## Supplementary Acknowledgement Table for "Rapid evaluation of COVID-19 vaccine effectiveness against VOC/VOIs by genetic mismatch"

We gratefully acknowledge the following Authors from the Originating laboratories responsible for obtaining the specimens, as well as the Submitting laboratories where the genome data were generated and shared via GISAID, on which this research is based.

All Submitters of data may be contacted directly via www.gisaid.org

Authors are sorted alphabetically.

| Accession ID | Originating Laboratory | Submitting Laboratory | Authors |
|---|---|---|---|
| EPI_ISL_1395777 | Laboratorio de Virología del Hospital de Niños Dr. Ricardo Gutierrez | Área de Secuenciación del Laboratorio de Virología del Hospital de Niños Dr. Ricardo Gutierrez on behalf of 'Proyecto Argentino Interinstitucional de genomica de SARS-CoV-2' (PAIS Consortium) | Nabaes Jodar, M; Goya, S; Lusso, S; Acuña, D; Alexay, S; Natale, MI; Valinotto, LE; Viegas, M. |
| EPI_ISL_1395778, EPI_ISL_1395779, EPI_ISL_1395780 | Laboratorio de Virología del Hospital de Niños Dr. Ricardo Gutierrez | Área de Secuenciación del Laboratorio de Virología del Hospital de Niños Dr. Ricardo Gutierrez on behalf of 'Proyecto Argentino Interinstitucional de genomica de SARS-CoV-2' (PAIS Consortium) | Alexay, S; Thomas, G; Medina, C; Labarta, N; Streitenberger, C; Villegas, E; Barreda Frank, M; Grandis, E; Acevedo, ME; Alvarez Lopez, C; Jacques, O; Mistchenko, A; Nabaes Jodar, M; Goya, S; Lusso, S; Acuña, D; Natale, MI; Valinotto, LE; Viegas, M. |
| EPI_ISL_1395790 | Laboratorio de salud pública, Facultad de Ciencias Exactas, UNLP | Área de Secuenciación del Laboratorio de Virología del Hospital de Niños Dr. Ricardo Gutierrez on behalf of 'Proyecto Argentino Interinstitucional de genomica de SARS-CoV-2' (PAIS Consortium) | Rosana Isabel Toro; Andrés Angelletti; Víctoria Cabassi; Victoria Nadalich; Andrés Cordero; Carina Tersigni; Laura Delaplace; Nabaes Jodar, M; Goya, S; Lusso, S; Acuña, D; Natale, MI; Valinotto, LE; Viegas, M. |
| EPI_ISL_1395791, EPI_ISL_1395793 | Laboratorio de Virología del Hospital de Niños Dr. Ricardo Gutierrez | Área de Secuenciación del Laboratorio de Virología del Hospital de Niños Dr. Ricardo Gutierrez on behalf of 'Proyecto Argentino Interinstitucional de genomica de SARS-CoV-2' (PAIS Consortium) | Alexay, S; Thomas, G; Medina, C; Labarta, N; Streitenberger, C; Villegas, E; Barreda Frank, M; Grandis, E; Acevedo, ME; Alvarez Lopez, C; Jacques, O; Mistchenko, A; Nabaes Jodar, M; Goya, S; Lusso, S; Acuña, D; Natale, MI; Valinotto, LE; Viegas, M. |
| EPI_ISL_1395808, EPI_ISL_1395809, EPI_ISL_1395810, EPI_ISL_1395811 | Hospital Pediatrico Dr Avelino Castelan, Hospital 4 de Junio Dr Ramon Carrillo | Grupo de Genómica y Bioinformática del Instituto de Investigación de la Cadena Láctea CONICET-INTA on behalf of 'Proyecto Argentino Interinstitucional de genomica de SARS-CoV-2' (PAIS Consortium) | Marino Goia, Lopez María Cecilia, Gili Andrea, Mayra Mendoza, Andrea Piedra Buena, Gabriela Jurasek, Antonieta Cayré, Laura Lescano, Eberhardt, MF, Irazoqui, Amadio, AF |
| EPI_ISL_1395813, EPI_ISL_1395814, EPI_ISL_1395815, EPI_ISL_1395816, EPI_ISL_1395819, EPI_ISL_1395820, EPI_ISL_1395821, EPI_ISL_1395822, EPI_ISL_1395823, EPI_ISL_1395827, EPI_ISL_1395828, EPI_ISL_1395829, EPI_ISL_1395830, EPI_ISL_1395831, EPI_ISL_1395832, EPI_ISL_1395833, EPI_ISL_1395837, EPI_ISL_1395838, EPI_ISL_1395839 | | | |
| see above | Inmunología del Hospital Perrando e Instituto de Medicina Regional de la UNNE | Grupo de Genómica y Bioinformática del Instituto de Investigación de la Cadena Láctea CONICET-INTA on behalf of 'Proyecto Argentino Interinstitucional de genomica de SARS-CoV-2' (PAIS Consortium) | María Delia Foussal, Gerardo Deluca, Natalia Andrea Ayala, María Verónica Gómez, Gustavo Giusiano, Horacio Lucero, Marcelo Marin, Antonieta Cayré, Laura Lescano, Eberhardt, MF, Irazoqui, Amadio, AF |
| EPI_ISL_1395840, EPI_ISL_1395841, EPI_ISL_1395843, EPI_ISL_1395844, EPI_ISL_1395845, EPI_ISL_1395846, EPI_ISL_1395847, EPI_ISL_1395848, EPI_ISL_1395849, EPI_ISL_1395850, EPI_ISL_1395852, EPI_ISL_1395853, EPI_ISL_1395854, EPI_ISL_1395855, EPI_ISL_1395856, EPI_ISL_1395857, EPI_ISL_1395858, EPI_ISL_1395859, EPI_ISL_1395860, EPI_ISL_1395861, EPI_ISL_1395862, EPI_ISL_1395863, EPI_ISL_1395864, EPI_ISL_1395865, EPI_ISL_1395866, EPI_ISL_1395867, EPI_ISL_1395868, EPI_ISL_1395869, EPI_ISL_1395870, EPI_ISL_1395872, EPI_ISL_1395873, EPI_ISL_1395874, EPI_ISL_1395875, EPI_ISL_1395876, EPI_ISL_1395877, EPI_ISL_1395878, EPI_ISL_1395879, EPI_ISL_1395880, EPI_ISL_1395881, EPI_ISL_1395882, EPI_ISL_1395883, EPI_ISL_1395884, EPI_ISL_1395885, EPI_ISL_1395886, EPI_ISL_1395887, EPI_ISL_1395888, EPI_ISL_1395889, EPI_ISL_1395890, EPI_ISL_1395891, EPI_ISL_1395892, EPI_ISL_1395893, EPI_ISL_1395894, EPI_ISL_1395895, EPI_ISL_1395896, EPI_ISL_1395897, EPI_ISL_1395898, EPI_ISL_1395899, EPI_ISL_1395900, EPI_ISL_1395901, EPI_ISL_1395902, EPI_ISL_1395903, EPI_ISL_1395904, EPI_ISL_1395905, EPI_ISL_1395906, EPI_ISL_1395907, EPI_ISL_1395908, EPI_ISL_1395909, EPI_ISL_1395910, EPI_ISL_1395911, EPI_ISL_1395912, EPI_ISL_1395913, EPI_ISL_1395914, EPI_ISL_1395915, EPI_ISL_1395916, EPI_ISL_1395917, EPI_ISL_1395918, EPI_ISL_1395919, EPI_ISL_1395920, EPI_ISL_1395921, EPI_ISL_1395922, EPI_ISL_1395923, EPI_ISL_1395924, EPI_ISL_1395925, EPI_ISL_1395926, EPI_ISL_1395927, EPI_ISL_1395928, EPI_ISL_1395929, EPI_ISL_1395930, EPI_ISL_1395931 | | | |
| see above | Laboratorio Central de la Ciudad de Santa Fe | Grupo de Genómica y Bioinformática del Instituto de Investigación de la Cadena Láctea CONICET-INTA on behalf of 'Proyecto Argentino Interinstitucional de genomica de SARS-CoV-2' (PAIS Consortium) | Eberhardt, MF; Irazoqui, JM; Ojeda, G; Rompato, G; Mugna, V; Pastor, C; Amadio, AF |
| EPI_ISL_1395932 | Laboratorio Central De Redes y Programas | Grupo de Genómica y Bioinformática del Instituto de Investigación de la Cadena Láctea CONICET-INTA on behalf of 'Proyecto Argentino Interinstitucional de genomica de SARS-CoV-2' (PAIS Consortium) | Natalia Ruiz Diaz, Gerardo Andino, Antonieta Cayré, Laura Lescano, Eberhardt, MF, Irazoqui, Amadio, AF |
| EPI_ISL_1395936, EPI_ISL_1395937, EPI_ISL_1395938, EPI_ISL_1395941, EPI_ISL_1395942, EPI_ISL_1395943, EPI_ISL_1395944, EPI_ISL_1395945, EPI_ISL_1395948, EPI_ISL_1395952, EPI_ISL_1395953, EPI_ISL_1395954, EPI_ISL_1395955, EPI_ISL_1395956, EPI_ISL_1395957, EPI_ISL_1395958 | | | |
| see above | Laboratorio Central de Salud Pública | Grupo de Genómica y Bioinformática del Instituto de Investigación de la Cadena Láctea CONICET-INTA on behalf of 'Proyecto Argentino Interinstitucional de genomica de SARS-CoV-2' (PAIS Consortium) | Natalia Andrea Ayala y María Verónica Gómez, Erica Struss, Esteban Paredes, Antonieta Cayré, Laura Lescano, Eberhardt, MF, Irazoqui, Amadio, AF |
| EPI_ISL_1395960, EPI_ISL_1395963, EPI_ISL_1395966 | Laboratorio Central de Salud Pública de la Provincia de Jujuy | Instituto de Patologia Vegetal (CIAP-INTA) on behalf of 'Proyecto Argentino Interinstitucional de genomica de SARS-CoV-2' (PAIS Consortium) | Fernández, FD, Marquez, N., Debat, HJ., Irazoqui, M., Amadio, A. Miguel Alejandro Charre, Ariel David Fridman, Claudia Mamani, Fabiana Vaca. |
| EPI_ISL_1395967, EPI_ISL_1395968, EPI_ISL_1395969, EPI_ISL_1395970, EPI_ISL_1395971, EPI_ISL_1395972, EPI_ISL_1395973, EPI_ISL_1395974, EPI_ISL_1395975, EPI_ISL_1395976, EPI_ISL_1395977, EPI_ISL_1395978, EPI_ISL_1395979, EPI_ISL_1395980, EPI_ISL_1395981, EPI_ISL_1395982, EPI_ISL_1395983, EPI_ISL_1395984, EPI_ISL_1395985, EPI_ISL_1395986, EPI_ISL_1395987, EPI_ISL_1395988, EPI_ISL_1395989, EPI_ISL_1395990, EPI_ISL_1395991, EPI_ISL_1395992, EPI_ISL_1395995, EPI_ISL_1395996, EPI_ISL_1395997, EPI_ISL_1395998, EPI_ISL_1395999, EPI_ISL_1396000, EPI_ISL_1396001, EPI_ISL_1396002, EPI_ISL_1396003, EPI_ISL_1396004, EPI_ISL_1396005, EPI_ISL_1396008, EPI_ISL_1396009, EPI_ISL_1396010, EPI_ISL_1396012, EPI_ISL_1396013, EPI_ISL_1396015, EPI_ISL_1396016, EPI_ISL_1396017, EPI_ISL_1396019, EPI_ISL_1396020, EPI_ISL_1396022, EPI_ISL_1396023, EPI_ISL_1396024, EPI_ISL_1396025, EPI_ISL_1396026, EPI_ISL_1396027, EPI_ISL_1396028, EPI_ISL_1396029, EPI_ISL_1396030, EPI_ISL_1396031, EPI_ISL_1396032, EPI_ISL_1396033, EPI_ISL_1396034, EPI_ISL_1396035, EPI_ISL_1396036, EPI_ISL_1396037, EPI_ISL_1396038, EPI_ISL_1396039, EPI_ISL_1396040, EPI_ISL_1396041, EPI_ISL_1396043, EPI_ISL_1396044, EPI_ISL_1396045, EPI_ISL_1396046, EPI_ISL_1396047, EPI_ISL_1396048, EPI_ISL_1396049, EPI_ISL_1396050, EPI_ISL_1396051, EPI_ISL_1396055, EPI_ISL_1396056, EPI_ISL_1396057, EPI_ISL_1396060 | | | |
| see above | Laboratorio Central, Ministerio de Salud Cordoba | Instituto de Patologia Vegetal (CIAP-INTA) on behalf of 'Proyecto Argentino Interinstitucional de genomica de SARS-CoV-2' (PAIS Consortium) | Fernández, FD; Marquez, N.; Debat, HJ.; Re, V.; Pisano, M.B.; Castro, G.; Barbas, G. |
| EPI_ISL_1396092, EPI_ISL_1396093, EPI_ISL_1396094, EPI_ISL_1396096, EPI_ISL_1396099, EPI_ISL_1396100, EPI_ISL_1396101, EPI_ISL_1396108, EPI_ISL_1396111, EPI_ISL_1396112, EPI_ISL_1396113, EPI_ISL_1396114, EPI_ISL_1396116, EPI_ISL_1396118, EPI_ISL_1396119, EPI_ISL_1396120, EPI_ISL_1396122, EPI_ISL_1396123 | | | |
| see above | Laboratorio de Virus Respiratorios y Neurovirosis. Hospital Señor del Milagro | Instituto de Patologia Vegetal (CIAP-INTA) on behalf of 'Proyecto Argentino Interinstitucional de genomica de SARS-CoV-2' (PAIS Consortium) | Fernández, FD, Marquez, N., Debat, HJ., Irazoqui, M., Amadio, A. Dra. Raskovsky Viviana, Dr. Lavaque Esteban, Dra. Veronica Lesser. Tecnica: Pamela Cajal, Fernanda Aguero. |
| EPI_ISL_1396124, EPI_ISL_1396127, EPI_ISL_1396128, EPI_ISL_1396129, EPI_ISL_1396130, EPI_ISL_1396131, EPI_ISL_1396140, EPI_ISL_1396141, EPI_ISL_1396142, EPI_ISL_1396143, EPI_ISL_1396144, EPI_ISL_1396145, EPI_ISL_1396146, EPI_ISL_1396147 | | | |
| see above | Laboratorio de la Dirección de Epidemiología | Área de Secuenciación del Laboratorio de Virología del Hospital de Niños Dr. Ricardo Gutierrez on behalf of 'Proyecto Argentino Interinstitucional de genomica de SARS-CoV-2' (PAIS Consortium) | Rechimont, C; Usero, JC; Villasana, M; Bertone, AG; Nabaes Jodar, M; Goya, S; Lusso, S; Acuña, D; Alexay, S; Natale, MI; Valinotto, LE; Viegas, M. |

| | | | |
|---|---|---|---|
| EPI_ISL_1396149, EPI_ISL_1396153, EPI_ISL_1396154, EPI_ISL_1396155, EPI_ISL_1396156, EPI_ISL_1396157, EPI_ISL_1396158, EPI_ISL_1396159, EPI_ISL_1396160, EPI_ISL_1396161, EPI_ISL_1396162, EPI_ISL_1396163, EPI_ISL_1396164, EPI_ISL_1396165, EPI_ISL_1396166, EPI_ISL_1396167, EPI_ISL_1396168, EPI_ISL_1396169, EPI_ISL_1396170, EPI_ISL_1396171, EPI_ISL_1396172, EPI_ISL_1396173, EPI_ISL_1396174, EPI_ISL_1396175, EPI_ISL_1396176, EPI_ISL_1396177, EPI_ISL_1396178, EPI_ISL_1396179, EPI_ISL_1396180, EPI_ISL_1396181, EPI_ISL_1396182, EPI_ISL_1396183, EPI_ISL_1396184, EPI_ISL_1396185, EPI_ISL_1396186, EPI_ISL_1396187, EPI_ISL_1396188, EPI_ISL_1396189, EPI_ISL_1396190, EPI_ISL_1396191, EPI_ISL_1396192, EPI_ISL_1396193, EPI_ISL_1396194, EPI_ISL_1396195, EPI_ISL_1396196, EPI_ISL_1396197, EPI_ISL_1396198, EPI_ISL_1396199, EPI_ISL_1396200, EPI_ISL_1396201, EPI_ISL_1396202, EPI_ISL_1396203, EPI_ISL_1396204, EPI_ISL_1396205, EPI_ISL_1396206, EPI_ISL_1396207, EPI_ISL_1396208, EPI_ISL_1396209, EPI_ISL_1396210, EPI_ISL_1396211, EPI_ISL_1396212, EPI_ISL_1396213, EPI_ISL_1396214, EPI_ISL_1396215, EPI_ISL_1396216, EPI_ISL_1396217, EPI_ISL_1396218, EPI_ISL_1396219, EPI_ISL_1396220, EPI_ISL_1396221, EPI_ISL_1396222, EPI_ISL_1396223, EPI_ISL_1396224, EPI_ISL_1396225 | | | |
| see above | Laboratorio de Virología del Hospital de Niños Dr. Ricardo Gutierrez | Área de Secuenciación del Laboratorio de Virología del Hospital de Niños Dr. Ricardo Gutierrez on behalf of 'Proyecto Argentino Interinstitucional de genomica de SARS-CoV-2' (PAIS Consortium) | Alexay, S; Thomas, G; Medina, C; Labarta, N; Streitenberger, C; Villegas, E; Barreda Frank, M; Grandis, E; Acevedo, ME; Alvarez Lopez, C; Jacques, O; Mistchenko, A; Nabaes Jodar, M; Goya, S; Lusso, S; Acuña, D; Natale, MI; Valinotto, LE; Viegas, M. |
| EPI_ISL_1396284, EPI_ISL_1396285, EPI_ISL_1396286, EPI_ISL_1396287, EPI_ISL_1396288, EPI_ISL_1396289, EPI_ISL_1396290, EPI_ISL_1396291, EPI_ISL_1396292, EPI_ISL_1396293, EPI_ISL_1396294, EPI_ISL_1396295, EPI_ISL_1396296, EPI_ISL_1396297, EPI_ISL_1396298, EPI_ISL_1396299, EPI_ISL_1396300, EPI_ISL_1396301, EPI_ISL_1396302, EPI_ISL_1396303, EPI_ISL_1396304, EPI_ISL_1396305, EPI_ISL_1396306, EPI_ISL_1396307, EPI_ISL_1396308, EPI_ISL_1396309, EPI_ISL_1396310, EPI_ISL_1396311, EPI_ISL_1396312, EPI_ISL_1396313, EPI_ISL_1396314, EPI_ISL_1396315, EPI_ISL_1396316, EPI_ISL_1396317, EPI_ISL_1396318, EPI_ISL_1396319, EPI_ISL_1396320, EPI_ISL_1396321, EPI_ISL_1396322, EPI_ISL_1396323, EPI_ISL_1396324, EPI_ISL_1396325, EPI_ISL_1396326, EPI_ISL_1396327, EPI_ISL_1396328, EPI_ISL_1396329, EPI_ISL_1396330, EPI_ISL_1396331 | | | |
| see above | Laboratorio de Virología del Hospital de Niños Dr. Ricardo Gutierrez | Biocódices SA. on behalf of 'Proyecto Argentino Interinstitucional de genomica de SARS-CoV-2' (PAIS Consortium) | Alexay, S; Thomas, G; Medina, C; Labarta, N; Streitenberger, C; Villegas, E; Barreda Frank, M; Grandis, E; Acevedo, ME; Alvarez Lopez, C; Jacques, O; Mistchenko, A; J, Zubrzycki J; Berros, JM; Dopazo, H. |
| EPI_ISL_1396340, EPI_ISL_1396341, EPI_ISL_1396342, EPI_ISL_1396343, EPI_ISL_1396344, EPI_ISL_1396345, EPI_ISL_1396346, EPI_ISL_1396347, EPI_ISL_1396348, EPI_ISL_1396349, EPI_ISL_1396350, EPI_ISL_1396351, EPI_ISL_1396352, EPI_ISL_1396353, EPI_ISL_1396354, EPI_ISL_1396355 | | | |
| see above | Laboratorio del Hospital Regional Ushuaia Gdor. Ernesto Campos | Nodo de Secuenciación Tierra del Fuego - Hospital Regional Ushuaia - Centro Austral De Investigaciones Científicas - Universidad Nacional De Tierra Del Fuego on behalf of 'Proyecto Argentino Interinstitucional de genomica de SARS-CoV-2' (PAIS Consortium) | Carina Andrea De Roccis, Gabriel Alejandro Castro, Silvana Beatriz Cáceres, Carolina Beatriz Yulan, Manuel Fabian Boutureira, Alejandro Ezequiel Rojas, Fernando Gallego, Santiago Guillermo Ceballos, Cristina Fernanda Nardi, Ivan Dario Gramundi |
| EPI_ISL_1396364, EPI_ISL_1396365, EPI_ISL_1396366, EPI_ISL_1396367, EPI_ISL_1396368, EPI_ISL_1396369, EPI_ISL_1396370, EPI_ISL_1396371, EPI_ISL_1396372, EPI_ISL_1396373 | Hospital Central Mendoza | Nodo de Secuenciación Tierra del Fuego - Hospital Regional Ushuaia - Centro Austral De Investigaciones Científicas - Universidad Nacional De Tierra Del Fuego on behalf of 'Proyecto Argentino Interinstitucional de genomica de SARS-CoV-2' (PAIS Consortium) | María Liliana Videla, Luciano Lima, Andrea Naser, Pablo Rico, Valeria Fontana, Héctor Horacio Cuello, Andrea Falaschi, Aguirre Carolina, Alberto Carena, Bosio Lia, Patricia Robledo, Maria Belen Peralta Roca, Carlos Espul, Alejandro Ezequiel Rojas, Fernando Gallego, Santiago Guillermo Ceballos, Cristina Fernanda Nardi, Ivan Dario Gramundi |
| EPI_ISL_1396383, EPI_ISL_1396384, EPI_ISL_1396385, EPI_ISL_1396386, EPI_ISL_1396387, EPI_ISL_1396388, EPI_ISL_1396389, EPI_ISL_1396390 | Laboratorio Salud Publica Mendoza | Nodo de Secuenciación Tierra del Fuego - Hospital Regional Ushuaia - Centro Austral De Investigaciones Científicas - Universidad Nacional De Tierra Del Fuego on behalf of 'Proyecto Argentino Interinstitucional de genomica de SARS-CoV-2' (PAIS Consortium) | Silvia Zerrer, Belén Ortiz, Silvina Denita, Luciana Martinez, Cristian Garay, Fernando Giuliani, Andrea Falaschi, Aguirre Carolina, Alberto Carena, Bosio Lia, Patricia Robledo, Maria Belen Peralta Roca, Carlos Espul, Alejandro Ezequiel Rojas, Fernando Gallego, Santiago Guillermo Ceballos, Cristina Fernanda Nardi, Ivan Dario Gramundi |
| EPI_ISL_1396412, EPI_ISL_1396413, EPI_ISL_1396414, EPI_ISL_1396415, EPI_ISL_1396416, EPI_ISL_1396417, EPI_ISL_1396418, EPI_ISL_1396419, EPI_ISL_1396420, EPI_ISL_1396421, EPI_ISL_1396422, EPI_ISL_1396423, EPI_ISL_1396424 | | | |
| see above | Centro de Tecnología en Salud Pública de la Universidad Nacional de Rosario | Laboratorio Mixto de Biotecnología Acuática (LMBA) on behalf of 'Proyecto Argentino Interinstitucional de genomica de SARS-CoV-2' (PAIS Consortium) | Joaquín Ezpeleta, Ignacio García Labari, Victoria Posner, Vanina Villanova, Pablo Casal, Pilar Bulacio, Sofía Lavista Llanos, Federico Remes Lenicov, Ana Paletta, Leandro Ciappina, Flavio Spetale, Agustina Cerri, Silvana Spinelli, Elisa Bolatti, Diego Chouhy, María Re, Gastón Viarengo, Ana Cavatorta, Julian Acosta, Javier Murillo, Laura Angelone, Adriana Giri, Silvia Arranz, Elizabeth Tapia (argenTAG) |
| EPI_ISL_1396425 | Hospital Español de Rosario | Laboratorio Mixto de Biotecnología Acuática (LMBA) on behalf of 'Proyecto Argentino Interinstitucional de genomica de SARS-CoV-2' (PAIS Consortium) | Joaquín Ezpeleta, Ignacio García Labari, Victoria Posner, Vanina Villanova, Pablo Casal, Pilar Bulacio, Sofía Lavista Llanos, Federico Remes Lenicov, Ana Paletta, Leandro Ciappina, Flavio Spetale, Agustina Cerri, Silvana Spinelli, Elisa Bolatti, Diego Chouhy, María Re, Gastón Viarengo, Ana Cavatorta, Julian Acosta, Javier Murillo, Laura Angelone, Adriana Giri, Silvia Arranz, Elizabeth Tapia (argenTAG) |
| EPI_ISL_1396426, EPI_ISL_1396427 | Centro de Tecnología en Salud Pública de la Universidad Nacional de Rosario | Laboratorio Mixto de Biotecnología Acuática (LMBA) on behalf of 'Proyecto Argentino Interinstitucional de genomica de SARS-CoV-2' (PAIS Consortium) | Joaquín Ezpeleta, Ignacio García Labari, Victoria Posner, Vanina Villanova, Pablo Casal, Pilar Bulacio, Sofía Lavista Llanos, Federico Remes Lenicov, Ana Paletta, Leandro Ciappina, Flavio Spetale, Agustina Cerri, Silvana Spinelli, Elisa Bolatti, Diego Chouhy, María Re, Gastón Viarengo, Ana Cavatorta, Julian Acosta, Javier Murillo, Laura Angelone, Adriana Giri, Silvia Arranz, Elizabeth Tapia (argenTAG) |
| EPI_ISL_1396428 | Hospital Español de Rosario | Laboratorio Mixto de Biotecnología Acuática (LMBA) on behalf of 'Proyecto Argentino Interinstitucional de genomica de SARS-CoV-2' (PAIS Consortium) | Joaquín Ezpeleta, Ignacio García Labari, Victoria Posner, Vanina Villanova, Pablo Casal, Pilar Bulacio, Sofía Lavista Llanos, Federico Remes Lenicov, Ana Paletta, Leandro Ciappina, Flavio Spetale, Agustina Cerri, Silvana Spinelli, Elisa Bolatti, Diego Chouhy, María Re, Gastón Viarengo, Ana Cavatorta, Julian Acosta, Javier Murillo, Laura Angelone, Adriana Giri, Silvia Arranz, Elizabeth Tapia (argenTAG) |
| EPI_ISL_1396429, EPI_ISL_1396430, EPI_ISL_1396431, EPI_ISL_1396432, EPI_ISL_1396433, EPI_ISL_1396434, EPI_ISL_1396435, EPI_ISL_1396436, EPI_ISL_1396437, EPI_ISL_1396438, EPI_ISL_1396439, EPI_ISL_1396440, EPI_ISL_1396441, EPI_ISL_1396442, EPI_ISL_1396443, EPI_ISL_1396444, EPI_ISL_1396445, EPI_ISL_1396446, EPI_ISL_1396447, EPI_ISL_1396448, EPI_ISL_1396449, EPI_ISL_1396450, EPI_ISL_1396451, EPI_ISL_1396452, EPI_ISL_1396453, EPI_ISL_1396454, EPI_ISL_1396455, EPI_ISL_1396456, EPI_ISL_1396457, EPI_ISL_1396458, EPI_ISL_1396459, EPI_ISL_1396460, EPI_ISL_1396461, EPI_ISL_1396462, EPI_ISL_1396463, EPI_ISL_1396464, EPI_ISL_1396465, EPI_ISL_1396466, EPI_ISL_1396467, EPI_ISL_1396468, EPI_ISL_1396469, EPI_ISL_1396470, EPI_ISL_1396471, EPI_ISL_1396472, EPI_ISL_1396473, EPI_ISL_1396474, EPI_ISL_1396475, EPI_ISL_1396476, EPI_ISL_1396477, EPI_ISL_1396478, EPI_ISL_1396479, EPI_ISL_1396480, EPI_ISL_1396481, EPI_ISL_1396482, EPI_ISL_1396483, EPI_ISL_1396484, EPI_ISL_1396485, EPI_ISL_1396486, EPI_ISL_1396487, EPI_ISL_1396488, EPI_ISL_1396489, EPI_ISL_1396490, EPI_ISL_1396491, EPI_ISL_1396492, EPI_ISL_1396493, EPI_ISL_1396494, EPI_ISL_1396495, EPI_ISL_1396496, EPI_ISL_1396497, EPI_ISL_1396498, EPI_ISL_1396499, EPI_ISL_1396500, EPI_ISL_1396501, EPI_ISL_1396502, EPI_ISL_1396503, EPI_ISL_1396504, EPI_ISL_1396505, EPI_ISL_1396506, EPI_ISL_1396507, EPI_ISL_1396508, EPI_ISL_1396509, EPI_ISL_1396510, EPI_ISL_1396511, EPI_ISL_1396512, EPI_ISL_1396513, EPI_ISL_1396514, EPI_ISL_1396515, EPI_ISL_1396516, EPI_ISL_1396517, EPI_ISL_1396518 | | | |
| see above | Centro de Tecnología en Salud Pública de la Universidad Nacional de Rosario | Laboratorio Mixto de Biotecnología Acuática (LMBA) on behalf of 'Proyecto Argentino Interinstitucional de genomica de SARS-CoV-2' (PAIS Consortium) | Joaquín Ezpeleta, Ignacio García Labari, Victoria Posner, Vanina Villanova, Pablo Casal, Pilar Bulacio, Sofía Lavista Llanos, Federico Remes Lenicov, Ana Paletta, Leandro Ciappina, Flavio Spetale, Agustina Cerri, Silvana Spinelli, Elisa Bolatti, Diego Chouhy, María Re, Gastón Viarengo, Ana Cavatorta, Julian Acosta, Javier Murillo, Laura Angelone, Adriana Giri, Silvia Arranz, Elizabeth Tapia (argenTAG) |
| EPI_ISL_2104757, EPI_ISL_2104758, EPI_ISL_2104759, EPI_ISL_2104760, EPI_ISL_2104761, EPI_ISL_2104762, EPI_ISL_2104763, EPI_ISL_2104764, EPI_ISL_2104765, EPI_ISL_2104766, EPI_ISL_2104767, EPI_ISL_2104768, EPI_ISL_2104769, EPI_ISL_2104770, EPI_ISL_2104771, EPI_ISL_2104772, EPI_ISL_2104773, EPI_ISL_2104774, EPI_ISL_2104775, EPI_ISL_2104776, EPI_ISL_2104777, EPI_ISL_2104778, EPI_ISL_2104779, EPI_ISL_2105566, EPI_ISL_2105567, EPI_ISL_2105568, EPI_ISL_2105569, EPI_ISL_2105570, EPI_ISL_2105571, EPI_ISL_2105572, EPI_ISL_2135134, EPI_ISL_2135135, EPI_ISL_778843 | | | |
| see above | Servicio Virosis Respiratorias-Departamento Virología-INEI | Instituto Nacional Enfermedades Infecciosas C.G.Malbran | Baumeister E., Avaro M., Benedetti E., Russo M., Dattero ME, Pontoriero A., Cisterna D., Molina V., Perandones C., Tuduri E., Lorenzo F., Poklepovich T., Campos J. |
| EPI_ISL_792522, EPI_ISL_792523, EPI_ISL_792524 | Laboratorio de Virología del Hospital de Niños Dr. Ricardo Gutierrez | Grupo de Genómica y Bioinformática del Instituto de Investigación de la Cadena Láctea CONICET-INTA on behalf of 'Proyecto Argentino Interinstitucional de genomica de SARS-CoV-2' (PAIS Consortium) | Amadio, AF, Eberhardt, MF; Irazoqui, M; Torres, C; Aulicino, P; König, G; Acevedo, ME; Alvarez Lopez, C; Alexay, S; Jacques, O; Mistchenko, AS, Goya, S; Nabaes Jodar, MS; Viegas, M. |
| EPI_ISL_792525 | Laboratorio del Hospital Interzonal General de Agudos Evita | Grupo de Genómica y Bioinformática del Instituto de Investigación de la Cadena Láctea CONICET-INTA on behalf of 'Proyecto Argentino Interinstitucional de genomica de SARS-CoV-2' (PAIS Consortium) | Amadio, AF, Eberhardt, MF; Irazoqui, M; Torres, C; Aulicino, P; König, G; Desimone, I; Luczac, E; Serrano, L; Grossi, O; Musto; Alexay, S; Goya, S; Nabaes Jodar, MS; Viegas, M. |
| EPI_ISL_842652 | Laboratorio de Biología Molecular Hospital Pedro de Elizalde | Grupo de Genómica y Bioinformática del Instituto de Investigación de la Cadena Láctea CONICET-INTA on behalf of 'Proyecto Argentino Interinstitucional de genomica de SARS-CoV-2' (PAIS Consortium) | Amadio, AF, Eberhardt, MF; Irazoqui, M; Indart, J; Rocovich, J; Montoto Piazza, L; Wenk, G; Martín, ME; Sanchez, MF; Marchetti, P; Morandi, F; Sueiro, ML; Claps, A; Bressan, L; Torres, FJ; Chamorro, J; Gondolessi, J; Gómez, ML; Diaz, B; Rosales, D; Alegre, F; Zamora, N; Osaba, E; Paez, E; Lorenzo, F; Torres, C; Aulicino, P; König, G; Alexay, S; Natale, M; Valinotto, L; Lusso, S; Goya, S; Nabaes Jodar, MS; Viegas, M. |
| EPI_ISL_849350, EPI_ISL_849351, EPI_ISL_849352, EPI_ISL_849353, EPI_ISL_849354, EPI_ISL_849355, EPI_ISL_849356, EPI_ISL_849357, EPI_ISL_849358, EPI_ISL_849359, EPI_ISL_849360, EPI_ISL_849361, EPI_ISL_849362, EPI_ISL_849363, EPI_ISL_849364, EPI_ISL_849365, EPI_ISL_849366, EPI_ISL_849367, EPI_ISL_849368, EPI_ISL_849369, EPI_ISL_849370, EPI_ISL_849371, EPI_ISL_849372, EPI_ISL_849373, EPI_ISL_849374, EPI_ISL_849375, EPI_ISL_849376, EPI_ISL_849377, EPI_ISL_849378, EPI_ISL_849379, EPI_ISL_849380, EPI_ISL_849381, EPI_ISL_849382, EPI_ISL_856756, EPI_ISL_856757, EPI_ISL_856758, EPI_ISL_856759, EPI_ISL_856760, EPI_ISL_856761, EPI_ISL_856762, EPI_ISL_856763, EPI_ISL_856764, EPI_ISL_856765, EPI_ISL_856766, EPI_ISL_856767, EPI_ISL_856768, EPI_ISL_856769, EPI_ISL_856770, EPI_ISL_856771, EPI_ISL_856772, EPI_ISL_856773, EPI_ISL_856774, EPI_ISL_856775, EPI_ISL_856776, EPI_ISL_856777, EPI_ISL_856778, EPI_ISL_856779, EPI_ISL_856780, EPI_ISL_856781, EPI_ISL_856782, EPI_ISL_856783, EPI_ISL_856784, EPI_ISL_856785, EPI_ISL_856786, EPI_ISL_856787, EPI_ISL_856788, EPI_ISL_856789, EPI_ISL_856790, EPI_ISL_856791, EPI_ISL_856792, EPI_ISL_856793, EPI_ISL_856794, EPI_ISL_856795, EPI_ISL_856796, EPI_ISL_856797, EPI_ISL_856798, EPI_ISL_856799, EPI_ISL_856800, EPI_ISL_856801 | | | |
| see above | Servicio Virosis Respiratorias-Departamento Virología-INEI | Instituto Nacional Enfermedades Infecciosas C.G.Malbran | Baumeister E., Avaro M., Benedetti E., Russo M., Dattero ME, Pontoriero A., Cisterna D., Molina V., Perandones C., Tuduri E., Lorenzo F., Poklepovich T., Campos J. |

| | | | |
|---|---|---|---|
| EPI_ISL_964906, EPI_ISL_964907, EPI_ISL_964908, EPI_ISL_964909, EPI_ISL_964910 | Laboratorio Central Mg. Luis Alfredo Pianciola on behalf of 'Proyecto Argentino Interinstitucional de genomica de SARS-CoV-2' (PAIS Consortium) | Laboratorio Central Mg. Luis Alfredo Pianciola on behalf of 'Proyecto Argentino Interinstitucional de genomica de SARS-CoV-2' (PAIS Consortium) | M Mazzeo, C Ziehm, C Pintos, M Fernandez, J Ousset, M Nabaes, L Pianciola. |
| EPI_ISL_964913 | Sistema Integrado de Emergencias de Neuquén (SIEN) | Laboratorio Central Mg. Luis Alfredo Pianciola on behalf of 'Proyecto Argentino Interinstitucional de genomica de SARS-CoV-2' (PAIS Consortium) | Liliana Fonseca, Marcela Nóbile, María Laura Álvarez, Yesica Espasandin, Patricia Valeria Blanco, Silvana Cecchi, Darío Fabián di Prátula, Antonela De Fino, M Mazzeo, C Ziehm, C Pintos, M Fernandez, J Ousset, M Nabaes, L Pianciola. |
| EPI_ISL_964914 | Laboratorio Central Mg. Luis Alfredo Pianciola on behalf of 'Proyecto Argentino Interinstitucional de genomica de SARS-CoV-2' (PAIS Consortium) | Laboratorio Central Mg. Luis Alfredo Pianciola on behalf of 'Proyecto Argentino Interinstitucional de genomica de SARS-CoV-2' (PAIS Consortium) | M Mazzeo, C Ziehm, C Pintos, M Fernandez, J Ousset, M Nabaes, L Pianciola. |
| EPI_ISL_981031, EPI_ISL_981033, EPI_ISL_981035, EPI_ISL_981037, EPI_ISL_981042, EPI_ISL_981045, EPI_ISL_981047, EPI_ISL_981048, EPI_ISL_981049, EPI_ISL_981051, EPI_ISL_981052 | | | |
| see above | Laboratorio Central Sección Virología. Hospital Regional Artemides Zatti (Viedma, Rio Negro); Laboratorio MICROBIOM (General Roca, Río Negro); Hospital Zonal Dr. Ramón Carrillo (San Carlos De Bariloche, Río Negro) | Laboratorio Central Mg. Luis Alfredo Pianciola on behalf of 'Proyecto Argentino Interinstitucional de genomica de SARS-CoV-2' (PAIS Consortium) | Liliana Fonseca, Marcela Nóbile, María Laura Álvarez, Yesica Espasandin, Patricia Valeria Blanco, Silvana Cecchi, Darío Fabián di Prátula, Antonela De Fino, M Mazzeo, C Ziehm, C Pintos, M Fernandez, J Ousset, M Nabaes, L Pianciola. |
| EPI_ISL_981053 | Laboratorio Central Mg. Luis Alfredo Pianciola on behalf of 'Proyecto Argentino Interinstitucional de genomica de SARS-CoV-2' (PAIS Consortium) | Laboratorio Central Mg. Luis Alfredo Pianciola on behalf of 'Proyecto Argentino Interinstitucional de genomica de SARS-CoV-2' (PAIS Consortium) | M Mazzeo, C Ziehm, C Pintos, M Fernandez, J Ousset, M Nabaes, L Pianciola. |
| EPI_ISL_981054, EPI_ISL_981055, EPI_ISL_981056 | Laboratorio Central Sección Virología. Hospital Regional Artemides Zatti (Viedma, Rio Negro); Laboratorio MICROBIOM (General Roca, Río Negro); Hospital Zonal Dr. Ramón Carrillo (San Carlos De Bariloche, Río Negro) | Laboratorio Central Mg. Luis Alfredo Pianciola on behalf of 'Proyecto Argentino Interinstitucional de genomica de SARS-CoV-2' (PAIS Consortium) | Liliana Fonseca, Marcela Nóbile, María Laura Álvarez, Yesica Espasandin, Patricia Valeria Blanco, Silvana Cecchi, Darío Fabián di Prátula, Antonela De Fino, M Mazzeo, C Ziehm, C Pintos, M Fernandez, J Ousset, M Nabaes, L Pianciola. |

We gratefully acknowledge the following Authors from the Originating laboratories responsible for obtaining the specimens, as well as the Submitting laboratories where the genome data were generated and shared via GISAID, on which this research is based.

All Submitters of data may be contacted directly via www.gisaid.org

Authors are sorted alphabetically.

| Accession ID | Originating Laboratory | Submitting Laboratory | Authors |
|---|---|---|---|
| EPI_ISL_1233891 | Communicable Disease Laboratory, Public Health Directorate | Communicable Disease Laboratory, Public Health Directorate | Alwasti,H., Altaif,Z., AlHujairi,Z., AlAbbas,Z., Marhoon,A. |
| EPI_ISL_1660421, EPI_ISL_1660422, EPI_ISL_1660423, EPI_ISL_1660424, EPI_ISL_1660425, EPI_ISL_1660426, EPI_ISL_1660427, EPI_ISL_1660428, EPI_ISL_1660429, EPI_ISL_1660430, EPI_ISL_1660431, EPI_ISL_1660432, EPI_ISL_1660433, EPI_ISL_1660434, EPI_ISL_1660435, EPI_ISL_1660436, EPI_ISL_1660437, EPI_ISL_1660438, EPI_ISL_1660439, EPI_ISL_1660440, EPI_ISL_1660441, EPI_ISL_1660442, EPI_ISL_1660443, EPI_ISL_1660444, EPI_ISL_1660445, EPI_ISL_1660446, EPI_ISL_1660447, EPI_ISL_1660448, EPI_ISL_1660449, EPI_ISL_1660450, EPI_ISL_1660451, EPI_ISL_1660452, EPI_ISL_1660453, EPI_ISL_1660454, EPI_ISL_1660455, EPI_ISL_1660456, EPI_ISL_1660457, EPI_ISL_1660458, EPI_ISL_1660459, EPI_ISL_1660460, EPI_ISL_1660461, EPI_ISL_1660462, EPI_ISL_1660463, EPI_ISL_1660464, EPI_ISL_1660465, EPI_ISL_1660466, EPI_ISL_1660467, EPI_ISL_1660468, EPI_ISL_1660469, EPI_ISL_1660470, EPI_ISL_1660471 | | | |
| see above | Communicable Disease Laboratory, Public Health Directorate | Communicable Disease Laboratory, Public Health Directorate | Alwasti,H., AlHujairi,Z., AlAbbas,Z., Marhoon,A., Almoamen,G. |
| EPI_ISL_632250, EPI_ISL_632251, EPI_ISL_632252, EPI_ISL_632253, EPI_ISL_632254, EPI_ISL_632255 | Communicable Disease Laboratory, Public Health Directorate | Communicable Disease Laboratory, Public Health Directorate | AlWasti,H., Altaif,Z., AlHujairi,Z., AlAbbas,Z. |
| EPI_ISL_632256, EPI_ISL_632257, EPI_ISL_632258 | Communicable Disease Laboratory, Public Health Directorate | Communicable Disease Laboratory, Public Health Directorate | AlHujairi,Z., Altaif,Z., AlWasti,H., AlAbbas,Z. |
| EPI_ISL_632259, EPI_ISL_632260 | Communicable Disease Laboratory, Public Health Directorate | Communicable Disease Laboratory, Public Health Directorate | AlTaif,Z., AlWasti,H., AlHujairi,Z., AlAbbas,Z. |
| EPI_ISL_632262, EPI_ISL_632263, EPI_ISL_632264, EPI_ISL_632267, EPI_ISL_632268, EPI_ISL_632269, EPI_ISL_632270, EPI_ISL_632271, EPI_ISL_632272, EPI_ISL_632273, EPI_ISL_632274, EPI_ISL_632275, EPI_ISL_632276, EPI_ISL_632277, EPI_ISL_632278, EPI_ISL_632279, EPI_ISL_632280, EPI_ISL_632281, EPI_ISL_632282, EPI_ISL_632283, EPI_ISL_632284 | | | |
| see above | Communicable Disease Laboratory, Public Health Directorate | Communicable Disease Laboratory, Public Health Directorate | AlWasti,H., AlTaif,Z., AlHujairi,Z., AlAbbas,Z. |
| EPI_ISL_632899, EPI_ISL_632900, EPI_ISL_632901, EPI_ISL_632902, EPI_ISL_632903 | Communicable Disease Laboratory, Public Health Directorate | Communicable Disease Laboratory, Public Health Directorate | AlAbbas,Z., Altaif,Z., AlWasti,H., Alhujairi,Z. |
| EPI_ISL_632904 | Communicable Disease Laboratory, Public Health Directorate | Communicable Disease Laboratory, Public Health Directorate | AlTaif,Z., AlHujairi,Z., AlWasti,H., AlAbbas,Z. |
| EPI_ISL_632905 | Communicable Disease Laboratory, Public Health Directorate | Communicable Disease Laboratory, Public Health Directorate | AlHujairi,Z., Altaif,Z., AlWasti,H., AlAbbas,Z. |
| EPI_ISL_632906, EPI_ISL_632907 | Communicable Disease Laboratory, Public Health Directorate | Communicable Disease Laboratory, Public Health Directorate | AlAbbas,Z., Altaif,Z., AlWasti,H., Alhujairi,Z. |
| EPI_ISL_636973 | Public Health Lab | Public Health Lab | Alwasti, H |
| EPI_ISL_678261, EPI_ISL_678262, EPI_ISL_678263, EPI_ISL_678264, EPI_ISL_678265, EPI_ISL_678266, EPI_ISL_678267, EPI_ISL_678268, EPI_ISL_678269, EPI_ISL_678270, EPI_ISL_678271, EPI_ISL_678272, EPI_ISL_681298, EPI_ISL_681299, EPI_ISL_681300, EPI_ISL_681301, EPI_ISL_681302, EPI_ISL_681303, EPI_ISL_681304, EPI_ISL_681305, EPI_ISL_681306, EPI_ISL_681307, EPI_ISL_681308, EPI_ISL_681309, EPI_ISL_681310, EPI_ISL_681311, EPI_ISL_681312, EPI_ISL_681313, EPI_ISL_681314, EPI_ISL_681315, EPI_ISL_681316, EPI_ISL_681317, EPI_ISL_681318, EPI_ISL_681319, EPI_ISL_682299, EPI_ISL_682300, EPI_ISL_682301, EPI_ISL_682302, EPI_ISL_682303, EPI_ISL_682304, EPI_ISL_682305, EPI_ISL_682306, EPI_ISL_682307, EPI_ISL_682308, EPI_ISL_682309, EPI_ISL_682310, EPI_ISL_682311, EPI_ISL_682312, EPI_ISL_682313, EPI_ISL_682314, EPI_ISL_682315, EPI_ISL_682316, EPI_ISL_682317, EPI_ISL_682318, EPI_ISL_682319, EPI_ISL_682320, EPI_ISL_682321, EPI_ISL_682322, EPI_ISL_684028, EPI_ISL_684029, EPI_ISL_684030, EPI_ISL_684031, EPI_ISL_684032, EPI_ISL_684033, EPI_ISL_684034, EPI_ISL_684035, EPI_ISL_684036 | | | |
| see above | Communicable Disease Laboratory, Public Health Directorate | Communicable Disease Laboratory, Public Health Directorate | Alwasti,H., Altaif,Z., AlHujairi,Z., AlAbbas,Z. |

We gratefully acknowledge the following Authors from the Originating laboratories responsible for obtaining the specimens, as well as the Submitting laboratories where the genome data were generated and shared via GISAID, on which this research is based.

All Submitters of data may be contacted directly via www.gisaid.org

Authors are sorted alphabetically.

| Accession ID | Originating Laboratory | Submitting Laboratory | Authors |
|---|---|---|---|
| EPI_ISL_1000668, EPI_ISL_1000670, EPI_ISL_1000671, EPI_ISL_1000673, EPI_ISL_1000675, EPI_ISL_1000677 | Instituto de Biotecnologia - UNESP-Botucatu-SP | Instituto de Biotecnologia - UNESP-Botucatu-SP | Leila Sabrina Ullmann; Fábio Sossai Possebon, Camila Dantas Malossi, Paula Rahal, Paulo Inacio da Costa, João Pessoa Araújo Jr. |
| EPI_ISL_1004233, EPI_ISL_1004234, EPI_ISL_1004235, EPI_ISL_1004236, EPI_ISL_1004237, EPI_ISL_1004238, EPI_ISL_1004239, EPI_ISL_1004240, EPI_ISL_1004241, EPI_ISL_1004242 | Laboratório de Virologia - Instituto de Medicina Tropical - Universidade de São Paulo | Laboratório de Parasitologia Médica - Instituto de Medicina Tropical - Universidade de São Paulo | Camila Malta Romano, Jaqueline Goes de Jesus, Giulia Magalhães Ferreira, Pamela dos Santos Andrade, Esmenia Coelho, Alvina Clara Felix, Anderson de Paula, Darlan Candido, Ingra Morales Claro, Franciane Mendes, Midiã Ferreira, Lucas A. Moyses Franco, Flavia Cristina Sales, Nuno Faria, Ester C. Sabino; Brazil-UK Centre for Arbovirus Discovery Diagnosis Genomics and Epidemiology (CADDE) Genomic Network - Instituto de Medicina Tropical |
| EPI_ISL_1009675, EPI_ISL_1009676, EPI_ISL_1009677, EPI_ISL_1009678 | Diagnósticos da América - DASA | Instituto de Medicina Tropical de Sao Paulo | Brazil-UK Centre for Arbovirus Discovery Diagnosis Genomics and Epidemiology (CADDE) Genomic Network - Instituto de Medicina Tropical |
| EPI_ISL_1023783, EPI_ISL_1023784, EPI_ISL_1023786, EPI_ISL_1023788, EPI_ISL_1023790, EPI_ISL_1023792, EPI_ISL_1023794, EPI_ISL_1023796, EPI_ISL_1023798, EPI_ISL_1023800, EPI_ISL_1023801, EPI_ISL_1023803, EPI_ISL_1023805, EPI_ISL_1023807, EPI_ISL_1023809, EPI_ISL_1023811, EPI_ISL_1023812, EPI_ISL_1023815, EPI_ISL_1023816, EPI_ISL_1023818, EPI_ISL_1023820, EPI_ISL_1023822, EPI_ISL_1023824, EPI_ISL_1023826, EPI_ISL_1023827, EPI_ISL_1023829, EPI_ISL_1023831, EPI_ISL_1023833, EPI_ISL_1023835, EPI_ISL_1023837, EPI_ISL_1023839, EPI_ISL_1023841, EPI_ISL_1023843, EPI_ISL_1023845 | | | |
| see above | Centro de Desenvolvimento Tecnológico em Saúde - CDTS | Centro de Desenvolvimento Tecnológico em Saúde - CDTS | Souza,T.M., Fintelman-Rodrigues,N., De Paula,A.D., Saraiva,F.B., Ferreira,M.A. and Sacramento,C.Q. |
| EPI_ISL_1034304, EPI_ISL_1034306 | Laboratorio de Ecologia de Doencas Transmissiveis na Amazonia, Instituto Leonidas e Maria Deane - Fiocruz Amazonia | Laboratorio de Ecologia de Doencas Transmissiveis na Amazonia, Instituto Leonidas e Maria Deane - Fiocruz Amazonia | Valdinete Nascimento, Victor Souza, André Corado, Fernanda Nascimento, George Silva, Ágatha Costa, Debora Duarte, Karina Pessoa, Matilde Mejía, Luciana Gonçalves, Maria Júlia Brandão, Michele Jesus, Felipe Naveca on behalf of the Fiocruz COVID-19 Genomic Surveillance Network |
| EPI_ISL_1039691, EPI_ISL_1039692, EPI_ISL_1039693, EPI_ISL_1039694, EPI_ISL_1039695 | LACEN do Estado de Goias | Instituto Adolfo Lutz, Interdiciplinary Procedures Center, Strategic Laboratory | Claudio Tavares Sacchi, Claudia Regina Gonçalves, Erica Valessa Ramos Gomes, Karoline Rodrigues Campos |
| EPI_ISL_1039696 | Instituto Adolfo Lutz - Regional de Presidente Prudente | Instituto Adolfo Lutz, Interdiciplinary Procedures Center, Strategic Laboratory | Claudio Tavares Sacchi, Claudia Regina Gonçalves, Erica Valessa Ramos Gomes, Karoline Rodrigues Campos |
| EPI_ISL_1039697 | Instituto Adolfo Lutz Central | Instituto Adolfo Lutz, Interdiciplinary Procedures Center, Strategic Laboratory | Claudio Tavares Sacchi, Claudia Regina Gonçalves, Erica Valessa Ramos Gomes, Karoline Rodrigues Campos |
| EPI_ISL_1039698 | Lab Loc - Itapecerica da Serra | Instituto Adolfo Lutz, Interdiciplinary Procedures Center, Strategic Laboratory | Claudio Tavares Sacchi, Claudia Regina Gonçalves, Erica Valessa Ramos Gomes, Karoline Rodrigues Campos |
| EPI_ISL_1039699 | Instituto Adolfo Lutz - Regional de Taubate | Instituto Adolfo Lutz, Interdiciplinary Procedures Center, Strategic Laboratory | Claudio Tavares Sacchi, Claudia Regina Gonçalves, Erica Valessa Ramos Gomes, Karoline Rodrigues Campos |
| EPI_ISL_1039700, EPI_ISL_1039701 | Instituto Adolfo Lutz Central | Instituto Adolfo Lutz, Interdiciplinary Procedures Center, Strategic Laboratory | Claudio Tavares Sacchi, Claudia Regina Gonçalves, Erica Valessa Ramos Gomes, Karoline Rodrigues Campos |
| EPI_ISL_1039702 | Instituto Adolfo Lutz - Regional de Aracatuba | Instituto Adolfo Lutz, Interdiciplinary Procedures Center, Strategic Laboratory | Claudio Tavares Sacchi, Claudia Regina Gonçalves, Erica Valessa Ramos Gomes, Karoline Rodrigues Campos |
| EPI_ISL_1039703 | Instituto Adolfo Lutz - Regional de Taubate | Instituto Adolfo Lutz, Interdiciplinary Procedures Center, Strategic Laboratory | Claudio Tavares Sacchi, Claudia Regina Gonçalves, Erica Valessa Ramos Gomes, Karoline Rodrigues Campos |
| EPI_ISL_1039704 | Lab Loc - Itapecerica da Serra | Instituto Adolfo Lutz, Interdiciplinary Procedures Center, Strategic Laboratory | Claudio Tavares Sacchi, Claudia Regina Gonçalves, Erica Valessa Ramos Gomes, Karoline Rodrigues Campos |
| EPI_ISL_1039705, EPI_ISL_1039706, EPI_ISL_1039707, EPI_ISL_1039708, EPI_ISL_1039709, EPI_ISL_1039710 | Instituto Adolfo Lutz Central | Instituto Adolfo Lutz, Interdiciplinary Procedures Center, Strategic Laboratory | Claudio Tavares Sacchi, Claudia Regina Gonçalves, Erica Valessa Ramos Gomes, Karoline Rodrigues Campos |
| EPI_ISL_1040823 | Secretaria Municipal de Saude de Piracaia | Instituto Adolfo Lutz, Interdiciplinary Procedures Center, Strategic Laboratory | Claudio Tavares Sacchi, Claudia Regina Gonçalves, Erica Valessa Ramos Gomes, Karoline Rodrigues Campos |
| EPI_ISL_1040824, EPI_ISL_1040825, EPI_ISL_1040826, EPI_ISL_1040827, EPI_ISL_1040828, EPI_ISL_1040829, EPI_ISL_1040830, EPI_ISL_1040831, EPI_ISL_1040832, EPI_ISL_1040833, EPI_ISL_1040834, EPI_ISL_1040835, EPI_ISL_1040836, EPI_ISL_1040837, EPI_ISL_1040838, EPI_ISL_1040839, EPI_ISL_1040840, EPI_ISL_1040841, EPI_ISL_1040842, EPI_ISL_1040843, EPI_ISL_1040844, EPI_ISL_1040845, EPI_ISL_1040846, EPI_ISL_1040847, EPI_ISL_1040848, EPI_ISL_1040849, EPI_ISL_1040850, EPI_ISL_1040851 | | | |
| see above | LACEN do Mato Grosso do Sul | Instituto Adolfo Lutz, Interdiciplinary Procedures Center, Strategic Laboratory | Claudio Tavares Sacchi, Claudia Regina Gonçalves, Erica Valessa Ramos Gomes, Karoline Rodrigues Campos |
| EPI_ISL_1041509 | LACEN do Estado de Goias | Instituto Adolfo Lutz, Interdiciplinary Procedures Center, Strategic Laboratory | Claudio Tavares Sacchi, Claudia Regina Gonçalves, Erica Valessa Ramos Gomes, Karoline Rodrigues Campos |
| EPI_ISL_1060876 | DB Diagnosticos do Brasil | Instituto de Medicina Tropical de Sao Paulo | Brazil-UK Centre for Arbovirus Discovery Diagnosis Genomics and Epidemiology (CADDE) Genomic Network - Instituto de Medicina Tropical |
| EPI_ISL_1060877, EPI_ISL_1060878, EPI_ISL_1060879, EPI_ISL_1060880, EPI_ISL_1060881, EPI_ISL_1060882, EPI_ISL_1060883 | CDL Laboratorio Santos e Vidal LTDA. | Instituto de Medicina Tropical de Sao Paulo | Brazil-UK Centre for Arbovirus Discovery Diagnosis Genomics and Epidemiology (CADDE) Genomic Network - Instituto de Medicina Tropical |
| EPI_ISL_1060884 | DB Diagnosticos do Brasil | Instituto de Medicina Tropical de Sao Paulo | Brazil-UK Centre for Arbovirus Discovery Diagnosis Genomics and Epidemiology (CADDE) Genomic Network - Instituto de Medicina Tropical |
| EPI_ISL_1060885 | CDL Laboratorio Santos e Vidal LTDA. | Instituto de Medicina Tropical de Sao Paulo | Brazil-UK Centre for Arbovirus Discovery Diagnosis Genomics and Epidemiology (CADDE) Genomic Network - Instituto de Medicina Tropical |
| EPI_ISL_1060886, EPI_ISL_1060887 | DB Diagnosticos do Brasil | Instituto de Medicina Tropical de Sao Paulo | Brazil-UK Centre for Arbovirus Discovery Diagnosis Genomics and Epidemiology (CADDE) Genomic Network - Instituto de Medicina Tropical |
| EPI_ISL_1060888, EPI_ISL_1060889, EPI_ISL_1060890 | CDL Laboratorio Santos e Vidal LTDA. | Instituto de Medicina Tropical de Sao Paulo | Brazil-UK Centre for Arbovirus Discovery Diagnosis Genomics and Epidemiology (CADDE) Genomic Network - Instituto de Medicina Tropical |
| EPI_ISL_1060891 | DB Diagnosticos do Brasil | Instituto de Medicina Tropical de Sao Paulo | Brazil-UK Centre for Arbovirus Discovery Diagnosis Genomics and Epidemiology (CADDE) Genomic Network - Instituto de Medicina Tropical |
| EPI_ISL_1060892, EPI_ISL_1060893, EPI_ISL_1060894, EPI_ISL_1060895, EPI_ISL_1060896, EPI_ISL_1060897, EPI_ISL_1060898, EPI_ISL_1060899 | CDL Laboratorio Santos e Vidal LTDA. | Instituto de Medicina Tropical de Sao Paulo | Brazil-UK Centre for Arbovirus Discovery Diagnosis Genomics and Epidemiology (CADDE) Genomic Network - Instituto de Medicina Tropical |

| | | | |
|---|---|---|---|
| EPI_ISL_1060900 | DB Diagnosticos do Brasil | Instituto de Medicina Tropical de Sao Paulo | Brazil-UK Centre for Arbovirus Discovery Diagnosis Genomics and Epidemiology (CADDE) Genomic Network - Instituto de Medicina Tropical |
| EPI_ISL_1060901 | CDL Laboratorio Santos e Vidal LTDA. | Instituto de Medicina Tropical de Sao Paulo | Brazil-UK Centre for Arbovirus Discovery Diagnosis Genomics and Epidemiology (CADDE) Genomic Network - Instituto de Medicina Tropical |
| EPI_ISL_1060902 | DB Diagnosticos do Brasil | Instituto de Medicina Tropical de Sao Paulo | Brazil-UK Centre for Arbovirus Discovery Diagnosis Genomics and Epidemiology (CADDE) Genomic Network - Instituto de Medicina Tropical |
| EPI_ISL_1060903 | CDL Laboratorio Santos e Vidal LTDA. | Instituto de Medicina Tropical de Sao Paulo | Brazil-UK Centre for Arbovirus Discovery Diagnosis Genomics and Epidemiology (CADDE) Genomic Network - Instituto de Medicina Tropical |
| EPI_ISL_1060904 | DB Diagnosticos do Brasil | Instituto de Medicina Tropical de Sao Paulo | Brazil-UK Centre for Arbovirus Discovery Diagnosis Genomics and Epidemiology (CADDE) Genomic Network - Instituto de Medicina Tropical |
| EPI_ISL_1060905, EPI_ISL_1060906, EPI_ISL_1060907, EPI_ISL_1060908, EPI_ISL_1060909 | CDL Laboratorio Santos e Vidal LTDA. | Instituto de Medicina Tropical de Sao Paulo | Brazil-UK Centre for Arbovirus Discovery Diagnosis Genomics and Epidemiology (CADDE) Genomic Network - Instituto de Medicina Tropical |
| EPI_ISL_1060910 | DB Diagnosticos do Brasil | Instituto de Medicina Tropical de Sao Paulo | Brazil-UK Centre for Arbovirus Discovery Diagnosis Genomics and Epidemiology (CADDE) Genomic Network - Instituto de Medicina Tropical |
| EPI_ISL_1060911, EPI_ISL_1060912, EPI_ISL_1060913 | CDL Laboratorio Santos e Vidal LTDA. | Instituto de Medicina Tropical de Sao Paulo | Brazil-UK Centre for Arbovirus Discovery Diagnosis Genomics and Epidemiology (CADDE) Genomic Network - Instituto de Medicina Tropical |
| EPI_ISL_1060914 | DB Diagnosticos do Brasil | Instituto de Medicina Tropical de Sao Paulo | Brazil-UK Centre for Arbovirus Discovery Diagnosis Genomics and Epidemiology (CADDE) Genomic Network - Instituto de Medicina Tropical |
| EPI_ISL_1060915, EPI_ISL_1060916, EPI_ISL_1060917 | CDL Laboratorio Santos e Vidal LTDA. | Instituto de Medicina Tropical de Sao Paulo | Brazil-UK Centre for Arbovirus Discovery Diagnosis Genomics and Epidemiology (CADDE) Genomic Network - Instituto de Medicina Tropical |
| EPI_ISL_1060918 | DB Diagnosticos do Brasil | Instituto de Medicina Tropical de Sao Paulo | Brazil-UK Centre for Arbovirus Discovery Diagnosis Genomics and Epidemiology (CADDE) Genomic Network - Instituto de Medicina Tropical |
| EPI_ISL_1060919 | CDL Laboratorio Santos e Vidal LTDA. | Instituto de Medicina Tropical de Sao Paulo | Brazil-UK Centre for Arbovirus Discovery Diagnosis Genomics and Epidemiology (CADDE) Genomic Network - Instituto de Medicina Tropical |
| EPI_ISL_1060920 | DB Diagnosticos do Brasil | Instituto de Medicina Tropical de Sao Paulo | Brazil-UK Centre for Arbovirus Discovery Diagnosis Genomics and Epidemiology (CADDE) Genomic Network - Instituto de Medicina Tropical |
| EPI_ISL_1060921, EPI_ISL_1060922 | CDL Laboratorio Santos e Vidal LTDA. | Instituto de Medicina Tropical de Sao Paulo | Brazil-UK Centre for Arbovirus Discovery Diagnosis Genomics and Epidemiology (CADDE) Genomic Network - Instituto de Medicina Tropical |
| EPI_ISL_1060923, EPI_ISL_1060924, EPI_ISL_1060925, EPI_ISL_1060926, EPI_ISL_1060927, EPI_ISL_1060928 | DB Diagnosticos do Brasil | Instituto de Medicina Tropical de Sao Paulo | Brazil-UK Centre for Arbovirus Discovery Diagnosis Genomics and Epidemiology (CADDE) Genomic Network - Instituto de Medicina Tropical |
| EPI_ISL_1060929, EPI_ISL_1060930, EPI_ISL_1060931, EPI_ISL_1060932, EPI_ISL_1060933, EPI_ISL_1060934, EPI_ISL_1060935, EPI_ISL_1060936, EPI_ISL_1060937, EPI_ISL_1060938, EPI_ISL_1060939, EPI_ISL_1060940, EPI_ISL_1060941, EPI_ISL_1060942 | | | |
| see above | CDL Laboratorio Santos e Vidal LTDA. | Instituto de Medicina Tropical de Sao Paulo | Brazil-UK Centre for Arbovirus Discovery Diagnosis Genomics and Epidemiology (CADDE) Genomic Network - Instituto de Medicina Tropical |
| EPI_ISL_1060943 | DB Diagnosticos do Brasil | Instituto de Medicina Tropical de Sao Paulo | Brazil-UK Centre for Arbovirus Discovery Diagnosis Genomics and Epidemiology (CADDE) Genomic Network - Instituto de Medicina Tropical |
| EPI_ISL_1060944, EPI_ISL_1060945, EPI_ISL_1060946 | CDL Laboratorio Santos e Vidal LTDA. | Instituto de Medicina Tropical de Sao Paulo | Brazil-UK Centre for Arbovirus Discovery Diagnosis Genomics and Epidemiology (CADDE) Genomic Network - Instituto de Medicina Tropical |
| EPI_ISL_1060947 | DB Diagnosticos do Brasil | Instituto de Medicina Tropical de Sao Paulo | Brazil-UK Centre for Arbovirus Discovery Diagnosis Genomics and Epidemiology (CADDE) Genomic Network - Instituto de Medicina Tropical |
| EPI_ISL_1060948 | CDL Laboratorio Santos e Vidal LTDA. | Instituto de Medicina Tropical de Sao Paulo | Brazil-UK Centre for Arbovirus Discovery Diagnosis Genomics and Epidemiology (CADDE) Genomic Network - Instituto de Medicina Tropical |
| EPI_ISL_1060949 | DB Diagnosticos do Brasil | Instituto de Medicina Tropical de Sao Paulo | Brazil-UK Centre for Arbovirus Discovery Diagnosis Genomics and Epidemiology (CADDE) Genomic Network - Instituto de Medicina Tropical |
| EPI_ISL_1060950, EPI_ISL_1060951, EPI_ISL_1060952, EPI_ISL_1060953 | CDL Laboratorio Santos e Vidal LTDA. | Instituto de Medicina Tropical de Sao Paulo | Brazil-UK Centre for Arbovirus Discovery Diagnosis Genomics and Epidemiology (CADDE) Genomic Network - Instituto de Medicina Tropical |
| EPI_ISL_1060954 | DB Diagnosticos do Brasil | Instituto de Medicina Tropical de Sao Paulo | Brazil-UK Centre for Arbovirus Discovery Diagnosis Genomics and Epidemiology (CADDE) Genomic Network - Instituto de Medicina Tropical |
| EPI_ISL_1060955, EPI_ISL_1060956, EPI_ISL_1060957, EPI_ISL_1060958, EPI_ISL_1060959, EPI_ISL_1060960, EPI_ISL_1060961, EPI_ISL_1060962 | CDL Laboratorio Santos e Vidal LTDA. | Instituto de Medicina Tropical de Sao Paulo | Brazil-UK Centre for Arbovirus Discovery Diagnosis Genomics and Epidemiology (CADDE) Genomic Network - Instituto de Medicina Tropical |
| EPI_ISL_1060963 | DB Diagnosticos do Brasil | Instituto de Medicina Tropical de Sao Paulo | Brazil-UK Centre for Arbovirus Discovery Diagnosis Genomics and Epidemiology (CADDE) Genomic Network - Instituto de Medicina Tropical |
| EPI_ISL_1060964, EPI_ISL_1060966, EPI_ISL_1060969, EPI_ISL_1060970, EPI_ISL_1060975, EPI_ISL_1060977 | CDL Laboratorio Santos e Vidal LTDA. | Instituto de Medicina Tropical de Sao Paulo | Brazil-UK Centre for Arbovirus Discovery Diagnosis Genomics and Epidemiology (CADDE) Genomic Network - Instituto de Medicina Tropical |
| EPI_ISL_1060978 | DB Diagnosticos do Brasil | Instituto de Medicina Tropical de Sao Paulo | Brazil-UK Centre for Arbovirus Discovery Diagnosis Genomics and Epidemiology (CADDE) Genomic Network - Instituto de Medicina Tropical |
| EPI_ISL_1060979, EPI_ISL_1060985, EPI_ISL_1060986, EPI_ISL_1060988, EPI_ISL_1060989, EPI_ISL_1060991, EPI_ISL_1060992, EPI_ISL_1060993 | CDL Laboratorio Santos e Vidal LTDA. | Instituto de Medicina Tropical de Sao Paulo | Brazil-UK Centre for Arbovirus Discovery Diagnosis Genomics and Epidemiology (CADDE) Genomic Network - Instituto de Medicina Tropical |
| EPI_ISL_1060995 | DB Diagnosticos do Brasil | Instituto de Medicina Tropical de Sao Paulo | Brazil-UK Centre for Arbovirus Discovery Diagnosis Genomics and Epidemiology (CADDE) Genomic Network - Instituto de Medicina Tropical |
| EPI_ISL_1060996 | CDL Laboratorio Santos e Vidal LTDA. | Instituto de Medicina Tropical de Sao Paulo | Brazil-UK Centre for Arbovirus Discovery Diagnosis Genomics and Epidemiology (CADDE) Genomic Network - Instituto de Medicina Tropical |
| EPI_ISL_1060997 | DB Diagnosticos do Brasil | Instituto de Medicina Tropical de Sao Paulo | Brazil-UK Centre for Arbovirus Discovery Diagnosis Genomics and Epidemiology (CADDE) Genomic Network - Instituto de Medicina Tropical |
| EPI_ISL_1060998, EPI_ISL_1060999, EPI_ISL_1061000, EPI_ISL_1061001, EPI_ISL_1061002, EPI_ISL_1061003, EPI_ISL_1061004, EPI_ISL_1061005 | CDL Laboratorio Santos e Vidal LTDA. | Instituto de Medicina Tropical de Sao Paulo | Brazil-UK Centre for Arbovirus Discovery Diagnosis Genomics and Epidemiology (CADDE) Genomic Network - Instituto de Medicina Tropical |
| EPI_ISL_1061006 | DB Diagnosticos do Brasil | Instituto de Medicina Tropical de Sao Paulo | Brazil-UK Centre for Arbovirus Discovery Diagnosis Genomics and Epidemiology (CADDE) Genomic Network - Instituto de Medicina Tropical |
| EPI_ISL_1061007, EPI_ISL_1061008, EPI_ISL_1061009, EPI_ISL_1061010, EPI_ISL_1061011, EPI_ISL_1061013, EPI_ISL_1061014, EPI_ISL_1061015, EPI_ISL_1061016, EPI_ISL_1061017, EPI_ISL_1061018, EPI_ISL_1061019, EPI_ISL_1061020, EPI_ISL_1061021, EPI_ISL_1061022, EPI_ISL_1061023, EPI_ISL_1061024 | | | |
| see above | CDL Laboratorio Santos e Vidal LTDA. | Instituto de Medicina Tropical de Sao Paulo | Brazil-UK Centre for Arbovirus Discovery Diagnosis Genomics and Epidemiology (CADDE) Genomic Network - Instituto de Medicina Tropical |
| EPI_ISL_1061025 | DB Diagnosticos do Brasil | Instituto de Medicina Tropical de Sao Paulo | Brazil-UK Centre for Arbovirus Discovery Diagnosis Genomics and Epidemiology (CADDE) Genomic Network - Instituto de Medicina Tropical |
| EPI_ISL_1061026 | CDL Laboratorio Santos e Vidal LTDA. | Instituto de Medicina Tropical de Sao Paulo | Brazil-UK Centre for Arbovirus Discovery Diagnosis Genomics and Epidemiology (CADDE) Genomic Network - Instituto de Medicina Tropical |
| EPI_ISL_1061027 | DB Diagnosticos do Brasil | Instituto de Medicina Tropical de Sao Paulo | Brazil-UK Centre for Arbovirus Discovery Diagnosis Genomics and Epidemiology (CADDE) Genomic Network - Instituto de Medicina Tropical |
| EPI_ISL_1061028 | CDL Laboratorio Santos e Vidal LTDA. | Instituto de Medicina Tropical de Sao Paulo | Brazil-UK Centre for Arbovirus Discovery Diagnosis Genomics and Epidemiology (CADDE) Genomic Network - Instituto de Medicina Tropical |
| EPI_ISL_1061029 | DB Diagnosticos do Brasil | Instituto de Medicina Tropical de Sao Paulo | Brazil-UK Centre for Arbovirus Discovery Diagnosis Genomics and Epidemiology (CADDE) Genomic Network - Instituto de Medicina Tropical |
| EPI_ISL_1061030, EPI_ISL_1061031, EPI_ISL_1061032 | CDL Laboratorio Santos e Vidal LTDA. | Instituto de Medicina Tropical de Sao Paulo | Brazil-UK Centre for Arbovirus Discovery Diagnosis Genomics and Epidemiology (CADDE) Genomic Network - Instituto de Medicina Tropical |
| EPI_ISL_1063789 | Evandro Chagas Institute | Evandro Chagas Institute Virology | Santos, M.C.; Silva, A.M.; Junior, W.D.C.; Barbagelata, L.S.; Ferreira, J.A.; Sousa, E.M.A.; da Silva, P.S.; Pinheiro, K.C.; L.C.; Sousa Junior, E.C. |
| EPI_ISL_1064736, EPI_ISL_1064737, EPI_ISL_1064738, EPI_ISL_1064739, EPI_ISL_1064740, EPI_ISL_1064741, EPI_ISL_1064742, EPI_ISL_1064743, EPI_ISL_1064744, EPI_ISL_1064745, EPI_ISL_1064746, EPI_ISL_1064747, EPI_ISL_1064748, EPI_ISL_1064749, EPI_ISL_1064750 | | | |
| see above | CDL Laboratorio Santos e Vidal LTDA. | Instituto de Medicina Tropical de Sao Paulo | Brazil-UK Centre for Arbovirus Discovery Diagnosis Genomics and Epidemiology (CADDE) Genomic Network - Instituto de Medicina Tropical |

| | | | |
|---|---|---|---|
| EPI_ISL_1067728 | Center for Biotechnology and Cell Therapy, São Rafael Hospital, Salvador, Brazil | Central Public Health Laboratory - LACEN -Bahia, Salvador, Brazil | Stephane Tosta, Luciana Oliveira, Vanessa Nardy,Patrícia Cajado,Marcela Gómez, Breno Dominguez, Jaqueline Gomes, Vagner Fonseca,Marta Giovanetti,Luiz Alcantara, Felicidade Pereira, Arabela Leal |
| EPI_ISL_1067729, EPI_ISL_1067730, EPI_ISL_1067731 | Central Public Health Laboratory - LACEN -Bahia, Salvador, Brazil | Central Public Health Laboratory - LACEN -Bahia, Salvador, Brazil | Stephane Tosta, Luciana Oliveira, Vanessa Nardy,Patrícia Cajado,Marcela Gómez, Breno Dominguez, Jaqueline Gomes, Vagner Fonseca,Marta Giovanetti,Luiz Alcantara, Felicidade Pereira, Arabela Leal |
| EPI_ISL_1067732 | Center for Biotechnology and Cell Therapy, São Rafael Hospital, Salvador, Brazil | Central Public Health Laboratory - LACEN -Bahia, Salvador, Brazil | Stephane Tosta, Luciana Oliveira, Vanessa Nardy,Patrícia Cajado,Marcela Gómez, Breno Dominguez, Jaqueline Gomes, Vagner Fonseca,Marta Giovanetti,Luiz Alcantara, Felicidade Pereira, Arabela Leal |
| EPI_ISL_1067733, EPI_ISL_1067734, EPI_ISL_1067735 | Central Public Health Laboratory - LACEN -Bahia, Salvador, Brazil | Central Public Health Laboratory - LACEN -Bahia, Salvador, Brazil | Stephane Tosta, Luciana Oliveira, Vanessa Nardy,Patrícia Cajado,Marcela Gómez, Breno Dominguez, Jaqueline Gomes, Vagner Fonseca,Marta Giovanetti,Luiz Alcantara, Felicidade Pereira, Arabela Leal |
| EPI_ISL_1067736 | Center for Biotechnology and Cell Therapy, São Rafael Hospital, Salvador, Brazil | Central Public Health Laboratory - LACEN -Bahia, Salvador, Brazil | Stephane Tosta, Luciana Oliveira, Vanessa Nardy,Patrícia Cajado,Marcela Gómez, Breno Dominguez, Jaqueline Gomes, Vagner Fonseca,Marta Giovanetti,Luiz Alcantara, Felicidade Pereira, Arabela Leal |
| EPI_ISL_1067737, EPI_ISL_1067738 | Central Public Health Laboratory - LACEN -Bahia, Salvador, Brazil | Central Public Health Laboratory - LACEN -Bahia, Salvador, Brazil | Stephane Tosta, Luciana Oliveira, Vanessa Nardy,Patrícia Cajado,Marcela Gómez, Breno Dominguez, Jaqueline Gomes, Vagner Fonseca,Marta Giovanetti,Luiz Alcantara, Felicidade Pereira, Arabela Leal |
| EPI_ISL_1068086, EPI_ISL_1068087, EPI_ISL_1068088, EPI_ISL_1068089, EPI_ISL_1068090, EPI_ISL_1068091, EPI_ISL_1068092, EPI_ISL_1068093, EPI_ISL_1068094, EPI_ISL_1068095, EPI_ISL_1068096, EPI_ISL_1068097, EPI_ISL_1068098, EPI_ISL_1068099, EPI_ISL_1068100, EPI_ISL_1068101, EPI_ISL_1068102, EPI_ISL_1068103, EPI_ISL_1068104, EPI_ISL_1068105, EPI_ISL_1068106, EPI_ISL_1068107, EPI_ISL_1068108, EPI_ISL_1068109, EPI_ISL_1068110, EPI_ISL_1068111, EPI_ISL_1068112, EPI_ISL_1068113, EPI_ISL_1068114, EPI_ISL_1068115, EPI_ISL_1068120, EPI_ISL_1068121, EPI_ISL_1068122, EPI_ISL_1068123, EPI_ISL_1068124, EPI_ISL_1068125, EPI_ISL_1068126, EPI_ISL_1068127, EPI_ISL_1068128, EPI_ISL_1068129, EPI_ISL_1068130, EPI_ISL_1068131, EPI_ISL_1068132, EPI_ISL_1068133, EPI_ISL_1068134, EPI_ISL_1068135, EPI_ISL_1068136, EPI_ISL_1068137, EPI_ISL_1068138, EPI_ISL_1068139, EPI_ISL_1068140, EPI_ISL_1068141, EPI_ISL_1068142, EPI_ISL_1068143, EPI_ISL_1068144, EPI_ISL_1068145, EPI_ISL_1068146, EPI_ISL_1068147, EPI_ISL_1068148, EPI_ISL_1068149, EPI_ISL_1068150, EPI_ISL_1068151, EPI_ISL_1068152, EPI_ISL_1068153, EPI_ISL_1068154, EPI_ISL_1068155, EPI_ISL_1068156, EPI_ISL_1068157, EPI_ISL_1068158, EPI_ISL_1068159, EPI_ISL_1068160, EPI_ISL_1068161, EPI_ISL_1068163, EPI_ISL_1068167, EPI_ISL_1068168, EPI_ISL_1068169, EPI_ISL_1068170, EPI_ISL_1068171, EPI_ISL_1068172, EPI_ISL_1068173, EPI_ISL_1068174, EPI_ISL_1068175, EPI_ISL_1068177, EPI_ISL_1068180, EPI_ISL_1068182, EPI_ISL_1068184, EPI_ISL_1068185, EPI_ISL_1068186, EPI_ISL_1068187, EPI_ISL_1068188, EPI_ISL_1068189, EPI_ISL_1068190, EPI_ISL_1068191, EPI_ISL_1068192, EPI_ISL_1068196, EPI_ISL_1068197, EPI_ISL_1068198, EPI_ISL_1068199, EPI_ISL_1068200, EPI_ISL_1068201, EPI_ISL_1068202, EPI_ISL_1068203, EPI_ISL_1068204, EPI_ISL_1068205, EPI_ISL_1068206, EPI_ISL_1068207, EPI_ISL_1068208, EPI_ISL_1068209, EPI_ISL_1068211, EPI_ISL_1068212, EPI_ISL_1068213, EPI_ISL_1068214, EPI_ISL_1068215, EPI_ISL_1068216, EPI_ISL_1068217, EPI_ISL_1068218, EPI_ISL_1068219, EPI_ISL_1068220, EPI_ISL_1068221, EPI_ISL_1068222, EPI_ISL_1068223, EPI_ISL_1068224, EPI_ISL_1068225, EPI_ISL_1068226, EPI_ISL_1068227, EPI_ISL_1068228, EPI_ISL_1068229, EPI_ISL_1068230, EPI_ISL_1068231, EPI_ISL_1068232, EPI_ISL_1068233, EPI_ISL_1068234, EPI_ISL_1068235, EPI_ISL_1068236, EPI_ISL_1068237, EPI_ISL_1068238, EPI_ISL_1068239, EPI_ISL_1068240, EPI_ISL_1068241, EPI_ISL_1068242, EPI_ISL_1068243, EPI_ISL_1068244, EPI_ISL_1068245, EPI_ISL_1068246, EPI_ISL_1068247, EPI_ISL_1068248, EPI_ISL_1068249, EPI_ISL_1068250, EPI_ISL_1068251, EPI_ISL_1068252, EPI_ISL_1068253, EPI_ISL_1068254, EPI_ISL_1068255, EPI_ISL_1068256, EPI_ISL_1068257, EPI_ISL_1068258, EPI_ISL_1068259, EPI_ISL_1068260, EPI_ISL_1068261, EPI_ISL_1068262, EPI_ISL_1068263, EPI_ISL_1068264, EPI_ISL_1068265, EPI_ISL_1068266, EPI_ISL_1068267, EPI_ISL_1068268, EPI_ISL_1068269, EPI_ISL_1068270, EPI_ISL_1068271, EPI_ISL_1068272, EPI_ISL_1068273, EPI_ISL_1068274, EPI_ISL_1068275, EPI_ISL_1068276, EPI_ISL_1068277, EPI_ISL_1068278, EPI_ISL_1068279, EPI_ISL_1068280, EPI_ISL_1068281, EPI_ISL_1068282, EPI_ISL_1068283, EPI_ISL_1068284, EPI_ISL_1068285, EPI_ISL_1068286, EPI_ISL_1068287, EPI_ISL_1068288, EPI_ISL_1068289, EPI_ISL_1068290, EPI_ISL_1068291, EPI_ISL_1068292 | | | |
| see above | Laboratorio de Ecologia de Doencas Transmissiveis na Amazonia, Instituto Leonidas e Maria Deane - Fiocruz Amazonia | Laboratorio de Ecologia de Doencas Transmissiveis na Amazonia, Instituto Leonidas e Maria Deane - Fiocruz Amazonia | Valdinete Nascimento, Victor Souza, André Corado, Fernanda Nascimento, George Silva, Ágatha Costa, Debora Duarte, Karina Pessoa, Matilde Mejía, Luciana Gonçalves, Maria Júlia Brandão, Michele Jesus, Felipe Naveca on behalf of the Fiocruz COVID-19 Genomic Surveillance Network |
| EPI_ISL_1068315, EPI_ISL_1068317, EPI_ISL_1068319, EPI_ISL_1068320, EPI_ISL_1068321, EPI_ISL_1068322, EPI_ISL_1068363, EPI_ISL_1068364, EPI_ISL_1068365, EPI_ISL_1068366, EPI_ISL_1068367, EPI_ISL_1068368, EPI_ISL_1068369, EPI_ISL_1068370, EPI_ISL_1068371, EPI_ISL_1068372, EPI_ISL_1068373, EPI_ISL_1068374, EPI_ISL_1068375, EPI_ISL_1068376, EPI_ISL_1068377, EPI_ISL_1068378, EPI_ISL_1068381, EPI_ISL_1068382, EPI_ISL_1068386, EPI_ISL_1068387, EPI_ISL_1068388, EPI_ISL_1068390, EPI_ISL_1068391 | | | |
| see above | Central Public Health Laboratory - LACEN -Bahia, Salvador, Brazil | Central Public Health Laboratory - LACEN -Bahia, Salvador, Brazil | Stephane Tosta, Luciana Oliveira, Vanessa Nardy,Patrícia Cajado,Marcela Gómez, Breno Dominguez, Jaqueline Gomes, Vagner Fonseca,Marta Giovanetti,Luiz Alcantara, Felicidade Pereira, Arabela Leal |
| EPI_ISL_1078981, EPI_ISL_1078983, EPI_ISL_1078984, EPI_ISL_1078986, EPI_ISL_1078987, EPI_ISL_1078988, EPI_ISL_1078989, EPI_ISL_1078991, EPI_ISL_1078992, EPI_ISL_1078993, EPI_ISL_1078995, EPI_ISL_1078996, EPI_ISL_1078997, EPI_ISL_1078999, EPI_ISL_1079000, EPI_ISL_1079002, EPI_ISL_1079003, EPI_ISL_1079004, EPI_ISL_1079006, EPI_ISL_1079007, EPI_ISL_1079008, EPI_ISL_1079158, EPI_ISL_1079159, EPI_ISL_1079161, EPI_ISL_1079162, EPI_ISL_1079163, EPI_ISL_1079165, EPI_ISL_1079166 | | | |
| see above | IAL Regional de Bauru | Instituto Adolfo Lutz, Interdiciplinary Procedures Center, Strategic Laboratory | Claudio Tavares Sacchi, Claudia Regina Gonçalves, Erica Valessa Ramos Gomes, Karoline Rodrigues Campos |
| EPI_ISL_1084723, EPI_ISL_1084724 | IMT_USP | Laboratório de Parasitologia Médica - Instituto de Medicina Tropical - Universidade de São Paulo | Brazil-UK Centre for Arbovirus Discovery Diagnosis Genomics and Epidemiology (CADDE) Genomic Network - Instituto de Medicina Tropical |
| EPI_ISL_1084726 | DB Diagnosticos do Brasil | Laboratório de Parasitologia Médica - Instituto de Medicina Tropical - Universidade de São Paulo | Brazil-UK Centre for Arbovirus Discovery Diagnosis Genomics and Epidemiology (CADDE) Genomic Network - Instituto de Medicina Tropical |
| EPI_ISL_1084727 | IMT_USP | Laboratório de Parasitologia Médica - Instituto de Medicina Tropical - Universidade de São Paulo | Brazil-UK Centre for Arbovirus Discovery Diagnosis Genomics and Epidemiology (CADDE) Genomic Network - Instituto de Medicina Tropical |
| EPI_ISL_1084730 | DB Diagnosticos do Brasil | Laboratório de Parasitologia Médica - Instituto de Medicina Tropical - Universidade de São Paulo | Brazil-UK Centre for Arbovirus Discovery Diagnosis Genomics and Epidemiology (CADDE) Genomic Network - Instituto de Medicina Tropical |
| EPI_ISL_1084734 | IMT_USP | Laboratório de Parasitologia Médica - Instituto de Medicina Tropical - Universidade de São Paulo | Brazil-UK Centre for Arbovirus Discovery Diagnosis Genomics and Epidemiology (CADDE) Genomic Network - Instituto de Medicina Tropical |
| EPI_ISL_1084737, EPI_ISL_1084738 | DB Diagnosticos do Brasil | Laboratório de Parasitologia Médica - Instituto de Medicina Tropical - Universidade de São Paulo | Brazil-UK Centre for Arbovirus Discovery Diagnosis Genomics and Epidemiology (CADDE) Genomic Network - Instituto de Medicina Tropical |
| EPI_ISL_1086034 | Diagnosticos da America - DASA | Instituto Adolfo Lutz, Interdiciplinary Procedures Center, Strategic Laboratory | Claudio Tavares Sacchi, Claudia Regina Gonçalves, Erica Valessa Ramos Gomes, Karoline Rodrigues Campos, Caio Vinicius Dias Lopes |
| EPI_ISL_1086035, EPI_ISL_1086036 | IAL Regional de Bauru | Instituto Adolfo Lutz, Interdiciplinary Procedures Center, Strategic Laboratory | Claudio Tavares Sacchi, Claudia Regina Gonçalves, Erica Valessa Ramos Gomes, Karoline Rodrigues Campos, Caio Vinicius Dias Lopes |
| EPI_ISL_1086037, EPI_ISL_1086038, EPI_ISL_1086039, EPI_ISL_1086040, EPI_ISL_1086041, EPI_ISL_1086042, EPI_ISL_1086043 | Diagnosticos da America - DASA | Instituto Adolfo Lutz, Interdiciplinary Procedures Center, Strategic Laboratory | Claudio Tavares Sacchi, Claudia Regina Gonçalves, Erica Valessa Ramos Gomes, Karoline Rodrigues Campos, Caio Vinicius Dias Lopes |
| EPI_ISL_1086044, EPI_ISL_1086045, EPI_ISL_1086046, EPI_ISL_1086047, EPI_ISL_1086048, EPI_ISL_1086049, EPI_ISL_1086050, EPI_ISL_1086051, EPI_ISL_1086052, EPI_ISL_1086053, EPI_ISL_1086054, EPI_ISL_1086055, EPI_ISL_1086056, EPI_ISL_1086057 | | | |
| see above | IAL Regional de Bauru | Instituto Adolfo Lutz, Interdiciplinary Procedures Center, Strategic Laboratory | Claudio Tavares Sacchi, Claudia Regina Gonçalves, Erica Valessa Ramos Gomes, Karoline Rodrigues Campos, Caio Vinicius Dias Lopes |
| EPI_ISL_1086373, EPI_ISL_1086374 | LACEN - Laboratório Central de Saúde Pública do Maranhao | Evandro Chagas Institute | Santos, M.C.; Silva, A.M.; Junior, W.D.C.; Barbagelata, L.S.; Ferreira, J.A.; Sousa, E.M.A.; da Silva, P.S.; Pinheiro, K.C.; L.C.; Sousa Junior, E.C. |
| EPI_ISL_1086375, EPI_ISL_1086376 | LACEN - Laboratório Central de Saúde Pública do Rio Grande do Norte | Evandro Chagas Institute | Santos, M.C.; Silva, A.M.; Junior, W.D.C.; Barbagelata, L.S.; Ferreira, J.A.; Sousa, E.M.A.; da Silva, P.S.; Pinheiro, K.C.; L.C.; Sousa Junior, E.C. |
| EPI_ISL_1086377 | LACEN - Laboratório Central de Saúde Pública do Paraiba | Evandro Chagas Institute | Santos, M.C.; Silva, A.M.; Junior, W.D.C.; Barbagelata, L.S.; Ferreira, J.A.; Sousa, E.M.A.; da Silva, P.S.; Pinheiro, K.C.; L.C.; Sousa Junior, E.C. |
| EPI_ISL_1092360 | IAL Regional de Bauru | Instituto Adolfo Lutz, Interdiciplinary Procedures Center, Strategic Laboratory | Claudio Tavares Sacchi, Claudia Regina Gonçalves, Erica Valessa Ramos Gomes, Karoline Rodrigues Campos |
| EPI_ISL_1092725 | Diagnosticos da America - DASA | Instituto Adolfo Lutz, Interdiciplinary Procedures Center, Strategic Laboratory | Claudio Tavares Sacchi, Claudia Regina Gonçalves, Erica Valessa Ramos Gomes, Karoline Rodrigues Campos |
| EPI_ISL_1095913 | IAL Regional de Bauru | Instituto Adolfo Lutz, Interdiciplinary Procedures Center, Strategic Laboratory | Claudio Tavares Sacchi, Claudia Regina Gonçalves, Erica Valessa Ramos Gomes, Karoline Rodrigues Campos |
| EPI_ISL_1096120 | IAL Regional de Bauru | Instituto Adolfo Lutz, Interdiciplinary Procedures Center, Strategic Laboratory | Claudio Tavares Sacchi, Claudia Regina Gonçalves, Erica Valessa Ramos Gomes, Karoline Rodrigues Campos, Caio Vinicius Dias Lopes |
| EPI_ISL_1096121 | Diagnosticos da America - DASA | Instituto Adolfo Lutz, Interdiciplinary Procedures Center, Strategic Laboratory | Claudio Tavares Sacchi, Claudia Regina Gonçalves, Erica Valessa Ramos Gomes, Karoline Rodrigues Campos, Caio Vinicius Dias Lopes |

| | | | |
|---|---|---|---|
| EPI_ISL_1096122, EPI_ISL_1096123, EPI_ISL_1096124, EPI_ISL_1096125, EPI_ISL_1096126, EPI_ISL_1096127, EPI_ISL_1096128, EPI_ISL_1096129, EPI_ISL_1096130, EPI_ISL_1096131, EPI_ISL_1096132, EPI_ISL_1096133, EPI_ISL_1096134 | | | |
| see above | IAL Regional de Bauru | Instituto Adolfo Lutz, Interdiciplinary Procedures Center, Strategic Laboratory | Claudio Tavares Sacchi, Claudia Regina Gonçalves, Erica Valessa Ramos Gomes, Karoline Rodrigues Campos, Caio Vinicius Dias Lopes |
| EPI_ISL_1096135 | Diagnosticos da America - DASA | Instituto Adolfo Lutz, Interdiciplinary Procedures Center, Strategic Laboratory | Claudio Tavares Sacchi, Claudia Regina Gonçalves, Erica Valessa Ramos Gomes, Karoline Rodrigues Campos, Caio Vinicius Dias Lopes |
| EPI_ISL_1096136 | IAL Regional de Bauru | Instituto Adolfo Lutz, Interdiciplinary Procedures Center, Strategic Laboratory | Claudio Tavares Sacchi, Claudia Regina Gonçalves, Erica Valessa Ramos Gomes, Karoline Rodrigues Campos, Caio Vinicius Dias Lopes |
| EPI_ISL_1109632 | Instituto de Biotecnologia - UNESP-Botucatu-SP | Instituto de Biotecnologia - UNESP-Botucatu-SP | Leila Sabrina Ullmann; Fábio Sossai Possebon; Cecília Artico Banho; Cíntia Bittar; Guilherme Campos; Helena Lage Ferreira; Jorge A. Petroli Marchesi; Lívia Sacchetto; Maisa C. Pereira Parra; Marília Moraes; Maurício L. Nogueira; Paula Rahal; Paulo Inacio da Costa; João Pessoa Araújo Jr. |
| EPI_ISL_1114151 | Laboratorio de Ecologia de Doencas Transmissiveis na Amazonia, Instituto Leonidas e Maria Deane - Fiocruz Amazonia | Laboratorio de Ecologia de Doencas Transmissiveis na Amazonia, Instituto Leonidas e Maria Deane - Fiocruz Amazonia | Valdinete Nascimento, Victor Souza, André Corado, Fernanda Nascimento, George Silva, Ágatha Costa, Debora Duarte, Karina Pessoa, Matilde Mejía, Luciana Gonçalves, Maria Júlia Brandão, Michele Jesus, Felipe Naveca on behalf of the Fiocruz COVID-19 Genomic Surveillance Network |
| EPI_ISL_1117379, EPI_ISL_1117380, EPI_ISL_1117381, EPI_ISL_1117382, EPI_ISL_1117383, EPI_ISL_1117384, EPI_ISL_1117385, EPI_ISL_1117386, EPI_ISL_1117387, EPI_ISL_1117388, EPI_ISL_1117389, EPI_ISL_1117390, EPI_ISL_1117391, EPI_ISL_1117392, EPI_ISL_1117393, EPI_ISL_1117394, EPI_ISL_1117395, EPI_ISL_1117396, EPI_ISL_1117397, EPI_ISL_1117398, EPI_ISL_1117399, EPI_ISL_1117400, EPI_ISL_1117401, EPI_ISL_1117402, EPI_ISL_1117403, EPI_ISL_1117404, EPI_ISL_1117405, EPI_ISL_1117406, EPI_ISL_1117407, EPI_ISL_1117408, EPI_ISL_1117409, EPI_ISL_1117410, EPI_ISL_1117411, EPI_ISL_1117412, EPI_ISL_1117413, EPI_ISL_1117414, EPI_ISL_1117415, EPI_ISL_1117416, EPI_ISL_1117417, EPI_ISL_1117418, EPI_ISL_1117419, EPI_ISL_1117420, EPI_ISL_1117421, EPI_ISL_1117422, EPI_ISL_1117423, EPI_ISL_1117424, EPI_ISL_1117425, EPI_ISL_1117426, EPI_ISL_1117427, EPI_ISL_1117428, EPI_ISL_1117429, EPI_ISL_1117430, EPI_ISL_1117431, EPI_ISL_1117432, EPI_ISL_1117433, EPI_ISL_1117434, EPI_ISL_1117435, EPI_ISL_1117436, EPI_ISL_1117437, EPI_ISL_1117438, EPI_ISL_1117439, EPI_ISL_1117440, EPI_ISL_1117441, EPI_ISL_1117442, EPI_ISL_1117443, EPI_ISL_1117444, EPI_ISL_1117445, EPI_ISL_1117446, EPI_ISL_1117447, EPI_ISL_1117448, EPI_ISL_1117449 | | | |
| see above | Nucleo de Pesquisa em Inovacao Terapeutica - UFPE | LABBE, Federal University of Pernambuco | Wilson Jose da Silva Junior, Marcos da Silveira Regueira Neto, Heidi Lacerda Alves da Cruz, Bruno Sampaio, Reginaldo Goncalves de Lima Neto, Maira Galdino da Rocha Pitta, Michelly Cristiny Pereira, Marco Katzenberger, Valdir de Queiroz Balbino |
| EPI_ISL_1121305 | Policlinica Maria Dirce | Instituto Adolfo Lutz, Interdiciplinary Procedures Center, Strategic Laboratory | Claudio Tavares Sacchi, Claudia Regina Gonçalves, Erica Valessa Ramos Gomes, Karoline Rodrigues Campos, Caio Vinicius Dias Lopes |
| EPI_ISL_1121306 | IAL Regional de Bauru | Instituto Adolfo Lutz, Interdiciplinary Procedures Center, Strategic Laboratory | Claudio Tavares Sacchi, Claudia Regina Gonçalves, Erica Valessa Ramos Gomes, Karoline Rodrigues Campos, Caio Vinicius Dias Lopes |
| EPI_ISL_1121307 | Hospital E Antonio Policarpo de Oliveira | Instituto Adolfo Lutz, Interdiciplinary Procedures Center, Strategic Laboratory | Claudio Tavares Sacchi, Claudia Regina Gonçalves, Erica Valessa Ramos Gomes, Karoline Rodrigues Campos, Caio Vinicius Dias Lopes |
| EPI_ISL_1121308 | Hospital e Pronto Socorro Portinari | Instituto Adolfo Lutz, Interdiciplinary Procedures Center, Strategic Laboratory | Claudio Tavares Sacchi, Claudia Regina Gonçalves, Erica Valessa Ramos Gomes, Karoline Rodrigues Campos, Caio Vinicius Dias Lopes |
| EPI_ISL_1121309 | UBS Jose Francisco Rezende | Instituto Adolfo Lutz, Interdiciplinary Procedures Center, Strategic Laboratory | Claudio Tavares Sacchi, Claudia Regina Gonçalves, Erica Valessa Ramos Gomes, Karoline Rodrigues Campos, Caio Vinicius Dias Lopes |
| EPI_ISL_1121310 | IAL Regional de Bauru | Instituto Adolfo Lutz, Interdiciplinary Procedures Center, Strategic Laboratory | Claudio Tavares Sacchi, Claudia Regina Gonçalves, Erica Valessa Ramos Gomes, Karoline Rodrigues Campos, Caio Vinicius Dias Lopes |
| EPI_ISL_1121311 | Centro de Saude II Dr. Jose Paione Mococa | Instituto Adolfo Lutz, Interdiciplinary Procedures Center, Strategic Laboratory | Claudio Tavares Sacchi, Claudia Regina Gonçalves, Erica Valessa Ramos Gomes, Karoline Rodrigues Campos, Caio Vinicius Dias Lopes |
| EPI_ISL_1121312, EPI_ISL_1121313, EPI_ISL_1121314, EPI_ISL_1121315 | IAL Regional de Bauru | Instituto Adolfo Lutz, Interdiciplinary Procedures Center, Strategic Laboratory | Claudio Tavares Sacchi, Claudia Regina Gonçalves, Erica Valessa Ramos Gomes, Karoline Rodrigues Campos, Caio Vinicius Dias Lopes |
| EPI_ISL_1121316 | LACEN do Rio Grande do Sul | Instituto Adolfo Lutz, Interdiciplinary Procedures Center, Strategic Laboratory | Claudio Tavares Sacchi, Claudia Regina Gonçalves, Erica Valessa Ramos Gomes, Karoline Rodrigues Campos, Caio Vinicius Dias Lopes |
| EPI_ISL_1121317 | IAL Regional de Bauru | Instituto Adolfo Lutz, Interdiciplinary Procedures Center, Strategic Laboratory | Claudio Tavares Sacchi, Claudia Regina Gonçalves, Erica Valessa Ramos Gomes, Karoline Rodrigues Campos, Caio Vinicius Dias Lopes |
| EPI_ISL_1121318, EPI_ISL_1121319 | Hospital de Campanha COVID 19 Caieiras | Instituto Adolfo Lutz, Interdiciplinary Procedures Center, Strategic Laboratory | Claudio Tavares Sacchi, Claudia Regina Gonçalves, Erica Valessa Ramos Gomes, Karoline Rodrigues Campos, Caio Vinicius Dias Lopes |
| EPI_ISL_1121320, EPI_ISL_1121321 | IAL Regional de Bauru | Instituto Adolfo Lutz, Interdiciplinary Procedures Center, Strategic Laboratory | Claudio Tavares Sacchi, Claudia Regina Gonçalves, Erica Valessa Ramos Gomes, Karoline Rodrigues Campos, Caio Vinicius Dias Lopes |
| EPI_ISL_1121322 | Santa Casa de Santa Isabel | Instituto Adolfo Lutz, Interdiciplinary Procedures Center, Strategic Laboratory | Claudio Tavares Sacchi, Claudia Regina Gonçalves, Erica Valessa Ramos Gomes, Karoline Rodrigues Campos, Caio Vinicius Dias Lopes |
| EPI_ISL_1121323, EPI_ISL_1121324 | Complexo Hospitalar Padre Bentode Guarulhos | Instituto Adolfo Lutz, Interdiciplinary Procedures Center, Strategic Laboratory | Claudio Tavares Sacchi, Claudia Regina Gonçalves, Erica Valessa Ramos Gomes, Karoline Rodrigues Campos, Caio Vinicius Dias Lopes |
| EPI_ISL_1121325 | IAL Regional de Santos | Instituto Adolfo Lutz, Interdiciplinary Procedures Center, Strategic Laboratory | Claudio Tavares Sacchi, Claudia Regina Gonçalves, Erica Valessa Ramos Gomes, Karoline Rodrigues Campos, Caio Vinicius Dias Lopes |
| EPI_ISL_1121326 | IAL Regional de Bauru | Instituto Adolfo Lutz, Interdiciplinary Procedures Center, Strategic Laboratory | Claudio Tavares Sacchi, Claudia Regina Gonçalves, Erica Valessa Ramos Gomes, Karoline Rodrigues Campos, Caio Vinicius Dias Lopes |
| EPI_ISL_1121327, EPI_ISL_1121328, EPI_ISL_1121329, EPI_ISL_1121330 | LACEN do Mato Grosso do Sul | Instituto Adolfo Lutz, Interdiciplinary Procedures Center, Strategic Laboratory | Claudio Tavares Sacchi, Claudia Regina Gonçalves, Erica Valessa Ramos Gomes, Karoline Rodrigues Campos, Caio Vinicius Dias Lopes |
| EPI_ISL_1123372 | UPA I Santa Isabel | Instituto Adolfo Lutz, Interdiciplinary Procedures Center, Strategic Laboratory | Claudio Tavares Sacchi, Claudia Regina Gonçalves, Erica Valessa Ramos Gomes, Karoline Rodrigues Campos, Caio Vinicius Dias Lopes |
| EPI_ISL_1123373 | Grupo Tecnico de Vigilancia Sanitaria e Epidemiologica | Instituto Adolfo Lutz, Interdiciplinary Procedures Center, Strategic Laboratory | Claudio Tavares Sacchi, Claudia Regina Gonçalves, Erica Valessa Ramos Gomes, Karoline Rodrigues Campos, Caio Vinicius Dias Lopes |
| EPI_ISL_1123374, EPI_ISL_1123375 | IAL Regional de Santos | Instituto Adolfo Lutz, Interdiciplinary Procedures Center, Strategic Laboratory | Claudio Tavares Sacchi, Claudia Regina Gonçalves, Erica Valessa Ramos Gomes, Karoline Rodrigues Campos, Caio Vinicius Dias Lopes |
| EPI_ISL_1133120, EPI_ISL_1133121, EPI_ISL_1133122, EPI_ISL_1133123, EPI_ISL_1133124, EPI_ISL_1133125, EPI_ISL_1133126, EPI_ISL_1133127, EPI_ISL_1133128, EPI_ISL_1133129, EPI_ISL_1133130, EPI_ISL_1133131, EPI_ISL_1133132, EPI_ISL_1133133, EPI_ISL_1133134, EPI_ISL_1133135, EPI_ISL_1133136, EPI_ISL_1133137, EPI_ISL_1133138, EPI_ISL_1133139, EPI_ISL_1133140, EPI_ISL_1133141, EPI_ISL_1133142, EPI_ISL_1133143, EPI_ISL_1133144 | | | |
| see above | LABCOVID_HCPA | LABRESIS_HCPA | Martins AF, Wink PL, Volpato F, Rosset C, de Paris F, Monteiro F, Barth AL |
| EPI_ISL_1133255, EPI_ISL_1133256, EPI_ISL_1133257, EPI_ISL_1133258, EPI_ISL_1133259, EPI_ISL_1133260, EPI_ISL_1133261, EPI_ISL_1133262, EPI_ISL_1133263, EPI_ISL_1133264, EPI_ISL_1133265, EPI_ISL_1133266, EPI_ISL_1133267, EPI_ISL_1133268, EPI_ISL_1133269, EPI_ISL_1133270, EPI_ISL_1133271, EPI_ISL_1133272, EPI_ISL_1133273, EPI_ISL_1133274, EPI_ISL_1133275, EPI_ISL_1133276, EPI_ISL_1133277, EPI_ISL_1133278, EPI_ISL_1133279 | | | |
| see above | Laboratório Hermes Pardini | Laboratório de Biologia Integrativa, Instituto de Ciências Biológicas, Universidade Federal de Minas Gerais | Filipe Romero Rebello Moreira, Diego Menezes Bonfim, Danielle Alves Gomes Zauli, Joice do Prado Silva, Aline Brito de Lima, Frederico Scott Varella Malta, Alessandro Clayton de Souza Ferreira, Victor Cavalcanti Pardini, Daniel Costa Queiroz, Rafael Marques de Souza, Victor Emmanuel Viana Geddes, Walyson Coelho Costa, Wagner Carlos Santos Magalhaes, Rennan Garcias Moreira, Carolina Moreira Voloch, Renan Pedra de Souza, Renato Santana Aguiar. |
| EPI_ISL_1139052, EPI_ISL_1139053, EPI_ISL_1139054, EPI_ISL_1139055, | LACEN do Mato Grosso do Sul | Instituto Adolfo Lutz, Interdiciplinary Procedures Center, Strategic Laboratory | Claudio Tavares Sacchi, Claudia Regina Gonçalves, Erica Valessa Ramos Gomes, Karoline Rodrigues Campos, Caio Vinicius Dias Lopes |

| | | | |
|---|---|---|---|
| EPI_ISL_1139056, EPI_ISL_1139057, EPI_ISL_1139059, EPI_ISL_1139063, EPI_ISL_1139064 | | | |
| EPI_ISL_1139069 | SMS Aruja | Instituto Adolfo Lutz, Interdiciplinary Procedures Center, Strategic Laboratory | Claudio Tavares Sacchi, Claudia Regina Gonçalves, Erica Valessa Ramos Gomes, Karoline Rodrigues Campos, Caio Vinicius Dias Lopes |
| EPI_ISL_1139070 | IAL Regional de Ribeirao Preto | Instituto Adolfo Lutz, Interdiciplinary Procedures Center, Strategic Laboratory | Claudio Tavares Sacchi, Claudia Regina Gonçalves, Erica Valessa Ramos Gomes, Karoline Rodrigues Campos, Caio Vinicius Dias Lopes |
| EPI_ISL_1139071, EPI_ISL_1139072, EPI_ISL_1139073, EPI_ISL_1139074 | Instituto Adolfo Lutz Central | Instituto Adolfo Lutz, Interdiciplinary Procedures Center, Strategic Laboratory | Claudio Tavares Sacchi, Claudia Regina Gonçalves, Erica Valessa Ramos Gomes, Karoline Rodrigues Campos, Caio Vinicius Dias Lopes |
| EPI_ISL_1139075, EPI_ISL_1139077 | Lab Loc - Itapecerica da Serra | Instituto Adolfo Lutz, Interdiciplinary Procedures Center, Strategic Laboratory | Claudio Tavares Sacchi, Claudia Regina Gonçalves, Erica Valessa Ramos Gomes, Karoline Rodrigues Campos, Caio Vinicius Dias Lopes |
| EPI_ISL_1161401, EPI_ISL_1161402, EPI_ISL_1161403, EPI_ISL_1161404, EPI_ISL_1161405, EPI_ISL_1161406, EPI_ISL_1161407, EPI_ISL_1161408, EPI_ISL_1161409, EPI_ISL_1161410, EPI_ISL_1161411, EPI_ISL_1161412, EPI_ISL_1161413, EPI_ISL_1161414, EPI_ISL_1161415, EPI_ISL_1163530, EPI_ISL_1163532, EPI_ISL_1163698, EPI_ISL_1163699, EPI_ISL_1163700, EPI_ISL_1163701, EPI_ISL_1163702, EPI_ISL_1163703, EPI_ISL_1163704, EPI_ISL_1163705, EPI_ISL_1163706, EPI_ISL_1163707, EPI_ISL_1163708, EPI_ISL_1163709, EPI_ISL_1163710, EPI_ISL_1163711, EPI_ISL_1163712, EPI_ISL_1163713, EPI_ISL_1163714, EPI_ISL_1163715, EPI_ISL_1163716 | | | |
| see above | LABCOVID_HCPA | LABRESIS_HCPA | Martins AF, Wink PL, Volpato F, Rosset C, de Paris F, Monteiro F, Zavascki AP, Barth AL |
| EPI_ISL_1163736 | LABCOVID_HCPA | LABCOVID_HCPA | Martins AF, Wink PL, Volpato F, Rosset C, de Paris F, Monteiro F, Zavascki AP, Barth AL |
| EPI_ISL_1163737 | LABCOVID_HCPA | LABRESIS_HCPA | Martins AF, Wink PL, Volpato F, Rosset C, de Paris F, Monteiro F, Zavascki AP, Barth AL |
| EPI_ISL_1164970, EPI_ISL_1164971 | LACEN - Laboratório Central de Saúde Pública do Ceará | Evandro Chagas Institute | Santos, M.C.; Silva, A.M.; Junior, W.D.C.; Barbagelata, L.S.; Ferreira, J.A.; Sousa, E.M.A.; da Silva, P.S.; Pinheiro, K.C.; L.C.; Sousa Junior, E.C. |
| EPI_ISL_1164972 | LACEN - Laboratório Central de Saúde Pública do Pará | Evandro Chagas Institute | Santos, M.C.; Silva, A.M.; Junior, W.D.C.; Barbagelata, L.S.; Ferreira, J.A.; Sousa, E.M.A.; da Silva, P.S.; Pinheiro, K.C.; L.C.; Sousa Junior, E.C. |
| EPI_ISL_1164973 | LACEN - Laboratório Central de Saúde Pública do Ceará | Evandro Chagas Institute | Santos, M.C.; Silva, A.M.; Junior, W.D.C.; Barbagelata, L.S.; Ferreira, J.A.; Sousa, E.M.A.; da Silva, P.S.; Pinheiro, K.C.; L.C.; Sousa Junior, E.C. |
| EPI_ISL_1164974, EPI_ISL_1164975 | LACEN - Laboratório Central de Saúde Pública do Pará | Evandro Chagas Institute | Santos, M.C.; Silva, A.M.; Junior, W.D.C.; Barbagelata, L.S.; Ferreira, J.A.; Sousa, E.M.A.; da Silva, P.S.; Pinheiro, K.C.; L.C.; Sousa Junior, E.C. |
| EPI_ISL_1164976 | LACEN - Laboratório Central de Saúde Pública do Amapá | Evandro Chagas Institute | Santos, M.C.; Silva, A.M.; Junior, W.D.C.; Barbagelata, L.S.; Ferreira, J.A.; Sousa, E.M.A.; da Silva, P.S.; Pinheiro, K.C.; L.C.; Sousa Junior, E.C. |
| EPI_ISL_1164977 | LACEN - Laboratório Central de Saúde Pública do Rio Grande do Norte | Evandro Chagas Institute | Santos, M.C.; Silva, A.M.; Junior, W.D.C.; Barbagelata, L.S.; Ferreira, J.A.; Sousa, E.M.A.; da Silva, P.S.; Pinheiro, K.C.; L.C.; Sousa Junior, E.C. |
| EPI_ISL_1164978 | LACEN - Laboratório Central de Saúde Pública do Pará | Evandro Chagas Institute | Santos, M.C.; Silva, A.M.; Junior, W.D.C.; Barbagelata, L.S.; Ferreira, J.A.; Sousa, E.M.A.; da Silva, P.S.; Pinheiro, K.C.; L.C.; Sousa Junior, E.C. |
| EPI_ISL_1164979 | LACEN - Laboratório Central de Saúde Pública do Maranhao | Evandro Chagas Institute | Santos, M.C.; Silva, A.M.; Junior, W.D.C.; Barbagelata, L.S.; Ferreira, J.A.; Sousa, E.M.A.; da Silva, P.S.; Pinheiro, K.C.; L.C.; Sousa Junior, E.C. |
| EPI_ISL_1164980 | LACEN - Laboratório Central de Saúde Pública do Ceará | Evandro Chagas Institute | Santos, M.C.; Silva, A.M.; Junior, W.D.C.; Barbagelata, L.S.; Ferreira, J.A.; Sousa, E.M.A.; da Silva, P.S.; Pinheiro, K.C.; L.C.; Sousa Junior, E.C. |
| EPI_ISL_1164981, EPI_ISL_1164982 | LACEN - Laboratório Central de Saúde Pública do Amapá | Evandro Chagas Institute | Santos, M.C.; Silva, A.M.; Junior, W.D.C.; Barbagelata, L.S.; Ferreira, J.A.; Sousa, E.M.A.; da Silva, P.S.; Pinheiro, K.C.; L.C.; Sousa Junior, E.C. |
| EPI_ISL_1164983 | LACEN - Laboratório Central de Saúde Pública do Pará | Evandro Chagas Institute | Santos, M.C.; Silva, A.M.; Junior, W.D.C.; Barbagelata, L.S.; Ferreira, J.A.; Sousa, E.M.A.; da Silva, P.S.; Pinheiro, K.C.; L.C.; Sousa Junior, E.C. |
| EPI_ISL_1164984, EPI_ISL_1164985 | LACEN - Laboratório Central de Saúde Pública do Amapá | Evandro Chagas Institute | Santos, M.C.; Silva, A.M.; Junior, W.D.C.; Barbagelata, L.S.; Ferreira, J.A.; Sousa, E.M.A.; da Silva, P.S.; Pinheiro, K.C.; L.C.; Sousa Junior, E.C. |
| EPI_ISL_1164986 | LACEN - Laboratório Central de Saúde Pública do Ceará | Evandro Chagas Institute | Santos, M.C.; Silva, A.M.; Junior, W.D.C.; Barbagelata, L.S.; Ferreira, J.A.; Sousa, E.M.A.; da Silva, P.S.; Pinheiro, K.C.; L.C.; Sousa Junior, E.C. |
| EPI_ISL_1164987, EPI_ISL_1164988 | LACEN - Laboratório Central de Saúde Pública do Rio Grande do Norte | Evandro Chagas Institute | Santos, M.C.; Silva, A.M.; Junior, W.D.C.; Barbagelata, L.S.; Ferreira, J.A.; Sousa, E.M.A.; da Silva, P.S.; Pinheiro, K.C.; L.C.; Sousa Junior, E.C. |
| EPI_ISL_1164989, EPI_ISL_1164990, EPI_ISL_1164991, EPI_ISL_1164992 | LACEN - Laboratório Central de Saúde Pública do Paraiba | Evandro Chagas Institute | Santos, M.C.; Silva, A.M.; Junior, W.D.C.; Barbagelata, L.S.; Ferreira, J.A.; Sousa, E.M.A.; da Silva, P.S.; Pinheiro, K.C.; L.C.; Sousa Junior, E.C. |
| EPI_ISL_1164993 | LACEN - Laboratório Central de Saúde Pública do Ceará | Evandro Chagas Institute | Santos, M.C.; Silva, A.M.; Junior, W.D.C.; Barbagelata, L.S.; Ferreira, J.A.; Sousa, E.M.A.; da Silva, P.S.; Pinheiro, K.C.; L.C.; Sousa Junior, E.C. |
| EPI_ISL_1164994 | LACEN - Laboratório Central de Saúde Pública do Paraiba | Evandro Chagas Institute | Santos, M.C.; Silva, A.M.; Junior, W.D.C.; Barbagelata, L.S.; Ferreira, J.A.; Sousa, E.M.A.; da Silva, P.S.; Pinheiro, K.C.; L.C.; Sousa Junior, E.C. |
| EPI_ISL_1164995 | LACEN - Laboratório Central de Saúde Pública do Ceará | Evandro Chagas Institute | Santos, M.C.; Silva, A.M.; Junior, W.D.C.; Barbagelata, L.S.; Ferreira, J.A.; Sousa, E.M.A.; da Silva, P.S.; Pinheiro, K.C.; L.C.; Sousa Junior, E.C. |
| EPI_ISL_1164996 | LACEN - Laboratório Central de Saúde Pública do Maranhao | Evandro Chagas Institute | Santos, M.C.; Silva, A.M.; Junior, W.D.C.; Barbagelata, L.S.; Ferreira, J.A.; Sousa, E.M.A.; da Silva, P.S.; Pinheiro, K.C.; L.C.; Sousa Junior, E.C. |
| EPI_ISL_1164997 | LACEN - Laboratório Central de Saúde Pública do Rio Grande do Norte | Evandro Chagas Institute | Santos, M.C.; Silva, A.M.; Junior, W.D.C.; Barbagelata, L.S.; Ferreira, J.A.; Sousa, E.M.A.; da Silva, P.S.; Pinheiro, K.C.; L.C.; Sousa Junior, E.C. |
| EPI_ISL_1164998 | LACEN - Laboratório Central de Saúde Pública do Maranhao | Evandro Chagas Institute | Santos, M.C.; Silva, A.M.; Junior, W.D.C.; Barbagelata, L.S.; Ferreira, J.A.; Sousa, E.M.A.; da Silva, P.S.; Pinheiro, K.C.; L.C.; Sousa Junior, E.C. |
| EPI_ISL_1164999 | LACEN - Laboratório Central de Saúde Pública do Paraiba | Evandro Chagas Institute | Santos, M.C.; Silva, A.M.; Junior, W.D.C.; Barbagelata, L.S.; Ferreira, J.A.; Sousa, E.M.A.; da Silva, P.S.; Pinheiro, K.C.; L.C.; Sousa Junior, E.C. |
| EPI_ISL_1166615 | LACEN - Laboratório Central de Saúde Pública do Rio Grande do Norte | Evandro Chagas Institute Virology | Santos, M.C.; Silva, A.M.; Junior, W.D.C.; Barbagelata, L.S.; Ferreira, J.A.; Sousa, E.M.A.; da Silva, P.S.; Pinheiro, K.C.; L.C.; Sousa Junior, E.C. |
| EPI_ISL_1171619 | IAL Regional de Presidente Prudente | Instituto Adolfo Lutz, Interdiciplinary Procedures Center, Strategic Laboratory | Claudio Tavares Sacchi, Claudia Regina Gonçalves, Erica Valessa Ramos Gomes, Karoline Rodrigues Campos |
| EPI_ISL_1171620 | Instituto Adolfo Lutz Central | Instituto Adolfo Lutz, Interdiciplinary Procedures Center, Strategic Laboratory | Claudio Tavares Sacchi, Claudia Regina Gonçalves, Erica Valessa Ramos Gomes, Karoline Rodrigues Campos, Caio Vinicius Dias Lopes |
| EPI_ISL_1171621 | LACEN do Mato Grosso do Sul | Instituto Adolfo Lutz, Interdiciplinary Procedures Center, Strategic Laboratory | Claudio Tavares Sacchi, Claudia Regina Gonçalves, Erica Valessa Ramos Gomes, Karoline Rodrigues Campos, Caio Vinicius Dias Lopes |
| EPI_ISL_1171622, EPI_ISL_1171623, EPI_ISL_1171624, EPI_ISL_1171625, EPI_ISL_1171626, EPI_ISL_1171627, EPI_ISL_1171628, EPI_ISL_1171629, EPI_ISL_1171630, EPI_ISL_1171631, EPI_ISL_1171632, EPI_ISL_1171633, EPI_ISL_1171634, EPI_ISL_1171635, EPI_ISL_1171636, EPI_ISL_1171637, EPI_ISL_1171638, EPI_ISL_1171639, EPI_ISL_1171640 | | | |
| see above | IAL Regional de Santos | Instituto Adolfo Lutz, Interdiciplinary Procedures Center, Strategic Laboratory | Claudio Tavares Sacchi, Claudia Regina Gonçalves, Erica Valessa Ramos Gomes, Karoline Rodrigues Campos, Caio Vinicius Dias Lopes |
| EPI_ISL_1171641, EPI_ISL_1171642, EPI_ISL_1171643, EPI_ISL_1171644, EPI_ISL_1171645, EPI_ISL_1171646, EPI_ISL_1171647 | IAL Regional de Marilia | Instituto Adolfo Lutz, Interdiciplinary Procedures Center, Strategic Laboratory | Claudio Tavares Sacchi, Claudia Regina Gonçalves, Erica Valessa Ramos Gomes, Karoline Rodrigues Campos, Caio Vinicius Dias Lopes |
| EPI_ISL_1171648, EPI_ISL_1171649, EPI_ISL_1171650 | Centro de Saude Dr. Jose Paione em Mococa | Instituto Adolfo Lutz, Interdiciplinary Procedures Center, Strategic Laboratory | Claudio Tavares Sacchi, Claudia Regina Gonçalves, Erica Valessa Ramos Gomes, Karoline Rodrigues Campos, Caio Vinicius Dias Lopes |
| EPI_ISL_1171651, EPI_ISL_1171652, EPI_ISL_1171653, EPI_ISL_1171654, EPI_ISL_1171655, EPI_ISL_1171656, EPI_ISL_1171657, EPI_ISL_1171658, EPI_ISL_1171659, EPI_ISL_1171660, EPI_ISL_1171661, EPI_ISL_1171662, EPI_ISL_1171663, EPI_ISL_1171664, EPI_ISL_1171665, EPI_ISL_1171666, EPI_ISL_1171667, EPI_ISL_1171668, EPI_ISL_1171669, EPI_ISL_1171670, EPI_ISL_1171671, EPI_ISL_1171672, EPI_ISL_1171673, EPI_ISL_1171674 | | | |
| see above | IAL Regional de Presidente Prudente | Instituto Adolfo Lutz, Interdiciplinary Procedures Center, Strategic Laboratory | Claudio Tavares Sacchi, Claudia Regina Gonçalves, Erica Valessa Ramos Gomes, Karoline Rodrigues Campos, Caio Vinicius Dias Lopes |
| EPI_ISL_1171871 | IMT_USP | Laboratório de Parasitologia Médica - Instituto de Medicina Tropical - Universidade de São Paulo | Brazil-UK Centre for Arbovirus Discovery Diagnosis Genomics and Epidemiology (CADDE) Genomic Network - Instituto de Medicina Tropical |

| | | | |
|---|---|---|---|
| EPI_ISL_1172015, EPI_ISL_1172016, EPI_ISL_1172017 | HC_FMUSP | Laboratório de Parasitologia Médica - Instituto de Medicina Tropical - Universidade de São Paulo | Brazil-UK Centre for Arbovirus Discovery Diagnosis Genomics and Epidemiology (CADDE) Genomic Network - Instituto de Medicina Tropical |
| EPI_ISL_1181353 | Laboratorio Central de Saude Publica do Estado do Rio de Janeiro (LACEN-RJ) | Laboratory of Respiratory Viruses and Measles, Oswaldo Cruz Institute, FIOCRUZ | Paola Resende, Luciana Appolinario, Fernando Motta, Anna Carolina Paixao, Ana Carolina Mendonca, Alice Sampaio Rocha, Renata Serrano Lopes, Andrea Cony Cavalcanti, Marilda Siqueira on behalf of the Fiocruz COVID-19 Genomic Surveillance Network |
| EPI_ISL_1181354, EPI_ISL_1181355 | Gonçalo Moniz Institute, FIOCRUZ, Bahia | Laboratory of Respiratory Viruses and Measles, Oswaldo Cruz Institute, FIOCRUZ | Paola Resende, Luciana Appolinario, Fernando Motta, Anna Carolina Paixao, Ana Carolina Mendonca, Alice Sampaio Rocha, Renata Serrano Lopes, Tiago Graf, Ricardo Khouri, Marilda Siqueira on behalf of the Fiocruz COVID-19 Genomic Surveillance Network |
| EPI_ISL_1181356 | Oswaldo Cruz Foundation, FIOCRUZ - Ceara (Fiocruz-CE) | Laboratory of Respiratory Viruses and Measles, Oswaldo Cruz Institute, FIOCRUZ | Paola Resende, Luciana Appolinario, Fernando Motta, Anna Carolina Paixao, Ana Carolina Mendonca, Alice Sampaio Rocha, Renata Serrano Lopes, Thais de Oliveira Costa, Joaquim César do Nascimento Sousa Júnior, Fabio Miyajima, Marilda Siqueira on behalf of the Fiocruz COVID-19 Genomic Surveillance Network |
| EPI_ISL_1181357, EPI_ISL_1181358, EPI_ISL_1181359, EPI_ISL_1181360, EPI_ISL_1181361 | Laboratory of Respiratory Viruses and Measles, Oswaldo Cruz Institute, FIOCRUZ | Laboratory of Respiratory Viruses and Measles, Oswaldo Cruz Institute, FIOCRUZ | Paola Resende, Luciana Appolinario, Fernando Motta, Anna Carolina Paixao, Ana Carolina Mendonca, Alice Sampaio Rocha, Renata Serrano Lopes, Marilda Siqueira on behalf of the Fiocruz COVID-19 Genomic Surveillance Network |
| EPI_ISL_1181362 | Instituto Aggeu Magalhães - Oswaldo Cruz Foundation, FIOCRUZ (FIOCRUZ-PE) | Laboratory of Respiratory Viruses and Measles, Oswaldo Cruz Institute, FIOCRUZ | Paola Resende, Luciana Appolinario, Fernando Motta, Anna Carolina Paixao, Ana Carolina Mendonca, Alice Sampaio Rocha, Renata Serrano Lopes, Gabriel Wallau, Marilda Siqueira on behalf of the Fiocruz COVID-19 Genomic Surveillance Network |
| EPI_ISL_1181363 | Laboratory of Respiratory Viruses and Measles, Oswaldo Cruz Institute, FIOCRUZ | Laboratory of Respiratory Viruses and Measles, Oswaldo Cruz Institute, FIOCRUZ | Paola Resende, Luciana Appolinario, Fernando Motta, Anna Carolina Paixao, Ana Carolina Mendonca, Alice Sampaio Rocha, Renata Serrano Lopes, Marilda Siqueira on behalf of the Fiocruz COVID-19 Genomic Surveillance Network |
| EPI_ISL_1181364 | Federal University of Southern Bahia (UFSB - Universidade Federal do Sul da Bahia) | Laboratory of Respiratory Viruses and Measles, Oswaldo Cruz Institute, FIOCRUZ | Paola Resende, Luciana Appolinario, Fernando Motta, Anna Carolina Paixao, Ana Carolina Mendonca, Alice Sampaio Rocha, Renata Serrano Lopes, Thiago Mafra, Marilda Siqueira on behalf of the Fiocruz COVID-19 Genomic Surveillance Network |
| EPI_ISL_1181365 | Laboratorio Central de Saude Publica do Estado do Alagoas (LACEN-AL) | Laboratory of Respiratory Viruses and Measles, Oswaldo Cruz Institute, FIOCRUZ | Paola Resende, Luciana Appolinario, Fernando Motta, Anna Carolina Paixao, Ana Carolina Mendonca, Alice Sampaio Rocha, Renata Serrano Lopes, Anderson Brandao Leite, Marilda Siqueira on behalf of the Fiocruz COVID-19 Genomic Surveillance Network |
| EPI_ISL_1181366, EPI_ISL_1181367 | Laboratory of Respiratory Viruses and Measles, Oswaldo Cruz Institute, FIOCRUZ | Laboratory of Respiratory Viruses and Measles, Oswaldo Cruz Institute, FIOCRUZ | Paola Resende, Luciana Appolinario, Fernando Motta, Anna Carolina Paixao, Ana Carolina Mendonca, Alice Sampaio Rocha, Renata Serrano Lopes, Marilda Siqueira on behalf of the Fiocruz COVID-19 Genomic Surveillance Network |
| EPI_ISL_1181368 | Laboratorio Central de Saude Publica do Estado do Rio Grande do Sul (LACEN-RS) | Laboratory of Respiratory Viruses and Measles, Oswaldo Cruz Institute, FIOCRUZ | Paola Resende, Luciana Appolinario, Fernando Motta, Anna Carolina Paixao, Ana Carolina Mendonca, Alice Sampaio Rocha, Renata Serrano Lopes, Tatiana Schaffer Gregianini, Marilda Siqueira on behalf of the Fiocruz COVID-19 Genomic Surveillance Network |
| EPI_ISL_1181369 | Laboratory of Respiratory Viruses and Measles, Oswaldo Cruz Institute, FIOCRUZ | Laboratory of Respiratory Viruses and Measles, Oswaldo Cruz Institute, FIOCRUZ | Paola Resende, Luciana Appolinario, Fernando Motta, Anna Carolina Paixao, Ana Carolina Mendonca, Alice Sampaio Rocha, Renata Serrano Lopes, Marilda Siqueira on behalf of the Fiocruz COVID-19 Genomic Surveillance Network |
| EPI_ISL_1181370, EPI_ISL_1181371 | Laboratorio Central de Saude Publica do Estado Maranhao (LACEN-MA) | Laboratory of Respiratory Viruses and Measles, Oswaldo Cruz Institute, FIOCRUZ | Paola Resende, Luciana Appolinario, Fernando Motta, Anna Carolina Paixao, Ana Carolina Mendonca, Alice Sampaio Rocha, Renata Serrano Lopes, Lidio Gonçalves Lima Neto, Marilda Siqueira on behalf of the Fiocruz COVID-19 Genomic Surveillance Network |
| EPI_ISL_1181372 | Laboratorio Central de Saude Publica do Estado do Rio de Janeiro (LACEN-RJ) | Laboratory of Respiratory Viruses and Measles, Oswaldo Cruz Institute, FIOCRUZ | Paola Resende, Luciana Appolinario, Fernando Motta, Anna Carolina Paixao, Ana Carolina Mendonca, Alice Sampaio Rocha, Renata Serrano Lopes, Andrea Cony Cavalcanti, Marilda Siqueira on behalf of the Fiocruz COVID-19 Genomic Surveillance Network |
| EPI_ISL_1181373, EPI_ISL_1181374, EPI_ISL_1181375, EPI_ISL_1181376, EPI_ISL_1181377 | Laboratory of Respiratory Viruses and Measles, Oswaldo Cruz Institute, FIOCRUZ | Laboratory of Respiratory Viruses and Measles, Oswaldo Cruz Institute, FIOCRUZ | Paola Resende, Luciana Appolinario, Fernando Motta, Anna Carolina Paixao, Ana Carolina Mendonca, Alice Sampaio Rocha, Renata Serrano Lopes, Marilda Siqueira on behalf of the Fiocruz COVID-19 Genomic Surveillance Network |
| EPI_ISL_1181378 | Laboratorio de Imunologia de Transplantes de Goias LTDA (HLAGYN) | Laboratory of Respiratory Viruses and Measles, Oswaldo Cruz Institute, FIOCRUZ | Paola Resende, Luciana Appolinario, Fernando Motta, Anna Carolina Paixao, Ana Carolina Mendonca, Alice Sampaio Rocha, Renata Serrano Lopes , Alessandro Leonardo Alvares Magalhaes, Fernando Antonio Vinhal dos Santos, Erika Lopes Rocha Batista, Marilda Siqueira on behalf of the Fiocruz COVID-19 Genomic Surveillance Network |
| EPI_ISL_1181379, EPI_ISL_1181380 | Gonçalo Moniz Institute, FIOCRUZ, Bahia | Laboratory of Respiratory Viruses and Measles, Oswaldo Cruz Institute, FIOCRUZ | Paola Resende, Luciana Appolinario, Fernando Motta, Anna Carolina Paixao, Ana Carolina Mendonca, Alice Sampaio Rocha, Renata Serrano Lopes, Tiago Graf, Ricardo Khouri, Marilda Siqueira on behalf of the Fiocruz COVID-19 Genomic Surveillance Network |
| EPI_ISL_1181381, EPI_ISL_1181382, EPI_ISL_1181383, EPI_ISL_1181384, EPI_ISL_1181385, EPI_ISL_1181386, EPI_ISL_1181387, EPI_ISL_1181388, EPI_ISL_1181389 | Laboratory of Respiratory Viruses and Measles, Oswaldo Cruz Institute, FIOCRUZ | Laboratory of Respiratory Viruses and Measles, Oswaldo Cruz Institute, FIOCRUZ | Paola Resende, Luciana Appolinario, Fernando Motta, Anna Carolina Paixao, Ana Carolina Mendonca, Alice Sampaio Rocha, Renata Serrano Lopes, Marilda Siqueira on behalf of the Fiocruz COVID-19 Genomic Surveillance Network |
| EPI_ISL_1181390 | Laboratorio Central de Saude Publica do Estado do Parana (LACEN-PR) | Laboratory of Respiratory Viruses and Measles, Oswaldo Cruz Institute, FIOCRUZ | Paola Resende, Luciana Appolinario, Fernando Motta, Anna Carolina Paixao, Ana Carolina Mendonca, Alice Sampaio Rocha, Renata Serrano Lopes, Maria do Carmo Debur, Irina Nastassja Riediger, Marilda Siqueira on behalf of the Fiocruz COVID-19 Genomic Surveillance Network |
| EPI_ISL_1181391 | Laboratory of Respiratory Viruses and Measles, Oswaldo Cruz Institute, FIOCRUZ | Laboratory of Respiratory Viruses and Measles, Oswaldo Cruz Institute, FIOCRUZ | Paola Resende, Luciana Appolinario, Fernando Motta, Anna Carolina Paixao, Ana Carolina Mendonca, Alice Sampaio Rocha, Renata Serrano Lopes, Marilda Siqueira on behalf of the Fiocruz COVID-19 Genomic Surveillance Network |
| EPI_ISL_1181392 | Laboratorio Central de Saude Publica do Estado do Parana (LACEN-PR) | Laboratory of Respiratory Viruses and Measles, Oswaldo Cruz Institute, FIOCRUZ | Paola Resende, Luciana Appolinario, Fernando Motta, Anna Carolina Paixao, Ana Carolina Mendonca, Alice Sampaio Rocha, Renata Serrano Lopes, Maria do Carmo Debur, Irina Nastassja Riediger, Marilda Siqueira on behalf of the Fiocruz COVID-19 Genomic Surveillance Network |
| EPI_ISL_1181393 | Laboratory of Respiratory Viruses and Measles, Oswaldo Cruz Institute, FIOCRUZ | Laboratory of Respiratory Viruses and Measles, Oswaldo Cruz Institute, FIOCRUZ | Paola Resende, Luciana Appolinario, Fernando Motta, Anna Carolina Paixao, Ana Carolina Mendonca, Alice Sampaio Rocha, Renata Serrano Lopes, Marilda Siqueira on behalf of the Fiocruz COVID-19 Genomic Surveillance Network |
| EPI_ISL_1181394 | Laboratorio Central de Saude Publica do Estado do Parana (LACEN-PR) | Laboratory of Respiratory Viruses and Measles, Oswaldo Cruz Institute, FIOCRUZ | Paola Resende, Luciana Appolinario, Fernando Motta, Anna Carolina Paixao, Ana Carolina Mendonca, Alice Sampaio Rocha, Renata Serrano Lopes, Maria do Carmo Debur, Irina Nastassja Riediger, Marilda Siqueira on behalf of the Fiocruz COVID-19 Genomic Surveillance Network |
| EPI_ISL_1181395 | Laboratorio Central de Saude Publica do Estado do Espitito Santo (LACEN-ES) | Laboratory of Respiratory Viruses and Measles, Oswaldo Cruz Institute, FIOCRUZ | Paola Resende, Luciana Appolinario, Fernando Motta, Anna Carolina Paixao, Ana Carolina Mendonca, Alice Sampaio Rocha, Renata Serrano Lopes, Rodrigo Ribeiro Rodrigues, Marilda Siqueira on behalf of the Fiocruz COVID-19 Genomic Surveillance Network |
| EPI_ISL_1181396 | Laboratorio Central de Saude Publica do Estado do Alagoas (LACEN-AL) | Laboratory of Respiratory Viruses and Measles, Oswaldo Cruz Institute, FIOCRUZ | Paola Resende, Luciana Appolinario, Fernando Motta, Anna Carolina Paixao, Ana Carolina Mendonca, Alice Sampaio Rocha, Renata Serrano Lopes, Anderson Brandao Leite, Marilda Siqueira on behalf of the Fiocruz COVID-19 Genomic Surveillance Network |
| EPI_ISL_1181397 | Laboratorio Central de Saude Publica do Estado do Espitito Santo (LACEN-ES) | Laboratory of Respiratory Viruses and Measles, Oswaldo Cruz Institute, FIOCRUZ | Paola Resende, Luciana Appolinario, Fernando Motta, Anna Carolina Paixao, Ana Carolina Mendonca, Alice Sampaio Rocha, Renata Serrano Lopes, Rodrigo Ribeiro Rodrigues, Marilda Siqueira on behalf of the Fiocruz COVID-19 Genomic Surveillance Network |
| EPI_ISL_1181398 | Laboratory of Respiratory Viruses and Measles, Oswaldo Cruz Institute, FIOCRUZ | Laboratory of Respiratory Viruses and Measles, Oswaldo Cruz Institute, FIOCRUZ | Paola Resende, Luciana Appolinario, Fernando Motta, Anna Carolina Paixao, Ana Carolina Mendonca, Alice Sampaio Rocha, Renata Serrano Lopes, Marilda Siqueira on behalf of the Fiocruz COVID-19 Genomic Surveillance Network |
| EPI_ISL_1181399 | Laboratorio Central de Saude Publica do Estado de Minas Gerais (LACEN-MG) | Laboratory of Respiratory Viruses and Measles, Oswaldo Cruz Institute, FIOCRUZ | Paola Resende, Luciana Appolinario, Fernando Motta, Anna Carolina Paixao, Ana Carolina Mendonca, Alice Sampaio Rocha, Renata Serrano Lopes, Felipe Iani, Marilda Siqueira on behalf of the Fiocruz COVID-19 Genomic Surveillance Network |
| EPI_ISL_1181400, EPI_ISL_1181401 | Laboratorio Central de Saude Publica do Estado Maranhao (LACEN-MA) | Laboratory of Respiratory Viruses and Measles, Oswaldo Cruz Institute, FIOCRUZ | Paola Resende, Luciana Appolinario, Fernando Motta, Anna Carolina Paixao, Ana Carolina Mendonca, Alice Sampaio Rocha, Renata Serrano Lopes, Lidio Gonçalves Lima Neto, Marilda Siqueira on behalf of the Fiocruz COVID-19 Genomic Surveillance Network |
| EPI_ISL_1181402 | Laboratory of Respiratory Viruses and Measles, Oswaldo Cruz Institute, FIOCRUZ | Laboratory of Respiratory Viruses and Measles, Oswaldo Cruz Institute, FIOCRUZ | Paola Resende, Luciana Appolinario, Fernando Motta, Anna Carolina Paixao, Ana Carolina Mendonca, Alice Sampaio Rocha, Renata Serrano Lopes, Marilda Siqueira on behalf of the Fiocruz COVID-19 Genomic Surveillance Network |
| EPI_ISL_1181403, EPI_ISL_1181404 | Laboratorio Central de Saude Publica do Estado de Santa Catarina (LACEN-SC) | Laboratory of Respiratory Viruses and Measles, Oswaldo Cruz Institute, FIOCRUZ | Paola Resende, Luciana Appolinario, Fernando Motta, Anna Carolina Paixao, Ana Carolina Mendonca, Alice Sampaio Rocha, Renata Serrano Lopes, Darcita Buerger Rovaris, Sandra Bianchini Fernandes, Marilda Siqueira on behalf of the Fiocruz COVID-19 Genomic Surveillance Network |
| EPI_ISL_1181405, EPI_ISL_1181406, EPI_ISL_1181407 | Laboratorio Central de Saude Publica do Estado de Sergipe (LACEN-SE) | Laboratory of Respiratory Viruses and Measles, Oswaldo Cruz Institute, FIOCRUZ | Paola Resende, Luciana Appolinario, Fernando Motta, Anna Carolina Paixao, Ana Carolina Mendonca, Alice Sampaio Rocha, Renata Serrano Lopes, Cliomar Alves dos Santos, Marilda Siqueira on behalf of the Fiocruz COVID-19 Genomic Surveillance Network |
| EPI_ISL_1181408 | Gonçalo Moniz Institute, FIOCRUZ, Bahia | Laboratory of Respiratory Viruses and Measles, Oswaldo Cruz | Paola Resende, Luciana Appolinario, Fernando Motta, Anna Carolina Paixao, Ana Carolina Mendonca, Alice Sampaio Rocha, Renata Serrano Lopes, |

| | | | |
|---|---|---|---|
| | | Institute, FIOCRUZ | Tiago Graf, Ricardo Khouri, Marilda Siqueira on behalf of the Fiocruz COVID-19 Genomic Surveillance Network |
| EPI_ISL_1181409 | Laboratorio Central de Saude Publica do Estado de Sergipe (LACEN-SE) | Laboratory of Respiratory Viruses and Measles, Oswaldo Cruz Institute, FIOCRUZ | Paola Resende, Luciana Appolinario, Fernando Motta, Anna Carolina Paixao, Ana Carolina Mendonca, Alice Sampaio Rocha, Renata Serrano Lopes, Cliomar Alves dos Santos, Marilda Siqueira on behalf of the Fiocruz COVID-19 Genomic Surveillance Network |
| EPI_ISL_1181410 | Laboratory of Respiratory Viruses and Measles, Oswaldo Cruz Institute, FIOCRUZ | Laboratory of Respiratory Viruses and Measles, Oswaldo Cruz Institute, FIOCRUZ | Paola Resende, Luciana Appolinario, Fernando Motta, Anna Carolina Paixao, Ana Carolina Mendonca, Alice Sampaio Rocha, Renata Serrano Lopes, Marilda Siqueira on behalf of the Fiocruz COVID-19 Genomic Surveillance Network |
| EPI_ISL_1181411 | Laboratorio Central de Saude Publica do Estado do Rio Grande do Sul (LACEN-RS) | Laboratory of Respiratory Viruses and Measles, Oswaldo Cruz Institute, FIOCRUZ | Paola Resende, Luciana Appolinario, Fernando Motta, Anna Carolina Paixao, Ana Carolina Mendonca, Alice Sampaio Rocha, Renata Serrano Lopes, Tatiana Schaffer Gregianini, Marilda Siqueira on behalf of the Fiocruz COVID-19 Genomic Surveillance Network |
| EPI_ISL_1181412 | Laboratorio Central de Saude Publica do Estado de Santa Catarina (LACEN-SC) | Laboratory of Respiratory Viruses and Measles, Oswaldo Cruz Institute, FIOCRUZ | Paola Resende, Luciana Appolinario, Fernando Motta, Anna Carolina Paixao, Ana Carolina Mendonca, Alice Sampaio Rocha, Renata Serrano Lopes, Darcita Buerger Rovaris, Sandra Bianchini Fernandes, Marilda Siqueira on behalf of the Fiocruz COVID-19 Genomic Surveillance Network |
| EPI_ISL_1181413, EPI_ISL_1181414 | Laboratorio Central de Saude Publica do Estado Maranhao (LACEN-MA) | Laboratory of Respiratory Viruses and Measles, Oswaldo Cruz Institute, FIOCRUZ | Paola Resende, Luciana Appolinario, Fernando Motta, Anna Carolina Paixao, Ana Carolina Mendonca, Alice Sampaio Rocha, Renata Serrano Lopes, Lidio Gonçalves Lima Neto, Marilda Siqueira on behalf of the Fiocruz COVID-19 Genomic Surveillance Network |
| EPI_ISL_1181415 | Laboratory of Respiratory Viruses and Measles, Oswaldo Cruz Institute, FIOCRUZ | Laboratory of Respiratory Viruses and Measles, Oswaldo Cruz Institute, FIOCRUZ | Paola Resende, Luciana Appolinario, Fernando Motta, Anna Carolina Paixao, Ana Carolina Mendonca, Alice Sampaio Rocha, Renata Serrano Lopes, Marilda Siqueira on behalf of the Fiocruz COVID-19 Genomic Surveillance Network |
| EPI_ISL_1181416 | Laboratorio Central de Saude Publica do Estado da Paraiba (LACEN-PB) | Laboratory of Respiratory Viruses and Measles, Oswaldo Cruz Institute, FIOCRUZ | Paola Resende, Luciana Appolinario, Fernando Motta, Anna Carolina Paixao, Ana Carolina Mendonca, Alice Sampaio Rocha, Renata Serrano Lopes, Joao Felipe Bezerra, Dalane Loudal Florentino Teixeira, Marilda Siqueira on behalf of the Fiocruz COVID-19 Genomic Surveillance Network |
| EPI_ISL_1181418 | Laboratorio Central de Saude Publica do Estado do Espitito Santo (LACEN-ES) | Laboratory of Respiratory Viruses and Measles, Oswaldo Cruz Institute, FIOCRUZ | Paola Resende, Luciana Appolinario, Fernando Motta, Anna Carolina Paixao, Ana Carolina Mendonca, Alice Sampaio Rocha, Renata Serrano Lopes, Rodrigo Ribeiro Rodrigues, Marilda Siqueira on behalf of the Fiocruz COVID-19 Genomic Surveillance Network |
| EPI_ISL_1181419 | Gonçalo Moniz Institute, FIOCRUZ, Bahia | Laboratory of Respiratory Viruses and Measles, Oswaldo Cruz Institute, FIOCRUZ | Paola Resende, Luciana Appolinario, Fernando Motta, Anna Carolina Paixao, Ana Carolina Mendonca, Alice Sampaio Rocha, Renata Serrano Lopes, Tiago Graf, Ricardo Khouri, Marilda Siqueira on behalf of the Fiocruz COVID-19 Genomic Surveillance Network |
| EPI_ISL_1181420, EPI_ISL_1181421 | Laboratorio Central de Saude Publica do Estado do Espitito Santo (LACEN-ES) | Laboratory of Respiratory Viruses and Measles, Oswaldo Cruz Institute, FIOCRUZ | Paola Resende, Luciana Appolinario, Fernando Motta, Anna Carolina Paixao, Ana Carolina Mendonca, Alice Sampaio Rocha, Renata Serrano Lopes, Rodrigo Ribeiro Rodrigues, Marilda Siqueira on behalf of the Fiocruz COVID-19 Genomic Surveillance Network |
| EPI_ISL_1181422 | Laboratorio Central de Saude Publica do Estado da Paraiba (LACEN-PB) | Laboratory of Respiratory Viruses and Measles, Oswaldo Cruz Institute, FIOCRUZ | Paola Resende, Luciana Appolinario, Fernando Motta, Anna Carolina Paixao, Ana Carolina Mendonca, Alice Sampaio Rocha, Renata Serrano Lopes, Joao Felipe Bezerra, Dalane Loudal Florentino Teixeira, Marilda Siqueira on behalf of the Fiocruz COVID-19 Genomic Surveillance Network |
| EPI_ISL_1181423 | Laboratorio Central de Saude Publica do Estado do Espitito Santo (LACEN-ES) | Laboratory of Respiratory Viruses and Measles, Oswaldo Cruz Institute, FIOCRUZ | Paola Resende, Luciana Appolinario, Fernando Motta, Anna Carolina Paixao, Ana Carolina Mendonca, Alice Sampaio Rocha, Renata Serrano Lopes, Rodrigo Ribeiro Rodrigues, Marilda Siqueira on behalf of the Fiocruz COVID-19 Genomic Surveillance Network |
| EPI_ISL_1181424 | Gonçalo Moniz Institute, FIOCRUZ, Bahia | Laboratory of Respiratory Viruses and Measles, Oswaldo Cruz Institute, FIOCRUZ | Paola Resende, Luciana Appolinario, Fernando Motta, Anna Carolina Paixao, Ana Carolina Mendonca, Alice Sampaio Rocha, Renata Serrano Lopes, Tiago Graf, Ricardo Khouri, Marilda Siqueira on behalf of the Fiocruz COVID-19 Genomic Surveillance Network |
| EPI_ISL_1181425 | Laboratorio Central de Saude Publica do Estado do Rio de Janeiro (LACEN-RJ) | Laboratory of Respiratory Viruses and Measles, Oswaldo Cruz Institute, FIOCRUZ | Paola Resende, Luciana Appolinario, Fernando Motta, Anna Carolina Paixao, Ana Carolina Mendonca, Alice Sampaio Rocha, Renata Serrano Lopes, Andrea Cony Cavalcanti, Marilda Siqueira on behalf of the Fiocruz COVID-19 Genomic Surveillance Network |
| EPI_ISL_1181426, EPI_ISL_1181427, EPI_ISL_1181428 | Laboratorio Central de Saude Publica do Estado de Minas Gerais (LACEN-MG) | Laboratory of Respiratory Viruses and Measles, Oswaldo Cruz Institute, FIOCRUZ | Paola Resende, Luciana Appolinario, Fernando Motta, Anna Carolina Paixao, Ana Carolina Mendonca, Alice Sampaio Rocha, Renata Serrano Lopes, Felipe Iani, Marilda Siqueira on behalf of the Fiocruz COVID-19 Genomic Surveillance Network |
| EPI_ISL_1181429 | Gonçalo Moniz Institute, FIOCRUZ, Bahia | Laboratory of Respiratory Viruses and Measles, Oswaldo Cruz Institute, FIOCRUZ | Paola Resende, Luciana Appolinario, Fernando Motta, Anna Carolina Paixao, Ana Carolina Mendonca, Alice Sampaio Rocha, Renata Serrano Lopes, Tiago Graf, Ricardo Khouri, Marilda Siqueira on behalf of the Fiocruz COVID-19 Genomic Surveillance Network |
| EPI_ISL_1181430 | Laboratory of Respiratory Viruses and Measles, Oswaldo Cruz Institute, FIOCRUZ | Laboratory of Respiratory Viruses and Measles, Oswaldo Cruz Institute, FIOCRUZ | Paola Resende, Luciana Appolinario, Fernando Motta, Anna Carolina Paixao, Ana Carolina Mendonca, Alice Sampaio Rocha, Renata Serrano Lopes, Marilda Siqueira on behalf of the Fiocruz COVID-19 Genomic Surveillance Network |
| EPI_ISL_1181431 | Laboratorio Central de Saude Publica do Estado do Rio de Janeiro (LACEN-RJ) | Laboratory of Respiratory Viruses and Measles, Oswaldo Cruz Institute, FIOCRUZ | Paola Resende, Luciana Appolinario, Fernando Motta, Anna Carolina Paixao, Ana Carolina Mendonca, Alice Sampaio Rocha, Renata Serrano Lopes, Andrea Cony Cavalcanti, Marilda Siqueira on behalf of the Fiocruz COVID-19 Genomic Surveillance Network |
| EPI_ISL_1181432 | Laboratorio Central de Saude Publica do Estado do Espitito Santo (LACEN-ES) | Laboratory of Respiratory Viruses and Measles, Oswaldo Cruz Institute, FIOCRUZ | Paola Resende, Luciana Appolinario, Fernando Motta, Anna Carolina Paixao, Ana Carolina Mendonca, Alice Sampaio Rocha, Renata Serrano Lopes, Rodrigo Ribeiro Rodrigues, Marilda Siqueira on behalf of the Fiocruz COVID-19 Genomic Surveillance Network |
| EPI_ISL_1181433, EPI_ISL_1181434 | Laboratorio Central de Saude Publica do Estado do Rio Grande do Sul (LACEN-RS) | Laboratory of Respiratory Viruses and Measles, Oswaldo Cruz Institute, FIOCRUZ | Paola Resende, Luciana Appolinario, Fernando Motta, Anna Carolina Paixao, Ana Carolina Mendonca, Alice Sampaio Rocha, Renata Serrano Lopes, Tatiana Schaffer Gregianini, Marilda Siqueira on behalf of the Fiocruz COVID-19 Genomic Surveillance Network |
| EPI_ISL_1181435, EPI_ISL_1181436 | Federal University of Southern Bahia (UFSB - Universidade Federal do Sul da Bahia) | Laboratory of Respiratory Viruses and Measles, Oswaldo Cruz Institute, FIOCRUZ | Paola Resende, Luciana Appolinario, Fernando Motta, Anna Carolina Paixao, Ana Carolina Mendonca, Alice Sampaio Rocha, Renata Serrano Lopes, Thiago Mafra, Marilda Siqueira on behalf of the Fiocruz COVID-19 Genomic Surveillance Network |
| EPI_ISL_1181437, EPI_ISL_1181438, EPI_ISL_1181439 | Laboratory of Respiratory Viruses and Measles, Oswaldo Cruz Institute, FIOCRUZ | Laboratory of Respiratory Viruses and Measles, Oswaldo Cruz Institute, FIOCRUZ | Paola Resende, Luciana Appolinario, Fernando Motta, Anna Carolina Paixao, Ana Carolina Mendonca, Alice Sampaio Rocha, Renata Serrano Lopes, Marilda Siqueira on behalf of the Fiocruz COVID-19 Genomic Surveillance Network |
| EPI_ISL_1181440, EPI_ISL_1181441 | Gonçalo Moniz Institute, FIOCRUZ, Bahia | Laboratory of Respiratory Viruses and Measles, Oswaldo Cruz Institute, FIOCRUZ | Paola Resende, Luciana Appolinario, Fernando Motta, Anna Carolina Paixao, Ana Carolina Mendonca, Alice Sampaio Rocha, Renata Serrano Lopes, Tiago Graf, Ricardo Khouri, Marilda Siqueira on behalf of the Fiocruz COVID-19 Genomic Surveillance Network |
| EPI_ISL_1181442 | Laboratory of Respiratory Viruses and Measles, Oswaldo Cruz Institute, FIOCRUZ | Laboratory of Respiratory Viruses and Measles, Oswaldo Cruz Institute, FIOCRUZ | Paola Resende, Luciana Appolinario, Fernando Motta, Anna Carolina Paixao, Ana Carolina Mendonca, Alice Sampaio Rocha, Renata Serrano Lopes, Marilda Siqueira on behalf of the Fiocruz COVID-19 Genomic Surveillance Network |
| EPI_ISL_1181443 | Federal University of Southern Bahia (UFSB - Universidade Federal do Sul da Bahia) | Laboratory of Respiratory Viruses and Measles, Oswaldo Cruz Institute, FIOCRUZ | Paola Resende, Luciana Appolinario, Fernando Motta, Anna Carolina Paixao, Ana Carolina Mendonca, Alice Sampaio Rocha, Renata Serrano Lopes, Thiago Mafra, Marilda Siqueira on behalf of the Fiocruz COVID-19 Genomic Surveillance Network |
| EPI_ISL_1181444, EPI_ISL_1181445 | Gonçalo Moniz Institute, FIOCRUZ, Bahia | Laboratory of Respiratory Viruses and Measles, Oswaldo Cruz Institute, FIOCRUZ | Paola Resende, Luciana Appolinario, Fernando Motta, Anna Carolina Paixao, Ana Carolina Mendonca, Alice Sampaio Rocha, Renata Serrano Lopes, Tiago Graf, Ricardo Khouri, Marilda Siqueira on behalf of the Fiocruz COVID-19 Genomic Surveillance Network |
| EPI_ISL_1181446, EPI_ISL_1181447, EPI_ISL_1181448 | Laboratory of Respiratory Viruses and Measles, Oswaldo Cruz Institute, FIOCRUZ | Laboratory of Respiratory Viruses and Measles, Oswaldo Cruz Institute, FIOCRUZ | Paola Resende, Luciana Appolinario, Fernando Motta, Anna Carolina Paixao, Ana Carolina Mendonca, Alice Sampaio Rocha, Renata Serrano Lopes, Marilda Siqueira on behalf of the Fiocruz COVID-19 Genomic Surveillance Network |
| EPI_ISL_1181449 | Gonçalo Moniz Institute, FIOCRUZ, Bahia | Laboratory of Respiratory Viruses and Measles, Oswaldo Cruz Institute, FIOCRUZ | Paola Resende, Luciana Appolinario, Fernando Motta, Anna Carolina Paixao, Ana Carolina Mendonca, Alice Sampaio Rocha, Renata Serrano Lopes, Tiago Graf, Ricardo Khouri, Marilda Siqueira on behalf of the Fiocruz COVID-19 Genomic Surveillance Network |
| EPI_ISL_1181450 | Federal University of Southern Bahia (UFSB - Universidade Federal do Sul da Bahia) | Laboratory of Respiratory Viruses and Measles, Oswaldo Cruz Institute, FIOCRUZ | Paola Resende, Luciana Appolinario, Fernando Motta, Anna Carolina Paixao, Ana Carolina Mendonca, Alice Sampaio Rocha, Renata Serrano Lopes, Thiago Mafra, Marilda Siqueira on behalf of the Fiocruz COVID-19 Genomic Surveillance Network |
| EPI_ISL_1181451 | Laboratorio Central de Saude Publica do Estado do Rio Grande do Sul (LACEN-RS) | Laboratory of Respiratory Viruses and Measles, Oswaldo Cruz Institute, FIOCRUZ | Paola Resende, Luciana Appolinario, Fernando Motta, Anna Carolina Paixao, Ana Carolina Mendonca, Alice Sampaio Rocha, Renata Serrano Lopes, Tatiana Schaffer Gregianini, Marilda Siqueira on behalf of the Fiocruz COVID-19 Genomic Surveillance Network |
| EPI_ISL_1181452 | Laboratorio de Imunologia de Transplantes de Goias LTDA (HLAGYN) | Laboratory of Respiratory Viruses and Measles, Oswaldo Cruz Institute, FIOCRUZ | Paola Resende, Luciana Appolinario, Fernando Motta, Anna Carolina Paixao, Ana Carolina Mendonca, Alice Sampaio Rocha, Renata Serrano Lopes , Alessandro Leonardo Alvares Magalhaes, Fernando Antonio Vinhal dos Santos, Erika Lopes Rocha Batista, Marilda Siqueira on behalf of the Fiocruz COVID-19 Genomic Surveillance Network |
| EPI_ISL_1181453 | Laboratorio Central de Saude Publica do Estado de Santa Catarina (LACEN-SC) | Laboratory of Respiratory Viruses and Measles, Oswaldo Cruz Institute, FIOCRUZ | Paola Resende, Luciana Appolinario, Fernando Motta, Anna Carolina Paixao, Ana Carolina Mendonca, Alice Sampaio Rocha, Renata Serrano Lopes, Darcita Buerger Rovaris, Sandra Bianchini Fernandes, Marilda Siqueira on behalf of the Fiocruz COVID-19 Genomic Surveillance Network |
| EPI_ISL_1181454, EPI_ISL_1181455, EPI_ISL_1181456, EPI_ISL_1181457, EPI_ISL_1181458, EPI_ISL_1181459, EPI_ISL_1181460, EPI_ISL_1181461, EPI_ISL_1181462, EPI_ISL_1181463, EPI_ISL_1181464, EPI_ISL_1181465, EPI_ISL_1181466, EPI_ISL_1181467 | | | |
| see above | Laboratory of Respiratory Viruses and Measles, Oswaldo Cruz Institute, FIOCRUZ | Laboratory of Respiratory Viruses and Measles, Oswaldo Cruz Institute, FIOCRUZ | Paola Resende, Luciana Appolinario, Fernando Motta, Anna Carolina Paixao, Ana Carolina Mendonca, Alice Sampaio Rocha, Renata Serrano Lopes, Marilda Siqueira on behalf of the Fiocruz COVID-19 Genomic Surveillance Network |
| EPI_ISL_1181468 | Laboratorio Central de Saude Publica do Estado do Rio | Laboratory of Respiratory Viruses and Measles, Oswaldo Cruz | Paola Resende, Luciana Appolinario, Fernando Motta, Anna Carolina Paixao, Ana Carolina Mendonca, Alice Sampaio Rocha, Renata Serrano Lopes, |

| | | | |
|---|---|---|---|
| | Grande do Sul (LACEN-RS) | Institute, FIOCRUZ | Tatiana Schaffer Gregianini, Marilda Siqueira on behalf of the Fiocruz COVID-19 Genomic Surveillance Network |
| EPI_ISL_1181469 | Federal University of Mato Grosso (UFMT) | Laboratory of Respiratory Viruses and Measles, Oswaldo Cruz Institute, FIOCRUZ | Paola Resende, Luciana Appolinario, Fernando Motta, Anna Carolina Paixao, Ana Carolina Mendonca, Alice Sampaio Rocha, Renata Serrano Lopes, Renata Dezengrini, Marilda Siqueira on behalf of the Fiocruz COVID-19 Genomic Surveillance Network |
| EPI_ISL_1181470 | Laboratory of Respiratory Viruses and Measles, Oswaldo Cruz Institute, FIOCRUZ | Laboratory of Respiratory Viruses and Measles, Oswaldo Cruz Institute, FIOCRUZ | Paola Resende, Luciana Appolinario, Fernando Motta, Anna Carolina Paixao, Ana Carolina Mendonca, Alice Sampaio Rocha, Renata Serrano Lopes, Marilda Siqueira on behalf of the Fiocruz COVID-19 Genomic Surveillance Network |
| EPI_ISL_1181471, EPI_ISL_1181472, EPI_ISL_1181473 | Laboratorio de Imunologia de Transplantes de Goias LTDA (HLAGYN) | Laboratory of Respiratory Viruses and Measles, Oswaldo Cruz Institute, FIOCRUZ | Paola Resende, Luciana Appolinario, Fernando Motta, Anna Carolina Paixao, Ana Carolina Mendonca, Alice Sampaio Rocha, Renata Serrano Lopes , Alessandro Leonardo Alvares Magalhaes, Fernando Antonio Vinhal dos Santos, Erika Lopes Rocha Batista, Marilda Siqueira on behalf of the Fiocruz COVID-19 Genomic Surveillance Network |
| EPI_ISL_1181474 | Laboratorio Central de Saude Publica do Estado do Parana (LACEN-PR) | Laboratory of Respiratory Viruses and Measles, Oswaldo Cruz Institute, FIOCRUZ | Paola Resende, Luciana Appolinario, Fernando Motta, Anna Carolina Paixao, Ana Carolina Mendonca, Alice Sampaio Rocha, Renata Serrano Lopes, Maria do Carmo Debur, Irina Nastassja Riediger, Marilda Siqueira on behalf of the Fiocruz COVID-19 Genomic Surveillance Network |
| EPI_ISL_1181475, EPI_ISL_1181476, EPI_ISL_1181477 | Laboratory of Respiratory Viruses and Measles, Oswaldo Cruz Institute, FIOCRUZ | Laboratory of Respiratory Viruses and Measles, Oswaldo Cruz Institute, FIOCRUZ | Paola Resende, Luciana Appolinario, Fernando Motta, Anna Carolina Paixao, Ana Carolina Mendonca, Alice Sampaio Rocha, Renata Serrano Lopes, Marilda Siqueira on behalf of the Fiocruz COVID-19 Genomic Surveillance Network |
| EPI_ISL_1181478, EPI_ISL_1181479, EPI_ISL_1181480 | Laboratorio de Imunologia de Transplantes de Goias LTDA (HLAGYN) | Laboratory of Respiratory Viruses and Measles, Oswaldo Cruz Institute, FIOCRUZ | Paola Resende, Luciana Appolinario, Fernando Motta, Anna Carolina Paixao, Ana Carolina Mendonca, Alice Sampaio Rocha, Renata Serrano Lopes , Alessandro Leonardo Alvares Magalhaes, Fernando Antonio Vinhal dos Santos, Erika Lopes Rocha Batista, Marilda Siqueira on behalf of the Fiocruz COVID-19 Genomic Surveillance Network |
| EPI_ISL_1181481, EPI_ISL_1181482, EPI_ISL_1181483 | Laboratory of Respiratory Viruses and Measles, Oswaldo Cruz Institute, FIOCRUZ | Laboratory of Respiratory Viruses and Measles, Oswaldo Cruz Institute, FIOCRUZ | Paola Resende, Luciana Appolinario, Fernando Motta, Anna Carolina Paixao, Ana Carolina Mendonca, Alice Sampaio Rocha, Renata Serrano Lopes, Marilda Siqueira on behalf of the Fiocruz COVID-19 Genomic Surveillance Network |
| EPI_ISL_1181484, EPI_ISL_1181485 | Gonçalo Moniz Institute, FIOCRUZ, Bahia | Laboratory of Respiratory Viruses and Measles, Oswaldo Cruz Institute, FIOCRUZ | Paola Resende, Luciana Appolinario, Fernando Motta, Anna Carolina Paixao, Ana Carolina Mendonca, Alice Sampaio Rocha, Renata Serrano Lopes, Tiago Graf, Ricardo Khouri, Marilda Siqueira on behalf of the Fiocruz COVID-19 Genomic Surveillance Network |
| EPI_ISL_1181486 | Laboratory of Respiratory Viruses and Measles, Oswaldo Cruz Institute, FIOCRUZ | Laboratory of Respiratory Viruses and Measles, Oswaldo Cruz Institute, FIOCRUZ | Paola Resende, Luciana Appolinario, Fernando Motta, Anna Carolina Paixao, Ana Carolina Mendonca, Alice Sampaio Rocha, Renata Serrano Lopes, Marilda Siqueira on behalf of the Fiocruz COVID-19 Genomic Surveillance Network |
| EPI_ISL_1181487 | Laboratorio de Imunologia de Transplantes de Goias LTDA (HLAGYN) | Laboratory of Respiratory Viruses and Measles, Oswaldo Cruz Institute, FIOCRUZ | Paola Resende, Luciana Appolinario, Fernando Motta, Anna Carolina Paixao, Ana Carolina Mendonca, Alice Sampaio Rocha, Renata Serrano Lopes , Alessandro Leonardo Alvares Magalhaes, Fernando Antonio Vinhal dos Santos, Erika Lopes Rocha Batista, Marilda Siqueira on behalf of the Fiocruz COVID-19 Genomic Surveillance Network |
| EPI_ISL_1181488, EPI_ISL_1181489, EPI_ISL_1181490, EPI_ISL_1181491 | Laboratory of Respiratory Viruses and Measles, Oswaldo Cruz Institute, FIOCRUZ | Laboratory of Respiratory Viruses and Measles, Oswaldo Cruz Institute, FIOCRUZ | Paola Resende, Luciana Appolinario, Fernando Motta, Anna Carolina Paixao, Ana Carolina Mendonca, Alice Sampaio Rocha, Renata Serrano Lopes, Marilda Siqueira on behalf of the Fiocruz COVID-19 Genomic Surveillance Network |
| EPI_ISL_1181492 | Laboratorio de Imunologia de Transplantes de Goias LTDA (HLAGYN) | Laboratory of Respiratory Viruses and Measles, Oswaldo Cruz Institute, FIOCRUZ | Paola Resende, Luciana Appolinario, Fernando Motta, Anna Carolina Paixao, Ana Carolina Mendonca, Alice Sampaio Rocha, Renata Serrano Lopes , Alessandro Leonardo Alvares Magalhaes, Fernando Antonio Vinhal dos Santos, Erika Lopes Rocha Batista, Marilda Siqueira on behalf of the Fiocruz COVID-19 Genomic Surveillance Network |
| EPI_ISL_1181493, EPI_ISL_1181494, EPI_ISL_1181495, EPI_ISL_1181496, EPI_ISL_1181497 | Laboratory of Respiratory Viruses and Measles, Oswaldo Cruz Institute, FIOCRUZ | Laboratory of Respiratory Viruses and Measles, Oswaldo Cruz Institute, FIOCRUZ | Paola Resende, Luciana Appolinario, Fernando Motta, Anna Carolina Paixao, Ana Carolina Mendonca, Alice Sampaio Rocha, Renata Serrano Lopes, Marilda Siqueira on behalf of the Fiocruz COVID-19 Genomic Surveillance Network |
| EPI_ISL_1181498 | Laboratorio Central de Saude Publica do Estado do Parana (LACEN-PR) | Laboratory of Respiratory Viruses and Measles, Oswaldo Cruz Institute, FIOCRUZ | Paola Resende, Luciana Appolinario, Fernando Motta, Anna Carolina Paixao, Ana Carolina Mendonca, Alice Sampaio Rocha, Renata Serrano Lopes, Maria do Carmo Debur, Irina Nastassja Riediger, Marilda Siqueira on behalf of the Fiocruz COVID-19 Genomic Surveillance Network |
| EPI_ISL_1181499, EPI_ISL_1181500, EPI_ISL_1181501, EPI_ISL_1181502, EPI_ISL_1181503, EPI_ISL_1181504, EPI_ISL_1181505, EPI_ISL_1181506, EPI_ISL_1181507, EPI_ISL_1181508, EPI_ISL_1181509, EPI_ISL_1181510, EPI_ISL_1181511, EPI_ISL_1181512, EPI_ISL_1181513, EPI_ISL_1181514, EPI_ISL_1181515 | | | |
| see above | Laboratory of Respiratory Viruses and Measles, Oswaldo Cruz Institute, FIOCRUZ | Laboratory of Respiratory Viruses and Measles, Oswaldo Cruz Institute, FIOCRUZ | Paola Resende, Luciana Appolinario, Fernando Motta, Anna Carolina Paixao, Ana Carolina Mendonca, Alice Sampaio Rocha, Renata Serrano Lopes, Marilda Siqueira on behalf of the Fiocruz COVID-19 Genomic Surveillance Network |
| EPI_ISL_1181516 | Laboratorio de Imunologia de Transplantes de Goias LTDA (HLAGYN) | Laboratory of Respiratory Viruses and Measles, Oswaldo Cruz Institute, FIOCRUZ | Paola Resende, Luciana Appolinario, Fernando Motta, Anna Carolina Paixao, Ana Carolina Mendonca, Alice Sampaio Rocha, Renata Serrano Lopes , Alessandro Leonardo Alvares Magalhaes, Fernando Antonio Vinhal dos Santos, Erika Lopes Rocha Batista, Marilda Siqueira on behalf of the Fiocruz COVID-19 Genomic Surveillance Network |
| EPI_ISL_1181517 | Laboratorio Central de Saude Publica do Estado do Parana (LACEN-PR) | Laboratory of Respiratory Viruses and Measles, Oswaldo Cruz Institute, FIOCRUZ | Paola Resende, Luciana Appolinario, Fernando Motta, Anna Carolina Paixao, Ana Carolina Mendonca, Alice Sampaio Rocha, Renata Serrano Lopes, Maria do Carmo Debur, Irina Nastassja Riediger, Marilda Siqueira on behalf of the Fiocruz COVID-19 Genomic Surveillance Network |
| EPI_ISL_1181518, EPI_ISL_1181519 | Laboratory of Respiratory Viruses and Measles, Oswaldo Cruz Institute, FIOCRUZ | Laboratory of Respiratory Viruses and Measles, Oswaldo Cruz Institute, FIOCRUZ | Paola Resende, Luciana Appolinario, Fernando Motta, Anna Carolina Paixao, Ana Carolina Mendonca, Alice Sampaio Rocha, Renata Serrano Lopes, Marilda Siqueira on behalf of the Fiocruz COVID-19 Genomic Surveillance Network |
| EPI_ISL_1181520 | Oswaldo Cruz Foundation, FIOCRUZ - Ceara (Fiocruz-CE) | Laboratory of Respiratory Viruses and Measles, Oswaldo Cruz Institute, FIOCRUZ | Paola Resende, Luciana Appolinario, Fernando Motta, Anna Carolina Paixao, Ana Carolina Mendonca, Alice Sampaio Rocha, Renata Serrano Lopes, Thais de Oliveira Costa, Joaquim César do Nascimento Sousa Júnior, Fabio Miyajima, Marilda Siqueira on behalf of the Fiocruz COVID-19 Genomic Surveillance Network |
| EPI_ISL_1181521 | Laboratory of Respiratory Viruses and Measles, Oswaldo Cruz Institute, FIOCRUZ | Laboratory of Respiratory Viruses and Measles, Oswaldo Cruz Institute, FIOCRUZ | Paola Resende, Luciana Appolinario, Fernando Motta, Anna Carolina Paixao, Ana Carolina Mendonca, Alice Sampaio Rocha, Renata Serrano Lopes, Marilda Siqueira on behalf of the Fiocruz COVID-19 Genomic Surveillance Network |
| EPI_ISL_1181522 | Oswaldo Cruz Foundation, FIOCRUZ - Ceara (Fiocruz-CE) | Laboratory of Respiratory Viruses and Measles, Oswaldo Cruz Institute, FIOCRUZ | Paola Resende, Luciana Appolinario, Fernando Motta, Anna Carolina Paixao, Ana Carolina Mendonca, Alice Sampaio Rocha, Renata Serrano Lopes, Thais de Oliveira Costa, Joaquim César do Nascimento Sousa Júnior, Fabio Miyajima, Marilda Siqueira on behalf of the Fiocruz COVID-19 Genomic Surveillance Network |
| EPI_ISL_1181523 | Laboratory of Respiratory Viruses and Measles, Oswaldo Cruz Institute, FIOCRUZ | Laboratory of Respiratory Viruses and Measles, Oswaldo Cruz Institute, FIOCRUZ | Paola Resende, Luciana Appolinario, Fernando Motta, Anna Carolina Paixao, Ana Carolina Mendonca, Alice Sampaio Rocha, Renata Serrano Lopes, Marilda Siqueira on behalf of the Fiocruz COVID-19 Genomic Surveillance Network |
| EPI_ISL_1181524 | Laboratorio Central de Saude Publica do Estado do Rio Grande do Sul (LACEN-RS) | Laboratory of Respiratory Viruses and Measles, Oswaldo Cruz Institute, FIOCRUZ | Paola Resende, Luciana Appolinario, Fernando Motta, Anna Carolina Paixao, Ana Carolina Mendonca, Alice Sampaio Rocha, Renata Serrano Lopes, Tatiana Schaffer Gregianini, Marilda Siqueira on behalf of the Fiocruz COVID-19 Genomic Surveillance Network |
| EPI_ISL_1181525 | Laboratorio de Imunologia de Transplantes de Goias LTDA (HLAGYN) | Laboratory of Respiratory Viruses and Measles, Oswaldo Cruz Institute, FIOCRUZ | Paola Resende, Luciana Appolinario, Fernando Motta, Anna Carolina Paixao, Ana Carolina Mendonca, Alice Sampaio Rocha, Renata Serrano Lopes , Alessandro Leonardo Alvares Magalhaes, Fernando Antonio Vinhal dos Santos, Erika Lopes Rocha Batista, Marilda Siqueira on behalf of the Fiocruz COVID-19 Genomic Surveillance Network |
| EPI_ISL_1181526, EPI_ISL_1181527, EPI_ISL_1181528 | Laboratorio Central de Saude Publica do Estado do Parana (LACEN-PR) | Laboratory of Respiratory Viruses and Measles, Oswaldo Cruz Institute, FIOCRUZ | Paola Resende, Luciana Appolinario, Fernando Motta, Anna Carolina Paixao, Ana Carolina Mendonca, Alice Sampaio Rocha, Renata Serrano Lopes, Maria do Carmo Debur, Irina Nastassja Riediger, Marilda Siqueira on behalf of the Fiocruz COVID-19 Genomic Surveillance Network |
| EPI_ISL_1181529 | Laboratory of Respiratory Viruses and Measles, Oswaldo Cruz Institute, FIOCRUZ | Laboratory of Respiratory Viruses and Measles, Oswaldo Cruz Institute, FIOCRUZ | Paola Resende, Luciana Appolinario, Fernando Motta, Anna Carolina Paixao, Ana Carolina Mendonca, Alice Sampaio Rocha, Renata Serrano Lopes, Marilda Siqueira on behalf of the Fiocruz COVID-19 Genomic Surveillance Network |
| EPI_ISL_1181530 | Laboratorio Central de Saude Publica do Estado do Parana (LACEN-PR) | Laboratory of Respiratory Viruses and Measles, Oswaldo Cruz Institute, FIOCRUZ | Paola Resende, Luciana Appolinario, Fernando Motta, Anna Carolina Paixao, Ana Carolina Mendonca, Alice Sampaio Rocha, Renata Serrano Lopes, Maria do Carmo Debur, Irina Nastassja Riediger, Marilda Siqueira on behalf of the Fiocruz COVID-19 Genomic Surveillance Network |
| EPI_ISL_1181531 | Laboratory of Respiratory Viruses and Measles, Oswaldo Cruz Institute, FIOCRUZ | Laboratory of Respiratory Viruses and Measles, Oswaldo Cruz Institute, FIOCRUZ | Paola Resende, Luciana Appolinario, Fernando Motta, Anna Carolina Paixao, Ana Carolina Mendonca, Alice Sampaio Rocha, Renata Serrano Lopes, Marilda Siqueira on behalf of the Fiocruz COVID-19 Genomic Surveillance Network |
| EPI_ISL_1181532 | Laboratorio de Imunologia de Transplantes de Goias LTDA (HLAGYN) | Laboratory of Respiratory Viruses and Measles, Oswaldo Cruz Institute, FIOCRUZ | Paola Resende, Luciana Appolinario, Fernando Motta, Anna Carolina Paixao, Ana Carolina Mendonca, Alice Sampaio Rocha, Renata Serrano Lopes , Alessandro Leonardo Alvares Magalhaes, Fernando Antonio Vinhal dos Santos, Erika Lopes Rocha Batista, Marilda Siqueira on behalf of the Fiocruz COVID-19 Genomic Surveillance Network |
| EPI_ISL_1181533, EPI_ISL_1181534 | Laboratorio Central de Saude Publica do Estado do Parana (LACEN-PR) | Laboratory of Respiratory Viruses and Measles, Oswaldo Cruz Institute, FIOCRUZ | Paola Resende, Luciana Appolinario, Fernando Motta, Anna Carolina Paixao, Ana Carolina Mendonca, Alice Sampaio Rocha, Renata Serrano Lopes, Maria do Carmo Debur, Irina Nastassja Riediger, Marilda Siqueira on behalf of the Fiocruz COVID-19 Genomic Surveillance Network |

| | | | |
|---|---|---|---|
| EPI_ISL_1181535 | Laboratorio de Imunologia de Transplantes de Goias LTDA (HLAGYN) | Laboratory of Respiratory Viruses and Measles, Oswaldo Cruz Institute, FIOCRUZ | Paola Resende, Luciana Appolinario, Fernando Motta, Anna Carolina Paixao, Ana Carolina Mendonca, Alice Sampaio Rocha, Renata Serrano Lopes , Alessandro Leonardo Alvares Magalhaes, Fernando Antonio Vinhal dos Santos, Erika Lopes Rocha Batista, Marilda Siqueira on behalf of the Fiocruz COVID-19 Genomic Surveillance Network |
| EPI_ISL_1181536, EPI_ISL_1181537, EPI_ISL_1181538, EPI_ISL_1181539, EPI_ISL_1181540, EPI_ISL_1181541, EPI_ISL_1181542, EPI_ISL_1181543, EPI_ISL_1181544, EPI_ISL_1181545 | Laboratorio Central de Saude Publica do Estado do Parana (LACEN-PR) | Laboratory of Respiratory Viruses and Measles, Oswaldo Cruz Institute, FIOCRUZ | Paola Resende, Luciana Appolinario, Fernando Motta, Anna Carolina Paixao, Ana Carolina Mendonca, Alice Sampaio Rocha, Renata Serrano Lopes, Maria do Carmo Debur, Irina Nastassja Riediger, Marilda Siqueira on behalf of the Fiocruz COVID-19 Genomic Surveillance Network |
| EPI_ISL_1181546, EPI_ISL_1181547, EPI_ISL_1181548, EPI_ISL_1181549, EPI_ISL_1181550 | Laboratorio de Imunologia de Transplantes de Goias LTDA (HLAGYN) | Laboratory of Respiratory Viruses and Measles, Oswaldo Cruz Institute, FIOCRUZ | Paola Resende, Luciana Appolinario, Fernando Motta, Anna Carolina Paixao, Ana Carolina Mendonca, Alice Sampaio Rocha, Renata Serrano Lopes , Alessandro Leonardo Alvares Magalhaes, Fernando Antonio Vinhal dos Santos, Erika Lopes Rocha Batista, Marilda Siqueira on behalf of the Fiocruz COVID-19 Genomic Surveillance Network |
| EPI_ISL_1181551, EPI_ISL_1181552, EPI_ISL_1181553, EPI_ISL_1181554, EPI_ISL_1181555, EPI_ISL_1181556, EPI_ISL_1181557, EPI_ISL_1181558, EPI_ISL_1181559 | Laboratorio Central de Saude Publica do Estado do Parana (LACEN-PR) | Laboratory of Respiratory Viruses and Measles, Oswaldo Cruz Institute, FIOCRUZ | Paola Resende, Luciana Appolinario, Fernando Motta, Anna Carolina Paixao, Ana Carolina Mendonca, Alice Sampaio Rocha, Renata Serrano Lopes, Maria do Carmo Debur, Irina Nastassja Riediger, Marilda Siqueira on behalf of the Fiocruz COVID-19 Genomic Surveillance Network |
| EPI_ISL_1181560, EPI_ISL_1181561, EPI_ISL_1181562 | Laboratory of Respiratory Viruses and Measles, Oswaldo Cruz Institute, FIOCRUZ | Laboratory of Respiratory Viruses and Measles, Oswaldo Cruz Institute, FIOCRUZ | Paola Resende, Luciana Appolinario, Fernando Motta, Anna Carolina Paixao, Ana Carolina Mendonca, Alice Sampaio Rocha, Renata Serrano Lopes, Marilda Siqueira on behalf of the Fiocruz COVID-19 Genomic Surveillance Network |
| EPI_ISL_1181563, EPI_ISL_1181564, EPI_ISL_1181565, EPI_ISL_1181566 | Laboratorio Central de Saude Publica do Estado do Parana (LACEN-PR) | Laboratory of Respiratory Viruses and Measles, Oswaldo Cruz Institute, FIOCRUZ | Paola Resende, Luciana Appolinario, Fernando Motta, Anna Carolina Paixao, Ana Carolina Mendonca, Alice Sampaio Rocha, Renata Serrano Lopes, Maria do Carmo Debur, Irina Nastassja Riediger, Marilda Siqueira on behalf of the Fiocruz COVID-19 Genomic Surveillance Network |
| EPI_ISL_1181567 | Laboratorio Central de Saude Publica do Estado do Rio Grande do Sul (LACEN-RS) | Laboratory of Respiratory Viruses and Measles, Oswaldo Cruz Institute, FIOCRUZ | Paola Resende, Luciana Appolinario, Fernando Motta, Anna Carolina Paixao, Ana Carolina Mendonca, Alice Sampaio Rocha, Renata Serrano Lopes, Tatiana Schaffer Gregianini, Marilda Siqueira on behalf of the Fiocruz COVID-19 Genomic Surveillance Network |
| EPI_ISL_1181568 | Laboratorio Central de Saude Publica do Estado do Parana (LACEN-PR) | Laboratory of Respiratory Viruses and Measles, Oswaldo Cruz Institute, FIOCRUZ | Paola Resende, Luciana Appolinario, Fernando Motta, Anna Carolina Paixao, Ana Carolina Mendonca, Alice Sampaio Rocha, Renata Serrano Lopes, Maria do Carmo Debur, Irina Nastassja Riediger, Marilda Siqueira on behalf of the Fiocruz COVID-19 Genomic Surveillance Network |
| EPI_ISL_1181569 | Laboratorio de Imunologia de Transplantes de Goias LTDA (HLAGYN) | Laboratory of Respiratory Viruses and Measles, Oswaldo Cruz Institute, FIOCRUZ | Paola Resende, Luciana Appolinario, Fernando Motta, Anna Carolina Paixao, Ana Carolina Mendonca, Alice Sampaio Rocha, Renata Serrano Lopes , Alessandro Leonardo Alvares Magalhaes, Fernando Antonio Vinhal dos Santos, Erika Lopes Rocha Batista, Marilda Siqueira on behalf of the Fiocruz COVID-19 Genomic Surveillance Network |
| EPI_ISL_1181570 | Laboratorio Central de Saude Publica do Estado do Parana (LACEN-PR) | Laboratory of Respiratory Viruses and Measles, Oswaldo Cruz Institute, FIOCRUZ | Paola Resende, Luciana Appolinario, Fernando Motta, Anna Carolina Paixao, Ana Carolina Mendonca, Alice Sampaio Rocha, Renata Serrano Lopes, Maria do Carmo Debur, Irina Nastassja Riediger, Marilda Siqueira on behalf of the Fiocruz COVID-19 Genomic Surveillance Network |
| EPI_ISL_1181571, EPI_ISL_1181572, EPI_ISL_1181573, EPI_ISL_1181574 | Laboratory of Respiratory Viruses and Measles, Oswaldo Cruz Institute, FIOCRUZ | Laboratory of Respiratory Viruses and Measles, Oswaldo Cruz Institute, FIOCRUZ | Paola Resende, Luciana Appolinario, Fernando Motta, Anna Carolina Paixao, Ana Carolina Mendonca, Alice Sampaio Rocha, Renata Serrano Lopes, Marilda Siqueira on behalf of the Fiocruz COVID-19 Genomic Surveillance Network |
| EPI_ISL_1181575 | Oswaldo Cruz Foundation, FIOCRUZ - Ceara (Fiocruz-CE) | Laboratory of Respiratory Viruses and Measles, Oswaldo Cruz Institute, FIOCRUZ | Paola Resende, Luciana Appolinario, Fernando Motta, Anna Carolina Paixao, Ana Carolina Mendonca, Alice Sampaio Rocha, Renata Serrano Lopes, Thais de Oliveira Costa, Joaquim César do Nascimento Sousa Júnior, Fabio Miyajima, Marilda Siqueira on behalf of the Fiocruz COVID-19 Genomic Surveillance Network |
| EPI_ISL_1181576 | Laboratorio Central de Saude Publica do Estado do Parana (LACEN-PR) | Laboratory of Respiratory Viruses and Measles, Oswaldo Cruz Institute, FIOCRUZ | Paola Resende, Luciana Appolinario, Fernando Motta, Anna Carolina Paixao, Ana Carolina Mendonca, Alice Sampaio Rocha, Renata Serrano Lopes, Maria do Carmo Debur, Irina Nastassja Riediger, Marilda Siqueira on behalf of the Fiocruz COVID-19 Genomic Surveillance Network |
| EPI_ISL_1181577 | Oswaldo Cruz Foundation, FIOCRUZ - Ceara (Fiocruz-CE) | Laboratory of Respiratory Viruses and Measles, Oswaldo Cruz Institute, FIOCRUZ | Paola Resende, Luciana Appolinario, Fernando Motta, Anna Carolina Paixao, Ana Carolina Mendonca, Alice Sampaio Rocha, Renata Serrano Lopes, Thais de Oliveira Costa, Joaquim César do Nascimento Sousa Júnior, Fabio Miyajima, Marilda Siqueira on behalf of the Fiocruz COVID-19 Genomic Surveillance Network |
| EPI_ISL_1181578, EPI_ISL_1181579 | Laboratory of Respiratory Viruses and Measles, Oswaldo Cruz Institute, FIOCRUZ | Laboratory of Respiratory Viruses and Measles, Oswaldo Cruz Institute, FIOCRUZ | Paola Resende, Luciana Appolinario, Fernando Motta, Anna Carolina Paixao, Ana Carolina Mendonca, Alice Sampaio Rocha, Renata Serrano Lopes, Marilda Siqueira on behalf of the Fiocruz COVID-19 Genomic Surveillance Network |
| EPI_ISL_1181580, EPI_ISL_1181581 | Laboratorio Central de Saude Publica do Estado do Parana (LACEN-PR) | Laboratory of Respiratory Viruses and Measles, Oswaldo Cruz Institute, FIOCRUZ | Paola Resende, Luciana Appolinario, Fernando Motta, Anna Carolina Paixao, Ana Carolina Mendonca, Alice Sampaio Rocha, Renata Serrano Lopes, Maria do Carmo Debur, Irina Nastassja Riediger, Marilda Siqueira on behalf of the Fiocruz COVID-19 Genomic Surveillance Network |
| EPI_ISL_1181582, EPI_ISL_1181583 | Laboratorio Central de Saude Publica do Estado do Espitito Santo (LACEN-ES) | Laboratory of Respiratory Viruses and Measles, Oswaldo Cruz Institute, FIOCRUZ | Paola Resende, Luciana Appolinario, Fernando Motta, Anna Carolina Paixao, Ana Carolina Mendonca, Alice Sampaio Rocha, Renata Serrano Lopes, Rodrigo Ribeiro Rodrigues, Marilda Siqueira on behalf of the Fiocruz COVID-19 Genomic Surveillance Network |
| EPI_ISL_1181584 | Laboratorio Central de Saude Publica do Estado do Parana (LACEN-PR) | Laboratory of Respiratory Viruses and Measles, Oswaldo Cruz Institute, FIOCRUZ | Paola Resende, Luciana Appolinario, Fernando Motta, Anna Carolina Paixao, Ana Carolina Mendonca, Alice Sampaio Rocha, Renata Serrano Lopes, Maria do Carmo Debur, Irina Nastassja Riediger, Marilda Siqueira on behalf of the Fiocruz COVID-19 Genomic Surveillance Network |
| EPI_ISL_1181585 | Laboratorio Central de Saude Publica do Estado do Espitito Santo (LACEN-ES) | Laboratory of Respiratory Viruses and Measles, Oswaldo Cruz Institute, FIOCRUZ | Paola Resende, Luciana Appolinario, Fernando Motta, Anna Carolina Paixao, Ana Carolina Mendonca, Alice Sampaio Rocha, Renata Serrano Lopes, Rodrigo Ribeiro Rodrigues, Marilda Siqueira on behalf of the Fiocruz COVID-19 Genomic Surveillance Network |
| EPI_ISL_1181586 | Oswaldo Cruz Foundation, FIOCRUZ - Ceara (Fiocruz-CE) | Laboratory of Respiratory Viruses and Measles, Oswaldo Cruz Institute, FIOCRUZ | Paola Resende, Luciana Appolinario, Fernando Motta, Anna Carolina Paixao, Ana Carolina Mendonca, Alice Sampaio Rocha, Renata Serrano Lopes, Thais de Oliveira Costa, Joaquim César do Nascimento Sousa Júnior, Fabio Miyajima, Marilda Siqueira on behalf of the Fiocruz COVID-19 Genomic Surveillance Network |
| EPI_ISL_1181587, EPI_ISL_1181589 | Laboratory of Respiratory Viruses and Measles, Oswaldo Cruz Institute, FIOCRUZ | Laboratory of Respiratory Viruses and Measles, Oswaldo Cruz Institute, FIOCRUZ | Paola Resende, Luciana Appolinario, Fernando Motta, Anna Carolina Paixao, Ana Carolina Mendonca, Alice Sampaio Rocha, Renata Serrano Lopes, Marilda Siqueira on behalf of the Fiocruz COVID-19 Genomic Surveillance Network |
| EPI_ISL_1181590 | Laboratorio Central de Saude Publica do Estado de Sergipe (LACEN-SE) | Laboratory of Respiratory Viruses and Measles, Oswaldo Cruz Institute, FIOCRUZ | Paola Resende, Luciana Appolinario, Fernando Motta, Anna Carolina Paixao, Ana Carolina Mendonca, Alice Sampaio Rocha, Renata Serrano Lopes, Cliomar Alves dos Santos, Marilda Siqueira on behalf of the Fiocruz COVID-19 Genomic Surveillance Network |
| EPI_ISL_1181591, EPI_ISL_1181593, EPI_ISL_1181596, EPI_ISL_1181598 | Oswaldo Cruz Foundation, FIOCRUZ - Ceara (Fiocruz-CE) | Laboratory of Respiratory Viruses and Measles, Oswaldo Cruz Institute, FIOCRUZ | Paola Resende, Luciana Appolinario, Fernando Motta, Anna Carolina Paixao, Ana Carolina Mendonca, Alice Sampaio Rocha, Renata Serrano Lopes, Thais de Oliveira Costa, Joaquim César do Nascimento Sousa Júnior, Fabio Miyajima, Marilda Siqueira on behalf of the Fiocruz COVID-19 Genomic Surveillance Network |
| EPI_ISL_1182541 | Fundação Ezequiel Dias (FUNED) | Coordenação Geral de Laboratórios de Saúde Pública (CGLAB/DAEVS/SVS/MS) | Vagner Fonseca, et al. |
| EPI_ISL_1182542 | Laboratório Central do Estado do Paraná | Coordenação Geral de Laboratórios de Saúde Pública (CGLAB/DAEVS/SVS/MS) | Vagner Fonseca, et al. |
| EPI_ISL_1182543, EPI_ISL_1182544, EPI_ISL_1182545 | Fundação Ezequiel Dias (FUNED) | Coordenação Geral de Laboratórios de Saúde Pública (CGLAB/DAEVS/SVS/MS) | Vagner Fonseca, et al. |
| EPI_ISL_1182546 | Laboratório Central do Estado do Paraná | Coordenação Geral de Laboratórios de Saúde Pública (CGLAB/DAEVS/SVS/MS) | Vagner Fonseca, et al. |
| EPI_ISL_1182547 | Laboratório Central de Saúde Pública do Rio Grande do Sul | Coordenação Geral de Laboratórios de Saúde Pública (CGLAB/DAEVS/SVS/MS) | Vagner Fonseca, et al. |
| EPI_ISL_1182548, EPI_ISL_1182549, EPI_ISL_1182550, EPI_ISL_1182551 | Fundação Ezequiel Dias (FUNED) | Coordenação Geral de Laboratórios de Saúde Pública (CGLAB/DAEVS/SVS/MS) | Vagner Fonseca, et al. |
| EPI_ISL_1182552, EPI_ISL_1182553, EPI_ISL_1182554 | Laboratório Central do Estado do Rio de Janeiro | Coordenação Geral de Laboratórios de Saúde Pública (CGLAB/DAEVS/SVS/MS) | Vagner Fonseca, et al. |

| | | | |
|---|---|---|---|
| EPI_ISL_1182555 | Fundação Ezequiel Dias (FUNED) | Coordenação Geral de Laboratórios de Saúde Pública (CGLAB/DAEVS/SVS/MS) | Vagner Fonseca, et al. |
| EPI_ISL_1182556, EPI_ISL_1182557, EPI_ISL_1182558 | Laboratório Central do Estado do Rio de Janeiro | Coordenação Geral de Laboratórios de Saúde Pública (CGLAB/DAEVS/SVS/MS) | Vagner Fonseca, et al. |
| EPI_ISL_1182559, EPI_ISL_1182560, EPI_ISL_1182561 | Fundação Ezequiel Dias (FUNED) | Coordenação Geral de Laboratórios de Saúde Pública (CGLAB/DAEVS/SVS/MS) | Vagner Fonseca, et al. |
| EPI_ISL_1182562, EPI_ISL_1182563 | Laboratório Central do Estado do Paraná | Coordenação Geral de Laboratórios de Saúde Pública (CGLAB/DAEVS/SVS/MS) | Vagner Fonseca, et al. |
| EPI_ISL_1182564 | Fundação Ezequiel Dias (FUNED) | Coordenação Geral de Laboratórios de Saúde Pública (CGLAB/DAEVS/SVS/MS) | Vagner Fonseca, et al. |
| EPI_ISL_1182565 | Laboratório Central do Estado do Paraná | Coordenação Geral de Laboratórios de Saúde Pública (CGLAB/DAEVS/SVS/MS) | Vagner Fonseca, et al. |
| EPI_ISL_1182566 | Fundação Ezequiel Dias (FUNED) | Coordenação Geral de Laboratórios de Saúde Pública (CGLAB/DAEVS/SVS/MS) | Vagner Fonseca, et al. |
| EPI_ISL_1182567, EPI_ISL_1182568 | Laboratório Central do Estado do Paraná | Coordenação Geral de Laboratórios de Saúde Pública (CGLAB/DAEVS/SVS/MS) | Vagner Fonseca, et al. |
| EPI_ISL_1182569, EPI_ISL_1182570 | Fundação Ezequiel Dias (FUNED) | Coordenação Geral de Laboratórios de Saúde Pública (CGLAB/DAEVS/SVS/MS) | Vagner Fonseca, et al. |
| EPI_ISL_1182571, EPI_ISL_1182572 | Laboratório Central do Estado do Paraná | Coordenação Geral de Laboratórios de Saúde Pública (CGLAB/DAEVS/SVS/MS) | Vagner Fonseca, et al. |
| EPI_ISL_1182573, EPI_ISL_1182574 | Fundação Ezequiel Dias (FUNED) | Coordenação Geral de Laboratórios de Saúde Pública (CGLAB/DAEVS/SVS/MS) | Vagner Fonseca, et al. |
| EPI_ISL_1182575, EPI_ISL_1182576 | Laboratório Central do Estado do Paraná | Coordenação Geral de Laboratórios de Saúde Pública (CGLAB/DAEVS/SVS/MS) | Vagner Fonseca, et al. |
| EPI_ISL_1182577, EPI_ISL_1182578, EPI_ISL_1182579 | Fundação Ezequiel Dias (FUNED) | Coordenação Geral de Laboratórios de Saúde Pública (CGLAB/DAEVS/SVS/MS) | Vagner Fonseca, et al. |
| EPI_ISL_1182580, EPI_ISL_1182581, EPI_ISL_1182582, EPI_ISL_1182583, EPI_ISL_1182584 | Laboratório Central do Estado do Paraná | Coordenação Geral de Laboratórios de Saúde Pública (CGLAB/DAEVS/SVS/MS) | Vagner Fonseca, et al. |
| EPI_ISL_1182585, EPI_ISL_1182586, EPI_ISL_1182587 | Fundação Ezequiel Dias (FUNED) | Coordenação Geral de Laboratórios de Saúde Pública (CGLAB/DAEVS/SVS/MS) | Vagner Fonseca, et al. |
| EPI_ISL_1182588, EPI_ISL_1182589 | Laboratório Central do Estado do Paraná | Coordenação Geral de Laboratórios de Saúde Pública (CGLAB/DAEVS/SVS/MS) | Vagner Fonseca, et al. |
| EPI_ISL_1182590, EPI_ISL_1182591 | Fundação Ezequiel Dias (FUNED) | Coordenação Geral de Laboratórios de Saúde Pública (CGLAB/DAEVS/SVS/MS) | Vagner Fonseca, et al. |
| EPI_ISL_1182592 | Laboratório Central do Estado do Paraná | Coordenação Geral de Laboratórios de Saúde Pública (CGLAB/DAEVS/SVS/MS) | Vagner Fonseca, et al. |
| EPI_ISL_1182593 | Fundação Ezequiel Dias (FUNED) | Coordenação Geral de Laboratórios de Saúde Pública (CGLAB/DAEVS/SVS/MS) | Vagner Fonseca, et al. |
| EPI_ISL_1182594, EPI_ISL_1182595, EPI_ISL_1182596 | Laboratório Central do Estado do Paraná | Coordenação Geral de Laboratórios de Saúde Pública (CGLAB/DAEVS/SVS/MS) | Vagner Fonseca, et al. |
| EPI_ISL_1182597 | Laboratório Central de Saúde Pública do Rio Grande do Sul | Coordenação Geral de Laboratórios de Saúde Pública (CGLAB/DAEVS/SVS/MS) | Vagner Fonseca, et al. |
| EPI_ISL_1182598 | Fundação Ezequiel Dias (FUNED) | Coordenação Geral de Laboratórios de Saúde Pública (CGLAB/DAEVS/SVS/MS) | Vagner Fonseca, et al. |
| EPI_ISL_1182599 | Laboratório Central de Saúde Pública do Rio Grande do Sul | Coordenação Geral de Laboratórios de Saúde Pública (CGLAB/DAEVS/SVS/MS) | Vagner Fonseca, et al. |
| EPI_ISL_1182600, EPI_ISL_1182601, EPI_ISL_1182602 | Fundação Ezequiel Dias (FUNED) | Coordenação Geral de Laboratórios de Saúde Pública (CGLAB/DAEVS/SVS/MS) | Vagner Fonseca, et al. |
| EPI_ISL_1182603, EPI_ISL_1182604 | Laboratório Central do Estado do Paraná | Coordenação Geral de Laboratórios de Saúde Pública (CGLAB/DAEVS/SVS/MS) | Vagner Fonseca, et al. |
| EPI_ISL_1182605, EPI_ISL_1182606 | Laboratório Central de Saúde Pública do Rio Grande do Sul | Coordenação Geral de Laboratórios de Saúde Pública (CGLAB/DAEVS/SVS/MS) | Vagner Fonseca, et al. |
| EPI_ISL_1182607 | Fundação Ezequiel Dias (FUNED) | Coordenação Geral de Laboratórios de Saúde Pública (CGLAB/DAEVS/SVS/MS) | Vagner Fonseca, et al. |
| EPI_ISL_1182608 | Laboratório Central de Saúde Pública do Rio Grande do Sul | Coordenação Geral de Laboratórios de Saúde Pública (CGLAB/DAEVS/SVS/MS) | Vagner Fonseca, et al. |
| EPI_ISL_1182609 | Fundação Ezequiel Dias (FUNED) | Coordenação Geral de Laboratórios de Saúde Pública (CGLAB/DAEVS/SVS/MS) | Vagner Fonseca, et al. |
| EPI_ISL_1182610 | Laboratório Central de Saúde Pública do Rio Grande do Sul | Coordenação Geral de Laboratórios de Saúde Pública (CGLAB/DAEVS/SVS/MS) | Vagner Fonseca, et al. |
| EPI_ISL_1182611, EPI_ISL_1182612 | Fundação Ezequiel Dias (FUNED) | Coordenação Geral de Laboratórios de Saúde Pública (CGLAB/DAEVS/SVS/MS) | Vagner Fonseca, et al. |
| EPI_ISL_1182613, EPI_ISL_1182614 | Laboratório Central do Estado do Paraná | Coordenação Geral de Laboratórios de Saúde Pública (CGLAB/DAEVS/SVS/MS) | Vagner Fonseca, et al. |
| EPI_ISL_1182615 | Fundação Ezequiel Dias (FUNED) | Coordenação Geral de Laboratórios de Saúde Pública (CGLAB/DAEVS/SVS/MS) | Vagner Fonseca, et al. |
| EPI_ISL_1182616 | Laboratório Central de Saúde Pública do Rio Grande do Sul | Coordenação Geral de Laboratórios de Saúde Pública (CGLAB/DAEVS/SVS/MS) | Vagner Fonseca, et al. |
| EPI_ISL_1182617 | Laboratório Central do Estado do Paraná | Coordenação Geral de Laboratórios de Saúde Pública (CGLAB/DAEVS/SVS/MS) | Vagner Fonseca, et al. |

| | | | |
|---|---|---|---|
| EPI_ISL_1182618 | Fundação Ezequiel Dias (FUNED) | Coordenação Geral de Laboratórios de Saúde Pública (CGLAB/DAEVS/SVS/MS) | Vagner Fonseca, et al. |
| EPI_ISL_1182619, EPI_ISL_1182620, EPI_ISL_1182621 | Laboratório Central de Saúde Pública do Rio Grande do Sul | Coordenação Geral de Laboratórios de Saúde Pública (CGLAB/DAEVS/SVS/MS) | Vagner Fonseca, et al. |
| EPI_ISL_1182622 | Laboratório Central do Estado do Paraná | Coordenação Geral de Laboratórios de Saúde Pública (CGLAB/DAEVS/SVS/MS) | Vagner Fonseca, et al. |
| EPI_ISL_1182623 | Laboratório Central de Saúde Pública do Rio Grande do Sul | Coordenação Geral de Laboratórios de Saúde Pública (CGLAB/DAEVS/SVS/MS) | Vagner Fonseca, et al. |
| EPI_ISL_1182624 | Laboratório Central do Estado do Paraná | Coordenação Geral de Laboratórios de Saúde Pública (CGLAB/DAEVS/SVS/MS) | Vagner Fonseca, et al. |
| EPI_ISL_1182625 | Fundação Ezequiel Dias (FUNED) | Coordenação Geral de Laboratórios de Saúde Pública (CGLAB/DAEVS/SVS/MS) | Vagner Fonseca, et al. |
| EPI_ISL_1195272 | FUNDACAO HOSPITALAR DE SAPUCAIA DO SUL | Epiclin | Fernando Hayashi Sant'Anna, Ana Paula Muterle, Janira Prichula, Juliana Comerlato, Carolina Comerlato, Eliana Márcia Da Ros Wendland |
| EPI_ISL_1195273 | FUNDACAO DE SAUDE PUBLICA SAO CAMILO DE ESTEIO | Epiclin | Fernando Hayashi Sant'Anna, Ana Paula Muterle, Janira Prichula, Juliana Comerlato, Carolina Comerlato, Eliana Márcia Da Ros Wendland |
| EPI_ISL_1195274 | SECRETARIA MUNICIPAL DE SAUDE DE TRES COROAS | Epiclin | Fernando Hayashi Sant'Anna, Ana Paula Muterle, Janira Prichula, Juliana Comerlato, Carolina Comerlato, Eliana Márcia Da Ros Wendland |
| EPI_ISL_1195275, EPI_ISL_1195276 | CENTRO DE REFERENCIA EM SINDROMES GRIPAIS | Epiclin | Fernando Hayashi Sant'Anna, Ana Paula Muterle, Janira Prichula, Juliana Comerlato, Carolina Comerlato, Eliana Márcia Da Ros Wendland |
| EPI_ISL_1195277 | Unidade de Atendimento DST AIDS TB e Han | Epiclin | Fernando Hayashi Sant'Anna, Ana Paula Muterle, Janira Prichula, Juliana Comerlato, Carolina Comerlato, Eliana Márcia Da Ros Wendland |
| EPI_ISL_1195278 | DIRETORIA DE VIGILANCIA EM SAUDE | Epiclin | Fernando Hayashi Sant'Anna, Ana Paula Muterle, Janira Prichula, Juliana Comerlato, Carolina Comerlato, Eliana Márcia Da Ros Wendland |
| EPI_ISL_1195279 | CENTRO DE REFERENCIA EM SINDROMES GRIPAIS | Epiclin | Fernando Hayashi Sant'Anna, Ana Paula Muterle, Janira Prichula, Juliana Comerlato, Carolina Comerlato, Eliana Márcia Da Ros Wendland |
| EPI_ISL_1195280 | SECRETARIA MUNICIPAL DE SAUDE DE ARARICA | Epiclin | Fernando Hayashi Sant'Anna, Ana Paula Muterle, Janira Prichula, Juliana Comerlato, Carolina Comerlato, Eliana Márcia Da Ros Wendland |
| EPI_ISL_1195281 | FUNDACAO DE SAUDE PUBLICA SAO CAMILO DE ESTEIO | Epiclin | Fernando Hayashi Sant'Anna, Ana Paula Muterle, Janira Prichula, Juliana Comerlato, Carolina Comerlato, Eliana Márcia Da Ros Wendland |
| EPI_ISL_1195282 | UNIDADE SANITARIA DE IGREJINHA | Epiclin | Fernando Hayashi Sant'Anna, Ana Paula Muterle, Janira Prichula, Juliana Comerlato, Carolina Comerlato, Eliana Márcia Da Ros Wendland |
| EPI_ISL_1195283 | SECRETARIA MUNICIPAL DE SAUDE DE TRES COROAS | Epiclin | Fernando Hayashi Sant'Anna, Ana Paula Muterle, Janira Prichula, Juliana Comerlato, Carolina Comerlato, Eliana Márcia Da Ros Wendland |
| EPI_ISL_1195284 | Centro de Especialidades Triunfo | Epiclin | Fernando Hayashi Sant'Anna, Ana Paula Muterle, Janira Prichula, Juliana Comerlato, Carolina Comerlato, Eliana Márcia Da Ros Wendland |
| EPI_ISL_1195285, EPI_ISL_1195286 | SECRETARIA MUNICIPAL DE SAUDE DE TRES COROAS | Epiclin | Fernando Hayashi Sant'Anna, Ana Paula Muterle, Janira Prichula, Juliana Comerlato, Carolina Comerlato, Eliana Márcia Da Ros Wendland |
| EPI_ISL_1195287 | SECRETARIA MUNICIPAL DE SAUDE DE SAO LEOPOLDO | Epiclin | Fernando Hayashi Sant'Anna, Ana Paula Muterle, Janira Prichula, Juliana Comerlato, Carolina Comerlato, Eliana Márcia Da Ros Wendland |
| EPI_ISL_1195288 | DIRETORIA DE VIGILANCIA EM SAUDE | Epiclin | Fernando Hayashi Sant'Anna, Ana Paula Muterle, Janira Prichula, Juliana Comerlato, Carolina Comerlato, Eliana Márcia Da Ros Wendland |
| EPI_ISL_1195289 | SECRETARIA MUNICIPAL DE SAUDE DE SAO LEOPOLDO | Epiclin | Fernando Hayashi Sant'Anna, Ana Paula Muterle, Janira Prichula, Juliana Comerlato, Carolina Comerlato, Eliana Márcia Da Ros Wendland |
| EPI_ISL_1195290 | SECRETARIA MUNICIPAL DE SAUDE DE TRES COROAS | Epiclin | Fernando Hayashi Sant'Anna, Ana Paula Muterle, Janira Prichula, Juliana Comerlato, Carolina Comerlato, Eliana Márcia Da Ros Wendland |
| EPI_ISL_1195291 | DIRETORIA DE VIGILANCIA EM SAUDE | Epiclin | Fernando Hayashi Sant'Anna, Ana Paula Muterle, Janira Prichula, Juliana Comerlato, Carolina Comerlato, Eliana Márcia Da Ros Wendland |
| EPI_ISL_1195292 | SECRETARIA MUNICIPAL DE SAUDE DE TRES COROAS | Epiclin | Fernando Hayashi Sant'Anna, Ana Paula Muterle, Janira Prichula, Juliana Comerlato, Carolina Comerlato, Eliana Márcia Da Ros Wendland |
| EPI_ISL_1195293 | SECRETARIA MUNICIPAL DE SAUDE DE TAQUARA | Epiclin | Fernando Hayashi Sant'Anna, Ana Paula Muterle, Janira Prichula, Juliana Comerlato, Carolina Comerlato, Eliana Márcia Da Ros Wendland |
| EPI_ISL_1196282, EPI_ISL_1196283, EPI_ISL_1196284 | LACEN do Mato Grosso do Sul | Instituto Adolfo Lutz, Interdiciplinary Procedures Center, Strategic Laboratory | Claudio Tavares Sacchi, Claudia Regina Gonçalves, Erica Valessa Ramos Gomes, Karoline Rodrigues Campos, Caio Vinicius Dias Lopes |
| EPI_ISL_1196285, EPI_ISL_1196286, EPI_ISL_1196287, EPI_ISL_1196288, EPI_ISL_1196289, EPI_ISL_1196290, EPI_ISL_1196291, EPI_ISL_1196292, EPI_ISL_1196293, EPI_ISL_1196294 | LACEN do Distrito Federal | Instituto Adolfo Lutz, Interdiciplinary Procedures Center, Strategic Laboratory | Claudio Tavares Sacchi, Claudia Regina Gonçalves, Erica Valessa Ramos Gomes, Karoline Rodrigues Campos, Caio Vinicius Dias Lopes |
| EPI_ISL_1196295 | UBS Otacilio Firmino Lopes | Instituto Adolfo Lutz, Interdiciplinary Procedures Center, Strategic Laboratory | Claudio Tavares Sacchi, Claudia Regina Gonçalves, Erica Valessa Ramos Gomes, Karoline Rodrigues Campos, Caio Vinicius Dias Lopes |
| EPI_ISL_1196296 | Centro de Saude II Dr. Jose Paione Mococa | Instituto Adolfo Lutz, Interdiciplinary Procedures Center, Strategic Laboratory | Claudio Tavares Sacchi, Claudia Regina Gonçalves, Erica Valessa Ramos Gomes, Karoline Rodrigues Campos, Caio Vinicius Dias Lopes |
| EPI_ISL_1196297, EPI_ISL_1196298, EPI_ISL_1196299, EPI_ISL_1196300, EPI_ISL_1196301, EPI_ISL_1196302 | IAL Regional de Marilia | Instituto Adolfo Lutz, Interdiciplinary Procedures Center, Strategic Laboratory | Claudio Tavares Sacchi, Claudia Regina Gonçalves, Erica Valessa Ramos Gomes, Karoline Rodrigues Campos, Caio Vinicius Dias Lopes |
| EPI_ISL_1201884, EPI_ISL_1201885, EPI_ISL_1201886, EPI_ISL_1201887, EPI_ISL_1201888 | Aeroporto Internacional de Guarulhos | Instituto Adolfo Lutz, Interdiciplinary Procedures Center, Strategic Laboratory | Claudio Tavares Sacchi, Claudia Regina Gonçalves, Erica Valessa Ramos Gomes, Karoline Rodrigues Campos, Caio Vinicius Dias Lopes |
| EPI_ISL_1201889, EPI_ISL_1201890, EPI_ISL_1201891, EPI_ISL_1201892 | IAL Regional de Marilia | Instituto Adolfo Lutz, Interdiciplinary Procedures Center, Strategic Laboratory | Claudio Tavares Sacchi, Claudia Regina Gonçalves, Erica Valessa Ramos Gomes, Karoline Rodrigues Campos, Caio Vinicius Dias Lopes |
| EPI_ISL_1201893 | IAL Regional de Sorocaba | Instituto Adolfo Lutz, Interdiciplinary Procedures Center, Strategic Laboratory | Claudio Tavares Sacchi, Claudia Regina Gonçalves, Erica Valessa Ramos Gomes, Karoline Rodrigues Campos, Caio Vinicius Dias Lopes |
| EPI_ISL_1201894, EPI_ISL_1201895, EPI_ISL_1201896 | Diagnosticos da America - DASA | Instituto Adolfo Lutz, Interdiciplinary Procedures Center, Strategic Laboratory | Claudio Tavares Sacchi, Claudia Regina Gonçalves, Erica Valessa Ramos Gomes, Karoline Rodrigues Campos, Caio Vinicius Dias Lopes |
| EPI_ISL_1213148, EPI_ISL_1213149, EPI_ISL_1213151 | Laboratório HLA/UERJ | Bioinformatics Laboratory / LNCC | Alessandra P Lamarca, Luiz G P de Almeida, Ronaldo da Silva Francisco Jr, Lucymara Fassarella Agnez Lima, Kátia Castanho Scortecci, Vinícius Pietta Perez, Otavio J. Brustolini, Eduardo Sérgio Soares Sousa, Danielle Angst Secco, Angela Maria Guimarães Santos, George Rego Albuquerque, Ana Paula Melo Mariano, Bianca Mendes Maciel, Alexandra L Gerber, Ana Paula de C Guimarães, Paulo Ricardo Nascimento, Francisco Paulo Freire Neto, Sandra Rocha Gadelha, Luís Cristóvão Porto, Eloiza Helena Campana, Selma Maria Bezerra Jeronimo, Ana Tereza R Vasconcelos |
| EPI_ISL_1213153 | LBM/UFPB | Bioinformatics Laboratory / LNCC | Alessandra P Lamarca, Luiz G P de Almeida, Ronaldo da Silva Francisco Jr, Lucymara Fassarella Agnez Lima, Kátia Castanho Scortecci, Vinícius Pietta Perez, Otavio J. Brustolini, Eduardo Sérgio Soares Sousa, Danielle Angst Secco, Angela Maria Guimarães Santos, George Rego Albuquerque, Ana Paula Melo Mariano, Bianca Mendes Maciel, Alexandra L Gerber, Ana Paula de C Guimarães, Paulo Ricardo Nascimento, Francisco Paulo Freire Neto, Sandra Rocha Gadelha, Luís Cristóvão Porto, Eloiza Helena Campana, Selma Maria Bezerra Jeronimo, Ana Tereza R Vasconcelos |
| EPI_ISL_1213155 | Laboratório HLA/UERJ | Bioinformatics Laboratory / LNCC | Alessandra P Lamarca, Luiz G P de Almeida, Ronaldo da Silva Francisco Jr, Lucymara Fassarella Agnez Lima, Kátia Castanho Scortecci, Vinícius Pietta Perez, Otavio J. Brustolini, Eduardo Sérgio Soares Sousa, Danielle Angst Secco, Angela Maria Guimarães Santos, George Rego Albuquerque, Ana Paula Melo Mariano, Bianca Mendes Maciel, Alexandra L Gerber, Ana Paula de C Guimarães, Paulo Ricardo Nascimento, Francisco Paulo Freire Neto, Sandra Rocha Gadelha, Luís Cristóvão Porto, Eloiza Helena Campana, Selma Maria Bezerra Jeronimo, Ana Tereza R Vasconcelos |
| EPI_ISL_1213156 | LBM/UFPB | Bioinformatics Laboratory / LNCC | Alessandra P Lamarca, Luiz G P de Almeida, Ronaldo da Silva Francisco Jr, Lucymara Fassarella Agnez Lima, Kátia Castanho Scortecci, Vinícius Pietta |

| | | | |
|---|---|---|---|
| | | | Perez, Otavio J. Brustolini, Eduardo Sérgio Soares Sousa, Danielle Angst Secco, Angela Maria Guimarães Santos, George Rego Albuquerque, Ana Paula Melo Mariano, Bianca Mendes Maciel, Alexandra L Gerber, Ana Paula de C Guimarães, Paulo Ricardo Nascimento, Francisco Paulo Freire Neto, Sandra Rocha Gadelha, Luís Cristóvão Porto, Eloiza Helena Campana, Selma Maria Bezerra Jeronimo, Ana Tereza R Vasconcelos |
| EPI_ISL_1213158 | Laboratório HLA/UERJ | Bioinformatics Laboratory / LNCC | Alessandra P Lamarca, Luiz G P de Almeida, Ronaldo da Silva Francisco Jr, Lucymara Fassarella Agnez Lima, Kátia Castanho Scortecci, Vinícius Pietta Perez, Otavio J. Brustolini, Eduardo Sérgio Soares Sousa, Danielle Angst Secco, Angela Maria Guimarães Santos, George Rego Albuquerque, Ana Paula Melo Mariano, Bianca Mendes Maciel, Alexandra L Gerber, Ana Paula de C Guimarães, Paulo Ricardo Nascimento, Francisco Paulo Freire Neto, Sandra Rocha Gadelha, Luís Cristóvão Porto, Eloiza Helena Campana, Selma Maria Bezerra Jeronimo, Ana Tereza R Vasconcelos |
| EPI_ISL_1213160, EPI_ISL_1213161 | LBM/UFPB | Bioinformatics Laboratory / LNCC | Alessandra P Lamarca, Luiz G P de Almeida, Ronaldo da Silva Francisco Jr, Lucymara Fassarella Agnez Lima, Kátia Castanho Scortecci, Vinícius Pietta Perez, Otavio J. Brustolini, Eduardo Sérgio Soares Sousa, Danielle Angst Secco, Angela Maria Guimarães Santos, George Rego Albuquerque, Ana Paula Melo Mariano, Bianca Mendes Maciel, Alexandra L Gerber, Ana Paula de C Guimarães, Paulo Ricardo Nascimento, Francisco Paulo Freire Neto, Sandra Rocha Gadelha, Luís Cristóvão Porto, Eloiza Helena Campana, Selma Maria Bezerra Jeronimo, Ana Tereza R Vasconcelos |
| EPI_ISL_1213163 | Laboratório HLA/UERJ | Bioinformatics Laboratory / LNCC | Alessandra P Lamarca, Luiz G P de Almeida, Ronaldo da Silva Francisco Jr, Lucymara Fassarella Agnez Lima, Kátia Castanho Scortecci, Vinícius Pietta Perez, Otavio J. Brustolini, Eduardo Sérgio Soares Sousa, Danielle Angst Secco, Angela Maria Guimarães Santos, George Rego Albuquerque, Ana Paula Melo Mariano, Bianca Mendes Maciel, Alexandra L Gerber, Ana Paula de C Guimarães, Paulo Ricardo Nascimento, Francisco Paulo Freire Neto, Sandra Rocha Gadelha, Luís Cristóvão Porto, Eloiza Helena Campana, Selma Maria Bezerra Jeronimo, Ana Tereza R Vasconcelos |
| EPI_ISL_1213165, EPI_ISL_1213166 | LBM/UFPB | Bioinformatics Laboratory / LNCC | Alessandra P Lamarca, Luiz G P de Almeida, Ronaldo da Silva Francisco Jr, Lucymara Fassarella Agnez Lima, Kátia Castanho Scortecci, Vinícius Pietta Perez, Otavio J. Brustolini, Eduardo Sérgio Soares Sousa, Danielle Angst Secco, Angela Maria Guimarães Santos, George Rego Albuquerque, Ana Paula Melo Mariano, Bianca Mendes Maciel, Alexandra L Gerber, Ana Paula de C Guimarães, Paulo Ricardo Nascimento, Francisco Paulo Freire Neto, Sandra Rocha Gadelha, Luís Cristóvão Porto, Eloiza Helena Campana, Selma Maria Bezerra Jeronimo, Ana Tereza R Vasconcelos |
| EPI_ISL_1213168, EPI_ISL_1213170 | LAFEM/UESC | Bioinformatics Laboratory / LNCC | Alessandra P Lamarca, Luiz G P de Almeida, Ronaldo da Silva Francisco Jr, Lucymara Fassarella Agnez Lima, Kátia Castanho Scortecci, Vinícius Pietta Perez, Otavio J. Brustolini, Eduardo Sérgio Soares Sousa, Danielle Angst Secco, Angela Maria Guimarães Santos, George Rego Albuquerque, Ana Paula Melo Mariano, Bianca Mendes Maciel, Alexandra L Gerber, Ana Paula de C Guimarães, Paulo Ricardo Nascimento, Francisco Paulo Freire Neto, Sandra Rocha Gadelha, Luís Cristóvão Porto, Eloiza Helena Campana, Selma Maria Bezerra Jeronimo, Ana Tereza R Vasconcelos |
| EPI_ISL_1213171 | Laboratório HLA/UERJ | Bioinformatics Laboratory / LNCC | Alessandra P Lamarca, Luiz G P de Almeida, Ronaldo da Silva Francisco Jr, Lucymara Fassarella Agnez Lima, Kátia Castanho Scortecci, Vinícius Pietta Perez, Otavio J. Brustolini, Eduardo Sérgio Soares Sousa, Danielle Angst Secco, Angela Maria Guimarães Santos, George Rego Albuquerque, Ana Paula Melo Mariano, Bianca Mendes Maciel, Alexandra L Gerber, Ana Paula de C Guimarães, Paulo Ricardo Nascimento, Francisco Paulo Freire Neto, Sandra Rocha Gadelha, Luís Cristóvão Porto, Eloiza Helena Campana, Selma Maria Bezerra Jeronimo, Ana Tereza R Vasconcelos |
| EPI_ISL_1213173, EPI_ISL_1213175, EPI_ISL_1213177, EPI_ISL_1213178, EPI_ISL_1213180, EPI_ISL_1213182, EPI_ISL_1213183, EPI_ISL_1213185, EPI_ISL_1213187 | IMT-UFRN/RN | Bioinformatics Laboratory / LNCC | Alessandra P Lamarca, Luiz G P de Almeida, Ronaldo da Silva Francisco Jr, Lucymara Fassarella Agnez Lima, Kátia Castanho Scortecci, Vinícius Pietta Perez, Otavio J. Brustolini, Eduardo Sérgio Soares Sousa, Danielle Angst Secco, Angela Maria Guimarães Santos, George Rego Albuquerque, Ana Paula Melo Mariano, Bianca Mendes Maciel, Alexandra L Gerber, Ana Paula de C Guimarães, Paulo Ricardo Nascimento, Francisco Paulo Freire Neto, Sandra Rocha Gadelha, Luís Cristóvão Porto, Eloiza Helena Campana, Selma Maria Bezerra Jeronimo, Ana Tereza R Vasconcelos |
| EPI_ISL_1213189, EPI_ISL_1213190 | LBM/UFPB | Bioinformatics Laboratory / LNCC | Alessandra P Lamarca, Luiz G P de Almeida, Ronaldo da Silva Francisco Jr, Lucymara Fassarella Agnez Lima, Kátia Castanho Scortecci, Vinícius Pietta Perez, Otavio J. Brustolini, Eduardo Sérgio Soares Sousa, Danielle Angst Secco, Angela Maria Guimarães Santos, George Rego Albuquerque, Ana Paula Melo Mariano, Bianca Mendes Maciel, Alexandra L Gerber, Ana Paula de C Guimarães, Paulo Ricardo Nascimento, Francisco Paulo Freire Neto, Sandra Rocha Gadelha, Luís Cristóvão Porto, Eloiza Helena Campana, Selma Maria Bezerra Jeronimo, Ana Tereza R Vasconcelos |
| EPI_ISL_1213192, EPI_ISL_1213194, EPI_ISL_1213196, EPI_ISL_1213197, EPI_ISL_1213199 | IMT-UFRN/RN | Bioinformatics Laboratory / LNCC | Alessandra P Lamarca, Luiz G P de Almeida, Ronaldo da Silva Francisco Jr, Lucymara Fassarella Agnez Lima, Kátia Castanho Scortecci, Vinícius Pietta Perez, Otavio J. Brustolini, Eduardo Sérgio Soares Sousa, Danielle Angst Secco, Angela Maria Guimarães Santos, George Rego Albuquerque, Ana Paula Melo Mariano, Bianca Mendes Maciel, Alexandra L Gerber, Ana Paula de C Guimarães, Paulo Ricardo Nascimento, Francisco Paulo Freire Neto, Sandra Rocha Gadelha, Luís Cristóvão Porto, Eloiza Helena Campana, Selma Maria Bezerra Jeronimo, Ana Tereza R Vasconcelos |
| EPI_ISL_1213201, EPI_ISL_1213202, EPI_ISL_1213204, EPI_ISL_1213206, EPI_ISL_1213207 | LBM/UFPB | Bioinformatics Laboratory / LNCC | Alessandra P Lamarca, Luiz G P de Almeida, Ronaldo da Silva Francisco Jr, Lucymara Fassarella Agnez Lima, Kátia Castanho Scortecci, Vinícius Pietta Perez, Otavio J. Brustolini, Eduardo Sérgio Soares Sousa, Danielle Angst Secco, Angela Maria Guimarães Santos, George Rego Albuquerque, Ana Paula Melo Mariano, Bianca Mendes Maciel, Alexandra L Gerber, Ana Paula de C Guimarães, Paulo Ricardo Nascimento, Francisco Paulo Freire Neto, Sandra Rocha Gadelha, Luís Cristóvão Porto, Eloiza Helena Campana, Selma Maria Bezerra Jeronimo, Ana Tereza R Vasconcelos |
| EPI_ISL_1213209 | IMT-UFRN/RN | Bioinformatics Laboratory / LNCC | Alessandra P Lamarca, Luiz G P de Almeida, Ronaldo da Silva Francisco Jr, Lucymara Fassarella Agnez Lima, Kátia Castanho Scortecci, Vinícius Pietta Perez, Otavio J. Brustolini, Eduardo Sérgio Soares Sousa, Danielle Angst Secco, Angela Maria Guimarães Santos, George Rego Albuquerque, Ana Paula Melo Mariano, Bianca Mendes Maciel, Alexandra L Gerber, Ana Paula de C Guimarães, Paulo Ricardo Nascimento, Francisco Paulo Freire Neto, Sandra Rocha Gadelha, Luís Cristóvão Porto, Eloiza Helena Campana, Selma Maria Bezerra Jeronimo, Ana Tereza R Vasconcelos |
| EPI_ISL_1213211 | LAFEM/UESC | Bioinformatics Laboratory / LNCC | Alessandra P Lamarca, Luiz G P de Almeida, Ronaldo da Silva Francisco Jr, Lucymara Fassarella Agnez Lima, Kátia Castanho Scortecci, Vinícius Pietta Perez, Otavio J. Brustolini, Eduardo Sérgio Soares Sousa, Danielle Angst Secco, Angela Maria Guimarães Santos, George Rego Albuquerque, Ana Paula Melo Mariano, Bianca Mendes Maciel, Alexandra L Gerber, Ana Paula de C Guimarães, Paulo Ricardo Nascimento, Francisco Paulo Freire Neto, Sandra Rocha Gadelha, Luís Cristóvão Porto, Eloiza Helena Campana, Selma Maria Bezerra Jeronimo, Ana Tereza R Vasconcelos |
| EPI_ISL_1213213, EPI_ISL_1213215 | LBM/UFPB | Bioinformatics Laboratory / LNCC | Alessandra P Lamarca, Luiz G P de Almeida, Ronaldo da Silva Francisco Jr, Lucymara Fassarella Agnez Lima, Kátia Castanho Scortecci, Vinícius Pietta Perez, Otavio J. Brustolini, Eduardo Sérgio Soares Sousa, Danielle Angst Secco, Angela Maria Guimarães Santos, George Rego Albuquerque, Ana Paula Melo Mariano, Bianca Mendes Maciel, Alexandra L Gerber, Ana Paula de C Guimarães, Paulo Ricardo Nascimento, Francisco Paulo Freire Neto, Sandra Rocha Gadelha, Luís Cristóvão Porto, Eloiza Helena Campana, Selma Maria Bezerra Jeronimo, Ana Tereza R Vasconcelos |
| EPI_ISL_1213216, EPI_ISL_1213218, EPI_ISL_1213220, EPI_ISL_1213222, EPI_ISL_1213224, EPI_ISL_1213226, EPI_ISL_1213227, EPI_ISL_1213229, EPI_ISL_1213230, EPI_ISL_1213232, EPI_ISL_1213234, EPI_ISL_1213236, EPI_ISL_1213237, EPI_ISL_1213239, EPI_ISL_1213241, EPI_ISL_1213242, EPI_ISL_1213244, EPI_ISL_1213245 | | | |
| see above | LAFEM/UESC | Bioinformatics Laboratory / LNCC | Alessandra P Lamarca, Luiz G P de Almeida, Ronaldo da Silva Francisco Jr, Lucymara Fassarella Agnez Lima, Kátia Castanho Scortecci, Vinícius Pietta Perez, Otavio J. Brustolini, Eduardo Sérgio Soares Sousa, Danielle Angst Secco, Angela Maria Guimarães Santos, George Rego Albuquerque, Ana Paula Melo Mariano, Bianca Mendes Maciel, Alexandra L Gerber, Ana Paula de C Guimarães, Paulo Ricardo Nascimento, Francisco Paulo Freire Neto, Sandra Rocha Gadelha, Luís Cristóvão Porto, Eloiza Helena Campana, Selma Maria Bezerra Jeronimo, Ana Tereza R Vasconcelos |
| EPI_ISL_1213248, EPI_ISL_1213249 | Laboratório HLA/UERJ | Bioinformatics Laboratory / LNCC | Alessandra P Lamarca, Luiz G P de Almeida, Ronaldo da Silva Francisco Jr, Lucymara Fassarella Agnez Lima, Kátia Castanho Scortecci, Vinícius Pietta Perez, Otavio J. Brustolini, Eduardo Sérgio Soares Sousa, Danielle Angst Secco, Angela Maria Guimarães Santos, George Rego Albuquerque, Ana Paula Melo Mariano, Bianca Mendes Maciel, Alexandra L Gerber, Ana Paula de C Guimarães, Paulo Ricardo Nascimento, Francisco Paulo Freire Neto, Sandra Rocha Gadelha, Luís Cristóvão Porto, Eloiza Helena Campana, Selma Maria Bezerra Jeronimo, Ana Tereza R Vasconcelos |
| EPI_ISL_1213251, EPI_ISL_1213252, EPI_ISL_1213254, EPI_ISL_1213256, EPI_ISL_1213258, EPI_ISL_1213259, EPI_ISL_1213261, EPI_ISL_1213263, EPI_ISL_1213265, EPI_ISL_1213267, EPI_ISL_1213269 | | | |
| see above | LAFEM/UESC | Bioinformatics Laboratory / LNCC | Alessandra P Lamarca, Luiz G P de Almeida, Ronaldo da Silva Francisco Jr, Lucymara Fassarella Agnez Lima, Kátia Castanho Scortecci, Vinícius Pietta Perez, Otavio J. Brustolini, Eduardo Sérgio Soares Sousa, Danielle Angst Secco, Angela Maria Guimarães Santos, George Rego Albuquerque, Ana Paula Melo Mariano, Bianca Mendes Maciel, Alexandra L Gerber, Ana Paula de C Guimarães, Paulo Ricardo Nascimento, Francisco Paulo Freire Neto, Sandra Rocha Gadelha, Luís Cristóvão Porto, Eloiza Helena Campana, Selma Maria Bezerra Jeronimo, Ana Tereza R Vasconcelos |
| EPI_ISL_1213270, EPI_ISL_1213272 | Laboratório HLA/UERJ | Bioinformatics Laboratory / LNCC | Alessandra P Lamarca, Luiz G P de Almeida, Ronaldo da Silva Francisco Jr, Lucymara Fassarella Agnez Lima, Kátia Castanho Scortecci, Vinícius Pietta Perez, Otavio J. Brustolini, Eduardo Sérgio Soares Sousa, Danielle Angst Secco, Angela Maria Guimarães Santos, George Rego Albuquerque, Ana Paula Melo Mariano, Bianca Mendes Maciel, Alexandra L Gerber, Ana Paula de C Guimarães, Paulo Ricardo Nascimento, Francisco Paulo Freire Neto, Sandra Rocha Gadelha, Luís Cristóvão Porto, Eloiza Helena Campana, Selma Maria Bezerra Jeronimo, Ana Tereza R Vasconcelos |
| EPI_ISL_1213274 | LAFEM/UESC | Bioinformatics Laboratory / LNCC | Alessandra P Lamarca, Luiz G P de Almeida, Ronaldo da Silva Francisco Jr, Lucymara Fassarella Agnez Lima, Kátia Castanho Scortecci, Vinícius Pietta Perez, Otavio J. Brustolini, Eduardo Sérgio Soares Sousa, Danielle Angst Secco, Angela Maria Guimarães Santos, George Rego Albuquerque, Ana Paula |

| | | | |
|---|---|---|---|
| | | | Melo Mariano, Bianca Mendes Maciel, Alexandra L Gerber, Ana Paula de C Guimarães, Paulo Ricardo Nascimento, Francisco Paulo Freire Neto, Sandra Rocha Gadelha, Luís Cristóvão Porto, Eloiza Helena Campana, Selma Maria Bezerra Jeronimo, Ana Tereza R Vasconcelos |
| EPI_ISL_1213275, EPI_ISL_1213277, EPI_ISL_1213279, EPI_ISL_1213281, EPI_ISL_1213282, EPI_ISL_1213284, EPI_ISL_1213286, EPI_ISL_1213288, EPI_ISL_1213289, EPI_ISL_1213291, EPI_ISL_1213293 | | | |
| see above | IMT-UFRN/RN | Bioinformatics Laboratory / LNCC | Alessandra P Lamarca, Luiz G P de Almeida, Ronaldo da Silva Francisco Jr, Lucymara Fassarella Agnez Lima, Kátia Castanho Scortecci, Vinícius Pietta Perez, Otavio J. Brustolini, Eduardo Sérgio Soares Sousa, Danielle Angst Secco, Angela Maria Guimarães Santos, George Rego Albuquerque, Ana Paula Melo Mariano, Bianca Mendes Maciel, Alexandra L Gerber, Ana Paula de C Guimarães, Paulo Ricardo Nascimento, Francisco Paulo Freire Neto, Sandra Rocha Gadelha, Luís Cristóvão Porto, Eloiza Helena Campana, Selma Maria Bezerra Jeronimo, Ana Tereza R Vasconcelos |
| EPI_ISL_1213294, EPI_ISL_1213297, EPI_ISL_1213298, EPI_ISL_1213300, EPI_ISL_1213302, EPI_ISL_1213303, EPI_ISL_1213305 | LAFEM/UESC | Bioinformatics Laboratory / LNCC | Alessandra P Lamarca, Luiz G P de Almeida, Ronaldo da Silva Francisco Jr, Lucymara Fassarella Agnez Lima, Kátia Castanho Scortecci, Vinícius Pietta Perez, Otavio J. Brustolini, Eduardo Sérgio Soares Sousa, Danielle Angst Secco, Angela Maria Guimarães Santos, George Rego Albuquerque, Ana Paula Melo Mariano, Bianca Mendes Maciel, Alexandra L Gerber, Ana Paula de C Guimarães, Paulo Ricardo Nascimento, Francisco Paulo Freire Neto, Sandra Rocha Gadelha, Luís Cristóvão Porto, Eloiza Helena Campana, Selma Maria Bezerra Jeronimo, Ana Tereza R Vasconcelos |
| EPI_ISL_1213307, EPI_ISL_1213309, EPI_ISL_1213310, EPI_ISL_1213312, EPI_ISL_1213314 | Laboratório HLA/UERJ | Bioinformatics Laboratory / LNCC | Alessandra P Lamarca, Luiz G P de Almeida, Ronaldo da Silva Francisco Jr, Lucymara Fassarella Agnez Lima, Kátia Castanho Scortecci, Vinícius Pietta Perez, Otavio J. Brustolini, Eduardo Sérgio Soares Sousa, Danielle Angst Secco, Angela Maria Guimarães Santos, George Rego Albuquerque, Ana Paula Melo Mariano, Bianca Mendes Maciel, Alexandra L Gerber, Ana Paula de C Guimarães, Paulo Ricardo Nascimento, Francisco Paulo Freire Neto, Sandra Rocha Gadelha, Luís Cristóvão Porto, Eloiza Helena Campana, Selma Maria Bezerra Jeronimo, Ana Tereza R Vasconcelos |
| EPI_ISL_1213315, EPI_ISL_1213317, EPI_ISL_1213319, EPI_ISL_1213320, EPI_ISL_1213322 | IMT-UFRN/RN | Bioinformatics Laboratory / LNCC | Alessandra P Lamarca, Luiz G P de Almeida, Ronaldo da Silva Francisco Jr, Lucymara Fassarella Agnez Lima, Kátia Castanho Scortecci, Vinícius Pietta Perez, Otavio J. Brustolini, Eduardo Sérgio Soares Sousa, Danielle Angst Secco, Angela Maria Guimarães Santos, George Rego Albuquerque, Ana Paula Melo Mariano, Bianca Mendes Maciel, Alexandra L Gerber, Ana Paula de C Guimarães, Paulo Ricardo Nascimento, Francisco Paulo Freire Neto, Sandra Rocha Gadelha, Luís Cristóvão Porto, Eloiza Helena Campana, Selma Maria Bezerra Jeronimo, Ana Tereza R Vasconcelos |
| EPI_ISL_1213324, EPI_ISL_1213326, EPI_ISL_1213328 | LAFEM/UESC | Bioinformatics Laboratory / LNCC | Alessandra P Lamarca, Luiz G P de Almeida, Ronaldo da Silva Francisco Jr, Lucymara Fassarella Agnez Lima, Kátia Castanho Scortecci, Vinícius Pietta Perez, Otavio J. Brustolini, Eduardo Sérgio Soares Sousa, Danielle Angst Secco, Angela Maria Guimarães Santos, George Rego Albuquerque, Ana Paula Melo Mariano, Bianca Mendes Maciel, Alexandra L Gerber, Ana Paula de C Guimarães, Paulo Ricardo Nascimento, Francisco Paulo Freire Neto, Sandra Rocha Gadelha, Luís Cristóvão Porto, Eloiza Helena Campana, Selma Maria Bezerra Jeronimo, Ana Tereza R Vasconcelos |
| EPI_ISL_1213329 | IMT-UFRN/RN | Bioinformatics Laboratory / LNCC | Alessandra P Lamarca, Luiz G P de Almeida, Ronaldo da Silva Francisco Jr, Lucymara Fassarella Agnez Lima, Kátia Castanho Scortecci, Vinícius Pietta Perez, Otavio J. Brustolini, Eduardo Sérgio Soares Sousa, Danielle Angst Secco, Angela Maria Guimarães Santos, George Rego Albuquerque, Ana Paula Melo Mariano, Bianca Mendes Maciel, Alexandra L Gerber, Ana Paula de C Guimarães, Paulo Ricardo Nascimento, Francisco Paulo Freire Neto, Sandra Rocha Gadelha, Luís Cristóvão Porto, Eloiza Helena Campana, Selma Maria Bezerra Jeronimo, Ana Tereza R Vasconcelos |
| EPI_ISL_1213331, EPI_ISL_1213333, EPI_ISL_1213335 | LAFEM/UESC | Bioinformatics Laboratory / LNCC | Alessandra P Lamarca, Luiz G P de Almeida, Ronaldo da Silva Francisco Jr, Lucymara Fassarella Agnez Lima, Kátia Castanho Scortecci, Vinícius Pietta Perez, Otavio J. Brustolini, Eduardo Sérgio Soares Sousa, Danielle Angst Secco, Angela Maria Guimarães Santos, George Rego Albuquerque, Ana Paula Melo Mariano, Bianca Mendes Maciel, Alexandra L Gerber, Ana Paula de C Guimarães, Paulo Ricardo Nascimento, Francisco Paulo Freire Neto, Sandra Rocha Gadelha, Luís Cristóvão Porto, Eloiza Helena Campana, Selma Maria Bezerra Jeronimo, Ana Tereza R Vasconcelos |
| EPI_ISL_1213336 | LBM/UFPB | Bioinformatics Laboratory / LNCC | Alessandra P Lamarca, Luiz G P de Almeida, Ronaldo da Silva Francisco Jr, Lucymara Fassarella Agnez Lima, Kátia Castanho Scortecci, Vinícius Pietta Perez, Otavio J. Brustolini, Eduardo Sérgio Soares Sousa, Danielle Angst Secco, Angela Maria Guimarães Santos, George Rego Albuquerque, Ana Paula Melo Mariano, Bianca Mendes Maciel, Alexandra L Gerber, Ana Paula de C Guimarães, Paulo Ricardo Nascimento, Francisco Paulo Freire Neto, Sandra Rocha Gadelha, Luís Cristóvão Porto, Eloiza Helena Campana, Selma Maria Bezerra Jeronimo, Ana Tereza R Vasconcelos |
| EPI_ISL_1213339, EPI_ISL_1213340 | Laboratório HLA/UERJ | Bioinformatics Laboratory / LNCC | Alessandra P Lamarca, Luiz G P de Almeida, Ronaldo da Silva Francisco Jr, Lucymara Fassarella Agnez Lima, Kátia Castanho Scortecci, Vinícius Pietta Perez, Otavio J. Brustolini, Eduardo Sérgio Soares Sousa, Danielle Angst Secco, Angela Maria Guimarães Santos, George Rego Albuquerque, Ana Paula Melo Mariano, Bianca Mendes Maciel, Alexandra L Gerber, Ana Paula de C Guimarães, Paulo Ricardo Nascimento, Francisco Paulo Freire Neto, Sandra Rocha Gadelha, Luís Cristóvão Porto, Eloiza Helena Campana, Selma Maria Bezerra Jeronimo, Ana Tereza R Vasconcelos |
| EPI_ISL_1213341, EPI_ISL_1213342 | LAFEM/UESC | Bioinformatics Laboratory / LNCC | Alessandra P Lamarca, Luiz G P de Almeida, Ronaldo da Silva Francisco Jr, Lucymara Fassarella Agnez Lima, Kátia Castanho Scortecci, Vinícius Pietta Perez, Otavio J. Brustolini, Eduardo Sérgio Soares Sousa, Danielle Angst Secco, Angela Maria Guimarães Santos, George Rego Albuquerque, Ana Paula Melo Mariano, Bianca Mendes Maciel, Alexandra L Gerber, Ana Paula de C Guimarães, Paulo Ricardo Nascimento, Francisco Paulo Freire Neto, Sandra Rocha Gadelha, Luís Cristóvão Porto, Eloiza Helena Campana, Selma Maria Bezerra Jeronimo, Ana Tereza R Vasconcelos |
| EPI_ISL_1213343, EPI_ISL_1213344 | LBM/UFPB | Bioinformatics Laboratory / LNCC | Alessandra P Lamarca, Luiz G P de Almeida, Ronaldo da Silva Francisco Jr, Lucymara Fassarella Agnez Lima, Kátia Castanho Scortecci, Vinícius Pietta Perez, Otavio J. Brustolini, Eduardo Sérgio Soares Sousa, Danielle Angst Secco, Angela Maria Guimarães Santos, George Rego Albuquerque, Ana Paula Melo Mariano, Bianca Mendes Maciel, Alexandra L Gerber, Ana Paula de C Guimarães, Paulo Ricardo Nascimento, Francisco Paulo Freire Neto, Sandra Rocha Gadelha, Luís Cristóvão Porto, Eloiza Helena Campana, Selma Maria Bezerra Jeronimo, Ana Tereza R Vasconcelos |
| EPI_ISL_1213345, EPI_ISL_1213346, EPI_ISL_1213348, EPI_ISL_1213350, EPI_ISL_1213352, EPI_ISL_1213353, EPI_ISL_1213355, EPI_ISL_1213357, EPI_ISL_1213358, EPI_ISL_1213360, EPI_ISL_1213362, EPI_ISL_1213364 | | | |
| see above | IMT-UFRN/RN | Bioinformatics Laboratory / LNCC | Alessandra P Lamarca, Luiz G P de Almeida, Ronaldo da Silva Francisco Jr, Lucymara Fassarella Agnez Lima, Kátia Castanho Scortecci, Vinícius Pietta Perez, Otavio J. Brustolini, Eduardo Sérgio Soares Sousa, Danielle Angst Secco, Angela Maria Guimarães Santos, George Rego Albuquerque, Ana Paula Melo Mariano, Bianca Mendes Maciel, Alexandra L Gerber, Ana Paula de C Guimarães, Paulo Ricardo Nascimento, Francisco Paulo Freire Neto, Sandra Rocha Gadelha, Luís Cristóvão Porto, Eloiza Helena Campana, Selma Maria Bezerra Jeronimo, Ana Tereza R Vasconcelos |
| EPI_ISL_1213365 | LAFEM/UESC | Bioinformatics Laboratory / LNCC | Alessandra P Lamarca, Luiz G P de Almeida, Ronaldo da Silva Francisco Jr, Lucymara Fassarella Agnez Lima, Kátia Castanho Scortecci, Vinícius Pietta Perez, Otavio J. Brustolini, Eduardo Sérgio Soares Sousa, Danielle Angst Secco, Angela Maria Guimarães Santos, George Rego Albuquerque, Ana Paula Melo Mariano, Bianca Mendes Maciel, Alexandra L Gerber, Ana Paula de C Guimarães, Paulo Ricardo Nascimento, Francisco Paulo Freire Neto, Sandra Rocha Gadelha, Luís Cristóvão Porto, Eloiza Helena Campana, Selma Maria Bezerra Jeronimo, Ana Tereza R Vasconcelos |
| EPI_ISL_1213367, EPI_ISL_1213369, EPI_ISL_1213370 | LBM/UFPB | Bioinformatics Laboratory / LNCC | Alessandra P Lamarca, Luiz G P de Almeida, Ronaldo da Silva Francisco Jr, Lucymara Fassarella Agnez Lima, Kátia Castanho Scortecci, Vinícius Pietta Perez, Otavio J. Brustolini, Eduardo Sérgio Soares Sousa, Danielle Angst Secco, Angela Maria Guimarães Santos, George Rego Albuquerque, Ana Paula Melo Mariano, Bianca Mendes Maciel, Alexandra L Gerber, Ana Paula de C Guimarães, Paulo Ricardo Nascimento, Francisco Paulo Freire Neto, Sandra Rocha Gadelha, Luís Cristóvão Porto, Eloiza Helena Campana, Selma Maria Bezerra Jeronimo, Ana Tereza R Vasconcelos |
| EPI_ISL_1213372, EPI_ISL_1213374, EPI_ISL_1213376, EPI_ISL_1213378, EPI_ISL_1213379, EPI_ISL_1213381, EPI_ISL_1213382 | Laboratório HLA/UERJ | Bioinformatics Laboratory / LNCC | Alessandra P Lamarca, Luiz G P de Almeida, Ronaldo da Silva Francisco Jr, Lucymara Fassarella Agnez Lima, Kátia Castanho Scortecci, Vinícius Pietta Perez, Otavio J. Brustolini, Eduardo Sérgio Soares Sousa, Danielle Angst Secco, Angela Maria Guimarães Santos, George Rego Albuquerque, Ana Paula Melo Mariano, Bianca Mendes Maciel, Alexandra L Gerber, Ana Paula de C Guimarães, Paulo Ricardo Nascimento, Francisco Paulo Freire Neto, Sandra Rocha Gadelha, Luís Cristóvão Porto, Eloiza Helena Campana, Selma Maria Bezerra Jeronimo, Ana Tereza R Vasconcelos |
| EPI_ISL_1213384, EPI_ISL_1213386 | LAFEM/UESC | Bioinformatics Laboratory / LNCC | Alessandra P Lamarca, Luiz G P de Almeida, Ronaldo da Silva Francisco Jr, Lucymara Fassarella Agnez Lima, Kátia Castanho Scortecci, Vinícius Pietta Perez, Otavio J. Brustolini, Eduardo Sérgio Soares Sousa, Danielle Angst Secco, Angela Maria Guimarães Santos, George Rego Albuquerque, Ana Paula Melo Mariano, Bianca Mendes Maciel, Alexandra L Gerber, Ana Paula de C Guimarães, Paulo Ricardo Nascimento, Francisco Paulo Freire Neto, Sandra Rocha Gadelha, Luís Cristóvão Porto, Eloiza Helena Campana, Selma Maria Bezerra Jeronimo, Ana Tereza R Vasconcelos |
| EPI_ISL_1213388 | LBM/UFPB | Bioinformatics Laboratory / LNCC | Alessandra P Lamarca, Luiz G P de Almeida, Ronaldo da Silva Francisco Jr, Lucymara Fassarella Agnez Lima, Kátia Castanho Scortecci, Vinícius Pietta Perez, Otavio J. Brustolini, Eduardo Sérgio Soares Sousa, Danielle Angst Secco, Angela Maria Guimarães Santos, George Rego Albuquerque, Ana Paula Melo Mariano, Bianca Mendes Maciel, Alexandra L Gerber, Ana Paula de C Guimarães, Paulo Ricardo Nascimento, Francisco Paulo Freire Neto, Sandra Rocha Gadelha, Luís Cristóvão Porto, Eloiza Helena Campana, Selma Maria Bezerra Jeronimo, Ana Tereza R Vasconcelos |
| EPI_ISL_1213390 | LAFEM/UESC | Bioinformatics Laboratory / LNCC | Alessandra P Lamarca, Luiz G P de Almeida, Ronaldo da Silva Francisco Jr, Lucymara Fassarella Agnez Lima, Kátia Castanho Scortecci, Vinícius Pietta Perez, Otavio J. Brustolini, Eduardo Sérgio Soares Sousa, Danielle Angst Secco, Angela Maria Guimarães Santos, George Rego Albuquerque, Ana Paula Melo Mariano, Bianca Mendes Maciel, Alexandra L Gerber, Ana Paula de C Guimarães, Paulo Ricardo Nascimento, Francisco Paulo Freire Neto, Sandra Rocha Gadelha, Luís Cristóvão Porto, Eloiza Helena Campana, Selma Maria Bezerra Jeronimo, Ana Tereza R Vasconcelos |
| EPI_ISL_1213392 | LBM/UFPB | Bioinformatics Laboratory / LNCC | Alessandra P Lamarca, Luiz G P de Almeida, Ronaldo da Silva Francisco Jr, Lucymara Fassarella Agnez Lima, Kátia Castanho Scortecci, Vinícius Pietta |

| | | | |
|---|---|---|---|
| | | | Perez, Otavio J. Brustolini, Eduardo Sérgio Soares Sousa, Danielle Angst Secco, Angela Maria Guimarães Santos, George Rego Albuquerque, Ana Paula Melo Mariano, Bianca Mendes Maciel, Alexandra L Gerber, Ana Paula de C Guimarães, Paulo Ricardo Nascimento, Francisco Paulo Freire Neto, Sandra Rocha Gadelha, Luís Cristóvão Porto, Eloiza Helena Campana, Selma Maria Bezerra Jeronimo, Ana Tereza R Vasconcelos |
| EPI_ISL_1213393, EPI_ISL_1213395, EPI_ISL_1213397 | Laboratório HLA/UERJ | Bioinformatics Laboratory / LNCC | Alessandra P Lamarca, Luiz G P de Almeida, Ronaldo da Silva Francisco Jr, Lucymara Fassarella Agnez Lima, Kátia Castanho Scortecci, Vinícius Pietta Perez, Otavio J. Brustolini, Eduardo Sérgio Soares Sousa, Danielle Angst Secco, Angela Maria Guimarães Santos, George Rego Albuquerque, Ana Paula Melo Mariano, Bianca Mendes Maciel, Alexandra L Gerber, Ana Paula de C Guimarães, Paulo Ricardo Nascimento, Francisco Paulo Freire Neto, Sandra Rocha Gadelha, Luís Cristóvão Porto, Eloiza Helena Campana, Selma Maria Bezerra Jeronimo, Ana Tereza R Vasconcelos |
| EPI_ISL_1213399, EPI_ISL_1213401, EPI_ISL_1213402 | LAFEM/UESC | Bioinformatics Laboratory / LNCC | Alessandra P Lamarca, Luiz G P de Almeida, Ronaldo da Silva Francisco Jr, Lucymara Fassarella Agnez Lima, Kátia Castanho Scortecci, Vinícius Pietta Perez, Otavio J. Brustolini, Eduardo Sérgio Soares Sousa, Danielle Angst Secco, Angela Maria Guimarães Santos, George Rego Albuquerque, Ana Paula Melo Mariano, Bianca Mendes Maciel, Alexandra L Gerber, Ana Paula de C Guimarães, Paulo Ricardo Nascimento, Francisco Paulo Freire Neto, Sandra Rocha Gadelha, Luís Cristóvão Porto, Eloiza Helena Campana, Selma Maria Bezerra Jeronimo, Ana Tereza R Vasconcelos |
| EPI_ISL_1213404, EPI_ISL_1213406 | LBM/UFPB | Bioinformatics Laboratory / LNCC | Alessandra P Lamarca, Luiz G P de Almeida, Ronaldo da Silva Francisco Jr, Lucymara Fassarella Agnez Lima, Kátia Castanho Scortecci, Vinícius Pietta Perez, Otavio J. Brustolini, Eduardo Sérgio Soares Sousa, Danielle Angst Secco, Angela Maria Guimarães Santos, George Rego Albuquerque, Ana Paula Melo Mariano, Bianca Mendes Maciel, Alexandra L Gerber, Ana Paula de C Guimarães, Paulo Ricardo Nascimento, Francisco Paulo Freire Neto, Sandra Rocha Gadelha, Luís Cristóvão Porto, Eloiza Helena Campana, Selma Maria Bezerra Jeronimo, Ana Tereza R Vasconcelos |
| EPI_ISL_1213408, EPI_ISL_1213410 | Laboratório HLA/UERJ | Bioinformatics Laboratory / LNCC | Alessandra P Lamarca, Luiz G P de Almeida, Ronaldo da Silva Francisco Jr, Lucymara Fassarella Agnez Lima, Kátia Castanho Scortecci, Vinícius Pietta Perez, Otavio J. Brustolini, Eduardo Sérgio Soares Sousa, Danielle Angst Secco, Angela Maria Guimarães Santos, George Rego Albuquerque, Ana Paula Melo Mariano, Bianca Mendes Maciel, Alexandra L Gerber, Ana Paula de C Guimarães, Paulo Ricardo Nascimento, Francisco Paulo Freire Neto, Sandra Rocha Gadelha, Luís Cristóvão Porto, Eloiza Helena Campana, Selma Maria Bezerra Jeronimo, Ana Tereza R Vasconcelos |
| EPI_ISL_1213411, EPI_ISL_1213413, EPI_ISL_1213415 | LAFEM/UESC | Bioinformatics Laboratory / LNCC | Alessandra P Lamarca, Luiz G P de Almeida, Ronaldo da Silva Francisco Jr, Lucymara Fassarella Agnez Lima, Kátia Castanho Scortecci, Vinícius Pietta Perez, Otavio J. Brustolini, Eduardo Sérgio Soares Sousa, Danielle Angst Secco, Angela Maria Guimarães Santos, George Rego Albuquerque, Ana Paula Melo Mariano, Bianca Mendes Maciel, Alexandra L Gerber, Ana Paula de C Guimarães, Paulo Ricardo Nascimento, Francisco Paulo Freire Neto, Sandra Rocha Gadelha, Luís Cristóvão Porto, Eloiza Helena Campana, Selma Maria Bezerra Jeronimo, Ana Tereza R Vasconcelos |
| EPI_ISL_1213417, EPI_ISL_1213418, EPI_ISL_1213420, EPI_ISL_1213422, EPI_ISL_1213424, EPI_ISL_1213425 | Laboratório HLA/UERJ | Bioinformatics Laboratory / LNCC | Alessandra P Lamarca, Luiz G P de Almeida, Ronaldo da Silva Francisco Jr, Lucymara Fassarella Agnez Lima, Kátia Castanho Scortecci, Vinícius Pietta Perez, Otavio J. Brustolini, Eduardo Sérgio Soares Sousa, Danielle Angst Secco, Angela Maria Guimarães Santos, George Rego Albuquerque, Ana Paula Melo Mariano, Bianca Mendes Maciel, Alexandra L Gerber, Ana Paula de C Guimarães, Paulo Ricardo Nascimento, Francisco Paulo Freire Neto, Sandra Rocha Gadelha, Luís Cristóvão Porto, Eloiza Helena Campana, Selma Maria Bezerra Jeronimo, Ana Tereza R Vasconcelos |
| EPI_ISL_1213427, EPI_ISL_1213429, EPI_ISL_1213431, EPI_ISL_1213433, EPI_ISL_1213434, EPI_ISL_1213436, EPI_ISL_1213438, EPI_ISL_1213439, EPI_ISL_1213441, EPI_ISL_1213443, EPI_ISL_1213444, EPI_ISL_1213446, EPI_ISL_1213448, EPI_ISL_1213449, EPI_ISL_1213451, EPI_ISL_1213453, EPI_ISL_1213454, EPI_ISL_1213456, EPI_ISL_1213458, EPI_ISL_1213459, EPI_ISL_1213461, EPI_ISL_1213463 | | | |
| see above | LBM/UFPB | Bioinformatics Laboratory / LNCC | Alessandra P Lamarca, Luiz G P de Almeida, Ronaldo da Silva Francisco Jr, Lucymara Fassarella Agnez Lima, Kátia Castanho Scortecci, Vinícius Pietta Perez, Otavio J. Brustolini, Eduardo Sérgio Soares Sousa, Danielle Angst Secco, Angela Maria Guimarães Santos, George Rego Albuquerque, Ana Paula Melo Mariano, Bianca Mendes Maciel, Alexandra L Gerber, Ana Paula de C Guimarães, Paulo Ricardo Nascimento, Francisco Paulo Freire Neto, Sandra Rocha Gadelha, Luís Cristóvão Porto, Eloiza Helena Campana, Selma Maria Bezerra Jeronimo, Ana Tereza R Vasconcelos |
| EPI_ISL_1219021 | Hospital Aviccena | Instituto Adolfo Lutz, Interdiciplinary Procedures Center, Strategic Laboratory | Claudio Tavares Sacchi, Claudia Regina Gonçalves, Erica Valessa Ramos Gomes, Karoline Rodrigues Campos, Caio Vinicius Dias Lopes |
| EPI_ISL_1219022, EPI_ISL_1219023, EPI_ISL_1219024, EPI_ISL_1219025, EPI_ISL_1219026 | IAL Regional de Sorocaba | Instituto Adolfo Lutz, Interdiciplinary Procedures Center, Strategic Laboratory | Claudio Tavares Sacchi, Claudia Regina Gonçalves, Erica Valessa Ramos Gomes, Karoline Rodrigues Campos, Caio Vinicius Dias Lopes |
| EPI_ISL_1219027 | IAL Regional de Presidente Prudente | Instituto Adolfo Lutz, Interdiciplinary Procedures Center, Strategic Laboratory | Claudio Tavares Sacchi, Claudia Regina Gonçalves, Erica Valessa Ramos Gomes, Karoline Rodrigues Campos, Caio Vinicius Dias Lopes |
| EPI_ISL_1219028, EPI_ISL_1219029, EPI_ISL_1219030, EPI_ISL_1219031, EPI_ISL_1219032, EPI_ISL_1219033, EPI_ISL_1219034, EPI_ISL_1219035, EPI_ISL_1219036 | Aeroporto Internacional de Guarulhos | Instituto Adolfo Lutz, Interdiciplinary Procedures Center, Strategic Laboratory | Claudio Tavares Sacchi, Claudia Regina Gonçalves, Erica Valessa Ramos Gomes, Karoline Rodrigues Campos, Caio Vinicius Dias Lopes |
| EPI_ISL_1219037 | IAL Regional de Presidente Prudente | Instituto Adolfo Lutz, Interdiciplinary Procedures Center, Strategic Laboratory | Claudio Tavares Sacchi, Claudia Regina Gonçalves, Erica Valessa Ramos Gomes, Karoline Rodrigues Campos, Caio Vinicius Dias Lopes |
| EPI_ISL_1219132, EPI_ISL_1219133 | Laboratorio Central de Saude Publica do Estado do Parana (LACEN-PR) | Laboratory of Respiratory Viruses and Measles, Oswaldo Cruz Institute, FIOCRUZ | Paola Resende, Luciana Appolinario, Fernando Motta, Anna Carolina Paixao, Ana Carolina Mendonca, Alice Sampaio Rocha, Renata Serrano Lopes, Maria do Carmo Debur, Irina Nastassja Riediger, Marilda Siqueira on behalf of the Fiocruz COVID-19 Genomic Surveillance Network |
| EPI_ISL_1219134, EPI_ISL_1219135 | Laboratorio Central de Saude Publica do Estado do Alagoas (LACEN-AL) | Laboratory of Respiratory Viruses and Measles, Oswaldo Cruz Institute, FIOCRUZ | Paola Resende, Luciana Appolinario, Fernando Motta, Anna Carolina Paixao, Ana Carolina Mendonca, Alice Sampaio Rocha, Renata Serrano Lopes, Anderson Brandao Leite, Marilda Siqueira on behalf of the Fiocruz COVID-19 Genomic Surveillance Network |
| EPI_ISL_1219136 | Gonçalo Moniz Institute, FIOCRUZ, Bahia | Laboratory of Respiratory Viruses and Measles, Oswaldo Cruz Institute, FIOCRUZ | Paola Resende, Luciana Appolinario, Fernando Motta, Anna Carolina Paixao, Ana Carolina Mendonca, Alice Sampaio Rocha, Renata Serrano Lopes, Tiago Graf, Ricardo Khouri, Marilda Siqueira on behalf of the Fiocruz COVID-19 Genomic Surveillance Network |
| EPI_ISL_1219137 | Laboratorio Central de Saude Publica do Estado de Minas Gerais (LACEN-MG) | Laboratory of Respiratory Viruses and Measles, Oswaldo Cruz Institute, FIOCRUZ | Paola Resende, Luciana Appolinario, Fernando Motta, Anna Carolina Paixao, Ana Carolina Mendonca, Alice Sampaio Rocha, Renata Serrano Lopes, Felipe Iani, Marilda Siqueira on behalf of the Fiocruz COVID-19 Genomic Surveillance Network |
| EPI_ISL_1219138 | Federal University of Mato Grosso (UFMT) | Laboratory of Respiratory Viruses and Measles, Oswaldo Cruz Institute, FIOCRUZ | Paola Resende, Luciana Appolinario, Fernando Motta, Anna Carolina Paixao, Ana Carolina Mendonca, Alice Sampaio Rocha, Renata Serrano Lopes, Renata Dezengrini, Marilda Siqueira on behalf of the Fiocruz COVID-19 Genomic Surveillance Network |
| EPI_ISL_1220095 | Hospital Municipal Gov. Mario Covas Jr. | Instituto Butantan (genome assembly and bioinformatics) and Mendelics (sequencing) | Maria Carolina Quartim Barbosa Elias Sabbaga, Jose Patane, Simone Haddad, Rafael dos Santos Bezerra, Sandra Coccuzzo Sampaio Vessoni, Antonio Jorge Martins, Dimas Tadeu Covas |
| EPI_ISL_1239012, EPI_ISL_1239013, EPI_ISL_1239014, EPI_ISL_1239015, EPI_ISL_1239016 | Laboratório Central de Saúde Pública do Estado de Pernambuco (LACEN-PE) | WallauLab, Aggeu Magalhaes Institute | Marcelo Henrique dos Santos Paiva, Duschinka Ribeiro Duarte Guedes, Cássia Docena, Matheus Filgueira Bezerra, Filipe Zimmer Dezordi, Laís Ceschini Machado, Larissa Krokovsky, Elisama Helvecio, Alexandre Freitas da Silva, Antonio Mauro Rezende, Sinval Pinto Brandão Filho, Constância Flávia Junqueira Ayres, Gabriel Luz Wallau on behalf of the Fiocruz COVID-19 Genomic Surveillance Network |
| EPI_ISL_1239111, EPI_ISL_1239112, EPI_ISL_1239113 | Laboratório Central de Saúde Pública Noel Nutels | Coordenação Geral de Laboratórios de Saúde Pública (CGLAB) | Vagner Fonseca et al, |
| EPI_ISL_1239114, EPI_ISL_1239115, EPI_ISL_1239116, EPI_ISL_1239117, EPI_ISL_1239118, EPI_ISL_1239119, EPI_ISL_1239120, EPI_ISL_1239121, EPI_ISL_1239122, EPI_ISL_1239123 | Laboratório Central de Saúde Pública do Espírito Santo | Coordenação Geral de Laboratórios de Saúde Pública (CGLAB) | Vagner Fonseca et al, |
| EPI_ISL_1239124 | Fundação Ezequiel Dias | Coordenação Geral de Laboratórios de Saúde Pública (CGLAB) | Vagner Fonseca et al, |
| EPI_ISL_1239125, EPI_ISL_1239126, EPI_ISL_1239127, EPI_ISL_1239128, EPI_ISL_1239129, EPI_ISL_1239130, EPI_ISL_1239131, EPI_ISL_1239132, EPI_ISL_1239133, EPI_ISL_1239134, EPI_ISL_1239135, EPI_ISL_1239136 | | | |
| see above | Laboratório Central de Saúde Pública do Espírito Santo | Coordenação Geral de Laboratórios de Saúde Pública (CGLAB) | Vagner Fonseca et al, |

| | | | |
|---|---|---|---|
| EPI_ISL_1239137, EPI_ISL_1239138, EPI_ISL_1239139 | Fundação Ezequiel Dias | Coordenação Geral de Laboratórios de Saúde Pública (CGLAB) | Vagner Fonseca et al, |
| EPI_ISL_1240639, EPI_ISL_1240640, EPI_ISL_1240641 | Laboratório Central de Saúde Pública do Espírito Santo | Coordenação Geral de Laboratórios de Saúde Pública (CGLAB) | Vagner Fonseca et al. |
| EPI_ISL_1240642 | Fundação Ezequiel Dias | Coordenação Geral de Laboratórios de Saúde Pública (CGLAB) | Vagner Fonseca et al. |
| EPI_ISL_1251212, EPI_ISL_1251213, EPI_ISL_1251214, EPI_ISL_1251215, EPI_ISL_1251216, EPI_ISL_1251217, EPI_ISL_1251218, EPI_ISL_1251219, EPI_ISL_1251220, EPI_ISL_1251221, EPI_ISL_1251222, EPI_ISL_1251223, EPI_ISL_1251224, EPI_ISL_1251225 | | | |
| see above | Raimundo Reginaldo de Almeida | MOA Lab | Vanessa C. Nicolete, Priscila T. Rodrigues, Marcelo U. Ferreira |
| EPI_ISL_1261122, EPI_ISL_1261123 | Laboratorio de Ecologia de Doencas Transmissiveis na Amazonia, Instituto Leonidas e Maria Deane - Fiocruz Amazonia | Laboratorio de Ecologia de Doencas Transmissiveis na Amazonia, Instituto Leonidas e Maria Deane - Fiocruz Amazonia | Valdinete Nascimento, Victor Souza, André Corado, Fernanda Nascimento, George Silva, Ágatha Costa, Debora Duarte, Karina Pessoa, Matilde Mejía, Luciana Gonçalves, Maria Júlia Brandão, Michele Jesus, Felipe Naveca |
| EPI_ISL_1261683 | LACEN - Laboratório Central de Saúde Pública do Amazonas | Evandro Chagas Institute | Santos, M.C.; Silva, A.M.; Junior, W.D.C.; Barbagelata, L.S.; Ferreira, J.A.; Sousa, E.M.A.; da Silva, P.S.; Pinheiro, K.C.; L.C.; Sousa Junior, E.C. |
| EPI_ISL_1261684 | LACEN - Laboratório Central de Saúde Pública do Ceará | Evandro Chagas Institute | Santos, M.C.; Silva, A.M.; Junior, W.D.C.; Barbagelata, L.S.; Ferreira, J.A.; Sousa, E.M.A.; da Silva, P.S.; Pinheiro, K.C.; L.C.; Sousa Junior, E.C. |
| EPI_ISL_1261685 | LACEN - Laboratório Central de Saúde Pública do Amazonas | Evandro Chagas Institute | Santos, M.C.; Silva, A.M.; Junior, W.D.C.; Barbagelata, L.S.; Ferreira, J.A.; Sousa, E.M.A.; da Silva, P.S.; Pinheiro, K.C.; L.C.; Sousa Junior, E.C. |
| EPI_ISL_1261686 | LACEN - Laboratório Central de Saúde Pública do Amapá | Evandro Chagas Institute | Santos, M.C.; Silva, A.M.; Junior, W.D.C.; Barbagelata, L.S.; Ferreira, J.A.; Sousa, E.M.A.; da Silva, P.S.; Pinheiro, K.C.; L.C.; Sousa Junior, E.C. |
| EPI_ISL_1261687 | LACEN - Laboratório Central de Saúde Pública de Roraima | Evandro Chagas Institute | Santos, M.C.; Silva, A.M.; Junior, W.D.C.; Barbagelata, L.S.; Ferreira, J.A.; Sousa, E.M.A.; da Silva, P.S.; Pinheiro, K.C.; L.C.; Sousa Junior, E.C. |
| EPI_ISL_1261688, EPI_ISL_1261689 | LACEN - Laboratório Central de Saúde Pública do Amapá | Evandro Chagas Institute | Santos, M.C.; Silva, A.M.; Junior, W.D.C.; Barbagelata, L.S.; Ferreira, J.A.; Sousa, E.M.A.; da Silva, P.S.; Pinheiro, K.C.; L.C.; Sousa Junior, E.C. |
| EPI_ISL_1261690 | LACEN - Laboratório Central de Saúde Pública do Amazonas | Evandro Chagas Institute | Santos, M.C.; Silva, A.M.; Junior, W.D.C.; Barbagelata, L.S.; Ferreira, J.A.; Sousa, E.M.A.; da Silva, P.S.; Pinheiro, K.C.; L.C.; Sousa Junior, E.C. |
| EPI_ISL_1261691 | Laboratório Paulo C. Azevedo | Evandro Chagas Institute | Santos, M.C.; Silva, A.M.; Junior, W.D.C.; Barbagelata, L.S.; Ferreira, J.A.; Sousa, E.M.A.; da Silva, P.S.; Pinheiro, K.C.; L.C.; Sousa Junior, E.C. |
| EPI_ISL_1261692 | LACEN - Laboratório Central de Saúde Pública do Amapá | Evandro Chagas Institute | Santos, M.C.; Silva, A.M.; Junior, W.D.C.; Barbagelata, L.S.; Ferreira, J.A.; Sousa, E.M.A.; da Silva, P.S.; Pinheiro, K.C.; L.C.; Sousa Junior, E.C. |
| EPI_ISL_1261693 | LACEN - Laboratório Central de Saúde Pública do Ceará | Evandro Chagas Institute | Santos, M.C.; Silva, A.M.; Junior, W.D.C.; Barbagelata, L.S.; Ferreira, J.A.; Sousa, E.M.A.; da Silva, P.S.; Pinheiro, K.C.; L.C.; Sousa Junior, E.C. |
| EPI_ISL_1261694 | LACEN - Laboratório Central de Saúde Pública do Amazonas | Evandro Chagas Institute | Santos, M.C.; Silva, A.M.; Junior, W.D.C.; Barbagelata, L.S.; Ferreira, J.A.; Sousa, E.M.A.; da Silva, P.S.; Pinheiro, K.C.; L.C.; Sousa Junior, E.C. |
| EPI_ISL_1261695, EPI_ISL_1261696 | LACEN - Laboratório Central de Saúde Pública do Amapá | Evandro Chagas Institute | Santos, M.C.; Silva, A.M.; Junior, W.D.C.; Barbagelata, L.S.; Ferreira, J.A.; Sousa, E.M.A.; da Silva, P.S.; Pinheiro, K.C.; L.C.; Sousa Junior, E.C. |
| EPI_ISL_1261697 | LACEN - Laboratório Central de Saúde Pública do Ceará | Evandro Chagas Institute | Santos, M.C.; Silva, A.M.; Junior, W.D.C.; Barbagelata, L.S.; Ferreira, J.A.; Sousa, E.M.A.; da Silva, P.S.; Pinheiro, K.C.; L.C.; Sousa Junior, E.C. |
| EPI_ISL_1261698, EPI_ISL_1261699 | LACEN - Laboratório Central de Saúde Pública de Pernambuco | Evandro Chagas Institute | Santos, M.C.; Silva, A.M.; Junior, W.D.C.; Barbagelata, L.S.; Ferreira, J.A.; Sousa, E.M.A.; da Silva, P.S.; Pinheiro, K.C.; L.C.; Sousa Junior, E.C. |
| EPI_ISL_1261700 | LACEN - Laboratório Central de Saúde Pública do Maranhao | Evandro Chagas Institute | Santos, M.C.; Silva, A.M.; Junior, W.D.C.; Barbagelata, L.S.; Ferreira, J.A.; Sousa, E.M.A.; da Silva, P.S.; Pinheiro, K.C.; L.C.; Sousa Junior, E.C. |
| EPI_ISL_1271944, EPI_ISL_1272074, EPI_ISL_1272076, EPI_ISL_1272188, EPI_ISL_1272236 | Universidade Federal do Norte do Tocantins (UFNT) | Laboratório de Bioinformática e Biotecnologia (Labinftec/UFT) | Ueric José Borges de Souza, Fabrício Souza Campos, Raíssa Nunes dos Santos, José Carlos Ribeiro Júnior, Rogério Fernandes Carvalho, Monike da Silva Oliveira, Bergmann Morais Ribeiro, Fernando Lucas Melo |
| EPI_ISL_1278274, EPI_ISL_1289956, EPI_ISL_1289957, EPI_ISL_1289958, EPI_ISL_1289959, EPI_ISL_1289960 | Laboratory of Virology, Ribeirão Preto General Hospital, Ribeirão Preto Medical School, University of São Paulo | Laboratory of Oncology, Blood Center of Ribeirão Preto, Ribeirão Preto School of Medicine, University of São Paulo | CAMPOS, MR; SANTOS, A.L.P.; YAMAMOTO, A.Y.; COLLI, L.M.; FONSECA, B.A.L.; BELLISSIMO-RODRIGUES, F. |
| EPI_ISL_1290802 | Genomic and molecular Biology Group, A.C.Camargo Cancer Center | Laboratory of Bioinformatics and Computational Biology, A.C.Camargo Cancer Center | Giovana Torrezan, Dirce Carraro, Israel Tojal, Alexandre Defelicibus |
| EPI_ISL_1293051 | LACEN do Distrito Federal | Instituto Adolfo Lutz, Interdiciplinary Procedures Center, Strategic Laboratory | Claudio Tavares Sacchi, Claudia Regina Gonçalves, Erica Valessa Ramos Gomes, Karoline Rodrigues Campos, Caio Vinicius Dias Lopes |
| EPI_ISL_1293052, EPI_ISL_1293053, EPI_ISL_1293054, EPI_ISL_1293055 | LACEN de Rondonia | Instituto Adolfo Lutz, Interdiciplinary Procedures Center, Strategic Laboratory | Claudio Tavares Sacchi, Claudia Regina Gonçalves, Erica Valessa Ramos Gomes, Karoline Rodrigues Campos, Caio Vinicius Dias Lopes |
| EPI_ISL_1293056, EPI_ISL_1293057, EPI_ISL_1293058, EPI_ISL_1293059, EPI_ISL_1293060, EPI_ISL_1293061, EPI_ISL_1293062, EPI_ISL_1293063, EPI_ISL_1293064, EPI_ISL_1293065, EPI_ISL_1293066, EPI_ISL_1293067, EPI_ISL_1293068, EPI_ISL_1293069, EPI_ISL_1293070, EPI_ISL_1293071, EPI_ISL_1293072, EPI_ISL_1293073, EPI_ISL_1293074, EPI_ISL_1293075, EPI_ISL_1293076, EPI_ISL_1293077, EPI_ISL_1293078, EPI_ISL_1293079, EPI_ISL_1293080, EPI_ISL_1293081 | | | |
| see above | IAL Regional de Sorocaba | Instituto Adolfo Lutz, Interdiciplinary Procedures Center, Strategic Laboratory | Claudio Tavares Sacchi, Claudia Regina Gonçalves, Erica Valessa Ramos Gomes, Karoline Rodrigues Campos, Caio Vinicius Dias Lopes |
| EPI_ISL_1303499, EPI_ISL_1303500, EPI_ISL_1303501, EPI_ISL_1303502, EPI_ISL_1303503, EPI_ISL_1303504, EPI_ISL_1303505 | LACEN de Rondonia | Instituto Adolfo Lutz, Interdiciplinary Procedures Center, Strategic Laboratory | Claudio Tavares Sacchi, Claudia Regina Gonçalves, Erica Valessa Ramos Gomes, Karoline Rodrigues Campos, Caio Vinicius Dias Lopes |
| EPI_ISL_1303506, EPI_ISL_1303507, EPI_ISL_1303508 | LACEN do Distrito Federal | Instituto Adolfo Lutz, Interdiciplinary Procedures Center, Strategic Laboratory | Claudio Tavares Sacchi, Claudia Regina Gonçalves, Erica Valessa Ramos Gomes, Karoline Rodrigues Campos, Caio Vinicius Dias Lopes |
| EPI_ISL_1303509 | LACEN do Estado de Tocantins | Instituto Adolfo Lutz, Interdiciplinary Procedures Center, Strategic Laboratory | Claudio Tavares Sacchi, Claudia Regina Gonçalves, Erica Valessa Ramos Gomes, Karoline Rodrigues Campos, Caio Vinicius Dias Lopes |
| EPI_ISL_1303510, EPI_ISL_1303511, EPI_ISL_1303512, EPI_ISL_1303513, EPI_ISL_1303514, EPI_ISL_1303515, EPI_ISL_1303516, EPI_ISL_1303517 | LACEN do Estado de Goias | Instituto Adolfo Lutz, Interdiciplinary Procedures Center, Strategic Laboratory | Claudio Tavares Sacchi, Claudia Regina Gonçalves, Erica Valessa Ramos Gomes, Karoline Rodrigues Campos, Caio Vinicius Dias Lopes |
| EPI_ISL_1303518, EPI_ISL_1303519, EPI_ISL_1303520, EPI_ISL_1303521, EPI_ISL_1303522, EPI_ISL_1303523, EPI_ISL_1303524, EPI_ISL_1303525, EPI_ISL_1303526, EPI_ISL_1303527, EPI_ISL_1303528, EPI_ISL_1303529, EPI_ISL_1303530, EPI_ISL_1303531, EPI_ISL_1303532, EPI_ISL_1303533, EPI_ISL_1303534 | | | |
| see above | IAL Regional de São Jose do Rio Preto | Instituto Adolfo Lutz, Interdiciplinary Procedures Center, Strategic Laboratory | Claudio Tavares Sacchi, Claudia Regina Gonçalves, Erica Valessa Ramos Gomes, Karoline Rodrigues Campos, Caio Vinicius Dias Lopes |
| EPI_ISL_1303535, EPI_ISL_1303536 | Hospital Heliopolis | Instituto Adolfo Lutz, Interdiciplinary Procedures Center, Strategic Laboratory | Claudio Tavares Sacchi, Claudia Regina Gonçalves, Erica Valessa Ramos Gomes, Karoline Rodrigues Campos, Caio Vinicius Dias Lopes |
| EPI_ISL_1303537 | Hospital Estadual de Vila Alpina | Instituto Adolfo Lutz, Interdiciplinary Procedures Center, Strategic Laboratory | Claudio Tavares Sacchi, Claudia Regina Gonçalves, Erica Valessa Ramos Gomes, Karoline Rodrigues Campos, Caio Vinicius Dias Lopes |
| EPI_ISL_1303538, EPI_ISL_1303539 | Hospital Presidente | Instituto Adolfo Lutz, Interdiciplinary Procedures Center, Strategic Laboratory | Claudio Tavares Sacchi, Claudia Regina Gonçalves, Erica Valessa Ramos Gomes, Karoline Rodrigues Campos, Caio Vinicius Dias Lopes |
| EPI_ISL_1303540, EPI_ISL_1303541 | Hospital Municipal Cidade Tiradentes Carmen Prudente | Instituto Adolfo Lutz, Interdiciplinary Procedures Center, Strategic Laboratory | Claudio Tavares Sacchi, Claudia Regina Gonçalves, Erica Valessa Ramos Gomes, Karoline Rodrigues Campos, Caio Vinicius Dias Lopes |
| EPI_ISL_1303542, EPI_ISL_1303543 | Hospital Estadual de Campanha Barradas | Instituto Adolfo Lutz, Interdiciplinary Procedures Center, Strategic Laboratory | Claudio Tavares Sacchi, Claudia Regina Gonçalves, Erica Valessa Ramos Gomes, Karoline Rodrigues Campos, Caio Vinicius Dias Lopes |

| | | | |
|---|---|---|---|
| EPI_ISL_1303544, EPI_ISL_1303545 | Hospital Municipal Cidade Tiradentes Carmen Prudente | Instituto Adolfo Lutz, Interdiciplinary Procedures Center, Strategic Laboratory | Claudio Tavares Sacchi, Claudia Regina Gonçalves, Erica Valessa Ramos Gomes, Karoline Rodrigues Campos, Caio Vinicius Dias Lopes |
| EPI_ISL_1303546, EPI_ISL_1303547, EPI_ISL_1303548, EPI_ISL_1303549 | UPA Vila Santa Catarina | Instituto Adolfo Lutz, Interdiciplinary Procedures Center, Strategic Laboratory | Claudio Tavares Sacchi, Claudia Regina Gonçalves, Erica Valessa Ramos Gomes, Karoline Rodrigues Campos, Caio Vinicius Dias Lopes |
| EPI_ISL_1324137, EPI_ISL_1324139, EPI_ISL_1324140, EPI_ISL_1324142, EPI_ISL_1324145, EPI_ISL_1324147, EPI_ISL_1324149 | UW Virology Lab | UW Virology Lab | Pavitra Roychoudhury, Hong Xie, Lasata Shrestha, Shah Mohamed Bakhash, Michelle Lin, Margaret Mills, Noah Baker, Sean Ellis, Saraswathi Sathees, Meei-Li Huang, Keith R Jerome, Alexander Greninger |
| EPI_ISL_1358285 | Centro de Treinamento e Referencia DST AIDS | Instituto Adolfo Lutz, Interdiciplinary Procedures Center, Strategic Laboratory | Claudio Tavares Sacchi, Claudia Regina Gonçalves, Erica Valessa Ramos Gomes, Karoline Rodrigues Campos, Caio Vinicius Dias Lopes |
| EPI_ISL_1358286 | Hospital Municipal Josanias Castanha Braga | Instituto Adolfo Lutz, Interdiciplinary Procedures Center, Strategic Laboratory | Claudio Tavares Sacchi, Claudia Regina Gonçalves, Erica Valessa Ramos Gomes, Karoline Rodrigues Campos, Caio Vinicius Dias Lopes |
| EPI_ISL_1358287 | Hospital Nipo Brasileiro | Instituto Adolfo Lutz, Interdiciplinary Procedures Center, Strategic Laboratory | Claudio Tavares Sacchi, Claudia Regina Gonçalves, Erica Valessa Ramos Gomes, Karoline Rodrigues Campos, Caio Vinicius Dias Lopes |
| EPI_ISL_1358288, EPI_ISL_1358289, EPI_ISL_1358290 | IAL Regional de Aracatuba | Instituto Adolfo Lutz, Interdiciplinary Procedures Center, Strategic Laboratory | Claudio Tavares Sacchi, Claudia Regina Gonçalves, Erica Valessa Ramos Gomes, Karoline Rodrigues Campos, Caio Vinicius Dias Lopes |
| EPI_ISL_1358291, EPI_ISL_1358292, EPI_ISL_1358293, EPI_ISL_1358294, EPI_ISL_1358295, EPI_ISL_1358296, EPI_ISL_1358297, EPI_ISL_1358298, EPI_ISL_1358299 | IAL Regional de Santo Andre | Instituto Adolfo Lutz, Interdiciplinary Procedures Center, Strategic Laboratory | Claudio Tavares Sacchi, Claudia Regina Gonçalves, Erica Valessa Ramos Gomes, Karoline Rodrigues Campos, Caio Vinicius Dias Lopes |
| EPI_ISL_1358300, EPI_ISL_1358301, EPI_ISL_1358302, EPI_ISL_1358303 | Lacen de Tocantins | Instituto Adolfo Lutz, Interdiciplinary Procedures Center, Strategic Laboratory | Claudio Tavares Sacchi, Claudia Regina Gonçalves, Erica Valessa Ramos Gomes, Karoline Rodrigues Campos, Caio Vinicius Dias Lopes |
| EPI_ISL_1358304, EPI_ISL_1358305, EPI_ISL_1358306, EPI_ISL_1358307, EPI_ISL_1358308, EPI_ISL_1358309, EPI_ISL_1358310, EPI_ISL_1358311, EPI_ISL_1358312, EPI_ISL_1358313, EPI_ISL_1358314, EPI_ISL_1358315, EPI_ISL_1358316, EPI_ISL_1358317 | | | |
| see above | LACEN do Mato Grosso do Sul | Instituto Adolfo Lutz, Interdiciplinary Procedures Center, Strategic Laboratory | Claudio Tavares Sacchi, Claudia Regina Gonçalves, Erica Valessa Ramos Gomes, Karoline Rodrigues Campos, Caio Vinicius Dias Lopes |
| EPI_ISL_1358318, EPI_ISL_1358319, EPI_ISL_1358320, EPI_ISL_1358321, EPI_ISL_1358322 | UPA Vila Santa Catarina | Instituto Adolfo Lutz, Interdiciplinary Procedures Center, Strategic Laboratory | Claudio Tavares Sacchi, Claudia Regina Gonçalves, Erica Valessa Ramos Gomes, Karoline Rodrigues Campos, Caio Vinicius Dias Lopes |
| EPI_ISL_1365747 | Associação Fundo de Incentivo a Pesquisa | Associação Fundo de Incentivo à Pesquisa (AFIP) | Priscila Farias Tempaku, Juliana Nogueira Martins Rodrigues, Erika Rodrigues de Oliveira, Soraya Sgambatti de Andrade, Debora Ribeiro Ramadan, Sergio Tufik |
| EPI_ISL_1381043, EPI_ISL_1381044, EPI_ISL_1381045, EPI_ISL_1381046, EPI_ISL_1381047, EPI_ISL_1381048, EPI_ISL_1381049, EPI_ISL_1381050, EPI_ISL_1381051, EPI_ISL_1381052, EPI_ISL_1381053, EPI_ISL_1381054, EPI_ISL_1381055, EPI_ISL_1381056, EPI_ISL_1381057, EPI_ISL_1381058, EPI_ISL_1381059, EPI_ISL_1381060, EPI_ISL_1381061, EPI_ISL_1381062, EPI_ISL_1381063, EPI_ISL_1381064, EPI_ISL_1381065 | | | |
| see above | IAL Regional de Santo Andre | Instituto Adolfo Lutz, Interdiciplinary Procedures Center, Strategic Laboratory | Claudio Tavares Sacchi, Claudia Regina Gonçalves, Erica Valessa Ramos Gomes, Karoline Rodrigues Campos, Caio Vinicius Dias Lopes |
| EPI_ISL_1381066, EPI_ISL_1381067 | LACEN do Mato Grosso do Sul | Instituto Adolfo Lutz, Interdiciplinary Procedures Center, Strategic Laboratory | Claudio Tavares Sacchi, Claudia Regina Gonçalves, Erica Valessa Ramos Gomes, Karoline Rodrigues Campos, Caio Vinicius Dias Lopes |
| EPI_ISL_1381068 | Conjunto Hospitalar do Mandaqui de Sao Paulo | Instituto Adolfo Lutz, Interdiciplinary Procedures Center, Strategic Laboratory | Claudio Tavares Sacchi, Claudia Regina Gonçalves, Erica Valessa Ramos Gomes, Karoline Rodrigues Campos, Caio Vinicius Dias Lopes |
| EPI_ISL_1381069 | Hospital Heliopolis | Instituto Adolfo Lutz, Interdiciplinary Procedures Center, Strategic Laboratory | Claudio Tavares Sacchi, Claudia Regina Gonçalves, Erica Valessa Ramos Gomes, Karoline Rodrigues Campos, Caio Vinicius Dias Lopes |
| EPI_ISL_1381070, EPI_ISL_1381071 | Hospital Municipal Cidade Tiradentes Carmem Prudente | Instituto Adolfo Lutz, Interdiciplinary Procedures Center, Strategic Laboratory | Claudio Tavares Sacchi, Claudia Regina Gonçalves, Erica Valessa Ramos Gomes, Karoline Rodrigues Campos, Caio Vinicius Dias Lopes |
| EPI_ISL_1402429, EPI_ISL_1402430 | Laboratory of Respiratory Viruses and Measles, Oswaldo Cruz Institute, FIOCRUZ | Laboratory of Respiratory Viruses and Measles, Oswaldo Cruz Institute, FIOCRUZ | Paola Resende, Alex Pauvolid-Correa, Mia Ferreira Araujo, Ana Beatriz Machado Lima, Luciana Appolinario, Fernando Motta, Anna Carolina Paixao, Ana Carolina Mendonca, Alice Sampaio Rocha, Renata Serrano Lopes, Marilda Siqueira on behalf of the Fiocruz COVID-19 Genomic Surveillance Network |
| EPI_ISL_1402431 | Laboratory of Respiratory Viruses and Measles, Oswaldo Cruz Institute, FIOCRUZ | Laboratory of Respiratory Viruses and Measles, Oswaldo Cruz Institute, FIOCRUZ | Paola Resende, Felipe Naveca, Alex Pauvolid-Correa, Mia Ferreira Araujo, Ana Beatriz Machado Lima, Luciana Appolinario, Fernando Motta, Anna Carolina Paixao, Ana Carolina Mendonca, Alice Sampaio Rocha, Renata Serrano Lopes, Marilda Siqueira on behalf of the Fiocruz COVID-19 Genomic Surveillance Network |
| EPI_ISL_1443196, EPI_ISL_1443197, EPI_ISL_1443198 | Hospital Aliança | Hospital São Rafael - IDOR | Isadora Cristina de Siqueira, Aquiles Assunção Camelier, Elves A.P. Maciel, Margarida Celia L. C. Neves, Carolina Kymie Vasques Nonaka, Karoline Almeida Félix de Sousa, Victor Costa Araujo, Yasmin Santos Freitas Macêdo, Aurea Angelica Paste, Bruno Solano de Freitas Souza, Tiago Gräf |
| EPI_ISL_1445065 | UNIDADE MISTA DE TAIACU | Instituto Butantan / Mendelics | Dimas Tadeu Covas, Sandra Coccuzzo Sampaio, Maria Carolina Elias, José Salvatore Leister Patané, Vincent Louis Viala, Antonio Jorge Martins, Ricardo Haddad, Claudia Renata dos Santos Barros, Elaine Cristina Marqueze, Raul Machado Neto, Debora Botequio Moretti, Bibiana Santos, João Paulo Kitajima, Erika Freitas, David Schlesinger, Simone Kashima, Evandra Strazza Rodrigues, Svetoslav Nanev Slavov, Elaine Vieira dos Santos, Rafael dos Santos Bezerra, Luiz Carlos Junior de Alcantara, Marta Giovanetti, Vagner Fonseca, Flavia Aburjaile, Rodrigo Tocantins Calado. |
| EPI_ISL_1445066 | SECRETARIA MUNICIPAL DA SAUDE DE GUARIBA | Instituto Butantan / Mendelics | Dimas Tadeu Covas, Sandra Coccuzzo Sampaio, Maria Carolina Elias, José Salvatore Leister Patané, Vincent Louis Viala, Antonio Jorge Martins, Ricardo Haddad, Claudia Renata dos Santos Barros, Elaine Cristina Marqueze, Raul Machado Neto, Debora Botequio Moretti, Bibiana Santos, João Paulo Kitajima, Erika Freitas, David Schlesinger, Simone Kashima, Evandra Strazza Rodrigues, Svetoslav Nanev Slavov, Elaine Vieira dos Santos, Rafael dos Santos Bezerra, Luiz Carlos Junior de Alcantara, Marta Giovanetti, Vagner Fonseca, Flavia Aburjaile, Rodrigo Tocantins Calado. |
| EPI_ISL_1445067 | USF TRES PONTES | Instituto Butantan / Mendelics | Dimas Tadeu Covas, Sandra Coccuzzo Sampaio, Maria Carolina Elias, José Salvatore Leister Patané, Vincent Louis Viala, Antonio Jorge Martins, Ricardo Haddad, Claudia Renata dos Santos Barros, Elaine Cristina Marqueze, Raul Machado Neto, Debora Botequio Moretti, Bibiana Santos, João Paulo Kitajima, Erika Freitas, David Schlesinger, Simone Kashima, Evandra Strazza Rodrigues, Svetoslav Nanev Slavov, Elaine Vieira dos Santos, Rafael dos Santos Bezerra, Luiz Carlos Junior de Alcantara, Marta Giovanetti, Vagner Fonseca, Flavia Aburjaile, Rodrigo Tocantins Calado. |
| EPI_ISL_1445068 | USF JARDIM DAS AVES MOREIRINHA | Instituto Butantan / Mendelics | Dimas Tadeu Covas, Sandra Coccuzzo Sampaio, Maria Carolina Elias, José Salvatore Leister Patané, Vincent Louis Viala, Antonio Jorge Martins, Ricardo Haddad, Claudia Renata dos Santos Barros, Elaine Cristina Marqueze, Raul Machado Neto, Debora Botequio Moretti, Bibiana Santos, João Paulo Kitajima, Erika Freitas, David Schlesinger, Simone Kashima, Evandra Strazza Rodrigues, Svetoslav Nanev Slavov, Elaine Vieira dos Santos, Rafael dos Santos Bezerra, Luiz Carlos Junior de Alcantara, Marta Giovanetti, Vagner Fonseca, Flavia Aburjaile, Rodrigo Tocantins Calado. |
| EPI_ISL_1445069 | USF JARDIM SAO DIMAS | Instituto Butantan / Mendelics | Dimas Tadeu Covas, Sandra Coccuzzo Sampaio, Maria Carolina Elias, José Salvatore Leister Patané, Vincent Louis Viala, Antonio Jorge Martins, Ricardo Haddad, Claudia Renata dos Santos Barros, Elaine Cristina Marqueze, Raul Machado Neto, Debora Botequio Moretti, Bibiana Santos, João Paulo Kitajima, Erika Freitas, David Schlesinger, Simone Kashima, Evandra Strazza Rodrigues, Svetoslav Nanev Slavov, Elaine Vieira dos Santos, Rafael dos Santos Bezerra, Luiz Carlos Junior de Alcantara, Marta Giovanetti, Vagner Fonseca, Flavia Aburjaile, Rodrigo Tocantins Calado. |
| EPI_ISL_1445070 | USF ARCADAS | Instituto Butantan / Mendelics | Dimas Tadeu Covas, Sandra Coccuzzo Sampaio, Maria Carolina Elias, José Salvatore Leister Patané, Vincent Louis Viala, Antonio Jorge Martins, Ricardo Haddad, Claudia Renata dos Santos Barros, Elaine Cristina Marqueze, Raul Machado Neto, Debora Botequio Moretti, Bibiana Santos, João Paulo Kitajima, Erika Freitas, David Schlesinger, Simone Kashima, Evandra Strazza Rodrigues, Svetoslav Nanev Slavov, Elaine Vieira dos Santos, Rafael dos Santos Bezerra, Luiz Carlos Junior de Alcantara, Marta Giovanetti, Vagner Fonseca, Flavia Aburjaile, Rodrigo Tocantins Calado. |

| | | | |
|---|---|---|---|
| EPI_ISL_1445071 | USF TRES PONTES | Instituto Butantan / Mendelics | Dimas Tadeu Covas, Sandra Coccuzzo Sampaio, Maria Carolina Elias, José Salvatore Leister Patané, Vincent Louis Viala, Antonio Jorge Martins, Ricardo Haddad, Claudia Renata dos Santos Barros, Elaine Cristina Marqueze, Raul Machado Neto, Debora Botequio Moretti, Bibiana Santos, João Paulo Kitajima, Erika Freitas, David Schlesinger, Simone Kashima, Evandra Strazza Rodrigues, Svetoslav Nanev Slavov, Elaine Vieira dos Santos, Rafael dos Santos Bezerra, Luiz Carlos Junior de Alcantara, Marta Giovanetti, Vagner Fonseca, Flavia Aburjaile, Rodrigo Tocantins Calado. |
| EPI_ISL_1445072 | USF JARDIM SAO DIMAS | Instituto Butantan / Mendelics | Dimas Tadeu Covas, Sandra Coccuzzo Sampaio, Maria Carolina Elias, José Salvatore Leister Patané, Vincent Louis Viala, Antonio Jorge Martins, Ricardo Haddad, Claudia Renata dos Santos Barros, Elaine Cristina Marqueze, Raul Machado Neto, Debora Botequio Moretti, Bibiana Santos, João Paulo Kitajima, Erika Freitas, David Schlesinger, Simone Kashima, Evandra Strazza Rodrigues, Svetoslav Nanev Slavov, Elaine Vieira dos Santos, Rafael dos Santos Bezerra, Luiz Carlos Junior de Alcantara, Marta Giovanetti, Vagner Fonseca, Flavia Aburjaile, Rodrigo Tocantins Calado. |
| EPI_ISL_1445073 | USF SANTA MARIA DO AMPARO | Instituto Butantan / Mendelics | Dimas Tadeu Covas, Sandra Coccuzzo Sampaio, Maria Carolina Elias, José Salvatore Leister Patané, Vincent Louis Viala, Antonio Jorge Martins, Ricardo Haddad, Claudia Renata dos Santos Barros, Elaine Cristina Marqueze, Raul Machado Neto, Debora Botequio Moretti, Bibiana Santos, João Paulo Kitajima, Erika Freitas, David Schlesinger, Simone Kashima, Evandra Strazza Rodrigues, Svetoslav Nanev Slavov, Elaine Vieira dos Santos, Rafael dos Santos Bezerra, Luiz Carlos Junior de Alcantara, Marta Giovanetti, Vagner Fonseca, Flavia Aburjaile, Rodrigo Tocantins Calado. |
| EPI_ISL_1445074 | USF TRES PONTES | Instituto Butantan / Mendelics | Dimas Tadeu Covas, Sandra Coccuzzo Sampaio, Maria Carolina Elias, José Salvatore Leister Patané, Vincent Louis Viala, Antonio Jorge Martins, Ricardo Haddad, Claudia Renata dos Santos Barros, Elaine Cristina Marqueze, Raul Machado Neto, Debora Botequio Moretti, Bibiana Santos, João Paulo Kitajima, Erika Freitas, David Schlesinger, Simone Kashima, Evandra Strazza Rodrigues, Svetoslav Nanev Slavov, Elaine Vieira dos Santos, Rafael dos Santos Bezerra, Luiz Carlos Junior de Alcantara, Marta Giovanetti, Vagner Fonseca, Flavia Aburjaile, Rodrigo Tocantins Calado. |
| EPI_ISL_1445075, EPI_ISL_1445076, EPI_ISL_1445077, EPI_ISL_1445078, EPI_ISL_1445079 | CENTRO DE SAUDE II DR GABRIEL MESQUITA VARGEM GDE DO SUL | Instituto Butantan / Mendelics | Dimas Tadeu Covas, Sandra Coccuzzo Sampaio, Maria Carolina Elias, José Salvatore Leister Patané, Vincent Louis Viala, Antonio Jorge Martins, Ricardo Haddad, Claudia Renata dos Santos Barros, Elaine Cristina Marqueze, Raul Machado Neto, Debora Botequio Moretti, Bibiana Santos, João Paulo Kitajima, Erika Freitas, David Schlesinger, Simone Kashima, Evandra Strazza Rodrigues, Svetoslav Nanev Slavov, Elaine Vieira dos Santos, Rafael dos Santos Bezerra, Luiz Carlos Junior de Alcantara, Marta Giovanetti, Vagner Fonseca, Flavia Aburjaile, Rodrigo Tocantins Calado. |
| EPI_ISL_1445080 | SAUDE COLETIVA CAPAO BONITO | Instituto Butantan / Mendelics | Dimas Tadeu Covas, Sandra Coccuzzo Sampaio, Maria Carolina Elias, José Salvatore Leister Patané, Vincent Louis Viala, Antonio Jorge Martins, Ricardo Haddad, Claudia Renata dos Santos Barros, Elaine Cristina Marqueze, Raul Machado Neto, Debora Botequio Moretti, Bibiana Santos, João Paulo Kitajima, Erika Freitas, David Schlesinger, Simone Kashima, Evandra Strazza Rodrigues, Svetoslav Nanev Slavov, Elaine Vieira dos Santos, Rafael dos Santos Bezerra, Luiz Carlos Junior de Alcantara, Marta Giovanetti, Vagner Fonseca, Flavia Aburjaile, Rodrigo Tocantins Calado. |
| EPI_ISL_1445081 | CENTRO DE SAUDE SAO ROQUE DR JOSE CARVALHO BRITO | Instituto Butantan / Mendelics | Dimas Tadeu Covas, Sandra Coccuzzo Sampaio, Maria Carolina Elias, José Salvatore Leister Patané, Vincent Louis Viala, Antonio Jorge Martins, Ricardo Haddad, Claudia Renata dos Santos Barros, Elaine Cristina Marqueze, Raul Machado Neto, Debora Botequio Moretti, Bibiana Santos, João Paulo Kitajima, Erika Freitas, David Schlesinger, Simone Kashima, Evandra Strazza Rodrigues, Svetoslav Nanev Slavov, Elaine Vieira dos Santos, Rafael dos Santos Bezerra, Luiz Carlos Junior de Alcantara, Marta Giovanetti, Vagner Fonseca, Flavia Aburjaile, Rodrigo Tocantins Calado. |
| EPI_ISL_1445082 | SECRETARIA MUNICIPAL DE SAUDE SOROCABA | Instituto Butantan / Mendelics | Dimas Tadeu Covas, Sandra Coccuzzo Sampaio, Maria Carolina Elias, José Salvatore Leister Patané, Vincent Louis Viala, Antonio Jorge Martins, Ricardo Haddad, Claudia Renata dos Santos Barros, Elaine Cristina Marqueze, Raul Machado Neto, Debora Botequio Moretti, Bibiana Santos, João Paulo Kitajima, Erika Freitas, David Schlesinger, Simone Kashima, Evandra Strazza Rodrigues, Svetoslav Nanev Slavov, Elaine Vieira dos Santos, Rafael dos Santos Bezerra, Luiz Carlos Junior de Alcantara, Marta Giovanetti, Vagner Fonseca, Flavia Aburjaile, Rodrigo Tocantins Calado. |
| EPI_ISL_1445083 | HOSPITAL REGIONAL DE ITAPETININGA | Instituto Butantan / Mendelics | Dimas Tadeu Covas, Sandra Coccuzzo Sampaio, Maria Carolina Elias, José Salvatore Leister Patané, Vincent Louis Viala, Antonio Jorge Martins, Ricardo Haddad, Claudia Renata dos Santos Barros, Elaine Cristina Marqueze, Raul Machado Neto, Debora Botequio Moretti, Bibiana Santos, João Paulo Kitajima, Erika Freitas, David Schlesinger, Simone Kashima, Evandra Strazza Rodrigues, Svetoslav Nanev Slavov, Elaine Vieira dos Santos, Rafael dos Santos Bezerra, Luiz Carlos Junior de Alcantara, Marta Giovanetti, Vagner Fonseca, Flavia Aburjaile, Rodrigo Tocantins Calado. |
| EPI_ISL_1445084 | SMS SECRETARIA MUNICIPAL DE SAUDE DE BOITUVA | Instituto Butantan / Mendelics | Dimas Tadeu Covas, Sandra Coccuzzo Sampaio, Maria Carolina Elias, José Salvatore Leister Patané, Vincent Louis Viala, Antonio Jorge Martins, Ricardo Haddad, Claudia Renata dos Santos Barros, Elaine Cristina Marqueze, Raul Machado Neto, Debora Botequio Moretti, Bibiana Santos, João Paulo Kitajima, Erika Freitas, David Schlesinger, Simone Kashima, Evandra Strazza Rodrigues, Svetoslav Nanev Slavov, Elaine Vieira dos Santos, Rafael dos Santos Bezerra, Luiz Carlos Junior de Alcantara, Marta Giovanetti, Vagner Fonseca, Flavia Aburjaile, Rodrigo Tocantins Calado. |
| EPI_ISL_1445085 | SAUDE COLETIVA CAPAO BONITO | Instituto Butantan / Mendelics | Dimas Tadeu Covas, Sandra Coccuzzo Sampaio, Maria Carolina Elias, José Salvatore Leister Patané, Vincent Louis Viala, Antonio Jorge Martins, Ricardo Haddad, Claudia Renata dos Santos Barros, Elaine Cristina Marqueze, Raul Machado Neto, Debora Botequio Moretti, Bibiana Santos, João Paulo Kitajima, Erika Freitas, David Schlesinger, Simone Kashima, Evandra Strazza Rodrigues, Svetoslav Nanev Slavov, Elaine Vieira dos Santos, Rafael dos Santos Bezerra, Luiz Carlos Junior de Alcantara, Marta Giovanetti, Vagner Fonseca, Flavia Aburjaile, Rodrigo Tocantins Calado. |
| EPI_ISL_1445086 | CENTRO DE SAUDE II MAIRINQUE MAIRINQUE | Instituto Butantan / Mendelics | Dimas Tadeu Covas, Sandra Coccuzzo Sampaio, Maria Carolina Elias, José Salvatore Leister Patané, Vincent Louis Viala, Antonio Jorge Martins, Ricardo Haddad, Claudia Renata dos Santos Barros, Elaine Cristina Marqueze, Raul Machado Neto, Debora Botequio Moretti, Bibiana Santos, João Paulo Kitajima, Erika Freitas, David Schlesinger, Simone Kashima, Evandra Strazza Rodrigues, Svetoslav Nanev Slavov, Elaine Vieira dos Santos, Rafael dos Santos Bezerra, Luiz Carlos Junior de Alcantara, Marta Giovanetti, Vagner Fonseca, Flavia Aburjaile, Rodrigo Tocantins Calado. |
| EPI_ISL_1445087, EPI_ISL_1445088 | SECRETARIA MUNICIPAL DE SAUDE SOROCABA | Instituto Butantan / Mendelics | Dimas Tadeu Covas, Sandra Coccuzzo Sampaio, Maria Carolina Elias, José Salvatore Leister Patané, Vincent Louis Viala, Antonio Jorge Martins, Ricardo Haddad, Claudia Renata dos Santos Barros, Elaine Cristina Marqueze, Raul Machado Neto, Debora Botequio Moretti, Bibiana Santos, João Paulo Kitajima, Erika Freitas, David Schlesinger, Simone Kashima, Evandra Strazza Rodrigues, Svetoslav Nanev Slavov, Elaine Vieira dos Santos, Rafael dos Santos Bezerra, Luiz Carlos Junior de Alcantara, Marta Giovanetti, Vagner Fonseca, Flavia Aburjaile, Rodrigo Tocantins Calado. |
| EPI_ISL_1445089 | CENTRO DE SAUDE II MAIRINQUE MAIRINQUE | Instituto Butantan / Mendelics | Dimas Tadeu Covas, Sandra Coccuzzo Sampaio, Maria Carolina Elias, José Salvatore Leister Patané, Vincent Louis Viala, Antonio Jorge Martins, Ricardo Haddad, Claudia Renata dos Santos Barros, Elaine Cristina Marqueze, Raul Machado Neto, Debora Botequio Moretti, Bibiana Santos, João Paulo Kitajima, Erika Freitas, David Schlesinger, Simone Kashima, Evandra Strazza Rodrigues, Svetoslav Nanev Slavov, Elaine Vieira dos Santos, Rafael dos Santos Bezerra, Luiz Carlos Junior de Alcantara, Marta Giovanetti, Vagner Fonseca, Flavia Aburjaile, Rodrigo Tocantins Calado. |
| EPI_ISL_1445090 | POLICLINICA COVID 19 ITAPETININGA | Instituto Butantan / Mendelics | Dimas Tadeu Covas, Sandra Coccuzzo Sampaio, Maria Carolina Elias, José Salvatore Leister Patané, Vincent Louis Viala, Antonio Jorge Martins, Ricardo Haddad, Claudia Renata dos Santos Barros, Elaine Cristina Marqueze, Raul Machado Neto, Debora Botequio Moretti, Bibiana Santos, João Paulo Kitajima, Erika Freitas, David Schlesinger, Simone Kashima, Evandra Strazza Rodrigues, Svetoslav Nanev Slavov, Elaine Vieira dos Santos, Rafael dos Santos Bezerra, Luiz Carlos Junior de Alcantara, Marta Giovanetti, Vagner Fonseca, Flavia Aburjaile, Rodrigo Tocantins Calado. |
| EPI_ISL_1445091, EPI_ISL_1445092, EPI_ISL_1445093, EPI_ISL_1445094, EPI_ISL_1445095 | SECRETARIA MUNICIPAL DE SAUDE DE PIRACAIA | Instituto Butantan / Mendelics | Dimas Tadeu Covas, Sandra Coccuzzo Sampaio, Maria Carolina Elias, José Salvatore Leister Patané, Vincent Louis Viala, Antonio Jorge Martins, Ricardo Haddad, Claudia Renata dos Santos Barros, Elaine Cristina Marqueze, Raul Machado Neto, Debora Botequio Moretti, Bibiana Santos, João Paulo Kitajima, Erika Freitas, David Schlesinger, Simone Kashima, Evandra Strazza Rodrigues, Svetoslav Nanev Slavov, Elaine Vieira dos Santos, Rafael dos Santos Bezerra, Luiz Carlos Junior de Alcantara, Marta Giovanetti, Vagner Fonseca, Flavia Aburjaile, Rodrigo Tocantins Calado. |
| EPI_ISL_1445096, EPI_ISL_1445097, EPI_ISL_1445098, EPI_ISL_1445099, EPI_ISL_1445100 | PRONTO ATENDIMENTO VILA PADRE ANCHIETA | Instituto Butantan / Mendelics | Dimas Tadeu Covas, Sandra Coccuzzo Sampaio, Maria Carolina Elias, José Salvatore Leister Patané, Vincent Louis Viala, Antonio Jorge Martins, Ricardo Haddad, Claudia Renata dos Santos Barros, Elaine Cristina Marqueze, Raul Machado Neto, Debora Botequio Moretti, Bibiana Santos, João Paulo Kitajima, Erika Freitas, David Schlesinger, Simone Kashima, Evandra Strazza Rodrigues, Svetoslav Nanev Slavov, Elaine Vieira dos Santos, Rafael dos Santos Bezerra, Luiz Carlos Junior de Alcantara, Marta Giovanetti, Vagner Fonseca, Flavia Aburjaile, Rodrigo Tocantins Calado. |
| EPI_ISL_1445101 | SECRETARIA MUNICIPAL DE SAUDE DE PIRACAIA | Instituto Butantan / Mendelics | Dimas Tadeu Covas, Sandra Coccuzzo Sampaio, Maria Carolina Elias, José Salvatore Leister Patané, Vincent Louis Viala, Antonio Jorge Martins, Ricardo Haddad, Claudia Renata dos Santos Barros, Elaine Cristina Marqueze, Raul Machado Neto, Debora Botequio Moretti, Bibiana Santos, João Paulo Kitajima, Erika Freitas, David Schlesinger, Simone Kashima, Evandra Strazza Rodrigues, Svetoslav Nanev Slavov, Elaine Vieira dos Santos, Rafael dos Santos Bezerra, Luiz Carlos Junior de Alcantara, Marta Giovanetti, Vagner Fonseca, Flavia Aburjaile, Rodrigo Tocantins Calado. |
| EPI_ISL_1445102, EPI_ISL_1445103, EPI_ISL_1445104, EPI_ISL_1445105, EPI_ISL_1445106, EPI_ISL_1445107, EPI_ISL_1445108 | PRONTO ATENDIMENTO VILA PADRE ANCHIETA | Instituto Butantan / Mendelics | Dimas Tadeu Covas, Sandra Coccuzzo Sampaio, Maria Carolina Elias, José Salvatore Leister Patané, Vincent Louis Viala, Antonio Jorge Martins, Ricardo Haddad, Claudia Renata dos Santos Barros, Elaine Cristina Marqueze, Raul Machado Neto, Debora Botequio Moretti, Bibiana Santos, João Paulo Kitajima, Erika Freitas, David Schlesinger, Simone Kashima, Evandra Strazza Rodrigues, Svetoslav Nanev Slavov, Elaine Vieira dos Santos, Rafael dos Santos Bezerra, Luiz Carlos Junior de Alcantara, Marta Giovanetti, Vagner Fonseca, Flavia Aburjaile, Rodrigo Tocantins Calado. |
| EPI_ISL_1445109, EPI_ISL_1445110 | COMPLEXO HOSPITALAR OURO VERDE DE CAMPINAS | Instituto Butantan / Mendelics | Dimas Tadeu Covas, Sandra Coccuzzo Sampaio, Maria Carolina Elias, José Salvatore Leister Patané, Vincent Louis Viala, Antonio Jorge Martins, Ricardo Haddad, Claudia Renata dos Santos Barros, Elaine Cristina Marqueze, Raul Machado Neto, Debora Botequio Moretti, Bibiana Santos, João Paulo |

| | | | |
|---|---|---|---|
| | | | Kitajima, Erika Freitas, David Schlesinger, Simone Kashima, Evandra Strazza Rodrigues, Svetoslav Nanev Slavov, Elaine Vieira dos Santos, Rafael dos Santos Bezerra, Luiz Carlos Junior de Alcantara, Marta Giovanetti, Vagner Fonseca, Flavia Aburjaile, Rodrigo Tocantins Calado. |
| EPI_ISL_1445111, EPI_ISL_1445112 | PRONTO ATENDIMENTO VILA PADRE ANCHIETA | Instituto Butantan / Mendelics | Dimas Tadeu Covas, Sandra Coccuzzo Sampaio, Maria Carolina Elias, José Salvatore Leister Patané, Vincent Louis Viala, Antonio Jorge Martins, Ricardo Haddad, Claudia Renata dos Santos Barros, Elaine Cristina Marqueze, Raul Machado Neto, Debora Botequio Moretti, Bibiana Santos, João Paulo Kitajima, Erika Freitas, David Schlesinger, Simone Kashima, Evandra Strazza Rodrigues, Svetoslav Nanev Slavov, Elaine Vieira dos Santos, Rafael dos Santos Bezerra, Luiz Carlos Junior de Alcantara, Marta Giovanetti, Vagner Fonseca, Flavia Aburjaile, Rodrigo Tocantins Calado. |
| EPI_ISL_1445113 | UNIDADE RESPIRATORIA NOVA HORTOLANDIA | Instituto Butantan / Mendelics | Dimas Tadeu Covas, Sandra Coccuzzo Sampaio, Maria Carolina Elias, José Salvatore Leister Patané, Vincent Louis Viala, Antonio Jorge Martins, Ricardo Haddad, Claudia Renata dos Santos Barros, Elaine Cristina Marqueze, Raul Machado Neto, Debora Botequio Moretti, Bibiana Santos, João Paulo Kitajima, Erika Freitas, David Schlesinger, Simone Kashima, Evandra Strazza Rodrigues, Svetoslav Nanev Slavov, Elaine Vieira dos Santos, Rafael dos Santos Bezerra, Luiz Carlos Junior de Alcantara, Marta Giovanetti, Vagner Fonseca, Flavia Aburjaile, Rodrigo Tocantins Calado. |
| EPI_ISL_1445114, EPI_ISL_1445115, EPI_ISL_1445116, EPI_ISL_1445117, EPI_ISL_1445118 | COMPLEXO HOSPITALAR OURO VERDE DE CAMPINAS | Instituto Butantan / Mendelics | Dimas Tadeu Covas, Sandra Coccuzzo Sampaio, Maria Carolina Elias, José Salvatore Leister Patané, Vincent Louis Viala, Antonio Jorge Martins, Ricardo Haddad, Claudia Renata dos Santos Barros, Elaine Cristina Marqueze, Raul Machado Neto, Debora Botequio Moretti, Bibiana Santos, João Paulo Kitajima, Erika Freitas, David Schlesinger, Simone Kashima, Evandra Strazza Rodrigues, Svetoslav Nanev Slavov, Elaine Vieira dos Santos, Rafael dos Santos Bezerra, Luiz Carlos Junior de Alcantara, Marta Giovanetti, Vagner Fonseca, Flavia Aburjaile, Rodrigo Tocantins Calado. |
| EPI_ISL_1445119 | UNIDADE RESPIRATORIA NOVA HORTOLANDIA | Instituto Butantan / Mendelics | Dimas Tadeu Covas, Sandra Coccuzzo Sampaio, Maria Carolina Elias, José Salvatore Leister Patané, Vincent Louis Viala, Antonio Jorge Martins, Ricardo Haddad, Claudia Renata dos Santos Barros, Elaine Cristina Marqueze, Raul Machado Neto, Debora Botequio Moretti, Bibiana Santos, João Paulo Kitajima, Erika Freitas, David Schlesinger, Simone Kashima, Evandra Strazza Rodrigues, Svetoslav Nanev Slavov, Elaine Vieira dos Santos, Rafael dos Santos Bezerra, Luiz Carlos Junior de Alcantara, Marta Giovanetti, Vagner Fonseca, Flavia Aburjaile, Rodrigo Tocantins Calado. |
| EPI_ISL_1445120, EPI_ISL_1445121, EPI_ISL_1445122, EPI_ISL_1445123, EPI_ISL_1445124, EPI_ISL_1445125, EPI_ISL_1445126 | UNIDADE DE PRONTO ATENDIMENTO UPA DRA ANA OLIVIA BENTIVOGLIO | Instituto Butantan / Mendelics | Dimas Tadeu Covas, Sandra Coccuzzo Sampaio, Maria Carolina Elias, José Salvatore Leister Patané, Vincent Louis Viala, Antonio Jorge Martins, Ricardo Haddad, Claudia Renata dos Santos Barros, Elaine Cristina Marqueze, Raul Machado Neto, Debora Botequio Moretti, Bibiana Santos, João Paulo Kitajima, Erika Freitas, David Schlesinger, Simone Kashima, Evandra Strazza Rodrigues, Svetoslav Nanev Slavov, Elaine Vieira dos Santos, Rafael dos Santos Bezerra, Luiz Carlos Junior de Alcantara, Marta Giovanetti, Vagner Fonseca, Flavia Aburjaile, Rodrigo Tocantins Calado. |
| EPI_ISL_1445127, EPI_ISL_1445128, EPI_ISL_1445129 | UNIDADE RESPIRATORIA NOVA HORTOLANDIA | Instituto Butantan / Mendelics | Dimas Tadeu Covas, Sandra Coccuzzo Sampaio, Maria Carolina Elias, José Salvatore Leister Patané, Vincent Louis Viala, Antonio Jorge Martins, Ricardo Haddad, Claudia Renata dos Santos Barros, Elaine Cristina Marqueze, Raul Machado Neto, Debora Botequio Moretti, Bibiana Santos, João Paulo Kitajima, Erika Freitas, David Schlesinger, Simone Kashima, Evandra Strazza Rodrigues, Svetoslav Nanev Slavov, Elaine Vieira dos Santos, Rafael dos Santos Bezerra, Luiz Carlos Junior de Alcantara, Marta Giovanetti, Vagner Fonseca, Flavia Aburjaile, Rodrigo Tocantins Calado. |
| EPI_ISL_1445130, EPI_ISL_1445131, EPI_ISL_1445132 | PRONTO ATENDIMENTO DA PONTE DE SAO JOAO | Instituto Butantan / Mendelics | Dimas Tadeu Covas, Sandra Coccuzzo Sampaio, Maria Carolina Elias, José Salvatore Leister Patané, Vincent Louis Viala, Antonio Jorge Martins, Ricardo Haddad, Claudia Renata dos Santos Barros, Elaine Cristina Marqueze, Raul Machado Neto, Debora Botequio Moretti, Bibiana Santos, João Paulo Kitajima, Erika Freitas, David Schlesinger, Simone Kashima, Evandra Strazza Rodrigues, Svetoslav Nanev Slavov, Elaine Vieira dos Santos, Rafael dos Santos Bezerra, Luiz Carlos Junior de Alcantara, Marta Giovanetti, Vagner Fonseca, Flavia Aburjaile, Rodrigo Tocantins Calado. |
| EPI_ISL_1445133, EPI_ISL_1445134, EPI_ISL_1445135 | SECRETARIA MUNICIPAL DE SAUDE SOROCABA | Instituto Butantan / Mendelics | Dimas Tadeu Covas, Sandra Coccuzzo Sampaio, Maria Carolina Elias, José Salvatore Leister Patané, Vincent Louis Viala, Antonio Jorge Martins, Ricardo Haddad, Claudia Renata dos Santos Barros, Elaine Cristina Marqueze, Raul Machado Neto, Debora Botequio Moretti, Bibiana Santos, João Paulo Kitajima, Erika Freitas, David Schlesinger, Simone Kashima, Evandra Strazza Rodrigues, Svetoslav Nanev Slavov, Elaine Vieira dos Santos, Rafael dos Santos Bezerra, Luiz Carlos Junior de Alcantara, Marta Giovanetti, Vagner Fonseca, Flavia Aburjaile, Rodrigo Tocantins Calado. |
| EPI_ISL_1445136 | SMS IPERO | Instituto Butantan / Mendelics | Dimas Tadeu Covas, Sandra Coccuzzo Sampaio, Maria Carolina Elias, José Salvatore Leister Patané, Vincent Louis Viala, Antonio Jorge Martins, Ricardo Haddad, Claudia Renata dos Santos Barros, Elaine Cristina Marqueze, Raul Machado Neto, Debora Botequio Moretti, Bibiana Santos, João Paulo Kitajima, Erika Freitas, David Schlesinger, Simone Kashima, Evandra Strazza Rodrigues, Svetoslav Nanev Slavov, Elaine Vieira dos Santos, Rafael dos Santos Bezerra, Luiz Carlos Junior de Alcantara, Marta Giovanetti, Vagner Fonseca, Flavia Aburjaile, Rodrigo Tocantins Calado. |
| EPI_ISL_1445137, EPI_ISL_1445138, EPI_ISL_1445139 | SECRETARIA MUNICIPAL DE SAUDE SOROCABA | Instituto Butantan / Mendelics | Dimas Tadeu Covas, Sandra Coccuzzo Sampaio, Maria Carolina Elias, José Salvatore Leister Patané, Vincent Louis Viala, Antonio Jorge Martins, Ricardo Haddad, Claudia Renata dos Santos Barros, Elaine Cristina Marqueze, Raul Machado Neto, Debora Botequio Moretti, Bibiana Santos, João Paulo Kitajima, Erika Freitas, David Schlesinger, Simone Kashima, Evandra Strazza Rodrigues, Svetoslav Nanev Slavov, Elaine Vieira dos Santos, Rafael dos Santos Bezerra, Luiz Carlos Junior de Alcantara, Marta Giovanetti, Vagner Fonseca, Flavia Aburjaile, Rodrigo Tocantins Calado. |
| EPI_ISL_1445140 | UNIDADE MISTA DE SAUDE CAPELA DO ALTO | Instituto Butantan / Mendelics | Dimas Tadeu Covas, Sandra Coccuzzo Sampaio, Maria Carolina Elias, José Salvatore Leister Patané, Vincent Louis Viala, Antonio Jorge Martins, Ricardo Haddad, Claudia Renata dos Santos Barros, Elaine Cristina Marqueze, Raul Machado Neto, Debora Botequio Moretti, Bibiana Santos, João Paulo Kitajima, Erika Freitas, David Schlesinger, Simone Kashima, Evandra Strazza Rodrigues, Svetoslav Nanev Slavov, Elaine Vieira dos Santos, Rafael dos Santos Bezerra, Luiz Carlos Junior de Alcantara, Marta Giovanetti, Vagner Fonseca, Flavia Aburjaile, Rodrigo Tocantins Calado. |
| EPI_ISL_1445141 | UBS SALTO DE SAO JOSE | Instituto Butantan / Mendelics | Dimas Tadeu Covas, Sandra Coccuzzo Sampaio, Maria Carolina Elias, José Salvatore Leister Patané, Vincent Louis Viala, Antonio Jorge Martins, Ricardo Haddad, Claudia Renata dos Santos Barros, Elaine Cristina Marqueze, Raul Machado Neto, Debora Botequio Moretti, Bibiana Santos, João Paulo Kitajima, Erika Freitas, David Schlesinger, Simone Kashima, Evandra Strazza Rodrigues, Svetoslav Nanev Slavov, Elaine Vieira dos Santos, Rafael dos Santos Bezerra, Luiz Carlos Junior de Alcantara, Marta Giovanetti, Vagner Fonseca, Flavia Aburjaile, Rodrigo Tocantins Calado. |
| EPI_ISL_1445142, EPI_ISL_1445143, EPI_ISL_1445144, EPI_ISL_1445145, EPI_ISL_1445146 | CS II EGIDIO BRUNHARA MORRO AGUDO | Instituto Butantan / Mendelics | Dimas Tadeu Covas, Sandra Coccuzzo Sampaio, Maria Carolina Elias, José Salvatore Leister Patané, Vincent Louis Viala, Antonio Jorge Martins, Ricardo Haddad, Claudia Renata dos Santos Barros, Elaine Cristina Marqueze, Raul Machado Neto, Debora Botequio Moretti, Bibiana Santos, João Paulo Kitajima, Erika Freitas, David Schlesinger, Simone Kashima, Evandra Strazza Rodrigues, Svetoslav Nanev Slavov, Elaine Vieira dos Santos, Rafael dos Santos Bezerra, Luiz Carlos Junior de Alcantara, Marta Giovanetti, Vagner Fonseca, Flavia Aburjaile, Rodrigo Tocantins Calado. |
| EPI_ISL_1445147, EPI_ISL_1445148, EPI_ISL_1445149, EPI_ISL_1445150, EPI_ISL_1445151, EPI_ISL_1445152 | CENTRO DE REFERENCIA DO IDOSO DR HUMBERTO MENDES DE CARVALHO | Instituto Butantan / Mendelics | Dimas Tadeu Covas, Sandra Coccuzzo Sampaio, Maria Carolina Elias, José Salvatore Leister Patané, Vincent Louis Viala, Antonio Jorge Martins, Ricardo Haddad, Claudia Renata dos Santos Barros, Elaine Cristina Marqueze, Raul Machado Neto, Debora Botequio Moretti, Bibiana Santos, João Paulo Kitajima, Erika Freitas, David Schlesinger, Simone Kashima, Evandra Strazza Rodrigues, Svetoslav Nanev Slavov, Elaine Vieira dos Santos, Rafael dos Santos Bezerra, Luiz Carlos Junior de Alcantara, Marta Giovanetti, Vagner Fonseca, Flavia Aburjaile, Rodrigo Tocantins Calado. |
| EPI_ISL_1445153, EPI_ISL_1445154 | SAE SERVICO DE ATENDIMENTO ESPECIALIZADO | Instituto Butantan / Mendelics | Dimas Tadeu Covas, Sandra Coccuzzo Sampaio, Maria Carolina Elias, José Salvatore Leister Patané, Vincent Louis Viala, Antonio Jorge Martins, Ricardo Haddad, Claudia Renata dos Santos Barros, Elaine Cristina Marqueze, Raul Machado Neto, Debora Botequio Moretti, Bibiana Santos, João Paulo Kitajima, Erika Freitas, David Schlesinger, Simone Kashima, Evandra Strazza Rodrigues, Svetoslav Nanev Slavov, Elaine Vieira dos Santos, Rafael dos Santos Bezerra, Luiz Carlos Junior de Alcantara, Marta Giovanetti, Vagner Fonseca, Flavia Aburjaile, Rodrigo Tocantins Calado. |
| EPI_ISL_1445155 | LABORATORIO DE FRANCA | Instituto Butantan / Mendelics | Dimas Tadeu Covas, Sandra Coccuzzo Sampaio, Maria Carolina Elias, José Salvatore Leister Patané, Vincent Louis Viala, Antonio Jorge Martins, Ricardo Haddad, Claudia Renata dos Santos Barros, Elaine Cristina Marqueze, Raul Machado Neto, Debora Botequio Moretti, Bibiana Santos, João Paulo Kitajima, Erika Freitas, David Schlesinger, Simone Kashima, Evandra Strazza Rodrigues, Svetoslav Nanev Slavov, Elaine Vieira dos Santos, Rafael dos Santos Bezerra, Luiz Carlos Junior de Alcantara, Marta Giovanetti, Vagner Fonseca, Flavia Aburjaile, Rodrigo Tocantins Calado. |
| EPI_ISL_1445156 | SAE SERVICO DE ATENDIMENTO ESPECIALIZADO | Instituto Butantan / Mendelics | Dimas Tadeu Covas, Sandra Coccuzzo Sampaio, Maria Carolina Elias, José Salvatore Leister Patané, Vincent Louis Viala, Antonio Jorge Martins, Ricardo Haddad, Claudia Renata dos Santos Barros, Elaine Cristina Marqueze, Raul Machado Neto, Debora Botequio Moretti, Bibiana Santos, João Paulo Kitajima, Erika Freitas, David Schlesinger, Simone Kashima, Evandra Strazza Rodrigues, Svetoslav Nanev Slavov, Elaine Vieira dos Santos, Rafael dos Santos Bezerra, Luiz Carlos Junior de Alcantara, Marta Giovanetti, Vagner Fonseca, Flavia Aburjaile, Rodrigo Tocantins Calado. |
| EPI_ISL_1445157, EPI_ISL_1445158, EPI_ISL_1445159, EPI_ISL_1445160, EPI_ISL_1445161 | SECAO CENTRO DE DIAGNOSTICO SECEDI | Instituto Butantan / Mendelics | Dimas Tadeu Covas, Sandra Coccuzzo Sampaio, Maria Carolina Elias, José Salvatore Leister Patané, Vincent Louis Viala, Antonio Jorge Martins, Ricardo Haddad, Claudia Renata dos Santos Barros, Elaine Cristina Marqueze, Raul Machado Neto, Debora Botequio Moretti, Bibiana Santos, João Paulo Kitajima, Erika Freitas, David Schlesinger, Simone Kashima, Evandra Strazza Rodrigues, Svetoslav Nanev Slavov, Elaine Vieira dos Santos, Rafael dos Santos Bezerra, Luiz Carlos Junior de Alcantara, Marta Giovanetti, Vagner Fonseca, Flavia Aburjaile, Rodrigo Tocantins Calado. |
| EPI_ISL_1445162, EPI_ISL_1445163, EPI_ISL_1445164, EPI_ISL_1445165, EPI_ISL_1445166, EPI_ISL_1445167 | SERV DE VIG SANITARIA EPIDEMIO E CTRL DE ZOONOZES GUARUJA | Instituto Butantan / Mendelics | Dimas Tadeu Covas, Sandra Coccuzzo Sampaio, Maria Carolina Elias, José Salvatore Leister Patané, Vincent Louis Viala, Antonio Jorge Martins, Ricardo Haddad, Claudia Renata dos Santos Barros, Elaine Cristina Marqueze, Raul Machado Neto, Debora Botequio Moretti, Bibiana Santos, João Paulo Kitajima, Erika Freitas, David Schlesinger, Simone Kashima, Evandra Strazza Rodrigues, Svetoslav Nanev Slavov, Elaine Vieira dos Santos, Rafael dos Santos Bezerra, Luiz Carlos Junior de Alcantara, Marta Giovanetti, Vagner Fonseca, Flavia Aburjaile, Rodrigo Tocantins Calado. |

| | | | |
|---|---|---|---|
| EPI_ISL_1445168, EPI_ISL_1445169, EPI_ISL_1445170 | SECAO CENTRO DE DIAGNOSTICO SECEDI | Instituto Butantan / Mendelics | Dimas Tadeu Covas, Sandra Coccuzzo Sampaio, Maria Carolina Elias, José Salvatore Leister Patané, Vincent Louis Viala, Antonio Jorge Martins, Ricardo Haddad, Claudia Renata dos Santos Barros, Elaine Cristina Marqueze, Raul Machado Neto, Debora Botequio Moretti, Bibiana Santos, João Paulo Kitajima, Erika Freitas, David Schlesinger, Simone Kashima, Evandra Strazza Rodrigues, Svetoslav Nanev Slavov, Elaine Vieira dos Santos, Rafael dos Santos Bezerra, Luiz Carlos Junior de Alcantara, Marta Giovanetti, Vagner Fonseca, Flavia Aburjaile, Rodrigo Tocantins Calado. |
| EPI_ISL_1445171 | SECRETARIA MUNICIPAL DE SAUDE SOROCABA | Instituto Butantan / Mendelics | Dimas Tadeu Covas, Sandra Coccuzzo Sampaio, Maria Carolina Elias, José Salvatore Leister Patané, Vincent Louis Viala, Antonio Jorge Martins, Ricardo Haddad, Claudia Renata dos Santos Barros, Elaine Cristina Marqueze, Raul Machado Neto, Debora Botequio Moretti, Bibiana Santos, João Paulo Kitajima, Erika Freitas, David Schlesinger, Simone Kashima, Evandra Strazza Rodrigues, Svetoslav Nanev Slavov, Elaine Vieira dos Santos, Rafael dos Santos Bezerra, Luiz Carlos Junior de Alcantara, Marta Giovanetti, Vagner Fonseca, Flavia Aburjaile, Rodrigo Tocantins Calado. |
| EPI_ISL_1445172 | UNIDADE MISTA ARACOIABA DA SERRA | Instituto Butantan / Mendelics | Dimas Tadeu Covas, Sandra Coccuzzo Sampaio, Maria Carolina Elias, José Salvatore Leister Patané, Vincent Louis Viala, Antonio Jorge Martins, Ricardo Haddad, Claudia Renata dos Santos Barros, Elaine Cristina Marqueze, Raul Machado Neto, Debora Botequio Moretti, Bibiana Santos, João Paulo Kitajima, Erika Freitas, David Schlesinger, Simone Kashima, Evandra Strazza Rodrigues, Svetoslav Nanev Slavov, Elaine Vieira dos Santos, Rafael dos Santos Bezerra, Luiz Carlos Junior de Alcantara, Marta Giovanetti, Vagner Fonseca, Flavia Aburjaile, Rodrigo Tocantins Calado. |
| EPI_ISL_1445173 | HOSPITAL MUNICIPAL DE IBIUNA IBIUNA SP | Instituto Butantan / Mendelics | Dimas Tadeu Covas, Sandra Coccuzzo Sampaio, Maria Carolina Elias, José Salvatore Leister Patané, Vincent Louis Viala, Antonio Jorge Martins, Ricardo Haddad, Claudia Renata dos Santos Barros, Elaine Cristina Marqueze, Raul Machado Neto, Debora Botequio Moretti, Bibiana Santos, João Paulo Kitajima, Erika Freitas, David Schlesinger, Simone Kashima, Evandra Strazza Rodrigues, Svetoslav Nanev Slavov, Elaine Vieira dos Santos, Rafael dos Santos Bezerra, Luiz Carlos Junior de Alcantara, Marta Giovanetti, Vagner Fonseca, Flavia Aburjaile, Rodrigo Tocantins Calado. |
| EPI_ISL_1445174 | PRONTO SOCORRO MUNICIPAL DE TATUI | Instituto Butantan / Mendelics | Dimas Tadeu Covas, Sandra Coccuzzo Sampaio, Maria Carolina Elias, José Salvatore Leister Patané, Vincent Louis Viala, Antonio Jorge Martins, Ricardo Haddad, Claudia Renata dos Santos Barros, Elaine Cristina Marqueze, Raul Machado Neto, Debora Botequio Moretti, Bibiana Santos, João Paulo Kitajima, Erika Freitas, David Schlesinger, Simone Kashima, Evandra Strazza Rodrigues, Svetoslav Nanev Slavov, Elaine Vieira dos Santos, Rafael dos Santos Bezerra, Luiz Carlos Junior de Alcantara, Marta Giovanetti, Vagner Fonseca, Flavia Aburjaile, Rodrigo Tocantins Calado. |
| EPI_ISL_1445175 | DEPARTAMENTO DE SAUDE COLETIVA | Instituto Butantan / Mendelics | Dimas Tadeu Covas, Sandra Coccuzzo Sampaio, Maria Carolina Elias, José Salvatore Leister Patané, Vincent Louis Viala, Antonio Jorge Martins, Ricardo Haddad, Claudia Renata dos Santos Barros, Elaine Cristina Marqueze, Raul Machado Neto, Debora Botequio Moretti, Bibiana Santos, João Paulo Kitajima, Erika Freitas, David Schlesinger, Simone Kashima, Evandra Strazza Rodrigues, Svetoslav Nanev Slavov, Elaine Vieira dos Santos, Rafael dos Santos Bezerra, Luiz Carlos Junior de Alcantara, Marta Giovanetti, Vagner Fonseca, Flavia Aburjaile, Rodrigo Tocantins Calado. |
| EPI_ISL_1445176 | SECRETARIA MUNICIPAL DE SAUDE SOROCABA | Instituto Butantan / Mendelics | Dimas Tadeu Covas, Sandra Coccuzzo Sampaio, Maria Carolina Elias, José Salvatore Leister Patané, Vincent Louis Viala, Antonio Jorge Martins, Ricardo Haddad, Claudia Renata dos Santos Barros, Elaine Cristina Marqueze, Raul Machado Neto, Debora Botequio Moretti, Bibiana Santos, João Paulo Kitajima, Erika Freitas, David Schlesinger, Simone Kashima, Evandra Strazza Rodrigues, Svetoslav Nanev Slavov, Elaine Vieira dos Santos, Rafael dos Santos Bezerra, Luiz Carlos Junior de Alcantara, Marta Giovanetti, Vagner Fonseca, Flavia Aburjaile, Rodrigo Tocantins Calado. |
| EPI_ISL_1445177 | SAUDE COLETIVA CAPAO BONITO | Instituto Butantan / Mendelics | Dimas Tadeu Covas, Sandra Coccuzzo Sampaio, Maria Carolina Elias, José Salvatore Leister Patané, Vincent Louis Viala, Antonio Jorge Martins, Ricardo Haddad, Claudia Renata dos Santos Barros, Elaine Cristina Marqueze, Raul Machado Neto, Debora Botequio Moretti, Bibiana Santos, João Paulo Kitajima, Erika Freitas, David Schlesinger, Simone Kashima, Evandra Strazza Rodrigues, Svetoslav Nanev Slavov, Elaine Vieira dos Santos, Rafael dos Santos Bezerra, Luiz Carlos Junior de Alcantara, Marta Giovanetti, Vagner Fonseca, Flavia Aburjaile, Rodrigo Tocantins Calado. |
| EPI_ISL_1445178, EPI_ISL_1445179, EPI_ISL_1445180 | SECRETARIA MUNICIPAL DE SAUDE SOROCABA | Instituto Butantan / Mendelics | Dimas Tadeu Covas, Sandra Coccuzzo Sampaio, Maria Carolina Elias, José Salvatore Leister Patané, Vincent Louis Viala, Antonio Jorge Martins, Ricardo Haddad, Claudia Renata dos Santos Barros, Elaine Cristina Marqueze, Raul Machado Neto, Debora Botequio Moretti, Bibiana Santos, João Paulo Kitajima, Erika Freitas, David Schlesinger, Simone Kashima, Evandra Strazza Rodrigues, Svetoslav Nanev Slavov, Elaine Vieira dos Santos, Rafael dos Santos Bezerra, Luiz Carlos Junior de Alcantara, Marta Giovanetti, Vagner Fonseca, Flavia Aburjaile, Rodrigo Tocantins Calado. |
| EPI_ISL_1445181 | HOSPITAL REGIONAL DE ITAPETININGA | Instituto Butantan / Mendelics | Dimas Tadeu Covas, Sandra Coccuzzo Sampaio, Maria Carolina Elias, José Salvatore Leister Patané, Vincent Louis Viala, Antonio Jorge Martins, Ricardo Haddad, Claudia Renata dos Santos Barros, Elaine Cristina Marqueze, Raul Machado Neto, Debora Botequio Moretti, Bibiana Santos, João Paulo Kitajima, Erika Freitas, David Schlesinger, Simone Kashima, Evandra Strazza Rodrigues, Svetoslav Nanev Slavov, Elaine Vieira dos Santos, Rafael dos Santos Bezerra, Luiz Carlos Junior de Alcantara, Marta Giovanetti, Vagner Fonseca, Flavia Aburjaile, Rodrigo Tocantins Calado. |
| EPI_ISL_1445182 | SECRETARIA MUNICIPAL DE SAUDE SOROCABA | Instituto Butantan / Mendelics | Dimas Tadeu Covas, Sandra Coccuzzo Sampaio, Maria Carolina Elias, José Salvatore Leister Patané, Vincent Louis Viala, Antonio Jorge Martins, Ricardo Haddad, Claudia Renata dos Santos Barros, Elaine Cristina Marqueze, Raul Machado Neto, Debora Botequio Moretti, Bibiana Santos, João Paulo Kitajima, Erika Freitas, David Schlesinger, Simone Kashima, Evandra Strazza Rodrigues, Svetoslav Nanev Slavov, Elaine Vieira dos Santos, Rafael dos Santos Bezerra, Luiz Carlos Junior de Alcantara, Marta Giovanetti, Vagner Fonseca, Flavia Aburjaile, Rodrigo Tocantins Calado. |
| EPI_ISL_1445183, EPI_ISL_1445184 | UNIDADE BASICA DE SAUDE DA FAMILIA BILAC | Instituto Butantan / Mendelics | Dimas Tadeu Covas, Sandra Coccuzzo Sampaio, Maria Carolina Elias, José Salvatore Leister Patané, Vincent Louis Viala, Antonio Jorge Martins, Ricardo Haddad, Claudia Renata dos Santos Barros, Elaine Cristina Marqueze, Raul Machado Neto, Debora Botequio Moretti, Bibiana Santos, João Paulo Kitajima, Erika Freitas, David Schlesinger, Simone Kashima, Evandra Strazza Rodrigues, Svetoslav Nanev Slavov, Elaine Vieira dos Santos, Rafael dos Santos Bezerra, Luiz Carlos Junior de Alcantara, Marta Giovanetti, Vagner Fonseca, Flavia Aburjaile, Rodrigo Tocantins Calado. |
| EPI_ISL_1445185 | PRONTO ATENDIMENTO SAO JOSE | Instituto Butantan / Mendelics | Dimas Tadeu Covas, Sandra Coccuzzo Sampaio, Maria Carolina Elias, José Salvatore Leister Patané, Vincent Louis Viala, Antonio Jorge Martins, Ricardo Haddad, Claudia Renata dos Santos Barros, Elaine Cristina Marqueze, Raul Machado Neto, Debora Botequio Moretti, Bibiana Santos, João Paulo Kitajima, Erika Freitas, David Schlesinger, Simone Kashima, Evandra Strazza Rodrigues, Svetoslav Nanev Slavov, Elaine Vieira dos Santos, Rafael dos Santos Bezerra, Luiz Carlos Junior de Alcantara, Marta Giovanetti, Vagner Fonseca, Flavia Aburjaile, Rodrigo Tocantins Calado. |
| EPI_ISL_1445186 | PRONTO ATENDIMENTO VILA PADRE ANCHIETA | Instituto Butantan / Mendelics | Dimas Tadeu Covas, Sandra Coccuzzo Sampaio, Maria Carolina Elias, José Salvatore Leister Patané, Vincent Louis Viala, Antonio Jorge Martins, Ricardo Haddad, Claudia Renata dos Santos Barros, Elaine Cristina Marqueze, Raul Machado Neto, Debora Botequio Moretti, Bibiana Santos, João Paulo Kitajima, Erika Freitas, David Schlesinger, Simone Kashima, Evandra Strazza Rodrigues, Svetoslav Nanev Slavov, Elaine Vieira dos Santos, Rafael dos Santos Bezerra, Luiz Carlos Junior de Alcantara, Marta Giovanetti, Vagner Fonseca, Flavia Aburjaile, Rodrigo Tocantins Calado. |
| EPI_ISL_1445187 | PRONTO ATENDIMENTO SAO JOSE | Instituto Butantan / Mendelics | Dimas Tadeu Covas, Sandra Coccuzzo Sampaio, Maria Carolina Elias, José Salvatore Leister Patané, Vincent Louis Viala, Antonio Jorge Martins, Ricardo Haddad, Claudia Renata dos Santos Barros, Elaine Cristina Marqueze, Raul Machado Neto, Debora Botequio Moretti, Bibiana Santos, João Paulo Kitajima, Erika Freitas, David Schlesinger, Simone Kashima, Evandra Strazza Rodrigues, Svetoslav Nanev Slavov, Elaine Vieira dos Santos, Rafael dos Santos Bezerra, Luiz Carlos Junior de Alcantara, Marta Giovanetti, Vagner Fonseca, Flavia Aburjaile, Rodrigo Tocantins Calado. |
| EPI_ISL_1445188, EPI_ISL_1445189, EPI_ISL_1445190, EPI_ISL_1445191, EPI_ISL_1445192 | PRONTO ATENDIMENTO VILA PADRE ANCHIETA | Instituto Butantan / Mendelics | Dimas Tadeu Covas, Sandra Coccuzzo Sampaio, Maria Carolina Elias, José Salvatore Leister Patané, Vincent Louis Viala, Antonio Jorge Martins, Ricardo Haddad, Claudia Renata dos Santos Barros, Elaine Cristina Marqueze, Raul Machado Neto, Debora Botequio Moretti, Bibiana Santos, João Paulo Kitajima, Erika Freitas, David Schlesinger, Simone Kashima, Evandra Strazza Rodrigues, Svetoslav Nanev Slavov, Elaine Vieira dos Santos, Rafael dos Santos Bezerra, Luiz Carlos Junior de Alcantara, Marta Giovanetti, Vagner Fonseca, Flavia Aburjaile, Rodrigo Tocantins Calado. |
| EPI_ISL_1445193, EPI_ISL_1445194 | PRONTO ATENDIMENTO SAO JOSE | Instituto Butantan / Mendelics | Dimas Tadeu Covas, Sandra Coccuzzo Sampaio, Maria Carolina Elias, José Salvatore Leister Patané, Vincent Louis Viala, Antonio Jorge Martins, Ricardo Haddad, Claudia Renata dos Santos Barros, Elaine Cristina Marqueze, Raul Machado Neto, Debora Botequio Moretti, Bibiana Santos, João Paulo Kitajima, Erika Freitas, David Schlesinger, Simone Kashima, Evandra Strazza Rodrigues, Svetoslav Nanev Slavov, Elaine Vieira dos Santos, Rafael dos Santos Bezerra, Luiz Carlos Junior de Alcantara, Marta Giovanetti, Vagner Fonseca, Flavia Aburjaile, Rodrigo Tocantins Calado. |
| EPI_ISL_1445195 | CENTRO DE SAUDE DE BORA | Instituto Butantan / Mendelics | Dimas Tadeu Covas, Sandra Coccuzzo Sampaio, Maria Carolina Elias, José Salvatore Leister Patané, Vincent Louis Viala, Antonio Jorge Martins, Ricardo Haddad, Claudia Renata dos Santos Barros, Elaine Cristina Marqueze, Raul Machado Neto, Debora Botequio Moretti, Bibiana Santos, João Paulo Kitajima, Erika Freitas, David Schlesinger, Simone Kashima, Evandra Strazza Rodrigues, Svetoslav Nanev Slavov, Elaine Vieira dos Santos, Rafael dos Santos Bezerra, Luiz Carlos Junior de Alcantara, Marta Giovanetti, Vagner Fonseca, Flavia Aburjaile, Rodrigo Tocantins Calado. |
| EPI_ISL_1445196 | UBS SAO JUDAS | Instituto Butantan / Mendelics | Dimas Tadeu Covas, Sandra Coccuzzo Sampaio, Maria Carolina Elias, José Salvatore Leister Patané, Vincent Louis Viala, Antonio Jorge Martins, Ricardo Haddad, Claudia Renata dos Santos Barros, Elaine Cristina Marqueze, Raul Machado Neto, Debora Botequio Moretti, Bibiana Santos, João Paulo Kitajima, Erika Freitas, David Schlesinger, Simone Kashima, Evandra Strazza Rodrigues, Svetoslav Nanev Slavov, Elaine Vieira dos Santos, Rafael dos Santos Bezerra, Luiz Carlos Junior de Alcantara, Marta Giovanetti, Vagner Fonseca, Flavia Aburjaile, Rodrigo Tocantins Calado. |
| EPI_ISL_1445197 | CENTRO DE SAUDE DE JULIO MESQUITA | Instituto Butantan / Mendelics | Dimas Tadeu Covas, Sandra Coccuzzo Sampaio, Maria Carolina Elias, José Salvatore Leister Patané, Vincent Louis Viala, Antonio Jorge Martins, Ricardo Haddad, Claudia Renata dos Santos Barros, Elaine Cristina Marqueze, Raul Machado Neto, Debora Botequio Moretti, Bibiana Santos, João Paulo |

| | | | |
|---|---|---|---|
| | | | Kitajima, Erika Freitas, David Schlesinger, Simone Kashima, Evandra Strazza Rodrigues, Svetoslav Nanev Slavov, Elaine Vieira dos Santos, Rafael dos Santos Bezerra, Luiz Carlos Junior de Alcantara, Marta Giovanetti, Vagner Fonseca, Flavia Aburjaile, Rodrigo Tocantins Calado. |
| EPI_ISL_1445198 | UNIDADE MISTA DE TAIACU | Instituto Butantan / Mendelics | Dimas Tadeu Covas, Sandra Coccuzzo Sampaio, Maria Carolina Elias, José Salvatore Leister Patané, Vincent Louis Viala, Antonio Jorge Martins, Ricardo Haddad, Claudia Renata dos Santos Barros, Elaine Cristina Marqueze, Raul Machado Neto, Debora Botequio Moretti, Bibiana Santos, João Paulo Kitajima, Erika Freitas, David Schlesinger, Simone Kashima, Evandra Strazza Rodrigues, Svetoslav Nanev Slavov, Elaine Vieira dos Santos, Rafael dos Santos Bezerra, Luiz Carlos Junior de Alcantara, Marta Giovanetti, Vagner Fonseca, Flavia Aburjaile, Rodrigo Tocantins Calado. |
| EPI_ISL_1445199 | CENTRO DE SAUDE III SALES OLIVEIRA | Instituto Butantan / Mendelics | Dimas Tadeu Covas, Sandra Coccuzzo Sampaio, Maria Carolina Elias, José Salvatore Leister Patané, Vincent Louis Viala, Antonio Jorge Martins, Ricardo Haddad, Claudia Renata dos Santos Barros, Elaine Cristina Marqueze, Raul Machado Neto, Debora Botequio Moretti, Bibiana Santos, João Paulo Kitajima, Erika Freitas, David Schlesinger, Simone Kashima, Evandra Strazza Rodrigues, Svetoslav Nanev Slavov, Elaine Vieira dos Santos, Rafael dos Santos Bezerra, Luiz Carlos Junior de Alcantara, Marta Giovanetti, Vagner Fonseca, Flavia Aburjaile, Rodrigo Tocantins Calado. |
| EPI_ISL_1445200 | HOSPITAL E MATERNIDADE NOSSA SENHORA DA AJUDA | Instituto Butantan / Mendelics | Dimas Tadeu Covas, Sandra Coccuzzo Sampaio, Maria Carolina Elias, José Salvatore Leister Patané, Vincent Louis Viala, Antonio Jorge Martins, Ricardo Haddad, Claudia Renata dos Santos Barros, Elaine Cristina Marqueze, Raul Machado Neto, Debora Botequio Moretti, Bibiana Santos, João Paulo Kitajima, Erika Freitas, David Schlesinger, Simone Kashima, Evandra Strazza Rodrigues, Svetoslav Nanev Slavov, Elaine Vieira dos Santos, Rafael dos Santos Bezerra, Luiz Carlos Junior de Alcantara, Marta Giovanetti, Vagner Fonseca, Flavia Aburjaile, Rodrigo Tocantins Calado. |
| EPI_ISL_1445201 | USAFA TUDE BASTOS | Instituto Butantan / Mendelics | Dimas Tadeu Covas, Sandra Coccuzzo Sampaio, Maria Carolina Elias, José Salvatore Leister Patané, Vincent Louis Viala, Antonio Jorge Martins, Ricardo Haddad, Claudia Renata dos Santos Barros, Elaine Cristina Marqueze, Raul Machado Neto, Debora Botequio Moretti, Bibiana Santos, João Paulo Kitajima, Erika Freitas, David Schlesinger, Simone Kashima, Evandra Strazza Rodrigues, Svetoslav Nanev Slavov, Elaine Vieira dos Santos, Rafael dos Santos Bezerra, Luiz Carlos Junior de Alcantara, Marta Giovanetti, Vagner Fonseca, Flavia Aburjaile, Rodrigo Tocantins Calado. |
| EPI_ISL_1445202 | USAFA MARACANA | Instituto Butantan / Mendelics | Dimas Tadeu Covas, Sandra Coccuzzo Sampaio, Maria Carolina Elias, José Salvatore Leister Patané, Vincent Louis Viala, Antonio Jorge Martins, Ricardo Haddad, Claudia Renata dos Santos Barros, Elaine Cristina Marqueze, Raul Machado Neto, Debora Botequio Moretti, Bibiana Santos, João Paulo Kitajima, Erika Freitas, David Schlesinger, Simone Kashima, Evandra Strazza Rodrigues, Svetoslav Nanev Slavov, Elaine Vieira dos Santos, Rafael dos Santos Bezerra, Luiz Carlos Junior de Alcantara, Marta Giovanetti, Vagner Fonseca, Flavia Aburjaile, Rodrigo Tocantins Calado. |
| EPI_ISL_1445203 | USAFA GUILHERMINA | Instituto Butantan / Mendelics | Dimas Tadeu Covas, Sandra Coccuzzo Sampaio, Maria Carolina Elias, José Salvatore Leister Patané, Vincent Louis Viala, Antonio Jorge Martins, Ricardo Haddad, Claudia Renata dos Santos Barros, Elaine Cristina Marqueze, Raul Machado Neto, Debora Botequio Moretti, Bibiana Santos, João Paulo Kitajima, Erika Freitas, David Schlesinger, Simone Kashima, Evandra Strazza Rodrigues, Svetoslav Nanev Slavov, Elaine Vieira dos Santos, Rafael dos Santos Bezerra, Luiz Carlos Junior de Alcantara, Marta Giovanetti, Vagner Fonseca, Flavia Aburjaile, Rodrigo Tocantins Calado. |
| EPI_ISL_1445204 | USAFA MARACANA | Instituto Butantan / Mendelics | Dimas Tadeu Covas, Sandra Coccuzzo Sampaio, Maria Carolina Elias, José Salvatore Leister Patané, Vincent Louis Viala, Antonio Jorge Martins, Ricardo Haddad, Claudia Renata dos Santos Barros, Elaine Cristina Marqueze, Raul Machado Neto, Debora Botequio Moretti, Bibiana Santos, João Paulo Kitajima, Erika Freitas, David Schlesinger, Simone Kashima, Evandra Strazza Rodrigues, Svetoslav Nanev Slavov, Elaine Vieira dos Santos, Rafael dos Santos Bezerra, Luiz Carlos Junior de Alcantara, Marta Giovanetti, Vagner Fonseca, Flavia Aburjaile, Rodrigo Tocantins Calado. |
| EPI_ISL_1445205 | UBS FRANCISCA LIMA DE LIRA | Instituto Butantan / Mendelics | Dimas Tadeu Covas, Sandra Coccuzzo Sampaio, Maria Carolina Elias, José Salvatore Leister Patané, Vincent Louis Viala, Antonio Jorge Martins, Ricardo Haddad, Claudia Renata dos Santos Barros, Elaine Cristina Marqueze, Raul Machado Neto, Debora Botequio Moretti, Bibiana Santos, João Paulo Kitajima, Erika Freitas, David Schlesinger, Simone Kashima, Evandra Strazza Rodrigues, Svetoslav Nanev Slavov, Elaine Vieira dos Santos, Rafael dos Santos Bezerra, Luiz Carlos Junior de Alcantara, Marta Giovanetti, Vagner Fonseca, Flavia Aburjaile, Rodrigo Tocantins Calado. |
| EPI_ISL_1445206 | UBS DARCY ALVES E ROBALINHO | Instituto Butantan / Mendelics | Dimas Tadeu Covas, Sandra Coccuzzo Sampaio, Maria Carolina Elias, José Salvatore Leister Patané, Vincent Louis Viala, Antonio Jorge Martins, Ricardo Haddad, Claudia Renata dos Santos Barros, Elaine Cristina Marqueze, Raul Machado Neto, Debora Botequio Moretti, Bibiana Santos, João Paulo Kitajima, Erika Freitas, David Schlesinger, Simone Kashima, Evandra Strazza Rodrigues, Svetoslav Nanev Slavov, Elaine Vieira dos Santos, Rafael dos Santos Bezerra, Luiz Carlos Junior de Alcantara, Marta Giovanetti, Vagner Fonseca, Flavia Aburjaile, Rodrigo Tocantins Calado. |
| EPI_ISL_1445207 | PPA MARCIO VALDEVINO BATISTA | Instituto Butantan / Mendelics | Dimas Tadeu Covas, Sandra Coccuzzo Sampaio, Maria Carolina Elias, José Salvatore Leister Patané, Vincent Louis Viala, Antonio Jorge Martins, Ricardo Haddad, Claudia Renata dos Santos Barros, Elaine Cristina Marqueze, Raul Machado Neto, Debora Botequio Moretti, Bibiana Santos, João Paulo Kitajima, Erika Freitas, David Schlesinger, Simone Kashima, Evandra Strazza Rodrigues, Svetoslav Nanev Slavov, Elaine Vieira dos Santos, Rafael dos Santos Bezerra, Luiz Carlos Junior de Alcantara, Marta Giovanetti, Vagner Fonseca, Flavia Aburjaile, Rodrigo Tocantins Calado. |
| EPI_ISL_1445208 | UBS DARCY ALVES E ROBALINHO | Instituto Butantan / Mendelics | Dimas Tadeu Covas, Sandra Coccuzzo Sampaio, Maria Carolina Elias, José Salvatore Leister Patané, Vincent Louis Viala, Antonio Jorge Martins, Ricardo Haddad, Claudia Renata dos Santos Barros, Elaine Cristina Marqueze, Raul Machado Neto, Debora Botequio Moretti, Bibiana Santos, João Paulo Kitajima, Erika Freitas, David Schlesinger, Simone Kashima, Evandra Strazza Rodrigues, Svetoslav Nanev Slavov, Elaine Vieira dos Santos, Rafael dos Santos Bezerra, Luiz Carlos Junior de Alcantara, Marta Giovanetti, Vagner Fonseca, Flavia Aburjaile, Rodrigo Tocantins Calado. |
| EPI_ISL_1445209 | UBS OTACILIO FIRMINO LOPES | Instituto Butantan / Mendelics | Dimas Tadeu Covas, Sandra Coccuzzo Sampaio, Maria Carolina Elias, José Salvatore Leister Patané, Vincent Louis Viala, Antonio Jorge Martins, Ricardo Haddad, Claudia Renata dos Santos Barros, Elaine Cristina Marqueze, Raul Machado Neto, Debora Botequio Moretti, Bibiana Santos, João Paulo Kitajima, Erika Freitas, David Schlesinger, Simone Kashima, Evandra Strazza Rodrigues, Svetoslav Nanev Slavov, Elaine Vieira dos Santos, Rafael dos Santos Bezerra, Luiz Carlos Junior de Alcantara, Marta Giovanetti, Vagner Fonseca, Flavia Aburjaile, Rodrigo Tocantins Calado. |
| EPI_ISL_1445210, EPI_ISL_1445211 | UBS JOSE SABINO FERREIRA | Instituto Butantan / Mendelics | Dimas Tadeu Covas, Sandra Coccuzzo Sampaio, Maria Carolina Elias, José Salvatore Leister Patané, Vincent Louis Viala, Antonio Jorge Martins, Ricardo Haddad, Claudia Renata dos Santos Barros, Elaine Cristina Marqueze, Raul Machado Neto, Debora Botequio Moretti, Bibiana Santos, João Paulo Kitajima, Erika Freitas, David Schlesinger, Simone Kashima, Evandra Strazza Rodrigues, Svetoslav Nanev Slavov, Elaine Vieira dos Santos, Rafael dos Santos Bezerra, Luiz Carlos Junior de Alcantara, Marta Giovanetti, Vagner Fonseca, Flavia Aburjaile, Rodrigo Tocantins Calado. |
| EPI_ISL_1445212 | UBS IRMA AGUEDA MARIA JAIME | Instituto Butantan / Mendelics | Dimas Tadeu Covas, Sandra Coccuzzo Sampaio, Maria Carolina Elias, José Salvatore Leister Patané, Vincent Louis Viala, Antonio Jorge Martins, Ricardo Haddad, Claudia Renata dos Santos Barros, Elaine Cristina Marqueze, Raul Machado Neto, Debora Botequio Moretti, Bibiana Santos, João Paulo Kitajima, Erika Freitas, David Schlesinger, Simone Kashima, Evandra Strazza Rodrigues, Svetoslav Nanev Slavov, Elaine Vieira dos Santos, Rafael dos Santos Bezerra, Luiz Carlos Junior de Alcantara, Marta Giovanetti, Vagner Fonseca, Flavia Aburjaile, Rodrigo Tocantins Calado. |
| EPI_ISL_1445213 | UBS OTACILIO FIRMINO LOPES | Instituto Butantan / Mendelics | Dimas Tadeu Covas, Sandra Coccuzzo Sampaio, Maria Carolina Elias, José Salvatore Leister Patané, Vincent Louis Viala, Antonio Jorge Martins, Ricardo Haddad, Claudia Renata dos Santos Barros, Elaine Cristina Marqueze, Raul Machado Neto, Debora Botequio Moretti, Bibiana Santos, João Paulo Kitajima, Erika Freitas, David Schlesinger, Simone Kashima, Evandra Strazza Rodrigues, Svetoslav Nanev Slavov, Elaine Vieira dos Santos, Rafael dos Santos Bezerra, Luiz Carlos Junior de Alcantara, Marta Giovanetti, Vagner Fonseca, Flavia Aburjaile, Rodrigo Tocantins Calado. |
| EPI_ISL_1445214 | UBS JOSE HILARIO DOS SANTOS | Instituto Butantan / Mendelics | Dimas Tadeu Covas, Sandra Coccuzzo Sampaio, Maria Carolina Elias, José Salvatore Leister Patané, Vincent Louis Viala, Antonio Jorge Martins, Ricardo Haddad, Claudia Renata dos Santos Barros, Elaine Cristina Marqueze, Raul Machado Neto, Debora Botequio Moretti, Bibiana Santos, João Paulo Kitajima, Erika Freitas, David Schlesinger, Simone Kashima, Evandra Strazza Rodrigues, Svetoslav Nanev Slavov, Elaine Vieira dos Santos, Rafael dos Santos Bezerra, Luiz Carlos Junior de Alcantara, Marta Giovanetti, Vagner Fonseca, Flavia Aburjaile, Rodrigo Tocantins Calado. |
| EPI_ISL_1445215, EPI_ISL_1445216 | UBS HELENA MARREY | Instituto Butantan / Mendelics | Dimas Tadeu Covas, Sandra Coccuzzo Sampaio, Maria Carolina Elias, José Salvatore Leister Patané, Vincent Louis Viala, Antonio Jorge Martins, Ricardo Haddad, Claudia Renata dos Santos Barros, Elaine Cristina Marqueze, Raul Machado Neto, Debora Botequio Moretti, Bibiana Santos, João Paulo Kitajima, Erika Freitas, David Schlesinger, Simone Kashima, Evandra Strazza Rodrigues, Svetoslav Nanev Slavov, Elaine Vieira dos Santos, Rafael dos Santos Bezerra, Luiz Carlos Junior de Alcantara, Marta Giovanetti, Vagner Fonseca, Flavia Aburjaile, Rodrigo Tocantins Calado. |
| EPI_ISL_1445217 | UBS EMILIA COSME CERQUEIRA | Instituto Butantan / Mendelics | Dimas Tadeu Covas, Sandra Coccuzzo Sampaio, Maria Carolina Elias, José Salvatore Leister Patané, Vincent Louis Viala, Antonio Jorge Martins, Ricardo Haddad, Claudia Renata dos Santos Barros, Elaine Cristina Marqueze, Raul Machado Neto, Debora Botequio Moretti, Bibiana Santos, João Paulo Kitajima, Erika Freitas, David Schlesinger, Simone Kashima, Evandra Strazza Rodrigues, Svetoslav Nanev Slavov, Elaine Vieira dos Santos, Rafael dos Santos Bezerra, Luiz Carlos Junior de Alcantara, Marta Giovanetti, Vagner Fonseca, Flavia Aburjaile, Rodrigo Tocantins Calado. |
| EPI_ISL_1445218 | UBS SYLVIO JOAO L DE LUCIA | Instituto Butantan / Mendelics | Dimas Tadeu Covas, Sandra Coccuzzo Sampaio, Maria Carolina Elias, José Salvatore Leister Patané, Vincent Louis Viala, Antonio Jorge Martins, Ricardo Haddad, Claudia Renata dos Santos Barros, Elaine Cristina Marqueze, Raul Machado Neto, Debora Botequio Moretti, Bibiana Santos, João Paulo Kitajima, Erika Freitas, David Schlesinger, Simone Kashima, Evandra Strazza Rodrigues, Svetoslav Nanev Slavov, Elaine Vieira dos Santos, Rafael dos Santos Bezerra, Luiz Carlos Junior de Alcantara, Marta Giovanetti, Vagner Fonseca, Flavia Aburjaile, Rodrigo Tocantins Calado. |

| | | | |
|---|---|---|---|
| EPI_ISL_1445219, EPI_ISL_1445220 | UBS EMILIA COSME CERQUEIRA | Instituto Butantan / Mendelics | Dimas Tadeu Covas, Sandra Coccuzzo Sampaio, Maria Carolina Elias, José Salvatore Leister Patané, Vincent Louis Viala, Antonio Jorge Martins, Ricardo Haddad, Claudia Renata dos Santos Barros, Elaine Cristina Marqueze, Raul Machado Neto, Debora Botequio Moretti, Bibiana Santos, João Paulo Kitajima, Erika Freitas, David Schlesinger, Simone Kashima, Evandra Strazza Rodrigues, Svetoslav Nanev Slavov, Elaine Vieira dos Santos, Rafael dos Santos Bezerra, Luiz Carlos Junior de Alcantara, Marta Giovanetti, Vagner Fonseca, Flavia Aburjaile, Rodrigo Tocantins Calado. |
| EPI_ISL_1445221, EPI_ISL_1445222, EPI_ISL_1445223 | UBS HELENA MARREY | Instituto Butantan / Mendelics | Dimas Tadeu Covas, Sandra Coccuzzo Sampaio, Maria Carolina Elias, José Salvatore Leister Patané, Vincent Louis Viala, Antonio Jorge Martins, Ricardo Haddad, Claudia Renata dos Santos Barros, Elaine Cristina Marqueze, Raul Machado Neto, Debora Botequio Moretti, Bibiana Santos, João Paulo Kitajima, Erika Freitas, David Schlesinger, Simone Kashima, Evandra Strazza Rodrigues, Svetoslav Nanev Slavov, Elaine Vieira dos Santos, Rafael dos Santos Bezerra, Luiz Carlos Junior de Alcantara, Marta Giovanetti, Vagner Fonseca, Flavia Aburjaile, Rodrigo Tocantins Calado. |
| EPI_ISL_1445224 | PPA FENELON GUEDES PEREIRA | Instituto Butantan / Mendelics | Dimas Tadeu Covas, Sandra Coccuzzo Sampaio, Maria Carolina Elias, José Salvatore Leister Patané, Vincent Louis Viala, Antonio Jorge Martins, Ricardo Haddad, Claudia Renata dos Santos Barros, Elaine Cristina Marqueze, Raul Machado Neto, Debora Botequio Moretti, Bibiana Santos, João Paulo Kitajima, Erika Freitas, David Schlesinger, Simone Kashima, Evandra Strazza Rodrigues, Svetoslav Nanev Slavov, Elaine Vieira dos Santos, Rafael dos Santos Bezerra, Luiz Carlos Junior de Alcantara, Marta Giovanetti, Vagner Fonseca, Flavia Aburjaile, Rodrigo Tocantins Calado. |
| EPI_ISL_1445225 | PRONTO SOCORRO DR ANTONIO FLAVIO FRANCA | Instituto Butantan / Mendelics | Dimas Tadeu Covas, Sandra Coccuzzo Sampaio, Maria Carolina Elias, José Salvatore Leister Patané, Vincent Louis Viala, Antonio Jorge Martins, Ricardo Haddad, Claudia Renata dos Santos Barros, Elaine Cristina Marqueze, Raul Machado Neto, Debora Botequio Moretti, Bibiana Santos, João Paulo Kitajima, Erika Freitas, David Schlesinger, Simone Kashima, Evandra Strazza Rodrigues, Svetoslav Nanev Slavov, Elaine Vieira dos Santos, Rafael dos Santos Bezerra, Luiz Carlos Junior de Alcantara, Marta Giovanetti, Vagner Fonseca, Flavia Aburjaile, Rodrigo Tocantins Calado. |
| EPI_ISL_1445226 | UBS SYLVIO JOAO L DE LUCIA | Instituto Butantan / Mendelics | Dimas Tadeu Covas, Sandra Coccuzzo Sampaio, Maria Carolina Elias, José Salvatore Leister Patané, Vincent Louis Viala, Antonio Jorge Martins, Ricardo Haddad, Claudia Renata dos Santos Barros, Elaine Cristina Marqueze, Raul Machado Neto, Debora Botequio Moretti, Bibiana Santos, João Paulo Kitajima, Erika Freitas, David Schlesinger, Simone Kashima, Evandra Strazza Rodrigues, Svetoslav Nanev Slavov, Elaine Vieira dos Santos, Rafael dos Santos Bezerra, Luiz Carlos Junior de Alcantara, Marta Giovanetti, Vagner Fonseca, Flavia Aburjaile, Rodrigo Tocantins Calado. |
| EPI_ISL_1445227 | UBS EMILIA COSME CERQUEIRA | Instituto Butantan / Mendelics | Dimas Tadeu Covas, Sandra Coccuzzo Sampaio, Maria Carolina Elias, José Salvatore Leister Patané, Vincent Louis Viala, Antonio Jorge Martins, Ricardo Haddad, Claudia Renata dos Santos Barros, Elaine Cristina Marqueze, Raul Machado Neto, Debora Botequio Moretti, Bibiana Santos, João Paulo Kitajima, Erika Freitas, David Schlesinger, Simone Kashima, Evandra Strazza Rodrigues, Svetoslav Nanev Slavov, Elaine Vieira dos Santos, Rafael dos Santos Bezerra, Luiz Carlos Junior de Alcantara, Marta Giovanetti, Vagner Fonseca, Flavia Aburjaile, Rodrigo Tocantins Calado. |
| EPI_ISL_1445228 | UBS IRMA AGUEDA MARIA JAIME | Instituto Butantan / Mendelics | Dimas Tadeu Covas, Sandra Coccuzzo Sampaio, Maria Carolina Elias, José Salvatore Leister Patané, Vincent Louis Viala, Antonio Jorge Martins, Ricardo Haddad, Claudia Renata dos Santos Barros, Elaine Cristina Marqueze, Raul Machado Neto, Debora Botequio Moretti, Bibiana Santos, João Paulo Kitajima, Erika Freitas, David Schlesinger, Simone Kashima, Evandra Strazza Rodrigues, Svetoslav Nanev Slavov, Elaine Vieira dos Santos, Rafael dos Santos Bezerra, Luiz Carlos Junior de Alcantara, Marta Giovanetti, Vagner Fonseca, Flavia Aburjaile, Rodrigo Tocantins Calado. |
| EPI_ISL_1445229, EPI_ISL_1445230, EPI_ISL_1445231, EPI_ISL_1445232, EPI_ISL_1445233, EPI_ISL_1445234, EPI_ISL_1445235, EPI_ISL_1445236, EPI_ISL_1445237 | VIGILANCIA EPIDEMIOLOGICA | Instituto Butantan / Mendelics | Dimas Tadeu Covas, Sandra Coccuzzo Sampaio, Maria Carolina Elias, José Salvatore Leister Patané, Vincent Louis Viala, Antonio Jorge Martins, Ricardo Haddad, Claudia Renata dos Santos Barros, Elaine Cristina Marqueze, Raul Machado Neto, Debora Botequio Moretti, Bibiana Santos, João Paulo Kitajima, Erika Freitas, David Schlesinger, Simone Kashima, Evandra Strazza Rodrigues, Svetoslav Nanev Slavov, Elaine Vieira dos Santos, Rafael dos Santos Bezerra, Luiz Carlos Junior de Alcantara, Marta Giovanetti, Vagner Fonseca, Flavia Aburjaile, Rodrigo Tocantins Calado. |
| EPI_ISL_1445238, EPI_ISL_1445239, EPI_ISL_1445240, EPI_ISL_1445241, EPI_ISL_1445242, EPI_ISL_1445243, EPI_ISL_1445244, EPI_ISL_1445245, EPI_ISL_1445246, EPI_ISL_1445247, EPI_ISL_1445248 | | | |
| see above | SECAO CENTRO DE DIAGNOSTICO SECEDI | Instituto Butantan / Mendelics | Dimas Tadeu Covas, Sandra Coccuzzo Sampaio, Maria Carolina Elias, José Salvatore Leister Patané, Vincent Louis Viala, Antonio Jorge Martins, Ricardo Haddad, Claudia Renata dos Santos Barros, Elaine Cristina Marqueze, Raul Machado Neto, Debora Botequio Moretti, Bibiana Santos, João Paulo Kitajima, Erika Freitas, David Schlesinger, Simone Kashima, Evandra Strazza Rodrigues, Svetoslav Nanev Slavov, Elaine Vieira dos Santos, Rafael dos Santos Bezerra, Luiz Carlos Junior de Alcantara, Marta Giovanetti, Vagner Fonseca, Flavia Aburjaile, Rodrigo Tocantins Calado. |
| EPI_ISL_1445249, EPI_ISL_1445250, EPI_ISL_1445251 | VIGILANCIA EPIDEMIOLOGICA | Instituto Butantan / Mendelics | Dimas Tadeu Covas, Sandra Coccuzzo Sampaio, Maria Carolina Elias, José Salvatore Leister Patané, Vincent Louis Viala, Antonio Jorge Martins, Ricardo Haddad, Claudia Renata dos Santos Barros, Elaine Cristina Marqueze, Raul Machado Neto, Debora Botequio Moretti, Bibiana Santos, João Paulo Kitajima, Erika Freitas, David Schlesinger, Simone Kashima, Evandra Strazza Rodrigues, Svetoslav Nanev Slavov, Elaine Vieira dos Santos, Rafael dos Santos Bezerra, Luiz Carlos Junior de Alcantara, Marta Giovanetti, Vagner Fonseca, Flavia Aburjaile, Rodrigo Tocantins Calado. |
| EPI_ISL_1445252, EPI_ISL_1445253 | SECAO CENTRO DE DIAGNOSTICO SECEDI | Instituto Butantan / Mendelics | Dimas Tadeu Covas, Sandra Coccuzzo Sampaio, Maria Carolina Elias, José Salvatore Leister Patané, Vincent Louis Viala, Antonio Jorge Martins, Ricardo Haddad, Claudia Renata dos Santos Barros, Elaine Cristina Marqueze, Raul Machado Neto, Debora Botequio Moretti, Bibiana Santos, João Paulo Kitajima, Erika Freitas, David Schlesinger, Simone Kashima, Evandra Strazza Rodrigues, Svetoslav Nanev Slavov, Elaine Vieira dos Santos, Rafael dos Santos Bezerra, Luiz Carlos Junior de Alcantara, Marta Giovanetti, Vagner Fonseca, Flavia Aburjaile, Rodrigo Tocantins Calado. |
| EPI_ISL_1445254, EPI_ISL_1445255, EPI_ISL_1445256, EPI_ISL_1445257, EPI_ISL_1445258, EPI_ISL_1445259, EPI_ISL_1445260, EPI_ISL_1445261 | VIGILANCIA EPIDEMIOLOGICA | Instituto Butantan / Mendelics | Dimas Tadeu Covas, Sandra Coccuzzo Sampaio, Maria Carolina Elias, José Salvatore Leister Patané, Vincent Louis Viala, Antonio Jorge Martins, Ricardo Haddad, Claudia Renata dos Santos Barros, Elaine Cristina Marqueze, Raul Machado Neto, Debora Botequio Moretti, Bibiana Santos, João Paulo Kitajima, Erika Freitas, David Schlesinger, Simone Kashima, Evandra Strazza Rodrigues, Svetoslav Nanev Slavov, Elaine Vieira dos Santos, Rafael dos Santos Bezerra, Luiz Carlos Junior de Alcantara, Marta Giovanetti, Vagner Fonseca, Flavia Aburjaile, Rodrigo Tocantins Calado. |
| EPI_ISL_1445262 | SECAO CENTRO DE DIAGNOSTICO SECEDI | Instituto Butantan / Mendelics | Dimas Tadeu Covas, Sandra Coccuzzo Sampaio, Maria Carolina Elias, José Salvatore Leister Patané, Vincent Louis Viala, Antonio Jorge Martins, Ricardo Haddad, Claudia Renata dos Santos Barros, Elaine Cristina Marqueze, Raul Machado Neto, Debora Botequio Moretti, Bibiana Santos, João Paulo Kitajima, Erika Freitas, David Schlesinger, Simone Kashima, Evandra Strazza Rodrigues, Svetoslav Nanev Slavov, Elaine Vieira dos Santos, Rafael dos Santos Bezerra, Luiz Carlos Junior de Alcantara, Marta Giovanetti, Vagner Fonseca, Flavia Aburjaile, Rodrigo Tocantins Calado. |
| EPI_ISL_1445263 | VIGILANCIA EPIDEMIOLOGICA | Instituto Butantan / Mendelics | Dimas Tadeu Covas, Sandra Coccuzzo Sampaio, Maria Carolina Elias, José Salvatore Leister Patané, Vincent Louis Viala, Antonio Jorge Martins, Ricardo Haddad, Claudia Renata dos Santos Barros, Elaine Cristina Marqueze, Raul Machado Neto, Debora Botequio Moretti, Bibiana Santos, João Paulo Kitajima, Erika Freitas, David Schlesinger, Simone Kashima, Evandra Strazza Rodrigues, Svetoslav Nanev Slavov, Elaine Vieira dos Santos, Rafael dos Santos Bezerra, Luiz Carlos Junior de Alcantara, Marta Giovanetti, Vagner Fonseca, Flavia Aburjaile, Rodrigo Tocantins Calado. |
| EPI_ISL_1445264 | AMBULATORIO MEDICO DE ESPECIALIDADES DE PERUIBE | Instituto Butantan / Mendelics | Dimas Tadeu Covas, Sandra Coccuzzo Sampaio, Maria Carolina Elias, José Salvatore Leister Patané, Vincent Louis Viala, Antonio Jorge Martins, Ricardo Haddad, Claudia Renata dos Santos Barros, Elaine Cristina Marqueze, Raul Machado Neto, Debora Botequio Moretti, Bibiana Santos, João Paulo Kitajima, Erika Freitas, David Schlesinger, Simone Kashima, Evandra Strazza Rodrigues, Svetoslav Nanev Slavov, Elaine Vieira dos Santos, Rafael dos Santos Bezerra, Luiz Carlos Junior de Alcantara, Marta Giovanetti, Vagner Fonseca, Flavia Aburjaile, Rodrigo Tocantins Calado. |
| EPI_ISL_1445265 | VIGILANCIA EPIDEMIOLOGICA | Instituto Butantan / Mendelics | Dimas Tadeu Covas, Sandra Coccuzzo Sampaio, Maria Carolina Elias, José Salvatore Leister Patané, Vincent Louis Viala, Antonio Jorge Martins, Ricardo Haddad, Claudia Renata dos Santos Barros, Elaine Cristina Marqueze, Raul Machado Neto, Debora Botequio Moretti, Bibiana Santos, João Paulo Kitajima, Erika Freitas, David Schlesinger, Simone Kashima, Evandra Strazza Rodrigues, Svetoslav Nanev Slavov, Elaine Vieira dos Santos, Rafael dos Santos Bezerra, Luiz Carlos Junior de Alcantara, Marta Giovanetti, Vagner Fonseca, Flavia Aburjaile, Rodrigo Tocantins Calado. |
| EPI_ISL_1445266, EPI_ISL_1445267 | AMBULATORIO MEDICO DE ESPECIALIDADES DE PERUIBE | Instituto Butantan / Mendelics | Dimas Tadeu Covas, Sandra Coccuzzo Sampaio, Maria Carolina Elias, José Salvatore Leister Patané, Vincent Louis Viala, Antonio Jorge Martins, Ricardo Haddad, Claudia Renata dos Santos Barros, Elaine Cristina Marqueze, Raul Machado Neto, Debora Botequio Moretti, Bibiana Santos, João Paulo Kitajima, Erika Freitas, David Schlesinger, Simone Kashima, Evandra Strazza Rodrigues, Svetoslav Nanev Slavov, Elaine Vieira dos Santos, Rafael dos Santos Bezerra, Luiz Carlos Junior de Alcantara, Marta Giovanetti, Vagner Fonseca, Flavia Aburjaile, Rodrigo Tocantins Calado. |
| EPI_ISL_1445268 | VIGILANCIA EPIDEMIOLOGICA | Instituto Butantan / Mendelics | Dimas Tadeu Covas, Sandra Coccuzzo Sampaio, Maria Carolina Elias, José Salvatore Leister Patané, Vincent Louis Viala, Antonio Jorge Martins, Ricardo Haddad, Claudia Renata dos Santos Barros, Elaine Cristina Marqueze, Raul Machado Neto, Debora Botequio Moretti, Bibiana Santos, João Paulo Kitajima, Erika Freitas, David Schlesinger, Simone Kashima, Evandra Strazza Rodrigues, Svetoslav Nanev Slavov, Elaine Vieira dos Santos, Rafael dos Santos Bezerra, Luiz Carlos Junior de Alcantara, Marta Giovanetti, Vagner Fonseca, Flavia Aburjaile, Rodrigo Tocantins Calado. |
| EPI_ISL_1445269 | SERV DE VIG SANITARIA EPIDEMIO E CTRL DE ZOONOZES GUARUJA | Instituto Butantan / Mendelics | Dimas Tadeu Covas, Sandra Coccuzzo Sampaio, Maria Carolina Elias, José Salvatore Leister Patané, Vincent Louis Viala, Antonio Jorge Martins, Ricardo Haddad, Claudia Renata dos Santos Barros, Elaine Cristina Marqueze, Raul Machado Neto, Debora Botequio Moretti, Bibiana Santos, João Paulo Kitajima, Erika Freitas, David Schlesinger, Simone Kashima, Evandra Strazza Rodrigues, Svetoslav Nanev Slavov, Elaine Vieira dos Santos, Rafael dos Santos Bezerra, Luiz Carlos Junior de Alcantara, Marta Giovanetti, Vagner Fonseca, Flavia Aburjaile, Rodrigo Tocantins Calado. |

| | | | |
|---|---|---|---|
| EPI_ISL_1445270, EPI_ISL_1445271 | VIGILANCIA EPIDEMIOLOGICA | Instituto Butantan / Mendelics | Dimas Tadeu Covas, Sandra Coccuzzo Sampaio, Maria Carolina Elias, José Salvatore Leister Patané, Vincent Louis Viala, Antonio Jorge Martins, Ricardo Haddad, Claudia Renata dos Santos Barros, Elaine Cristina Marqueze, Raul Machado Neto, Debora Botequio Moretti, Bibiana Santos, João Paulo Kitajima, Erika Freitas, David Schlesinger, Simone Kashima, Evandra Strazza Rodrigues, Svetoslav Nanev Slavov, Elaine Vieira dos Santos, Rafael dos Santos Bezerra, Luiz Carlos Junior de Alcantara, Marta Giovanetti, Vagner Fonseca, Flavia Aburjaile, Rodrigo Tocantins Calado. |
| EPI_ISL_1445272 | SERV DE VIG SANITARIA EPIDEMIO E CTRL DE ZOONOZES GUARUJA | Instituto Butantan / Mendelics | Dimas Tadeu Covas, Sandra Coccuzzo Sampaio, Maria Carolina Elias, José Salvatore Leister Patané, Vincent Louis Viala, Antonio Jorge Martins, Ricardo Haddad, Claudia Renata dos Santos Barros, Elaine Cristina Marqueze, Raul Machado Neto, Debora Botequio Moretti, Bibiana Santos, João Paulo Kitajima, Erika Freitas, David Schlesinger, Simone Kashima, Evandra Strazza Rodrigues, Svetoslav Nanev Slavov, Elaine Vieira dos Santos, Rafael dos Santos Bezerra, Luiz Carlos Junior de Alcantara, Marta Giovanetti, Vagner Fonseca, Flavia Aburjaile, Rodrigo Tocantins Calado. |
| EPI_ISL_1445273, EPI_ISL_1445274 | AMBULATORIO MEDICO DE ESPECIALIDADES DE PERUIBE | Instituto Butantan / Mendelics | Dimas Tadeu Covas, Sandra Coccuzzo Sampaio, Maria Carolina Elias, José Salvatore Leister Patané, Vincent Louis Viala, Antonio Jorge Martins, Ricardo Haddad, Claudia Renata dos Santos Barros, Elaine Cristina Marqueze, Raul Machado Neto, Debora Botequio Moretti, Bibiana Santos, João Paulo Kitajima, Erika Freitas, David Schlesinger, Simone Kashima, Evandra Strazza Rodrigues, Svetoslav Nanev Slavov, Elaine Vieira dos Santos, Rafael dos Santos Bezerra, Luiz Carlos Junior de Alcantara, Marta Giovanetti, Vagner Fonseca, Flavia Aburjaile, Rodrigo Tocantins Calado. |
| EPI_ISL_1446194 | LATE - Laboratório de Técnicas Especiais - Hospital Israelita Albert Einstein | LATE - Laboratório de Técnicas Especiais - Hospital Israelita Albert Einstein | Deyvid Amgarten, Fernanda de Mello Malta, Raquel Riyuzo, Ana Paula Moreira Salles, Pedro Henrique Sebe Rodrigues, João Renato Rebello Pinho |
| EPI_ISL_1464626, EPI_ISL_1464627, EPI_ISL_1464628, EPI_ISL_1464629, EPI_ISL_1464630, EPI_ISL_1464631, EPI_ISL_1464632, EPI_ISL_1464633, EPI_ISL_1464634, EPI_ISL_1464635, EPI_ISL_1464636, EPI_ISL_1464637, EPI_ISL_1464638, EPI_ISL_1464639, EPI_ISL_1464640, EPI_ISL_1464641, EPI_ISL_1464642, EPI_ISL_1464643, EPI_ISL_1464644, EPI_ISL_1464645, EPI_ISL_1464646, EPI_ISL_1464647, EPI_ISL_1464648, EPI_ISL_1464649, EPI_ISL_1464650, EPI_ISL_1464651, EPI_ISL_1464652, EPI_ISL_1464653, EPI_ISL_1464654, EPI_ISL_1464655, EPI_ISL_1464656, EPI_ISL_1464657, EPI_ISL_1464658, EPI_ISL_1464659, EPI_ISL_1464660, EPI_ISL_1464661, EPI_ISL_1464662, EPI_ISL_1464663, EPI_ISL_1464664, EPI_ISL_1464665, EPI_ISL_1464666, EPI_ISL_1464667, EPI_ISL_1464668, EPI_ISL_1464669, EPI_ISL_1464670, EPI_ISL_1464671, EPI_ISL_1464672, EPI_ISL_1464673, EPI_ISL_1464674, EPI_ISL_1464675, EPI_ISL_1464676, EPI_ISL_1464677 | | | |
| see above | Laboratório de Virologia - UNIFESP | Laboratory of Respiratory Viruses and Measles, Oswaldo Cruz Institute, FIOCRUZ | Paola Resende, Nancy Belei, Luciana Appolinario, Fernando Motta, Anna Carolina Paixao, Ana Carolina Mendonca, Alice Sampaio Rocha, Renata Serrano Lopes, Marilda Siqueira on behalf of the Fiocruz COVID-19 Genomic Surveillance Network |
| EPI_ISL_1465185, EPI_ISL_1465186, EPI_ISL_1465188, EPI_ISL_1465189, EPI_ISL_1465191, EPI_ISL_1465192, EPI_ISL_1465194, EPI_ISL_1465195, EPI_ISL_1465196, EPI_ISL_1465198, EPI_ISL_1465199, EPI_ISL_1465201, EPI_ISL_1465202, EPI_ISL_1465203, EPI_ISL_1465205, EPI_ISL_1465206, EPI_ISL_1465208, EPI_ISL_1465209, EPI_ISL_1465210, EPI_ISL_1465212, EPI_ISL_1465213, EPI_ISL_1465215, EPI_ISL_1465216, EPI_ISL_1465217, EPI_ISL_1465219, EPI_ISL_1465220, EPI_ISL_1465221, EPI_ISL_1465222, EPI_ISL_1465224, EPI_ISL_1465225, EPI_ISL_1465226, EPI_ISL_1465228, EPI_ISL_1465229, EPI_ISL_1465231, EPI_ISL_1465232, EPI_ISL_1465234, EPI_ISL_1465235, EPI_ISL_1465236, EPI_ISL_1465238, EPI_ISL_1465239, EPI_ISL_1465241, EPI_ISL_1465242, EPI_ISL_1465243, EPI_ISL_1465245, EPI_ISL_1465246, EPI_ISL_1465248, EPI_ISL_1465249, EPI_ISL_1465250, EPI_ISL_1465252, EPI_ISL_1465253, EPI_ISL_1465254, EPI_ISL_1465255, EPI_ISL_1465257, EPI_ISL_1465258, EPI_ISL_1465259, EPI_ISL_1465261, EPI_ISL_1465262, EPI_ISL_1465264, EPI_ISL_1465265, EPI_ISL_1465267, EPI_ISL_1465268, EPI_ISL_1465270, EPI_ISL_1465271, EPI_ISL_1465273, EPI_ISL_1465274, EPI_ISL_1465275, EPI_ISL_1465277, EPI_ISL_1465278 | | | |
| see above | Laboratorio Central de Saude Publica do Estado do Maranhao (LACEN-MA) | Laboratory of Respiratory Viruses and Measles, Oswaldo Cruz Institute, FIOCRUZ | Paola Resende, Luciana Appolinario, Fernando Motta, Anna Carolina Paixao, Ana Carolina Mendonca, Alice Sampaio Rocha, Renata Serrano Lopes, Lidio Gonçalves Lima Neto, Marilda Siqueira on behalf of the Fiocruz COVID-19 Genomic Surveillance Network |
| EPI_ISL_1468411 | Centro de Saude II Matao | Instituto Adolfo Lutz, Interdiciplinary Procedures Center, Strategic Laboratory | Claudio Tavares Sacchi, Claudia Regina Gonçalves, Erica Valessa Ramos Gomes, Karoline Rodrigues Campos, Caio Vinicius Dias Lopes |
| EPI_ISL_1468412 | Hospital Estadual de Mirandopolis | Instituto Adolfo Lutz, Interdiciplinary Procedures Center, Strategic Laboratory | Claudio Tavares Sacchi, Claudia Regina Gonçalves, Erica Valessa Ramos Gomes, Karoline Rodrigues Campos, Caio Vinicius Dias Lopes |
| EPI_ISL_1468413, EPI_ISL_1468414, EPI_ISL_1468415 | LACEN do Estado de Goias | Instituto Adolfo Lutz, Interdiciplinary Procedures Center, Strategic Laboratory | Claudio Tavares Sacchi, Claudia Regina Gonçalves, Erica Valessa Ramos Gomes, Karoline Rodrigues Campos, Caio Vinicius Dias Lopes |
| EPI_ISL_1468416 | Secretaria Municipal de Saude de Andradina | Instituto Adolfo Lutz, Interdiciplinary Procedures Center, Strategic Laboratory | Claudio Tavares Sacchi, Claudia Regina Gonçalves, Erica Valessa Ramos Gomes, Karoline Rodrigues Campos, Caio Vinicius Dias Lopes |
| EPI_ISL_1468417 | Santa Casa de Aracatuba Hospital Sagrado Coracao de Jesus | Instituto Adolfo Lutz, Interdiciplinary Procedures Center, Strategic Laboratory | Claudio Tavares Sacchi, Claudia Regina Gonçalves, Erica Valessa Ramos Gomes, Karoline Rodrigues Campos, Caio Vinicius Dias Lopes |
| EPI_ISL_1468418 | Secretaria Municipal de Saude de Birigui | Instituto Adolfo Lutz, Interdiciplinary Procedures Center, Strategic Laboratory | Claudio Tavares Sacchi, Claudia Regina Gonçalves, Erica Valessa Ramos Gomes, Karoline Rodrigues Campos, Caio Vinicius Dias Lopes |
| EPI_ISL_1468419 | Santa Casa de Aracatuba Hospital Sagrado Coracao de Jesus | Instituto Adolfo Lutz, Interdiciplinary Procedures Center, Strategic Laboratory | Claudio Tavares Sacchi, Claudia Regina Gonçalves, Erica Valessa Ramos Gomes, Karoline Rodrigues Campos, Caio Vinicius Dias Lopes |
| EPI_ISL_1468420, EPI_ISL_1468421 | Santa Casa de Birigui | Instituto Adolfo Lutz, Interdiciplinary Procedures Center, Strategic Laboratory | Claudio Tavares Sacchi, Claudia Regina Gonçalves, Erica Valessa Ramos Gomes, Karoline Rodrigues Campos, Caio Vinicius Dias Lopes |
| EPI_ISL_1468422, EPI_ISL_1468423 | Penitenciaria Compacta de Avanhandava | Instituto Adolfo Lutz, Interdiciplinary Procedures Center, Strategic Laboratory | Claudio Tavares Sacchi, Claudia Regina Gonçalves, Erica Valessa Ramos Gomes, Karoline Rodrigues Campos, Caio Vinicius Dias Lopes |
| EPI_ISL_1468424 | Santa Casa de Aracatuba Hospital Sagrado Coracao de Jesus | Instituto Adolfo Lutz, Interdiciplinary Procedures Center, Strategic Laboratory | Claudio Tavares Sacchi, Claudia Regina Gonçalves, Erica Valessa Ramos Gomes, Karoline Rodrigues Campos, Caio Vinicius Dias Lopes |
| EPI_ISL_1468425 | Santa Casa de Birigui | Instituto Adolfo Lutz, Interdiciplinary Procedures Center, Strategic Laboratory | Claudio Tavares Sacchi, Claudia Regina Gonçalves, Erica Valessa Ramos Gomes, Karoline Rodrigues Campos, Caio Vinicius Dias Lopes |
| EPI_ISL_1468426, EPI_ISL_1468427 | Centro de Saude II Matao | Instituto Adolfo Lutz, Interdiciplinary Procedures Center, Strategic Laboratory | Claudio Tavares Sacchi, Claudia Regina Gonçalves, Erica Valessa Ramos Gomes, Karoline Rodrigues Campos, Caio Vinicius Dias Lopes |
| EPI_ISL_1468428 | SAE Servico de Atendimento Especializado | Instituto Adolfo Lutz, Interdiciplinary Procedures Center, Strategic Laboratory | Claudio Tavares Sacchi, Claudia Regina Gonçalves, Erica Valessa Ramos Gomes, Karoline Rodrigues Campos, Caio Vinicius Dias Lopes |
| EPI_ISL_1468429, EPI_ISL_1468430 | Centro de Saude II Matao | Instituto Adolfo Lutz, Interdiciplinary Procedures Center, Strategic Laboratory | Claudio Tavares Sacchi, Claudia Regina Gonçalves, Erica Valessa Ramos Gomes, Karoline Rodrigues Campos, Caio Vinicius Dias Lopes |
| EPI_ISL_1468431 | LACEN do Estado de Goias | Instituto Adolfo Lutz, Interdiciplinary Procedures Center, Strategic Laboratory | Claudio Tavares Sacchi, Claudia Regina Gonçalves, Erica Valessa Ramos Gomes, Karoline Rodrigues Campos, Caio Vinicius Dias Lopes |
| EPI_ISL_1468432, EPI_ISL_1468433, EPI_ISL_1468434, EPI_ISL_1468435, EPI_ISL_1468436, EPI_ISL_1468437, EPI_ISL_1468438, EPI_ISL_1468439, EPI_ISL_1468440, EPI_ISL_1468441 | LACEN do Mato Grosso do Sul | Instituto Adolfo Lutz, Interdiciplinary Procedures Center, Strategic Laboratory | Claudio Tavares Sacchi, Claudia Regina Gonçalves, Erica Valessa Ramos Gomes, Karoline Rodrigues Campos, Caio Vinicius Dias Lopes |
| EPI_ISL_1468442 | Santa Casa de Birigui | Instituto Adolfo Lutz, Interdiciplinary Procedures Center, Strategic Laboratory | Claudio Tavares Sacchi, Claudia Regina Gonçalves, Erica Valessa Ramos Gomes, Karoline Rodrigues Campos, Caio Vinicius Dias Lopes |
| EPI_ISL_1468443 | Hospital Estadual de Mirandopolis | Instituto Adolfo Lutz, Interdiciplinary Procedures Center, Strategic Laboratory | Claudio Tavares Sacchi, Claudia Regina Gonçalves, Erica Valessa Ramos Gomes, Karoline Rodrigues Campos, Caio Vinicius Dias Lopes |
| EPI_ISL_1468444 | Secretaria Municipal de Saude de Birigui | Instituto Adolfo Lutz, Interdiciplinary Procedures Center, Strategic Laboratory | Claudio Tavares Sacchi, Claudia Regina Gonçalves, Erica Valessa Ramos Gomes, Karoline Rodrigues Campos, Caio Vinicius Dias Lopes |
| EPI_ISL_1468445 | Santa Casa de Misericordia de Pereira Barreto | Instituto Adolfo Lutz, Interdiciplinary Procedures Center, Strategic Laboratory | Claudio Tavares Sacchi, Claudia Regina Gonçalves, Erica Valessa Ramos Gomes, Karoline Rodrigues Campos, Caio Vinicius Dias Lopes |
| EPI_ISL_1468446 | Secretaria Municipal de Saude de Birigui | Instituto Adolfo Lutz, Interdiciplinary Procedures Center, Strategic Laboratory | Claudio Tavares Sacchi, Claudia Regina Gonçalves, Erica Valessa Ramos Gomes, Karoline Rodrigues Campos, Caio Vinicius Dias Lopes |
| EPI_ISL_1468447 | Secretaria Municipal de Saude de Andradina | Instituto Adolfo Lutz, Interdiciplinary Procedures Center, Strategic Laboratory | Claudio Tavares Sacchi, Claudia Regina Gonçalves, Erica Valessa Ramos Gomes, Karoline Rodrigues Campos, Caio Vinicius Dias Lopes |

| | | | |
|---|---|---|---|
| EPI_ISL_1468448 | Santa Casa de Birigui | Instituto Adolfo Lutz, Interdiciplinary Procedures Center, Strategic Laboratory | Claudio Tavares Sacchi, Claudia Regina Gonçalves, Erica Valessa Ramos Gomes, Karoline Rodrigues Campos, Caio Vinicius Dias Lopes |
| EPI_ISL_1468449, EPI_ISL_1468450 | Santa Casa de Aracatuba Hospital Sagrado Coracao de Jesus | Instituto Adolfo Lutz, Interdiciplinary Procedures Center, Strategic Laboratory | Claudio Tavares Sacchi, Claudia Regina Gonçalves, Erica Valessa Ramos Gomes, Karoline Rodrigues Campos, Caio Vinicius Dias Lopes |
| EPI_ISL_1468451 | UBS IV Guararapes | Instituto Adolfo Lutz, Interdiciplinary Procedures Center, Strategic Laboratory | Claudio Tavares Sacchi, Claudia Regina Gonçalves, Erica Valessa Ramos Gomes, Karoline Rodrigues Campos, Caio Vinicius Dias Lopes |
| EPI_ISL_1468452 | Centro de Atendimento COVID | Instituto Adolfo Lutz, Interdiciplinary Procedures Center, Strategic Laboratory | Claudio Tavares Sacchi, Claudia Regina Gonçalves, Erica Valessa Ramos Gomes, Karoline Rodrigues Campos, Caio Vinicius Dias Lopes |
| EPI_ISL_1468453 | Santa Casa de Misericordia de Pereira Barreto | Instituto Adolfo Lutz, Interdiciplinary Procedures Center, Strategic Laboratory | Claudio Tavares Sacchi, Claudia Regina Gonçalves, Erica Valessa Ramos Gomes, Karoline Rodrigues Campos, Caio Vinicius Dias Lopes |
| EPI_ISL_1468454 | Santa Casa de Andradina | Instituto Adolfo Lutz, Interdiciplinary Procedures Center, Strategic Laboratory | Claudio Tavares Sacchi, Claudia Regina Gonçalves, Erica Valessa Ramos Gomes, Karoline Rodrigues Campos, Caio Vinicius Dias Lopes |
| EPI_ISL_1468455 | UBS 02 Jardim Toselar Birigui | Instituto Adolfo Lutz, Interdiciplinary Procedures Center, Strategic Laboratory | Claudio Tavares Sacchi, Claudia Regina Gonçalves, Erica Valessa Ramos Gomes, Karoline Rodrigues Campos, Caio Vinicius Dias Lopes |
| EPI_ISL_1468456, EPI_ISL_1468457 | UBS Dr Alfredo Dantas de Souza Umuarama | Instituto Adolfo Lutz, Interdiciplinary Procedures Center, Strategic Laboratory | Claudio Tavares Sacchi, Claudia Regina Gonçalves, Erica Valessa Ramos Gomes, Karoline Rodrigues Campos, Caio Vinicius Dias Lopes |
| EPI_ISL_1468458, EPI_ISL_1468459 | Secretaria Municipal de Saude de Valparaiso SP | Instituto Adolfo Lutz, Interdiciplinary Procedures Center, Strategic Laboratory | Claudio Tavares Sacchi, Claudia Regina Gonçalves, Erica Valessa Ramos Gomes, Karoline Rodrigues Campos, Caio Vinicius Dias Lopes |
| EPI_ISL_1468460 | Santa Casa de Birigui | Instituto Adolfo Lutz, Interdiciplinary Procedures Center, Strategic Laboratory | Claudio Tavares Sacchi, Claudia Regina Gonçalves, Erica Valessa Ramos Gomes, Karoline Rodrigues Campos, Caio Vinicius Dias Lopes |
| EPI_ISL_1468461 | Santa Casa de Aracatuba Hospital Sagrado Coracao de Jesus | Instituto Adolfo Lutz, Interdiciplinary Procedures Center, Strategic Laboratory | Claudio Tavares Sacchi, Claudia Regina Gonçalves, Erica Valessa Ramos Gomes, Karoline Rodrigues Campos, Caio Vinicius Dias Lopes |
| EPI_ISL_1468462 | Servico Especial de Saude de Araraquara Sesa Araraquara | Instituto Adolfo Lutz, Interdiciplinary Procedures Center, Strategic Laboratory | Claudio Tavares Sacchi, Claudia Regina Gonçalves, Erica Valessa Ramos Gomes, Karoline Rodrigues Campos, Caio Vinicius Dias Lopes |
| EPI_ISL_1468463 | Centro de Saude II Matao | Instituto Adolfo Lutz, Interdiciplinary Procedures Center, Strategic Laboratory | Claudio Tavares Sacchi, Claudia Regina Gonçalves, Erica Valessa Ramos Gomes, Karoline Rodrigues Campos, Caio Vinicius Dias Lopes |
| EPI_ISL_1468464, EPI_ISL_1468465 | Sae servico de Atendimento Especializado | Instituto Adolfo Lutz, Interdiciplinary Procedures Center, Strategic Laboratory | Claudio Tavares Sacchi, Claudia Regina Gonçalves, Erica Valessa Ramos Gomes, Karoline Rodrigues Campos, Caio Vinicius Dias Lopes |
| EPI_ISL_1468466 | Santa Casa de Sao Carlos | Instituto Adolfo Lutz, Interdiciplinary Procedures Center, Strategic Laboratory | Claudio Tavares Sacchi, Claudia Regina Gonçalves, Erica Valessa Ramos Gomes, Karoline Rodrigues Campos, Caio Vinicius Dias Lopes |
| EPI_ISL_1468467 | Secretaria Municipal de Saude Descalvado | Instituto Adolfo Lutz, Interdiciplinary Procedures Center, Strategic Laboratory | Claudio Tavares Sacchi, Claudia Regina Gonçalves, Erica Valessa Ramos Gomes, Karoline Rodrigues Campos, Caio Vinicius Dias Lopes |
| EPI_ISL_1468468 | Secretaria municipal de saude de Itapolis | Instituto Adolfo Lutz, Interdiciplinary Procedures Center, Strategic Laboratory | Claudio Tavares Sacchi, Claudia Regina Gonçalves, Erica Valessa Ramos Gomes, Karoline Rodrigues Campos, Caio Vinicius Dias Lopes |
| EPI_ISL_1468469 | Secretaria Municipal de Saude Porto Ferreira | Instituto Adolfo Lutz, Interdiciplinary Procedures Center, Strategic Laboratory | Claudio Tavares Sacchi, Claudia Regina Gonçalves, Erica Valessa Ramos Gomes, Karoline Rodrigues Campos, Caio Vinicius Dias Lopes |
| EPI_ISL_1468470, EPI_ISL_1468471 | Secretaria Municipal de Saude Descalvado | Instituto Adolfo Lutz, Interdiciplinary Procedures Center, Strategic Laboratory | Claudio Tavares Sacchi, Claudia Regina Gonçalves, Erica Valessa Ramos Gomes, Karoline Rodrigues Campos, Caio Vinicius Dias Lopes |
| EPI_ISL_1468472, EPI_ISL_1468473 | Centro de Saude II Matao | Instituto Adolfo Lutz, Interdiciplinary Procedures Center, Strategic Laboratory | Claudio Tavares Sacchi, Claudia Regina Gonçalves, Erica Valessa Ramos Gomes, Karoline Rodrigues Campos, Caio Vinicius Dias Lopes |
| EPI_ISL_1468474 | Sae servico de Atendimento Especializado | Instituto Adolfo Lutz, Interdiciplinary Procedures Center, Strategic Laboratory | Claudio Tavares Sacchi, Claudia Regina Gonçalves, Erica Valessa Ramos Gomes, Karoline Rodrigues Campos, Caio Vinicius Dias Lopes |
| EPI_ISL_1469549 | FUNDACAO HOSPITALAR DE SAPUCAIA DO SUL | Epiclin | Fernando Hayashi Sant'Anna, Ana Paula Muterle, Janira Prichula, Juliana Comerlato, Carolina Comerlato, Eliana Márcia Da Ros Wendland |
| EPI_ISL_1469550 | Vigilância em Saúde de Sapucaia do Sul | Epiclin | Fernando Hayashi Sant'Anna, Ana Paula Muterle, Janira Prichula, Juliana Comerlato, Carolina Comerlato, Eliana Márcia Da Ros Wendland |
| EPI_ISL_1469551 | Secretaria Municipal de Saúde de São Leopoldo | Epiclin | Fernando Hayashi Sant'Anna, Ana Paula Muterle, Janira Prichula, Juliana Comerlato, Carolina Comerlato, Eliana Márcia Da Ros Wendland |
| EPI_ISL_1469552 | Unidade Sanitária de Igrejinha | Epiclin | Fernando Hayashi Sant'Anna, Ana Paula Muterle, Janira Prichula, Juliana Comerlato, Carolina Comerlato, Eliana Márcia Da Ros Wendland |
| EPI_ISL_1469553 | Diretoria de Vigilância em Saúde | Epiclin | Fernando Hayashi Sant'Anna, Ana Paula Muterle, Janira Prichula, Juliana Comerlato, Carolina Comerlato, Eliana Márcia Da Ros Wendland |
| EPI_ISL_1469554 | Fundação de Saúde Pública São Camilo de Esteio | Epiclin | Fernando Hayashi Sant'Anna, Ana Paula Muterle, Janira Prichula, Juliana Comerlato, Carolina Comerlato, Eliana Márcia Da Ros Wendland |
| EPI_ISL_1469555 | FUNDACAO DE SAUDE PUBLICA DE NOVO HAMBURGO FSNH | Epiclin | Fernando Hayashi Sant'Anna, Ana Paula Muterle, Janira Prichula, Juliana Comerlato, Carolina Comerlato, Eliana Márcia Da Ros Wendland |
| EPI_ISL_1469556 | Secretaria Municipal de Saúde de São Leopoldo | Epiclin | Fernando Hayashi Sant'Anna, Ana Paula Muterle, Janira Prichula, Juliana Comerlato, Carolina Comerlato, Eliana Márcia Da Ros Wendland |
| EPI_ISL_1469557 | Centro de Serviços Especializados Santa Rita | Epiclin | Fernando Hayashi Sant'Anna, Ana Paula Muterle, Janira Prichula, Juliana Comerlato, Carolina Comerlato, Eliana Márcia Da Ros Wendland |
| EPI_ISL_1469558 | DIRETORIA DE VIGILANCIA EM SAUDE | Epiclin | Fernando Hayashi Sant'Anna, Ana Paula Muterle, Janira Prichula, Juliana Comerlato, Carolina Comerlato, Eliana Márcia Da Ros Wendland |
| EPI_ISL_1469559 | Secretaria Municipal de Saúde de Três Coroas | Epiclin | Fernando Hayashi Sant'Anna, Ana Paula Muterle, Janira Prichula, Juliana Comerlato, Carolina Comerlato, Eliana Márcia Da Ros Wendland |
| EPI_ISL_1469560 | Diretoria de Vigilância em Saúde | Epiclin | Fernando Hayashi Sant'Anna, Ana Paula Muterle, Janira Prichula, Juliana Comerlato, Carolina Comerlato, Eliana Márcia Da Ros Wendland |
| EPI_ISL_1469561 | Fundação de Saúde Pública São Camilo de Esteio | Epiclin | Fernando Hayashi Sant'Anna, Ana Paula Muterle, Janira Prichula, Juliana Comerlato, Carolina Comerlato, Eliana Márcia Da Ros Wendland |
| EPI_ISL_1469562 | Hospital Dia e Pronto Atendimento | Epiclin | Fernando Hayashi Sant'Anna, Ana Paula Muterle, Janira Prichula, Juliana Comerlato, Carolina Comerlato, Eliana Márcia Da Ros Wendland |
| EPI_ISL_1469563 | Unidade Básica de Saúde de Riozinho | Epiclin | Fernando Hayashi Sant'Anna, Ana Paula Muterle, Janira Prichula, Juliana Comerlato, Carolina Comerlato, Eliana Márcia Da Ros Wendland |
| EPI_ISL_1469564 | Pronto Atendimento Campo Bom | Epiclin | Fernando Hayashi Sant'Anna, Ana Paula Muterle, Janira Prichula, Juliana Comerlato, Carolina Comerlato, Eliana Márcia Da Ros Wendland |
| EPI_ISL_1469565 | Unidade de Pronto Atendimento de Sapucaia do Sul | Epiclin | Fernando Hayashi Sant'Anna, Ana Paula Muterle, Janira Prichula, Juliana Comerlato, Carolina Comerlato, Eliana Márcia Da Ros Wendland |
| EPI_ISL_1469566 | DIRETORIA DE VIGILANCIA EM SAUDE | Epiclin | Fernando Hayashi Sant'Anna, Ana Paula Muterle, Janira Prichula, Juliana Comerlato, Carolina Comerlato, Eliana Márcia Da Ros Wendland |
| EPI_ISL_1469567 | Unidade Sanitária de Igrejinha | Epiclin | Fernando Hayashi Sant'Anna, Ana Paula Muterle, Janira Prichula, Juliana Comerlato, Carolina Comerlato, Eliana Márcia Da Ros Wendland |
| EPI_ISL_1469568 | SECRETARIA MUNICIPAL DE SAUDE DE SAO LEOPOLDO | Epiclin | Fernando Hayashi Sant'Anna, Ana Paula Muterle, Janira Prichula, Juliana Comerlato, Carolina Comerlato, Eliana Márcia Da Ros Wendland |
| EPI_ISL_1469569 | Fundação de Saúde Pública São Camilo de Esteio | Epiclin | Fernando Hayashi Sant'Anna, Ana Paula Muterle, Janira Prichula, Juliana Comerlato, Carolina Comerlato, Eliana Márcia Da Ros Wendland |
| EPI_ISL_1469570 | Hospital Universitário | Epiclin | Fernando Hayashi Sant'Anna, Ana Paula Muterle, Janira Prichula, Juliana Comerlato, Carolina Comerlato, Eliana Márcia Da Ros Wendland |
| EPI_ISL_1469571 | Unidade de Pronto Atendimento de Sapucaia do Sul | Epiclin | Fernando Hayashi Sant'Anna, Ana Paula Muterle, Janira Prichula, Juliana Comerlato, Carolina Comerlato, Eliana Márcia Da Ros Wendland |

| | | | |
|---|---|---|---|
| EPI_ISL_1469572 | FUNDACAO DE SAUDE PUBLICA DE NOVO HAMBURGO FSNH | Epiclin | Fernando Hayashi Sant'Anna, Ana Paula Muterle, Janira Prichula, Juliana Comerlato, Carolina Comerlato, Eliana Márcia Da Ros Wendland |
| EPI_ISL_1469573 | VIGILANCIA EM SAUDE NH | Epiclin | Fernando Hayashi Sant'Anna, Ana Paula Muterle, Janira Prichula, Juliana Comerlato, Carolina Comerlato, Eliana Márcia Da Ros Wendland |
| EPI_ISL_1469574 | CENTRO DE SERVICOS ESPECIALIZADOS SANTA RITA DE CASSIA | Epiclin | Fernando Hayashi Sant'Anna, Ana Paula Muterle, Janira Prichula, Juliana Comerlato, Carolina Comerlato, Eliana Márcia Da Ros Wendland |
| EPI_ISL_1469575 | HOSPITAL SAO FRANCISCO DE ASSIS | Epiclin | Fernando Hayashi Sant'Anna, Ana Paula Muterle, Janira Prichula, Juliana Comerlato, Carolina Comerlato, Eliana Márcia Da Ros Wendland |
| EPI_ISL_1469576 | Unidade Sanitária de Igrejinha | Epiclin | Fernando Hayashi Sant'Anna, Ana Paula Muterle, Janira Prichula, Juliana Comerlato, Carolina Comerlato, Eliana Márcia Da Ros Wendland |
| EPI_ISL_1469577 | Unidade de Pronto Atendimento de Sapucaia do Sul | Epiclin | Fernando Hayashi Sant'Anna, Ana Paula Muterle, Janira Prichula, Juliana Comerlato, Carolina Comerlato, Eliana Márcia Da Ros Wendland |
| EPI_ISL_1469578 | HOSPITAL SAO FRANCISCO DE ASSIS | Epiclin | Fernando Hayashi Sant'Anna, Ana Paula Muterle, Janira Prichula, Juliana Comerlato, Carolina Comerlato, Eliana Márcia Da Ros Wendland |
| EPI_ISL_1469579 | DIRETORIA DE VIGILANCIA EM SAUDE | Epiclin | Fernando Hayashi Sant'Anna, Ana Paula Muterle, Janira Prichula, Juliana Comerlato, Carolina Comerlato, Eliana Márcia Da Ros Wendland |
| EPI_ISL_1469580 | Unidade de Pronto Atendimento de Sapucaia do Sul | Epiclin | Fernando Hayashi Sant'Anna, Ana Paula Muterle, Janira Prichula, Juliana Comerlato, Carolina Comerlato, Eliana Márcia Da Ros Wendland |
| EPI_ISL_1469581 | Hospital Dia e Pronto Atendimento | Epiclin | Fernando Hayashi Sant'Anna, Ana Paula Muterle, Janira Prichula, Juliana Comerlato, Carolina Comerlato, Eliana Márcia Da Ros Wendland |
| EPI_ISL_1469582 | Unidade de Pronto Atendimento de Sapucaia do Sul | Epiclin | Fernando Hayashi Sant'Anna, Ana Paula Muterle, Janira Prichula, Juliana Comerlato, Carolina Comerlato, Eliana Márcia Da Ros Wendland |
| EPI_ISL_1469583 | UNIDADE SANITARIA DE IGREJINHA | Epiclin | Fernando Hayashi Sant'Anna, Ana Paula Muterle, Janira Prichula, Juliana Comerlato, Carolina Comerlato, Eliana Márcia Da Ros Wendland |
| EPI_ISL_1469584 | CENTRO MUNICIPAL DE SAUDE DE ROLANTE | Epiclin | Fernando Hayashi Sant'Anna, Ana Paula Muterle, Janira Prichula, Juliana Comerlato, Carolina Comerlato, Eliana Márcia Da Ros Wendland |
| EPI_ISL_1469585 | Fundação Hospitalar de Sapucaia do Sul | Epiclin | Fernando Hayashi Sant'Anna, Ana Paula Muterle, Janira Prichula, Juliana Comerlato, Carolina Comerlato, Eliana Márcia Da Ros Wendland |
| EPI_ISL_1469586 | FUNDACAO HOSPITALAR SAO JOSE | Epiclin | Fernando Hayashi Sant'Anna, Ana Paula Muterle, Janira Prichula, Juliana Comerlato, Carolina Comerlato, Eliana Márcia Da Ros Wendland |
| EPI_ISL_1469587 | Hospital Sapiranga | Epiclin | Fernando Hayashi Sant'Anna, Ana Paula Muterle, Janira Prichula, Juliana Comerlato, Carolina Comerlato, Eliana Márcia Da Ros Wendland |
| EPI_ISL_1469588 | Secretaria Municipal de Saúde de Taquara | Epiclin | Fernando Hayashi Sant'Anna, Ana Paula Muterle, Janira Prichula, Juliana Comerlato, Carolina Comerlato, Eliana Márcia Da Ros Wendland |
| EPI_ISL_1469589 | UNIDADE DE PRONTO ATENDIMENTO DE SAPUCAIA DO SUL UPA | Epiclin | Fernando Hayashi Sant'Anna, Ana Paula Muterle, Janira Prichula, Juliana Comerlato, Carolina Comerlato, Eliana Márcia Da Ros Wendland |
| EPI_ISL_1469590 | DIRETORIA DE VIGILANCIA EM SAUDE | Epiclin | Fernando Hayashi Sant'Anna, Ana Paula Muterle, Janira Prichula, Juliana Comerlato, Carolina Comerlato, Eliana Márcia Da Ros Wendland |
| EPI_ISL_1469591 | Hospital Nossa Senhora das Graças | Epiclin | Fernando Hayashi Sant'Anna, Ana Paula Muterle, Janira Prichula, Juliana Comerlato, Carolina Comerlato, Eliana Márcia Da Ros Wendland |
| EPI_ISL_1469592 | Diretoria de Vigilância em Saúde | Epiclin | Fernando Hayashi Sant'Anna, Ana Paula Muterle, Janira Prichula, Juliana Comerlato, Carolina Comerlato, Eliana Márcia Da Ros Wendland |
| EPI_ISL_1469593 | Hospital Sapiranga | Epiclin | Fernando Hayashi Sant'Anna, Ana Paula Muterle, Janira Prichula, Juliana Comerlato, Carolina Comerlato, Eliana Márcia Da Ros Wendland |
| EPI_ISL_1469594 | DIRETORIA DE VIGILANCIA EM SAUDE | Epiclin | Fernando Hayashi Sant'Anna, Ana Paula Muterle, Janira Prichula, Juliana Comerlato, Carolina Comerlato, Eliana Márcia Da Ros Wendland |
| EPI_ISL_1469595 | Fundação de Saúde Pública de Novo Hamburgo | Epiclin | Fernando Hayashi Sant'Anna, Ana Paula Muterle, Janira Prichula, Juliana Comerlato, Carolina Comerlato, Eliana Márcia Da Ros Wendland |
| EPI_ISL_1469596 | CENTRO DE REFERENCIA EM SINDROMES GRIPAIS | Epiclin | Fernando Hayashi Sant'Anna, Ana Paula Muterle, Janira Prichula, Juliana Comerlato, Carolina Comerlato, Eliana Márcia Da Ros Wendland |
| EPI_ISL_1469597 | Fundação de Saúde Pública de Novo Hamburgo | Epiclin | Fernando Hayashi Sant'Anna, Ana Paula Muterle, Janira Prichula, Juliana Comerlato, Carolina Comerlato, Eliana Márcia Da Ros Wendland |
| EPI_ISL_1469598 | Diretoria de Vigilância em Saúde | Epiclin | Fernando Hayashi Sant'Anna, Ana Paula Muterle, Janira Prichula, Juliana Comerlato, Carolina Comerlato, Eliana Márcia Da Ros Wendland |
| EPI_ISL_1469599 | Hospital Montenegro | Epiclin | Fernando Hayashi Sant'Anna, Ana Paula Muterle, Janira Prichula, Juliana Comerlato, Carolina Comerlato, Eliana Márcia Da Ros Wendland |
| EPI_ISL_1469600 | Hospital de Portão | Epiclin | Fernando Hayashi Sant'Anna, Ana Paula Muterle, Janira Prichula, Juliana Comerlato, Carolina Comerlato, Eliana Márcia Da Ros Wendland |
| EPI_ISL_1469601 | SECRETARIA MUNICIPAL DE SAUDE DE TAQUARA | Epiclin | Fernando Hayashi Sant'Anna, Ana Paula Muterle, Janira Prichula, Juliana Comerlato, Carolina Comerlato, Eliana Márcia Da Ros Wendland |
| EPI_ISL_1469602 | Pronto Atendimento Campo Bom | Epiclin | Fernando Hayashi Sant'Anna, Ana Paula Muterle, Janira Prichula, Juliana Comerlato, Carolina Comerlato, Eliana Márcia Da Ros Wendland |
| EPI_ISL_1469603 | Centro de Referência em Síndromes Gripais | Epiclin | Fernando Hayashi Sant'Anna, Ana Paula Muterle, Janira Prichula, Juliana Comerlato, Carolina Comerlato, Eliana Márcia Da Ros Wendland |
| EPI_ISL_1469604 | COORDENADORIA GERAL DE VIGILANCIA EM SAUDE | Epiclin | Fernando Hayashi Sant'Anna, Ana Paula Muterle, Janira Prichula, Juliana Comerlato, Carolina Comerlato, Eliana Márcia Da Ros Wendland |
| EPI_ISL_1469605 | Diretoria de Vigilância em Saúde | Epiclin | Fernando Hayashi Sant'Anna, Ana Paula Muterle, Janira Prichula, Juliana Comerlato, Carolina Comerlato, Eliana Márcia Da Ros Wendland |
| EPI_ISL_1469607 | Secretaria Municipal de Saúde de Três Coroas | Epiclin | Fernando Hayashi Sant'Anna, Ana Paula Muterle, Janira Prichula, Juliana Comerlato, Carolina Comerlato, Eliana Márcia Da Ros Wendland |
| EPI_ISL_1469608 | COORDENADORIA GERAL DE VIGILANCIA EM SAUDE | Epiclin | Fernando Hayashi Sant'Anna, Ana Paula Muterle, Janira Prichula, Juliana Comerlato, Carolina Comerlato, Eliana Márcia Da Ros Wendland |
| EPI_ISL_1469609, EPI_ISL_1469610 | Diretoria de Vigilância em Saúde | Epiclin | Fernando Hayashi Sant'Anna, Ana Paula Muterle, Janira Prichula, Juliana Comerlato, Carolina Comerlato, Eliana Márcia Da Ros Wendland |
| EPI_ISL_1469611 | DIRETORIA DE VIGILANCIA EM SAUDE | Epiclin | Fernando Hayashi Sant'Anna, Ana Paula Muterle, Janira Prichula, Juliana Comerlato, Carolina Comerlato, Eliana Márcia Da Ros Wendland |
| EPI_ISL_1469612 | FUNDACAO DE SAUDE PUBLICA DE NOVO HAMBURGO FSNH | Epiclin | Fernando Hayashi Sant'Anna, Ana Paula Muterle, Janira Prichula, Juliana Comerlato, Carolina Comerlato, Eliana Márcia Da Ros Wendland |
| EPI_ISL_1469613 | Fundação de Saúde Pública de Novo Hamburgo | Epiclin | Fernando Hayashi Sant'Anna, Ana Paula Muterle, Janira Prichula, Juliana Comerlato, Carolina Comerlato, Eliana Márcia Da Ros Wendland |
| EPI_ISL_1469614 | Unidade Sanitária de Igrejinha | Epiclin | Fernando Hayashi Sant'Anna, Ana Paula Muterle, Janira Prichula, Juliana Comerlato, Carolina Comerlato, Eliana Márcia Da Ros Wendland |
| EPI_ISL_1469615 | Pronto Atendimento Campo Bom | Epiclin | Fernando Hayashi Sant'Anna, Ana Paula Muterle, Janira Prichula, Juliana Comerlato, Carolina Comerlato, Eliana Márcia Da Ros Wendland |
| EPI_ISL_1469616 | Diretoria de Vigilância em Saúde | Epiclin | Fernando Hayashi Sant'Anna, Ana Paula Muterle, Janira Prichula, Juliana Comerlato, Carolina Comerlato, Eliana Márcia Da Ros Wendland |
| EPI_ISL_1469617 | Fundação Hospitalar de Sapucaia do Sul | Epiclin | Fernando Hayashi Sant'Anna, Ana Paula Muterle, Janira Prichula, Juliana Comerlato, Carolina Comerlato, Eliana Márcia Da Ros Wendland |
| EPI_ISL_1469618 | Diretoria de Vigilância em Saúde | Epiclin | Fernando Hayashi Sant'Anna, Ana Paula Muterle, Janira Prichula, Juliana Comerlato, Carolina Comerlato, Eliana Márcia Da Ros Wendland |
| EPI_ISL_1469619 | Fundação Hospitalar de Sapucaia do Sul | Epiclin | Fernando Hayashi Sant'Anna, Ana Paula Muterle, Janira Prichula, Juliana Comerlato, Carolina Comerlato, Eliana Márcia Da Ros Wendland |
| EPI_ISL_1469620 | FUNDACAO DE SAUDE PUBLICA DE NOVO HAMBURGO FSNH | Epiclin | Fernando Hayashi Sant'Anna, Ana Paula Muterle, Janira Prichula, Juliana Comerlato, Carolina Comerlato, Eliana Márcia Da Ros Wendland |
| EPI_ISL_1469621 | Pronto Atendimento Campo Bom | Epiclin | Fernando Hayashi Sant'Anna, Ana Paula Muterle, Janira Prichula, Juliana Comerlato, Carolina Comerlato, Eliana Márcia Da Ros Wendland |
| EPI_ISL_1469622 | Fundação de Saúde Pública de Novo Hamburgo | Epiclin | Fernando Hayashi Sant'Anna, Ana Paula Muterle, Janira Prichula, Juliana Comerlato, Carolina Comerlato, Eliana Márcia Da Ros Wendland |
| EPI_ISL_1469623 | Diretoria de Vigilância em Saúde | Epiclin | Fernando Hayashi Sant'Anna, Ana Paula Muterle, Janira Prichula, Juliana Comerlato, Carolina Comerlato, Eliana Márcia Da Ros Wendland |
| EPI_ISL_1469624 | HOSPITAL MUNICIPAL GETULIO VARGAS | Epiclin | Fernando Hayashi Sant'Anna, Ana Paula Muterle, Janira Prichula, Juliana Comerlato, Carolina Comerlato, Eliana Márcia Da Ros Wendland |
| EPI_ISL_1469625 | Diretoria de Vigilância em Saúde | Epiclin | Fernando Hayashi Sant'Anna, Ana Paula Muterle, Janira Prichula, Juliana Comerlato, Carolina Comerlato, Eliana Márcia Da Ros Wendland |
| EPI_ISL_1469626 | Fundação de Saúde Pública de Novo Hamburgo | Epiclin | Fernando Hayashi Sant'Anna, Ana Paula Muterle, Janira Prichula, Juliana Comerlato, Carolina Comerlato, Eliana Márcia Da Ros Wendland |
| EPI_ISL_1469627 | Diretoria de Vigilância em Saúde | Epiclin | Fernando Hayashi Sant'Anna, Ana Paula Muterle, Janira Prichula, Juliana Comerlato, Carolina Comerlato, Eliana Márcia Da Ros Wendland |
| EPI_ISL_1469628 | CENTRO DE SERVICOS ESPECIALIZADOS SANTA RITA DE CASSIA | Epiclin | Fernando Hayashi Sant'Anna, Ana Paula Muterle, Janira Prichula, Juliana Comerlato, Carolina Comerlato, Eliana Márcia Da Ros Wendland |

| | | | |
|---|---|---|---|
| EPI_ISL_1469629 | CENTRO MUNICIPAL DE SAUDE DE ROLANTE | Epiclin | Fernando Hayashi Sant'Anna, Ana Paula Muterle, Janira Prichula, Juliana Comerlato, Carolina Comerlato, Eliana Márcia Da Ros Wendland |
| EPI_ISL_1469630 | FUNDACAO DE SAUDE PUBLICA DE NOVO HAMBURGO FSNH | Epiclin | Fernando Hayashi Sant'Anna, Ana Paula Muterle, Janira Prichula, Juliana Comerlato, Carolina Comerlato, Eliana Márcia Da Ros Wendland |
| EPI_ISL_1469631 | HOSPITAL MUNICIPAL GETULIO VARGAS | Epiclin | Fernando Hayashi Sant'Anna, Ana Paula Muterle, Janira Prichula, Juliana Comerlato, Carolina Comerlato, Eliana Márcia Da Ros Wendland |
| EPI_ISL_1469632 | Diretoria de Vigilância em Saúde | Epiclin | Fernando Hayashi Sant'Anna, Ana Paula Muterle, Janira Prichula, Juliana Comerlato, Carolina Comerlato, Eliana Márcia Da Ros Wendland |
| EPI_ISL_1469633 | DIRETORIA DE VIGILANCIA EM SAUDE | Epiclin | Fernando Hayashi Sant'Anna, Ana Paula Muterle, Janira Prichula, Juliana Comerlato, Carolina Comerlato, Eliana Márcia Da Ros Wendland |
| EPI_ISL_1469634 | Diretoria de Vigilância em Saúde | Epiclin | Fernando Hayashi Sant'Anna, Ana Paula Muterle, Janira Prichula, Juliana Comerlato, Carolina Comerlato, Eliana Márcia Da Ros Wendland |
| EPI_ISL_1469635, EPI_ISL_1469636 | COORDENADORIA GERAL DE VIGILANCIA EM SAUDE | Epiclin | Fernando Hayashi Sant'Anna, Ana Paula Muterle, Janira Prichula, Juliana Comerlato, Carolina Comerlato, Eliana Márcia Da Ros Wendland |
| EPI_ISL_1469637, EPI_ISL_1469638 | DIRETORIA DE VIGILANCIA EM SAUDE | Epiclin | Fernando Hayashi Sant'Anna, Ana Paula Muterle, Janira Prichula, Juliana Comerlato, Carolina Comerlato, Eliana Márcia Da Ros Wendland |
| EPI_ISL_1469639 | SECRETARIA MUNICIPAL DE SAUDE DE SAO LEOPOLDO | Epiclin | Fernando Hayashi Sant'Anna, Ana Paula Muterle, Janira Prichula, Juliana Comerlato, Carolina Comerlato, Eliana Márcia Da Ros Wendland |
| EPI_ISL_1469640 | Fundação de Saúde Pública São Camilo de Esteio | Epiclin | Fernando Hayashi Sant'Anna, Ana Paula Muterle, Janira Prichula, Juliana Comerlato, Carolina Comerlato, Eliana Márcia Da Ros Wendland |
| EPI_ISL_1469641 | COORDENADORIA GERAL DE VIGILANCIA EM SAUDE | Epiclin | Fernando Hayashi Sant'Anna, Ana Paula Muterle, Janira Prichula, Juliana Comerlato, Carolina Comerlato, Eliana Márcia Da Ros Wendland |
| EPI_ISL_1469642 | CENTRO DE REFERENCIA EM SINDROMES GRIPAIS | Epiclin | Fernando Hayashi Sant'Anna, Ana Paula Muterle, Janira Prichula, Juliana Comerlato, Carolina Comerlato, Eliana Márcia Da Ros Wendland |
| EPI_ISL_1469643 | HOSPITAL SAO FRANCISCO DE ASSIS | Epiclin | Fernando Hayashi Sant'Anna, Ana Paula Muterle, Janira Prichula, Juliana Comerlato, Carolina Comerlato, Eliana Márcia Da Ros Wendland |
| EPI_ISL_1469644 | Diretoria de Vigilância em Saúde | Epiclin | Fernando Hayashi Sant'Anna, Ana Paula Muterle, Janira Prichula, Juliana Comerlato, Carolina Comerlato, Eliana Márcia Da Ros Wendland |
| EPI_ISL_1469645 | SECRETARIA MUNICIPAL DE SAUDE DE TRES COROAS | Epiclin | Fernando Hayashi Sant'Anna, Ana Paula Muterle, Janira Prichula, Juliana Comerlato, Carolina Comerlato, Eliana Márcia Da Ros Wendland |
| EPI_ISL_1469646 | UNIDADE DE PRONTO ATENDIMENTO DE SAPUCAIA DO SUL UPA | Epiclin | Fernando Hayashi Sant'Anna, Ana Paula Muterle, Janira Prichula, Juliana Comerlato, Carolina Comerlato, Eliana Márcia Da Ros Wendland |
| EPI_ISL_1469647 | Secretaria Municipal de Saúde de Taquara | Epiclin | Fernando Hayashi Sant'Anna, Ana Paula Muterle, Janira Prichula, Juliana Comerlato, Carolina Comerlato, Eliana Márcia Da Ros Wendland |
| EPI_ISL_1469648 | HOSPITAL SAO FRANCISCO DE ASSIS | Epiclin | Fernando Hayashi Sant'Anna, Ana Paula Muterle, Janira Prichula, Juliana Comerlato, Carolina Comerlato, Eliana Márcia Da Ros Wendland |
| EPI_ISL_1469649 | Unidade de Atendimento DST AIDS TB e Han | Epiclin | Fernando Hayashi Sant'Anna, Ana Paula Muterle, Janira Prichula, Juliana Comerlato, Carolina Comerlato, Eliana Márcia Da Ros Wendland |
| EPI_ISL_1469650 | Diretoria de Vigilância em Saúde | Epiclin | Fernando Hayashi Sant'Anna, Ana Paula Muterle, Janira Prichula, Juliana Comerlato, Carolina Comerlato, Eliana Márcia Da Ros Wendland |
| EPI_ISL_1469651 | FUNDACAO DE SAUDE PUBLICA DE NOVO HAMBURGO FSNH | Epiclin | Fernando Hayashi Sant'Anna, Ana Paula Muterle, Janira Prichula, Juliana Comerlato, Carolina Comerlato, Eliana Márcia Da Ros Wendland |
| EPI_ISL_1469652 | DIRETORIA DE VIGILANCIA EM SAUDE | Epiclin | Fernando Hayashi Sant'Anna, Ana Paula Muterle, Janira Prichula, Juliana Comerlato, Carolina Comerlato, Eliana Márcia Da Ros Wendland |
| EPI_ISL_1469653 | Fundação de Saúde Pública de Novo Hamburgo | Epiclin | Fernando Hayashi Sant'Anna, Ana Paula Muterle, Janira Prichula, Juliana Comerlato, Carolina Comerlato, Eliana Márcia Da Ros Wendland |
| EPI_ISL_1469654 | Fundação Hospitalar de Sapucaia do Sul | Epiclin | Fernando Hayashi Sant'Anna, Ana Paula Muterle, Janira Prichula, Juliana Comerlato, Carolina Comerlato, Eliana Márcia Da Ros Wendland |
| EPI_ISL_1469655 | Centro Municipal de Saúde de Rolante | Epiclin | Fernando Hayashi Sant'Anna, Ana Paula Muterle, Janira Prichula, Juliana Comerlato, Carolina Comerlato, Eliana Márcia Da Ros Wendland |
| EPI_ISL_1469656 | Diretoria de Vigilância em Saúde | Epiclin | Fernando Hayashi Sant'Anna, Ana Paula Muterle, Janira Prichula, Juliana Comerlato, Carolina Comerlato, Eliana Márcia Da Ros Wendland |
| EPI_ISL_1469657 | Pronto Atendimento Campo Bom | Epiclin | Fernando Hayashi Sant'Anna, Ana Paula Muterle, Janira Prichula, Juliana Comerlato, Carolina Comerlato, Eliana Márcia Da Ros Wendland |
| EPI_ISL_1469658 | CENTRO DE ESPECIALIDADES TRIUNFO | Epiclin | Fernando Hayashi Sant'Anna, Ana Paula Muterle, Janira Prichula, Juliana Comerlato, Carolina Comerlato, Eliana Márcia Da Ros Wendland |
| EPI_ISL_1469659 | DIRETORIA DE VIGILANCIA EM SAUDE | Epiclin | Fernando Hayashi Sant'Anna, Ana Paula Muterle, Janira Prichula, Juliana Comerlato, Carolina Comerlato, Eliana Márcia Da Ros Wendland |
| EPI_ISL_1469660 | UNIDADE BASICA DE SAUDE DE RIOZINHO | Epiclin | Fernando Hayashi Sant'Anna, Ana Paula Muterle, Janira Prichula, Juliana Comerlato, Carolina Comerlato, Eliana Márcia Da Ros Wendland |
| EPI_ISL_1469661 | FUNDACAO DE SAUDE PUBLICA DE NOVO HAMBURGO FSNH | Epiclin | Fernando Hayashi Sant'Anna, Ana Paula Muterle, Janira Prichula, Juliana Comerlato, Carolina Comerlato, Eliana Márcia Da Ros Wendland |
| EPI_ISL_1469662 | CENTRO DE SAUDE DR BRUNO CASSEL | Epiclin | Fernando Hayashi Sant'Anna, Ana Paula Muterle, Janira Prichula, Juliana Comerlato, Carolina Comerlato, Eliana Márcia Da Ros Wendland |
| EPI_ISL_1469663 | DIRETORIA DE VIGILANCIA EM SAUDE | Epiclin | Fernando Hayashi Sant'Anna, Ana Paula Muterle, Janira Prichula, Juliana Comerlato, Carolina Comerlato, Eliana Márcia Da Ros Wendland |
| EPI_ISL_1469664 | Unidade de Atendimento DST AIDS TB e Han | Epiclin | Fernando Hayashi Sant'Anna, Ana Paula Muterle, Janira Prichula, Juliana Comerlato, Carolina Comerlato, Eliana Márcia Da Ros Wendland |
| EPI_ISL_1469665 | FUNDACAO DE SAUDE PUBLICA SAO CAMILO DE ESTEIO | Epiclin | Fernando Hayashi Sant'Anna, Ana Paula Muterle, Janira Prichula, Juliana Comerlato, Carolina Comerlato, Eliana Márcia Da Ros Wendland |
| EPI_ISL_1469666 | Secretaria Municipal de Saúde de Montenegro | Epiclin | Fernando Hayashi Sant'Anna, Ana Paula Muterle, Janira Prichula, Juliana Comerlato, Carolina Comerlato, Eliana Márcia Da Ros Wendland |
| EPI_ISL_1469667 | Centro de Referência em Síndromes Gripais | Epiclin | Fernando Hayashi Sant'Anna, Ana Paula Muterle, Janira Prichula, Juliana Comerlato, Carolina Comerlato, Eliana Márcia Da Ros Wendland |
| EPI_ISL_1469668 | Hospital Unimed Vale do Caí | Epiclin | Fernando Hayashi Sant'Anna, Ana Paula Muterle, Janira Prichula, Juliana Comerlato, Carolina Comerlato, Eliana Márcia Da Ros Wendland |
| EPI_ISL_1469669 | Diretoria de Vigilância em Saúde | Epiclin | Fernando Hayashi Sant'Anna, Ana Paula Muterle, Janira Prichula, Juliana Comerlato, Carolina Comerlato, Eliana Márcia Da Ros Wendland |
| EPI_ISL_1469670 | Hospital São Francisco de Assis | Epiclin | Fernando Hayashi Sant'Anna, Ana Paula Muterle, Janira Prichula, Juliana Comerlato, Carolina Comerlato, Eliana Márcia Da Ros Wendland |
| EPI_ISL_1469671 | Vigilância em Saúde de Sapucaia do Sul | Epiclin | Fernando Hayashi Sant'Anna, Ana Paula Muterle, Janira Prichula, Juliana Comerlato, Carolina Comerlato, Eliana Márcia Da Ros Wendland |
| EPI_ISL_1469672 | Diretoria de Vigilância em Saúde | Epiclin | Fernando Hayashi Sant'Anna, Ana Paula Muterle, Janira Prichula, Juliana Comerlato, Carolina Comerlato, Eliana Márcia Da Ros Wendland |
| EPI_ISL_1469673 | Fundação de Saúde Pública de Novo Hamburgo | Epiclin | Fernando Hayashi Sant'Anna, Ana Paula Muterle, Janira Prichula, Juliana Comerlato, Carolina Comerlato, Eliana Márcia Da Ros Wendland |
| EPI_ISL_1469674, EPI_ISL_1469675 | DIRETORIA DE VIGILANCIA EM SAUDE | Epiclin | Fernando Hayashi Sant'Anna, Ana Paula Muterle, Janira Prichula, Juliana Comerlato, Carolina Comerlato, Eliana Márcia Da Ros Wendland |
| EPI_ISL_1469676 | HOSPITAL SAO FRANCISCO DE ASSIS | Epiclin | Fernando Hayashi Sant'Anna, Ana Paula Muterle, Janira Prichula, Juliana Comerlato, Carolina Comerlato, Eliana Márcia Da Ros Wendland |
| EPI_ISL_1469677 | DIRETORIA DE VIGILANCIA EM SAUDE | Epiclin | Fernando Hayashi Sant'Anna, Ana Paula Muterle, Janira Prichula, Juliana Comerlato, Carolina Comerlato, Eliana Márcia Da Ros Wendland |
| EPI_ISL_1469678 | SECRETARIA MUNICIPAL DE SAUDE DE TRES COROAS | Epiclin | Fernando Hayashi Sant'Anna, Ana Paula Muterle, Janira Prichula, Juliana Comerlato, Carolina Comerlato, Eliana Márcia Da Ros Wendland |
| EPI_ISL_1469679 | DIRETORIA DE VIGILANCIA EM SAUDE | Epiclin | Fernando Hayashi Sant'Anna, Ana Paula Muterle, Janira Prichula, Juliana Comerlato, Carolina Comerlato, Eliana Márcia Da Ros Wendland |
| EPI_ISL_1469680 | UNIDADE DE ATENDIMENTO DST AIDS TB E HAN | Epiclin | Fernando Hayashi Sant'Anna, Ana Paula Muterle, Janira Prichula, Juliana Comerlato, Carolina Comerlato, Eliana Márcia Da Ros Wendland |
| EPI_ISL_1469681 | DIRETORIA DE VIGILANCIA EM SAUDE | Epiclin | Fernando Hayashi Sant'Anna, Ana Paula Muterle, Janira Prichula, Juliana Comerlato, Carolina Comerlato, Eliana Márcia Da Ros Wendland |
| EPI_ISL_1469682 | CENTRO DE SERVICOS ESPECIALIZADOS SANTA RITA DE CASSIA | Epiclin | Fernando Hayashi Sant'Anna, Ana Paula Muterle, Janira Prichula, Juliana Comerlato, Carolina Comerlato, Eliana Márcia Da Ros Wendland |
| EPI_ISL_1469683 | SECRETARIA MUNICIPAL DE SAUDE DE TAQUARA | Epiclin | Fernando Hayashi Sant'Anna, Ana Paula Muterle, Janira Prichula, Juliana Comerlato, Carolina Comerlato, Eliana Márcia Da Ros Wendland |
| EPI_ISL_1469684 | DIRETORIA DE VIGILANCIA EM SAUDE | Epiclin | Fernando Hayashi Sant'Anna, Ana Paula Muterle, Janira Prichula, Juliana Comerlato, Carolina Comerlato, Eliana Márcia Da Ros Wendland |
| EPI_ISL_1469685 | HOSPITAL SAO FRANCISCO DE ASSIS | Epiclin | Fernando Hayashi Sant'Anna, Ana Paula Muterle, Janira Prichula, Juliana Comerlato, Carolina Comerlato, Eliana Márcia Da Ros Wendland |
| EPI_ISL_1469686 | DIRETORIA DE VIGILANCIA EM SAUDE | Epiclin | Fernando Hayashi Sant'Anna, Ana Paula Muterle, Janira Prichula, Juliana Comerlato, Carolina Comerlato, Eliana Márcia Da Ros Wendland |

| | | | |
|---|---|---|---|
| EPI_ISL_1469687 | SECRETARIA MUNICIPAL DE SAUDE DE TAQUARA | Epiclin | Fernando Hayashi Sant'Anna, Ana Paula Muterle, Janira Prichula, Juliana Comerlato, Carolina Comerlato, Eliana Márcia Da Ros Wendland |
| EPI_ISL_1469688 | DIRETORIA DE VIGILANCIA EM SAUDE | Epiclin | Fernando Hayashi Sant'Anna, Ana Paula Muterle, Janira Prichula, Juliana Comerlato, Carolina Comerlato, Eliana Márcia Da Ros Wendland |
| EPI_ISL_1469689 | FUNDACAO DE SAUDE PUBLICA SAO CAMILO DE ESTEIO | Epiclin | Fernando Hayashi Sant'Anna, Ana Paula Muterle, Janira Prichula, Juliana Comerlato, Carolina Comerlato, Eliana Márcia Da Ros Wendland |
| EPI_ISL_1469690 | SECRETARIA MUNICIPAL DE SAUDE DE SAO LEOPOLDO | Epiclin | Fernando Hayashi Sant'Anna, Ana Paula Muterle, Janira Prichula, Juliana Comerlato, Carolina Comerlato, Eliana Márcia Da Ros Wendland |
| EPI_ISL_1469691 | Fundação de Saúde Pública São Camilo de Esteio | Epiclin | Fernando Hayashi Sant'Anna, Ana Paula Muterle, Janira Prichula, Juliana Comerlato, Carolina Comerlato, Eliana Márcia Da Ros Wendland |
| EPI_ISL_1469692 | DIRETORIA DE VIGILANCIA EM SAUDE | Epiclin | Fernando Hayashi Sant'Anna, Ana Paula Muterle, Janira Prichula, Juliana Comerlato, Carolina Comerlato, Eliana Márcia Da Ros Wendland |
| EPI_ISL_1469693 | FUNDACAO DE SAUDE PUBLICA SAO CAMILO DE ESTEIO | Epiclin | Fernando Hayashi Sant'Anna, Ana Paula Muterle, Janira Prichula, Juliana Comerlato, Carolina Comerlato, Eliana Márcia Da Ros Wendland |
| EPI_ISL_1469694 | UNIDADE BASICA DE SAUDE PARECI NOVO | Epiclin | Fernando Hayashi Sant'Anna, Ana Paula Muterle, Janira Prichula, Juliana Comerlato, Carolina Comerlato, Eliana Márcia Da Ros Wendland |
| EPI_ISL_1469695 | Unidade de Pronto Atendimento de Sapucaia do Sul | Epiclin | Fernando Hayashi Sant'Anna, Ana Paula Muterle, Janira Prichula, Juliana Comerlato, Carolina Comerlato, Eliana Márcia Da Ros Wendland |
| EPI_ISL_1469696 | Vigilância em Saúde de Sapucaia do Sul | Epiclin | Fernando Hayashi Sant'Anna, Ana Paula Muterle, Janira Prichula, Juliana Comerlato, Carolina Comerlato, Eliana Márcia Da Ros Wendland |
| EPI_ISL_1469697 | Hospital Regina | Epiclin | Fernando Hayashi Sant'Anna, Ana Paula Muterle, Janira Prichula, Juliana Comerlato, Carolina Comerlato, Eliana Márcia Da Ros Wendland |
| EPI_ISL_1469698 | CENTRO DE SERVICOS ESPECIALIZADOS SANTA RITA DE CASSIA | Epiclin | Fernando Hayashi Sant'Anna, Ana Paula Muterle, Janira Prichula, Juliana Comerlato, Carolina Comerlato, Eliana Márcia Da Ros Wendland |
| EPI_ISL_1469699 | HOSPITAL MUNICIPAL GETULIO VARGAS | Epiclin | Fernando Hayashi Sant'Anna, Ana Paula Muterle, Janira Prichula, Juliana Comerlato, Carolina Comerlato, Eliana Márcia Da Ros Wendland |
| EPI_ISL_1469700 | Diretoria de Vigilância em Saúde | Epiclin | Fernando Hayashi Sant'Anna, Ana Paula Muterle, Janira Prichula, Juliana Comerlato, Carolina Comerlato, Eliana Márcia Da Ros Wendland |
| EPI_ISL_1469701 | Hospital Sapiranga | Epiclin | Fernando Hayashi Sant'Anna, Ana Paula Muterle, Janira Prichula, Juliana Comerlato, Carolina Comerlato, Eliana Márcia Da Ros Wendland |
| EPI_ISL_1469702 | Diretoria de Vigilância em Saúde | Epiclin | Fernando Hayashi Sant'Anna, Ana Paula Muterle, Janira Prichula, Juliana Comerlato, Carolina Comerlato, Eliana Márcia Da Ros Wendland |
| EPI_ISL_1469704 | FUNDACAO DE SAUDE PUBLICA SAO CAMILO DE ESTEIO | Epiclin | Fernando Hayashi Sant'Anna, Ana Paula Muterle, Janira Prichula, Juliana Comerlato, Carolina Comerlato, Eliana Márcia Da Ros Wendland |
| EPI_ISL_1469705 | HOSPITAL DE CAMPO BOM DR LAURO REUS | Epiclin | Fernando Hayashi Sant'Anna, Ana Paula Muterle, Janira Prichula, Juliana Comerlato, Carolina Comerlato, Eliana Márcia Da Ros Wendland |
| EPI_ISL_1469706 | CENTRO DE SAUDE DR BRUNO CASSEL | Epiclin | Fernando Hayashi Sant'Anna, Ana Paula Muterle, Janira Prichula, Juliana Comerlato, Carolina Comerlato, Eliana Márcia Da Ros Wendland |
| EPI_ISL_1469707 | Hospital Bom Jesus | Epiclin | Fernando Hayashi Sant'Anna, Ana Paula Muterle, Janira Prichula, Juliana Comerlato, Carolina Comerlato, Eliana Márcia Da Ros Wendland |
| EPI_ISL_1469708 | Diretoria de Vigilância em Saúde | Epiclin | Fernando Hayashi Sant'Anna, Ana Paula Muterle, Janira Prichula, Juliana Comerlato, Carolina Comerlato, Eliana Márcia Da Ros Wendland |
| EPI_ISL_1469709 | SECRETARIA MUNICIPAL DE SAUDE DE TAQUARA | Epiclin | Fernando Hayashi Sant'Anna, Ana Paula Muterle, Janira Prichula, Juliana Comerlato, Carolina Comerlato, Eliana Márcia Da Ros Wendland |
| EPI_ISL_1469710 | DIRETORIA DE VIGILANCIA EM SAUDE | Epiclin | Fernando Hayashi Sant'Anna, Ana Paula Muterle, Janira Prichula, Juliana Comerlato, Carolina Comerlato, Eliana Márcia Da Ros Wendland |
| EPI_ISL_1469711 | Hospital Dia e Pronto Atendimento | Epiclin | Fernando Hayashi Sant'Anna, Ana Paula Muterle, Janira Prichula, Juliana Comerlato, Carolina Comerlato, Eliana Márcia Da Ros Wendland |
| EPI_ISL_1469712 | Pronto Atendimento Campo Bom | Epiclin | Fernando Hayashi Sant'Anna, Ana Paula Muterle, Janira Prichula, Juliana Comerlato, Carolina Comerlato, Eliana Márcia Da Ros Wendland |
| EPI_ISL_1469713 | Fundação Hospitalar de Sapucaia do Sul | Epiclin | Fernando Hayashi Sant'Anna, Ana Paula Muterle, Janira Prichula, Juliana Comerlato, Carolina Comerlato, Eliana Márcia Da Ros Wendland |
| EPI_ISL_1469714 | Unidade de Atendimento DST AIDS TB e Han | Epiclin | Fernando Hayashi Sant'Anna, Ana Paula Muterle, Janira Prichula, Juliana Comerlato, Carolina Comerlato, Eliana Márcia Da Ros Wendland |
| EPI_ISL_1469715 | COORDENADORIA GERAL DE VIGILANCIA EM SAUDE | Epiclin | Fernando Hayashi Sant'Anna, Ana Paula Muterle, Janira Prichula, Juliana Comerlato, Carolina Comerlato, Eliana Márcia Da Ros Wendland |
| EPI_ISL_1469716 | Unidade Sanitária de Igrejinha | Epiclin | Fernando Hayashi Sant'Anna, Ana Paula Muterle, Janira Prichula, Juliana Comerlato, Carolina Comerlato, Eliana Márcia Da Ros Wendland |
| EPI_ISL_1469717 | Hospital São Francisco de Assis | Epiclin | Fernando Hayashi Sant'Anna, Ana Paula Muterle, Janira Prichula, Juliana Comerlato, Carolina Comerlato, Eliana Márcia Da Ros Wendland |
| EPI_ISL_1469718 | Unidade de Pronto Atendimento de Sapucaia do Sul | Epiclin | Fernando Hayashi Sant'Anna, Ana Paula Muterle, Janira Prichula, Juliana Comerlato, Carolina Comerlato, Eliana Márcia Da Ros Wendland |
| EPI_ISL_1469719 | UNIDADE DE PRONTO ATENDIMENTO DE SAPUCAIA DO SUL UPA | Epiclin | Fernando Hayashi Sant'Anna, Ana Paula Muterle, Janira Prichula, Juliana Comerlato, Carolina Comerlato, Eliana Márcia Da Ros Wendland |
| EPI_ISL_1469720 | UNIDADE SANITARIA DE IGREJINHA | Epiclin | Fernando Hayashi Sant'Anna, Ana Paula Muterle, Janira Prichula, Juliana Comerlato, Carolina Comerlato, Eliana Márcia Da Ros Wendland |
| EPI_ISL_1469721 | Pronto Atendimento Cruzeiro do Sul | Epiclin | Fernando Hayashi Sant'Anna, Ana Paula Muterle, Janira Prichula, Juliana Comerlato, Carolina Comerlato, Eliana Márcia Da Ros Wendland |
| EPI_ISL_1469722 | UNIDADE SANITARIA DE IGREJINHA | Epiclin | Fernando Hayashi Sant'Anna, Ana Paula Muterle, Janira Prichula, Juliana Comerlato, Carolina Comerlato, Eliana Márcia Da Ros Wendland |
| EPI_ISL_1469723 | FUNDACAO DE SAUDE PUBLICA SAO CAMILO DE ESTEIO | Epiclin | Fernando Hayashi Sant'Anna, Ana Paula Muterle, Janira Prichula, Juliana Comerlato, Carolina Comerlato, Eliana Márcia Da Ros Wendland |
| EPI_ISL_1469724 | Fundação Hospitalar de Sapucaia do Sul | Epiclin | Fernando Hayashi Sant'Anna, Ana Paula Muterle, Janira Prichula, Juliana Comerlato, Carolina Comerlato, Eliana Márcia Da Ros Wendland |
| EPI_ISL_1469725 | DIRETORIA DE VIGILANCIA EM SAUDE | Epiclin | Fernando Hayashi Sant'Anna, Ana Paula Muterle, Janira Prichula, Juliana Comerlato, Carolina Comerlato, Eliana Márcia Da Ros Wendland |
| EPI_ISL_1469726 | HOSPITAL SAPIRANGA | Epiclin | Fernando Hayashi Sant'Anna, Ana Paula Muterle, Janira Prichula, Juliana Comerlato, Carolina Comerlato, Eliana Márcia Da Ros Wendland |
| EPI_ISL_1469727 | Secretaria Municipal de Saúde de São Leopoldo | Epiclin | Fernando Hayashi Sant'Anna, Ana Paula Muterle, Janira Prichula, Juliana Comerlato, Carolina Comerlato, Eliana Márcia Da Ros Wendland |
| EPI_ISL_1469728 | Diretoria de Vigilância em Saúde | Epiclin | Fernando Hayashi Sant'Anna, Ana Paula Muterle, Janira Prichula, Juliana Comerlato, Carolina Comerlato, Eliana Márcia Da Ros Wendland |
| EPI_ISL_1469729 | Centro de Especialidades Triunfo | Epiclin | Fernando Hayashi Sant'Anna, Ana Paula Muterle, Janira Prichula, Juliana Comerlato, Carolina Comerlato, Eliana Márcia Da Ros Wendland |
| EPI_ISL_1469730 | CENTRO DE REFERENCIA EM SINDROMES GRIPAIS | Epiclin | Fernando Hayashi Sant'Anna, Ana Paula Muterle, Janira Prichula, Juliana Comerlato, Carolina Comerlato, Eliana Márcia Da Ros Wendland |
| EPI_ISL_1469731 | DIRETORIA DE VIGILANCIA EM SAUDE | Epiclin | Fernando Hayashi Sant'Anna, Ana Paula Muterle, Janira Prichula, Juliana Comerlato, Carolina Comerlato, Eliana Márcia Da Ros Wendland |
| EPI_ISL_1469732 | CENTRO DE REFERENCIA EM SINDROMES GRIPAIS | Epiclin | Fernando Hayashi Sant'Anna, Ana Paula Muterle, Janira Prichula, Juliana Comerlato, Carolina Comerlato, Eliana Márcia Da Ros Wendland |
| EPI_ISL_1469733 | UNIDADE DE PRONTO ATENDIMENTO DE SAPUCAIA DO SUL UPA | Epiclin | Fernando Hayashi Sant'Anna, Ana Paula Muterle, Janira Prichula, Juliana Comerlato, Carolina Comerlato, Eliana Márcia Da Ros Wendland |
| EPI_ISL_1469734 | Hospital Dia e Pronto Atendimento | Epiclin | Fernando Hayashi Sant'Anna, Ana Paula Muterle, Janira Prichula, Juliana Comerlato, Carolina Comerlato, Eliana Márcia Da Ros Wendland |
| EPI_ISL_1469735 | COORDENADORIA GERAL DE VIGILANCIA EM SAUDE | Epiclin | Fernando Hayashi Sant'Anna, Ana Paula Muterle, Janira Prichula, Juliana Comerlato, Carolina Comerlato, Eliana Márcia Da Ros Wendland |
| EPI_ISL_1469736 | SECRETARIA MUNICIPAL DE SAUDE DE SAO LEOPOLDO | Epiclin | Fernando Hayashi Sant'Anna, Ana Paula Muterle, Janira Prichula, Juliana Comerlato, Carolina Comerlato, Eliana Márcia Da Ros Wendland |
| EPI_ISL_1469737 | CENTRO DE REFERENCIA EM SINDROMES GRIPAIS | Epiclin | Fernando Hayashi Sant'Anna, Ana Paula Muterle, Janira Prichula, Juliana Comerlato, Carolina Comerlato, Eliana Márcia Da Ros Wendland |
| EPI_ISL_1469738 | HOSPITAL SAO FRANCISCO DE ASSIS | Epiclin | Fernando Hayashi Sant'Anna, Ana Paula Muterle, Janira Prichula, Juliana Comerlato, Carolina Comerlato, Eliana Márcia Da Ros Wendland |
| EPI_ISL_1469739 | Hospital São Francisco de Assis | Epiclin | Fernando Hayashi Sant'Anna, Ana Paula Muterle, Janira Prichula, Juliana Comerlato, Carolina Comerlato, Eliana Márcia Da Ros Wendland |
| EPI_ISL_1469740 | SECRETARIA MUNICIPAL DE SAUDE DE TRES COROAS | Epiclin | Fernando Hayashi Sant'Anna, Ana Paula Muterle, Janira Prichula, Juliana Comerlato, Carolina Comerlato, Eliana Márcia Da Ros Wendland |
| EPI_ISL_1469741 | Centro de Referência em Síndromes Gripais | Epiclin | Fernando Hayashi Sant'Anna, Ana Paula Muterle, Janira Prichula, Juliana Comerlato, Carolina Comerlato, Eliana Márcia Da Ros Wendland |
| EPI_ISL_1469742 | CENTRO DE REFERENCIA EM SINDROMES GRIPAIS | Epiclin | Fernando Hayashi Sant'Anna, Ana Paula Muterle, Janira Prichula, Juliana Comerlato, Carolina Comerlato, Eliana Márcia Da Ros Wendland |
| EPI_ISL_1469743 | DIRETORIA DE VIGILANCIA EM SAUDE | Epiclin | Fernando Hayashi Sant'Anna, Ana Paula Muterle, Janira Prichula, Juliana Comerlato, Carolina Comerlato, Eliana Márcia Da Ros Wendland |
| EPI_ISL_1469744 | HOSPITAL SAO FRANCISCO DE ASSIS | Epiclin | Fernando Hayashi Sant'Anna, Ana Paula Muterle, Janira Prichula, Juliana Comerlato, Carolina Comerlato, Eliana Márcia Da Ros Wendland |

| | | | |
|---|---|---|---|
| EPI_ISL_1469745 | UNIDADE DE PRONTO ATENDIMENTO DE SAPUCAIA DO SUL UPA | Epiclin | Fernando Hayashi Sant'Anna, Ana Paula Muterle, Janira Prichula, Juliana Comerlato, Carolina Comerlato, Eliana Márcia Da Ros Wendland |
| EPI_ISL_1469746 | HOSPITAL MONTENEGRO | Epiclin | Fernando Hayashi Sant'Anna, Ana Paula Muterle, Janira Prichula, Juliana Comerlato, Carolina Comerlato, Eliana Márcia Da Ros Wendland |
| EPI_ISL_1469747 | Unidade de Pronto Atendimento de Sapucaia do Sul | Epiclin | Fernando Hayashi Sant'Anna, Ana Paula Muterle, Janira Prichula, Juliana Comerlato, Carolina Comerlato, Eliana Márcia Da Ros Wendland |
| EPI_ISL_1469748 | Diretoria de Vigilância em Saúde | Epiclin | Fernando Hayashi Sant'Anna, Ana Paula Muterle, Janira Prichula, Juliana Comerlato, Carolina Comerlato, Eliana Márcia Da Ros Wendland |
| EPI_ISL_1469749 | Centro de Referência em Síndromes Gripais | Epiclin | Fernando Hayashi Sant'Anna, Ana Paula Muterle, Janira Prichula, Juliana Comerlato, Carolina Comerlato, Eliana Márcia Da Ros Wendland |
| EPI_ISL_1469750 | Hospital São Francisco de Assis | Epiclin | Fernando Hayashi Sant'Anna, Ana Paula Muterle, Janira Prichula, Juliana Comerlato, Carolina Comerlato, Eliana Márcia Da Ros Wendland |
| EPI_ISL_1469751 | UNIDADE BASICA DE SAUDE DE RIOZINHO | Epiclin | Fernando Hayashi Sant'Anna, Ana Paula Muterle, Janira Prichula, Juliana Comerlato, Carolina Comerlato, Eliana Márcia Da Ros Wendland |
| EPI_ISL_1469752 | Fundação Hospitalar de Sapucaia do Sul | Epiclin | Fernando Hayashi Sant'Anna, Ana Paula Muterle, Janira Prichula, Juliana Comerlato, Carolina Comerlato, Eliana Márcia Da Ros Wendland |
| EPI_ISL_1469753 | Diretoria de Vigilância em Saúde | Epiclin | Fernando Hayashi Sant'Anna, Ana Paula Muterle, Janira Prichula, Juliana Comerlato, Carolina Comerlato, Eliana Márcia Da Ros Wendland |
| EPI_ISL_1469754 | Hospital Municipal Getúlio Vargas | Epiclin | Fernando Hayashi Sant'Anna, Ana Paula Muterle, Janira Prichula, Juliana Comerlato, Carolina Comerlato, Eliana Márcia Da Ros Wendland |
| EPI_ISL_1469755 | Hospital São Francisco de Assis | Epiclin | Fernando Hayashi Sant'Anna, Ana Paula Muterle, Janira Prichula, Juliana Comerlato, Carolina Comerlato, Eliana Márcia Da Ros Wendland |
| EPI_ISL_1469756 | Diretoria de Vigilância em Saúde | Epiclin | Fernando Hayashi Sant'Anna, Ana Paula Muterle, Janira Prichula, Juliana Comerlato, Carolina Comerlato, Eliana Márcia Da Ros Wendland |
| EPI_ISL_1469757 | Secretaria Municipal de Saúde de São Leopoldo | Epiclin | Fernando Hayashi Sant'Anna, Ana Paula Muterle, Janira Prichula, Juliana Comerlato, Carolina Comerlato, Eliana Márcia Da Ros Wendland |
| EPI_ISL_1469758 | Secretaria Municipal de Saúde de Três Coroas | Epiclin | Fernando Hayashi Sant'Anna, Ana Paula Muterle, Janira Prichula, Juliana Comerlato, Carolina Comerlato, Eliana Márcia Da Ros Wendland |
| EPI_ISL_1469759 | HOSPITAL MUNICIPAL GETULIO VARGAS | Epiclin | Fernando Hayashi Sant'Anna, Ana Paula Muterle, Janira Prichula, Juliana Comerlato, Carolina Comerlato, Eliana Márcia Da Ros Wendland |
| EPI_ISL_1469760 | DIRETORIA DE VIGILANCIA EM SAUDE | Epiclin | Fernando Hayashi Sant'Anna, Ana Paula Muterle, Janira Prichula, Juliana Comerlato, Carolina Comerlato, Eliana Márcia Da Ros Wendland |
| EPI_ISL_1469761 | Diretoria de Vigilância em Saúde | Epiclin | Fernando Hayashi Sant'Anna, Ana Paula Muterle, Janira Prichula, Juliana Comerlato, Carolina Comerlato, Eliana Márcia Da Ros Wendland |
| EPI_ISL_1469762 | Hospital Nossa Senhora das Graças | Epiclin | Fernando Hayashi Sant'Anna, Ana Paula Muterle, Janira Prichula, Juliana Comerlato, Carolina Comerlato, Eliana Márcia Da Ros Wendland |
| EPI_ISL_1469763 | Unidade Básica de Saúde de Riozinho | Epiclin | Fernando Hayashi Sant'Anna, Ana Paula Muterle, Janira Prichula, Juliana Comerlato, Carolina Comerlato, Eliana Márcia Da Ros Wendland |
| EPI_ISL_1469764 | DIRETORIA DE VIGILANCIA EM SAUDE | Epiclin | Fernando Hayashi Sant'Anna, Ana Paula Muterle, Janira Prichula, Juliana Comerlato, Carolina Comerlato, Eliana Márcia Da Ros Wendland |
| EPI_ISL_1469765 | UNIDADE SANITARIA DE IGREJINHA | Epiclin | Fernando Hayashi Sant'Anna, Ana Paula Muterle, Janira Prichula, Juliana Comerlato, Carolina Comerlato, Eliana Márcia Da Ros Wendland |
| EPI_ISL_1469766 | Coordenadoria Geral de Vigilância em Saúde · Vigilância em Saúde | Epiclin | Fernando Hayashi Sant'Anna, Ana Paula Muterle, Janira Prichula, Juliana Comerlato, Carolina Comerlato, Eliana Márcia Da Ros Wendland |
| EPI_ISL_1469767 | Fundação de Saúde Pública de Novo Hamburgo | Epiclin | Fernando Hayashi Sant'Anna, Ana Paula Muterle, Janira Prichula, Juliana Comerlato, Carolina Comerlato, Eliana Márcia Da Ros Wendland |
| EPI_ISL_1469768 | UNIDADE DE PRONTO ATENDIMENTO DE SAPUCAIA DO SUL UPA | Epiclin | Fernando Hayashi Sant'Anna, Ana Paula Muterle, Janira Prichula, Juliana Comerlato, Carolina Comerlato, Eliana Márcia Da Ros Wendland |
| EPI_ISL_1469769 | Diretoria de Vigilância em Saúde | Epiclin | Fernando Hayashi Sant'Anna, Ana Paula Muterle, Janira Prichula, Juliana Comerlato, Carolina Comerlato, Eliana Márcia Da Ros Wendland |
| EPI_ISL_1469770 | Fundação de Saúde Pública de Novo Hamburgo | Epiclin | Fernando Hayashi Sant'Anna, Ana Paula Muterle, Janira Prichula, Juliana Comerlato, Carolina Comerlato, Eliana Márcia Da Ros Wendland |
| EPI_ISL_1469771 | Hospital de Portão | Epiclin | Fernando Hayashi Sant'Anna, Ana Paula Muterle, Janira Prichula, Juliana Comerlato, Carolina Comerlato, Eliana Márcia Da Ros Wendland |
| EPI_ISL_1469772 | DIRETORIA DE VIGILANCIA EM SAUDE | Epiclin | Fernando Hayashi Sant'Anna, Ana Paula Muterle, Janira Prichula, Juliana Comerlato, Carolina Comerlato, Eliana Márcia Da Ros Wendland |
| EPI_ISL_1469773 | Hospital Universitário | Epiclin | Fernando Hayashi Sant'Anna, Ana Paula Muterle, Janira Prichula, Juliana Comerlato, Carolina Comerlato, Eliana Márcia Da Ros Wendland |
| EPI_ISL_1469774 | Hospital Universitário de Canoas | Epiclin | Fernando Hayashi Sant'Anna, Ana Paula Muterle, Janira Prichula, Juliana Comerlato, Carolina Comerlato, Eliana Márcia Da Ros Wendland |
| EPI_ISL_1469775 | Diretoria de Vigilância em Saúde | Epiclin | Fernando Hayashi Sant'Anna, Ana Paula Muterle, Janira Prichula, Juliana Comerlato, Carolina Comerlato, Eliana Márcia Da Ros Wendland |
| EPI_ISL_1469776 | Centro Municipal de Saúde de Rolante | Epiclin | Fernando Hayashi Sant'Anna, Ana Paula Muterle, Janira Prichula, Juliana Comerlato, Carolina Comerlato, Eliana Márcia Da Ros Wendland |
| EPI_ISL_1469777 | Centro de Referência em Síndromes Gripais | Epiclin | Fernando Hayashi Sant'Anna, Ana Paula Muterle, Janira Prichula, Juliana Comerlato, Carolina Comerlato, Eliana Márcia Da Ros Wendland |
| EPI_ISL_1469778 | Hospital Municipal Getúlio Vargas | Epiclin | Fernando Hayashi Sant'Anna, Ana Paula Muterle, Janira Prichula, Juliana Comerlato, Carolina Comerlato, Eliana Márcia Da Ros Wendland |
| EPI_ISL_1469779 | Centro de Especialidades Triunfo | Epiclin | Fernando Hayashi Sant'Anna, Ana Paula Muterle, Janira Prichula, Juliana Comerlato, Carolina Comerlato, Eliana Márcia Da Ros Wendland |
| EPI_ISL_1469780 | Secretaria Municipal de Saúde de Taquara | Epiclin | Fernando Hayashi Sant'Anna, Ana Paula Muterle, Janira Prichula, Juliana Comerlato, Carolina Comerlato, Eliana Márcia Da Ros Wendland |
| EPI_ISL_1469781 | DIRETORIA DE VIGILANCIA EM SAUDE | Epiclin | Fernando Hayashi Sant'Anna, Ana Paula Muterle, Janira Prichula, Juliana Comerlato, Carolina Comerlato, Eliana Márcia Da Ros Wendland |
| EPI_ISL_1469782 | COORDENADORIA GERAL DE VIGILANCIA EM SAUDE | Epiclin | Fernando Hayashi Sant'Anna, Ana Paula Muterle, Janira Prichula, Juliana Comerlato, Carolina Comerlato, Eliana Márcia Da Ros Wendland |
| EPI_ISL_1469783 | Diretoria de Vigilância em Saúde | Epiclin | Fernando Hayashi Sant'Anna, Ana Paula Muterle, Janira Prichula, Juliana Comerlato, Carolina Comerlato, Eliana Márcia Da Ros Wendland |
| EPI_ISL_1469784 | DIRETORIA DE VIGILANCIA EM SAUDE | Epiclin | Fernando Hayashi Sant'Anna, Ana Paula Muterle, Janira Prichula, Juliana Comerlato, Carolina Comerlato, Eliana Márcia Da Ros Wendland |
| EPI_ISL_1469785 | FUNDACAO DE SAUDE PUBLICA DE NOVO HAMBURGO FSNH | Epiclin | Fernando Hayashi Sant'Anna, Ana Paula Muterle, Janira Prichula, Juliana Comerlato, Carolina Comerlato, Eliana Márcia Da Ros Wendland |
| EPI_ISL_1469786 | Diretoria de Vigilância em Saúde | Epiclin | Fernando Hayashi Sant'Anna, Ana Paula Muterle, Janira Prichula, Juliana Comerlato, Carolina Comerlato, Eliana Márcia Da Ros Wendland |
| EPI_ISL_1469787 | Fundação Hospitalar de Sapucaia do Sul | Epiclin | Fernando Hayashi Sant'Anna, Ana Paula Muterle, Janira Prichula, Juliana Comerlato, Carolina Comerlato, Eliana Márcia Da Ros Wendland |
| EPI_ISL_1469788 | SECRETARIA MUNICIPAL DE SAUDE DE SAO LEOPOLDO | Epiclin | Fernando Hayashi Sant'Anna, Ana Paula Muterle, Janira Prichula, Juliana Comerlato, Carolina Comerlato, Eliana Márcia Da Ros Wendland |
| EPI_ISL_1469789 | Hospital São Francisco de Assis | Epiclin | Fernando Hayashi Sant'Anna, Ana Paula Muterle, Janira Prichula, Juliana Comerlato, Carolina Comerlato, Eliana Márcia Da Ros Wendland |
| EPI_ISL_1469790 | Unidade de Pronto Atendimento de Sapucaia do Sul | Epiclin | Fernando Hayashi Sant'Anna, Ana Paula Muterle, Janira Prichula, Juliana Comerlato, Carolina Comerlato, Eliana Márcia Da Ros Wendland |
| EPI_ISL_1469791 | DIRETORIA DE VIGILANCIA EM SAUDE | Epiclin | Fernando Hayashi Sant'Anna, Ana Paula Muterle, Janira Prichula, Juliana Comerlato, Carolina Comerlato, Eliana Márcia Da Ros Wendland |
| EPI_ISL_1469792 | Diretoria de Vigilância em Saúde | Epiclin | Fernando Hayashi Sant'Anna, Ana Paula Muterle, Janira Prichula, Juliana Comerlato, Carolina Comerlato, Eliana Márcia Da Ros Wendland |
| EPI_ISL_1469793 | Hospital Universitário de Canoas | Epiclin | Fernando Hayashi Sant'Anna, Ana Paula Muterle, Janira Prichula, Juliana Comerlato, Carolina Comerlato, Eliana Márcia Da Ros Wendland |
| EPI_ISL_1469794, EPI_ISL_1469795 | Hospital Dia e Pronto Atendimento | Epiclin | Fernando Hayashi Sant'Anna, Ana Paula Muterle, Janira Prichula, Juliana Comerlato, Carolina Comerlato, Eliana Márcia Da Ros Wendland |
| EPI_ISL_1469796, EPI_ISL_1469797 | Diretoria de Vigilância em Saúde | Epiclin | Fernando Hayashi Sant'Anna, Ana Paula Muterle, Janira Prichula, Juliana Comerlato, Carolina Comerlato, Eliana Márcia Da Ros Wendland |
| EPI_ISL_1469798 | DIRETORIA DE VIGILANCIA EM SAUDE | Epiclin | Fernando Hayashi Sant'Anna, Ana Paula Muterle, Janira Prichula, Juliana Comerlato, Carolina Comerlato, Eliana Márcia Da Ros Wendland |
| EPI_ISL_1469799 | Hospital Regina | Epiclin | Fernando Hayashi Sant'Anna, Ana Paula Muterle, Janira Prichula, Juliana Comerlato, Carolina Comerlato, Eliana Márcia Da Ros Wendland |
| EPI_ISL_1469800 | Fundação de Saúde Pública de Novo Hamburgo | Epiclin | Fernando Hayashi Sant'Anna, Ana Paula Muterle, Janira Prichula, Juliana Comerlato, Carolina Comerlato, Eliana Márcia Da Ros Wendland |
| EPI_ISL_1469801 | Hospital Sapiranga | Epiclin | Fernando Hayashi Sant'Anna, Ana Paula Muterle, Janira Prichula, Juliana Comerlato, Carolina Comerlato, Eliana Márcia Da Ros Wendland |
| EPI_ISL_1469802 | Diretoria de Vigilância em Saúde | Epiclin | Fernando Hayashi Sant'Anna, Ana Paula Muterle, Janira Prichula, Juliana Comerlato, Carolina Comerlato, Eliana Márcia Da Ros Wendland |

| | | | |
|---|---|---|---|
| EPI_ISL_1469803 | Secretaria Municipal de Saúde de Três Coroas | Epiclin | Fernando Hayashi Sant'Anna, Ana Paula Muterle, Janira Prichula, Juliana Comerlato, Carolina Comerlato, Eliana Márcia Da Ros Wendland |
| EPI_ISL_1469804 | UNIDADE DE PRONTO ATENDIMENTO DE SAPUCAIA DO SUL UPA | Epiclin | Fernando Hayashi Sant'Anna, Ana Paula Muterle, Janira Prichula, Juliana Comerlato, Carolina Comerlato, Eliana Márcia Da Ros Wendland |
| EPI_ISL_1469806 | Centro de Referência em Síndromes Gripais | Epiclin | Fernando Hayashi Sant'Anna, Ana Paula Muterle, Janira Prichula, Juliana Comerlato, Carolina Comerlato, Eliana Márcia Da Ros Wendland |
| EPI_ISL_1469807 | Hospital São Francisco de Assis | Epiclin | Fernando Hayashi Sant'Anna, Ana Paula Muterle, Janira Prichula, Juliana Comerlato, Carolina Comerlato, Eliana Márcia Da Ros Wendland |
| EPI_ISL_1469808 | Diretoria de Vigilância em Saúde | Epiclin | Fernando Hayashi Sant'Anna, Ana Paula Muterle, Janira Prichula, Juliana Comerlato, Carolina Comerlato, Eliana Márcia Da Ros Wendland |
| EPI_ISL_1469809 | Fundação Hospitalar de Sapucaia do Sul | Epiclin | Fernando Hayashi Sant'Anna, Ana Paula Muterle, Janira Prichula, Juliana Comerlato, Carolina Comerlato, Eliana Márcia Da Ros Wendland |
| EPI_ISL_1469810 | Unidade de Pronto Atendimento de Sapucaia do Sul | Epiclin | Fernando Hayashi Sant'Anna, Ana Paula Muterle, Janira Prichula, Juliana Comerlato, Carolina Comerlato, Eliana Márcia Da Ros Wendland |
| EPI_ISL_1469811 | Hospital Municipal Getúlio Vargas | Epiclin | Fernando Hayashi Sant'Anna, Ana Paula Muterle, Janira Prichula, Juliana Comerlato, Carolina Comerlato, Eliana Márcia Da Ros Wendland |
| EPI_ISL_1469812 | Posto de Saúde Cambará do Sul | Epiclin | Fernando Hayashi Sant'Anna, Ana Paula Muterle, Janira Prichula, Juliana Comerlato, Carolina Comerlato, Eliana Márcia Da Ros Wendland |
| EPI_ISL_1469813 | HOSPITAL BOM JESUS | Epiclin | Fernando Hayashi Sant'Anna, Ana Paula Muterle, Janira Prichula, Juliana Comerlato, Carolina Comerlato, Eliana Márcia Da Ros Wendland |
| EPI_ISL_1469814 | SECRETARIA MUNICIPAL DE SAUDE DE SAO LEOPOLDO | Epiclin | Fernando Hayashi Sant'Anna, Ana Paula Muterle, Janira Prichula, Juliana Comerlato, Carolina Comerlato, Eliana Márcia Da Ros Wendland |
| EPI_ISL_1469815 | Hospital Universitário | Epiclin | Fernando Hayashi Sant'Anna, Ana Paula Muterle, Janira Prichula, Juliana Comerlato, Carolina Comerlato, Eliana Márcia Da Ros Wendland |
| EPI_ISL_1469816 | Diretoria de Vigilância em Saúde | Epiclin | Fernando Hayashi Sant'Anna, Ana Paula Muterle, Janira Prichula, Juliana Comerlato, Carolina Comerlato, Eliana Márcia Da Ros Wendland |
| EPI_ISL_1469817 | Fundação Hospitalar de Sapucaia do Sul | Epiclin | Fernando Hayashi Sant'Anna, Ana Paula Muterle, Janira Prichula, Juliana Comerlato, Carolina Comerlato, Eliana Márcia Da Ros Wendland |
| EPI_ISL_1469818 | Hospital São Francisco de Assis | Epiclin | Fernando Hayashi Sant'Anna, Ana Paula Muterle, Janira Prichula, Juliana Comerlato, Carolina Comerlato, Eliana Márcia Da Ros Wendland |
| EPI_ISL_1469819 | Diretoria de Vigilância em Saúde | Epiclin | Fernando Hayashi Sant'Anna, Ana Paula Muterle, Janira Prichula, Juliana Comerlato, Carolina Comerlato, Eliana Márcia Da Ros Wendland |
| EPI_ISL_1469820 | Hospital Sapiranga | Epiclin | Fernando Hayashi Sant'Anna, Ana Paula Muterle, Janira Prichula, Juliana Comerlato, Carolina Comerlato, Eliana Márcia Da Ros Wendland |
| EPI_ISL_1469821 | Centro de Saúde Bruno Cassel | Epiclin | Fernando Hayashi Sant'Anna, Ana Paula Muterle, Janira Prichula, Juliana Comerlato, Carolina Comerlato, Eliana Márcia Da Ros Wendland |
| EPI_ISL_1469822 | Hospital São Francisco de Assis | Epiclin | Fernando Hayashi Sant'Anna, Ana Paula Muterle, Janira Prichula, Juliana Comerlato, Carolina Comerlato, Eliana Márcia Da Ros Wendland |
| EPI_ISL_1469823 | Diretoria de Vigilância em Saúde | Epiclin | Fernando Hayashi Sant'Anna, Ana Paula Muterle, Janira Prichula, Juliana Comerlato, Carolina Comerlato, Eliana Márcia Da Ros Wendland |
| EPI_ISL_1469824 | Hospital Regina | Epiclin | Fernando Hayashi Sant'Anna, Ana Paula Muterle, Janira Prichula, Juliana Comerlato, Carolina Comerlato, Eliana Márcia Da Ros Wendland |
| EPI_ISL_1469825 | Pronto Atendimento Campo Bom | Epiclin | Fernando Hayashi Sant'Anna, Ana Paula Muterle, Janira Prichula, Juliana Comerlato, Carolina Comerlato, Eliana Márcia Da Ros Wendland |
| EPI_ISL_1469826 | Fundação de Saúde Pública de Novo Hamburgo | Epiclin | Fernando Hayashi Sant'Anna, Ana Paula Muterle, Janira Prichula, Juliana Comerlato, Carolina Comerlato, Eliana Márcia Da Ros Wendland |
| EPI_ISL_1469827 | Hospital de Portão | Epiclin | Fernando Hayashi Sant'Anna, Ana Paula Muterle, Janira Prichula, Juliana Comerlato, Carolina Comerlato, Eliana Márcia Da Ros Wendland |
| EPI_ISL_1469828 | Fundação de Saúde Pública de Novo Hamburgo | Epiclin | Fernando Hayashi Sant'Anna, Ana Paula Muterle, Janira Prichula, Juliana Comerlato, Carolina Comerlato, Eliana Márcia Da Ros Wendland |
| EPI_ISL_1469829 | Diretoria de Vigilância em Saúde | Epiclin | Fernando Hayashi Sant'Anna, Ana Paula Muterle, Janira Prichula, Juliana Comerlato, Carolina Comerlato, Eliana Márcia Da Ros Wendland |
| EPI_ISL_1469830 | Hospital São Francisco de Assis | Epiclin | Fernando Hayashi Sant'Anna, Ana Paula Muterle, Janira Prichula, Juliana Comerlato, Carolina Comerlato, Eliana Márcia Da Ros Wendland |
| EPI_ISL_1469831 | Fundação Hospitalar de Sapucaia do Sul | Epiclin | Fernando Hayashi Sant'Anna, Ana Paula Muterle, Janira Prichula, Juliana Comerlato, Carolina Comerlato, Eliana Márcia Da Ros Wendland |
| EPI_ISL_1469832 | Diretoria de Vigilância em Saúde | Epiclin | Fernando Hayashi Sant'Anna, Ana Paula Muterle, Janira Prichula, Juliana Comerlato, Carolina Comerlato, Eliana Márcia Da Ros Wendland |
| EPI_ISL_1469833 | Hospital Nossa Senhora das Graças | Epiclin | Fernando Hayashi Sant'Anna, Ana Paula Muterle, Janira Prichula, Juliana Comerlato, Carolina Comerlato, Eliana Márcia Da Ros Wendland |
| EPI_ISL_1469834 | HOSPITAL SAO FRANCISCO DE ASSIS | Epiclin | Fernando Hayashi Sant'Anna, Ana Paula Muterle, Janira Prichula, Juliana Comerlato, Carolina Comerlato, Eliana Márcia Da Ros Wendland |
| EPI_ISL_1469835, EPI_ISL_1469836 | Diretoria de Vigilância em Saúde | Epiclin | Fernando Hayashi Sant'Anna, Ana Paula Muterle, Janira Prichula, Juliana Comerlato, Carolina Comerlato, Eliana Márcia Da Ros Wendland |
| EPI_ISL_1469837 | Fundação de Saúde Pública São Camilo de Esteio | Epiclin | Fernando Hayashi Sant'Anna, Ana Paula Muterle, Janira Prichula, Juliana Comerlato, Carolina Comerlato, Eliana Márcia Da Ros Wendland |
| EPI_ISL_1469838 | Secretaria Municipal de Saúde de Três Coroas | Epiclin | Fernando Hayashi Sant'Anna, Ana Paula Muterle, Janira Prichula, Juliana Comerlato, Carolina Comerlato, Eliana Márcia Da Ros Wendland |
| EPI_ISL_1469839 | HOSPITAL DE CAMPO BOM DR LAURO REUS | Epiclin | Fernando Hayashi Sant'Anna, Ana Paula Muterle, Janira Prichula, Juliana Comerlato, Carolina Comerlato, Eliana Márcia Da Ros Wendland |
| EPI_ISL_1469840 | Hospital Sapiranga | Epiclin | Fernando Hayashi Sant'Anna, Ana Paula Muterle, Janira Prichula, Juliana Comerlato, Carolina Comerlato, Eliana Márcia Da Ros Wendland |
| EPI_ISL_1469841 | Diretoria de Vigilância em Saúde | Epiclin | Fernando Hayashi Sant'Anna, Ana Paula Muterle, Janira Prichula, Juliana Comerlato, Carolina Comerlato, Eliana Márcia Da Ros Wendland |
| EPI_ISL_1469842 | Hospital São Francisco de Assis | Epiclin | Fernando Hayashi Sant'Anna, Ana Paula Muterle, Janira Prichula, Juliana Comerlato, Carolina Comerlato, Eliana Márcia Da Ros Wendland |
| EPI_ISL_1469843 | Hospital Sapiranga | Epiclin | Fernando Hayashi Sant'Anna, Ana Paula Muterle, Janira Prichula, Juliana Comerlato, Carolina Comerlato, Eliana Márcia Da Ros Wendland |
| EPI_ISL_1469844 | Hospital Universitário de Canoas | Epiclin | Fernando Hayashi Sant'Anna, Ana Paula Muterle, Janira Prichula, Juliana Comerlato, Carolina Comerlato, Eliana Márcia Da Ros Wendland |
| EPI_ISL_1469845 | Fundação Hospitalar de Sapucaia do Sul | Epiclin | Fernando Hayashi Sant'Anna, Ana Paula Muterle, Janira Prichula, Juliana Comerlato, Carolina Comerlato, Eliana Márcia Da Ros Wendland |
| EPI_ISL_1469846 | Hospital Nossa Senhora das Graças | Epiclin | Fernando Hayashi Sant'Anna, Ana Paula Muterle, Janira Prichula, Juliana Comerlato, Carolina Comerlato, Eliana Márcia Da Ros Wendland |
| EPI_ISL_1469847 | Secretaria Municipal de Saúde de Três Coroas | Epiclin | Fernando Hayashi Sant'Anna, Ana Paula Muterle, Janira Prichula, Juliana Comerlato, Carolina Comerlato, Eliana Márcia Da Ros Wendland |
| EPI_ISL_1469848 | Diretoria de Vigilância em Saúde | Epiclin | Fernando Hayashi Sant'Anna, Ana Paula Muterle, Janira Prichula, Juliana Comerlato, Carolina Comerlato, Eliana Márcia Da Ros Wendland |
| EPI_ISL_1469849 | Hospital Universitário de Canoas | Epiclin | Fernando Hayashi Sant'Anna, Ana Paula Muterle, Janira Prichula, Juliana Comerlato, Carolina Comerlato, Eliana Márcia Da Ros Wendland |
| EPI_ISL_1469850 | Fundação de Saúde Pública São Camilo de Esteio | Epiclin | Fernando Hayashi Sant'Anna, Ana Paula Muterle, Janira Prichula, Juliana Comerlato, Carolina Comerlato, Eliana Márcia Da Ros Wendland |
| EPI_ISL_1469851 | Hospital Nossa Senhora das Graças | Epiclin | Fernando Hayashi Sant'Anna, Ana Paula Muterle, Janira Prichula, Juliana Comerlato, Carolina Comerlato, Eliana Márcia Da Ros Wendland |
| EPI_ISL_1479119 | DIRETORIA DE VIGILANCIA EM SAUDE | Epiclin | Fernando Hayashi Sant'Anna, Ana Paula Muterle, Janira Prichula, Juliana Comerlato, Carolina Comerlato, Eliana Márcia Da Ros Wendland |
| EPI_ISL_1479120 | Diretoria de Vigilância em Saúde | Epiclin | Fernando Hayashi Sant'Anna, Ana Paula Muterle, Janira Prichula, Juliana Comerlato, Carolina Comerlato, Eliana Márcia Da Ros Wendland |
| EPI_ISL_1479121 | DIRETORIA DE VIGILANCIA EM SAUDE | Epiclin | Fernando Hayashi Sant'Anna, Ana Paula Muterle, Janira Prichula, Juliana Comerlato, Carolina Comerlato, Eliana Márcia Da Ros Wendland |
| EPI_ISL_1479122 | Fundação Hospitalar São José | Epiclin | Fernando Hayashi Sant'Anna, Ana Paula Muterle, Janira Prichula, Juliana Comerlato, Carolina Comerlato, Eliana Márcia Da Ros Wendland |
| EPI_ISL_1479123 | Fundação de Saúde Pública de Novo Hamburgo | Epiclin | Fernando Hayashi Sant'Anna, Ana Paula Muterle, Janira Prichula, Juliana Comerlato, Carolina Comerlato, Eliana Márcia Da Ros Wendland |
| EPI_ISL_1479124 | UNIDADE DE SAUDE NOVA HARTZ | Epiclin | Fernando Hayashi Sant'Anna, Ana Paula Muterle, Janira Prichula, Juliana Comerlato, Carolina Comerlato, Eliana Márcia Da Ros Wendland |
| EPI_ISL_1479125 | CENTRO DE REFERENCIA EM SINDROMES GRIPAIS | Epiclin | Fernando Hayashi Sant'Anna, Ana Paula Muterle, Janira Prichula, Juliana Comerlato, Carolina Comerlato, Eliana Márcia Da Ros Wendland |
| EPI_ISL_1479126 | UNIDADE DE ATENDIMENTO DST AIDS TB E HAN | Epiclin | Fernando Hayashi Sant'Anna, Ana Paula Muterle, Janira Prichula, Juliana Comerlato, Carolina Comerlato, Eliana Márcia Da Ros Wendland |
| EPI_ISL_1479127 | Hospital Regina | Epiclin | Fernando Hayashi Sant'Anna, Ana Paula Muterle, Janira Prichula, Juliana Comerlato, Carolina Comerlato, Eliana Márcia Da Ros Wendland |
| EPI_ISL_1479128 | HOSPITAL SAO FRANCISCO DE ASSIS | Epiclin | Fernando Hayashi Sant'Anna, Ana Paula Muterle, Janira Prichula, Juliana Comerlato, Carolina Comerlato, Eliana Márcia Da Ros Wendland |
| EPI_ISL_1479129 | Pronto Atendimento Campo Bom | Epiclin | Fernando Hayashi Sant'Anna, Ana Paula Muterle, Janira Prichula, Juliana Comerlato, Carolina Comerlato, Eliana Márcia Da Ros Wendland |

| | | | |
|---|---|---|---|
| EPI_ISL_1479130 | SECRETARIA MUNICIPAL DE SAUDE DE TAQUARA | Epiclin | Fernando Hayashi Sant'Anna, Ana Paula Muterle, Janira Prichula, Juliana Comerlato, Carolina Comerlato, Eliana Márcia Da Ros Wendland |
| EPI_ISL_1479131, EPI_ISL_1479132 | Fundação de Saúde Pública de Novo Hamburgo | Epiclin | Fernando Hayashi Sant'Anna, Ana Paula Muterle, Janira Prichula, Juliana Comerlato, Carolina Comerlato, Eliana Márcia Da Ros Wendland |
| EPI_ISL_1479133 | HOSPITAL SAO FRANCISCO DE ASSIS | Epiclin | Fernando Hayashi Sant'Anna, Ana Paula Muterle, Janira Prichula, Juliana Comerlato, Carolina Comerlato, Eliana Márcia Da Ros Wendland |
| EPI_ISL_1493572 | Centro de Saude II Dr Alcides Facundo Arroyo | Instituto Adolfo Lutz, Interdiciplinary Procedures Center, Strategic Laboratory | Claudio Tavares Sacchi, Claudia Regina Gonçalves, Erica Valessa Ramos Gomes, Karoline Rodrigues Campos, Caio Vinicius Dias Lopes |
| EPI_ISL_1493573, EPI_ISL_1493574, EPI_ISL_1493575, EPI_ISL_1493576, EPI_ISL_1493577 | LACEN do Estado de Goias | Instituto Adolfo Lutz, Interdiciplinary Procedures Center, Strategic Laboratory | Claudio Tavares Sacchi, Claudia Regina Gonçalves, Erica Valessa Ramos Gomes, Karoline Rodrigues Campos, Caio Vinicius Dias Lopes |
| EPI_ISL_1493578, EPI_ISL_1493579 | LACEN do Estado de Rondonia | Instituto Adolfo Lutz, Interdiciplinary Procedures Center, Strategic Laboratory | Claudio Tavares Sacchi, Claudia Regina Gonçalves, Erica Valessa Ramos Gomes, Karoline Rodrigues Campos, Caio Vinicius Dias Lopes |
| EPI_ISL_1493580 | CS II Dr Miguel Vitaliano Orlandia | Instituto Adolfo Lutz, Interdiciplinary Procedures Center, Strategic Laboratory | Claudio Tavares Sacchi, Claudia Regina Gonçalves, Erica Valessa Ramos Gomes, Karoline Rodrigues Campos, Caio Vinicius Dias Lopes |
| EPI_ISL_1493581 | Sae servico de Atendimento Especializado | Instituto Adolfo Lutz, Interdiciplinary Procedures Center, Strategic Laboratory | Claudio Tavares Sacchi, Claudia Regina Gonçalves, Erica Valessa Ramos Gomes, Karoline Rodrigues Campos, Caio Vinicius Dias Lopes |
| EPI_ISL_1493582 | CS II Dr Miguel Vitaliano Orlandia | Instituto Adolfo Lutz, Interdiciplinary Procedures Center, Strategic Laboratory | Claudio Tavares Sacchi, Claudia Regina Gonçalves, Erica Valessa Ramos Gomes, Karoline Rodrigues Campos, Caio Vinicius Dias Lopes |
| EPI_ISL_1493583, EPI_ISL_1493584 | LACEN do Estado de Rondonia | Instituto Adolfo Lutz, Interdiciplinary Procedures Center, Strategic Laboratory | Claudio Tavares Sacchi, Claudia Regina Gonçalves, Erica Valessa Ramos Gomes, Karoline Rodrigues Campos, Caio Vinicius Dias Lopes |
| EPI_ISL_1493585 | Unidade de Saude Dr Phebo de Oliveira Roge Ferreira | Instituto Adolfo Lutz, Interdiciplinary Procedures Center, Strategic Laboratory | Claudio Tavares Sacchi, Claudia Regina Gonçalves, Erica Valessa Ramos Gomes, Karoline Rodrigues Campos, Caio Vinicius Dias Lopes |
| EPI_ISL_1493586 | Hospital Sao Marcos da Samamorro Agudo | Instituto Adolfo Lutz, Interdiciplinary Procedures Center, Strategic Laboratory | Claudio Tavares Sacchi, Claudia Regina Gonçalves, Erica Valessa Ramos Gomes, Karoline Rodrigues Campos, Caio Vinicius Dias Lopes |
| EPI_ISL_1493587, EPI_ISL_1493588 | CS III de Patrocinio Paulista | Instituto Adolfo Lutz, Interdiciplinary Procedures Center, Strategic Laboratory | Claudio Tavares Sacchi, Claudia Regina Gonçalves, Erica Valessa Ramos Gomes, Karoline Rodrigues Campos, Caio Vinicius Dias Lopes |
| EPI_ISL_1493589 | CS II Dr Jose Ferreira Telles | Instituto Adolfo Lutz, Interdiciplinary Procedures Center, Strategic Laboratory | Claudio Tavares Sacchi, Claudia Regina Gonçalves, Erica Valessa Ramos Gomes, Karoline Rodrigues Campos, Caio Vinicius Dias Lopes |
| EPI_ISL_1493590 | Hospital Sao Marcos da Samamorro Agudo | Instituto Adolfo Lutz, Interdiciplinary Procedures Center, Strategic Laboratory | Claudio Tavares Sacchi, Claudia Regina Gonçalves, Erica Valessa Ramos Gomes, Karoline Rodrigues Campos, Caio Vinicius Dias Lopes |
| EPI_ISL_1493591 | Centro de Saude II Dr Alcides Facundo Arroyo | Instituto Adolfo Lutz, Interdiciplinary Procedures Center, Strategic Laboratory | Claudio Tavares Sacchi, Claudia Regina Gonçalves, Erica Valessa Ramos Gomes, Karoline Rodrigues Campos, Caio Vinicius Dias Lopes |
| EPI_ISL_1493592 | Santa Casa de Guaira | Instituto Adolfo Lutz, Interdiciplinary Procedures Center, Strategic Laboratory | Claudio Tavares Sacchi, Claudia Regina Gonçalves, Erica Valessa Ramos Gomes, Karoline Rodrigues Campos, Caio Vinicius Dias Lopes |
| EPI_ISL_1493593 | CS II Dr Jahyr de Paula Ribeiro Guara | Instituto Adolfo Lutz, Interdiciplinary Procedures Center, Strategic Laboratory | Claudio Tavares Sacchi, Claudia Regina Gonçalves, Erica Valessa Ramos Gomes, Karoline Rodrigues Campos, Caio Vinicius Dias Lopes |
| EPI_ISL_1493594 | Unidade de Saude Dr Phebo de Oliveira Roge Ferreira | Instituto Adolfo Lutz, Interdiciplinary Procedures Center, Strategic Laboratory | Claudio Tavares Sacchi, Claudia Regina Gonçalves, Erica Valessa Ramos Gomes, Karoline Rodrigues Campos, Caio Vinicius Dias Lopes |
| EPI_ISL_1493595, EPI_ISL_1493596, EPI_ISL_1493597, EPI_ISL_1493598, EPI_ISL_1493599, EPI_ISL_1493600 | LACEN do Estado de Rondonia | Instituto Adolfo Lutz, Interdiciplinary Procedures Center, Strategic Laboratory | Claudio Tavares Sacchi, Claudia Regina Gonçalves, Erica Valessa Ramos Gomes, Karoline Rodrigues Campos, Caio Vinicius Dias Lopes |
| EPI_ISL_1494923 | CS II Dr Jose Ferreira Telles | Instituto Adolfo Lutz, Interdiciplinary Procedures Center, Strategic Laboratory | Claudio Tavares Sacchi, Claudia Regina Gonçalves, Erica Valessa Ramos Gomes, Karoline Rodrigues Campos, Caio Vinicius Dias Lopes |
| EPI_ISL_1494924 | LACEN do Estado de Rondonia | Instituto Adolfo Lutz, Interdiciplinary Procedures Center, Strategic Laboratory | Claudio Tavares Sacchi, Claudia Regina Gonçalves, Erica Valessa Ramos Gomes, Karoline Rodrigues Campos, Caio Vinicius Dias Lopes |
| EPI_ISL_1494960, EPI_ISL_1494961, EPI_ISL_1494962, EPI_ISL_1494963, EPI_ISL_1494964, EPI_ISL_1494965, EPI_ISL_1494966, EPI_ISL_1494967, EPI_ISL_1494968, EPI_ISL_1494969, EPI_ISL_1494970, EPI_ISL_1494971, EPI_ISL_1494972, EPI_ISL_1494973, EPI_ISL_1494974, EPI_ISL_1494975, EPI_ISL_1494976, EPI_ISL_1494977, EPI_ISL_1494978, EPI_ISL_1494979, EPI_ISL_1494980, EPI_ISL_1494981, EPI_ISL_1494982, EPI_ISL_1494983, EPI_ISL_1494984, EPI_ISL_1494985, EPI_ISL_1494986, EPI_ISL_1494987, EPI_ISL_1494988, EPI_ISL_1494989, EPI_ISL_1494990, EPI_ISL_1494991, EPI_ISL_1494992, EPI_ISL_1494993, EPI_ISL_1494994, EPI_ISL_1494995, EPI_ISL_1494996, EPI_ISL_1494997, EPI_ISL_1494998, EPI_ISL_1494999, EPI_ISL_1495000, EPI_ISL_1495001, EPI_ISL_1495002, EPI_ISL_1495003, EPI_ISL_1495004, EPI_ISL_1495005, EPI_ISL_1495006, EPI_ISL_1495007, EPI_ISL_1495008, EPI_ISL_1495009, EPI_ISL_1495010, EPI_ISL_1495011, EPI_ISL_1495012, EPI_ISL_1495013, EPI_ISL_1495014, EPI_ISL_1495015, EPI_ISL_1495016, EPI_ISL_1495017, EPI_ISL_1495018, EPI_ISL_1495019, EPI_ISL_1495020, EPI_ISL_1495021, EPI_ISL_1495022, EPI_ISL_1495023, EPI_ISL_1495024, EPI_ISL_1495025, EPI_ISL_1495026, EPI_ISL_1495027, EPI_ISL_1495028, EPI_ISL_1495029, EPI_ISL_1495030, EPI_ISL_1495031, EPI_ISL_1495032, EPI_ISL_1495033, EPI_ISL_1495034, EPI_ISL_1495035, EPI_ISL_1495036, EPI_ISL_1495037, EPI_ISL_1495038, EPI_ISL_1495039, EPI_ISL_1495040, EPI_ISL_1495041, EPI_ISL_1495042, EPI_ISL_1497548, EPI_ISL_1497549 | | | |
| see above | Laboratório de Biologia Integrativa | Laboratório de Biologia Integrativa | Filipe Romero Rebello Moreira, Diego Menezes Bonfim, Victor Emmanuel Viana Geddes, Danielle Alves Gomes Zauli, Joice do Prado Silva, Aline Brito de Lima, Frederico Scott Varella Malta, Alessandro Clayton de Souza Ferreira, Victor Cavalcanti Pardini, Daniel Costa Queiroz, Rafael Marques de Souza, Lucyene Miguita Luiz, Paula Luize Camargos Fonseca, Rennan Garcias Moreira, Nuno Rodrigues Faria, Carolina Moreira Voloch, Renan Pedra de Souza, Renato Santana Aguiar |
| EPI_ISL_1498263 | Associação Fundo de Incentivo à Pesquisa (AFIP) | Associação Fundo de Incentivo à Pesquisa (AFIP) | Priscila Farias Tempaku, Juliana Nogueira Martins Rodrigues, Erika Rodrigues de Oliveira, Debora R. Ramadan, Soraya Sgambatti de Andrade, Sergio Tufik. |
| EPI_ISL_1498380 | Associação Fundo de Incentivo à Pesquisa (AFIP). | Associação Fundo de Incentivo à Pesquisa (AFIP) | Priscila Farias Tempaku, Juliana Nogueira Martins Rodrigues, Erika Rodrigues de Oliveira, Debora R. Ramadan, Soraya Sgambatti de Andrade, Sergio Tufik. |
| EPI_ISL_1498585, EPI_ISL_1498821, EPI_ISL_1498822, EPI_ISL_1498823, EPI_ISL_1498824, EPI_ISL_1498825, EPI_ISL_1498916 | Associação Fundo de Incentivo à Pesquisa (AFIP) | Associação Fundo de Incentivo à Pesquisa (AFIP) | Priscila Farias Tempaku, Juliana Nogueira Martins Rodrigues, Erika Rodrigues de Oliveira, Debora R. Ramadan, Soraya Sgambatti de Andrade, Sergio Tufik. |
| EPI_ISL_1498917 | Hospital Estadual de Mirandopolis | Instituto Adolfo Lutz, Interdiciplinary Procedures Center, Strategic Laboratory | Claudio Tavares Sacchi, Claudia Regina Gonçalves, Erica Valessa Ramos Gomes, Karoline Rodrigues Campos, Caio Vinicius Dias Lopes |
| EPI_ISL_1498919, EPI_ISL_1499020, EPI_ISL_1499105, EPI_ISL_1499114, EPI_ISL_1499201, EPI_ISL_1499202, EPI_ISL_1499203, EPI_ISL_1499204, EPI_ISL_1499205, EPI_ISL_1499297, EPI_ISL_1499298, EPI_ISL_1499299, EPI_ISL_1499300 | | | |
| see above | Associação Fundo de Incentivo à Pesquisa (AFIP) | Associação Fundo de Incentivo à Pesquisa (AFIP) | Priscila Farias Tempaku, Juliana Nogueira Martins Rodrigues, Erika Rodrigues de Oliveira, Debora R. Ramadan, Soraya Sgambatti de Andrade, Sergio Tufik. |
| EPI_ISL_1509639 | UBS de Auriflama | Instituto Adolfo Lutz, Interdiciplinary Procedures Center, Strategic Laboratory | Claudio Tavares Sacchi, Claudia Regina Gonçalves, Erica Valessa Ramos Gomes, Karoline Rodrigues Campos, Caio Vinicius Dias Lopes, Leonardo Jose Tadeu de Araujo |
| EPI_ISL_1509720 | UBS de Auriflama | Instituto Adolfo Lutz, Interdiciplinary Procedures Center, Strategic Laboratory | Claudio Tavares Sacchi, Claudia Regina Gonçalves, Erica Valessa Ramos Gomes, Karoline Rodrigues Campos, Caio Vinicius Dias Lopes |
| EPI_ISL_1511399, EPI_ISL_1511603 | Laboratorio de Patologia Clinica - UNICAMP | Laboratorio de Estudos de Virus Emergentes - UNICAMP | Mariene R. Amorim, William M. Souza, Antonio C. G. Carlos Jr, Daniel A. Toledo-Teixeira, Karina Bispo-dos-Santos, Camila L. Simeoni, Pierina L. Parise, Aline Vieira, Julia Forato, Ingra M. Claro, Luciana S. Mofatto, Natalia S. Brunetti, Emerson S.S. França, Gisele A. Pedroso, Barbara F. N. Carvalho, Tania R. Zaccariotto, Kamila C. S. Krywacz, André S. Vieira, Marcelo A. Mori, Alessandro S. Farias, Maria H. P. Pavan, Luís Felipe Bachur, Luís G. O. Cardoso, |

| | | | |
|---|---|---|---|
| | | | Fernando R. Spilki, Ester C. Sabino, Nuno R. Faria, Magnun N. N. Santos, Rodrigo Angerami, Patricia A. F. Leme, Angelica Schreiber, Maria L. Moretti, Fabiana Granja, José Luiz Proenca-Modena |
| EPI_ISL_1511643 | Genomic and molecular Biology Group, A.C.Camargo Cancer Center | Laboratory of Bioinformatics and Computational Biology, A.C.Camargo Cancer Center | Giovana Torrezan, Dirce Carraro, Israel Tojal, Alexandre Defelicibus |
| EPI_ISL_1511644 | Laboratorio de Patologia Clinica - UNICAMP | Laboratorio de Estudos de Virus Emergentes | Mariene R. Amorim, William M. Souza, Antonio C. G. Carlos Jr, Daniel A. Toledo-Teixeira, Karina Bispo-dos-Santos, Camila L. Simeoni, Pierina L. Parise, Aline Vieira, Julia Forato, Ingra M. Claro, Luciana S. Mofatto, Natalia S. Brunetti, Emerson S.S. França, Gisele A. Pedroso, Barbara F. N. Carvalho, Tania R. Zaccariotto, Kamila C. S. Krywacz, André S. Vieira, Marcelo A. Mori, Alessandro S. Farias, Maria H. P. Pavan, Luís Felipe Bachur, Luís G. O. Cardoso, Fernando R. Spilki, Ester C. Sabino, Nuno R. Faria, Magnun N. N. Santos, Rodrigo Angerami, Patricia A. F. Leme, Angelica Schreiber, Maria L. Moretti, Fabiana Granja, José Luiz Proenca-Modena |
| EPI_ISL_1520107, EPI_ISL_1520108, EPI_ISL_1520109 | LACEN do Estado de Rondonia | Instituto Adolfo Lutz, Interdiciplinary Procedures Center, Strategic Laboratory | Claudio Tavares Sacchi, Claudia Regina Gonçalves, Erica Valessa Ramos Gomes, Karoline Rodrigues Campos, Caio Vinicius Dias Lopes |
| EPI_ISL_1520110, EPI_ISL_1520111, EPI_ISL_1520112 | Hospital Municipal Reynaldo Guerra Cajati | Instituto Adolfo Lutz, Interdiciplinary Procedures Center, Strategic Laboratory | Claudio Tavares Sacchi, Claudia Regina Gonçalves, Erica Valessa Ramos Gomes, Karoline Rodrigues Campos, Caio Vinicius Dias Lopes |
| EPI_ISL_1520113 | Hospital Santo Antonio de Juquia Juquia | Instituto Adolfo Lutz, Interdiciplinary Procedures Center, Strategic Laboratory | Claudio Tavares Sacchi, Claudia Regina Gonçalves, Erica Valessa Ramos Gomes, Karoline Rodrigues Campos, Caio Vinicius Dias Lopes |
| EPI_ISL_1520114 | UBS III de Pariquera Acu Pariquera Acu | Instituto Adolfo Lutz, Interdiciplinary Procedures Center, Strategic Laboratory | Claudio Tavares Sacchi, Claudia Regina Gonçalves, Erica Valessa Ramos Gomes, Karoline Rodrigues Campos, Caio Vinicius Dias Lopes |
| EPI_ISL_1520115, EPI_ISL_1520116 | Unidade de Pronto Atendimento UPA | Instituto Adolfo Lutz, Interdiciplinary Procedures Center, Strategic Laboratory | Claudio Tavares Sacchi, Claudia Regina Gonçalves, Erica Valessa Ramos Gomes, Karoline Rodrigues Campos, Caio Vinicius Dias Lopes |
| EPI_ISL_1520117, EPI_ISL_1520118, EPI_ISL_1520119, EPI_ISL_1520120, EPI_ISL_1520121, EPI_ISL_1520122, EPI_ISL_1520123, EPI_ISL_1520124, EPI_ISL_1520125, EPI_ISL_1520126, EPI_ISL_1520127, EPI_ISL_1520128 | | | |
| see above | Centro de Saude II Dr Jose de Felipe Espito Santo do Pinhal SP | Instituto Adolfo Lutz, Interdiciplinary Procedures Center, Strategic Laboratory | Claudio Tavares Sacchi, Claudia Regina Gonçalves, Erica Valessa Ramos Gomes, Karoline Rodrigues Campos, Caio Vinicius Dias Lopes |
| EPI_ISL_1520129, EPI_ISL_1520130, EPI_ISL_1520131, EPI_ISL_1520132, EPI_ISL_1520133, EPI_ISL_1520134, EPI_ISL_1520135, EPI_ISL_1520136, EPI_ISL_1520137 | Centro de Saude II Dr Jose Paione Mococa | Instituto Adolfo Lutz, Interdiciplinary Procedures Center, Strategic Laboratory | Claudio Tavares Sacchi, Claudia Regina Gonçalves, Erica Valessa Ramos Gomes, Karoline Rodrigues Campos, Caio Vinicius Dias Lopes |
| EPI_ISL_1533609 | Laboratorio de Ecologia de Doencas Transmissiveis na Amazonia, Instituto Leonidas e Maria Deane - Fiocruz Amazonia | Laboratorio de Ecologia de Doencas Transmissiveis na Amazonia, Instituto Leonidas e Maria Deane - Fiocruz Amazonia | Valdinete Nascimento, Victor Souza, André Corado, Fernanda Nascimento, George Silva, Ágatha Costa, Debora Duarte, Karina Pessoa, Matilde Mejía, Luciana Gonçalves, Maria Júlia Brandão, Michele Jesus, Felipe Naveca |
| EPI_ISL_1533689, EPI_ISL_1533690, EPI_ISL_1533691 | Centro de Saude II Dr. Jose Paione Mococa | Instituto Adolfo Lutz, Interdiciplinary Procedures Center, Strategic Laboratory | Claudio Tavares Sacchi, Claudia Regina Gonçalves, Erica Valessa Ramos Gomes, Karoline Rodrigues Campos, Caio Vinicius Dias Lopes, Leonardo Jose Tadeu de Araujo |
| EPI_ISL_1533692 | Santa Casa de Sao Paulo | Instituto Adolfo Lutz, Interdiciplinary Procedures Center, Strategic Laboratory | Claudio Tavares Sacchi, Claudia Regina Gonçalves, Erica Valessa Ramos Gomes, Karoline Rodrigues Campos, Caio Vinicius Dias Lopes, Leonardo Jose Tadeu de Araujo |
| EPI_ISL_1533693 | Secretaria Municipal de Saude de Ubatuba | Instituto Adolfo Lutz, Interdiciplinary Procedures Center, Strategic Laboratory | Claudio Tavares Sacchi, Claudia Regina Gonçalves, Erica Valessa Ramos Gomes, Karoline Rodrigues Campos, Caio Vinicius Dias Lopes, Leonardo Jose Tadeu de Araujo |
| EPI_ISL_1533694 | Santa Casa de Birigui | Instituto Adolfo Lutz, Interdiciplinary Procedures Center, Strategic Laboratory | Claudio Tavares Sacchi, Claudia Regina Gonçalves, Erica Valessa Ramos Gomes, Karoline Rodrigues Campos, Caio Vinicius Dias Lopes, Leonardo Jose Tadeu de Araujo |
| EPI_ISL_1533695 | Hospital Municipal Dr Mario Gatti | Instituto Adolfo Lutz, Interdiciplinary Procedures Center, Strategic Laboratory | Claudio Tavares Sacchi, Claudia Regina Gonçalves, Erica Valessa Ramos Gomes, Karoline Rodrigues Campos, Caio Vinicius Dias Lopes, Leonardo Jose Tadeu de Araujo |
| EPI_ISL_1533696 | Hospital Municipal de Pedreira | Instituto Adolfo Lutz, Interdiciplinary Procedures Center, Strategic Laboratory | Claudio Tavares Sacchi, Claudia Regina Gonçalves, Erica Valessa Ramos Gomes, Karoline Rodrigues Campos, Caio Vinicius Dias Lopes, Leonardo Jose Tadeu de Araujo |
| EPI_ISL_1533697 | Hospital Estadual de Vila Alpina | Instituto Adolfo Lutz, Interdiciplinary Procedures Center, Strategic Laboratory | Claudio Tavares Sacchi, Claudia Regina Gonçalves, Erica Valessa Ramos Gomes, Karoline Rodrigues Campos, Caio Vinicius Dias Lopes, Leonardo Jose Tadeu de Araujo |
| EPI_ISL_1533698 | Secretaria Municipal de Saude de Birigui | Instituto Adolfo Lutz, Interdiciplinary Procedures Center, Strategic Laboratory | Claudio Tavares Sacchi, Claudia Regina Gonçalves, Erica Valessa Ramos Gomes, Karoline Rodrigues Campos, Caio Vinicius Dias Lopes, Leonardo Jose Tadeu de Araujo |
| EPI_ISL_1533699 | Hospital das Clinicas Luzia de Pinho Melo | Instituto Adolfo Lutz, Interdiciplinary Procedures Center, Strategic Laboratory | Claudio Tavares Sacchi, Claudia Regina Gonçalves, Erica Valessa Ramos Gomes, Karoline Rodrigues Campos, Caio Vinicius Dias Lopes, Leonardo Jose Tadeu de Araujo |
| EPI_ISL_1533700 | Hospital Estadual de Sapopemba Sao Paulo | Instituto Adolfo Lutz, Interdiciplinary Procedures Center, Strategic Laboratory | Claudio Tavares Sacchi, Claudia Regina Gonçalves, Erica Valessa Ramos Gomes, Karoline Rodrigues Campos, Caio Vinicius Dias Lopes, Leonardo Jose Tadeu de Araujo |
| EPI_ISL_1533701 | Hospital Estadual de Vila Alpina Org Social Seconci Sao Paulo | Instituto Adolfo Lutz, Interdiciplinary Procedures Center, Strategic Laboratory | Claudio Tavares Sacchi, Claudia Regina Gonçalves, Erica Valessa Ramos Gomes, Karoline Rodrigues Campos, Caio Vinicius Dias Lopes, Leonardo Jose Tadeu de Araujo |
| EPI_ISL_1533702 | Hospital Universitario da USP Sao Paulo | Instituto Adolfo Lutz, Interdiciplinary Procedures Center, Strategic Laboratory | Claudio Tavares Sacchi, Claudia Regina Gonçalves, Erica Valessa Ramos Gomes, Karoline Rodrigues Campos, Caio Vinicius Dias Lopes, Leonardo Jose Tadeu de Araujo |
| EPI_ISL_1533703 | Pronto Atendimento Sao Jose | Instituto Adolfo Lutz, Interdiciplinary Procedures Center, Strategic Laboratory | Claudio Tavares Sacchi, Claudia Regina Gonçalves, Erica Valessa Ramos Gomes, Karoline Rodrigues Campos, Caio Vinicius Dias Lopes, Leonardo Jose Tadeu de Araujo |
| EPI_ISL_1533704 | Santa Casa de Atibaia Pro Saude | Instituto Adolfo Lutz, Interdiciplinary Procedures Center, Strategic Laboratory | Claudio Tavares Sacchi, Claudia Regina Gonçalves, Erica Valessa Ramos Gomes, Karoline Rodrigues Campos, Caio Vinicius Dias Lopes, Leonardo Jose Tadeu de Araujo |
| EPI_ISL_1533705 | Vigilancia em Saude | Instituto Adolfo Lutz, Interdiciplinary Procedures Center, Strategic Laboratory | Claudio Tavares Sacchi, Claudia Regina Gonçalves, Erica Valessa Ramos Gomes, Karoline Rodrigues Campos, Caio Vinicius Dias Lopes, Leonardo Jose Tadeu de Araujo |
| EPI_ISL_1533706 | PS Mun Santana Lauro Ribas Braga | Instituto Adolfo Lutz, Interdiciplinary Procedures Center, Strategic Laboratory | Claudio Tavares Sacchi, Claudia Regina Gonçalves, Erica Valessa Ramos Gomes, Karoline Rodrigues Campos, Caio Vinicius Dias Lopes, Leonardo Jose Tadeu de Araujo |
| EPI_ISL_1533707 | Hosp Mun Planalto Waldomiro de Paula | Instituto Adolfo Lutz, Interdiciplinary Procedures Center, Strategic Laboratory | Claudio Tavares Sacchi, Claudia Regina Gonçalves, Erica Valessa Ramos Gomes, Karoline Rodrigues Campos, Caio Vinicius Dias Lopes, Leonardo Jose Tadeu de Araujo |
| EPI_ISL_1533708 | Hospital Municipal Dr Mario Gatti Campinas | Instituto Adolfo Lutz, Interdiciplinary Procedures Center, Strategic Laboratory | Claudio Tavares Sacchi, Claudia Regina Gonçalves, Erica Valessa Ramos Gomes, Karoline Rodrigues Campos, Caio Vinicius Dias Lopes, Leonardo Jose Tadeu de Araujo |
| EPI_ISL_1533709, EPI_ISL_1533710 | Hospital Estadual de Vila Alpina Org Social Seconci Sao Paulo | Instituto Adolfo Lutz, Interdiciplinary Procedures Center, Strategic Laboratory | Claudio Tavares Sacchi, Claudia Regina Gonçalves, Erica Valessa Ramos Gomes, Karoline Rodrigues Campos, Caio Vinicius Dias Lopes, Leonardo Jose Tadeu de Araujo |
| EPI_ISL_1533711 | Hospital Santa Clara | Instituto Adolfo Lutz, Interdiciplinary Procedures Center, Strategic Laboratory | Claudio Tavares Sacchi, Claudia Regina Gonçalves, Erica Valessa Ramos Gomes, Karoline Rodrigues Campos, Caio Vinicius Dias Lopes, Leonardo Jose Tadeu de Araujo |
| EPI_ISL_1533712 | Hospital Universitario da USP Sao Paulo | Instituto Adolfo Lutz, Interdiciplinary Procedures Center, Strategic Laboratory | Claudio Tavares Sacchi, Claudia Regina Gonçalves, Erica Valessa Ramos Gomes, Karoline Rodrigues Campos, Caio Vinicius Dias Lopes, Leonardo Jose Tadeu de Araujo |

| | | | |
|---|---|---|---|
| EPI_ISL_1533713 | Unidade de Pronto Atendimento Jd Amanda | Instituto Adolfo Lutz, Interdiciplinary Procedures Center, Strategic Laboratory | Claudio Tavares Sacchi, Claudia Regina Gonçalves, Erica Valessa Ramos Gomes, Karoline Rodrigues Campos, Caio Vinicius Dias Lopes, Leonardo Jose Tadeu de Araujo |
| EPI_ISL_1533714 | Pronto Socorro Dr Osmar Mesquita | Instituto Adolfo Lutz, Interdiciplinary Procedures Center, Strategic Laboratory | Claudio Tavares Sacchi, Claudia Regina Gonçalves, Erica Valessa Ramos Gomes, Karoline Rodrigues Campos, Caio Vinicius Dias Lopes, Leonardo Jose Tadeu de Araujo |
| EPI_ISL_1533715 | Santa Casa de Aracatuba Hospital Sagrado Coracao de Jesus | Instituto Adolfo Lutz, Interdiciplinary Procedures Center, Strategic Laboratory | Claudio Tavares Sacchi, Claudia Regina Gonçalves, Erica Valessa Ramos Gomes, Karoline Rodrigues Campos, Caio Vinicius Dias Lopes, Leonardo Jose Tadeu de Araujo |
| EPI_ISL_1533716 | Vigilancia Em Saude | Instituto Adolfo Lutz, Interdiciplinary Procedures Center, Strategic Laboratory | Claudio Tavares Sacchi, Claudia Regina Gonçalves, Erica Valessa Ramos Gomes, Karoline Rodrigues Campos, Caio Vinicius Dias Lopes, Leonardo Jose Tadeu de Araujo |
| EPI_ISL_1533717 | Unidade de Pronto Atendimento Jd Amanda | Instituto Adolfo Lutz, Interdiciplinary Procedures Center, Strategic Laboratory | Claudio Tavares Sacchi, Claudia Regina Gonçalves, Erica Valessa Ramos Gomes, Karoline Rodrigues Campos, Caio Vinicius Dias Lopes, Leonardo Jose Tadeu de Araujo |
| EPI_ISL_1533718 | Servico de Verificacao de Obitos Svo Guarulhos | Instituto Adolfo Lutz, Interdiciplinary Procedures Center, Strategic Laboratory | Claudio Tavares Sacchi, Claudia Regina Gonçalves, Erica Valessa Ramos Gomes, Karoline Rodrigues Campos, Caio Vinicius Dias Lopes, Leonardo Jose Tadeu de Araujo |
| EPI_ISL_1533719 | Policlinica Munic da Est Turistica de Holambra | Instituto Adolfo Lutz, Interdiciplinary Procedures Center, Strategic Laboratory | Claudio Tavares Sacchi, Claudia Regina Gonçalves, Erica Valessa Ramos Gomes, Karoline Rodrigues Campos, Caio Vinicius Dias Lopes, Leonardo Jose Tadeu de Araujo |
| EPI_ISL_1533720 | Santa Casa de Penapolis | Instituto Adolfo Lutz, Interdiciplinary Procedures Center, Strategic Laboratory | Claudio Tavares Sacchi, Claudia Regina Gonçalves, Erica Valessa Ramos Gomes, Karoline Rodrigues Campos, Caio Vinicius Dias Lopes, Leonardo Jose Tadeu de Araujo |
| EPI_ISL_1533721 | Santa Casa de Aracatuba Hospital Sagrado Coracao De Jesus | Instituto Adolfo Lutz, Interdiciplinary Procedures Center, Strategic Laboratory | Claudio Tavares Sacchi, Claudia Regina Gonçalves, Erica Valessa Ramos Gomes, Karoline Rodrigues Campos, Caio Vinicius Dias Lopes, Leonardo Jose Tadeu de Araujo |
| EPI_ISL_1533722 | Secretaria Municipal de Saude De Piracaia | Instituto Adolfo Lutz, Interdiciplinary Procedures Center, Strategic Laboratory | Claudio Tavares Sacchi, Claudia Regina Gonçalves, Erica Valessa Ramos Gomes, Karoline Rodrigues Campos, Caio Vinicius Dias Lopes, Leonardo Jose Tadeu de Araujo |
| EPI_ISL_1533723 | Hospital Geral de Itaquaquecetuba | Instituto Adolfo Lutz, Interdiciplinary Procedures Center, Strategic Laboratory | Claudio Tavares Sacchi, Claudia Regina Gonçalves, Erica Valessa Ramos Gomes, Karoline Rodrigues Campos, Caio Vinicius Dias Lopes, Leonardo Jose Tadeu de Araujo |
| EPI_ISL_1533724 | Diretoria Municipal de Saude | Instituto Adolfo Lutz, Interdiciplinary Procedures Center, Strategic Laboratory | Claudio Tavares Sacchi, Claudia Regina Gonçalves, Erica Valessa Ramos Gomes, Karoline Rodrigues Campos, Caio Vinicius Dias Lopes, Leonardo Jose Tadeu de Araujo |
| EPI_ISL_1533725 | Hospital Geral de Guarulhos | Instituto Adolfo Lutz, Interdiciplinary Procedures Center, Strategic Laboratory | Claudio Tavares Sacchi, Claudia Regina Gonçalves, Erica Valessa Ramos Gomes, Karoline Rodrigues Campos, Caio Vinicius Dias Lopes, Leonardo Jose Tadeu de Araujo |
| EPI_ISL_1533726 | Hospital Estadual de Campanha Covid 19 Barradas | Instituto Adolfo Lutz, Interdiciplinary Procedures Center, Strategic Laboratory | Claudio Tavares Sacchi, Claudia Regina Gonçalves, Erica Valessa Ramos Gomes, Karoline Rodrigues Campos, Caio Vinicius Dias Lopes, Leonardo Jose Tadeu de Araujo |
| EPI_ISL_1533727 | Hospital Universitario da USP Sao Paulo | Instituto Adolfo Lutz, Interdiciplinary Procedures Center, Strategic Laboratory | Claudio Tavares Sacchi, Claudia Regina Gonçalves, Erica Valessa Ramos Gomes, Karoline Rodrigues Campos, Caio Vinicius Dias Lopes, Leonardo Jose Tadeu de Araujo |
| EPI_ISL_1533977 | Laboratorio de Virologia Molecular / UFRJ | Laboratory of Respiratory Viruses and Measles, Oswaldo Cruz Institute, FIOCRUZ | Paola Resende, Carolina M Voloch, Luciana Appolinario, Fernando Motta, Anna Carolina Paixao, Ana Carolina Mendonca, Alice Sampaio Rocha, Renata Serrano Lopes, Amilcar Tanuri, Marilda Siqueira on behalf of the Fiocruz COVID-19 Genomic Surveillance Network |
| EPI_ISL_1533978 | Laboratorio Central de Saude Publica do Estado do Parana (LACEN-PR) | Laboratory of Respiratory Viruses and Measles, Oswaldo Cruz Institute, FIOCRUZ | Paola Resende, Luciana Appolinario, Fernando Motta, Anna Carolina Paixao, Ana Carolina Mendonca, Alice Sampaio Rocha, Renata Serrano Lopes, Maria do Carmo Debur, Irina Nastassja Riediger, Marilda Siqueira on behalf of the Fiocruz COVID-19 Genomic Surveillance Network |
| EPI_ISL_1533979 | Laboratorio Central de Saude Publica do Estado do Rio Grande do Sul (LACEN-RS) | Laboratory of Respiratory Viruses and Measles, Oswaldo Cruz Institute, FIOCRUZ | Paola Resende, Luciana Appolinario, Fernando Motta, Anna Carolina Paixao, Ana Carolina Mendonca, Alice Sampaio Rocha, Renata Serrano Lopes, Richard Salvato, Tatiana Schaffer Gregianini, Marilda Siqueira on behalf of the Fiocruz COVID-19 Genomic Surveillance Network |
| EPI_ISL_1533980, EPI_ISL_1533983 | Laboratory of Respiratory Viruses and Measles, Oswaldo Cruz Institute, FIOCRUZ | Laboratory of Respiratory Viruses and Measles, Oswaldo Cruz Institute, FIOCRUZ | Paola Resende, Luciana Appolinario, Fernando Motta, Anna Carolina Paixao, Ana Carolina Mendonca, Alice Sampaio Rocha, Renata Serrano Lopes, Marilda Siqueira on behalf of the Fiocruz COVID-19 Genomic Surveillance Network |
| EPI_ISL_1533990 | Laboratorio Central de Saude Publica do Estado de Santa Catarina (LACEN-SC) | Laboratory of Respiratory Viruses and Measles, Oswaldo Cruz Institute, FIOCRUZ | Paola Resende, Luciana Appolinario, Fernando Motta, Anna Carolina Paixao, Ana Carolina Mendonca, Alice Sampaio Rocha, Renata Serrano Lopes, Darcita Buerger Rovaris, Sandra Bianchini Fernandes, Marilda Siqueira on behalf of the Fiocruz COVID-19 Genomic Surveillance Network |
| EPI_ISL_1533991 | Laboratorio de Virologia Molecular / UFRJ | Laboratory of Respiratory Viruses and Measles, Oswaldo Cruz Institute, FIOCRUZ | Paola Resende, Carolina M Voloch, Luciana Appolinario, Fernando Motta, Anna Carolina Paixao, Ana Carolina Mendonca, Alice Sampaio Rocha, Renata Serrano Lopes, Amilcar Tanuri, Marilda Siqueira on behalf of the Fiocruz COVID-19 Genomic Surveillance Network |
| EPI_ISL_1533992, EPI_ISL_1533993, EPI_ISL_1533994 | Laboratorio Central de Saude Publica do Estado de Santa Catarina (LACEN-SC) | Laboratory of Respiratory Viruses and Measles, Oswaldo Cruz Institute, FIOCRUZ | Paola Resende, Luciana Appolinario, Fernando Motta, Anna Carolina Paixao, Ana Carolina Mendonca, Alice Sampaio Rocha, Renata Serrano Lopes, Darcita Buerger Rovaris, Sandra Bianchini Fernandes, Marilda Siqueira on behalf of the Fiocruz COVID-19 Genomic Surveillance Network |
| EPI_ISL_1533995 | Laboratorio de Virologia Molecular / UFRJ | Laboratory of Respiratory Viruses and Measles, Oswaldo Cruz Institute, FIOCRUZ | Paola Resende, Carolina M Voloch, Luciana Appolinario, Fernando Motta, Anna Carolina Paixao, Ana Carolina Mendonca, Alice Sampaio Rocha, Renata Serrano Lopes, Amilcar Tanuri, Marilda Siqueira on behalf of the Fiocruz COVID-19 Genomic Surveillance Network |
| EPI_ISL_1533996, EPI_ISL_1533997 | Laboratorio Central de Saude Publica do Estado de Santa Catarina (LACEN-SC) | Laboratory of Respiratory Viruses and Measles, Oswaldo Cruz Institute, FIOCRUZ | Paola Resende, Luciana Appolinario, Fernando Motta, Anna Carolina Paixao, Ana Carolina Mendonca, Alice Sampaio Rocha, Renata Serrano Lopes, Darcita Buerger Rovaris, Sandra Bianchini Fernandes, Marilda Siqueira on behalf of the Fiocruz COVID-19 Genomic Surveillance Network |
| EPI_ISL_1533998 | Laboratorio de Virologia Molecular / UFRJ | Laboratory of Respiratory Viruses and Measles, Oswaldo Cruz Institute, FIOCRUZ | Paola Resende, Carolina M Voloch, Luciana Appolinario, Fernando Motta, Anna Carolina Paixao, Ana Carolina Mendonca, Alice Sampaio Rocha, Renata Serrano Lopes, Amilcar Tanuri, Marilda Siqueira on behalf of the Fiocruz COVID-19 Genomic Surveillance Network |
| EPI_ISL_1533999, EPI_ISL_1534000, EPI_ISL_1534001 | Laboratorio Central de Saude Publica do Estado do Parana (LACEN-PR) | Laboratory of Respiratory Viruses and Measles, Oswaldo Cruz Institute, FIOCRUZ | Paola Resende, Luciana Appolinario, Fernando Motta, Anna Carolina Paixao, Ana Carolina Mendonca, Alice Sampaio Rocha, Renata Serrano Lopes, Maria do Carmo Debur, Irina Nastassja Riediger, Marilda Siqueira on behalf of the Fiocruz COVID-19 Genomic Surveillance Network |
| EPI_ISL_1534002, EPI_ISL_1534003 | Laboratorio Central de Saude Publica do Estado de Santa Catarina (LACEN-SC) | Laboratory of Respiratory Viruses and Measles, Oswaldo Cruz Institute, FIOCRUZ | Paola Resende, Luciana Appolinario, Fernando Motta, Anna Carolina Paixao, Ana Carolina Mendonca, Alice Sampaio Rocha, Renata Serrano Lopes, Darcita Buerger Rovaris, Sandra Bianchini Fernandes, Marilda Siqueira on behalf of the Fiocruz COVID-19 Genomic Surveillance Network |
| EPI_ISL_1534004 | Laboratorio Central de Saude Publica do Estado de Sergipe (LACEN-SE) | Laboratory of Respiratory Viruses and Measles, Oswaldo Cruz Institute, FIOCRUZ | Paola Resende, Luciana Appolinario, Fernando Motta, Anna Carolina Paixao, Ana Carolina Mendonca, Alice Sampaio Rocha, Renata Serrano Lopes, Cliomar Alves dos Santos, Marilda Siqueira on behalf of the Fiocruz COVID-19 Genomic Surveillance Network |
| EPI_ISL_1534005 | Laboratory of Respiratory Viruses and Measles, Oswaldo Cruz Institute, FIOCRUZ | Laboratory of Respiratory Viruses and Measles, Oswaldo Cruz Institute, FIOCRUZ | Paola Resende, Luciana Appolinario, Fernando Motta, Anna Carolina Paixao, Ana Carolina Mendonca, Alice Sampaio Rocha, Renata Serrano Lopes, Marilda Siqueira on behalf of the Fiocruz COVID-19 Genomic Surveillance Network |
| EPI_ISL_1534006, EPI_ISL_1534007, EPI_ISL_1534008, EPI_ISL_1534009, EPI_ISL_1534010 | Laboratorio Central de Saude Publica do Estado de Santa Catarina (LACEN-SC) | Laboratory of Respiratory Viruses and Measles, Oswaldo Cruz Institute, FIOCRUZ | Paola Resende, Luciana Appolinario, Fernando Motta, Anna Carolina Paixao, Ana Carolina Mendonca, Alice Sampaio Rocha, Renata Serrano Lopes, Darcita Buerger Rovaris, Sandra Bianchini Fernandes, Marilda Siqueira on behalf of the Fiocruz COVID-19 Genomic Surveillance Network |
| EPI_ISL_1534011 | Laboratory of Respiratory Viruses and Measles, Oswaldo Cruz Institute, FIOCRUZ | Laboratory of Respiratory Viruses and Measles, Oswaldo Cruz Institute, FIOCRUZ | Paola Resende, Luciana Appolinario, Fernando Motta, Anna Carolina Paixao, Ana Carolina Mendonca, Alice Sampaio Rocha, Renata Serrano Lopes, Marilda Siqueira on behalf of the Fiocruz COVID-19 Genomic Surveillance Network |
| EPI_ISL_1534012 | Laboratorio Central de Saude Publica do Estado do Rio de Janeiro (LACEN-RJ) | Laboratory of Respiratory Viruses and Measles, Oswaldo Cruz Institute, FIOCRUZ | Paola Resende, Luciana Appolinario, Fernando Motta, Anna Carolina Paixao, Ana Carolina Mendonca, Alice Sampaio Rocha, Renata Serrano Lopes, Andrea Cony Cavalcanti, Marilda Siqueira on behalf of the Fiocruz COVID-19 Genomic Surveillance Network |
| EPI_ISL_1534013 | Laboratorio Central de Saude Publica do Estado do Rio Grande do Sul (LACEN-RS) | Laboratory of Respiratory Viruses and Measles, Oswaldo Cruz Institute, FIOCRUZ | Paola Resende, Luciana Appolinario, Fernando Motta, Anna Carolina Paixao, Ana Carolina Mendonca, Alice Sampaio Rocha, Renata Serrano Lopes, Richard Salvato, Tatiana Schaffer Gregianini, Marilda Siqueira on behalf of the Fiocruz COVID-19 Genomic Surveillance Network |
| EPI_ISL_1534014, EPI_ISL_1534015, EPI_ISL_1534016 | Laboratory of Respiratory Viruses and Measles, Oswaldo Cruz Institute, FIOCRUZ | Laboratory of Respiratory Viruses and Measles, Oswaldo Cruz Institute, FIOCRUZ | Paola Resende, Luciana Appolinario, Fernando Motta, Anna Carolina Paixao, Ana Carolina Mendonca, Alice Sampaio Rocha, Renata Serrano Lopes, Marilda Siqueira on behalf of the Fiocruz COVID-19 Genomic Surveillance Network |
| EPI_ISL_1550388 | Instituto Oswaldo Cruz | Laboratorio de Virologia, Faculdade de Medicina, | María de Fátima Ferreira, Renata Dezengrini Slhessarenko, Janeth Aracely Ramirez Pavon, Luciano Nakazato, Valeria Dutra, Rosane Christiane Hahn, |

| | | | |
|---|---|---|---|
| | | Universidade Federal de Mato Grosso, campus Cuiabá | Paola Cristina Resende |
| EPI_ISL_1557222 | Instituto de Medicina Tropical de Sao Paulo | Instituto de Medicina Tropical de Sao Paulo | Camila Malta Romano, Lucy Santos Vilas Boas, Maria Cassia J Mendes-Correa |
| EPI_ISL_1580504 | Laboratório de Biologia Molecular do Hospital das Clínicas da Faculdade de Medicina de Botucatu/SP | Laboratórios de Genômica Funcional (FCA/UNESP) e Biologia Molecular (FMB-HC/UNESP) - Rede de Vigilância Genômica (Vigenômica)/UNESP | Patrícia Akemi Assato; Felipe Allan da Silva da Costa; Bianca Cechetto Carlos; Flavia Hebner Barbosa Trovão; Guilherme Targino Valente; Rejane Maria Tommasini Grotto; Jayme A. Souza-Neto. |
| EPI_ISL_1583640, EPI_ISL_1583641, EPI_ISL_1583642, EPI_ISL_1583643, EPI_ISL_1583644, EPI_ISL_1583645, EPI_ISL_1583646, EPI_ISL_1583647, EPI_ISL_1583648, EPI_ISL_1583649, EPI_ISL_1583650, EPI_ISL_1583651, EPI_ISL_1583652, EPI_ISL_1583653, EPI_ISL_1583654, EPI_ISL_1583655, EPI_ISL_1583656, EPI_ISL_1583657, EPI_ISL_1583658, EPI_ISL_1583659, EPI_ISL_1583660, EPI_ISL_1583661, EPI_ISL_1583662, EPI_ISL_1583663, EPI_ISL_1583664, EPI_ISL_1583665, EPI_ISL_1583666, EPI_ISL_1583667, EPI_ISL_1583668, EPI_ISL_1583669, EPI_ISL_1583670, EPI_ISL_1583671, EPI_ISL_1583672, EPI_ISL_1583673, EPI_ISL_1583674 | | | |
| see above | Central Public Health Laboratory - LACEN -Bahia, Salvador, Brazil | Central Public Health Laboratory - LACEN -Bahia, Salvador, Brazil | Stephane Tosta, Luciana Oliveira, Vanessa Nardy,Patrícia Cajado,Marcela Gómez, Breno Dominguez, Jaqueline Gomes, Vagner Fonseca,Marta Giovanetti,Luiz Alcantara, Felicidade Pereira, Arabela Leal |
| EPI_ISL_1583675 | DNA Laboratory | Central Public Health Laboratory - LACEN -Bahia, Salvador, Brazil | Stephane Tosta, Luciana Oliveira, Vanessa Nardy,Patrícia Cajado,Marcela Gómez, Breno Dominguez, Jaqueline Gomes, Vagner Fonseca,Marta Giovanetti,Luiz Alcantara, Felicidade Pereira, Arabela Leal |
| EPI_ISL_1583676, EPI_ISL_1583677, EPI_ISL_1583678, EPI_ISL_1583679, EPI_ISL_1583680, EPI_ISL_1583681, EPI_ISL_1583682, EPI_ISL_1583683, EPI_ISL_1583686, EPI_ISL_1583689, EPI_ISL_1583691, EPI_ISL_1583694, EPI_ISL_1583697, EPI_ISL_1583700, EPI_ISL_1583703, EPI_ISL_1583705, EPI_ISL_1583708, EPI_ISL_1583710, EPI_ISL_1583714, EPI_ISL_1583716, EPI_ISL_1583719, EPI_ISL_1583722, EPI_ISL_1583725, EPI_ISL_1583727, EPI_ISL_1583730, EPI_ISL_1583733 | | | |
| see above | Central Public Health Laboratory - LACEN -Bahia, Salvador, Brazil | Central Public Health Laboratory - LACEN -Bahia, Salvador, Brazil | Stephane Tosta, Luciana Oliveira, Vanessa Nardy,Patrícia Cajado,Marcela Gómez, Breno Dominguez, Jaqueline Gomes, Vagner Fonseca,Marta Giovanetti,Luiz Alcantara, Felicidade Pereira, Arabela Leal |
| EPI_ISL_1592227, EPI_ISL_1592228, EPI_ISL_1592229, EPI_ISL_1592230 | Laboratório Hermes Pardini | Laboratório de Biologia Integrativa, Instituto de Ciências Biológicas, Universidade Federal de Minas Gerais | Filipe Romero Rebello Moreira, Diego Menezes Bonfim, Danielle Alves Gomes Zauli, Joice do Prado Silva, Aline Brito de Lima, Frederico Scott Varella Malta, Alessandro Clayton de Souza Ferreira, Victor Cavalcanti Pardini, Daniel Costa Queiroz, Rafael Marques de Souza, Victor Emmanuel Viana Geddes, Walyson Coelho Costa, Wagner Carlos Santos Magalhaes, Rennan Garcias Moreira, Carolina Moreira Voloch, Renan Pedra de Souza, Renato Santana Aguiar. |
| EPI_ISL_1608161, EPI_ISL_1608162, EPI_ISL_1608163, EPI_ISL_1608164, EPI_ISL_1608165, EPI_ISL_1608166, EPI_ISL_1608167, EPI_ISL_1608168, EPI_ISL_1608169, EPI_ISL_1608170, EPI_ISL_1608171 | | | |
| see above | Hospital São Rafael - IDOR | Hospital São Rafael - IDOR | Carolina Kymie Vasques Nonaka, Tiago Gräf, Camila Araújo de Lorenzo Barcia, Vanessa Ferreira Costa, Janderson Lopes de Oliveira, Rogério da Hora Passos, Clarissa Araújo Gurgel Rocha, Iasmin Nogueira Bastos, Maria Clara Brito de Santana, Ian Marinho Santos, Karoline Almeida Felix de Sousa, Isadora Cristina de Siqueira, Thamires Gomes Lopes Weber, Ana Verena Almeida Mendes, Bruno Solano de Freitas Souza |
| EPI_ISL_1620638 | Hospital Municipal Dr. Ignacio Proença de Gouvea | Instituto Butantan / Mendelics | Alexander Roberto Precioso, Dimas Tadeu Covas, Sandra Coccuzzo Sampaio, Maria Carolina Elias, José Salvatore Leister Patané, Vincent Louis Viala, Antonio Jorge Martins, Ricardo Haddad, Claudia Renata dos Santos Barros, Elaine Cristina Marqueze, Raul Machado Neto, Debora Botequio Moretti, Bibiana Santos, João Paulo Kitajima, Erika Freitas, David Schlesinger, Simone Kashima, Evandra Strazza Rodrigues, Svetoslav Nanev Slavov, Elaine Vieira dos Santos, Rafael dos Santos Bezerra, Luiz Carlos Junior de Alcantara, Marta Giovanetti, Vagner Fonseca, Flavia Aburjaile, Rodrigo Tocantins Calado. |
| EPI_ISL_1620639 | Fleury | Instituto Butantan / Mendelics | Alexander Roberto Precioso, Dimas Tadeu Covas, Sandra Coccuzzo Sampaio, Maria Carolina Elias, José Salvatore Leister Patané, Vincent Louis Viala, Antonio Jorge Martins, Ricardo Haddad, Claudia Renata dos Santos Barros, Elaine Cristina Marqueze, Raul Machado Neto, Debora Botequio Moretti, Bibiana Santos, João Paulo Kitajima, Erika Freitas, David Schlesinger, Simone Kashima, Evandra Strazza Rodrigues, Svetoslav Nanev Slavov, Elaine Vieira dos Santos, Rafael dos Santos Bezerra, Luiz Carlos Junior de Alcantara, Marta Giovanetti, Vagner Fonseca, Flavia Aburjaile, Rodrigo Tocantins Calado. |
| EPI_ISL_1625972 | Ama J Angela | Instituto Adolfo Lutz, Interdiciplinary Procedures Center, Strategic Laboratory | Claudio Tavares Sacchi, Claudia Regina Gonçalves, Erica Valessa Ramos Gomes, Karoline Rodrigues Campos, Caio Vinicius Dias Lopes, Leonardo Jose Tadeu de Araujo, Katia Correa de Oliveira Santos |
| EPI_ISL_1625973, EPI_ISL_1625974 | Santa Casa de Aracatuba Hospital Sagrado Coracao de Jesus | Instituto Adolfo Lutz, Interdiciplinary Procedures Center, Strategic Laboratory | Claudio Tavares Sacchi, Claudia Regina Gonçalves, Erica Valessa Ramos Gomes, Karoline Rodrigues Campos, Caio Vinicius Dias Lopes, Leonardo Jose Tadeu de Araujo, Katia Correa de Oliveira Santos |
| EPI_ISL_1625975 | CS II Dr Antonio Vicoso Moreira De Rezende Sumare | Instituto Adolfo Lutz, Interdiciplinary Procedures Center, Strategic Laboratory | Claudio Tavares Sacchi, Claudia Regina Gonçalves, Erica Valessa Ramos Gomes, Karoline Rodrigues Campos, Caio Vinicius Dias Lopes, Leonardo Jose Tadeu de Araujo, Katia Correa de Oliveira Santos |
| EPI_ISL_1625976 | Centro de Saude III de Divinolandia | Instituto Adolfo Lutz, Interdiciplinary Procedures Center, Strategic Laboratory | Claudio Tavares Sacchi, Claudia Regina Gonçalves, Erica Valessa Ramos Gomes, Karoline Rodrigues Campos, Caio Vinicius Dias Lopes, Leonardo Jose Tadeu de Araujo, Katia Correa de Oliveira Santos |
| EPI_ISL_1625977, EPI_ISL_1625978, EPI_ISL_1625979, EPI_ISL_1625980, EPI_ISL_1625981, EPI_ISL_1625982, EPI_ISL_1625983, EPI_ISL_1625984, EPI_ISL_1625985, EPI_ISL_1625986, EPI_ISL_1625987, EPI_ISL_1625988, EPI_ISL_1625989, EPI_ISL_1625990, EPI_ISL_1625991, EPI_ISL_1625992, EPI_ISL_1625993, EPI_ISL_1625994, EPI_ISL_1625995, EPI_ISL_1625996, EPI_ISL_1625997, EPI_ISL_1625998, EPI_ISL_1625999, EPI_ISL_1626000, EPI_ISL_1626001, EPI_ISL_1626002, EPI_ISL_1626003, EPI_ISL_1626004, EPI_ISL_1626005, EPI_ISL_1626006, EPI_ISL_1626007, EPI_ISL_1626008, EPI_ISL_1626009, EPI_ISL_1626606 | | | |
| see above | Instituto Adolfo Lutz - Regional de Rio Claro | Instituto Adolfo Lutz, Interdiciplinary Procedures Center, Strategic Laboratory | Claudio Tavares Sacchi, Claudia Regina Gonçalves, Erica Valessa Ramos Gomes, Karoline Rodrigues Campos, Caio Vinicius Dias Lopes, Leonardo Jose Tadeu de Araujo, Katia Correa de Oliveira Santos |
| EPI_ISL_1628344 | Hospital Geral de Pedreira | Instituto Adolfo Lutz, Interdiciplinary Procedures Center, Strategic Laboratory | Claudio Tavares Sacchi, Claudia Regina Gonçalves, Erica Valessa Ramos Gomes, Karoline Rodrigues Campos, Caio Vinicius Dias Lopes, Leonardo Jose Tadeu de Araujo, Katia Correa de Oliveira Santos |
| EPI_ISL_1628345, EPI_ISL_1628346 | Instituto Adolfo Lutz - Regional de Rio Claro | Instituto Adolfo Lutz, Interdiciplinary Procedures Center, Strategic Laboratory | Claudio Tavares Sacchi, Claudia Regina Gonçalves, Erica Valessa Ramos Gomes, Karoline Rodrigues Campos, Caio Vinicius Dias Lopes, Leonardo Jose Tadeu de Araujo, Katia Correa de Oliveira Santos |
| EPI_ISL_1628347, EPI_ISL_1628348, EPI_ISL_1628349, EPI_ISL_1628350, EPI_ISL_1628351, EPI_ISL_1628352, EPI_ISL_1628353, EPI_ISL_1628354, EPI_ISL_1628355, EPI_ISL_1628356, EPI_ISL_1628357, EPI_ISL_1628358, EPI_ISL_1628359, EPI_ISL_1628360, EPI_ISL_1628361, EPI_ISL_1628362 | | | |
| see above | Instituto Adolfo Lutz Central | Instituto Adolfo Lutz, Interdiciplinary Procedures Center, Strategic Laboratory | Claudio Tavares Sacchi, Claudia Regina Gonçalves, Erica Valessa Ramos Gomes, Karoline Rodrigues Campos, Caio Vinicius Dias Lopes, Leonardo Jose Tadeu de Araujo, Katia Correa de Oliveira Santos |
| EPI_ISL_1628363, EPI_ISL_1628364, EPI_ISL_1628365 | LACEN do Estado de Goias | Instituto Adolfo Lutz, Interdiciplinary Procedures Center, Strategic Laboratory | Claudio Tavares Sacchi, Claudia Regina Gonçalves, Erica Valessa Ramos Gomes, Karoline Rodrigues Campos, Caio Vinicius Dias Lopes, Leonardo Jose Tadeu de Araujo, Katia Correa de Oliveira Santos |
| EPI_ISL_1628366 | CS III de Patrocinio Paulista | Instituto Adolfo Lutz, Interdiciplinary Procedures Center, Strategic Laboratory | Claudio Tavares Sacchi, Claudia Regina Gonçalves, Erica Valessa Ramos Gomes, Karoline Rodrigues Campos, Caio Vinicius Dias Lopes, Leonardo Jose Tadeu de Araujo, Katia Correa de Oliveira Santos |
| EPI_ISL_1628367 | Santa Casa de Guaira | Instituto Adolfo Lutz, Interdiciplinary Procedures Center, Strategic Laboratory | Claudio Tavares Sacchi, Claudia Regina Gonçalves, Erica Valessa Ramos Gomes, Karoline Rodrigues Campos, Caio Vinicius Dias Lopes, Leonardo Jose Tadeu de Araujo, Katia Correa de Oliveira Santos |
| EPI_ISL_1628368 | Centro Medico Social Comunitario Januario Teodoro de Souza | Instituto Adolfo Lutz, Interdiciplinary Procedures Center, Strategic Laboratory | Claudio Tavares Sacchi, Claudia Regina Gonçalves, Erica Valessa Ramos Gomes, Karoline Rodrigues Campos, Caio Vinicius Dias Lopes, Leonardo Jose Tadeu de Araujo, Katia Correa de Oliveira Santos |
| EPI_ISL_1628369 | UPA Dr Luis Atilio Losi Viana Ribeirao Preto | Instituto Adolfo Lutz, Interdiciplinary Procedures Center, Strategic Laboratory | Claudio Tavares Sacchi, Claudia Regina Gonçalves, Erica Valessa Ramos Gomes, Karoline Rodrigues Campos, Caio Vinicius Dias Lopes, Leonardo Jose Tadeu de Araujo, Katia Correa de Oliveira Santos |
| EPI_ISL_1628370 | Sae Servico De Atendimento Especializado | Instituto Adolfo Lutz, Interdiciplinary Procedures Center, Strategic Laboratory | Claudio Tavares Sacchi, Claudia Regina Gonçalves, Erica Valessa Ramos Gomes, Karoline Rodrigues Campos, Caio Vinicius Dias Lopes, Leonardo Jose Tadeu de Araujo, Katia Correa de Oliveira Santos |
| EPI_ISL_1628371 | Centro De Saude II Ibitinga | Instituto Adolfo Lutz, Interdiciplinary Procedures Center, Strategic Laboratory | Claudio Tavares Sacchi, Claudia Regina Gonçalves, Erica Valessa Ramos Gomes, Karoline Rodrigues Campos, Caio Vinicius Dias Lopes, Leonardo Jose Tadeu de Araujo, Katia Correa de Oliveira Santos |
| EPI_ISL_1628372 | Hospital Sao Marcos da Sama Morro Agudo | Instituto Adolfo Lutz, Interdiciplinary Procedures Center, Strategic Laboratory | Claudio Tavares Sacchi, Claudia Regina Gonçalves, Erica Valessa Ramos Gomes, Karoline Rodrigues Campos, Caio Vinicius Dias Lopes, Leonardo Jose Tadeu de Araujo, Katia Correa de Oliveira Santos |
| EPI_ISL_1628373 | UPA De Bebedouro | Instituto Adolfo Lutz, Interdiciplinary Procedures Center, Strategic Laboratory | Claudio Tavares Sacchi, Claudia Regina Gonçalves, Erica Valessa Ramos Gomes, Karoline Rodrigues Campos, Caio Vinicius Dias Lopes, Leonardo Jose Tadeu de Araujo, Katia Correa de Oliveira Santos |
| EPI_ISL_1628374 | Santa Casa De Cravinhos | Instituto Adolfo Lutz, Interdiciplinary Procedures Center, | Claudio Tavares Sacchi, Claudia Regina Gonçalves, Erica Valessa Ramos Gomes, Karoline Rodrigues Campos, Caio Vinicius Dias Lopes, Leonardo Jose |

| | | | |
|---|---|---|---|
| | | Strategic Laboratory | Tadeu de Araujo, Katia Correa de Oliveira Santos |
| EPI_ISL_1628375, EPI_ISL_1628376 | Secretaria Municipal de Saude de Guariba | Instituto Adolfo Lutz, Interdiciplinary Procedures Center, Strategic Laboratory | Claudio Tavares Sacchi, Claudia Regina Gonçalves, Erica Valessa Ramos Gomes, Karoline Rodrigues Campos, Caio Vinicius Dias Lopes, Leonardo Jose Tadeu de Araujo, Katia Correa de Oliveira Santos |
| EPI_ISL_1628377 | Centro De Saude II Ibitinga | Instituto Adolfo Lutz, Interdiciplinary Procedures Center, Strategic Laboratory | Claudio Tavares Sacchi, Claudia Regina Gonçalves, Erica Valessa Ramos Gomes, Karoline Rodrigues Campos, Caio Vinicius Dias Lopes, Leonardo Jose Tadeu de Araujo, Katia Correa de Oliveira Santos |
| EPI_ISL_1628378 | Vigilancia Epidemiologica Jardinopolis | Instituto Adolfo Lutz, Interdiciplinary Procedures Center, Strategic Laboratory | Claudio Tavares Sacchi, Claudia Regina Gonçalves, Erica Valessa Ramos Gomes, Karoline Rodrigues Campos, Caio Vinicius Dias Lopes, Leonardo Jose Tadeu de Araujo, Katia Correa de Oliveira Santos |
| EPI_ISL_1628379 | UPA Dr Luis Atilio Losi Viana Ribeirao Preto | Instituto Adolfo Lutz, Interdiciplinary Procedures Center, Strategic Laboratory | Claudio Tavares Sacchi, Claudia Regina Gonçalves, Erica Valessa Ramos Gomes, Karoline Rodrigues Campos, Caio Vinicius Dias Lopes, Leonardo Jose Tadeu de Araujo, Katia Correa de Oliveira Santos |
| EPI_ISL_1628380 | Santa Casa De Pitangueiras | Instituto Adolfo Lutz, Interdiciplinary Procedures Center, Strategic Laboratory | Claudio Tavares Sacchi, Claudia Regina Gonçalves, Erica Valessa Ramos Gomes, Karoline Rodrigues Campos, Caio Vinicius Dias Lopes, Leonardo Jose Tadeu de Araujo, Katia Correa de Oliveira Santos |
| EPI_ISL_1629809 | Laboratório de Microbiologia Molecular - Universidade FEEVALE | Molecular Microbiology Laboratory | Alana Witt Hansen, Fágner Henrique Heldt, Fernando Rosado Spilki, Flávio Silveira, Juliana Schons Gulart, Juliane Deise Fleck, Mariana Soares da Silva, Meriane Demoliner, Matheus Nunes Weber, Paula Rodrigues de Almeida, Michele Filippi. |
| EPI_ISL_1630158 | Laboratório de Microbiologia Molecular - Universidade FEEVALE | Laboratório de Microbiologia Molecular - Universidade FEEVALE | Alana Witt Hansen, Fágner Henrique Heldt, Fernando Rosado Spilki, Flávio Silveira, Juliana Schons Gularte, Juliane Deise Fleck, Mariana Soares da Silva, Meriane Demoliner, Matheus Nunes Weber, Paula Rodrigues de Almeida, Micheli Filippi. |
| EPI_ISL_1652237 | Fleury | Instituto Butantan / Mendelics | Alexander Roberto Precioso, Dimas Tadeu Covas, Sandra Coccuzzo Sampaio, Maria Carolina Elias, José Salvatore Leister Patané, Vincent Louis Viala, Antonio Jorge Martins, Ricardo Haddad, Claudia Renata dos Santos Barros, Elaine Cristina Marqueze, Raul Machado Neto, Debora Botequio Moretti, Bibiana Santos, João Paulo Kitajima, Erika Freitas, David Schlesinger, Simone Kashima, Evandra Strazza Rodrigues, Svetoslav Nanev Slavov, Elaine Vieira dos Santos, Rafael dos Santos Bezerra, Luiz Carlos Junior de Alcantara, Marta Giovanetti, Vagner Fonseca, Flavia Aburjaile, Rodrigo Tocantins Calado. |
| EPI_ISL_1661250, EPI_ISL_1661251, EPI_ISL_1661252 | Laboratorio de Ecologia de Doencas Transmissiveis na Amazonia, Instituto Leonidas e Maria Deane - Fiocruz Amazonia | Laboratorio de Ecologia de Doencas Transmissiveis na Amazonia, Instituto Leonidas e Maria Deane - Fiocruz Amazonia | Valdinete Nascimento, Victor Souza, André Corado, Fernanda Nascimento, George Silva, Ágatha Costa, Debora Duarte, Karina Pessoa, Matilde Mejía, Luciana Gonçalves, Maria Júlia Brandão, Michele Jesus, Felipe Naveca |
| EPI_ISL_1664113, EPI_ISL_1664114, EPI_ISL_1664115, EPI_ISL_1664116, EPI_ISL_1664117, EPI_ISL_1664118, EPI_ISL_1664119, EPI_ISL_1664120, EPI_ISL_1664121, EPI_ISL_1664122, EPI_ISL_1664149, EPI_ISL_1664150, EPI_ISL_1664151, EPI_ISL_1664152, EPI_ISL_1664153, EPI_ISL_1664154, EPI_ISL_1664155, EPI_ISL_1664156, EPI_ISL_1664157, EPI_ISL_1664164, EPI_ISL_1664165, EPI_ISL_1664166, EPI_ISL_1664167, EPI_ISL_1664168, EPI_ISL_1664169, EPI_ISL_1664170, EPI_ISL_1664171, EPI_ISL_1664172, EPI_ISL_1664173 | | | |
| see above | Laboratorio Central Noel Nutels | Bioinformatics Laboratory / LNCC | Luiz G P de Almeida, Alessandra P Lamarca, Ronaldo da Silva F Jr, Liliane Cavalcante, Alexandra L Gerber, Ana Paula de C Guimarães, Douglas Terra Machado, Cassia Alves, Diana Mariani, Thais Felix Cruz, Mario Sergio Ribeiro, Silvia Carvalho, Flávio Dias da Silva, Marcio Henrique de Oliveira Garcia, Leandro Magalhães de Souza, Cristiane Gomes da Silva, Caio Luiz Pereira Ribeiro, Andréa Cony Cavalcanti, Claudia Maria Braga de Mello, Amilcar Tanuri, Ana Tereza R Vasconcelos |
| EPI_ISL_1694628, EPI_ISL_1694631, EPI_ISL_1694632, EPI_ISL_1694634, EPI_ISL_1695909 | Instituto Estadual do Cérebro Paulo Niemayer (IECPN) | Laboratório de Imunofarmacologia | Thiago Moreno Lopes Souza |
| EPI_ISL_1701311 | Laboratório de diagnóstico molecular da COVID-19 - Bahia, Santo Antônio de Jesus, Brazil | Laboratório Baculovírus | Josilene R Pinheiro, Jaime H Amorim, Aline Belmok, Fernando L Melo, Bergmann M Ribeiro |
| EPI_ISL_1707688 | UBDS Dr Marco Antonio Sahao Vila Virginia | Instituto Adolfo Lutz, Interdiciplinary Procedures Center, Strategic Laboratory | Claudio Tavares Sacchi, Claudia Regina Gonçalves, Erica Valessa Ramos Gomes, Karoline Rodrigues Campos, Caio Vinicius Dias Lopes, Leonardo Jose Tadeu de Araujo, Katia Correa de Oliveira Santos |
| EPI_ISL_1708317 | Fundação Hospitalar de Hematologia e Hemoterapia do Amazonas | Laboratório de Estudos de Vírus Emergentes | William M. Souza; Mariene R. Amorim, Renata Sesti-Costa, Lais D. Coimbra, Daniel A. Toledo-Teixeira, Pierina L. Parise, , Priscilla P. Barbosa, Karina Bispo-dos-Santos, Luciana S. Mofatto, Camila L. Simeoni, Natalia S. Brunetti, Ingra M. Claro, Adriana S. S. Duarte, Thais M. Coletti, Audrey B. Zangirolami, Carolina Costa-Lima, Arilson Bernardo S. P. Gomes, Lucas I. Buscaratti, Flavia C. Sales, Vitor A. Costa, Lucas A.M. Franco, Darlan S. Candido, Oliver G. Pybus, Jaqueline G. de Jesus, Camila A.M. Silva, Mariana S. Ramundo, Giulia M. Ferreira, Mariana C. Pinho, Leandro M. Souza, Esmenia C. Rocha, Pamela S. Andrade, Myuki A.E. Crispim, Grazielle C. Maktura, Erika R. Manuli, Magnun N.N. Santos, Cecilia C. Camilo, Rodrigo N. Angerami, Maria L. Moretti, Fernando R. Spilki, Clarice W. Arns, Marcelo Addas-Carvalho, Bruno D. Benites, Marcelo A.S. Mori, Nelson Gaburo, Christopher Dye, Chieh-Hsi Wu, Henrique Marques-Souza, Rafael E. Marques, Alessandro S. Farias, Michael S. Diamond, Nuno R. Faria, Ester C. Sabino, Fab |
| EPI_ISL_1708318 | Fundação Hospitalar de Hematologia e Hemoterapia do Amazonas | Laboratório de Estudos de Vírus Emergentes | William M. Souza; Mariene R. Amorim, Renata Sesti-Costa, Lais D. Coimbra, Daniel A. Toledo-Teixeira, Pierina L. Parise, Priscilla P. Barbosa, Karina Bispo-dos-Santos, Luciana S. Mofatto, Camila L. Simeoni, Natalia S. Brunetti, Ingra M. Claro, Adriana S. S. Duarte, Thais M. Coletti, Audrey B. Zangirolami, Carolina Costa-Lima, Arilson Bernardo S. P. Gomes, Lucas I. Buscaratti, Flavia C. Sales, Vitor A. Costa, Lucas A.M. Franco, Darlan S. Candido, Oliver G. Pybus, Jaqueline G. de Jesus, Camila A.M. Silva, Mariana S. Ramundo, Giulia M. Ferreira, Mariana C. Pinho, Leandro M. Souza, Esmenia C. Rocha, Pamela S. Andrade, Myuki A.E. Crispim, Grazielle C. Maktura, Erika R. Manuli, Magnun N.N. Santos, Cecilia C. Camilo, Rodrigo N. Angerami, Maria L. Moretti, Fernando R. Spilki, Clarice W. Arns, Marcelo Addas-Carvalho, Bruno D. Benites, Marcelo A.S. Mori, Nelson Gaburo, Christopher Dye, Chieh-Hsi Wu, Henrique Marques-Souza, Rafael E. Marques, Alessandro S. Farias, Michael S. Diamond, Nuno R. Faria, Ester C. Sabino, Fabia |
| EPI_ISL_1715134 | Centro de Saude II Ibitinga | Instituto Adolfo Lutz, Interdiciplinary Procedures Center, Strategic Laboratory | Claudio Tavares Sacchi, Claudia Regina Gonçalves, Erica Valessa Ramos Gomes, Karoline Rodrigues Campos, Caio Vinicius Dias Lopes, Leonardo Jose Tadeu de Araujo, Katia Correa de Oliveira Santos |
| EPI_ISL_1715135, EPI_ISL_1715136 | CS II Dr Jose Ferreira Telles | Instituto Adolfo Lutz, Interdiciplinary Procedures Center, Strategic Laboratory | Claudio Tavares Sacchi, Claudia Regina Gonçalves, Erica Valessa Ramos Gomes, Karoline Rodrigues Campos, Caio Vinicius Dias Lopes, Leonardo Jose Tadeu de Araujo, Katia Correa de Oliveira Santos |
| EPI_ISL_1715137 | Secretaria Municipal De Saude | Instituto Adolfo Lutz, Interdiciplinary Procedures Center, Strategic Laboratory | Claudio Tavares Sacchi, Claudia Regina Gonçalves, Erica Valessa Ramos Gomes, Karoline Rodrigues Campos, Caio Vinicius Dias Lopes, Leonardo Jose Tadeu de Araujo, Katia Correa de Oliveira Santos |
| EPI_ISL_1715138 | Santa Casa De Cravinhos | Instituto Adolfo Lutz, Interdiciplinary Procedures Center, Strategic Laboratory | Claudio Tavares Sacchi, Claudia Regina Gonçalves, Erica Valessa Ramos Gomes, Karoline Rodrigues Campos, Caio Vinicius Dias Lopes, Leonardo Jose Tadeu de Araujo, Katia Correa de Oliveira Santos |
| EPI_ISL_1715139 | Sae Servico De Atendimento Especializado | Instituto Adolfo Lutz, Interdiciplinary Procedures Center, Strategic Laboratory | Claudio Tavares Sacchi, Claudia Regina Gonçalves, Erica Valessa Ramos Gomes, Karoline Rodrigues Campos, Caio Vinicius Dias Lopes, Leonardo Jose Tadeu de Araujo, Katia Correa de Oliveira Santos |
| EPI_ISL_1715140 | UPA Dr Luis Atilio Losi Viana Ribeirao Preto | Instituto Adolfo Lutz, Interdiciplinary Procedures Center, Strategic Laboratory | Claudio Tavares Sacchi, Claudia Regina Gonçalves, Erica Valessa Ramos Gomes, Karoline Rodrigues Campos, Caio Vinicius Dias Lopes, Leonardo Jose Tadeu de Araujo, Katia Correa de Oliveira Santos |
| EPI_ISL_1715141 | Centro De Saude II Ibitinga | Instituto Adolfo Lutz, Interdiciplinary Procedures Center, Strategic Laboratory | Claudio Tavares Sacchi, Claudia Regina Gonçalves, Erica Valessa Ramos Gomes, Karoline Rodrigues Campos, Caio Vinicius Dias Lopes, Leonardo Jose Tadeu de Araujo, Katia Correa de Oliveira Santos |
| EPI_ISL_1715142 | CS III de Patrocinio Paulista | Instituto Adolfo Lutz, Interdiciplinary Procedures Center, Strategic Laboratory | Claudio Tavares Sacchi, Claudia Regina Gonçalves, Erica Valessa Ramos Gomes, Karoline Rodrigues Campos, Caio Vinicius Dias Lopes, Leonardo Jose Tadeu de Araujo, Katia Correa de Oliveira Santos |
| EPI_ISL_1715143 | Secretaria Municipal de Saude De Guariba | Instituto Adolfo Lutz, Interdiciplinary Procedures Center, Strategic Laboratory | Claudio Tavares Sacchi, Claudia Regina Gonçalves, Erica Valessa Ramos Gomes, Karoline Rodrigues Campos, Caio Vinicius Dias Lopes, Leonardo Jose Tadeu de Araujo, Katia Correa de Oliveira Santos |
| EPI_ISL_1715144 | UBDS Dr Joao Baptista Quartim Central | Instituto Adolfo Lutz, Interdiciplinary Procedures Center, Strategic Laboratory | Claudio Tavares Sacchi, Claudia Regina Gonçalves, Erica Valessa Ramos Gomes, Karoline Rodrigues Campos, Caio Vinicius Dias Lopes, Leonardo Jose Tadeu de Araujo, Katia Correa de Oliveira Santos |
| EPI_ISL_1715145 | UPA de Bebedouro | Instituto Adolfo Lutz, Interdiciplinary Procedures Center, Strategic Laboratory | Claudio Tavares Sacchi, Claudia Regina Gonçalves, Erica Valessa Ramos Gomes, Karoline Rodrigues Campos, Caio Vinicius Dias Lopes, Leonardo Jose Tadeu de Araujo, Katia Correa de Oliveira Santos |

| | | | |
|---|---|---|---|
| EPI_ISL_1715159 | Instituto Adolfo Lutz Central | Instituto Adolfo Lutz, Interdiciplinary Procedures Center, Strategic Laboratory | Claudio Tavares Sacchi, Claudia Regina Gonçalves, Erica Valessa Ramos Gomes, Karoline Rodrigues Campos, Caio Vinicius Dias Lopes, Leonardo Jose Tadeu de Araujo, Katia Correa de Oliveira Santos |
| EPI_ISL_1716397, EPI_ISL_1716398, EPI_ISL_1716399, EPI_ISL_1716400, EPI_ISL_1716401, EPI_ISL_1716402, EPI_ISL_1716403, EPI_ISL_1716404, EPI_ISL_1716405, EPI_ISL_1716406, EPI_ISL_1716407, EPI_ISL_1716408, EPI_ISL_1716409, EPI_ISL_1716410, EPI_ISL_1716411, EPI_ISL_1716412, EPI_ISL_1716413, EPI_ISL_1716414, EPI_ISL_1716415, EPI_ISL_1716416, EPI_ISL_1716417, EPI_ISL_1716418, EPI_ISL_1716419, EPI_ISL_1716420, EPI_ISL_1716421, EPI_ISL_1716422, EPI_ISL_1716423, EPI_ISL_1716424, EPI_ISL_1716425, EPI_ISL_1716426, EPI_ISL_1716427, EPI_ISL_1716428, EPI_ISL_1716429, EPI_ISL_1716430, EPI_ISL_1716431, EPI_ISL_1716432, EPI_ISL_1716433, EPI_ISL_1716434, EPI_ISL_1716435, EPI_ISL_1716436, EPI_ISL_1716437, EPI_ISL_1716438, EPI_ISL_1716439, EPI_ISL_1716440, EPI_ISL_1716441, EPI_ISL_1716442, EPI_ISL_1716443, EPI_ISL_1716444, EPI_ISL_1716445, EPI_ISL_1716446, EPI_ISL_1716447, EPI_ISL_1716448, EPI_ISL_1716449, EPI_ISL_1716450, EPI_ISL_1716451, EPI_ISL_1716452, EPI_ISL_1716453, EPI_ISL_1716454, EPI_ISL_1716455, EPI_ISL_1716456 | | | |
| see above | Laboratorio Saude | LGBio (Laboratorio de Genetica & Biodiversidade) | Mariana Pires de Campos Telles, Daniela de Melo e Silva, Elisangela de Paula Silveira Lacerda, Renata de Oliveira Dias, Rhewter Nunes, Cintia Pelegrineti Targueta de Azevedo Brito, Ramilla dos Santos Braga, Thais Guimarães Castro, Thays Millena Alves Pedroso, Amanda Alves de Melo, Francyelli Mello Andrade |
| EPI_ISL_1716457, EPI_ISL_1716458, EPI_ISL_1716459, EPI_ISL_1716460 | LACEN (Laboratorio de Saude Publica Dr. Giovanni Cysneiros) | LGBio (Laboratorio de Genetica & Biodiversidade) | Mariana Pires de Campos Telles, Daniela de Melo e Silva, Elisangela de Paula Silveira Lacerda, Renata de Oliveira Dias, Rhewter Nunes, Cintia Pelegrineti Targueta de Azevedo Brito, Ramilla dos Santos Braga, Thais Guimarães Castro, Thays Millena Alves Pedroso, Amanda Alves de Melo, Francyelli Mello Andrade |
| EPI_ISL_1716461, EPI_ISL_1716462, EPI_ISL_1716463, EPI_ISL_1716464, EPI_ISL_1716465, EPI_ISL_1716466, EPI_ISL_1716467, EPI_ISL_1716468, EPI_ISL_1716469, EPI_ISL_1716470, EPI_ISL_1716471, EPI_ISL_1716472, EPI_ISL_1716473, EPI_ISL_1716474, EPI_ISL_1716475, EPI_ISL_1716476, EPI_ISL_1716477, EPI_ISL_1716478, EPI_ISL_1716479, EPI_ISL_1716480, EPI_ISL_1716481, EPI_ISL_1716482, EPI_ISL_1716483, EPI_ISL_1716484, EPI_ISL_1716485, EPI_ISL_1716486, EPI_ISL_1716487, EPI_ISL_1716488, EPI_ISL_1716489, EPI_ISL_1716490, EPI_ISL_1716491, EPI_ISL_1716492, EPI_ISL_1716493, EPI_ISL_1716494, EPI_ISL_1716495, EPI_ISL_1716496, EPI_ISL_1716497, EPI_ISL_1716498, EPI_ISL_1716499, EPI_ISL_1716500, EPI_ISL_1716501, EPI_ISL_1716502, EPI_ISL_1716503, EPI_ISL_1716504, EPI_ISL_1716505, EPI_ISL_1716506, EPI_ISL_1716507, EPI_ISL_1716508, EPI_ISL_1716509, EPI_ISL_1716510, EPI_ISL_1716511, EPI_ISL_1716512, EPI_ISL_1716513, EPI_ISL_1716514 | | | |
| see above | LACEN (Laboratorio de Saude Publica Dr. Giovanni Cysneiros) | LGBio (Laboratorio de Genetica & Biodiversidade) | Mariana Pires de Campos Telles, Daniela de Melo e Silva, Elisangela de Paula Silveira Lacerda, Renata de Oliveira Dias, Rhewter Nunes, Cintia Pelegrineti Targueta de Azevedo Brito, Ramilla dos Santos Braga, Thais Guimarães Castro, Thays Millena Alves Pedroso, Amanda Alves de Melo, Aparecido Divino da Cruz, Luiz Augusto Pereira, Thais Cidália Vieira Gigonzac, Marc Alexandre Duarte Gigonzac, Alex Honda Bernardes, Francyelli Mello Andrade |
| EPI_ISL_1716877 | SECRETARIA MUNICIPAL DE SAUDE SOROCABA | Instituto Butantan / Mendelics | Instituto Butantan: Alexander Roberto Precioso, Dimas Tadeu Covas, Sandra Coccuzzo Sampaio, Maria Carolina Elias, José Salvatore Leister Patané, Vincent Louis Viala, Antonio Jorge Martins, Ricardo Haddad, Claudia Renata dos Santos Barros, Elaine Cristina Marqueze, Raul Machado Neto, Debora Botequio Moretti. Centro de Genômica Funcional da ESALQ: Luiz Lehmann Coutinho, Ricardo Augusto Brassaloti, Raquel de Lello Rocha Campos Cassano. NGS Soluções Genômicas: Pilar Drummond Sampaio Corrêa Mariani. FZEA-USP Pirassununga: Mirele Daiana Poleti, Jessika Cristina Chagas Lesbon, Elisangela Chicaroni Mattos, Heidge Fukumasu. USP-Botucatu: Rejane Maria Tommasini Grotto, Jayme A. Souza-Neto, Guilherme Targino Valente, Patricia Akemi Assato, Felipe Allan da Silva da Costa, Bianca Cechetto Carlos. Mendelics: Bibiana Santos, João Paulo Kitajima, Erika Freitas, David Schlesinger. Hemocentro Ribeirão Preto: Simone Kashima, Evandra Strazza Rodrigues, Svetoslav Nanev Slavov, Elaine Vieira dos Santos, Rafael dos Santos Bezerra, Luiz Carlos Junior de Alcantara, Marta Giovanetti, Vagner Fonseca, Flavia Aburjaile, Rodrigo Tocantins Calado. |
| EPI_ISL_1716878 | VIGILANCIA EPIDEMIOLOGICA JARDINOPOLIS SP | Instituto Butantan / ESALQ-USP | Instituto Butantan: Alexander Roberto Precioso, Dimas Tadeu Covas, Sandra Coccuzzo Sampaio, Maria Carolina Elias, José Salvatore Leister Patané, Vincent Louis Viala, Antonio Jorge Martins, Ricardo Haddad, Claudia Renata dos Santos Barros, Elaine Cristina Marqueze, Raul Machado Neto, Debora Botequio Moretti. Centro de Genômica Funcional da ESALQ: Luiz Lehmann Coutinho, Ricardo Augusto Brassaloti, Raquel de Lello Rocha Campos Cassano. NGS Soluções Genômicas: Pilar Drummond Sampaio Corrêa Mariani. FZEA-USP Pirassununga: Mirele Daiana Poleti, Jessika Cristina Chagas Lesbon, Elisangela Chicaroni Mattos, Heidge Fukumasu. USP-Botucatu: Rejane Maria Tommasini Grotto, Jayme A. Souza-Neto, Guilherme Targino Valente, Patricia Akemi Assato, Felipe Allan da Silva da Costa, Bianca Cechetto Carlos. Mendelics: Bibiana Santos, João Paulo Kitajima, Erika Freitas, David Schlesinger. Hemocentro Ribeirão Preto: Simone Kashima, Evandra Strazza Rodrigues, Svetoslav Nanev Slavov, Elaine Vieira dos Santos, Rafael dos Santos Bezerra, Luiz Carlos Junior de Alcantara, Marta Giovanetti, Vagner Fonseca, Flavia Aburjaile, Rodrigo Tocantins Calado. |
| EPI_ISL_1716879 | CENTRO MEDICO PMESP | Instituto Butantan / ESALQ-USP | Instituto Butantan: Alexander Roberto Precioso, Dimas Tadeu Covas, Sandra Coccuzzo Sampaio, Maria Carolina Elias, José Salvatore Leister Patané, Vincent Louis Viala, Antonio Jorge Martins, Ricardo Haddad, Claudia Renata dos Santos Barros, Elaine Cristina Marqueze, Raul Machado Neto, Debora Botequio Moretti. Centro de Genômica Funcional da ESALQ: Luiz Lehmann Coutinho, Ricardo Augusto Brassaloti, Raquel de Lello Rocha Campos Cassano. NGS Soluções Genômicas: Pilar Drummond Sampaio Corrêa Mariani. FZEA-USP Pirassununga: Mirele Daiana Poleti, Jessika Cristina Chagas Lesbon, Elisangela Chicaroni Mattos, Heidge Fukumasu. USP-Botucatu: Rejane Maria Tommasini Grotto, Jayme A. Souza-Neto, Guilherme Targino Valente, Patricia Akemi Assato, Felipe Allan da Silva da Costa, Bianca Cechetto Carlos. Mendelics: Bibiana Santos, João Paulo Kitajima, Erika Freitas, David Schlesinger. Hemocentro Ribeirão Preto: Simone Kashima, Evandra Strazza Rodrigues, Svetoslav Nanev Slavov, Elaine Vieira dos Santos, Rafael dos Santos Bezerra, Luiz Carlos Junior de Alcantara, Marta Giovanetti, Vagner Fonseca, Flavia Aburjaile, Rodrigo Tocantins Calado. |
| EPI_ISL_1731574 | UBDS Dr Marco Antonio Sahao Vila Virginia | Instituto Adolfo Lutz, Interdiciplinary Procedures Center, Strategic Laboratory | Claudio Tavares Sacchi, Claudia Regina Gonçalves, Erica Valessa Ramos Gomes, Karoline Rodrigues Campos, Caio Vinicius Dias Lopes, Leonardo Jose Tadeu de Araujo, Katia Correa de Oliveira Santos |
| EPI_ISL_1731575 | Casa de Caridade Sao Vicente de Paulo Cajuru | Instituto Adolfo Lutz, Interdiciplinary Procedures Center, Strategic Laboratory | Claudio Tavares Sacchi, Claudia Regina Gonçalves, Erica Valessa Ramos Gomes, Karoline Rodrigues Campos, Caio Vinicius Dias Lopes, Leonardo Jose Tadeu de Araujo, Katia Correa de Oliveira Santos |
| EPI_ISL_1731576 | UPA Dr Luis Atilio Losi Viana Ribeirao Preto | Instituto Adolfo Lutz, Interdiciplinary Procedures Center, Strategic Laboratory | Claudio Tavares Sacchi, Claudia Regina Gonçalves, Erica Valessa Ramos Gomes, Karoline Rodrigues Campos, Caio Vinicius Dias Lopes, Leonardo Jose Tadeu de Araujo, Katia Correa de Oliveira Santos |
| EPI_ISL_1731581 | Instituto Adolfo Lutz - Regional de Ribeirao Preto | Instituto Adolfo Lutz, Interdiciplinary Procedures Center, Strategic Laboratory | Claudio Tavares Sacchi, Claudia Regina Gonçalves, Erica Valessa Ramos Gomes, Karoline Rodrigues Campos, Caio Vinicius Dias Lopes, Leonardo Jose Tadeu de Araujo, Katia Correa de Oliveira Santos |
| EPI_ISL_1731598, EPI_ISL_1731599, EPI_ISL_1731600, EPI_ISL_1731601, EPI_ISL_1731602, EPI_ISL_1731603 | Instituto Adolfo Lutz Central | Instituto Adolfo Lutz, Interdiciplinary Procedures Center, Strategic Laboratory | Claudio Tavares Sacchi, Claudia Regina Gonçalves, Erica Valessa Ramos Gomes, Karoline Rodrigues Campos, Caio Vinicius Dias Lopes, Leonardo Jose Tadeu de Araujo, Katia Correa de Oliveira Santos |
| EPI_ISL_1734841 | Instituto de Biotecnologia - UNESP-Botucatu-SP | Instituto de Biotecnologia - UNESP-Botucatu-SP | Fábio Sossai Possebon; Leila Sabrina Ullmann; Cecília Artico Banho; Cíntia Bittar; Guilherme Campos; Helena Lage Ferreira; Jorge A. Petroli Marchesi; Lívia Sacchetto; Maisa C. Pereira Parra; Marília Moraes; Maurício L. Nogueira; Paula Rahal; Paulo Inacio da Costa; João Pessoa Araújo Jr. |
| EPI_ISL_1752637 | UBDS Dr Marco Antonio Sahao Vila Virginia | Instituto Adolfo Lutz, Interdiciplinary Procedures Center, Strategic Laboratory | Claudio Tavares Sacchi, Claudia Regina Gonçalves, Erica Valessa Ramos Gomes, Karoline Rodrigues Campos, Caio Vinicius Dias Lopes, Leonardo Jose Tadeu de Araujo, Katia Correa de Oliveira Santos |
| EPI_ISL_1752648, EPI_ISL_1752649, EPI_ISL_1752650, EPI_ISL_1752651, EPI_ISL_1752652, EPI_ISL_1752653, EPI_ISL_1752654, EPI_ISL_1752655, EPI_ISL_1752656, EPI_ISL_1752657, EPI_ISL_1752658, EPI_ISL_1752659, EPI_ISL_1752660, EPI_ISL_1752661, EPI_ISL_1752662, EPI_ISL_1752663, EPI_ISL_1752664, EPI_ISL_1752665, EPI_ISL_1752666, EPI_ISL_1752667, EPI_ISL_1752668 | | | |
| see above | Instituto Adolfo Lutz Central | Instituto Adolfo Lutz, Interdiciplinary Procedures Center, Strategic Laboratory | Claudio Tavares Sacchi, Claudia Regina Gonçalves, Erica Valessa Ramos Gomes, Karoline Rodrigues Campos, Caio Vinicius Dias Lopes, Leonardo Jose Tadeu de Araujo, Katia Correa de Oliveira Santos |
| EPI_ISL_1754186 | Laboratório de Pesquisa em Virologia, FAMERP, SJRP | Laboratório de Pesquisa em Virologia, FAMERP, SJRP | Cecília Artico Banho; Fábio Sossai Possebon; Leila Sabrina Ullmann; Cíntia Bittar; Guilherme Campos; Helena Lage Ferreira; Jorge A. Petroli Marchesi; Lívia Sacchetto; Maisa C. Pereira Parra; Marília Moraes; Paula Rahal; Paulo Inacio da Costa; João Pessoa Araújo Jr.; Maurício L. Nogueira |
| EPI_ISL_1785609, EPI_ISL_1785610, EPI_ISL_1785611, EPI_ISL_1785612, EPI_ISL_1785613, EPI_ISL_1785614 | Laboratório de Pesquisa em Virologia, FAMERP, SJRP | Laboratório de Pesquisa em Virologia, FAMERP, SJRP | Fábio Sossai Possebon; Leila Sabrina Ullmann; Cecília Artico Banho; Cíntia Bittar; Guilherme Campos; Helena Lage Ferreira; Jorge A. Petroli Marchesi; Lívia Sacchetto; Maisa C. Pereira Parra; Marília Moraes; Maurício L. Nogueira; Paula Rahal; Paulo Inacio da Costa; João Pessoa Araújo Jr. |
| EPI_ISL_1786560, EPI_ISL_1786561, EPI_ISL_1786562, EPI_ISL_1786563, EPI_ISL_1786564, EPI_ISL_1786565, EPI_ISL_1786566, EPI_ISL_1786567, EPI_ISL_1786568, EPI_ISL_1786569, EPI_ISL_1786570 | | | |
| see above | Laboratorio de Virologia Clinica do HCFMRP-USP | Hocentro de Ribeirao Preto - Laboratório de Biologia Molecular | Svetoslav Nanev Slavov, Marta Giovanetti, Rafael dos Santos Bezerra, Vagner Fonseca, Elaine Vieira Santos, Evandra Strazza Rodrigues, Talita Adelino, Joilson Xavier, Josiane Serrano Borges, Mariane Evaristo, Glauco de Carvalho Pereira, Aparecida Yulie Yamamoto, Diego Villa Clé, Rodrigo Tocantins Calado, Dimas Tadeu Covas, Luiz Carlos Junior Alcantara, Simone Kashima |
| EPI_ISL_1795061 | POLICLINICA HORTOLANDIA | Instituto Butantan / ESALQ-Piracicaba | Instituto Butantan: Alexander Roberto Precioso, Dimas Tadeu Covas, Sandra Coccuzzo Sampaio, Maria Carolina Elias, José Salvatore Leister Patané, Vincent Louis Viala, Antonio Jorge Martins, Ricardo Haddad, Claudia Renata dos Santos Barros, Elaine Cristina Marqueze, Raul Machado Neto, Debora Botequio Moretti. Centro de Genômica Funcional da ESALQ: Luiz Lehmann Coutinho, Ricardo Augusto Brassaloti, Raquel de Lello Rocha Campos Cassano. NGS Soluções Genômicas: Pilar Drummond Sampaio Corrêa Mariani. FZEA-USP Pirassununga: Mirele Daiana Poleti, Jessika Cristina Chagas |

| | | | |
|---|---|---|---|
| | | | Lesbon, Elisangela Chicaroni Mattos, Heidge Fukumasu. USP-Botucatu: Rejane Maria Tommasini Grotto, Jayme A. Souza-Neto, Guilherme Targino Valente, Patricia Akemi Assato, Felipe Allan da Silva da Costa, Bianca Cechetto Carlos. Mendelics: Bibiana Santos, João Paulo Kitajima, Erika Freitas, David Schlesinger. Hemocentro Ribeirão Preto: Simone Kashima, Evandra Strazza Rodrigues, Svetoslav Nanev Slavov, Elaine Vieira dos Santos, Rafael dos Santos Bezerra, Luiz Carlos Junior de Alcantara, Marta Giovanetti, Vagner Fonseca, Flavia Aburjaile, Rodrigo Tocantins Calado. |
| EPI_ISL_1795062 | AMBULATORIO DE ESPECIALIDADE V E MOGI MIRIM | Instituto Butantan / ESALQ-Piracicaba | Instituto Butantan: Alexander Roberto Precioso, Dimas Tadeu Covas, Sandra Coccuzzo Sampaio, Maria Carolina Elias, José Salvatore Leister Patané, Vincent Louis Viala, Antonio Jorge Martins, Ricardo Haddad, Claudia Renata dos Santos Barros, Elaine Cristina Marqueze, Raul Machado Neto, Debora Botequio Moretti. Centro de Genômica Funcional da ESALQ: Luiz Lehmann Coutinho, Ricardo Augusto Brassaloti, Raquel de Lello Rocha Campos Cassano. NGS Soluções Genômicas: Pilar Drummond Sampaio Corrêa Mariani. FZEA-USP Pirassununga: Mirele Daiana Poleti, Jessika Cristina Chagas Lesbon, Elisangela Chicaroni Mattos, Heidge Fukumasu. USP-Botucatu: Rejane Maria Tommasini Grotto, Jayme A. Souza-Neto, Guilherme Targino Valente, Patricia Akemi Assato, Felipe Allan da Silva da Costa, Bianca Cechetto Carlos. Mendelics: Bibiana Santos, João Paulo Kitajima, Erika Freitas, David Schlesinger. Hemocentro Ribeirão Preto: Simone Kashima, Evandra Strazza Rodrigues, Svetoslav Nanev Slavov, Elaine Vieira dos Santos, Rafael dos Santos Bezerra, Luiz Carlos Junior de Alcantara, Marta Giovanetti, Vagner Fonseca, Flavia Aburjaile, Rodrigo Tocantins Calado. |
| EPI_ISL_1795063 | UNIDADE DE VIGILANCIA EM SAUDE | Instituto Butantan / ESALQ-Piracicaba | Instituto Butantan: Alexander Roberto Precioso, Dimas Tadeu Covas, Sandra Coccuzzo Sampaio, Maria Carolina Elias, José Salvatore Leister Patané, Vincent Louis Viala, Antonio Jorge Martins, Ricardo Haddad, Claudia Renata dos Santos Barros, Elaine Cristina Marqueze, Raul Machado Neto, Debora Botequio Moretti. Centro de Genômica Funcional da ESALQ: Luiz Lehmann Coutinho, Ricardo Augusto Brassaloti, Raquel de Lello Rocha Campos Cassano. NGS Soluções Genômicas: Pilar Drummond Sampaio Corrêa Mariani. FZEA-USP Pirassununga: Mirele Daiana Poleti, Jessika Cristina Chagas Lesbon, Elisangela Chicaroni Mattos, Heidge Fukumasu. USP-Botucatu: Rejane Maria Tommasini Grotto, Jayme A. Souza-Neto, Guilherme Targino Valente, Patricia Akemi Assato, Felipe Allan da Silva da Costa, Bianca Cechetto Carlos. Mendelics: Bibiana Santos, João Paulo Kitajima, Erika Freitas, David Schlesinger. Hemocentro Ribeirão Preto: Simone Kashima, Evandra Strazza Rodrigues, Svetoslav Nanev Slavov, Elaine Vieira dos Santos, Rafael dos Santos Bezerra, Luiz Carlos Junior de Alcantara, Marta Giovanetti, Vagner Fonseca, Flavia Aburjaile, Rodrigo Tocantins Calado. |
| EPI_ISL_1795064 | USF SALERNO | Instituto Butantan / ESALQ-Piracicaba | Instituto Butantan: Alexander Roberto Precioso, Dimas Tadeu Covas, Sandra Coccuzzo Sampaio, Maria Carolina Elias, José Salvatore Leister Patané, Vincent Louis Viala, Antonio Jorge Martins, Ricardo Haddad, Claudia Renata dos Santos Barros, Elaine Cristina Marqueze, Raul Machado Neto, Debora Botequio Moretti. Centro de Genômica Funcional da ESALQ: Luiz Lehmann Coutinho, Ricardo Augusto Brassaloti, Raquel de Lello Rocha Campos Cassano. NGS Soluções Genômicas: Pilar Drummond Sampaio Corrêa Mariani. FZEA-USP Pirassununga: Mirele Daiana Poleti, Jessika Cristina Chagas Lesbon, Elisangela Chicaroni Mattos, Heidge Fukumasu. USP-Botucatu: Rejane Maria Tommasini Grotto, Jayme A. Souza-Neto, Guilherme Targino Valente, Patricia Akemi Assato, Felipe Allan da Silva da Costa, Bianca Cechetto Carlos. Mendelics: Bibiana Santos, João Paulo Kitajima, Erika Freitas, David Schlesinger. Hemocentro Ribeirão Preto: Simone Kashima, Evandra Strazza Rodrigues, Svetoslav Nanev Slavov, Elaine Vieira dos Santos, Rafael dos Santos Bezerra, Luiz Carlos Junior de Alcantara, Marta Giovanetti, Vagner Fonseca, Flavia Aburjaile, Rodrigo Tocantins Calado. |
| EPI_ISL_1795065 | UNIDADE DE SAUDE DA FAMILIA JOSE ADALBERTO LELLIS GARCIA | Instituto Butantan / ESALQ-Piracicaba | Instituto Butantan: Alexander Roberto Precioso, Dimas Tadeu Covas, Sandra Coccuzzo Sampaio, Maria Carolina Elias, José Salvatore Leister Patané, Vincent Louis Viala, Antonio Jorge Martins, Ricardo Haddad, Claudia Renata dos Santos Barros, Elaine Cristina Marqueze, Raul Machado Neto, Debora Botequio Moretti. Centro de Genômica Funcional da ESALQ: Luiz Lehmann Coutinho, Ricardo Augusto Brassaloti, Raquel de Lello Rocha Campos Cassano. NGS Soluções Genômicas: Pilar Drummond Sampaio Corrêa Mariani. FZEA-USP Pirassununga: Mirele Daiana Poleti, Jessika Cristina Chagas Lesbon, Elisangela Chicaroni Mattos, Heidge Fukumasu. USP-Botucatu: Rejane Maria Tommasini Grotto, Jayme A. Souza-Neto, Guilherme Targino Valente, Patricia Akemi Assato, Felipe Allan da Silva da Costa, Bianca Cechetto Carlos. Mendelics: Bibiana Santos, João Paulo Kitajima, Erika Freitas, David Schlesinger. Hemocentro Ribeirão Preto: Simone Kashima, Evandra Strazza Rodrigues, Svetoslav Nanev Slavov, Elaine Vieira dos Santos, Rafael dos Santos Bezerra, Luiz Carlos Junior de Alcantara, Marta Giovanetti, Vagner Fonseca, Flavia Aburjaile, Rodrigo Tocantins Calado. |
| EPI_ISL_1795066, EPI_ISL_1795067, EPI_ISL_1795068, EPI_ISL_1795069, EPI_ISL_1795070, EPI_ISL_1795071 | LABORATORIO DE FRANCA | Instituto Butantan / ESALQ-Piracicaba | Instituto Butantan: Alexander Roberto Precioso, Dimas Tadeu Covas, Sandra Coccuzzo Sampaio, Maria Carolina Elias, José Salvatore Leister Patané, Vincent Louis Viala, Antonio Jorge Martins, Ricardo Haddad, Claudia Renata dos Santos Barros, Elaine Cristina Marqueze, Raul Machado Neto, Debora Botequio Moretti. Centro de Genômica Funcional da ESALQ: Luiz Lehmann Coutinho, Ricardo Augusto Brassaloti, Raquel de Lello Rocha Campos Cassano. NGS Soluções Genômicas: Pilar Drummond Sampaio Corrêa Mariani. FZEA-USP Pirassununga: Mirele Daiana Poleti, Jessika Cristina Chagas Lesbon, Elisangela Chicaroni Mattos, Heidge Fukumasu. USP-Botucatu: Rejane Maria Tommasini Grotto, Jayme A. Souza-Neto, Guilherme Targino Valente, Patricia Akemi Assato, Felipe Allan da Silva da Costa, Bianca Cechetto Carlos. Mendelics: Bibiana Santos, João Paulo Kitajima, Erika Freitas, David Schlesinger. Hemocentro Ribeirão Preto: Simone Kashima, Evandra Strazza Rodrigues, Svetoslav Nanev Slavov, Elaine Vieira dos Santos, Rafael dos Santos Bezerra, Luiz Carlos Junior de Alcantara, Marta Giovanetti, Vagner Fonseca, Flavia Aburjaile, Rodrigo Tocantins Calado. |
| EPI_ISL_1795072, EPI_ISL_1795073, EPI_ISL_1795074 | LABORATORIO DR PAULO EMILIO DALESSANDRO PINDAMONHANGABA | Instituto Butantan / ESALQ-Piracicaba | Instituto Butantan: Alexander Roberto Precioso, Dimas Tadeu Covas, Sandra Coccuzzo Sampaio, Maria Carolina Elias, José Salvatore Leister Patané, Vincent Louis Viala, Antonio Jorge Martins, Ricardo Haddad, Claudia Renata dos Santos Barros, Elaine Cristina Marqueze, Raul Machado Neto, Debora Botequio Moretti. Centro de Genômica Funcional da ESALQ: Luiz Lehmann Coutinho, Ricardo Augusto Brassaloti, Raquel de Lello Rocha Campos Cassano. NGS Soluções Genômicas: Pilar Drummond Sampaio Corrêa Mariani. FZEA-USP Pirassununga: Mirele Daiana Poleti, Jessika Cristina Chagas Lesbon, Elisangela Chicaroni Mattos, Heidge Fukumasu. USP-Botucatu: Rejane Maria Tommasini Grotto, Jayme A. Souza-Neto, Guilherme Targino Valente, Patricia Akemi Assato, Felipe Allan da Silva da Costa, Bianca Cechetto Carlos. Mendelics: Bibiana Santos, João Paulo Kitajima, Erika Freitas, David Schlesinger. Hemocentro Ribeirão Preto: Simone Kashima, Evandra Strazza Rodrigues, Svetoslav Nanev Slavov, Elaine Vieira dos Santos, Rafael dos Santos Bezerra, Luiz Carlos Junior de Alcantara, Marta Giovanetti, Vagner Fonseca, Flavia Aburjaile, Rodrigo Tocantins Calado. |
| EPI_ISL_1795075, EPI_ISL_1795076 | NUCLEO SAUDE III N HABITACIONAL PRESIDENTE GEISEL BAURU | Instituto Butantan / ESALQ-Piracicaba | Instituto Butantan: Alexander Roberto Precioso, Dimas Tadeu Covas, Sandra Coccuzzo Sampaio, Maria Carolina Elias, José Salvatore Leister Patané, Vincent Louis Viala, Antonio Jorge Martins, Ricardo Haddad, Claudia Renata dos Santos Barros, Elaine Cristina Marqueze, Raul Machado Neto, Debora Botequio Moretti. Centro de Genômica Funcional da ESALQ: Luiz Lehmann Coutinho, Ricardo Augusto Brassaloti, Raquel de Lello Rocha Campos Cassano. NGS Soluções Genômicas: Pilar Drummond Sampaio Corrêa Mariani. FZEA-USP Pirassununga: Mirele Daiana Poleti, Jessika Cristina Chagas Lesbon, Elisangela Chicaroni Mattos, Heidge Fukumasu. USP-Botucatu: Rejane Maria Tommasini Grotto, Jayme A. Souza-Neto, Guilherme Targino Valente, Patricia Akemi Assato, Felipe Allan da Silva da Costa, Bianca Cechetto Carlos. Mendelics: Bibiana Santos, João Paulo Kitajima, Erika Freitas, David Schlesinger. Hemocentro Ribeirão Preto: Simone Kashima, Evandra Strazza Rodrigues, Svetoslav Nanev Slavov, Elaine Vieira dos Santos, Rafael dos Santos Bezerra, Luiz Carlos Junior de Alcantara, Marta Giovanetti, Vagner Fonseca, Flavia Aburjaile, Rodrigo Tocantins Calado. |
| EPI_ISL_1795077 | MARIA TEREZINHA BOGNAR CHECHETO | Instituto Butantan / ESALQ-Piracicaba | Instituto Butantan: Alexander Roberto Precioso, Dimas Tadeu Covas, Sandra Coccuzzo Sampaio, Maria Carolina Elias, José Salvatore Leister Patané, Vincent Louis Viala, Antonio Jorge Martins, Ricardo Haddad, Claudia Renata dos Santos Barros, Elaine Cristina Marqueze, Raul Machado Neto, Debora Botequio Moretti. Centro de Genômica Funcional da ESALQ: Luiz Lehmann Coutinho, Ricardo Augusto Brassaloti, Raquel de Lello Rocha Campos Cassano. NGS Soluções Genômicas: Pilar Drummond Sampaio Corrêa Mariani. FZEA-USP Pirassununga: Mirele Daiana Poleti, Jessika Cristina Chagas Lesbon, Elisangela Chicaroni Mattos, Heidge Fukumasu. USP-Botucatu: Rejane Maria Tommasini Grotto, Jayme A. Souza-Neto, Guilherme Targino Valente, Patricia Akemi Assato, Felipe Allan da Silva da Costa, Bianca Cechetto Carlos. Mendelics: Bibiana Santos, João Paulo Kitajima, Erika Freitas, David Schlesinger. Hemocentro Ribeirão Preto: Simone Kashima, Evandra Strazza Rodrigues, Svetoslav Nanev Slavov, Elaine Vieira dos Santos, Rafael dos Santos Bezerra, Luiz Carlos Junior de Alcantara, Marta Giovanetti, Vagner Fonseca, Flavia Aburjaile, Rodrigo Tocantins Calado. |
| EPI_ISL_1795078 | LABORATORIO DE FRANCA | Instituto Butantan / ESALQ-Piracicaba | Instituto Butantan: Alexander Roberto Precioso, Dimas Tadeu Covas, Sandra Coccuzzo Sampaio, Maria Carolina Elias, José Salvatore Leister Patané, Vincent Louis Viala, Antonio Jorge Martins, Ricardo Haddad, Claudia Renata dos Santos Barros, Elaine Cristina Marqueze, Raul Machado Neto, Debora Botequio Moretti. Centro de Genômica Funcional da ESALQ: Luiz Lehmann Coutinho, Ricardo Augusto Brassaloti, Raquel de Lello Rocha Campos Cassano. NGS Soluções Genômicas: Pilar Drummond Sampaio Corrêa Mariani. FZEA-USP Pirassununga: Mirele Daiana Poleti, Jessika Cristina Chagas Lesbon, Elisangela Chicaroni Mattos, Heidge Fukumasu. USP-Botucatu: Rejane Maria Tommasini Grotto, Jayme A. Souza-Neto, Guilherme Targino Valente, Patricia Akemi Assato, Felipe Allan da Silva da Costa, Bianca Cechetto Carlos. Mendelics: Bibiana Santos, João Paulo Kitajima, Erika Freitas, David Schlesinger. Hemocentro Ribeirão Preto: Simone Kashima, Evandra Strazza Rodrigues, Svetoslav Nanev Slavov, Elaine Vieira dos Santos, Rafael dos Santos Bezerra, Luiz Carlos Junior de Alcantara, Marta Giovanetti, Vagner Fonseca, Flavia Aburjaile, Rodrigo Tocantins Calado. |
| EPI_ISL_1795079 | CS DE PARANAPUA | Instituto Butantan / ESALQ-Piracicaba | Instituto Butantan: Alexander Roberto Precioso, Dimas Tadeu Covas, Sandra Coccuzzo Sampaio, Maria Carolina Elias, José Salvatore Leister Patané, Vincent Louis Viala, Antonio Jorge Martins, Ricardo Haddad, Claudia Renata dos Santos Barros, Elaine Cristina Marqueze, Raul Machado Neto, Debora Botequio Moretti. Centro de Genômica Funcional da ESALQ: Luiz Lehmann Coutinho, Ricardo Augusto Brassaloti, Raquel de Lello Rocha Campos Cassano. NGS Soluções Genômicas: Pilar Drummond Sampaio Corrêa Mariani. FZEA-USP Pirassununga: Mirele Daiana Poleti, Jessika Cristina Chagas Lesbon, Elisangela Chicaroni Mattos, Heidge Fukumasu. USP-Botucatu: Rejane Maria Tommasini Grotto, Jayme A. Souza-Neto, Guilherme Targino Valente, Patricia Akemi Assato, Felipe Allan da Silva da Costa, Bianca Cechetto Carlos. Mendelics: Bibiana Santos, João Paulo Kitajima, Erika Freitas, |

| | | | |
|---|---|---|---|
| | | | David Schlesinger. Hemocentro Ribeirão Preto: Simone Kashima, Evandra Strazza Rodrigues, Svetoslav Nanev Slavov, Elaine Vieira dos Santos, Rafael dos Santos Bezerra, Luiz Carlos Junior de Alcantara, Marta Giovanetti, Vagner Fonseca, Flavia Aburjaile, Rodrigo Tocantins Calado. |
| EPI_ISL_1795080 | CENTRO MEDICO PMESP | Instituto Butantan / ESALQ-Piracicaba | Instituto Butantan: Alexander Roberto Precioso, Dimas Tadeu Covas, Sandra Coccuzzo Sampaio, Maria Carolina Elias, José Salvatore Leister Patané, Vincent Louis Viala, Antonio Jorge Martins, Ricardo Haddad, Claudia Renata dos Santos Barros, Elaine Cristina Marqueze, Raul Machado Neto, Debora Botequio Moretti. Centro de Genômica Funcional da ESALQ: Luiz Lehmann Coutinho, Ricardo Augusto Brassaloti, Raquel de Lello Rocha Campos Cassano. NGS Soluções Genômicas: Pilar Drummond Sampaio Corrêa Mariani. FZEA-USP Pirassununga: Mirele Daiana Poleti, Jessika Cristina Chagas Lesbon, Elisangela Chicaroni Mattos, Heidge Fukumasu. USP-Botucatu: Rejane Maria Tommasini Grotto, Jayme A. Souza-Neto, Guilherme Targino Valente, Patricia Akemi Assato, Felipe Allan da Silva da Costa, Bianca Cechetto Carlos. Mendelics: Bibiana Santos, João Paulo Kitajima, Erika Freitas, David Schlesinger. Hemocentro Ribeirão Preto: Simone Kashima, Evandra Strazza Rodrigues, Svetoslav Nanev Slavov, Elaine Vieira dos Santos, Rafael dos Santos Bezerra, Luiz Carlos Junior de Alcantara, Marta Giovanetti, Vagner Fonseca, Flavia Aburjaile, Rodrigo Tocantins Calado. |
| EPI_ISL_1795081 | CENTRO DE SAUDE III AFFONSO LUZZI SANTA CRUZ DAS PALMEIRAS | Instituto Butantan / ESALQ-Piracicaba | Instituto Butantan: Alexander Roberto Precioso, Dimas Tadeu Covas, Sandra Coccuzzo Sampaio, Maria Carolina Elias, José Salvatore Leister Patané, Vincent Louis Viala, Antonio Jorge Martins, Ricardo Haddad, Claudia Renata dos Santos Barros, Elaine Cristina Marqueze, Raul Machado Neto, Debora Botequio Moretti. Centro de Genômica Funcional da ESALQ: Luiz Lehmann Coutinho, Ricardo Augusto Brassaloti, Raquel de Lello Rocha Campos Cassano. NGS Soluções Genômicas: Pilar Drummond Sampaio Corrêa Mariani. FZEA-USP Pirassununga: Mirele Daiana Poleti, Jessika Cristina Chagas Lesbon, Elisangela Chicaroni Mattos, Heidge Fukumasu. USP-Botucatu: Rejane Maria Tommasini Grotto, Jayme A. Souza-Neto, Guilherme Targino Valente, Patricia Akemi Assato, Felipe Allan da Silva da Costa, Bianca Cechetto Carlos. Mendelics: Bibiana Santos, João Paulo Kitajima, Erika Freitas, David Schlesinger. Hemocentro Ribeirão Preto: Simone Kashima, Evandra Strazza Rodrigues, Svetoslav Nanev Slavov, Elaine Vieira dos Santos, Rafael dos Santos Bezerra, Luiz Carlos Junior de Alcantara, Marta Giovanetti, Vagner Fonseca, Flavia Aburjaile, Rodrigo Tocantins Calado. |
| EPI_ISL_1795082, EPI_ISL_1795083 | POLICLINICA HORTOLANDIA | Instituto Butantan / ESALQ-Piracicaba | Instituto Butantan: Alexander Roberto Precioso, Dimas Tadeu Covas, Sandra Coccuzzo Sampaio, Maria Carolina Elias, José Salvatore Leister Patané, Vincent Louis Viala, Antonio Jorge Martins, Ricardo Haddad, Claudia Renata dos Santos Barros, Elaine Cristina Marqueze, Raul Machado Neto, Debora Botequio Moretti. Centro de Genômica Funcional da ESALQ: Luiz Lehmann Coutinho, Ricardo Augusto Brassaloti, Raquel de Lello Rocha Campos Cassano. NGS Soluções Genômicas: Pilar Drummond Sampaio Corrêa Mariani. FZEA-USP Pirassununga: Mirele Daiana Poleti, Jessika Cristina Chagas Lesbon, Elisangela Chicaroni Mattos, Heidge Fukumasu. USP-Botucatu: Rejane Maria Tommasini Grotto, Jayme A. Souza-Neto, Guilherme Targino Valente, Patricia Akemi Assato, Felipe Allan da Silva da Costa, Bianca Cechetto Carlos. Mendelics: Bibiana Santos, João Paulo Kitajima, Erika Freitas, David Schlesinger. Hemocentro Ribeirão Preto: Simone Kashima, Evandra Strazza Rodrigues, Svetoslav Nanev Slavov, Elaine Vieira dos Santos, Rafael dos Santos Bezerra, Luiz Carlos Junior de Alcantara, Marta Giovanetti, Vagner Fonseca, Flavia Aburjaile, Rodrigo Tocantins Calado. |
| EPI_ISL_1795084, EPI_ISL_1795085, EPI_ISL_1795086 | PRONTO SOCORRO MUNICIPAL TAMBAU | Instituto Butantan / ESALQ-Piracicaba | Instituto Butantan: Alexander Roberto Precioso, Dimas Tadeu Covas, Sandra Coccuzzo Sampaio, Maria Carolina Elias, José Salvatore Leister Patané, Vincent Louis Viala, Antonio Jorge Martins, Ricardo Haddad, Claudia Renata dos Santos Barros, Elaine Cristina Marqueze, Raul Machado Neto, Debora Botequio Moretti. Centro de Genômica Funcional da ESALQ: Luiz Lehmann Coutinho, Ricardo Augusto Brassaloti, Raquel de Lello Rocha Campos Cassano. NGS Soluções Genômicas: Pilar Drummond Sampaio Corrêa Mariani. FZEA-USP Pirassununga: Mirele Daiana Poleti, Jessika Cristina Chagas Lesbon, Elisangela Chicaroni Mattos, Heidge Fukumasu. USP-Botucatu: Rejane Maria Tommasini Grotto, Jayme A. Souza-Neto, Guilherme Targino Valente, Patricia Akemi Assato, Felipe Allan da Silva da Costa, Bianca Cechetto Carlos. Mendelics: Bibiana Santos, João Paulo Kitajima, Erika Freitas, David Schlesinger. Hemocentro Ribeirão Preto: Simone Kashima, Evandra Strazza Rodrigues, Svetoslav Nanev Slavov, Elaine Vieira dos Santos, Rafael dos Santos Bezerra, Luiz Carlos Junior de Alcantara, Marta Giovanetti, Vagner Fonseca, Flavia Aburjaile, Rodrigo Tocantins Calado. |
| EPI_ISL_1795087, EPI_ISL_1795088, EPI_ISL_1795089, EPI_ISL_1795090 | CENTRO DE SAUDE III AFFONSO LUZZI SANTA CRUZ DAS PALMEIRAS | Instituto Butantan / ESALQ-Piracicaba | Instituto Butantan: Alexander Roberto Precioso, Dimas Tadeu Covas, Sandra Coccuzzo Sampaio, Maria Carolina Elias, José Salvatore Leister Patané, Vincent Louis Viala, Antonio Jorge Martins, Ricardo Haddad, Claudia Renata dos Santos Barros, Elaine Cristina Marqueze, Raul Machado Neto, Debora Botequio Moretti. Centro de Genômica Funcional da ESALQ: Luiz Lehmann Coutinho, Ricardo Augusto Brassaloti, Raquel de Lello Rocha Campos Cassano. NGS Soluções Genômicas: Pilar Drummond Sampaio Corrêa Mariani. FZEA-USP Pirassununga: Mirele Daiana Poleti, Jessika Cristina Chagas Lesbon, Elisangela Chicaroni Mattos, Heidge Fukumasu. USP-Botucatu: Rejane Maria Tommasini Grotto, Jayme A. Souza-Neto, Guilherme Targino Valente, Patricia Akemi Assato, Felipe Allan da Silva da Costa, Bianca Cechetto Carlos. Mendelics: Bibiana Santos, João Paulo Kitajima, Erika Freitas, David Schlesinger. Hemocentro Ribeirão Preto: Simone Kashima, Evandra Strazza Rodrigues, Svetoslav Nanev Slavov, Elaine Vieira dos Santos, Rafael dos Santos Bezerra, Luiz Carlos Junior de Alcantara, Marta Giovanetti, Vagner Fonseca, Flavia Aburjaile, Rodrigo Tocantins Calado. |
| EPI_ISL_1795091, EPI_ISL_1795092 | PRONTO SOCORRO MUNICIPAL TAMBAU | Instituto Butantan / ESALQ-Piracicaba | Instituto Butantan: Alexander Roberto Precioso, Dimas Tadeu Covas, Sandra Coccuzzo Sampaio, Maria Carolina Elias, José Salvatore Leister Patané, Vincent Louis Viala, Antonio Jorge Martins, Ricardo Haddad, Claudia Renata dos Santos Barros, Elaine Cristina Marqueze, Raul Machado Neto, Debora Botequio Moretti. Centro de Genômica Funcional da ESALQ: Luiz Lehmann Coutinho, Ricardo Augusto Brassaloti, Raquel de Lello Rocha Campos Cassano. NGS Soluções Genômicas: Pilar Drummond Sampaio Corrêa Mariani. FZEA-USP Pirassununga: Mirele Daiana Poleti, Jessika Cristina Chagas Lesbon, Elisangela Chicaroni Mattos, Heidge Fukumasu. USP-Botucatu: Rejane Maria Tommasini Grotto, Jayme A. Souza-Neto, Guilherme Targino Valente, Patricia Akemi Assato, Felipe Allan da Silva da Costa, Bianca Cechetto Carlos. Mendelics: Bibiana Santos, João Paulo Kitajima, Erika Freitas, David Schlesinger. Hemocentro Ribeirão Preto: Simone Kashima, Evandra Strazza Rodrigues, Svetoslav Nanev Slavov, Elaine Vieira dos Santos, Rafael dos Santos Bezerra, Luiz Carlos Junior de Alcantara, Marta Giovanetti, Vagner Fonseca, Flavia Aburjaile, Rodrigo Tocantins Calado. |
| EPI_ISL_1795093 | CENTRO INTEGRADO DE SAUDE | Instituto Butantan / ESALQ-Piracicaba | Instituto Butantan: Alexander Roberto Precioso, Dimas Tadeu Covas, Sandra Coccuzzo Sampaio, Maria Carolina Elias, José Salvatore Leister Patané, Vincent Louis Viala, Antonio Jorge Martins, Ricardo Haddad, Claudia Renata dos Santos Barros, Elaine Cristina Marqueze, Raul Machado Neto, Debora Botequio Moretti. Centro de Genômica Funcional da ESALQ: Luiz Lehmann Coutinho, Ricardo Augusto Brassaloti, Raquel de Lello Rocha Campos Cassano. NGS Soluções Genômicas: Pilar Drummond Sampaio Corrêa Mariani. FZEA-USP Pirassununga: Mirele Daiana Poleti, Jessika Cristina Chagas Lesbon, Elisangela Chicaroni Mattos, Heidge Fukumasu. USP-Botucatu: Rejane Maria Tommasini Grotto, Jayme A. Souza-Neto, Guilherme Targino Valente, Patricia Akemi Assato, Felipe Allan da Silva da Costa, Bianca Cechetto Carlos. Mendelics: Bibiana Santos, João Paulo Kitajima, Erika Freitas, David Schlesinger. Hemocentro Ribeirão Preto: Simone Kashima, Evandra Strazza Rodrigues, Svetoslav Nanev Slavov, Elaine Vieira dos Santos, Rafael dos Santos Bezerra, Luiz Carlos Junior de Alcantara, Marta Giovanetti, Vagner Fonseca, Flavia Aburjaile, Rodrigo Tocantins Calado. |
| EPI_ISL_1795094 | VIGILANCIA EPIDEMIOLOGICA DE LEME | Instituto Butantan / ESALQ-Piracicaba | Instituto Butantan: Alexander Roberto Precioso, Dimas Tadeu Covas, Sandra Coccuzzo Sampaio, Maria Carolina Elias, José Salvatore Leister Patané, Vincent Louis Viala, Antonio Jorge Martins, Ricardo Haddad, Claudia Renata dos Santos Barros, Elaine Cristina Marqueze, Raul Machado Neto, Debora Botequio Moretti. Centro de Genômica Funcional da ESALQ: Luiz Lehmann Coutinho, Ricardo Augusto Brassaloti, Raquel de Lello Rocha Campos Cassano. NGS Soluções Genômicas: Pilar Drummond Sampaio Corrêa Mariani. FZEA-USP Pirassununga: Mirele Daiana Poleti, Jessika Cristina Chagas Lesbon, Elisangela Chicaroni Mattos, Heidge Fukumasu. USP-Botucatu: Rejane Maria Tommasini Grotto, Jayme A. Souza-Neto, Guilherme Targino Valente, Patricia Akemi Assato, Felipe Allan da Silva da Costa, Bianca Cechetto Carlos. Mendelics: Bibiana Santos, João Paulo Kitajima, Erika Freitas, David Schlesinger. Hemocentro Ribeirão Preto: Simone Kashima, Evandra Strazza Rodrigues, Svetoslav Nanev Slavov, Elaine Vieira dos Santos, Rafael dos Santos Bezerra, Luiz Carlos Junior de Alcantara, Marta Giovanetti, Vagner Fonseca, Flavia Aburjaile, Rodrigo Tocantins Calado. |
| EPI_ISL_1795096 | USF SALERNO | Instituto Butantan / ESALQ-Piracicaba | Instituto Butantan: Alexander Roberto Precioso, Dimas Tadeu Covas, Sandra Coccuzzo Sampaio, Maria Carolina Elias, José Salvatore Leister Patané, Vincent Louis Viala, Antonio Jorge Martins, Ricardo Haddad, Claudia Renata dos Santos Barros, Elaine Cristina Marqueze, Raul Machado Neto, Debora Botequio Moretti. Centro de Genômica Funcional da ESALQ: Luiz Lehmann Coutinho, Ricardo Augusto Brassaloti, Raquel de Lello Rocha Campos Cassano. NGS Soluções Genômicas: Pilar Drummond Sampaio Corrêa Mariani. FZEA-USP Pirassununga: Mirele Daiana Poleti, Jessika Cristina Chagas Lesbon, Elisangela Chicaroni Mattos, Heidge Fukumasu. USP-Botucatu: Rejane Maria Tommasini Grotto, Jayme A. Souza-Neto, Guilherme Targino Valente, Patricia Akemi Assato, Felipe Allan da Silva da Costa, Bianca Cechetto Carlos. Mendelics: Bibiana Santos, João Paulo Kitajima, Erika Freitas, David Schlesinger. Hemocentro Ribeirão Preto: Simone Kashima, Evandra Strazza Rodrigues, Svetoslav Nanev Slavov, Elaine Vieira dos Santos, Rafael dos Santos Bezerra, Luiz Carlos Junior de Alcantara, Marta Giovanetti, Vagner Fonseca, Flavia Aburjaile, Rodrigo Tocantins Calado. |
| EPI_ISL_1795097 | LABORATORIO DR PAULO EMILIO DALESSANDRO PINDAMONHANGABA | Instituto Butantan / ESALQ-Piracicaba | Instituto Butantan: Alexander Roberto Precioso, Dimas Tadeu Covas, Sandra Coccuzzo Sampaio, Maria Carolina Elias, José Salvatore Leister Patané, Vincent Louis Viala, Antonio Jorge Martins, Ricardo Haddad, Claudia Renata dos Santos Barros, Elaine Cristina Marqueze, Raul Machado Neto, Debora Botequio Moretti. Centro de Genômica Funcional da ESALQ: Luiz Lehmann Coutinho, Ricardo Augusto Brassaloti, Raquel de Lello Rocha Campos Cassano. NGS Soluções Genômicas: Pilar Drummond Sampaio Corrêa Mariani. FZEA-USP Pirassununga: Mirele Daiana Poleti, Jessika Cristina Chagas Lesbon, Elisangela Chicaroni Mattos, Heidge Fukumasu. USP-Botucatu: Rejane Maria Tommasini Grotto, Jayme A. Souza-Neto, Guilherme Targino Valente, Patricia Akemi Assato, Felipe Allan da Silva da Costa, Bianca Cechetto Carlos. Mendelics: Bibiana Santos, João Paulo Kitajima, Erika Freitas, David Schlesinger. Hemocentro Ribeirão Preto: Simone Kashima, Evandra Strazza Rodrigues, Svetoslav Nanev Slavov, Elaine Vieira dos Santos, Rafael dos Santos Bezerra, Luiz Carlos Junior de Alcantara, Marta Giovanetti, Vagner Fonseca, Flavia Aburjaile, Rodrigo Tocantins Calado. |

| | | | |
|---|---|---|---|
| EPI_ISL_1795098 | ESALQ | Instituto Butantan / ESALQ-Piracicaba | Instituto Butantan: Alexander Roberto Precioso, Dimas Tadeu Covas, Sandra Coccuzzo Sampaio, Maria Carolina Elias, José Salvatore Leister Patané, Vincent Louis Viala, Antonio Jorge Martins, Ricardo Haddad, Claudia Renata dos Santos Barros, Elaine Cristina Marqueze, Raul Machado Neto, Debora Botequio Moretti. Centro de Genômica Funcional da ESALQ: Luiz Lehmann Coutinho, Ricardo Augusto Brassaloti, Raquel de Lello Rocha Campos Cassano. NGS Soluções Genômicas: Pilar Drummond Sampaio Corrêa Mariani. FZEA-USP Pirassununga: Mirele Daiana Poleti, Jessika Cristina Chagas Lesbon, Elisangela Chicaroni Mattos, Heidge Fukumasu. USP-Botucatu: Rejane Maria Tommasini Grotto, Jayme A. Souza-Neto, Guilherme Targino Valente, Patricia Akemi Assato, Felipe Allan da Silva da Costa, Bianca Cechetto Carlos. Mendelics: Bibiana Santos, João Paulo Kitajima, Erika Freitas, David Schlesinger. Hemocentro Ribeirão Preto: Simone Kashima, Evandra Strazza Rodrigues, Svetoslav Nanev Slavov, Elaine Vieira dos Santos, Rafael dos Santos Bezerra, Luiz Carlos Junior de Alcantara, Marta Giovanetti, Vagner Fonseca, Flavia Aburjaile, Rodrigo Tocantins Calado. |
| EPI_ISL_1795107 | COORDENADORIA MUNICIPAL DE SAUDE DE IRACEMAPOLIS | Instituto Butantan / ESALQ-Piracicaba | Instituto Butantan: Alexander Roberto Precioso, Dimas Tadeu Covas, Sandra Coccuzzo Sampaio, Maria Carolina Elias, José Salvatore Leister Patané, Vincent Louis Viala, Antonio Jorge Martins, Ricardo Haddad, Claudia Renata dos Santos Barros, Elaine Cristina Marqueze, Raul Machado Neto, Debora Botequio Moretti. Centro de Genômica Funcional da ESALQ: Luiz Lehmann Coutinho, Ricardo Augusto Brassaloti, Raquel de Lello Rocha Campos Cassano. NGS Soluções Genômicas: Pilar Drummond Sampaio Corrêa Mariani. FZEA-USP Pirassununga: Mirele Daiana Poleti, Jessika Cristina Chagas Lesbon, Elisangela Chicaroni Mattos, Heidge Fukumasu. USP-Botucatu: Rejane Maria Tommasini Grotto, Jayme A. Souza-Neto, Guilherme Targino Valente, Patricia Akemi Assato, Felipe Allan da Silva da Costa, Bianca Cechetto Carlos. Mendelics: Bibiana Santos, João Paulo Kitajima, Erika Freitas, David Schlesinger. Hemocentro Ribeirão Preto: Simone Kashima, Evandra Strazza Rodrigues, Svetoslav Nanev Slavov, Elaine Vieira dos Santos, Rafael dos Santos Bezerra, Luiz Carlos Junior de Alcantara, Marta Giovanetti, Vagner Fonseca, Flavia Aburjaile, Rodrigo Tocantins Calado. |
| EPI_ISL_1795108 | SECRETARIA DE SAUDE DE SAO PEDRO | Instituto Butantan / ESALQ-Piracicaba | Instituto Butantan: Alexander Roberto Precioso, Dimas Tadeu Covas, Sandra Coccuzzo Sampaio, Maria Carolina Elias, José Salvatore Leister Patané, Vincent Louis Viala, Antonio Jorge Martins, Ricardo Haddad, Claudia Renata dos Santos Barros, Elaine Cristina Marqueze, Raul Machado Neto, Debora Botequio Moretti. Centro de Genômica Funcional da ESALQ: Luiz Lehmann Coutinho, Ricardo Augusto Brassaloti, Raquel de Lello Rocha Campos Cassano. NGS Soluções Genômicas: Pilar Drummond Sampaio Corrêa Mariani. FZEA-USP Pirassununga: Mirele Daiana Poleti, Jessika Cristina Chagas Lesbon, Elisangela Chicaroni Mattos, Heidge Fukumasu. USP-Botucatu: Rejane Maria Tommasini Grotto, Jayme A. Souza-Neto, Guilherme Targino Valente, Patricia Akemi Assato, Felipe Allan da Silva da Costa, Bianca Cechetto Carlos. Mendelics: Bibiana Santos, João Paulo Kitajima, Erika Freitas, David Schlesinger. Hemocentro Ribeirão Preto: Simone Kashima, Evandra Strazza Rodrigues, Svetoslav Nanev Slavov, Elaine Vieira dos Santos, Rafael dos Santos Bezerra, Luiz Carlos Junior de Alcantara, Marta Giovanetti, Vagner Fonseca, Flavia Aburjaile, Rodrigo Tocantins Calado. |
| EPI_ISL_1795110 | UNIDADE ESF DR LUIZ SPINA | Instituto Butantan / ESALQ-Piracicaba | Instituto Butantan: Alexander Roberto Precioso, Dimas Tadeu Covas, Sandra Coccuzzo Sampaio, Maria Carolina Elias, José Salvatore Leister Patané, Vincent Louis Viala, Antonio Jorge Martins, Ricardo Haddad, Claudia Renata dos Santos Barros, Elaine Cristina Marqueze, Raul Machado Neto, Debora Botequio Moretti. Centro de Genômica Funcional da ESALQ: Luiz Lehmann Coutinho, Ricardo Augusto Brassaloti, Raquel de Lello Rocha Campos Cassano. NGS Soluções Genômicas: Pilar Drummond Sampaio Corrêa Mariani. FZEA-USP Pirassununga: Mirele Daiana Poleti, Jessika Cristina Chagas Lesbon, Elisangela Chicaroni Mattos, Heidge Fukumasu. USP-Botucatu: Rejane Maria Tommasini Grotto, Jayme A. Souza-Neto, Guilherme Targino Valente, Patricia Akemi Assato, Felipe Allan da Silva da Costa, Bianca Cechetto Carlos. Mendelics: Bibiana Santos, João Paulo Kitajima, Erika Freitas, David Schlesinger. Hemocentro Ribeirão Preto: Simone Kashima, Evandra Strazza Rodrigues, Svetoslav Nanev Slavov, Elaine Vieira dos Santos, Rafael dos Santos Bezerra, Luiz Carlos Junior de Alcantara, Marta Giovanetti, Vagner Fonseca, Flavia Aburjaile, Rodrigo Tocantins Calado. |
| EPI_ISL_1795111, EPI_ISL_1795112 | SECRETARIA MUNICIPAL DA SAUDE DE GUARIBA | Instituto Butantan / ESALQ-Piracicaba | Instituto Butantan: Alexander Roberto Precioso, Dimas Tadeu Covas, Sandra Coccuzzo Sampaio, Maria Carolina Elias, José Salvatore Leister Patané, Vincent Louis Viala, Antonio Jorge Martins, Ricardo Haddad, Claudia Renata dos Santos Barros, Elaine Cristina Marqueze, Raul Machado Neto, Debora Botequio Moretti. Centro de Genômica Funcional da ESALQ: Luiz Lehmann Coutinho, Ricardo Augusto Brassaloti, Raquel de Lello Rocha Campos Cassano. NGS Soluções Genômicas: Pilar Drummond Sampaio Corrêa Mariani. FZEA-USP Pirassununga: Mirele Daiana Poleti, Jessika Cristina Chagas Lesbon, Elisangela Chicaroni Mattos, Heidge Fukumasu. USP-Botucatu: Rejane Maria Tommasini Grotto, Jayme A. Souza-Neto, Guilherme Targino Valente, Patricia Akemi Assato, Felipe Allan da Silva da Costa, Bianca Cechetto Carlos. Mendelics: Bibiana Santos, João Paulo Kitajima, Erika Freitas, David Schlesinger. Hemocentro Ribeirão Preto: Simone Kashima, Evandra Strazza Rodrigues, Svetoslav Nanev Slavov, Elaine Vieira dos Santos, Rafael dos Santos Bezerra, Luiz Carlos Junior de Alcantara, Marta Giovanetti, Vagner Fonseca, Flavia Aburjaile, Rodrigo Tocantins Calado. |
| EPI_ISL_1795113 | AMBULATORIO DE ATENDIMENTO DE DST DE GUARIBA | Instituto Butantan / ESALQ-Piracicaba | Instituto Butantan: Alexander Roberto Precioso, Dimas Tadeu Covas, Sandra Coccuzzo Sampaio, Maria Carolina Elias, José Salvatore Leister Patané, Vincent Louis Viala, Antonio Jorge Martins, Ricardo Haddad, Claudia Renata dos Santos Barros, Elaine Cristina Marqueze, Raul Machado Neto, Debora Botequio Moretti. Centro de Genômica Funcional da ESALQ: Luiz Lehmann Coutinho, Ricardo Augusto Brassaloti, Raquel de Lello Rocha Campos Cassano. NGS Soluções Genômicas: Pilar Drummond Sampaio Corrêa Mariani. FZEA-USP Pirassununga: Mirele Daiana Poleti, Jessika Cristina Chagas Lesbon, Elisangela Chicaroni Mattos, Heidge Fukumasu. USP-Botucatu: Rejane Maria Tommasini Grotto, Jayme A. Souza-Neto, Guilherme Targino Valente, Patricia Akemi Assato, Felipe Allan da Silva da Costa, Bianca Cechetto Carlos. Mendelics: Bibiana Santos, João Paulo Kitajima, Erika Freitas, David Schlesinger. Hemocentro Ribeirão Preto: Simone Kashima, Evandra Strazza Rodrigues, Svetoslav Nanev Slavov, Elaine Vieira dos Santos, Rafael dos Santos Bezerra, Luiz Carlos Junior de Alcantara, Marta Giovanetti, Vagner Fonseca, Flavia Aburjaile, Rodrigo Tocantins Calado. |
| EPI_ISL_1795114, EPI_ISL_1795115 | CENTRO DE SAUDE III BORBOREMA | Instituto Butantan / ESALQ-Piracicaba | Instituto Butantan: Alexander Roberto Precioso, Dimas Tadeu Covas, Sandra Coccuzzo Sampaio, Maria Carolina Elias, José Salvatore Leister Patané, Vincent Louis Viala, Antonio Jorge Martins, Ricardo Haddad, Claudia Renata dos Santos Barros, Elaine Cristina Marqueze, Raul Machado Neto, Debora Botequio Moretti. Centro de Genômica Funcional da ESALQ: Luiz Lehmann Coutinho, Ricardo Augusto Brassaloti, Raquel de Lello Rocha Campos Cassano. NGS Soluções Genômicas: Pilar Drummond Sampaio Corrêa Mariani. FZEA-USP Pirassununga: Mirele Daiana Poleti, Jessika Cristina Chagas Lesbon, Elisangela Chicaroni Mattos, Heidge Fukumasu. USP-Botucatu: Rejane Maria Tommasini Grotto, Jayme A. Souza-Neto, Guilherme Targino Valente, Patricia Akemi Assato, Felipe Allan da Silva da Costa, Bianca Cechetto Carlos. Mendelics: Bibiana Santos, João Paulo Kitajima, Erika Freitas, David Schlesinger. Hemocentro Ribeirão Preto: Simone Kashima, Evandra Strazza Rodrigues, Svetoslav Nanev Slavov, Elaine Vieira dos Santos, Rafael dos Santos Bezerra, Luiz Carlos Junior de Alcantara, Marta Giovanetti, Vagner Fonseca, Flavia Aburjaile, Rodrigo Tocantins Calado. |
| EPI_ISL_1795116, EPI_ISL_1795117, EPI_ISL_1795118, EPI_ISL_1795119 | UNIDADE MISTA DE LUIZ ANTONIO | Instituto Butantan / ESALQ-Piracicaba | Instituto Butantan: Alexander Roberto Precioso, Dimas Tadeu Covas, Sandra Coccuzzo Sampaio, Maria Carolina Elias, José Salvatore Leister Patané, Vincent Louis Viala, Antonio Jorge Martins, Ricardo Haddad, Claudia Renata dos Santos Barros, Elaine Cristina Marqueze, Raul Machado Neto, Debora Botequio Moretti. Centro de Genômica Funcional da ESALQ: Luiz Lehmann Coutinho, Ricardo Augusto Brassaloti, Raquel de Lello Rocha Campos Cassano. NGS Soluções Genômicas: Pilar Drummond Sampaio Corrêa Mariani. FZEA-USP Pirassununga: Mirele Daiana Poleti, Jessika Cristina Chagas Lesbon, Elisangela Chicaroni Mattos, Heidge Fukumasu. USP-Botucatu: Rejane Maria Tommasini Grotto, Jayme A. Souza-Neto, Guilherme Targino Valente, Patricia Akemi Assato, Felipe Allan da Silva da Costa, Bianca Cechetto Carlos. Mendelics: Bibiana Santos, João Paulo Kitajima, Erika Freitas, David Schlesinger. Hemocentro Ribeirão Preto: Simone Kashima, Evandra Strazza Rodrigues, Svetoslav Nanev Slavov, Elaine Vieira dos Santos, Rafael dos Santos Bezerra, Luiz Carlos Junior de Alcantara, Marta Giovanetti, Vagner Fonseca, Flavia Aburjaile, Rodrigo Tocantins Calado. |
| EPI_ISL_1795120, EPI_ISL_1795121 | CENTRO DE SAUDE DE CAJOBI | Instituto Butantan / ESALQ-Piracicaba | Instituto Butantan: Alexander Roberto Precioso, Dimas Tadeu Covas, Sandra Coccuzzo Sampaio, Maria Carolina Elias, José Salvatore Leister Patané, Vincent Louis Viala, Antonio Jorge Martins, Ricardo Haddad, Claudia Renata dos Santos Barros, Elaine Cristina Marqueze, Raul Machado Neto, Debora Botequio Moretti. Centro de Genômica Funcional da ESALQ: Luiz Lehmann Coutinho, Ricardo Augusto Brassaloti, Raquel de Lello Rocha Campos Cassano. NGS Soluções Genômicas: Pilar Drummond Sampaio Corrêa Mariani. FZEA-USP Pirassununga: Mirele Daiana Poleti, Jessika Cristina Chagas Lesbon, Elisangela Chicaroni Mattos, Heidge Fukumasu. USP-Botucatu: Rejane Maria Tommasini Grotto, Jayme A. Souza-Neto, Guilherme Targino Valente, Patricia Akemi Assato, Felipe Allan da Silva da Costa, Bianca Cechetto Carlos. Mendelics: Bibiana Santos, João Paulo Kitajima, Erika Freitas, David Schlesinger. Hemocentro Ribeirão Preto: Simone Kashima, Evandra Strazza Rodrigues, Svetoslav Nanev Slavov, Elaine Vieira dos Santos, Rafael dos Santos Bezerra, Luiz Carlos Junior de Alcantara, Marta Giovanetti, Vagner Fonseca, Flavia Aburjaile, Rodrigo Tocantins Calado. |
| EPI_ISL_1795122, EPI_ISL_1795123, EPI_ISL_1795124 | UNIDADE DE VIGILANCIA EM SAUDE | Instituto Butantan / ESALQ-Piracicaba | Instituto Butantan: Alexander Roberto Precioso, Dimas Tadeu Covas, Sandra Coccuzzo Sampaio, Maria Carolina Elias, José Salvatore Leister Patané, Vincent Louis Viala, Antonio Jorge Martins, Ricardo Haddad, Claudia Renata dos Santos Barros, Elaine Cristina Marqueze, Raul Machado Neto, Debora Botequio Moretti. Centro de Genômica Funcional da ESALQ: Luiz Lehmann Coutinho, Ricardo Augusto Brassaloti, Raquel de Lello Rocha Campos Cassano. NGS Soluções Genômicas: Pilar Drummond Sampaio Corrêa Mariani. FZEA-USP Pirassununga: Mirele Daiana Poleti, Jessika Cristina Chagas Lesbon, Elisangela Chicaroni Mattos, Heidge Fukumasu. USP-Botucatu: Rejane Maria Tommasini Grotto, Jayme A. Souza-Neto, Guilherme Targino Valente, Patricia Akemi Assato, Felipe Allan da Silva da Costa, Bianca Cechetto Carlos. Mendelics: Bibiana Santos, João Paulo Kitajima, Erika Freitas, David Schlesinger. Hemocentro Ribeirão Preto: Simone Kashima, Evandra Strazza Rodrigues, Svetoslav Nanev Slavov, Elaine Vieira dos Santos, Rafael dos Santos Bezerra, Luiz Carlos Junior de Alcantara, Marta Giovanetti, Vagner Fonseca, Flavia Aburjaile, Rodrigo Tocantins Calado. |
| EPI_ISL_1795125 | CENTRO DE SAUDE III CANDIDO RODRIGUES | Instituto Butantan / ESALQ-Piracicaba | Instituto Butantan: Alexander Roberto Precioso, Dimas Tadeu Covas, Sandra Coccuzzo Sampaio, Maria Carolina Elias, José Salvatore Leister Patané, Vincent Louis Viala, Antonio Jorge Martins, Ricardo Haddad, Claudia Renata dos Santos Barros, Elaine Cristina Marqueze, Raul Machado Neto, Debora |

| | | | |
|---|---|---|---|
| | | | Botequio Moretti. Centro de Genômica Funcional da ESALQ: Luiz Lehmann Coutinho, Ricardo Augusto Brassaloti, Raquel de Lello Rocha Campos Cassano. NGS Soluções Genômicas: Pilar Drummond Sampaio Corrêa Mariani. FZEA-USP Pirassununga: Mirele Daiana Poleti, Jessika Cristina Chagas Lesbon, Elisangela Chicaroni Mattos, Heidge Fukumasu. USP-Botucatu: Rejane Maria Tommasini Grotto, Jayme A. Souza-Neto, Guilherme Targino Valente, Patricia Akemi Assato, Felipe Allan da Silva da Costa, Bianca Cechetto Carlos. Mendelics: Bibiana Santos, João Paulo Kitajima, Erika Freitas, David Schlesinger. Hemocentro Ribeirão Preto: Simone Kashima, Evandra Strazza Rodrigues, Svetoslav Nanev Slavov, Elaine Vieira dos Santos, Rafael dos Santos Bezerra, Luiz Carlos Junior de Alcantara, Marta Giovanetti, Vagner Fonseca, Flavia Aburjaile, Rodrigo Tocantins Calado. |
| EPI_ISL_1795126 | CENTRO DE SAUDE III SALES OLIVEIRA | Instituto Butantan / ESALQ-Piracicaba | Instituto Butantan: Alexander Roberto Precioso, Dimas Tadeu Covas, Sandra Coccuzzo Sampaio, Maria Carolina Elias, José Salvatore Leister Patané, Vincent Louis Viala, Antonio Jorge Martins, Ricardo Haddad, Claudia Renata dos Santos Barros, Elaine Cristina Marqueze, Raul Machado Neto, Debora Botequio Moretti. Centro de Genômica Funcional da ESALQ: Luiz Lehmann Coutinho, Ricardo Augusto Brassaloti, Raquel de Lello Rocha Campos Cassano. NGS Soluções Genômicas: Pilar Drummond Sampaio Corrêa Mariani. FZEA-USP Pirassununga: Mirele Daiana Poleti, Jessika Cristina Chagas Lesbon, Elisangela Chicaroni Mattos, Heidge Fukumasu. USP-Botucatu: Rejane Maria Tommasini Grotto, Jayme A. Souza-Neto, Guilherme Targino Valente, Patricia Akemi Assato, Felipe Allan da Silva da Costa, Bianca Cechetto Carlos. Mendelics: Bibiana Santos, João Paulo Kitajima, Erika Freitas, David Schlesinger. Hemocentro Ribeirão Preto: Simone Kashima, Evandra Strazza Rodrigues, Svetoslav Nanev Slavov, Elaine Vieira dos Santos, Rafael dos Santos Bezerra, Luiz Carlos Junior de Alcantara, Marta Giovanetti, Vagner Fonseca, Flavia Aburjaile, Rodrigo Tocantins Calado. |
| EPI_ISL_1795127 | CENTRO DE SAUDE III CANDIDO RODRIGUES | Instituto Butantan / ESALQ-Piracicaba | Instituto Butantan: Alexander Roberto Precioso, Dimas Tadeu Covas, Sandra Coccuzzo Sampaio, Maria Carolina Elias, José Salvatore Leister Patané, Vincent Louis Viala, Antonio Jorge Martins, Ricardo Haddad, Claudia Renata dos Santos Barros, Elaine Cristina Marqueze, Raul Machado Neto, Debora Botequio Moretti. Centro de Genômica Funcional da ESALQ: Luiz Lehmann Coutinho, Ricardo Augusto Brassaloti, Raquel de Lello Rocha Campos Cassano. NGS Soluções Genômicas: Pilar Drummond Sampaio Corrêa Mariani. FZEA-USP Pirassununga: Mirele Daiana Poleti, Jessika Cristina Chagas Lesbon, Elisangela Chicaroni Mattos, Heidge Fukumasu. USP-Botucatu: Rejane Maria Tommasini Grotto, Jayme A. Souza-Neto, Guilherme Targino Valente, Patricia Akemi Assato, Felipe Allan da Silva da Costa, Bianca Cechetto Carlos. Mendelics: Bibiana Santos, João Paulo Kitajima, Erika Freitas, David Schlesinger. Hemocentro Ribeirão Preto: Simone Kashima, Evandra Strazza Rodrigues, Svetoslav Nanev Slavov, Elaine Vieira dos Santos, Rafael dos Santos Bezerra, Luiz Carlos Junior de Alcantara, Marta Giovanetti, Vagner Fonseca, Flavia Aburjaile, Rodrigo Tocantins Calado. |
| EPI_ISL_1795128, EPI_ISL_1795129, EPI_ISL_1795130, EPI_ISL_1795131, EPI_ISL_1795132, EPI_ISL_1795133, EPI_ISL_1795134, EPI_ISL_1795135 | UNIDADE DE VIGILANCIA EM SAUDE | Instituto Butantan / ESALQ-Piracicaba | Instituto Butantan: Alexander Roberto Precioso, Dimas Tadeu Covas, Sandra Coccuzzo Sampaio, Maria Carolina Elias, José Salvatore Leister Patané, Vincent Louis Viala, Antonio Jorge Martins, Ricardo Haddad, Claudia Renata dos Santos Barros, Elaine Cristina Marqueze, Raul Machado Neto, Debora Botequio Moretti. Centro de Genômica Funcional da ESALQ: Luiz Lehmann Coutinho, Ricardo Augusto Brassaloti, Raquel de Lello Rocha Campos Cassano. NGS Soluções Genômicas: Pilar Drummond Sampaio Corrêa Mariani. FZEA-USP Pirassununga: Mirele Daiana Poleti, Jessika Cristina Chagas Lesbon, Elisangela Chicaroni Mattos, Heidge Fukumasu. USP-Botucatu: Rejane Maria Tommasini Grotto, Jayme A. Souza-Neto, Guilherme Targino Valente, Patricia Akemi Assato, Felipe Allan da Silva da Costa, Bianca Cechetto Carlos. Mendelics: Bibiana Santos, João Paulo Kitajima, Erika Freitas, David Schlesinger. Hemocentro Ribeirão Preto: Simone Kashima, Evandra Strazza Rodrigues, Svetoslav Nanev Slavov, Elaine Vieira dos Santos, Rafael dos Santos Bezerra, Luiz Carlos Junior de Alcantara, Marta Giovanetti, Vagner Fonseca, Flavia Aburjaile, Rodrigo Tocantins Calado. |
| EPI_ISL_1795136, EPI_ISL_1795137, EPI_ISL_1795138, EPI_ISL_1795139, EPI_ISL_1795140 | UNIDADE MISTA DE VISTA ALEGRE DO ALTO VISTA ALEGRE DO ALTO | Instituto Butantan / ESALQ-Piracicaba | Instituto Butantan: Alexander Roberto Precioso, Dimas Tadeu Covas, Sandra Coccuzzo Sampaio, Maria Carolina Elias, José Salvatore Leister Patané, Vincent Louis Viala, Antonio Jorge Martins, Ricardo Haddad, Claudia Renata dos Santos Barros, Elaine Cristina Marqueze, Raul Machado Neto, Debora Botequio Moretti. Centro de Genômica Funcional da ESALQ: Luiz Lehmann Coutinho, Ricardo Augusto Brassaloti, Raquel de Lello Rocha Campos Cassano. NGS Soluções Genômicas: Pilar Drummond Sampaio Corrêa Mariani. FZEA-USP Pirassununga: Mirele Daiana Poleti, Jessika Cristina Chagas Lesbon, Elisangela Chicaroni Mattos, Heidge Fukumasu. USP-Botucatu: Rejane Maria Tommasini Grotto, Jayme A. Souza-Neto, Guilherme Targino Valente, Patricia Akemi Assato, Felipe Allan da Silva da Costa, Bianca Cechetto Carlos. Mendelics: Bibiana Santos, João Paulo Kitajima, Erika Freitas, David Schlesinger. Hemocentro Ribeirão Preto: Simone Kashima, Evandra Strazza Rodrigues, Svetoslav Nanev Slavov, Elaine Vieira dos Santos, Rafael dos Santos Bezerra, Luiz Carlos Junior de Alcantara, Marta Giovanetti, Vagner Fonseca, Flavia Aburjaile, Rodrigo Tocantins Calado. |
| EPI_ISL_1795141, EPI_ISL_1795142, EPI_ISL_1795143, EPI_ISL_1795144, EPI_ISL_1795145, EPI_ISL_1795146, EPI_ISL_1795147, EPI_ISL_1795148 | POLICLINICA HORTOLANDIA | Instituto Butantan / ESALQ-Piracicaba | Instituto Butantan: Alexander Roberto Precioso, Dimas Tadeu Covas, Sandra Coccuzzo Sampaio, Maria Carolina Elias, José Salvatore Leister Patané, Vincent Louis Viala, Antonio Jorge Martins, Ricardo Haddad, Claudia Renata dos Santos Barros, Elaine Cristina Marqueze, Raul Machado Neto, Debora Botequio Moretti. Centro de Genômica Funcional da ESALQ: Luiz Lehmann Coutinho, Ricardo Augusto Brassaloti, Raquel de Lello Rocha Campos Cassano. NGS Soluções Genômicas: Pilar Drummond Sampaio Corrêa Mariani. FZEA-USP Pirassununga: Mirele Daiana Poleti, Jessika Cristina Chagas Lesbon, Elisangela Chicaroni Mattos, Heidge Fukumasu. USP-Botucatu: Rejane Maria Tommasini Grotto, Jayme A. Souza-Neto, Guilherme Targino Valente, Patricia Akemi Assato, Felipe Allan da Silva da Costa, Bianca Cechetto Carlos. Mendelics: Bibiana Santos, João Paulo Kitajima, Erika Freitas, David Schlesinger. Hemocentro Ribeirão Preto: Simone Kashima, Evandra Strazza Rodrigues, Svetoslav Nanev Slavov, Elaine Vieira dos Santos, Rafael dos Santos Bezerra, Luiz Carlos Junior de Alcantara, Marta Giovanetti, Vagner Fonseca, Flavia Aburjaile, Rodrigo Tocantins Calado. |
| EPI_ISL_1795149, EPI_ISL_1795150 | UNIDADE MISTA DE VISTA ALEGRE DO ALTO VISTA ALEGRE DO ALTO | Instituto Butantan / ESALQ-Piracicaba | Instituto Butantan: Alexander Roberto Precioso, Dimas Tadeu Covas, Sandra Coccuzzo Sampaio, Maria Carolina Elias, José Salvatore Leister Patané, Vincent Louis Viala, Antonio Jorge Martins, Ricardo Haddad, Claudia Renata dos Santos Barros, Elaine Cristina Marqueze, Raul Machado Neto, Debora Botequio Moretti. Centro de Genômica Funcional da ESALQ: Luiz Lehmann Coutinho, Ricardo Augusto Brassaloti, Raquel de Lello Rocha Campos Cassano. NGS Soluções Genômicas: Pilar Drummond Sampaio Corrêa Mariani. FZEA-USP Pirassununga: Mirele Daiana Poleti, Jessika Cristina Chagas Lesbon, Elisangela Chicaroni Mattos, Heidge Fukumasu. USP-Botucatu: Rejane Maria Tommasini Grotto, Jayme A. Souza-Neto, Guilherme Targino Valente, Patricia Akemi Assato, Felipe Allan da Silva da Costa, Bianca Cechetto Carlos. Mendelics: Bibiana Santos, João Paulo Kitajima, Erika Freitas, David Schlesinger. Hemocentro Ribeirão Preto: Simone Kashima, Evandra Strazza Rodrigues, Svetoslav Nanev Slavov, Elaine Vieira dos Santos, Rafael dos Santos Bezerra, Luiz Carlos Junior de Alcantara, Marta Giovanetti, Vagner Fonseca, Flavia Aburjaile, Rodrigo Tocantins Calado. |
| EPI_ISL_1795151 | CENTRO DE SAUDE DE CAJOBI | Instituto Butantan / ESALQ-Piracicaba | Instituto Butantan: Alexander Roberto Precioso, Dimas Tadeu Covas, Sandra Coccuzzo Sampaio, Maria Carolina Elias, José Salvatore Leister Patané, Vincent Louis Viala, Antonio Jorge Martins, Ricardo Haddad, Claudia Renata dos Santos Barros, Elaine Cristina Marqueze, Raul Machado Neto, Debora Botequio Moretti. Centro de Genômica Funcional da ESALQ: Luiz Lehmann Coutinho, Ricardo Augusto Brassaloti, Raquel de Lello Rocha Campos Cassano. NGS Soluções Genômicas: Pilar Drummond Sampaio Corrêa Mariani. FZEA-USP Pirassununga: Mirele Daiana Poleti, Jessika Cristina Chagas Lesbon, Elisangela Chicaroni Mattos, Heidge Fukumasu. USP-Botucatu: Rejane Maria Tommasini Grotto, Jayme A. Souza-Neto, Guilherme Targino Valente, Patricia Akemi Assato, Felipe Allan da Silva da Costa, Bianca Cechetto Carlos. Mendelics: Bibiana Santos, João Paulo Kitajima, Erika Freitas, David Schlesinger. Hemocentro Ribeirão Preto: Simone Kashima, Evandra Strazza Rodrigues, Svetoslav Nanev Slavov, Elaine Vieira dos Santos, Rafael dos Santos Bezerra, Luiz Carlos Junior de Alcantara, Marta Giovanetti, Vagner Fonseca, Flavia Aburjaile, Rodrigo Tocantins Calado. |
| EPI_ISL_1795152, EPI_ISL_1795153, EPI_ISL_1795154, EPI_ISL_1795155, EPI_ISL_1795156, EPI_ISL_1795157, EPI_ISL_1795158, EPI_ISL_1795159 | POLICLINICA HORTOLANDIA | Instituto Butantan / ESALQ-Piracicaba | Instituto Butantan: Alexander Roberto Precioso, Dimas Tadeu Covas, Sandra Coccuzzo Sampaio, Maria Carolina Elias, José Salvatore Leister Patané, Vincent Louis Viala, Antonio Jorge Martins, Ricardo Haddad, Claudia Renata dos Santos Barros, Elaine Cristina Marqueze, Raul Machado Neto, Debora Botequio Moretti. Centro de Genômica Funcional da ESALQ: Luiz Lehmann Coutinho, Ricardo Augusto Brassaloti, Raquel de Lello Rocha Campos Cassano. NGS Soluções Genômicas: Pilar Drummond Sampaio Corrêa Mariani. FZEA-USP Pirassununga: Mirele Daiana Poleti, Jessika Cristina Chagas Lesbon, Elisangela Chicaroni Mattos, Heidge Fukumasu. USP-Botucatu: Rejane Maria Tommasini Grotto, Jayme A. Souza-Neto, Guilherme Targino Valente, Patricia Akemi Assato, Felipe Allan da Silva da Costa, Bianca Cechetto Carlos. Mendelics: Bibiana Santos, João Paulo Kitajima, Erika Freitas, David Schlesinger. Hemocentro Ribeirão Preto: Simone Kashima, Evandra Strazza Rodrigues, Svetoslav Nanev Slavov, Elaine Vieira dos Santos, Rafael dos Santos Bezerra, Luiz Carlos Junior de Alcantara, Marta Giovanetti, Vagner Fonseca, Flavia Aburjaile, Rodrigo Tocantins Calado. |
| EPI_ISL_1795160 | UBS JARDIM ITAMARATY | Instituto Butantan / ESALQ-Piracicaba | Instituto Butantan: Alexander Roberto Precioso, Dimas Tadeu Covas, Sandra Coccuzzo Sampaio, Maria Carolina Elias, José Salvatore Leister Patané, Vincent Louis Viala, Antonio Jorge Martins, Ricardo Haddad, Claudia Renata dos Santos Barros, Elaine Cristina Marqueze, Raul Machado Neto, Debora Botequio Moretti. Centro de Genômica Funcional da ESALQ: Luiz Lehmann Coutinho, Ricardo Augusto Brassaloti, Raquel de Lello Rocha Campos Cassano. NGS Soluções Genômicas: Pilar Drummond Sampaio Corrêa Mariani. FZEA-USP Pirassununga: Mirele Daiana Poleti, Jessika Cristina Chagas Lesbon, Elisangela Chicaroni Mattos, Heidge Fukumasu. USP-Botucatu: Rejane Maria Tommasini Grotto, Jayme A. Souza-Neto, Guilherme Targino Valente, Patricia Akemi Assato, Felipe Allan da Silva da Costa, Bianca Cechetto Carlos. Mendelics: Bibiana Santos, João Paulo Kitajima, Erika Freitas, David Schlesinger. Hemocentro Ribeirão Preto: Simone Kashima, Evandra Strazza Rodrigues, Svetoslav Nanev Slavov, Elaine Vieira dos Santos, Rafael dos Santos Bezerra, Luiz Carlos Junior de Alcantara, Marta Giovanetti, Vagner Fonseca, Flavia Aburjaile, Rodrigo Tocantins Calado. |
| EPI_ISL_1795161 | CS III VILA ODILON | Instituto Butantan / ESALQ-Piracicaba | Instituto Butantan: Alexander Roberto Precioso, Dimas Tadeu Covas, Sandra Coccuzzo Sampaio, Maria Carolina Elias, José Salvatore Leister Patané, Vincent Louis Viala, Antonio Jorge Martins, Ricardo Haddad, Claudia Renata dos Santos Barros, Elaine Cristina Marqueze, Raul Machado Neto, Debora Botequio Moretti. Centro de Genômica Funcional da ESALQ: Luiz Lehmann Coutinho, Ricardo Augusto Brassaloti, Raquel de Lello Rocha Campos Cassano. NGS Soluções Genômicas: Pilar Drummond Sampaio Corrêa Mariani. FZEA-USP Pirassununga: Mirele Daiana Poleti, Jessika Cristina Chagas |

| | | | |
|---|---|---|---|
| | | | Lesbon, Elisangela Chicaroni Mattos, Heidge Fukumasu. USP-Botucatu: Rejane Maria Tommasini Grotto, Jayme A. Souza-Neto, Guilherme Targino Valente, Patricia Akemi Assato, Felipe Allan da Silva da Costa, Bianca Cechetto Carlos. Mendelics: Bibiana Santos, João Paulo Kitajima, Erika Freitas, David Schlesinger. Hemocentro Ribeirão Preto: Simone Kashima, Evandra Strazza Rodrigues, Svetoslav Nanev Slavov, Elaine Vieira dos Santos, Rafael dos Santos Bezerra, Luiz Carlos Junior de Alcantara, Marta Giovanetti, Vagner Fonseca, Flavia Aburjaile, Rodrigo Tocantins Calado. |
| EPI_ISL_1795162, EPI_ISL_1795163, EPI_ISL_1795164 | CENTRO DE SAUDE DR PLINIO ALBERS DE GALIA | Instituto Butantan / ESALQ-Piracicaba | Instituto Butantan: Alexander Roberto Precioso, Dimas Tadeu Covas, Sandra Coccuzzo Sampaio, Maria Carolina Elias, José Salvatore Leister Patané, Vincent Louis Viala, Antonio Jorge Martins, Ricardo Haddad, Claudia Renata dos Santos Barros, Elaine Cristina Marqueze, Raul Machado Neto, Debora Botequio Moretti. Centro de Genômica Funcional da ESALQ: Luiz Lehmann Coutinho, Ricardo Augusto Brassaloti, Raquel de Lello Rocha Campos Cassano. NGS Soluções Genômicas: Pilar Drummond Sampaio Corrêa Mariani. FZEA-USP Pirassununga: Mirele Daiana Poleti, Jessika Cristina Chagas Lesbon, Elisangela Chicaroni Mattos, Heidge Fukumasu. USP-Botucatu: Rejane Maria Tommasini Grotto, Jayme A. Souza-Neto, Guilherme Targino Valente, Patricia Akemi Assato, Felipe Allan da Silva da Costa, Bianca Cechetto Carlos. Mendelics: Bibiana Santos, João Paulo Kitajima, Erika Freitas, David Schlesinger. Hemocentro Ribeirão Preto: Simone Kashima, Evandra Strazza Rodrigues, Svetoslav Nanev Slavov, Elaine Vieira dos Santos, Rafael dos Santos Bezerra, Luiz Carlos Junior de Alcantara, Marta Giovanetti, Vagner Fonseca, Flavia Aburjaile, Rodrigo Tocantins Calado. |
| EPI_ISL_1795165 | CS DE ALVARO CARVALHO | Instituto Butantan / ESALQ-Piracicaba | Instituto Butantan: Alexander Roberto Precioso, Dimas Tadeu Covas, Sandra Coccuzzo Sampaio, Maria Carolina Elias, José Salvatore Leister Patané, Vincent Louis Viala, Antonio Jorge Martins, Ricardo Haddad, Claudia Renata dos Santos Barros, Elaine Cristina Marqueze, Raul Machado Neto, Debora Botequio Moretti. Centro de Genômica Funcional da ESALQ: Luiz Lehmann Coutinho, Ricardo Augusto Brassaloti, Raquel de Lello Rocha Campos Cassano. NGS Soluções Genômicas: Pilar Drummond Sampaio Corrêa Mariani. FZEA-USP Pirassununga: Mirele Daiana Poleti, Jessika Cristina Chagas Lesbon, Elisangela Chicaroni Mattos, Heidge Fukumasu. USP-Botucatu: Rejane Maria Tommasini Grotto, Jayme A. Souza-Neto, Guilherme Targino Valente, Patricia Akemi Assato, Felipe Allan da Silva da Costa, Bianca Cechetto Carlos. Mendelics: Bibiana Santos, João Paulo Kitajima, Erika Freitas, David Schlesinger. Hemocentro Ribeirão Preto: Simone Kashima, Evandra Strazza Rodrigues, Svetoslav Nanev Slavov, Elaine Vieira dos Santos, Rafael dos Santos Bezerra, Luiz Carlos Junior de Alcantara, Marta Giovanetti, Vagner Fonseca, Flavia Aburjaile, Rodrigo Tocantins Calado. |
| EPI_ISL_1795166 | CENTRO DE SAUDE DR PLINIO ALBERS DE GALIA | Instituto Butantan / ESALQ-Piracicaba | Instituto Butantan: Alexander Roberto Precioso, Dimas Tadeu Covas, Sandra Coccuzzo Sampaio, Maria Carolina Elias, José Salvatore Leister Patané, Vincent Louis Viala, Antonio Jorge Martins, Ricardo Haddad, Claudia Renata dos Santos Barros, Elaine Cristina Marqueze, Raul Machado Neto, Debora Botequio Moretti. Centro de Genômica Funcional da ESALQ: Luiz Lehmann Coutinho, Ricardo Augusto Brassaloti, Raquel de Lello Rocha Campos Cassano. NGS Soluções Genômicas: Pilar Drummond Sampaio Corrêa Mariani. FZEA-USP Pirassununga: Mirele Daiana Poleti, Jessika Cristina Chagas Lesbon, Elisangela Chicaroni Mattos, Heidge Fukumasu. USP-Botucatu: Rejane Maria Tommasini Grotto, Jayme A. Souza-Neto, Guilherme Targino Valente, Patricia Akemi Assato, Felipe Allan da Silva da Costa, Bianca Cechetto Carlos. Mendelics: Bibiana Santos, João Paulo Kitajima, Erika Freitas, David Schlesinger. Hemocentro Ribeirão Preto: Simone Kashima, Evandra Strazza Rodrigues, Svetoslav Nanev Slavov, Elaine Vieira dos Santos, Rafael dos Santos Bezerra, Luiz Carlos Junior de Alcantara, Marta Giovanetti, Vagner Fonseca, Flavia Aburjaile, Rodrigo Tocantins Calado. |
| EPI_ISL_1795167, EPI_ISL_1795168, EPI_ISL_1795169 | CENTRO DE SAUDE DE FLORINEA | Instituto Butantan / ESALQ-Piracicaba | Instituto Butantan: Alexander Roberto Precioso, Dimas Tadeu Covas, Sandra Coccuzzo Sampaio, Maria Carolina Elias, José Salvatore Leister Patané, Vincent Louis Viala, Antonio Jorge Martins, Ricardo Haddad, Claudia Renata dos Santos Barros, Elaine Cristina Marqueze, Raul Machado Neto, Debora Botequio Moretti. Centro de Genômica Funcional da ESALQ: Luiz Lehmann Coutinho, Ricardo Augusto Brassaloti, Raquel de Lello Rocha Campos Cassano. NGS Soluções Genômicas: Pilar Drummond Sampaio Corrêa Mariani. FZEA-USP Pirassununga: Mirele Daiana Poleti, Jessika Cristina Chagas Lesbon, Elisangela Chicaroni Mattos, Heidge Fukumasu. USP-Botucatu: Rejane Maria Tommasini Grotto, Jayme A. Souza-Neto, Guilherme Targino Valente, Patricia Akemi Assato, Felipe Allan da Silva da Costa, Bianca Cechetto Carlos. Mendelics: Bibiana Santos, João Paulo Kitajima, Erika Freitas, David Schlesinger. Hemocentro Ribeirão Preto: Simone Kashima, Evandra Strazza Rodrigues, Svetoslav Nanev Slavov, Elaine Vieira dos Santos, Rafael dos Santos Bezerra, Luiz Carlos Junior de Alcantara, Marta Giovanetti, Vagner Fonseca, Flavia Aburjaile, Rodrigo Tocantins Calado. |
| EPI_ISL_1795170 | CENTRO DE SAUDE DE MARACAI | Instituto Butantan / ESALQ-Piracicaba | Instituto Butantan: Alexander Roberto Precioso, Dimas Tadeu Covas, Sandra Coccuzzo Sampaio, Maria Carolina Elias, José Salvatore Leister Patané, Vincent Louis Viala, Antonio Jorge Martins, Ricardo Haddad, Claudia Renata dos Santos Barros, Elaine Cristina Marqueze, Raul Machado Neto, Debora Botequio Moretti. Centro de Genômica Funcional da ESALQ: Luiz Lehmann Coutinho, Ricardo Augusto Brassaloti, Raquel de Lello Rocha Campos Cassano. NGS Soluções Genômicas: Pilar Drummond Sampaio Corrêa Mariani. FZEA-USP Pirassununga: Mirele Daiana Poleti, Jessika Cristina Chagas Lesbon, Elisangela Chicaroni Mattos, Heidge Fukumasu. USP-Botucatu: Rejane Maria Tommasini Grotto, Jayme A. Souza-Neto, Guilherme Targino Valente, Patricia Akemi Assato, Felipe Allan da Silva da Costa, Bianca Cechetto Carlos. Mendelics: Bibiana Santos, João Paulo Kitajima, Erika Freitas, David Schlesinger. Hemocentro Ribeirão Preto: Simone Kashima, Evandra Strazza Rodrigues, Svetoslav Nanev Slavov, Elaine Vieira dos Santos, Rafael dos Santos Bezerra, Luiz Carlos Junior de Alcantara, Marta Giovanetti, Vagner Fonseca, Flavia Aburjaile, Rodrigo Tocantins Calado. |
| EPI_ISL_1795171 | CS DE OSCAR BRESSANE PSF | Instituto Butantan / ESALQ-Piracicaba | Instituto Butantan: Alexander Roberto Precioso, Dimas Tadeu Covas, Sandra Coccuzzo Sampaio, Maria Carolina Elias, José Salvatore Leister Patané, Vincent Louis Viala, Antonio Jorge Martins, Ricardo Haddad, Claudia Renata dos Santos Barros, Elaine Cristina Marqueze, Raul Machado Neto, Debora Botequio Moretti. Centro de Genômica Funcional da ESALQ: Luiz Lehmann Coutinho, Ricardo Augusto Brassaloti, Raquel de Lello Rocha Campos Cassano. NGS Soluções Genômicas: Pilar Drummond Sampaio Corrêa Mariani. FZEA-USP Pirassununga: Mirele Daiana Poleti, Jessika Cristina Chagas Lesbon, Elisangela Chicaroni Mattos, Heidge Fukumasu. USP-Botucatu: Rejane Maria Tommasini Grotto, Jayme A. Souza-Neto, Guilherme Targino Valente, Patricia Akemi Assato, Felipe Allan da Silva da Costa, Bianca Cechetto Carlos. Mendelics: Bibiana Santos, João Paulo Kitajima, Erika Freitas, David Schlesinger. Hemocentro Ribeirão Preto: Simone Kashima, Evandra Strazza Rodrigues, Svetoslav Nanev Slavov, Elaine Vieira dos Santos, Rafael dos Santos Bezerra, Luiz Carlos Junior de Alcantara, Marta Giovanetti, Vagner Fonseca, Flavia Aburjaile, Rodrigo Tocantins Calado. |
| EPI_ISL_1795172 | CENTRO DE SAUDE DE MARACAI | Instituto Butantan / ESALQ-Piracicaba | Instituto Butantan: Alexander Roberto Precioso, Dimas Tadeu Covas, Sandra Coccuzzo Sampaio, Maria Carolina Elias, José Salvatore Leister Patané, Vincent Louis Viala, Antonio Jorge Martins, Ricardo Haddad, Claudia Renata dos Santos Barros, Elaine Cristina Marqueze, Raul Machado Neto, Debora Botequio Moretti. Centro de Genômica Funcional da ESALQ: Luiz Lehmann Coutinho, Ricardo Augusto Brassaloti, Raquel de Lello Rocha Campos Cassano. NGS Soluções Genômicas: Pilar Drummond Sampaio Corrêa Mariani. FZEA-USP Pirassununga: Mirele Daiana Poleti, Jessika Cristina Chagas Lesbon, Elisangela Chicaroni Mattos, Heidge Fukumasu. USP-Botucatu: Rejane Maria Tommasini Grotto, Jayme A. Souza-Neto, Guilherme Targino Valente, Patricia Akemi Assato, Felipe Allan da Silva da Costa, Bianca Cechetto Carlos. Mendelics: Bibiana Santos, João Paulo Kitajima, Erika Freitas, David Schlesinger. Hemocentro Ribeirão Preto: Simone Kashima, Evandra Strazza Rodrigues, Svetoslav Nanev Slavov, Elaine Vieira dos Santos, Rafael dos Santos Bezerra, Luiz Carlos Junior de Alcantara, Marta Giovanetti, Vagner Fonseca, Flavia Aburjaile, Rodrigo Tocantins Calado. |
| EPI_ISL_1795173 | CS DE OSCAR BRESSANE PSF | Instituto Butantan / ESALQ-Piracicaba | Instituto Butantan: Alexander Roberto Precioso, Dimas Tadeu Covas, Sandra Coccuzzo Sampaio, Maria Carolina Elias, José Salvatore Leister Patané, Vincent Louis Viala, Antonio Jorge Martins, Ricardo Haddad, Claudia Renata dos Santos Barros, Elaine Cristina Marqueze, Raul Machado Neto, Debora Botequio Moretti. Centro de Genômica Funcional da ESALQ: Luiz Lehmann Coutinho, Ricardo Augusto Brassaloti, Raquel de Lello Rocha Campos Cassano. NGS Soluções Genômicas: Pilar Drummond Sampaio Corrêa Mariani. FZEA-USP Pirassununga: Mirele Daiana Poleti, Jessika Cristina Chagas Lesbon, Elisangela Chicaroni Mattos, Heidge Fukumasu. USP-Botucatu: Rejane Maria Tommasini Grotto, Jayme A. Souza-Neto, Guilherme Targino Valente, Patricia Akemi Assato, Felipe Allan da Silva da Costa, Bianca Cechetto Carlos. Mendelics: Bibiana Santos, João Paulo Kitajima, Erika Freitas, David Schlesinger. Hemocentro Ribeirão Preto: Simone Kashima, Evandra Strazza Rodrigues, Svetoslav Nanev Slavov, Elaine Vieira dos Santos, Rafael dos Santos Bezerra, Luiz Carlos Junior de Alcantara, Marta Giovanetti, Vagner Fonseca, Flavia Aburjaile, Rodrigo Tocantins Calado. |
| EPI_ISL_1795174, EPI_ISL_1795175, EPI_ISL_1795176 | USF SALERNO | Instituto Butantan / ESALQ-Piracicaba | Instituto Butantan: Alexander Roberto Precioso, Dimas Tadeu Covas, Sandra Coccuzzo Sampaio, Maria Carolina Elias, José Salvatore Leister Patané, Vincent Louis Viala, Antonio Jorge Martins, Ricardo Haddad, Claudia Renata dos Santos Barros, Elaine Cristina Marqueze, Raul Machado Neto, Debora Botequio Moretti. Centro de Genômica Funcional da ESALQ: Luiz Lehmann Coutinho, Ricardo Augusto Brassaloti, Raquel de Lello Rocha Campos Cassano. NGS Soluções Genômicas: Pilar Drummond Sampaio Corrêa Mariani. FZEA-USP Pirassununga: Mirele Daiana Poleti, Jessika Cristina Chagas Lesbon, Elisangela Chicaroni Mattos, Heidge Fukumasu. USP-Botucatu: Rejane Maria Tommasini Grotto, Jayme A. Souza-Neto, Guilherme Targino Valente, Patricia Akemi Assato, Felipe Allan da Silva da Costa, Bianca Cechetto Carlos. Mendelics: Bibiana Santos, João Paulo Kitajima, Erika Freitas, David Schlesinger. Hemocentro Ribeirão Preto: Simone Kashima, Evandra Strazza Rodrigues, Svetoslav Nanev Slavov, Elaine Vieira dos Santos, Rafael dos Santos Bezerra, Luiz Carlos Junior de Alcantara, Marta Giovanetti, Vagner Fonseca, Flavia Aburjaile, Rodrigo Tocantins Calado. |
| EPI_ISL_1795177, EPI_ISL_1795178, EPI_ISL_1795179, EPI_ISL_1795180, EPI_ISL_1795181 | POLICLINICA HORTOLANDIA | Instituto Butantan / ESALQ-Piracicaba | Instituto Butantan: Alexander Roberto Precioso, Dimas Tadeu Covas, Sandra Coccuzzo Sampaio, Maria Carolina Elias, José Salvatore Leister Patané, Vincent Louis Viala, Antonio Jorge Martins, Ricardo Haddad, Claudia Renata dos Santos Barros, Elaine Cristina Marqueze, Raul Machado Neto, Debora Botequio Moretti. Centro de Genômica Funcional da ESALQ: Luiz Lehmann Coutinho, Ricardo Augusto Brassaloti, Raquel de Lello Rocha Campos Cassano. NGS Soluções Genômicas: Pilar Drummond Sampaio Corrêa Mariani. FZEA-USP Pirassununga: Mirele Daiana Poleti, Jessika Cristina Chagas Lesbon, Elisangela Chicaroni Mattos, Heidge Fukumasu. USP-Botucatu: Rejane Maria Tommasini Grotto, Jayme A. Souza-Neto, Guilherme Targino Valente, Patricia Akemi Assato, Felipe Allan da Silva da Costa, Bianca Cechetto Carlos. Mendelics: Bibiana Santos, João Paulo Kitajima, Erika Freitas, |

| | | | |
|---|---|---|---|
| | | | David Schlesinger. Hemocentro Ribeirão Preto: Simone Kashima, Evandra Strazza Rodrigues, Svetoslav Nanev Slavov, Elaine Vieira dos Santos, Rafael dos Santos Bezerra, Luiz Carlos Junior de Alcantara, Marta Giovanetti, Vagner Fonseca, Flavia Aburjaile, Rodrigo Tocantins Calado. |
| EPI_ISL_1795182 | DEPARTAMENTO DE SAUDE MUNICIPAL SOCORRO SP | Instituto Butantan / ESALQ-Piracicaba | Instituto Butantan: Alexander Roberto Precioso, Dimas Tadeu Covas, Sandra Coccuzzo Sampaio, Maria Carolina Elias, José Salvatore Leister Patané, Vincent Louis Viala, Antonio Jorge Martins, Ricardo Haddad, Claudia Renata dos Santos Barros, Elaine Cristina Marqueze, Raul Machado Neto, Debora Botequio Moretti. Centro de Genômica Funcional da ESALQ: Luiz Lehmann Coutinho, Ricardo Augusto Brassaloti, Raquel de Lello Rocha Campos Cassano. NGS Soluções Genômicas: Pilar Drummond Sampaio Corrêa Mariani. FZEA-USP Pirassununga: Mirele Daiana Poleti, Jessika Cristina Chagas Lesbon, Elisangela Chicaroni Mattos, Heidge Fukumasu. USP-Botucatu: Rejane Maria Tommasini Grotto, Jayme A. Souza-Neto, Guilherme Targino Valente, Patricia Akemi Assato, Felipe Allan da Silva da Costa, Bianca Cechetto Carlos. Mendelics: Bibiana Santos, João Paulo Kitajima, Erika Freitas, David Schlesinger. Hemocentro Ribeirão Preto: Simone Kashima, Evandra Strazza Rodrigues, Svetoslav Nanev Slavov, Elaine Vieira dos Santos, Rafael dos Santos Bezerra, Luiz Carlos Junior de Alcantara, Marta Giovanetti, Vagner Fonseca, Flavia Aburjaile, Rodrigo Tocantins Calado. |
| EPI_ISL_1795183, EPI_ISL_1795184, EPI_ISL_1795185, EPI_ISL_1795186 | POLICLINICA HORTOLANDIA | Instituto Butantan / ESALQ-Piracicaba | Instituto Butantan: Alexander Roberto Precioso, Dimas Tadeu Covas, Sandra Coccuzzo Sampaio, Maria Carolina Elias, José Salvatore Leister Patané, Vincent Louis Viala, Antonio Jorge Martins, Ricardo Haddad, Claudia Renata dos Santos Barros, Elaine Cristina Marqueze, Raul Machado Neto, Debora Botequio Moretti. Centro de Genômica Funcional da ESALQ: Luiz Lehmann Coutinho, Ricardo Augusto Brassaloti, Raquel de Lello Rocha Campos Cassano. NGS Soluções Genômicas: Pilar Drummond Sampaio Corrêa Mariani. FZEA-USP Pirassununga: Mirele Daiana Poleti, Jessika Cristina Chagas Lesbon, Elisangela Chicaroni Mattos, Heidge Fukumasu. USP-Botucatu: Rejane Maria Tommasini Grotto, Jayme A. Souza-Neto, Guilherme Targino Valente, Patricia Akemi Assato, Felipe Allan da Silva da Costa, Bianca Cechetto Carlos. Mendelics: Bibiana Santos, João Paulo Kitajima, Erika Freitas, David Schlesinger. Hemocentro Ribeirão Preto: Simone Kashima, Evandra Strazza Rodrigues, Svetoslav Nanev Slavov, Elaine Vieira dos Santos, Rafael dos Santos Bezerra, Luiz Carlos Junior de Alcantara, Marta Giovanetti, Vagner Fonseca, Flavia Aburjaile, Rodrigo Tocantins Calado. |
| EPI_ISL_1795187 | USF SALERNO | Instituto Butantan / ESALQ-Piracicaba | Instituto Butantan: Alexander Roberto Precioso, Dimas Tadeu Covas, Sandra Coccuzzo Sampaio, Maria Carolina Elias, José Salvatore Leister Patané, Vincent Louis Viala, Antonio Jorge Martins, Ricardo Haddad, Claudia Renata dos Santos Barros, Elaine Cristina Marqueze, Raul Machado Neto, Debora Botequio Moretti. Centro de Genômica Funcional da ESALQ: Luiz Lehmann Coutinho, Ricardo Augusto Brassaloti, Raquel de Lello Rocha Campos Cassano. NGS Soluções Genômicas: Pilar Drummond Sampaio Corrêa Mariani. FZEA-USP Pirassununga: Mirele Daiana Poleti, Jessika Cristina Chagas Lesbon, Elisangela Chicaroni Mattos, Heidge Fukumasu. USP-Botucatu: Rejane Maria Tommasini Grotto, Jayme A. Souza-Neto, Guilherme Targino Valente, Patricia Akemi Assato, Felipe Allan da Silva da Costa, Bianca Cechetto Carlos. Mendelics: Bibiana Santos, João Paulo Kitajima, Erika Freitas, David Schlesinger. Hemocentro Ribeirão Preto: Simone Kashima, Evandra Strazza Rodrigues, Svetoslav Nanev Slavov, Elaine Vieira dos Santos, Rafael dos Santos Bezerra, Luiz Carlos Junior de Alcantara, Marta Giovanetti, Vagner Fonseca, Flavia Aburjaile, Rodrigo Tocantins Calado. |
| EPI_ISL_1795188 | POLICLINICA HORTOLANDIA | Instituto Butantan / ESALQ-Piracicaba | Instituto Butantan: Alexander Roberto Precioso, Dimas Tadeu Covas, Sandra Coccuzzo Sampaio, Maria Carolina Elias, José Salvatore Leister Patané, Vincent Louis Viala, Antonio Jorge Martins, Ricardo Haddad, Claudia Renata dos Santos Barros, Elaine Cristina Marqueze, Raul Machado Neto, Debora Botequio Moretti. Centro de Genômica Funcional da ESALQ: Luiz Lehmann Coutinho, Ricardo Augusto Brassaloti, Raquel de Lello Rocha Campos Cassano. NGS Soluções Genômicas: Pilar Drummond Sampaio Corrêa Mariani. FZEA-USP Pirassununga: Mirele Daiana Poleti, Jessika Cristina Chagas Lesbon, Elisangela Chicaroni Mattos, Heidge Fukumasu. USP-Botucatu: Rejane Maria Tommasini Grotto, Jayme A. Souza-Neto, Guilherme Targino Valente, Patricia Akemi Assato, Felipe Allan da Silva da Costa, Bianca Cechetto Carlos. Mendelics: Bibiana Santos, João Paulo Kitajima, Erika Freitas, David Schlesinger. Hemocentro Ribeirão Preto: Simone Kashima, Evandra Strazza Rodrigues, Svetoslav Nanev Slavov, Elaine Vieira dos Santos, Rafael dos Santos Bezerra, Luiz Carlos Junior de Alcantara, Marta Giovanetti, Vagner Fonseca, Flavia Aburjaile, Rodrigo Tocantins Calado. |
| EPI_ISL_1795189 | USF SALERNO | Instituto Butantan / ESALQ-Piracicaba | Instituto Butantan: Alexander Roberto Precioso, Dimas Tadeu Covas, Sandra Coccuzzo Sampaio, Maria Carolina Elias, José Salvatore Leister Patané, Vincent Louis Viala, Antonio Jorge Martins, Ricardo Haddad, Claudia Renata dos Santos Barros, Elaine Cristina Marqueze, Raul Machado Neto, Debora Botequio Moretti. Centro de Genômica Funcional da ESALQ: Luiz Lehmann Coutinho, Ricardo Augusto Brassaloti, Raquel de Lello Rocha Campos Cassano. NGS Soluções Genômicas: Pilar Drummond Sampaio Corrêa Mariani. FZEA-USP Pirassununga: Mirele Daiana Poleti, Jessika Cristina Chagas Lesbon, Elisangela Chicaroni Mattos, Heidge Fukumasu. USP-Botucatu: Rejane Maria Tommasini Grotto, Jayme A. Souza-Neto, Guilherme Targino Valente, Patricia Akemi Assato, Felipe Allan da Silva da Costa, Bianca Cechetto Carlos. Mendelics: Bibiana Santos, João Paulo Kitajima, Erika Freitas, David Schlesinger. Hemocentro Ribeirão Preto: Simone Kashima, Evandra Strazza Rodrigues, Svetoslav Nanev Slavov, Elaine Vieira dos Santos, Rafael dos Santos Bezerra, Luiz Carlos Junior de Alcantara, Marta Giovanetti, Vagner Fonseca, Flavia Aburjaile, Rodrigo Tocantins Calado. |
| EPI_ISL_1795190, EPI_ISL_1795191 | POLICLINICA HORTOLANDIA | Instituto Butantan / ESALQ-Piracicaba | Instituto Butantan: Alexander Roberto Precioso, Dimas Tadeu Covas, Sandra Coccuzzo Sampaio, Maria Carolina Elias, José Salvatore Leister Patané, Vincent Louis Viala, Antonio Jorge Martins, Ricardo Haddad, Claudia Renata dos Santos Barros, Elaine Cristina Marqueze, Raul Machado Neto, Debora Botequio Moretti. Centro de Genômica Funcional da ESALQ: Luiz Lehmann Coutinho, Ricardo Augusto Brassaloti, Raquel de Lello Rocha Campos Cassano. NGS Soluções Genômicas: Pilar Drummond Sampaio Corrêa Mariani. FZEA-USP Pirassununga: Mirele Daiana Poleti, Jessika Cristina Chagas Lesbon, Elisangela Chicaroni Mattos, Heidge Fukumasu. USP-Botucatu: Rejane Maria Tommasini Grotto, Jayme A. Souza-Neto, Guilherme Targino Valente, Patricia Akemi Assato, Felipe Allan da Silva da Costa, Bianca Cechetto Carlos. Mendelics: Bibiana Santos, João Paulo Kitajima, Erika Freitas, David Schlesinger. Hemocentro Ribeirão Preto: Simone Kashima, Evandra Strazza Rodrigues, Svetoslav Nanev Slavov, Elaine Vieira dos Santos, Rafael dos Santos Bezerra, Luiz Carlos Junior de Alcantara, Marta Giovanetti, Vagner Fonseca, Flavia Aburjaile, Rodrigo Tocantins Calado. |
| EPI_ISL_1795192, EPI_ISL_1795193 | USF SALERNO | Instituto Butantan / ESALQ-Piracicaba | Instituto Butantan: Alexander Roberto Precioso, Dimas Tadeu Covas, Sandra Coccuzzo Sampaio, Maria Carolina Elias, José Salvatore Leister Patané, Vincent Louis Viala, Antonio Jorge Martins, Ricardo Haddad, Claudia Renata dos Santos Barros, Elaine Cristina Marqueze, Raul Machado Neto, Debora Botequio Moretti. Centro de Genômica Funcional da ESALQ: Luiz Lehmann Coutinho, Ricardo Augusto Brassaloti, Raquel de Lello Rocha Campos Cassano. NGS Soluções Genômicas: Pilar Drummond Sampaio Corrêa Mariani. FZEA-USP Pirassununga: Mirele Daiana Poleti, Jessika Cristina Chagas Lesbon, Elisangela Chicaroni Mattos, Heidge Fukumasu. USP-Botucatu: Rejane Maria Tommasini Grotto, Jayme A. Souza-Neto, Guilherme Targino Valente, Patricia Akemi Assato, Felipe Allan da Silva da Costa, Bianca Cechetto Carlos. Mendelics: Bibiana Santos, João Paulo Kitajima, Erika Freitas, David Schlesinger. Hemocentro Ribeirão Preto: Simone Kashima, Evandra Strazza Rodrigues, Svetoslav Nanev Slavov, Elaine Vieira dos Santos, Rafael dos Santos Bezerra, Luiz Carlos Junior de Alcantara, Marta Giovanetti, Vagner Fonseca, Flavia Aburjaile, Rodrigo Tocantins Calado. |
| EPI_ISL_1795194, EPI_ISL_1795195, EPI_ISL_1795196, EPI_ISL_1795197, EPI_ISL_1795198, EPI_ISL_1795199, EPI_ISL_1795200, EPI_ISL_1795201, EPI_ISL_1795202, EPI_ISL_1795203, EPI_ISL_1795204, EPI_ISL_1795205, EPI_ISL_1795206, EPI_ISL_1795207 | | | |
| see above | AMBULATORIO DE ESPECIALIDADE V E MOGI MIRIM | Instituto Butantan / ESALQ-Piracicaba | Instituto Butantan: Alexander Roberto Precioso, Dimas Tadeu Covas, Sandra Coccuzzo Sampaio, Maria Carolina Elias, José Salvatore Leister Patané, Vincent Louis Viala, Antonio Jorge Martins, Ricardo Haddad, Claudia Renata dos Santos Barros, Elaine Cristina Marqueze, Raul Machado Neto, Debora Botequio Moretti. Centro de Genômica Funcional da ESALQ: Luiz Lehmann Coutinho, Ricardo Augusto Brassaloti, Raquel de Lello Rocha Campos Cassano. NGS Soluções Genômicas: Pilar Drummond Sampaio Corrêa Mariani. FZEA-USP Pirassununga: Mirele Daiana Poleti, Jessika Cristina Chagas Lesbon, Elisangela Chicaroni Mattos, Heidge Fukumasu. USP-Botucatu: Rejane Maria Tommasini Grotto, Jayme A. Souza-Neto, Guilherme Targino Valente, Patricia Akemi Assato, Felipe Allan da Silva da Costa, Bianca Cechetto Carlos. Mendelics: Bibiana Santos, João Paulo Kitajima, Erika Freitas, David Schlesinger. Hemocentro Ribeirão Preto: Simone Kashima, Evandra Strazza Rodrigues, Svetoslav Nanev Slavov, Elaine Vieira dos Santos, Rafael dos Santos Bezerra, Luiz Carlos Junior de Alcantara, Marta Giovanetti, Vagner Fonseca, Flavia Aburjaile, Rodrigo Tocantins Calado. |
| EPI_ISL_1795208, EPI_ISL_1795209 | PRONTO SOCORRO MUNICIPAL TAMBAU | Instituto Butantan / ESALQ-Piracicaba | Instituto Butantan: Alexander Roberto Precioso, Dimas Tadeu Covas, Sandra Coccuzzo Sampaio, Maria Carolina Elias, José Salvatore Leister Patané, Vincent Louis Viala, Antonio Jorge Martins, Ricardo Haddad, Claudia Renata dos Santos Barros, Elaine Cristina Marqueze, Raul Machado Neto, Debora Botequio Moretti. Centro de Genômica Funcional da ESALQ: Luiz Lehmann Coutinho, Ricardo Augusto Brassaloti, Raquel de Lello Rocha Campos Cassano. NGS Soluções Genômicas: Pilar Drummond Sampaio Corrêa Mariani. FZEA-USP Pirassununga: Mirele Daiana Poleti, Jessika Cristina Chagas Lesbon, Elisangela Chicaroni Mattos, Heidge Fukumasu. USP-Botucatu: Rejane Maria Tommasini Grotto, Jayme A. Souza-Neto, Guilherme Targino Valente, Patricia Akemi Assato, Felipe Allan da Silva da Costa, Bianca Cechetto Carlos. Mendelics: Bibiana Santos, João Paulo Kitajima, Erika Freitas, David Schlesinger. Hemocentro Ribeirão Preto: Simone Kashima, Evandra Strazza Rodrigues, Svetoslav Nanev Slavov, Elaine Vieira dos Santos, Rafael dos Santos Bezerra, Luiz Carlos Junior de Alcantara, Marta Giovanetti, Vagner Fonseca, Flavia Aburjaile, Rodrigo Tocantins Calado. |
| EPI_ISL_1795210 | AMBULATORIO DE ESPECIALIDADE V E MOGI MIRIM | Instituto Butantan / ESALQ-Piracicaba | Instituto Butantan: Alexander Roberto Precioso, Dimas Tadeu Covas, Sandra Coccuzzo Sampaio, Maria Carolina Elias, José Salvatore Leister Patané, Vincent Louis Viala, Antonio Jorge Martins, Ricardo Haddad, Claudia Renata dos Santos Barros, Elaine Cristina Marqueze, Raul Machado Neto, Debora Botequio Moretti. Centro de Genômica Funcional da ESALQ: Luiz Lehmann Coutinho, Ricardo Augusto Brassaloti, Raquel de Lello Rocha Campos Cassano. NGS Soluções Genômicas: Pilar Drummond Sampaio Corrêa Mariani. FZEA-USP Pirassununga: Mirele Daiana Poleti, Jessika Cristina Chagas Lesbon, Elisangela Chicaroni Mattos, Heidge Fukumasu. USP-Botucatu: Rejane Maria Tommasini Grotto, Jayme A. Souza-Neto, Guilherme Targino |

| | | | |
|---|---|---|---|
| | | | Valente, Patricia Akemi Assato, Felipe Allan da Silva da Costa, Bianca Cechetto Carlos. Mendelics: Bibiana Santos, João Paulo Kitajima, Erika Freitas, David Schlesinger. Hemocentro Ribeirão Preto: Simone Kashima, Evandra Strazza Rodrigues, Svetoslav Nanev Slavov, Elaine Vieira dos Santos, Rafael dos Santos Bezerra, Luiz Carlos Junior de Alcantara, Marta Giovanetti, Vagner Fonseca, Flavia Aburjaile, Rodrigo Tocantins Calado. |
| EPI_ISL_1795211 | HOSPITAL MUNICIPAL DR MARIO GATTI CAMPINAS | Instituto Butantan / ESALQ-Piracicaba | Instituto Butantan: Alexander Roberto Precioso, Dimas Tadeu Covas, Sandra Coccuzzo Sampaio, Maria Carolina Elias, José Salvatore Leister Patané, Vincent Louis Viala, Antonio Jorge Martins, Ricardo Haddad, Claudia Renata dos Santos Barros, Elaine Cristina Marqueze, Raul Machado Neto, Debora Botequio Moretti. Centro de Genômica Funcional da ESALQ: Luiz Lehmann Coutinho, Ricardo Augusto Brassaloti, Raquel de Lello Rocha Campos Cassano. NGS Soluções Genômicas: Pilar Drummond Sampaio Corrêa Mariani. FZEA-USP Pirassununga: Mirele Daiana Poleti, Jessika Cristina Chagas Lesbon, Elisangela Chicaroni Mattos, Heidge Fukumasu. USP-Botucatu: Rejane Maria Tommasini Grotto, Jayme A. Souza-Neto, Guilherme Targino Valente, Patricia Akemi Assato, Felipe Allan da Silva da Costa, Bianca Cechetto Carlos. Mendelics: Bibiana Santos, João Paulo Kitajima, Erika Freitas, David Schlesinger. Hemocentro Ribeirão Preto: Simone Kashima, Evandra Strazza Rodrigues, Svetoslav Nanev Slavov, Elaine Vieira dos Santos, Rafael dos Santos Bezerra, Luiz Carlos Junior de Alcantara, Marta Giovanetti, Vagner Fonseca, Flavia Aburjaile, Rodrigo Tocantins Calado. |
| EPI_ISL_1795212 | CENTRO DE SAUDE II DR ALCIDES FACUNDO ARROYO | Instituto Butantan / ESALQ-Piracicaba | Instituto Butantan: Alexander Roberto Precioso, Dimas Tadeu Covas, Sandra Coccuzzo Sampaio, Maria Carolina Elias, José Salvatore Leister Patané, Vincent Louis Viala, Antonio Jorge Martins, Ricardo Haddad, Claudia Renata dos Santos Barros, Elaine Cristina Marqueze, Raul Machado Neto, Debora Botequio Moretti. Centro de Genômica Funcional da ESALQ: Luiz Lehmann Coutinho, Ricardo Augusto Brassaloti, Raquel de Lello Rocha Campos Cassano. NGS Soluções Genômicas: Pilar Drummond Sampaio Corrêa Mariani. FZEA-USP Pirassununga: Mirele Daiana Poleti, Jessika Cristina Chagas Lesbon, Elisangela Chicaroni Mattos, Heidge Fukumasu. USP-Botucatu: Rejane Maria Tommasini Grotto, Jayme A. Souza-Neto, Guilherme Targino Valente, Patricia Akemi Assato, Felipe Allan da Silva da Costa, Bianca Cechetto Carlos. Mendelics: Bibiana Santos, João Paulo Kitajima, Erika Freitas, David Schlesinger. Hemocentro Ribeirão Preto: Simone Kashima, Evandra Strazza Rodrigues, Svetoslav Nanev Slavov, Elaine Vieira dos Santos, Rafael dos Santos Bezerra, Luiz Carlos Junior de Alcantara, Marta Giovanetti, Vagner Fonseca, Flavia Aburjaile, Rodrigo Tocantins Calado. |
| EPI_ISL_1795213, EPI_ISL_1795214, EPI_ISL_1795215, EPI_ISL_1795216 | PRONTO SOCORRO MUNICIPAL DE SEVERINIA | Instituto Butantan / ESALQ-Piracicaba | Instituto Butantan: Alexander Roberto Precioso, Dimas Tadeu Covas, Sandra Coccuzzo Sampaio, Maria Carolina Elias, José Salvatore Leister Patané, Vincent Louis Viala, Antonio Jorge Martins, Ricardo Haddad, Claudia Renata dos Santos Barros, Elaine Cristina Marqueze, Raul Machado Neto, Debora Botequio Moretti. Centro de Genômica Funcional da ESALQ: Luiz Lehmann Coutinho, Ricardo Augusto Brassaloti, Raquel de Lello Rocha Campos Cassano. NGS Soluções Genômicas: Pilar Drummond Sampaio Corrêa Mariani. FZEA-USP Pirassununga: Mirele Daiana Poleti, Jessika Cristina Chagas Lesbon, Elisangela Chicaroni Mattos, Heidge Fukumasu. USP-Botucatu: Rejane Maria Tommasini Grotto, Jayme A. Souza-Neto, Guilherme Targino Valente, Patricia Akemi Assato, Felipe Allan da Silva da Costa, Bianca Cechetto Carlos. Mendelics: Bibiana Santos, João Paulo Kitajima, Erika Freitas, David Schlesinger. Hemocentro Ribeirão Preto: Simone Kashima, Evandra Strazza Rodrigues, Svetoslav Nanev Slavov, Elaine Vieira dos Santos, Rafael dos Santos Bezerra, Luiz Carlos Junior de Alcantara, Marta Giovanetti, Vagner Fonseca, Flavia Aburjaile, Rodrigo Tocantins Calado. |
| EPI_ISL_1795217 | VIGILANCIA EPIDEMIOLOGICA JARDINOPOLIS SP | Instituto Butantan / ESALQ-Piracicaba | Instituto Butantan: Alexander Roberto Precioso, Dimas Tadeu Covas, Sandra Coccuzzo Sampaio, Maria Carolina Elias, José Salvatore Leister Patané, Vincent Louis Viala, Antonio Jorge Martins, Ricardo Haddad, Claudia Renata dos Santos Barros, Elaine Cristina Marqueze, Raul Machado Neto, Debora Botequio Moretti. Centro de Genômica Funcional da ESALQ: Luiz Lehmann Coutinho, Ricardo Augusto Brassaloti, Raquel de Lello Rocha Campos Cassano. NGS Soluções Genômicas: Pilar Drummond Sampaio Corrêa Mariani. FZEA-USP Pirassununga: Mirele Daiana Poleti, Jessika Cristina Chagas Lesbon, Elisangela Chicaroni Mattos, Heidge Fukumasu. USP-Botucatu: Rejane Maria Tommasini Grotto, Jayme A. Souza-Neto, Guilherme Targino Valente, Patricia Akemi Assato, Felipe Allan da Silva da Costa, Bianca Cechetto Carlos. Mendelics: Bibiana Santos, João Paulo Kitajima, Erika Freitas, David Schlesinger. Hemocentro Ribeirão Preto: Simone Kashima, Evandra Strazza Rodrigues, Svetoslav Nanev Slavov, Elaine Vieira dos Santos, Rafael dos Santos Bezerra, Luiz Carlos Junior de Alcantara, Marta Giovanetti, Vagner Fonseca, Flavia Aburjaile, Rodrigo Tocantins Calado. |
| EPI_ISL_1795218 | VIGILANCIA EPIDIMIOLOGICA DE ARACARIGUAMA | Instituto Butantan / ESALQ-Piracicaba | Instituto Butantan: Alexander Roberto Precioso, Dimas Tadeu Covas, Sandra Coccuzzo Sampaio, Maria Carolina Elias, José Salvatore Leister Patané, Vincent Louis Viala, Antonio Jorge Martins, Ricardo Haddad, Claudia Renata dos Santos Barros, Elaine Cristina Marqueze, Raul Machado Neto, Debora Botequio Moretti. Centro de Genômica Funcional da ESALQ: Luiz Lehmann Coutinho, Ricardo Augusto Brassaloti, Raquel de Lello Rocha Campos Cassano. NGS Soluções Genômicas: Pilar Drummond Sampaio Corrêa Mariani. FZEA-USP Pirassununga: Mirele Daiana Poleti, Jessika Cristina Chagas Lesbon, Elisangela Chicaroni Mattos, Heidge Fukumasu. USP-Botucatu: Rejane Maria Tommasini Grotto, Jayme A. Souza-Neto, Guilherme Targino Valente, Patricia Akemi Assato, Felipe Allan da Silva da Costa, Bianca Cechetto Carlos. Mendelics: Bibiana Santos, João Paulo Kitajima, Erika Freitas, David Schlesinger. Hemocentro Ribeirão Preto: Simone Kashima, Evandra Strazza Rodrigues, Svetoslav Nanev Slavov, Elaine Vieira dos Santos, Rafael dos Santos Bezerra, Luiz Carlos Junior de Alcantara, Marta Giovanetti, Vagner Fonseca, Flavia Aburjaile, Rodrigo Tocantins Calado. |
| EPI_ISL_1795219 | PRONTO ATENDIMENTO UNIDADE SAUDE ADALBERTO ROCHA GUAREI | Instituto Butantan / ESALQ-Piracicaba | Instituto Butantan: Alexander Roberto Precioso, Dimas Tadeu Covas, Sandra Coccuzzo Sampaio, Maria Carolina Elias, José Salvatore Leister Patané, Vincent Louis Viala, Antonio Jorge Martins, Ricardo Haddad, Claudia Renata dos Santos Barros, Elaine Cristina Marqueze, Raul Machado Neto, Debora Botequio Moretti. Centro de Genômica Funcional da ESALQ: Luiz Lehmann Coutinho, Ricardo Augusto Brassaloti, Raquel de Lello Rocha Campos Cassano. NGS Soluções Genômicas: Pilar Drummond Sampaio Corrêa Mariani. FZEA-USP Pirassununga: Mirele Daiana Poleti, Jessika Cristina Chagas Lesbon, Elisangela Chicaroni Mattos, Heidge Fukumasu. USP-Botucatu: Rejane Maria Tommasini Grotto, Jayme A. Souza-Neto, Guilherme Targino Valente, Patricia Akemi Assato, Felipe Allan da Silva da Costa, Bianca Cechetto Carlos. Mendelics: Bibiana Santos, João Paulo Kitajima, Erika Freitas, David Schlesinger. Hemocentro Ribeirão Preto: Simone Kashima, Evandra Strazza Rodrigues, Svetoslav Nanev Slavov, Elaine Vieira dos Santos, Rafael dos Santos Bezerra, Luiz Carlos Junior de Alcantara, Marta Giovanetti, Vagner Fonseca, Flavia Aburjaile, Rodrigo Tocantins Calado. |
| EPI_ISL_1795220 | UNIDADE SENTINELA COVID19 | Instituto Butantan / ESALQ-Piracicaba | Instituto Butantan: Alexander Roberto Precioso, Dimas Tadeu Covas, Sandra Coccuzzo Sampaio, Maria Carolina Elias, José Salvatore Leister Patané, Vincent Louis Viala, Antonio Jorge Martins, Ricardo Haddad, Claudia Renata dos Santos Barros, Elaine Cristina Marqueze, Raul Machado Neto, Debora Botequio Moretti. Centro de Genômica Funcional da ESALQ: Luiz Lehmann Coutinho, Ricardo Augusto Brassaloti, Raquel de Lello Rocha Campos Cassano. NGS Soluções Genômicas: Pilar Drummond Sampaio Corrêa Mariani. FZEA-USP Pirassununga: Mirele Daiana Poleti, Jessika Cristina Chagas Lesbon, Elisangela Chicaroni Mattos, Heidge Fukumasu. USP-Botucatu: Rejane Maria Tommasini Grotto, Jayme A. Souza-Neto, Guilherme Targino Valente, Patricia Akemi Assato, Felipe Allan da Silva da Costa, Bianca Cechetto Carlos. Mendelics: Bibiana Santos, João Paulo Kitajima, Erika Freitas, David Schlesinger. Hemocentro Ribeirão Preto: Simone Kashima, Evandra Strazza Rodrigues, Svetoslav Nanev Slavov, Elaine Vieira dos Santos, Rafael dos Santos Bezerra, Luiz Carlos Junior de Alcantara, Marta Giovanetti, Vagner Fonseca, Flavia Aburjaile, Rodrigo Tocantins Calado. |
| EPI_ISL_1795221 | HOSPITAL REGIONAL DE ITAPETININGA | Instituto Butantan / ESALQ-Piracicaba | Instituto Butantan: Alexander Roberto Precioso, Dimas Tadeu Covas, Sandra Coccuzzo Sampaio, Maria Carolina Elias, José Salvatore Leister Patané, Vincent Louis Viala, Antonio Jorge Martins, Ricardo Haddad, Claudia Renata dos Santos Barros, Elaine Cristina Marqueze, Raul Machado Neto, Debora Botequio Moretti. Centro de Genômica Funcional da ESALQ: Luiz Lehmann Coutinho, Ricardo Augusto Brassaloti, Raquel de Lello Rocha Campos Cassano. NGS Soluções Genômicas: Pilar Drummond Sampaio Corrêa Mariani. FZEA-USP Pirassununga: Mirele Daiana Poleti, Jessika Cristina Chagas Lesbon, Elisangela Chicaroni Mattos, Heidge Fukumasu. USP-Botucatu: Rejane Maria Tommasini Grotto, Jayme A. Souza-Neto, Guilherme Targino Valente, Patricia Akemi Assato, Felipe Allan da Silva da Costa, Bianca Cechetto Carlos. Mendelics: Bibiana Santos, João Paulo Kitajima, Erika Freitas, David Schlesinger. Hemocentro Ribeirão Preto: Simone Kashima, Evandra Strazza Rodrigues, Svetoslav Nanev Slavov, Elaine Vieira dos Santos, Rafael dos Santos Bezerra, Luiz Carlos Junior de Alcantara, Marta Giovanetti, Vagner Fonseca, Flavia Aburjaile, Rodrigo Tocantins Calado. |
| EPI_ISL_1795222 | UNIDADE MISTA ARACOIABA DA SERRA | Instituto Butantan / ESALQ-Piracicaba | Instituto Butantan: Alexander Roberto Precioso, Dimas Tadeu Covas, Sandra Coccuzzo Sampaio, Maria Carolina Elias, José Salvatore Leister Patané, Vincent Louis Viala, Antonio Jorge Martins, Ricardo Haddad, Claudia Renata dos Santos Barros, Elaine Cristina Marqueze, Raul Machado Neto, Debora Botequio Moretti. Centro de Genômica Funcional da ESALQ: Luiz Lehmann Coutinho, Ricardo Augusto Brassaloti, Raquel de Lello Rocha Campos Cassano. NGS Soluções Genômicas: Pilar Drummond Sampaio Corrêa Mariani. FZEA-USP Pirassununga: Mirele Daiana Poleti, Jessika Cristina Chagas Lesbon, Elisangela Chicaroni Mattos, Heidge Fukumasu. USP-Botucatu: Rejane Maria Tommasini Grotto, Jayme A. Souza-Neto, Guilherme Targino Valente, Patricia Akemi Assato, Felipe Allan da Silva da Costa, Bianca Cechetto Carlos. Mendelics: Bibiana Santos, João Paulo Kitajima, Erika Freitas, David Schlesinger. Hemocentro Ribeirão Preto: Simone Kashima, Evandra Strazza Rodrigues, Svetoslav Nanev Slavov, Elaine Vieira dos Santos, Rafael dos Santos Bezerra, Luiz Carlos Junior de Alcantara, Marta Giovanetti, Vagner Fonseca, Flavia Aburjaile, Rodrigo Tocantins Calado. |
| EPI_ISL_1795223 | HOSPITAL REGIONAL DE ITAPETININGA | Instituto Butantan / ESALQ-Piracicaba | Instituto Butantan: Alexander Roberto Precioso, Dimas Tadeu Covas, Sandra Coccuzzo Sampaio, Maria Carolina Elias, José Salvatore Leister Patané, Vincent Louis Viala, Antonio Jorge Martins, Ricardo Haddad, Claudia Renata dos Santos Barros, Elaine Cristina Marqueze, Raul Machado Neto, Debora Botequio Moretti. Centro de Genômica Funcional da ESALQ: Luiz Lehmann Coutinho, Ricardo Augusto Brassaloti, Raquel de Lello Rocha Campos Cassano. NGS Soluções Genômicas: Pilar Drummond Sampaio Corrêa Mariani. FZEA-USP Pirassununga: Mirele Daiana Poleti, Jessika Cristina Chagas Lesbon, Elisangela Chicaroni Mattos, Heidge Fukumasu. USP-Botucatu: Rejane Maria Tommasini Grotto, Jayme A. Souza-Neto, Guilherme Targino |

| | | | |
|---|---|---|---|
| | | | Valente, Patricia Akemi Assato, Felipe Allan da Silva da Costa, Bianca Cechetto Carlos. Mendelics: Bibiana Santos, João Paulo Kitajima, Erika Freitas, David Schlesinger. Hemocentro Ribeirão Preto: Simone Kashima, Evandra Strazza Rodrigues, Svetoslav Nanev Slavov, Elaine Vieira dos Santos, Rafael dos Santos Bezerra, Luiz Carlos Junior de Alcantara, Marta Giovanetti, Vagner Fonseca, Flavia Aburjaile, Rodrigo Tocantins Calado. |
| EPI_ISL_1795224 | UNIDADE BASICA DE SAUDE DR APOLONIO MORAES E SOUZA | Instituto Butantan / ESALQ-Piracicaba | Instituto Butantan: Alexander Roberto Precioso, Dimas Tadeu Covas, Sandra Coccuzzo Sampaio, Maria Carolina Elias, José Salvatore Leister Patané, Vincent Louis Viala, Antonio Jorge Martins, Ricardo Haddad, Claudia Renata dos Santos Barros, Elaine Cristina Marqueze, Raul Machado Neto, Debora Botequio Moretti. Centro de Genômica Funcional da ESALQ: Luiz Lehmann Coutinho, Ricardo Augusto Brassaloti, Raquel de Lello Rocha Campos Cassano. NGS Soluções Genômicas: Pilar Drummond Sampaio Corrêa Mariani. FZEA-USP Pirassununga: Mirele Daiana Poleti, Jessika Cristina Chagas Lesbon, Elisangela Chicaroni Mattos, Heidge Fukumasu. USP-Botucatu: Rejane Maria Tommasini Grotto, Jayme A. Souza-Neto, Guilherme Targino Valente, Patricia Akemi Assato, Felipe Allan da Silva da Costa, Bianca Cechetto Carlos. Mendelics: Bibiana Santos, João Paulo Kitajima, Erika Freitas, David Schlesinger. Hemocentro Ribeirão Preto: Simone Kashima, Evandra Strazza Rodrigues, Svetoslav Nanev Slavov, Elaine Vieira dos Santos, Rafael dos Santos Bezerra, Luiz Carlos Junior de Alcantara, Marta Giovanetti, Vagner Fonseca, Flavia Aburjaile, Rodrigo Tocantins Calado. |
| EPI_ISL_1795225 | UNIDADE BASICA DE SAUDE DR WILSON HAYEK SAIHG | Instituto Butantan / ESALQ-Piracicaba | Instituto Butantan: Alexander Roberto Precioso, Dimas Tadeu Covas, Sandra Coccuzzo Sampaio, Maria Carolina Elias, José Salvatore Leister Patané, Vincent Louis Viala, Antonio Jorge Martins, Ricardo Haddad, Claudia Renata dos Santos Barros, Elaine Cristina Marqueze, Raul Machado Neto, Debora Botequio Moretti. Centro de Genômica Funcional da ESALQ: Luiz Lehmann Coutinho, Ricardo Augusto Brassaloti, Raquel de Lello Rocha Campos Cassano. NGS Soluções Genômicas: Pilar Drummond Sampaio Corrêa Mariani. FZEA-USP Pirassununga: Mirele Daiana Poleti, Jessika Cristina Chagas Lesbon, Elisangela Chicaroni Mattos, Heidge Fukumasu. USP-Botucatu: Rejane Maria Tommasini Grotto, Jayme A. Souza-Neto, Guilherme Targino Valente, Patricia Akemi Assato, Felipe Allan da Silva da Costa, Bianca Cechetto Carlos. Mendelics: Bibiana Santos, João Paulo Kitajima, Erika Freitas, David Schlesinger. Hemocentro Ribeirão Preto: Simone Kashima, Evandra Strazza Rodrigues, Svetoslav Nanev Slavov, Elaine Vieira dos Santos, Rafael dos Santos Bezerra, Luiz Carlos Junior de Alcantara, Marta Giovanetti, Vagner Fonseca, Flavia Aburjaile, Rodrigo Tocantins Calado. |
| EPI_ISL_1795226 | UNIDADE BASICA DR JOSE PARASSU CARVALHO | Instituto Butantan / ESALQ-Piracicaba | Instituto Butantan: Alexander Roberto Precioso, Dimas Tadeu Covas, Sandra Coccuzzo Sampaio, Maria Carolina Elias, José Salvatore Leister Patané, Vincent Louis Viala, Antonio Jorge Martins, Ricardo Haddad, Claudia Renata dos Santos Barros, Elaine Cristina Marqueze, Raul Machado Neto, Debora Botequio Moretti. Centro de Genômica Funcional da ESALQ: Luiz Lehmann Coutinho, Ricardo Augusto Brassaloti, Raquel de Lello Rocha Campos Cassano. NGS Soluções Genômicas: Pilar Drummond Sampaio Corrêa Mariani. FZEA-USP Pirassununga: Mirele Daiana Poleti, Jessika Cristina Chagas Lesbon, Elisangela Chicaroni Mattos, Heidge Fukumasu. USP-Botucatu: Rejane Maria Tommasini Grotto, Jayme A. Souza-Neto, Guilherme Targino Valente, Patricia Akemi Assato, Felipe Allan da Silva da Costa, Bianca Cechetto Carlos. Mendelics: Bibiana Santos, João Paulo Kitajima, Erika Freitas, David Schlesinger. Hemocentro Ribeirão Preto: Simone Kashima, Evandra Strazza Rodrigues, Svetoslav Nanev Slavov, Elaine Vieira dos Santos, Rafael dos Santos Bezerra, Luiz Carlos Junior de Alcantara, Marta Giovanetti, Vagner Fonseca, Flavia Aburjaile, Rodrigo Tocantins Calado. |
| EPI_ISL_1795227 | UNIDADE BASICA DE SAUDE DR LOTFALLAH MIZIARA | Instituto Butantan / ESALQ-Piracicaba | Instituto Butantan: Alexander Roberto Precioso, Dimas Tadeu Covas, Sandra Coccuzzo Sampaio, Maria Carolina Elias, José Salvatore Leister Patané, Vincent Louis Viala, Antonio Jorge Martins, Ricardo Haddad, Claudia Renata dos Santos Barros, Elaine Cristina Marqueze, Raul Machado Neto, Debora Botequio Moretti. Centro de Genômica Funcional da ESALQ: Luiz Lehmann Coutinho, Ricardo Augusto Brassaloti, Raquel de Lello Rocha Campos Cassano. NGS Soluções Genômicas: Pilar Drummond Sampaio Corrêa Mariani. FZEA-USP Pirassununga: Mirele Daiana Poleti, Jessika Cristina Chagas Lesbon, Elisangela Chicaroni Mattos, Heidge Fukumasu. USP-Botucatu: Rejane Maria Tommasini Grotto, Jayme A. Souza-Neto, Guilherme Targino Valente, Patricia Akemi Assato, Felipe Allan da Silva da Costa, Bianca Cechetto Carlos. Mendelics: Bibiana Santos, João Paulo Kitajima, Erika Freitas, David Schlesinger. Hemocentro Ribeirão Preto: Simone Kashima, Evandra Strazza Rodrigues, Svetoslav Nanev Slavov, Elaine Vieira dos Santos, Rafael dos Santos Bezerra, Luiz Carlos Junior de Alcantara, Marta Giovanetti, Vagner Fonseca, Flavia Aburjaile, Rodrigo Tocantins Calado. |
| EPI_ISL_1795228, EPI_ISL_1795229 | CENTRO DE REFERENCIA DO IDOSO DR HUMBERTO MENDES DE CARVALHO | Instituto Butantan / ESALQ-Piracicaba | Instituto Butantan: Alexander Roberto Precioso, Dimas Tadeu Covas, Sandra Coccuzzo Sampaio, Maria Carolina Elias, José Salvatore Leister Patané, Vincent Louis Viala, Antonio Jorge Martins, Ricardo Haddad, Claudia Renata dos Santos Barros, Elaine Cristina Marqueze, Raul Machado Neto, Debora Botequio Moretti. Centro de Genômica Funcional da ESALQ: Luiz Lehmann Coutinho, Ricardo Augusto Brassaloti, Raquel de Lello Rocha Campos Cassano. NGS Soluções Genômicas: Pilar Drummond Sampaio Corrêa Mariani. FZEA-USP Pirassununga: Mirele Daiana Poleti, Jessika Cristina Chagas Lesbon, Elisangela Chicaroni Mattos, Heidge Fukumasu. USP-Botucatu: Rejane Maria Tommasini Grotto, Jayme A. Souza-Neto, Guilherme Targino Valente, Patricia Akemi Assato, Felipe Allan da Silva da Costa, Bianca Cechetto Carlos. Mendelics: Bibiana Santos, João Paulo Kitajima, Erika Freitas, David Schlesinger. Hemocentro Ribeirão Preto: Simone Kashima, Evandra Strazza Rodrigues, Svetoslav Nanev Slavov, Elaine Vieira dos Santos, Rafael dos Santos Bezerra, Luiz Carlos Junior de Alcantara, Marta Giovanetti, Vagner Fonseca, Flavia Aburjaile, Rodrigo Tocantins Calado. |
| EPI_ISL_1795230, EPI_ISL_1795231 | UNIDADE DE SAUDE DA FAMILIA JOSE ADALBERTO LELLIS GARCIA | Instituto Butantan / ESALQ-Piracicaba | Instituto Butantan: Alexander Roberto Precioso, Dimas Tadeu Covas, Sandra Coccuzzo Sampaio, Maria Carolina Elias, José Salvatore Leister Patané, Vincent Louis Viala, Antonio Jorge Martins, Ricardo Haddad, Claudia Renata dos Santos Barros, Elaine Cristina Marqueze, Raul Machado Neto, Debora Botequio Moretti. Centro de Genômica Funcional da ESALQ: Luiz Lehmann Coutinho, Ricardo Augusto Brassaloti, Raquel de Lello Rocha Campos Cassano. NGS Soluções Genômicas: Pilar Drummond Sampaio Corrêa Mariani. FZEA-USP Pirassununga: Mirele Daiana Poleti, Jessika Cristina Chagas Lesbon, Elisangela Chicaroni Mattos, Heidge Fukumasu. USP-Botucatu: Rejane Maria Tommasini Grotto, Jayme A. Souza-Neto, Guilherme Targino Valente, Patricia Akemi Assato, Felipe Allan da Silva da Costa, Bianca Cechetto Carlos. Mendelics: Bibiana Santos, João Paulo Kitajima, Erika Freitas, David Schlesinger. Hemocentro Ribeirão Preto: Simone Kashima, Evandra Strazza Rodrigues, Svetoslav Nanev Slavov, Elaine Vieira dos Santos, Rafael dos Santos Bezerra, Luiz Carlos Junior de Alcantara, Marta Giovanetti, Vagner Fonseca, Flavia Aburjaile, Rodrigo Tocantins Calado. |
| EPI_ISL_1795232, EPI_ISL_1795233 | CS DE URUPES | Instituto Butantan / ESALQ-Piracicaba | Instituto Butantan: Alexander Roberto Precioso, Dimas Tadeu Covas, Sandra Coccuzzo Sampaio, Maria Carolina Elias, José Salvatore Leister Patané, Vincent Louis Viala, Antonio Jorge Martins, Ricardo Haddad, Claudia Renata dos Santos Barros, Elaine Cristina Marqueze, Raul Machado Neto, Debora Botequio Moretti. Centro de Genômica Funcional da ESALQ: Luiz Lehmann Coutinho, Ricardo Augusto Brassaloti, Raquel de Lello Rocha Campos Cassano. NGS Soluções Genômicas: Pilar Drummond Sampaio Corrêa Mariani. FZEA-USP Pirassununga: Mirele Daiana Poleti, Jessika Cristina Chagas Lesbon, Elisangela Chicaroni Mattos, Heidge Fukumasu. USP-Botucatu: Rejane Maria Tommasini Grotto, Jayme A. Souza-Neto, Guilherme Targino Valente, Patricia Akemi Assato, Felipe Allan da Silva da Costa, Bianca Cechetto Carlos. Mendelics: Bibiana Santos, João Paulo Kitajima, Erika Freitas, David Schlesinger. Hemocentro Ribeirão Preto: Simone Kashima, Evandra Strazza Rodrigues, Svetoslav Nanev Slavov, Elaine Vieira dos Santos, Rafael dos Santos Bezerra, Luiz Carlos Junior de Alcantara, Marta Giovanetti, Vagner Fonseca, Flavia Aburjaile, Rodrigo Tocantins Calado. |
| EPI_ISL_1795234, EPI_ISL_1795235, EPI_ISL_1795236, EPI_ISL_1795237, EPI_ISL_1795238 | CS DE BALSAMO | Instituto Butantan / ESALQ-Piracicaba | Instituto Butantan: Alexander Roberto Precioso, Dimas Tadeu Covas, Sandra Coccuzzo Sampaio, Maria Carolina Elias, José Salvatore Leister Patané, Vincent Louis Viala, Antonio Jorge Martins, Ricardo Haddad, Claudia Renata dos Santos Barros, Elaine Cristina Marqueze, Raul Machado Neto, Debora Botequio Moretti. Centro de Genômica Funcional da ESALQ: Luiz Lehmann Coutinho, Ricardo Augusto Brassaloti, Raquel de Lello Rocha Campos Cassano. NGS Soluções Genômicas: Pilar Drummond Sampaio Corrêa Mariani. FZEA-USP Pirassununga: Mirele Daiana Poleti, Jessika Cristina Chagas Lesbon, Elisangela Chicaroni Mattos, Heidge Fukumasu. USP-Botucatu: Rejane Maria Tommasini Grotto, Jayme A. Souza-Neto, Guilherme Targino Valente, Patricia Akemi Assato, Felipe Allan da Silva da Costa, Bianca Cechetto Carlos. Mendelics: Bibiana Santos, João Paulo Kitajima, Erika Freitas, David Schlesinger. Hemocentro Ribeirão Preto: Simone Kashima, Evandra Strazza Rodrigues, Svetoslav Nanev Slavov, Elaine Vieira dos Santos, Rafael dos Santos Bezerra, Luiz Carlos Junior de Alcantara, Marta Giovanetti, Vagner Fonseca, Flavia Aburjaile, Rodrigo Tocantins Calado. |
| EPI_ISL_1795239 | CS DE URUPES | Instituto Butantan / ESALQ-Piracicaba | Instituto Butantan: Alexander Roberto Precioso, Dimas Tadeu Covas, Sandra Coccuzzo Sampaio, Maria Carolina Elias, José Salvatore Leister Patané, Vincent Louis Viala, Antonio Jorge Martins, Ricardo Haddad, Claudia Renata dos Santos Barros, Elaine Cristina Marqueze, Raul Machado Neto, Debora Botequio Moretti. Centro de Genômica Funcional da ESALQ: Luiz Lehmann Coutinho, Ricardo Augusto Brassaloti, Raquel de Lello Rocha Campos Cassano. NGS Soluções Genômicas: Pilar Drummond Sampaio Corrêa Mariani. FZEA-USP Pirassununga: Mirele Daiana Poleti, Jessika Cristina Chagas Lesbon, Elisangela Chicaroni Mattos, Heidge Fukumasu. USP-Botucatu: Rejane Maria Tommasini Grotto, Jayme A. Souza-Neto, Guilherme Targino Valente, Patricia Akemi Assato, Felipe Allan da Silva da Costa, Bianca Cechetto Carlos. Mendelics: Bibiana Santos, João Paulo Kitajima, Erika Freitas, David Schlesinger. Hemocentro Ribeirão Preto: Simone Kashima, Evandra Strazza Rodrigues, Svetoslav Nanev Slavov, Elaine Vieira dos Santos, Rafael dos Santos Bezerra, Luiz Carlos Junior de Alcantara, Marta Giovanetti, Vagner Fonseca, Flavia Aburjaile, Rodrigo Tocantins Calado. |
| EPI_ISL_1795240 | CS DE SEBASTIANOPOLIS DO SUL | Instituto Butantan / ESALQ-Piracicaba | Instituto Butantan: Alexander Roberto Precioso, Dimas Tadeu Covas, Sandra Coccuzzo Sampaio, Maria Carolina Elias, José Salvatore Leister Patané, Vincent Louis Viala, Antonio Jorge Martins, Ricardo Haddad, Claudia Renata dos Santos Barros, Elaine Cristina Marqueze, Raul Machado Neto, Debora Botequio Moretti. Centro de Genômica Funcional da ESALQ: Luiz Lehmann Coutinho, Ricardo Augusto Brassaloti, Raquel de Lello Rocha Campos Cassano. NGS Soluções Genômicas: Pilar Drummond Sampaio Corrêa Mariani. FZEA-USP Pirassununga: Mirele Daiana Poleti, Jessika Cristina Chagas Lesbon, Elisangela Chicaroni Mattos, Heidge Fukumasu. USP-Botucatu: Rejane Maria Tommasini Grotto, Jayme A. Souza-Neto, Guilherme Targino |

| | | | |
|---|---|---|---|
| | | | Valente, Patricia Akemi Assato, Felipe Allan da Silva da Costa, Bianca Cechetto Carlos. Mendelics: Bibiana Santos, João Paulo Kitajima, Erika Freitas, David Schlesinger. Hemocentro Ribeirão Preto: Simone Kashima, Evandra Strazza Rodrigues, Svetoslav Nanev Slavov, Elaine Vieira dos Santos, Rafael dos Santos Bezerra, Luiz Carlos Junior de Alcantara, Marta Giovanetti, Vagner Fonseca, Flavia Aburjaile, Rodrigo Tocantins Calado. |
| EPI_ISL_1795241 | CS DE BALSAMO | Instituto Butantan / ESALQ-Piracicaba | Instituto Butantan: Alexander Roberto Precioso, Dimas Tadeu Covas, Sandra Coccuzzo Sampaio, Maria Carolina Elias, José Salvatore Leister Patané, Vincent Louis Viala, Antonio Jorge Martins, Ricardo Haddad, Claudia Renata dos Santos Barros, Elaine Cristina Marqueze, Raul Machado Neto, Debora Botequio Moretti. Centro de Genômica Funcional da ESALQ: Luiz Lehmann Coutinho, Ricardo Augusto Brassaloti, Raquel de Lello Rocha Campos Cassano. NGS Soluções Genômicas: Pilar Drummond Sampaio Corrêa Mariani. FZEA-USP Pirassununga: Mirele Daiana Poleti, Jessika Cristina Chagas Lesbon, Elisangela Chicaroni Mattos, Heidge Fukumasu. USP-Botucatu: Rejane Maria Tommasini Grotto, Jayme A. Souza-Neto, Guilherme Targino Valente, Patricia Akemi Assato, Felipe Allan da Silva da Costa, Bianca Cechetto Carlos. Mendelics: Bibiana Santos, João Paulo Kitajima, Erika Freitas, David Schlesinger. Hemocentro Ribeirão Preto: Simone Kashima, Evandra Strazza Rodrigues, Svetoslav Nanev Slavov, Elaine Vieira dos Santos, Rafael dos Santos Bezerra, Luiz Carlos Junior de Alcantara, Marta Giovanetti, Vagner Fonseca, Flavia Aburjaile, Rodrigo Tocantins Calado. |
| EPI_ISL_1795242, EPI_ISL_1795243, EPI_ISL_1795244 | CS DE URUPES | Instituto Butantan / ESALQ-Piracicaba | Instituto Butantan: Alexander Roberto Precioso, Dimas Tadeu Covas, Sandra Coccuzzo Sampaio, Maria Carolina Elias, José Salvatore Leister Patané, Vincent Louis Viala, Antonio Jorge Martins, Ricardo Haddad, Claudia Renata dos Santos Barros, Elaine Cristina Marqueze, Raul Machado Neto, Debora Botequio Moretti. Centro de Genômica Funcional da ESALQ: Luiz Lehmann Coutinho, Ricardo Augusto Brassaloti, Raquel de Lello Rocha Campos Cassano. NGS Soluções Genômicas: Pilar Drummond Sampaio Corrêa Mariani. FZEA-USP Pirassununga: Mirele Daiana Poleti, Jessika Cristina Chagas Lesbon, Elisangela Chicaroni Mattos, Heidge Fukumasu. USP-Botucatu: Rejane Maria Tommasini Grotto, Jayme A. Souza-Neto, Guilherme Targino Valente, Patricia Akemi Assato, Felipe Allan da Silva da Costa, Bianca Cechetto Carlos. Mendelics: Bibiana Santos, João Paulo Kitajima, Erika Freitas, David Schlesinger. Hemocentro Ribeirão Preto: Simone Kashima, Evandra Strazza Rodrigues, Svetoslav Nanev Slavov, Elaine Vieira dos Santos, Rafael dos Santos Bezerra, Luiz Carlos Junior de Alcantara, Marta Giovanetti, Vagner Fonseca, Flavia Aburjaile, Rodrigo Tocantins Calado. |
| EPI_ISL_1795245, EPI_ISL_1795246, EPI_ISL_1795247 | EMERGENCIA RESPIRATORIA DE NOVA GRANADA | Instituto Butantan / ESALQ-Piracicaba | Instituto Butantan: Alexander Roberto Precioso, Dimas Tadeu Covas, Sandra Coccuzzo Sampaio, Maria Carolina Elias, José Salvatore Leister Patané, Vincent Louis Viala, Antonio Jorge Martins, Ricardo Haddad, Claudia Renata dos Santos Barros, Elaine Cristina Marqueze, Raul Machado Neto, Debora Botequio Moretti. Centro de Genômica Funcional da ESALQ: Luiz Lehmann Coutinho, Ricardo Augusto Brassaloti, Raquel de Lello Rocha Campos Cassano. NGS Soluções Genômicas: Pilar Drummond Sampaio Corrêa Mariani. FZEA-USP Pirassununga: Mirele Daiana Poleti, Jessika Cristina Chagas Lesbon, Elisangela Chicaroni Mattos, Heidge Fukumasu. USP-Botucatu: Rejane Maria Tommasini Grotto, Jayme A. Souza-Neto, Guilherme Targino Valente, Patricia Akemi Assato, Felipe Allan da Silva da Costa, Bianca Cechetto Carlos. Mendelics: Bibiana Santos, João Paulo Kitajima, Erika Freitas, David Schlesinger. Hemocentro Ribeirão Preto: Simone Kashima, Evandra Strazza Rodrigues, Svetoslav Nanev Slavov, Elaine Vieira dos Santos, Rafael dos Santos Bezerra, Luiz Carlos Junior de Alcantara, Marta Giovanetti, Vagner Fonseca, Flavia Aburjaile, Rodrigo Tocantins Calado. |
| EPI_ISL_1795248, EPI_ISL_1795249, EPI_ISL_1795250 | ESF NOVA TANABI II | Instituto Butantan / ESALQ-Piracicaba | Instituto Butantan: Alexander Roberto Precioso, Dimas Tadeu Covas, Sandra Coccuzzo Sampaio, Maria Carolina Elias, José Salvatore Leister Patané, Vincent Louis Viala, Antonio Jorge Martins, Ricardo Haddad, Claudia Renata dos Santos Barros, Elaine Cristina Marqueze, Raul Machado Neto, Debora Botequio Moretti. Centro de Genômica Funcional da ESALQ: Luiz Lehmann Coutinho, Ricardo Augusto Brassaloti, Raquel de Lello Rocha Campos Cassano. NGS Soluções Genômicas: Pilar Drummond Sampaio Corrêa Mariani. FZEA-USP Pirassununga: Mirele Daiana Poleti, Jessika Cristina Chagas Lesbon, Elisangela Chicaroni Mattos, Heidge Fukumasu. USP-Botucatu: Rejane Maria Tommasini Grotto, Jayme A. Souza-Neto, Guilherme Targino Valente, Patricia Akemi Assato, Felipe Allan da Silva da Costa, Bianca Cechetto Carlos. Mendelics: Bibiana Santos, João Paulo Kitajima, Erika Freitas, David Schlesinger. Hemocentro Ribeirão Preto: Simone Kashima, Evandra Strazza Rodrigues, Svetoslav Nanev Slavov, Elaine Vieira dos Santos, Rafael dos Santos Bezerra, Luiz Carlos Junior de Alcantara, Marta Giovanetti, Vagner Fonseca, Flavia Aburjaile, Rodrigo Tocantins Calado. |
| EPI_ISL_1795251, EPI_ISL_1795252, EPI_ISL_1795253, EPI_ISL_1795254, EPI_ISL_1795255, EPI_ISL_1795256, EPI_ISL_1795257 | UBS II DE TANABI MILTON MARTINS PERCHES | Instituto Butantan / ESALQ-Piracicaba | Instituto Butantan: Alexander Roberto Precioso, Dimas Tadeu Covas, Sandra Coccuzzo Sampaio, Maria Carolina Elias, José Salvatore Leister Patané, Vincent Louis Viala, Antonio Jorge Martins, Ricardo Haddad, Claudia Renata dos Santos Barros, Elaine Cristina Marqueze, Raul Machado Neto, Debora Botequio Moretti. Centro de Genômica Funcional da ESALQ: Luiz Lehmann Coutinho, Ricardo Augusto Brassaloti, Raquel de Lello Rocha Campos Cassano. NGS Soluções Genômicas: Pilar Drummond Sampaio Corrêa Mariani. FZEA-USP Pirassununga: Mirele Daiana Poleti, Jessika Cristina Chagas Lesbon, Elisangela Chicaroni Mattos, Heidge Fukumasu. USP-Botucatu: Rejane Maria Tommasini Grotto, Jayme A. Souza-Neto, Guilherme Targino Valente, Patricia Akemi Assato, Felipe Allan da Silva da Costa, Bianca Cechetto Carlos. Mendelics: Bibiana Santos, João Paulo Kitajima, Erika Freitas, David Schlesinger. Hemocentro Ribeirão Preto: Simone Kashima, Evandra Strazza Rodrigues, Svetoslav Nanev Slavov, Elaine Vieira dos Santos, Rafael dos Santos Bezerra, Luiz Carlos Junior de Alcantara, Marta Giovanetti, Vagner Fonseca, Flavia Aburjaile, Rodrigo Tocantins Calado. |
| EPI_ISL_1795258 | CS DE PLANALTO | Instituto Butantan / ESALQ-Piracicaba | Instituto Butantan: Alexander Roberto Precioso, Dimas Tadeu Covas, Sandra Coccuzzo Sampaio, Maria Carolina Elias, José Salvatore Leister Patané, Vincent Louis Viala, Antonio Jorge Martins, Ricardo Haddad, Claudia Renata dos Santos Barros, Elaine Cristina Marqueze, Raul Machado Neto, Debora Botequio Moretti. Centro de Genômica Funcional da ESALQ: Luiz Lehmann Coutinho, Ricardo Augusto Brassaloti, Raquel de Lello Rocha Campos Cassano. NGS Soluções Genômicas: Pilar Drummond Sampaio Corrêa Mariani. FZEA-USP Pirassununga: Mirele Daiana Poleti, Jessika Cristina Chagas Lesbon, Elisangela Chicaroni Mattos, Heidge Fukumasu. USP-Botucatu: Rejane Maria Tommasini Grotto, Jayme A. Souza-Neto, Guilherme Targino Valente, Patricia Akemi Assato, Felipe Allan da Silva da Costa, Bianca Cechetto Carlos. Mendelics: Bibiana Santos, João Paulo Kitajima, Erika Freitas, David Schlesinger. Hemocentro Ribeirão Preto: Simone Kashima, Evandra Strazza Rodrigues, Svetoslav Nanev Slavov, Elaine Vieira dos Santos, Rafael dos Santos Bezerra, Luiz Carlos Junior de Alcantara, Marta Giovanetti, Vagner Fonseca, Flavia Aburjaile, Rodrigo Tocantins Calado. |
| EPI_ISL_1795259, EPI_ISL_1795260, EPI_ISL_1795261, EPI_ISL_1795262, EPI_ISL_1795263 | CS DE ICEM | Instituto Butantan / ESALQ-Piracicaba | Instituto Butantan: Alexander Roberto Precioso, Dimas Tadeu Covas, Sandra Coccuzzo Sampaio, Maria Carolina Elias, José Salvatore Leister Patané, Vincent Louis Viala, Antonio Jorge Martins, Ricardo Haddad, Claudia Renata dos Santos Barros, Elaine Cristina Marqueze, Raul Machado Neto, Debora Botequio Moretti. Centro de Genômica Funcional da ESALQ: Luiz Lehmann Coutinho, Ricardo Augusto Brassaloti, Raquel de Lello Rocha Campos Cassano. NGS Soluções Genômicas: Pilar Drummond Sampaio Corrêa Mariani. FZEA-USP Pirassununga: Mirele Daiana Poleti, Jessika Cristina Chagas Lesbon, Elisangela Chicaroni Mattos, Heidge Fukumasu. USP-Botucatu: Rejane Maria Tommasini Grotto, Jayme A. Souza-Neto, Guilherme Targino Valente, Patricia Akemi Assato, Felipe Allan da Silva da Costa, Bianca Cechetto Carlos. Mendelics: Bibiana Santos, João Paulo Kitajima, Erika Freitas, David Schlesinger. Hemocentro Ribeirão Preto: Simone Kashima, Evandra Strazza Rodrigues, Svetoslav Nanev Slavov, Elaine Vieira dos Santos, Rafael dos Santos Bezerra, Luiz Carlos Junior de Alcantara, Marta Giovanetti, Vagner Fonseca, Flavia Aburjaile, Rodrigo Tocantins Calado. |
| EPI_ISL_1795264 | CS DE PARANAPUA | Instituto Butantan / ESALQ-Piracicaba | Instituto Butantan: Alexander Roberto Precioso, Dimas Tadeu Covas, Sandra Coccuzzo Sampaio, Maria Carolina Elias, José Salvatore Leister Patané, Vincent Louis Viala, Antonio Jorge Martins, Ricardo Haddad, Claudia Renata dos Santos Barros, Elaine Cristina Marqueze, Raul Machado Neto, Debora Botequio Moretti. Centro de Genômica Funcional da ESALQ: Luiz Lehmann Coutinho, Ricardo Augusto Brassaloti, Raquel de Lello Rocha Campos Cassano. NGS Soluções Genômicas: Pilar Drummond Sampaio Corrêa Mariani. FZEA-USP Pirassununga: Mirele Daiana Poleti, Jessika Cristina Chagas Lesbon, Elisangela Chicaroni Mattos, Heidge Fukumasu. USP-Botucatu: Rejane Maria Tommasini Grotto, Jayme A. Souza-Neto, Guilherme Targino Valente, Patricia Akemi Assato, Felipe Allan da Silva da Costa, Bianca Cechetto Carlos. Mendelics: Bibiana Santos, João Paulo Kitajima, Erika Freitas, David Schlesinger. Hemocentro Ribeirão Preto: Simone Kashima, Evandra Strazza Rodrigues, Svetoslav Nanev Slavov, Elaine Vieira dos Santos, Rafael dos Santos Bezerra, Luiz Carlos Junior de Alcantara, Marta Giovanetti, Vagner Fonseca, Flavia Aburjaile, Rodrigo Tocantins Calado. |
| EPI_ISL_1795265 | CS DE ICEM | Instituto Butantan / ESALQ-Piracicaba | Instituto Butantan: Alexander Roberto Precioso, Dimas Tadeu Covas, Sandra Coccuzzo Sampaio, Maria Carolina Elias, José Salvatore Leister Patané, Vincent Louis Viala, Antonio Jorge Martins, Ricardo Haddad, Claudia Renata dos Santos Barros, Elaine Cristina Marqueze, Raul Machado Neto, Debora Botequio Moretti. Centro de Genômica Funcional da ESALQ: Luiz Lehmann Coutinho, Ricardo Augusto Brassaloti, Raquel de Lello Rocha Campos Cassano. NGS Soluções Genômicas: Pilar Drummond Sampaio Corrêa Mariani. FZEA-USP Pirassununga: Mirele Daiana Poleti, Jessika Cristina Chagas Lesbon, Elisangela Chicaroni Mattos, Heidge Fukumasu. USP-Botucatu: Rejane Maria Tommasini Grotto, Jayme A. Souza-Neto, Guilherme Targino Valente, Patricia Akemi Assato, Felipe Allan da Silva da Costa, Bianca Cechetto Carlos. Mendelics: Bibiana Santos, João Paulo Kitajima, Erika Freitas, David Schlesinger. Hemocentro Ribeirão Preto: Simone Kashima, Evandra Strazza Rodrigues, Svetoslav Nanev Slavov, Elaine Vieira dos Santos, Rafael dos Santos Bezerra, Luiz Carlos Junior de Alcantara, Marta Giovanetti, Vagner Fonseca, Flavia Aburjaile, Rodrigo Tocantins Calado. |
| EPI_ISL_1795266, EPI_ISL_1795267, EPI_ISL_1795268 | UBS LUIS FACHIN IPIGUA | Instituto Butantan / ESALQ-Piracicaba | Instituto Butantan: Alexander Roberto Precioso, Dimas Tadeu Covas, Sandra Coccuzzo Sampaio, Maria Carolina Elias, José Salvatore Leister Patané, Vincent Louis Viala, Antonio Jorge Martins, Ricardo Haddad, Claudia Renata dos Santos Barros, Elaine Cristina Marqueze, Raul Machado Neto, Debora Botequio Moretti. Centro de Genômica Funcional da ESALQ: Luiz Lehmann Coutinho, Ricardo Augusto Brassaloti, Raquel de Lello Rocha Campos Cassano. NGS Soluções Genômicas: Pilar Drummond Sampaio Corrêa Mariani. FZEA-USP Pirassununga: Mirele Daiana Poleti, Jessika Cristina Chagas Lesbon, Elisangela Chicaroni Mattos, Heidge Fukumasu. USP-Botucatu: Rejane Maria Tommasini Grotto, Jayme A. Souza-Neto, Guilherme Targino |

| | | | |
|---|---|---|---|
| | | | Valente, Patricia Akemi Assato, Felipe Allan da Silva da Costa, Bianca Cechetto Carlos. Mendelics: Bibiana Santos, João Paulo Kitajima, Erika Freitas, David Schlesinger. Hemocentro Ribeirão Preto: Simone Kashima, Evandra Strazza Rodrigues, Svetoslav Nanev Slavov, Elaine Vieira dos Santos, Rafael dos Santos Bezerra, Luiz Carlos Junior de Alcantara, Marta Giovanetti, Vagner Fonseca, Flavia Aburjaile, Rodrigo Tocantins Calado. |
| EPI_ISL_1795269, EPI_ISL_1795270 | UBS TEREZA GALLO UCHOA | Instituto Butantan / ESALQ-Piracicaba | Instituto Butantan: Alexander Roberto Precioso, Dimas Tadeu Covas, Sandra Coccuzzo Sampaio, Maria Carolina Elias, José Salvatore Leister Patané, Vincent Louis Viala, Antonio Jorge Martins, Ricardo Haddad, Claudia Renata dos Santos Barros, Elaine Cristina Marqueze, Raul Machado Neto, Debora Botequio Moretti. Centro de Genômica Funcional da ESALQ: Luiz Lehmann Coutinho, Ricardo Augusto Brassaloti, Raquel de Lello Rocha Campos Cassano. NGS Soluções Genômicas: Pilar Drummond Sampaio Corrêa Mariani. FZEA-USP Pirassununga: Mirele Daiana Poleti, Jessika Cristina Chagas Lesbon, Elisangela Chicaroni Mattos, Heidge Fukumasu. USP-Botucatu: Rejane Maria Tommasini Grotto, Jayme A. Souza-Neto, Guilherme Targino Valente, Patricia Akemi Assato, Felipe Allan da Silva da Costa, Bianca Cechetto Carlos. Mendelics: Bibiana Santos, João Paulo Kitajima, Erika Freitas, David Schlesinger. Hemocentro Ribeirão Preto: Simone Kashima, Evandra Strazza Rodrigues, Svetoslav Nanev Slavov, Elaine Vieira dos Santos, Rafael dos Santos Bezerra, Luiz Carlos Junior de Alcantara, Marta Giovanetti, Vagner Fonseca, Flavia Aburjaile, Rodrigo Tocantins Calado. |
| EPI_ISL_1795271 | CS DE PARANAPUA | Instituto Butantan / ESALQ-Piracicaba | Instituto Butantan: Alexander Roberto Precioso, Dimas Tadeu Covas, Sandra Coccuzzo Sampaio, Maria Carolina Elias, José Salvatore Leister Patané, Vincent Louis Viala, Antonio Jorge Martins, Ricardo Haddad, Claudia Renata dos Santos Barros, Elaine Cristina Marqueze, Raul Machado Neto, Debora Botequio Moretti. Centro de Genômica Funcional da ESALQ: Luiz Lehmann Coutinho, Ricardo Augusto Brassaloti, Raquel de Lello Rocha Campos Cassano. NGS Soluções Genômicas: Pilar Drummond Sampaio Corrêa Mariani. FZEA-USP Pirassununga: Mirele Daiana Poleti, Jessika Cristina Chagas Lesbon, Elisangela Chicaroni Mattos, Heidge Fukumasu. USP-Botucatu: Rejane Maria Tommasini Grotto, Jayme A. Souza-Neto, Guilherme Targino Valente, Patricia Akemi Assato, Felipe Allan da Silva da Costa, Bianca Cechetto Carlos. Mendelics: Bibiana Santos, João Paulo Kitajima, Erika Freitas, David Schlesinger. Hemocentro Ribeirão Preto: Simone Kashima, Evandra Strazza Rodrigues, Svetoslav Nanev Slavov, Elaine Vieira dos Santos, Rafael dos Santos Bezerra, Luiz Carlos Junior de Alcantara, Marta Giovanetti, Vagner Fonseca, Flavia Aburjaile, Rodrigo Tocantins Calado. |
| EPI_ISL_1795272, EPI_ISL_1795273 | UBS TEREZA GALLO UCHOA | Instituto Butantan / ESALQ-Piracicaba | Instituto Butantan: Alexander Roberto Precioso, Dimas Tadeu Covas, Sandra Coccuzzo Sampaio, Maria Carolina Elias, José Salvatore Leister Patané, Vincent Louis Viala, Antonio Jorge Martins, Ricardo Haddad, Claudia Renata dos Santos Barros, Elaine Cristina Marqueze, Raul Machado Neto, Debora Botequio Moretti. Centro de Genômica Funcional da ESALQ: Luiz Lehmann Coutinho, Ricardo Augusto Brassaloti, Raquel de Lello Rocha Campos Cassano. NGS Soluções Genômicas: Pilar Drummond Sampaio Corrêa Mariani. FZEA-USP Pirassununga: Mirele Daiana Poleti, Jessika Cristina Chagas Lesbon, Elisangela Chicaroni Mattos, Heidge Fukumasu. USP-Botucatu: Rejane Maria Tommasini Grotto, Jayme A. Souza-Neto, Guilherme Targino Valente, Patricia Akemi Assato, Felipe Allan da Silva da Costa, Bianca Cechetto Carlos. Mendelics: Bibiana Santos, João Paulo Kitajima, Erika Freitas, David Schlesinger. Hemocentro Ribeirão Preto: Simone Kashima, Evandra Strazza Rodrigues, Svetoslav Nanev Slavov, Elaine Vieira dos Santos, Rafael dos Santos Bezerra, Luiz Carlos Junior de Alcantara, Marta Giovanetti, Vagner Fonseca, Flavia Aburjaile, Rodrigo Tocantins Calado. |
| EPI_ISL_1795274, EPI_ISL_1795275 | CENTRO MEDICO PMESP | Instituto Butantan / ESALQ-Piracicaba | Instituto Butantan: Alexander Roberto Precioso, Dimas Tadeu Covas, Sandra Coccuzzo Sampaio, Maria Carolina Elias, José Salvatore Leister Patané, Vincent Louis Viala, Antonio Jorge Martins, Ricardo Haddad, Claudia Renata dos Santos Barros, Elaine Cristina Marqueze, Raul Machado Neto, Debora Botequio Moretti. Centro de Genômica Funcional da ESALQ: Luiz Lehmann Coutinho, Ricardo Augusto Brassaloti, Raquel de Lello Rocha Campos Cassano. NGS Soluções Genômicas: Pilar Drummond Sampaio Corrêa Mariani. FZEA-USP Pirassununga: Mirele Daiana Poleti, Jessika Cristina Chagas Lesbon, Elisangela Chicaroni Mattos, Heidge Fukumasu. USP-Botucatu: Rejane Maria Tommasini Grotto, Jayme A. Souza-Neto, Guilherme Targino Valente, Patricia Akemi Assato, Felipe Allan da Silva da Costa, Bianca Cechetto Carlos. Mendelics: Bibiana Santos, João Paulo Kitajima, Erika Freitas, David Schlesinger. Hemocentro Ribeirão Preto: Simone Kashima, Evandra Strazza Rodrigues, Svetoslav Nanev Slavov, Elaine Vieira dos Santos, Rafael dos Santos Bezerra, Luiz Carlos Junior de Alcantara, Marta Giovanetti, Vagner Fonseca, Flavia Aburjaile, Rodrigo Tocantins Calado. |
| EPI_ISL_1795276, EPI_ISL_1795277, EPI_ISL_1795278, EPI_ISL_1795279, EPI_ISL_1795280, EPI_ISL_1795281, EPI_ISL_1795282, EPI_ISL_1795283, EPI_ISL_1795284, EPI_ISL_1795285, EPI_ISL_1795286 | | | |
| see above | CS DE PALESTINA | Instituto Butantan / ESALQ-Piracicaba | Instituto Butantan: Alexander Roberto Precioso, Dimas Tadeu Covas, Sandra Coccuzzo Sampaio, Maria Carolina Elias, José Salvatore Leister Patané, Vincent Louis Viala, Antonio Jorge Martins, Ricardo Haddad, Claudia Renata dos Santos Barros, Elaine Cristina Marqueze, Raul Machado Neto, Debora Botequio Moretti. Centro de Genômica Funcional da ESALQ: Luiz Lehmann Coutinho, Ricardo Augusto Brassaloti, Raquel de Lello Rocha Campos Cassano. NGS Soluções Genômicas: Pilar Drummond Sampaio Corrêa Mariani. FZEA-USP Pirassununga: Mirele Daiana Poleti, Jessika Cristina Chagas Lesbon, Elisangela Chicaroni Mattos, Heidge Fukumasu. USP-Botucatu: Rejane Maria Tommasini Grotto, Jayme A. Souza-Neto, Guilherme Targino Valente, Patricia Akemi Assato, Felipe Allan da Silva da Costa, Bianca Cechetto Carlos. Mendelics: Bibiana Santos, João Paulo Kitajima, Erika Freitas, David Schlesinger. Hemocentro Ribeirão Preto: Simone Kashima, Evandra Strazza Rodrigues, Svetoslav Nanev Slavov, Elaine Vieira dos Santos, Rafael dos Santos Bezerra, Luiz Carlos Junior de Alcantara, Marta Giovanetti, Vagner Fonseca, Flavia Aburjaile, Rodrigo Tocantins Calado. |
| EPI_ISL_1795287 | UBS ALCIMINIO DE ASSIS LOURENCO BADY BASSITT | Instituto Butantan / ESALQ-Piracicaba | Instituto Butantan: Alexander Roberto Precioso, Dimas Tadeu Covas, Sandra Coccuzzo Sampaio, Maria Carolina Elias, José Salvatore Leister Patané, Vincent Louis Viala, Antonio Jorge Martins, Ricardo Haddad, Claudia Renata dos Santos Barros, Elaine Cristina Marqueze, Raul Machado Neto, Debora Botequio Moretti. Centro de Genômica Funcional da ESALQ: Luiz Lehmann Coutinho, Ricardo Augusto Brassaloti, Raquel de Lello Rocha Campos Cassano. NGS Soluções Genômicas: Pilar Drummond Sampaio Corrêa Mariani. FZEA-USP Pirassununga: Mirele Daiana Poleti, Jessika Cristina Chagas Lesbon, Elisangela Chicaroni Mattos, Heidge Fukumasu. USP-Botucatu: Rejane Maria Tommasini Grotto, Jayme A. Souza-Neto, Guilherme Targino Valente, Patricia Akemi Assato, Felipe Allan da Silva da Costa, Bianca Cechetto Carlos. Mendelics: Bibiana Santos, João Paulo Kitajima, Erika Freitas, David Schlesinger. Hemocentro Ribeirão Preto: Simone Kashima, Evandra Strazza Rodrigues, Svetoslav Nanev Slavov, Elaine Vieira dos Santos, Rafael dos Santos Bezerra, Luiz Carlos Junior de Alcantara, Marta Giovanetti, Vagner Fonseca, Flavia Aburjaile, Rodrigo Tocantins Calado. |
| EPI_ISL_1795288 | CS DE NIPOA | Instituto Butantan / ESALQ-Piracicaba | Instituto Butantan: Alexander Roberto Precioso, Dimas Tadeu Covas, Sandra Coccuzzo Sampaio, Maria Carolina Elias, José Salvatore Leister Patané, Vincent Louis Viala, Antonio Jorge Martins, Ricardo Haddad, Claudia Renata dos Santos Barros, Elaine Cristina Marqueze, Raul Machado Neto, Debora Botequio Moretti. Centro de Genômica Funcional da ESALQ: Luiz Lehmann Coutinho, Ricardo Augusto Brassaloti, Raquel de Lello Rocha Campos Cassano. NGS Soluções Genômicas: Pilar Drummond Sampaio Corrêa Mariani. FZEA-USP Pirassununga: Mirele Daiana Poleti, Jessika Cristina Chagas Lesbon, Elisangela Chicaroni Mattos, Heidge Fukumasu. USP-Botucatu: Rejane Maria Tommasini Grotto, Jayme A. Souza-Neto, Guilherme Targino Valente, Patricia Akemi Assato, Felipe Allan da Silva da Costa, Bianca Cechetto Carlos. Mendelics: Bibiana Santos, João Paulo Kitajima, Erika Freitas, David Schlesinger. Hemocentro Ribeirão Preto: Simone Kashima, Evandra Strazza Rodrigues, Svetoslav Nanev Slavov, Elaine Vieira dos Santos, Rafael dos Santos Bezerra, Luiz Carlos Junior de Alcantara, Marta Giovanetti, Vagner Fonseca, Flavia Aburjaile, Rodrigo Tocantins Calado. |
| EPI_ISL_1795289 | UBS ALCIMINIO DE ASSIS LOURENCO BADY BASSITT | Instituto Butantan / ESALQ-Piracicaba | Instituto Butantan: Alexander Roberto Precioso, Dimas Tadeu Covas, Sandra Coccuzzo Sampaio, Maria Carolina Elias, José Salvatore Leister Patané, Vincent Louis Viala, Antonio Jorge Martins, Ricardo Haddad, Claudia Renata dos Santos Barros, Elaine Cristina Marqueze, Raul Machado Neto, Debora Botequio Moretti. Centro de Genômica Funcional da ESALQ: Luiz Lehmann Coutinho, Ricardo Augusto Brassaloti, Raquel de Lello Rocha Campos Cassano. NGS Soluções Genômicas: Pilar Drummond Sampaio Corrêa Mariani. FZEA-USP Pirassununga: Mirele Daiana Poleti, Jessika Cristina Chagas Lesbon, Elisangela Chicaroni Mattos, Heidge Fukumasu. USP-Botucatu: Rejane Maria Tommasini Grotto, Jayme A. Souza-Neto, Guilherme Targino Valente, Patricia Akemi Assato, Felipe Allan da Silva da Costa, Bianca Cechetto Carlos. Mendelics: Bibiana Santos, João Paulo Kitajima, Erika Freitas, David Schlesinger. Hemocentro Ribeirão Preto: Simone Kashima, Evandra Strazza Rodrigues, Svetoslav Nanev Slavov, Elaine Vieira dos Santos, Rafael dos Santos Bezerra, Luiz Carlos Junior de Alcantara, Marta Giovanetti, Vagner Fonseca, Flavia Aburjaile, Rodrigo Tocantins Calado. |
| EPI_ISL_1795290 | USF EUCALIPTOS | Instituto Butantan / ESALQ-Piracicaba | Instituto Butantan: Alexander Roberto Precioso, Dimas Tadeu Covas, Sandra Coccuzzo Sampaio, Maria Carolina Elias, José Salvatore Leister Patané, Vincent Louis Viala, Antonio Jorge Martins, Ricardo Haddad, Claudia Renata dos Santos Barros, Elaine Cristina Marqueze, Raul Machado Neto, Debora Botequio Moretti. Centro de Genômica Funcional da ESALQ: Luiz Lehmann Coutinho, Ricardo Augusto Brassaloti, Raquel de Lello Rocha Campos Cassano. NGS Soluções Genômicas: Pilar Drummond Sampaio Corrêa Mariani. FZEA-USP Pirassununga: Mirele Daiana Poleti, Jessika Cristina Chagas Lesbon, Elisangela Chicaroni Mattos, Heidge Fukumasu. USP-Botucatu: Rejane Maria Tommasini Grotto, Jayme A. Souza-Neto, Guilherme Targino Valente, Patricia Akemi Assato, Felipe Allan da Silva da Costa, Bianca Cechetto Carlos. Mendelics: Bibiana Santos, João Paulo Kitajima, Erika Freitas, David Schlesinger. Hemocentro Ribeirão Preto: Simone Kashima, Evandra Strazza Rodrigues, Svetoslav Nanev Slavov, Elaine Vieira dos Santos, Rafael dos Santos Bezerra, Luiz Carlos Junior de Alcantara, Marta Giovanetti, Vagner Fonseca, Flavia Aburjaile, Rodrigo Tocantins Calado. |
| EPI_ISL_1795291, EPI_ISL_1795292 | AMBULATORIO DE ESPECIALIDADE V E MOGI MIRIM | Instituto Butantan / ESALQ-Piracicaba | Instituto Butantan: Alexander Roberto Precioso, Dimas Tadeu Covas, Sandra Coccuzzo Sampaio, Maria Carolina Elias, José Salvatore Leister Patané, Vincent Louis Viala, Antonio Jorge Martins, Ricardo Haddad, Claudia Renata dos Santos Barros, Elaine Cristina Marqueze, Raul Machado Neto, Debora Botequio Moretti. Centro de Genômica Funcional da ESALQ: Luiz Lehmann Coutinho, Ricardo Augusto Brassaloti, Raquel de Lello Rocha Campos Cassano. NGS Soluções Genômicas: Pilar Drummond Sampaio Corrêa Mariani. FZEA-USP Pirassununga: Mirele Daiana Poleti, Jessika Cristina Chagas Lesbon, Elisangela Chicaroni Mattos, Heidge Fukumasu. USP-Botucatu: Rejane Maria Tommasini Grotto, Jayme A. Souza-Neto, Guilherme Targino Valente, Patricia Akemi Assato, Felipe Allan da Silva da Costa, Bianca Cechetto Carlos. Mendelics: Bibiana Santos, João Paulo Kitajima, Erika Freitas, |

| | | | |
|---|---|---|---|
| | | | David Schlesinger. Hemocentro Ribeirão Preto: Simone Kashima, Evandra Strazza Rodrigues, Svetoslav Nanev Slavov, Elaine Vieira dos Santos, Rafael dos Santos Bezerra, Luiz Carlos Junior de Alcantara, Marta Giovanetti, Vagner Fonseca, Flavia Aburjaile, Rodrigo Tocantins Calado. |
| EPI_ISL_1795293, EPI_ISL_1795294 | UNIDADE DE SAUDE DE ITOBI ALCIBIADES PIRES | Instituto Butantan / ESALQ-Piracicaba | Instituto Butantan: Alexander Roberto Precioso, Dimas Tadeu Covas, Sandra Coccuzzo Sampaio, Maria Carolina Elias, José Salvatore Leister Patané, Vincent Louis Viala, Antonio Jorge Martins, Ricardo Haddad, Claudia Renata dos Santos Barros, Elaine Cristina Marqueze, Raul Machado Neto, Debora Botequio Moretti. Centro de Genômica Funcional da ESALQ: Luiz Lehmann Coutinho, Ricardo Augusto Brassaloti, Raquel de Lello Rocha Campos Cassano. NGS Soluções Genômicas: Pilar Drummond Sampaio Corrêa Mariani. FZEA-USP Pirassununga: Mirele Daiana Poleti, Jessika Cristina Chagas Lesbon, Elisangela Chicaroni Mattos, Heidge Fukumasu. USP-Botucatu: Rejane Maria Tommasini Grotto, Jayme A. Souza-Neto, Guilherme Targino Valente, Patricia Akemi Assato, Felipe Allan da Silva da Costa, Bianca Cechetto Carlos. Mendelics: Bibiana Santos, João Paulo Kitajima, Erika Freitas, David Schlesinger. Hemocentro Ribeirão Preto: Simone Kashima, Evandra Strazza Rodrigues, Svetoslav Nanev Slavov, Elaine Vieira dos Santos, Rafael dos Santos Bezerra, Luiz Carlos Junior de Alcantara, Marta Giovanetti, Vagner Fonseca, Flavia Aburjaile, Rodrigo Tocantins Calado. |
| EPI_ISL_1795295 | USF GUACUANO | Instituto Butantan / ESALQ-Piracicaba | Instituto Butantan: Alexander Roberto Precioso, Dimas Tadeu Covas, Sandra Coccuzzo Sampaio, Maria Carolina Elias, José Salvatore Leister Patané, Vincent Louis Viala, Antonio Jorge Martins, Ricardo Haddad, Claudia Renata dos Santos Barros, Elaine Cristina Marqueze, Raul Machado Neto, Debora Botequio Moretti. Centro de Genômica Funcional da ESALQ: Luiz Lehmann Coutinho, Ricardo Augusto Brassaloti, Raquel de Lello Rocha Campos Cassano. NGS Soluções Genômicas: Pilar Drummond Sampaio Corrêa Mariani. FZEA-USP Pirassununga: Mirele Daiana Poleti, Jessika Cristina Chagas Lesbon, Elisangela Chicaroni Mattos, Heidge Fukumasu. USP-Botucatu: Rejane Maria Tommasini Grotto, Jayme A. Souza-Neto, Guilherme Targino Valente, Patricia Akemi Assato, Felipe Allan da Silva da Costa, Bianca Cechetto Carlos. Mendelics: Bibiana Santos, João Paulo Kitajima, Erika Freitas, David Schlesinger. Hemocentro Ribeirão Preto: Simone Kashima, Evandra Strazza Rodrigues, Svetoslav Nanev Slavov, Elaine Vieira dos Santos, Rafael dos Santos Bezerra, Luiz Carlos Junior de Alcantara, Marta Giovanetti, Vagner Fonseca, Flavia Aburjaile, Rodrigo Tocantins Calado. |
| EPI_ISL_1795296, EPI_ISL_1795297 | CENTRO DE SAUDE III AFFONSO LUZZI SANTA CRUZ DAS PALMEIRAS | Instituto Butantan / ESALQ-Piracicaba | Instituto Butantan: Alexander Roberto Precioso, Dimas Tadeu Covas, Sandra Coccuzzo Sampaio, Maria Carolina Elias, José Salvatore Leister Patané, Vincent Louis Viala, Antonio Jorge Martins, Ricardo Haddad, Claudia Renata dos Santos Barros, Elaine Cristina Marqueze, Raul Machado Neto, Debora Botequio Moretti. Centro de Genômica Funcional da ESALQ: Luiz Lehmann Coutinho, Ricardo Augusto Brassaloti, Raquel de Lello Rocha Campos Cassano. NGS Soluções Genômicas: Pilar Drummond Sampaio Corrêa Mariani. FZEA-USP Pirassununga: Mirele Daiana Poleti, Jessika Cristina Chagas Lesbon, Elisangela Chicaroni Mattos, Heidge Fukumasu. USP-Botucatu: Rejane Maria Tommasini Grotto, Jayme A. Souza-Neto, Guilherme Targino Valente, Patricia Akemi Assato, Felipe Allan da Silva da Costa, Bianca Cechetto Carlos. Mendelics: Bibiana Santos, João Paulo Kitajima, Erika Freitas, David Schlesinger. Hemocentro Ribeirão Preto: Simone Kashima, Evandra Strazza Rodrigues, Svetoslav Nanev Slavov, Elaine Vieira dos Santos, Rafael dos Santos Bezerra, Luiz Carlos Junior de Alcantara, Marta Giovanetti, Vagner Fonseca, Flavia Aburjaile, Rodrigo Tocantins Calado. |
| EPI_ISL_1795298 | UNIDADE DE SAUDE DE ITOBI ALCIBIADES PIRES | Instituto Butantan / ESALQ-Piracicaba | Instituto Butantan: Alexander Roberto Precioso, Dimas Tadeu Covas, Sandra Coccuzzo Sampaio, Maria Carolina Elias, José Salvatore Leister Patané, Vincent Louis Viala, Antonio Jorge Martins, Ricardo Haddad, Claudia Renata dos Santos Barros, Elaine Cristina Marqueze, Raul Machado Neto, Debora Botequio Moretti. Centro de Genômica Funcional da ESALQ: Luiz Lehmann Coutinho, Ricardo Augusto Brassaloti, Raquel de Lello Rocha Campos Cassano. NGS Soluções Genômicas: Pilar Drummond Sampaio Corrêa Mariani. FZEA-USP Pirassununga: Mirele Daiana Poleti, Jessika Cristina Chagas Lesbon, Elisangela Chicaroni Mattos, Heidge Fukumasu. USP-Botucatu: Rejane Maria Tommasini Grotto, Jayme A. Souza-Neto, Guilherme Targino Valente, Patricia Akemi Assato, Felipe Allan da Silva da Costa, Bianca Cechetto Carlos. Mendelics: Bibiana Santos, João Paulo Kitajima, Erika Freitas, David Schlesinger. Hemocentro Ribeirão Preto: Simone Kashima, Evandra Strazza Rodrigues, Svetoslav Nanev Slavov, Elaine Vieira dos Santos, Rafael dos Santos Bezerra, Luiz Carlos Junior de Alcantara, Marta Giovanetti, Vagner Fonseca, Flavia Aburjaile, Rodrigo Tocantins Calado. |
| EPI_ISL_1795299 | CENTRO DE SAUDE III AFFONSO LUZZI SANTA CRUZ DAS PALMEIRAS | Instituto Butantan / ESALQ-Piracicaba | Instituto Butantan: Alexander Roberto Precioso, Dimas Tadeu Covas, Sandra Coccuzzo Sampaio, Maria Carolina Elias, José Salvatore Leister Patané, Vincent Louis Viala, Antonio Jorge Martins, Ricardo Haddad, Claudia Renata dos Santos Barros, Elaine Cristina Marqueze, Raul Machado Neto, Debora Botequio Moretti. Centro de Genômica Funcional da ESALQ: Luiz Lehmann Coutinho, Ricardo Augusto Brassaloti, Raquel de Lello Rocha Campos Cassano. NGS Soluções Genômicas: Pilar Drummond Sampaio Corrêa Mariani. FZEA-USP Pirassununga: Mirele Daiana Poleti, Jessika Cristina Chagas Lesbon, Elisangela Chicaroni Mattos, Heidge Fukumasu. USP-Botucatu: Rejane Maria Tommasini Grotto, Jayme A. Souza-Neto, Guilherme Targino Valente, Patricia Akemi Assato, Felipe Allan da Silva da Costa, Bianca Cechetto Carlos. Mendelics: Bibiana Santos, João Paulo Kitajima, Erika Freitas, David Schlesinger. Hemocentro Ribeirão Preto: Simone Kashima, Evandra Strazza Rodrigues, Svetoslav Nanev Slavov, Elaine Vieira dos Santos, Rafael dos Santos Bezerra, Luiz Carlos Junior de Alcantara, Marta Giovanetti, Vagner Fonseca, Flavia Aburjaile, Rodrigo Tocantins Calado. |
| EPI_ISL_1795300, EPI_ISL_1795301, EPI_ISL_1795302 | PRONTO SOCORRO MUNICIPAL TAMBAU | Instituto Butantan / ESALQ-Piracicaba | Instituto Butantan: Alexander Roberto Precioso, Dimas Tadeu Covas, Sandra Coccuzzo Sampaio, Maria Carolina Elias, José Salvatore Leister Patané, Vincent Louis Viala, Antonio Jorge Martins, Ricardo Haddad, Claudia Renata dos Santos Barros, Elaine Cristina Marqueze, Raul Machado Neto, Debora Botequio Moretti. Centro de Genômica Funcional da ESALQ: Luiz Lehmann Coutinho, Ricardo Augusto Brassaloti, Raquel de Lello Rocha Campos Cassano. NGS Soluções Genômicas: Pilar Drummond Sampaio Corrêa Mariani. FZEA-USP Pirassununga: Mirele Daiana Poleti, Jessika Cristina Chagas Lesbon, Elisangela Chicaroni Mattos, Heidge Fukumasu. USP-Botucatu: Rejane Maria Tommasini Grotto, Jayme A. Souza-Neto, Guilherme Targino Valente, Patricia Akemi Assato, Felipe Allan da Silva da Costa, Bianca Cechetto Carlos. Mendelics: Bibiana Santos, João Paulo Kitajima, Erika Freitas, David Schlesinger. Hemocentro Ribeirão Preto: Simone Kashima, Evandra Strazza Rodrigues, Svetoslav Nanev Slavov, Elaine Vieira dos Santos, Rafael dos Santos Bezerra, Luiz Carlos Junior de Alcantara, Marta Giovanetti, Vagner Fonseca, Flavia Aburjaile, Rodrigo Tocantins Calado. |
| EPI_ISL_1795303 | LABORATORIO DE FRANCA | Instituto Butantan / ESALQ-Piracicaba | Instituto Butantan: Alexander Roberto Precioso, Dimas Tadeu Covas, Sandra Coccuzzo Sampaio, Maria Carolina Elias, José Salvatore Leister Patané, Vincent Louis Viala, Antonio Jorge Martins, Ricardo Haddad, Claudia Renata dos Santos Barros, Elaine Cristina Marqueze, Raul Machado Neto, Debora Botequio Moretti. Centro de Genômica Funcional da ESALQ: Luiz Lehmann Coutinho, Ricardo Augusto Brassaloti, Raquel de Lello Rocha Campos Cassano. NGS Soluções Genômicas: Pilar Drummond Sampaio Corrêa Mariani. FZEA-USP Pirassununga: Mirele Daiana Poleti, Jessika Cristina Chagas Lesbon, Elisangela Chicaroni Mattos, Heidge Fukumasu. USP-Botucatu: Rejane Maria Tommasini Grotto, Jayme A. Souza-Neto, Guilherme Targino Valente, Patricia Akemi Assato, Felipe Allan da Silva da Costa, Bianca Cechetto Carlos. Mendelics: Bibiana Santos, João Paulo Kitajima, Erika Freitas, David Schlesinger. Hemocentro Ribeirão Preto: Simone Kashima, Evandra Strazza Rodrigues, Svetoslav Nanev Slavov, Elaine Vieira dos Santos, Rafael dos Santos Bezerra, Luiz Carlos Junior de Alcantara, Marta Giovanetti, Vagner Fonseca, Flavia Aburjaile, Rodrigo Tocantins Calado. |
| EPI_ISL_1795304 | PRONTO SOCORRO MUNICIPAL TAMBAU | Instituto Butantan / ESALQ-Piracicaba | Instituto Butantan: Alexander Roberto Precioso, Dimas Tadeu Covas, Sandra Coccuzzo Sampaio, Maria Carolina Elias, José Salvatore Leister Patané, Vincent Louis Viala, Antonio Jorge Martins, Ricardo Haddad, Claudia Renata dos Santos Barros, Elaine Cristina Marqueze, Raul Machado Neto, Debora Botequio Moretti. Centro de Genômica Funcional da ESALQ: Luiz Lehmann Coutinho, Ricardo Augusto Brassaloti, Raquel de Lello Rocha Campos Cassano. NGS Soluções Genômicas: Pilar Drummond Sampaio Corrêa Mariani. FZEA-USP Pirassununga: Mirele Daiana Poleti, Jessika Cristina Chagas Lesbon, Elisangela Chicaroni Mattos, Heidge Fukumasu. USP-Botucatu: Rejane Maria Tommasini Grotto, Jayme A. Souza-Neto, Guilherme Targino Valente, Patricia Akemi Assato, Felipe Allan da Silva da Costa, Bianca Cechetto Carlos. Mendelics: Bibiana Santos, João Paulo Kitajima, Erika Freitas, David Schlesinger. Hemocentro Ribeirão Preto: Simone Kashima, Evandra Strazza Rodrigues, Svetoslav Nanev Slavov, Elaine Vieira dos Santos, Rafael dos Santos Bezerra, Luiz Carlos Junior de Alcantara, Marta Giovanetti, Vagner Fonseca, Flavia Aburjaile, Rodrigo Tocantins Calado. |
| EPI_ISL_1795305 | UNIDADE DA SAUDE DO ADULTO CASA BRANCA PREFEITURA | Instituto Butantan / ESALQ-Piracicaba | Instituto Butantan: Alexander Roberto Precioso, Dimas Tadeu Covas, Sandra Coccuzzo Sampaio, Maria Carolina Elias, José Salvatore Leister Patané, Vincent Louis Viala, Antonio Jorge Martins, Ricardo Haddad, Claudia Renata dos Santos Barros, Elaine Cristina Marqueze, Raul Machado Neto, Debora Botequio Moretti. Centro de Genômica Funcional da ESALQ: Luiz Lehmann Coutinho, Ricardo Augusto Brassaloti, Raquel de Lello Rocha Campos Cassano. NGS Soluções Genômicas: Pilar Drummond Sampaio Corrêa Mariani. FZEA-USP Pirassununga: Mirele Daiana Poleti, Jessika Cristina Chagas Lesbon, Elisangela Chicaroni Mattos, Heidge Fukumasu. USP-Botucatu: Rejane Maria Tommasini Grotto, Jayme A. Souza-Neto, Guilherme Targino Valente, Patricia Akemi Assato, Felipe Allan da Silva da Costa, Bianca Cechetto Carlos. Mendelics: Bibiana Santos, João Paulo Kitajima, Erika Freitas, David Schlesinger. Hemocentro Ribeirão Preto: Simone Kashima, Evandra Strazza Rodrigues, Svetoslav Nanev Slavov, Elaine Vieira dos Santos, Rafael dos Santos Bezerra, Luiz Carlos Junior de Alcantara, Marta Giovanetti, Vagner Fonseca, Flavia Aburjaile, Rodrigo Tocantins Calado. |
| EPI_ISL_1795306, EPI_ISL_1795307, EPI_ISL_1795308, EPI_ISL_1795309, EPI_ISL_1795310, EPI_ISL_1795311, EPI_ISL_1795312, EPI_ISL_1795313, EPI_ISL_1795314, EPI_ISL_1795315, EPI_ISL_1795316, EPI_ISL_1795317, EPI_ISL_1795318, EPI_ISL_1795319, EPI_ISL_1795320, EPI_ISL_1795321, EPI_ISL_1795322, EPI_ISL_1795323 | | | |
| see above | LABORATORIO DE FRANCA | Instituto Butantan / ESALQ-Piracicaba | Instituto Butantan: Alexander Roberto Precioso, Dimas Tadeu Covas, Sandra Coccuzzo Sampaio, Maria Carolina Elias, José Salvatore Leister Patané, Vincent Louis Viala, Antonio Jorge Martins, Ricardo Haddad, Claudia Renata dos Santos Barros, Elaine Cristina Marqueze, Raul Machado Neto, Debora Botequio Moretti. Centro de Genômica Funcional da ESALQ: Luiz Lehmann Coutinho, Ricardo Augusto Brassaloti, Raquel de Lello Rocha Campos Cassano. NGS Soluções Genômicas: Pilar Drummond Sampaio Corrêa Mariani. FZEA-USP Pirassununga: Mirele Daiana Poleti, Jessika Cristina Chagas Lesbon, Elisangela Chicaroni Mattos, Heidge Fukumasu. USP-Botucatu: Rejane Maria Tommasini Grotto, Jayme A. Souza-Neto, Guilherme Targino Valente, Patricia Akemi Assato, Felipe Allan da Silva da Costa, Bianca Cechetto Carlos. Mendelics: Bibiana Santos, João Paulo Kitajima, Erika Freitas, |

| | | | |
|---|---|---|---|
| | | | David Schlesinger. Hemocentro Ribeirão Preto: Simone Kashima, Evandra Strazza Rodrigues, Svetoslav Nanev Slavov, Elaine Vieira dos Santos, Rafael dos Santos Bezerra, Luiz Carlos Junior de Alcantara, Marta Giovanetti, Vagner Fonseca, Flavia Aburjaile, Rodrigo Tocantins Calado. |
| EPI_ISL_1795324 | UBS II DE NARANDIBA | Instituto Butantan / ESALQ-Piracicaba | Instituto Butantan: Alexander Roberto Precioso, Dimas Tadeu Covas, Sandra Coccuzzo Sampaio, Maria Carolina Elias, José Salvatore Leister Patané, Vincent Louis Viala, Antonio Jorge Martins, Ricardo Haddad, Claudia Renata dos Santos Barros, Elaine Cristina Marqueze, Raul Machado Neto, Debora Botequio Moretti. Centro de Genômica Funcional da ESALQ: Luiz Lehmann Coutinho, Ricardo Augusto Brassaloti, Raquel de Lello Rocha Campos Cassano. NGS Soluções Genômicas: Pilar Drummond Sampaio Corrêa Mariani. FZEA-USP Pirassununga: Mirele Daiana Poleti, Jessika Cristina Chagas Lesbon, Elisangela Chicaroni Mattos, Heidge Fukumasu. USP-Botucatu: Rejane Maria Tommasini Grotto, Jayme A. Souza-Neto, Guilherme Targino Valente, Patricia Akemi Assato, Felipe Allan da Silva da Costa, Bianca Cechetto Carlos. Mendelics: Bibiana Santos, João Paulo Kitajima, Erika Freitas, David Schlesinger. Hemocentro Ribeirão Preto: Simone Kashima, Evandra Strazza Rodrigues, Svetoslav Nanev Slavov, Elaine Vieira dos Santos, Rafael dos Santos Bezerra, Luiz Carlos Junior de Alcantara, Marta Giovanetti, Vagner Fonseca, Flavia Aburjaile, Rodrigo Tocantins Calado. |
| EPI_ISL_1795325 | CENTRO DE ESPECIALIDADES DE PRIMAVERA | Instituto Butantan / ESALQ-Piracicaba | Instituto Butantan: Alexander Roberto Precioso, Dimas Tadeu Covas, Sandra Coccuzzo Sampaio, Maria Carolina Elias, José Salvatore Leister Patané, Vincent Louis Viala, Antonio Jorge Martins, Ricardo Haddad, Claudia Renata dos Santos Barros, Elaine Cristina Marqueze, Raul Machado Neto, Debora Botequio Moretti. Centro de Genômica Funcional da ESALQ: Luiz Lehmann Coutinho, Ricardo Augusto Brassaloti, Raquel de Lello Rocha Campos Cassano. NGS Soluções Genômicas: Pilar Drummond Sampaio Corrêa Mariani. FZEA-USP Pirassununga: Mirele Daiana Poleti, Jessika Cristina Chagas Lesbon, Elisangela Chicaroni Mattos, Heidge Fukumasu. USP-Botucatu: Rejane Maria Tommasini Grotto, Jayme A. Souza-Neto, Guilherme Targino Valente, Patricia Akemi Assato, Felipe Allan da Silva da Costa, Bianca Cechetto Carlos. Mendelics: Bibiana Santos, João Paulo Kitajima, Erika Freitas, David Schlesinger. Hemocentro Ribeirão Preto: Simone Kashima, Evandra Strazza Rodrigues, Svetoslav Nanev Slavov, Elaine Vieira dos Santos, Rafael dos Santos Bezerra, Luiz Carlos Junior de Alcantara, Marta Giovanetti, Vagner Fonseca, Flavia Aburjaile, Rodrigo Tocantins Calado. |
| EPI_ISL_1795326 | UBS II DE REGENTE FEIJO | Instituto Butantan / ESALQ-Piracicaba | Instituto Butantan: Alexander Roberto Precioso, Dimas Tadeu Covas, Sandra Coccuzzo Sampaio, Maria Carolina Elias, José Salvatore Leister Patané, Vincent Louis Viala, Antonio Jorge Martins, Ricardo Haddad, Claudia Renata dos Santos Barros, Elaine Cristina Marqueze, Raul Machado Neto, Debora Botequio Moretti. Centro de Genômica Funcional da ESALQ: Luiz Lehmann Coutinho, Ricardo Augusto Brassaloti, Raquel de Lello Rocha Campos Cassano. NGS Soluções Genômicas: Pilar Drummond Sampaio Corrêa Mariani. FZEA-USP Pirassununga: Mirele Daiana Poleti, Jessika Cristina Chagas Lesbon, Elisangela Chicaroni Mattos, Heidge Fukumasu. USP-Botucatu: Rejane Maria Tommasini Grotto, Jayme A. Souza-Neto, Guilherme Targino Valente, Patricia Akemi Assato, Felipe Allan da Silva da Costa, Bianca Cechetto Carlos. Mendelics: Bibiana Santos, João Paulo Kitajima, Erika Freitas, David Schlesinger. Hemocentro Ribeirão Preto: Simone Kashima, Evandra Strazza Rodrigues, Svetoslav Nanev Slavov, Elaine Vieira dos Santos, Rafael dos Santos Bezerra, Luiz Carlos Junior de Alcantara, Marta Giovanetti, Vagner Fonseca, Flavia Aburjaile, Rodrigo Tocantins Calado. |
| EPI_ISL_1795327 | CENTRO DE SAUDE II JUNQUEIROPOLIS | Instituto Butantan / ESALQ-Piracicaba | Instituto Butantan: Alexander Roberto Precioso, Dimas Tadeu Covas, Sandra Coccuzzo Sampaio, Maria Carolina Elias, José Salvatore Leister Patané, Vincent Louis Viala, Antonio Jorge Martins, Ricardo Haddad, Claudia Renata dos Santos Barros, Elaine Cristina Marqueze, Raul Machado Neto, Debora Botequio Moretti. Centro de Genômica Funcional da ESALQ: Luiz Lehmann Coutinho, Ricardo Augusto Brassaloti, Raquel de Lello Rocha Campos Cassano. NGS Soluções Genômicas: Pilar Drummond Sampaio Corrêa Mariani. FZEA-USP Pirassununga: Mirele Daiana Poleti, Jessika Cristina Chagas Lesbon, Elisangela Chicaroni Mattos, Heidge Fukumasu. USP-Botucatu: Rejane Maria Tommasini Grotto, Jayme A. Souza-Neto, Guilherme Targino Valente, Patricia Akemi Assato, Felipe Allan da Silva da Costa, Bianca Cechetto Carlos. Mendelics: Bibiana Santos, João Paulo Kitajima, Erika Freitas, David Schlesinger. Hemocentro Ribeirão Preto: Simone Kashima, Evandra Strazza Rodrigues, Svetoslav Nanev Slavov, Elaine Vieira dos Santos, Rafael dos Santos Bezerra, Luiz Carlos Junior de Alcantara, Marta Giovanetti, Vagner Fonseca, Flavia Aburjaile, Rodrigo Tocantins Calado. |
| EPI_ISL_1795328 | UBS II DR EXPEDITO SHIZUO KUROCE | Instituto Butantan / ESALQ-Piracicaba | Instituto Butantan: Alexander Roberto Precioso, Dimas Tadeu Covas, Sandra Coccuzzo Sampaio, Maria Carolina Elias, José Salvatore Leister Patané, Vincent Louis Viala, Antonio Jorge Martins, Ricardo Haddad, Claudia Renata dos Santos Barros, Elaine Cristina Marqueze, Raul Machado Neto, Debora Botequio Moretti. Centro de Genômica Funcional da ESALQ: Luiz Lehmann Coutinho, Ricardo Augusto Brassaloti, Raquel de Lello Rocha Campos Cassano. NGS Soluções Genômicas: Pilar Drummond Sampaio Corrêa Mariani. FZEA-USP Pirassununga: Mirele Daiana Poleti, Jessika Cristina Chagas Lesbon, Elisangela Chicaroni Mattos, Heidge Fukumasu. USP-Botucatu: Rejane Maria Tommasini Grotto, Jayme A. Souza-Neto, Guilherme Targino Valente, Patricia Akemi Assato, Felipe Allan da Silva da Costa, Bianca Cechetto Carlos. Mendelics: Bibiana Santos, João Paulo Kitajima, Erika Freitas, David Schlesinger. Hemocentro Ribeirão Preto: Simone Kashima, Evandra Strazza Rodrigues, Svetoslav Nanev Slavov, Elaine Vieira dos Santos, Rafael dos Santos Bezerra, Luiz Carlos Junior de Alcantara, Marta Giovanetti, Vagner Fonseca, Flavia Aburjaile, Rodrigo Tocantins Calado. |
| EPI_ISL_1795329 | CENTRO DE ESPECIALIDADES DE PRIMAVERA | Instituto Butantan / ESALQ-Piracicaba | Instituto Butantan: Alexander Roberto Precioso, Dimas Tadeu Covas, Sandra Coccuzzo Sampaio, Maria Carolina Elias, José Salvatore Leister Patané, Vincent Louis Viala, Antonio Jorge Martins, Ricardo Haddad, Claudia Renata dos Santos Barros, Elaine Cristina Marqueze, Raul Machado Neto, Debora Botequio Moretti. Centro de Genômica Funcional da ESALQ: Luiz Lehmann Coutinho, Ricardo Augusto Brassaloti, Raquel de Lello Rocha Campos Cassano. NGS Soluções Genômicas: Pilar Drummond Sampaio Corrêa Mariani. FZEA-USP Pirassununga: Mirele Daiana Poleti, Jessika Cristina Chagas Lesbon, Elisangela Chicaroni Mattos, Heidge Fukumasu. USP-Botucatu: Rejane Maria Tommasini Grotto, Jayme A. Souza-Neto, Guilherme Targino Valente, Patricia Akemi Assato, Felipe Allan da Silva da Costa, Bianca Cechetto Carlos. Mendelics: Bibiana Santos, João Paulo Kitajima, Erika Freitas, David Schlesinger. Hemocentro Ribeirão Preto: Simone Kashima, Evandra Strazza Rodrigues, Svetoslav Nanev Slavov, Elaine Vieira dos Santos, Rafael dos Santos Bezerra, Luiz Carlos Junior de Alcantara, Marta Giovanetti, Vagner Fonseca, Flavia Aburjaile, Rodrigo Tocantins Calado. |
| EPI_ISL_1795330 | UBS II DR EXPEDITO SHIZUO KUROCE | Instituto Butantan / ESALQ-Piracicaba | Instituto Butantan: Alexander Roberto Precioso, Dimas Tadeu Covas, Sandra Coccuzzo Sampaio, Maria Carolina Elias, José Salvatore Leister Patané, Vincent Louis Viala, Antonio Jorge Martins, Ricardo Haddad, Claudia Renata dos Santos Barros, Elaine Cristina Marqueze, Raul Machado Neto, Debora Botequio Moretti. Centro de Genômica Funcional da ESALQ: Luiz Lehmann Coutinho, Ricardo Augusto Brassaloti, Raquel de Lello Rocha Campos Cassano. NGS Soluções Genômicas: Pilar Drummond Sampaio Corrêa Mariani. FZEA-USP Pirassununga: Mirele Daiana Poleti, Jessika Cristina Chagas Lesbon, Elisangela Chicaroni Mattos, Heidge Fukumasu. USP-Botucatu: Rejane Maria Tommasini Grotto, Jayme A. Souza-Neto, Guilherme Targino Valente, Patricia Akemi Assato, Felipe Allan da Silva da Costa, Bianca Cechetto Carlos. Mendelics: Bibiana Santos, João Paulo Kitajima, Erika Freitas, David Schlesinger. Hemocentro Ribeirão Preto: Simone Kashima, Evandra Strazza Rodrigues, Svetoslav Nanev Slavov, Elaine Vieira dos Santos, Rafael dos Santos Bezerra, Luiz Carlos Junior de Alcantara, Marta Giovanetti, Vagner Fonseca, Flavia Aburjaile, Rodrigo Tocantins Calado. |
| EPI_ISL_1795331 | CENTRO DE SAUDE II JUNQUEIROPOLIS | Instituto Butantan / ESALQ-Piracicaba | Instituto Butantan: Alexander Roberto Precioso, Dimas Tadeu Covas, Sandra Coccuzzo Sampaio, Maria Carolina Elias, José Salvatore Leister Patané, Vincent Louis Viala, Antonio Jorge Martins, Ricardo Haddad, Claudia Renata dos Santos Barros, Elaine Cristina Marqueze, Raul Machado Neto, Debora Botequio Moretti. Centro de Genômica Funcional da ESALQ: Luiz Lehmann Coutinho, Ricardo Augusto Brassaloti, Raquel de Lello Rocha Campos Cassano. NGS Soluções Genômicas: Pilar Drummond Sampaio Corrêa Mariani. FZEA-USP Pirassununga: Mirele Daiana Poleti, Jessika Cristina Chagas Lesbon, Elisangela Chicaroni Mattos, Heidge Fukumasu. USP-Botucatu: Rejane Maria Tommasini Grotto, Jayme A. Souza-Neto, Guilherme Targino Valente, Patricia Akemi Assato, Felipe Allan da Silva da Costa, Bianca Cechetto Carlos. Mendelics: Bibiana Santos, João Paulo Kitajima, Erika Freitas, David Schlesinger. Hemocentro Ribeirão Preto: Simone Kashima, Evandra Strazza Rodrigues, Svetoslav Nanev Slavov, Elaine Vieira dos Santos, Rafael dos Santos Bezerra, Luiz Carlos Junior de Alcantara, Marta Giovanetti, Vagner Fonseca, Flavia Aburjaile, Rodrigo Tocantins Calado. |
| EPI_ISL_1795332 | UBS III DE RANCHARIA | Instituto Butantan / ESALQ-Piracicaba | Instituto Butantan: Alexander Roberto Precioso, Dimas Tadeu Covas, Sandra Coccuzzo Sampaio, Maria Carolina Elias, José Salvatore Leister Patané, Vincent Louis Viala, Antonio Jorge Martins, Ricardo Haddad, Claudia Renata dos Santos Barros, Elaine Cristina Marqueze, Raul Machado Neto, Debora Botequio Moretti. Centro de Genômica Funcional da ESALQ: Luiz Lehmann Coutinho, Ricardo Augusto Brassaloti, Raquel de Lello Rocha Campos Cassano. NGS Soluções Genômicas: Pilar Drummond Sampaio Corrêa Mariani. FZEA-USP Pirassununga: Mirele Daiana Poleti, Jessika Cristina Chagas Lesbon, Elisangela Chicaroni Mattos, Heidge Fukumasu. USP-Botucatu: Rejane Maria Tommasini Grotto, Jayme A. Souza-Neto, Guilherme Targino Valente, Patricia Akemi Assato, Felipe Allan da Silva da Costa, Bianca Cechetto Carlos. Mendelics: Bibiana Santos, João Paulo Kitajima, Erika Freitas, David Schlesinger. Hemocentro Ribeirão Preto: Simone Kashima, Evandra Strazza Rodrigues, Svetoslav Nanev Slavov, Elaine Vieira dos Santos, Rafael dos Santos Bezerra, Luiz Carlos Junior de Alcantara, Marta Giovanetti, Vagner Fonseca, Flavia Aburjaile, Rodrigo Tocantins Calado. |
| EPI_ISL_1795333 | CENTRO DE ESPECIALIDADES DE PRIMAVERA | Instituto Butantan / ESALQ-Piracicaba | Instituto Butantan: Alexander Roberto Precioso, Dimas Tadeu Covas, Sandra Coccuzzo Sampaio, Maria Carolina Elias, José Salvatore Leister Patané, Vincent Louis Viala, Antonio Jorge Martins, Ricardo Haddad, Claudia Renata dos Santos Barros, Elaine Cristina Marqueze, Raul Machado Neto, Debora Botequio Moretti. Centro de Genômica Funcional da ESALQ: Luiz Lehmann Coutinho, Ricardo Augusto Brassaloti, Raquel de Lello Rocha Campos Cassano. NGS Soluções Genômicas: Pilar Drummond Sampaio Corrêa Mariani. FZEA-USP Pirassununga: Mirele Daiana Poleti, Jessika Cristina Chagas Lesbon, Elisangela Chicaroni Mattos, Heidge Fukumasu. USP-Botucatu: Rejane Maria Tommasini Grotto, Jayme A. Souza-Neto, Guilherme Targino Valente, Patricia Akemi Assato, Felipe Allan da Silva da Costa, Bianca Cechetto Carlos. Mendelics: Bibiana Santos, João Paulo Kitajima, Erika Freitas, David Schlesinger. Hemocentro Ribeirão Preto: Simone Kashima, Evandra Strazza Rodrigues, Svetoslav Nanev Slavov, Elaine Vieira dos Santos, Rafael dos Santos Bezerra, Luiz Carlos Junior de Alcantara, Marta Giovanetti, Vagner Fonseca, Flavia Aburjaile, Rodrigo Tocantins Calado. |

| | | | |
|---|---|---|---|
| EPI_ISL_1795334 | UBS MARCIA CRISTIANE DA SILVA DE OURO VERDE | Instituto Butantan / ESALQ-Piracicaba | Instituto Butantan: Alexander Roberto Precioso, Dimas Tadeu Covas, Sandra Coccuzzo Sampaio, Maria Carolina Elias, José Salvatore Leister Patané, Vincent Louis Viala, Antonio Jorge Martins, Ricardo Haddad, Claudia Renata dos Santos Barros, Elaine Cristina Marqueze, Raul Machado Neto, Debora Botequio Moretti. Centro de Genômica Funcional da ESALQ: Luiz Lehmann Coutinho, Ricardo Augusto Brassaloti, Raquel de Lello Rocha Campos Cassano. NGS Soluções Genômicas: Pilar Drummond Sampaio Corrêa Mariani. FZEA-USP Pirassununga: Mirele Daiana Poleti, Jessika Cristina Chagas Lesbon, Elisangela Chicaroni Mattos, Heidge Fukumasu. USP-Botucatu: Rejane Maria Tommasini Grotto, Jayme A. Souza-Neto, Guilherme Targino Valente, Patricia Akemi Assato, Felipe Allan da Silva da Costa, Bianca Cechetto Carlos. Mendelics: Bibiana Santos, João Paulo Kitajima, Erika Freitas, David Schlesinger. Hemocentro Ribeirão Preto: Simone Kashima, Evandra Strazza Rodrigues, Svetoslav Nanev Slavov, Elaine Vieira dos Santos, Rafael dos Santos Bezerra, Luiz Carlos Junior de Alcantara, Marta Giovanetti, Vagner Fonseca, Flavia Aburjaile, Rodrigo Tocantins Calado. |
| EPI_ISL_1795335 | UBS III DE RANCHARIA | Instituto Butantan / ESALQ-Piracicaba | Instituto Butantan: Alexander Roberto Precioso, Dimas Tadeu Covas, Sandra Coccuzzo Sampaio, Maria Carolina Elias, José Salvatore Leister Patané, Vincent Louis Viala, Antonio Jorge Martins, Ricardo Haddad, Claudia Renata dos Santos Barros, Elaine Cristina Marqueze, Raul Machado Neto, Debora Botequio Moretti. Centro de Genômica Funcional da ESALQ: Luiz Lehmann Coutinho, Ricardo Augusto Brassaloti, Raquel de Lello Rocha Campos Cassano. NGS Soluções Genômicas: Pilar Drummond Sampaio Corrêa Mariani. FZEA-USP Pirassununga: Mirele Daiana Poleti, Jessika Cristina Chagas Lesbon, Elisangela Chicaroni Mattos, Heidge Fukumasu. USP-Botucatu: Rejane Maria Tommasini Grotto, Jayme A. Souza-Neto, Guilherme Targino Valente, Patricia Akemi Assato, Felipe Allan da Silva da Costa, Bianca Cechetto Carlos. Mendelics: Bibiana Santos, João Paulo Kitajima, Erika Freitas, David Schlesinger. Hemocentro Ribeirão Preto: Simone Kashima, Evandra Strazza Rodrigues, Svetoslav Nanev Slavov, Elaine Vieira dos Santos, Rafael dos Santos Bezerra, Luiz Carlos Junior de Alcantara, Marta Giovanetti, Vagner Fonseca, Flavia Aburjaile, Rodrigo Tocantins Calado. |
| EPI_ISL_1795336 | PAS JOAO ANTONIO DO NASCIMENTO | Instituto Butantan / ESALQ-Piracicaba | Instituto Butantan: Alexander Roberto Precioso, Dimas Tadeu Covas, Sandra Coccuzzo Sampaio, Maria Carolina Elias, José Salvatore Leister Patané, Vincent Louis Viala, Antonio Jorge Martins, Ricardo Haddad, Claudia Renata dos Santos Barros, Elaine Cristina Marqueze, Raul Machado Neto, Debora Botequio Moretti. Centro de Genômica Funcional da ESALQ: Luiz Lehmann Coutinho, Ricardo Augusto Brassaloti, Raquel de Lello Rocha Campos Cassano. NGS Soluções Genômicas: Pilar Drummond Sampaio Corrêa Mariani. FZEA-USP Pirassununga: Mirele Daiana Poleti, Jessika Cristina Chagas Lesbon, Elisangela Chicaroni Mattos, Heidge Fukumasu. USP-Botucatu: Rejane Maria Tommasini Grotto, Jayme A. Souza-Neto, Guilherme Targino Valente, Patricia Akemi Assato, Felipe Allan da Silva da Costa, Bianca Cechetto Carlos. Mendelics: Bibiana Santos, João Paulo Kitajima, Erika Freitas, David Schlesinger. Hemocentro Ribeirão Preto: Simone Kashima, Evandra Strazza Rodrigues, Svetoslav Nanev Slavov, Elaine Vieira dos Santos, Rafael dos Santos Bezerra, Luiz Carlos Junior de Alcantara, Marta Giovanetti, Vagner Fonseca, Flavia Aburjaile, Rodrigo Tocantins Calado. |
| EPI_ISL_1795344 | HOSPITAL E MATERNIDADE NOSSA SENHORA DA AJUDA | Instituto Butantan / ESALQ-Piracicaba | Instituto Butantan: Alexander Roberto Precioso, Dimas Tadeu Covas, Sandra Coccuzzo Sampaio, Maria Carolina Elias, José Salvatore Leister Patané, Vincent Louis Viala, Antonio Jorge Martins, Ricardo Haddad, Claudia Renata dos Santos Barros, Elaine Cristina Marqueze, Raul Machado Neto, Debora Botequio Moretti. Centro de Genômica Funcional da ESALQ: Luiz Lehmann Coutinho, Ricardo Augusto Brassaloti, Raquel de Lello Rocha Campos Cassano. NGS Soluções Genômicas: Pilar Drummond Sampaio Corrêa Mariani. FZEA-USP Pirassununga: Mirele Daiana Poleti, Jessika Cristina Chagas Lesbon, Elisangela Chicaroni Mattos, Heidge Fukumasu. USP-Botucatu: Rejane Maria Tommasini Grotto, Jayme A. Souza-Neto, Guilherme Targino Valente, Patricia Akemi Assato, Felipe Allan da Silva da Costa, Bianca Cechetto Carlos. Mendelics: Bibiana Santos, João Paulo Kitajima, Erika Freitas, David Schlesinger. Hemocentro Ribeirão Preto: Simone Kashima, Evandra Strazza Rodrigues, Svetoslav Nanev Slavov, Elaine Vieira dos Santos, Rafael dos Santos Bezerra, Luiz Carlos Junior de Alcantara, Marta Giovanetti, Vagner Fonseca, Flavia Aburjaile, Rodrigo Tocantins Calado. |
| EPI_ISL_1795345 | HOSPITAL DE CAMPANHA COVID 19 MUNICIPIO DE TAUBATE | Instituto Butantan / ESALQ-Piracicaba | Instituto Butantan: Alexander Roberto Precioso, Dimas Tadeu Covas, Sandra Coccuzzo Sampaio, Maria Carolina Elias, José Salvatore Leister Patané, Vincent Louis Viala, Antonio Jorge Martins, Ricardo Haddad, Claudia Renata dos Santos Barros, Elaine Cristina Marqueze, Raul Machado Neto, Debora Botequio Moretti. Centro de Genômica Funcional da ESALQ: Luiz Lehmann Coutinho, Ricardo Augusto Brassaloti, Raquel de Lello Rocha Campos Cassano. NGS Soluções Genômicas: Pilar Drummond Sampaio Corrêa Mariani. FZEA-USP Pirassununga: Mirele Daiana Poleti, Jessika Cristina Chagas Lesbon, Elisangela Chicaroni Mattos, Heidge Fukumasu. USP-Botucatu: Rejane Maria Tommasini Grotto, Jayme A. Souza-Neto, Guilherme Targino Valente, Patricia Akemi Assato, Felipe Allan da Silva da Costa, Bianca Cechetto Carlos. Mendelics: Bibiana Santos, João Paulo Kitajima, Erika Freitas, David Schlesinger. Hemocentro Ribeirão Preto: Simone Kashima, Evandra Strazza Rodrigues, Svetoslav Nanev Slavov, Elaine Vieira dos Santos, Rafael dos Santos Bezerra, Luiz Carlos Junior de Alcantara, Marta Giovanetti, Vagner Fonseca, Flavia Aburjaile, Rodrigo Tocantins Calado. |
| EPI_ISL_1795346, EPI_ISL_1795347, EPI_ISL_1795348, EPI_ISL_1795349, EPI_ISL_1795350, EPI_ISL_1795351, EPI_ISL_1795352, EPI_ISL_1795353, EPI_ISL_1795354, EPI_ISL_1795355, EPI_ISL_1795356, EPI_ISL_1795357, EPI_ISL_1795358, EPI_ISL_1795359, EPI_ISL_1795360 | | | |
| see above | LABORATORIO DR PAULO EMILIO DALESSANDRO PINDAMONHANGABA | Instituto Butantan / ESALQ-Piracicaba | Instituto Butantan: Alexander Roberto Precioso, Dimas Tadeu Covas, Sandra Coccuzzo Sampaio, Maria Carolina Elias, José Salvatore Leister Patané, Vincent Louis Viala, Antonio Jorge Martins, Ricardo Haddad, Claudia Renata dos Santos Barros, Elaine Cristina Marqueze, Raul Machado Neto, Debora Botequio Moretti. Centro de Genômica Funcional da ESALQ: Luiz Lehmann Coutinho, Ricardo Augusto Brassaloti, Raquel de Lello Rocha Campos Cassano. NGS Soluções Genômicas: Pilar Drummond Sampaio Corrêa Mariani. FZEA-USP Pirassununga: Mirele Daiana Poleti, Jessika Cristina Chagas Lesbon, Elisangela Chicaroni Mattos, Heidge Fukumasu. USP-Botucatu: Rejane Maria Tommasini Grotto, Jayme A. Souza-Neto, Guilherme Targino Valente, Patricia Akemi Assato, Felipe Allan da Silva da Costa, Bianca Cechetto Carlos. Mendelics: Bibiana Santos, João Paulo Kitajima, Erika Freitas, David Schlesinger. Hemocentro Ribeirão Preto: Simone Kashima, Evandra Strazza Rodrigues, Svetoslav Nanev Slavov, Elaine Vieira dos Santos, Rafael dos Santos Bezerra, Luiz Carlos Junior de Alcantara, Marta Giovanetti, Vagner Fonseca, Flavia Aburjaile, Rodrigo Tocantins Calado. |
| EPI_ISL_1795362, EPI_ISL_1795363 | UNIDADE BASICA DE SAUDE DE PIQUETE | Instituto Butantan / ESALQ-Piracicaba | Instituto Butantan: Alexander Roberto Precioso, Dimas Tadeu Covas, Sandra Coccuzzo Sampaio, Maria Carolina Elias, José Salvatore Leister Patané, Vincent Louis Viala, Antonio Jorge Martins, Ricardo Haddad, Claudia Renata dos Santos Barros, Elaine Cristina Marqueze, Raul Machado Neto, Debora Botequio Moretti. Centro de Genômica Funcional da ESALQ: Luiz Lehmann Coutinho, Ricardo Augusto Brassaloti, Raquel de Lello Rocha Campos Cassano. NGS Soluções Genômicas: Pilar Drummond Sampaio Corrêa Mariani. FZEA-USP Pirassununga: Mirele Daiana Poleti, Jessika Cristina Chagas Lesbon, Elisangela Chicaroni Mattos, Heidge Fukumasu. USP-Botucatu: Rejane Maria Tommasini Grotto, Jayme A. Souza-Neto, Guilherme Targino Valente, Patricia Akemi Assato, Felipe Allan da Silva da Costa, Bianca Cechetto Carlos. Mendelics: Bibiana Santos, João Paulo Kitajima, Erika Freitas, David Schlesinger. Hemocentro Ribeirão Preto: Simone Kashima, Evandra Strazza Rodrigues, Svetoslav Nanev Slavov, Elaine Vieira dos Santos, Rafael dos Santos Bezerra, Luiz Carlos Junior de Alcantara, Marta Giovanetti, Vagner Fonseca, Flavia Aburjaile, Rodrigo Tocantins Calado. |
| EPI_ISL_1795364 | SECRETARIA DE SAUDE | Instituto Butantan / ESALQ-Piracicaba | Instituto Butantan: Alexander Roberto Precioso, Dimas Tadeu Covas, Sandra Coccuzzo Sampaio, Maria Carolina Elias, José Salvatore Leister Patané, Vincent Louis Viala, Antonio Jorge Martins, Ricardo Haddad, Claudia Renata dos Santos Barros, Elaine Cristina Marqueze, Raul Machado Neto, Debora Botequio Moretti. Centro de Genômica Funcional da ESALQ: Luiz Lehmann Coutinho, Ricardo Augusto Brassaloti, Raquel de Lello Rocha Campos Cassano. NGS Soluções Genômicas: Pilar Drummond Sampaio Corrêa Mariani. FZEA-USP Pirassununga: Mirele Daiana Poleti, Jessika Cristina Chagas Lesbon, Elisangela Chicaroni Mattos, Heidge Fukumasu. USP-Botucatu: Rejane Maria Tommasini Grotto, Jayme A. Souza-Neto, Guilherme Targino Valente, Patricia Akemi Assato, Felipe Allan da Silva da Costa, Bianca Cechetto Carlos. Mendelics: Bibiana Santos, João Paulo Kitajima, Erika Freitas, David Schlesinger. Hemocentro Ribeirão Preto: Simone Kashima, Evandra Strazza Rodrigues, Svetoslav Nanev Slavov, Elaine Vieira dos Santos, Rafael dos Santos Bezerra, Luiz Carlos Junior de Alcantara, Marta Giovanetti, Vagner Fonseca, Flavia Aburjaile, Rodrigo Tocantins Calado. |
| EPI_ISL_1795365, EPI_ISL_1795366 | LABORATORIO DR PAULO EMILIO DALESSANDRO PINDAMONHANGABA | Instituto Butantan / ESALQ-Piracicaba | Instituto Butantan: Alexander Roberto Precioso, Dimas Tadeu Covas, Sandra Coccuzzo Sampaio, Maria Carolina Elias, José Salvatore Leister Patané, Vincent Louis Viala, Antonio Jorge Martins, Ricardo Haddad, Claudia Renata dos Santos Barros, Elaine Cristina Marqueze, Raul Machado Neto, Debora Botequio Moretti. Centro de Genômica Funcional da ESALQ: Luiz Lehmann Coutinho, Ricardo Augusto Brassaloti, Raquel de Lello Rocha Campos Cassano. NGS Soluções Genômicas: Pilar Drummond Sampaio Corrêa Mariani. FZEA-USP Pirassununga: Mirele Daiana Poleti, Jessika Cristina Chagas Lesbon, Elisangela Chicaroni Mattos, Heidge Fukumasu. USP-Botucatu: Rejane Maria Tommasini Grotto, Jayme A. Souza-Neto, Guilherme Targino Valente, Patricia Akemi Assato, Felipe Allan da Silva da Costa, Bianca Cechetto Carlos. Mendelics: Bibiana Santos, João Paulo Kitajima, Erika Freitas, David Schlesinger. Hemocentro Ribeirão Preto: Simone Kashima, Evandra Strazza Rodrigues, Svetoslav Nanev Slavov, Elaine Vieira dos Santos, Rafael dos Santos Bezerra, Luiz Carlos Junior de Alcantara, Marta Giovanetti, Vagner Fonseca, Flavia Aburjaile, Rodrigo Tocantins Calado. |
| EPI_ISL_1795367 | AMBULATORIO MEDICO MUNICIPAL DE AGUDOS | Instituto Butantan / ESALQ-Piracicaba | Instituto Butantan: Alexander Roberto Precioso, Dimas Tadeu Covas, Sandra Coccuzzo Sampaio, Maria Carolina Elias, José Salvatore Leister Patané, Vincent Louis Viala, Antonio Jorge Martins, Ricardo Haddad, Claudia Renata dos Santos Barros, Elaine Cristina Marqueze, Raul Machado Neto, Debora Botequio Moretti. Centro de Genômica Funcional da ESALQ: Luiz Lehmann Coutinho, Ricardo Augusto Brassaloti, Raquel de Lello Rocha Campos Cassano. NGS Soluções Genômicas: Pilar Drummond Sampaio Corrêa Mariani. FZEA-USP Pirassununga: Mirele Daiana Poleti, Jessika Cristina Chagas Lesbon, Elisangela Chicaroni Mattos, Heidge Fukumasu. USP-Botucatu: Rejane Maria Tommasini Grotto, Jayme A. Souza-Neto, Guilherme Targino Valente, Patricia Akemi Assato, Felipe Allan da Silva da Costa, Bianca Cechetto Carlos. Mendelics: Bibiana Santos, João Paulo Kitajima, Erika Freitas, David Schlesinger. Hemocentro Ribeirão Preto: Simone Kashima, Evandra Strazza Rodrigues, Svetoslav Nanev Slavov, Elaine Vieira dos Santos, Rafael dos Santos Bezerra, Luiz Carlos Junior de Alcantara, Marta Giovanetti, Vagner Fonseca, Flavia Aburjaile, Rodrigo Tocantins Calado. |
| EPI_ISL_1795368 | CENTRO INTEGRADO DE SAUDE | Instituto Butantan / ESALQ-Piracicaba | Instituto Butantan: Alexander Roberto Precioso, Dimas Tadeu Covas, Sandra Coccuzzo Sampaio, Maria Carolina Elias, José Salvatore Leister Patané, |

| | | | |
|---|---|---|---|
| | | | Vincent Louis Viala, Antonio Jorge Martins, Ricardo Haddad, Claudia Renata dos Santos Barros, Elaine Cristina Marqueze, Raul Machado Neto, Debora Botequio Moretti. Centro de Genômica Funcional da ESALQ: Luiz Lehmann Coutinho, Ricardo Augusto Brassaloti, Raquel de Lello Rocha Campos Cassano. NGS Soluções Genômicas: Pilar Drummond Sampaio Corrêa Mariani. FZEA-USP Pirassununga: Mirele Daiana Poleti, Jessika Cristina Chagas Lesbon, Elisangela Chicaroni Mattos, Heidge Fukumasu. USP-Botucatu: Rejane Maria Tommasini Grotto, Jayme A. Souza-Neto, Guilherme Targino Valente, Patricia Akemi Assato, Felipe Allan da Silva da Costa, Bianca Cechetto Carlos. Mendelics: Bibiana Santos, João Paulo Kitajima, Erika Freitas, David Schlesinger. Hemocentro Ribeirão Preto: Simone Kashima, Evandra Strazza Rodrigues, Svetoslav Nanev Slavov, Elaine Vieira dos Santos, Rafael dos Santos Bezerra, Luiz Carlos Junior de Alcantara, Marta Giovanetti, Vagner Fonseca, Flavia Aburjaile, Rodrigo Tocantins Calado. |
| EPI_ISL_1795369 | AMBULATORIO MEDICO MUNICIPAL DE AGUDOS | Instituto Butantan / ESALQ-Piracicaba | Instituto Butantan: Alexander Roberto Precioso, Dimas Tadeu Covas, Sandra Coccuzzo Sampaio, Maria Carolina Elias, José Salvatore Leister Patané, Vincent Louis Viala, Antonio Jorge Martins, Ricardo Haddad, Claudia Renata dos Santos Barros, Elaine Cristina Marqueze, Raul Machado Neto, Debora Botequio Moretti. Centro de Genômica Funcional da ESALQ: Luiz Lehmann Coutinho, Ricardo Augusto Brassaloti, Raquel de Lello Rocha Campos Cassano. NGS Soluções Genômicas: Pilar Drummond Sampaio Corrêa Mariani. FZEA-USP Pirassununga: Mirele Daiana Poleti, Jessika Cristina Chagas Lesbon, Elisangela Chicaroni Mattos, Heidge Fukumasu. USP-Botucatu: Rejane Maria Tommasini Grotto, Jayme A. Souza-Neto, Guilherme Targino Valente, Patricia Akemi Assato, Felipe Allan da Silva da Costa, Bianca Cechetto Carlos. Mendelics: Bibiana Santos, João Paulo Kitajima, Erika Freitas, David Schlesinger. Hemocentro Ribeirão Preto: Simone Kashima, Evandra Strazza Rodrigues, Svetoslav Nanev Slavov, Elaine Vieira dos Santos, Rafael dos Santos Bezerra, Luiz Carlos Junior de Alcantara, Marta Giovanetti, Vagner Fonseca, Flavia Aburjaile, Rodrigo Tocantins Calado. |
| EPI_ISL_1795370 | CENTRO INTEGRADO DE SAUDE | Instituto Butantan / ESALQ-Piracicaba | Instituto Butantan: Alexander Roberto Precioso, Dimas Tadeu Covas, Sandra Coccuzzo Sampaio, Maria Carolina Elias, José Salvatore Leister Patané, Vincent Louis Viala, Antonio Jorge Martins, Ricardo Haddad, Claudia Renata dos Santos Barros, Elaine Cristina Marqueze, Raul Machado Neto, Debora Botequio Moretti. Centro de Genômica Funcional da ESALQ: Luiz Lehmann Coutinho, Ricardo Augusto Brassaloti, Raquel de Lello Rocha Campos Cassano. NGS Soluções Genômicas: Pilar Drummond Sampaio Corrêa Mariani. FZEA-USP Pirassununga: Mirele Daiana Poleti, Jessika Cristina Chagas Lesbon, Elisangela Chicaroni Mattos, Heidge Fukumasu. USP-Botucatu: Rejane Maria Tommasini Grotto, Jayme A. Souza-Neto, Guilherme Targino Valente, Patricia Akemi Assato, Felipe Allan da Silva da Costa, Bianca Cechetto Carlos. Mendelics: Bibiana Santos, João Paulo Kitajima, Erika Freitas, David Schlesinger. Hemocentro Ribeirão Preto: Simone Kashima, Evandra Strazza Rodrigues, Svetoslav Nanev Slavov, Elaine Vieira dos Santos, Rafael dos Santos Bezerra, Luiz Carlos Junior de Alcantara, Marta Giovanetti, Vagner Fonseca, Flavia Aburjaile, Rodrigo Tocantins Calado. |
| EPI_ISL_1795371, EPI_ISL_1795372 | HOSPITAL SANTA THEREZINHA BROTAS | Instituto Butantan / ESALQ-Piracicaba | Instituto Butantan: Alexander Roberto Precioso, Dimas Tadeu Covas, Sandra Coccuzzo Sampaio, Maria Carolina Elias, José Salvatore Leister Patané, Vincent Louis Viala, Antonio Jorge Martins, Ricardo Haddad, Claudia Renata dos Santos Barros, Elaine Cristina Marqueze, Raul Machado Neto, Debora Botequio Moretti. Centro de Genômica Funcional da ESALQ: Luiz Lehmann Coutinho, Ricardo Augusto Brassaloti, Raquel de Lello Rocha Campos Cassano. NGS Soluções Genômicas: Pilar Drummond Sampaio Corrêa Mariani. FZEA-USP Pirassununga: Mirele Daiana Poleti, Jessika Cristina Chagas Lesbon, Elisangela Chicaroni Mattos, Heidge Fukumasu. USP-Botucatu: Rejane Maria Tommasini Grotto, Jayme A. Souza-Neto, Guilherme Targino Valente, Patricia Akemi Assato, Felipe Allan da Silva da Costa, Bianca Cechetto Carlos. Mendelics: Bibiana Santos, João Paulo Kitajima, Erika Freitas, David Schlesinger. Hemocentro Ribeirão Preto: Simone Kashima, Evandra Strazza Rodrigues, Svetoslav Nanev Slavov, Elaine Vieira dos Santos, Rafael dos Santos Bezerra, Luiz Carlos Junior de Alcantara, Marta Giovanetti, Vagner Fonseca, Flavia Aburjaile, Rodrigo Tocantins Calado. |
| EPI_ISL_1795373 | SECRETARIA MUNICIPAL DE SAUDE DE PEDERNEIRAS | Instituto Butantan / ESALQ-Piracicaba | Instituto Butantan: Alexander Roberto Precioso, Dimas Tadeu Covas, Sandra Coccuzzo Sampaio, Maria Carolina Elias, José Salvatore Leister Patané, Vincent Louis Viala, Antonio Jorge Martins, Ricardo Haddad, Claudia Renata dos Santos Barros, Elaine Cristina Marqueze, Raul Machado Neto, Debora Botequio Moretti. Centro de Genômica Funcional da ESALQ: Luiz Lehmann Coutinho, Ricardo Augusto Brassaloti, Raquel de Lello Rocha Campos Cassano. NGS Soluções Genômicas: Pilar Drummond Sampaio Corrêa Mariani. FZEA-USP Pirassununga: Mirele Daiana Poleti, Jessika Cristina Chagas Lesbon, Elisangela Chicaroni Mattos, Heidge Fukumasu. USP-Botucatu: Rejane Maria Tommasini Grotto, Jayme A. Souza-Neto, Guilherme Targino Valente, Patricia Akemi Assato, Felipe Allan da Silva da Costa, Bianca Cechetto Carlos. Mendelics: Bibiana Santos, João Paulo Kitajima, Erika Freitas, David Schlesinger. Hemocentro Ribeirão Preto: Simone Kashima, Evandra Strazza Rodrigues, Svetoslav Nanev Slavov, Elaine Vieira dos Santos, Rafael dos Santos Bezerra, Luiz Carlos Junior de Alcantara, Marta Giovanetti, Vagner Fonseca, Flavia Aburjaile, Rodrigo Tocantins Calado. |
| EPI_ISL_1795374, EPI_ISL_1795375 | SANTA CASA DE DOIS CORREGOS | Instituto Butantan / ESALQ-Piracicaba | Instituto Butantan: Alexander Roberto Precioso, Dimas Tadeu Covas, Sandra Coccuzzo Sampaio, Maria Carolina Elias, José Salvatore Leister Patané, Vincent Louis Viala, Antonio Jorge Martins, Ricardo Haddad, Claudia Renata dos Santos Barros, Elaine Cristina Marqueze, Raul Machado Neto, Debora Botequio Moretti. Centro de Genômica Funcional da ESALQ: Luiz Lehmann Coutinho, Ricardo Augusto Brassaloti, Raquel de Lello Rocha Campos Cassano. NGS Soluções Genômicas: Pilar Drummond Sampaio Corrêa Mariani. FZEA-USP Pirassununga: Mirele Daiana Poleti, Jessika Cristina Chagas Lesbon, Elisangela Chicaroni Mattos, Heidge Fukumasu. USP-Botucatu: Rejane Maria Tommasini Grotto, Jayme A. Souza-Neto, Guilherme Targino Valente, Patricia Akemi Assato, Felipe Allan da Silva da Costa, Bianca Cechetto Carlos. Mendelics: Bibiana Santos, João Paulo Kitajima, Erika Freitas, David Schlesinger. Hemocentro Ribeirão Preto: Simone Kashima, Evandra Strazza Rodrigues, Svetoslav Nanev Slavov, Elaine Vieira dos Santos, Rafael dos Santos Bezerra, Luiz Carlos Junior de Alcantara, Marta Giovanetti, Vagner Fonseca, Flavia Aburjaile, Rodrigo Tocantins Calado. |
| EPI_ISL_1795376, EPI_ISL_1795377, EPI_ISL_1795378 | ESF MINEIROS DO TIETE | Instituto Butantan / ESALQ-Piracicaba | Instituto Butantan: Alexander Roberto Precioso, Dimas Tadeu Covas, Sandra Coccuzzo Sampaio, Maria Carolina Elias, José Salvatore Leister Patané, Vincent Louis Viala, Antonio Jorge Martins, Ricardo Haddad, Claudia Renata dos Santos Barros, Elaine Cristina Marqueze, Raul Machado Neto, Debora Botequio Moretti. Centro de Genômica Funcional da ESALQ: Luiz Lehmann Coutinho, Ricardo Augusto Brassaloti, Raquel de Lello Rocha Campos Cassano. NGS Soluções Genômicas: Pilar Drummond Sampaio Corrêa Mariani. FZEA-USP Pirassununga: Mirele Daiana Poleti, Jessika Cristina Chagas Lesbon, Elisangela Chicaroni Mattos, Heidge Fukumasu. USP-Botucatu: Rejane Maria Tommasini Grotto, Jayme A. Souza-Neto, Guilherme Targino Valente, Patricia Akemi Assato, Felipe Allan da Silva da Costa, Bianca Cechetto Carlos. Mendelics: Bibiana Santos, João Paulo Kitajima, Erika Freitas, David Schlesinger. Hemocentro Ribeirão Preto: Simone Kashima, Evandra Strazza Rodrigues, Svetoslav Nanev Slavov, Elaine Vieira dos Santos, Rafael dos Santos Bezerra, Luiz Carlos Junior de Alcantara, Marta Giovanetti, Vagner Fonseca, Flavia Aburjaile, Rodrigo Tocantins Calado. |
| EPI_ISL_1795379 | NUCLEO DE SAUDE VILA FALCAO DE BAURU | Instituto Butantan / ESALQ-Piracicaba | Instituto Butantan: Alexander Roberto Precioso, Dimas Tadeu Covas, Sandra Coccuzzo Sampaio, Maria Carolina Elias, José Salvatore Leister Patané, Vincent Louis Viala, Antonio Jorge Martins, Ricardo Haddad, Claudia Renata dos Santos Barros, Elaine Cristina Marqueze, Raul Machado Neto, Debora Botequio Moretti. Centro de Genômica Funcional da ESALQ: Luiz Lehmann Coutinho, Ricardo Augusto Brassaloti, Raquel de Lello Rocha Campos Cassano. NGS Soluções Genômicas: Pilar Drummond Sampaio Corrêa Mariani. FZEA-USP Pirassununga: Mirele Daiana Poleti, Jessika Cristina Chagas Lesbon, Elisangela Chicaroni Mattos, Heidge Fukumasu. USP-Botucatu: Rejane Maria Tommasini Grotto, Jayme A. Souza-Neto, Guilherme Targino Valente, Patricia Akemi Assato, Felipe Allan da Silva da Costa, Bianca Cechetto Carlos. Mendelics: Bibiana Santos, João Paulo Kitajima, Erika Freitas, David Schlesinger. Hemocentro Ribeirão Preto: Simone Kashima, Evandra Strazza Rodrigues, Svetoslav Nanev Slavov, Elaine Vieira dos Santos, Rafael dos Santos Bezerra, Luiz Carlos Junior de Alcantara, Marta Giovanetti, Vagner Fonseca, Flavia Aburjaile, Rodrigo Tocantins Calado. |
| EPI_ISL_1795380 | HOSPITAL MATERNIDADE SAO JOSE ITAPUI | Instituto Butantan / ESALQ-Piracicaba | Instituto Butantan: Alexander Roberto Precioso, Dimas Tadeu Covas, Sandra Coccuzzo Sampaio, Maria Carolina Elias, José Salvatore Leister Patané, Vincent Louis Viala, Antonio Jorge Martins, Ricardo Haddad, Claudia Renata dos Santos Barros, Elaine Cristina Marqueze, Raul Machado Neto, Debora Botequio Moretti. Centro de Genômica Funcional da ESALQ: Luiz Lehmann Coutinho, Ricardo Augusto Brassaloti, Raquel de Lello Rocha Campos Cassano. NGS Soluções Genômicas: Pilar Drummond Sampaio Corrêa Mariani. FZEA-USP Pirassununga: Mirele Daiana Poleti, Jessika Cristina Chagas Lesbon, Elisangela Chicaroni Mattos, Heidge Fukumasu. USP-Botucatu: Rejane Maria Tommasini Grotto, Jayme A. Souza-Neto, Guilherme Targino Valente, Patricia Akemi Assato, Felipe Allan da Silva da Costa, Bianca Cechetto Carlos. Mendelics: Bibiana Santos, João Paulo Kitajima, Erika Freitas, David Schlesinger. Hemocentro Ribeirão Preto: Simone Kashima, Evandra Strazza Rodrigues, Svetoslav Nanev Slavov, Elaine Vieira dos Santos, Rafael dos Santos Bezerra, Luiz Carlos Junior de Alcantara, Marta Giovanetti, Vagner Fonseca, Flavia Aburjaile, Rodrigo Tocantins Calado. |
| EPI_ISL_1795381 | SECRETARIA MUNICIPAL DE SAUDE DE MACATUBA | Instituto Butantan / ESALQ-Piracicaba | Instituto Butantan: Alexander Roberto Precioso, Dimas Tadeu Covas, Sandra Coccuzzo Sampaio, Maria Carolina Elias, José Salvatore Leister Patané, Vincent Louis Viala, Antonio Jorge Martins, Ricardo Haddad, Claudia Renata dos Santos Barros, Elaine Cristina Marqueze, Raul Machado Neto, Debora Botequio Moretti. Centro de Genômica Funcional da ESALQ: Luiz Lehmann Coutinho, Ricardo Augusto Brassaloti, Raquel de Lello Rocha Campos Cassano. NGS Soluções Genômicas: Pilar Drummond Sampaio Corrêa Mariani. FZEA-USP Pirassununga: Mirele Daiana Poleti, Jessika Cristina Chagas Lesbon, Elisangela Chicaroni Mattos, Heidge Fukumasu. USP-Botucatu: Rejane Maria Tommasini Grotto, Jayme A. Souza-Neto, Guilherme Targino Valente, Patricia Akemi Assato, Felipe Allan da Silva da Costa, Bianca Cechetto Carlos. Mendelics: Bibiana Santos, João Paulo Kitajima, Erika Freitas, David Schlesinger. Hemocentro Ribeirão Preto: Simone Kashima, Evandra Strazza Rodrigues, Svetoslav Nanev Slavov, Elaine Vieira dos Santos, Rafael dos Santos Bezerra, Luiz Carlos Junior de Alcantara, Marta Giovanetti, Vagner Fonseca, Flavia Aburjaile, Rodrigo Tocantins Calado. |
| EPI_ISL_1795382, EPI_ISL_1795383 | LABORATORIO DR PAULO EMILIO DALESSANDRO PINDAMONHANGABA | Instituto Butantan / ESALQ-Piracicaba | Instituto Butantan: Alexander Roberto Precioso, Dimas Tadeu Covas, Sandra Coccuzzo Sampaio, Maria Carolina Elias, José Salvatore Leister Patané, Vincent Louis Viala, Antonio Jorge Martins, Ricardo Haddad, Claudia Renata dos Santos Barros, Elaine Cristina Marqueze, Raul Machado Neto, Debora Botequio Moretti. Centro de Genômica Funcional da ESALQ: Luiz Lehmann Coutinho, Ricardo Augusto Brassaloti, Raquel de Lello Rocha Campos |

| | | | |
|---|---|---|---|
| | | | Cassano. NGS Soluções Genômicas: Pilar Drummond Sampaio Corrêa Mariani. FZEA-USP Pirassununga: Mirele Daiana Poleti, Jessika Cristina Chagas Lesbon, Elisangela Chicaroni Mattos, Heidge Fukumasu. USP-Botucatu: Rejane Maria Tommasini Grotto, Jayme A. Souza-Neto, Guilherme Targino Valente, Patricia Akemi Assato, Felipe Allan da Silva da Costa, Bianca Cechetto Carlos. Mendelics: Bibiana Santos, João Paulo Kitajima, Erika Freitas, David Schlesinger. Hemocentro Ribeirão Preto: Simone Kashima, Evandra Strazza Rodrigues, Svetoslav Nanev Slavov, Elaine Vieira dos Santos, Rafael dos Santos Bezerra, Luiz Carlos Junior de Alcantara, Marta Giovanetti, Vagner Fonseca, Flavia Aburjaile, Rodrigo Tocantins Calado. |
| EPI_ISL_1795384 | USF SALERNO | Instituto Butantan / ESALQ-Piracicaba | Instituto Butantan: Alexander Roberto Precioso, Dimas Tadeu Covas, Sandra Coccuzzo Sampaio, Maria Carolina Elias, José Salvatore Leister Patané, Vincent Louis Viala, Antonio Jorge Martins, Ricardo Haddad, Claudia Renata dos Santos Barros, Elaine Cristina Marqueze, Raul Machado Neto, Debora Botequio Moretti. Centro de Genômica Funcional da ESALQ: Luiz Lehmann Coutinho, Ricardo Augusto Brassaloti, Raquel de Lello Rocha Campos Cassano. NGS Soluções Genômicas: Pilar Drummond Sampaio Corrêa Mariani. FZEA-USP Pirassununga: Mirele Daiana Poleti, Jessika Cristina Chagas Lesbon, Elisangela Chicaroni Mattos, Heidge Fukumasu. USP-Botucatu: Rejane Maria Tommasini Grotto, Jayme A. Souza-Neto, Guilherme Targino Valente, Patricia Akemi Assato, Felipe Allan da Silva da Costa, Bianca Cechetto Carlos. Mendelics: Bibiana Santos, João Paulo Kitajima, Erika Freitas, David Schlesinger. Hemocentro Ribeirão Preto: Simone Kashima, Evandra Strazza Rodrigues, Svetoslav Nanev Slavov, Elaine Vieira dos Santos, Rafael dos Santos Bezerra, Luiz Carlos Junior de Alcantara, Marta Giovanetti, Vagner Fonseca, Flavia Aburjaile, Rodrigo Tocantins Calado. |
| EPI_ISL_1795385, EPI_ISL_1795386 | UNIDADE DE VIGILANCIA EM SAUDE | Instituto Butantan / ESALQ-Piracicaba | Instituto Butantan: Alexander Roberto Precioso, Dimas Tadeu Covas, Sandra Coccuzzo Sampaio, Maria Carolina Elias, José Salvatore Leister Patané, Vincent Louis Viala, Antonio Jorge Martins, Ricardo Haddad, Claudia Renata dos Santos Barros, Elaine Cristina Marqueze, Raul Machado Neto, Debora Botequio Moretti. Centro de Genômica Funcional da ESALQ: Luiz Lehmann Coutinho, Ricardo Augusto Brassaloti, Raquel de Lello Rocha Campos Cassano. NGS Soluções Genômicas: Pilar Drummond Sampaio Corrêa Mariani. FZEA-USP Pirassununga: Mirele Daiana Poleti, Jessika Cristina Chagas Lesbon, Elisangela Chicaroni Mattos, Heidge Fukumasu. USP-Botucatu: Rejane Maria Tommasini Grotto, Jayme A. Souza-Neto, Guilherme Targino Valente, Patricia Akemi Assato, Felipe Allan da Silva da Costa, Bianca Cechetto Carlos. Mendelics: Bibiana Santos, João Paulo Kitajima, Erika Freitas, David Schlesinger. Hemocentro Ribeirão Preto: Simone Kashima, Evandra Strazza Rodrigues, Svetoslav Nanev Slavov, Elaine Vieira dos Santos, Rafael dos Santos Bezerra, Luiz Carlos Junior de Alcantara, Marta Giovanetti, Vagner Fonseca, Flavia Aburjaile, Rodrigo Tocantins Calado. |
| EPI_ISL_1795387, EPI_ISL_1795388 | POLICLINICA HORTOLANDIA | Instituto Butantan / ESALQ-Piracicaba | Instituto Butantan: Alexander Roberto Precioso, Dimas Tadeu Covas, Sandra Coccuzzo Sampaio, Maria Carolina Elias, José Salvatore Leister Patané, Vincent Louis Viala, Antonio Jorge Martins, Ricardo Haddad, Claudia Renata dos Santos Barros, Elaine Cristina Marqueze, Raul Machado Neto, Debora Botequio Moretti. Centro de Genômica Funcional da ESALQ: Luiz Lehmann Coutinho, Ricardo Augusto Brassaloti, Raquel de Lello Rocha Campos Cassano. NGS Soluções Genômicas: Pilar Drummond Sampaio Corrêa Mariani. FZEA-USP Pirassununga: Mirele Daiana Poleti, Jessika Cristina Chagas Lesbon, Elisangela Chicaroni Mattos, Heidge Fukumasu. USP-Botucatu: Rejane Maria Tommasini Grotto, Jayme A. Souza-Neto, Guilherme Targino Valente, Patricia Akemi Assato, Felipe Allan da Silva da Costa, Bianca Cechetto Carlos. Mendelics: Bibiana Santos, João Paulo Kitajima, Erika Freitas, David Schlesinger. Hemocentro Ribeirão Preto: Simone Kashima, Evandra Strazza Rodrigues, Svetoslav Nanev Slavov, Elaine Vieira dos Santos, Rafael dos Santos Bezerra, Luiz Carlos Junior de Alcantara, Marta Giovanetti, Vagner Fonseca, Flavia Aburjaile, Rodrigo Tocantins Calado. |
| EPI_ISL_1795389 | CS DE BALSAMO | Instituto Butantan / ESALQ-Piracicaba | Instituto Butantan: Alexander Roberto Precioso, Dimas Tadeu Covas, Sandra Coccuzzo Sampaio, Maria Carolina Elias, José Salvatore Leister Patané, Vincent Louis Viala, Antonio Jorge Martins, Ricardo Haddad, Claudia Renata dos Santos Barros, Elaine Cristina Marqueze, Raul Machado Neto, Debora Botequio Moretti. Centro de Genômica Funcional da ESALQ: Luiz Lehmann Coutinho, Ricardo Augusto Brassaloti, Raquel de Lello Rocha Campos Cassano. NGS Soluções Genômicas: Pilar Drummond Sampaio Corrêa Mariani. FZEA-USP Pirassununga: Mirele Daiana Poleti, Jessika Cristina Chagas Lesbon, Elisangela Chicaroni Mattos, Heidge Fukumasu. USP-Botucatu: Rejane Maria Tommasini Grotto, Jayme A. Souza-Neto, Guilherme Targino Valente, Patricia Akemi Assato, Felipe Allan da Silva da Costa, Bianca Cechetto Carlos. Mendelics: Bibiana Santos, João Paulo Kitajima, Erika Freitas, David Schlesinger. Hemocentro Ribeirão Preto: Simone Kashima, Evandra Strazza Rodrigues, Svetoslav Nanev Slavov, Elaine Vieira dos Santos, Rafael dos Santos Bezerra, Luiz Carlos Junior de Alcantara, Marta Giovanetti, Vagner Fonseca, Flavia Aburjaile, Rodrigo Tocantins Calado. |
| EPI_ISL_1795390 | CS DE OSCAR BRESSANE PSF | Instituto Butantan / ESALQ-Piracicaba | Instituto Butantan: Alexander Roberto Precioso, Dimas Tadeu Covas, Sandra Coccuzzo Sampaio, Maria Carolina Elias, José Salvatore Leister Patané, Vincent Louis Viala, Antonio Jorge Martins, Ricardo Haddad, Claudia Renata dos Santos Barros, Elaine Cristina Marqueze, Raul Machado Neto, Debora Botequio Moretti. Centro de Genômica Funcional da ESALQ: Luiz Lehmann Coutinho, Ricardo Augusto Brassaloti, Raquel de Lello Rocha Campos Cassano. NGS Soluções Genômicas: Pilar Drummond Sampaio Corrêa Mariani. FZEA-USP Pirassununga: Mirele Daiana Poleti, Jessika Cristina Chagas Lesbon, Elisangela Chicaroni Mattos, Heidge Fukumasu. USP-Botucatu: Rejane Maria Tommasini Grotto, Jayme A. Souza-Neto, Guilherme Targino Valente, Patricia Akemi Assato, Felipe Allan da Silva da Costa, Bianca Cechetto Carlos. Mendelics: Bibiana Santos, João Paulo Kitajima, Erika Freitas, David Schlesinger. Hemocentro Ribeirão Preto: Simone Kashima, Evandra Strazza Rodrigues, Svetoslav Nanev Slavov, Elaine Vieira dos Santos, Rafael dos Santos Bezerra, Luiz Carlos Junior de Alcantara, Marta Giovanetti, Vagner Fonseca, Flavia Aburjaile, Rodrigo Tocantins Calado. |
| EPI_ISL_1795391 | USF ROSA CRUZ | Instituto Butantan / ESALQ-Piracicaba | Instituto Butantan: Alexander Roberto Precioso, Dimas Tadeu Covas, Sandra Coccuzzo Sampaio, Maria Carolina Elias, José Salvatore Leister Patané, Vincent Louis Viala, Antonio Jorge Martins, Ricardo Haddad, Claudia Renata dos Santos Barros, Elaine Cristina Marqueze, Raul Machado Neto, Debora Botequio Moretti. Centro de Genômica Funcional da ESALQ: Luiz Lehmann Coutinho, Ricardo Augusto Brassaloti, Raquel de Lello Rocha Campos Cassano. NGS Soluções Genômicas: Pilar Drummond Sampaio Corrêa Mariani. FZEA-USP Pirassununga: Mirele Daiana Poleti, Jessika Cristina Chagas Lesbon, Elisangela Chicaroni Mattos, Heidge Fukumasu. USP-Botucatu: Rejane Maria Tommasini Grotto, Jayme A. Souza-Neto, Guilherme Targino Valente, Patricia Akemi Assato, Felipe Allan da Silva da Costa, Bianca Cechetto Carlos. Mendelics: Bibiana Santos, João Paulo Kitajima, Erika Freitas, David Schlesinger. Hemocentro Ribeirão Preto: Simone Kashima, Evandra Strazza Rodrigues, Svetoslav Nanev Slavov, Elaine Vieira dos Santos, Rafael dos Santos Bezerra, Luiz Carlos Junior de Alcantara, Marta Giovanetti, Vagner Fonseca, Flavia Aburjaile, Rodrigo Tocantins Calado. |
| EPI_ISL_1795392 | POLICLINICA HORTOLANDIA | Instituto Butantan / ESALQ-Piracicaba | Instituto Butantan: Alexander Roberto Precioso, Dimas Tadeu Covas, Sandra Coccuzzo Sampaio, Maria Carolina Elias, José Salvatore Leister Patané, Vincent Louis Viala, Antonio Jorge Martins, Ricardo Haddad, Claudia Renata dos Santos Barros, Elaine Cristina Marqueze, Raul Machado Neto, Debora Botequio Moretti. Centro de Genômica Funcional da ESALQ: Luiz Lehmann Coutinho, Ricardo Augusto Brassaloti, Raquel de Lello Rocha Campos Cassano. NGS Soluções Genômicas: Pilar Drummond Sampaio Corrêa Mariani. FZEA-USP Pirassununga: Mirele Daiana Poleti, Jessika Cristina Chagas Lesbon, Elisangela Chicaroni Mattos, Heidge Fukumasu. USP-Botucatu: Rejane Maria Tommasini Grotto, Jayme A. Souza-Neto, Guilherme Targino Valente, Patricia Akemi Assato, Felipe Allan da Silva da Costa, Bianca Cechetto Carlos. Mendelics: Bibiana Santos, João Paulo Kitajima, Erika Freitas, David Schlesinger. Hemocentro Ribeirão Preto: Simone Kashima, Evandra Strazza Rodrigues, Svetoslav Nanev Slavov, Elaine Vieira dos Santos, Rafael dos Santos Bezerra, Luiz Carlos Junior de Alcantara, Marta Giovanetti, Vagner Fonseca, Flavia Aburjaile, Rodrigo Tocantins Calado. |
| EPI_ISL_1795393, EPI_ISL_1795394 | LABORATORIO DE FRANCA | Instituto Butantan / ESALQ-Piracicaba | Instituto Butantan: Alexander Roberto Precioso, Dimas Tadeu Covas, Sandra Coccuzzo Sampaio, Maria Carolina Elias, José Salvatore Leister Patané, Vincent Louis Viala, Antonio Jorge Martins, Ricardo Haddad, Claudia Renata dos Santos Barros, Elaine Cristina Marqueze, Raul Machado Neto, Debora Botequio Moretti. Centro de Genômica Funcional da ESALQ: Luiz Lehmann Coutinho, Ricardo Augusto Brassaloti, Raquel de Lello Rocha Campos Cassano. NGS Soluções Genômicas: Pilar Drummond Sampaio Corrêa Mariani. FZEA-USP Pirassununga: Mirele Daiana Poleti, Jessika Cristina Chagas Lesbon, Elisangela Chicaroni Mattos, Heidge Fukumasu. USP-Botucatu: Rejane Maria Tommasini Grotto, Jayme A. Souza-Neto, Guilherme Targino Valente, Patricia Akemi Assato, Felipe Allan da Silva da Costa, Bianca Cechetto Carlos. Mendelics: Bibiana Santos, João Paulo Kitajima, Erika Freitas, David Schlesinger. Hemocentro Ribeirão Preto: Simone Kashima, Evandra Strazza Rodrigues, Svetoslav Nanev Slavov, Elaine Vieira dos Santos, Rafael dos Santos Bezerra, Luiz Carlos Junior de Alcantara, Marta Giovanetti, Vagner Fonseca, Flavia Aburjaile, Rodrigo Tocantins Calado. |
| EPI_ISL_1795395, EPI_ISL_1795396, EPI_ISL_1795397 | UBS III DE RANCHARIA | Instituto Butantan / ESALQ-Piracicaba | Instituto Butantan: Alexander Roberto Precioso, Dimas Tadeu Covas, Sandra Coccuzzo Sampaio, Maria Carolina Elias, José Salvatore Leister Patané, Vincent Louis Viala, Antonio Jorge Martins, Ricardo Haddad, Claudia Renata dos Santos Barros, Elaine Cristina Marqueze, Raul Machado Neto, Debora Botequio Moretti. Centro de Genômica Funcional da ESALQ: Luiz Lehmann Coutinho, Ricardo Augusto Brassaloti, Raquel de Lello Rocha Campos Cassano. NGS Soluções Genômicas: Pilar Drummond Sampaio Corrêa Mariani. FZEA-USP Pirassununga: Mirele Daiana Poleti, Jessika Cristina Chagas Lesbon, Elisangela Chicaroni Mattos, Heidge Fukumasu. USP-Botucatu: Rejane Maria Tommasini Grotto, Jayme A. Souza-Neto, Guilherme Targino Valente, Patricia Akemi Assato, Felipe Allan da Silva da Costa, Bianca Cechetto Carlos. Mendelics: Bibiana Santos, João Paulo Kitajima, Erika Freitas, David Schlesinger. Hemocentro Ribeirão Preto: Simone Kashima, Evandra Strazza Rodrigues, Svetoslav Nanev Slavov, Elaine Vieira dos Santos, Rafael dos Santos Bezerra, Luiz Carlos Junior de Alcantara, Marta Giovanetti, Vagner Fonseca, Flavia Aburjaile, Rodrigo Tocantins Calado. |
| EPI_ISL_1795398 | SMS SECRETARIA MUNICIPAL DE SAUDE DE BOITUVA | Instituto Butantan / ESALQ-Piracicaba | Instituto Butantan: Alexander Roberto Precioso, Dimas Tadeu Covas, Sandra Coccuzzo Sampaio, Maria Carolina Elias, José Salvatore Leister Patané, Vincent Louis Viala, Antonio Jorge Martins, Ricardo Haddad, Claudia Renata dos Santos Barros, Elaine Cristina Marqueze, Raul Machado Neto, Debora Botequio Moretti. Centro de Genômica Funcional da ESALQ: Luiz Lehmann Coutinho, Ricardo Augusto Brassaloti, Raquel de Lello Rocha Campos Cassano. NGS Soluções Genômicas: Pilar Drummond Sampaio Corrêa Mariani. FZEA-USP Pirassununga: Mirele Daiana Poleti, Jessika Cristina Chagas Lesbon, Elisangela Chicaroni Mattos, Heidge Fukumasu. USP-Botucatu: Rejane Maria Tommasini Grotto, Jayme A. Souza-Neto, Guilherme Targino |

| | | | |
|---|---|---|---|
| | | | Valente, Patricia Akemi Assato, Felipe Allan da Silva da Costa, Bianca Cechetto Carlos. Mendelics: Bibiana Santos, João Paulo Kitajima, Erika Freitas, David Schlesinger. Hemocentro Ribeirão Preto: Simone Kashima, Evandra Strazza Rodrigues, Svetoslav Nanev Slavov, Elaine Vieira dos Santos, Rafael dos Santos Bezerra, Luiz Carlos Junior de Alcantara, Marta Giovanetti, Vagner Fonseca, Flavia Aburjaile, Rodrigo Tocantins Calado. |
| EPI_ISL_1795399 | POLICLINICA HORTOLANDIA | Instituto Butantan / ESALQ-Piracicaba | Instituto Butantan: Alexander Roberto Precioso, Dimas Tadeu Covas, Sandra Coccuzzo Sampaio, Maria Carolina Elias, José Salvatore Leister Patané, Vincent Louis Viala, Antonio Jorge Martins, Ricardo Haddad, Claudia Renata dos Santos Barros, Elaine Cristina Marqueze, Raul Machado Neto, Debora Botequio Moretti. Centro de Genômica Funcional da ESALQ: Luiz Lehmann Coutinho, Ricardo Augusto Brassaloti, Raquel de Lello Rocha Campos Cassano. NGS Soluções Genômicas: Pilar Drummond Sampaio Corrêa Mariani. FZEA-USP Pirassununga: Mirele Daiana Poleti, Jessika Cristina Chagas Lesbon, Elisangela Chicaroni Mattos, Heidge Fukumasu. USP-Botucatu: Rejane Maria Tommasini Grotto, Jayme A. Souza-Neto, Guilherme Targino Valente, Patricia Akemi Assato, Felipe Allan da Silva da Costa, Bianca Cechetto Carlos. Mendelics: Bibiana Santos, João Paulo Kitajima, Erika Freitas, David Schlesinger. Hemocentro Ribeirão Preto: Simone Kashima, Evandra Strazza Rodrigues, Svetoslav Nanev Slavov, Elaine Vieira dos Santos, Rafael dos Santos Bezerra, Luiz Carlos Junior de Alcantara, Marta Giovanetti, Vagner Fonseca, Flavia Aburjaile, Rodrigo Tocantins Calado. |
| EPI_ISL_1795400, EPI_ISL_1795401 | LABORATORIO MUNICIPAL DE ANALISES CLINICAS DE RIO CLARO | Instituto Butantan / ESALQ-Piracicaba | Instituto Butantan: Alexander Roberto Precioso, Dimas Tadeu Covas, Sandra Coccuzzo Sampaio, Maria Carolina Elias, José Salvatore Leister Patané, Vincent Louis Viala, Antonio Jorge Martins, Ricardo Haddad, Claudia Renata dos Santos Barros, Elaine Cristina Marqueze, Raul Machado Neto, Debora Botequio Moretti. Centro de Genômica Funcional da ESALQ: Luiz Lehmann Coutinho, Ricardo Augusto Brassaloti, Raquel de Lello Rocha Campos Cassano. NGS Soluções Genômicas: Pilar Drummond Sampaio Corrêa Mariani. FZEA-USP Pirassununga: Mirele Daiana Poleti, Jessika Cristina Chagas Lesbon, Elisangela Chicaroni Mattos, Heidge Fukumasu. USP-Botucatu: Rejane Maria Tommasini Grotto, Jayme A. Souza-Neto, Guilherme Targino Valente, Patricia Akemi Assato, Felipe Allan da Silva da Costa, Bianca Cechetto Carlos. Mendelics: Bibiana Santos, João Paulo Kitajima, Erika Freitas, David Schlesinger. Hemocentro Ribeirão Preto: Simone Kashima, Evandra Strazza Rodrigues, Svetoslav Nanev Slavov, Elaine Vieira dos Santos, Rafael dos Santos Bezerra, Luiz Carlos Junior de Alcantara, Marta Giovanetti, Vagner Fonseca, Flavia Aburjaile, Rodrigo Tocantins Calado. |
| EPI_ISL_1795402 | SECRETARIA MUNICIPAL DE SAUDE DE CORDEIROPOLIS | Instituto Butantan / ESALQ-Piracicaba | Instituto Butantan: Alexander Roberto Precioso, Dimas Tadeu Covas, Sandra Coccuzzo Sampaio, Maria Carolina Elias, José Salvatore Leister Patané, Vincent Louis Viala, Antonio Jorge Martins, Ricardo Haddad, Claudia Renata dos Santos Barros, Elaine Cristina Marqueze, Raul Machado Neto, Debora Botequio Moretti. Centro de Genômica Funcional da ESALQ: Luiz Lehmann Coutinho, Ricardo Augusto Brassaloti, Raquel de Lello Rocha Campos Cassano. NGS Soluções Genômicas: Pilar Drummond Sampaio Corrêa Mariani. FZEA-USP Pirassununga: Mirele Daiana Poleti, Jessika Cristina Chagas Lesbon, Elisangela Chicaroni Mattos, Heidge Fukumasu. USP-Botucatu: Rejane Maria Tommasini Grotto, Jayme A. Souza-Neto, Guilherme Targino Valente, Patricia Akemi Assato, Felipe Allan da Silva da Costa, Bianca Cechetto Carlos. Mendelics: Bibiana Santos, João Paulo Kitajima, Erika Freitas, David Schlesinger. Hemocentro Ribeirão Preto: Simone Kashima, Evandra Strazza Rodrigues, Svetoslav Nanev Slavov, Elaine Vieira dos Santos, Rafael dos Santos Bezerra, Luiz Carlos Junior de Alcantara, Marta Giovanetti, Vagner Fonseca, Flavia Aburjaile, Rodrigo Tocantins Calado. |
| EPI_ISL_1795403 | UNIDADE MISTA DE LUIZ ANTONIO | Instituto Butantan / ESALQ-Piracicaba | Instituto Butantan: Alexander Roberto Precioso, Dimas Tadeu Covas, Sandra Coccuzzo Sampaio, Maria Carolina Elias, José Salvatore Leister Patané, Vincent Louis Viala, Antonio Jorge Martins, Ricardo Haddad, Claudia Renata dos Santos Barros, Elaine Cristina Marqueze, Raul Machado Neto, Debora Botequio Moretti. Centro de Genômica Funcional da ESALQ: Luiz Lehmann Coutinho, Ricardo Augusto Brassaloti, Raquel de Lello Rocha Campos Cassano. NGS Soluções Genômicas: Pilar Drummond Sampaio Corrêa Mariani. FZEA-USP Pirassununga: Mirele Daiana Poleti, Jessika Cristina Chagas Lesbon, Elisangela Chicaroni Mattos, Heidge Fukumasu. USP-Botucatu: Rejane Maria Tommasini Grotto, Jayme A. Souza-Neto, Guilherme Targino Valente, Patricia Akemi Assato, Felipe Allan da Silva da Costa, Bianca Cechetto Carlos. Mendelics: Bibiana Santos, João Paulo Kitajima, Erika Freitas, David Schlesinger. Hemocentro Ribeirão Preto: Simone Kashima, Evandra Strazza Rodrigues, Svetoslav Nanev Slavov, Elaine Vieira dos Santos, Rafael dos Santos Bezerra, Luiz Carlos Junior de Alcantara, Marta Giovanetti, Vagner Fonseca, Flavia Aburjaile, Rodrigo Tocantins Calado. |
| EPI_ISL_1795404 | POLICLINICA HORTOLANDIA | Instituto Butantan / ESALQ-Piracicaba | Instituto Butantan: Alexander Roberto Precioso, Dimas Tadeu Covas, Sandra Coccuzzo Sampaio, Maria Carolina Elias, José Salvatore Leister Patané, Vincent Louis Viala, Antonio Jorge Martins, Ricardo Haddad, Claudia Renata dos Santos Barros, Elaine Cristina Marqueze, Raul Machado Neto, Debora Botequio Moretti. Centro de Genômica Funcional da ESALQ: Luiz Lehmann Coutinho, Ricardo Augusto Brassaloti, Raquel de Lello Rocha Campos Cassano. NGS Soluções Genômicas: Pilar Drummond Sampaio Corrêa Mariani. FZEA-USP Pirassununga: Mirele Daiana Poleti, Jessika Cristina Chagas Lesbon, Elisangela Chicaroni Mattos, Heidge Fukumasu. USP-Botucatu: Rejane Maria Tommasini Grotto, Jayme A. Souza-Neto, Guilherme Targino Valente, Patricia Akemi Assato, Felipe Allan da Silva da Costa, Bianca Cechetto Carlos. Mendelics: Bibiana Santos, João Paulo Kitajima, Erika Freitas, David Schlesinger. Hemocentro Ribeirão Preto: Simone Kashima, Evandra Strazza Rodrigues, Svetoslav Nanev Slavov, Elaine Vieira dos Santos, Rafael dos Santos Bezerra, Luiz Carlos Junior de Alcantara, Marta Giovanetti, Vagner Fonseca, Flavia Aburjaile, Rodrigo Tocantins Calado. |
| EPI_ISL_1795405 | UBS CAIC OURINHOS | Instituto Butantan / ESALQ-Piracicaba | Instituto Butantan: Alexander Roberto Precioso, Dimas Tadeu Covas, Sandra Coccuzzo Sampaio, Maria Carolina Elias, José Salvatore Leister Patané, Vincent Louis Viala, Antonio Jorge Martins, Ricardo Haddad, Claudia Renata dos Santos Barros, Elaine Cristina Marqueze, Raul Machado Neto, Debora Botequio Moretti. Centro de Genômica Funcional da ESALQ: Luiz Lehmann Coutinho, Ricardo Augusto Brassaloti, Raquel de Lello Rocha Campos Cassano. NGS Soluções Genômicas: Pilar Drummond Sampaio Corrêa Mariani. FZEA-USP Pirassununga: Mirele Daiana Poleti, Jessika Cristina Chagas Lesbon, Elisangela Chicaroni Mattos, Heidge Fukumasu. USP-Botucatu: Rejane Maria Tommasini Grotto, Jayme A. Souza-Neto, Guilherme Targino Valente, Patricia Akemi Assato, Felipe Allan da Silva da Costa, Bianca Cechetto Carlos. Mendelics: Bibiana Santos, João Paulo Kitajima, Erika Freitas, David Schlesinger. Hemocentro Ribeirão Preto: Simone Kashima, Evandra Strazza Rodrigues, Svetoslav Nanev Slavov, Elaine Vieira dos Santos, Rafael dos Santos Bezerra, Luiz Carlos Junior de Alcantara, Marta Giovanetti, Vagner Fonseca, Flavia Aburjaile, Rodrigo Tocantins Calado. |
| EPI_ISL_1795406 | CS DE OSCAR BRESSANE PSF | Instituto Butantan / ESALQ-Piracicaba | Instituto Butantan: Alexander Roberto Precioso, Dimas Tadeu Covas, Sandra Coccuzzo Sampaio, Maria Carolina Elias, José Salvatore Leister Patané, Vincent Louis Viala, Antonio Jorge Martins, Ricardo Haddad, Claudia Renata dos Santos Barros, Elaine Cristina Marqueze, Raul Machado Neto, Debora Botequio Moretti. Centro de Genômica Funcional da ESALQ: Luiz Lehmann Coutinho, Ricardo Augusto Brassaloti, Raquel de Lello Rocha Campos Cassano. NGS Soluções Genômicas: Pilar Drummond Sampaio Corrêa Mariani. FZEA-USP Pirassununga: Mirele Daiana Poleti, Jessika Cristina Chagas Lesbon, Elisangela Chicaroni Mattos, Heidge Fukumasu. USP-Botucatu: Rejane Maria Tommasini Grotto, Jayme A. Souza-Neto, Guilherme Targino Valente, Patricia Akemi Assato, Felipe Allan da Silva da Costa, Bianca Cechetto Carlos. Mendelics: Bibiana Santos, João Paulo Kitajima, Erika Freitas, David Schlesinger. Hemocentro Ribeirão Preto: Simone Kashima, Evandra Strazza Rodrigues, Svetoslav Nanev Slavov, Elaine Vieira dos Santos, Rafael dos Santos Bezerra, Luiz Carlos Junior de Alcantara, Marta Giovanetti, Vagner Fonseca, Flavia Aburjaile, Rodrigo Tocantins Calado. |
| EPI_ISL_1795407 | CENTRO DE SAUDE DE RIBEIRAO DO SUL | Instituto Butantan / ESALQ-Piracicaba | Instituto Butantan: Alexander Roberto Precioso, Dimas Tadeu Covas, Sandra Coccuzzo Sampaio, Maria Carolina Elias, José Salvatore Leister Patané, Vincent Louis Viala, Antonio Jorge Martins, Ricardo Haddad, Claudia Renata dos Santos Barros, Elaine Cristina Marqueze, Raul Machado Neto, Debora Botequio Moretti. Centro de Genômica Funcional da ESALQ: Luiz Lehmann Coutinho, Ricardo Augusto Brassaloti, Raquel de Lello Rocha Campos Cassano. NGS Soluções Genômicas: Pilar Drummond Sampaio Corrêa Mariani. FZEA-USP Pirassununga: Mirele Daiana Poleti, Jessika Cristina Chagas Lesbon, Elisangela Chicaroni Mattos, Heidge Fukumasu. USP-Botucatu: Rejane Maria Tommasini Grotto, Jayme A. Souza-Neto, Guilherme Targino Valente, Patricia Akemi Assato, Felipe Allan da Silva da Costa, Bianca Cechetto Carlos. Mendelics: Bibiana Santos, João Paulo Kitajima, Erika Freitas, David Schlesinger. Hemocentro Ribeirão Preto: Simone Kashima, Evandra Strazza Rodrigues, Svetoslav Nanev Slavov, Elaine Vieira dos Santos, Rafael dos Santos Bezerra, Luiz Carlos Junior de Alcantara, Marta Giovanetti, Vagner Fonseca, Flavia Aburjaile, Rodrigo Tocantins Calado. |
| EPI_ISL_1795408 | CS DE OSCAR BRESSANE PSF | Instituto Butantan / ESALQ-Piracicaba | Instituto Butantan: Alexander Roberto Precioso, Dimas Tadeu Covas, Sandra Coccuzzo Sampaio, Maria Carolina Elias, José Salvatore Leister Patané, Vincent Louis Viala, Antonio Jorge Martins, Ricardo Haddad, Claudia Renata dos Santos Barros, Elaine Cristina Marqueze, Raul Machado Neto, Debora Botequio Moretti. Centro de Genômica Funcional da ESALQ: Luiz Lehmann Coutinho, Ricardo Augusto Brassaloti, Raquel de Lello Rocha Campos Cassano. NGS Soluções Genômicas: Pilar Drummond Sampaio Corrêa Mariani. FZEA-USP Pirassununga: Mirele Daiana Poleti, Jessika Cristina Chagas Lesbon, Elisangela Chicaroni Mattos, Heidge Fukumasu. USP-Botucatu: Rejane Maria Tommasini Grotto, Jayme A. Souza-Neto, Guilherme Targino Valente, Patricia Akemi Assato, Felipe Allan da Silva da Costa, Bianca Cechetto Carlos. Mendelics: Bibiana Santos, João Paulo Kitajima, Erika Freitas, David Schlesinger. Hemocentro Ribeirão Preto: Simone Kashima, Evandra Strazza Rodrigues, Svetoslav Nanev Slavov, Elaine Vieira dos Santos, Rafael dos Santos Bezerra, Luiz Carlos Junior de Alcantara, Marta Giovanetti, Vagner Fonseca, Flavia Aburjaile, Rodrigo Tocantins Calado. |
| EPI_ISL_1795409, EPI_ISL_1795410 | CENTRO DE SAUDE DE MARACAI | Instituto Butantan / ESALQ-Piracicaba | Instituto Butantan: Alexander Roberto Precioso, Dimas Tadeu Covas, Sandra Coccuzzo Sampaio, Maria Carolina Elias, José Salvatore Leister Patané, Vincent Louis Viala, Antonio Jorge Martins, Ricardo Haddad, Claudia Renata dos Santos Barros, Elaine Cristina Marqueze, Raul Machado Neto, Debora Botequio Moretti. Centro de Genômica Funcional da ESALQ: Luiz Lehmann Coutinho, Ricardo Augusto Brassaloti, Raquel de Lello Rocha Campos Cassano. NGS Soluções Genômicas: Pilar Drummond Sampaio Corrêa Mariani. FZEA-USP Pirassununga: Mirele Daiana Poleti, Jessika Cristina Chagas Lesbon, Elisangela Chicaroni Mattos, Heidge Fukumasu. USP-Botucatu: Rejane Maria Tommasini Grotto, Jayme A. Souza-Neto, Guilherme Targino |

| | | | |
|---|---|---|---|
| | | | Valente, Patricia Akemi Assato, Felipe Allan da Silva da Costa, Bianca Cechetto Carlos. Mendelics: Bibiana Santos, João Paulo Kitajima, Erika Freitas, David Schlesinger. Hemocentro Ribeirão Preto: Simone Kashima, Evandra Strazza Rodrigues, Svetoslav Nanev Slavov, Elaine Vieira dos Santos, Rafael dos Santos Bezerra, Luiz Carlos Junior de Alcantara, Marta Giovanetti, Vagner Fonseca, Flavia Aburjaile, Rodrigo Tocantins Calado. |
| EPI_ISL_1795411 | POLICLINICA HORTOLANDIA | Instituto Butantan / ESALQ-Piracicaba | Instituto Butantan: Alexander Roberto Precioso, Dimas Tadeu Covas, Sandra Coccuzzo Sampaio, Maria Carolina Elias, José Salvatore Leister Patané, Vincent Louis Viala, Antonio Jorge Martins, Ricardo Haddad, Claudia Renata dos Santos Barros, Elaine Cristina Marqueze, Raul Machado Neto, Debora Botequio Moretti. Centro de Genômica Funcional da ESALQ: Luiz Lehmann Coutinho, Ricardo Augusto Brassaloti, Raquel de Lello Rocha Campos Cassano. NGS Soluções Genômicas: Pilar Drummond Sampaio Corrêa Mariani. FZEA-USP Pirassununga: Mirele Daiana Poleti, Jessika Cristina Chagas Lesbon, Elisangela Chicaroni Mattos, Heidge Fukumasu. USP-Botucatu: Rejane Maria Tommasini Grotto, Jayme A. Souza-Neto, Guilherme Targino Valente, Patricia Akemi Assato, Felipe Allan da Silva da Costa, Bianca Cechetto Carlos. Mendelics: Bibiana Santos, João Paulo Kitajima, Erika Freitas, David Schlesinger. Hemocentro Ribeirão Preto: Simone Kashima, Evandra Strazza Rodrigues, Svetoslav Nanev Slavov, Elaine Vieira dos Santos, Rafael dos Santos Bezerra, Luiz Carlos Junior de Alcantara, Marta Giovanetti, Vagner Fonseca, Flavia Aburjaile, Rodrigo Tocantins Calado. |
| EPI_ISL_1795412 | USF SALERNO | Instituto Butantan / ESALQ-Piracicaba | Instituto Butantan: Alexander Roberto Precioso, Dimas Tadeu Covas, Sandra Coccuzzo Sampaio, Maria Carolina Elias, José Salvatore Leister Patané, Vincent Louis Viala, Antonio Jorge Martins, Ricardo Haddad, Claudia Renata dos Santos Barros, Elaine Cristina Marqueze, Raul Machado Neto, Debora Botequio Moretti. Centro de Genômica Funcional da ESALQ: Luiz Lehmann Coutinho, Ricardo Augusto Brassaloti, Raquel de Lello Rocha Campos Cassano. NGS Soluções Genômicas: Pilar Drummond Sampaio Corrêa Mariani. FZEA-USP Pirassununga: Mirele Daiana Poleti, Jessika Cristina Chagas Lesbon, Elisangela Chicaroni Mattos, Heidge Fukumasu. USP-Botucatu: Rejane Maria Tommasini Grotto, Jayme A. Souza-Neto, Guilherme Targino Valente, Patricia Akemi Assato, Felipe Allan da Silva da Costa, Bianca Cechetto Carlos. Mendelics: Bibiana Santos, João Paulo Kitajima, Erika Freitas, David Schlesinger. Hemocentro Ribeirão Preto: Simone Kashima, Evandra Strazza Rodrigues, Svetoslav Nanev Slavov, Elaine Vieira dos Santos, Rafael dos Santos Bezerra, Luiz Carlos Junior de Alcantara, Marta Giovanetti, Vagner Fonseca, Flavia Aburjaile, Rodrigo Tocantins Calado. |
| EPI_ISL_1795413 | PRONTO SOCORRO MUNICIPAL TAMBAU | Instituto Butantan / ESALQ-Piracicaba | Instituto Butantan: Alexander Roberto Precioso, Dimas Tadeu Covas, Sandra Coccuzzo Sampaio, Maria Carolina Elias, José Salvatore Leister Patané, Vincent Louis Viala, Antonio Jorge Martins, Ricardo Haddad, Claudia Renata dos Santos Barros, Elaine Cristina Marqueze, Raul Machado Neto, Debora Botequio Moretti. Centro de Genômica Funcional da ESALQ: Luiz Lehmann Coutinho, Ricardo Augusto Brassaloti, Raquel de Lello Rocha Campos Cassano. NGS Soluções Genômicas: Pilar Drummond Sampaio Corrêa Mariani. FZEA-USP Pirassununga: Mirele Daiana Poleti, Jessika Cristina Chagas Lesbon, Elisangela Chicaroni Mattos, Heidge Fukumasu. USP-Botucatu: Rejane Maria Tommasini Grotto, Jayme A. Souza-Neto, Guilherme Targino Valente, Patricia Akemi Assato, Felipe Allan da Silva da Costa, Bianca Cechetto Carlos. Mendelics: Bibiana Santos, João Paulo Kitajima, Erika Freitas, David Schlesinger. Hemocentro Ribeirão Preto: Simone Kashima, Evandra Strazza Rodrigues, Svetoslav Nanev Slavov, Elaine Vieira dos Santos, Rafael dos Santos Bezerra, Luiz Carlos Junior de Alcantara, Marta Giovanetti, Vagner Fonseca, Flavia Aburjaile, Rodrigo Tocantins Calado. |
| EPI_ISL_1795414 | HOSPITAL MUNICIPAL DE IBIUNA IBIUNA SP | Instituto Butantan / ESALQ-Piracicaba | Instituto Butantan: Alexander Roberto Precioso, Dimas Tadeu Covas, Sandra Coccuzzo Sampaio, Maria Carolina Elias, José Salvatore Leister Patané, Vincent Louis Viala, Antonio Jorge Martins, Ricardo Haddad, Claudia Renata dos Santos Barros, Elaine Cristina Marqueze, Raul Machado Neto, Debora Botequio Moretti. Centro de Genômica Funcional da ESALQ: Luiz Lehmann Coutinho, Ricardo Augusto Brassaloti, Raquel de Lello Rocha Campos Cassano. NGS Soluções Genômicas: Pilar Drummond Sampaio Corrêa Mariani. FZEA-USP Pirassununga: Mirele Daiana Poleti, Jessika Cristina Chagas Lesbon, Elisangela Chicaroni Mattos, Heidge Fukumasu. USP-Botucatu: Rejane Maria Tommasini Grotto, Jayme A. Souza-Neto, Guilherme Targino Valente, Patricia Akemi Assato, Felipe Allan da Silva da Costa, Bianca Cechetto Carlos. Mendelics: Bibiana Santos, João Paulo Kitajima, Erika Freitas, David Schlesinger. Hemocentro Ribeirão Preto: Simone Kashima, Evandra Strazza Rodrigues, Svetoslav Nanev Slavov, Elaine Vieira dos Santos, Rafael dos Santos Bezerra, Luiz Carlos Junior de Alcantara, Marta Giovanetti, Vagner Fonseca, Flavia Aburjaile, Rodrigo Tocantins Calado. |
| EPI_ISL_1795415 | UBS DE SANTA SALETE | Instituto Butantan / ESALQ-Piracicaba | Instituto Butantan: Alexander Roberto Precioso, Dimas Tadeu Covas, Sandra Coccuzzo Sampaio, Maria Carolina Elias, José Salvatore Leister Patané, Vincent Louis Viala, Antonio Jorge Martins, Ricardo Haddad, Claudia Renata dos Santos Barros, Elaine Cristina Marqueze, Raul Machado Neto, Debora Botequio Moretti. Centro de Genômica Funcional da ESALQ: Luiz Lehmann Coutinho, Ricardo Augusto Brassaloti, Raquel de Lello Rocha Campos Cassano. NGS Soluções Genômicas: Pilar Drummond Sampaio Corrêa Mariani. FZEA-USP Pirassununga: Mirele Daiana Poleti, Jessika Cristina Chagas Lesbon, Elisangela Chicaroni Mattos, Heidge Fukumasu. USP-Botucatu: Rejane Maria Tommasini Grotto, Jayme A. Souza-Neto, Guilherme Targino Valente, Patricia Akemi Assato, Felipe Allan da Silva da Costa, Bianca Cechetto Carlos. Mendelics: Bibiana Santos, João Paulo Kitajima, Erika Freitas, David Schlesinger. Hemocentro Ribeirão Preto: Simone Kashima, Evandra Strazza Rodrigues, Svetoslav Nanev Slavov, Elaine Vieira dos Santos, Rafael dos Santos Bezerra, Luiz Carlos Junior de Alcantara, Marta Giovanetti, Vagner Fonseca, Flavia Aburjaile, Rodrigo Tocantins Calado. |
| EPI_ISL_1795416 | CS DE MARAPOAMA | Instituto Butantan / ESALQ-Piracicaba | Instituto Butantan: Alexander Roberto Precioso, Dimas Tadeu Covas, Sandra Coccuzzo Sampaio, Maria Carolina Elias, José Salvatore Leister Patané, Vincent Louis Viala, Antonio Jorge Martins, Ricardo Haddad, Claudia Renata dos Santos Barros, Elaine Cristina Marqueze, Raul Machado Neto, Debora Botequio Moretti. Centro de Genômica Funcional da ESALQ: Luiz Lehmann Coutinho, Ricardo Augusto Brassaloti, Raquel de Lello Rocha Campos Cassano. NGS Soluções Genômicas: Pilar Drummond Sampaio Corrêa Mariani. FZEA-USP Pirassununga: Mirele Daiana Poleti, Jessika Cristina Chagas Lesbon, Elisangela Chicaroni Mattos, Heidge Fukumasu. USP-Botucatu: Rejane Maria Tommasini Grotto, Jayme A. Souza-Neto, Guilherme Targino Valente, Patricia Akemi Assato, Felipe Allan da Silva da Costa, Bianca Cechetto Carlos. Mendelics: Bibiana Santos, João Paulo Kitajima, Erika Freitas, David Schlesinger. Hemocentro Ribeirão Preto: Simone Kashima, Evandra Strazza Rodrigues, Svetoslav Nanev Slavov, Elaine Vieira dos Santos, Rafael dos Santos Bezerra, Luiz Carlos Junior de Alcantara, Marta Giovanetti, Vagner Fonseca, Flavia Aburjaile, Rodrigo Tocantins Calado. |
| EPI_ISL_1795417 | AMBULATORIO DE ESPECIALIDADE V E MOGI MIRIM | Instituto Butantan / ESALQ-Piracicaba | Instituto Butantan: Alexander Roberto Precioso, Dimas Tadeu Covas, Sandra Coccuzzo Sampaio, Maria Carolina Elias, José Salvatore Leister Patané, Vincent Louis Viala, Antonio Jorge Martins, Ricardo Haddad, Claudia Renata dos Santos Barros, Elaine Cristina Marqueze, Raul Machado Neto, Debora Botequio Moretti. Centro de Genômica Funcional da ESALQ: Luiz Lehmann Coutinho, Ricardo Augusto Brassaloti, Raquel de Lello Rocha Campos Cassano. NGS Soluções Genômicas: Pilar Drummond Sampaio Corrêa Mariani. FZEA-USP Pirassununga: Mirele Daiana Poleti, Jessika Cristina Chagas Lesbon, Elisangela Chicaroni Mattos, Heidge Fukumasu. USP-Botucatu: Rejane Maria Tommasini Grotto, Jayme A. Souza-Neto, Guilherme Targino Valente, Patricia Akemi Assato, Felipe Allan da Silva da Costa, Bianca Cechetto Carlos. Mendelics: Bibiana Santos, João Paulo Kitajima, Erika Freitas, David Schlesinger. Hemocentro Ribeirão Preto: Simone Kashima, Evandra Strazza Rodrigues, Svetoslav Nanev Slavov, Elaine Vieira dos Santos, Rafael dos Santos Bezerra, Luiz Carlos Junior de Alcantara, Marta Giovanetti, Vagner Fonseca, Flavia Aburjaile, Rodrigo Tocantins Calado. |
| EPI_ISL_1795418, EPI_ISL_1795419, EPI_ISL_1795420, EPI_ISL_1795421, EPI_ISL_1795422, EPI_ISL_1795423 | LABORATORIO DE FRANCA | Instituto Butantan / ESALQ-Piracicaba | Instituto Butantan: Alexander Roberto Precioso, Dimas Tadeu Covas, Sandra Coccuzzo Sampaio, Maria Carolina Elias, José Salvatore Leister Patané, Vincent Louis Viala, Antonio Jorge Martins, Ricardo Haddad, Claudia Renata dos Santos Barros, Elaine Cristina Marqueze, Raul Machado Neto, Debora Botequio Moretti. Centro de Genômica Funcional da ESALQ: Luiz Lehmann Coutinho, Ricardo Augusto Brassaloti, Raquel de Lello Rocha Campos Cassano. NGS Soluções Genômicas: Pilar Drummond Sampaio Corrêa Mariani. FZEA-USP Pirassununga: Mirele Daiana Poleti, Jessika Cristina Chagas Lesbon, Elisangela Chicaroni Mattos, Heidge Fukumasu. USP-Botucatu: Rejane Maria Tommasini Grotto, Jayme A. Souza-Neto, Guilherme Targino Valente, Patricia Akemi Assato, Felipe Allan da Silva da Costa, Bianca Cechetto Carlos. Mendelics: Bibiana Santos, João Paulo Kitajima, Erika Freitas, David Schlesinger. Hemocentro Ribeirão Preto: Simone Kashima, Evandra Strazza Rodrigues, Svetoslav Nanev Slavov, Elaine Vieira dos Santos, Rafael dos Santos Bezerra, Luiz Carlos Junior de Alcantara, Marta Giovanetti, Vagner Fonseca, Flavia Aburjaile, Rodrigo Tocantins Calado. |
| EPI_ISL_1795424 | UBS II DE REGENTE FEIJO | Instituto Butantan / ESALQ-Piracicaba | Instituto Butantan: Alexander Roberto Precioso, Dimas Tadeu Covas, Sandra Coccuzzo Sampaio, Maria Carolina Elias, José Salvatore Leister Patané, Vincent Louis Viala, Antonio Jorge Martins, Ricardo Haddad, Claudia Renata dos Santos Barros, Elaine Cristina Marqueze, Raul Machado Neto, Debora Botequio Moretti. Centro de Genômica Funcional da ESALQ: Luiz Lehmann Coutinho, Ricardo Augusto Brassaloti, Raquel de Lello Rocha Campos Cassano. NGS Soluções Genômicas: Pilar Drummond Sampaio Corrêa Mariani. FZEA-USP Pirassununga: Mirele Daiana Poleti, Jessika Cristina Chagas Lesbon, Elisangela Chicaroni Mattos, Heidge Fukumasu. USP-Botucatu: Rejane Maria Tommasini Grotto, Jayme A. Souza-Neto, Guilherme Targino Valente, Patricia Akemi Assato, Felipe Allan da Silva da Costa, Bianca Cechetto Carlos. Mendelics: Bibiana Santos, João Paulo Kitajima, Erika Freitas, David Schlesinger. Hemocentro Ribeirão Preto: Simone Kashima, Evandra Strazza Rodrigues, Svetoslav Nanev Slavov, Elaine Vieira dos Santos, Rafael dos Santos Bezerra, Luiz Carlos Junior de Alcantara, Marta Giovanetti, Vagner Fonseca, Flavia Aburjaile, Rodrigo Tocantins Calado. |
| EPI_ISL_1795425 | UBS III DE RANCHARIA | Instituto Butantan / ESALQ-Piracicaba | Instituto Butantan: Alexander Roberto Precioso, Dimas Tadeu Covas, Sandra Coccuzzo Sampaio, Maria Carolina Elias, José Salvatore Leister Patané, Vincent Louis Viala, Antonio Jorge Martins, Ricardo Haddad, Claudia Renata dos Santos Barros, Elaine Cristina Marqueze, Raul Machado Neto, Debora Botequio Moretti. Centro de Genômica Funcional da ESALQ: Luiz Lehmann Coutinho, Ricardo Augusto Brassaloti, Raquel de Lello Rocha Campos Cassano. NGS Soluções Genômicas: Pilar Drummond Sampaio Corrêa Mariani. FZEA-USP Pirassununga: Mirele Daiana Poleti, Jessika Cristina Chagas Lesbon, Elisangela Chicaroni Mattos, Heidge Fukumasu. USP-Botucatu: Rejane Maria Tommasini Grotto, Jayme A. Souza-Neto, Guilherme Targino |

| | | | |
|---|---|---|---|
| | | | Valente, Patricia Akemi Assato, Felipe Allan da Silva da Costa, Bianca Cechetto Carlos. Mendelics: Bibiana Santos, João Paulo Kitajima, Erika Freitas, David Schlesinger. Hemocentro Ribeirão Preto: Simone Kashima, Evandra Strazza Rodrigues, Svetoslav Nanev Slavov, Elaine Vieira dos Santos, Rafael dos Santos Bezerra, Luiz Carlos Junior de Alcantara, Marta Giovanetti, Vagner Fonseca, Flavia Aburjaile, Rodrigo Tocantins Calado. |
| EPI_ISL_1795426 | PRONTO SOCORRO MUNICIPAL DE PARAIBUNA | Instituto Butantan / ESALQ-Piracicaba | Instituto Butantan: Alexander Roberto Precioso, Dimas Tadeu Covas, Sandra Coccuzzo Sampaio, Maria Carolina Elias, José Salvatore Leister Patané, Vincent Louis Viala, Antonio Jorge Martins, Ricardo Haddad, Claudia Renata dos Santos Barros, Elaine Cristina Marqueze, Raul Machado Neto, Debora Botequio Moretti. Centro de Genômica Funcional da ESALQ: Luiz Lehmann Coutinho, Ricardo Augusto Brassaloti, Raquel de Lello Rocha Campos Cassano. NGS Soluções Genômicas: Pilar Drummond Sampaio Corrêa Mariani. FZEA-USP Pirassununga: Mirele Daiana Poleti, Jessika Cristina Chagas Lesbon, Elisangela Chicaroni Mattos, Heidge Fukumasu. USP-Botucatu: Rejane Maria Tommasini Grotto, Jayme A. Souza-Neto, Guilherme Targino Valente, Patricia Akemi Assato, Felipe Allan da Silva da Costa, Bianca Cechetto Carlos. Mendelics: Bibiana Santos, João Paulo Kitajima, Erika Freitas, David Schlesinger. Hemocentro Ribeirão Preto: Simone Kashima, Evandra Strazza Rodrigues, Svetoslav Nanev Slavov, Elaine Vieira dos Santos, Rafael dos Santos Bezerra, Luiz Carlos Junior de Alcantara, Marta Giovanetti, Vagner Fonseca, Flavia Aburjaile, Rodrigo Tocantins Calado. |
| EPI_ISL_1795427 | CENTRO DE SAUDE DE NATIVIDADE DA SERRA | Instituto Butantan / ESALQ-Piracicaba | Instituto Butantan: Alexander Roberto Precioso, Dimas Tadeu Covas, Sandra Coccuzzo Sampaio, Maria Carolina Elias, José Salvatore Leister Patané, Vincent Louis Viala, Antonio Jorge Martins, Ricardo Haddad, Claudia Renata dos Santos Barros, Elaine Cristina Marqueze, Raul Machado Neto, Debora Botequio Moretti. Centro de Genômica Funcional da ESALQ: Luiz Lehmann Coutinho, Ricardo Augusto Brassaloti, Raquel de Lello Rocha Campos Cassano. NGS Soluções Genômicas: Pilar Drummond Sampaio Corrêa Mariani. FZEA-USP Pirassununga: Mirele Daiana Poleti, Jessika Cristina Chagas Lesbon, Elisangela Chicaroni Mattos, Heidge Fukumasu. USP-Botucatu: Rejane Maria Tommasini Grotto, Jayme A. Souza-Neto, Guilherme Targino Valente, Patricia Akemi Assato, Felipe Allan da Silva da Costa, Bianca Cechetto Carlos. Mendelics: Bibiana Santos, João Paulo Kitajima, Erika Freitas, David Schlesinger. Hemocentro Ribeirão Preto: Simone Kashima, Evandra Strazza Rodrigues, Svetoslav Nanev Slavov, Elaine Vieira dos Santos, Rafael dos Santos Bezerra, Luiz Carlos Junior de Alcantara, Marta Giovanetti, Vagner Fonseca, Flavia Aburjaile, Rodrigo Tocantins Calado. |
| EPI_ISL_1795428 | LABORATORIO DR PAULO EMILIO DALESSANDRO PINDAMONHANGABA | Instituto Butantan / ESALQ-Piracicaba | Instituto Butantan: Alexander Roberto Precioso, Dimas Tadeu Covas, Sandra Coccuzzo Sampaio, Maria Carolina Elias, José Salvatore Leister Patané, Vincent Louis Viala, Antonio Jorge Martins, Ricardo Haddad, Claudia Renata dos Santos Barros, Elaine Cristina Marqueze, Raul Machado Neto, Debora Botequio Moretti. Centro de Genômica Funcional da ESALQ: Luiz Lehmann Coutinho, Ricardo Augusto Brassaloti, Raquel de Lello Rocha Campos Cassano. NGS Soluções Genômicas: Pilar Drummond Sampaio Corrêa Mariani. FZEA-USP Pirassununga: Mirele Daiana Poleti, Jessika Cristina Chagas Lesbon, Elisangela Chicaroni Mattos, Heidge Fukumasu. USP-Botucatu: Rejane Maria Tommasini Grotto, Jayme A. Souza-Neto, Guilherme Targino Valente, Patricia Akemi Assato, Felipe Allan da Silva da Costa, Bianca Cechetto Carlos. Mendelics: Bibiana Santos, João Paulo Kitajima, Erika Freitas, David Schlesinger. Hemocentro Ribeirão Preto: Simone Kashima, Evandra Strazza Rodrigues, Svetoslav Nanev Slavov, Elaine Vieira dos Santos, Rafael dos Santos Bezerra, Luiz Carlos Junior de Alcantara, Marta Giovanetti, Vagner Fonseca, Flavia Aburjaile, Rodrigo Tocantins Calado. |
| EPI_ISL_1795429 | CENTRO INTEGRADO DE SAUDE | Instituto Butantan / ESALQ-Piracicaba | Instituto Butantan: Alexander Roberto Precioso, Dimas Tadeu Covas, Sandra Coccuzzo Sampaio, Maria Carolina Elias, José Salvatore Leister Patané, Vincent Louis Viala, Antonio Jorge Martins, Ricardo Haddad, Claudia Renata dos Santos Barros, Elaine Cristina Marqueze, Raul Machado Neto, Debora Botequio Moretti. Centro de Genômica Funcional da ESALQ: Luiz Lehmann Coutinho, Ricardo Augusto Brassaloti, Raquel de Lello Rocha Campos Cassano. NGS Soluções Genômicas: Pilar Drummond Sampaio Corrêa Mariani. FZEA-USP Pirassununga: Mirele Daiana Poleti, Jessika Cristina Chagas Lesbon, Elisangela Chicaroni Mattos, Heidge Fukumasu. USP-Botucatu: Rejane Maria Tommasini Grotto, Jayme A. Souza-Neto, Guilherme Targino Valente, Patricia Akemi Assato, Felipe Allan da Silva da Costa, Bianca Cechetto Carlos. Mendelics: Bibiana Santos, João Paulo Kitajima, Erika Freitas, David Schlesinger. Hemocentro Ribeirão Preto: Simone Kashima, Evandra Strazza Rodrigues, Svetoslav Nanev Slavov, Elaine Vieira dos Santos, Rafael dos Santos Bezerra, Luiz Carlos Junior de Alcantara, Marta Giovanetti, Vagner Fonseca, Flavia Aburjaile, Rodrigo Tocantins Calado. |
| EPI_ISL_1799498 | Laboratório de Microbiologia Molecular - Universidade FEEVALE | Molecular Microbiology Laboratory | Alana Witt Hansen, Fágner Henrique Heldt, Fernando Rosado Spilki, Flávio Silveira, Juliana Schons Gularte, Juliane Deise Fleck, Mariana Soares da Silva, Meriane Demoliner, Matheus Nunes Weber, Paula Rodrigues de Almeida, Micheli Filippi. |
| EPI_ISL_1799499, EPI_ISL_1799501, EPI_ISL_1799502, EPI_ISL_1799504, EPI_ISL_1799505, EPI_ISL_1799507 | Laboratório de Microbiologia Molecular - Universidade FEEVALE | Molecular Microbiology Laboratory | Alana Witt Hansen, Fágner Henrique Heldt, Fernando Rosado Spilki, Flávio Silveira, Juliana Schons Gulart, Juliane Deise Fleck, Mariana Soares da Silva, Meriane Demoliner, Matheus Nunes Weber, Paula Rodrigues de Almeida, Michele Filippi. |
| EPI_ISL_1821204 | Santa Casa de Cravinhos | Instituto Adolfo Lutz, Interdiciplinary Procedures Center, Strategic Laboratory | Claudio Tavares Sacchi, Claudia Regina Gonçalves, Erica Valessa Ramos Gomes, Karoline Rodrigues Campos, Caio Vinicius Dias Lopes, Leonardo Jose Tadeu de Araujo |
| EPI_ISL_1821205 | UPA Dr Luis Atilio Losi Viana Ribeirao Preto | Instituto Adolfo Lutz, Interdiciplinary Procedures Center, Strategic Laboratory | Claudio Tavares Sacchi, Claudia Regina Gonçalves, Erica Valessa Ramos Gomes, Karoline Rodrigues Campos, Caio Vinicius Dias Lopes, Leonardo Jose Tadeu de Araujo |
| EPI_ISL_1821206 | Sae Servico de Atendimento Especializado | Instituto Adolfo Lutz, Interdiciplinary Procedures Center, Strategic Laboratory | Claudio Tavares Sacchi, Claudia Regina Gonçalves, Erica Valessa Ramos Gomes, Karoline Rodrigues Campos, Caio Vinicius Dias Lopes, Leonardo Jose Tadeu de Araujo |
| EPI_ISL_1821207 | Secretaria Municipal de Saúde de Lins | Instituto Adolfo Lutz, Interdiciplinary Procedures Center, Strategic Laboratory | Claudio Tavares Sacchi, Claudia Regina Gonçalves, Erica Valessa Ramos Gomes, Karoline Rodrigues Campos, Caio Vinicius Dias Lopes, Leonardo Jose Tadeu de Araujo |
| EPI_ISL_1821208 | Hospital de Base de Bauru Hospital de Base Setima Regiao | Instituto Adolfo Lutz, Interdiciplinary Procedures Center, Strategic Laboratory | Claudio Tavares Sacchi, Claudia Regina Gonçalves, Erica Valessa Ramos Gomes, Karoline Rodrigues Campos, Caio Vinicius Dias Lopes, Leonardo Jose Tadeu de Araujo |
| EPI_ISL_1821209 | CS de Paulo de Faria | Instituto Adolfo Lutz, Interdiciplinary Procedures Center, Strategic Laboratory | Claudio Tavares Sacchi, Claudia Regina Gonçalves, Erica Valessa Ramos Gomes, Karoline Rodrigues Campos, Caio Vinicius Dias Lopes, Leonardo Jose Tadeu de Araujo |
| EPI_ISL_1821210 | Hospital Universitario de Marilia | Instituto Adolfo Lutz, Interdiciplinary Procedures Center, Strategic Laboratory | Claudio Tavares Sacchi, Claudia Regina Gonçalves, Erica Valessa Ramos Gomes, Karoline Rodrigues Campos, Caio Vinicius Dias Lopes, Leonardo Jose Tadeu de Araujo |
| EPI_ISL_1821211, EPI_ISL_1821212, EPI_ISL_1821213 | Diagnóstico da America S/A | Instituto Adolfo Lutz, Interdiciplinary Procedures Center, Strategic Laboratory | Claudio Tavares Sacchi, Claudia Regina Gonçalves, Erica Valessa Ramos Gomes, Karoline Rodrigues Campos, Caio Vinicius Dias Lopes, Leonardo Jose Tadeu de Araujo |
| EPI_ISL_1821225, EPI_ISL_1821226, EPI_ISL_1821227, EPI_ISL_1821228, EPI_ISL_1821229, EPI_ISL_1821230, EPI_ISL_1821231, EPI_ISL_1821232, EPI_ISL_1821233, EPI_ISL_1821234, EPI_ISL_1821235, EPI_ISL_1821236, EPI_ISL_1821237, EPI_ISL_1821238, EPI_ISL_1821239, EPI_ISL_1821240, EPI_ISL_1821241, EPI_ISL_1821242, EPI_ISL_1821243, EPI_ISL_1821244, EPI_ISL_1821245 | | | |
| see above | Instituto Adolfo Lutz - Regional de Marilia | Instituto Adolfo Lutz, Interdiciplinary Procedures Center, Strategic Laboratory | Claudio Tavares Sacchi, Claudia Regina Gonçalves, Erica Valessa Ramos Gomes, Karoline Rodrigues Campos, Caio Vinicius Dias Lopes, Leonardo Jose Tadeu de Araujo |
| EPI_ISL_1821250, EPI_ISL_1821253, EPI_ISL_1821256, EPI_ISL_1821260, EPI_ISL_1821263, EPI_ISL_1821265 | Instituto Adolfo Lutz - Regional de Ribeirao Preto | Instituto Adolfo Lutz, Interdiciplinary Procedures Center, Strategic Laboratory | Claudio Tavares Sacchi, Claudia Regina Gonçalves, Erica Valessa Ramos Gomes, Karoline Rodrigues Campos, Caio Vinicius Dias Lopes, Leonardo Jose Tadeu de Araujo |
| EPI_ISL_1857094, EPI_ISL_1857095, EPI_ISL_1857096, EPI_ISL_1857097, EPI_ISL_1857098 | Virology Lab- IMTSP | Virology Laboratory, Institute of Tropical Medicine of Sao Paulo- USP | Flavia Salles; Ingra Morales Claro, Ester C Sabino, Maria Cassia J M Correa |
| EPI_ISL_1858758, EPI_ISL_1858761, EPI_ISL_1858763, EPI_ISL_1858765, EPI_ISL_1858767, EPI_ISL_1858769, EPI_ISL_1858771, EPI_ISL_1858773, EPI_ISL_1858775, EPI_ISL_1858777, EPI_ISL_1858779, EPI_ISL_1858781, EPI_ISL_1858783, EPI_ISL_1858786, EPI_ISL_1858788, EPI_ISL_1858790, EPI_ISL_1858792, EPI_ISL_1858794, EPI_ISL_1858796, EPI_ISL_1858798, EPI_ISL_1858800, EPI_ISL_1858802, EPI_ISL_1858807, EPI_ISL_1858809, EPI_ISL_1858811, EPI_ISL_1858814, EPI_ISL_1858816, EPI_ISL_1858819, EPI_ISL_1858822, EPI_ISL_1858824, EPI_ISL_1858826, EPI_ISL_1858828, EPI_ISL_1858830, EPI_ISL_1858832, EPI_ISL_1858834, EPI_ISL_1858836, EPI_ISL_1858838, EPI_ISL_1858840, EPI_ISL_1858842, EPI_ISL_1858843, EPI_ISL_1858846, EPI_ISL_1858847, EPI_ISL_1858850, EPI_ISL_1858852, EPI_ISL_1858853, EPI_ISL_1858856, EPI_ISL_1858858, EPI_ISL_1858860, EPI_ISL_1858862, EPI_ISL_1858864, EPI_ISL_1858866, EPI_ISL_1858868, EPI_ISL_1858870, EPI_ISL_1858872, EPI_ISL_1858880, EPI_ISL_1858882, EPI_ISL_1858884, EPI_ISL_1858886, EPI_ISL_1858888, EPI_ISL_1858890, EPI_ISL_1858892, EPI_ISL_1858894, EPI_ISL_1858896, EPI_ISL_1858898, EPI_ISL_1858900, EPI_ISL_1858902, EPI_ISL_1858904, EPI_ISL_1858918, EPI_ISL_1858920, EPI_ISL_1858922, EPI_ISL_1858926, EPI_ISL_1858928, EPI_ISL_1858930, EPI_ISL_1858933, EPI_ISL_1858934, EPI_ISL_1858936, EPI_ISL_1858940, EPI_ISL_1858944, EPI_ISL_1858950, EPI_ISL_1858952, EPI_ISL_1858954, EPI_ISL_1858971, EPI_ISL_1858973, EPI_ISL_1858975, EPI_ISL_1858977, EPI_ISL_1858979, EPI_ISL_1858981, EPI_ISL_1858984, EPI_ISL_1858986, EPI_ISL_1858988, EPI_ISL_1858990, EPI_ISL_1858992, EPI_ISL_1858994, EPI_ISL_1858996, EPI_ISL_1858998, EPI_ISL_1859000, EPI_ISL_1859002, EPI_ISL_1859005, EPI_ISL_1859007, EPI_ISL_1859008 | | | |
| see above | Unidade de apoio ao diagnóstico da COVID - UNADIG | Bioinformatics Laboratory / LNCC | Luiz G P de Almeida, Alessandra P Lamarca, Ronaldo da Silva F Jr, Liliane Cavalcante, Alexandra L Gerber, Ana Paula de C Guimarães, Douglas Terra Machado, Cassia Alves, Diana Mariani, Thais Felix Cruz, Mario Sergio Ribeiro, Silvia Carvalho, Flávio Dias da Silva, Marcio Henrique de Oliveira Garcia, Leandro Magalhães de Souza, Cristiane Gomes da Silva, Caio Luiz Pereira Ribeiro, Andréa Cony Cavalcanti, Claudia Maria Braga de Mello, Amilcar Tanuri, Ana Tereza R Vasconcelos |
| EPI_ISL_1941583 | Laboratorio de Pesquisa em Virologia | Laboratorio de Pesquisa em Virologia | Cecília Artico Banho, Lívia Sacchetto, Fábio Sossai Possebon, Leila Sabrina Ullmann, Cíntia Bittar, Guilherme Campos, Helena Lage Ferreira, Jorge A. |

| | | | |
|---|---|---|---|
| | | | Petroli Marchesi, Maisa C. Pereira Parra, Marília Moraes, Paula Rahal, Paulo Inacio da Costa, João Pessoa Araújo Jr., Maurício Lacerda Nogueira. |
| EPI_ISL_1966059, EPI_ISL_1966060, EPI_ISL_1966061, EPI_ISL_1966062, EPI_ISL_1966063, EPI_ISL_1966064, EPI_ISL_1966065, EPI_ISL_1966066, EPI_ISL_1966067, EPI_ISL_1966068, EPI_ISL_1966069 | | | |
| see above | VIGILANCIA EPIDEMIOLOGICA | Instituto Butantan / Mendelics | Instituto Butantan: Dimas Tadeu Covas, Sandra Coccuzzo Sampaio, Maria Carolina Elias, José Salvatore Leister Patané, Vincent Louis Viala, Antonio Jorge Martins, Ricardo Haddad, Claudia Renata dos Santos Barros, Elaine Cristina Marqueze, Raul Machado Neto, Debora Botequio Moretti, Jardelina de Souza Todao Bernardino, Loyze Paola Oliveira de Lima, Luiz Aurelio de Campos Crispin. Centro de Genômica Funcional da ESALQ: Luiz Lehmann Coutinho, Ricardo Augusto Brassaloti, Raquel de Lello Rocha Campos Cassano. NGS Soluções Genômicas: Pilar Drummond Sampaio Corrêa Mariani. FZEA-USP Pirassununga: Mirele Daiana Poleti, Jessika Cristina Chagas Lesbon, Elisangela Chicaroni Mattos, Heidge Fukumasu. USP-Botucatu: Rejane Maria Tommasini Grotto, Jayme A. Souza-Neto, Guilherme Targino Valente, Patricia Akemi Assato, Felipe Allan da Silva da Costa, Bianca Cechetto Carlos. Mendelics: Bibiana Santos, João Paulo Kitajima, Erika Freitas, David Schlesinger. Hemocentro Ribeirão Preto: Simone Kashima, Evandra Strazza Rodrigues, Svetoslav Nanev Slavov, Elaine Vieira dos Santos, Rafael dos Santos Bezerra, Luiz Carlos Junior de Alcantara, Marta Giovanetti, Vagner Fonseca, Flavia Aburjaile, Rodrigo Tocantins Calado. FAMERP-SJRP: Cecília Artico Banho, Lívia Sacchetto, Fábio Sossai Possebon, Leila Sabrina Ullmann, Cíntia Bittar, Guilherme Campos, Helena Lage Ferreira, Jorge A. Petroli Marchesi, Maisa C. Pereira Parra, Marília Moraes, Paula Rahal, Paulo Inacio da Costa, João Pessoa Araújo Jr., Maurício Lacerda Nogueira. Prefeitura de Sao Paulo: Melissa Palmieri. |
| EPI_ISL_1966070, EPI_ISL_1966071, EPI_ISL_1966072, EPI_ISL_1966073, EPI_ISL_1966074, EPI_ISL_1966075, EPI_ISL_1966076, EPI_ISL_1966077, EPI_ISL_1966078, EPI_ISL_1966079 | SECAO CENTRO DE DIAGNOSTICO SECEDI | Instituto Butantan / Mendelics | Instituto Butantan: Dimas Tadeu Covas, Sandra Coccuzzo Sampaio, Maria Carolina Elias, José Salvatore Leister Patané, Vincent Louis Viala, Antonio Jorge Martins, Ricardo Haddad, Claudia Renata dos Santos Barros, Elaine Cristina Marqueze, Raul Machado Neto, Debora Botequio Moretti, Jardelina de Souza Todao Bernardino, Loyze Paola Oliveira de Lima, Luiz Aurelio de Campos Crispin. Centro de Genômica Funcional da ESALQ: Luiz Lehmann Coutinho, Ricardo Augusto Brassaloti, Raquel de Lello Rocha Campos Cassano. NGS Soluções Genômicas: Pilar Drummond Sampaio Corrêa Mariani. FZEA-USP Pirassununga: Mirele Daiana Poleti, Jessika Cristina Chagas Lesbon, Elisangela Chicaroni Mattos, Heidge Fukumasu. USP-Botucatu: Rejane Maria Tommasini Grotto, Jayme A. Souza-Neto, Guilherme Targino Valente, Patricia Akemi Assato, Felipe Allan da Silva da Costa, Bianca Cechetto Carlos. Mendelics: Bibiana Santos, João Paulo Kitajima, Erika Freitas, David Schlesinger. Hemocentro Ribeirão Preto: Simone Kashima, Evandra Strazza Rodrigues, Svetoslav Nanev Slavov, Elaine Vieira dos Santos, Rafael dos Santos Bezerra, Luiz Carlos Junior de Alcantara, Marta Giovanetti, Vagner Fonseca, Flavia Aburjaile, Rodrigo Tocantins Calado. FAMERP-SJRP: Cecília Artico Banho, Lívia Sacchetto, Fábio Sossai Possebon, Leila Sabrina Ullmann, Cíntia Bittar, Guilherme Campos, Helena Lage Ferreira, Jorge A. Petroli Marchesi, Maisa C. Pereira Parra, Marília Moraes, Paula Rahal, Paulo Inacio da Costa, João Pessoa Araújo Jr., Maurício Lacerda Nogueira. Prefeitura de Sao Paulo: Melissa Palmieri. |
| EPI_ISL_1966080, EPI_ISL_1966081, EPI_ISL_1966082, EPI_ISL_1966083, EPI_ISL_1966084, EPI_ISL_1966085, EPI_ISL_1966086, EPI_ISL_1966087, EPI_ISL_1966088, EPI_ISL_1966089, EPI_ISL_1966090, EPI_ISL_1966091, EPI_ISL_1966092 | | | |
| see above | VIGILANCIA EPIDEMIOLOGICA | Instituto Butantan / Mendelics | Instituto Butantan: Dimas Tadeu Covas, Sandra Coccuzzo Sampaio, Maria Carolina Elias, José Salvatore Leister Patané, Vincent Louis Viala, Antonio Jorge Martins, Ricardo Haddad, Claudia Renata dos Santos Barros, Elaine Cristina Marqueze, Raul Machado Neto, Debora Botequio Moretti, Jardelina de Souza Todao Bernardino, Loyze Paola Oliveira de Lima, Luiz Aurelio de Campos Crispin. Centro de Genômica Funcional da ESALQ: Luiz Lehmann Coutinho, Ricardo Augusto Brassaloti, Raquel de Lello Rocha Campos Cassano. NGS Soluções Genômicas: Pilar Drummond Sampaio Corrêa Mariani. FZEA-USP Pirassununga: Mirele Daiana Poleti, Jessika Cristina Chagas Lesbon, Elisangela Chicaroni Mattos, Heidge Fukumasu. USP-Botucatu: Rejane Maria Tommasini Grotto, Jayme A. Souza-Neto, Guilherme Targino Valente, Patricia Akemi Assato, Felipe Allan da Silva da Costa, Bianca Cechetto Carlos. Mendelics: Bibiana Santos, João Paulo Kitajima, Erika Freitas, David Schlesinger. Hemocentro Ribeirão Preto: Simone Kashima, Evandra Strazza Rodrigues, Svetoslav Nanev Slavov, Elaine Vieira dos Santos, Rafael dos Santos Bezerra, Luiz Carlos Junior de Alcantara, Marta Giovanetti, Vagner Fonseca, Flavia Aburjaile, Rodrigo Tocantins Calado. FAMERP-SJRP: Cecília Artico Banho, Lívia Sacchetto, Fábio Sossai Possebon, Leila Sabrina Ullmann, Cíntia Bittar, Guilherme Campos, Helena Lage Ferreira, Jorge A. Petroli Marchesi, Maisa C. Pereira Parra, Marília Moraes, Paula Rahal, Paulo Inacio da Costa, João Pessoa Araújo Jr., Maurício Lacerda Nogueira. Prefeitura de Sao Paulo: Melissa Palmieri. |
| EPI_ISL_1966093, EPI_ISL_1966094 | SERV DE VIG SANITARIA EPIDEMIO E CTRL DE ZOONOZES GUARUJA | Instituto Butantan / Mendelics | Instituto Butantan: Dimas Tadeu Covas, Sandra Coccuzzo Sampaio, Maria Carolina Elias, José Salvatore Leister Patané, Vincent Louis Viala, Antonio Jorge Martins, Ricardo Haddad, Claudia Renata dos Santos Barros, Elaine Cristina Marqueze, Raul Machado Neto, Debora Botequio Moretti, Jardelina de Souza Todao Bernardino, Loyze Paola Oliveira de Lima, Luiz Aurelio de Campos Crispin. Centro de Genômica Funcional da ESALQ: Luiz Lehmann Coutinho, Ricardo Augusto Brassaloti, Raquel de Lello Rocha Campos Cassano. NGS Soluções Genômicas: Pilar Drummond Sampaio Corrêa Mariani. FZEA-USP Pirassununga: Mirele Daiana Poleti, Jessika Cristina Chagas Lesbon, Elisangela Chicaroni Mattos, Heidge Fukumasu. USP-Botucatu: Rejane Maria Tommasini Grotto, Jayme A. Souza-Neto, Guilherme Targino Valente, Patricia Akemi Assato, Felipe Allan da Silva da Costa, Bianca Cechetto Carlos. Mendelics: Bibiana Santos, João Paulo Kitajima, Erika Freitas, David Schlesinger. Hemocentro Ribeirão Preto: Simone Kashima, Evandra Strazza Rodrigues, Svetoslav Nanev Slavov, Elaine Vieira dos Santos, Rafael dos Santos Bezerra, Luiz Carlos Junior de Alcantara, Marta Giovanetti, Vagner Fonseca, Flavia Aburjaile, Rodrigo Tocantins Calado. FAMERP-SJRP: Cecília Artico Banho, Lívia Sacchetto, Fábio Sossai Possebon, Leila Sabrina Ullmann, Cíntia Bittar, Guilherme Campos, Helena Lage Ferreira, Jorge A. Petroli Marchesi, Maisa C. Pereira Parra, Marília Moraes, Paula Rahal, Paulo Inacio da Costa, João Pessoa Araújo Jr., Maurício Lacerda Nogueira. Prefeitura de Sao Paulo: Melissa Palmieri. |
| EPI_ISL_1966095, EPI_ISL_1966096, EPI_ISL_1966097, EPI_ISL_1966098 | AMBULATORIO MEDICO DE ESPECIALIDADES DE PERUIBE | Instituto Butantan / Mendelics | Instituto Butantan: Dimas Tadeu Covas, Sandra Coccuzzo Sampaio, Maria Carolina Elias, José Salvatore Leister Patané, Vincent Louis Viala, Antonio Jorge Martins, Ricardo Haddad, Claudia Renata dos Santos Barros, Elaine Cristina Marqueze, Raul Machado Neto, Debora Botequio Moretti, Jardelina de Souza Todao Bernardino, Loyze Paola Oliveira de Lima, Luiz Aurelio de Campos Crispin. Centro de Genômica Funcional da ESALQ: Luiz Lehmann Coutinho, Ricardo Augusto Brassaloti, Raquel de Lello Rocha Campos Cassano. NGS Soluções Genômicas: Pilar Drummond Sampaio Corrêa Mariani. FZEA-USP Pirassununga: Mirele Daiana Poleti, Jessika Cristina Chagas Lesbon, Elisangela Chicaroni Mattos, Heidge Fukumasu. USP-Botucatu: Rejane Maria Tommasini Grotto, Jayme A. Souza-Neto, Guilherme Targino Valente, Patricia Akemi Assato, Felipe Allan da Silva da Costa, Bianca Cechetto Carlos. Mendelics: Bibiana Santos, João Paulo Kitajima, Erika Freitas, David Schlesinger. Hemocentro Ribeirão Preto: Simone Kashima, Evandra Strazza Rodrigues, Svetoslav Nanev Slavov, Elaine Vieira dos Santos, Rafael dos Santos Bezerra, Luiz Carlos Junior de Alcantara, Marta Giovanetti, Vagner Fonseca, Flavia Aburjaile, Rodrigo Tocantins Calado. FAMERP-SJRP: Cecília Artico Banho, Lívia Sacchetto, Fábio Sossai Possebon, Leila Sabrina Ullmann, Cíntia Bittar, Guilherme Campos, Helena Lage Ferreira, Jorge A. Petroli Marchesi, Maisa C. Pereira Parra, Marília Moraes, Paula Rahal, Paulo Inacio da Costa, João Pessoa Araújo Jr., Maurício Lacerda Nogueira. Prefeitura de Sao Paulo: Melissa Palmieri. |
| EPI_ISL_1966099, EPI_ISL_1966100, EPI_ISL_1966101 | SECAO CENTRO DE DIAGNOSTICO SECEDI | Instituto Butantan / Mendelics | Instituto Butantan: Dimas Tadeu Covas, Sandra Coccuzzo Sampaio, Maria Carolina Elias, José Salvatore Leister Patané, Vincent Louis Viala, Antonio Jorge Martins, Ricardo Haddad, Claudia Renata dos Santos Barros, Elaine Cristina Marqueze, Raul Machado Neto, Debora Botequio Moretti, Jardelina de Souza Todao Bernardino, Loyze Paola Oliveira de Lima, Luiz Aurelio de Campos Crispin. Centro de Genômica Funcional da ESALQ: Luiz Lehmann Coutinho, Ricardo Augusto Brassaloti, Raquel de Lello Rocha Campos Cassano. NGS Soluções Genômicas: Pilar Drummond Sampaio Corrêa Mariani. FZEA-USP Pirassununga: Mirele Daiana Poleti, Jessika Cristina Chagas Lesbon, Elisangela Chicaroni Mattos, Heidge Fukumasu. USP-Botucatu: Rejane Maria Tommasini Grotto, Jayme A. Souza-Neto, Guilherme Targino Valente, Patricia Akemi Assato, Felipe Allan da Silva da Costa, Bianca Cechetto Carlos. Mendelics: Bibiana Santos, João Paulo Kitajima, Erika Freitas, David Schlesinger. Hemocentro Ribeirão Preto: Simone Kashima, Evandra Strazza Rodrigues, Svetoslav Nanev Slavov, Elaine Vieira dos Santos, Rafael dos Santos Bezerra, Luiz Carlos Junior de Alcantara, Marta Giovanetti, Vagner Fonseca, Flavia Aburjaile, Rodrigo Tocantins Calado. FAMERP-SJRP: Cecília Artico Banho, Lívia Sacchetto, Fábio Sossai Possebon, Leila Sabrina Ullmann, Cíntia Bittar, Guilherme Campos, Helena Lage Ferreira, Jorge A. Petroli Marchesi, Maisa C. Pereira Parra, Marília Moraes, Paula Rahal, Paulo Inacio da Costa, João Pessoa Araújo Jr., Maurício Lacerda Nogueira. Prefeitura de Sao Paulo: Melissa Palmieri. |
| EPI_ISL_1966102 | USF TRES PONTES | Instituto Butantan / Mendelics | Instituto Butantan: Dimas Tadeu Covas, Sandra Coccuzzo Sampaio, Maria Carolina Elias, José Salvatore Leister Patané, Vincent Louis Viala, Antonio Jorge Martins, Ricardo Haddad, Claudia Renata dos Santos Barros, Elaine Cristina Marqueze, Raul Machado Neto, Debora Botequio Moretti, Jardelina de Souza Todao Bernardino, Loyze Paola Oliveira de Lima, Luiz Aurelio de Campos Crispin. Centro de Genômica Funcional da ESALQ: Luiz Lehmann Coutinho, Ricardo Augusto Brassaloti, Raquel de Lello Rocha Campos Cassano. NGS Soluções Genômicas: Pilar Drummond Sampaio Corrêa Mariani. FZEA-USP Pirassununga: Mirele Daiana Poleti, Jessika Cristina Chagas Lesbon, Elisangela Chicaroni Mattos, Heidge Fukumasu. USP-Botucatu: Rejane Maria Tommasini Grotto, Jayme A. Souza-Neto, Guilherme Targino Valente, Patricia Akemi Assato, Felipe Allan da Silva da Costa, Bianca Cechetto Carlos. Mendelics: Bibiana Santos, João Paulo Kitajima, Erika Freitas, David Schlesinger. Hemocentro Ribeirão Preto: Simone Kashima, Evandra Strazza Rodrigues, Svetoslav Nanev Slavov, Elaine Vieira dos Santos, Rafael dos Santos Bezerra, Luiz Carlos Junior de Alcantara, Marta Giovanetti, Vagner Fonseca, Flavia Aburjaile, Rodrigo Tocantins Calado. FAMERP-SJRP: Cecília Artico Banho, Lívia Sacchetto, Fábio Sossai Possebon, Leila Sabrina Ullmann, Cíntia Bittar, Guilherme Campos, Helena Lage Ferreira, Jorge A. Petroli Marchesi, Maisa C. Pereira Parra, Marília Moraes, Paula Rahal, Paulo Inacio da Costa, João Pessoa Araújo Jr., Maurício Lacerda Nogueira. Prefeitura de Sao Paulo: Melissa Palmieri. |
| EPI_ISL_1966103 | SMS SECRETARIA MUNICIPAL DE SAUDE DE BOITUVA | Instituto Butantan / Mendelics | Instituto Butantan: Dimas Tadeu Covas, Sandra Coccuzzo Sampaio, Maria Carolina Elias, José Salvatore Leister Patané, Vincent Louis Viala, Antonio Jorge Martins, Ricardo Haddad, Claudia Renata dos Santos Barros, Elaine Cristina Marqueze, Raul Machado Neto, Debora Botequio Moretti, Jardelina de |

| | | | |
|---|---|---|---|
| | | | Souza Todao Bernardino, Loyze Paola Oliveira de Lima, Luiz Aurelio de Campos Crispin. Centro de Genômica Funcional da ESALQ: Luiz Lehmann Coutinho, Ricardo Augusto Brassaloti, Raquel de Lello Rocha Campos Cassano. NGS Soluções Genômicas: Pilar Drummond Sampaio Corrêa Mariani. FZEA-USP Pirassununga: Mirele Daiana Poleti, Jessika Cristina Chagas Lesbon, Elisangela Chicaroni Mattos, Heidge Fukumasu. USP-Botucatu: Rejane Maria Tommasini Grotto, Jayme A. Souza-Neto, Guilherme Targino Valente, Patricia Akemi Assato, Felipe Allan da Silva da Costa, Bianca Cechetto Carlos. Mendelics: Bibiana Santos, João Paulo Kitajima, Erika Freitas, David Schlesinger. Hemocentro Ribeirão Preto: Simone Kashima, Evandra Strazza Rodrigues, Svetoslav Nanev Slavov, Elaine Vieira dos Santos, Rafael dos Santos Bezerra, Luiz Carlos Junior de Alcantara, Marta Giovanetti, Vagner Fonseca, Flavia Aburjaile, Rodrigo Tocantins Calado. FAMERP-SJRP: Cecília Artico Banho, Lívia Sacchetto, Fábio Sossai Possebon, Leila Sabrina Ullmann, Cíntia Bittar, Guilherme Campos, Helena Lage Ferreira, Jorge A. Petroli Marchesi, Maisa C. Pereira Parra, Marília Moraes, Paula Rahal, Paulo Inacio da Costa, João Pessoa Araújo Jr., Maurício Lacerda Nogueira. Prefeitura de Sao Paulo: Melissa Palmieri. |
| EPI_ISL_1966104 | POLICLINICA COVID 19 ITAPETININGA | Instituto Butantan / Mendelics | Instituto Butantan: Dimas Tadeu Covas, Sandra Coccuzzo Sampaio, Maria Carolina Elias, José Salvatore Leister Patané, Vincent Louis Viala, Antonio Jorge Martins, Ricardo Haddad, Claudia Renata dos Santos Barros, Elaine Cristina Marqueze, Raul Machado Neto, Debora Botequio Moretti, Jardelina de Souza Todao Bernardino, Loyze Paola Oliveira de Lima, Luiz Aurelio de Campos Crispin. Centro de Genômica Funcional da ESALQ: Luiz Lehmann Coutinho, Ricardo Augusto Brassaloti, Raquel de Lello Rocha Campos Cassano. NGS Soluções Genômicas: Pilar Drummond Sampaio Corrêa Mariani. FZEA-USP Pirassununga: Mirele Daiana Poleti, Jessika Cristina Chagas Lesbon, Elisangela Chicaroni Mattos, Heidge Fukumasu. USP-Botucatu: Rejane Maria Tommasini Grotto, Jayme A. Souza-Neto, Guilherme Targino Valente, Patricia Akemi Assato, Felipe Allan da Silva da Costa, Bianca Cechetto Carlos. Mendelics: Bibiana Santos, João Paulo Kitajima, Erika Freitas, David Schlesinger. Hemocentro Ribeirão Preto: Simone Kashima, Evandra Strazza Rodrigues, Svetoslav Nanev Slavov, Elaine Vieira dos Santos, Rafael dos Santos Bezerra, Luiz Carlos Junior de Alcantara, Marta Giovanetti, Vagner Fonseca, Flavia Aburjaile, Rodrigo Tocantins Calado. FAMERP-SJRP: Cecília Artico Banho, Lívia Sacchetto, Fábio Sossai Possebon, Leila Sabrina Ullmann, Cíntia Bittar, Guilherme Campos, Helena Lage Ferreira, Jorge A. Petroli Marchesi, Maisa C. Pereira Parra, Marília Moraes, Paula Rahal, Paulo Inacio da Costa, João Pessoa Araújo Jr., Maurício Lacerda Nogueira. Prefeitura de Sao Paulo: Melissa Palmieri. |
| EPI_ISL_1966105 | HOSPITAL REGIONAL DE ITAPETININGA | Instituto Butantan / Mendelics | Instituto Butantan: Dimas Tadeu Covas, Sandra Coccuzzo Sampaio, Maria Carolina Elias, José Salvatore Leister Patané, Vincent Louis Viala, Antonio Jorge Martins, Ricardo Haddad, Claudia Renata dos Santos Barros, Elaine Cristina Marqueze, Raul Machado Neto, Debora Botequio Moretti, Jardelina de Souza Todao Bernardino, Loyze Paola Oliveira de Lima, Luiz Aurelio de Campos Crispin. Centro de Genômica Funcional da ESALQ: Luiz Lehmann Coutinho, Ricardo Augusto Brassaloti, Raquel de Lello Rocha Campos Cassano. NGS Soluções Genômicas: Pilar Drummond Sampaio Corrêa Mariani. FZEA-USP Pirassununga: Mirele Daiana Poleti, Jessika Cristina Chagas Lesbon, Elisangela Chicaroni Mattos, Heidge Fukumasu. USP-Botucatu: Rejane Maria Tommasini Grotto, Jayme A. Souza-Neto, Guilherme Targino Valente, Patricia Akemi Assato, Felipe Allan da Silva da Costa, Bianca Cechetto Carlos. Mendelics: Bibiana Santos, João Paulo Kitajima, Erika Freitas, David Schlesinger. Hemocentro Ribeirão Preto: Simone Kashima, Evandra Strazza Rodrigues, Svetoslav Nanev Slavov, Elaine Vieira dos Santos, Rafael dos Santos Bezerra, Luiz Carlos Junior de Alcantara, Marta Giovanetti, Vagner Fonseca, Flavia Aburjaile, Rodrigo Tocantins Calado. FAMERP-SJRP: Cecília Artico Banho, Lívia Sacchetto, Fábio Sossai Possebon, Leila Sabrina Ullmann, Cíntia Bittar, Guilherme Campos, Helena Lage Ferreira, Jorge A. Petroli Marchesi, Maisa C. Pereira Parra, Marília Moraes, Paula Rahal, Paulo Inacio da Costa, João Pessoa Araújo Jr., Maurício Lacerda Nogueira. Prefeitura de Sao Paulo: Melissa Palmieri. |
| EPI_ISL_1966106 | CENTRO DE SAUDE II MAIRINQUE MAIRINQUE | Instituto Butantan / Mendelics | Instituto Butantan: Dimas Tadeu Covas, Sandra Coccuzzo Sampaio, Maria Carolina Elias, José Salvatore Leister Patané, Vincent Louis Viala, Antonio Jorge Martins, Ricardo Haddad, Claudia Renata dos Santos Barros, Elaine Cristina Marqueze, Raul Machado Neto, Debora Botequio Moretti, Jardelina de Souza Todao Bernardino, Loyze Paola Oliveira de Lima, Luiz Aurelio de Campos Crispin. Centro de Genômica Funcional da ESALQ: Luiz Lehmann Coutinho, Ricardo Augusto Brassaloti, Raquel de Lello Rocha Campos Cassano. NGS Soluções Genômicas: Pilar Drummond Sampaio Corrêa Mariani. FZEA-USP Pirassununga: Mirele Daiana Poleti, Jessika Cristina Chagas Lesbon, Elisangela Chicaroni Mattos, Heidge Fukumasu. USP-Botucatu: Rejane Maria Tommasini Grotto, Jayme A. Souza-Neto, Guilherme Targino Valente, Patricia Akemi Assato, Felipe Allan da Silva da Costa, Bianca Cechetto Carlos. Mendelics: Bibiana Santos, João Paulo Kitajima, Erika Freitas, David Schlesinger. Hemocentro Ribeirão Preto: Simone Kashima, Evandra Strazza Rodrigues, Svetoslav Nanev Slavov, Elaine Vieira dos Santos, Rafael dos Santos Bezerra, Luiz Carlos Junior de Alcantara, Marta Giovanetti, Vagner Fonseca, Flavia Aburjaile, Rodrigo Tocantins Calado. FAMERP-SJRP: Cecília Artico Banho, Lívia Sacchetto, Fábio Sossai Possebon, Leila Sabrina Ullmann, Cíntia Bittar, Guilherme Campos, Helena Lage Ferreira, Jorge A. Petroli Marchesi, Maisa C. Pereira Parra, Marília Moraes, Paula Rahal, Paulo Inacio da Costa, João Pessoa Araújo Jr., Maurício Lacerda Nogueira. Prefeitura de Sao Paulo: Melissa Palmieri. |
| EPI_ISL_1966107 | SECRETARIA MUNICIPAL DE SAUDE SOROCABA | Instituto Butantan / Mendelics | Instituto Butantan: Dimas Tadeu Covas, Sandra Coccuzzo Sampaio, Maria Carolina Elias, José Salvatore Leister Patané, Vincent Louis Viala, Antonio Jorge Martins, Ricardo Haddad, Claudia Renata dos Santos Barros, Elaine Cristina Marqueze, Raul Machado Neto, Debora Botequio Moretti, Jardelina de Souza Todao Bernardino, Loyze Paola Oliveira de Lima, Luiz Aurelio de Campos Crispin. Centro de Genômica Funcional da ESALQ: Luiz Lehmann Coutinho, Ricardo Augusto Brassaloti, Raquel de Lello Rocha Campos Cassano. NGS Soluções Genômicas: Pilar Drummond Sampaio Corrêa Mariani. FZEA-USP Pirassununga: Mirele Daiana Poleti, Jessika Cristina Chagas Lesbon, Elisangela Chicaroni Mattos, Heidge Fukumasu. USP-Botucatu: Rejane Maria Tommasini Grotto, Jayme A. Souza-Neto, Guilherme Targino Valente, Patricia Akemi Assato, Felipe Allan da Silva da Costa, Bianca Cechetto Carlos. Mendelics: Bibiana Santos, João Paulo Kitajima, Erika Freitas, David Schlesinger. Hemocentro Ribeirão Preto: Simone Kashima, Evandra Strazza Rodrigues, Svetoslav Nanev Slavov, Elaine Vieira dos Santos, Rafael dos Santos Bezerra, Luiz Carlos Junior de Alcantara, Marta Giovanetti, Vagner Fonseca, Flavia Aburjaile, Rodrigo Tocantins Calado. FAMERP-SJRP: Cecília Artico Banho, Lívia Sacchetto, Fábio Sossai Possebon, Leila Sabrina Ullmann, Cíntia Bittar, Guilherme Campos, Helena Lage Ferreira, Jorge A. Petroli Marchesi, Maisa C. Pereira Parra, Marília Moraes, Paula Rahal, Paulo Inacio da Costa, João Pessoa Araújo Jr., Maurício Lacerda Nogueira. Prefeitura de Sao Paulo: Melissa Palmieri. |
| EPI_ISL_1966108 | CENTRO DE SAUDE II DR GABRIEL MESQUITA VARGEM GDE DO SUL | Instituto Butantan / Mendelics | Dimas Tadeu Covas, Antonio Jorge Martins, Claudia Renata dos Santos Barros, David Schlesinger, Debora Botequio Moretti, Elaine Cristina Marqueze, Elaine Vieira Santos, Evandra Strazza Rodrigues, Heidge Fukumasu, Jayme Augusto de Souza-Neto, José Salvatore Leister Patané, Luiz Alcantara, Luiz Lehmann Coutinho, Maria Carolina Elias, Maurício Lacerda Nogueira, Rafael dos Santos Bezerra, Raul Machado Neto, Rejane Maria Tommasini Grotto, Ricardo Haddad, Sandra Coccuzzo Sampaio Vessoni, Simone Kashima, Svetoslav Nanev Slavov, Vincent Louis Viala |
| EPI_ISL_1966109, EPI_ISL_1966110 | CENTRO DE SAUDE II DR GABRIEL MESQUITA VARGEM GDE DO SUL | Instituto Butantan / Mendelics | Instituto Butantan: Dimas Tadeu Covas, Sandra Coccuzzo Sampaio, Maria Carolina Elias, José Salvatore Leister Patané, Vincent Louis Viala, Antonio Jorge Martins, Ricardo Haddad, Claudia Renata dos Santos Barros, Elaine Cristina Marqueze, Raul Machado Neto, Debora Botequio Moretti, Jardelina de Souza Todao Bernardino, Loyze Paola Oliveira de Lima, Luiz Aurelio de Campos Crispin. Centro de Genômica Funcional da ESALQ: Luiz Lehmann Coutinho, Ricardo Augusto Brassaloti, Raquel de Lello Rocha Campos Cassano. NGS Soluções Genômicas: Pilar Drummond Sampaio Corrêa Mariani. FZEA-USP Pirassununga: Mirele Daiana Poleti, Jessika Cristina Chagas Lesbon, Elisangela Chicaroni Mattos, Heidge Fukumasu. USP-Botucatu: Rejane Maria Tommasini Grotto, Jayme A. Souza-Neto, Guilherme Targino Valente, Patricia Akemi Assato, Felipe Allan da Silva da Costa, Bianca Cechetto Carlos. Mendelics: Bibiana Santos, João Paulo Kitajima, Erika Freitas, David Schlesinger. Hemocentro Ribeirão Preto: Simone Kashima, Evandra Strazza Rodrigues, Svetoslav Nanev Slavov, Elaine Vieira dos Santos, Rafael dos Santos Bezerra, Luiz Carlos Junior de Alcantara, Marta Giovanetti, Vagner Fonseca, Flavia Aburjaile, Rodrigo Tocantins Calado. FAMERP-SJRP: Cecília Artico Banho, Lívia Sacchetto, Fábio Sossai Possebon, Leila Sabrina Ullmann, Cíntia Bittar, Guilherme Campos, Helena Lage Ferreira, Jorge A. Petroli Marchesi, Maisa C. Pereira Parra, Marília Moraes, Paula Rahal, Paulo Inacio da Costa, João Pessoa Araújo Jr., Maurício Lacerda Nogueira. Prefeitura de Sao Paulo: Melissa Palmieri. |
| EPI_ISL_1966111 | UNIDADE MISTA ARACOIABA DA SERRA | Instituto Butantan / Mendelics | Dimas Tadeu Covas, Antonio Jorge Martins, Claudia Renata dos Santos Barros, David Schlesinger, Debora Botequio Moretti, Elaine Cristina Marqueze, Elaine Vieira Santos, Evandra Strazza Rodrigues, Heidge Fukumasu, Jayme Augusto de Souza-Neto, José Salvatore Leister Patané, Luiz Alcantara, Luiz Lehmann Coutinho, Maria Carolina Elias, Maurício Lacerda Nogueira, Rafael dos Santos Bezerra, Raul Machado Neto, Rejane Maria Tommasini Grotto, Ricardo Haddad, Sandra Coccuzzo Sampaio Vessoni, Simone Kashima, Svetoslav Nanev Slavov, Vincent Louis Viala |
| EPI_ISL_1966112 | SAUDE COLETIVA CAPAO BONITO | Instituto Butantan / Mendelics | Instituto Butantan: Dimas Tadeu Covas, Sandra Coccuzzo Sampaio, Maria Carolina Elias, José Salvatore Leister Patané, Vincent Louis Viala, Antonio Jorge Martins, Ricardo Haddad, Claudia Renata dos Santos Barros, Elaine Cristina Marqueze, Raul Machado Neto, Debora Botequio Moretti, Jardelina de Souza Todao Bernardino, Loyze Paola Oliveira de Lima, Luiz Aurelio de Campos Crispin. Centro de Genômica Funcional da ESALQ: Luiz Lehmann Coutinho, Ricardo Augusto Brassaloti, Raquel de Lello Rocha Campos Cassano. NGS Soluções Genômicas: Pilar Drummond Sampaio Corrêa Mariani. FZEA-USP Pirassununga: Mirele Daiana Poleti, Jessika Cristina Chagas Lesbon, Elisangela Chicaroni Mattos, Heidge Fukumasu. USP-Botucatu: Rejane Maria Tommasini Grotto, Jayme A. Souza-Neto, Guilherme Targino Valente, Patricia Akemi Assato, Felipe Allan da Silva da Costa, Bianca Cechetto Carlos. Mendelics: Bibiana Santos, João Paulo Kitajima, Erika Freitas, David Schlesinger. Hemocentro Ribeirão Preto: Simone Kashima, Evandra Strazza Rodrigues, Svetoslav Nanev Slavov, Elaine Vieira dos Santos, Rafael dos Santos Bezerra, Luiz Carlos Junior de Alcantara, Marta Giovanetti, Vagner Fonseca, Flavia Aburjaile, Rodrigo Tocantins Calado. FAMERP-SJRP: Cecília Artico Banho, Lívia Sacchetto, Fábio Sossai Possebon, Leila Sabrina Ullmann, Cíntia Bittar, Guilherme Campos, Helena Lage Ferreira, Jorge A. Petroli Marchesi, Maisa C. Pereira Parra, Marília Moraes, Paula Rahal, Paulo Inacio da Costa, João Pessoa Araújo Jr., Maurício Lacerda Nogueira. Prefeitura de Sao Paulo: Melissa Palmieri. |

| | | | |
|---|---|---|---|
| EPI_ISL_1966113, EPI_ISL_1966114, EPI_ISL_1966115, EPI_ISL_1966116, EPI_ISL_1966117 | CS II EGIDIO BRUNHARA MORRO AGUDO | Instituto Butantan / Mendelics | Instituto Butantan: Dimas Tadeu Covas, Sandra Coccuzzo Sampaio, Maria Carolina Elias, José Salvatore Leister Patané, Vincent Louis Viala, Antonio Jorge Martins, Ricardo Haddad, Claudia Renata dos Santos Barros, Elaine Cristina Marqueze, Raul Machado Neto, Debora Botequio Moretti, Jardelina de Souza Todao Bernardino, Loyze Paola Oliveira de Lima, Luiz Aurelio de Campos Crispin. Centro de Genômica Funcional da ESALQ: Luiz Lehmann Coutinho, Ricardo Augusto Brassaloti, Raquel de Lello Rocha Campos Cassano. NGS Soluções Genômicas: Pilar Drummond Sampaio Corrêa Mariani. FZEA-USP Pirassununga: Mirele Daiana Poleti, Jessika Cristina Chagas Lesbon, Elisangela Chicaroni Mattos, Heidge Fukumasu. USP-Botucatu: Rejane Maria Tommasini Grotto, Jayme A. Souza-Neto, Guilherme Targino Valente, Patricia Akemi Assato, Felipe Allan da Silva da Costa, Bianca Cechetto Carlos. Mendelics: Bibiana Santos, João Paulo Kitajima, Erika Freitas, David Schlesinger. Hemocentro Ribeirão Preto: Simone Kashima, Evandra Strazza Rodrigues, Svetoslav Nanev Slavov, Elaine Vieira dos Santos, Rafael dos Santos Bezerra, Luiz Carlos Junior de Alcantara, Marta Giovanetti, Vagner Fonseca, Flavia Aburjaile, Rodrigo Tocantins Calado. FAMERP-SJRP: Cecília Artico Banho, Lívia Sacchetto, Fábio Sossai Possebon, Leila Sabrina Ullmann, Cíntia Bittar, Guilherme Campos, Helena Lage Ferreira, Jorge A. Petroli Marchesi, Maisa C. Pereira Parra, Marília Moraes, Paula Rahal, Paulo Inacio da Costa, João Pessoa Araújo Jr., Maurício Lacerda Nogueira. Prefeitura de Sao Paulo: Melissa Palmieri. |
| EPI_ISL_1966118, EPI_ISL_1966119, EPI_ISL_1966120, EPI_ISL_1966121, EPI_ISL_1966122, EPI_ISL_1966123 | CENTRO DE REFERENCIA DO IDOSO DR HUMBERTO MENDES DE CARVALHO | Instituto Butantan / Mendelics | Instituto Butantan: Dimas Tadeu Covas, Sandra Coccuzzo Sampaio, Maria Carolina Elias, José Salvatore Leister Patané, Vincent Louis Viala, Antonio Jorge Martins, Ricardo Haddad, Claudia Renata dos Santos Barros, Elaine Cristina Marqueze, Raul Machado Neto, Debora Botequio Moretti, Jardelina de Souza Todao Bernardino, Loyze Paola Oliveira de Lima, Luiz Aurelio de Campos Crispin. Centro de Genômica Funcional da ESALQ: Luiz Lehmann Coutinho, Ricardo Augusto Brassaloti, Raquel de Lello Rocha Campos Cassano. NGS Soluções Genômicas: Pilar Drummond Sampaio Corrêa Mariani. FZEA-USP Pirassununga: Mirele Daiana Poleti, Jessika Cristina Chagas Lesbon, Elisangela Chicaroni Mattos, Heidge Fukumasu. USP-Botucatu: Rejane Maria Tommasini Grotto, Jayme A. Souza-Neto, Guilherme Targino Valente, Patricia Akemi Assato, Felipe Allan da Silva da Costa, Bianca Cechetto Carlos. Mendelics: Bibiana Santos, João Paulo Kitajima, Erika Freitas, David Schlesinger. Hemocentro Ribeirão Preto: Simone Kashima, Evandra Strazza Rodrigues, Svetoslav Nanev Slavov, Elaine Vieira dos Santos, Rafael dos Santos Bezerra, Luiz Carlos Junior de Alcantara, Marta Giovanetti, Vagner Fonseca, Flavia Aburjaile, Rodrigo Tocantins Calado. FAMERP-SJRP: Cecília Artico Banho, Lívia Sacchetto, Fábio Sossai Possebon, Leila Sabrina Ullmann, Cíntia Bittar, Guilherme Campos, Helena Lage Ferreira, Jorge A. Petroli Marchesi, Maisa C. Pereira Parra, Marília Moraes, Paula Rahal, Paulo Inacio da Costa, João Pessoa Araújo Jr., Maurício Lacerda Nogueira. Prefeitura de Sao Paulo: Melissa Palmieri. |
| EPI_ISL_1966124, EPI_ISL_1966125 | SECRETARIA MUNICIPAL DE SAUDE SOROCABA | Instituto Butantan / Mendelics | Instituto Butantan: Dimas Tadeu Covas, Sandra Coccuzzo Sampaio, Maria Carolina Elias, José Salvatore Leister Patané, Vincent Louis Viala, Antonio Jorge Martins, Ricardo Haddad, Claudia Renata dos Santos Barros, Elaine Cristina Marqueze, Raul Machado Neto, Debora Botequio Moretti, Jardelina de Souza Todao Bernardino, Loyze Paola Oliveira de Lima, Luiz Aurelio de Campos Crispin. Centro de Genômica Funcional da ESALQ: Luiz Lehmann Coutinho, Ricardo Augusto Brassaloti, Raquel de Lello Rocha Campos Cassano. NGS Soluções Genômicas: Pilar Drummond Sampaio Corrêa Mariani. FZEA-USP Pirassununga: Mirele Daiana Poleti, Jessika Cristina Chagas Lesbon, Elisangela Chicaroni Mattos, Heidge Fukumasu. USP-Botucatu: Rejane Maria Tommasini Grotto, Jayme A. Souza-Neto, Guilherme Targino Valente, Patricia Akemi Assato, Felipe Allan da Silva da Costa, Bianca Cechetto Carlos. Mendelics: Bibiana Santos, João Paulo Kitajima, Erika Freitas, David Schlesinger. Hemocentro Ribeirão Preto: Simone Kashima, Evandra Strazza Rodrigues, Svetoslav Nanev Slavov, Elaine Vieira dos Santos, Rafael dos Santos Bezerra, Luiz Carlos Junior de Alcantara, Marta Giovanetti, Vagner Fonseca, Flavia Aburjaile, Rodrigo Tocantins Calado. FAMERP-SJRP: Cecília Artico Banho, Lívia Sacchetto, Fábio Sossai Possebon, Leila Sabrina Ullmann, Cíntia Bittar, Guilherme Campos, Helena Lage Ferreira, Jorge A. Petroli Marchesi, Maisa C. Pereira Parra, Marília Moraes, Paula Rahal, Paulo Inacio da Costa, João Pessoa Araújo Jr., Maurício Lacerda Nogueira. Prefeitura de Sao Paulo: Melissa Palmieri. |
| EPI_ISL_1966126, EPI_ISL_1966127, EPI_ISL_1966128 | SAE SERVICO DE ATENDIMENTO ESPECIALIZADO | Instituto Butantan / Mendelics | Instituto Butantan: Dimas Tadeu Covas, Sandra Coccuzzo Sampaio, Maria Carolina Elias, José Salvatore Leister Patané, Vincent Louis Viala, Antonio Jorge Martins, Ricardo Haddad, Claudia Renata dos Santos Barros, Elaine Cristina Marqueze, Raul Machado Neto, Debora Botequio Moretti, Jardelina de Souza Todao Bernardino, Loyze Paola Oliveira de Lima, Luiz Aurelio de Campos Crispin. Centro de Genômica Funcional da ESALQ: Luiz Lehmann Coutinho, Ricardo Augusto Brassaloti, Raquel de Lello Rocha Campos Cassano. NGS Soluções Genômicas: Pilar Drummond Sampaio Corrêa Mariani. FZEA-USP Pirassununga: Mirele Daiana Poleti, Jessika Cristina Chagas Lesbon, Elisangela Chicaroni Mattos, Heidge Fukumasu. USP-Botucatu: Rejane Maria Tommasini Grotto, Jayme A. Souza-Neto, Guilherme Targino Valente, Patricia Akemi Assato, Felipe Allan da Silva da Costa, Bianca Cechetto Carlos. Mendelics: Bibiana Santos, João Paulo Kitajima, Erika Freitas, David Schlesinger. Hemocentro Ribeirão Preto: Simone Kashima, Evandra Strazza Rodrigues, Svetoslav Nanev Slavov, Elaine Vieira dos Santos, Rafael dos Santos Bezerra, Luiz Carlos Junior de Alcantara, Marta Giovanetti, Vagner Fonseca, Flavia Aburjaile, Rodrigo Tocantins Calado. FAMERP-SJRP: Cecília Artico Banho, Lívia Sacchetto, Fábio Sossai Possebon, Leila Sabrina Ullmann, Cíntia Bittar, Guilherme Campos, Helena Lage Ferreira, Jorge A. Petroli Marchesi, Maisa C. Pereira Parra, Marília Moraes, Paula Rahal, Paulo Inacio da Costa, João Pessoa Araújo Jr., Maurício Lacerda Nogueira. Prefeitura de Sao Paulo: Melissa Palmieri. |
| EPI_ISL_1966129 | PRONTO SOCORRO MUNICIPAL DE TATUI | Instituto Butantan / Mendelics | Instituto Butantan: Dimas Tadeu Covas, Sandra Coccuzzo Sampaio, Maria Carolina Elias, José Salvatore Leister Patané, Vincent Louis Viala, Antonio Jorge Martins, Ricardo Haddad, Claudia Renata dos Santos Barros, Elaine Cristina Marqueze, Raul Machado Neto, Debora Botequio Moretti, Jardelina de Souza Todao Bernardino, Loyze Paola Oliveira de Lima, Luiz Aurelio de Campos Crispin. Centro de Genômica Funcional da ESALQ: Luiz Lehmann Coutinho, Ricardo Augusto Brassaloti, Raquel de Lello Rocha Campos Cassano. NGS Soluções Genômicas: Pilar Drummond Sampaio Corrêa Mariani. FZEA-USP Pirassununga: Mirele Daiana Poleti, Jessika Cristina Chagas Lesbon, Elisangela Chicaroni Mattos, Heidge Fukumasu. USP-Botucatu: Rejane Maria Tommasini Grotto, Jayme A. Souza-Neto, Guilherme Targino Valente, Patricia Akemi Assato, Felipe Allan da Silva da Costa, Bianca Cechetto Carlos. Mendelics: Bibiana Santos, João Paulo Kitajima, Erika Freitas, David Schlesinger. Hemocentro Ribeirão Preto: Simone Kashima, Evandra Strazza Rodrigues, Svetoslav Nanev Slavov, Elaine Vieira dos Santos, Rafael dos Santos Bezerra, Luiz Carlos Junior de Alcantara, Marta Giovanetti, Vagner Fonseca, Flavia Aburjaile, Rodrigo Tocantins Calado. FAMERP-SJRP: Cecília Artico Banho, Lívia Sacchetto, Fábio Sossai Possebon, Leila Sabrina Ullmann, Cíntia Bittar, Guilherme Campos, Helena Lage Ferreira, Jorge A. Petroli Marchesi, Maisa C. Pereira Parra, Marília Moraes, Paula Rahal, Paulo Inacio da Costa, João Pessoa Araújo Jr., Maurício Lacerda Nogueira. Prefeitura de Sao Paulo: Melissa Palmieri. |
| EPI_ISL_1966130 | DEPARTAMENTO DE SAUDE COLETIVA | Instituto Butantan / Mendelics | Instituto Butantan: Dimas Tadeu Covas, Sandra Coccuzzo Sampaio, Maria Carolina Elias, José Salvatore Leister Patané, Vincent Louis Viala, Antonio Jorge Martins, Ricardo Haddad, Claudia Renata dos Santos Barros, Elaine Cristina Marqueze, Raul Machado Neto, Debora Botequio Moretti, Jardelina de Souza Todao Bernardino, Loyze Paola Oliveira de Lima, Luiz Aurelio de Campos Crispin. Centro de Genômica Funcional da ESALQ: Luiz Lehmann Coutinho, Ricardo Augusto Brassaloti, Raquel de Lello Rocha Campos Cassano. NGS Soluções Genômicas: Pilar Drummond Sampaio Corrêa Mariani. FZEA-USP Pirassununga: Mirele Daiana Poleti, Jessika Cristina Chagas Lesbon, Elisangela Chicaroni Mattos, Heidge Fukumasu. USP-Botucatu: Rejane Maria Tommasini Grotto, Jayme A. Souza-Neto, Guilherme Targino Valente, Patricia Akemi Assato, Felipe Allan da Silva da Costa, Bianca Cechetto Carlos. Mendelics: Bibiana Santos, João Paulo Kitajima, Erika Freitas, David Schlesinger. Hemocentro Ribeirão Preto: Simone Kashima, Evandra Strazza Rodrigues, Svetoslav Nanev Slavov, Elaine Vieira dos Santos, Rafael dos Santos Bezerra, Luiz Carlos Junior de Alcantara, Marta Giovanetti, Vagner Fonseca, Flavia Aburjaile, Rodrigo Tocantins Calado. FAMERP-SJRP: Cecília Artico Banho, Lívia Sacchetto, Fábio Sossai Possebon, Leila Sabrina Ullmann, Cíntia Bittar, Guilherme Campos, Helena Lage Ferreira, Jorge A. Petroli Marchesi, Maisa C. Pereira Parra, Marília Moraes, Paula Rahal, Paulo Inacio da Costa, João Pessoa Araújo Jr., Maurício Lacerda Nogueira. Prefeitura de Sao Paulo: Melissa Palmieri. |
| EPI_ISL_1966131 | CENTRO DE SAUDE II MAIRINQUE MAIRINQUE | Instituto Butantan / Mendelics | Instituto Butantan: Dimas Tadeu Covas, Sandra Coccuzzo Sampaio, Maria Carolina Elias, José Salvatore Leister Patané, Vincent Louis Viala, Antonio Jorge Martins, Ricardo Haddad, Claudia Renata dos Santos Barros, Elaine Cristina Marqueze, Raul Machado Neto, Debora Botequio Moretti, Jardelina de Souza Todao Bernardino, Loyze Paola Oliveira de Lima, Luiz Aurelio de Campos Crispin. Centro de Genômica Funcional da ESALQ: Luiz Lehmann Coutinho, Ricardo Augusto Brassaloti, Raquel de Lello Rocha Campos Cassano. NGS Soluções Genômicas: Pilar Drummond Sampaio Corrêa Mariani. FZEA-USP Pirassununga: Mirele Daiana Poleti, Jessika Cristina Chagas Lesbon, Elisangela Chicaroni Mattos, Heidge Fukumasu. USP-Botucatu: Rejane Maria Tommasini Grotto, Jayme A. Souza-Neto, Guilherme Targino Valente, Patricia Akemi Assato, Felipe Allan da Silva da Costa, Bianca Cechetto Carlos. Mendelics: Bibiana Santos, João Paulo Kitajima, Erika Freitas, David Schlesinger. Hemocentro Ribeirão Preto: Simone Kashima, Evandra Strazza Rodrigues, Svetoslav Nanev Slavov, Elaine Vieira dos Santos, Rafael dos Santos Bezerra, Luiz Carlos Junior de Alcantara, Marta Giovanetti, Vagner Fonseca, Flavia Aburjaile, Rodrigo Tocantins Calado. FAMERP-SJRP: Cecília Artico Banho, Lívia Sacchetto, Fábio Sossai Possebon, Leila Sabrina Ullmann, Cíntia Bittar, Guilherme Campos, Helena Lage Ferreira, Jorge A. Petroli Marchesi, Maisa C. Pereira Parra, Marília Moraes, Paula Rahal, Paulo Inacio da Costa, João Pessoa Araújo Jr., Maurício Lacerda Nogueira. Prefeitura de Sao Paulo: Melissa Palmieri. |
| EPI_ISL_1966132 | UNIDADE MISTA DE TAIACU | Instituto Butantan / Mendelics | Instituto Butantan: Dimas Tadeu Covas, Sandra Coccuzzo Sampaio, Maria Carolina Elias, José Salvatore Leister Patané, Vincent Louis Viala, Antonio Jorge Martins, Ricardo Haddad, Claudia Renata dos Santos Barros, Elaine Cristina Marqueze, Raul Machado Neto, Debora Botequio Moretti, Jardelina de Souza Todao Bernardino, Loyze Paola Oliveira de Lima, Luiz Aurelio de Campos Crispin. Centro de Genômica Funcional da ESALQ: Luiz Lehmann Coutinho, Ricardo Augusto Brassaloti, Raquel de Lello Rocha Campos Cassano. NGS Soluções Genômicas: Pilar Drummond Sampaio Corrêa Mariani. FZEA-USP Pirassununga: Mirele Daiana Poleti, Jessika Cristina Chagas Lesbon, Elisangela Chicaroni Mattos, Heidge Fukumasu. USP-Botucatu: Rejane Maria Tommasini Grotto, Jayme A. Souza-Neto, Guilherme Targino Valente, Patricia Akemi Assato, Felipe Allan da Silva da Costa, Bianca Cechetto Carlos. Mendelics: Bibiana Santos, João Paulo Kitajima, Erika Freitas, David Schlesinger. Hemocentro Ribeirão Preto: Simone Kashima, Evandra Strazza |

| | | | |
|---|---|---|---|
| | | | Rodrigues, Svetoslav Nanev Slavov, Elaine Vieira dos Santos, Rafael dos Santos Bezerra, Luiz Carlos Junior de Alcantara, Marta Giovanetti, Vagner Fonseca, Flavia Aburjaile, Rodrigo Tocantins Calado. FAMERP-SJRP: Cecília Artico Banho, Lívia Sacchetto, Fábio Sossai Possebon, Leila Sabrina Ullmann, Cíntia Bittar, Guilherme Campos, Helena Lage Ferreira, Jorge A. Petroli Marchesi, Maisa C. Pereira Parra, Marília Moraes, Paula Rahal, Paulo Inacio da Costa, João Pessoa Araújo Jr., Maurício Lacerda Nogueira. Prefeitura de Sao Paulo: Melissa Palmieri. |
| EPI_ISL_1966133, EPI_ISL_1966134, EPI_ISL_1966135, EPI_ISL_1966136, EPI_ISL_1966137, EPI_ISL_1966138, EPI_ISL_1966139, EPI_ISL_1966140, EPI_ISL_1966141, EPI_ISL_1966142, EPI_ISL_1966143, EPI_ISL_1966144 | | | |
| see above | PRONTO ATENDIMENTO VILA PADRE ANCHIETA | Instituto Butantan / Mendelics | Instituto Butantan: Dimas Tadeu Covas, Sandra Coccuzzo Sampaio, Maria Carolina Elias, José Salvatore Leister Patané, Vincent Louis Viala, Antonio Jorge Martins, Ricardo Haddad, Claudia Renata dos Santos Barros, Elaine Cristina Marqueze, Raul Machado Neto, Debora Botequio Moretti, Jardelina de Souza Todao Bernardino, Loyze Paola Oliveira de Lima, Luiz Aurelio de Campos Crispin. Centro de Genômica Funcional da ESALQ: Luiz Lehmann Coutinho, Ricardo Augusto Brassaloti, Raquel de Lello Rocha Campos Cassano. NGS Soluções Genômicas: Pilar Drummond Sampaio Corrêa Mariani. FZEA-USP Pirassununga: Mirele Daiana Poleti, Jessika Cristina Chagas Lesbon, Elisangela Chicaroni Mattos, Heidge Fukumasu. USP-Botucatu: Rejane Maria Tommasini Grotto, Jayme A. Souza-Neto, Guilherme Targino Valente, Patricia Akemi Assato, Felipe Allan da Silva da Costa, Bianca Cechetto Carlos. Mendelics: Bibiana Santos, João Paulo Kitajima, Erika Freitas, David Schlesinger. Hemocentro Ribeirão Preto: Simone Kashima, Evandra Strazza Rodrigues, Svetoslav Nanev Slavov, Elaine Vieira dos Santos, Rafael dos Santos Bezerra, Luiz Carlos Junior de Alcantara, Marta Giovanetti, Vagner Fonseca, Flavia Aburjaile, Rodrigo Tocantins Calado. FAMERP-SJRP: Cecília Artico Banho, Lívia Sacchetto, Fábio Sossai Possebon, Leila Sabrina Ullmann, Cíntia Bittar, Guilherme Campos, Helena Lage Ferreira, Jorge A. Petroli Marchesi, Maisa C. Pereira Parra, Marília Moraes, Paula Rahal, Paulo Inacio da Costa, João Pessoa Araújo Jr., Maurício Lacerda Nogueira. Prefeitura de Sao Paulo: Melissa Palmieri. |
| EPI_ISL_1966145, EPI_ISL_1966146 | COMPLEXO HOSPITALAR OURO VERDE DE CAMPINAS | Instituto Butantan / Mendelics | Instituto Butantan: Dimas Tadeu Covas, Sandra Coccuzzo Sampaio, Maria Carolina Elias, José Salvatore Leister Patané, Vincent Louis Viala, Antonio Jorge Martins, Ricardo Haddad, Claudia Renata dos Santos Barros, Elaine Cristina Marqueze, Raul Machado Neto, Debora Botequio Moretti, Jardelina de Souza Todao Bernardino, Loyze Paola Oliveira de Lima, Luiz Aurelio de Campos Crispin. Centro de Genômica Funcional da ESALQ: Luiz Lehmann Coutinho, Ricardo Augusto Brassaloti, Raquel de Lello Rocha Campos Cassano. NGS Soluções Genômicas: Pilar Drummond Sampaio Corrêa Mariani. FZEA-USP Pirassununga: Mirele Daiana Poleti, Jessika Cristina Chagas Lesbon, Elisangela Chicaroni Mattos, Heidge Fukumasu. USP-Botucatu: Rejane Maria Tommasini Grotto, Jayme A. Souza-Neto, Guilherme Targino Valente, Patricia Akemi Assato, Felipe Allan da Silva da Costa, Bianca Cechetto Carlos. Mendelics: Bibiana Santos, João Paulo Kitajima, Erika Freitas, David Schlesinger. Hemocentro Ribeirão Preto: Simone Kashima, Evandra Strazza Rodrigues, Svetoslav Nanev Slavov, Elaine Vieira dos Santos, Rafael dos Santos Bezerra, Luiz Carlos Junior de Alcantara, Marta Giovanetti, Vagner Fonseca, Flavia Aburjaile, Rodrigo Tocantins Calado. FAMERP-SJRP: Cecília Artico Banho, Lívia Sacchetto, Fábio Sossai Possebon, Leila Sabrina Ullmann, Cíntia Bittar, Guilherme Campos, Helena Lage Ferreira, Jorge A. Petroli Marchesi, Maisa C. Pereira Parra, Marília Moraes, Paula Rahal, Paulo Inacio da Costa, João Pessoa Araújo Jr., Maurício Lacerda Nogueira. Prefeitura de Sao Paulo: Melissa Palmieri. |
| EPI_ISL_1966147 | HOSPITAL REGIONAL DE ITAPETININGA | Instituto Butantan / Mendelics | Instituto Butantan: Dimas Tadeu Covas, Sandra Coccuzzo Sampaio, Maria Carolina Elias, José Salvatore Leister Patané, Vincent Louis Viala, Antonio Jorge Martins, Ricardo Haddad, Claudia Renata dos Santos Barros, Elaine Cristina Marqueze, Raul Machado Neto, Debora Botequio Moretti, Jardelina de Souza Todao Bernardino, Loyze Paola Oliveira de Lima, Luiz Aurelio de Campos Crispin. Centro de Genômica Funcional da ESALQ: Luiz Lehmann Coutinho, Ricardo Augusto Brassaloti, Raquel de Lello Rocha Campos Cassano. NGS Soluções Genômicas: Pilar Drummond Sampaio Corrêa Mariani. FZEA-USP Pirassununga: Mirele Daiana Poleti, Jessika Cristina Chagas Lesbon, Elisangela Chicaroni Mattos, Heidge Fukumasu. USP-Botucatu: Rejane Maria Tommasini Grotto, Jayme A. Souza-Neto, Guilherme Targino Valente, Patricia Akemi Assato, Felipe Allan da Silva da Costa, Bianca Cechetto Carlos. Mendelics: Bibiana Santos, João Paulo Kitajima, Erika Freitas, David Schlesinger. Hemocentro Ribeirão Preto: Simone Kashima, Evandra Strazza Rodrigues, Svetoslav Nanev Slavov, Elaine Vieira dos Santos, Rafael dos Santos Bezerra, Luiz Carlos Junior de Alcantara, Marta Giovanetti, Vagner Fonseca, Flavia Aburjaile, Rodrigo Tocantins Calado. FAMERP-SJRP: Cecília Artico Banho, Lívia Sacchetto, Fábio Sossai Possebon, Leila Sabrina Ullmann, Cíntia Bittar, Guilherme Campos, Helena Lage Ferreira, Jorge A. Petroli Marchesi, Maisa C. Pereira Parra, Marília Moraes, Paula Rahal, Paulo Inacio da Costa, João Pessoa Araújo Jr., Maurício Lacerda Nogueira. Prefeitura de Sao Paulo: Melissa Palmieri. |
| EPI_ISL_1966148, EPI_ISL_1966149, EPI_ISL_1966150, EPI_ISL_1966151, EPI_ISL_1966152, EPI_ISL_1966153 | SECRETARIA MUNICIPAL DE SAUDE DE PIRACAIA | Instituto Butantan / Mendelics | Instituto Butantan: Dimas Tadeu Covas, Sandra Coccuzzo Sampaio, Maria Carolina Elias, José Salvatore Leister Patané, Vincent Louis Viala, Antonio Jorge Martins, Ricardo Haddad, Claudia Renata dos Santos Barros, Elaine Cristina Marqueze, Raul Machado Neto, Debora Botequio Moretti, Jardelina de Souza Todao Bernardino, Loyze Paola Oliveira de Lima, Luiz Aurelio de Campos Crispin. Centro de Genômica Funcional da ESALQ: Luiz Lehmann Coutinho, Ricardo Augusto Brassaloti, Raquel de Lello Rocha Campos Cassano. NGS Soluções Genômicas: Pilar Drummond Sampaio Corrêa Mariani. FZEA-USP Pirassununga: Mirele Daiana Poleti, Jessika Cristina Chagas Lesbon, Elisangela Chicaroni Mattos, Heidge Fukumasu. USP-Botucatu: Rejane Maria Tommasini Grotto, Jayme A. Souza-Neto, Guilherme Targino Valente, Patricia Akemi Assato, Felipe Allan da Silva da Costa, Bianca Cechetto Carlos. Mendelics: Bibiana Santos, João Paulo Kitajima, Erika Freitas, David Schlesinger. Hemocentro Ribeirão Preto: Simone Kashima, Evandra Strazza Rodrigues, Svetoslav Nanev Slavov, Elaine Vieira dos Santos, Rafael dos Santos Bezerra, Luiz Carlos Junior de Alcantara, Marta Giovanetti, Vagner Fonseca, Flavia Aburjaile, Rodrigo Tocantins Calado. FAMERP-SJRP: Cecília Artico Banho, Lívia Sacchetto, Fábio Sossai Possebon, Leila Sabrina Ullmann, Cíntia Bittar, Guilherme Campos, Helena Lage Ferreira, Jorge A. Petroli Marchesi, Maisa C. Pereira Parra, Marília Moraes, Paula Rahal, Paulo Inacio da Costa, João Pessoa Araújo Jr., Maurício Lacerda Nogueira. Prefeitura de Sao Paulo: Melissa Palmieri. |
| EPI_ISL_1966154, EPI_ISL_1966155 | SECRETARIA MUNICIPAL DE SAUDE SOROCABA | Instituto Butantan / Mendelics | Instituto Butantan: Dimas Tadeu Covas, Sandra Coccuzzo Sampaio, Maria Carolina Elias, José Salvatore Leister Patané, Vincent Louis Viala, Antonio Jorge Martins, Ricardo Haddad, Claudia Renata dos Santos Barros, Elaine Cristina Marqueze, Raul Machado Neto, Debora Botequio Moretti, Jardelina de Souza Todao Bernardino, Loyze Paola Oliveira de Lima, Luiz Aurelio de Campos Crispin. Centro de Genômica Funcional da ESALQ: Luiz Lehmann Coutinho, Ricardo Augusto Brassaloti, Raquel de Lello Rocha Campos Cassano. NGS Soluções Genômicas: Pilar Drummond Sampaio Corrêa Mariani. FZEA-USP Pirassununga: Mirele Daiana Poleti, Jessika Cristina Chagas Lesbon, Elisangela Chicaroni Mattos, Heidge Fukumasu. USP-Botucatu: Rejane Maria Tommasini Grotto, Jayme A. Souza-Neto, Guilherme Targino Valente, Patricia Akemi Assato, Felipe Allan da Silva da Costa, Bianca Cechetto Carlos. Mendelics: Bibiana Santos, João Paulo Kitajima, Erika Freitas, David Schlesinger. Hemocentro Ribeirão Preto: Simone Kashima, Evandra Strazza Rodrigues, Svetoslav Nanev Slavov, Elaine Vieira dos Santos, Rafael dos Santos Bezerra, Luiz Carlos Junior de Alcantara, Marta Giovanetti, Vagner Fonseca, Flavia Aburjaile, Rodrigo Tocantins Calado. FAMERP-SJRP: Cecília Artico Banho, Lívia Sacchetto, Fábio Sossai Possebon, Leila Sabrina Ullmann, Cíntia Bittar, Guilherme Campos, Helena Lage Ferreira, Jorge A. Petroli Marchesi, Maisa C. Pereira Parra, Marília Moraes, Paula Rahal, Paulo Inacio da Costa, João Pessoa Araújo Jr., Maurício Lacerda Nogueira. Prefeitura de Sao Paulo: Melissa Palmieri. |
| EPI_ISL_1966156 | PRONTO ATENDIMENTO VILA PADRE ANCHIETA | Instituto Butantan / Mendelics | Instituto Butantan: Dimas Tadeu Covas, Sandra Coccuzzo Sampaio, Maria Carolina Elias, José Salvatore Leister Patané, Vincent Louis Viala, Antonio Jorge Martins, Ricardo Haddad, Claudia Renata dos Santos Barros, Elaine Cristina Marqueze, Raul Machado Neto, Debora Botequio Moretti, Jardelina de Souza Todao Bernardino, Loyze Paola Oliveira de Lima, Luiz Aurelio de Campos Crispin. Centro de Genômica Funcional da ESALQ: Luiz Lehmann Coutinho, Ricardo Augusto Brassaloti, Raquel de Lello Rocha Campos Cassano. NGS Soluções Genômicas: Pilar Drummond Sampaio Corrêa Mariani. FZEA-USP Pirassununga: Mirele Daiana Poleti, Jessika Cristina Chagas Lesbon, Elisangela Chicaroni Mattos, Heidge Fukumasu. USP-Botucatu: Rejane Maria Tommasini Grotto, Jayme A. Souza-Neto, Guilherme Targino Valente, Patricia Akemi Assato, Felipe Allan da Silva da Costa, Bianca Cechetto Carlos. Mendelics: Bibiana Santos, João Paulo Kitajima, Erika Freitas, David Schlesinger. Hemocentro Ribeirão Preto: Simone Kashima, Evandra Strazza Rodrigues, Svetoslav Nanev Slavov, Elaine Vieira dos Santos, Rafael dos Santos Bezerra, Luiz Carlos Junior de Alcantara, Marta Giovanetti, Vagner Fonseca, Flavia Aburjaile, Rodrigo Tocantins Calado. FAMERP-SJRP: Cecília Artico Banho, Lívia Sacchetto, Fábio Sossai Possebon, Leila Sabrina Ullmann, Cíntia Bittar, Guilherme Campos, Helena Lage Ferreira, Jorge A. Petroli Marchesi, Maisa C. Pereira Parra, Marília Moraes, Paula Rahal, Paulo Inacio da Costa, João Pessoa Araújo Jr., Maurício Lacerda Nogueira. Prefeitura de Sao Paulo: Melissa Palmieri. |
| EPI_ISL_1966157, EPI_ISL_1966158, EPI_ISL_1966159, EPI_ISL_1966160, EPI_ISL_1966161 | COMPLEXO HOSPITALAR OURO VERDE DE CAMPINAS | Instituto Butantan / Mendelics | Instituto Butantan: Dimas Tadeu Covas, Sandra Coccuzzo Sampaio, Maria Carolina Elias, José Salvatore Leister Patané, Vincent Louis Viala, Antonio Jorge Martins, Ricardo Haddad, Claudia Renata dos Santos Barros, Elaine Cristina Marqueze, Raul Machado Neto, Debora Botequio Moretti, Jardelina de Souza Todao Bernardino, Loyze Paola Oliveira de Lima, Luiz Aurelio de Campos Crispin. Centro de Genômica Funcional da ESALQ: Luiz Lehmann Coutinho, Ricardo Augusto Brassaloti, Raquel de Lello Rocha Campos Cassano. NGS Soluções Genômicas: Pilar Drummond Sampaio Corrêa Mariani. FZEA-USP Pirassununga: Mirele Daiana Poleti, Jessika Cristina Chagas Lesbon, Elisangela Chicaroni Mattos, Heidge Fukumasu. USP-Botucatu: Rejane Maria Tommasini Grotto, Jayme A. Souza-Neto, Guilherme Targino Valente, Patricia Akemi Assato, Felipe Allan da Silva da Costa, Bianca Cechetto Carlos. Mendelics: Bibiana Santos, João Paulo Kitajima, Erika Freitas, David Schlesinger. Hemocentro Ribeirão Preto: Simone Kashima, Evandra Strazza Rodrigues, Svetoslav Nanev Slavov, Elaine Vieira dos Santos, Rafael dos Santos Bezerra, Luiz Carlos Junior de Alcantara, Marta Giovanetti, Vagner Fonseca, Flavia Aburjaile, Rodrigo Tocantins Calado. FAMERP-SJRP: Cecília Artico Banho, Lívia Sacchetto, Fábio Sossai Possebon, Leila Sabrina Ullmann, Cíntia Bittar, Guilherme Campos, Helena Lage Ferreira, Jorge A. Petroli Marchesi, Maisa C. Pereira Parra, Marília Moraes, Paula Rahal, Paulo Inacio da Costa, João Pessoa Araújo Jr., Maurício Lacerda Nogueira. Prefeitura de Sao Paulo: Melissa Palmieri. |
| EPI_ISL_1966162, EPI_ISL_1966163, | UNIDADE RESPIRATORIA NOVA HORTOLANDIA | Instituto Butantan / Mendelics | Instituto Butantan: Dimas Tadeu Covas, Sandra Coccuzzo Sampaio, Maria Carolina Elias, José Salvatore Leister Patané, Vincent Louis Viala, Antonio |

| | | | |
|---|---|---|---|
| EPI_ISL_1966164, EPI_ISL_1966165, EPI_ISL_1966166 | | | Jorge Martins, Ricardo Haddad, Claudia Renata dos Santos Barros, Elaine Cristina Marqueze, Raul Machado Neto, Debora Botequio Moretti, Jardelina de Souza Todao Bernardino, Loyze Paola Oliveira de Lima, Luiz Aurelio de Campos Crispin. Centro de Genômica Funcional da ESALQ: Luiz Lehmann Coutinho, Ricardo Augusto Brassaloti, Raquel de Lello Rocha Campos Cassano. NGS Soluções Genômicas: Pilar Drummond Sampaio Corrêa Mariani. FZEA-USP Pirassununga: Mirele Daiana Poleti, Jessika Cristina Chagas Lesbon, Elisangela Chicaroni Mattos, Heidge Fukumasu. USP-Botucatu: Rejane Maria Tommasini Grotto, Jayme A. Souza-Neto, Guilherme Targino Valente, Patricia Akemi Assato, Felipe Allan da Silva da Costa, Bianca Cechetto Carlos. Mendelics: Bibiana Santos, João Paulo Kitajima, Erika Freitas, David Schlesinger. Hemocentro Ribeirão Preto: Simone Kashima, Evandra Strazza Rodrigues, Svetoslav Nanev Slavov, Elaine Vieira dos Santos, Rafael dos Santos Bezerra, Luiz Carlos Junior de Alcantara, Marta Giovanetti, Vagner Fonseca, Flavia Aburjaile, Rodrigo Tocantins Calado. FAMERP-SJRP: Cecília Artico Banho, Lívia Sacchetto, Fábio Sossai Possebon, Leila Sabrina Ullmann, Cíntia Bittar, Guilherme Campos, Helena Lage Ferreira, Jorge A. Petroli Marchesi, Maisa C. Pereira Parra, Marília Moraes, Paula Rahal, Paulo Inacio da Costa, João Pessoa Araújo Jr., Maurício Lacerda Nogueira. Prefeitura de Sao Paulo: Melissa Palmieri. |
| EPI_ISL_1966167, EPI_ISL_1966168, EPI_ISL_1966169, EPI_ISL_1966170, EPI_ISL_1966171, EPI_ISL_1966172, EPI_ISL_1966173 | UNIDADE DE PRONTO ATENDIMENTO UPA DRA ANA OLIVIA BENTIVOGLIO | Instituto Butantan / Mendelics | Instituto Butantan: Dimas Tadeu Covas, Sandra Coccuzzo Sampaio, Maria Carolina Elias, José Salvatore Leister Patané, Vincent Louis Viala, Antonio Jorge Martins, Ricardo Haddad, Claudia Renata dos Santos Barros, Elaine Cristina Marqueze, Raul Machado Neto, Debora Botequio Moretti, Jardelina de Souza Todao Bernardino, Loyze Paola Oliveira de Lima, Luiz Aurelio de Campos Crispin. Centro de Genômica Funcional da ESALQ: Luiz Lehmann Coutinho, Ricardo Augusto Brassaloti, Raquel de Lello Rocha Campos Cassano. NGS Soluções Genômicas: Pilar Drummond Sampaio Corrêa Mariani. FZEA-USP Pirassununga: Mirele Daiana Poleti, Jessika Cristina Chagas Lesbon, Elisangela Chicaroni Mattos, Heidge Fukumasu. USP-Botucatu: Rejane Maria Tommasini Grotto, Jayme A. Souza-Neto, Guilherme Targino Valente, Patricia Akemi Assato, Felipe Allan da Silva da Costa, Bianca Cechetto Carlos. Mendelics: Bibiana Santos, João Paulo Kitajima, Erika Freitas, David Schlesinger. Hemocentro Ribeirão Preto: Simone Kashima, Evandra Strazza Rodrigues, Svetoslav Nanev Slavov, Elaine Vieira dos Santos, Rafael dos Santos Bezerra, Luiz Carlos Junior de Alcantara, Marta Giovanetti, Vagner Fonseca, Flavia Aburjaile, Rodrigo Tocantins Calado. FAMERP-SJRP: Cecília Artico Banho, Lívia Sacchetto, Fábio Sossai Possebon, Leila Sabrina Ullmann, Cíntia Bittar, Guilherme Campos, Helena Lage Ferreira, Jorge A. Petroli Marchesi, Maisa C. Pereira Parra, Marília Moraes, Paula Rahal, Paulo Inacio da Costa, João Pessoa Araújo Jr., Maurício Lacerda Nogueira. Prefeitura de Sao Paulo: Melissa Palmieri. |
| EPI_ISL_1966174, EPI_ISL_1966175, EPI_ISL_1966176 | PRONTO ATENDIMENTO DA PONTE DE SAO JOAO | Instituto Butantan / Mendelics | Instituto Butantan: Dimas Tadeu Covas, Sandra Coccuzzo Sampaio, Maria Carolina Elias, José Salvatore Leister Patané, Vincent Louis Viala, Antonio Jorge Martins, Ricardo Haddad, Claudia Renata dos Santos Barros, Elaine Cristina Marqueze, Raul Machado Neto, Debora Botequio Moretti, Jardelina de Souza Todao Bernardino, Loyze Paola Oliveira de Lima, Luiz Aurelio de Campos Crispin. Centro de Genômica Funcional da ESALQ: Luiz Lehmann Coutinho, Ricardo Augusto Brassaloti, Raquel de Lello Rocha Campos Cassano. NGS Soluções Genômicas: Pilar Drummond Sampaio Corrêa Mariani. FZEA-USP Pirassununga: Mirele Daiana Poleti, Jessika Cristina Chagas Lesbon, Elisangela Chicaroni Mattos, Heidge Fukumasu. USP-Botucatu: Rejane Maria Tommasini Grotto, Jayme A. Souza-Neto, Guilherme Targino Valente, Patricia Akemi Assato, Felipe Allan da Silva da Costa, Bianca Cechetto Carlos. Mendelics: Bibiana Santos, João Paulo Kitajima, Erika Freitas, David Schlesinger. Hemocentro Ribeirão Preto: Simone Kashima, Evandra Strazza Rodrigues, Svetoslav Nanev Slavov, Elaine Vieira dos Santos, Rafael dos Santos Bezerra, Luiz Carlos Junior de Alcantara, Marta Giovanetti, Vagner Fonseca, Flavia Aburjaile, Rodrigo Tocantins Calado. FAMERP-SJRP: Cecília Artico Banho, Lívia Sacchetto, Fábio Sossai Possebon, Leila Sabrina Ullmann, Cíntia Bittar, Guilherme Campos, Helena Lage Ferreira, Jorge A. Petroli Marchesi, Maisa C. Pereira Parra, Marília Moraes, Paula Rahal, Paulo Inacio da Costa, João Pessoa Araújo Jr., Maurício Lacerda Nogueira. Prefeitura de Sao Paulo: Melissa Palmieri. |
| EPI_ISL_1966177, EPI_ISL_1966178 | SECRETARIA MUNICIPAL DE SAUDE SOROCABA | Instituto Butantan / Mendelics | Instituto Butantan: Dimas Tadeu Covas, Sandra Coccuzzo Sampaio, Maria Carolina Elias, José Salvatore Leister Patané, Vincent Louis Viala, Antonio Jorge Martins, Ricardo Haddad, Claudia Renata dos Santos Barros, Elaine Cristina Marqueze, Raul Machado Neto, Debora Botequio Moretti, Jardelina de Souza Todao Bernardino, Loyze Paola Oliveira de Lima, Luiz Aurelio de Campos Crispin. Centro de Genômica Funcional da ESALQ: Luiz Lehmann Coutinho, Ricardo Augusto Brassaloti, Raquel de Lello Rocha Campos Cassano. NGS Soluções Genômicas: Pilar Drummond Sampaio Corrêa Mariani. FZEA-USP Pirassununga: Mirele Daiana Poleti, Jessika Cristina Chagas Lesbon, Elisangela Chicaroni Mattos, Heidge Fukumasu. USP-Botucatu: Rejane Maria Tommasini Grotto, Jayme A. Souza-Neto, Guilherme Targino Valente, Patricia Akemi Assato, Felipe Allan da Silva da Costa, Bianca Cechetto Carlos. Mendelics: Bibiana Santos, João Paulo Kitajima, Erika Freitas, David Schlesinger. Hemocentro Ribeirão Preto: Simone Kashima, Evandra Strazza Rodrigues, Svetoslav Nanev Slavov, Elaine Vieira dos Santos, Rafael dos Santos Bezerra, Luiz Carlos Junior de Alcantara, Marta Giovanetti, Vagner Fonseca, Flavia Aburjaile, Rodrigo Tocantins Calado. FAMERP-SJRP: Cecília Artico Banho, Lívia Sacchetto, Fábio Sossai Possebon, Leila Sabrina Ullmann, Cíntia Bittar, Guilherme Campos, Helena Lage Ferreira, Jorge A. Petroli Marchesi, Maisa C. Pereira Parra, Marília Moraes, Paula Rahal, Paulo Inacio da Costa, João Pessoa Araújo Jr., Maurício Lacerda Nogueira. Prefeitura de Sao Paulo: Melissa Palmieri. |
| EPI_ISL_1966179 | PRONTO ATENDIMENTO VILA PADRE ANCHIETA | Instituto Butantan / Mendelics | Instituto Butantan: Dimas Tadeu Covas, Sandra Coccuzzo Sampaio, Maria Carolina Elias, José Salvatore Leister Patané, Vincent Louis Viala, Antonio Jorge Martins, Ricardo Haddad, Claudia Renata dos Santos Barros, Elaine Cristina Marqueze, Raul Machado Neto, Debora Botequio Moretti, Jardelina de Souza Todao Bernardino, Loyze Paola Oliveira de Lima, Luiz Aurelio de Campos Crispin. Centro de Genômica Funcional da ESALQ: Luiz Lehmann Coutinho, Ricardo Augusto Brassaloti, Raquel de Lello Rocha Campos Cassano. NGS Soluções Genômicas: Pilar Drummond Sampaio Corrêa Mariani. FZEA-USP Pirassununga: Mirele Daiana Poleti, Jessika Cristina Chagas Lesbon, Elisangela Chicaroni Mattos, Heidge Fukumasu. USP-Botucatu: Rejane Maria Tommasini Grotto, Jayme A. Souza-Neto, Guilherme Targino Valente, Patricia Akemi Assato, Felipe Allan da Silva da Costa, Bianca Cechetto Carlos. Mendelics: Bibiana Santos, João Paulo Kitajima, Erika Freitas, David Schlesinger. Hemocentro Ribeirão Preto: Simone Kashima, Evandra Strazza Rodrigues, Svetoslav Nanev Slavov, Elaine Vieira dos Santos, Rafael dos Santos Bezerra, Luiz Carlos Junior de Alcantara, Marta Giovanetti, Vagner Fonseca, Flavia Aburjaile, Rodrigo Tocantins Calado. FAMERP-SJRP: Cecília Artico Banho, Lívia Sacchetto, Fábio Sossai Possebon, Leila Sabrina Ullmann, Cíntia Bittar, Guilherme Campos, Helena Lage Ferreira, Jorge A. Petroli Marchesi, Maisa C. Pereira Parra, Marília Moraes, Paula Rahal, Paulo Inacio da Costa, João Pessoa Araújo Jr., Maurício Lacerda Nogueira. Prefeitura de Sao Paulo: Melissa Palmieri. |
| EPI_ISL_1966180 | UNIDADE MISTA DE SAUDE CAPELA DO ALTO | Instituto Butantan / Mendelics | Instituto Butantan: Dimas Tadeu Covas, Sandra Coccuzzo Sampaio, Maria Carolina Elias, José Salvatore Leister Patané, Vincent Louis Viala, Antonio Jorge Martins, Ricardo Haddad, Claudia Renata dos Santos Barros, Elaine Cristina Marqueze, Raul Machado Neto, Debora Botequio Moretti, Jardelina de Souza Todao Bernardino, Loyze Paola Oliveira de Lima, Luiz Aurelio de Campos Crispin. Centro de Genômica Funcional da ESALQ: Luiz Lehmann Coutinho, Ricardo Augusto Brassaloti, Raquel de Lello Rocha Campos Cassano. NGS Soluções Genômicas: Pilar Drummond Sampaio Corrêa Mariani. FZEA-USP Pirassununga: Mirele Daiana Poleti, Jessika Cristina Chagas Lesbon, Elisangela Chicaroni Mattos, Heidge Fukumasu. USP-Botucatu: Rejane Maria Tommasini Grotto, Jayme A. Souza-Neto, Guilherme Targino Valente, Patricia Akemi Assato, Felipe Allan da Silva da Costa, Bianca Cechetto Carlos. Mendelics: Bibiana Santos, João Paulo Kitajima, Erika Freitas, David Schlesinger. Hemocentro Ribeirão Preto: Simone Kashima, Evandra Strazza Rodrigues, Svetoslav Nanev Slavov, Elaine Vieira dos Santos, Rafael dos Santos Bezerra, Luiz Carlos Junior de Alcantara, Marta Giovanetti, Vagner Fonseca, Flavia Aburjaile, Rodrigo Tocantins Calado. FAMERP-SJRP: Cecília Artico Banho, Lívia Sacchetto, Fábio Sossai Possebon, Leila Sabrina Ullmann, Cíntia Bittar, Guilherme Campos, Helena Lage Ferreira, Jorge A. Petroli Marchesi, Maisa C. Pereira Parra, Marília Moraes, Paula Rahal, Paulo Inacio da Costa, João Pessoa Araújo Jr., Maurício Lacerda Nogueira. Prefeitura de Sao Paulo: Melissa Palmieri. |
| EPI_ISL_1966181, EPI_ISL_1966182, EPI_ISL_1966183, EPI_ISL_1966184, EPI_ISL_1966185, EPI_ISL_1966186 | SERV DE VIG SANITARIA EPIDEMIO E CTRL DE ZOONOZES GUARUJA | Instituto Butantan / Mendelics | Instituto Butantan: Dimas Tadeu Covas, Sandra Coccuzzo Sampaio, Maria Carolina Elias, José Salvatore Leister Patané, Vincent Louis Viala, Antonio Jorge Martins, Ricardo Haddad, Claudia Renata dos Santos Barros, Elaine Cristina Marqueze, Raul Machado Neto, Debora Botequio Moretti, Jardelina de Souza Todao Bernardino, Loyze Paola Oliveira de Lima, Luiz Aurelio de Campos Crispin. Centro de Genômica Funcional da ESALQ: Luiz Lehmann Coutinho, Ricardo Augusto Brassaloti, Raquel de Lello Rocha Campos Cassano. NGS Soluções Genômicas: Pilar Drummond Sampaio Corrêa Mariani. FZEA-USP Pirassununga: Mirele Daiana Poleti, Jessika Cristina Chagas Lesbon, Elisangela Chicaroni Mattos, Heidge Fukumasu. USP-Botucatu: Rejane Maria Tommasini Grotto, Jayme A. Souza-Neto, Guilherme Targino Valente, Patricia Akemi Assato, Felipe Allan da Silva da Costa, Bianca Cechetto Carlos. Mendelics: Bibiana Santos, João Paulo Kitajima, Erika Freitas, David Schlesinger. Hemocentro Ribeirão Preto: Simone Kashima, Evandra Strazza Rodrigues, Svetoslav Nanev Slavov, Elaine Vieira dos Santos, Rafael dos Santos Bezerra, Luiz Carlos Junior de Alcantara, Marta Giovanetti, Vagner Fonseca, Flavia Aburjaile, Rodrigo Tocantins Calado. FAMERP-SJRP: Cecília Artico Banho, Lívia Sacchetto, Fábio Sossai Possebon, Leila Sabrina Ullmann, Cíntia Bittar, Guilherme Campos, Helena Lage Ferreira, Jorge A. Petroli Marchesi, Maisa C. Pereira Parra, Marília Moraes, Paula Rahal, Paulo Inacio da Costa, João Pessoa Araújo Jr., Maurício Lacerda Nogueira. Prefeitura de Sao Paulo: Melissa Palmieri. |
| EPI_ISL_1966187 | SMS IPERO | Instituto Butantan / Mendelics | Instituto Butantan: Dimas Tadeu Covas, Sandra Coccuzzo Sampaio, Maria Carolina Elias, José Salvatore Leister Patané, Vincent Louis Viala, Antonio Jorge Martins, Ricardo Haddad, Claudia Renata dos Santos Barros, Elaine Cristina Marqueze, Raul Machado Neto, Debora Botequio Moretti, Jardelina de Souza Todao Bernardino, Loyze Paola Oliveira de Lima, Luiz Aurelio de Campos Crispin. Centro de Genômica Funcional da ESALQ: Luiz Lehmann Coutinho, Ricardo Augusto Brassaloti, Raquel de Lello Rocha Campos Cassano. NGS Soluções Genômicas: Pilar Drummond Sampaio Corrêa Mariani. FZEA-USP Pirassununga: Mirele Daiana Poleti, Jessika Cristina Chagas Lesbon, Elisangela Chicaroni Mattos, Heidge Fukumasu. USP-Botucatu: Rejane Maria Tommasini Grotto, Jayme A. Souza-Neto, Guilherme Targino Valente, Patricia Akemi Assato, Felipe Allan da Silva da Costa, Bianca Cechetto Carlos. Mendelics: Bibiana Santos, João Paulo Kitajima, Erika Freitas, David Schlesinger. Hemocentro Ribeirão Preto: Simone Kashima, Evandra Strazza Rodrigues, Svetoslav Nanev Slavov, Elaine Vieira dos Santos, Rafael dos Santos Bezerra, Luiz Carlos Junior de Alcantara, Marta Giovanetti, Vagner |

| | | | |
|---|---|---|---|
| | | | Fonseca, Flavia Aburjaile, Rodrigo Tocantins Calado. FAMERP-SJRP: Cecília Artico Banho, Lívia Sacchetto, Fábio Sossai Possebon, Leila Sabrina Ullmann, Cíntia Bittar, Guilherme Campos, Helena Lage Ferreira, Jorge A. Petroli Marchesi, Maisa C. Pereira Parra, Marília Moraes, Paula Rahal, Paulo Inacio da Costa, João Pessoa Araújo Jr., Maurício Lacerda Nogueira. Prefeitura de Sao Paulo: Melissa Palmieri. |
| EPI_ISL_1966188 | UBS SALTO DE SAO JOSE | Instituto Butantan / Mendelics | Instituto Butantan: Dimas Tadeu Covas, Sandra Coccuzzo Sampaio, Maria Carolina Elias, José Salvatore Leister Patané, Vincent Louis Viala, Antonio Jorge Martins, Ricardo Haddad, Claudia Renata dos Santos Barros, Elaine Cristina Marqueze, Raul Machado Neto, Debora Botequio Moretti, Jardelina de Souza Todao Bernardino, Loyze Paola Oliveira de Lima, Luiz Aurelio de Campos Crispin. Centro de Genômica Funcional da ESALQ: Luiz Lehmann Coutinho, Ricardo Augusto Brassaloti, Raquel de Lello Rocha Campos Cassano. NGS Soluções Genômicas: Pilar Drummond Sampaio Corrêa Mariani. FZEA-USP Pirassununga: Mirele Daiana Poleti, Jessika Cristina Chagas Lesbon, Elisangela Chicaroni Mattos, Heidge Fukumasu. USP-Botucatu: Rejane Maria Tommasini Grotto, Jayme A. Souza-Neto, Guilherme Targino Valente, Patricia Akemi Assato, Felipe Allan da Silva da Costa, Bianca Cechetto Carlos. Mendelics: Bibiana Santos, João Paulo Kitajima, Erika Freitas, David Schlesinger. Hemocentro Ribeirão Preto: Simone Kashima, Evandra Strazza Rodrigues, Svetoslav Nanev Slavov, Elaine Vieira dos Santos, Rafael dos Santos Bezerra, Luiz Carlos Junior de Alcantara, Marta Giovanetti, Vagner Fonseca, Flavia Aburjaile, Rodrigo Tocantins Calado. FAMERP-SJRP: Cecília Artico Banho, Lívia Sacchetto, Fábio Sossai Possebon, Leila Sabrina Ullmann, Cíntia Bittar, Guilherme Campos, Helena Lage Ferreira, Jorge A. Petroli Marchesi, Maisa C. Pereira Parra, Marília Moraes, Paula Rahal, Paulo Inacio da Costa, João Pessoa Araújo Jr., Maurício Lacerda Nogueira. Prefeitura de Sao Paulo: Melissa Palmieri. |
| EPI_ISL_1966189, EPI_ISL_1966190, EPI_ISL_1966191, EPI_ISL_1966192, EPI_ISL_1966193, EPI_ISL_1966194, EPI_ISL_1966195, EPI_ISL_1966196 | SECAO CENTRO DE DIAGNOSTICO SECEDI | Instituto Butantan / Mendelics | Instituto Butantan: Dimas Tadeu Covas, Sandra Coccuzzo Sampaio, Maria Carolina Elias, José Salvatore Leister Patané, Vincent Louis Viala, Antonio Jorge Martins, Ricardo Haddad, Claudia Renata dos Santos Barros, Elaine Cristina Marqueze, Raul Machado Neto, Debora Botequio Moretti, Jardelina de Souza Todao Bernardino, Loyze Paola Oliveira de Lima, Luiz Aurelio de Campos Crispin. Centro de Genômica Funcional da ESALQ: Luiz Lehmann Coutinho, Ricardo Augusto Brassaloti, Raquel de Lello Rocha Campos Cassano. NGS Soluções Genômicas: Pilar Drummond Sampaio Corrêa Mariani. FZEA-USP Pirassununga: Mirele Daiana Poleti, Jessika Cristina Chagas Lesbon, Elisangela Chicaroni Mattos, Heidge Fukumasu. USP-Botucatu: Rejane Maria Tommasini Grotto, Jayme A. Souza-Neto, Guilherme Targino Valente, Patricia Akemi Assato, Felipe Allan da Silva da Costa, Bianca Cechetto Carlos. Mendelics: Bibiana Santos, João Paulo Kitajima, Erika Freitas, David Schlesinger. Hemocentro Ribeirão Preto: Simone Kashima, Evandra Strazza Rodrigues, Svetoslav Nanev Slavov, Elaine Vieira dos Santos, Rafael dos Santos Bezerra, Luiz Carlos Junior de Alcantara, Marta Giovanetti, Vagner Fonseca, Flavia Aburjaile, Rodrigo Tocantins Calado. FAMERP-SJRP: Cecília Artico Banho, Lívia Sacchetto, Fábio Sossai Possebon, Leila Sabrina Ullmann, Cíntia Bittar, Guilherme Campos, Helena Lage Ferreira, Jorge A. Petroli Marchesi, Maisa C. Pereira Parra, Marília Moraes, Paula Rahal, Paulo Inacio da Costa, João Pessoa Araújo Jr., Maurício Lacerda Nogueira. Prefeitura de Sao Paulo: Melissa Palmieri. |
| EPI_ISL_1966197, EPI_ISL_1966198, EPI_ISL_1966199, EPI_ISL_1966200 | SECRETARIA MUNICIPAL DE SAUDE SOROCABA | Instituto Butantan / Mendelics | Instituto Butantan: Dimas Tadeu Covas, Sandra Coccuzzo Sampaio, Maria Carolina Elias, José Salvatore Leister Patané, Vincent Louis Viala, Antonio Jorge Martins, Ricardo Haddad, Claudia Renata dos Santos Barros, Elaine Cristina Marqueze, Raul Machado Neto, Debora Botequio Moretti, Jardelina de Souza Todao Bernardino, Loyze Paola Oliveira de Lima, Luiz Aurelio de Campos Crispin. Centro de Genômica Funcional da ESALQ: Luiz Lehmann Coutinho, Ricardo Augusto Brassaloti, Raquel de Lello Rocha Campos Cassano. NGS Soluções Genômicas: Pilar Drummond Sampaio Corrêa Mariani. FZEA-USP Pirassununga: Mirele Daiana Poleti, Jessika Cristina Chagas Lesbon, Elisangela Chicaroni Mattos, Heidge Fukumasu. USP-Botucatu: Rejane Maria Tommasini Grotto, Jayme A. Souza-Neto, Guilherme Targino Valente, Patricia Akemi Assato, Felipe Allan da Silva da Costa, Bianca Cechetto Carlos. Mendelics: Bibiana Santos, João Paulo Kitajima, Erika Freitas, David Schlesinger. Hemocentro Ribeirão Preto: Simone Kashima, Evandra Strazza Rodrigues, Svetoslav Nanev Slavov, Elaine Vieira dos Santos, Rafael dos Santos Bezerra, Luiz Carlos Junior de Alcantara, Marta Giovanetti, Vagner Fonseca, Flavia Aburjaile, Rodrigo Tocantins Calado. FAMERP-SJRP: Cecília Artico Banho, Lívia Sacchetto, Fábio Sossai Possebon, Leila Sabrina Ullmann, Cíntia Bittar, Guilherme Campos, Helena Lage Ferreira, Jorge A. Petroli Marchesi, Maisa C. Pereira Parra, Marília Moraes, Paula Rahal, Paulo Inacio da Costa, João Pessoa Araújo Jr., Maurício Lacerda Nogueira. Prefeitura de Sao Paulo: Melissa Palmieri. |
| EPI_ISL_1966201 | UNIDADE BASICA DE SAUDE DA FAMILIA BILAC | Instituto Butantan / Mendelics | Instituto Butantan: Dimas Tadeu Covas, Sandra Coccuzzo Sampaio, Maria Carolina Elias, José Salvatore Leister Patané, Vincent Louis Viala, Antonio Jorge Martins, Ricardo Haddad, Claudia Renata dos Santos Barros, Elaine Cristina Marqueze, Raul Machado Neto, Debora Botequio Moretti, Jardelina de Souza Todao Bernardino, Loyze Paola Oliveira de Lima, Luiz Aurelio de Campos Crispin. Centro de Genômica Funcional da ESALQ: Luiz Lehmann Coutinho, Ricardo Augusto Brassaloti, Raquel de Lello Rocha Campos Cassano. NGS Soluções Genômicas: Pilar Drummond Sampaio Corrêa Mariani. FZEA-USP Pirassununga: Mirele Daiana Poleti, Jessika Cristina Chagas Lesbon, Elisangela Chicaroni Mattos, Heidge Fukumasu. USP-Botucatu: Rejane Maria Tommasini Grotto, Jayme A. Souza-Neto, Guilherme Targino Valente, Patricia Akemi Assato, Felipe Allan da Silva da Costa, Bianca Cechetto Carlos. Mendelics: Bibiana Santos, João Paulo Kitajima, Erika Freitas, David Schlesinger. Hemocentro Ribeirão Preto: Simone Kashima, Evandra Strazza Rodrigues, Svetoslav Nanev Slavov, Elaine Vieira dos Santos, Rafael dos Santos Bezerra, Luiz Carlos Junior de Alcantara, Marta Giovanetti, Vagner Fonseca, Flavia Aburjaile, Rodrigo Tocantins Calado. FAMERP-SJRP: Cecília Artico Banho, Lívia Sacchetto, Fábio Sossai Possebon, Leila Sabrina Ullmann, Cíntia Bittar, Guilherme Campos, Helena Lage Ferreira, Jorge A. Petroli Marchesi, Maisa C. Pereira Parra, Marília Moraes, Paula Rahal, Paulo Inacio da Costa, João Pessoa Araújo Jr., Maurício Lacerda Nogueira. Prefeitura de Sao Paulo: Melissa Palmieri. |
| EPI_ISL_1966202 | UBS JOSE SABINO FERREIRA | Instituto Butantan / Mendelics | Instituto Butantan: Dimas Tadeu Covas, Sandra Coccuzzo Sampaio, Maria Carolina Elias, José Salvatore Leister Patané, Vincent Louis Viala, Antonio Jorge Martins, Ricardo Haddad, Claudia Renata dos Santos Barros, Elaine Cristina Marqueze, Raul Machado Neto, Debora Botequio Moretti, Jardelina de Souza Todao Bernardino, Loyze Paola Oliveira de Lima, Luiz Aurelio de Campos Crispin. Centro de Genômica Funcional da ESALQ: Luiz Lehmann Coutinho, Ricardo Augusto Brassaloti, Raquel de Lello Rocha Campos Cassano. NGS Soluções Genômicas: Pilar Drummond Sampaio Corrêa Mariani. FZEA-USP Pirassununga: Mirele Daiana Poleti, Jessika Cristina Chagas Lesbon, Elisangela Chicaroni Mattos, Heidge Fukumasu. USP-Botucatu: Rejane Maria Tommasini Grotto, Jayme A. Souza-Neto, Guilherme Targino Valente, Patricia Akemi Assato, Felipe Allan da Silva da Costa, Bianca Cechetto Carlos. Mendelics: Bibiana Santos, João Paulo Kitajima, Erika Freitas, David Schlesinger. Hemocentro Ribeirão Preto: Simone Kashima, Evandra Strazza Rodrigues, Svetoslav Nanev Slavov, Elaine Vieira dos Santos, Rafael dos Santos Bezerra, Luiz Carlos Junior de Alcantara, Marta Giovanetti, Vagner Fonseca, Flavia Aburjaile, Rodrigo Tocantins Calado. FAMERP-SJRP: Cecília Artico Banho, Lívia Sacchetto, Fábio Sossai Possebon, Leila Sabrina Ullmann, Cíntia Bittar, Guilherme Campos, Helena Lage Ferreira, Jorge A. Petroli Marchesi, Maisa C. Pereira Parra, Marília Moraes, Paula Rahal, Paulo Inacio da Costa, João Pessoa Araújo Jr., Maurício Lacerda Nogueira. Prefeitura de Sao Paulo: Melissa Palmieri. |
| EPI_ISL_1966203 | UBS IRMA AGUEDA MARIA JAIME | Instituto Butantan / Mendelics | Instituto Butantan: Dimas Tadeu Covas, Sandra Coccuzzo Sampaio, Maria Carolina Elias, José Salvatore Leister Patané, Vincent Louis Viala, Antonio Jorge Martins, Ricardo Haddad, Claudia Renata dos Santos Barros, Elaine Cristina Marqueze, Raul Machado Neto, Debora Botequio Moretti, Jardelina de Souza Todao Bernardino, Loyze Paola Oliveira de Lima, Luiz Aurelio de Campos Crispin. Centro de Genômica Funcional da ESALQ: Luiz Lehmann Coutinho, Ricardo Augusto Brassaloti, Raquel de Lello Rocha Campos Cassano. NGS Soluções Genômicas: Pilar Drummond Sampaio Corrêa Mariani. FZEA-USP Pirassununga: Mirele Daiana Poleti, Jessika Cristina Chagas Lesbon, Elisangela Chicaroni Mattos, Heidge Fukumasu. USP-Botucatu: Rejane Maria Tommasini Grotto, Jayme A. Souza-Neto, Guilherme Targino Valente, Patricia Akemi Assato, Felipe Allan da Silva da Costa, Bianca Cechetto Carlos. Mendelics: Bibiana Santos, João Paulo Kitajima, Erika Freitas, David Schlesinger. Hemocentro Ribeirão Preto: Simone Kashima, Evandra Strazza Rodrigues, Svetoslav Nanev Slavov, Elaine Vieira dos Santos, Rafael dos Santos Bezerra, Luiz Carlos Junior de Alcantara, Marta Giovanetti, Vagner Fonseca, Flavia Aburjaile, Rodrigo Tocantins Calado. FAMERP-SJRP: Cecília Artico Banho, Lívia Sacchetto, Fábio Sossai Possebon, Leila Sabrina Ullmann, Cíntia Bittar, Guilherme Campos, Helena Lage Ferreira, Jorge A. Petroli Marchesi, Maisa C. Pereira Parra, Marília Moraes, Paula Rahal, Paulo Inacio da Costa, João Pessoa Araújo Jr., Maurício Lacerda Nogueira. Prefeitura de Sao Paulo: Melissa Palmieri. |
| EPI_ISL_1966204 | UBS JOSE HILARIO DOS SANTOS | Instituto Butantan / Mendelics | Instituto Butantan: Dimas Tadeu Covas, Sandra Coccuzzo Sampaio, Maria Carolina Elias, José Salvatore Leister Patané, Vincent Louis Viala, Antonio Jorge Martins, Ricardo Haddad, Claudia Renata dos Santos Barros, Elaine Cristina Marqueze, Raul Machado Neto, Debora Botequio Moretti, Jardelina de Souza Todao Bernardino, Loyze Paola Oliveira de Lima, Luiz Aurelio de Campos Crispin. Centro de Genômica Funcional da ESALQ: Luiz Lehmann Coutinho, Ricardo Augusto Brassaloti, Raquel de Lello Rocha Campos Cassano. NGS Soluções Genômicas: Pilar Drummond Sampaio Corrêa Mariani. FZEA-USP Pirassununga: Mirele Daiana Poleti, Jessika Cristina Chagas Lesbon, Elisangela Chicaroni Mattos, Heidge Fukumasu. USP-Botucatu: Rejane Maria Tommasini Grotto, Jayme A. Souza-Neto, Guilherme Targino Valente, Patricia Akemi Assato, Felipe Allan da Silva da Costa, Bianca Cechetto Carlos. Mendelics: Bibiana Santos, João Paulo Kitajima, Erika Freitas, David Schlesinger. Hemocentro Ribeirão Preto: Simone Kashima, Evandra Strazza Rodrigues, Svetoslav Nanev Slavov, Elaine Vieira dos Santos, Rafael dos Santos Bezerra, Luiz Carlos Junior de Alcantara, Marta Giovanetti, Vagner Fonseca, Flavia Aburjaile, Rodrigo Tocantins Calado. FAMERP-SJRP: Cecília Artico Banho, Lívia Sacchetto, Fábio Sossai Possebon, Leila Sabrina Ullmann, Cíntia Bittar, Guilherme Campos, Helena Lage Ferreira, Jorge A. Petroli Marchesi, Maisa C. Pereira Parra, Marília Moraes, Paula Rahal, Paulo Inacio da Costa, João Pessoa Araújo Jr., Maurício Lacerda Nogueira. Prefeitura de Sao Paulo: Melissa Palmieri. |
| EPI_ISL_1966205 | UBS DARCY ALVES E ROBALINHO | Instituto Butantan / Mendelics | Instituto Butantan: Dimas Tadeu Covas, Sandra Coccuzzo Sampaio, Maria Carolina Elias, José Salvatore Leister Patané, Vincent Louis Viala, Antonio Jorge Martins, Ricardo Haddad, Claudia Renata dos Santos Barros, Elaine Cristina Marqueze, Raul Machado Neto, Debora Botequio Moretti, Jardelina de Souza Todao Bernardino, Loyze Paola Oliveira de Lima, Luiz Aurelio de Campos Crispin. Centro de Genômica Funcional da ESALQ: Luiz Lehmann |

| | | | |
|---|---|---|---|
| | | | Coutinho, Ricardo Augusto Brassaloti, Raquel de Lello Rocha Campos Cassano. NGS Soluções Genômicas: Pilar Drummond Sampaio Corrêa Mariani. FZEA-USP Pirassununga: Mirele Daiana Poleti, Jessika Cristina Chagas Lesbon, Elisangela Chicaroni Mattos, Heidge Fukumasu. USP-Botucatu: Rejane Maria Tommasini Grotto, Jayme A. Souza-Neto, Guilherme Targino Valente, Patricia Akemi Assato, Felipe Allan da Silva da Costa, Bianca Cechetto Carlos. Mendelics: Bibiana Santos, João Paulo Kitajima, Erika Freitas, David Schlesinger. Hemocentro Ribeirão Preto: Simone Kashima, Evandra Strazza Rodrigues, Svetoslav Nanev Slavov, Elaine Vieira dos Santos, Rafael dos Santos Bezerra, Luiz Carlos Junior de Alcantara, Marta Giovanetti, Vagner Fonseca, Flavia Aburjaile, Rodrigo Tocantins Calado. FAMERP-SJRP: Cecília Artico Banho, Lívia Sacchetto, Fábio Sossai Possebon, Leila Sabrina Ullmann, Cíntia Bittar, Guilherme Campos, Helena Lage Ferreira, Jorge A. Petroli Marchesi, Maisa C. Pereira Parra, Marília Moraes, Paula Rahal, Paulo Inacio da Costa, João Pessoa Araújo Jr., Maurício Lacerda Nogueira. Prefeitura de Sao Paulo: Melissa Palmieri. |
| EPI_ISL_1966206 | UBS OTACILIO FIRMINO LOPES | Instituto Butantan / Mendelics | Instituto Butantan: Dimas Tadeu Covas, Sandra Coccuzzo Sampaio, Maria Carolina Elias, José Salvatore Leister Patané, Vincent Louis Viala, Antonio Jorge Martins, Ricardo Haddad, Claudia Renata dos Santos Barros, Elaine Cristina Marqueze, Raul Machado Neto, Debora Botequio Moretti, Jardelina de Souza Todao Bernardino, Loyze Paola Oliveira de Lima, Luiz Aurelio de Campos Crispin. Centro de Genômica Funcional da ESALQ: Luiz Lehmann Coutinho, Ricardo Augusto Brassaloti, Raquel de Lello Rocha Campos Cassano. NGS Soluções Genômicas: Pilar Drummond Sampaio Corrêa Mariani. FZEA-USP Pirassununga: Mirele Daiana Poleti, Jessika Cristina Chagas Lesbon, Elisangela Chicaroni Mattos, Heidge Fukumasu. USP-Botucatu: Rejane Maria Tommasini Grotto, Jayme A. Souza-Neto, Guilherme Targino Valente, Patricia Akemi Assato, Felipe Allan da Silva da Costa, Bianca Cechetto Carlos. Mendelics: Bibiana Santos, João Paulo Kitajima, Erika Freitas, David Schlesinger. Hemocentro Ribeirão Preto: Simone Kashima, Evandra Strazza Rodrigues, Svetoslav Nanev Slavov, Elaine Vieira dos Santos, Rafael dos Santos Bezerra, Luiz Carlos Junior de Alcantara, Marta Giovanetti, Vagner Fonseca, Flavia Aburjaile, Rodrigo Tocantins Calado. FAMERP-SJRP: Cecília Artico Banho, Lívia Sacchetto, Fábio Sossai Possebon, Leila Sabrina Ullmann, Cíntia Bittar, Guilherme Campos, Helena Lage Ferreira, Jorge A. Petroli Marchesi, Maisa C. Pereira Parra, Marília Moraes, Paula Rahal, Paulo Inacio da Costa, João Pessoa Araújo Jr., Maurício Lacerda Nogueira. Prefeitura de Sao Paulo: Melissa Palmieri. |
| EPI_ISL_1966207 | UBS FRANCISCA LIMA DE LIRA | Instituto Butantan / Mendelics | Instituto Butantan: Dimas Tadeu Covas, Sandra Coccuzzo Sampaio, Maria Carolina Elias, José Salvatore Leister Patané, Vincent Louis Viala, Antonio Jorge Martins, Ricardo Haddad, Claudia Renata dos Santos Barros, Elaine Cristina Marqueze, Raul Machado Neto, Debora Botequio Moretti, Jardelina de Souza Todao Bernardino, Loyze Paola Oliveira de Lima, Luiz Aurelio de Campos Crispin. Centro de Genômica Funcional da ESALQ: Luiz Lehmann Coutinho, Ricardo Augusto Brassaloti, Raquel de Lello Rocha Campos Cassano. NGS Soluções Genômicas: Pilar Drummond Sampaio Corrêa Mariani. FZEA-USP Pirassununga: Mirele Daiana Poleti, Jessika Cristina Chagas Lesbon, Elisangela Chicaroni Mattos, Heidge Fukumasu. USP-Botucatu: Rejane Maria Tommasini Grotto, Jayme A. Souza-Neto, Guilherme Targino Valente, Patricia Akemi Assato, Felipe Allan da Silva da Costa, Bianca Cechetto Carlos. Mendelics: Bibiana Santos, João Paulo Kitajima, Erika Freitas, David Schlesinger. Hemocentro Ribeirão Preto: Simone Kashima, Evandra Strazza Rodrigues, Svetoslav Nanev Slavov, Elaine Vieira dos Santos, Rafael dos Santos Bezerra, Luiz Carlos Junior de Alcantara, Marta Giovanetti, Vagner Fonseca, Flavia Aburjaile, Rodrigo Tocantins Calado. FAMERP-SJRP: Cecília Artico Banho, Lívia Sacchetto, Fábio Sossai Possebon, Leila Sabrina Ullmann, Cíntia Bittar, Guilherme Campos, Helena Lage Ferreira, Jorge A. Petroli Marchesi, Maisa C. Pereira Parra, Marília Moraes, Paula Rahal, Paulo Inacio da Costa, João Pessoa Araújo Jr., Maurício Lacerda Nogueira. Prefeitura de Sao Paulo: Melissa Palmieri. |
| EPI_ISL_1966208 | UBS OTACILIO FIRMINO LOPES | Instituto Butantan / Mendelics | Instituto Butantan: Dimas Tadeu Covas, Sandra Coccuzzo Sampaio, Maria Carolina Elias, José Salvatore Leister Patané, Vincent Louis Viala, Antonio Jorge Martins, Ricardo Haddad, Claudia Renata dos Santos Barros, Elaine Cristina Marqueze, Raul Machado Neto, Debora Botequio Moretti, Jardelina de Souza Todao Bernardino, Loyze Paola Oliveira de Lima, Luiz Aurelio de Campos Crispin. Centro de Genômica Funcional da ESALQ: Luiz Lehmann Coutinho, Ricardo Augusto Brassaloti, Raquel de Lello Rocha Campos Cassano. NGS Soluções Genômicas: Pilar Drummond Sampaio Corrêa Mariani. FZEA-USP Pirassununga: Mirele Daiana Poleti, Jessika Cristina Chagas Lesbon, Elisangela Chicaroni Mattos, Heidge Fukumasu. USP-Botucatu: Rejane Maria Tommasini Grotto, Jayme A. Souza-Neto, Guilherme Targino Valente, Patricia Akemi Assato, Felipe Allan da Silva da Costa, Bianca Cechetto Carlos. Mendelics: Bibiana Santos, João Paulo Kitajima, Erika Freitas, David Schlesinger. Hemocentro Ribeirão Preto: Simone Kashima, Evandra Strazza Rodrigues, Svetoslav Nanev Slavov, Elaine Vieira dos Santos, Rafael dos Santos Bezerra, Luiz Carlos Junior de Alcantara, Marta Giovanetti, Vagner Fonseca, Flavia Aburjaile, Rodrigo Tocantins Calado. FAMERP-SJRP: Cecília Artico Banho, Lívia Sacchetto, Fábio Sossai Possebon, Leila Sabrina Ullmann, Cíntia Bittar, Guilherme Campos, Helena Lage Ferreira, Jorge A. Petroli Marchesi, Maisa C. Pereira Parra, Marília Moraes, Paula Rahal, Paulo Inacio da Costa, João Pessoa Araújo Jr., Maurício Lacerda Nogueira. Prefeitura de Sao Paulo: Melissa Palmieri. |
| EPI_ISL_1966209, EPI_ISL_1966210 | UBS HELENA MARREY | Instituto Butantan / Mendelics | Instituto Butantan: Dimas Tadeu Covas, Sandra Coccuzzo Sampaio, Maria Carolina Elias, José Salvatore Leister Patané, Vincent Louis Viala, Antonio Jorge Martins, Ricardo Haddad, Claudia Renata dos Santos Barros, Elaine Cristina Marqueze, Raul Machado Neto, Debora Botequio Moretti, Jardelina de Souza Todao Bernardino, Loyze Paola Oliveira de Lima, Luiz Aurelio de Campos Crispin. Centro de Genômica Funcional da ESALQ: Luiz Lehmann Coutinho, Ricardo Augusto Brassaloti, Raquel de Lello Rocha Campos Cassano. NGS Soluções Genômicas: Pilar Drummond Sampaio Corrêa Mariani. FZEA-USP Pirassununga: Mirele Daiana Poleti, Jessika Cristina Chagas Lesbon, Elisangela Chicaroni Mattos, Heidge Fukumasu. USP-Botucatu: Rejane Maria Tommasini Grotto, Jayme A. Souza-Neto, Guilherme Targino Valente, Patricia Akemi Assato, Felipe Allan da Silva da Costa, Bianca Cechetto Carlos. Mendelics: Bibiana Santos, João Paulo Kitajima, Erika Freitas, David Schlesinger. Hemocentro Ribeirão Preto: Simone Kashima, Evandra Strazza Rodrigues, Svetoslav Nanev Slavov, Elaine Vieira dos Santos, Rafael dos Santos Bezerra, Luiz Carlos Junior de Alcantara, Marta Giovanetti, Vagner Fonseca, Flavia Aburjaile, Rodrigo Tocantins Calado. FAMERP-SJRP: Cecília Artico Banho, Lívia Sacchetto, Fábio Sossai Possebon, Leila Sabrina Ullmann, Cíntia Bittar, Guilherme Campos, Helena Lage Ferreira, Jorge A. Petroli Marchesi, Maisa C. Pereira Parra, Marília Moraes, Paula Rahal, Paulo Inacio da Costa, João Pessoa Araújo Jr., Maurício Lacerda Nogueira. Prefeitura de Sao Paulo: Melissa Palmieri. |
| EPI_ISL_1966211 | UBS EMILIA COSME CERQUEIRA | Instituto Butantan / Mendelics | Instituto Butantan: Dimas Tadeu Covas, Sandra Coccuzzo Sampaio, Maria Carolina Elias, José Salvatore Leister Patané, Vincent Louis Viala, Antonio Jorge Martins, Ricardo Haddad, Claudia Renata dos Santos Barros, Elaine Cristina Marqueze, Raul Machado Neto, Debora Botequio Moretti, Jardelina de Souza Todao Bernardino, Loyze Paola Oliveira de Lima, Luiz Aurelio de Campos Crispin. Centro de Genômica Funcional da ESALQ: Luiz Lehmann Coutinho, Ricardo Augusto Brassaloti, Raquel de Lello Rocha Campos Cassano. NGS Soluções Genômicas: Pilar Drummond Sampaio Corrêa Mariani. FZEA-USP Pirassununga: Mirele Daiana Poleti, Jessika Cristina Chagas Lesbon, Elisangela Chicaroni Mattos, Heidge Fukumasu. USP-Botucatu: Rejane Maria Tommasini Grotto, Jayme A. Souza-Neto, Guilherme Targino Valente, Patricia Akemi Assato, Felipe Allan da Silva da Costa, Bianca Cechetto Carlos. Mendelics: Bibiana Santos, João Paulo Kitajima, Erika Freitas, David Schlesinger. Hemocentro Ribeirão Preto: Simone Kashima, Evandra Strazza Rodrigues, Svetoslav Nanev Slavov, Elaine Vieira dos Santos, Rafael dos Santos Bezerra, Luiz Carlos Junior de Alcantara, Marta Giovanetti, Vagner Fonseca, Flavia Aburjaile, Rodrigo Tocantins Calado. FAMERP-SJRP: Cecília Artico Banho, Lívia Sacchetto, Fábio Sossai Possebon, Leila Sabrina Ullmann, Cíntia Bittar, Guilherme Campos, Helena Lage Ferreira, Jorge A. Petroli Marchesi, Maisa C. Pereira Parra, Marília Moraes, Paula Rahal, Paulo Inacio da Costa, João Pessoa Araújo Jr., Maurício Lacerda Nogueira. Prefeitura de Sao Paulo: Melissa Palmieri. |
| EPI_ISL_1966212 | UBS SYLVIO JOAO L DE LUCIA | Instituto Butantan / Mendelics | Instituto Butantan: Dimas Tadeu Covas, Sandra Coccuzzo Sampaio, Maria Carolina Elias, José Salvatore Leister Patané, Vincent Louis Viala, Antonio Jorge Martins, Ricardo Haddad, Claudia Renata dos Santos Barros, Elaine Cristina Marqueze, Raul Machado Neto, Debora Botequio Moretti, Jardelina de Souza Todao Bernardino, Loyze Paola Oliveira de Lima, Luiz Aurelio de Campos Crispin. Centro de Genômica Funcional da ESALQ: Luiz Lehmann Coutinho, Ricardo Augusto Brassaloti, Raquel de Lello Rocha Campos Cassano. NGS Soluções Genômicas: Pilar Drummond Sampaio Corrêa Mariani. FZEA-USP Pirassununga: Mirele Daiana Poleti, Jessika Cristina Chagas Lesbon, Elisangela Chicaroni Mattos, Heidge Fukumasu. USP-Botucatu: Rejane Maria Tommasini Grotto, Jayme A. Souza-Neto, Guilherme Targino Valente, Patricia Akemi Assato, Felipe Allan da Silva da Costa, Bianca Cechetto Carlos. Mendelics: Bibiana Santos, João Paulo Kitajima, Erika Freitas, David Schlesinger. Hemocentro Ribeirão Preto: Simone Kashima, Evandra Strazza Rodrigues, Svetoslav Nanev Slavov, Elaine Vieira dos Santos, Rafael dos Santos Bezerra, Luiz Carlos Junior de Alcantara, Marta Giovanetti, Vagner Fonseca, Flavia Aburjaile, Rodrigo Tocantins Calado. FAMERP-SJRP: Cecília Artico Banho, Lívia Sacchetto, Fábio Sossai Possebon, Leila Sabrina Ullmann, Cíntia Bittar, Guilherme Campos, Helena Lage Ferreira, Jorge A. Petroli Marchesi, Maisa C. Pereira Parra, Marília Moraes, Paula Rahal, Paulo Inacio da Costa, João Pessoa Araújo Jr., Maurício Lacerda Nogueira. Prefeitura de Sao Paulo: Melissa Palmieri. |
| EPI_ISL_1966213, EPI_ISL_1966214 | UBS EMILIA COSME CERQUEIRA | Instituto Butantan / Mendelics | Instituto Butantan: Dimas Tadeu Covas, Sandra Coccuzzo Sampaio, Maria Carolina Elias, José Salvatore Leister Patané, Vincent Louis Viala, Antonio Jorge Martins, Ricardo Haddad, Claudia Renata dos Santos Barros, Elaine Cristina Marqueze, Raul Machado Neto, Debora Botequio Moretti, Jardelina de Souza Todao Bernardino, Loyze Paola Oliveira de Lima, Luiz Aurelio de Campos Crispin. Centro de Genômica Funcional da ESALQ: Luiz Lehmann Coutinho, Ricardo Augusto Brassaloti, Raquel de Lello Rocha Campos Cassano. NGS Soluções Genômicas: Pilar Drummond Sampaio Corrêa Mariani. FZEA-USP Pirassununga: Mirele Daiana Poleti, Jessika Cristina Chagas Lesbon, Elisangela Chicaroni Mattos, Heidge Fukumasu. USP-Botucatu: Rejane Maria Tommasini Grotto, Jayme A. Souza-Neto, Guilherme Targino Valente, Patricia Akemi Assato, Felipe Allan da Silva da Costa, Bianca Cechetto Carlos. Mendelics: Bibiana Santos, João Paulo Kitajima, Erika Freitas, David Schlesinger. Hemocentro Ribeirão Preto: Simone Kashima, Evandra Strazza Rodrigues, Svetoslav Nanev Slavov, Elaine Vieira dos Santos, Rafael dos Santos Bezerra, Luiz Carlos Junior de Alcantara, Marta Giovanetti, Vagner |

| | | | |
|---|---|---|---|
| | | | Fonseca, Flavia Aburjaile, Rodrigo Tocantins Calado. FAMERP-SJRP: Cecília Artico Banho, Lívia Sacchetto, Fábio Sossai Possebon, Leila Sabrina Ullmann, Cíntia Bittar, Guilherme Campos, Helena Lage Ferreira, Jorge A. Petroli Marchesi, Maisa C. Pereira Parra, Marília Moraes, Paula Rahal, Paulo Inacio da Costa, João Pessoa Araújo Jr., Maurício Lacerda Nogueira. Prefeitura de Sao Paulo: Melissa Palmieri. |
| EPI_ISL_1966215, EPI_ISL_1966216, EPI_ISL_1966217 | UBS HELENA MARREY | Instituto Butantan / Mendelics | Instituto Butantan: Dimas Tadeu Covas, Sandra Coccuzzo Sampaio, Maria Carolina Elias, José Salvatore Leister Patané, Vincent Louis Viala, Antonio Jorge Martins, Ricardo Haddad, Claudia Renata dos Santos Barros, Elaine Cristina Marqueze, Raul Machado Neto, Debora Botequio Moretti, Jardelina de Souza Todao Bernardino, Loyze Paola Oliveira de Lima, Luiz Aurelio de Campos Crispin. Centro de Genômica Funcional da ESALQ: Luiz Lehmann Coutinho, Ricardo Augusto Brassaloti, Raquel de Lello Rocha Campos Cassano. NGS Soluções Genômicas: Pilar Drummond Sampaio Corrêa Mariani. FZEA-USP Pirassununga: Mirele Daiana Poleti, Jessika Cristina Chagas Lesbon, Elisangela Chicaroni Mattos, Heidge Fukumasu. USP-Botucatu: Rejane Maria Tommasini Grotto, Jayme A. Souza-Neto, Guilherme Targino Valente, Patricia Akemi Assato, Felipe Allan da Silva da Costa, Bianca Cechetto Carlos. Mendelics: Bibiana Santos, João Paulo Kitajima, Erika Freitas, David Schlesinger. Hemocentro Ribeirão Preto: Simone Kashima, Evandra Strazza Rodrigues, Svetoslav Nanev Slavov, Elaine Vieira dos Santos, Rafael dos Santos Bezerra, Luiz Carlos Junior de Alcantara, Marta Giovanetti, Vagner Fonseca, Flavia Aburjaile, Rodrigo Tocantins Calado. FAMERP-SJRP: Cecília Artico Banho, Lívia Sacchetto, Fábio Sossai Possebon, Leila Sabrina Ullmann, Cíntia Bittar, Guilherme Campos, Helena Lage Ferreira, Jorge A. Petroli Marchesi, Maisa C. Pereira Parra, Marília Moraes, Paula Rahal, Paulo Inacio da Costa, João Pessoa Araújo Jr., Maurício Lacerda Nogueira. Prefeitura de Sao Paulo: Melissa Palmieri. |
| EPI_ISL_1966218 | PPA FENELON GUEDES PEREIRA | Instituto Butantan / Mendelics | Instituto Butantan: Dimas Tadeu Covas, Sandra Coccuzzo Sampaio, Maria Carolina Elias, José Salvatore Leister Patané, Vincent Louis Viala, Antonio Jorge Martins, Ricardo Haddad, Claudia Renata dos Santos Barros, Elaine Cristina Marqueze, Raul Machado Neto, Debora Botequio Moretti, Jardelina de Souza Todao Bernardino, Loyze Paola Oliveira de Lima, Luiz Aurelio de Campos Crispin. Centro de Genômica Funcional da ESALQ: Luiz Lehmann Coutinho, Ricardo Augusto Brassaloti, Raquel de Lello Rocha Campos Cassano. NGS Soluções Genômicas: Pilar Drummond Sampaio Corrêa Mariani. FZEA-USP Pirassununga: Mirele Daiana Poleti, Jessika Cristina Chagas Lesbon, Elisangela Chicaroni Mattos, Heidge Fukumasu. USP-Botucatu: Rejane Maria Tommasini Grotto, Jayme A. Souza-Neto, Guilherme Targino Valente, Patricia Akemi Assato, Felipe Allan da Silva da Costa, Bianca Cechetto Carlos. Mendelics: Bibiana Santos, João Paulo Kitajima, Erika Freitas, David Schlesinger. Hemocentro Ribeirão Preto: Simone Kashima, Evandra Strazza Rodrigues, Svetoslav Nanev Slavov, Elaine Vieira dos Santos, Rafael dos Santos Bezerra, Luiz Carlos Junior de Alcantara, Marta Giovanetti, Vagner Fonseca, Flavia Aburjaile, Rodrigo Tocantins Calado. FAMERP-SJRP: Cecília Artico Banho, Lívia Sacchetto, Fábio Sossai Possebon, Leila Sabrina Ullmann, Cíntia Bittar, Guilherme Campos, Helena Lage Ferreira, Jorge A. Petroli Marchesi, Maisa C. Pereira Parra, Marília Moraes, Paula Rahal, Paulo Inacio da Costa, João Pessoa Araújo Jr., Maurício Lacerda Nogueira. Prefeitura de Sao Paulo: Melissa Palmieri. |
| EPI_ISL_1966219 | CENTRO DE SAUDE DE BORA | Instituto Butantan / Mendelics | Instituto Butantan: Dimas Tadeu Covas, Sandra Coccuzzo Sampaio, Maria Carolina Elias, José Salvatore Leister Patané, Vincent Louis Viala, Antonio Jorge Martins, Ricardo Haddad, Claudia Renata dos Santos Barros, Elaine Cristina Marqueze, Raul Machado Neto, Debora Botequio Moretti, Jardelina de Souza Todao Bernardino, Loyze Paola Oliveira de Lima, Luiz Aurelio de Campos Crispin. Centro de Genômica Funcional da ESALQ: Luiz Lehmann Coutinho, Ricardo Augusto Brassaloti, Raquel de Lello Rocha Campos Cassano. NGS Soluções Genômicas: Pilar Drummond Sampaio Corrêa Mariani. FZEA-USP Pirassununga: Mirele Daiana Poleti, Jessika Cristina Chagas Lesbon, Elisangela Chicaroni Mattos, Heidge Fukumasu. USP-Botucatu: Rejane Maria Tommasini Grotto, Jayme A. Souza-Neto, Guilherme Targino Valente, Patricia Akemi Assato, Felipe Allan da Silva da Costa, Bianca Cechetto Carlos. Mendelics: Bibiana Santos, João Paulo Kitajima, Erika Freitas, David Schlesinger. Hemocentro Ribeirão Preto: Simone Kashima, Evandra Strazza Rodrigues, Svetoslav Nanev Slavov, Elaine Vieira dos Santos, Rafael dos Santos Bezerra, Luiz Carlos Junior de Alcantara, Marta Giovanetti, Vagner Fonseca, Flavia Aburjaile, Rodrigo Tocantins Calado. FAMERP-SJRP: Cecília Artico Banho, Lívia Sacchetto, Fábio Sossai Possebon, Leila Sabrina Ullmann, Cíntia Bittar, Guilherme Campos, Helena Lage Ferreira, Jorge A. Petroli Marchesi, Maisa C. Pereira Parra, Marília Moraes, Paula Rahal, Paulo Inacio da Costa, João Pessoa Araújo Jr., Maurício Lacerda Nogueira. Prefeitura de Sao Paulo: Melissa Palmieri. |
| EPI_ISL_1966220 | PRONTO ATENDIMENTO SAO JOSE | Instituto Butantan / Mendelics | Instituto Butantan: Dimas Tadeu Covas, Sandra Coccuzzo Sampaio, Maria Carolina Elias, José Salvatore Leister Patané, Vincent Louis Viala, Antonio Jorge Martins, Ricardo Haddad, Claudia Renata dos Santos Barros, Elaine Cristina Marqueze, Raul Machado Neto, Debora Botequio Moretti, Jardelina de Souza Todao Bernardino, Loyze Paola Oliveira de Lima, Luiz Aurelio de Campos Crispin. Centro de Genômica Funcional da ESALQ: Luiz Lehmann Coutinho, Ricardo Augusto Brassaloti, Raquel de Lello Rocha Campos Cassano. NGS Soluções Genômicas: Pilar Drummond Sampaio Corrêa Mariani. FZEA-USP Pirassununga: Mirele Daiana Poleti, Jessika Cristina Chagas Lesbon, Elisangela Chicaroni Mattos, Heidge Fukumasu. USP-Botucatu: Rejane Maria Tommasini Grotto, Jayme A. Souza-Neto, Guilherme Targino Valente, Patricia Akemi Assato, Felipe Allan da Silva da Costa, Bianca Cechetto Carlos. Mendelics: Bibiana Santos, João Paulo Kitajima, Erika Freitas, David Schlesinger. Hemocentro Ribeirão Preto: Simone Kashima, Evandra Strazza Rodrigues, Svetoslav Nanev Slavov, Elaine Vieira dos Santos, Rafael dos Santos Bezerra, Luiz Carlos Junior de Alcantara, Marta Giovanetti, Vagner Fonseca, Flavia Aburjaile, Rodrigo Tocantins Calado. FAMERP-SJRP: Cecília Artico Banho, Lívia Sacchetto, Fábio Sossai Possebon, Leila Sabrina Ullmann, Cíntia Bittar, Guilherme Campos, Helena Lage Ferreira, Jorge A. Petroli Marchesi, Maisa C. Pereira Parra, Marília Moraes, Paula Rahal, Paulo Inacio da Costa, João Pessoa Araújo Jr., Maurício Lacerda Nogueira. Prefeitura de Sao Paulo: Melissa Palmieri. |
| EPI_ISL_1966221 | PRONTO ATENDIMENTO VILA PADRE ANCHIETA | Instituto Butantan / Mendelics | Instituto Butantan: Dimas Tadeu Covas, Sandra Coccuzzo Sampaio, Maria Carolina Elias, José Salvatore Leister Patané, Vincent Louis Viala, Antonio Jorge Martins, Ricardo Haddad, Claudia Renata dos Santos Barros, Elaine Cristina Marqueze, Raul Machado Neto, Debora Botequio Moretti, Jardelina de Souza Todao Bernardino, Loyze Paola Oliveira de Lima, Luiz Aurelio de Campos Crispin. Centro de Genômica Funcional da ESALQ: Luiz Lehmann Coutinho, Ricardo Augusto Brassaloti, Raquel de Lello Rocha Campos Cassano. NGS Soluções Genômicas: Pilar Drummond Sampaio Corrêa Mariani. FZEA-USP Pirassununga: Mirele Daiana Poleti, Jessika Cristina Chagas Lesbon, Elisangela Chicaroni Mattos, Heidge Fukumasu. USP-Botucatu: Rejane Maria Tommasini Grotto, Jayme A. Souza-Neto, Guilherme Targino Valente, Patricia Akemi Assato, Felipe Allan da Silva da Costa, Bianca Cechetto Carlos. Mendelics: Bibiana Santos, João Paulo Kitajima, Erika Freitas, David Schlesinger. Hemocentro Ribeirão Preto: Simone Kashima, Evandra Strazza Rodrigues, Svetoslav Nanev Slavov, Elaine Vieira dos Santos, Rafael dos Santos Bezerra, Luiz Carlos Junior de Alcantara, Marta Giovanetti, Vagner Fonseca, Flavia Aburjaile, Rodrigo Tocantins Calado. FAMERP-SJRP: Cecília Artico Banho, Lívia Sacchetto, Fábio Sossai Possebon, Leila Sabrina Ullmann, Cíntia Bittar, Guilherme Campos, Helena Lage Ferreira, Jorge A. Petroli Marchesi, Maisa C. Pereira Parra, Marília Moraes, Paula Rahal, Paulo Inacio da Costa, João Pessoa Araújo Jr., Maurício Lacerda Nogueira. Prefeitura de Sao Paulo: Melissa Palmieri. |
| EPI_ISL_1966222 | PRONTO ATENDIMENTO SAO JOSE | Instituto Butantan / Mendelics | Instituto Butantan: Dimas Tadeu Covas, Sandra Coccuzzo Sampaio, Maria Carolina Elias, José Salvatore Leister Patané, Vincent Louis Viala, Antonio Jorge Martins, Ricardo Haddad, Claudia Renata dos Santos Barros, Elaine Cristina Marqueze, Raul Machado Neto, Debora Botequio Moretti, Jardelina de Souza Todao Bernardino, Loyze Paola Oliveira de Lima, Luiz Aurelio de Campos Crispin. Centro de Genômica Funcional da ESALQ: Luiz Lehmann Coutinho, Ricardo Augusto Brassaloti, Raquel de Lello Rocha Campos Cassano. NGS Soluções Genômicas: Pilar Drummond Sampaio Corrêa Mariani. FZEA-USP Pirassununga: Mirele Daiana Poleti, Jessika Cristina Chagas Lesbon, Elisangela Chicaroni Mattos, Heidge Fukumasu. USP-Botucatu: Rejane Maria Tommasini Grotto, Jayme A. Souza-Neto, Guilherme Targino Valente, Patricia Akemi Assato, Felipe Allan da Silva da Costa, Bianca Cechetto Carlos. Mendelics: Bibiana Santos, João Paulo Kitajima, Erika Freitas, David Schlesinger. Hemocentro Ribeirão Preto: Simone Kashima, Evandra Strazza Rodrigues, Svetoslav Nanev Slavov, Elaine Vieira dos Santos, Rafael dos Santos Bezerra, Luiz Carlos Junior de Alcantara, Marta Giovanetti, Vagner Fonseca, Flavia Aburjaile, Rodrigo Tocantins Calado. FAMERP-SJRP: Cecília Artico Banho, Lívia Sacchetto, Fábio Sossai Possebon, Leila Sabrina Ullmann, Cíntia Bittar, Guilherme Campos, Helena Lage Ferreira, Jorge A. Petroli Marchesi, Maisa C. Pereira Parra, Marília Moraes, Paula Rahal, Paulo Inacio da Costa, João Pessoa Araújo Jr., Maurício Lacerda Nogueira. Prefeitura de Sao Paulo: Melissa Palmieri. |
| EPI_ISL_1966223, EPI_ISL_1966224, EPI_ISL_1966225 | PRONTO ATENDIMENTO VILA PADRE ANCHIETA | Instituto Butantan / Mendelics | Instituto Butantan: Dimas Tadeu Covas, Sandra Coccuzzo Sampaio, Maria Carolina Elias, José Salvatore Leister Patané, Vincent Louis Viala, Antonio Jorge Martins, Ricardo Haddad, Claudia Renata dos Santos Barros, Elaine Cristina Marqueze, Raul Machado Neto, Debora Botequio Moretti, Jardelina de Souza Todao Bernardino, Loyze Paola Oliveira de Lima, Luiz Aurelio de Campos Crispin. Centro de Genômica Funcional da ESALQ: Luiz Lehmann Coutinho, Ricardo Augusto Brassaloti, Raquel de Lello Rocha Campos Cassano. NGS Soluções Genômicas: Pilar Drummond Sampaio Corrêa Mariani. FZEA-USP Pirassununga: Mirele Daiana Poleti, Jessika Cristina Chagas Lesbon, Elisangela Chicaroni Mattos, Heidge Fukumasu. USP-Botucatu: Rejane Maria Tommasini Grotto, Jayme A. Souza-Neto, Guilherme Targino Valente, Patricia Akemi Assato, Felipe Allan da Silva da Costa, Bianca Cechetto Carlos. Mendelics: Bibiana Santos, João Paulo Kitajima, Erika Freitas, David Schlesinger. Hemocentro Ribeirão Preto: Simone Kashima, Evandra Strazza Rodrigues, Svetoslav Nanev Slavov, Elaine Vieira dos Santos, Rafael dos Santos Bezerra, Luiz Carlos Junior de Alcantara, Marta Giovanetti, Vagner Fonseca, Flavia Aburjaile, Rodrigo Tocantins Calado. FAMERP-SJRP: Cecília Artico Banho, Lívia Sacchetto, Fábio Sossai Possebon, Leila Sabrina Ullmann, Cíntia Bittar, Guilherme Campos, Helena Lage Ferreira, Jorge A. Petroli Marchesi, Maisa C. Pereira Parra, Marília Moraes, Paula Rahal, Paulo Inacio da Costa, João Pessoa Araújo Jr., Maurício Lacerda Nogueira. Prefeitura de Sao Paulo: Melissa Palmieri. |
| EPI_ISL_1966226 | PRONTO ATENDIMENTO SAO JOSE | Instituto Butantan / Mendelics | Instituto Butantan: Dimas Tadeu Covas, Sandra Coccuzzo Sampaio, Maria Carolina Elias, José Salvatore Leister Patané, Vincent Louis Viala, Antonio Jorge Martins, Ricardo Haddad, Claudia Renata dos Santos Barros, Elaine Cristina Marqueze, Raul Machado Neto, Debora Botequio Moretti, Jardelina de Souza Todao Bernardino, Loyze Paola Oliveira de Lima, Luiz Aurelio de Campos Crispin. Centro de Genômica Funcional da ESALQ: Luiz Lehmann |

| | | | |
|---|---|---|---|
| | | | Coutinho, Ricardo Augusto Brassaloti, Raquel de Lello Rocha Campos Cassano. NGS Soluções Genômicas: Pilar Drummond Sampaio Corrêa Mariani. FZEA-USP Pirassununga: Mirele Daiana Poleti, Jessika Cristina Chagas Lesbon, Elisangela Chicaroni Mattos, Heidge Fukumasu. USP-Botucatu: Rejane Maria Tommasini Grotto, Jayme A. Souza-Neto, Guilherme Targino Valente, Patricia Akemi Assato, Felipe Allan da Silva da Costa, Bianca Cechetto Carlos. Mendelics: Bibiana Santos, João Paulo Kitajima, Erika Freitas, David Schlesinger. Hemocentro Ribeirão Preto: Simone Kashima, Evandra Strazza Rodrigues, Svetoslav Nanev Slavov, Elaine Vieira dos Santos, Rafael dos Santos Bezerra, Luiz Carlos Junior de Alcantara, Marta Giovanetti, Vagner Fonseca, Flavia Aburjaile, Rodrigo Tocantins Calado. FAMERP-SJRP: Cecília Artico Banho, Lívia Sacchetto, Fábio Sossai Possebon, Leila Sabrina Ullmann, Cíntia Bittar, Guilherme Campos, Helena Lage Ferreira, Jorge A. Petroli Marchesi, Maisa C. Pereira Parra, Marília Moraes, Paula Rahal, Paulo Inacio da Costa, João Pessoa Araújo Jr., Maurício Lacerda Nogueira. Prefeitura de Sao Paulo: Melissa Palmieri. |
| EPI_ISL_1966227 | UBS SAO JUDAS | Instituto Butantan / Mendelics | Instituto Butantan: Dimas Tadeu Covas, Sandra Coccuzzo Sampaio, Maria Carolina Elias, José Salvatore Leister Patané, Vincent Louis Viala, Antonio Jorge Martins, Ricardo Haddad, Claudia Renata dos Santos Barros, Elaine Cristina Marqueze, Raul Machado Neto, Debora Botequio Moretti, Jardelina de Souza Todao Bernardino, Loyze Paola Oliveira de Lima, Luiz Aurelio de Campos Crispin. Centro de Genômica Funcional da ESALQ: Luiz Lehmann Coutinho, Ricardo Augusto Brassaloti, Raquel de Lello Rocha Campos Cassano. NGS Soluções Genômicas: Pilar Drummond Sampaio Corrêa Mariani. FZEA-USP Pirassununga: Mirele Daiana Poleti, Jessika Cristina Chagas Lesbon, Elisangela Chicaroni Mattos, Heidge Fukumasu. USP-Botucatu: Rejane Maria Tommasini Grotto, Jayme A. Souza-Neto, Guilherme Targino Valente, Patricia Akemi Assato, Felipe Allan da Silva da Costa, Bianca Cechetto Carlos. Mendelics: Bibiana Santos, João Paulo Kitajima, Erika Freitas, David Schlesinger. Hemocentro Ribeirão Preto: Simone Kashima, Evandra Strazza Rodrigues, Svetoslav Nanev Slavov, Elaine Vieira dos Santos, Rafael dos Santos Bezerra, Luiz Carlos Junior de Alcantara, Marta Giovanetti, Vagner Fonseca, Flavia Aburjaile, Rodrigo Tocantins Calado. FAMERP-SJRP: Cecília Artico Banho, Lívia Sacchetto, Fábio Sossai Possebon, Leila Sabrina Ullmann, Cíntia Bittar, Guilherme Campos, Helena Lage Ferreira, Jorge A. Petroli Marchesi, Maisa C. Pereira Parra, Marília Moraes, Paula Rahal, Paulo Inacio da Costa, João Pessoa Araújo Jr., Maurício Lacerda Nogueira. Prefeitura de Sao Paulo: Melissa Palmieri. |
| EPI_ISL_1966228 | CENTRO DE SAUDE DE JULIO MESQUITA | Instituto Butantan / Mendelics | Instituto Butantan: Dimas Tadeu Covas, Sandra Coccuzzo Sampaio, Maria Carolina Elias, José Salvatore Leister Patané, Vincent Louis Viala, Antonio Jorge Martins, Ricardo Haddad, Claudia Renata dos Santos Barros, Elaine Cristina Marqueze, Raul Machado Neto, Debora Botequio Moretti, Jardelina de Souza Todao Bernardino, Loyze Paola Oliveira de Lima, Luiz Aurelio de Campos Crispin. Centro de Genômica Funcional da ESALQ: Luiz Lehmann Coutinho, Ricardo Augusto Brassaloti, Raquel de Lello Rocha Campos Cassano. NGS Soluções Genômicas: Pilar Drummond Sampaio Corrêa Mariani. FZEA-USP Pirassununga: Mirele Daiana Poleti, Jessika Cristina Chagas Lesbon, Elisangela Chicaroni Mattos, Heidge Fukumasu. USP-Botucatu: Rejane Maria Tommasini Grotto, Jayme A. Souza-Neto, Guilherme Targino Valente, Patricia Akemi Assato, Felipe Allan da Silva da Costa, Bianca Cechetto Carlos. Mendelics: Bibiana Santos, João Paulo Kitajima, Erika Freitas, David Schlesinger. Hemocentro Ribeirão Preto: Simone Kashima, Evandra Strazza Rodrigues, Svetoslav Nanev Slavov, Elaine Vieira dos Santos, Rafael dos Santos Bezerra, Luiz Carlos Junior de Alcantara, Marta Giovanetti, Vagner Fonseca, Flavia Aburjaile, Rodrigo Tocantins Calado. FAMERP-SJRP: Cecília Artico Banho, Lívia Sacchetto, Fábio Sossai Possebon, Leila Sabrina Ullmann, Cíntia Bittar, Guilherme Campos, Helena Lage Ferreira, Jorge A. Petroli Marchesi, Maisa C. Pereira Parra, Marília Moraes, Paula Rahal, Paulo Inacio da Costa, João Pessoa Araújo Jr., Maurício Lacerda Nogueira. Prefeitura de Sao Paulo: Melissa Palmieri. |
| EPI_ISL_1966229 | UBS SYLVIO JOAO L DE LUCIA | Instituto Butantan / Mendelics | Instituto Butantan: Dimas Tadeu Covas, Sandra Coccuzzo Sampaio, Maria Carolina Elias, José Salvatore Leister Patané, Vincent Louis Viala, Antonio Jorge Martins, Ricardo Haddad, Claudia Renata dos Santos Barros, Elaine Cristina Marqueze, Raul Machado Neto, Debora Botequio Moretti, Jardelina de Souza Todao Bernardino, Loyze Paola Oliveira de Lima, Luiz Aurelio de Campos Crispin. Centro de Genômica Funcional da ESALQ: Luiz Lehmann Coutinho, Ricardo Augusto Brassaloti, Raquel de Lello Rocha Campos Cassano. NGS Soluções Genômicas: Pilar Drummond Sampaio Corrêa Mariani. FZEA-USP Pirassununga: Mirele Daiana Poleti, Jessika Cristina Chagas Lesbon, Elisangela Chicaroni Mattos, Heidge Fukumasu. USP-Botucatu: Rejane Maria Tommasini Grotto, Jayme A. Souza-Neto, Guilherme Targino Valente, Patricia Akemi Assato, Felipe Allan da Silva da Costa, Bianca Cechetto Carlos. Mendelics: Bibiana Santos, João Paulo Kitajima, Erika Freitas, David Schlesinger. Hemocentro Ribeirão Preto: Simone Kashima, Evandra Strazza Rodrigues, Svetoslav Nanev Slavov, Elaine Vieira dos Santos, Rafael dos Santos Bezerra, Luiz Carlos Junior de Alcantara, Marta Giovanetti, Vagner Fonseca, Flavia Aburjaile, Rodrigo Tocantins Calado. FAMERP-SJRP: Cecília Artico Banho, Lívia Sacchetto, Fábio Sossai Possebon, Leila Sabrina Ullmann, Cíntia Bittar, Guilherme Campos, Helena Lage Ferreira, Jorge A. Petroli Marchesi, Maisa C. Pereira Parra, Marília Moraes, Paula Rahal, Paulo Inacio da Costa, João Pessoa Araújo Jr., Maurício Lacerda Nogueira. Prefeitura de Sao Paulo: Melissa Palmieri. |
| EPI_ISL_1966230 | UBS EMILIA COSME CERQUEIRA | Instituto Butantan / Mendelics | Instituto Butantan: Dimas Tadeu Covas, Sandra Coccuzzo Sampaio, Maria Carolina Elias, José Salvatore Leister Patané, Vincent Louis Viala, Antonio Jorge Martins, Ricardo Haddad, Claudia Renata dos Santos Barros, Elaine Cristina Marqueze, Raul Machado Neto, Debora Botequio Moretti, Jardelina de Souza Todao Bernardino, Loyze Paola Oliveira de Lima, Luiz Aurelio de Campos Crispin. Centro de Genômica Funcional da ESALQ: Luiz Lehmann Coutinho, Ricardo Augusto Brassaloti, Raquel de Lello Rocha Campos Cassano. NGS Soluções Genômicas: Pilar Drummond Sampaio Corrêa Mariani. FZEA-USP Pirassununga: Mirele Daiana Poleti, Jessika Cristina Chagas Lesbon, Elisangela Chicaroni Mattos, Heidge Fukumasu. USP-Botucatu: Rejane Maria Tommasini Grotto, Jayme A. Souza-Neto, Guilherme Targino Valente, Patricia Akemi Assato, Felipe Allan da Silva da Costa, Bianca Cechetto Carlos. Mendelics: Bibiana Santos, João Paulo Kitajima, Erika Freitas, David Schlesinger. Hemocentro Ribeirão Preto: Simone Kashima, Evandra Strazza Rodrigues, Svetoslav Nanev Slavov, Elaine Vieira dos Santos, Rafael dos Santos Bezerra, Luiz Carlos Junior de Alcantara, Marta Giovanetti, Vagner Fonseca, Flavia Aburjaile, Rodrigo Tocantins Calado. FAMERP-SJRP: Cecília Artico Banho, Lívia Sacchetto, Fábio Sossai Possebon, Leila Sabrina Ullmann, Cíntia Bittar, Guilherme Campos, Helena Lage Ferreira, Jorge A. Petroli Marchesi, Maisa C. Pereira Parra, Marília Moraes, Paula Rahal, Paulo Inacio da Costa, João Pessoa Araújo Jr., Maurício Lacerda Nogueira. Prefeitura de Sao Paulo: Melissa Palmieri. |
| EPI_ISL_1966231 | UBS IRMA AGUEDA MARIA JAIME | Instituto Butantan / Mendelics | Instituto Butantan: Dimas Tadeu Covas, Sandra Coccuzzo Sampaio, Maria Carolina Elias, José Salvatore Leister Patané, Vincent Louis Viala, Antonio Jorge Martins, Ricardo Haddad, Claudia Renata dos Santos Barros, Elaine Cristina Marqueze, Raul Machado Neto, Debora Botequio Moretti, Jardelina de Souza Todao Bernardino, Loyze Paola Oliveira de Lima, Luiz Aurelio de Campos Crispin. Centro de Genômica Funcional da ESALQ: Luiz Lehmann Coutinho, Ricardo Augusto Brassaloti, Raquel de Lello Rocha Campos Cassano. NGS Soluções Genômicas: Pilar Drummond Sampaio Corrêa Mariani. FZEA-USP Pirassununga: Mirele Daiana Poleti, Jessika Cristina Chagas Lesbon, Elisangela Chicaroni Mattos, Heidge Fukumasu. USP-Botucatu: Rejane Maria Tommasini Grotto, Jayme A. Souza-Neto, Guilherme Targino Valente, Patricia Akemi Assato, Felipe Allan da Silva da Costa, Bianca Cechetto Carlos. Mendelics: Bibiana Santos, João Paulo Kitajima, Erika Freitas, David Schlesinger. Hemocentro Ribeirão Preto: Simone Kashima, Evandra Strazza Rodrigues, Svetoslav Nanev Slavov, Elaine Vieira dos Santos, Rafael dos Santos Bezerra, Luiz Carlos Junior de Alcantara, Marta Giovanetti, Vagner Fonseca, Flavia Aburjaile, Rodrigo Tocantins Calado. FAMERP-SJRP: Cecília Artico Banho, Lívia Sacchetto, Fábio Sossai Possebon, Leila Sabrina Ullmann, Cíntia Bittar, Guilherme Campos, Helena Lage Ferreira, Jorge A. Petroli Marchesi, Maisa C. Pereira Parra, Marília Moraes, Paula Rahal, Paulo Inacio da Costa, João Pessoa Araújo Jr., Maurício Lacerda Nogueira. Prefeitura de Sao Paulo: Melissa Palmieri. |
| EPI_ISL_1966232 | USAFA GUILHERMINA | Instituto Butantan / Mendelics | Instituto Butantan: Dimas Tadeu Covas, Sandra Coccuzzo Sampaio, Maria Carolina Elias, José Salvatore Leister Patané, Vincent Louis Viala, Antonio Jorge Martins, Ricardo Haddad, Claudia Renata dos Santos Barros, Elaine Cristina Marqueze, Raul Machado Neto, Debora Botequio Moretti, Jardelina de Souza Todao Bernardino, Loyze Paola Oliveira de Lima, Luiz Aurelio de Campos Crispin. Centro de Genômica Funcional da ESALQ: Luiz Lehmann Coutinho, Ricardo Augusto Brassaloti, Raquel de Lello Rocha Campos Cassano. NGS Soluções Genômicas: Pilar Drummond Sampaio Corrêa Mariani. FZEA-USP Pirassununga: Mirele Daiana Poleti, Jessika Cristina Chagas Lesbon, Elisangela Chicaroni Mattos, Heidge Fukumasu. USP-Botucatu: Rejane Maria Tommasini Grotto, Jayme A. Souza-Neto, Guilherme Targino Valente, Patricia Akemi Assato, Felipe Allan da Silva da Costa, Bianca Cechetto Carlos. Mendelics: Bibiana Santos, João Paulo Kitajima, Erika Freitas, David Schlesinger. Hemocentro Ribeirão Preto: Simone Kashima, Evandra Strazza Rodrigues, Svetoslav Nanev Slavov, Elaine Vieira dos Santos, Rafael dos Santos Bezerra, Luiz Carlos Junior de Alcantara, Marta Giovanetti, Vagner Fonseca, Flavia Aburjaile, Rodrigo Tocantins Calado. FAMERP-SJRP: Cecília Artico Banho, Lívia Sacchetto, Fábio Sossai Possebon, Leila Sabrina Ullmann, Cíntia Bittar, Guilherme Campos, Helena Lage Ferreira, Jorge A. Petroli Marchesi, Maisa C. Pereira Parra, Marília Moraes, Paula Rahal, Paulo Inacio da Costa, João Pessoa Araújo Jr., Maurício Lacerda Nogueira. Prefeitura de Sao Paulo: Melissa Palmieri. |
| EPI_ISL_1966233 | USAFA MARACANA | Instituto Butantan / Mendelics | Instituto Butantan: Dimas Tadeu Covas, Sandra Coccuzzo Sampaio, Maria Carolina Elias, José Salvatore Leister Patané, Vincent Louis Viala, Antonio Jorge Martins, Ricardo Haddad, Claudia Renata dos Santos Barros, Elaine Cristina Marqueze, Raul Machado Neto, Debora Botequio Moretti, Jardelina de Souza Todao Bernardino, Loyze Paola Oliveira de Lima, Luiz Aurelio de Campos Crispin. Centro de Genômica Funcional da ESALQ: Luiz Lehmann Coutinho, Ricardo Augusto Brassaloti, Raquel de Lello Rocha Campos Cassano. NGS Soluções Genômicas: Pilar Drummond Sampaio Corrêa Mariani. FZEA-USP Pirassununga: Mirele Daiana Poleti, Jessika Cristina Chagas Lesbon, Elisangela Chicaroni Mattos, Heidge Fukumasu. USP-Botucatu: Rejane Maria Tommasini Grotto, Jayme A. Souza-Neto, Guilherme Targino Valente, Patricia Akemi Assato, Felipe Allan da Silva da Costa, Bianca Cechetto Carlos. Mendelics: Bibiana Santos, João Paulo Kitajima, Erika Freitas, David Schlesinger. Hemocentro Ribeirão Preto: Simone Kashima, Evandra Strazza Rodrigues, Svetoslav Nanev Slavov, Elaine Vieira dos Santos, Rafael dos Santos Bezerra, Luiz Carlos Junior de Alcantara, Marta Giovanetti, Vagner |

| | | | |
|---|---|---|---|
| | | | Fonseca, Flavia Aburjaile, Rodrigo Tocantins Calado. FAMERP-SJRP: Cecília Artico Banho, Lívia Sacchetto, Fábio Sossai Possebon, Leila Sabrina Ullmann, Cíntia Bittar, Guilherme Campos, Helena Lage Ferreira, Jorge A. Petroli Marchesi, Maisa C. Pereira Parra, Marília Moraes, Paula Rahal, Paulo Inacio da Costa, João Pessoa Araújo Jr., Maurício Lacerda Nogueira. Prefeitura de Sao Paulo: Melissa Palmieri. |
| EPI_ISL_1966234 | USAFA TUDE BASTOS | Instituto Butantan / Mendelics | Instituto Butantan: Dimas Tadeu Covas, Sandra Coccuzzo Sampaio, Maria Carolina Elias, José Salvatore Leister Patané, Vincent Louis Viala, Antonio Jorge Martins, Ricardo Haddad, Claudia Renata dos Santos Barros, Elaine Cristina Marqueze, Raul Machado Neto, Debora Botequio Moretti, Jardelina de Souza Todao Bernardino, Loyze Paola Oliveira de Lima, Luiz Aurelio de Campos Crispin. Centro de Genômica Funcional da ESALQ: Luiz Lehmann Coutinho, Ricardo Augusto Brassaloti, Raquel de Lello Rocha Campos Cassano. NGS Soluções Genômicas: Pilar Drummond Sampaio Corrêa Mariani. FZEA-USP Pirassununga: Mirele Daiana Poleti, Jessika Cristina Chagas Lesbon, Elisangela Chicaroni Mattos, Heidge Fukumasu. USP-Botucatu: Rejane Maria Tommasini Grotto, Jayme A. Souza-Neto, Guilherme Targino Valente, Patricia Akemi Assato, Felipe Allan da Silva da Costa, Bianca Cechetto Carlos. Mendelics: Bibiana Santos, João Paulo Kitajima, Erika Freitas, David Schlesinger. Hemocentro Ribeirão Preto: Simone Kashima, Evandra Strazza Rodrigues, Svetoslav Nanev Slavov, Elaine Vieira dos Santos, Rafael dos Santos Bezerra, Luiz Carlos Junior de Alcantara, Marta Giovanetti, Vagner Fonseca, Flavia Aburjaile, Rodrigo Tocantins Calado. FAMERP-SJRP: Cecília Artico Banho, Lívia Sacchetto, Fábio Sossai Possebon, Leila Sabrina Ullmann, Cíntia Bittar, Guilherme Campos, Helena Lage Ferreira, Jorge A. Petroli Marchesi, Maisa C. Pereira Parra, Marília Moraes, Paula Rahal, Paulo Inacio da Costa, João Pessoa Araújo Jr., Maurício Lacerda Nogueira. Prefeitura de Sao Paulo: Melissa Palmieri. |
| EPI_ISL_1966235 | CENTRO DE SAUDE III SALES OLIVEIRA | Instituto Butantan / Mendelics | Instituto Butantan: Dimas Tadeu Covas, Sandra Coccuzzo Sampaio, Maria Carolina Elias, José Salvatore Leister Patané, Vincent Louis Viala, Antonio Jorge Martins, Ricardo Haddad, Claudia Renata dos Santos Barros, Elaine Cristina Marqueze, Raul Machado Neto, Debora Botequio Moretti, Jardelina de Souza Todao Bernardino, Loyze Paola Oliveira de Lima, Luiz Aurelio de Campos Crispin. Centro de Genômica Funcional da ESALQ: Luiz Lehmann Coutinho, Ricardo Augusto Brassaloti, Raquel de Lello Rocha Campos Cassano. NGS Soluções Genômicas: Pilar Drummond Sampaio Corrêa Mariani. FZEA-USP Pirassununga: Mirele Daiana Poleti, Jessika Cristina Chagas Lesbon, Elisangela Chicaroni Mattos, Heidge Fukumasu. USP-Botucatu: Rejane Maria Tommasini Grotto, Jayme A. Souza-Neto, Guilherme Targino Valente, Patricia Akemi Assato, Felipe Allan da Silva da Costa, Bianca Cechetto Carlos. Mendelics: Bibiana Santos, João Paulo Kitajima, Erika Freitas, David Schlesinger. Hemocentro Ribeirão Preto: Simone Kashima, Evandra Strazza Rodrigues, Svetoslav Nanev Slavov, Elaine Vieira dos Santos, Rafael dos Santos Bezerra, Luiz Carlos Junior de Alcantara, Marta Giovanetti, Vagner Fonseca, Flavia Aburjaile, Rodrigo Tocantins Calado. FAMERP-SJRP: Cecília Artico Banho, Lívia Sacchetto, Fábio Sossai Possebon, Leila Sabrina Ullmann, Cíntia Bittar, Guilherme Campos, Helena Lage Ferreira, Jorge A. Petroli Marchesi, Maisa C. Pereira Parra, Marília Moraes, Paula Rahal, Paulo Inacio da Costa, João Pessoa Araújo Jr., Maurício Lacerda Nogueira. Prefeitura de Sao Paulo: Melissa Palmieri. |
| EPI_ISL_1966236 | UNIDADE MISTA DE TAIACU | Instituto Butantan / Mendelics | Instituto Butantan: Dimas Tadeu Covas, Sandra Coccuzzo Sampaio, Maria Carolina Elias, José Salvatore Leister Patané, Vincent Louis Viala, Antonio Jorge Martins, Ricardo Haddad, Claudia Renata dos Santos Barros, Elaine Cristina Marqueze, Raul Machado Neto, Debora Botequio Moretti, Jardelina de Souza Todao Bernardino, Loyze Paola Oliveira de Lima, Luiz Aurelio de Campos Crispin. Centro de Genômica Funcional da ESALQ: Luiz Lehmann Coutinho, Ricardo Augusto Brassaloti, Raquel de Lello Rocha Campos Cassano. NGS Soluções Genômicas: Pilar Drummond Sampaio Corrêa Mariani. FZEA-USP Pirassununga: Mirele Daiana Poleti, Jessika Cristina Chagas Lesbon, Elisangela Chicaroni Mattos, Heidge Fukumasu. USP-Botucatu: Rejane Maria Tommasini Grotto, Jayme A. Souza-Neto, Guilherme Targino Valente, Patricia Akemi Assato, Felipe Allan da Silva da Costa, Bianca Cechetto Carlos. Mendelics: Bibiana Santos, João Paulo Kitajima, Erika Freitas, David Schlesinger. Hemocentro Ribeirão Preto: Simone Kashima, Evandra Strazza Rodrigues, Svetoslav Nanev Slavov, Elaine Vieira dos Santos, Rafael dos Santos Bezerra, Luiz Carlos Junior de Alcantara, Marta Giovanetti, Vagner Fonseca, Flavia Aburjaile, Rodrigo Tocantins Calado. FAMERP-SJRP: Cecília Artico Banho, Lívia Sacchetto, Fábio Sossai Possebon, Leila Sabrina Ullmann, Cíntia Bittar, Guilherme Campos, Helena Lage Ferreira, Jorge A. Petroli Marchesi, Maisa C. Pereira Parra, Marília Moraes, Paula Rahal, Paulo Inacio da Costa, João Pessoa Araújo Jr., Maurício Lacerda Nogueira. Prefeitura de Sao Paulo: Melissa Palmieri. |
| EPI_ISL_1966237 | UPA DE BEBEDOURO | Instituto Butantan / Mendelics | Instituto Butantan: Dimas Tadeu Covas, Sandra Coccuzzo Sampaio, Maria Carolina Elias, José Salvatore Leister Patané, Vincent Louis Viala, Antonio Jorge Martins, Ricardo Haddad, Claudia Renata dos Santos Barros, Elaine Cristina Marqueze, Raul Machado Neto, Debora Botequio Moretti, Jardelina de Souza Todao Bernardino, Loyze Paola Oliveira de Lima, Luiz Aurelio de Campos Crispin. Centro de Genômica Funcional da ESALQ: Luiz Lehmann Coutinho, Ricardo Augusto Brassaloti, Raquel de Lello Rocha Campos Cassano. NGS Soluções Genômicas: Pilar Drummond Sampaio Corrêa Mariani. FZEA-USP Pirassununga: Mirele Daiana Poleti, Jessika Cristina Chagas Lesbon, Elisangela Chicaroni Mattos, Heidge Fukumasu. USP-Botucatu: Rejane Maria Tommasini Grotto, Jayme A. Souza-Neto, Guilherme Targino Valente, Patricia Akemi Assato, Felipe Allan da Silva da Costa, Bianca Cechetto Carlos. Mendelics: Bibiana Santos, João Paulo Kitajima, Erika Freitas, David Schlesinger. Hemocentro Ribeirão Preto: Simone Kashima, Evandra Strazza Rodrigues, Svetoslav Nanev Slavov, Elaine Vieira dos Santos, Rafael dos Santos Bezerra, Luiz Carlos Junior de Alcantara, Marta Giovanetti, Vagner Fonseca, Flavia Aburjaile, Rodrigo Tocantins Calado. FAMERP-SJRP: Cecília Artico Banho, Lívia Sacchetto, Fábio Sossai Possebon, Leila Sabrina Ullmann, Cíntia Bittar, Guilherme Campos, Helena Lage Ferreira, Jorge A. Petroli Marchesi, Maisa C. Pereira Parra, Marília Moraes, Paula Rahal, Paulo Inacio da Costa, João Pessoa Araújo Jr., Maurício Lacerda Nogueira. Prefeitura de Sao Paulo: Melissa Palmieri. |
| EPI_ISL_1966238 | CS II DR SEBASTIAO RIBEIRO DO VALLE | Instituto Butantan / Mendelics | Instituto Butantan: Dimas Tadeu Covas, Sandra Coccuzzo Sampaio, Maria Carolina Elias, José Salvatore Leister Patané, Vincent Louis Viala, Antonio Jorge Martins, Ricardo Haddad, Claudia Renata dos Santos Barros, Elaine Cristina Marqueze, Raul Machado Neto, Debora Botequio Moretti, Jardelina de Souza Todao Bernardino, Loyze Paola Oliveira de Lima, Luiz Aurelio de Campos Crispin. Centro de Genômica Funcional da ESALQ: Luiz Lehmann Coutinho, Ricardo Augusto Brassaloti, Raquel de Lello Rocha Campos Cassano. NGS Soluções Genômicas: Pilar Drummond Sampaio Corrêa Mariani. FZEA-USP Pirassununga: Mirele Daiana Poleti, Jessika Cristina Chagas Lesbon, Elisangela Chicaroni Mattos, Heidge Fukumasu. USP-Botucatu: Rejane Maria Tommasini Grotto, Jayme A. Souza-Neto, Guilherme Targino Valente, Patricia Akemi Assato, Felipe Allan da Silva da Costa, Bianca Cechetto Carlos. Mendelics: Bibiana Santos, João Paulo Kitajima, Erika Freitas, David Schlesinger. Hemocentro Ribeirão Preto: Simone Kashima, Evandra Strazza Rodrigues, Svetoslav Nanev Slavov, Elaine Vieira dos Santos, Rafael dos Santos Bezerra, Luiz Carlos Junior de Alcantara, Marta Giovanetti, Vagner Fonseca, Flavia Aburjaile, Rodrigo Tocantins Calado. FAMERP-SJRP: Cecília Artico Banho, Lívia Sacchetto, Fábio Sossai Possebon, Leila Sabrina Ullmann, Cíntia Bittar, Guilherme Campos, Helena Lage Ferreira, Jorge A. Petroli Marchesi, Maisa C. Pereira Parra, Marília Moraes, Paula Rahal, Paulo Inacio da Costa, João Pessoa Araújo Jr., Maurício Lacerda Nogueira. Prefeitura de Sao Paulo: Melissa Palmieri. |
| EPI_ISL_1966239, EPI_ISL_1966240, EPI_ISL_1966241 | CENTRO DE SAUDE DR MARIO DIAS DE AGUIAR CAPIVARI | Instituto Butantan / Mendelics | Instituto Butantan: Dimas Tadeu Covas, Sandra Coccuzzo Sampaio, Maria Carolina Elias, José Salvatore Leister Patané, Vincent Louis Viala, Antonio Jorge Martins, Ricardo Haddad, Claudia Renata dos Santos Barros, Elaine Cristina Marqueze, Raul Machado Neto, Debora Botequio Moretti, Jardelina de Souza Todao Bernardino, Loyze Paola Oliveira de Lima, Luiz Aurelio de Campos Crispin. Centro de Genômica Funcional da ESALQ: Luiz Lehmann Coutinho, Ricardo Augusto Brassaloti, Raquel de Lello Rocha Campos Cassano. NGS Soluções Genômicas: Pilar Drummond Sampaio Corrêa Mariani. FZEA-USP Pirassununga: Mirele Daiana Poleti, Jessika Cristina Chagas Lesbon, Elisangela Chicaroni Mattos, Heidge Fukumasu. USP-Botucatu: Rejane Maria Tommasini Grotto, Jayme A. Souza-Neto, Guilherme Targino Valente, Patricia Akemi Assato, Felipe Allan da Silva da Costa, Bianca Cechetto Carlos. Mendelics: Bibiana Santos, João Paulo Kitajima, Erika Freitas, David Schlesinger. Hemocentro Ribeirão Preto: Simone Kashima, Evandra Strazza Rodrigues, Svetoslav Nanev Slavov, Elaine Vieira dos Santos, Rafael dos Santos Bezerra, Luiz Carlos Junior de Alcantara, Marta Giovanetti, Vagner Fonseca, Flavia Aburjaile, Rodrigo Tocantins Calado. FAMERP-SJRP: Cecília Artico Banho, Lívia Sacchetto, Fábio Sossai Possebon, Leila Sabrina Ullmann, Cíntia Bittar, Guilherme Campos, Helena Lage Ferreira, Jorge A. Petroli Marchesi, Maisa C. Pereira Parra, Marília Moraes, Paula Rahal, Paulo Inacio da Costa, João Pessoa Araújo Jr., Maurício Lacerda Nogueira. Prefeitura de Sao Paulo: Melissa Palmieri. |
| EPI_ISL_1966242 | SANTA CASA DE MIS E MATERNIDADE N SRA DA CONCEICAO CUNHA | Instituto Butantan / Mendelics | Instituto Butantan: Dimas Tadeu Covas, Sandra Coccuzzo Sampaio, Maria Carolina Elias, José Salvatore Leister Patané, Vincent Louis Viala, Antonio Jorge Martins, Ricardo Haddad, Claudia Renata dos Santos Barros, Elaine Cristina Marqueze, Raul Machado Neto, Debora Botequio Moretti, Jardelina de Souza Todao Bernardino, Loyze Paola Oliveira de Lima, Luiz Aurelio de Campos Crispin. Centro de Genômica Funcional da ESALQ: Luiz Lehmann Coutinho, Ricardo Augusto Brassaloti, Raquel de Lello Rocha Campos Cassano. NGS Soluções Genômicas: Pilar Drummond Sampaio Corrêa Mariani. FZEA-USP Pirassununga: Mirele Daiana Poleti, Jessika Cristina Chagas Lesbon, Elisangela Chicaroni Mattos, Heidge Fukumasu. USP-Botucatu: Rejane Maria Tommasini Grotto, Jayme A. Souza-Neto, Guilherme Targino Valente, Patricia Akemi Assato, Felipe Allan da Silva da Costa, Bianca Cechetto Carlos. Mendelics: Bibiana Santos, João Paulo Kitajima, Erika Freitas, David Schlesinger. Hemocentro Ribeirão Preto: Simone Kashima, Evandra Strazza Rodrigues, Svetoslav Nanev Slavov, Elaine Vieira dos Santos, Rafael dos Santos Bezerra, Luiz Carlos Junior de Alcantara, Marta Giovanetti, Vagner Fonseca, Flavia Aburjaile, Rodrigo Tocantins Calado. FAMERP-SJRP: Cecília Artico Banho, Lívia Sacchetto, Fábio Sossai Possebon, Leila Sabrina Ullmann, Cíntia Bittar, Guilherme Campos, Helena Lage Ferreira, Jorge A. Petroli Marchesi, Maisa C. Pereira Parra, Marília Moraes, Paula Rahal, Paulo Inacio da Costa, João Pessoa Araújo Jr., Maurício Lacerda Nogueira. Prefeitura de Sao Paulo: Melissa Palmieri. |
| EPI_ISL_1966243 | UNIDADE MISTA DE SAUDE DE DUMONT | Instituto Butantan / Mendelics | Instituto Butantan: Dimas Tadeu Covas, Sandra Coccuzzo Sampaio, Maria Carolina Elias, José Salvatore Leister Patané, Vincent Louis Viala, Antonio Jorge Martins, Ricardo Haddad, Claudia Renata dos Santos Barros, Elaine Cristina Marqueze, Raul Machado Neto, Debora Botequio Moretti, Jardelina de Souza Todao Bernardino, Loyze Paola Oliveira de Lima, Luiz Aurelio de Campos Crispin. Centro de Genômica Funcional da ESALQ: Luiz Lehmann |

| | | | |
|---|---|---|---|
| | | | Coutinho, Ricardo Augusto Brassaloti, Raquel de Lello Rocha Campos Cassano. NGS Soluções Genômicas: Pilar Drummond Sampaio Corrêa Mariani. FZEA-USP Pirassununga: Mirele Daiana Poleti, Jessika Cristina Chagas Lesbon, Elisangela Chicaroni Mattos, Heidge Fukumasu. USP-Botucatu: Rejane Maria Tommasini Grotto, Jayme A. Souza-Neto, Guilherme Targino Valente, Patricia Akemi Assato, Felipe Allan da Silva da Costa, Bianca Cechetto Carlos. Mendelics: Bibiana Santos, João Paulo Kitajima, Erika Freitas, David Schlesinger. Hemocentro Ribeirão Preto: Simone Kashima, Evandra Strazza Rodrigues, Svetoslav Nanev Slavov, Elaine Vieira dos Santos, Rafael dos Santos Bezerra, Luiz Carlos Junior de Alcantara, Marta Giovanetti, Vagner Fonseca, Flavia Aburjaile, Rodrigo Tocantins Calado. FAMERP-SJRP: Cecília Artico Banho, Lívia Sacchetto, Fábio Sossai Possebon, Leila Sabrina Ullmann, Cíntia Bittar, Guilherme Campos, Helena Lage Ferreira, Jorge A. Petroli Marchesi, Maisa C. Pereira Parra, Marília Moraes, Paula Rahal, Paulo Inacio da Costa, João Pessoa Araújo Jr., Maurício Lacerda Nogueira. Prefeitura de Sao Paulo: Melissa Palmieri. |
| EPI_ISL_1966244 | POSTO MEDICO NATAL DIEGUES DE ESTIVA GERBI | Instituto Butantan / Mendelics | Instituto Butantan: Dimas Tadeu Covas, Sandra Coccuzzo Sampaio, Maria Carolina Elias, José Salvatore Leister Patané, Vincent Louis Viala, Antonio Jorge Martins, Ricardo Haddad, Claudia Renata dos Santos Barros, Elaine Cristina Marqueze, Raul Machado Neto, Debora Botequio Moretti, Jardelina de Souza Todao Bernardino, Loyze Paola Oliveira de Lima, Luiz Aurelio de Campos Crispin. Centro de Genômica Funcional da ESALQ: Luiz Lehmann Coutinho, Ricardo Augusto Brassaloti, Raquel de Lello Rocha Campos Cassano. NGS Soluções Genômicas: Pilar Drummond Sampaio Corrêa Mariani. FZEA-USP Pirassununga: Mirele Daiana Poleti, Jessika Cristina Chagas Lesbon, Elisangela Chicaroni Mattos, Heidge Fukumasu. USP-Botucatu: Rejane Maria Tommasini Grotto, Jayme A. Souza-Neto, Guilherme Targino Valente, Patricia Akemi Assato, Felipe Allan da Silva da Costa, Bianca Cechetto Carlos. Mendelics: Bibiana Santos, João Paulo Kitajima, Erika Freitas, David Schlesinger. Hemocentro Ribeirão Preto: Simone Kashima, Evandra Strazza Rodrigues, Svetoslav Nanev Slavov, Elaine Vieira dos Santos, Rafael dos Santos Bezerra, Luiz Carlos Junior de Alcantara, Marta Giovanetti, Vagner Fonseca, Flavia Aburjaile, Rodrigo Tocantins Calado. FAMERP-SJRP: Cecília Artico Banho, Lívia Sacchetto, Fábio Sossai Possebon, Leila Sabrina Ullmann, Cíntia Bittar, Guilherme Campos, Helena Lage Ferreira, Jorge A. Petroli Marchesi, Maisa C. Pereira Parra, Marília Moraes, Paula Rahal, Paulo Inacio da Costa, João Pessoa Araújo Jr., Maurício Lacerda Nogueira. Prefeitura de Sao Paulo: Melissa Palmieri. |
| EPI_ISL_1966245 | UNIDADE BASICA DE SAUDE DR ORESTES DE MOURA PINTO | Instituto Butantan / Mendelics | Instituto Butantan: Dimas Tadeu Covas, Sandra Coccuzzo Sampaio, Maria Carolina Elias, José Salvatore Leister Patané, Vincent Louis Viala, Antonio Jorge Martins, Ricardo Haddad, Claudia Renata dos Santos Barros, Elaine Cristina Marqueze, Raul Machado Neto, Debora Botequio Moretti, Jardelina de Souza Todao Bernardino, Loyze Paola Oliveira de Lima, Luiz Aurelio de Campos Crispin. Centro de Genômica Funcional da ESALQ: Luiz Lehmann Coutinho, Ricardo Augusto Brassaloti, Raquel de Lello Rocha Campos Cassano. NGS Soluções Genômicas: Pilar Drummond Sampaio Corrêa Mariani. FZEA-USP Pirassununga: Mirele Daiana Poleti, Jessika Cristina Chagas Lesbon, Elisangela Chicaroni Mattos, Heidge Fukumasu. USP-Botucatu: Rejane Maria Tommasini Grotto, Jayme A. Souza-Neto, Guilherme Targino Valente, Patricia Akemi Assato, Felipe Allan da Silva da Costa, Bianca Cechetto Carlos. Mendelics: Bibiana Santos, João Paulo Kitajima, Erika Freitas, David Schlesinger. Hemocentro Ribeirão Preto: Simone Kashima, Evandra Strazza Rodrigues, Svetoslav Nanev Slavov, Elaine Vieira dos Santos, Rafael dos Santos Bezerra, Luiz Carlos Junior de Alcantara, Marta Giovanetti, Vagner Fonseca, Flavia Aburjaile, Rodrigo Tocantins Calado. FAMERP-SJRP: Cecília Artico Banho, Lívia Sacchetto, Fábio Sossai Possebon, Leila Sabrina Ullmann, Cíntia Bittar, Guilherme Campos, Helena Lage Ferreira, Jorge A. Petroli Marchesi, Maisa C. Pereira Parra, Marília Moraes, Paula Rahal, Paulo Inacio da Costa, João Pessoa Araújo Jr., Maurício Lacerda Nogueira. Prefeitura de Sao Paulo: Melissa Palmieri. |
| EPI_ISL_1966246 | UNIDADE MISTA DE IGUAPE | Instituto Butantan / Mendelics | Instituto Butantan: Dimas Tadeu Covas, Sandra Coccuzzo Sampaio, Maria Carolina Elias, José Salvatore Leister Patané, Vincent Louis Viala, Antonio Jorge Martins, Ricardo Haddad, Claudia Renata dos Santos Barros, Elaine Cristina Marqueze, Raul Machado Neto, Debora Botequio Moretti, Jardelina de Souza Todao Bernardino, Loyze Paola Oliveira de Lima, Luiz Aurelio de Campos Crispin. Centro de Genômica Funcional da ESALQ: Luiz Lehmann Coutinho, Ricardo Augusto Brassaloti, Raquel de Lello Rocha Campos Cassano. NGS Soluções Genômicas: Pilar Drummond Sampaio Corrêa Mariani. FZEA-USP Pirassununga: Mirele Daiana Poleti, Jessika Cristina Chagas Lesbon, Elisangela Chicaroni Mattos, Heidge Fukumasu. USP-Botucatu: Rejane Maria Tommasini Grotto, Jayme A. Souza-Neto, Guilherme Targino Valente, Patricia Akemi Assato, Felipe Allan da Silva da Costa, Bianca Cechetto Carlos. Mendelics: Bibiana Santos, João Paulo Kitajima, Erika Freitas, David Schlesinger. Hemocentro Ribeirão Preto: Simone Kashima, Evandra Strazza Rodrigues, Svetoslav Nanev Slavov, Elaine Vieira dos Santos, Rafael dos Santos Bezerra, Luiz Carlos Junior de Alcantara, Marta Giovanetti, Vagner Fonseca, Flavia Aburjaile, Rodrigo Tocantins Calado. FAMERP-SJRP: Cecília Artico Banho, Lívia Sacchetto, Fábio Sossai Possebon, Leila Sabrina Ullmann, Cíntia Bittar, Guilherme Campos, Helena Lage Ferreira, Jorge A. Petroli Marchesi, Maisa C. Pereira Parra, Marília Moraes, Paula Rahal, Paulo Inacio da Costa, João Pessoa Araújo Jr., Maurício Lacerda Nogueira. Prefeitura de Sao Paulo: Melissa Palmieri. |
| EPI_ISL_1966247 | SECRETARIA DE SAUDE DO MUNICIPIO DE IPUA | Instituto Butantan / Mendelics | Instituto Butantan: Dimas Tadeu Covas, Sandra Coccuzzo Sampaio, Maria Carolina Elias, José Salvatore Leister Patané, Vincent Louis Viala, Antonio Jorge Martins, Ricardo Haddad, Claudia Renata dos Santos Barros, Elaine Cristina Marqueze, Raul Machado Neto, Debora Botequio Moretti, Jardelina de Souza Todao Bernardino, Loyze Paola Oliveira de Lima, Luiz Aurelio de Campos Crispin. Centro de Genômica Funcional da ESALQ: Luiz Lehmann Coutinho, Ricardo Augusto Brassaloti, Raquel de Lello Rocha Campos Cassano. NGS Soluções Genômicas: Pilar Drummond Sampaio Corrêa Mariani. FZEA-USP Pirassununga: Mirele Daiana Poleti, Jessika Cristina Chagas Lesbon, Elisangela Chicaroni Mattos, Heidge Fukumasu. USP-Botucatu: Rejane Maria Tommasini Grotto, Jayme A. Souza-Neto, Guilherme Targino Valente, Patricia Akemi Assato, Felipe Allan da Silva da Costa, Bianca Cechetto Carlos. Mendelics: Bibiana Santos, João Paulo Kitajima, Erika Freitas, David Schlesinger. Hemocentro Ribeirão Preto: Simone Kashima, Evandra Strazza Rodrigues, Svetoslav Nanev Slavov, Elaine Vieira dos Santos, Rafael dos Santos Bezerra, Luiz Carlos Junior de Alcantara, Marta Giovanetti, Vagner Fonseca, Flavia Aburjaile, Rodrigo Tocantins Calado. FAMERP-SJRP: Cecília Artico Banho, Lívia Sacchetto, Fábio Sossai Possebon, Leila Sabrina Ullmann, Cíntia Bittar, Guilherme Campos, Helena Lage Ferreira, Jorge A. Petroli Marchesi, Maisa C. Pereira Parra, Marília Moraes, Paula Rahal, Paulo Inacio da Costa, João Pessoa Araújo Jr., Maurício Lacerda Nogueira. Prefeitura de Sao Paulo: Melissa Palmieri. |
| EPI_ISL_1966248, EPI_ISL_1966249 | UNIDADE DE PRONTO ATENDIMENTO DE JALES | Instituto Butantan / Mendelics | Instituto Butantan: Dimas Tadeu Covas, Sandra Coccuzzo Sampaio, Maria Carolina Elias, José Salvatore Leister Patané, Vincent Louis Viala, Antonio Jorge Martins, Ricardo Haddad, Claudia Renata dos Santos Barros, Elaine Cristina Marqueze, Raul Machado Neto, Debora Botequio Moretti, Jardelina de Souza Todao Bernardino, Loyze Paola Oliveira de Lima, Luiz Aurelio de Campos Crispin. Centro de Genômica Funcional da ESALQ: Luiz Lehmann Coutinho, Ricardo Augusto Brassaloti, Raquel de Lello Rocha Campos Cassano. NGS Soluções Genômicas: Pilar Drummond Sampaio Corrêa Mariani. FZEA-USP Pirassununga: Mirele Daiana Poleti, Jessika Cristina Chagas Lesbon, Elisangela Chicaroni Mattos, Heidge Fukumasu. USP-Botucatu: Rejane Maria Tommasini Grotto, Jayme A. Souza-Neto, Guilherme Targino Valente, Patricia Akemi Assato, Felipe Allan da Silva da Costa, Bianca Cechetto Carlos. Mendelics: Bibiana Santos, João Paulo Kitajima, Erika Freitas, David Schlesinger. Hemocentro Ribeirão Preto: Simone Kashima, Evandra Strazza Rodrigues, Svetoslav Nanev Slavov, Elaine Vieira dos Santos, Rafael dos Santos Bezerra, Luiz Carlos Junior de Alcantara, Marta Giovanetti, Vagner Fonseca, Flavia Aburjaile, Rodrigo Tocantins Calado. FAMERP-SJRP: Cecília Artico Banho, Lívia Sacchetto, Fábio Sossai Possebon, Leila Sabrina Ullmann, Cíntia Bittar, Guilherme Campos, Helena Lage Ferreira, Jorge A. Petroli Marchesi, Maisa C. Pereira Parra, Marília Moraes, Paula Rahal, Paulo Inacio da Costa, João Pessoa Araújo Jr., Maurício Lacerda Nogueira. Prefeitura de Sao Paulo: Melissa Palmieri. |
| EPI_ISL_1966250 | UNIDADE SAUDE DA FAMILIA DRA ZILDA ARNS NEUMANN JALES | Instituto Butantan / Mendelics | Instituto Butantan: Dimas Tadeu Covas, Sandra Coccuzzo Sampaio, Maria Carolina Elias, José Salvatore Leister Patané, Vincent Louis Viala, Antonio Jorge Martins, Ricardo Haddad, Claudia Renata dos Santos Barros, Elaine Cristina Marqueze, Raul Machado Neto, Debora Botequio Moretti, Jardelina de Souza Todao Bernardino, Loyze Paola Oliveira de Lima, Luiz Aurelio de Campos Crispin. Centro de Genômica Funcional da ESALQ: Luiz Lehmann Coutinho, Ricardo Augusto Brassaloti, Raquel de Lello Rocha Campos Cassano. NGS Soluções Genômicas: Pilar Drummond Sampaio Corrêa Mariani. FZEA-USP Pirassununga: Mirele Daiana Poleti, Jessika Cristina Chagas Lesbon, Elisangela Chicaroni Mattos, Heidge Fukumasu. USP-Botucatu: Rejane Maria Tommasini Grotto, Jayme A. Souza-Neto, Guilherme Targino Valente, Patricia Akemi Assato, Felipe Allan da Silva da Costa, Bianca Cechetto Carlos. Mendelics: Bibiana Santos, João Paulo Kitajima, Erika Freitas, David Schlesinger. Hemocentro Ribeirão Preto: Simone Kashima, Evandra Strazza Rodrigues, Svetoslav Nanev Slavov, Elaine Vieira dos Santos, Rafael dos Santos Bezerra, Luiz Carlos Junior de Alcantara, Marta Giovanetti, Vagner Fonseca, Flavia Aburjaile, Rodrigo Tocantins Calado. FAMERP-SJRP: Cecília Artico Banho, Lívia Sacchetto, Fábio Sossai Possebon, Leila Sabrina Ullmann, Cíntia Bittar, Guilherme Campos, Helena Lage Ferreira, Jorge A. Petroli Marchesi, Maisa C. Pereira Parra, Marília Moraes, Paula Rahal, Paulo Inacio da Costa, João Pessoa Araújo Jr., Maurício Lacerda Nogueira. Prefeitura de Sao Paulo: Melissa Palmieri. |
| EPI_ISL_1966251 | ESF DR LUIS ERNESTO SANDI MORI JALES | Instituto Butantan / Mendelics | Instituto Butantan: Dimas Tadeu Covas, Sandra Coccuzzo Sampaio, Maria Carolina Elias, José Salvatore Leister Patané, Vincent Louis Viala, Antonio Jorge Martins, Ricardo Haddad, Claudia Renata dos Santos Barros, Elaine Cristina Marqueze, Raul Machado Neto, Debora Botequio Moretti, Jardelina de Souza Todao Bernardino, Loyze Paola Oliveira de Lima, Luiz Aurelio de Campos Crispin. Centro de Genômica Funcional da ESALQ: Luiz Lehmann Coutinho, Ricardo Augusto Brassaloti, Raquel de Lello Rocha Campos Cassano. NGS Soluções Genômicas: Pilar Drummond Sampaio Corrêa Mariani. FZEA-USP Pirassununga: Mirele Daiana Poleti, Jessika Cristina Chagas Lesbon, Elisangela Chicaroni Mattos, Heidge Fukumasu. USP-Botucatu: Rejane Maria Tommasini Grotto, Jayme A. Souza-Neto, Guilherme Targino Valente, Patricia Akemi Assato, Felipe Allan da Silva da Costa, Bianca Cechetto Carlos. Mendelics: Bibiana Santos, João Paulo Kitajima, Erika Freitas, David Schlesinger. Hemocentro Ribeirão Preto: Simone Kashima, Evandra Strazza Rodrigues, Svetoslav Nanev Slavov, Elaine Vieira dos Santos, Rafael dos Santos Bezerra, Luiz Carlos Junior de Alcantara, Marta Giovanetti, Vagner |

| | | | |
|---|---|---|---|
| | | | Fonseca, Flavia Aburjaile, Rodrigo Tocantins Calado. FAMERP-SJRP: Cecília Artico Banho, Lívia Sacchetto, Fábio Sossai Possebon, Leila Sabrina Ullmann, Cíntia Bittar, Guilherme Campos, Helena Lage Ferreira, Jorge A. Petroli Marchesi, Maisa C. Pereira Parra, Marília Moraes, Paula Rahal, Paulo Inacio da Costa, João Pessoa Araújo Jr., Maurício Lacerda Nogueira. Prefeitura de Sao Paulo: Melissa Palmieri. |
| EPI_ISL_1966252, EPI_ISL_1966253, EPI_ISL_1966254, EPI_ISL_1966255 | UBS DR MOHANNA ADAS | Instituto Butantan / Mendelics | Instituto Butantan: Dimas Tadeu Covas, Sandra Coccuzzo Sampaio, Maria Carolina Elias, José Salvatore Leister Patané, Vincent Louis Viala, Antonio Jorge Martins, Ricardo Haddad, Claudia Renata dos Santos Barros, Elaine Cristina Marqueze, Raul Machado Neto, Debora Botequio Moretti, Jardelina de Souza Todao Bernardino, Loyze Paola Oliveira de Lima, Luiz Aurelio de Campos Crispin. Centro de Genômica Funcional da ESALQ: Luiz Lehmann Coutinho, Ricardo Augusto Brassaloti, Raquel de Lello Rocha Campos Cassano. NGS Soluções Genômicas: Pilar Drummond Sampaio Corrêa Mariani. FZEA-USP Pirassununga: Mirele Daiana Poleti, Jessika Cristina Chagas Lesbon, Elisangela Chicaroni Mattos, Heidge Fukumasu. USP-Botucatu: Rejane Maria Tommasini Grotto, Jayme A. Souza-Neto, Guilherme Targino Valente, Patricia Akemi Assato, Felipe Allan da Silva da Costa, Bianca Cechetto Carlos. Mendelics: Bibiana Santos, João Paulo Kitajima, Erika Freitas, David Schlesinger. Hemocentro Ribeirão Preto: Simone Kashima, Evandra Strazza Rodrigues, Svetoslav Nanev Slavov, Elaine Vieira dos Santos, Rafael dos Santos Bezerra, Luiz Carlos Junior de Alcantara, Marta Giovanetti, Vagner Fonseca, Flavia Aburjaile, Rodrigo Tocantins Calado. FAMERP-SJRP: Cecília Artico Banho, Lívia Sacchetto, Fábio Sossai Possebon, Leila Sabrina Ullmann, Cíntia Bittar, Guilherme Campos, Helena Lage Ferreira, Jorge A. Petroli Marchesi, Maisa C. Pereira Parra, Marília Moraes, Paula Rahal, Paulo Inacio da Costa, João Pessoa Araújo Jr., Maurício Lacerda Nogueira. Prefeitura de Sao Paulo: Melissa Palmieri. |
| EPI_ISL_1966256 | CENTRO INTEGRADO DE SAUDE | Instituto Butantan / Mendelics | Instituto Butantan: Dimas Tadeu Covas, Sandra Coccuzzo Sampaio, Maria Carolina Elias, José Salvatore Leister Patané, Vincent Louis Viala, Antonio Jorge Martins, Ricardo Haddad, Claudia Renata dos Santos Barros, Elaine Cristina Marqueze, Raul Machado Neto, Debora Botequio Moretti, Jardelina de Souza Todao Bernardino, Loyze Paola Oliveira de Lima, Luiz Aurelio de Campos Crispin. Centro de Genômica Funcional da ESALQ: Luiz Lehmann Coutinho, Ricardo Augusto Brassaloti, Raquel de Lello Rocha Campos Cassano. NGS Soluções Genômicas: Pilar Drummond Sampaio Corrêa Mariani. FZEA-USP Pirassununga: Mirele Daiana Poleti, Jessika Cristina Chagas Lesbon, Elisangela Chicaroni Mattos, Heidge Fukumasu. USP-Botucatu: Rejane Maria Tommasini Grotto, Jayme A. Souza-Neto, Guilherme Targino Valente, Patricia Akemi Assato, Felipe Allan da Silva da Costa, Bianca Cechetto Carlos. Mendelics: Bibiana Santos, João Paulo Kitajima, Erika Freitas, David Schlesinger. Hemocentro Ribeirão Preto: Simone Kashima, Evandra Strazza Rodrigues, Svetoslav Nanev Slavov, Elaine Vieira dos Santos, Rafael dos Santos Bezerra, Luiz Carlos Junior de Alcantara, Marta Giovanetti, Vagner Fonseca, Flavia Aburjaile, Rodrigo Tocantins Calado. FAMERP-SJRP: Cecília Artico Banho, Lívia Sacchetto, Fábio Sossai Possebon, Leila Sabrina Ullmann, Cíntia Bittar, Guilherme Campos, Helena Lage Ferreira, Jorge A. Petroli Marchesi, Maisa C. Pereira Parra, Marília Moraes, Paula Rahal, Paulo Inacio da Costa, João Pessoa Araújo Jr., Maurício Lacerda Nogueira. Prefeitura de Sao Paulo: Melissa Palmieri. |
| EPI_ISL_1966257, EPI_ISL_1966258 | CS DE RUBINEIA | Instituto Butantan / Mendelics | Instituto Butantan: Dimas Tadeu Covas, Sandra Coccuzzo Sampaio, Maria Carolina Elias, José Salvatore Leister Patané, Vincent Louis Viala, Antonio Jorge Martins, Ricardo Haddad, Claudia Renata dos Santos Barros, Elaine Cristina Marqueze, Raul Machado Neto, Debora Botequio Moretti, Jardelina de Souza Todao Bernardino, Loyze Paola Oliveira de Lima, Luiz Aurelio de Campos Crispin. Centro de Genômica Funcional da ESALQ: Luiz Lehmann Coutinho, Ricardo Augusto Brassaloti, Raquel de Lello Rocha Campos Cassano. NGS Soluções Genômicas: Pilar Drummond Sampaio Corrêa Mariani. FZEA-USP Pirassununga: Mirele Daiana Poleti, Jessika Cristina Chagas Lesbon, Elisangela Chicaroni Mattos, Heidge Fukumasu. USP-Botucatu: Rejane Maria Tommasini Grotto, Jayme A. Souza-Neto, Guilherme Targino Valente, Patricia Akemi Assato, Felipe Allan da Silva da Costa, Bianca Cechetto Carlos. Mendelics: Bibiana Santos, João Paulo Kitajima, Erika Freitas, David Schlesinger. Hemocentro Ribeirão Preto: Simone Kashima, Evandra Strazza Rodrigues, Svetoslav Nanev Slavov, Elaine Vieira dos Santos, Rafael dos Santos Bezerra, Luiz Carlos Junior de Alcantara, Marta Giovanetti, Vagner Fonseca, Flavia Aburjaile, Rodrigo Tocantins Calado. FAMERP-SJRP: Cecília Artico Banho, Lívia Sacchetto, Fábio Sossai Possebon, Leila Sabrina Ullmann, Cíntia Bittar, Guilherme Campos, Helena Lage Ferreira, Jorge A. Petroli Marchesi, Maisa C. Pereira Parra, Marília Moraes, Paula Rahal, Paulo Inacio da Costa, João Pessoa Araújo Jr., Maurício Lacerda Nogueira. Prefeitura de Sao Paulo: Melissa Palmieri. |
| EPI_ISL_1966259 | SANTA CASA DE MISERICORDIA DE UBATUBA | Instituto Butantan / Mendelics | Instituto Butantan: Dimas Tadeu Covas, Sandra Coccuzzo Sampaio, Maria Carolina Elias, José Salvatore Leister Patané, Vincent Louis Viala, Antonio Jorge Martins, Ricardo Haddad, Claudia Renata dos Santos Barros, Elaine Cristina Marqueze, Raul Machado Neto, Debora Botequio Moretti, Jardelina de Souza Todao Bernardino, Loyze Paola Oliveira de Lima, Luiz Aurelio de Campos Crispin. Centro de Genômica Funcional da ESALQ: Luiz Lehmann Coutinho, Ricardo Augusto Brassaloti, Raquel de Lello Rocha Campos Cassano. NGS Soluções Genômicas: Pilar Drummond Sampaio Corrêa Mariani. FZEA-USP Pirassununga: Mirele Daiana Poleti, Jessika Cristina Chagas Lesbon, Elisangela Chicaroni Mattos, Heidge Fukumasu. USP-Botucatu: Rejane Maria Tommasini Grotto, Jayme A. Souza-Neto, Guilherme Targino Valente, Patricia Akemi Assato, Felipe Allan da Silva da Costa, Bianca Cechetto Carlos. Mendelics: Bibiana Santos, João Paulo Kitajima, Erika Freitas, David Schlesinger. Hemocentro Ribeirão Preto: Simone Kashima, Evandra Strazza Rodrigues, Svetoslav Nanev Slavov, Elaine Vieira dos Santos, Rafael dos Santos Bezerra, Luiz Carlos Junior de Alcantara, Marta Giovanetti, Vagner Fonseca, Flavia Aburjaile, Rodrigo Tocantins Calado. FAMERP-SJRP: Cecília Artico Banho, Lívia Sacchetto, Fábio Sossai Possebon, Leila Sabrina Ullmann, Cíntia Bittar, Guilherme Campos, Helena Lage Ferreira, Jorge A. Petroli Marchesi, Maisa C. Pereira Parra, Marília Moraes, Paula Rahal, Paulo Inacio da Costa, João Pessoa Araújo Jr., Maurício Lacerda Nogueira. Prefeitura de Sao Paulo: Melissa Palmieri. |
| EPI_ISL_1966260, EPI_ISL_1966261, EPI_ISL_1966262, EPI_ISL_1966263, EPI_ISL_1966264 | SMS SECRETARIA MUNICIPAL DE SAUDE DE BOITUVA | Instituto Butantan / Mendelics | Instituto Butantan: Dimas Tadeu Covas, Sandra Coccuzzo Sampaio, Maria Carolina Elias, José Salvatore Leister Patané, Vincent Louis Viala, Antonio Jorge Martins, Ricardo Haddad, Claudia Renata dos Santos Barros, Elaine Cristina Marqueze, Raul Machado Neto, Debora Botequio Moretti, Jardelina de Souza Todao Bernardino, Loyze Paola Oliveira de Lima, Luiz Aurelio de Campos Crispin. Centro de Genômica Funcional da ESALQ: Luiz Lehmann Coutinho, Ricardo Augusto Brassaloti, Raquel de Lello Rocha Campos Cassano. NGS Soluções Genômicas: Pilar Drummond Sampaio Corrêa Mariani. FZEA-USP Pirassununga: Mirele Daiana Poleti, Jessika Cristina Chagas Lesbon, Elisangela Chicaroni Mattos, Heidge Fukumasu. USP-Botucatu: Rejane Maria Tommasini Grotto, Jayme A. Souza-Neto, Guilherme Targino Valente, Patricia Akemi Assato, Felipe Allan da Silva da Costa, Bianca Cechetto Carlos. Mendelics: Bibiana Santos, João Paulo Kitajima, Erika Freitas, David Schlesinger. Hemocentro Ribeirão Preto: Simone Kashima, Evandra Strazza Rodrigues, Svetoslav Nanev Slavov, Elaine Vieira dos Santos, Rafael dos Santos Bezerra, Luiz Carlos Junior de Alcantara, Marta Giovanetti, Vagner Fonseca, Flavia Aburjaile, Rodrigo Tocantins Calado. FAMERP-SJRP: Cecília Artico Banho, Lívia Sacchetto, Fábio Sossai Possebon, Leila Sabrina Ullmann, Cíntia Bittar, Guilherme Campos, Helena Lage Ferreira, Jorge A. Petroli Marchesi, Maisa C. Pereira Parra, Marília Moraes, Paula Rahal, Paulo Inacio da Costa, João Pessoa Araújo Jr., Maurício Lacerda Nogueira. Prefeitura de Sao Paulo: Melissa Palmieri. |
| EPI_ISL_1966265 | UBS II DE NARANDIBA | Instituto Butantan / Mendelics | Instituto Butantan: Dimas Tadeu Covas, Sandra Coccuzzo Sampaio, Maria Carolina Elias, José Salvatore Leister Patané, Vincent Louis Viala, Antonio Jorge Martins, Ricardo Haddad, Claudia Renata dos Santos Barros, Elaine Cristina Marqueze, Raul Machado Neto, Debora Botequio Moretti, Jardelina de Souza Todao Bernardino, Loyze Paola Oliveira de Lima, Luiz Aurelio de Campos Crispin. Centro de Genômica Funcional da ESALQ: Luiz Lehmann Coutinho, Ricardo Augusto Brassaloti, Raquel de Lello Rocha Campos Cassano. NGS Soluções Genômicas: Pilar Drummond Sampaio Corrêa Mariani. FZEA-USP Pirassununga: Mirele Daiana Poleti, Jessika Cristina Chagas Lesbon, Elisangela Chicaroni Mattos, Heidge Fukumasu. USP-Botucatu: Rejane Maria Tommasini Grotto, Jayme A. Souza-Neto, Guilherme Targino Valente, Patricia Akemi Assato, Felipe Allan da Silva da Costa, Bianca Cechetto Carlos. Mendelics: Bibiana Santos, João Paulo Kitajima, Erika Freitas, David Schlesinger. Hemocentro Ribeirão Preto: Simone Kashima, Evandra Strazza Rodrigues, Svetoslav Nanev Slavov, Elaine Vieira dos Santos, Rafael dos Santos Bezerra, Luiz Carlos Junior de Alcantara, Marta Giovanetti, Vagner Fonseca, Flavia Aburjaile, Rodrigo Tocantins Calado. FAMERP-SJRP: Cecília Artico Banho, Lívia Sacchetto, Fábio Sossai Possebon, Leila Sabrina Ullmann, Cíntia Bittar, Guilherme Campos, Helena Lage Ferreira, Jorge A. Petroli Marchesi, Maisa C. Pereira Parra, Marília Moraes, Paula Rahal, Paulo Inacio da Costa, João Pessoa Araújo Jr., Maurício Lacerda Nogueira. Prefeitura de Sao Paulo: Melissa Palmieri. |
| EPI_ISL_1966266 | UBS IRMA AGUEDA MARIA JAIME | Instituto Butantan / Mendelics | Instituto Butantan: Dimas Tadeu Covas, Sandra Coccuzzo Sampaio, Maria Carolina Elias, José Salvatore Leister Patané, Vincent Louis Viala, Antonio Jorge Martins, Ricardo Haddad, Claudia Renata dos Santos Barros, Elaine Cristina Marqueze, Raul Machado Neto, Debora Botequio Moretti, Jardelina de Souza Todao Bernardino, Loyze Paola Oliveira de Lima, Luiz Aurelio de Campos Crispin. Centro de Genômica Funcional da ESALQ: Luiz Lehmann Coutinho, Ricardo Augusto Brassaloti, Raquel de Lello Rocha Campos Cassano. NGS Soluções Genômicas: Pilar Drummond Sampaio Corrêa Mariani. FZEA-USP Pirassununga: Mirele Daiana Poleti, Jessika Cristina Chagas Lesbon, Elisangela Chicaroni Mattos, Heidge Fukumasu. USP-Botucatu: Rejane Maria Tommasini Grotto, Jayme A. Souza-Neto, Guilherme Targino Valente, Patricia Akemi Assato, Felipe Allan da Silva da Costa, Bianca Cechetto Carlos. Mendelics: Bibiana Santos, João Paulo Kitajima, Erika Freitas, David Schlesinger. Hemocentro Ribeirão Preto: Simone Kashima, Evandra Strazza Rodrigues, Svetoslav Nanev Slavov, Elaine Vieira dos Santos, Rafael dos Santos Bezerra, Luiz Carlos Junior de Alcantara, Marta Giovanetti, Vagner Fonseca, Flavia Aburjaile, Rodrigo Tocantins Calado. FAMERP-SJRP: Cecília Artico Banho, Lívia Sacchetto, Fábio Sossai Possebon, Leila Sabrina Ullmann, Cíntia Bittar, Guilherme Campos, Helena Lage Ferreira, Jorge A. Petroli Marchesi, Maisa C. Pereira Parra, Marília Moraes, Paula Rahal, Paulo Inacio da Costa, João Pessoa Araújo Jr., Maurício Lacerda Nogueira. Prefeitura de Sao Paulo: Melissa Palmieri. |
| EPI_ISL_1966267, EPI_ISL_1966268, EPI_ISL_1966269 | VIGILANCIA EM SAUDE SAE VIGILANCIA EPIDEMIOLOGICA | Instituto Butantan / Mendelics | Instituto Butantan: Dimas Tadeu Covas, Sandra Coccuzzo Sampaio, Maria Carolina Elias, José Salvatore Leister Patané, Vincent Louis Viala, Antonio Jorge Martins, Ricardo Haddad, Claudia Renata dos Santos Barros, Elaine Cristina Marqueze, Raul Machado Neto, Debora Botequio Moretti, Jardelina de Souza Todao Bernardino, Loyze Paola Oliveira de Lima, Luiz Aurelio de Campos Crispin. Centro de Genômica Funcional da ESALQ: Luiz Lehmann |

| | | | |
|---|---|---|---|
| | | | Coutinho, Ricardo Augusto Brassaloti, Raquel de Lello Rocha Campos Cassano. NGS Soluções Genômicas: Pilar Drummond Sampaio Corrêa Mariani. FZEA-USP Pirassununga: Mirele Daiana Poleti, Jessika Cristina Chagas Lesbon, Elisangela Chicaroni Mattos, Heidge Fukumasu. USP-Botucatu: Rejane Maria Tommasini Grotto, Jayme A. Souza-Neto, Guilherme Targino Valente, Patricia Akemi Assato, Felipe Allan da Silva da Costa, Bianca Cechetto Carlos. Mendelics: Bibiana Santos, João Paulo Kitajima, Erika Freitas, David Schlesinger. Hemocentro Ribeirão Preto: Simone Kashima, Evandra Strazza Rodrigues, Svetoslav Nanev Slavov, Elaine Vieira dos Santos, Rafael dos Santos Bezerra, Luiz Carlos Junior de Alcantara, Marta Giovanetti, Vagner Fonseca, Flavia Aburjaile, Rodrigo Tocantins Calado. FAMERP-SJRP: Cecília Artico Banho, Lívia Sacchetto, Fábio Sossai Possebon, Leila Sabrina Ullmann, Cíntia Bittar, Guilherme Campos, Helena Lage Ferreira, Jorge A. Petroli Marchesi, Maisa C. Pereira Parra, Marília Moraes, Paula Rahal, Paulo Inacio da Costa, João Pessoa Araújo Jr., Maurício Lacerda Nogueira. Prefeitura de Sao Paulo: Melissa Palmieri. |
| EPI_ISL_1966270, EPI_ISL_1966271, EPI_ISL_1966272, EPI_ISL_1966273, EPI_ISL_1966274, EPI_ISL_1966275, EPI_ISL_1966276, EPI_ISL_1966277 | SECRETARIA MUNICIPAL DE SAUDE DE ANDRADINA | Instituto Butantan / Mendelics | Instituto Butantan: Dimas Tadeu Covas, Sandra Coccuzzo Sampaio, Maria Carolina Elias, José Salvatore Leister Patané, Vincent Louis Viala, Antonio Jorge Martins, Ricardo Haddad, Claudia Renata dos Santos Barros, Elaine Cristina Marqueze, Raul Machado Neto, Debora Botequio Moretti, Jardelina de Souza Todao Bernardino, Loyze Paola Oliveira de Lima, Luiz Aurelio de Campos Crispin. Centro de Genômica Funcional da ESALQ: Luiz Lehmann Coutinho, Ricardo Augusto Brassaloti, Raquel de Lello Rocha Campos Cassano. NGS Soluções Genômicas: Pilar Drummond Sampaio Corrêa Mariani. FZEA-USP Pirassununga: Mirele Daiana Poleti, Jessika Cristina Chagas Lesbon, Elisangela Chicaroni Mattos, Heidge Fukumasu. USP-Botucatu: Rejane Maria Tommasini Grotto, Jayme A. Souza-Neto, Guilherme Targino Valente, Patricia Akemi Assato, Felipe Allan da Silva da Costa, Bianca Cechetto Carlos. Mendelics: Bibiana Santos, João Paulo Kitajima, Erika Freitas, David Schlesinger. Hemocentro Ribeirão Preto: Simone Kashima, Evandra Strazza Rodrigues, Svetoslav Nanev Slavov, Elaine Vieira dos Santos, Rafael dos Santos Bezerra, Luiz Carlos Junior de Alcantara, Marta Giovanetti, Vagner Fonseca, Flavia Aburjaile, Rodrigo Tocantins Calado. FAMERP-SJRP: Cecília Artico Banho, Lívia Sacchetto, Fábio Sossai Possebon, Leila Sabrina Ullmann, Cíntia Bittar, Guilherme Campos, Helena Lage Ferreira, Jorge A. Petroli Marchesi, Maisa C. Pereira Parra, Marília Moraes, Paula Rahal, Paulo Inacio da Costa, João Pessoa Araújo Jr., Maurício Lacerda Nogueira. Prefeitura de Sao Paulo: Melissa Palmieri. |
| EPI_ISL_1966278, EPI_ISL_1966279, EPI_ISL_1966280, EPI_ISL_1966281, EPI_ISL_1966282, EPI_ISL_1966283, EPI_ISL_1966284, EPI_ISL_1966285, EPI_ISL_1966286 | UBS DR ALFREDO DANTAS DE SOUZA UMUARAMA | Instituto Butantan / Mendelics | Instituto Butantan: Dimas Tadeu Covas, Sandra Coccuzzo Sampaio, Maria Carolina Elias, José Salvatore Leister Patané, Vincent Louis Viala, Antonio Jorge Martins, Ricardo Haddad, Claudia Renata dos Santos Barros, Elaine Cristina Marqueze, Raul Machado Neto, Debora Botequio Moretti, Jardelina de Souza Todao Bernardino, Loyze Paola Oliveira de Lima, Luiz Aurelio de Campos Crispin. Centro de Genômica Funcional da ESALQ: Luiz Lehmann Coutinho, Ricardo Augusto Brassaloti, Raquel de Lello Rocha Campos Cassano. NGS Soluções Genômicas: Pilar Drummond Sampaio Corrêa Mariani. FZEA-USP Pirassununga: Mirele Daiana Poleti, Jessika Cristina Chagas Lesbon, Elisangela Chicaroni Mattos, Heidge Fukumasu. USP-Botucatu: Rejane Maria Tommasini Grotto, Jayme A. Souza-Neto, Guilherme Targino Valente, Patricia Akemi Assato, Felipe Allan da Silva da Costa, Bianca Cechetto Carlos. Mendelics: Bibiana Santos, João Paulo Kitajima, Erika Freitas, David Schlesinger. Hemocentro Ribeirão Preto: Simone Kashima, Evandra Strazza Rodrigues, Svetoslav Nanev Slavov, Elaine Vieira dos Santos, Rafael dos Santos Bezerra, Luiz Carlos Junior de Alcantara, Marta Giovanetti, Vagner Fonseca, Flavia Aburjaile, Rodrigo Tocantins Calado. FAMERP-SJRP: Cecília Artico Banho, Lívia Sacchetto, Fábio Sossai Possebon, Leila Sabrina Ullmann, Cíntia Bittar, Guilherme Campos, Helena Lage Ferreira, Jorge A. Petroli Marchesi, Maisa C. Pereira Parra, Marília Moraes, Paula Rahal, Paulo Inacio da Costa, João Pessoa Araújo Jr., Maurício Lacerda Nogueira. Prefeitura de Sao Paulo: Melissa Palmieri. |
| EPI_ISL_1966287 | UBS DE AURIFLAMA | Instituto Butantan / Mendelics | Instituto Butantan: Dimas Tadeu Covas, Sandra Coccuzzo Sampaio, Maria Carolina Elias, José Salvatore Leister Patané, Vincent Louis Viala, Antonio Jorge Martins, Ricardo Haddad, Claudia Renata dos Santos Barros, Elaine Cristina Marqueze, Raul Machado Neto, Debora Botequio Moretti, Jardelina de Souza Todao Bernardino, Loyze Paola Oliveira de Lima, Luiz Aurelio de Campos Crispin. Centro de Genômica Funcional da ESALQ: Luiz Lehmann Coutinho, Ricardo Augusto Brassaloti, Raquel de Lello Rocha Campos Cassano. NGS Soluções Genômicas: Pilar Drummond Sampaio Corrêa Mariani. FZEA-USP Pirassununga: Mirele Daiana Poleti, Jessika Cristina Chagas Lesbon, Elisangela Chicaroni Mattos, Heidge Fukumasu. USP-Botucatu: Rejane Maria Tommasini Grotto, Jayme A. Souza-Neto, Guilherme Targino Valente, Patricia Akemi Assato, Felipe Allan da Silva da Costa, Bianca Cechetto Carlos. Mendelics: Bibiana Santos, João Paulo Kitajima, Erika Freitas, David Schlesinger. Hemocentro Ribeirão Preto: Simone Kashima, Evandra Strazza Rodrigues, Svetoslav Nanev Slavov, Elaine Vieira dos Santos, Rafael dos Santos Bezerra, Luiz Carlos Junior de Alcantara, Marta Giovanetti, Vagner Fonseca, Flavia Aburjaile, Rodrigo Tocantins Calado. FAMERP-SJRP: Cecília Artico Banho, Lívia Sacchetto, Fábio Sossai Possebon, Leila Sabrina Ullmann, Cíntia Bittar, Guilherme Campos, Helena Lage Ferreira, Jorge A. Petroli Marchesi, Maisa C. Pereira Parra, Marília Moraes, Paula Rahal, Paulo Inacio da Costa, João Pessoa Araújo Jr., Maurício Lacerda Nogueira. Prefeitura de Sao Paulo: Melissa Palmieri. |
| EPI_ISL_1966288 | UNIDADE BASICA DE SAUDE DR BARTOLOMEU MARAGLIANO VENERE | Instituto Butantan / Mendelics | Instituto Butantan: Dimas Tadeu Covas, Sandra Coccuzzo Sampaio, Maria Carolina Elias, José Salvatore Leister Patané, Vincent Louis Viala, Antonio Jorge Martins, Ricardo Haddad, Claudia Renata dos Santos Barros, Elaine Cristina Marqueze, Raul Machado Neto, Debora Botequio Moretti, Jardelina de Souza Todao Bernardino, Loyze Paola Oliveira de Lima, Luiz Aurelio de Campos Crispin. Centro de Genômica Funcional da ESALQ: Luiz Lehmann Coutinho, Ricardo Augusto Brassaloti, Raquel de Lello Rocha Campos Cassano. NGS Soluções Genômicas: Pilar Drummond Sampaio Corrêa Mariani. FZEA-USP Pirassununga: Mirele Daiana Poleti, Jessika Cristina Chagas Lesbon, Elisangela Chicaroni Mattos, Heidge Fukumasu. USP-Botucatu: Rejane Maria Tommasini Grotto, Jayme A. Souza-Neto, Guilherme Targino Valente, Patricia Akemi Assato, Felipe Allan da Silva da Costa, Bianca Cechetto Carlos. Mendelics: Bibiana Santos, João Paulo Kitajima, Erika Freitas, David Schlesinger. Hemocentro Ribeirão Preto: Simone Kashima, Evandra Strazza Rodrigues, Svetoslav Nanev Slavov, Elaine Vieira dos Santos, Rafael dos Santos Bezerra, Luiz Carlos Junior de Alcantara, Marta Giovanetti, Vagner Fonseca, Flavia Aburjaile, Rodrigo Tocantins Calado. FAMERP-SJRP: Cecília Artico Banho, Lívia Sacchetto, Fábio Sossai Possebon, Leila Sabrina Ullmann, Cíntia Bittar, Guilherme Campos, Helena Lage Ferreira, Jorge A. Petroli Marchesi, Maisa C. Pereira Parra, Marília Moraes, Paula Rahal, Paulo Inacio da Costa, João Pessoa Araújo Jr., Maurício Lacerda Nogueira. Prefeitura de Sao Paulo: Melissa Palmieri. |
| EPI_ISL_1966289 | UBS DR ALLY ALAHMAR DE BARRETOS | Instituto Butantan / Mendelics | Instituto Butantan: Dimas Tadeu Covas, Sandra Coccuzzo Sampaio, Maria Carolina Elias, José Salvatore Leister Patané, Vincent Louis Viala, Antonio Jorge Martins, Ricardo Haddad, Claudia Renata dos Santos Barros, Elaine Cristina Marqueze, Raul Machado Neto, Debora Botequio Moretti, Jardelina de Souza Todao Bernardino, Loyze Paola Oliveira de Lima, Luiz Aurelio de Campos Crispin. Centro de Genômica Funcional da ESALQ: Luiz Lehmann Coutinho, Ricardo Augusto Brassaloti, Raquel de Lello Rocha Campos Cassano. NGS Soluções Genômicas: Pilar Drummond Sampaio Corrêa Mariani. FZEA-USP Pirassununga: Mirele Daiana Poleti, Jessika Cristina Chagas Lesbon, Elisangela Chicaroni Mattos, Heidge Fukumasu. USP-Botucatu: Rejane Maria Tommasini Grotto, Jayme A. Souza-Neto, Guilherme Targino Valente, Patricia Akemi Assato, Felipe Allan da Silva da Costa, Bianca Cechetto Carlos. Mendelics: Bibiana Santos, João Paulo Kitajima, Erika Freitas, David Schlesinger. Hemocentro Ribeirão Preto: Simone Kashima, Evandra Strazza Rodrigues, Svetoslav Nanev Slavov, Elaine Vieira dos Santos, Rafael dos Santos Bezerra, Luiz Carlos Junior de Alcantara, Marta Giovanetti, Vagner Fonseca, Flavia Aburjaile, Rodrigo Tocantins Calado. FAMERP-SJRP: Cecília Artico Banho, Lívia Sacchetto, Fábio Sossai Possebon, Leila Sabrina Ullmann, Cíntia Bittar, Guilherme Campos, Helena Lage Ferreira, Jorge A. Petroli Marchesi, Maisa C. Pereira Parra, Marília Moraes, Paula Rahal, Paulo Inacio da Costa, João Pessoa Araújo Jr., Maurício Lacerda Nogueira. Prefeitura de Sao Paulo: Melissa Palmieri. |
| EPI_ISL_1966290, EPI_ISL_1966291 | UNIDADE BASICA DE SAUDE DA FAMILIA BILAC | Instituto Butantan / Mendelics | Instituto Butantan: Dimas Tadeu Covas, Sandra Coccuzzo Sampaio, Maria Carolina Elias, José Salvatore Leister Patané, Vincent Louis Viala, Antonio Jorge Martins, Ricardo Haddad, Claudia Renata dos Santos Barros, Elaine Cristina Marqueze, Raul Machado Neto, Debora Botequio Moretti, Jardelina de Souza Todao Bernardino, Loyze Paola Oliveira de Lima, Luiz Aurelio de Campos Crispin. Centro de Genômica Funcional da ESALQ: Luiz Lehmann Coutinho, Ricardo Augusto Brassaloti, Raquel de Lello Rocha Campos Cassano. NGS Soluções Genômicas: Pilar Drummond Sampaio Corrêa Mariani. FZEA-USP Pirassununga: Mirele Daiana Poleti, Jessika Cristina Chagas Lesbon, Elisangela Chicaroni Mattos, Heidge Fukumasu. USP-Botucatu: Rejane Maria Tommasini Grotto, Jayme A. Souza-Neto, Guilherme Targino Valente, Patricia Akemi Assato, Felipe Allan da Silva da Costa, Bianca Cechetto Carlos. Mendelics: Bibiana Santos, João Paulo Kitajima, Erika Freitas, David Schlesinger. Hemocentro Ribeirão Preto: Simone Kashima, Evandra Strazza Rodrigues, Svetoslav Nanev Slavov, Elaine Vieira dos Santos, Rafael dos Santos Bezerra, Luiz Carlos Junior de Alcantara, Marta Giovanetti, Vagner Fonseca, Flavia Aburjaile, Rodrigo Tocantins Calado. FAMERP-SJRP: Cecília Artico Banho, Lívia Sacchetto, Fábio Sossai Possebon, Leila Sabrina Ullmann, Cíntia Bittar, Guilherme Campos, Helena Lage Ferreira, Jorge A. Petroli Marchesi, Maisa C. Pereira Parra, Marília Moraes, Paula Rahal, Paulo Inacio da Costa, João Pessoa Araújo Jr., Maurício Lacerda Nogueira. Prefeitura de Sao Paulo: Melissa Palmieri. |
| EPI_ISL_1966292 | CENTRO DE SAUDE DE FLORINEA | Instituto Butantan / Mendelics | Instituto Butantan: Dimas Tadeu Covas, Sandra Coccuzzo Sampaio, Maria Carolina Elias, José Salvatore Leister Patané, Vincent Louis Viala, Antonio Jorge Martins, Ricardo Haddad, Claudia Renata dos Santos Barros, Elaine Cristina Marqueze, Raul Machado Neto, Debora Botequio Moretti, Jardelina de Souza Todao Bernardino, Loyze Paola Oliveira de Lima, Luiz Aurelio de Campos Crispin. Centro de Genômica Funcional da ESALQ: Luiz Lehmann Coutinho, Ricardo Augusto Brassaloti, Raquel de Lello Rocha Campos Cassano. NGS Soluções Genômicas: Pilar Drummond Sampaio Corrêa Mariani. FZEA-USP Pirassununga: Mirele Daiana Poleti, Jessika Cristina Chagas Lesbon, Elisangela Chicaroni Mattos, Heidge Fukumasu. USP-Botucatu: Rejane Maria Tommasini Grotto, Jayme A. Souza-Neto, Guilherme Targino Valente, Patricia Akemi Assato, Felipe Allan da Silva da Costa, Bianca Cechetto Carlos. Mendelics: Bibiana Santos, João Paulo Kitajima, Erika Freitas, David Schlesinger. Hemocentro Ribeirão Preto: Simone Kashima, Evandra Strazza Rodrigues, Svetoslav Nanev Slavov, Elaine Vieira dos Santos, Rafael dos Santos Bezerra, Luiz Carlos Junior de Alcantara, Marta Giovanetti, Vagner |

| | | | |
|---|---|---|---|
| | | | Fonseca, Flavia Aburjaile, Rodrigo Tocantins Calado. FAMERP-SJRP: Cecília Artico Banho, Lívia Sacchetto, Fábio Sossai Possebon, Leila Sabrina Ullmann, Cíntia Bittar, Guilherme Campos, Helena Lage Ferreira, Jorge A. Petroli Marchesi, Maisa C. Pereira Parra, Marília Moraes, Paula Rahal, Paulo Inacio da Costa, João Pessoa Araújo Jr., Maurício Lacerda Nogueira. Prefeitura de Sao Paulo: Melissa Palmieri. |
| EPI_ISL_1966293 | CENTRO DE SAUDE III GLICERIO GIMAIEL ELIAS GIMAIEL | Instituto Butantan / Mendelics | Instituto Butantan: Dimas Tadeu Covas, Sandra Coccuzzo Sampaio, Maria Carolina Elias, José Salvatore Leister Patané, Vincent Louis Viala, Antonio Jorge Martins, Ricardo Haddad, Claudia Renata dos Santos Barros, Elaine Cristina Marqueze, Raul Machado Neto, Debora Botequio Moretti, Jardelina de Souza Todao Bernardino, Loyze Paola Oliveira de Lima, Luiz Aurelio de Campos Crispin. Centro de Genômica Funcional da ESALQ: Luiz Lehmann Coutinho, Ricardo Augusto Brassaloti, Raquel de Lello Rocha Campos Cassano. NGS Soluções Genômicas: Pilar Drummond Sampaio Corrêa Mariani. FZEA-USP Pirassununga: Mirele Daiana Poleti, Jessika Cristina Chagas Lesbon, Elisangela Chicaroni Mattos, Heidge Fukumasu. USP-Botucatu: Rejane Maria Tommasini Grotto, Jayme A. Souza-Neto, Guilherme Targino Valente, Patricia Akemi Assato, Felipe Allan da Silva da Costa, Bianca Cechetto Carlos. Mendelics: Bibiana Santos, João Paulo Kitajima, Erika Freitas, David Schlesinger. Hemocentro Ribeirão Preto: Simone Kashima, Evandra Strazza Rodrigues, Svetoslav Nanev Slavov, Elaine Vieira dos Santos, Rafael dos Santos Bezerra, Luiz Carlos Junior de Alcantara, Marta Giovanetti, Vagner Fonseca, Flavia Aburjaile, Rodrigo Tocantins Calado. FAMERP-SJRP: Cecília Artico Banho, Lívia Sacchetto, Fábio Sossai Possebon, Leila Sabrina Ullmann, Cíntia Bittar, Guilherme Campos, Helena Lage Ferreira, Jorge A. Petroli Marchesi, Maisa C. Pereira Parra, Marília Moraes, Paula Rahal, Paulo Inacio da Costa, João Pessoa Araújo Jr., Maurício Lacerda Nogueira. Prefeitura de Sao Paulo: Melissa Palmieri. |
| EPI_ISL_1966294, EPI_ISL_1966295, EPI_ISL_1966296, EPI_ISL_1966297, EPI_ISL_1966298, EPI_ISL_1966299, EPI_ISL_1966300, EPI_ISL_1966301 | VIGILANCIA EPIDEMIOLOGICA DE IBATE | Instituto Butantan / Mendelics | Instituto Butantan: Dimas Tadeu Covas, Sandra Coccuzzo Sampaio, Maria Carolina Elias, José Salvatore Leister Patané, Vincent Louis Viala, Antonio Jorge Martins, Ricardo Haddad, Claudia Renata dos Santos Barros, Elaine Cristina Marqueze, Raul Machado Neto, Debora Botequio Moretti, Jardelina de Souza Todao Bernardino, Loyze Paola Oliveira de Lima, Luiz Aurelio de Campos Crispin. Centro de Genômica Funcional da ESALQ: Luiz Lehmann Coutinho, Ricardo Augusto Brassaloti, Raquel de Lello Rocha Campos Cassano. NGS Soluções Genômicas: Pilar Drummond Sampaio Corrêa Mariani. FZEA-USP Pirassununga: Mirele Daiana Poleti, Jessika Cristina Chagas Lesbon, Elisangela Chicaroni Mattos, Heidge Fukumasu. USP-Botucatu: Rejane Maria Tommasini Grotto, Jayme A. Souza-Neto, Guilherme Targino Valente, Patricia Akemi Assato, Felipe Allan da Silva da Costa, Bianca Cechetto Carlos. Mendelics: Bibiana Santos, João Paulo Kitajima, Erika Freitas, David Schlesinger. Hemocentro Ribeirão Preto: Simone Kashima, Evandra Strazza Rodrigues, Svetoslav Nanev Slavov, Elaine Vieira dos Santos, Rafael dos Santos Bezerra, Luiz Carlos Junior de Alcantara, Marta Giovanetti, Vagner Fonseca, Flavia Aburjaile, Rodrigo Tocantins Calado. FAMERP-SJRP: Cecília Artico Banho, Lívia Sacchetto, Fábio Sossai Possebon, Leila Sabrina Ullmann, Cíntia Bittar, Guilherme Campos, Helena Lage Ferreira, Jorge A. Petroli Marchesi, Maisa C. Pereira Parra, Marília Moraes, Paula Rahal, Paulo Inacio da Costa, João Pessoa Araújo Jr., Maurício Lacerda Nogueira. Prefeitura de Sao Paulo: Melissa Palmieri. |
| EPI_ISL_1966302, EPI_ISL_1966303, EPI_ISL_1966304, EPI_ISL_1966305 | PRONTO SOCORRO DE AGENOR DE CAMPOS MONGAGUA | Instituto Butantan / Mendelics | Instituto Butantan: Dimas Tadeu Covas, Sandra Coccuzzo Sampaio, Maria Carolina Elias, José Salvatore Leister Patané, Vincent Louis Viala, Antonio Jorge Martins, Ricardo Haddad, Claudia Renata dos Santos Barros, Elaine Cristina Marqueze, Raul Machado Neto, Debora Botequio Moretti, Jardelina de Souza Todao Bernardino, Loyze Paola Oliveira de Lima, Luiz Aurelio de Campos Crispin. Centro de Genômica Funcional da ESALQ: Luiz Lehmann Coutinho, Ricardo Augusto Brassaloti, Raquel de Lello Rocha Campos Cassano. NGS Soluções Genômicas: Pilar Drummond Sampaio Corrêa Mariani. FZEA-USP Pirassununga: Mirele Daiana Poleti, Jessika Cristina Chagas Lesbon, Elisangela Chicaroni Mattos, Heidge Fukumasu. USP-Botucatu: Rejane Maria Tommasini Grotto, Jayme A. Souza-Neto, Guilherme Targino Valente, Patricia Akemi Assato, Felipe Allan da Silva da Costa, Bianca Cechetto Carlos. Mendelics: Bibiana Santos, João Paulo Kitajima, Erika Freitas, David Schlesinger. Hemocentro Ribeirão Preto: Simone Kashima, Evandra Strazza Rodrigues, Svetoslav Nanev Slavov, Elaine Vieira dos Santos, Rafael dos Santos Bezerra, Luiz Carlos Junior de Alcantara, Marta Giovanetti, Vagner Fonseca, Flavia Aburjaile, Rodrigo Tocantins Calado. FAMERP-SJRP: Cecília Artico Banho, Lívia Sacchetto, Fábio Sossai Possebon, Leila Sabrina Ullmann, Cíntia Bittar, Guilherme Campos, Helena Lage Ferreira, Jorge A. Petroli Marchesi, Maisa C. Pereira Parra, Marília Moraes, Paula Rahal, Paulo Inacio da Costa, João Pessoa Araújo Jr., Maurício Lacerda Nogueira. Prefeitura de Sao Paulo: Melissa Palmieri. |
| EPI_ISL_1966306, EPI_ISL_1966307 | UNIDADE DE PRONTO ATENDIMENTO DE AGENOR DE CAMPOS | Instituto Butantan / Mendelics | Instituto Butantan: Dimas Tadeu Covas, Sandra Coccuzzo Sampaio, Maria Carolina Elias, José Salvatore Leister Patané, Vincent Louis Viala, Antonio Jorge Martins, Ricardo Haddad, Claudia Renata dos Santos Barros, Elaine Cristina Marqueze, Raul Machado Neto, Debora Botequio Moretti, Jardelina de Souza Todao Bernardino, Loyze Paola Oliveira de Lima, Luiz Aurelio de Campos Crispin. Centro de Genômica Funcional da ESALQ: Luiz Lehmann Coutinho, Ricardo Augusto Brassaloti, Raquel de Lello Rocha Campos Cassano. NGS Soluções Genômicas: Pilar Drummond Sampaio Corrêa Mariani. FZEA-USP Pirassununga: Mirele Daiana Poleti, Jessika Cristina Chagas Lesbon, Elisangela Chicaroni Mattos, Heidge Fukumasu. USP-Botucatu: Rejane Maria Tommasini Grotto, Jayme A. Souza-Neto, Guilherme Targino Valente, Patricia Akemi Assato, Felipe Allan da Silva da Costa, Bianca Cechetto Carlos. Mendelics: Bibiana Santos, João Paulo Kitajima, Erika Freitas, David Schlesinger. Hemocentro Ribeirão Preto: Simone Kashima, Evandra Strazza Rodrigues, Svetoslav Nanev Slavov, Elaine Vieira dos Santos, Rafael dos Santos Bezerra, Luiz Carlos Junior de Alcantara, Marta Giovanetti, Vagner Fonseca, Flavia Aburjaile, Rodrigo Tocantins Calado. FAMERP-SJRP: Cecília Artico Banho, Lívia Sacchetto, Fábio Sossai Possebon, Leila Sabrina Ullmann, Cíntia Bittar, Guilherme Campos, Helena Lage Ferreira, Jorge A. Petroli Marchesi, Maisa C. Pereira Parra, Marília Moraes, Paula Rahal, Paulo Inacio da Costa, João Pessoa Araújo Jr., Maurício Lacerda Nogueira. Prefeitura de Sao Paulo: Melissa Palmieri. |
| EPI_ISL_1966308 | PRONTO SOCORRO DE AGENOR DE CAMPOS MONGAGUA | Instituto Butantan / Mendelics | Instituto Butantan: Dimas Tadeu Covas, Sandra Coccuzzo Sampaio, Maria Carolina Elias, José Salvatore Leister Patané, Vincent Louis Viala, Antonio Jorge Martins, Ricardo Haddad, Claudia Renata dos Santos Barros, Elaine Cristina Marqueze, Raul Machado Neto, Debora Botequio Moretti, Jardelina de Souza Todao Bernardino, Loyze Paola Oliveira de Lima, Luiz Aurelio de Campos Crispin. Centro de Genômica Funcional da ESALQ: Luiz Lehmann Coutinho, Ricardo Augusto Brassaloti, Raquel de Lello Rocha Campos Cassano. NGS Soluções Genômicas: Pilar Drummond Sampaio Corrêa Mariani. FZEA-USP Pirassununga: Mirele Daiana Poleti, Jessika Cristina Chagas Lesbon, Elisangela Chicaroni Mattos, Heidge Fukumasu. USP-Botucatu: Rejane Maria Tommasini Grotto, Jayme A. Souza-Neto, Guilherme Targino Valente, Patricia Akemi Assato, Felipe Allan da Silva da Costa, Bianca Cechetto Carlos. Mendelics: Bibiana Santos, João Paulo Kitajima, Erika Freitas, David Schlesinger. Hemocentro Ribeirão Preto: Simone Kashima, Evandra Strazza Rodrigues, Svetoslav Nanev Slavov, Elaine Vieira dos Santos, Rafael dos Santos Bezerra, Luiz Carlos Junior de Alcantara, Marta Giovanetti, Vagner Fonseca, Flavia Aburjaile, Rodrigo Tocantins Calado. FAMERP-SJRP: Cecília Artico Banho, Lívia Sacchetto, Fábio Sossai Possebon, Leila Sabrina Ullmann, Cíntia Bittar, Guilherme Campos, Helena Lage Ferreira, Jorge A. Petroli Marchesi, Maisa C. Pereira Parra, Marília Moraes, Paula Rahal, Paulo Inacio da Costa, João Pessoa Araújo Jr., Maurício Lacerda Nogueira. Prefeitura de Sao Paulo: Melissa Palmieri. |
| EPI_ISL_1966309, EPI_ISL_1966310 | UNIDADE DE PRONTO ATENDIMENTO DE AGENOR DE CAMPOS | Instituto Butantan / Mendelics | Instituto Butantan: Dimas Tadeu Covas, Sandra Coccuzzo Sampaio, Maria Carolina Elias, José Salvatore Leister Patané, Vincent Louis Viala, Antonio Jorge Martins, Ricardo Haddad, Claudia Renata dos Santos Barros, Elaine Cristina Marqueze, Raul Machado Neto, Debora Botequio Moretti, Jardelina de Souza Todao Bernardino, Loyze Paola Oliveira de Lima, Luiz Aurelio de Campos Crispin. Centro de Genômica Funcional da ESALQ: Luiz Lehmann Coutinho, Ricardo Augusto Brassaloti, Raquel de Lello Rocha Campos Cassano. NGS Soluções Genômicas: Pilar Drummond Sampaio Corrêa Mariani. FZEA-USP Pirassununga: Mirele Daiana Poleti, Jessika Cristina Chagas Lesbon, Elisangela Chicaroni Mattos, Heidge Fukumasu. USP-Botucatu: Rejane Maria Tommasini Grotto, Jayme A. Souza-Neto, Guilherme Targino Valente, Patricia Akemi Assato, Felipe Allan da Silva da Costa, Bianca Cechetto Carlos. Mendelics: Bibiana Santos, João Paulo Kitajima, Erika Freitas, David Schlesinger. Hemocentro Ribeirão Preto: Simone Kashima, Evandra Strazza Rodrigues, Svetoslav Nanev Slavov, Elaine Vieira dos Santos, Rafael dos Santos Bezerra, Luiz Carlos Junior de Alcantara, Marta Giovanetti, Vagner Fonseca, Flavia Aburjaile, Rodrigo Tocantins Calado. FAMERP-SJRP: Cecília Artico Banho, Lívia Sacchetto, Fábio Sossai Possebon, Leila Sabrina Ullmann, Cíntia Bittar, Guilherme Campos, Helena Lage Ferreira, Jorge A. Petroli Marchesi, Maisa C. Pereira Parra, Marília Moraes, Paula Rahal, Paulo Inacio da Costa, João Pessoa Araújo Jr., Maurício Lacerda Nogueira. Prefeitura de Sao Paulo: Melissa Palmieri. |
| EPI_ISL_1966311, EPI_ISL_1966312, EPI_ISL_1966313, EPI_ISL_1966314, EPI_ISL_1966315 | PRONTO SOCORRO DE AGENOR DE CAMPOS MONGAGUA | Instituto Butantan / Mendelics | Instituto Butantan: Dimas Tadeu Covas, Sandra Coccuzzo Sampaio, Maria Carolina Elias, José Salvatore Leister Patané, Vincent Louis Viala, Antonio Jorge Martins, Ricardo Haddad, Claudia Renata dos Santos Barros, Elaine Cristina Marqueze, Raul Machado Neto, Debora Botequio Moretti, Jardelina de Souza Todao Bernardino, Loyze Paola Oliveira de Lima, Luiz Aurelio de Campos Crispin. Centro de Genômica Funcional da ESALQ: Luiz Lehmann Coutinho, Ricardo Augusto Brassaloti, Raquel de Lello Rocha Campos Cassano. NGS Soluções Genômicas: Pilar Drummond Sampaio Corrêa Mariani. FZEA-USP Pirassununga: Mirele Daiana Poleti, Jessika Cristina Chagas Lesbon, Elisangela Chicaroni Mattos, Heidge Fukumasu. USP-Botucatu: Rejane Maria Tommasini Grotto, Jayme A. Souza-Neto, Guilherme Targino Valente, Patricia Akemi Assato, Felipe Allan da Silva da Costa, Bianca Cechetto Carlos. Mendelics: Bibiana Santos, João Paulo Kitajima, Erika Freitas, David Schlesinger. Hemocentro Ribeirão Preto: Simone Kashima, Evandra Strazza Rodrigues, Svetoslav Nanev Slavov, Elaine Vieira dos Santos, Rafael dos Santos Bezerra, Luiz Carlos Junior de Alcantara, Marta Giovanetti, Vagner Fonseca, Flavia Aburjaile, Rodrigo Tocantins Calado. FAMERP-SJRP: Cecília Artico Banho, Lívia Sacchetto, Fábio Sossai Possebon, Leila Sabrina Ullmann, Cíntia Bittar, Guilherme Campos, Helena Lage Ferreira, Jorge A. Petroli Marchesi, Maisa C. Pereira Parra, Marília Moraes, Paula Rahal, Paulo Inacio da Costa, João Pessoa Araújo Jr., Maurício Lacerda Nogueira. Prefeitura de Sao Paulo: Melissa Palmieri. |
| EPI_ISL_1966316 | UNIDADE DE PRONTO ATENDIMENTO DE AGENOR DE CAMPOS | Instituto Butantan / Mendelics | Instituto Butantan: Dimas Tadeu Covas, Sandra Coccuzzo Sampaio, Maria Carolina Elias, José Salvatore Leister Patané, Vincent Louis Viala, Antonio Jorge Martins, Ricardo Haddad, Claudia Renata dos Santos Barros, Elaine Cristina Marqueze, Raul Machado Neto, Debora Botequio Moretti, Jardelina de Souza Todao Bernardino, Loyze Paola Oliveira de Lima, Luiz Aurelio de Campos Crispin. Centro de Genômica Funcional da ESALQ: Luiz Lehmann |

| | | | |
|---|---|---|---|
| | | | Coutinho, Ricardo Augusto Brassaloti, Raquel de Lello Rocha Campos Cassano. NGS Soluções Genômicas: Pilar Drummond Sampaio Corrêa Mariani. FZEA-USP Pirassununga: Mirele Daiana Poleti, Jessika Cristina Chagas Lesbon, Elisangela Chicaroni Mattos, Heidge Fukumasu. USP-Botucatu: Rejane Maria Tommasini Grotto, Jayme A. Souza-Neto, Guilherme Targino Valente, Patricia Akemi Assato, Felipe Allan da Silva da Costa, Bianca Cechetto Carlos. Mendelics: Bibiana Santos, João Paulo Kitajima, Erika Freitas, David Schlesinger. Hemocentro Ribeirão Preto: Simone Kashima, Evandra Strazza Rodrigues, Svetoslav Nanev Slavov, Elaine Vieira dos Santos, Rafael dos Santos Bezerra, Luiz Carlos Junior de Alcantara, Marta Giovanetti, Vagner Fonseca, Flavia Aburjaile, Rodrigo Tocantins Calado. FAMERP-SJRP: Cecília Artico Banho, Lívia Sacchetto, Fábio Sossai Possebon, Leila Sabrina Ullmann, Cíntia Bittar, Guilherme Campos, Helena Lage Ferreira, Jorge A. Petroli Marchesi, Maisa C. Pereira Parra, Marília Moraes, Paula Rahal, Paulo Inacio da Costa, João Pessoa Araújo Jr., Maurício Lacerda Nogueira. Prefeitura de Sao Paulo: Melissa Palmieri. |
| EPI_ISL_1966317, EPI_ISL_1966318 | PRONTO SOCORRO DE AGENOR DE CAMPOS MONGAGUA | Instituto Butantan / Mendelics | Instituto Butantan: Dimas Tadeu Covas, Sandra Coccuzzo Sampaio, Maria Carolina Elias, José Salvatore Leister Patané, Vincent Louis Viala, Antonio Jorge Martins, Ricardo Haddad, Claudia Renata dos Santos Barros, Elaine Cristina Marqueze, Raul Machado Neto, Debora Botequio Moretti, Jardelina de Souza Todao Bernardino, Loyze Paola Oliveira de Lima, Luiz Aurelio de Campos Crispin. Centro de Genômica Funcional da ESALQ: Luiz Lehmann Coutinho, Ricardo Augusto Brassaloti, Raquel de Lello Rocha Campos Cassano. NGS Soluções Genômicas: Pilar Drummond Sampaio Corrêa Mariani. FZEA-USP Pirassununga: Mirele Daiana Poleti, Jessika Cristina Chagas Lesbon, Elisangela Chicaroni Mattos, Heidge Fukumasu. USP-Botucatu: Rejane Maria Tommasini Grotto, Jayme A. Souza-Neto, Guilherme Targino Valente, Patricia Akemi Assato, Felipe Allan da Silva da Costa, Bianca Cechetto Carlos. Mendelics: Bibiana Santos, João Paulo Kitajima, Erika Freitas, David Schlesinger. Hemocentro Ribeirão Preto: Simone Kashima, Evandra Strazza Rodrigues, Svetoslav Nanev Slavov, Elaine Vieira dos Santos, Rafael dos Santos Bezerra, Luiz Carlos Junior de Alcantara, Marta Giovanetti, Vagner Fonseca, Flavia Aburjaile, Rodrigo Tocantins Calado. FAMERP-SJRP: Cecília Artico Banho, Lívia Sacchetto, Fábio Sossai Possebon, Leila Sabrina Ullmann, Cíntia Bittar, Guilherme Campos, Helena Lage Ferreira, Jorge A. Petroli Marchesi, Maisa C. Pereira Parra, Marília Moraes, Paula Rahal, Paulo Inacio da Costa, João Pessoa Araújo Jr., Maurício Lacerda Nogueira. Prefeitura de Sao Paulo: Melissa Palmieri. |
| EPI_ISL_1966319 | UNIDADE DE PRONTO ATENDIMENTO DE AGENOR DE CAMPOS | Instituto Butantan / Mendelics | Instituto Butantan: Dimas Tadeu Covas, Sandra Coccuzzo Sampaio, Maria Carolina Elias, José Salvatore Leister Patané, Vincent Louis Viala, Antonio Jorge Martins, Ricardo Haddad, Claudia Renata dos Santos Barros, Elaine Cristina Marqueze, Raul Machado Neto, Debora Botequio Moretti, Jardelina de Souza Todao Bernardino, Loyze Paola Oliveira de Lima, Luiz Aurelio de Campos Crispin. Centro de Genômica Funcional da ESALQ: Luiz Lehmann Coutinho, Ricardo Augusto Brassaloti, Raquel de Lello Rocha Campos Cassano. NGS Soluções Genômicas: Pilar Drummond Sampaio Corrêa Mariani. FZEA-USP Pirassununga: Mirele Daiana Poleti, Jessika Cristina Chagas Lesbon, Elisangela Chicaroni Mattos, Heidge Fukumasu. USP-Botucatu: Rejane Maria Tommasini Grotto, Jayme A. Souza-Neto, Guilherme Targino Valente, Patricia Akemi Assato, Felipe Allan da Silva da Costa, Bianca Cechetto Carlos. Mendelics: Bibiana Santos, João Paulo Kitajima, Erika Freitas, David Schlesinger. Hemocentro Ribeirão Preto: Simone Kashima, Evandra Strazza Rodrigues, Svetoslav Nanev Slavov, Elaine Vieira dos Santos, Rafael dos Santos Bezerra, Luiz Carlos Junior de Alcantara, Marta Giovanetti, Vagner Fonseca, Flavia Aburjaile, Rodrigo Tocantins Calado. FAMERP-SJRP: Cecília Artico Banho, Lívia Sacchetto, Fábio Sossai Possebon, Leila Sabrina Ullmann, Cíntia Bittar, Guilherme Campos, Helena Lage Ferreira, Jorge A. Petroli Marchesi, Maisa C. Pereira Parra, Marília Moraes, Paula Rahal, Paulo Inacio da Costa, João Pessoa Araújo Jr., Maurício Lacerda Nogueira. Prefeitura de Sao Paulo: Melissa Palmieri. |
| EPI_ISL_1966320 | PRONTO SOCORRO DE AGENOR DE CAMPOS MONGAGUA | Instituto Butantan / Mendelics | Instituto Butantan: Dimas Tadeu Covas, Sandra Coccuzzo Sampaio, Maria Carolina Elias, José Salvatore Leister Patané, Vincent Louis Viala, Antonio Jorge Martins, Ricardo Haddad, Claudia Renata dos Santos Barros, Elaine Cristina Marqueze, Raul Machado Neto, Debora Botequio Moretti, Jardelina de Souza Todao Bernardino, Loyze Paola Oliveira de Lima, Luiz Aurelio de Campos Crispin. Centro de Genômica Funcional da ESALQ: Luiz Lehmann Coutinho, Ricardo Augusto Brassaloti, Raquel de Lello Rocha Campos Cassano. NGS Soluções Genômicas: Pilar Drummond Sampaio Corrêa Mariani. FZEA-USP Pirassununga: Mirele Daiana Poleti, Jessika Cristina Chagas Lesbon, Elisangela Chicaroni Mattos, Heidge Fukumasu. USP-Botucatu: Rejane Maria Tommasini Grotto, Jayme A. Souza-Neto, Guilherme Targino Valente, Patricia Akemi Assato, Felipe Allan da Silva da Costa, Bianca Cechetto Carlos. Mendelics: Bibiana Santos, João Paulo Kitajima, Erika Freitas, David Schlesinger. Hemocentro Ribeirão Preto: Simone Kashima, Evandra Strazza Rodrigues, Svetoslav Nanev Slavov, Elaine Vieira dos Santos, Rafael dos Santos Bezerra, Luiz Carlos Junior de Alcantara, Marta Giovanetti, Vagner Fonseca, Flavia Aburjaile, Rodrigo Tocantins Calado. FAMERP-SJRP: Cecília Artico Banho, Lívia Sacchetto, Fábio Sossai Possebon, Leila Sabrina Ullmann, Cíntia Bittar, Guilherme Campos, Helena Lage Ferreira, Jorge A. Petroli Marchesi, Maisa C. Pereira Parra, Marília Moraes, Paula Rahal, Paulo Inacio da Costa, João Pessoa Araújo Jr., Maurício Lacerda Nogueira. Prefeitura de Sao Paulo: Melissa Palmieri. |
| EPI_ISL_1966321, EPI_ISL_1966322 | UPA SILVINA | Instituto Butantan / Mendelics | Instituto Butantan: Dimas Tadeu Covas, Sandra Coccuzzo Sampaio, Maria Carolina Elias, José Salvatore Leister Patané, Vincent Louis Viala, Antonio Jorge Martins, Ricardo Haddad, Claudia Renata dos Santos Barros, Elaine Cristina Marqueze, Raul Machado Neto, Debora Botequio Moretti, Jardelina de Souza Todao Bernardino, Loyze Paola Oliveira de Lima, Luiz Aurelio de Campos Crispin. Centro de Genômica Funcional da ESALQ: Luiz Lehmann Coutinho, Ricardo Augusto Brassaloti, Raquel de Lello Rocha Campos Cassano. NGS Soluções Genômicas: Pilar Drummond Sampaio Corrêa Mariani. FZEA-USP Pirassununga: Mirele Daiana Poleti, Jessika Cristina Chagas Lesbon, Elisangela Chicaroni Mattos, Heidge Fukumasu. USP-Botucatu: Rejane Maria Tommasini Grotto, Jayme A. Souza-Neto, Guilherme Targino Valente, Patricia Akemi Assato, Felipe Allan da Silva da Costa, Bianca Cechetto Carlos. Mendelics: Bibiana Santos, João Paulo Kitajima, Erika Freitas, David Schlesinger. Hemocentro Ribeirão Preto: Simone Kashima, Evandra Strazza Rodrigues, Svetoslav Nanev Slavov, Elaine Vieira dos Santos, Rafael dos Santos Bezerra, Luiz Carlos Junior de Alcantara, Marta Giovanetti, Vagner Fonseca, Flavia Aburjaile, Rodrigo Tocantins Calado. FAMERP-SJRP: Cecília Artico Banho, Lívia Sacchetto, Fábio Sossai Possebon, Leila Sabrina Ullmann, Cíntia Bittar, Guilherme Campos, Helena Lage Ferreira, Jorge A. Petroli Marchesi, Maisa C. Pereira Parra, Marília Moraes, Paula Rahal, Paulo Inacio da Costa, João Pessoa Araújo Jr., Maurício Lacerda Nogueira. Prefeitura de Sao Paulo: Melissa Palmieri. |
| EPI_ISL_1966323, EPI_ISL_1966324, EPI_ISL_1966326, EPI_ISL_1966327, EPI_ISL_1966328, EPI_ISL_1966329, EPI_ISL_1966330, EPI_ISL_1966331, EPI_ISL_1966332, EPI_ISL_1966334, EPI_ISL_1966335, EPI_ISL_1966337, EPI_ISL_1966338, EPI_ISL_1966340, EPI_ISL_1966341 | | | |
| see above | Ambulatório IB | Instituto Butantan / Mendelics | Instituto Butantan: Dimas Tadeu Covas, Sandra Coccuzzo Sampaio, Maria Carolina Elias, José Salvatore Leister Patané, Vincent Louis Viala, Antonio Jorge Martins, Ricardo Haddad, Claudia Renata dos Santos Barros, Elaine Cristina Marqueze, Raul Machado Neto, Debora Botequio Moretti, Jardelina de Souza Todao Bernardino, Loyze Paola Oliveira de Lima, Luiz Aurelio de Campos Crispin. Centro de Genômica Funcional da ESALQ: Luiz Lehmann Coutinho, Ricardo Augusto Brassaloti, Raquel de Lello Rocha Campos Cassano. NGS Soluções Genômicas: Pilar Drummond Sampaio Corrêa Mariani. FZEA-USP Pirassununga: Mirele Daiana Poleti, Jessika Cristina Chagas Lesbon, Elisangela Chicaroni Mattos, Heidge Fukumasu. USP-Botucatu: Rejane Maria Tommasini Grotto, Jayme A. Souza-Neto, Guilherme Targino Valente, Patricia Akemi Assato, Felipe Allan da Silva da Costa, Bianca Cechetto Carlos. Mendelics: Bibiana Santos, João Paulo Kitajima, Erika Freitas, David Schlesinger. Hemocentro Ribeirão Preto: Simone Kashima, Evandra Strazza Rodrigues, Svetoslav Nanev Slavov, Elaine Vieira dos Santos, Rafael dos Santos Bezerra, Luiz Carlos Junior de Alcantara, Marta Giovanetti, Vagner Fonseca, Flavia Aburjaile, Rodrigo Tocantins Calado. FAMERP-SJRP: Cecília Artico Banho, Lívia Sacchetto, Fábio Sossai Possebon, Leila Sabrina Ullmann, Cíntia Bittar, Guilherme Campos, Helena Lage Ferreira, Jorge A. Petroli Marchesi, Maisa C. Pereira Parra, Marília Moraes, Paula Rahal, Paulo Inacio da Costa, João Pessoa Araújo Jr., Maurício Lacerda Nogueira. Prefeitura de Sao Paulo: Melissa Palmieri. |
| EPI_ISL_1966342 | UPA 24 HORAS CENTRO | Instituto Butantan / Mendelics | Instituto Butantan: Dimas Tadeu Covas, Sandra Coccuzzo Sampaio, Maria Carolina Elias, José Salvatore Leister Patané, Vincent Louis Viala, Antonio Jorge Martins, Ricardo Haddad, Claudia Renata dos Santos Barros, Elaine Cristina Marqueze, Raul Machado Neto, Debora Botequio Moretti, Jardelina de Souza Todao Bernardino, Loyze Paola Oliveira de Lima, Luiz Aurelio de Campos Crispin. Centro de Genômica Funcional da ESALQ: Luiz Lehmann Coutinho, Ricardo Augusto Brassaloti, Raquel de Lello Rocha Campos Cassano. NGS Soluções Genômicas: Pilar Drummond Sampaio Corrêa Mariani. FZEA-USP Pirassununga: Mirele Daiana Poleti, Jessika Cristina Chagas Lesbon, Elisangela Chicaroni Mattos, Heidge Fukumasu. USP-Botucatu: Rejane Maria Tommasini Grotto, Jayme A. Souza-Neto, Guilherme Targino Valente, Patricia Akemi Assato, Felipe Allan da Silva da Costa, Bianca Cechetto Carlos. Mendelics: Bibiana Santos, João Paulo Kitajima, Erika Freitas, David Schlesinger. Hemocentro Ribeirão Preto: Simone Kashima, Evandra Strazza Rodrigues, Svetoslav Nanev Slavov, Elaine Vieira dos Santos, Rafael dos Santos Bezerra, Luiz Carlos Junior de Alcantara, Marta Giovanetti, Vagner Fonseca, Flavia Aburjaile, Rodrigo Tocantins Calado. FAMERP-SJRP: Cecília Artico Banho, Lívia Sacchetto, Fábio Sossai Possebon, Leila Sabrina Ullmann, Cíntia Bittar, Guilherme Campos, Helena Lage Ferreira, Jorge A. Petroli Marchesi, Maisa C. Pereira Parra, Marília Moraes, Paula Rahal, Paulo Inacio da Costa, João Pessoa Araújo Jr., Maurício Lacerda Nogueira. Prefeitura de Sao Paulo: Melissa Palmieri. |
| EPI_ISL_1966343, EPI_ISL_1966344, EPI_ISL_1966345 | CENTRO DE SAUDE III TABATINGA | Instituto Butantan / Mendelics | Instituto Butantan: Dimas Tadeu Covas, Sandra Coccuzzo Sampaio, Maria Carolina Elias, José Salvatore Leister Patané, Vincent Louis Viala, Antonio Jorge Martins, Ricardo Haddad, Claudia Renata dos Santos Barros, Elaine Cristina Marqueze, Raul Machado Neto, Debora Botequio Moretti, Jardelina de Souza Todao Bernardino, Loyze Paola Oliveira de Lima, Luiz Aurelio de Campos Crispin. Centro de Genômica Funcional da ESALQ: Luiz Lehmann Coutinho, Ricardo Augusto Brassaloti, Raquel de Lello Rocha Campos Cassano. NGS Soluções Genômicas: Pilar Drummond Sampaio Corrêa Mariani. FZEA-USP Pirassununga: Mirele Daiana Poleti, Jessika Cristina Chagas Lesbon, Elisangela Chicaroni Mattos, Heidge Fukumasu. USP-Botucatu: Rejane Maria Tommasini Grotto, Jayme A. Souza-Neto, Guilherme Targino Valente, Patricia Akemi Assato, Felipe Allan da Silva da Costa, Bianca Cechetto Carlos. Mendelics: Bibiana Santos, João Paulo Kitajima, Erika Freitas, David Schlesinger. Hemocentro Ribeirão Preto: Simone Kashima, Evandra Strazza Rodrigues, Svetoslav Nanev Slavov, Elaine Vieira dos Santos, Rafael dos Santos Bezerra, Luiz Carlos Junior de Alcantara, Marta Giovanetti, Vagner |

| | | | |
|---|---|---|---|
| | | | Fonseca, Flavia Aburjaile, Rodrigo Tocantins Calado. FAMERP-SJRP: Cecília Artico Banho, Lívia Sacchetto, Fábio Sossai Possebon, Leila Sabrina Ullmann, Cíntia Bittar, Guilherme Campos, Helena Lage Ferreira, Jorge A. Petroli Marchesi, Maisa C. Pereira Parra, Marília Moraes, Paula Rahal, Paulo Inacio da Costa, João Pessoa Araújo Jr., Maurício Lacerda Nogueira. Prefeitura de Sao Paulo: Melissa Palmieri. |
| EPI_ISL_1966346, EPI_ISL_1966347, EPI_ISL_1966348, EPI_ISL_1966349 | SECRETARIA MUNICIPAL DE SAUDE DE VALPARAISO SP | Instituto Butantan / Mendelics | Instituto Butantan: Dimas Tadeu Covas, Sandra Coccuzzo Sampaio, Maria Carolina Elias, José Salvatore Leister Patané, Vincent Louis Viala, Antonio Jorge Martins, Ricardo Haddad, Claudia Renata dos Santos Barros, Elaine Cristina Marqueze, Raul Machado Neto, Debora Botequio Moretti, Jardelina de Souza Todao Bernardino, Loyze Paola Oliveira de Lima, Luiz Aurelio de Campos Crispin. Centro de Genômica Funcional da ESALQ: Luiz Lehmann Coutinho, Ricardo Augusto Brassaloti, Raquel de Lello Rocha Campos Cassano. NGS Soluções Genômicas: Pilar Drummond Sampaio Corrêa Mariani. FZEA-USP Pirassununga: Mirele Daiana Poleti, Jessika Cristina Chagas Lesbon, Elisangela Chicaroni Mattos, Heidge Fukumasu. USP-Botucatu: Rejane Maria Tommasini Grotto, Jayme A. Souza-Neto, Guilherme Targino Valente, Patricia Akemi Assato, Felipe Allan da Silva da Costa, Bianca Cechetto Carlos. Mendelics: Bibiana Santos, João Paulo Kitajima, Erika Freitas, David Schlesinger. Hemocentro Ribeirão Preto: Simone Kashima, Evandra Strazza Rodrigues, Svetoslav Nanev Slavov, Elaine Vieira dos Santos, Rafael dos Santos Bezerra, Luiz Carlos Junior de Alcantara, Marta Giovanetti, Vagner Fonseca, Flavia Aburjaile, Rodrigo Tocantins Calado. FAMERP-SJRP: Cecília Artico Banho, Lívia Sacchetto, Fábio Sossai Possebon, Leila Sabrina Ullmann, Cíntia Bittar, Guilherme Campos, Helena Lage Ferreira, Jorge A. Petroli Marchesi, Maisa C. Pereira Parra, Marília Moraes, Paula Rahal, Paulo Inacio da Costa, João Pessoa Araújo Jr., Maurício Lacerda Nogueira. Prefeitura de Sao Paulo: Melissa Palmieri. |
| EPI_ISL_2003111 | Pronto Socorro Central de Bauru | Instituto Adolfo Lutz, Interdiciplinary Procedures Center, Strategic Laboratory | Claudio Tavares Sacchi, Claudia Regina Gonçalves, Erica Valessa Ramos Gomes, Karoline Rodrigues Campos, Caio Vinicius Dias Lopes, Leonardo Jose Tadeu de Araujo |
| EPI_ISL_2003112 | CS de Paulo de Faria | Instituto Adolfo Lutz, Interdiciplinary Procedures Center, Strategic Laboratory | Claudio Tavares Sacchi, Claudia Regina Gonçalves, Erica Valessa Ramos Gomes, Karoline Rodrigues Campos, Caio Vinicius Dias Lopes, Leonardo Jose Tadeu de Araujo |
| EPI_ISL_2003113 | Hospital Municipal Dr Mario Gatti Campinas | Instituto Adolfo Lutz, Interdiciplinary Procedures Center, Strategic Laboratory | Claudio Tavares Sacchi, Claudia Regina Gonçalves, Erica Valessa Ramos Gomes, Karoline Rodrigues Campos, Caio Vinicius Dias Lopes, Leonardo Jose Tadeu de Araujo |
| EPI_ISL_2003114, EPI_ISL_2003116, EPI_ISL_2003117, EPI_ISL_2003131 | Instituto Adolfo Lutz Central | Instituto Adolfo Lutz, Interdiciplinary Procedures Center, Strategic Laboratory | Claudio Tavares Sacchi, Claudia Regina Gonçalves, Erica Valessa Ramos Gomes, Karoline Rodrigues Campos, Caio Vinicius Dias Lopes, Leonardo Jose Tadeu de Araujo |
| EPI_ISL_2003134, EPI_ISL_2003135, EPI_ISL_2003136, EPI_ISL_2003137, EPI_ISL_2003138, EPI_ISL_2003139, EPI_ISL_2003140, EPI_ISL_2003147, EPI_ISL_2003148, EPI_ISL_2003149 | Instituto Adolfo Lutz - Regional de Santos | Instituto Adolfo Lutz, Interdiciplinary Procedures Center, Strategic Laboratory | Claudio Tavares Sacchi, Claudia Regina Gonçalves, Erica Valessa Ramos Gomes, Karoline Rodrigues Campos, Caio Vinicius Dias Lopes, Leonardo Jose Tadeu de Araujo |
| EPI_ISL_2003150, EPI_ISL_2003151 | Instituto Adolfo Lutz - Regional de Bauru | Instituto Adolfo Lutz, Interdiciplinary Procedures Center, Strategic Laboratory | Claudio Tavares Sacchi, Claudia Regina Gonçalves, Erica Valessa Ramos Gomes, Karoline Rodrigues Campos, Caio Vinicius Dias Lopes, Leonardo Jose Tadeu de Araujo |
| EPI_ISL_2003152 | Instituto Adolfo Lutz - Regional de Marilia | Instituto Adolfo Lutz, Interdiciplinary Procedures Center, Strategic Laboratory | Claudio Tavares Sacchi, Claudia Regina Gonçalves, Erica Valessa Ramos Gomes, Karoline Rodrigues Campos, Caio Vinicius Dias Lopes, Leonardo Jose Tadeu de Araujo |
| EPI_ISL_2003153, EPI_ISL_2003154, EPI_ISL_2003155, EPI_ISL_2003156, EPI_ISL_2003157, EPI_ISL_2003158, EPI_ISL_2003159, EPI_ISL_2003160, EPI_ISL_2003161 | Instituto Adolfo Lutz Central | Instituto Adolfo Lutz, Interdiciplinary Procedures Center, Strategic Laboratory | Claudio Tavares Sacchi, Claudia Regina Gonçalves, Erica Valessa Ramos Gomes, Karoline Rodrigues Campos, Caio Vinicius Dias Lopes, Leonardo Jose Tadeu de Araujo |
| EPI_ISL_2003169, EPI_ISL_2003170 | Instituto Adolfo Lutz - Regional de Campinas | Instituto Adolfo Lutz, Interdiciplinary Procedures Center, Strategic Laboratory | Claudio Tavares Sacchi, Claudia Regina Gonçalves, Erica Valessa Ramos Gomes, Karoline Rodrigues Campos, Caio Vinicius Dias Lopes, Leonardo Jose Tadeu de Araujo |
| EPI_ISL_2008936, EPI_ISL_2008937, EPI_ISL_2008938, EPI_ISL_2008939, EPI_ISL_2008940, EPI_ISL_2008941, EPI_ISL_2008942, EPI_ISL_2008943, EPI_ISL_2013854, EPI_ISL_2013855 | Laboratorio de Pesquisa em Virologia, FAMERP, SJRP | Laboratorio de Pesquisa em Virologia, FAMERP, SJRP | Cecília Artico Banho; Lívia Sacchetto; Fábio Sossai Possebon; Leila Sabrina Ullmann; Cíntia Bittar; Guilherme Campos; Helena Lage Ferreira; Jorge A. Petroli Marchesi; Maisa C. Pereira Parra; Marília Moraes; Paula Rahal; Paulo Inacio da Costa; João Pessoa Araújo Jr.; Maurício L. Nogueira. |
| EPI_ISL_2017241, EPI_ISL_2017242, EPI_ISL_2017243, EPI_ISL_2017244, EPI_ISL_2017245, EPI_ISL_2017246, EPI_ISL_2017247, EPI_ISL_2017248, EPI_ISL_2017249, EPI_ISL_2017250, EPI_ISL_2017251, EPI_ISL_2017252, EPI_ISL_2017253, EPI_ISL_2017254, EPI_ISL_2017255, EPI_ISL_2017256, EPI_ISL_2017257, EPI_ISL_2017258, EPI_ISL_2017259, EPI_ISL_2017260, EPI_ISL_2017261, EPI_ISL_2017262, EPI_ISL_2017263, EPI_ISL_2017264, EPI_ISL_2017265, EPI_ISL_2017266, EPI_ISL_2017267, EPI_ISL_2017268, EPI_ISL_2017269, EPI_ISL_2017270, EPI_ISL_2017271, EPI_ISL_2017272, EPI_ISL_2017273, EPI_ISL_2017274, EPI_ISL_2017275, EPI_ISL_2017276, EPI_ISL_2017277, EPI_ISL_2017278, EPI_ISL_2017279, EPI_ISL_2017280, EPI_ISL_2017281, EPI_ISL_2017283, EPI_ISL_2017284, EPI_ISL_2017285, EPI_ISL_2017286, EPI_ISL_2017287, EPI_ISL_2017288, EPI_ISL_2017289, EPI_ISL_2017290, EPI_ISL_2017291, EPI_ISL_2017292, EPI_ISL_2017293, EPI_ISL_2017294, EPI_ISL_2017295, EPI_ISL_2017296, EPI_ISL_2017297, EPI_ISL_2017298, EPI_ISL_2017299, EPI_ISL_2017300, EPI_ISL_2017301, EPI_ISL_2017302, EPI_ISL_2017303, EPI_ISL_2017304, EPI_ISL_2017305, EPI_ISL_2017306, EPI_ISL_2017307, EPI_ISL_2017308, EPI_ISL_2017309, EPI_ISL_2017310, EPI_ISL_2017311, EPI_ISL_2017312, EPI_ISL_2017313, EPI_ISL_2017314, EPI_ISL_2017315, EPI_ISL_2017316, EPI_ISL_2017317, EPI_ISL_2017318, EPI_ISL_2017319, EPI_ISL_2017320, EPI_ISL_2017321, EPI_ISL_2017322, EPI_ISL_2017323, EPI_ISL_2017324 | | | |
| see above | HLAGYN - Laboratorio de Imunologia de Transplantes de Goias | HLAGYN - Laboratorio de Imunologia de Transplantes de Goias | Fernando Antonio Vinhal dos Santos, Erika Lopes Rocha Batista, Alessandro Leonardo Alvares Magalhaes, Raphael Bessa Parmigiane, Frederico Rodrigues Vinhal, Sabrina Sara Moreira Duarte, Danielle de Paiva Rezende, Lucas Carlos Gomes Pereira, Paola Cristina Resende Silva |
| EPI_ISL_2017412, EPI_ISL_2017413, EPI_ISL_2017414, EPI_ISL_2017415, EPI_ISL_2017416, EPI_ISL_2017417, EPI_ISL_2017418, EPI_ISL_2017419, EPI_ISL_2017420, EPI_ISL_2017421, EPI_ISL_2017422, EPI_ISL_2017423, EPI_ISL_2017426, EPI_ISL_2017427, EPI_ISL_2017428, EPI_ISL_2017429, EPI_ISL_2017447, EPI_ISL_2017449, EPI_ISL_2017450, EPI_ISL_2017452, EPI_ISL_2017457, EPI_ISL_2017458, EPI_ISL_2017459, EPI_ISL_2017460, EPI_ISL_2017461, EPI_ISL_2017465, EPI_ISL_2017466, EPI_ISL_2017467, EPI_ISL_2017468, EPI_ISL_2017469, EPI_ISL_2017470, EPI_ISL_2017471, EPI_ISL_2017472, EPI_ISL_2017476, EPI_ISL_2017478, EPI_ISL_2017479, EPI_ISL_2017480, EPI_ISL_2017481, EPI_ISL_2017483 | | | |
| see above | HLAGYN - Laboratorio de Imunologia de Transplantes de Goias | HLAGYN - Laboratorio de Imunologia de Transplantes de Goias | Fernando Antonio Vinhal dos Santos, Erika Lopes Rocha Batista, Alessandro Leonardo Alvares Magalhaes, Frederico Rodrigues Vinhal, Sabrina Sara Moreira Duarte, Danielle de Paiva Rezende, Lucas Carlos Gomes Pereira, Paola Cristina Resende Silva |
| EPI_ISL_2038926, EPI_ISL_2038927, EPI_ISL_2038928, EPI_ISL_2038929, EPI_ISL_2038930, EPI_ISL_2038931, EPI_ISL_2038932, EPI_ISL_2038933, EPI_ISL_2038934, EPI_ISL_2038935, EPI_ISL_2038936, EPI_ISL_2038937, EPI_ISL_2038938, EPI_ISL_2038939, EPI_ISL_2038940, EPI_ISL_2038941, EPI_ISL_2038942, EPI_ISL_2038943, EPI_ISL_2038945, EPI_ISL_2038946, EPI_ISL_2038947, EPI_ISL_2038948, EPI_ISL_2038949, EPI_ISL_2038950, EPI_ISL_2038951, EPI_ISL_2038952, EPI_ISL_2038953, EPI_ISL_2038954 | | | |
| see above | Laboratorio Central de Saude Publica do Estado de Santa Catarina (LACEN-SC) | Laboratory of Respiratory Viruses and Measles, Oswaldo Cruz Institute, FIOCRUZ | Paola Resende, Luciana Appolinario, Fernando Motta, Anna Carolina Paixao, Ana Carolina Mendonca, Alice Sampaio Rocha, Taina Venas, Elisa Cavalcante Pereira, Renata Serrano Lopes, Darcita Buerger Rovaris, Sandra Bianchini Fernandes, Marilda Siqueira on behalf of the Fiocruz COVID-19 Genomic Surveillance Network |
| EPI_ISL_2038955 | Laboratorio Central de Saude Publica do Estado do Parana (LACEN-PR) | Laboratory of Respiratory Viruses and Measles, Oswaldo Cruz Institute, FIOCRUZ | Paola Resende, Luciana Appolinario, Fernando Motta, Anna Carolina Paixao, Ana Carolina Mendonca, Alice Sampaio Rocha, Taina Venas, Elisa Cavalcante Pereira, Renata Serrano Lopes, Irina Riediger, Marilda Siqueira on behalf of the Fiocruz COVID-19 Genomic Surveillance Network |
| EPI_ISL_2038956 | Laboratorio Central de Saude Publica do Estado do Rio de Janeiro (LACEN-RJ) | Laboratory of Respiratory Viruses and Measles, Oswaldo Cruz Institute, FIOCRUZ | Paola Resende, Luciana Appolinario, Fernando Motta, Anna Carolina Paixao, Ana Carolina Mendonca, Alice Sampaio Rocha, Taina Venas, Elisa Cavalcante Pereira, Renata Serrano Lopes, Andrea Cony Cavalcanti, Marilda Siqueira on behalf of the Fiocruz COVID-19 Genomic Surveillance Network |
| EPI_ISL_2038957, EPI_ISL_2038958, EPI_ISL_2038959, EPI_ISL_2038960 | Laboratorio Central de Saude Publica do Estado do Rio Grande do Sul (LACEN-RS) | Laboratory of Respiratory Viruses and Measles, Oswaldo Cruz Institute, FIOCRUZ | Paola Resende, Luciana Appolinario, Fernando Motta, Anna Carolina Paixao, Ana Carolina Mendonca, Alice Sampaio Rocha, Taina Venas, Elisa Cavalcante Pereira, Renata Serrano Lopes, Tatiana Schaffer Gregianini, Richard Salvato, Marilda Siqueira on behalf of the Fiocruz COVID-19 Genomic Surveillance Network |
| EPI_ISL_2038961, EPI_ISL_2038962, EPI_ISL_2038963 | Laboratory of Respiratory Viruses and Measles, Oswaldo Cruz Institute, FIOCRUZ | Laboratory of Respiratory Viruses and Measles, Oswaldo Cruz Institute, FIOCRUZ | Paola Resende, Luciana Appolinario, Fernando Motta, Anna Carolina Paixao, Ana Carolina Mendonca, Alice Sampaio Rocha, Taina Venas, Elisa Cavalcante Pereira, Renata Serrano Lopes, Marilda Siqueira on behalf of the Fiocruz COVID-19 Genomic Surveillance Network |
| EPI_ISL_2038964 | Laboratorio Central de Saude Publica do Estado do Alagoas (LACEN-AL) | Laboratory of Respiratory Viruses and Measles, Oswaldo Cruz Institute, FIOCRUZ | Paola Resende, Luciana Appolinario, Fernando Motta, Anna Carolina Paixao, Ana Carolina Mendonca, Alice Sampaio Rocha, Taina Venas, Elisa Cavalcante Pereira, Renata Serrano Lopes, Anderson Brandao Leite, Marilda Siqueira on behalf of the Fiocruz COVID-19 Genomic Surveillance Network |
| EPI_ISL_2038965 | Laboratorio Central de Saude Publica do Estado de Minas Gerais (LACEN-MG) | Laboratory of Respiratory Viruses and Measles, Oswaldo Cruz Institute, FIOCRUZ | Paola Resende, Luciana Appolinario, Fernando Motta, Anna Carolina Paixao, Ana Carolina Mendonca, Alice Sampaio Rocha, Taina Venas, Elisa Cavalcante Pereira, Renata Serrano Lopes, Andre Felipe Leal Bernardes, Marilda Siqueira on behalf of the Fiocruz COVID-19 Genomic Surveillance Network |
| EPI_ISL_2038966, EPI_ISL_2038967 | Laboratorio Central de Saude Publica do Estado de Santa | Laboratory of Respiratory Viruses and Measles, Oswaldo Cruz | Paola Resende, Luciana Appolinario, Fernando Motta, Anna Carolina Paixao, Ana Carolina Mendonca, Alice Sampaio Rocha, Taina Venas, Elisa |

| | | | |
|---|---|---|---|
| | Catarina (LACEN-SC) | Institute, FIOCRUZ | Cavalcante Pereira, Renata Serrano Lopes, Darcita Buerger Rovaris, Sandra Bianchini Fernandes, Marilda Siqueira on behalf of the Fiocruz COVID-19 Genomic Surveillance Network |
| EPI_ISL_2038968 | Laboratorio Central de Saude Publica do Estado do Espitito Santo (LACEN-ES) | Laboratory of Respiratory Viruses and Measles, Oswaldo Cruz Institute, FIOCRUZ | Paola Resende, Luciana Appolinario, Fernando Motta, Anna Carolina Paixao, Ana Carolina Mendonca, Alice Sampaio Rocha, Renata Serrano Lopes, Rodrigo Ribeiro Rodrigues, Marilda Siqueira on behalf of the Fiocruz COVID-19 Genomic Surveillance Network |
| EPI_ISL_2086268 | Laboratory of Molecular Biology of Flavivirus, Oswaldo Cruz Institute, FIOCRUZ | Laboratory of Molecular Biology of Flavivirus, Oswaldo Cruz Institute, FIOCRUZ | Fernando Lopez Tort, Ieda Ribeiro, Lidiane Menezes, Alexandre dos Santos, Marta Pereira, Myrna Bonaldo |
| EPI_ISL_2086585, EPI_ISL_2086586, EPI_ISL_2086587, EPI_ISL_2086588, EPI_ISL_2086589, EPI_ISL_2086590, EPI_ISL_2086591, EPI_ISL_2086592, EPI_ISL_2086593, EPI_ISL_2086594, EPI_ISL_2086595, EPI_ISL_2086596, EPI_ISL_2086597, EPI_ISL_2086598, EPI_ISL_2086599, EPI_ISL_2086600, EPI_ISL_2086601, EPI_ISL_2086602, EPI_ISL_2086603, EPI_ISL_2086604, EPI_ISL_2086605, EPI_ISL_2086606, EPI_ISL_2086607, EPI_ISL_2086608, EPI_ISL_2086609, EPI_ISL_2086763, EPI_ISL_2086764 | | | |
| see above | LABCOVID_HCPA | LABRESIS_HCPA | Wink PL, Martins AF, Volpato F, Monteiro F, Zavascki AP, Barth AL |
| EPI_ISL_2098666 | LABCOVID HCPA | LABRESIS | Wink PL, Martins AF, Volpato F, Monteiro F, Zavascki AP, Barth AL |
| EPI_ISL_2101541 | Laboratorio Central Noel Nutels | Bioinformatics Laboratory / LNCC | Luiz G P de Almeida, Alessandra P Lamarca, Ronaldo da Silva F Jr, Liliane Cavalcante, Alexandra L Gerber, Ana Paula de C Guimaraes, Douglas Terra Machado, Cassia Alves, Diana Mariani, Cintia Policarpo, Gleidson da Silva de Oliveira, Mario Sergio Ribeiro, Silvia Carvalho, Flavio Dias da Silva, Marcio Henrique de Oliveira Garcia, Leandro Magalhaes de Souza, Cristiane Gomes da Silva, Caio Luiz Pereira Ribeiro, Andrea Cony Cavalcanti, Claudia Maria Braga de Mello, Amilcar Tanuri, Ana Tereza R Vasconcelos |
| EPI_ISL_2102018, EPI_ISL_2102063 | Laboratório Central de Saúde Pública do Estado do Amazonas (LACEN-AM) | Laboratorio de Ecologia de Doencas Transmissiveis na Amazonia, Instituto Leonidas e Maria Deane - Fiocruz Amazonia | Valdinete Nascimento, Victor Souza, André Corado, Fernanda Nascimento, George Silva, Ágatha Costa, Debora Duarte, Karina Pessoa, Matilde Mejía, Luciana Gonçalves, Maria Júlia Brandão, Felipe Naveca |
| EPI_ISL_2102517, EPI_ISL_2102518, EPI_ISL_2102519, EPI_ISL_2102520 | HLAGYN - Laboratorio de Imunologia de Transplantes de Goias | HLAGYN - Laboratorio de Imunologia de Transplantes de Goias | Fernando Antonio Vinhal dos Santos, Erika Lopes Rocha Batista, Alessandro Leonardo Alvares Magalhaes, Frederico Rodrigues Vinhal, Sabrina Sara Moreira Duarte, Danielle de Paiva Rezende, Lucas Carlos Gomes Pereira, Paola Cristina Resende Silva |
| EPI_ISL_2103421 | Universidade Federal do Norte do Tocantins (UFNT) | Laboratório de Bioinformática e Biotecnologia (Labinftec/UFT) | Ueric José Borges de Souza, Fabrício Souza Campos, Raíssa Nunes dos Santos, José Carlos Ribeiro Júnior, Rogério Fernandes Carvalho, Monike daSilva Oliveira, Bergmann Morais Ribeiro, Fernando Lucas Melo |
| EPI_ISL_2106183 | Biology, UFLA, Universidade Federal de Lavras | Biology, UFLA, Universidade Federal de Lavras | Pylro,V., Barcante,J., Cherem,J., Luciano,P., Fernandes,G. and Melo,D. |
| EPI_ISL_2106184, EPI_ISL_2106185, EPI_ISL_2106186, EPI_ISL_2106187, EPI_ISL_2106188, EPI_ISL_2106189, EPI_ISL_2106190 | Biology, UFLA, Universidade Federal de Lavras | Biology, UFLA, Universidade Federal de Lavras | Pylro,V., Barcante,J., Cherem,J., Luciano,P., Fernandes,G., Melo,D. |
| EPI_ISL_2107299, EPI_ISL_2107300, EPI_ISL_2107304 | Laboratorio de Pesquisa em Virologia, FAMERP, SJRP | Laboratorio de Pesquisa em Virologia, FAMERP, SJRP | Cecília Artico Banho; Lívia Sacchetto; Fábio Sossai Possebon; Leila Sabrina Ullmann; Cíntia Bittar; Guilherme Campos; Helena Lage Ferreira; Jorge A. Petroli Marchesi; Maisa C. Pereira Parra; Marília Moraes; Paula Rahal; Paulo Inacio da Costa; João Pessoa Araújo Jr.; Maurício L. Nogueira. |
| EPI_ISL_2134821, EPI_ISL_2134822 | HospitalIsraelita Albert Eistein | Virology Laboratory- Institute of Tropical Medicine SP | Fernando Lucas Melo, Camila Malta Romano |
| EPI_ISL_2139494, EPI_ISL_2139495, EPI_ISL_2139496, EPI_ISL_2139497, EPI_ISL_2139498, EPI_ISL_2139499, EPI_ISL_2139500, EPI_ISL_2139501, EPI_ISL_2139502, EPI_ISL_2139503, EPI_ISL_2139504, EPI_ISL_2139505, EPI_ISL_2139506, EPI_ISL_2139507, EPI_ISL_2139508, EPI_ISL_2139509, EPI_ISL_2139510, EPI_ISL_2139511, EPI_ISL_2139512, EPI_ISL_2139513, EPI_ISL_2139514, EPI_ISL_2139515, EPI_ISL_2139516, EPI_ISL_2139517, EPI_ISL_2139518, EPI_ISL_2139519, EPI_ISL_2139520, EPI_ISL_2139521, EPI_ISL_2139522, EPI_ISL_2139523, EPI_ISL_2139524, EPI_ISL_2139525, EPI_ISL_2139526, EPI_ISL_2139527, EPI_ISL_2139528, EPI_ISL_2139529, EPI_ISL_2139530, EPI_ISL_2139531, EPI_ISL_2139532, EPI_ISL_2139533, EPI_ISL_2139534, EPI_ISL_2139535, EPI_ISL_2139536, EPI_ISL_2139537, EPI_ISL_2139538, EPI_ISL_2139539, EPI_ISL_2139540, EPI_ISL_2139541, EPI_ISL_2139542, EPI_ISL_2139543, EPI_ISL_2139544, EPI_ISL_2139545, EPI_ISL_2139546, EPI_ISL_2139547, EPI_ISL_2139548, EPI_ISL_2139549 | | | |
| see above | Laboratorio Exame | Universidade Federal de Ciencias da Saude de Porto Alegre | Vinícius Bonetti Franceschi, Gabriel Dickin Caldana et al. |
| EPI_ISL_458141, EPI_ISL_458146, EPI_ISL_458147, EPI_ISL_458148 | Evandro Chagas Institute | Evandro Chagas Institute | Santos, M.C.; Silva, A.M.; Junior, W.D.C.; Barbagelata, L.S.; Ferreira, J.A.; Sousa, E.M.A.; da Silva, P.S.; Resque, H.R; Martins, L.C.; Sousa Junior, E.C.;Viana, G.M.R |
| EPI_ISL_467353, EPI_ISL_467354, EPI_ISL_467355, EPI_ISL_467356, EPI_ISL_467357, EPI_ISL_467358, EPI_ISL_467359, EPI_ISL_467360, EPI_ISL_467361, EPI_ISL_467362, EPI_ISL_467363, EPI_ISL_467364, EPI_ISL_467365, EPI_ISL_467366, EPI_ISL_467367, EPI_ISL_467368, EPI_ISL_467369, EPI_ISL_467370, EPI_ISL_467371 | | | |
| see above | Laboratory of Respiratory Viruses and Measles, Oswaldo Cruz Institute, FIOCRUZ | Laboratory of Respiratory Viruses and Measles, Oswaldo Cruz Institute, FIOCRUZ | Paola Resende, Luciana Appolinario, Fernando Motta, Anna Carolina Paixão, Ana Carolina Mendonça, Aline Mattos, Milene Miranda, Cristiana Garcia, Braulia Caetano, Maria Ogrzewalska, Jonathan Lopes, Marilda Siqueira on behalf of the Fiocruz COVID-19 Genomic Surveillance Network |
| EPI_ISL_470646, EPI_ISL_470647, EPI_ISL_470648, EPI_ISL_470649, EPI_ISL_470650 | Laboratorio de Virologia Molecular / UFRJ | Bioinformatics Laboratory / LNCC | Alexandra Gerber, Ana Paula Guimarães, Luiz Gonzaga Paula de Almeida, Ronaldo da Silva Francisco Junior, Mariane Talon, Filipe Romero, Átila Duque Rossi, Terezinha Marta Pereira, working group UFRJ, Jaqueline Goes de Jesus, Ingra Morales Claro, Ester Cerdeira Sabino, Nuno Rodrigues Faria, CADDE-group, Laboratorio Hermes Pardini, Laboratorio Simile, working group UFMG, Amilcar Tanuri, Carolina Voloch, Renato Santana Aguiar e Ana Tereza Vasconcelos |
| EPI_ISL_471562, EPI_ISL_471581, EPI_ISL_471582 | Hosp. Municipal Prof. Dr. Alípio Corrêa Netto | Instituto Adolfo Lutz, Interdiciplinary Procedures Center, Strategic Laboratory | Claudio Tavares Sacchi, Claudia Regina Gonçalves, Erica Valessa Ramos Gomes |
| EPI_ISL_471647 | Hospital Municipal de Barueri Dr. Francisco Moran | Instituto Adolfo Lutz, Interdiciplinary Procedures Center, Strategic Laboratory | Claudio Tavares Sacchi, Claudia Regina Gonçalves, Erica Valessa Ramos Gomes |
| EPI_ISL_471648 | UBS e Pronto Socorro Jd. Jacira | Instituto Adolfo Lutz, Interdiciplinary Procedures Center, Strategic Laboratory | Claudio Tavares Sacchi, Claudia Regina Gonçalves, Erica Valessa Ramos Gomes |
| EPI_ISL_476346, EPI_ISL_476347, EPI_ISL_476348, EPI_ISL_476349, EPI_ISL_476408, EPI_ISL_476409, EPI_ISL_476410, EPI_ISL_476411, EPI_ISL_476412, EPI_ISL_476413, EPI_ISL_476414, EPI_ISL_476415, EPI_ISL_476416, EPI_ISL_476417, EPI_ISL_476418, EPI_ISL_476419, EPI_ISL_476420, EPI_ISL_476421, EPI_ISL_476422, EPI_ISL_476423, EPI_ISL_476424, EPI_ISL_476425 | | | |
| see above | Laboratório de Patologia Clínica - UNICAMP | Laboratório de Estudos de Vírus Emergentes - UNICAMP | José Luiz Proença-Modena, Magnun Nueldo Nunes dos Santos, Angelica Schreiber, Julia Forato,Camila Simeoni, Marcilio Jorge Fumagalli, Mariene Ribeiro Amorim, Darlan da Silva Candido, Nuno Rodrigues Faria, Julien Theze, Luiz Gonzaga,Jaqueline Goes Jesus e William Marciel de Souza |
| EPI_ISL_492033 | Instituto de Biologia do Exército | Laboratório Metabolismo Macromolecular FirminoTorres de Castro, Instituto de Biofísica Carlos Chagas Filho, Universidade Federal do Rio de Janeiro | Bianca Catarina Azevedo Cabral, Aline Rosa Vianna de Souza, Caleb GM Santos, Marcos Dornelas-Ribeiro, Tatiana LS Nogueira, Nádia Vaez Gonçalves da Cruz, Elizabeth Valentin, Marcio da Costa Cipitelli, Virginia Sara Grancieri do Amaral, Rodrigo Soares de Moura Neto, Clarissa Damaso, Rosane Silva |
| EPI_ISL_492035 | Instituto de Biologia do Exército | Laboratório Metabolismo Macromolecular FirminoTorres de Castro, Instituto de Biofísica Carlos Chagas Filho, Universidade Federal do Rio de Janeiro | Bianca Catarina Azevedo Cabral, Aline Rosa Vianna de Souza, Tatiana LS Nogueira, Nádia Vaez Gonçalves da Cruz, Caleb GM Santos, Marcos Dornelas-Ribeiro, Elizabeth Valentin, Marcio da Costa Cipitelli, Virginia Sara Grancieri do Amaral, Rodrigo Soares de Moura Neto, Clarissa Damaso, Rosane Silva |
| EPI_ISL_492036 | Instituto de Biologia do Exército | Laboratório Metabolismo Macromolecular FirminoTorres de Castro, Instituto de Biofísica Carlos Chagas Filho, Universidade Federal do Rio de Janeiro | Bianca Catarina Azevedo Cabral, Aline Rosa Vianna de Souza , Marcos Dornelas-Ribeiro, Tatiana LS Nogueira, Nádia Vaez Gonçalves da Cruz, Caleb GM Santos, Elizabeth Valentin, Marcio da Costa Cipitelli, Virginia Sara Grancieri do Amaral, Rodrigo Soares de Moura Neto, Clarissa Damaso, Rosane Silva |
| EPI_ISL_492037 | Instituto de Biologia do Exército | Laboratório Metabolismo Macromolecular FirminoTorres de Castro, Instituto de Biofísica Carlos Chagas Filho, Universidade Federal do Rio de Janeiro | Bianca Catarina Azevedo Cabral, Aline Rosa Vianna de Souza, Caleb GM Santos, Marcos Dornelas-Ribeiro, Tatiana LS Nogueira, Nádia Vaez Gonçalves da Cruz, Elizabeth Valentin, Marcio da Costa Cipitelli, Virginia Sara Grancieri do Amaral, Rodrigo Soares de Moura Neto, Clarissa Damaso, Rosane Silva |
| EPI_ISL_492038 | Instituto de Biologia do Exército | Laboratório Metabolismo Macromolecular FirminoTorres de Castro, Instituto de Biofísica Carlos Chagas Filho, Universidade Federal do Rio de Janeiro | Bianca Catarina Azevedo Cabral, Aline Rosa Vianna de Souza, Nádia Vaez Gonçalves da Cruz, Caleb GM Santos, Marcos Dornelas-Ribeiro, Tatiana LS Nogueira, Elizabeth Valentin, Marcio da Costa Cipitelli, Virginia Sara Grancieri do Amaral, Rodrigo Soares de Moura Neto, Clarissa Damaso, Rosane Silva |
| EPI_ISL_492039 | Instituto de Biologia do Exército | Laboratório Metabolismo Macromolecular FirminoTorres de Castro, Instituto de Biofísica Carlos Chagas Filho, Universidade Federal do Rio de Janeiro | Bianca Catarina Azevedo Cabral, Aline Rosa Vianna de Souza, Tatiana LS Nogueira, Nádia Vaez Gonçalves da Cruz, Caleb GM Santos, Marcos Dornelas-Ribeiro, Elizabeth Valentin, Marcio da Costa Cipitelli, Virginia Sara Grancieri do Amaral, Rodrigo Soares de Moura Neto, Clarissa Damaso, Rosane Silva |

| | | | |
|---|---|---|---|
| EPI_ISL_492040 | Instituto de Biologia do Exército | Laboratório Metabolismo Macromolecular FirminoTorres de Castro, Instituto de Biofísica Carlos Chagas Filho, Universidade Federal do Rio de Janeiro | Bianca Catarina Azevedo Cabral, Aline Rosa Vianna de Souza , Marcos Dornelas-Ribeiro, Tatiana LS Nogueira, Nádia Vaez Gonçalves da Cruz, Caleb GM Santos, Elizabeth Valentin, Marcio da Costa Cipitelli, Virginia Sara Grancieri do Amaral, Rodrigo Soares de Moura Neto, Clarissa Damaso, Rosane Silva |
| EPI_ISL_492041 | Instituto de Biologia do Exército | Laboratório Metabolismo Macromolecular FirminoTorres de Castro, Instituto de Biofísica Carlos Chagas Filho, Universidade Federal do Rio de Janeiro | Bianca Catarina Azevedo Cabral, Aline Rosa Vianna de Souza, Caleb GM Santos, Marcos Dornelas-Ribeiro, Tatiana LS Nogueira, Nádia Vaez Gonçalves da Cruz, Elizabeth Valentin, Marcio da Costa Cipitelli, Virginia Sara Grancieri do Amaral, Rodrigo Soares de Moura Neto, Clarissa Damaso, Rosane Silva |
| EPI_ISL_492043 | Instituto de Biologia do Exército | Laboratório Metabolismo Macromolecular FirminoTorres de Castro, Instituto de Biofísica Carlos Chagas Filho, Universidade Federal do Rio de Janeiro | Bianca Catarina Azevedo Cabral, Aline Rosa Vianna de Souza, Tatiana LS Nogueira, Nádia Vaez Gonçalves da Cruz, Caleb GM Santos, Marcos Dornelas-Ribeiro, Elizabeth Valentin, Marcio da Costa Cipitelli, Virginia Sara Grancieri do Amaral, Rodrigo Soares de Moura Neto, Clarissa Damaso, Rosane Silva |
| EPI_ISL_492044 | Instituto de Biologia do Exército | Laboratório Metabolismo Macromolecular FirminoTorres de Castro, Instituto de Biofísica Carlos Chagas Filho, Universidade Federal do Rio de Janeiro | Bianca Catarina Azevedo Cabral, Aline Rosa Vianna de Souza , Marcos Dornelas-Ribeiro, Tatiana LS Nogueira, Nádia Vaez Gonçalves da Cruz, Caleb GM Santos, Elizabeth Valentin, Marcio da Costa Cipitelli, Virginia Sara Grancieri do Amaral, Rodrigo Soares de Moura Neto, Clarissa Damaso, Rosane Silva |
| EPI_ISL_492045 | Instituto de Biologia do Exército | Laboratório Metabolismo Macromolecular FirminoTorres de Castro, Instituto de Biofísica Carlos Chagas Filho, Universidade Federal do Rio de Janeiro | Bianca Catarina Azevedo Cabral, Aline Rosa Vianna de Souza, Caleb GM Santos, Marcos Dornelas-Ribeiro, Tatiana LS Nogueira, Nádia Vaez Gonçalves da Cruz, Elizabeth Valentin, Marcio da Costa Cipitelli, Virginia Sara Grancieri do Amaral, Rodrigo Soares de Moura Neto, Clarissa Damaso, Rosane Silva |
| EPI_ISL_492046 | Instituto de Biologia do Exército | Laboratório Metabolismo Macromolecular FirminoTorres de Castro, Instituto de Biofísica Carlos Chagas Filho, Universidade Federal do Rio de Janeiro | Bianca Catarina Azevedo Cabral, Aline Rosa Vianna de Souza, Nádia Vaez Gonçalves da Cruz, Caleb GM Santos, Marcos Dornelas-Ribeiro, Tatiana LS Nogueira, Elizabeth Valentin, Marcio da Costa Cipitelli, Virginia Sara Grancieri do Amaral, Rodrigo Soares de Moura Neto, Clarissa Damaso, Rosane Silva |
| EPI_ISL_492047 | Instituto de Biologia do Exército | Laboratório Metabolismo Macromolecular FirminoTorres de Castro, Instituto de Biofísica Carlos Chagas Filho, Universidade Federal do Rio de Janeiro | Bianca Catarina Azevedo Cabral, Aline Rosa Vianna de Souza, Tatiana LS Nogueira, Nádia Vaez Gonçalves da Cruz, Caleb GM Santos, Marcos Dornelas-Ribeiro, Elizabeth Valentin, Marcio da Costa Cipitelli, Virginia Sara Grancieri do Amaral, Rodrigo Soares de Moura Neto, Clarissa Damaso, Rosane Silva |
| EPI_ISL_492048 | Instituto de Biologia do Exército | Laboratório Metabolismo Macromolecular FirminoTorres de Castro, Instituto de Biofísica Carlos Chagas Filho, Universidade Federal do Rio de Janeiro | Bianca Catarina Azevedo Cabral, Aline Rosa Vianna de Souza , Marcos Dornelas-Ribeiro, Tatiana LS Nogueira, Nádia Vaez Gonçalves da Cruz, Caleb GM Santos, Elizabeth Valentin, Marcio da Costa Cipitelli, Virginia Sara Grancieri do Amaral, Rodrigo Soares de Moura Neto, Clarissa Damaso, Rosane Silva |
| EPI_ISL_500864 | Instituto de Pesquisas Biomédicas, Hospital Naval Marcílio Dias | Bioinformatics Laboratory / LNCC | Andressa Rangel de Oliveira Lima, Luiz Gonzaga Paula de Almeida, Alexandra Lehmkuhl Gerber, Marlon Daniel Lima Tonin, Mônica Gomes Monteiro da R. Santos, Claudio Alberto Mule Monteiro, Átila Duque Rossi, Isabel de Medeiros Magalhaes de As, Luana Ferreira Martins de Toledo, Amilcar Tanuri, Albina Luciana da Silva Freitas, Fernanda Conceição Silva do Amorim, Shana Priscila Coutinho Barroso, Cynthia Chester Cardoso, Carolina Moreira Voloch, Ana Tereza Vasconcelos |
| EPI_ISL_502779 | LACEN/PE | LABBE, Federal University of Pernambuco | WILSON JOSE DA SILVA JUNIOR, HEIDI LACERDA ALVES DA CRUZ, MARCOS DA SILVEIRA REGUEIRA NETO, BRUNO SAMPAIO, SERGIO DE SA LEITAO PAIVA JUNIOR, ZILDENE DE SOUSA SILVEIRA, MAIRA GALDINO DA ROCHA PITTA, MICHELLY CRISTINY PEREIRA, MARCOS ANTONIO DE MORAIS JUNIOR, ANTONIO CARLOS DE FREITAS, VALDIR DE QUEIROZ BALBINO. |
| EPI_ISL_502875 | LACEN/PE | LABBE, Federal University of Pernambuco | WILSON JOSE DA SILVA JUNIOR, HEIDI LACERDA ALVES DA CRUZ, MARCOS DA SILVEIRA REGUEIRA NETO, BRUNO SAMPAIO, SERGIO DE SA LEITAO PAIVA JUNIOR, ZILDENE DE SOUSA SILVEIRA, MAIRA GALDINO DA ROCHA PITTA, MICHELLY CRISTINY PEREIRA, REGINALDO GONCALVES DE LIMA NETO, MARCOS ANTONIO DE MORAIS JUNIOR, ANTONIO CARLOS DE FREITAS, VALDIR DE QUEIROZ BALBINO. |
| EPI_ISL_509430, EPI_ISL_509431, EPI_ISL_509432, EPI_ISL_509433, EPI_ISL_509435 | Centro de Desenvolvimento Tecnologico em Saude, Fundacao Oswaldo Cruz | Centro de Desenvolvimento Tecnologico em Saude, Fundacao Oswaldo Cruz | Souza,T.M., Fintelman-Rodrigues,N., De Paula,A.D., Saraiva,F.B., Ferreira,M.A., Sacramento,C.Q., Medeiros,M.A. |
| EPI_ISL_510535 | Molecular Virology, Instituto Carlos Chagas / Fiocruz Paraná | Universidade Federal do Parana (UFPR) | Suzukawa,A., Tscha,M., Zanluca,C., Raboni,S., Duarte dos Santos,C. |
| EPI_ISL_510536 | Centro de Desenvolvimento Tecnologico em Saude, Fundacao Oswaldo Cruz | Centro de Desenvolvimento Tecnologico em Saude, Fundacao Oswaldo Cruz | Souza,T.M., Fintelman-Rodrigues,N., De Paula,A.D., Saraiva,F.B., Ferreira,M.A., Sacramento,C.Q. and Medeiros,M.A. |
| EPI_ISL_510541 | Centro de Desenvolvimento Tecnologico em Saude, Fundacao Oswaldo Cruz | Centro de Desenvolvimento Tecnologico em Saude, Fundacao Oswaldo Cruz | Souza,T.M., Fintelman-Rodrigues,N., De Paula,A.D., Saraiva,F.B., Ferreira,M.A., Sacramento,C.Q. and Medeiros,M.A. |
| EPI_ISL_513519, EPI_ISL_513520, EPI_ISL_513521, EPI_ISL_513522, EPI_ISL_513523, EPI_ISL_513524, EPI_ISL_513525, EPI_ISL_513526, EPI_ISL_513527, EPI_ISL_513528, EPI_ISL_513529, EPI_ISL_513563, EPI_ISL_513564, EPI_ISL_513565, EPI_ISL_513566, EPI_ISL_513567, EPI_ISL_513568, EPI_ISL_513569, EPI_ISL_513570, EPI_ISL_513571, EPI_ISL_513572, EPI_ISL_513573, EPI_ISL_513574, EPI_ISL_513575, EPI_ISL_513576, EPI_ISL_513577, EPI_ISL_513578, EPI_ISL_513579, EPI_ISL_513580, EPI_ISL_513581, EPI_ISL_513582, EPI_ISL_513583 | | | |
| see above | Programa de Oncovirologia, Instituto Nacional de Câncer | Programa de Oncovirologia, Instituto Nacional de Câncer | Juliana D. Siqueira, Livia R. Goes, Brunna M. Alves, Claudia Cicala,James Arthos, João P.B. Viola, Andreia C. de Melo, Marcelo A. Soares |
| EPI_ISL_514131 | Rondônia Central Public Health Laboratory (LACEN/RO), vinculated to State Health Secretariat of Rondônia (SESAU/RO) | Molecular Virology Laboratory of Oswaldo Cruz Foundation of Rondônia | Luan Felipo Botelho-Souza, Felipe Souza Nogueira-Lima, Tárcio Peixoto Roca, Alcione de Oliveira dos Santos, Felipe Gomes Naveca, Adriana Cristina Salvador Maia, Cicileia Correia da Silva, Aline Linhares Ferreira de Melo Mendonça, Celina Aparecida Bertoni Lugtenburg, Camila Flávia Gomes Azzi, Juliana Loca Furtado, Suelen Cavalcante, Rita de Cássia Pontello Rampazzo, Caio Henrique Nemeth Santos, Alice Paula Di Sabatino Guimarães, Jansen Fernandes de Medeiros, Fernando Rodrigues Máximo, Juan Miguel Vilallobos-Salcedo and Deusilene Souza Vieira1 |
| EPI_ISL_514132 | Rondônia Central Public Health Laboratory (LACEN/RO), vinculated to State Health Secretariat of Rondônia (SESAU/RO) | Molecular Virology Laboratory of Oswaldo Cruz Foundation of Rondônia | Luan Felipo Botelho-Souza, Felipe Souza Nogueira-Lima, Tárcio Peixoto Roca, Alcione de Oliveira dos Santos, Felipe Gomes Naveca, Adriana Cristina Salvador Maia, Cicileia Correia da Silva, Aline Linhares Ferreira de Melo Mendonça, Celina Aparecida Bertoni Lugtenburg, Camila Flávia Gomes Azzi, Juliana Loca Furtado, Suelen Cavalcante, Rita de Cássia Pontello Rampazzo, Caio Henrique Nemeth Santos, Alice Paula Di Sabatino Guimarães, Jansen Fernandes de Medeiros, Fernando Rodrigues Máximo, Juan Miguel Vilallobos-Salcedo and Deusilene Souza Vieira. |
| EPI_ISL_514133, EPI_ISL_514134, EPI_ISL_514135, EPI_ISL_514136, EPI_ISL_514137, EPI_ISL_514138 | Rondônia Central Public Health Laboratory (LACEN/RO), vinculated to State Health Secretariat of Rondônia (SESAU/RO) | Molecular Virology Laboratory of Oswaldo Cruz Foundation of Rondônia | Luan Felipo Botelho-Souza, Felipe Souza Nogueira-Lima, Tárcio Peixoto Roca, Alcione de Oliveira dos Santos, Felipe Gomes Naveca, Adriana Cristina Salvador Maia, Cicileia Correia da Silva, Aline Linhares Ferreira de Melo Mendonça, Celina Aparecida Bertoni Lugtenburg, Camila Flávia Gomes Azzi, Juliana Loca Furtado, Suelen Cavalcante, Rita de Cássia Pontello Rampazzo, Caio Henrique Nemeth Santos, Alice Paula Di Sabatino Guimarães, Jansen Fernandes de Medeiros, Fernando Rodrigues Máximo, Juan Miguel Vilallobos-Salcedo and Deusilene Souza Vieira |
| EPI_ISL_515525 | National Influenza Center - Instituto Adolfo Lutz | Instituto Adolfo Lutz, Interdiciplinary Procedures Center, Strategic Laboratory | Claudio Tavares Sacchi, Claudia Regina Gonçalves, Erica Valessa Ramos Gomes |
| EPI_ISL_515566 | PS Municipal Dr Lauro Ribas Braga | Instituto Adolfo Lutz, Interdiciplinary Procedures Center, Strategic Laboratory | Claudio Tavares Sacchi, Claudia Regina Gonçalves, Erica Valessa Ramos Gomes |
| EPI_ISL_523959 | Pronto Socorro Municipal de Perus | Instituto Adolfo Lutz, Interdiciplinary Procedures Center, Strategic Laboratory | Claudio Tavares Sacchi, Claudia Regina Gonçalves, Erica Valessa Ramos Gomes |
| EPI_ISL_523986 | Ama Dr Jose Soares Hungria | Instituto Adolfo Lutz, Interdiciplinary Procedures Center, Strategic Laboratory | Claudio Tavares Sacchi, Claudia Regina Gonçalves, Erica Valessa Ramos Gomes |
| EPI_ISL_523992 | Hospital Municipal Carmen Prudente | Instituto Adolfo Lutz, Interdiciplinary Procedures Center, Strategic Laboratory | Claudio Tavares Sacchi, Claudia Regina Gonçalves, Erica Valessa Ramos Gomes |
| EPI_ISL_524468 | Hospital Municipal Vereador Jose Storopolli | Instituto Adolfo Lutz, Interdiciplinary Procedures Center, Strategic Laboratory | Claudio Tavares Sacchi, Claudia Regina Gonçalves, Erica Valessa Ramos Gomes |
| EPI_ISL_524469 | Santa Casa de Misericordia de Sao Paulo | Instituto Adolfo Lutz, Interdiciplinary Procedures Center, Strategic Laboratory | Claudio Tavares Sacchi, Claudia Regina Gonçalves, Erica Valessa Ramos Gomes |
| EPI_ISL_524470 | Hospital do Servidor Público Estadual Francisco Morato de Oliveira | Instituto Adolfo Lutz, Interdiciplinary Procedures Center, Strategic Laboratory | Claudio Tavares Sacchi, Claudia Regina Gonçalves, Erica Valessa Ramos Gomes |

| | | | |
|---|---|---|---|
| EPI_ISL_524783, EPI_ISL_524784, EPI_ISL_524785, EPI_ISL_524786, EPI_ISL_524788, EPI_ISL_524789, EPI_ISL_524790, EPI_ISL_524791, EPI_ISL_524793, EPI_ISL_524794, EPI_ISL_524796, EPI_ISL_524799, EPI_ISL_524800 | | | |
| see above | Evandro Chagas Institute | Evandro Chagas Institute | Santos, M.C.; Silva, A.M.; Junior, W.D.C.; Barbagelata, L.S.; Ferreira, J.A.; Sousa, E.M.A.; da Silva, P.S.; Resque, H.R; Martins, L.C.; Sousa Junior, E.C.;Viana, G.M.R |
| EPI_ISL_527864 | Hospital e Pronto Socorro Comunitário Vila Iolanda | Instituto Adolfo Lutz, Interdiciplinary Procedures Center, Strategic Laboratory | Claudio Tavares Sacchi, Claudia Regina Gonçalves, Erica Valessa Ramos Gomes |
| EPI_ISL_527870 | Hospital Municipal Mário Gatti | Instituto Adolfo Lutz, Interdiciplinary Procedures Center, Strategic Laboratory | Claudio Tavares Sacchi, Claudia Regina Gonçalves, Erica Valessa Ramos Gomes |
| EPI_ISL_529139 | Centro de Desenvolvimento Tecnologico em Saude, Fundacao Oswaldo Cruz | Centro de Desenvolvimento Tecnologico em Saude, Fundacao Oswaldo Cruz | Souza,T.M., Fintelman-Rodrigues,N., De Paula,A.D., Saraiva,F.B., Ferreira,M.A., Sacramento,C.Q., Medeiros,M.A. |
| EPI_ISL_529140 | Centro de Desenvolvimento Tecnologico em Saude, Fundacao Oswaldo Cruz | Centro de Desenvolvimento Tecnologico em Saude, Fundacao Oswaldo Cruz | Souza,T.M., Fintelman-Rodrigues,N., De Paula,A.D., Saraiva,F.B., Ferreira,M.A., Sacramento,C.Q., Medeiros,M.A. |
| EPI_ISL_534311 | UPA III 26 de Agosto | Instituto Adolfo Lutz, Interdiciplinary Procedures Center, Strategic Laboratory | Claudio Tavares Sacchi, Claudia Regina Gonçalves, Erica Valessa Ramos Gomes |
| EPI_ISL_534312 | Distrito Sanitario Sul | Instituto Adolfo Lutz, Interdiciplinary Procedures Center, Strategic Laboratory | Claudio Tavares Sacchi, Claudia Regina Gonçalves, Erica Valessa Ramos Gomes |
| EPI_ISL_534314 | Hospital Universitario da USP de SP | Instituto Adolfo Lutz, Interdiciplinary Procedures Center, Strategic Laboratory | Claudio Tavares Sacchi, Claudia Regina Gonçalves, Erica Valessa Ramos Gomes |
| EPI_ISL_534315 | Serviço de Verificação de Óbitos SVO Guaruulhos | Instituto Adolfo Lutz, Interdiciplinary Procedures Center, Strategic Laboratory | Claudio Tavares Sacchi, Claudia Regina Gonçalves, Erica Valessa Ramos Gomes |
| EPI_ISL_534316 | OS Mun Santana Lauro Ribas Braga | Instituto Adolfo Lutz, Interdiciplinary Procedures Center, Strategic Laboratory | Claudio Tavares Sacchi, Claudia Regina Gonçalves, Erica Valessa Ramos Gomes |
| EPI_ISL_534317 | Hospital Geral de Itapevi | Instituto Adolfo Lutz, Interdiciplinary Procedures Center, Strategic Laboratory | Claudio Tavares Sacchi, Claudia Regina Gonçalves, Erica Valessa Ramos Gomes |
| EPI_ISL_534318 | Hospital Municipal Antonio Giglio | Instituto Adolfo Lutz, Interdiciplinary Procedures Center, Strategic Laboratory | Claudio Tavares Sacchi, Claudia Regina Gonçalves, Erica Valessa Ramos Gomes |
| EPI_ISL_534319, EPI_ISL_534320 | Hospital do Serv Pub ESTAFCO Morato de Oliveira | Instituto Adolfo Lutz, Interdiciplinary Procedures Center, Strategic Laboratory | Claudio Tavares Sacchi, Claudia Regina Gonçalves, Erica Valessa Ramos Gomes |
| EPI_ISL_534321 | PS e Maternidade Nair Fonseca Leitao Arantes | Instituto Adolfo Lutz, Interdiciplinary Procedures Center, Strategic Laboratory | Claudio Tavares Sacchi, Claudia Regina Gonçalves, Erica Valessa Ramos Gomes |
| EPI_ISL_534322 | PS Mun Julio Tupy | Instituto Adolfo Lutz, Interdiciplinary Procedures Center, Strategic Laboratory | Claudio Tavares Sacchi, Claudia Regina Gonçalves, Erica Valessa Ramos Gomes |
| EPI_ISL_534323 | Hospital e Pronto Socorro Comunitario Vila Yolanda | Instituto Adolfo Lutz, Interdiciplinary Procedures Center, Strategic Laboratory | Claudio Tavares Sacchi, Claudia Regina Gonçalves, Erica Valessa Ramos Gomes |
| EPI_ISL_534324 | Hospital Mun Ver Jose Storopolli | Instituto Adolfo Lutz, Interdiciplinary Procedures Center, Strategic Laboratory | Claudio Tavares Sacchi, Claudia Regina Gonçalves, Erica Valessa Ramos Gomes |
| EPI_ISL_534325 | Unidade de Vigilancia em Saude de Guarulhos | Instituto Adolfo Lutz, Interdiciplinary Procedures Center, Strategic Laboratory | Claudio Tavares Sacchi, Claudia Regina Gonçalves, Erica Valessa Ramos Gomes |
| EPI_ISL_534326 | Notre Dame Intermedica Saude AS | Instituto Adolfo Lutz, Interdiciplinary Procedures Center, Strategic Laboratory | Claudio Tavares Sacchi, Claudia Regina Gonçalves, Erica Valessa Ramos Gomes |
| EPI_ISL_541362, EPI_ISL_541363 | Laboratory of Respiratory Viruses and Measles, Oswaldo Cruz Institute, FIOCRUZ | Laboratory of Respiratory Viruses and Measles, Oswaldo Cruz Institute, FIOCRUZ | Paola Resende, Luciana Appolinario, Fernando Motta, Anna Carolina Paixão, Ana Carolina Mendonça, Jonathan Lopes, Marilda Siqueira on behalf of the Fiocruz COVID-19 Genomic Surveillance Network |
| EPI_ISL_541364, EPI_ISL_541365, EPI_ISL_541366 | Laboratory of Respiratory Viruses and Measles, Oswaldo Cruz Institute, FIOCRUZ | Laboratory of Respiratory Viruses and Measles, Oswaldo Cruz Institute, FIOCRUZ | Paola Resende, Roxana Loayza, Cinthia Avila, Luciana Appolinario, Fernando Motta, Anna Carolina Paixao, Ana Carolina Mendonca, Marilda Siqueira on behalf of the Fiocruz COVID-19 Genomic Surveillance Network |
| EPI_ISL_541367 | Laboratory of Respiratory Viruses and Measles, Oswaldo Cruz Institute, FIOCRUZ | Laboratory of Respiratory Viruses and Measles, Oswaldo Cruz Institute, FIOCRUZ | Paola Resende, Luciana Appolinario, Fernando Motta, Anna Carolina Paixão, Ana Carolina Mendonça, Jonathan Lopes, Marilda Siqueira on behalf of the Fiocruz COVID-19 Genomic Surveillance Network |
| EPI_ISL_541368, EPI_ISL_541369 | Laboratory of Respiratory Viruses and Measles, Oswaldo Cruz Institute, FIOCRUZ | Laboratory of Respiratory Viruses and Measles, Oswaldo Cruz Institute, FIOCRUZ | Paola Resende, Roxana Loayza, Cinthia Avila, Luciana Appolinario, Fernando Motta, Anna Carolina Paixao, Ana Carolina Mendonca, Marilda Siqueira on behalf of the Fiocruz COVID-19 Genomic Surveillance Network |
| EPI_ISL_541377, EPI_ISL_541378, EPI_ISL_541379, EPI_ISL_541380, EPI_ISL_541381, EPI_ISL_541382, EPI_ISL_541383, EPI_ISL_541384, EPI_ISL_541385, EPI_ISL_541386, EPI_ISL_541387, EPI_ISL_541390, EPI_ISL_541391 | | | |
| see above | Laboratório Central de Saúde Pública do Estado de Sergipe (LACEN-SE) | Laboratory of Respiratory Viruses and Measles, Oswaldo Cruz Institute, FIOCRUZ | Paola Resende, Luciana Appolinario, Fernando Motta, Anna Carolina Paixão, Ana Carolina Mendonça, Jonathan Lopes, Clioma Santos, Marilda Siqueira on behalf of the Fiocruz COVID-19 Genomic Surveillance Network |
| EPI_ISL_547570 | PS Municipal Dr Augusto Gomes de Mattos | Instituto Adolfo Lutz, Interdiciplinary Procedures Center, Strategic Laboratory | Claudio Tavares Sacchi, Claudia Regina Gonçalves, Erica Valessa Ramos Gomes, Karoline Rodrigues Campos |
| EPI_ISL_547571 | Hospital Municipal Antônio Giglio | Instituto Adolfo Lutz, Interdiciplinary Procedures Center, Strategic Laboratory | Claudio Tavares Sacchi, Claudia Regina Gonçalves, Erica Valessa Ramos Gomes, Karoline Rodrigues Campos |
| EPI_ISL_547573 | Vigilância em Saúde de Cajamar | Instituto Adolfo Lutz, Interdiciplinary Procedures Center, Strategic Laboratory | Claudio Tavares Sacchi, Claudia Regina Gonçalves, Erica Valessa Ramos Gomes, Karoline Rodrigues Campos |
| EPI_ISL_547574 | Hospital Universitario da USP | Instituto Adolfo Lutz, Interdiciplinary Procedures Center, Strategic Laboratory | Claudio Tavares Sacchi, Claudia Regina Gonçalves, Erica Valessa Ramos Gomes, Karoline Rodrigues Campos |
| EPI_ISL_547575 | SVO Jundiaí | Instituto Adolfo Lutz, Interdiciplinary Procedures Center, Strategic Laboratory | Claudio Tavares Sacchi, Claudia Regina Gonçalves, Erica Valessa Ramos Gomes, Karoline Rodrigues Campos |
| EPI_ISL_547576 | Secretaria Municipal de Saúde | Instituto Adolfo Lutz, Interdiciplinary Procedures Center, Strategic Laboratory | Claudio Tavares Sacchi, Claudia Regina Gonçalves, Erica Valessa Ramos Gomes, Karoline Rodrigues Campos |
| EPI_ISL_547577 | Hospital e Maternidade Nossa Senhora das Graças | Instituto Adolfo Lutz, Interdiciplinary Procedures Center, Strategic Laboratory | Claudio Tavares Sacchi, Claudia Regina Gonçalves, Erica Valessa Ramos Gomes, Karoline Rodrigues Campos |
| EPI_ISL_547578 | Hospital Doutor Domingos Leonardo Cerávolo | Instituto Adolfo Lutz, Interdiciplinary Procedures Center, Strategic Laboratory | Claudio Tavares Sacchi, Claudia Regina Gonçalves, Erica Valessa Ramos Gomes, Karoline Rodrigues Campos |
| EPI_ISL_547579 | Santa Casa de Misericórdia de Araçatuba | Instituto Adolfo Lutz, Interdiciplinary Procedures Center, Strategic Laboratory | Claudio Tavares Sacchi, Claudia Regina Gonçalves, Erica Valessa Ramos Gomes, Karoline Rodrigues Campos |
| EPI_ISL_547580 | Santa Casa da Misericórdia de Presidente Prudente | Instituto Adolfo Lutz, Interdiciplinary Procedures Center, Strategic Laboratory | Claudio Tavares Sacchi, Claudia Regina Gonçalves, Erica Valessa Ramos Gomes, Karoline Rodrigues Campos |

| | | | |
|---|---|---|---|
| EPI_ISL_572339, EPI_ISL_572350, EPI_ISL_572351, EPI_ISL_572352, EPI_ISL_572353, EPI_ISL_572354, EPI_ISL_572355, EPI_ISL_572356, EPI_ISL_572357, EPI_ISL_572358, EPI_ISL_572359, EPI_ISL_572360, EPI_ISL_572361, EPI_ISL_572362, EPI_ISL_572363, EPI_ISL_572364, EPI_ISL_572365, EPI_ISL_572366, EPI_ISL_572367, EPI_ISL_572368, EPI_ISL_572369, EPI_ISL_572370, EPI_ISL_572371, EPI_ISL_572372, EPI_ISL_572373, EPI_ISL_572374, EPI_ISL_572375, EPI_ISL_572376, EPI_ISL_572377, EPI_ISL_572378, EPI_ISL_572379, EPI_ISL_572380, EPI_ISL_572381, EPI_ISL_572382, EPI_ISL_572383, EPI_ISL_572384, EPI_ISL_572385, EPI_ISL_572386, EPI_ISL_572387, EPI_ISL_572388, EPI_ISL_572389, EPI_ISL_572390, EPI_ISL_572391, EPI_ISL_572392, EPI_ISL_572393, EPI_ISL_572394, EPI_ISL_572395, EPI_ISL_572396 | | | |
| see above | Laboratório Central de Saúde Pública do Estado de Pernambuco (LACEN-PE) | WallauLab, Aggeu Magalhaes Institute | Marcelo Henrique Santos Paiva, Duschinka Ribeiro Duarte Guedes, Cássia Docena, Matheus Filgueira Bezerra, Filipe Zimmer Dezordi, Laís Ceschini Machado, Larissa Krokovsky, Elisama Helvecio, Alexandre Freitas da Silva, Luydson Richardson Silva Vasconcelos, Antonio Mauro Rezende, Severino Jefferson Ribeiro da Silva, Kamila Gaudêncio da Silva Sales, Bruna Santos Lima Figueiredo de Sá, Derciliano Lopes da Cruz, Claudio Eduardo Cavalcanti, Armando de Menezes Neto, Caroline Targino Alves da Silva, Renata Pessôa Germano Mendes, Maria Almerice Lopes da Silva, Tiago Gräf, Paola Cristina Resende, Gonzalo Bello, Michelle da Silva Barros, Wheverton Ricardo Correia do Nascimento, Rodrigo Moraes Loyo Arcoverde, Luciane Caroline Albuquerque Bezerra, Sinval Pinto Brandão Filho, Constância Flávia Junqueira Ayres, Gabriel Luz Wallau on behalf of the Fiocruz COVID-19 Genomic Surveillance Network |
| EPI_ISL_574578 | Hospital Municipal Mário Gatti | Instituto Adolfo Lutz, Interdiciplinary Procedures Center, Strategic Laboratory | Claudio Tavares Sacchi, Claudia Regina Gonçalves, Erica Valessa Ramos Gomes, Karoline Rodrigues Campos |
| EPI_ISL_574579 | Hospital Municipal Dr. Ignacio Proença de Gouvea | Instituto Adolfo Lutz, Interdiciplinary Procedures Center, Strategic Laboratory | Claudio Tavares Sacchi, Claudia Regina Gonçalves, Erica Valessa Ramos Gomes, Karoline Rodrigues Campos |
| EPI_ISL_574582 | Hospital Municipal Dr. Jose Soares Hungria | Instituto Adolfo Lutz, Interdiciplinary Procedures Center, Strategic Laboratory | Claudio Tavares Sacchi, Claudia Regina Gonçalves, Erica Valessa Ramos Gomes, Karoline Rodrigues Campos |
| EPI_ISL_574583 | Secretaria Municipal de Saude de Jandira | Instituto Adolfo Lutz, Interdiciplinary Procedures Center, Strategic Laboratory | Claudio Tavares Sacchi, Claudia Regina Gonçalves, Erica Valessa Ramos Gomes, Karoline Rodrigues Campos |
| EPI_ISL_574588 | Hospital Estadual Sumare | Instituto Adolfo Lutz, Interdiciplinary Procedures Center, Strategic Laboratory | Claudio Tavares Sacchi, Claudia Regina Gonçalves, Erica Valessa Ramos Gomes, Karoline Rodrigues Campos |
| EPI_ISL_574589 | Hospital Municipal Dr. Jose Soares Hungria | Instituto Adolfo Lutz, Interdiciplinary Procedures Center, Strategic Laboratory | Claudio Tavares Sacchi, Claudia Regina Gonçalves, Erica Valessa Ramos Gomes, Karoline Rodrigues Campos |
| EPI_ISL_574590 | Unidade de Printo Atendimento UPA I Santa Isabel | Instituto Adolfo Lutz, Interdiciplinary Procedures Center, Strategic Laboratory | Claudio Tavares Sacchi, Claudia Regina Gonçalves, Erica Valessa Ramos Gomes, Karoline Rodrigues Campos |
| EPI_ISL_574593 | CS II Dr. Antonio Vicoso Moreira de Rezende Sumare | Instituto Adolfo Lutz, Interdiciplinary Procedures Center, Strategic Laboratory | Claudio Tavares Sacchi, Claudia Regina Gonçalves, Erica Valessa Ramos Gomes, Karoline Rodrigues Campos |
| EPI_ISL_574594 | Hospital Escola da Universidade de Taubate | Instituto Adolfo Lutz, Interdiciplinary Procedures Center, Strategic Laboratory | Claudio Tavares Sacchi, Claudia Regina Gonçalves, Erica Valessa Ramos Gomes, Karoline Rodrigues Campos |
| EPI_ISL_574596 | CS II Dr. Antonio Vicoso Moreira de Rezende Sumare | Instituto Adolfo Lutz, Interdiciplinary Procedures Center, Strategic Laboratory | Claudio Tavares Sacchi, Claudia Regina Gonçalves, Erica Valessa Ramos Gomes, Karoline Rodrigues Campos |
| EPI_ISL_574597 | Secretaria Municipal de Saude de Jarinu | Instituto Adolfo Lutz, Interdiciplinary Procedures Center, Strategic Laboratory | Claudio Tavares Sacchi, Claudia Regina Gonçalves, Erica Valessa Ramos Gomes, Karoline Rodrigues Campos |
| EPI_ISL_574598 | Servico de Verificacao de Obito SVO | Instituto Adolfo Lutz, Interdiciplinary Procedures Center, Strategic Laboratory | Claudio Tavares Sacchi, Claudia Regina Gonçalves, Erica Valessa Ramos Gomes, Karoline Rodrigues Campos |
| EPI_ISL_583493 | Vigilância em Saúde de Cajamar | Instituto Adolfo Lutz, Interdiciplinary Procedures Center, Strategic Laboratory | Claudio Tavares Sacchi, Claudia Regina Gonçalves, Erica Valessa Ramos Gomes, Karoline Rodrigues Campos |
| EPI_ISL_583494 | CS II Dr. Antonio Vicoso Moreira de Rezende Sumare | Instituto Adolfo Lutz, Interdiciplinary Procedures Center, Strategic Laboratory | Claudio Tavares Sacchi, Claudia Regina Gonçalves, Erica Valessa Ramos Gomes, Karoline Rodrigues Campos |
| EPI_ISL_583495 | Serviço de Verificação de Óbitos SVO Guaruulhos | Instituto Adolfo Lutz, Interdiciplinary Procedures Center, Strategic Laboratory | Claudio Tavares Sacchi, Claudia Regina Gonçalves, Erica Valessa Ramos Gomes, Karoline Rodrigues Campos |
| EPI_ISL_583496 | UPA Jandira | Instituto Adolfo Lutz, Interdiciplinary Procedures Center, Strategic Laboratory | Claudio Tavares Sacchi, Claudia Regina Gonçalves, Erica Valessa Ramos Gomes, Karoline Rodrigues Campos |
| EPI_ISL_583497 | Complexo Hospitalar Ouro Verde de Campinas | Instituto Adolfo Lutz, Interdiciplinary Procedures Center, Strategic Laboratory | Claudio Tavares Sacchi, Claudia Regina Gonçalves, Erica Valessa Ramos Gomes, Karoline Rodrigues Campos |
| EPI_ISL_583498 | Hospital Municipal Dr. Waldemar Tebaldi | Instituto Adolfo Lutz, Interdiciplinary Procedures Center, Strategic Laboratory | Claudio Tavares Sacchi, Claudia Regina Gonçalves, Erica Valessa Ramos Gomes, Karoline Rodrigues Campos |
| EPI_ISL_583499 | Distrito Sanitario Sul Campinas | Instituto Adolfo Lutz, Interdiciplinary Procedures Center, Strategic Laboratory | Claudio Tavares Sacchi, Claudia Regina Gonçalves, Erica Valessa Ramos Gomes, Karoline Rodrigues Campos |
| EPI_ISL_583500 | Centro de Saude I Tacito Leite de Carvalho e Silva | Instituto Adolfo Lutz, Interdiciplinary Procedures Center, Strategic Laboratory | Claudio Tavares Sacchi, Claudia Regina Gonçalves, Erica Valessa Ramos Gomes, Karoline Rodrigues Campos |
| EPI_ISL_583501 | Hospital Estadual de CampanhaCOVID 19 Barradas | Instituto Adolfo Lutz, Interdiciplinary Procedures Center, Strategic Laboratory | Claudio Tavares Sacchi, Claudia Regina Gonçalves, Erica Valessa Ramos Gomes, Karoline Rodrigues Campos |
| EPI_ISL_583502 | Serv de Vig Sanitaria Epidemio e CTRL de Zoonoses Guaruja | Instituto Adolfo Lutz, Interdiciplinary Procedures Center, Strategic Laboratory | Claudio Tavares Sacchi, Claudia Regina Gonçalves, Erica Valessa Ramos Gomes, Karoline Rodrigues Campos |
| EPI_ISL_583503 | CTA Centro de Testagem e Aconselhamento | Instituto Adolfo Lutz, Interdiciplinary Procedures Center, Strategic Laboratory | Claudio Tavares Sacchi, Claudia Regina Gonçalves, Erica Valessa Ramos Gomes, Karoline Rodrigues Campos |
| EPI_ISL_583504, EPI_ISL_583505 | Casa de Saude Stella Maris | Instituto Adolfo Lutz, Interdiciplinary Procedures Center, Strategic Laboratory | Claudio Tavares Sacchi, Claudia Regina Gonçalves, Erica Valessa Ramos Gomes, Karoline Rodrigues Campos |
| EPI_ISL_603021 | Pronto Socorro Dr. Conrado Cesarino Nuvolini | Instituto Adolfo Lutz, Interdiciplinary Procedures Center, Strategic Laboratory | Claudio Tavares Sacchi, Claudia Regina Gonçalves, Erica Valessa Ramos Gomes, Karoline Rodrigues Campos |
| EPI_ISL_603022 | Departamento de Vigilância à Saúde | Instituto Adolfo Lutz, Interdiciplinary Procedures Center, Strategic Laboratory | Claudio Tavares Sacchi, Claudia Regina Gonçalves, Erica Valessa Ramos Gomes, Karoline Rodrigues Campos |
| EPI_ISL_603023 | Vigilância em Saúde Visa Sul | Instituto Adolfo Lutz, Interdiciplinary Procedures Center, Strategic Laboratory | Claudio Tavares Sacchi, Claudia Regina Gonçalves, Erica Valessa Ramos Gomes, Karoline Rodrigues Campos |
| EPI_ISL_603024 | Santa Casa de Misericordia de Araçatuba | Instituto Adolfo Lutz, Interdiciplinary Procedures Center, Strategic Laboratory | Claudio Tavares Sacchi, Claudia Regina Gonçalves, Erica Valessa Ramos Gomes, Karoline Rodrigues Campos |
| EPI_ISL_603025 | UPA Central de Caraguatatuba | Instituto Adolfo Lutz, Interdiciplinary Procedures Center, Strategic Laboratory | Claudio Tavares Sacchi, Claudia Regina Gonçalves, Erica Valessa Ramos Gomes, Karoline Rodrigues Campos |
| EPI_ISL_603026 | Santa Casa da Misericórdia de Presidente Prudente | Instituto Adolfo Lutz, Interdiciplinary Procedures Center, Strategic Laboratory | Claudio Tavares Sacchi, Claudia Regina Gonçalves, Erica Valessa Ramos Gomes, Karoline Rodrigues Campos |
| EPI_ISL_603027 | Santa Casa de Misericordia de Araçatuba | Instituto Adolfo Lutz, Interdiciplinary Procedures Center, Strategic Laboratory | Claudio Tavares Sacchi, Claudia Regina Gonçalves, Erica Valessa Ramos Gomes, Karoline Rodrigues Campos |

| | | | |
|---|---|---|---|
| EPI_ISL_603028 | Hospital Municipal Santa Ana | Instituto Adolfo Lutz, Interdiciplinary Procedures Center, Strategic Laboratory | Claudio Tavares Sacchi, Claudia Regina Gonçalves, Erica Valessa Ramos Gomes, Karoline Rodrigues Campos |
| EPI_ISL_603029 | Hospital Municipal Mário Gatti | Instituto Adolfo Lutz, Interdiciplinary Procedures Center, Strategic Laboratory | Claudio Tavares Sacchi, Claudia Regina Gonçalves, Erica Valessa Ramos Gomes, Karoline Rodrigues Campos |
| EPI_ISL_603030 | Hospital Domingos Leonardo Ceravolo Presidente Prudente | Instituto Adolfo Lutz, Interdiciplinary Procedures Center, Strategic Laboratory | Claudio Tavares Sacchi, Claudia Regina Gonçalves, Erica Valessa Ramos Gomes, Karoline Rodrigues Campos |
| EPI_ISL_603031 | Santa Casa de Presidente Epitácio | Instituto Adolfo Lutz, Interdiciplinary Procedures Center, Strategic Laboratory | Claudio Tavares Sacchi, Claudia Regina Gonçalves, Erica Valessa Ramos Gomes, Karoline Rodrigues Campos |
| EPI_ISL_603032 | Santa Casa da Misericórdia de Presidente Prudente | Instituto Adolfo Lutz, Interdiciplinary Procedures Center, Strategic Laboratory | Claudio Tavares Sacchi, Claudia Regina Gonçalves, Erica Valessa Ramos Gomes, Karoline Rodrigues Campos |
| EPI_ISL_603033 | Vigilancia Epidemiologica de São Bernardo do Campo | Instituto Adolfo Lutz, Interdiciplinary Procedures Center, Strategic Laboratory | Claudio Tavares Sacchi, Claudia Regina Gonçalves, Erica Valessa Ramos Gomes, Karoline Rodrigues Campos |
| EPI_ISL_603034 | Departamento de Vigilância à Saúde | Instituto Adolfo Lutz, Interdiciplinary Procedures Center, Strategic Laboratory | Claudio Tavares Sacchi, Claudia Regina Gonçalves, Erica Valessa Ramos Gomes, Karoline Rodrigues Campos |
| EPI_ISL_603035 | Secretaria Municipal de Saúde | Instituto Adolfo Lutz, Interdiciplinary Procedures Center, Strategic Laboratory | Claudio Tavares Sacchi, Claudia Regina Gonçalves, Erica Valessa Ramos Gomes, Karoline Rodrigues Campos |
| EPI_ISL_603036 | Hospital Santa Ana | Instituto Adolfo Lutz, Interdiciplinary Procedures Center, Strategic Laboratory | Claudio Tavares Sacchi, Claudia Regina Gonçalves, Erica Valessa Ramos Gomes, Karoline Rodrigues Campos |
| EPI_ISL_603037 | Hospital Geral de Pedreira | Instituto Adolfo Lutz, Interdiciplinary Procedures Center, Strategic Laboratory | Claudio Tavares Sacchi, Claudia Regina Gonçalves, Erica Valessa Ramos Gomes, Karoline Rodrigues Campos |
| EPI_ISL_603038 | Santa Casa de Misericordia de Araçatuba | Instituto Adolfo Lutz, Interdiciplinary Procedures Center, Strategic Laboratory | Claudio Tavares Sacchi, Claudia Regina Gonçalves, Erica Valessa Ramos Gomes, Karoline Rodrigues Campos |
| EPI_ISL_603039 | Hospital Municipal Mário Gatti | Instituto Adolfo Lutz, Interdiciplinary Procedures Center, Strategic Laboratory | Claudio Tavares Sacchi, Claudia Regina Gonçalves, Erica Valessa Ramos Gomes, Karoline Rodrigues Campos |
| EPI_ISL_623106, EPI_ISL_623107, EPI_ISL_623109, EPI_ISL_623110, EPI_ISL_623111, EPI_ISL_623113, EPI_ISL_623114, EPI_ISL_623115, EPI_ISL_623117, EPI_ISL_623118, EPI_ISL_623119, EPI_ISL_623121, EPI_ISL_623123, EPI_ISL_623125, EPI_ISL_623129, EPI_ISL_623131, EPI_ISL_623133, EPI_ISL_623134, EPI_ISL_623135, EPI_ISL_623136, EPI_ISL_623137, EPI_ISL_623141, EPI_ISL_623143, EPI_ISL_623144, EPI_ISL_623145, EPI_ISL_623146, EPI_ISL_623147, EPI_ISL_623148, EPI_ISL_623149, EPI_ISL_623150, EPI_ISL_623151, EPI_ISL_623153, EPI_ISL_623154, EPI_ISL_623155, EPI_ISL_623157, EPI_ISL_623158, EPI_ISL_623159, EPI_ISL_623161, EPI_ISL_623162, EPI_ISL_623163, EPI_ISL_623164, EPI_ISL_623165, EPI_ISL_623167, EPI_ISL_623169 | | | |
| see above | Laboratorio de Virologia Molecular / UFRJ | Bioinformatics Laboratory / LNCC | Carolina M Voloch, Ronaldo S Francisco Jr, Luiz G P de Almeida, Otavio J. Brustolini, Cynthia C Cardoso, Alexandra L Gerber, Ana Paula de C Guimarães, Diana Mariani, Covid19-UFRJ Workgroup, Luís Cristóvão Pôrto, Renato S Aguiar, Terezinha M P P Castiñeiras, Orlando C. Ferreira, Amilcar Tanuri, Ana Tereza R de Vasconcelos |
| EPI_ISL_636737, EPI_ISL_636835, EPI_ISL_636837 | Laboratório de Imunofarmacologia - Instituto Oswaldo Cruz | Laboratório de Imunofarmacologia - Instituto Oswaldo Cruz | Souza,T.M., Fintelman-Rodrigues,N., De Paula,A.D., Saraiva,F.B., Ferreira,M.A. and Sacramento,C.Q. |
| EPI_ISL_672674, EPI_ISL_672678, EPI_ISL_672679 | DB Diagnosticos do Brasil | Laboratório de Parasitologia Médica - Instituto de Medicina Tropical - Universidade de São Paulo | Brazil-UK Centre for Arbovirus Discovery Diagnosis Genomics and Epidemiology (CADDE) Genomic Network - Instituto de Medicina Tropical |
| EPI_ISL_672738, EPI_ISL_672743, EPI_ISL_672744, EPI_ISL_672745 | Hospital das Clínicas da Faculdade de Medicina da Universidade de São Paulo (HC-FMUSP) | Laboratório de Parasitologia Médica - Instituto de Medicina Tropical - Universidade de São Paulo | Brazil-UK Centre for Arbovirus Discovery Diagnosis Genomics and Epidemiology (CADDE) Genomic Network - Instituto de Medicina Tropical |
| EPI_ISL_693196 | Hospital Santa Clara | Instituto Adolfo Lutz, Interdiciplinary Procedures Center, Strategic Laboratory | Claudio Tavares Sacchi, Claudia Regina Gonçalves, Erica Valessa Ramos Gomes, Karoline Rodrigues Campos |
| EPI_ISL_693197 | Central de Rede de Frio Municipal | Instituto Adolfo Lutz, Interdiciplinary Procedures Center, Strategic Laboratory | Claudio Tavares Sacchi, Claudia Regina Gonçalves, Erica Valessa Ramos Gomes, Karoline Rodrigues Campos |
| EPI_ISL_693198 | Santa Casa de Misericordia de Sao Paulo - Hospital Central | Instituto Adolfo Lutz, Interdiciplinary Procedures Center, Strategic Laboratory | Claudio Tavares Sacchi, Claudia Regina Gonçalves, Erica Valessa Ramos Gomes, Karoline Rodrigues Campos |
| EPI_ISL_693199 | Hospital do Servidor Publico Estadual Francisco Morato de Oliveira | Instituto Adolfo Lutz, Interdiciplinary Procedures Center, Strategic Laboratory | Claudio Tavares Sacchi, Claudia Regina Gonçalves, Erica Valessa Ramos Gomes, Karoline Rodrigues Campos |
| EPI_ISL_693200 | Hospital e Maternidade Mairipora | Instituto Adolfo Lutz, Interdiciplinary Procedures Center, Strategic Laboratory | Claudio Tavares Sacchi, Claudia Regina Gonçalves, Erica Valessa Ramos Gomes, Karoline Rodrigues Campos |
| EPI_ISL_693204 | Pronto Socorro Dr. Conrado Cesarino Nuvolini | Instituto Adolfo Lutz, Interdiciplinary Procedures Center, Strategic Laboratory | Claudio Tavares Sacchi, Claudia Regina Gonçalves, Erica Valessa Ramos Gomes, Karoline Rodrigues Campos |
| EPI_ISL_693205 | Hospital de Campanha Covid-19 Assis | Instituto Adolfo Lutz, Interdiciplinary Procedures Center, Strategic Laboratory | Claudio Tavares Sacchi, Claudia Regina Gonçalves, Erica Valessa Ramos Gomes, Karoline Rodrigues Campos |
| EPI_ISL_693206 | Hospital Municipal Mario Gatti | Instituto Adolfo Lutz, Interdiciplinary Procedures Center, Strategic Laboratory | Claudio Tavares Sacchi, Claudia Regina Gonçalves, Erica Valessa Ramos Gomes, Karoline Rodrigues Campos |
| EPI_ISL_693207 | Cs II Doutor Antonio Vicoso Moreira de Rezende | Instituto Adolfo Lutz, Interdiciplinary Procedures Center, Strategic Laboratory | Claudio Tavares Sacchi, Claudia Regina Gonçalves, Erica Valessa Ramos Gomes, Karoline Rodrigues Campos |
| EPI_ISL_693208, EPI_ISL_693209 | Hospital Municipal Antonio Giglio | Instituto Adolfo Lutz, Interdiciplinary Procedures Center, Strategic Laboratory | Claudio Tavares Sacchi, Claudia Regina Gonçalves, Erica Valessa Ramos Gomes, Karoline Rodrigues Campos |
| EPI_ISL_693210 | Pronto-Socorro Dr. Osmar Mesquita | Instituto Adolfo Lutz, Interdiciplinary Procedures Center, Strategic Laboratory | Claudio Tavares Sacchi, Claudia Regina Gonçalves, Erica Valessa Ramos Gomes, Karoline Rodrigues Campos |
| EPI_ISL_693211 | Santa Casa de Misericordia e Maternidade | Instituto Adolfo Lutz, Interdiciplinary Procedures Center, Strategic Laboratory | Claudio Tavares Sacchi, Claudia Regina Gonçalves, Erica Valessa Ramos Gomes, Karoline Rodrigues Campos |
| EPI_ISL_693212 | Santa Casa de Misericordia de Braganca Paulista | Instituto Adolfo Lutz, Interdiciplinary Procedures Center, Strategic Laboratory | Claudio Tavares Sacchi, Claudia Regina Gonçalves, Erica Valessa Ramos Gomes, Karoline Rodrigues Campos |
| EPI_ISL_693213 | Hospital E Maternidade Municipal Governador Mario Covas | Instituto Adolfo Lutz, Interdiciplinary Procedures Center, Strategic Laboratory | Claudio Tavares Sacchi, Claudia Regina Gonçalves, Erica Valessa Ramos Gomes, Karoline Rodrigues Campos |
| EPI_ISL_693214 | Unidade de Pronto Atendimento Central de Caraguatatuba | Instituto Adolfo Lutz, Interdiciplinary Procedures Center, Strategic Laboratory | Claudio Tavares Sacchi, Claudia Regina Gonçalves, Erica Valessa Ramos Gomes, Karoline Rodrigues Campos |
| EPI_ISL_693215 | Secretaria Municipal de Saúde de Iracemapolis | Instituto Adolfo Lutz, Interdiciplinary Procedures Center, Strategic Laboratory | Claudio Tavares Sacchi, Claudia Regina Gonçalves, Erica Valessa Ramos Gomes, Karoline Rodrigues Campos |
| EPI_ISL_693216, EPI_ISL_693217 | Unidade de Vigilância Epiudemiológica de Araras | Instituto Adolfo Lutz, Interdiciplinary Procedures Center, Strategic Laboratory | Claudio Tavares Sacchi, Claudia Regina Gonçalves, Erica Valessa Ramos Gomes, Karoline Rodrigues Campos |

| | | | |
|---|---|---|---|
| EPI_ISL_693218 | Hospital Domingos Leonardo Ceravolo Presidente Prudente | Instituto Adolfo Lutz, Interdiciplinary Procedures Center, Strategic Laboratory | Claudio Tavares Sacchi, Claudia Regina Gonçalves, Erica Valessa Ramos Gomes, Karoline Rodrigues Campos |
| EPI_ISL_693219 | Santa Casa da Misericórdia de Presidente Prudente | Instituto Adolfo Lutz, Interdiciplinary Procedures Center, Strategic Laboratory | Claudio Tavares Sacchi, Claudia Regina Gonçalves, Erica Valessa Ramos Gomes, Karoline Rodrigues Campos |
| EPI_ISL_693220 | Laboratório Municipal de Piracicaba | Instituto Adolfo Lutz, Interdiciplinary Procedures Center, Strategic Laboratory | Claudio Tavares Sacchi, Claudia Regina Gonçalves, Erica Valessa Ramos Gomes, Karoline Rodrigues Campos |
| EPI_ISL_693221, EPI_ISL_693222 | Secretaria Municipal de Saúde de Birigui | Instituto Adolfo Lutz, Interdiciplinary Procedures Center, Strategic Laboratory | Claudio Tavares Sacchi, Claudia Regina Gonçalves, Erica Valessa Ramos Gomes, Karoline Rodrigues Campos |
| EPI_ISL_693223, EPI_ISL_693224 | Laboratório Municipal de Piracicaba | Instituto Adolfo Lutz, Interdiciplinary Procedures Center, Strategic Laboratory | Claudio Tavares Sacchi, Claudia Regina Gonçalves, Erica Valessa Ramos Gomes, Karoline Rodrigues Campos |
| EPI_ISL_693225 | Ubs Vila Rosa - Olímpia Gomes De Almeida | Instituto Adolfo Lutz, Interdiciplinary Procedures Center, Strategic Laboratory | Claudio Tavares Sacchi, Claudia Regina Gonçalves, Erica Valessa Ramos Gomes, Karoline Rodrigues Campos |
| EPI_ISL_693226 | Unidade de Pronto Atendimento Sao José | Instituto Adolfo Lutz, Interdiciplinary Procedures Center, Strategic Laboratory | Claudio Tavares Sacchi, Claudia Regina Gonçalves, Erica Valessa Ramos Gomes, Karoline Rodrigues Campos |
| EPI_ISL_693227 | UBS Vila Marchi | Instituto Adolfo Lutz, Interdiciplinary Procedures Center, Strategic Laboratory | Claudio Tavares Sacchi, Claudia Regina Gonçalves, Erica Valessa Ramos Gomes, Karoline Rodrigues Campos |
| EPI_ISL_693228 | Secretaria Municipal de Sorocaba | Instituto Adolfo Lutz, Interdiciplinary Procedures Center, Strategic Laboratory | Claudio Tavares Sacchi, Claudia Regina Gonçalves, Erica Valessa Ramos Gomes, Karoline Rodrigues Campos |
| EPI_ISL_693229 | Hospital 8 de Maio | Instituto Adolfo Lutz, Interdiciplinary Procedures Center, Strategic Laboratory | Claudio Tavares Sacchi, Claudia Regina Gonçalves, Erica Valessa Ramos Gomes, Karoline Rodrigues Campos |
| EPI_ISL_693230 | Hospital e Pronto Socorro Portinari | Instituto Adolfo Lutz, Interdiciplinary Procedures Center, Strategic Laboratory | Claudio Tavares Sacchi, Claudia Regina Gonçalves, Erica Valessa Ramos Gomes, Karoline Rodrigues Campos |
| EPI_ISL_693231 | Pronto Socorro Municipal de Santa Branca | Instituto Adolfo Lutz, Interdiciplinary Procedures Center, Strategic Laboratory | Claudio Tavares Sacchi, Claudia Regina Gonçalves, Erica Valessa Ramos Gomes, Karoline Rodrigues Campos |
| EPI_ISL_693232 | Hospital e Pronto Socorro Portinari | Instituto Adolfo Lutz, Interdiciplinary Procedures Center, Strategic Laboratory | Claudio Tavares Sacchi, Claudia Regina Gonçalves, Erica Valessa Ramos Gomes, Karoline Rodrigues Campos |
| EPI_ISL_693233 | Hospital Santa Cruz | Instituto Adolfo Lutz, Interdiciplinary Procedures Center, Strategic Laboratory | Claudio Tavares Sacchi, Claudia Regina Gonçalves, Erica Valessa Ramos Gomes, Karoline Rodrigues Campos |
| EPI_ISL_693234 | Upa Vereador Jose Da Rocha Goncalves | Instituto Adolfo Lutz, Interdiciplinary Procedures Center, Strategic Laboratory | Claudio Tavares Sacchi, Claudia Regina Gonçalves, Erica Valessa Ramos Gomes, Karoline Rodrigues Campos |
| EPI_ISL_693235 | Casmi Centro Atendimento Saude da Mulher e Infancia | Instituto Adolfo Lutz, Interdiciplinary Procedures Center, Strategic Laboratory | Claudio Tavares Sacchi, Claudia Regina Gonçalves, Erica Valessa Ramos Gomes, Karoline Rodrigues Campos |
| EPI_ISL_693236 | Hospital Santa Marcelina Sao Paulo | Instituto Adolfo Lutz, Interdiciplinary Procedures Center, Strategic Laboratory | Claudio Tavares Sacchi, Claudia Regina Gonçalves, Erica Valessa Ramos Gomes, Karoline Rodrigues Campos |
| EPI_ISL_693237 | UPA Santa Isabel | Instituto Adolfo Lutz, Interdiciplinary Procedures Center, Strategic Laboratory | Claudio Tavares Sacchi, Claudia Regina Gonçalves, Erica Valessa Ramos Gomes, Karoline Rodrigues Campos |
| EPI_ISL_693238, EPI_ISL_693239 | Secao Centro de Diagnostico Secedi | Instituto Adolfo Lutz, Interdiciplinary Procedures Center, Strategic Laboratory | Claudio Tavares Sacchi, Claudia Regina Gonçalves, Erica Valessa Ramos Gomes, Karoline Rodrigues Campos |
| EPI_ISL_693240 | Centro de Vigilância a Saude de Diadema | Instituto Adolfo Lutz, Interdiciplinary Procedures Center, Strategic Laboratory | Claudio Tavares Sacchi, Claudia Regina Gonçalves, Erica Valessa Ramos Gomes, Karoline Rodrigues Campos |
| EPI_ISL_693241 | Hospital e Maternidade Sao Lucas | Instituto Adolfo Lutz, Interdiciplinary Procedures Center, Strategic Laboratory | Claudio Tavares Sacchi, Claudia Regina Gonçalves, Erica Valessa Ramos Gomes, Karoline Rodrigues Campos |
| EPI_ISL_693242 | Centro de Vigilância a Saude de Diadema | Instituto Adolfo Lutz, Interdiciplinary Procedures Center, Strategic Laboratory | Claudio Tavares Sacchi, Claudia Regina Gonçalves, Erica Valessa Ramos Gomes, Karoline Rodrigues Campos |
| EPI_ISL_693243 | Laboratório Municipal de Piracicaba | Instituto Adolfo Lutz, Interdiciplinary Procedures Center, Strategic Laboratory | Claudio Tavares Sacchi, Claudia Regina Gonçalves, Erica Valessa Ramos Gomes, Karoline Rodrigues Campos |
| EPI_ISL_693244 | Centro Médico da Polícia Militar do Estado de Sao Paulo | Instituto Adolfo Lutz, Interdiciplinary Procedures Center, Strategic Laboratory | Claudio Tavares Sacchi, Claudia Regina Gonçalves, Erica Valessa Ramos Gomes, Karoline Rodrigues Campos |
| EPI_ISL_693245 | UPA Santa Isabel | Instituto Adolfo Lutz, Interdiciplinary Procedures Center, Strategic Laboratory | Claudio Tavares Sacchi, Claudia Regina Gonçalves, Erica Valessa Ramos Gomes, Karoline Rodrigues Campos |
| EPI_ISL_693246 | Laboratorio Municipal de Rio Grande da Serra | Instituto Adolfo Lutz, Interdiciplinary Procedures Center, Strategic Laboratory | Claudio Tavares Sacchi, Claudia Regina Gonçalves, Erica Valessa Ramos Gomes, Karoline Rodrigues Campos |
| EPI_ISL_693247 | Secao Centro de Diagnostico Secedi | Instituto Adolfo Lutz, Interdiciplinary Procedures Center, Strategic Laboratory | Claudio Tavares Sacchi, Claudia Regina Gonçalves, Erica Valessa Ramos Gomes, Karoline Rodrigues Campos |
| EPI_ISL_693248 | Centro Municipal de Epidemiologia e Imunizações | Instituto Adolfo Lutz, Interdiciplinary Procedures Center, Strategic Laboratory | Claudio Tavares Sacchi, Claudia Regina Gonçalves, Erica Valessa Ramos Gomes, Karoline Rodrigues Campos |
| EPI_ISL_708529 | Secretária Municipal de Saude de Fernandópolis | Instituto Adolfo Lutz, Interdiciplinary Procedures Center, Strategic Laboratory | Claudio Tavares Sacchi, Claudia Regina Gonçalves, Erica Valessa Ramos Gomes, Carlos Henrique Camargo, Karoline Rodrigues Campos, Fernanda Modesto Tolentino Binhardi, Maricelia Navarro Pinheiro Flores, Marcia Maria Costa Nunes Soares, Janaina Other Martins Montanha |
| EPI_ISL_708530 | Secretaria Municipal de Saude de Fernandópolis | Instituto Adolfo Lutz, Interdiciplinary Procedures Center, Strategic Laboratory | Claudio Tavares Sacchi, Claudia Regina Gonçalves, Erica Valessa Ramos Gomes, Carlos Henrique Camargo, Karoline Rodrigues Campos, Fernanda Modesto Tolentino Binhardi, Maricelia Navarro Pinheiro Flores, Marcia Maria Costa Nunes Soares, Janaina Other Martins Montanha |
| EPI_ISL_717785, EPI_ISL_717786, EPI_ISL_717787, EPI_ISL_717788, EPI_ISL_717789, EPI_ISL_717790 | LACEN RJ - Noel Nutels | Bioinformatics Laboratory / LNCC | Carolina M Voloch, Ronaldo da Silva F Jr, Luiz G P de Almeida, Cynthia C Cardoso, Otavio Bustrolini, Alexandra L Gerber, Ana Paula de C Guimarães, Diana Mariani, Andréa Cony Cavalcanti, Claudia dos Santos Rodrigues, Terezinha M P P Castiñeira, Amilcar Tanuri, Ana Tereza R de Vasconcelos |
| EPI_ISL_717791 | Laboratorio de Virologia Molecular / UFRJ | Bioinformatics Laboratory / LNCC | Carolina M Voloch, Ronaldo da Silva F Jr, Luiz G P de Almeida, Cynthia C Cardoso, Otavio Bustrolini, Alexandra L Gerber, Ana Paula de C Guimarães, Diana Mariani, Andréa Cony Cavalcanti, Claudia dos Santos Rodrigues, Terezinha M P P Castiñeira, Amilcar Tanuri, Ana Tereza R de Vasconcelos |
| EPI_ISL_717792 | LACEN RJ - Noel Nutels | Bioinformatics Laboratory / LNCC | Carolina M Voloch, Ronaldo da Silva F Jr, Luiz G P de Almeida, Cynthia C Cardoso, Otavio Bustrolini, Alexandra L Gerber, Ana Paula de C Guimarães, Diana Mariani, Andréa Cony Cavalcanti, Claudia dos Santos Rodrigues, Terezinha M P P Castiñeira, Amilcar Tanuri, Ana Tereza R de Vasconcelos |
| EPI_ISL_717793 | Laboratorio de Virologia Molecular / UFRJ | Bioinformatics Laboratory / LNCC | Carolina M Voloch, Ronaldo da Silva F Jr, Luiz G P de Almeida, Cynthia C Cardoso, Otavio Bustrolini, Alexandra L Gerber, Ana Paula de C Guimarães, Diana Mariani, Andréa Cony Cavalcanti, Claudia dos Santos Rodrigues, Terezinha M P P Castiñeira, Amilcar Tanuri, Ana Tereza R de Vasconcelos |
| EPI_ISL_717794, EPI_ISL_717795, EPI_ISL_717796, EPI_ISL_717797, | LACEN RJ - Noel Nutels | Bioinformatics Laboratory / LNCC | Carolina M Voloch, Ronaldo da Silva F Jr, Luiz G P de Almeida, Cynthia C Cardoso, Otavio Bustrolini, Alexandra L Gerber, Ana Paula de C Guimarães, Diana Mariani, Andréa Cony Cavalcanti, Claudia dos Santos Rodrigues, Terezinha M P P Castiñeira, Amilcar Tanuri, Ana Tereza R de Vasconcelos |

| | | | |
|---|---|---|---|
| EPI_ISL_717798, EPI_ISL_717799, EPI_ISL_717800 | | | |
| EPI_ISL_717801, EPI_ISL_717802 | Laboratorio de Virologia Molecular / UFRJ | Bioinformatics Laboratory / LNCC | Carolina M Voloch, Ronaldo da Silva F Jr, Luiz G P de Almeida, Cynthia C Cardoso, Otavio Bustrolini, Alexandra L Gerber, Ana Paula de C Guimarães, Diana Mariani, Andréa Cony Cavalcanti, Claudia dos Santos Rodrigues, Terezinha M P P Castiñeira, Amilcar Tanuri, Ana Tereza R de Vasconcelos |
| EPI_ISL_717803, EPI_ISL_717804 | LACEN RJ - Noel Nutels | Bioinformatics Laboratory / LNCC | Carolina M Voloch, Ronaldo da Silva F Jr, Luiz G P de Almeida, Cynthia C Cardoso, Otavio Bustrolini, Alexandra L Gerber, Ana Paula de C Guimarães, Diana Mariani, Andréa Cony Cavalcanti, Claudia dos Santos Rodrigues, Terezinha M P P Castiñeira, Amilcar Tanuri, Ana Tereza R de Vasconcelos |
| EPI_ISL_717805, EPI_ISL_717806, EPI_ISL_717807, EPI_ISL_717808 | Laboratorio de Virologia Molecular / UFRJ | Bioinformatics Laboratory / LNCC | Carolina M Voloch, Ronaldo da Silva F Jr, Luiz G P de Almeida, Cynthia C Cardoso, Otavio Bustrolini, Alexandra L Gerber, Ana Paula de C Guimarães, Diana Mariani, Andréa Cony Cavalcanti, Claudia dos Santos Rodrigues, Terezinha M P P Castiñeira, Amilcar Tanuri, Ana Tereza R de Vasconcelos |
| EPI_ISL_717809 | LACEN Dr. Francisco Rimolo Neto | Bioinformatics Laboratory / LNCC | Carolina M Voloch, Ronaldo da Silva F Jr, Luiz G P de Almeida, Cynthia C Cardoso, Otavio Bustrolini, Alexandra L Gerber, Ana Paula de C Guimarães, Diana Mariani, Andréa Cony Cavalcanti, Claudia dos Santos Rodrigues, Terezinha M P P Castiñeira, Amilcar Tanuri, Ana Tereza R de Vasconcelos |
| EPI_ISL_717812, EPI_ISL_717813, EPI_ISL_717814, EPI_ISL_717815, EPI_ISL_717816, EPI_ISL_717817, EPI_ISL_717818, EPI_ISL_717819, EPI_ISL_717820, EPI_ISL_717821, EPI_ISL_717822, EPI_ISL_717823, EPI_ISL_717824, EPI_ISL_717825, EPI_ISL_717826, EPI_ISL_717827, EPI_ISL_717828, EPI_ISL_717829, EPI_ISL_717830, EPI_ISL_717831 | | | |
| see above | Laboratorio de Virologia Molecular / UFRJ | Bioinformatics Laboratory / LNCC | Carolina M Voloch, Ronaldo da Silva F Jr, Luiz G P de Almeida, Cynthia C Cardoso, Otavio Bustrolini, Alexandra L Gerber, Ana Paula de C Guimarães, Diana Mariani, Andréa Cony Cavalcanti, Claudia dos Santos Rodrigues, Terezinha M P P Castiñeira, Amilcar Tanuri, Ana Tereza R de Vasconcelos |
| EPI_ISL_717832, EPI_ISL_717833, EPI_ISL_717834, EPI_ISL_717835, EPI_ISL_717836, EPI_ISL_717841 | LACEN Dr. Francisco Rimolo Neto | Bioinformatics Laboratory / LNCC | Carolina M Voloch, Ronaldo da Silva F Jr, Luiz G P de Almeida, Cynthia C Cardoso, Otavio Bustrolini, Alexandra L Gerber, Ana Paula de C Guimarães, Diana Mariani, Andréa Cony Cavalcanti, Claudia dos Santos Rodrigues, Terezinha M P P Castiñeira, Amilcar Tanuri, Ana Tereza R de Vasconcelos |
| EPI_ISL_717844, EPI_ISL_717845, EPI_ISL_717846, EPI_ISL_717847, EPI_ISL_717848, EPI_ISL_717849, EPI_ISL_717850, EPI_ISL_717851, EPI_ISL_717852, EPI_ISL_717853, EPI_ISL_717854, EPI_ISL_717855, EPI_ISL_717856, EPI_ISL_717857, EPI_ISL_717858, EPI_ISL_717859, EPI_ISL_717860, EPI_ISL_717861, EPI_ISL_717862, EPI_ISL_717863, EPI_ISL_717864, EPI_ISL_717865, EPI_ISL_717866, EPI_ISL_717867, EPI_ISL_717868, EPI_ISL_717869, EPI_ISL_717870, EPI_ISL_717871, EPI_ISL_717872, EPI_ISL_717873, EPI_ISL_717874, EPI_ISL_717875, EPI_ISL_717876, EPI_ISL_717877, EPI_ISL_717878, EPI_ISL_717879, EPI_ISL_717880, EPI_ISL_717881, EPI_ISL_717882, EPI_ISL_717883, EPI_ISL_717884, EPI_ISL_717885, EPI_ISL_717886, EPI_ISL_717887, EPI_ISL_717888, EPI_ISL_717889, EPI_ISL_717890, EPI_ISL_717891, EPI_ISL_717892, EPI_ISL_717893, EPI_ISL_717894, EPI_ISL_717895, EPI_ISL_717896, EPI_ISL_717897, EPI_ISL_717898 | | | |
| see above | Laboratorio de Virologia Molecular / UFRJ | Bioinformatics Laboratory / LNCC | Carolina M Voloch, Ronaldo da Silva F Jr, Luiz G P de Almeida, Cynthia C Cardoso, Otavio Bustrolini, Alexandra L Gerber, Ana Paula de C Guimarães, Diana Mariani, Andréa Cony Cavalcanti, Claudia dos Santos Rodrigues, Terezinha M P P Castiñeira, Amilcar Tanuri, Ana Tereza R de Vasconcelos |
| EPI_ISL_717899, EPI_ISL_717900, EPI_ISL_717901, EPI_ISL_717902, EPI_ISL_717903, EPI_ISL_717904, EPI_ISL_717905, EPI_ISL_717906, EPI_ISL_717907, EPI_ISL_717908, EPI_ISL_717909 | | | |
| see above | LACEN RJ - Noel Nutels | Bioinformatics Laboratory / LNCC | Carolina M Voloch, Ronaldo da Silva F Jr, Luiz G P de Almeida, Cynthia C Cardoso, Otavio Bustrolini, Alexandra L Gerber, Ana Paula de C Guimarães, Diana Mariani, Andréa Cony Cavalcanti, Claudia dos Santos Rodrigues, Terezinha M P P Castiñeira, Amilcar Tanuri, Ana Tereza R de Vasconcelos |
| EPI_ISL_717910, EPI_ISL_717911, EPI_ISL_717912, EPI_ISL_717913, EPI_ISL_717914, EPI_ISL_717915, EPI_ISL_717916, EPI_ISL_717917, EPI_ISL_717918, EPI_ISL_717919 | LACEN Dr. Francisco Rimolo Neto | Bioinformatics Laboratory / LNCC | Carolina M Voloch, Ronaldo da Silva F Jr, Luiz G P de Almeida, Cynthia C Cardoso, Otavio Bustrolini, Alexandra L Gerber, Ana Paula de C Guimarães, Diana Mariani, Andréa Cony Cavalcanti, Claudia dos Santos Rodrigues, Terezinha M P P Castiñeira, Amilcar Tanuri, Ana Tereza R de Vasconcelos |
| EPI_ISL_717920, EPI_ISL_717921, EPI_ISL_717922, EPI_ISL_717923, EPI_ISL_717924, EPI_ISL_717925, EPI_ISL_717926, EPI_ISL_717927, EPI_ISL_717928, EPI_ISL_717929, EPI_ISL_717930, EPI_ISL_717931, EPI_ISL_717932, EPI_ISL_717933, EPI_ISL_717934, EPI_ISL_717935, EPI_ISL_717936, EPI_ISL_717937, EPI_ISL_717938, EPI_ISL_717939, EPI_ISL_717940, EPI_ISL_717941, EPI_ISL_717942, EPI_ISL_717943, EPI_ISL_717944, EPI_ISL_717945, EPI_ISL_717946, EPI_ISL_717947, EPI_ISL_717948, EPI_ISL_717949, EPI_ISL_717950, EPI_ISL_717951, EPI_ISL_717952, EPI_ISL_717953, EPI_ISL_717954, EPI_ISL_717955, EPI_ISL_717956, EPI_ISL_717957 | | | |
| see above | Laboratorio de Virologia Molecular / UFRJ | Bioinformatics Laboratory / LNCC | Carolina M Voloch, Ronaldo da Silva F Jr, Luiz G P de Almeida, Cynthia C Cardoso, Otavio Bustrolini, Alexandra L Gerber, Ana Paula de C Guimarães, Diana Mariani, Andréa Cony Cavalcanti, Claudia dos Santos Rodrigues, Terezinha M P P Castiñeira, Amilcar Tanuri, Ana Tereza R de Vasconcelos |
| EPI_ISL_717958 | LACEN Dr. Francisco Rimolo Neto | Bioinformatics Laboratory / LNCC | Carolina M Voloch, Ronaldo da Silva F Jr, Luiz G P de Almeida, Cynthia C Cardoso, Otavio Bustrolini, Alexandra L Gerber, Ana Paula de C Guimarães, Diana Mariani, Andréa Cony Cavalcanti, Claudia dos Santos Rodrigues, Terezinha M P P Castiñeira, Amilcar Tanuri, Ana Tereza R de Vasconcelos |
| EPI_ISL_717959, EPI_ISL_717960, EPI_ISL_717961 | Laboratorio de Virologia Molecular / UFRJ | Bioinformatics Laboratory / LNCC | Carolina M Voloch, Ronaldo da Silva F Jr, Luiz G P de Almeida, Cynthia C Cardoso, Otavio Bustrolini, Alexandra L Gerber, Ana Paula de C Guimarães, Diana Mariani, Andréa Cony Cavalcanti, Claudia dos Santos Rodrigues, Terezinha M P P Castiñeira, Amilcar Tanuri, Ana Tereza R de Vasconcelos |
| EPI_ISL_717962 | LACEN RJ - Noel Nutels | Bioinformatics Laboratory / LNCC | Carolina M Voloch, Ronaldo da Silva F Jr, Luiz G P de Almeida, Cynthia C Cardoso, Otavio Bustrolini, Alexandra L Gerber, Ana Paula de C Guimarães, Diana Mariani, Andréa Cony Cavalcanti, Claudia dos Santos Rodrigues, Terezinha M P P Castiñeira, Amilcar Tanuri, Ana Tereza R de Vasconcelos |
| EPI_ISL_717963, EPI_ISL_717964 | LACEN Dr. Francisco Rimolo Neto | Bioinformatics Laboratory / LNCC | Carolina M Voloch, Ronaldo da Silva F Jr, Luiz G P de Almeida, Cynthia C Cardoso, Otavio Bustrolini, Alexandra L Gerber, Ana Paula de C Guimarães, Diana Mariani, Andréa Cony Cavalcanti, Claudia dos Santos Rodrigues, Terezinha M P P Castiñeira, Amilcar Tanuri, Ana Tereza R de Vasconcelos |
| EPI_ISL_721988, EPI_ISL_721989, EPI_ISL_721990, EPI_ISL_721991, EPI_ISL_721993, EPI_ISL_721994, EPI_ISL_721995, EPI_ISL_721996, EPI_ISL_721997, EPI_ISL_721998, EPI_ISL_721999, EPI_ISL_722006, EPI_ISL_722007, EPI_ISL_722008, EPI_ISL_722010, EPI_ISL_722011, EPI_ISL_722016, EPI_ISL_722017, EPI_ISL_722019, EPI_ISL_722020, EPI_ISL_722021, EPI_ISL_722022, EPI_ISL_722027, EPI_ISL_722028, EPI_ISL_722037, EPI_ISL_722038, EPI_ISL_722039, EPI_ISL_722040, EPI_ISL_722041, EPI_ISL_722044, EPI_ISL_722046, EPI_ISL_722047, EPI_ISL_722048, EPI_ISL_722049, EPI_ISL_722050, EPI_ISL_722051, EPI_ISL_722058, EPI_ISL_722060, EPI_ISL_722061, EPI_ISL_722062, EPI_ISL_722064, EPI_ISL_722066, EPI_ISL_722069, EPI_ISL_722070, EPI_ISL_722071, EPI_ISL_722074, EPI_ISL_722076, EPI_ISL_722077, EPI_ISL_722081, EPI_ISL_722086, EPI_ISL_722087, EPI_ISL_722089, EPI_ISL_722091, EPI_ISL_722092, EPI_ISL_722093, EPI_ISL_722094, EPI_ISL_722095, EPI_ISL_722096, EPI_ISL_722097, EPI_ISL_722098, EPI_ISL_722100, EPI_ISL_722102, EPI_ISL_722103, EPI_ISL_722104, EPI_ISL_722105, EPI_ISL_722106, EPI_ISL_722107, EPI_ISL_722109, EPI_ISL_722110, EPI_ISL_722111, EPI_ISL_722113, EPI_ISL_722115, EPI_ISL_722116, EPI_ISL_722117, EPI_ISL_722120, EPI_ISL_722121, EPI_ISL_722123, EPI_ISL_722124, EPI_ISL_722125, EPI_ISL_722126, EPI_ISL_722128, EPI_ISL_722129 | | | |
| see above | Hospital das Clínicas Universidade de São Paulo Medical School | Laboratório de Parasitologia Médica - Instituto de Medicina Tropical - Universidade de São Paulo | Brazil-UK Centre for Arbovirus Discovery Diagnosis Genomics and Epidemiology (CADDE) Genomic Network - Instituto de Medicina Tropical |
| EPI_ISL_729794, EPI_ISL_729795, EPI_ISL_729796, EPI_ISL_729797, EPI_ISL_729798, EPI_ISL_729799, EPI_ISL_729800, EPI_ISL_729801, EPI_ISL_729802, EPI_ISL_729803, EPI_ISL_729820, EPI_ISL_729821, EPI_ISL_729822, EPI_ISL_729823, EPI_ISL_729824, EPI_ISL_729825, EPI_ISL_729826, EPI_ISL_729827, EPI_ISL_729828, EPI_ISL_729829, EPI_ISL_729830, EPI_ISL_729831, EPI_ISL_729832, EPI_ISL_729833, EPI_ISL_729835, EPI_ISL_729836, EPI_ISL_729837, EPI_ISL_729838, EPI_ISL_729839, EPI_ISL_729840, EPI_ISL_729841, EPI_ISL_729842, EPI_ISL_729843, EPI_ISL_729844, EPI_ISL_729845, EPI_ISL_729846, EPI_ISL_729847, EPI_ISL_729848, EPI_ISL_729849, EPI_ISL_729850, EPI_ISL_729851, EPI_ISL_729852, EPI_ISL_729853, EPI_ISL_729854, EPI_ISL_729855, EPI_ISL_729856, EPI_ISL_729857, EPI_ISL_729858, EPI_ISL_729859, EPI_ISL_729860, EPI_ISL_729861 | | | |
| see above | Laboratório Central de Saúde Pública do Estado do Rio Grande do Sul (LACEN-RS) | Laboratory of Respiratory Viruses and Measles, Oswaldo Cruz Institute, FIOCRUZ | Paola Resende, Luciana Appolinario, Fernando Motta, Anna Carolina Paixão, Ana Carolina Mendonça, Tatiana Schaffer Gregianini, Marilda Tereza Mar da Rosa, Marilda Siqueira on behalf of the Fiocruz COVID-19 Genomic Surveillance Network |
| EPI_ISL_735396 | Hospital de Camplanha COVID 19 SER | Instituto Adolfo Lutz, Interdiciplinary Procedures Center, Strategic Laboratory | Claudio Tavares Sacchi, Claudia Regina Gonçalves, Erica Valessa Ramos Gomes, Karoline Rodrigues Campos |
| EPI_ISL_735397 | Unidade Respiratória Nova Hortolandia | Instituto Adolfo Lutz, Interdiciplinary Procedures Center, Strategic Laboratory | Claudio Tavares Sacchi, Claudia Regina Gonçalves, Erica Valessa Ramos Gomes, Karoline Rodrigues Campos |
| EPI_ISL_735398 | Laboratorio Fleury | Instituto Adolfo Lutz, Interdiciplinary Procedures Center, Strategic Laboratory | Claudio Tavares Sacchi, Claudia Regina Gonçalves, Erica Valessa Ramos Gomes, Karoline Rodrigues Campos |
| EPI_ISL_735400 | Instituto Adolfo Lutz - Regional de Santos | Instituto Adolfo Lutz, Interdiciplinary Procedures Center, Strategic Laboratory | Claudio Tavares Sacchi, Claudia Regina Gonçalves, Erica Valessa Ramos Gomes, Karoline Rodrigues Campos |
| EPI_ISL_735401, EPI_ISL_735402, EPI_ISL_735403, EPI_ISL_735404 | Instituto Adolfo Lutz - Regional de Rio Claro | Instituto Adolfo Lutz, Interdiciplinary Procedures Center, Strategic Laboratory | Claudio Tavares Sacchi, Claudia Regina Gonçalves, Erica Valessa Ramos Gomes, Karoline Rodrigues Campos |
| EPI_ISL_735405 | Secretaria Minucipal de Saude de Birigui | Instituto Adolfo Lutz, Interdiciplinary Procedures Center, Strategic Laboratory | Claudio Tavares Sacchi, Claudia Regina Gonçalves, Erica Valessa Ramos Gomes, Karoline Rodrigues Campos |
| EPI_ISL_735406 | Unidade de Pronto Atendimento UPA I Sta Isabel | Instituto Adolfo Lutz, Interdiciplinary Procedures Center, Strategic Laboratory | Claudio Tavares Sacchi, Claudia Regina Gonçalves, Erica Valessa Ramos Gomes, Karoline Rodrigues Campos |
| EPI_ISL_735407 | Santa Casa de Marilia | Instituto Adolfo Lutz, Interdiciplinary Procedures Center, Strategic Laboratory | Claudio Tavares Sacchi, Claudia Regina Gonçalves, Erica Valessa Ramos Gomes, Karoline Rodrigues Campos |

| | | | |
|---|---|---|---|
| EPI_ISL_735408 | COVID 19 Centro de Combate ao Coronavirus CCC Jandira | Instituto Adolfo Lutz, Interdiciplinary Procedures Center, Strategic Laboratory | Claudio Tavares Sacchi, Claudia Regina Gonçalves, Erica Valessa Ramos Gomes, Karoline Rodrigues Campos |
| EPI_ISL_735409 | Unidade de Pronto Atendimento Carlos Lourenco | Instituto Adolfo Lutz, Interdiciplinary Procedures Center, Strategic Laboratory | Claudio Tavares Sacchi, Claudia Regina Gonçalves, Erica Valessa Ramos Gomes, Karoline Rodrigues Campos |
| EPI_ISL_735410 | Instituto Adolfo Lutz - Regional de Rio Claro | Instituto Adolfo Lutz, Interdiciplinary Procedures Center, Strategic Laboratory | Claudio Tavares Sacchi, Claudia Regina Gonçalves, Erica Valessa Ramos Gomes, Karoline Rodrigues Campos |
| EPI_ISL_735411 | Centro de Vigilancia a Saude de Diadema | Instituto Adolfo Lutz, Interdiciplinary Procedures Center, Strategic Laboratory | Claudio Tavares Sacchi, Claudia Regina Gonçalves, Erica Valessa Ramos Gomes, Karoline Rodrigues Campos |
| EPI_ISL_735412 | Hospital e Pronto Socorro Portinari | Instituto Adolfo Lutz, Interdiciplinary Procedures Center, Strategic Laboratory | Claudio Tavares Sacchi, Claudia Regina Gonçalves, Erica Valessa Ramos Gomes, Karoline Rodrigues Campos |
| EPI_ISL_735413 | Militello Centro de Diagnosticos e Biopesquisa Clinica | Instituto Adolfo Lutz, Interdiciplinary Procedures Center, Strategic Laboratory | Claudio Tavares Sacchi, Claudia Regina Gonçalves, Erica Valessa Ramos Gomes, Karoline Rodrigues Campos |
| EPI_ISL_735414, EPI_ISL_735415 | Unidade de Pronto Atendimento de Agenor de Campos | Instituto Adolfo Lutz, Interdiciplinary Procedures Center, Strategic Laboratory | Claudio Tavares Sacchi, Claudia Regina Gonçalves, Erica Valessa Ramos Gomes, Karoline Rodrigues Campos |
| EPI_ISL_735416 | Centro de Saude II Dr Jose Paione Mococa | Instituto Adolfo Lutz, Interdiciplinary Procedures Center, Strategic Laboratory | Claudio Tavares Sacchi, Claudia Regina Gonçalves, Erica Valessa Ramos Gomes, Karoline Rodrigues Campos |
| EPI_ISL_735417 | Unidade de Pronto Atendimento de Agenor de Campos | Instituto Adolfo Lutz, Interdiciplinary Procedures Center, Strategic Laboratory | Claudio Tavares Sacchi, Claudia Regina Gonçalves, Erica Valessa Ramos Gomes, Karoline Rodrigues Campos |
| EPI_ISL_735418 | Hospital Regional do Vale do Paraiba | Instituto Adolfo Lutz, Interdiciplinary Procedures Center, Strategic Laboratory | Claudio Tavares Sacchi, Claudia Regina Gonçalves, Erica Valessa Ramos Gomes, Karoline Rodrigues Campos |
| EPI_ISL_735419 | UBS Alvarenga | Instituto Adolfo Lutz, Interdiciplinary Procedures Center, Strategic Laboratory | Claudio Tavares Sacchi, Claudia Regina Gonçalves, Erica Valessa Ramos Gomes, Karoline Rodrigues Campos |
| EPI_ISL_735420 | UBS Riacho Grande | Instituto Adolfo Lutz, Interdiciplinary Procedures Center, Strategic Laboratory | Claudio Tavares Sacchi, Claudia Regina Gonçalves, Erica Valessa Ramos Gomes, Karoline Rodrigues Campos |
| EPI_ISL_735421 | UBS Sta Terezinha | Instituto Adolfo Lutz, Interdiciplinary Procedures Center, Strategic Laboratory | Claudio Tavares Sacchi, Claudia Regina Gonçalves, Erica Valessa Ramos Gomes, Karoline Rodrigues Campos |
| EPI_ISL_735422 | UBS Dematchi | Instituto Adolfo Lutz, Interdiciplinary Procedures Center, Strategic Laboratory | Claudio Tavares Sacchi, Claudia Regina Gonçalves, Erica Valessa Ramos Gomes, Karoline Rodrigues Campos |
| EPI_ISL_735423, EPI_ISL_735424 | Centro de Vigilancia a Saude de Diadema | Instituto Adolfo Lutz, Interdiciplinary Procedures Center, Strategic Laboratory | Claudio Tavares Sacchi, Claudia Regina Gonçalves, Erica Valessa Ramos Gomes, Karoline Rodrigues Campos |
| EPI_ISL_735425 | Hospital e Maternidade Sao Lucas | Instituto Adolfo Lutz, Interdiciplinary Procedures Center, Strategic Laboratory | Claudio Tavares Sacchi, Claudia Regina Gonçalves, Erica Valessa Ramos Gomes, Karoline Rodrigues Campos |
| EPI_ISL_735426 | Centro de Vigilancia a Saude de Diadema | Instituto Adolfo Lutz, Interdiciplinary Procedures Center, Strategic Laboratory | Claudio Tavares Sacchi, Claudia Regina Gonçalves, Erica Valessa Ramos Gomes, Karoline Rodrigues Campos |
| EPI_ISL_735427 | Instituto Adolfo Lutz - Regional de Santos | Instituto Adolfo Lutz, Interdiciplinary Procedures Center, Strategic Laboratory | Claudio Tavares Sacchi, Claudia Regina Gonçalves, Erica Valessa Ramos Gomes, Karoline Rodrigues Campos |
| EPI_ISL_735428, EPI_ISL_735429 | Hospital Nipo Brasileiro | Instituto Adolfo Lutz, Interdiciplinary Procedures Center, Strategic Laboratory | Claudio Tavares Sacchi, Claudia Regina Gonçalves, Erica Valessa Ramos Gomes, Karoline Rodrigues Campos |
| EPI_ISL_735430 | Instituto Adolfo Lutz - Regional de Santos | Instituto Adolfo Lutz, Interdiciplinary Procedures Center, Strategic Laboratory | Claudio Tavares Sacchi, Claudia Regina Gonçalves, Erica Valessa Ramos Gomes, Karoline Rodrigues Campos |
| EPI_ISL_735431, EPI_ISL_735432 | Hospital Nipo Brasileiro | Instituto Adolfo Lutz, Interdiciplinary Procedures Center, Strategic Laboratory | Claudio Tavares Sacchi, Claudia Regina Gonçalves, Erica Valessa Ramos Gomes, Karoline Rodrigues Campos |
| EPI_ISL_735433 | Posto de Atendimento Saude Cidade Pasc Cajati | Instituto Adolfo Lutz, Interdiciplinary Procedures Center, Strategic Laboratory | Claudio Tavares Sacchi, Claudia Regina Gonçalves, Erica Valessa Ramos Gomes, Karoline Rodrigues Campos |
| EPI_ISL_754236 | Diagnósticos da América - DASA | Instituto de Medicina Tropical Universidade de São Paulo | Brazil-UK Centre for Arbovirus Discovery Diagnosis Genomics and Epidemiology (CADDE) Genomic Network - Instituto de Medicina Tropical |
| EPI_ISL_754237 | Diagnósticos da América - DASA | Instituto de Medicina Tropical de Sao Paulo | Brazil-UK Centre for Arbovirus Discovery Diagnosis Genomics and Epidemiology (CADDE) Genomic Network - Instituto de Medicina Tropical |
| EPI_ISL_755640 | Instituto Adolfo Lutz - Central | Instituto Adolfo Lutz, Interdiciplinary Procedures Center, Strategic Laboratory | Claudio Tavares Sacchi, Claudia Regina Gonçalves, Erica Valessa Ramos Gomes, Karoline Rodrigues Campos |
| EPI_ISL_755641 | Instituto Adolfo Lutz - Regional de Santo Andre | Instituto Adolfo Lutz, Interdiciplinary Procedures Center, Strategic Laboratory | Claudio Tavares Sacchi, Claudia Regina Gonçalves, Erica Valessa Ramos Gomes, Karoline Rodrigues Campos |
| EPI_ISL_755642, EPI_ISL_755643 | Instituto Adolfo Lutz - Central | Instituto Adolfo Lutz, Interdiciplinary Procedures Center, Strategic Laboratory | Claudio Tavares Sacchi, Claudia Regina Gonçalves, Erica Valessa Ramos Gomes, Karoline Rodrigues Campos |
| EPI_ISL_755644, EPI_ISL_755645 | Lab LOC - Itapecerica da Serra | Instituto Adolfo Lutz, Interdiciplinary Procedures Center, Strategic Laboratory | Claudio Tavares Sacchi, Claudia Regina Gonçalves, Erica Valessa Ramos Gomes, Karoline Rodrigues Campos |
| EPI_ISL_755646, EPI_ISL_755647 | Instituto Adolfo Lutz - Regional de Santo Andre | Instituto Adolfo Lutz, Interdiciplinary Procedures Center, Strategic Laboratory | Claudio Tavares Sacchi, Claudia Regina Gonçalves, Erica Valessa Ramos Gomes, Karoline Rodrigues Campos |
| EPI_ISL_755648 | Instituto Adolfo Lutz - Regional de Taubate | Instituto Adolfo Lutz, Interdiciplinary Procedures Center, Strategic Laboratory | Claudio Tavares Sacchi, Claudia Regina Gonçalves, Erica Valessa Ramos Gomes, Karoline Rodrigues Campos |
| EPI_ISL_755649 | Instituto Adolfo Lutz - Regional de Santo Andre | Instituto Adolfo Lutz, Interdiciplinary Procedures Center, Strategic Laboratory | Claudio Tavares Sacchi, Claudia Regina Gonçalves, Erica Valessa Ramos Gomes, Karoline Rodrigues Campos |
| EPI_ISL_755650 | Instituto Adolfo Lutz - Regional de Taubate | Instituto Adolfo Lutz, Interdiciplinary Procedures Center, Strategic Laboratory | Claudio Tavares Sacchi, Claudia Regina Gonçalves, Erica Valessa Ramos Gomes, Karoline Rodrigues Campos |
| EPI_ISL_755651 | Instituto Adolfo Lutz - Central | Instituto Adolfo Lutz, Interdiciplinary Procedures Center, Strategic Laboratory | Claudio Tavares Sacchi, Claudia Regina Gonçalves, Erica Valessa Ramos Gomes, Karoline Rodrigues Campos |
| EPI_ISL_755652 | Lab LOC - Itapecerica da Serra | Instituto Adolfo Lutz, Interdiciplinary Procedures Center, Strategic Laboratory | Claudio Tavares Sacchi, Claudia Regina Gonçalves, Erica Valessa Ramos Gomes, Karoline Rodrigues Campos |
| EPI_ISL_755653, EPI_ISL_755654 | Instituto Adolfo Lutz - Central | Instituto Adolfo Lutz, Interdiciplinary Procedures Center, Strategic Laboratory | Claudio Tavares Sacchi, Claudia Regina Gonçalves, Erica Valessa Ramos Gomes, Karoline Rodrigues Campos |
| EPI_ISL_755655 | Instituto Adolfo Lutz - Regional de Campinas | Instituto Adolfo Lutz, Interdiciplinary Procedures Center, Strategic Laboratory | Claudio Tavares Sacchi, Claudia Regina Gonçalves, Erica Valessa Ramos Gomes, Karoline Rodrigues Campos |

| | | | |
|---|---|---|---|
| EPI_ISL_756293, EPI_ISL_756294 | Center for Biotechnology and Cell Therapy, São Rafael Hospital, Salvador, Brazil | Center for Biotechnology and Cell Therapy, São Rafael Hospital, Salvador, Brazil | Carolina Kymie Vasques Nonaka, Marília Miranda Franco, Tiago Gräf, Ana Verena Almeida Mendes, Renato Santana de Aguiar, Marta Giovanetti, Bruno Solano de Freitas Souza |
| EPI_ISL_763074, EPI_ISL_763075 | Diagnosticos da America - DASA | Instituto Adolfo Lutz, Interdiciplinary Procedures Center, Strategic Laboratory | Claudio Tavares Sacchi, Claudia Regina Gonçalves, Erica Valessa Ramos Gomes, Karoline Rodrigues Campos |
| EPI_ISL_770551, EPI_ISL_770552, EPI_ISL_770553, EPI_ISL_770554, EPI_ISL_770555, EPI_ISL_770556, EPI_ISL_770557, EPI_ISL_770558, EPI_ISL_770559, EPI_ISL_770560, EPI_ISL_770561, EPI_ISL_770562, EPI_ISL_770563, EPI_ISL_770564, EPI_ISL_770565, EPI_ISL_770566, EPI_ISL_770567, EPI_ISL_770568, EPI_ISL_770569, EPI_ISL_770570, EPI_ISL_770571, EPI_ISL_770572, EPI_ISL_770573, EPI_ISL_770574, EPI_ISL_770575, EPI_ISL_770576, EPI_ISL_770577, EPI_ISL_770578, EPI_ISL_770579, EPI_ISL_770580, EPI_ISL_770581, EPI_ISL_770582, EPI_ISL_770583, EPI_ISL_770584, EPI_ISL_770585, EPI_ISL_770586, EPI_ISL_770587, EPI_ISL_770588, EPI_ISL_770589, EPI_ISL_770590, EPI_ISL_770591, EPI_ISL_770592, EPI_ISL_770593, EPI_ISL_770594, EPI_ISL_770595, EPI_ISL_770596, EPI_ISL_770597, EPI_ISL_770598, EPI_ISL_770599, EPI_ISL_770600, EPI_ISL_770601, EPI_ISL_770602, EPI_ISL_770603, EPI_ISL_770604, EPI_ISL_770605, EPI_ISL_770606, EPI_ISL_770607, EPI_ISL_770608, EPI_ISL_770609, EPI_ISL_770610, EPI_ISL_770611, EPI_ISL_770612, EPI_ISL_770613, EPI_ISL_770614, EPI_ISL_770615, EPI_ISL_770616, EPI_ISL_770617, EPI_ISL_770618, EPI_ISL_770619, EPI_ISL_770620, EPI_ISL_770621, EPI_ISL_770622, EPI_ISL_770623, EPI_ISL_770624, EPI_ISL_770625, EPI_ISL_770626, EPI_ISL_770627, EPI_ISL_770628, EPI_ISL_770629, EPI_ISL_770630 | | | |
| see above | Laboratório de Microbiologia Molecular - Universidade FEEVALE | Bioinformatics Laboratory / LNCC | Felipe Benites, Fernando Rosado Spilki, Alana Witt Hansen, Juliane Deise Fleck, Juliana Schons, Meriane Demoliner, Ana Karolina Eisen Antunes, Fagner Henrique Heldt, Larissa Mallmann, Bruna Hermann, Ana Luiza Ziulkoski, Vyctoria Goes, Karoline Schallenberger, Matheus Nunes Weber, Paula Rodrigues de Almeida, Alessandra Pavan Lamarca da Silva, Ronaldo da Silva F Jr , Luiz G P de Almeida, Alexandra L Gerber , Ana Paula de C Guimarães,Ana Tereza R de Vasconcelos |
| EPI_ISL_776755, EPI_ISL_776756 | Instituto Adolfo Lutz - Central | Instituto Adolfo Lutz, Interdiciplinary Procedures Center, Strategic Laboratory | Claudio Tavares Sacchi, Claudia Regina Gonçalves, Erica Valessa Ramos Gomes, Karoline Rodrigues Campos |
| EPI_ISL_776757, EPI_ISL_776758 | Instituto Adolfo Lutz - Regional de Marilia | Instituto Adolfo Lutz, Interdiciplinary Procedures Center, Strategic Laboratory | Claudio Tavares Sacchi, Claudia Regina Gonçalves, Erica Valessa Ramos Gomes, Karoline Rodrigues Campos |
| EPI_ISL_776759 | Instituto Adolfo Lutz - Central | Instituto Adolfo Lutz, Interdiciplinary Procedures Center, Strategic Laboratory | Claudio Tavares Sacchi, Claudia Regina Gonçalves, Erica Valessa Ramos Gomes, Karoline Rodrigues Campos |
| EPI_ISL_776764, EPI_ISL_776765, EPI_ISL_776766 | Instituto Adolfo Lutz - Regional de Santo Andre | Instituto Adolfo Lutz, Interdiciplinary Procedures Center, Strategic Laboratory | Claudio Tavares Sacchi, Claudia Regina Gonçalves, Erica Valessa Ramos Gomes, Karoline Rodrigues Campos |
| EPI_ISL_776767 | Instituto Adolfo Lutz - Regional de Marilia | Instituto Adolfo Lutz, Interdiciplinary Procedures Center, Strategic Laboratory | Claudio Tavares Sacchi, Claudia Regina Gonçalves, Erica Valessa Ramos Gomes, Karoline Rodrigues Campos |
| EPI_ISL_776768 | Instituto Adolfo Lutz - Regional de Aracatuba | Instituto Adolfo Lutz, Interdiciplinary Procedures Center, Strategic Laboratory | Claudio Tavares Sacchi, Claudia Regina Gonçalves, Erica Valessa Ramos Gomes, Karoline Rodrigues Campos |
| EPI_ISL_776769 | Instituto Adolfo Lutz - Regional de Santo Andre | Instituto Adolfo Lutz, Interdiciplinary Procedures Center, Strategic Laboratory | Claudio Tavares Sacchi, Claudia Regina Gonçalves, Erica Valessa Ramos Gomes, Karoline Rodrigues Campos |
| EPI_ISL_779155, EPI_ISL_779156, EPI_ISL_779157, EPI_ISL_779158, EPI_ISL_779159, EPI_ISL_779160, EPI_ISL_779161, EPI_ISL_779162, EPI_ISL_779163, EPI_ISL_779164, EPI_ISL_779165, EPI_ISL_779166, EPI_ISL_779167, EPI_ISL_779168 | | | |
| see above | Laboratório de Microbiologia Molecular - Universidade FEEVALE | Bioinformatics Laboratory / LNCC | Felipe Benites, Fernando Rosado Spilki, Alana Witt Hansen, Juliane Deise Fleck, Juliana Schons, Meriane Demoliner, Ana Karolina Eisen Antunes, Fagner Henrique Heldt, Larissa Mallmann, Bruna Hermann, Ana Luiza Ziulkoski, Vyctoria Goes, Karoline Schallenberger, Matheus Nunes Weber, Paula Rodrigues de Almeida, Alessandra Pavan Lamarca da Silva, Ronaldo da Silva F Jr , Luiz G P de Almeida, Alexandra L Gerber , Ana Paula de C Guimarães,Ana Tereza R de Vasconcelos |
| EPI_ISL_792112, EPI_ISL_792113, EPI_ISL_792114 | Instituto Adolfo Lutz - Central | Instituto Adolfo Lutz, Interdiciplinary Procedures Center, Strategic Laboratory | Claudio Tavares Sacchi, Claudia Regina Gonçalves, Erica Valessa Ramos Gomes, Karoline Rodrigues Campos |
| EPI_ISL_792115, EPI_ISL_792116 | Instituto Adolfo Lutz - Regional de Taubate | Instituto Adolfo Lutz, Interdiciplinary Procedures Center, Strategic Laboratory | Claudio Tavares Sacchi, Claudia Regina Gonçalves, Erica Valessa Ramos Gomes, Karoline Rodrigues Campos |
| EPI_ISL_792560 | Laboratorio de Ecologia de Doencas Transmissiveis na Amazonia, Instituto Leonidas e Maria Deane - Fiocruz Amazonia | Laboratorio de Ecologia de Doencas Transmissiveis na Amazonia, Instituto Leonidas e Maria Deane - Fiocruz Amazonia | Valdinete Nascimento, Victor Souza, André Corado, Fernanda Nascimento, George Silva, Ágatha Costa, Karina Pessoa, Debora Duarte, Luciana Gonçalves, Maria Júlia Brandão, Michele Jesus, Felipe Naveca on behalf of the Fiocruz COVID-19 Genomic Surveillance Network |
| EPI_ISL_792561, EPI_ISL_792562, EPI_ISL_792563, EPI_ISL_792564, EPI_ISL_792565, EPI_ISL_792566, EPI_ISL_792567, EPI_ISL_792568, EPI_ISL_792569, EPI_ISL_792570, EPI_ISL_792571, EPI_ISL_792573, EPI_ISL_792574, EPI_ISL_792575, EPI_ISL_792586, EPI_ISL_792587, EPI_ISL_792588, EPI_ISL_792589, EPI_ISL_792593, EPI_ISL_792594, EPI_ISL_792595, EPI_ISL_792596, EPI_ISL_792597, EPI_ISL_792598, EPI_ISL_792599, EPI_ISL_792600, EPI_ISL_792601, EPI_ISL_792602, EPI_ISL_792603, EPI_ISL_792604, EPI_ISL_792605, EPI_ISL_792606, EPI_ISL_792607, EPI_ISL_792608, EPI_ISL_792609, EPI_ISL_792610, EPI_ISL_792611, EPI_ISL_792612, EPI_ISL_792613, EPI_ISL_792614, EPI_ISL_792615, EPI_ISL_792616, EPI_ISL_792617, EPI_ISL_792618, EPI_ISL_792619, EPI_ISL_792620, EPI_ISL_792621, EPI_ISL_792622, EPI_ISL_792623, EPI_ISL_792624, EPI_ISL_792625, EPI_ISL_792626, EPI_ISL_792627, EPI_ISL_792628, EPI_ISL_792629, EPI_ISL_792630, EPI_ISL_792631, EPI_ISL_792632, EPI_ISL_792633, EPI_ISL_792634, EPI_ISL_792635, EPI_ISL_792636, EPI_ISL_792637, EPI_ISL_792638 | | | |
| see above | Laboratório Central de Saúde Pública do Estado da Paraíba (LACEN-PB) | Laboratory of Respiratory Viruses and Measles, Oswaldo Cruz Institute, FIOCRUZ | Paola Resende, Luciana Appolinario, Fernando Motta, Anna Carolina Paixao, Ana Carolina Mendonca, João Felipe Bezerra, Romero Henrique Teixeira de Vasconcelos, Dalane Loudal Florentino Teixeira, Thiago Franco de Oliveira Carneiro, Marilda Siqueira on behalf of the Fiocruz COVID-19 Genomic Surveillance Network |
| EPI_ISL_792639, EPI_ISL_792640, EPI_ISL_792641, EPI_ISL_792642, EPI_ISL_792643, EPI_ISL_792644 | Laboratório Central de Saúde Pública do Estado de Alagoas (LACEN-AL) | Laboratory of Respiratory Viruses and Measles, Oswaldo Cruz Institute, FIOCRUZ | Paola Resende, Luciana Appolinario, Fernando Motta, Anna Carolina Paixao, Ana Carolina Mendonca, Anderson Brandao Leite, Marilda Siqueira on behalf of the Fiocruz COVID-19 Genomic Surveillance Network |
| EPI_ISL_792645, EPI_ISL_792646, EPI_ISL_792647, EPI_ISL_792648, EPI_ISL_792649, EPI_ISL_792650, EPI_ISL_792651, EPI_ISL_792652, EPI_ISL_792653, EPI_ISL_792654 | Laboratório Central de Saúde Pública do Estado do Paraná (LACEN-PR) | Laboratory of Respiratory Viruses and Measles, Oswaldo Cruz Institute, FIOCRUZ | Paola Resende, Luciana Appolinario, Fernando Motta, Anna Carolina Paixao, Ana Carolina Mendonca, Maria do Carmo Debur, Irina Nastassja Riediger, Marilda Siqueira on behalf of the Fiocruz COVID-19 Genomic Surveillance Network |
| EPI_ISL_801386, EPI_ISL_801387, EPI_ISL_801388, EPI_ISL_801389, EPI_ISL_801390, EPI_ISL_801391, EPI_ISL_801392, EPI_ISL_801393, EPI_ISL_801394, EPI_ISL_801395, EPI_ISL_801396 | | | |
| see above | Laboratorio de Ecologia de Doencas Transmissiveis na Amazonia, Instituto Leonidas e Maria Deane - Fiocruz Amazonia | Laboratorio de Ecologia de Doencas Transmissiveis na Amazonia, Instituto Leonidas e Maria Deane - Fiocruz Amazonia | Valdinete Nascimento, Victor Souza, André Corado, Fernanda Nascimento, George Silva, Ágatha Costa, Debora Duarte, Luciana Gonçalves, Maria Júlia Brandão, Michele Jesus, Felipe Naveca on behalf of the Fiocruz COVID-19 Genomic Surveillance Network |
| EPI_ISL_801397, EPI_ISL_801398, EPI_ISL_801399, EPI_ISL_801400, EPI_ISL_801401, EPI_ISL_801402, EPI_ISL_801403 | Laboratório Central de Saúde Pública do Estado do Amazonas (LACEN-AM) | Laboratorio de Ecologia de Doencas Transmissiveis na Amazonia, Instituto Leonidas e Maria Deane - Fiocruz Amazonia | Valdinete Nascimento, Victor Souza, André Corado, Fernanda Nascimento, George Silva, Ágatha Costa, Debora Duarte, Luciana Gonçalves, Maria Júlia Brandão, Michele Jesus, Felipe Naveca on behalf of the Fiocruz COVID-19 Genomic Surveillance Network |
| EPI_ISL_804814, EPI_ISL_804815, EPI_ISL_804816, EPI_ISL_804817, EPI_ISL_804818, EPI_ISL_804819, EPI_ISL_804820, EPI_ISL_804821, EPI_ISL_804822, EPI_ISL_804823, EPI_ISL_804824, EPI_ISL_804825, EPI_ISL_804826, EPI_ISL_804827, EPI_ISL_804828, EPI_ISL_804829, EPI_ISL_804830, EPI_ISL_804831, EPI_ISL_804832, EPI_ISL_804833, EPI_ISL_804834, EPI_ISL_804835, EPI_ISL_804836, EPI_ISL_804839, EPI_ISL_804840, EPI_ISL_804841, EPI_ISL_804842, EPI_ISL_804843, EPI_ISL_804844 | | | |
| see above | DB Diagnosticos do Brasil | Laboratório de Parasitologia Médica - Instituto de Medicina Tropical - Universidade de São Paulo | Nuno Faria, Ingra Morales Claro, Darlan Candido, Lucas A. Moyses Franco, Pamela dos Santos Andrade, Thais de Moura Coletti, Camila A. Maia da Silva, Flavia Cristina Sales, Erika Regina Manuli, Renato A. Santana, Nelson Gaburo, Cecília da Cunha Camilo, Nelson Abrahim Fraiji, Myuki Alfaia Esashika Crispim, Maria do Perpétuo Socorro Sampaio Carvalho, Andrew Rambaut, Nick Loman, Oliver G. Pybus, Ester C. Sabino; DB; HEMOAM; CDL; CADDE Genomic Network. |
| EPI_ISL_811149 | Laboratorio de Ecologia de Doencas Transmissiveis na Amazonia, Instituto Leonidas e Maria Deane - Fiocruz Amazonia | Laboratorio de Ecologia de Doencas Transmissiveis na Amazonia, Instituto Leonidas e Maria Deane - Fiocruz Amazonia | Valdinete Nascimento, Victor Souza, André Corado, Fernanda Nascimento, George Silva, Ágatha Costa, Debora Duarte, Luciana Gonçalves, Matilde Mejía, Karina Pessoa, Maria Júlia Brandão, Michele Jesus, Felipe Naveca on behalf of the Fiocruz COVID-19 Genomic Surveillance Network |
| EPI_ISL_831474, EPI_ISL_831645, EPI_ISL_831646, EPI_ISL_831660, EPI_ISL_831678, EPI_ISL_831681, EPI_ISL_831683, EPI_ISL_831685, EPI_ISL_831688, EPI_ISL_831689, EPI_ISL_831892, EPI_ISL_831898, EPI_ISL_831913, EPI_ISL_831938, EPI_ISL_831939, EPI_ISL_831940, EPI_ISL_832009, EPI_ISL_832010, EPI_ISL_832011, EPI_ISL_832012, EPI_ISL_832013 | | | |
| see above | Laboratório de Microbiologia Molecular - Universidade | Universidade Federal de Ciências da Saúde de Porto Alegre | Vinícius Bonetti Franceschi, Amanda de Menezes Mayer, Gabriel Dickin Caldana, Carla Andretta Moreira Neves, Patrícia Aline Gröhs Ferrareze, Gabriela |

| | | | |
|---|---|---|---|
| | FEEVALE | | Bettella Cybis, Ricardo Ariel Zimerman, Lívia Kmetzsch, Fernando Rosado Spilki, Claudia Elizabeth Thompson |
| EPI_ISL_833131, EPI_ISL_833132, EPI_ISL_833133, EPI_ISL_833134, EPI_ISL_833135, EPI_ISL_833136, EPI_ISL_833137, EPI_ISL_833138, EPI_ISL_833139, EPI_ISL_833140 | Laboratorio de Ecologia de Doencas Transmissiveis na Amazonia, Instituto Leonidas e Maria Deane - Fiocruz Amazonia | Laboratorio de Ecologia de Doencas Transmissiveis na Amazonia, Instituto Leonidas e Maria Deane - Fiocruz Amazonia | Valdinete Nascimento, Victor Souza, André Corado, Fernanda Nascimento, George Silva, Ágatha Costa, Debora Duarte, Karina Pessoa, Matilde Mejía, Luciana Gonçalves, Maria Júlia Brandão, Michele Jesus, Felipe Naveca on behalf of the Fiocruz COVID-19 Genomic Surveillance Network |
| EPI_ISL_833155 | Instituto Adolfo Lutz - Central | Instituto Adolfo Lutz, Interdiciplinary Procedures Center, Strategic Laboratory | Claudio Tavares Sacchi, Claudia Regina Gonçalves, Erica Valessa Ramos Gomes, Karoline Rodrigues Campos |
| EPI_ISL_833156 | Instituto Adolfo Lutz - Regional de Sorocaba | Instituto Adolfo Lutz, Interdiciplinary Procedures Center, Strategic Laboratory | Claudio Tavares Sacchi, Claudia Regina Gonçalves, Erica Valessa Ramos Gomes, Karoline Rodrigues Campos |
| EPI_ISL_833157, EPI_ISL_833158 | Instituto Adolfo Lutz - Regional de Santo Andre | Instituto Adolfo Lutz, Interdiciplinary Procedures Center, Strategic Laboratory | Claudio Tavares Sacchi, Claudia Regina Gonçalves, Erica Valessa Ramos Gomes, Karoline Rodrigues Campos |
| EPI_ISL_833159 | Instituto Adolfo Lutz - Central | Instituto Adolfo Lutz, Interdiciplinary Procedures Center, Strategic Laboratory | Claudio Tavares Sacchi, Claudia Regina Gonçalves, Erica Valessa Ramos Gomes, Karoline Rodrigues Campos |
| EPI_ISL_833160 | Instituto Adolfo Lutz - Regional de Santo Andre | Instituto Adolfo Lutz, Interdiciplinary Procedures Center, Strategic Laboratory | Claudio Tavares Sacchi, Claudia Regina Gonçalves, Erica Valessa Ramos Gomes, Karoline Rodrigues Campos |
| EPI_ISL_833161 | Instituto Adolfo Lutz - Central | Instituto Adolfo Lutz, Interdiciplinary Procedures Center, Strategic Laboratory | Claudio Tavares Sacchi, Claudia Regina Gonçalves, Erica Valessa Ramos Gomes, Karoline Rodrigues Campos |
| EPI_ISL_833162 | Lab LOC - Itapecerica da Serra | Instituto Adolfo Lutz, Interdiciplinary Procedures Center, Strategic Laboratory | Claudio Tavares Sacchi, Claudia Regina Gonçalves, Erica Valessa Ramos Gomes, Karoline Rodrigues Campos |
| EPI_ISL_833163 | Instituto Adolfo Lutz - Regional de Marilia | Instituto Adolfo Lutz, Interdiciplinary Procedures Center, Strategic Laboratory | Claudio Tavares Sacchi, Claudia Regina Gonçalves, Erica Valessa Ramos Gomes, Karoline Rodrigues Campos |
| EPI_ISL_833164 | Secretaria Municipal de Saude de Santa Barbara d'oeste | Instituto Adolfo Lutz, Interdiciplinary Procedures Center, Strategic Laboratory | Claudio Tavares Sacchi, Claudia Regina Gonçalves, Erica Valessa Ramos Gomes, Karoline Rodrigues Campos |
| EPI_ISL_833165, EPI_ISL_833166 | Hospital Samaritano | Instituto Adolfo Lutz, Interdiciplinary Procedures Center, Strategic Laboratory | Claudio Tavares Sacchi, Claudia Regina Gonçalves, Erica Valessa Ramos Gomes, Karoline Rodrigues Campos |
| EPI_ISL_833167, EPI_ISL_833168, EPI_ISL_833169, EPI_ISL_833170, EPI_ISL_833171, EPI_ISL_833172, EPI_ISL_833173, EPI_ISL_833174, EPI_ISL_833175, EPI_ISL_833176 | DB Diagnosticos do Brasil | Instituto Adolfo Lutz, Interdiciplinary Procedures Center, Strategic Laboratory | Claudio Tavares Sacchi, Claudia Regina Gonçalves, Erica Valessa Ramos Gomes, Karoline Rodrigues Campos |
| EPI_ISL_836143 | Hospital de Campanha COVID-19 de Mairipora | Instituto Adolfo Lutz, Interdiciplinary Procedures Center, Strategic Laboratory | Claudio Tavares Sacchi, Claudia Regina Gonçalves, Erica Valessa Ramos Gomes, Karoline Rodrigues Campos |
| EPI_ISL_836977 | Hospital Municiplal Dr. Jose de Carvalho Florence | Instituto Adolfo Lutz, Interdiciplinary Procedures Center, Strategic Laboratory | Claudio Tavares Sacchi, Claudia Regina Gonçalves, Erica Valessa Ramos Gomes, Karoline Rodrigues Campos |
| EPI_ISL_836978 | Irmandade da Santa Casa de Misericordia de Lorena | Instituto Adolfo Lutz, Interdiciplinary Procedures Center, Strategic Laboratory | Claudio Tavares Sacchi, Claudia Regina Gonçalves, Erica Valessa Ramos Gomes, Karoline Rodrigues Campos |
| EPI_ISL_837053 | UBS Darcy Alves e Robalinho | Instituto Adolfo Lutz, Interdiciplinary Procedures Center, Strategic Laboratory | Claudio Tavares Sacchi, Claudia Regina Gonçalves, Erica Valessa Ramos Gomes, Karoline Rodrigues Campos |
| EPI_ISL_837054 | UBS Jose Sabino Ferreira | Instituto Adolfo Lutz, Interdiciplinary Procedures Center, Strategic Laboratory | Claudio Tavares Sacchi, Claudia Regina Gonçalves, Erica Valessa Ramos Gomes, Karoline Rodrigues Campos |
| EPI_ISL_848555, EPI_ISL_848556, EPI_ISL_848557, EPI_ISL_848558, EPI_ISL_848559, EPI_ISL_848560, EPI_ISL_848561, EPI_ISL_848562, EPI_ISL_848563, EPI_ISL_848564, EPI_ISL_848565, EPI_ISL_848566, EPI_ISL_848567, EPI_ISL_848568, EPI_ISL_848569, EPI_ISL_848570, EPI_ISL_848571, EPI_ISL_848572, EPI_ISL_848573, EPI_ISL_848574, EPI_ISL_848575, EPI_ISL_848576, EPI_ISL_848577, EPI_ISL_848578, EPI_ISL_848579, EPI_ISL_848580, EPI_ISL_848581, EPI_ISL_848582, EPI_ISL_848583, EPI_ISL_848584, EPI_ISL_848585, EPI_ISL_848586, EPI_ISL_848587, EPI_ISL_848588, EPI_ISL_848606, EPI_ISL_848607, EPI_ISL_848608, EPI_ISL_848609, EPI_ISL_848610, EPI_ISL_848611, EPI_ISL_848612, EPI_ISL_848613, EPI_ISL_848614, EPI_ISL_848615, EPI_ISL_848616, EPI_ISL_848617, EPI_ISL_848618, EPI_ISL_848619, EPI_ISL_848620 | | | |
| see above | Evandro Chagas Institute | Evandro Chagas Institute | Santos, M.C.; Silva, A.M.; Junior, W.D.C.; Barbagelata, L.S.; Ferreira, J.A.; Sousa, E.M.A.; da Silva, P.S.; Pinheiro, K.C.; L.C.; Sousa Junior, E.C. |
| EPI_ISL_860317, EPI_ISL_860633 | Instituto Adolfo Lutz - Regional de Sorocaba | Instituto Adolfo Lutz, Interdiciplinary Procedures Center, Strategic Laboratory | Claudio Tavares Sacchi, Claudia Regina Gonçalves, Erica Valessa Ramos Gomes, Karoline Rodrigues Campos |
| EPI_ISL_861242 | Instituto de Biotecnologia - UNESP-Botucatu-SP | Instituto de Biotecnologia - UNESP-Botucatu-SP | Leila Sabrina Ullmann; Fábio Sossai Possebon, Camila Dantas Malossi, Paula Rahal, Paulo Inacio da Costa, João Pessoa Araújo Jr. |
| EPI_ISL_861639 | Hospital Sao Paulo de Ensino da Unifesp | Instituto Adolfo Lutz, Interdiciplinary Procedures Center, Strategic Laboratory | Claudio Tavares Sacchi, Claudia Regina Gonçalves, Erica Valessa Ramos Gomes, Karoline Rodrigues Campos |
| EPI_ISL_861642, EPI_ISL_861643 | Instituto Adolfo Lutz - Central | Instituto Adolfo Lutz, Interdiciplinary Procedures Center, Strategic Laboratory | Claudio Tavares Sacchi, Claudia Regina Gonçalves, Erica Valessa Ramos Gomes, Karoline Rodrigues Campos |
| EPI_ISL_861644 | Hospital Santa Virginia | Instituto Adolfo Lutz, Interdiciplinary Procedures Center, Strategic Laboratory | Claudio Tavares Sacchi, Claudia Regina Gonçalves, Erica Valessa Ramos Gomes, Karoline Rodrigues Campos |
| EPI_ISL_861645 | Hospital e Pronto Socorro Comunitario Vila Iolanda | Instituto Adolfo Lutz, Interdiciplinary Procedures Center, Strategic Laboratory | Claudio Tavares Sacchi, Claudia Regina Gonçalves, Erica Valessa Ramos Gomes, Karoline Rodrigues Campos |
| EPI_ISL_861646, EPI_ISL_861647 | Hospital Santa Marcelina Sao Paulo | Instituto Adolfo Lutz, Interdiciplinary Procedures Center, Strategic Laboratory | Claudio Tavares Sacchi, Claudia Regina Gonçalves, Erica Valessa Ramos Gomes, Karoline Rodrigues Campos |
| EPI_ISL_861648 | Hospital e Pronto Socorro Portinari | Instituto Adolfo Lutz, Interdiciplinary Procedures Center, Strategic Laboratory | Claudio Tavares Sacchi, Claudia Regina Gonçalves, Erica Valessa Ramos Gomes, Karoline Rodrigues Campos |
| EPI_ISL_861649 | Hospital Renascença Campinas | Instituto Adolfo Lutz, Interdiciplinary Procedures Center, Strategic Laboratory | Claudio Tavares Sacchi, Claudia Regina Gonçalves, Erica Valessa Ramos Gomes, Karoline Rodrigues Campos |
| EPI_ISL_861650 | Hospital Santa Marcelina Sao Paulo | Instituto Adolfo Lutz, Interdiciplinary Procedures Center, Strategic Laboratory | Claudio Tavares Sacchi, Claudia Regina Gonçalves, Erica Valessa Ramos Gomes, Karoline Rodrigues Campos |
| EPI_ISL_861651 | AMA Jardim Brasil | Instituto Adolfo Lutz, Interdiciplinary Procedures Center, Strategic Laboratory | Claudio Tavares Sacchi, Claudia Regina Gonçalves, Erica Valessa Ramos Gomes, Karoline Rodrigues Campos |
| EPI_ISL_861652 | AMA Wamberto Dias da Costa | Instituto Adolfo Lutz, Interdiciplinary Procedures Center, Strategic Laboratory | Claudio Tavares Sacchi, Claudia Regina Gonçalves, Erica Valessa Ramos Gomes, Karoline Rodrigues Campos |
| EPI_ISL_861653 | Hospital Santa Virgínia | Instituto Adolfo Lutz, Interdiciplinary Procedures Center, Strategic Laboratory | Claudio Tavares Sacchi, Claudia Regina Gonçalves, Erica Valessa Ramos Gomes, Karoline Rodrigues Campos |
| EPI_ISL_861654, EPI_ISL_861655 | Hospital Santa Marcelina Sao Paulo | Instituto Adolfo Lutz, Interdiciplinary Procedures Center, | Claudio Tavares Sacchi, Claudia Regina Gonçalves, Erica Valessa Ramos Gomes, Karoline Rodrigues Campos |

| | | | |
|---|---|---|---|
| | | Strategic Laboratory | |
| EPI_ISL_861656 | UPA de Jandira | Instituto Adolfo Lutz, Interdiciplinary Procedures Center, Strategic Laboratory | Claudio Tavares Sacchi, Claudia Regina Gonçalves, Erica Valessa Ramos Gomes, Karoline Rodrigues Campos |
| EPI_ISL_861657 | Hospital e Maternidade Sino Brasileiro | Instituto Adolfo Lutz, Interdiciplinary Procedures Center, Strategic Laboratory | Claudio Tavares Sacchi, Claudia Regina Gonçalves, Erica Valessa Ramos Gomes, Karoline Rodrigues Campos |
| EPI_ISL_861658 | Hospital Municipal Antônio Giglio | Instituto Adolfo Lutz, Interdiciplinary Procedures Center, Strategic Laboratory | Claudio Tavares Sacchi, Claudia Regina Gonçalves, Erica Valessa Ramos Gomes, Karoline Rodrigues Campos |
| EPI_ISL_861659, EPI_ISL_861660, EPI_ISL_861661 | PS e Maternidade Nair Fonseca Leitao Arantes | Instituto Adolfo Lutz, Interdiciplinary Procedures Center, Strategic Laboratory | Claudio Tavares Sacchi, Claudia Regina Gonçalves, Erica Valessa Ramos Gomes, Karoline Rodrigues Campos |
| EPI_ISL_861662 | CS I Tacito Leite de Carvalho e Silva | Instituto Adolfo Lutz, Interdiciplinary Procedures Center, Strategic Laboratory | Claudio Tavares Sacchi, Claudia Regina Gonçalves, Erica Valessa Ramos Gomes, Karoline Rodrigues Campos |
| EPI_ISL_861664 | Instituto Adolfo Lutz - Regional de Campinas | Instituto Adolfo Lutz, Interdiciplinary Procedures Center, Strategic Laboratory | Claudio Tavares Sacchi, Claudia Regina Gonçalves, Erica Valessa Ramos Gomes, Karoline Rodrigues Campos |
| EPI_ISL_861665 | Instituto Adolfo Lutz - Regional de Taubate | Instituto Adolfo Lutz, Interdiciplinary Procedures Center, Strategic Laboratory | Claudio Tavares Sacchi, Claudia Regina Gonçalves, Erica Valessa Ramos Gomes, Karoline Rodrigues Campos |
| EPI_ISL_861666 | PSF Dr. Antonio Pires de Almeida | Instituto Adolfo Lutz, Interdiciplinary Procedures Center, Strategic Laboratory | Claudio Tavares Sacchi, Claudia Regina Gonçalves, Erica Valessa Ramos Gomes, Karoline Rodrigues Campos |
| EPI_ISL_861667 | Instituto Adolfo Lutz - Regional de Rio Claro | Instituto Adolfo Lutz, Interdiciplinary Procedures Center, Strategic Laboratory | Claudio Tavares Sacchi, Claudia Regina Gonçalves, Erica Valessa Ramos Gomes, Karoline Rodrigues Campos |
| EPI_ISL_861668 | Centro de Triagem Covid19 | Instituto Adolfo Lutz, Interdiciplinary Procedures Center, Strategic Laboratory | Claudio Tavares Sacchi, Claudia Regina Gonçalves, Erica Valessa Ramos Gomes, Karoline Rodrigues Campos |
| EPI_ISL_861669 | Lab LOC - Itapecerica da Serra | Instituto Adolfo Lutz, Interdiciplinary Procedures Center, Strategic Laboratory | Claudio Tavares Sacchi, Claudia Regina Gonçalves, Erica Valessa Ramos Gomes, Karoline Rodrigues Campos |
| EPI_ISL_861670 | Instituto Adolfo Lutz - Regional de Taubate | Instituto Adolfo Lutz, Interdiciplinary Procedures Center, Strategic Laboratory | Claudio Tavares Sacchi, Claudia Regina Gonçalves, Erica Valessa Ramos Gomes, Karoline Rodrigues Campos |
| EPI_ISL_861671 | Hospital Municipal Prefeito Waldemar Costa Filho | Instituto Adolfo Lutz, Interdiciplinary Procedures Center, Strategic Laboratory | Claudio Tavares Sacchi, Claudia Regina Gonçalves, Erica Valessa Ramos Gomes, Karoline Rodrigues Campos |
| EPI_ISL_861672 | Day Hospital de Ermelino Matarazzo | Instituto Adolfo Lutz, Interdiciplinary Procedures Center, Strategic Laboratory | Claudio Tavares Sacchi, Claudia Regina Gonçalves, Erica Valessa Ramos Gomes, Karoline Rodrigues Campos |
| EPI_ISL_861673 | PA Novo Osasco | Instituto Adolfo Lutz, Interdiciplinary Procedures Center, Strategic Laboratory | Claudio Tavares Sacchi, Claudia Regina Gonçalves, Erica Valessa Ramos Gomes, Karoline Rodrigues Campos |
| EPI_ISL_861674, EPI_ISL_861675 | UPA Central de Caraguatatuba | Instituto Adolfo Lutz, Interdiciplinary Procedures Center, Strategic Laboratory | Claudio Tavares Sacchi, Claudia Regina Gonçalves, Erica Valessa Ramos Gomes, Karoline Rodrigues Campos |
| EPI_ISL_861676 | UPA Vila Santa Catarina | Instituto Adolfo Lutz, Interdiciplinary Procedures Center, Strategic Laboratory | Claudio Tavares Sacchi, Claudia Regina Gonçalves, Erica Valessa Ramos Gomes, Karoline Rodrigues Campos |
| EPI_ISL_861677 | Instituto Adolfo Lutz - Central | Instituto Adolfo Lutz, Interdiciplinary Procedures Center, Strategic Laboratory | Claudio Tavares Sacchi, Claudia Regina Gonçalves, Erica Valessa Ramos Gomes, Karoline Rodrigues Campos |
| EPI_ISL_861678 | Laboratorio Municipal de Guarulhos | Instituto Adolfo Lutz, Interdiciplinary Procedures Center, Strategic Laboratory | Claudio Tavares Sacchi, Claudia Regina Gonçalves, Erica Valessa Ramos Gomes, Karoline Rodrigues Campos |
| EPI_ISL_861679 | Instituto Adolfo Lutz - Regional de Taubate | Instituto Adolfo Lutz, Interdiciplinary Procedures Center, Strategic Laboratory | Claudio Tavares Sacchi, Claudia Regina Gonçalves, Erica Valessa Ramos Gomes, Karoline Rodrigues Campos |
| EPI_ISL_861680 | Hospital e Pronto Socorro Portinari | Instituto Adolfo Lutz, Interdiciplinary Procedures Center, Strategic Laboratory | Claudio Tavares Sacchi, Claudia Regina Gonçalves, Erica Valessa Ramos Gomes, Karoline Rodrigues Campos |
| EPI_ISL_861681 | Hospital Municipal Dr. Waldemar Tebaldi | Instituto Adolfo Lutz, Interdiciplinary Procedures Center, Strategic Laboratory | Claudio Tavares Sacchi, Claudia Regina Gonçalves, Erica Valessa Ramos Gomes, Karoline Rodrigues Campos |
| EPI_ISL_861682 | UPA Vila Santa Catarina | Instituto Adolfo Lutz, Interdiciplinary Procedures Center, Strategic Laboratory | Claudio Tavares Sacchi, Claudia Regina Gonçalves, Erica Valessa Ramos Gomes, Karoline Rodrigues Campos |
| EPI_ISL_861683 | Complexo Hospitalar Padre Bento de Guarulhos | Instituto Adolfo Lutz, Interdiciplinary Procedures Center, Strategic Laboratory | Claudio Tavares Sacchi, Claudia Regina Gonçalves, Erica Valessa Ramos Gomes, Karoline Rodrigues Campos |
| EPI_ISL_861684, EPI_ISL_861685 | Hospital Pronto Socorro Itaquera | Instituto Adolfo Lutz, Interdiciplinary Procedures Center, Strategic Laboratory | Claudio Tavares Sacchi, Claudia Regina Gonçalves, Erica Valessa Ramos Gomes, Karoline Rodrigues Campos |
| EPI_ISL_861867, EPI_ISL_861868, EPI_ISL_861870, EPI_ISL_861871, EPI_ISL_861872, EPI_ISL_861873, EPI_ISL_861875, EPI_ISL_861876, EPI_ISL_861877, EPI_ISL_861878, EPI_ISL_861880, EPI_ISL_861881, EPI_ISL_861882, EPI_ISL_861883, EPI_ISL_861884, EPI_ISL_861885, EPI_ISL_861886, EPI_ISL_861887, EPI_ISL_861888, EPI_ISL_861891, EPI_ISL_861892, EPI_ISL_861894, EPI_ISL_861895, EPI_ISL_861897, EPI_ISL_861898, EPI_ISL_861899, EPI_ISL_861901, EPI_ISL_861903, EPI_ISL_861904, EPI_ISL_861908, EPI_ISL_861909, EPI_ISL_861910 | | | |
| see above | LATE - Laboratório de Técnicas Especiais - Hospital Israelita Albert Einstein | LATE - Laboratório de Técnicas Especiais - Hospital Israelita Albert Einstein | Deyvid Amgarten, Fernanda de Mello Malta, Raquel Riyuzo, Ana Paula Moreira Salles, Pedro Henrique Sebe Rodrigues, João Renato Rebello Pinho |
| EPI_ISL_872191, EPI_ISL_872192 | Conjunto Hospitalar do Mandaqui de Sao Paulo | Instituto Adolfo Lutz, Interdiciplinary Procedures Center, Strategic Laboratory | Claudio Tavares Sacchi, Claudia Regina Gonçalves, Erica Valessa Ramos Gomes, Karoline Rodrigues Campos, Katia Correa de Oliveira Santos, Ana Lucia de Carvalho Avelino, Fabiana Cristina Pereira dos Santos |
| EPI_ISL_875540, EPI_ISL_875542, EPI_ISL_875543, EPI_ISL_875546, EPI_ISL_875547, EPI_ISL_875548, EPI_ISL_875549 | Instituto de Biotecnologia - UNESP-Botucatu-SP | Instituto de Biotecnologia - UNESP-Botucatu-SP | Leila Sabrina Ullmann; Fábio Sossai Possebon, Camila Dantas Malossi, Paula Rahal, Paulo Inacio da Costa, João Pessoa Araújo Jr. |
| EPI_ISL_875688 | National Influenza Center - Instituto Adolfo Lutz | Instituto Adolfo Lutz, Interdiciplinary Procedures Center, Strategic Laboratory | Claudio Tavares Sacchi, Claudia Regina Gonçalves, Erica Valessa Ramos Gomes, Karoline Rodrigues Campos, Katia Correa de Oliveira Santos, Ana Lucia de Carvalho Avelino, Clovis Roberto Abe Constantino |
| EPI_ISL_875689 | Hospital do Servidor Publico | Instituto Adolfo Lutz, Interdiciplinary Procedures Center, Strategic Laboratory | Claudio Tavares Sacchi, Claudia Regina Gonçalves, Erica Valessa Ramos Gomes, Karoline Rodrigues Campos |
| EPI_ISL_882657 | LACEN do Estado do Piaui, Dr. Costa Alvarenga | Instituto Adolfo Lutz, Interdiciplinary Procedures Center, Strategic Laboratory | Claudio Tavares Sacchi, Claudia Regina Gonçalves, Erica Valessa Ramos Gomes, Karoline Rodrigues Campos |
| EPI_ISL_882658 | Secretaria Municipal de Saude | Instituto Adolfo Lutz, Interdiciplinary Procedures Center, Strategic Laboratory | Claudio Tavares Sacchi, Claudia Regina Gonçalves, Erica Valessa Ramos Gomes, Karoline Rodrigues Campos |
| EPI_ISL_882659 | Centro de Triagem Covid19 | Instituto Adolfo Lutz, Interdiciplinary Procedures Center, Strategic Laboratory | Claudio Tavares Sacchi, Claudia Regina Gonçalves, Erica Valessa Ramos Gomes, Karoline Rodrigues Campos |

| | | | |
|---|---|---|---|
| EPI_ISL_882660 | Hospital Municipal Prefeito Waldemar Costa Filho | Instituto Adolfo Lutz, Interdiciplinary Procedures Center, Strategic Laboratory | Claudio Tavares Sacchi, Claudia Regina Gonçalves, Erica Valessa Ramos Gomes, Karoline Rodrigues Campos |
| EPI_ISL_882661, EPI_ISL_882662 | Hospital de Santa Barbara de Goias | Instituto Adolfo Lutz, Interdiciplinary Procedures Center, Strategic Laboratory | Claudio Tavares Sacchi, Claudia Regina Gonçalves, Erica Valessa Ramos Gomes, Karoline Rodrigues Campos |
| EPI_ISL_882663, EPI_ISL_882664 | LACEN do Distrito Federal | Instituto Adolfo Lutz, Interdiciplinary Procedures Center, Strategic Laboratory | Claudio Tavares Sacchi, Claudia Regina Gonçalves, Erica Valessa Ramos Gomes, Karoline Rodrigues Campos |
| EPI_ISL_882665 | Unidade de Pronto Atendimento Dra Zilda Arns | Instituto Adolfo Lutz, Interdiciplinary Procedures Center, Strategic Laboratory | Claudio Tavares Sacchi, Claudia Regina Gonçalves, Erica Valessa Ramos Gomes, Karoline Rodrigues Campos |
| EPI_ISL_882666 | UMS de Juquitiba | Instituto Adolfo Lutz, Interdiciplinary Procedures Center, Strategic Laboratory | Claudio Tavares Sacchi, Claudia Regina Gonçalves, Erica Valessa Ramos Gomes, Karoline Rodrigues Campos |
| EPI_ISL_882667 | Hospital de Clinicas Caieiras | Instituto Adolfo Lutz, Interdiciplinary Procedures Center, Strategic Laboratory | Claudio Tavares Sacchi, Claudia Regina Gonçalves, Erica Valessa Ramos Gomes, Karoline Rodrigues Campos |
| EPI_ISL_882668 | UMS de Juquitiba | Instituto Adolfo Lutz, Interdiciplinary Procedures Center, Strategic Laboratory | Claudio Tavares Sacchi, Claudia Regina Gonçalves, Erica Valessa Ramos Gomes, Karoline Rodrigues Campos |
| EPI_ISL_882669 | UPA Vila Santa Catarina | Instituto Adolfo Lutz, Interdiciplinary Procedures Center, Strategic Laboratory | Claudio Tavares Sacchi, Claudia Regina Gonçalves, Erica Valessa Ramos Gomes, Karoline Rodrigues Campos |
| EPI_ISL_882670 | Hospital Samaritano Paulista | Instituto Adolfo Lutz, Interdiciplinary Procedures Center, Strategic Laboratory | Claudio Tavares Sacchi, Claudia Regina Gonçalves, Erica Valessa Ramos Gomes, Karoline Rodrigues Campos |
| EPI_ISL_882671, EPI_ISL_882672, EPI_ISL_882673 | Hospital Municipal Dr. Guido Guida | Instituto Adolfo Lutz, Interdiciplinary Procedures Center, Strategic Laboratory | Claudio Tavares Sacchi, Claudia Regina Gonçalves, Erica Valessa Ramos Gomes, Karoline Rodrigues Campos |
| EPI_ISL_884249, EPI_ISL_884250 | LATE - Laboratório de Técnicas Especiais - Hospital Israelita Albert Einstein | LATE - Laboratório de Técnicas Especiais - Hospital Israelita Albert Einstein | Deyvid Amgarten, Fernanda de Mello Malta, Raquel Riyuzo, Ana Paula Moreira Salles, Pedro Henrique Sebe Rodrigues, João Renato Rebello Pinho |
| EPI_ISL_888672 | Instituto de Biotecnologia - UNESP-Botucatu-SP | Instituto de Biotecnologia - UNESP-Botucatu-SP | Leila Sabrina Ullmann; Fábio Sossai Possebon, Camila Dantas Malossi, Paula Rahal, Paulo Inacio da Costa, João Pessoa Araújo Jr. |
| EPI_ISL_904027, EPI_ISL_904028, EPI_ISL_904029, EPI_ISL_904030 | DB Diagnosticos do Brasil | Laboratório de Parasitologia Médica - Instituto de Medicina Tropical - Universidade de São Paulo | Brazil-UK Centre for Arbovirus Discovery Diagnosis Genomics and Epidemiology (CADDE) Genomic Network - Instituto de Medicina Tropical |
| EPI_ISL_904120, EPI_ISL_904121 | LACEN - Laboratório Central de Saúde Pública do Pará | Evandro Chagas Institute | Santos, M.C.; Silva, A.M.; Junior, W.D.C.; Barbagelata, L.S.; Ferreira, J.A.; Sousa, E.M.A.; da Silva, P.S.; Pinheiro, K.C.; L.C.; Sousa Junior, E.C. |
| EPI_ISL_906067 | PS e Maternidade Nair Fonseca Leitao Arantes | Instituto Adolfo Lutz, Interdiciplinary Procedures Center, Strategic Laboratory | Claudio Tavares Sacchi, Claudia Regina Gonçalves, Erica Valessa Ramos Gomes, Karoline Rodrigues Campos |
| EPI_ISL_906068, EPI_ISL_906069 | Instituto Adolfo Lutz - Regional de Campinas | Instituto Adolfo Lutz, Interdiciplinary Procedures Center, Strategic Laboratory | Claudio Tavares Sacchi, Claudia Regina Gonçalves, Erica Valessa Ramos Gomes, Karoline Rodrigues Campos |
| EPI_ISL_906070 | UPA Dr. Akira Tada | Instituto Adolfo Lutz, Interdiciplinary Procedures Center, Strategic Laboratory | Claudio Tavares Sacchi, Claudia Regina Gonçalves, Erica Valessa Ramos Gomes, Karoline Rodrigues Campos |
| EPI_ISL_906071 | LACEN-PI DR. Costa Alvarenga | Instituto Adolfo Lutz, Interdiciplinary Procedures Center, Strategic Laboratory | Claudio Tavares Sacchi, Claudia Regina Gonçalves, Erica Valessa Ramos Gomes, Karoline Rodrigues Campos |
| EPI_ISL_906072 | UPA Dr. Akira Tada | Instituto Adolfo Lutz, Interdiciplinary Procedures Center, Strategic Laboratory | Claudio Tavares Sacchi, Claudia Regina Gonçalves, Erica Valessa Ramos Gomes, Karoline Rodrigues Campos |
| EPI_ISL_906073 | Hospital Vila Lobos | Instituto Adolfo Lutz, Interdiciplinary Procedures Center, Strategic Laboratory | Claudio Tavares Sacchi, Claudia Regina Gonçalves, Erica Valessa Ramos Gomes, Karoline Rodrigues Campos |
| EPI_ISL_906074 | Searom Laboratorio Diagnostico | Instituto Adolfo Lutz, Interdiciplinary Procedures Center, Strategic Laboratory | Claudio Tavares Sacchi, Claudia Regina Gonçalves, Erica Valessa Ramos Gomes, Karoline Rodrigues Campos |
| EPI_ISL_906075 | Hospital Geral de Vila Penteado Dr Jose Pangella Sao Paulo | Instituto Adolfo Lutz, Interdiciplinary Procedures Center, Strategic Laboratory | Claudio Tavares Sacchi, Claudia Regina Gonçalves, Erica Valessa Ramos Gomes, Karoline Rodrigues Campos |
| EPI_ISL_906076, EPI_ISL_906077 | Hospital Sao Luiz Sao Caetano | Instituto Adolfo Lutz, Interdiciplinary Procedures Center, Strategic Laboratory | Claudio Tavares Sacchi, Claudia Regina Gonçalves, Erica Valessa Ramos Gomes, Karoline Rodrigues Campos |
| EPI_ISL_906078 | Hospital Carlos Chagas | Instituto Adolfo Lutz, Interdiciplinary Procedures Center, Strategic Laboratory | Claudio Tavares Sacchi, Claudia Regina Gonçalves, Erica Valessa Ramos Gomes, Karoline Rodrigues Campos |
| EPI_ISL_906079 | Hospital Paulistano Paulista | Instituto Adolfo Lutz, Interdiciplinary Procedures Center, Strategic Laboratory | Claudio Tavares Sacchi, Claudia Regina Gonçalves, Erica Valessa Ramos Gomes, Karoline Rodrigues Campos |
| EPI_ISL_906080, EPI_ISL_906081 | Hospital Beneficiencia Portuguesa | Instituto Adolfo Lutz, Interdiciplinary Procedures Center, Strategic Laboratory | Claudio Tavares Sacchi, Claudia Regina Gonçalves, Erica Valessa Ramos Gomes, Karoline Rodrigues Campos |
| EPI_ISL_918499, EPI_ISL_918500, EPI_ISL_918501, EPI_ISL_918502, EPI_ISL_918503, EPI_ISL_918504, EPI_ISL_918505, EPI_ISL_918506, EPI_ISL_918507, EPI_ISL_918508, EPI_ISL_918509, EPI_ISL_918510, EPI_ISL_918511, EPI_ISL_918512 | | | |
| see above | LACEN - Laboratório Central de Saúde Pública do Amazonas | Evandro Chagas Institute | Santos, M.C.; Silva, A.M.; Junior, W.D.C.; Barbagelata, L.S.; Ferreira, J.A.; Sousa, E.M.A.; da Silva, P.S.; Pinheiro, K.C.; L.C.; Sousa Junior, E.C. |
| EPI_ISL_918513 | LACEN - Laboratório Central de Saúde Pública do Roraima | Evandro Chagas Institute | Santos, M.C.; Silva, A.M.; Junior, W.D.C.; Barbagelata, L.S.; Ferreira, J.A.; Sousa, E.M.A.; da Silva, P.S.; Pinheiro, K.C.; L.C.; Sousa Junior, E.C. |
| EPI_ISL_918514 | Evandro Chagas Institute | Evandro Chagas Institute | Santos, M.C.; Silva, A.M.; Junior, W.D.C.; Barbagelata, L.S.; Ferreira, J.A.; Sousa, E.M.A.; da Silva, P.S.; Pinheiro, K.C.; L.C.; Sousa Junior, E.C. |
| EPI_ISL_918515, EPI_ISL_918516, EPI_ISL_918517 | LACEN - Laboratório Central de Saúde Pública do Para | Evandro Chagas Institute | Santos, M.C.; Silva, A.M.; Junior, W.D.C.; Barbagelata, L.S.; Ferreira, J.A.; Sousa, E.M.A.; da Silva, P.S.; Pinheiro, K.C.; L.C.; Sousa Junior, E.C. |
| EPI_ISL_918518, EPI_ISL_918519, EPI_ISL_918520, EPI_ISL_918521 | Evandro Chagas Institute | Evandro Chagas Institute | Santos, M.C.; Silva, A.M.; Junior, W.D.C.; Barbagelata, L.S.; Ferreira, J.A.; Sousa, E.M.A.; da Silva, P.S.; Pinheiro, K.C.; L.C.; Sousa Junior, E.C. |
| EPI_ISL_918522, EPI_ISL_918523, EPI_ISL_918524, EPI_ISL_918525, EPI_ISL_918526, EPI_ISL_918527, EPI_ISL_918528, EPI_ISL_918529, EPI_ISL_918530 | LACEN - Laboratório Central de Saúde Pública do Para | Evandro Chagas Institute | Santos, M.C.; Silva, A.M.; Junior, W.D.C.; Barbagelata, L.S.; Ferreira, J.A.; Sousa, E.M.A.; da Silva, P.S.; Pinheiro, K.C.; L.C.; Sousa Junior, E.C. |
| EPI_ISL_918531, EPI_ISL_918532, EPI_ISL_918533, EPI_ISL_918534, EPI_ISL_918535 | LACEN - Laboratório Central de Saúde Pública do Amazonas | Evandro Chagas Institute | Santos, M.C.; Silva, A.M.; Junior, W.D.C.; Barbagelata, L.S.; Ferreira, J.A.; Sousa, E.M.A.; da Silva, P.S.; Pinheiro, K.C.; L.C.; Sousa Junior, E.C. |
| EPI_ISL_918537, EPI_ISL_918538, EPI_ISL_918539, EPI_ISL_918540, EPI_ISL_918541, EPI_ISL_918542, EPI_ISL_918543, EPI_ISL_918544 | LACEN - Laboratório Central de Saúde Pública do Ceara | Evandro Chagas Institute | Santos, M.C.; Silva, A.M.; Junior, W.D.C.; Barbagelata, L.S.; Ferreira, J.A.; Sousa, E.M.A.; da Silva, P.S.; Pinheiro, K.C.; L.C.; Sousa Junior, E.C. |

| | | | |
|---|---|---|---|
| EPI_ISL_918545, EPI_ISL_918546, EPI_ISL_918547, EPI_ISL_918548, EPI_ISL_918549, EPI_ISL_918550 | LACEN - Laboratório Central de Saúde Pública do Para | Evandro Chagas Institute | Santos, M.C.; Silva, A.M.; Junior, W.D.C.; Barbagelata, L.S.; Ferreira, J.A.; Sousa, E.M.A.; da Silva, P.S.; Pinheiro, K.C.; L.C.; Sousa Junior, E.C. |
| EPI_ISL_918551 | LACEN - Laboratório Central de Saúde Pública do Amapa | Evandro Chagas Institute | Santos, M.C.; Silva, A.M.; Junior, W.D.C.; Barbagelata, L.S.; Ferreira, J.A.; Sousa, E.M.A.; da Silva, P.S.; Pinheiro, K.C.; L.C.; Sousa Junior, E.C. |
| EPI_ISL_918552 | LACEN - Laboratório Central de Saúde Pública do Para | Evandro Chagas Institute | Santos, M.C.; Silva, A.M.; Junior, W.D.C.; Barbagelata, L.S.; Ferreira, J.A.; Sousa, E.M.A.; da Silva, P.S.; Pinheiro, K.C.; L.C.; Sousa Junior, E.C. |
| EPI_ISL_918553, EPI_ISL_918554, EPI_ISL_918555, EPI_ISL_918556, EPI_ISL_918557, EPI_ISL_918558, EPI_ISL_918559, EPI_ISL_918560, EPI_ISL_918561 | LACEN - Laboratório Central de Saúde Pública do Amapa | Evandro Chagas Institute | Santos, M.C.; Silva, A.M.; Junior, W.D.C.; Barbagelata, L.S.; Ferreira, J.A.; Sousa, E.M.A.; da Silva, P.S.; Pinheiro, K.C.; L.C.; Sousa Junior, E.C. |
| EPI_ISL_925846, EPI_ISL_925916, EPI_ISL_926446 | LACEN - Laboratório Central de Saúde Pública do Amazonas | Evandro Chagas Institute Virology | Santos, M.C.; Silva, A.M.; Junior, W.D.C.; Barbagelata, L.S.; Ferreira, J.A.; Sousa, E.M.A.; da Silva, P.S.; Pinheiro, K.C.; L.C.; Sousa Junior, E.C. |
| EPI_ISL_930854, EPI_ISL_930855, EPI_ISL_930856, EPI_ISL_930857, EPI_ISL_930858 | Central Laboratory of Public Health of Rio Grande do Sul (Lacen-RS) | State Center for Health Surveillance of the Health Department of the State of Rio Grande do Sul (CEVS/SES-RS) | Barcellos R, Campos A, Dornelles C, Godinho F, Gonzalez A, Gregianini T, Molina C, Salvato R, Schaurich A, |
| EPI_ISL_940608 | Laboratório Sao Lucas | Instituto Adolfo Lutz, Interdiciplinary Procedures Center, Strategic Laboratory | Claudio Tavares Sacchi, Claudia Regina Gonçalves, Erica Valessa Ramos Gomes, Karoline Rodrigues Campos |
| EPI_ISL_940609 | Hospital e Maternidade Celso Pierro | Instituto Adolfo Lutz, Interdiciplinary Procedures Center, Strategic Laboratory | Claudio Tavares Sacchi, Claudia Regina Gonçalves, Erica Valessa Ramos Gomes, Karoline Rodrigues Campos |
| EPI_ISL_940610, EPI_ISL_940611 | Hospital Paulistano Paulista | Instituto Adolfo Lutz, Interdiciplinary Procedures Center, Strategic Laboratory | Claudio Tavares Sacchi, Claudia Regina Gonçalves, Erica Valessa Ramos Gomes, Karoline Rodrigues Campos |
| EPI_ISL_940612 | Hospital de Campanha para Enfrentamento do Coronavirus - Goiania | Instituto Adolfo Lutz, Interdiciplinary Procedures Center, Strategic Laboratory | Claudio Tavares Sacchi, Claudia Regina Gonçalves, Erica Valessa Ramos Gomes, Karoline Rodrigues Campos |
| EPI_ISL_940613, EPI_ISL_940614, EPI_ISL_940615, EPI_ISL_940616, EPI_ISL_940617, EPI_ISL_940618 | LACEN-PI DR. Costa Alvarenga | Instituto Adolfo Lutz, Interdiciplinary Procedures Center, Strategic Laboratory | Claudio Tavares Sacchi, Claudia Regina Gonçalves, Erica Valessa Ramos Gomes, Karoline Rodrigues Campos |
| EPI_ISL_940619, EPI_ISL_940620, EPI_ISL_940621, EPI_ISL_940622, EPI_ISL_940623, EPI_ISL_940624, EPI_ISL_940625 | Hospital Sao Joaquim - Beneficiencia Portuguesa | Instituto Adolfo Lutz, Interdiciplinary Procedures Center, Strategic Laboratory | Claudio Tavares Sacchi, Claudia Regina Gonçalves, Erica Valessa Ramos Gomes, Karoline Rodrigues Campos |
| EPI_ISL_940626, EPI_ISL_940627 | Hospital Central Sao Caetano do Sul | Instituto Adolfo Lutz, Interdiciplinary Procedures Center, Strategic Laboratory | Claudio Tavares Sacchi, Claudia Regina Gonçalves, Erica Valessa Ramos Gomes, Karoline Rodrigues Campos |
| EPI_ISL_940628 | Unidade Mista de Iguape | Instituto Adolfo Lutz, Interdiciplinary Procedures Center, Strategic Laboratory | Claudio Tavares Sacchi, Claudia Regina Gonçalves, Erica Valessa Ramos Gomes, Karoline Rodrigues Campos |
| EPI_ISL_940629 | Hospital Municipal Josanias Castanha Braga | Instituto Adolfo Lutz, Interdiciplinary Procedures Center, Strategic Laboratory | Claudio Tavares Sacchi, Claudia Regina Gonçalves, Erica Valessa Ramos Gomes, Karoline Rodrigues Campos |
| EPI_ISL_940630 | Hospital Geral de Sao Paulo | Instituto Adolfo Lutz, Interdiciplinary Procedures Center, Strategic Laboratory | Claudio Tavares Sacchi, Claudia Regina Gonçalves, Erica Valessa Ramos Gomes, Karoline Rodrigues Campos |
| EPI_ISL_942374, EPI_ISL_942375, EPI_ISL_942407, EPI_ISL_942896, EPI_ISL_942897, EPI_ISL_942898, EPI_ISL_942899, EPI_ISL_942930, EPI_ISL_942931 | Central Laboratory of Public Health of Rio Grande do Sul (Lacen-RS) | State Center for Health Surveillance of the Health Department of the State of Rio Grande do Sul (CEVS/SES-RS) | Barcellos R, Campos A, Crescente L, Da Silva A, Dornelles C, Fonseca V, Garay L, Godinho F, Gonzalez A, Gregianini T, Molina C, Salvato R, Schaurich A |
| EPI_ISL_943574, EPI_ISL_943575, EPI_ISL_943576, EPI_ISL_943577, EPI_ISL_943578, EPI_ISL_943579, EPI_ISL_943580, EPI_ISL_943581, EPI_ISL_943582, EPI_ISL_943583, EPI_ISL_943584, EPI_ISL_943585, EPI_ISL_943586, EPI_ISL_943587, EPI_ISL_943588, EPI_ISL_943589, EPI_ISL_943590, EPI_ISL_943591, EPI_ISL_943593, EPI_ISL_943594, EPI_ISL_943595, EPI_ISL_943596, EPI_ISL_943597, EPI_ISL_943598, EPI_ISL_943599, EPI_ISL_943600, EPI_ISL_943601, EPI_ISL_943602, EPI_ISL_943603, EPI_ISL_943604, EPI_ISL_943605, EPI_ISL_943606, EPI_ISL_943607, EPI_ISL_943608, EPI_ISL_943609, EPI_ISL_943610, EPI_ISL_943611, EPI_ISL_943612, EPI_ISL_943613 | | | |
| see above | Central Laboratory of Public Health of Rio Grande do Sul (Lacen-RS) | State Center for Health Surveillance of the Health Department of the State of Rio Grande do Sul (CEVS/SES-RS) | Aline Campos, Amanda da Silva, Anelise Schaurich, Claudia Dornelles, Cynthia Molina, Fernanda Godinho, Lara Crescente, Leticia Garay, Regina Barcellos, Richard Salvato, Tatiana Gregianini, Vagner Fonseca |
| EPI_ISL_943967, EPI_ISL_943968, EPI_ISL_943969, EPI_ISL_943970, EPI_ISL_943971, EPI_ISL_943972 | Hospital Geral de Sao Paulo | Instituto Adolfo Lutz, Interdiciplinary Procedures Center, Strategic Laboratory | Claudio Tavares Sacchi, Claudia Regina Gonçalves, Erica Valessa Ramos Gomes, Karoline Rodrigues Campos |
| EPI_ISL_943973, EPI_ISL_943974, EPI_ISL_943975, EPI_ISL_943976, EPI_ISL_943977, EPI_ISL_943978, EPI_ISL_943979, EPI_ISL_943980, EPI_ISL_943981, EPI_ISL_943982, EPI_ISL_943983, EPI_ISL_943984, EPI_ISL_943985, EPI_ISL_943986, EPI_ISL_943987 | | | |
| see above | LACEN do Estado de Tocantins | Instituto Adolfo Lutz, Interdiciplinary Procedures Center, Strategic Laboratory | Claudio Tavares Sacchi, Claudia Regina Gonçalves, Erica Valessa Ramos Gomes, Karoline Rodrigues Campos |
| EPI_ISL_943988, EPI_ISL_943989, EPI_ISL_943990 | LACEN do Estado de Goias | Instituto Adolfo Lutz, Interdiciplinary Procedures Center, Strategic Laboratory | Claudio Tavares Sacchi, Claudia Regina Gonçalves, Erica Valessa Ramos Gomes, Karoline Rodrigues Campos |
| EPI_ISL_943991 | LACEN do Estado de Tocantins | Instituto Adolfo Lutz, Interdiciplinary Procedures Center, Strategic Laboratory | Claudio Tavares Sacchi, Claudia Regina Gonçalves, Erica Valessa Ramos Gomes, Karoline Rodrigues Campos |
| EPI_ISL_977471 | Instituto Adolfo Lutz - Regional de Presidente Prudente | Instituto Adolfo Lutz, Interdiciplinary Procedures Center, Strategic Laboratory | Claudio Tavares Sacchi, Claudia Regina Gonçalves, Erica Valessa Ramos Gomes, Karoline Rodrigues Campos |
| EPI_ISL_977472, EPI_ISL_977473, EPI_ISL_977474 | Instituto Adolfo Lutz Central | Instituto Adolfo Lutz, Interdiciplinary Procedures Center, Strategic Laboratory | Claudio Tavares Sacchi, Claudia Regina Gonçalves, Erica Valessa Ramos Gomes, Karoline Rodrigues Campos |
| EPI_ISL_977475 | Instituto Adolfo Lutz - Regional de Presidente Prudente | Instituto Adolfo Lutz, Interdiciplinary Procedures Center, Strategic Laboratory | Claudio Tavares Sacchi, Claudia Regina Gonçalves, Erica Valessa Ramos Gomes, Karoline Rodrigues Campos |
| EPI_ISL_977476, EPI_ISL_977477 | Instituto Adolfo Lutz Central | Instituto Adolfo Lutz, Interdiciplinary Procedures Center, Strategic Laboratory | Claudio Tavares Sacchi, Claudia Regina Gonçalves, Erica Valessa Ramos Gomes, Karoline Rodrigues Campos |
| EPI_ISL_977478 | Instituto Adolfo Lutz - Regional de Presidente Prudente | Instituto Adolfo Lutz, Interdiciplinary Procedures Center, Strategic Laboratory | Claudio Tavares Sacchi, Claudia Regina Gonçalves, Erica Valessa Ramos Gomes, Karoline Rodrigues Campos |
| EPI_ISL_977479 | Lab Loc - Itapecerica da Serra | Instituto Adolfo Lutz, Interdiciplinary Procedures Center, Strategic Laboratory | Claudio Tavares Sacchi, Claudia Regina Gonçalves, Erica Valessa Ramos Gomes, Karoline Rodrigues Campos |
| EPI_ISL_977480, EPI_ISL_977481 | Instituto Adolfo Lutz - Regional de Presidente Prudente | Instituto Adolfo Lutz, Interdiciplinary Procedures Center, Strategic Laboratory | Claudio Tavares Sacchi, Claudia Regina Gonçalves, Erica Valessa Ramos Gomes, Karoline Rodrigues Campos |

| | | | |
|---|---|---|---|
| EPI_ISL_977482 | Instituto Adolfo Lutz - Regional de Aracatuba | Instituto Adolfo Lutz, Interdiciplinary Procedures Center, Strategic Laboratory | Claudio Tavares Sacchi, Claudia Regina Gonçalves, Erica Valessa Ramos Gomes, Karoline Rodrigues Campos |
| EPI_ISL_977483, EPI_ISL_977484 | Instituto Adolfo Lutz Central | Instituto Adolfo Lutz, Interdiciplinary Procedures Center, Strategic Laboratory | Claudio Tavares Sacchi, Claudia Regina Gonçalves, Erica Valessa Ramos Gomes, Karoline Rodrigues Campos |
| EPI_ISL_977485 | Instituto Adolfo Lutz - Regional de Presidente Prudente | Instituto Adolfo Lutz, Interdiciplinary Procedures Center, Strategic Laboratory | Claudio Tavares Sacchi, Claudia Regina Gonçalves, Erica Valessa Ramos Gomes, Karoline Rodrigues Campos |
| EPI_ISL_977486 | Instituto Adolfo Lutz - Regional de Santo Andre | Instituto Adolfo Lutz, Interdiciplinary Procedures Center, Strategic Laboratory | Claudio Tavares Sacchi, Claudia Regina Gonçalves, Erica Valessa Ramos Gomes, Karoline Rodrigues Campos |
| EPI_ISL_977487 | Instituto Adolfo Lutz Central | Instituto Adolfo Lutz, Interdiciplinary Procedures Center, Strategic Laboratory | Claudio Tavares Sacchi, Claudia Regina Gonçalves, Erica Valessa Ramos Gomes, Karoline Rodrigues Campos |
| EPI_ISL_977488 | Instituto Adolfo Lutz - Regional de Presidente Prudente | Instituto Adolfo Lutz, Interdiciplinary Procedures Center, Strategic Laboratory | Claudio Tavares Sacchi, Claudia Regina Gonçalves, Erica Valessa Ramos Gomes, Karoline Rodrigues Campos |
| EPI_ISL_977489 | UPA Dr. Akira Tada | Instituto Adolfo Lutz, Interdiciplinary Procedures Center, Strategic Laboratory | Claudio Tavares Sacchi, Claudia Regina Gonçalves, Erica Valessa Ramos Gomes, Karoline Rodrigues Campos |
| EPI_ISL_977490 | Hospital Municipal Guido Guida | Instituto Adolfo Lutz, Interdiciplinary Procedures Center, Strategic Laboratory | Claudio Tavares Sacchi, Claudia Regina Gonçalves, Erica Valessa Ramos Gomes, Karoline Rodrigues Campos |
| EPI_ISL_977500 | LATE - Laboratório de Técnicas Especiais - Hospital Israelita Albert Einstein | LATE - Laboratório de Técnicas Especiais - Hospital Israelita Albert Einstein | Deyvid Amgarten, Fernanda de Mello Malta, Raquel Riyuzo, Ana Paula Moreira Salles, Pedro Henrique Sebe Rodrigues, João Renato Rebello Pinho |
| EPI_ISL_978495, EPI_ISL_978496, EPI_ISL_978497, EPI_ISL_978498, EPI_ISL_978499, EPI_ISL_978500, EPI_ISL_978501, EPI_ISL_978522, EPI_ISL_978528, EPI_ISL_978529, EPI_ISL_978531, EPI_ISL_978532 | | | |
| see above | Central Public Health Laboratory - LACEN -Bahia, Salvador, Brazil | Central Public Health Laboratory - LACEN -Bahia, Salvador, Brazil | Stephane Tosta, Luciana Oliveira, Vanessa Nardy,Patrícia Cajado,Marcela Gómez, Breno Dominguez, Jaqueline Gomes, Vagner Fonseca,Marta Giovanetti,Luiz Alcantara, Felicidade Pereira, Arabela Leal |
| EPI_ISL_981383, EPI_ISL_981385, EPI_ISL_981387 | IAL Regional de Bauru | Instituto Adolfo Lutz, Interdiciplinary Procedures Center, Strategic Laboratory | Claudio Tavares Sacchi, Claudia Regina Gonçalves, Erica Valessa Ramos Gomes, Karoline Rodrigues Campos |
| EPI_ISL_983863, EPI_ISL_983864, EPI_ISL_983865, EPI_ISL_983866, EPI_ISL_983867, EPI_ISL_983868, EPI_ISL_983869 | Central Laboratory of Public Health of Rio Grande do Sul (Lacen-RS) | State Center for Health Surveillance of the Health Department of the State of Rio Grande do Sul (CEVS/SES-RS) | Aline Campos, Cynthia Molina, Lara Crescente, Leticia Garay, Ludmila Fiorenzano Baethgen, Richard Salvato, Tatiana Gregianini |
| EPI_ISL_984242 | Instituto Adolfo Lutz Central | Instituto Adolfo Lutz, Interdiciplinary Procedures Center, Strategic Laboratory | Claudio Tavares Sacchi, Claudia Regina Gonçalves, Erica Valessa Ramos Gomes, Karoline Rodrigues Campos |
| EPI_ISL_984243, EPI_ISL_984244 | Instituto Adolfo Lutz - Regional de Marilia | Instituto Adolfo Lutz, Interdiciplinary Procedures Center, Strategic Laboratory | Claudio Tavares Sacchi, Claudia Regina Gonçalves, Erica Valessa Ramos Gomes, Karoline Rodrigues Campos |
| EPI_ISL_984245, EPI_ISL_984246 | Instituto Adolfo Lutz Central | Instituto Adolfo Lutz, Interdiciplinary Procedures Center, Strategic Laboratory | Claudio Tavares Sacchi, Claudia Regina Gonçalves, Erica Valessa Ramos Gomes, Karoline Rodrigues Campos |
| EPI_ISL_984247, EPI_ISL_984248, EPI_ISL_984249, EPI_ISL_984250, EPI_ISL_984251, EPI_ISL_984252, EPI_ISL_984253, EPI_ISL_984254, EPI_ISL_984255, EPI_ISL_984256, EPI_ISL_984257, EPI_ISL_984258, EPI_ISL_984259, EPI_ISL_984260, EPI_ISL_984261, EPI_ISL_984262 | | | |
| see above | IAL Regional de Marilia | Instituto Adolfo Lutz, Interdiciplinary Procedures Center, Strategic Laboratory | Claudio Tavares Sacchi, Claudia Regina Gonçalves, Erica Valessa Ramos Gomes, Karoline Rodrigues Campos |
| EPI_ISL_984263 | IAL Regional de Bauru | Instituto Adolfo Lutz, Interdiciplinary Procedures Center, Strategic Laboratory | Claudio Tavares Sacchi, Claudia Regina Gonçalves, Erica Valessa Ramos Gomes, Karoline Rodrigues Campos |
| EPI_ISL_984619, EPI_ISL_984620, EPI_ISL_984621 | Central Laboratory of Public Health of Rio Grande do Sul (Lacen-RS) | State Center for Health Surveillance of the Health Department of the State of Rio Grande do Sul (CEVS/SES-RS) | Aline Campos, Cynthia Molina, Lara Crescente, Leticia Garay, Ludmila Fiorenzano Baethgen, Richard Salvato, Tatiana Gregianini |
| EPI_ISL_985170 | Instituto Adolfo Lutz - Regional de Presidente Prudente | Instituto Adolfo Lutz, Interdiciplinary Procedures Center, Strategic Laboratory | Claudio Tavares Sacchi, Claudia Regina Gonçalves, Erica Valessa Ramos Gomes, Karoline Rodrigues Campos |
| EPI_ISL_985171, EPI_ISL_985172, EPI_ISL_985173, EPI_ISL_985174 | Instituto Adolfo Lutz - Regional de Taubate | Instituto Adolfo Lutz, Interdiciplinary Procedures Center, Strategic Laboratory | Claudio Tavares Sacchi, Claudia Regina Gonçalves, Erica Valessa Ramos Gomes, Karoline Rodrigues Campos |
| EPI_ISL_985175, EPI_ISL_985176, EPI_ISL_985177 | Instituto Adolfo Lutz Central | Instituto Adolfo Lutz, Interdiciplinary Procedures Center, Strategic Laboratory | Claudio Tavares Sacchi, Claudia Regina Gonçalves, Erica Valessa Ramos Gomes, Karoline Rodrigues Campos |
| EPI_ISL_985178 | Lab Loc - Itapecerica da Serra | Instituto Adolfo Lutz, Interdiciplinary Procedures Center, Strategic Laboratory | Claudio Tavares Sacchi, Claudia Regina Gonçalves, Erica Valessa Ramos Gomes, Karoline Rodrigues Campos |
| EPI_ISL_985303, EPI_ISL_985304, EPI_ISL_985305, EPI_ISL_985306, EPI_ISL_985307, EPI_ISL_985308, EPI_ISL_985309, EPI_ISL_985310, EPI_ISL_985311, EPI_ISL_985312, EPI_ISL_985313, EPI_ISL_985314, EPI_ISL_985315, EPI_ISL_985316, EPI_ISL_985317 | | | |
| see above | LACEN do Estado de Goias | Instituto Adolfo Lutz, Interdiciplinary Procedures Center, Strategic Laboratory | Claudio Tavares Sacchi, Claudia Regina Gonçalves, Erica Valessa Ramos Gomes, Karoline Rodrigues Campos |
| EPI_ISL_985318, EPI_ISL_985319 | LACEN de Santa Catarina | Instituto Adolfo Lutz, Interdiciplinary Procedures Center, Strategic Laboratory | Claudio Tavares Sacchi, Claudia Regina Gonçalves, Erica Valessa Ramos Gomes, Karoline Rodrigues Campos |

We gratefully acknowledge the following Authors from the Originating laboratories responsible for obtaining the specimens, as well as the Submitting laboratories where the genome data were generated and shared via GISAID, on which this research is based.

All Submitters of data may be contacted directly via www.gisaid.org

Authors are sorted alphabetically.

| Accession ID | Originating Laboratory | Submitting Laboratory | Authors |
|---|---|---|---|
| EPI_ISL_1225963, EPI_ISL_1225964, EPI_ISL_1225965, EPI_ISL_1225966 | "CDPH, Viral and Rickettsial Disease Laboratory" | Genomics and Discovery, Respiratory Viruses Branch, Division of Viral Diseases, Centers for Disease Control and Prevention | Anna Montmayeur; Anna Uehara; Ben L. Rambo-Martin; Clinton R. Paden; Dhwani Batra; Haibin Wang; Jasmine Padilla; Jing Zhang; Justin Lee; Katie Dillon; Krista Queen; Kristen Knipe; Kristine Lacek; Lori Rowe; Mark Burroughs; Matthew Schmerer; Mili Sheth; Peter W. Cook; Rachel Marine; Sam Shepard; Sarah Nobles; Shoshona Le; Suxiang Tong; Yan Li; Ying Tao |
| EPI_ISL_1093547, EPI_ISL_1093548, EPI_ISL_1093550, EPI_ISL_1093576, EPI_ISL_1093577, EPI_ISL_1093578, EPI_ISL_1093579, EPI_ISL_1093580, EPI_ISL_1093581, EPI_ISL_1093582, EPI_ISL_1093583, EPI_ISL_1093584, EPI_ISL_1093585, EPI_ISL_1093586, EPI_ISL_1093587, EPI_ISL_1093588, EPI_ISL_1093589, EPI_ISL_1093590, EPI_ISL_1093591, EPI_ISL_1093592 | | | |
| see above | Alameda County Public Health Department | Alameda County Public Health Department | Kevin Libuit; Kristna Hsieh; Maria Elma |
| EPI_ISL_625522, EPI_ISL_625523, EPI_ISL_625524, EPI_ISL_625525, EPI_ISL_625526, EPI_ISL_625527, EPI_ISL_625528, EPI_ISL_625529, EPI_ISL_625530, EPI_ISL_625531, EPI_ISL_625532, EPI_ISL_625533, EPI_ISL_625534, EPI_ISL_625535, EPI_ISL_625536, EPI_ISL_625537, EPI_ISL_625538, EPI_ISL_625539, EPI_ISL_625540, EPI_ISL_625541, EPI_ISL_625542, EPI_ISL_625543, EPI_ISL_625544, EPI_ISL_625545, EPI_ISL_625546, EPI_ISL_625611, EPI_ISL_625612, EPI_ISL_625613, EPI_ISL_625614, EPI_ISL_625615, EPI_ISL_625616, EPI_ISL_625617, EPI_ISL_625618, EPI_ISL_625619, EPI_ISL_625620, EPI_ISL_625621, EPI_ISL_625622, EPI_ISL_672024, EPI_ISL_672029, EPI_ISL_672036, EPI_ISL_672045, EPI_ISL_672062, EPI_ISL_672076, EPI_ISL_672086, EPI_ISL_672091, EPI_ISL_672113, EPI_ISL_672454, EPI_ISL_672455, EPI_ISL_672456, EPI_ISL_672457, EPI_ISL_672458, EPI_ISL_672459, EPI_ISL_672460, EPI_ISL_672461, EPI_ISL_672462, EPI_ISL_672463, EPI_ISL_672464, EPI_ISL_672465, EPI_ISL_672466, EPI_ISL_672467, EPI_ISL_672468, EPI_ISL_672469, EPI_ISL_672470, EPI_ISL_672471, EPI_ISL_672472, EPI_ISL_672473, EPI_ISL_672474, EPI_ISL_672475, EPI_ISL_672476, EPI_ISL_738510, EPI_ISL_738513, EPI_ISL_738523, EPI_ISL_738527, EPI_ISL_738531, EPI_ISL_738537, EPI_ISL_738538, EPI_ISL_738540, EPI_ISL_738545, EPI_ISL_738547, EPI_ISL_738553, EPI_ISL_738554, EPI_ISL_738561, EPI_ISL_738562, EPI_ISL_738564, EPI_ISL_738572, EPI_ISL_738573, EPI_ISL_738592, EPI_ISL_738604, EPI_ISL_738609, EPI_ISL_738613, EPI_ISL_738615, EPI_ISL_738619, EPI_ISL_738623, EPI_ISL_738625, EPI_ISL_738627, EPI_ISL_738629, EPI_ISL_738633, EPI_ISL_738642, EPI_ISL_738654, EPI_ISL_738674, EPI_ISL_738679, EPI_ISL_738680, EPI_ISL_738692, EPI_ISL_738711, EPI_ISL_738721, EPI_ISL_738743, EPI_ISL_738744, EPI_ISL_738745, EPI_ISL_738748, EPI_ISL_738766, EPI_ISL_738767, EPI_ISL_738774, EPI_ISL_738776, EPI_ISL_738783, EPI_ISL_738792, EPI_ISL_738793, EPI_ISL_738796, EPI_ISL_738797, EPI_ISL_738800, EPI_ISL_738802, EPI_ISL_738813, EPI_ISL_738814, EPI_ISL_738819, EPI_ISL_738825, EPI_ISL_738831, EPI_ISL_738833, EPI_ISL_738835, EPI_ISL_738846, EPI_ISL_738848, EPI_ISL_738868, EPI_ISL_738895, EPI_ISL_738919, EPI_ISL_738925, EPI_ISL_738932, EPI_ISL_738940, EPI_ISL_738963, EPI_ISL_738974, EPI_ISL_738982, EPI_ISL_739023, EPI_ISL_739033, EPI_ISL_739046, EPI_ISL_739049, EPI_ISL_739059, EPI_ISL_739062, EPI_ISL_739073, EPI_ISL_739082, EPI_ISL_739088, EPI_ISL_739090, EPI_ISL_739093, EPI_ISL_739096, EPI_ISL_739109, EPI_ISL_739110, EPI_ISL_739113, EPI_ISL_739115, EPI_ISL_739127, EPI_ISL_739136, EPI_ISL_739137, EPI_ISL_739150, EPI_ISL_739158, EPI_ISL_739163, EPI_ISL_739173, EPI_ISL_739179, EPI_ISL_739202, EPI_ISL_739209, EPI_ISL_739213, EPI_ISL_739215, EPI_ISL_739218, EPI_ISL_739220, EPI_ISL_739226, EPI_ISL_739231, EPI_ISL_739244, EPI_ISL_739261, EPI_ISL_739262, EPI_ISL_739280, EPI_ISL_739283, EPI_ISL_739285, EPI_ISL_739288, EPI_ISL_739300, EPI_ISL_739302, EPI_ISL_739303, EPI_ISL_739308, EPI_ISL_739312, EPI_ISL_739326, EPI_ISL_739328, EPI_ISL_739351, EPI_ISL_739356, EPI_ISL_739361, EPI_ISL_739368, EPI_ISL_739387, EPI_ISL_739406, EPI_ISL_739426, EPI_ISL_739427, EPI_ISL_739444, EPI_ISL_739448, EPI_ISL_739449, EPI_ISL_739470, EPI_ISL_739472, EPI_ISL_739473, EPI_ISL_739483, EPI_ISL_739507, EPI_ISL_739515, EPI_ISL_739518, EPI_ISL_739519, EPI_ISL_739523, EPI_ISL_739526, EPI_ISL_739535, EPI_ISL_739541, EPI_ISL_739555, EPI_ISL_739563, EPI_ISL_739565, EPI_ISL_739572, EPI_ISL_739575, EPI_ISL_739583, EPI_ISL_739604, EPI_ISL_739606, EPI_ISL_739610, EPI_ISL_739616, EPI_ISL_739623, EPI_ISL_739631, EPI_ISL_739634, EPI_ISL_955386, EPI_ISL_955389, EPI_ISL_1027296, EPI_ISL_1027303, EPI_ISL_1027307, EPI_ISL_1027308, EPI_ISL_1027309, EPI_ISL_1027323, EPI_ISL_1027324, EPI_ISL_1027330, EPI_ISL_1027333, EPI_ISL_1027336, EPI_ISL_1027341, EPI_ISL_1027345, EPI_ISL_1027348, EPI_ISL_1027361, EPI_ISL_1027363, EPI_ISL_1027365, EPI_ISL_1027445, EPI_ISL_1027446, EPI_ISL_1027447, EPI_ISL_1027448, EPI_ISL_1027449, EPI_ISL_1027450, EPI_ISL_1027451, EPI_ISL_1027452, EPI_ISL_1027453, EPI_ISL_1027454, EPI_ISL_1027455, EPI_ISL_1027456, EPI_ISL_1027457, EPI_ISL_1027458, EPI_ISL_1027459, EPI_ISL_1027460, EPI_ISL_1027461, EPI_ISL_1027462, EPI_ISL_1027463, EPI_ISL_1027464, EPI_ISL_1027465, EPI_ISL_1027466, EPI_ISL_1027467, EPI_ISL_1027468, EPI_ISL_1027469, EPI_ISL_1027470, EPI_ISL_1027471, EPI_ISL_1027472, EPI_ISL_1027473, EPI_ISL_1027543, EPI_ISL_1137085, EPI_ISL_1137086, EPI_ISL_1137087, EPI_ISL_1137088, EPI_ISL_1137089, EPI_ISL_1184624, EPI_ISL_1184625, EPI_ISL_1184626, EPI_ISL_1184627, EPI_ISL_1184628, EPI_ISL_1184629, EPI_ISL_1184630, EPI_ISL_1184631, EPI_ISL_1184632, EPI_ISL_1184633, EPI_ISL_1184634, EPI_ISL_1184635, EPI_ISL_1184636, EPI_ISL_1184637, EPI_ISL_1184638, EPI_ISL_1184639, EPI_ISL_1184640, EPI_ISL_1184641, EPI_ISL_1184642, EPI_ISL_1184643, EPI_ISL_1184644, EPI_ISL_1184645, EPI_ISL_1184646, EPI_ISL_1184647, EPI_ISL_1184648, EPI_ISL_1184649, EPI_ISL_1184650, EPI_ISL_1184651, EPI_ISL_1184652, EPI_ISL_1184653, EPI_ISL_1184654, EPI_ISL_1184655, EPI_ISL_1184656, EPI_ISL_1184657, EPI_ISL_1184658, EPI_ISL_1184659, EPI_ISL_1184660, EPI_ISL_1184661, EPI_ISL_1184662, EPI_ISL_1184663, EPI_ISL_1184664, EPI_ISL_1184665, EPI_ISL_1184666, EPI_ISL_1184667, EPI_ISL_1184668, EPI_ISL_1184669, EPI_ISL_1184670, EPI_ISL_1184671, EPI_ISL_1184672, EPI_ISL_1184673, EPI_ISL_1184674, EPI_ISL_1184675, EPI_ISL_1184676, EPI_ISL_1184677, EPI_ISL_1184678, EPI_ISL_1184679, EPI_ISL_1184680, EPI_ISL_1184861, EPI_ISL_1184862, EPI_ISL_1184863, EPI_ISL_1184864, EPI_ISL_1184865, EPI_ISL_1184866, EPI_ISL_1184867, EPI_ISL_1184868, EPI_ISL_1184869, EPI_ISL_1184870, EPI_ISL_1184871, EPI_ISL_1184872, EPI_ISL_1184873, EPI_ISL_1184874, EPI_ISL_1184875, EPI_ISL_1184876, EPI_ISL_1184877, EPI_ISL_1184878, EPI_ISL_1184879, EPI_ISL_1184880, EPI_ISL_1184881, EPI_ISL_1184882, EPI_ISL_1184883, EPI_ISL_1184884, EPI_ISL_1184885, EPI_ISL_1184886, EPI_ISL_1184887, EPI_ISL_1184888, EPI_ISL_1184889, EPI_ISL_1184890, EPI_ISL_1184891, EPI_ISL_1184892, EPI_ISL_1184893, EPI_ISL_1184894, EPI_ISL_1184895, EPI_ISL_1184896, EPI_ISL_1184897, EPI_ISL_1184898, EPI_ISL_1184899, EPI_ISL_1184900, EPI_ISL_1184901, EPI_ISL_1184902, EPI_ISL_1184903, EPI_ISL_1184904, EPI_ISL_1184905, EPI_ISL_1184906, EPI_ISL_1184907, EPI_ISL_1184908, EPI_ISL_1184909, EPI_ISL_1184910, EPI_ISL_1184911, EPI_ISL_1184912, EPI_ISL_1184913, EPI_ISL_1184914, EPI_ISL_1184915, EPI_ISL_1184916, EPI_ISL_1184917, EPI_ISL_1184918, EPI_ISL_1184919, EPI_ISL_1184920, EPI_ISL_1184921, EPI_ISL_1184922, EPI_ISL_1234542, EPI_ISL_1234543, EPI_ISL_1234562, EPI_ISL_1234564, EPI_ISL_1234565, EPI_ISL_1234566, EPI_ISL_1234567, EPI_ISL_1234568, EPI_ISL_1234574, EPI_ISL_1234597, EPI_ISL_1234600, EPI_ISL_1234603, EPI_ISL_1234604, EPI_ISL_1234611, EPI_ISL_1234612, EPI_ISL_1234617, EPI_ISL_1234621, EPI_ISL_1234637, EPI_ISL_1234640, EPI_ISL_1234646, EPI_ISL_1234648, EPI_ISL_1234661, EPI_ISL_1234665, EPI_ISL_1234666, EPI_ISL_1234670, EPI_ISL_1234672, EPI_ISL_1234676, EPI_ISL_1234677, EPI_ISL_1234679, EPI_ISL_1234683, EPI_ISL_1234698, EPI_ISL_1234701, EPI_ISL_1234716, EPI_ISL_1234718, EPI_ISL_1234720, EPI_ISL_1234723, EPI_ISL_1234732, EPI_ISL_1234739, EPI_ISL_1234742, EPI_ISL_1234743, EPI_ISL_1234751, EPI_ISL_1234755, EPI_ISL_1234758, EPI_ISL_1234762, EPI_ISL_1234765, EPI_ISL_1234767, EPI_ISL_1234773, EPI_ISL_1234776, EPI_ISL_1234779, EPI_ISL_1234782, EPI_ISL_1234785, EPI_ISL_1234790, EPI_ISL_1234795, EPI_ISL_1234802, EPI_ISL_1234808, EPI_ISL_1234809, EPI_ISL_1234815, EPI_ISL_1234819, EPI_ISL_1234820, EPI_ISL_1234827, EPI_ISL_1234829, EPI_ISL_1234830, EPI_ISL_1234832, EPI_ISL_1234834, EPI_ISL_1234835, EPI_ISL_1234839, EPI_ISL_1234844, EPI_ISL_1234848, EPI_ISL_1234858, EPI_ISL_1234860, EPI_ISL_1234862, EPI_ISL_1234870, EPI_ISL_1234872, EPI_ISL_1234873, EPI_ISL_1234877, EPI_ISL_1234892, EPI_ISL_1234894, EPI_ISL_1234895, EPI_ISL_1234900, EPI_ISL_1234901, EPI_ISL_1234904, EPI_ISL_1234907, EPI_ISL_1234920, EPI_ISL_1234921, EPI_ISL_1234924, EPI_ISL_1234932, EPI_ISL_1234933, EPI_ISL_1234939, EPI_ISL_1234941, EPI_ISL_1234950, EPI_ISL_1234952, EPI_ISL_1234953, EPI_ISL_1234956, EPI_ISL_1234959, EPI_ISL_1234968, EPI_ISL_1234969, EPI_ISL_1234979, EPI_ISL_1234982, EPI_ISL_1234986, EPI_ISL_1234987, EPI_ISL_1234990, EPI_ISL_1234995, EPI_ISL_1235006, EPI_ISL_1235013, EPI_ISL_1235021, EPI_ISL_1235022, EPI_ISL_1235025, EPI_ISL_1235028, EPI_ISL_1235032, EPI_ISL_1235042, EPI_ISL_1235048, EPI_ISL_1235054, EPI_ISL_1235059, EPI_ISL_1235075, EPI_ISL_1235077, EPI_ISL_1235083, EPI_ISL_1235091, EPI_ISL_1235093, EPI_ISL_1235094, EPI_ISL_1235098, EPI_ISL_1235099, EPI_ISL_1235102, EPI_ISL_1235111, EPI_ISL_1235115, EPI_ISL_1235116, EPI_ISL_1235122, EPI_ISL_1235124, EPI_ISL_1235141, EPI_ISL_1235142, EPI_ISL_1235147, EPI_ISL_1235149, EPI_ISL_1235158, EPI_ISL_1235165, EPI_ISL_1235171, EPI_ISL_1235175, EPI_ISL_1235178, EPI_ISL_1235181, EPI_ISL_1235183, EPI_ISL_1235185, EPI_ISL_1235188, EPI_ISL_1235194, EPI_ISL_1235198, EPI_ISL_1235210, EPI_ISL_1235213, EPI_ISL_1235217, EPI_ISL_1235219, EPI_ISL_1235223, EPI_ISL_1235230, EPI_ISL_1235235, EPI_ISL_1235241, EPI_ISL_1235243, EPI_ISL_1235244, EPI_ISL_1235249, EPI_ISL_1235255, EPI_ISL_1235256, EPI_ISL_1235263, EPI_ISL_1235270, EPI_ISL_1235276, EPI_ISL_1235279, EPI_ISL_1235281, EPI_ISL_1235289, EPI_ISL_1235292, EPI_ISL_1235294, EPI_ISL_1235299, EPI_ISL_1235303, EPI_ISL_1235308, EPI_ISL_1235309, EPI_ISL_1235312, EPI_ISL_1235313, EPI_ISL_1235314, EPI_ISL_1235318, EPI_ISL_1235335, EPI_ISL_1235338, EPI_ISL_1235339, EPI_ISL_1235359, EPI_ISL_1235368, EPI_ISL_1235369, EPI_ISL_1235380, EPI_ISL_1235387, EPI_ISL_1235388, EPI_ISL_1235391, EPI_ISL_1235392, EPI_ISL_1235404, EPI_ISL_1235405, EPI_ISL_1235406, EPI_ISL_1235407, EPI_ISL_1235414, EPI_ISL_1235440, EPI_ISL_1235448, EPI_ISL_1235459, EPI_ISL_1235471, EPI_ISL_1235477, EPI_ISL_1235480, EPI_ISL_1235482, EPI_ISL_1235485, EPI_ISL_1235486, EPI_ISL_1235492, EPI_ISL_1235495, EPI_ISL_1235497, EPI_ISL_1235499, EPI_ISL_1235503, EPI_ISL_1235504, EPI_ISL_1235505, EPI_ISL_1235512, EPI_ISL_1235516, EPI_ISL_1235520, EPI_ISL_1235524, EPI_ISL_1235529, EPI_ISL_1235532, EPI_ISL_1235537, EPI_ISL_1235543, EPI_ISL_1235550, EPI_ISL_1235552, EPI_ISL_1235554, EPI_ISL_1235557, EPI_ISL_1235559, EPI_ISL_1235562, EPI_ISL_1235564, EPI_ISL_1235566, EPI_ISL_1235567, EPI_ISL_1235573, EPI_ISL_1235574, EPI_ISL_1235575, EPI_ISL_1235579, EPI_ISL_1235580, EPI_ISL_1235586, EPI_ISL_1235589, EPI_ISL_1235590, EPI_ISL_1235592, EPI_ISL_1235597, EPI_ISL_1235598, EPI_ISL_1235599, EPI_ISL_1235605, EPI_ISL_1235610, EPI_ISL_1477170, EPI_ISL_1477175, EPI_ISL_1477178, EPI_ISL_1477199, EPI_ISL_1477216, EPI_ISL_1477228, EPI_ISL_1477243, EPI_ISL_1477244, EPI_ISL_1477251, EPI_ISL_1477252, EPI_ISL_1477259, EPI_ISL_1477260, EPI_ISL_1477262, EPI_ISL_1477268, EPI_ISL_1477269, EPI_ISL_1477270, EPI_ISL_1477271, EPI_ISL_1477279, EPI_ISL_1477287, EPI_ISL_1477288, EPI_ISL_1477292, EPI_ISL_1477293, EPI_ISL_1477295, EPI_ISL_1477301, EPI_ISL_1477308, EPI_ISL_1477309, EPI_ISL_1477313, EPI_ISL_1477318, EPI_ISL_1477322, EPI_ISL_1477328, EPI_ISL_1477329, EPI_ISL_1477335, EPI_ISL_1477337, EPI_ISL_1477343, EPI_ISL_1477352, EPI_ISL_1477359, EPI_ISL_1477364, EPI_ISL_1477367, EPI_ISL_1477371, EPI_ISL_1477373, EPI_ISL_1477377, EPI_ISL_1477380, EPI_ISL_1477382, EPI_ISL_1477385, EPI_ISL_1477388, EPI_ISL_1477405, EPI_ISL_1477406, EPI_ISL_1477410, EPI_ISL_1477411, EPI_ISL_1477413, EPI_ISL_1477414, EPI_ISL_1477415, EPI_ISL_1477432, EPI_ISL_1477433, EPI_ISL_1477435, EPI_ISL_1477446, EPI_ISL_1477447, EPI_ISL_1477462, EPI_ISL_1477466, EPI_ISL_1477468, EPI_ISL_1477483, EPI_ISL_1477487, EPI_ISL_1477489, EPI_ISL_1477491, EPI_ISL_1477497, EPI_ISL_1477499, EPI_ISL_1477520, EPI_ISL_1477545, EPI_ISL_1477546, EPI_ISL_1477547, EPI_ISL_1477554, EPI_ISL_1477557, EPI_ISL_1477565, EPI_ISL_1477569, EPI_ISL_1477584, EPI_ISL_1477594, EPI_ISL_1477595, EPI_ISL_1477605, EPI_ISL_1477617, EPI_ISL_1477627, EPI_ISL_1477630, EPI_ISL_1477632, EPI_ISL_1477639, EPI_ISL_1477647, EPI_ISL_1477649, EPI_ISL_1477657, EPI_ISL_1477668, EPI_ISL_1477669, EPI_ISL_1477671, EPI_ISL_1477672, EPI_ISL_1477674, EPI_ISL_1477683, EPI_ISL_1477684, EPI_ISL_1477686, EPI_ISL_1477688, EPI_ISL_1477691, EPI_ISL_1477696, EPI_ISL_1477701, EPI_ISL_1477702, EPI_ISL_1477703, EPI_ISL_1477708, EPI_ISL_1477710, EPI_ISL_1477712, EPI_ISL_1477720, EPI_ISL_1477733, EPI_ISL_1477735, EPI_ISL_1477741, EPI_ISL_1477747, EPI_ISL_1477751, EPI_ISL_1477753, EPI_ISL_1477754, EPI_ISL_1477757, EPI_ISL_1477758, EPI_ISL_1477766, EPI_ISL_1477771, EPI_ISL_1477772, EPI_ISL_1477773, EPI_ISL_1477775, EPI_ISL_1477779, EPI_ISL_1477783, EPI_ISL_1477784, EPI_ISL_1477791, EPI_ISL_1477801, EPI_ISL_1477804, EPI_ISL_1477805, EPI_ISL_1477807, EPI_ISL_1477809, EPI_ISL_1477820, EPI_ISL_1477823, EPI_ISL_1477824, EPI_ISL_1477825, EPI_ISL_1477828, EPI_ISL_1477832, EPI_ISL_1477841, EPI_ISL_1477847, EPI_ISL_1477857, EPI_ISL_1477858, EPI_ISL_1477862, EPI_ISL_1477865, EPI_ISL_1477880, EPI_ISL_1477886, EPI_ISL_1477887, EPI_ISL_1477889, EPI_ISL_1477896, EPI_ISL_1477899, EPI_ISL_1477900, EPI_ISL_1477904, EPI_ISL_1477905, EPI_ISL_1477906, EPI_ISL_1477914, EPI_ISL_1477915, EPI_ISL_1477919, EPI_ISL_1477922, EPI_ISL_1477928, EPI_ISL_1477932, EPI_ISL_1477933, EPI_ISL_1477936, EPI_ISL_1477941, EPI_ISL_1477942, EPI_ISL_1477945, EPI_ISL_1477948, EPI_ISL_1663927, EPI_ISL_1663928, EPI_ISL_1663935, EPI_ISL_1663938, EPI_ISL_1663947, EPI_ISL_1663956, EPI_ISL_1663958, EPI_ISL_1663959, EPI_ISL_1663960, EPI_ISL_1663969, EPI_ISL_1663972, EPI_ISL_1663986, EPI_ISL_1663990, EPI_ISL_1664003, EPI_ISL_1664006, EPI_ISL_1664010, EPI_ISL_1664015, EPI_ISL_1664021, EPI_ISL_1664037, EPI_ISL_1664039, EPI_ISL_1664040, EPI_ISL_1664051, EPI_ISL_1664052, EPI_ISL_1664056, EPI_ISL_1664065, EPI_ISL_1664090, EPI_ISL_1664101, EPI_ISL_1664106, EPI_ISL_1701046, EPI_ISL_1701057, EPI_ISL_1701065, EPI_ISL_1701071, EPI_ISL_1701072, EPI_ISL_1701074, EPI_ISL_1701078, EPI_ISL_1701080, EPI_ISL_1701081, EPI_ISL_1701082, EPI_ISL_1701083, EPI_ISL_1701084, EPI_ISL_1701087, EPI_ISL_1701090, EPI_ISL_1701091, EPI_ISL_1701092, EPI_ISL_1701099, EPI_ISL_1701104, EPI_ISL_1701106, EPI_ISL_1701115, EPI_ISL_1701119, EPI_ISL_1701127, EPI_ISL_1701135, EPI_ISL_1701137, EPI_ISL_1701147, EPI_ISL_1701148, EPI_ISL_1701150, EPI_ISL_1701155, EPI_ISL_1701156, EPI_ISL_1701157, EPI_ISL_1701161, EPI_ISL_1701162, EPI_ISL_1701167, EPI_ISL_1701171, EPI_ISL_1701172, EPI_ISL_1701174, EPI_ISL_1701175, EPI_ISL_1701176, EPI_ISL_1701177, EPI_ISL_1701178, EPI_ISL_1701179, EPI_ISL_1701180, EPI_ISL_1701194, EPI_ISL_1701205, EPI_ISL_1701210, EPI_ISL_1701212, EPI_ISL_1701214, EPI_ISL_1701217, EPI_ISL_1701218, EPI_ISL_1701222, EPI_ISL_1701228, EPI_ISL_1701234, EPI_ISL_1701249, EPI_ISL_1701250, EPI_ISL_1701267, EPI_ISL_1701273, EPI_ISL_1701274, EPI_ISL_1701277, EPI_ISL_1701285, EPI_ISL_1701289, EPI_ISL_1701291, EPI_ISL_1701293, EPI_ISL_1701295, EPI_ISL_1701299, EPI_ISL_1701301, EPI_ISL_1701303, EPI_ISL_1701306, EPI_ISL_1701307, EPI_ISL_1701409, EPI_ISL_1701415, EPI_ISL_1701417, EPI_ISL_1701418, EPI_ISL_1701421, EPI_ISL_1701424, EPI_ISL_1701432, EPI_ISL_1701434, EPI_ISL_1701442, EPI_ISL_1701449, EPI_ISL_1701452, EPI_ISL_1701470, EPI_ISL_1701485, EPI_ISL_1701506, EPI_ISL_1701512, EPI_ISL_1701524, EPI_ISL_1701529, EPI_ISL_1701535, EPI_ISL_1701546, EPI_ISL_1701552, EPI_ISL_1701553, EPI_ISL_1701556, EPI_ISL_1701576, EPI_ISL_1701592, EPI_ISL_1701610, EPI_ISL_1701622, EPI_ISL_1701632, EPI_ISL_1701633, EPI_ISL_1701635, EPI_ISL_1701642, EPI_ISL_1701650, EPI_ISL_1701651, EPI_ISL_1701652 | | | |
| see above | Alameda County Public Health Lab | Chan-Zuckerberg Biohub | CZB Cliahub Consortium |
| EPI_ISL_884866, EPI_ISL_884867, EPI_ISL_884868, EPI_ISL_884869, EPI_ISL_884870, EPI_ISL_1184809, EPI_ISL_1184810, EPI_ISL_1184811, EPI_ISL_1184812, EPI_ISL_1184813, EPI_ISL_1184817, EPI_ISL_1185289, EPI_ISL_1185290, EPI_ISL_1185293, EPI_ISL_1185295, EPI_ISL_1185296, EPI_ISL_1185297, EPI_ISL_1185299, EPI_ISL_1185303, EPI_ISL_1185307, EPI_ISL_1185310, EPI_ISL_1185311, EPI_ISL_1185314, EPI_ISL_1185316, EPI_ISL_1185317, EPI_ISL_1185320, EPI_ISL_1185321, EPI_ISL_1185327, EPI_ISL_1185330, EPI_ISL_1185331, EPI_ISL_1185332, EPI_ISL_1185333, EPI_ISL_1185334, EPI_ISL_1185335, EPI_ISL_1185336, EPI_ISL_1185337, EPI_ISL_1185338, EPI_ISL_1185339, EPI_ISL_1185341, EPI_ISL_1185342, EPI_ISL_1185343, EPI_ISL_1185344, EPI_ISL_1185347, EPI_ISL_1185348, EPI_ISL_1185349, EPI_ISL_1185350, EPI_ISL_1185354, EPI_ISL_1185366, EPI_ISL_1185370, EPI_ISL_1185371, EPI_ISL_1185373, EPI_ISL_1185374, EPI_ISL_1185375, EPI_ISL_1185377, EPI_ISL_1185378, EPI_ISL_1185379, EPI_ISL_1185380, EPI_ISL_1185381, EPI_ISL_1185384, EPI_ISL_1185386, EPI_ISL_1185389, EPI_ISL_1185390, EPI_ISL_1185391, EPI_ISL_1185392, EPI_ISL_1185396, EPI_ISL_1185397, EPI_ISL_1185398, EPI_ISL_1185400, EPI_ISL_1185402, EPI_ISL_1185403, EPI_ISL_1185404, EPI_ISL_1185405, EPI_ISL_1185406, EPI_ISL_1185411, EPI_ISL_1185415, EPI_ISL_1185416, EPI_ISL_1185418, EPI_ISL_1185419, EPI_ISL_1185420, EPI_ISL_1185421, EPI_ISL_1185423, EPI_ISL_1185426, EPI_ISL_1185429, EPI_ISL_1185433, EPI_ISL_1185434, EPI_ISL_1185435, EPI_ISL_1185437, EPI_ISL_1185441, EPI_ISL_1185442, EPI_ISL_1185444, EPI_ISL_1185445, EPI_ISL_1185446, EPI_ISL_1185450, EPI_ISL_1185451, EPI_ISL_1185453, EPI_ISL_1185458, EPI_ISL_1185462, EPI_ISL_1185464, EPI_ISL_1185465, EPI_ISL_1185467, EPI_ISL_1185470, EPI_ISL_1185472, EPI_ISL_1185473, EPI_ISL_1185477, EPI_ISL_1185478, EPI_ISL_1185482, EPI_ISL_1185483, EPI_ISL_1185484, EPI_ISL_1185485, EPI_ISL_1185490, EPI_ISL_1185492, EPI_ISL_1185494, EPI_ISL_1185495, EPI_ISL_1185500, EPI_ISL_1185506, EPI_ISL_1185508, EPI_ISL_1185510, EPI_ISL_1185511, EPI_ISL_1185512, EPI_ISL_1185513, EPI_ISL_1185514, EPI_ISL_1185515, EPI_ISL_1185520, EPI_ISL_1185522, EPI_ISL_1185523, EPI_ISL_1185524, EPI_ISL_1185525, EPI_ISL_1185527, EPI_ISL_1185530, EPI_ISL_1185531, EPI_ISL_1185532, EPI_ISL_1924583, EPI_ISL_1924584, EPI_ISL_1924585, EPI_ISL_1924586, EPI_ISL_1924587, EPI_ISL_1924588, EPI_ISL_1924589, EPI_ISL_1924591, EPI_ISL_1924594, EPI_ISL_1924595, EPI_ISL_1924597, EPI_ISL_1924598, EPI_ISL_1924599, EPI_ISL_1924601, EPI_ISL_1924603, EPI_ISL_1924606, EPI_ISL_1924607, EPI_ISL_1924608, EPI_ISL_1924609, EPI_ISL_1924610, EPI_ISL_1924611, EPI_ISL_1924612, EPI_ISL_1924613, EPI_ISL_1924614, EPI_ISL_1924615, EPI_ISL_1924617, EPI_ISL_1924620, EPI_ISL_1924621, EPI_ISL_1924622, EPI_ISL_1924623, EPI_ISL_1924624, EPI_ISL_1924625, EPI_ISL_1924626, EPI_ISL_1924627, EPI_ISL_1924628 | | | |
| see above | CA DPH Viral and Rickettsial Disease Laboratory | Chan-Zuckerberg Biohub | CZB Cliahub Consortium |
| EPI_ISL_754914, EPI_ISL_754915, EPI_ISL_754916, EPI_ISL_754917, EPI_ISL_754918, EPI_ISL_754919, EPI_ISL_754921, EPI_ISL_754922, EPI_ISL_754923, EPI_ISL_754924, EPI_ISL_754925, EPI_ISL_754926, EPI_ISL_754928, EPI_ISL_754929, EPI_ISL_754931, EPI_ISL_754934, EPI_ISL_754936, EPI_ISL_754937, EPI_ISL_754938, EPI_ISL_754939, EPI_ISL_754940, EPI_ISL_754941, EPI_ISL_754942, EPI_ISL_754943, EPI_ISL_754948, EPI_ISL_754949, EPI_ISL_754951, EPI_ISL_754952, EPI_ISL_754954, EPI_ISL_754955, EPI_ISL_754957, EPI_ISL_754958, EPI_ISL_754959, EPI_ISL_754960, EPI_ISL_754963, EPI_ISL_754964, EPI_ISL_754965, EPI_ISL_754966, EPI_ISL_754967, EPI_ISL_754971, EPI_ISL_754972, EPI_ISL_754973, EPI_ISL_754974, EPI_ISL_754975, EPI_ISL_754978, EPI_ISL_754979, EPI_ISL_754981, EPI_ISL_754982, EPI_ISL_754983, EPI_ISL_754985, EPI_ISL_754987, EPI_ISL_754989, EPI_ISL_754990, EPI_ISL_754993, EPI_ISL_754994, EPI_ISL_754995, EPI_ISL_754997, EPI_ISL_754999, EPI_ISL_755000, EPI_ISL_755001, EPI_ISL_755002, EPI_ISL_755003, EPI_ISL_755005, EPI_ISL_755007, EPI_ISL_755008, EPI_ISL_755009, EPI_ISL_755010, EPI_ISL_755011, EPI_ISL_755012, EPI_ISL_755013, EPI_ISL_755014, EPI_ISL_755015, EPI_ISL_755016, EPI_ISL_755017, EPI_ISL_755020, EPI_ISL_755021, EPI_ISL_755023, EPI_ISL_755024, EPI_ISL_755026, EPI_ISL_755027, EPI_ISL_755029, EPI_ISL_755031, EPI_ISL_755032, EPI_ISL_755044, EPI_ISL_755045, EPI_ISL_755046, EPI_ISL_755047, EPI_ISL_755048, EPI_ISL_755049, EPI_ISL_755050, EPI_ISL_755051, EPI_ISL_755052, EPI_ISL_755053, EPI_ISL_755057, EPI_ISL_755058, EPI_ISL_755059, EPI_ISL_755062, EPI_ISL_755063, EPI_ISL_755064, EPI_ISL_755065, EPI_ISL_755066, EPI_ISL_755067, EPI_ISL_755068, EPI_ISL_755069, EPI_ISL_755070, EPI_ISL_755071, EPI_ISL_755072, EPI_ISL_755073, EPI_ISL_755074, EPI_ISL_755075, EPI_ISL_755076, EPI_ISL_755077, EPI_ISL_755078, EPI_ISL_755079, EPI_ISL_755080, EPI_ISL_755082, EPI_ISL_755083, EPI_ISL_755084, EPI_ISL_755085, EPI_ISL_755086, EPI_ISL_755087, EPI_ISL_755088, EPI_ISL_755089, EPI_ISL_755090, EPI_ISL_755091, EPI_ISL_755092, EPI_ISL_755093, EPI_ISL_755094, EPI_ISL_755095, EPI_ISL_755096, EPI_ISL_755097, EPI_ISL_755098, EPI_ISL_755099, EPI_ISL_755100, EPI_ISL_755101, EPI_ISL_755102, EPI_ISL_755103, EPI_ISL_755104, EPI_ISL_755105, EPI_ISL_755106, EPI_ISL_755107, EPI_ISL_755108, EPI_ISL_755109, EPI_ISL_755110, EPI_ISL_755111, EPI_ISL_755112, EPI_ISL_755113, EPI_ISL_755114, EPI_ISL_824390, EPI_ISL_824391, EPI_ISL_824392, EPI_ISL_824393, EPI_ISL_824394, EPI_ISL_824395, EPI_ISL_824396, EPI_ISL_824397, EPI_ISL_824398, EPI_ISL_824399, EPI_ISL_824400, EPI_ISL_824401, EPI_ISL_824402, EPI_ISL_824403, EPI_ISL_824404, EPI_ISL_824405, EPI_ISL_824406 | | | |
| see above | California Department of Public Health | California Department of Public Health | CDPH IDLB COVIDNet |
| EPI_ISL_755940, EPI_ISL_755941, EPI_ISL_755942, EPI_ISL_847516, EPI_ISL_847518, EPI_ISL_847519, EPI_ISL_847520, EPI_ISL_847521, EPI_ISL_847522, EPI_ISL_847523, EPI_ISL_847524, EPI_ISL_847525, EPI_ISL_847526, EPI_ISL_847527, EPI_ISL_847528, EPI_ISL_847529, EPI_ISL_847530, EPI_ISL_847531, EPI_ISL_847532, EPI_ISL_847533, EPI_ISL_847534, EPI_ISL_847535, EPI_ISL_847537, EPI_ISL_847538, EPI_ISL_847539, EPI_ISL_847541, EPI_ISL_847542, EPI_ISL_847543, EPI_ISL_847544, EPI_ISL_847547, EPI_ISL_847548, EPI_ISL_847549, EPI_ISL_847550, EPI_ISL_847551, EPI_ISL_847552, EPI_ISL_847554, EPI_ISL_847555, EPI_ISL_847556, EPI_ISL_847557, EPI_ISL_847558, EPI_ISL_847559, EPI_ISL_847560, EPI_ISL_847561, EPI_ISL_847562, EPI_ISL_847565, EPI_ISL_847566, EPI_ISL_847567, EPI_ISL_847569, EPI_ISL_847570, EPI_ISL_847571, EPI_ISL_847573, EPI_ISL_847575, EPI_ISL_847576, EPI_ISL_847577, EPI_ISL_847578, EPI_ISL_847580, EPI_ISL_847581, EPI_ISL_847582, EPI_ISL_847583, EPI_ISL_847584, EPI_ISL_847585, EPI_ISL_847586, EPI_ISL_847587, EPI_ISL_847588, EPI_ISL_847604, EPI_ISL_847605, EPI_ISL_847606, EPI_ISL_847607, EPI_ISL_847608, EPI_ISL_847609, EPI_ISL_847610, EPI_ISL_847611, EPI_ISL_847612, EPI_ISL_847613, EPI_ISL_847614, EPI_ISL_847615, EPI_ISL_847616, EPI_ISL_847617, EPI_ISL_847618, EPI_ISL_847623, EPI_ISL_847624, EPI_ISL_847625, EPI_ISL_847626, EPI_ISL_847627, EPI_ISL_847628, EPI_ISL_847629, EPI_ISL_847630, EPI_ISL_847631, EPI_ISL_847632, EPI_ISL_847633, EPI_ISL_847634, EPI_ISL_847635, EPI_ISL_847636, EPI_ISL_847637, EPI_ISL_847638, EPI_ISL_847639, EPI_ISL_847645, EPI_ISL_847646, EPI_ISL_847647, EPI_ISL_847648, EPI_ISL_847649, EPI_ISL_847650, EPI_ISL_847651, EPI_ISL_847652, EPI_ISL_847653, EPI_ISL_847654, EPI_ISL_847655, EPI_ISL_847656, EPI_ISL_847657, EPI_ISL_847658, EPI_ISL_847659, EPI_ISL_847660, EPI_ISL_847661, EPI_ISL_847662, EPI_ISL_847663, EPI_ISL_847664, EPI_ISL_847665, EPI_ISL_847666, EPI_ISL_847667, EPI_ISL_847668, EPI_ISL_847669, EPI_ISL_847670, EPI_ISL_847671, EPI_ISL_847672, EPI_ISL_847673, EPI_ISL_847674, EPI_ISL_847675, EPI_ISL_847676, EPI_ISL_847677, EPI_ISL_847678, EPI_ISL_847679, EPI_ISL_847680, EPI_ISL_847681, EPI_ISL_847682, EPI_ISL_847683, EPI_ISL_847684, EPI_ISL_847685, EPI_ISL_847686, EPI_ISL_847687, EPI_ISL_847688, EPI_ISL_847689, EPI_ISL_847690, EPI_ISL_847691, EPI_ISL_847692, EPI_ISL_847693, EPI_ISL_847694, EPI_ISL_847695, EPI_ISL_847696, EPI_ISL_847697, EPI_ISL_847698, EPI_ISL_847699, EPI_ISL_847700, EPI_ISL_847701, EPI_ISL_847702, EPI_ISL_847703, EPI_ISL_847704, EPI_ISL_847705, EPI_ISL_847706, EPI_ISL_847707, EPI_ISL_847708, EPI_ISL_847709, EPI_ISL_847710, EPI_ISL_847711, EPI_ISL_847712, EPI_ISL_847713, EPI_ISL_847714, EPI_ISL_847715, EPI_ISL_847717, EPI_ISL_847718, EPI_ISL_847719, EPI_ISL_847721, EPI_ISL_847722, EPI_ISL_847723, EPI_ISL_847724, EPI_ISL_847725, EPI_ISL_847726, EPI_ISL_847727, EPI_ISL_847728, EPI_ISL_847729, EPI_ISL_847732, EPI_ISL_847733, EPI_ISL_847734, EPI_ISL_847735, EPI_ISL_847737, EPI_ISL_847739, EPI_ISL_847740, EPI_ISL_847745, EPI_ISL_847748, EPI_ISL_847749, EPI_ISL_847750, EPI_ISL_847751, EPI_ISL_847752, EPI_ISL_847753, EPI_ISL_847754, EPI_ISL_847755, EPI_ISL_847769, EPI_ISL_847770, EPI_ISL_847771, EPI_ISL_847772, EPI_ISL_847773, EPI_ISL_847774, EPI_ISL_847775, EPI_ISL_847776, EPI_ISL_847777, EPI_ISL_847778, EPI_ISL_847779, EPI_ISL_847780, EPI_ISL_847781, EPI_ISL_847783, EPI_ISL_847784, EPI_ISL_847785, EPI_ISL_847786, EPI_ISL_847787, EPI_ISL_847788, EPI_ISL_847789, EPI_ISL_847790, EPI_ISL_847791, EPI_ISL_847792, EPI_ISL_847793, EPI_ISL_847794, EPI_ISL_847795, EPI_ISL_847796, EPI_ISL_847797, EPI_ISL_847798, EPI_ISL_847799, EPI_ISL_847800, EPI_ISL_847801, EPI_ISL_847802, EPI_ISL_847803, EPI_ISL_847804, EPI_ISL_847805, EPI_ISL_847806, EPI_ISL_847807, EPI_ISL_847808, EPI_ISL_847809, EPI_ISL_847810, EPI_ISL_847811, EPI_ISL_847812, EPI_ISL_847813, EPI_ISL_847814, EPI_ISL_847815, EPI_ISL_847816, EPI_ISL_847817, EPI_ISL_847818, EPI_ISL_855389, EPI_ISL_855390, EPI_ISL_855391, EPI_ISL_855392, EPI_ISL_855395, EPI_ISL_855396, EPI_ISL_855397, EPI_ISL_855398, EPI_ISL_977721, EPI_ISL_977722, EPI_ISL_977723, EPI_ISL_977724, EPI_ISL_977725, EPI_ISL_977726, EPI_ISL_977727, EPI_ISL_977728, EPI_ISL_977729, EPI_ISL_977730, EPI_ISL_977731, EPI_ISL_977732, EPI_ISL_977733, EPI_ISL_977734, EPI_ISL_977735, EPI_ISL_977736, EPI_ISL_977737, EPI_ISL_977738, EPI_ISL_977739, EPI_ISL_977740, EPI_ISL_977741, EPI_ISL_977742, EPI_ISL_977745, EPI_ISL_977746, EPI_ISL_977747, EPI_ISL_977748, EPI_ISL_977749, EPI_ISL_977750, EPI_ISL_977751, EPI_ISL_977752, EPI_ISL_977753, EPI_ISL_977754, EPI_ISL_977755, EPI_ISL_977757, EPI_ISL_977758, EPI_ISL_977759, EPI_ISL_977760, EPI_ISL_977761, EPI_ISL_977762, EPI_ISL_977763, EPI_ISL_977764, EPI_ISL_977765, EPI_ISL_977766, EPI_ISL_977767, EPI_ISL_977768, EPI_ISL_977769, EPI_ISL_977770, EPI_ISL_977771, EPI_ISL_977772, EPI_ISL_977773, EPI_ISL_977774, EPI_ISL_977775, EPI_ISL_977776, EPI_ISL_977777, EPI_ISL_977778, EPI_ISL_977779, EPI_ISL_977780, EPI_ISL_977781, EPI_ISL_977782, EPI_ISL_977783, EPI_ISL_977784, EPI_ISL_977785, EPI_ISL_977786, EPI_ISL_977787, EPI_ISL_977788, EPI_ISL_977789, EPI_ISL_977790, EPI_ISL_977791, EPI_ISL_977792, EPI_ISL_977793, EPI_ISL_977794, EPI_ISL_977795, EPI_ISL_977796, EPI_ISL_977797, EPI_ISL_977798, EPI_ISL_977799, EPI_ISL_977800, EPI_ISL_977801, EPI_ISL_977802, EPI_ISL_977803, EPI_ISL_977804, EPI_ISL_977805, EPI_ISL_977806, EPI_ISL_977807, EPI_ISL_977808, EPI_ISL_977809, EPI_ISL_977810, EPI_ISL_977811, EPI_ISL_977812, EPI_ISL_977813, EPI_ISL_977814, EPI_ISL_977815, EPI_ISL_977816, EPI_ISL_977817, EPI_ISL_977818, EPI_ISL_977819, EPI_ISL_977820, EPI_ISL_977821, EPI_ISL_977822, EPI_ISL_977823, EPI_ISL_977824, EPI_ISL_977825, EPI_ISL_977826, EPI_ISL_977827, EPI_ISL_977828, EPI_ISL_977829, EPI_ISL_977830, EPI_ISL_977831, EPI_ISL_977832, EPI_ISL_977833, EPI_ISL_977834, EPI_ISL_977835, EPI_ISL_977836, EPI_ISL_977837, EPI_ISL_977838, EPI_ISL_977839, EPI_ISL_977840, EPI_ISL_977841, EPI_ISL_977842, EPI_ISL_977843, EPI_ISL_977844, EPI_ISL_977845, EPI_ISL_977846, EPI_ISL_977847, EPI_ISL_977848, EPI_ISL_977849, EPI_ISL_977850, EPI_ISL_977851, EPI_ISL_977864, EPI_ISL_977865, EPI_ISL_977866, EPI_ISL_977867, EPI_ISL_977869, EPI_ISL_977878, EPI_ISL_978976, EPI_ISL_978977, EPI_ISL_978978, EPI_ISL_978979, EPI_ISL_978980, EPI_ISL_978981, EPI_ISL_978982, EPI_ISL_978983, EPI_ISL_984318, EPI_ISL_984319, EPI_ISL_984320, EPI_ISL_984321, EPI_ISL_984322, EPI_ISL_984323, EPI_ISL_984324, EPI_ISL_984325, EPI_ISL_984326, EPI_ISL_984327, EPI_ISL_984328, EPI_ISL_984329, EPI_ISL_984330, EPI_ISL_984331, EPI_ISL_984333, EPI_ISL_984334, EPI_ISL_984335, EPI_ISL_984336, EPI_ISL_984337, EPI_ISL_984338, EPI_ISL_984339, EPI_ISL_984340, EPI_ISL_984341, EPI_ISL_984342, EPI_ISL_984343, EPI_ISL_984344, EPI_ISL_984345, EPI_ISL_984346, EPI_ISL_984348, EPI_ISL_984349, EPI_ISL_984350, EPI_ISL_984351, EPI_ISL_984352, EPI_ISL_984353, EPI_ISL_984355, EPI_ISL_984356, EPI_ISL_984357, EPI_ISL_984359, EPI_ISL_984361, EPI_ISL_984362, EPI_ISL_984365, EPI_ISL_984366, EPI_ISL_984367, EPI_ISL_984368, EPI_ISL_984369, EPI_ISL_984370, EPI_ISL_984375, EPI_ISL_984376, EPI_ISL_984377, EPI_ISL_984378, EPI_ISL_984380, EPI_ISL_984385, EPI_ISL_984386, EPI_ISL_984389, EPI_ISL_984390, EPI_ISL_984391, EPI_ISL_984393, EPI_ISL_984394, EPI_ISL_984395, EPI_ISL_984396, EPI_ISL_984397, EPI_ISL_984398, EPI_ISL_984399, EPI_ISL_984400, EPI_ISL_984401, EPI_ISL_984402, EPI_ISL_984403, EPI_ISL_984404, EPI_ISL_984405, EPI_ISL_984406, EPI_ISL_984407, EPI_ISL_984408, EPI_ISL_984409, EPI_ISL_984410, EPI_ISL_984411, EPI_ISL_984412, EPI_ISL_984413, EPI_ISL_984414, EPI_ISL_984415, EPI_ISL_984416, EPI_ISL_984417, EPI_ISL_984418, EPI_ISL_984420, EPI_ISL_984421, EPI_ISL_984422, EPI_ISL_984424, EPI_ISL_984425, EPI_ISL_984426, EPI_ISL_984427, EPI_ISL_984428, EPI_ISL_984429, EPI_ISL_984430, EPI_ISL_984431, EPI_ISL_984432, EPI_ISL_984433, EPI_ISL_984434, EPI_ISL_984435, EPI_ISL_984437, EPI_ISL_984490, EPI_ISL_984491, EPI_ISL_984492, EPI_ISL_984493, EPI_ISL_984494, EPI_ISL_984495, EPI_ISL_984496, EPI_ISL_984497, EPI_ISL_984498, EPI_ISL_984499, EPI_ISL_984500, EPI_ISL_984501, EPI_ISL_984502, EPI_ISL_984503, EPI_ISL_984504, EPI_ISL_984505, EPI_ISL_984506, EPI_ISL_984507, EPI_ISL_984508, EPI_ISL_984509, EPI_ISL_984510, EPI_ISL_984511, EPI_ISL_984512, EPI_ISL_984513, EPI_ISL_984514, EPI_ISL_984515, EPI_ISL_984516, EPI_ISL_984517, EPI_ISL_984518, EPI_ISL_984519, EPI_ISL_984520, EPI_ISL_984521, EPI_ISL_984522, EPI_ISL_984523, EPI_ISL_984524, EPI_ISL_984525, EPI_ISL_984526, EPI_ISL_984527, EPI_ISL_984528, EPI_ISL_984529, EPI_ISL_984530, EPI_ISL_984531, EPI_ISL_984532, EPI_ISL_984533, EPI_ISL_984534, EPI_ISL_984535, EPI_ISL_984536, EPI_ISL_984537, EPI_ISL_984538, EPI_ISL_984539, EPI_ISL_984540, EPI_ISL_984541, EPI_ISL_984542, EPI_ISL_984543, EPI_ISL_984544, EPI_ISL_984545, EPI_ISL_984546, EPI_ISL_984547, EPI_ISL_984548, EPI_ISL_984549, EPI_ISL_984550, EPI_ISL_984551, EPI_ISL_984552, EPI_ISL_984553, EPI_ISL_984554, EPI_ISL_984555, EPI_ISL_984556, EPI_ISL_984557, EPI_ISL_984558, EPI_ISL_984559, EPI_ISL_984560, EPI_ISL_984561, EPI_ISL_984562, EPI_ISL_984563, EPI_ISL_984564, EPI_ISL_984565, EPI_ISL_984566, EPI_ISL_984567, EPI_ISL_984568, EPI_ISL_984569, EPI_ISL_984570, EPI_ISL_984571, EPI_ISL_984572, EPI_ISL_984573, EPI_ISL_984574, EPI_ISL_984575, EPI_ISL_984576, EPI_ISL_984577, EPI_ISL_984578, EPI_ISL_984579, EPI_ISL_984580, EPI_ISL_984581, EPI_ISL_984582, EPI_ISL_984583, EPI_ISL_984584, EPI_ISL_984585, EPI_ISL_984586, EPI_ISL_984587, EPI_ISL_984588, EPI_ISL_984589, EPI_ISL_984590, EPI_ISL_984591, EPI_ISL_984592, EPI_ISL_984593, EPI_ISL_984594, EPI_ISL_984595, EPI_ISL_984596, EPI_ISL_984597, EPI_ISL_984598, EPI_ISL_984599, EPI_ISL_984600, EPI_ISL_984601, EPI_ISL_984602, EPI_ISL_984603, EPI_ISL_984604, EPI_ISL_984605, EPI_ISL_984606, EPI_ISL_984607, EPI_ISL_984608, EPI_ISL_984609, EPI_ISL_984610, EPI_ISL_984611, EPI_ISL_984612, EPI_ISL_984613 | | | |
| see above | California Department of Public Health | Chiu Laboratory, University of California, San Francisco | Brian Bushnell; Candace Wang; Charles Chiu; Debra Wadford; Jill Hacker; Scot Federman; Venice Servellita; Xianding (Wayne) Deng |

| | | | |
|---|---|---|---|
| EPI_ISL_885128, EPI_ISL_885129, EPI_ISL_885130, EPI_ISL_885131, EPI_ISL_885132, EPI_ISL_885133, EPI_ISL_885134, EPI_ISL_885135, EPI_ISL_885136, EPI_ISL_885137, EPI_ISL_885138, EPI_ISL_912187, EPI_ISL_912188, EPI_ISL_912189, EPI_ISL_912190, EPI_ISL_912191, EPI_ISL_912192, EPI_ISL_912193, EPI_ISL_912194, EPI_ISL_1234947, EPI_ISL_1234951, EPI_ISL_1234964, EPI_ISL_1235014, EPI_ISL_1235027, EPI_ISL_1235035, EPI_ISL_1235056, EPI_ISL_1701619, EPI_ISL_1701638 | | | |
| see above | California Institute of Technology | Chan-Zuckerberg Biohub | CZB Cliahub Consortium |
| EPI_ISL_824555, EPI_ISL_824556, EPI_ISL_824557, EPI_ISL_824558, EPI_ISL_824559, EPI_ISL_824560, EPI_ISL_824561, EPI_ISL_824562, EPI_ISL_824563, EPI_ISL_824564, EPI_ISL_824565, EPI_ISL_824566, EPI_ISL_824567, EPI_ISL_824568, EPI_ISL_824569, EPI_ISL_824570, EPI_ISL_824571, EPI_ISL_824572, EPI_ISL_824573, EPI_ISL_824574, EPI_ISL_824575, EPI_ISL_824576, EPI_ISL_824577, EPI_ISL_824578, EPI_ISL_824579, EPI_ISL_824580, EPI_ISL_824581, EPI_ISL_824582, EPI_ISL_824583, EPI_ISL_824585, EPI_ISL_824587, EPI_ISL_824588, EPI_ISL_824589, EPI_ISL_824590, EPI_ISL_824591, EPI_ISL_824592, EPI_ISL_824593, EPI_ISL_824594, EPI_ISL_824595, EPI_ISL_824596, EPI_ISL_824598, EPI_ISL_824599, EPI_ISL_824600, EPI_ISL_824601, EPI_ISL_824602, EPI_ISL_824603, EPI_ISL_824604, EPI_ISL_824605, EPI_ISL_824606, EPI_ISL_824607, EPI_ISL_824608, EPI_ISL_824609, EPI_ISL_824610, EPI_ISL_824611, EPI_ISL_824612, EPI_ISL_824613, EPI_ISL_824614, EPI_ISL_824615, EPI_ISL_824616, EPI_ISL_824618, EPI_ISL_824619, EPI_ISL_824620, EPI_ISL_824621, EPI_ISL_824622, EPI_ISL_824623, EPI_ISL_824624, EPI_ISL_824626, EPI_ISL_824627, EPI_ISL_824628, EPI_ISL_824629, EPI_ISL_824630, EPI_ISL_824631, EPI_ISL_824632, EPI_ISL_824633, EPI_ISL_824634, EPI_ISL_824635, EPI_ISL_824636, EPI_ISL_824637, EPI_ISL_824638, EPI_ISL_824639, EPI_ISL_824640, EPI_ISL_824641, EPI_ISL_824642, EPI_ISL_824643, EPI_ISL_824644, EPI_ISL_824645, EPI_ISL_824646, EPI_ISL_824647, EPI_ISL_824648, EPI_ISL_824649, EPI_ISL_824650, EPI_ISL_824651, EPI_ISL_824652, EPI_ISL_824653, EPI_ISL_824654, EPI_ISL_824655, EPI_ISL_824656, EPI_ISL_824658, EPI_ISL_824659, EPI_ISL_824660, EPI_ISL_824661, EPI_ISL_824662, EPI_ISL_824663, EPI_ISL_824664, EPI_ISL_824665, EPI_ISL_824666, EPI_ISL_824667, EPI_ISL_824668, EPI_ISL_824669, EPI_ISL_824670, EPI_ISL_824671, EPI_ISL_824672, EPI_ISL_824673, EPI_ISL_824674, EPI_ISL_824675, EPI_ISL_824676, EPI_ISL_824677, EPI_ISL_824678, EPI_ISL_824679, EPI_ISL_824680, EPI_ISL_824681, EPI_ISL_824682, EPI_ISL_824683, EPI_ISL_824684, EPI_ISL_824685, EPI_ISL_824686, EPI_ISL_824687, EPI_ISL_824688, EPI_ISL_824689, EPI_ISL_824690, EPI_ISL_824691, EPI_ISL_824692, EPI_ISL_824693, EPI_ISL_824694, EPI_ISL_824695, EPI_ISL_824696, EPI_ISL_824697, EPI_ISL_824698, EPI_ISL_824699, EPI_ISL_824700, EPI_ISL_824701, EPI_ISL_824702, EPI_ISL_824703, EPI_ISL_824704, EPI_ISL_824705, EPI_ISL_824706, EPI_ISL_824707, EPI_ISL_824708, EPI_ISL_824709, EPI_ISL_824710, EPI_ISL_824711, EPI_ISL_824712, EPI_ISL_824713, EPI_ISL_824714, EPI_ISL_824715, EPI_ISL_824716, EPI_ISL_824717, EPI_ISL_824718, EPI_ISL_824719, EPI_ISL_824720, EPI_ISL_824721, EPI_ISL_824722, EPI_ISL_824723, EPI_ISL_824724, EPI_ISL_824725, EPI_ISL_824726, EPI_ISL_824727, EPI_ISL_824728, EPI_ISL_824729, EPI_ISL_824730, EPI_ISL_824731, EPI_ISL_824732, EPI_ISL_824733, EPI_ISL_824734, EPI_ISL_824735, EPI_ISL_824736, EPI_ISL_824737, EPI_ISL_824738, EPI_ISL_824739, EPI_ISL_824740, EPI_ISL_824741 | | | |
| see above | Cedars-Sinai Medical Center, Department of Pathology & Laboratory Medicine, Molecular Pathology Laboratory | Cedars-Sinai Medical Center, Molecular Pathology Laboratory of Department of Pathology & Laboratory Medicine and Genomic Core | Brian Davis; Eric Vail; Jasmine T Plummer; Jorge Mario Sincuir Martinez; Stephanie Chen; Wenjuan Zhang |
| EPI_ISL_1531823, EPI_ISL_1531827, EPI_ISL_1531828, EPI_ISL_1531829, EPI_ISL_1531830, EPI_ISL_1531831, EPI_ISL_1531832, EPI_ISL_1531833, EPI_ISL_1531834, EPI_ISL_1531835, EPI_ISL_1531836, EPI_ISL_1531837, EPI_ISL_1531838, EPI_ISL_1531839, EPI_ISL_1531840, EPI_ISL_1531841, EPI_ISL_1531842, EPI_ISL_1531843, EPI_ISL_1531844, EPI_ISL_1531845, EPI_ISL_1531846, EPI_ISL_1531847, EPI_ISL_1531848, EPI_ISL_1531849, EPI_ISL_1531850, EPI_ISL_1531851, EPI_ISL_1531852, EPI_ISL_1531853, EPI_ISL_1531854, EPI_ISL_1531855, EPI_ISL_1531856, EPI_ISL_1531857, EPI_ISL_1531858, EPI_ISL_1531859, EPI_ISL_1531860, EPI_ISL_1531861, EPI_ISL_1531862, EPI_ISL_1531863, EPI_ISL_1531864, EPI_ISL_1531865, EPI_ISL_1531866, EPI_ISL_1531867, EPI_ISL_1531868, EPI_ISL_1531869, EPI_ISL_1531870, EPI_ISL_1531871, EPI_ISL_1531872, EPI_ISL_1531873, EPI_ISL_1531874, EPI_ISL_1531875, EPI_ISL_1531876, EPI_ISL_1531877, EPI_ISL_1531878, EPI_ISL_1531879, EPI_ISL_1531880, EPI_ISL_1531881, EPI_ISL_1531882, EPI_ISL_1531883, EPI_ISL_1531884, EPI_ISL_1531885, EPI_ISL_1531886, EPI_ISL_1531887, EPI_ISL_1531888, EPI_ISL_1531889, EPI_ISL_1531890, EPI_ISL_1531891, EPI_ISL_1531892, EPI_ISL_1531893, EPI_ISL_1531894, EPI_ISL_1531895, EPI_ISL_1531896, EPI_ISL_1531897, EPI_ISL_1531898, EPI_ISL_1531899, EPI_ISL_1531900, EPI_ISL_1531901, EPI_ISL_1531902, EPI_ISL_1531903, EPI_ISL_1531904, EPI_ISL_1531905, EPI_ISL_1531906, EPI_ISL_1531907, EPI_ISL_1531908, EPI_ISL_1531909, EPI_ISL_1531910, EPI_ISL_1531911, EPI_ISL_1531912, EPI_ISL_1531913, EPI_ISL_1531914, EPI_ISL_1531915, EPI_ISL_1531916, EPI_ISL_1531917, EPI_ISL_1531918, EPI_ISL_1531919, EPI_ISL_1531920, EPI_ISL_1531921, EPI_ISL_1531922, EPI_ISL_1531923, EPI_ISL_1531924, EPI_ISL_1531925, EPI_ISL_1531926, EPI_ISL_1531927, EPI_ISL_1531928, EPI_ISL_1531929, EPI_ISL_1531930, EPI_ISL_1531931, EPI_ISL_1531932, EPI_ISL_1531933, EPI_ISL_1531934, EPI_ISL_1531935, EPI_ISL_1531936, EPI_ISL_1531937, EPI_ISL_1531938, EPI_ISL_1531939, EPI_ISL_1531940, EPI_ISL_1531941 | | | |
| see above | Centro de Diagnostico COVID-19 UABC Tijuana | Andersen lab at Scripps Research | German Ibarra; Jonathan Vincent Baena; Jorge Luis Jimenez Niebla; Manuel Sanchez Alavez; Oscar Efren Zazueta Fierro; SEARCH Alliance San Diego with Idanya Rubi Serafin Higuera |
| EPI_ISL_847515, EPI_ISL_847517, EPI_ISL_847540, EPI_ISL_847545, EPI_ISL_847546, EPI_ISL_847553, EPI_ISL_847563, EPI_ISL_847579, EPI_ISL_847619, EPI_ISL_847620, EPI_ISL_847621, EPI_ISL_847622, EPI_ISL_847640, EPI_ISL_847641, EPI_ISL_847642, EPI_ISL_847643, EPI_ISL_847644, EPI_ISL_847716, EPI_ISL_847720, EPI_ISL_847731, EPI_ISL_847738, EPI_ISL_847742, EPI_ISL_847744, EPI_ISL_847746, EPI_ISL_847747, EPI_ISL_847782, EPI_ISL_855399, EPI_ISL_855400, EPI_ISL_977885, EPI_ISL_977886, EPI_ISL_977887, EPI_ISL_977888, EPI_ISL_977889, EPI_ISL_977890, EPI_ISL_977891, EPI_ISL_977892, EPI_ISL_977893, EPI_ISL_977894, EPI_ISL_977895, EPI_ISL_977896, EPI_ISL_977897, EPI_ISL_977898, EPI_ISL_977899, EPI_ISL_977900, EPI_ISL_977901, EPI_ISL_977902, EPI_ISL_977903, EPI_ISL_977904, EPI_ISL_977905, EPI_ISL_977906, EPI_ISL_977907, EPI_ISL_977908, EPI_ISL_977909, EPI_ISL_977910, EPI_ISL_977911, EPI_ISL_977912, EPI_ISL_977913, EPI_ISL_977914, EPI_ISL_977915, EPI_ISL_977916, EPI_ISL_977917, EPI_ISL_977981, EPI_ISL_977982, EPI_ISL_977983, EPI_ISL_977985, EPI_ISL_977986, EPI_ISL_977987, EPI_ISL_977988, EPI_ISL_977989, EPI_ISL_977990, EPI_ISL_977991, EPI_ISL_977992, EPI_ISL_977993, EPI_ISL_977994, EPI_ISL_977995, EPI_ISL_977996, EPI_ISL_977997, EPI_ISL_977998, EPI_ISL_977999, EPI_ISL_978000, EPI_ISL_978001, EPI_ISL_978002, EPI_ISL_978003, EPI_ISL_978004, EPI_ISL_978005, EPI_ISL_978006, EPI_ISL_978007, EPI_ISL_978008, EPI_ISL_978009, EPI_ISL_978010, EPI_ISL_978011, EPI_ISL_978012, EPI_ISL_978013, EPI_ISL_978014, EPI_ISL_978015, EPI_ISL_978016, EPI_ISL_978017, EPI_ISL_978018, EPI_ISL_978019, EPI_ISL_978020, EPI_ISL_978021, EPI_ISL_978022, EPI_ISL_978023, EPI_ISL_978024, EPI_ISL_978025, EPI_ISL_978026, EPI_ISL_978027, EPI_ISL_978028, EPI_ISL_978029, EPI_ISL_978030, EPI_ISL_978031, EPI_ISL_978032, EPI_ISL_978033, EPI_ISL_978034, EPI_ISL_978035, EPI_ISL_978036, EPI_ISL_978037, EPI_ISL_978038, EPI_ISL_978039, EPI_ISL_978040, EPI_ISL_978041, EPI_ISL_978042, EPI_ISL_978043, EPI_ISL_978044, EPI_ISL_978045, EPI_ISL_978046, EPI_ISL_978047, EPI_ISL_978048, EPI_ISL_978049, EPI_ISL_978050, EPI_ISL_978051, EPI_ISL_978052, EPI_ISL_978053, EPI_ISL_978054, EPI_ISL_978055, EPI_ISL_978056, EPI_ISL_978057, EPI_ISL_978058, EPI_ISL_978059, EPI_ISL_978060, EPI_ISL_978061, EPI_ISL_978062, EPI_ISL_978072, EPI_ISL_978073, EPI_ISL_978074, EPI_ISL_978131, EPI_ISL_978160, EPI_ISL_978161, EPI_ISL_978162, EPI_ISL_978163, EPI_ISL_978164, EPI_ISL_978165, EPI_ISL_978166, EPI_ISL_978167, EPI_ISL_978168, EPI_ISL_978955, EPI_ISL_978956, EPI_ISL_978957, EPI_ISL_978958, EPI_ISL_978959, EPI_ISL_978960, EPI_ISL_978967, EPI_ISL_978968, EPI_ISL_978969, EPI_ISL_978970, EPI_ISL_978971, EPI_ISL_978972 | | | |
| see above | Chiu Laboratory, University of California, San Francisco | Chiu Laboratory, University of California, San Francisco | Brian Bushnell; Candace Wang; Charles Chiu; Debra Wadford; Jill Hacker; Scot Federman; Venice Servellita; Xianding (Wayne) Deng |
| EPI_ISL_753262, EPI_ISL_753263, EPI_ISL_753264, EPI_ISL_753265, EPI_ISL_753266, EPI_ISL_753267, EPI_ISL_753268, EPI_ISL_753269, EPI_ISL_753270, EPI_ISL_753271, EPI_ISL_753272, EPI_ISL_753273, EPI_ISL_753274, EPI_ISL_753275, EPI_ISL_753276, EPI_ISL_753277, EPI_ISL_753278, EPI_ISL_753279, EPI_ISL_753280, EPI_ISL_753281, EPI_ISL_753282, EPI_ISL_753283, EPI_ISL_753284, EPI_ISL_753285, EPI_ISL_753286, EPI_ISL_753287, EPI_ISL_753288, EPI_ISL_753289, EPI_ISL_753290, EPI_ISL_753291, EPI_ISL_753292, EPI_ISL_753293, EPI_ISL_753294, EPI_ISL_753295, EPI_ISL_753296, EPI_ISL_753297, EPI_ISL_753298, EPI_ISL_753299, EPI_ISL_753300, EPI_ISL_753301, EPI_ISL_753302, EPI_ISL_753303, EPI_ISL_753304, EPI_ISL_753305, EPI_ISL_753306, EPI_ISL_753307, EPI_ISL_753308, EPI_ISL_753309, EPI_ISL_753310, EPI_ISL_753311, EPI_ISL_753312, EPI_ISL_753313, EPI_ISL_753314, EPI_ISL_753315, EPI_ISL_753316, EPI_ISL_753317, EPI_ISL_753318, EPI_ISL_753319, EPI_ISL_753320, EPI_ISL_753321, EPI_ISL_753322, EPI_ISL_753323, EPI_ISL_753324, EPI_ISL_753325, EPI_ISL_753326, EPI_ISL_753327, EPI_ISL_753328, EPI_ISL_753329, EPI_ISL_753330, EPI_ISL_753331, EPI_ISL_753332, EPI_ISL_753333, EPI_ISL_753334, EPI_ISL_753335, EPI_ISL_753336, EPI_ISL_753337, EPI_ISL_753338, EPI_ISL_753339, EPI_ISL_753340, EPI_ISL_753341, EPI_ISL_753342, EPI_ISL_753343, EPI_ISL_753344, EPI_ISL_753345, EPI_ISL_753346, EPI_ISL_753347, EPI_ISL_753348, EPI_ISL_753349, EPI_ISL_753350, EPI_ISL_753351, EPI_ISL_753352, EPI_ISL_753353, EPI_ISL_753354, EPI_ISL_753355, EPI_ISL_753356, EPI_ISL_753357, EPI_ISL_753358, EPI_ISL_753359, EPI_ISL_753360, EPI_ISL_753361, EPI_ISL_753362, EPI_ISL_753363, EPI_ISL_753364, EPI_ISL_753365, EPI_ISL_753366, EPI_ISL_753367, EPI_ISL_753368, EPI_ISL_753369, EPI_ISL_753370, EPI_ISL_753371, EPI_ISL_753372, EPI_ISL_753373, EPI_ISL_753374, EPI_ISL_753375, EPI_ISL_753376, EPI_ISL_753377, EPI_ISL_753378, EPI_ISL_753379, EPI_ISL_753380, EPI_ISL_753381, EPI_ISL_753382, EPI_ISL_753383, EPI_ISL_753384, EPI_ISL_753385, EPI_ISL_753386, EPI_ISL_753387, EPI_ISL_753388, EPI_ISL_753389, EPI_ISL_753390, EPI_ISL_753391, EPI_ISL_753392, EPI_ISL_753393, EPI_ISL_753394, EPI_ISL_753395, EPI_ISL_753396, EPI_ISL_753397, EPI_ISL_753398, EPI_ISL_753399, EPI_ISL_753400, EPI_ISL_753401, EPI_ISL_753402, EPI_ISL_753403, EPI_ISL_753404, EPI_ISL_753405, EPI_ISL_753406, EPI_ISL_753407, EPI_ISL_753408, EPI_ISL_753409, EPI_ISL_753410, EPI_ISL_753411, EPI_ISL_753412, EPI_ISL_753413, EPI_ISL_753414, EPI_ISL_753415, EPI_ISL_753416, EPI_ISL_753417, EPI_ISL_753418, EPI_ISL_753419, EPI_ISL_753420, EPI_ISL_753421, EPI_ISL_753422, EPI_ISL_753423, EPI_ISL_753424, EPI_ISL_753425, EPI_ISL_753426, EPI_ISL_753427, EPI_ISL_753428, EPI_ISL_753429, EPI_ISL_753430, EPI_ISL_753431, EPI_ISL_753432, EPI_ISL_753433, EPI_ISL_753434, EPI_ISL_753435, EPI_ISL_753436, EPI_ISL_753437, EPI_ISL_753438, EPI_ISL_753439, EPI_ISL_753440, EPI_ISL_753441, EPI_ISL_753442, EPI_ISL_753443, EPI_ISL_753444, EPI_ISL_753445, EPI_ISL_753446, EPI_ISL_753447, EPI_ISL_753448, EPI_ISL_753449, EPI_ISL_753450, EPI_ISL_753451, EPI_ISL_753452, EPI_ISL_753453, EPI_ISL_753454, EPI_ISL_753455, EPI_ISL_753456, EPI_ISL_753457, EPI_ISL_753458, EPI_ISL_753459, EPI_ISL_753460, EPI_ISL_753461, EPI_ISL_753462, EPI_ISL_753463, EPI_ISL_753464, EPI_ISL_753465, EPI_ISL_753466, EPI_ISL_753467, EPI_ISL_753468, EPI_ISL_753469, EPI_ISL_753470, EPI_ISL_753471, EPI_ISL_753472, EPI_ISL_753473, EPI_ISL_753474, EPI_ISL_753475, EPI_ISL_753476, EPI_ISL_753477, EPI_ISL_753478, EPI_ISL_753479, EPI_ISL_753480, EPI_ISL_753481, EPI_ISL_753482, EPI_ISL_753483, EPI_ISL_753484, EPI_ISL_753485, EPI_ISL_753486, EPI_ISL_753487, EPI_ISL_753488, EPI_ISL_753489, EPI_ISL_753490, EPI_ISL_753491, EPI_ISL_753492, EPI_ISL_753493, EPI_ISL_753494, EPI_ISL_753495, EPI_ISL_753496, EPI_ISL_753497, EPI_ISL_753498, EPI_ISL_753499, EPI_ISL_753500, EPI_ISL_753501, EPI_ISL_753502, EPI_ISL_753503, EPI_ISL_753504, EPI_ISL_753505, EPI_ISL_753506, EPI_ISL_753507, EPI_ISL_753508, EPI_ISL_753509, EPI_ISL_753510, EPI_ISL_753511, EPI_ISL_753512, EPI_ISL_753513, EPI_ISL_753514, EPI_ISL_753515, EPI_ISL_753516, EPI_ISL_753517, EPI_ISL_753518, EPI_ISL_753519, EPI_ISL_753520, EPI_ISL_753521, EPI_ISL_753522, EPI_ISL_753523, EPI_ISL_753524, EPI_ISL_753525, EPI_ISL_753526, EPI_ISL_753527, EPI_ISL_753528, EPI_ISL_753529, EPI_ISL_753530, EPI_ISL_753531, EPI_ISL_753532, EPI_ISL_753533, EPI_ISL_753534, EPI_ISL_753535, EPI_ISL_753536, EPI_ISL_753537, EPI_ISL_753538, EPI_ISL_753539, EPI_ISL_753540, EPI_ISL_753541, EPI_ISL_753542, EPI_ISL_753543, EPI_ISL_753544, EPI_ISL_753545, EPI_ISL_753546, EPI_ISL_753547, EPI_ISL_753548, EPI_ISL_753549, EPI_ISL_753550, EPI_ISL_753551, EPI_ISL_753552, EPI_ISL_753553, EPI_ISL_753554, EPI_ISL_753555, EPI_ISL_753556, EPI_ISL_753557, EPI_ISL_753558, EPI_ISL_753559, EPI_ISL_753560, EPI_ISL_753561, EPI_ISL_753562, EPI_ISL_753563, EPI_ISL_753564, EPI_ISL_753565, EPI_ISL_753566, EPI_ISL_753567, EPI_ISL_753568, EPI_ISL_753569, EPI_ISL_753570, EPI_ISL_753571, EPI_ISL_753572, EPI_ISL_753573, EPI_ISL_753574, EPI_ISL_753575, EPI_ISL_753576, EPI_ISL_753577, EPI_ISL_753578, EPI_ISL_753579, EPI_ISL_753580, EPI_ISL_753581, EPI_ISL_753582, EPI_ISL_753583, EPI_ISL_753584, EPI_ISL_753585, EPI_ISL_753586, EPI_ISL_753587, EPI_ISL_753588, EPI_ISL_753589, EPI_ISL_753590, EPI_ISL_753591, EPI_ISL_753592, EPI_ISL_753593, EPI_ISL_753594, EPI_ISL_753595, EPI_ISL_753596, EPI_ISL_753597, EPI_ISL_753598, EPI_ISL_753599, EPI_ISL_753600, EPI_ISL_753601, EPI_ISL_753602, EPI_ISL_753603, EPI_ISL_753604, EPI_ISL_753605, EPI_ISL_753606, EPI_ISL_753607, EPI_ISL_753608, EPI_ISL_753609, EPI_ISL_753610, EPI_ISL_753611, EPI_ISL_753612, EPI_ISL_753613, EPI_ISL_753614, EPI_ISL_753615, EPI_ISL_753616, EPI_ISL_753617, EPI_ISL_753618, EPI_ISL_753619, EPI_ISL_753620, EPI_ISL_753621, EPI_ISL_753622, EPI_ISL_753623, EPI_ISL_753624, EPI_ISL_753625, EPI_ISL_753626, EPI_ISL_753627, EPI_ISL_753628, EPI_ISL_753629, EPI_ISL_753630, EPI_ISL_753631, EPI_ISL_753632, EPI_ISL_753633, EPI_ISL_753634, EPI_ISL_753635, EPI_ISL_753636, EPI_ISL_753637, EPI_ISL_753638, EPI_ISL_753639, EPI_ISL_753640, EPI_ISL_753641, EPI_ISL_753642, EPI_ISL_753643, EPI_ISL_753644, EPI_ISL_753645, EPI_ISL_753646, EPI_ISL_753647, EPI_ISL_753648, EPI_ISL_753649, EPI_ISL_753650, EPI_ISL_753651, EPI_ISL_753652, EPI_ISL_753653, EPI_ISL_753654, EPI_ISL_753655, EPI_ISL_753656, EPI_ISL_753657, EPI_ISL_753658, EPI_ISL_753659, EPI_ISL_753660, EPI_ISL_753661, EPI_ISL_753662, EPI_ISL_753663, EPI_ISL_753664, EPI_ISL_753665, EPI_ISL_753666, EPI_ISL_753667, EPI_ISL_753668, EPI_ISL_753669, EPI_ISL_753670, EPI_ISL_753671, EPI_ISL_753672, EPI_ISL_753673, EPI_ISL_753674, EPI_ISL_753675, EPI_ISL_753676, EPI_ISL_753677, EPI_ISL_753678, EPI_ISL_753679, EPI_ISL_753680, EPI_ISL_753681, EPI_ISL_753682, EPI_ISL_753683, EPI_ISL_753684, EPI_ISL_753685, EPI_ISL_753686, EPI_ISL_753687, EPI_ISL_753688, EPI_ISL_753689, EPI_ISL_753690, EPI_ISL_753691, EPI_ISL_753692, EPI_ISL_1295564, EPI_ISL_1295565, EPI_ISL_1295568, EPI_ISL_1295569, EPI_ISL_1295574, EPI_ISL_1295575, EPI_ISL_1295576, EPI_ISL_1295581, EPI_ISL_1295583, EPI_ISL_1295584 | | | |
| see above | Clinical virology Laboratory, Children's Hospital Los Angeles | Center for Personalized Medicine, Children's Hospital Los Angeles | Alex Ryutov; Alexander R. Judkins; Ananthanarayanan Govindarajan; David Ruble; Dejerianne Ostrow; Dennis Maglinte; Gai et al; Jaclyn A. Biegel; Jennifer Dien Bard; Jennifer Han; Lishuang Shen; Moiz Bootwalla; Timothy J. Triche; Utsav Pandey; Xiaowu Gai |
| EPI_ISL_672488, EPI_ISL_672489 | Contra Costa County Public Health Lab | Chan-Zuckerberg Biohub | CZB Cliahub Consortium |
| EPI_ISL_548491, EPI_ISL_548492, EPI_ISL_548506, EPI_ISL_548507, EPI_ISL_548508, EPI_ISL_548510, EPI_ISL_548515, EPI_ISL_548516, EPI_ISL_548520, EPI_ISL_548531, EPI_ISL_548534, EPI_ISL_548552, EPI_ISL_548554, EPI_ISL_548556, EPI_ISL_548558, EPI_ISL_548566, EPI_ISL_548569, EPI_ISL_548573, EPI_ISL_548574, EPI_ISL_548577 | | | |
| see above | Contra Costa Public Health Lab | Chan-Zuckerberg Biohub | CZB Cliahub Consortium |
| EPI_ISL_548347, EPI_ISL_548360, EPI_ISL_548365, EPI_ISL_548366, EPI_ISL_548370, EPI_ISL_548502, EPI_ISL_548504, EPI_ISL_548532, EPI_ISL_548544, EPI_ISL_672028, EPI_ISL_738890, EPI_ISL_738990, EPI_ISL_739017, EPI_ISL_739038, EPI_ISL_739095, EPI_ISL_739363, EPI_ISL_739399, EPI_ISL_739480, EPI_ISL_739492, EPI_ISL_739588, EPI_ISL_1477965, EPI_ISL_1478006, EPI_ISL_1478018, EPI_ISL_1478025, EPI_ISL_1478044, EPI_ISL_1478045, EPI_ISL_1478061, EPI_ISL_1478070, EPI_ISL_1478074, EPI_ISL_1478080, EPI_ISL_1478118, EPI_ISL_1478136, EPI_ISL_1478179, EPI_ISL_1478195, EPI_ISL_1478198, EPI_ISL_1529147, EPI_ISL_1529168, EPI_ISL_1529225, EPI_ISL_1529237, EPI_ISL_1529246, EPI_ISL_1529248, EPI_ISL_1529255, EPI_ISL_1529256, EPI_ISL_1529268, EPI_ISL_1529270, EPI_ISL_1529279, EPI_ISL_1529285, EPI_ISL_1529292, EPI_ISL_1529293, EPI_ISL_1529311, EPI_ISL_1529343, EPI_ISL_1701066, EPI_ISL_1701073, EPI_ISL_1701093, EPI_ISL_1701168, EPI_ISL_1701189, EPI_ISL_1701219, EPI_ISL_1701264, EPI_ISL_1701268, EPI_ISL_1701408, EPI_ISL_1701419, EPI_ISL_1701460, EPI_ISL_1701461, EPI_ISL_1701464, EPI_ISL_1701468, EPI_ISL_1701476, EPI_ISL_1701490, EPI_ISL_1701519, EPI_ISL_1701531, EPI_ISL_1701539, EPI_ISL_1701540, EPI_ISL_1701557, EPI_ISL_1701562, EPI_ISL_1701569, EPI_ISL_1701582, EPI_ISL_2102802, EPI_ISL_2102870, EPI_ISL_2102889, EPI_ISL_2102925, EPI_ISL_2102926, EPI_ISL_2102938, EPI_ISL_2102953, EPI_ISL_2102971, EPI_ISL_2103096 | | | |
| see above | County of San Luis Obispo Public Health Laboratory | Chan-Zuckerberg Biohub | CZB Cliahub Consortium |
| EPI_ISL_548337, EPI_ISL_548338, EPI_ISL_548339, EPI_ISL_548340, EPI_ISL_548341, EPI_ISL_548342, EPI_ISL_548343, EPI_ISL_548344, EPI_ISL_548420, EPI_ISL_548434, EPI_ISL_548470, EPI_ISL_548582, EPI_ISL_548583, EPI_ISL_548584, EPI_ISL_548588, EPI_ISL_548593, EPI_ISL_548595, EPI_ISL_548596, EPI_ISL_548597, EPI_ISL_548598, EPI_ISL_548600, EPI_ISL_548602, EPI_ISL_548604, EPI_ISL_548605, EPI_ISL_548606, EPI_ISL_548607, EPI_ISL_548610, EPI_ISL_548612, EPI_ISL_548616, EPI_ISL_548617, EPI_ISL_548618, EPI_ISL_548619, EPI_ISL_548621, EPI_ISL_548622, EPI_ISL_548624, EPI_ISL_548629, EPI_ISL_548630, EPI_ISL_548632, EPI_ISL_548633, EPI_ISL_548636, EPI_ISL_548639, EPI_ISL_548642, EPI_ISL_548643, EPI_ISL_548645, EPI_ISL_548647, EPI_ISL_548649, EPI_ISL_548650, EPI_ISL_548652, EPI_ISL_548655, EPI_ISL_548656, EPI_ISL_548657, EPI_ISL_548658, EPI_ISL_548659, EPI_ISL_548661, EPI_ISL_548664, EPI_ISL_548669, EPI_ISL_548670, EPI_ISL_548672, EPI_ISL_548673, EPI_ISL_548676, EPI_ISL_548678, EPI_ISL_548679, EPI_ISL_582887, EPI_ISL_582888, EPI_ISL_582889, EPI_ISL_582890, EPI_ISL_582891, EPI_ISL_582892, EPI_ISL_582893, EPI_ISL_582894, EPI_ISL_582895, EPI_ISL_582896, EPI_ISL_582897, EPI_ISL_582898, EPI_ISL_582899, EPI_ISL_582900, EPI_ISL_582901, EPI_ISL_582902, EPI_ISL_582903, EPI_ISL_582904, EPI_ISL_582905, EPI_ISL_582906, EPI_ISL_582907, EPI_ISL_582908, EPI_ISL_582909, EPI_ISL_582910, EPI_ISL_582911, EPI_ISL_582912, EPI_ISL_582913, EPI_ISL_582914, EPI_ISL_582915, EPI_ISL_582916, EPI_ISL_582917, EPI_ISL_582918, EPI_ISL_582919, EPI_ISL_582920, EPI_ISL_582921, EPI_ISL_582922, EPI_ISL_582923, EPI_ISL_582924, EPI_ISL_582925, EPI_ISL_582926, EPI_ISL_582927, EPI_ISL_582928, EPI_ISL_582929, EPI_ISL_582930, EPI_ISL_582931, EPI_ISL_582932, EPI_ISL_582933, EPI_ISL_582934, EPI_ISL_582935, EPI_ISL_582936, EPI_ISL_582937, EPI_ISL_582938, EPI_ISL_582939, EPI_ISL_582940, EPI_ISL_582941, EPI_ISL_582942, EPI_ISL_582943, EPI_ISL_582944, EPI_ISL_582945, EPI_ISL_582946, EPI_ISL_582947, EPI_ISL_582948, EPI_ISL_582949, EPI_ISL_582950, EPI_ISL_582951, EPI_ISL_582952, EPI_ISL_582953, EPI_ISL_582954, EPI_ISL_582955, EPI_ISL_582956, EPI_ISL_582957, EPI_ISL_582958 | | | |
| see above | County of Santa Clara Public Health Department | Chan-Zuckerberg Biohub | CZB Cliahub Consortium |
| EPI_ISL_1811644, EPI_ISL_1811645, EPI_ISL_1811648, EPI_ISL_1811655, EPI_ISL_1811657, EPI_ISL_1811660, EPI_ISL_1811663, EPI_ISL_1811664, EPI_ISL_1811667, EPI_ISL_1811668, EPI_ISL_1811672, EPI_ISL_1811673, EPI_ISL_1811677, EPI_ISL_1811684, EPI_ISL_1811709, EPI_ISL_1811713, EPI_ISL_1811716, EPI_ISL_1811723, EPI_ISL_1811734, EPI_ISL_1811790, EPI_ISL_1811791, EPI_ISL_1811792, EPI_ISL_1811793, EPI_ISL_1811794, EPI_ISL_1811812, EPI_ISL_1811824, EPI_ISL_1811835, EPI_ISL_1811851, EPI_ISL_1811867, EPI_ISL_1811869, EPI_ISL_1811940, EPI_ISL_1811966, EPI_ISL_1812007, EPI_ISL_1812013, EPI_ISL_1812039, EPI_ISL_1812073, EPI_ISL_1812139, EPI_ISL_1812159, EPI_ISL_1812178, EPI_ISL_1812199, EPI_ISL_1812217, EPI_ISL_1812230, EPI_ISL_1812261, EPI_ISL_1812271, EPI_ISL_1812281, EPI_ISL_1812282, EPI_ISL_1812291, EPI_ISL_1812361, EPI_ISL_1812363, EPI_ISL_1812367, EPI_ISL_1812381, EPI_ISL_1812412, EPI_ISL_1812448, EPI_ISL_1812449, EPI_ISL_1812492, EPI_ISL_1812495, EPI_ISL_1812520, EPI_ISL_1812553, EPI_ISL_1812558, EPI_ISL_1812590, EPI_ISL_1812638, EPI_ISL_1812646, EPI_ISL_1812671, EPI_ISL_1812683, EPI_ISL_1812757, EPI_ISL_1812758, EPI_ISL_1812777, EPI_ISL_1812784, EPI_ISL_1812799, EPI_ISL_1812810, EPI_ISL_1812826, EPI_ISL_1812854, EPI_ISL_1812905, EPI_ISL_1812910, EPI_ISL_1812917, EPI_ISL_1812925, EPI_ISL_1812929, EPI_ISL_1812963, EPI_ISL_1812981, EPI_ISL_1812987, EPI_ISL_1812994, EPI_ISL_1813002, EPI_ISL_1813040, EPI_ISL_1813054, EPI_ISL_1813059, EPI_ISL_1813063, EPI_ISL_1813079, EPI_ISL_1813093, EPI_ISL_1813097, EPI_ISL_1813113, EPI_ISL_1813134, EPI_ISL_1813145, EPI_ISL_1813177, EPI_ISL_1813304, EPI_ISL_1813306, EPI_ISL_1813340, EPI_ISL_1813342, EPI_ISL_1813377, EPI_ISL_1813405, EPI_ISL_1813409, EPI_ISL_1813425, EPI_ISL_1813430, EPI_ISL_1813475, EPI_ISL_1813497, EPI_ISL_1813500, EPI_ISL_1813508, EPI_ISL_1813512, EPI_ISL_1813600, EPI_ISL_1813711, EPI_ISL_1813783, EPI_ISL_1813806, EPI_ISL_1813925, EPI_ISL_1813975, EPI_ISL_1814049, EPI_ISL_1814055, EPI_ISL_1814237, EPI_ISL_1814460, EPI_ISL_1814601, EPI_ISL_1814637, EPI_ISL_1814649, EPI_ISL_1814744, EPI_ISL_1814766, EPI_ISL_1815209, EPI_ISL_1815351, EPI_ISL_1815380, EPI_ISL_1815495, EPI_ISL_1815515, EPI_ISL_1815519, EPI_ISL_1815594, EPI_ISL_1815677, EPI_ISL_1815748, EPI_ISL_1815764, EPI_ISL_1815789, EPI_ISL_1816066, EPI_ISL_1816079, EPI_ISL_1816081, EPI_ISL_1816127, EPI_ISL_1816197, EPI_ISL_1816220, EPI_ISL_1816264, EPI_ISL_1816265, EPI_ISL_1816270, EPI_ISL_1816277, EPI_ISL_1816290, EPI_ISL_1816297, EPI_ISL_1816405, EPI_ISL_1816489, EPI_ISL_1816556, EPI_ISL_1816583, EPI_ISL_1816633, EPI_ISL_1816659, EPI_ISL_1816674, EPI_ISL_1816742, EPI_ISL_1816750, EPI_ISL_1816760, EPI_ISL_1816761, EPI_ISL_1816764, EPI_ISL_1816792, EPI_ISL_1816808, EPI_ISL_1816821, EPI_ISL_1816825, EPI_ISL_1816830, EPI_ISL_1816831, EPI_ISL_1816832, EPI_ISL_1816833, EPI_ISL_1816834, EPI_ISL_1816852, EPI_ISL_1816856, EPI_ISL_1816859, EPI_ISL_1816860, EPI_ISL_1816865, EPI_ISL_1816871, EPI_ISL_1816873, EPI_ISL_1816876, EPI_ISL_1816878, EPI_ISL_1816880, EPI_ISL_1816882, EPI_ISL_1816885, EPI_ISL_1816895, EPI_ISL_1816897, EPI_ISL_1816899, EPI_ISL_1816901, EPI_ISL_1816908, EPI_ISL_1816912 | | | |
| see above | EXCITE Lab | Andersen lab at Scripps Research | Abigail Schnapper; Angela Scioscia; Chip Schooley; David Pride; Helena Tubb; Natasha Martin Cheryl Anderson; Sawyer Farmer; Sharon Reed; Sydney Morgan + Marni Jacobs + Celestine Magallanes + Holly Valentine + Louise C. Laurent + SEARCH; Tommy Valles + SEARCH |
| EPI_ISL_672423, EPI_ISL_672424, EPI_ISL_672425, EPI_ISL_1235371, EPI_ISL_1235408, EPI_ISL_1235413, EPI_ISL_1235422, EPI_ISL_1235560, EPI_ISL_1235607 | | | |
| see above | Fresno County Public Health Laboratory | Chan-Zuckerberg Biohub | CZB Cliahub Consortium |
| EPI_ISL_1005636, EPI_ISL_1027224, EPI_ISL_1027229 | Fulgent Genetics | Fulgent Genetics | Becky Tsai; Benafsh Sapra; Doreen Ng; Harry Gao; James Xie; John Gao; Joseph Fierro; Mickey Li; Yan Meng |
| EPI_ISL_747243 | General Hospital Laboratory | Pathogen Laboratory (BSL3), Biomedical Innovation Department, Experimental and Applied Biology Division, Scientific Research Center and High Education from Ensenada (CICESE) | Cervantes-Luevano K.; Galindo C. and Licea-Navarro A.; Martinez M.A.; Saavedra-Flores A. |
| EPI_ISL_730209, EPI_ISL_730210, EPI_ISL_730211, EPI_ISL_730212, EPI_ISL_730213, EPI_ISL_730214, EPI_ISL_730215, EPI_ISL_730216, EPI_ISL_730217, EPI_ISL_730218, EPI_ISL_730219, EPI_ISL_730220, EPI_ISL_730221, EPI_ISL_730222, EPI_ISL_730223, EPI_ISL_730224, EPI_ISL_730225, EPI_ISL_730226, EPI_ISL_730227, EPI_ISL_730228, EPI_ISL_1081423, EPI_ISL_1081424, EPI_ISL_1081425, EPI_ISL_1081426, EPI_ISL_1081427, EPI_ISL_1081428, EPI_ISL_1081429, EPI_ISL_1081430, EPI_ISL_1081431, EPI_ISL_1081432, EPI_ISL_1081433, EPI_ISL_1081434, EPI_ISL_1081435, EPI_ISL_1081436, EPI_ISL_1081437, EPI_ISL_1081438, EPI_ISL_1081439, EPI_ISL_1081440, EPI_ISL_1081441, EPI_ISL_1081442, EPI_ISL_1081443, EPI_ISL_1081444, EPI_ISL_1081445, EPI_ISL_1081446, EPI_ISL_1081447, EPI_ISL_1081448, EPI_ISL_1081449, EPI_ISL_1081451, EPI_ISL_1081473, EPI_ISL_1081474, EPI_ISL_1081475, EPI_ISL_1081476, EPI_ISL_1081477, EPI_ISL_1081478, EPI_ISL_1081479, EPI_ISL_1081480, EPI_ISL_1081481, EPI_ISL_1081482, EPI_ISL_1081483, EPI_ISL_1081484, EPI_ISL_1081485, EPI_ISL_1081486, EPI_ISL_1081487, EPI_ISL_1081488, EPI_ISL_1081489, EPI_ISL_1081490, EPI_ISL_1081491, EPI_ISL_1081492, EPI_ISL_1081493, EPI_ISL_1081494, EPI_ISL_1081495 | | | |
| see above | Genomica Lab Molecular, M√©xico | Andersen lab at Scripps Research | Jose Horacio Reyna Verdugo; Jose Roman Chavez Mendez; Luis Alberto Rangel Gonzalez; Martin Gonzalez Ibarra; SEARCH Alliance San Diego with Jonathan Gonzalez Garcia |
| EPI_ISL_755594, EPI_ISL_755595, EPI_ISL_755596, EPI_ISL_755597, EPI_ISL_755602, EPI_ISL_755613, EPI_ISL_802610, EPI_ISL_802611, EPI_ISL_802612, EPI_ISL_802613, EPI_ISL_802614, EPI_ISL_802615, EPI_ISL_802616, EPI_ISL_802617, EPI_ISL_802659, EPI_ISL_802660, EPI_ISL_802661, EPI_ISL_802720 | | | |
| see above | Helix/Illumina | Genomics and Discovery, Respiratory Viruses Branch, Division of Viral Diseases, Centers for Disease Control and Prevention | Alexandre Bolze; Ary Ascencio; Ben L. Rambo-Martin Eileen de Feo; Brad Sickler; Charlotte Rivera-Garcia; Christine Tran; Clinton R. Paden; David Becker; Dhwani Batra; Duncan MacCannell; Efren Sandoval; Eileen de Feo; Elizabeth Cirulli; Eric Allen; Geraint Levan; James Lu; Jan Antico; Jason Nguyen; Jimmy Ramirez; Jingtao Liu; Kelly Schiabor Barrett; Kim Gietzen; Magnus Isaksson; Marc Laurent; Matthew Tolentino; Nicole L. Washington; Peter W. Cook; Phil Febbo; Ryan Cho; Shannon Wickline; Sherry Wang; Simon White; Summer Galloway; Suxiang Tong; Tyler Cassens; William Lee |
| EPI_ISL_583093, EPI_ISL_583094, EPI_ISL_583095, EPI_ISL_583096, EPI_ISL_583097, EPI_ISL_583098, EPI_ISL_583099, EPI_ISL_583100, EPI_ISL_583101, EPI_ISL_583102, EPI_ISL_583103, EPI_ISL_583104, EPI_ISL_583105, EPI_ISL_583106, EPI_ISL_583107, EPI_ISL_583108, EPI_ISL_583109, EPI_ISL_583110, EPI_ISL_583111, EPI_ISL_583112, EPI_ISL_583113, EPI_ISL_583114, EPI_ISL_583115, EPI_ISL_583116, | | | |

| | | | |
|---|---|---|---|
| EPI_ISL_583117, EPI_ISL_583118, EPI_ISL_583119, EPI_ISL_583120, EPI_ISL_583121, EPI_ISL_583122, EPI_ISL_583123, EPI_ISL_583124, EPI_ISL_583125, EPI_ISL_583126, EPI_ISL_583127, EPI_ISL_583128, EPI_ISL_583129, EPI_ISL_583130, EPI_ISL_583131, EPI_ISL_583132, EPI_ISL_583133, EPI_ISL_583134, EPI_ISL_583135, EPI_ISL_583136, EPI_ISL_583137, EPI_ISL_583138, EPI_ISL_583139, EPI_ISL_583140, EPI_ISL_583141, EPI_ISL_583142, EPI_ISL_583143, EPI_ISL_583144, EPI_ISL_583145, EPI_ISL_583146, EPI_ISL_583147, EPI_ISL_583148, EPI_ISL_583149, EPI_ISL_583150, EPI_ISL_583151, EPI_ISL_583152, EPI_ISL_583153, EPI_ISL_583154, EPI_ISL_583155, EPI_ISL_583156, EPI_ISL_583157, EPI_ISL_583158, EPI_ISL_583159, EPI_ISL_583160, EPI_ISL_583161, EPI_ISL_583162, EPI_ISL_583163, EPI_ISL_583164, EPI_ISL_583165, EPI_ISL_583166, EPI_ISL_583167, EPI_ISL_583168, EPI_ISL_583169, EPI_ISL_583170, EPI_ISL_738504, EPI_ISL_738505, EPI_ISL_738530, EPI_ISL_738539, EPI_ISL_738563, EPI_ISL_738600, EPI_ISL_738647, EPI_ISL_738649, EPI_ISL_738651, EPI_ISL_738742, EPI_ISL_738756, EPI_ISL_738759, EPI_ISL_738778, EPI_ISL_738779, EPI_ISL_738785, EPI_ISL_738786, EPI_ISL_738795, EPI_ISL_738801, EPI_ISL_738806, EPI_ISL_738807, EPI_ISL_738809, EPI_ISL_738810, EPI_ISL_738827, EPI_ISL_738832, EPI_ISL_738844, EPI_ISL_738849, EPI_ISL_738852, EPI_ISL_738854, EPI_ISL_738865, EPI_ISL_738875, EPI_ISL_738879, EPI_ISL_738902, EPI_ISL_738912, EPI_ISL_738913, EPI_ISL_738917, EPI_ISL_738918, EPI_ISL_738923, EPI_ISL_738929, EPI_ISL_738938, EPI_ISL_738947, EPI_ISL_738951, EPI_ISL_738953, EPI_ISL_738958, EPI_ISL_738961, EPI_ISL_738969, EPI_ISL_738980, EPI_ISL_738985, EPI_ISL_738988, EPI_ISL_738999, EPI_ISL_739000, EPI_ISL_739001, EPI_ISL_739007, EPI_ISL_739011, EPI_ISL_739032, EPI_ISL_739043, EPI_ISL_739051, EPI_ISL_739052, EPI_ISL_739079, EPI_ISL_739086, EPI_ISL_739106, EPI_ISL_739112, EPI_ISL_739114, EPI_ISL_739123, EPI_ISL_739126, EPI_ISL_739131, EPI_ISL_739148, EPI_ISL_739155, EPI_ISL_739159, EPI_ISL_739165, EPI_ISL_739170, EPI_ISL_739177, EPI_ISL_739183, EPI_ISL_739194, EPI_ISL_739198, EPI_ISL_739210, EPI_ISL_739225, EPI_ISL_739228, EPI_ISL_739235, EPI_ISL_739240, EPI_ISL_739242, EPI_ISL_739250, EPI_ISL_739264, EPI_ISL_739269, EPI_ISL_739272, EPI_ISL_739274, EPI_ISL_739287, EPI_ISL_739292, EPI_ISL_739297, EPI_ISL_739299, EPI_ISL_739311, EPI_ISL_739313, EPI_ISL_739318, EPI_ISL_739330, EPI_ISL_739366, EPI_ISL_739367, EPI_ISL_739389, EPI_ISL_739403, EPI_ISL_739405, EPI_ISL_739411, EPI_ISL_739417, EPI_ISL_739418, EPI_ISL_739425, EPI_ISL_739430, EPI_ISL_739433, EPI_ISL_739436, EPI_ISL_739442, EPI_ISL_739447, EPI_ISL_739452, EPI_ISL_739461, EPI_ISL_739475, EPI_ISL_739489, EPI_ISL_739499, EPI_ISL_739525, EPI_ISL_739527, EPI_ISL_739530, EPI_ISL_739531, EPI_ISL_739533, EPI_ISL_739542, EPI_ISL_739553, EPI_ISL_739558, EPI_ISL_739571, EPI_ISL_739585, EPI_ISL_739608, EPI_ISL_739611, EPI_ISL_739621, EPI_ISL_739651, EPI_ISL_955496, EPI_ISL_955497, EPI_ISL_955498, EPI_ISL_955499, EPI_ISL_955500, EPI_ISL_955501, EPI_ISL_955502, EPI_ISL_955503, EPI_ISL_955504, EPI_ISL_955505, EPI_ISL_955506, EPI_ISL_955507, EPI_ISL_955508, EPI_ISL_955509, EPI_ISL_955510, EPI_ISL_955511, EPI_ISL_955512, EPI_ISL_955513, EPI_ISL_955514, EPI_ISL_955515, EPI_ISL_955516, EPI_ISL_955517, EPI_ISL_955518, EPI_ISL_955519, EPI_ISL_955520, EPI_ISL_955521, EPI_ISL_955522, EPI_ISL_955523, EPI_ISL_955524, EPI_ISL_955525, EPI_ISL_955526, EPI_ISL_955527, EPI_ISL_955528, EPI_ISL_955529, EPI_ISL_955530, EPI_ISL_955531, EPI_ISL_955532, EPI_ISL_955533, EPI_ISL_955534, EPI_ISL_955535, EPI_ISL_955536, EPI_ISL_955537, EPI_ISL_955538, EPI_ISL_955539, EPI_ISL_955540, EPI_ISL_955541, EPI_ISL_955542, EPI_ISL_955543, EPI_ISL_955544, EPI_ISL_955545, EPI_ISL_955546, EPI_ISL_955547, EPI_ISL_955548, EPI_ISL_955549, EPI_ISL_955550, EPI_ISL_955551, EPI_ISL_955552, EPI_ISL_955553, EPI_ISL_955554, EPI_ISL_955555, EPI_ISL_955556, EPI_ISL_955557, EPI_ISL_955558, EPI_ISL_955559, EPI_ISL_955560, EPI_ISL_955693, EPI_ISL_955694, EPI_ISL_955695, EPI_ISL_955696, EPI_ISL_955697, EPI_ISL_955698, EPI_ISL_955699, EPI_ISL_955701, EPI_ISL_955702, EPI_ISL_955703, EPI_ISL_955705, EPI_ISL_955707, EPI_ISL_979162, EPI_ISL_979163, EPI_ISL_979164, EPI_ISL_979165, EPI_ISL_979166, EPI_ISL_979167, EPI_ISL_979168, EPI_ISL_979169, EPI_ISL_979170, EPI_ISL_979171, EPI_ISL_979172, EPI_ISL_979173, EPI_ISL_979174, EPI_ISL_979175, EPI_ISL_979176, EPI_ISL_979177, EPI_ISL_979178, EPI_ISL_979179, EPI_ISL_979180, EPI_ISL_979181, EPI_ISL_979182, EPI_ISL_979183, EPI_ISL_979184, EPI_ISL_979185, EPI_ISL_979186, EPI_ISL_979187, EPI_ISL_979188, EPI_ISL_979189, EPI_ISL_979190, EPI_ISL_979191, EPI_ISL_979192, EPI_ISL_979193, EPI_ISL_979194, EPI_ISL_979195, EPI_ISL_979196, EPI_ISL_979197, EPI_ISL_979198, EPI_ISL_979199, EPI_ISL_979200, EPI_ISL_979201, EPI_ISL_979202, EPI_ISL_979203, EPI_ISL_979204, EPI_ISL_979205, EPI_ISL_979206, EPI_ISL_979207, EPI_ISL_979208, EPI_ISL_979209, EPI_ISL_979210, EPI_ISL_979211, EPI_ISL_979212, EPI_ISL_979213, EPI_ISL_979214, EPI_ISL_979215, EPI_ISL_979216, EPI_ISL_979217, EPI_ISL_979218, EPI_ISL_979219, EPI_ISL_979220, EPI_ISL_979221, EPI_ISL_979222, EPI_ISL_979223, EPI_ISL_979224, EPI_ISL_979225, EPI_ISL_979226, EPI_ISL_979227, EPI_ISL_979228, EPI_ISL_979229, EPI_ISL_979230, EPI_ISL_979231, EPI_ISL_979232, EPI_ISL_979233, EPI_ISL_979234, EPI_ISL_979235, EPI_ISL_979236, EPI_ISL_979237, EPI_ISL_979238, EPI_ISL_979239, EPI_ISL_979240, EPI_ISL_979241, EPI_ISL_979242, EPI_ISL_979243, EPI_ISL_979244, EPI_ISL_979245, EPI_ISL_979710, EPI_ISL_979711, EPI_ISL_979712, EPI_ISL_979713, EPI_ISL_979714, EPI_ISL_979715, EPI_ISL_979716, EPI_ISL_979717, EPI_ISL_979718, EPI_ISL_979719, EPI_ISL_979720, EPI_ISL_979721, EPI_ISL_979722, EPI_ISL_979723, EPI_ISL_979724, EPI_ISL_979725, EPI_ISL_979726, EPI_ISL_979727, EPI_ISL_979728, EPI_ISL_979729, EPI_ISL_979730, EPI_ISL_979731, EPI_ISL_979732, EPI_ISL_979733, EPI_ISL_979734, EPI_ISL_979735, EPI_ISL_979736, EPI_ISL_979737, EPI_ISL_979738, EPI_ISL_979739, EPI_ISL_979740, EPI_ISL_979741, EPI_ISL_979742, EPI_ISL_979743, EPI_ISL_979744, EPI_ISL_979745, EPI_ISL_979746, EPI_ISL_979747, EPI_ISL_979748, EPI_ISL_979749, EPI_ISL_979750, EPI_ISL_979751, EPI_ISL_979752, EPI_ISL_979753, EPI_ISL_979754, EPI_ISL_979755, EPI_ISL_979756, EPI_ISL_979757, EPI_ISL_979758, EPI_ISL_979759, EPI_ISL_979760, EPI_ISL_979761, EPI_ISL_979762, EPI_ISL_979763, EPI_ISL_979764, EPI_ISL_979765, EPI_ISL_979766, EPI_ISL_979767, EPI_ISL_979768, EPI_ISL_979769, EPI_ISL_979770, EPI_ISL_979771, EPI_ISL_979772, EPI_ISL_979773, EPI_ISL_979774, EPI_ISL_979775, EPI_ISL_979776, EPI_ISL_979777, EPI_ISL_979778, EPI_ISL_979779, EPI_ISL_979780, EPI_ISL_979781, EPI_ISL_979782, EPI_ISL_979783, EPI_ISL_979784, EPI_ISL_979785, EPI_ISL_979786, EPI_ISL_979787, EPI_ISL_979788, EPI_ISL_979789, EPI_ISL_979790, EPI_ISL_979791, EPI_ISL_979792, EPI_ISL_979793, EPI_ISL_979794, EPI_ISL_979795, EPI_ISL_979796, EPI_ISL_979797, EPI_ISL_1027535, EPI_ISL_1027536, EPI_ISL_1027537, EPI_ISL_1027538, EPI_ISL_1027539, EPI_ISL_1027540, EPI_ISL_1027541, EPI_ISL_1030499 | | | |
| see above | Humboldt County Public Health Laboratory | Chan-Zuckerberg Biohub | CZB Cliahub Consortium |
| EPI_ISL_1081170 | IEH Laboratories & Consulting Group | ID Genomics | Curtis Fowler; Evgeni Sokurenko; Hao Geng; Hemantha Kulakesara; Kai Deng; Kirthi Kutumbaka; Lydia Larson; Mansour Samadpour; Veronika Tchesnokova |
| EPI_ISL_1233893, EPI_ISL_1233894, EPI_ISL_2253173 | Infectious Diseases, Chan-Zuckerberg Biohub Covidtracker | Infectious Diseases, Chan-Zuckerberg Biohub Covidtracker | CZB Cliahub Consortium |
| EPI_ISL_884308, EPI_ISL_884309, EPI_ISL_884310, EPI_ISL_884311, EPI_ISL_884312, EPI_ISL_884313, EPI_ISL_884314, EPI_ISL_884315 | | | |
| see above | Infectious Diseases, Quest Diagnostics | Infectious Diseases, Quest Diagnostics | Anderson, B.; Bernstein; D.F.; Gerasimova, A.; Hua, M.; K.E.; Kagan; L.E.; Lacbawan, F.; Liu, Y.; Livingston; Owen, R.; R.M.; Rosenthal; S.H.; Shalhout |
| EPI_ISL_1512783, EPI_ISL_1512788, EPI_ISL_1512859, EPI_ISL_1512861, EPI_ISL_1512878 | Infinity Biologix | Centers for Disease Control and Prevention Division of Viral Diseases, Pathogen Discovery | Adrian Paskey; Benjamin Rambo-Martin; Chirayu Goswami; Christian Bixby; Christopher Gulvick; Clinton R. Paden; Dakota Howard; Darlene Wagner; Dhwani Batra; Duncan MacCannell; Jason Caravas; Jonathan Schultz; Kara Moser; Matthew Schmerer; Peter W. Cook; Robin Grimwood; Russ Hager; Scott Sammons; Shatavia Morrison; Yihe Wang; Yvette Unoarumhi |
| EPI_ISL_677826, EPI_ISL_677827, EPI_ISL_677828, EPI_ISL_677829, EPI_ISL_677830, EPI_ISL_677831, EPI_ISL_677832, EPI_ISL_677833, EPI_ISL_677834, EPI_ISL_677835, EPI_ISL_677836, EPI_ISL_677837, EPI_ISL_677838, EPI_ISL_677839, EPI_ISL_677840, EPI_ISL_677841, EPI_ISL_677842, EPI_ISL_677843, EPI_ISL_677844, EPI_ISL_677845, EPI_ISL_677846, EPI_ISL_677847, EPI_ISL_677848, EPI_ISL_677849, EPI_ISL_677850, EPI_ISL_677851, EPI_ISL_677852, EPI_ISL_677853, EPI_ISL_677854, EPI_ISL_677855, EPI_ISL_677856, EPI_ISL_677857, EPI_ISL_677858, EPI_ISL_677859, EPI_ISL_677860, EPI_ISL_677861, EPI_ISL_677862, EPI_ISL_677864, EPI_ISL_677865, EPI_ISL_677866, EPI_ISL_677867, EPI_ISL_677868, EPI_ISL_677869, EPI_ISL_677870, EPI_ISL_677871, EPI_ISL_677872, EPI_ISL_677873, EPI_ISL_677874, EPI_ISL_677875, EPI_ISL_677876, EPI_ISL_677878, EPI_ISL_677879, EPI_ISL_677880, EPI_ISL_677881, EPI_ISL_677882, EPI_ISL_677883, EPI_ISL_677884, EPI_ISL_677885, EPI_ISL_677886, EPI_ISL_677887, EPI_ISL_677888, EPI_ISL_677895, EPI_ISL_677904, EPI_ISL_677906, EPI_ISL_756331, EPI_ISL_756332, EPI_ISL_756337, EPI_ISL_918178, EPI_ISL_918184, EPI_ISL_918186, EPI_ISL_918197, EPI_ISL_918202, EPI_ISL_918205, EPI_ISL_918206, EPI_ISL_918207, EPI_ISL_918208, EPI_ISL_918209, EPI_ISL_918210, EPI_ISL_918211, EPI_ISL_918212, EPI_ISL_918213, EPI_ISL_918214, EPI_ISL_918215, EPI_ISL_918216, EPI_ISL_918217, EPI_ISL_918218, EPI_ISL_918219, EPI_ISL_918220, EPI_ISL_918221, EPI_ISL_918222, EPI_ISL_918223, EPI_ISL_1041034, EPI_ISL_1041035, EPI_ISL_1041036, EPI_ISL_1041037, EPI_ISL_1041038, EPI_ISL_1041039, EPI_ISL_1041040, EPI_ISL_1041041, EPI_ISL_1041042, EPI_ISL_1041043, EPI_ISL_1041044, EPI_ISL_1041045, EPI_ISL_1041046, EPI_ISL_1041047, EPI_ISL_1041048, EPI_ISL_1041049, EPI_ISL_1041050, EPI_ISL_1041051, EPI_ISL_1041052, EPI_ISL_1041053, EPI_ISL_1041055, EPI_ISL_1041056, EPI_ISL_1041057, EPI_ISL_1041058, EPI_ISL_1041059, EPI_ISL_1041060, EPI_ISL_1041061, EPI_ISL_1041062, EPI_ISL_1041063, EPI_ISL_1041064, EPI_ISL_1041065, EPI_ISL_1041066, EPI_ISL_1041067, EPI_ISL_1041068, EPI_ISL_1041069, EPI_ISL_1041070, EPI_ISL_1041071, EPI_ISL_1041072, EPI_ISL_1041073, EPI_ISL_1041074, EPI_ISL_1041075, EPI_ISL_1041076, EPI_ISL_1041077, EPI_ISL_1041078, EPI_ISL_1041079, EPI_ISL_1041080, EPI_ISL_1041081, EPI_ISL_1041082, EPI_ISL_1041083, EPI_ISL_1041084, EPI_ISL_1041085, EPI_ISL_1041086, EPI_ISL_1041087, EPI_ISL_1041088, EPI_ISL_1041089, EPI_ISL_1041090, EPI_ISL_1041091, EPI_ISL_1041092, EPI_ISL_1041093, EPI_ISL_1041094, EPI_ISL_1041095, EPI_ISL_1041096, EPI_ISL_1041097, EPI_ISL_1041098, EPI_ISL_1041099, EPI_ISL_1041100, EPI_ISL_1041101, EPI_ISL_1041102, EPI_ISL_1041103, EPI_ISL_1041104, EPI_ISL_1041105, EPI_ISL_1041106, EPI_ISL_1041107, EPI_ISL_1041181, EPI_ISL_1041182, EPI_ISL_1041183, EPI_ISL_1041184, EPI_ISL_1041185, EPI_ISL_1041186, EPI_ISL_1041187, EPI_ISL_1041188, EPI_ISL_1041189, EPI_ISL_1041190, EPI_ISL_1041191, EPI_ISL_1041192, EPI_ISL_1041193, EPI_ISL_1041194, EPI_ISL_1049262, EPI_ISL_1049263, EPI_ISL_1049264, EPI_ISL_1049265, EPI_ISL_1049266, EPI_ISL_1049267, EPI_ISL_1049268, EPI_ISL_1049269, EPI_ISL_1049270, EPI_ISL_1049271, EPI_ISL_1049272, EPI_ISL_1049273, EPI_ISL_1049274, EPI_ISL_1049275, EPI_ISL_1049276, EPI_ISL_1049277, EPI_ISL_1049278, EPI_ISL_1049279, EPI_ISL_1049280, EPI_ISL_1049281, EPI_ISL_1049282, EPI_ISL_1049283, EPI_ISL_1049284, EPI_ISL_1049285, EPI_ISL_1049286, EPI_ISL_1049287, EPI_ISL_1049288, EPI_ISL_1049289, EPI_ISL_1049290, EPI_ISL_1049291, EPI_ISL_1049292, EPI_ISL_1049293, EPI_ISL_1049294, EPI_ISL_1049295, EPI_ISL_1049296, EPI_ISL_1049297, EPI_ISL_1049298, EPI_ISL_1049299, EPI_ISL_1049300, EPI_ISL_1049301, EPI_ISL_1049302, EPI_ISL_1049303, EPI_ISL_1049304, EPI_ISL_1049305, EPI_ISL_1049306, EPI_ISL_1049307, EPI_ISL_1049308, EPI_ISL_1049309, EPI_ISL_1049310, EPI_ISL_1049311, EPI_ISL_1049312, EPI_ISL_1049313, EPI_ISL_1049314, EPI_ISL_1049315, EPI_ISL_1049316, EPI_ISL_1049317, EPI_ISL_1049318, EPI_ISL_1049319, EPI_ISL_1049320, EPI_ISL_1049321, EPI_ISL_1049322, EPI_ISL_1049323, EPI_ISL_1049324, EPI_ISL_1049325, EPI_ISL_1049326, EPI_ISL_1049327, EPI_ISL_1049328, EPI_ISL_1049329, EPI_ISL_1049330, EPI_ISL_1049331, EPI_ISL_1049332, EPI_ISL_1049333, EPI_ISL_1049334, EPI_ISL_1049335, EPI_ISL_1049336, EPI_ISL_1049337, EPI_ISL_1712457, EPI_ISL_1712458, EPI_ISL_1712459, EPI_ISL_1712460, EPI_ISL_1712461, EPI_ISL_1712480, EPI_ISL_1712481, EPI_ISL_1712482, EPI_ISL_1712483, EPI_ISL_1712484, EPI_ISL_1712485, EPI_ISL_1712487, EPI_ISL_1712488, EPI_ISL_1712489, EPI_ISL_1712490, EPI_ISL_1712491, EPI_ISL_1712492, EPI_ISL_1712494, EPI_ISL_1712495, EPI_ISL_1712496, EPI_ISL_1712497, EPI_ISL_1712498, EPI_ISL_1712499, EPI_ISL_1712500, EPI_ISL_1712501, EPI_ISL_1712502, EPI_ISL_1712503, EPI_ISL_1712504, EPI_ISL_1712505, EPI_ISL_1712506, EPI_ISL_1712508, EPI_ISL_1712510, EPI_ISL_1712511, EPI_ISL_1712513, EPI_ISL_1712514, EPI_ISL_1712515, EPI_ISL_1712516, EPI_ISL_1712520, EPI_ISL_1712521, EPI_ISL_1712522 | | | |
| see above | Innovative Genomics Institute, UC Berkeley | Innovative Genomics Institute, UC Berkeley | Alison Ciling; Haridha Shivram; Holly Gildea; John Boyle; Justin Choi; Liana Lareau; Netravathi Krishnappa; Phil Frankino; Shana McDevitt; Stacia Wyman |
| EPI_ISL_1060681, EPI_ISL_1060703, EPI_ISL_1060705, EPI_ISL_1060720, EPI_ISL_1060724, EPI_ISL_1060726, EPI_ISL_1060747, EPI_ISL_1060748, EPI_ISL_1060749, EPI_ISL_1060751 | | | |
| see above | Instituto de Diagnostico y Referencia Epidemiologicos (INDRE)_RNLSP | Instituto de Diagnostico y Referencia Epidemiologicos (INDRE) | Abril Rodriguez-Maldonado; Adnan Araiza-Rodriguez; Claudia Wong-Arambula; David Fragoso-Fonseca; Ernesto Ramirez-Gonzalez.; Fabiola Garces-Ayala; Gisela Barrera-Badillo; Irma Lopez-Martinez; Lucia Hernandez-Rivas; Mayra Jimenez-Morales; Nancy Munoz-Hernandez; Natividad Cruz-Ortiz; Sergio Rangel-Guerrero; Tatiana Nunez-Garcia |
| EPI_ISL_913936, EPI_ISL_913937, EPI_ISL_913942, EPI_ISL_913943, EPI_ISL_933678, EPI_ISL_1054984 | Instituto de Diagnostico y Referencia Epidemiologicos INDRE_RNLSP | Instituto de Diagnostico y Referencia Epidemiologicos (INDRE) | Abril Rodriguez-Maldonado; Adnan Araiza-Rodriguez; Claudia Wong-Arambula; David Fragoso-Fonseca; Ernesto Ramirez-Gonzalez.; Fabiola Garces-Ayala; Gisela Barrera-Badillo; Irma Lopez-Martinez; Lucia Hernandez-Rivas; Mayra Jimenez-Morales; Nancy Munoz-Hernandez; Natividad Cruz-Ortiz; Sergio Rangel-Guerrero; Tatiana Nunez-Garcia |
| EPI_ISL_1302182 | Instituto de Diagnostico y Referencia Epidemiologicos InDRE_RNLSP | Instituto de Biotecnología de la UNAM | Alejandra Hernández-Terán; Alejandro Sanchez-Flores; Alma Rincón-Rubio; Andrea Santos Coy-Arechavaleta; Authors from IBT; Blanca Taboada; Celia Boukadida; Clara Esperanza Santacruz-Tinoco; Edgar Mendieta-Condado; Eduardo Becerril-Vargas; Fidencio Mejía-Nepomuceno; Francisco Pulido; Gisela Barrera-Badillo; Gloria Vazquez; Hector Esteban Paz-Juárez; IMSS; InDRE and INER (in alphabetical order): Carlos F. Arias; Irma Lopez-Martinez; Jerome Jean Verleyen; Joel Armando Vázquez-Pérez; Jorge Salas-Hernández; José Arturo Martínez-Orozco; José Ernesto Ramírez-González; José Esteban Muñoz-Medina; Larissa Fernandes-Matano; Lucia Hernandez-Rivas; Luis Alberto Ochoa-Carrera; Margarita Matías-Florentino; Mario Mújica-Sánchez; Natividad Cruz-Ortiz; Pavel Isa; Ricardo Grande; Santiago Ávila-Ríos; Tatiana Nunez-Garcia; Teresita Rojas-Mendoza |
| EPI_ISL_1113761, EPI_ISL_1121043 | Kashi Clinical Laboratory | Kashi Clinical Laboratory | Jenefer DeKoning; Zahra Kashi |
| EPI_ISL_887678, EPI_ISL_887696, EPI_ISL_887698, EPI_ISL_887762, EPI_ISL_887766, EPI_ISL_887779, EPI_ISL_887831, EPI_ISL_887859, EPI_ISL_887890, EPI_ISL_887891, EPI_ISL_887901, EPI_ISL_887903, EPI_ISL_887953, EPI_ISL_887960, EPI_ISL_887971, EPI_ISL_887994, EPI_ISL_888015, EPI_ISL_888018, EPI_ISL_888019, EPI_ISL_888024, EPI_ISL_888030, EPI_ISL_888057, EPI_ISL_888066, EPI_ISL_888116, EPI_ISL_888117, EPI_ISL_888122, EPI_ISL_888139, EPI_ISL_888151, EPI_ISL_888163, EPI_ISL_888199, EPI_ISL_888221, EPI_ISL_888225, EPI_ISL_888241, EPI_ISL_888272, EPI_ISL_888274, EPI_ISL_888280, EPI_ISL_888325, EPI_ISL_888330, EPI_ISL_888334, EPI_ISL_888373, EPI_ISL_888382, EPI_ISL_888454, EPI_ISL_888486, EPI_ISL_888547, EPI_ISL_888567, EPI_ISL_888582 | | | |
| see above | Labcorp | Genomics and Discovery, Respiratory Viruses Branch, Division of Viral Diseases, Centers for Disease Control and Prevention | ; Amanda Douglas; Amanda Suchanek; Andrea Throop; Ayla Burns; Ben L. Rambo-Martin; Bobbi Croy; Brian Krueger; Brian Norvell; Christos Petropoulos; Clinton R. Paden; Craig Lukasik; Debbie Boles; Dhwani Batra; Duncan MacCannell; Eyad Almasri; Goran Stevovic; Howard Engler; Hrushikesh Deshmukh; Jake Humphrey; Jana Schroth; Joe Voshell; John Pruitt; Jonathan Meltzer; Jonathan Williams; Kimberly Wagner; Lax Iyer; Lyndon Tilson; Manoj Jain; Marcia Eisenberg; Mary Ann Cristobal; Mary Williamson; Michael Levandoski; Mike Sapeta; Mindy Nye; Minoo Agarwal; Mohan Kolli; Nuthawin Charoensri; Oren Cohen; Peter W. Cook; Prashant Gupta; Qian Zeng; Rama Ghatti; Scott Parker; Scott Ryan; Stanley Letovsky; Steven Ragan; Summer Galloway; Suresh Babu Selvaraju; Susan Countryman; Susan Hicks; Suxiang Tong; Suzanne Dale; Thomas Urban; Tim Kuphal; Tricia Zwiefelhofer; Vincent Drouillon |
| EPI_ISL_1027773, EPI_ISL_1027789, EPI_ISL_1027838, EPI_ISL_1027839, EPI_ISL_1027912, EPI_ISL_1027916, EPI_ISL_1027917, EPI_ISL_1027936, EPI_ISL_1028029, EPI_ISL_1028053, EPI_ISL_1028145, EPI_ISL_1028202, EPI_ISL_1028241, EPI_ISL_1028294, EPI_ISL_1028347, EPI_ISL_1028348, EPI_ISL_1028381, EPI_ISL_1028412, EPI_ISL_1028448, EPI_ISL_1028516, EPI_ISL_1028580, EPI_ISL_1028610, EPI_ISL_1028679, EPI_ISL_1028731, EPI_ISL_1028800, EPI_ISL_1028822, EPI_ISL_1028825, EPI_ISL_1028826, EPI_ISL_1028864, EPI_ISL_1028866, EPI_ISL_1028867, EPI_ISL_1028873, EPI_ISL_1028874, EPI_ISL_1028886, EPI_ISL_1028899, EPI_ISL_1028941, EPI_ISL_1028956, EPI_ISL_1028957, EPI_ISL_1028965, EPI_ISL_1028998, EPI_ISL_1029035, EPI_ISL_1029068, EPI_ISL_1029069, EPI_ISL_1029079, EPI_ISL_1029274, EPI_ISL_1029285, EPI_ISL_1029300, EPI_ISL_1029301, EPI_ISL_1029302, EPI_ISL_1029303, EPI_ISL_1029305, EPI_ISL_1029306, EPI_ISL_1029307, EPI_ISL_1029308, EPI_ISL_1029309, EPI_ISL_1029310, EPI_ISL_1029311, EPI_ISL_1029312, EPI_ISL_1029313, EPI_ISL_1029314, EPI_ISL_1029315, EPI_ISL_1029316, EPI_ISL_1029317, EPI_ISL_1029318, EPI_ISL_1029319, EPI_ISL_1029320, EPI_ISL_1029321, EPI_ISL_1029322, EPI_ISL_1029323, EPI_ISL_1029324, EPI_ISL_1029325, EPI_ISL_1029326, EPI_ISL_1029414, EPI_ISL_1029415, EPI_ISL_1029416, EPI_ISL_1029417, EPI_ISL_1029418, EPI_ISL_1029419, EPI_ISL_1029420, EPI_ISL_1029421, EPI_ISL_1029422, EPI_ISL_1029423, EPI_ISL_1029532, EPI_ISL_1029579, EPI_ISL_1029580, EPI_ISL_1029581, EPI_ISL_1029721 | | | |
| see above | Laboratory Corporation of America | Respiratory Viruses Branch, Division of Viral Diseases, Centers for Disease Control and Prevention | Ben L. Rambo-Martin; Clinton R. Paden; Dakota Howard; Dhwani Batra; Duncan MacCannell; Peter W. Cook; Suxiang Tong |
| EPI_ISL_708182, EPI_ISL_708183, EPI_ISL_708528, EPI_ISL_710099, EPI_ISL_710100, EPI_ISL_710101, EPI_ISL_710102, EPI_ISL_710103, EPI_ISL_710104, EPI_ISL_710105, EPI_ISL_710106, EPI_ISL_710107, EPI_ISL_710108, EPI_ISL_710109, EPI_ISL_710110, EPI_ISL_710111, EPI_ISL_710112, EPI_ISL_710113, EPI_ISL_710114, EPI_ISL_710115, EPI_ISL_710116, EPI_ISL_710117, EPI_ISL_710118, EPI_ISL_710416, EPI_ISL_710417, EPI_ISL_710418, EPI_ISL_710419, EPI_ISL_710421, EPI_ISL_710422, EPI_ISL_710423, EPI_ISL_710424, EPI_ISL_710425, EPI_ISL_710426, EPI_ISL_710427, EPI_ISL_710428, EPI_ISL_710429, EPI_ISL_710430, EPI_ISL_710431, EPI_ISL_710432, EPI_ISL_737236, EPI_ISL_737237, EPI_ISL_737238, EPI_ISL_737239, EPI_ISL_737240, EPI_ISL_737241, EPI_ISL_737242, EPI_ISL_737243, EPI_ISL_737244, EPI_ISL_737245, EPI_ISL_737246, EPI_ISL_737247, EPI_ISL_737248, EPI_ISL_737249, EPI_ISL_737250, EPI_ISL_737251, EPI_ISL_737252, EPI_ISL_737253, EPI_ISL_737254, EPI_ISL_737255, EPI_ISL_737256, EPI_ISL_737257, EPI_ISL_737258, EPI_ISL_737259, EPI_ISL_737260, EPI_ISL_737261, EPI_ISL_754130, EPI_ISL_754131, EPI_ISL_754132, EPI_ISL_765998, EPI_ISL_765999, EPI_ISL_766000, EPI_ISL_766001, EPI_ISL_766002, EPI_ISL_766003, EPI_ISL_766004, EPI_ISL_766005, EPI_ISL_766006, EPI_ISL_766007, EPI_ISL_766008, EPI_ISL_766009, EPI_ISL_766010, EPI_ISL_766011, EPI_ISL_766012, EPI_ISL_766013, EPI_ISL_766014, EPI_ISL_792655, EPI_ISL_792656, EPI_ISL_792657, EPI_ISL_792658, EPI_ISL_792659, EPI_ISL_792660, EPI_ISL_792661, EPI_ISL_792662, EPI_ISL_792663, EPI_ISL_792664, EPI_ISL_792665, EPI_ISL_792666, EPI_ISL_792667, EPI_ISL_792676, EPI_ISL_792677, EPI_ISL_792678, EPI_ISL_861698, EPI_ISL_903367, EPI_ISL_903368, EPI_ISL_903369, EPI_ISL_903370, EPI_ISL_915438, EPI_ISL_1503992, EPI_ISL_1503995, EPI_ISL_1707677, EPI_ISL_1707678, EPI_ISL_1707679, EPI_ISL_2031979, EPI_ISL_2031980, EPI_ISL_2031981, EPI_ISL_2031982, EPI_ISL_2031983, EPI_ISL_2031984, EPI_ISL_2031985, EPI_ISL_2031986, EPI_ISL_2031987, EPI_ISL_2031988, EPI_ISL_2031989, EPI_ISL_2031990, EPI_ISL_2031991, EPI_ISL_2031992, EPI_ISL_2031994, EPI_ISL_2031995, EPI_ISL_2031996, EPI_ISL_2031997 | | | |
| see above | Los Angeles County PHL | Los Angeles County PHL | P. Hemarajata et al. |
| EPI_ISL_2106182 | Los Angeles County Public Health Laboratories, Los Angeles County Public Health Lab microbial pathogen submission group | Los Angeles County Public Health Laboratories, Los Angeles County Public Health Lab microbial pathogen submission group | Hemarajata; P. and Green, N. |
| EPI_ISL_694046, EPI_ISL_707694, EPI_ISL_707695, EPI_ISL_707883, EPI_ISL_707884, EPI_ISL_707885, EPI_ISL_707886, EPI_ISL_707887, EPI_ISL_707888, EPI_ISL_707889, EPI_ISL_708507, EPI_ISL_708724, EPI_ISL_708727, EPI_ISL_708728, EPI_ISL_708729, EPI_ISL_710433 | | | |
| see above | Los Angeles County Public Health Laboratory | Los Angeles County Public Health Laboratory | P. Hemarajata et al.; Peera Hemarajata et al. |
| EPI_ISL_832105, EPI_ISL_832106, EPI_ISL_832107, EPI_ISL_832108, EPI_ISL_832109, EPI_ISL_832110, EPI_ISL_832111, EPI_ISL_832112, EPI_ISL_832113, EPI_ISL_832114, EPI_ISL_832115, EPI_ISL_832117, EPI_ISL_832118, EPI_ISL_832119, EPI_ISL_832120 | | | |
| see above | MD Laboratories | Los Angeles County PHL | P. Hemarajata et al. |
| EPI_ISL_648545, EPI_ISL_648546, EPI_ISL_648547, EPI_ISL_648548, EPI_ISL_648549, EPI_ISL_648550, EPI_ISL_648551, EPI_ISL_648552, EPI_ISL_648553, EPI_ISL_648554, EPI_ISL_672019, EPI_ISL_672022, EPI_ISL_672026, EPI_ISL_672037, EPI_ISL_672041, EPI_ISL_672046, EPI_ISL_672048, EPI_ISL_672059, EPI_ISL_672067, EPI_ISL_672081, EPI_ISL_672098, EPI_ISL_672099, EPI_ISL_672100, EPI_ISL_672101, EPI_ISL_672102, EPI_ISL_672394, EPI_ISL_672395, EPI_ISL_672396, EPI_ISL_672397, EPI_ISL_672398, EPI_ISL_672399, EPI_ISL_672400, EPI_ISL_672401, EPI_ISL_672402, EPI_ISL_672403, EPI_ISL_672404, EPI_ISL_672405, EPI_ISL_672406, EPI_ISL_672407, EPI_ISL_672408, EPI_ISL_672409, EPI_ISL_672410, EPI_ISL_672411, EPI_ISL_672412, EPI_ISL_672413, EPI_ISL_672414, EPI_ISL_672415, EPI_ISL_672416, EPI_ISL_672417, EPI_ISL_672418, EPI_ISL_672419, EPI_ISL_672420, EPI_ISL_672421, EPI_ISL_672422, EPI_ISL_672504, EPI_ISL_672505, EPI_ISL_672506, EPI_ISL_672507, EPI_ISL_672508, EPI_ISL_672509, EPI_ISL_672510, EPI_ISL_672511, EPI_ISL_672512, EPI_ISL_672513, EPI_ISL_672514, EPI_ISL_672515, EPI_ISL_672516, EPI_ISL_672517, EPI_ISL_672518, EPI_ISL_672519, EPI_ISL_672520, EPI_ISL_672521, EPI_ISL_672522, EPI_ISL_672523, EPI_ISL_672524, EPI_ISL_672525, EPI_ISL_672526, EPI_ISL_672527, EPI_ISL_672528, EPI_ISL_672529, EPI_ISL_672530, EPI_ISL_738666, EPI_ISL_738707, EPI_ISL_738708, EPI_ISL_738916, EPI_ISL_738964, EPI_ISL_739061, EPI_ISL_739133, EPI_ISL_739172, EPI_ISL_739197, EPI_ISL_739390, EPI_ISL_739393, EPI_ISL_739429, EPI_ISL_739539, EPI_ISL_739567 | | | |
| see above | Madera County Department of Public Health | Chan-Zuckerberg Biohub | CZB Cliahub Consortium |
| EPI_ISL_583198 | Marin County Public Health Department | Chan-Zuckerberg Biohub | CZB Cliahub Consortium |
| EPI_ISL_1093144 | Maya Juarez, Div. of Pulmonary, Critical Care and Sleep Medicine, UC Davis Medical Center | Díaz-Muñoz Lab, Department of Microbiology and Molecular Genetics, University of California, Davis | A.J. Campbell; Alexandre Tremeau-Bravard; Ana M. M. Stoian; Angela Haczku; Brandt A Robinson; Christian Sandrock; Daniel G Tompkins; David A. Coil; Greg Brennan; Ivy R. Jose; Jonathan A. Eisen; Maya Juarez; Samuel L Díaz-Muñoz; Satya Dandekar; Shefali Banerjee; Stefan Rothenburg; Stuart H. Cohen; Timothy Albertson; Tracey Goldstein |
| EPI_ISL_1810946, EPI_ISL_2006728, EPI_ISL_2006747, EPI_ISL_2006753, EPI_ISL_2006754, EPI_ISL_2006758, EPI_ISL_2006761, EPI_ISL_2006774, EPI_ISL_2006779, EPI_ISL_2006780, EPI_ISL_2006789, EPI_ISL_2006790, EPI_ISL_2009412, EPI_ISL_2009448, EPI_ISL_2009450, EPI_ISL_2009495, EPI_ISL_2009496, EPI_ISL_2009505, EPI_ISL_2011702, EPI_ISL_2019737, EPI_ISL_2019738, EPI_ISL_2019765, | | | |

| | | | |
|---|---|---|---|
| EPI_ISL_2019773, EPI_ISL_2019776, EPI_ISL_2019786, EPI_ISL_2019793, EPI_ISL_2019798, EPI_ISL_2019810, EPI_ISL_2019844, EPI_ISL_2019847, EPI_ISL_2019848, EPI_ISL_2019872, EPI_ISL_2019902, EPI_ISL_2019939, EPI_ISL_2019947, EPI_ISL_2019950, EPI_ISL_2019951, EPI_ISL_2019971, EPI_ISL_2019975, EPI_ISL_2019989, EPI_ISL_2019990 | | | |
| see above | Moderna Inc. | Moderna Inc. | Groves Dixon; Rolando Pajon; Yamuna D. Paila; Yamuna Paila |
| EPI_ISL_738658, EPI_ISL_739336, EPI_ISL_739372, EPI_ISL_739546 | Monterey County Public Health Lab | Chan-Zuckerberg Biohub | CZB Cliahub Consortium |
| EPI_ISL_1263207, EPI_ISL_1263208, EPI_ISL_1263209, EPI_ISL_1263210, EPI_ISL_1263211, EPI_ISL_1263212, EPI_ISL_1263213, EPI_ISL_1263214, EPI_ISL_1263215, EPI_ISL_1263216, EPI_ISL_1263217, EPI_ISL_1263218, EPI_ISL_1263219, EPI_ISL_1263220, EPI_ISL_1263221, EPI_ISL_1263222, EPI_ISL_1263223, EPI_ISL_1263224, EPI_ISL_1263225, EPI_ISL_1263226 | | | |
| see above | Monterey County Public Health Laboratory | Monterey County Public Health Laboratory | Monterey County Public Health Laboratory |
| EPI_ISL_625579, EPI_ISL_625580, EPI_ISL_625581, EPI_ISL_625582, EPI_ISL_625583, EPI_ISL_625584, EPI_ISL_625585, EPI_ISL_625586, EPI_ISL_625587, EPI_ISL_625588, EPI_ISL_625589, EPI_ISL_625590, EPI_ISL_625591, EPI_ISL_625592, EPI_ISL_625593 | | | |
| see above | NaN | Chan-Zuckerberg Biohub | CZB Cliahub Consortium |
| EPI_ISL_738512, EPI_ISL_738541, EPI_ISL_738754, EPI_ISL_738836, EPI_ISL_738870, EPI_ISL_738911, EPI_ISL_738952, EPI_ISL_738959, EPI_ISL_739169, EPI_ISL_739181, EPI_ISL_739223, EPI_ISL_739315, EPI_ISL_739325, EPI_ISL_739339, EPI_ISL_739469, EPI_ISL_739471, EPI_ISL_739474, EPI_ISL_739557, EPI_ISL_739628, EPI_ISL_739645 | | | |
| see above | Napa-Solano-Yolo- Marin County (NSYM) Public Health Laboratories | Chan-Zuckerberg Biohub | CZB Cliahub Consortium |
| EPI_ISL_1171051 | OHSU Lab Services Molecular Microbiology Lab | Ginkgo Bioworks Clinical Laboratory | Alec J. Hirsch; Alex Plocik; Amanda Riccardi; Andrew C. Adey; Becky Schilling; Benjamin N. Bimber; Birgitte Simen; Brendan L. O'Connell; Brian J. O'Roak; Daniel N. Streblow; Donna Hansel; Guang Fan; James McGann; Jim Griffin; Kayla Carter; Keith Robison; Malaika Mckenzie-Bennett; Martha Pierson; Michelle Spencer; Rebecca Littlefield; Ruth V. Nichols; Sally Grindstaff; William B. Messer; Xuan Qin |
| EPI_ISL_667088, EPI_ISL_667300, EPI_ISL_667417, EPI_ISL_667424, EPI_ISL_832675, EPI_ISL_832758 | OHSU Lab Services Molecular Microbiology Lab | Oregon SARS-CoV-2 Genome Sequencing Center | Alec J. Hirsch; Andrew C. Adey; Benjamin N. Bimber; Brendan L. O'Connell; Brian J. O'Roak; Daniel N. Streblow; Donna Hansel; Guang Fan; Ruth V. Nichols; Sally Grindstaff; William B. Messer |
| EPI_ISL_792679 | Olive View Medical Center | Los Angeles County PHL | P. Hemarajata et al. |
| EPI_ISL_1714787, EPI_ISL_1714790 | Operational Infectious Diseases, Naval Health Research Center | Operational Infectious Diseases, Naval Health Research Center | A.E.; Balansay-Ames; C.A.; Fierro, M.; Iniguez-Stevens, E.; Kriner, P.; M.S.; Moser, K.; Myers; Pan; Phippard; R.S.; R.W.; Underwood; Vo, S. |
| EPI_ISL_582975, EPI_ISL_582976, EPI_ISL_582977, EPI_ISL_582978, EPI_ISL_582979, EPI_ISL_582980, EPI_ISL_582981, EPI_ISL_582982, EPI_ISL_582983, EPI_ISL_582984, EPI_ISL_582985, EPI_ISL_582986, EPI_ISL_582987, EPI_ISL_582988, EPI_ISL_582989, EPI_ISL_582990, EPI_ISL_582991, EPI_ISL_582992, EPI_ISL_582993, EPI_ISL_582994, EPI_ISL_582995, EPI_ISL_582996, EPI_ISL_582997, EPI_ISL_582998, EPI_ISL_582999, EPI_ISL_583000, EPI_ISL_583001, EPI_ISL_583002, EPI_ISL_583003, EPI_ISL_583004, EPI_ISL_583005, EPI_ISL_583006, EPI_ISL_583007, EPI_ISL_583008, EPI_ISL_583009, EPI_ISL_583010, EPI_ISL_583011, EPI_ISL_583012, EPI_ISL_583013, EPI_ISL_583014, EPI_ISL_583015, EPI_ISL_583016, EPI_ISL_583017, EPI_ISL_583018, EPI_ISL_583019, EPI_ISL_583020, EPI_ISL_583021, EPI_ISL_583022, EPI_ISL_583023, EPI_ISL_583024, EPI_ISL_583025, EPI_ISL_583026, EPI_ISL_583027, EPI_ISL_583028, EPI_ISL_583029, EPI_ISL_583030, EPI_ISL_583031, EPI_ISL_583032, EPI_ISL_583033, EPI_ISL_583034, EPI_ISL_583035, EPI_ISL_583036, EPI_ISL_583037, EPI_ISL_583038, EPI_ISL_583039, EPI_ISL_583040, EPI_ISL_583041, EPI_ISL_583042, EPI_ISL_583043, EPI_ISL_583044, EPI_ISL_583045, EPI_ISL_625627, EPI_ISL_625628, EPI_ISL_625629, EPI_ISL_625630, EPI_ISL_625631, EPI_ISL_625632, EPI_ISL_625633, EPI_ISL_625634, EPI_ISL_625635, EPI_ISL_625636, EPI_ISL_625637, EPI_ISL_625638, EPI_ISL_625639, EPI_ISL_625640, EPI_ISL_625641, EPI_ISL_625642, EPI_ISL_625643, EPI_ISL_625644, EPI_ISL_625645, EPI_ISL_625646, EPI_ISL_625647, EPI_ISL_625648, EPI_ISL_625649, EPI_ISL_625650, EPI_ISL_625651, EPI_ISL_625652, EPI_ISL_625653, EPI_ISL_625654, EPI_ISL_625655, EPI_ISL_625656, EPI_ISL_625657, EPI_ISL_625658, EPI_ISL_625659, EPI_ISL_648496, EPI_ISL_648497, EPI_ISL_648498, EPI_ISL_648499, EPI_ISL_648500, EPI_ISL_648501, EPI_ISL_648502, EPI_ISL_648503, EPI_ISL_648504, EPI_ISL_648505, EPI_ISL_648506, EPI_ISL_648507, EPI_ISL_648508, EPI_ISL_648509, EPI_ISL_648510, EPI_ISL_648511, EPI_ISL_648512, EPI_ISL_648513, EPI_ISL_648514, EPI_ISL_648515, EPI_ISL_648516, EPI_ISL_648517, EPI_ISL_648518, EPI_ISL_648519, EPI_ISL_648520, EPI_ISL_648521, EPI_ISL_648522, EPI_ISL_648523, EPI_ISL_648524, EPI_ISL_648525, EPI_ISL_648526, EPI_ISL_648527, EPI_ISL_648528, EPI_ISL_648529, EPI_ISL_648530, EPI_ISL_672075, EPI_ISL_672084, EPI_ISL_672103, EPI_ISL_672109, EPI_ISL_672110, EPI_ISL_672361, EPI_ISL_672362, EPI_ISL_672363, EPI_ISL_672364, EPI_ISL_672365, EPI_ISL_672366, EPI_ISL_672367, EPI_ISL_672368, EPI_ISL_672369, EPI_ISL_672370, EPI_ISL_672371, EPI_ISL_672372, EPI_ISL_672373, EPI_ISL_672374, EPI_ISL_672477, EPI_ISL_672478, EPI_ISL_672479, EPI_ISL_672480, EPI_ISL_672481, EPI_ISL_672482, EPI_ISL_672483, EPI_ISL_672484, EPI_ISL_672485, EPI_ISL_672486, EPI_ISL_672487, EPI_ISL_738503, EPI_ISL_738507, EPI_ISL_738515, EPI_ISL_738518, EPI_ISL_738529, EPI_ISL_738548, EPI_ISL_738559, EPI_ISL_738569, EPI_ISL_738570, EPI_ISL_738571, EPI_ISL_738588, EPI_ISL_738594, EPI_ISL_738599, EPI_ISL_738602, EPI_ISL_738603, EPI_ISL_738620, EPI_ISL_738621, EPI_ISL_738624, EPI_ISL_738630, EPI_ISL_738632, EPI_ISL_738634, EPI_ISL_738635, EPI_ISL_738636, EPI_ISL_738637, EPI_ISL_738638, EPI_ISL_738639, EPI_ISL_738640, EPI_ISL_738641, EPI_ISL_738646, EPI_ISL_738655, EPI_ISL_738664, EPI_ISL_738669, EPI_ISL_738688, EPI_ISL_738693, EPI_ISL_738694, EPI_ISL_738703, EPI_ISL_738704, EPI_ISL_738709, EPI_ISL_738722, EPI_ISL_738729, EPI_ISL_738731, EPI_ISL_738732, EPI_ISL_738733, EPI_ISL_738734, EPI_ISL_738735, EPI_ISL_738736, EPI_ISL_738737, EPI_ISL_738738, EPI_ISL_738739, EPI_ISL_738740, EPI_ISL_738741, EPI_ISL_738752, EPI_ISL_738755, EPI_ISL_738757, EPI_ISL_738758, EPI_ISL_738761, EPI_ISL_738762, EPI_ISL_738782, EPI_ISL_738784, EPI_ISL_738787, EPI_ISL_738790, EPI_ISL_738799, EPI_ISL_738808, EPI_ISL_738811, EPI_ISL_738815, EPI_ISL_738816, EPI_ISL_738818, EPI_ISL_738823, EPI_ISL_738824, EPI_ISL_738826, EPI_ISL_738829, EPI_ISL_738830, EPI_ISL_738840, EPI_ISL_738841, EPI_ISL_738845, EPI_ISL_738850, EPI_ISL_738856, EPI_ISL_738857, EPI_ISL_738860, EPI_ISL_738862, EPI_ISL_738863, EPI_ISL_738866, EPI_ISL_738869, EPI_ISL_738871, EPI_ISL_738872, EPI_ISL_738873, EPI_ISL_738874, EPI_ISL_738878, EPI_ISL_738880, EPI_ISL_738882, EPI_ISL_738884, EPI_ISL_738885, EPI_ISL_738888, EPI_ISL_738893, EPI_ISL_738899, EPI_ISL_738900, EPI_ISL_738905, EPI_ISL_738907, EPI_ISL_738908, EPI_ISL_738921, EPI_ISL_738928, EPI_ISL_738930, EPI_ISL_738935, EPI_ISL_738937, EPI_ISL_738939, EPI_ISL_738944, EPI_ISL_738946, EPI_ISL_738949, EPI_ISL_738954, EPI_ISL_738966, EPI_ISL_738968, EPI_ISL_738970, EPI_ISL_738973, EPI_ISL_738975, EPI_ISL_738978, EPI_ISL_738981, EPI_ISL_738989, EPI_ISL_738991, EPI_ISL_738993, EPI_ISL_738995, EPI_ISL_739002, EPI_ISL_739013, EPI_ISL_739016, EPI_ISL_739028, EPI_ISL_739034, EPI_ISL_739042, EPI_ISL_739044, EPI_ISL_739048, EPI_ISL_739055, EPI_ISL_739063, EPI_ISL_739069, EPI_ISL_739071, EPI_ISL_739072, EPI_ISL_739078, EPI_ISL_739081, EPI_ISL_739089, EPI_ISL_739092, EPI_ISL_739094, EPI_ISL_739097, EPI_ISL_739099, EPI_ISL_739105, EPI_ISL_739108, EPI_ISL_739116, EPI_ISL_739125, EPI_ISL_739132, EPI_ISL_739139, EPI_ISL_739142, EPI_ISL_739143, EPI_ISL_739151, EPI_ISL_739154, EPI_ISL_739156, EPI_ISL_739162, EPI_ISL_739175, EPI_ISL_739176, EPI_ISL_739182, EPI_ISL_739185, EPI_ISL_739191, EPI_ISL_739199, EPI_ISL_739205, EPI_ISL_739211, EPI_ISL_739216, EPI_ISL_739222, EPI_ISL_739227, EPI_ISL_739230, EPI_ISL_739238, EPI_ISL_739239, EPI_ISL_739245, EPI_ISL_739255, EPI_ISL_739258, EPI_ISL_739259, EPI_ISL_739265, EPI_ISL_739266, EPI_ISL_739267, EPI_ISL_739271, EPI_ISL_739276, EPI_ISL_739279, EPI_ISL_739289, EPI_ISL_739291, EPI_ISL_739296, EPI_ISL_739304, EPI_ISL_739314, EPI_ISL_739322, EPI_ISL_739331, EPI_ISL_739334, EPI_ISL_739341, EPI_ISL_739342, EPI_ISL_739345, EPI_ISL_739347, EPI_ISL_739350, EPI_ISL_739353, EPI_ISL_739359, EPI_ISL_739362, EPI_ISL_739369, EPI_ISL_739373, EPI_ISL_739375, EPI_ISL_739379, EPI_ISL_739384, EPI_ISL_739386, EPI_ISL_739396, EPI_ISL_739397, EPI_ISL_739401, EPI_ISL_739410, EPI_ISL_739412, EPI_ISL_739420, EPI_ISL_739421, EPI_ISL_739423, EPI_ISL_739428, EPI_ISL_739432, EPI_ISL_739437, EPI_ISL_739438, EPI_ISL_739441, EPI_ISL_739445, EPI_ISL_739450, EPI_ISL_739455, EPI_ISL_739460, EPI_ISL_739462, EPI_ISL_739464, EPI_ISL_739465, EPI_ISL_739477, EPI_ISL_739482, EPI_ISL_739486, EPI_ISL_739488, EPI_ISL_739490, EPI_ISL_739493, EPI_ISL_739494, EPI_ISL_739496, EPI_ISL_739497, EPI_ISL_739500, EPI_ISL_739513, EPI_ISL_739517, EPI_ISL_739524, EPI_ISL_739529, EPI_ISL_739532, EPI_ISL_739534, EPI_ISL_739540, EPI_ISL_739545, EPI_ISL_739547, EPI_ISL_739556, EPI_ISL_739559, EPI_ISL_739566, EPI_ISL_739568, EPI_ISL_739570, EPI_ISL_739573, EPI_ISL_739577, EPI_ISL_739581, EPI_ISL_739582, EPI_ISL_739586, EPI_ISL_739591, EPI_ISL_739592, EPI_ISL_739595, EPI_ISL_739599, EPI_ISL_739607, EPI_ISL_739609, EPI_ISL_739614, EPI_ISL_739620, EPI_ISL_739625, EPI_ISL_739627, EPI_ISL_739636, EPI_ISL_739637, EPI_ISL_739641, EPI_ISL_739646, EPI_ISL_739654, EPI_ISL_739658, EPI_ISL_885052, EPI_ISL_885053, EPI_ISL_885054, EPI_ISL_885055, EPI_ISL_885056, EPI_ISL_885057, EPI_ISL_885058, EPI_ISL_885059, EPI_ISL_885060, EPI_ISL_885061, EPI_ISL_885062, EPI_ISL_885063, EPI_ISL_885064, EPI_ISL_885065, EPI_ISL_885066, EPI_ISL_885067, EPI_ISL_885068, EPI_ISL_885069, EPI_ISL_885070, EPI_ISL_885071, EPI_ISL_885072, EPI_ISL_885073, EPI_ISL_885074, EPI_ISL_885075, EPI_ISL_885076, EPI_ISL_885077, EPI_ISL_885078, EPI_ISL_885079, EPI_ISL_885080, EPI_ISL_885081, EPI_ISL_885082, EPI_ISL_885083, EPI_ISL_885084, EPI_ISL_885085, EPI_ISL_885086, EPI_ISL_885087, EPI_ISL_885088, EPI_ISL_885089, EPI_ISL_885090, EPI_ISL_885091, EPI_ISL_885092, EPI_ISL_885093, EPI_ISL_885094, EPI_ISL_885095, EPI_ISL_885096, EPI_ISL_885097, EPI_ISL_885098, EPI_ISL_885099, EPI_ISL_885100, EPI_ISL_885101, EPI_ISL_885102, EPI_ISL_885103, EPI_ISL_885104, EPI_ISL_885105, EPI_ISL_885106, EPI_ISL_885107, EPI_ISL_885108, EPI_ISL_885109, EPI_ISL_885110, EPI_ISL_885111, EPI_ISL_885112, EPI_ISL_885119, EPI_ISL_955628, EPI_ISL_955629, EPI_ISL_955630, EPI_ISL_955631, EPI_ISL_955632, EPI_ISL_955633, EPI_ISL_955634, EPI_ISL_955635, EPI_ISL_955636, EPI_ISL_955637, EPI_ISL_955638, EPI_ISL_955639, EPI_ISL_955640, EPI_ISL_955641, EPI_ISL_955642, EPI_ISL_955643, EPI_ISL_955644, EPI_ISL_955645, EPI_ISL_955646, EPI_ISL_955647, EPI_ISL_955648, EPI_ISL_955649, EPI_ISL_955650, EPI_ISL_955651, EPI_ISL_955652, EPI_ISL_955653, EPI_ISL_955654, EPI_ISL_955655, EPI_ISL_955656, EPI_ISL_955657, EPI_ISL_955658, EPI_ISL_955659, EPI_ISL_955660, EPI_ISL_955661, EPI_ISL_955662, EPI_ISL_955663, EPI_ISL_955664, EPI_ISL_955665, EPI_ISL_955666, EPI_ISL_955667, EPI_ISL_955668, EPI_ISL_955669, EPI_ISL_955670, EPI_ISL_955671, EPI_ISL_955672, EPI_ISL_955673, EPI_ISL_955674, EPI_ISL_955675, EPI_ISL_955676, EPI_ISL_955677, EPI_ISL_955678, EPI_ISL_955679, EPI_ISL_955680, EPI_ISL_955681, EPI_ISL_955682, EPI_ISL_955683, EPI_ISL_955684, EPI_ISL_955685, EPI_ISL_955686, EPI_ISL_955687, EPI_ISL_955688, EPI_ISL_955689, EPI_ISL_955690, EPI_ISL_955691, EPI_ISL_955692, EPI_ISL_1027350, EPI_ISL_1027436, EPI_ISL_1030290, EPI_ISL_1030291, EPI_ISL_1030292, EPI_ISL_1030293, EPI_ISL_1030294, EPI_ISL_1030295, EPI_ISL_1030296, EPI_ISL_1030297, EPI_ISL_1030298, EPI_ISL_1030299, EPI_ISL_1030308, EPI_ISL_1030312, EPI_ISL_1030444, EPI_ISL_1030445, EPI_ISL_1030446, EPI_ISL_1030447, EPI_ISL_1030448, EPI_ISL_1030449, EPI_ISL_1030450, EPI_ISL_1030451, EPI_ISL_1030452, EPI_ISL_1030453, EPI_ISL_1030454, EPI_ISL_1030455, EPI_ISL_1030456, EPI_ISL_1030457, EPI_ISL_1030458, EPI_ISL_1030459, EPI_ISL_1030460, EPI_ISL_1030461, EPI_ISL_1030462, EPI_ISL_1030463, EPI_ISL_1030464, EPI_ISL_1030465, EPI_ISL_1030466, EPI_ISL_1030467, EPI_ISL_1030468, EPI_ISL_1030469, EPI_ISL_1030470, EPI_ISL_1030471, EPI_ISL_1030472, EPI_ISL_1030473, EPI_ISL_1030474, EPI_ISL_1030475, EPI_ISL_1030476, EPI_ISL_1030477, EPI_ISL_1030478, EPI_ISL_1030479, EPI_ISL_1030480, EPI_ISL_1030481, EPI_ISL_1030482, EPI_ISL_1030483, EPI_ISL_1030484, EPI_ISL_1030485, EPI_ISL_1030486, EPI_ISL_1030487, EPI_ISL_1030488, EPI_ISL_1030489, EPI_ISL_1030490, EPI_ISL_1030491, EPI_ISL_1030492, EPI_ISL_1030493, EPI_ISL_1030494, EPI_ISL_1030495, EPI_ISL_1030496, EPI_ISL_1030497, EPI_ISL_1030498, EPI_ISL_1184681, EPI_ISL_1184682, EPI_ISL_1184683, EPI_ISL_1184684, EPI_ISL_1184685, EPI_ISL_1184686, EPI_ISL_1184687, EPI_ISL_1184688, EPI_ISL_1184689, EPI_ISL_1184690, EPI_ISL_1184691, EPI_ISL_1184692, EPI_ISL_1184693, EPI_ISL_1184694, EPI_ISL_1184695, EPI_ISL_1184696, EPI_ISL_1184697, EPI_ISL_1184698, EPI_ISL_1184699, EPI_ISL_1184700, EPI_ISL_1184701, EPI_ISL_1184702, EPI_ISL_1184703, EPI_ISL_1184704, EPI_ISL_1184705, EPI_ISL_1184706, EPI_ISL_1184707, EPI_ISL_1184708, EPI_ISL_1184709, EPI_ISL_1184710, EPI_ISL_1184711, EPI_ISL_1184712, EPI_ISL_1184713, EPI_ISL_1184714, EPI_ISL_1184715, EPI_ISL_1184716, EPI_ISL_1184717, EPI_ISL_1184718, EPI_ISL_1184719, EPI_ISL_1184720, EPI_ISL_1184721, EPI_ISL_1184722, EPI_ISL_1184723, EPI_ISL_1184724, EPI_ISL_1184725, EPI_ISL_1184726, EPI_ISL_1184727, EPI_ISL_1184728, EPI_ISL_1184729, EPI_ISL_1184730, EPI_ISL_1184731, EPI_ISL_1184732, EPI_ISL_1184733, EPI_ISL_1184734, EPI_ISL_1184735, EPI_ISL_1184736, EPI_ISL_1184737, EPI_ISL_1184738, EPI_ISL_1184739, EPI_ISL_1184740, EPI_ISL_1184741, EPI_ISL_1184742, EPI_ISL_1184743, EPI_ISL_1184744, EPI_ISL_1184745, EPI_ISL_1184746, EPI_ISL_1184747, EPI_ISL_1184748, EPI_ISL_1184749, EPI_ISL_1184750, EPI_ISL_1184751, EPI_ISL_1184752, EPI_ISL_1184753, EPI_ISL_1184754, EPI_ISL_1184755, EPI_ISL_1184756, EPI_ISL_1184757, EPI_ISL_1184758, EPI_ISL_1184759, EPI_ISL_1184760, EPI_ISL_1184761, EPI_ISL_1184762, EPI_ISL_1184763, EPI_ISL_1184764, EPI_ISL_1184765, EPI_ISL_1184766, EPI_ISL_1184767, EPI_ISL_1184768, EPI_ISL_1184769, EPI_ISL_1184770, EPI_ISL_1184771, EPI_ISL_1184772, EPI_ISL_1184773, EPI_ISL_1184774, EPI_ISL_1184775, EPI_ISL_1184776, EPI_ISL_1184777, EPI_ISL_1184778, EPI_ISL_1184779, EPI_ISL_1184780, EPI_ISL_1184781, EPI_ISL_1184782, EPI_ISL_1184783, EPI_ISL_1184784, EPI_ISL_1184785, EPI_ISL_1184786, EPI_ISL_1184787, EPI_ISL_1184788, EPI_ISL_1184789, EPI_ISL_1184790, EPI_ISL_1184791, EPI_ISL_1184792, EPI_ISL_1184793, EPI_ISL_1184794, EPI_ISL_1184795, EPI_ISL_1184796, EPI_ISL_1184797, EPI_ISL_1184798, EPI_ISL_1184799, EPI_ISL_1184800, EPI_ISL_1184801, EPI_ISL_1184802, EPI_ISL_1184803, EPI_ISL_1184804, EPI_ISL_1184805, EPI_ISL_1184806, EPI_ISL_1184807, EPI_ISL_1184808, EPI_ISL_1184967, EPI_ISL_1184968, EPI_ISL_1184969, EPI_ISL_1184970, EPI_ISL_1184971, EPI_ISL_1184972, EPI_ISL_1184973, EPI_ISL_1184974, EPI_ISL_1184975, EPI_ISL_1184976, EPI_ISL_1184977, EPI_ISL_1184978, EPI_ISL_1184979, EPI_ISL_1184980, EPI_ISL_1184981, EPI_ISL_1184982, EPI_ISL_1184983, EPI_ISL_1184984, EPI_ISL_1184985, EPI_ISL_1184986, EPI_ISL_1184987, EPI_ISL_1184988, EPI_ISL_1184989, EPI_ISL_1184990, EPI_ISL_1184991, EPI_ISL_1184992, EPI_ISL_1184993, EPI_ISL_1184994, EPI_ISL_1184995, EPI_ISL_1184996, EPI_ISL_1184997, EPI_ISL_1184998, EPI_ISL_1184999, EPI_ISL_1185000, EPI_ISL_1185001, EPI_ISL_1185002, EPI_ISL_1185003, EPI_ISL_1185004, EPI_ISL_1185005, EPI_ISL_1185006, EPI_ISL_1185007, EPI_ISL_1185008, EPI_ISL_1185009, EPI_ISL_1185010, EPI_ISL_1185011, EPI_ISL_1185012, EPI_ISL_1185013, EPI_ISL_1185014, EPI_ISL_1185015, EPI_ISL_1701400, EPI_ISL_1701426, EPI_ISL_1701431, EPI_ISL_1701450, EPI_ISL_1701465, EPI_ISL_1701478, EPI_ISL_1701483, EPI_ISL_1701491, EPI_ISL_1701511, EPI_ISL_1701513, EPI_ISL_1701533, EPI_ISL_1701537, EPI_ISL_1701543, EPI_ISL_1701545, EPI_ISL_1701586, EPI_ISL_1701599, EPI_ISL_1701611, EPI_ISL_1701616, EPI_ISL_1701623, EPI_ISL_1701629, EPI_ISL_1701657, EPI_ISL_2103006, EPI_ISL_2103025, EPI_ISL_2103028, EPI_ISL_2103029, EPI_ISL_2103035, EPI_ISL_2103036, EPI_ISL_2103067, EPI_ISL_2103081, EPI_ISL_2103085, EPI_ISL_2103086, EPI_ISL_2103094, EPI_ISL_2103100, EPI_ISL_2103114, EPI_ISL_2103122, EPI_ISL_2103134, EPI_ISL_2103152, EPI_ISL_2103158, EPI_ISL_2103159, EPI_ISL_2103178, EPI_ISL_2103187 | | | |
| see above | Orange County Public Health Lab | Chan-Zuckerberg Biohub | CZB Cliahub Consortium |
| EPI_ISL_548375, EPI_ISL_548381, EPI_ISL_548383, EPI_ISL_548387, EPI_ISL_548389, EPI_ISL_548405, EPI_ISL_548411, EPI_ISL_548412, EPI_ISL_548413, EPI_ISL_548440, EPI_ISL_548441, EPI_ISL_548465, EPI_ISL_548467, EPI_ISL_548469, EPI_ISL_548484, EPI_ISL_548580, EPI_ISL_548581, EPI_ISL_548585, EPI_ISL_548586, EPI_ISL_548587, EPI_ISL_548589, EPI_ISL_548590, EPI_ISL_548591, EPI_ISL_548592, EPI_ISL_548594, EPI_ISL_548601, EPI_ISL_548608, EPI_ISL_548609, EPI_ISL_548611, EPI_ISL_548613, EPI_ISL_548614, EPI_ISL_548615, EPI_ISL_548620, EPI_ISL_548623, EPI_ISL_548625, EPI_ISL_548627, EPI_ISL_548628, EPI_ISL_548634, EPI_ISL_548635, EPI_ISL_548637, EPI_ISL_548638, EPI_ISL_548640, EPI_ISL_548641, EPI_ISL_548646, EPI_ISL_548648, EPI_ISL_548651, EPI_ISL_548653, EPI_ISL_548654, EPI_ISL_548660, EPI_ISL_548662, EPI_ISL_548665, EPI_ISL_548666, EPI_ISL_548667, EPI_ISL_548668, EPI_ISL_548674, EPI_ISL_548675, EPI_ISL_548677 | | | |
| see above | Orange County Public Health Laboratory | Chan-Zuckerberg Biohub | CZB Cliahub Consortium |
| EPI_ISL_747242 | Pathogen Lab (BSL3), Biomedical Innovation Department, Applied and Experimental Biology Division, Scientific Research Center and High Education from Ensenada (CICESE) | Pathogen Laboratory (BSL3), Biomedical Innovation Department, Experimental and Applied Biology Division, Scientific Research Center and High Education from Ensenada (CICESE) | Cervantes-Luevano K; Galindo C and Licea-Navarro A; Martinez M; Saavedra A |
| EPI_ISL_1017051, EPI_ISL_1017059, EPI_ISL_1017060, EPI_ISL_1017061, EPI_ISL_1017062, EPI_ISL_1017063, EPI_ISL_1017065, EPI_ISL_1017097, EPI_ISL_1017098, EPI_ISL_1017099, EPI_ISL_1017108, EPI_ISL_1017109, EPI_ISL_1017110, EPI_ISL_1017111, EPI_ISL_1017116, EPI_ISL_1017117, EPI_ISL_1017126, EPI_ISL_1017127, EPI_ISL_1017128, EPI_ISL_1017129, EPI_ISL_1017130, EPI_ISL_1017131, EPI_ISL_1017132, EPI_ISL_1017147, EPI_ISL_1017148, EPI_ISL_1017149, EPI_ISL_1017150, EPI_ISL_1017151, EPI_ISL_1017152, EPI_ISL_1017153, EPI_ISL_1017154, EPI_ISL_1017155, EPI_ISL_1017156, EPI_ISL_1017157, EPI_ISL_1017158, EPI_ISL_1017159, EPI_ISL_1017160 | | | |
| see above | Providence Saint Joseph Medical Center - Burbank | Providence St. Joseph Health Molecular Genomics Laboratory | Alexa K Dowdell; Brian D Piening; Carlo B Bifulco; Fred L Robinson; Mary Campbell |
| EPI_ISL_1017106, EPI_ISL_1017118, EPI_ISL_1017119, EPI_ISL_1017133, EPI_ISL_1017134, EPI_ISL_1017135, EPI_ISL_1017136, EPI_ISL_1017137, EPI_ISL_1017138, EPI_ISL_1017139, EPI_ISL_1017140, EPI_ISL_1017141, EPI_ISL_1017142, EPI_ISL_1017143, EPI_ISL_1017144, EPI_ISL_1017145, EPI_ISL_1017146 | | | |
| see above | Providence St. Joseph Health Orange - Chapman | Providence St. Joseph Health Molecular Genomics Laboratory | Alexa K Dowdell; Brian D Piening; Carlo B Bifulco; Fred L Robinson; Mary Campbell |
| EPI_ISL_802996, EPI_ISL_802997, EPI_ISL_802998, EPI_ISL_802999, EPI_ISL_803000, EPI_ISL_803001, EPI_ISL_803002, EPI_ISL_803003, EPI_ISL_803004, EPI_ISL_803005, EPI_ISL_803006, EPI_ISL_803007, EPI_ISL_803008, EPI_ISL_803009, EPI_ISL_803010, EPI_ISL_803011, EPI_ISL_803012, EPI_ISL_803013, EPI_ISL_803014, EPI_ISL_803015, EPI_ISL_803016, EPI_ISL_803017, EPI_ISL_803018, EPI_ISL_803019, EPI_ISL_803020, EPI_ISL_803021, EPI_ISL_803022, EPI_ISL_803023, EPI_ISL_803024, EPI_ISL_803025, EPI_ISL_803026, EPI_ISL_803027, EPI_ISL_803028, EPI_ISL_803029, EPI_ISL_803030, EPI_ISL_803031, EPI_ISL_803032, EPI_ISL_803033, EPI_ISL_803034, EPI_ISL_803035, EPI_ISL_803036, EPI_ISL_803037, EPI_ISL_803038, EPI_ISL_803039, EPI_ISL_803040, EPI_ISL_803041, EPI_ISL_803042, EPI_ISL_803043, EPI_ISL_803044, EPI_ISL_803045, EPI_ISL_803046, EPI_ISL_803047, EPI_ISL_803048, EPI_ISL_803049, EPI_ISL_803050, EPI_ISL_803051, EPI_ISL_803052, EPI_ISL_803053, EPI_ISL_803054, EPI_ISL_803055, EPI_ISL_803056, EPI_ISL_803057, EPI_ISL_803058, EPI_ISL_803059, EPI_ISL_803060, EPI_ISL_803061, EPI_ISL_803062, EPI_ISL_803063, EPI_ISL_803064, EPI_ISL_803065, EPI_ISL_803066, EPI_ISL_803067, EPI_ISL_803068, EPI_ISL_803069, EPI_ISL_803070, EPI_ISL_803071, EPI_ISL_803072, EPI_ISL_803073, EPI_ISL_803074, EPI_ISL_803075, EPI_ISL_803076, EPI_ISL_803077, EPI_ISL_803078, EPI_ISL_803079, EPI_ISL_803080, EPI_ISL_803081, EPI_ISL_803082, EPI_ISL_803083, EPI_ISL_803084, EPI_ISL_803085, EPI_ISL_803086, EPI_ISL_803087, EPI_ISL_803088, EPI_ISL_803089, EPI_ISL_803090, EPI_ISL_803091, EPI_ISL_803092, EPI_ISL_803093, EPI_ISL_803094, EPI_ISL_803095, EPI_ISL_803096, EPI_ISL_803097, EPI_ISL_845657, EPI_ISL_845659, EPI_ISL_845660, EPI_ISL_845661, EPI_ISL_876827, EPI_ISL_876828, EPI_ISL_876882, EPI_ISL_876887, EPI_ISL_876888, EPI_ISL_876894, EPI_ISL_876905, EPI_ISL_876910, EPI_ISL_876913, EPI_ISL_876926, EPI_ISL_876930, EPI_ISL_876936, EPI_ISL_876950, EPI_ISL_937027, EPI_ISL_937028, EPI_ISL_937029, EPI_ISL_937030, EPI_ISL_937031, EPI_ISL_937032, EPI_ISL_937033, EPI_ISL_937034 | | | |
| see above | Quest Diagnostics | Quest Diagnostics | Anderson, B.; Bernstein; D.F.; Gerasimova, A.; Hua, M.; I.A.; K.E.; Kagan; L.E.; Lacbawan, F.; Liu Y.; Livingston; Owen, R.; Perez, A.; R.M.; Rosenthal; S.H.; Shalhout; Shlyakhter; Tanpaiboon, P. |
| EPI_ISL_878637, EPI_ISL_878640, EPI_ISL_878643, EPI_ISL_878645, EPI_ISL_878648, EPI_ISL_878651, EPI_ISL_878653, EPI_ISL_878656, EPI_ISL_878658, EPI_ISL_878661, EPI_ISL_878664, EPI_ISL_878666, EPI_ISL_878669, EPI_ISL_878671, EPI_ISL_878674, EPI_ISL_878677, EPI_ISL_878679, EPI_ISL_878682, EPI_ISL_878684, EPI_ISL_878687, EPI_ISL_878690, EPI_ISL_878693, EPI_ISL_878697, EPI_ISL_878700, EPI_ISL_878702, EPI_ISL_878705, EPI_ISL_878707, EPI_ISL_878710, EPI_ISL_878713, EPI_ISL_878716, EPI_ISL_878719, EPI_ISL_878721, EPI_ISL_878724, EPI_ISL_878726, EPI_ISL_878729, EPI_ISL_878732, EPI_ISL_878736, EPI_ISL_878739, EPI_ISL_878742, EPI_ISL_878747, EPI_ISL_878755, EPI_ISL_878757, EPI_ISL_878760, EPI_ISL_878763, EPI_ISL_878765, EPI_ISL_878768, EPI_ISL_878776, EPI_ISL_878781, EPI_ISL_878786, EPI_ISL_878791, EPI_ISL_878797, EPI_ISL_880172, EPI_ISL_880174, EPI_ISL_880175, EPI_ISL_880180, EPI_ISL_880181, EPI_ISL_880184, EPI_ISL_880185, EPI_ISL_880186, EPI_ISL_880187, EPI_ISL_880188, EPI_ISL_880189, EPI_ISL_880190, EPI_ISL_880191, EPI_ISL_880193, EPI_ISL_880194, EPI_ISL_880196, EPI_ISL_880197, EPI_ISL_880198, EPI_ISL_880199, EPI_ISL_880200 | | | |
| see above | Rady's Childrens Hospital | Andersen lab at Scripps Research | Christina Clarke; David Dimmock; Denise Malicki; Kathryn Bouic; Linda Luo; SEARCH Alliance San Diego with Nanda Radamchar; Teresa Mueller |
| EPI_ISL_738567, EPI_ISL_738574, EPI_ISL_738575, EPI_ISL_738576, EPI_ISL_738591, EPI_ISL_738660, EPI_ISL_738662, EPI_ISL_738712, EPI_ISL_738713, EPI_ISL_738714, EPI_ISL_738723, EPI_ISL_738725, EPI_ISL_738727, EPI_ISL_738770, EPI_ISL_738851, EPI_ISL_738877, EPI_ISL_738901, EPI_ISL_738977, EPI_ISL_739014, EPI_ISL_739022, EPI_ISL_739316, EPI_ISL_739348, EPI_ISL_739355, EPI_ISL_739394, EPI_ISL_739466, EPI_ISL_739467, EPI_ISL_739478, EPI_ISL_739564, EPI_ISL_739630, EPI_ISL_739655, EPI_ISL_850019, EPI_ISL_850020, EPI_ISL_850021, EPI_ISL_850022, EPI_ISL_850023, EPI_ISL_850024, EPI_ISL_850025, EPI_ISL_850026, EPI_ISL_850027, EPI_ISL_850028, EPI_ISL_850029, EPI_ISL_850030, EPI_ISL_850031, EPI_ISL_850032, EPI_ISL_850033, EPI_ISL_850034, EPI_ISL_850035, EPI_ISL_850036, EPI_ISL_850037, EPI_ISL_850038, EPI_ISL_850039, EPI_ISL_850040, EPI_ISL_850041, EPI_ISL_850042, EPI_ISL_850043, EPI_ISL_850044, EPI_ISL_850045, EPI_ISL_850046, EPI_ISL_850047, EPI_ISL_850048, EPI_ISL_850049, EPI_ISL_850050, EPI_ISL_850051, EPI_ISL_850052, EPI_ISL_850053, EPI_ISL_850054, EPI_ISL_850055, EPI_ISL_850056, EPI_ISL_850057, EPI_ISL_850058, EPI_ISL_850059, EPI_ISL_850060, EPI_ISL_850061, EPI_ISL_850062, EPI_ISL_850063, EPI_ISL_850064, EPI_ISL_850065, EPI_ISL_850066, EPI_ISL_850067, EPI_ISL_850068, EPI_ISL_850069, EPI_ISL_850070, EPI_ISL_850071, EPI_ISL_850072, EPI_ISL_850073, EPI_ISL_850074, EPI_ISL_850075, EPI_ISL_850076, EPI_ISL_850077, EPI_ISL_850078, EPI_ISL_850079, EPI_ISL_850080, EPI_ISL_850081, EPI_ISL_850082, EPI_ISL_850083, EPI_ISL_850084, EPI_ISL_850085, EPI_ISL_850086, EPI_ISL_850087, EPI_ISL_850088, EPI_ISL_850089, EPI_ISL_850090, EPI_ISL_850091, EPI_ISL_850092, EPI_ISL_850093, EPI_ISL_850094, EPI_ISL_850095, EPI_ISL_850096, EPI_ISL_850097, EPI_ISL_850098, EPI_ISL_850099, EPI_ISL_850100, EPI_ISL_850101, EPI_ISL_850102, EPI_ISL_850103, EPI_ISL_850104, EPI_ISL_850105, EPI_ISL_850106, EPI_ISL_850107, EPI_ISL_850108, EPI_ISL_850109, EPI_ISL_850110, EPI_ISL_850111, EPI_ISL_850112, EPI_ISL_850113, EPI_ISL_850114, EPI_ISL_850115, EPI_ISL_850116, EPI_ISL_850117, EPI_ISL_850118, EPI_ISL_850119, EPI_ISL_850120, EPI_ISL_850121, EPI_ISL_850122, EPI_ISL_850123, EPI_ISL_850124, EPI_ISL_850125, EPI_ISL_850126, EPI_ISL_850127, EPI_ISL_850128, EPI_ISL_850129, EPI_ISL_850130, EPI_ISL_850131, EPI_ISL_850132, EPI_ISL_850133, EPI_ISL_850134, EPI_ISL_850135, EPI_ISL_850136, EPI_ISL_850137, EPI_ISL_850138, EPI_ISL_850139, EPI_ISL_850140, EPI_ISL_850141, EPI_ISL_850142, EPI_ISL_850143, EPI_ISL_850144, EPI_ISL_850145, EPI_ISL_850146, EPI_ISL_850147, EPI_ISL_850148, EPI_ISL_850149, EPI_ISL_850150, EPI_ISL_850151, EPI_ISL_850152, EPI_ISL_850153, EPI_ISL_850154, EPI_ISL_850155, EPI_ISL_850156, EPI_ISL_850157, EPI_ISL_850158, EPI_ISL_850159, EPI_ISL_850160, EPI_ISL_850161, EPI_ISL_850162, EPI_ISL_850163, EPI_ISL_850164, EPI_ISL_850165, EPI_ISL_850166, EPI_ISL_850167, EPI_ISL_850168, EPI_ISL_850169, EPI_ISL_850170, EPI_ISL_850171, EPI_ISL_850172, EPI_ISL_850173, EPI_ISL_850174, EPI_ISL_850175, EPI_ISL_850176, EPI_ISL_850177, EPI_ISL_850178, EPI_ISL_850179 | | | |
| see above | Renegade | Chan-Zuckerberg Biohub | CZB Cliahub Consortium |

EPI_ISL_635681, EPI_ISL_635688, EPI_ISL_635701, EPI_ISL_635705, EPI_ISL_635707, EPI_ISL_635708, EPI_ISL_635709, EPI_ISL_635711, EPI_ISL_635713, EPI_ISL_635714, EPI_ISL_635716, EPI_ISL_635717, EPI_ISL_635718, EPI_ISL_635719, EPI_ISL_635720, EPI_ISL_635721, EPI_ISL_635731, EPI_ISL_635733, EPI_ISL_635735, EPI_ISL_635749, EPI_ISL_635750, EPI_ISL_635751, EPI_ISL_635752, EPI_ISL_635753, EPI_ISL_635754, EPI_ISL_635755, EPI_ISL_635756, EPI_ISL_635757, EPI_ISL_635758, EPI_ISL_635759, EPI_ISL_635760, EPI_ISL_635761, EPI_ISL_635762, EPI_ISL_635763, EPI_ISL_635764, EPI_ISL_635765, EPI_ISL_635766, EPI_ISL_635767, EPI_ISL_635768, EPI_ISL_635769, EPI_ISL_635770, EPI_ISL_635771, EPI_ISL_635772, EPI_ISL_635773, EPI_ISL_635774, EPI_ISL_635775, EPI_ISL_635776, EPI_ISL_635789, EPI_ISL_635790, EPI_ISL_635791, EPI_ISL_635792, EPI_ISL_635793, EPI_ISL_635794, EPI_ISL_635795, EPI_ISL_635796, EPI_ISL_635797, EPI_ISL_635798, EPI_ISL_635799, EPI_ISL_635800, EPI_ISL_635801, EPI_ISL_635802, EPI_ISL_635803, EPI_ISL_635804, EPI_ISL_635805, EPI_ISL_635806, EPI_ISL_635807, EPI_ISL_635808, EPI_ISL_635809, EPI_ISL_635810, EPI_ISL_635811, EPI_ISL_635812, EPI_ISL_635813, EPI_ISL_635814, EPI_ISL_635815, EPI_ISL_635816, EPI_ISL_635817, EPI_ISL_635818, EPI_ISL_635819, EPI_ISL_635820, EPI_ISL_635821, EPI_ISL_635822, EPI_ISL_635823, EPI_ISL_635824, EPI_ISL_635825, EPI_ISL_635826, EPI_ISL_635827, EPI_ISL_635828, EPI_ISL_635829, EPI_ISL_635830, EPI_ISL_635831, EPI_ISL_635832, EPI_ISL_635833, EPI_ISL_635834, EPI_ISL_635835, EPI_ISL_635836, EPI_ISL_635837, EPI_ISL_635838, EPI_ISL_635839, EPI_ISL_635840, EPI_ISL_635841, EPI_ISL_635842, EPI_ISL_635843, EPI_ISL_635844, EPI_ISL_635847, EPI_ISL_635854, EPI_ISL_635859, EPI_ISL_635860, EPI_ISL_635861, EPI_ISL_635862, EPI_ISL_635863, EPI_ISL_635864, EPI_ISL_635865, EPI_ISL_635866, EPI_ISL_635867, EPI_ISL_635868, EPI_ISL_635869, EPI_ISL_635870, EPI_ISL_635871, EPI_ISL_635873, EPI_ISL_635875, EPI_ISL_635876, EPI_ISL_635877, EPI_ISL_635878, EPI_ISL_635879, EPI_ISL_635880, EPI_ISL_635881, EPI_ISL_635882, EPI_ISL_635884, EPI_ISL_635885, EPI_ISL_635886, EPI_ISL_635887, EPI_ISL_635888, EPI_ISL_635891, EPI_ISL_635892, EPI_ISL_635893, EPI_ISL_635894, EPI_ISL_635910, EPI_ISL_635911, EPI_ISL_635913, EPI_ISL_635914, EPI_ISL_635915, EPI_ISL_635916, EPI_ISL_635917, EPI_ISL_635918, EPI_ISL_635919, EPI_ISL_635922, EPI_ISL_635924, EPI_ISL_635925, EPI_ISL_635926, EPI_ISL_635927, EPI_ISL_635928, EPI_ISL_635929, EPI_ISL_635930, EPI_ISL_635932, EPI_ISL_635933, EPI_ISL_635934, EPI_ISL_635935, EPI_ISL_635936, EPI_ISL_635937, EPI_ISL_635938, EPI_ISL_635939, EPI_ISL_635940, EPI_ISL_635941, EPI_ISL_635942, EPI_ISL_635943, EPI_ISL_635944, EPI_ISL_635945, EPI_ISL_635946, EPI_ISL_635947, EPI_ISL_635948, EPI_ISL_635949, EPI_ISL_635950, EPI_ISL_635951, EPI_ISL_635952, EPI_ISL_635953, EPI_ISL_635954, EPI_ISL_635955, EPI_ISL_635956, EPI_ISL_635957, EPI_ISL_635958, EPI_ISL_635959, EPI_ISL_635960, EPI_ISL_635961, EPI_ISL_635962, EPI_ISL_635963, EPI_ISL_635964, EPI_ISL_635965, EPI_ISL_635966, EPI_ISL_635967, EPI_ISL_635968, EPI_ISL_635969, EPI_ISL_635970, EPI_ISL_635971, EPI_ISL_635972, EPI_ISL_635973, EPI_ISL_635974, EPI_ISL_635975, EPI_ISL_635976, EPI_ISL_635977, EPI_ISL_635978, EPI_ISL_635979, EPI_ISL_635980, EPI_ISL_635981, EPI_ISL_635982, EPI_ISL_635985, EPI_ISL_635986, EPI_ISL_635987, EPI_ISL_635994, EPI_ISL_636002, EPI_ISL_636031, EPI_ISL_636032, EPI_ISL_636033, EPI_ISL_636034, EPI_ISL_636035, EPI_ISL_636036, EPI_ISL_636037, EPI_ISL_636038, EPI_ISL_636039, EPI_ISL_636040, EPI_ISL_636041, EPI_ISL_636042, EPI_ISL_636043, EPI_ISL_636044, EPI_ISL_636046, EPI_ISL_636047, EPI_ISL_636048, EPI_ISL_636049, EPI_ISL_636050, EPI_ISL_636051, EPI_ISL_636052, EPI_ISL_636053, EPI_ISL_636054, EPI_ISL_636055, EPI_ISL_636056, EPI_ISL_636057, EPI_ISL_636059, EPI_ISL_636060, EPI_ISL_636061, EPI_ISL_636062, EPI_ISL_636063, EPI_ISL_636066, EPI_ISL_636067, EPI_ISL_636068, EPI_ISL_636069, EPI_ISL_636070, EPI_ISL_636071, EPI_ISL_636121, EPI_ISL_636122, EPI_ISL_636123, EPI_ISL_636124, EPI_ISL_636125, EPI_ISL_636126, EPI_ISL_636127, EPI_ISL_636128, EPI_ISL_636129, EPI_ISL_636130, EPI_ISL_636134, EPI_ISL_636135, EPI_ISL_636136, EPI_ISL_636140, EPI_ISL_636141, EPI_ISL_636142, EPI_ISL_636144, EPI_ISL_636145, EPI_ISL_636146, EPI_ISL_636147, EPI_ISL_636148, EPI_ISL_636150, EPI_ISL_636151, EPI_ISL_636153, EPI_ISL_636154, EPI_ISL_636155, EPI_ISL_636156, EPI_ISL_636157, EPI_ISL_636158, EPI_ISL_636159, EPI_ISL_636160, EPI_ISL_636161, EPI_ISL_636162, EPI_ISL_648868, EPI_ISL_648869, EPI_ISL_648870, EPI_ISL_648871, EPI_ISL_648872, EPI_ISL_648873, EPI_ISL_648876, EPI_ISL_648877, EPI_ISL_648878, EPI_ISL_648881, EPI_ISL_648882, EPI_ISL_648883, EPI_ISL_648885, EPI_ISL_648886, EPI_ISL_648887, EPI_ISL_648888, EPI_ISL_648890, EPI_ISL_648891, EPI_ISL_648892, EPI_ISL_648894, EPI_ISL_648895, EPI_ISL_648898, EPI_ISL_648899, EPI_ISL_648900, EPI_ISL_648901, EPI_ISL_648902, EPI_ISL_648903, EPI_ISL_648904, EPI_ISL_648906, EPI_ISL_648907, EPI_ISL_648908, EPI_ISL_648909, EPI_ISL_648910, EPI_ISL_648911, EPI_ISL_648913, EPI_ISL_648914, EPI_ISL_648915, EPI_ISL_648916, EPI_ISL_648917, EPI_ISL_648919, EPI_ISL_648920, EPI_ISL_648921, EPI_ISL_648922, EPI_ISL_648923, EPI_ISL_648924, EPI_ISL_648925, EPI_ISL_648926, EPI_ISL_648927, EPI_ISL_648928, EPI_ISL_648929, EPI_ISL_648930, EPI_ISL_648931, EPI_ISL_648932, EPI_ISL_648933, EPI_ISL_648934, EPI_ISL_648935, EPI_ISL_648936, EPI_ISL_648937, EPI_ISL_648938, EPI_ISL_648939, EPI_ISL_648940, EPI_ISL_648941, EPI_ISL_648942, EPI_ISL_648943, EPI_ISL_648944, EPI_ISL_648945, EPI_ISL_648946, EPI_ISL_648947, EPI_ISL_648948, EPI_ISL_648949, EPI_ISL_648950, EPI_ISL_648951, EPI_ISL_648952, EPI_ISL_648953, EPI_ISL_648954, EPI_ISL_648955, EPI_ISL_648956, EPI_ISL_648957, EPI_ISL_648958, EPI_ISL_648959, EPI_ISL_648960, EPI_ISL_648961, EPI_ISL_648962, EPI_ISL_648963, EPI_ISL_648964, EPI_ISL_648965, EPI_ISL_648966, EPI_ISL_648967, EPI_ISL_648968, EPI_ISL_648969, EPI_ISL_648970, EPI_ISL_648971, EPI_ISL_648972, EPI_ISL_648973, EPI_ISL_648974, EPI_ISL_648975, EPI_ISL_648976, EPI_ISL_648977, EPI_ISL_648978, EPI_ISL_648979, EPI_ISL_648980, EPI_ISL_648985, EPI_ISL_648988, EPI_ISL_648989, EPI_ISL_648990, EPI_ISL_648993, EPI_ISL_648994, EPI_ISL_649000, EPI_ISL_649001, EPI_ISL_649002, EPI_ISL_649003, EPI_ISL_649005, EPI_ISL_649006, EPI_ISL_649007, EPI_ISL_649009, EPI_ISL_649010, EPI_ISL_649011, EPI_ISL_649012, EPI_ISL_649013, EPI_ISL_649014, EPI_ISL_649015, EPI_ISL_649018, EPI_ISL_649020, EPI_ISL_649021, EPI_ISL_649022, EPI_ISL_649024, EPI_ISL_649025, EPI_ISL_649026, EPI_ISL_649027, EPI_ISL_649028, EPI_ISL_649029, EPI_ISL_649030, EPI_ISL_649031, EPI_ISL_649032, EPI_ISL_649034, EPI_ISL_649035, EPI_ISL_649038, EPI_ISL_649039, EPI_ISL_649040, EPI_ISL_649041, EPI_ISL_649042, EPI_ISL_649044, EPI_ISL_649045, EPI_ISL_649046, EPI_ISL_649048, EPI_ISL_649049, EPI_ISL_649050, EPI_ISL_649051, EPI_ISL_649053, EPI_ISL_649054, EPI_ISL_649056, EPI_ISL_666965, EPI_ISL_666966, EPI_ISL_666967, EPI_ISL_666968, EPI_ISL_666969, EPI_ISL_666970, EPI_ISL_666971, EPI_ISL_666972, EPI_ISL_666973, EPI_ISL_666974, EPI_ISL_666975, EPI_ISL_666976, EPI_ISL_666977, EPI_ISL_666978, EPI_ISL_666979, EPI_ISL_666980, EPI_ISL_666981, EPI_ISL_666982, EPI_ISL_666983, EPI_ISL_666984, EPI_ISL_666985, EPI_ISL_666986, EPI_ISL_666987, EPI_ISL_666988, EPI_ISL_666989, EPI_ISL_666990, EPI_ISL_666991, EPI_ISL_666993, EPI_ISL_666994, EPI_ISL_666995, EPI_ISL_666996, EPI_ISL_666997, EPI_ISL_666998, EPI_ISL_667042, EPI_ISL_667043, EPI_ISL_667044, EPI_ISL_667045, EPI_ISL_667046, EPI_ISL_667047, EPI_ISL_667048, EPI_ISL_667049, EPI_ISL_667050, EPI_ISL_667051, EPI_ISL_667052, EPI_ISL_667053, EPI_ISL_667054, EPI_ISL_667055, EPI_ISL_667056, EPI_ISL_667057, EPI_ISL_667058, EPI_ISL_667059, EPI_ISL_667060, EPI_ISL_667061, EPI_ISL_667062, EPI_ISL_667063, EPI_ISL_667064, EPI_ISL_667065, EPI_ISL_667066, EPI_ISL_667067, EPI_ISL_667068, EPI_ISL_667069, EPI_ISL_667070, EPI_ISL_667071, EPI_ISL_667072, EPI_ISL_667074, EPI_ISL_667075, EPI_ISL_667076, EPI_ISL_667077, EPI_ISL_667078, EPI_ISL_730085, EPI_ISL_730086, EPI_ISL_730087, EPI_ISL_730089, EPI_ISL_730090, EPI_ISL_730091, EPI_ISL_730092, EPI_ISL_730093, EPI_ISL_730094, EPI_ISL_730095, EPI_ISL_730097, EPI_ISL_730098, EPI_ISL_730099, EPI_ISL_730100, EPI_ISL_730101, EPI_ISL_730102, EPI_ISL_730105, EPI_ISL_730106, EPI_ISL_730107, EPI_ISL_730108, EPI_ISL_730109, EPI_ISL_730110, EPI_ISL_730111, EPI_ISL_730112, EPI_ISL_730113, EPI_ISL_730114, EPI_ISL_730115, EPI_ISL_730116, EPI_ISL_730117, EPI_ISL_730120, EPI_ISL_730121, EPI_ISL_730122, EPI_ISL_730123, EPI_ISL_730124, EPI_ISL_730125, EPI_ISL_730161, EPI_ISL_730162, EPI_ISL_730163, EPI_ISL_730164, EPI_ISL_730165, EPI_ISL_730166, EPI_ISL_730167, EPI_ISL_730168, EPI_ISL_730169, EPI_ISL_730170, EPI_ISL_730171, EPI_ISL_730172, EPI_ISL_730173, EPI_ISL_730174, EPI_ISL_730175, EPI_ISL_730176, EPI_ISL_730177, EPI_ISL_730178, EPI_ISL_730179, EPI_ISL_730180, EPI_ISL_730181, EPI_ISL_730182, EPI_ISL_730183, EPI_ISL_730184, EPI_ISL_730185, EPI_ISL_730186, EPI_ISL_730187, EPI_ISL_730188, EPI_ISL_730189, EPI_ISL_730190, EPI_ISL_730191, EPI_ISL_730192, EPI_ISL_730193, EPI_ISL_730194, EPI_ISL_730195, EPI_ISL_730196, EPI_ISL_730291, EPI_ISL_730292, EPI_ISL_730293, EPI_ISL_730294, EPI_ISL_730295, EPI_ISL_730296, EPI_ISL_730297, EPI_ISL_730298, EPI_ISL_730299, EPI_ISL_730300, EPI_ISL_730301, EPI_ISL_730302, EPI_ISL_730303, EPI_ISL_730304, EPI_ISL_730305, EPI_ISL_730306, EPI_ISL_730307, EPI_ISL_730308, EPI_ISL_730309, EPI_ISL_730310, EPI_ISL_730311, EPI_ISL_730312, EPI_ISL_730313, EPI_ISL_730314, EPI_ISL_730315, EPI_ISL_730316, EPI_ISL_730317, EPI_ISL_730318, EPI_ISL_730319, EPI_ISL_730320, EPI_ISL_730322, EPI_ISL_730323, EPI_ISL_730324, EPI_ISL_730325, EPI_ISL_730326, EPI_ISL_730328, EPI_ISL_730329, EPI_ISL_730332, EPI_ISL_730333, EPI_ISL_730335, EPI_ISL_730336, EPI_ISL_730337, EPI_ISL_730338, EPI_ISL_730340, EPI_ISL_730341, EPI_ISL_730342, EPI_ISL_730345, EPI_ISL_730346, EPI_ISL_730348, EPI_ISL_730349, EPI_ISL_730350, EPI_ISL_730351, EPI_ISL_730352, EPI_ISL_730353, EPI_ISL_730354, EPI_ISL_730355, EPI_ISL_730356, EPI_ISL_730357, EPI_ISL_730358, EPI_ISL_730359, EPI_ISL_730360, EPI_ISL_730361, EPI_ISL_730362, EPI_ISL_730363, EPI_ISL_730364, EPI_ISL_730365, EPI_ISL_730366, EPI_ISL_730367, EPI_ISL_730368, EPI_ISL_730369, EPI_ISL_730370, EPI_ISL_730371, EPI_ISL_755115, EPI_ISL_755116, EPI_ISL_755117, EPI_ISL_755119, EPI_ISL_755130, EPI_ISL_755132, EPI_ISL_755134, EPI_ISL_755135, EPI_ISL_755136, EPI_ISL_755138, EPI_ISL_755139, EPI_ISL_755269, EPI_ISL_755270, EPI_ISL_755271, EPI_ISL_755272, EPI_ISL_755273, EPI_ISL_755274, EPI_ISL_755275, EPI_ISL_755276, EPI_ISL_755277, EPI_ISL_755278, EPI_ISL_755279, EPI_ISL_755280, EPI_ISL_755281, EPI_ISL_755282, EPI_ISL_755283, EPI_ISL_755284, EPI_ISL_755285, EPI_ISL_755286, EPI_ISL_755287, EPI_ISL_755288, EPI_ISL_755289, EPI_ISL_755290, EPI_ISL_755291, EPI_ISL_755292, EPI_ISL_755293, EPI_ISL_755294, EPI_ISL_755295, EPI_ISL_755296, EPI_ISL_755297, EPI_ISL_755298, EPI_ISL_755299, EPI_ISL_755300, EPI_ISL_878152, EPI_ISL_878155, EPI_ISL_878157, EPI_ISL_878159, EPI_ISL_878162, EPI_ISL_878165, EPI_ISL_878167, EPI_ISL_878170, EPI_ISL_878172, EPI_ISL_878175, EPI_ISL_878178, EPI_ISL_878180, EPI_ISL_878183, EPI_ISL_878186, EPI_ISL_878189, EPI_ISL_878191, EPI_ISL_878194, EPI_ISL_878197, EPI_ISL_878199, EPI_ISL_878202, EPI_ISL_878205, EPI_ISL_878209, EPI_ISL_878212, EPI_ISL_878215, EPI_ISL_878217, EPI_ISL_878221, EPI_ISL_878223, EPI_ISL_878226, EPI_ISL_878229, EPI_ISL_878232, EPI_ISL_878234, EPI_ISL_878237, EPI_ISL_878239, EPI_ISL_878242, EPI_ISL_878245, EPI_ISL_878248, EPI_ISL_878250, EPI_ISL_878253, EPI_ISL_878256, EPI_ISL_878258, EPI_ISL_878260, EPI_ISL_878263, EPI_ISL_878266, EPI_ISL_878268, EPI_ISL_878271, EPI_ISL_878274, EPI_ISL_878276, EPI_ISL_878279, EPI_ISL_878281, EPI_ISL_878284, EPI_ISL_878287, EPI_ISL_878290, EPI_ISL_878292, EPI_ISL_878295, EPI_ISL_878298, EPI_ISL_878300, EPI_ISL_878303, EPI_ISL_878305, EPI_ISL_878308, EPI_ISL_878311, EPI_ISL_878314, EPI_ISL_878316, EPI_ISL_878319, EPI_ISL_878322, EPI_ISL_878325, EPI_ISL_878327, EPI_ISL_878330, EPI_ISL_878333, EPI_ISL_878336, EPI_ISL_878338, EPI_ISL_878341, EPI_ISL_878342, EPI_ISL_878343, EPI_ISL_878344, EPI_ISL_878345, EPI_ISL_878346, EPI_ISL_878347, EPI_ISL_878348, EPI_ISL_878349, EPI_ISL_878350, EPI_ISL_878351, EPI_ISL_878352, EPI_ISL_878353, EPI_ISL_878354, EPI_ISL_878355, EPI_ISL_878356, EPI_ISL_878357, EPI_ISL_878358, EPI_ISL_878359, EPI_ISL_878360, EPI_ISL_878361, EPI_ISL_878362, EPI_ISL_878363, EPI_ISL_878364, EPI_ISL_878365, EPI_ISL_878366, EPI_ISL_878367, EPI_ISL_878368, EPI_ISL_878369, EPI_ISL_878370, EPI_ISL_878371, EPI_ISL_878372, EPI_ISL_878373, EPI_ISL_878374, EPI_ISL_878375, EPI_ISL_878376, EPI_ISL_878377, EPI_ISL_878378, EPI_ISL_878379, EPI_ISL_878380, EPI_ISL_878381, EPI_ISL_878382, EPI_ISL_878383, EPI_ISL_878384, EPI_ISL_878385, EPI_ISL_878386, EPI_ISL_878387, EPI_ISL_878388, EPI_ISL_878389, EPI_ISL_878390, EPI_ISL_878391, EPI_ISL_878392, EPI_ISL_878393, EPI_ISL_878394, EPI_ISL_878395, EPI_ISL_878396, EPI_ISL_878397, EPI_ISL_878398, EPI_ISL_878399, EPI_ISL_878400, EPI_ISL_878401, EPI_ISL_878402, EPI_ISL_878403, EPI_ISL_878404, EPI_ISL_878405, EPI_ISL_878406, EPI_ISL_878407, EPI_ISL_878408, EPI_ISL_878411, EPI_ISL_878413, EPI_ISL_878416, EPI_ISL_878418, EPI_ISL_878421, EPI_ISL_878423, EPI_ISL_878426, EPI_ISL_878429, EPI_ISL_878431, EPI_ISL_878434, EPI_ISL_878437, EPI_ISL_878584, EPI_ISL_878587, EPI_ISL_878589, EPI_ISL_878592, EPI_ISL_878595, EPI_ISL_878597, EPI_ISL_878600, EPI_ISL_878603, EPI_ISL_878605, EPI_ISL_878608, EPI_ISL_878611, EPI_ISL_878613, EPI_ISL_878616, EPI_ISL_878619, EPI_ISL_878624, EPI_ISL_878629, EPI_ISL_878632, EPI_ISL_878635, EPI_ISL_879924, EPI_ISL_879926, EPI_ISL_879929, EPI_ISL_879932, EPI_ISL_879934, EPI_ISL_879937, EPI_ISL_879939, EPI_ISL_879942, EPI_ISL_879945, EPI_ISL_879947, EPI_ISL_879950, EPI_ISL_879953, EPI_ISL_879956, EPI_ISL_879959, EPI_ISL_879961, EPI_ISL_879964, EPI_ISL_879967, EPI_ISL_879969, EPI_ISL_879972, EPI_ISL_879975, EPI_ISL_879977, EPI_ISL_879980, EPI_ISL_879982, EPI_ISL_879985, EPI_ISL_879987, EPI_ISL_879990, EPI_ISL_879993, EPI_ISL_879995, EPI_ISL_879998, EPI_ISL_880000, EPI_ISL_880003, EPI_ISL_880006, EPI_ISL_880009, EPI_ISL_880011, EPI_ISL_880014, EPI_ISL_880017, EPI_ISL_880019, EPI_ISL_880022, EPI_ISL_880024, EPI_ISL_880027, EPI_ISL_880029, EPI_ISL_880032, EPI_ISL_880034, EPI_ISL_962609, EPI_ISL_962610, EPI_ISL_962612, EPI_ISL_962614, EPI_ISL_962615, EPI_ISL_962616, EPI_ISL_962617, EPI_ISL_962766, EPI_ISL_962767, EPI_ISL_962768, EPI_ISL_962769, EPI_ISL_962770, EPI_ISL_962771, EPI_ISL_962772, EPI_ISL_962773, EPI_ISL_962774, EPI_ISL_962775, EPI_ISL_962776, EPI_ISL_962777, EPI_ISL_962778, EPI_ISL_962779, EPI_ISL_962780, EPI_ISL_962781, EPI_ISL_962782, EPI_ISL_962783, EPI_ISL_962784, EPI_ISL_962785, EPI_ISL_962786, EPI_ISL_962788, EPI_ISL_962789, EPI_ISL_962790, EPI_ISL_962791, EPI_ISL_962792, EPI_ISL_962793, EPI_ISL_962794, EPI_ISL_962795, EPI_ISL_962796, EPI_ISL_962797, EPI_ISL_962798, EPI_ISL_962799, EPI_ISL_962800, EPI_ISL_962801, EPI_ISL_962802, EPI_ISL_962803, EPI_ISL_962804, EPI_ISL_962805, EPI_ISL_962806, EPI_ISL_962807, EPI_ISL_962808, EPI_ISL_962809, EPI_ISL_962810, EPI_ISL_962811, EPI_ISL_1081457, EPI_ISL_1081458, EPI_ISL_1366747, EPI_ISL_1531942, EPI_ISL_1531943, EPI_ISL_1531944, EPI_ISL_1531945, EPI_ISL_1531946, EPI_ISL_1531947, EPI_ISL_1531948, EPI_ISL_1531949, EPI_ISL_1531950, EPI_ISL_1531951, EPI_ISL_1531952, EPI_ISL_1531953, EPI_ISL_1531954, EPI_ISL_1531955, EPI_ISL_1531956, EPI_ISL_1531957, EPI_ISL_1531958, EPI_ISL_1531959, EPI_ISL_1531960, EPI_ISL_1531961, EPI_ISL_1531962, EPI_ISL_1531963, EPI_ISL_1531964, EPI_ISL_1531965, EPI_ISL_1531966, EPI_ISL_1531967, EPI_ISL_1531968, EPI_ISL_1531969, EPI_ISL_1531970, EPI_ISL_1531971, EPI_ISL_1531972, EPI_ISL_1531973, EPI_ISL_1531974, EPI_ISL_1531975, EPI_ISL_1531976, EPI_ISL_1531977, EPI_ISL_1531978, EPI_ISL_1531979, EPI_ISL_1531980, EPI_ISL_1531981, EPI_ISL_1531982, EPI_ISL_1531983, EPI_ISL_1531984, EPI_ISL_1531985, EPI_ISL_1531986, EPI_ISL_1531987, EPI_ISL_1531988, EPI_ISL_1531989, EPI_ISL_1531990, EPI_ISL_1531991, EPI_ISL_1531992, EPI_ISL_1531993, EPI_ISL_1531994, EPI_ISL_1531995, EPI_ISL_1531996, EPI_ISL_1531997, EPI_ISL_1531998, EPI_ISL_1531999, EPI_ISL_1532000, EPI_ISL_1532001, EPI_ISL_1532002, EPI_ISL_1532003, EPI_ISL_1532004, EPI_ISL_1532005, EPI_ISL_1532006, EPI_ISL_1532007, EPI_ISL_1532008, EPI_ISL_1532009, EPI_ISL_1532010, EPI_ISL_1532011, EPI_ISL_1532012, EPI_ISL_1532013, EPI_ISL_1532014, EPI_ISL_1532015, EPI_ISL_1532016, EPI_ISL_1532017, EPI_ISL_1532018, EPI_ISL_1532019, EPI_ISL_1532020, EPI_ISL_1532021, EPI_ISL_1532022, EPI_ISL_1532023, EPI_ISL_1532024, EPI_ISL_1532025, EPI_ISL_1532026, EPI_ISL_1532027, EPI_ISL_1532028, EPI_ISL_1532029, EPI_ISL_1532030, EPI_ISL_1532031, EPI_ISL_1532032, EPI_ISL_1532033, EPI_ISL_1532034, EPI_ISL_1532035, EPI_ISL_1532036, EPI_ISL_1532037, EPI_ISL_1532038, EPI_ISL_1532039, EPI_ISL_1532040, EPI_ISL_1532041, EPI_ISL_1532042, EPI_ISL_1532043, EPI_ISL_1532044, EPI_ISL_1532045, EPI_ISL_1532046, EPI_ISL_1532047, EPI_ISL_1532048, EPI_ISL_1532049, EPI_ISL_1532050, EPI_ISL_1532051, EPI_ISL_1532052, EPI_ISL_1532053, EPI_ISL_1532054, EPI_ISL_1532055, EPI_ISL_1532056, EPI_ISL_1532057, EPI_ISL_1532058, EPI_ISL_1532059, EPI_ISL_1532060, EPI_ISL_1532061, EPI_ISL_1532062, EPI_ISL_1532063, EPI_ISL_1532064, EPI_ISL_1794620, EPI_ISL_1794627, EPI_ISL_1794629, EPI_ISL_1794634, EPI_ISL_1794636, EPI_ISL_1794641, EPI_ISL_1794645, EPI_ISL_1794650, EPI_ISL_1794656, EPI_ISL_1794657, EPI_ISL_1794658, EPI_ISL_1794659, EPI_ISL_1794660, EPI_ISL_1794661, EPI_ISL_1794662, EPI_ISL_1794663, EPI_ISL_1794664, EPI_ISL_1794665, EPI_ISL_1794666, EPI_ISL_1794667, EPI_ISL_1794669, EPI_ISL_1794671, EPI_ISL_1794672, EPI_ISL_1794673, EPI_ISL_1794677, EPI_ISL_1794678, EPI_ISL_1794680, EPI_ISL_1794682, EPI_ISL_1794683, EPI_ISL_1794684, EPI_ISL_1794939, EPI_ISL_1794940, EPI_ISL_1794941, EPI_ISL_1794943, EPI_ISL_1794945, EPI_ISL_1794946, EPI_ISL_1794947, EPI_ISL_1794951, EPI_ISL_1794955, EPI_ISL_1794956, EPI_ISL_1794960, EPI_ISL_1794962, EPI_ISL_1794963, EPI_ISL_1794964, EPI_ISL_1794967, EPI_ISL_1794968, EPI_ISL_1794971, EPI_ISL_1794972, EPI_ISL_1794976, EPI_ISL_1794977, EPI_ISL_1794978, EPI_ISL_1794980, EPI_ISL_1794981, EPI_ISL_1794982, EPI_ISL_1969282, EPI_ISL_1969284, EPI_ISL_1969285, EPI_ISL_1969286, EPI_ISL_1969287, EPI_ISL_1969289, EPI_ISL_1969293, EPI_ISL_1969295, EPI_ISL_1969300, EPI_ISL_1969301, EPI_ISL_1969303, EPI_ISL_1969305, EPI_ISL_1969306, EPI_ISL_1969308, EPI_ISL_1969319, EPI_ISL_1969320, EPI_ISL_1969321, EPI_ISL_1969322, EPI_ISL_1969323, EPI_ISL_1969372, EPI_ISL_1969379, EPI_ISL_1969428, EPI_ISL_1969433, EPI_ISL_1969451, EPI_ISL_1969452, EPI_ISL_1969453, EPI_ISL_1969454, EPI_ISL_1969455, EPI_ISL_1969456, EPI_ISL_1969457, EPI_ISL_1969458, EPI_ISL_1969459, EPI_ISL_1969460, EPI_ISL_1969461, EPI_ISL_1969462, EPI_ISL_1969463, EPI_ISL_1969464, EPI_ISL_1969465, EPI_ISL_1969466, EPI_ISL_1969467, EPI_ISL_1969468, EPI_ISL_1969469, EPI_ISL_1969470, EPI_ISL_1969471, EPI_ISL_1969472, EPI_ISL_1969473, EPI_ISL_1969474, EPI_ISL_1969475, EPI_ISL_1969476, EPI_ISL_1969477, EPI_ISL_1969478, EPI_ISL_1969479, EPI_ISL_1969480, EPI_ISL_1969481, EPI_ISL_1969482, EPI_ISL_1969483, EPI_ISL_1969484, EPI_ISL_1969485, EPI_ISL_1969486, EPI_ISL_1969487, EPI_ISL_1969488, EPI_ISL_1969489, EPI_ISL_1969490, EPI_ISL_1969491, EPI_ISL_1969492, EPI_ISL_1969493, EPI_ISL_1969494, EPI_ISL_1969495, EPI_ISL_1969496, EPI_ISL_1969497, EPI_ISL_1969498, EPI_ISL_1969501, EPI_ISL_1969502, EPI_ISL_1969503, EPI_ISL_1969504, EPI_ISL_1969505, EPI_ISL_1969506

| | | | |
|---|---|---|---|
| see above | San Diego County Public Health Laboratory | Andersen lab at Scripps Research | Brett Austin; Jovan Shephard; SEARCH Alliance San Diego with Ashleigh Murphy; SEARCH Alliance San Diego with Tracy Basler |

EPI_ISL_583190, EPI_ISL_583191, EPI_ISL_583192, EPI_ISL_583193, EPI_ISL_583194, EPI_ISL_583195, EPI_ISL_583196, EPI_ISL_583197, EPI_ISL_672378, EPI_ISL_672380, EPI_ISL_672381, EPI_ISL_739603

| | | | |
|---|---|---|---|
| see above | San Francisco Public Health Laboratory | Chan-Zuckerberg Biohub | CZB Cliahub Consortium |
| EPI_ISL_625660 | San Joaquin County Public Health Lab | Chan-Zuckerberg Biohub | CZB Cliahub Consortium |
| EPI_ISL_582974 | San Luis Obispo Public Health Department | Chan-Zuckerberg Biohub | CZB Cliahub Consortium |

EPI_ISL_625478, EPI_ISL_625479, EPI_ISL_625480, EPI_ISL_625481, EPI_ISL_625482, EPI_ISL_625483, EPI_ISL_625484, EPI_ISL_625485, EPI_ISL_625486, EPI_ISL_625487, EPI_ISL_625488, EPI_ISL_625489, EPI_ISL_625490, EPI_ISL_625491, EPI_ISL_625492, EPI_ISL_625493, EPI_ISL_625494, EPI_ISL_625495, EPI_ISL_625496, EPI_ISL_625497, EPI_ISL_625498, EPI_ISL_625499, EPI_ISL_625600, EPI_ISL_625601, EPI_ISL_625602, EPI_ISL_625603, EPI_ISL_625604, EPI_ISL_625605, EPI_ISL_625606, EPI_ISL_625607, EPI_ISL_625608, EPI_ISL_625609, EPI_ISL_625610, EPI_ISL_648381, EPI_ISL_648382, EPI_ISL_648383, EPI_ISL_648384, EPI_ISL_648385, EPI_ISL_648386, EPI_ISL_648387, EPI_ISL_648388, EPI_ISL_648389, EPI_ISL_648390, EPI_ISL_648391, EPI_ISL_648392, EPI_ISL_648393, EPI_ISL_648394, EPI_ISL_648395, EPI_ISL_648396, EPI_ISL_648397, EPI_ISL_648398, EPI_ISL_648399, EPI_ISL_648400, EPI_ISL_648401, EPI_ISL_648402, EPI_ISL_648403, EPI_ISL_648404, EPI_ISL_648405, EPI_ISL_648406, EPI_ISL_648407, EPI_ISL_648408, EPI_ISL_648409, EPI_ISL_648410, EPI_ISL_648411, EPI_ISL_648412, EPI_ISL_648413, EPI_ISL_648414, EPI_ISL_648415, EPI_ISL_648416, EPI_ISL_648417, EPI_ISL_648418, EPI_ISL_648419, EPI_ISL_648420, EPI_ISL_648421, EPI_ISL_648422, EPI_ISL_648423, EPI_ISL_648424, EPI_ISL_648425, EPI_ISL_648426, EPI_ISL_648427, EPI_ISL_648428, EPI_ISL_648429, EPI_ISL_648430, EPI_ISL_648431, EPI_ISL_648432, EPI_ISL_648433, EPI_ISL_648434, EPI_ISL_648435, EPI_ISL_648436, EPI_ISL_648437, EPI_ISL_648438, EPI_ISL_648439, EPI_ISL_648440, EPI_ISL_648441, EPI_ISL_648442, EPI_ISL_648443, EPI_ISL_648444, EPI_ISL_648445, EPI_ISL_648446, EPI_ISL_648447, EPI_ISL_648448, EPI_ISL_648449, EPI_ISL_648450, EPI_ISL_648451, EPI_ISL_648452, EPI_ISL_648453, EPI_ISL_648454, EPI_ISL_648455, EPI_ISL_648456, EPI_ISL_648457, EPI_ISL_648458, EPI_ISL_648459, EPI_ISL_648460, EPI_ISL_648461, EPI_ISL_648462, EPI_ISL_648463, EPI_ISL_648464, EPI_ISL_648465, EPI_ISL_648466, EPI_ISL_648467, EPI_ISL_648468, EPI_ISL_648469, EPI_ISL_648470, EPI_ISL_648471, EPI_ISL_648472, EPI_ISL_648473, EPI_ISL_648474, EPI_ISL_648475, EPI_ISL_648476, EPI_ISL_648477, EPI_ISL_648478, EPI_ISL_648479, EPI_ISL_648480, EPI_ISL_648481, EPI_ISL_648482, EPI_ISL_648483, EPI_ISL_648484, EPI_ISL_648485, EPI_ISL_648486, EPI_ISL_648487, EPI_ISL_648488, EPI_ISL_648489, EPI_ISL_648490, EPI_ISL_648491, EPI_ISL_648492, EPI_ISL_648493, EPI_ISL_648494, EPI_ISL_648495, EPI_ISL_672013, EPI_ISL_672031, EPI_ISL_672033, EPI_ISL_672035, EPI_ISL_672043, EPI_ISL_672044, EPI_ISL_672052, EPI_ISL_672057, EPI_ISL_672065, EPI_ISL_672069, EPI_ISL_672070, EPI_ISL_672072, EPI_ISL_672073, EPI_ISL_672080, EPI_ISL_672085, EPI_ISL_672088, EPI_ISL_672089, EPI_ISL_672104, EPI_ISL_672106, EPI_ISL_672107, EPI_ISL_672108, EPI_ISL_672111, EPI_ISL_672277, EPI_ISL_672278, EPI_ISL_672279, EPI_ISL_672280, EPI_ISL_672281, EPI_ISL_672282, EPI_ISL_672283, EPI_ISL_672284, EPI_ISL_672285, EPI_ISL_672286, EPI_ISL_672287, EPI_ISL_672288, EPI_ISL_672289, EPI_ISL_672290, EPI_ISL_672291, EPI_ISL_672292, EPI_ISL_672293, EPI_ISL_672294, EPI_ISL_672295, EPI_ISL_672296, EPI_ISL_672297, EPI_ISL_672298, EPI_ISL_672299, EPI_ISL_672300, EPI_ISL_672301, EPI_ISL_672302, EPI_ISL_672304, EPI_ISL_672429, EPI_ISL_672430, EPI_ISL_672431, EPI_ISL_672432, EPI_ISL_672433, EPI_ISL_672434, EPI_ISL_672435, EPI_ISL_672436, EPI_ISL_672437, EPI_ISL_672438, EPI_ISL_738500, EPI_ISL_738501, EPI_ISL_738502, EPI_ISL_738508, EPI_ISL_738511, EPI_ISL_738516, EPI_ISL_738517, EPI_ISL_738520, EPI_ISL_738521, EPI_ISL_738525, EPI_ISL_738526, EPI_ISL_738532, EPI_ISL_738533, EPI_ISL_738534, EPI_ISL_738536, EPI_ISL_738542, EPI_ISL_738543, EPI_ISL_738546, EPI_ISL_738549, EPI_ISL_738550, EPI_ISL_738551, EPI_ISL_738552, EPI_ISL_738555, EPI_ISL_738556, EPI_ISL_738557, EPI_ISL_738558, EPI_ISL_738560, EPI_ISL_738566, EPI_ISL_738568, EPI_ISL_738578, EPI_ISL_738579, EPI_ISL_738581, EPI_ISL_738582, EPI_ISL_738583, EPI_ISL_738586, EPI_ISL_738589, EPI_ISL_738590, EPI_ISL_738593, EPI_ISL_738595, EPI_ISL_738596, EPI_ISL_738598, EPI_ISL_738601, EPI_ISL_738610, EPI_ISL_738611, EPI_ISL_738612, EPI_ISL_738617, EPI_ISL_738618, EPI_ISL_738628, EPI_ISL_738631, EPI_ISL_738643, EPI_ISL_738644, EPI_ISL_738645, EPI_ISL_738650, EPI_ISL_738652, EPI_ISL_738653, EPI_ISL_738657, EPI_ISL_738661, EPI_ISL_738663, EPI_ISL_738667, EPI_ISL_738670, EPI_ISL_738672, EPI_ISL_738673, EPI_ISL_738681, EPI_ISL_738682, EPI_ISL_738683, EPI_ISL_738684, EPI_ISL_738685, EPI_ISL_738689, EPI_ISL_738690, EPI_ISL_738691, EPI_ISL_738695, EPI_ISL_738696, EPI_ISL_738697, EPI_ISL_738698, EPI_ISL_738700, EPI_ISL_738701, EPI_ISL_738702, EPI_ISL_738705, EPI_ISL_738706, EPI_ISL_738710, EPI_ISL_738716, EPI_ISL_738717, EPI_ISL_738718, EPI_ISL_738719, EPI_ISL_738720, EPI_ISL_738726, EPI_ISL_738728, EPI_ISL_738730, EPI_ISL_738746, EPI_ISL_738747, EPI_ISL_738750, EPI_ISL_738763, EPI_ISL_738764, EPI_ISL_738765, EPI_ISL_738769, EPI_ISL_738771, EPI_ISL_738772, EPI_ISL_738780, EPI_ISL_738789, EPI_ISL_738791, EPI_ISL_738794, EPI_ISL_738798, EPI_ISL_738803, EPI_ISL_738805, EPI_ISL_738812, EPI_ISL_738834, EPI_ISL_738839, EPI_ISL_738842, EPI_ISL_738853, EPI_ISL_738855, EPI_ISL_738859, EPI_ISL_738861, EPI_ISL_738864, EPI_ISL_738867, EPI_ISL_738883, EPI_ISL_738887, EPI_ISL_738889, EPI_ISL_738894, EPI_ISL_738898, EPI_ISL_738903, EPI_ISL_738906, EPI_ISL_738909, EPI_ISL_738910, EPI_ISL_738915, EPI_ISL_738920, EPI_ISL_738922, EPI_ISL_738924, EPI_ISL_738926, EPI_ISL_738927, EPI_ISL_738931, EPI_ISL_738933, EPI_ISL_738936, EPI_ISL_738941, EPI_ISL_738942, EPI_ISL_738943, EPI_ISL_738948, EPI_ISL_738955, EPI_ISL_738956, EPI_ISL_738960, EPI_ISL_738962, EPI_ISL_738965, EPI_ISL_738967, EPI_ISL_738971, EPI_ISL_738972, EPI_ISL_738979, EPI_ISL_738984, EPI_ISL_738994, EPI_ISL_738996, EPI_ISL_738997, EPI_ISL_738998, EPI_ISL_739005, EPI_ISL_739006, EPI_ISL_739008, EPI_ISL_739009, EPI_ISL_739010, EPI_ISL_739012, EPI_ISL_739015, EPI_ISL_739019, EPI_ISL_739020, EPI_ISL_739021, EPI_ISL_739025, EPI_ISL_739026, EPI_ISL_739027, EPI_ISL_739030, EPI_ISL_739031, EPI_ISL_739036, EPI_ISL_739037, EPI_ISL_739039, EPI_ISL_739040, EPI_ISL_739041, EPI_ISL_739045, EPI_ISL_739050, EPI_ISL_739053, EPI_ISL_739058, EPI_ISL_739060, EPI_ISL_739065, EPI_ISL_739066, EPI_ISL_739067, EPI_ISL_739068, EPI_ISL_739070, EPI_ISL_739074, EPI_ISL_739076, EPI_ISL_739080, EPI_ISL_739085, EPI_ISL_739087, EPI_ISL_739091, EPI_ISL_739098, EPI_ISL_739104, EPI_ISL_739107, EPI_ISL_739117, EPI_ISL_739121, EPI_ISL_739122, EPI_ISL_739128, EPI_ISL_739129, EPI_ISL_739130, EPI_ISL_739138, EPI_ISL_739141, EPI_ISL_739145, EPI_ISL_739146, EPI_ISL_739147, EPI_ISL_739152, EPI_ISL_739153, EPI_ISL_739157, EPI_ISL_739166, EPI_ISL_739167, EPI_ISL_739168, EPI_ISL_739180, EPI_ISL_739184, EPI_ISL_739186, EPI_ISL_739187, EPI_ISL_739188, EPI_ISL_739195, EPI_ISL_739204, EPI_ISL_739214, EPI_ISL_739219, EPI_ISL_739221, EPI_ISL_739224, EPI_ISL_739232, EPI_ISL_739234, EPI_ISL_739237, EPI_ISL_739246, EPI_ISL_739247, EPI_ISL_739248, EPI_ISL_739252, EPI_ISL_739253, EPI_ISL_739254, EPI_ISL_739256, EPI_ISL_739260, EPI_ISL_739263, EPI_ISL_739273, EPI_ISL_739275, EPI_ISL_739277, EPI_ISL_739278, EPI_ISL_739281, EPI_ISL_739282, EPI_ISL_739284, EPI_ISL_739293, EPI_ISL_739298, EPI_ISL_739305, EPI_ISL_739310, EPI_ISL_739317, EPI_ISL_739319, EPI_ISL_739320, EPI_ISL_739321, EPI_ISL_739327, EPI_ISL_739332, EPI_ISL_739335, EPI_ISL_739343, EPI_ISL_739344, EPI_ISL_739346, EPI_ISL_739352, EPI_ISL_739354, EPI_ISL_739357, EPI_ISL_739360, EPI_ISL_739365, EPI_ISL_739370, EPI_ISL_739371, EPI_ISL_739374, EPI_ISL_739376, EPI_ISL_739377, EPI_ISL_739380, EPI_ISL_739381, EPI_ISL_739382, EPI_ISL_739388, EPI_ISL_739392, EPI_ISL_739398, EPI_ISL_739400, EPI_ISL_739402, EPI_ISL_739404, EPI_ISL_739409, EPI_ISL_739413, EPI_ISL_739415, EPI_ISL_739416, EPI_ISL_739419, EPI_ISL_739431, EPI_ISL_739440, EPI_ISL_739443, EPI_ISL_739451, EPI_ISL_739453, EPI_ISL_739456, EPI_ISL_739457, EPI_ISL_739458, EPI_ISL_739479, EPI_ISL_739481, EPI_ISL_739486, EPI_ISL_739487, EPI_ISL_739491, EPI_ISL_739498, EPI_ISL_739501, EPI_ISL_739502, EPI_ISL_739504, EPI_ISL_739505, EPI_ISL_739508, EPI_ISL_739509, EPI_ISL_739510, EPI_ISL_739512, EPI_ISL_739520, EPI_ISL_739521, EPI_ISL_739536, EPI_ISL_739537, EPI_ISL_739543, EPI_ISL_739548, EPI_ISL_739549, EPI_ISL_739550, EPI_ISL_739560, EPI_ISL_739561, EPI_ISL_739562, EPI_ISL_739574, EPI_ISL_739579, EPI_ISL_739580, EPI_ISL_739584, EPI_ISL_739593, EPI_ISL_739594, EPI_ISL_739596, EPI_ISL_739597, EPI_ISL_739602, EPI_ISL_739605, EPI_ISL_739612, EPI_ISL_739617, EPI_ISL_739618, EPI_ISL_739624, EPI_ISL_739626, EPI_ISL_739629, EPI_ISL_739642, EPI_ISL_739647, EPI_ISL_739649, EPI_ISL_739650, EPI_ISL_739653, EPI_ISL_739657, EPI_ISL_849953, EPI_ISL_849954, EPI_ISL_849955, EPI_ISL_849956, EPI_ISL_849957, EPI_ISL_849958, EPI_ISL_849959, EPI_ISL_849960, EPI_ISL_849961, EPI_ISL_849962, EPI_ISL_849963, EPI_ISL_849964, EPI_ISL_849965, EPI_ISL_849966, EPI_ISL_849967, EPI_ISL_849968, EPI_ISL_849969, EPI_ISL_849970, EPI_ISL_849971, EPI_ISL_849972, EPI_ISL_849973, EPI_ISL_849974, EPI_ISL_849975, EPI_ISL_849976, EPI_ISL_849977, EPI_ISL_849978, EPI_ISL_849979, EPI_ISL_849980, EPI_ISL_849981, EPI_ISL_849982, EPI_ISL_849983, EPI_ISL_849984, EPI_ISL_849985, EPI_ISL_849986, EPI_ISL_849987, EPI_ISL_849988, EPI_ISL_849989, EPI_ISL_849990, EPI_ISL_849991, EPI_ISL_849992, EPI_ISL_849993, EPI_ISL_849994, EPI_ISL_849995, EPI_ISL_849996, EPI_ISL_849997, EPI_ISL_849998, EPI_ISL_849999, EPI_ISL_850000, EPI_ISL_850001, EPI_ISL_850002, EPI_ISL_850003, EPI_ISL_850004, EPI_ISL_850005, EPI_ISL_850007, EPI_ISL_850008, EPI_ISL_850009, EPI_ISL_850010, EPI_ISL_850011, EPI_ISL_850012, EPI_ISL_850013, EPI_ISL_850014, EPI_ISL_850015, EPI_ISL_850016, EPI_ISL_850017, EPI_ISL_850018, EPI_ISL_853417, EPI_ISL_853418, EPI_ISL_853419, EPI_ISL_853420, EPI_ISL_853421, EPI_ISL_853422, EPI_ISL_853423, EPI_ISL_853424, EPI_ISL_853425, EPI_ISL_853426, EPI_ISL_853427, EPI_ISL_853428, EPI_ISL_853429, EPI_ISL_853430, EPI_ISL_853431, EPI_ISL_853432, EPI_ISL_853433, EPI_ISL_853434, EPI_ISL_853435, EPI_ISL_853436, EPI_ISL_853437, EPI_ISL_853438, EPI_ISL_853439, EPI_ISL_853440, EPI_ISL_853441, EPI_ISL_853442, EPI_ISL_853443, EPI_ISL_853444, EPI_ISL_853445, EPI_ISL_853446, EPI_ISL_853447, EPI_ISL_853448, EPI_ISL_853449, EPI_ISL_853450, EPI_ISL_853451, EPI_ISL_853452, EPI_ISL_853453, EPI_ISL_853454, EPI_ISL_853455, EPI_ISL_853456, EPI_ISL_853457, EPI_ISL_853458, EPI_ISL_853459, EPI_ISL_853460, EPI_ISL_853461, EPI_ISL_853462, EPI_ISL_853463, EPI_ISL_853464, EPI_ISL_853465, EPI_ISL_853466, EPI_ISL_853467, EPI_ISL_853468, EPI_ISL_853469, EPI_ISL_853470, EPI_ISL_853471, EPI_ISL_853472, EPI_ISL_853473, EPI_ISL_853474, EPI_ISL_853475, EPI_ISL_853476, EPI_ISL_853477, EPI_ISL_853478, EPI_ISL_853479, EPI_ISL_853480, EPI_ISL_853481, EPI_ISL_853482, EPI_ISL_853483, EPI_ISL_853484, EPI_ISL_853485, EPI_ISL_853486, EPI_ISL_853487, EPI_ISL_853488, EPI_ISL_853489, EPI_ISL_853490, EPI_ISL_853491, EPI_ISL_853492, EPI_ISL_853493, EPI_ISL_853494, EPI_ISL_853495, EPI_ISL_853496, EPI_ISL_853497, EPI_ISL_853498, EPI_ISL_853499, EPI_ISL_853500, EPI_ISL_853501, EPI_ISL_853502, EPI_ISL_853503, EPI_ISL_853504, EPI_ISL_853505, EPI_ISL_853506, EPI_ISL_853507, EPI_ISL_853508, EPI_ISL_853509, EPI_ISL_853510, EPI_ISL_853511, EPI_ISL_853512, EPI_ISL_853513, EPI_ISL_853514, EPI_ISL_853515, EPI_ISL_853516, EPI_ISL_853517, EPI_ISL_853518, EPI_ISL_853519, EPI_ISL_853520, EPI_ISL_853521, EPI_ISL_853522, EPI_ISL_853523, EPI_ISL_853524, EPI_ISL_853525, EPI_ISL_853526, EPI_ISL_853527, EPI_ISL_853528, EPI_ISL_853529, EPI_ISL_853530, EPI_ISL_853531, EPI_ISL_853532, EPI_ISL_853533, EPI_ISL_853534, EPI_ISL_853535, EPI_ISL_853536, EPI_ISL_853537, EPI_ISL_853538, EPI_ISL_853539, EPI_ISL_853540, EPI_ISL_853541, EPI_ISL_853542, EPI_ISL_853543, EPI_ISL_853544, EPI_ISL_853545, EPI_ISL_853560, EPI_ISL_853561, EPI_ISL_853562, EPI_ISL_853563, EPI_ISL_853564, EPI_ISL_853565, EPI_ISL_884871, EPI_ISL_884872, EPI_ISL_884873, EPI_ISL_884874, EPI_ISL_884875, EPI_ISL_884876, EPI_ISL_884877, EPI_ISL_884878, EPI_ISL_884879, EPI_ISL_884880, EPI_ISL_884881, EPI_ISL_884882, EPI_ISL_884883, EPI_ISL_884884, EPI_ISL_884885, EPI_ISL_884886, EPI_ISL_884887, EPI_ISL_884888, EPI_ISL_884889, EPI_ISL_884890, EPI_ISL_884891, EPI_ISL_884892, EPI_ISL_884893, EPI_ISL_884894, EPI_ISL_884895, EPI_ISL_884896, EPI_ISL_884897, EPI_ISL_884898, EPI_ISL_884899, EPI_ISL_884900, EPI_ISL_884901, EPI_ISL_884902, EPI_ISL_884903, EPI_ISL_884904, EPI_ISL_884905, EPI_ISL_884906, EPI_ISL_884907, EPI_ISL_884908, EPI_ISL_884909, EPI_ISL_884910, EPI_ISL_884911, EPI_ISL_884912, EPI_ISL_884913, EPI_ISL_884914, EPI_ISL_884915, EPI_ISL_884916, EPI_ISL_884917, EPI_ISL_884918, EPI_ISL_884919, EPI_ISL_884920, EPI_ISL_884921, EPI_ISL_884922, EPI_ISL_884923, EPI_ISL_884924, EPI_ISL_884925, EPI_ISL_884926, EPI_ISL_884927, EPI_ISL_884928, EPI_ISL_884929, EPI_ISL_884930, EPI_ISL_884931, EPI_ISL_884932, EPI_ISL_884933, EPI_ISL_884934, EPI_ISL_884935, EPI_ISL_884936, EPI_ISL_884937, EPI_ISL_884938, EPI_ISL_884939, EPI_ISL_884940, EPI_ISL_884941, EPI_ISL_884942, EPI_ISL_884943, EPI_ISL_884944, EPI_ISL_884945, EPI_ISL_884946, EPI_ISL_884947, EPI_ISL_884948, EPI_ISL_884949, EPI_ISL_884950, EPI_ISL_884951, EPI_ISL_884952, EPI_ISL_884953, EPI_ISL_884954, EPI_ISL_884955, EPI_ISL_884956, EPI_ISL_884957, EPI_ISL_884958, EPI_ISL_884959, EPI_ISL_884960, EPI_ISL_884961, EPI_ISL_884962, EPI_ISL_884963, EPI_ISL_884964, EPI_ISL_884965, EPI_ISL_884966, EPI_ISL_884967, EPI_ISL_884968, EPI_ISL_884969, EPI_ISL_884970, EPI_ISL_884971, EPI_ISL_884972, EPI_ISL_884973, EPI_ISL_884974, EPI_ISL_884975, EPI_ISL_884976, EPI_ISL_884977, EPI_ISL_884978, EPI_ISL_884979, EPI_ISL_884980, EPI_ISL_884981, EPI_ISL_884982, EPI_ISL_884995, EPI_ISL_884996, EPI_ISL_884997, EPI_ISL_884998, EPI_ISL_884999, EPI_ISL_885000, EPI_ISL_885001, EPI_ISL_885002, EPI_ISL_885003, EPI_ISL_885004, EPI_ISL_885005, EPI_ISL_885006, EPI_ISL_885007, EPI_ISL_885008, EPI_ISL_885009, EPI_ISL_885010, EPI_ISL_885011, EPI_ISL_885012, EPI_ISL_885051, EPI_ISL_955353, EPI_ISL_955354, EPI_ISL_955355, EPI_ISL_955356, EPI_ISL_955357, EPI_ISL_955358, EPI_ISL_955359, EPI_ISL_955360, EPI_ISL_955361, EPI_ISL_955362, EPI_ISL_955363, EPI_ISL_955364, EPI_ISL_955365, EPI_ISL_955366, EPI_ISL_955367, EPI_ISL_955368, EPI_ISL_955369, EPI_ISL_955370, EPI_ISL_955371, EPI_ISL_955372, EPI_ISL_955373, EPI_ISL_955374, EPI_ISL_955375, EPI_ISL_955376, EPI_ISL_955377, EPI_ISL_955378, EPI_ISL_955379, EPI_ISL_955380, EPI_ISL_955381, EPI_ISL_955382, EPI_ISL_955383, EPI_ISL_955384, EPI_ISL_955385, EPI_ISL_955561, EPI_ISL_955562, EPI_ISL_955563, EPI_ISL_955564, EPI_ISL_955565, EPI_ISL_955566, EPI_ISL_955567, EPI_ISL_955568, EPI_ISL_955569, EPI_ISL_955570, EPI_ISL_955571, EPI_ISL_955572, EPI_ISL_955573, EPI_ISL_955574, EPI_ISL_955575, EPI_ISL_955576, EPI_ISL_955577, EPI_ISL_955578, EPI_ISL_955579, EPI_ISL_955580, EPI_ISL_955581, EPI_ISL_955582, EPI_ISL_955583, EPI_ISL_955584, EPI_ISL_955585, EPI_ISL_955586, EPI_ISL_955587, EPI_ISL_955588, EPI_ISL_955589, EPI_ISL_955590, EPI_ISL_955591, EPI_ISL_955592, EPI_ISL_955593, EPI_ISL_955594, EPI_ISL_955595, EPI_ISL_955596, EPI_ISL_955597, EPI_ISL_955598, EPI_ISL_955599, EPI_ISL_955600, EPI_ISL_955601, EPI_ISL_955602, EPI_ISL_955603, EPI_ISL_955604, EPI_ISL_955605, EPI_ISL_955606, EPI_ISL_955607, EPI_ISL_955608, EPI_ISL_955609, EPI_ISL_955610, EPI_ISL_955611, EPI_ISL_955612, EPI_ISL_955613, EPI_ISL_955614, EPI_ISL_955615, EPI_ISL_955616, EPI_ISL_955617, EPI_ISL_955618, EPI_ISL_955619, EPI_ISL_955620, EPI_ISL_955621, EPI_ISL_955622, EPI_ISL_955623, EPI_ISL_955624, EPI_ISL_955625, EPI_ISL_955626, EPI_ISL_955627, EPI_ISL_978989, EPI_ISL_978990, EPI_ISL_978991, EPI_ISL_978992, EPI_ISL_978993, EPI_ISL_978994, EPI_ISL_978995, EPI_ISL_978996, EPI_ISL_978997, EPI_ISL_978998, EPI_ISL_978999, EPI_ISL_979000, EPI_ISL_979001, EPI_ISL_979002, EPI_ISL_979003, EPI_ISL_979004, EPI_ISL_979005, EPI_ISL_979006, EPI_ISL_979007, EPI_ISL_979008, EPI_ISL_979009, EPI_ISL_979010, EPI_ISL_979011, EPI_ISL_979012, EPI_ISL_979013, EPI_ISL_979014, EPI_ISL_979015, EPI_ISL_979016, EPI_ISL_979017, EPI_ISL_979018, EPI_ISL_979019, EPI_ISL_979020, EPI_ISL_979021, EPI_ISL_979022, EPI_ISL_979023, EPI_ISL_979024, EPI_ISL_979025, EPI_ISL_979026, EPI_ISL_979027, EPI_ISL_979028, EPI_ISL_979029, EPI_ISL_979030, EPI_ISL_979031, EPI_ISL_979032, EPI_ISL_979033, EPI_ISL_979034, EPI_ISL_979035, EPI_ISL_979036, EPI_ISL_979037, EPI_ISL_979038, EPI_ISL_979039, EPI_ISL_979040, EPI_ISL_979041, EPI_ISL_979042, EPI_ISL_979043, EPI_ISL_979044, EPI_ISL_979045, EPI_ISL_979046, EPI_ISL_979047, EPI_ISL_979049, EPI_ISL_979050, EPI_ISL_979051, EPI_ISL_979052, EPI_ISL_979053, EPI_ISL_979054, EPI_ISL_979055, EPI_ISL_979545, EPI_ISL_979546, EPI_ISL_979547, EPI_ISL_979548, EPI_ISL_979549, EPI_ISL_979550, EPI_ISL_979551, EPI_ISL_979552, EPI_ISL_979553, EPI_ISL_979554, EPI_ISL_979555, EPI_ISL_979556, EPI_ISL_979557, EPI_ISL_979558, EPI_ISL_979559, EPI_ISL_979560, EPI_ISL_979561, EPI_ISL_979562, EPI_ISL_979563, EPI_ISL_979564, EPI_ISL_979565, EPI_ISL_979566, EPI_ISL_979567, EPI_ISL_979568, EPI_ISL_979569, EPI_ISL_979570, EPI_ISL_979571, EPI_ISL_979572, EPI_ISL_979573, EPI_ISL_979574, EPI_ISL_979575, EPI_ISL_979576, EPI_ISL_979577, EPI_ISL_979578, EPI_ISL_979579, EPI_ISL_979580, EPI_ISL_979581, EPI_ISL_979582, EPI_ISL_979583, EPI_ISL_979584, EPI_ISL_979585, EPI_ISL_979586, EPI_ISL_979587, EPI_ISL_979588, EPI_ISL_979589, EPI_ISL_979590, EPI_ISL_979591, EPI_ISL_979592, EPI_ISL_979593, EPI_ISL_979594, EPI_ISL_979595, EPI_ISL_979598, EPI_ISL_979606, EPI_ISL_979607, EPI_ISL_979608, EPI_ISL_979609, EPI_ISL_979610, EPI_ISL_979611, EPI_ISL_979613, EPI_ISL_979614, EPI_ISL_979615, EPI_ISL_979616, EPI_ISL_979617, EPI_ISL_979618, EPI_ISL_979619, EPI_ISL_979620, EPI_ISL_979621, EPI_ISL_979622, EPI_ISL_1027293, EPI_ISL_1027306, EPI_ISL_1027312, EPI_ISL_1027314, EPI_ISL_1027315, EPI_ISL_1027317, EPI_ISL_1027319, EPI_ISL_1027320, EPI_ISL_1027325, EPI_ISL_1027326, EPI_ISL_1027327, EPI_ISL_1027332, EPI_ISL_1027337, EPI_ISL_1027338, EPI_ISL_1027342, EPI_ISL_1027343, EPI_ISL_1027347, EPI_ISL_1027355, EPI_ISL_1027357, EPI_ISL_1027362, EPI_ISL_1027364, EPI_ISL_1027437, EPI_ISL_1027438, EPI_ISL_1027439, EPI_ISL_1027440, EPI_ISL_1027441, EPI_ISL_1027442, EPI_ISL_1027443, EPI_ISL_1027444, EPI_ISL_1027474, EPI_ISL_1027477, EPI_ISL_1027478, EPI_ISL_1027481, EPI_ISL_1027483, EPI_ISL_1027485, EPI_ISL_1027488, EPI_ISL_1027492, EPI_ISL_1027493, EPI_ISL_1027495, EPI_ISL_1027497, EPI_ISL_1027500, EPI_ISL_1027508, EPI_ISL_1027509, EPI_ISL_1027510, EPI_ISL_1027511, EPI_ISL_1027512, EPI_ISL_1027514, EPI_ISL_1027516, EPI_ISL_1027519, EPI_ISL_1027522, EPI_ISL_1027524, EPI_ISL_1027527, EPI_ISL_1027529, EPI_ISL_1027533, EPI_ISL_1027534, EPI_ISL_1027542, EPI_ISL_1027544, EPI_ISL_1027545, EPI_ISL_1027546, EPI_ISL_1027547, EPI_ISL_1027548, EPI_ISL_1027549, EPI_ISL_1027550, EPI_ISL_1027551, EPI_ISL_1027552, EPI_ISL_1027553, EPI_ISL_1027554, EPI_ISL_1027555, EPI_ISL_1027556, EPI_ISL_1027557, EPI_ISL_1027558, EPI_ISL_1027559, EPI_ISL_1030252, EPI_ISL_1030253, EPI_ISL_1030254, EPI_ISL_1030255, EPI_ISL_1030256, EPI_ISL_1030257, EPI_ISL_1030258, EPI_ISL_1030259, EPI_ISL_1030260, EPI_ISL_1030261, EPI_ISL_1030263, EPI_ISL_1030264, EPI_ISL_1030265, EPI_ISL_1030266, EPI_ISL_1030268, EPI_ISL_1030270, EPI_ISL_1030271, EPI_ISL_1030273, EPI_ISL_1030274, EPI_ISL_1030275, EPI_ISL_1030277, EPI_ISL_1030278, EPI_ISL_1030279, EPI_ISL_1030280, EPI_ISL_1030284, EPI_ISL_1030285, EPI_ISL_1030287, EPI_ISL_1030288, EPI_ISL_1030289, EPI_ISL_1030300, EPI_ISL_1030301,

| | | | |
|---|---|---|---|
| EPI_ISL_1030304, EPI_ISL_1030305, EPI_ISL_1030398, EPI_ISL_1030399, EPI_ISL_1030400, EPI_ISL_1030401, EPI_ISL_1030402, EPI_ISL_1030403, EPI_ISL_1030404, EPI_ISL_1030405, EPI_ISL_1030406, EPI_ISL_1030407, EPI_ISL_1030408, EPI_ISL_1030409, EPI_ISL_1030410, EPI_ISL_1030411, EPI_ISL_1030412, EPI_ISL_1030413, EPI_ISL_1030414, EPI_ISL_1030415, EPI_ISL_1030416, EPI_ISL_1030417, EPI_ISL_1030418, EPI_ISL_1030419, EPI_ISL_1030420, EPI_ISL_1030421, EPI_ISL_1030422, EPI_ISL_1030423, EPI_ISL_1030424, EPI_ISL_1030425, EPI_ISL_1030426, EPI_ISL_1030427, EPI_ISL_1030428, EPI_ISL_1030429, EPI_ISL_1030430, EPI_ISL_1030431, EPI_ISL_1030432, EPI_ISL_1030433, EPI_ISL_1030434, EPI_ISL_1030435, EPI_ISL_1030436, EPI_ISL_1030437, EPI_ISL_1030438, EPI_ISL_1030439, EPI_ISL_1030440, EPI_ISL_1030441, EPI_ISL_1030442, EPI_ISL_1030443, EPI_ISL_1030500, EPI_ISL_1030501, EPI_ISL_1030502, EPI_ISL_1184583, EPI_ISL_1184584, EPI_ISL_1184585, EPI_ISL_1184586, EPI_ISL_1184587, EPI_ISL_1184588, EPI_ISL_1184589, EPI_ISL_1184590, EPI_ISL_1184591, EPI_ISL_1184592, EPI_ISL_1184593, EPI_ISL_1184594, EPI_ISL_1184595, EPI_ISL_1184596, EPI_ISL_1184597, EPI_ISL_1184598, EPI_ISL_1184599, EPI_ISL_1184600, EPI_ISL_1184601, EPI_ISL_1184602, EPI_ISL_1184603, EPI_ISL_1184604, EPI_ISL_1184605, EPI_ISL_1184606, EPI_ISL_1184607, EPI_ISL_1184608, EPI_ISL_1184609, EPI_ISL_1184610, EPI_ISL_1184611, EPI_ISL_1184612, EPI_ISL_1184613, EPI_ISL_1184614, EPI_ISL_1184615, EPI_ISL_1184616, EPI_ISL_1184617, EPI_ISL_1184618, EPI_ISL_1184619, EPI_ISL_1184620, EPI_ISL_1184621, EPI_ISL_1184622, EPI_ISL_1184623, EPI_ISL_1184827, EPI_ISL_1184828, EPI_ISL_1184829, EPI_ISL_1184830, EPI_ISL_1184831, EPI_ISL_1184832, EPI_ISL_1184833, EPI_ISL_1184834, EPI_ISL_1184835, EPI_ISL_1184836, EPI_ISL_1184837, EPI_ISL_1184838, EPI_ISL_1184839, EPI_ISL_1184840, EPI_ISL_1184841, EPI_ISL_1184842, EPI_ISL_1184843, EPI_ISL_1184844, EPI_ISL_1184845, EPI_ISL_1184846, EPI_ISL_1184847, EPI_ISL_1184848, EPI_ISL_1184849, EPI_ISL_1184850, EPI_ISL_1184851, EPI_ISL_1184852, EPI_ISL_1184853, EPI_ISL_1184854, EPI_ISL_1184855, EPI_ISL_1184856, EPI_ISL_1184857, EPI_ISL_1184858, EPI_ISL_1184859, EPI_ISL_1184860 | | | |
| see above | Santa Clara County Public Health Laboratory | Chan-Zuckerberg Biohub | CZB Cliahub Consortium |
| EPI_ISL_806543, EPI_ISL_806545, EPI_ISL_812115, EPI_ISL_812116, EPI_ISL_812119, EPI_ISL_812120, EPI_ISL_812121, EPI_ISL_812350, EPI_ISL_831648, EPI_ISL_831662, EPI_ISL_831687, EPI_ISL_831893, EPI_ISL_831894, EPI_ISL_831895, EPI_ISL_831896, EPI_ISL_831897, EPI_ISL_831899, EPI_ISL_832066, EPI_ISL_832068, EPI_ISL_832075, EPI_ISL_832076, EPI_ISL_832375, EPI_ISL_832376, EPI_ISL_832377, EPI_ISL_832379, EPI_ISL_832380, EPI_ISL_832381, EPI_ISL_832382, EPI_ISL_832383, EPI_ISL_832388, EPI_ISL_832391, EPI_ISL_832392, EPI_ISL_832394, EPI_ISL_832395, EPI_ISL_832396, EPI_ISL_832398, EPI_ISL_832399, EPI_ISL_832400, EPI_ISL_832401, EPI_ISL_832402, EPI_ISL_832403, EPI_ISL_832404, EPI_ISL_914646, EPI_ISL_1081192, EPI_ISL_1137637, EPI_ISL_1169590, EPI_ISL_1169591, EPI_ISL_1203818, EPI_ISL_1920833, EPI_ISL_1920834, EPI_ISL_1920835, EPI_ISL_1920838, EPI_ISL_1920839, EPI_ISL_1920840, EPI_ISL_1920841, EPI_ISL_1920842, EPI_ISL_1920845, EPI_ISL_1920846, EPI_ISL_1920848, EPI_ISL_1920850, EPI_ISL_1920851, EPI_ISL_1920852, EPI_ISL_1920854, EPI_ISL_1920855, EPI_ISL_1920856, EPI_ISL_1920857, EPI_ISL_1920858, EPI_ISL_1920859, EPI_ISL_1920860, EPI_ISL_1920861, EPI_ISL_1920862, EPI_ISL_1920863, EPI_ISL_1920865, EPI_ISL_1920866, EPI_ISL_1920867, EPI_ISL_1920868, EPI_ISL_1920869, EPI_ISL_1920871, EPI_ISL_1920872, EPI_ISL_1920873, EPI_ISL_1920874, EPI_ISL_1920876, EPI_ISL_1920877, EPI_ISL_1920879, EPI_ISL_1920880, EPI_ISL_1920881, EPI_ISL_1920882, EPI_ISL_1920884, EPI_ISL_1920885, EPI_ISL_1920886, EPI_ISL_1920887, EPI_ISL_1920888, EPI_ISL_1920889, EPI_ISL_1920890, EPI_ISL_1920891, EPI_ISL_1920892, EPI_ISL_1920893, EPI_ISL_1920894, EPI_ISL_1920895, EPI_ISL_1920896, EPI_ISL_1920897, EPI_ISL_1920898, EPI_ISL_1920899, EPI_ISL_1920900, EPI_ISL_1920901 | | | |
| see above | Santa Clara County Public Health Laboratory | Santa Clara County Public Health Laboratory | Santa CLara County Public Health Department; Santa Clara Co Public Health Department; Santa Clara County Public Health Department |
| EPI_ISL_1794577, EPI_ISL_1794578, EPI_ISL_1794579, EPI_ISL_1794580, EPI_ISL_1794581, EPI_ISL_1794582, EPI_ISL_1794583, EPI_ISL_1794584, EPI_ISL_1794585, EPI_ISL_1794586, EPI_ISL_1794587, EPI_ISL_1794588, EPI_ISL_1794589, EPI_ISL_1794590, EPI_ISL_1794591, EPI_ISL_1794592, EPI_ISL_1794593, EPI_ISL_1794594, EPI_ISL_1794595, EPI_ISL_1794596, EPI_ISL_1794597, EPI_ISL_1794598, EPI_ISL_1794599, EPI_ISL_1794600, EPI_ISL_1794601, EPI_ISL_1794602, EPI_ISL_1794603, EPI_ISL_1794604, EPI_ISL_1794605, EPI_ISL_1794606, EPI_ISL_1794607, EPI_ISL_1794608, EPI_ISL_1794609, EPI_ISL_1794610, EPI_ISL_1794611, EPI_ISL_1794612, EPI_ISL_1794613, EPI_ISL_1794614, EPI_ISL_1794615, EPI_ISL_1794787, EPI_ISL_1794788, EPI_ISL_1794789, EPI_ISL_1794790, EPI_ISL_1794791, EPI_ISL_1794792, EPI_ISL_1794793, EPI_ISL_1794794, EPI_ISL_1794795, EPI_ISL_1794796, EPI_ISL_1794797, EPI_ISL_1794798, EPI_ISL_1794799, EPI_ISL_1794800, EPI_ISL_1794801, EPI_ISL_1794802, EPI_ISL_1794803, EPI_ISL_1794804, EPI_ISL_1794805, EPI_ISL_1794806, EPI_ISL_1794807, EPI_ISL_1794808, EPI_ISL_1794809, EPI_ISL_1794810, EPI_ISL_1794811, EPI_ISL_1794812, EPI_ISL_1794813, EPI_ISL_1794814, EPI_ISL_1794815, EPI_ISL_1794816, EPI_ISL_1794817, EPI_ISL_1794818, EPI_ISL_1794819, EPI_ISL_1794820, EPI_ISL_1794821, EPI_ISL_1794822, EPI_ISL_1794823, EPI_ISL_1794824, EPI_ISL_1794825, EPI_ISL_1794826, EPI_ISL_1794827, EPI_ISL_1794828, EPI_ISL_1794829, EPI_ISL_1794830, EPI_ISL_1794831, EPI_ISL_1794832, EPI_ISL_1794833, EPI_ISL_1794834, EPI_ISL_1794835, EPI_ISL_1794836, EPI_ISL_1794837, EPI_ISL_1794838, EPI_ISL_1969271, EPI_ISL_1969272, EPI_ISL_1969273, EPI_ISL_1969274, EPI_ISL_1969275, EPI_ISL_1969276, EPI_ISL_1969277, EPI_ISL_1969278, EPI_ISL_1969279, EPI_ISL_1969280, EPI_ISL_1969281, EPI_ISL_1969314, EPI_ISL_1969327, EPI_ISL_1969328, EPI_ISL_1969329, EPI_ISL_1969330, EPI_ISL_1969331, EPI_ISL_1969332, EPI_ISL_1969333, EPI_ISL_1969334, EPI_ISL_1969335, EPI_ISL_1969336, EPI_ISL_1969337, EPI_ISL_1969338, EPI_ISL_1969339, EPI_ISL_1969340, EPI_ISL_1969341, EPI_ISL_1969342, EPI_ISL_1969343, EPI_ISL_1969344, EPI_ISL_1969345, EPI_ISL_1969346, EPI_ISL_1969347, EPI_ISL_1969348, EPI_ISL_1969349, EPI_ISL_1969350, EPI_ISL_1969351, EPI_ISL_1969352, EPI_ISL_1969353, EPI_ISL_1969354, EPI_ISL_1969355, EPI_ISL_1969356, EPI_ISL_1969357, EPI_ISL_1969358, EPI_ISL_1969359, EPI_ISL_1969360, EPI_ISL_1969361, EPI_ISL_1969362, EPI_ISL_1969363, EPI_ISL_1969364, EPI_ISL_1969365, EPI_ISL_1969366, EPI_ISL_1969367, EPI_ISL_1969368, EPI_ISL_1969369, EPI_ISL_1969370 | | | |
| see above | Scripps Medical Laboratory | Andersen lab at Scripps Research | Ellen Stefanski; Ian Mchardy; SEARCH Alliance San Diego with Michael Quigley |
| EPI_ISL_730127, EPI_ISL_730128, EPI_ISL_730129, EPI_ISL_730130, EPI_ISL_730131, EPI_ISL_730132, EPI_ISL_730133, EPI_ISL_730134, EPI_ISL_730135, EPI_ISL_730136, EPI_ISL_1969317, EPI_ISL_1969554, EPI_ISL_1969555, EPI_ISL_1969556, EPI_ISL_1969557, EPI_ISL_1969558, EPI_ISL_1969559, EPI_ISL_1969560, EPI_ISL_1969561, EPI_ISL_1969562, EPI_ISL_1969602, EPI_ISL_1969603, EPI_ISL_1969604, EPI_ISL_1969605, EPI_ISL_1969607, EPI_ISL_1969611, EPI_ISL_1969612, EPI_ISL_1969613, EPI_ISL_1969614, EPI_ISL_1969615, EPI_ISL_1969616, EPI_ISL_1969617, EPI_ISL_1969618, EPI_ISL_1969619, EPI_ISL_1969620, EPI_ISL_1969621, EPI_ISL_1969622, EPI_ISL_1969623, EPI_ISL_1969624, EPI_ISL_1969625, EPI_ISL_1969626, EPI_ISL_1969627, EPI_ISL_1969628, EPI_ISL_1969629, EPI_ISL_1969630, EPI_ISL_1969631, EPI_ISL_1969632, EPI_ISL_1969633, EPI_ISL_1969634, EPI_ISL_1969635, EPI_ISL_1969636, EPI_ISL_1969637, EPI_ISL_1969638, EPI_ISL_1969639, EPI_ISL_1969640, EPI_ISL_1969641, EPI_ISL_1969642, EPI_ISL_1969643, EPI_ISL_1969644, EPI_ISL_1969645, EPI_ISL_1969646, EPI_ISL_1969647, EPI_ISL_1969648, EPI_ISL_1969649, EPI_ISL_1969650, EPI_ISL_1969651, EPI_ISL_1969652, EPI_ISL_1969653, EPI_ISL_1969654, EPI_ISL_1969655, EPI_ISL_1969656, EPI_ISL_1969657, EPI_ISL_1969658, EPI_ISL_1969659, EPI_ISL_1969660, EPI_ISL_1969661, EPI_ISL_1969662, EPI_ISL_1969663, EPI_ISL_1969664, EPI_ISL_1969665, EPI_ISL_1969666 | | | |
| see above | Sharp HealthCare Laboratory | Andersen lab at Scripps Research | Art Mendoza; Cathy Woerle; Jacquelyn Berumen; Liam McGinnis; Omid Bakhtar; SEARCH Alliance San Diego with Aaron Harding |
| EPI_ISL_1159373 | Stanford Health Care | Clinical virology Laboratory, Stanford University School of Medicine | ChunHong Huang; Daniel Solis; Fumiko Yamamoto; James Zehnder; Malaya K. Sahoo; Mamdouh Sibai; Michelle Verghese; and Benjamin A. Pinsky |
| EPI_ISL_1379719, EPI_ISL_1379720, EPI_ISL_1379721, EPI_ISL_1379722, EPI_ISL_1379723, EPI_ISL_1379724, EPI_ISL_1379725, EPI_ISL_1379726, EPI_ISL_1379731, EPI_ISL_1379733, EPI_ISL_1379751, EPI_ISL_1379829, EPI_ISL_1379832, EPI_ISL_1379833, EPI_ISL_1379841, EPI_ISL_1379845, EPI_ISL_1379846, EPI_ISL_1379861, EPI_ISL_1424067, EPI_ISL_1424068, EPI_ISL_1424069, EPI_ISL_1424070, EPI_ISL_1424071, EPI_ISL_1424095, EPI_ISL_2009050, EPI_ISL_2009051, EPI_ISL_2009070, EPI_ISL_2009071 | | | |
| see above | Stanford Health Care | Stanford University School of Medicine, Clinical Virology Laboratory | ChunHong Huang; Daniel Solis; Fumiko Yamamoto; James Zehnder; Malaya K. Sahoo; Mamdouh Sibai; Michelle Vergese; Michelle Verghese; and Benjamin A. Pinsky |
| EPI_ISL_583171, EPI_ISL_583172, EPI_ISL_648537, EPI_ISL_648538, EPI_ISL_648539, EPI_ISL_648540, EPI_ISL_648541, EPI_ISL_739119, EPI_ISL_739193, EPI_ISL_739639 | | | |
| see above | Tulare County Public Health Lab | Chan-Zuckerberg Biohub | CZB Cliahub Consortium |
| EPI_ISL_548599, EPI_ISL_548626 | Tuolumne County Public Health | Chan-Zuckerberg Biohub | CZB Cliahub Consortium |
| EPI_ISL_849939, EPI_ISL_849940, EPI_ISL_849941, EPI_ISL_849942, EPI_ISL_849943, EPI_ISL_849944, EPI_ISL_849945, EPI_ISL_849946, EPI_ISL_849948, EPI_ISL_849950, EPI_ISL_849951, EPI_ISL_849952, EPI_ISL_955337, EPI_ISL_955338, EPI_ISL_955339, EPI_ISL_955340, EPI_ISL_955341, EPI_ISL_955342, EPI_ISL_955343, EPI_ISL_955344, EPI_ISL_955345, EPI_ISL_955346, EPI_ISL_955347, EPI_ISL_955348, EPI_ISL_955349, EPI_ISL_955350, EPI_ISL_955351, EPI_ISL_955352 | | | |
| see above | UC Davis- Department of Pathology and Laboratory Medicine | Chan-Zuckerberg Biohub | CZB Cliahub Consortium |
| EPI_ISL_754110, EPI_ISL_754111, EPI_ISL_754112, EPI_ISL_754113, EPI_ISL_754114, EPI_ISL_754115, EPI_ISL_754116, EPI_ISL_754117, EPI_ISL_754118, EPI_ISL_754119, EPI_ISL_754120, EPI_ISL_754121, EPI_ISL_754122, EPI_ISL_754123, EPI_ISL_765987, EPI_ISL_765988, EPI_ISL_765989, EPI_ISL_765990, EPI_ISL_765991, EPI_ISL_765992, EPI_ISL_765993, EPI_ISL_962506, EPI_ISL_962507, EPI_ISL_962508, EPI_ISL_962514 | | | |
| see above | UCLA Clinical Micro Lab | Los Angeles County PHL | P. Hemarajata et al. |
| EPI_ISL_1469854, EPI_ISL_1469855, EPI_ISL_1469856, EPI_ISL_1469857, EPI_ISL_1469858, EPI_ISL_1469859, EPI_ISL_1469860, EPI_ISL_1469861, EPI_ISL_1469862, EPI_ISL_1469863, EPI_ISL_1469864, EPI_ISL_1469865, EPI_ISL_1469866, EPI_ISL_1469867, EPI_ISL_1469868, EPI_ISL_1469869, EPI_ISL_1469870, EPI_ISL_1469871, EPI_ISL_1469872, EPI_ISL_1469873, EPI_ISL_1469874, EPI_ISL_1469884, EPI_ISL_1469885, EPI_ISL_1469886, EPI_ISL_1469887, EPI_ISL_1469888, EPI_ISL_1469889, EPI_ISL_1469890, EPI_ISL_1469891, EPI_ISL_1469892, EPI_ISL_1469893, EPI_ISL_1469894, EPI_ISL_1469895, EPI_ISL_1469896, EPI_ISL_1469897, EPI_ISL_1469898, EPI_ISL_1469899, EPI_ISL_1469900, EPI_ISL_1469901, EPI_ISL_1469902, EPI_ISL_1469903, EPI_ISL_1469904, EPI_ISL_1469905, EPI_ISL_1469906, EPI_ISL_1469907, EPI_ISL_1469909, EPI_ISL_1469910, EPI_ISL_1469911, EPI_ISL_1469912, EPI_ISL_1469913, EPI_ISL_1469914, EPI_ISL_1469915, EPI_ISL_1469916, EPI_ISL_1469917, EPI_ISL_1469918, EPI_ISL_1469919, EPI_ISL_1469920, EPI_ISL_1469921, EPI_ISL_1469922, EPI_ISL_1469923, EPI_ISL_1469924, EPI_ISL_1469925, EPI_ISL_1469926, EPI_ISL_1469927, EPI_ISL_1469928, EPI_ISL_1469929, EPI_ISL_1469930, EPI_ISL_1469931, EPI_ISL_1469932, EPI_ISL_1469933, EPI_ISL_1469934, EPI_ISL_1469935, EPI_ISL_1469936, EPI_ISL_1921085, EPI_ISL_1921086, EPI_ISL_1921087, EPI_ISL_1921089, EPI_ISL_1921090, EPI_ISL_1921091, EPI_ISL_1921092, EPI_ISL_1921093, EPI_ISL_1921094, EPI_ISL_1921095, EPI_ISL_1921098, EPI_ISL_1921099, EPI_ISL_1921100, EPI_ISL_1921101, EPI_ISL_1921102, EPI_ISL_1921103, EPI_ISL_1921104, EPI_ISL_1921105, EPI_ISL_1921106, EPI_ISL_1921107, EPI_ISL_1921108, EPI_ISL_1921109, EPI_ISL_1921110, EPI_ISL_1921111, EPI_ISL_1921116, EPI_ISL_1921117, EPI_ISL_1921118, EPI_ISL_1921119, EPI_ISL_1921120, EPI_ISL_1921121, EPI_ISL_1921122, EPI_ISL_1921123, EPI_ISL_1921124, EPI_ISL_1921125, EPI_ISL_1921126, EPI_ISL_1921127, EPI_ISL_1921129, EPI_ISL_1921130, EPI_ISL_1921131, EPI_ISL_1921132, EPI_ISL_1921133, EPI_ISL_1921134, EPI_ISL_1921135, EPI_ISL_1921136, EPI_ISL_1921137, EPI_ISL_1921138, EPI_ISL_1921139, EPI_ISL_1921140, EPI_ISL_1921141, EPI_ISL_1921142, EPI_ISL_1921143, EPI_ISL_1921144, EPI_ISL_1921145, EPI_ISL_1921147, EPI_ISL_1921148, EPI_ISL_1921149, EPI_ISL_1921150, EPI_ISL_1921151, EPI_ISL_1921152, EPI_ISL_1921153, EPI_ISL_1921154, EPI_ISL_1921155, EPI_ISL_1921156, EPI_ISL_1921157, EPI_ISL_1921158, EPI_ISL_1921159, EPI_ISL_1921160, EPI_ISL_1921161, EPI_ISL_1921162, EPI_ISL_1921163, EPI_ISL_1921164, EPI_ISL_2031787, EPI_ISL_2031788, EPI_ISL_2031789, EPI_ISL_2031790, EPI_ISL_2031791, EPI_ISL_2031792, EPI_ISL_2031793, EPI_ISL_2031794, EPI_ISL_2031795, EPI_ISL_2031796, EPI_ISL_2031797, EPI_ISL_2031798, EPI_ISL_2031799, EPI_ISL_2031800, EPI_ISL_2031801, EPI_ISL_2031802, EPI_ISL_2031803, EPI_ISL_2031804, EPI_ISL_2031805, EPI_ISL_2031806, EPI_ISL_2031807, EPI_ISL_2031808, EPI_ISL_2031809, EPI_ISL_2031810, EPI_ISL_2031811, EPI_ISL_2031812, EPI_ISL_2031813, EPI_ISL_2031814, EPI_ISL_2031815, EPI_ISL_2031816, EPI_ISL_2031817, EPI_ISL_2031818, EPI_ISL_2031819, EPI_ISL_2031820, EPI_ISL_2031821, EPI_ISL_2031822, EPI_ISL_2031823, EPI_ISL_2031824, EPI_ISL_2031825, EPI_ISL_2031826, EPI_ISL_2031827, EPI_ISL_2031828, EPI_ISL_2031830, EPI_ISL_2031831, EPI_ISL_2031832, EPI_ISL_2031833, EPI_ISL_2031834, EPI_ISL_2031835, EPI_ISL_2031836, EPI_ISL_2031837, EPI_ISL_2031838, EPI_ISL_2031840, EPI_ISL_2031841, EPI_ISL_2031842, EPI_ISL_2031843, EPI_ISL_2031844, EPI_ISL_2031845, EPI_ISL_2031846, EPI_ISL_2031847, EPI_ISL_2031848, EPI_ISL_2031849, EPI_ISL_2031850, EPI_ISL_2031851, EPI_ISL_2031852, EPI_ISL_2031854, EPI_ISL_2031855, EPI_ISL_2031856, EPI_ISL_2031857, EPI_ISL_2081230, EPI_ISL_2081235, EPI_ISL_2081236, EPI_ISL_2081240, EPI_ISL_2081243, EPI_ISL_2081244, EPI_ISL_2081245, EPI_ISL_2081247, EPI_ISL_2081248, EPI_ISL_2081249, EPI_ISL_2081250, EPI_ISL_2081251, EPI_ISL_2081252, EPI_ISL_2081253, EPI_ISL_2081254, EPI_ISL_2081255, EPI_ISL_2081256, EPI_ISL_2081257, EPI_ISL_2081258, EPI_ISL_2081259, EPI_ISL_2081260, EPI_ISL_2081261, EPI_ISL_2081262, EPI_ISL_2081263, EPI_ISL_2081264, EPI_ISL_2081265, EPI_ISL_2081266, EPI_ISL_2081267, EPI_ISL_2081268, EPI_ISL_2081269, EPI_ISL_2081270, EPI_ISL_2081271, EPI_ISL_2081272, EPI_ISL_2081273, EPI_ISL_2081274, EPI_ISL_2081275, EPI_ISL_2081276, EPI_ISL_2081277, EPI_ISL_2081278, EPI_ISL_2081280, EPI_ISL_2081282, EPI_ISL_2081283, EPI_ISL_2081284, EPI_ISL_2081285, EPI_ISL_2081286, EPI_ISL_2081287, EPI_ISL_2081288, EPI_ISL_2081290, EPI_ISL_2081291, EPI_ISL_2081293, EPI_ISL_2081294, EPI_ISL_2081295, EPI_ISL_2081296, EPI_ISL_2081297, EPI_ISL_2081298, EPI_ISL_2081299, EPI_ISL_2081300, EPI_ISL_2081301, EPI_ISL_2081302, EPI_ISL_2081303, EPI_ISL_2081304, EPI_ISL_2081307, EPI_ISL_2081308, EPI_ISL_2081309, EPI_ISL_2081530, EPI_ISL_2081531, EPI_ISL_2081532, EPI_ISL_2081534, EPI_ISL_2081535, EPI_ISL_2081536, EPI_ISL_2081538, EPI_ISL_2081539, EPI_ISL_2081542, EPI_ISL_2081543, EPI_ISL_2081545, EPI_ISL_2081546, EPI_ISL_2081548, EPI_ISL_2081549, EPI_ISL_2081550, EPI_ISL_2081553, EPI_ISL_2081554, EPI_ISL_2081555, EPI_ISL_2081556, EPI_ISL_2081557, EPI_ISL_2081558, EPI_ISL_2081559, EPI_ISL_2081560, EPI_ISL_2081561, EPI_ISL_2081562, EPI_ISL_2081563, EPI_ISL_2081564, EPI_ISL_2081565, EPI_ISL_2081566, EPI_ISL_2081567, EPI_ISL_2081568, EPI_ISL_2081569, EPI_ISL_2081570, EPI_ISL_2081571, EPI_ISL_2081572, EPI_ISL_2081573, EPI_ISL_2081574, EPI_ISL_2081575, EPI_ISL_2081576, EPI_ISL_2081577, EPI_ISL_2081578, EPI_ISL_2081579, EPI_ISL_2081580, EPI_ISL_2081581, EPI_ISL_2081582, EPI_ISL_2081583, EPI_ISL_2081584, EPI_ISL_2081585, EPI_ISL_2081586, EPI_ISL_2081587, EPI_ISL_2081588, EPI_ISL_2081589, EPI_ISL_2081590, EPI_ISL_2081591, EPI_ISL_2081592, EPI_ISL_2081593, EPI_ISL_2081594, EPI_ISL_2081595, EPI_ISL_2081596, EPI_ISL_2081597, EPI_ISL_2081598, EPI_ISL_2081599, EPI_ISL_2081600, EPI_ISL_2081601, EPI_ISL_2081602, EPI_ISL_2081603, EPI_ISL_2081604, EPI_ISL_2081605, EPI_ISL_2081606, EPI_ISL_2081607, EPI_ISL_2081608, EPI_ISL_2081609, EPI_ISL_2081611, EPI_ISL_2081612, EPI_ISL_2081613, EPI_ISL_2082936, EPI_ISL_2082937, EPI_ISL_2082938, EPI_ISL_2082939, EPI_ISL_2082940, EPI_ISL_2082941, EPI_ISL_2082943, EPI_ISL_2082944, EPI_ISL_2082945, EPI_ISL_2082946, EPI_ISL_2082947, EPI_ISL_2082948, EPI_ISL_2082949, EPI_ISL_2082950, EPI_ISL_2082951, EPI_ISL_2082952, EPI_ISL_2082953, EPI_ISL_2082954, EPI_ISL_2082955, EPI_ISL_2082956, EPI_ISL_2082957, EPI_ISL_2082958, EPI_ISL_2082959, EPI_ISL_2082960, EPI_ISL_2082961, EPI_ISL_2082962, EPI_ISL_2082963, EPI_ISL_2082964, EPI_ISL_2082965, EPI_ISL_2082966, EPI_ISL_2082967, EPI_ISL_2082968, EPI_ISL_2082969, EPI_ISL_2082970, EPI_ISL_2082971, EPI_ISL_2082972, EPI_ISL_2082973, EPI_ISL_2082974, EPI_ISL_2082975, EPI_ISL_2082976, EPI_ISL_2082977, EPI_ISL_2082978, EPI_ISL_2082979, EPI_ISL_2082980, EPI_ISL_2082981, EPI_ISL_2082982, EPI_ISL_2082983, EPI_ISL_2082986, EPI_ISL_2082987, EPI_ISL_2082988, EPI_ISL_2082989, EPI_ISL_2082990, EPI_ISL_2082991, EPI_ISL_2082992, EPI_ISL_2082993, EPI_ISL_2082994, EPI_ISL_2082996, EPI_ISL_2082997, EPI_ISL_2082998, EPI_ISL_2082999, EPI_ISL_2083000, EPI_ISL_2083001, EPI_ISL_2083004, EPI_ISL_2083005, EPI_ISL_2083006, EPI_ISL_2083007, EPI_ISL_2083008, EPI_ISL_2083009, EPI_ISL_2083011, EPI_ISL_2083012, EPI_ISL_2083013, EPI_ISL_2083014, EPI_ISL_2083015 | | | |
| see above | UCSC Genomics Institute | UCSC Genomics Institute | A. Marm Kilpatrick; Angie Hinrichs; Beth Shapiro; Bryan Thornlow; Ciara Wanket; David Haussler; Eric Beraut; Hugh Olsen; Ikenna Anigbogu; Isabel Bjork; Jakob McBroome; Jeremy Sanford; Joshua Kapp; Mark Akeson; Maximilian Haeussler; Michael Stone; Miten Jain; Molly Cassatt-Johnstone; Namrita Dhillon; Russell Corbett-Detig; Terren Chang; Yatish Turakhia |
| EPI_ISL_755133, EPI_ISL_755137, EPI_ISL_755140, EPI_ISL_755146, EPI_ISL_755147, EPI_ISL_755148, EPI_ISL_755149, EPI_ISL_755150, EPI_ISL_755151, EPI_ISL_755152, EPI_ISL_755153, EPI_ISL_755154, EPI_ISL_755155, EPI_ISL_755156, EPI_ISL_755157, EPI_ISL_755158, EPI_ISL_755159, EPI_ISL_755160, EPI_ISL_755161, EPI_ISL_755162, EPI_ISL_755163, EPI_ISL_755164, EPI_ISL_755165, EPI_ISL_755166, EPI_ISL_755167, EPI_ISL_755168, EPI_ISL_755169, EPI_ISL_755170, EPI_ISL_755171, EPI_ISL_755172, EPI_ISL_755173, EPI_ISL_755174, EPI_ISL_755175, EPI_ISL_755176, EPI_ISL_755177, EPI_ISL_755178, EPI_ISL_755179, EPI_ISL_755180, EPI_ISL_755181, EPI_ISL_755182, EPI_ISL_755183, EPI_ISL_755184, EPI_ISL_755185, EPI_ISL_755186, EPI_ISL_755187, EPI_ISL_755188, EPI_ISL_755189, EPI_ISL_755190, EPI_ISL_755191, EPI_ISL_755192, EPI_ISL_755193, EPI_ISL_755194, EPI_ISL_755195, EPI_ISL_755196, EPI_ISL_755197, EPI_ISL_755198, EPI_ISL_755199, EPI_ISL_755200, EPI_ISL_755201, EPI_ISL_755202, EPI_ISL_755203, EPI_ISL_755204, EPI_ISL_755205, EPI_ISL_755206, EPI_ISL_755207, EPI_ISL_755208, EPI_ISL_755209, EPI_ISL_755215, EPI_ISL_755216, EPI_ISL_755217, EPI_ISL_755218, EPI_ISL_755219, EPI_ISL_755220, EPI_ISL_755221, EPI_ISL_755222, EPI_ISL_755223, EPI_ISL_755224, EPI_ISL_755225, EPI_ISL_755226, EPI_ISL_755227, EPI_ISL_755228, EPI_ISL_755229, EPI_ISL_755230, EPI_ISL_755231 | | | |
| see above | UCSD EXCITE lab | Andersen lab at Scripps Research | SEARCH Alliance San Diego |
| EPI_ISL_516745, EPI_ISL_516746, EPI_ISL_516747, EPI_ISL_548505, EPI_ISL_548513, EPI_ISL_548522, EPI_ISL_548555, EPI_ISL_548571, EPI_ISL_583199, EPI_ISL_583200, EPI_ISL_583201, EPI_ISL_583202, EPI_ISL_583203, EPI_ISL_583204, EPI_ISL_583205, EPI_ISL_583206, EPI_ISL_583207, EPI_ISL_583208, EPI_ISL_583209, EPI_ISL_583216, EPI_ISL_583217, EPI_ISL_583218, EPI_ISL_583219, EPI_ISL_583220, EPI_ISL_583221, EPI_ISL_583222, EPI_ISL_583223, EPI_ISL_583224, EPI_ISL_583225, EPI_ISL_583226, EPI_ISL_583227, EPI_ISL_583228, EPI_ISL_583229, EPI_ISL_583230, EPI_ISL_583231, EPI_ISL_625661, EPI_ISL_625662, EPI_ISL_625663, EPI_ISL_625664, EPI_ISL_625665, EPI_ISL_625666, EPI_ISL_625667, EPI_ISL_625668, EPI_ISL_625669, EPI_ISL_625670, EPI_ISL_648543, EPI_ISL_648544, EPI_ISL_672018, EPI_ISL_672021, EPI_ISL_672388, EPI_ISL_672393, EPI_ISL_672426, EPI_ISL_672427, EPI_ISL_672428, EPI_ISL_672501, EPI_ISL_738544, EPI_ISL_739004, EPI_ISL_739203, EPI_ISL_739295, EPI_ISL_739613, EPI_ISL_739632, EPI_ISL_739633, EPI_ISL_849926 | | | |
| see above | UCSF Clinical Microbiology Laboratory | Chan-Zuckerberg Biohub | CZB Cliahub Consortium |
| EPI_ISL_754124, EPI_ISL_754125, EPI_ISL_754126, EPI_ISL_754127, EPI_ISL_754128, EPI_ISL_754129, EPI_ISL_754133, EPI_ISL_754134, EPI_ISL_754135, EPI_ISL_754390, EPI_ISL_939619, EPI_ISL_939620, EPI_ISL_939621, EPI_ISL_939622, EPI_ISL_939623, EPI_ISL_939624, EPI_ISL_939625, EPI_ISL_1005491, EPI_ISL_1005492, EPI_ISL_1005493, EPI_ISL_1005494, EPI_ISL_1040172, EPI_ISL_1040173, EPI_ISL_1040174, EPI_ISL_1040175, EPI_ISL_1040176, EPI_ISL_1040177, EPI_ISL_1040178, EPI_ISL_1040179, EPI_ISL_1040180, EPI_ISL_1040427, EPI_ISL_1040428, EPI_ISL_1040429, EPI_ISL_1040430, EPI_ISL_1040431, EPI_ISL_1040432, EPI_ISL_1040433, EPI_ISL_1040434, EPI_ISL_1040435, EPI_ISL_1040436, EPI_ISL_1040437, EPI_ISL_1192367, EPI_ISL_1201558 | | | |
| see above | USC Clinical Lab | Los Angeles County PHL | P. Hemarajata et al. |
| EPI_ISL_812687, EPI_ISL_831700, EPI_ISL_831737, EPI_ISL_831784, EPI_ISL_831816, EPI_ISL_831840, EPI_ISL_831849, EPI_ISL_831864, EPI_ISL_831880 | | | |
| see above | United States Air Force School of Aerospace Medicine | United States Air Force School of Aerospace Medicine | Amanda Javorina; Anthony Fries; Clarise Starr; Elizabeth Macias; Jennifer Meyer; Sarah Purves; William Gruner |
| EPI_ISL_536668, EPI_ISL_536678 | University of Wisconsin-Madison AIDS Vaccine Research Laboratories | University of Wisconsin-Madison AIDS Vaccine Research Laboratories | Gage Moreno; Katarina Braun; et al. AIDS Vaccine Research Laboratories |
| EPI_ISL_2101781, EPI_ISL_2101782, EPI_ISL_2101799, EPI_ISL_2101859, EPI_ISL_2101860, EPI_ISL_2101861, EPI_ISL_2101862, EPI_ISL_2101863, EPI_ISL_2101864, EPI_ISL_2101865, EPI_ISL_2101866, EPI_ISL_2101867, EPI_ISL_2101868 | | | |
| see above | WLA VAMC | Pathogen Discovery, Respiratory Viruses Branch, Division of Viral Diseases, Centers for Disease Control and Prevention | Adam Retchless; Anna Kelleher; Anna Uehara; Brian Lynch; Clinton R. Paden; Dhwani Batra; Haibin Wang; Han Jia Justin Ng; Jasmine Padilla; Jing Zhang; Justin Lee; Krista Queen; Mark Burroughs; Mili Sheth; Morgan Davis; Peter Cook; Rachel Marine; Sarah Nobles; Suxiang Tong; Tara Coalter; Yan Li; Ying Tao |
| EPI_ISL_849577 | Washington State Department of Health | Seattle Flu Study | Amanda Adler; Barry R. Lutz; Benjamin Pelle; Brian Hiatt; Caitlin R. Wolf; Chris D. Frazar; Deborah A. Nickerson; Elisabeth Brandstetter; Geoff Melly; Helen Y. Chu; Janet A. Englund; Jay Shendure; Jover Lee; Kairsten Fay; Kirsten Lacombe; Lea M. Starita; Mark J. Rieder; Matthew Richardson; Matthew Thompson; Melissa Truong; Michael Boeckh; Michael Famulare; Misja Ilcisin; Peter D. Han; Philip Dykema; Romesh Gautom; Scott Lindquist; Thomas R. Sibley; Trevor Bedford |
| EPI_ISL_977984 | ` | Chiu Laboratory, University of California, San Francisco | Candace Wang; Charles Chiu; Debra Wadford; Jill Hacker; Venice Servellita; Xianding (Wayne) Deng |

We gratefully acknowledge the following Authors from the Originating laboratories responsible for obtaining the specimens, as well as the Submitting laboratories where the genome data were generated and shared via GISAID, on which this research is based.

All Submitters of data may be contacted directly via www.gisaid.org

Authors are sorted alphabetically.

| Accession ID | Originating Laboratory | Submitting Laboratory | Authors |
|---|---|---|---|
| EPI_ISL_1225958, EPI_ISL_1225959, EPI_ISL_1225960, EPI_ISL_1225961, EPI_ISL_1225962, EPI_ISL_1225967, EPI_ISL_1225968, EPI_ISL_1225969, EPI_ISL_1225970, EPI_ISL_1225971, EPI_ISL_1225972, EPI_ISL_1225973, EPI_ISL_1225974, EPI_ISL_1226237, EPI_ISL_1226238, EPI_ISL_1226274 | | | |
| see above | "CDPH, Viral and Rickettsial Disease Laboratory" | Genomics and Discovery, Respiratory Viruses Branch, Division of Viral Diseases, Centers for Disease Control and Prevention | Anna Montmayeur; Anna Uehara; Ben L. Rambo-Martin; Clinton R. Paden; Dhwani Batra; Haibin Wang; Jasmine Padilla; Jing Zhang; Justin Lee; Katie Dillon; Krista Queen; Kristen Knipe; Kristine Lacek; Lori Rowe; Mark Burroughs; Matthew Schmerer; Mili Sheth; Peter W. Cook; Rachel Marine; Sam Shepard; Sarah Nobles; Shoshona Le; Suxiang Tong; Yan Li; Ying Tao |
| EPI_ISL_1093549, EPI_ISL_1093553, EPI_ISL_1093554, EPI_ISL_1093555, EPI_ISL_1093556, EPI_ISL_1093563, EPI_ISL_1093564, EPI_ISL_1093565, EPI_ISL_1093566, EPI_ISL_1093567, EPI_ISL_1093568, EPI_ISL_1093570, EPI_ISL_1093571, EPI_ISL_1093572, EPI_ISL_1093573, EPI_ISL_1093574, EPI_ISL_1093575, EPI_ISL_1373135, EPI_ISL_1373136, EPI_ISL_1373137, EPI_ISL_1373138, EPI_ISL_1373139, EPI_ISL_1373140, EPI_ISL_1373141, EPI_ISL_1373142, EPI_ISL_1373143, EPI_ISL_1373144, EPI_ISL_1373146, EPI_ISL_1373147, EPI_ISL_1373148, EPI_ISL_1373149, EPI_ISL_1373150, EPI_ISL_1373151, EPI_ISL_1373152, EPI_ISL_1373153, EPI_ISL_1373154, EPI_ISL_1373155, EPI_ISL_1373156, EPI_ISL_1373157, EPI_ISL_1373158, EPI_ISL_1373159, EPI_ISL_1576992, EPI_ISL_1576993, EPI_ISL_1576994, EPI_ISL_1576995, EPI_ISL_1576996, EPI_ISL_1576997 | | | |
| see above | Alameda County Public Health Department | Alameda County Public Health Department | Kevin Libuit; Kristna Hsieh; Maria Elma |
| EPI_ISL_955387, EPI_ISL_955388, EPI_ISL_955390, EPI_ISL_955391, EPI_ISL_955392, EPI_ISL_955393, EPI_ISL_955394, EPI_ISL_955395, EPI_ISL_955396, EPI_ISL_955397, EPI_ISL_955398, EPI_ISL_955399, EPI_ISL_955400, EPI_ISL_955401, EPI_ISL_955402, EPI_ISL_955403, EPI_ISL_955404, EPI_ISL_955405, EPI_ISL_955406, EPI_ISL_955407, EPI_ISL_955408, EPI_ISL_955409, EPI_ISL_955410, EPI_ISL_955411, EPI_ISL_955412, EPI_ISL_955413, EPI_ISL_955414, EPI_ISL_955415, EPI_ISL_955416, EPI_ISL_955417, EPI_ISL_955418, EPI_ISL_955419, EPI_ISL_955420, EPI_ISL_955421, EPI_ISL_955422, EPI_ISL_955423, EPI_ISL_955424, EPI_ISL_955425, EPI_ISL_955426, EPI_ISL_1027479, EPI_ISL_1027560, EPI_ISL_1027568, EPI_ISL_1027578, EPI_ISL_1027582, EPI_ISL_1184923, EPI_ISL_1184924, EPI_ISL_1184925, EPI_ISL_1184926, EPI_ISL_1184927, EPI_ISL_1184928, EPI_ISL_1184929, EPI_ISL_1184930, EPI_ISL_1184931, EPI_ISL_1184932, EPI_ISL_1184933, EPI_ISL_1184934, EPI_ISL_1184935, EPI_ISL_1184936, EPI_ISL_1184937, EPI_ISL_1184938, EPI_ISL_1184939, EPI_ISL_1184940, EPI_ISL_1184941, EPI_ISL_1184942, EPI_ISL_1184943, EPI_ISL_1184944, EPI_ISL_1184945, EPI_ISL_1184946, EPI_ISL_1184947, EPI_ISL_1184948, EPI_ISL_1184949, EPI_ISL_1184950, EPI_ISL_1184951, EPI_ISL_1184952, EPI_ISL_1184953, EPI_ISL_1184954, EPI_ISL_1184955, EPI_ISL_1184956, EPI_ISL_1184957, EPI_ISL_1184958, EPI_ISL_1184959, EPI_ISL_1184960, EPI_ISL_1184961, EPI_ISL_1184962, EPI_ISL_1184963, EPI_ISL_1184964, EPI_ISL_1184965, EPI_ISL_1184966, EPI_ISL_1185027, EPI_ISL_1185028, EPI_ISL_1185031, EPI_ISL_1185033, EPI_ISL_1185034, EPI_ISL_1185035, EPI_ISL_1185036, EPI_ISL_1185037, EPI_ISL_1185038, EPI_ISL_1185041, EPI_ISL_1185047, EPI_ISL_1185054, EPI_ISL_1185056, EPI_ISL_1185057, EPI_ISL_1185066, EPI_ISL_1185071, EPI_ISL_1185073, EPI_ISL_1185082, EPI_ISL_1185085, EPI_ISL_1185087, EPI_ISL_1185094, EPI_ISL_1185095, EPI_ISL_1185100, EPI_ISL_1185102, EPI_ISL_1185105, EPI_ISL_1185109, EPI_ISL_1185117, EPI_ISL_1185121, EPI_ISL_1185124, EPI_ISL_1185126, EPI_ISL_1185128, EPI_ISL_1185130, EPI_ISL_1185136, EPI_ISL_1185142, EPI_ISL_1185157, EPI_ISL_1185161, EPI_ISL_1185162, EPI_ISL_1185164, EPI_ISL_1185172, EPI_ISL_1185176, EPI_ISL_1185179, EPI_ISL_1185187, EPI_ISL_1185198, EPI_ISL_1185199, EPI_ISL_1185204, EPI_ISL_1185207, EPI_ISL_1185215, EPI_ISL_1185222, EPI_ISL_1185229, EPI_ISL_1185232, EPI_ISL_1185243, EPI_ISL_1185250, EPI_ISL_1185258, EPI_ISL_1185259, EPI_ISL_1185262, EPI_ISL_1185271, EPI_ISL_1185281, EPI_ISL_1185283, EPI_ISL_1185286, EPI_ISL_1185305, EPI_ISL_1185313, EPI_ISL_1185315, EPI_ISL_1185322, EPI_ISL_1185351, EPI_ISL_1185356, EPI_ISL_1185357, EPI_ISL_1185358, EPI_ISL_1185362, EPI_ISL_1185363, EPI_ISL_1185364, EPI_ISL_1185367, EPI_ISL_1185368, EPI_ISL_1185372, EPI_ISL_1185376, EPI_ISL_1185383, EPI_ISL_1185388, EPI_ISL_1185393, EPI_ISL_1185401, EPI_ISL_1185414, EPI_ISL_1185422, EPI_ISL_1185427, EPI_ISL_1185430, EPI_ISL_1185431, EPI_ISL_1185447, EPI_ISL_1185449, EPI_ISL_1185452, EPI_ISL_1185456, EPI_ISL_1185457, EPI_ISL_1185459, EPI_ISL_1185461, EPI_ISL_1185466, EPI_ISL_1185475, EPI_ISL_1185480, EPI_ISL_1185491, EPI_ISL_1185493, EPI_ISL_1185496, EPI_ISL_1185499, EPI_ISL_1185501, EPI_ISL_1185502, EPI_ISL_1185503, EPI_ISL_1185517, EPI_ISL_1185529, EPI_ISL_1185533, EPI_ISL_1185534, EPI_ISL_1235361, EPI_ISL_1235363, EPI_ISL_1235364, EPI_ISL_1235365, EPI_ISL_1235366, EPI_ISL_1235374, EPI_ISL_1235376, EPI_ISL_1235385, EPI_ISL_1235386, EPI_ISL_1235389, EPI_ISL_1235390, EPI_ISL_1235393, EPI_ISL_1235397, EPI_ISL_1235398, EPI_ISL_1235399, EPI_ISL_1235402, EPI_ISL_1235421, EPI_ISL_1235423, EPI_ISL_1235425, EPI_ISL_1235426, EPI_ISL_1235432, EPI_ISL_1235433, EPI_ISL_1235437, EPI_ISL_1235443, EPI_ISL_1235446, EPI_ISL_1235447, EPI_ISL_1235456, EPI_ISL_1235457, EPI_ISL_1235458, EPI_ISL_1235460, EPI_ISL_1235462, EPI_ISL_1235463, EPI_ISL_1235464, EPI_ISL_1235465, EPI_ISL_1235475, EPI_ISL_1235476, EPI_ISL_1235481, EPI_ISL_1235489, EPI_ISL_1235493, EPI_ISL_1235494, EPI_ISL_1235500, EPI_ISL_1235510, EPI_ISL_1235513, EPI_ISL_1235514, EPI_ISL_1235517, EPI_ISL_1235523, EPI_ISL_1235527, EPI_ISL_1235538, EPI_ISL_1235548, EPI_ISL_1235553, EPI_ISL_1235555, EPI_ISL_1235561, EPI_ISL_1235565, EPI_ISL_1235568, EPI_ISL_1235569, EPI_ISL_1235571, EPI_ISL_1235576, EPI_ISL_1235577, EPI_ISL_1235584, EPI_ISL_1235585, EPI_ISL_1235587, EPI_ISL_1235594, EPI_ISL_1235595, EPI_ISL_1235603, EPI_ISL_1235609, EPI_ISL_1235611, EPI_ISL_1701224, EPI_ISL_1701397, EPI_ISL_1701403, EPI_ISL_1701407, EPI_ISL_1701416, EPI_ISL_1701427, EPI_ISL_1701436, EPI_ISL_1701443, EPI_ISL_1701453, EPI_ISL_1701455, EPI_ISL_1701459, EPI_ISL_1701463, EPI_ISL_1701466, EPI_ISL_1701475, EPI_ISL_1701487, EPI_ISL_1701488, EPI_ISL_1701489, EPI_ISL_1701492, EPI_ISL_1701499, EPI_ISL_1701503, EPI_ISL_1701508, EPI_ISL_1701514, EPI_ISL_1701515, EPI_ISL_1701521, EPI_ISL_1701522, EPI_ISL_1701547, EPI_ISL_1701551, EPI_ISL_1701561, EPI_ISL_1701563, EPI_ISL_1701570, EPI_ISL_1701572, EPI_ISL_1701578, EPI_ISL_1701580, EPI_ISL_1701585, EPI_ISL_1701606, EPI_ISL_1701613, EPI_ISL_1701621, EPI_ISL_1701644, EPI_ISL_1701647, EPI_ISL_1701649, EPI_ISL_1701655 | | | |
| see above | Alameda County Public Health Lab | Chan-Zuckerberg Biohub | CZB Cliahub Consortium |
| EPI_ISL_1015897 | Altius Institute for Biomedical Sciences | Seattle Flu Study | Alex Nguyen; Amanda Adler; Andrew Meuser; Barry R. Lutz; Benjamin Pelle; Caitlin R. Wolf; Chris D. Frazar; Clem Green; Daniel Bates; Deborah A. Nickerson; Elisabeth Brandstetter; Erica Ryke; Hannah Petersen; Helen Y. Chu; Jacob Rodriguez; Janet A. Englund; Jay Shendure; Jessica Halow; John Stamatoyannopoulos; Joshua Richards; Jover Lee; Julia Wald; Kairsten Fay; Kirsten Lacombe; Kneshay Harper; Lea M. Starita; Mark J. Rieder; Matt Hartman; Matthew Richardson; Matthew Thompson; Melissa Truong; Michael Boeckh; Michael Famulare; Misja Ilcisin; Muhammad Halimun; Olivia Waltner; Peter D. Han; Rebecca Bruders; Ryan Alexander; Sadie Patraw; Sofia Olsson; Stephanie DeBaun; Thomas R. Sibley; Tobias Ragoczy; Trevor Bedford; Truong Nguyen |
| EPI_ISL_833405, EPI_ISL_833406, EPI_ISL_833407, EPI_ISL_833408, EPI_ISL_833409, EPI_ISL_833410, EPI_ISL_833411, EPI_ISL_833412 | | | |
| see above | Beverly Hospital | Los Angeles County PHL | P. Hemarajata et al. |
| EPI_ISL_1184814, EPI_ISL_1184815, EPI_ISL_1184816, EPI_ISL_1184818, EPI_ISL_1184819, EPI_ISL_1184820, EPI_ISL_1184821, EPI_ISL_1184822, EPI_ISL_1184823, EPI_ISL_1184824, EPI_ISL_1185016, EPI_ISL_1185017, EPI_ISL_1185018, EPI_ISL_1185019, EPI_ISL_1185020, EPI_ISL_1185021, EPI_ISL_1185022, EPI_ISL_1185023, EPI_ISL_1185024, EPI_ISL_1185025, EPI_ISL_1185030, EPI_ISL_1185042, EPI_ISL_1185050, EPI_ISL_1185052, EPI_ISL_1185059, EPI_ISL_1185060, EPI_ISL_1185061, EPI_ISL_1185063, EPI_ISL_1185064, EPI_ISL_1185067, EPI_ISL_1185068, EPI_ISL_1185069, EPI_ISL_1185077, EPI_ISL_1185078, EPI_ISL_1185079, EPI_ISL_1185080, EPI_ISL_1185084, EPI_ISL_1185088, EPI_ISL_1185089, EPI_ISL_1185090, EPI_ISL_1185096, EPI_ISL_1185099, EPI_ISL_1185103, EPI_ISL_1185119, EPI_ISL_1185122, EPI_ISL_1185132, EPI_ISL_1185133, EPI_ISL_1185135, EPI_ISL_1185138, EPI_ISL_1185139, EPI_ISL_1185144, EPI_ISL_1185147, EPI_ISL_1185150, EPI_ISL_1185153, EPI_ISL_1185155, EPI_ISL_1185156, EPI_ISL_1185158, EPI_ISL_1185166, EPI_ISL_1185168, EPI_ISL_1185169, EPI_ISL_1185173, EPI_ISL_1185177, EPI_ISL_1185178, EPI_ISL_1185182, EPI_ISL_1185184, EPI_ISL_1185186, EPI_ISL_1185188, EPI_ISL_1185190, EPI_ISL_1185192, EPI_ISL_1185193, EPI_ISL_1185195, EPI_ISL_1185196, EPI_ISL_1185202, EPI_ISL_1185203, EPI_ISL_1185205, EPI_ISL_1185208, EPI_ISL_1185210, EPI_ISL_1185211, EPI_ISL_1185213, EPI_ISL_1185220, EPI_ISL_1185223, EPI_ISL_1185226, EPI_ISL_1185230, EPI_ISL_1185244, EPI_ISL_1185246, EPI_ISL_1185247, EPI_ISL_1185248, EPI_ISL_1185249, EPI_ISL_1185253, EPI_ISL_1185254, EPI_ISL_1185256, EPI_ISL_1185260, EPI_ISL_1185264, EPI_ISL_1185265, EPI_ISL_1185268, EPI_ISL_1185270, EPI_ISL_1185275, EPI_ISL_1185276, EPI_ISL_1185279, EPI_ISL_1185282, EPI_ISL_1234544, EPI_ISL_1234545, EPI_ISL_1234550, EPI_ISL_1234551, EPI_ISL_1234554, EPI_ISL_1234557, EPI_ISL_1234571, EPI_ISL_1234575, EPI_ISL_1234576, EPI_ISL_1234581, EPI_ISL_1234582, EPI_ISL_1234589, EPI_ISL_1234591, EPI_ISL_1234592, EPI_ISL_1234593, EPI_ISL_1234594, EPI_ISL_1234595, EPI_ISL_1234599, EPI_ISL_1234601, EPI_ISL_1234605, EPI_ISL_1234608, EPI_ISL_1234619, EPI_ISL_1234620, EPI_ISL_1234623, EPI_ISL_1234624, EPI_ISL_1234625, EPI_ISL_1234627, EPI_ISL_1234633, EPI_ISL_1234634, EPI_ISL_1234635, EPI_ISL_1234638, EPI_ISL_1234639, EPI_ISL_1234644, EPI_ISL_1234649, EPI_ISL_1234650, EPI_ISL_1234651, EPI_ISL_1234652, EPI_ISL_1234656, EPI_ISL_1234658, EPI_ISL_1234663, EPI_ISL_1234668, EPI_ISL_1234669, EPI_ISL_1234673, EPI_ISL_1234681, EPI_ISL_1234695, EPI_ISL_1234699, EPI_ISL_1234700, EPI_ISL_1234703, EPI_ISL_1234704, EPI_ISL_1234712, EPI_ISL_1234713, EPI_ISL_1234714, EPI_ISL_1234715, EPI_ISL_1234717, EPI_ISL_1234719, EPI_ISL_1234722, EPI_ISL_1234730, EPI_ISL_1234731, EPI_ISL_1234733, EPI_ISL_1234734, EPI_ISL_1234735, EPI_ISL_1234740, EPI_ISL_1234747, EPI_ISL_1234748, EPI_ISL_1234750, EPI_ISL_1234752, EPI_ISL_1234753, EPI_ISL_1234754, EPI_ISL_1234761, EPI_ISL_1234766, EPI_ISL_1234771, EPI_ISL_1234774, EPI_ISL_1234778, EPI_ISL_1234780, EPI_ISL_1234781, EPI_ISL_1234784, EPI_ISL_1235060, EPI_ISL_1235063, EPI_ISL_1235068, EPI_ISL_1235070, EPI_ISL_1235074, EPI_ISL_1235079, EPI_ISL_1235084, EPI_ISL_1235092, EPI_ISL_1235095, EPI_ISL_1235097, EPI_ISL_1235104, EPI_ISL_1235107, EPI_ISL_1235112, EPI_ISL_1235118, EPI_ISL_1235119, EPI_ISL_1235121, EPI_ISL_1235123, EPI_ISL_1235125, EPI_ISL_1235126, EPI_ISL_1235127, EPI_ISL_1235128, EPI_ISL_1235132, EPI_ISL_1235133, EPI_ISL_1235135, EPI_ISL_1235136, EPI_ISL_1235138, EPI_ISL_1235140, EPI_ISL_1235144, EPI_ISL_1235148, EPI_ISL_1235150, EPI_ISL_1235152, EPI_ISL_1235162, EPI_ISL_1235176, EPI_ISL_1235182, EPI_ISL_1235186, EPI_ISL_1235190, EPI_ISL_1235192, EPI_ISL_1235193, EPI_ISL_1235195, EPI_ISL_1235197, EPI_ISL_1235199, EPI_ISL_1235203, EPI_ISL_1235206, EPI_ISL_1235212, EPI_ISL_1235220, EPI_ISL_1235222, EPI_ISL_1235226, EPI_ISL_1235229, EPI_ISL_1235232, EPI_ISL_1235234, EPI_ISL_1235236, EPI_ISL_1235238, EPI_ISL_1235242, EPI_ISL_1235248, EPI_ISL_1235251, EPI_ISL_1235252, EPI_ISL_1235254, EPI_ISL_1235269, EPI_ISL_1235272, EPI_ISL_1235273, EPI_ISL_1235274, EPI_ISL_1235282, EPI_ISL_1235285, EPI_ISL_1235287, EPI_ISL_1235290, EPI_ISL_1235291, EPI_ISL_1235295, EPI_ISL_1235298, EPI_ISL_1235307, EPI_ISL_1235310, EPI_ISL_1235317, EPI_ISL_1235322, EPI_ISL_1235346, EPI_ISL_1235351, EPI_ISL_1235354, EPI_ISL_1477438, EPI_ISL_1477439, EPI_ISL_1477444, EPI_ISL_1477449, EPI_ISL_1477452, EPI_ISL_1477453, EPI_ISL_1477457, EPI_ISL_1477458, EPI_ISL_1477459, EPI_ISL_1477460, EPI_ISL_1477463, EPI_ISL_1477465, EPI_ISL_1477471, EPI_ISL_1477473, EPI_ISL_1477476, EPI_ISL_1477477, EPI_ISL_1477478, EPI_ISL_1477481, EPI_ISL_1477482, EPI_ISL_1477488, EPI_ISL_1477493, EPI_ISL_1477500, EPI_ISL_1477502, EPI_ISL_1477503, EPI_ISL_1477504, EPI_ISL_1477506, EPI_ISL_1477508, EPI_ISL_1477511, EPI_ISL_1477514, EPI_ISL_1477515, EPI_ISL_1477517, EPI_ISL_1477521, EPI_ISL_1477525, EPI_ISL_1477531, EPI_ISL_1477532, EPI_ISL_1477533, EPI_ISL_1477536, EPI_ISL_1477537, EPI_ISL_1477539, EPI_ISL_1477543, EPI_ISL_1477544, EPI_ISL_1477551, EPI_ISL_1477555, EPI_ISL_1477561, EPI_ISL_1477562, EPI_ISL_1477571, EPI_ISL_1477577, EPI_ISL_1477587, EPI_ISL_1477588, EPI_ISL_1477589, EPI_ISL_1477593, EPI_ISL_1477597, EPI_ISL_1477600, EPI_ISL_1477601, EPI_ISL_1477604, EPI_ISL_1477610, EPI_ISL_1477612, EPI_ISL_1477613, EPI_ISL_1477614, EPI_ISL_1477615, EPI_ISL_1477619, EPI_ISL_1477623, EPI_ISL_1477624, EPI_ISL_1477631, EPI_ISL_1477635, EPI_ISL_1477641, EPI_ISL_1477645, EPI_ISL_1477646, EPI_ISL_1477650, EPI_ISL_1477656, EPI_ISL_1477658, EPI_ISL_1477659, EPI_ISL_1477660, EPI_ISL_1477665, EPI_ISL_1477666, EPI_ISL_1477667, EPI_ISL_1477673, EPI_ISL_1477678, EPI_ISL_1477680, EPI_ISL_1477687, EPI_ISL_1477692, EPI_ISL_1477693, EPI_ISL_1477694, EPI_ISL_1477700, EPI_ISL_1477705, EPI_ISL_1477707, EPI_ISL_1477709, EPI_ISL_1477711, EPI_ISL_1477715, EPI_ISL_1477717, EPI_ISL_1477718, EPI_ISL_1477719, EPI_ISL_1477721, EPI_ISL_1477723, EPI_ISL_1477725, EPI_ISL_1477728, EPI_ISL_1477736, EPI_ISL_1477740, EPI_ISL_1477742, EPI_ISL_1477756, EPI_ISL_1477759, EPI_ISL_1477763, EPI_ISL_1477769, EPI_ISL_1477777, EPI_ISL_1477778, EPI_ISL_1477786, EPI_ISL_1477787, EPI_ISL_1477792, EPI_ISL_1477796, EPI_ISL_1477798, EPI_ISL_1477803, EPI_ISL_1477808, EPI_ISL_1477811, EPI_ISL_1477812, EPI_ISL_1477819, EPI_ISL_1477830, EPI_ISL_1477831, EPI_ISL_1477837, EPI_ISL_1477838, EPI_ISL_1477840, EPI_ISL_1477848, EPI_ISL_1477852, EPI_ISL_1477853, EPI_ISL_1477855, EPI_ISL_1477868, EPI_ISL_1477873, EPI_ISL_1477875, EPI_ISL_1477877, EPI_ISL_1477883, EPI_ISL_1477884, EPI_ISL_1477890, EPI_ISL_1477891, EPI_ISL_1477897, EPI_ISL_1477901, EPI_ISL_1477903, EPI_ISL_1477908, EPI_ISL_1477913, EPI_ISL_1477917, EPI_ISL_1477920, EPI_ISL_1477921, EPI_ISL_1477924, EPI_ISL_1477929, EPI_ISL_1477934, EPI_ISL_1477938, EPI_ISL_1477940, EPI_ISL_1477949 | | | |
| see above | CA DPH Viral and Rickettsial Disease Laboratory | Chan-Zuckerberg Biohub | CZB Cliahub Consortium |
| EPI_ISL_1225949, EPI_ISL_1225950, EPI_ISL_1225951, EPI_ISL_1225952, EPI_ISL_1225953, EPI_ISL_1225954, EPI_ISL_1225955, EPI_ISL_1225956, EPI_ISL_1225957 | | | |
| see above | CA-Los Angeles County Public Health Laboratory | Genomics and Discovery, Respiratory Viruses Branch, Division of Viral Diseases, Centers for Disease Control and Prevention | Anna Montmayeur; Anna Uehara; Ben L. Rambo-Martin; Clinton R. Paden; Dhwani Batra; Haibin Wang; Jasmine Padilla; Jing Zhang; Justin Lee; Katie Dillon; Krista Queen; Kristen Knipe; Kristine Lacek; Lori Rowe; Mark Burroughs; Matthew Schmerer; Mili Sheth; Peter W. Cook; Rachel Marine; Sam Shepard; Sarah Nobles; Shoshona Le; Suxiang Tong; Yan Li; Ying Tao |
| EPI_ISL_1094325, EPI_ISL_1094361, EPI_ISL_1094362, EPI_ISL_1094363, EPI_ISL_1094559, EPI_ISL_1094560, EPI_ISL_1094561, EPI_ISL_1094562, EPI_ISL_1094563, EPI_ISL_1094564, EPI_ISL_1094565, EPI_ISL_1094566, EPI_ISL_1094567, EPI_ISL_1094568, EPI_ISL_1094569, EPI_ISL_1094570, EPI_ISL_1094571 | | | |
| see above | CA-Los Angeles County Public Health Laboratory | Respiratory Viruses Branch, Division of Viral Diseases, Centers for Disease Control and Prevention | Anna Montmayeur; Anna Uehara; Ben L. Rambo-Martin; Clinton R. Paden; Dhwani Batra; Haibin Wang; Jasmine Padilla; Jing Zhang; Justin Lee; Krista Queen; Lori Rowe; Mark Burroughs; Mili Sheth; Peter W. Cook; Rachel Marine; Sarah Nobles; Suxiang Tong; Yan Li; Ying Tao |
| EPI_ISL_1094536 | CDPH, Microbial Diseases Laboratory | Respiratory Viruses Branch, Division of Viral Diseases, Centers for Disease Control and Prevention | Anna Montmayeur; Anna Uehara; Ben L. Rambo-Martin; Clinton R. Paden; Dhwani Batra; Haibin Wang; Jasmine Padilla; Jing Zhang; Justin Lee; Krista Queen; Lori Rowe; Mark Burroughs; Mili Sheth; Peter W. Cook; Rachel Marine; Sarah Nobles; Suxiang Tong; Yan Li; Ying Tao |
| EPI_ISL_1798774, EPI_ISL_1798785, EPI_ISL_1798795 | CDPH, Viral and Rickettsial Disease Laboratory | Centers for Disease Control and Prevention Division of Viral Diseases, Pathogen Discovery | Alison Laufer Halpin; Ben L. Rambo-Martin; Clinton R. Paden; Dakota Howard; Darlene Wagner; Dave Wentworth; Dhwani Batra; Jasmine Padilla; Justin Lee; Katie Dillon; Krista Queen; Kristen Knipe; Kristine Lacek; Mark Burroughs; Matthew Schmerer; Mili Sheth; Peter Cook; Sam Shepard; Sarah Nobles; Shoshona Le; Suxiang Tong; Vivien Dugan; Yvette Unoarumhi |
| EPI_ISL_1373857 | CDPH, Viral and Rickettsial Disease Laboratory | Genomics and Discovery, Respiratory Viruses Branch, Division of Viral Diseases, Centers for Disease Control and Prevention | Anna Kelleher; Anna Uehara; Brian Lynch; Clinton R. Paden; Haibin Wang; Han Jia Justin Ng; Jing Zhang; Krista Queen; Peter Cook; Suxiang Tong; Yan Li; Ying Tao |
| EPI_ISL_1094216, EPI_ISL_1094224, EPI_ISL_1094233, EPI_ISL_1094268, EPI_ISL_1094348, EPI_ISL_1094577, EPI_ISL_1094579, EPI_ISL_1094580, EPI_ISL_1094581, EPI_ISL_1094583, EPI_ISL_1094584, EPI_ISL_1094585, EPI_ISL_1094586, EPI_ISL_1094587, EPI_ISL_1094588, EPI_ISL_1094589, EPI_ISL_1095190, EPI_ISL_1095191, EPI_ISL_1095192, EPI_ISL_1095193, EPI_ISL_1095194, EPI_ISL_1095195, EPI_ISL_1095196, EPI_ISL_1095197, EPI_ISL_1095198, EPI_ISL_1095199, EPI_ISL_1095200, EPI_ISL_1095201, EPI_ISL_1095203 | | | |
| see above | CDPH, Viral and Rickettsial Disease Laboratory | Respiratory Viruses Branch, Division of Viral Diseases, Centers for Disease Control and Prevention | Anna Montmayeur; Anna Uehara; Ben L. Rambo-Martin; Clinton R. Paden; Dhwani Batra; Haibin Wang; Jasmine Padilla; Jing Zhang; Justin Lee; Krista Queen; Lori Rowe; Mark Burroughs; Mili Sheth; Peter W. Cook; Rachel Marine; Sarah Nobles; Suxiang Tong; Yan Li; Ying Tao |
| EPI_ISL_847564, EPI_ISL_847568, EPI_ISL_847572, EPI_ISL_847574, EPI_ISL_847589, EPI_ISL_847590, EPI_ISL_847591, EPI_ISL_847592, EPI_ISL_847593, EPI_ISL_847594, EPI_ISL_847595, EPI_ISL_847596, EPI_ISL_847597, EPI_ISL_847598, EPI_ISL_847599, EPI_ISL_847600, EPI_ISL_847601, EPI_ISL_847602, EPI_ISL_847603, EPI_ISL_847756, EPI_ISL_847757, EPI_ISL_847758, EPI_ISL_847759, EPI_ISL_847760, EPI_ISL_847761, EPI_ISL_847762, EPI_ISL_847763, EPI_ISL_847764, EPI_ISL_847765, EPI_ISL_847766, EPI_ISL_847767, EPI_ISL_847768, EPI_ISL_855393, EPI_ISL_855394, EPI_ISL_984332, EPI_ISL_984347, EPI_ISL_984354, EPI_ISL_984358, EPI_ISL_984360, EPI_ISL_984364, EPI_ISL_984371, EPI_ISL_984372, EPI_ISL_984373, EPI_ISL_984374, EPI_ISL_984379, EPI_ISL_984381, EPI_ISL_984382, EPI_ISL_984383, EPI_ISL_984384, EPI_ISL_984387, EPI_ISL_984388, EPI_ISL_984392, EPI_ISL_984419, EPI_ISL_984423, EPI_ISL_984436, EPI_ISL_984438, EPI_ISL_984439, EPI_ISL_984440, EPI_ISL_984441, EPI_ISL_984442, EPI_ISL_984443, EPI_ISL_984444, EPI_ISL_984445, EPI_ISL_984446, EPI_ISL_984447, EPI_ISL_984448, EPI_ISL_984449, EPI_ISL_984450, EPI_ISL_984451, EPI_ISL_984452, EPI_ISL_984453, EPI_ISL_984454, EPI_ISL_984455, EPI_ISL_984456, EPI_ISL_984457, EPI_ISL_984458, EPI_ISL_984459, EPI_ISL_984460, EPI_ISL_984461, EPI_ISL_984462, EPI_ISL_984463, EPI_ISL_984464, EPI_ISL_984465, EPI_ISL_984466, EPI_ISL_984467, EPI_ISL_984468, EPI_ISL_984469, EPI_ISL_984470, EPI_ISL_984471, EPI_ISL_984472, EPI_ISL_984473, EPI_ISL_984474, EPI_ISL_984475, EPI_ISL_984476, EPI_ISL_984477, EPI_ISL_984478, EPI_ISL_984479, EPI_ISL_984480, EPI_ISL_984481, EPI_ISL_984482, EPI_ISL_984483, EPI_ISL_984484, EPI_ISL_984485, EPI_ISL_984486, EPI_ISL_984487, EPI_ISL_984488, EPI_ISL_984489 | | | |
| see above | California Department of Public Health | Chiu Laboratory, University of California, San Francisco | Brian Bushnell; Candace Wang; Charles Chiu; Debra Wadford; Jill Hacker; Scot Federman; Venice Servellita; Xianding (Wayne) Deng |
| EPI_ISL_1234845, EPI_ISL_1234867, EPI_ISL_1234885, EPI_ISL_1234915, EPI_ISL_1234978, EPI_ISL_1234992, EPI_ISL_1235026, EPI_ISL_1235036, EPI_ISL_1235055, EPI_ISL_1701456 | | | |
| see above | California Institute of Technology | Chan-Zuckerberg Biohub | CZB Cliahub Consortium |
| EPI_ISL_824584, EPI_ISL_824586, EPI_ISL_824597, EPI_ISL_824625 | Cedars-Sinai Medical Center, Department of Pathology & Laboratory Medicine, Molecular Pathology Laboratory | Cedars-Sinai Medical Center, Molecular Pathology Laboratory of Department of Pathology & Laboratory Medicine and Genomic | Brian Davis; Eric Vail; Jasmine T Plummer; Jorge Mario Sincuir Martinez; Stephanie Chen; Wenjuan Zhang |

| | | | |
|---|---|---|---|
| | | Core | |
| EPI_ISL_847536, EPI_ISL_847730, EPI_ISL_847736, EPI_ISL_847741, EPI_ISL_847743, EPI_ISL_855401, EPI_ISL_977918, EPI_ISL_977919, EPI_ISL_977920, EPI_ISL_977921, EPI_ISL_977922, EPI_ISL_977923, EPI_ISL_977924, EPI_ISL_977925, EPI_ISL_977926, EPI_ISL_977927, EPI_ISL_977928, EPI_ISL_977929, EPI_ISL_977930, EPI_ISL_977931, EPI_ISL_977932, EPI_ISL_977933, EPI_ISL_977934, EPI_ISL_977935, EPI_ISL_977936, EPI_ISL_977937, EPI_ISL_977938, EPI_ISL_977939, EPI_ISL_977940, EPI_ISL_977941, EPI_ISL_977942, EPI_ISL_977943, EPI_ISL_977944, EPI_ISL_977945, EPI_ISL_977946, EPI_ISL_977947, EPI_ISL_977948, EPI_ISL_977949, EPI_ISL_977950, EPI_ISL_977951, EPI_ISL_977952, EPI_ISL_977953, EPI_ISL_977954, EPI_ISL_977955, EPI_ISL_977956, EPI_ISL_977957, EPI_ISL_977958, EPI_ISL_977959, EPI_ISL_977960, EPI_ISL_977961, EPI_ISL_977962, EPI_ISL_977963, EPI_ISL_977964, EPI_ISL_977965, EPI_ISL_977966, EPI_ISL_977967, EPI_ISL_977968, EPI_ISL_977969, EPI_ISL_977970, EPI_ISL_977971, EPI_ISL_977972, EPI_ISL_977973, EPI_ISL_977974, EPI_ISL_977975, EPI_ISL_977976, EPI_ISL_977977, EPI_ISL_977978, EPI_ISL_977979, EPI_ISL_977980, EPI_ISL_978063, EPI_ISL_978064, EPI_ISL_978065, EPI_ISL_978066, EPI_ISL_978067, EPI_ISL_978068, EPI_ISL_978069, EPI_ISL_978070, EPI_ISL_978071, EPI_ISL_978075, EPI_ISL_978076, EPI_ISL_978077, EPI_ISL_978078, EPI_ISL_978079, EPI_ISL_978080, EPI_ISL_978081, EPI_ISL_978082, EPI_ISL_978083, EPI_ISL_978084, EPI_ISL_978085, EPI_ISL_978086, EPI_ISL_978087, EPI_ISL_978088, EPI_ISL_978089, EPI_ISL_978090, EPI_ISL_978091, EPI_ISL_978092, EPI_ISL_978093, EPI_ISL_978094, EPI_ISL_978095, EPI_ISL_978096, EPI_ISL_978097, EPI_ISL_978098, EPI_ISL_978099, EPI_ISL_978100, EPI_ISL_978101, EPI_ISL_978102, EPI_ISL_978103, EPI_ISL_978104, EPI_ISL_978105, EPI_ISL_978106, EPI_ISL_978107, EPI_ISL_978108, EPI_ISL_978109, EPI_ISL_978110, EPI_ISL_978111, EPI_ISL_978112, EPI_ISL_978113, EPI_ISL_978114, EPI_ISL_978115, EPI_ISL_978116, EPI_ISL_978117, EPI_ISL_978118, EPI_ISL_978119, EPI_ISL_978120, EPI_ISL_978121, EPI_ISL_978122, EPI_ISL_978123, EPI_ISL_978124, EPI_ISL_978125, EPI_ISL_978126, EPI_ISL_978127, EPI_ISL_978128, EPI_ISL_978129, EPI_ISL_978130, EPI_ISL_978132, EPI_ISL_978133, EPI_ISL_978134, EPI_ISL_978135, EPI_ISL_978136, EPI_ISL_978137, EPI_ISL_978138, EPI_ISL_978139, EPI_ISL_978140, EPI_ISL_978141, EPI_ISL_978142, EPI_ISL_978143, EPI_ISL_978144, EPI_ISL_978145, EPI_ISL_978146, EPI_ISL_978147, EPI_ISL_978148, EPI_ISL_978149, EPI_ISL_978150, EPI_ISL_978151, EPI_ISL_978152, EPI_ISL_978153, EPI_ISL_978154, EPI_ISL_978155, EPI_ISL_978156, EPI_ISL_978157, EPI_ISL_978158, EPI_ISL_978159, EPI_ISL_978961, EPI_ISL_978962, EPI_ISL_978963, EPI_ISL_978964, EPI_ISL_978965, EPI_ISL_978966, EPI_ISL_978973, EPI_ISL_1824296, EPI_ISL_1824297, EPI_ISL_1824298 | | | |
| see above | Chiu Laboratory, University of California, San Francisco | Chiu Laboratory, University of California, San Francisco | Alicia Sotomayor-Gonzalez; Brian Bushnell; Candace Wang; Charles Chiu; Daniel Wong; Debra Wadford; Jill Hacker; Scot Federman; Venice Servellita; Xianding (Wayne) Deng; Yueyuan Zhang |
| EPI_ISL_833413 | City of Hope Comprehensive Cancer Center | Los Angeles County PHL | P. Hemarajata et al. |
| EPI_ISL_1824720, EPI_ISL_1824721, EPI_ISL_1824722, EPI_ISL_1824723, EPI_ISL_1824724, EPI_ISL_1824725, EPI_ISL_1824726, EPI_ISL_1824727, EPI_ISL_1824728, EPI_ISL_1824729, EPI_ISL_1824730, EPI_ISL_1824731, EPI_ISL_1824732 | | | |
| see above | Color Genomics | Chiu Laboratory, University of California, San Francisco | Alicia Sotomayor-Gonzalez; Alicia Zhou; Candace Wang; Charles Chiu; Daniel Wong; Erika Torres; Madyson Wanner; Scott Topper; Venice Servellita; Yueyuan Zhang |
| EPI_ISL_1030269, EPI_ISL_1030281, EPI_ISL_1030282, EPI_ISL_1030302, EPI_ISL_1030303, EPI_ISL_1030306, EPI_ISL_1030307, EPI_ISL_1030309, EPI_ISL_1030310, EPI_ISL_1030311, EPI_ISL_1030313, EPI_ISL_1030314, EPI_ISL_1030315, EPI_ISL_1030316, EPI_ISL_1030317, EPI_ISL_1030318, EPI_ISL_1030319, EPI_ISL_1030320, EPI_ISL_1030321, EPI_ISL_1030322, EPI_ISL_1030323, EPI_ISL_1030324, EPI_ISL_1030325, EPI_ISL_1030326, EPI_ISL_1030327, EPI_ISL_1030328, EPI_ISL_1030329, EPI_ISL_1030331, EPI_ISL_1030332, EPI_ISL_1030333, EPI_ISL_1030334, EPI_ISL_1030335, EPI_ISL_1030336, EPI_ISL_1030338, EPI_ISL_1030339, EPI_ISL_1030340, EPI_ISL_1030341, EPI_ISL_1030342, EPI_ISL_1030343, EPI_ISL_1030344, EPI_ISL_1030345, EPI_ISL_1030346, EPI_ISL_1030347, EPI_ISL_1030348, EPI_ISL_1030349, EPI_ISL_1030350, EPI_ISL_1030351, EPI_ISL_1030352, EPI_ISL_1030353, EPI_ISL_1030354, EPI_ISL_1030355, EPI_ISL_1030356, EPI_ISL_1030357, EPI_ISL_1030358, EPI_ISL_1030359, EPI_ISL_1030360, EPI_ISL_1030361, EPI_ISL_1030362, EPI_ISL_1030363, EPI_ISL_1030364, EPI_ISL_1030365, EPI_ISL_1030366, EPI_ISL_1030367, EPI_ISL_1030368, EPI_ISL_1030369, EPI_ISL_1030370, EPI_ISL_1030371, EPI_ISL_1030372, EPI_ISL_1030373, EPI_ISL_1030374, EPI_ISL_1030375, EPI_ISL_1030376, EPI_ISL_1030377, EPI_ISL_1030378, EPI_ISL_1030379, EPI_ISL_1030380, EPI_ISL_1030381, EPI_ISL_1030382, EPI_ISL_1030383, EPI_ISL_1030384, EPI_ISL_1030385, EPI_ISL_1030386, EPI_ISL_1030387, EPI_ISL_1030388, EPI_ISL_1030389, EPI_ISL_1030390, EPI_ISL_1234538, EPI_ISL_1234540, EPI_ISL_1234546, EPI_ISL_1234552, EPI_ISL_1234553, EPI_ISL_1234555, EPI_ISL_1234556, EPI_ISL_1234558, EPI_ISL_1234559, EPI_ISL_1234563, EPI_ISL_1234569, EPI_ISL_1234570, EPI_ISL_1234572, EPI_ISL_1234573, EPI_ISL_1234577, EPI_ISL_1234579, EPI_ISL_1234580, EPI_ISL_1234583, EPI_ISL_1234585, EPI_ISL_1234587, EPI_ISL_1234590, EPI_ISL_1234596, EPI_ISL_1234598, EPI_ISL_1234602, EPI_ISL_1234607, EPI_ISL_1234610, EPI_ISL_1234614, EPI_ISL_1234615, EPI_ISL_1234616, EPI_ISL_1234618, EPI_ISL_1234622, EPI_ISL_1234626, EPI_ISL_1234628, EPI_ISL_1234629, EPI_ISL_1234630, EPI_ISL_1234631, EPI_ISL_1234632, EPI_ISL_1234636, EPI_ISL_1234642, EPI_ISL_1234643, EPI_ISL_1234647, EPI_ISL_1234654, EPI_ISL_1234655, EPI_ISL_1234659, EPI_ISL_1234662, EPI_ISL_1234664, EPI_ISL_1234674, EPI_ISL_1234675, EPI_ISL_1234680, EPI_ISL_1234682, EPI_ISL_1234684, EPI_ISL_1234685, EPI_ISL_1234686, EPI_ISL_1234688, EPI_ISL_1234690, EPI_ISL_1234691, EPI_ISL_1234692, EPI_ISL_1234693, EPI_ISL_1234696, EPI_ISL_1234697, EPI_ISL_1234702, EPI_ISL_1234705, EPI_ISL_1234707, EPI_ISL_1234708, EPI_ISL_1234709, EPI_ISL_1234710, EPI_ISL_1234726, EPI_ISL_1234727, EPI_ISL_1234736, EPI_ISL_1234738, EPI_ISL_1234741, EPI_ISL_1234745, EPI_ISL_1234746, EPI_ISL_1234749, EPI_ISL_1234756, EPI_ISL_1234757, EPI_ISL_1234763, EPI_ISL_1234764, EPI_ISL_1234768, EPI_ISL_1234770, EPI_ISL_1234772, EPI_ISL_1234775, EPI_ISL_1234777, EPI_ISL_1234783, EPI_ISL_1235061, EPI_ISL_1235064, EPI_ISL_1235067, EPI_ISL_1235069, EPI_ISL_1235071, EPI_ISL_1235072, EPI_ISL_1235078, EPI_ISL_1235080, EPI_ISL_1235081, EPI_ISL_1235082, EPI_ISL_1235085, EPI_ISL_1235088, EPI_ISL_1235090, EPI_ISL_1235105, EPI_ISL_1235106, EPI_ISL_1235108, EPI_ISL_1235117, EPI_ISL_1235120, EPI_ISL_1235130, EPI_ISL_1235134, EPI_ISL_1235137, EPI_ISL_1235139, EPI_ISL_1235143, EPI_ISL_1235145, EPI_ISL_1235146, EPI_ISL_1235153, EPI_ISL_1235154, EPI_ISL_1235155, EPI_ISL_1235157, EPI_ISL_1235159, EPI_ISL_1235161, EPI_ISL_1235164, EPI_ISL_1235166, EPI_ISL_1235167, EPI_ISL_1235170, EPI_ISL_1235172, EPI_ISL_1235173, EPI_ISL_1235177, EPI_ISL_1235179, EPI_ISL_1235180, EPI_ISL_1235184, EPI_ISL_1235187, EPI_ISL_1235189, EPI_ISL_1235191, EPI_ISL_1235200, EPI_ISL_1235202, EPI_ISL_1235204, EPI_ISL_1235208, EPI_ISL_1235209, EPI_ISL_1235214, EPI_ISL_1235221, EPI_ISL_1235224, EPI_ISL_1235225, EPI_ISL_1235228, EPI_ISL_1235237, EPI_ISL_1235240, EPI_ISL_1235245, EPI_ISL_1235246, EPI_ISL_1235247, EPI_ISL_1235258, EPI_ISL_1235259, EPI_ISL_1235260, EPI_ISL_1235261, EPI_ISL_1235262, EPI_ISL_1235264, EPI_ISL_1235271, EPI_ISL_1235277, EPI_ISL_1235278, EPI_ISL_1235283, EPI_ISL_1235284, EPI_ISL_1235286, EPI_ISL_1235288, EPI_ISL_1235293, EPI_ISL_1235301, EPI_ISL_1235302, EPI_ISL_1235304, EPI_ISL_1235306, EPI_ISL_1235311, EPI_ISL_1235315, EPI_ISL_1235316, EPI_ISL_1235319, EPI_ISL_1235321, EPI_ISL_1235323, EPI_ISL_1235325, EPI_ISL_1235326, EPI_ISL_1235328, EPI_ISL_1235329, EPI_ISL_1235330, EPI_ISL_1235331, EPI_ISL_1235332, EPI_ISL_1235334, EPI_ISL_1235342, EPI_ISL_1235345, EPI_ISL_1235348, EPI_ISL_1235352, EPI_ISL_1235353, EPI_ISL_1235357, EPI_ISL_1235358, EPI_ISL_1477737, EPI_ISL_1477746, EPI_ISL_1477760, EPI_ISL_1477794, EPI_ISL_1477821, EPI_ISL_1477833, EPI_ISL_1477844, EPI_ISL_1477867, EPI_ISL_1477930, EPI_ISL_1477950, EPI_ISL_1477952, EPI_ISL_1477953, EPI_ISL_1477960, EPI_ISL_1477972, EPI_ISL_1477977, EPI_ISL_1477990, EPI_ISL_1477993, EPI_ISL_1478002, EPI_ISL_1478003, EPI_ISL_1478008, EPI_ISL_1478011, EPI_ISL_1478017, EPI_ISL_1478022, EPI_ISL_1478032, EPI_ISL_1478034, EPI_ISL_1478035, EPI_ISL_1478036, EPI_ISL_1478037, EPI_ISL_1478039, EPI_ISL_1478040, EPI_ISL_1478043, EPI_ISL_1478053, EPI_ISL_1478057, EPI_ISL_1478072, EPI_ISL_1478078, EPI_ISL_1478088, EPI_ISL_1478099, EPI_ISL_1478108, EPI_ISL_1478111, EPI_ISL_1478122, EPI_ISL_1478125, EPI_ISL_1478126, EPI_ISL_1478131, EPI_ISL_1478147, EPI_ISL_1478154, EPI_ISL_1478180, EPI_ISL_1478187, EPI_ISL_1478188, EPI_ISL_1478193, EPI_ISL_1478196, EPI_ISL_1478197, EPI_ISL_1478199, EPI_ISL_1529103, EPI_ISL_1529149, EPI_ISL_1529163, EPI_ISL_1529165, EPI_ISL_1529171, EPI_ISL_1529177, EPI_ISL_1529181, EPI_ISL_1529186, EPI_ISL_1529188, EPI_ISL_1529194, EPI_ISL_1529195, EPI_ISL_1529196, EPI_ISL_1529210, EPI_ISL_1529211, EPI_ISL_1529212, EPI_ISL_1529216, EPI_ISL_1529227, EPI_ISL_1529228, EPI_ISL_1529229, EPI_ISL_1529236, EPI_ISL_1529239, EPI_ISL_1529250, EPI_ISL_1529253, EPI_ISL_1529261, EPI_ISL_1529271, EPI_ISL_1529272, EPI_ISL_1529282, EPI_ISL_1529297, EPI_ISL_1529298, EPI_ISL_1529301, EPI_ISL_1529302, EPI_ISL_1529304, EPI_ISL_1529305, EPI_ISL_1529310, EPI_ISL_1529320, EPI_ISL_1529328, EPI_ISL_1529330, EPI_ISL_1529331, EPI_ISL_1529334, EPI_ISL_1529346, EPI_ISL_1529348, EPI_ISL_1529352, EPI_ISL_1529353, EPI_ISL_1529354, EPI_ISL_1529365, EPI_ISL_1663923, EPI_ISL_1663926, EPI_ISL_1663930, EPI_ISL_1663934, EPI_ISL_1663936, EPI_ISL_1663937, EPI_ISL_1663940, EPI_ISL_1663943, EPI_ISL_1663946, EPI_ISL_1663948, EPI_ISL_1663950, EPI_ISL_1663951, EPI_ISL_1663953, EPI_ISL_1663954, EPI_ISL_1663955, EPI_ISL_1663957, EPI_ISL_1663962, EPI_ISL_1663964, EPI_ISL_1663968, EPI_ISL_1663970, EPI_ISL_1663971, EPI_ISL_1663974, EPI_ISL_1663975, EPI_ISL_1663976, EPI_ISL_1663977, EPI_ISL_1663983, EPI_ISL_1663985, EPI_ISL_1663987, EPI_ISL_1663989, EPI_ISL_1663992, EPI_ISL_1663994, EPI_ISL_1663995, EPI_ISL_1663996, EPI_ISL_1663999, EPI_ISL_1664005, EPI_ISL_1664008, EPI_ISL_1664014, EPI_ISL_1664024, EPI_ISL_1664026, EPI_ISL_1664030, EPI_ISL_1664032, EPI_ISL_1664035, EPI_ISL_1664038, EPI_ISL_1664045, EPI_ISL_1664046, EPI_ISL_1664047, EPI_ISL_1664049, EPI_ISL_1664050, EPI_ISL_1664053, EPI_ISL_1664054, EPI_ISL_1664057, EPI_ISL_1664058, EPI_ISL_1664060, EPI_ISL_1664063, EPI_ISL_1664068, EPI_ISL_1664069, EPI_ISL_1664071, EPI_ISL_1664072, EPI_ISL_1664081, EPI_ISL_1664082, EPI_ISL_1664084, EPI_ISL_1664087, EPI_ISL_1664088, EPI_ISL_1664096, EPI_ISL_1664097, EPI_ISL_1664098, EPI_ISL_1664100, EPI_ISL_1664102, EPI_ISL_1664103, EPI_ISL_1664104, EPI_ISL_1664105, EPI_ISL_1664108, EPI_ISL_1664109, EPI_ISL_1664110, EPI_ISL_1664111, EPI_ISL_1664112, EPI_ISL_2102522, EPI_ISL_2102524, EPI_ISL_2102526, EPI_ISL_2102534, EPI_ISL_2102538, EPI_ISL_2102541, EPI_ISL_2102543, EPI_ISL_2102555, EPI_ISL_2102569, EPI_ISL_2102579, EPI_ISL_2102588, EPI_ISL_2102590, EPI_ISL_2102591, EPI_ISL_2102746, EPI_ISL_2102758, EPI_ISL_2102759, EPI_ISL_2102760, EPI_ISL_2102761, EPI_ISL_2102762, EPI_ISL_2102764, EPI_ISL_2102767, EPI_ISL_2102769, EPI_ISL_2102770, EPI_ISL_2102782, EPI_ISL_2102788, EPI_ISL_2102792, EPI_ISL_2102795, EPI_ISL_2102796, EPI_ISL_2102800, EPI_ISL_2102801, EPI_ISL_2102804, EPI_ISL_2102814, EPI_ISL_2102815, EPI_ISL_2102817, EPI_ISL_2102828, EPI_ISL_2102836, EPI_ISL_2102840, EPI_ISL_2102841, EPI_ISL_2102858, EPI_ISL_2102863, EPI_ISL_2102868, EPI_ISL_2102880, EPI_ISL_2102885, EPI_ISL_2102894, EPI_ISL_2102896, EPI_ISL_2102901, EPI_ISL_2102907, EPI_ISL_2102908, EPI_ISL_2102909, EPI_ISL_2102916, EPI_ISL_2102917, EPI_ISL_2102918, EPI_ISL_2102919, EPI_ISL_2102922, EPI_ISL_2102927, EPI_ISL_2102930, EPI_ISL_2102931, EPI_ISL_2102936, EPI_ISL_2102939, EPI_ISL_2102949, EPI_ISL_2102951, EPI_ISL_2102952, EPI_ISL_2102970, EPI_ISL_2102973, EPI_ISL_2102975, EPI_ISL_2102981, EPI_ISL_2102985, EPI_ISL_2102990, EPI_ISL_2103049, EPI_ISL_2103195 | | | |
| see above | Contra Costa County Public Health Lab | Chan-Zuckerberg Biohub | CZB Cliahub Consortium |
| EPI_ISL_979159, EPI_ISL_979160, EPI_ISL_979161, EPI_ISL_1701604, EPI_ISL_2103044 | County of San Luis Obispo Public Health Laboratory | Chan-Zuckerberg Biohub | CZB Cliahub Consortium |
| EPI_ISL_1195946, EPI_ISL_1255102 | Curative Inc | Curative Inc | Katie Bond; Vladimir Slepnev |
| EPI_ISL_1093602, EPI_ISL_1093603, EPI_ISL_1093605, EPI_ISL_1093606, EPI_ISL_1093607, EPI_ISL_1093608, EPI_ISL_1093609, EPI_ISL_1093610, EPI_ISL_1093611, EPI_ISL_1093612, EPI_ISL_1093613, EPI_ISL_1093614, EPI_ISL_1093615, EPI_ISL_1093616, EPI_ISL_1093617, EPI_ISL_1093618, EPI_ISL_1093619, EPI_ISL_1093621, EPI_ISL_1093622, EPI_ISL_1093623, EPI_ISL_1093625, EPI_ISL_1093626, EPI_ISL_1093627, EPI_ISL_1093628, EPI_ISL_1093630, EPI_ISL_1093631, EPI_ISL_1093632, EPI_ISL_1093633, EPI_ISL_1093635, EPI_ISL_1093636, EPI_ISL_1093637, EPI_ISL_1093638, EPI_ISL_1093639, EPI_ISL_1093640, EPI_ISL_1093641, EPI_ISL_1093643, EPI_ISL_1093644, EPI_ISL_1093645, EPI_ISL_1093646, EPI_ISL_1093647, EPI_ISL_1093648, EPI_ISL_1093649, EPI_ISL_1093650, EPI_ISL_1093651, EPI_ISL_1093652, EPI_ISL_1093654, EPI_ISL_1093655, EPI_ISL_1093656, EPI_ISL_1093657, EPI_ISL_1093658, EPI_ISL_1093659, EPI_ISL_1093660, EPI_ISL_1093661, EPI_ISL_1093662, EPI_ISL_1093663, EPI_ISL_1093664, EPI_ISL_1093665, EPI_ISL_1093666, EPI_ISL_1093667, EPI_ISL_1093668, EPI_ISL_1093669, EPI_ISL_1093670, EPI_ISL_1093671, EPI_ISL_1093672, EPI_ISL_1093673, EPI_ISL_1093674, EPI_ISL_1093675, EPI_ISL_1093676, EPI_ISL_1093677, EPI_ISL_1093678, EPI_ISL_1093679, EPI_ISL_1093680, EPI_ISL_1093681, EPI_ISL_1093682, EPI_ISL_1093684, EPI_ISL_1093685, EPI_ISL_1093686, EPI_ISL_1093687, EPI_ISL_1093688, EPI_ISL_1093691, EPI_ISL_1093693, EPI_ISL_1093695, EPI_ISL_1093696, EPI_ISL_1093697, EPI_ISL_1093698, EPI_ISL_1093699, EPI_ISL_1093700, EPI_ISL_1093701, EPI_ISL_1093702, EPI_ISL_1093703, EPI_ISL_1093704, EPI_ISL_1093705, EPI_ISL_1093706, EPI_ISL_1093707, EPI_ISL_1093708, EPI_ISL_1093709, EPI_ISL_1093710, EPI_ISL_1093711, EPI_ISL_1093713, EPI_ISL_1093714, EPI_ISL_1093715, EPI_ISL_1093716, EPI_ISL_1093717, EPI_ISL_1093718, EPI_ISL_1093719, EPI_ISL_1093720, EPI_ISL_1093721, EPI_ISL_1093722, EPI_ISL_1093723, EPI_ISL_1093724, EPI_ISL_1093725, EPI_ISL_1093726, EPI_ISL_1093727, EPI_ISL_1093728, EPI_ISL_1093729, EPI_ISL_1093730, EPI_ISL_1093732, EPI_ISL_1093733, EPI_ISL_1093734, EPI_ISL_1093735, EPI_ISL_1093736, EPI_ISL_1093737, EPI_ISL_1093738, EPI_ISL_1093739, EPI_ISL_1093740, EPI_ISL_1093741, EPI_ISL_1093742, EPI_ISL_1093743, EPI_ISL_1093744, EPI_ISL_1093745, EPI_ISL_1093746, EPI_ISL_1093747, EPI_ISL_1093748, EPI_ISL_1093749, EPI_ISL_1093750, EPI_ISL_1093751, EPI_ISL_1093752, EPI_ISL_1093753, EPI_ISL_1093754, EPI_ISL_1093755, EPI_ISL_1093756, EPI_ISL_1093757, EPI_ISL_1093758, EPI_ISL_1093760, EPI_ISL_1093761, EPI_ISL_1093762, EPI_ISL_1093763, EPI_ISL_1093764, EPI_ISL_1093765, EPI_ISL_1093766, EPI_ISL_1093767, EPI_ISL_1093768, EPI_ISL_1093769, EPI_ISL_1093771, EPI_ISL_1093772, EPI_ISL_1093773, EPI_ISL_1093775, EPI_ISL_1093776, EPI_ISL_1093777, EPI_ISL_1093778, EPI_ISL_1093779, EPI_ISL_1093780, EPI_ISL_1093781, EPI_ISL_1093782, EPI_ISL_1093783, EPI_ISL_1093784, EPI_ISL_1093785, EPI_ISL_1093786, EPI_ISL_1093787, EPI_ISL_1093788, EPI_ISL_1093789, EPI_ISL_1093790, EPI_ISL_1093791, EPI_ISL_1093792, EPI_ISL_1093793, EPI_ISL_1093794, EPI_ISL_1093795, EPI_ISL_1093796, EPI_ISL_1093797, EPI_ISL_1093798, EPI_ISL_1093799, EPI_ISL_1093800, EPI_ISL_1093801, EPI_ISL_1093802, EPI_ISL_1093803, EPI_ISL_1093805, EPI_ISL_1093806, EPI_ISL_1093807, EPI_ISL_1093808, EPI_ISL_1093809, EPI_ISL_1093810, EPI_ISL_1093812, EPI_ISL_1093813, EPI_ISL_1093815, EPI_ISL_1093816, EPI_ISL_1093817, EPI_ISL_1093818, EPI_ISL_1093819, EPI_ISL_1093820, EPI_ISL_1093821, EPI_ISL_1093822, EPI_ISL_1093823, EPI_ISL_1093824, EPI_ISL_1093825, EPI_ISL_1093826, EPI_ISL_1093827, EPI_ISL_1093828, EPI_ISL_1093829, EPI_ISL_1093830, EPI_ISL_1093832, EPI_ISL_1093833, EPI_ISL_1093834, EPI_ISL_1093835, EPI_ISL_1093837, EPI_ISL_1093838, EPI_ISL_1093839, EPI_ISL_1093840, EPI_ISL_1093841, EPI_ISL_1093842, EPI_ISL_1093843, EPI_ISL_1093844, EPI_ISL_1093845, EPI_ISL_1093846, EPI_ISL_1093847, EPI_ISL_1093848, EPI_ISL_1093849, EPI_ISL_1093850, EPI_ISL_1093851, EPI_ISL_1093852, EPI_ISL_1093853, EPI_ISL_1093854, EPI_ISL_1093855, EPI_ISL_1093856, EPI_ISL_1093857, EPI_ISL_1093858, EPI_ISL_1093859, EPI_ISL_1093860, EPI_ISL_1093861, EPI_ISL_1093862, EPI_ISL_1093863, EPI_ISL_1093864, EPI_ISL_1093865, EPI_ISL_1093866, EPI_ISL_1093867, EPI_ISL_1093868, EPI_ISL_1093869, EPI_ISL_1093870, EPI_ISL_1093871, EPI_ISL_1093873, EPI_ISL_1093874, EPI_ISL_1093875, EPI_ISL_1093876, EPI_ISL_1093877, EPI_ISL_1093878, EPI_ISL_1093879, EPI_ISL_1093880, EPI_ISL_1093881, EPI_ISL_1093882, EPI_ISL_1093883, EPI_ISL_1093884, EPI_ISL_1093885, EPI_ISL_1093886, EPI_ISL_1093887, EPI_ISL_1093888, EPI_ISL_1093889, EPI_ISL_1093890, EPI_ISL_1093891, EPI_ISL_1093892, EPI_ISL_1093893, EPI_ISL_1093894, EPI_ISL_1093895, EPI_ISL_1093896, EPI_ISL_1093897, EPI_ISL_1093898, EPI_ISL_1093899, EPI_ISL_1093900, EPI_ISL_1093901, EPI_ISL_1093902, EPI_ISL_1093903, EPI_ISL_1093904, EPI_ISL_1093905, EPI_ISL_1093906, EPI_ISL_1093908, EPI_ISL_1093909, EPI_ISL_1093910, EPI_ISL_1093911, EPI_ISL_1093912, EPI_ISL_1093913, EPI_ISL_1093916, EPI_ISL_1093917, EPI_ISL_1093918, EPI_ISL_1093919, EPI_ISL_1093920, EPI_ISL_1093921, EPI_ISL_1093922, EPI_ISL_1093923, EPI_ISL_1093924, EPI_ISL_1093925, EPI_ISL_1093926, EPI_ISL_1093927, EPI_ISL_1093928, EPI_ISL_1093929, EPI_ISL_1093930, EPI_ISL_1093931, EPI_ISL_1093932, EPI_ISL_1093933, EPI_ISL_1093934, EPI_ISL_1093935, EPI_ISL_1093936, EPI_ISL_1093937, EPI_ISL_1093938, EPI_ISL_1093939, EPI_ISL_1093940, EPI_ISL_1093942, EPI_ISL_1093943, EPI_ISL_1093944, EPI_ISL_1093945, EPI_ISL_1093946, EPI_ISL_1093947, EPI_ISL_1093948, EPI_ISL_1093949, EPI_ISL_1093950, EPI_ISL_1093951, EPI_ISL_1093952, EPI_ISL_1093953, EPI_ISL_1093955, EPI_ISL_1093956, EPI_ISL_1093957, EPI_ISL_1093958, EPI_ISL_1093959, EPI_ISL_1093960, EPI_ISL_1093961, EPI_ISL_1093962, EPI_ISL_1093963, EPI_ISL_1093964, EPI_ISL_1093965, EPI_ISL_1093966, EPI_ISL_1093967, EPI_ISL_1093968, EPI_ISL_1093969, EPI_ISL_1093970, EPI_ISL_1093971, EPI_ISL_1093972, EPI_ISL_1093973, EPI_ISL_1093974, EPI_ISL_1093976, EPI_ISL_1093978, EPI_ISL_1093979, EPI_ISL_1093980, EPI_ISL_1093981, EPI_ISL_1093982, EPI_ISL_1093983, EPI_ISL_1093984, EPI_ISL_1093985, EPI_ISL_1093986, EPI_ISL_1093988, EPI_ISL_1093989, EPI_ISL_1093991, EPI_ISL_1093992, EPI_ISL_1093993, EPI_ISL_1093994, EPI_ISL_1093995, EPI_ISL_1093996, EPI_ISL_1093997, EPI_ISL_1093998, EPI_ISL_1093999, EPI_ISL_1094000, EPI_ISL_1094001, EPI_ISL_1094002, EPI_ISL_1094003, EPI_ISL_1094004, EPI_ISL_1094005, EPI_ISL_1094006, EPI_ISL_1094007, EPI_ISL_1094008, EPI_ISL_1094009, EPI_ISL_1094010, EPI_ISL_1094011, EPI_ISL_1094012, EPI_ISL_1094013, EPI_ISL_1094014, EPI_ISL_1094015, EPI_ISL_1094016, EPI_ISL_1094017, EPI_ISL_1094018, EPI_ISL_1094019, EPI_ISL_1094020, EPI_ISL_1094021, EPI_ISL_1094022, EPI_ISL_1094023, EPI_ISL_1094024, EPI_ISL_1094025, EPI_ISL_1094026, EPI_ISL_1094027, EPI_ISL_1094028, EPI_ISL_1094029, EPI_ISL_1094030, EPI_ISL_1094032, EPI_ISL_1094033, EPI_ISL_1094034, EPI_ISL_1094035, EPI_ISL_1094036, EPI_ISL_1094037, EPI_ISL_1094038, EPI_ISL_1094039, EPI_ISL_1094040, EPI_ISL_1094042, EPI_ISL_1094043, EPI_ISL_1094044, EPI_ISL_1094045, EPI_ISL_1094046, EPI_ISL_1094047, EPI_ISL_1094048, EPI_ISL_1094049, EPI_ISL_1094050, EPI_ISL_1094051, EPI_ISL_1094052, EPI_ISL_1094053, EPI_ISL_1094054, EPI_ISL_1094055, EPI_ISL_1094056, EPI_ISL_1094057, EPI_ISL_1094058, EPI_ISL_1094059, EPI_ISL_1094060, EPI_ISL_1094061, EPI_ISL_1094062, EPI_ISL_1094063, EPI_ISL_1094064, EPI_ISL_1094065, EPI_ISL_1094066, EPI_ISL_1094067, EPI_ISL_1094068, EPI_ISL_1094069, EPI_ISL_1094070, EPI_ISL_1094071, EPI_ISL_1094072, EPI_ISL_1094073, EPI_ISL_1094074, EPI_ISL_1094075, EPI_ISL_1094076, EPI_ISL_1094078, EPI_ISL_1094080, EPI_ISL_1094081, EPI_ISL_1094082, EPI_ISL_1094083, EPI_ISL_1094084, EPI_ISL_1094086, EPI_ISL_1094088, EPI_ISL_1094089, EPI_ISL_1094090, EPI_ISL_1094091, EPI_ISL_1094092, EPI_ISL_1094093, EPI_ISL_1094094, EPI_ISL_1094095, EPI_ISL_1094096, EPI_ISL_1094097, EPI_ISL_1094098, EPI_ISL_1094099, EPI_ISL_1094100, EPI_ISL_1094102, EPI_ISL_1094103, EPI_ISL_1094104, EPI_ISL_1094106, EPI_ISL_1094107, EPI_ISL_1094108, EPI_ISL_1094109, EPI_ISL_1094111, EPI_ISL_1094112, EPI_ISL_1094113, EPI_ISL_1094115, EPI_ISL_1094116, EPI_ISL_1094117, EPI_ISL_1094118, EPI_ISL_1094119, EPI_ISL_1094120, EPI_ISL_1094121, EPI_ISL_1094122, EPI_ISL_1094123, EPI_ISL_1094124, EPI_ISL_1094125, EPI_ISL_1094126, EPI_ISL_1094127, EPI_ISL_1094128, EPI_ISL_1094129, EPI_ISL_1094130, EPI_ISL_1094131, EPI_ISL_1094132, EPI_ISL_1094133, EPI_ISL_1094134, EPI_ISL_1094135, EPI_ISL_1094136, EPI_ISL_1094137, EPI_ISL_1094138, EPI_ISL_1094140, EPI_ISL_1094142, EPI_ISL_1094143, EPI_ISL_1094144, EPI_ISL_1094145, EPI_ISL_1094146, EPI_ISL_1094147, EPI_ISL_1094148, EPI_ISL_1094149, EPI_ISL_1094150, EPI_ISL_1094151, EPI_ISL_1094152, EPI_ISL_1094153, EPI_ISL_1094154, EPI_ISL_1094155, EPI_ISL_1094156, EPI_ISL_1094157, EPI_ISL_1094158, EPI_ISL_1094159, EPI_ISL_1094160, EPI_ISL_1094161, EPI_ISL_1094162, EPI_ISL_1094163, EPI_ISL_1094164, EPI_ISL_1094165, EPI_ISL_1094166, EPI_ISL_1094167, EPI_ISL_1094168, EPI_ISL_1094170, EPI_ISL_1094171, EPI_ISL_1094172, EPI_ISL_1094173, EPI_ISL_1094174, EPI_ISL_1094175, EPI_ISL_1094176, EPI_ISL_1094177, EPI_ISL_1094178, EPI_ISL_1094179, EPI_ISL_1094180, EPI_ISL_1094181, EPI_ISL_1094183, EPI_ISL_1094184, EPI_ISL_1094185, EPI_ISL_1094186, EPI_ISL_1094187, EPI_ISL_1094188, EPI_ISL_1094189, EPI_ISL_1094190, EPI_ISL_1094191, EPI_ISL_1098800, EPI_ISL_1098801, EPI_ISL_1098802, EPI_ISL_1098803, EPI_ISL_1098804, EPI_ISL_1098805, EPI_ISL_1098806, EPI_ISL_1098807, EPI_ISL_1098808, EPI_ISL_1098809, EPI_ISL_1098810, EPI_ISL_1098811, EPI_ISL_1098812, EPI_ISL_1098813, EPI_ISL_1098814, EPI_ISL_1098815, EPI_ISL_1098817, EPI_ISL_1098818, EPI_ISL_1098819, EPI_ISL_1098820, EPI_ISL_1098821, EPI_ISL_1098822, EPI_ISL_1098823, EPI_ISL_1098824, EPI_ISL_1098825, EPI_ISL_1098827, EPI_ISL_1098828, EPI_ISL_1098829, EPI_ISL_1318698, EPI_ISL_1318700, EPI_ISL_1318703, EPI_ISL_1318706, EPI_ISL_1318707, EPI_ISL_1318710, EPI_ISL_1318717, EPI_ISL_1318719, EPI_ISL_1318723, EPI_ISL_1318724, EPI_ISL_1318725, EPI_ISL_1318729, EPI_ISL_1318732, EPI_ISL_1318734, EPI_ISL_1318735, EPI_ISL_1318737, EPI_ISL_1318747, EPI_ISL_1318749, EPI_ISL_1318752, EPI_ISL_1318756, EPI_ISL_1318762, EPI_ISL_1318765, EPI_ISL_1318766, EPI_ISL_1318768, EPI_ISL_1318771, EPI_ISL_1318772, EPI_ISL_1318774, EPI_ISL_1318778, EPI_ISL_1318784, EPI_ISL_1318785, EPI_ISL_1318788, EPI_ISL_1318790, EPI_ISL_1318792, EPI_ISL_1318795, EPI_ISL_1318797, EPI_ISL_1318800, EPI_ISL_1318801, EPI_ISL_1318807, EPI_ISL_1318809, EPI_ISL_1318813, EPI_ISL_1318814, EPI_ISL_1318815, EPI_ISL_1318816, EPI_ISL_1318822, EPI_ISL_1318824, EPI_ISL_1318825, EPI_ISL_1318835, EPI_ISL_1318837, EPI_ISL_1318838, EPI_ISL_1318839, EPI_ISL_1318843, EPI_ISL_1318844, EPI_ISL_1318849, EPI_ISL_1318854, EPI_ISL_1318859, EPI_ISL_1318860, EPI_ISL_1318864, EPI_ISL_1318865, EPI_ISL_1318870, EPI_ISL_1318872, EPI_ISL_1318877, EPI_ISL_1318881, EPI_ISL_1318882, EPI_ISL_1318883, EPI_ISL_1318887, EPI_ISL_1318891, EPI_ISL_1318892, EPI_ISL_1318894, EPI_ISL_1318895, EPI_ISL_1318897, EPI_ISL_1318899, EPI_ISL_1318903, EPI_ISL_1318909, EPI_ISL_1318910, EPI_ISL_1318912, EPI_ISL_1318913, EPI_ISL_1318914, EPI_ISL_1318922, EPI_ISL_1318923, EPI_ISL_1318931, EPI_ISL_1318932, EPI_ISL_1318939, EPI_ISL_1318940, EPI_ISL_1318941, EPI_ISL_1318942, EPI_ISL_1318944, EPI_ISL_1318947, EPI_ISL_1318949, EPI_ISL_1318950, EPI_ISL_1318959, EPI_ISL_1318960, EPI_ISL_1318961, EPI_ISL_1318964, EPI_ISL_1318965, EPI_ISL_1318966, EPI_ISL_1318967, EPI_ISL_1318968, EPI_ISL_1318973, EPI_ISL_1318974, EPI_ISL_1318975, EPI_ISL_1318977, EPI_ISL_1318980, EPI_ISL_1318983, EPI_ISL_1318984, EPI_ISL_1318985, EPI_ISL_1347778, EPI_ISL_1347779, EPI_ISL_1347780, EPI_ISL_1347781, EPI_ISL_1347782, EPI_ISL_1347784, EPI_ISL_1349077, EPI_ISL_1349087 | | | |
| see above | DeRisi Lab, University of California, San Francisco | DeRisi Lab, University of California, San Francisco | Amy Kistler; Anthea Mitchell; Carina Marquez; Diane Havlir; Diane Jones; Douglas Black; Eric Chow; Gabriel Chamie; Genay Pilarowski; IDseq Team; Jackie Martinez; James Peng; Jamin Liu; Jon Jacobo; Lucy Li; Luis Rubio; Manu Vanaerschot; Matthew Laurie; Maya Petersen; Patrick Ayscue; Sabrina Mann; Sara Sunshine; Susana Rojas; Susy Rojas; Valerie Tulier-Laiwa; and Joseph DeRisi |
| EPI_ISL_1811676, EPI_ISL_1811687, EPI_ISL_1811691, EPI_ISL_1811694, EPI_ISL_1811699, EPI_ISL_1811700, EPI_ISL_1811704, EPI_ISL_1811707, EPI_ISL_1811728, EPI_ISL_1811731, EPI_ISL_1811735, EPI_ISL_1811752, EPI_ISL_1811756, EPI_ISL_1811757, EPI_ISL_1811767, EPI_ISL_1811768, EPI_ISL_1811774, EPI_ISL_1811781, EPI_ISL_1811783, EPI_ISL_1811798, EPI_ISL_1811800, EPI_ISL_1811823, EPI_ISL_1811834, EPI_ISL_1811838, EPI_ISL_1811839, EPI_ISL_1811841, EPI_ISL_1811842, EPI_ISL_1811844, EPI_ISL_1811846, EPI_ISL_1811856, EPI_ISL_1811857, EPI_ISL_1811859, EPI_ISL_1811860, EPI_ISL_1811861, EPI_ISL_1811863, EPI_ISL_1811864, EPI_ISL_1811865, EPI_ISL_1811866, EPI_ISL_1811868, EPI_ISL_1811870, EPI_ISL_1811871, EPI_ISL_1811874, EPI_ISL_1811876, EPI_ISL_1811877, EPI_ISL_1811878, EPI_ISL_1811879, EPI_ISL_1811880, EPI_ISL_1811881, EPI_ISL_1811882, EPI_ISL_1811883, EPI_ISL_1811884, EPI_ISL_1811886, EPI_ISL_1811888, EPI_ISL_1811890, EPI_ISL_1811891, EPI_ISL_1811892, EPI_ISL_1811893, EPI_ISL_1811894, EPI_ISL_1811895, EPI_ISL_1811896, EPI_ISL_1811898, EPI_ISL_1811899, EPI_ISL_1811900, EPI_ISL_1811901, EPI_ISL_1811903, EPI_ISL_1811904, EPI_ISL_1811905, EPI_ISL_1811906, EPI_ISL_1811907, EPI_ISL_1811908, EPI_ISL_1811909, EPI_ISL_1811910, EPI_ISL_1811911, EPI_ISL_1811912, EPI_ISL_1811913, EPI_ISL_1811915, EPI_ISL_1811916, EPI_ISL_1811917, EPI_ISL_1811918, EPI_ISL_1811919, EPI_ISL_1811920, EPI_ISL_1811921, EPI_ISL_1811922, EPI_ISL_1811923, EPI_ISL_1811925, EPI_ISL_1811926, EPI_ISL_1811928, EPI_ISL_1811930, EPI_ISL_1811933, EPI_ISL_1811934, EPI_ISL_1811935, EPI_ISL_1811937, EPI_ISL_1811938, EPI_ISL_1811939, EPI_ISL_1811941, EPI_ISL_1811942, EPI_ISL_1811943, EPI_ISL_1811944, EPI_ISL_1811945, EPI_ISL_1811946, EPI_ISL_1811947, EPI_ISL_1811948, EPI_ISL_1811949, EPI_ISL_1811950, EPI_ISL_1811951, EPI_ISL_1811952, EPI_ISL_1811953, EPI_ISL_1811954, EPI_ISL_1811955, EPI_ISL_1811956, EPI_ISL_1811957, EPI_ISL_1811958, EPI_ISL_1811959, EPI_ISL_1811960, EPI_ISL_1811961, EPI_ISL_1811962, EPI_ISL_1811963, EPI_ISL_1811964, EPI_ISL_1811967, EPI_ISL_1811968, EPI_ISL_1811970, EPI_ISL_1811971, EPI_ISL_1811972, EPI_ISL_1811974, EPI_ISL_1811975, EPI_ISL_1811976, EPI_ISL_1811977, EPI_ISL_1811978, EPI_ISL_1811979, EPI_ISL_1811981, EPI_ISL_1811982, EPI_ISL_1811983, EPI_ISL_1811984, EPI_ISL_1811985, EPI_ISL_1811986, EPI_ISL_1811987, EPI_ISL_1811988, EPI_ISL_1811989, EPI_ISL_1811990, EPI_ISL_1811991, EPI_ISL_1811993, EPI_ISL_1811995, EPI_ISL_1811997, EPI_ISL_1811998, EPI_ISL_1811999, EPI_ISL_1812002, EPI_ISL_1812003, EPI_ISL_1812004, EPI_ISL_1812005, EPI_ISL_1812006, EPI_ISL_1812008, EPI_ISL_1812009, EPI_ISL_1812010, EPI_ISL_1812014, EPI_ISL_1812018, EPI_ISL_1812020, EPI_ISL_1812022, EPI_ISL_1812023, EPI_ISL_1812024, EPI_ISL_1812025, EPI_ISL_1812026, EPI_ISL_1812027, EPI_ISL_1812028, EPI_ISL_1812030, EPI_ISL_1812032, EPI_ISL_1812033, EPI_ISL_1812034, EPI_ISL_1812035, EPI_ISL_1812037, EPI_ISL_1812038, EPI_ISL_1812040, EPI_ISL_1812041, EPI_ISL_1812046, EPI_ISL_1812047, EPI_ISL_1812048, EPI_ISL_1812053, EPI_ISL_1812054, EPI_ISL_1812055, EPI_ISL_1812056, EPI_ISL_1812057, EPI_ISL_1812058, EPI_ISL_1812059, EPI_ISL_1812063, EPI_ISL_1812064, EPI_ISL_1812066, EPI_ISL_1812067, EPI_ISL_1812069, EPI_ISL_1812070, EPI_ISL_1812072, EPI_ISL_1812078, EPI_ISL_1812080, EPI_ISL_1812081, EPI_ISL_1812082, EPI_ISL_1812083, EPI_ISL_1812084, EPI_ISL_1812085, EPI_ISL_1812088, EPI_ISL_1812089, EPI_ISL_1812091, EPI_ISL_1812092, EPI_ISL_1812093, EPI_ISL_1812094, EPI_ISL_1812095, EPI_ISL_1812096, EPI_ISL_1812097, EPI_ISL_1812101, EPI_ISL_1812103, EPI_ISL_1812105, EPI_ISL_1812107, EPI_ISL_1812108, EPI_ISL_1812109, EPI_ISL_1812110, EPI_ISL_1812111, EPI_ISL_1812112, EPI_ISL_1812113, EPI_ISL_1812115, EPI_ISL_1812118, EPI_ISL_1812124, EPI_ISL_1812125, EPI_ISL_1812126, EPI_ISL_1812128, EPI_ISL_1812129, EPI_ISL_1812132, EPI_ISL_1812133, EPI_ISL_1812135, EPI_ISL_1812136, EPI_ISL_1812138, EPI_ISL_1812140, EPI_ISL_1812141, EPI_ISL_1812142, EPI_ISL_1812143, EPI_ISL_1812144, EPI_ISL_1812146, EPI_ISL_1812147, EPI_ISL_1812148, EPI_ISL_1812149, EPI_ISL_1812151, EPI_ISL_1812152, EPI_ISL_1812153, EPI_ISL_1812155, EPI_ISL_1812156, EPI_ISL_1812158, EPI_ISL_1812161, EPI_ISL_1812162, EPI_ISL_1812164, EPI_ISL_1812165, EPI_ISL_1812166, EPI_ISL_1812167, EPI_ISL_1812169, EPI_ISL_1812172, EPI_ISL_1812173, EPI_ISL_1812174, EPI_ISL_1812175, EPI_ISL_1812176, EPI_ISL_1812177, EPI_ISL_1812179, EPI_ISL_1812182, EPI_ISL_1812184, EPI_ISL_1812186, EPI_ISL_1812189, EPI_ISL_1812190, EPI_ISL_1812194, EPI_ISL_1812195, EPI_ISL_1812196, EPI_ISL_1812200, EPI_ISL_1812202, EPI_ISL_1812205, EPI_ISL_1812207, EPI_ISL_1812208, EPI_ISL_1812209, EPI_ISL_1812210, EPI_ISL_1812212, EPI_ISL_1812213, EPI_ISL_1812214, EPI_ISL_1812218, EPI_ISL_1812220, EPI_ISL_1812221, EPI_ISL_1812222, EPI_ISL_1812223, EPI_ISL_1812224, EPI_ISL_1812225, EPI_ISL_1812226, EPI_ISL_1812232, EPI_ISL_1812233, EPI_ISL_1812234, EPI_ISL_1812235, EPI_ISL_1812236, EPI_ISL_1812237, EPI_ISL_1812240, EPI_ISL_1812241, EPI_ISL_1812243, EPI_ISL_1812244, EPI_ISL_1812245, EPI_ISL_1812246, EPI_ISL_1812247, EPI_ISL_1812248, EPI_ISL_1812249, EPI_ISL_1812251, EPI_ISL_1812253, EPI_ISL_1812254, EPI_ISL_1812255, EPI_ISL_1812256, EPI_ISL_1812258, EPI_ISL_1812260, EPI_ISL_1812262, EPI_ISL_1812263, EPI_ISL_1812264, EPI_ISL_1812266, EPI_ISL_1812267, EPI_ISL_1812269, EPI_ISL_1812270, EPI_ISL_1812272, EPI_ISL_1812273, EPI_ISL_1812274, EPI_ISL_1812277, EPI_ISL_1812278, EPI_ISL_1812279, EPI_ISL_1812280, EPI_ISL_1812283, EPI_ISL_1812285, EPI_ISL_1812288, EPI_ISL_1812289, EPI_ISL_1812293, EPI_ISL_1812296, EPI_ISL_1812297, EPI_ISL_1812298, EPI_ISL_1812299, EPI_ISL_1812300, EPI_ISL_1812301, EPI_ISL_1812302, EPI_ISL_1812303, EPI_ISL_1812304, EPI_ISL_1812306, EPI_ISL_1812307, EPI_ISL_1812308, EPI_ISL_1812309, EPI_ISL_1812313, EPI_ISL_1812315, EPI_ISL_1812318, EPI_ISL_1812319, EPI_ISL_1812320, EPI_ISL_1812321, EPI_ISL_1812322, EPI_ISL_1812325, EPI_ISL_1812330, EPI_ISL_1812331, EPI_ISL_1812332, EPI_ISL_1812333, EPI_ISL_1812334, EPI_ISL_1812335, EPI_ISL_1812337, EPI_ISL_1812339, EPI_ISL_1812340, EPI_ISL_1812342, EPI_ISL_1812344, EPI_ISL_1812345, EPI_ISL_1812346, EPI_ISL_1812347, EPI_ISL_1812350, EPI_ISL_1812351, EPI_ISL_1812353, EPI_ISL_1812355, EPI_ISL_1812356, EPI_ISL_1812358, EPI_ISL_1812359, EPI_ISL_1812362, EPI_ISL_1812366, EPI_ISL_1812369, EPI_ISL_1812370, EPI_ISL_1812371, EPI_ISL_1812374, EPI_ISL_1812375, EPI_ISL_1812376, EPI_ISL_1812377, EPI_ISL_1812380, EPI_ISL_1812383, EPI_ISL_1812386, EPI_ISL_1812394, EPI_ISL_1812396, EPI_ISL_1812397, EPI_ISL_1812399, EPI_ISL_1812401, EPI_ISL_1812403, EPI_ISL_1812405, EPI_ISL_1812409, EPI_ISL_1812410, EPI_ISL_1812413, EPI_ISL_1812415, EPI_ISL_1812418, EPI_ISL_1812419, EPI_ISL_1812421, EPI_ISL_1812422, EPI_ISL_1812425, EPI_ISL_1812426, EPI_ISL_1812428, EPI_ISL_1812429, EPI_ISL_1812431, EPI_ISL_1812432, EPI_ISL_1812435, EPI_ISL_1812437, EPI_ISL_1812438, EPI_ISL_1812439, EPI_ISL_1812440, EPI_ISL_1812442, EPI_ISL_1812443, EPI_ISL_1812444, EPI_ISL_1812450, EPI_ISL_1812451, EPI_ISL_1812452, EPI_ISL_1812454, EPI_ISL_1812455, EPI_ISL_1812456, EPI_ISL_1812457, EPI_ISL_1812459, EPI_ISL_1812461, EPI_ISL_1812462, EPI_ISL_1812463, EPI_ISL_1812466, EPI_ISL_1812467, EPI_ISL_1812469, EPI_ISL_1812470, EPI_ISL_1812471, EPI_ISL_1812473, EPI_ISL_1812474, EPI_ISL_1812475, EPI_ISL_1812476, EPI_ISL_1812478, EPI_ISL_1812479, EPI_ISL_1812483, EPI_ISL_1812484, EPI_ISL_1812485, EPI_ISL_1812488, EPI_ISL_1812490, EPI_ISL_1812493, EPI_ISL_1812494, EPI_ISL_1812496, EPI_ISL_1812498, EPI_ISL_1812500, EPI_ISL_1812501, EPI_ISL_1812502, EPI_ISL_1812503, EPI_ISL_1812504, EPI_ISL_1812505, EPI_ISL_1812507, EPI_ISL_1812508, EPI_ISL_1812510, EPI_ISL_1812511, EPI_ISL_1812513, EPI_ISL_1812514, EPI_ISL_1812515, EPI_ISL_1812516, EPI_ISL_1812519, EPI_ISL_1812521, EPI_ISL_1812523, EPI_ISL_1812527, EPI_ISL_1812528, EPI_ISL_1812529, EPI_ISL_1812532, EPI_ISL_1812539, EPI_ISL_1812543, EPI_ISL_1812544, EPI_ISL_1812545, EPI_ISL_1812546, EPI_ISL_1812547, EPI_ISL_1812551, EPI_ISL_1812554, EPI_ISL_1812555, EPI_ISL_1812557, EPI_ISL_1812560, EPI_ISL_1812561, EPI_ISL_1812563, EPI_ISL_1812565, EPI_ISL_1812567, EPI_ISL_1812570, EPI_ISL_1812572, EPI_ISL_1812573, EPI_ISL_1812575, EPI_ISL_1812580, | | | |

EPI_ISL_1812581, EPI_ISL_1812583, EPI_ISL_1812584, EPI_ISL_1812585, EPI_ISL_1812588, EPI_ISL_1812589, EPI_ISL_1812591, EPI_ISL_1812594, EPI_ISL_1812595, EPI_ISL_1812596, EPI_ISL_1812600, EPI_ISL_1812602, EPI_ISL_1812605, EPI_ISL_1812606, EPI_ISL_1812607, EPI_ISL_1812609, EPI_ISL_1812610, EPI_ISL_1812613, EPI_ISL_1812616, EPI_ISL_1812617, EPI_ISL_1812618, EPI_ISL_1812619, EPI_ISL_1812620, EPI_ISL_1812622, EPI_ISL_1812624, EPI_ISL_1812630, EPI_ISL_1812631, EPI_ISL_1812633, EPI_ISL_1812635, EPI_ISL_1812636, EPI_ISL_1812643, EPI_ISL_1812645, EPI_ISL_1812649, EPI_ISL_1812652, EPI_ISL_1812654, EPI_ISL_1812656, EPI_ISL_1812658, EPI_ISL_1812659, EPI_ISL_1812662, EPI_ISL_1812663, EPI_ISL_1812664, EPI_ISL_1812667, EPI_ISL_1812669, EPI_ISL_1812672, EPI_ISL_1812678, EPI_ISL_1812680, EPI_ISL_1812681, EPI_ISL_1812685, EPI_ISL_1812690, EPI_ISL_1812693, EPI_ISL_1812694, EPI_ISL_1812697, EPI_ISL_1812700, EPI_ISL_1812701, EPI_ISL_1812704, EPI_ISL_1812706, EPI_ISL_1812707, EPI_ISL_1812708, EPI_ISL_1812709, EPI_ISL_1812710, EPI_ISL_1812711, EPI_ISL_1812713, EPI_ISL_1812714, EPI_ISL_1812715, EPI_ISL_1812717, EPI_ISL_1812718, EPI_ISL_1812719, EPI_ISL_1812720, EPI_ISL_1812721, EPI_ISL_1812723, EPI_ISL_1812725, EPI_ISL_1812726, EPI_ISL_1812730, EPI_ISL_1812734, EPI_ISL_1812735, EPI_ISL_1812738, EPI_ISL_1812740, EPI_ISL_1812741, EPI_ISL_1812742, EPI_ISL_1812744, EPI_ISL_1812747, EPI_ISL_1812748, EPI_ISL_1812754, EPI_ISL_1812759, EPI_ISL_1812761, EPI_ISL_1812762, EPI_ISL_1812766, EPI_ISL_1812770, EPI_ISL_1812772, EPI_ISL_1812773, EPI_ISL_1812774, EPI_ISL_1812776, EPI_ISL_1812778, EPI_ISL_1812779, EPI_ISL_1812782, EPI_ISL_1812783, EPI_ISL_1812785, EPI_ISL_1812786, EPI_ISL_1812787, EPI_ISL_1812789, EPI_ISL_1812791, EPI_ISL_1812793, EPI_ISL_1812794, EPI_ISL_1812798, EPI_ISL_1812802, EPI_ISL_1812805, EPI_ISL_1812806, EPI_ISL_1812807, EPI_ISL_1812808, EPI_ISL_1812809, EPI_ISL_1812813, EPI_ISL_1812814, EPI_ISL_1812818, EPI_ISL_1812819, EPI_ISL_1812821, EPI_ISL_1812822, EPI_ISL_1812823, EPI_ISL_1812824, EPI_ISL_1812830, EPI_ISL_1812832, EPI_ISL_1812834, EPI_ISL_1812835, EPI_ISL_1812836, EPI_ISL_1812839, EPI_ISL_1812843, EPI_ISL_1812845, EPI_ISL_1812847, EPI_ISL_1812848, EPI_ISL_1812849, EPI_ISL_1812851, EPI_ISL_1812855, EPI_ISL_1812856, EPI_ISL_1812859, EPI_ISL_1812862, EPI_ISL_1812863, EPI_ISL_1812867, EPI_ISL_1812870, EPI_ISL_1812871, EPI_ISL_1812875, EPI_ISL_1812883, EPI_ISL_1812884, EPI_ISL_1812885, EPI_ISL_1812888, EPI_ISL_1812889, EPI_ISL_1812894, EPI_ISL_1812898, EPI_ISL_1812899, EPI_ISL_1812900, EPI_ISL_1812902, EPI_ISL_1812903, EPI_ISL_1812906, EPI_ISL_1812907, EPI_ISL_1812908, EPI_ISL_1812909, EPI_ISL_1812911, EPI_ISL_1812912, EPI_ISL_1812916, EPI_ISL_1812920, EPI_ISL_1812923, EPI_ISL_1812924, EPI_ISL_1812927, EPI_ISL_1812928, EPI_ISL_1812930, EPI_ISL_1812932, EPI_ISL_1812933, EPI_ISL_1812939, EPI_ISL_1812941, EPI_ISL_1812943, EPI_ISL_1812945, EPI_ISL_1812947, EPI_ISL_1812948, EPI_ISL_1812949, EPI_ISL_1812950, EPI_ISL_1812951, EPI_ISL_1812952, EPI_ISL_1812953, EPI_ISL_1812954, EPI_ISL_1812955, EPI_ISL_1812957, EPI_ISL_1812958, EPI_ISL_1812961, EPI_ISL_1812962, EPI_ISL_1812965, EPI_ISL_1812966, EPI_ISL_1812967, EPI_ISL_1812968, EPI_ISL_1812971, EPI_ISL_1812972, EPI_ISL_1812973, EPI_ISL_1812975, EPI_ISL_1812976, EPI_ISL_1812977, EPI_ISL_1812979, EPI_ISL_1812980, EPI_ISL_1812982, EPI_ISL_1812984, EPI_ISL_1812988, EPI_ISL_1812989, EPI_ISL_1812991, EPI_ISL_1812995, EPI_ISL_1813000, EPI_ISL_1813003, EPI_ISL_1813006, EPI_ISL_1813007, EPI_ISL_1813011, EPI_ISL_1813012, EPI_ISL_1813013, EPI_ISL_1813014, EPI_ISL_1813015, EPI_ISL_1813017, EPI_ISL_1813021, EPI_ISL_1813023, EPI_ISL_1813024, EPI_ISL_1813027, EPI_ISL_1813029, EPI_ISL_1813031, EPI_ISL_1813033, EPI_ISL_1813034, EPI_ISL_1813036, EPI_ISL_1813037, EPI_ISL_1813039, EPI_ISL_1813042, EPI_ISL_1813044, EPI_ISL_1813046, EPI_ISL_1813049, EPI_ISL_1813050, EPI_ISL_1813052, EPI_ISL_1813053, EPI_ISL_1813055, EPI_ISL_1813056, EPI_ISL_1813057, EPI_ISL_1813061, EPI_ISL_1813062, EPI_ISL_1813065, EPI_ISL_1813066, EPI_ISL_1813069, EPI_ISL_1813073, EPI_ISL_1813077, EPI_ISL_1813080, EPI_ISL_1813082, EPI_ISL_1813084, EPI_ISL_1813085, EPI_ISL_1813086, EPI_ISL_1813087, EPI_ISL_1813088, EPI_ISL_1813089, EPI_ISL_1813090, EPI_ISL_1813091, EPI_ISL_1813092, EPI_ISL_1813095, EPI_ISL_1813099, EPI_ISL_1813100, EPI_ISL_1813103, EPI_ISL_1813105, EPI_ISL_1813106, EPI_ISL_1813108, EPI_ISL_1813110, EPI_ISL_1813111, EPI_ISL_1813112, EPI_ISL_1813114, EPI_ISL_1813115, EPI_ISL_1813118, EPI_ISL_1813121, EPI_ISL_1813122, EPI_ISL_1813124, EPI_ISL_1813125, EPI_ISL_1813128, EPI_ISL_1813129, EPI_ISL_1813131, EPI_ISL_1813132, EPI_ISL_1813135, EPI_ISL_1813136, EPI_ISL_1813139, EPI_ISL_1813140, EPI_ISL_1813141, EPI_ISL_1813142, EPI_ISL_1813143, EPI_ISL_1813147, EPI_ISL_1813150, EPI_ISL_1813152, EPI_ISL_1813154, EPI_ISL_1813155, EPI_ISL_1813156, EPI_ISL_1813159, EPI_ISL_1813160, EPI_ISL_1813161, EPI_ISL_1813163, EPI_ISL_1813164, EPI_ISL_1813167, EPI_ISL_1813169, EPI_ISL_1813170, EPI_ISL_1813171, EPI_ISL_1813173, EPI_ISL_1813174, EPI_ISL_1813180, EPI_ISL_1813183, EPI_ISL_1813184, EPI_ISL_1813186, EPI_ISL_1813187, EPI_ISL_1813188, EPI_ISL_1813190, EPI_ISL_1813191, EPI_ISL_1813194, EPI_ISL_1813196, EPI_ISL_1813197, EPI_ISL_1813200, EPI_ISL_1813203, EPI_ISL_1813205, EPI_ISL_1813206, EPI_ISL_1813207, EPI_ISL_1813209, EPI_ISL_1813210, EPI_ISL_1813213, EPI_ISL_1813216, EPI_ISL_1813221, EPI_ISL_1813225, EPI_ISL_1813226, EPI_ISL_1813227, EPI_ISL_1813228, EPI_ISL_1813229, EPI_ISL_1813230, EPI_ISL_1813232, EPI_ISL_1813233, EPI_ISL_1813234, EPI_ISL_1813235, EPI_ISL_1813239, EPI_ISL_1813242, EPI_ISL_1813243, EPI_ISL_1813244, EPI_ISL_1813247, EPI_ISL_1813250, EPI_ISL_1813251, EPI_ISL_1813252, EPI_ISL_1813256, EPI_ISL_1813258, EPI_ISL_1813259, EPI_ISL_1813262, EPI_ISL_1813263, EPI_ISL_1813265, EPI_ISL_1813266, EPI_ISL_1813267, EPI_ISL_1813268, EPI_ISL_1813270, EPI_ISL_1813273, EPI_ISL_1813276, EPI_ISL_1813279, EPI_ISL_1813280, EPI_ISL_1813281, EPI_ISL_1813282, EPI_ISL_1813287, EPI_ISL_1813288, EPI_ISL_1813291, EPI_ISL_1813292, EPI_ISL_1813293, EPI_ISL_1813298, EPI_ISL_1813300, EPI_ISL_1813302, EPI_ISL_1813303, EPI_ISL_1813310, EPI_ISL_1813312, EPI_ISL_1813313, EPI_ISL_1813315, EPI_ISL_1813318, EPI_ISL_1813322, EPI_ISL_1813323, EPI_ISL_1813324, EPI_ISL_1813325, EPI_ISL_1813329, EPI_ISL_1813330, EPI_ISL_1813331, EPI_ISL_1813332, EPI_ISL_1813335, EPI_ISL_1813337, EPI_ISL_1813338, EPI_ISL_1813339, EPI_ISL_1813343, EPI_ISL_1813345, EPI_ISL_1813347, EPI_ISL_1813350, EPI_ISL_1813353, EPI_ISL_1813359, EPI_ISL_1813360, EPI_ISL_1813361, EPI_ISL_1813362, EPI_ISL_1813363, EPI_ISL_1813369, EPI_ISL_1813375, EPI_ISL_1813378, EPI_ISL_1813381, EPI_ISL_1813382, EPI_ISL_1813386, EPI_ISL_1813389, EPI_ISL_1813390, EPI_ISL_1813392, EPI_ISL_1813393, EPI_ISL_1813394, EPI_ISL_1813395, EPI_ISL_1813396, EPI_ISL_1813398, EPI_ISL_1813399, EPI_ISL_1813401, EPI_ISL_1813402, EPI_ISL_1813403, EPI_ISL_1813404, EPI_ISL_1813408, EPI_ISL_1813412, EPI_ISL_1813413, EPI_ISL_1813414, EPI_ISL_1813416, EPI_ISL_1813420, EPI_ISL_1813421, EPI_ISL_1813426, EPI_ISL_1813427, EPI_ISL_1813434, EPI_ISL_1813435, EPI_ISL_1813436, EPI_ISL_1813439, EPI_ISL_1813442, EPI_ISL_1813444, EPI_ISL_1813446, EPI_ISL_1813449, EPI_ISL_1813450, EPI_ISL_1813451, EPI_ISL_1813452, EPI_ISL_1813454, EPI_ISL_1813456, EPI_ISL_1813457, EPI_ISL_1813458, EPI_ISL_1813460, EPI_ISL_1813462, EPI_ISL_1813465, EPI_ISL_1813467, EPI_ISL_1813470, EPI_ISL_1813473, EPI_ISL_1813474, EPI_ISL_1813483, EPI_ISL_1813486, EPI_ISL_1813492, EPI_ISL_1813495, EPI_ISL_1813501, EPI_ISL_1813504, EPI_ISL_1813506, EPI_ISL_1813511, EPI_ISL_1813513, EPI_ISL_1813516, EPI_ISL_1813517, EPI_ISL_1813518, EPI_ISL_1813520, EPI_ISL_1813525, EPI_ISL_1813526, EPI_ISL_1813532, EPI_ISL_1813534, EPI_ISL_1813535, EPI_ISL_1813536, EPI_ISL_1813537, EPI_ISL_1813538, EPI_ISL_1813539, EPI_ISL_1813540, EPI_ISL_1813544, EPI_ISL_1813545, EPI_ISL_1813548, EPI_ISL_1813553, EPI_ISL_1813554, EPI_ISL_1813559, EPI_ISL_1813560, EPI_ISL_1813562, EPI_ISL_1813564, EPI_ISL_1813565, EPI_ISL_1813567, EPI_ISL_1813568, EPI_ISL_1813571, EPI_ISL_1813573, EPI_ISL_1813574, EPI_ISL_1813583, EPI_ISL_1813584, EPI_ISL_1813586, EPI_ISL_1813587, EPI_ISL_1813588, EPI_ISL_1813589, EPI_ISL_1813590, EPI_ISL_1813592, EPI_ISL_1813593, EPI_ISL_1813595, EPI_ISL_1813596, EPI_ISL_1813597, EPI_ISL_1813599, EPI_ISL_1813605, EPI_ISL_1813608, EPI_ISL_1813610, EPI_ISL_1813611, EPI_ISL_1813615, EPI_ISL_1813616, EPI_ISL_1813624, EPI_ISL_1813626, EPI_ISL_1813629, EPI_ISL_1813632, EPI_ISL_1813633, EPI_ISL_1813638, EPI_ISL_1813644, EPI_ISL_1813647, EPI_ISL_1813650, EPI_ISL_1813653, EPI_ISL_1813654, EPI_ISL_1813657, EPI_ISL_1813660, EPI_ISL_1813663, EPI_ISL_1813665, EPI_ISL_1813676, EPI_ISL_1813677, EPI_ISL_1813682, EPI_ISL_1813684, EPI_ISL_1813687, EPI_ISL_1813690, EPI_ISL_1813691, EPI_ISL_1813696, EPI_ISL_1813697, EPI_ISL_1813699, EPI_ISL_1813700, EPI_ISL_1813702, EPI_ISL_1813704, EPI_ISL_1813710, EPI_ISL_1813713, EPI_ISL_1813716, EPI_ISL_1813720, EPI_ISL_1813721, EPI_ISL_1813723, EPI_ISL_1813727, EPI_ISL_1813728, EPI_ISL_1813731, EPI_ISL_1813732, EPI_ISL_1813737, EPI_ISL_1813738, EPI_ISL_1813739, EPI_ISL_1813743, EPI_ISL_1813744, EPI_ISL_1813745, EPI_ISL_1813746, EPI_ISL_1813752, EPI_ISL_1813753, EPI_ISL_1813755, EPI_ISL_1813758, EPI_ISL_1813759, EPI_ISL_1813764, EPI_ISL_1813765, EPI_ISL_1813766, EPI_ISL_1813767, EPI_ISL_1813770, EPI_ISL_1813773, EPI_ISL_1813774, EPI_ISL_1813776, EPI_ISL_1813777, EPI_ISL_1813780, EPI_ISL_1813782, EPI_ISL_1813784, EPI_ISL_1813788, EPI_ISL_1813789, EPI_ISL_1813791, EPI_ISL_1813793, EPI_ISL_1813794, EPI_ISL_1813798, EPI_ISL_1813800, EPI_ISL_1813801, EPI_ISL_1813804, EPI_ISL_1813807, EPI_ISL_1813808, EPI_ISL_1813809, EPI_ISL_1813811, EPI_ISL_1813815, EPI_ISL_1813817, EPI_ISL_1813823, EPI_ISL_1813824, EPI_ISL_1813825, EPI_ISL_1813831, EPI_ISL_1813832, EPI_ISL_1813833, EPI_ISL_1813834, EPI_ISL_1813841, EPI_ISL_1813843, EPI_ISL_1813846, EPI_ISL_1813847, EPI_ISL_1813851, EPI_ISL_1813853, EPI_ISL_1813856, EPI_ISL_1813858, EPI_ISL_1813860, EPI_ISL_1813861, EPI_ISL_1813862, EPI_ISL_1813868, EPI_ISL_1813871, EPI_ISL_1813873, EPI_ISL_1813874, EPI_ISL_1813875, EPI_ISL_1813876, EPI_ISL_1813881, EPI_ISL_1813886, EPI_ISL_1813887, EPI_ISL_1813888, EPI_ISL_1813892, EPI_ISL_1813894, EPI_ISL_1813897, EPI_ISL_1813898, EPI_ISL_1813903, EPI_ISL_1813908, EPI_ISL_1813910, EPI_ISL_1813919, EPI_ISL_1813928, EPI_ISL_1813930, EPI_ISL_1813933, EPI_ISL_1813934, EPI_ISL_1813938, EPI_ISL_1813939, EPI_ISL_1813940, EPI_ISL_1813946, EPI_ISL_1813947, EPI_ISL_1813949, EPI_ISL_1813950, EPI_ISL_1813953, EPI_ISL_1813954, EPI_ISL_1813956, EPI_ISL_1813963, EPI_ISL_1813964, EPI_ISL_1813965, EPI_ISL_1813968, EPI_ISL_1813971, EPI_ISL_1813972, EPI_ISL_1813977, EPI_ISL_1813979, EPI_ISL_1813980, EPI_ISL_1813981, EPI_ISL_1813983, EPI_ISL_1813985, EPI_ISL_1813988, EPI_ISL_1813990, EPI_ISL_1813991, EPI_ISL_1813992, EPI_ISL_1813995, EPI_ISL_1814000, EPI_ISL_1814006, EPI_ISL_1814007, EPI_ISL_1814008, EPI_ISL_1814012, EPI_ISL_1814015, EPI_ISL_1814018, EPI_ISL_1814020, EPI_ISL_1814022, EPI_ISL_1814026, EPI_ISL_1814031, EPI_ISL_1814039, EPI_ISL_1814045, EPI_ISL_1814050, EPI_ISL_1814052, EPI_ISL_1814054, EPI_ISL_1814059, EPI_ISL_1814061, EPI_ISL_1814064, EPI_ISL_1814072, EPI_ISL_1814075, EPI_ISL_1814077, EPI_ISL_1814078, EPI_ISL_1814081, EPI_ISL_1814083, EPI_ISL_1814088, EPI_ISL_1814091, EPI_ISL_1814093, EPI_ISL_1814094, EPI_ISL_1814097, EPI_ISL_1814099, EPI_ISL_1814107, EPI_ISL_1814108, EPI_ISL_1814111, EPI_ISL_1814112, EPI_ISL_1814113, EPI_ISL_1814114, EPI_ISL_1814115, EPI_ISL_1814116, EPI_ISL_1814118, EPI_ISL_1814120, EPI_ISL_1814125, EPI_ISL_1814126, EPI_ISL_1814127, EPI_ISL_1814128, EPI_ISL_1814132, EPI_ISL_1814137, EPI_ISL_1814141, EPI_ISL_1814144, EPI_ISL_1814146, EPI_ISL_1814148, EPI_ISL_1814151, EPI_ISL_1814155, EPI_ISL_1814159, EPI_ISL_1814160, EPI_ISL_1814162, EPI_ISL_1814163, EPI_ISL_1814175, EPI_ISL_1814179, EPI_ISL_1814180, EPI_ISL_1814181, EPI_ISL_1814183, EPI_ISL_1814186, EPI_ISL_1814193, EPI_ISL_1814195, EPI_ISL_1814198, EPI_ISL_1814200, EPI_ISL_1814201, EPI_ISL_1814202, EPI_ISL_1814209, EPI_ISL_1814211, EPI_ISL_1814213, EPI_ISL_1814225, EPI_ISL_1814235, EPI_ISL_1814238, EPI_ISL_1814239, EPI_ISL_1814240, EPI_ISL_1814242, EPI_ISL_1814245, EPI_ISL_1814246, EPI_ISL_1814247, EPI_ISL_1814249, EPI_ISL_1814250, EPI_ISL_1814258, EPI_ISL_1814265, EPI_ISL_1814266, EPI_ISL_1814267, EPI_ISL_1814273, EPI_ISL_1814274, EPI_ISL_1814277, EPI_ISL_1814280, EPI_ISL_1814284, EPI_ISL_1814285, EPI_ISL_1814289, EPI_ISL_1814292, EPI_ISL_1814293, EPI_ISL_1814294, EPI_ISL_1814295, EPI_ISL_1814303, EPI_ISL_1814304, EPI_ISL_1814308, EPI_ISL_1814311, EPI_ISL_1814319, EPI_ISL_1814331, EPI_ISL_1814337, EPI_ISL_1814347, EPI_ISL_1814348, EPI_ISL_1814350, EPI_ISL_1814351, EPI_ISL_1814355, EPI_ISL_1814358, EPI_ISL_1814363, EPI_ISL_1814365, EPI_ISL_1814366, EPI_ISL_1814367, EPI_ISL_1814370, EPI_ISL_1814372, EPI_ISL_1814375, EPI_ISL_1814376, EPI_ISL_1814379, EPI_ISL_1814380, EPI_ISL_1814387, EPI_ISL_1814389, EPI_ISL_1814390, EPI_ISL_1814393, EPI_ISL_1814400, EPI_ISL_1814403, EPI_ISL_1814405, EPI_ISL_1814406, EPI_ISL_1814408, EPI_ISL_1814409, EPI_ISL_1814418, EPI_ISL_1814422, EPI_ISL_1814428, EPI_ISL_1814431, EPI_ISL_1814436, EPI_ISL_1814438, EPI_ISL_1814445, EPI_ISL_1814448, EPI_ISL_1814450, EPI_ISL_1814454, EPI_ISL_1814456, EPI_ISL_1814458, EPI_ISL_1814466, EPI_ISL_1814469, EPI_ISL_1814470, EPI_ISL_1814477, EPI_ISL_1814480, EPI_ISL_1814486, EPI_ISL_1814487, EPI_ISL_1814489, EPI_ISL_1814491, EPI_ISL_1814492, EPI_ISL_1814496, EPI_ISL_1814499, EPI_ISL_1814501, EPI_ISL_1814503, EPI_ISL_1814511, EPI_ISL_1814513, EPI_ISL_1814520, EPI_ISL_1814521, EPI_ISL_1814526, EPI_ISL_1814528, EPI_ISL_1814529, EPI_ISL_1814531, EPI_ISL_1814535, EPI_ISL_1814542, EPI_ISL_1814550, EPI_ISL_1814554, EPI_ISL_1814560, EPI_ISL_1814562, EPI_ISL_1814564, EPI_ISL_1814565, EPI_ISL_1814567, EPI_ISL_1814571, EPI_ISL_1814576, EPI_ISL_1814580, EPI_ISL_1814581, EPI_ISL_1814585, EPI_ISL_1814587, EPI_ISL_1814595, EPI_ISL_1814599, EPI_ISL_1814603, EPI_ISL_1814608, EPI_ISL_1814609, EPI_ISL_1814610, EPI_ISL_1814615, EPI_ISL_1814616, EPI_ISL_1814618, EPI_ISL_1814620, EPI_ISL_1814622, EPI_ISL_1814627, EPI_ISL_1814640, EPI_ISL_1814644, EPI_ISL_1814647, EPI_ISL_1814651, EPI_ISL_1814653, EPI_ISL_1814660, EPI_ISL_1814661, EPI_ISL_1814664, EPI_ISL_1814665, EPI_ISL_1814671, EPI_ISL_1814678, EPI_ISL_1814679, EPI_ISL_1814681, EPI_ISL_1814690, EPI_ISL_1814691, EPI_ISL_1814693, EPI_ISL_1814694, EPI_ISL_1814696, EPI_ISL_1814700, EPI_ISL_1814701, EPI_ISL_1814703, EPI_ISL_1814707, EPI_ISL_1814722, EPI_ISL_1814724, EPI_ISL_1814728, EPI_ISL_1814729, EPI_ISL_1814735, EPI_ISL_1814750, EPI_ISL_1814753, EPI_ISL_1814760, EPI_ISL_1814767, EPI_ISL_1814768, EPI_ISL_1814769, EPI_ISL_1814770, EPI_ISL_1814776, EPI_ISL_1814777, EPI_ISL_1814778, EPI_ISL_1814784, EPI_ISL_1814785, EPI_ISL_1814786, EPI_ISL_1814790, EPI_ISL_1814796, EPI_ISL_1814803, EPI_ISL_1814807, EPI_ISL_1814813, EPI_ISL_1814815, EPI_ISL_1814819, EPI_ISL_1814821, EPI_ISL_1814828, EPI_ISL_1814838, EPI_ISL_1814841, EPI_ISL_1814849, EPI_ISL_1814854, EPI_ISL_1814857, EPI_ISL_1814858, EPI_ISL_1814860, EPI_ISL_1814862, EPI_ISL_1814864, EPI_ISL_1814867, EPI_ISL_1814873, EPI_ISL_1814880, EPI_ISL_1814885, EPI_ISL_1814886, EPI_ISL_1814889, EPI_ISL_1814899, EPI_ISL_1814901, EPI_ISL_1814902, EPI_ISL_1814903, EPI_ISL_1814909, EPI_ISL_1814910, EPI_ISL_1814912, EPI_ISL_1814919, EPI_ISL_1814923, EPI_ISL_1814927, EPI_ISL_1814928, EPI_ISL_1814935, EPI_ISL_1814940, EPI_ISL_1814943, EPI_ISL_1814946, EPI_ISL_1814949, EPI_ISL_1814952, EPI_ISL_1814955, EPI_ISL_1814956, EPI_ISL_1814958, EPI_ISL_1814961, EPI_ISL_1814963, EPI_ISL_1814965, EPI_ISL_1814967, EPI_ISL_1814968, EPI_ISL_1814969, EPI_ISL_1814973, EPI_ISL_1814974, EPI_ISL_1814976, EPI_ISL_1814980, EPI_ISL_1814987, EPI_ISL_1814991, EPI_ISL_1814993, EPI_ISL_1815001, EPI_ISL_1815005, EPI_ISL_1815010, EPI_ISL_1815011, EPI_ISL_1815012, EPI_ISL_1815019, EPI_ISL_1815020, EPI_ISL_1815022, EPI_ISL_1815027, EPI_ISL_1815028, EPI_ISL_1815030, EPI_ISL_1815033, EPI_ISL_1815034, EPI_ISL_1815036, EPI_ISL_1815038, EPI_ISL_1815041, EPI_ISL_1815045, EPI_ISL_1815047, EPI_ISL_1815053, EPI_ISL_1815056, EPI_ISL_1815057, EPI_ISL_1815064, EPI_ISL_1815067, EPI_ISL_1815079, EPI_ISL_1815086, EPI_ISL_1815090, EPI_ISL_1815094, EPI_ISL_1815095, EPI_ISL_1815097, EPI_ISL_1815099, EPI_ISL_1815100, EPI_ISL_1815101, EPI_ISL_1815107, EPI_ISL_1815111, EPI_ISL_1815113, EPI_ISL_1815116, EPI_ISL_1815119, EPI_ISL_1815122, EPI_ISL_1815125, EPI_ISL_1815134, EPI_ISL_1815137, EPI_ISL_1815144, EPI_ISL_1815145, EPI_ISL_1815149, EPI_ISL_1815150, EPI_ISL_1815156, EPI_ISL_1815163, EPI_ISL_1815167, EPI_ISL_1815176, EPI_ISL_1815179, EPI_ISL_1815181, EPI_ISL_1815184, EPI_ISL_1815186, EPI_ISL_1815202, EPI_ISL_1815205, EPI_ISL_1815208, EPI_ISL_1815211, EPI_ISL_1815217, EPI_ISL_1815219, EPI_ISL_1815221, EPI_ISL_1815222, EPI_ISL_1815223, EPI_ISL_1815224, EPI_ISL_1815225, EPI_ISL_1815232, EPI_ISL_1815233, EPI_ISL_1815234, EPI_ISL_1815236, EPI_ISL_1815238, EPI_ISL_1815239, EPI_ISL_1815240, EPI_ISL_1815241, EPI_ISL_1815245, EPI_ISL_1815246, EPI_ISL_1815247, EPI_ISL_1815248, EPI_ISL_1815260, EPI_ISL_1815261, EPI_ISL_1815263, EPI_ISL_1815268, EPI_ISL_1815275, EPI_ISL_1815276, EPI_ISL_1815277, EPI_ISL_1815280, EPI_ISL_1815282, EPI_ISL_1815284, EPI_ISL_1815285, EPI_ISL_1815297, EPI_ISL_1815299, EPI_ISL_1815306, EPI_ISL_1815312, EPI_ISL_1815314, EPI_ISL_1815316, EPI_ISL_1815317, EPI_ISL_1815321, EPI_ISL_1815324, EPI_ISL_1815331, EPI_ISL_1815332, EPI_ISL_1815334, EPI_ISL_1815338, EPI_ISL_1815342, EPI_ISL_1815344, EPI_ISL_1815345, EPI_ISL_1815349, EPI_ISL_1815352, EPI_ISL_1815355, EPI_ISL_1815364, EPI_ISL_1815365, EPI_ISL_1815369, EPI_ISL_1815370, EPI_ISL_1815372, EPI_ISL_1815374, EPI_ISL_1815377, EPI_ISL_1815381, EPI_ISL_1815397, EPI_ISL_1815401, EPI_ISL_1815407, EPI_ISL_1815411, EPI_ISL_1815412, EPI_ISL_1815414, EPI_ISL_1815416, EPI_ISL_1815418, EPI_ISL_1815419, EPI_ISL_1815421, EPI_ISL_1815423, EPI_ISL_1815426, EPI_ISL_1815433, EPI_ISL_1815436, EPI_ISL_1815438, EPI_ISL_1815440, EPI_ISL_1815456, EPI_ISL_1815461, EPI_ISL_1815468, EPI_ISL_1815482, EPI_ISL_1815484, EPI_ISL_1815500, EPI_ISL_1815511, EPI_ISL_1815512, EPI_ISL_1815516, EPI_ISL_1815527, EPI_ISL_1815532, EPI_ISL_1815543, EPI_ISL_1815545, EPI_ISL_1815547, EPI_ISL_1815549, EPI_ISL_1815561, EPI_ISL_1815566, EPI_ISL_1815568, EPI_ISL_1815569, EPI_ISL_1815581, EPI_ISL_1815583, EPI_ISL_1815586, EPI_ISL_1815592, EPI_ISL_1815595, EPI_ISL_1815608, EPI_ISL_1815609, EPI_ISL_1815611, EPI_ISL_1815612, EPI_ISL_1815621, EPI_ISL_1815625, EPI_ISL_1815627, EPI_ISL_1815630, EPI_ISL_1815645, EPI_ISL_1815646, EPI_ISL_1815648, EPI_ISL_1815649, EPI_ISL_1815651, EPI_ISL_1815652, EPI_ISL_1815659, EPI_ISL_1815661, EPI_ISL_1815666, EPI_ISL_1815675, EPI_ISL_1815678, EPI_ISL_1815684, EPI_ISL_1815688, EPI_ISL_1815693, EPI_ISL_1815696, EPI_ISL_1815697, EPI_ISL_1815699, EPI_ISL_1815704, EPI_ISL_1815707, EPI_ISL_1815710, EPI_ISL_1815711, EPI_ISL_1815716, EPI_ISL_1815717, EPI_ISL_1815719, EPI_ISL_1815724, EPI_ISL_1815730, EPI_ISL_1815734, EPI_ISL_1815737, EPI_ISL_1815738, EPI_ISL_1815742, EPI_ISL_1815750, EPI_ISL_1815756, EPI_ISL_1815766, EPI_ISL_1815777, EPI_ISL_1815791, EPI_ISL_1815795, EPI_ISL_1815796, EPI_ISL_1815800, EPI_ISL_1815801, EPI_ISL_1815803, EPI_ISL_1815804, EPI_ISL_1815807, EPI_ISL_1815814, EPI_ISL_1815816, EPI_ISL_1815817, EPI_ISL_1815819, EPI_ISL_1815820, EPI_ISL_1815826, EPI_ISL_1815834, EPI_ISL_1815840, EPI_ISL_1815841, EPI_ISL_1815844, EPI_ISL_1815859, EPI_ISL_1815877, EPI_ISL_1815883, EPI_ISL_1815886, EPI_ISL_1815888, EPI_ISL_1815889, EPI_ISL_1815896, EPI_ISL_1815898, EPI_ISL_1815903, EPI_ISL_1815906, EPI_ISL_1815909, EPI_ISL_1815918, EPI_ISL_1815919, EPI_ISL_1815920, EPI_ISL_1815925, EPI_ISL_1815927, EPI_ISL_1815928, EPI_ISL_1815929, EPI_ISL_1815930, EPI_ISL_1815934, EPI_ISL_1815939, EPI_ISL_1815941, EPI_ISL_1815944, EPI_ISL_1815947, EPI_ISL_1815956, EPI_ISL_1815958, EPI_ISL_1815960, EPI_ISL_1815965, EPI_ISL_1815968, EPI_ISL_1815969, EPI_ISL_1815974, EPI_ISL_1815975, EPI_ISL_1815977, EPI_ISL_1815978, EPI_ISL_1815981, EPI_ISL_1815983, EPI_ISL_1815984, EPI_ISL_1815986, EPI_ISL_1815997, EPI_ISL_1816004, EPI_ISL_1816006, EPI_ISL_1816009, EPI_ISL_1816015, EPI_ISL_1816017, EPI_ISL_1816018, EPI_ISL_1816021, EPI_ISL_1816026, EPI_ISL_1816033, EPI_ISL_1816036, EPI_ISL_1816039, EPI_ISL_1816041, EPI_ISL_1816045, EPI_ISL_1816052, EPI_ISL_1816059, EPI_ISL_1816063, EPI_ISL_1816068, EPI_ISL_1816072, EPI_ISL_1816074, EPI_ISL_1816076, EPI_ISL_1816082, EPI_ISL_1816083, EPI_ISL_1816086, EPI_ISL_1816088, EPI_ISL_1816094, EPI_ISL_1816108, EPI_ISL_1816118, EPI_ISL_1816123, EPI_ISL_1816125, EPI_ISL_1816129, EPI_ISL_1816132, EPI_ISL_1816134, EPI_ISL_1816135, EPI_ISL_1816137, EPI_ISL_1816142, EPI_ISL_1816147, EPI_ISL_1816152, EPI_ISL_1816153, EPI_ISL_1816155, EPI_ISL_1816156, EPI_ISL_1816161, EPI_ISL_1816165, EPI_ISL_1816168, EPI_ISL_1816172, EPI_ISL_1816175, EPI_ISL_1816179, EPI_ISL_1816181, EPI_ISL_1816182, EPI_ISL_1816195, EPI_ISL_1816198, EPI_ISL_1816199, EPI_ISL_1816208, EPI_ISL_1816216, EPI_ISL_1816218, EPI_ISL_1816230, EPI_ISL_1816231, EPI_ISL_1816237, EPI_ISL_1816242, EPI_ISL_1816243, EPI_ISL_1816244, EPI_ISL_1816248, EPI_ISL_1816249, EPI_ISL_1816251, EPI_ISL_1816252, EPI_ISL_1816254, EPI_ISL_1816255, EPI_ISL_1816262, EPI_ISL_1816272, EPI_ISL_1816276, EPI_ISL_1816281, EPI_ISL_1816282, EPI_ISL_1816283, EPI_ISL_1816286, EPI_ISL_1816287, EPI_ISL_1816291, EPI_ISL_1816296, EPI_ISL_1816298, EPI_ISL_1816303, EPI_ISL_1816313, EPI_ISL_1816317, EPI_ISL_1816318, EPI_ISL_1816323, EPI_ISL_1816332, EPI_ISL_1816336, EPI_ISL_1816340, EPI_ISL_1816344, EPI_ISL_1816347, EPI_ISL_1816352, EPI_ISL_1816358, EPI_ISL_1816359, EPI_ISL_1816362, EPI_ISL_1816365, EPI_ISL_1816367, EPI_ISL_1816368, EPI_ISL_1816370, EPI_ISL_1816372, EPI_ISL_1816374, EPI_ISL_1816376, EPI_ISL_1816378, EPI_ISL_1816379, EPI_ISL_1816382, EPI_ISL_1816386, EPI_ISL_1816387, EPI_ISL_1816389, EPI_ISL_1816390, EPI_ISL_1816397, EPI_ISL_1816406, EPI_ISL_1816407, EPI_ISL_1816408, EPI_ISL_1816410, EPI_ISL_1816413, EPI_ISL_1816417, EPI_ISL_1816419, EPI_ISL_1816421, EPI_ISL_1816422, EPI_ISL_1816424, EPI_ISL_1816427, EPI_ISL_1816431, EPI_ISL_1816434, EPI_ISL_1816439, EPI_ISL_1816441, EPI_ISL_1816445, EPI_ISL_1816452, EPI_ISL_1816455, EPI_ISL_1816456, EPI_ISL_1816457, EPI_ISL_1816460, EPI_ISL_1816462, EPI_ISL_1816468, EPI_ISL_1816474, EPI_ISL_1816477, EPI_ISL_1816479, EPI_ISL_1816480, EPI_ISL_1816485, EPI_ISL_1816490, EPI_ISL_1816491, EPI_ISL_1816492, EPI_ISL_1816497, EPI_ISL_1816501, EPI_ISL_1816504, EPI_ISL_1816513, EPI_ISL_1816514, EPI_ISL_1816517, EPI_ISL_1816518, EPI_ISL_1816536, EPI_ISL_1816538, EPI_ISL_1816539, EPI_ISL_1816543, EPI_ISL_1816549, EPI_ISL_1816555, EPI_ISL_1816558, EPI_ISL_1816561, EPI_ISL_1816562, EPI_ISL_1816564, EPI_ISL_1816565, EPI_ISL_1816566, EPI_ISL_1816570, EPI_ISL_1816580, EPI_ISL_1816581, EPI_ISL_1816582, EPI_ISL_1816585, EPI_ISL_1816586, EPI_ISL_1816588, EPI_ISL_1816589, EPI_ISL_1816590, EPI_ISL_1816591, EPI_ISL_1816595, EPI_ISL_1816596, EPI_ISL_1816597, EPI_ISL_1816600, EPI_ISL_1816601, EPI_ISL_1816606, EPI_ISL_1816607, EPI_ISL_1816617, EPI_ISL_1816620, EPI_ISL_1816626, EPI_ISL_1816631, EPI_ISL_1816635, EPI_ISL_1816645, EPI_ISL_1816646, EPI_ISL_1816647, EPI_ISL_1816651, EPI_ISL_1816654, EPI_ISL_1816656, EPI_ISL_1816657, EPI_ISL_1816658, EPI_ISL_1816660, EPI_ISL_1816662, EPI_ISL_1816663, EPI_ISL_1816664, EPI_ISL_1816667, EPI_ISL_1816668, EPI_ISL_1816672, EPI_ISL_1816673, EPI_ISL_1816676, EPI_ISL_1816677, EPI_ISL_1816682, EPI_ISL_1816683, EPI_ISL_1816685, EPI_ISL_1816691, EPI_ISL_1816692, EPI_ISL_1816696, EPI_ISL_1816697, EPI_ISL_1816698, EPI_ISL_1816701, EPI_ISL_1816703, EPI_ISL_1816706, EPI_ISL_1816709, EPI_ISL_1816710, EPI_ISL_1816713, EPI_ISL_1816715, EPI_ISL_1816720, EPI_ISL_1816721, EPI_ISL_1816724, EPI_ISL_1816728, EPI_ISL_1816729, EPI_ISL_1816730, EPI_ISL_1816732, EPI_ISL_1816735, EPI_ISL_1816738, EPI_ISL_1816739, EPI_ISL_1816740, EPI_ISL_1816743, EPI_ISL_1816745, EPI_ISL_1816746, EPI_ISL_1816747, EPI_ISL_1816749, EPI_ISL_1816751, EPI_ISL_1816757, EPI_ISL_1816758, EPI_ISL_1816765, EPI_ISL_1816766, EPI_ISL_1816769, EPI_ISL_1816771, EPI_ISL_1816775, EPI_ISL_1816777, EPI_ISL_1816778, EPI_ISL_1816782, EPI_ISL_1816784, EPI_ISL_1816788, EPI_ISL_1816789, EPI_ISL_1816790, EPI_ISL_1816791, EPI_ISL_1816801, EPI_ISL_1816803, EPI_ISL_1816804, EPI_ISL_1816810, EPI_ISL_1816814, EPI_ISL_1816816, EPI_ISL_1816817, EPI_ISL_1816820, EPI_ISL_1816822, EPI_ISL_1816823, EPI_ISL_1816826, EPI_ISL_1816836, EPI_ISL_1816837, EPI_ISL_1816844, EPI_ISL_1816845, EPI_ISL_1816846, EPI_ISL_1816847, EPI_ISL_1816850, EPI_ISL_1816851, EPI_ISL_1816857, EPI_ISL_1816862, EPI_ISL_1816863, EPI_ISL_1816866, EPI_ISL_1816868, EPI_ISL_1816869, EPI_ISL_1816870, EPI_ISL_1816888, EPI_ISL_1816889, EPI_ISL_1816890, EPI_ISL_1816896, EPI_ISL_1816898, EPI_ISL_1816906, EPI_ISL_1816910, EPI_ISL_1816914

| | | | |
|---|---|---|---|
| see above | EXCITE Lab | Andersen lab at Scripps Research | Abigail Schnapper; Alexandre Bolze; Alice Summerfield; Angela Scioscia; Araz Majnoonian; Ashkan Hassani; Brent Brainard+ SEARCH; Celena Andrade; Charlotte Rivera-Garcia; Cheryl Anderson; Chip Schooley; Christine Mitchell + SEARCH; David Becker; David Pride; Efren Sandoval; Elizabeth Cirulli; Fitri Carrissa Wijaya; Francisco Tanudjaja; Fred Wu; Geraint Levan; Greg Humphrey; Helena Tubb; J. Joelle Donofrio-Odmann; James Lu + SEARCH; Jason Nguyen; Jimmy Ramirez; Kelly Schiabor Barrett; Laura Kohn; Magnus Isaksson; Marc Laurent; Marlene Flores; Megan Nguyen; Natasha Martin; Natasha Martin Cheryl Anderson; Nicole L Washington; Patricia Gonzalez-Zuniga; Rebecca Fielding-Miller; Rodolfo Salido; Ryan Cho; Sawyer Farmer; Sharon Reed; Sherry Wang; Simon White; Smruthi Karthikeyan; Sydney Morgan + Marni Jacobs + Celestine Magallanes + Holly Valentine + Louise C. Laurent + SEARCH; Tommy Valles + SEARCH; Tyler Cassens; Victor Cantu; Vinton Omaleki; William Lee |
| EPI_ISL_917943, EPI_ISL_1097739 | Flow Diagnostics Lab | Los Angeles County PHL | P. Hemarajata et al. |

EPI_ISL_977120, EPI_ISL_977121, EPI_ISL_977122, EPI_ISL_977123, EPI_ISL_977124, EPI_ISL_977125, EPI_ISL_977126, EPI_ISL_977127, EPI_ISL_977128, EPI_ISL_977129, EPI_ISL_977130, EPI_ISL_977131, EPI_ISL_977132, EPI_ISL_977133, EPI_ISL_977134, EPI_ISL_977135, EPI_ISL_977136, EPI_ISL_977137, EPI_ISL_977138, EPI_ISL_977139, EPI_ISL_977140, EPI_ISL_977141, EPI_ISL_977142, EPI_ISL_977143, EPI_ISL_977144, EPI_ISL_1253713, EPI_ISL_1253714, EPI_ISL_1253715, EPI_ISL_1253716, EPI_ISL_1253717, EPI_ISL_1253718

| | | | |
|---|---|---|---|
| see above | Flow Health | Infectious Disease Program, Broad Institute of Harvard and MIT | Adams, G.; B.L.; B.W.; Bauer, M.; Birren; Carter, A.; Chaluvadi, S.; D.J.; DeRuff, K.; Gallagher, G.; Gladden-Young, A.; J.E.; K.J.; Lagerborg, K.; Lemieux; Loreth, C.; MacInnis; Normandin, E.; P.C.; Park; Reilly, S.; Rudy, M.; Siddle; Smole, S.; Tomkins-Tinch, C.; and Sabeti |

EPI_ISL_955261, EPI_ISL_955263, EPI_ISL_984754, EPI_ISL_1005583, EPI_ISL_1005584, EPI_ISL_1005585, EPI_ISL_1005586, EPI_ISL_1005587, EPI_ISL_1005588, EPI_ISL_1005589, EPI_ISL_1005590, EPI_ISL_1005591, EPI_ISL_1005592, EPI_ISL_1005593, EPI_ISL_1005594, EPI_ISL_1005595, EPI_ISL_1005596, EPI_ISL_1005597, EPI_ISL_1005598, EPI_ISL_1005599, EPI_ISL_1005600, EPI_ISL_1005601, EPI_ISL_1005602, EPI_ISL_1005603, EPI_ISL_1005604, EPI_ISL_1005605, EPI_ISL_1005606, EPI_ISL_1005607, EPI_ISL_1005608, EPI_ISL_1005609, EPI_ISL_1005610, EPI_ISL_1005611, EPI_ISL_1005612, EPI_ISL_1005613, EPI_ISL_1005614, EPI_ISL_1005615, EPI_ISL_1005616, EPI_ISL_1005617, EPI_ISL_1005618, EPI_ISL_1005619, EPI_ISL_1005620, EPI_ISL_1005621, EPI_ISL_1005622, EPI_ISL_1005623, EPI_ISL_1005624, EPI_ISL_1005625, EPI_ISL_1005626, EPI_ISL_1005627, EPI_ISL_1005628, EPI_ISL_1005629, EPI_ISL_1005630, EPI_ISL_1005631, EPI_ISL_1005632, EPI_ISL_1005633, EPI_ISL_1005634, EPI_ISL_1005635, EPI_ISL_1005636, EPI_ISL_1005637, EPI_ISL_1005638, EPI_ISL_1005639, EPI_ISL_1005640, EPI_ISL_1005641, EPI_ISL_1005642, EPI_ISL_1005643, EPI_ISL_1005644, EPI_ISL_1005645, EPI_ISL_1005646, EPI_ISL_1005647, EPI_ISL_1005648, EPI_ISL_1005649, EPI_ISL_1005650, EPI_ISL_1005651, EPI_ISL_1005652, EPI_ISL_1005653, EPI_ISL_1005654, EPI_ISL_1009868, EPI_ISL_1009869, EPI_ISL_1009870, EPI_ISL_1009871, EPI_ISL_1009872, EPI_ISL_1009873, EPI_ISL_1009874, EPI_ISL_1009875, EPI_ISL_1009876, EPI_ISL_1009877, EPI_ISL_1009878, EPI_ISL_1009879, EPI_ISL_1009880, EPI_ISL_1009881, EPI_ISL_1009882, EPI_ISL_1009883, EPI_ISL_1009884, EPI_ISL_1009885, EPI_ISL_1009886, EPI_ISL_1009887, EPI_ISL_1009888, EPI_ISL_1009889, EPI_ISL_1009890, EPI_ISL_1009891, EPI_ISL_1009892, EPI_ISL_1009893, EPI_ISL_1009894, EPI_ISL_1009895, EPI_ISL_1009896, EPI_ISL_1009897, EPI_ISL_1009898, EPI_ISL_1009899, EPI_ISL_1009900, EPI_ISL_1009901, EPI_ISL_1009902, EPI_ISL_1009903, EPI_ISL_1009904, EPI_ISL_1009905, EPI_ISL_1009906, EPI_ISL_1009907, EPI_ISL_1009908, EPI_ISL_1009909, EPI_ISL_1009910, EPI_ISL_1009911, EPI_ISL_1009912, EPI_ISL_1009913, EPI_ISL_1009914, EPI_ISL_1009915, EPI_ISL_1009916, EPI_ISL_1009917, EPI_ISL_1009918, EPI_ISL_1009919, EPI_ISL_1009920, EPI_ISL_1009921, EPI_ISL_1009922, EPI_ISL_1009923, EPI_ISL_1009924, EPI_ISL_1009925, EPI_ISL_1009926, EPI_ISL_1009927, EPI_ISL_1009928, EPI_ISL_1009929, EPI_ISL_1009930, EPI_ISL_1009931, EPI_ISL_1009932, EPI_ISL_1009933, EPI_ISL_1009934, EPI_ISL_1009935, EPI_ISL_1009936, EPI_ISL_1009937, EPI_ISL_1009938, EPI_ISL_1009939, EPI_ISL_1009940, EPI_ISL_1009941, EPI_ISL_1009942, EPI_ISL_1009943, EPI_ISL_1009944, EPI_ISL_1009945, EPI_ISL_1009946, EPI_ISL_1009947, EPI_ISL_1009948, EPI_ISL_1009949, EPI_ISL_1009950, EPI_ISL_1009951, EPI_ISL_1009952, EPI_ISL_1009953, EPI_ISL_1009954, EPI_ISL_1009955, EPI_ISL_1009956, EPI_ISL_1009957, EPI_ISL_1009958, EPI_ISL_1009959, EPI_ISL_1009960, EPI_ISL_1009961, EPI_ISL_1009962, EPI_ISL_1009963, EPI_ISL_1009964, EPI_ISL_1009965, EPI_ISL_1009966, EPI_ISL_1009967, EPI_ISL_1009968, EPI_ISL_1009969, EPI_ISL_1009970, EPI_ISL_1009971, EPI_ISL_1009972, EPI_ISL_1009973, EPI_ISL_1009974, EPI_ISL_1009975, EPI_ISL_1009976, EPI_ISL_1009977, EPI_ISL_1009978, EPI_ISL_1009979, EPI_ISL_1009980, EPI_ISL_1009981, EPI_ISL_1009982, EPI_ISL_1009983, EPI_ISL_1009984, EPI_ISL_1009985, EPI_ISL_1009986, EPI_ISL_1009987, EPI_ISL_1009988, EPI_ISL_1009989, EPI_ISL_1009990, EPI_ISL_1009991, EPI_ISL_1009992, EPI_ISL_1009993, EPI_ISL_1009994, EPI_ISL_1009995, EPI_ISL_1009996, EPI_ISL_1009997, EPI_ISL_1009998, EPI_ISL_1009999, EPI_ISL_1010000, EPI_ISL_1010001, EPI_ISL_1010002, EPI_ISL_1010003, EPI_ISL_1010004, EPI_ISL_1010005, EPI_ISL_1010006, EPI_ISL_1010007, EPI_ISL_1010008, EPI_ISL_1010009, EPI_ISL_1010010, EPI_ISL_1010011, EPI_ISL_1010012, EPI_ISL_1010013, EPI_ISL_1010014, EPI_ISL_1010015, EPI_ISL_1010016, EPI_ISL_1010017, EPI_ISL_1010018, EPI_ISL_1010019, EPI_ISL_1010020, EPI_ISL_1010021, EPI_ISL_1010022, EPI_ISL_1010023, EPI_ISL_1010024, EPI_ISL_1010025, EPI_ISL_1010026, EPI_ISL_1010027, EPI_ISL_1010028, EPI_ISL_1010029, EPI_ISL_1010030, EPI_ISL_1010031, EPI_ISL_1010032, EPI_ISL_1010033, EPI_ISL_1010034, EPI_ISL_1010035, EPI_ISL_1010036, EPI_ISL_1010037, EPI_ISL_1010038, EPI_ISL_1010039, EPI_ISL_1010040, EPI_ISL_1010041, EPI_ISL_1010042, EPI_ISL_1010043, EPI_ISL_1010044, EPI_ISL_1010045, EPI_ISL_1010046, EPI_ISL_1010047, EPI_ISL_1010048, EPI_ISL_1010049, EPI_ISL_1010050, EPI_ISL_1010051, EPI_ISL_1010052, EPI_ISL_1010053, EPI_ISL_1010054, EPI_ISL_1010055, EPI_ISL_1010056, EPI_ISL_1010057, EPI_ISL_1010058, EPI_ISL_1010059, EPI_ISL_1010060, EPI_ISL_1010061, EPI_ISL_1010062, EPI_ISL_1010063, EPI_ISL_1010064, EPI_ISL_1010065, EPI_ISL_1010066, EPI_ISL_1010067, EPI_ISL_1010068, EPI_ISL_1010069, EPI_ISL_1010070, EPI_ISL_1010071, EPI_ISL_1010072, EPI_ISL_1010073, EPI_ISL_1010074, EPI_ISL_1010075, EPI_ISL_1010076, EPI_ISL_1010077, EPI_ISL_1010078, EPI_ISL_1010079, EPI_ISL_1010080, EPI_ISL_1010081, EPI_ISL_1010082, EPI_ISL_1010083, EPI_ISL_1010084, EPI_ISL_1010085, EPI_ISL_1010086, EPI_ISL_1010087, EPI_ISL_1010088, EPI_ISL_1010089, EPI_ISL_1010090, EPI_ISL_1010091, EPI_ISL_1010092, EPI_ISL_1010093, EPI_ISL_1010094, EPI_ISL_1010095, EPI_ISL_1010096, EPI_ISL_1010097, EPI_ISL_1010098, EPI_ISL_1010099, EPI_ISL_1010100, EPI_ISL_1010101, EPI_ISL_1010102, EPI_ISL_1010103, EPI_ISL_1010104, EPI_ISL_1010105, EPI_ISL_1010106, EPI_ISL_1010107, EPI_ISL_1010108, EPI_ISL_1010109, EPI_ISL_1010110, EPI_ISL_1010111, EPI_ISL_1010112, EPI_ISL_1010113, EPI_ISL_1010114, EPI_ISL_1010115, EPI_ISL_1010116, EPI_ISL_1010117, EPI_ISL_1010118, EPI_ISL_1010119, EPI_ISL_1010120, EPI_ISL_1010121, EPI_ISL_1010122, EPI_ISL_1010123, EPI_ISL_1010124, EPI_ISL_1010125, EPI_ISL_1010126, EPI_ISL_1010127, EPI_ISL_1010128, EPI_ISL_1010129, EPI_ISL_1010130, EPI_ISL_1010131, EPI_ISL_1010132, EPI_ISL_1010133, EPI_ISL_1010134, EPI_ISL_1010135, EPI_ISL_1010136, EPI_ISL_1010137, EPI_ISL_1010138, EPI_ISL_1010139, EPI_ISL_1010140, EPI_ISL_1010141, EPI_ISL_1010142, EPI_ISL_1010143, EPI_ISL_1010144, EPI_ISL_1010145, EPI_ISL_1010146, EPI_ISL_1010147, EPI_ISL_1010148, EPI_ISL_1010149, EPI_ISL_1010150, EPI_ISL_1010151, EPI_ISL_1010152, EPI_ISL_1010153, EPI_ISL_1010154, EPI_ISL_1010155, EPI_ISL_1010156, EPI_ISL_1010157, EPI_ISL_1010158, EPI_ISL_1010159, EPI_ISL_1010160, EPI_ISL_1010161, EPI_ISL_1010162, EPI_ISL_1010163, EPI_ISL_1010164, EPI_ISL_1010165, EPI_ISL_1010166, EPI_ISL_1010167, EPI_ISL_1010168, EPI_ISL_1010169, EPI_ISL_1010170, EPI_ISL_1010171, EPI_ISL_1010172, EPI_ISL_1010173, EPI_ISL_1010174, EPI_ISL_1010175, EPI_ISL_1010176, EPI_ISL_1010177, EPI_ISL_1010178, EPI_ISL_1010179, EPI_ISL_1010180, EPI_ISL_1010181, EPI_ISL_1010182, EPI_ISL_1010183, EPI_ISL_1010184, EPI_ISL_1010185, EPI_ISL_1010186, EPI_ISL_1010187, EPI_ISL_1010188, EPI_ISL_1010189, EPI_ISL_1010190, EPI_ISL_1010191, EPI_ISL_1010192, EPI_ISL_1010193, EPI_ISL_1010194, EPI_ISL_1010195, EPI_ISL_1010196, EPI_ISL_1010197, EPI_ISL_1010198, EPI_ISL_1010199, EPI_ISL_1010200, EPI_ISL_1010201, EPI_ISL_1010202, EPI_ISL_1010203, EPI_ISL_1010204, EPI_ISL_1010205, EPI_ISL_1010206, EPI_ISL_1010207, EPI_ISL_1010208, EPI_ISL_1010209, EPI_ISL_1010210, EPI_ISL_1010211, EPI_ISL_1010212, EPI_ISL_1010213, EPI_ISL_1010214, EPI_ISL_1010215, EPI_ISL_1010216, EPI_ISL_1010217, EPI_ISL_1010218, EPI_ISL_1010219, EPI_ISL_1010220, EPI_ISL_1010221, EPI_ISL_1010222, EPI_ISL_1010223, EPI_ISL_1010224, EPI_ISL_1010225, EPI_ISL_1010226, EPI_ISL_1010227, EPI_ISL_1010228, EPI_ISL_1010229, EPI_ISL_1010230, EPI_ISL_1010231, EPI_ISL_1010232, EPI_ISL_1010233, EPI_ISL_1010234, EPI_ISL_1010235, EPI_ISL_1010236, EPI_ISL_1010237, EPI_ISL_1010238, EPI_ISL_1010239, EPI_ISL_1010240, EPI_ISL_1010241, EPI_ISL_1010242, EPI_ISL_1010243, EPI_ISL_1010244, EPI_ISL_1010245, EPI_ISL_1010246, EPI_ISL_1010247, EPI_ISL_1010248, EPI_ISL_1010249, EPI_ISL_1010250, EPI_ISL_1010251, EPI_ISL_1010252, EPI_ISL_1010253, EPI_ISL_1010254, EPI_ISL_1010255, EPI_ISL_1010256, EPI_ISL_1010257, EPI_ISL_1010258, EPI_ISL_1010259, EPI_ISL_1010260, EPI_ISL_1010261, EPI_ISL_1010262, EPI_ISL_1010263, EPI_ISL_1010264, EPI_ISL_1010265, EPI_ISL_1010266, EPI_ISL_1010267, EPI_ISL_1010268, EPI_ISL_1010269, EPI_ISL_1010270, EPI_ISL_1010271, EPI_ISL_1010272, EPI_ISL_1010273, EPI_ISL_1010274, EPI_ISL_1010275, EPI_ISL_1010276, EPI_ISL_1010277, EPI_ISL_1010278, EPI_ISL_1010279, EPI_ISL_1010280, EPI_ISL_1010281, EPI_ISL_1010282, EPI_ISL_1010283, EPI_ISL_1010284, EPI_ISL_1010285, EPI_ISL_1010286, EPI_ISL_1010287, EPI_ISL_1010288, EPI_ISL_1010289, EPI_ISL_1010290, EPI_ISL_1010291, EPI_ISL_1010292, EPI_ISL_1010293, EPI_ISL_1010294, EPI_ISL_1010295, EPI_ISL_1010296, EPI_ISL_1010297, EPI_ISL_1010298, EPI_ISL_1010299, EPI_ISL_1010300, EPI_ISL_1010301, EPI_ISL_1010302, EPI_ISL_1010303, EPI_ISL_1010305, EPI_ISL_1010306, EPI_ISL_1010307, EPI_ISL_1010309, EPI_ISL_1010310, EPI_ISL_1010311, EPI_ISL_1010312, EPI_ISL_1010313, EPI_ISL_1010314, EPI_ISL_1010315, EPI_ISL_1010316, EPI_ISL_1010317, EPI_ISL_1010318, EPI_ISL_1010319, EPI_ISL_1010320, EPI_ISL_1010321, EPI_ISL_1010322, EPI_ISL_1010323, EPI_ISL_1010324, EPI_ISL_1010325, EPI_ISL_1010326, EPI_ISL_1010327, EPI_ISL_1010328, EPI_ISL_1010329, EPI_ISL_1010330, EPI_ISL_1010331, EPI_ISL_1010332, EPI_ISL_1010333, EPI_ISL_1010334, EPI_ISL_1010335, EPI_ISL_1010336, EPI_ISL_1010337, EPI_ISL_1010338, EPI_ISL_1010339, EPI_ISL_1010340, EPI_ISL_1010341, EPI_ISL_1010342, EPI_ISL_1010343, EPI_ISL_1010344, EPI_ISL_1010345, EPI_ISL_1010346, EPI_ISL_1010347, EPI_ISL_1010348, EPI_ISL_1010349, EPI_ISL_1010350, EPI_ISL_1010351, EPI_ISL_1010352, EPI_ISL_1010353, EPI_ISL_1010354, EPI_ISL_1010355, EPI_ISL_1010356, EPI_ISL_1010357, EPI_ISL_1010358, EPI_ISL_1010359, EPI_ISL_1010360, EPI_ISL_1010361, EPI_ISL_1010362, EPI_ISL_1010363, EPI_ISL_1010364, EPI_ISL_1010365, EPI_ISL_1010366, EPI_ISL_1010367, EPI_ISL_1010368, EPI_ISL_1010369, EPI_ISL_1010370, EPI_ISL_1010371, EPI_ISL_1010372, EPI_ISL_1010373, EPI_ISL_1010374, EPI_ISL_1010375, EPI_ISL_1010376, EPI_ISL_1010377, EPI_ISL_1010378, EPI_ISL_1010379, EPI_ISL_1010380, EPI_ISL_1010381, EPI_ISL_1010382, EPI_ISL_1010383, EPI_ISL_1010384, EPI_ISL_1010385, EPI_ISL_1010386, EPI_ISL_1010387, EPI_ISL_1010388, EPI_ISL_1010389, EPI_ISL_1010390, EPI_ISL_1010391, EPI_ISL_1010392, EPI_ISL_1010393, EPI_ISL_1010394, EPI_ISL_1010395, EPI_ISL_1010396, EPI_ISL_1010397, EPI_ISL_1010398, EPI_ISL_1010399, EPI_ISL_1010400, EPI_ISL_1010401, EPI_ISL_1010402, EPI_ISL_1010403, EPI_ISL_1010404, EPI_ISL_1010405, EPI_ISL_1010406, EPI_ISL_1010407, EPI_ISL_1010553, EPI_ISL_1010554, EPI_ISL_1010555, EPI_ISL_1010556, EPI_ISL_1010557, EPI_ISL_1010558, EPI_ISL_1010559, EPI_ISL_1010560, EPI_ISL_1010561, EPI_ISL_1010562, EPI_ISL_1010563, EPI_ISL_1010564, EPI_ISL_1010565, EPI_ISL_1010566, EPI_ISL_1010567, EPI_ISL_1010568, EPI_ISL_1010569, EPI_ISL_1010570, EPI_ISL_1010571, EPI_ISL_1010572, EPI_ISL_1010573, EPI_ISL_1010574, EPI_ISL_1010575, EPI_ISL_1010576, EPI_ISL_1010577, EPI_ISL_1010578, EPI_ISL_1010579, EPI_ISL_1010580, EPI_ISL_1010581, EPI_ISL_1010583, EPI_ISL_1010584, EPI_ISL_1010585, EPI_ISL_1010586, EPI_ISL_1010587, EPI_ISL_1010588, EPI_ISL_1010589, EPI_ISL_1010590, EPI_ISL_1010591, EPI_ISL_1010592, EPI_ISL_1010593, EPI_ISL_1010594, EPI_ISL_1010595, EPI_ISL_1010596, EPI_ISL_1010597, EPI_ISL_1010598, EPI_ISL_1010599, EPI_ISL_1010600, EPI_ISL_1010601, EPI_ISL_1010602, EPI_ISL_1010603, EPI_ISL_1010604, EPI_ISL_1010605, EPI_ISL_1010606, EPI_ISL_1010607, EPI_ISL_1010608, EPI_ISL_1010609, EPI_ISL_1010610, EPI_ISL_1010611, EPI_ISL_1010612, EPI_ISL_1010613, EPI_ISL_1010614, EPI_ISL_1010615, EPI_ISL_1010616, EPI_ISL_1010617, EPI_ISL_1010618, EPI_ISL_1010619, EPI_ISL_1010620,

| | | | |
|---|---|---|---|
| EPI_ISL_1010621, EPI_ISL_1010622, EPI_ISL_1010623, EPI_ISL_1010624, EPI_ISL_1010625, EPI_ISL_1010626, EPI_ISL_1010627, EPI_ISL_1010628, EPI_ISL_1010629, EPI_ISL_1010630, EPI_ISL_1010631, EPI_ISL_1010632, EPI_ISL_1010633, EPI_ISL_1010634, EPI_ISL_1010635, EPI_ISL_1010636, EPI_ISL_1010637, EPI_ISL_1010638, EPI_ISL_1010639, EPI_ISL_1010640, EPI_ISL_1010641, EPI_ISL_1010642, EPI_ISL_1010643, EPI_ISL_1010644, EPI_ISL_1010645, EPI_ISL_1010646, EPI_ISL_1010647, EPI_ISL_1010648, EPI_ISL_1010649, EPI_ISL_1010650, EPI_ISL_1010651, EPI_ISL_1010652, EPI_ISL_1010653, EPI_ISL_1010654, EPI_ISL_1010655, EPI_ISL_1010656, EPI_ISL_1010657, EPI_ISL_1010658, EPI_ISL_1010659, EPI_ISL_1010660, EPI_ISL_1010661, EPI_ISL_1010662, EPI_ISL_1010663, EPI_ISL_1010664, EPI_ISL_1010665, EPI_ISL_1010666, EPI_ISL_1010667, EPI_ISL_1010668, EPI_ISL_1010669, EPI_ISL_1010670, EPI_ISL_1010671, EPI_ISL_1010672, EPI_ISL_1010673, EPI_ISL_1010674, EPI_ISL_1010675, EPI_ISL_1010676, EPI_ISL_1010677, EPI_ISL_1010678, EPI_ISL_1010679, EPI_ISL_1010680, EPI_ISL_1010681, EPI_ISL_1010682, EPI_ISL_1010683, EPI_ISL_1010684, EPI_ISL_1010685, EPI_ISL_1010686, EPI_ISL_1010687, EPI_ISL_1010688, EPI_ISL_1010689, EPI_ISL_1010690, EPI_ISL_1010691, EPI_ISL_1010692, EPI_ISL_1010693, EPI_ISL_1010694, EPI_ISL_1010695, EPI_ISL_1010696, EPI_ISL_1010697, EPI_ISL_1010698, EPI_ISL_1027178, EPI_ISL_1027179, EPI_ISL_1027180, EPI_ISL_1027181, EPI_ISL_1027182, EPI_ISL_1027183, EPI_ISL_1027184, EPI_ISL_1027185, EPI_ISL_1027186, EPI_ISL_1027187, EPI_ISL_1027188, EPI_ISL_1027189, EPI_ISL_1027190, EPI_ISL_1027191, EPI_ISL_1027192, EPI_ISL_1027193, EPI_ISL_1027194, EPI_ISL_1027195, EPI_ISL_1027196, EPI_ISL_1027197, EPI_ISL_1027198, EPI_ISL_1027199, EPI_ISL_1027200, EPI_ISL_1027201, EPI_ISL_1027202, EPI_ISL_1027203, EPI_ISL_1027204, EPI_ISL_1027205, EPI_ISL_1027206, EPI_ISL_1027207, EPI_ISL_1027208, EPI_ISL_1027209, EPI_ISL_1027210, EPI_ISL_1027211, EPI_ISL_1027212, EPI_ISL_1027213, EPI_ISL_1027214, EPI_ISL_1027215, EPI_ISL_1027216, EPI_ISL_1027217, EPI_ISL_1027218, EPI_ISL_1027219, EPI_ISL_1027220, EPI_ISL_1027221, EPI_ISL_1027222, EPI_ISL_1027223, EPI_ISL_1027225, EPI_ISL_1027226, EPI_ISL_1027227, EPI_ISL_1027228, EPI_ISL_1027230, EPI_ISL_1027231, EPI_ISL_1027232, EPI_ISL_1027233, EPI_ISL_1027234, EPI_ISL_1027235, EPI_ISL_1027236, EPI_ISL_1027237, EPI_ISL_1027238, EPI_ISL_1027239, EPI_ISL_1027240, EPI_ISL_1027241, EPI_ISL_1027242, EPI_ISL_1027243, EPI_ISL_1027244, EPI_ISL_1027245, EPI_ISL_1027246, EPI_ISL_1027247, EPI_ISL_1032946, EPI_ISL_1032948, EPI_ISL_1032949 | | | |
| see above | Fulgent Genetics | Fulgent Genetics | Becky Tsai; Benafsh Sapra; Doreen Ng; Harry Gao; James Xie; John Gao; Joseph Fierro; Mickey Li; Yan Meng |
| EPI_ISL_1366326, EPI_ISL_1366327, EPI_ISL_1366328, EPI_ISL_1366329, EPI_ISL_1366330, EPI_ISL_1366331, EPI_ISL_1366332 | | | |
| see above | Genomica Lab Molecular, Mexico | Andersen lab at Scripps Research | Jose Horacio Reyna Verdugo; Jose Roman Chavez Mendez; Luis Alberto Rangel Gonzalez; Martin Gonzalez Ibarra; SEARCH Alliance San Diego with Jonathan Gonzalez Garcia |
| EPI_ISL_1081450, EPI_ISL_1081452, EPI_ISL_1081453, EPI_ISL_1081454, EPI_ISL_1081455, EPI_ISL_1081456 | Genomica Lab Molecular, M√©xico | Andersen lab at Scripps Research | Jose Horacio Reyna Verdugo; Jose Roman Chavez Mendez; Luis Alberto Rangel Gonzalez; Martin Gonzalez Ibarra; SEARCH Alliance San Diego with Jonathan Gonzalez Garcia |
| EPI_ISL_850690, EPI_ISL_850691, EPI_ISL_850693, EPI_ISL_850695, EPI_ISL_850696, EPI_ISL_850697, EPI_ISL_850698, EPI_ISL_850699, EPI_ISL_850700, EPI_ISL_850701, EPI_ISL_850702, EPI_ISL_850703, EPI_ISL_850704, EPI_ISL_850710, EPI_ISL_850735, EPI_ISL_850765, EPI_ISL_850781, EPI_ISL_850784, EPI_ISL_850788, EPI_ISL_850790, EPI_ISL_850791, EPI_ISL_850792, EPI_ISL_850793, EPI_ISL_850794, EPI_ISL_850795, EPI_ISL_850796, EPI_ISL_850797, EPI_ISL_850798, EPI_ISL_850799, EPI_ISL_850800, EPI_ISL_850801, EPI_ISL_850802, EPI_ISL_850803, EPI_ISL_850804, EPI_ISL_850805, EPI_ISL_850806, EPI_ISL_850807, EPI_ISL_850808, EPI_ISL_850809, EPI_ISL_850811, EPI_ISL_850812, EPI_ISL_850813, EPI_ISL_850814, EPI_ISL_850815, EPI_ISL_850816 | | | |
| see above | Helix / Illumina | Genomics and Discovery, Respiratory Viruses Branch, Division of Viral Diseases, Centers for Disease Control and Prevention | Alexandre Bolze; Ary Ascencio; Ben L. Rambo-Martin Eileen de Feo; Brad Sickler; Charlotte Rivera-Garcia; Christine Tran; Clinton R. Paden; David Becker; Dhwani Batra; Duncan MacCannell; Efren Sandoval; Elizabeth Cirulli; Eric Allen; Geraint Levan; James Lu; Jan Antico; Jason Nguyen; Jimmy Ramirez; Jingtao Liu; Kelly Schiabor Barrett; Kim Gietzen; Magnus Isaksson; Marc Laurent; Matthew Tolentino; Nicole L. Washington; Peter W. Cook; Phil Febbo; Ryan Cho; Shannon Wickline; Sherry Wang; Simon White; Summer Galloway; Suxiang Tong; Tyler Cassens; William Lee |
| EPI_ISL_802601, EPI_ISL_802604, EPI_ISL_802605, EPI_ISL_802607, EPI_ISL_802608, EPI_ISL_802609, EPI_ISL_802618, EPI_ISL_802619, EPI_ISL_802620, EPI_ISL_802621, EPI_ISL_802622, EPI_ISL_802623, EPI_ISL_802624, EPI_ISL_802625, EPI_ISL_802626, EPI_ISL_802627, EPI_ISL_802628, EPI_ISL_802652, EPI_ISL_802657, EPI_ISL_802658, EPI_ISL_802662, EPI_ISL_802663, EPI_ISL_802664, EPI_ISL_802665, EPI_ISL_802666, EPI_ISL_802667, EPI_ISL_802668, EPI_ISL_850517, EPI_ISL_850518, EPI_ISL_850519, EPI_ISL_850520, EPI_ISL_850521, EPI_ISL_850522, EPI_ISL_850523, EPI_ISL_850524, EPI_ISL_850525, EPI_ISL_850527, EPI_ISL_850528, EPI_ISL_850588, EPI_ISL_850631, EPI_ISL_850632, EPI_ISL_850633, EPI_ISL_850634, EPI_ISL_850635, EPI_ISL_850637, EPI_ISL_850638, EPI_ISL_850953, EPI_ISL_850954, EPI_ISL_850956, EPI_ISL_850957, EPI_ISL_850958, EPI_ISL_850959, EPI_ISL_851000, EPI_ISL_851001, EPI_ISL_851002, EPI_ISL_851003, EPI_ISL_851004, EPI_ISL_851005, EPI_ISL_851006, EPI_ISL_851011, EPI_ISL_851013, EPI_ISL_851021, EPI_ISL_851022, EPI_ISL_851023, EPI_ISL_851025, EPI_ISL_851032, EPI_ISL_876634, EPI_ISL_876635, EPI_ISL_876641, EPI_ISL_876644, EPI_ISL_876645, EPI_ISL_876658, EPI_ISL_876661, EPI_ISL_876670, EPI_ISL_876671, EPI_ISL_876672, EPI_ISL_876673, EPI_ISL_876674, EPI_ISL_876675, EPI_ISL_876676 | | | |
| see above | Helix/Illumina | Genomics and Discovery, Respiratory Viruses Branch, Division of Viral Diseases, Centers for Disease Control and Prevention | ; Alexandre Bolze; Ary Ascencio; Ben L. Rambo-Martin; Ben L. Rambo-Martin Eileen de Feo; Brad Sickler; Charlotte Rivera-Garcia; Christine Tran; Clinton R. Paden; David Becker; Dhwani Batra; Duncan MacCannell; Efren Sandoval; Eileen de Feo; Elizabeth Cirulli; Eric Allen; Geraint Levan; James Lu; Jan Antico; Jason Nguyen; Jimmy Ramirez; Jingtao Liu; Kelly Schiabor Barrett; Kim Gietzen; Magnus Isaksson; Marc Laurent; Matthew Tolentino; Nicole L. Washington; Peter W. Cook; Phil Febbo; Ryan Cho; Shannon Wickline; Sherry Wang; Simon White; Summer Galloway; Suxiang Tong; Tyler Cassens; William Lee |
| EPI_ISL_966497, EPI_ISL_966498, EPI_ISL_966499, EPI_ISL_966500, EPI_ISL_966501, EPI_ISL_966505, EPI_ISL_966532, EPI_ISL_966534, EPI_ISL_966535, EPI_ISL_966536, EPI_ISL_966537, EPI_ISL_966538, EPI_ISL_966545, EPI_ISL_966556, EPI_ISL_966561, EPI_ISL_966562, EPI_ISL_966563, EPI_ISL_966567, EPI_ISL_966568, EPI_ISL_966570, EPI_ISL_966571, EPI_ISL_966583, EPI_ISL_966584, EPI_ISL_966585, EPI_ISL_966586, EPI_ISL_966587, EPI_ISL_966588, EPI_ISL_966590, EPI_ISL_966603, EPI_ISL_966608, EPI_ISL_966615, EPI_ISL_966622, EPI_ISL_966624, EPI_ISL_966625, EPI_ISL_966626, EPI_ISL_966627, EPI_ISL_966628, EPI_ISL_966630, EPI_ISL_966632, EPI_ISL_966633, EPI_ISL_966636, EPI_ISL_966637, EPI_ISL_966638, EPI_ISL_966640, EPI_ISL_966657, EPI_ISL_966665, EPI_ISL_966670, EPI_ISL_966672, EPI_ISL_966680, EPI_ISL_966683, EPI_ISL_966686, EPI_ISL_966694, EPI_ISL_966702, EPI_ISL_966720, EPI_ISL_966725, EPI_ISL_966729, EPI_ISL_966910, EPI_ISL_966912, EPI_ISL_966921, EPI_ISL_966922, EPI_ISL_966923, EPI_ISL_966924, EPI_ISL_966925, EPI_ISL_966926, EPI_ISL_966927, EPI_ISL_966928, EPI_ISL_966942, EPI_ISL_966974, EPI_ISL_966984, EPI_ISL_966985, EPI_ISL_966986, EPI_ISL_966991, EPI_ISL_966992, EPI_ISL_966993, EPI_ISL_966995, EPI_ISL_967000, EPI_ISL_967046, EPI_ISL_967047, EPI_ISL_967048, EPI_ISL_967057, EPI_ISL_967058, EPI_ISL_967060, EPI_ISL_967061, EPI_ISL_967062, EPI_ISL_967065, EPI_ISL_967072, EPI_ISL_967073, EPI_ISL_967081, EPI_ISL_967099, EPI_ISL_967104, EPI_ISL_967107, EPI_ISL_967113, EPI_ISL_967116, EPI_ISL_967117, EPI_ISL_967175, EPI_ISL_967193, EPI_ISL_967199, EPI_ISL_967211, EPI_ISL_967214, EPI_ISL_967218, EPI_ISL_967243, EPI_ISL_967257, EPI_ISL_967259, EPI_ISL_967305, EPI_ISL_967318, EPI_ISL_967324, EPI_ISL_967346, EPI_ISL_967347, EPI_ISL_967354, EPI_ISL_967355, EPI_ISL_967361, EPI_ISL_967376, EPI_ISL_967390, EPI_ISL_967392, EPI_ISL_967403, EPI_ISL_967407, EPI_ISL_967413, EPI_ISL_967418, EPI_ISL_967425, EPI_ISL_967426, EPI_ISL_967427, EPI_ISL_967428, EPI_ISL_967429, EPI_ISL_967430, EPI_ISL_967431, EPI_ISL_967432, EPI_ISL_967433, EPI_ISL_967437, EPI_ISL_967439, EPI_ISL_967451, EPI_ISL_967454, EPI_ISL_967456, EPI_ISL_967460, EPI_ISL_967464, EPI_ISL_967465, EPI_ISL_967487, EPI_ISL_967488, EPI_ISL_967489, EPI_ISL_967490, EPI_ISL_967492, EPI_ISL_967493, EPI_ISL_967494, EPI_ISL_967495, EPI_ISL_967505, EPI_ISL_967506, EPI_ISL_967514, EPI_ISL_967517, EPI_ISL_967700, EPI_ISL_967710, EPI_ISL_967719, EPI_ISL_967726, EPI_ISL_967736, EPI_ISL_967748, EPI_ISL_967775, EPI_ISL_967777, EPI_ISL_967779, EPI_ISL_967780, EPI_ISL_967813, EPI_ISL_967817, EPI_ISL_967818, EPI_ISL_967819, EPI_ISL_967820, EPI_ISL_967849, EPI_ISL_978618, EPI_ISL_978622, EPI_ISL_978623, EPI_ISL_978624, EPI_ISL_978630, EPI_ISL_978640, EPI_ISL_978642, EPI_ISL_978643, EPI_ISL_978644, EPI_ISL_978646, EPI_ISL_978648, EPI_ISL_978718, EPI_ISL_978727, EPI_ISL_978730, EPI_ISL_978733, EPI_ISL_978734, EPI_ISL_1016746, EPI_ISL_1016804, EPI_ISL_1016833, EPI_ISL_1016834, EPI_ISL_1036687, EPI_ISL_1036698, EPI_ISL_1036699, EPI_ISL_1036700, EPI_ISL_1036707, EPI_ISL_1036709, EPI_ISL_1036712, EPI_ISL_1036714, EPI_ISL_1036716, EPI_ISL_1036719 | | | |
| see above | Helix/Illumina | Respiratory Viruses Branch, Division of Viral Diseases, Centers for Disease Control and Prevention | ; Alexandre Bolze; Ary Ascencio; Ben L. Rambo-Martin; Brad Sickler; Charlotte Rivera-Garcia; Christine Tran; Clinton R. Paden; Dakota Howard; David Becker; Dhwani Batra; Duncan MacCannell; Efren Sandoval; Eileen de Feo; Elizabeth Cirulli; Eric Allen; Geraint Levan; James Lu; Jan Antico; Jason Nguyen; Jimmy Ramirez; Jingtao Liu; Kelly Schiabor Barrett; Kim Gietzen; Magnus Isaksson; Marc Laurent; Matthew Tolentino; Nicole L. Washington; Peter W. Cook; Phil Febbo; Ryan Cho; Shannon Wickline; Sherry Wang; Simon White; Summer Galloway; Suxiang Tong; Tyler Cassens; William Lee |
| EPI_ISL_955700, EPI_ISL_955704, EPI_ISL_955706, EPI_ISL_955708, EPI_ISL_955709, EPI_ISL_955710, EPI_ISL_955711, EPI_ISL_955712, EPI_ISL_955713, EPI_ISL_955714, EPI_ISL_955715, EPI_ISL_955716, EPI_ISL_955717, EPI_ISL_955718, EPI_ISL_955719, EPI_ISL_955720, EPI_ISL_955721, EPI_ISL_955722, EPI_ISL_955723, EPI_ISL_955724, EPI_ISL_955725, EPI_ISL_955726, EPI_ISL_955727, EPI_ISL_955728, EPI_ISL_955729, EPI_ISL_955730, EPI_ISL_955731, EPI_ISL_955732, EPI_ISL_955733, EPI_ISL_955734, EPI_ISL_955735, EPI_ISL_955736, EPI_ISL_955737, EPI_ISL_955738, EPI_ISL_955739, EPI_ISL_955740, EPI_ISL_955741, EPI_ISL_955742, EPI_ISL_955743, EPI_ISL_955744, EPI_ISL_955745, EPI_ISL_955746, EPI_ISL_955747, EPI_ISL_955748, EPI_ISL_955749, EPI_ISL_955750, EPI_ISL_955751, EPI_ISL_955752, EPI_ISL_955753, EPI_ISL_955754, EPI_ISL_955755, EPI_ISL_955756, EPI_ISL_955757, EPI_ISL_955758, EPI_ISL_955759, EPI_ISL_955760, EPI_ISL_955761, EPI_ISL_955762, EPI_ISL_955763, EPI_ISL_955764, EPI_ISL_955765, EPI_ISL_955766, EPI_ISL_955767, EPI_ISL_955768, EPI_ISL_955769, EPI_ISL_955770, EPI_ISL_955771, EPI_ISL_955772, EPI_ISL_955773, EPI_ISL_955774, EPI_ISL_955775, EPI_ISL_955776, EPI_ISL_955777, EPI_ISL_1027476, EPI_ISL_1027517, EPI_ISL_1027561, EPI_ISL_1185287, EPI_ISL_1185291, EPI_ISL_1185292, EPI_ISL_1185294, EPI_ISL_1185298, EPI_ISL_1185300, EPI_ISL_1185301, EPI_ISL_1185302, EPI_ISL_1185304, EPI_ISL_1185306, EPI_ISL_1185308, EPI_ISL_1185309, EPI_ISL_1185312, EPI_ISL_1185318, EPI_ISL_1185319, EPI_ISL_1185323, EPI_ISL_1185324, EPI_ISL_1185325, EPI_ISL_1185326, EPI_ISL_1185328, EPI_ISL_1185329, EPI_ISL_1185340, EPI_ISL_1185345, EPI_ISL_1185346, EPI_ISL_1185352, EPI_ISL_1185353, EPI_ISL_1185355, EPI_ISL_1185359, EPI_ISL_1185360, EPI_ISL_1185361, EPI_ISL_1185365, EPI_ISL_1185369, EPI_ISL_1185382, EPI_ISL_1185385, EPI_ISL_1185394, EPI_ISL_1185395, EPI_ISL_1185399, EPI_ISL_1185407, EPI_ISL_1185408, EPI_ISL_1185409, EPI_ISL_1185410, EPI_ISL_1185412, EPI_ISL_1185413, EPI_ISL_1185417, EPI_ISL_1185424, EPI_ISL_1185425, EPI_ISL_1185428, EPI_ISL_1185432, EPI_ISL_1185436, EPI_ISL_1185438, EPI_ISL_1185439, EPI_ISL_1185440, EPI_ISL_1185448, EPI_ISL_1185454, EPI_ISL_1185455, EPI_ISL_1185460, EPI_ISL_1185463, EPI_ISL_1185468, EPI_ISL_1185469, EPI_ISL_1185471, EPI_ISL_1185474, EPI_ISL_1185476, EPI_ISL_1185479, EPI_ISL_1185481, EPI_ISL_1185486, EPI_ISL_1185487, EPI_ISL_1185488, EPI_ISL_1185489, EPI_ISL_1185497, EPI_ISL_1185498, EPI_ISL_1185504, EPI_ISL_1185505, EPI_ISL_1185507, EPI_ISL_1185509, EPI_ISL_1185516, EPI_ISL_1185518, EPI_ISL_1185519, EPI_ISL_1185521, EPI_ISL_1185526, EPI_ISL_1185528, EPI_ISL_1234786, EPI_ISL_1234788, EPI_ISL_1234789, EPI_ISL_1234792, EPI_ISL_1234793, EPI_ISL_1234794, EPI_ISL_1234799, EPI_ISL_1234801, EPI_ISL_1234803, EPI_ISL_1234805, EPI_ISL_1234807, EPI_ISL_1234811, EPI_ISL_1234814, EPI_ISL_1234816, EPI_ISL_1234817, EPI_ISL_1234821, EPI_ISL_1234822, EPI_ISL_1234823, EPI_ISL_1234824, EPI_ISL_1234826, EPI_ISL_1234831, EPI_ISL_1234833, EPI_ISL_1234836, EPI_ISL_1234852, EPI_ISL_1234853, EPI_ISL_1234856, EPI_ISL_1234859, EPI_ISL_1234861, EPI_ISL_1234863, EPI_ISL_1234868, EPI_ISL_1234871, EPI_ISL_1234874, EPI_ISL_1234882, EPI_ISL_1234888, EPI_ISL_1234889, EPI_ISL_1234890, EPI_ISL_1234893, EPI_ISL_1234896, EPI_ISL_1234898, EPI_ISL_1234903, EPI_ISL_1234905, EPI_ISL_1234906, EPI_ISL_1234908, EPI_ISL_1234910, EPI_ISL_1234911, EPI_ISL_1234912, EPI_ISL_1234914, EPI_ISL_1234916, EPI_ISL_1234922, EPI_ISL_1234927, EPI_ISL_1234928, EPI_ISL_1234930, EPI_ISL_1234931, EPI_ISL_1234934, EPI_ISL_1234936, EPI_ISL_1234948, EPI_ISL_1234949, EPI_ISL_1234955, EPI_ISL_1234960, EPI_ISL_1234963, EPI_ISL_1234966, EPI_ISL_1234971, EPI_ISL_1234974, EPI_ISL_1234980, EPI_ISL_1234981, EPI_ISL_1234984, EPI_ISL_1234989, EPI_ISL_1234998, EPI_ISL_1235001, EPI_ISL_1235002, EPI_ISL_1235003, EPI_ISL_1235004, EPI_ISL_1235005, EPI_ISL_1235011, EPI_ISL_1235012, EPI_ISL_1235023, EPI_ISL_1235030, EPI_ISL_1235033, EPI_ISL_1235037, EPI_ISL_1235039, EPI_ISL_1235040, EPI_ISL_1235045, EPI_ISL_1235046, EPI_ISL_1235053 | | | |
| see above | Humboldt County Public Health Laboratory | Chan-Zuckerberg Biohub | CZB Cliahub Consortium |
| EPI_ISL_1081337, EPI_ISL_1081339, EPI_ISL_1081341, EPI_ISL_1081346, EPI_ISL_1081347 | IEH Laboratories and Consulting Group | IEH and ID Genomics | Curtis Fowler; Evgeni Sokurenko; Hao Geng; Hemantha Kulakesara; Kai Deng; Kirthi Kutumbaka; Lydia Larson; Mansour Samadpour; Veronika Tchesnokova |
| EPI_ISL_2253171 | Infectious Diseases, Chan-Zuckerberg Biohub Covidtracker | Infectious Diseases, Chan-Zuckerberg Biohub Covidtracker | CZB Cliahub Consortium |
| EPI_ISL_906920, EPI_ISL_906921, EPI_ISL_906922, EPI_ISL_906923, EPI_ISL_906924, EPI_ISL_906925, EPI_ISL_906926, EPI_ISL_906927, EPI_ISL_906928, EPI_ISL_906929, EPI_ISL_906930, EPI_ISL_906931, EPI_ISL_906932, EPI_ISL_906933, EPI_ISL_906934, EPI_ISL_906935, EPI_ISL_906936, EPI_ISL_906937, EPI_ISL_906938, EPI_ISL_906939, EPI_ISL_906940, EPI_ISL_906941, EPI_ISL_906942, EPI_ISL_906943, EPI_ISL_906944, EPI_ISL_906945, EPI_ISL_906946, EPI_ISL_906947, EPI_ISL_906948, EPI_ISL_906949, EPI_ISL_906950, EPI_ISL_906951, EPI_ISL_906952, EPI_ISL_906953, EPI_ISL_906954, EPI_ISL_906955, EPI_ISL_906956, EPI_ISL_906957, EPI_ISL_906958, EPI_ISL_906959, EPI_ISL_906960, EPI_ISL_906961, EPI_ISL_906962, EPI_ISL_906963, EPI_ISL_906964, EPI_ISL_906965, EPI_ISL_906966, EPI_ISL_906967, EPI_ISL_906968, EPI_ISL_906969, EPI_ISL_906970, EPI_ISL_906971, EPI_ISL_906972, EPI_ISL_906973, EPI_ISL_906974, EPI_ISL_906975, EPI_ISL_906976, EPI_ISL_906977, EPI_ISL_906978, EPI_ISL_906979, EPI_ISL_906980, EPI_ISL_906981, EPI_ISL_906982, EPI_ISL_906983, EPI_ISL_906984, EPI_ISL_906985, EPI_ISL_906986, EPI_ISL_906987, EPI_ISL_906988, EPI_ISL_906989, EPI_ISL_906990, EPI_ISL_906991, EPI_ISL_906992, EPI_ISL_906993, EPI_ISL_906994, EPI_ISL_906995, EPI_ISL_906996, EPI_ISL_906997, EPI_ISL_906998, EPI_ISL_906999, EPI_ISL_907000, EPI_ISL_907001, EPI_ISL_907002, EPI_ISL_907003, EPI_ISL_907004, EPI_ISL_907005, EPI_ISL_907006, EPI_ISL_907007, EPI_ISL_907008, EPI_ISL_907009, EPI_ISL_907010, EPI_ISL_907011, EPI_ISL_907012, EPI_ISL_907013, EPI_ISL_907014, EPI_ISL_907015, EPI_ISL_907016, EPI_ISL_907017, EPI_ISL_907018, EPI_ISL_907019, EPI_ISL_907020, EPI_ISL_907021, EPI_ISL_907022, EPI_ISL_907023, EPI_ISL_907024, EPI_ISL_907025, EPI_ISL_907026, EPI_ISL_907027, EPI_ISL_907028, EPI_ISL_907029, EPI_ISL_907030, EPI_ISL_907031, EPI_ISL_907032, EPI_ISL_907033, EPI_ISL_907034, EPI_ISL_907035, EPI_ISL_907036, EPI_ISL_907037, EPI_ISL_907038, EPI_ISL_907039, EPI_ISL_907040, EPI_ISL_907041, EPI_ISL_907042, EPI_ISL_907043, EPI_ISL_907044, EPI_ISL_907045, EPI_ISL_907046, EPI_ISL_907047, EPI_ISL_907048, EPI_ISL_907049, EPI_ISL_907050, EPI_ISL_907051, EPI_ISL_907052, EPI_ISL_907053, EPI_ISL_907054, EPI_ISL_907055, EPI_ISL_907056, EPI_ISL_907057, EPI_ISL_907058, EPI_ISL_907059, EPI_ISL_907060, EPI_ISL_907061, EPI_ISL_907062, EPI_ISL_907063, EPI_ISL_1233901, EPI_ISL_1233902, EPI_ISL_1233903, EPI_ISL_1233919, EPI_ISL_1233920 | | | |
| see above | Infectious Diseases, Quest Diagnostics | Infectious Diseases, Quest Diagnostics | Anderson, B.; Bernstein; D.F.; Gerasimova, A.; Hua, M.; I.A.; K.E.; Kagan; L.E.; Lacbawan, F.; Liu, Y.; Livingston; Owen, R.; Perez, A.; R.M.; Rosenthal; S.H.; Shalhout; Shlyakhter; Tanpaiboon, P. |
| EPI_ISL_918177, EPI_ISL_918179, EPI_ISL_918180, EPI_ISL_918181, EPI_ISL_918182, EPI_ISL_918183, EPI_ISL_918185, EPI_ISL_918187, EPI_ISL_918188, EPI_ISL_918189, EPI_ISL_918190, EPI_ISL_918191, EPI_ISL_918192, EPI_ISL_918193, EPI_ISL_918194, EPI_ISL_918195, EPI_ISL_918196, EPI_ISL_918198, EPI_ISL_918199, EPI_ISL_918200, EPI_ISL_918201, EPI_ISL_918203, EPI_ISL_918204, EPI_ISL_918224, EPI_ISL_918225, EPI_ISL_918226, EPI_ISL_918227, EPI_ISL_918228, EPI_ISL_918229, EPI_ISL_918230, EPI_ISL_918231, EPI_ISL_918232, EPI_ISL_918233, EPI_ISL_918234, EPI_ISL_918235, EPI_ISL_918236, EPI_ISL_918237, EPI_ISL_918238, EPI_ISL_918239, EPI_ISL_918240, EPI_ISL_918241, EPI_ISL_918242, EPI_ISL_918243, EPI_ISL_918244, EPI_ISL_918245, EPI_ISL_918246, EPI_ISL_918247, EPI_ISL_918248, EPI_ISL_918249, EPI_ISL_918250, EPI_ISL_918251, EPI_ISL_918252, EPI_ISL_918253, EPI_ISL_918254, EPI_ISL_918255, EPI_ISL_918256, EPI_ISL_918257, EPI_ISL_918258, EPI_ISL_918259, EPI_ISL_918260, EPI_ISL_918261, EPI_ISL_918262, EPI_ISL_955126, EPI_ISL_955127, EPI_ISL_955128, EPI_ISL_955129, EPI_ISL_980840, EPI_ISL_980841, EPI_ISL_980842, EPI_ISL_980843, EPI_ISL_980844, EPI_ISL_980845, EPI_ISL_980846, EPI_ISL_980847, EPI_ISL_980848, EPI_ISL_980849, EPI_ISL_980850, EPI_ISL_980851, EPI_ISL_980852, EPI_ISL_980853, EPI_ISL_980854, EPI_ISL_980855, EPI_ISL_980856, EPI_ISL_980859, EPI_ISL_980860, EPI_ISL_980861, EPI_ISL_980862, EPI_ISL_980863, EPI_ISL_980864, EPI_ISL_980865, EPI_ISL_980866, EPI_ISL_980867, EPI_ISL_980868, EPI_ISL_980869, EPI_ISL_980870, EPI_ISL_980871, EPI_ISL_980872, EPI_ISL_980873, EPI_ISL_980874, EPI_ISL_980875, EPI_ISL_980876, EPI_ISL_980877, EPI_ISL_980878, EPI_ISL_980879, EPI_ISL_980880, EPI_ISL_980881, EPI_ISL_980882, EPI_ISL_980883, EPI_ISL_980884, EPI_ISL_980886, EPI_ISL_980887, EPI_ISL_980888, EPI_ISL_980889, EPI_ISL_980890, EPI_ISL_980891, EPI_ISL_980892, EPI_ISL_980893, EPI_ISL_980894, EPI_ISL_980895, EPI_ISL_980896, EPI_ISL_980897, EPI_ISL_980898, EPI_ISL_980899, EPI_ISL_980900, EPI_ISL_980901, EPI_ISL_980902, EPI_ISL_980903, EPI_ISL_980904, EPI_ISL_980905, EPI_ISL_980906, EPI_ISL_980907, EPI_ISL_980908, EPI_ISL_980909, EPI_ISL_980910, EPI_ISL_980911, EPI_ISL_980912, EPI_ISL_980913, EPI_ISL_980914, EPI_ISL_980915, EPI_ISL_980916, EPI_ISL_980917, EPI_ISL_980918, EPI_ISL_980919, EPI_ISL_980920, EPI_ISL_980921, EPI_ISL_980922, EPI_ISL_980923, EPI_ISL_980924, EPI_ISL_980925, EPI_ISL_980926, EPI_ISL_980927, EPI_ISL_980928, EPI_ISL_980929, EPI_ISL_980930, EPI_ISL_980931, EPI_ISL_980932, EPI_ISL_980933, EPI_ISL_980934, EPI_ISL_980935, EPI_ISL_980936, EPI_ISL_980937, EPI_ISL_980938, EPI_ISL_980939, EPI_ISL_980940, EPI_ISL_980941, EPI_ISL_980942, EPI_ISL_980943, EPI_ISL_980944, EPI_ISL_980945, EPI_ISL_980946, EPI_ISL_980947, EPI_ISL_980948, EPI_ISL_980949, EPI_ISL_980950, EPI_ISL_980951, EPI_ISL_980952, EPI_ISL_980953, EPI_ISL_980954, EPI_ISL_980955, EPI_ISL_980956, EPI_ISL_980957, EPI_ISL_980958, EPI_ISL_980959, EPI_ISL_980960, EPI_ISL_980961, EPI_ISL_980962, EPI_ISL_980963, EPI_ISL_980964, EPI_ISL_980965, EPI_ISL_980966, EPI_ISL_980967, EPI_ISL_980968, EPI_ISL_980969, EPI_ISL_980970, EPI_ISL_980971, EPI_ISL_980972, EPI_ISL_980973, EPI_ISL_980974, EPI_ISL_980975, EPI_ISL_980976, EPI_ISL_980977, EPI_ISL_980978, EPI_ISL_980979, EPI_ISL_980980, EPI_ISL_980981, EPI_ISL_980982, EPI_ISL_980983, EPI_ISL_980984, EPI_ISL_980985, EPI_ISL_980986, EPI_ISL_980987, EPI_ISL_980988, EPI_ISL_980989, EPI_ISL_980990, EPI_ISL_980991, EPI_ISL_980992, EPI_ISL_980993, EPI_ISL_980994, EPI_ISL_980995, EPI_ISL_980996, EPI_ISL_981003, EPI_ISL_984659, EPI_ISL_984660, EPI_ISL_984661, EPI_ISL_984662, EPI_ISL_1712442, EPI_ISL_1712443, EPI_ISL_1712444, EPI_ISL_1712445, EPI_ISL_1712446, EPI_ISL_1712447, EPI_ISL_1712448, EPI_ISL_1712449, EPI_ISL_1712450, EPI_ISL_1712451, EPI_ISL_1712452, EPI_ISL_1712453, EPI_ISL_1712454, EPI_ISL_1712455, EPI_ISL_1712456 | | | |
| see above | Innovative Genomics Institute, UC Berkeley | Innovative Genomics Institute, UC Berkeley | Haridha Shivram; Holly Gildea; John Boyle; Justin Choi; Liana Lareau; Phil Frankino; Shana McDevitt; Stacia Wyman |
| EPI_ISL_1168525, EPI_ISL_1168538, EPI_ISL_1168539, EPI_ISL_1168540, EPI_ISL_1168541, EPI_ISL_1168542 | Instituto de Diagnostico y Referencia Epidemiologicos INDRE_RNLSP | Instituto de Diagnostico y Referencia Epidemiologicos (INDRE) | Abril Rodriguez-Maldonado; Ariadna Medina-Benitez; Claudia Wong-Arambula; David Fragoso-Fonseca; Ernesto Ramirez-Gonzalez.; Gisela Barrera-Badillo; Irma Lopez-Martinez; Joaquin Quiroz-Mercado; Lucia Hernandez-Rivas; Natividad Cruz-Ortiz; Sergio Rangel-Guerrero; Tatiana Nunez-Garcia; Vanessa Rivero-Arredondo |
| EPI_ISL_1532141, EPI_ISL_1532142, EPI_ISL_1532143 | Kaiser San Jose | Santa Clara County Public Health Laboratory | Santa Clara County Public Health Department |
| EPI_ISL_1664228, EPI_ISL_1664229, EPI_ISL_1664232, EPI_ISL_1664250, EPI_ISL_1664251, EPI_ISL_1664252, EPI_ISL_1664253 | | | |
| see above | Kaiser San Jose | Santa Clara County public Health Laboratory | Santa Clara County Public Health Department |
| EPI_ISL_886178, EPI_ISL_886303, EPI_ISL_886330, EPI_ISL_886348, EPI_ISL_886351, EPI_ISL_886352, EPI_ISL_886354, EPI_ISL_886358, EPI_ISL_886463, EPI_ISL_886465, EPI_ISL_886557, EPI_ISL_886720, EPI_ISL_886730, EPI_ISL_886865, EPI_ISL_886907, EPI_ISL_887035, EPI_ISL_887056, EPI_ISL_887067, EPI_ISL_887641, EPI_ISL_887729, EPI_ISL_887917, EPI_ISL_887923, EPI_ISL_887995, EPI_ISL_888059, EPI_ISL_888522, EPI_ISL_888583 | | | |
| see above | Labcorp | Genomics and Discovery, Respiratory | ; Amanda Douglas; Amanda Suchanek; Andrea Throop; Ayla Burns; Ben L. Rambo-Martin; Bobbi Croy; Brian Krueger; Brian Norvell; Christos Petropoulos; Clinton R. Paden; Craig Lukasik; Debbie Boles; Dhwani Batra; Duncan MacCannell; Eyad Almasri; Goran Stevovic; Howard Engler; Hrushikesh |

| | | | |
|---|---|---|---|
| | | Viruses Branch, Division of Viral Diseases, Centers for Disease Control and Prevention | Deshmukh; Jake Humphrey; Jana Schroth; Joe Voshell; John Pruitt; Jonathan Meltzer; Jonathan Williams; Kimberly Wagner; Lax Iyer; Lyndon Tilson; Manoj Jain; Marcia Eisenberg; Mary Ann Cristobal; Mary Williamson; Michael Levandoski; Mike Sapeta; Mindy Nye; Minoo Agarwal; Mohan Kolli; Nuthawin Charoensri; Oren Cohen; Peter W. Cook; Prashant Gupta; Qian Zeng; Rama Ghatti; Scott Parker; Scott Ryan; Stanley Letovsky; Steven Ragan; Summer Galloway; Suresh Babu Selvaraju; Susan Countryman; Susan Hicks; Suxiang Tong; Suzanne Dale; Thomas Urban; Tim Kuphal; Tricia Zwiefelhofer; Vincent Drouillon |
| EPI_ISL_1020770, EPI_ISL_1020784, EPI_ISL_1020807, EPI_ISL_1020853, EPI_ISL_1020857, EPI_ISL_1020859, EPI_ISL_1020877, EPI_ISL_1020882, EPI_ISL_1020885, EPI_ISL_1020890, EPI_ISL_1020899, EPI_ISL_1020944, EPI_ISL_1020966, EPI_ISL_1020967, EPI_ISL_1020977, EPI_ISL_1021008, EPI_ISL_1021037, EPI_ISL_1021045, EPI_ISL_1021049, EPI_ISL_1021060, EPI_ISL_1021063, EPI_ISL_1021086, EPI_ISL_1021095, EPI_ISL_1021099, EPI_ISL_1021101, EPI_ISL_1021129, EPI_ISL_1021130, EPI_ISL_1021131, EPI_ISL_1021139, EPI_ISL_1021173, EPI_ISL_1021245, EPI_ISL_1021246, EPI_ISL_1021280, EPI_ISL_1021308, EPI_ISL_1021309, EPI_ISL_1021310, EPI_ISL_1021311, EPI_ISL_1021312, EPI_ISL_1021313, EPI_ISL_1021314, EPI_ISL_1021341, EPI_ISL_1021343, EPI_ISL_1021367, EPI_ISL_1021373, EPI_ISL_1021374, EPI_ISL_1021380, EPI_ISL_1021382, EPI_ISL_1021395, EPI_ISL_1021397, EPI_ISL_1021402, EPI_ISL_1021403, EPI_ISL_1021415, EPI_ISL_1021416, EPI_ISL_1021445, EPI_ISL_1021448, EPI_ISL_1021462, EPI_ISL_1021468, EPI_ISL_1021526, EPI_ISL_1021529, EPI_ISL_1021554, EPI_ISL_1021559, EPI_ISL_1021579, EPI_ISL_1021612, EPI_ISL_1021616, EPI_ISL_1021617, EPI_ISL_1021622, EPI_ISL_1021623, EPI_ISL_1021624, EPI_ISL_1021646, EPI_ISL_1021647, EPI_ISL_1021648, EPI_ISL_1021651, EPI_ISL_1021652, EPI_ISL_1021665, EPI_ISL_1021671, EPI_ISL_1021680, EPI_ISL_1021696, EPI_ISL_1021697, EPI_ISL_1021698, EPI_ISL_1021723, EPI_ISL_1021724, EPI_ISL_1021794, EPI_ISL_1021795, EPI_ISL_1021810, EPI_ISL_1021814, EPI_ISL_1021815, EPI_ISL_1021823, EPI_ISL_1021826, EPI_ISL_1021827, EPI_ISL_1021830, EPI_ISL_1021834, EPI_ISL_1021837, EPI_ISL_1021838, EPI_ISL_1021841, EPI_ISL_1021853, EPI_ISL_1021860, EPI_ISL_1021861, EPI_ISL_1021862, EPI_ISL_1021871, EPI_ISL_1021877, EPI_ISL_1021892, EPI_ISL_1021897, EPI_ISL_1021899, EPI_ISL_1021906, EPI_ISL_1021907, EPI_ISL_1021908, EPI_ISL_1021914, EPI_ISL_1021915, EPI_ISL_1021923, EPI_ISL_1021926, EPI_ISL_1021932, EPI_ISL_1021933, EPI_ISL_1021934, EPI_ISL_1021935, EPI_ISL_1021936, EPI_ISL_1021937, EPI_ISL_1021961, EPI_ISL_1021964, EPI_ISL_1021975, EPI_ISL_1021978, EPI_ISL_1021987, EPI_ISL_1021998, EPI_ISL_1022003, EPI_ISL_1022007, EPI_ISL_1022009, EPI_ISL_1022018, EPI_ISL_1022019, EPI_ISL_1022020, EPI_ISL_1022025, EPI_ISL_1022028, EPI_ISL_1022029, EPI_ISL_1022033, EPI_ISL_1022036, EPI_ISL_1022040, EPI_ISL_1022045, EPI_ISL_1022052, EPI_ISL_1022055, EPI_ISL_1022057, EPI_ISL_1022060, EPI_ISL_1022064, EPI_ISL_1022065, EPI_ISL_1022067, EPI_ISL_1022068, EPI_ISL_1022070, EPI_ISL_1022072, EPI_ISL_1022073, EPI_ISL_1022079, EPI_ISL_1022083, EPI_ISL_1022084, EPI_ISL_1022086, EPI_ISL_1022090, EPI_ISL_1022091, EPI_ISL_1022093, EPI_ISL_1022094, EPI_ISL_1022095, EPI_ISL_1022099, EPI_ISL_1022101, EPI_ISL_1022103, EPI_ISL_1022104, EPI_ISL_1022105, EPI_ISL_1022106, EPI_ISL_1022109, EPI_ISL_1022110, EPI_ISL_1022116, EPI_ISL_1022120, EPI_ISL_1022124, EPI_ISL_1022125, EPI_ISL_1022126, EPI_ISL_1022127, EPI_ISL_1022128, EPI_ISL_1022138, EPI_ISL_1022139, EPI_ISL_1022140, EPI_ISL_1022141, EPI_ISL_1022159, EPI_ISL_1022160, EPI_ISL_1022161, EPI_ISL_1022162, EPI_ISL_1022163, EPI_ISL_1022164, EPI_ISL_1022169, EPI_ISL_1022174, EPI_ISL_1022188, EPI_ISL_1022221, EPI_ISL_1022222, EPI_ISL_1022225, EPI_ISL_1022226, EPI_ISL_1022232, EPI_ISL_1022250, EPI_ISL_1022255, EPI_ISL_1022267, EPI_ISL_1022295, EPI_ISL_1022326, EPI_ISL_1022369, EPI_ISL_1022375, EPI_ISL_1022380, EPI_ISL_1022393, EPI_ISL_1022394, EPI_ISL_1022407, EPI_ISL_1022419, EPI_ISL_1025966, EPI_ISL_1025999, EPI_ISL_1026019, EPI_ISL_1026025, EPI_ISL_1026056, EPI_ISL_1026151, EPI_ISL_1026171, EPI_ISL_1026183, EPI_ISL_1026340, EPI_ISL_1026355, EPI_ISL_1026469, EPI_ISL_1026495, EPI_ISL_1026564, EPI_ISL_1026638, EPI_ISL_1026728, EPI_ISL_1026760, EPI_ISL_1026809, EPI_ISL_1027752, EPI_ISL_1027888, EPI_ISL_1028135, EPI_ISL_1028205, EPI_ISL_1028413, EPI_ISL_1028426, EPI_ISL_1028552, EPI_ISL_1028634, EPI_ISL_1028641, EPI_ISL_1028751, EPI_ISL_1029767, EPI_ISL_1029768, EPI_ISL_1029774, EPI_ISL_1029775, EPI_ISL_1029801, EPI_ISL_1029866, EPI_ISL_1030654, EPI_ISL_1030691, EPI_ISL_1030709, EPI_ISL_1030972, EPI_ISL_1031067, EPI_ISL_1031072, EPI_ISL_1031132, EPI_ISL_1031536, EPI_ISL_1031697, EPI_ISL_1031798, EPI_ISL_1031799, EPI_ISL_1031803, EPI_ISL_1032409, EPI_ISL_1032411, EPI_ISL_1032412, EPI_ISL_1032413, EPI_ISL_1032527, EPI_ISL_1032528, EPI_ISL_1032542, EPI_ISL_1037515, EPI_ISL_1037540, EPI_ISL_1037760, EPI_ISL_1037875, EPI_ISL_1038246, EPI_ISL_1038351, EPI_ISL_1038362, EPI_ISL_1038488, EPI_ISL_1081267 | | | |
| see above | Laboratory Corporation of America | Respiratory Viruses Branch, Division of Viral Diseases, Centers for Disease Control and Prevention | Alexandre Bolze; Ary Ascencio; Ben L. Rambo-Martin; Brad Sickler; Charlotte Rivera-Garcia; Christine Tran; Clinton R. Paden; Dakota Howard; David Becker; Dhwani Batra; Duncan MacCannell; Efren Sandoval; Eileen de Feo; Elizabeth Cirulli; Eric Allen; Geraint Levan; James Lu; Jan Antico; Jason Nguyen; Jimmy Ramirez; Jingtao Liu; Kelly Schiabor Barrett; Kim Gietzen; Magnus Isaksson; Marc Laurent; Matthew Tolentino; Nicole L. Washington; Peter W. Cook; Phil Febbo; Ryan Cho; Shannon Wickline; Sherry Wang; Simon White; Summer Galloway; Suxiang Tong; Tyler Cassens; William Lee |
| EPI_ISL_792668, EPI_ISL_792669, EPI_ISL_792670, EPI_ISL_792671, EPI_ISL_792672, EPI_ISL_792673, EPI_ISL_792674, EPI_ISL_792675, EPI_ISL_861686, EPI_ISL_861687, EPI_ISL_861688, EPI_ISL_861689, EPI_ISL_861690, EPI_ISL_861691, EPI_ISL_861692, EPI_ISL_861693, EPI_ISL_861694, EPI_ISL_861695, EPI_ISL_861696, EPI_ISL_861697, EPI_ISL_861699, EPI_ISL_903371, EPI_ISL_903372, EPI_ISL_917936, EPI_ISL_917938, EPI_ISL_917941, EPI_ISL_917951, EPI_ISL_917952, EPI_ISL_917953, EPI_ISL_917955, EPI_ISL_939609, EPI_ISL_939610, EPI_ISL_939611, EPI_ISL_939612, EPI_ISL_939613, EPI_ISL_939614, EPI_ISL_939615, EPI_ISL_939616, EPI_ISL_939617, EPI_ISL_939618, EPI_ISL_939652, EPI_ISL_962848, EPI_ISL_962849, EPI_ISL_962850, EPI_ISL_962851, EPI_ISL_962852, EPI_ISL_962853, EPI_ISL_962854, EPI_ISL_962855, EPI_ISL_962856, EPI_ISL_962857, EPI_ISL_962858, EPI_ISL_962859, EPI_ISL_962860, EPI_ISL_962861, EPI_ISL_962862, EPI_ISL_962863, EPI_ISL_962864, EPI_ISL_962865, EPI_ISL_962866, EPI_ISL_962867, EPI_ISL_1005483, EPI_ISL_1005484, EPI_ISL_1005486, EPI_ISL_1005487, EPI_ISL_1005488, EPI_ISL_1005489, EPI_ISL_1005490, EPI_ISL_1005509, EPI_ISL_1005510, EPI_ISL_1040052, EPI_ISL_1040425, EPI_ISL_1040426, EPI_ISL_2031993 | | | |
| see above | Los Angeles County PHL | Los Angeles County PHL | P. Hemarajata et al. |
| EPI_ISL_1018060, EPI_ISL_1018061, EPI_ISL_1018062 | Los Angeles County Public Health Laboratories, Los Angeles County Public Health Lab microbial pathogen submission group | Los Angeles County Public Health Laboratories, Los Angeles County Public Health Lab microbial pathogen submission group | Green, N.; Hemarajata, P. |
| EPI_ISL_832116, EPI_ISL_833417, EPI_ISL_833418, EPI_ISL_833419, EPI_ISL_833420, EPI_ISL_833421, EPI_ISL_833422, EPI_ISL_833423, EPI_ISL_833424, EPI_ISL_833425, EPI_ISL_833426, EPI_ISL_833427, EPI_ISL_833490, EPI_ISL_833491, EPI_ISL_861700, EPI_ISL_861701, EPI_ISL_861702, EPI_ISL_861703, EPI_ISL_862121, EPI_ISL_882963, EPI_ISL_882964, EPI_ISL_882965, EPI_ISL_882966, EPI_ISL_882985, EPI_ISL_882986, EPI_ISL_882987, EPI_ISL_882988, EPI_ISL_882989, EPI_ISL_882990, EPI_ISL_882991, EPI_ISL_882992, EPI_ISL_882993, EPI_ISL_882994, EPI_ISL_882995, EPI_ISL_882996, EPI_ISL_892249, EPI_ISL_892250, EPI_ISL_892251, EPI_ISL_892252, EPI_ISL_892253, EPI_ISL_892254, EPI_ISL_892255, EPI_ISL_892256, EPI_ISL_892257, EPI_ISL_892258, EPI_ISL_892259, EPI_ISL_892260, EPI_ISL_892261, EPI_ISL_892262, EPI_ISL_892263, EPI_ISL_892264, EPI_ISL_892265, EPI_ISL_892266, EPI_ISL_902925, EPI_ISL_903373, EPI_ISL_903374, EPI_ISL_905765, EPI_ISL_905766, EPI_ISL_905767, EPI_ISL_905768, EPI_ISL_905769, EPI_ISL_905770, EPI_ISL_905771, EPI_ISL_905772, EPI_ISL_915434, EPI_ISL_915435, EPI_ISL_915436, EPI_ISL_915437, EPI_ISL_915439, EPI_ISL_917944, EPI_ISL_917945, EPI_ISL_917946, EPI_ISL_917947, EPI_ISL_917948, EPI_ISL_917949, EPI_ISL_917950, EPI_ISL_939651, EPI_ISL_939653, EPI_ISL_962515, EPI_ISL_962516, EPI_ISL_962847, EPI_ISL_1005555, EPI_ISL_1098628, EPI_ISL_1098646, EPI_ISL_1303483 | | | |
| see above | MD Laboratories | Los Angeles County PHL | P. Hemarajata et al. |
| EPI_ISL_906047, EPI_ISL_906048 | MD Laboratories | Los Angeles County Public Health Laboratory | P. Hemarajata et al. |
| EPI_ISL_1015733 | Medical diagnostic laboratories (MD Laboratories) | Los Angeles County Public Health Laboratory | P. Hemarajata et al. |
| EPI_ISL_2009465, EPI_ISL_2009467, EPI_ISL_2019597, EPI_ISL_2019615, EPI_ISL_2019617, EPI_ISL_2019625, EPI_ISL_2019647, EPI_ISL_2019685, EPI_ISL_2019687, EPI_ISL_2019704, EPI_ISL_2019725, EPI_ISL_2019729, EPI_ISL_2019730 | | | |
| see above | Moderna Inc. | Moderna Inc. | Groves Dixon; Rolando Pajon; Yamuna Paila |
| EPI_ISL_930788, EPI_ISL_930789, EPI_ISL_930791, EPI_ISL_930792, EPI_ISL_930794, EPI_ISL_930796, EPI_ISL_930797, EPI_ISL_930800, EPI_ISL_930804, EPI_ISL_930805, EPI_ISL_930830, EPI_ISL_930833, EPI_ISL_930845, EPI_ISL_930847, EPI_ISL_930848, EPI_ISL_930851, EPI_ISL_930852, EPI_ISL_968069, EPI_ISL_968070, EPI_ISL_968071, EPI_ISL_968072, EPI_ISL_968073, EPI_ISL_968074, EPI_ISL_968075, EPI_ISL_968076, EPI_ISL_968077, EPI_ISL_968079, EPI_ISL_1263227, EPI_ISL_1263229, EPI_ISL_1263230, EPI_ISL_1664599, EPI_ISL_1664600, EPI_ISL_1664601, EPI_ISL_1664602, EPI_ISL_1664603, EPI_ISL_1664604, EPI_ISL_1664605, EPI_ISL_1664606, EPI_ISL_1664607, EPI_ISL_1664608, EPI_ISL_1664609, EPI_ISL_1664610, EPI_ISL_1664611, EPI_ISL_1664612, EPI_ISL_1664613, EPI_ISL_1664614, EPI_ISL_1664615, EPI_ISL_1664616, EPI_ISL_1700696, EPI_ISL_1700697, EPI_ISL_1700698, EPI_ISL_1700699 | | | |
| see above | Monterey County Public Health Laboratory | Monterey County Public Health Laboratory | ; C. Anaya et al.; Monterey County Public Health Laboratory |
| EPI_ISL_882997, EPI_ISL_882998 | Motion Picture Television Fund Hospital Lab | Los Angeles County PHL | P. Hemarajata et al. |
| EPI_ISL_832204 | Northwest Pathology | UW Virology Lab | Alexander Greninger; Hong Xie; Keith R Jerome; Lasata Shrestha; Meei-Li Huang; Michelle Lin; Pavitra Roychoudhury |
| EPI_ISL_936990 | Northwestern Memorial Hospital | Ozer Lab | Chad J. Achenbach; Egon A. Ozer; Judd F. Hultquist; Lacy M. Simons; Lawrence J. Jennings; Michael G. Ison; Ramon Lorenzo-Redondo |
| EPI_ISL_1122131, EPI_ISL_1541297, EPI_ISL_1541732, EPI_ISL_1541768, EPI_ISL_1541889 | OHSU Lab Services Molecular Microbiology Lab | Oregon SARS-CoV-2 Genome Sequencing Center | Alec J. Hirsch; Andrew C. Adey; Benjamin N. Bimber; Brendan L. O'Connell; Brian J. O'Roak; Daniel N. Streblow; Donna Hansel; Guang Fan; Kayla Carter; Qin; Ruth V. Nichols; Sally Grindstaff; Sonia Acharya; William B. Messer; Xuan; Xuan Qin |
| EPI_ISL_1005541 | Olive View Medical Center | Los Angeles County PHL | P. Hemarajata et al. |
| EPI_ISL_849922, EPI_ISL_885113, EPI_ISL_885114, EPI_ISL_885115, EPI_ISL_885116, EPI_ISL_885117, EPI_ISL_885118, EPI_ISL_885120, EPI_ISL_885121, EPI_ISL_885122, EPI_ISL_885123, EPI_ISL_885124, EPI_ISL_885125, EPI_ISL_885126, EPI_ISL_885127, EPI_ISL_955427, EPI_ISL_955428, EPI_ISL_955429, EPI_ISL_955430, EPI_ISL_955431, EPI_ISL_955432, EPI_ISL_955433, EPI_ISL_955434, EPI_ISL_955435, EPI_ISL_955436, EPI_ISL_955437, EPI_ISL_955438, EPI_ISL_955439, EPI_ISL_955440, EPI_ISL_955441, EPI_ISL_955442, EPI_ISL_955443, EPI_ISL_955444, EPI_ISL_955445, EPI_ISL_955446, EPI_ISL_955447, EPI_ISL_955448, EPI_ISL_955449, EPI_ISL_955450, EPI_ISL_955451, EPI_ISL_955452, EPI_ISL_955453, EPI_ISL_955454, EPI_ISL_955455, EPI_ISL_955456, EPI_ISL_955457, EPI_ISL_955458, EPI_ISL_955459, EPI_ISL_955460, EPI_ISL_955461, EPI_ISL_955462, EPI_ISL_955463, EPI_ISL_955464, EPI_ISL_955465, EPI_ISL_955466, EPI_ISL_955467, EPI_ISL_955468, EPI_ISL_955469, EPI_ISL_955470, EPI_ISL_955471, EPI_ISL_955472, EPI_ISL_955473, EPI_ISL_955474, EPI_ISL_955475, EPI_ISL_955476, EPI_ISL_955477, EPI_ISL_955478, EPI_ISL_955479, EPI_ISL_955480, EPI_ISL_955481, EPI_ISL_955482, EPI_ISL_955483, EPI_ISL_955484, EPI_ISL_955485, EPI_ISL_955486, EPI_ISL_955487, EPI_ISL_955488, EPI_ISL_955489, EPI_ISL_955490, EPI_ISL_955491, EPI_ISL_955492, EPI_ISL_955493, EPI_ISL_955494, EPI_ISL_955495, EPI_ISL_979643, EPI_ISL_979644, EPI_ISL_979645, EPI_ISL_979646, EPI_ISL_979647, EPI_ISL_979648, EPI_ISL_979649, EPI_ISL_979650, EPI_ISL_979651, EPI_ISL_979652, EPI_ISL_979653, EPI_ISL_979654, EPI_ISL_979655, EPI_ISL_979656, EPI_ISL_979657, EPI_ISL_979658, EPI_ISL_979659, EPI_ISL_979660, EPI_ISL_979661, EPI_ISL_979662, EPI_ISL_979663, EPI_ISL_979664, EPI_ISL_979665, EPI_ISL_979666, EPI_ISL_979667, EPI_ISL_979668, EPI_ISL_979669, EPI_ISL_979670, EPI_ISL_979671, EPI_ISL_979672, EPI_ISL_979673, EPI_ISL_979674, EPI_ISL_979675, EPI_ISL_979676, EPI_ISL_979677, EPI_ISL_979678, EPI_ISL_979679, EPI_ISL_979680, EPI_ISL_979681, EPI_ISL_979682, EPI_ISL_979683, EPI_ISL_979684, EPI_ISL_979685, EPI_ISL_979686, EPI_ISL_979687, EPI_ISL_979688, EPI_ISL_979689, EPI_ISL_979690, EPI_ISL_979691, EPI_ISL_979692, EPI_ISL_979693, EPI_ISL_979694, EPI_ISL_979695, EPI_ISL_979696, EPI_ISL_979697, EPI_ISL_979698, EPI_ISL_979699, EPI_ISL_979700, EPI_ISL_979701, EPI_ISL_979702, EPI_ISL_979703, EPI_ISL_979704, EPI_ISL_979705, EPI_ISL_979706, EPI_ISL_979707, EPI_ISL_979708, EPI_ISL_979709, EPI_ISL_1027294, EPI_ISL_1027321, EPI_ISL_1027329, EPI_ISL_1027331, EPI_ISL_1027335, EPI_ISL_1027339, EPI_ISL_1027340, EPI_ISL_1027344, EPI_ISL_1027351, EPI_ISL_1027359, EPI_ISL_1027378, EPI_ISL_1027379, EPI_ISL_1027380, EPI_ISL_1027381, EPI_ISL_1027382, EPI_ISL_1027383, EPI_ISL_1027384, EPI_ISL_1027385, EPI_ISL_1027386, EPI_ISL_1027387, EPI_ISL_1027388, EPI_ISL_1027389, EPI_ISL_1027390, EPI_ISL_1027391, EPI_ISL_1027392, EPI_ISL_1027393, EPI_ISL_1027394, EPI_ISL_1027395, EPI_ISL_1027396, EPI_ISL_1027397, EPI_ISL_1027398, EPI_ISL_1027399, EPI_ISL_1027400, EPI_ISL_1027401, EPI_ISL_1027402, EPI_ISL_1027403, EPI_ISL_1027404, EPI_ISL_1027405, EPI_ISL_1027406, EPI_ISL_1027407, EPI_ISL_1027408, EPI_ISL_1027409, EPI_ISL_1027410, EPI_ISL_1027411, EPI_ISL_1027412, EPI_ISL_1027413, EPI_ISL_1027414, EPI_ISL_1027415, EPI_ISL_1027416, EPI_ISL_1027417, EPI_ISL_1027418, EPI_ISL_1027419, EPI_ISL_1027420, EPI_ISL_1027421, EPI_ISL_1027422, EPI_ISL_1027423, EPI_ISL_1027424, EPI_ISL_1027425, EPI_ISL_1027426, EPI_ISL_1027427, EPI_ISL_1027428, EPI_ISL_1027429, EPI_ISL_1027430, EPI_ISL_1027431, EPI_ISL_1027432, EPI_ISL_1027433, EPI_ISL_1027434, EPI_ISL_1027435, EPI_ISL_1027487, EPI_ISL_1027503, EPI_ISL_1027504, EPI_ISL_1027507, EPI_ISL_1027530, EPI_ISL_1027531, EPI_ISL_1027532, EPI_ISL_1027573, EPI_ISL_1027583, EPI_ISL_1027584, EPI_ISL_1027587, EPI_ISL_1027588, EPI_ISL_1027589, EPI_ISL_1027590, EPI_ISL_1027591, EPI_ISL_1027592, EPI_ISL_1027593, EPI_ISL_1027594, EPI_ISL_1027595, EPI_ISL_1027596, EPI_ISL_1027597, EPI_ISL_1027598, EPI_ISL_1027599, EPI_ISL_1027600, EPI_ISL_1027601, EPI_ISL_1027602, EPI_ISL_1027603, EPI_ISL_1027604, EPI_ISL_1027605, EPI_ISL_1027606, EPI_ISL_1027607, EPI_ISL_1027608, EPI_ISL_1027609, EPI_ISL_1027610, EPI_ISL_1027611, EPI_ISL_1027612, EPI_ISL_1027613, EPI_ISL_1027614, EPI_ISL_1027615, EPI_ISL_1027616, EPI_ISL_1027617, EPI_ISL_1027618, EPI_ISL_1027619, EPI_ISL_1027620, EPI_ISL_1027621, EPI_ISL_1027622, EPI_ISL_1027623, EPI_ISL_1027624, EPI_ISL_1027625, EPI_ISL_1027626, EPI_ISL_1027627, EPI_ISL_1185032, EPI_ISL_1185039, EPI_ISL_1185040, EPI_ISL_1185043, EPI_ISL_1185045, EPI_ISL_1185048, EPI_ISL_1185049, EPI_ISL_1185055, EPI_ISL_1185058, EPI_ISL_1185062, EPI_ISL_1185070, EPI_ISL_1185075, EPI_ISL_1185093, EPI_ISL_1185098, EPI_ISL_1185101, EPI_ISL_1185104, EPI_ISL_1185106, EPI_ISL_1185107, EPI_ISL_1185108, EPI_ISL_1185112, EPI_ISL_1185113, EPI_ISL_1185115, EPI_ISL_1185118, EPI_ISL_1185123, EPI_ISL_1185125, EPI_ISL_1185127, EPI_ISL_1185131, EPI_ISL_1185134, EPI_ISL_1185140, EPI_ISL_1185143, EPI_ISL_1185145, EPI_ISL_1185146, EPI_ISL_1185148, EPI_ISL_1185149, EPI_ISL_1185152, EPI_ISL_1185154, EPI_ISL_1185159, EPI_ISL_1185160, EPI_ISL_1185163, EPI_ISL_1185165, EPI_ISL_1185167, EPI_ISL_1185171, EPI_ISL_1185174, EPI_ISL_1185175, EPI_ISL_1185180, EPI_ISL_1185189, EPI_ISL_1185194, EPI_ISL_1185201, EPI_ISL_1185206, EPI_ISL_1185209, EPI_ISL_1185212, EPI_ISL_1185214, EPI_ISL_1185216, EPI_ISL_1185217, EPI_ISL_1185221, EPI_ISL_1185224, EPI_ISL_1185225, EPI_ISL_1185228, EPI_ISL_1185231, EPI_ISL_1185233, EPI_ISL_1185235, EPI_ISL_1185236, EPI_ISL_1185237, EPI_ISL_1185238, EPI_ISL_1185240, EPI_ISL_1185241, EPI_ISL_1185242, EPI_ISL_1185257, EPI_ISL_1185261, EPI_ISL_1185263, EPI_ISL_1185266, EPI_ISL_1185269, EPI_ISL_1185272, EPI_ISL_1185274, EPI_ISL_1185278, EPI_ISL_1185280, EPI_ISL_1234804, EPI_ISL_1234825, EPI_ISL_1234869, EPI_ISL_1234923, EPI_ISL_1234942, EPI_ISL_1234944, EPI_ISL_1235041, EPI_ISL_1235052, EPI_ISL_1701566, EPI_ISL_2103061, EPI_ISL_2103110, EPI_ISL_2103115 | | | |
| see above | Orange County Public Health Lab | Chan-Zuckerberg Biohub | CZB Cliahub Consortium |
| EPI_ISL_917954 | Pomona Valley Hospital | Los Angeles County PHL | P. Hemarajata et al. |
| EPI_ISL_1017052, EPI_ISL_1017058, EPI_ISL_1017075, EPI_ISL_1017076, EPI_ISL_1017077, EPI_ISL_1017078, EPI_ISL_1017079, EPI_ISL_1017080, EPI_ISL_1017081, EPI_ISL_1017082, EPI_ISL_1017083, EPI_ISL_1017084, EPI_ISL_1017085, EPI_ISL_1017086, EPI_ISL_1017087, EPI_ISL_1017088, EPI_ISL_1017089, EPI_ISL_1017091, EPI_ISL_1017092, EPI_ISL_1017093, EPI_ISL_1017094, EPI_ISL_1017095 | | | |
| see above | Providence St. Joseph Health Orange - Chapman | Providence St. Joseph Health Molecular Genomics Laboratory | Alexa K Dowdell; Brian D Piening; Carlo B Bifulco; Fred L Robinson; Mary Campbell |
| EPI_ISL_854810, EPI_ISL_854811, EPI_ISL_854812, EPI_ISL_854813, EPI_ISL_854814, EPI_ISL_854815, EPI_ISL_854816, EPI_ISL_854817, EPI_ISL_854818, EPI_ISL_854819, EPI_ISL_854820, EPI_ISL_854821, EPI_ISL_854822, EPI_ISL_854823, EPI_ISL_854824, EPI_ISL_854825, EPI_ISL_854826, EPI_ISL_854827, EPI_ISL_854828, EPI_ISL_854829, EPI_ISL_854830, EPI_ISL_854831, EPI_ISL_854832, EPI_ISL_854833, EPI_ISL_854834, EPI_ISL_854835, EPI_ISL_854836, EPI_ISL_854837, EPI_ISL_854838, EPI_ISL_854839, EPI_ISL_854840, EPI_ISL_854841, EPI_ISL_855015, EPI_ISL_855016, EPI_ISL_855017, EPI_ISL_855018, EPI_ISL_855019, EPI_ISL_855020, EPI_ISL_855021, EPI_ISL_855022, EPI_ISL_855023, EPI_ISL_855024, EPI_ISL_855025, EPI_ISL_855026, EPI_ISL_855027, EPI_ISL_855028, EPI_ISL_855029, EPI_ISL_855030, EPI_ISL_855031, EPI_ISL_855032, EPI_ISL_855033, EPI_ISL_855034, EPI_ISL_855035, EPI_ISL_855036, EPI_ISL_855037, EPI_ISL_855038, EPI_ISL_855039, EPI_ISL_855040, EPI_ISL_855041, EPI_ISL_855042, EPI_ISL_855043, EPI_ISL_855044, EPI_ISL_855045, EPI_ISL_855046, EPI_ISL_855047, EPI_ISL_855048, EPI_ISL_855049, EPI_ISL_855050, EPI_ISL_855051, EPI_ISL_855052, EPI_ISL_855053, EPI_ISL_855054, EPI_ISL_855055, EPI_ISL_855056, EPI_ISL_855057, EPI_ISL_855058, EPI_ISL_855059, EPI_ISL_855060, EPI_ISL_855061, EPI_ISL_855062, EPI_ISL_855063, EPI_ISL_855064, EPI_ISL_855065, EPI_ISL_855066, EPI_ISL_855067, EPI_ISL_855068, EPI_ISL_855069, EPI_ISL_855070, EPI_ISL_855071, EPI_ISL_855072, EPI_ISL_855073, EPI_ISL_855074, EPI_ISL_855075, EPI_ISL_855076, EPI_ISL_855077, EPI_ISL_855078, EPI_ISL_855079, EPI_ISL_855080, EPI_ISL_877037, EPI_ISL_877038, EPI_ISL_877039, EPI_ISL_877040, EPI_ISL_877041, EPI_ISL_877042, EPI_ISL_877043, EPI_ISL_877044, EPI_ISL_877045, EPI_ISL_877046, EPI_ISL_877047, EPI_ISL_877048, EPI_ISL_877049, EPI_ISL_877050, EPI_ISL_877053, EPI_ISL_877054, EPI_ISL_877055, EPI_ISL_877056, EPI_ISL_877057, EPI_ISL_877058, EPI_ISL_877059, EPI_ISL_877060, EPI_ISL_877061, EPI_ISL_877062, EPI_ISL_877063, EPI_ISL_877064, EPI_ISL_877065, EPI_ISL_877066, EPI_ISL_877067, EPI_ISL_877068, EPI_ISL_877069, EPI_ISL_877070, EPI_ISL_877071, EPI_ISL_877072, EPI_ISL_877073, EPI_ISL_877074, EPI_ISL_877075, EPI_ISL_877076, EPI_ISL_877077, EPI_ISL_877078, EPI_ISL_877079, EPI_ISL_877080, EPI_ISL_877081, EPI_ISL_877082, EPI_ISL_877083, EPI_ISL_877084, EPI_ISL_877085, EPI_ISL_877086, EPI_ISL_877087, EPI_ISL_877088, EPI_ISL_877089, EPI_ISL_877090, EPI_ISL_877091, EPI_ISL_877092, EPI_ISL_877093, EPI_ISL_877094, EPI_ISL_877095, EPI_ISL_877096, EPI_ISL_877097, EPI_ISL_877098, EPI_ISL_877099, EPI_ISL_877100, EPI_ISL_877101, EPI_ISL_877102, EPI_ISL_877103, EPI_ISL_877104, EPI_ISL_877105, EPI_ISL_877106, EPI_ISL_877107, EPI_ISL_877108, EPI_ISL_877109, EPI_ISL_877110, EPI_ISL_877111, EPI_ISL_877112, EPI_ISL_877113, EPI_ISL_877114, EPI_ISL_877115, EPI_ISL_877116, EPI_ISL_877117, EPI_ISL_877118, EPI_ISL_877186, EPI_ISL_877187, EPI_ISL_877188, EPI_ISL_877197, EPI_ISL_903390, EPI_ISL_903391, EPI_ISL_903392, EPI_ISL_903394, EPI_ISL_903395, EPI_ISL_903396, EPI_ISL_903397, EPI_ISL_903398, EPI_ISL_903399, EPI_ISL_903400, EPI_ISL_903401, EPI_ISL_903402, EPI_ISL_903403, EPI_ISL_903404, EPI_ISL_903405, EPI_ISL_903407, EPI_ISL_903408, EPI_ISL_903409, EPI_ISL_903410, EPI_ISL_903411, EPI_ISL_903412, EPI_ISL_903413, EPI_ISL_903414, EPI_ISL_903415, EPI_ISL_903416, EPI_ISL_903417, EPI_ISL_903418, EPI_ISL_903419, EPI_ISL_903420, EPI_ISL_903421, EPI_ISL_903422, EPI_ISL_903423, EPI_ISL_903424, EPI_ISL_903425, EPI_ISL_903426, EPI_ISL_903427, EPI_ISL_903428, EPI_ISL_903429, EPI_ISL_903430, EPI_ISL_903431, EPI_ISL_903432, EPI_ISL_903433, EPI_ISL_903434, EPI_ISL_903435, EPI_ISL_903436, EPI_ISL_903437, EPI_ISL_903438, EPI_ISL_903439, EPI_ISL_903440, EPI_ISL_903441, EPI_ISL_903442, EPI_ISL_903443, EPI_ISL_903444, EPI_ISL_903445, EPI_ISL_903446, EPI_ISL_903447, EPI_ISL_903448, EPI_ISL_903449, EPI_ISL_903450, EPI_ISL_903451, EPI_ISL_903452, EPI_ISL_903453, EPI_ISL_903454, EPI_ISL_903455, EPI_ISL_903456, EPI_ISL_903457, EPI_ISL_903458, EPI_ISL_903459, EPI_ISL_903460, EPI_ISL_903461, EPI_ISL_903462, EPI_ISL_903463, EPI_ISL_903464, EPI_ISL_903465, EPI_ISL_903466, EPI_ISL_903467, EPI_ISL_903468, EPI_ISL_903469, EPI_ISL_903470, EPI_ISL_903471, EPI_ISL_903472, EPI_ISL_903473, EPI_ISL_903474, EPI_ISL_903475, EPI_ISL_903476, EPI_ISL_903477, EPI_ISL_903478, EPI_ISL_903479, EPI_ISL_903480, EPI_ISL_903481, EPI_ISL_903482, EPI_ISL_903483, EPI_ISL_903484, EPI_ISL_903485, EPI_ISL_903486, EPI_ISL_903487, EPI_ISL_903488, EPI_ISL_903489, EPI_ISL_903490, EPI_ISL_903491, EPI_ISL_903492, EPI_ISL_903493, EPI_ISL_903494, EPI_ISL_903495, EPI_ISL_903496, EPI_ISL_903497, EPI_ISL_903498, EPI_ISL_903499, EPI_ISL_903500, EPI_ISL_903501, EPI_ISL_903502, EPI_ISL_903503, EPI_ISL_903504, EPI_ISL_903505, EPI_ISL_903506, EPI_ISL_903507, EPI_ISL_903508, EPI_ISL_903509, EPI_ISL_903510, EPI_ISL_903511, EPI_ISL_903512, EPI_ISL_903513, EPI_ISL_903514, EPI_ISL_903515, EPI_ISL_903516, EPI_ISL_903517, EPI_ISL_903518, EPI_ISL_903519, EPI_ISL_903520, EPI_ISL_903521, EPI_ISL_903522, EPI_ISL_903523, EPI_ISL_903524, EPI_ISL_903525, EPI_ISL_904127, EPI_ISL_904128, EPI_ISL_904129, EPI_ISL_904130, EPI_ISL_906308, EPI_ISL_906309, EPI_ISL_906310, EPI_ISL_906311, EPI_ISL_906312, EPI_ISL_906313, EPI_ISL_906314, EPI_ISL_906315, EPI_ISL_906316, EPI_ISL_906317, EPI_ISL_906318, EPI_ISL_906319, EPI_ISL_906320, EPI_ISL_906321, EPI_ISL_906322, EPI_ISL_906323, EPI_ISL_906324, EPI_ISL_906325, EPI_ISL_906326, EPI_ISL_906327, EPI_ISL_906328, EPI_ISL_906329, EPI_ISL_906330, EPI_ISL_906331, EPI_ISL_906332, EPI_ISL_906333, EPI_ISL_906334, EPI_ISL_906335, EPI_ISL_906336, EPI_ISL_906337, EPI_ISL_906338, EPI_ISL_906339, EPI_ISL_906340, EPI_ISL_906341, EPI_ISL_906342, EPI_ISL_906343, EPI_ISL_906344, EPI_ISL_906345, EPI_ISL_906346, EPI_ISL_906347, EPI_ISL_906348, EPI_ISL_906349, EPI_ISL_906350, EPI_ISL_906351, EPI_ISL_906352, EPI_ISL_906353, EPI_ISL_906354, EPI_ISL_906355, EPI_ISL_906356, EPI_ISL_906357, EPI_ISL_906358, EPI_ISL_906359, EPI_ISL_906360, EPI_ISL_906361, EPI_ISL_906362, EPI_ISL_906363, EPI_ISL_906364, EPI_ISL_906365, EPI_ISL_906366, EPI_ISL_906367, EPI_ISL_906368, EPI_ISL_906369, EPI_ISL_906370, EPI_ISL_906371, EPI_ISL_906372, EPI_ISL_906373, EPI_ISL_906374, EPI_ISL_906375, EPI_ISL_906376, EPI_ISL_906377, EPI_ISL_906378, EPI_ISL_906379, EPI_ISL_906380, EPI_ISL_906381, EPI_ISL_906382, EPI_ISL_906383, EPI_ISL_906384, EPI_ISL_906385, EPI_ISL_906386, EPI_ISL_906387, EPI_ISL_906388, EPI_ISL_906389, EPI_ISL_906390, EPI_ISL_906391, EPI_ISL_906392, EPI_ISL_906393, EPI_ISL_906394, EPI_ISL_906395, EPI_ISL_906396, EPI_ISL_906397, EPI_ISL_906398, EPI_ISL_906399, EPI_ISL_906400, EPI_ISL_906401, EPI_ISL_906402, EPI_ISL_906403, EPI_ISL_906404, EPI_ISL_906405, EPI_ISL_906406, EPI_ISL_906407, EPI_ISL_906408, EPI_ISL_906409, EPI_ISL_906410, EPI_ISL_906411, EPI_ISL_906412, EPI_ISL_906413, EPI_ISL_906414, EPI_ISL_906415, EPI_ISL_906416, EPI_ISL_906417, EPI_ISL_906418, EPI_ISL_906419, EPI_ISL_906420, EPI_ISL_906421, EPI_ISL_906422, EPI_ISL_906423, EPI_ISL_911704, EPI_ISL_911705, EPI_ISL_911706, EPI_ISL_914890, EPI_ISL_914891, EPI_ISL_914892, EPI_ISL_914893, EPI_ISL_914894, EPI_ISL_914895, EPI_ISL_914896, EPI_ISL_914897, EPI_ISL_914898, EPI_ISL_914899, EPI_ISL_914900, EPI_ISL_914901, EPI_ISL_914902, EPI_ISL_914903, EPI_ISL_914904, EPI_ISL_914905, EPI_ISL_914906, EPI_ISL_914907, EPI_ISL_914908, EPI_ISL_914909, EPI_ISL_914910, EPI_ISL_914911, EPI_ISL_914912, EPI_ISL_914913, EPI_ISL_914914, EPI_ISL_914915, EPI_ISL_914916, EPI_ISL_914917, EPI_ISL_914918, EPI_ISL_914919, EPI_ISL_914920, EPI_ISL_914921, EPI_ISL_914922, EPI_ISL_914923, EPI_ISL_914924, EPI_ISL_914925, EPI_ISL_914926, EPI_ISL_914927, EPI_ISL_914928, EPI_ISL_914929, EPI_ISL_914930, EPI_ISL_914931, EPI_ISL_914932, EPI_ISL_914933, EPI_ISL_914934, EPI_ISL_914935, EPI_ISL_914936, EPI_ISL_914937, EPI_ISL_914938, EPI_ISL_914939, EPI_ISL_914940, EPI_ISL_914941, EPI_ISL_914942, EPI_ISL_914943, EPI_ISL_914944, EPI_ISL_914945, EPI_ISL_914946, EPI_ISL_914947, EPI_ISL_914948, EPI_ISL_914949, EPI_ISL_914950, EPI_ISL_914951, EPI_ISL_914952, EPI_ISL_914953, EPI_ISL_914954, EPI_ISL_914955, EPI_ISL_914956, EPI_ISL_914957, EPI_ISL_914958, EPI_ISL_914959, EPI_ISL_914960, EPI_ISL_914961, EPI_ISL_914962, EPI_ISL_914963, EPI_ISL_914964, EPI_ISL_914965, EPI_ISL_914966, EPI_ISL_914967, EPI_ISL_914968, EPI_ISL_914969, EPI_ISL_915197, EPI_ISL_915198, EPI_ISL_915199, EPI_ISL_915200, EPI_ISL_915201, EPI_ISL_915202, EPI_ISL_915203, EPI_ISL_915204, EPI_ISL_915205, EPI_ISL_915206, EPI_ISL_915207, EPI_ISL_915208, EPI_ISL_915209, EPI_ISL_915210, EPI_ISL_915211, EPI_ISL_915212, EPI_ISL_915213, EPI_ISL_915214, EPI_ISL_915215, EPI_ISL_915216, EPI_ISL_915217, EPI_ISL_915218, EPI_ISL_915219, EPI_ISL_915220, EPI_ISL_915221, EPI_ISL_915222, EPI_ISL_915223, EPI_ISL_915224, EPI_ISL_915225, EPI_ISL_915226, EPI_ISL_915227, EPI_ISL_915228, EPI_ISL_915229, EPI_ISL_915230, EPI_ISL_915231, EPI_ISL_915232, EPI_ISL_915233, EPI_ISL_915234, EPI_ISL_915235, EPI_ISL_915236, EPI_ISL_915237, EPI_ISL_915238, EPI_ISL_915239, EPI_ISL_915240, EPI_ISL_915241, EPI_ISL_915242, EPI_ISL_915243, EPI_ISL_915244, EPI_ISL_915245, EPI_ISL_915246, EPI_ISL_915247, EPI_ISL_915248, EPI_ISL_915249, EPI_ISL_915250, EPI_ISL_915251, EPI_ISL_915252, EPI_ISL_915253, EPI_ISL_915254, EPI_ISL_915255, EPI_ISL_915256, EPI_ISL_915257, EPI_ISL_915258, EPI_ISL_915259, EPI_ISL_915260, EPI_ISL_915261, EPI_ISL_915262, EPI_ISL_915263, EPI_ISL_915264, EPI_ISL_915265, EPI_ISL_915266, EPI_ISL_915267, EPI_ISL_915268, EPI_ISL_915269, EPI_ISL_915270, EPI_ISL_915271, EPI_ISL_915272, EPI_ISL_915273, EPI_ISL_915274, EPI_ISL_915275, EPI_ISL_915276, EPI_ISL_915277, EPI_ISL_915278, EPI_ISL_915279, EPI_ISL_915280, EPI_ISL_915281, EPI_ISL_915282, EPI_ISL_915283, EPI_ISL_915284, EPI_ISL_915285, EPI_ISL_915286, EPI_ISL_915287, EPI_ISL_915288, EPI_ISL_915289, EPI_ISL_915290, EPI_ISL_915291, EPI_ISL_915292, EPI_ISL_915293, EPI_ISL_915294, EPI_ISL_915295, EPI_ISL_915296, EPI_ISL_915297, EPI_ISL_915298, EPI_ISL_915299, EPI_ISL_915300, EPI_ISL_915301, | | | |

| | | | |
|---|---|---|---|
| EPI_ISL_915302, EPI_ISL_915303, EPI_ISL_915304, EPI_ISL_915305, EPI_ISL_915306, EPI_ISL_915307, EPI_ISL_925137, EPI_ISL_925138, EPI_ISL_925139, EPI_ISL_925140, EPI_ISL_937121, EPI_ISL_937122, EPI_ISL_937123, EPI_ISL_937124, EPI_ISL_937125 | | | |
| see above | Quest Diagnostics | Quest Diagnostics | Anderson, B.; Bernstein; D.F.; Gerasimova, A.; Hua, M.; I.A.; K.E.; Kagan; L.E.; Lacbawan, F.; Liu Y.; Livingston; Owen, R.; Perez, A.; R.M.; Rosenthal; S.H.; Shalhout; Shlyakhter; Tanpaiboon, P. |
| EPI_ISL_1086225, EPI_ISL_1086232, EPI_ISL_1086236, EPI_ISL_1086244, EPI_ISL_1086245, EPI_ISL_1086252, EPI_ISL_1086270, EPI_ISL_1086277, EPI_ISL_1086280, EPI_ISL_1086292, EPI_ISL_1086301, EPI_ISL_1086331, EPI_ISL_1086368, EPI_ISL_1086494, EPI_ISL_1086500, EPI_ISL_1086503, EPI_ISL_1086506, EPI_ISL_1086508, EPI_ISL_1086510, EPI_ISL_1086512, EPI_ISL_1086518, EPI_ISL_1086523, EPI_ISL_1086525, EPI_ISL_1086526, EPI_ISL_1086537, EPI_ISL_1086539, EPI_ISL_1086541, EPI_ISL_1086542, EPI_ISL_1086543, EPI_ISL_1086546, EPI_ISL_1086548, EPI_ISL_1086586, EPI_ISL_1086667, EPI_ISL_1086668, EPI_ISL_1086670, EPI_ISL_1086671, EPI_ISL_1086674, EPI_ISL_1086677, EPI_ISL_1086679, EPI_ISL_1086681, EPI_ISL_1086682, EPI_ISL_1086684, EPI_ISL_1086685, EPI_ISL_1086688, EPI_ISL_1086689, EPI_ISL_1086690, EPI_ISL_1086693, EPI_ISL_1086694, EPI_ISL_1086697, EPI_ISL_1086699, EPI_ISL_1086700, EPI_ISL_1086701, EPI_ISL_1086703, EPI_ISL_1086709, EPI_ISL_1086710, EPI_ISL_1086712, EPI_ISL_1086715, EPI_ISL_1086716, EPI_ISL_1086719, EPI_ISL_1086721, EPI_ISL_1086722, EPI_ISL_1086725, EPI_ISL_1086726, EPI_ISL_1086728, EPI_ISL_1086729, EPI_ISL_1086730, EPI_ISL_1086731, EPI_ISL_1086733, EPI_ISL_1086735, EPI_ISL_1086736, EPI_ISL_1086738, EPI_ISL_1086739, EPI_ISL_1086741, EPI_ISL_1086742, EPI_ISL_1086745, EPI_ISL_1086747, EPI_ISL_1086749, EPI_ISL_1086750, EPI_ISL_1086752, EPI_ISL_1086756, EPI_ISL_1086759, EPI_ISL_1086760, EPI_ISL_1086761, EPI_ISL_1086762, EPI_ISL_1086765, EPI_ISL_1086767, EPI_ISL_1086769, EPI_ISL_1086770, EPI_ISL_1086771, EPI_ISL_1086772, EPI_ISL_1086774, EPI_ISL_1086775, EPI_ISL_1086777, EPI_ISL_1086778, EPI_ISL_1086779, EPI_ISL_1086780, EPI_ISL_1086783, EPI_ISL_1086784, EPI_ISL_1086785, EPI_ISL_1086787, EPI_ISL_1086792, EPI_ISL_1086793, EPI_ISL_1086796, EPI_ISL_1086797, EPI_ISL_1086799, EPI_ISL_1086801, EPI_ISL_1086802, EPI_ISL_1086803, EPI_ISL_1086804, EPI_ISL_1086805, EPI_ISL_1086806, EPI_ISL_1086808, EPI_ISL_1086809, EPI_ISL_1086810, EPI_ISL_1086811, EPI_ISL_1086813, EPI_ISL_1086814, EPI_ISL_1086815, EPI_ISL_1086816, EPI_ISL_1086819, EPI_ISL_1086820, EPI_ISL_1086822, EPI_ISL_1091089 | | | |
| see above | Quest Diagnostics Incorporated | Respiratory Viruses Branch, Division of Viral Diseases, Centers for Disease Control and Prevention | A. Gerasimova; A. Perez; B. Anderson; Ben L. Rambo-Martin; Clinton R. Paden; Dakota Howard; Dhwani Batra; Duncan MacCannell; F. Lacbawan; I. A. Shlyakhter; K.E. Livingston; L.E. Bernstein; M. Hua; P. Tanpaiboon; Peter W. Cook; R. M. Kagan; R. Owen; R. V. Rolando; S. H. Rosenthal; Suxiang Tong; Y. Liu |
| EPI_ISL_1295796, EPI_ISL_1295797, EPI_ISL_1295801, EPI_ISL_1295805, EPI_ISL_1295806, EPI_ISL_1295808, EPI_ISL_1295810, EPI_ISL_1295814, EPI_ISL_1794771, EPI_ISL_1794774, EPI_ISL_1794775, EPI_ISL_1794780 | | | |
| see above | Rady Children's Hospital - San Diego | Andersen lab at Scripps Research | Christina Clarke; David Dimmock; Denise Malicki; Kathryn Bouic; Linda Luo; SEARCH Alliance San Diego with Nanda Radamchar; Teresa Mueller |
| EPI_ISL_878695, EPI_ISL_878734, EPI_ISL_878744, EPI_ISL_878750, EPI_ISL_878752, EPI_ISL_878771, EPI_ISL_878773, EPI_ISL_878779, EPI_ISL_878784, EPI_ISL_878789, EPI_ISL_878794, EPI_ISL_880171, EPI_ISL_880173, EPI_ISL_880176, EPI_ISL_880177, EPI_ISL_880178, EPI_ISL_880179, EPI_ISL_880182, EPI_ISL_880183, EPI_ISL_880192, EPI_ISL_880195, EPI_ISL_1081571, EPI_ISL_1081572, EPI_ISL_1081573, EPI_ISL_1081574, EPI_ISL_1081575, EPI_ISL_1081576, EPI_ISL_1081577, EPI_ISL_1081578, EPI_ISL_1081579, EPI_ISL_1081580, EPI_ISL_1081581, EPI_ISL_1081582, EPI_ISL_1081583, EPI_ISL_1081584, EPI_ISL_1081585, EPI_ISL_1081586, EPI_ISL_1081587, EPI_ISL_1081588, EPI_ISL_1081589, EPI_ISL_1081590, EPI_ISL_1081591, EPI_ISL_1081592, EPI_ISL_1081593, EPI_ISL_1081594, EPI_ISL_1081595, EPI_ISL_1081596, EPI_ISL_1081597 | | | |
| see above | Rady's Childrens Hospital | Andersen lab at Scripps Research | Christina Clarke; David Dimmock; Denise Malicki; Kathryn Bouic; Linda Luo; SEARCH Alliance San Diego with Nanda Radamchar; Teresa Mueller |
| EPI_ISL_884582, EPI_ISL_884584, EPI_ISL_884586, EPI_ISL_884590, EPI_ISL_884638, EPI_ISL_884641, EPI_ISL_884642, EPI_ISL_884643, EPI_ISL_884644, EPI_ISL_884645, EPI_ISL_884646, EPI_ISL_884647, EPI_ISL_884648, EPI_ISL_884650, EPI_ISL_884653, EPI_ISL_884656, EPI_ISL_884658, EPI_ISL_884663, EPI_ISL_884665, EPI_ISL_884690, EPI_ISL_884695, EPI_ISL_884704, EPI_ISL_884729, EPI_ISL_884730, EPI_ISL_884733, EPI_ISL_884735, EPI_ISL_884737, EPI_ISL_884739, EPI_ISL_884741, EPI_ISL_884742, EPI_ISL_884743, EPI_ISL_884746, EPI_ISL_884823, EPI_ISL_1233929 | | | |
| see above | Respiratory Viruses Branch, Centers for Disease Control and Prevention | Respiratory Viruses Branch, Centers for Disease Control and Prevention | Allen, E.; Antico, J.; Ascencio, A.; B.L.; Barret; Batra, D.; Becker, D.; Bolze, A.; C.R.; Cassens, T.; Cho, R.; Cirulli, E.; Cook; Febbo, P.; Galloway, S.; Gietzen, K.; Isaksson, M.; K.S.; Laurent, M.; Lee, W.; Levan, G.; Liu, J.; Lu, J.; MacCannell, D.; N.L.; Nguyen, J.; P.W.; Paden; Rambo-Martin; Ramirez, J.; Rivera-Garcia, C.; Sandoval, E.; Sickler, B.; Tolentino, M.; Tong, S.; Tran, C.; Wang, S.; Washington; White, S.; Wickline, S.; de Feo, E. |
| EPI_ISL_862722, EPI_ISL_862723, EPI_ISL_878622, EPI_ISL_878627, EPI_ISL_962611, EPI_ISL_962613, EPI_ISL_1081459, EPI_ISL_1081460, EPI_ISL_1081461, EPI_ISL_1081462, EPI_ISL_1081463, EPI_ISL_1081464, EPI_ISL_1081465, EPI_ISL_1081466, EPI_ISL_1081467, EPI_ISL_1081468, EPI_ISL_1081469, EPI_ISL_1081470, EPI_ISL_1081471, EPI_ISL_1081472, EPI_ISL_1081619, EPI_ISL_1081620, EPI_ISL_1081621, EPI_ISL_1081622, EPI_ISL_1081623, EPI_ISL_1081624, EPI_ISL_1081625, EPI_ISL_1081626, EPI_ISL_1081627, EPI_ISL_1081742, EPI_ISL_1081743, EPI_ISL_1081744, EPI_ISL_1081745, EPI_ISL_1081746, EPI_ISL_1081747, EPI_ISL_1081748, EPI_ISL_1081749, EPI_ISL_1081750, EPI_ISL_1081751, EPI_ISL_1081752, EPI_ISL_1081753, EPI_ISL_1081754, EPI_ISL_1081755, EPI_ISL_1081756, EPI_ISL_1081757, EPI_ISL_1081758, EPI_ISL_1081759, EPI_ISL_1081760, EPI_ISL_1081761, EPI_ISL_1081762, EPI_ISL_1081763, EPI_ISL_1081764, EPI_ISL_1081765, EPI_ISL_1081766, EPI_ISL_1081767, EPI_ISL_1081768, EPI_ISL_1081769, EPI_ISL_1081770, EPI_ISL_1081771, EPI_ISL_1081772, EPI_ISL_1081773, EPI_ISL_1081774, EPI_ISL_1081775, EPI_ISL_1081776, EPI_ISL_1081777, EPI_ISL_1081778, EPI_ISL_1081779, EPI_ISL_1081780, EPI_ISL_1081781, EPI_ISL_1081782, EPI_ISL_1081783, EPI_ISL_1081784, EPI_ISL_1081785, EPI_ISL_1081786, EPI_ISL_1081787, EPI_ISL_1081788, EPI_ISL_1081789, EPI_ISL_1081790, EPI_ISL_1081791, EPI_ISL_1366377, EPI_ISL_1366378, EPI_ISL_1366748, EPI_ISL_1468837, EPI_ISL_1468959, EPI_ISL_1794621, EPI_ISL_1794668, EPI_ISL_1794670, EPI_ISL_1794674, EPI_ISL_1794675, EPI_ISL_1794676, EPI_ISL_1794679, EPI_ISL_1794681, EPI_ISL_1794983, EPI_ISL_1794984, EPI_ISL_1794985, EPI_ISL_1794986, EPI_ISL_1794988, EPI_ISL_1794989, EPI_ISL_1794990, EPI_ISL_1794991, EPI_ISL_1794992, EPI_ISL_1794993, EPI_ISL_1794994, EPI_ISL_1794995, EPI_ISL_1794998, EPI_ISL_1794999, EPI_ISL_1795000, EPI_ISL_1795003, EPI_ISL_1795004, EPI_ISL_1795005, EPI_ISL_1795008, EPI_ISL_1795009, EPI_ISL_1795010, EPI_ISL_1795013, EPI_ISL_1795014, EPI_ISL_1795015, EPI_ISL_1795016, EPI_ISL_1795018, EPI_ISL_1795019, EPI_ISL_1795020, EPI_ISL_1795021, EPI_ISL_1795024, EPI_ISL_1795027, EPI_ISL_1795028, EPI_ISL_1795029, EPI_ISL_1795030, EPI_ISL_1795031, EPI_ISL_1795035, EPI_ISL_1795036, EPI_ISL_1795037, EPI_ISL_1795039, EPI_ISL_1795040, EPI_ISL_1795041, EPI_ISL_1795042, EPI_ISL_1969324, EPI_ISL_1969325, EPI_ISL_1969326 | | | |
| see above | San Diego County Public Health Laboratory | Andersen lab at Scripps Research | Brett Austin; Jovan Shephard; SEARCH Alliance San Diego with Tracy Basler |
| EPI_ISL_1701040, EPI_ISL_1701069, EPI_ISL_1701117, EPI_ISL_1701208, EPI_ISL_1701280 | San Joaquin County Public Health Lab | Chan-Zuckerberg Biohub | CZB Cliahub Consortium |
| EPI_ISL_884983, EPI_ISL_884984, EPI_ISL_884985, EPI_ISL_884986, EPI_ISL_884987, EPI_ISL_884988, EPI_ISL_884989, EPI_ISL_884990, EPI_ISL_884991, EPI_ISL_884992, EPI_ISL_884993, EPI_ISL_884994, EPI_ISL_885013, EPI_ISL_885014, EPI_ISL_885015, EPI_ISL_885016, EPI_ISL_885017, EPI_ISL_885018, EPI_ISL_885019, EPI_ISL_885020, EPI_ISL_885021, EPI_ISL_885022, EPI_ISL_885023, EPI_ISL_885024, EPI_ISL_885025, EPI_ISL_885026, EPI_ISL_885027, EPI_ISL_885028, EPI_ISL_885029, EPI_ISL_885030, EPI_ISL_885031, EPI_ISL_885032, EPI_ISL_885033, EPI_ISL_885034, EPI_ISL_885035, EPI_ISL_885036, EPI_ISL_885037, EPI_ISL_885038, EPI_ISL_885039, EPI_ISL_885040, EPI_ISL_885041, EPI_ISL_885042, EPI_ISL_885043, EPI_ISL_885044, EPI_ISL_885045, EPI_ISL_885046, EPI_ISL_885047, EPI_ISL_885048, EPI_ISL_885049, EPI_ISL_885050, EPI_ISL_979056, EPI_ISL_979057, EPI_ISL_979058, EPI_ISL_979059, EPI_ISL_979060, EPI_ISL_979061, EPI_ISL_979062, EPI_ISL_979063, EPI_ISL_979064, EPI_ISL_979065, EPI_ISL_979066, EPI_ISL_979067, EPI_ISL_979068, EPI_ISL_979069, EPI_ISL_979070, EPI_ISL_979071, EPI_ISL_979072, EPI_ISL_979073, EPI_ISL_979074, EPI_ISL_979075, EPI_ISL_979076, EPI_ISL_979077, EPI_ISL_979078, EPI_ISL_979079, EPI_ISL_979080, EPI_ISL_979081, EPI_ISL_979082, EPI_ISL_979083, EPI_ISL_979084, EPI_ISL_979085, EPI_ISL_979086, EPI_ISL_979087, EPI_ISL_979088, EPI_ISL_979089, EPI_ISL_979090, EPI_ISL_979091, EPI_ISL_979092, EPI_ISL_979093, EPI_ISL_979094, EPI_ISL_979095, EPI_ISL_979096, EPI_ISL_979097, EPI_ISL_979098, EPI_ISL_979099, EPI_ISL_979100, EPI_ISL_979101, EPI_ISL_979102, EPI_ISL_979103, EPI_ISL_979104, EPI_ISL_979105, EPI_ISL_979106, EPI_ISL_979107, EPI_ISL_979108, EPI_ISL_979109, EPI_ISL_979110, EPI_ISL_979111, EPI_ISL_979112, EPI_ISL_979113, EPI_ISL_979114, EPI_ISL_979115, EPI_ISL_979116, EPI_ISL_979117, EPI_ISL_979118, EPI_ISL_979119, EPI_ISL_979120, EPI_ISL_979121, EPI_ISL_979122, EPI_ISL_979123, EPI_ISL_979124, EPI_ISL_979125, EPI_ISL_979126, EPI_ISL_979127, EPI_ISL_979128, EPI_ISL_979129, EPI_ISL_979130, EPI_ISL_979131, EPI_ISL_979132, EPI_ISL_979133, EPI_ISL_979134, EPI_ISL_979135, EPI_ISL_979136, EPI_ISL_979137, EPI_ISL_979138, EPI_ISL_979139, EPI_ISL_979140, EPI_ISL_979141, EPI_ISL_979142, EPI_ISL_979143, EPI_ISL_979144, EPI_ISL_979596, EPI_ISL_979597, EPI_ISL_979599, EPI_ISL_979600, EPI_ISL_979601, EPI_ISL_979602, EPI_ISL_979603, EPI_ISL_979604, EPI_ISL_979605, EPI_ISL_979623, EPI_ISL_979624, EPI_ISL_979625, EPI_ISL_979626, EPI_ISL_979627, EPI_ISL_979628, EPI_ISL_979629, EPI_ISL_979630, EPI_ISL_979631, EPI_ISL_979632, EPI_ISL_979633, EPI_ISL_979634, EPI_ISL_979635, EPI_ISL_979636, EPI_ISL_979637, EPI_ISL_979638, EPI_ISL_979639, EPI_ISL_979640, EPI_ISL_979641, EPI_ISL_979642, EPI_ISL_1027295, EPI_ISL_1027297, EPI_ISL_1027298, EPI_ISL_1027299, EPI_ISL_1027300, EPI_ISL_1027301, EPI_ISL_1027302, EPI_ISL_1027304, EPI_ISL_1027305, EPI_ISL_1027310, EPI_ISL_1027311, EPI_ISL_1027313, EPI_ISL_1027316, EPI_ISL_1027318, EPI_ISL_1027322, EPI_ISL_1027328, EPI_ISL_1027334, EPI_ISL_1027346, EPI_ISL_1027349, EPI_ISL_1027352, EPI_ISL_1027353, EPI_ISL_1027354, EPI_ISL_1027356, EPI_ISL_1027358, EPI_ISL_1027360, EPI_ISL_1027368, EPI_ISL_1027369, EPI_ISL_1027370, EPI_ISL_1027371, EPI_ISL_1027372, EPI_ISL_1027373, EPI_ISL_1027374, EPI_ISL_1027375, EPI_ISL_1027376, EPI_ISL_1027377, EPI_ISL_1027475, EPI_ISL_1027480, EPI_ISL_1027482, EPI_ISL_1027484, EPI_ISL_1027486, EPI_ISL_1027489, EPI_ISL_1027490, EPI_ISL_1027491, EPI_ISL_1027494, EPI_ISL_1027496, EPI_ISL_1027498, EPI_ISL_1027499, EPI_ISL_1027501, EPI_ISL_1027502, EPI_ISL_1027505, EPI_ISL_1027506, EPI_ISL_1027513, EPI_ISL_1027515, EPI_ISL_1027518, EPI_ISL_1027520, EPI_ISL_1027521, EPI_ISL_1027523, EPI_ISL_1027525, EPI_ISL_1027526, EPI_ISL_1027528, EPI_ISL_1027562, EPI_ISL_1027563, EPI_ISL_1027564, EPI_ISL_1027565, EPI_ISL_1027566, EPI_ISL_1027567, EPI_ISL_1027569, EPI_ISL_1027570, EPI_ISL_1027571, EPI_ISL_1027572, EPI_ISL_1027574, EPI_ISL_1027575, EPI_ISL_1027576, EPI_ISL_1027577, EPI_ISL_1027579, EPI_ISL_1027580, EPI_ISL_1027581, EPI_ISL_1027585, EPI_ISL_1027586, EPI_ISL_1030262, EPI_ISL_1030267, EPI_ISL_1030272, EPI_ISL_1030276, EPI_ISL_1030283, EPI_ISL_1030286, EPI_ISL_1030330, EPI_ISL_1030391, EPI_ISL_1030392, EPI_ISL_1030393, EPI_ISL_1030394, EPI_ISL_1030395, EPI_ISL_1030396, EPI_ISL_1030397, EPI_ISL_1185029, EPI_ISL_1185044, EPI_ISL_1185046, EPI_ISL_1185051, EPI_ISL_1185053, EPI_ISL_1185065, EPI_ISL_1185072, EPI_ISL_1185074, EPI_ISL_1185081, EPI_ISL_1185083, EPI_ISL_1185086, EPI_ISL_1185091, EPI_ISL_1185092, EPI_ISL_1185097, EPI_ISL_1185110, EPI_ISL_1185111, EPI_ISL_1185116, EPI_ISL_1185120, EPI_ISL_1185129, EPI_ISL_1185137, EPI_ISL_1185141, EPI_ISL_1185151, EPI_ISL_1185170, EPI_ISL_1185181, EPI_ISL_1185183, EPI_ISL_1185185, EPI_ISL_1185191, EPI_ISL_1185197, EPI_ISL_1185200, EPI_ISL_1185218, EPI_ISL_1185219, EPI_ISL_1185234, EPI_ISL_1185239, EPI_ISL_1185251, EPI_ISL_1185252, EPI_ISL_1185255, EPI_ISL_1185267, EPI_ISL_1185273, EPI_ISL_1185277, EPI_ISL_1185284, EPI_ISL_1234537, EPI_ISL_1234539, EPI_ISL_1234547, EPI_ISL_1234548, EPI_ISL_1234549, EPI_ISL_1234560, EPI_ISL_1234586, EPI_ISL_1234588, EPI_ISL_1234606, EPI_ISL_1234609, EPI_ISL_1234613, EPI_ISL_1234645, EPI_ISL_1234653, EPI_ISL_1234660, EPI_ISL_1234667, EPI_ISL_1234671, EPI_ISL_1234678, EPI_ISL_1234689, EPI_ISL_1234721, EPI_ISL_1234724, EPI_ISL_1234725, EPI_ISL_1234728, EPI_ISL_1234729, EPI_ISL_1234744, EPI_ISL_1234759, EPI_ISL_1234760, EPI_ISL_1235018, EPI_ISL_1477961, EPI_ISL_1477963, EPI_ISL_1477983, EPI_ISL_1478013, EPI_ISL_1478038, EPI_ISL_1478084, EPI_ISL_1478092, EPI_ISL_1478129, EPI_ISL_1478153, EPI_ISL_1478170, EPI_ISL_1701405, EPI_ISL_1701406, EPI_ISL_1701410, EPI_ISL_1701414, EPI_ISL_1701467, EPI_ISL_1701469, EPI_ISL_1701482, EPI_ISL_1701502, EPI_ISL_1701504, EPI_ISL_1701505, EPI_ISL_1701517, EPI_ISL_1701520, EPI_ISL_1701525, EPI_ISL_1701530, EPI_ISL_1701536, EPI_ISL_1701541, EPI_ISL_1701542, EPI_ISL_1701559, EPI_ISL_1701574, EPI_ISL_1701579, EPI_ISL_1701581, EPI_ISL_1701602, EPI_ISL_1701628, EPI_ISL_1701636, EPI_ISL_1701639, EPI_ISL_1701654, EPI_ISL_1701656 | | | |
| see above | Santa Clara County Public Health Laboratory | Chan-Zuckerberg Biohub | CZB Cliahub Consortium |
| EPI_ISL_832079, EPI_ISL_832080, EPI_ISL_832081, EPI_ISL_832090, EPI_ISL_832095, EPI_ISL_832097, EPI_ISL_845544, EPI_ISL_884297, EPI_ISL_884298, EPI_ISL_884299, EPI_ISL_884300, EPI_ISL_884301, EPI_ISL_884302, EPI_ISL_902730, EPI_ISL_902731, EPI_ISL_902732, EPI_ISL_902733, EPI_ISL_902734, EPI_ISL_914638, EPI_ISL_914639, EPI_ISL_914640, EPI_ISL_914641, EPI_ISL_914642, EPI_ISL_914643, EPI_ISL_914644, EPI_ISL_936493, EPI_ISL_1001011, EPI_ISL_1001012, EPI_ISL_1001023, EPI_ISL_1060545, EPI_ISL_1060547, EPI_ISL_1060549, EPI_ISL_1060554, EPI_ISL_1060555, EPI_ISL_1060556, EPI_ISL_1137631, EPI_ISL_1137632, EPI_ISL_1137633, EPI_ISL_1137634, EPI_ISL_1137635, EPI_ISL_1137636, EPI_ISL_1137638, EPI_ISL_1137639, EPI_ISL_1137640, EPI_ISL_1137641, EPI_ISL_1137642, EPI_ISL_1137643, EPI_ISL_1137644, EPI_ISL_1137645, EPI_ISL_1137648, EPI_ISL_1169579, EPI_ISL_1169580, EPI_ISL_1169581, EPI_ISL_1169582, EPI_ISL_1169583, EPI_ISL_1169584, EPI_ISL_1169585, EPI_ISL_1169586, EPI_ISL_1169587, EPI_ISL_1169588, EPI_ISL_1169589, EPI_ISL_1169592, EPI_ISL_1169593, EPI_ISL_1169594, EPI_ISL_1169595, EPI_ISL_1169596, EPI_ISL_1169597, EPI_ISL_1169598, EPI_ISL_1169599, EPI_ISL_1222758, EPI_ISL_1235639, EPI_ISL_1235641, EPI_ISL_1335836, EPI_ISL_1335839, EPI_ISL_1335841, EPI_ISL_1335842, EPI_ISL_1335843, EPI_ISL_1335844, EPI_ISL_1335845, EPI_ISL_1335846, EPI_ISL_1335847, EPI_ISL_1335848, EPI_ISL_1335849, EPI_ISL_1335850, EPI_ISL_1335851, EPI_ISL_1335852, EPI_ISL_1335859, EPI_ISL_1335860, EPI_ISL_1406156, EPI_ISL_1406157, EPI_ISL_1406158, EPI_ISL_1406159, EPI_ISL_1406160, EPI_ISL_1406164, EPI_ISL_1406167, EPI_ISL_1406171, EPI_ISL_1406172, EPI_ISL_1406173, EPI_ISL_1406174, EPI_ISL_1406175, EPI_ISL_1406176, EPI_ISL_1920836, EPI_ISL_1920843, EPI_ISL_1920847, EPI_ISL_1920875, EPI_ISL_1920883 | | | |
| see above | Santa Clara County Public Health Laboratory | Santa Clara County Public Health Laboratory | Santa Clara County Public Health Department |
| EPI_ISL_880201, EPI_ISL_880202, EPI_ISL_880203, EPI_ISL_880204, EPI_ISL_880205, EPI_ISL_880206, EPI_ISL_880207, EPI_ISL_880208, EPI_ISL_880209, EPI_ISL_880212, EPI_ISL_880215, EPI_ISL_880218, EPI_ISL_880220, EPI_ISL_880223, EPI_ISL_880225, EPI_ISL_880228, EPI_ISL_880231, EPI_ISL_880233, EPI_ISL_880236, EPI_ISL_880239, EPI_ISL_880241, EPI_ISL_880244, EPI_ISL_880246, EPI_ISL_880249, EPI_ISL_880251, EPI_ISL_880253, EPI_ISL_880256, EPI_ISL_880259, EPI_ISL_880261, EPI_ISL_880263, EPI_ISL_880266, EPI_ISL_880268, EPI_ISL_880271, EPI_ISL_880274, EPI_ISL_880277, EPI_ISL_880279, EPI_ISL_880282, EPI_ISL_880284, EPI_ISL_880287, EPI_ISL_880290, EPI_ISL_880292, EPI_ISL_880294, EPI_ISL_880297, EPI_ISL_880300, EPI_ISL_880303, EPI_ISL_880305, EPI_ISL_880308, EPI_ISL_880310, EPI_ISL_880313, EPI_ISL_880316, EPI_ISL_880319, EPI_ISL_880321, EPI_ISL_880324, EPI_ISL_880326, EPI_ISL_880329, EPI_ISL_962585, EPI_ISL_962586, EPI_ISL_962587, EPI_ISL_962588, EPI_ISL_962589, EPI_ISL_962590, EPI_ISL_962591, EPI_ISL_962592, EPI_ISL_962593, EPI_ISL_962594, EPI_ISL_962595, EPI_ISL_962596, EPI_ISL_962597, EPI_ISL_962598, EPI_ISL_962599, EPI_ISL_962600, EPI_ISL_962601, EPI_ISL_962618, EPI_ISL_962619, EPI_ISL_962620, EPI_ISL_962621, EPI_ISL_962622, EPI_ISL_962623, EPI_ISL_962624, EPI_ISL_962625, EPI_ISL_962626, EPI_ISL_962627, EPI_ISL_962628, EPI_ISL_962629, EPI_ISL_962630, EPI_ISL_962631, EPI_ISL_962632, EPI_ISL_962633, EPI_ISL_962634, EPI_ISL_962635, EPI_ISL_962636, EPI_ISL_962637, EPI_ISL_962638, EPI_ISL_962639, EPI_ISL_962640, EPI_ISL_962641, EPI_ISL_962642, EPI_ISL_962643, EPI_ISL_962644, EPI_ISL_962645, EPI_ISL_962646, EPI_ISL_962647, EPI_ISL_962648, EPI_ISL_962649, EPI_ISL_1081551, EPI_ISL_1081552, EPI_ISL_1081553, EPI_ISL_1081554, EPI_ISL_1081555, EPI_ISL_1081557, EPI_ISL_1081558, EPI_ISL_1081559, EPI_ISL_1081560, EPI_ISL_1081561, EPI_ISL_1081562, EPI_ISL_1081563, EPI_ISL_1081564, EPI_ISL_1081565, EPI_ISL_1081566, EPI_ISL_1081567, EPI_ISL_1081598, EPI_ISL_1081599, EPI_ISL_1081600, EPI_ISL_1081601, EPI_ISL_1081602, EPI_ISL_1081603, EPI_ISL_1081604, EPI_ISL_1081605, EPI_ISL_1081606, EPI_ISL_1081607, EPI_ISL_1081608, EPI_ISL_1081609, EPI_ISL_1081610, EPI_ISL_1081611, EPI_ISL_1081612, EPI_ISL_1081613, EPI_ISL_1081614, EPI_ISL_1081615, EPI_ISL_1081616, EPI_ISL_1081617, EPI_ISL_1081618, EPI_ISL_1081628, EPI_ISL_1081629, EPI_ISL_1081630, EPI_ISL_1081631, EPI_ISL_1081632, EPI_ISL_1081633, EPI_ISL_1081634, EPI_ISL_1081635, EPI_ISL_1081636, EPI_ISL_1081637, EPI_ISL_1081638, EPI_ISL_1081639, EPI_ISL_1081640, EPI_ISL_1081641, EPI_ISL_1081642, EPI_ISL_1081643, EPI_ISL_1081644, EPI_ISL_1081645, EPI_ISL_1081646, EPI_ISL_1081647, EPI_ISL_1081648, EPI_ISL_1081649, EPI_ISL_1081650, EPI_ISL_1081651, EPI_ISL_1081652, EPI_ISL_1081653, EPI_ISL_1081654, EPI_ISL_1081655, EPI_ISL_1081656, EPI_ISL_1081657, EPI_ISL_1081658, EPI_ISL_1081659, EPI_ISL_1081660, EPI_ISL_1081661, EPI_ISL_1081662, EPI_ISL_1081663, EPI_ISL_1081664, EPI_ISL_1081665, EPI_ISL_1081666, EPI_ISL_1081667, EPI_ISL_1081668, EPI_ISL_1081669, EPI_ISL_1081670, EPI_ISL_1081671, EPI_ISL_1081672, EPI_ISL_1081673, EPI_ISL_1081674, EPI_ISL_1081675, EPI_ISL_1081676, EPI_ISL_1081677, EPI_ISL_1081678, EPI_ISL_1081679, EPI_ISL_1081680, EPI_ISL_1081681, EPI_ISL_1081682, EPI_ISL_1081683, EPI_ISL_1081684, EPI_ISL_1081685, EPI_ISL_1081686, EPI_ISL_1081687, EPI_ISL_1081688, EPI_ISL_1081689, EPI_ISL_1081690, EPI_ISL_1081691, EPI_ISL_1081692, EPI_ISL_1081693, EPI_ISL_1081694, EPI_ISL_1081695, EPI_ISL_1081696, EPI_ISL_1081697, EPI_ISL_1081698, EPI_ISL_1081699, EPI_ISL_1081700, EPI_ISL_1081701, EPI_ISL_1081702, EPI_ISL_1081703, EPI_ISL_1081704, EPI_ISL_1081705, EPI_ISL_1081706, EPI_ISL_1081707, EPI_ISL_1081708, EPI_ISL_1081709, EPI_ISL_1081710, EPI_ISL_1081711, EPI_ISL_1081712, EPI_ISL_1081713, EPI_ISL_1081714, EPI_ISL_1081715, EPI_ISL_1081716, EPI_ISL_1081717, EPI_ISL_1081718, EPI_ISL_1081719, EPI_ISL_1081720, EPI_ISL_1081721, EPI_ISL_1081722, EPI_ISL_1081723, EPI_ISL_1081724, EPI_ISL_1081725, EPI_ISL_1081726, EPI_ISL_1081727, EPI_ISL_1081728, EPI_ISL_1081729, EPI_ISL_1081730, EPI_ISL_1081731, EPI_ISL_1081732, EPI_ISL_1081733, EPI_ISL_1081734, EPI_ISL_1081735, EPI_ISL_1081736, EPI_ISL_1081737, EPI_ISL_1081738, EPI_ISL_1081739, EPI_ISL_1081740, EPI_ISL_1081741, EPI_ISL_1185854, EPI_ISL_1185855, EPI_ISL_1185856, EPI_ISL_1185857, EPI_ISL_1185858, EPI_ISL_1185859, EPI_ISL_1185860, EPI_ISL_1185861, EPI_ISL_1185862, EPI_ISL_1185863, EPI_ISL_1185864, EPI_ISL_1185865, EPI_ISL_1185866, EPI_ISL_1185867, EPI_ISL_1185868, EPI_ISL_1185869, EPI_ISL_1185870, EPI_ISL_1185871, EPI_ISL_1185872, EPI_ISL_1185873, EPI_ISL_1185874, EPI_ISL_1185875, EPI_ISL_1185876, EPI_ISL_1185877, EPI_ISL_1185878, EPI_ISL_1185879, EPI_ISL_1185880, EPI_ISL_1185881, EPI_ISL_1185882, EPI_ISL_1185883, EPI_ISL_1185884, EPI_ISL_1185885, EPI_ISL_1185886, EPI_ISL_1185887, EPI_ISL_1185888, EPI_ISL_1185889, EPI_ISL_1185890, EPI_ISL_1469043, EPI_ISL_1469044 | | | |
| see above | Scripps Medical Laboratory | Andersen lab at Scripps Research | Ellen Stefanski; Ian Mchardy; SEARCH Alliance San Diego with Michael Quigley |
| EPI_ISL_880037, EPI_ISL_880040, EPI_ISL_880042, EPI_ISL_880045, EPI_ISL_880048, EPI_ISL_880050, EPI_ISL_880053, EPI_ISL_880056, EPI_ISL_880058, EPI_ISL_880061, EPI_ISL_880063, EPI_ISL_880066, EPI_ISL_880069, EPI_ISL_880071, EPI_ISL_880074, EPI_ISL_880077, EPI_ISL_880079, EPI_ISL_880082, EPI_ISL_880085, EPI_ISL_880087, EPI_ISL_880090, EPI_ISL_880092, EPI_ISL_880095, EPI_ISL_880098, EPI_ISL_880101, EPI_ISL_880103, EPI_ISL_880106, EPI_ISL_880109, EPI_ISL_880111, EPI_ISL_880114, EPI_ISL_880117, EPI_ISL_880119, EPI_ISL_880122, EPI_ISL_880125, EPI_ISL_880128, EPI_ISL_880130, EPI_ISL_880133, EPI_ISL_880135, EPI_ISL_880138, EPI_ISL_880140, EPI_ISL_880141, EPI_ISL_880142, EPI_ISL_880143, EPI_ISL_880144, EPI_ISL_880145, EPI_ISL_880146, EPI_ISL_880147, EPI_ISL_880148, EPI_ISL_880149, EPI_ISL_880150, EPI_ISL_880151, EPI_ISL_880152, EPI_ISL_880153, EPI_ISL_880154, EPI_ISL_880155, EPI_ISL_880156, EPI_ISL_880157, EPI_ISL_880158, EPI_ISL_880159, EPI_ISL_880160, EPI_ISL_880161, EPI_ISL_880162, EPI_ISL_880163, EPI_ISL_880164, EPI_ISL_880165, EPI_ISL_880166, EPI_ISL_880167, EPI_ISL_880168, EPI_ISL_880169, EPI_ISL_880170, EPI_ISL_962602, EPI_ISL_962603, EPI_ISL_962604, EPI_ISL_962605, EPI_ISL_962606, EPI_ISL_962607, EPI_ISL_962608, EPI_ISL_962650, EPI_ISL_962651, EPI_ISL_962652, EPI_ISL_962653, EPI_ISL_962654, EPI_ISL_962655, EPI_ISL_962656, EPI_ISL_962657, EPI_ISL_962658, EPI_ISL_962659, EPI_ISL_962660, EPI_ISL_962661, EPI_ISL_962662, EPI_ISL_962663, EPI_ISL_962664, EPI_ISL_962665, EPI_ISL_962666, EPI_ISL_962667, EPI_ISL_962668, EPI_ISL_962669, EPI_ISL_962670, EPI_ISL_962671, EPI_ISL_962672, EPI_ISL_962673, EPI_ISL_962674, EPI_ISL_962675, EPI_ISL_962676, EPI_ISL_962677, EPI_ISL_962678, EPI_ISL_962679, EPI_ISL_962680, EPI_ISL_962681, EPI_ISL_962682, EPI_ISL_962683, EPI_ISL_962684, EPI_ISL_962685, EPI_ISL_962686, EPI_ISL_962687, EPI_ISL_962688, EPI_ISL_962689, EPI_ISL_962690, EPI_ISL_962691, EPI_ISL_962692, EPI_ISL_962693, EPI_ISL_962694, EPI_ISL_962695, EPI_ISL_962696, EPI_ISL_962697, EPI_ISL_962698, EPI_ISL_962699, EPI_ISL_962700, EPI_ISL_962701, EPI_ISL_962702, EPI_ISL_962703, EPI_ISL_962704, EPI_ISL_962705, EPI_ISL_962706, EPI_ISL_962707, EPI_ISL_962708, EPI_ISL_962709, EPI_ISL_962710, EPI_ISL_962711, EPI_ISL_962712, EPI_ISL_962713, EPI_ISL_962714, EPI_ISL_962715, EPI_ISL_962716, EPI_ISL_962717, EPI_ISL_962718, EPI_ISL_962719, EPI_ISL_962720, EPI_ISL_962721, EPI_ISL_962722, EPI_ISL_962723, EPI_ISL_962724, EPI_ISL_962725, EPI_ISL_962726, EPI_ISL_962727, EPI_ISL_962728, EPI_ISL_962729, EPI_ISL_962730, EPI_ISL_962731, EPI_ISL_962732, EPI_ISL_962733, EPI_ISL_962734, EPI_ISL_962735, EPI_ISL_962736, EPI_ISL_962737, EPI_ISL_962738, EPI_ISL_962739, EPI_ISL_962740, EPI_ISL_962741, EPI_ISL_962742, EPI_ISL_962743, EPI_ISL_962744, EPI_ISL_962745, EPI_ISL_962746, EPI_ISL_962747, EPI_ISL_962748, EPI_ISL_962749, EPI_ISL_962750, EPI_ISL_962751, EPI_ISL_962752, EPI_ISL_962753, EPI_ISL_962754, EPI_ISL_962755, EPI_ISL_962756, EPI_ISL_962757, EPI_ISL_962758, EPI_ISL_962759, EPI_ISL_962760, EPI_ISL_962761, EPI_ISL_962762, EPI_ISL_962763, EPI_ISL_962764, EPI_ISL_962765, EPI_ISL_1081373, EPI_ISL_1081374, EPI_ISL_1081375, EPI_ISL_1081376, EPI_ISL_1081377, EPI_ISL_1081378, EPI_ISL_1081379, EPI_ISL_1081380, EPI_ISL_1081381, EPI_ISL_1081382, EPI_ISL_1081383, EPI_ISL_1081384, EPI_ISL_1081385, EPI_ISL_1081386, EPI_ISL_1081387, EPI_ISL_1081388, EPI_ISL_1081389, EPI_ISL_1081390, EPI_ISL_1081391, EPI_ISL_1081392, EPI_ISL_1081393, EPI_ISL_1081394, EPI_ISL_1081395, EPI_ISL_1081396, EPI_ISL_1081397, EPI_ISL_1081398, EPI_ISL_1081399, EPI_ISL_1081400, EPI_ISL_1081401, EPI_ISL_1081402, EPI_ISL_1081403, EPI_ISL_1081404, EPI_ISL_1081405, EPI_ISL_1081406, EPI_ISL_1081407, EPI_ISL_1081408, EPI_ISL_1081409, EPI_ISL_1081410, EPI_ISL_1081411, EPI_ISL_1081412, EPI_ISL_1081413, EPI_ISL_1081414, EPI_ISL_1081415, EPI_ISL_1081416, EPI_ISL_1081417, EPI_ISL_1081418, EPI_ISL_1081419, EPI_ISL_1081420, EPI_ISL_1081421, EPI_ISL_1081422, EPI_ISL_1081496, EPI_ISL_1081497, EPI_ISL_1081498, EPI_ISL_1081499, EPI_ISL_1081500, EPI_ISL_1081501, EPI_ISL_1081502, EPI_ISL_1081503, EPI_ISL_1081504, EPI_ISL_1081505, EPI_ISL_1081506, EPI_ISL_1081507, EPI_ISL_1081508, EPI_ISL_1081509, EPI_ISL_1081510, EPI_ISL_1081511, EPI_ISL_1081512, EPI_ISL_1081513, EPI_ISL_1081514, EPI_ISL_1081515, EPI_ISL_1081516, EPI_ISL_1081517, EPI_ISL_1081518, EPI_ISL_1081519, EPI_ISL_1081520, EPI_ISL_1081521, EPI_ISL_1081522, EPI_ISL_1081523, EPI_ISL_1081524, EPI_ISL_1081525, EPI_ISL_1081526, EPI_ISL_1081527, EPI_ISL_1081528, EPI_ISL_1081529, EPI_ISL_1081530, EPI_ISL_1081531, EPI_ISL_1081532, EPI_ISL_1081533, EPI_ISL_1081534, EPI_ISL_1081535, EPI_ISL_1081536, EPI_ISL_1081537, EPI_ISL_1081538, EPI_ISL_1081539, EPI_ISL_1081540, EPI_ISL_1081541, EPI_ISL_1081542, EPI_ISL_1081543, EPI_ISL_1081544, EPI_ISL_1081545, EPI_ISL_1081546, EPI_ISL_1081547, EPI_ISL_1081548, EPI_ISL_1081549, EPI_ISL_1081568, EPI_ISL_1081569, EPI_ISL_1081570, EPI_ISL_1185891, EPI_ISL_1185892, EPI_ISL_1185893, EPI_ISL_1185894, EPI_ISL_1185895, EPI_ISL_1185896, EPI_ISL_1185897, EPI_ISL_1185898, EPI_ISL_1185899, EPI_ISL_1185900, EPI_ISL_1185901, EPI_ISL_1185902, EPI_ISL_1185903, EPI_ISL_1185904, EPI_ISL_1185905, EPI_ISL_1185906, EPI_ISL_1185907, EPI_ISL_1185908, EPI_ISL_1185909, EPI_ISL_1185910, EPI_ISL_1185911, EPI_ISL_1185912, EPI_ISL_1185913, EPI_ISL_1185914, EPI_ISL_1185915, EPI_ISL_1185916, EPI_ISL_1185917, EPI_ISL_1185918, EPI_ISL_1185919, EPI_ISL_1185920, EPI_ISL_1185921, EPI_ISL_1185922, EPI_ISL_1185923, EPI_ISL_1185924, EPI_ISL_1185925, EPI_ISL_1185926, EPI_ISL_1185927, EPI_ISL_1185928, EPI_ISL_1185929, EPI_ISL_1185930, EPI_ISL_1185931, EPI_ISL_1185932, EPI_ISL_1185933, EPI_ISL_1185934, EPI_ISL_1185935, EPI_ISL_1185936, EPI_ISL_1185937, EPI_ISL_1185938, EPI_ISL_1185939, EPI_ISL_1185940, EPI_ISL_1185941, EPI_ISL_1185942, EPI_ISL_1185943, EPI_ISL_1185944, EPI_ISL_1185945, EPI_ISL_1185946, EPI_ISL_1185947, EPI_ISL_1185948, EPI_ISL_1185949, EPI_ISL_1185950, EPI_ISL_1185951, EPI_ISL_1185952, EPI_ISL_1185953, EPI_ISL_1185954, EPI_ISL_1185955, EPI_ISL_1185956, EPI_ISL_1185957, EPI_ISL_1185958, EPI_ISL_1185959, EPI_ISL_1366280, EPI_ISL_1366293, EPI_ISL_1366294, EPI_ISL_1366295, EPI_ISL_1366296, EPI_ISL_1366316, EPI_ISL_1366324, EPI_ISL_1366335, EPI_ISL_1366336, EPI_ISL_1366337, EPI_ISL_1366338, EPI_ISL_1366339, EPI_ISL_1366340, EPI_ISL_1366341, EPI_ISL_1366342, EPI_ISL_1366343, EPI_ISL_1366344, EPI_ISL_1366345, EPI_ISL_1366346, EPI_ISL_1366347, EPI_ISL_1366348, EPI_ISL_1366349, EPI_ISL_1366350, EPI_ISL_1366351, EPI_ISL_1366352, EPI_ISL_1366353, EPI_ISL_1366354, EPI_ISL_1366355, EPI_ISL_1366356, EPI_ISL_1366357, EPI_ISL_1366358, EPI_ISL_1366359, EPI_ISL_1366360, EPI_ISL_1366361, EPI_ISL_1366362, EPI_ISL_1366363, EPI_ISL_1366364, EPI_ISL_1366365, EPI_ISL_1366366, EPI_ISL_1366367, EPI_ISL_1366368, EPI_ISL_1366369, EPI_ISL_1366370, EPI_ISL_1366371, EPI_ISL_1366372, EPI_ISL_1366373, EPI_ISL_1366374, EPI_ISL_1366375, EPI_ISL_1794854, EPI_ISL_1794855, EPI_ISL_1794856, EPI_ISL_1794857, EPI_ISL_1794858, EPI_ISL_1794859, EPI_ISL_1794860, EPI_ISL_1794861, EPI_ISL_1794862, EPI_ISL_1794863, EPI_ISL_1794864, EPI_ISL_1794865, EPI_ISL_1794866, EPI_ISL_1794867, EPI_ISL_1794868, EPI_ISL_1794869, EPI_ISL_1794870, EPI_ISL_1794871, EPI_ISL_1794872, EPI_ISL_1794873, EPI_ISL_1794874, EPI_ISL_1794875, EPI_ISL_1794876, EPI_ISL_1794877, EPI_ISL_1794878, EPI_ISL_1794879, EPI_ISL_1794880, EPI_ISL_1794881, EPI_ISL_1794882, EPI_ISL_1794883, EPI_ISL_1794884, EPI_ISL_1794919, EPI_ISL_1794920, EPI_ISL_1794921, EPI_ISL_1794922, EPI_ISL_1794923, EPI_ISL_1794924, EPI_ISL_1794925, EPI_ISL_1794926, EPI_ISL_1794927, EPI_ISL_1794928, EPI_ISL_1794929, EPI_ISL_1794930, EPI_ISL_1794931, EPI_ISL_1794932, EPI_ISL_1794933, EPI_ISL_1969315, EPI_ISL_1969316, EPI_ISL_1969318 | | | |
| see above | Sharp HealthCare Laboratory | Andersen lab at Scripps Research | Art Mendoza; Cathy Woerle; Jacquelyn Berumen; Liam McGinnis; Omid Bakhtar; SEARCH Alliance San Diego with Aaron Harding |
| EPI_ISL_833394, EPI_ISL_833395, EPI_ISL_833396, EPI_ISL_833397, EPI_ISL_833398, EPI_ISL_833399, EPI_ISL_833400, EPI_ISL_833401, EPI_ISL_833402, EPI_ISL_833403, EPI_ISL_833404, EPI_ISL_833489 | | | |
| see above | St. Francis Medical Center | Los Angeles County PHL | P. Hemarajata et al. |
| EPI_ISL_1508963 | Stanford Clinical Virology | Santa Clara County Public Health | Santa Clara County Public Health Department |

| | | | |
|---|---|---|---|
| | Laboratory | Laboratory | |
| EPI_ISL_1335871, EPI_ISL_1335872 | Stanford Health Care | Clinical Virology Laboratory, Stanford University School or Medicine | ChunHong Huang; Daniel Solis; Fumiko Yamamoto; James Zehnder; Malaya K. Sahoo; Mamdouh Sibai; Michelle Vergese; and Benjamin A. Pinsky |
| EPI_ISL_1364488, EPI_ISL_1364489, EPI_ISL_1364490, EPI_ISL_1364491, EPI_ISL_1364492, EPI_ISL_1364493, EPI_ISL_1364494, EPI_ISL_1364495, EPI_ISL_1364496, EPI_ISL_1364497, EPI_ISL_1364498, EPI_ISL_1364499, EPI_ISL_1364500, EPI_ISL_1364501, EPI_ISL_1364502, EPI_ISL_1364503, EPI_ISL_1364504, EPI_ISL_1364505, EPI_ISL_1364506, EPI_ISL_1364508, EPI_ISL_1364509, EPI_ISL_1364510, EPI_ISL_1364511, EPI_ISL_1364512, EPI_ISL_1364515, EPI_ISL_1364516, EPI_ISL_1364518, EPI_ISL_1364520, EPI_ISL_1379727, EPI_ISL_1379728, EPI_ISL_1379729, EPI_ISL_1379730, EPI_ISL_1379732, EPI_ISL_1379734, EPI_ISL_1379735, EPI_ISL_1379736, EPI_ISL_1379737, EPI_ISL_1379738, EPI_ISL_1379739, EPI_ISL_1379740, EPI_ISL_1379741, EPI_ISL_1379742, EPI_ISL_1379743, EPI_ISL_1379744, EPI_ISL_1379745, EPI_ISL_1379746, EPI_ISL_1379747, EPI_ISL_1379748, EPI_ISL_1379749, EPI_ISL_1379750, EPI_ISL_1379752, EPI_ISL_1379753, EPI_ISL_1379754, EPI_ISL_1379755, EPI_ISL_1379756, EPI_ISL_1379757, EPI_ISL_1379758, EPI_ISL_1379759, EPI_ISL_1379760, EPI_ISL_1379761, EPI_ISL_1379762, EPI_ISL_1379763, EPI_ISL_1379764, EPI_ISL_1379765, EPI_ISL_1379766, EPI_ISL_1379767, EPI_ISL_1379768, EPI_ISL_1379769, EPI_ISL_1379770, EPI_ISL_1379771, EPI_ISL_1379772, EPI_ISL_1379773, EPI_ISL_1379774, EPI_ISL_1379775, EPI_ISL_1379776, EPI_ISL_1379777, EPI_ISL_1379778, EPI_ISL_1379779, EPI_ISL_1379780, EPI_ISL_1379781, EPI_ISL_1379782, EPI_ISL_1379783, EPI_ISL_1379784, EPI_ISL_1379785, EPI_ISL_1379786, EPI_ISL_1379787, EPI_ISL_1379788, EPI_ISL_1379789, EPI_ISL_1379790, EPI_ISL_1379791, EPI_ISL_1379792, EPI_ISL_1379793, EPI_ISL_1379794, EPI_ISL_1379795, EPI_ISL_1379796, EPI_ISL_1379797, EPI_ISL_1379798, EPI_ISL_1379799, EPI_ISL_1379800, EPI_ISL_1379801, EPI_ISL_1379802, EPI_ISL_1379803, EPI_ISL_1379804, EPI_ISL_1379805, EPI_ISL_1379806, EPI_ISL_1379807, EPI_ISL_1379808, EPI_ISL_1379809, EPI_ISL_1379810, EPI_ISL_1379811, EPI_ISL_1379812, EPI_ISL_1379813, EPI_ISL_1379814, EPI_ISL_1379815, EPI_ISL_1379816, EPI_ISL_1379817, EPI_ISL_1379818, EPI_ISL_1379819, EPI_ISL_1379820, EPI_ISL_1379821, EPI_ISL_1379822, EPI_ISL_1379823, EPI_ISL_1379824, EPI_ISL_1379825, EPI_ISL_1379826, EPI_ISL_1379827, EPI_ISL_1379828, EPI_ISL_1379830, EPI_ISL_1379831, EPI_ISL_1379834, EPI_ISL_1379836, EPI_ISL_1379837, EPI_ISL_1379838, EPI_ISL_1379839, EPI_ISL_1379840, EPI_ISL_1379842, EPI_ISL_1379843, EPI_ISL_1379844, EPI_ISL_1379849, EPI_ISL_1379850, EPI_ISL_1379851, EPI_ISL_1379852, EPI_ISL_1379855, EPI_ISL_1379856, EPI_ISL_1379860, EPI_ISL_1379864, EPI_ISL_1379867, EPI_ISL_1379868, EPI_ISL_1379871, EPI_ISL_1379872, EPI_ISL_1379873, EPI_ISL_1379874, EPI_ISL_1379875, EPI_ISL_1379876, EPI_ISL_1379877, EPI_ISL_1379878, EPI_ISL_1379879, EPI_ISL_1379880, EPI_ISL_1379881, EPI_ISL_1379882, EPI_ISL_1379884, EPI_ISL_1379885, EPI_ISL_1379886, EPI_ISL_1424066, EPI_ISL_1424072, EPI_ISL_1424073, EPI_ISL_1424074, EPI_ISL_1424075, EPI_ISL_1424076, EPI_ISL_1424077, EPI_ISL_1424078, EPI_ISL_1424079, EPI_ISL_1424080, EPI_ISL_1424081, EPI_ISL_1424082, EPI_ISL_1424083, EPI_ISL_1424084, EPI_ISL_1424085, EPI_ISL_1424086, EPI_ISL_1424087, EPI_ISL_1424088, EPI_ISL_1424089, EPI_ISL_1424092, EPI_ISL_1424093, EPI_ISL_1424094, EPI_ISL_1424096, EPI_ISL_1424097, EPI_ISL_1424100, EPI_ISL_1424101, EPI_ISL_1424102, EPI_ISL_1424103, EPI_ISL_1424104, EPI_ISL_1424106, EPI_ISL_1424107, EPI_ISL_1424108, EPI_ISL_1424109, EPI_ISL_1424110, EPI_ISL_1424111, EPI_ISL_1424112, EPI_ISL_1424113, EPI_ISL_1424114, EPI_ISL_1424115, EPI_ISL_1424126, EPI_ISL_2009049, EPI_ISL_2009052, EPI_ISL_2009053, EPI_ISL_2009054, EPI_ISL_2009056, EPI_ISL_2027140, EPI_ISL_2027147, EPI_ISL_2027153, EPI_ISL_2027157, EPI_ISL_2027160, EPI_ISL_2027164, EPI_ISL_2027202, EPI_ISL_2027205, EPI_ISL_2027208, EPI_ISL_2027212, EPI_ISL_2027215, EPI_ISL_2027218, EPI_ISL_2027221, EPI_ISL_2027225, EPI_ISL_2027228 | | | |
| see above | Stanford Health Care | Stanford University School of Medicine, Clinical Virology Laboratory | Becky Jiang; Bernadette Troung; ChunHong Huang; Daniel Solis; Fumiko Yamamoto; James Zehnder; Malaya K. Sahoo; Mamdouh Sibai; Michelle Vergese; Michelle Verghese; and Benjamin A. Pinsky |
| EPI_ISL_845836 | TGen North | TGen North | Ashlyn Pfeiffer; Chris French; Darrin Lemmer; Dave Engelthaler; Hayley Yaglom; Jolene Bowers; Megan Folkerts; The Arizona COVID Genomics Union (ACGU) |
| EPI_ISL_833432 | Torrance Memorial Medical Center Microbiology | Los Angeles County PHL | P. Hemarajata et al. |
| EPI_ISL_1009813, EPI_ISL_1009827 | UC Davis Genome Center and Healthy Davis Together | UC Davis Genome Center | Sam Diaz-Muñoz and AJ Campbell |
| EPI_ISL_794641, EPI_ISL_794642, EPI_ISL_794643, EPI_ISL_794644, EPI_ISL_794645, EPI_ISL_794646, EPI_ISL_794647, EPI_ISL_794648, EPI_ISL_794670, EPI_ISL_833428, EPI_ISL_833429, EPI_ISL_833430, EPI_ISL_833431, EPI_ISL_861704, EPI_ISL_861705, EPI_ISL_861706, EPI_ISL_862122, EPI_ISL_862123, EPI_ISL_882967, EPI_ISL_882968, EPI_ISL_882969, EPI_ISL_882970, EPI_ISL_882971, EPI_ISL_882972, EPI_ISL_882973, EPI_ISL_882974, EPI_ISL_882975, EPI_ISL_882976, EPI_ISL_882977, EPI_ISL_882978, EPI_ISL_882979, EPI_ISL_882980, EPI_ISL_882981, EPI_ISL_882982, EPI_ISL_882983, EPI_ISL_882984, EPI_ISL_892267, EPI_ISL_892268, EPI_ISL_905773, EPI_ISL_905774, EPI_ISL_905775, EPI_ISL_905776, EPI_ISL_905777, EPI_ISL_905778, EPI_ISL_905779, EPI_ISL_905780, EPI_ISL_905781, EPI_ISL_905782, EPI_ISL_905783, EPI_ISL_905784, EPI_ISL_905785, EPI_ISL_905786, EPI_ISL_905787, EPI_ISL_905788, EPI_ISL_905789, EPI_ISL_905790, EPI_ISL_905791, EPI_ISL_962505, EPI_ISL_962509, EPI_ISL_962510, EPI_ISL_962511, EPI_ISL_962512, EPI_ISL_962513, EPI_ISL_962521, EPI_ISL_962833, EPI_ISL_962834, EPI_ISL_962835, EPI_ISL_962836, EPI_ISL_962837, EPI_ISL_962838, EPI_ISL_962839, EPI_ISL_962840, EPI_ISL_962841, EPI_ISL_962842, EPI_ISL_962843, EPI_ISL_962844, EPI_ISL_962845, EPI_ISL_962846, EPI_ISL_962876, EPI_ISL_962879, EPI_ISL_1005511, EPI_ISL_1005512, EPI_ISL_1005513, EPI_ISL_1005514, EPI_ISL_1040053, EPI_ISL_1098629, EPI_ISL_1191129, EPI_ISL_1706603, EPI_ISL_1706604 | | | |
| see above | UCLA Clinical Micro Lab | Los Angeles County PHL | P. Hemarajata et al. |
| EPI_ISL_1405599, EPI_ISL_1405600, EPI_ISL_1405601, EPI_ISL_1405602, EPI_ISL_1405603, EPI_ISL_1405604, EPI_ISL_1405605, EPI_ISL_1405606, EPI_ISL_1405607, EPI_ISL_1405608, EPI_ISL_1405609, EPI_ISL_1405610, EPI_ISL_1405611, EPI_ISL_1405612, EPI_ISL_1405613, EPI_ISL_1405614, EPI_ISL_1405615, EPI_ISL_1405616, EPI_ISL_1405617, EPI_ISL_1405618, EPI_ISL_1405619, EPI_ISL_1405620, EPI_ISL_1405621, EPI_ISL_1405622, EPI_ISL_1405623, EPI_ISL_1405624, EPI_ISL_1405625, EPI_ISL_1405626, EPI_ISL_1405627, EPI_ISL_1405628, EPI_ISL_1405629, EPI_ISL_1405630, EPI_ISL_1405631, EPI_ISL_1405632, EPI_ISL_1405633, EPI_ISL_1405634, EPI_ISL_1405635, EPI_ISL_1405636, EPI_ISL_1405637, EPI_ISL_1405638, EPI_ISL_1405639, EPI_ISL_1405640, EPI_ISL_1405641, EPI_ISL_1405642, EPI_ISL_1405643, EPI_ISL_1405644, EPI_ISL_1405645, EPI_ISL_1405646, EPI_ISL_1405647, EPI_ISL_1405648, EPI_ISL_1405649, EPI_ISL_1405650, EPI_ISL_1405651, EPI_ISL_1405652, EPI_ISL_1405653, EPI_ISL_1405654, EPI_ISL_1405655, EPI_ISL_1405656, EPI_ISL_1405657, EPI_ISL_1405658, EPI_ISL_1405659, EPI_ISL_1405660, EPI_ISL_1405661, EPI_ISL_1405662, EPI_ISL_1405663, EPI_ISL_1405664, EPI_ISL_1405814, EPI_ISL_1405815, EPI_ISL_1405816, EPI_ISL_1405817, EPI_ISL_1405818, EPI_ISL_1405819, EPI_ISL_1405820, EPI_ISL_1405821, EPI_ISL_1405822, EPI_ISL_1405823, EPI_ISL_1405824, EPI_ISL_1405825, EPI_ISL_1405826, EPI_ISL_1405827, EPI_ISL_1405828, EPI_ISL_1405829, EPI_ISL_1405830, EPI_ISL_1405831, EPI_ISL_1405832, EPI_ISL_1405833, EPI_ISL_1405834, EPI_ISL_1405835, EPI_ISL_1405836, EPI_ISL_1405837, EPI_ISL_1405838, EPI_ISL_1405839, EPI_ISL_1405840, EPI_ISL_1405841, EPI_ISL_1405842, EPI_ISL_1405843, EPI_ISL_1405844, EPI_ISL_1405845, EPI_ISL_1405846, EPI_ISL_1405847, EPI_ISL_1405848, EPI_ISL_1405849, EPI_ISL_1405850, EPI_ISL_1405851, EPI_ISL_1405852, EPI_ISL_1405853, EPI_ISL_1405854, EPI_ISL_1405855, EPI_ISL_1405856, EPI_ISL_1405857, EPI_ISL_1405858, EPI_ISL_1405859, EPI_ISL_1405860, EPI_ISL_1405861, EPI_ISL_1405862, EPI_ISL_1405863, EPI_ISL_1405864, EPI_ISL_1405865, EPI_ISL_1405866, EPI_ISL_1405867, EPI_ISL_1405868, EPI_ISL_1405869, EPI_ISL_1405870, EPI_ISL_1405871, EPI_ISL_1405872, EPI_ISL_1405873, EPI_ISL_1405874, EPI_ISL_1405875, EPI_ISL_1405876, EPI_ISL_1405877, EPI_ISL_1405878, EPI_ISL_1405879, EPI_ISL_1405880, EPI_ISL_1405881, EPI_ISL_1405882, EPI_ISL_1405883, EPI_ISL_1405884, EPI_ISL_1405885, EPI_ISL_1405886, EPI_ISL_1405887, EPI_ISL_1405888, EPI_ISL_1405889, EPI_ISL_1405890, EPI_ISL_1405891, EPI_ISL_1405892, EPI_ISL_1405893, EPI_ISL_1405894, EPI_ISL_1405895, EPI_ISL_1405896, EPI_ISL_1405897, EPI_ISL_1469120, EPI_ISL_1469121, EPI_ISL_1469908, EPI_ISL_1921088, EPI_ISL_1921096, EPI_ISL_1921097, EPI_ISL_2031829, EPI_ISL_2031839, EPI_ISL_2031853, EPI_ISL_2081241, EPI_ISL_2081242, EPI_ISL_2081279, EPI_ISL_2081281, EPI_ISL_2081289, EPI_ISL_2081292, EPI_ISL_2081305, EPI_ISL_2081306, EPI_ISL_2081544, EPI_ISL_2081610, EPI_ISL_2082984, EPI_ISL_2082985, EPI_ISL_2082995, EPI_ISL_2083003 | | | |
| see above | UCSC Genomics Institute | UCSC Genomics Institute | A. Marm Kilpatrick; Angie Hinrichs; Beth Shapiro; Bryan Thornlow; Ciara Wanket; David Haussler; Eric Beraut; Hugh Olsen; Ikenna Anigbogu; Isabel Bjork; Jakob McBroome; Jeremy Sanford; Joshua Kapp; Mark Akeson; Maximilian Haeussler; Michael Stone; Miten Jain; Molly Cassatt-Johnstone; Namrita Dhillon; Russell Corbett-Detig; Terren Chang; Yatish Turakhia |
| EPI_ISL_751801 | UCSD EXCITE | Andersen lab at Scripps Research | SEARCH Alliance San Diego |
| EPI_ISL_1301442, EPI_ISL_1301443 | UCSD EXCITE | NBACC | Aaron Phillips; Adam Bazinet; Amanda Graham; Brian Holland; David Miller; Denise Freeburger; Elyse Abbott; Gregory Williams; Jason Yolitz; Jennifer Biryukov; Jordan Bohannon; Karla Garcia; Katie Beck; M.J. Rosovitz; Melissa Krause; Michael Schuit; Paul Dabisch; Sean Lovett; Shanna Ratnesar-Shumate; Stewart Wood; Victoria Wahl; Wade Weaver |
| EPI_ISL_849923, EPI_ISL_849924, EPI_ISL_849925, EPI_ISL_849927 | UCSF Clinical Microbiology Laboratory | Chan-Zuckerberg Biohub | CZB Cliahub Consortium |
| EPI_ISL_794635, EPI_ISL_794636, EPI_ISL_794637, EPI_ISL_794638, EPI_ISL_794639, EPI_ISL_794640, EPI_ISL_794669, EPI_ISL_833414, EPI_ISL_833415, EPI_ISL_833416, EPI_ISL_1508930 | | | |
| see above | USC Clinical Lab | Los Angeles County PHL | P. Hemarajata et al. |
| EPI_ISL_962517, EPI_ISL_962518, EPI_ISL_962519, EPI_ISL_962520 | VA Medical Center | Los Angeles County PHL | P. Hemarajata et al. |

We gratefully acknowledge the following Authors from the Originating laboratories responsible for obtaining the specimens, as well as the Submitting laboratories where the genome data were generated and shared via GISAID, on which this research is based.

All Submitters of data may be contacted directly via www.gisaid.org

Authors are sorted alphabetically.

| Accession ID | Originating Laboratory | Submitting Laboratory | Authors |
|---|---|---|---|
| EPI_ISL_1318634, EPI_ISL_1318635, EPI_ISL_1318636, EPI_ISL_1318637, EPI_ISL_1318638, EPI_ISL_1318639, EPI_ISL_1318640, EPI_ISL_1318641, EPI_ISL_1318643, EPI_ISL_1318644, EPI_ISL_1318645, EPI_ISL_1318646, EPI_ISL_1318647, EPI_ISL_1318648, EPI_ISL_1318649, EPI_ISL_1318650, EPI_ISL_1318651, EPI_ISL_1318652, EPI_ISL_1318653, EPI_ISL_1318654, EPI_ISL_1318655, EPI_ISL_1318656, EPI_ISL_1318657, EPI_ISL_1318659, EPI_ISL_1318660, EPI_ISL_1318661, EPI_ISL_1318662, EPI_ISL_1318663, EPI_ISL_1318664, EPI_ISL_1318665, EPI_ISL_1318666, EPI_ISL_1318667, EPI_ISL_1446781, EPI_ISL_1446784, EPI_ISL_1446791, EPI_ISL_1446792, EPI_ISL_1446794, EPI_ISL_1446801, EPI_ISL_1447018, EPI_ISL_1678147, EPI_ISL_1678155 | | | |
| see above | "CDPH, Viral and Rickettsial Disease Laboratory" | Centers for Disease Control and Prevention Division of Viral Diseases, Pathogen Discovery | Alison Laufer Halpin; Anna Montmayeur; Anna Uehara; Ben L. Rambo-Martin; Clinton R. Paden; Dakota Howard; Darlene Wagner; Dave Wentworth; Dhwani Batra; Haibin Wang; Jasmine Padilla; Jing Zhang; Justin Lee; Katie Dillon; Krista Queen; Kristen Knipe; Kristine Lacek; Lori Rowe; Mark Burroughs; Matthew Schmerer; Mili Sheth; Peter Cook; Peter W. Cook; Rachel Marine; Sam Shepard; Sarah Nobles; Shoshona Le; Suxiang Tong; Vivien Dugan; Yan Li; Ying Tao; Yvette Unoarumhi |
| EPI_ISL_1226239, EPI_ISL_1226240, EPI_ISL_1226241, EPI_ISL_1226242, EPI_ISL_1226243, EPI_ISL_1226244, EPI_ISL_1226245, EPI_ISL_1226246, EPI_ISL_1226247, EPI_ISL_1226248, EPI_ISL_1226249, EPI_ISL_1226250, EPI_ISL_1226251, EPI_ISL_1226252, EPI_ISL_1226253, EPI_ISL_1226254, EPI_ISL_1226255, EPI_ISL_1226256, EPI_ISL_1226257, EPI_ISL_1226258, EPI_ISL_1226259, EPI_ISL_1226260, EPI_ISL_1226261, EPI_ISL_1226262, EPI_ISL_1226263, EPI_ISL_1226264, EPI_ISL_1226265, EPI_ISL_1226266, EPI_ISL_1226267, EPI_ISL_1226268, EPI_ISL_1226269, EPI_ISL_1226270, EPI_ISL_1226271, EPI_ISL_1226272, EPI_ISL_1226273, EPI_ISL_1226275, EPI_ISL_1491516, EPI_ISL_1491517, EPI_ISL_1491522 | | | |
| see above | "CDPH, Viral and Rickettsial Disease Laboratory" | Genomics and Discovery, Respiratory Viruses Branch, Division of Viral Diseases, Centers for Disease Control and Prevention | Anna Kelleher; Anna Montmayeur; Anna Uehara; Ben L. Rambo-Martin; Brian Lynch; Clinton R. Paden; Dhwani Batra; Haibin Wang; Han Jia Justin Ng; Jasmine Padilla; Jing Zhang; Justin Lee; Katie Dillon; Krista Queen; Kristen Knipe; Kristine Lacek; Lori Rowe; Mark Burroughs; Matthew Schmerer; Mili Sheth; Peter Cook; Peter W. Cook; Rachel Marine; Sam Shepard; Sarah Nobles; Shoshona Le; Suxiang Tong; Yan Li; Ying Tao |
| EPI_ISL_1093546, EPI_ISL_1093551, EPI_ISL_1093552, EPI_ISL_1093557, EPI_ISL_1093558, EPI_ISL_1093559, EPI_ISL_1093560, EPI_ISL_1093561, EPI_ISL_1093562, EPI_ISL_1093569, EPI_ISL_1093593, EPI_ISL_1093594, EPI_ISL_1093595, EPI_ISL_1093596, EPI_ISL_1093597, EPI_ISL_1093598, EPI_ISL_1093599, EPI_ISL_1093600, EPI_ISL_1093601, EPI_ISL_1196775, EPI_ISL_1196776, EPI_ISL_1196777, EPI_ISL_1196778, EPI_ISL_1196779, EPI_ISL_1196780, EPI_ISL_1196781, EPI_ISL_1196782, EPI_ISL_1196783, EPI_ISL_1196784, EPI_ISL_1196785, EPI_ISL_1196786, EPI_ISL_1196787, EPI_ISL_1196788, EPI_ISL_1196789, EPI_ISL_1196790, EPI_ISL_1196791, EPI_ISL_1205471, EPI_ISL_1205472, EPI_ISL_1205473, EPI_ISL_1205475, EPI_ISL_1205476, EPI_ISL_1205477, EPI_ISL_1205478, EPI_ISL_1205479, EPI_ISL_1205480, EPI_ISL_1205481, EPI_ISL_1205482, EPI_ISL_1373145, EPI_ISL_1373160, EPI_ISL_1576998, EPI_ISL_1576999 | | | |
| see above | Alameda County Public Health Department | Alameda County Public Health Department | Kevin Libuit; Kristna Hsieh; Maria Elma |
| EPI_ISL_1114336 | Altius Institute for Biomedical Sciences | Seattle Flu Study | Alex Nguyen; Amanda Adler; Andrew Meuser; Barry R. Lutz; Benjamin Pelle; Caitlin R. Wolf; Chris D. Frazar; Clem Green; Daniel Bates; Deborah A. Nickerson; Elisabeth Brandstetter; Erica Ryke; Hannah Petersen; Helen Y. Chu; Jacob Rodriguez; Janet A. Englund; Jay Shendure; Jessica Halow; John Stamatoyannopoulos; Joshua Richards; Jover Lee; Julia Wald; Kairsten Fay; Kirsten Lacombe; Kneshay Harper; Lea M. Starita; Mark J. Rieder; Matt Hartman; Matthew Richardson; Matthew Thompson; Melissa Truong; Michael Boeckh; Michael Famulare; Misja Ilcisin; Muhammad Halimun; Olivia Waltner; Peter D. Han; Rebecca Bruders; Ryan Alexander; Sadie Patraw; Sofia Olsson; Stephanie DeBaun; Thomas R. Sibley; Tobias Ragoczy; Trevor Bedford; Truong Nguyen |
| EPI_ISL_1364729 | Arizona State University | Arizona State University | Efrem S. Lim; Emily A. Kaelin; Joshua LaBaer; Joy M. Blain; Kristina Buss; LaRinda A. Holland; Neal W Woodbury; Nicholas J. Mellor; Peter T. Skidmore; Rabia Maqsood; Valerie Harris; Vel Murugan |
| EPI_ISL_1235360, EPI_ISL_1235367, EPI_ISL_1235372, EPI_ISL_1235373, EPI_ISL_1235375, EPI_ISL_1235377, EPI_ISL_1235378, EPI_ISL_1235381, EPI_ISL_1235383, EPI_ISL_1235384, EPI_ISL_1235400, EPI_ISL_1235401, EPI_ISL_1235403, EPI_ISL_1235410, EPI_ISL_1235416, EPI_ISL_1235417, EPI_ISL_1235418, EPI_ISL_1235419, EPI_ISL_1235424, EPI_ISL_1235427, EPI_ISL_1235434, EPI_ISL_1235435, EPI_ISL_1235438, EPI_ISL_1235441, EPI_ISL_1235442, EPI_ISL_1235450, EPI_ISL_1235451, EPI_ISL_1235455, EPI_ISL_1235468, EPI_ISL_1235469, EPI_ISL_1235472, EPI_ISL_1235473, EPI_ISL_1235478, EPI_ISL_1235488, EPI_ISL_1235490, EPI_ISL_1235496, EPI_ISL_1235498, EPI_ISL_1235501, EPI_ISL_1235502, EPI_ISL_1235506, EPI_ISL_1235507, EPI_ISL_1235509, EPI_ISL_1235511, EPI_ISL_1235515, EPI_ISL_1235521, EPI_ISL_1235522, EPI_ISL_1235525, EPI_ISL_1235526, EPI_ISL_1235528, EPI_ISL_1235539, EPI_ISL_1235540, EPI_ISL_1235544, EPI_ISL_1235545, EPI_ISL_1235546, EPI_ISL_1235547, EPI_ISL_1235549, EPI_ISL_1235551, EPI_ISL_1235558, EPI_ISL_1235570, EPI_ISL_1235572, EPI_ISL_1235578, EPI_ISL_1235582, EPI_ISL_1235588, EPI_ISL_1235591, EPI_ISL_1235596, EPI_ISL_1235600, EPI_ISL_1235602, EPI_ISL_1235604, EPI_ISL_1235606, EPI_ISL_1235608, EPI_ISL_1477167, EPI_ISL_1477168, EPI_ISL_1477171, EPI_ISL_1477177, EPI_ISL_1477179, EPI_ISL_1477181, EPI_ISL_1477182, EPI_ISL_1477184, EPI_ISL_1477185, EPI_ISL_1477186, EPI_ISL_1477187, EPI_ISL_1477188, EPI_ISL_1477190, EPI_ISL_1477191, EPI_ISL_1477192, EPI_ISL_1477193, EPI_ISL_1477194, EPI_ISL_1477196, EPI_ISL_1477197, EPI_ISL_1477198, EPI_ISL_1477200, EPI_ISL_1477201, EPI_ISL_1477202, EPI_ISL_1477203, EPI_ISL_1477204, EPI_ISL_1477205, EPI_ISL_1477209, EPI_ISL_1477210, EPI_ISL_1477211, EPI_ISL_1477212, EPI_ISL_1477213, EPI_ISL_1477214, EPI_ISL_1477215, EPI_ISL_1477217, EPI_ISL_1477219, EPI_ISL_1477221, EPI_ISL_1477222, EPI_ISL_1477223, EPI_ISL_1477225, EPI_ISL_1477229, EPI_ISL_1477230, EPI_ISL_1477231, EPI_ISL_1477232, EPI_ISL_1477233, EPI_ISL_1477235, EPI_ISL_1477237, EPI_ISL_1477238, EPI_ISL_1477240, EPI_ISL_1477241, EPI_ISL_1477242, EPI_ISL_1477245, EPI_ISL_1477246, EPI_ISL_1477248, EPI_ISL_1477250, EPI_ISL_1477253, EPI_ISL_1477254, EPI_ISL_1477256, EPI_ISL_1477258, EPI_ISL_1477261, EPI_ISL_1477263, EPI_ISL_1477265, EPI_ISL_1477266, EPI_ISL_1477267, EPI_ISL_1477273, EPI_ISL_1477274, EPI_ISL_1477275, EPI_ISL_1477278, EPI_ISL_1477281, EPI_ISL_1477283, EPI_ISL_1477285, EPI_ISL_1477286, EPI_ISL_1477289, EPI_ISL_1477291, EPI_ISL_1477297, EPI_ISL_1477298, EPI_ISL_1477300, EPI_ISL_1477302, EPI_ISL_1477303, EPI_ISL_1477304, EPI_ISL_1477305, EPI_ISL_1477307, EPI_ISL_1477310, EPI_ISL_1477311, EPI_ISL_1477314, EPI_ISL_1477315, EPI_ISL_1477316, EPI_ISL_1477317, EPI_ISL_1477319, EPI_ISL_1477320, EPI_ISL_1477321, EPI_ISL_1477323, EPI_ISL_1477324, EPI_ISL_1477327, EPI_ISL_1477330, EPI_ISL_1477331, EPI_ISL_1477333, EPI_ISL_1477336, EPI_ISL_1477338, EPI_ISL_1477342, EPI_ISL_1477344, EPI_ISL_1477345, EPI_ISL_1477347, EPI_ISL_1477349, EPI_ISL_1477350, EPI_ISL_1477351, EPI_ISL_1477353, EPI_ISL_1477356, EPI_ISL_1477357, EPI_ISL_1477358, EPI_ISL_1477360, EPI_ISL_1477361, EPI_ISL_1477362, EPI_ISL_1477363, EPI_ISL_1477365, EPI_ISL_1477366, EPI_ISL_1477368, EPI_ISL_1477369, EPI_ISL_1477370, EPI_ISL_1477372, EPI_ISL_1477374, EPI_ISL_1477378, EPI_ISL_1477383, EPI_ISL_1477384, EPI_ISL_1477386, EPI_ISL_1477387, EPI_ISL_1477390, EPI_ISL_1477391, EPI_ISL_1477392, EPI_ISL_1477393, EPI_ISL_1477395, EPI_ISL_1477398, EPI_ISL_1477400, EPI_ISL_1477402, EPI_ISL_1477403, EPI_ISL_1477404, EPI_ISL_1477407, EPI_ISL_1477409, EPI_ISL_1477416, EPI_ISL_1477418, EPI_ISL_1477419, EPI_ISL_1477420, EPI_ISL_1477421, EPI_ISL_1477422, EPI_ISL_1477423, EPI_ISL_1477424, EPI_ISL_1477425, EPI_ISL_1477426, EPI_ISL_1477427, EPI_ISL_1477428, EPI_ISL_1477429, EPI_ISL_1477431, EPI_ISL_1477434, EPI_ISL_1477437, EPI_ISL_1477440, EPI_ISL_1477442, EPI_ISL_1477448, EPI_ISL_1477450, EPI_ISL_1477451, EPI_ISL_1477456, EPI_ISL_1477464, EPI_ISL_1477467, EPI_ISL_1477469, EPI_ISL_1477470, EPI_ISL_1477472, EPI_ISL_1477474, EPI_ISL_1477475, EPI_ISL_1477479, EPI_ISL_1477480, EPI_ISL_1477485, EPI_ISL_1477490, EPI_ISL_1477492, EPI_ISL_1477505, EPI_ISL_1477509, EPI_ISL_1477512, EPI_ISL_1477513, EPI_ISL_1477516, EPI_ISL_1477522, EPI_ISL_1477523, EPI_ISL_1477524, EPI_ISL_1477526, EPI_ISL_1477527, EPI_ISL_1477528, EPI_ISL_1477538, EPI_ISL_1477542, EPI_ISL_1477548, EPI_ISL_1477549, EPI_ISL_1477553, EPI_ISL_1477556, EPI_ISL_1477559, EPI_ISL_1477560, EPI_ISL_1477563, EPI_ISL_1477567, EPI_ISL_1477568, EPI_ISL_1477570, EPI_ISL_1477573, EPI_ISL_1477574, EPI_ISL_1477576, EPI_ISL_1477579, EPI_ISL_1477580, EPI_ISL_1477583, EPI_ISL_1477585, EPI_ISL_1477586, EPI_ISL_1477591, EPI_ISL_1477592, EPI_ISL_1477596, EPI_ISL_1477599, EPI_ISL_1477602, EPI_ISL_1477606, EPI_ISL_1477607, EPI_ISL_1477608, EPI_ISL_1477609, EPI_ISL_1477616, EPI_ISL_1477618, EPI_ISL_1477620, EPI_ISL_1477621, EPI_ISL_1477625, EPI_ISL_1477628, EPI_ISL_1477629, EPI_ISL_1477633, EPI_ISL_1477636, EPI_ISL_1477638, EPI_ISL_1477640, EPI_ISL_1477642, EPI_ISL_1477643, EPI_ISL_1477648, EPI_ISL_1477651, EPI_ISL_1477652, EPI_ISL_1477653, EPI_ISL_1477654, EPI_ISL_1477661, EPI_ISL_1477662, EPI_ISL_1477959, EPI_ISL_1477964, EPI_ISL_1477968, EPI_ISL_1477971, EPI_ISL_1477978, EPI_ISL_1477984, EPI_ISL_1477985, EPI_ISL_1477988, EPI_ISL_1477989, EPI_ISL_1477994, EPI_ISL_1477996, EPI_ISL_1478001, EPI_ISL_1478004, EPI_ISL_1478014, EPI_ISL_1478016, EPI_ISL_1478021, EPI_ISL_1478027, EPI_ISL_1478029, EPI_ISL_1478033, EPI_ISL_1478042, EPI_ISL_1478051, EPI_ISL_1478056, EPI_ISL_1478060, EPI_ISL_1478064, EPI_ISL_1478068, EPI_ISL_1478071, EPI_ISL_1478073, EPI_ISL_1478076, EPI_ISL_1478079, EPI_ISL_1478085, EPI_ISL_1478100, EPI_ISL_1478104, EPI_ISL_1478109, EPI_ISL_1478112, EPI_ISL_1478113, EPI_ISL_1478114, EPI_ISL_1478115, EPI_ISL_1478116, EPI_ISL_1478119, EPI_ISL_1478127, EPI_ISL_1478130, EPI_ISL_1478132, EPI_ISL_1478142, EPI_ISL_1478144, EPI_ISL_1478146, EPI_ISL_1478148, EPI_ISL_1478151, EPI_ISL_1478158, EPI_ISL_1478162, EPI_ISL_1478166, EPI_ISL_1478167, EPI_ISL_1478168, EPI_ISL_1478173, EPI_ISL_1478178, EPI_ISL_1478183, EPI_ISL_1478184, EPI_ISL_1478185, EPI_ISL_1478191, EPI_ISL_1478194, EPI_ISL_1529100, EPI_ISL_1529104, EPI_ISL_1529105, EPI_ISL_1529111, EPI_ISL_1529112, EPI_ISL_1529114, EPI_ISL_1529115, EPI_ISL_1529116, EPI_ISL_1529117, EPI_ISL_1529120, EPI_ISL_1529125, EPI_ISL_1529126, EPI_ISL_1529130, EPI_ISL_1529139, EPI_ISL_1529140, EPI_ISL_1529143, EPI_ISL_1529145, EPI_ISL_1529146, EPI_ISL_1529150, EPI_ISL_1529153, EPI_ISL_1529157, EPI_ISL_1529159, EPI_ISL_1529160, EPI_ISL_1529167, EPI_ISL_1529170, EPI_ISL_1529172, EPI_ISL_1529174, EPI_ISL_1529178, EPI_ISL_1529182, EPI_ISL_1529187, EPI_ISL_1529189, EPI_ISL_1529192, EPI_ISL_1529197, EPI_ISL_1529198, EPI_ISL_1529199, EPI_ISL_1529200, EPI_ISL_1529201, EPI_ISL_1529203, EPI_ISL_1529214, EPI_ISL_1529215, EPI_ISL_1529219, EPI_ISL_1529222, EPI_ISL_1529223, EPI_ISL_1529230, EPI_ISL_1529231, EPI_ISL_1529233, EPI_ISL_1529242, EPI_ISL_1529243, EPI_ISL_1529244, EPI_ISL_1529249, EPI_ISL_1529254, EPI_ISL_1529257, EPI_ISL_1529264, EPI_ISL_1529265, EPI_ISL_1529266, EPI_ISL_1529274, EPI_ISL_1529276, EPI_ISL_1529277, EPI_ISL_1529280, EPI_ISL_1529281, EPI_ISL_1529286, EPI_ISL_1529288, EPI_ISL_1529290, EPI_ISL_1529291, EPI_ISL_1529294, EPI_ISL_1529299, EPI_ISL_1529306, EPI_ISL_1529312, EPI_ISL_1529313, EPI_ISL_1529314, EPI_ISL_1529319, EPI_ISL_1529321, EPI_ISL_1529324, EPI_ISL_1529325, EPI_ISL_1529326, EPI_ISL_1529332, EPI_ISL_1529333, EPI_ISL_1529335, EPI_ISL_1529337, EPI_ISL_1529357, EPI_ISL_1529359, EPI_ISL_1529360, EPI_ISL_1529361, EPI_ISL_1529362, EPI_ISL_1529363, EPI_ISL_1529366, EPI_ISL_1529369, EPI_ISL_1663921, EPI_ISL_1663922, EPI_ISL_1663924, EPI_ISL_1663929, EPI_ISL_1663932, EPI_ISL_1663941, EPI_ISL_1663942, EPI_ISL_1663944, EPI_ISL_1663949, EPI_ISL_1663961, EPI_ISL_1663965, EPI_ISL_1663966, EPI_ISL_1663967, EPI_ISL_1663973, EPI_ISL_1663978, EPI_ISL_1663981, EPI_ISL_1663988, EPI_ISL_1663991, EPI_ISL_1663997, EPI_ISL_1663998, EPI_ISL_1664000, EPI_ISL_1664001, EPI_ISL_1664002, EPI_ISL_1664007, EPI_ISL_1664011, EPI_ISL_1664012, EPI_ISL_1664017, EPI_ISL_1664018, EPI_ISL_1664019, EPI_ISL_1664022, EPI_ISL_1664023, EPI_ISL_1664027, EPI_ISL_1664029, EPI_ISL_1664031, EPI_ISL_1664033, EPI_ISL_1664034, EPI_ISL_1664041, EPI_ISL_1664042, EPI_ISL_1664044, EPI_ISL_1664055, EPI_ISL_1664059, EPI_ISL_1664061, EPI_ISL_1664066, EPI_ISL_1664067, EPI_ISL_1664074, EPI_ISL_1664075, EPI_ISL_1664076, EPI_ISL_1664077, EPI_ISL_1664078, EPI_ISL_1664092, EPI_ISL_1664099, EPI_ISL_1701103, EPI_ISL_1701263, EPI_ISL_1821316, EPI_ISL_1821318, EPI_ISL_1821321, EPI_ISL_1821322, EPI_ISL_1821325, EPI_ISL_1821334, EPI_ISL_1821335, EPI_ISL_1821336, EPI_ISL_1821343, EPI_ISL_1821347, EPI_ISL_1821348, EPI_ISL_1821349, EPI_ISL_1821351, EPI_ISL_1821352, EPI_ISL_1821353, EPI_ISL_1821370, EPI_ISL_1821393, EPI_ISL_1821395, EPI_ISL_1821400, EPI_ISL_1821401, EPI_ISL_1821406, EPI_ISL_1821407, EPI_ISL_1821409, EPI_ISL_1821411, EPI_ISL_1821415, EPI_ISL_1821416, EPI_ISL_1821417, EPI_ISL_1821418, EPI_ISL_1821420, EPI_ISL_1821421, EPI_ISL_1821422, EPI_ISL_1821423, EPI_ISL_1821425, EPI_ISL_1821429, EPI_ISL_1821430, EPI_ISL_1821431, EPI_ISL_1821432, EPI_ISL_1821433, EPI_ISL_1821435, EPI_ISL_1821436, EPI_ISL_1821438, EPI_ISL_1821439, EPI_ISL_1821441, EPI_ISL_1821444, EPI_ISL_1821446, EPI_ISL_1821448, EPI_ISL_1821449, EPI_ISL_1821450, EPI_ISL_1821451, EPI_ISL_1821452, EPI_ISL_1821453, EPI_ISL_1821456, EPI_ISL_1821457, EPI_ISL_1821459, EPI_ISL_1821462, EPI_ISL_1821464, EPI_ISL_1821465, EPI_ISL_1821467, EPI_ISL_1821469, EPI_ISL_1821471, EPI_ISL_1821472, EPI_ISL_2102523, EPI_ISL_2102529, EPI_ISL_2102530, EPI_ISL_2102531, EPI_ISL_2102532, EPI_ISL_2102535, EPI_ISL_2102536, EPI_ISL_2102544, EPI_ISL_2102545, EPI_ISL_2102547, EPI_ISL_2102548, EPI_ISL_2102549, EPI_ISL_2102553, EPI_ISL_2102554, EPI_ISL_2102556, EPI_ISL_2102557, EPI_ISL_2102558, EPI_ISL_2102559, EPI_ISL_2102560, EPI_ISL_2102563, EPI_ISL_2102565, EPI_ISL_2102567, EPI_ISL_2102571, EPI_ISL_2102574, EPI_ISL_2102576, EPI_ISL_2102577, EPI_ISL_2102578, EPI_ISL_2102581, EPI_ISL_2102582, EPI_ISL_2102583, EPI_ISL_2102589, EPI_ISL_2102593, EPI_ISL_2102594, EPI_ISL_2102596, EPI_ISL_2102598, EPI_ISL_2103149 | | | |
| see above | CA DPH Viral and Rickettsial Disease Laboratory | Chan-Zuckerberg Biohub | CZB Cliahub Consortium |
| EPI_ISL_1272873, EPI_ISL_1272874, EPI_ISL_1272875, EPI_ISL_1272876, EPI_ISL_1272877, EPI_ISL_1272879, EPI_ISL_1272880, EPI_ISL_1272881, EPI_ISL_1272882, EPI_ISL_1272883, EPI_ISL_1272884, EPI_ISL_1272885, EPI_ISL_1272886, EPI_ISL_1272887, EPI_ISL_1272889, EPI_ISL_1272890, EPI_ISL_1272892, EPI_ISL_1272893, EPI_ISL_1272894, EPI_ISL_1272895, EPI_ISL_1447049 | | | |
| see above | CA-Los Angeles County Public Health Laboratory | Centers for Disease Control and Prevention Division of Viral Diseases, Pathogen Discovery | Alison Laufer Halpin; Anna Montmayeur; Anna Uehara; Ben L. Rambo-Martin; Clinton R. Paden; Dakota Howard; Darlene Wagner; Dave Wentworth; Dhwani Batra; Haibin Wang; Jasmine Padilla; Jing Zhang; Justin Lee; Katie Dillon; Krista Queen; Kristen Knipe; Kristine Lacek; Lori Rowe; Mark Burroughs; Matthew Schmerer; Mili Sheth; Peter Cook; Peter W. Cook; Rachel Marine; Sam Shepard; Sarah Nobles; Shoshona Le; Suxiang Tong; Vivien Dugan; Yan Li; Ying Tao; Yvette Unoarumhi |
| EPI_ISL_1226169, EPI_ISL_1226170, EPI_ISL_1226171, EPI_ISL_1226172, EPI_ISL_1226173, EPI_ISL_1226174, EPI_ISL_1226175, EPI_ISL_1226176 | | | |
| see above | CA-Los Angeles County Public Health Laboratory | Genomics and Discovery, Respiratory Viruses Branch, Division of Viral Diseases, Centers for Disease Control and Prevention | Anna Montmayeur; Anna Uehara; Ben L. Rambo-Martin; Clinton R. Paden; Dhwani Batra; Haibin Wang; Jasmine Padilla; Jing Zhang; Justin Lee; Katie Dillon; Krista Queen; Kristen Knipe; Kristine Lacek; Lori Rowe; Mark Burroughs; Matthew Schmerer; Mili Sheth; Peter W. Cook; Rachel Marine; Sam Shepard; Sarah Nobles; Shoshona Le; Suxiang Tong; Yan Li; Ying Tao |
| EPI_ISL_1373726, EPI_ISL_1373729, EPI_ISL_1373741, EPI_ISL_1373751, EPI_ISL_1373770, EPI_ISL_1373813, EPI_ISL_1373814, EPI_ISL_1373816, EPI_ISL_1373837, EPI_ISL_1373839, EPI_ISL_1373840, EPI_ISL_1373841, EPI_ISL_1373842, EPI_ISL_1373843, EPI_ISL_1373844, EPI_ISL_1373845, EPI_ISL_1373846, EPI_ISL_1373848, EPI_ISL_1373849, EPI_ISL_1373851, EPI_ISL_1373852, EPI_ISL_1373855, EPI_ISL_1373856, EPI_ISL_1373859, EPI_ISL_1373860, EPI_ISL_1373861, EPI_ISL_1373864, EPI_ISL_1373866, EPI_ISL_1373867, EPI_ISL_1373868, EPI_ISL_1373869, EPI_ISL_1373870, EPI_ISL_1373871, EPI_ISL_1373873, EPI_ISL_1373874, EPI_ISL_1373875, EPI_ISL_1373876, EPI_ISL_1373878 | | | |
| see above | CDPH, Viral and Rickettsial Disease Laboratory | Genomics and Discovery, Respiratory Viruses Branch, Division of Viral Diseases, Centers for Disease Control and Prevention | Anna Kelleher; Anna Uehara; Brian Lynch; Clinton R. Paden; Haibin Wang; Han Jia Justin Ng; Jing Zhang; Krista Queen; Peter Cook; Suxiang Tong; Yan Li; Ying Tao |
| EPI_ISL_1483240 | CDPH, Viral and Rickettsial Disease Laboratory | Pathogen Discovery, Respiratory Viruses Branch, Division of Viral Diseases, Centers for Disease Control and Prevention | Anna Kelleher; Anna Uehara; Brian Lynch; Clinton R. Paden; Haibin Wang; Han Jia Justin Ng; Jing Zhang; Krista Queen; Peter Cook; Suxiang Tong; Yan Li; Ying Tao |
| EPI_ISL_1234813, EPI_ISL_1234891, EPI_ISL_1234967, EPI_ISL_1234994, EPI_ISL_1701473, EPI_ISL_1701620 | California Institute of Technology | Chan-Zuckerberg Biohub | CZB Cliahub Consortium |
| EPI_ISL_1595603 | Centro de Investigación Biomédica de Occidente (CIBO) | Instituto Nacional de Enfermedades Respiratorias (INER): Centro de Investigación en Enfermedades Infecciosas (CIENI) | Alejandro Sanchez-Flores; Alfredo Herrera-Estrella; Alicia Ocaña-Mondragón; Angel Gustavo Salas-Lais; Bernardo Martínez-Miguel; Blanca Taboada; Brenda Irasema Maldonado-Meza; Carla Ivón Herrera-Najera; Carlos F. Arias; Celia Boukadida; Clara Esperanza Santacruz-Tinoco; Concepción Grajales-Muñiz; Consorcio Mexicano de Vigilancia Genómica (CoViGen-Mex). Authors (in alphabetical order): Julio Elias Alvarado-Yaah; Célida Duque Molina; Fernando Fontove-Herrera; Francisco Pulido; Gloria Elena Espinosa-Ayala; Gloria María Molina-Salinas; Gloria Vazquez; Hector Esteban Paz-Juárez; Hector Montoya-Fuentes; Helen Haydee Fernanda Ramírez-Plascencia; José Antonio Enciso-Moreno; José Esteban Muñoz-Medina; José de Jesús Nuñez-Contreras; Juan Bautista Chale-Dzul; Luis Alberto Ochoa-Carrera; Margarita Matías-Florentino; María Guadalupe Santiago-Mauricio; María Guadalupe de Jesús Mireles-Rivera; Nelly Sélem-Mojica; Pavel Isa; Ricardo Grande; Santiago Ávila-Ríos; Victor Eduardo Garcia-Arias; Víctor Hugo Borja-Aburto |
| EPI_ISL_1824299, EPI_ISL_1824300, EPI_ISL_1824301, EPI_ISL_1824302, EPI_ISL_1824303, EPI_ISL_1824304, EPI_ISL_1824305, EPI_ISL_1824306, EPI_ISL_1824307, EPI_ISL_1824308, EPI_ISL_1824309, EPI_ISL_1824310, EPI_ISL_1824311, EPI_ISL_1824312, EPI_ISL_1824313, EPI_ISL_1824314, EPI_ISL_1824315, EPI_ISL_1824316, EPI_ISL_1824317, EPI_ISL_1824318, EPI_ISL_1824319, EPI_ISL_1824320, EPI_ISL_1824321, EPI_ISL_1824322, EPI_ISL_1824323, EPI_ISL_1824324, EPI_ISL_1824325, EPI_ISL_1824326, EPI_ISL_1824327, EPI_ISL_1824328, EPI_ISL_1824329, EPI_ISL_1824330, EPI_ISL_1824331, EPI_ISL_1824332, EPI_ISL_1824333, EPI_ISL_1824334, EPI_ISL_1824335, EPI_ISL_1824336, EPI_ISL_1824337, EPI_ISL_1824338, EPI_ISL_1824339, EPI_ISL_1824340, EPI_ISL_1824341, EPI_ISL_1824342, EPI_ISL_1824343, EPI_ISL_1824344, EPI_ISL_1824345, EPI_ISL_1824346, EPI_ISL_1824347, EPI_ISL_1824348, EPI_ISL_1824349, EPI_ISL_1824351, EPI_ISL_1824353, EPI_ISL_1824354, EPI_ISL_1824355, EPI_ISL_1824356, EPI_ISL_1824363, EPI_ISL_1824365, EPI_ISL_1824366 | | | |
| see above | Chiu Laboratory, University of California, San Francisco | Chiu Laboratory, University of California, San Francisco | Alicia Sotomayor-Gonzalez; Candace Wang; Charles Chiu; Daniel Wong; Venice Servellita; Yueyuan Zhang |

EPI_ISL_1824733, EPI_ISL_1824734, EPI_ISL_1824735, EPI_ISL_1824736, EPI_ISL_1824737, EPI_ISL_1824738, EPI_ISL_1824739, EPI_ISL_1824740, EPI_ISL_1824741, EPI_ISL_1824742, EPI_ISL_1824743, EPI_ISL_1824744, EPI_ISL_1824745, EPI_ISL_1824746, EPI_ISL_1824747, EPI_ISL_1824748, EPI_ISL_1824749, EPI_ISL_1824750, EPI_ISL_1824751, EPI_ISL_1824752, EPI_ISL_1824753, EPI_ISL_1824754, EPI_ISL_1824755, EPI_ISL_1824756, EPI_ISL_1824757, EPI_ISL_1824758, EPI_ISL_1824759, EPI_ISL_1824760, EPI_ISL_1824761, EPI_ISL_1824762, EPI_ISL_1824763, EPI_ISL_1824764, EPI_ISL_1824765, EPI_ISL_1824766, EPI_ISL_1824767, EPI_ISL_1824768, EPI_ISL_1824769, EPI_ISL_1824770, EPI_ISL_1824771, EPI_ISL_1824772, EPI_ISL_1824773, EPI_ISL_1824774, EPI_ISL_1824775, EPI_ISL_1824776, EPI_ISL_1824777, EPI_ISL_1824778, EPI_ISL_1824779, EPI_ISL_1824780, EPI_ISL_1824781, EPI_ISL_1824782, EPI_ISL_1824783, EPI_ISL_1824784, EPI_ISL_1824785, EPI_ISL_1824786, EPI_ISL_1824787, EPI_ISL_1824788, EPI_ISL_1824789, EPI_ISL_1824790, EPI_ISL_1824791, EPI_ISL_1824792, EPI_ISL_1824793, EPI_ISL_1824794, EPI_ISL_1824795, EPI_ISL_1824796, EPI_ISL_1824797, EPI_ISL_1824798, EPI_ISL_1824799, EPI_ISL_1824800, EPI_ISL_1824801, EPI_ISL_1824802, EPI_ISL_1824803, EPI_ISL_1824804, EPI_ISL_1824805, EPI_ISL_1824806, EPI_ISL_1824807, EPI_ISL_1824808, EPI_ISL_1824809, EPI_ISL_1824810, EPI_ISL_1824811, EPI_ISL_1824812, EPI_ISL_1824813, EPI_ISL_1824814, EPI_ISL_1824815, EPI_ISL_1824816, EPI_ISL_1824817, EPI_ISL_1824818, EPI_ISL_1824819, EPI_ISL_1824820, EPI_ISL_1824821, EPI_ISL_1824822, EPI_ISL_1824823, EPI_ISL_1824824, EPI_ISL_1824825, EPI_ISL_1824826, EPI_ISL_1824827, EPI_ISL_1824828, EPI_ISL_1824829, EPI_ISL_1824830, EPI_ISL_1824831, EPI_ISL_1824832, EPI_ISL_1824833, EPI_ISL_1824834, EPI_ISL_1824835, EPI_ISL_1824836, EPI_ISL_1824837, EPI_ISL_1824838, EPI_ISL_1824839, EPI_ISL_1824840, EPI_ISL_1824841, EPI_ISL_1824842, EPI_ISL_1824843, EPI_ISL_1824844, EPI_ISL_1824845, EPI_ISL_1824846, EPI_ISL_1824847, EPI_ISL_1824848, EPI_ISL_1824849, EPI_ISL_1824850, EPI_ISL_1824851, EPI_ISL_1824852, EPI_ISL_1824853, EPI_ISL_1824854, EPI_ISL_1824855, EPI_ISL_1824856, EPI_ISL_1824857, EPI_ISL_1824858, EPI_ISL_1824859, EPI_ISL_1824860, EPI_ISL_1824861, EPI_ISL_1824862, EPI_ISL_1824863, EPI_ISL_1824864, EPI_ISL_1824865, EPI_ISL_1824866, EPI_ISL_1824867, EPI_ISL_1824868, EPI_ISL_1824869, EPI_ISL_1824870, EPI_ISL_1824871, EPI_ISL_1824872, EPI_ISL_1824873, EPI_ISL_1824874, EPI_ISL_1824875, EPI_ISL_1824876, EPI_ISL_1824877, EPI_ISL_1824878, EPI_ISL_1824879, EPI_ISL_1824880, EPI_ISL_1824881, EPI_ISL_1824882, EPI_ISL_1824883, EPI_ISL_1824884, EPI_ISL_1824885, EPI_ISL_1824886, EPI_ISL_1824887, EPI_ISL_1824888, EPI_ISL_1824889, EPI_ISL_1824890, EPI_ISL_1824891, EPI_ISL_1824892, EPI_ISL_1824893, EPI_ISL_1824894, EPI_ISL_1824895, EPI_ISL_1824896, EPI_ISL_1824897, EPI_ISL_1824898, EPI_ISL_1824899, EPI_ISL_1824900, EPI_ISL_1824901, EPI_ISL_1824902, EPI_ISL_1824903, EPI_ISL_1824904, EPI_ISL_1824905, EPI_ISL_1824906, EPI_ISL_1824907, EPI_ISL_1824908, EPI_ISL_1824909, EPI_ISL_1824910, EPI_ISL_1824911, EPI_ISL_1824912, EPI_ISL_1824913, EPI_ISL_1824914, EPI_ISL_1824915, EPI_ISL_1824916, EPI_ISL_1824917, EPI_ISL_1824918, EPI_ISL_1824919, EPI_ISL_1824920, EPI_ISL_1824921, EPI_ISL_1824922, EPI_ISL_1824923, EPI_ISL_1824924, EPI_ISL_1824925, EPI_ISL_1824926, EPI_ISL_1824927, EPI_ISL_1824928, EPI_ISL_1824929, EPI_ISL_1824930, EPI_ISL_1824931, EPI_ISL_1824932, EPI_ISL_1824933, EPI_ISL_1824934, EPI_ISL_1824935, EPI_ISL_1824936, EPI_ISL_1824937, EPI_ISL_1824938, EPI_ISL_1824939, EPI_ISL_1824940, EPI_ISL_1824941, EPI_ISL_1824942, EPI_ISL_1824943, EPI_ISL_1824944, EPI_ISL_1824945, EPI_ISL_1824946, EPI_ISL_1824947, EPI_ISL_1824948, EPI_ISL_1824949, EPI_ISL_1824950, EPI_ISL_1824951, EPI_ISL_1824952, EPI_ISL_1824953, EPI_ISL_1824954, EPI_ISL_1824955, EPI_ISL_1824956, EPI_ISL_1824957, EPI_ISL_1824958, EPI_ISL_1824959, EPI_ISL_1824960, EPI_ISL_1824961, EPI_ISL_1824962, EPI_ISL_1824963, EPI_ISL_1824964, EPI_ISL_1824965, EPI_ISL_1824966, EPI_ISL_1824967, EPI_ISL_1824968, EPI_ISL_1824969, EPI_ISL_1824970, EPI_ISL_1824971, EPI_ISL_1824972, EPI_ISL_1824973, EPI_ISL_1824974, EPI_ISL_1824975, EPI_ISL_1824976, EPI_ISL_1824977, EPI_ISL_1824978, EPI_ISL_1824979

| | | | |
|---|---|---|---|
| see above | Color Genomics | Chiu Laboratory, University of California, San Francisco | Alicia Sotomayor-Gonzalez; Alicia Zhou; Candace Wang; Charles Chiu; Daniel Wong; Erika Torres; Madyson Wanner; Scott Topper; Venice Servellita; Yueyuan Zhang |

EPI_ISL_1235062, EPI_ISL_1235065, EPI_ISL_1235066, EPI_ISL_1235073, EPI_ISL_1235076, EPI_ISL_1235086, EPI_ISL_1235087, EPI_ISL_1235089, EPI_ISL_1235096, EPI_ISL_1235100, EPI_ISL_1235101, EPI_ISL_1235103, EPI_ISL_1235109, EPI_ISL_1235110, EPI_ISL_1235113, EPI_ISL_1235114, EPI_ISL_1235129, EPI_ISL_1235131, EPI_ISL_1235151, EPI_ISL_1235156, EPI_ISL_1235160, EPI_ISL_1235163, EPI_ISL_1235168, EPI_ISL_1235169, EPI_ISL_1235174, EPI_ISL_1235196, EPI_ISL_1235201, EPI_ISL_1235205, EPI_ISL_1235207, EPI_ISL_1235211, EPI_ISL_1235215, EPI_ISL_1235216, EPI_ISL_1235218, EPI_ISL_1235227, EPI_ISL_1235231, EPI_ISL_1235233, EPI_ISL_1235239, EPI_ISL_1235250, EPI_ISL_1235253, EPI_ISL_1235257, EPI_ISL_1235265, EPI_ISL_1235266, EPI_ISL_1235267, EPI_ISL_1235268, EPI_ISL_1235275, EPI_ISL_1235280, EPI_ISL_1235296, EPI_ISL_1235297, EPI_ISL_1235300, EPI_ISL_1235305, EPI_ISL_1235320, EPI_ISL_1235324, EPI_ISL_1235327, EPI_ISL_1235333, EPI_ISL_1235336, EPI_ISL_1235337, EPI_ISL_1235340, EPI_ISL_1235341, EPI_ISL_1235343, EPI_ISL_1235344, EPI_ISL_1235347, EPI_ISL_1235349, EPI_ISL_1235350, EPI_ISL_1235355, EPI_ISL_1235356, EPI_ISL_1477675, EPI_ISL_1477697, EPI_ISL_1477716, EPI_ISL_1477732, EPI_ISL_1477738, EPI_ISL_1477743, EPI_ISL_1477764, EPI_ISL_1477774, EPI_ISL_1477781, EPI_ISL_1477785, EPI_ISL_1477788, EPI_ISL_1477802, EPI_ISL_1477810, EPI_ISL_1477813, EPI_ISL_1477826, EPI_ISL_1477827, EPI_ISL_1477829, EPI_ISL_1477834, EPI_ISL_1477835, EPI_ISL_1477849, EPI_ISL_1477851, EPI_ISL_1477861, EPI_ISL_1477866, EPI_ISL_1477869, EPI_ISL_1477870, EPI_ISL_1477894, EPI_ISL_1477910, EPI_ISL_1477918, EPI_ISL_1477927, EPI_ISL_1477939, EPI_ISL_1477946, EPI_ISL_1477956, EPI_ISL_1477957, EPI_ISL_1477966, EPI_ISL_1477980, EPI_ISL_1477995, EPI_ISL_1477998, EPI_ISL_1477999, EPI_ISL_1478000, EPI_ISL_1478005, EPI_ISL_1478007, EPI_ISL_1478020, EPI_ISL_1478023, EPI_ISL_1478047, EPI_ISL_1478049, EPI_ISL_1478052, EPI_ISL_1478069, EPI_ISL_1478086, EPI_ISL_1478090, EPI_ISL_1478091, EPI_ISL_1478098, EPI_ISL_1478101, EPI_ISL_1478120, EPI_ISL_1478128, EPI_ISL_1478133, EPI_ISL_1478139, EPI_ISL_1478141, EPI_ISL_1478145, EPI_ISL_1478155, EPI_ISL_1478157, EPI_ISL_1478160, EPI_ISL_1478181, EPI_ISL_1478182, EPI_ISL_1478189, EPI_ISL_1529144, EPI_ISL_1529164, EPI_ISL_1529166, EPI_ISL_1529204, EPI_ISL_1529217, EPI_ISL_1529269, EPI_ISL_1529275, EPI_ISL_1529296, EPI_ISL_1529340, EPI_ISL_1529350, EPI_ISL_1529355, EPI_ISL_2102745, EPI_ISL_2102757, EPI_ISL_2102821, EPI_ISL_2102833, EPI_ISL_2102856, EPI_ISL_2102886, EPI_ISL_2102937, EPI_ISL_2102961, EPI_ISL_2102996, EPI_ISL_2103003, EPI_ISL_2103005, EPI_ISL_2103007, EPI_ISL_2103010, EPI_ISL_2103011, EPI_ISL_2103012, EPI_ISL_2103013, EPI_ISL_2103016, EPI_ISL_2103017, EPI_ISL_2103022, EPI_ISL_2103024, EPI_ISL_2103030, EPI_ISL_2103037, EPI_ISL_2103039, EPI_ISL_2103040, EPI_ISL_2103041, EPI_ISL_2103043, EPI_ISL_2103048, EPI_ISL_2103052, EPI_ISL_2103054, EPI_ISL_2103055, EPI_ISL_2103056, EPI_ISL_2103057, EPI_ISL_2103060, EPI_ISL_2103063, EPI_ISL_2103065, EPI_ISL_2103068, EPI_ISL_2103069, EPI_ISL_2103070, EPI_ISL_2103072, EPI_ISL_2103076, EPI_ISL_2103077, EPI_ISL_2103078, EPI_ISL_2103079, EPI_ISL_2103080, EPI_ISL_2103082, EPI_ISL_2103084, EPI_ISL_2103087, EPI_ISL_2103099, EPI_ISL_2103101, EPI_ISL_2103102, EPI_ISL_2103104, EPI_ISL_2103105, EPI_ISL_2103108, EPI_ISL_2103111, EPI_ISL_2103113, EPI_ISL_2103117, EPI_ISL_2103125, EPI_ISL_2103127, EPI_ISL_2103131, EPI_ISL_2103136, EPI_ISL_2103141, EPI_ISL_2103143, EPI_ISL_2103145, EPI_ISL_2103150, EPI_ISL_2103154, EPI_ISL_2103156, EPI_ISL_2103157, EPI_ISL_2103164, EPI_ISL_2103168, EPI_ISL_2103170, EPI_ISL_2103171, EPI_ISL_2103174, EPI_ISL_2103175, EPI_ISL_2103179, EPI_ISL_2103182, EPI_ISL_2103183, EPI_ISL_2103184, EPI_ISL_2103185, EPI_ISL_2103186, EPI_ISL_2103191, EPI_ISL_2103196, EPI_ISL_2103198

| | | | |
|---|---|---|---|
| see above | Contra Costa County Public Health Lab | Chan-Zuckerberg Biohub | CZB Cliahub Consortium |

EPI_ISL_1027366, EPI_ISL_1027367, EPI_ISL_1185288, EPI_ISL_1185387, EPI_ISL_1185443, EPI_ISL_1234806, EPI_ISL_1234938, EPI_ISL_1529232, EPI_ISL_1701088, EPI_ISL_1701201, EPI_ISL_1701471, EPI_ISL_1701548, EPI_ISL_1701660, EPI_ISL_1821326, EPI_ISL_2102855, EPI_ISL_2102943, EPI_ISL_2103091, EPI_ISL_2103188

| | | | |
|---|---|---|---|
| see above | County of San Luis Obispo Public Health Laboratory | Chan-Zuckerberg Biohub | CZB Cliahub Consortium |

EPI_ISL_1093604, EPI_ISL_1093620, EPI_ISL_1093624, EPI_ISL_1093629, EPI_ISL_1093634, EPI_ISL_1093642, EPI_ISL_1093653, EPI_ISL_1093683, EPI_ISL_1093689, EPI_ISL_1093690, EPI_ISL_1093692, EPI_ISL_1093694, EPI_ISL_1093712, EPI_ISL_1093731, EPI_ISL_1093759, EPI_ISL_1093770, EPI_ISL_1093774, EPI_ISL_1093811, EPI_ISL_1093814, EPI_ISL_1093831, EPI_ISL_1093836, EPI_ISL_1093872, EPI_ISL_1093907, EPI_ISL_1093914, EPI_ISL_1093915, EPI_ISL_1093941, EPI_ISL_1093954, EPI_ISL_1093975, EPI_ISL_1093977, EPI_ISL_1093990, EPI_ISL_1094031, EPI_ISL_1094041, EPI_ISL_1094077, EPI_ISL_1094079, EPI_ISL_1094085, EPI_ISL_1094087, EPI_ISL_1094101, EPI_ISL_1094105, EPI_ISL_1094110, EPI_ISL_1094139, EPI_ISL_1094141, EPI_ISL_1094169, EPI_ISL_1094182, EPI_ISL_1098816, EPI_ISL_1098826, EPI_ISL_1318699, EPI_ISL_1318701, EPI_ISL_1318702, EPI_ISL_1318704, EPI_ISL_1318705, EPI_ISL_1318709, EPI_ISL_1318712, EPI_ISL_1318713, EPI_ISL_1318714, EPI_ISL_1318715, EPI_ISL_1318716, EPI_ISL_1318720, EPI_ISL_1318721, EPI_ISL_1318722, EPI_ISL_1318730, EPI_ISL_1318731, EPI_ISL_1318733, EPI_ISL_1318736, EPI_ISL_1318738, EPI_ISL_1318739, EPI_ISL_1318741, EPI_ISL_1318742, EPI_ISL_1318743, EPI_ISL_1318744, EPI_ISL_1318745, EPI_ISL_1318746, EPI_ISL_1318750, EPI_ISL_1318751, EPI_ISL_1318753, EPI_ISL_1318754, EPI_ISL_1318755, EPI_ISL_1318757, EPI_ISL_1318758, EPI_ISL_1318759, EPI_ISL_1318761, EPI_ISL_1318763, EPI_ISL_1318764, EPI_ISL_1318769, EPI_ISL_1318770, EPI_ISL_1318773, EPI_ISL_1318776, EPI_ISL_1318779, EPI_ISL_1318780, EPI_ISL_1318781, EPI_ISL_1318782, EPI_ISL_1318783, EPI_ISL_1318787, EPI_ISL_1318789, EPI_ISL_1318791, EPI_ISL_1318793, EPI_ISL_1318794, EPI_ISL_1318798, EPI_ISL_1318799, EPI_ISL_1318804, EPI_ISL_1318806, EPI_ISL_1318808, EPI_ISL_1318810, EPI_ISL_1318811, EPI_ISL_1318812, EPI_ISL_1318817, EPI_ISL_1318818, EPI_ISL_1318819, EPI_ISL_1318821, EPI_ISL_1318826, EPI_ISL_1318827, EPI_ISL_1318828, EPI_ISL_1318829, EPI_ISL_1318830, EPI_ISL_1318831, EPI_ISL_1318833, EPI_ISL_1318834, EPI_ISL_1318836, EPI_ISL_1318840, EPI_ISL_1318841, EPI_ISL_1318842, EPI_ISL_1318846, EPI_ISL_1318847, EPI_ISL_1318848, EPI_ISL_1318850, EPI_ISL_1318851, EPI_ISL_1318852, EPI_ISL_1318853, EPI_ISL_1318855, EPI_ISL_1318858, EPI_ISL_1318861, EPI_ISL_1318862, EPI_ISL_1318863, EPI_ISL_1318866, EPI_ISL_1318867, EPI_ISL_1318868, EPI_ISL_1318869, EPI_ISL_1318873, EPI_ISL_1318874, EPI_ISL_1318875, EPI_ISL_1318876, EPI_ISL_1318878, EPI_ISL_1318879, EPI_ISL_1318880, EPI_ISL_1318884, EPI_ISL_1318885, EPI_ISL_1318888, EPI_ISL_1318889, EPI_ISL_1318890, EPI_ISL_1318893, EPI_ISL_1318901, EPI_ISL_1318902, EPI_ISL_1318904, EPI_ISL_1318905, EPI_ISL_1318907, EPI_ISL_1318911, EPI_ISL_1318915, EPI_ISL_1318916, EPI_ISL_1318918, EPI_ISL_1318919, EPI_ISL_1318920, EPI_ISL_1318921, EPI_ISL_1318924, EPI_ISL_1318925, EPI_ISL_1318926, EPI_ISL_1318927, EPI_ISL_1318928, EPI_ISL_1318929, EPI_ISL_1318930, EPI_ISL_1318933, EPI_ISL_1318935, EPI_ISL_1318936, EPI_ISL_1318938, EPI_ISL_1318946, EPI_ISL_1318948, EPI_ISL_1318954, EPI_ISL_1318955, EPI_ISL_1318956, EPI_ISL_1318957, EPI_ISL_1318958, EPI_ISL_1318962, EPI_ISL_1318963, EPI_ISL_1318969, EPI_ISL_1318971, EPI_ISL_1318972, EPI_ISL_1318978, EPI_ISL_1318981, EPI_ISL_1318982, EPI_ISL_1318986, EPI_ISL_1318987, EPI_ISL_1318988, EPI_ISL_1347783, EPI_ISL_1347785, EPI_ISL_1347786, EPI_ISL_1347787, EPI_ISL_1347788, EPI_ISL_1347789, EPI_ISL_1349075, EPI_ISL_1349080, EPI_ISL_1349082, EPI_ISL_1349084, EPI_ISL_1349085, EPI_ISL_1349089, EPI_ISL_1349091

| | | | |
|---|---|---|---|
| see above | DeRisi Lab, University of California, San Francisco | DeRisi Lab, University of California, San Francisco | Amy Kistler; Anthea Mitchell; Carina Marquez; Diane Havlir; Diane Jones; Douglas Black; Eric Chow; Gabriel Chamie; Genay Pilarowski; IDseq Team; Jackie Martinez; James Peng; Jamin Liu; Jon Jacobo; Lucy Li; Luis Rubio; Manu Vanaerschot; Matthew Laurie; Maya Petersen; Patrick Ayscue; Sabrina Mann; Sara Sunshine; Susana Rojas; Susy Rojas; Valerie Tulier-Laiwa; and Joseph DeRisi |

EPI_ISL_1811642, EPI_ISL_1811643, EPI_ISL_1811646, EPI_ISL_1811647, EPI_ISL_1811649, EPI_ISL_1811650, EPI_ISL_1811651, EPI_ISL_1811652, EPI_ISL_1811654, EPI_ISL_1811656, EPI_ISL_1811658, EPI_ISL_1811659, EPI_ISL_1811665, EPI_ISL_1811666, EPI_ISL_1811670, EPI_ISL_1811671, EPI_ISL_1811674, EPI_ISL_1811675, EPI_ISL_1811678, EPI_ISL_1811681, EPI_ISL_1811682, EPI_ISL_1811683, EPI_ISL_1811686, EPI_ISL_1811688, EPI_ISL_1811690, EPI_ISL_1811692, EPI_ISL_1811695, EPI_ISL_1811696, EPI_ISL_1811698, EPI_ISL_1811701, EPI_ISL_1811703, EPI_ISL_1811705, EPI_ISL_1811708, EPI_ISL_1811710, EPI_ISL_1811711, EPI_ISL_1811712, EPI_ISL_1811714, EPI_ISL_1811715, EPI_ISL_1811717, EPI_ISL_1811718, EPI_ISL_1811719, EPI_ISL_1811720, EPI_ISL_1811721, EPI_ISL_1811722, EPI_ISL_1811724, EPI_ISL_1811725, EPI_ISL_1811726, EPI_ISL_1811727, EPI_ISL_1811730, EPI_ISL_1811733, EPI_ISL_1811736, EPI_ISL_1811737, EPI_ISL_1811738, EPI_ISL_1811739, EPI_ISL_1811741, EPI_ISL_1811742, EPI_ISL_1811743, EPI_ISL_1811744, EPI_ISL_1811745, EPI_ISL_1811746, EPI_ISL_1811747, EPI_ISL_1811748, EPI_ISL_1811749, EPI_ISL_1811750, EPI_ISL_1811751, EPI_ISL_1811753, EPI_ISL_1811754, EPI_ISL_1811755, EPI_ISL_1811758, EPI_ISL_1811759, EPI_ISL_1811760, EPI_ISL_1811761, EPI_ISL_1811762, EPI_ISL_1811764, EPI_ISL_1811765, EPI_ISL_1811766, EPI_ISL_1811770, EPI_ISL_1811772, EPI_ISL_1811773, EPI_ISL_1811775, EPI_ISL_1811776, EPI_ISL_1811777, EPI_ISL_1811778, EPI_ISL_1811779, EPI_ISL_1811780, EPI_ISL_1811782, EPI_ISL_1811784, EPI_ISL_1811785, EPI_ISL_1811786, EPI_ISL_1811787, EPI_ISL_1811788, EPI_ISL_1811789, EPI_ISL_1811795, EPI_ISL_1811796, EPI_ISL_1811797, EPI_ISL_1811799, EPI_ISL_1811801, EPI_ISL_1811802, EPI_ISL_1811803, EPI_ISL_1811804, EPI_ISL_1811805, EPI_ISL_1811806, EPI_ISL_1811808, EPI_ISL_1811809, EPI_ISL_1811810, EPI_ISL_1811811, EPI_ISL_1811813, EPI_ISL_1811814, EPI_ISL_1811815, EPI_ISL_1811816, EPI_ISL_1811818, EPI_ISL_1811819, EPI_ISL_1811820, EPI_ISL_1811822, EPI_ISL_1811826, EPI_ISL_1811827, EPI_ISL_1811829, EPI_ISL_1811830, EPI_ISL_1811832, EPI_ISL_1811833, EPI_ISL_1811837, EPI_ISL_1811840, EPI_ISL_1811843, EPI_ISL_1811845, EPI_ISL_1811848, EPI_ISL_1811849, EPI_ISL_1811850, EPI_ISL_1811852, EPI_ISL_1811854, EPI_ISL_1811855, EPI_ISL_1811862, EPI_ISL_1811872, EPI_ISL_1811873, EPI_ISL_1811875, EPI_ISL_1811885, EPI_ISL_1811887, EPI_ISL_1811889, EPI_ISL_1811897, EPI_ISL_1811902, EPI_ISL_1811927, EPI_ISL_1811929, EPI_ISL_1811931, EPI_ISL_1811936, EPI_ISL_1811965, EPI_ISL_1811969, EPI_ISL_1811992, EPI_ISL_1811996, EPI_ISL_1812000, EPI_ISL_1812001, EPI_ISL_1812011, EPI_ISL_1812012, EPI_ISL_1812016, EPI_ISL_1812017, EPI_ISL_1812019, EPI_ISL_1812021, EPI_ISL_1812031, EPI_ISL_1812036, EPI_ISL_1812042, EPI_ISL_1812043, EPI_ISL_1812044, EPI_ISL_1812045, EPI_ISL_1812049, EPI_ISL_1812050, EPI_ISL_1812051, EPI_ISL_1812052, EPI_ISL_1812060, EPI_ISL_1812061, EPI_ISL_1812062, EPI_ISL_1812065, EPI_ISL_1812068, EPI_ISL_1812071, EPI_ISL_1812074, EPI_ISL_1812075, EPI_ISL_1812076, EPI_ISL_1812077, EPI_ISL_1812086, EPI_ISL_1812087, EPI_ISL_1812090, EPI_ISL_1812098, EPI_ISL_1812099, EPI_ISL_1812100, EPI_ISL_1812102, EPI_ISL_1812104, EPI_ISL_1812116, EPI_ISL_1812120, EPI_ISL_1812121, EPI_ISL_1812122, EPI_ISL_1812123, EPI_ISL_1812127, EPI_ISL_1812130, EPI_ISL_1812131, EPI_ISL_1812134, EPI_ISL_1812137, EPI_ISL_1812145, EPI_ISL_1812150, EPI_ISL_1812154, EPI_ISL_1812157, EPI_ISL_1812163, EPI_ISL_1812168, EPI_ISL_1812170, EPI_ISL_1812180, EPI_ISL_1812181, EPI_ISL_1812183, EPI_ISL_1812185, EPI_ISL_1812187, EPI_ISL_1812188, EPI_ISL_1812191, EPI_ISL_1812192, EPI_ISL_1812193, EPI_ISL_1812197, EPI_ISL_1812198, EPI_ISL_1812201, EPI_ISL_1812203, EPI_ISL_1812204, EPI_ISL_1812206, EPI_ISL_1812211, EPI_ISL_1812216, EPI_ISL_1812219, EPI_ISL_1812227, EPI_ISL_1812228, EPI_ISL_1812231, EPI_ISL_1812238, EPI_ISL_1812239, EPI_ISL_1812242, EPI_ISL_1812252, EPI_ISL_1812257, EPI_ISL_1812265, EPI_ISL_1812268, EPI_ISL_1812275, EPI_ISL_1812276, EPI_ISL_1812284, EPI_ISL_1812286, EPI_ISL_1812287, EPI_ISL_1812290, EPI_ISL_1812292, EPI_ISL_1812294, EPI_ISL_1812295, EPI_ISL_1812310, EPI_ISL_1812311, EPI_ISL_1812314, EPI_ISL_1812323, EPI_ISL_1812326, EPI_ISL_1812327, EPI_ISL_1812328, EPI_ISL_1812329, EPI_ISL_1812336, EPI_ISL_1812338, EPI_ISL_1812341, EPI_ISL_1812343, EPI_ISL_1812348, EPI_ISL_1812349, EPI_ISL_1812352, EPI_ISL_1812357, EPI_ISL_1812360, EPI_ISL_1812364, EPI_ISL_1812368, EPI_ISL_1812372, EPI_ISL_1812373, EPI_ISL_1812378, EPI_ISL_1812379, EPI_ISL_1812382, EPI_ISL_1812384, EPI_ISL_1812387, EPI_ISL_1812388, EPI_ISL_1812389, EPI_ISL_1812390, EPI_ISL_1812391, EPI_ISL_1812392, EPI_ISL_1812393, EPI_ISL_1812395, EPI_ISL_1812398, EPI_ISL_1812400, EPI_ISL_1812402, EPI_ISL_1812404, EPI_ISL_1812407, EPI_ISL_1812408, EPI_ISL_1812411, EPI_ISL_1812414, EPI_ISL_1812420, EPI_ISL_1812423, EPI_ISL_1812424, EPI_ISL_1812427, EPI_ISL_1812430, EPI_ISL_1812433, EPI_ISL_1812434, EPI_ISL_1812436, EPI_ISL_1812445, EPI_ISL_1812446, EPI_ISL_1812447, EPI_ISL_1812453, EPI_ISL_1812458, EPI_ISL_1812460, EPI_ISL_1812464, EPI_ISL_1812465, EPI_ISL_1812468, EPI_ISL_1812472, EPI_ISL_1812477, EPI_ISL_1812480, EPI_ISL_1812481, EPI_ISL_1812482, EPI_ISL_1812486, EPI_ISL_1812487, EPI_ISL_1812489, EPI_ISL_1812491, EPI_ISL_1812499, EPI_ISL_1812506, EPI_ISL_1812509, EPI_ISL_1812512, EPI_ISL_1812517, EPI_ISL_1812518, EPI_ISL_1812522, EPI_ISL_1812524, EPI_ISL_1812525, EPI_ISL_1812526, EPI_ISL_1812530, EPI_ISL_1812531, EPI_ISL_1812533, EPI_ISL_1812534, EPI_ISL_1812535, EPI_ISL_1812536, EPI_ISL_1812537, EPI_ISL_1812538, EPI_ISL_1812541, EPI_ISL_1812548, EPI_ISL_1812549, EPI_ISL_1812550, EPI_ISL_1812556, EPI_ISL_1812559, EPI_ISL_1812566, EPI_ISL_1812568, EPI_ISL_1812569, EPI_ISL_1812574, EPI_ISL_1812577, EPI_ISL_1812578, EPI_ISL_1812579, EPI_ISL_1812582, EPI_ISL_1812586, EPI_ISL_1812593, EPI_ISL_1812597, EPI_ISL_1812598, EPI_ISL_1812599, EPI_ISL_1812601, EPI_ISL_1812604, EPI_ISL_1812608, EPI_ISL_1812611, EPI_ISL_1812614, EPI_ISL_1812615, EPI_ISL_1812621, EPI_ISL_1812623, EPI_ISL_1812625, EPI_ISL_1812626, EPI_ISL_1812627, EPI_ISL_1812628, EPI_ISL_1812629, EPI_ISL_1812632, EPI_ISL_1812634, EPI_ISL_1812637, EPI_ISL_1812639, EPI_ISL_1812640, EPI_ISL_1812641, EPI_ISL_1812642, EPI_ISL_1812644, EPI_ISL_1812647, EPI_ISL_1812648, EPI_ISL_1812650, EPI_ISL_1812651, EPI_ISL_1812653, EPI_ISL_1812657, EPI_ISL_1812660, EPI_ISL_1812661, EPI_ISL_1812665, EPI_ISL_1812666, EPI_ISL_1812673, EPI_ISL_1812674, EPI_ISL_1812676, EPI_ISL_1812677, EPI_ISL_1812679, EPI_ISL_1812682, EPI_ISL_1812684, EPI_ISL_1812686, EPI_ISL_1812687, EPI_ISL_1812688, EPI_ISL_1812691, EPI_ISL_1812692, EPI_ISL_1812696, EPI_ISL_1812698, EPI_ISL_1812699, EPI_ISL_1812702, EPI_ISL_1812703, EPI_ISL_1812705, EPI_ISL_1812712, EPI_ISL_1812716, EPI_ISL_1812724, EPI_ISL_1812727, EPI_ISL_1812728, EPI_ISL_1812729, EPI_ISL_1812731, EPI_ISL_1812732, EPI_ISL_1812733, EPI_ISL_1812736, EPI_ISL_1812737, EPI_ISL_1812739, EPI_ISL_1812743, EPI_ISL_1812745, EPI_ISL_1812746, EPI_ISL_1812749, EPI_ISL_1812750, EPI_ISL_1812751, EPI_ISL_1812752, EPI_ISL_1812753, EPI_ISL_1812755, EPI_ISL_1812764, EPI_ISL_1812767, EPI_ISL_1812768, EPI_ISL_1812769, EPI_ISL_1812771, EPI_ISL_1812775, EPI_ISL_1812780, EPI_ISL_1812790, EPI_ISL_1812795, EPI_ISL_1812796, EPI_ISL_1812797, EPI_ISL_1812800, EPI_ISL_1812801, EPI_ISL_1812803, EPI_ISL_1812804, EPI_ISL_1812811, EPI_ISL_1812812, EPI_ISL_1812815, EPI_ISL_1812816, EPI_ISL_1812820, EPI_ISL_1812825, EPI_ISL_1812827, EPI_ISL_1812829, EPI_ISL_1812831, EPI_ISL_1812833, EPI_ISL_1812837, EPI_ISL_1812838, EPI_ISL_1812840, EPI_ISL_1812844, EPI_ISL_1812846, EPI_ISL_1812850, EPI_ISL_1812853, EPI_ISL_1812857, EPI_ISL_1812860, EPI_ISL_1812864, EPI_ISL_1812865, EPI_ISL_1812866, EPI_ISL_1812868, EPI_ISL_1812869, EPI_ISL_1812872, EPI_ISL_1812873, EPI_ISL_1812876, EPI_ISL_1812880, EPI_ISL_1812881, EPI_ISL_1812886, EPI_ISL_1812887, EPI_ISL_1812890, EPI_ISL_1812891, EPI_ISL_1812892, EPI_ISL_1812893, EPI_ISL_1812895, EPI_ISL_1812896, EPI_ISL_1812901, EPI_ISL_1812913, EPI_ISL_1812914, EPI_ISL_1812915, EPI_ISL_1812918, EPI_ISL_1812919, EPI_ISL_1812921, EPI_ISL_1812922, EPI_ISL_1812926, EPI_ISL_1812931, EPI_ISL_1812934, EPI_ISL_1812935, EPI_ISL_1812936, EPI_ISL_1812937, EPI_ISL_1812938, EPI_ISL_1812940, EPI_ISL_1812942, EPI_ISL_1812944, EPI_ISL_1812946, EPI_ISL_1812956, EPI_ISL_1812959, EPI_ISL_1812964, EPI_ISL_1812969, EPI_ISL_1812983, EPI_ISL_1812986, EPI_ISL_1812992, EPI_ISL_1812993, EPI_ISL_1812996, EPI_ISL_1812997, EPI_ISL_1812998, EPI_ISL_1812999, EPI_ISL_1813001, EPI_ISL_1813004, EPI_ISL_1813005, EPI_ISL_1813010, EPI_ISL_1813016, EPI_ISL_1813018, EPI_ISL_1813019, EPI_ISL_1813020, EPI_ISL_1813022, EPI_ISL_1813025, EPI_ISL_1813026, EPI_ISL_1813028, EPI_ISL_1813030, EPI_ISL_1813032, EPI_ISL_1813035, EPI_ISL_1813041, EPI_ISL_1813047, EPI_ISL_1813048, EPI_ISL_1813058, EPI_ISL_1813060, EPI_ISL_1813064, EPI_ISL_1813067, EPI_ISL_1813068, EPI_ISL_1813071, EPI_ISL_1813072, EPI_ISL_1813074, EPI_ISL_1813076, EPI_ISL_1813078, EPI_ISL_1813083, EPI_ISL_1813094, EPI_ISL_1813096, EPI_ISL_1813098, EPI_ISL_1813102, EPI_ISL_1813107, EPI_ISL_1813109, EPI_ISL_1813116, EPI_ISL_1813117, EPI_ISL_1813120, EPI_ISL_1813123, EPI_ISL_1813133, EPI_ISL_1813137, EPI_ISL_1813138, EPI_ISL_1813146, EPI_ISL_1813148, EPI_ISL_1813165, EPI_ISL_1813168, EPI_ISL_1813172, EPI_ISL_1813175, EPI_ISL_1813176, EPI_ISL_1813178, EPI_ISL_1813179, EPI_ISL_1813181, EPI_ISL_1813182, EPI_ISL_1813185, EPI_ISL_1813189, EPI_ISL_1813193, EPI_ISL_1813195, EPI_ISL_1813199, EPI_ISL_1813201, EPI_ISL_1813202, EPI_ISL_1813204, EPI_ISL_1813208, EPI_ISL_1813212, EPI_ISL_1813214, EPI_ISL_1813215, EPI_ISL_1813218, EPI_ISL_1813219, EPI_ISL_1813220, EPI_ISL_1813222, EPI_ISL_1813224, EPI_ISL_1813231, EPI_ISL_1813240, EPI_ISL_1813245, EPI_ISL_1813249, EPI_ISL_1813253, EPI_ISL_1813254, EPI_ISL_1813260, EPI_ISL_1813269, EPI_ISL_1813271, EPI_ISL_1813272, EPI_ISL_1813274, EPI_ISL_1813275, EPI_ISL_1813277, EPI_ISL_1813278, EPI_ISL_1813283, EPI_ISL_1813284, EPI_ISL_1813285, EPI_ISL_1813286, EPI_ISL_1813289, EPI_ISL_1813294, EPI_ISL_1813295, EPI_ISL_1813297, EPI_ISL_1813299, EPI_ISL_1813301, EPI_ISL_1813305, EPI_ISL_1813307, EPI_ISL_1813308, EPI_ISL_1813309, EPI_ISL_1813311, EPI_ISL_1813314, EPI_ISL_1813316, EPI_ISL_1813317, EPI_ISL_1813319, EPI_ISL_1813320, EPI_ISL_1813327, EPI_ISL_1813328, EPI_ISL_1813334, EPI_ISL_1813344, EPI_ISL_1813346, EPI_ISL_1813348, EPI_ISL_1813351, EPI_ISL_1813352, EPI_ISL_1813354, EPI_ISL_1813355, EPI_ISL_1813357, EPI_ISL_1813358, EPI_ISL_1813365, EPI_ISL_1813366, EPI_ISL_1813368, EPI_ISL_1813370, EPI_ISL_1813372, EPI_ISL_1813373, EPI_ISL_1813374, EPI_ISL_1813376, EPI_ISL_1813380, EPI_ISL_1813383, EPI_ISL_1813384, EPI_ISL_1813388, EPI_ISL_1813400, EPI_ISL_1813406, EPI_ISL_1813407, EPI_ISL_1813410, EPI_ISL_1813411, EPI_ISL_1813415, EPI_ISL_1813418, EPI_ISL_1813422, EPI_ISL_1813423, EPI_ISL_1813424, EPI_ISL_1813429, EPI_ISL_1813431, EPI_ISL_1813433, EPI_ISL_1813437, EPI_ISL_1813438, EPI_ISL_1813441, EPI_ISL_1813443, EPI_ISL_1813448, EPI_ISL_1813453, EPI_ISL_1813455, EPI_ISL_1813459, EPI_ISL_1813461, EPI_ISL_1813463, EPI_ISL_1813464, EPI_ISL_1813466, EPI_ISL_1813468, EPI_ISL_1813469, EPI_ISL_1813472, EPI_ISL_1813479, EPI_ISL_1813481, EPI_ISL_1813484, EPI_ISL_1813487, EPI_ISL_1813488, EPI_ISL_1813489, EPI_ISL_1813491, EPI_ISL_1813494, EPI_ISL_1813496, EPI_ISL_1813499, EPI_ISL_1813502, EPI_ISL_1813505, EPI_ISL_1813507, EPI_ISL_1813509, EPI_ISL_1813510, EPI_ISL_1813514, EPI_ISL_1813515, EPI_ISL_1813519, EPI_ISL_1813522, EPI_ISL_1813523, EPI_ISL_1813524, EPI_ISL_1813529, EPI_ISL_1813541, EPI_ISL_1813546, EPI_ISL_1813549, EPI_ISL_1813550, EPI_ISL_1813552, EPI_ISL_1813556, EPI_ISL_1813557, EPI_ISL_1813563, EPI_ISL_1813566, EPI_ISL_1813569, EPI_ISL_1813570, EPI_ISL_1813572, EPI_ISL_1813575, EPI_ISL_1813576, EPI_ISL_1813577, EPI_ISL_1813578, EPI_ISL_1813580, EPI_ISL_1813585, EPI_ISL_1813598, EPI_ISL_1813602, EPI_ISL_1813603, EPI_ISL_1813607, EPI_ISL_1813609, EPI_ISL_1813612, EPI_ISL_1813613, EPI_ISL_1813614, EPI_ISL_1813619, EPI_ISL_1813620, EPI_ISL_1813621, EPI_ISL_1813622, EPI_ISL_1813623, EPI_ISL_1813625, EPI_ISL_1813627, EPI_ISL_1813631, EPI_ISL_1813635, EPI_ISL_1813637, EPI_ISL_1813639, EPI_ISL_1813640, EPI_ISL_1813641, EPI_ISL_1813642, EPI_ISL_1813643, EPI_ISL_1813645, EPI_ISL_1813646, EPI_ISL_1813649, EPI_ISL_1813651, EPI_ISL_1813652, EPI_ISL_1813656, EPI_ISL_1813658, EPI_ISL_1813659, EPI_ISL_1813661, EPI_ISL_1813664, EPI_ISL_1813667, EPI_ISL_1813670, EPI_ISL_1813672, EPI_ISL_1813674, EPI_ISL_1813675, EPI_ISL_1813679, EPI_ISL_1813683, EPI_ISL_1813685, EPI_ISL_1813686, EPI_ISL_1813688, EPI_ISL_1813689, EPI_ISL_1813692, EPI_ISL_1813694, EPI_ISL_1813695, EPI_ISL_1813698, EPI_ISL_1813708, EPI_ISL_1813709, EPI_ISL_1813712, EPI_ISL_1813715, EPI_ISL_1813717, EPI_ISL_1813718, EPI_ISL_1813719, EPI_ISL_1813724, EPI_ISL_1813729, EPI_ISL_1813730, EPI_ISL_1813733, EPI_ISL_1813736, EPI_ISL_1813740, EPI_ISL_1813741, EPI_ISL_1813742, EPI_ISL_1813747, EPI_ISL_1813748, EPI_ISL_1813749, EPI_ISL_1813754, EPI_ISL_1813757, EPI_ISL_1813762, EPI_ISL_1813763, EPI_ISL_1813768, EPI_ISL_1813769, EPI_ISL_1813772, EPI_ISL_1813775, EPI_ISL_1813781, EPI_ISL_1813785, EPI_ISL_1813796, EPI_ISL_1813797, EPI_ISL_1813799, EPI_ISL_1813803, EPI_ISL_1813805, EPI_ISL_1813810, EPI_ISL_1813812, EPI_ISL_1813813, EPI_ISL_1813816, EPI_ISL_1813819, EPI_ISL_1813820, EPI_ISL_1813826, EPI_ISL_1813828, EPI_ISL_1813830, EPI_ISL_1813835, EPI_ISL_1813836, EPI_ISL_1813838, EPI_ISL_1813839, EPI_ISL_1813844, EPI_ISL_1813848, EPI_ISL_1813852, EPI_ISL_1813854, EPI_ISL_1813867, EPI_ISL_1813869, EPI_ISL_1813872, EPI_ISL_1813879, EPI_ISL_1813880, EPI_ISL_1813882, EPI_ISL_1813883, EPI_ISL_1813885, EPI_ISL_1813889, EPI_ISL_1813890, EPI_ISL_1813895, EPI_ISL_1813900, EPI_ISL_1813902, EPI_ISL_1813907, EPI_ISL_1813909, EPI_ISL_1813911, EPI_ISL_1813913, EPI_ISL_1813914, EPI_ISL_1813915, EPI_ISL_1813916, EPI_ISL_1813917, EPI_ISL_1813918, EPI_ISL_1813920, EPI_ISL_1813923, EPI_ISL_1813926, EPI_ISL_1813927, EPI_ISL_1813929, EPI_ISL_1813931, EPI_ISL_1813932, EPI_ISL_1813935, EPI_ISL_1813936, EPI_ISL_1813937, EPI_ISL_1813941, EPI_ISL_1813942, EPI_ISL_1813944, EPI_ISL_1813951, EPI_ISL_1813955, EPI_ISL_1813958, EPI_ISL_1813959, EPI_ISL_1813960, EPI_ISL_1813961, EPI_ISL_1813966, EPI_ISL_1813967, EPI_ISL_1813969, EPI_ISL_1813970, EPI_ISL_1813978, EPI_ISL_1813989, EPI_ISL_1813993, EPI_ISL_1813996, EPI_ISL_1813998, EPI_ISL_1813999, EPI_ISL_1814001, EPI_ISL_1814002, EPI_ISL_1814004, EPI_ISL_1814005, EPI_ISL_1814010, EPI_ISL_1814011, EPI_ISL_1814013, EPI_ISL_1814016, EPI_ISL_1814017, EPI_ISL_1814021, EPI_ISL_1814023, EPI_ISL_1814024, EPI_ISL_1814025, EPI_ISL_1814027, EPI_ISL_1814028, EPI_ISL_1814029, EPI_ISL_1814030, EPI_ISL_1814032, EPI_ISL_1814033, EPI_ISL_1814034, EPI_ISL_1814035, EPI_ISL_1814037, EPI_ISL_1814038, EPI_ISL_1814040, EPI_ISL_1814041, EPI_ISL_1814042, EPI_ISL_1814044, EPI_ISL_1814047, EPI_ISL_1814048, EPI_ISL_1814051, EPI_ISL_1814056, EPI_ISL_1814057, EPI_ISL_1814058, EPI_ISL_1814062, EPI_ISL_1814067, EPI_ISL_1814068, EPI_ISL_1814069, EPI_ISL_1814070, EPI_ISL_1814073, EPI_ISL_1814074, EPI_ISL_1814079, EPI_ISL_1814080, EPI_ISL_1814084, EPI_ISL_1814085, EPI_ISL_1814086, EPI_ISL_1814087, EPI_ISL_1814090, EPI_ISL_1814092, EPI_ISL_1814095, EPI_ISL_1814096, EPI_ISL_1814100, EPI_ISL_1814102, EPI_ISL_1814103, EPI_ISL_1814104, EPI_ISL_1814105, EPI_ISL_1814106, EPI_ISL_1814109, EPI_ISL_1814110, EPI_ISL_1814119, EPI_ISL_1814124, EPI_ISL_1814129, EPI_ISL_1814133, EPI_ISL_1814134, EPI_ISL_1814136, EPI_ISL_1814138, EPI_ISL_1814139, EPI_ISL_1814142, EPI_ISL_1814143, EPI_ISL_1814149, EPI_ISL_1814152, EPI_ISL_1814156, EPI_ISL_1814157, EPI_ISL_1814158, EPI_ISL_1814161, EPI_ISL_1814164, EPI_ISL_1814165, EPI_ISL_1814166, EPI_ISL_1814167, EPI_ISL_1814168, EPI_ISL_1814169, EPI_ISL_1814171, EPI_ISL_1814172, EPI_ISL_1814173, EPI_ISL_1814174, EPI_ISL_1814176, EPI_ISL_1814177, EPI_ISL_1814178, EPI_ISL_1814182, EPI_ISL_1814184, EPI_ISL_1814185, EPI_ISL_1814187, EPI_ISL_1814188, EPI_ISL_1814189, EPI_ISL_1814190, EPI_ISL_1814191, EPI_ISL_1814192, EPI_ISL_1814194, EPI_ISL_1814196, EPI_ISL_1814203, EPI_ISL_1814204, EPI_ISL_1814205, EPI_ISL_1814206, EPI_ISL_1814208, EPI_ISL_1814215, EPI_ISL_1814216, EPI_ISL_1814217, EPI_ISL_1814219, EPI_ISL_1814220, EPI_ISL_1814221, EPI_ISL_1814222, EPI_ISL_1814223, EPI_ISL_1814224, EPI_ISL_1814228, EPI_ISL_1814229, EPI_ISL_1814230, EPI_ISL_1814231, EPI_ISL_1814232, EPI_ISL_1814233, EPI_ISL_1814236, EPI_ISL_1814241, EPI_ISL_1814244, EPI_ISL_1814248, EPI_ISL_1814251, EPI_ISL_1814253, EPI_ISL_1814254, EPI_ISL_1814255, EPI_ISL_1814256, EPI_ISL_1814257, EPI_ISL_1814262, EPI_ISL_1814263, EPI_ISL_1814264, EPI_ISL_1814268, EPI_ISL_1814270, EPI_ISL_1814271, EPI_ISL_1814272, EPI_ISL_1814275, EPI_ISL_1814281, EPI_ISL_1814282, EPI_ISL_1814283, EPI_ISL_1814288, EPI_ISL_1814290, EPI_ISL_1814291, EPI_ISL_1814296, EPI_ISL_1814299, EPI_ISL_1814300, EPI_ISL_1814302, EPI_ISL_1814305, EPI_ISL_1814307, EPI_ISL_1814309, EPI_ISL_1814316, EPI_ISL_1814317, EPI_ISL_1814321, EPI_ISL_1814322, EPI_ISL_1814323, EPI_ISL_1814327, EPI_ISL_1814328, EPI_ISL_1814332, EPI_ISL_1814339, EPI_ISL_1814340, EPI_ISL_1814342, EPI_ISL_1814343, EPI_ISL_1814344, EPI_ISL_1814349, EPI_ISL_1814352, EPI_ISL_1814356, EPI_ISL_1814362, EPI_ISL_1814368, EPI_ISL_1814369, EPI_ISL_1814371, EPI_ISL_1814373, EPI_ISL_1814374, EPI_ISL_1814377, EPI_ISL_1814382, EPI_ISL_1814383, EPI_ISL_1814384, EPI_ISL_1814388, EPI_ISL_1814391, EPI_ISL_1814394, EPI_ISL_1814395, EPI_ISL_1814399, EPI_ISL_1814404, EPI_ISL_1814407, EPI_ISL_1814410, EPI_ISL_1814411, EPI_ISL_1814412, EPI_ISL_1814413, EPI_ISL_1814416, EPI_ISL_1814419, EPI_ISL_1814420, EPI_ISL_1814421, EPI_ISL_1814423, EPI_ISL_1814424, EPI_ISL_1814425, EPI_ISL_1814426, EPI_ISL_1814429, EPI_ISL_1814430, EPI_ISL_1814432, EPI_ISL_1814433, EPI_ISL_1814435, EPI_ISL_1814437, EPI_ISL_1814440, EPI_ISL_1814441, EPI_ISL_1814442, EPI_ISL_1814444, EPI_ISL_1814447, EPI_ISL_1814451, EPI_ISL_1814452, EPI_ISL_1814453, EPI_ISL_1814455, EPI_ISL_1814459, EPI_ISL_1814464, EPI_ISL_1814467, EPI_ISL_1814468, EPI_ISL_1814471, EPI_ISL_1814472, EPI_ISL_1814473, EPI_ISL_1814474, EPI_ISL_1814475, EPI_ISL_1814476, EPI_ISL_1814478, EPI_ISL_1814479, EPI_ISL_1814481, EPI_ISL_1814482, EPI_ISL_1814485, EPI_ISL_1814490, EPI_ISL_1814495, EPI_ISL_1814497, EPI_ISL_1814500, EPI_ISL_1814502, EPI_ISL_1814505, EPI_ISL_1814506, EPI_ISL_1814507, EPI_ISL_1814508, EPI_ISL_1814509, EPI_ISL_1814510, EPI_ISL_1814512, EPI_ISL_1814514, EPI_ISL_1814516, EPI_ISL_1814517, EPI_ISL_1814518, EPI_ISL_1814523, EPI_ISL_1814530, EPI_ISL_1814532, EPI_ISL_1814533, EPI_ISL_1814534, EPI_ISL_1814538, EPI_ISL_1814543, EPI_ISL_1814544, EPI_ISL_1814546, EPI_ISL_1814548, EPI_ISL_1814551, EPI_ISL_1814555, EPI_ISL_1814559, EPI_ISL_1814563, EPI_ISL_1814566, EPI_ISL_1814568, EPI_ISL_1814573, EPI_ISL_1814575, EPI_ISL_1814577, EPI_ISL_1814578, EPI_ISL_1814579, EPI_ISL_1814584, EPI_ISL_1814586, EPI_ISL_1814588, EPI_ISL_1814589, EPI_ISL_1814590, EPI_ISL_1814591, EPI_ISL_1814592, EPI_ISL_1814593, EPI_ISL_1814594, EPI_ISL_1814596, EPI_ISL_1814597, EPI_ISL_1814604, EPI_ISL_1814606, EPI_ISL_1814607, EPI_ISL_1814611, EPI_ISL_1814613, EPI_ISL_1814614, EPI_ISL_1814617, EPI_ISL_1814619, EPI_ISL_1814621, EPI_ISL_1814624, EPI_ISL_1814626, EPI_ISL_1814629, EPI_ISL_1814633, EPI_ISL_1814636, EPI_ISL_1814638, EPI_ISL_1814639, EPI_ISL_1814641, EPI_ISL_1814643, EPI_ISL_1814645, EPI_ISL_1814646, EPI_ISL_1814648, EPI_ISL_1814650, EPI_ISL_1814654, EPI_ISL_1814657, EPI_ISL_1814659, EPI_ISL_1814663, EPI_ISL_1814666, EPI_ISL_1814667, EPI_ISL_1814668, EPI_ISL_1814672, EPI_ISL_1814675, EPI_ISL_1814676, EPI_ISL_1814677, EPI_ISL_1814680, EPI_ISL_1814685, EPI_ISL_1814688, EPI_ISL_1814689, EPI_ISL_1814692, EPI_ISL_1814695, EPI_ISL_1814698, EPI_ISL_1814699, EPI_ISL_1814702, EPI_ISL_1814705, EPI_ISL_1814706, EPI_ISL_1814708, EPI_ISL_1814709, EPI_ISL_1814711, EPI_ISL_1814712, EPI_ISL_1814713, EPI_ISL_1814714, EPI_ISL_1814716, EPI_ISL_1814719, EPI_ISL_1814723, EPI_ISL_1814725, EPI_ISL_1814726, EPI_ISL_1814727, EPI_ISL_1814730, EPI_ISL_1814731, EPI_ISL_1814734, EPI_ISL_1814737, EPI_ISL_1814738, EPI_ISL_1814740, EPI_ISL_1814741, EPI_ISL_1814748, EPI_ISL_1814749, EPI_ISL_1814752, EPI_ISL_1814754, EPI_ISL_1814755, EPI_ISL_1814756, EPI_ISL_1814758, EPI_ISL_1814759, EPI_ISL_1814761, EPI_ISL_1814764, EPI_ISL_1814772, EPI_ISL_1814774, EPI_ISL_1814779, EPI_ISL_1814780, EPI_ISL_1814782, EPI_ISL_1814787, EPI_ISL_1814789, EPI_ISL_1814791, EPI_ISL_1814793, EPI_ISL_1814794, EPI_ISL_1814797, EPI_ISL_1814799, EPI_ISL_1814800, EPI_ISL_1814801, EPI_ISL_1814802, EPI_ISL_1814804, EPI_ISL_1814805, EPI_ISL_1814808, EPI_ISL_1814809, EPI_ISL_1814810, EPI_ISL_1814811, EPI_ISL_1814817, EPI_ISL_1814818, EPI_ISL_1814820, EPI_ISL_1814822, EPI_ISL_1814823, EPI_ISL_1814824, EPI_ISL_1814827, EPI_ISL_1814829, EPI_ISL_1814831, EPI_ISL_1814832, EPI_ISL_1814835, EPI_ISL_1814837, EPI_ISL_1814842, EPI_ISL_1814843, EPI_ISL_1814852, EPI_ISL_1814853, EPI_ISL_1814855, EPI_ISL_1814863, EPI_ISL_1814865, EPI_ISL_1814866, EPI_ISL_1814869, EPI_ISL_1814870, EPI_ISL_1814871, EPI_ISL_1814874, EPI_ISL_1814875, EPI_ISL_1814876, EPI_ISL_1814882, EPI_ISL_1814884, EPI_ISL_1814887, EPI_ISL_1814888, EPI_ISL_1814890, EPI_ISL_1814897, EPI_ISL_1814898, EPI_ISL_1814900, EPI_ISL_1814904, EPI_ISL_1814907, EPI_ISL_1814908, EPI_ISL_1814911, EPI_ISL_1814913, EPI_ISL_1814915, EPI_ISL_1814916, EPI_ISL_1814920, EPI_ISL_1814921, EPI_ISL_1814922, EPI_ISL_1814925, EPI_ISL_1814929, EPI_ISL_1814931, EPI_ISL_1814932, EPI_ISL_1814933, EPI_ISL_1814934, EPI_ISL_1814936, EPI_ISL_1814937, EPI_ISL_1814938, EPI_ISL_1814941, EPI_ISL_1814944, EPI_ISL_1814945, EPI_ISL_1814947, EPI_ISL_1814948, EPI_ISL_1814951, EPI_ISL_1814953, EPI_ISL_1814959, EPI_ISL_1814960, EPI_ISL_1814964, EPI_ISL_1814966, EPI_ISL_1814970, EPI_ISL_1814972, EPI_ISL_1814975, EPI_ISL_1814977, EPI_ISL_1814978, EPI_ISL_1814979, EPI_ISL_1814982, EPI_ISL_1814983, EPI_ISL_1814985, EPI_ISL_1814988, EPI_ISL_1814990, EPI_ISL_1814992, EPI_ISL_1814994, EPI_ISL_1814995, EPI_ISL_1814996, EPI_ISL_1814997, EPI_ISL_1814999, EPI_ISL_1815003, EPI_ISL_1815004, EPI_ISL_1815006, EPI_ISL_1815007, EPI_ISL_1815009, EPI_ISL_1815013, EPI_ISL_1815014, EPI_ISL_1815015, EPI_ISL_1815016, EPI_ISL_1815021, EPI_ISL_1815023, EPI_ISL_1815024, EPI_ISL_1815025, EPI_ISL_1815029, EPI_ISL_1815032, EPI_ISL_1815037, EPI_ISL_1815042, EPI_ISL_1815043, EPI_ISL_1815044, EPI_ISL_1815046, EPI_ISL_1815049, EPI_ISL_1815050, EPI_ISL_1815051, EPI_ISL_1815052, EPI_ISL_1815054, EPI_ISL_1815055, EPI_ISL_1815058, EPI_ISL_1815059, EPI_ISL_1815061, EPI_ISL_1815065, EPI_ISL_1815066, EPI_ISL_1815068, EPI_ISL_1815069, EPI_ISL_1815071, EPI_ISL_1815072, EPI_ISL_1815073, EPI_ISL_1815076, EPI_ISL_1815077, EPI_ISL_1815078, EPI_ISL_1815080, EPI_ISL_1815081, EPI_ISL_1815082, EPI_ISL_1815083, EPI_ISL_1815085, EPI_ISL_1815087, EPI_ISL_1815089, EPI_ISL_1815091, EPI_ISL_1815093, EPI_ISL_1815096, EPI_ISL_1815098, EPI_ISL_1815102, EPI_ISL_1815104, EPI_ISL_1815105, EPI_ISL_1815106, EPI_ISL_1815117, EPI_ISL_1815120, EPI_ISL_1815123, EPI_ISL_1815124, EPI_ISL_1815127, EPI_ISL_1815128, EPI_ISL_1815129, EPI_ISL_1815130, EPI_ISL_1815131, EPI_ISL_1815132, EPI_ISL_1815133, EPI_ISL_1815136, EPI_ISL_1815138, EPI_ISL_1815141, EPI_ISL_1815143, EPI_ISL_1815146, EPI_ISL_1815148, EPI_ISL_1815152, EPI_ISL_1815153, EPI_ISL_1815154, EPI_ISL_1815159, EPI_ISL_1815168, EPI_ISL_1815170, EPI_ISL_1815171,

EPI_ISL_1815172, EPI_ISL_1815173, EPI_ISL_1815174, EPI_ISL_1815177, EPI_ISL_1815182, EPI_ISL_1815185, EPI_ISL_1815187, EPI_ISL_1815188, EPI_ISL_1815189, EPI_ISL_1815190, EPI_ISL_1815192, EPI_ISL_1815193, EPI_ISL_1815194, EPI_ISL_1815195, EPI_ISL_1815196, EPI_ISL_1815199, EPI_ISL_1815204, EPI_ISL_1815210, EPI_ISL_1815214, EPI_ISL_1815215, EPI_ISL_1815218, EPI_ISL_1815226,
EPI_ISL_1815227, EPI_ISL_1815228, EPI_ISL_1815229, EPI_ISL_1815242, EPI_ISL_1815243, EPI_ISL_1815244, EPI_ISL_1815249, EPI_ISL_1815250, EPI_ISL_1815252, EPI_ISL_1815254, EPI_ISL_1815256, EPI_ISL_1815258, EPI_ISL_1815259, EPI_ISL_1815262, EPI_ISL_1815265, EPI_ISL_1815266, EPI_ISL_1815269, EPI_ISL_1815271, EPI_ISL_1815272, EPI_ISL_1815279, EPI_ISL_1815281, EPI_ISL_1815286,
EPI_ISL_1815289, EPI_ISL_1815291, EPI_ISL_1815292, EPI_ISL_1815294, EPI_ISL_1815295, EPI_ISL_1815300, EPI_ISL_1815302, EPI_ISL_1815303, EPI_ISL_1815304, EPI_ISL_1815308, EPI_ISL_1815310, EPI_ISL_1815311, EPI_ISL_1815313, EPI_ISL_1815319, EPI_ISL_1815322, EPI_ISL_1815323, EPI_ISL_1815325, EPI_ISL_1815327, EPI_ISL_1815328, EPI_ISL_1815330, EPI_ISL_1815339, EPI_ISL_1815343,
EPI_ISL_1815347, EPI_ISL_1815348, EPI_ISL_1815353, EPI_ISL_1815356, EPI_ISL_1815358, EPI_ISL_1815360, EPI_ISL_1815361, EPI_ISL_1815362, EPI_ISL_1815366, EPI_ISL_1815367, EPI_ISL_1815376, EPI_ISL_1815379, EPI_ISL_1815382, EPI_ISL_1815384, EPI_ISL_1815389, EPI_ISL_1815394, EPI_ISL_1815395, EPI_ISL_1815398, EPI_ISL_1815400, EPI_ISL_1815403, EPI_ISL_1815406, EPI_ISL_1815408,
EPI_ISL_1815409, EPI_ISL_1815410, EPI_ISL_1815413, EPI_ISL_1815417, EPI_ISL_1815424, EPI_ISL_1815425, EPI_ISL_1815427, EPI_ISL_1815428, EPI_ISL_1815430, EPI_ISL_1815431, EPI_ISL_1815432, EPI_ISL_1815434, EPI_ISL_1815437, EPI_ISL_1815439, EPI_ISL_1815442, EPI_ISL_1815443, EPI_ISL_1815444, EPI_ISL_1815445, EPI_ISL_1815447, EPI_ISL_1815448, EPI_ISL_1815458, EPI_ISL_1815459,
EPI_ISL_1815460, EPI_ISL_1815462, EPI_ISL_1815465, EPI_ISL_1815467, EPI_ISL_1815469, EPI_ISL_1815470, EPI_ISL_1815472, EPI_ISL_1815476, EPI_ISL_1815477, EPI_ISL_1815478, EPI_ISL_1815479, EPI_ISL_1815480, EPI_ISL_1815481, EPI_ISL_1815483, EPI_ISL_1815485, EPI_ISL_1815487, EPI_ISL_1815488, EPI_ISL_1815489, EPI_ISL_1815490, EPI_ISL_1815492, EPI_ISL_1815494, EPI_ISL_1815497,
EPI_ISL_1815498, EPI_ISL_1815499, EPI_ISL_1815503, EPI_ISL_1815504, EPI_ISL_1815505, EPI_ISL_1815508, EPI_ISL_1815510, EPI_ISL_1815517, EPI_ISL_1815518, EPI_ISL_1815520, EPI_ISL_1815522, EPI_ISL_1815523, EPI_ISL_1815524, EPI_ISL_1815525, EPI_ISL_1815528, EPI_ISL_1815529, EPI_ISL_1815530, EPI_ISL_1815531, EPI_ISL_1815536, EPI_ISL_1815537, EPI_ISL_1815538, EPI_ISL_1815539,
EPI_ISL_1815540, EPI_ISL_1815541, EPI_ISL_1815542, EPI_ISL_1815551, EPI_ISL_1815552, EPI_ISL_1815553, EPI_ISL_1815555, EPI_ISL_1815556, EPI_ISL_1815557, EPI_ISL_1815558, EPI_ISL_1815560, EPI_ISL_1815562, EPI_ISL_1815563, EPI_ISL_1815565, EPI_ISL_1815567, EPI_ISL_1815570, EPI_ISL_1815571, EPI_ISL_1815572, EPI_ISL_1815580, EPI_ISL_1815585, EPI_ISL_1815587, EPI_ISL_1815589,
EPI_ISL_1815591, EPI_ISL_1815593, EPI_ISL_1815596, EPI_ISL_1815597, EPI_ISL_1815598, EPI_ISL_1815599, EPI_ISL_1815600, EPI_ISL_1815601, EPI_ISL_1815602, EPI_ISL_1815610, EPI_ISL_1815613, EPI_ISL_1815614, EPI_ISL_1815615, EPI_ISL_1815620, EPI_ISL_1815626, EPI_ISL_1815629, EPI_ISL_1815632, EPI_ISL_1815633, EPI_ISL_1815636, EPI_ISL_1815637, EPI_ISL_1815640, EPI_ISL_1815642,
EPI_ISL_1815643, EPI_ISL_1815644, EPI_ISL_1815650, EPI_ISL_1815655, EPI_ISL_1815658, EPI_ISL_1815662, EPI_ISL_1815668, EPI_ISL_1815674, EPI_ISL_1815676, EPI_ISL_1815679, EPI_ISL_1815680, EPI_ISL_1815682, EPI_ISL_1815685, EPI_ISL_1815686, EPI_ISL_1815687, EPI_ISL_1815689, EPI_ISL_1815690, EPI_ISL_1815692, EPI_ISL_1815694, EPI_ISL_1815695, EPI_ISL_1815696, EPI_ISL_1815701,
EPI_ISL_1815703, EPI_ISL_1815709, EPI_ISL_1815715, EPI_ISL_1815718, EPI_ISL_1815721, EPI_ISL_1815722, EPI_ISL_1815723, EPI_ISL_1815726, EPI_ISL_1815729, EPI_ISL_1815731, EPI_ISL_1815732, EPI_ISL_1815733, EPI_ISL_1815736, EPI_ISL_1815739, EPI_ISL_1815740, EPI_ISL_1815745, EPI_ISL_1815746, EPI_ISL_1815752, EPI_ISL_1815755, EPI_ISL_1815758, EPI_ISL_1815763, EPI_ISL_1815767,
EPI_ISL_1815769, EPI_ISL_1815771, EPI_ISL_1815772, EPI_ISL_1815773, EPI_ISL_1815779, EPI_ISL_1815780, EPI_ISL_1815783, EPI_ISL_1815787, EPI_ISL_1815790, EPI_ISL_1815793, EPI_ISL_1815797, EPI_ISL_1815798, EPI_ISL_1815799, EPI_ISL_1815802, EPI_ISL_1815805, EPI_ISL_1815810, EPI_ISL_1815811, EPI_ISL_1815815, EPI_ISL_1815822, EPI_ISL_1815824, EPI_ISL_1815827, EPI_ISL_1815828,
EPI_ISL_1815829, EPI_ISL_1815830, EPI_ISL_1815831, EPI_ISL_1815835, EPI_ISL_1815836, EPI_ISL_1815837, EPI_ISL_1815839, EPI_ISL_1815842, EPI_ISL_1815843, EPI_ISL_1815845, EPI_ISL_1815846, EPI_ISL_1815850, EPI_ISL_1815853, EPI_ISL_1815854, EPI_ISL_1815858, EPI_ISL_1815860, EPI_ISL_1815862, EPI_ISL_1815864, EPI_ISL_1815865, EPI_ISL_1815866, EPI_ISL_1815867, EPI_ISL_1815871,
EPI_ISL_1815872, EPI_ISL_1815874, EPI_ISL_1815876, EPI_ISL_1815880, EPI_ISL_1815881, EPI_ISL_1815882, EPI_ISL_1815884, EPI_ISL_1815885, EPI_ISL_1815891, EPI_ISL_1815892, EPI_ISL_1815894, EPI_ISL_1815895, EPI_ISL_1815900, EPI_ISL_1815907, EPI_ISL_1815908, EPI_ISL_1815911, EPI_ISL_1815912, EPI_ISL_1815913, EPI_ISL_1815916, EPI_ISL_1815917, EPI_ISL_1815922, EPI_ISL_1815923,
EPI_ISL_1815926, EPI_ISL_1815931, EPI_ISL_1815933, EPI_ISL_1815935, EPI_ISL_1815936, EPI_ISL_1815940, EPI_ISL_1815943, EPI_ISL_1815945, EPI_ISL_1815949, EPI_ISL_1815950, EPI_ISL_1815952, EPI_ISL_1815953, EPI_ISL_1815954, EPI_ISL_1815955, EPI_ISL_1815961, EPI_ISL_1815962, EPI_ISL_1815964, EPI_ISL_1815966, EPI_ISL_1815967, EPI_ISL_1815971, EPI_ISL_1815979, EPI_ISL_1815982,
EPI_ISL_1815987, EPI_ISL_1815989, EPI_ISL_1815990, EPI_ISL_1815992, EPI_ISL_1815994, EPI_ISL_1815995, EPI_ISL_1815996, EPI_ISL_1815998, EPI_ISL_1815999, EPI_ISL_1816000, EPI_ISL_1816005, EPI_ISL_1816007, EPI_ISL_1816008, EPI_ISL_1816011, EPI_ISL_1816012, EPI_ISL_1816013, EPI_ISL_1816014, EPI_ISL_1816024, EPI_ISL_1816025, EPI_ISL_1816027, EPI_ISL_1816028, EPI_ISL_1816029,
EPI_ISL_1816031, EPI_ISL_1816032, EPI_ISL_1816034, EPI_ISL_1816035, EPI_ISL_1816037, EPI_ISL_1816038, EPI_ISL_1816040, EPI_ISL_1816042, EPI_ISL_1816044, EPI_ISL_1816048, EPI_ISL_1816049, EPI_ISL_1816050, EPI_ISL_1816056, EPI_ISL_1816057, EPI_ISL_1816061, EPI_ISL_1816067, EPI_ISL_1816069, EPI_ISL_1816073, EPI_ISL_1816077, EPI_ISL_1816078, EPI_ISL_1816080, EPI_ISL_1816089,
EPI_ISL_1816097, EPI_ISL_1816102, EPI_ISL_1816103, EPI_ISL_1816107, EPI_ISL_1816110, EPI_ISL_1816111, EPI_ISL_1816115, EPI_ISL_1816119, EPI_ISL_1816120, EPI_ISL_1816124, EPI_ISL_1816128, EPI_ISL_1816130, EPI_ISL_1816131, EPI_ISL_1816138, EPI_ISL_1816139, EPI_ISL_1816140, EPI_ISL_1816145, EPI_ISL_1816148, EPI_ISL_1816149, EPI_ISL_1816151, EPI_ISL_1816157, EPI_ISL_1816158,
EPI_ISL_1816159, EPI_ISL_1816160, EPI_ISL_1816164, EPI_ISL_1816167, EPI_ISL_1816171, EPI_ISL_1816173, EPI_ISL_1816177, EPI_ISL_1816178, EPI_ISL_1816184, EPI_ISL_1816185, EPI_ISL_1816187, EPI_ISL_1816188, EPI_ISL_1816189, EPI_ISL_1816190, EPI_ISL_1816191, EPI_ISL_1816193, EPI_ISL_1816194, EPI_ISL_1816200, EPI_ISL_1816201, EPI_ISL_1816207, EPI_ISL_1816215, EPI_ISL_1816217,
EPI_ISL_1816219, EPI_ISL_1816221, EPI_ISL_1816223, EPI_ISL_1816224, EPI_ISL_1816225, EPI_ISL_1816227, EPI_ISL_1816229, EPI_ISL_1816232, EPI_ISL_1816233, EPI_ISL_1816235, EPI_ISL_1816236, EPI_ISL_1816238, EPI_ISL_1816240, EPI_ISL_1816250, EPI_ISL_1816257, EPI_ISL_1816259, EPI_ISL_1816268, EPI_ISL_1816273, EPI_ISL_1816279, EPI_ISL_1816288, EPI_ISL_1816289, EPI_ISL_1816292,
EPI_ISL_1816294, EPI_ISL_1816295, EPI_ISL_1816301, EPI_ISL_1816304, EPI_ISL_1816305, EPI_ISL_1816306, EPI_ISL_1816310, EPI_ISL_1816315, EPI_ISL_1816321, EPI_ISL_1816325, EPI_ISL_1816327, EPI_ISL_1816328, EPI_ISL_1816329, EPI_ISL_1816330, EPI_ISL_1816333, EPI_ISL_1816334, EPI_ISL_1816337, EPI_ISL_1816338, EPI_ISL_1816341, EPI_ISL_1816342, EPI_ISL_1816343, EPI_ISL_1816345,
EPI_ISL_1816350, EPI_ISL_1816353, EPI_ISL_1816357, EPI_ISL_1816360, EPI_ISL_1816363, EPI_ISL_1816364, EPI_ISL_1816380, EPI_ISL_1816383, EPI_ISL_1816393, EPI_ISL_1816395, EPI_ISL_1816396, EPI_ISL_1816403, EPI_ISL_1816404, EPI_ISL_1816412, EPI_ISL_1816416, EPI_ISL_1816418, EPI_ISL_1816423, EPI_ISL_1816425, EPI_ISL_1816426, EPI_ISL_1816429, EPI_ISL_1816430, EPI_ISL_1816432,
EPI_ISL_1816435, EPI_ISL_1816437, EPI_ISL_1816438, EPI_ISL_1816442, EPI_ISL_1816443, EPI_ISL_1816446, EPI_ISL_1816448, EPI_ISL_1816449, EPI_ISL_1816451, EPI_ISL_1816453, EPI_ISL_1816463, EPI_ISL_1816464, EPI_ISL_1816466, EPI_ISL_1816471, EPI_ISL_1816475, EPI_ISL_1816476, EPI_ISL_1816478, EPI_ISL_1816481, EPI_ISL_1816482, EPI_ISL_1816483, EPI_ISL_1816486, EPI_ISL_1816488,
EPI_ISL_1816494, EPI_ISL_1816495, EPI_ISL_1816496, EPI_ISL_1816499, EPI_ISL_1816502, EPI_ISL_1816506, EPI_ISL_1816507, EPI_ISL_1816509, EPI_ISL_1816510, EPI_ISL_1816520, EPI_ISL_1816524, EPI_ISL_1816527, EPI_ISL_1816528, EPI_ISL_1816529, EPI_ISL_1816531, EPI_ISL_1816532, EPI_ISL_1816534, EPI_ISL_1816544, EPI_ISL_1816546, EPI_ISL_1816547, EPI_ISL_1816550, EPI_ISL_1816554,
EPI_ISL_1816557, EPI_ISL_1816563, EPI_ISL_1816571, EPI_ISL_1816572, EPI_ISL_1816573, EPI_ISL_1816575, EPI_ISL_1816576, EPI_ISL_1816577, EPI_ISL_1816578, EPI_ISL_1816579, EPI_ISL_1816592, EPI_ISL_1816599, EPI_ISL_1816602, EPI_ISL_1816604, EPI_ISL_1816605, EPI_ISL_1816612, EPI_ISL_1816614, EPI_ISL_1816618, EPI_ISL_1816621, EPI_ISL_1816622, EPI_ISL_1816623, EPI_ISL_1816625,
EPI_ISL_1816627, EPI_ISL_1816628, EPI_ISL_1816638, EPI_ISL_1816641, EPI_ISL_1816642, EPI_ISL_1816643, EPI_ISL_1816649, EPI_ISL_1816653, EPI_ISL_1816655, EPI_ISL_1816661, EPI_ISL_1816670, EPI_ISL_1816679, EPI_ISL_1816684, EPI_ISL_1816686, EPI_ISL_1816688, EPI_ISL_1816689, EPI_ISL_1816690, EPI_ISL_1816693, EPI_ISL_1816705, EPI_ISL_1816707, EPI_ISL_1816711, EPI_ISL_1816712,
EPI_ISL_1816714, EPI_ISL_1816717, EPI_ISL_1816726, EPI_ISL_1816731, EPI_ISL_1816734, EPI_ISL_1816744, EPI_ISL_1816753, EPI_ISL_1816755, EPI_ISL_1816762, EPI_ISL_1816763, EPI_ISL_1816768, EPI_ISL_1816776, EPI_ISL_1816779, EPI_ISL_1816781, EPI_ISL_1816798, EPI_ISL_1816806, EPI_ISL_1816809, EPI_ISL_1816812, EPI_ISL_1816828, EPI_ISL_1816854, EPI_ISL_1816877, EPI_ISL_1816883

see above | EXCITE Lab | Andersen lab at Scripps Research | Abigail Schnapper; Alexandre Bolze; Alice Summerfield; Angela Scioscia; Araz Majnoonian; Ashkan Hassani; Brent Brainard+ SEARCH; Celena Andrade; Charlotte Rivera-Garcia; Cheryl Anderson; Chip Schooley; Christine Mitchell + SEARCH; David Becker; David Pride; Efren Sandoval; Elizabeth Cirulli; Fitri Carrissa Wijaya; Francisco Tanudjaja; Fred Wu; Geraint Levan; Greg Humphrey; Helena Tubb; J. Joelle Donofrio-Odmann; James Lu + SEARCH; Jason Nguyen; Jimmy Ramirez; Kelly Schiabor Barrett; Laura Kohn; Magnus Isaksson; Marc Laurent; Marlene Flores; Megan Nguyen; Natasha Martin; Natasha Martin Cheryl Anderson; Nicole L Washington; Patricia Gonzalez-Zuniga; Rebecca Fielding-Miller; Rodolfo Salido; Ryan Cho; Sawyer Farmer; Sharon Reed; Sherry Wang; Simon White; Smruthi Karthikeyan; Sydney Morgan + Marni Jacobs + Celestine Magallanes + Holly Valentine + Louise C. Laurent + SEARCH; Tommy Valles + SEARCH; Tyler Cassens; Victor Cantu; Vinton Omaleki; William Lee

EPI_ISL_1098636, EPI_ISL_1098641, EPI_ISL_1192366, EPI_ISL_1192369, EPI_ISL_1192370, EPI_ISL_1192371, EPI_ISL_1197059, EPI_ISL_1197060, EPI_ISL_1197061, EPI_ISL_1198005

see above | Flow Diagnostics Lab | Los Angeles County PHL | P. Hemarajata et al.

EPI_ISL_1253710, EPI_ISL_1253711, EPI_ISL_1253712, EPI_ISL_1254009, EPI_ISL_1254010, EPI_ISL_1254011, EPI_ISL_1254012

see above | Flow Health | Infectious Disease Program, Broad Institute of Harvard and MIT | Adams, G.; B.L.; B.W.; Bauer, M.; Birren; Carter, A.; Chaluvadi, S.; D.J.; DeRuff, K.; Gallagher, G.; Gladden-Young, A.; J.E.; K.J.; Lagerborg, K.; Lemieux; Loreth, C.; MacInnis; Normandin, E.; P.C.; Park; Reilly, S.; Rudy, M.; Siddle; Smole, S.; Tomkins-Tinch, C.; and Sabeti

EPI_ISL_984753, EPI_ISL_984948, EPI_ISL_1010408, EPI_ISL_1010409, EPI_ISL_1010410, EPI_ISL_1010411, EPI_ISL_1010412, EPI_ISL_1010413, EPI_ISL_1010414, EPI_ISL_1010415, EPI_ISL_1010416, EPI_ISL_1010417, EPI_ISL_1010418, EPI_ISL_1010419, EPI_ISL_1010420, EPI_ISL_1010421, EPI_ISL_1010422, EPI_ISL_1010423, EPI_ISL_1010424, EPI_ISL_1010425, EPI_ISL_1010426, EPI_ISL_1010427,
EPI_ISL_1010428, EPI_ISL_1010429, EPI_ISL_1010430, EPI_ISL_1010431, EPI_ISL_1010432, EPI_ISL_1010433, EPI_ISL_1010434, EPI_ISL_1010435, EPI_ISL_1010436, EPI_ISL_1010437, EPI_ISL_1010438, EPI_ISL_1010439, EPI_ISL_1010440, EPI_ISL_1010441, EPI_ISL_1010442, EPI_ISL_1010443, EPI_ISL_1010444, EPI_ISL_1010445, EPI_ISL_1010446, EPI_ISL_1010447, EPI_ISL_1010448, EPI_ISL_1010449,
EPI_ISL_1010450, EPI_ISL_1010451, EPI_ISL_1010452, EPI_ISL_1010453, EPI_ISL_1010454, EPI_ISL_1010455, EPI_ISL_1010456, EPI_ISL_1010457, EPI_ISL_1010458, EPI_ISL_1010459, EPI_ISL_1010460, EPI_ISL_1010461, EPI_ISL_1010462, EPI_ISL_1010463, EPI_ISL_1010464, EPI_ISL_1010465, EPI_ISL_1010466, EPI_ISL_1010467, EPI_ISL_1010468, EPI_ISL_1010469, EPI_ISL_1010470, EPI_ISL_1010471,
EPI_ISL_1010472, EPI_ISL_1010473, EPI_ISL_1010474, EPI_ISL_1010475, EPI_ISL_1010476, EPI_ISL_1010477, EPI_ISL_1010478, EPI_ISL_1010479, EPI_ISL_1010480, EPI_ISL_1010481, EPI_ISL_1010482, EPI_ISL_1010483, EPI_ISL_1010484, EPI_ISL_1010485, EPI_ISL_1010486, EPI_ISL_1010487, EPI_ISL_1010488, EPI_ISL_1010489, EPI_ISL_1010490, EPI_ISL_1010491, EPI_ISL_1010492, EPI_ISL_1010493,
EPI_ISL_1010494, EPI_ISL_1010495, EPI_ISL_1010496, EPI_ISL_1010497, EPI_ISL_1010498, EPI_ISL_1010499, EPI_ISL_1010500, EPI_ISL_1010501, EPI_ISL_1010502, EPI_ISL_1010503, EPI_ISL_1010504, EPI_ISL_1010505, EPI_ISL_1010506, EPI_ISL_1010507, EPI_ISL_1010508, EPI_ISL_1010509, EPI_ISL_1010510, EPI_ISL_1010511, EPI_ISL_1010512, EPI_ISL_1010513, EPI_ISL_1010514, EPI_ISL_1010515,
EPI_ISL_1010516, EPI_ISL_1010517, EPI_ISL_1010518, EPI_ISL_1010519, EPI_ISL_1010520, EPI_ISL_1010521, EPI_ISL_1010522, EPI_ISL_1010523, EPI_ISL_1010524, EPI_ISL_1010525, EPI_ISL_1010526, EPI_ISL_1010527, EPI_ISL_1010528, EPI_ISL_1010529, EPI_ISL_1010530, EPI_ISL_1010531, EPI_ISL_1010532, EPI_ISL_1010533, EPI_ISL_1010534, EPI_ISL_1010535, EPI_ISL_1010536, EPI_ISL_1010537,
EPI_ISL_1010538, EPI_ISL_1010539, EPI_ISL_1010540, EPI_ISL_1010541, EPI_ISL_1010542, EPI_ISL_1010543, EPI_ISL_1010544, EPI_ISL_1010545, EPI_ISL_1010546, EPI_ISL_1010547, EPI_ISL_1010548, EPI_ISL_1010549, EPI_ISL_1010550, EPI_ISL_1010551, EPI_ISL_1010737, EPI_ISL_1010738, EPI_ISL_1010739, EPI_ISL_1010740, EPI_ISL_1010741, EPI_ISL_1010742, EPI_ISL_1010743, EPI_ISL_1010744,
EPI_ISL_1010745, EPI_ISL_1010746, EPI_ISL_1010747, EPI_ISL_1010748, EPI_ISL_1010749, EPI_ISL_1010750, EPI_ISL_1010751, EPI_ISL_1010752, EPI_ISL_1010753, EPI_ISL_1010754, EPI_ISL_1010755, EPI_ISL_1010756, EPI_ISL_1010757, EPI_ISL_1010758, EPI_ISL_1010759, EPI_ISL_1010760, EPI_ISL_1010761, EPI_ISL_1010762, EPI_ISL_1010763, EPI_ISL_1010764, EPI_ISL_1010765, EPI_ISL_1010766,
EPI_ISL_1010767, EPI_ISL_1010768, EPI_ISL_1010769, EPI_ISL_1010770, EPI_ISL_1010771, EPI_ISL_1010772, EPI_ISL_1010773, EPI_ISL_1010774, EPI_ISL_1010775, EPI_ISL_1010776, EPI_ISL_1010777, EPI_ISL_1010778, EPI_ISL_1010779, EPI_ISL_1010780, EPI_ISL_1010781, EPI_ISL_1010782, EPI_ISL_1010783, EPI_ISL_1010784, EPI_ISL_1010785, EPI_ISL_1010786, EPI_ISL_1010787, EPI_ISL_1010788,
EPI_ISL_1010789, EPI_ISL_1010790, EPI_ISL_1010791, EPI_ISL_1010792, EPI_ISL_1010793, EPI_ISL_1010794, EPI_ISL_1010795, EPI_ISL_1010796, EPI_ISL_1010797, EPI_ISL_1010798, EPI_ISL_1010799, EPI_ISL_1010800, EPI_ISL_1010801, EPI_ISL_1010802, EPI_ISL_1010803, EPI_ISL_1010804, EPI_ISL_1010805, EPI_ISL_1010806, EPI_ISL_1010807, EPI_ISL_1010808, EPI_ISL_1010809, EPI_ISL_1010810,
EPI_ISL_1010811, EPI_ISL_1010812, EPI_ISL_1010813, EPI_ISL_1010814, EPI_ISL_1010815, EPI_ISL_1010816, EPI_ISL_1010817, EPI_ISL_1010818, EPI_ISL_1010819, EPI_ISL_1010820, EPI_ISL_1010821, EPI_ISL_1010822, EPI_ISL_1010823, EPI_ISL_1010824, EPI_ISL_1010825, EPI_ISL_1010826, EPI_ISL_1010827, EPI_ISL_1010828, EPI_ISL_1010829, EPI_ISL_1010830, EPI_ISL_1010831, EPI_ISL_1010832,
EPI_ISL_1010833, EPI_ISL_1010834, EPI_ISL_1010835, EPI_ISL_1010836, EPI_ISL_1010837, EPI_ISL_1010838, EPI_ISL_1010839, EPI_ISL_1010840, EPI_ISL_1010841, EPI_ISL_1010842, EPI_ISL_1010843, EPI_ISL_1010844, EPI_ISL_1010845, EPI_ISL_1010846, EPI_ISL_1010847, EPI_ISL_1010848, EPI_ISL_1010849, EPI_ISL_1010850, EPI_ISL_1010851, EPI_ISL_1010852, EPI_ISL_1010853, EPI_ISL_1010854,
EPI_ISL_1010855, EPI_ISL_1010856, EPI_ISL_1010857, EPI_ISL_1010858, EPI_ISL_1010859, EPI_ISL_1010860, EPI_ISL_1010861, EPI_ISL_1010862, EPI_ISL_1010863, EPI_ISL_1010864, EPI_ISL_1010865, EPI_ISL_1010866, EPI_ISL_1010867, EPI_ISL_1010868, EPI_ISL_1010869, EPI_ISL_1010870, EPI_ISL_1010871, EPI_ISL_1010872, EPI_ISL_1010873, EPI_ISL_1010874, EPI_ISL_1010875, EPI_ISL_1010876,
EPI_ISL_1010877, EPI_ISL_1010878, EPI_ISL_1010879, EPI_ISL_1010880, EPI_ISL_1010881, EPI_ISL_1010882, EPI_ISL_1010883, EPI_ISL_1010884, EPI_ISL_1010885, EPI_ISL_1010886, EPI_ISL_1010887, EPI_ISL_1010888, EPI_ISL_1010889, EPI_ISL_1010890, EPI_ISL_1010891, EPI_ISL_1010892, EPI_ISL_1010893, EPI_ISL_1010894, EPI_ISL_1010895, EPI_ISL_1010896, EPI_ISL_1010897, EPI_ISL_1010898,
EPI_ISL_1010899, EPI_ISL_1010900, EPI_ISL_1010901, EPI_ISL_1010902, EPI_ISL_1010903, EPI_ISL_1010904, EPI_ISL_1010905, EPI_ISL_1010906, EPI_ISL_1010907, EPI_ISL_1010908, EPI_ISL_1010909, EPI_ISL_1010910, EPI_ISL_1010911, EPI_ISL_1010912, EPI_ISL_1010913, EPI_ISL_1010914, EPI_ISL_1010915, EPI_ISL_1010916, EPI_ISL_1010917, EPI_ISL_1010918, EPI_ISL_1010919, EPI_ISL_1010920,
EPI_ISL_1010921, EPI_ISL_1010922, EPI_ISL_1010923, EPI_ISL_1010924, EPI_ISL_1010925, EPI_ISL_1010926, EPI_ISL_1010927, EPI_ISL_1010928, EPI_ISL_1010929, EPI_ISL_1010930, EPI_ISL_1010931, EPI_ISL_1010932, EPI_ISL_1010933, EPI_ISL_1010934, EPI_ISL_1010935, EPI_ISL_1010936, EPI_ISL_1010937, EPI_ISL_1010938, EPI_ISL_1010939, EPI_ISL_1010940, EPI_ISL_1010941, EPI_ISL_1010942,
EPI_ISL_1010943, EPI_ISL_1010944, EPI_ISL_1010945, EPI_ISL_1010946, EPI_ISL_1010947, EPI_ISL_1010948, EPI_ISL_1010949, EPI_ISL_1010950, EPI_ISL_1010951, EPI_ISL_1010952, EPI_ISL_1010953, EPI_ISL_1010954, EPI_ISL_1010955, EPI_ISL_1010956, EPI_ISL_1010957, EPI_ISL_1010958, EPI_ISL_1010959, EPI_ISL_1010960, EPI_ISL_1010961, EPI_ISL_1010962, EPI_ISL_1010963, EPI_ISL_1010964,
EPI_ISL_1010965, EPI_ISL_1010966, EPI_ISL_1010967, EPI_ISL_1010968, EPI_ISL_1010969, EPI_ISL_1010970, EPI_ISL_1010971, EPI_ISL_1010972, EPI_ISL_1010973, EPI_ISL_1010974, EPI_ISL_1010975, EPI_ISL_1010976, EPI_ISL_1010977, EPI_ISL_1010978, EPI_ISL_1010979, EPI_ISL_1010980, EPI_ISL_1010981, EPI_ISL_1010982, EPI_ISL_1010983, EPI_ISL_1010984, EPI_ISL_1010985, EPI_ISL_1010986,
EPI_ISL_1010987, EPI_ISL_1010988, EPI_ISL_1010989, EPI_ISL_1010990, EPI_ISL_1010991, EPI_ISL_1010992, EPI_ISL_1010993, EPI_ISL_1010994, EPI_ISL_1010995, EPI_ISL_1010996, EPI_ISL_1010997, EPI_ISL_1010998, EPI_ISL_1010999, EPI_ISL_1011000, EPI_ISL_1011001, EPI_ISL_1011002, EPI_ISL_1011003, EPI_ISL_1011004, EPI_ISL_1011005, EPI_ISL_1011006, EPI_ISL_1011007, EPI_ISL_1011008,
EPI_ISL_1011009, EPI_ISL_1011010, EPI_ISL_1011011, EPI_ISL_1011012, EPI_ISL_1011013, EPI_ISL_1011014, EPI_ISL_1011015, EPI_ISL_1011016, EPI_ISL_1011017, EPI_ISL_1011018, EPI_ISL_1011019, EPI_ISL_1011020, EPI_ISL_1011021, EPI_ISL_1011022, EPI_ISL_1011023, EPI_ISL_1011024, EPI_ISL_1011025, EPI_ISL_1011026, EPI_ISL_1011027, EPI_ISL_1011028, EPI_ISL_1011029, EPI_ISL_1011030,
EPI_ISL_1011031, EPI_ISL_1011032, EPI_ISL_1011033, EPI_ISL_1011034, EPI_ISL_1011035, EPI_ISL_1011036, EPI_ISL_1011037, EPI_ISL_1011038, EPI_ISL_1011039, EPI_ISL_1011040, EPI_ISL_1011041, EPI_ISL_1011042, EPI_ISL_1011043, EPI_ISL_1011044, EPI_ISL_1011045, EPI_ISL_1011046, EPI_ISL_1011047, EPI_ISL_1011048, EPI_ISL_1011049, EPI_ISL_1011050, EPI_ISL_1011051, EPI_ISL_1011052,
EPI_ISL_1011053, EPI_ISL_1011054, EPI_ISL_1011055, EPI_ISL_1011056, EPI_ISL_1011057, EPI_ISL_1011058, EPI_ISL_1011059, EPI_ISL_1011060, EPI_ISL_1011061, EPI_ISL_1011062, EPI_ISL_1011063, EPI_ISL_1011064, EPI_ISL_1011065, EPI_ISL_1011066, EPI_ISL_1011067, EPI_ISL_1011068, EPI_ISL_1011069, EPI_ISL_1011070, EPI_ISL_1011071, EPI_ISL_1011072, EPI_ISL_1011073, EPI_ISL_1011074,
EPI_ISL_1011075, EPI_ISL_1011076, EPI_ISL_1011077, EPI_ISL_1011078, EPI_ISL_1011079, EPI_ISL_1011080, EPI_ISL_1011081, EPI_ISL_1011082, EPI_ISL_1011083, EPI_ISL_1011084, EPI_ISL_1011085, EPI_ISL_1011086, EPI_ISL_1011087, EPI_ISL_1011088, EPI_ISL_1011089, EPI_ISL_1011090, EPI_ISL_1011091, EPI_ISL_1011092, EPI_ISL_1011093, EPI_ISL_1011094, EPI_ISL_1011095, EPI_ISL_1011096,
EPI_ISL_1011097, EPI_ISL_1011098, EPI_ISL_1011099, EPI_ISL_1011100, EPI_ISL_1011101, EPI_ISL_1011102, EPI_ISL_1011103, EPI_ISL_1011104, EPI_ISL_1011105, EPI_ISL_1011106, EPI_ISL_1011107, EPI_ISL_1011108, EPI_ISL_1011109, EPI_ISL_1011110, EPI_ISL_1011111, EPI_ISL_1011112, EPI_ISL_1011113, EPI_ISL_1011114, EPI_ISL_1011115, EPI_ISL_1011116, EPI_ISL_1011117, EPI_ISL_1011118,
EPI_ISL_1011119, EPI_ISL_1011120, EPI_ISL_1011121, EPI_ISL_1011122, EPI_ISL_1011123, EPI_ISL_1011124, EPI_ISL_1011125, EPI_ISL_1011126, EPI_ISL_1011127, EPI_ISL_1011128, EPI_ISL_1011129, EPI_ISL_1011130, EPI_ISL_1011131, EPI_ISL_1011132, EPI_ISL_1011133, EPI_ISL_1011134, EPI_ISL_1011135, EPI_ISL_1011136, EPI_ISL_1011137, EPI_ISL_1011138, EPI_ISL_1011139, EPI_ISL_1011140,
EPI_ISL_1011141, EPI_ISL_1011142, EPI_ISL_1011143, EPI_ISL_1011144, EPI_ISL_1011145, EPI_ISL_1011146, EPI_ISL_1011147, EPI_ISL_1011148, EPI_ISL_1011149, EPI_ISL_1011150, EPI_ISL_1011151, EPI_ISL_1011152, EPI_ISL_1011153, EPI_ISL_1011154, EPI_ISL_1011155, EPI_ISL_1011156, EPI_ISL_1011157, EPI_ISL_1011158, EPI_ISL_1011159, EPI_ISL_1011160, EPI_ISL_1011161, EPI_ISL_1011162,
EPI_ISL_1011163, EPI_ISL_1011164, EPI_ISL_1011165, EPI_ISL_1011166, EPI_ISL_1011167, EPI_ISL_1011168, EPI_ISL_1011169, EPI_ISL_1011170, EPI_ISL_1011171, EPI_ISL_1011172, EPI_ISL_1011173, EPI_ISL_1011174, EPI_ISL_1011175, EPI_ISL_1011176, EPI_ISL_1011177, EPI_ISL_1011178, EPI_ISL_1011179, EPI_ISL_1011180, EPI_ISL_1011181, EPI_ISL_1011182, EPI_ISL_1011183, EPI_ISL_1011184,
EPI_ISL_1011185, EPI_ISL_1011186, EPI_ISL_1011187, EPI_ISL_1011188, EPI_ISL_1011189, EPI_ISL_1011190, EPI_ISL_1011191, EPI_ISL_1011192, EPI_ISL_1011193, EPI_ISL_1011194, EPI_ISL_1011195, EPI_ISL_1011196, EPI_ISL_1011197, EPI_ISL_1011198, EPI_ISL_1011199, EPI_ISL_1011200, EPI_ISL_1011201, EPI_ISL_1011202, EPI_ISL_1011203, EPI_ISL_1011204, EPI_ISL_1011205, EPI_ISL_1011206,
EPI_ISL_1011207, EPI_ISL_1011208, EPI_ISL_1011209, EPI_ISL_1011210, EPI_ISL_1011211, EPI_ISL_1011212, EPI_ISL_1011213, EPI_ISL_1011214, EPI_ISL_1011215, EPI_ISL_1011216, EPI_ISL_1011217, EPI_ISL_1011218, EPI_ISL_1011219, EPI_ISL_1011220, EPI_ISL_1011221, EPI_ISL_1011222, EPI_ISL_1011223, EPI_ISL_1011224, EPI_ISL_1011225, EPI_ISL_1011226, EPI_ISL_1011227, EPI_ISL_1011228,
EPI_ISL_1011229, EPI_ISL_1011230, EPI_ISL_1011231, EPI_ISL_1011232, EPI_ISL_1011233, EPI_ISL_1011234, EPI_ISL_1011235, EPI_ISL_1011236, EPI_ISL_1011237, EPI_ISL_1011238, EPI_ISL_1011239, EPI_ISL_1011240, EPI_ISL_1011241, EPI_ISL_1011242, EPI_ISL_1011243, EPI_ISL_1011244, EPI_ISL_1011245, EPI_ISL_1011246, EPI_ISL_1011247, EPI_ISL_1011248, EPI_ISL_1011249, EPI_ISL_1011250,
EPI_ISL_1011251, EPI_ISL_1011252, EPI_ISL_1011253, EPI_ISL_1011254, EPI_ISL_1011255, EPI_ISL_1011256, EPI_ISL_1011257, EPI_ISL_1011258, EPI_ISL_1011259, EPI_ISL_1011260, EPI_ISL_1011261, EPI_ISL_1011262, EPI_ISL_1011263, EPI_ISL_1011264, EPI_ISL_1011265, EPI_ISL_1011266, EPI_ISL_1011267, EPI_ISL_1011268, EPI_ISL_1011269, EPI_ISL_1011270, EPI_ISL_1011271, EPI_ISL_1011272,
EPI_ISL_1011273, EPI_ISL_1011274, EPI_ISL_1011275, EPI_ISL_1011276, EPI_ISL_1011277, EPI_ISL_1011278, EPI_ISL_1011279, EPI_ISL_1011280, EPI_ISL_1011281, EPI_ISL_1011282, EPI_ISL_1011283, EPI_ISL_1011284, EPI_ISL_1011285, EPI_ISL_1011286, EPI_ISL_1011287, EPI_ISL_1011288, EPI_ISL_1011289, EPI_ISL_1011290, EPI_ISL_1011291, EPI_ISL_1011292, EPI_ISL_1011293, EPI_ISL_1011294,
EPI_ISL_1011295, EPI_ISL_1011296, EPI_ISL_1011297, EPI_ISL_1011298, EPI_ISL_1011299, EPI_ISL_1011300, EPI_ISL_1011301, EPI_ISL_1011302, EPI_ISL_1011303, EPI_ISL_1011304, EPI_ISL_1011305, EPI_ISL_1011306, EPI_ISL_1011307, EPI_ISL_1011308, EPI_ISL_1011309, EPI_ISL_1011310, EPI_ISL_1011311, EPI_ISL_1011312, EPI_ISL_1011313, EPI_ISL_1011314, EPI_ISL_1011315, EPI_ISL_1011316,
EPI_ISL_1011317, EPI_ISL_1011318, EPI_ISL_1011319, EPI_ISL_1011320, EPI_ISL_1011321, EPI_ISL_1011322, EPI_ISL_1011323, EPI_ISL_1011324, EPI_ISL_1011325, EPI_ISL_1011326, EPI_ISL_1011327, EPI_ISL_1011328, EPI_ISL_1011329, EPI_ISL_1011330, EPI_ISL_1011331, EPI_ISL_1011332, EPI_ISL_1011333, EPI_ISL_1011334, EPI_ISL_1011335, EPI_ISL_1011336, EPI_ISL_1011337, EPI_ISL_1011338,
EPI_ISL_1011339, EPI_ISL_1011340, EPI_ISL_1011341, EPI_ISL_1011342, EPI_ISL_1011343, EPI_ISL_1011344, EPI_ISL_1011345, EPI_ISL_1011346, EPI_ISL_1011347, EPI_ISL_1011348, EPI_ISL_1011349, EPI_ISL_1011350, EPI_ISL_1011351, EPI_ISL_1011352, EPI_ISL_1011353, EPI_ISL_1011354, EPI_ISL_1011355, EPI_ISL_1011356, EPI_ISL_1011357, EPI_ISL_1011358, EPI_ISL_1011359, EPI_ISL_1011360,
EPI_ISL_1011361, EPI_ISL_1011362, EPI_ISL_1011363, EPI_ISL_1011364, EPI_ISL_1011365, EPI_ISL_1011366, EPI_ISL_1011367, EPI_ISL_1011368, EPI_ISL_1011369, EPI_ISL_1011370, EPI_ISL_1011371, EPI_ISL_1011372, EPI_ISL_1011373, EPI_ISL_1011374, EPI_ISL_1011375, EPI_ISL_1011376, EPI_ISL_1011377, EPI_ISL_1011378, EPI_ISL_1011379, EPI_ISL_1011380, EPI_ISL_1011381, EPI_ISL_1011382,
EPI_ISL_1011383, EPI_ISL_1011384, EPI_ISL_1011385, EPI_ISL_1011386, EPI_ISL_1011387, EPI_ISL_1011388, EPI_ISL_1011389, EPI_ISL_1011390, EPI_ISL_1011391, EPI_ISL_1011392, EPI_ISL_1011393, EPI_ISL_1011394, EPI_ISL_1011395, EPI_ISL_1011396, EPI_ISL_1011397, EPI_ISL_1011398, EPI_ISL_1011399, EPI_ISL_1011400, EPI_ISL_1011401, EPI_ISL_1011402, EPI_ISL_1011403, EPI_ISL_1011404,
EPI_ISL_1011405, EPI_ISL_1011406, EPI_ISL_1011407, EPI_ISL_1011408, EPI_ISL_1011409, EPI_ISL_1011410, EPI_ISL_1011411, EPI_ISL_1011412, EPI_ISL_1011413, EPI_ISL_1011414, EPI_ISL_1011415, EPI_ISL_1011416, EPI_ISL_1011417, EPI_ISL_1011418, EPI_ISL_1011419, EPI_ISL_1011420, EPI_ISL_1011421, EPI_ISL_1011422, EPI_ISL_1011423, EPI_ISL_1011424, EPI_ISL_1011425, EPI_ISL_1011426,
EPI_ISL_1011427, EPI_ISL_1011428, EPI_ISL_1011429, EPI_ISL_1011430, EPI_ISL_1011431, EPI_ISL_1011432, EPI_ISL_1011433, EPI_ISL_1011434, EPI_ISL_1011435, EPI_ISL_1011436, EPI_ISL_1011437, EPI_ISL_1011438, EPI_ISL_1011439, EPI_ISL_1011440, EPI_ISL_1011441, EPI_ISL_1011442, EPI_ISL_1011443, EPI_ISL_1011444, EPI_ISL_1011445, EPI_ISL_1011446, EPI_ISL_1011447, EPI_ISL_1011448,
EPI_ISL_1011449, EPI_ISL_1011450, EPI_ISL_1011451, EPI_ISL_1011452, EPI_ISL_1011453, EPI_ISL_1011454, EPI_ISL_1011455, EPI_ISL_1011456, EPI_ISL_1011457, EPI_ISL_1011458, EPI_ISL_1011459, EPI_ISL_1011460, EPI_ISL_1011461, EPI_ISL_1011462, EPI_ISL_1011463, EPI_ISL_1011464, EPI_ISL_1011465, EPI_ISL_1011466, EPI_ISL_1011467, EPI_ISL_1011468, EPI_ISL_1011469, EPI_ISL_1011470,
EPI_ISL_1011471, EPI_ISL_1011472, EPI_ISL_1011473, EPI_ISL_1011474, EPI_ISL_1011475, EPI_ISL_1011476, EPI_ISL_1011477, EPI_ISL_1011478, EPI_ISL_1011479, EPI_ISL_1011480, EPI_ISL_1011481, EPI_ISL_1011482, EPI_ISL_1011483, EPI_ISL_1011484, EPI_ISL_1011485, EPI_ISL_1011486, EPI_ISL_1011487, EPI_ISL_1011488, EPI_ISL_1011489, EPI_ISL_1011490, EPI_ISL_1011491, EPI_ISL_1011492,
EPI_ISL_1011493, EPI_ISL_1011494, EPI_ISL_1011495, EPI_ISL_1011496, EPI_ISL_1011497, EPI_ISL_1011498, EPI_ISL_1011499, EPI_ISL_1011500, EPI_ISL_1011501, EPI_ISL_1011502, EPI_ISL_1011503, EPI_ISL_1011504, EPI_ISL_1011505, EPI_ISL_1011506, EPI_ISL_1011507, EPI_ISL_1011508, EPI_ISL_1026866, EPI_ISL_1026867, EPI_ISL_1026868, EPI_ISL_1026869, EPI_ISL_1026870, EPI_ISL_1026871,
EPI_ISL_1026872, EPI_ISL_1026873, EPI_ISL_1026874, EPI_ISL_1026875, EPI_ISL_1026876, EPI_ISL_1026877, EPI_ISL_1026878, EPI_ISL_1026879, EPI_ISL_1026880, EPI_ISL_1026881, EPI_ISL_1026882, EPI_ISL_1026883, EPI_ISL_1026884, EPI_ISL_1026885, EPI_ISL_1026886, EPI_ISL_1026887, EPI_ISL_1026888, EPI_ISL_1026889, EPI_ISL_1026890, EPI_ISL_1026891, EPI_ISL_1026892, EPI_ISL_1026893,
EPI_ISL_1026894, EPI_ISL_1026895, EPI_ISL_1026896, EPI_ISL_1026897, EPI_ISL_1026898, EPI_ISL_1026899, EPI_ISL_1026900, EPI_ISL_1026901, EPI_ISL_1026902, EPI_ISL_1026903, EPI_ISL_1026904, EPI_ISL_1026905, EPI_ISL_1026906, EPI_ISL_1026907, EPI_ISL_1026908, EPI_ISL_1026909, EPI_ISL_1026910, EPI_ISL_1026911, EPI_ISL_1026912, EPI_ISL_1026913, EPI_ISL_1026914, EPI_ISL_1026915,
EPI_ISL_1026916, EPI_ISL_1026917, EPI_ISL_1026918, EPI_ISL_1026919, EPI_ISL_1026920, EPI_ISL_1026921, EPI_ISL_1026922, EPI_ISL_1026923, EPI_ISL_1026924, EPI_ISL_1026925, EPI_ISL_1026926, EPI_ISL_1026927, EPI_ISL_1026928, EPI_ISL_1026929, EPI_ISL_1026930, EPI_ISL_1026931, EPI_ISL_1026932, EPI_ISL_1026933, EPI_ISL_1026934, EPI_ISL_1026935, EPI_ISL_1026936, EPI_ISL_1026937,
EPI_ISL_1026938, EPI_ISL_1026939, EPI_ISL_1026940, EPI_ISL_1026941, EPI_ISL_1026942, EPI_ISL_1026943, EPI_ISL_1026944, EPI_ISL_1026945, EPI_ISL_1026946, EPI_ISL_1026947, EPI_ISL_1026948, EPI_ISL_1026949, EPI_ISL_1026950, EPI_ISL_1026951, EPI_ISL_1026952, EPI_ISL_1026953, EPI_ISL_1026954, EPI_ISL_1026955, EPI_ISL_1026956, EPI_ISL_1026957, EPI_ISL_1026958, EPI_ISL_1026959,
EPI_ISL_1026960, EPI_ISL_1026961, EPI_ISL_1026962, EPI_ISL_1026963, EPI_ISL_1026964, EPI_ISL_1026965, EPI_ISL_1026966, EPI_ISL_1026967, EPI_ISL_1026968, EPI_ISL_1026969, EPI_ISL_1026970, EPI_ISL_1026971, EPI_ISL_1026972, EPI_ISL_1026973, EPI_ISL_1026974, EPI_ISL_1026975, EPI_ISL_1026976, EPI_ISL_1026977, EPI_ISL_1026978, EPI_ISL_1026979, EPI_ISL_1026980, EPI_ISL_1026981,
EPI_ISL_1026982, EPI_ISL_1026983, EPI_ISL_1026984, EPI_ISL_1026985, EPI_ISL_1026986, EPI_ISL_1026987, EPI_ISL_1026988, EPI_ISL_1026989, EPI_ISL_1026990, EPI_ISL_1026991, EPI_ISL_1026992, EPI_ISL_1026993, EPI_ISL_1026994, EPI_ISL_1026995, EPI_ISL_1026996, EPI_ISL_1026997, EPI_ISL_1026998, EPI_ISL_1026999, EPI_ISL_1027000, EPI_ISL_1027001, EPI_ISL_1027002, EPI_ISL_1027003,
EPI_ISL_1027004, EPI_ISL_1027005, EPI_ISL_1027006, EPI_ISL_1027007, EPI_ISL_1027008, EPI_ISL_1027009, EPI_ISL_1027010, EPI_ISL_1027011, EPI_ISL_1027012, EPI_ISL_1027013, EPI_ISL_1027014, EPI_ISL_1027015, EPI_ISL_1027016, EPI_ISL_1027017, EPI_ISL_1027018, EPI_ISL_1027019, EPI_ISL_1027020, EPI_ISL_1027021, EPI_ISL_1027022, EPI_ISL_1027023, EPI_ISL_1027024, EPI_ISL_1027025,
EPI_ISL_1027026, EPI_ISL_1027028, EPI_ISL_1027029, EPI_ISL_1027030, EPI_ISL_1027031, EPI_ISL_1027032, EPI_ISL_1027033, EPI_ISL_1027034, EPI_ISL_1027035, EPI_ISL_1027037, EPI_ISL_1027038, EPI_ISL_1027039, EPI_ISL_1027040, EPI_ISL_1027041, EPI_ISL_1027042, EPI_ISL_1027043, EPI_ISL_1027044, EPI_ISL_1027045, EPI_ISL_1027046, EPI_ISL_1027047, EPI_ISL_1027048, EPI_ISL_1027049,
EPI_ISL_1027050, EPI_ISL_1027051, EPI_ISL_1027052, EPI_ISL_1027053, EPI_ISL_1027054, EPI_ISL_1027055, EPI_ISL_1027056, EPI_ISL_1027057, EPI_ISL_1027058, EPI_ISL_1027059, EPI_ISL_1027060, EPI_ISL_1027061, EPI_ISL_1027062, EPI_ISL_1027063, EPI_ISL_1027064, EPI_ISL_1027065, EPI_ISL_1027066, EPI_ISL_1027067, EPI_ISL_1027068, EPI_ISL_1027069, EPI_ISL_1027070, EPI_ISL_1027071,
EPI_ISL_1027072, EPI_ISL_1027073, EPI_ISL_1027074, EPI_ISL_1027075, EPI_ISL_1027076, EPI_ISL_1027077, EPI_ISL_1027078, EPI_ISL_1027079, EPI_ISL_1027080, EPI_ISL_1027081, EPI_ISL_1027082, EPI_ISL_1027083, EPI_ISL_1027084, EPI_ISL_1027086, EPI_ISL_1027087, EPI_ISL_1027088, EPI_ISL_1027089, EPI_ISL_1027090, EPI_ISL_1027091, EPI_ISL_1027092, EPI_ISL_1027093, EPI_ISL_1027094,
EPI_ISL_1027095, EPI_ISL_1027096, EPI_ISL_1027097, EPI_ISL_1027098, EPI_ISL_1027099, EPI_ISL_1027100, EPI_ISL_1027101, EPI_ISL_1027102, EPI_ISL_1027103, EPI_ISL_1027104, EPI_ISL_1027105, EPI_ISL_1027106, EPI_ISL_1027107, EPI_ISL_1027108, EPI_ISL_1027109, EPI_ISL_1027110, EPI_ISL_1027111, EPI_ISL_1027112, EPI_ISL_1027113, EPI_ISL_1027114, EPI_ISL_1027115, EPI_ISL_1027116,
EPI_ISL_1027117, EPI_ISL_1027118, EPI_ISL_1027119, EPI_ISL_1027120, EPI_ISL_1027121, EPI_ISL_1027122, EPI_ISL_1027123, EPI_ISL_1027124, EPI_ISL_1027125, EPI_ISL_1027126, EPI_ISL_1027127, EPI_ISL_1027128, EPI_ISL_1027129, EPI_ISL_1027130, EPI_ISL_1027131, EPI_ISL_1027132, EPI_ISL_1027133, EPI_ISL_1027134, EPI_ISL_1027135, EPI_ISL_1027136, EPI_ISL_1027137, EPI_ISL_1027138,
EPI_ISL_1027139, EPI_ISL_1027140, EPI_ISL_1027141, EPI_ISL_1027142, EPI_ISL_1027143, EPI_ISL_1027144, EPI_ISL_1027145, EPI_ISL_1027146, EPI_ISL_1027147, EPI_ISL_1027148, EPI_ISL_1027149, EPI_ISL_1027150, EPI_ISL_1027151, EPI_ISL_1027152, EPI_ISL_1027153, EPI_ISL_1027154, EPI_ISL_1027155, EPI_ISL_1027156, EPI_ISL_1027157, EPI_ISL_1027158, EPI_ISL_1027159, EPI_ISL_1027160,
EPI_ISL_1027161, EPI_ISL_1027162, EPI_ISL_1027163, EPI_ISL_1027164, EPI_ISL_1027165, EPI_ISL_1027166, EPI_ISL_1027167, EPI_ISL_1027168, EPI_ISL_1027169, EPI_ISL_1027170, EPI_ISL_1027171, EPI_ISL_1027172, EPI_ISL_1027173, EPI_ISL_1027174, EPI_ISL_1027175, EPI_ISL_1027176, EPI_ISL_1027177, EPI_ISL_1032947, EPI_ISL_1032950, EPI_ISL_1032952, EPI_ISL_1032953, EPI_ISL_1032954,
EPI_ISL_1032955, EPI_ISL_1032956, EPI_ISL_1032957, EPI_ISL_1032958, EPI_ISL_1032959, EPI_ISL_1032960, EPI_ISL_1032961, EPI_ISL_1032962, EPI_ISL_1032963, EPI_ISL_1032964, EPI_ISL_1032965, EPI_ISL_1032966, EPI_ISL_1032967, EPI_ISL_1032968, EPI_ISL_1032969, EPI_ISL_1032970, EPI_ISL_1032971, EPI_ISL_1032972, EPI_ISL_1032973, EPI_ISL_1032974, EPI_ISL_1032975, EPI_ISL_1032976,
EPI_ISL_1032977, EPI_ISL_1032978, EPI_ISL_1032979, EPI_ISL_1032980, EPI_ISL_1032981, EPI_ISL_1032982, EPI_ISL_1032983, EPI_ISL_1032984, EPI_ISL_1032985, EPI_ISL_1032986, EPI_ISL_1032987, EPI_ISL_1032988, EPI_ISL_1032989, EPI_ISL_1032990, EPI_ISL_1032991, EPI_ISL_1032992, EPI_ISL_1032993, EPI_ISL_1032994, EPI_ISL_1032995, EPI_ISL_1032996, EPI_ISL_1032997, EPI_ISL_1032998,
EPI_ISL_1032999, EPI_ISL_1033000, EPI_ISL_1033001, EPI_ISL_1033002, EPI_ISL_1033003, EPI_ISL_1033004, EPI_ISL_1033005, EPI_ISL_1033006, EPI_ISL_1033007, EPI_ISL_1033008, EPI_ISL_1033009, EPI_ISL_1033010, EPI_ISL_1033011, EPI_ISL_1033012, EPI_ISL_1033013, EPI_ISL_1033014, EPI_ISL_1033015, EPI_ISL_1033016, EPI_ISL_1033017, EPI_ISL_1033018, EPI_ISL_1033019, EPI_ISL_1033020,
EPI_ISL_1033021, EPI_ISL_1033022, EPI_ISL_1033023, EPI_ISL_1068395, EPI_ISL_1068396, EPI_ISL_1068397, EPI_ISL_1068398, EPI_ISL_1068399, EPI_ISL_1068400, EPI_ISL_1068401, EPI_ISL_1068402, EPI_ISL_1068403, EPI_ISL_1068404, EPI_ISL_1068405, EPI_ISL_1068406, EPI_ISL_1068407, EPI_ISL_1068408, EPI_ISL_1068409, EPI_ISL_1068410, EPI_ISL_1068411, EPI_ISL_1068412, EPI_ISL_1068413,
EPI_ISL_1068414, EPI_ISL_1068415, EPI_ISL_1068416, EPI_ISL_1068417, EPI_ISL_1068418, EPI_ISL_1068419, EPI_ISL_1068420, EPI_ISL_1068421, EPI_ISL_1068422, EPI_ISL_1068423, EPI_ISL_1068424, EPI_ISL_1068425, EPI_ISL_1068426, EPI_ISL_1068427, EPI_ISL_1068428, EPI_ISL_1068429, EPI_ISL_1068430, EPI_ISL_1068431, EPI_ISL_1068432, EPI_ISL_1068433, EPI_ISL_1068434, EPI_ISL_1068435,
EPI_ISL_1068436, EPI_ISL_1068437, EPI_ISL_1068438, EPI_ISL_1068439, EPI_ISL_1068440, EPI_ISL_1068441, EPI_ISL_1068442, EPI_ISL_1068443, EPI_ISL_1068444, EPI_ISL_1068445, EPI_ISL_1068446, EPI_ISL_1068447, EPI_ISL_1068448, EPI_ISL_1068449, EPI_ISL_1068450, EPI_ISL_1068451, EPI_ISL_1068452, EPI_ISL_1068453, EPI_ISL_1068454, EPI_ISL_1068455, EPI_ISL_1068456, EPI_ISL_1068457,
EPI_ISL_1068458, EPI_ISL_1068459, EPI_ISL_1068460, EPI_ISL_1068461, EPI_ISL_1068462, EPI_ISL_1068463, EPI_ISL_1068464, EPI_ISL_1068465, EPI_ISL_1068466, EPI_ISL_1068467, EPI_ISL_1068468, EPI_ISL_1068469, EPI_ISL_1068470, EPI_ISL_1068471, EPI_ISL_1068472, EPI_ISL_1068473, EPI_ISL_1068474, EPI_ISL_1068475, EPI_ISL_1068476, EPI_ISL_1068477, EPI_ISL_1068478, EPI_ISL_1068479,
EPI_ISL_1068480, EPI_ISL_1068481, EPI_ISL_1068482, EPI_ISL_1068483, EPI_ISL_1068484, EPI_ISL_1068485, EPI_ISL_1068486, EPI_ISL_1068487, EPI_ISL_1068488, EPI_ISL_1068489, EPI_ISL_1068490, EPI_ISL_1068491, EPI_ISL_1068492, EPI_ISL_1068493, EPI_ISL_1068494, EPI_ISL_1068495, EPI_ISL_1068496, EPI_ISL_1068497, EPI_ISL_1068498, EPI_ISL_1068499, EPI_ISL_1068500, EPI_ISL_1068501,
EPI_ISL_1068502, EPI_ISL_1068503, EPI_ISL_1068504, EPI_ISL_1068505, EPI_ISL_1068506, EPI_ISL_1068507, EPI_ISL_1068508, EPI_ISL_1068509, EPI_ISL_1068510, EPI_ISL_1068511, EPI_ISL_1068512, EPI_ISL_1068513, EPI_ISL_1068514, EPI_ISL_1068515, EPI_ISL_1068516, EPI_ISL_1068517, EPI_ISL_1068518, EPI_ISL_1068519, EPI_ISL_1068520, EPI_ISL_1068521, EPI_ISL_1068522, EPI_ISL_1068523,
EPI_ISL_1068524, EPI_ISL_1068525, EPI_ISL_1068526, EPI_ISL_1068527, EPI_ISL_1068528, EPI_ISL_1068529, EPI_ISL_1068530, EPI_ISL_1068531, EPI_ISL_1068532, EPI_ISL_1068533, EPI_ISL_1068534, EPI_ISL_1068535, EPI_ISL_1068536, EPI_ISL_1068537, EPI_ISL_1068538, EPI_ISL_1068539, EPI_ISL_1068540, EPI_ISL_1068541, EPI_ISL_1068542, EPI_ISL_1068543, EPI_ISL_1068544, EPI_ISL_1068545,
EPI_ISL_1068546, EPI_ISL_1068547, EPI_ISL_1068548, EPI_ISL_1068549, EPI_ISL_1068550, EPI_ISL_1068551, EPI_ISL_1068552, EPI_ISL_1068553, EPI_ISL_1068554, EPI_ISL_1068555, EPI_ISL_1068556, EPI_ISL_1068557, EPI_ISL_1068558, EPI_ISL_1068559, EPI_ISL_1068560, EPI_ISL_1068561, EPI_ISL_1068562, EPI_ISL_1068563, EPI_ISL_1068564, EPI_ISL_1068565, EPI_ISL_1068566, EPI_ISL_1068567,
EPI_ISL_1068568, EPI_ISL_1068569, EPI_ISL_1068570, EPI_ISL_1068571, EPI_ISL_1068572, EPI_ISL_1068573, EPI_ISL_1068574, EPI_ISL_1068575, EPI_ISL_1068576, EPI_ISL_1068577, EPI_ISL_1068578, EPI_ISL_1068579, EPI_ISL_1068580, EPI_ISL_1068581, EPI_ISL_1068582, EPI_ISL_1068583, EPI_ISL_1068584, EPI_ISL_1068585, EPI_ISL_1068586, EPI_ISL_1068587, EPI_ISL_1068588, EPI_ISL_1068589,
EPI_ISL_1068590, EPI_ISL_1068591, EPI_ISL_1068592, EPI_ISL_1068594, EPI_ISL_1068595, EPI_ISL_1068596, EPI_ISL_1068597, EPI_ISL_1068598, EPI_ISL_1068599, EPI_ISL_1068600, EPI_ISL_1068601, EPI_ISL_1068602, EPI_ISL_1068603, EPI_ISL_1068604, EPI_ISL_1068605, EPI_ISL_1068606, EPI_ISL_1068607, EPI_ISL_1068608, EPI_ISL_1068609, EPI_ISL_1068610, EPI_ISL_1068611, EPI_ISL_1068612,
EPI_ISL_1068613, EPI_ISL_1068614, EPI_ISL_1068615, EPI_ISL_1068616, EPI_ISL_1068617, EPI_ISL_1068618, EPI_ISL_1068619, EPI_ISL_1068620, EPI_ISL_1068621, EPI_ISL_1068622, EPI_ISL_1068623, EPI_ISL_1068624, EPI_ISL_1068625, EPI_ISL_1068626, EPI_ISL_1068627, EPI_ISL_1068628, EPI_ISL_1068629, EPI_ISL_1068630, EPI_ISL_1068631, EPI_ISL_1068632, EPI_ISL_1068633, EPI_ISL_1068634,
EPI_ISL_1068635, EPI_ISL_1068636, EPI_ISL_1068637, EPI_ISL_1068638, EPI_ISL_1068639, EPI_ISL_1068641, EPI_ISL_1068642, EPI_ISL_1068643, EPI_ISL_1068644, EPI_ISL_1068645, EPI_ISL_1068646, EPI_ISL_1068647, EPI_ISL_1068648, EPI_ISL_1068649, EPI_ISL_1068650, EPI_ISL_1068651, EPI_ISL_1068652, EPI_ISL_1068653, EPI_ISL_1068654, EPI_ISL_1068655, EPI_ISL_1068656, EPI_ISL_1068657,
EPI_ISL_1068658, EPI_ISL_1068659, EPI_ISL_1068660, EPI_ISL_1068661, EPI_ISL_1068662, EPI_ISL_1068663, EPI_ISL_1068664, EPI_ISL_1068665, EPI_ISL_1068666, EPI_ISL_1068667, EPI_ISL_1068668, EPI_ISL_1068669, EPI_ISL_1068670, EPI_ISL_1068671, EPI_ISL_1068672, EPI_ISL_1068673, EPI_ISL_1068674, EPI_ISL_1068675, EPI_ISL_1068676, EPI_ISL_1068677, EPI_ISL_1068678, EPI_ISL_1068679,

| | | | |
|---|---|---|---|
| EPI_ISL_1068680, EPI_ISL_1068681, EPI_ISL_1068683, EPI_ISL_1068684, EPI_ISL_1068685, EPI_ISL_1068686, EPI_ISL_1068687, EPI_ISL_1068688, EPI_ISL_1068689, EPI_ISL_1068690, EPI_ISL_1068691, EPI_ISL_1068692, EPI_ISL_1068693, EPI_ISL_1068694, EPI_ISL_1068695, EPI_ISL_1068696, EPI_ISL_1068697, EPI_ISL_1068698, EPI_ISL_1068699, EPI_ISL_1068700, EPI_ISL_1068701, EPI_ISL_1068702, EPI_ISL_1068703, EPI_ISL_1068704, EPI_ISL_1068705, EPI_ISL_1068706, EPI_ISL_1068707, EPI_ISL_1068708, EPI_ISL_1068709, EPI_ISL_1068710, EPI_ISL_1068711, EPI_ISL_1068712, EPI_ISL_1068713, EPI_ISL_1068714, EPI_ISL_1068715, EPI_ISL_1068716, EPI_ISL_1068717, EPI_ISL_1068718, EPI_ISL_1068719, EPI_ISL_1068720, EPI_ISL_1068721, EPI_ISL_1068722, EPI_ISL_1068723, EPI_ISL_1068724, EPI_ISL_1068725, EPI_ISL_1068726, EPI_ISL_1068727, EPI_ISL_1068728, EPI_ISL_1068729, EPI_ISL_1068730, EPI_ISL_1068731, EPI_ISL_1068732, EPI_ISL_1068733, EPI_ISL_1068734, EPI_ISL_1068735, EPI_ISL_1068736, EPI_ISL_1068737, EPI_ISL_1068738, EPI_ISL_1068739, EPI_ISL_1068740, EPI_ISL_1068741, EPI_ISL_1068742, EPI_ISL_1068743, EPI_ISL_1068744, EPI_ISL_1068745, EPI_ISL_1068746, EPI_ISL_1068747, EPI_ISL_1068748, EPI_ISL_1068749, EPI_ISL_1068750, EPI_ISL_1068751, EPI_ISL_1068752, EPI_ISL_1068753, EPI_ISL_1068754, EPI_ISL_1068755, EPI_ISL_1068756, EPI_ISL_1068757, EPI_ISL_1068758, EPI_ISL_1068759, EPI_ISL_1068760, EPI_ISL_1068761, EPI_ISL_1068762, EPI_ISL_1068763, EPI_ISL_1068764, EPI_ISL_1068765, EPI_ISL_1068766, EPI_ISL_1068767, EPI_ISL_1068768, EPI_ISL_1068769, EPI_ISL_1068770, EPI_ISL_1068771, EPI_ISL_1068772, EPI_ISL_1068773, EPI_ISL_1068774, EPI_ISL_1068775, EPI_ISL_1068776, EPI_ISL_1068777, EPI_ISL_1068778, EPI_ISL_1068779, EPI_ISL_1068780, EPI_ISL_1068781, EPI_ISL_1068782, EPI_ISL_1068783, EPI_ISL_1068784, EPI_ISL_1068785, EPI_ISL_1068786, EPI_ISL_1068787, EPI_ISL_1068788, EPI_ISL_1068789, EPI_ISL_1068790, EPI_ISL_1068791, EPI_ISL_1068792, EPI_ISL_1068793, EPI_ISL_1068794, EPI_ISL_1068795, EPI_ISL_1068796, EPI_ISL_1068797, EPI_ISL_1068798, EPI_ISL_1068799, EPI_ISL_1068800, EPI_ISL_1068801, EPI_ISL_1068802, EPI_ISL_1068803, EPI_ISL_1068804, EPI_ISL_1068805, EPI_ISL_1068806, EPI_ISL_1068807, EPI_ISL_1068808, EPI_ISL_1068809, EPI_ISL_1068810, EPI_ISL_1068811, EPI_ISL_1068812, EPI_ISL_1068813, EPI_ISL_1068814, EPI_ISL_1068815, EPI_ISL_1068816, EPI_ISL_1068817, EPI_ISL_1068818, EPI_ISL_1068819, EPI_ISL_1068820, EPI_ISL_1068821, EPI_ISL_1068822, EPI_ISL_1068823, EPI_ISL_1068824, EPI_ISL_1068825, EPI_ISL_1068826, EPI_ISL_1068827, EPI_ISL_1068828, EPI_ISL_1068829, EPI_ISL_1068830, EPI_ISL_1068831, EPI_ISL_1068832, EPI_ISL_1068833, EPI_ISL_1068834, EPI_ISL_1068835, EPI_ISL_1068836, EPI_ISL_1068837, EPI_ISL_1068838, EPI_ISL_1068839, EPI_ISL_1068840, EPI_ISL_1068841, EPI_ISL_1068842, EPI_ISL_1068843, EPI_ISL_1068844, EPI_ISL_1068845, EPI_ISL_1068846, EPI_ISL_1068847, EPI_ISL_1068848, EPI_ISL_1068852, EPI_ISL_1068853, EPI_ISL_1068854, EPI_ISL_1068855, EPI_ISL_1068856, EPI_ISL_1068857, EPI_ISL_1068858, EPI_ISL_1068859, EPI_ISL_1068860, EPI_ISL_1068861, EPI_ISL_1068862, EPI_ISL_1068863, EPI_ISL_1068864, EPI_ISL_1068865, EPI_ISL_1068866, EPI_ISL_1068867, EPI_ISL_1068868, EPI_ISL_1068869, EPI_ISL_1068870, EPI_ISL_1068871, EPI_ISL_1068872, EPI_ISL_1068873, EPI_ISL_1068874, EPI_ISL_1068875, EPI_ISL_1068876, EPI_ISL_1068877, EPI_ISL_1068878, EPI_ISL_1068879, EPI_ISL_1068880, EPI_ISL_1068881, EPI_ISL_1068882, EPI_ISL_1068883, EPI_ISL_1068884, EPI_ISL_1068885, EPI_ISL_1068886, EPI_ISL_1068887, EPI_ISL_1068888, EPI_ISL_1068889, EPI_ISL_1068890, EPI_ISL_1068891, EPI_ISL_1068892, EPI_ISL_1068893, EPI_ISL_1068894, EPI_ISL_1068895, EPI_ISL_1068896, EPI_ISL_1068897, EPI_ISL_1068898, EPI_ISL_1068899, EPI_ISL_1068900, EPI_ISL_1068901, EPI_ISL_1068902, EPI_ISL_1068903, EPI_ISL_1068904, EPI_ISL_1068905, EPI_ISL_1068906, EPI_ISL_1068907, EPI_ISL_1068908, EPI_ISL_1068909, EPI_ISL_1068910, EPI_ISL_1068911, EPI_ISL_1068912, EPI_ISL_1068913, EPI_ISL_1068914, EPI_ISL_1068915, EPI_ISL_1068916, EPI_ISL_1068917, EPI_ISL_1068918, EPI_ISL_1068919, EPI_ISL_1068920, EPI_ISL_1068921, EPI_ISL_1068922, EPI_ISL_1068923, EPI_ISL_1068924, EPI_ISL_1068925, EPI_ISL_1068926, EPI_ISL_1068930, EPI_ISL_1068931, EPI_ISL_1068932, EPI_ISL_1068933, EPI_ISL_1068934, EPI_ISL_1068935, EPI_ISL_1068936, EPI_ISL_1068937, EPI_ISL_1068938, EPI_ISL_1068939, EPI_ISL_1068940, EPI_ISL_1068941, EPI_ISL_1068942, EPI_ISL_1068943, EPI_ISL_1068944, EPI_ISL_1068945, EPI_ISL_1068946, EPI_ISL_1068947, EPI_ISL_1068948, EPI_ISL_1068949, EPI_ISL_1068950, EPI_ISL_1068951, EPI_ISL_1068952, EPI_ISL_1068953, EPI_ISL_1068954, EPI_ISL_1068955, EPI_ISL_1068956, EPI_ISL_1068957, EPI_ISL_1068958, EPI_ISL_1068959, EPI_ISL_1068960, EPI_ISL_1068961, EPI_ISL_1068962, EPI_ISL_1068963, EPI_ISL_1068964, EPI_ISL_1068965, EPI_ISL_1068966, EPI_ISL_1068967, EPI_ISL_1068968, EPI_ISL_1068969, EPI_ISL_1068970, EPI_ISL_1068971, EPI_ISL_1068972, EPI_ISL_1068973, EPI_ISL_1068974, EPI_ISL_1068975, EPI_ISL_1068976, EPI_ISL_1068977, EPI_ISL_1068978, EPI_ISL_1068979, EPI_ISL_1068980, EPI_ISL_1068981, EPI_ISL_1068982, EPI_ISL_1068983, EPI_ISL_1068984, EPI_ISL_1068985, EPI_ISL_1068986, EPI_ISL_1068987, EPI_ISL_1068988, EPI_ISL_1068989, EPI_ISL_1068990, EPI_ISL_1068991, EPI_ISL_1068992, EPI_ISL_1068993, EPI_ISL_1068994, EPI_ISL_1068995, EPI_ISL_1069000, EPI_ISL_1069001, EPI_ISL_1069002, EPI_ISL_1069003, EPI_ISL_1069004, EPI_ISL_1069005, EPI_ISL_1069006, EPI_ISL_1069007, EPI_ISL_1069008, EPI_ISL_1069009, EPI_ISL_1069010, EPI_ISL_1069011, EPI_ISL_1069012, EPI_ISL_1069013, EPI_ISL_1069014, EPI_ISL_1069015, EPI_ISL_1069016, EPI_ISL_1069017, EPI_ISL_1069018, EPI_ISL_1069019, EPI_ISL_1069020, EPI_ISL_1069021, EPI_ISL_1069022, EPI_ISL_1069023, EPI_ISL_1069024, EPI_ISL_1069025, EPI_ISL_1069026, EPI_ISL_1069027, EPI_ISL_1069028, EPI_ISL_1069029, EPI_ISL_1069030, EPI_ISL_1069031, EPI_ISL_1069032, EPI_ISL_1069033, EPI_ISL_1069034, EPI_ISL_1069035, EPI_ISL_1069036, EPI_ISL_1069037, EPI_ISL_1069038, EPI_ISL_1069039, EPI_ISL_1069040, EPI_ISL_1069041, EPI_ISL_1069042, EPI_ISL_1069043, EPI_ISL_1069044, EPI_ISL_1069045, EPI_ISL_1069046, EPI_ISL_1069047, EPI_ISL_1069048, EPI_ISL_1069049, EPI_ISL_1069050, EPI_ISL_1069051, EPI_ISL_1069052, EPI_ISL_1069053, EPI_ISL_1069054, EPI_ISL_1069055, EPI_ISL_1069056, EPI_ISL_1069057, EPI_ISL_1069058, EPI_ISL_1069059, EPI_ISL_1069060, EPI_ISL_1069061, EPI_ISL_1069062, EPI_ISL_1069063, EPI_ISL_1069064, EPI_ISL_1069065, EPI_ISL_1069066, EPI_ISL_1069067, EPI_ISL_1069068, EPI_ISL_1069069, EPI_ISL_1069070, EPI_ISL_1069071, EPI_ISL_1069072, EPI_ISL_1069073, EPI_ISL_1069074, EPI_ISL_1069075, EPI_ISL_1069085, EPI_ISL_1069086, EPI_ISL_1069087, EPI_ISL_1069088, EPI_ISL_1069089, EPI_ISL_1069090, EPI_ISL_1069091, EPI_ISL_1069092, EPI_ISL_1069093, EPI_ISL_1069094, EPI_ISL_1069095, EPI_ISL_1069096, EPI_ISL_1069097, EPI_ISL_1069098, EPI_ISL_1069099, EPI_ISL_1069100, EPI_ISL_1069101, EPI_ISL_1069102, EPI_ISL_1069103, EPI_ISL_1069104, EPI_ISL_1069105, EPI_ISL_1069106, EPI_ISL_1069107, EPI_ISL_1069108, EPI_ISL_1069109, EPI_ISL_1069110, EPI_ISL_1069111, EPI_ISL_1069112, EPI_ISL_1069113, EPI_ISL_1069114, EPI_ISL_1069115, EPI_ISL_1069116, EPI_ISL_1069117, EPI_ISL_1069118, EPI_ISL_1069119, EPI_ISL_1069120, EPI_ISL_1069121, EPI_ISL_1069122, EPI_ISL_1069123, EPI_ISL_1069124, EPI_ISL_1069125, EPI_ISL_1069126, EPI_ISL_1069127, EPI_ISL_1069128, EPI_ISL_1069129, EPI_ISL_1069130, EPI_ISL_1069131, EPI_ISL_1069132, EPI_ISL_1069133, EPI_ISL_1069134, EPI_ISL_1069135, EPI_ISL_1069136, EPI_ISL_1069137, EPI_ISL_1069138, EPI_ISL_1069139, EPI_ISL_1069140, EPI_ISL_1069141, EPI_ISL_1069142, EPI_ISL_1069143, EPI_ISL_1069144, EPI_ISL_1069145, EPI_ISL_1069146, EPI_ISL_1069147, EPI_ISL_1069148, EPI_ISL_1069149, EPI_ISL_1069150, EPI_ISL_1069151, EPI_ISL_1069152, EPI_ISL_1095645, EPI_ISL_1095646, EPI_ISL_1095647, EPI_ISL_1095648, EPI_ISL_1095649, EPI_ISL_1095650, EPI_ISL_1095651, EPI_ISL_1095652, EPI_ISL_1095653, EPI_ISL_1095654, EPI_ISL_1095655, EPI_ISL_1095656, EPI_ISL_1095657, EPI_ISL_1095658, EPI_ISL_1095659, EPI_ISL_1095660, EPI_ISL_1095661, EPI_ISL_1095662, EPI_ISL_1095663, EPI_ISL_1095664, EPI_ISL_1095665, EPI_ISL_1095666, EPI_ISL_1095667, EPI_ISL_1095668, EPI_ISL_1095669, EPI_ISL_1095670, EPI_ISL_1095671, EPI_ISL_1095672, EPI_ISL_1095673, EPI_ISL_1095674, EPI_ISL_1095675, EPI_ISL_1095676, EPI_ISL_1095677, EPI_ISL_1095678, EPI_ISL_1095679, EPI_ISL_1095680, EPI_ISL_1095681, EPI_ISL_1095682, EPI_ISL_1095683, EPI_ISL_1095684, EPI_ISL_1095685, EPI_ISL_1095686, EPI_ISL_1095687, EPI_ISL_1095688, EPI_ISL_1095689, EPI_ISL_1095690, EPI_ISL_1095691, EPI_ISL_1095692, EPI_ISL_1095693, EPI_ISL_1095694, EPI_ISL_1095695, EPI_ISL_1095696, EPI_ISL_1095697, EPI_ISL_1095698, EPI_ISL_1095699, EPI_ISL_1095700, EPI_ISL_1095701, EPI_ISL_1095702, EPI_ISL_1095703, EPI_ISL_1095704, EPI_ISL_1095705, EPI_ISL_1095706, EPI_ISL_1095707, EPI_ISL_1095708, EPI_ISL_1095709, EPI_ISL_1095710, EPI_ISL_1095711, EPI_ISL_1095712, EPI_ISL_1095713, EPI_ISL_1095714, EPI_ISL_1095715, EPI_ISL_1095716, EPI_ISL_1095717, EPI_ISL_1095718, EPI_ISL_1095719, EPI_ISL_1095720, EPI_ISL_1095721, EPI_ISL_1095722, EPI_ISL_1095723, EPI_ISL_1095724, EPI_ISL_1095725, EPI_ISL_1095726, EPI_ISL_1095727, EPI_ISL_1095728, EPI_ISL_1095729, EPI_ISL_1095730, EPI_ISL_1095731, EPI_ISL_1095732, EPI_ISL_1095733, EPI_ISL_1095734, EPI_ISL_1095735, EPI_ISL_1095736, EPI_ISL_1095737, EPI_ISL_1095738, EPI_ISL_1095739, EPI_ISL_1095740, EPI_ISL_1095741, EPI_ISL_1095742, EPI_ISL_1095743, EPI_ISL_1095744, EPI_ISL_1095745, EPI_ISL_1095746, EPI_ISL_1095747, EPI_ISL_1095748, EPI_ISL_1095749, EPI_ISL_1095750, EPI_ISL_1095751, EPI_ISL_1095752, EPI_ISL_1095753, EPI_ISL_1095754, EPI_ISL_1095755, EPI_ISL_1095756, EPI_ISL_1095757, EPI_ISL_1095758, EPI_ISL_1095759, EPI_ISL_1095760, EPI_ISL_1095761, EPI_ISL_1095762, EPI_ISL_1095763, EPI_ISL_1095764, EPI_ISL_1095765, EPI_ISL_1095766, EPI_ISL_1095767, EPI_ISL_1095768, EPI_ISL_1095769, EPI_ISL_1095770, EPI_ISL_1095771, EPI_ISL_1095772, EPI_ISL_1095773, EPI_ISL_1095774, EPI_ISL_1095775, EPI_ISL_1095776, EPI_ISL_1095777, EPI_ISL_1095778, EPI_ISL_1095779, EPI_ISL_1095780, EPI_ISL_1095781, EPI_ISL_1095782, EPI_ISL_1095783, EPI_ISL_1095784, EPI_ISL_1095785, EPI_ISL_1095786, EPI_ISL_1095787, EPI_ISL_1095788, EPI_ISL_1095789, EPI_ISL_1095790, EPI_ISL_1095791, EPI_ISL_1095792, EPI_ISL_1095793, EPI_ISL_1095794, EPI_ISL_1095795, EPI_ISL_1095796, EPI_ISL_1095797, EPI_ISL_1095798, EPI_ISL_1095799, EPI_ISL_1095800, EPI_ISL_1095801, EPI_ISL_1095802, EPI_ISL_1095803, EPI_ISL_1095804, EPI_ISL_1095805, EPI_ISL_1095806, EPI_ISL_1095807, EPI_ISL_1095808, EPI_ISL_1095809, EPI_ISL_1095810, EPI_ISL_1095811, EPI_ISL_1095812, EPI_ISL_1095813, EPI_ISL_1095814, EPI_ISL_1095815, EPI_ISL_1095816, EPI_ISL_1095817, EPI_ISL_1095818, EPI_ISL_1095819, EPI_ISL_1095820, EPI_ISL_1095821, EPI_ISL_1095822, EPI_ISL_1095823, EPI_ISL_1095824, EPI_ISL_1095825, EPI_ISL_1095826, EPI_ISL_1095827, EPI_ISL_1095828, EPI_ISL_1095829, EPI_ISL_1095830, EPI_ISL_1095831, EPI_ISL_1095832, EPI_ISL_1095833, EPI_ISL_1095834, EPI_ISL_1095835, EPI_ISL_1095836, EPI_ISL_1095837, EPI_ISL_1095838, EPI_ISL_1095839, EPI_ISL_1095840, EPI_ISL_1095841, EPI_ISL_1095842, EPI_ISL_1095843, EPI_ISL_1095844, EPI_ISL_1095845, EPI_ISL_1095846, EPI_ISL_1095847, EPI_ISL_1095848, EPI_ISL_1095849, EPI_ISL_1095850, EPI_ISL_1095851, EPI_ISL_1095852, EPI_ISL_1095853, EPI_ISL_1095854, EPI_ISL_1095855, EPI_ISL_1095856, EPI_ISL_1095857, EPI_ISL_1095858, EPI_ISL_1095859, EPI_ISL_1095860, EPI_ISL_1095861, EPI_ISL_1095862, EPI_ISL_1095863, EPI_ISL_1095864, EPI_ISL_1095865, EPI_ISL_1095866, EPI_ISL_1095867, EPI_ISL_1095868, EPI_ISL_1095869, EPI_ISL_1095870, EPI_ISL_1095871, EPI_ISL_1095872, EPI_ISL_1095873, EPI_ISL_1095874, EPI_ISL_1095875, EPI_ISL_1095876, EPI_ISL_1095877, EPI_ISL_1095878, EPI_ISL_1095879, EPI_ISL_1095880, EPI_ISL_1095881, EPI_ISL_1095882, EPI_ISL_1095883, EPI_ISL_1095884, EPI_ISL_1095885, EPI_ISL_1095886, EPI_ISL_1095887, EPI_ISL_1095888, EPI_ISL_1095889, EPI_ISL_1095890, EPI_ISL_1095891, EPI_ISL_1095892, EPI_ISL_1095893, EPI_ISL_1095894, EPI_ISL_1095895, EPI_ISL_1095896, EPI_ISL_1095897, EPI_ISL_1095898, EPI_ISL_1095899, EPI_ISL_1095900, EPI_ISL_1095901, EPI_ISL_1095902, EPI_ISL_1095903, EPI_ISL_1095904, EPI_ISL_1095905, EPI_ISL_1095906, EPI_ISL_1095907, EPI_ISL_1095908, EPI_ISL_1095909, EPI_ISL_1095910, EPI_ISL_1095911, EPI_ISL_1095912, EPI_ISL_1095913, EPI_ISL_1095914, EPI_ISL_1095915, EPI_ISL_1095916, EPI_ISL_1095917, EPI_ISL_1095918, EPI_ISL_1095919, EPI_ISL_1095920, EPI_ISL_1095921, EPI_ISL_1095922, EPI_ISL_1095923, EPI_ISL_1095924, EPI_ISL_1095925, EPI_ISL_1095926, EPI_ISL_1095927, EPI_ISL_1095928, EPI_ISL_1095929, EPI_ISL_1095930, EPI_ISL_1095931, EPI_ISL_1095932, EPI_ISL_1095933, EPI_ISL_1095934, EPI_ISL_1095935, EPI_ISL_1095936, EPI_ISL_1095937, EPI_ISL_1095938, EPI_ISL_1095939, EPI_ISL_1095940, EPI_ISL_1095941, EPI_ISL_1095942, EPI_ISL_1095943, EPI_ISL_1095944, EPI_ISL_1095945, EPI_ISL_1095946, EPI_ISL_1095947, EPI_ISL_1095948, EPI_ISL_1095949, EPI_ISL_1095950, EPI_ISL_1095951, EPI_ISL_1095952, EPI_ISL_1095953, EPI_ISL_1095954, EPI_ISL_1095955, EPI_ISL_1095956, EPI_ISL_1095957, EPI_ISL_1095958, EPI_ISL_1095959, EPI_ISL_1095960, EPI_ISL_1095961, EPI_ISL_1095962, EPI_ISL_1095963, EPI_ISL_1095964, EPI_ISL_1095965, EPI_ISL_1095966, EPI_ISL_1095967, EPI_ISL_1095968, EPI_ISL_1095969, EPI_ISL_1095970, EPI_ISL_1095971, EPI_ISL_1095972, EPI_ISL_1095973, EPI_ISL_1095974, EPI_ISL_1095975, EPI_ISL_1095976, EPI_ISL_1095977, EPI_ISL_1095978, EPI_ISL_1095979, EPI_ISL_1095980, EPI_ISL_1095981, EPI_ISL_1095982, EPI_ISL_1095983, EPI_ISL_1095984, EPI_ISL_1095985, EPI_ISL_1095986, EPI_ISL_1095987, EPI_ISL_1095988, EPI_ISL_1095989, EPI_ISL_1095990, EPI_ISL_1095991, EPI_ISL_1095992, EPI_ISL_1095993, EPI_ISL_1095994, EPI_ISL_1095995, EPI_ISL_1095996, EPI_ISL_1095997, EPI_ISL_1095998, EPI_ISL_1095999, EPI_ISL_1096000, EPI_ISL_1096001, EPI_ISL_1096002, EPI_ISL_1096003, EPI_ISL_1096004, EPI_ISL_1096005, EPI_ISL_1096006, EPI_ISL_1096007, EPI_ISL_1096008, EPI_ISL_1096009, EPI_ISL_1096010, EPI_ISL_1096011, EPI_ISL_1096012, EPI_ISL_1096013, EPI_ISL_1096014, EPI_ISL_1096015, EPI_ISL_1096016, EPI_ISL_1096017, EPI_ISL_1096018, EPI_ISL_1096019, EPI_ISL_1096020, EPI_ISL_1096021, EPI_ISL_1096022, EPI_ISL_1096023, EPI_ISL_1096024, EPI_ISL_1096025, EPI_ISL_1096026, EPI_ISL_1096027, EPI_ISL_1096028, EPI_ISL_1096029, EPI_ISL_1132845, EPI_ISL_1132846, EPI_ISL_1132847, EPI_ISL_1132848, EPI_ISL_1132849, EPI_ISL_1132850, EPI_ISL_1132851, EPI_ISL_1132852, EPI_ISL_1132853, EPI_ISL_1132854, EPI_ISL_1132855, EPI_ISL_1132856, EPI_ISL_1132857, EPI_ISL_1132858, EPI_ISL_1132859, EPI_ISL_1132860, EPI_ISL_1132861, EPI_ISL_1132862, EPI_ISL_1132863, EPI_ISL_1132864, EPI_ISL_1132865, EPI_ISL_1132866, EPI_ISL_1132867, EPI_ISL_1132868, EPI_ISL_1132869, EPI_ISL_1132870, EPI_ISL_1132871, EPI_ISL_1132872, EPI_ISL_1132873, EPI_ISL_1132874, EPI_ISL_1132875, EPI_ISL_1132876, EPI_ISL_1132877, EPI_ISL_1132878, EPI_ISL_1132879, EPI_ISL_1132880, EPI_ISL_1132881, EPI_ISL_1132882, EPI_ISL_1132883, EPI_ISL_1132884, EPI_ISL_1132885, EPI_ISL_1132886, EPI_ISL_1132887, EPI_ISL_1132888, EPI_ISL_1132889, EPI_ISL_1132890, EPI_ISL_1132891, EPI_ISL_1132892, EPI_ISL_1132893, EPI_ISL_1132894, EPI_ISL_1132895, EPI_ISL_1132896, EPI_ISL_1132897, EPI_ISL_1132898, EPI_ISL_1132899, EPI_ISL_1132900, EPI_ISL_1132901, EPI_ISL_1132902, EPI_ISL_1132903, EPI_ISL_1132904, EPI_ISL_1132905, EPI_ISL_1132906, EPI_ISL_1132907, EPI_ISL_1132908, EPI_ISL_1132909, EPI_ISL_1132910, EPI_ISL_1132911, EPI_ISL_1132912, EPI_ISL_1132913, EPI_ISL_1132914, EPI_ISL_1132915, EPI_ISL_1132916, EPI_ISL_1132917, EPI_ISL_1132918, EPI_ISL_1132919, EPI_ISL_1132920, EPI_ISL_1132921, EPI_ISL_1132922, EPI_ISL_1132923, EPI_ISL_1132924, EPI_ISL_1132925, EPI_ISL_1132926, EPI_ISL_1132927, EPI_ISL_1132928, EPI_ISL_1132929, EPI_ISL_1132930, EPI_ISL_1132931, EPI_ISL_1132932, EPI_ISL_1132933, EPI_ISL_1132934, EPI_ISL_1132935, EPI_ISL_1132936, EPI_ISL_1132937, EPI_ISL_1132938, EPI_ISL_1132939, EPI_ISL_1132940, EPI_ISL_1132941, EPI_ISL_1132942, EPI_ISL_1132943, EPI_ISL_1132944, EPI_ISL_1132945, EPI_ISL_1132946, EPI_ISL_1132947, EPI_ISL_1132948, EPI_ISL_1132949, EPI_ISL_1132950, EPI_ISL_1132951, EPI_ISL_1132952, EPI_ISL_1132953, EPI_ISL_1132954, EPI_ISL_1132955, EPI_ISL_1132956, EPI_ISL_1132957, EPI_ISL_1132958, EPI_ISL_1132959, EPI_ISL_1132960, EPI_ISL_1132961, EPI_ISL_1132962, EPI_ISL_1132963, EPI_ISL_1132964, EPI_ISL_1132965, EPI_ISL_1132966, EPI_ISL_1132967, EPI_ISL_1132968, EPI_ISL_1132969, EPI_ISL_1132970, EPI_ISL_1134840, EPI_ISL_1134841, EPI_ISL_1134842, EPI_ISL_1134843, EPI_ISL_1134844, EPI_ISL_1134845, EPI_ISL_1134846, EPI_ISL_1134847, EPI_ISL_1134848, EPI_ISL_1134849, EPI_ISL_1134850, EPI_ISL_1134851, EPI_ISL_1134852, EPI_ISL_1134853, EPI_ISL_1134854, EPI_ISL_1134855, EPI_ISL_1134856, EPI_ISL_1134857, EPI_ISL_1134858, EPI_ISL_1134859, EPI_ISL_1134860, EPI_ISL_1134861, EPI_ISL_1134862, EPI_ISL_1134863, EPI_ISL_1134864, EPI_ISL_1134865, EPI_ISL_1134866, EPI_ISL_1134867, EPI_ISL_1134868, EPI_ISL_1134869, EPI_ISL_1134870, EPI_ISL_1134871, EPI_ISL_1134872, EPI_ISL_1134873, EPI_ISL_1134874, EPI_ISL_1134875, EPI_ISL_1134876, EPI_ISL_1134877, EPI_ISL_1134878, EPI_ISL_1134879, EPI_ISL_1134880, EPI_ISL_1134881, EPI_ISL_1134882, EPI_ISL_1134883, EPI_ISL_1134884, EPI_ISL_1134885, EPI_ISL_1134886, EPI_ISL_1134887, EPI_ISL_1134888, EPI_ISL_1134889, EPI_ISL_1134890, EPI_ISL_1134891, EPI_ISL_1134892, EPI_ISL_1134893, EPI_ISL_1134894, EPI_ISL_1134895, EPI_ISL_1134896, EPI_ISL_1134897, EPI_ISL_1134898, EPI_ISL_1134899, EPI_ISL_1134900, EPI_ISL_1134901, EPI_ISL_1134902, EPI_ISL_1134903, EPI_ISL_1164360, EPI_ISL_1164361, EPI_ISL_1164362, EPI_ISL_1164363, EPI_ISL_1164364, EPI_ISL_1164365, EPI_ISL_1164366, EPI_ISL_1164367, EPI_ISL_1164368, EPI_ISL_1164369, EPI_ISL_1164370, EPI_ISL_1164371, EPI_ISL_1164372, EPI_ISL_1164373, EPI_ISL_1164374, EPI_ISL_1164375, EPI_ISL_1164376, EPI_ISL_1164377, EPI_ISL_1164378, EPI_ISL_1164379, EPI_ISL_1164380, EPI_ISL_1164381, EPI_ISL_1164382, EPI_ISL_1164383, EPI_ISL_1164384, EPI_ISL_1164385, EPI_ISL_1164386, EPI_ISL_1164387, EPI_ISL_1164388, EPI_ISL_1164389, EPI_ISL_1164390, EPI_ISL_1164391, EPI_ISL_1164392, EPI_ISL_1164393, EPI_ISL_1164394, EPI_ISL_1164395, EPI_ISL_1164396, EPI_ISL_1164397, EPI_ISL_1164398, EPI_ISL_1164399, EPI_ISL_1164400, EPI_ISL_1164401, EPI_ISL_1164402, EPI_ISL_1164403, EPI_ISL_1164404, EPI_ISL_1164405, EPI_ISL_1164406, EPI_ISL_1164407, EPI_ISL_1164408, EPI_ISL_1164409, EPI_ISL_1164410, EPI_ISL_1164411, EPI_ISL_1164412, EPI_ISL_1164413, EPI_ISL_1164414, EPI_ISL_1164415, EPI_ISL_1164416, EPI_ISL_1164417, EPI_ISL_1164418, EPI_ISL_1164419, EPI_ISL_1164420, EPI_ISL_1164421, EPI_ISL_1164422, EPI_ISL_1164423, EPI_ISL_1164424, EPI_ISL_1164425, EPI_ISL_1164426, EPI_ISL_1164427, EPI_ISL_1164428, EPI_ISL_1164429, EPI_ISL_1164430, EPI_ISL_1164431, EPI_ISL_1164432, EPI_ISL_1164433, EPI_ISL_1164434, EPI_ISL_1164435, EPI_ISL_1164436, EPI_ISL_1164437, EPI_ISL_1164438, EPI_ISL_1172046, EPI_ISL_1172047, EPI_ISL_1172048, EPI_ISL_1172049, EPI_ISL_1172050, EPI_ISL_1172051, EPI_ISL_1172052, EPI_ISL_1172053, EPI_ISL_1172054, EPI_ISL_1172055, EPI_ISL_1172056, EPI_ISL_1172057, EPI_ISL_1172058, EPI_ISL_1172059, EPI_ISL_1172060, EPI_ISL_1172061, EPI_ISL_1172062, EPI_ISL_1172063, EPI_ISL_1172064, EPI_ISL_1172065, EPI_ISL_1172066, EPI_ISL_1172067, EPI_ISL_1172068, EPI_ISL_1172069, EPI_ISL_1172070, EPI_ISL_1172071, EPI_ISL_1172072, EPI_ISL_1172073, EPI_ISL_1172074, EPI_ISL_1172075, EPI_ISL_1172076, EPI_ISL_1172077, EPI_ISL_1172078, EPI_ISL_1172079, EPI_ISL_1172080, EPI_ISL_1172081, EPI_ISL_1172082, EPI_ISL_1172083, EPI_ISL_1172084, EPI_ISL_1172085, EPI_ISL_1172086, EPI_ISL_1172087, EPI_ISL_1172088, EPI_ISL_1172089, EPI_ISL_1172090, EPI_ISL_1172091, EPI_ISL_1172092, EPI_ISL_1172093, EPI_ISL_1172094, EPI_ISL_1172095, EPI_ISL_1172096, EPI_ISL_1172097, EPI_ISL_1172098, EPI_ISL_1172099, EPI_ISL_1172100, EPI_ISL_1172101, EPI_ISL_1172102, EPI_ISL_1172103, EPI_ISL_1172104, EPI_ISL_1172105, EPI_ISL_1172106, EPI_ISL_1172107, EPI_ISL_1172108, EPI_ISL_1172109, EPI_ISL_1195947, EPI_ISL_1195948, EPI_ISL_1195949, EPI_ISL_1251260, EPI_ISL_1252050 | | | |
| see above | Fulgent Genetics | Fulgent Genetics | Becky Tsai; Benafsh Sapra; Doreen Ng; Harry Gao; James Xie; John Gao; Joseph Fierro; Mickey Li; Yan Meng |
| EPI_ISL_1366333, EPI_ISL_1366334 | Genomica Lab Molecular, Mexico | Andersen lab at Scripps Research | Jose Horacio Reyna Verdugo; Jose Roman Chavez Mendez; Luis Alberto Rangel Gonzalez; Martin Gonzalez Ibarra; SEARCH Alliance San Diego with Jonathan Gonzalez Garcia |
| EPI_ISL_1508978, EPI_ISL_1508979, EPI_ISL_1508980, EPI_ISL_1508981, EPI_ISL_1508982, EPI_ISL_1508983, EPI_ISL_1508987, EPI_ISL_1532118, EPI_ISL_1532120, EPI_ISL_1532121, EPI_ISL_1532122 | | | |
| see above | Good Samaritan Hospital | Santa Clara County Public Health Laboratory | Santa Clara County Public Health Department |
| EPI_ISL_1088585, EPI_ISL_1088591, EPI_ISL_1088593, EPI_ISL_1088595, EPI_ISL_1088599, EPI_ISL_1088600, EPI_ISL_1088601, EPI_ISL_1088603, EPI_ISL_1088605, EPI_ISL_1088606, EPI_ISL_1088610, EPI_ISL_1088611, EPI_ISL_1088616, EPI_ISL_1088617, EPI_ISL_1088625, EPI_ISL_1088627, EPI_ISL_1088628, EPI_ISL_1088630, EPI_ISL_1088633, EPI_ISL_1088635, EPI_ISL_1088640, EPI_ISL_1088652, EPI_ISL_1088654, EPI_ISL_1088655, EPI_ISL_1088661, EPI_ISL_1088663, EPI_ISL_1088666, EPI_ISL_1088677, EPI_ISL_1088678, EPI_ISL_1088681, EPI_ISL_1088684, EPI_ISL_1088689, EPI_ISL_1088690, EPI_ISL_1088696, EPI_ISL_1088698, EPI_ISL_1088703, EPI_ISL_1088706, EPI_ISL_1088715, EPI_ISL_1088720, EPI_ISL_1088725, EPI_ISL_1088726, EPI_ISL_1088727, EPI_ISL_1088730, EPI_ISL_1088751, EPI_ISL_1088755, EPI_ISL_1088762, EPI_ISL_1088763, EPI_ISL_1088765, EPI_ISL_1088772, EPI_ISL_1088774, EPI_ISL_1088777, EPI_ISL_1088791, EPI_ISL_1088793, EPI_ISL_1088816, EPI_ISL_1088819, EPI_ISL_1088820, EPI_ISL_1088822, EPI_ISL_1088828, EPI_ISL_1088833, EPI_ISL_1088835, EPI_ISL_1088841, EPI_ISL_1088850, EPI_ISL_1088851, EPI_ISL_1088853, EPI_ISL_1088865, EPI_ISL_1088867, EPI_ISL_1088871, EPI_ISL_1088874, EPI_ISL_1088880, EPI_ISL_1088887, EPI_ISL_1088891, EPI_ISL_1088900, EPI_ISL_1088901, EPI_ISL_1088903, EPI_ISL_1088904, EPI_ISL_1088907, EPI_ISL_1088913, EPI_ISL_1088914, EPI_ISL_1088933, EPI_ISL_1088956, EPI_ISL_1088965, EPI_ISL_1088967, EPI_ISL_1088974, EPI_ISL_1088979, EPI_ISL_1088995, EPI_ISL_1088997, EPI_ISL_1088999, EPI_ISL_1089000, EPI_ISL_1089001, EPI_ISL_1089004, EPI_ISL_1089007, EPI_ISL_1089012, EPI_ISL_1089027, EPI_ISL_1089050, EPI_ISL_1089053, EPI_ISL_1089054, EPI_ISL_1089062, EPI_ISL_1089063, EPI_ISL_1089069, EPI_ISL_1089074, EPI_ISL_1089075, EPI_ISL_1089080, EPI_ISL_1089086, EPI_ISL_1089090, EPI_ISL_1089101, EPI_ISL_1089108, EPI_ISL_1089117, EPI_ISL_1089118, EPI_ISL_1089127, EPI_ISL_1089136, EPI_ISL_1089138, EPI_ISL_1089148, EPI_ISL_1089152, EPI_ISL_1089153, EPI_ISL_1089154, EPI_ISL_1089155, EPI_ISL_1089157, EPI_ISL_1089159, EPI_ISL_1089171, EPI_ISL_1089173, EPI_ISL_1089185, EPI_ISL_1089197, EPI_ISL_1089198, EPI_ISL_1089207, EPI_ISL_1089212, EPI_ISL_1089214, EPI_ISL_1089215, EPI_ISL_1089216, EPI_ISL_1089221, EPI_ISL_1089222, EPI_ISL_1089239, EPI_ISL_1089244, EPI_ISL_1089258, EPI_ISL_1089260, EPI_ISL_1089262, EPI_ISL_1089266, EPI_ISL_1089267, EPI_ISL_1089269, EPI_ISL_1089272, EPI_ISL_1089273, EPI_ISL_1089274, EPI_ISL_1089275, EPI_ISL_1089279, EPI_ISL_1089280, EPI_ISL_1089284, EPI_ISL_1089285, EPI_ISL_1089288, EPI_ISL_1089289, EPI_ISL_1089290, EPI_ISL_1089291, EPI_ISL_1089293, EPI_ISL_1089297, EPI_ISL_1089298, EPI_ISL_1089299, EPI_ISL_1089302, EPI_ISL_1089304, EPI_ISL_1089305, EPI_ISL_1089308, EPI_ISL_1089310, EPI_ISL_1089311, EPI_ISL_1089312, EPI_ISL_1089314, EPI_ISL_1089316, EPI_ISL_1089318, EPI_ISL_1089319, EPI_ISL_1089320, EPI_ISL_1089321, EPI_ISL_1089429, EPI_ISL_1089430, EPI_ISL_1089433 | | | |
| see above | Helix / Illumina | Respiratory Viruses Branch, Division of Viral Diseases, Centers for Disease Control and Prevention | Alexandre Bolze; Ary Ascencio; Ben L. Rambo-Martin; Brad Sickler; Charlotte Rivera-Garcia; Christine Tran; Clinton R. Paden; Dakota Howard; David Becker; Dhwani Batra; Duncan MacCannell; Efren Sandoval; Eileen de Feo; Elizabeth Cirulli; Eric Allen; Geraint Levan; James Lu; Jan Antico; Jason Nguyen; Jimmy Ramirez; Jingtao Liu; Kelly Schiabor Barrett; Kim Gietzen; Magnus Isaksson; Marc Laurent; Matthew Tolentino; Nicole L. Washington; Peter W. Cook; Phil Febbo; Ryan Cho; Shannon Wickline; Sherry Wang; Simon White; Summer Galloway; Suxiang Tong; Tyler Cassens; William Lee |
| EPI_ISL_1194743, EPI_ISL_1194746, EPI_ISL_1194785, EPI_ISL_1194808, EPI_ISL_1194818, EPI_ISL_1194828, EPI_ISL_1262278, EPI_ISL_1262280, EPI_ISL_1262284, EPI_ISL_1262291, EPI_ISL_1262300, EPI_ISL_1262306, EPI_ISL_1262336, EPI_ISL_1262338, EPI_ISL_1262339, EPI_ISL_1262347, EPI_ISL_1262353, EPI_ISL_1262360, EPI_ISL_1262379, EPI_ISL_1262381, EPI_ISL_1262385, EPI_ISL_1262393, EPI_ISL_1262404, EPI_ISL_1262414, EPI_ISL_1267505, EPI_ISL_1267513, EPI_ISL_1267530, EPI_ISL_1267537, EPI_ISL_1267538, EPI_ISL_1267542, EPI_ISL_1267545, EPI_ISL_1267548, EPI_ISL_1267551, EPI_ISL_1267556, EPI_ISL_1267557, EPI_ISL_1267558, EPI_ISL_1267559, EPI_ISL_1267568, EPI_ISL_1267569, EPI_ISL_1267575, EPI_ISL_1267582, EPI_ISL_1267589, EPI_ISL_1267621, EPI_ISL_1267627, EPI_ISL_1267629, EPI_ISL_1267631, EPI_ISL_1267635, EPI_ISL_1267636, EPI_ISL_1267640, EPI_ISL_1267642, EPI_ISL_1267652, EPI_ISL_1267657, EPI_ISL_1267661, EPI_ISL_1267667, EPI_ISL_1267680, EPI_ISL_1267681, EPI_ISL_1267689, EPI_ISL_1267695, EPI_ISL_1267704, EPI_ISL_1267746, EPI_ISL_1267758, EPI_ISL_1267767, EPI_ISL_1267776, EPI_ISL_1267790, EPI_ISL_1267797, EPI_ISL_1267798, EPI_ISL_1267800, EPI_ISL_1267809, EPI_ISL_1267815, EPI_ISL_1267820, EPI_ISL_1267821, EPI_ISL_1267833, EPI_ISL_1267839, EPI_ISL_1267844, EPI_ISL_1267845, EPI_ISL_1267851, EPI_ISL_1267876, EPI_ISL_1267892, EPI_ISL_1267897, EPI_ISL_1267898, EPI_ISL_1267901, EPI_ISL_1267903, EPI_ISL_1267904, EPI_ISL_1267906, EPI_ISL_1267911, EPI_ISL_1267912, EPI_ISL_1267913, EPI_ISL_1267916, EPI_ISL_1267920, EPI_ISL_1267971, EPI_ISL_1267978, EPI_ISL_1267993, EPI_ISL_1268000, EPI_ISL_1268012, EPI_ISL_1268017, EPI_ISL_1268038, EPI_ISL_1268039, EPI_ISL_1268048, EPI_ISL_1268049, EPI_ISL_1268073, EPI_ISL_1268075, EPI_ISL_1268085, EPI_ISL_1268086, EPI_ISL_1268096, EPI_ISL_1268098, EPI_ISL_1268099, EPI_ISL_1268110, EPI_ISL_1268111, EPI_ISL_1268113, EPI_ISL_1268119, EPI_ISL_1268138, EPI_ISL_1268146, EPI_ISL_1268147, EPI_ISL_1268150, EPI_ISL_1268173, EPI_ISL_1268174, EPI_ISL_1268180, EPI_ISL_1268181, EPI_ISL_1268185, EPI_ISL_1268186, EPI_ISL_1268190, EPI_ISL_1268197, EPI_ISL_1268199, EPI_ISL_1268201, EPI_ISL_1268231, EPI_ISL_1268260, EPI_ISL_1268272, EPI_ISL_1268273, EPI_ISL_1268276, EPI_ISL_1268286, EPI_ISL_1268312, EPI_ISL_1268339, EPI_ISL_1268344, EPI_ISL_1268363, EPI_ISL_1268378, EPI_ISL_1268404, EPI_ISL_1268413, EPI_ISL_1268434, EPI_ISL_1268436, EPI_ISL_1268440, EPI_ISL_1268442, EPI_ISL_1268443, EPI_ISL_1268473, EPI_ISL_1268485, EPI_ISL_1268509, EPI_ISL_1268534, EPI_ISL_1268544, EPI_ISL_1268549, EPI_ISL_1268554, EPI_ISL_1268575, EPI_ISL_1268599, EPI_ISL_1268603, EPI_ISL_1268604, EPI_ISL_1268606, EPI_ISL_1268634, EPI_ISL_1268640, EPI_ISL_1268744, EPI_ISL_1268748, EPI_ISL_1268842, EPI_ISL_1268866, EPI_ISL_1268870, EPI_ISL_1268897, EPI_ISL_1268901, EPI_ISL_1268909, EPI_ISL_1268915, EPI_ISL_1268917, EPI_ISL_1268931, EPI_ISL_1268935, EPI_ISL_1268953, EPI_ISL_1268955, EPI_ISL_1268991, EPI_ISL_1269016, EPI_ISL_1269023, EPI_ISL_1269039, EPI_ISL_1269059, EPI_ISL_1269094, EPI_ISL_1269109, EPI_ISL_1269132, EPI_ISL_1269135, EPI_ISL_1269139, EPI_ISL_1269141, EPI_ISL_1269148, EPI_ISL_1269151, EPI_ISL_1269152, EPI_ISL_1269194, EPI_ISL_1269196, EPI_ISL_1269198, EPI_ISL_1269205, EPI_ISL_1269207, EPI_ISL_1269219, EPI_ISL_1269230, EPI_ISL_1269239, EPI_ISL_1269270, EPI_ISL_1269278, EPI_ISL_1269312, EPI_ISL_1269313, EPI_ISL_1269332, EPI_ISL_1269532, EPI_ISL_1269554, EPI_ISL_1269557, EPI_ISL_1269576, EPI_ISL_1269588, EPI_ISL_1269593, EPI_ISL_1269596, EPI_ISL_1269606, EPI_ISL_1269609, EPI_ISL_1270145, EPI_ISL_1270147, EPI_ISL_1270161, EPI_ISL_1270174, EPI_ISL_1270207, EPI_ISL_1270246, EPI_ISL_1298206, EPI_ISL_1336965, EPI_ISL_1336973, EPI_ISL_1336975, EPI_ISL_1337010, EPI_ISL_1337017, EPI_ISL_1337027, EPI_ISL_1337029, EPI_ISL_1337039, EPI_ISL_1337040, EPI_ISL_1337062, EPI_ISL_1337068, EPI_ISL_1337071, EPI_ISL_1337073, EPI_ISL_1337095, EPI_ISL_1337109, EPI_ISL_1337132, EPI_ISL_1337138, EPI_ISL_1337188, EPI_ISL_1337211, EPI_ISL_1391777, EPI_ISL_1391778, EPI_ISL_1391782, EPI_ISL_1391808, EPI_ISL_1391814, EPI_ISL_1391982, EPI_ISL_1391991, EPI_ISL_1391995, EPI_ISL_1493792 | | | |
| see above | Helix/Illumina | Centers for Disease Control and Prevention Division of Viral Diseases, Pathogen Discovery | Adrian Paskey; Alexandre Bolze; Ary Ascencio; Ben L. Rambo-Martin; Benjamin Rambo-Martin; Brad Sickler; Charlotte Rivera-Garcia; Christine Tran; Christopher Gulvick; Clinton R. Paden; Dakota Howard; Darlene Wagner; David Becker; Dhwani Batra; Duncan MacCannell; Efren Sandoval; Eileen de Feo; Elizabeth Cirulli; Eric Allen; Geraint Levan; James Lu; Jan Antico; Jason Caravas; Jason Nguyen; Jimmy Ramirez; Jingtao Liu; Kara Moser; Kelly Schiabor Barrett; Kim Gietzen; Magnus Isaksson; Marc Laurent; Matthew Schmerer; Matthew Tolentino; Nicole L. Washington; Peter W. Cook; Phil Febbo; Ryan Cho; Scott Sammons; Shannon Wickline; Shatavia Morrison; Sherry Wang; Simon White; Summer Galloway; Suxiang Tong; Tyler Cassens; William Lee; Yvette Unoarumhi |
| EPI_ISL_966852, EPI_ISL_966853, EPI_ISL_966855, EPI_ISL_966857, EPI_ISL_966858, EPI_ISL_966860, EPI_ISL_966861, EPI_ISL_966862, EPI_ISL_978557, EPI_ISL_978561, EPI_ISL_978562, EPI_ISL_978564, EPI_ISL_978565, EPI_ISL_978568, EPI_ISL_978569, EPI_ISL_978581, EPI_ISL_978585, EPI_ISL_978586, EPI_ISL_978587, EPI_ISL_978589, EPI_ISL_978597, EPI_ISL_978609, EPI_ISL_1016510, EPI_ISL_1016513, EPI_ISL_1016518, EPI_ISL_1016528, EPI_ISL_1016532, EPI_ISL_1016538, EPI_ISL_1016545, EPI_ISL_1016549, EPI_ISL_1016554, EPI_ISL_1016555, EPI_ISL_1016575, EPI_ISL_1016578, EPI_ISL_1016592, EPI_ISL_1016599, EPI_ISL_1016603, EPI_ISL_1016604, EPI_ISL_1016606, EPI_ISL_1016608, EPI_ISL_1016609, EPI_ISL_1016610, EPI_ISL_1016611, EPI_ISL_1016612, EPI_ISL_1016618, EPI_ISL_1016621, EPI_ISL_1016622, EPI_ISL_1016623, EPI_ISL_1016647, EPI_ISL_1016652, EPI_ISL_1016654, EPI_ISL_1016657, EPI_ISL_1016658, EPI_ISL_1016786, EPI_ISL_1081049, EPI_ISL_1081059, EPI_ISL_1081063, EPI_ISL_1081083, EPI_ISL_1110663, EPI_ISL_1133793, EPI_ISL_1133796, EPI_ISL_1133802, EPI_ISL_1133805, EPI_ISL_1133822, EPI_ISL_1133824, EPI_ISL_1133832, EPI_ISL_1133838, EPI_ISL_1133842, EPI_ISL_1133858, EPI_ISL_1133859, EPI_ISL_1133862, EPI_ISL_1133866, EPI_ISL_1133876, EPI_ISL_1133878, EPI_ISL_1133884, EPI_ISL_1133887, EPI_ISL_1133891, EPI_ISL_1133892, EPI_ISL_1133894, EPI_ISL_1133895, EPI_ISL_1133896, EPI_ISL_1133899, EPI_ISL_1133901, EPI_ISL_1133912, EPI_ISL_1133915, EPI_ISL_1133916, EPI_ISL_1133917, EPI_ISL_1133929, EPI_ISL_1133937, EPI_ISL_1133944, EPI_ISL_1133945, EPI_ISL_1133954, EPI_ISL_1133960, EPI_ISL_1133964, EPI_ISL_1133965, EPI_ISL_1133969, EPI_ISL_1133972, EPI_ISL_1133975, EPI_ISL_1133980, EPI_ISL_1133981, EPI_ISL_1133984, EPI_ISL_1133985, EPI_ISL_1133987, EPI_ISL_1133996, EPI_ISL_1134002, EPI_ISL_1134005, EPI_ISL_1134011, EPI_ISL_1134019, EPI_ISL_1134041, EPI_ISL_1134048, EPI_ISL_1134056, EPI_ISL_1134058, EPI_ISL_1134063, EPI_ISL_1134065, EPI_ISL_1134070, EPI_ISL_1134073, EPI_ISL_1134085, EPI_ISL_1134099, EPI_ISL_1134109, EPI_ISL_1134114, EPI_ISL_1134121, EPI_ISL_1134127, EPI_ISL_1134145, EPI_ISL_1134146, EPI_ISL_1134147, EPI_ISL_1134148, EPI_ISL_1134154, EPI_ISL_1134157, EPI_ISL_1134180, EPI_ISL_1134182, EPI_ISL_1134193, EPI_ISL_1134260, EPI_ISL_1134272, EPI_ISL_1134276, EPI_ISL_1134279, EPI_ISL_1134280, EPI_ISL_1134283, EPI_ISL_1134291, EPI_ISL_1134292, EPI_ISL_1134297, EPI_ISL_1134299, EPI_ISL_1134305, EPI_ISL_1134306, EPI_ISL_1134307, EPI_ISL_1134312, EPI_ISL_1134314, EPI_ISL_1134315, EPI_ISL_1134323, EPI_ISL_1134326, EPI_ISL_1134332, EPI_ISL_1134338, EPI_ISL_1134345, EPI_ISL_1134351, EPI_ISL_1134369, EPI_ISL_1134395, EPI_ISL_1134404, EPI_ISL_1134407, EPI_ISL_1134412, EPI_ISL_1134432, EPI_ISL_1134458, EPI_ISL_1134463, EPI_ISL_1134497, EPI_ISL_1134504, EPI_ISL_1134531, EPI_ISL_1134539, EPI_ISL_1134557, EPI_ISL_1134563, EPI_ISL_1134568, EPI_ISL_1134569, EPI_ISL_1134570, EPI_ISL_1134574, EPI_ISL_1134580, EPI_ISL_1134582, EPI_ISL_1134585, EPI_ISL_1134648, EPI_ISL_1134649, EPI_ISL_1134654, EPI_ISL_1134656, EPI_ISL_1134664, EPI_ISL_1134670, EPI_ISL_1134673, EPI_ISL_1134685, EPI_ISL_1134687, EPI_ISL_1134695, EPI_ISL_1134714, EPI_ISL_1134729, EPI_ISL_1134732, EPI_ISL_1134740, EPI_ISL_1134751, EPI_ISL_1134754, EPI_ISL_1134767, EPI_ISL_1134785, EPI_ISL_1134818, EPI_ISL_1134829 | | | |
| see above | Helix/Illumina | Respiratory Viruses Branch, Division of Viral Diseases, Centers for Disease Control and Prevention | ; Alexandre Bolze; Ary Ascencio; Ben L. Rambo-Martin; Brad Sickler; Charlotte Rivera-Garcia; Christine Tran; Clinton R. Paden; Dakota Howard; David Becker; Dhwani Batra; Duncan MacCannell; Efren Sandoval; Eileen de Feo; Elizabeth Cirulli; Eric Allen; Geraint Levan; James Lu; Jan Antico; Jason Nguyen; Jimmy Ramirez; Jingtao Liu; Kelly Schiabor Barrett; Kim Gietzen; Magnus Isaksson; Marc Laurent; Matthew Tolentino; Nicole L. Washington; Peter W. Cook; Phil Febbo; Ryan Cho; Shannon Wickline; Sherry Wang; Simon White; Summer Galloway; Suxiang Tong; Tyler Cassens; William Lee |
| EPI_ISL_1477169, EPI_ISL_1477172, EPI_ISL_1477173, EPI_ISL_1477174, EPI_ISL_1477176, EPI_ISL_1477180, EPI_ISL_1477189, EPI_ISL_1477206, EPI_ISL_1477207, EPI_ISL_1477208, EPI_ISL_1477218, EPI_ISL_1477220, EPI_ISL_1477224, EPI_ISL_1477226, EPI_ISL_1477227, EPI_ISL_1477234, EPI_ISL_1477236, EPI_ISL_1477239, EPI_ISL_1477249, EPI_ISL_1477255, EPI_ISL_1477257, EPI_ISL_1477264, EPI_ISL_1477272, EPI_ISL_1477276, EPI_ISL_1477277, EPI_ISL_1477280, EPI_ISL_1477284, EPI_ISL_1477290, EPI_ISL_1477294, EPI_ISL_1477299, EPI_ISL_1477306, EPI_ISL_1477312, EPI_ISL_1477325, EPI_ISL_1477326, EPI_ISL_1477332, EPI_ISL_1477334, EPI_ISL_1477339, EPI_ISL_1477340, EPI_ISL_1477341, EPI_ISL_1477348, EPI_ISL_1477354, EPI_ISL_1477355, EPI_ISL_1477375, EPI_ISL_1477376, EPI_ISL_1477381, EPI_ISL_1477389, EPI_ISL_1477394, EPI_ISL_1477396, EPI_ISL_1477397, EPI_ISL_1477399, EPI_ISL_1477401, EPI_ISL_1477412, EPI_ISL_1477417, EPI_ISL_1477430 | | | |
| see above | Humboldt County Public Health Laboratory | Chan-Zuckerberg Biohub | CZB Cliahub Consortium |
| EPI_ISL_1192364 | Huntington Memorial | Los Angeles County PHL | P. Hemarajata et al. |

| | | | |
|---|---|---|---|
| | Hospital | | |
| EPI_ISL_2253166, EPI_ISL_2253168 | Infectious Diseases, Chan-Zuckerberg Biohub Covidtracker | Infectious Diseases, Chan-Zuckerberg Biohub Covidtracker | CZB Cliahub Consortium |
| EPI_ISL_980857, EPI_ISL_980858, EPI_ISL_980885, EPI_ISL_980997, EPI_ISL_980998, EPI_ISL_980999, EPI_ISL_981000, EPI_ISL_981001, EPI_ISL_981002, EPI_ISL_1060040, EPI_ISL_1060041, EPI_ISL_1060042, EPI_ISL_1060043, EPI_ISL_1060044, EPI_ISL_1060045, EPI_ISL_1060046, EPI_ISL_1060047, EPI_ISL_1060048, EPI_ISL_1060049, EPI_ISL_1060050, EPI_ISL_1060051, EPI_ISL_1060052, EPI_ISL_1060053, EPI_ISL_1060054, EPI_ISL_1060055, EPI_ISL_1060056, EPI_ISL_1060057, EPI_ISL_1060058, EPI_ISL_1060059, EPI_ISL_1060060, EPI_ISL_1060061, EPI_ISL_1060062, EPI_ISL_1060063, EPI_ISL_1060064, EPI_ISL_1060065, EPI_ISL_1060066, EPI_ISL_1060067, EPI_ISL_1060068, EPI_ISL_1060069, EPI_ISL_1060070, EPI_ISL_1060071, EPI_ISL_1060072, EPI_ISL_1060073, EPI_ISL_1060074, EPI_ISL_1060075, EPI_ISL_1060076, EPI_ISL_1060077, EPI_ISL_1060078, EPI_ISL_1060079, EPI_ISL_1060080, EPI_ISL_1060081, EPI_ISL_1060082, EPI_ISL_1060083, EPI_ISL_1060084, EPI_ISL_1060085, EPI_ISL_1060086, EPI_ISL_1060087, EPI_ISL_1060088, EPI_ISL_1060089, EPI_ISL_1060090, EPI_ISL_1060091, EPI_ISL_1060092, EPI_ISL_1060093, EPI_ISL_1060094, EPI_ISL_1060095, EPI_ISL_1060096, EPI_ISL_1060097, EPI_ISL_1060098, EPI_ISL_1060099, EPI_ISL_1060100, EPI_ISL_1060101, EPI_ISL_1060102, EPI_ISL_1060103, EPI_ISL_1060104, EPI_ISL_1060105, EPI_ISL_1060106, EPI_ISL_1060107, EPI_ISL_1060108, EPI_ISL_1060109, EPI_ISL_1060110, EPI_ISL_1060111, EPI_ISL_1060112, EPI_ISL_1060113, EPI_ISL_1060114, EPI_ISL_1060115, EPI_ISL_1060116, EPI_ISL_1060117, EPI_ISL_1060118, EPI_ISL_1060119, EPI_ISL_1060120, EPI_ISL_1060121, EPI_ISL_1060122, EPI_ISL_1060123, EPI_ISL_1060124, EPI_ISL_1060125, EPI_ISL_1060126, EPI_ISL_1060127, EPI_ISL_1060128, EPI_ISL_1060129, EPI_ISL_1060130, EPI_ISL_1060131, EPI_ISL_1060132, EPI_ISL_1060133, EPI_ISL_1060134, EPI_ISL_1060135, EPI_ISL_1060136, EPI_ISL_1060137, EPI_ISL_1060138, EPI_ISL_1060139, EPI_ISL_1060140, EPI_ISL_1060141, EPI_ISL_1060142, EPI_ISL_1060143, EPI_ISL_1060144, EPI_ISL_1060145, EPI_ISL_1060146, EPI_ISL_1060147, EPI_ISL_1060148, EPI_ISL_1060149, EPI_ISL_1060150, EPI_ISL_1060151, EPI_ISL_1060152, EPI_ISL_1060153, EPI_ISL_1060154, EPI_ISL_1060155, EPI_ISL_1060156, EPI_ISL_1060157, EPI_ISL_1060158, EPI_ISL_1060159, EPI_ISL_1060160, EPI_ISL_1060161, EPI_ISL_1060162, EPI_ISL_1060163, EPI_ISL_1060164, EPI_ISL_1060165, EPI_ISL_1060166, EPI_ISL_1060167, EPI_ISL_1060168, EPI_ISL_1060169, EPI_ISL_1060170, EPI_ISL_1060171, EPI_ISL_1060172, EPI_ISL_1060173, EPI_ISL_1060174, EPI_ISL_1060175, EPI_ISL_1060176, EPI_ISL_1060177, EPI_ISL_1060178, EPI_ISL_1060179, EPI_ISL_1060180, EPI_ISL_1060181, EPI_ISL_1060182, EPI_ISL_1060183, EPI_ISL_1060184, EPI_ISL_1060185, EPI_ISL_1060186, EPI_ISL_1060187, EPI_ISL_1060188, EPI_ISL_1060189, EPI_ISL_1060190, EPI_ISL_1060191, EPI_ISL_1081284, EPI_ISL_1081285, EPI_ISL_1081286, EPI_ISL_1081287, EPI_ISL_1081288, EPI_ISL_1081289, EPI_ISL_1081290, EPI_ISL_1712439, EPI_ISL_1712440, EPI_ISL_1712441 | | | |
| see above | Innovative Genomics Institute, UC Berkeley | Innovative Genomics Institute, UC Berkeley | Alison Ciling; Haridha Shivram; Holly Gildea; John Boyle; Liana Lareau; Netravathi Krishnappa; Phil Frankino; Stacia Wyman |
| EPI_ISL_1664230, EPI_ISL_1664231, EPI_ISL_1664233, EPI_ISL_1664234, EPI_ISL_1664235, EPI_ISL_1664236, EPI_ISL_1664237, EPI_ISL_1664238, EPI_ISL_1664239, EPI_ISL_1664240, EPI_ISL_1664241, EPI_ISL_1664242, EPI_ISL_1664243, EPI_ISL_1664244, EPI_ISL_1664245, EPI_ISL_1664246, EPI_ISL_1664247, EPI_ISL_1664248, EPI_ISL_1664249 | | | |
| see above | Kaiser San Jose | Santa Clara County public Health Laboratory | Santa Clara County Public Health Department |
| EPI_ISL_1508971, EPI_ISL_1508976, EPI_ISL_1508977 | Kaiser Santa Clara | Santa Clara County Public Health Laboratory | Santa Clara County Public Health Department |
| EPI_ISL_1365658, EPI_ISL_1365659, EPI_ISL_1365660 | LESP Baja California | Instituto de Diagnostico y Referencia Epidemiologicos (INDRE) | Abril Rodriguez-Maldonado; Ariadna Medina-Benitez; Claudia Wong-Arambula; Ernesto Ramirez-Gonzalez.; Gisela Barrera-Badillo; Irma Lopez-Martinez; Joaquin Quiroz-Mercado; Lucia Hernandez-Rivas; Natividad Cruz-Ortiz; Sergio Rangel-Guerrero; Tatiana Nunez-Garcia; Vanessa Rivero-Arredondo |
| EPI_ISL_1337370, EPI_ISL_1337371, EPI_ISL_1337373, EPI_ISL_1337374 | LESP Baja California Sur | Instituto de Diagnostico y Referencia Epidemiologicos (INDRE) | Abril Rodriguez-Maldonado; Ariadna Medina-Benitez; Claudia Wong-Arambula; Ernesto Ramirez-Gonzalez.; Gisela Barrera-Badillo; Irma Lopez-Martinez; Joaquin Quiroz-Mercado; Lucia Hernandez-Rivas; Natividad Cruz-Ortiz; Sergio Rangel-Guerrero; Tatiana Nunez-Garcia; Vanessa Rivero-Arredondo |
| EPI_ISL_1288179, EPI_ISL_1288180, EPI_ISL_1288181, EPI_ISL_1288182, EPI_ISL_1288308, EPI_ISL_1288309, EPI_ISL_1288310, EPI_ISL_1288312, EPI_ISL_1288313, EPI_ISL_1288314, EPI_ISL_1288316, EPI_ISL_1288317, EPI_ISL_1288318, EPI_ISL_1288423, EPI_ISL_1288424, EPI_ISL_1288425, EPI_ISL_1288426, EPI_ISL_1288427, EPI_ISL_1288428, EPI_ISL_1288429, EPI_ISL_1288430, EPI_ISL_1288431, EPI_ISL_1288432, EPI_ISL_1288433, EPI_ISL_1288434, EPI_ISL_1288435, EPI_ISL_1288436, EPI_ISL_1288437, EPI_ISL_1288438, EPI_ISL_1288439, EPI_ISL_1288440, EPI_ISL_1288491, EPI_ISL_1288492, EPI_ISL_1288493 | | | |
| see above | Laboratorio Central de Epidemiología (LCE) | Instituto de Biotecnología de la UNAM | Alejandro Sanchez-Flores; Alfredo Herrera-Estrella; Alicia Ocaña-Mondragón; Angel Gustavo Salas-Lais; Bernardo Martínez-Miguel; Blanca Taboada; Brenda Irasema Maldonado-Meza; Carla Ivón Herrera-Najera; Carlos F. Arias; Celia Boukadida; Clara Esperanza Santacruz-Tinoco; Concepción Grajales-Muñiz; Consorcio Mexicano de Vigilancia Genómica (CoViGen-Mex). Authors (in alphabetical order): Julio Elias Alvarado-Yaah; Fernando Fontove-Herrera; Francisco Pulido; Gloria Elena Espinoza-Ayala; Gloria María Molina-Salinas; Gloria Vazquez; Hector Esteban Paz-Juárez; Hector Montoya-Fuentes; Helen Haydee Fernanda Ramírez-Plascencia; Jorge Ivan Salinal-Nevarez; José Antonio Enciso-Moreno; José Esteban Muñoz-Medina; José de Jesús Nuñez-Contreras; Juan Bautista Chale-Dzul; Luis Alberto Ochoa-Carrera; Margarita Matías-Florentino; María Guadalupe Santiago-Mauricio; María Guadalupe de Jesús Mireles-Rivera; Nelly Sélem-Mojica; Pavel Isa; Ricardo Grande; Santiago Ávila-Ríos; Víctor Hugo Borja-Aburto |
| EPI_ISL_1302220 | Laboratorio Central de Epidemiología IMSS | Instituto de Biotecnología de la UNAM | Alejandra Hernández-Terán; Alejandro Sanchez-Flores; Alma Rincón-Rubio; Andrea Santos Coy-Arechavaleta; Authors from IBT; Blanca Taboada; Celia Boukadida; Clara Esperanza Santacruz-Tinoco; Edgar Mendieta-Condado; Eduardo Becerril-Vargas; Fidencio Mejía-Nepomuceno; Francisco Pulido; Gisela Barrera-Badillo; Gloria Vazquez; Hector Esteban Paz-Juárez; IMSS; InDRE and INER (in alphabetical order): Carlos F. Arias; Irma Lopez-Martinez; Jerome Jean Verleyen; Joel Armando Vázquez-Pérez; Jorge Salas-Hernández; José Arturo Martínez-Orozco; José Ernesto Ramírez-González; José Esteban Muñoz-Medina; Larissa Fernandes-Matano; Lucia Hernandez-Rivas; Luis Alberto Ochoa-Carrera; Margarita Matías-Florentino; Mario Mújica-Sánchez; Natividad Cruz-Ortiz; Pavel Isa; Ricardo Grande; Santiago Ávila-Ríos; Tatiana Nunez-Garcia; Teresita Rojas-Mendoza |
| EPI_ISL_1193289, EPI_ISL_1193290, EPI_ISL_1220991, EPI_ISL_1221355, EPI_ISL_1221386, EPI_ISL_1221388, EPI_ISL_1221420, EPI_ISL_1221660, EPI_ISL_1221810, EPI_ISL_1221811, EPI_ISL_1221830, EPI_ISL_1221831, EPI_ISL_1221832, EPI_ISL_1221833, EPI_ISL_1221863, EPI_ISL_1221864, EPI_ISL_1221865, EPI_ISL_1222032, EPI_ISL_1222033, EPI_ISL_1291297, EPI_ISL_1291298, EPI_ISL_1298296, EPI_ISL_1320038, EPI_ISL_1320039, EPI_ISL_1320040, EPI_ISL_1320041, EPI_ISL_1320042, EPI_ISL_1320043, EPI_ISL_1320044, EPI_ISL_1320045, EPI_ISL_1320046, EPI_ISL_1320047, EPI_ISL_1320048, EPI_ISL_1320049, EPI_ISL_1320050, EPI_ISL_1320051, EPI_ISL_1320052, EPI_ISL_1320053, EPI_ISL_1320054, EPI_ISL_1320055, EPI_ISL_1320056, EPI_ISL_1320057, EPI_ISL_1320058, EPI_ISL_1320059, EPI_ISL_1320060, EPI_ISL_1320061, EPI_ISL_1320062, EPI_ISL_1320063, EPI_ISL_1320064, EPI_ISL_1320065, EPI_ISL_1320066, EPI_ISL_1320067, EPI_ISL_1320068, EPI_ISL_1320069, EPI_ISL_1320070, EPI_ISL_1320071, EPI_ISL_1320072, EPI_ISL_1320073, EPI_ISL_1320074, EPI_ISL_1320075, EPI_ISL_1320076, EPI_ISL_1320077, EPI_ISL_1320078, EPI_ISL_1320079, EPI_ISL_1320080, EPI_ISL_1320081, EPI_ISL_1320082, EPI_ISL_1320083, EPI_ISL_1320084, EPI_ISL_1320085, EPI_ISL_1320086, EPI_ISL_1320087, EPI_ISL_1320088, EPI_ISL_1320089, EPI_ISL_1320090, EPI_ISL_1320091, EPI_ISL_1320092, EPI_ISL_1320093, EPI_ISL_1320094, EPI_ISL_1320095, EPI_ISL_1320096, EPI_ISL_1320097, EPI_ISL_1320098, EPI_ISL_1320099, EPI_ISL_1320100, EPI_ISL_1321010, EPI_ISL_1321011, EPI_ISL_1321012, EPI_ISL_1321013, EPI_ISL_1321014, EPI_ISL_1321015, EPI_ISL_1321016, EPI_ISL_1321017, EPI_ISL_1321018, EPI_ISL_1321019, EPI_ISL_1321020, EPI_ISL_1321021, EPI_ISL_1321022, EPI_ISL_1321023, EPI_ISL_1321024, EPI_ISL_1321025, EPI_ISL_1321026, EPI_ISL_1321027, EPI_ISL_1321028, EPI_ISL_1321029, EPI_ISL_1321030, EPI_ISL_1321031, EPI_ISL_1321032, EPI_ISL_1321033, EPI_ISL_1321034, EPI_ISL_1321035, EPI_ISL_1321036, EPI_ISL_1321037, EPI_ISL_1321038, EPI_ISL_1321039, EPI_ISL_1321040, EPI_ISL_1321041, EPI_ISL_1321042, EPI_ISL_1321043, EPI_ISL_1321044, EPI_ISL_1321045, EPI_ISL_1321046, EPI_ISL_1321047, EPI_ISL_1321048, EPI_ISL_1321049, EPI_ISL_1321050, EPI_ISL_1321051, EPI_ISL_1321052, EPI_ISL_1321053, EPI_ISL_1321055, EPI_ISL_1321056, EPI_ISL_1321057, EPI_ISL_1321058, EPI_ISL_1321059, EPI_ISL_1321060, EPI_ISL_1321061, EPI_ISL_1321062, EPI_ISL_1321063, EPI_ISL_1321064, EPI_ISL_1321065, EPI_ISL_1321066, EPI_ISL_1321067, EPI_ISL_1321068, EPI_ISL_1321069, EPI_ISL_1321070, EPI_ISL_1321071, EPI_ISL_1321072, EPI_ISL_1321073, EPI_ISL_1321074, EPI_ISL_1421979, EPI_ISL_1422164, EPI_ISL_1422165, EPI_ISL_1422166, EPI_ISL_1422167, EPI_ISL_1422168 | | | |
| see above | Laboratory Corporation of America | Centers for Disease Control and Prevention Division of Viral Diseases, Pathogen Discovery | Amanda Douglas; Amanda Suchanek; Andrea Throop; Ayla Burns; Ben L. Rambo-Martin; Bobbi Croy; Brian Krueger; Brian Norvell; Christos Petropoulos; Clinton R. Paden; Craig Lukasik; Dakota Howard; Debbie Boles; Dhwani Batra; Duncan MacCannell; Eyad Almasri; Eyad Almasri Debbie Boles; Goran Stevovic; Howard Engler; Hrushikesh Deshmukh; Jake Humphrey; Jana Schroth; Joe Voshell; John Pruitt; Jonathan Meltzer; Jonathan Williams; Kimberly Wagner; Lax Iyer; Lyndon Tilson; Manoj Jain; Marcia Eisenberg; Mary Ann Cristobal; Mary Williamson; Michael Levandoski; Mike Sapeta; Mindy Nye; Minoo Agarwal; Mohan Kolli; Nuthawin Charoensri; Oren Cohen; Peter W. Cook; Prashant Gupta; Qian Zeng; Rama Ghatti; Scott Parker; Scott Ryan; Stanley Letovsky; Steven Ragan; Suresh Babu Selvaraju; Susan Countryman; Susan Hicks; Suxiang Tong; Suzanne Dale; Thomas Urban; Tim Kuphal; Tricia Zwiefelhofer; Vincent Drouillon |
| EPI_ISL_1036992, EPI_ISL_1037031, EPI_ISL_1037106, EPI_ISL_1037109, EPI_ISL_1037112, EPI_ISL_1037115, EPI_ISL_1037126, EPI_ISL_1037144, EPI_ISL_1037147, EPI_ISL_1037180, EPI_ISL_1037222, EPI_ISL_1037249, EPI_ISL_1037316, EPI_ISL_1037373, EPI_ISL_1037404, EPI_ISL_1037417, EPI_ISL_1037423, EPI_ISL_1037448, EPI_ISL_1037473, EPI_ISL_1037559, EPI_ISL_1037576, EPI_ISL_1037578, EPI_ISL_1037612, EPI_ISL_1037639, EPI_ISL_1037651, EPI_ISL_1037660, EPI_ISL_1037674, EPI_ISL_1037682, EPI_ISL_1037709, EPI_ISL_1037740, EPI_ISL_1037747, EPI_ISL_1037757, EPI_ISL_1037762, EPI_ISL_1037763, EPI_ISL_1037781, EPI_ISL_1037782, EPI_ISL_1037783, EPI_ISL_1037806, EPI_ISL_1037874, EPI_ISL_1037886, EPI_ISL_1037888, EPI_ISL_1037918, EPI_ISL_1037938, EPI_ISL_1037993, EPI_ISL_1038066, EPI_ISL_1038085, EPI_ISL_1038148, EPI_ISL_1038150, EPI_ISL_1038181, EPI_ISL_1038243, EPI_ISL_1038244, EPI_ISL_1038245, EPI_ISL_1038275, EPI_ISL_1038283, EPI_ISL_1038489, EPI_ISL_1038673, EPI_ISL_1038674, EPI_ISL_1038675, EPI_ISL_1038676, EPI_ISL_1038677, EPI_ISL_1038678, EPI_ISL_1038679, EPI_ISL_1038680, EPI_ISL_1038681, EPI_ISL_1038682, EPI_ISL_1038683, EPI_ISL_1038684, EPI_ISL_1038685, EPI_ISL_1038686, EPI_ISL_1038687, EPI_ISL_1038689, EPI_ISL_1038690, EPI_ISL_1038691, EPI_ISL_1038692, EPI_ISL_1038693, EPI_ISL_1038694, EPI_ISL_1038695, EPI_ISL_1038769, EPI_ISL_1160013, EPI_ISL_1160036, EPI_ISL_1160061, EPI_ISL_1160078, EPI_ISL_1160082, EPI_ISL_1160113, EPI_ISL_1160210, EPI_ISL_1160222, EPI_ISL_1160278, EPI_ISL_1160329, EPI_ISL_1160354, EPI_ISL_1160367, EPI_ISL_1160373, EPI_ISL_1160390, EPI_ISL_1160411, EPI_ISL_1160445, EPI_ISL_1160453, EPI_ISL_1160497, EPI_ISL_1160508, EPI_ISL_1160511, EPI_ISL_1160527, EPI_ISL_1160560, EPI_ISL_1160583, EPI_ISL_1160660, EPI_ISL_1160799, EPI_ISL_1160807, EPI_ISL_1160886, EPI_ISL_1160923, EPI_ISL_1160924, EPI_ISL_1161290, EPI_ISL_1161291, EPI_ISL_1161335, EPI_ISL_1161343, EPI_ISL_1161344, EPI_ISL_1161345, EPI_ISL_1161346, EPI_ISL_1161347, EPI_ISL_1161351, EPI_ISL_1161352, EPI_ISL_1161353, EPI_ISL_1161358, EPI_ISL_1161359, EPI_ISL_1161360, EPI_ISL_1161419, EPI_ISL_1161420, EPI_ISL_1161422, EPI_ISL_1161426, EPI_ISL_1161598, EPI_ISL_1161599, EPI_ISL_1161600, EPI_ISL_1161601, EPI_ISL_1161602, EPI_ISL_1161603, EPI_ISL_1161691, EPI_ISL_1161709, EPI_ISL_1161710, EPI_ISL_1161711, EPI_ISL_1161735, EPI_ISL_1161761, EPI_ISL_1161923, EPI_ISL_1161943, EPI_ISL_1161950, EPI_ISL_1161960, EPI_ISL_1161970, EPI_ISL_1161997, EPI_ISL_1161998, EPI_ISL_1162011, EPI_ISL_1162020, EPI_ISL_1162050, EPI_ISL_1162096, EPI_ISL_1162130, EPI_ISL_1162136, EPI_ISL_1162293, EPI_ISL_1162312, EPI_ISL_1162342, EPI_ISL_1162494, EPI_ISL_1162497, EPI_ISL_1162539, EPI_ISL_1162542, EPI_ISL_1162553, EPI_ISL_1162561, EPI_ISL_1162562, EPI_ISL_1162604, EPI_ISL_1162624, EPI_ISL_1162640, EPI_ISL_1162641, EPI_ISL_1162687, EPI_ISL_1162754, EPI_ISL_1162783, EPI_ISL_1162839, EPI_ISL_1162937, EPI_ISL_1162938, EPI_ISL_1162939, EPI_ISL_1162940, EPI_ISL_1162976, EPI_ISL_1162984, EPI_ISL_1163192, EPI_ISL_1163423, EPI_ISL_1163424, EPI_ISL_1163425, EPI_ISL_1163426, EPI_ISL_1163427, EPI_ISL_1163428, EPI_ISL_1163429, EPI_ISL_1163430, EPI_ISL_1163431, EPI_ISL_1163432, EPI_ISL_1163433, EPI_ISL_1163434, EPI_ISL_1163435, EPI_ISL_1163436, EPI_ISL_1163437, EPI_ISL_1163438 | | | |
| see above | Laboratory Corporation of America | Respiratory Viruses Branch, Division of Viral Diseases, Centers for Disease Control and Prevention | Amanda Douglas; Amanda Suchanek; Andrea Throop; Ayla Burns; Ben L. Rambo-Martin; Bobbi Croy; Brian Krueger; Brian Norvell; Christos Petropoulos; Clinton R. Paden; Craig Lukasik; Dakota Howard; Dhwani Batra; Duncan MacCannell; Eyad Almasri Debbie Boles; Goran Stevovic; Howard Engler; Hrushikesh Deshmukh; Jake Humphrey; Jana Schroth; Joe Voshell; John Pruitt; Jonathan Meltzer; Jonathan Williams; Kimberly Wagner; Lax Iyer; Lyndon Tilson; Manoj Jain; Marcia Eisenberg; Mary Ann Cristobal; Mary Williamson; Michael Levandoski; Mike Sapeta; Mindy Nye; Minoo Agarwal; Mohan Kolli; Nuthawin Charoensri; Oren Cohen; Peter W. Cook; Prashant Gupta; Qian Zeng; Rama Ghatti; Scott Parker; Scott Ryan; Stanley Letovsky; Steven Ragan; Suresh Babu Selvaraju; Susan Countryman; Susan Hicks; Suxiang Tong; Suzanne Dale; Thomas Urban; Tim Kuphal; Tricia Zwiefelhofer; Vincent Drouillon |
| EPI_ISL_1005557 | Los Angeles Community Hospital | Los Angeles County PHL | P. Hemarajata et al. |
| EPI_ISL_1005485, EPI_ISL_1005508, EPI_ISL_1097724, EPI_ISL_1097725, EPI_ISL_1097726, EPI_ISL_1097727, EPI_ISL_1097728, EPI_ISL_1097729, EPI_ISL_1097730, EPI_ISL_1097731, EPI_ISL_1097732, EPI_ISL_1097733, EPI_ISL_1097734, EPI_ISL_1097735, EPI_ISL_1097736, EPI_ISL_1097737, EPI_ISL_1097740, EPI_ISL_1097741, EPI_ISL_1097743, EPI_ISL_1098632, EPI_ISL_1098633, EPI_ISL_1098639, EPI_ISL_1098640, EPI_ISL_1191109 | | | |
| see above | Los Angeles County PHL | Los Angeles County PHL | P. Hemarajata et al. |
| EPI_ISL_1303416, EPI_ISL_1303417, EPI_ISL_1303418, EPI_ISL_1303419 | MD Laboratories | Los Angeles County PHL | P. Hemarajata et al. |
| EPI_ISL_1197043, EPI_ISL_1197045, EPI_ISL_1197048, EPI_ISL_1197050, EPI_ISL_1197052, EPI_ISL_1197055, EPI_ISL_1197056, EPI_ISL_1197057, EPI_ISL_1303439, EPI_ISL_1303447 | | | |
| see above | Martin Luther King Jr. Community Hospital | Los Angeles County PHL | P. Hemarajata et al. |
| EPI_ISL_968078, EPI_ISL_1263228, EPI_ISL_1263231, EPI_ISL_1263232, EPI_ISL_1263233, EPI_ISL_1263234, EPI_ISL_1263235, EPI_ISL_1664617, EPI_ISL_1700695, EPI_ISL_1921880, EPI_ISL_1921881, EPI_ISL_1921882, EPI_ISL_1921883, EPI_ISL_1921884, EPI_ISL_1921885, EPI_ISL_1921886, EPI_ISL_1921887, EPI_ISL_1921888, EPI_ISL_1921889 | | | |
| see above | Monterey County Public Health Laboratory | Monterey County Public Health Laboratory | Monterey County Public Health Laboratory |
| EPI_ISL_1541555 | OHSU Lab Services Molecular Microbiology Lab | Oregon SARS-CoV-2 Genome Sequencing Center | Alec J. Hirsch; Andrew C. Adey; Benjamin N. Bimber; Brendan L. O'Connell; Brian J. O'Roak; Daniel N. Streblow; Donna Hansel; Guang Fan; Kayla Carter; Ruth V. Nichols; Sally Grindstaff; Sonia Acharya; William B. Messer; Xuan Qin |
| EPI_ISL_1005556 | Olive View Medical Center | Los Angeles County PHL | P. Hemarajata et al. |
| EPI_ISL_1234791, EPI_ISL_1234796, EPI_ISL_1234797, EPI_ISL_1234828, EPI_ISL_1234838, EPI_ISL_1234842, EPI_ISL_1234847, EPI_ISL_1234855, EPI_ISL_1234864, EPI_ISL_1234887, EPI_ISL_1234909, EPI_ISL_1234918, EPI_ISL_1234926, EPI_ISL_1234929, EPI_ISL_1234937, EPI_ISL_1234940, EPI_ISL_1234943, EPI_ISL_1234945, EPI_ISL_1234954, EPI_ISL_1234958, EPI_ISL_1234965, EPI_ISL_1234970, EPI_ISL_1234977, EPI_ISL_1234997, EPI_ISL_1235007, EPI_ISL_1235008, EPI_ISL_1235009, EPI_ISL_1235016, EPI_ISL_1235044, EPI_ISL_1235057, EPI_ISL_1235058, EPI_ISL_1477443, EPI_ISL_1477611, EPI_ISL_1477663, EPI_ISL_1701438, EPI_ISL_1701565, EPI_ISL_1701571, EPI_ISL_1701589, EPI_ISL_1701646 | | | |
| see above | Orange County Public Health Lab | Chan-Zuckerberg Biohub | CZB Cliahub Consortium |
| EPI_ISL_1912975 | Poplar Healthcare | Colleen B. Jonsson | Colleen Jonsson; Jacob Nichols; Joythi Parvathareddy; Mariah Taylor; Surekha Surendranathan; Walter Reichard |
| EPI_ISL_1159233, EPI_ISL_1159244, EPI_ISL_1159250, EPI_ISL_1159262, EPI_ISL_1159268, EPI_ISL_1159269, EPI_ISL_1159274, EPI_ISL_1159275, EPI_ISL_1159287, EPI_ISL_1159288, EPI_ISL_1159291, EPI_ISL_1159293, EPI_ISL_1159294, EPI_ISL_1159295, EPI_ISL_1159308, EPI_ISL_1159311, EPI_ISL_1159319, EPI_ISL_1159321, EPI_ISL_1159324, EPI_ISL_1159325, EPI_ISL_1159332, EPI_ISL_1193374, EPI_ISL_1193375, EPI_ISL_1193379, EPI_ISL_1193380, EPI_ISL_1193381, EPI_ISL_1193382, EPI_ISL_1193383, EPI_ISL_1193385, EPI_ISL_1193386, EPI_ISL_1193388, EPI_ISL_1193390, EPI_ISL_1193392, EPI_ISL_1193394, EPI_ISL_1193395, EPI_ISL_1193405, EPI_ISL_1193411, EPI_ISL_1193413, EPI_ISL_1193414, EPI_ISL_1193415, EPI_ISL_1193416, EPI_ISL_1193417, EPI_ISL_1193419, EPI_ISL_1193421, EPI_ISL_1193422, EPI_ISL_1193423, EPI_ISL_1193426, EPI_ISL_1193430, EPI_ISL_1193431, EPI_ISL_1193433, EPI_ISL_1193434, EPI_ISL_1193435, EPI_ISL_1193436, EPI_ISL_1193437, EPI_ISL_1193438, EPI_ISL_1193439, EPI_ISL_1193442, EPI_ISL_1193443, EPI_ISL_1193444, EPI_ISL_1193460, EPI_ISL_1193464, EPI_ISL_1193466, EPI_ISL_1193471, EPI_ISL_1193472, EPI_ISL_1193479, EPI_ISL_1193481, EPI_ISL_1193482, EPI_ISL_1193483, EPI_ISL_1193484, EPI_ISL_1193486, EPI_ISL_1193487, EPI_ISL_1193490, EPI_ISL_1193491, EPI_ISL_1193492, EPI_ISL_1193493, EPI_ISL_1193494, EPI_ISL_1193495, EPI_ISL_1193497, EPI_ISL_1193498, EPI_ISL_1193501, EPI_ISL_1193503, EPI_ISL_1193504, EPI_ISL_1193505, EPI_ISL_1193506, EPI_ISL_1193511, EPI_ISL_1193512, EPI_ISL_1193514, EPI_ISL_1193515, EPI_ISL_1193518, EPI_ISL_1193520, EPI_ISL_1193522, EPI_ISL_1193523, EPI_ISL_1193524, EPI_ISL_1193525, EPI_ISL_1193526, EPI_ISL_1193527, EPI_ISL_1193530, EPI_ISL_1193532, EPI_ISL_1193533, EPI_ISL_1193534, EPI_ISL_1193535, EPI_ISL_1193536, EPI_ISL_1193538, EPI_ISL_1193546, EPI_ISL_1193548, EPI_ISL_1193552, EPI_ISL_1193557, EPI_ISL_1193559, EPI_ISL_1193562, EPI_ISL_1193570, EPI_ISL_1193571, EPI_ISL_1193576, EPI_ISL_1193585, EPI_ISL_1193588, EPI_ISL_1193601, EPI_ISL_1193607, EPI_ISL_1193628, EPI_ISL_1193630, EPI_ISL_1193647, EPI_ISL_1193653, EPI_ISL_1193780, EPI_ISL_1193784, EPI_ISL_1193799, EPI_ISL_1193803, EPI_ISL_1193810, EPI_ISL_1193831, EPI_ISL_1193862, EPI_ISL_1193863, EPI_ISL_1193865, EPI_ISL_1193877, EPI_ISL_1193892, EPI_ISL_1193899, EPI_ISL_1193913, EPI_ISL_1193914, EPI_ISL_1193916, EPI_ISL_1193917, EPI_ISL_1193918, EPI_ISL_1193919, EPI_ISL_1193920, EPI_ISL_1193921, EPI_ISL_1193923, EPI_ISL_1193924, EPI_ISL_1193925, EPI_ISL_1193926, EPI_ISL_1193927, EPI_ISL_1193928, EPI_ISL_1193929, EPI_ISL_1193931, EPI_ISL_1193932, EPI_ISL_1193933, EPI_ISL_1193934, EPI_ISL_1193935, EPI_ISL_1193936, EPI_ISL_1193939, EPI_ISL_1193942, EPI_ISL_1193943, EPI_ISL_1193944, EPI_ISL_1193945, EPI_ISL_1193946, EPI_ISL_1193947, EPI_ISL_1193950, EPI_ISL_1193951, EPI_ISL_1193952, EPI_ISL_1193953, EPI_ISL_1193954, EPI_ISL_1193956, EPI_ISL_1193957, EPI_ISL_1193958, EPI_ISL_1193959, EPI_ISL_1193960, EPI_ISL_1193961, EPI_ISL_1193962, EPI_ISL_1193963, EPI_ISL_1193964, EPI_ISL_1193966, EPI_ISL_1193967, EPI_ISL_1193968, EPI_ISL_1193969, EPI_ISL_1193970, EPI_ISL_1193971, EPI_ISL_1193972, EPI_ISL_1193976, EPI_ISL_1193977, EPI_ISL_1193978, EPI_ISL_1193979, EPI_ISL_1193981, EPI_ISL_1193982, EPI_ISL_1193984, EPI_ISL_1193986, EPI_ISL_1193989, EPI_ISL_1193991, EPI_ISL_1193992, EPI_ISL_1193994, EPI_ISL_1193995, EPI_ISL_1193996, EPI_ISL_1193998, EPI_ISL_1194000, EPI_ISL_1194002, EPI_ISL_1194003, EPI_ISL_1194004, EPI_ISL_1194005, EPI_ISL_1194006, EPI_ISL_1194007, EPI_ISL_1194008, EPI_ISL_1194009, EPI_ISL_1194010, EPI_ISL_1194012, EPI_ISL_1194013, EPI_ISL_1194014, EPI_ISL_1194016, EPI_ISL_1194017, EPI_ISL_1194018, EPI_ISL_1194020, EPI_ISL_1194021, EPI_ISL_1194022, EPI_ISL_1194023, EPI_ISL_1194024, EPI_ISL_1194025, EPI_ISL_1194026, EPI_ISL_1194027, | | | |

| | | | |
|---|---|---|---|
| EPI_ISL_1194028, EPI_ISL_1194029, EPI_ISL_1194030, EPI_ISL_1194031, EPI_ISL_1194032, EPI_ISL_1194033, EPI_ISL_1194034, EPI_ISL_1194035, EPI_ISL_1194036, EPI_ISL_1194037, EPI_ISL_1194038, EPI_ISL_1194039, EPI_ISL_1194040, EPI_ISL_1194041, EPI_ISL_1194042, EPI_ISL_1194074, EPI_ISL_1194113, EPI_ISL_1194125, EPI_ISL_1194154, EPI_ISL_1194172, EPI_ISL_1194173, EPI_ISL_1194175, EPI_ISL_1194186, EPI_ISL_1194195, EPI_ISL_1194196, EPI_ISL_1194198, EPI_ISL_1194200, EPI_ISL_1194202, EPI_ISL_1194207, EPI_ISL_1194214, EPI_ISL_1194227, EPI_ISL_1194229, EPI_ISL_1194236, EPI_ISL_1194237, EPI_ISL_1194244, EPI_ISL_1194245, EPI_ISL_1194247, EPI_ISL_1194258, EPI_ISL_1194260, EPI_ISL_1194263, EPI_ISL_1194264, EPI_ISL_1194270, EPI_ISL_1194275, EPI_ISL_1194276, EPI_ISL_1194281, EPI_ISL_1194283, EPI_ISL_1194284, EPI_ISL_1194287, EPI_ISL_1194289, EPI_ISL_1194291, EPI_ISL_1194295, EPI_ISL_1194297, EPI_ISL_1194298, EPI_ISL_1194299, EPI_ISL_1194304, EPI_ISL_1194305, EPI_ISL_1194307, EPI_ISL_1194311, EPI_ISL_1194315, EPI_ISL_1194316, EPI_ISL_1194320, EPI_ISL_1194323, EPI_ISL_1194324, EPI_ISL_1194329, EPI_ISL_1194330, EPI_ISL_1194333, EPI_ISL_1194343, EPI_ISL_1194345, EPI_ISL_1194354, EPI_ISL_1194355, EPI_ISL_1194359, EPI_ISL_1194362, EPI_ISL_1194381, EPI_ISL_1194383, EPI_ISL_1194402, EPI_ISL_1194403, EPI_ISL_1194404, EPI_ISL_1194420, EPI_ISL_1194488, EPI_ISL_1220375, EPI_ISL_1220429, EPI_ISL_1220430, EPI_ISL_1220472, EPI_ISL_1220507, EPI_ISL_1220520, EPI_ISL_1220547, EPI_ISL_1220549, EPI_ISL_1220596, EPI_ISL_1220601, EPI_ISL_1220611, EPI_ISL_1220629, EPI_ISL_1220636, EPI_ISL_1220660, EPI_ISL_1220661, EPI_ISL_1220662, EPI_ISL_1220672, EPI_ISL_1220687, EPI_ISL_1220695, EPI_ISL_1220715, EPI_ISL_1220723, EPI_ISL_1220724, EPI_ISL_1220730, EPI_ISL_1220737, EPI_ISL_1220739, EPI_ISL_1220740, EPI_ISL_1220741, EPI_ISL_1220743, EPI_ISL_1220745, EPI_ISL_1220747, EPI_ISL_1220749, EPI_ISL_1220751, EPI_ISL_1220753, EPI_ISL_1220754, EPI_ISL_1220757, EPI_ISL_1220761, EPI_ISL_1220766, EPI_ISL_1220767, EPI_ISL_1220776, EPI_ISL_1220778, EPI_ISL_1220781, EPI_ISL_1220783, EPI_ISL_1220784, EPI_ISL_1220786, EPI_ISL_1220791, EPI_ISL_1220792, EPI_ISL_1220794, EPI_ISL_1220796, EPI_ISL_1220797, EPI_ISL_1220798, EPI_ISL_1220801, EPI_ISL_1220804, EPI_ISL_1220806, EPI_ISL_1220808, EPI_ISL_1220811, EPI_ISL_1220817, EPI_ISL_1220818, EPI_ISL_1220820, EPI_ISL_1220827, EPI_ISL_1220831, EPI_ISL_1220832, EPI_ISL_1220833, EPI_ISL_1220834, EPI_ISL_1220835, EPI_ISL_1220837, EPI_ISL_1220838, EPI_ISL_1220839, EPI_ISL_1220840, EPI_ISL_1220841, EPI_ISL_1220843, EPI_ISL_1220845, EPI_ISL_1220857, EPI_ISL_1220859, EPI_ISL_1220860, EPI_ISL_1220861, EPI_ISL_1252944, EPI_ISL_1252945, EPI_ISL_1252979, EPI_ISL_1253007, EPI_ISL_1253018, EPI_ISL_1253019, EPI_ISL_1253136, EPI_ISL_1253141, EPI_ISL_1253142, EPI_ISL_1253152, EPI_ISL_1253185, EPI_ISL_1253190, EPI_ISL_1253193, EPI_ISL_1253239, EPI_ISL_1253253, EPI_ISL_1253390, EPI_ISL_1253395, EPI_ISL_1253397, EPI_ISL_1253404, EPI_ISL_1253420, EPI_ISL_1253423, EPI_ISL_1253424, EPI_ISL_1253427, EPI_ISL_1253429, EPI_ISL_1253431, EPI_ISL_1253436, EPI_ISL_1253441, EPI_ISL_1253446, EPI_ISL_1253448, EPI_ISL_1253453, EPI_ISL_1253455, EPI_ISL_1253464, EPI_ISL_1253465, EPI_ISL_1253471, EPI_ISL_1253473, EPI_ISL_1253476, EPI_ISL_1253477, EPI_ISL_1253480, EPI_ISL_1253482, EPI_ISL_1253484, EPI_ISL_1253485, EPI_ISL_1253486, EPI_ISL_1253497, EPI_ISL_1253506, EPI_ISL_1253512, EPI_ISL_1253519, EPI_ISL_1253525, EPI_ISL_1253529, EPI_ISL_1253536, EPI_ISL_1254022, EPI_ISL_1254027, EPI_ISL_1254035, EPI_ISL_1254041, EPI_ISL_1254066, EPI_ISL_1254071, EPI_ISL_1254073, EPI_ISL_1254079, EPI_ISL_1254080, EPI_ISL_1254082, EPI_ISL_1254089, EPI_ISL_1254095, EPI_ISL_1254096, EPI_ISL_1254104, EPI_ISL_1254109, EPI_ISL_1254129, EPI_ISL_1254133, EPI_ISL_1254143, EPI_ISL_1254151, EPI_ISL_1254158, EPI_ISL_1254159, EPI_ISL_1254160, EPI_ISL_1254162, EPI_ISL_1254181, EPI_ISL_1254192, EPI_ISL_1254198, EPI_ISL_1254216, EPI_ISL_1254220, EPI_ISL_1254222, EPI_ISL_1254224, EPI_ISL_1254267, EPI_ISL_1254275, EPI_ISL_1254301, EPI_ISL_1254302, EPI_ISL_1254308, EPI_ISL_1291400, EPI_ISL_1291420, EPI_ISL_1291433, EPI_ISL_1291438, EPI_ISL_1291451, EPI_ISL_1291480, EPI_ISL_1291482, EPI_ISL_1291541, EPI_ISL_1298235, EPI_ISL_1335915, EPI_ISL_1335973, EPI_ISL_1335987, EPI_ISL_1335995, EPI_ISL_1336104 | | | |
| see above | Quest Diagnostics Incorporated | Centers for Disease Control and Prevention Division of Viral Diseases, Pathogen Discovery | A. Gerasimova; A. Perez; B. Anderson; Ben L. Rambo-Martin; Clinton R. Paden; Dakota Howard; Dhwani Batra; Duncan MacCannell; F. Lacbawan; I. A. Shlyakhter; K.E. Livingston; L.E. Bernstein; M. Hua; P. Tanpaiboon; Peter W. Cook; R. M. Kagan; R. Owen; R. V. Rolando; S. H. Rosenthal; Suxiang Tong; Y. Liu |
| EPI_ISL_1086378, EPI_ISL_1086380, EPI_ISL_1086382, EPI_ISL_1086383, EPI_ISL_1086384, EPI_ISL_1086402, EPI_ISL_1086549, EPI_ISL_1086557, EPI_ISL_1086558, EPI_ISL_1086566, EPI_ISL_1086568, EPI_ISL_1086571, EPI_ISL_1086576, EPI_ISL_1086579, EPI_ISL_1086585, EPI_ISL_1086587, EPI_ISL_1086589, EPI_ISL_1086598, EPI_ISL_1086605, EPI_ISL_1086607, EPI_ISL_1086609, EPI_ISL_1086615, EPI_ISL_1086620, EPI_ISL_1086621, EPI_ISL_1086622, EPI_ISL_1086628, EPI_ISL_1086632, EPI_ISL_1086633, EPI_ISL_1086637, EPI_ISL_1086639, EPI_ISL_1086644, EPI_ISL_1086648, EPI_ISL_1086649, EPI_ISL_1086650, EPI_ISL_1086657, EPI_ISL_1086658, EPI_ISL_1086665, EPI_ISL_1086666, EPI_ISL_1087439, EPI_ISL_1087517, EPI_ISL_1087525, EPI_ISL_1087541, EPI_ISL_1087552, EPI_ISL_1087628, EPI_ISL_1087633, EPI_ISL_1087634, EPI_ISL_1087636, EPI_ISL_1087651, EPI_ISL_1087655, EPI_ISL_1087656, EPI_ISL_1087659, EPI_ISL_1087661, EPI_ISL_1087663, EPI_ISL_1087667, EPI_ISL_1087669, EPI_ISL_1087675, EPI_ISL_1087679, EPI_ISL_1087684, EPI_ISL_1087693, EPI_ISL_1087727, EPI_ISL_1087755, EPI_ISL_1087760, EPI_ISL_1087768, EPI_ISL_1087782, EPI_ISL_1087786, EPI_ISL_1087788, EPI_ISL_1087789, EPI_ISL_1087793, EPI_ISL_1087794, EPI_ISL_1087799, EPI_ISL_1087800, EPI_ISL_1087813, EPI_ISL_1087814, EPI_ISL_1087817, EPI_ISL_1087819, EPI_ISL_1087824, EPI_ISL_1087825, EPI_ISL_1087826, EPI_ISL_1087827, EPI_ISL_1087828, EPI_ISL_1087829, EPI_ISL_1087831, EPI_ISL_1087834, EPI_ISL_1087835, EPI_ISL_1087837, EPI_ISL_1087838, EPI_ISL_1087840, EPI_ISL_1087842, EPI_ISL_1087845, EPI_ISL_1087847, EPI_ISL_1087850, EPI_ISL_1087852, EPI_ISL_1087853, EPI_ISL_1087854, EPI_ISL_1087855, EPI_ISL_1087856, EPI_ISL_1087857, EPI_ISL_1087858, EPI_ISL_1087859, EPI_ISL_1087860, EPI_ISL_1087895, EPI_ISL_1087896, EPI_ISL_1087897, EPI_ISL_1087898, EPI_ISL_1087900, EPI_ISL_1087903, EPI_ISL_1087905, EPI_ISL_1087906, EPI_ISL_1087909, EPI_ISL_1087912, EPI_ISL_1087917, EPI_ISL_1087918, EPI_ISL_1087919, EPI_ISL_1087921, EPI_ISL_1087925, EPI_ISL_1087930, EPI_ISL_1087932, EPI_ISL_1087933, EPI_ISL_1087934, EPI_ISL_1087935, EPI_ISL_1087943, EPI_ISL_1087945, EPI_ISL_1087948, EPI_ISL_1087949, EPI_ISL_1087950, EPI_ISL_1087951, EPI_ISL_1087952, EPI_ISL_1087954, EPI_ISL_1087957, EPI_ISL_1087959, EPI_ISL_1087976, EPI_ISL_1088079, EPI_ISL_1088080, EPI_ISL_1088083, EPI_ISL_1088087, EPI_ISL_1088094, EPI_ISL_1088107, EPI_ISL_1088111, EPI_ISL_1088115, EPI_ISL_1088117, EPI_ISL_1088123, EPI_ISL_1088124, EPI_ISL_1088140, EPI_ISL_1088142, EPI_ISL_1088143, EPI_ISL_1088152, EPI_ISL_1088170, EPI_ISL_1088188, EPI_ISL_1088192, EPI_ISL_1088196, EPI_ISL_1088210, EPI_ISL_1088211, EPI_ISL_1088216, EPI_ISL_1088231, EPI_ISL_1088232, EPI_ISL_1088239, EPI_ISL_1088271, EPI_ISL_1088299, EPI_ISL_1088333, EPI_ISL_1088354, EPI_ISL_1088381, EPI_ISL_1088382, EPI_ISL_1088383, EPI_ISL_1088385, EPI_ISL_1088386, EPI_ISL_1088390, EPI_ISL_1088393, EPI_ISL_1088395, EPI_ISL_1088398, EPI_ISL_1088399, EPI_ISL_1088400, EPI_ISL_1088402, EPI_ISL_1088403, EPI_ISL_1088404, EPI_ISL_1088409, EPI_ISL_1088410, EPI_ISL_1088411, EPI_ISL_1088412, EPI_ISL_1088413, EPI_ISL_1088414, EPI_ISL_1088415, EPI_ISL_1088416, EPI_ISL_1088417, EPI_ISL_1088419, EPI_ISL_1088420, EPI_ISL_1088422, EPI_ISL_1088423, EPI_ISL_1088425, EPI_ISL_1088426, EPI_ISL_1088428, EPI_ISL_1088429, EPI_ISL_1088430, EPI_ISL_1088431, EPI_ISL_1088432, EPI_ISL_1088434, EPI_ISL_1088437, EPI_ISL_1088438, EPI_ISL_1088439, EPI_ISL_1088440, EPI_ISL_1088441, EPI_ISL_1088442, EPI_ISL_1088443, EPI_ISL_1088444, EPI_ISL_1088445, EPI_ISL_1088447, EPI_ISL_1088448, EPI_ISL_1088453, EPI_ISL_1088455, EPI_ISL_1088456, EPI_ISL_1088457, EPI_ISL_1088458, EPI_ISL_1088460, EPI_ISL_1088462, EPI_ISL_1088464, EPI_ISL_1088466, EPI_ISL_1088467, EPI_ISL_1088469, EPI_ISL_1088471, EPI_ISL_1088472, EPI_ISL_1088474, EPI_ISL_1088475, EPI_ISL_1088477, EPI_ISL_1088478, EPI_ISL_1088479, EPI_ISL_1088480, EPI_ISL_1088481, EPI_ISL_1088482, EPI_ISL_1088483, EPI_ISL_1088488, EPI_ISL_1088489, EPI_ISL_1088490, EPI_ISL_1088491, EPI_ISL_1088493, EPI_ISL_1088494, EPI_ISL_1088495, EPI_ISL_1088496, EPI_ISL_1088497, EPI_ISL_1088499, EPI_ISL_1088501, EPI_ISL_1088502, EPI_ISL_1088503, EPI_ISL_1088505, EPI_ISL_1088506, EPI_ISL_1088508, EPI_ISL_1088509, EPI_ISL_1088511, EPI_ISL_1088512, EPI_ISL_1088513, EPI_ISL_1088514, EPI_ISL_1088516, EPI_ISL_1088517, EPI_ISL_1088518, EPI_ISL_1088520, EPI_ISL_1088521, EPI_ISL_1088522, EPI_ISL_1088523, EPI_ISL_1088525, EPI_ISL_1088528, EPI_ISL_1088529, EPI_ISL_1088530, EPI_ISL_1088532, EPI_ISL_1088533, EPI_ISL_1088534, EPI_ISL_1088537, EPI_ISL_1088538, EPI_ISL_1088539, EPI_ISL_1088541, EPI_ISL_1088542, EPI_ISL_1088543, EPI_ISL_1088545, EPI_ISL_1088547, EPI_ISL_1088548, EPI_ISL_1088549, EPI_ISL_1090291, EPI_ISL_1090327, EPI_ISL_1090351, EPI_ISL_1090370, EPI_ISL_1090397, EPI_ISL_1090426, EPI_ISL_1090496, EPI_ISL_1090520, EPI_ISL_1090522, EPI_ISL_1090559, EPI_ISL_1090562, EPI_ISL_1090565, EPI_ISL_1090568, EPI_ISL_1090595, EPI_ISL_1090618, EPI_ISL_1090653, EPI_ISL_1090672, EPI_ISL_1090674, EPI_ISL_1090679, EPI_ISL_1090684, EPI_ISL_1090694, EPI_ISL_1090697, EPI_ISL_1090702, EPI_ISL_1090713, EPI_ISL_1090714, EPI_ISL_1090715, EPI_ISL_1090716, EPI_ISL_1090717, EPI_ISL_1090718, EPI_ISL_1090719, EPI_ISL_1090721, EPI_ISL_1090723, EPI_ISL_1090724, EPI_ISL_1090725, EPI_ISL_1090726, EPI_ISL_1090727, EPI_ISL_1090728, EPI_ISL_1090730, EPI_ISL_1090731, EPI_ISL_1090733, EPI_ISL_1090758, EPI_ISL_1090782, EPI_ISL_1090842, EPI_ISL_1090847, EPI_ISL_1090849, EPI_ISL_1090865, EPI_ISL_1090880, EPI_ISL_1090881, EPI_ISL_1090950, EPI_ISL_1090951, EPI_ISL_1090952, EPI_ISL_1090953, EPI_ISL_1090955, EPI_ISL_1090956, EPI_ISL_1090957, EPI_ISL_1090959, EPI_ISL_1090960, EPI_ISL_1090961, EPI_ISL_1090962, EPI_ISL_1090963, EPI_ISL_1090964, EPI_ISL_1090965, EPI_ISL_1090981, EPI_ISL_1090982, EPI_ISL_1090983, EPI_ISL_1090984, EPI_ISL_1090985, EPI_ISL_1090986, EPI_ISL_1090987, EPI_ISL_1090988, EPI_ISL_1090990, EPI_ISL_1090991, EPI_ISL_1090992, EPI_ISL_1090993, EPI_ISL_1090994, EPI_ISL_1090995, EPI_ISL_1090997, EPI_ISL_1090998, EPI_ISL_1090999, EPI_ISL_1091000, EPI_ISL_1091001, EPI_ISL_1091002, EPI_ISL_1091003, EPI_ISL_1091004, EPI_ISL_1091005, EPI_ISL_1091011, EPI_ISL_1091012, EPI_ISL_1091037, EPI_ISL_1091040, EPI_ISL_1091073, EPI_ISL_1091078, EPI_ISL_1113902, EPI_ISL_1114075, EPI_ISL_1114137, EPI_ISL_1139180, EPI_ISL_1139223, EPI_ISL_1139297, EPI_ISL_1139322, EPI_ISL_1139381, EPI_ISL_1139383, EPI_ISL_1139384, EPI_ISL_1139385, EPI_ISL_1139387, EPI_ISL_1139388, EPI_ISL_1139389, EPI_ISL_1139390, EPI_ISL_1139391, EPI_ISL_1139392, EPI_ISL_1139393, EPI_ISL_1139394, EPI_ISL_1139395, EPI_ISL_1139396, EPI_ISL_1139397, EPI_ISL_1139398, EPI_ISL_1139399, EPI_ISL_1139400, EPI_ISL_1139401, EPI_ISL_1139404, EPI_ISL_1139405, EPI_ISL_1139407, EPI_ISL_1139408, EPI_ISL_1139409, EPI_ISL_1139410, EPI_ISL_1139413, EPI_ISL_1139415, EPI_ISL_1139416, EPI_ISL_1139417, EPI_ISL_1139419, EPI_ISL_1139420, EPI_ISL_1139421, EPI_ISL_1139425, EPI_ISL_1139428, EPI_ISL_1139429, EPI_ISL_1139480, EPI_ISL_1139491, EPI_ISL_1139503, EPI_ISL_1139505, EPI_ISL_1139506, EPI_ISL_1139811, EPI_ISL_1139812, EPI_ISL_1139817, EPI_ISL_1139819, EPI_ISL_1139820, EPI_ISL_1139821, EPI_ISL_1139822, EPI_ISL_1139823, EPI_ISL_1139824, EPI_ISL_1139825, EPI_ISL_1139826, EPI_ISL_1139827, EPI_ISL_1139828, EPI_ISL_1139831, EPI_ISL_1139832, EPI_ISL_1139833, EPI_ISL_1139834, EPI_ISL_1139835, EPI_ISL_1139836, EPI_ISL_1139837, EPI_ISL_1139838, EPI_ISL_1139839, EPI_ISL_1139840, EPI_ISL_1139841, EPI_ISL_1139842, EPI_ISL_1139843, EPI_ISL_1139844, EPI_ISL_1139845, EPI_ISL_1139846, EPI_ISL_1139847, EPI_ISL_1139848, EPI_ISL_1139849, EPI_ISL_1139850, EPI_ISL_1139851, EPI_ISL_1139852, EPI_ISL_1139853, EPI_ISL_1139855, EPI_ISL_1139856, EPI_ISL_1139857, EPI_ISL_1139858, EPI_ISL_1139859, EPI_ISL_1139860, EPI_ISL_1139861, EPI_ISL_1139862, EPI_ISL_1139863, EPI_ISL_1139864, EPI_ISL_1139865, EPI_ISL_1139866, EPI_ISL_1139867, EPI_ISL_1139868, EPI_ISL_1139869, EPI_ISL_1139870, EPI_ISL_1139871, EPI_ISL_1139872, EPI_ISL_1139873, EPI_ISL_1139874, EPI_ISL_1139875, EPI_ISL_1139876, EPI_ISL_1139877, EPI_ISL_1139878, EPI_ISL_1139879, EPI_ISL_1139880, EPI_ISL_1139881, EPI_ISL_1139882, EPI_ISL_1139883, EPI_ISL_1139884, EPI_ISL_1139885, EPI_ISL_1139886, EPI_ISL_1139887, EPI_ISL_1139888, EPI_ISL_1139889, EPI_ISL_1139890, EPI_ISL_1139891, EPI_ISL_1139892, EPI_ISL_1139893, EPI_ISL_1139894, EPI_ISL_1139895, EPI_ISL_1139896, EPI_ISL_1139898, EPI_ISL_1139899, EPI_ISL_1139900, EPI_ISL_1139901, EPI_ISL_1139902, EPI_ISL_1139903, EPI_ISL_1139904, EPI_ISL_1139905, EPI_ISL_1139906, EPI_ISL_1139907, EPI_ISL_1139908, EPI_ISL_1139909, EPI_ISL_1139910, EPI_ISL_1139911, EPI_ISL_1139912, EPI_ISL_1139913, EPI_ISL_1139914, EPI_ISL_1139915, EPI_ISL_1139916, EPI_ISL_1139917, EPI_ISL_1139918, EPI_ISL_1139919, EPI_ISL_1139920, EPI_ISL_1139921, EPI_ISL_1139922, EPI_ISL_1139923, EPI_ISL_1139924, EPI_ISL_1139925, EPI_ISL_1139926, EPI_ISL_1139927, EPI_ISL_1139928, EPI_ISL_1139929, EPI_ISL_1139930, EPI_ISL_1139931, EPI_ISL_1139932, EPI_ISL_1139933, EPI_ISL_1139934, EPI_ISL_1139935, EPI_ISL_1139936, EPI_ISL_1139937, EPI_ISL_1139938, EPI_ISL_1139939, EPI_ISL_1139940, EPI_ISL_1139941, EPI_ISL_1139942, EPI_ISL_1139943, EPI_ISL_1139944, EPI_ISL_1139945, EPI_ISL_1139946, EPI_ISL_1139947, EPI_ISL_1139948, EPI_ISL_1139949, EPI_ISL_1139950, EPI_ISL_1139951, EPI_ISL_1139952, EPI_ISL_1139953, EPI_ISL_1139954, EPI_ISL_1139955, EPI_ISL_1139956, EPI_ISL_1139957, EPI_ISL_1139958, EPI_ISL_1139959, EPI_ISL_1139960, EPI_ISL_1139961, EPI_ISL_1139962, EPI_ISL_1139963, EPI_ISL_1139964, EPI_ISL_1139965, EPI_ISL_1139966, EPI_ISL_1139967, EPI_ISL_1139968, EPI_ISL_1139969, EPI_ISL_1139970, EPI_ISL_1139971, EPI_ISL_1139972, EPI_ISL_1139973, EPI_ISL_1139974, EPI_ISL_1139975, EPI_ISL_1139976, EPI_ISL_1139977, EPI_ISL_1139978, EPI_ISL_1139979, EPI_ISL_1139980, EPI_ISL_1139981, EPI_ISL_1139982, EPI_ISL_1139983, EPI_ISL_1139984, EPI_ISL_1139985, EPI_ISL_1139986, EPI_ISL_1139987, EPI_ISL_1139988, EPI_ISL_1139989, EPI_ISL_1139990, EPI_ISL_1139991, EPI_ISL_1139992, EPI_ISL_1139993, EPI_ISL_1139994, EPI_ISL_1139995, EPI_ISL_1139996, EPI_ISL_1139997, EPI_ISL_1139998, EPI_ISL_1139999, EPI_ISL_1140000, EPI_ISL_1140001, EPI_ISL_1140002, EPI_ISL_1140003, EPI_ISL_1140004, EPI_ISL_1140005, EPI_ISL_1140006, EPI_ISL_1140007, EPI_ISL_1140008, EPI_ISL_1140009, EPI_ISL_1140010, EPI_ISL_1140011, EPI_ISL_1140012, EPI_ISL_1140013, EPI_ISL_1140014, EPI_ISL_1140015, EPI_ISL_1140016, EPI_ISL_1140017, EPI_ISL_1140018, EPI_ISL_1140019, EPI_ISL_1140020, EPI_ISL_1140021, EPI_ISL_1140022, EPI_ISL_1140023, EPI_ISL_1140024, EPI_ISL_1140025, EPI_ISL_1140026, EPI_ISL_1140027, EPI_ISL_1140028, EPI_ISL_1140029, EPI_ISL_1140030, EPI_ISL_1140031, EPI_ISL_1140032, EPI_ISL_1140033, EPI_ISL_1140034, EPI_ISL_1140035, EPI_ISL_1140036, EPI_ISL_1140037, EPI_ISL_1140038, EPI_ISL_1140039, EPI_ISL_1140040, EPI_ISL_1140041, EPI_ISL_1140042, EPI_ISL_1140043, EPI_ISL_1140044, EPI_ISL_1140045, EPI_ISL_1140046, EPI_ISL_1158931, EPI_ISL_1158988, EPI_ISL_1159006, EPI_ISL_1159009, EPI_ISL_1159067, EPI_ISL_1159409, EPI_ISL_1159437, EPI_ISL_1159549, EPI_ISL_1159579, EPI_ISL_1159591, EPI_ISL_1159600, EPI_ISL_1159603, EPI_ISL_1159620, EPI_ISL_1159623, EPI_ISL_1159643, EPI_ISL_1159646, EPI_ISL_1159653, EPI_ISL_1159659, EPI_ISL_1159674, EPI_ISL_1293334, EPI_ISL_1293335, EPI_ISL_1293340 | | | |
| see above | Quest Diagnostics Incorporated | Respiratory Viruses Branch, Division of Viral Diseases, Centers for Disease Control and Prevention | A. Gerasimova; A. Perez; B. Anderson; Ben L. Rambo-Martin; Clinton R. Paden; Dakota Howard; Dhwani Batra; Duncan MacCannell; F. Lacbawan; I. A. Shlyakhter; K.E. Livingston; L.E. Bernstein; M. Hua; P. Tanpaiboon; Peter W. Cook; R. M. Kagan; R. Owen; R. V. Rolando; S. H. Rosenthal; Suxiang Tong; Y. Liu |
| EPI_ISL_1295798, EPI_ISL_1295799, EPI_ISL_1295800, EPI_ISL_1295802, EPI_ISL_1295803, EPI_ISL_1295804, EPI_ISL_1295807, EPI_ISL_1295809, EPI_ISL_1295811, EPI_ISL_1295812, EPI_ISL_1295813, EPI_ISL_1794773 | | | |
| see above | Rady Children's Hospital - San Diego | Andersen lab at Scripps Research | Christina Clarke; David Dimmock; Denise Malicki; Kathryn Bouic; Linda Luo; SEARCH Alliance San Diego with Nanda Radamchar; Teresa Mueller |
| EPI_ISL_1233880, EPI_ISL_1233889 | Respiratory Viruses Branch, Centers for Disease Control and Prevention | Respiratory Viruses Branch, Centers for Disease Control and Prevention | Allen, E.; Antico, J.; Ascencio, A.; B.L.; Barrett; Batra, D.; Becker, D.; Bolze, A.; C.R.; Cassens, T.; Cho, R.; Cirulli, E.; Cook; D.T.; Febbo, P.; Galloway, S.; Gietzen, K.; Howard; Isaksson, M.; K.S.; Laurent, M.; Lee, W.; Levan, G.; Liu, J.; Lu, J.; MacCannell, D.; N.L.; Nguyen, J.; P.W.; Paden; Rambo-Martin; Ramirez, J.; Rivera-Garcia, C.; Sandoval, E.; Sickler, B.; Tolentino, M.; Tong, S.; Tran, C.; Wang, S.; Washington; White, S.; Wickline, S.; de Feo, E. |
| EPI_ISL_1185960, EPI_ISL_1185961, EPI_ISL_1185962, EPI_ISL_1185963, EPI_ISL_1185964, EPI_ISL_1185965, EPI_ISL_1185966, EPI_ISL_1185967, EPI_ISL_1185968, EPI_ISL_1185969, EPI_ISL_1185970, EPI_ISL_1185971, EPI_ISL_1185972, EPI_ISL_1185973, EPI_ISL_1185974, EPI_ISL_1185975, EPI_ISL_1185976, EPI_ISL_1185977, EPI_ISL_1185978, EPI_ISL_1185979, EPI_ISL_1185980, EPI_ISL_1185981, EPI_ISL_1185982, EPI_ISL_1185983, EPI_ISL_1185984, EPI_ISL_1185985, EPI_ISL_1185986, EPI_ISL_1185987, EPI_ISL_1185988, EPI_ISL_1185989, EPI_ISL_1185990, EPI_ISL_1185991, EPI_ISL_1185992, EPI_ISL_1185993, EPI_ISL_1185994, EPI_ISL_1185995, EPI_ISL_1185996, EPI_ISL_1185997, EPI_ISL_1185998, EPI_ISL_1185999, EPI_ISL_1186000, EPI_ISL_1186001, EPI_ISL_1186002, EPI_ISL_1186003, EPI_ISL_1186004, EPI_ISL_1186005, EPI_ISL_1186006, EPI_ISL_1186007, EPI_ISL_1186008, EPI_ISL_1186009, EPI_ISL_1186010, EPI_ISL_1186011, EPI_ISL_1186012, EPI_ISL_1186013, EPI_ISL_1186014, EPI_ISL_1186015, EPI_ISL_1186016, EPI_ISL_1186017, EPI_ISL_1186018, EPI_ISL_1186019, EPI_ISL_1186020, EPI_ISL_1186021, EPI_ISL_1186022, EPI_ISL_1295815, EPI_ISL_1468841, EPI_ISL_1468847, EPI_ISL_1468854, EPI_ISL_1468875, EPI_ISL_1468876, EPI_ISL_1468906, EPI_ISL_1468907, EPI_ISL_1468908, EPI_ISL_1468909, EPI_ISL_1468950, EPI_ISL_1468953, EPI_ISL_1468979, EPI_ISL_1468980, EPI_ISL_1468983, EPI_ISL_1468984, EPI_ISL_1468986, EPI_ISL_1468987, EPI_ISL_1468989, EPI_ISL_1468990, EPI_ISL_1468991, EPI_ISL_1468994, EPI_ISL_1468995, EPI_ISL_1468996, EPI_ISL_1468999, EPI_ISL_1469000, EPI_ISL_1469001, EPI_ISL_1469002, EPI_ISL_1469004, EPI_ISL_1469005, EPI_ISL_1469008, EPI_ISL_1469009, EPI_ISL_1469012, EPI_ISL_1469013, EPI_ISL_1469016, EPI_ISL_1469020, EPI_ISL_1469021, EPI_ISL_1469022, EPI_ISL_1794618, EPI_ISL_1794628, EPI_ISL_1794635, EPI_ISL_1794638, EPI_ISL_1794639, EPI_ISL_1794643, EPI_ISL_1794648, EPI_ISL_1794653, EPI_ISL_1969255, EPI_ISL_1969257, EPI_ISL_1969258, EPI_ISL_1969260, EPI_ISL_1969262, EPI_ISL_1969263, EPI_ISL_1969265, EPI_ISL_1969266, EPI_ISL_1969268, EPI_ISL_1969270 | | | |
| see above | San Diego County Public Health Laboratory | Andersen lab at Scripps Research | Brett Austin; Jovan Shephard; SEARCH Alliance San Diego with Tracy Basler |
| EPI_ISL_1701041, EPI_ISL_1701050, EPI_ISL_1701052, EPI_ISL_1701061, EPI_ISL_1701079, EPI_ISL_1701086, EPI_ISL_1701096, EPI_ISL_1701142, EPI_ISL_1701204, EPI_ISL_1701211, EPI_ISL_1701236, EPI_ISL_1701262, EPI_ISL_1701281, EPI_ISL_1701290 | | | |
| see above | San Joaquin County Public Health Lab | Chan-Zuckerberg Biohub | CZB Cliahub Consortium |
| EPI_ISL_1234541, EPI_ISL_1234561, EPI_ISL_1234578, EPI_ISL_1234584, EPI_ISL_1234641, EPI_ISL_1234657, EPI_ISL_1234687, EPI_ISL_1234694, EPI_ISL_1234706, EPI_ISL_1234711, EPI_ISL_1234737, EPI_ISL_1234769, EPI_ISL_1234787, EPI_ISL_1234798, EPI_ISL_1234800, EPI_ISL_1234810, EPI_ISL_1234812, EPI_ISL_1234818, EPI_ISL_1234837, EPI_ISL_1234840, EPI_ISL_1234841, EPI_ISL_1234843, EPI_ISL_1234846, EPI_ISL_1234849, EPI_ISL_1234850, EPI_ISL_1234851, EPI_ISL_1234854, EPI_ISL_1234857, EPI_ISL_1234865, EPI_ISL_1234866, EPI_ISL_1234875, EPI_ISL_1234876, EPI_ISL_1234878, EPI_ISL_1234879, EPI_ISL_1234880, EPI_ISL_1234881, EPI_ISL_1234883, EPI_ISL_1234884, EPI_ISL_1234886, EPI_ISL_1234897, EPI_ISL_1234899, EPI_ISL_1234902, EPI_ISL_1234913, EPI_ISL_1234917, EPI_ISL_1234919, EPI_ISL_1234925, EPI_ISL_1234935, EPI_ISL_1234946, EPI_ISL_1234957, EPI_ISL_1234961, EPI_ISL_1234962, EPI_ISL_1234972, EPI_ISL_1234973, EPI_ISL_1234975, EPI_ISL_1234976, EPI_ISL_1234983, EPI_ISL_1234985, EPI_ISL_1234988, EPI_ISL_1234991, EPI_ISL_1234993, EPI_ISL_1234996, EPI_ISL_1234999, EPI_ISL_1235000, EPI_ISL_1235010, EPI_ISL_1235015, EPI_ISL_1235017, EPI_ISL_1235019, EPI_ISL_1235020, EPI_ISL_1235024, EPI_ISL_1235031, EPI_ISL_1235034, EPI_ISL_1235038, EPI_ISL_1235043, EPI_ISL_1235047, EPI_ISL_1235049, EPI_ISL_1235050, EPI_ISL_1235051, EPI_ISL_1235362, EPI_ISL_1235370, EPI_ISL_1235379, EPI_ISL_1235382, EPI_ISL_1235394, EPI_ISL_1235395, EPI_ISL_1235396, EPI_ISL_1235409, EPI_ISL_1235411, EPI_ISL_1235412, EPI_ISL_1235415, EPI_ISL_1235420, EPI_ISL_1235428, EPI_ISL_1235429, EPI_ISL_1235430, EPI_ISL_1235431, EPI_ISL_1235436, EPI_ISL_1235439, EPI_ISL_1235444, EPI_ISL_1235445, EPI_ISL_1235449, EPI_ISL_1235452, EPI_ISL_1235453, EPI_ISL_1235454, EPI_ISL_1235461, EPI_ISL_1235466, EPI_ISL_1235467, EPI_ISL_1235470, EPI_ISL_1235474, EPI_ISL_1235479, EPI_ISL_1235483, EPI_ISL_1235484, EPI_ISL_1235487, EPI_ISL_1235491, EPI_ISL_1235508, EPI_ISL_1235518, EPI_ISL_1235519, EPI_ISL_1235530, EPI_ISL_1235531, EPI_ISL_1235533, EPI_ISL_1235534, EPI_ISL_1235535, EPI_ISL_1235536, EPI_ISL_1235541, EPI_ISL_1235542, EPI_ISL_1235556, EPI_ISL_1235563, EPI_ISL_1235581, EPI_ISL_1235583, EPI_ISL_1235593, EPI_ISL_1235601, EPI_ISL_1235612, EPI_ISL_1477441, EPI_ISL_1477445, EPI_ISL_1477455, EPI_ISL_1477461, EPI_ISL_1477484, EPI_ISL_1477486, EPI_ISL_1477494, EPI_ISL_1477496, EPI_ISL_1477501, EPI_ISL_1477507, EPI_ISL_1477510, EPI_ISL_1477518, EPI_ISL_1477519, EPI_ISL_1477529, EPI_ISL_1477534, EPI_ISL_1477535, EPI_ISL_1477540, EPI_ISL_1477541, EPI_ISL_1477552, EPI_ISL_1477558, EPI_ISL_1477566, EPI_ISL_1477575, EPI_ISL_1477578, EPI_ISL_1477581, EPI_ISL_1477590, EPI_ISL_1477598, EPI_ISL_1477603, EPI_ISL_1477622, EPI_ISL_1477637, EPI_ISL_1477644, EPI_ISL_1477655, EPI_ISL_1477664, EPI_ISL_1477670, EPI_ISL_1477677, EPI_ISL_1477681, EPI_ISL_1477682, EPI_ISL_1477690, EPI_ISL_1477695, EPI_ISL_1477699, EPI_ISL_1477706, EPI_ISL_1477722, EPI_ISL_1477726, EPI_ISL_1477727, EPI_ISL_1477729, EPI_ISL_1477730, EPI_ISL_1477731, EPI_ISL_1477734, EPI_ISL_1477745, EPI_ISL_1477749, EPI_ISL_1477750, EPI_ISL_1477752, EPI_ISL_1477755, EPI_ISL_1477762, EPI_ISL_1477767, EPI_ISL_1477768, EPI_ISL_1477776, EPI_ISL_1477780, EPI_ISL_1477790, EPI_ISL_1477793, EPI_ISL_1477797, EPI_ISL_1477799, EPI_ISL_1477800, EPI_ISL_1477814, EPI_ISL_1477817, EPI_ISL_1477818, EPI_ISL_1477822, EPI_ISL_1477836, EPI_ISL_1477839, EPI_ISL_1477842, EPI_ISL_1477845, EPI_ISL_1477846, EPI_ISL_1477850, EPI_ISL_1477854, EPI_ISL_1477856, EPI_ISL_1477859, EPI_ISL_1477860, EPI_ISL_1477863, EPI_ISL_1477871, EPI_ISL_1477874, EPI_ISL_1477878, EPI_ISL_1477879, EPI_ISL_1477881, EPI_ISL_1477882, EPI_ISL_1477885, EPI_ISL_1477888, EPI_ISL_1477892, EPI_ISL_1477893, EPI_ISL_1477895, EPI_ISL_1477902, EPI_ISL_1477907, EPI_ISL_1477909, EPI_ISL_1477912, EPI_ISL_1477923, EPI_ISL_1477925, EPI_ISL_1477926, EPI_ISL_1477935, EPI_ISL_1477937, EPI_ISL_1477943, EPI_ISL_1477958, EPI_ISL_1477967, EPI_ISL_1477974, EPI_ISL_1477979, EPI_ISL_1477982, EPI_ISL_1478012, EPI_ISL_1478019, EPI_ISL_1478028, EPI_ISL_1478031, EPI_ISL_1478046, EPI_ISL_1478048, EPI_ISL_1478050, EPI_ISL_1478059, EPI_ISL_1478081, EPI_ISL_1478087, EPI_ISL_1478089, EPI_ISL_1478095, EPI_ISL_1478097, EPI_ISL_1478102, EPI_ISL_1478103, EPI_ISL_1478123, EPI_ISL_1478124, EPI_ISL_1478149, EPI_ISL_1478152, EPI_ISL_1478161, EPI_ISL_1478171, EPI_ISL_1478192, EPI_ISL_1478200, EPI_ISL_1529101, EPI_ISL_1529102, EPI_ISL_1529106, EPI_ISL_1529107, EPI_ISL_1529108, EPI_ISL_1529109, EPI_ISL_1529110, EPI_ISL_1529113, EPI_ISL_1529119, EPI_ISL_1529121, EPI_ISL_1529138, EPI_ISL_1529151, EPI_ISL_1529152, EPI_ISL_1529162, EPI_ISL_1529173, EPI_ISL_1529183, EPI_ISL_1529206, EPI_ISL_1529207, EPI_ISL_1529218, EPI_ISL_1529221, EPI_ISL_1529224, EPI_ISL_1529226, EPI_ISL_1529238, EPI_ISL_1529241, EPI_ISL_1529252, EPI_ISL_1529258, EPI_ISL_1529262, EPI_ISL_1529273, EPI_ISL_1529283, EPI_ISL_1529287, EPI_ISL_1529295, EPI_ISL_1529309, EPI_ISL_1529316, EPI_ISL_1529344, EPI_ISL_1529347, EPI_ISL_1529349, EPI_ISL_1529351, EPI_ISL_1663980, EPI_ISL_1821317, EPI_ISL_1821328, EPI_ISL_1821344, EPI_ISL_1821346, EPI_ISL_1821428, EPI_ISL_2102742, EPI_ISL_2102768, EPI_ISL_2102805, EPI_ISL_2102812, EPI_ISL_2102813, EPI_ISL_2102859, EPI_ISL_2102878, EPI_ISL_2102883, EPI_ISL_2102892, EPI_ISL_2102895, EPI_ISL_2102898, EPI_ISL_2102899, EPI_ISL_2102904, EPI_ISL_2102913, EPI_ISL_2102914, EPI_ISL_2102923, EPI_ISL_2102967 | | | |
| see above | Santa Clara County Public Health Laboratory | Chan-Zuckerberg Biohub | CZB Cliahub Consortium |
| EPI_ISL_1001005, EPI_ISL_1001006, EPI_ISL_1001007, EPI_ISL_1001008, EPI_ISL_1001009, EPI_ISL_1001010, EPI_ISL_1001013, EPI_ISL_1001014, EPI_ISL_1001015, EPI_ISL_1001016, EPI_ISL_1001017, EPI_ISL_1001018, EPI_ISL_1001019, EPI_ISL_1001020, EPI_ISL_1001021, EPI_ISL_1001022, EPI_ISL_1001024, EPI_ISL_1001025, EPI_ISL_1001026, EPI_ISL_1001027, EPI_ISL_1001028, EPI_ISL_1001029, EPI_ISL_1001030, EPI_ISL_1001031, EPI_ISL_1001032, EPI_ISL_1001033, EPI_ISL_1001034, EPI_ISL_1001035, EPI_ISL_1001036, EPI_ISL_1001037, EPI_ISL_1001038, EPI_ISL_1001039, EPI_ISL_1001040, EPI_ISL_1009655, EPI_ISL_1009657, EPI_ISL_1009659, EPI_ISL_1009660, EPI_ISL_1009661, EPI_ISL_1009662, EPI_ISL_1009663, EPI_ISL_1009664, EPI_ISL_1060543, EPI_ISL_1060544, EPI_ISL_1060546, EPI_ISL_1060548, EPI_ISL_1060550, EPI_ISL_1060551, EPI_ISL_1060552, EPI_ISL_1060553, EPI_ISL_1137646, EPI_ISL_1137647, EPI_ISL_1235636, EPI_ISL_1235638, EPI_ISL_1235640, EPI_ISL_1291743, EPI_ISL_1291745, EPI_ISL_1335804, EPI_ISL_1335805, EPI_ISL_1335806, EPI_ISL_1335807, EPI_ISL_1335808, EPI_ISL_1335809, EPI_ISL_1335810, EPI_ISL_1335811, EPI_ISL_1335812, EPI_ISL_1335813, EPI_ISL_1335814, EPI_ISL_1335815, EPI_ISL_1335816, EPI_ISL_1335817, EPI_ISL_1335818, EPI_ISL_1335819, EPI_ISL_1335820, EPI_ISL_1335821, EPI_ISL_1335822, EPI_ISL_1335823, EPI_ISL_1335824, EPI_ISL_1335825, EPI_ISL_1335826, EPI_ISL_1335827, EPI_ISL_1335828, EPI_ISL_1335829, EPI_ISL_1335830, EPI_ISL_1335831, EPI_ISL_1335832, EPI_ISL_1335833, EPI_ISL_1335834, EPI_ISL_1335835, EPI_ISL_1335837, EPI_ISL_1335838, EPI_ISL_1335840, EPI_ISL_1335853, EPI_ISL_1335854, EPI_ISL_1335855, EPI_ISL_1335856, EPI_ISL_1335857, EPI_ISL_1335858, EPI_ISL_1335861, EPI_ISL_1335862, EPI_ISL_1335863, EPI_ISL_1335864, EPI_ISL_1335865, EPI_ISL_1335866, EPI_ISL_1335867, EPI_ISL_1406161, EPI_ISL_1406162, EPI_ISL_1406163, EPI_ISL_1406170 | | | |
| see above | Santa Clara County Public Health Laboratory | Santa Clara County Public Health Laboratory | Santa Clara County Public Health Department |
| EPI_ISL_1081550, EPI_ISL_1081556, EPI_ISL_1366275, EPI_ISL_1366278, EPI_ISL_1366282, EPI_ISL_1366285, EPI_ISL_1366297, EPI_ISL_1366318, EPI_ISL_1366320, EPI_ISL_1366322, EPI_ISL_1366379, EPI_ISL_1366380, EPI_ISL_1366381, EPI_ISL_1366382, EPI_ISL_1366383, EPI_ISL_1366384, EPI_ISL_1366385, EPI_ISL_1366386, EPI_ISL_1366387, EPI_ISL_1366388, EPI_ISL_1366389, EPI_ISL_1366390, EPI_ISL_1366391, EPI_ISL_1366392, EPI_ISL_1366393, EPI_ISL_1366394, EPI_ISL_1366395, EPI_ISL_1366396, EPI_ISL_1366397, EPI_ISL_1366398, EPI_ISL_1366399, EPI_ISL_1366400, EPI_ISL_1366401, EPI_ISL_1366402, EPI_ISL_1366403, EPI_ISL_1366404, EPI_ISL_1366405, EPI_ISL_1366406, EPI_ISL_1366407, EPI_ISL_1366408, EPI_ISL_1366409, EPI_ISL_1366410, EPI_ISL_1366411, EPI_ISL_1366412, EPI_ISL_1366413, EPI_ISL_1366414, EPI_ISL_1366415, EPI_ISL_1366416, EPI_ISL_1366417, EPI_ISL_1366418, EPI_ISL_1366419, EPI_ISL_1366420, EPI_ISL_1366421, EPI_ISL_1366422, EPI_ISL_1366423, EPI_ISL_1468849, EPI_ISL_1468887, EPI_ISL_1468888, EPI_ISL_1468947, EPI_ISL_1469047, EPI_ISL_1469048, EPI_ISL_1469049, EPI_ISL_1469050, EPI_ISL_1469058, EPI_ISL_1469059, EPI_ISL_1469060 | | | |
| see above | Scripps Medical Laboratory | Andersen lab at Scripps Research | Ellen Stefanski; Ian Mchardy; SEARCH Alliance San Diego with Michael Quigley |
| EPI_ISL_1295646, EPI_ISL_1295647, EPI_ISL_1295648, EPI_ISL_1295649, EPI_ISL_1295650, EPI_ISL_1295651, EPI_ISL_1295652, EPI_ISL_1295653, EPI_ISL_1295654, EPI_ISL_1295655, EPI_ISL_1295656, EPI_ISL_1295657, EPI_ISL_1295658, EPI_ISL_1295659, EPI_ISL_1295660, EPI_ISL_1295661, EPI_ISL_1295662, EPI_ISL_1295663, EPI_ISL_1295664, EPI_ISL_1295665, EPI_ISL_1295666, EPI_ISL_1295667, EPI_ISL_1295668, EPI_ISL_1295669, EPI_ISL_1295670, EPI_ISL_1295671, EPI_ISL_1295672, EPI_ISL_1295673, EPI_ISL_1295674, EPI_ISL_1295675, EPI_ISL_1295676, EPI_ISL_1295677, EPI_ISL_1295678, EPI_ISL_1295679, EPI_ISL_1295680, EPI_ISL_1295681, EPI_ISL_1295682, EPI_ISL_1295683, EPI_ISL_1295684, EPI_ISL_1295685, EPI_ISL_1295686, EPI_ISL_1295687, EPI_ISL_1295688, EPI_ISL_1295689, EPI_ISL_1295690, EPI_ISL_1295691, EPI_ISL_1295692, EPI_ISL_1295693, EPI_ISL_1295694, EPI_ISL_1295695, EPI_ISL_1295696, EPI_ISL_1295697, EPI_ISL_1295698, EPI_ISL_1295699, EPI_ISL_1295700, EPI_ISL_1295701, EPI_ISL_1295702, EPI_ISL_1295703, EPI_ISL_1295704, EPI_ISL_1295705, EPI_ISL_1295706, EPI_ISL_1295707, EPI_ISL_1295708, EPI_ISL_1295709, EPI_ISL_1295710, EPI_ISL_1295711, EPI_ISL_1295712, EPI_ISL_1295713, EPI_ISL_1295714, EPI_ISL_1295715, EPI_ISL_1295716, EPI_ISL_1295717, EPI_ISL_1295718, EPI_ISL_1295719, EPI_ISL_1295720, EPI_ISL_1295721, EPI_ISL_1295722, EPI_ISL_1295723, EPI_ISL_1295724, EPI_ISL_1295725, EPI_ISL_1295726, EPI_ISL_1295727, EPI_ISL_1295728, EPI_ISL_1295729, EPI_ISL_1295730, EPI_ISL_1295731, EPI_ISL_1295732, EPI_ISL_1295733, | | | |

| | | | |
|---|---|---|---|
| EPI_ISL_1295734, EPI_ISL_1295735, EPI_ISL_1295736, EPI_ISL_1295737, EPI_ISL_1295738, EPI_ISL_1295739, EPI_ISL_1295740, EPI_ISL_1295741, EPI_ISL_1295742, EPI_ISL_1295743, EPI_ISL_1295744, EPI_ISL_1295745, EPI_ISL_1295746, EPI_ISL_1295747, EPI_ISL_1295748, EPI_ISL_1295749, EPI_ISL_1295750, EPI_ISL_1295751, EPI_ISL_1295752, EPI_ISL_1295753, EPI_ISL_1295754, EPI_ISL_1295755, EPI_ISL_1295756, EPI_ISL_1295757, EPI_ISL_1295758, EPI_ISL_1295759, EPI_ISL_1295760, EPI_ISL_1295761, EPI_ISL_1295762, EPI_ISL_1295763, EPI_ISL_1295764, EPI_ISL_1295765, EPI_ISL_1295766, EPI_ISL_1295767, EPI_ISL_1295768, EPI_ISL_1295769, EPI_ISL_1295770, EPI_ISL_1295771, EPI_ISL_1295772, EPI_ISL_1295773, EPI_ISL_1295774, EPI_ISL_1295775, EPI_ISL_1295778, EPI_ISL_1295782, EPI_ISL_1295787, EPI_ISL_1295789, EPI_ISL_1295790, EPI_ISL_1295818, EPI_ISL_1295819, EPI_ISL_1295820, EPI_ISL_1295821, EPI_ISL_1295822, EPI_ISL_1295823, EPI_ISL_1295824, EPI_ISL_1295825, EPI_ISL_1295826, EPI_ISL_1295827, EPI_ISL_1295828, EPI_ISL_1295829, EPI_ISL_1295830, EPI_ISL_1295831, EPI_ISL_1295832, EPI_ISL_1295833, EPI_ISL_1295834, EPI_ISL_1295835, EPI_ISL_1295836, EPI_ISL_1295837, EPI_ISL_1295838, EPI_ISL_1295839, EPI_ISL_1295840, EPI_ISL_1295841, EPI_ISL_1295842, EPI_ISL_1295843, EPI_ISL_1295844, EPI_ISL_1295845, EPI_ISL_1295846, EPI_ISL_1295847, EPI_ISL_1295848, EPI_ISL_1295849, EPI_ISL_1295850, EPI_ISL_1295851, EPI_ISL_1295852, EPI_ISL_1295853, EPI_ISL_1295854, EPI_ISL_1295855, EPI_ISL_1295856, EPI_ISL_1295857, EPI_ISL_1295858, EPI_ISL_1295859, EPI_ISL_1295860, EPI_ISL_1295861, EPI_ISL_1295862, EPI_ISL_1295863, EPI_ISL_1295864, EPI_ISL_1295865, EPI_ISL_1295866, EPI_ISL_1295867, EPI_ISL_1295868, EPI_ISL_1295869, EPI_ISL_1295870, EPI_ISL_1295871, EPI_ISL_1295872, EPI_ISL_1295873, EPI_ISL_1295874, EPI_ISL_1295875, EPI_ISL_1295876, EPI_ISL_1295877, EPI_ISL_1295878, EPI_ISL_1295879, EPI_ISL_1295880, EPI_ISL_1295881, EPI_ISL_1295882, EPI_ISL_1295883, EPI_ISL_1295884, EPI_ISL_1295885, EPI_ISL_1295886, EPI_ISL_1295887, EPI_ISL_1295888, EPI_ISL_1295889, EPI_ISL_1295890, EPI_ISL_1295891, EPI_ISL_1295892, EPI_ISL_1295893, EPI_ISL_1295894, EPI_ISL_1295895, EPI_ISL_1295896, EPI_ISL_1295897, EPI_ISL_1295898, EPI_ISL_1295899, EPI_ISL_1295900, EPI_ISL_1366284, EPI_ISL_1366287, EPI_ISL_1366288, EPI_ISL_1366289, EPI_ISL_1366290, EPI_ISL_1366291, EPI_ISL_1366292, EPI_ISL_1366298, EPI_ISL_1366299, EPI_ISL_1366300, EPI_ISL_1366303, EPI_ISL_1366306, EPI_ISL_1366307, EPI_ISL_1366310, EPI_ISL_1366317, EPI_ISL_1366323, EPI_ISL_1366325, EPI_ISL_1366376, EPI_ISL_1366424, EPI_ISL_1366425, EPI_ISL_1366426, EPI_ISL_1366427, EPI_ISL_1366428, EPI_ISL_1366429, EPI_ISL_1366430, EPI_ISL_1366431, EPI_ISL_1366432, EPI_ISL_1366433, EPI_ISL_1366434, EPI_ISL_1366435, EPI_ISL_1366436, EPI_ISL_1366437, EPI_ISL_1366438, EPI_ISL_1366439, EPI_ISL_1366440, EPI_ISL_1366441, EPI_ISL_1366442, EPI_ISL_1366443, EPI_ISL_1366444, EPI_ISL_1366445, EPI_ISL_1366446, EPI_ISL_1366447, EPI_ISL_1366448, EPI_ISL_1366449, EPI_ISL_1366450, EPI_ISL_1366451, EPI_ISL_1366452, EPI_ISL_1366453, EPI_ISL_1366454, EPI_ISL_1366455, EPI_ISL_1366456, EPI_ISL_1366457, EPI_ISL_1366458, EPI_ISL_1366459, EPI_ISL_1366460, EPI_ISL_1366461, EPI_ISL_1366463, EPI_ISL_1366464, EPI_ISL_1366465, EPI_ISL_1366469, EPI_ISL_1366470, EPI_ISL_1366471, EPI_ISL_1366475, EPI_ISL_1366477, EPI_ISL_1366479, EPI_ISL_1366481, EPI_ISL_1366484, EPI_ISL_1366495, EPI_ISL_1366496 | | | |
| see above | Sharp HealthCare Laboratory | Andersen lab at Scripps Research | Art Mendoza; Cathy Woerle; Jacquelyn Berumen; Liam McGinnis; Omid Bakhtar; SEARCH Alliance San Diego with Aaron Harding |
| EPI_ISL_1483795, EPI_ISL_1483796, EPI_ISL_1483797, EPI_ISL_1483798 | Stanford Clinical Virology Laboratory | Santa Clara County Public Health Laboratory | Santa Clara County Public Health Department |
| EPI_ISL_1364507, EPI_ISL_1364513, EPI_ISL_1364514, EPI_ISL_1364517, EPI_ISL_1364519, EPI_ISL_1364521, EPI_ISL_1364522, EPI_ISL_1364523, EPI_ISL_1364524, EPI_ISL_1364525, EPI_ISL_1364526, EPI_ISL_1379835, EPI_ISL_1379847, EPI_ISL_1379848, EPI_ISL_1379853, EPI_ISL_1379854, EPI_ISL_1379857, EPI_ISL_1379858, EPI_ISL_1379859, EPI_ISL_1379862, EPI_ISL_1379863, EPI_ISL_1379865, EPI_ISL_1379866, EPI_ISL_1379869, EPI_ISL_1379870, EPI_ISL_1379883, EPI_ISL_1379887, EPI_ISL_1379888, EPI_ISL_1424090, EPI_ISL_1424091, EPI_ISL_1424098, EPI_ISL_1424099, EPI_ISL_1424105, EPI_ISL_1424118, EPI_ISL_1424119, EPI_ISL_1424120, EPI_ISL_1424121, EPI_ISL_1424122, EPI_ISL_1424123, EPI_ISL_1424124, EPI_ISL_1424127, EPI_ISL_1424128, EPI_ISL_1424129, EPI_ISL_2009055, EPI_ISL_2009057, EPI_ISL_2027144, EPI_ISL_2027150, EPI_ISL_2027171, EPI_ISL_2027174, EPI_ISL_2027177, EPI_ISL_2027178, EPI_ISL_2027183, EPI_ISL_2027185, EPI_ISL_2027187, EPI_ISL_2027191, EPI_ISL_2027195, EPI_ISL_2027198 | | | |
| see above | Stanford Health Care | Stanford University School of Medicine, Clinical Virology Laboratory | Becky Jiang; ChunHong Huang; Daniel Solis; Fumiko Yamamoto; James Zehnder; Malaya K. Sahoo; Mamdouh Sibai; Michelle Verghese; and Benjamin A. Pinsky |
| EPI_ISL_1005515, EPI_ISL_1005516, EPI_ISL_1005517, EPI_ISL_1005518, EPI_ISL_1005519, EPI_ISL_1005520, EPI_ISL_1005521, EPI_ISL_1005542, EPI_ISL_1005543, EPI_ISL_1005544, EPI_ISL_1005545, EPI_ISL_1005546, EPI_ISL_1005547, EPI_ISL_1005548, EPI_ISL_1005549, EPI_ISL_1005550, EPI_ISL_1005551, EPI_ISL_1005552, EPI_ISL_1005553, EPI_ISL_1005554, EPI_ISL_1040054, EPI_ISL_1040055, EPI_ISL_1040056, EPI_ISL_1040057, EPI_ISL_1040058, EPI_ISL_1040059, EPI_ISL_1040060, EPI_ISL_1040061, EPI_ISL_1040062, EPI_ISL_1040063, EPI_ISL_1040064, EPI_ISL_1040065, EPI_ISL_1040066, EPI_ISL_1040067, EPI_ISL_1040068, EPI_ISL_1040069, EPI_ISL_1040070, EPI_ISL_1040071, EPI_ISL_1097742, EPI_ISL_1098609, EPI_ISL_1098610, EPI_ISL_1098611, EPI_ISL_1098612, EPI_ISL_1098613, EPI_ISL_1098614, EPI_ISL_1098615, EPI_ISL_1098616, EPI_ISL_1098618, EPI_ISL_1098619, EPI_ISL_1098620, EPI_ISL_1098621, EPI_ISL_1098622, EPI_ISL_1098623, EPI_ISL_1098624, EPI_ISL_1098625, EPI_ISL_1098626, EPI_ISL_1098627, EPI_ISL_1098631, EPI_ISL_1191110, EPI_ISL_1191111, EPI_ISL_1191112, EPI_ISL_1191113, EPI_ISL_1191114, EPI_ISL_1191115, EPI_ISL_1191116, EPI_ISL_1191117, EPI_ISL_1191118, EPI_ISL_1191119, EPI_ISL_1191120, EPI_ISL_1191121, EPI_ISL_1191122, EPI_ISL_1191123, EPI_ISL_1191124, EPI_ISL_1191125, EPI_ISL_1191126, EPI_ISL_1191127, EPI_ISL_1191128, EPI_ISL_1191130, EPI_ISL_1191131, EPI_ISL_1197149, EPI_ISL_1303450, EPI_ISL_1303451, EPI_ISL_1303452 | | | |
| see above | UCLA Clinical Micro Lab | Los Angeles County PHL | P. Hemarajata et al. |
| EPI_ISL_1402586, EPI_ISL_1402587, EPI_ISL_1402588, EPI_ISL_1402589, EPI_ISL_1402590, EPI_ISL_1402591, EPI_ISL_1402592, EPI_ISL_1402593, EPI_ISL_1402594, EPI_ISL_1402595, EPI_ISL_1402596, EPI_ISL_1402597, EPI_ISL_1402598, EPI_ISL_1402599, EPI_ISL_1402600, EPI_ISL_1402601, EPI_ISL_1402602, EPI_ISL_1402603, EPI_ISL_1402604, EPI_ISL_1402605, EPI_ISL_1402606, EPI_ISL_1402607, EPI_ISL_1405665, EPI_ISL_1405666, EPI_ISL_1405667, EPI_ISL_1405668, EPI_ISL_1405669, EPI_ISL_1405670, EPI_ISL_1405671, EPI_ISL_1405672, EPI_ISL_1405673, EPI_ISL_1405674, EPI_ISL_1405675, EPI_ISL_1405676, EPI_ISL_1405677, EPI_ISL_1405678, EPI_ISL_1405679 | | | |
| see above | UCSC Genomics Institute | UCSC Genomics Institute | A. Marm Kilpatrick; Angie Hinrichs; Beth Shapiro; Bryan Thornlow; David Haussler; Eric Beraut; Hugh Olsen; Ikenna Anigbogu; Isabel Bjork; Jakob McBroome; Jeremy Sanford; Joshua Kapp; Mark Akeson; Maximilian Haeussler; Michael Stone; Miten Jain; Molly Cassatt-Johnstone; Namrita Dhillon; Russell Corbett-Detig; Terren Chang; Yatish Turakhia |
| EPI_ISL_1005495, EPI_ISL_1005496, EPI_ISL_1005497, EPI_ISL_1005498, EPI_ISL_1005499, EPI_ISL_1005500, EPI_ISL_1005501, EPI_ISL_1005502, EPI_ISL_1005503, EPI_ISL_1005504, EPI_ISL_1005505, EPI_ISL_1005506, EPI_ISL_1005507, EPI_ISL_1005522, EPI_ISL_1005523, EPI_ISL_1005524, EPI_ISL_1005525, EPI_ISL_1005526, EPI_ISL_1005527, EPI_ISL_1005528, EPI_ISL_1005529, EPI_ISL_1005530, EPI_ISL_1005531, EPI_ISL_1005532, EPI_ISL_1005533, EPI_ISL_1005534, EPI_ISL_1005535, EPI_ISL_1040051, EPI_ISL_1192368, EPI_ISL_1197042, EPI_ISL_1197044, EPI_ISL_1197046, EPI_ISL_1197047, EPI_ISL_1197049, EPI_ISL_1197051, EPI_ISL_1201559, EPI_ISL_1201560, EPI_ISL_1303408, EPI_ISL_1303424, EPI_ISL_1303438 | | | |
| see above | USC Clinical Lab | Los Angeles County PHL | P. Hemarajata et al. |
| EPI_ISL_1307638 | VA Medical Center | Los Angeles County PHL | P. Hemarajata et al. |

We gratefully acknowledge the following Authors from the Originating laboratories responsible for obtaining the specimens, as well as the Submitting laboratories where the genome data were generated and shared via GISAID, on which this research is based.

All Submitters of data may be contacted directly via www.gisaid.org

Authors are sorted alphabetically.

| Accession ID | Originating Laboratory | Submitting Laboratory | Authors |
|---|---|---|---|
| EPI_ISL_1318642, EPI_ISL_1446773, EPI_ISL_1446774, EPI_ISL_1446775, EPI_ISL_1446776, EPI_ISL_1446777, EPI_ISL_1446778, EPI_ISL_1446779, EPI_ISL_1446780, EPI_ISL_1446782, EPI_ISL_1446783, EPI_ISL_1446785, EPI_ISL_1446786, EPI_ISL_1446788, EPI_ISL_1446789, EPI_ISL_1446793, EPI_ISL_1446795, EPI_ISL_1446796, EPI_ISL_1446797, EPI_ISL_1446798, EPI_ISL_1446799, EPI_ISL_1446800, EPI_ISL_1446802, EPI_ISL_1446840, EPI_ISL_1446841, EPI_ISL_1446842, EPI_ISL_1446843, EPI_ISL_1446844, EPI_ISL_1446845, EPI_ISL_1678143, EPI_ISL_1678144, EPI_ISL_1678145, EPI_ISL_1678146, EPI_ISL_1678148, EPI_ISL_1678149, EPI_ISL_1678150, EPI_ISL_1678151, EPI_ISL_1678152, EPI_ISL_1678153, EPI_ISL_1678154, EPI_ISL_1678156, EPI_ISL_1678157, EPI_ISL_1678158, EPI_ISL_1678159, EPI_ISL_1678160, EPI_ISL_1678161, EPI_ISL_1678162, EPI_ISL_1678313, EPI_ISL_1678314, EPI_ISL_1678315 | | | |
| see above | "CDPH, Viral and Rickettsial Disease Laboratory" | Centers for Disease Control and Prevention Division of Viral Diseases, Pathogen Discovery | Alison Laufer Halpin; Anna Montmayeur; Anna Uehara; Ben L. Rambo-Martin; Clinton R. Paden; Dakota Howard; Darlene Wagner; Dave Wentworth; Dhwani Batra; Haibin Wang; Jasmine Padilla; Jing Zhang; Justin Lee; Katie Dillon; Krista Queen; Kristen Knipe; Kristine Lacek; Lori Rowe; Mark Burroughs; Matthew Schmerer; Mili Sheth; Peter Cook; Peter W. Cook; Rachel Marine; Sam Shepard; Sarah Nobles; Shoshona Le; Suxiang Tong; Vivien Dugan; Yan Li; Ying Tao; Yvette Unoarumhi |
| EPI_ISL_1445279, EPI_ISL_1445280, EPI_ISL_1445289, EPI_ISL_1445292, EPI_ISL_1445293, EPI_ISL_1445294, EPI_ISL_1445384, EPI_ISL_1445425, EPI_ISL_1445426, EPI_ISL_1445427, EPI_ISL_1445429, EPI_ISL_1445430, EPI_ISL_1445431, EPI_ISL_1445434, EPI_ISL_1445454, EPI_ISL_1445455, EPI_ISL_1445461, EPI_ISL_1445462, EPI_ISL_1445468, EPI_ISL_1445472, EPI_ISL_1445491, EPI_ISL_1445506, EPI_ISL_1445508, EPI_ISL_1445509, EPI_ISL_1445511, EPI_ISL_1445537, EPI_ISL_1445538, EPI_ISL_1445539, EPI_ISL_1445540, EPI_ISL_1445541, EPI_ISL_1445542, EPI_ISL_1445544, EPI_ISL_1445545, EPI_ISL_1445546, EPI_ISL_1445547, EPI_ISL_1445548, EPI_ISL_1445550, EPI_ISL_1445551, EPI_ISL_1445552, EPI_ISL_1445554, EPI_ISL_1445555, EPI_ISL_1445556, EPI_ISL_1445557, EPI_ISL_1445558, EPI_ISL_1445559, EPI_ISL_1445560, EPI_ISL_1445561, EPI_ISL_1445562, EPI_ISL_1445565, EPI_ISL_1445566, EPI_ISL_1445568, EPI_ISL_1445569, EPI_ISL_1445573, EPI_ISL_1445584, EPI_ISL_1445593, EPI_ISL_1445595, EPI_ISL_1445598, EPI_ISL_1445605, EPI_ISL_1445608, EPI_ISL_1445609, EPI_ISL_1445610, EPI_ISL_1445613, EPI_ISL_1445615, EPI_ISL_1445616, EPI_ISL_1445620, EPI_ISL_1445622, EPI_ISL_1445627, EPI_ISL_1445651, EPI_ISL_1445652, EPI_ISL_1445653, EPI_ISL_1445654, EPI_ISL_1445655, EPI_ISL_1445656, EPI_ISL_1445657, EPI_ISL_1445658, EPI_ISL_1445659, EPI_ISL_1445660, EPI_ISL_1445671, EPI_ISL_1445673, EPI_ISL_1445675, EPI_ISL_1445685, EPI_ISL_1445690, EPI_ISL_1445691, EPI_ISL_1445694, EPI_ISL_1445695, EPI_ISL_1445703, EPI_ISL_1445709, EPI_ISL_1445720, EPI_ISL_1445725, EPI_ISL_1445731, EPI_ISL_1445735, EPI_ISL_1445739, EPI_ISL_1445754, EPI_ISL_1445764, EPI_ISL_1445769, EPI_ISL_1445774, EPI_ISL_1445775, EPI_ISL_1445776, EPI_ISL_1445789, EPI_ISL_1445790, EPI_ISL_1445791, EPI_ISL_1445796, EPI_ISL_1445797, EPI_ISL_1445798, EPI_ISL_1445799, EPI_ISL_1445800, EPI_ISL_1445801, EPI_ISL_1445802, EPI_ISL_1445803, EPI_ISL_1445805, EPI_ISL_1445806, EPI_ISL_1445809, EPI_ISL_1445815, EPI_ISL_1445816, EPI_ISL_1445817, EPI_ISL_1445818, EPI_ISL_1445819, EPI_ISL_1445820, EPI_ISL_1445821, EPI_ISL_1445835, EPI_ISL_1445843, EPI_ISL_1445845, EPI_ISL_1445858, EPI_ISL_1445871, EPI_ISL_1445904, EPI_ISL_1445913, EPI_ISL_1445931, EPI_ISL_1445932, EPI_ISL_1445940, EPI_ISL_1445941, EPI_ISL_1445942, EPI_ISL_1445950, EPI_ISL_1445959, EPI_ISL_1446032, EPI_ISL_1446035, EPI_ISL_1446066, EPI_ISL_1446083, EPI_ISL_1446084, EPI_ISL_1446090, EPI_ISL_1446091, EPI_ISL_1446092, EPI_ISL_1446096, EPI_ISL_1446097, EPI_ISL_1446110, EPI_ISL_1446116, EPI_ISL_1446118, EPI_ISL_1446122, EPI_ISL_1446123, EPI_ISL_1446132, EPI_ISL_1446147, EPI_ISL_1446162, EPI_ISL_1446179, EPI_ISL_1446181, EPI_ISL_1446192, EPI_ISL_1479498, EPI_ISL_1479512, EPI_ISL_1479518, EPI_ISL_1479587, EPI_ISL_1479590, EPI_ISL_1479591, EPI_ISL_1479608, EPI_ISL_1479613, EPI_ISL_1479656, EPI_ISL_1479805, EPI_ISL_1479825, EPI_ISL_1479828, EPI_ISL_1479830, EPI_ISL_1479834, EPI_ISL_1479842, EPI_ISL_1491734, EPI_ISL_1491744, EPI_ISL_1491786, EPI_ISL_1491787, EPI_ISL_1491788, EPI_ISL_1491814, EPI_ISL_1491817, EPI_ISL_1491829, EPI_ISL_1491842, EPI_ISL_1491878, EPI_ISL_1491884, EPI_ISL_1491885, EPI_ISL_1491888, EPI_ISL_1491898, EPI_ISL_1491899, EPI_ISL_1491917, EPI_ISL_1491918, EPI_ISL_1491929, EPI_ISL_1491939, EPI_ISL_1491940, EPI_ISL_1493502, EPI_ISL_1493503, EPI_ISL_1493535, EPI_ISL_1493550, EPI_ISL_1493570, EPI_ISL_1493571, EPI_ISL_1513139, EPI_ISL_1513152, EPI_ISL_1513153, EPI_ISL_1513160, EPI_ISL_1513172, EPI_ISL_1513183, EPI_ISL_1513185, EPI_ISL_1513186, EPI_ISL_1513190, EPI_ISL_1513199, EPI_ISL_1513200, EPI_ISL_1513220, EPI_ISL_1513222, EPI_ISL_1513236, EPI_ISL_1513239, EPI_ISL_1513240, EPI_ISL_1513242, EPI_ISL_1513262, EPI_ISL_1513263, EPI_ISL_1513294, EPI_ISL_1513296, EPI_ISL_1513297, EPI_ISL_1513301, EPI_ISL_1513303, EPI_ISL_1513306, EPI_ISL_1513307, EPI_ISL_1513309, EPI_ISL_1513323, EPI_ISL_1513324, EPI_ISL_1513325, EPI_ISL_1513352, EPI_ISL_1513359, EPI_ISL_1513360, EPI_ISL_1513361, EPI_ISL_1513362, EPI_ISL_1513386, EPI_ISL_1513387, EPI_ISL_1513410, EPI_ISL_1513487, EPI_ISL_1513488, EPI_ISL_1513489, EPI_ISL_1513513, EPI_ISL_1513518, EPI_ISL_1513524, EPI_ISL_1513541, EPI_ISL_1513542, EPI_ISL_1513548, EPI_ISL_1513570, EPI_ISL_1513583, EPI_ISL_1513622, EPI_ISL_1513625, EPI_ISL_1513627, EPI_ISL_1525033, EPI_ISL_1525034, EPI_ISL_1525035, EPI_ISL_1525040, EPI_ISL_1525041, EPI_ISL_1525042, EPI_ISL_1525048, EPI_ISL_1525049, EPI_ISL_1525066, EPI_ISL_1525075, EPI_ISL_1525123, EPI_ISL_1525146, EPI_ISL_1525147, EPI_ISL_1525152, EPI_ISL_1525153, EPI_ISL_1525155, EPI_ISL_1525161, EPI_ISL_1525162, EPI_ISL_1525163, EPI_ISL_1525176, EPI_ISL_1525186, EPI_ISL_1525187, EPI_ISL_1525220, EPI_ISL_1550455, EPI_ISL_1550465, EPI_ISL_1550466, EPI_ISL_1550492, EPI_ISL_1550516, EPI_ISL_1550519, EPI_ISL_1550521, EPI_ISL_1550522, EPI_ISL_1550550, EPI_ISL_1550565, EPI_ISL_1550669, EPI_ISL_1550670, EPI_ISL_1550690, EPI_ISL_1550739, EPI_ISL_1550742, EPI_ISL_1550743, EPI_ISL_1550794, EPI_ISL_1550845, EPI_ISL_1550972, EPI_ISL_1550975, EPI_ISL_1550976, EPI_ISL_1550982, EPI_ISL_1550984, EPI_ISL_1551127, EPI_ISL_1551128, EPI_ISL_1551220, EPI_ISL_1551266, EPI_ISL_1551393, EPI_ISL_1551422, EPI_ISL_1551543, EPI_ISL_1551556, EPI_ISL_1551585, EPI_ISL_1551636, EPI_ISL_1559656, EPI_ISL_1559657, EPI_ISL_1559658, EPI_ISL_1559659, EPI_ISL_1559660, EPI_ISL_1559661, EPI_ISL_1559662, EPI_ISL_1559663, EPI_ISL_1559664, EPI_ISL_1559665, EPI_ISL_1559667, EPI_ISL_1559668, EPI_ISL_1559669, EPI_ISL_1559670, EPI_ISL_1559672, EPI_ISL_1559740, EPI_ISL_1559741, EPI_ISL_1559758, EPI_ISL_1559759, EPI_ISL_1559760, EPI_ISL_1559761, EPI_ISL_1559806, EPI_ISL_1559807, EPI_ISL_1559808, EPI_ISL_1559809, EPI_ISL_1559828, EPI_ISL_1559829, EPI_ISL_1559847, EPI_ISL_1559877, EPI_ISL_1559880, EPI_ISL_1559881, EPI_ISL_1559883, EPI_ISL_1559887, EPI_ISL_1559903, EPI_ISL_1560009, EPI_ISL_1560010, EPI_ISL_1560011, EPI_ISL_1560018, EPI_ISL_1560037, EPI_ISL_1560106, EPI_ISL_1560123, EPI_ISL_1560137, EPI_ISL_1560142, EPI_ISL_1560150, EPI_ISL_1560177, EPI_ISL_1560179, EPI_ISL_1560255, EPI_ISL_1560350, EPI_ISL_1560382, EPI_ISL_1560383, EPI_ISL_1560491, EPI_ISL_1560496, EPI_ISL_1560519, EPI_ISL_1560527, EPI_ISL_1560603, EPI_ISL_1560606, EPI_ISL_1560644, EPI_ISL_1560645, EPI_ISL_1560672, EPI_ISL_1560679, EPI_ISL_1560681, EPI_ISL_1560702, EPI_ISL_1560707, EPI_ISL_1560717, EPI_ISL_1560720, EPI_ISL_1560726, EPI_ISL_1560740, EPI_ISL_1560787, EPI_ISL_1560788, EPI_ISL_1560942, EPI_ISL_1560949, EPI_ISL_1561002, EPI_ISL_1561020, EPI_ISL_1561022, EPI_ISL_1561030, EPI_ISL_1561043, EPI_ISL_1561202, EPI_ISL_1561222, EPI_ISL_1561223, EPI_ISL_1561224, EPI_ISL_1561225, EPI_ISL_1561226, EPI_ISL_1561237, EPI_ISL_1561248, EPI_ISL_1561249, EPI_ISL_1561260, EPI_ISL_1561289, EPI_ISL_1561298, EPI_ISL_1561328, EPI_ISL_1561330, EPI_ISL_1561335, EPI_ISL_1561342, EPI_ISL_1561345, EPI_ISL_1561366, EPI_ISL_1561374, EPI_ISL_1561400, EPI_ISL_1561404, EPI_ISL_1561449, EPI_ISL_1561496, EPI_ISL_1561525, EPI_ISL_1561541, EPI_ISL_1561551, EPI_ISL_1561553, EPI_ISL_1561563, EPI_ISL_1561580, EPI_ISL_1561588, EPI_ISL_1561595, EPI_ISL_1561596, EPI_ISL_1561614, EPI_ISL_1561615, EPI_ISL_1561622, EPI_ISL_1561637, EPI_ISL_1561638, EPI_ISL_1561647, EPI_ISL_1561682, EPI_ISL_1561693, EPI_ISL_1561717, EPI_ISL_1561719, EPI_ISL_1561788, EPI_ISL_1561794, EPI_ISL_1561802, EPI_ISL_1561803, EPI_ISL_1561804, EPI_ISL_1561805, EPI_ISL_1561806, EPI_ISL_1561809, EPI_ISL_1561810, EPI_ISL_1561811, EPI_ISL_1561817, EPI_ISL_1561818, EPI_ISL_1561819, EPI_ISL_1561828, EPI_ISL_1561866, EPI_ISL_1561870, EPI_ISL_1561891, EPI_ISL_1561893, EPI_ISL_1561902, EPI_ISL_1561904, EPI_ISL_1561907, EPI_ISL_1561918, EPI_ISL_1561926, EPI_ISL_1561927, EPI_ISL_1561975, EPI_ISL_1561976, EPI_ISL_1561983, EPI_ISL_1561987, EPI_ISL_1561998, EPI_ISL_1562015, EPI_ISL_1562017, EPI_ISL_1562018, EPI_ISL_1562025, EPI_ISL_1562043, EPI_ISL_1562046, EPI_ISL_1562054, EPI_ISL_1562073, EPI_ISL_1562096, EPI_ISL_1562102, EPI_ISL_1562103, EPI_ISL_1562104, EPI_ISL_1562145, EPI_ISL_1562146, EPI_ISL_1562152, EPI_ISL_1562156, EPI_ISL_1562181, EPI_ISL_1562182, EPI_ISL_1562183, EPI_ISL_1562184, EPI_ISL_1562198, EPI_ISL_1562219, EPI_ISL_1562225, EPI_ISL_1562254, EPI_ISL_1562255, EPI_ISL_1562267, EPI_ISL_1562268, EPI_ISL_1562343, EPI_ISL_1562352, EPI_ISL_1562353, EPI_ISL_1562364, EPI_ISL_1562377, EPI_ISL_1562378, EPI_ISL_1562380, EPI_ISL_1562381, EPI_ISL_1562384, EPI_ISL_1562387, EPI_ISL_1562391, EPI_ISL_1562392, EPI_ISL_1562400, EPI_ISL_1562411, EPI_ISL_1562412, EPI_ISL_1562416, EPI_ISL_1562418, EPI_ISL_1562426, EPI_ISL_1562450, EPI_ISL_1562451, EPI_ISL_1562454, EPI_ISL_1562456, EPI_ISL_1562503, EPI_ISL_1562506, EPI_ISL_1562554, EPI_ISL_1562567, EPI_ISL_1562601, EPI_ISL_1562631, EPI_ISL_1562791, EPI_ISL_1562812, EPI_ISL_1562820, EPI_ISL_1562823, EPI_ISL_1562827, EPI_ISL_1562831, EPI_ISL_1562836, EPI_ISL_1562848, EPI_ISL_1562945, EPI_ISL_1562950, EPI_ISL_1562987, EPI_ISL_1562990, EPI_ISL_1563006, EPI_ISL_1563044, EPI_ISL_1563048, EPI_ISL_1563055, EPI_ISL_1563071, EPI_ISL_1563072, EPI_ISL_1563128, EPI_ISL_1563132, EPI_ISL_1563154, EPI_ISL_1563224, EPI_ISL_1563225, EPI_ISL_1563226, EPI_ISL_1563229, EPI_ISL_1563239, EPI_ISL_1563281, EPI_ISL_1563329, EPI_ISL_1563350, EPI_ISL_1563353, EPI_ISL_1563398, EPI_ISL_1563409, EPI_ISL_1563410, EPI_ISL_1563411, EPI_ISL_1563417, EPI_ISL_1563418, EPI_ISL_1563454, EPI_ISL_1563549, EPI_ISL_1563550, EPI_ISL_1563558, EPI_ISL_1563573, EPI_ISL_1563607, EPI_ISL_1563608, EPI_ISL_1648968, EPI_ISL_1649028, EPI_ISL_1649029, EPI_ISL_1649030, EPI_ISL_1649031, EPI_ISL_1649048, EPI_ISL_1649126, EPI_ISL_1649150, EPI_ISL_1649159, EPI_ISL_1649210, EPI_ISL_1649211, EPI_ISL_1649263, EPI_ISL_1649264, EPI_ISL_1649336, EPI_ISL_1649381, EPI_ISL_1649382, EPI_ISL_1649414, EPI_ISL_1649447, EPI_ISL_1649448, EPI_ISL_1649471, EPI_ISL_1649525, EPI_ISL_1649546, EPI_ISL_1649547, EPI_ISL_1649548, EPI_ISL_1649550, EPI_ISL_1649556, EPI_ISL_1649566, EPI_ISL_1649598, EPI_ISL_1649599, EPI_ISL_1649622, EPI_ISL_1649652, EPI_ISL_1649694, EPI_ISL_1649732, EPI_ISL_1649734, EPI_ISL_1649737, EPI_ISL_1649738, EPI_ISL_1649757, EPI_ISL_1649764, EPI_ISL_1649766, EPI_ISL_1649767, EPI_ISL_1649769, EPI_ISL_1649777, EPI_ISL_1649787, EPI_ISL_1649796, EPI_ISL_1649804, EPI_ISL_1649858, EPI_ISL_1649894, EPI_ISL_1649897, EPI_ISL_1649907, EPI_ISL_1649944, EPI_ISL_1650002, EPI_ISL_1650041, EPI_ISL_1650088, EPI_ISL_1650205, EPI_ISL_1650271, EPI_ISL_1650278, EPI_ISL_1650292, EPI_ISL_1650409, EPI_ISL_1650412, EPI_ISL_1650472, EPI_ISL_1650534, EPI_ISL_1650538, EPI_ISL_1650632, EPI_ISL_1650638, EPI_ISL_1650659, EPI_ISL_1650672, EPI_ISL_1650693, EPI_ISL_1650695, EPI_ISL_1650703, EPI_ISL_1650729, EPI_ISL_1650730, EPI_ISL_1650944, EPI_ISL_1650954, EPI_ISL_1650966, EPI_ISL_1650967, EPI_ISL_1651036, EPI_ISL_1651045, EPI_ISL_1651061, EPI_ISL_1651075, EPI_ISL_1651094, EPI_ISL_1651099, EPI_ISL_1651133, EPI_ISL_1651152, EPI_ISL_1651169, EPI_ISL_1651180, EPI_ISL_1651182, EPI_ISL_1685472, EPI_ISL_1685852, EPI_ISL_1686708, EPI_ISL_1687815, EPI_ISL_1695152, EPI_ISL_1695225, EPI_ISL_1695275, EPI_ISL_1695302, EPI_ISL_1695422, EPI_ISL_1695448, EPI_ISL_1925479 | | | |
| see above | Aegis Sciences Corporation | Centers for Disease Control and Prevention Division of Viral Diseases, Pathogen Discovery | Adrian Paskey; Alec Vest; Benjamin Rambo-Martin; Christopher Gulvick; Clinton R. Paden; Cyndi Clark; Dakota Howard; Darlene Wagner; Dhwani Batra; Dillon Nall; Duncan MacCannell; Ethan Sanders; Holly Houdeshell; Jason Caravas; Kara Moser; Matthew Hardison; Matthew Schmerer; Ola Kvalvaag; Patrick Campbell; Peter W. Cook; Rob Case; Scott Sammons; Shatavia Morrison; Shaun Westlund; Vikramsinha Ghorpade; Yvette Unoarumhi |
| EPI_ISL_1205474, EPI_ISL_1205483, EPI_ISL_1205484, EPI_ISL_1575114, EPI_ISL_1575115, EPI_ISL_1575116, EPI_ISL_1575117, EPI_ISL_1575118, EPI_ISL_1575119, EPI_ISL_1575120, EPI_ISL_1575121, EPI_ISL_1575122, EPI_ISL_1575123, EPI_ISL_1575124, EPI_ISL_1575125, EPI_ISL_1575126, EPI_ISL_1576991, EPI_ISL_1577000, EPI_ISL_1577001, EPI_ISL_1577002 | | | |
| see above | Alameda County Public Health Department | Alameda County Public Health Department | Kevin Libuit; Kristna Hsieh; Maria Elma |
| EPI_ISL_1222640 | Altius Institute | Seattle Flu Study | Alex Nguyen; Amanda Adler; Andrew Meuser; Barry R. Lutz; Benjamin Pelle; Caitlin R. Wolf; Chris D. Frazar; Clem Green; Daniel Bates; Deborah A. Nickerson; Elisabeth Brandstetter; Erica Ryke; Hannah Petersen; Helen Y. Chu; Jacob Rodriguez; Janet A. Englund; Jay Shendure; Jessica Halow; John Stamatoyannopoulos; Joshua Richards; Jover Lee; Julia Wald; Kairsten Fay; Kirsten Lacombe; Kneshay Harper; Lea M. Starita; Mark J. Rieder; Matt Hartman; Matthew Richardson; Matthew Thompson; Melissa Truong; Michael Boeckh; Michael Famulare; Misja Ilcisin; Muhammad Halimun; Olivia Waltner; Peter D. Han; Rebecca Bruders; Ryan Alexander; Sadie Patraw; Sofia Olsson; Stephanie DeBaun; Thomas R. Sibley; Tobias Ragoczy; Trevor Bedford; Truong Nguyen |
| EPI_ISL_1853453, EPI_ISL_1853470, EPI_ISL_1853565 | Avellino Labs | Avellino Labs | Milos Pjanic |
| EPI_ISL_1477969, EPI_ISL_1478137, EPI_ISL_1478201, EPI_ISL_1529118, EPI_ISL_1529132, EPI_ISL_1529136, EPI_ISL_1529141, EPI_ISL_1529142, EPI_ISL_1529156, EPI_ISL_1529169, EPI_ISL_1529175, EPI_ISL_1529180, EPI_ISL_1529184, EPI_ISL_1529220, EPI_ISL_1529260, EPI_ISL_1529278, EPI_ISL_1529289, EPI_ISL_1529303, EPI_ISL_1529307, EPI_ISL_1529315, EPI_ISL_1529338, EPI_ISL_1529339, EPI_ISL_1529356, EPI_ISL_1701042, EPI_ISL_1701043, EPI_ISL_1701044, EPI_ISL_1701045, EPI_ISL_1701047, EPI_ISL_1701048, EPI_ISL_1701049, EPI_ISL_1701051, EPI_ISL_1701053, EPI_ISL_1701054, EPI_ISL_1701055, EPI_ISL_1701056, EPI_ISL_1701058, EPI_ISL_1701059, EPI_ISL_1701060, EPI_ISL_1701062, EPI_ISL_1701063, EPI_ISL_1701064, EPI_ISL_1701067, EPI_ISL_1701068, EPI_ISL_1701070, EPI_ISL_1701075, EPI_ISL_1701076, EPI_ISL_1701077, EPI_ISL_1701085, EPI_ISL_1701089, EPI_ISL_1701094, EPI_ISL_1701095, EPI_ISL_1701097, EPI_ISL_1701098, EPI_ISL_1701100, EPI_ISL_1701101, EPI_ISL_1701102, EPI_ISL_1701105, EPI_ISL_1701107, EPI_ISL_1701108, EPI_ISL_1701109, EPI_ISL_1701110, EPI_ISL_1701111, EPI_ISL_1701112, EPI_ISL_1701113, EPI_ISL_1701114, EPI_ISL_1701116, EPI_ISL_1701118, EPI_ISL_1701121, EPI_ISL_1701122, EPI_ISL_1701125, EPI_ISL_1701126, EPI_ISL_1701128, EPI_ISL_1701129, EPI_ISL_1701130, EPI_ISL_1701131, EPI_ISL_1701133, EPI_ISL_1701134, EPI_ISL_1701138, EPI_ISL_1701139, EPI_ISL_1701140, EPI_ISL_1701141, EPI_ISL_1701143, EPI_ISL_1701144, EPI_ISL_1701145, EPI_ISL_1701146, EPI_ISL_1701149, EPI_ISL_1701151, EPI_ISL_1701152, EPI_ISL_1701153, EPI_ISL_1701154, EPI_ISL_1701158, EPI_ISL_1701159, EPI_ISL_1701160, EPI_ISL_1701163, EPI_ISL_1701164, EPI_ISL_1701165, EPI_ISL_1701166, EPI_ISL_1701169, EPI_ISL_1701170, EPI_ISL_1701173, EPI_ISL_1701181, EPI_ISL_1701182, EPI_ISL_1701183, EPI_ISL_1701184, EPI_ISL_1701185, EPI_ISL_1701186, EPI_ISL_1701187, EPI_ISL_1701190, EPI_ISL_1701191, EPI_ISL_1701192, EPI_ISL_1701193, EPI_ISL_1701195, EPI_ISL_1701196, EPI_ISL_1701197, EPI_ISL_1701198, EPI_ISL_1701199, EPI_ISL_1701200, EPI_ISL_1701202, EPI_ISL_1701203, EPI_ISL_1701206, EPI_ISL_1701207, EPI_ISL_1701209, EPI_ISL_1701215, EPI_ISL_1701216, EPI_ISL_1701220, EPI_ISL_1701221, EPI_ISL_1701223, EPI_ISL_1701225, EPI_ISL_1701226, EPI_ISL_1701227, EPI_ISL_1701229, EPI_ISL_1701231, EPI_ISL_1701232, EPI_ISL_1701237, EPI_ISL_1701238, EPI_ISL_1701239, EPI_ISL_1701240, EPI_ISL_1701241, EPI_ISL_1701242, EPI_ISL_1701243, EPI_ISL_1701244, EPI_ISL_1701246, EPI_ISL_1701248, EPI_ISL_1701252, EPI_ISL_1701253, EPI_ISL_1701254, EPI_ISL_1701255, EPI_ISL_1701256, EPI_ISL_1701257, EPI_ISL_1701258, EPI_ISL_1701259, EPI_ISL_1701260, EPI_ISL_1701261, EPI_ISL_1701265, EPI_ISL_1701269, EPI_ISL_1701270, EPI_ISL_1701271, EPI_ISL_1701272, EPI_ISL_1701275, EPI_ISL_1701276, EPI_ISL_1701278, EPI_ISL_1701279, EPI_ISL_1701282, EPI_ISL_1701283, EPI_ISL_1701284, EPI_ISL_1701286, EPI_ISL_1701287, EPI_ISL_1701288, EPI_ISL_1701292, EPI_ISL_1701294, EPI_ISL_1701296, EPI_ISL_1701298, EPI_ISL_1701300, EPI_ISL_1701304, EPI_ISL_1701305, EPI_ISL_1701308, EPI_ISL_1701309, EPI_ISL_1701310, EPI_ISL_1821315, EPI_ISL_1821340, EPI_ISL_1821345, EPI_ISL_1821356, EPI_ISL_1821357, EPI_ISL_1821360, EPI_ISL_1821361, EPI_ISL_1821364, EPI_ISL_1821369, EPI_ISL_1821379, EPI_ISL_1821382, EPI_ISL_1821403, EPI_ISL_1821410, EPI_ISL_1821412, EPI_ISL_1821424, EPI_ISL_1821440, EPI_ISL_1821442, EPI_ISL_1821466, EPI_ISL_2102525, EPI_ISL_2102527, EPI_ISL_2102539, EPI_ISL_2102540, EPI_ISL_2102542, EPI_ISL_2102546, EPI_ISL_2102551, EPI_ISL_2102552, EPI_ISL_2102562, EPI_ISL_2102564, EPI_ISL_2102566, EPI_ISL_2102568, EPI_ISL_2102570, EPI_ISL_2102572, EPI_ISL_2102573, EPI_ISL_2102575, EPI_ISL_2102580, EPI_ISL_2102584, EPI_ISL_2102586, EPI_ISL_2102587, EPI_ISL_2102592, EPI_ISL_2102595, EPI_ISL_2102597, EPI_ISL_2102741, EPI_ISL_2102743, EPI_ISL_2102750, EPI_ISL_2102754, EPI_ISL_2102756, EPI_ISL_2102765, EPI_ISL_2102771, EPI_ISL_2102773, EPI_ISL_2102775, EPI_ISL_2102777, EPI_ISL_2102778, EPI_ISL_2102780, EPI_ISL_2102783, EPI_ISL_2102786, EPI_ISL_2102787, EPI_ISL_2102789, EPI_ISL_2102793, EPI_ISL_2102794, EPI_ISL_2102799, EPI_ISL_2102807, EPI_ISL_2102816, EPI_ISL_2102818, EPI_ISL_2102819, EPI_ISL_2102829, EPI_ISL_2102834, EPI_ISL_2102835, EPI_ISL_2102839, EPI_ISL_2102846, EPI_ISL_2102854, EPI_ISL_2102857, EPI_ISL_2102860, EPI_ISL_2102864, EPI_ISL_2102867, EPI_ISL_2102873, EPI_ISL_2102874, EPI_ISL_2102877, EPI_ISL_2102879, EPI_ISL_2102881, EPI_ISL_2102884, EPI_ISL_2102897, EPI_ISL_2102900, EPI_ISL_2102910, EPI_ISL_2102915, EPI_ISL_2102921, EPI_ISL_2102924, EPI_ISL_2102928, EPI_ISL_2102929, EPI_ISL_2102934, EPI_ISL_2102940, EPI_ISL_2102942, EPI_ISL_2102944, EPI_ISL_2102957, EPI_ISL_2102962, EPI_ISL_2102965, EPI_ISL_2102968, EPI_ISL_2102974, EPI_ISL_2102977, EPI_ISL_2102980, EPI_ISL_2102983, EPI_ISL_2102984, EPI_ISL_2102987, EPI_ISL_2102989, EPI_ISL_2102991, EPI_ISL_2102993, EPI_ISL_2103000, EPI_ISL_2103004, EPI_ISL_2103015, EPI_ISL_2103027, EPI_ISL_2103047, EPI_ISL_2103050, EPI_ISL_2103053, EPI_ISL_2103066, EPI_ISL_2103075, EPI_ISL_2103095, EPI_ISL_2103098, EPI_ISL_2103103, EPI_ISL_2103116, EPI_ISL_2103118, EPI_ISL_2103120, EPI_ISL_2103124, EPI_ISL_2103135, EPI_ISL_2103139, EPI_ISL_2103142, EPI_ISL_2103155, EPI_ISL_2103160, EPI_ISL_2103163, EPI_ISL_2103166, EPI_ISL_2103180, EPI_ISL_2103181, EPI_ISL_2103193 | | | |
| see above | CA DPH Viral and Rickettsial Disease Laboratory | Chan-Zuckerberg Biohub | CZB Cliahub Consortium |
| EPI_ISL_1272878, EPI_ISL_1272888, EPI_ISL_1272891, EPI_ISL_1318377, EPI_ISL_1318378, EPI_ISL_1318379, EPI_ISL_1318380, EPI_ISL_1446714, EPI_ISL_1446715, EPI_ISL_1447050, EPI_ISL_1447051, EPI_ISL_1516361, EPI_ISL_1678085, EPI_ISL_1678086, EPI_ISL_1678088, EPI_ISL_1678089, EPI_ISL_1678090, EPI_ISL_1678091, EPI_ISL_1678092, EPI_ISL_1678276, EPI_ISL_1678277, EPI_ISL_1678278, EPI_ISL_1678279, EPI_ISL_1678280, EPI_ISL_1678281, EPI_ISL_1678283, EPI_ISL_1678284, EPI_ISL_1678285, EPI_ISL_1678286, EPI_ISL_1678287, EPI_ISL_1798830, EPI_ISL_1798832, EPI_ISL_1798836, EPI_ISL_1798839, EPI_ISL_1798843, EPI_ISL_1798844, EPI_ISL_1798846, EPI_ISL_1798848, EPI_ISL_1798850, EPI_ISL_1798858, EPI_ISL_1798862, EPI_ISL_1798864, EPI_ISL_1798866, EPI_ISL_1823714, EPI_ISL_1823721, EPI_ISL_1823723, EPI_ISL_1823731, EPI_ISL_1823735, EPI_ISL_1823740, EPI_ISL_1823742, EPI_ISL_1823746 | | | |
| see above | CA-Los Angeles County Public Health Laboratory | Centers for Disease Control and Prevention Division of Viral Diseases, Pathogen Discovery | Alison Laufer Halpin; Anna Montmayeur; Anna Uehara; Ben L. Rambo-Martin; Clinton R. Paden; Dakota Howard; Darlene Wagner; Dave Wentworth; Dhwani Batra; Haibin Wang; Jasmine Padilla; Jing Zhang; Justin Lee; Katie Dillon; Krista Queen; Kristen Knipe; Kristine Lacek; Lori Rowe; Mark Burroughs; Matthew Schmerer; Mili Sheth; Peter Cook; Peter W. Cook; Rachel Marine; Sam Shepard; Sarah Nobles; Shoshona Le; Suxiang Tong; Vivien Dugan; Yan Li; Ying Tao; Yvette Unoarumhi |
| EPI_ISL_1798789, EPI_ISL_1798797, EPI_ISL_1798799, EPI_ISL_1798801 | CDPH, Viral and Rickettsial Disease Laboratory | Centers for Disease Control and Prevention Division of Viral Diseases, Pathogen Discovery | Alison Laufer Halpin; Ben L. Rambo-Martin; Clinton R. Paden; Dakota Howard; Darlene Wagner; Dave Wentworth; Dhwani Batra; Jasmine Padilla; Justin Lee; Katie Dillon; Krista Queen; Kristen Knipe; Kristine Lacek; Mark Burroughs; Matthew Schmerer; Mili Sheth; Peter Cook; Sam Shepard; Sarah Nobles; Shoshona Le; Suxiang Tong; Vivien Dugan; Yvette Unoarumhi |
| EPI_ISL_1373737, EPI_ISL_1373739, EPI_ISL_1373776, EPI_ISL_1373778, EPI_ISL_1373780, EPI_ISL_1373786, EPI_ISL_1373787, EPI_ISL_1373793, EPI_ISL_1373801, EPI_ISL_1373804, EPI_ISL_1373828 | | | |
| see above | CDPH, Viral and Rickettsial Disease Laboratory | Genomics and Discovery, Respiratory Viruses Branch, Division of Viral Diseases, Centers for Disease Control and Prevention | Anna Kelleher; Anna Uehara; Brian Lynch; Clinton R. Paden; Haibin Wang; Han Jia Justin Ng; Jing Zhang; Krista Queen; Peter Cook; Suxiang Tong; Yan Li; Ying Tao |
| EPI_ISL_1591240, EPI_ISL_1591241, EPI_ISL_1591242, EPI_ISL_1623836, EPI_ISL_1911349, EPI_ISL_1911350, EPI_ISL_1911351, EPI_ISL_1911352, EPI_ISL_1911353, EPI_ISL_1911354, EPI_ISL_1911355, EPI_ISL_1911356, EPI_ISL_1911357, EPI_ISL_1911358, EPI_ISL_1911359, EPI_ISL_1911360, EPI_ISL_1911361, EPI_ISL_1911362, EPI_ISL_1911363, EPI_ISL_1911364, EPI_ISL_1911365, EPI_ISL_1911366, EPI_ISL_1911367, EPI_ISL_1911368 | | | |
| see above | CDPH, Viral and Rickettsial Disease Laboratory | Pathogen Discovery, Respiratory Viruses Branch, Division of Viral Diseases, Centers for Disease Control and Prevention | Adam Retchless; Anna Kelleher; Anna Montmayeur; Anna Uehara; Brian Lynch; Clinton R. Paden; Dhwani Batra; Haibin Wang; Han Jia Justin Ng; Jasmine Padilla; Jing Zhang; Justin Lee; Krista Queen; Mark Burroughs; Mili Sheth; Morgan Davis; Peter Cook; Rachel Marine; Sarah Nobles; Suxiang Tong; Tara Coalter; Yan Li; Ying Tao |

| | | | |
|---|---|---|---|
| EPI_ISL_1701494, EPI_ISL_1701495 | California Institute of Technology | Chan-Zuckerberg Biohub | CZB Cliahub Consortium |
| EPI_ISL_1661979 | Centro de Investigacion Biomedica de Occidente (CIBO) | Unidad de Genomica Avanzada | Alejandro Sanchez-Flores; Alfredo Herrera-Estrella; Alicia Ocana-Mondragon; Angel Gustavo Salas-Lais; Bernardo Martinez-Miguel; Blanca Taboada; Brenda Irasema Maldonado-Meza; Carla Ivon Herrera-Najera; Carlos F. Arias; Celia Boukadida; Celida Duque Molina; Clara Esperanza Santacruz-Tinoco; Concepcion Grajales-Muniz; Consorcio Mexicano de Vigilancia Genomica (CoViGen-Mex). Authors (in alphabetical order): Julio Elias Alvarado-Yaah; Fernando Fontove-Herrera; Francisco Pulido; Gloria Elena Espinosa-Ayala; Gloria Maria Molina-Salinas; Gloria Vazquez; Hector Esteban Paz-Juarez; Hector Montoya-Fuentes; Helen Haydee Fernanda Ramirez-Plascencia; Jose Antonio Enciso-Moreno; Jose Esteban Munoz-Medina; Jose de Jesus Nunez-Contreras; Juan Bautista Chale-Dzul; Luis Alberto Ochoa-Carrera; Margarita Matias-Florentino; Maria Guadalupe Santiago-Mauricio; Maria Guadalupe de Jesus Mireles-Rivera; Nelly Selem-Mojica; Pavel Isa; Ricardo Grande; Santiago Ávila-Rios; Victor Eduardo Garcia-Arias; Victor Hugo Borja-Aburto |
| EPI_ISL_1824350, EPI_ISL_1824352, EPI_ISL_1824357, EPI_ISL_1824358, EPI_ISL_1824359, EPI_ISL_1824360, EPI_ISL_1824361, EPI_ISL_1824362, EPI_ISL_1824364, EPI_ISL_1824367, EPI_ISL_1824368, EPI_ISL_1824369, EPI_ISL_1824370, EPI_ISL_1824371, EPI_ISL_1824372, EPI_ISL_1824373, EPI_ISL_1824374, EPI_ISL_1824375, EPI_ISL_1824376, EPI_ISL_1824377, EPI_ISL_1824378, EPI_ISL_1824379, EPI_ISL_1824380, EPI_ISL_1824381, EPI_ISL_1824382, EPI_ISL_1824383, EPI_ISL_1824384, EPI_ISL_1824385, EPI_ISL_1824386, EPI_ISL_1824387, EPI_ISL_1824388, EPI_ISL_1824389, EPI_ISL_1824390, EPI_ISL_1824391, EPI_ISL_1824392, EPI_ISL_1824393, EPI_ISL_1824394, EPI_ISL_1824395, EPI_ISL_1824396, EPI_ISL_1824397, EPI_ISL_1824398, EPI_ISL_1824399, EPI_ISL_1824400, EPI_ISL_1824401, EPI_ISL_1824402, EPI_ISL_1824403, EPI_ISL_1824404, EPI_ISL_1824405, EPI_ISL_1824406, EPI_ISL_1824407, EPI_ISL_1824408, EPI_ISL_1824413, EPI_ISL_1824416, EPI_ISL_1824417 | | | |
| see above | Chiu Laboratory, University of California, San Francisco | Chiu Laboratory, University of California, San Francisco | Alicia Sotomayor-Gonzalez; Candace Wang; Charles Chiu; Daniel Wong; Venice Servellita; Yueyuan Zhang |
| EPI_ISL_1192363 | City of Hope Comprehensive Cancer Center | Los Angeles County PHL | P. Hemarajata et al. |
| EPI_ISL_1824980, EPI_ISL_1824981, EPI_ISL_1824982, EPI_ISL_1824983, EPI_ISL_1824984, EPI_ISL_1824985, EPI_ISL_1824986, EPI_ISL_1824987, EPI_ISL_1824988, EPI_ISL_1824989, EPI_ISL_1824990, EPI_ISL_1824991, EPI_ISL_1824992, EPI_ISL_1824993, EPI_ISL_1824994, EPI_ISL_1824995, EPI_ISL_1824996, EPI_ISL_1824997, EPI_ISL_1824998, EPI_ISL_1824999, EPI_ISL_1825000, EPI_ISL_1825001, EPI_ISL_1825002, EPI_ISL_1825003, EPI_ISL_1825004, EPI_ISL_1825005, EPI_ISL_1825006, EPI_ISL_1825007, EPI_ISL_1825008, EPI_ISL_1825009, EPI_ISL_1825010, EPI_ISL_1825011, EPI_ISL_1825012, EPI_ISL_1825013, EPI_ISL_1825014, EPI_ISL_1825015, EPI_ISL_1825016, EPI_ISL_1825017, EPI_ISL_1825018, EPI_ISL_1825019, EPI_ISL_1825020, EPI_ISL_1825021, EPI_ISL_1825022, EPI_ISL_1825023, EPI_ISL_1825024, EPI_ISL_1825025, EPI_ISL_1825026, EPI_ISL_1825027, EPI_ISL_1825028, EPI_ISL_1825029, EPI_ISL_1825030, EPI_ISL_1825031, EPI_ISL_1825032, EPI_ISL_1825033, EPI_ISL_1825034, EPI_ISL_1825035, EPI_ISL_1825036, EPI_ISL_1825037, EPI_ISL_1825038, EPI_ISL_1825039, EPI_ISL_1825040, EPI_ISL_1825041, EPI_ISL_1825042, EPI_ISL_1825043, EPI_ISL_1825044, EPI_ISL_1825045, EPI_ISL_1825046, EPI_ISL_1825047, EPI_ISL_1825048, EPI_ISL_1825049, EPI_ISL_1825050, EPI_ISL_1825051, EPI_ISL_1825052, EPI_ISL_1825053, EPI_ISL_1825054, EPI_ISL_1825055, EPI_ISL_1825056, EPI_ISL_1825057, EPI_ISL_1825058, EPI_ISL_1825059, EPI_ISL_1825060, EPI_ISL_1825061, EPI_ISL_1825062, EPI_ISL_1825063, EPI_ISL_1825064, EPI_ISL_1825065, EPI_ISL_1825066, EPI_ISL_1825067, EPI_ISL_1825068, EPI_ISL_1825069, EPI_ISL_1825070, EPI_ISL_1825071, EPI_ISL_1825072, EPI_ISL_1825073, EPI_ISL_1825074, EPI_ISL_1825075, EPI_ISL_1825076, EPI_ISL_1825077, EPI_ISL_1825078 | | | |
| see above | Color Genomics | Chiu Laboratory, University of California, San Francisco | Alicia Sotomayor-Gonzalez; Alicia Zhou; Candace Wang; Charles Chiu; Daniel Wong; Erika Torres; Madyson Wanner; Scott Topper; Venice Servellita; Yueyuan Zhang |
| EPI_ISL_1477676, EPI_ISL_1477679, EPI_ISL_1477685, EPI_ISL_1477689, EPI_ISL_1477698, EPI_ISL_1477704, EPI_ISL_1477713, EPI_ISL_1477714, EPI_ISL_1477724, EPI_ISL_1477739, EPI_ISL_1477744, EPI_ISL_1477748, EPI_ISL_1477761, EPI_ISL_1477765, EPI_ISL_1477770, EPI_ISL_1477782, EPI_ISL_1477789, EPI_ISL_1477795, EPI_ISL_1477806, EPI_ISL_1477815, EPI_ISL_1477816, EPI_ISL_1477843, EPI_ISL_1477864, EPI_ISL_1477872, EPI_ISL_1477876, EPI_ISL_1477898, EPI_ISL_1477911, EPI_ISL_1477916, EPI_ISL_1477931, EPI_ISL_1477944, EPI_ISL_1477947, EPI_ISL_1477962, EPI_ISL_1478117, EPI_ISL_1529122, EPI_ISL_1529123, EPI_ISL_1529124, EPI_ISL_1529131, EPI_ISL_1529133, EPI_ISL_1529134, EPI_ISL_1529135, EPI_ISL_1529155, EPI_ISL_1529176, EPI_ISL_1529179, EPI_ISL_1529185, EPI_ISL_1529190, EPI_ISL_1529202, EPI_ISL_1529209, EPI_ISL_1529235, EPI_ISL_1529247, EPI_ISL_1529251, EPI_ISL_1529300, EPI_ISL_1529308, EPI_ISL_1529318, EPI_ISL_1529322, EPI_ISL_1529323, EPI_ISL_1529327, EPI_ISL_1529345, EPI_ISL_1529358, EPI_ISL_1529364, EPI_ISL_1529367, EPI_ISL_1529368, EPI_ISL_1664020, EPI_ISL_1664070, EPI_ISL_1664086, EPI_ISL_1664091, EPI_ISL_1664093, EPI_ISL_2102751, EPI_ISL_2102824, EPI_ISL_2102843, EPI_ISL_2102866, EPI_ISL_2102945, EPI_ISL_2102946, EPI_ISL_2102982 | | | |
| see above | Contra Costa County Public Health Lab | Chan-Zuckerberg Biohub | CZB Cliahub Consortium |
| EPI_ISL_1805719, EPI_ISL_1805720, EPI_ISL_1805721, EPI_ISL_1805722, EPI_ISL_1805723, EPI_ISL_1805727, EPI_ISL_1805728, EPI_ISL_1805729, EPI_ISL_1910451, EPI_ISL_1910452, EPI_ISL_1910453, EPI_ISL_1910454, EPI_ISL_1910455, EPI_ISL_1910456, EPI_ISL_1910457, EPI_ISL_1910458, EPI_ISL_1910459, EPI_ISL_1910460, EPI_ISL_1910461, EPI_ISL_1910463 | | | |
| see above | County of San Diego Health and Human Services Agency | Pathogen Discovery, Respiratory Viruses Branch, Division of Viral Diseases, Centers for Disease Control and Prevention | Adam Retchless; Anna Kelleher; Anna Montmayeur; Anna Uehara; Brian Lynch; Clinton R. Paden; Dhwani Batra; Haibin Wang; Han Jia Justin Ng; Jasmine Padilla; Jing Zhang; Justin Lee; Krista Queen; Mark Burroughs; Mili Sheth; Morgan Davis; Peter Cook; Rachel Marine; Sarah Nobles; Suxiang Tong; Tara Coalter; Yan Li; Ying Tao |
| EPI_ISL_1477550, EPI_ISL_1477626, EPI_ISL_1477992, EPI_ISL_1478174, EPI_ISL_1529127, EPI_ISL_1529128, EPI_ISL_1529129, EPI_ISL_1529161, EPI_ISL_1529208, EPI_ISL_1529213, EPI_ISL_1529329, EPI_ISL_1529341, EPI_ISL_1529342, EPI_ISL_1701136, EPI_ISL_1701266, EPI_ISL_1701297, EPI_ISL_1821359, EPI_ISL_1821363, EPI_ISL_1821367, EPI_ISL_1821378, EPI_ISL_1821404, EPI_ISL_1821408, EPI_ISL_1821447, EPI_ISL_1821468, EPI_ISL_2102998, EPI_ISL_2103073, EPI_ISL_2103172 | | | |
| see above | County of San Luis Obispo Public Health Laboratory | Chan-Zuckerberg Biohub | CZB Cliahub Consortium |
| EPI_ISL_1503991 | Curative Labs | Los Angeles County PHL | P. Hemarajata et al. |
| EPI_ISL_1318697, EPI_ISL_1318708, EPI_ISL_1318711, EPI_ISL_1318718, EPI_ISL_1318726, EPI_ISL_1318727, EPI_ISL_1318728, EPI_ISL_1318740, EPI_ISL_1318748, EPI_ISL_1318760, EPI_ISL_1318767, EPI_ISL_1318775, EPI_ISL_1318777, EPI_ISL_1318786, EPI_ISL_1318796, EPI_ISL_1318802, EPI_ISL_1318803, EPI_ISL_1318805, EPI_ISL_1318820, EPI_ISL_1318823, EPI_ISL_1318832, EPI_ISL_1318845, EPI_ISL_1318856, EPI_ISL_1318857, EPI_ISL_1318871, EPI_ISL_1318886, EPI_ISL_1318896, EPI_ISL_1318898, EPI_ISL_1318900, EPI_ISL_1318906, EPI_ISL_1318908, EPI_ISL_1318917, EPI_ISL_1318934, EPI_ISL_1318937, EPI_ISL_1318943, EPI_ISL_1318945, EPI_ISL_1318951, EPI_ISL_1318952, EPI_ISL_1318953, EPI_ISL_1318970, EPI_ISL_1318976, EPI_ISL_1318979, EPI_ISL_1349078, EPI_ISL_1349093 | | | |
| see above | DeRisi Lab, University of California, San Francisco | DeRisi Lab, University of California, San Francisco | Amy Kistler; Anthea Mitchell; Carina Marquez; Diane Havlir; Diane Jones; Douglas Black; Eric Chow; Gabriel Chamie; Genay Pilarowski; IDseq Team; Jackie Martinez; James Peng; Jamin Liu; Jon Jacobo; Lucy Li; Luis Rubio; Manu Vanaerschot; Matthew Laurie; Maya Petersen; Patrick Ayscue; Sabrina Mann; Sara Sunshine; Susana Rojas; Susy Rojas; Valerie Tulier-Laiwa; and Joseph DeRisi |
| EPI_ISL_1811653, EPI_ISL_1811662, EPI_ISL_1811669, EPI_ISL_1811685, EPI_ISL_1811689, EPI_ISL_1811693, EPI_ISL_1811697, EPI_ISL_1811702, EPI_ISL_1811706, EPI_ISL_1811729, EPI_ISL_1811732, EPI_ISL_1811740, EPI_ISL_1811763, EPI_ISL_1811769, EPI_ISL_1811771, EPI_ISL_1811807, EPI_ISL_1811817, EPI_ISL_1811825, EPI_ISL_1811828, EPI_ISL_1811831, EPI_ISL_1811836, EPI_ISL_1811847, EPI_ISL_1811853, EPI_ISL_1811858, EPI_ISL_1811914, EPI_ISL_1811924, EPI_ISL_1811973, EPI_ISL_1811980, EPI_ISL_1811994, EPI_ISL_1812015, EPI_ISL_1812029, EPI_ISL_1812079, EPI_ISL_1812114, EPI_ISL_1812160, EPI_ISL_1812171, EPI_ISL_1812229, EPI_ISL_1812312, EPI_ISL_1812317, EPI_ISL_1812324, EPI_ISL_1812354, EPI_ISL_1812365, EPI_ISL_1812385, EPI_ISL_1812406, EPI_ISL_1812417, EPI_ISL_1812441, EPI_ISL_1812497, EPI_ISL_1812540, EPI_ISL_1812542, EPI_ISL_1812552, EPI_ISL_1812562, EPI_ISL_1812564, EPI_ISL_1812571, EPI_ISL_1812576, EPI_ISL_1812587, EPI_ISL_1812592, EPI_ISL_1812603, EPI_ISL_1812612, EPI_ISL_1812655, EPI_ISL_1812670, EPI_ISL_1812675, EPI_ISL_1812689, EPI_ISL_1812695, EPI_ISL_1812722, EPI_ISL_1812756, EPI_ISL_1812760, EPI_ISL_1812763, EPI_ISL_1812765, EPI_ISL_1812781, EPI_ISL_1812792, EPI_ISL_1812817, EPI_ISL_1812828, EPI_ISL_1812841, EPI_ISL_1812842, EPI_ISL_1812852, EPI_ISL_1812858, EPI_ISL_1812861, EPI_ISL_1812874, EPI_ISL_1812877, EPI_ISL_1812878, EPI_ISL_1812879, EPI_ISL_1812882, EPI_ISL_1812897, EPI_ISL_1812960, EPI_ISL_1812970, EPI_ISL_1812974, EPI_ISL_1812978, EPI_ISL_1812985, EPI_ISL_1812990, EPI_ISL_1813008, EPI_ISL_1813009, EPI_ISL_1813038, EPI_ISL_1813043, EPI_ISL_1813045, EPI_ISL_1813051, EPI_ISL_1813070, EPI_ISL_1813075, EPI_ISL_1813081, EPI_ISL_1813101, EPI_ISL_1813104, EPI_ISL_1813119, EPI_ISL_1813126, EPI_ISL_1813127, EPI_ISL_1813130, EPI_ISL_1813144, EPI_ISL_1813149, EPI_ISL_1813151, EPI_ISL_1813153, EPI_ISL_1813157, EPI_ISL_1813158, EPI_ISL_1813162, EPI_ISL_1813166, EPI_ISL_1813192, EPI_ISL_1813198, EPI_ISL_1813211, EPI_ISL_1813217, EPI_ISL_1813223, EPI_ISL_1813236, EPI_ISL_1813237, EPI_ISL_1813238, EPI_ISL_1813241, EPI_ISL_1813246, EPI_ISL_1813248, EPI_ISL_1813255, EPI_ISL_1813257, EPI_ISL_1813261, EPI_ISL_1813264, EPI_ISL_1813290, EPI_ISL_1813296, EPI_ISL_1813321, EPI_ISL_1813326, EPI_ISL_1813333, EPI_ISL_1813336, EPI_ISL_1813341, EPI_ISL_1813349, EPI_ISL_1813356, EPI_ISL_1813364, EPI_ISL_1813367, EPI_ISL_1813371, EPI_ISL_1813379, EPI_ISL_1813385, EPI_ISL_1813387, EPI_ISL_1813391, EPI_ISL_1813397, EPI_ISL_1813417, EPI_ISL_1813419, EPI_ISL_1813428, EPI_ISL_1813432, EPI_ISL_1813440, EPI_ISL_1813445, EPI_ISL_1813471, EPI_ISL_1813476, EPI_ISL_1813477, EPI_ISL_1813478, EPI_ISL_1813480, EPI_ISL_1813482, EPI_ISL_1813485, EPI_ISL_1813490, EPI_ISL_1813493, EPI_ISL_1813498, EPI_ISL_1813503, EPI_ISL_1813521, EPI_ISL_1813527, EPI_ISL_1813528, EPI_ISL_1813530, EPI_ISL_1813531, EPI_ISL_1813533, EPI_ISL_1813542, EPI_ISL_1813543, EPI_ISL_1813547, EPI_ISL_1813551, EPI_ISL_1813555, EPI_ISL_1813558, EPI_ISL_1813561, EPI_ISL_1813579, EPI_ISL_1813581, EPI_ISL_1813582, EPI_ISL_1813591, EPI_ISL_1813594, EPI_ISL_1813601, EPI_ISL_1813604, EPI_ISL_1813606, EPI_ISL_1813617, EPI_ISL_1813618, EPI_ISL_1813628, EPI_ISL_1813634, EPI_ISL_1813636, EPI_ISL_1813648, EPI_ISL_1813655, EPI_ISL_1813662, EPI_ISL_1813666, EPI_ISL_1813668, EPI_ISL_1813671, EPI_ISL_1813673, EPI_ISL_1813678, EPI_ISL_1813680, EPI_ISL_1813681, EPI_ISL_1813693, EPI_ISL_1813701, EPI_ISL_1813703, EPI_ISL_1813705, EPI_ISL_1813706, EPI_ISL_1813707, EPI_ISL_1813714, EPI_ISL_1813722, EPI_ISL_1813725, EPI_ISL_1813726, EPI_ISL_1813734, EPI_ISL_1813735, EPI_ISL_1813750, EPI_ISL_1813751, EPI_ISL_1813756, EPI_ISL_1813760, EPI_ISL_1813761, EPI_ISL_1813771, EPI_ISL_1813778, EPI_ISL_1813779, EPI_ISL_1813786, EPI_ISL_1813787, EPI_ISL_1813790, EPI_ISL_1813792, EPI_ISL_1813795, EPI_ISL_1813802, EPI_ISL_1813814, EPI_ISL_1813818, EPI_ISL_1813821, EPI_ISL_1813822, EPI_ISL_1813827, EPI_ISL_1813829, EPI_ISL_1813837, EPI_ISL_1813840, EPI_ISL_1813842, EPI_ISL_1813845, EPI_ISL_1813849, EPI_ISL_1813850, EPI_ISL_1813855, EPI_ISL_1813857, EPI_ISL_1813859, EPI_ISL_1813863, EPI_ISL_1813864, EPI_ISL_1813865, EPI_ISL_1813866, EPI_ISL_1813870, EPI_ISL_1813877, EPI_ISL_1813878, EPI_ISL_1813884, EPI_ISL_1813891, EPI_ISL_1813893, EPI_ISL_1813896, EPI_ISL_1813899, EPI_ISL_1813901, EPI_ISL_1813904, EPI_ISL_1813905, EPI_ISL_1813906, EPI_ISL_1813912, EPI_ISL_1813921, EPI_ISL_1813922, EPI_ISL_1813924, EPI_ISL_1813943, EPI_ISL_1813945, EPI_ISL_1813948, EPI_ISL_1813952, EPI_ISL_1813956, EPI_ISL_1813957, EPI_ISL_1813973, EPI_ISL_1813974, EPI_ISL_1813976, EPI_ISL_1813982, EPI_ISL_1813984, EPI_ISL_1813986, EPI_ISL_1813987, EPI_ISL_1813994, EPI_ISL_1813997, EPI_ISL_1814003, EPI_ISL_1814009, EPI_ISL_1814014, EPI_ISL_1814019, EPI_ISL_1814036, EPI_ISL_1814043, EPI_ISL_1814046, EPI_ISL_1814053, EPI_ISL_1814060, EPI_ISL_1814063, EPI_ISL_1814065, EPI_ISL_1814066, EPI_ISL_1814071, EPI_ISL_1814076, EPI_ISL_1814082, EPI_ISL_1814089, EPI_ISL_1814098, EPI_ISL_1814101, EPI_ISL_1814117, EPI_ISL_1814121, EPI_ISL_1814122, EPI_ISL_1814123, EPI_ISL_1814130, EPI_ISL_1814131, EPI_ISL_1814135, EPI_ISL_1814140, EPI_ISL_1814145, EPI_ISL_1814147, EPI_ISL_1814150, EPI_ISL_1814153, EPI_ISL_1814154, EPI_ISL_1814170, EPI_ISL_1814197, EPI_ISL_1814199, EPI_ISL_1814207, EPI_ISL_1814210, EPI_ISL_1814212, EPI_ISL_1814214, EPI_ISL_1814218, EPI_ISL_1814226, EPI_ISL_1814227, EPI_ISL_1814234, EPI_ISL_1814252, EPI_ISL_1814259, EPI_ISL_1814260, EPI_ISL_1814261, EPI_ISL_1814269, EPI_ISL_1814276, EPI_ISL_1814278, EPI_ISL_1814279, EPI_ISL_1814287, EPI_ISL_1814297, EPI_ISL_1814298, EPI_ISL_1814301, EPI_ISL_1814306, EPI_ISL_1814310, EPI_ISL_1814312, EPI_ISL_1814313, EPI_ISL_1814314, EPI_ISL_1814315, EPI_ISL_1814318, EPI_ISL_1814320, EPI_ISL_1814324, EPI_ISL_1814325, EPI_ISL_1814326, EPI_ISL_1814329, EPI_ISL_1814330, EPI_ISL_1814333, EPI_ISL_1814334, EPI_ISL_1814335, EPI_ISL_1814336, EPI_ISL_1814338, EPI_ISL_1814341, EPI_ISL_1814345, EPI_ISL_1814346, EPI_ISL_1814353, EPI_ISL_1814354, EPI_ISL_1814357, EPI_ISL_1814359, EPI_ISL_1814360, EPI_ISL_1814361, EPI_ISL_1814364, EPI_ISL_1814378, EPI_ISL_1814381, EPI_ISL_1814385, EPI_ISL_1814386, EPI_ISL_1814392, EPI_ISL_1814397, EPI_ISL_1814398, EPI_ISL_1814401, EPI_ISL_1814402, EPI_ISL_1814414, EPI_ISL_1814415, EPI_ISL_1814427, EPI_ISL_1814434, EPI_ISL_1814439, EPI_ISL_1814443, EPI_ISL_1814446, EPI_ISL_1814449, EPI_ISL_1814457, EPI_ISL_1814461, EPI_ISL_1814462, EPI_ISL_1814463, EPI_ISL_1814465, EPI_ISL_1814483, EPI_ISL_1814484, EPI_ISL_1814488, EPI_ISL_1814493, EPI_ISL_1814494, EPI_ISL_1814498, EPI_ISL_1814504, EPI_ISL_1814515, EPI_ISL_1814519, EPI_ISL_1814522, EPI_ISL_1814524, EPI_ISL_1814525, EPI_ISL_1814527, EPI_ISL_1814536, EPI_ISL_1814537, EPI_ISL_1814539, EPI_ISL_1814540, EPI_ISL_1814541, EPI_ISL_1814545, EPI_ISL_1814547, EPI_ISL_1814549, EPI_ISL_1814552, EPI_ISL_1814553, EPI_ISL_1814556, EPI_ISL_1814557, EPI_ISL_1814558, EPI_ISL_1814561, EPI_ISL_1814569, EPI_ISL_1814570, EPI_ISL_1814572, EPI_ISL_1814574, EPI_ISL_1814582, EPI_ISL_1814583, EPI_ISL_1814598, EPI_ISL_1814600, EPI_ISL_1814602, EPI_ISL_1814605, EPI_ISL_1814612, EPI_ISL_1814623, EPI_ISL_1814625, EPI_ISL_1814628, EPI_ISL_1814630, EPI_ISL_1814631, EPI_ISL_1814632, EPI_ISL_1814634, EPI_ISL_1814635, EPI_ISL_1814642, EPI_ISL_1814652, EPI_ISL_1814655, EPI_ISL_1814656, EPI_ISL_1814658, EPI_ISL_1814662, EPI_ISL_1814669, EPI_ISL_1814670, EPI_ISL_1814673, EPI_ISL_1814674, EPI_ISL_1814682, EPI_ISL_1814683, EPI_ISL_1814684, EPI_ISL_1814686, EPI_ISL_1814687, EPI_ISL_1814697, EPI_ISL_1814704, EPI_ISL_1814710, EPI_ISL_1814715, EPI_ISL_1814717, EPI_ISL_1814718, EPI_ISL_1814720, EPI_ISL_1814721, EPI_ISL_1814732, EPI_ISL_1814733, EPI_ISL_1814736, EPI_ISL_1814739, EPI_ISL_1814742, EPI_ISL_1814743, EPI_ISL_1814745, EPI_ISL_1814746, EPI_ISL_1814747, EPI_ISL_1814751, EPI_ISL_1814757, EPI_ISL_1814762, EPI_ISL_1814763, EPI_ISL_1814765, EPI_ISL_1814771, EPI_ISL_1814773, EPI_ISL_1814775, EPI_ISL_1814781, EPI_ISL_1814783, EPI_ISL_1814788, EPI_ISL_1814792, EPI_ISL_1814795, EPI_ISL_1814798, EPI_ISL_1814806, EPI_ISL_1814812, EPI_ISL_1814814, EPI_ISL_1814816, EPI_ISL_1814825, EPI_ISL_1814826, EPI_ISL_1814830, EPI_ISL_1814833, EPI_ISL_1814834, EPI_ISL_1814836, EPI_ISL_1814839, EPI_ISL_1814840, EPI_ISL_1814844, EPI_ISL_1814845, EPI_ISL_1814846, EPI_ISL_1814847, EPI_ISL_1814848, EPI_ISL_1814850, EPI_ISL_1814851, EPI_ISL_1814856, EPI_ISL_1814859, EPI_ISL_1814861, EPI_ISL_1814868, EPI_ISL_1814872, EPI_ISL_1814877, EPI_ISL_1814878, EPI_ISL_1814879, EPI_ISL_1814881, EPI_ISL_1814883, EPI_ISL_1814891, EPI_ISL_1814892, EPI_ISL_1814893, EPI_ISL_1814894, EPI_ISL_1814895, EPI_ISL_1814896, EPI_ISL_1814905, EPI_ISL_1814906, EPI_ISL_1814914, EPI_ISL_1814917, EPI_ISL_1814918, EPI_ISL_1814924, EPI_ISL_1814926, EPI_ISL_1814930, EPI_ISL_1814939, EPI_ISL_1814942, EPI_ISL_1814950, EPI_ISL_1814954, EPI_ISL_1814957, EPI_ISL_1814962, EPI_ISL_1814971, EPI_ISL_1814981, EPI_ISL_1814984, EPI_ISL_1814986, EPI_ISL_1814989, EPI_ISL_1814998, EPI_ISL_1815000, EPI_ISL_1815002, EPI_ISL_1815008, EPI_ISL_1815017, EPI_ISL_1815018, EPI_ISL_1815026, EPI_ISL_1815031, EPI_ISL_1815035, EPI_ISL_1815040, EPI_ISL_1815048, EPI_ISL_1815060, EPI_ISL_1815062, EPI_ISL_1815063, EPI_ISL_1815070, EPI_ISL_1815074, EPI_ISL_1815075, EPI_ISL_1815084, EPI_ISL_1815088, EPI_ISL_1815092, EPI_ISL_1815103, EPI_ISL_1815108, EPI_ISL_1815109, EPI_ISL_1815110, EPI_ISL_1815112, EPI_ISL_1815114, EPI_ISL_1815115, EPI_ISL_1815118, EPI_ISL_1815121, EPI_ISL_1815126, EPI_ISL_1815135, EPI_ISL_1815139, EPI_ISL_1815140, EPI_ISL_1815142, EPI_ISL_1815147, EPI_ISL_1815151, EPI_ISL_1815155, EPI_ISL_1815157, EPI_ISL_1815158, EPI_ISL_1815160, EPI_ISL_1815161, EPI_ISL_1815162, EPI_ISL_1815164, EPI_ISL_1815165, EPI_ISL_1815166, EPI_ISL_1815169, EPI_ISL_1815175, EPI_ISL_1815178, EPI_ISL_1815180, EPI_ISL_1815183, EPI_ISL_1815191, EPI_ISL_1815197, EPI_ISL_1815198, EPI_ISL_1815200, EPI_ISL_1815201, EPI_ISL_1815203, EPI_ISL_1815206, EPI_ISL_1815207, EPI_ISL_1815212, EPI_ISL_1815213, EPI_ISL_1815216, EPI_ISL_1815220, EPI_ISL_1815230, EPI_ISL_1815231, EPI_ISL_1815235, EPI_ISL_1815237, EPI_ISL_1815251, EPI_ISL_1815253, EPI_ISL_1815255, EPI_ISL_1815257, EPI_ISL_1815264, EPI_ISL_1815267, EPI_ISL_1815270, EPI_ISL_1815273, EPI_ISL_1815274, EPI_ISL_1815278, EPI_ISL_1815283, EPI_ISL_1815287, EPI_ISL_1815288, EPI_ISL_1815290, EPI_ISL_1815293, EPI_ISL_1815296, EPI_ISL_1815298, EPI_ISL_1815301, EPI_ISL_1815305, EPI_ISL_1815307, EPI_ISL_1815309, EPI_ISL_1815315, EPI_ISL_1815318, EPI_ISL_1815320, EPI_ISL_1815326, EPI_ISL_1815329, EPI_ISL_1815333, EPI_ISL_1815335, EPI_ISL_1815336, EPI_ISL_1815337, EPI_ISL_1815340, EPI_ISL_1815341, EPI_ISL_1815346, EPI_ISL_1815350, EPI_ISL_1815354, EPI_ISL_1815357, EPI_ISL_1815359, EPI_ISL_1815363, EPI_ISL_1815368, EPI_ISL_1815371, EPI_ISL_1815373, EPI_ISL_1815375, EPI_ISL_1815378, EPI_ISL_1815383, EPI_ISL_1815385, EPI_ISL_1815386, EPI_ISL_1815387, EPI_ISL_1815388, EPI_ISL_1815390, EPI_ISL_1815391, EPI_ISL_1815392, EPI_ISL_1815393, EPI_ISL_1815396, EPI_ISL_1815399, EPI_ISL_1815402, EPI_ISL_1815404, EPI_ISL_1815405, EPI_ISL_1815415, EPI_ISL_1815420, EPI_ISL_1815422, EPI_ISL_1815429, EPI_ISL_1815435, EPI_ISL_1815441, EPI_ISL_1815446, EPI_ISL_1815449, EPI_ISL_1815450, EPI_ISL_1815451, EPI_ISL_1815452, EPI_ISL_1815453, EPI_ISL_1815454, EPI_ISL_1815455, EPI_ISL_1815457, EPI_ISL_1815463, EPI_ISL_1815464, EPI_ISL_1815466, EPI_ISL_1815471, EPI_ISL_1815473, EPI_ISL_1815474, EPI_ISL_1815475, EPI_ISL_1815486, EPI_ISL_1815491, EPI_ISL_1815493, EPI_ISL_1815496, EPI_ISL_1815501, EPI_ISL_1815502, EPI_ISL_1815506, EPI_ISL_1815507, EPI_ISL_1815509, EPI_ISL_1815513, EPI_ISL_1815514, EPI_ISL_1815521, EPI_ISL_1815526, EPI_ISL_1815533, EPI_ISL_1815534, EPI_ISL_1815535, EPI_ISL_1815544, EPI_ISL_1815546, EPI_ISL_1815548, EPI_ISL_1815550, EPI_ISL_1815554, EPI_ISL_1815559, EPI_ISL_1815564, EPI_ISL_1815573, EPI_ISL_1815574, EPI_ISL_1815575, EPI_ISL_1815576, EPI_ISL_1815577, EPI_ISL_1815578, EPI_ISL_1815579, EPI_ISL_1815582, EPI_ISL_1815584, EPI_ISL_1815588, EPI_ISL_1815590, EPI_ISL_1815603, EPI_ISL_1815604, EPI_ISL_1815605, EPI_ISL_1815606, EPI_ISL_1815607, EPI_ISL_1815616, EPI_ISL_1815617, EPI_ISL_1815618, EPI_ISL_1815619, EPI_ISL_1815622, EPI_ISL_1815623, EPI_ISL_1815624, EPI_ISL_1815631, EPI_ISL_1815634, EPI_ISL_1815635, EPI_ISL_1815638, EPI_ISL_1815639, EPI_ISL_1815641, EPI_ISL_1815647, EPI_ISL_1815653, EPI_ISL_1815654, EPI_ISL_1815656, EPI_ISL_1815657, EPI_ISL_1815660, EPI_ISL_1815663, EPI_ISL_1815664, EPI_ISL_1815665, EPI_ISL_1815667, EPI_ISL_1815669, EPI_ISL_1815670, EPI_ISL_1815671, EPI_ISL_1815672, EPI_ISL_1815673, EPI_ISL_1815681, EPI_ISL_1815683, EPI_ISL_1815691, EPI_ISL_1815698, EPI_ISL_1815700, EPI_ISL_1815702, EPI_ISL_1815705, EPI_ISL_1815706, EPI_ISL_1815708, EPI_ISL_1815712, EPI_ISL_1815713, EPI_ISL_1815714, EPI_ISL_1815720, EPI_ISL_1815725, EPI_ISL_1815727, EPI_ISL_1815728, EPI_ISL_1815735, EPI_ISL_1815741, EPI_ISL_1815743, EPI_ISL_1815744, EPI_ISL_1815747, EPI_ISL_1815749, EPI_ISL_1815751, EPI_ISL_1815753, EPI_ISL_1815754, EPI_ISL_1815757, EPI_ISL_1815759, EPI_ISL_1815760, EPI_ISL_1815761, EPI_ISL_1815762, EPI_ISL_1815765, EPI_ISL_1815768, EPI_ISL_1815770, EPI_ISL_1815774, EPI_ISL_1815775, EPI_ISL_1815776, EPI_ISL_1815778, EPI_ISL_1815781, EPI_ISL_1815782, EPI_ISL_1815784, EPI_ISL_1815785, EPI_ISL_1815786, EPI_ISL_1815788, EPI_ISL_1815792, EPI_ISL_1815794, EPI_ISL_1815806, EPI_ISL_1815808, EPI_ISL_1815809, EPI_ISL_1815812, EPI_ISL_1815813, EPI_ISL_1815818, EPI_ISL_1815821, EPI_ISL_1815823, EPI_ISL_1815825, EPI_ISL_1815832, EPI_ISL_1815833, EPI_ISL_1815838, EPI_ISL_1815847, EPI_ISL_1815848, EPI_ISL_1815849, EPI_ISL_1815851, EPI_ISL_1815852, EPI_ISL_1815855, EPI_ISL_1815856, EPI_ISL_1815857, EPI_ISL_1815861, EPI_ISL_1815863, EPI_ISL_1815868, EPI_ISL_1815869, EPI_ISL_1815870, EPI_ISL_1815873, EPI_ISL_1815875, EPI_ISL_1815878, EPI_ISL_1815879, EPI_ISL_1815887, EPI_ISL_1815890, EPI_ISL_1815893, EPI_ISL_1815897, EPI_ISL_1815901, EPI_ISL_1815902, EPI_ISL_1815904, EPI_ISL_1815905, EPI_ISL_1815910, EPI_ISL_1815914, EPI_ISL_1815915, EPI_ISL_1815921, EPI_ISL_1815924, EPI_ISL_1815932, EPI_ISL_1815937, EPI_ISL_1815938, EPI_ISL_1815942, EPI_ISL_1815946, EPI_ISL_1815948, EPI_ISL_1815951, EPI_ISL_1815957, EPI_ISL_1815959, EPI_ISL_1815963, EPI_ISL_1815970, EPI_ISL_1815972, EPI_ISL_1815973, EPI_ISL_1815976, EPI_ISL_1815980, EPI_ISL_1815985, EPI_ISL_1815988, EPI_ISL_1815991, EPI_ISL_1815993, EPI_ISL_1816001, EPI_ISL_1816002, EPI_ISL_1816003, EPI_ISL_1816010, EPI_ISL_1816016, EPI_ISL_1816019, EPI_ISL_1816020, EPI_ISL_1816022, EPI_ISL_1816023, EPI_ISL_1816043, EPI_ISL_1816046, EPI_ISL_1816047, EPI_ISL_1816051, EPI_ISL_1816053, EPI_ISL_1816054, EPI_ISL_1816055, EPI_ISL_1816058, EPI_ISL_1816060, EPI_ISL_1816062, EPI_ISL_1816064, EPI_ISL_1816065, EPI_ISL_1816070, EPI_ISL_1816071, EPI_ISL_1816075, EPI_ISL_1816084, EPI_ISL_1816085, EPI_ISL_1816087, EPI_ISL_1816090, EPI_ISL_1816091, EPI_ISL_1816092, EPI_ISL_1816093, EPI_ISL_1816095, EPI_ISL_1816096, EPI_ISL_1816098, EPI_ISL_1816099, EPI_ISL_1816100, EPI_ISL_1816101, EPI_ISL_1816104, EPI_ISL_1816105, EPI_ISL_1816106, EPI_ISL_1816109, EPI_ISL_1816112, EPI_ISL_1816113, EPI_ISL_1816114, EPI_ISL_1816116, EPI_ISL_1816117, EPI_ISL_1816121, EPI_ISL_1816122, EPI_ISL_1816126, EPI_ISL_1816133, EPI_ISL_1816136, EPI_ISL_1816141, EPI_ISL_1816143, EPI_ISL_1816144, EPI_ISL_1816146, EPI_ISL_1816150, EPI_ISL_1816154, EPI_ISL_1816162, EPI_ISL_1816163, EPI_ISL_1816166, EPI_ISL_1816169, EPI_ISL_1816170, EPI_ISL_1816174, EPI_ISL_1816176, EPI_ISL_1816180, EPI_ISL_1816183, EPI_ISL_1816186, EPI_ISL_1816192, EPI_ISL_1816196, EPI_ISL_1816202, EPI_ISL_1816203, EPI_ISL_1816204, EPI_ISL_1816205, EPI_ISL_1816206, EPI_ISL_1816209, EPI_ISL_1816210, EPI_ISL_1816211, EPI_ISL_1816212, EPI_ISL_1816213, EPI_ISL_1816214, EPI_ISL_1816222, EPI_ISL_1816226, EPI_ISL_1816228, EPI_ISL_1816234, EPI_ISL_1816239, EPI_ISL_1816241, EPI_ISL_1816245, EPI_ISL_1816246, EPI_ISL_1816247, EPI_ISL_1816253, EPI_ISL_1816256, EPI_ISL_1816258, EPI_ISL_1816260, EPI_ISL_1816261, EPI_ISL_1816263, EPI_ISL_1816266, EPI_ISL_1816267, EPI_ISL_1816269, EPI_ISL_1816271, EPI_ISL_1816274, EPI_ISL_1816275, EPI_ISL_1816278, EPI_ISL_1816280, EPI_ISL_1816284, EPI_ISL_1816285, EPI_ISL_1816293, EPI_ISL_1816299, EPI_ISL_1816300, EPI_ISL_1816302, EPI_ISL_1816307, EPI_ISL_1816308, EPI_ISL_1816309, EPI_ISL_1816311, EPI_ISL_1816312, EPI_ISL_1816314, EPI_ISL_1816316, EPI_ISL_1816319, EPI_ISL_1816320, EPI_ISL_1816322, EPI_ISL_1816324, EPI_ISL_1816326, EPI_ISL_1816331, EPI_ISL_1816335, EPI_ISL_1816339, EPI_ISL_1816346, EPI_ISL_1816348, EPI_ISL_1816349, EPI_ISL_1816351, EPI_ISL_1816354, EPI_ISL_1816355, EPI_ISL_1816356, EPI_ISL_1816361, EPI_ISL_1816366, EPI_ISL_1816369, EPI_ISL_1816371, EPI_ISL_1816373, EPI_ISL_1816375, EPI_ISL_1816377, EPI_ISL_1816381, EPI_ISL_1816384, EPI_ISL_1816385, EPI_ISL_1816388, EPI_ISL_1816391, EPI_ISL_1816392, EPI_ISL_1816394, EPI_ISL_1816398, EPI_ISL_1816399, EPI_ISL_1816400, EPI_ISL_1816401, EPI_ISL_1816402, EPI_ISL_1816409, EPI_ISL_1816411, EPI_ISL_1816414, EPI_ISL_1816415, EPI_ISL_1816420, EPI_ISL_1816428, EPI_ISL_1816433, EPI_ISL_1816436, EPI_ISL_1816440, EPI_ISL_1816444, EPI_ISL_1816447, EPI_ISL_1816450, EPI_ISL_1816454, EPI_ISL_1816458, EPI_ISL_1816459, EPI_ISL_1816461, EPI_ISL_1816465, EPI_ISL_1816467, EPI_ISL_1816469, EPI_ISL_1816470, EPI_ISL_1816472, EPI_ISL_1816473, EPI_ISL_1816484, EPI_ISL_1816487, EPI_ISL_1816493, EPI_ISL_1816498, EPI_ISL_1816500, EPI_ISL_1816503, EPI_ISL_1816505, EPI_ISL_1816508, EPI_ISL_1816511, EPI_ISL_1816512, EPI_ISL_1816515, EPI_ISL_1816516, EPI_ISL_1816519, EPI_ISL_1816521, EPI_ISL_1816522, EPI_ISL_1816523, EPI_ISL_1816525, EPI_ISL_1816526, EPI_ISL_1816530, EPI_ISL_1816533, EPI_ISL_1816535, EPI_ISL_1816537, EPI_ISL_1816540, EPI_ISL_1816541, EPI_ISL_1816542, EPI_ISL_1816548, EPI_ISL_1816551, EPI_ISL_1816552, EPI_ISL_1816553, EPI_ISL_1816559, EPI_ISL_1816560, EPI_ISL_1816567, EPI_ISL_1816568, EPI_ISL_1816569, EPI_ISL_1816574, EPI_ISL_1816584, EPI_ISL_1816587, EPI_ISL_1816594, EPI_ISL_1816598, EPI_ISL_1816603, EPI_ISL_1816608, EPI_ISL_1816609, EPI_ISL_1816610, EPI_ISL_1816611, EPI_ISL_1816613, EPI_ISL_1816615, EPI_ISL_1816616, EPI_ISL_1816619, EPI_ISL_1816624, EPI_ISL_1816629, EPI_ISL_1816632, EPI_ISL_1816634, EPI_ISL_1816636, EPI_ISL_1816637, EPI_ISL_1816639, EPI_ISL_1816640, EPI_ISL_1816644, EPI_ISL_1816648, EPI_ISL_1816650, EPI_ISL_1816652, EPI_ISL_1816665, EPI_ISL_1816666, EPI_ISL_1816669, EPI_ISL_1816671, EPI_ISL_1816675, EPI_ISL_1816678, EPI_ISL_1816680, EPI_ISL_1816681, EPI_ISL_1816687, EPI_ISL_1816694, EPI_ISL_1816695, EPI_ISL_1816699, EPI_ISL_1816700, EPI_ISL_1816704, EPI_ISL_1816708, EPI_ISL_1816716, EPI_ISL_1816718, EPI_ISL_1816719, EPI_ISL_1816722, EPI_ISL_1816723, EPI_ISL_1816725, EPI_ISL_1816727, EPI_ISL_1816733, EPI_ISL_1816736, EPI_ISL_1816737, EPI_ISL_1816741, EPI_ISL_1816748, EPI_ISL_1816752, EPI_ISL_1816754, EPI_ISL_1816756, EPI_ISL_1816759, EPI_ISL_1816767, EPI_ISL_1816770, EPI_ISL_1816772, EPI_ISL_1816773, EPI_ISL_1816774, EPI_ISL_1816780, EPI_ISL_1816783, EPI_ISL_1816785, EPI_ISL_1816786, EPI_ISL_1816787, EPI_ISL_1816793, EPI_ISL_1816794, EPI_ISL_1816795, EPI_ISL_1816796, EPI_ISL_1816797, EPI_ISL_1816799, EPI_ISL_1816802, EPI_ISL_1816805, EPI_ISL_1816807, EPI_ISL_1816813, EPI_ISL_1816818, EPI_ISL_1816819, EPI_ISL_1816824, EPI_ISL_1816827, EPI_ISL_1816829, EPI_ISL_1816835, EPI_ISL_1816838, EPI_ISL_1816839, EPI_ISL_1816840, EPI_ISL_1816841, EPI_ISL_1816842, EPI_ISL_1816843, EPI_ISL_1816848, EPI_ISL_1816849, EPI_ISL_1816855, EPI_ISL_1816861, EPI_ISL_1816864, EPI_ISL_1816867, EPI_ISL_1816875, EPI_ISL_1816879, EPI_ISL_1816881, | | | |

EPI_ISL_1816884, EPI_ISL_1816887, EPI_ISL_1816891, EPI_ISL_1816892, EPI_ISL_1816893, EPI_ISL_1816894, EPI_ISL_1816902, EPI_ISL_1816903, EPI_ISL_1816904, EPI_ISL_1816905, EPI_ISL_1816907, EPI_ISL_1816909, EPI_ISL_1816911, EPI_ISL_1816913, EPI_ISL_1816915

see above | EXCITE Lab | Andersen lab at Scripps Research | Abigail Schnapper; Alexandre Bolze; Alice Summerfield; Angela Scioscia; Araz Majnoonian; Ashkan Hassani; Celena Andrade; Charlotte Rivera-Garcia; Cheryl Anderson; Chip Schooley; Christine Mitchell + SEARCH; David Becker; David Pride; Efren Sandoval; Elizabeth Cirulli; Fitri Carrissa Wijaya; Francisco Tanudjaja; Fred Wu; Geraint Levan; Greg Humphrey; Helena Tubb; James Lu + SEARCH; Jason Nguyen; Jimmy Ramirez; Kelly Schiabor Barrett; Laura Kohn; Magnus Isaksson; Marc Laurent; Marlene Flores; Megan Nguyen; Natasha Martin; Natasha Martin Cheryl Anderson; Nicole L Washington; Patricia Gonzalez-Zuniga; Rebecca Fielding-Miller; Rodolfo Salido; Ryan Cho; Sawyer Farmer; Sharon Reed; Sherry Wang; Simon White; Smruthi Karthikeyan; Tommy Valles + SEARCH; Tyler Cassens; Victor Cantu; Vinton Omaleki; William Lee

EPI_ISL_1483789, EPI_ISL_1483790, EPI_ISL_1483791, EPI_ISL_1483792, EPI_ISL_1483793, EPI_ISL_1483794, EPI_ISL_1483800, EPI_ISL_1483801

see above | El Camino Hospital | Santa Clara County Public Health Laboratory | Santa Clara County Public Health Department

EPI_ISL_1303426, EPI_ISL_1303474, EPI_ISL_1303475, EPI_ISL_1303476, EPI_ISL_1303477, EPI_ISL_1303478, EPI_ISL_1303479, EPI_ISL_1303484, EPI_ISL_1303485, EPI_ISL_1303486, EPI_ISL_1303487, EPI_ISL_1303488, EPI_ISL_1307636, EPI_ISL_1307637, EPI_ISL_1379978, EPI_ISL_1497542, EPI_ISL_1669341, EPI_ISL_1669342, EPI_ISL_1669343, EPI_ISL_1669344, EPI_ISL_1669345, EPI_ISL_1669346, EPI_ISL_1669347, EPI_ISL_1697372, EPI_ISL_1706605, EPI_ISL_1706606, EPI_ISL_1706607, EPI_ISL_1707675, EPI_ISL_1707676

see above | Flow Diagnostics Lab | Los Angeles County PHL | P. Hemarajata et al.

EPI_ISL_1413763, EPI_ISL_1413766, EPI_ISL_1516746, EPI_ISL_1578215, EPI_ISL_1578327, EPI_ISL_1578328, EPI_ISL_1578329, EPI_ISL_1578331, EPI_ISL_1578339, EPI_ISL_1578451

see above | Flow Health | Infectious Disease Program, Broad Institute of Harvard and MIT | Adams, G.; B.L.; B.W.; Bauer, M.; Birren; Carter, A.; Chaluvadi, S.; D.J.; DeRuff, K.; Gallagher, G.; Gladden-Young, A.; J.E.; K.J.; Lagerborg, K.; Lemieux; Loreth, C.; MacInnis; Normandin, E.; P.C.; Park; Reilly, S.; Rudy, M.; Siddle; Smole, S.; Tomkins-Tinch, C.; and Sabeti

EPI_ISL_1457792, EPI_ISL_1457793, EPI_ISL_1457794, EPI_ISL_1457795, EPI_ISL_1457796, EPI_ISL_1457797, EPI_ISL_1457798, EPI_ISL_1457799, EPI_ISL_1457800, EPI_ISL_1457801, EPI_ISL_1457802, EPI_ISL_1457803, EPI_ISL_1457804, EPI_ISL_1457805, EPI_ISL_1457806, EPI_ISL_1457807, EPI_ISL_1457808, EPI_ISL_1457809, EPI_ISL_1457810, EPI_ISL_1457811, EPI_ISL_1457812, EPI_ISL_1457813,
EPI_ISL_1457814, EPI_ISL_1457815, EPI_ISL_1457816, EPI_ISL_1457817, EPI_ISL_1457818, EPI_ISL_1457819, EPI_ISL_1457820, EPI_ISL_1457821, EPI_ISL_1457822, EPI_ISL_1457823, EPI_ISL_1457824, EPI_ISL_1457825, EPI_ISL_1457826, EPI_ISL_1457827, EPI_ISL_1457828, EPI_ISL_1457829, EPI_ISL_1457830, EPI_ISL_1457831, EPI_ISL_1457832, EPI_ISL_1457833, EPI_ISL_1457834, EPI_ISL_1457835,
EPI_ISL_1457836, EPI_ISL_1457837, EPI_ISL_1457838, EPI_ISL_1457839, EPI_ISL_1457840, EPI_ISL_1457841, EPI_ISL_1457842, EPI_ISL_1457843, EPI_ISL_1457844, EPI_ISL_1457845, EPI_ISL_1457846, EPI_ISL_1457847, EPI_ISL_1457848, EPI_ISL_1457849, EPI_ISL_1457850, EPI_ISL_1457851, EPI_ISL_1457852, EPI_ISL_1457853, EPI_ISL_1457854, EPI_ISL_1457855, EPI_ISL_1457856, EPI_ISL_1457857,
EPI_ISL_1457858, EPI_ISL_1457859, EPI_ISL_1457861, EPI_ISL_1457862, EPI_ISL_1457863, EPI_ISL_1457864, EPI_ISL_1457865, EPI_ISL_1457866, EPI_ISL_1457867, EPI_ISL_1457868, EPI_ISL_1457869, EPI_ISL_1457870, EPI_ISL_1457871, EPI_ISL_1457872, EPI_ISL_1457873, EPI_ISL_1457874, EPI_ISL_1457875, EPI_ISL_1457876, EPI_ISL_1457877, EPI_ISL_1457878, EPI_ISL_1457879, EPI_ISL_1457880,
EPI_ISL_1457881, EPI_ISL_1457882, EPI_ISL_1457883, EPI_ISL_1457884, EPI_ISL_1457885, EPI_ISL_1457886, EPI_ISL_1457887, EPI_ISL_1457888, EPI_ISL_1457889, EPI_ISL_1457890, EPI_ISL_1457891, EPI_ISL_1457892, EPI_ISL_1457893, EPI_ISL_1457894, EPI_ISL_1457895, EPI_ISL_1457896, EPI_ISL_1457897, EPI_ISL_1457898, EPI_ISL_1457899, EPI_ISL_1457900, EPI_ISL_1457901, EPI_ISL_1457902,
EPI_ISL_1457903, EPI_ISL_1457904, EPI_ISL_1457905, EPI_ISL_1457906, EPI_ISL_1457907, EPI_ISL_1457908, EPI_ISL_1457909, EPI_ISL_1457910, EPI_ISL_1457911, EPI_ISL_1457912, EPI_ISL_1457913, EPI_ISL_1457914, EPI_ISL_1457915, EPI_ISL_1457916, EPI_ISL_1457917, EPI_ISL_1457918, EPI_ISL_1457919, EPI_ISL_1457920, EPI_ISL_1457921, EPI_ISL_1457922, EPI_ISL_1457923, EPI_ISL_1457924,
EPI_ISL_1457925, EPI_ISL_1457926, EPI_ISL_1457927, EPI_ISL_1457928, EPI_ISL_1457929, EPI_ISL_1457930, EPI_ISL_1457931, EPI_ISL_1457932, EPI_ISL_1457933, EPI_ISL_1457934, EPI_ISL_1457935, EPI_ISL_1457936, EPI_ISL_1457937, EPI_ISL_1457938, EPI_ISL_1457939, EPI_ISL_1457940, EPI_ISL_1457941, EPI_ISL_1457942, EPI_ISL_1457943, EPI_ISL_1457944, EPI_ISL_1457945, EPI_ISL_1457946,
EPI_ISL_1457947, EPI_ISL_1457948, EPI_ISL_1457949, EPI_ISL_1457950, EPI_ISL_1457951, EPI_ISL_1457952, EPI_ISL_1457953, EPI_ISL_1457954, EPI_ISL_1457955, EPI_ISL_1457956, EPI_ISL_1457957, EPI_ISL_1457958, EPI_ISL_1457959, EPI_ISL_1457960, EPI_ISL_1457961, EPI_ISL_1457962, EPI_ISL_1457963, EPI_ISL_1457964, EPI_ISL_1457965, EPI_ISL_1457966, EPI_ISL_1457968, EPI_ISL_1457969,
EPI_ISL_1457970, EPI_ISL_1457971, EPI_ISL_1457972, EPI_ISL_1457973, EPI_ISL_1457974, EPI_ISL_1457975, EPI_ISL_1457976, EPI_ISL_1457977, EPI_ISL_1457978, EPI_ISL_1457980, EPI_ISL_1457981, EPI_ISL_1457982, EPI_ISL_1457983, EPI_ISL_1457984, EPI_ISL_1457985, EPI_ISL_1457986, EPI_ISL_1457987, EPI_ISL_1457988, EPI_ISL_1457989, EPI_ISL_1457990, EPI_ISL_1457991, EPI_ISL_1457992,
EPI_ISL_1457993, EPI_ISL_1457994, EPI_ISL_1457995, EPI_ISL_1457996, EPI_ISL_1457997, EPI_ISL_1457998, EPI_ISL_1457999, EPI_ISL_1458000, EPI_ISL_1458001, EPI_ISL_1458002, EPI_ISL_1458003, EPI_ISL_1458004, EPI_ISL_1458005, EPI_ISL_1458006, EPI_ISL_1458007, EPI_ISL_1458008, EPI_ISL_1458009, EPI_ISL_1458010, EPI_ISL_1458011, EPI_ISL_1458012, EPI_ISL_1458013, EPI_ISL_1458014,
EPI_ISL_1458015, EPI_ISL_1458016, EPI_ISL_1458017, EPI_ISL_1458018, EPI_ISL_1458019, EPI_ISL_1458020, EPI_ISL_1458021, EPI_ISL_1458022, EPI_ISL_1458023, EPI_ISL_1458024, EPI_ISL_1458025, EPI_ISL_1458026, EPI_ISL_1458027, EPI_ISL_1458028, EPI_ISL_1458029, EPI_ISL_1458030, EPI_ISL_1458031, EPI_ISL_1458032, EPI_ISL_1458033, EPI_ISL_1458034, EPI_ISL_1458035, EPI_ISL_1458036,
EPI_ISL_1458037, EPI_ISL_1458038, EPI_ISL_1458039, EPI_ISL_1458040, EPI_ISL_1458041, EPI_ISL_1458042, EPI_ISL_1458043, EPI_ISL_1458044, EPI_ISL_1458045, EPI_ISL_1458046, EPI_ISL_1458047, EPI_ISL_1458048, EPI_ISL_1458049, EPI_ISL_1458050, EPI_ISL_1458051, EPI_ISL_1458052, EPI_ISL_1458053, EPI_ISL_1458054, EPI_ISL_1458055, EPI_ISL_1458056, EPI_ISL_1458057, EPI_ISL_1458058,
EPI_ISL_1458059, EPI_ISL_1458060, EPI_ISL_1458061, EPI_ISL_1458062, EPI_ISL_1458063, EPI_ISL_1458064, EPI_ISL_1458065, EPI_ISL_1458066, EPI_ISL_1458067, EPI_ISL_1458068, EPI_ISL_1458069, EPI_ISL_1458070, EPI_ISL_1458071, EPI_ISL_1458072, EPI_ISL_1458073, EPI_ISL_1458074, EPI_ISL_1458075, EPI_ISL_1458076, EPI_ISL_1458077, EPI_ISL_1458078, EPI_ISL_1458079, EPI_ISL_1458080,
EPI_ISL_1458081, EPI_ISL_1458082, EPI_ISL_1458083, EPI_ISL_1458084, EPI_ISL_1458086, EPI_ISL_1458087, EPI_ISL_1458088, EPI_ISL_1458089, EPI_ISL_1458090, EPI_ISL_1458091, EPI_ISL_1458092, EPI_ISL_1458093, EPI_ISL_1458094, EPI_ISL_1458095, EPI_ISL_1458096, EPI_ISL_1458097, EPI_ISL_1458098, EPI_ISL_1458099, EPI_ISL_1458100, EPI_ISL_1458101, EPI_ISL_1458102, EPI_ISL_1458103,
EPI_ISL_1458104, EPI_ISL_1458105, EPI_ISL_1458106, EPI_ISL_1458107, EPI_ISL_1458108, EPI_ISL_1458109, EPI_ISL_1458110, EPI_ISL_1458111, EPI_ISL_1458112, EPI_ISL_1458113, EPI_ISL_1458114, EPI_ISL_1458115, EPI_ISL_1458116, EPI_ISL_1458117, EPI_ISL_1458118, EPI_ISL_1458119, EPI_ISL_1458120, EPI_ISL_1458121, EPI_ISL_1458122, EPI_ISL_1458123, EPI_ISL_1458124, EPI_ISL_1458125,
EPI_ISL_1458126, EPI_ISL_1458127, EPI_ISL_1458128, EPI_ISL_1458129, EPI_ISL_1458130, EPI_ISL_1458131, EPI_ISL_1458132, EPI_ISL_1458133, EPI_ISL_1458134, EPI_ISL_1458135, EPI_ISL_1458136, EPI_ISL_1458137, EPI_ISL_1458138, EPI_ISL_1458139, EPI_ISL_1458140, EPI_ISL_1458141, EPI_ISL_1458142, EPI_ISL_1458143, EPI_ISL_1458144, EPI_ISL_1458145, EPI_ISL_1458146, EPI_ISL_1458147,
EPI_ISL_1458148, EPI_ISL_1458149, EPI_ISL_1458150, EPI_ISL_1458151, EPI_ISL_1458152, EPI_ISL_1458153, EPI_ISL_1458154, EPI_ISL_1458155, EPI_ISL_1458156, EPI_ISL_1458157, EPI_ISL_1458158, EPI_ISL_1458159, EPI_ISL_1458160, EPI_ISL_1458161, EPI_ISL_1458162, EPI_ISL_1458163, EPI_ISL_1458164, EPI_ISL_1458165, EPI_ISL_1458166, EPI_ISL_1458167, EPI_ISL_1458168, EPI_ISL_1458169,
EPI_ISL_1458170, EPI_ISL_1458171, EPI_ISL_1458172, EPI_ISL_1458173, EPI_ISL_1458174, EPI_ISL_1458175, EPI_ISL_1458176, EPI_ISL_1458177, EPI_ISL_1458178, EPI_ISL_1458179, EPI_ISL_1458180, EPI_ISL_1458181, EPI_ISL_1458182, EPI_ISL_1458183, EPI_ISL_1458184, EPI_ISL_1458185, EPI_ISL_1458186, EPI_ISL_1458187, EPI_ISL_1458188, EPI_ISL_1458189, EPI_ISL_1458190, EPI_ISL_1458191,
EPI_ISL_1458192, EPI_ISL_1458193, EPI_ISL_1458194, EPI_ISL_1458195, EPI_ISL_1458196, EPI_ISL_1458197, EPI_ISL_1458198, EPI_ISL_1458199, EPI_ISL_1458200, EPI_ISL_1458201, EPI_ISL_1458202, EPI_ISL_1458203, EPI_ISL_1458204, EPI_ISL_1458205, EPI_ISL_1458206, EPI_ISL_1458207, EPI_ISL_1458208, EPI_ISL_1458209, EPI_ISL_1458210, EPI_ISL_1458211, EPI_ISL_1458212, EPI_ISL_1458213,
EPI_ISL_1458214, EPI_ISL_1458215, EPI_ISL_1458216, EPI_ISL_1458217, EPI_ISL_1458218, EPI_ISL_1458219, EPI_ISL_1458220, EPI_ISL_1458221, EPI_ISL_1458222, EPI_ISL_1458223, EPI_ISL_1458224, EPI_ISL_1458225, EPI_ISL_1458226, EPI_ISL_1458227, EPI_ISL_1458228, EPI_ISL_1458229, EPI_ISL_1458230, EPI_ISL_1458231, EPI_ISL_1458232, EPI_ISL_1458233, EPI_ISL_1458234, EPI_ISL_1458235,
EPI_ISL_1458236, EPI_ISL_1458237, EPI_ISL_1458238, EPI_ISL_1458239, EPI_ISL_1458240, EPI_ISL_1458241, EPI_ISL_1458242, EPI_ISL_1458243, EPI_ISL_1458244, EPI_ISL_1458245, EPI_ISL_1458246, EPI_ISL_1458247, EPI_ISL_1458248, EPI_ISL_1458249, EPI_ISL_1458250, EPI_ISL_1458251, EPI_ISL_1458252, EPI_ISL_1458253, EPI_ISL_1458254, EPI_ISL_1458255, EPI_ISL_1458256, EPI_ISL_1458257,
EPI_ISL_1458258, EPI_ISL_1458259, EPI_ISL_1458260, EPI_ISL_1458261, EPI_ISL_1458262, EPI_ISL_1458263, EPI_ISL_1458264, EPI_ISL_1458265, EPI_ISL_1458266, EPI_ISL_1458267, EPI_ISL_1458268, EPI_ISL_1458269, EPI_ISL_1458270, EPI_ISL_1458271, EPI_ISL_1458272, EPI_ISL_1458273, EPI_ISL_1458274, EPI_ISL_1458275, EPI_ISL_1458276, EPI_ISL_1458277, EPI_ISL_1458278, EPI_ISL_1458279,
EPI_ISL_1458280, EPI_ISL_1458281, EPI_ISL_1458282, EPI_ISL_1458283, EPI_ISL_1458285, EPI_ISL_1458286, EPI_ISL_1458289, EPI_ISL_1458290, EPI_ISL_1458291, EPI_ISL_1458292, EPI_ISL_1458293, EPI_ISL_1458294, EPI_ISL_1458295, EPI_ISL_1458296, EPI_ISL_1458297, EPI_ISL_1458298, EPI_ISL_1458299, EPI_ISL_1458300, EPI_ISL_1458301, EPI_ISL_1458302, EPI_ISL_1458303, EPI_ISL_1458304,
EPI_ISL_1458305, EPI_ISL_1458306, EPI_ISL_1458307, EPI_ISL_1458308, EPI_ISL_1458309, EPI_ISL_1458310, EPI_ISL_1458311, EPI_ISL_1458312, EPI_ISL_1458313, EPI_ISL_1458314, EPI_ISL_1458315, EPI_ISL_1458316, EPI_ISL_1458317, EPI_ISL_1458318, EPI_ISL_1458319, EPI_ISL_1458320, EPI_ISL_1458321, EPI_ISL_1458322, EPI_ISL_1458323, EPI_ISL_1458324, EPI_ISL_1458325, EPI_ISL_1458326,
EPI_ISL_1458327, EPI_ISL_1458328, EPI_ISL_1458329, EPI_ISL_1458330, EPI_ISL_1458331, EPI_ISL_1458332, EPI_ISL_1458333, EPI_ISL_1458334, EPI_ISL_1458335, EPI_ISL_1458336, EPI_ISL_1458337, EPI_ISL_1458338, EPI_ISL_1458339, EPI_ISL_1458340, EPI_ISL_1458341, EPI_ISL_1458342, EPI_ISL_1458343, EPI_ISL_1458344, EPI_ISL_1458345, EPI_ISL_1458346, EPI_ISL_1458347, EPI_ISL_1458348,
EPI_ISL_1458349, EPI_ISL_1458350, EPI_ISL_1458351, EPI_ISL_1458352, EPI_ISL_1458353, EPI_ISL_1458354, EPI_ISL_1458355, EPI_ISL_1458356, EPI_ISL_1458357, EPI_ISL_1458358, EPI_ISL_1458359, EPI_ISL_1458360, EPI_ISL_1458361, EPI_ISL_1458362, EPI_ISL_1458363, EPI_ISL_1458364, EPI_ISL_1458365, EPI_ISL_1458366, EPI_ISL_1458367, EPI_ISL_1458368, EPI_ISL_1458369, EPI_ISL_1458370,
EPI_ISL_1458371, EPI_ISL_1458372, EPI_ISL_1458373, EPI_ISL_1458374, EPI_ISL_1458375, EPI_ISL_1458376, EPI_ISL_1458377, EPI_ISL_1458378, EPI_ISL_1458379, EPI_ISL_1458380, EPI_ISL_1458381, EPI_ISL_1458382, EPI_ISL_1458383, EPI_ISL_1458384, EPI_ISL_1458385, EPI_ISL_1458386, EPI_ISL_1458387, EPI_ISL_1458388, EPI_ISL_1458389, EPI_ISL_1458390, EPI_ISL_1458391, EPI_ISL_1458392,
EPI_ISL_1458393, EPI_ISL_1458394, EPI_ISL_1458395, EPI_ISL_1458396, EPI_ISL_1458397, EPI_ISL_1458398, EPI_ISL_1458399, EPI_ISL_1458400, EPI_ISL_1458401, EPI_ISL_1458402, EPI_ISL_1458403, EPI_ISL_1458404, EPI_ISL_1458405, EPI_ISL_1458406, EPI_ISL_1458407, EPI_ISL_1458408, EPI_ISL_1458409, EPI_ISL_1458410, EPI_ISL_1458411, EPI_ISL_1458412, EPI_ISL_1458413, EPI_ISL_1458414,
EPI_ISL_1458415, EPI_ISL_1458416, EPI_ISL_1458417, EPI_ISL_1458418, EPI_ISL_1458481, EPI_ISL_1458482, EPI_ISL_1458483, EPI_ISL_1458484, EPI_ISL_1458485, EPI_ISL_1458486, EPI_ISL_1458487, EPI_ISL_1458488, EPI_ISL_1458489, EPI_ISL_1458490, EPI_ISL_1458491, EPI_ISL_1458492, EPI_ISL_1458493, EPI_ISL_1458494, EPI_ISL_1458495, EPI_ISL_1458496, EPI_ISL_1458497, EPI_ISL_1458498,
EPI_ISL_1458499, EPI_ISL_1458500, EPI_ISL_1458501, EPI_ISL_1458502, EPI_ISL_1458503, EPI_ISL_1458504, EPI_ISL_1458505, EPI_ISL_1458506, EPI_ISL_1458507, EPI_ISL_1458508, EPI_ISL_1458509, EPI_ISL_1458510, EPI_ISL_1458511, EPI_ISL_1458512, EPI_ISL_1458513, EPI_ISL_1458514, EPI_ISL_1458515, EPI_ISL_1458516, EPI_ISL_1458517, EPI_ISL_1458518, EPI_ISL_1458519, EPI_ISL_1458520,
EPI_ISL_1458521, EPI_ISL_1458522, EPI_ISL_1458523, EPI_ISL_1458524, EPI_ISL_1458525, EPI_ISL_1458526, EPI_ISL_1458527, EPI_ISL_1458528, EPI_ISL_1458529, EPI_ISL_1458530, EPI_ISL_1458531, EPI_ISL_1458532, EPI_ISL_1458533, EPI_ISL_1458534, EPI_ISL_1458535, EPI_ISL_1458536, EPI_ISL_1458537, EPI_ISL_1458538, EPI_ISL_1458539, EPI_ISL_1458540, EPI_ISL_1458541, EPI_ISL_1458542,
EPI_ISL_1458543, EPI_ISL_1458544, EPI_ISL_1458545, EPI_ISL_1458546, EPI_ISL_1458547, EPI_ISL_1458548, EPI_ISL_1458549, EPI_ISL_1458550, EPI_ISL_1458551, EPI_ISL_1458552, EPI_ISL_1458553, EPI_ISL_1458554, EPI_ISL_1458555, EPI_ISL_1458556, EPI_ISL_1458557, EPI_ISL_1458558, EPI_ISL_1458559, EPI_ISL_1458560, EPI_ISL_1458561, EPI_ISL_1458562, EPI_ISL_1458563, EPI_ISL_1458564,
EPI_ISL_1458565, EPI_ISL_1458566, EPI_ISL_1458567, EPI_ISL_1458568, EPI_ISL_1458569, EPI_ISL_1458570, EPI_ISL_1458571, EPI_ISL_1458572, EPI_ISL_1458573, EPI_ISL_1458574, EPI_ISL_1458575, EPI_ISL_1458576, EPI_ISL_1458577, EPI_ISL_1458578, EPI_ISL_1458579, EPI_ISL_1458580, EPI_ISL_1458581, EPI_ISL_1458582, EPI_ISL_1458583, EPI_ISL_1458584, EPI_ISL_1458585, EPI_ISL_1458586,
EPI_ISL_1458587, EPI_ISL_1458588, EPI_ISL_1458589, EPI_ISL_1458590, EPI_ISL_1458591, EPI_ISL_1458592, EPI_ISL_1458593, EPI_ISL_1458594, EPI_ISL_1458595, EPI_ISL_1458596, EPI_ISL_1458597, EPI_ISL_1458598, EPI_ISL_1458599, EPI_ISL_1458600, EPI_ISL_1458601, EPI_ISL_1458602, EPI_ISL_1458603, EPI_ISL_1458604, EPI_ISL_1458605, EPI_ISL_1458606, EPI_ISL_1458607, EPI_ISL_1458608,
EPI_ISL_1458609, EPI_ISL_1458610, EPI_ISL_1458611, EPI_ISL_1458612, EPI_ISL_1458614, EPI_ISL_1458615, EPI_ISL_1458616, EPI_ISL_1458617, EPI_ISL_1458618, EPI_ISL_1458619, EPI_ISL_1458620, EPI_ISL_1458621, EPI_ISL_1458622, EPI_ISL_1458623, EPI_ISL_1458624, EPI_ISL_1458625, EPI_ISL_1458626, EPI_ISL_1458627, EPI_ISL_1458628, EPI_ISL_1458629, EPI_ISL_1458630, EPI_ISL_1458631,
EPI_ISL_1458632, EPI_ISL_1458633, EPI_ISL_1458634, EPI_ISL_1458635, EPI_ISL_1458636, EPI_ISL_1458637, EPI_ISL_1458638, EPI_ISL_1458639, EPI_ISL_1458640, EPI_ISL_1458641, EPI_ISL_1458642, EPI_ISL_1458643, EPI_ISL_1458644, EPI_ISL_1458645, EPI_ISL_1458646, EPI_ISL_1458647, EPI_ISL_1458648, EPI_ISL_1458649, EPI_ISL_1458650, EPI_ISL_1458651, EPI_ISL_1458652, EPI_ISL_1458653,
EPI_ISL_1458654, EPI_ISL_1458655, EPI_ISL_1458656, EPI_ISL_1458657, EPI_ISL_1458658, EPI_ISL_1458659, EPI_ISL_1458660, EPI_ISL_1458661, EPI_ISL_1458662, EPI_ISL_1458663, EPI_ISL_1458664, EPI_ISL_1458665, EPI_ISL_1458666, EPI_ISL_1458667, EPI_ISL_1458668, EPI_ISL_1458669, EPI_ISL_1458670, EPI_ISL_1458671, EPI_ISL_1458672, EPI_ISL_1458673, EPI_ISL_1458674, EPI_ISL_1458675,
EPI_ISL_1458676, EPI_ISL_1458677, EPI_ISL_1458678, EPI_ISL_1458679, EPI_ISL_1458680, EPI_ISL_1458681, EPI_ISL_1458682, EPI_ISL_1458683, EPI_ISL_1458684, EPI_ISL_1458685, EPI_ISL_1458686, EPI_ISL_1458687, EPI_ISL_1458688, EPI_ISL_1458689, EPI_ISL_1458690, EPI_ISL_1458691, EPI_ISL_1458692, EPI_ISL_1458693, EPI_ISL_1458694, EPI_ISL_1458695, EPI_ISL_1458696, EPI_ISL_1458697,
EPI_ISL_1458698, EPI_ISL_1458699, EPI_ISL_1458700, EPI_ISL_1458701, EPI_ISL_1458702, EPI_ISL_1458703, EPI_ISL_1458704, EPI_ISL_1458705, EPI_ISL_1458706, EPI_ISL_1458707, EPI_ISL_1458708, EPI_ISL_1458709, EPI_ISL_1458710, EPI_ISL_1458711, EPI_ISL_1458714, EPI_ISL_1458715, EPI_ISL_1458716, EPI_ISL_1458717, EPI_ISL_1458718, EPI_ISL_1458719, EPI_ISL_1458720, EPI_ISL_1458721,
EPI_ISL_1458722, EPI_ISL_1458723, EPI_ISL_1458724, EPI_ISL_1458725, EPI_ISL_1458726, EPI_ISL_1458727, EPI_ISL_1458728, EPI_ISL_1458729, EPI_ISL_1458730, EPI_ISL_1458731, EPI_ISL_1458732, EPI_ISL_1458733, EPI_ISL_1458734, EPI_ISL_1458735, EPI_ISL_1458736, EPI_ISL_1458737, EPI_ISL_1458738, EPI_ISL_1458739, EPI_ISL_1458740, EPI_ISL_1458741, EPI_ISL_1458742, EPI_ISL_1458743,
EPI_ISL_1458744, EPI_ISL_1458745, EPI_ISL_1458746, EPI_ISL_1458747, EPI_ISL_1458748, EPI_ISL_1458749, EPI_ISL_1458750, EPI_ISL_1458751, EPI_ISL_1458752, EPI_ISL_1458753, EPI_ISL_1458754, EPI_ISL_1458755, EPI_ISL_1458756, EPI_ISL_1458757, EPI_ISL_1458758, EPI_ISL_1458759, EPI_ISL_1458760, EPI_ISL_1458761, EPI_ISL_1458762, EPI_ISL_1458763, EPI_ISL_1458764, EPI_ISL_1458765,
EPI_ISL_1458766, EPI_ISL_1458767, EPI_ISL_1458768, EPI_ISL_1458769, EPI_ISL_1458770, EPI_ISL_1458771, EPI_ISL_1458772, EPI_ISL_1458773, EPI_ISL_1458774, EPI_ISL_1458775, EPI_ISL_1458776, EPI_ISL_1458777, EPI_ISL_1458778, EPI_ISL_1458779, EPI_ISL_1458780, EPI_ISL_1458781, EPI_ISL_1458782, EPI_ISL_1458783, EPI_ISL_1458784, EPI_ISL_1458785, EPI_ISL_1458786, EPI_ISL_1458787,
EPI_ISL_1458788, EPI_ISL_1458789, EPI_ISL_1458790, EPI_ISL_1458791, EPI_ISL_1458792, EPI_ISL_1458793, EPI_ISL_1458794, EPI_ISL_1458795, EPI_ISL_1458796, EPI_ISL_1458797, EPI_ISL_1458798, EPI_ISL_1458799, EPI_ISL_1458800, EPI_ISL_1458801, EPI_ISL_1458802, EPI_ISL_1458803, EPI_ISL_1458804, EPI_ISL_1458805, EPI_ISL_1458806, EPI_ISL_1458807, EPI_ISL_1458808, EPI_ISL_1458809,
EPI_ISL_1458810, EPI_ISL_1458811, EPI_ISL_1458812, EPI_ISL_1458813, EPI_ISL_1458814, EPI_ISL_1458815, EPI_ISL_1458816, EPI_ISL_1458817, EPI_ISL_1458818, EPI_ISL_1458819, EPI_ISL_1458844, EPI_ISL_1458845, EPI_ISL_1458847, EPI_ISL_1458850, EPI_ISL_1458851, EPI_ISL_1458852, EPI_ISL_1458853, EPI_ISL_1458854, EPI_ISL_1458855, EPI_ISL_1458856, EPI_ISL_1458857, EPI_ISL_1458858,
EPI_ISL_1458859, EPI_ISL_1458900, EPI_ISL_1458901, EPI_ISL_1458902, EPI_ISL_1458903, EPI_ISL_1458904, EPI_ISL_1458905, EPI_ISL_1458906, EPI_ISL_1458907, EPI_ISL_1458908, EPI_ISL_1458909, EPI_ISL_1458910, EPI_ISL_1458911, EPI_ISL_1458912, EPI_ISL_1458913, EPI_ISL_1458914, EPI_ISL_1458915, EPI_ISL_1458916, EPI_ISL_1458917, EPI_ISL_1458918, EPI_ISL_1458919, EPI_ISL_1458920,
EPI_ISL_1458921, EPI_ISL_1458922, EPI_ISL_1458923, EPI_ISL_1458924, EPI_ISL_1458925, EPI_ISL_1458926, EPI_ISL_1458927, EPI_ISL_1458928, EPI_ISL_1458929, EPI_ISL_1458930, EPI_ISL_1458931, EPI_ISL_1458932, EPI_ISL_1458933, EPI_ISL_1458934, EPI_ISL_1458935, EPI_ISL_1458936, EPI_ISL_1458937, EPI_ISL_1458938, EPI_ISL_1458939, EPI_ISL_1458965, EPI_ISL_1458968, EPI_ISL_1458970,
EPI_ISL_1458971, EPI_ISL_1458972, EPI_ISL_1458973, EPI_ISL_1458974, EPI_ISL_1458975, EPI_ISL_1458976, EPI_ISL_1458977, EPI_ISL_1458978, EPI_ISL_1458979, EPI_ISL_1459207, EPI_ISL_1459208, EPI_ISL_1459209, EPI_ISL_1459256, EPI_ISL_1459257, EPI_ISL_1459258, EPI_ISL_1459259, EPI_ISL_1459260, EPI_ISL_1459261, EPI_ISL_1459262, EPI_ISL_1459263, EPI_ISL_1459264, EPI_ISL_1459265,
EPI_ISL_1459266, EPI_ISL_1459267, EPI_ISL_1459268, EPI_ISL_1459269, EPI_ISL_1459292, EPI_ISL_1459293, EPI_ISL_1459294, EPI_ISL_1459295, EPI_ISL_1459296, EPI_ISL_1459297, EPI_ISL_1459298, EPI_ISL_1459299, EPI_ISL_1459300, EPI_ISL_1459301, EPI_ISL_1459302, EPI_ISL_1459303, EPI_ISL_1459304, EPI_ISL_1459305, EPI_ISL_1459306, EPI_ISL_1459307, EPI_ISL_1459308, EPI_ISL_1459309,
EPI_ISL_1459310, EPI_ISL_1459311, EPI_ISL_1459312, EPI_ISL_1459313, EPI_ISL_1459314, EPI_ISL_1459315, EPI_ISL_1459316, EPI_ISL_1459317, EPI_ISL_1459318, EPI_ISL_1459319, EPI_ISL_1459320, EPI_ISL_1459321, EPI_ISL_1459322, EPI_ISL_1459323, EPI_ISL_1459324, EPI_ISL_1459325, EPI_ISL_1459326, EPI_ISL_1459327, EPI_ISL_1459328, EPI_ISL_1459329, EPI_ISL_1459330, EPI_ISL_1459331,
EPI_ISL_1459332, EPI_ISL_1459333, EPI_ISL_1459334, EPI_ISL_1459335, EPI_ISL_1459336, EPI_ISL_1459337, EPI_ISL_1459338, EPI_ISL_1459339, EPI_ISL_1459340, EPI_ISL_1459341, EPI_ISL_1459342, EPI_ISL_1459343, EPI_ISL_1459344, EPI_ISL_1459345, EPI_ISL_1459346, EPI_ISL_1459347, EPI_ISL_1459348, EPI_ISL_1459349, EPI_ISL_1459350, EPI_ISL_1459351, EPI_ISL_1459352, EPI_ISL_1459353,
EPI_ISL_1459354, EPI_ISL_1459355, EPI_ISL_1459356, EPI_ISL_1459357, EPI_ISL_1459358, EPI_ISL_1459359, EPI_ISL_1459360, EPI_ISL_1459361, EPI_ISL_1459362, EPI_ISL_1459363, EPI_ISL_1459364, EPI_ISL_1459365, EPI_ISL_1459366, EPI_ISL_1459367, EPI_ISL_1459368, EPI_ISL_1459369, EPI_ISL_1459370, EPI_ISL_1459371, EPI_ISL_1459372, EPI_ISL_1459373, EPI_ISL_1459374, EPI_ISL_1459375,
EPI_ISL_1459376, EPI_ISL_1459377, EPI_ISL_1459378, EPI_ISL_1459379, EPI_ISL_1459381, EPI_ISL_1459382, EPI_ISL_1459383, EPI_ISL_1459384, EPI_ISL_1459385, EPI_ISL_1459386, EPI_ISL_1459387, EPI_ISL_1459388, EPI_ISL_1459389, EPI_ISL_1459390, EPI_ISL_1459391, EPI_ISL_1459392, EPI_ISL_1459393, EPI_ISL_1459394, EPI_ISL_1459395, EPI_ISL_1459396, EPI_ISL_1459397, EPI_ISL_1459398,
EPI_ISL_1459399, EPI_ISL_1459400, EPI_ISL_1459401, EPI_ISL_1459402, EPI_ISL_1459403, EPI_ISL_1459404, EPI_ISL_1459405, EPI_ISL_1459406, EPI_ISL_1459407, EPI_ISL_1459408, EPI_ISL_1459409, EPI_ISL_1459410, EPI_ISL_1459411, EPI_ISL_1459412, EPI_ISL_1459413, EPI_ISL_1459414, EPI_ISL_1459415, EPI_ISL_1459416, EPI_ISL_1459417, EPI_ISL_1459418, EPI_ISL_1459419, EPI_ISL_1459420,
EPI_ISL_1459421, EPI_ISL_1459422, EPI_ISL_1459423, EPI_ISL_1459424, EPI_ISL_1459425, EPI_ISL_1459426, EPI_ISL_1459427, EPI_ISL_1459428, EPI_ISL_1459429, EPI_ISL_1459430, EPI_ISL_1459431, EPI_ISL_1459432, EPI_ISL_1459433, EPI_ISL_1459434, EPI_ISL_1459435, EPI_ISL_1459436, EPI_ISL_1459437, EPI_ISL_1459438, EPI_ISL_1459439, EPI_ISL_1459440, EPI_ISL_1459441, EPI_ISL_1459442,
EPI_ISL_1459443, EPI_ISL_1459444, EPI_ISL_1459445, EPI_ISL_1459446, EPI_ISL_1459447, EPI_ISL_1459448, EPI_ISL_1459449, EPI_ISL_1459450, EPI_ISL_1459451, EPI_ISL_1459452, EPI_ISL_1459453, EPI_ISL_1459454, EPI_ISL_1459455, EPI_ISL_1459456, EPI_ISL_1459457, EPI_ISL_1459458, EPI_ISL_1459459, EPI_ISL_1459460, EPI_ISL_1459461, EPI_ISL_1459462, EPI_ISL_1459463, EPI_ISL_1459464,
EPI_ISL_1459465, EPI_ISL_1459466, EPI_ISL_1459467, EPI_ISL_1459468, EPI_ISL_1459469, EPI_ISL_1459470, EPI_ISL_1459471, EPI_ISL_1459472, EPI_ISL_1459473, EPI_ISL_1459474, EPI_ISL_1459475, EPI_ISL_1459476, EPI_ISL_1459477, EPI_ISL_1459478, EPI_ISL_1459479, EPI_ISL_1459480, EPI_ISL_1459481, EPI_ISL_1459482, EPI_ISL_1459483, EPI_ISL_1459484, EPI_ISL_1459485, EPI_ISL_1459486,
EPI_ISL_1459487, EPI_ISL_1459488, EPI_ISL_1459489, EPI_ISL_1459490, EPI_ISL_1459491, EPI_ISL_1459492, EPI_ISL_1459493, EPI_ISL_1459494, EPI_ISL_1459495, EPI_ISL_1459496, EPI_ISL_1459497, EPI_ISL_1459498, EPI_ISL_1459499, EPI_ISL_1459500, EPI_ISL_1459501, EPI_ISL_1459502, EPI_ISL_1459503, EPI_ISL_1459504, EPI_ISL_1459505, EPI_ISL_1459506, EPI_ISL_1459507, EPI_ISL_1459508,
EPI_ISL_1459529, EPI_ISL_1459530, EPI_ISL_1459531, EPI_ISL_1459532, EPI_ISL_1459533, EPI_ISL_1459534, EPI_ISL_1459535, EPI_ISL_1459536, EPI_ISL_1459537, EPI_ISL_1459538, EPI_ISL_1459539, EPI_ISL_1459540, EPI_ISL_1459541, EPI_ISL_1459542, EPI_ISL_1459543, EPI_ISL_1459544, EPI_ISL_1459545, EPI_ISL_1459546, EPI_ISL_1459547, EPI_ISL_1459548, EPI_ISL_1459549, EPI_ISL_1459550,
EPI_ISL_1459551, EPI_ISL_1459552, EPI_ISL_1459553, EPI_ISL_1459554, EPI_ISL_1459555, EPI_ISL_1459556, EPI_ISL_1459557, EPI_ISL_1459558, EPI_ISL_1459559, EPI_ISL_1459560, EPI_ISL_1459561, EPI_ISL_1459562, EPI_ISL_1459563, EPI_ISL_1459564, EPI_ISL_1459565, EPI_ISL_1459566, EPI_ISL_1459567, EPI_ISL_1459568, EPI_ISL_1459569, EPI_ISL_1459570, EPI_ISL_1459571, EPI_ISL_1459572,
EPI_ISL_1459573, EPI_ISL_1459574, EPI_ISL_1459575, EPI_ISL_1459576, EPI_ISL_1459577, EPI_ISL_1459578, EPI_ISL_1459579, EPI_ISL_1459580, EPI_ISL_1459581, EPI_ISL_1459582, EPI_ISL_1459583, EPI_ISL_1459584, EPI_ISL_1459585, EPI_ISL_1459586, EPI_ISL_1459587, EPI_ISL_1459588, EPI_ISL_1459589, EPI_ISL_1459590, EPI_ISL_1459591, EPI_ISL_1459592, EPI_ISL_1459593, EPI_ISL_1459594,
EPI_ISL_1459595, EPI_ISL_1459596, EPI_ISL_1459597, EPI_ISL_1459598, EPI_ISL_1459599, EPI_ISL_1459600, EPI_ISL_1459601, EPI_ISL_1459602, EPI_ISL_1459603, EPI_ISL_1459604, EPI_ISL_1459605, EPI_ISL_1459606, EPI_ISL_1459607, EPI_ISL_1459608, EPI_ISL_1459609, EPI_ISL_1459610, EPI_ISL_1459611, EPI_ISL_1459612, EPI_ISL_1459613, EPI_ISL_1459614, EPI_ISL_1459615, EPI_ISL_1459616,
EPI_ISL_1459617, EPI_ISL_1459618, EPI_ISL_1459619, EPI_ISL_1459620, EPI_ISL_1459621, EPI_ISL_1459622, EPI_ISL_1459623, EPI_ISL_1459624, EPI_ISL_1459625, EPI_ISL_1459626, EPI_ISL_1459627, EPI_ISL_1459628, EPI_ISL_1459629, EPI_ISL_1459630, EPI_ISL_1459631, EPI_ISL_1459632, EPI_ISL_1459633, EPI_ISL_1459634, EPI_ISL_1459635, EPI_ISL_1459636, EPI_ISL_1459637, EPI_ISL_1459638,
EPI_ISL_1459639, EPI_ISL_1459640, EPI_ISL_1459641, EPI_ISL_1459642, EPI_ISL_1459643, EPI_ISL_1459644, EPI_ISL_1459645, EPI_ISL_1459646, EPI_ISL_1459647, EPI_ISL_1459648, EPI_ISL_1459649, EPI_ISL_1459650, EPI_ISL_1459651, EPI_ISL_1459652, EPI_ISL_1459653, EPI_ISL_1459654, EPI_ISL_1459655, EPI_ISL_1459656, EPI_ISL_1459657, EPI_ISL_1459658, EPI_ISL_1459659, EPI_ISL_1459660,
EPI_ISL_1459661, EPI_ISL_1459662, EPI_ISL_1459663, EPI_ISL_1459664, EPI_ISL_1459665, EPI_ISL_1459666, EPI_ISL_1459667, EPI_ISL_1459668, EPI_ISL_1459669, EPI_ISL_1459670, EPI_ISL_1459671, EPI_ISL_1459672, EPI_ISL_1459673, EPI_ISL_1459674, EPI_ISL_1459675, EPI_ISL_1459676, EPI_ISL_1459677, EPI_ISL_1459678, EPI_ISL_1459679, EPI_ISL_1459680, EPI_ISL_1459681, EPI_ISL_1459682,
EPI_ISL_1459683, EPI_ISL_1459684, EPI_ISL_1459685, EPI_ISL_1459686, EPI_ISL_1459687, EPI_ISL_1459688, EPI_ISL_1459689, EPI_ISL_1459690, EPI_ISL_1459691, EPI_ISL_1459692, EPI_ISL_1459693, EPI_ISL_1459694, EPI_ISL_1459695, EPI_ISL_1459696, EPI_ISL_1459697, EPI_ISL_1459698, EPI_ISL_1459699, EPI_ISL_1459700, EPI_ISL_1459701, EPI_ISL_1459702, EPI_ISL_1459703, EPI_ISL_1459704,
EPI_ISL_1459705, EPI_ISL_1459706, EPI_ISL_1459707, EPI_ISL_1459708, EPI_ISL_1459709, EPI_ISL_1459710, EPI_ISL_1459711, EPI_ISL_1459712, EPI_ISL_1459713, EPI_ISL_1459714, EPI_ISL_1459715, EPI_ISL_1459716, EPI_ISL_1459717, EPI_ISL_1459718, EPI_ISL_1459719, EPI_ISL_1459720, EPI_ISL_1459721, EPI_ISL_1459722, EPI_ISL_1459723, EPI_ISL_1459724, EPI_ISL_1459725, EPI_ISL_1459726,
EPI_ISL_1459727, EPI_ISL_1459728, EPI_ISL_1459729, EPI_ISL_1459730, EPI_ISL_1459731, EPI_ISL_1459732, EPI_ISL_1459733, EPI_ISL_1459734, EPI_ISL_1459735, EPI_ISL_1459736, EPI_ISL_1459737, EPI_ISL_1459738, EPI_ISL_1459739, EPI_ISL_1459740, EPI_ISL_1459741, EPI_ISL_1459742, EPI_ISL_1459743, EPI_ISL_1459744, EPI_ISL_1459745, EPI_ISL_1459746, EPI_ISL_1459747, EPI_ISL_1459748,
EPI_ISL_1459749, EPI_ISL_1459750, EPI_ISL_1459751, EPI_ISL_1459752, EPI_ISL_1459753, EPI_ISL_1459754, EPI_ISL_1459755, EPI_ISL_1459756, EPI_ISL_1459757, EPI_ISL_1459758, EPI_ISL_1459759, EPI_ISL_1459760, EPI_ISL_1459761, EPI_ISL_1459762, EPI_ISL_1459763, EPI_ISL_1459764, EPI_ISL_1459765, EPI_ISL_1459766, EPI_ISL_1459767, EPI_ISL_1459768, EPI_ISL_1459769, EPI_ISL_1459771,
EPI_ISL_1459772, EPI_ISL_1459773, EPI_ISL_1459774, EPI_ISL_1459775, EPI_ISL_1459776, EPI_ISL_1459777, EPI_ISL_1459778, EPI_ISL_1459779, EPI_ISL_1459780, EPI_ISL_1459781, EPI_ISL_1459782, EPI_ISL_1459783, EPI_ISL_1459784, EPI_ISL_1459785, EPI_ISL_1459786, EPI_ISL_1459787, EPI_ISL_1459788, EPI_ISL_1459789, EPI_ISL_1459790, EPI_ISL_1459791, EPI_ISL_1459792, EPI_ISL_1459793,
EPI_ISL_1459794, EPI_ISL_1459795, EPI_ISL_1459796, EPI_ISL_1459797, EPI_ISL_1459798, EPI_ISL_1459799, EPI_ISL_1459800, EPI_ISL_1459801, EPI_ISL_1459802, EPI_ISL_1459803, EPI_ISL_1459804, EPI_ISL_1459805, EPI_ISL_1459806, EPI_ISL_1459807, EPI_ISL_1459808, EPI_ISL_1459809, EPI_ISL_1459810, EPI_ISL_1459811, EPI_ISL_1459812, EPI_ISL_1459813, EPI_ISL_1459814, EPI_ISL_1459815,
EPI_ISL_1459816, EPI_ISL_1459817, EPI_ISL_1459818, EPI_ISL_1459819, EPI_ISL_1459820, EPI_ISL_1459821, EPI_ISL_1459822, EPI_ISL_1459823, EPI_ISL_1459824, EPI_ISL_1459825, EPI_ISL_1459826, EPI_ISL_1459827, EPI_ISL_1459828, EPI_ISL_1459829, EPI_ISL_1459830, EPI_ISL_1459831, EPI_ISL_1459832, EPI_ISL_1459833, EPI_ISL_1459834, EPI_ISL_1459835, EPI_ISL_1459836, EPI_ISL_1459837,
EPI_ISL_1459838, EPI_ISL_1459839, EPI_ISL_1459840, EPI_ISL_1459842, EPI_ISL_1459843, EPI_ISL_1459844, EPI_ISL_1459845, EPI_ISL_1459846, EPI_ISL_1459848, EPI_ISL_1459849, EPI_ISL_1459850, EPI_ISL_1459851, EPI_ISL_1459852, EPI_ISL_1459853, EPI_ISL_1459854, EPI_ISL_1459855, EPI_ISL_1459856, EPI_ISL_1459857, EPI_ISL_1459858, EPI_ISL_1459859, EPI_ISL_1459860, EPI_ISL_1459861,
EPI_ISL_1459862, EPI_ISL_1459863, EPI_ISL_1459864, EPI_ISL_1459865, EPI_ISL_1459866, EPI_ISL_1459867, EPI_ISL_1459868, EPI_ISL_1459869, EPI_ISL_1459870, EPI_ISL_1459871, EPI_ISL_1459872, EPI_ISL_1459873, EPI_ISL_1459874, EPI_ISL_1459875, EPI_ISL_1459876, EPI_ISL_1459877, EPI_ISL_1459878, EPI_ISL_1459879, EPI_ISL_1459880, EPI_ISL_1459881, EPI_ISL_1459882, EPI_ISL_1459883,
EPI_ISL_1459884, EPI_ISL_1459885, EPI_ISL_1459886, EPI_ISL_1459887, EPI_ISL_1459888, EPI_ISL_1459889, EPI_ISL_1459890, EPI_ISL_1459891, EPI_ISL_1459892, EPI_ISL_1459893, EPI_ISL_1459894, EPI_ISL_1459895, EPI_ISL_1459896, EPI_ISL_1459897, EPI_ISL_1459898, EPI_ISL_1459899, EPI_ISL_1459900, EPI_ISL_1459901, EPI_ISL_1459902, EPI_ISL_1459903, EPI_ISL_1459904, EPI_ISL_1459905,
EPI_ISL_1459906, EPI_ISL_1459907, EPI_ISL_1459908, EPI_ISL_1459909, EPI_ISL_1459910, EPI_ISL_1459911, EPI_ISL_1459912, EPI_ISL_1459913, EPI_ISL_1459914, EPI_ISL_1459915, EPI_ISL_1459916, EPI_ISL_1459917, EPI_ISL_1459918, EPI_ISL_1459919, EPI_ISL_1459920, EPI_ISL_1459921, EPI_ISL_1459922, EPI_ISL_1459923, EPI_ISL_1459924, EPI_ISL_1459925, EPI_ISL_1459926, EPI_ISL_1459927,
EPI_ISL_1459928, EPI_ISL_1459929, EPI_ISL_1459930, EPI_ISL_1459931, EPI_ISL_1459932, EPI_ISL_1459933, EPI_ISL_1459934, EPI_ISL_1459935, EPI_ISL_1459936, EPI_ISL_1459937, EPI_ISL_1459938, EPI_ISL_1459939, EPI_ISL_1459940, EPI_ISL_1459941, EPI_ISL_1459942, EPI_ISL_1459943, EPI_ISL_1459944, EPI_ISL_1459945, EPI_ISL_1459946, EPI_ISL_1459947, EPI_ISL_1459948, EPI_ISL_1459949,
EPI_ISL_1459950, EPI_ISL_1459951, EPI_ISL_1459952, EPI_ISL_1459953, EPI_ISL_1459954, EPI_ISL_1459955, EPI_ISL_1459956, EPI_ISL_1459957, EPI_ISL_1459958, EPI_ISL_1459959, EPI_ISL_1459960, EPI_ISL_1459961, EPI_ISL_1459962, EPI_ISL_1459963, EPI_ISL_1459964, EPI_ISL_1459965, EPI_ISL_1459966, EPI_ISL_1459967, EPI_ISL_1459968, EPI_ISL_1459969, EPI_ISL_1459970, EPI_ISL_1459971,
EPI_ISL_1459972, EPI_ISL_1459973, EPI_ISL_1459974, EPI_ISL_1459975, EPI_ISL_1459976, EPI_ISL_1459977, EPI_ISL_1459978, EPI_ISL_1459979, EPI_ISL_1459980, EPI_ISL_1459981, EPI_ISL_1459982, EPI_ISL_1459983, EPI_ISL_1459984, EPI_ISL_1459985, EPI_ISL_1459986, EPI_ISL_1459987, EPI_ISL_1459988, EPI_ISL_1459989, EPI_ISL_1459990, EPI_ISL_1459991, EPI_ISL_1459992, EPI_ISL_1459993,
EPI_ISL_1459994, EPI_ISL_1459995, EPI_ISL_1459996, EPI_ISL_1459997, EPI_ISL_1459998, EPI_ISL_1459999, EPI_ISL_1460000, EPI_ISL_1460001, EPI_ISL_1460002, EPI_ISL_1460003, EPI_ISL_1460004, EPI_ISL_1460005, EPI_ISL_1460006, EPI_ISL_1460007, EPI_ISL_1460008, EPI_ISL_1460009, EPI_ISL_1460010, EPI_ISL_1460011, EPI_ISL_1460012, EPI_ISL_1460013, EPI_ISL_1460014, EPI_ISL_1460015,
EPI_ISL_1460016, EPI_ISL_1460017, EPI_ISL_1460018, EPI_ISL_1460019, EPI_ISL_1460020, EPI_ISL_1460021, EPI_ISL_1460022, EPI_ISL_1460023, EPI_ISL_1460024, EPI_ISL_1460025, EPI_ISL_1460026, EPI_ISL_1460027, EPI_ISL_1460028, EPI_ISL_1460029, EPI_ISL_1460030, EPI_ISL_1460031, EPI_ISL_1460032, EPI_ISL_1460033, EPI_ISL_1460034, EPI_ISL_1460035, EPI_ISL_1460036, EPI_ISL_1460037,
EPI_ISL_1460038, EPI_ISL_1460039, EPI_ISL_1460040, EPI_ISL_1460041, EPI_ISL_1460042, EPI_ISL_1460043, EPI_ISL_1460044, EPI_ISL_1460045, EPI_ISL_1460046, EPI_ISL_1460047, EPI_ISL_1460048, EPI_ISL_1460049, EPI_ISL_1460050, EPI_ISL_1460051, EPI_ISL_1460052, EPI_ISL_1460053, EPI_ISL_1460054, EPI_ISL_1460055, EPI_ISL_1460056, EPI_ISL_1460057, EPI_ISL_1460058, EPI_ISL_1460059,
EPI_ISL_1460060, EPI_ISL_1460061, EPI_ISL_1460062, EPI_ISL_1460063, EPI_ISL_1460064, EPI_ISL_1460065, EPI_ISL_1460066, EPI_ISL_1460067, EPI_ISL_1460068, EPI_ISL_1460069, EPI_ISL_1460070, EPI_ISL_1460071, EPI_ISL_1460072, EPI_ISL_1460073, EPI_ISL_1460074, EPI_ISL_1460075, EPI_ISL_1460076, EPI_ISL_1460077, EPI_ISL_1460078, EPI_ISL_1460079, EPI_ISL_1460080, EPI_ISL_1460081,
EPI_ISL_1460082, EPI_ISL_1460083, EPI_ISL_1460084, EPI_ISL_1460085, EPI_ISL_1460086, EPI_ISL_1460087, EPI_ISL_1460088, EPI_ISL_1460089, EPI_ISL_1460090, EPI_ISL_1460091, EPI_ISL_1460092, EPI_ISL_1460093, EPI_ISL_1460094, EPI_ISL_1460095, EPI_ISL_1460096, EPI_ISL_1460097, EPI_ISL_1460098, EPI_ISL_1460099, EPI_ISL_1460100, EPI_ISL_1460101, EPI_ISL_1460102, EPI_ISL_1460103,
EPI_ISL_1460104, EPI_ISL_1460105, EPI_ISL_1460106, EPI_ISL_1460107, EPI_ISL_1460108, EPI_ISL_1460109, EPI_ISL_1460110, EPI_ISL_1460111, EPI_ISL_1460112, EPI_ISL_1460113, EPI_ISL_1460114, EPI_ISL_1460115, EPI_ISL_1460116, EPI_ISL_1460117, EPI_ISL_1460118, EPI_ISL_1460119, EPI_ISL_1460120, EPI_ISL_1460121, EPI_ISL_1460123, EPI_ISL_1460124, EPI_ISL_1460125, EPI_ISL_1460126,
EPI_ISL_1460127, EPI_ISL_1460128, EPI_ISL_1460129, EPI_ISL_1460130, EPI_ISL_1460131, EPI_ISL_1460132, EPI_ISL_1460133, EPI_ISL_1460134, EPI_ISL_1460135, EPI_ISL_1460136, EPI_ISL_1460137, EPI_ISL_1460138, EPI_ISL_1460139, EPI_ISL_1460140, EPI_ISL_1460141, EPI_ISL_1460142, EPI_ISL_1460143, EPI_ISL_1460144, EPI_ISL_1460145, EPI_ISL_1460146, EPI_ISL_1460147, EPI_ISL_1460148,
EPI_ISL_1460149, EPI_ISL_1460150, EPI_ISL_1460151, EPI_ISL_1460152, EPI_ISL_1460153, EPI_ISL_1460154, EPI_ISL_1460155, EPI_ISL_1460156, EPI_ISL_1460157, EPI_ISL_1460158, EPI_ISL_1460159, EPI_ISL_1460160, EPI_ISL_1460161, EPI_ISL_1460162, EPI_ISL_1460163, EPI_ISL_1460164, EPI_ISL_1460165, EPI_ISL_1460166, EPI_ISL_1460167, EPI_ISL_1460168, EPI_ISL_1460169, EPI_ISL_1460170,
EPI_ISL_1460171, EPI_ISL_1460172, EPI_ISL_1460173, EPI_ISL_1460174, EPI_ISL_1460175, EPI_ISL_1460176, EPI_ISL_1460177, EPI_ISL_1460178, EPI_ISL_1460179, EPI_ISL_1460180, EPI_ISL_1460181, EPI_ISL_1460182, EPI_ISL_1460183, EPI_ISL_1460184, EPI_ISL_1460185, EPI_ISL_1460186, EPI_ISL_1460187, EPI_ISL_1460188, EPI_ISL_1460189, EPI_ISL_1460190, EPI_ISL_1460191, EPI_ISL_1460192,
EPI_ISL_1460193, EPI_ISL_1460194, EPI_ISL_1460195, EPI_ISL_1460196, EPI_ISL_1460197, EPI_ISL_1460198, EPI_ISL_1460199, EPI_ISL_1460200, EPI_ISL_1460201, EPI_ISL_1460202, EPI_ISL_1460203, EPI_ISL_1460204, EPI_ISL_1460205, EPI_ISL_1460206, EPI_ISL_1460207, EPI_ISL_1460208, EPI_ISL_1460209, EPI_ISL_1460210, EPI_ISL_1460211, EPI_ISL_1460212, EPI_ISL_1460213, EPI_ISL_1460214,
EPI_ISL_1460263, EPI_ISL_1460295, EPI_ISL_1460296, EPI_ISL_1460297, EPI_ISL_1460298, EPI_ISL_1460299, EPI_ISL_1460300, EPI_ISL_1460301, EPI_ISL_1460302, EPI_ISL_1460303, EPI_ISL_1460304, EPI_ISL_1460305, EPI_ISL_1460306, EPI_ISL_1460307, EPI_ISL_1460308, EPI_ISL_1460309, EPI_ISL_1460310, EPI_ISL_1460311, EPI_ISL_1460312, EPI_ISL_1460313, EPI_ISL_1460314, EPI_ISL_1460315,
EPI_ISL_1460316, EPI_ISL_1460317, EPI_ISL_1460318, EPI_ISL_1460319, EPI_ISL_1460320, EPI_ISL_1460321, EPI_ISL_1460322, EPI_ISL_1460323, EPI_ISL_1460324, EPI_ISL_1460325, EPI_ISL_1460326, EPI_ISL_1460327, EPI_ISL_1460328, EPI_ISL_1460329, EPI_ISL_1460330, EPI_ISL_1460331, EPI_ISL_1460332, EPI_ISL_1460333, EPI_ISL_1460334, EPI_ISL_1460335, EPI_ISL_1460336, EPI_ISL_1460337,
EPI_ISL_1460338, EPI_ISL_1460339, EPI_ISL_1460340, EPI_ISL_1460341, EPI_ISL_1460342, EPI_ISL_1460343, EPI_ISL_1460344, EPI_ISL_1460345, EPI_ISL_1460346, EPI_ISL_1460347, EPI_ISL_1460348, EPI_ISL_1460349, EPI_ISL_1460350, EPI_ISL_1460351, EPI_ISL_1460352, EPI_ISL_1460353, EPI_ISL_1460356, EPI_ISL_1460357, EPI_ISL_1460358, EPI_ISL_1460359, EPI_ISL_1460360, EPI_ISL_1460361,
EPI_ISL_1460362, EPI_ISL_1460363, EPI_ISL_1460364, EPI_ISL_1460365, EPI_ISL_1460366, EPI_ISL_1460367, EPI_ISL_1460368, EPI_ISL_1460369, EPI_ISL_1460370, EPI_ISL_1460371, EPI_ISL_1460372, EPI_ISL_1460373, EPI_ISL_1460374, EPI_ISL_1460375, EPI_ISL_1460376, EPI_ISL_1460377, EPI_ISL_1460378, EPI_ISL_1460379, EPI_ISL_1460380, EPI_ISL_1460381, EPI_ISL_1460382, EPI_ISL_1460383,
EPI_ISL_1460384, EPI_ISL_1460385, EPI_ISL_1460386, EPI_ISL_1460387, EPI_ISL_1460388, EPI_ISL_1460389, EPI_ISL_1460390, EPI_ISL_1460391, EPI_ISL_1460392, EPI_ISL_1460393, EPI_ISL_1460394, EPI_ISL_1460395, EPI_ISL_1460396, EPI_ISL_1460397, EPI_ISL_1460398, EPI_ISL_1460399, EPI_ISL_1460400, EPI_ISL_1460401, EPI_ISL_1460402, EPI_ISL_1460403, EPI_ISL_1460404, EPI_ISL_1460405,

EPI_ISL_1460406, EPI_ISL_1460407, EPI_ISL_1460408, EPI_ISL_1460409, EPI_ISL_1460410, EPI_ISL_1460411, EPI_ISL_1460412, EPI_ISL_1460413, EPI_ISL_1460414, EPI_ISL_1460415, EPI_ISL_1460416, EPI_ISL_1460417, EPI_ISL_1460418, EPI_ISL_1460419, EPI_ISL_1460420, EPI_ISL_1460421, EPI_ISL_1460422, EPI_ISL_1460423, EPI_ISL_1460424, EPI_ISL_1460425, EPI_ISL_1460426, EPI_ISL_1460427, EPI_ISL_1460428, EPI_ISL_1460429, EPI_ISL_1460430, EPI_ISL_1460431, EPI_ISL_1460432, EPI_ISL_1460433, EPI_ISL_1460434, EPI_ISL_1460435, EPI_ISL_1460436, EPI_ISL_1460437, EPI_ISL_1460438, EPI_ISL_1460439, EPI_ISL_1460440, EPI_ISL_1460442, EPI_ISL_1460443, EPI_ISL_1460444, EPI_ISL_1460445, EPI_ISL_1460446, EPI_ISL_1460447, EPI_ISL_1460448, EPI_ISL_1460449, EPI_ISL_1460450, EPI_ISL_1460451, EPI_ISL_1460452, EPI_ISL_1460453, EPI_ISL_1460454, EPI_ISL_1460455, EPI_ISL_1460456, EPI_ISL_1460457, EPI_ISL_1460458, EPI_ISL_1460459, EPI_ISL_1460460, EPI_ISL_1460461, EPI_ISL_1460462, EPI_ISL_1460463, EPI_ISL_1460464, EPI_ISL_1460465, EPI_ISL_1460466, EPI_ISL_1460467, EPI_ISL_1460468, EPI_ISL_1460469, EPI_ISL_1460470, EPI_ISL_1460471, EPI_ISL_1460472, EPI_ISL_1460473, EPI_ISL_1460474, EPI_ISL_1460475, EPI_ISL_1460476, EPI_ISL_1460477, EPI_ISL_1460478, EPI_ISL_1460479, EPI_ISL_1460480, EPI_ISL_1460481, EPI_ISL_1460482, EPI_ISL_1460483, EPI_ISL_1460484, EPI_ISL_1460485, EPI_ISL_1460486, EPI_ISL_1460487, EPI_ISL_1460488, EPI_ISL_1460489, EPI_ISL_1460490, EPI_ISL_1460491, EPI_ISL_1460492, EPI_ISL_1460493, EPI_ISL_1460494, EPI_ISL_1460495, EPI_ISL_1460496, EPI_ISL_1460497, EPI_ISL_1460498, EPI_ISL_1460499, EPI_ISL_1460500, EPI_ISL_1460501, EPI_ISL_1460502, EPI_ISL_1460503, EPI_ISL_1460504, EPI_ISL_1460505, EPI_ISL_1460506, EPI_ISL_1460507, EPI_ISL_1460508, EPI_ISL_1460509, EPI_ISL_1460510, EPI_ISL_1460511, EPI_ISL_1460512, EPI_ISL_1460513, EPI_ISL_1460514, EPI_ISL_1460515, EPI_ISL_1460516, EPI_ISL_1460517, EPI_ISL_1460518, EPI_ISL_1460519, EPI_ISL_1460520, EPI_ISL_1460521, EPI_ISL_1460522, EPI_ISL_1460523, EPI_ISL_1460524, EPI_ISL_1460525, EPI_ISL_1460526, EPI_ISL_1460527, EPI_ISL_1460528, EPI_ISL_1460529, EPI_ISL_1460530, EPI_ISL_1460531, EPI_ISL_1460532, EPI_ISL_1460533, EPI_ISL_1460534, EPI_ISL_1460535, EPI_ISL_1460536, EPI_ISL_1460537, EPI_ISL_1460538, EPI_ISL_1460539, EPI_ISL_1460540, EPI_ISL_1460541, EPI_ISL_1460542, EPI_ISL_1460543, EPI_ISL_1460544, EPI_ISL_1460545, EPI_ISL_1460546, EPI_ISL_1460547, EPI_ISL_1460548, EPI_ISL_1460549, EPI_ISL_1460550, EPI_ISL_1460551, EPI_ISL_1460552, EPI_ISL_1460553, EPI_ISL_1460554, EPI_ISL_1460555, EPI_ISL_1460556, EPI_ISL_1460557, EPI_ISL_1460558, EPI_ISL_1460559, EPI_ISL_1460560, EPI_ISL_1460561, EPI_ISL_1460562, EPI_ISL_1460563, EPI_ISL_1460564, EPI_ISL_1460565, EPI_ISL_1460566, EPI_ISL_1460567, EPI_ISL_1460568, EPI_ISL_1460569, EPI_ISL_1460570, EPI_ISL_1460571, EPI_ISL_1460572, EPI_ISL_1460573, EPI_ISL_1460574, EPI_ISL_1460575, EPI_ISL_1460576, EPI_ISL_1460577, EPI_ISL_1460578, EPI_ISL_1460580, EPI_ISL_1460581, EPI_ISL_1460582, EPI_ISL_1460583, EPI_ISL_1460584, EPI_ISL_1460585, EPI_ISL_1460586, EPI_ISL_1460587, EPI_ISL_1460588, EPI_ISL_1460589, EPI_ISL_1460590, EPI_ISL_1460591, EPI_ISL_1460592, EPI_ISL_1460593, EPI_ISL_1460594, EPI_ISL_1460595, EPI_ISL_1460596, EPI_ISL_1460597, EPI_ISL_1460598, EPI_ISL_1460599, EPI_ISL_1460600, EPI_ISL_1460601, EPI_ISL_1460602, EPI_ISL_1460603, EPI_ISL_1460604, EPI_ISL_1460605, EPI_ISL_1460606, EPI_ISL_1460607, EPI_ISL_1460608, EPI_ISL_1460609, EPI_ISL_1460610, EPI_ISL_1460611, EPI_ISL_1460612, EPI_ISL_1460613, EPI_ISL_1460614, EPI_ISL_1460615, EPI_ISL_1460616, EPI_ISL_1460617, EPI_ISL_1460618, EPI_ISL_1460619, EPI_ISL_1460620, EPI_ISL_1460621, EPI_ISL_1460622, EPI_ISL_1460623, EPI_ISL_1460624, EPI_ISL_1460625, EPI_ISL_1460626, EPI_ISL_1460627, EPI_ISL_1460628, EPI_ISL_1460629, EPI_ISL_1460630, EPI_ISL_1460631, EPI_ISL_1460632, EPI_ISL_1460633, EPI_ISL_1460634, EPI_ISL_1460635, EPI_ISL_1460636, EPI_ISL_1460637, EPI_ISL_1460638, EPI_ISL_1460639, EPI_ISL_1460640, EPI_ISL_1460641, EPI_ISL_1460642, EPI_ISL_1460643, EPI_ISL_1460644, EPI_ISL_1460645, EPI_ISL_1460646, EPI_ISL_1460647, EPI_ISL_1460648, EPI_ISL_1460649, EPI_ISL_1460650, EPI_ISL_1460651, EPI_ISL_1460652, EPI_ISL_1460653, EPI_ISL_1460654, EPI_ISL_1460655, EPI_ISL_1460656, EPI_ISL_1460657, EPI_ISL_1460658, EPI_ISL_1460659, EPI_ISL_1460660, EPI_ISL_1460661, EPI_ISL_1460662, EPI_ISL_1460663, EPI_ISL_1460664, EPI_ISL_1460665, EPI_ISL_1460666, EPI_ISL_1460667, EPI_ISL_1460668, EPI_ISL_1460669, EPI_ISL_1460670, EPI_ISL_1460671, EPI_ISL_1460672, EPI_ISL_1460673, EPI_ISL_1460674, EPI_ISL_1460675, EPI_ISL_1460676, EPI_ISL_1460677, EPI_ISL_1460678, EPI_ISL_1460679, EPI_ISL_1460680, EPI_ISL_1460681, EPI_ISL_1460682, EPI_ISL_1460683, EPI_ISL_1460684, EPI_ISL_1460685, EPI_ISL_1460686, EPI_ISL_1460687, EPI_ISL_1460688, EPI_ISL_1460689, EPI_ISL_1460690, EPI_ISL_1460691, EPI_ISL_1460692, EPI_ISL_1460693, EPI_ISL_1460694, EPI_ISL_1460695, EPI_ISL_1460696, EPI_ISL_1460697, EPI_ISL_1460698, EPI_ISL_1460699, EPI_ISL_1460700, EPI_ISL_1460701, EPI_ISL_1460702, EPI_ISL_1460703, EPI_ISL_1460704, EPI_ISL_1460705, EPI_ISL_1460706, EPI_ISL_1460707, EPI_ISL_1460708, EPI_ISL_1460709, EPI_ISL_1460710, EPI_ISL_1460711, EPI_ISL_1460712, EPI_ISL_1460713, EPI_ISL_1460714, EPI_ISL_1460715, EPI_ISL_1460716, EPI_ISL_1460717, EPI_ISL_1460718, EPI_ISL_1460719, EPI_ISL_1460720, EPI_ISL_1460721, EPI_ISL_1460722, EPI_ISL_1460723, EPI_ISL_1460724, EPI_ISL_1460725, EPI_ISL_1460726, EPI_ISL_1460727, EPI_ISL_1460728, EPI_ISL_1460729, EPI_ISL_1481550, EPI_ISL_1481551, EPI_ISL_1481552, EPI_ISL_1481553, EPI_ISL_1481555, EPI_ISL_1481556, EPI_ISL_1481557, EPI_ISL_1481558, EPI_ISL_1481559, EPI_ISL_1481560, EPI_ISL_1481561, EPI_ISL_1481562, EPI_ISL_1481563, EPI_ISL_1481564, EPI_ISL_1481565, EPI_ISL_1481566, EPI_ISL_1481567, EPI_ISL_1481568, EPI_ISL_1481569, EPI_ISL_1481570, EPI_ISL_1481571, EPI_ISL_1481572, EPI_ISL_1481573, EPI_ISL_1481574, EPI_ISL_1481575, EPI_ISL_1481576, EPI_ISL_1481577, EPI_ISL_1481578, EPI_ISL_1481579, EPI_ISL_1481580, EPI_ISL_1481581, EPI_ISL_1481582, EPI_ISL_1481583, EPI_ISL_1481584, EPI_ISL_1481585, EPI_ISL_1481586, EPI_ISL_1481587, EPI_ISL_1481588, EPI_ISL_1481589, EPI_ISL_1481590, EPI_ISL_1481591, EPI_ISL_1481592, EPI_ISL_1481593, EPI_ISL_1481594, EPI_ISL_1481595, EPI_ISL_1481596, EPI_ISL_1481597, EPI_ISL_1481679, EPI_ISL_1481716, EPI_ISL_1481717, EPI_ISL_1481718, EPI_ISL_1481719, EPI_ISL_1481720, EPI_ISL_1481721, EPI_ISL_1481722, EPI_ISL_1481723, EPI_ISL_1481724, EPI_ISL_1481725, EPI_ISL_1481726, EPI_ISL_1481727, EPI_ISL_1481728, EPI_ISL_1481729, EPI_ISL_1481730, EPI_ISL_1481731, EPI_ISL_1481732, EPI_ISL_1481733, EPI_ISL_1481734, EPI_ISL_1481735, EPI_ISL_1481736, EPI_ISL_1481737, EPI_ISL_1481738, EPI_ISL_1481739, EPI_ISL_1481740, EPI_ISL_1481741, EPI_ISL_1481742, EPI_ISL_1481743, EPI_ISL_1481744, EPI_ISL_1481745, EPI_ISL_1481746, EPI_ISL_1481747, EPI_ISL_1481748, EPI_ISL_1481749, EPI_ISL_1481750, EPI_ISL_1481751, EPI_ISL_1481752, EPI_ISL_1481753, EPI_ISL_1481754, EPI_ISL_1481755, EPI_ISL_1481756, EPI_ISL_1481757, EPI_ISL_1481758, EPI_ISL_1481759, EPI_ISL_1481760, EPI_ISL_1481761, EPI_ISL_1481762, EPI_ISL_1481763, EPI_ISL_1481764, EPI_ISL_1481765, EPI_ISL_1481766, EPI_ISL_1481767, EPI_ISL_1481768, EPI_ISL_1481769, EPI_ISL_1481770, EPI_ISL_1481771, EPI_ISL_1481772, EPI_ISL_1481773, EPI_ISL_1481774, EPI_ISL_1481775, EPI_ISL_1481776, EPI_ISL_1481777, EPI_ISL_1481778, EPI_ISL_1481779, EPI_ISL_1481780, EPI_ISL_1481781, EPI_ISL_1481782, EPI_ISL_1481783, EPI_ISL_1481784, EPI_ISL_1481785, EPI_ISL_1481786, EPI_ISL_1481787, EPI_ISL_1481788, EPI_ISL_1481789, EPI_ISL_1481790, EPI_ISL_1481791, EPI_ISL_1481792, EPI_ISL_1481793, EPI_ISL_1481794, EPI_ISL_1481795, EPI_ISL_1481796, EPI_ISL_1481797, EPI_ISL_1481798, EPI_ISL_1481799, EPI_ISL_1481800, EPI_ISL_1481801, EPI_ISL_1481802, EPI_ISL_1481803, EPI_ISL_1481804, EPI_ISL_1481805, EPI_ISL_1481806, EPI_ISL_1481807, EPI_ISL_1481808, EPI_ISL_1481809, EPI_ISL_1481810, EPI_ISL_1481811, EPI_ISL_1481812, EPI_ISL_1481813, EPI_ISL_1481814, EPI_ISL_1481815, EPI_ISL_1481816, EPI_ISL_1481817, EPI_ISL_1481818, EPI_ISL_1481819, EPI_ISL_1481837, EPI_ISL_1481901, EPI_ISL_1481904, EPI_ISL_1481905, EPI_ISL_1481906, EPI_ISL_1481907, EPI_ISL_1481908, EPI_ISL_1481909, EPI_ISL_1481910, EPI_ISL_1481911, EPI_ISL_1481912, EPI_ISL_1481913, EPI_ISL_1481914, EPI_ISL_1481915, EPI_ISL_1481916, EPI_ISL_1481917, EPI_ISL_1481918, EPI_ISL_1481919, EPI_ISL_1481920, EPI_ISL_1481921, EPI_ISL_1481922, EPI_ISL_1481923, EPI_ISL_1481924, EPI_ISL_1481925, EPI_ISL_1481926, EPI_ISL_1481927, EPI_ISL_1481928, EPI_ISL_1481929, EPI_ISL_1481930, EPI_ISL_1481931, EPI_ISL_1481932, EPI_ISL_1481933, EPI_ISL_1481934, EPI_ISL_1481935, EPI_ISL_1481936, EPI_ISL_1481937, EPI_ISL_1481938, EPI_ISL_1481939, EPI_ISL_1481940, EPI_ISL_1481941, EPI_ISL_1481942, EPI_ISL_1481943, EPI_ISL_1481944, EPI_ISL_1481945, EPI_ISL_1481946, EPI_ISL_1481947, EPI_ISL_1481948, EPI_ISL_1481949, EPI_ISL_1481950, EPI_ISL_1481951, EPI_ISL_1481952, EPI_ISL_1481953, EPI_ISL_1481954, EPI_ISL_1481955, EPI_ISL_1481956, EPI_ISL_1481957, EPI_ISL_1481958, EPI_ISL_1481959, EPI_ISL_1481960, EPI_ISL_1481961, EPI_ISL_1481962, EPI_ISL_1481963, EPI_ISL_1481964, EPI_ISL_1481965, EPI_ISL_1481966, EPI_ISL_1481967, EPI_ISL_1481968, EPI_ISL_1481969, EPI_ISL_1481970, EPI_ISL_1481971, EPI_ISL_1481972, EPI_ISL_1481973, EPI_ISL_1481974, EPI_ISL_1481975, EPI_ISL_1481976, EPI_ISL_1481977, EPI_ISL_1481978, EPI_ISL_1481979, EPI_ISL_1481980, EPI_ISL_1481981, EPI_ISL_1481982, EPI_ISL_1481983, EPI_ISL_1481984, EPI_ISL_1481985, EPI_ISL_1481986, EPI_ISL_1481987, EPI_ISL_1481988, EPI_ISL_1481989, EPI_ISL_1481990, EPI_ISL_1481991, EPI_ISL_1481992, EPI_ISL_1481993, EPI_ISL_1481994, EPI_ISL_1481995, EPI_ISL_1481996, EPI_ISL_1481997, EPI_ISL_1482000, EPI_ISL_1482001, EPI_ISL_1482002, EPI_ISL_1482003, EPI_ISL_1482004, EPI_ISL_1482005, EPI_ISL_1482006, EPI_ISL_1482007, EPI_ISL_1482008, EPI_ISL_1482009, EPI_ISL_1482010, EPI_ISL_1482011, EPI_ISL_1482012, EPI_ISL_1482013, EPI_ISL_1482014, EPI_ISL_1482015, EPI_ISL_1482016, EPI_ISL_1482017, EPI_ISL_1482018, EPI_ISL_1482019, EPI_ISL_1482020, EPI_ISL_1482021, EPI_ISL_1482022, EPI_ISL_1482023, EPI_ISL_1482024, EPI_ISL_1482025, EPI_ISL_1482026, EPI_ISL_1482027, EPI_ISL_1482028, EPI_ISL_1482029, EPI_ISL_1482030, EPI_ISL_1482031, EPI_ISL_1482032, EPI_ISL_1482033, EPI_ISL_1482034, EPI_ISL_1482035, EPI_ISL_1482042, EPI_ISL_1482043, EPI_ISL_1482044, EPI_ISL_1482045, EPI_ISL_1482046, EPI_ISL_1482047, EPI_ISL_1482048, EPI_ISL_1482050, EPI_ISL_1482051, EPI_ISL_1482052, EPI_ISL_1482053, EPI_ISL_1482054, EPI_ISL_1482055, EPI_ISL_1482056, EPI_ISL_1482057, EPI_ISL_1482058, EPI_ISL_1482059, EPI_ISL_1482060, EPI_ISL_1482061, EPI_ISL_1482062, EPI_ISL_1482063, EPI_ISL_1482064, EPI_ISL_1482065, EPI_ISL_1482066, EPI_ISL_1482067, EPI_ISL_1482068, EPI_ISL_1482069, EPI_ISL_1482070, EPI_ISL_1482071, EPI_ISL_1482072, EPI_ISL_1482073, EPI_ISL_1482074, EPI_ISL_1482075, EPI_ISL_1482076, EPI_ISL_1482077, EPI_ISL_1482078, EPI_ISL_1482079, EPI_ISL_1482080, EPI_ISL_1482081, EPI_ISL_1482082, EPI_ISL_1482083, EPI_ISL_1482084, EPI_ISL_1482085, EPI_ISL_1482086, EPI_ISL_1482087, EPI_ISL_1482088, EPI_ISL_1482089, EPI_ISL_1482090, EPI_ISL_1482091, EPI_ISL_1482092, EPI_ISL_1482093, EPI_ISL_1482094, EPI_ISL_1482095, EPI_ISL_1482096, EPI_ISL_1482097, EPI_ISL_1482098, EPI_ISL_1482099, EPI_ISL_1482101, EPI_ISL_1482102, EPI_ISL_1482103, EPI_ISL_1482104, EPI_ISL_1482105, EPI_ISL_1482106, EPI_ISL_1482107, EPI_ISL_1482108, EPI_ISL_1482109, EPI_ISL_1482110, EPI_ISL_1482111, EPI_ISL_1482112, EPI_ISL_1482113, EPI_ISL_1482114, EPI_ISL_1482115, EPI_ISL_1482116, EPI_ISL_1482117, EPI_ISL_1482118, EPI_ISL_1482119, EPI_ISL_1482120, EPI_ISL_1482121, EPI_ISL_1482122, EPI_ISL_1482123, EPI_ISL_1482124, EPI_ISL_1482125, EPI_ISL_1482126, EPI_ISL_1482127, EPI_ISL_1482128, EPI_ISL_1482129, EPI_ISL_1482130, EPI_ISL_1482131, EPI_ISL_1482132, EPI_ISL_1482133, EPI_ISL_1482134, EPI_ISL_1482135, EPI_ISL_1482136, EPI_ISL_1482137, EPI_ISL_1482138, EPI_ISL_1482139, EPI_ISL_1482140, EPI_ISL_1482141, EPI_ISL_1482142, EPI_ISL_1482143, EPI_ISL_1482144, EPI_ISL_1482145, EPI_ISL_1482146, EPI_ISL_1482147, EPI_ISL_1482148, EPI_ISL_1482149, EPI_ISL_1482150, EPI_ISL_1482151, EPI_ISL_1482152, EPI_ISL_1482153, EPI_ISL_1482154, EPI_ISL_1482155, EPI_ISL_1482156, EPI_ISL_1482157, EPI_ISL_1482158, EPI_ISL_1482159, EPI_ISL_1482160, EPI_ISL_1482161, EPI_ISL_1482162, EPI_ISL_1482163, EPI_ISL_1482164, EPI_ISL_1482165, EPI_ISL_1482166, EPI_ISL_1482167, EPI_ISL_1482168, EPI_ISL_1482169, EPI_ISL_1482170, EPI_ISL_1482171, EPI_ISL_1482172, EPI_ISL_1482173, EPI_ISL_1482174, EPI_ISL_1482175, EPI_ISL_1482176, EPI_ISL_1482194, EPI_ISL_1525545, EPI_ISL_1525546, EPI_ISL_1525547, EPI_ISL_1525548, EPI_ISL_1525549, EPI_ISL_1525550, EPI_ISL_1525551, EPI_ISL_1525552, EPI_ISL_1525553, EPI_ISL_1525554, EPI_ISL_1525555, EPI_ISL_1525556, EPI_ISL_1525557, EPI_ISL_1525558, EPI_ISL_1525559, EPI_ISL_1525560, EPI_ISL_1525561, EPI_ISL_1525562, EPI_ISL_1525563, EPI_ISL_1525564, EPI_ISL_1525565, EPI_ISL_1525566, EPI_ISL_1525567, EPI_ISL_1525568, EPI_ISL_1525569, EPI_ISL_1525570, EPI_ISL_1525571, EPI_ISL_1525572, EPI_ISL_1525573, EPI_ISL_1525574, EPI_ISL_1525575, EPI_ISL_1525576, EPI_ISL_1525577, EPI_ISL_1525578, EPI_ISL_1525579, EPI_ISL_1525580, EPI_ISL_1525581, EPI_ISL_1525582, EPI_ISL_1525583, EPI_ISL_1525584, EPI_ISL_1525585, EPI_ISL_1525586, EPI_ISL_1525587, EPI_ISL_1525588, EPI_ISL_1525589, EPI_ISL_1525590, EPI_ISL_1525591, EPI_ISL_1525592, EPI_ISL_1525593, EPI_ISL_1525594, EPI_ISL_1525595, EPI_ISL_1525596, EPI_ISL_1525597, EPI_ISL_1525598, EPI_ISL_1525599, EPI_ISL_1525600, EPI_ISL_1525601, EPI_ISL_1525602, EPI_ISL_1525603, EPI_ISL_1525604, EPI_ISL_1525605, EPI_ISL_1525606, EPI_ISL_1525607, EPI_ISL_1525608, EPI_ISL_1525609, EPI_ISL_1525610, EPI_ISL_1525611, EPI_ISL_1525612, EPI_ISL_1525613, EPI_ISL_1525614, EPI_ISL_1525615, EPI_ISL_1525616, EPI_ISL_1525617, EPI_ISL_1525618, EPI_ISL_1525619, EPI_ISL_1525620, EPI_ISL_1525621, EPI_ISL_1525622, EPI_ISL_1525623, EPI_ISL_1525624, EPI_ISL_1525625, EPI_ISL_1525626, EPI_ISL_1525627, EPI_ISL_1525635, EPI_ISL_1525641, EPI_ISL_1525654, EPI_ISL_1525657, EPI_ISL_1525664, EPI_ISL_1525665, EPI_ISL_1525693, EPI_ISL_1525715, EPI_ISL_1554950, EPI_ISL_1554951, EPI_ISL_1554953, EPI_ISL_1554954, EPI_ISL_1554956, EPI_ISL_1554957, EPI_ISL_1554959, EPI_ISL_1554962, EPI_ISL_1554963, EPI_ISL_1554964, EPI_ISL_1554965, EPI_ISL_1554966, EPI_ISL_1554967, EPI_ISL_1554968, EPI_ISL_1554969, EPI_ISL_1554970, EPI_ISL_1554971, EPI_ISL_1554972, EPI_ISL_1554973, EPI_ISL_1554974, EPI_ISL_1554975, EPI_ISL_1554976, EPI_ISL_1554977, EPI_ISL_1554978, EPI_ISL_1554979, EPI_ISL_1554980, EPI_ISL_1554982, EPI_ISL_1554983, EPI_ISL_1554984, EPI_ISL_1554985, EPI_ISL_1554986, EPI_ISL_1554987, EPI_ISL_1554988, EPI_ISL_1554989, EPI_ISL_1554990, EPI_ISL_1554991, EPI_ISL_1554992, EPI_ISL_1554993, EPI_ISL_1554994, EPI_ISL_1554995, EPI_ISL_1554996, EPI_ISL_1554997, EPI_ISL_1554998, EPI_ISL_1554999, EPI_ISL_1555000, EPI_ISL_1555001, EPI_ISL_1555002, EPI_ISL_1555003, EPI_ISL_1555004, EPI_ISL_1555005, EPI_ISL_1555006, EPI_ISL_1555007, EPI_ISL_1555010, EPI_ISL_1555011, EPI_ISL_1555012, EPI_ISL_1555013, EPI_ISL_1555014, EPI_ISL_1555015, EPI_ISL_1555016, EPI_ISL_1555017, EPI_ISL_1555018, EPI_ISL_1555019, EPI_ISL_1555020, EPI_ISL_1555021, EPI_ISL_1555022, EPI_ISL_1555023, EPI_ISL_1555024, EPI_ISL_1555025, EPI_ISL_1555026, EPI_ISL_1555027, EPI_ISL_1555028, EPI_ISL_1555029, EPI_ISL_1555030, EPI_ISL_1555031, EPI_ISL_1555032, EPI_ISL_1555033, EPI_ISL_1555034, EPI_ISL_1555035, EPI_ISL_1555036, EPI_ISL_1555037, EPI_ISL_1555042, EPI_ISL_1555043, EPI_ISL_1555044, EPI_ISL_1555045, EPI_ISL_1555046, EPI_ISL_1555047, EPI_ISL_1555048, EPI_ISL_1555049, EPI_ISL_1555050, EPI_ISL_1555052, EPI_ISL_1555053, EPI_ISL_1555054, EPI_ISL_1555055, EPI_ISL_1555056, EPI_ISL_1555057, EPI_ISL_1555058, EPI_ISL_1555059, EPI_ISL_1555060, EPI_ISL_1555061, EPI_ISL_1555062, EPI_ISL_1555063, EPI_ISL_1555064, EPI_ISL_1555065, EPI_ISL_1555066, EPI_ISL_1555067, EPI_ISL_1555068, EPI_ISL_1555069, EPI_ISL_1555070, EPI_ISL_1555071, EPI_ISL_1555072, EPI_ISL_1555073, EPI_ISL_1555074, EPI_ISL_1555075, EPI_ISL_1555076, EPI_ISL_1555077, EPI_ISL_1555078, EPI_ISL_1555079, EPI_ISL_1555080, EPI_ISL_1555081, EPI_ISL_1555082, EPI_ISL_1555083, EPI_ISL_1555084, EPI_ISL_1555085, EPI_ISL_1555086, EPI_ISL_1555087, EPI_ISL_1555088, EPI_ISL_1555089, EPI_ISL_1555090, EPI_ISL_1555091, EPI_ISL_1555092, EPI_ISL_1555093, EPI_ISL_1555094, EPI_ISL_1555095, EPI_ISL_1555096, EPI_ISL_1555097, EPI_ISL_1555098, EPI_ISL_1555099, EPI_ISL_1555100, EPI_ISL_1555101, EPI_ISL_1555102, EPI_ISL_1555103, EPI_ISL_1555104, EPI_ISL_1555105, EPI_ISL_1555106, EPI_ISL_1555107, EPI_ISL_1555108, EPI_ISL_1555109, EPI_ISL_1555110, EPI_ISL_1555112, EPI_ISL_1555113, EPI_ISL_1555114, EPI_ISL_1555117, EPI_ISL_1555118, EPI_ISL_1555119, EPI_ISL_1555120, EPI_ISL_1555121, EPI_ISL_1555122, EPI_ISL_1555123, EPI_ISL_1555124, EPI_ISL_1555125, EPI_ISL_1555126, EPI_ISL_1555127, EPI_ISL_1555128, EPI_ISL_1555129, EPI_ISL_1555130, EPI_ISL_1555131, EPI_ISL_1555132, EPI_ISL_1555133, EPI_ISL_1555134, EPI_ISL_1555135, EPI_ISL_1555136, EPI_ISL_1555137, EPI_ISL_1555138, EPI_ISL_1555139, EPI_ISL_1555140, EPI_ISL_1555141, EPI_ISL_1555142, EPI_ISL_1555143, EPI_ISL_1555144, EPI_ISL_1555145, EPI_ISL_1555146, EPI_ISL_1555147, EPI_ISL_1555148, EPI_ISL_1555149, EPI_ISL_1555150, EPI_ISL_1555151, EPI_ISL_1555152, EPI_ISL_1555153, EPI_ISL_1555154, EPI_ISL_1555155, EPI_ISL_1555156, EPI_ISL_1555157, EPI_ISL_1555158, EPI_ISL_1555159, EPI_ISL_1555264, EPI_ISL_1555265, EPI_ISL_1555266, EPI_ISL_1555267, EPI_ISL_1555268, EPI_ISL_1555269, EPI_ISL_1555270, EPI_ISL_1555271, EPI_ISL_1555272, EPI_ISL_1555273, EPI_ISL_1555274, EPI_ISL_1555275, EPI_ISL_1555276, EPI_ISL_1555277, EPI_ISL_1555278, EPI_ISL_1555279, EPI_ISL_1555280, EPI_ISL_1555281, EPI_ISL_1555282, EPI_ISL_1555283, EPI_ISL_1555284, EPI_ISL_1555285, EPI_ISL_1555286, EPI_ISL_1555287, EPI_ISL_1555289, EPI_ISL_1555290, EPI_ISL_1555291, EPI_ISL_1555292, EPI_ISL_1555293, EPI_ISL_1555294, EPI_ISL_1555295, EPI_ISL_1555296, EPI_ISL_1555297, EPI_ISL_1555298, EPI_ISL_1555299, EPI_ISL_1555300, EPI_ISL_1555301, EPI_ISL_1555302, EPI_ISL_1555303, EPI_ISL_1555306, EPI_ISL_1555307, EPI_ISL_1555308, EPI_ISL_1555309, EPI_ISL_1555310, EPI_ISL_1555311, EPI_ISL_1555312, EPI_ISL_1555313, EPI_ISL_1555314, EPI_ISL_1555315, EPI_ISL_1555316, EPI_ISL_1555317, EPI_ISL_1555318, EPI_ISL_1555319, EPI_ISL_1555320, EPI_ISL_1555321, EPI_ISL_1555322, EPI_ISL_1555323, EPI_ISL_1555324, EPI_ISL_1555325, EPI_ISL_1555326, EPI_ISL_1555327, EPI_ISL_1555328, EPI_ISL_1555329, EPI_ISL_1555330, EPI_ISL_1555331, EPI_ISL_1555332, EPI_ISL_1555333, EPI_ISL_1555334, EPI_ISL_1555335, EPI_ISL_1555336, EPI_ISL_1555337, EPI_ISL_1555338, EPI_ISL_1555339, EPI_ISL_1555340, EPI_ISL_1555539, EPI_ISL_1555540, EPI_ISL_1555563, EPI_ISL_1555566, EPI_ISL_1555567, EPI_ISL_1555571, EPI_ISL_1555572, EPI_ISL_1555573, EPI_ISL_1555574, EPI_ISL_1555575, EPI_ISL_1555577, EPI_ISL_1555579, EPI_ISL_1555580, EPI_ISL_1555581, EPI_ISL_1555582, EPI_ISL_1555583, EPI_ISL_1555584, EPI_ISL_1555585, EPI_ISL_1555586, EPI_ISL_1555587, EPI_ISL_1555588, EPI_ISL_1555589, EPI_ISL_1555590, EPI_ISL_1555591, EPI_ISL_1555592, EPI_ISL_1555593, EPI_ISL_1555594, EPI_ISL_1555595, EPI_ISL_1555596, EPI_ISL_1555597, EPI_ISL_1555598, EPI_ISL_1555599, EPI_ISL_1555600, EPI_ISL_1555601, EPI_ISL_1555602, EPI_ISL_1555603, EPI_ISL_1555604, EPI_ISL_1555605, EPI_ISL_1555606, EPI_ISL_1555607, EPI_ISL_1555608, EPI_ISL_1555609, EPI_ISL_1555610, EPI_ISL_1555611, EPI_ISL_1555612, EPI_ISL_1555613, EPI_ISL_1555614, EPI_ISL_1555615, EPI_ISL_1555616, EPI_ISL_1555617, EPI_ISL_1555618, EPI_ISL_1555619, EPI_ISL_1555620, EPI_ISL_1555621, EPI_ISL_1555622, EPI_ISL_1555623, EPI_ISL_1555624, EPI_ISL_1555625, EPI_ISL_1555626, EPI_ISL_1555627, EPI_ISL_1555628, EPI_ISL_1555629, EPI_ISL_1555630, EPI_ISL_1555631, EPI_ISL_1555632, EPI_ISL_1555633, EPI_ISL_1555635, EPI_ISL_1555636, EPI_ISL_1555637, EPI_ISL_1555638, EPI_ISL_1555639, EPI_ISL_1555640, EPI_ISL_1555641, EPI_ISL_1555642, EPI_ISL_1555643, EPI_ISL_1555644, EPI_ISL_1555645, EPI_ISL_1555646, EPI_ISL_1555647, EPI_ISL_1555648, EPI_ISL_1555649, EPI_ISL_1555650, EPI_ISL_1555651, EPI_ISL_1555652, EPI_ISL_1555653, EPI_ISL_1555654, EPI_ISL_1555655, EPI_ISL_1555656, EPI_ISL_1555657, EPI_ISL_1555658, EPI_ISL_1555660, EPI_ISL_1555661, EPI_ISL_1555662, EPI_ISL_1555663, EPI_ISL_1555664, EPI_ISL_1555665, EPI_ISL_1555666, EPI_ISL_1555667, EPI_ISL_1555668, EPI_ISL_1555669, EPI_ISL_1555670, EPI_ISL_1555671, EPI_ISL_1555672, EPI_ISL_1555673, EPI_ISL_1555674, EPI_ISL_1555675, EPI_ISL_1555676, EPI_ISL_1555677, EPI_ISL_1555678, EPI_ISL_1555679, EPI_ISL_1555680, EPI_ISL_1555681, EPI_ISL_1555682, EPI_ISL_1555683, EPI_ISL_1555684, EPI_ISL_1555685, EPI_ISL_1555686, EPI_ISL_1555687, EPI_ISL_1555688, EPI_ISL_1555689, EPI_ISL_1555690, EPI_ISL_1555691, EPI_ISL_1555692, EPI_ISL_1555693, EPI_ISL_1555694, EPI_ISL_1555699, EPI_ISL_1555700, EPI_ISL_1555701, EPI_ISL_1555702, EPI_ISL_1555703, EPI_ISL_1555704, EPI_ISL_1555705, EPI_ISL_1555706, EPI_ISL_1555707, EPI_ISL_1555708, EPI_ISL_1555709, EPI_ISL_1555710, EPI_ISL_1555711, EPI_ISL_1555712, EPI_ISL_1555713, EPI_ISL_1555714, EPI_ISL_1555715, EPI_ISL_1555716, EPI_ISL_1555717, EPI_ISL_1555718, EPI_ISL_1555719, EPI_ISL_1555720, EPI_ISL_1555721, EPI_ISL_1555722, EPI_ISL_1555723, EPI_ISL_1555724, EPI_ISL_1555725, EPI_ISL_1555726, EPI_ISL_1555727, EPI_ISL_1555728, EPI_ISL_1555729, EPI_ISL_1555730, EPI_ISL_1555731, EPI_ISL_1555732, EPI_ISL_1555733, EPI_ISL_1555734, EPI_ISL_1555735, EPI_ISL_1555736, EPI_ISL_1555737, EPI_ISL_1555738, EPI_ISL_1555739, EPI_ISL_1555740, EPI_ISL_1555741, EPI_ISL_1555742, EPI_ISL_1555743, EPI_ISL_1555744, EPI_ISL_1555745, EPI_ISL_1555746, EPI_ISL_1555747, EPI_ISL_1555748, EPI_ISL_1555749, EPI_ISL_1555877, EPI_ISL_1555878, EPI_ISL_1555879, EPI_ISL_1555880, EPI_ISL_1555881, EPI_ISL_1555882, EPI_ISL_1555883, EPI_ISL_1555884, EPI_ISL_1555886, EPI_ISL_1555887, EPI_ISL_1555888, EPI_ISL_1555889, EPI_ISL_1555890, EPI_ISL_1555891, EPI_ISL_1555892, EPI_ISL_1555894, EPI_ISL_1555895, EPI_ISL_1555896, EPI_ISL_1555897, EPI_ISL_1555898, EPI_ISL_1555899, EPI_ISL_1555900, EPI_ISL_1555901, EPI_ISL_1555902, EPI_ISL_1555903, EPI_ISL_1555904, EPI_ISL_1555905, EPI_ISL_1555906, EPI_ISL_1555907, EPI_ISL_1555908, EPI_ISL_1555909, EPI_ISL_1555910, EPI_ISL_1555911, EPI_ISL_1555912, EPI_ISL_1555913, EPI_ISL_1555914, EPI_ISL_1555915, EPI_ISL_1555916, EPI_ISL_1555917, EPI_ISL_1555918, EPI_ISL_1555919, EPI_ISL_1555920, EPI_ISL_1555921, EPI_ISL_1555922, EPI_ISL_1555923, EPI_ISL_1555924, EPI_ISL_1555925, EPI_ISL_1555926, EPI_ISL_1555927, EPI_ISL_1555928, EPI_ISL_1555929, EPI_ISL_1555930, EPI_ISL_1555931, EPI_ISL_1555932, EPI_ISL_1555933, EPI_ISL_1555934, EPI_ISL_1555935, EPI_ISL_1555936, EPI_ISL_1555937, EPI_ISL_1555938, EPI_ISL_1555939, EPI_ISL_1555940, EPI_ISL_1555941, EPI_ISL_1555942, EPI_ISL_1555943, EPI_ISL_1555944, EPI_ISL_1555945, EPI_ISL_1555946, EPI_ISL_1555947, EPI_ISL_1555948, EPI_ISL_1555949, EPI_ISL_1555950, EPI_ISL_1555951, EPI_ISL_1555952, EPI_ISL_1555953, EPI_ISL_1555954, EPI_ISL_1555955, EPI_ISL_1555956, EPI_ISL_1555957, EPI_ISL_1555958, EPI_ISL_1555959, EPI_ISL_1555960, EPI_ISL_1555961, EPI_ISL_1555962, EPI_ISL_1555963, EPI_ISL_1555964, EPI_ISL_1555965, EPI_ISL_1555966, EPI_ISL_1555967, EPI_ISL_1555968, EPI_ISL_1556078, EPI_ISL_1556079, EPI_ISL_1556080, EPI_ISL_1556081, EPI_ISL_1556082, EPI_ISL_1556083, EPI_ISL_1556084, EPI_ISL_1556085, EPI_ISL_1556086, EPI_ISL_1556087, EPI_ISL_1556088, EPI_ISL_1556089, EPI_ISL_1556090, EPI_ISL_1556091, EPI_ISL_1556092, EPI_ISL_1556093, EPI_ISL_1556094, EPI_ISL_1556095, EPI_ISL_1556096, EPI_ISL_1556097, EPI_ISL_1556098, EPI_ISL_1556099, EPI_ISL_1556100, EPI_ISL_1556101, EPI_ISL_1556102, EPI_ISL_1556103, EPI_ISL_1556104, EPI_ISL_1556105, EPI_ISL_1556106, EPI_ISL_1556107, EPI_ISL_1556108, EPI_ISL_1556109, EPI_ISL_1556110, EPI_ISL_1556111, EPI_ISL_1556112, EPI_ISL_1556113, EPI_ISL_1556114, EPI_ISL_1556115, EPI_ISL_1556116, EPI_ISL_1556117, EPI_ISL_1556119, EPI_ISL_1556197, EPI_ISL_1556198, EPI_ISL_1556199, EPI_ISL_1556200, EPI_ISL_1556202, EPI_ISL_1556203, EPI_ISL_1556204, EPI_ISL_1556205, EPI_ISL_1556206, EPI_ISL_1556207, EPI_ISL_1556208, EPI_ISL_1556209, EPI_ISL_1556210, EPI_ISL_1556211, EPI_ISL_1556212, EPI_ISL_1556213, EPI_ISL_1556214, EPI_ISL_1556215, EPI_ISL_1556216, EPI_ISL_1556217, EPI_ISL_1556218, EPI_ISL_1556219, EPI_ISL_1556220, EPI_ISL_1556221, EPI_ISL_1556222, EPI_ISL_1556223, EPI_ISL_1556224, EPI_ISL_1556225, EPI_ISL_1556230, EPI_ISL_1556231, EPI_ISL_1556232, EPI_ISL_1556233, EPI_ISL_1556243, EPI_ISL_1556244, EPI_ISL_1556245, EPI_ISL_1556246, EPI_ISL_1556247, EPI_ISL_1556248, EPI_ISL_1556249, EPI_ISL_1556250, EPI_ISL_1556251, EPI_ISL_1556254, EPI_ISL_1556255, EPI_ISL_1556256, EPI_ISL_1556257, EPI_ISL_1556281, EPI_ISL_1556282, EPI_ISL_1556875, EPI_ISL_1556876, EPI_ISL_1556877, EPI_ISL_1556878, EPI_ISL_1556879, EPI_ISL_1556880, EPI_ISL_1556881, EPI_ISL_1556882, EPI_ISL_1556883, EPI_ISL_1556884, EPI_ISL_1556885, EPI_ISL_1556886, EPI_ISL_1556887, EPI_ISL_1556888, EPI_ISL_1556889, EPI_ISL_1556890, EPI_ISL_1556891, EPI_ISL_1556892, EPI_ISL_1556893, EPI_ISL_1556894, EPI_ISL_1556895, EPI_ISL_1556896, EPI_ISL_1556897, EPI_ISL_1556898, EPI_ISL_1556899, EPI_ISL_1556900, EPI_ISL_1556901, EPI_ISL_1556902, EPI_ISL_1556903, EPI_ISL_1556904, EPI_ISL_1556905, EPI_ISL_1556906, EPI_ISL_1556907, EPI_ISL_1556908, EPI_ISL_1556909, EPI_ISL_1556910, EPI_ISL_1556911, EPI_ISL_1556912, EPI_ISL_1556913, EPI_ISL_1556914, EPI_ISL_1556915, EPI_ISL_1556916, EPI_ISL_1556917, EPI_ISL_1556918, EPI_ISL_1556919, EPI_ISL_1556920, EPI_ISL_1556921, EPI_ISL_1556939, EPI_ISL_1557186, EPI_ISL_1667103, EPI_ISL_1667105, EPI_ISL_1667106, EPI_ISL_1667107, EPI_ISL_1694957, EPI_ISL_1694960, EPI_ISL_1694961, EPI_ISL_1694962, EPI_ISL_1694964, EPI_ISL_1694965, EPI_ISL_1694966, EPI_ISL_1694967, EPI_ISL_1694968, EPI_ISL_1694973

| | | | |
|---|---|---|---|
| see above | Fulgent Genetics | Centers for Disease Control and Prevention Division of Viral Diseases, Pathogen Discovery | Adrian Paskey; Becky Tsai; Benafsh Sapra; Benjamin Rambo-Martin; Christopher Gulvick; Clinton R. Paden; Dakota Howard; Darlene Wagner; Dhwani Batra; Doreen Ng; Duncan MacCannell; Harry Gao; James Xie; Jason Caravas; John Gao; Joseph Fierro; Kara Moser; Matthew Schmerer; Mickey Li; Peter W. Cook; Scott Sammons; Shatavia Morrison; Yan Meng; Yvette Unoarumhi |

EPI_ISL_1231437, EPI_ISL_1231438, EPI_ISL_1231439, EPI_ISL_1231440, EPI_ISL_1231441, EPI_ISL_1231442, EPI_ISL_1231443, EPI_ISL_1231444, EPI_ISL_1231445, EPI_ISL_1231446, EPI_ISL_1231447, EPI_ISL_1231448, EPI_ISL_1231449, EPI_ISL_1231450, EPI_ISL_1231451, EPI_ISL_1231452, EPI_ISL_1231453, EPI_ISL_1231454, EPI_ISL_1231455, EPI_ISL_1231456, EPI_ISL_1231457, EPI_ISL_1231458, EPI_ISL_1231459, EPI_ISL_1231460, EPI_ISL_1231461, EPI_ISL_1231462, EPI_ISL_1231463, EPI_ISL_1231464, EPI_ISL_1231465, EPI_ISL_1231466, EPI_ISL_1231467, EPI_ISL_1231468, EPI_ISL_1231469, EPI_ISL_1231470, EPI_ISL_1231471, EPI_ISL_1231472, EPI_ISL_1231473, EPI_ISL_1231474, EPI_ISL_1231475, EPI_ISL_1231476, EPI_ISL_1231477, EPI_ISL_1231478, EPI_ISL_1231479, EPI_ISL_1231480, EPI_ISL_1231481, EPI_ISL_1231482, EPI_ISL_1231483, EPI_ISL_1231484, EPI_ISL_1231485, EPI_ISL_1231486, EPI_ISL_1231487, EPI_ISL_1231488, EPI_ISL_1231489, EPI_ISL_1231490, EPI_ISL_1231491, EPI_ISL_1231492, EPI_ISL_1231493, EPI_ISL_1231494, EPI_ISL_1231495, EPI_ISL_1231496, EPI_ISL_1231497, EPI_ISL_1231498, EPI_ISL_1231499, EPI_ISL_1231500, EPI_ISL_1231501, EPI_ISL_1231502, EPI_ISL_1231503, EPI_ISL_1231504, EPI_ISL_1231505, EPI_ISL_1231506, EPI_ISL_1231507, EPI_ISL_1231508, EPI_ISL_1231509, EPI_ISL_1231510, EPI_ISL_1231512, EPI_ISL_1231513, EPI_ISL_1231514, EPI_ISL_1231515, EPI_ISL_1231516, EPI_ISL_1231517, EPI_ISL_1231518, EPI_ISL_1231519, EPI_ISL_1231520, EPI_ISL_1231521, EPI_ISL_1231522, EPI_ISL_1231523, EPI_ISL_1231524, EPI_ISL_1231525, EPI_ISL_1231526, EPI_ISL_1231527, EPI_ISL_1231528, EPI_ISL_1231529, EPI_ISL_1231530, EPI_ISL_1231533, EPI_ISL_1231537, EPI_ISL_1231541, EPI_ISL_1231545, EPI_ISL_1231549, EPI_ISL_1231553, EPI_ISL_1231557, EPI_ISL_1231561, EPI_ISL_1231565, EPI_ISL_1231569, EPI_ISL_1231573, EPI_ISL_1231577, EPI_ISL_1231581, EPI_ISL_1231585, EPI_ISL_1231590, EPI_ISL_1231594, EPI_ISL_1231598, EPI_ISL_1231602, EPI_ISL_1231606, EPI_ISL_1231610, EPI_ISL_1231615, EPI_ISL_1231618, EPI_ISL_1231623, EPI_ISL_1231626, EPI_ISL_1231630, EPI_ISL_1231634, EPI_ISL_1231638, EPI_ISL_1231642, EPI_ISL_1231646, EPI_ISL_1231651, EPI_ISL_1231655, EPI_ISL_1231659, EPI_ISL_1231663, EPI_ISL_1231666, EPI_ISL_1231670, EPI_ISL_1231675, EPI_ISL_1231678, EPI_ISL_1231682, EPI_ISL_1231686, EPI_ISL_1231690, EPI_ISL_1231694, EPI_ISL_1231698, EPI_ISL_1231702, EPI_ISL_1231706, EPI_ISL_1231710, EPI_ISL_1231714, EPI_ISL_1231719, EPI_ISL_1231723, EPI_ISL_1231727, EPI_ISL_1231730, EPI_ISL_1231735, EPI_ISL_1231739, EPI_ISL_1231743, EPI_ISL_1231747, EPI_ISL_1231751, EPI_ISL_1231755, EPI_ISL_1231758, EPI_ISL_1231762, EPI_ISL_1231767, EPI_ISL_1231771, EPI_ISL_1231774, EPI_ISL_1231775, EPI_ISL_1231776, EPI_ISL_1231777, EPI_ISL_1231778, EPI_ISL_1231779, EPI_ISL_1231780, EPI_ISL_1231781, EPI_ISL_1231782, EPI_ISL_1231783, EPI_ISL_1231784, EPI_ISL_1231785, EPI_ISL_1231786, EPI_ISL_1231787, EPI_ISL_1231788, EPI_ISL_1231789, EPI_ISL_1231790, EPI_ISL_1231791, EPI_ISL_1231793, EPI_ISL_1231794, EPI_ISL_1231795, EPI_ISL_1231796, EPI_ISL_1231797, EPI_ISL_1231798, EPI_ISL_1231799, EPI_ISL_1231800, EPI_ISL_1231802, EPI_ISL_1231803, EPI_ISL_1231804, EPI_ISL_1231805, EPI_ISL_1231806, EPI_ISL_1231807, EPI_ISL_1231808, EPI_ISL_1231809, EPI_ISL_1231810, EPI_ISL_1231811, EPI_ISL_1231812, EPI_ISL_1231813, EPI_ISL_1231814, EPI_ISL_1231815, EPI_ISL_1231816, EPI_ISL_1231817, EPI_ISL_1231818, EPI_ISL_1231819, EPI_ISL_1231820, EPI_ISL_1231821, EPI_ISL_1231822, EPI_ISL_1231823, EPI_ISL_1231824, EPI_ISL_1231825, EPI_ISL_1231826, EPI_ISL_1231827, EPI_ISL_1231828, EPI_ISL_1231829, EPI_ISL_1231830, EPI_ISL_1231831, EPI_ISL_1231832, EPI_ISL_1231833, EPI_ISL_1231834, EPI_ISL_1231835, EPI_ISL_1231836, EPI_ISL_1231837, EPI_ISL_1231838, EPI_ISL_1231839, EPI_ISL_1231840, EPI_ISL_1231841, EPI_ISL_1231842, EPI_ISL_1231843, EPI_ISL_1231844, EPI_ISL_1231845, EPI_ISL_1231846, EPI_ISL_1231847, EPI_ISL_1231848, EPI_ISL_1231849, EPI_ISL_1231850, EPI_ISL_1231851, EPI_ISL_1231852, EPI_ISL_1231853, EPI_ISL_1231854, EPI_ISL_1231855, EPI_ISL_1231856, EPI_ISL_1231857, EPI_ISL_1231858, EPI_ISL_1231859, EPI_ISL_1231860, EPI_ISL_1231861, EPI_ISL_1231862, EPI_ISL_1231863, EPI_ISL_1231864, EPI_ISL_1231865, EPI_ISL_1231866, EPI_ISL_1231867, EPI_ISL_1231868, EPI_ISL_1231869, EPI_ISL_1231870, EPI_ISL_1231871, EPI_ISL_1231872, EPI_ISL_1231873, EPI_ISL_1231874, EPI_ISL_1231875, EPI_ISL_1231876, EPI_ISL_1231877, EPI_ISL_1231878, EPI_ISL_1231879, EPI_ISL_1231880, EPI_ISL_1231881, EPI_ISL_1231882, EPI_ISL_1231883, EPI_ISL_1231884, EPI_ISL_1231885, EPI_ISL_1231886, EPI_ISL_1231887, EPI_ISL_1231888, EPI_ISL_1231889, EPI_ISL_1231890, EPI_ISL_1231891, EPI_ISL_1231892, EPI_ISL_1231893, EPI_ISL_1231894, EPI_ISL_1231895, EPI_ISL_1231896, EPI_ISL_1231897, EPI_ISL_1231898, EPI_ISL_1231899, EPI_ISL_1231900, EPI_ISL_1231901, EPI_ISL_1231902, EPI_ISL_1231903, EPI_ISL_1231904, EPI_ISL_1231905, EPI_ISL_1231906, EPI_ISL_1231907, EPI_ISL_1231908, EPI_ISL_1231909, EPI_ISL_1231910, EPI_ISL_1231911, EPI_ISL_1231912, EPI_ISL_1231913, EPI_ISL_1232029, EPI_ISL_1232030, EPI_ISL_1232031, EPI_ISL_1232032, EPI_ISL_1232033, EPI_ISL_1232034, EPI_ISL_1232035, EPI_ISL_1232036, EPI_ISL_1232037, EPI_ISL_1232038, EPI_ISL_1232039, EPI_ISL_1232040, EPI_ISL_1232041, EPI_ISL_1232042, EPI_ISL_1232043, EPI_ISL_1232044, EPI_ISL_1232045, EPI_ISL_1232046, EPI_ISL_1232047, EPI_ISL_1232048, EPI_ISL_1232049, EPI_ISL_1232050, EPI_ISL_1232051, EPI_ISL_1232052, EPI_ISL_1232053, EPI_ISL_1232054, EPI_ISL_1232055, EPI_ISL_1232056, EPI_ISL_1232057, EPI_ISL_1232058, EPI_ISL_1232059, EPI_ISL_1232060, EPI_ISL_1232061, EPI_ISL_1232062, EPI_ISL_1232063, EPI_ISL_1232064, EPI_ISL_1232065, EPI_ISL_1232066, EPI_ISL_1232067, EPI_ISL_1232068, EPI_ISL_1232069, EPI_ISL_1232070, EPI_ISL_1232071, EPI_ISL_1232072, EPI_ISL_1232073, EPI_ISL_1232074, EPI_ISL_1232075, EPI_ISL_1232076, EPI_ISL_1232077, EPI_ISL_1232078, EPI_ISL_1232079, EPI_ISL_1232080, EPI_ISL_1232081, EPI_ISL_1232082, EPI_ISL_1232083, EPI_ISL_1232084, EPI_ISL_1232085, EPI_ISL_1232086, EPI_ISL_1232087, EPI_ISL_1232088, EPI_ISL_1232089, EPI_ISL_1232090, EPI_ISL_1232091, EPI_ISL_1232092, EPI_ISL_1232093, EPI_ISL_1232094, EPI_ISL_1232095, EPI_ISL_1232096, EPI_ISL_1232099, EPI_ISL_1232100, EPI_ISL_1232101, EPI_ISL_1232102, EPI_ISL_1232103, EPI_ISL_1232104, EPI_ISL_1232105, EPI_ISL_1232106, EPI_ISL_1232107, EPI_ISL_1232108, EPI_ISL_1232109, EPI_ISL_1232110, EPI_ISL_1232111, EPI_ISL_1232112, EPI_ISL_1232113, EPI_ISL_1232114, EPI_ISL_1232115, EPI_ISL_1232116, EPI_ISL_1232117, EPI_ISL_1232118, EPI_ISL_1232119, EPI_ISL_1232120, EPI_ISL_1232121, EPI_ISL_1232122, EPI_ISL_1232123, EPI_ISL_1232124, EPI_ISL_1232125, EPI_ISL_1232126, EPI_ISL_1232127, EPI_ISL_1232128, EPI_ISL_1232129, EPI_ISL_1232130, EPI_ISL_1232131, EPI_ISL_1232132, EPI_ISL_1232133, EPI_ISL_1232134, EPI_ISL_1232135, EPI_ISL_1232136, EPI_ISL_1232137, EPI_ISL_1232138, EPI_ISL_1232139, EPI_ISL_1232140, EPI_ISL_1232141, EPI_ISL_1232142, EPI_ISL_1232143, EPI_ISL_1232144, EPI_ISL_1232145, EPI_ISL_1232146, EPI_ISL_1232147, EPI_ISL_1232148, EPI_ISL_1232149, EPI_ISL_1232150, EPI_ISL_1232151, EPI_ISL_1232152, EPI_ISL_1232153, EPI_ISL_1232154, EPI_ISL_1232155, EPI_ISL_1232156, EPI_ISL_1232157, EPI_ISL_1232158, EPI_ISL_1232159, EPI_ISL_1232160, EPI_ISL_1232161, EPI_ISL_1232162, EPI_ISL_1232163, EPI_ISL_1232164, EPI_ISL_1232165, EPI_ISL_1232166, EPI_ISL_1232167, EPI_ISL_1232168, EPI_ISL_1232169, EPI_ISL_1232170, EPI_ISL_1232171, EPI_ISL_1232172, EPI_ISL_1232173, EPI_ISL_1232174, EPI_ISL_1232175, EPI_ISL_1232176, EPI_ISL_1232177, EPI_ISL_1232178, EPI_ISL_1232179, EPI_ISL_1232180, EPI_ISL_1232181, EPI_ISL_1232182, EPI_ISL_1232183, EPI_ISL_1232184, EPI_ISL_1232185, EPI_ISL_1232186, EPI_ISL_1232187, EPI_ISL_1232188, EPI_ISL_1232189, EPI_ISL_1232190, EPI_ISL_1232191, EPI_ISL_1232192, EPI_ISL_1251273

| | | | |
|---|---|---|---|
| see above | Fulgent Genetics | Fulgent Genetics | Becky Tsai; Benafsh Sapra; Doreen Ng; Harry Gao; James Xie; John Gao; Joseph Fierro; Mickey Li; Yan Meng |
| EPI_ISL_1532119, EPI_ISL_1532123 | Good Samaritan Hospital | Santa Clara County Public Health Laboratory | Santa Clara County Public Health Department |

EPI_ISL_1261900, EPI_ISL_1261904, EPI_ISL_1261907, EPI_ISL_1261921, EPI_ISL_1261932, EPI_ISL_1261951, EPI_ISL_1261969, EPI_ISL_1262005, EPI_ISL_1262219, EPI_ISL_1269214, EPI_ISL_1269621, EPI_ISL_1269630, EPI_ISL_1269648, EPI_ISL_1269673, EPI_ISL_1269676, EPI_ISL_1269697, EPI_ISL_1269716, EPI_ISL_1269727, EPI_ISL_1269730, EPI_ISL_1270144, EPI_ISL_1270184, EPI_ISL_1270253, EPI_ISL_1270273, EPI_ISL_1270279, EPI_ISL_1270442, EPI_ISL_1270443, EPI_ISL_1270448, EPI_ISL_1270451, EPI_ISL_1270459, EPI_ISL_1270461, EPI_ISL_1270462, EPI_ISL_1270464, EPI_ISL_1270469, EPI_ISL_1270470, EPI_ISL_1270471, EPI_ISL_1270473, EPI_ISL_1270474, EPI_ISL_1270475, EPI_ISL_1270478, EPI_ISL_1270480, EPI_ISL_1270504, EPI_ISL_1270663, EPI_ISL_1270668, EPI_ISL_1270669, EPI_ISL_1270703, EPI_ISL_1270706, EPI_ISL_1270708, EPI_ISL_1270727, EPI_ISL_1270746, EPI_ISL_1270749, EPI_ISL_1270761, EPI_ISL_1270767, EPI_ISL_1270770, EPI_ISL_1283377, EPI_ISL_1283443, EPI_ISL_1283456, EPI_ISL_1283639, EPI_ISL_1283692, EPI_ISL_1283707, EPI_ISL_1283742, EPI_ISL_1283831, EPI_ISL_1283862, EPI_ISL_1283864, EPI_ISL_1283882, EPI_ISL_1283906, EPI_ISL_1283935, EPI_ISL_1283952, EPI_ISL_1283990, EPI_ISL_1284006, EPI_ISL_1284051, EPI_ISL_1284264, EPI_ISL_1284385, EPI_ISL_1284410, EPI_ISL_1297927, EPI_ISL_1297941, EPI_ISL_1297944, EPI_ISL_1297951, EPI_ISL_1297955, EPI_ISL_1297959, EPI_ISL_1297975, EPI_ISL_1298016, EPI_ISL_1298067, EPI_ISL_1298069, EPI_ISL_1298074, EPI_ISL_1298077, EPI_ISL_1298080, EPI_ISL_1298084, EPI_ISL_1298098, EPI_ISL_1298111, EPI_ISL_1298114, EPI_ISL_1298122, EPI_ISL_1298168, EPI_ISL_1298171, EPI_ISL_1298172, EPI_ISL_1298182, EPI_ISL_1337399, EPI_ISL_1338057, EPI_ISL_1338070, EPI_ISL_1338072, EPI_ISL_1338074, EPI_ISL_1338085, EPI_ISL_1338087, EPI_ISL_1338091, EPI_ISL_1338097, EPI_ISL_1338101, EPI_ISL_1338122, EPI_ISL_1338124, EPI_ISL_1338130, EPI_ISL_1338133, EPI_ISL_1338140, EPI_ISL_1338156, EPI_ISL_1338166, EPI_ISL_1338170, EPI_ISL_1338194, EPI_ISL_1338204, EPI_ISL_1338210, EPI_ISL_1338212, EPI_ISL_1338226, EPI_ISL_1338230, EPI_ISL_1338233, EPI_ISL_1338245, EPI_ISL_1338289, EPI_ISL_1338307, EPI_ISL_1338333, EPI_ISL_1338394, EPI_ISL_1338397, EPI_ISL_1340060, EPI_ISL_1340061, EPI_ISL_1340068, EPI_ISL_1340083, EPI_ISL_1340084, EPI_ISL_1340088, EPI_ISL_1340090, EPI_ISL_1340096, EPI_ISL_1340124, EPI_ISL_1340129, EPI_ISL_1340225, EPI_ISL_1340249, EPI_ISL_1340251, EPI_ISL_1340254, EPI_ISL_1340258, EPI_ISL_1340260, EPI_ISL_1340264, EPI_ISL_1340265, EPI_ISL_1340275, EPI_ISL_1340301, EPI_ISL_1340305, EPI_ISL_1340308, EPI_ISL_1340337, EPI_ISL_1340339, EPI_ISL_1340368, EPI_ISL_1340373, EPI_ISL_1340377, EPI_ISL_1340384, EPI_ISL_1340447, EPI_ISL_1391729, EPI_ISL_1391750, EPI_ISL_1391839, EPI_ISL_1391843, EPI_ISL_1391848, EPI_ISL_1391853, EPI_ISL_1391864, EPI_ISL_1391865, EPI_ISL_1391888, EPI_ISL_1391895, EPI_ISL_1391905, EPI_ISL_1391919, EPI_ISL_1391929, EPI_ISL_1391934, EPI_ISL_1391939, EPI_ISL_1391943, EPI_ISL_1391949, EPI_ISL_1391962, EPI_ISL_1420835, EPI_ISL_1420925, EPI_ISL_1421655, EPI_ISL_1443819, EPI_ISL_1443827, EPI_ISL_1443831, EPI_ISL_1443835, EPI_ISL_1443842, EPI_ISL_1443846, EPI_ISL_1443865, EPI_ISL_1443918, EPI_ISL_1443932, EPI_ISL_1443938, EPI_ISL_1443950, EPI_ISL_1443965, EPI_ISL_1443968, EPI_ISL_1443973, EPI_ISL_1443981, EPI_ISL_1443984, EPI_ISL_1443997, EPI_ISL_1444004, EPI_ISL_1444005, EPI_ISL_1444006, EPI_ISL_1444015, EPI_ISL_1444022, EPI_ISL_1444024, EPI_ISL_1444030, EPI_ISL_1444041, EPI_ISL_1444046, EPI_ISL_1444064, EPI_ISL_1444074, EPI_ISL_1444092, EPI_ISL_1444098, EPI_ISL_1444100, EPI_ISL_1444188, EPI_ISL_1444190, EPI_ISL_1444192, EPI_ISL_1444200, EPI_ISL_1444227, EPI_ISL_1444242, EPI_ISL_1460798, EPI_ISL_1460801, EPI_ISL_1460802, EPI_ISL_1460831, EPI_ISL_1460835, EPI_ISL_1460853, EPI_ISL_1460917,

| | | | |
|---|---|---|---|
| EPI_ISL_1460959, EPI_ISL_1460975, EPI_ISL_1460981, EPI_ISL_1460985, EPI_ISL_1460997, EPI_ISL_1460998, EPI_ISL_1479226, EPI_ISL_1479271, EPI_ISL_1479275, EPI_ISL_1479283, EPI_ISL_1479285, EPI_ISL_1479333, EPI_ISL_1479353, EPI_ISL_1479425, EPI_ISL_1480009, EPI_ISL_1480066, EPI_ISL_1480190, EPI_ISL_1480238, EPI_ISL_1480260, EPI_ISL_1480271, EPI_ISL_1480341, EPI_ISL_1480451, EPI_ISL_1480513, EPI_ISL_1493758, EPI_ISL_1493886, EPI_ISL_1493898, EPI_ISL_1493921, EPI_ISL_1511861, EPI_ISL_1511862, EPI_ISL_1511867, EPI_ISL_1511871, EPI_ISL_1511872, EPI_ISL_1511875, EPI_ISL_1511879, EPI_ISL_1511883, EPI_ISL_1511888, EPI_ISL_1511889, EPI_ISL_1511897, EPI_ISL_1511903, EPI_ISL_1511905, EPI_ISL_1511986, EPI_ISL_1511989, EPI_ISL_1512001, EPI_ISL_1512003, EPI_ISL_1512005, EPI_ISL_1512011, EPI_ISL_1512027, EPI_ISL_1512028, EPI_ISL_1512034, EPI_ISL_1512044, EPI_ISL_1512048, EPI_ISL_1512049, EPI_ISL_1512083, EPI_ISL_1512089, EPI_ISL_1512090, EPI_ISL_1512093, EPI_ISL_1512095, EPI_ISL_1512096, EPI_ISL_1512098, EPI_ISL_1512100, EPI_ISL_1512101, EPI_ISL_1512102, EPI_ISL_1512105, EPI_ISL_1512107, EPI_ISL_1512109, EPI_ISL_1512111, EPI_ISL_1512112, EPI_ISL_1512113, EPI_ISL_1512115, EPI_ISL_1512116, EPI_ISL_1512119, EPI_ISL_1512122, EPI_ISL_1512125, EPI_ISL_1512148, EPI_ISL_1512203, EPI_ISL_1512233, EPI_ISL_1512247, EPI_ISL_1512248, EPI_ISL_1512249, EPI_ISL_1512252, EPI_ISL_1512253, EPI_ISL_1512255, EPI_ISL_1512259, EPI_ISL_1512266, EPI_ISL_1512267, EPI_ISL_1512268, EPI_ISL_1512314, EPI_ISL_1512317, EPI_ISL_1512319, EPI_ISL_1512321, EPI_ISL_1512322, EPI_ISL_1512324, EPI_ISL_1512326, EPI_ISL_1512329, EPI_ISL_1512331, EPI_ISL_1512335, EPI_ISL_1512349, EPI_ISL_1512351, EPI_ISL_1512352, EPI_ISL_1512358, EPI_ISL_1512366, EPI_ISL_1512376, EPI_ISL_1512377, EPI_ISL_1512378, EPI_ISL_1512383, EPI_ISL_1512386, EPI_ISL_1512395, EPI_ISL_1512402, EPI_ISL_1512404, EPI_ISL_1512419, EPI_ISL_1512420, EPI_ISL_1512423, EPI_ISL_1512424, EPI_ISL_1512429, EPI_ISL_1512431, EPI_ISL_1512432, EPI_ISL_1512436, EPI_ISL_1512438, EPI_ISL_1512442, EPI_ISL_1512443, EPI_ISL_1512451, EPI_ISL_1512459, EPI_ISL_1512463, EPI_ISL_1512464, EPI_ISL_1512466, EPI_ISL_1512469, EPI_ISL_1512471, EPI_ISL_1512475, EPI_ISL_1512478, EPI_ISL_1512479, EPI_ISL_1512484, EPI_ISL_1512489, EPI_ISL_1512492, EPI_ISL_1512519, EPI_ISL_1512520, EPI_ISL_1512554, EPI_ISL_1512562, EPI_ISL_1512584, EPI_ISL_1512589, EPI_ISL_1512593, EPI_ISL_1512599, EPI_ISL_1512634, EPI_ISL_1512642, EPI_ISL_1512652, EPI_ISL_1512664, EPI_ISL_1512667, EPI_ISL_1512672, EPI_ISL_1512686, EPI_ISL_1512691, EPI_ISL_1512707, EPI_ISL_1512718, EPI_ISL_1512734, EPI_ISL_1512735, EPI_ISL_1512736, EPI_ISL_1512742, EPI_ISL_1512744, EPI_ISL_1553291, EPI_ISL_1553301, EPI_ISL_1553418, EPI_ISL_1553432, EPI_ISL_1553447, EPI_ISL_1553474, EPI_ISL_1553500, EPI_ISL_1553518, EPI_ISL_1553595, EPI_ISL_1553931, EPI_ISL_1553965, EPI_ISL_1554030, EPI_ISL_1554035, EPI_ISL_1554197, EPI_ISL_1554200, EPI_ISL_1554204, EPI_ISL_1554216, EPI_ISL_1554217, EPI_ISL_1554224, EPI_ISL_1554304, EPI_ISL_1554343, EPI_ISL_1554361, EPI_ISL_1554367, EPI_ISL_1554375, EPI_ISL_1554401, EPI_ISL_1554406, EPI_ISL_1554419, EPI_ISL_1554422, EPI_ISL_1554425, EPI_ISL_1554465, EPI_ISL_1554466, EPI_ISL_1554467, EPI_ISL_1554468, EPI_ISL_1554469, EPI_ISL_1554470, EPI_ISL_1554471, EPI_ISL_1554472, EPI_ISL_1554475, EPI_ISL_1554476, EPI_ISL_1554477, EPI_ISL_1554479, EPI_ISL_1554480, EPI_ISL_1554481, EPI_ISL_1554486, EPI_ISL_1554487, EPI_ISL_1554488, EPI_ISL_1554490, EPI_ISL_1554493, EPI_ISL_1554494, EPI_ISL_1554495, EPI_ISL_1554499, EPI_ISL_1554500, EPI_ISL_1554501, EPI_ISL_1554502, EPI_ISL_1554506, EPI_ISL_1554507, EPI_ISL_1554508, EPI_ISL_1554523, EPI_ISL_1554525, EPI_ISL_1554526, EPI_ISL_1554528, EPI_ISL_1554537, EPI_ISL_1554545, EPI_ISL_1554570, EPI_ISL_1554621, EPI_ISL_1554645, EPI_ISL_1554649, EPI_ISL_1554653, EPI_ISL_1554659, EPI_ISL_1554662, EPI_ISL_1554663, EPI_ISL_1554664, EPI_ISL_1554685, EPI_ISL_1554686, EPI_ISL_1554687, EPI_ISL_1554688, EPI_ISL_1554689, EPI_ISL_1554701, EPI_ISL_1554715, EPI_ISL_1554720, EPI_ISL_1554725, EPI_ISL_1554728, EPI_ISL_1554744, EPI_ISL_1554747, EPI_ISL_1554769, EPI_ISL_1554770, EPI_ISL_1554771, EPI_ISL_1554776, EPI_ISL_1554778, EPI_ISL_1554780, EPI_ISL_1554784, EPI_ISL_1554790, EPI_ISL_1554791, EPI_ISL_1554794, EPI_ISL_1554801, EPI_ISL_1554806, EPI_ISL_1554818, EPI_ISL_1554819, EPI_ISL_1554820, EPI_ISL_1554824, EPI_ISL_1554827, EPI_ISL_1554829, EPI_ISL_1554831, EPI_ISL_1554834, EPI_ISL_1554839, EPI_ISL_1554844, EPI_ISL_1554845, EPI_ISL_1554849, EPI_ISL_1554850, EPI_ISL_1554857, EPI_ISL_1554859, EPI_ISL_1554860, EPI_ISL_1554865, EPI_ISL_1554866, EPI_ISL_1554872, EPI_ISL_1575494, EPI_ISL_1575499, EPI_ISL_1575506, EPI_ISL_1575512, EPI_ISL_1575521, EPI_ISL_1575524, EPI_ISL_1575527, EPI_ISL_1575534, EPI_ISL_1575537, EPI_ISL_1575546, EPI_ISL_1575549, EPI_ISL_1575551, EPI_ISL_1575552, EPI_ISL_1575573, EPI_ISL_1575581, EPI_ISL_1575582, EPI_ISL_1575595, EPI_ISL_1575606, EPI_ISL_1575617, EPI_ISL_1575623, EPI_ISL_1575628, EPI_ISL_1575631, EPI_ISL_1575640, EPI_ISL_1575653, EPI_ISL_1575665, EPI_ISL_1575836, EPI_ISL_1575837, EPI_ISL_1575847, EPI_ISL_1575848, EPI_ISL_1575851, EPI_ISL_1575863, EPI_ISL_1575882, EPI_ISL_1575895, EPI_ISL_1575900, EPI_ISL_1575909, EPI_ISL_1575910, EPI_ISL_1575919, EPI_ISL_1575933, EPI_ISL_1575954, EPI_ISL_1575975, EPI_ISL_1575977, EPI_ISL_1575992, EPI_ISL_1575997, EPI_ISL_1576016, EPI_ISL_1576032, EPI_ISL_1576041, EPI_ISL_1576042, EPI_ISL_1576046, EPI_ISL_1576047, EPI_ISL_1576048, EPI_ISL_1576049, EPI_ISL_1576051, EPI_ISL_1576053, EPI_ISL_1576058, EPI_ISL_1576059, EPI_ISL_1576060, EPI_ISL_1576062, EPI_ISL_1576066, EPI_ISL_1576070, EPI_ISL_1576072, EPI_ISL_1576076, EPI_ISL_1576078, EPI_ISL_1576082, EPI_ISL_1576084, EPI_ISL_1576086, EPI_ISL_1576087, EPI_ISL_1576094, EPI_ISL_1576102, EPI_ISL_1576104, EPI_ISL_1576115, EPI_ISL_1576122, EPI_ISL_1576132, EPI_ISL_1576168, EPI_ISL_1576171, EPI_ISL_1576183, EPI_ISL_1576278, EPI_ISL_1576279, EPI_ISL_1576302, EPI_ISL_1576309, EPI_ISL_1576320, EPI_ISL_1576328, EPI_ISL_1576330, EPI_ISL_1576334, EPI_ISL_1576340, EPI_ISL_1576351, EPI_ISL_1576352, EPI_ISL_1576354, EPI_ISL_1576357, EPI_ISL_1576366, EPI_ISL_1576374, EPI_ISL_1576376, EPI_ISL_1576378, EPI_ISL_1576391, EPI_ISL_1576395, EPI_ISL_1576587, EPI_ISL_1576615, EPI_ISL_1576637, EPI_ISL_1576658, EPI_ISL_1576666, EPI_ISL_1581154, EPI_ISL_1581158, EPI_ISL_1581169, EPI_ISL_1581175, EPI_ISL_1581190, EPI_ISL_1581193, EPI_ISL_1581213, EPI_ISL_1581297, EPI_ISL_1581309, EPI_ISL_1581704, EPI_ISL_1581740, EPI_ISL_1581751, EPI_ISL_1694635, EPI_ISL_1694637, EPI_ISL_1694638, EPI_ISL_1694641, EPI_ISL_1694664 | | | |
| see above | Helix/Illumina | Centers for Disease Control and Prevention Division of Viral Diseases, Pathogen Discovery | Adrian Paskey; Alexandre Bolze; Ary Ascencio; Ben L. Rambo-Martin; Benjamin Rambo-Martin; Brad Sickler; Charlotte Rivera-Garcia; Christine Tran; Christopher Gulvick; Clinton R. Paden; Dakota Howard; Darlene Wagner; David Becker; Dhwani Batra; Duncan MacCannell; Efren Sandoval; Eileen de Feo; Elizabeth Cirulli; Eric Allen; Geraint Levan; James Lu; Jan Antico; Jason Caravas; Jason Nguyen; Jimmy Ramirez; Jingtao Liu; Kara Moser; Kelly Schiabor Barrett; Kim Gietzen; Magnus Isaksson; Marc Laurent; Matthew Schmerer; Matthew Tolentino; Nicole L. Washington; Peter W. Cook; Phil Febbo; Ryan Cho; Scott Sammons; Shannon Wickline; Shatavia Morrison; Sherry Wang; Simon White; Summer Galloway; Suxiang Tong; Tyler Cassens; William Lee; Yvette Unoarumhi |
| EPI_ISL_1303471 | Henry Mayo Newhall Hospital | Los Angeles County PHL | P. Hemarajata et al. |
| EPI_ISL_1477183, EPI_ISL_1477195, EPI_ISL_1477247, EPI_ISL_1477282, EPI_ISL_1477296, EPI_ISL_1477346, EPI_ISL_1477379, EPI_ISL_1477408, EPI_ISL_1477951, EPI_ISL_1477954, EPI_ISL_1477955, EPI_ISL_1477970, EPI_ISL_1477973, EPI_ISL_1477975, EPI_ISL_1477976, EPI_ISL_1477981, EPI_ISL_1477987, EPI_ISL_1477991, EPI_ISL_1477997, EPI_ISL_1478009, EPI_ISL_1478010, EPI_ISL_1478015, EPI_ISL_1478026, EPI_ISL_1478041, EPI_ISL_1478054, EPI_ISL_1478055, EPI_ISL_1478058, EPI_ISL_1478062, EPI_ISL_1478063, EPI_ISL_1478065, EPI_ISL_1478066, EPI_ISL_1478067, EPI_ISL_1478075, EPI_ISL_1478077, EPI_ISL_1478082, EPI_ISL_1478083, EPI_ISL_1478093, EPI_ISL_1478094, EPI_ISL_1478096, EPI_ISL_1478105, EPI_ISL_1478107, EPI_ISL_1478121, EPI_ISL_1478134, EPI_ISL_1478135, EPI_ISL_1478138, EPI_ISL_1478140, EPI_ISL_1478143, EPI_ISL_1478150, EPI_ISL_1478156, EPI_ISL_1478159, EPI_ISL_1478163, EPI_ISL_1478164, EPI_ISL_1478165, EPI_ISL_1478169, EPI_ISL_1478172, EPI_ISL_1478176, EPI_ISL_1478177, EPI_ISL_1478186, EPI_ISL_1478190, EPI_ISL_1701396, EPI_ISL_1701398, EPI_ISL_1701402, EPI_ISL_1701404, EPI_ISL_1701412, EPI_ISL_1701420, EPI_ISL_1701422, EPI_ISL_1701428, EPI_ISL_1701429, EPI_ISL_1701433, EPI_ISL_1701435, EPI_ISL_1701441, EPI_ISL_1701446, EPI_ISL_1701448, EPI_ISL_1701451, EPI_ISL_1701454, EPI_ISL_1701458, EPI_ISL_1701472, EPI_ISL_1701474, EPI_ISL_1701484, EPI_ISL_1701500, EPI_ISL_1701507, EPI_ISL_1701509, EPI_ISL_1701527, EPI_ISL_1701534, EPI_ISL_1701538, EPI_ISL_1701549, EPI_ISL_1701558, EPI_ISL_1701568, EPI_ISL_1701577, EPI_ISL_1701584, EPI_ISL_1701588, EPI_ISL_1701590, EPI_ISL_1701595, EPI_ISL_1701597, EPI_ISL_1701600, EPI_ISL_1701603, EPI_ISL_1701608, EPI_ISL_1701617, EPI_ISL_1701618, EPI_ISL_1701624, EPI_ISL_1701627, EPI_ISL_1701637, EPI_ISL_1701643, EPI_ISL_1701658 | | | |
| see above | Humboldt County Public Health Laboratory | Chan-Zuckerberg Biohub | CZB Cliahub Consortium |
| EPI_ISL_1192365, EPI_ISL_1497544, EPI_ISL_1497545, EPI_ISL_1497546, EPI_ISL_1497547, EPI_ISL_1506116 | Huntington Memorial Hospital | Los Angeles County PHL | P. Hemarajata et al. |
| EPI_ISL_2253167, EPI_ISL_2253169, EPI_ISL_2253170, EPI_ISL_2253172, EPI_ISL_2253174, EPI_ISL_2253175 | Infectious Diseases, Chan-Zuckerberg Biohub Covidtracker | Infectious Diseases, Chan-Zuckerberg Biohub Covidtracker | CZB Cliahub Consortium |
| EPI_ISL_1512824, EPI_ISL_1512945, EPI_ISL_1691713, EPI_ISL_1692004, EPI_ISL_1692184, EPI_ISL_1692231, EPI_ISL_1692270, EPI_ISL_1692487, EPI_ISL_1702257 | | | |
| see above | Infinity Biologix | Centers for Disease Control and Prevention Division of Viral Diseases, Pathogen Discovery | Adrian Paskey; Benjamin Rambo-Martin; Chirayu Goswami; Christian Bixby; Christopher Gulvick; Clinton R. Paden; Dakota Howard; Darlene Wagner; Dhwani Batra; Duncan MacCannell; Jason Caravas; Jonathan Schultz; Kara Moser; Matthew Schmerer; Peter W. Cook; Robin Grimwood; Russ Hager; Scott Sammons; Shatavia Morrison; Yihe Wang; Yvette Unoarumhi |
| EPI_ISL_1712422, EPI_ISL_1712423, EPI_ISL_1712424, EPI_ISL_1712425, EPI_ISL_1712426, EPI_ISL_1712427, EPI_ISL_1712428, EPI_ISL_1712429, EPI_ISL_1712430, EPI_ISL_1712431, EPI_ISL_1712432, EPI_ISL_1712433, EPI_ISL_1712434, EPI_ISL_1712435, EPI_ISL_1712436, EPI_ISL_1712437, EPI_ISL_1712438, EPI_ISL_1712462, EPI_ISL_1712463, EPI_ISL_1712464, EPI_ISL_1712465, EPI_ISL_1712466, EPI_ISL_1712467, EPI_ISL_1712468, EPI_ISL_1712469, EPI_ISL_1712470, EPI_ISL_1712471, EPI_ISL_1712472, EPI_ISL_1712473, EPI_ISL_1712474, EPI_ISL_1712475, EPI_ISL_1712476, EPI_ISL_1712477, EPI_ISL_1712478, EPI_ISL_1712479, EPI_ISL_1712542, EPI_ISL_1712543, EPI_ISL_1712544, EPI_ISL_1712545, EPI_ISL_1712546, EPI_ISL_1712547, EPI_ISL_1712548, EPI_ISL_1712549 | | | |
| see above | Innovative Genomics Institute, UC Berkeley | Innovative Genomics Institute, UC Berkeley | Holly Gildea; John Boyle; Phil Frankino; Stacia Wyman |
| EPI_ISL_1508918 | Kaiser Permanente Downey Medical Center | Los Angeles County PHL | P. Hemarajata et al. |
| EPI_ISL_1508902, EPI_ISL_1508917 | Kaiser Permanente West Los Angeles Medical Center | Los Angeles County PHL | P. Hemarajata et al. |
| EPI_ISL_1508984, EPI_ISL_1508985, EPI_ISL_1508986 | Kaiser Regional | Santa Clara County Public Health Laboratory | Santa Clara County Public Health Department |
| EPI_ISL_1508966, EPI_ISL_1508967, EPI_ISL_1508969, EPI_ISL_1508970, EPI_ISL_1508972, EPI_ISL_1508973, EPI_ISL_1508975 | | | |
| see above | Kaiser Santa Clara | Santa Clara County Public Health Laboratory | Santa Clara County Public Health Department |
| EPI_ISL_1337390, EPI_ISL_1399280, EPI_ISL_1424050, EPI_ISL_1504108, EPI_ISL_1532264, EPI_ISL_1558798, EPI_ISL_1661550, EPI_ISL_1700837, EPI_ISL_1710971, EPI_ISL_1710972 | | | |
| see above | LESP Baja California | Instituto de Diagnostico y Referencia Epidemiologicos (INDRE) | Abril Rodriguez-Maldonado; Ariadna Medina-Benitez; Claudia Wong-Arambula; Ernesto Ramirez-Gonzalez.; Gisela Barrera-Badillo; Irma Lopez-Martinez; Joaquin Quiroz-Mercado; Lucia Hernandez-Rivas; Natividad Cruz-Ortiz; Sergio Rangel-Guerrero; Tatiana Nunez-Garcia; Vanessa Rivero-Arredondo |
| EPI_ISL_1337392, EPI_ISL_1337393, EPI_ISL_1504105, EPI_ISL_1532243, EPI_ISL_1532244, EPI_ISL_1532245, EPI_ISL_1558801, EPI_ISL_1626811, EPI_ISL_1626812, EPI_ISL_1626813, EPI_ISL_1626814, EPI_ISL_1626815, EPI_ISL_1651890, EPI_ISL_1651891, EPI_ISL_1651892, EPI_ISL_1651893, EPI_ISL_1651894, EPI_ISL_1651895, EPI_ISL_1651900, EPI_ISL_1700819, EPI_ISL_1700822 | | | |
| see above | LESP Baja California Sur | Instituto de Diagnostico y Referencia Epidemiologicos (INDRE) | Abril Rodriguez-Maldonado; Ariadna Medina-Benitez; Claudia Wong-Arambula; Ernesto Ramirez-Gonzalez.; Gisela Barrera-Badillo; Irma Lopez-Martinez; Joaquin Quiroz-Mercado; Lucia Hernandez-Rivas; Natividad Cruz-Ortiz; Sergio Rangel-Guerrero; Tatiana Nunez-Garcia; Vanessa Rivero-Arredondo |
| EPI_ISL_1661980, EPI_ISL_1661981 | Laboratorio Central de Epidemiologia (LCE) | Unidad de Genomica Avanzada | Alejandro Sanchez-Flores; Alfredo Herrera-Estrella; Alicia Ocana-Mondragon; Angel Gustavo Salas-Lais; Bernardo Martinez-Miguel; Blanca Taboada; Brenda Irasema Maldonado-Meza; Carla Ivon Herrera-Najera; Carlos F. Arias; Celia Boukadida; Celida Duque Molina; Clara Esperanza Santacruz-Tinoco; Concepcion Grajales-Muniz; Consorcio Mexicano de Vigilancia Genomica (CoViGen-Mex). Authors (in alphabetical order): Julio Elias Alvarado-Yaah; Fernando Fontove-Herrera; Francisco Pulido; Gloria Elena Espinoza-Ayala; Gloria Maria Molina-Salinas; Gloria Vazquez; Hector Esteban Paz-Juarez; Hector Montoya-Fuentes; Helen Haydee Fernanda Ramirez-Plascencia; Jose Antonio Enciso-Moreno; Jose Esteban Munoz-Medina; Jose de Jesus Nunez-Contreras; Juan Bautista Chale-Dzul; Luis Alberto Ochoa-Carrera; Margarita Matias-Florentino; Maria Guadalupe Santiago-Mauricio; Maria Guadalupe de Jesus Mireles-Rivera; Nelly Selem-Mojica; Pavel Isa; Ricardo Grande; Santiago Ávila-Rios; Victor Eduardo Garcia-Arias; Victor Hugo Borja-Aburto |
| EPI_ISL_1351981, EPI_ISL_1351984, EPI_ISL_1351987, EPI_ISL_1351990, EPI_ISL_1352048 | Laboratorio Central de EpidemiologÃa (LCE) | Unidad de Genomica Avanzada | Alejandro Sanchez-Flores; Alfredo Herrera-Estrella; Alicia Ocana-Mondragon; Angel Gustavo Salas-Lais; Bernardo Martinez-Miguel; Blanca Taboada; Brenda Irasema Maldonado-Meza; Carla Ivon Herrera-Najera; Carlos F. Arias; Celia Boukadida; Clara Esperanza Santacruz-Tinoco; Concepcion Grajales-Muniz; Consorcio Mexicano de Vigilancia Genomica (CoViGen-Mex). Authors (in alphabetical order): Julio Elias Alvarado-Yaah; Fernando Fontove-Herrera; Francisco Pulido; Gloria Elena Espinoza-Ayala; Gloria Maria Molina-Salinas; Gloria Vazquez; Hector Esteban Paz-Juarez; Hector Montoya-Fuentes; Helen Haydee Fernanda Ramirez-Plascencia; Jorge Ivan Salinal-Nevarez; Jose Antonio Enciso-Moreno; Jose Esteban Munoz-Medina; Jose de Jesus Nunez-Contreras; Juan Bautista Chale-Dzul; Luis Alberto Ochoa-Carrera; Margarita Matias-Florentino; Maria Guadalupe Santiago-Mauricio; Maria Guadalupe de Jesus Mireles-Rivera; Nelly Selem-Mojica; Pavel Isa; Ricardo Grande; Santiago Avila-Rios; Victor Hugo Borja-Aburto |
| EPI_ISL_1585658 | Laboratorio Central de Epidemiología (LCE) | Instituto Nacional de Enfermedades Respiratorias (INER): Centro de Investigación en Enfermedades Infecciosas (CIENI) | Alejandro Sanchez-Flores; Alfredo Herrera-Estrella; Alicia Ocaña-Mondragón; Angel Gustavo Salas-Lais; Bernardo Martínez-Miguel; Blanca Taboada; Brenda Irasema Maldonado-Meza; Carla Ivón Herrera-Najera; Carlos F. Arias; Celia Boukadida; Clara Esperanza Santacruz-Tinoco; Concepción Grajales-Muñiz; Consorcio Mexicano de Vigilancia Genómica (CoViGen-Mex). Authors (in alphabetical order): Julio Elias Alvarado-Yaah; Célida Duque Molina; Fernando Fontove-Herrera; Francisco Pulido; Gloria Elena Espinoza-Ayala; Gloria María Molina-Salinas; Gloria Vazquez; Hector Esteban Paz-Juárez; Hector Montoya-Fuentes; Helen Haydee Fernanda Ramírez-Plascencia; José Antonio Enciso-Moreno; José Esteban Muñoz-Medina; José de Jesús Nuñez-Contreras; Juan Bautista Chale-Dzul; Luis Alberto Ochoa-Carrera; Margarita Matías-Florentino; María Guadalupe Santiago-Mauricio; María Guadalupe de Jesús Mireles-Rivera; Nelly Sélem-Mojica; Pavel Isa; Ricardo Grande; Santiago Ávila-Ríos; Victor Eduardo Garcia-Arias; Víctor Hugo Borja-Aburto |
| EPI_ISL_1416468, EPI_ISL_1416469, EPI_ISL_1416470, EPI_ISL_1416471, EPI_ISL_1416472, EPI_ISL_1416473, EPI_ISL_1416474, EPI_ISL_1416547, EPI_ISL_1416548, EPI_ISL_1416549, EPI_ISL_1416594, EPI_ISL_1416599 | | | |
| see above | Laboratorio Central de Epidemiología (LCE) | Instituto de Biotecnología de la UNAM | Alejandro Sanchez-Flores; Alfredo Herrera-Estrella; Alicia Ocaña-Mondragón; Angel Gustavo Salas-Lais; Bernardo Martínez-Miguel; Blanca Taboada; Brenda Irasema Maldonado-Meza; Carla Ivón Herrera-Najera; Carlos F. Arias; Celia Boukadida; Clara Esperanza Santacruz-Tinoco; Concepción Grajales-Muñiz; Consorcio Mexicano de Vigilancia Genómica (CoViGen-Mex). Authors (in alphabetical order): Julio Elias Alvarado-Yaah; Fernando Fontove-Herrera; Francisco Pulido; Gloria Elena Espinoza-Ayala; Gloria María Molina-Salinas; Gloria Vazquez; Hector Esteban Paz-Juárez; Hector Montoya-Fuentes; Helen Haydee Fernanda Ramírez-Plascencia; Jorge Ivan Salinal-Nevarez; José Antonio Enciso-Moreno; José Esteban Muñoz-Medina; José de Jesús Nuñez-Contreras; Juan Bautista Chale-Dzul; Luis Alberto Ochoa-Carrera; Margarita Matías-Florentino; María Guadalupe Santiago-Mauricio; María Guadalupe de Jesús Mireles-Rivera; Nelly Sélem-Mojica; Pavel Isa; Ricardo Grande; Santiago Ávila-Ríos; Víctor Hugo Borja-Aburto |
| EPI_ISL_1221302, EPI_ISL_1221702, EPI_ISL_1221739, EPI_ISL_1221740, EPI_ISL_1221741, EPI_ISL_1320008, EPI_ISL_1320145, EPI_ISL_1320146, EPI_ISL_1320147, EPI_ISL_1320148, EPI_ISL_1320149, EPI_ISL_1320236, EPI_ISL_1320237, EPI_ISL_1320244, EPI_ISL_1320274, EPI_ISL_1320275, EPI_ISL_1320276, EPI_ISL_1320277, EPI_ISL_1320278, EPI_ISL_1320279, EPI_ISL_1320280, EPI_ISL_1320281, EPI_ISL_1320283, EPI_ISL_1320284, EPI_ISL_1320285, EPI_ISL_1320286, EPI_ISL_1320287, EPI_ISL_1320288, EPI_ISL_1320289, EPI_ISL_1320290, EPI_ISL_1320291, EPI_ISL_1320292, EPI_ISL_1320293, EPI_ISL_1320294, EPI_ISL_1320295, EPI_ISL_1320296, EPI_ISL_1320297, EPI_ISL_1320298, EPI_ISL_1320299, EPI_ISL_1320300, EPI_ISL_1320301, EPI_ISL_1320302, EPI_ISL_1320303, EPI_ISL_1320304, EPI_ISL_1320305, EPI_ISL_1320306, EPI_ISL_1320307, EPI_ISL_1320308, EPI_ISL_1320309, EPI_ISL_1320310, EPI_ISL_1320311, EPI_ISL_1320312, EPI_ISL_1320313, EPI_ISL_1320314, EPI_ISL_1320315, EPI_ISL_1320316, EPI_ISL_1320317, EPI_ISL_1320318, EPI_ISL_1320319, EPI_ISL_1320320, EPI_ISL_1320321, EPI_ISL_1320322, EPI_ISL_1320323, EPI_ISL_1320324, EPI_ISL_1320325, EPI_ISL_1320326, EPI_ISL_1320327, EPI_ISL_1320328, EPI_ISL_1320329, EPI_ISL_1320330, EPI_ISL_1320585, EPI_ISL_1321132, EPI_ISL_1321133, EPI_ISL_1321134, EPI_ISL_1321135, EPI_ISL_1321136, EPI_ISL_1321137, EPI_ISL_1321138, EPI_ISL_1321139, EPI_ISL_1321141, EPI_ISL_1321142, EPI_ISL_1321143, EPI_ISL_1321144, EPI_ISL_1321145, EPI_ISL_1321146, EPI_ISL_1321147, EPI_ISL_1321148, EPI_ISL_1321166, | | | |

| | | | |
|---|---|---|---|
| EPI_ISL_1321167, EPI_ISL_1321168, EPI_ISL_1321176, EPI_ISL_1321177, EPI_ISL_1321178, EPI_ISL_1321179, EPI_ISL_1321180, EPI_ISL_1321181, EPI_ISL_1321188, EPI_ISL_1321190, EPI_ISL_1321191, EPI_ISL_1321192, EPI_ISL_1321193, EPI_ISL_1321194, EPI_ISL_1321195, EPI_ISL_1321196, EPI_ISL_1321197, EPI_ISL_1321198, EPI_ISL_1321199, EPI_ISL_1321200, EPI_ISL_1321201, EPI_ISL_1321202, EPI_ISL_1321203, EPI_ISL_1321204, EPI_ISL_1321205, EPI_ISL_1321206, EPI_ISL_1321207, EPI_ISL_1321208, EPI_ISL_1321209, EPI_ISL_1321210, EPI_ISL_1321211, EPI_ISL_1321212, EPI_ISL_1321213, EPI_ISL_1321214, EPI_ISL_1321215, EPI_ISL_1321216, EPI_ISL_1321217, EPI_ISL_1321218, EPI_ISL_1321228, EPI_ISL_1321229, EPI_ISL_1321230, EPI_ISL_1321231, EPI_ISL_1321232, EPI_ISL_1321233, EPI_ISL_1321234, EPI_ISL_1321235, EPI_ISL_1321236, EPI_ISL_1321237, EPI_ISL_1321238, EPI_ISL_1321239, EPI_ISL_1321240, EPI_ISL_1321241, EPI_ISL_1321242, EPI_ISL_1321243, EPI_ISL_1321244, EPI_ISL_1321245, EPI_ISL_1321246, EPI_ISL_1321247, EPI_ISL_1321248, EPI_ISL_1321249, EPI_ISL_1321250, EPI_ISL_1321251, EPI_ISL_1321252, EPI_ISL_1321253, EPI_ISL_1321254, EPI_ISL_1321255, EPI_ISL_1321256, EPI_ISL_1321259, EPI_ISL_1321260, EPI_ISL_1321261, EPI_ISL_1321262, EPI_ISL_1321263, EPI_ISL_1321264, EPI_ISL_1321265, EPI_ISL_1321266, EPI_ISL_1321267, EPI_ISL_1321268, EPI_ISL_1321269, EPI_ISL_1321274, EPI_ISL_1321275, EPI_ISL_1321276, EPI_ISL_1321277, EPI_ISL_1321278, EPI_ISL_1321279, EPI_ISL_1321280, EPI_ISL_1321281, EPI_ISL_1321282, EPI_ISL_1321283, EPI_ISL_1321284, EPI_ISL_1321285, EPI_ISL_1321286, EPI_ISL_1321287, EPI_ISL_1321288, EPI_ISL_1321289, EPI_ISL_1321290, EPI_ISL_1321291, EPI_ISL_1321292, EPI_ISL_1321293, EPI_ISL_1321294, EPI_ISL_1321295, EPI_ISL_1321296, EPI_ISL_1321297, EPI_ISL_1321298, EPI_ISL_1321299, EPI_ISL_1321300, EPI_ISL_1321301, EPI_ISL_1321302, EPI_ISL_1321303, EPI_ISL_1321304, EPI_ISL_1321305, EPI_ISL_1321306, EPI_ISL_1321307, EPI_ISL_1321316, EPI_ISL_1321317, EPI_ISL_1321318, EPI_ISL_1321319, EPI_ISL_1321320, EPI_ISL_1321321, EPI_ISL_1321322, EPI_ISL_1321323, EPI_ISL_1321324, EPI_ISL_1321325, EPI_ISL_1321326, EPI_ISL_1321327, EPI_ISL_1321328, EPI_ISL_1321329, EPI_ISL_1321330, EPI_ISL_1321331, EPI_ISL_1321332, EPI_ISL_1321333, EPI_ISL_1321334, EPI_ISL_1321335, EPI_ISL_1321336, EPI_ISL_1321337, EPI_ISL_1321345, EPI_ISL_1321347, EPI_ISL_1321354, EPI_ISL_1338888, EPI_ISL_1338896, EPI_ISL_1338897, EPI_ISL_1338898, EPI_ISL_1338899, EPI_ISL_1338900, EPI_ISL_1338901, EPI_ISL_1338902, EPI_ISL_1338903, EPI_ISL_1338904, EPI_ISL_1338906, EPI_ISL_1338907, EPI_ISL_1338908, EPI_ISL_1338909, EPI_ISL_1338910, EPI_ISL_1338912, EPI_ISL_1338921, EPI_ISL_1338923, EPI_ISL_1338924, EPI_ISL_1338925, EPI_ISL_1338926, EPI_ISL_1338927, EPI_ISL_1338928, EPI_ISL_1338930, EPI_ISL_1338931, EPI_ISL_1338932, EPI_ISL_1338933, EPI_ISL_1338934, EPI_ISL_1338935, EPI_ISL_1338937, EPI_ISL_1338938, EPI_ISL_1338939, EPI_ISL_1338948, EPI_ISL_1338949, EPI_ISL_1338950, EPI_ISL_1338951, EPI_ISL_1338952, EPI_ISL_1338953, EPI_ISL_1338954, EPI_ISL_1338955, EPI_ISL_1338956, EPI_ISL_1338957, EPI_ISL_1338958, EPI_ISL_1338959, EPI_ISL_1338991, EPI_ISL_1338992, EPI_ISL_1339004, EPI_ISL_1339005, EPI_ISL_1339006, EPI_ISL_1339038, EPI_ISL_1339039, EPI_ISL_1339051, EPI_ISL_1339052, EPI_ISL_1339053, EPI_ISL_1339054, EPI_ISL_1339055, EPI_ISL_1339056, EPI_ISL_1339057, EPI_ISL_1339090, EPI_ISL_1339091, EPI_ISL_1339092, EPI_ISL_1339093, EPI_ISL_1339094, EPI_ISL_1339095, EPI_ISL_1339149, EPI_ISL_1339158, EPI_ISL_1339159, EPI_ISL_1339160, EPI_ISL_1339161, EPI_ISL_1339162, EPI_ISL_1339163, EPI_ISL_1339182, EPI_ISL_1339183, EPI_ISL_1339184, EPI_ISL_1339185, EPI_ISL_1339186, EPI_ISL_1339187, EPI_ISL_1339188, EPI_ISL_1339207, EPI_ISL_1339234, EPI_ISL_1339251, EPI_ISL_1339252, EPI_ISL_1339253, EPI_ISL_1421988, EPI_ISL_1421989, EPI_ISL_1421990, EPI_ISL_1421991, EPI_ISL_1421992, EPI_ISL_1422037, EPI_ISL_1422174, EPI_ISL_1422175, EPI_ISL_1422176, EPI_ISL_1422177, EPI_ISL_1422178, EPI_ISL_1422183, EPI_ISL_1422184, EPI_ISL_1422190, EPI_ISL_1422191, EPI_ISL_1422194, EPI_ISL_1422195, EPI_ISL_1422196, EPI_ISL_1422199, EPI_ISL_1422200, EPI_ISL_1422201, EPI_ISL_1422203, EPI_ISL_1422204, EPI_ISL_1422205, EPI_ISL_1422206, EPI_ISL_1461710, EPI_ISL_1461711, EPI_ISL_1461712, EPI_ISL_1461713, EPI_ISL_1461719, EPI_ISL_1461720, EPI_ISL_1461721, EPI_ISL_1461722, EPI_ISL_1461723, EPI_ISL_1461724, EPI_ISL_1461725, EPI_ISL_1461726, EPI_ISL_1461727, EPI_ISL_1461728, EPI_ISL_1461729, EPI_ISL_1461730, EPI_ISL_1461731, EPI_ISL_1461741, EPI_ISL_1461742, EPI_ISL_1461743, EPI_ISL_1461744, EPI_ISL_1461745, EPI_ISL_1461746, EPI_ISL_1461750, EPI_ISL_1461751, EPI_ISL_1461753, EPI_ISL_1461754, EPI_ISL_1461755, EPI_ISL_1461756, EPI_ISL_1461757, EPI_ISL_1461759, EPI_ISL_1461760, EPI_ISL_1461761, EPI_ISL_1461786, EPI_ISL_1461787, EPI_ISL_1461788, EPI_ISL_1461789, EPI_ISL_1461790, EPI_ISL_1461791, EPI_ISL_1461792, EPI_ISL_1461793, EPI_ISL_1461794, EPI_ISL_1461795, EPI_ISL_1461796, EPI_ISL_1461797, EPI_ISL_1461798, EPI_ISL_1461799, EPI_ISL_1461800, EPI_ISL_1461801, EPI_ISL_1461802, EPI_ISL_1461803, EPI_ISL_1461804, EPI_ISL_1461805, EPI_ISL_1461806, EPI_ISL_1461807, EPI_ISL_1461808, EPI_ISL_1461809, EPI_ISL_1461810, EPI_ISL_1461811, EPI_ISL_1461827, EPI_ISL_1461859, EPI_ISL_1461877, EPI_ISL_1461878, EPI_ISL_1461879, EPI_ISL_1481185, EPI_ISL_1481186, EPI_ISL_1481189, EPI_ISL_1481190, EPI_ISL_1481191, EPI_ISL_1481192, EPI_ISL_1481194, EPI_ISL_1481202, EPI_ISL_1481214, EPI_ISL_1481265, EPI_ISL_1481270, EPI_ISL_1513757, EPI_ISL_1513762, EPI_ISL_1513768, EPI_ISL_1513769, EPI_ISL_1513801, EPI_ISL_1513802, EPI_ISL_1513803, EPI_ISL_1513804, EPI_ISL_1513805, EPI_ISL_1513818, EPI_ISL_1513819, EPI_ISL_1513820, EPI_ISL_1513821, EPI_ISL_1513822, EPI_ISL_1513823, EPI_ISL_1513824, EPI_ISL_1513825, EPI_ISL_1513826, EPI_ISL_1513827, EPI_ISL_1513833, EPI_ISL_1513859, EPI_ISL_1513860, EPI_ISL_1513862, EPI_ISL_1513863, EPI_ISL_1513864, EPI_ISL_1513865, EPI_ISL_1513866, EPI_ISL_1513867, EPI_ISL_1513868, EPI_ISL_1513869, EPI_ISL_1513870, EPI_ISL_1513871, EPI_ISL_1513872, EPI_ISL_1513873, EPI_ISL_1513874, EPI_ISL_1513875, EPI_ISL_1513876, EPI_ISL_1513877, EPI_ISL_1513878, EPI_ISL_1513879, EPI_ISL_1513880, EPI_ISL_1513881, EPI_ISL_1513882, EPI_ISL_1513883, EPI_ISL_1513884, EPI_ISL_1513885, EPI_ISL_1513886, EPI_ISL_1513887, EPI_ISL_1513888, EPI_ISL_1513889, EPI_ISL_1513890, EPI_ISL_1513891, EPI_ISL_1513892, EPI_ISL_1513893, EPI_ISL_1513894, EPI_ISL_1513895, EPI_ISL_1513896, EPI_ISL_1513897, EPI_ISL_1513898, EPI_ISL_1513899, EPI_ISL_1513900, EPI_ISL_1513901, EPI_ISL_1513902, EPI_ISL_1513903, EPI_ISL_1513904, EPI_ISL_1513905, EPI_ISL_1513906, EPI_ISL_1513907, EPI_ISL_1513908, EPI_ISL_1513909, EPI_ISL_1514723, EPI_ISL_1514724, EPI_ISL_1514725, EPI_ISL_1514726, EPI_ISL_1514727, EPI_ISL_1514731, EPI_ISL_1514752, EPI_ISL_1514753, EPI_ISL_1514754, EPI_ISL_1514755, EPI_ISL_1514756, EPI_ISL_1514758, EPI_ISL_1514759, EPI_ISL_1514760, EPI_ISL_1514771, EPI_ISL_1514772, EPI_ISL_1514773, EPI_ISL_1514774, EPI_ISL_1514813, EPI_ISL_1514825, EPI_ISL_1514826, EPI_ISL_1514827, EPI_ISL_1514828, EPI_ISL_1514832, EPI_ISL_1514838, EPI_ISL_1514839, EPI_ISL_1514840, EPI_ISL_1514841, EPI_ISL_1514842, EPI_ISL_1514843, EPI_ISL_1514844, EPI_ISL_1514845, EPI_ISL_1514846, EPI_ISL_1514847, EPI_ISL_1514848, EPI_ISL_1514849, EPI_ISL_1514850, EPI_ISL_1514851, EPI_ISL_1514852, EPI_ISL_1514853, EPI_ISL_1514854, EPI_ISL_1514855, EPI_ISL_1514856, EPI_ISL_1514857, EPI_ISL_1514858, EPI_ISL_1514859, EPI_ISL_1514860, EPI_ISL_1514861, EPI_ISL_1514862, EPI_ISL_1514863, EPI_ISL_1514864, EPI_ISL_1514865, EPI_ISL_1514866, EPI_ISL_1514867, EPI_ISL_1514868, EPI_ISL_1514871, EPI_ISL_1514884, EPI_ISL_1514895, EPI_ISL_1514896, EPI_ISL_1514897, EPI_ISL_1514898, EPI_ISL_1514899, EPI_ISL_1514927, EPI_ISL_1514928, EPI_ISL_1514929, EPI_ISL_1514930, EPI_ISL_1514931, EPI_ISL_1514939, EPI_ISL_1514940, EPI_ISL_1514941, EPI_ISL_1514942, EPI_ISL_1514943, EPI_ISL_1514961, EPI_ISL_1514974, EPI_ISL_1514975, EPI_ISL_1514976, EPI_ISL_1514977, EPI_ISL_1514978, EPI_ISL_1514979, EPI_ISL_1514980, EPI_ISL_1514982, EPI_ISL_1514997, EPI_ISL_1514998, EPI_ISL_1514999, EPI_ISL_1515000, EPI_ISL_1515001, EPI_ISL_1515002, EPI_ISL_1515003, EPI_ISL_1515004, EPI_ISL_1515005, EPI_ISL_1515006, EPI_ISL_1515007, EPI_ISL_1515008, EPI_ISL_1515009, EPI_ISL_1515010, EPI_ISL_1515011, EPI_ISL_1515012, EPI_ISL_1515013, EPI_ISL_1515014, EPI_ISL_1515015, EPI_ISL_1515016, EPI_ISL_1515017, EPI_ISL_1515018, EPI_ISL_1515019, EPI_ISL_1515020, EPI_ISL_1515021, EPI_ISL_1515022, EPI_ISL_1515051, EPI_ISL_1515052, EPI_ISL_1515053, EPI_ISL_1515054, EPI_ISL_1515055, EPI_ISL_1515091, EPI_ISL_1515117, EPI_ISL_1515118, EPI_ISL_1515119, EPI_ISL_1515120, EPI_ISL_1515121, EPI_ISL_1515139, EPI_ISL_1515140, EPI_ISL_1515141, EPI_ISL_1515142, EPI_ISL_1515143, EPI_ISL_1515144, EPI_ISL_1515145, EPI_ISL_1515146, EPI_ISL_1515147, EPI_ISL_1515148, EPI_ISL_1515149, EPI_ISL_1515157, EPI_ISL_1515158, EPI_ISL_1515159, EPI_ISL_1515160, EPI_ISL_1515161, EPI_ISL_1515162, EPI_ISL_1515163, EPI_ISL_1515164, EPI_ISL_1515165, EPI_ISL_1515166, EPI_ISL_1515167, EPI_ISL_1515168, EPI_ISL_1515169, EPI_ISL_1515170, EPI_ISL_1515171, EPI_ISL_1515350, EPI_ISL_1515365, EPI_ISL_1515374, EPI_ISL_1515375, EPI_ISL_1515389, EPI_ISL_1515390, EPI_ISL_1515391, EPI_ISL_1515392, EPI_ISL_1515393, EPI_ISL_1515394, EPI_ISL_1515395, EPI_ISL_1515405, EPI_ISL_1515406, EPI_ISL_1515407, EPI_ISL_1515408, EPI_ISL_1515409, EPI_ISL_1515410, EPI_ISL_1515411, EPI_ISL_1515412, EPI_ISL_1515413, EPI_ISL_1515414, EPI_ISL_1515415, EPI_ISL_1515416, EPI_ISL_1515417, EPI_ISL_1515418, EPI_ISL_1515419, EPI_ISL_1515420, EPI_ISL_1515421, EPI_ISL_1515422, EPI_ISL_1515423, EPI_ISL_1515424, EPI_ISL_1515425, EPI_ISL_1515426, EPI_ISL_1515427, EPI_ISL_1515428, EPI_ISL_1515429, EPI_ISL_1515430, EPI_ISL_1515431, EPI_ISL_1515432, EPI_ISL_1515433, EPI_ISL_1515586, EPI_ISL_1515615, EPI_ISL_1515618, EPI_ISL_1515619, EPI_ISL_1515620, EPI_ISL_1515621, EPI_ISL_1515622, EPI_ISL_1515623, EPI_ISL_1515631, EPI_ISL_1515632, EPI_ISL_1515634, EPI_ISL_1515635, EPI_ISL_1515682, EPI_ISL_1515733, EPI_ISL_1548117, EPI_ISL_1548118, EPI_ISL_1548125, EPI_ISL_1548126, EPI_ISL_1548139, EPI_ISL_1548140, EPI_ISL_1549203, EPI_ISL_1549205, EPI_ISL_1549245, EPI_ISL_1549246, EPI_ISL_1549247, EPI_ISL_1549248, EPI_ISL_1549809, EPI_ISL_1549811, EPI_ISL_1549825, EPI_ISL_1549826, EPI_ISL_1549827, EPI_ISL_1549828, EPI_ISL_1549829, EPI_ISL_1549830, EPI_ISL_1549831, EPI_ISL_1549849, EPI_ISL_1549858, EPI_ISL_1549859, EPI_ISL_1549877, EPI_ISL_1549883, EPI_ISL_1549884, EPI_ISL_1549885, EPI_ISL_1549886, EPI_ISL_1549887, EPI_ISL_1549888, EPI_ISL_1549889, EPI_ISL_1549890, EPI_ISL_1549903, EPI_ISL_1549904, EPI_ISL_1549905, EPI_ISL_1549934, EPI_ISL_1549935, EPI_ISL_1549944, EPI_ISL_1549945, EPI_ISL_1549952, EPI_ISL_1550057, EPI_ISL_1550058, EPI_ISL_1550068, EPI_ISL_1550093, EPI_ISL_1550094, EPI_ISL_1550095, EPI_ISL_1550096, EPI_ISL_1550097, EPI_ISL_1550098, EPI_ISL_1550099, EPI_ISL_1550101, EPI_ISL_1550102, EPI_ISL_1550103, EPI_ISL_1550104, EPI_ISL_1550105, EPI_ISL_1550184, EPI_ISL_1550204, EPI_ISL_1550205, EPI_ISL_1550224, EPI_ISL_1550232, EPI_ISL_1550246, EPI_ISL_1550247, EPI_ISL_1695016, EPI_ISL_1695018 | | | |
| see above | Laboratory Corporation of America | Centers for Disease Control and Prevention Division of Viral Diseases, Pathogen Discovery | Adrian Paskey; Amanda Douglas; Amanda Suchanek; Andrea Throop; Ayla Burns; Ben L. Rambo-Martin; Benjamin Rambo-Martin; Bobbi Croy; Brian Krueger; Brian Norvell; Christopher Gulvick; Christos Petropoulos; Clinton R. Paden; Craig Lukasik; Dakota Howard; Darlene Wagner; Debbie Boles; Dhwani Batra; Duncan MacCannell; Eyad Almasri; Goran Stevovic; Howard Engler; Hrushikesh Deshmukh; Jake Humphrey; Jana Schroth; Jason Caravas; Joe Voshell; John Pruitt; Jonathan Meltzer; Jonathan Williams; Kara Moser; Kimberly Wagner; Lax Iyer; Lyndon Tilson; Manoj Jain; Marcia Eisenberg; Mary Ann Cristobal; Mary Williamson; Matthew Schmerer; Michael Levandoski; Mike Sapeta; Mindy Nye; Minoo Agarwal; Mohan Kolli; Nuthawin Charoensri; Oren Cohen; Peter W. Cook; Prashant Gupta; Qian Zeng; Rama Ghatti; Scott Parker; Scott Ryan; Scott Sammons; Shatavia Morrison; Stanley Letovsky; Steven Ragan; Suresh Babu Selvaraju; Susan Countryman; Susan Hicks; Suxiang Tong; Suzanne Dale; Thomas Urban; Tim Kuphal; Tricia Zwiefelhofer; Vincent Drouillon; Yvette Unoarumhi |
| EPI_ISL_1504027, EPI_ISL_1508909 | Los Angeles County PHL | Los Angeles County PHL | P. Hemarajata et al. |
| EPI_ISL_1303420, EPI_ISL_1303421, EPI_ISL_1303422, EPI_ISL_1303423, EPI_ISL_1379976, EPI_ISL_1502804, EPI_ISL_1502805, EPI_ISL_1502806, EPI_ISL_1502807, EPI_ISL_1502808, EPI_ISL_1502809, EPI_ISL_1506117, EPI_ISL_1506118, EPI_ISL_1506119 | | | |
| see above | MD Laboratories | Los Angeles County PHL | P. Hemarajata et al. |
| EPI_ISL_1197054, EPI_ISL_1303414, EPI_ISL_1303482 | Martin Luther King Jr. Community Hospital | Los Angeles County PHL | P. Hemarajata et al. |
| EPI_ISL_1664594, EPI_ISL_1664595, EPI_ISL_1664596, EPI_ISL_1664597, EPI_ISL_1664598, EPI_ISL_1664618, EPI_ISL_1664619, EPI_ISL_1669862, EPI_ISL_1921890, EPI_ISL_1921891 | | | |
| see above | Monterey County Public Health Laboratory | Monterey County Public Health Laboratory | Monterey County Public Health Laboratory |
| EPI_ISL_1195800, EPI_ISL_1195863 | NYU Langone Health | Departments of Pathology and Medicine, New York University School of Medicine | Adriana Heguy; Christian Marier; Dacia Dimartino; Emily Guzman; Gael Westby; Guiqing Wang; Paolo Cotzia; Paul Zappile; Peter Meyn; Sitharam Ramaswami; Yutong Zhang |
| EPI_ISL_1477495, EPI_ISL_1477498, EPI_ISL_1477572, EPI_ISL_1477582, EPI_ISL_1477986, EPI_ISL_1478024, EPI_ISL_1478030, EPI_ISL_1478106, EPI_ISL_1478110, EPI_ISL_1478175, EPI_ISL_1529158, EPI_ISL_1529336, EPI_ISL_1701251, EPI_ISL_1701399, EPI_ISL_1701430 | | | |
| see above | Orange County Public Health Lab | Chan-Zuckerberg Biohub | CZB Cliahub Consortium |
| EPI_ISL_1303404, EPI_ISL_1303405, EPI_ISL_1303406, EPI_ISL_1303472, EPI_ISL_1303473, EPI_ISL_1497525, EPI_ISL_1506115, EPI_ISL_1508898, EPI_ISL_1508899, EPI_ISL_1508900, EPI_ISL_1508901 | | | |
| see above | Pomona Valley Hospital Medical Center | Los Angeles County PHL | P. Hemarajata et al. |
| EPI_ISL_1252919, EPI_ISL_1252923, EPI_ISL_1252924, EPI_ISL_1252928, EPI_ISL_1252929, EPI_ISL_1252937, EPI_ISL_1252946, EPI_ISL_1252958, EPI_ISL_1252967, EPI_ISL_1252969, EPI_ISL_1252971, EPI_ISL_1252978, EPI_ISL_1252980, EPI_ISL_1252985, EPI_ISL_1252987, EPI_ISL_1252988, EPI_ISL_1252989, EPI_ISL_1252992, EPI_ISL_1252993, EPI_ISL_1253001, EPI_ISL_1253008, EPI_ISL_1253020, EPI_ISL_1253021, EPI_ISL_1253022, EPI_ISL_1253025, EPI_ISL_1253028, EPI_ISL_1253029, EPI_ISL_1253030, EPI_ISL_1253032, EPI_ISL_1253033, EPI_ISL_1253135, EPI_ISL_1253140, EPI_ISL_1253147, EPI_ISL_1253154, EPI_ISL_1253164, EPI_ISL_1253172, EPI_ISL_1253192, EPI_ISL_1253232, EPI_ISL_1253233, EPI_ISL_1253243, EPI_ISL_1253259, EPI_ISL_1253368, EPI_ISL_1253391, EPI_ISL_1253407, EPI_ISL_1253409, EPI_ISL_1253413, EPI_ISL_1253435, EPI_ISL_1253475, EPI_ISL_1253487, EPI_ISL_1253513, EPI_ISL_1253523, EPI_ISL_1253527, EPI_ISL_1253528, EPI_ISL_1253533, EPI_ISL_1253535, EPI_ISL_1254015, EPI_ISL_1254019, EPI_ISL_1254020, EPI_ISL_1254021, EPI_ISL_1254023, EPI_ISL_1254024, EPI_ISL_1254025, EPI_ISL_1254029, EPI_ISL_1254030, EPI_ISL_1254031, EPI_ISL_1254032, EPI_ISL_1254034, EPI_ISL_1254038, EPI_ISL_1254042, EPI_ISL_1254043, EPI_ISL_1254046, EPI_ISL_1254047, EPI_ISL_1254053, EPI_ISL_1254055, EPI_ISL_1254062, EPI_ISL_1254063, EPI_ISL_1254064, EPI_ISL_1254068, EPI_ISL_1254069, EPI_ISL_1254077, EPI_ISL_1254078, EPI_ISL_1254081, EPI_ISL_1254084, EPI_ISL_1254092, EPI_ISL_1254093, EPI_ISL_1254097, EPI_ISL_1254101, EPI_ISL_1254106, EPI_ISL_1254107, EPI_ISL_1254110, EPI_ISL_1254117, EPI_ISL_1254123, EPI_ISL_1254130, EPI_ISL_1254136, EPI_ISL_1254138, EPI_ISL_1254140, EPI_ISL_1254141, EPI_ISL_1254144, EPI_ISL_1254149, EPI_ISL_1254150, EPI_ISL_1254152, EPI_ISL_1254167, EPI_ISL_1254169, EPI_ISL_1254173, EPI_ISL_1254174, EPI_ISL_1254176, EPI_ISL_1254180, EPI_ISL_1254183, EPI_ISL_1254186, EPI_ISL_1254188, EPI_ISL_1254193, EPI_ISL_1254194, EPI_ISL_1254196, EPI_ISL_1254200, EPI_ISL_1254206, EPI_ISL_1254209, EPI_ISL_1254210, EPI_ISL_1254213, EPI_ISL_1254217, EPI_ISL_1254218, EPI_ISL_1254221, EPI_ISL_1254227, EPI_ISL_1254241, EPI_ISL_1254262, EPI_ISL_1254270, EPI_ISL_1254273, EPI_ISL_1254278, EPI_ISL_1254281, EPI_ISL_1254286, EPI_ISL_1254292, EPI_ISL_1254294, EPI_ISL_1254298, EPI_ISL_1254300, EPI_ISL_1254304, EPI_ISL_1254306, EPI_ISL_1254307, EPI_ISL_1254313, EPI_ISL_1254316, EPI_ISL_1254320, EPI_ISL_1254344, EPI_ISL_1254360, EPI_ISL_1254379, EPI_ISL_1254432, EPI_ISL_1254433, EPI_ISL_1254464, EPI_ISL_1254465, EPI_ISL_1254470, EPI_ISL_1254477, EPI_ISL_1254491, EPI_ISL_1265918, EPI_ISL_1265924, EPI_ISL_1265930, EPI_ISL_1265954, EPI_ISL_1265963, EPI_ISL_1265966, EPI_ISL_1265979, EPI_ISL_1266036, EPI_ISL_1266101, EPI_ISL_1266106, EPI_ISL_1266107, EPI_ISL_1266205, EPI_ISL_1266216, EPI_ISL_1266485, EPI_ISL_1266524, EPI_ISL_1266667, EPI_ISL_1266697, EPI_ISL_1266734, EPI_ISL_1266862, EPI_ISL_1267283, EPI_ISL_1267342, EPI_ISL_1267351, EPI_ISL_1267357, EPI_ISL_1267390, EPI_ISL_1267391, EPI_ISL_1267411, EPI_ISL_1267412, EPI_ISL_1267422, EPI_ISL_1267430, EPI_ISL_1267432, EPI_ISL_1267434, EPI_ISL_1267445, EPI_ISL_1267479, EPI_ISL_1267481, EPI_ISL_1291002, EPI_ISL_1291005, EPI_ISL_1291015, EPI_ISL_1291028, EPI_ISL_1291032, EPI_ISL_1291042, EPI_ISL_1291091, EPI_ISL_1291092, EPI_ISL_1291094, EPI_ISL_1291095, EPI_ISL_1291096, EPI_ISL_1291099, EPI_ISL_1291100, EPI_ISL_1291105, EPI_ISL_1291106, EPI_ISL_1291109, EPI_ISL_1291110, EPI_ISL_1291111, EPI_ISL_1291112, EPI_ISL_1291114, EPI_ISL_1291115, EPI_ISL_1291116, EPI_ISL_1291117, EPI_ISL_1291118, EPI_ISL_1291121, EPI_ISL_1291123, EPI_ISL_1291124, EPI_ISL_1291125, EPI_ISL_1291127, EPI_ISL_1291128, EPI_ISL_1291129, EPI_ISL_1291130, EPI_ISL_1291131, EPI_ISL_1291132, EPI_ISL_1291134, EPI_ISL_1291135, EPI_ISL_1291137, EPI_ISL_1291138, EPI_ISL_1291140, EPI_ISL_1291141, EPI_ISL_1291142, EPI_ISL_1291143, EPI_ISL_1291144, EPI_ISL_1291145, EPI_ISL_1291146, EPI_ISL_1291147, EPI_ISL_1291149, EPI_ISL_1291150, EPI_ISL_1291151, EPI_ISL_1291152, EPI_ISL_1291153, EPI_ISL_1291154, EPI_ISL_1291155, EPI_ISL_1291156, EPI_ISL_1291157, EPI_ISL_1291159, EPI_ISL_1291160, EPI_ISL_1291161, EPI_ISL_1291162, EPI_ISL_1291163, EPI_ISL_1291164, EPI_ISL_1291165, EPI_ISL_1291166, EPI_ISL_1291167, EPI_ISL_1291170, EPI_ISL_1291171, EPI_ISL_1291172, EPI_ISL_1291173, EPI_ISL_1291179, EPI_ISL_1291180, EPI_ISL_1291182, EPI_ISL_1291186, EPI_ISL_1291187, EPI_ISL_1291188, EPI_ISL_1297683, EPI_ISL_1297688, EPI_ISL_1297689, EPI_ISL_1297691, EPI_ISL_1297701, EPI_ISL_1297705, EPI_ISL_1297708, EPI_ISL_1297719, EPI_ISL_1297735, EPI_ISL_1297737, EPI_ISL_1297738, EPI_ISL_1297741, EPI_ISL_1297744, EPI_ISL_1297745, EPI_ISL_1297747, EPI_ISL_1297751, EPI_ISL_1297754, EPI_ISL_1297757, EPI_ISL_1297758, EPI_ISL_1297760, EPI_ISL_1314735, EPI_ISL_1335993, EPI_ISL_1335996, EPI_ISL_1336068, EPI_ISL_1336087, EPI_ISL_1336092, EPI_ISL_1336094, EPI_ISL_1336105, EPI_ISL_1336106, EPI_ISL_1336157, EPI_ISL_1339830, EPI_ISL_1339837, EPI_ISL_1339839, EPI_ISL_1339850, EPI_ISL_1339866, EPI_ISL_1339876, EPI_ISL_1339877, EPI_ISL_1339881, EPI_ISL_1339892, EPI_ISL_1339893, EPI_ISL_1339896, EPI_ISL_1339910, EPI_ISL_1339913, EPI_ISL_1339916, EPI_ISL_1339917, EPI_ISL_1339928, EPI_ISL_1339935, EPI_ISL_1339949, EPI_ISL_1339964, EPI_ISL_1340679, EPI_ISL_1367239, EPI_ISL_1367241, EPI_ISL_1367242, EPI_ISL_1367249, EPI_ISL_1367252, EPI_ISL_1367253, EPI_ISL_1367257, EPI_ISL_1367261, EPI_ISL_1367262, EPI_ISL_1367263, EPI_ISL_1367267, EPI_ISL_1367273, EPI_ISL_1367277, EPI_ISL_1367278, EPI_ISL_1367281, EPI_ISL_1367282, EPI_ISL_1367283, EPI_ISL_1367286, EPI_ISL_1367288, EPI_ISL_1367290, EPI_ISL_1367293, EPI_ISL_1367298, EPI_ISL_1367300, EPI_ISL_1367302, EPI_ISL_1367307, EPI_ISL_1367311, EPI_ISL_1367312, EPI_ISL_1367314, EPI_ISL_1367315, EPI_ISL_1367319, EPI_ISL_1367321, EPI_ISL_1367324, EPI_ISL_1367326, EPI_ISL_1367328, EPI_ISL_1391459, EPI_ISL_1391464, EPI_ISL_1391608, EPI_ISL_1391672, EPI_ISL_1391692, EPI_ISL_1422428, EPI_ISL_1422585, EPI_ISL_1422623, EPI_ISL_1422635, EPI_ISL_1422642, EPI_ISL_1422653, EPI_ISL_1422655, EPI_ISL_1422663, EPI_ISL_1422669, EPI_ISL_1422671, EPI_ISL_1422674, EPI_ISL_1422675, EPI_ISL_1422679, EPI_ISL_1422680, EPI_ISL_1422682, EPI_ISL_1422683, EPI_ISL_1422684, EPI_ISL_1422685, EPI_ISL_1422687, EPI_ISL_1422688, EPI_ISL_1422692, EPI_ISL_1422693, EPI_ISL_1422694, EPI_ISL_1422697, EPI_ISL_1422699, EPI_ISL_1422702, EPI_ISL_1422705, EPI_ISL_1422706, EPI_ISL_1422707, EPI_ISL_1422708, EPI_ISL_1422710, EPI_ISL_1422787, EPI_ISL_1422795, EPI_ISL_1422798, EPI_ISL_1444408, EPI_ISL_1444409, EPI_ISL_1444410, EPI_ISL_1444411, EPI_ISL_1444412, EPI_ISL_1444414, EPI_ISL_1444417, EPI_ISL_1444419, EPI_ISL_1444421, EPI_ISL_1444422, EPI_ISL_1444426, EPI_ISL_1444427, EPI_ISL_1444428, EPI_ISL_1444429, EPI_ISL_1444430, EPI_ISL_1444431, EPI_ISL_1444433, EPI_ISL_1444434, EPI_ISL_1444435, EPI_ISL_1444436, EPI_ISL_1444437, EPI_ISL_1444438, EPI_ISL_1444439, EPI_ISL_1444440, EPI_ISL_1444441, EPI_ISL_1444442, EPI_ISL_1444443, EPI_ISL_1444444, EPI_ISL_1444445, EPI_ISL_1444446, EPI_ISL_1444447, EPI_ISL_1444448, EPI_ISL_1444449, EPI_ISL_1444452, EPI_ISL_1444453, EPI_ISL_1444454, EPI_ISL_1444455, EPI_ISL_1444456, EPI_ISL_1444457, EPI_ISL_1444459, EPI_ISL_1444461, EPI_ISL_1444462, EPI_ISL_1444463, EPI_ISL_1444465, EPI_ISL_1444466, EPI_ISL_1444468, EPI_ISL_1444469, EPI_ISL_1444470, EPI_ISL_1444473, EPI_ISL_1444474, EPI_ISL_1444475, EPI_ISL_1444476, EPI_ISL_1444477, EPI_ISL_1444478, EPI_ISL_1444479, EPI_ISL_1444481, EPI_ISL_1444482, EPI_ISL_1444483, EPI_ISL_1444484, EPI_ISL_1444485, EPI_ISL_1444486, EPI_ISL_1444487, EPI_ISL_1444489, EPI_ISL_1444490, EPI_ISL_1444491, EPI_ISL_1444493, EPI_ISL_1444494, EPI_ISL_1444495, EPI_ISL_1444496, EPI_ISL_1444497, EPI_ISL_1444499, EPI_ISL_1444523, EPI_ISL_1444527, EPI_ISL_1444566, EPI_ISL_1444568, EPI_ISL_1444569, EPI_ISL_1444571, EPI_ISL_1444573, EPI_ISL_1444574, EPI_ISL_1444576, EPI_ISL_1444578, EPI_ISL_1444579, EPI_ISL_1444580, EPI_ISL_1444581, EPI_ISL_1444583, EPI_ISL_1444585, EPI_ISL_1444588, EPI_ISL_1444589, EPI_ISL_1444590, EPI_ISL_1444592, EPI_ISL_1444593, EPI_ISL_1444594, EPI_ISL_1444595, EPI_ISL_1444596, EPI_ISL_1444597, EPI_ISL_1444601, EPI_ISL_1444602, EPI_ISL_1444604, EPI_ISL_1444605, EPI_ISL_1444606, EPI_ISL_1444608, EPI_ISL_1444609, EPI_ISL_1444611, EPI_ISL_1444612, EPI_ISL_1444613, EPI_ISL_1444614, EPI_ISL_1444615, EPI_ISL_1444617, EPI_ISL_1444618, EPI_ISL_1444619, EPI_ISL_1444620, EPI_ISL_1444621, EPI_ISL_1444622, EPI_ISL_1444623, EPI_ISL_1444624, EPI_ISL_1444625, EPI_ISL_1444626, EPI_ISL_1444627, EPI_ISL_1444628, EPI_ISL_1444631, EPI_ISL_1444632, EPI_ISL_1444633, EPI_ISL_1444638, EPI_ISL_1444639, EPI_ISL_1444640, EPI_ISL_1444642, EPI_ISL_1444643, EPI_ISL_1444644, EPI_ISL_1444645, EPI_ISL_1444646, EPI_ISL_1444647, EPI_ISL_1444648, EPI_ISL_1444649, EPI_ISL_1444650, EPI_ISL_1444651, EPI_ISL_1444652, EPI_ISL_1444653, EPI_ISL_1444654, EPI_ISL_1444655, EPI_ISL_1444656, EPI_ISL_1444657, EPI_ISL_1444658, EPI_ISL_1444659, EPI_ISL_1444660, EPI_ISL_1444661, EPI_ISL_1444662, EPI_ISL_1444663, EPI_ISL_1444664, EPI_ISL_1444665, EPI_ISL_1444666, EPI_ISL_1444667, EPI_ISL_1444669, EPI_ISL_1444714, EPI_ISL_1460730, EPI_ISL_1460731, EPI_ISL_1460733, EPI_ISL_1460735, EPI_ISL_1460737, EPI_ISL_1460739, EPI_ISL_1460740, EPI_ISL_1460741, EPI_ISL_1460742, EPI_ISL_1460748, EPI_ISL_1460751, EPI_ISL_1460752, EPI_ISL_1460753, EPI_ISL_1460754, EPI_ISL_1460755, EPI_ISL_1460756, EPI_ISL_1460757, EPI_ISL_1460760, EPI_ISL_1460761, EPI_ISL_1460762, EPI_ISL_1460763, EPI_ISL_1460764, EPI_ISL_1460765, EPI_ISL_1460766, EPI_ISL_1460767, EPI_ISL_1460768, EPI_ISL_1460769, EPI_ISL_1460770, EPI_ISL_1460771, EPI_ISL_1460772, EPI_ISL_1460774, EPI_ISL_1460776, EPI_ISL_1460777, EPI_ISL_1460778, EPI_ISL_1460779, EPI_ISL_1460782, EPI_ISL_1460783, EPI_ISL_1460784, EPI_ISL_1479739, EPI_ISL_1479757, EPI_ISL_1479758, EPI_ISL_1479759, EPI_ISL_1479767, EPI_ISL_1479768, EPI_ISL_1479769, EPI_ISL_1479770, EPI_ISL_1479774, EPI_ISL_1479776, EPI_ISL_1479779, EPI_ISL_1479782, EPI_ISL_1479783, EPI_ISL_1479784, EPI_ISL_1479789, EPI_ISL_1479793, EPI_ISL_1479798, EPI_ISL_1479801, EPI_ISL_1479950, EPI_ISL_1480817, EPI_ISL_1480838, EPI_ISL_1480839, EPI_ISL_1480842, EPI_ISL_1480848, EPI_ISL_1480858, EPI_ISL_1480862, EPI_ISL_1491950, EPI_ISL_1493939, EPI_ISL_1493941, EPI_ISL_1493949, EPI_ISL_1493951, EPI_ISL_1493960, EPI_ISL_1493962, EPI_ISL_1493966, EPI_ISL_1493976, EPI_ISL_1493978, EPI_ISL_1493983, EPI_ISL_1494011, EPI_ISL_1494021, EPI_ISL_1511667, EPI_ISL_1511706, EPI_ISL_1511711, EPI_ISL_1511721, EPI_ISL_1511735, EPI_ISL_1525910, EPI_ISL_1525912, EPI_ISL_1525939, EPI_ISL_1525955, EPI_ISL_1525969, EPI_ISL_1525971, EPI_ISL_1525977, EPI_ISL_1525978, EPI_ISL_1525979, EPI_ISL_1525980, EPI_ISL_1525981, EPI_ISL_1525983, EPI_ISL_1525984, EPI_ISL_1525986, EPI_ISL_1525987, EPI_ISL_1525988, EPI_ISL_1525989, EPI_ISL_1525992, EPI_ISL_1525993, EPI_ISL_1525994, EPI_ISL_1525996, EPI_ISL_1525998, EPI_ISL_1525999, EPI_ISL_1526000, EPI_ISL_1526002, EPI_ISL_1526003, EPI_ISL_1526004, EPI_ISL_1526005, EPI_ISL_1526008, EPI_ISL_1526009, EPI_ISL_1526010, EPI_ISL_1526013, EPI_ISL_1526016, EPI_ISL_1526017, EPI_ISL_1526018, EPI_ISL_1526019, EPI_ISL_1526020, EPI_ISL_1526021, EPI_ISL_1526022, EPI_ISL_1526024, EPI_ISL_1526025, EPI_ISL_1526026, EPI_ISL_1526027, EPI_ISL_1526028, EPI_ISL_1526032, EPI_ISL_1526033, EPI_ISL_1526034, EPI_ISL_1526035, EPI_ISL_1526037, EPI_ISL_1526038, EPI_ISL_1526039, EPI_ISL_1526040, EPI_ISL_1526041, EPI_ISL_1526042, EPI_ISL_1526043, EPI_ISL_1526044, EPI_ISL_1526045, EPI_ISL_1526046, EPI_ISL_1526049, EPI_ISL_1526050, EPI_ISL_1526051, EPI_ISL_1526052, EPI_ISL_1526054, EPI_ISL_1526056, EPI_ISL_1526057, EPI_ISL_1526058, EPI_ISL_1526061, EPI_ISL_1526062, EPI_ISL_1526063, EPI_ISL_1526064, EPI_ISL_1526065, EPI_ISL_1526068, EPI_ISL_1526070, EPI_ISL_1551776, EPI_ISL_1551778, EPI_ISL_1551780, EPI_ISL_1551781, EPI_ISL_1551782, EPI_ISL_1551783, EPI_ISL_1551784, EPI_ISL_1551785, EPI_ISL_1551788, EPI_ISL_1551791, EPI_ISL_1551792, EPI_ISL_1551793, EPI_ISL_1551794, EPI_ISL_1551795, EPI_ISL_1551799, EPI_ISL_1551801, EPI_ISL_1551802, EPI_ISL_1551803, EPI_ISL_1551805, EPI_ISL_1551806, EPI_ISL_1551807, EPI_ISL_1551808, EPI_ISL_1551809, EPI_ISL_1551810, EPI_ISL_1551813, EPI_ISL_1551814, EPI_ISL_1551816, EPI_ISL_1551817, EPI_ISL_1551820, EPI_ISL_1551881, EPI_ISL_1551883, EPI_ISL_1551885, EPI_ISL_1551886, EPI_ISL_1551887, EPI_ISL_1551890, EPI_ISL_1551891, EPI_ISL_1551892, EPI_ISL_1551895, EPI_ISL_1551899, EPI_ISL_1551900, EPI_ISL_1551901, EPI_ISL_1551903, EPI_ISL_1551905, EPI_ISL_1551906, EPI_ISL_1551908, EPI_ISL_1551911, EPI_ISL_1551912, EPI_ISL_1551913, EPI_ISL_1551916, EPI_ISL_1551917, EPI_ISL_1551919, EPI_ISL_1551920, EPI_ISL_1551922, EPI_ISL_1551923, EPI_ISL_1552010, EPI_ISL_1552012, EPI_ISL_1552013, EPI_ISL_1552020, EPI_ISL_1552021, EPI_ISL_1552025, EPI_ISL_1552028, EPI_ISL_1552029, EPI_ISL_1552031, EPI_ISL_1552032, EPI_ISL_1552033, EPI_ISL_1552037, EPI_ISL_1552038, EPI_ISL_1552040, EPI_ISL_1552047, EPI_ISL_1552051, EPI_ISL_1552055, EPI_ISL_1552057, EPI_ISL_1552062, EPI_ISL_1552063, EPI_ISL_1552064, EPI_ISL_1552070, EPI_ISL_1552074, EPI_ISL_1552174, EPI_ISL_1552202, EPI_ISL_1552214, EPI_ISL_1552721, EPI_ISL_1552738, EPI_ISL_1552762, EPI_ISL_1552763, EPI_ISL_1552765, EPI_ISL_1552767, EPI_ISL_1552768, EPI_ISL_1552769, EPI_ISL_1552774, EPI_ISL_1552781, EPI_ISL_1552785, EPI_ISL_1552787, EPI_ISL_1552788, EPI_ISL_1552789, EPI_ISL_1552791, EPI_ISL_1552798, EPI_ISL_1552799, EPI_ISL_1552800, EPI_ISL_1552801, EPI_ISL_1552802, EPI_ISL_1552803, EPI_ISL_1552804, EPI_ISL_1552806, EPI_ISL_1552809, EPI_ISL_1552813, EPI_ISL_1552815, EPI_ISL_1552816, EPI_ISL_1559413, EPI_ISL_1559416, EPI_ISL_1559418, EPI_ISL_1559425, EPI_ISL_1559431, EPI_ISL_1559432, EPI_ISL_1559437, EPI_ISL_1559443, EPI_ISL_1559450, EPI_ISL_1559454, EPI_ISL_1559455, EPI_ISL_1559457, EPI_ISL_1559461, EPI_ISL_1559463, EPI_ISL_1559464, EPI_ISL_1559480, EPI_ISL_1559485, EPI_ISL_1559486, EPI_ISL_1559488, EPI_ISL_1559491, EPI_ISL_1559495, EPI_ISL_1559496, EPI_ISL_1559498, EPI_ISL_1559506, EPI_ISL_1559510, EPI_ISL_1559512, EPI_ISL_1559515, EPI_ISL_1559517, EPI_ISL_1559518, EPI_ISL_1559534, EPI_ISL_1559535, EPI_ISL_1559536, EPI_ISL_1559537, EPI_ISL_1559538, EPI_ISL_1559540, EPI_ISL_1559541, EPI_ISL_1559542, EPI_ISL_1559543, EPI_ISL_1559545, EPI_ISL_1559549, EPI_ISL_1559550, EPI_ISL_1559551, EPI_ISL_1559553, EPI_ISL_1559555, EPI_ISL_1559559, EPI_ISL_1559562, EPI_ISL_1559563, EPI_ISL_1559565, EPI_ISL_1559569, EPI_ISL_1559577, EPI_ISL_1559579, EPI_ISL_1559582, EPI_ISL_1559583, EPI_ISL_1559586, EPI_ISL_1582093, EPI_ISL_1582122, EPI_ISL_1582123, EPI_ISL_1582135, EPI_ISL_1582136, EPI_ISL_1582139, EPI_ISL_1582140, EPI_ISL_1582146, EPI_ISL_1582163, EPI_ISL_1694666, EPI_ISL_1694667, EPI_ISL_1694668, EPI_ISL_1694692, EPI_ISL_1694694 | | | |
| see above | Quest Diagnostics Incorporated | Centers for Disease Control and Prevention Division of Viral Diseases, Pathogen Discovery | A. Gerasimova; A. Perez; Adrian Paskey; B. Anderson; Ben L. Rambo-Martin; Benjamin Rambo-Martin; Christopher Gulvick; Clinton R. Paden; Dakota Howard; Darlene Wagner; Dhwani Batra; Duncan MacCannell; F. Lacbawan; I. A. Shlyakhter; Jason Caravas; K.E. Livingston; Kara Moser; L.E. Bernstein; M. Hua; Matthew Schmerer; P. Tanpaiboon; Peter W. Cook; R. M. Kagan; R. Owen; R. V. Rolando; S. H. Rosenthal; Scott Sammons; Shatavia Morrison; Suxiang Tong; Y. Liu; Yvette Unoarumhi |
| EPI_ISL_1794769, EPI_ISL_1794770, EPI_ISL_1794772, EPI_ISL_1794776, EPI_ISL_1794777, EPI_ISL_1794778, EPI_ISL_1794779, EPI_ISL_1794781 | | | |
| see above | Rady Children's | Andersen lab at Scripps Research | Christina Clarke; David Dimmock; Denise Malicki; Kathryn Bouic; Linda Luo; SEARCH Alliance San Diego with Nanda Radamchar; Teresa Mueller |

| | | | |
|---|---|---|---|
| | Hospital - San Diego | | |
| EPI_ISL_1295816, EPI_ISL_1295817, EPI_ISL_1315073, EPI_ISL_1366749, EPI_ISL_1366750, EPI_ISL_1366751, EPI_ISL_1468832, EPI_ISL_1468839, EPI_ISL_1468843, EPI_ISL_1468844, EPI_ISL_1468874, EPI_ISL_1468877, EPI_ISL_1468878, EPI_ISL_1468879, EPI_ISL_1468880, EPI_ISL_1468881, EPI_ISL_1468914, EPI_ISL_1468915, EPI_ISL_1468916, EPI_ISL_1468922, EPI_ISL_1468923, EPI_ISL_1468924, EPI_ISL_1468925, EPI_ISL_1468926, EPI_ISL_1468927, EPI_ISL_1468928, EPI_ISL_1468929, EPI_ISL_1468930, EPI_ISL_1468931, EPI_ISL_1468932, EPI_ISL_1468933, EPI_ISL_1468934, EPI_ISL_1468935, EPI_ISL_1468936, EPI_ISL_1468937, EPI_ISL_1468938, EPI_ISL_1468939, EPI_ISL_1468940, EPI_ISL_1468941, EPI_ISL_1468942, EPI_ISL_1468943, EPI_ISL_1468944, EPI_ISL_1468945, EPI_ISL_1468946, EPI_ISL_1468948, EPI_ISL_1468949, EPI_ISL_1468951, EPI_ISL_1468981, EPI_ISL_1468982, EPI_ISL_1468985, EPI_ISL_1468988, EPI_ISL_1468992, EPI_ISL_1468993, EPI_ISL_1468997, EPI_ISL_1468998, EPI_ISL_1469003, EPI_ISL_1469006, EPI_ISL_1469007, EPI_ISL_1469010, EPI_ISL_1469011, EPI_ISL_1469014, EPI_ISL_1469015, EPI_ISL_1469017, EPI_ISL_1469018, EPI_ISL_1469019, EPI_ISL_1469023, EPI_ISL_1469024, EPI_ISL_1469025, EPI_ISL_1469026, EPI_ISL_1469027, EPI_ISL_1469028, EPI_ISL_1469029, EPI_ISL_1469030, EPI_ISL_1469031, EPI_ISL_1469032, EPI_ISL_1469033, EPI_ISL_1469034, EPI_ISL_1469035, EPI_ISL_1469036, EPI_ISL_1469037, EPI_ISL_1469038, EPI_ISL_1469039, EPI_ISL_1469040, EPI_ISL_1469041, EPI_ISL_1469042, EPI_ISL_1791403, EPI_ISL_1791404, EPI_ISL_1791405, EPI_ISL_1791406, EPI_ISL_1791407, EPI_ISL_1791408, EPI_ISL_1791409, EPI_ISL_1791410, EPI_ISL_1791411, EPI_ISL_1791412, EPI_ISL_1791413, EPI_ISL_1791414, EPI_ISL_1791415, EPI_ISL_1791416, EPI_ISL_1791417, EPI_ISL_1791418, EPI_ISL_1791419, EPI_ISL_1791420, EPI_ISL_1791421, EPI_ISL_1791422, EPI_ISL_1791423, EPI_ISL_1791424, EPI_ISL_1791425, EPI_ISL_1791426, EPI_ISL_1791427, EPI_ISL_1791428, EPI_ISL_1794619, EPI_ISL_1794649, EPI_ISL_1794654, EPI_ISL_1794997, EPI_ISL_1795012, EPI_ISL_1795023, EPI_ISL_1795034, EPI_ISL_1969251, EPI_ISL_1969252, EPI_ISL_1969253, EPI_ISL_1969254, EPI_ISL_1969256, EPI_ISL_1969259, EPI_ISL_1969261, EPI_ISL_1969264, EPI_ISL_1969267, EPI_ISL_1969269 | | | |
| see above | San Diego County Public Health Laboratory | Andersen lab at Scripps Research | Brett Austin; Jovan Shephard; SEARCH Alliance San Diego with Ashleigh Murphy; SEARCH Alliance San Diego with Tracy Basler |
| EPI_ISL_1529137, EPI_ISL_1529148, EPI_ISL_1529154, EPI_ISL_1529191, EPI_ISL_1529193, EPI_ISL_1529205, EPI_ISL_1529234, EPI_ISL_1529240, EPI_ISL_1529245, EPI_ISL_1529259, EPI_ISL_1529263, EPI_ISL_1529267, EPI_ISL_1529284, EPI_ISL_1529317, EPI_ISL_1663931, EPI_ISL_1663933, EPI_ISL_1663939, EPI_ISL_1663963, EPI_ISL_1663979, EPI_ISL_1663982, EPI_ISL_1663993, EPI_ISL_1664004, EPI_ISL_1664016, EPI_ISL_1664025, EPI_ISL_1664036, EPI_ISL_1664043, EPI_ISL_1664062, EPI_ISL_1664073, EPI_ISL_1664079, EPI_ISL_1664080, EPI_ISL_1664083, EPI_ISL_1664089, EPI_ISL_1664094, EPI_ISL_1664095, EPI_ISL_1664107, EPI_ISL_1701401, EPI_ISL_1701413, EPI_ISL_1701437, EPI_ISL_1701439, EPI_ISL_1701440, EPI_ISL_1701445, EPI_ISL_1701457, EPI_ISL_1701462, EPI_ISL_1701477, EPI_ISL_1701480, EPI_ISL_1701481, EPI_ISL_1701486, EPI_ISL_1701496, EPI_ISL_1701498, EPI_ISL_1701501, EPI_ISL_1701510, EPI_ISL_1701516, EPI_ISL_1701518, EPI_ISL_1701523, EPI_ISL_1701526, EPI_ISL_1701528, EPI_ISL_1701532, EPI_ISL_1701544, EPI_ISL_1701550, EPI_ISL_1701554, EPI_ISL_1701564, EPI_ISL_1701567, EPI_ISL_1701573, EPI_ISL_1701575, EPI_ISL_1701583, EPI_ISL_1701593, EPI_ISL_1701596, EPI_ISL_1701598, EPI_ISL_1701609, EPI_ISL_1701612, EPI_ISL_1701625, EPI_ISL_1701630, EPI_ISL_1701631, EPI_ISL_1701640, EPI_ISL_1701645, EPI_ISL_1701653, EPI_ISL_1821319, EPI_ISL_1821320, EPI_ISL_1821324, EPI_ISL_1821327, EPI_ISL_1821329, EPI_ISL_1821330, EPI_ISL_1821331, EPI_ISL_1821333, EPI_ISL_1821337, EPI_ISL_1821338, EPI_ISL_1821339, EPI_ISL_1821341, EPI_ISL_1821342, EPI_ISL_1821350, EPI_ISL_1821355, EPI_ISL_1821392, EPI_ISL_1821396, EPI_ISL_1821397, EPI_ISL_1821398, EPI_ISL_1821402, EPI_ISL_1821405, EPI_ISL_1821413, EPI_ISL_1821414, EPI_ISL_1821419, EPI_ISL_1821426, EPI_ISL_1821427, EPI_ISL_1821437, EPI_ISL_1821443, EPI_ISL_1821445, EPI_ISL_1821455, EPI_ISL_1821458, EPI_ISL_1821460, EPI_ISL_1821461, EPI_ISL_1821470, EPI_ISL_2102744, EPI_ISL_2102766, EPI_ISL_2102774, EPI_ISL_2102779, EPI_ISL_2102781, EPI_ISL_2102784, EPI_ISL_2102785, EPI_ISL_2102803, EPI_ISL_2102810, EPI_ISL_2102823, EPI_ISL_2102825, EPI_ISL_2102827, EPI_ISL_2102831, EPI_ISL_2102851, EPI_ISL_2102862, EPI_ISL_2102865, EPI_ISL_2102871, EPI_ISL_2102875, EPI_ISL_2102876, EPI_ISL_2102882, EPI_ISL_2102911, EPI_ISL_2102920, EPI_ISL_2102941, EPI_ISL_2102950, EPI_ISL_2102956, EPI_ISL_2102959, EPI_ISL_2102963, EPI_ISL_2102966, EPI_ISL_2102969, EPI_ISL_2103014, EPI_ISL_2103021, EPI_ISL_2103042, EPI_ISL_2103059, EPI_ISL_2103083, EPI_ISL_2103089, EPI_ISL_2103123, EPI_ISL_2103144, EPI_ISL_2103148, EPI_ISL_2103190 | | | |
| see above | Santa Clara County Public Health Laboratory | Chan-Zuckerberg Biohub | CZB Cliahub Consortium |
| EPI_ISL_1169600, EPI_ISL_1203817, EPI_ISL_1222757, EPI_ISL_1235633, EPI_ISL_1235634, EPI_ISL_1235635, EPI_ISL_1235637, EPI_ISL_1335868, EPI_ISL_1335869, EPI_ISL_1335870, EPI_ISL_1372415, EPI_ISL_1406155, EPI_ISL_1406165, EPI_ISL_1406166, EPI_ISL_1406168, EPI_ISL_1406169, EPI_ISL_1483802, EPI_ISL_1920832 | | | |
| see above | Santa Clara County Public Health Laboratory | Santa Clara County Public Health Laboratory | Santa Clara County Public Health Department |
| EPI_ISL_1483799, EPI_ISL_1532131, EPI_ISL_1532133, EPI_ISL_1532134, EPI_ISL_1532135, EPI_ISL_1532136, EPI_ISL_1532137, EPI_ISL_1532138 | | | |
| see above | Santa Clara Valley Medical Center | Santa Clara County Public Health Laboratory | Santa Clara County Public Health Department |
| EPI_ISL_1664288 | Santa Clara Valley Medical Center | Santa Clara County public Health Laboratory | Santa Clara County Public Health Department |
| EPI_ISL_1468842, EPI_ISL_1468882, EPI_ISL_1468883, EPI_ISL_1468884, EPI_ISL_1468885, EPI_ISL_1468886, EPI_ISL_1468955, EPI_ISL_1468956, EPI_ISL_1469045, EPI_ISL_1469046, EPI_ISL_1469051, EPI_ISL_1469052, EPI_ISL_1469053, EPI_ISL_1469054, EPI_ISL_1469055, EPI_ISL_1469056, EPI_ISL_1469057 | | | |
| see above | Scripps Medical Laboratory | Andersen lab at Scripps Research | Ellen Stefanski; Ian Mchardy; SEARCH Alliance San Diego with Michael Quigley |
| EPI_ISL_1295776, EPI_ISL_1295777, EPI_ISL_1295779, EPI_ISL_1295780, EPI_ISL_1295781, EPI_ISL_1295783, EPI_ISL_1295784, EPI_ISL_1295785, EPI_ISL_1295786, EPI_ISL_1295788, EPI_ISL_1295791, EPI_ISL_1295792, EPI_ISL_1295793, EPI_ISL_1295794, EPI_ISL_1295795, EPI_ISL_1366276, EPI_ISL_1366277, EPI_ISL_1366279, EPI_ISL_1366281, EPI_ISL_1366283, EPI_ISL_1366286, EPI_ISL_1366301, EPI_ISL_1366302, EPI_ISL_1366304, EPI_ISL_1366305, EPI_ISL_1366308, EPI_ISL_1366309, EPI_ISL_1366311, EPI_ISL_1366312, EPI_ISL_1366313, EPI_ISL_1366314, EPI_ISL_1366315, EPI_ISL_1366319, EPI_ISL_1366321, EPI_ISL_1366462, EPI_ISL_1366466, EPI_ISL_1366467, EPI_ISL_1366468, EPI_ISL_1366472, EPI_ISL_1366473, EPI_ISL_1366474, EPI_ISL_1366476, EPI_ISL_1366478, EPI_ISL_1366480, EPI_ISL_1366482, EPI_ISL_1366483, EPI_ISL_1366485, EPI_ISL_1366486, EPI_ISL_1366487, EPI_ISL_1366488, EPI_ISL_1366489, EPI_ISL_1366490, EPI_ISL_1366491, EPI_ISL_1366492, EPI_ISL_1366493, EPI_ISL_1366494, EPI_ISL_1468829, EPI_ISL_1468830, EPI_ISL_1468831, EPI_ISL_1468833, EPI_ISL_1468834, EPI_ISL_1468835, EPI_ISL_1468836, EPI_ISL_1468838, EPI_ISL_1468840, EPI_ISL_1468845, EPI_ISL_1468846, EPI_ISL_1468848, EPI_ISL_1468850, EPI_ISL_1468851, EPI_ISL_1468852, EPI_ISL_1468853, EPI_ISL_1468855, EPI_ISL_1468856, EPI_ISL_1468857, EPI_ISL_1468858, EPI_ISL_1468859, EPI_ISL_1468860, EPI_ISL_1468861, EPI_ISL_1468862, EPI_ISL_1468863, EPI_ISL_1468864, EPI_ISL_1468865, EPI_ISL_1468866, EPI_ISL_1468867, EPI_ISL_1468868, EPI_ISL_1468869, EPI_ISL_1468870, EPI_ISL_1468871, EPI_ISL_1468872, EPI_ISL_1468873, EPI_ISL_1468889, EPI_ISL_1468890, EPI_ISL_1468891, EPI_ISL_1468892, EPI_ISL_1468893, EPI_ISL_1468894, EPI_ISL_1468895, EPI_ISL_1468896, EPI_ISL_1468897, EPI_ISL_1468898, EPI_ISL_1468899, EPI_ISL_1468900, EPI_ISL_1468901, EPI_ISL_1468902, EPI_ISL_1468903, EPI_ISL_1468904, EPI_ISL_1468905, EPI_ISL_1468910, EPI_ISL_1468911, EPI_ISL_1468912, EPI_ISL_1468913, EPI_ISL_1468917, EPI_ISL_1468918, EPI_ISL_1468919, EPI_ISL_1468920, EPI_ISL_1468921, EPI_ISL_1468952, EPI_ISL_1468954, EPI_ISL_1468957, EPI_ISL_1468958, EPI_ISL_1468960, EPI_ISL_1468961, EPI_ISL_1468962, EPI_ISL_1468963, EPI_ISL_1468964, EPI_ISL_1468965, EPI_ISL_1468966, EPI_ISL_1468967, EPI_ISL_1468968, EPI_ISL_1468969, EPI_ISL_1468970, EPI_ISL_1468971, EPI_ISL_1468972, EPI_ISL_1468973, EPI_ISL_1468974, EPI_ISL_1468975, EPI_ISL_1468976, EPI_ISL_1468977, EPI_ISL_1468978, EPI_ISL_1469061, EPI_ISL_1469062, EPI_ISL_1469063, EPI_ISL_1469064, EPI_ISL_1469065, EPI_ISL_1469066, EPI_ISL_1469067, EPI_ISL_1469068, EPI_ISL_1469069, EPI_ISL_1469070, EPI_ISL_1469071, EPI_ISL_1469072, EPI_ISL_1469073, EPI_ISL_1469074, EPI_ISL_1469075, EPI_ISL_1469076, EPI_ISL_1469077, EPI_ISL_1469078, EPI_ISL_1469079, EPI_ISL_1469080, EPI_ISL_1469081, EPI_ISL_1469082, EPI_ISL_1469083, EPI_ISL_1469084, EPI_ISL_1469085, EPI_ISL_1469086, EPI_ISL_1469087, EPI_ISL_1531777, EPI_ISL_1531778, EPI_ISL_1531779, EPI_ISL_1531780, EPI_ISL_1531781, EPI_ISL_1531782, EPI_ISL_1531783, EPI_ISL_1531784, EPI_ISL_1531785, EPI_ISL_1531786, EPI_ISL_1531787, EPI_ISL_1531788, EPI_ISL_1531789, EPI_ISL_1531790, EPI_ISL_1531791, EPI_ISL_1531792, EPI_ISL_1531793, EPI_ISL_1531794, EPI_ISL_1531795, EPI_ISL_1531796, EPI_ISL_1531797, EPI_ISL_1531798, EPI_ISL_1531799, EPI_ISL_1531800, EPI_ISL_1531801, EPI_ISL_1531802, EPI_ISL_1531803, EPI_ISL_1531804, EPI_ISL_1531805, EPI_ISL_1531806, EPI_ISL_1794685, EPI_ISL_1794686, EPI_ISL_1794687, EPI_ISL_1794688, EPI_ISL_1794689, EPI_ISL_1794690, EPI_ISL_1794691, EPI_ISL_1794692, EPI_ISL_1794693, EPI_ISL_1794694, EPI_ISL_1794695, EPI_ISL_1794696, EPI_ISL_1794697, EPI_ISL_1794698, EPI_ISL_1794699, EPI_ISL_1794703, EPI_ISL_1794704, EPI_ISL_1794713, EPI_ISL_1794714, EPI_ISL_1794715, EPI_ISL_1794716, EPI_ISL_1794717, EPI_ISL_1794718, EPI_ISL_1794719, EPI_ISL_1794720, EPI_ISL_1969312, EPI_ISL_1969313 | | | |
| see above | Sharp HealthCare Laboratory | Andersen lab at Scripps Research | Art Mendoza; Cathy Woerle; Jacquelyn Berumen; Liam McGinnis; Omid Bakhtar; SEARCH Alliance San Diego with Aaron Harding |
| EPI_ISL_1379889, EPI_ISL_1424116, EPI_ISL_1424117, EPI_ISL_1424125, EPI_ISL_1675223, EPI_ISL_1675224, EPI_ISL_1675225, EPI_ISL_1701679, EPI_ISL_2027133, EPI_ISL_2027136, EPI_ISL_2027138, EPI_ISL_2027167 | | | |
| see above | Stanford Health Care | Stanford University School of Medicine, Clinical Virology Laboratory | Becky Jiang; Bernadette Troung; Bernadette Truong; ChunHong Huang; Daniel Solis; Fumiko Yamamoto; Hannah Wang; Jacob Miller; James Zehnder; Kenji O. Mfuh; Malaya K. Sahoo; Mamdouh Sibai; Marilyn Mar; Michelle Verghese; Naomi Iwai; and Benjamin A. Pinsky |
| EPI_ISL_1303432, EPI_ISL_1303433, EPI_ISL_1303434, EPI_ISL_1303435, EPI_ISL_1303436, EPI_ISL_1303437, EPI_ISL_1303449, EPI_ISL_1303453, EPI_ISL_1303454, EPI_ISL_1303455, EPI_ISL_1303456, EPI_ISL_1303457, EPI_ISL_1303458, EPI_ISL_1303459, EPI_ISL_1303460, EPI_ISL_1303461, EPI_ISL_1379958, EPI_ISL_1379959, EPI_ISL_1379960, EPI_ISL_1379961, EPI_ISL_1379962, EPI_ISL_1379963, EPI_ISL_1379964, EPI_ISL_1379965, EPI_ISL_1379966, EPI_ISL_1379967, EPI_ISL_1379968, EPI_ISL_1379969, EPI_ISL_1379970, EPI_ISL_1379971, EPI_ISL_1379972, EPI_ISL_1379973, EPI_ISL_1379974, EPI_ISL_1379975, EPI_ISL_1497541, EPI_ISL_1506114, EPI_ISL_1707043, EPI_ISL_1707049 | | | |
| see above | UCLA Clinical Micro Lab | Los Angeles County PHL | P. Hemarajata et al. |
| EPI_ISL_1402608, EPI_ISL_1402609, EPI_ISL_1402610, EPI_ISL_1402611, EPI_ISL_1402612, EPI_ISL_1402613, EPI_ISL_1402614, EPI_ISL_1402615, EPI_ISL_1402616, EPI_ISL_1402617, EPI_ISL_1402618, EPI_ISL_1402619, EPI_ISL_1402620, EPI_ISL_1402621, EPI_ISL_1402622, EPI_ISL_1402623, EPI_ISL_1402624, EPI_ISL_1402625, EPI_ISL_1402626, EPI_ISL_1402627, EPI_ISL_1402628, EPI_ISL_1402629, EPI_ISL_1402630, EPI_ISL_1402631, EPI_ISL_1402632, EPI_ISL_1402633, EPI_ISL_1402634, EPI_ISL_1402635, EPI_ISL_1402636, EPI_ISL_1402637, EPI_ISL_1402638, EPI_ISL_1402639, EPI_ISL_1402640, EPI_ISL_1402641, EPI_ISL_1402642, EPI_ISL_1402643, EPI_ISL_1402644, EPI_ISL_1402645, EPI_ISL_1402646, EPI_ISL_1402647, EPI_ISL_1402648, EPI_ISL_1402649, EPI_ISL_1402650, EPI_ISL_1402651, EPI_ISL_1402652, EPI_ISL_1402653, EPI_ISL_1402654, EPI_ISL_1405050, EPI_ISL_1405051, EPI_ISL_1405052, EPI_ISL_1405053, EPI_ISL_1405054, EPI_ISL_1405055, EPI_ISL_1405056, EPI_ISL_1405057, EPI_ISL_1405058, EPI_ISL_1405059, EPI_ISL_1405060, EPI_ISL_1469853, EPI_ISL_1469877, EPI_ISL_1469882 | | | |
| see above | UCSC Genomics Institute | UCSC Genomics Institute | A. Marm Kilpatrick; Angie Hinrichs; Beth Shapiro; Bryan Thornlow; David Haussler; Eric Beraut; Hugh Olsen; Ikenna Anigbogu; Isabel Bjork; Jakob McBroome; Jeremy Sanford; Joshua Kapp; Mark Akeson; Maximilian Haeussler; Michael Stone; Miten Jain; Molly Cassatt-Johnstone; Namrita Dhillon; Russell Corbett-Detig; Terren Chang; Yatish Turakhia |
| EPI_ISL_1197053, EPI_ISL_1197131, EPI_ISL_1197132, EPI_ISL_1197133, EPI_ISL_1197134, EPI_ISL_1197135, EPI_ISL_1197136, EPI_ISL_1197137, EPI_ISL_1197138, EPI_ISL_1197139, EPI_ISL_1197140, EPI_ISL_1197141, EPI_ISL_1197142, EPI_ISL_1197143, EPI_ISL_1197144, EPI_ISL_1197145, EPI_ISL_1197146, EPI_ISL_1197147, EPI_ISL_1197148, EPI_ISL_1303407, EPI_ISL_1303409, EPI_ISL_1303410, EPI_ISL_1303411, EPI_ISL_1303412, EPI_ISL_1303413, EPI_ISL_1303415, EPI_ISL_1303425, EPI_ISL_1303440, EPI_ISL_1303441, EPI_ISL_1303442, EPI_ISL_1303443, EPI_ISL_1303444, EPI_ISL_1303445, EPI_ISL_1303446, EPI_ISL_1303448, EPI_ISL_1303462, EPI_ISL_1303464, EPI_ISL_1303465, EPI_ISL_1303466, EPI_ISL_1303467, EPI_ISL_1303468, EPI_ISL_1303469, EPI_ISL_1303470, EPI_ISL_1303480, EPI_ISL_1303481, EPI_ISL_1303489, EPI_ISL_1303490, EPI_ISL_1303491, EPI_ISL_1303492, EPI_ISL_1303493, EPI_ISL_1303494, EPI_ISL_1303495, EPI_ISL_1303496, EPI_ISL_1303497, EPI_ISL_1303498, EPI_ISL_1379977, EPI_ISL_1497526, EPI_ISL_1497527, EPI_ISL_1497528, EPI_ISL_1497529, EPI_ISL_1497530, EPI_ISL_1497531, EPI_ISL_1497532, EPI_ISL_1497533, EPI_ISL_1497534, EPI_ISL_1497535, EPI_ISL_1497536, EPI_ISL_1497537, EPI_ISL_1497538, EPI_ISL_1497539, EPI_ISL_1497540, EPI_ISL_1502810, EPI_ISL_1502811, EPI_ISL_1502812, EPI_ISL_1503989, EPI_ISL_1503990, EPI_ISL_1503993, EPI_ISL_1503994, EPI_ISL_1503996, EPI_ISL_1503997, EPI_ISL_1504025, EPI_ISL_1504028, EPI_ISL_1504029, EPI_ISL_1504030, EPI_ISL_1504031, EPI_ISL_1504032, EPI_ISL_1504033, EPI_ISL_1504034, EPI_ISL_1504035, EPI_ISL_1504036, EPI_ISL_1504037, EPI_ISL_1504038, EPI_ISL_1504039, EPI_ISL_1504040, EPI_ISL_1504041, EPI_ISL_1506120, EPI_ISL_1506121, EPI_ISL_1506122, EPI_ISL_1506123, EPI_ISL_1506124, EPI_ISL_1506125, EPI_ISL_1506126, EPI_ISL_1508903, EPI_ISL_1508904, EPI_ISL_1508905, EPI_ISL_1508906, EPI_ISL_1508907, EPI_ISL_1508908, EPI_ISL_1508910, EPI_ISL_1508911, EPI_ISL_1508912, EPI_ISL_1508913, EPI_ISL_1508914, EPI_ISL_1508919, EPI_ISL_1508920, EPI_ISL_1508929, EPI_ISL_1508931, EPI_ISL_1508932, EPI_ISL_1508933, EPI_ISL_1508934, EPI_ISL_1508935, EPI_ISL_1508936, EPI_ISL_1508937, EPI_ISL_1508938, EPI_ISL_1508939, EPI_ISL_1509274, EPI_ISL_1509275, EPI_ISL_1509276, EPI_ISL_1509277, EPI_ISL_1509278, EPI_ISL_1509279, EPI_ISL_1509280, EPI_ISL_1509281, EPI_ISL_1509282, EPI_ISL_1509283, EPI_ISL_1509284, EPI_ISL_1509285, EPI_ISL_1509286, EPI_ISL_1509287, EPI_ISL_1509288, EPI_ISL_1509289, EPI_ISL_1509290, EPI_ISL_1509291, EPI_ISL_1509292, EPI_ISL_1509293, EPI_ISL_1509294, EPI_ISL_1509295, EPI_ISL_1509296, EPI_ISL_1509297, EPI_ISL_1515800, EPI_ISL_1706608 | | | |
| see above | USC Clinical Lab | Los Angeles County PHL | P. Hemarajata et al. |
| EPI_ISL_1504026, EPI_ISL_1506127 | VA Medical Center | Los Angeles County PHL | P. Hemarajata et al. |

We gratefully acknowledge the following Authors from the Originating laboratories responsible for obtaining the specimens, as well as the Submitting laboratories where the genome data were generated and shared via GISAID, on which this research is based.

All Submitters of data may be contacted directly via www.gisaid.org

Authors are sorted alphabetically.

| Accession ID | Originating Laboratory | Submitting Laboratory | Authors |
|---|---|---|---|
| EPI_ISL_1650051, EPI_ISL_1650058, EPI_ISL_1650388, EPI_ISL_1650403, EPI_ISL_1650417, EPI_ISL_1650449, EPI_ISL_1650453, EPI_ISL_1650454, EPI_ISL_1650455, EPI_ISL_1650457, EPI_ISL_1650459, EPI_ISL_1650460, EPI_ISL_1650471, EPI_ISL_1667437, EPI_ISL_1667497, EPI_ISL_1667498, EPI_ISL_1667525, EPI_ISL_1667542, EPI_ISL_1667573, EPI_ISL_1667576, EPI_ISL_1667579, EPI_ISL_1667580, EPI_ISL_1667586, EPI_ISL_1667604, EPI_ISL_1667605, EPI_ISL_1667623, EPI_ISL_1667647, EPI_ISL_1667677, EPI_ISL_1667764, EPI_ISL_1667768, EPI_ISL_1667770, EPI_ISL_1667779, EPI_ISL_1667788, EPI_ISL_1667861, EPI_ISL_1667862, EPI_ISL_1667863, EPI_ISL_1667866, EPI_ISL_1667919, EPI_ISL_1667965, EPI_ISL_1667975, EPI_ISL_1667976, EPI_ISL_1667977, EPI_ISL_1667986, EPI_ISL_1667987, EPI_ISL_1668046, EPI_ISL_1668058, EPI_ISL_1668073, EPI_ISL_1668075, EPI_ISL_1668080, EPI_ISL_1668082, EPI_ISL_1668084, EPI_ISL_1668087, EPI_ISL_1668090, EPI_ISL_1668096, EPI_ISL_1668099, EPI_ISL_1668136, EPI_ISL_1668139, EPI_ISL_1668150, EPI_ISL_1668155, EPI_ISL_1668158, EPI_ISL_1668159, EPI_ISL_1668163, EPI_ISL_1668165, EPI_ISL_1668175, EPI_ISL_1668177, EPI_ISL_1668179, EPI_ISL_1668225, EPI_ISL_1668257, EPI_ISL_1668280, EPI_ISL_1668320, EPI_ISL_1668324, EPI_ISL_1668335, EPI_ISL_1685474, EPI_ISL_1685487, EPI_ISL_1685548, EPI_ISL_1685552, EPI_ISL_1685553, EPI_ISL_1685555, EPI_ISL_1685556, EPI_ISL_1685566, EPI_ISL_1685619, EPI_ISL_1685621, EPI_ISL_1685625, EPI_ISL_1685634, EPI_ISL_1685680, EPI_ISL_1685681, EPI_ISL_1685762, EPI_ISL_1685764, EPI_ISL_1685770, EPI_ISL_1685786, EPI_ISL_1685843, EPI_ISL_1685860, EPI_ISL_1685942, EPI_ISL_1685943, EPI_ISL_1686023, EPI_ISL_1686079, EPI_ISL_1686204, EPI_ISL_1686233, EPI_ISL_1686290, EPI_ISL_1686462, EPI_ISL_1686508, EPI_ISL_1686542, EPI_ISL_1686543, EPI_ISL_1686573, EPI_ISL_1686576, EPI_ISL_1686587, EPI_ISL_1686589, EPI_ISL_1686627, EPI_ISL_1686635, EPI_ISL_1686699, EPI_ISL_1686700, EPI_ISL_1686768, EPI_ISL_1686901, EPI_ISL_1686907, EPI_ISL_1686926, EPI_ISL_1686938, EPI_ISL_1686939, EPI_ISL_1686940, EPI_ISL_1686941, EPI_ISL_1686991, EPI_ISL_1686998, EPI_ISL_1687070, EPI_ISL_1687113, EPI_ISL_1687157, EPI_ISL_1687159, EPI_ISL_1687160, EPI_ISL_1687161, EPI_ISL_1687162, EPI_ISL_1687164, EPI_ISL_1687172, EPI_ISL_1687180, EPI_ISL_1687181, EPI_ISL_1687190, EPI_ISL_1687241, EPI_ISL_1687242, EPI_ISL_1687269, EPI_ISL_1687309, EPI_ISL_1687332, EPI_ISL_1687346, EPI_ISL_1687416, EPI_ISL_1687435, EPI_ISL_1687446, EPI_ISL_1687454, EPI_ISL_1687460, EPI_ISL_1687470, EPI_ISL_1687474, EPI_ISL_1687481, EPI_ISL_1687484, EPI_ISL_1687485, EPI_ISL_1687486, EPI_ISL_1687510, EPI_ISL_1687512, EPI_ISL_1687517, EPI_ISL_1687633, EPI_ISL_1687701, EPI_ISL_1687702, EPI_ISL_1687731, EPI_ISL_1687734, EPI_ISL_1687758, EPI_ISL_1687764, EPI_ISL_1687770, EPI_ISL_1687827, EPI_ISL_1687837, EPI_ISL_1687898, EPI_ISL_1687942, EPI_ISL_1687964, EPI_ISL_1687971, EPI_ISL_1687980, EPI_ISL_1687981, EPI_ISL_1687983, EPI_ISL_1687988, EPI_ISL_1687994, EPI_ISL_1688003, EPI_ISL_1688017, EPI_ISL_1688028, EPI_ISL_1688045, EPI_ISL_1688074, EPI_ISL_1688077, EPI_ISL_1688078, EPI_ISL_1688097, EPI_ISL_1688104, EPI_ISL_1688111, EPI_ISL_1688117, EPI_ISL_1688118, EPI_ISL_1688203, EPI_ISL_1688274, EPI_ISL_1688361, EPI_ISL_1688370, EPI_ISL_1688460, EPI_ISL_1688512, EPI_ISL_1688587, EPI_ISL_1688590, EPI_ISL_1688593, EPI_ISL_1688705, EPI_ISL_1688709, EPI_ISL_1688768, EPI_ISL_1688859, EPI_ISL_1688862, EPI_ISL_1688866, EPI_ISL_1689033, EPI_ISL_1689061, EPI_ISL_1689075, EPI_ISL_1689194, EPI_ISL_1689333, EPI_ISL_1689494, EPI_ISL_1689560, EPI_ISL_1689590, EPI_ISL_1689592, EPI_ISL_1689633, EPI_ISL_1689634, EPI_ISL_1689636, EPI_ISL_1689637, EPI_ISL_1689638, EPI_ISL_1689640, EPI_ISL_1689649, EPI_ISL_1689655, EPI_ISL_1689684, EPI_ISL_1689738, EPI_ISL_1689754, EPI_ISL_1689757, EPI_ISL_1689760, EPI_ISL_1689813, EPI_ISL_1689827, EPI_ISL_1689829, EPI_ISL_1689832, EPI_ISL_1689858, EPI_ISL_1689992, EPI_ISL_1690028, EPI_ISL_1690106, EPI_ISL_1690127, EPI_ISL_1690178, EPI_ISL_1690179, EPI_ISL_1690180, EPI_ISL_1690190, EPI_ISL_1690229, EPI_ISL_1690235, EPI_ISL_1690238, EPI_ISL_1690241, EPI_ISL_1695389, EPI_ISL_1695506, EPI_ISL_1695560, EPI_ISL_1701908, EPI_ISL_1701914, EPI_ISL_1701921, EPI_ISL_1701924, EPI_ISL_1701979, EPI_ISL_1701993, EPI_ISL_1702008, EPI_ISL_1702047, EPI_ISL_1702143, EPI_ISL_1702725, EPI_ISL_1702726, EPI_ISL_1702738, EPI_ISL_1702745, EPI_ISL_1702759, EPI_ISL_1702769, EPI_ISL_1702792, EPI_ISL_1702797, EPI_ISL_1702798, EPI_ISL_1702799, EPI_ISL_1702800, EPI_ISL_1702803, EPI_ISL_1702812, EPI_ISL_1702813, EPI_ISL_1702952, EPI_ISL_1735687, EPI_ISL_1735689, EPI_ISL_1735765, EPI_ISL_1735767, EPI_ISL_1735789, EPI_ISL_1735804, EPI_ISL_1735805, EPI_ISL_1735822, EPI_ISL_1735846, EPI_ISL_1735873, EPI_ISL_1735874, EPI_ISL_1735877, EPI_ISL_1735908, EPI_ISL_1735956, EPI_ISL_1735958, EPI_ISL_1735967, EPI_ISL_1735977, EPI_ISL_1736013, EPI_ISL_1736085, EPI_ISL_1736088, EPI_ISL_1736101, EPI_ISL_1736102, EPI_ISL_1736103, EPI_ISL_1736137, EPI_ISL_1736141, EPI_ISL_1736150, EPI_ISL_1736185, EPI_ISL_1736302, EPI_ISL_1736355, EPI_ISL_1736404, EPI_ISL_1736427, EPI_ISL_1736428, EPI_ISL_1736440, EPI_ISL_1736451, EPI_ISL_1736458, EPI_ISL_1736461, EPI_ISL_1736482, EPI_ISL_1736489, EPI_ISL_1736498, EPI_ISL_1736499, EPI_ISL_1736504, EPI_ISL_1736505, EPI_ISL_1736543, EPI_ISL_1736544, EPI_ISL_1736561, EPI_ISL_1736566, EPI_ISL_1736567, EPI_ISL_1736572, EPI_ISL_1736576, EPI_ISL_1736645, EPI_ISL_1736648, EPI_ISL_1736681, EPI_ISL_1736753, EPI_ISL_1736787, EPI_ISL_1736790, EPI_ISL_1736813, EPI_ISL_1736823, EPI_ISL_1736870, EPI_ISL_1736873, EPI_ISL_1736875, EPI_ISL_1736877, EPI_ISL_1736878, EPI_ISL_1737032, EPI_ISL_1737064, EPI_ISL_1737071, EPI_ISL_1737072, EPI_ISL_1737073, EPI_ISL_1737075, EPI_ISL_1737086, EPI_ISL_1737087, EPI_ISL_1737154, EPI_ISL_1737182, EPI_ISL_1737199, EPI_ISL_1737217, EPI_ISL_1737218, EPI_ISL_1737224, EPI_ISL_1737241, EPI_ISL_1737252, EPI_ISL_1737256, EPI_ISL_1737258, EPI_ISL_1737264, EPI_ISL_1737265, EPI_ISL_1737266, EPI_ISL_1737267, EPI_ISL_1737268, EPI_ISL_1737322, EPI_ISL_1737323, EPI_ISL_1737434, EPI_ISL_1737438, EPI_ISL_1737446, EPI_ISL_1737454, EPI_ISL_1737474, EPI_ISL_1737479, EPI_ISL_1737490, EPI_ISL_1737500, EPI_ISL_1737511, EPI_ISL_1737514, EPI_ISL_1737553, EPI_ISL_1737592, EPI_ISL_1737609, EPI_ISL_1737653, EPI_ISL_1737654, EPI_ISL_1737655, EPI_ISL_1737679, EPI_ISL_1737693, EPI_ISL_1737694, EPI_ISL_1737696, EPI_ISL_1737715, EPI_ISL_1737723, EPI_ISL_1737739, EPI_ISL_1737740, EPI_ISL_1737742, EPI_ISL_1737750, EPI_ISL_1737755, EPI_ISL_1737777, EPI_ISL_1737778, EPI_ISL_1737782, EPI_ISL_1737783, EPI_ISL_1737787, EPI_ISL_1737788, EPI_ISL_1737798, EPI_ISL_1737805, EPI_ISL_1737831, EPI_ISL_1737833, EPI_ISL_1737834, EPI_ISL_1737838, EPI_ISL_1737840, EPI_ISL_1738763, EPI_ISL_1753554, EPI_ISL_1753593, EPI_ISL_1753597, EPI_ISL_1753605, EPI_ISL_1753609, EPI_ISL_1834453, EPI_ISL_1834455, EPI_ISL_1834494, EPI_ISL_1834499, EPI_ISL_1834515, EPI_ISL_1834516, EPI_ISL_1834522, EPI_ISL_1834581, EPI_ISL_1834602, EPI_ISL_1834616, EPI_ISL_1834650, EPI_ISL_1834733, EPI_ISL_1834743, EPI_ISL_1834762, EPI_ISL_1834765, EPI_ISL_1834778, EPI_ISL_1834809, EPI_ISL_1834824, EPI_ISL_1834831, EPI_ISL_1834880, EPI_ISL_1834918, EPI_ISL_1834945, EPI_ISL_1834971, EPI_ISL_1835057, EPI_ISL_1835058, EPI_ISL_1835059, EPI_ISL_1835084, EPI_ISL_1835085, EPI_ISL_1835156, EPI_ISL_1835183, EPI_ISL_1835223, EPI_ISL_1835232, EPI_ISL_1835273, EPI_ISL_1835332, EPI_ISL_1835350, EPI_ISL_1835372, EPI_ISL_1835392, EPI_ISL_1835404, EPI_ISL_1835451, EPI_ISL_1835455, EPI_ISL_1835478, EPI_ISL_1835495, EPI_ISL_1835513, EPI_ISL_1835517, EPI_ISL_1835545, EPI_ISL_1835546, EPI_ISL_1835556, EPI_ISL_1835561, EPI_ISL_1835605, EPI_ISL_1835643, EPI_ISL_1835655, EPI_ISL_1835676, EPI_ISL_1835694, EPI_ISL_1835702, EPI_ISL_1835733, EPI_ISL_1835734, EPI_ISL_1835791, EPI_ISL_1835792, EPI_ISL_1835793, EPI_ISL_1835796, EPI_ISL_1835807, EPI_ISL_1835809, EPI_ISL_1835829, EPI_ISL_1835830, EPI_ISL_1835850, EPI_ISL_1835878, EPI_ISL_1835896, EPI_ISL_1835945, EPI_ISL_1835946, EPI_ISL_1835964, EPI_ISL_1835966, EPI_ISL_1836010, EPI_ISL_1836032, EPI_ISL_1836085, EPI_ISL_1836087, EPI_ISL_1836095, EPI_ISL_1836098, EPI_ISL_1836100, EPI_ISL_1836105, EPI_ISL_1836107, EPI_ISL_1836119, EPI_ISL_1836335, EPI_ISL_1836338, EPI_ISL_1836339, EPI_ISL_1836354, EPI_ISL_1836426, EPI_ISL_1836428, EPI_ISL_1836429, EPI_ISL_1836430, EPI_ISL_1836464, EPI_ISL_1836465, EPI_ISL_1836477, EPI_ISL_1836483, EPI_ISL_1836490, EPI_ISL_1836493, EPI_ISL_1836499, EPI_ISL_1836518, EPI_ISL_1836521, EPI_ISL_1836536, EPI_ISL_1836580, EPI_ISL_1836610, EPI_ISL_1836611, EPI_ISL_1836612, EPI_ISL_1836639, EPI_ISL_1836640, EPI_ISL_1836644, EPI_ISL_1836668, EPI_ISL_1836687, EPI_ISL_1836826, EPI_ISL_1836840, EPI_ISL_1836853, EPI_ISL_1836886, EPI_ISL_1836909, EPI_ISL_1836913, EPI_ISL_1836914, EPI_ISL_1836916, EPI_ISL_1836918, EPI_ISL_1836952, EPI_ISL_1836958, EPI_ISL_1836986, EPI_ISL_1837008, EPI_ISL_1837039, EPI_ISL_1837068, EPI_ISL_1837097, EPI_ISL_1837098, EPI_ISL_1837112, EPI_ISL_1837113, EPI_ISL_1837115, EPI_ISL_1837116, EPI_ISL_1837118, EPI_ISL_1837119, EPI_ISL_1837120, EPI_ISL_1837176, EPI_ISL_1837314, EPI_ISL_1837385, EPI_ISL_1837417, EPI_ISL_1837432, EPI_ISL_1837448, EPI_ISL_1837449, EPI_ISL_1837452, EPI_ISL_1837455, EPI_ISL_1837617, EPI_ISL_1837619, EPI_ISL_1837639, EPI_ISL_1837644, EPI_ISL_1837664, EPI_ISL_1837670, EPI_ISL_1837701, EPI_ISL_1837702, EPI_ISL_1837703, EPI_ISL_1837717, EPI_ISL_1837758, EPI_ISL_1837957, EPI_ISL_1837959, EPI_ISL_1837970, EPI_ISL_1837971, EPI_ISL_1838211, EPI_ISL_1838407, EPI_ISL_1900357, EPI_ISL_1900378, EPI_ISL_1900408, EPI_ISL_1922910, EPI_ISL_1922912, EPI_ISL_1922913, EPI_ISL_1922917, EPI_ISL_1922934, EPI_ISL_1922968, EPI_ISL_1922970, EPI_ISL_1923038, EPI_ISL_1923154, EPI_ISL_1923155, EPI_ISL_1923156, EPI_ISL_1925142, EPI_ISL_1925194, EPI_ISL_1925208, EPI_ISL_1925211, EPI_ISL_1925229, EPI_ISL_1925233, EPI_ISL_1925286, EPI_ISL_1925375, EPI_ISL_1925386, EPI_ISL_1925434, EPI_ISL_1925443, EPI_ISL_1925444, EPI_ISL_1925449, EPI_ISL_1925469, EPI_ISL_1925472, EPI_ISL_1925587, EPI_ISL_1925588, EPI_ISL_1995006, EPI_ISL_1995061, EPI_ISL_1995067, EPI_ISL_1995107, EPI_ISL_1995113, EPI_ISL_1995116, EPI_ISL_1995136, EPI_ISL_1995177, EPI_ISL_1995188, EPI_ISL_1995226, EPI_ISL_1995230, EPI_ISL_1995235, EPI_ISL_1995244, EPI_ISL_1995259, EPI_ISL_1995260, EPI_ISL_1995261, EPI_ISL_1995264, EPI_ISL_1995322, EPI_ISL_1995323, EPI_ISL_1995341, EPI_ISL_1995348, EPI_ISL_1995410, EPI_ISL_1995455, EPI_ISL_1995476, EPI_ISL_1995483, EPI_ISL_1995490, EPI_ISL_1995491, EPI_ISL_1995507, EPI_ISL_1995541, EPI_ISL_1995552, EPI_ISL_1995570, EPI_ISL_1995582, EPI_ISL_1995599, EPI_ISL_1995722, EPI_ISL_1995831, EPI_ISL_1995863, EPI_ISL_1995872, EPI_ISL_1995942, EPI_ISL_1995964, EPI_ISL_1995990, EPI_ISL_1995991, EPI_ISL_1996059, EPI_ISL_1996083, EPI_ISL_1996085, EPI_ISL_1996086, EPI_ISL_1996088, EPI_ISL_1996089, EPI_ISL_1996090, EPI_ISL_1996092, EPI_ISL_1996093, EPI_ISL_1996188, EPI_ISL_1996203, EPI_ISL_1996264, EPI_ISL_1996267, EPI_ISL_1996311, EPI_ISL_1996440, EPI_ISL_1996486, EPI_ISL_1996523, EPI_ISL_1996594, EPI_ISL_1996596, EPI_ISL_1996598, EPI_ISL_1996600, EPI_ISL_1996626, EPI_ISL_1996657, EPI_ISL_1996661, EPI_ISL_1996664, EPI_ISL_1996665, EPI_ISL_1996667, EPI_ISL_1996671, EPI_ISL_1996672, EPI_ISL_1996673, EPI_ISL_1996681, EPI_ISL_1996707, EPI_ISL_1996719, EPI_ISL_1996730, EPI_ISL_1996731, EPI_ISL_1996732, EPI_ISL_1996745, EPI_ISL_1996746, EPI_ISL_1996855, EPI_ISL_1996857, EPI_ISL_1996861, EPI_ISL_1996872, EPI_ISL_1996874, EPI_ISL_1996882, EPI_ISL_1996892, EPI_ISL_1996944, EPI_ISL_1996949, EPI_ISL_1996973, EPI_ISL_1996987, EPI_ISL_1996990, EPI_ISL_1997023, EPI_ISL_1997063, EPI_ISL_1997079, EPI_ISL_1997088, EPI_ISL_1997101, EPI_ISL_1997107, EPI_ISL_1997108, EPI_ISL_1997122, EPI_ISL_1997123, EPI_ISL_1997124, EPI_ISL_1997126, EPI_ISL_1997130, EPI_ISL_1997132, EPI_ISL_1997133, EPI_ISL_1997134, EPI_ISL_1997135, EPI_ISL_1997136, EPI_ISL_1997138, EPI_ISL_1997156, EPI_ISL_1997159, EPI_ISL_1997168, EPI_ISL_1997247, EPI_ISL_1997262, EPI_ISL_1997314, EPI_ISL_1997362, EPI_ISL_1997363, EPI_ISL_1997399, EPI_ISL_1997400, EPI_ISL_1997404, EPI_ISL_1997416, EPI_ISL_1997426, EPI_ISL_1997427, EPI_ISL_1997428, EPI_ISL_1997431, EPI_ISL_1997432, EPI_ISL_1997435, EPI_ISL_1997436, EPI_ISL_1997439, EPI_ISL_1997440, EPI_ISL_1997496, EPI_ISL_1997497, EPI_ISL_1997498, EPI_ISL_1997528, EPI_ISL_1997613, EPI_ISL_1997614, EPI_ISL_1997615, EPI_ISL_1997616, EPI_ISL_1997617, EPI_ISL_1997618, EPI_ISL_1997620, EPI_ISL_1997621, EPI_ISL_1997622, EPI_ISL_1997628, EPI_ISL_1997629, EPI_ISL_1997630, EPI_ISL_1997641, EPI_ISL_1997722, EPI_ISL_1997739, EPI_ISL_1997740, EPI_ISL_1997745, EPI_ISL_1997747, EPI_ISL_1997751, EPI_ISL_1997754, EPI_ISL_1997756, EPI_ISL_1997762, EPI_ISL_1997763, EPI_ISL_1997769, EPI_ISL_1997805, EPI_ISL_1997806, EPI_ISL_1997824, EPI_ISL_1997851, EPI_ISL_1997869, EPI_ISL_1997922, EPI_ISL_1997935, EPI_ISL_1997949, EPI_ISL_1997950, EPI_ISL_1997986, EPI_ISL_1998013, EPI_ISL_1998025, EPI_ISL_1998026, EPI_ISL_1998054, EPI_ISL_1998061, EPI_ISL_1998088, EPI_ISL_1998111, EPI_ISL_1998112, EPI_ISL_1998120, EPI_ISL_1998122, EPI_ISL_1998123, EPI_ISL_1998187, EPI_ISL_1998212, EPI_ISL_1998217, EPI_ISL_1998237, EPI_ISL_1998238, EPI_ISL_1998240, EPI_ISL_1998268, EPI_ISL_1998269, EPI_ISL_1998283, EPI_ISL_1998285, EPI_ISL_1998307, EPI_ISL_1998375, EPI_ISL_1998377, EPI_ISL_1998381, EPI_ISL_1998407, EPI_ISL_1998408, EPI_ISL_1998410, EPI_ISL_1998411, EPI_ISL_1998412, EPI_ISL_1998419, EPI_ISL_1998432, EPI_ISL_1998439, EPI_ISL_1998467, EPI_ISL_1998513, EPI_ISL_1998514, EPI_ISL_1998518, EPI_ISL_1998537, EPI_ISL_1998538, EPI_ISL_1998620, EPI_ISL_1998668, EPI_ISL_1998672, EPI_ISL_1998722, EPI_ISL_1998755, EPI_ISL_1998774, EPI_ISL_1998789, EPI_ISL_1998799, EPI_ISL_1998831, EPI_ISL_1998848, EPI_ISL_1998859, EPI_ISL_1998860, EPI_ISL_1998862, EPI_ISL_1998863, EPI_ISL_1998882, EPI_ISL_1998896, EPI_ISL_1998965, EPI_ISL_1998975, EPI_ISL_1999023, EPI_ISL_1999024, EPI_ISL_1999034, EPI_ISL_1999040, EPI_ISL_1999098, EPI_ISL_1999132, EPI_ISL_1999141, EPI_ISL_1999162, EPI_ISL_1999189, EPI_ISL_1999195, EPI_ISL_1999196, EPI_ISL_1999200, EPI_ISL_1999211, EPI_ISL_1999212, EPI_ISL_1999227, EPI_ISL_1999231, EPI_ISL_1999248, EPI_ISL_1999293, EPI_ISL_1999299, EPI_ISL_1999302, EPI_ISL_1999309, EPI_ISL_1999311, EPI_ISL_1999366, EPI_ISL_1999368, EPI_ISL_1999375, EPI_ISL_1999446, EPI_ISL_1999491, EPI_ISL_1999569, EPI_ISL_1999600, EPI_ISL_1999716, EPI_ISL_1999717, EPI_ISL_1999914, EPI_ISL_2000222, EPI_ISL_2000296, EPI_ISL_2000333, EPI_ISL_2000343, EPI_ISL_2000393, EPI_ISL_2000407, EPI_ISL_2000410, EPI_ISL_2000412, EPI_ISL_2000417, EPI_ISL_2000483, EPI_ISL_2000484, EPI_ISL_2000509, EPI_ISL_2000512, EPI_ISL_2000520, EPI_ISL_2035046, EPI_ISL_2035069, EPI_ISL_2035110, EPI_ISL_2035138, EPI_ISL_2035143, EPI_ISL_2035151, EPI_ISL_2035184, EPI_ISL_2035189, EPI_ISL_2035223, EPI_ISL_2035225, EPI_ISL_2040477, EPI_ISL_2040478, EPI_ISL_2040996 | | | |
| see above | Aegis Sciences Corporation | Centers for Disease Control and Prevention Division of Viral Diseases, Pathogen Discovery | Adrian Paskey; Alec Vest; Benjamin Rambo-Martin; Christopher Gulvick; Clinton R. Paden; Cyndi Clark; Dakota Howard; Darlene Wagner; Dhwani Batra; Dillon Nall; Duncan MacCannell; Ethan Sanders; Holly Houdeshell; Jason Caravas; Kara Moser; Matthew Hardison; Matthew Schmerer; Ola Kvalvaag; Patrick Campbell; Peter W. Cook; Rob Case; Scott Sammons; Shatavia Morrison; Shaun Westlund; Vikramsinha Ghorpade; Yvette Unoarumhi |
| EPI_ISL_1585085 | Arizona State University | Arizona State University | Efrem S. Lim; Joshua LaBaer; Joy M. Blain; LaRinda A. Holland; Nicholas J. Mellor; Peter T. Skidmore; Rabia Maqsood; Valerie Harris; Vel Murugan |
| EPI_ISL_2103032, EPI_ISL_2103064, EPI_ISL_2103128, EPI_ISL_2103129, EPI_ISL_2103194 | CA DPH Viral and Rickettsial Disease Laboratory | Chan-Zuckerberg Biohub | CZB Cliahub Consortium |
| EPI_ISL_1678087, EPI_ISL_1678093, EPI_ISL_1678094, EPI_ISL_1678095, EPI_ISL_1678282, EPI_ISL_1798834, EPI_ISL_1798841, EPI_ISL_1798853, EPI_ISL_1798855, EPI_ISL_1798857, EPI_ISL_1798860, EPI_ISL_1798868, EPI_ISL_1823713, EPI_ISL_1823715, EPI_ISL_1823716, EPI_ISL_1823717, EPI_ISL_1823718, EPI_ISL_1823719, EPI_ISL_1823720, EPI_ISL_1823722, EPI_ISL_1823724, EPI_ISL_1823725, EPI_ISL_1823726, EPI_ISL_1823727, EPI_ISL_1823728, EPI_ISL_1823729, EPI_ISL_1823730, EPI_ISL_1823732, EPI_ISL_1823733, EPI_ISL_1823734, EPI_ISL_1823736, EPI_ISL_1823737, EPI_ISL_1823738, EPI_ISL_1823739, EPI_ISL_1823741, EPI_ISL_1823743, EPI_ISL_1823744, EPI_ISL_1823745, EPI_ISL_1823747, EPI_ISL_1937812, EPI_ISL_1937813, EPI_ISL_1937814, EPI_ISL_1937815, EPI_ISL_1937816, EPI_ISL_1937817, EPI_ISL_1937818, EPI_ISL_1937819, EPI_ISL_1937820, EPI_ISL_1937821, EPI_ISL_1937822, EPI_ISL_1937823, EPI_ISL_1937824, EPI_ISL_1937825, EPI_ISL_1937826, EPI_ISL_1937827, EPI_ISL_1937828, EPI_ISL_1937829, EPI_ISL_2105128 | | | |
| see above | CA-Los Angeles County Public Health Laboratory | Centers for Disease Control and Prevention Division of Viral Diseases, Pathogen Discovery | Alison Laufer Halpin; Ben L. Rambo-Martin; Clinton R. Paden; Dakota Howard; Darlene Wagner; Dave Wentworth; Dhwani Batra; Jasmine Padilla; Justin Lee; Katie Dillon; Krista Queen; Kristen Knipe; Kristine Lacek; Mark Burroughs; Matthew Schmerer; Mili Sheth; Peter Cook; Sam Shepard; Sarah Nobles; Shoshona Le; Suxiang Tong; Vivien Dugan; Yvette Unoarumhi |
| EPI_ISL_1798766, EPI_ISL_1798768, EPI_ISL_1798770, EPI_ISL_1798772, EPI_ISL_1798777, EPI_ISL_1798779, EPI_ISL_1798781, EPI_ISL_1798783, EPI_ISL_1798787, EPI_ISL_1798791, EPI_ISL_1798793, EPI_ISL_2018313, EPI_ISL_2018315, EPI_ISL_2018316, EPI_ISL_2018318, EPI_ISL_2018319, EPI_ISL_2018320, EPI_ISL_2018321, EPI_ISL_2018324, EPI_ISL_2018325 | | | |
| see above | CDPH, Viral and Rickettsial Disease Laboratory | Centers for Disease Control and Prevention Division of Viral Diseases, Pathogen Discovery | Alison Laufer Halpin; Ben L. Rambo-Martin; Clinton R. Paden; Dakota Howard; Darlene Wagner; Dave Wentworth; Dhwani Batra; Jasmine Padilla; Justin Lee; Katie Dillon; Krista Queen; Kristen Knipe; Kristine Lacek; Mark Burroughs; Matthew Schmerer; Mili Sheth; Peter Cook; Sam Shepard; Sarah Nobles; Shoshona Le; Suxiang Tong; Vivien Dugan; Yvette Unoarumhi |
| EPI_ISL_1824409, EPI_ISL_1824410, EPI_ISL_1824411, EPI_ISL_1824412, EPI_ISL_1824414, EPI_ISL_1824415 | Chiu Laboratory, University of California, San Francisco | Chiu Laboratory, University of California, San Francisco | Alicia Sotomayor-Gonzalez; Candace Wang; Charles Chiu; Daniel Wong; Venice Servellita; Yueyuan Zhang |
| EPI_ISL_1825079, EPI_ISL_1825080, EPI_ISL_1825081, EPI_ISL_1825082, EPI_ISL_1825083 | Color Genomics | Chiu Laboratory, University of California, San Francisco | Alicia Sotomayor-Gonzalez; Alicia Zhou; Candace Wang; Charles Chiu; Daniel Wong; Erika Torres; Madyson Wanner; Scott Topper; Venice Servellita; Yueyuan Zhang |
| EPI_ISL_2102749, EPI_ISL_2102763, EPI_ISL_2102772, EPI_ISL_2102798, EPI_ISL_2102826, EPI_ISL_2102861, EPI_ISL_2102887, EPI_ISL_2102891, EPI_ISL_2102932, EPI_ISL_2102954, EPI_ISL_2102960, EPI_ISL_2102972, EPI_ISL_2102988, EPI_ISL_2102997, EPI_ISL_2103009, EPI_ISL_2103031, EPI_ISL_2103062, EPI_ISL_2103097, EPI_ISL_2103106, EPI_ISL_2103138, EPI_ISL_2103169, EPI_ISL_2103173 | | | |
| see above | Contra Costa County Public Health Lab | Chan-Zuckerberg Biohub | CZB Cliahub Consortium |
| EPI_ISL_1805724, EPI_ISL_1805725, EPI_ISL_1805726, EPI_ISL_1805730, EPI_ISL_1805731, EPI_ISL_1805732, EPI_ISL_1805733, EPI_ISL_1805734, EPI_ISL_1805735, EPI_ISL_1805736, EPI_ISL_1805737, EPI_ISL_1805738, EPI_ISL_1805739, EPI_ISL_1841392, EPI_ISL_1910462, EPI_ISL_1910464, EPI_ISL_1910465, EPI_ISL_1910466, EPI_ISL_1968661 | | | |
| see above | County of San Diego Health and Human Services Agency | Pathogen Discovery, Respiratory Viruses Branch, Division of Viral Diseases, Centers for Disease Control and Prevention | Adam Retchless; Anna Kelleher; Anna Montmayeur; Anna Uehara; Brian Lynch; Clinton R. Paden; Dhwani Batra; Haibin Wang; Han Jia Justin Ng; Jasmine Padilla; Jing Zhang; Justin Lee; Krista Queen; Mark Burroughs; Mili Sheth; Morgan Davis; Peter Cook; Rachel Marine; Sarah Nobles; Suxiang Tong; Tara Coalter; Yan Li; Ying Tao |
| EPI_ISL_1701423, EPI_ISL_1701591, EPI_ISL_1701607, EPI_ISL_1821323, EPI_ISL_1821332, EPI_ISL_1821354, EPI_ISL_1821358, EPI_ISL_1821362, EPI_ISL_1821365, EPI_ISL_1821366, EPI_ISL_1821368, EPI_ISL_1821371, EPI_ISL_1821372, EPI_ISL_1821373, EPI_ISL_1821374, EPI_ISL_1821375, EPI_ISL_1821376, EPI_ISL_1821377, EPI_ISL_1821380, EPI_ISL_1821381, EPI_ISL_1821383, EPI_ISL_1821384, EPI_ISL_1821385, EPI_ISL_1821386, EPI_ISL_1821387, EPI_ISL_1821388, EPI_ISL_1821389, EPI_ISL_1821390, EPI_ISL_1821391, EPI_ISL_1821394, EPI_ISL_1821399, EPI_ISL_1821434, EPI_ISL_1821454, EPI_ISL_1821463, EPI_ISL_2102747, EPI_ISL_2102748, EPI_ISL_2102755, EPI_ISL_2102776, EPI_ISL_2102790, EPI_ISL_2102809, EPI_ISL_2102811, EPI_ISL_2102820, EPI_ISL_2102830, EPI_ISL_2102837, EPI_ISL_2102842, EPI_ISL_2102844, EPI_ISL_2102845, EPI_ISL_2102848, EPI_ISL_2102849, EPI_ISL_2102850, EPI_ISL_2102869, EPI_ISL_2102890, EPI_ISL_2102893, EPI_ISL_2102902, EPI_ISL_2102905, EPI_ISL_2102912, EPI_ISL_2102933, EPI_ISL_2102947, EPI_ISL_2102955, EPI_ISL_2102958, EPI_ISL_2102976, EPI_ISL_2102978, EPI_ISL_2102994, EPI_ISL_2103045, EPI_ISL_2103092, EPI_ISL_2103126, EPI_ISL_2103130, EPI_ISL_2103153 | | | |
| see above | County of San Luis Obispo Public Health Laboratory | Chan-Zuckerberg Biohub | CZB Cliahub Consortium |
| EPI_ISL_1913219, EPI_ISL_1913220, EPI_ISL_1913221, EPI_ISL_1913222, EPI_ISL_1913223, | EL CAMINO HOSPITAL | Santa Clara County public Health Laboratory | Santa Clara County Public Health Department |

| | | | |
|---|---|---|---|
| EPI_ISL_1913224 | | | |
| EPI_ISL_1812119, EPI_ISL_1813447, EPI_ISL_1813669, EPI_ISL_1814286, EPI_ISL_1814396, EPI_ISL_1816811 | EXCITE Lab | Andersen lab at Scripps Research | Abigail Schnapper; Angela Scioscia; Cheryl Anderson; Chip Schooley; Greg Humphrey; Helena Tubb; Natasha Martin; Sawyer Farmer; Smruthi Karthikeyan; Tommy Valles + SEARCH |
| EPI_ISL_1532124, EPI_ISL_1532125, EPI_ISL_1532126 | El Camino Hospital | Santa Clara County Public Health Laboratory | Santa Clara County Public Health Department |
| EPI_ISL_1664270, EPI_ISL_1664271, EPI_ISL_1664272, EPI_ISL_1664273, EPI_ISL_1664274 | El Camino Hospital | Santa Clara County public Health Laboratory | Santa Clara County Public Health Department |
| EPI_ISL_2031859, EPI_ISL_2031860, EPI_ISL_2031861 | Flow Diagnostics Lab | Los Angeles County PHL | P. Hemarajata et al. |

EPI_ISL_1578136, EPI_ISL_1578139, EPI_ISL_1578144, EPI_ISL_1578153, EPI_ISL_1578157, EPI_ISL_1578162, EPI_ISL_1578176, EPI_ISL_1578180, EPI_ISL_1578299, EPI_ISL_1578330, EPI_ISL_1578333, EPI_ISL_1578336, EPI_ISL_1578338, EPI_ISL_1918991, EPI_ISL_1918992

| | | | |
|---|---|---|---|
| see above | Flow Health | Infectious Disease Program, Broad Institute of Harvard and MIT | Adams, G.; B.L.; B.W.; Bauer, M.; Birren; Carter, A.; Chaluvadi, S.; D.J.; DeRuff, K.; Gallagher, G.; Gladden-Young, A.; J.E.; K.J.; Lagerborg, K.; Lemieux; Loreth, C.; MacInnis; Normandin, E.; P.C.; Park; Reilly, S.; Rudy, M.; Siddle; Smole, S.; Tomkins-Tinch, C.; and Sabeti |

EPI_ISL_1525267, EPI_ISL_1525268, EPI_ISL_1525269, EPI_ISL_1525270, EPI_ISL_1525271, EPI_ISL_1525272, EPI_ISL_1525273, EPI_ISL_1525274, EPI_ISL_1525275, EPI_ISL_1525276, EPI_ISL_1525277, EPI_ISL_1525278, EPI_ISL_1525279, EPI_ISL_1525280, EPI_ISL_1525281, EPI_ISL_1525282, EPI_ISL_1525283, EPI_ISL_1525284, EPI_ISL_1525285, EPI_ISL_1525286, EPI_ISL_1525287, EPI_ISL_1525288,
EPI_ISL_1525289, EPI_ISL_1525290, EPI_ISL_1525291, EPI_ISL_1525292, EPI_ISL_1525293, EPI_ISL_1525294, EPI_ISL_1525295, EPI_ISL_1525296, EPI_ISL_1525297, EPI_ISL_1525298, EPI_ISL_1525299, EPI_ISL_1525300, EPI_ISL_1525301, EPI_ISL_1525302, EPI_ISL_1525303, EPI_ISL_1525304, EPI_ISL_1525305, EPI_ISL_1525306, EPI_ISL_1525307, EPI_ISL_1525308, EPI_ISL_1525309, EPI_ISL_1525310,
EPI_ISL_1525311, EPI_ISL_1525312, EPI_ISL_1525313, EPI_ISL_1525314, EPI_ISL_1525628, EPI_ISL_1525629, EPI_ISL_1525630, EPI_ISL_1525631, EPI_ISL_1525632, EPI_ISL_1525633, EPI_ISL_1525634, EPI_ISL_1525636, EPI_ISL_1525637, EPI_ISL_1525638, EPI_ISL_1525639, EPI_ISL_1525640, EPI_ISL_1525642, EPI_ISL_1525643, EPI_ISL_1525644, EPI_ISL_1525645, EPI_ISL_1525646, EPI_ISL_1525647,
EPI_ISL_1525648, EPI_ISL_1525649, EPI_ISL_1525650, EPI_ISL_1525651, EPI_ISL_1525652, EPI_ISL_1525653, EPI_ISL_1525655, EPI_ISL_1525656, EPI_ISL_1525658, EPI_ISL_1525659, EPI_ISL_1525660, EPI_ISL_1525661, EPI_ISL_1525662, EPI_ISL_1525663, EPI_ISL_1525666, EPI_ISL_1525667, EPI_ISL_1525668, EPI_ISL_1525669, EPI_ISL_1525670, EPI_ISL_1525671, EPI_ISL_1525672, EPI_ISL_1525673,
EPI_ISL_1525674, EPI_ISL_1525675, EPI_ISL_1525676, EPI_ISL_1525677, EPI_ISL_1525678, EPI_ISL_1525679, EPI_ISL_1525680, EPI_ISL_1525681, EPI_ISL_1525682, EPI_ISL_1525683, EPI_ISL_1525684, EPI_ISL_1525685, EPI_ISL_1525686, EPI_ISL_1525687, EPI_ISL_1525688, EPI_ISL_1525689, EPI_ISL_1525690, EPI_ISL_1525691, EPI_ISL_1525692, EPI_ISL_1525694, EPI_ISL_1525695, EPI_ISL_1525696,
EPI_ISL_1525697, EPI_ISL_1525698, EPI_ISL_1525699, EPI_ISL_1525700, EPI_ISL_1525701, EPI_ISL_1525702, EPI_ISL_1525703, EPI_ISL_1525704, EPI_ISL_1525705, EPI_ISL_1525706, EPI_ISL_1525707, EPI_ISL_1525708, EPI_ISL_1525709, EPI_ISL_1525710, EPI_ISL_1525711, EPI_ISL_1525712, EPI_ISL_1525713, EPI_ISL_1525714, EPI_ISL_1525716, EPI_ISL_1525717, EPI_ISL_1525718, EPI_ISL_1525719,
EPI_ISL_1525720, EPI_ISL_1525721, EPI_ISL_1525722, EPI_ISL_1525723, EPI_ISL_1525724, EPI_ISL_1525725, EPI_ISL_1525726, EPI_ISL_1525727, EPI_ISL_1525728, EPI_ISL_1525729, EPI_ISL_1525730, EPI_ISL_1525731, EPI_ISL_1525732, EPI_ISL_1525733, EPI_ISL_1525734, EPI_ISL_1525735, EPI_ISL_1525736, EPI_ISL_1525737, EPI_ISL_1525738, EPI_ISL_1525739, EPI_ISL_1525740, EPI_ISL_1525741,
EPI_ISL_1525742, EPI_ISL_1525743, EPI_ISL_1525744, EPI_ISL_1525745, EPI_ISL_1525746, EPI_ISL_1525747, EPI_ISL_1525748, EPI_ISL_1525749, EPI_ISL_1525750, EPI_ISL_1525751, EPI_ISL_1525752, EPI_ISL_1525753, EPI_ISL_1525754, EPI_ISL_1525755, EPI_ISL_1525756, EPI_ISL_1525757, EPI_ISL_1525758, EPI_ISL_1525759, EPI_ISL_1525760, EPI_ISL_1525761, EPI_ISL_1525762, EPI_ISL_1525763,
EPI_ISL_1525764, EPI_ISL_1525765, EPI_ISL_1525766, EPI_ISL_1525767, EPI_ISL_1525768, EPI_ISL_1525769, EPI_ISL_1525770, EPI_ISL_1525771, EPI_ISL_1525772, EPI_ISL_1525773, EPI_ISL_1525774, EPI_ISL_1525775, EPI_ISL_1525776, EPI_ISL_1525777, EPI_ISL_1525778, EPI_ISL_1525779, EPI_ISL_1525780, EPI_ISL_1525781, EPI_ISL_1525782, EPI_ISL_1525783, EPI_ISL_1525784, EPI_ISL_1525785,
EPI_ISL_1525786, EPI_ISL_1525787, EPI_ISL_1525788, EPI_ISL_1525789, EPI_ISL_1525790, EPI_ISL_1525791, EPI_ISL_1525792, EPI_ISL_1525793, EPI_ISL_1525794, EPI_ISL_1525795, EPI_ISL_1525796, EPI_ISL_1525797, EPI_ISL_1525798, EPI_ISL_1525799, EPI_ISL_1556394, EPI_ISL_1556395, EPI_ISL_1556396, EPI_ISL_1556397, EPI_ISL_1556398, EPI_ISL_1556399, EPI_ISL_1556400, EPI_ISL_1556401,
EPI_ISL_1556402, EPI_ISL_1556403, EPI_ISL_1556404, EPI_ISL_1556405, EPI_ISL_1556406, EPI_ISL_1556407, EPI_ISL_1556408, EPI_ISL_1556409, EPI_ISL_1556410, EPI_ISL_1556411, EPI_ISL_1556412, EPI_ISL_1556413, EPI_ISL_1556414, EPI_ISL_1556415, EPI_ISL_1556416, EPI_ISL_1556417, EPI_ISL_1556418, EPI_ISL_1556419, EPI_ISL_1556922, EPI_ISL_1556923, EPI_ISL_1556924, EPI_ISL_1556925,
EPI_ISL_1556926, EPI_ISL_1556927, EPI_ISL_1556928, EPI_ISL_1556929, EPI_ISL_1556930, EPI_ISL_1556931, EPI_ISL_1556932, EPI_ISL_1556933, EPI_ISL_1556934, EPI_ISL_1556935, EPI_ISL_1556936, EPI_ISL_1556937, EPI_ISL_1556938, EPI_ISL_1556940, EPI_ISL_1556941, EPI_ISL_1556942, EPI_ISL_1556943, EPI_ISL_1556944, EPI_ISL_1556945, EPI_ISL_1556946, EPI_ISL_1556947, EPI_ISL_1556948,
EPI_ISL_1556949, EPI_ISL_1556950, EPI_ISL_1556951, EPI_ISL_1556952, EPI_ISL_1556953, EPI_ISL_1556954, EPI_ISL_1556955, EPI_ISL_1556956, EPI_ISL_1556957, EPI_ISL_1556958, EPI_ISL_1556959, EPI_ISL_1556960, EPI_ISL_1556961, EPI_ISL_1556962, EPI_ISL_1556963, EPI_ISL_1556964, EPI_ISL_1556965, EPI_ISL_1556966, EPI_ISL_1556967, EPI_ISL_1556968, EPI_ISL_1556969, EPI_ISL_1556970,
EPI_ISL_1556971, EPI_ISL_1556972, EPI_ISL_1556973, EPI_ISL_1556974, EPI_ISL_1556976, EPI_ISL_1556977, EPI_ISL_1556978, EPI_ISL_1556979, EPI_ISL_1556980, EPI_ISL_1556981, EPI_ISL_1556982, EPI_ISL_1556983, EPI_ISL_1556984, EPI_ISL_1556985, EPI_ISL_1556986, EPI_ISL_1556987, EPI_ISL_1556988, EPI_ISL_1556989, EPI_ISL_1556990, EPI_ISL_1556991, EPI_ISL_1556992, EPI_ISL_1556993,
EPI_ISL_1556994, EPI_ISL_1556995, EPI_ISL_1556996, EPI_ISL_1556997, EPI_ISL_1556998, EPI_ISL_1556999, EPI_ISL_1557000, EPI_ISL_1557001, EPI_ISL_1557002, EPI_ISL_1557003, EPI_ISL_1557004, EPI_ISL_1557005, EPI_ISL_1557006, EPI_ISL_1557007, EPI_ISL_1557008, EPI_ISL_1557009, EPI_ISL_1557010, EPI_ISL_1557011, EPI_ISL_1557012, EPI_ISL_1557013, EPI_ISL_1557014, EPI_ISL_1557015,
EPI_ISL_1557016, EPI_ISL_1557017, EPI_ISL_1557018, EPI_ISL_1557019, EPI_ISL_1557020, EPI_ISL_1557021, EPI_ISL_1557022, EPI_ISL_1557023, EPI_ISL_1557024, EPI_ISL_1557025, EPI_ISL_1557026, EPI_ISL_1557027, EPI_ISL_1557028, EPI_ISL_1557029, EPI_ISL_1557030, EPI_ISL_1557031, EPI_ISL_1557032, EPI_ISL_1612850, EPI_ISL_1612953, EPI_ISL_1612954, EPI_ISL_1612955, EPI_ISL_1612956,
EPI_ISL_1612957, EPI_ISL_1612958, EPI_ISL_1612959, EPI_ISL_1612960, EPI_ISL_1612961, EPI_ISL_1612962, EPI_ISL_1612963, EPI_ISL_1612964, EPI_ISL_1612965, EPI_ISL_1612966, EPI_ISL_1612967, EPI_ISL_1612968, EPI_ISL_1612969, EPI_ISL_1612970, EPI_ISL_1612971, EPI_ISL_1612972, EPI_ISL_1612973, EPI_ISL_1612974, EPI_ISL_1612975, EPI_ISL_1612976, EPI_ISL_1612977, EPI_ISL_1612978,
EPI_ISL_1612979, EPI_ISL_1612980, EPI_ISL_1612981, EPI_ISL_1612982, EPI_ISL_1612983, EPI_ISL_1612984, EPI_ISL_1612985, EPI_ISL_1612986, EPI_ISL_1612987, EPI_ISL_1612988, EPI_ISL_1612989, EPI_ISL_1612990, EPI_ISL_1612991, EPI_ISL_1612992, EPI_ISL_1612993, EPI_ISL_1612994, EPI_ISL_1612995, EPI_ISL_1612996, EPI_ISL_1612997, EPI_ISL_1612998, EPI_ISL_1612999, EPI_ISL_1613000,
EPI_ISL_1613001, EPI_ISL_1613002, EPI_ISL_1613003, EPI_ISL_1613004, EPI_ISL_1613005, EPI_ISL_1613006, EPI_ISL_1613007, EPI_ISL_1613008, EPI_ISL_1613009, EPI_ISL_1613010, EPI_ISL_1613011, EPI_ISL_1613012, EPI_ISL_1613013, EPI_ISL_1613014, EPI_ISL_1613015, EPI_ISL_1613016, EPI_ISL_1613017, EPI_ISL_1613018, EPI_ISL_1613019, EPI_ISL_1613020, EPI_ISL_1613021, EPI_ISL_1613022,
EPI_ISL_1613023, EPI_ISL_1613024, EPI_ISL_1613025, EPI_ISL_1613026, EPI_ISL_1613027, EPI_ISL_1613028, EPI_ISL_1613029, EPI_ISL_1613030, EPI_ISL_1613031, EPI_ISL_1613032, EPI_ISL_1613033, EPI_ISL_1613034, EPI_ISL_1613035, EPI_ISL_1613036, EPI_ISL_1613037, EPI_ISL_1613038, EPI_ISL_1613039, EPI_ISL_1613040, EPI_ISL_1613041, EPI_ISL_1613042, EPI_ISL_1613043, EPI_ISL_1613044,
EPI_ISL_1613045, EPI_ISL_1613046, EPI_ISL_1613047, EPI_ISL_1613048, EPI_ISL_1613049, EPI_ISL_1613050, EPI_ISL_1613051, EPI_ISL_1613052, EPI_ISL_1613053, EPI_ISL_1613054, EPI_ISL_1613055, EPI_ISL_1613056, EPI_ISL_1613057, EPI_ISL_1613058, EPI_ISL_1613059, EPI_ISL_1613060, EPI_ISL_1613061, EPI_ISL_1613062, EPI_ISL_1613063, EPI_ISL_1613064, EPI_ISL_1613065, EPI_ISL_1613066,
EPI_ISL_1613067, EPI_ISL_1613068, EPI_ISL_1613069, EPI_ISL_1613070, EPI_ISL_1613071, EPI_ISL_1613072, EPI_ISL_1613073, EPI_ISL_1613074, EPI_ISL_1613075, EPI_ISL_1613076, EPI_ISL_1613077, EPI_ISL_1613078, EPI_ISL_1613079, EPI_ISL_1613182, EPI_ISL_1613185, EPI_ISL_1613188, EPI_ISL_1613191, EPI_ISL_1613194, EPI_ISL_1613197, EPI_ISL_1613200, EPI_ISL_1613203, EPI_ISL_1613206,
EPI_ISL_1613209, EPI_ISL_1613212, EPI_ISL_1613215, EPI_ISL_1613218, EPI_ISL_1613221, EPI_ISL_1613224, EPI_ISL_1613226, EPI_ISL_1613229, EPI_ISL_1613235, EPI_ISL_1613238, EPI_ISL_1613241, EPI_ISL_1613245, EPI_ISL_1613248, EPI_ISL_1613251, EPI_ISL_1613254, EPI_ISL_1613257, EPI_ISL_1613260, EPI_ISL_1613263, EPI_ISL_1613266, EPI_ISL_1613269, EPI_ISL_1613272, EPI_ISL_1613275,
EPI_ISL_1613277, EPI_ISL_1613280, EPI_ISL_1613284, EPI_ISL_1613287, EPI_ISL_1613290, EPI_ISL_1613292, EPI_ISL_1613295, EPI_ISL_1613298, EPI_ISL_1613301, EPI_ISL_1613304, EPI_ISL_1613307, EPI_ISL_1613310, EPI_ISL_1613313, EPI_ISL_1613316, EPI_ISL_1613319, EPI_ISL_1613321, EPI_ISL_1613324, EPI_ISL_1613327, EPI_ISL_1613331, EPI_ISL_1613334, EPI_ISL_1613339, EPI_ISL_1613342,
EPI_ISL_1613345, EPI_ISL_1613348, EPI_ISL_1613351, EPI_ISL_1613354, EPI_ISL_1613357, EPI_ISL_1613360, EPI_ISL_1613363, EPI_ISL_1613366, EPI_ISL_1613369, EPI_ISL_1613372, EPI_ISL_1613375, EPI_ISL_1613378, EPI_ISL_1613381, EPI_ISL_1613384, EPI_ISL_1613386, EPI_ISL_1613389, EPI_ISL_1613392, EPI_ISL_1613401, EPI_ISL_1613404, EPI_ISL_1613407, EPI_ISL_1613410, EPI_ISL_1613413,
EPI_ISL_1613416, EPI_ISL_1613419, EPI_ISL_1613422, EPI_ISL_1613425, EPI_ISL_1613428, EPI_ISL_1613431, EPI_ISL_1613433, EPI_ISL_1613436, EPI_ISL_1613439, EPI_ISL_1613442, EPI_ISL_1613445, EPI_ISL_1613448, EPI_ISL_1613451, EPI_ISL_1613454, EPI_ISL_1613458, EPI_ISL_1613461, EPI_ISL_1613464, EPI_ISL_1613467, EPI_ISL_1613469, EPI_ISL_1613472, EPI_ISL_1613475, EPI_ISL_1613478,
EPI_ISL_1613481, EPI_ISL_1613485, EPI_ISL_1613487, EPI_ISL_1613490, EPI_ISL_1613493, EPI_ISL_1613496, EPI_ISL_1613499, EPI_ISL_1613502, EPI_ISL_1613505, EPI_ISL_1613508, EPI_ISL_1613511, EPI_ISL_1613514, EPI_ISL_1613517, EPI_ISL_1613520, EPI_ISL_1613522, EPI_ISL_1613529, EPI_ISL_1613532, EPI_ISL_1613534, EPI_ISL_1613538, EPI_ISL_1613540, EPI_ISL_1613543, EPI_ISL_1613546,
EPI_ISL_1613549, EPI_ISL_1613552, EPI_ISL_1613555, EPI_ISL_1613558, EPI_ISL_1613562, EPI_ISL_1613565, EPI_ISL_1613567, EPI_ISL_1613570, EPI_ISL_1613573, EPI_ISL_1613576, EPI_ISL_1613579, EPI_ISL_1613582, EPI_ISL_1613585, EPI_ISL_1613588, EPI_ISL_1613591, EPI_ISL_1613593, EPI_ISL_1613596, EPI_ISL_1613599, EPI_ISL_1613602, EPI_ISL_1613605, EPI_ISL_1613608, EPI_ISL_1613805,
EPI_ISL_1613808, EPI_ISL_1613810, EPI_ISL_1613813, EPI_ISL_1613816, EPI_ISL_1613819, EPI_ISL_1613822, EPI_ISL_1613825, EPI_ISL_1613828, EPI_ISL_1613831, EPI_ISL_1613834, EPI_ISL_1613836, EPI_ISL_1613839, EPI_ISL_1613843, EPI_ISL_1613846, EPI_ISL_1613849, EPI_ISL_1613852, EPI_ISL_1613854, EPI_ISL_1613857, EPI_ISL_1613859, EPI_ISL_1613862, EPI_ISL_1613865, EPI_ISL_1613868,
EPI_ISL_1613871, EPI_ISL_1613874, EPI_ISL_1613877, EPI_ISL_1613880, EPI_ISL_1613883, EPI_ISL_1613886, EPI_ISL_1613889, EPI_ISL_1613891, EPI_ISL_1613894, EPI_ISL_1613897, EPI_ISL_1613900, EPI_ISL_1613902, EPI_ISL_1613905, EPI_ISL_1613908, EPI_ISL_1613911, EPI_ISL_1613914, EPI_ISL_1613916, EPI_ISL_1613919, EPI_ISL_1613922, EPI_ISL_1613925, EPI_ISL_1613928, EPI_ISL_1613931,
EPI_ISL_1613934, EPI_ISL_1613937, EPI_ISL_1613938, EPI_ISL_1613939, EPI_ISL_1613940, EPI_ISL_1613941, EPI_ISL_1613942, EPI_ISL_1613943, EPI_ISL_1613944, EPI_ISL_1613945, EPI_ISL_1613946, EPI_ISL_1613947, EPI_ISL_1613948, EPI_ISL_1613949, EPI_ISL_1613950, EPI_ISL_1613951, EPI_ISL_1613952, EPI_ISL_1613953, EPI_ISL_1613954, EPI_ISL_1613955, EPI_ISL_1613956, EPI_ISL_1613957,
EPI_ISL_1613958, EPI_ISL_1613959, EPI_ISL_1613960, EPI_ISL_1613961, EPI_ISL_1613962, EPI_ISL_1613963, EPI_ISL_1613964, EPI_ISL_1613965, EPI_ISL_1613966, EPI_ISL_1613967, EPI_ISL_1613968, EPI_ISL_1613969, EPI_ISL_1613970, EPI_ISL_1613973, EPI_ISL_1613974, EPI_ISL_1613975, EPI_ISL_1613976, EPI_ISL_1613977, EPI_ISL_1613978, EPI_ISL_1613979, EPI_ISL_1613980, EPI_ISL_1613981,
EPI_ISL_1613982, EPI_ISL_1613983, EPI_ISL_1613984, EPI_ISL_1613985, EPI_ISL_1613986, EPI_ISL_1613987, EPI_ISL_1613988, EPI_ISL_1613989, EPI_ISL_1613990, EPI_ISL_1613991, EPI_ISL_1613992, EPI_ISL_1613993, EPI_ISL_1613994, EPI_ISL_1613995, EPI_ISL_1613996, EPI_ISL_1613997, EPI_ISL_1613998, EPI_ISL_1613999, EPI_ISL_1614000, EPI_ISL_1614001, EPI_ISL_1614002, EPI_ISL_1614003,
EPI_ISL_1614004, EPI_ISL_1614005, EPI_ISL_1614006, EPI_ISL_1614007, EPI_ISL_1614008, EPI_ISL_1614009, EPI_ISL_1614010, EPI_ISL_1614011, EPI_ISL_1614012, EPI_ISL_1614013, EPI_ISL_1614014, EPI_ISL_1614015, EPI_ISL_1614016, EPI_ISL_1614017, EPI_ISL_1614018, EPI_ISL_1614019, EPI_ISL_1614020, EPI_ISL_1614021, EPI_ISL_1614022, EPI_ISL_1614023, EPI_ISL_1614024, EPI_ISL_1614025,
EPI_ISL_1614026, EPI_ISL_1614027, EPI_ISL_1614028, EPI_ISL_1614029, EPI_ISL_1614030, EPI_ISL_1614031, EPI_ISL_1614032, EPI_ISL_1614033, EPI_ISL_1614034, EPI_ISL_1614035, EPI_ISL_1614036, EPI_ISL_1614037, EPI_ISL_1614038, EPI_ISL_1614039, EPI_ISL_1614040, EPI_ISL_1614041, EPI_ISL_1614042, EPI_ISL_1614043, EPI_ISL_1614044, EPI_ISL_1614045, EPI_ISL_1614046, EPI_ISL_1614047,
EPI_ISL_1614048, EPI_ISL_1614049, EPI_ISL_1614050, EPI_ISL_1614051, EPI_ISL_1614052, EPI_ISL_1614053, EPI_ISL_1614054, EPI_ISL_1614055, EPI_ISL_1614056, EPI_ISL_1614057, EPI_ISL_1614058, EPI_ISL_1614059, EPI_ISL_1614060, EPI_ISL_1614061, EPI_ISL_1614062, EPI_ISL_1614063, EPI_ISL_1614064, EPI_ISL_1614065, EPI_ISL_1614066, EPI_ISL_1614067, EPI_ISL_1614068, EPI_ISL_1614069,
EPI_ISL_1614070, EPI_ISL_1614071, EPI_ISL_1614072, EPI_ISL_1614073, EPI_ISL_1614074, EPI_ISL_1614075, EPI_ISL_1614077, EPI_ISL_1614078, EPI_ISL_1614080, EPI_ISL_1614081, EPI_ISL_1614082, EPI_ISL_1614083, EPI_ISL_1614084, EPI_ISL_1614085, EPI_ISL_1614086, EPI_ISL_1614087, EPI_ISL_1614088, EPI_ISL_1614089, EPI_ISL_1614090, EPI_ISL_1614091, EPI_ISL_1614092, EPI_ISL_1614093,
EPI_ISL_1614094, EPI_ISL_1614095, EPI_ISL_1614096, EPI_ISL_1614097, EPI_ISL_1614098, EPI_ISL_1614099, EPI_ISL_1614100, EPI_ISL_1614101, EPI_ISL_1614102, EPI_ISL_1614103, EPI_ISL_1614104, EPI_ISL_1614105, EPI_ISL_1614106, EPI_ISL_1614107, EPI_ISL_1614108, EPI_ISL_1614109, EPI_ISL_1614110, EPI_ISL_1614111, EPI_ISL_1614112, EPI_ISL_1614113, EPI_ISL_1614114, EPI_ISL_1614115,
EPI_ISL_1614116, EPI_ISL_1614117, EPI_ISL_1614118, EPI_ISL_1614119, EPI_ISL_1614120, EPI_ISL_1614121, EPI_ISL_1614122, EPI_ISL_1614123, EPI_ISL_1614124, EPI_ISL_1614125, EPI_ISL_1614126, EPI_ISL_1614127, EPI_ISL_1614128, EPI_ISL_1614129, EPI_ISL_1614130, EPI_ISL_1614131, EPI_ISL_1614132, EPI_ISL_1614133, EPI_ISL_1614134, EPI_ISL_1614135, EPI_ISL_1614136, EPI_ISL_1614137,
EPI_ISL_1614138, EPI_ISL_1614139, EPI_ISL_1614140, EPI_ISL_1614141, EPI_ISL_1614142, EPI_ISL_1614143, EPI_ISL_1614144, EPI_ISL_1614145, EPI_ISL_1614146, EPI_ISL_1614147, EPI_ISL_1614148, EPI_ISL_1614149, EPI_ISL_1614150, EPI_ISL_1614151, EPI_ISL_1614152, EPI_ISL_1614153, EPI_ISL_1614154, EPI_ISL_1614155, EPI_ISL_1614156, EPI_ISL_1614157, EPI_ISL_1614158, EPI_ISL_1614159,
EPI_ISL_1614160, EPI_ISL_1614161, EPI_ISL_1614162, EPI_ISL_1614163, EPI_ISL_1614164, EPI_ISL_1614165, EPI_ISL_1614166, EPI_ISL_1614167, EPI_ISL_1614168, EPI_ISL_1614169, EPI_ISL_1614170, EPI_ISL_1614226, EPI_ISL_1614348, EPI_ISL_1614460, EPI_ISL_1614462, EPI_ISL_1614463, EPI_ISL_1614464, EPI_ISL_1614465, EPI_ISL_1614466, EPI_ISL_1614467, EPI_ISL_1614468, EPI_ISL_1614469,
EPI_ISL_1614470, EPI_ISL_1614471, EPI_ISL_1614472, EPI_ISL_1614473, EPI_ISL_1614474, EPI_ISL_1614475, EPI_ISL_1614476, EPI_ISL_1614477, EPI_ISL_1614478, EPI_ISL_1614479, EPI_ISL_1614480, EPI_ISL_1614481, EPI_ISL_1614482, EPI_ISL_1614483, EPI_ISL_1614484, EPI_ISL_1614485, EPI_ISL_1614486, EPI_ISL_1614487, EPI_ISL_1614488, EPI_ISL_1614489, EPI_ISL_1614490, EPI_ISL_1614491,
EPI_ISL_1614492, EPI_ISL_1614493, EPI_ISL_1614494, EPI_ISL_1614495, EPI_ISL_1614496, EPI_ISL_1614497, EPI_ISL_1614498, EPI_ISL_1614499, EPI_ISL_1614500, EPI_ISL_1614501, EPI_ISL_1614502, EPI_ISL_1614503, EPI_ISL_1614504, EPI_ISL_1614505, EPI_ISL_1614506, EPI_ISL_1614507, EPI_ISL_1614508, EPI_ISL_1614509, EPI_ISL_1614510, EPI_ISL_1614511, EPI_ISL_1614512, EPI_ISL_1614513,
EPI_ISL_1614514, EPI_ISL_1614515, EPI_ISL_1614516, EPI_ISL_1614517, EPI_ISL_1614518, EPI_ISL_1614519, EPI_ISL_1614520, EPI_ISL_1614521, EPI_ISL_1614522, EPI_ISL_1614523, EPI_ISL_1614524, EPI_ISL_1614525, EPI_ISL_1614526, EPI_ISL_1614527, EPI_ISL_1614528, EPI_ISL_1614529, EPI_ISL_1614530, EPI_ISL_1614531, EPI_ISL_1614532, EPI_ISL_1614533, EPI_ISL_1614534, EPI_ISL_1614536,
EPI_ISL_1614537, EPI_ISL_1614538, EPI_ISL_1614539, EPI_ISL_1614540, EPI_ISL_1614541, EPI_ISL_1614542, EPI_ISL_1614543, EPI_ISL_1614544, EPI_ISL_1614545, EPI_ISL_1614546, EPI_ISL_1614547, EPI_ISL_1614548, EPI_ISL_1614549, EPI_ISL_1614550, EPI_ISL_1614551, EPI_ISL_1614552, EPI_ISL_1614553, EPI_ISL_1614554, EPI_ISL_1614555, EPI_ISL_1614556, EPI_ISL_1614557, EPI_ISL_1614558,
EPI_ISL_1614559, EPI_ISL_1614560, EPI_ISL_1614561, EPI_ISL_1614562, EPI_ISL_1614563, EPI_ISL_1614564, EPI_ISL_1614565, EPI_ISL_1614566, EPI_ISL_1614567, EPI_ISL_1614568, EPI_ISL_1614569, EPI_ISL_1614570, EPI_ISL_1614571, EPI_ISL_1614572, EPI_ISL_1614573, EPI_ISL_1614574, EPI_ISL_1614575, EPI_ISL_1614576, EPI_ISL_1614577, EPI_ISL_1614578, EPI_ISL_1614579, EPI_ISL_1614580,
EPI_ISL_1614581, EPI_ISL_1614582, EPI_ISL_1614583, EPI_ISL_1614584, EPI_ISL_1614585, EPI_ISL_1614586, EPI_ISL_1614587, EPI_ISL_1614588, EPI_ISL_1614589, EPI_ISL_1614590, EPI_ISL_1614591, EPI_ISL_1614592, EPI_ISL_1614593, EPI_ISL_1614594, EPI_ISL_1614595, EPI_ISL_1614596, EPI_ISL_1614597, EPI_ISL_1614598, EPI_ISL_1614599, EPI_ISL_1614600, EPI_ISL_1614601, EPI_ISL_1614602,
EPI_ISL_1614603, EPI_ISL_1614604, EPI_ISL_1614605, EPI_ISL_1614606, EPI_ISL_1614607, EPI_ISL_1614608, EPI_ISL_1614609, EPI_ISL_1614610, EPI_ISL_1614611, EPI_ISL_1614612, EPI_ISL_1614613, EPI_ISL_1614614, EPI_ISL_1614615, EPI_ISL_1614616, EPI_ISL_1614617, EPI_ISL_1614618, EPI_ISL_1614619, EPI_ISL_1614620, EPI_ISL_1614621, EPI_ISL_1614622, EPI_ISL_1614623, EPI_ISL_1614624,
EPI_ISL_1614625, EPI_ISL_1614626, EPI_ISL_1614627, EPI_ISL_1614628, EPI_ISL_1614629, EPI_ISL_1614630, EPI_ISL_1614631, EPI_ISL_1614632, EPI_ISL_1614633, EPI_ISL_1614634, EPI_ISL_1614635, EPI_ISL_1614636, EPI_ISL_1614637, EPI_ISL_1614638, EPI_ISL_1614639, EPI_ISL_1614640, EPI_ISL_1614641, EPI_ISL_1614642, EPI_ISL_1614643, EPI_ISL_1614644, EPI_ISL_1614645, EPI_ISL_1614646,
EPI_ISL_1614647, EPI_ISL_1614648, EPI_ISL_1614649, EPI_ISL_1614650, EPI_ISL_1614651, EPI_ISL_1614652, EPI_ISL_1614653, EPI_ISL_1614654, EPI_ISL_1614655, EPI_ISL_1614656, EPI_ISL_1614657, EPI_ISL_1614658, EPI_ISL_1614659, EPI_ISL_1614660, EPI_ISL_1614661, EPI_ISL_1614662, EPI_ISL_1614663, EPI_ISL_1614664, EPI_ISL_1614665, EPI_ISL_1614666, EPI_ISL_1614667, EPI_ISL_1614668,
EPI_ISL_1614669, EPI_ISL_1614670, EPI_ISL_1614671, EPI_ISL_1614672, EPI_ISL_1614673, EPI_ISL_1614674, EPI_ISL_1614675, EPI_ISL_1614676, EPI_ISL_1614677, EPI_ISL_1614678, EPI_ISL_1614679, EPI_ISL_1614680, EPI_ISL_1614681, EPI_ISL_1614682, EPI_ISL_1614683, EPI_ISL_1614684, EPI_ISL_1614685, EPI_ISL_1614686, EPI_ISL_1614687, EPI_ISL_1614688, EPI_ISL_1614689, EPI_ISL_1614690,
EPI_ISL_1614691, EPI_ISL_1614692, EPI_ISL_1614693, EPI_ISL_1614694, EPI_ISL_1614695, EPI_ISL_1614696, EPI_ISL_1614697, EPI_ISL_1614698, EPI_ISL_1614699, EPI_ISL_1614700, EPI_ISL_1614702, EPI_ISL_1614703, EPI_ISL_1614704, EPI_ISL_1614705, EPI_ISL_1614706, EPI_ISL_1614707, EPI_ISL_1614708, EPI_ISL_1614709, EPI_ISL_1614710, EPI_ISL_1614711, EPI_ISL_1614712, EPI_ISL_1614713,
EPI_ISL_1614714, EPI_ISL_1614715, EPI_ISL_1614716, EPI_ISL_1614717, EPI_ISL_1614718, EPI_ISL_1614719, EPI_ISL_1614720, EPI_ISL_1614721, EPI_ISL_1614722, EPI_ISL_1614723, EPI_ISL_1614724, EPI_ISL_1614725, EPI_ISL_1614726, EPI_ISL_1614727, EPI_ISL_1614728, EPI_ISL_1614729, EPI_ISL_1614730, EPI_ISL_1614731, EPI_ISL_1614732, EPI_ISL_1614733, EPI_ISL_1614734, EPI_ISL_1614735,
EPI_ISL_1614736, EPI_ISL_1614737, EPI_ISL_1614738, EPI_ISL_1614739, EPI_ISL_1614740, EPI_ISL_1614741, EPI_ISL_1614742, EPI_ISL_1614743, EPI_ISL_1614745, EPI_ISL_1614746, EPI_ISL_1614747, EPI_ISL_1614748, EPI_ISL_1614749, EPI_ISL_1614750, EPI_ISL_1614751, EPI_ISL_1614752, EPI_ISL_1614753, EPI_ISL_1614754, EPI_ISL_1614755, EPI_ISL_1614756, EPI_ISL_1614757, EPI_ISL_1614760,
EPI_ISL_1614761, EPI_ISL_1614762, EPI_ISL_1614763, EPI_ISL_1614764, EPI_ISL_1614765, EPI_ISL_1614766, EPI_ISL_1614767, EPI_ISL_1614768, EPI_ISL_1614769, EPI_ISL_1614770, EPI_ISL_1614771, EPI_ISL_1614772, EPI_ISL_1614773, EPI_ISL_1614774, EPI_ISL_1614776, EPI_ISL_1614777, EPI_ISL_1614778, EPI_ISL_1614779, EPI_ISL_1614780, EPI_ISL_1614781, EPI_ISL_1614782, EPI_ISL_1614783,
EPI_ISL_1614784, EPI_ISL_1614785, EPI_ISL_1614786, EPI_ISL_1614787, EPI_ISL_1614788, EPI_ISL_1614789, EPI_ISL_1614790, EPI_ISL_1614791, EPI_ISL_1614792, EPI_ISL_1614793, EPI_ISL_1614794, EPI_ISL_1614795, EPI_ISL_1614796, EPI_ISL_1614797, EPI_ISL_1614798, EPI_ISL_1614799, EPI_ISL_1614800, EPI_ISL_1614801, EPI_ISL_1614802, EPI_ISL_1614803, EPI_ISL_1614804, EPI_ISL_1614805,
EPI_ISL_1614806, EPI_ISL_1614807, EPI_ISL_1614808, EPI_ISL_1614809, EPI_ISL_1614810, EPI_ISL_1614811, EPI_ISL_1614812, EPI_ISL_1614813, EPI_ISL_1614814, EPI_ISL_1614815, EPI_ISL_1614816, EPI_ISL_1614817, EPI_ISL_1614818, EPI_ISL_1614819, EPI_ISL_1614820, EPI_ISL_1614821, EPI_ISL_1614822, EPI_ISL_1614823, EPI_ISL_1614824, EPI_ISL_1614825, EPI_ISL_1614826, EPI_ISL_1614827,
EPI_ISL_1614828, EPI_ISL_1614829, EPI_ISL_1614830, EPI_ISL_1614831, EPI_ISL_1614832, EPI_ISL_1614833, EPI_ISL_1614834, EPI_ISL_1614835, EPI_ISL_1614836, EPI_ISL_1614837, EPI_ISL_1614838, EPI_ISL_1614839, EPI_ISL_1614840, EPI_ISL_1614841, EPI_ISL_1614842, EPI_ISL_1614843, EPI_ISL_1614844, EPI_ISL_1614845, EPI_ISL_1614846, EPI_ISL_1614847, EPI_ISL_1614848, EPI_ISL_1614849,
EPI_ISL_1614850, EPI_ISL_1614851, EPI_ISL_1614852, EPI_ISL_1614853, EPI_ISL_1614854, EPI_ISL_1614855, EPI_ISL_1614856, EPI_ISL_1614857, EPI_ISL_1614858, EPI_ISL_1614859, EPI_ISL_1614860, EPI_ISL_1614861, EPI_ISL_1614862, EPI_ISL_1614863, EPI_ISL_1614864, EPI_ISL_1614865, EPI_ISL_1614866, EPI_ISL_1614867, EPI_ISL_1614868, EPI_ISL_1614869, EPI_ISL_1614870, EPI_ISL_1614871,
EPI_ISL_1614872, EPI_ISL_1614873, EPI_ISL_1614874, EPI_ISL_1614875, EPI_ISL_1614876, EPI_ISL_1614877, EPI_ISL_1614878, EPI_ISL_1614879, EPI_ISL_1614880, EPI_ISL_1614881, EPI_ISL_1614882, EPI_ISL_1614883, EPI_ISL_1614884, EPI_ISL_1614885, EPI_ISL_1614886, EPI_ISL_1614887, EPI_ISL_1614888, EPI_ISL_1614889, EPI_ISL_1614890, EPI_ISL_1614891, EPI_ISL_1614893, EPI_ISL_1614895,
EPI_ISL_1614898, EPI_ISL_1614900, EPI_ISL_1614901, EPI_ISL_1614902, EPI_ISL_1614903, EPI_ISL_1614904, EPI_ISL_1665940, EPI_ISL_1666000, EPI_ISL_1666001, EPI_ISL_1666002, EPI_ISL_1666003, EPI_ISL_1666004, EPI_ISL_1666005, EPI_ISL_1666006, EPI_ISL_1666007, EPI_ISL_1666008, EPI_ISL_1666009, EPI_ISL_1666010, EPI_ISL_1666011, EPI_ISL_1666012, EPI_ISL_1666013, EPI_ISL_1666014,
EPI_ISL_1666015, EPI_ISL_1666016, EPI_ISL_1666017, EPI_ISL_1666018, EPI_ISL_1666019, EPI_ISL_1666020, EPI_ISL_1666021, EPI_ISL_1666022, EPI_ISL_1666023, EPI_ISL_1666024, EPI_ISL_1666025, EPI_ISL_1666026, EPI_ISL_1666027, EPI_ISL_1666028, EPI_ISL_1666029, EPI_ISL_1666030, EPI_ISL_1666031, EPI_ISL_1666032, EPI_ISL_1666033, EPI_ISL_1666034, EPI_ISL_1666035, EPI_ISL_1666036,
EPI_ISL_1666037, EPI_ISL_1666038, EPI_ISL_1666039, EPI_ISL_1666040, EPI_ISL_1666041, EPI_ISL_1666042, EPI_ISL_1666043, EPI_ISL_1666044, EPI_ISL_1666045, EPI_ISL_1666046, EPI_ISL_1666047, EPI_ISL_1666048, EPI_ISL_1666049, EPI_ISL_1666050, EPI_ISL_1666051, EPI_ISL_1666052, EPI_ISL_1666053, EPI_ISL_1666054, EPI_ISL_1666055, EPI_ISL_1666056, EPI_ISL_1666060, EPI_ISL_1666061,
EPI_ISL_1666062, EPI_ISL_1666063, EPI_ISL_1666064, EPI_ISL_1666065, EPI_ISL_1666066, EPI_ISL_1666067, EPI_ISL_1666068, EPI_ISL_1666069, EPI_ISL_1666070, EPI_ISL_1666071, EPI_ISL_1666072, EPI_ISL_1666073, EPI_ISL_1666074, EPI_ISL_1666075, EPI_ISL_1666076, EPI_ISL_1666077, EPI_ISL_1666078, EPI_ISL_1666079, EPI_ISL_1666080, EPI_ISL_1666081, EPI_ISL_1666082, EPI_ISL_1666083,
EPI_ISL_1666084, EPI_ISL_1666085, EPI_ISL_1666086, EPI_ISL_1666087, EPI_ISL_1666088, EPI_ISL_1666089, EPI_ISL_1666090, EPI_ISL_1666091, EPI_ISL_1666092, EPI_ISL_1666093, EPI_ISL_1666094, EPI_ISL_1666095, EPI_ISL_1666096, EPI_ISL_1666097, EPI_ISL_1666098, EPI_ISL_1666099, EPI_ISL_1666100, EPI_ISL_1666101, EPI_ISL_1666102, EPI_ISL_1666103, EPI_ISL_1666104, EPI_ISL_1666105,
EPI_ISL_1666106, EPI_ISL_1666107, EPI_ISL_1666108, EPI_ISL_1666109, EPI_ISL_1666110, EPI_ISL_1666111, EPI_ISL_1666112, EPI_ISL_1666113, EPI_ISL_1666114, EPI_ISL_1666115, EPI_ISL_1666116, EPI_ISL_1666117, EPI_ISL_1666118, EPI_ISL_1666119, EPI_ISL_1666120, EPI_ISL_1666121, EPI_ISL_1666122, EPI_ISL_1666123, EPI_ISL_1666124, EPI_ISL_1666125, EPI_ISL_1666126, EPI_ISL_1666127,
EPI_ISL_1666128, EPI_ISL_1666129, EPI_ISL_1666130, EPI_ISL_1666131, EPI_ISL_1666132, EPI_ISL_1666133, EPI_ISL_1666134, EPI_ISL_1666135, EPI_ISL_1666136, EPI_ISL_1666137, EPI_ISL_1666138, EPI_ISL_1666139, EPI_ISL_1666140, EPI_ISL_1666141, EPI_ISL_1666142, EPI_ISL_1666143, EPI_ISL_1666144, EPI_ISL_1666145, EPI_ISL_1666146, EPI_ISL_1666147, EPI_ISL_1666148, EPI_ISL_1666149,
EPI_ISL_1666150, EPI_ISL_1666151, EPI_ISL_1666152, EPI_ISL_1666153, EPI_ISL_1666154, EPI_ISL_1666155, EPI_ISL_1666156, EPI_ISL_1666157, EPI_ISL_1666158, EPI_ISL_1666159, EPI_ISL_1666160, EPI_ISL_1666161, EPI_ISL_1666162, EPI_ISL_1666163, EPI_ISL_1666164, EPI_ISL_1666165, EPI_ISL_1666166, EPI_ISL_1666167, EPI_ISL_1666168, EPI_ISL_1666169, EPI_ISL_1666170, EPI_ISL_1666171,
EPI_ISL_1666172, EPI_ISL_1666173, EPI_ISL_1666174, EPI_ISL_1666176, EPI_ISL_1666177, EPI_ISL_1666178, EPI_ISL_1666179, EPI_ISL_1666180, EPI_ISL_1666181, EPI_ISL_1666182, EPI_ISL_1666183, EPI_ISL_1666184, EPI_ISL_1666185, EPI_ISL_1666187, EPI_ISL_1666188, EPI_ISL_1666189, EPI_ISL_1666190, EPI_ISL_1666191, EPI_ISL_1666192, EPI_ISL_1666193, EPI_ISL_1666194, EPI_ISL_1666195,
EPI_ISL_1666196, EPI_ISL_1666197, EPI_ISL_1666198, EPI_ISL_1666199, EPI_ISL_1666200, EPI_ISL_1666201, EPI_ISL_1666202, EPI_ISL_1666203, EPI_ISL_1666204, EPI_ISL_1666205, EPI_ISL_1666206, EPI_ISL_1666207, EPI_ISL_1666208, EPI_ISL_1666209, EPI_ISL_1666210, EPI_ISL_1666211, EPI_ISL_1666212, EPI_ISL_1666213, EPI_ISL_1666214, EPI_ISL_1666215, EPI_ISL_1666216, EPI_ISL_1666217,
EPI_ISL_1666218, EPI_ISL_1666219, EPI_ISL_1666221, EPI_ISL_1666222, EPI_ISL_1666223, EPI_ISL_1666224, EPI_ISL_1666225, EPI_ISL_1666226, EPI_ISL_1666227, EPI_ISL_1666228, EPI_ISL_1666229, EPI_ISL_1666230, EPI_ISL_1666231, EPI_ISL_1666232, EPI_ISL_1666233, EPI_ISL_1666234, EPI_ISL_1666235, EPI_ISL_1666236, EPI_ISL_1666237, EPI_ISL_1666238, EPI_ISL_1666239, EPI_ISL_1666240,
EPI_ISL_1666241, EPI_ISL_1666242, EPI_ISL_1666243, EPI_ISL_1666244, EPI_ISL_1666245, EPI_ISL_1666246, EPI_ISL_1666247, EPI_ISL_1666248, EPI_ISL_1666249, EPI_ISL_1666250, EPI_ISL_1666251, EPI_ISL_1666252, EPI_ISL_1666253, EPI_ISL_1666254, EPI_ISL_1666255, EPI_ISL_1666256, EPI_ISL_1666257, EPI_ISL_1666258, EPI_ISL_1666259, EPI_ISL_1666260, EPI_ISL_1666261, EPI_ISL_1666262,
EPI_ISL_1666263, EPI_ISL_1666264, EPI_ISL_1666265, EPI_ISL_1666266, EPI_ISL_1666267, EPI_ISL_1666268, EPI_ISL_1666269, EPI_ISL_1666270, EPI_ISL_1666271, EPI_ISL_1666272, EPI_ISL_1666273, EPI_ISL_1666274, EPI_ISL_1666275, EPI_ISL_1666276, EPI_ISL_1666277, EPI_ISL_1666278, EPI_ISL_1666279, EPI_ISL_1666280, EPI_ISL_1666281, EPI_ISL_1666282, EPI_ISL_1666283, EPI_ISL_1666284,
EPI_ISL_1666285, EPI_ISL_1666286, EPI_ISL_1666287, EPI_ISL_1666288, EPI_ISL_1666289, EPI_ISL_1666290, EPI_ISL_1666291, EPI_ISL_1666292, EPI_ISL_1666293, EPI_ISL_1666294, EPI_ISL_1666295, EPI_ISL_1666296, EPI_ISL_1666297, EPI_ISL_1666298, EPI_ISL_1666299, EPI_ISL_1666300, EPI_ISL_1666301, EPI_ISL_1666302, EPI_ISL_1666303, EPI_ISL_1666304, EPI_ISL_1666305, EPI_ISL_1666306,
EPI_ISL_1666307, EPI_ISL_1666308, EPI_ISL_1666309, EPI_ISL_1666310, EPI_ISL_1666311, EPI_ISL_1666312, EPI_ISL_1666313, EPI_ISL_1666314, EPI_ISL_1666315, EPI_ISL_1666316, EPI_ISL_1666317, EPI_ISL_1666318, EPI_ISL_1666319, EPI_ISL_1666320, EPI_ISL_1666321, EPI_ISL_1666322, EPI_ISL_1666323, EPI_ISL_1666324, EPI_ISL_1666325, EPI_ISL_1666326, EPI_ISL_1666327, EPI_ISL_1666329,
EPI_ISL_1666330, EPI_ISL_1666331, EPI_ISL_1666332, EPI_ISL_1666333, EPI_ISL_1666334, EPI_ISL_1666335, EPI_ISL_1666336, EPI_ISL_1666337, EPI_ISL_1666338, EPI_ISL_1666339, EPI_ISL_1666340, EPI_ISL_1666341, EPI_ISL_1666342, EPI_ISL_1666343, EPI_ISL_1666344, EPI_ISL_1666345, EPI_ISL_1666346, EPI_ISL_1666347, EPI_ISL_1666348, EPI_ISL_1666349, EPI_ISL_1666350, EPI_ISL_1666351,
EPI_ISL_1666352, EPI_ISL_1666353, EPI_ISL_1666354, EPI_ISL_1666355, EPI_ISL_1666356, EPI_ISL_1666357, EPI_ISL_1666358, EPI_ISL_1666359, EPI_ISL_1666360, EPI_ISL_1666361, EPI_ISL_1666362, EPI_ISL_1666363, EPI_ISL_1666365, EPI_ISL_1666366, EPI_ISL_1666367, EPI_ISL_1666368, EPI_ISL_1666369, EPI_ISL_1666370, EPI_ISL_1666371, EPI_ISL_1666372, EPI_ISL_1666373, EPI_ISL_1666374,
EPI_ISL_1666375, EPI_ISL_1666376, EPI_ISL_1666377, EPI_ISL_1666378, EPI_ISL_1666379, EPI_ISL_1666380, EPI_ISL_1666381, EPI_ISL_1666382, EPI_ISL_1666383, EPI_ISL_1666384, EPI_ISL_1666385, EPI_ISL_1666386, EPI_ISL_1666387, EPI_ISL_1666388, EPI_ISL_1666389, EPI_ISL_1666390, EPI_ISL_1666391, EPI_ISL_1666392, EPI_ISL_1666393, EPI_ISL_1666394, EPI_ISL_1666395, EPI_ISL_1666396,
EPI_ISL_1666397, EPI_ISL_1666398, EPI_ISL_1666399, EPI_ISL_1666400, EPI_ISL_1666401, EPI_ISL_1666402, EPI_ISL_1666404, EPI_ISL_1666405, EPI_ISL_1666406, EPI_ISL_1666407, EPI_ISL_1666408, EPI_ISL_1666409, EPI_ISL_1666410, EPI_ISL_1666411, EPI_ISL_1666412, EPI_ISL_1666415, EPI_ISL_1666416, EPI_ISL_1666417, EPI_ISL_1666418, EPI_ISL_1666419, EPI_ISL_1666420, EPI_ISL_1666421,
EPI_ISL_1666422, EPI_ISL_1666423, EPI_ISL_1666424, EPI_ISL_1666425, EPI_ISL_1666426, EPI_ISL_1666427, EPI_ISL_1666428, EPI_ISL_1666429, EPI_ISL_1666430, EPI_ISL_1666431, EPI_ISL_1666432, EPI_ISL_1666433, EPI_ISL_1666434, EPI_ISL_1666435, EPI_ISL_1666436, EPI_ISL_1666437, EPI_ISL_1666438, EPI_ISL_1666439, EPI_ISL_1666440, EPI_ISL_1666441, EPI_ISL_1666442, EPI_ISL_1666443,
EPI_ISL_1666444, EPI_ISL_1666445, EPI_ISL_1666446, EPI_ISL_1666447, EPI_ISL_1666448, EPI_ISL_1666449, EPI_ISL_1666450, EPI_ISL_1666451, EPI_ISL_1666452, EPI_ISL_1666453, EPI_ISL_1666454, EPI_ISL_1666455, EPI_ISL_1666456, EPI_ISL_1666457, EPI_ISL_1666458, EPI_ISL_1666459, EPI_ISL_1666460, EPI_ISL_1666461, EPI_ISL_1666462, EPI_ISL_1666463, EPI_ISL_1666464, EPI_ISL_1666465,
EPI_ISL_1666466, EPI_ISL_1666467, EPI_ISL_1666468, EPI_ISL_1666469, EPI_ISL_1666470, EPI_ISL_1666471, EPI_ISL_1666472, EPI_ISL_1666473, EPI_ISL_1666474, EPI_ISL_1666475, EPI_ISL_1666476, EPI_ISL_1666478, EPI_ISL_1666479, EPI_ISL_1666480, EPI_ISL_1666481, EPI_ISL_1666482, EPI_ISL_1666483, EPI_ISL_1666484, EPI_ISL_1666485, EPI_ISL_1666486, EPI_ISL_1666487, EPI_ISL_1666488,
EPI_ISL_1666489, EPI_ISL_1666490, EPI_ISL_1666491, EPI_ISL_1666492, EPI_ISL_1666493, EPI_ISL_1666494, EPI_ISL_1666495, EPI_ISL_1666496, EPI_ISL_1666497, EPI_ISL_1666498, EPI_ISL_1666499, EPI_ISL_1666500, EPI_ISL_1666501, EPI_ISL_1666502, EPI_ISL_1666503, EPI_ISL_1666504, EPI_ISL_1666505, EPI_ISL_1666507, EPI_ISL_1666508, EPI_ISL_1666509, EPI_ISL_1666510, EPI_ISL_1666511,
EPI_ISL_1666512, EPI_ISL_1666513, EPI_ISL_1666514, EPI_ISL_1666515, EPI_ISL_1666516, EPI_ISL_1666517, EPI_ISL_1666518, EPI_ISL_1666519, EPI_ISL_1666520, EPI_ISL_1666521, EPI_ISL_1666522, EPI_ISL_1666523, EPI_ISL_1666524, EPI_ISL_1666525, EPI_ISL_1666526, EPI_ISL_1666527, EPI_ISL_1666528, EPI_ISL_1666529, EPI_ISL_1666530, EPI_ISL_1666531, EPI_ISL_1666532, EPI_ISL_1666533,
EPI_ISL_1666534, EPI_ISL_1666535, EPI_ISL_1666536, EPI_ISL_1666537, EPI_ISL_1666538, EPI_ISL_1666539, EPI_ISL_1666540, EPI_ISL_1666541, EPI_ISL_1666542, EPI_ISL_1666655, EPI_ISL_1666658, EPI_ISL_1666702, EPI_ISL_1666724, EPI_ISL_1666934, EPI_ISL_1667018, EPI_ISL_1667077, EPI_ISL_1667078, EPI_ISL_1667083, EPI_ISL_1667084, EPI_ISL_1667085, EPI_ISL_1667086, EPI_ISL_1667087,
EPI_ISL_1667088, EPI_ISL_1667089, EPI_ISL_1667090, EPI_ISL_1667091, EPI_ISL_1667092, EPI_ISL_1667093, EPI_ISL_1667094, EPI_ISL_1667095, EPI_ISL_1667096, EPI_ISL_1667097, EPI_ISL_1667098, EPI_ISL_1667099, EPI_ISL_1667100, EPI_ISL_1667101, EPI_ISL_1667102, EPI_ISL_1667104, EPI_ISL_1667108, EPI_ISL_1667109, EPI_ISL_1667110, EPI_ISL_1667111, EPI_ISL_1667112, EPI_ISL_1667113,

| | | | |
|---|---|---|---|
| EPI_ISL_1667114, EPI_ISL_1667115, EPI_ISL_1667116, EPI_ISL_1667117, EPI_ISL_1667118, EPI_ISL_1667119, EPI_ISL_1667120, EPI_ISL_1667121, EPI_ISL_1667122, EPI_ISL_1667123, EPI_ISL_1667124, EPI_ISL_1667125, EPI_ISL_1667126, EPI_ISL_1667127, EPI_ISL_1667128, EPI_ISL_1667129, EPI_ISL_1667130, EPI_ISL_1667131, EPI_ISL_1667132, EPI_ISL_1667133, EPI_ISL_1667134, EPI_ISL_1667135, EPI_ISL_1667136, EPI_ISL_1667137, EPI_ISL_1667138, EPI_ISL_1667139, EPI_ISL_1667140, EPI_ISL_1667141, EPI_ISL_1667142, EPI_ISL_1667143, EPI_ISL_1667144, EPI_ISL_1667145, EPI_ISL_1667146, EPI_ISL_1667147, EPI_ISL_1667148, EPI_ISL_1667149, EPI_ISL_1667150, EPI_ISL_1667151, EPI_ISL_1667152, EPI_ISL_1667153, EPI_ISL_1667154, EPI_ISL_1667155, EPI_ISL_1667156, EPI_ISL_1667157, EPI_ISL_1667158, EPI_ISL_1667159, EPI_ISL_1667160, EPI_ISL_1667161, EPI_ISL_1667162, EPI_ISL_1667163, EPI_ISL_1667164, EPI_ISL_1667165, EPI_ISL_1667166, EPI_ISL_1667167, EPI_ISL_1667168, EPI_ISL_1667169, EPI_ISL_1667170, EPI_ISL_1667171, EPI_ISL_1667172, EPI_ISL_1667174, EPI_ISL_1667175, EPI_ISL_1667176, EPI_ISL_1667177, EPI_ISL_1667178, EPI_ISL_1667179, EPI_ISL_1667180, EPI_ISL_1667181, EPI_ISL_1667182, EPI_ISL_1667183, EPI_ISL_1667184, EPI_ISL_1667185, EPI_ISL_1667186, EPI_ISL_1667187, EPI_ISL_1667188, EPI_ISL_1667189, EPI_ISL_1667190, EPI_ISL_1667191, EPI_ISL_1667192, EPI_ISL_1667193, EPI_ISL_1667194, EPI_ISL_1667195, EPI_ISL_1667196, EPI_ISL_1667197, EPI_ISL_1667198, EPI_ISL_1667199, EPI_ISL_1667200, EPI_ISL_1667202, EPI_ISL_1667203, EPI_ISL_1667204, EPI_ISL_1667205, EPI_ISL_1667206, EPI_ISL_1667207, EPI_ISL_1667208, EPI_ISL_1667209, EPI_ISL_1667210, EPI_ISL_1667211, EPI_ISL_1667212, EPI_ISL_1667213, EPI_ISL_1667214, EPI_ISL_1667215, EPI_ISL_1667216, EPI_ISL_1667217, EPI_ISL_1667218, EPI_ISL_1667219, EPI_ISL_1667220, EPI_ISL_1667221, EPI_ISL_1667222, EPI_ISL_1667223, EPI_ISL_1667224, EPI_ISL_1667225, EPI_ISL_1667226, EPI_ISL_1667227, EPI_ISL_1667228, EPI_ISL_1667229, EPI_ISL_1667230, EPI_ISL_1667231, EPI_ISL_1667232, EPI_ISL_1667233, EPI_ISL_1667234, EPI_ISL_1667235, EPI_ISL_1667236, EPI_ISL_1667237, EPI_ISL_1667238, EPI_ISL_1667239, EPI_ISL_1667240, EPI_ISL_1667241, EPI_ISL_1667242, EPI_ISL_1667243, EPI_ISL_1667244, EPI_ISL_1667245, EPI_ISL_1667246, EPI_ISL_1667247, EPI_ISL_1667248, EPI_ISL_1667249, EPI_ISL_1667250, EPI_ISL_1667251, EPI_ISL_1667252, EPI_ISL_1667253, EPI_ISL_1667254, EPI_ISL_1667255, EPI_ISL_1667256, EPI_ISL_1667257, EPI_ISL_1667258, EPI_ISL_1667259, EPI_ISL_1667260, EPI_ISL_1667261, EPI_ISL_1667262, EPI_ISL_1667263, EPI_ISL_1667264, EPI_ISL_1667265, EPI_ISL_1667266, EPI_ISL_1667267, EPI_ISL_1667268, EPI_ISL_1667269, EPI_ISL_1667270, EPI_ISL_1667271, EPI_ISL_1667272, EPI_ISL_1667273, EPI_ISL_1667274, EPI_ISL_1667275, EPI_ISL_1667276, EPI_ISL_1667277, EPI_ISL_1667278, EPI_ISL_1667279, EPI_ISL_1667280, EPI_ISL_1667281, EPI_ISL_1667282, EPI_ISL_1667283, EPI_ISL_1667284, EPI_ISL_1667285, EPI_ISL_1667286, EPI_ISL_1667287, EPI_ISL_1667288, EPI_ISL_1667289, EPI_ISL_1667290, EPI_ISL_1667291, EPI_ISL_1667292, EPI_ISL_1667293, EPI_ISL_1667294, EPI_ISL_1667295, EPI_ISL_1667296, EPI_ISL_1667297, EPI_ISL_1667298, EPI_ISL_1667299, EPI_ISL_1667300, EPI_ISL_1667302, EPI_ISL_1667303, EPI_ISL_1667304, EPI_ISL_1667305, EPI_ISL_1667306, EPI_ISL_1667307, EPI_ISL_1667308, EPI_ISL_1667309, EPI_ISL_1667310, EPI_ISL_1667311, EPI_ISL_1667312, EPI_ISL_1667313, EPI_ISL_1667314, EPI_ISL_1667315, EPI_ISL_1667316, EPI_ISL_1667317, EPI_ISL_1667318, EPI_ISL_1667319, EPI_ISL_1667320, EPI_ISL_1667321, EPI_ISL_1667322, EPI_ISL_1667323, EPI_ISL_1667324, EPI_ISL_1667325, EPI_ISL_1667326, EPI_ISL_1667327, EPI_ISL_1667328, EPI_ISL_1667329, EPI_ISL_1667330, EPI_ISL_1667331, EPI_ISL_1667332, EPI_ISL_1667333, EPI_ISL_1667334, EPI_ISL_1667335, EPI_ISL_1667336, EPI_ISL_1667337, EPI_ISL_1667338, EPI_ISL_1667339, EPI_ISL_1667340, EPI_ISL_1667341, EPI_ISL_1667342, EPI_ISL_1667343, EPI_ISL_1667344, EPI_ISL_1667345, EPI_ISL_1667346, EPI_ISL_1667347, EPI_ISL_1667348, EPI_ISL_1667349, EPI_ISL_1667350, EPI_ISL_1667351, EPI_ISL_1667352, EPI_ISL_1667353, EPI_ISL_1667354, EPI_ISL_1667355, EPI_ISL_1667356, EPI_ISL_1667357, EPI_ISL_1667358, EPI_ISL_1667359, EPI_ISL_1667360, EPI_ISL_1667361, EPI_ISL_1667362, EPI_ISL_1667363, EPI_ISL_1667364, EPI_ISL_1667365, EPI_ISL_1667366, EPI_ISL_1667367, EPI_ISL_1667368, EPI_ISL_1667369, EPI_ISL_1667370, EPI_ISL_1667371, EPI_ISL_1667372, EPI_ISL_1667373, EPI_ISL_1667374, EPI_ISL_1667375, EPI_ISL_1667376, EPI_ISL_1667377, EPI_ISL_1667378, EPI_ISL_1667379, EPI_ISL_1667380, EPI_ISL_1667381, EPI_ISL_1694974, EPI_ISL_1695033, EPI_ISL_1695036, EPI_ISL_1733806, EPI_ISL_1733839, EPI_ISL_1733840, EPI_ISL_1733841, EPI_ISL_1733842, EPI_ISL_1733843, EPI_ISL_1733844, EPI_ISL_1733847, EPI_ISL_1733848, EPI_ISL_1733849, EPI_ISL_1733850, EPI_ISL_1733851, EPI_ISL_1733852, EPI_ISL_1733853, EPI_ISL_1733854, EPI_ISL_1733855, EPI_ISL_1733856, EPI_ISL_1733857, EPI_ISL_1733858, EPI_ISL_1733859, EPI_ISL_1733860, EPI_ISL_1733861, EPI_ISL_1733862, EPI_ISL_1733863, EPI_ISL_1733864, EPI_ISL_1733865, EPI_ISL_1733866, EPI_ISL_1733867, EPI_ISL_1733868, EPI_ISL_1733869, EPI_ISL_1733870, EPI_ISL_1733874, EPI_ISL_1733875, EPI_ISL_1733876, EPI_ISL_1733877, EPI_ISL_1733878, EPI_ISL_1733879, EPI_ISL_1733880, EPI_ISL_1733881, EPI_ISL_1733882, EPI_ISL_1733883, EPI_ISL_1733884, EPI_ISL_1733885, EPI_ISL_1733887, EPI_ISL_1733888, EPI_ISL_1733889, EPI_ISL_1733890, EPI_ISL_1733891, EPI_ISL_1733892, EPI_ISL_1733893, EPI_ISL_1733894, EPI_ISL_1733895, EPI_ISL_1733896, EPI_ISL_1733897, EPI_ISL_1733898, EPI_ISL_1733899, EPI_ISL_1733900, EPI_ISL_1733901, EPI_ISL_1733902, EPI_ISL_1733903, EPI_ISL_1733906, EPI_ISL_1733909, EPI_ISL_1733910, EPI_ISL_1733912, EPI_ISL_1733916, EPI_ISL_1733922, EPI_ISL_1733928, EPI_ISL_1733939, EPI_ISL_1733942, EPI_ISL_1733943, EPI_ISL_1733944, EPI_ISL_1733945, EPI_ISL_1733946, EPI_ISL_1733949, EPI_ISL_1733951, EPI_ISL_1733954, EPI_ISL_1733955, EPI_ISL_1733957, EPI_ISL_1733962, EPI_ISL_1733968, EPI_ISL_1733969, EPI_ISL_1733970, EPI_ISL_1734413, EPI_ISL_2037981, EPI_ISL_2038260, EPI_ISL_2038261, EPI_ISL_2038262, EPI_ISL_2038263, EPI_ISL_2038266, EPI_ISL_2038267, EPI_ISL_2038268, EPI_ISL_2038269, EPI_ISL_2038270, EPI_ISL_2038271, EPI_ISL_2038272, EPI_ISL_2038273, EPI_ISL_2038274, EPI_ISL_2038275, EPI_ISL_2038276, EPI_ISL_2038277, EPI_ISL_2038278, EPI_ISL_2038279, EPI_ISL_2038280, EPI_ISL_2038281, EPI_ISL_2038282, EPI_ISL_2038283, EPI_ISL_2038284, EPI_ISL_2038285, EPI_ISL_2038286, EPI_ISL_2038287, EPI_ISL_2038288, EPI_ISL_2038289, EPI_ISL_2038290, EPI_ISL_2038291, EPI_ISL_2038292, EPI_ISL_2038293, EPI_ISL_2038294, EPI_ISL_2038295, EPI_ISL_2038296, EPI_ISL_2038297, EPI_ISL_2038298, EPI_ISL_2038299, EPI_ISL_2038300, EPI_ISL_2038301, EPI_ISL_2038303, EPI_ISL_2038304, EPI_ISL_2038305, EPI_ISL_2038306, EPI_ISL_2038307, EPI_ISL_2038309, EPI_ISL_2038310, EPI_ISL_2038311, EPI_ISL_2038312, EPI_ISL_2038313, EPI_ISL_2038314, EPI_ISL_2038315, EPI_ISL_2038316, EPI_ISL_2038317, EPI_ISL_2038318, EPI_ISL_2038319, EPI_ISL_2038320, EPI_ISL_2038321, EPI_ISL_2038322, EPI_ISL_2038323, EPI_ISL_2038324, EPI_ISL_2038325, EPI_ISL_2038326, EPI_ISL_2038327, EPI_ISL_2038328, EPI_ISL_2038329, EPI_ISL_2038515, EPI_ISL_2038548, EPI_ISL_2038627 | | | |
| see above | Fulgent Genetics | Centers for Disease Control and Prevention Division of Viral Diseases, Pathogen Discovery | Adrian Paskey; Becky Tsai; Benafsh Sapra; Benjamin Rambo-Martin; Christopher Gulvick; Clinton R. Paden; Dakota Howard; Darlene Wagner; Dhwani Batra; Doreen Ng; Duncan MacCannell; Harry Gao; James Xie; Jason Caravas; John Gao; Joseph Fierro; Kara Moser; Matthew Schmerer; Mickey Li; Peter W. Cook; Scott Sammons; Shatavia Morrison; Yan Meng; Yvette Unoarumhi |
| EPI_ISL_1707680, EPI_ISL_1707681, EPI_ISL_1707682, EPI_ISL_1707683, EPI_ISL_1707684, EPI_ISL_1707685 | Fulgent Genetics | Los Angeles County PHL | P. Hemarajata et al. |
| EPI_ISL_1664267 | Good Samaritan Hospital | Santa Clara County public Health Laboratory | Santa Clara County Public Health Department |
| EPI_ISL_1575872, EPI_ISL_1576607, EPI_ISL_1576614, EPI_ISL_1576619, EPI_ISL_1576629, EPI_ISL_1576636, EPI_ISL_1576641, EPI_ISL_1576657, EPI_ISL_1576669, EPI_ISL_1576678, EPI_ISL_1576681, EPI_ISL_1576683, EPI_ISL_1576689, EPI_ISL_1576692, EPI_ISL_1576701, EPI_ISL_1576704, EPI_ISL_1576713, EPI_ISL_1576721, EPI_ISL_1576723, EPI_ISL_1576734, EPI_ISL_1576735, EPI_ISL_1576744, EPI_ISL_1576746, EPI_ISL_1576747, EPI_ISL_1576754, EPI_ISL_1576756, EPI_ISL_1576757, EPI_ISL_1580871, EPI_ISL_1580873, EPI_ISL_1580880, EPI_ISL_1580885, EPI_ISL_1580887, EPI_ISL_1580895, EPI_ISL_1580904, EPI_ISL_1580907, EPI_ISL_1580908, EPI_ISL_1580919, EPI_ISL_1580925, EPI_ISL_1580935, EPI_ISL_1580949, EPI_ISL_1581240, EPI_ISL_1581321, EPI_ISL_1581334, EPI_ISL_1581357, EPI_ISL_1581366, EPI_ISL_1581368, EPI_ISL_1581369, EPI_ISL_1581378, EPI_ISL_1581379, EPI_ISL_1581402, EPI_ISL_1581561, EPI_ISL_1581817, EPI_ISL_1592405, EPI_ISL_1592408, EPI_ISL_1592409, EPI_ISL_1592413, EPI_ISL_1592415, EPI_ISL_1592422, EPI_ISL_1592423, EPI_ISL_1592424, EPI_ISL_1592432, EPI_ISL_1592435, EPI_ISL_1592436, EPI_ISL_1592440, EPI_ISL_1592441, EPI_ISL_1592448, EPI_ISL_1592461, EPI_ISL_1592463, EPI_ISL_1592471, EPI_ISL_1592477, EPI_ISL_1592479, EPI_ISL_1592483, EPI_ISL_1592484, EPI_ISL_1592487, EPI_ISL_1592489, EPI_ISL_1592496, EPI_ISL_1592497, EPI_ISL_1592573, EPI_ISL_1592593, EPI_ISL_1592594, EPI_ISL_1592595, EPI_ISL_1592596, EPI_ISL_1592597, EPI_ISL_1592604, EPI_ISL_1592605, EPI_ISL_1592606, EPI_ISL_1592608, EPI_ISL_1592609, EPI_ISL_1592614, EPI_ISL_1592615, EPI_ISL_1592616, EPI_ISL_1592617, EPI_ISL_1592618, EPI_ISL_1592619, EPI_ISL_1592620, EPI_ISL_1592626, EPI_ISL_1592627, EPI_ISL_1592628, EPI_ISL_1592629, EPI_ISL_1592630, EPI_ISL_1592631, EPI_ISL_1592638, EPI_ISL_1592639, EPI_ISL_1592642, EPI_ISL_1592643, EPI_ISL_1592644, EPI_ISL_1592650, EPI_ISL_1592652, EPI_ISL_1592653, EPI_ISL_1592654, EPI_ISL_1592662, EPI_ISL_1592664, EPI_ISL_1592665, EPI_ISL_1592667, EPI_ISL_1592674, EPI_ISL_1592677, EPI_ISL_1592678, EPI_ISL_1592896, EPI_ISL_1592940, EPI_ISL_1593021, EPI_ISL_1593054, EPI_ISL_1593063, EPI_ISL_1593074, EPI_ISL_1593162, EPI_ISL_1593166, EPI_ISL_1593182, EPI_ISL_1593197, EPI_ISL_1615101, EPI_ISL_1615136, EPI_ISL_1615233, EPI_ISL_1615234, EPI_ISL_1615240, EPI_ISL_1615241, EPI_ISL_1615295, EPI_ISL_1615322, EPI_ISL_1615323, EPI_ISL_1615370, EPI_ISL_1615372, EPI_ISL_1615393, EPI_ISL_1615409, EPI_ISL_1615430, EPI_ISL_1615436, EPI_ISL_1615441, EPI_ISL_1615450, EPI_ISL_1615478, EPI_ISL_1615531, EPI_ISL_1615537, EPI_ISL_1615555, EPI_ISL_1678894, EPI_ISL_1678901, EPI_ISL_1678928, EPI_ISL_1678936, EPI_ISL_1678974, EPI_ISL_1678983, EPI_ISL_1678984, EPI_ISL_1679020, EPI_ISL_1679021, EPI_ISL_1679159, EPI_ISL_1679214, EPI_ISL_1679218, EPI_ISL_1679220, EPI_ISL_1679236, EPI_ISL_1679239, EPI_ISL_1679247, EPI_ISL_1679249, EPI_ISL_1679258, EPI_ISL_1679267, EPI_ISL_1679288, EPI_ISL_1679301, EPI_ISL_1679303, EPI_ISL_1679309, EPI_ISL_1679363, EPI_ISL_1679370, EPI_ISL_1679384, EPI_ISL_1679399, EPI_ISL_1679450, EPI_ISL_1679451, EPI_ISL_1679472, EPI_ISL_1679523, EPI_ISL_1679529, EPI_ISL_1679613, EPI_ISL_1679616, EPI_ISL_1679677, EPI_ISL_1679702, EPI_ISL_1679713, EPI_ISL_1679720, EPI_ISL_1679738, EPI_ISL_1679771, EPI_ISL_1679775, EPI_ISL_1679776, EPI_ISL_1679781, EPI_ISL_1679791, EPI_ISL_1679792, EPI_ISL_1679841, EPI_ISL_1679862, EPI_ISL_1679878, EPI_ISL_1679900, EPI_ISL_1679908, EPI_ISL_1679911, EPI_ISL_1679915, EPI_ISL_1679922, EPI_ISL_1680073, EPI_ISL_1680144, EPI_ISL_1680178, EPI_ISL_1690283, EPI_ISL_1690290, EPI_ISL_1690295, EPI_ISL_1690301, EPI_ISL_1690311, EPI_ISL_1690312, EPI_ISL_1690330, EPI_ISL_1690342, EPI_ISL_1690435, EPI_ISL_1690452, EPI_ISL_1690457, EPI_ISL_1690459, EPI_ISL_1690498, EPI_ISL_1690584, EPI_ISL_1690600, EPI_ISL_1690617, EPI_ISL_1690649, EPI_ISL_1690708, EPI_ISL_1690772, EPI_ISL_1690817, EPI_ISL_1690833, EPI_ISL_1690899, EPI_ISL_1690984, EPI_ISL_1691022, EPI_ISL_1691032, EPI_ISL_1691062, EPI_ISL_1691063, EPI_ISL_1691082, EPI_ISL_1691133, EPI_ISL_1691155, EPI_ISL_1691162, EPI_ISL_1691163, EPI_ISL_1691190, EPI_ISL_1691192, EPI_ISL_1691199, EPI_ISL_1691201, EPI_ISL_1691216, EPI_ISL_1691218, EPI_ISL_1691222, EPI_ISL_1691310, EPI_ISL_1691312, EPI_ISL_1691340, EPI_ISL_1691343, EPI_ISL_1695048, EPI_ISL_1701848, EPI_ISL_1701852, EPI_ISL_1701854, EPI_ISL_1702158, EPI_ISL_1734681, EPI_ISL_1734703, EPI_ISL_1734784, EPI_ISL_1734798, EPI_ISL_1734832, EPI_ISL_1734848, EPI_ISL_1734876, EPI_ISL_1734894, EPI_ISL_1734905, EPI_ISL_1734906, EPI_ISL_1734911, EPI_ISL_1734915, EPI_ISL_1734935, EPI_ISL_1734962, EPI_ISL_1734980, EPI_ISL_1734992, EPI_ISL_1735013, EPI_ISL_1735015, EPI_ISL_1735018, EPI_ISL_1735174, EPI_ISL_1735217, EPI_ISL_1735227, EPI_ISL_1735228, EPI_ISL_1735238, EPI_ISL_1735291, EPI_ISL_1735300, EPI_ISL_1735323, EPI_ISL_1735416, EPI_ISL_1735427, EPI_ISL_1735429, EPI_ISL_1735438, EPI_ISL_1735604, EPI_ISL_1735621, EPI_ISL_1735631, EPI_ISL_1735674, EPI_ISL_1753503, EPI_ISL_1753510, EPI_ISL_1796139, EPI_ISL_1796218, EPI_ISL_1796229, EPI_ISL_1796231, EPI_ISL_1796237, EPI_ISL_1796241, EPI_ISL_1796250, EPI_ISL_1796251, EPI_ISL_1796259, EPI_ISL_1796275, EPI_ISL_1796285, EPI_ISL_1796341, EPI_ISL_1796428, EPI_ISL_1796435, EPI_ISL_1796454, EPI_ISL_1796474, EPI_ISL_1796487, EPI_ISL_1796500, EPI_ISL_1796595, EPI_ISL_1796613, EPI_ISL_1796621, EPI_ISL_1796630, EPI_ISL_1796639, EPI_ISL_1796641, EPI_ISL_1796708, EPI_ISL_1796718, EPI_ISL_1796853, EPI_ISL_1796869, EPI_ISL_1796882, EPI_ISL_1796894, EPI_ISL_1796897, EPI_ISL_1796911, EPI_ISL_1796922, EPI_ISL_1796977, EPI_ISL_1796992, EPI_ISL_1796993, EPI_ISL_1797102, EPI_ISL_1797124, EPI_ISL_1797134, EPI_ISL_1797265, EPI_ISL_1797276, EPI_ISL_1797277, EPI_ISL_1797299, EPI_ISL_1797335, EPI_ISL_1797371, EPI_ISL_1797385, EPI_ISL_1797392, EPI_ISL_1797395, EPI_ISL_1797416, EPI_ISL_1797425, EPI_ISL_1803526, EPI_ISL_1803535, EPI_ISL_1803554, EPI_ISL_1803663, EPI_ISL_1803669, EPI_ISL_1803673, EPI_ISL_1803674, EPI_ISL_1803676, EPI_ISL_1803694, EPI_ISL_1803697, EPI_ISL_1803704, EPI_ISL_1803709, EPI_ISL_1803722, EPI_ISL_1803729, EPI_ISL_1803815, EPI_ISL_1803820, EPI_ISL_1803964, EPI_ISL_1803987, EPI_ISL_1803990, EPI_ISL_1803997, EPI_ISL_1804000, EPI_ISL_1804083, EPI_ISL_1804094, EPI_ISL_1804095, EPI_ISL_1804103, EPI_ISL_1804108, EPI_ISL_1804122, EPI_ISL_1804135, EPI_ISL_1804143, EPI_ISL_1804169, EPI_ISL_1804179, EPI_ISL_1804191, EPI_ISL_1804202, EPI_ISL_1804213, EPI_ISL_1804256, EPI_ISL_1804518, EPI_ISL_1804524, EPI_ISL_1804534, EPI_ISL_1804535, EPI_ISL_1804554, EPI_ISL_1804564, EPI_ISL_1804568, EPI_ISL_1804583, EPI_ISL_1804589, EPI_ISL_1804601, EPI_ISL_1804611, EPI_ISL_1804625, EPI_ISL_1804643, EPI_ISL_1804676, EPI_ISL_1804681, EPI_ISL_1804702, EPI_ISL_1804713, EPI_ISL_1804722, EPI_ISL_1804729, EPI_ISL_1818008, EPI_ISL_1818009, EPI_ISL_1818028, EPI_ISL_1906970, EPI_ISL_1907014, EPI_ISL_1907023, EPI_ISL_1907031, EPI_ISL_1907068, EPI_ISL_1907098, EPI_ISL_1907130, EPI_ISL_1907133, EPI_ISL_1907464, EPI_ISL_1907469, EPI_ISL_1907475, EPI_ISL_1907480, EPI_ISL_1907492, EPI_ISL_1907493, EPI_ISL_1907513, EPI_ISL_1907521, EPI_ISL_1907531, EPI_ISL_1907544, EPI_ISL_1907546, EPI_ISL_1907679, EPI_ISL_1907692, EPI_ISL_1907702, EPI_ISL_1907745, EPI_ISL_1907797, EPI_ISL_1907860, EPI_ISL_1908068, EPI_ISL_1908103, EPI_ISL_1908114, EPI_ISL_1908138, EPI_ISL_1923926, EPI_ISL_1923936 | | | |
| see above | Helix/Illumina | Centers for Disease Control and Prevention Division of Viral Diseases, Pathogen Discovery | Adrian Paskey; Alexandre Bolze; Ary Ascencio; Benjamin Rambo-Martin; Brad Sickler; Charlotte Rivera-Garcia; Christine Tran; Christopher Gulvick; Clinton R. Paden; Dakota Howard; Darlene Wagner; David Becker; Dhwani Batra; Duncan MacCannell; Efren Sandoval; Eileen de Feo; Elizabeth Cirulli; Eric Allen; Geraint Levan; James Lu; Jan Antico; Jason Caravas; Jason Nguyen; Jimmy Ramirez; Jingtao Liu; Kara Moser; Kelly Schiabor Barrett; Kim Gietzen; Magnus Isaksson; Marc Laurent; Matthew Schmerer; Matthew Tolentino; Nicole L. Washington; Peter W. Cook; Phil Febbo; Ryan Cho; Scott Sammons; Shannon Wickline; Shatavia Morrison; Sherry Wang; Simon White; Tyler Cassens; William Lee; Yvette Unoarumhi |
| EPI_ISL_1701411, EPI_ISL_1701444, EPI_ISL_1701447, EPI_ISL_1701479, EPI_ISL_1701493, EPI_ISL_1701497, EPI_ISL_1701555, EPI_ISL_1701560, EPI_ISL_1701587, EPI_ISL_1701594, EPI_ISL_1701601, EPI_ISL_1701605, EPI_ISL_1701614, EPI_ISL_2102528, EPI_ISL_2102533, EPI_ISL_2102537, EPI_ISL_2102550, EPI_ISL_2102561, EPI_ISL_2102585, EPI_ISL_2102752, EPI_ISL_2102753, EPI_ISL_2102791, EPI_ISL_2102797, EPI_ISL_2102806, EPI_ISL_2102808, EPI_ISL_2102832, EPI_ISL_2102838, EPI_ISL_2102847, EPI_ISL_2102872, EPI_ISL_2102888, EPI_ISL_2102903, EPI_ISL_2102964 | | | |
| see above | Humboldt County Public Health Laboratory | Chan-Zuckerberg Biohub | CZB Cliahub Consortium |
| EPI_ISL_1512777, EPI_ISL_1512805, EPI_ISL_1512823, EPI_ISL_1512827, EPI_ISL_1512844, EPI_ISL_1512880, EPI_ISL_1512898, EPI_ISL_1513116, EPI_ISL_1513119, EPI_ISL_1513120, EPI_ISL_1513127, EPI_ISL_1513134, EPI_ISL_1513137, EPI_ISL_1513138, EPI_ISL_1587592, EPI_ISL_1587898, EPI_ISL_1587916, EPI_ISL_1587932, EPI_ISL_1587948, EPI_ISL_1588416, EPI_ISL_1692123, EPI_ISL_1693196, EPI_ISL_1693463, EPI_ISL_1693826, EPI_ISL_1694297, EPI_ISL_1694349, EPI_ISL_1694535, EPI_ISL_1795476, EPI_ISL_1795480, EPI_ISL_1795494, EPI_ISL_1795606, EPI_ISL_1802878, EPI_ISL_1818093, EPI_ISL_2013193, EPI_ISL_2013277, EPI_ISL_2013332, EPI_ISL_2014190 | | | |
| see above | Infinity Biologix | Centers for Disease Control and Prevention Division of Viral Diseases, Pathogen Discovery | Adrian Paskey; Benjamin Rambo-Martin; Chirayu Goswami; Christian Bixby; Christopher Gulvick; Clinton R. Paden; Dakota Howard; Darlene Wagner; Dhwani Batra; Duncan MacCannell; Jason Caravas; Jonathan Schultz; Kara Moser; Matthew Schmerer; Peter W. Cook; Robin Grimwood; Russ Hager; Scott Sammons; Shatavia Morrison; Yihe Wang; Yvette Unoarumhi |
| EPI_ISL_1712523, EPI_ISL_1712524, EPI_ISL_1712525, EPI_ISL_1712526, EPI_ISL_1712527, EPI_ISL_1712528, EPI_ISL_1712529, EPI_ISL_1712530, EPI_ISL_1712531, EPI_ISL_1712532, EPI_ISL_1712533, EPI_ISL_1712534, EPI_ISL_1712535, EPI_ISL_1712536, EPI_ISL_1712537, EPI_ISL_1712538, EPI_ISL_1712539, EPI_ISL_1712540, EPI_ISL_1712541, EPI_ISL_1712550, EPI_ISL_1712551, EPI_ISL_1712552, EPI_ISL_1712553, EPI_ISL_1712554, EPI_ISL_1712555, EPI_ISL_1712556, EPI_ISL_1712557, EPI_ISL_1712558, EPI_ISL_1712559, EPI_ISL_1712560, EPI_ISL_1712561, EPI_ISL_1712562 | | | |
| see above | Innovative Genomics Institute, UC Berkeley | Innovative Genomics Institute, UC Berkeley | Holly Gildea; John Boyle; Phil Frankino; Stacia Wyman |
| EPI_ISL_1913217, EPI_ISL_1913225 | KAISER SAN JOSE | Santa Clara County public Health Laboratory | Santa Clara County Public Health Department |
| EPI_ISL_1508968, EPI_ISL_1508974 | Kaiser Santa Clara | Santa Clara County Public Health Laboratory | Santa Clara County Public Health Department |
| EPI_ISL_1664268, EPI_ISL_1664269 | Kaiser Santa Clara | Santa Clara County public Health Laboratory | Santa Clara County Public Health Department |
| EPI_ISL_1700838, EPI_ISL_1754141, EPI_ISL_1805493, EPI_ISL_1821201, EPI_ISL_1821202, EPI_ISL_1821203, EPI_ISL_1857271, EPI_ISL_2139908, EPI_ISL_2139909 | | | |
| see above | LESP Baja California | Instituto de Diagnostico y Referencia Epidemiologicos (INDRE) | Abril Rodriguez-Maldonado; Ariadna Medina-Benitez; Claudia Wong-Arambula; Ernesto Ramirez-Gonzalez.; Gisela Barrera-Badillo; Irma Lopez-Martinez; Joaquin Quiroz-Mercado; Lucia Hernandez-Rivas; Natividad Cruz-Ortiz; Sergio Rangel-Guerrero; Tatiana Nunez-Garcia; Vanessa Rivero-Arredondo |
| EPI_ISL_1805487, EPI_ISL_1821104, EPI_ISL_1821105, EPI_ISL_1821180, EPI_ISL_1821181, EPI_ISL_1821182, EPI_ISL_1821183, EPI_ISL_1821184, EPI_ISL_1857224, EPI_ISL_1857225, EPI_ISL_1857226, EPI_ISL_1857227, EPI_ISL_1857235, EPI_ISL_1857266, EPI_ISL_1857267 | | | |
| see above | LESP Baja California Sur | Instituto de Diagnostico y Referencia Epidemiologicos (INDRE) | Abril Rodriguez-Maldonado; Ariadna Medina-Benitez; Claudia Wong-Arambula; Ernesto Ramirez-Gonzalez.; Gisela Barrera-Badillo; Irma Lopez-Martinez; Joaquin Quiroz-Mercado; Lucia Hernandez-Rivas; Natividad Cruz-Ortiz; Sergio Rangel-Guerrero; Tatiana Nunez-Garcia; Vanessa Rivero-Arredondo |
| EPI_ISL_1811524, EPI_ISL_1811531 | Laboratorio Central de Epidemiología (LCE) | Instituto de Biotecnología de la UNAM | Alejandro Sanchez-Flores; Alfredo Herrera-Estrella; Alicia Ocaña-Mondragón; Angel Gustavo Salas-Lais; Bernardo Martínez-Miguel; Blanca Taboada; Brenda Irasema Maldonado-Meza; Carla Ivón Herrera-Najera; Carlos F. Arias; Celia Boukadida; Clara Esperanza Santacruz-Tinoco; Concepción Grajales-Muñiz; Célida Duque-Molina; Fernando Fontove-Herrera; Francisco Pulido; Gloria Elena Espinosa-Ayala; Gloria María Molina-Salinas; Gloria Vazquez; Hector Esteban Paz-Juárez; Hector Montoya-Fuentes; Helen Haydee Fernanda Ramírez-Plascencia; José Antonio Enciso-Moreno; José Esteban Muñoz-Medina; José de Jesús Nuñez-Contreras; Juan Bautista Chale-Dzul; Julio Elias Alvarado-Yaah; Luis Alberto Ochoa-Carrera; Margarita Matías-Florentino; María Guadalupe Santiago-Mauricio; María Guadalupe de Jesús Mireles-Rivera; Nelly Sélem-Mojica; Pavel Isa; Ricardo Grande; Santiago Ávila-Ríos; Victor Eduardo Garcia-Arias; Víctor Hugo Borja-Aburto |
| EPI_ISL_1515636, EPI_ISL_1515637, EPI_ISL_1515638, EPI_ISL_1515640, EPI_ISL_1515744, EPI_ISL_1550248, EPI_ISL_1550361, EPI_ISL_1612132, EPI_ISL_1612231, EPI_ISL_1612243, EPI_ISL_1612244, EPI_ISL_1612245, EPI_ISL_1612259, EPI_ISL_1612263, EPI_ISL_1612266, EPI_ISL_1612394, EPI_ISL_1612395, EPI_ISL_1612396, EPI_ISL_1612397, EPI_ISL_1612398, EPI_ISL_1612399, EPI_ISL_1612400, EPI_ISL_1612401, EPI_ISL_1612406, EPI_ISL_1612441, EPI_ISL_1612442, EPI_ISL_1612443, EPI_ISL_1612444, EPI_ISL_1612445, EPI_ISL_1612446, EPI_ISL_1612447, EPI_ISL_1612448, EPI_ISL_1612449, EPI_ISL_1612450, EPI_ISL_1612451, EPI_ISL_1612452, EPI_ISL_1612453, EPI_ISL_1612454, EPI_ISL_1612455, EPI_ISL_1612456, EPI_ISL_1612457, EPI_ISL_1612458, EPI_ISL_1612459, EPI_ISL_1612510, EPI_ISL_1612616, EPI_ISL_1612617, EPI_ISL_1612618, EPI_ISL_1612619, EPI_ISL_1612620, EPI_ISL_1612621, EPI_ISL_1612622, EPI_ISL_1612623, EPI_ISL_1612624, EPI_ISL_1612625, EPI_ISL_1612626, EPI_ISL_1612627, EPI_ISL_1612628, EPI_ISL_1612629, EPI_ISL_1612671, EPI_ISL_1612672, EPI_ISL_1612673, EPI_ISL_1612677, EPI_ISL_1612678, EPI_ISL_1612679, EPI_ISL_1612680, EPI_ISL_1612681, EPI_ISL_1612682, EPI_ISL_1612683, EPI_ISL_1612684, EPI_ISL_1612685, EPI_ISL_1612686, EPI_ISL_1612687, EPI_ISL_1612688, EPI_ISL_1612689, EPI_ISL_1612690, EPI_ISL_1612691, EPI_ISL_1612692, EPI_ISL_1612694, EPI_ISL_1612695, EPI_ISL_1612696, EPI_ISL_1612697, EPI_ISL_1612698, EPI_ISL_1612699, EPI_ISL_1612700, EPI_ISL_1612701, EPI_ISL_1612702, EPI_ISL_1612703, EPI_ISL_1612704, EPI_ISL_1612705, EPI_ISL_1612706, EPI_ISL_1612707, EPI_ISL_1612708, EPI_ISL_1612709, EPI_ISL_1612710, EPI_ISL_1612711, EPI_ISL_1612712, EPI_ISL_1612713, EPI_ISL_1612714, EPI_ISL_1612715, EPI_ISL_1612716, EPI_ISL_1612717, EPI_ISL_1612718, EPI_ISL_1612719, EPI_ISL_1612720, EPI_ISL_1612721, EPI_ISL_1612722, EPI_ISL_1612723, EPI_ISL_1612724, EPI_ISL_1612728, EPI_ISL_1612729, EPI_ISL_1612731, EPI_ISL_1612732, EPI_ISL_1612733, EPI_ISL_1612734, EPI_ISL_1612735, EPI_ISL_1612736, EPI_ISL_1612737, EPI_ISL_1612738, EPI_ISL_1612739, EPI_ISL_1612740, EPI_ISL_1612741, EPI_ISL_1612742, EPI_ISL_1612743, EPI_ISL_1612744, EPI_ISL_1612745, EPI_ISL_1612746, EPI_ISL_1612747, EPI_ISL_1612748, EPI_ISL_1612749, EPI_ISL_1612750, EPI_ISL_1612751, EPI_ISL_1612752, EPI_ISL_1612753, EPI_ISL_1612759, EPI_ISL_1612760, EPI_ISL_1612769, EPI_ISL_1612770, EPI_ISL_1612771, EPI_ISL_1612772, EPI_ISL_1612773, EPI_ISL_1612781, EPI_ISL_1612782, EPI_ISL_1612783, EPI_ISL_1612784, EPI_ISL_1612785, EPI_ISL_1612786, EPI_ISL_1612794, EPI_ISL_1612795, EPI_ISL_1612796, EPI_ISL_1612797, EPI_ISL_1612798, EPI_ISL_1612799, EPI_ISL_1612800, EPI_ISL_1612801, EPI_ISL_1612802, EPI_ISL_1612803, EPI_ISL_1612804, EPI_ISL_1612805, EPI_ISL_1681623, EPI_ISL_1684343, EPI_ISL_1684344, EPI_ISL_1684345, EPI_ISL_1684346, EPI_ISL_1684347, EPI_ISL_1684348, EPI_ISL_1684354, EPI_ISL_1684358, EPI_ISL_1684359, EPI_ISL_1684360, EPI_ISL_1684364, EPI_ISL_1684365, EPI_ISL_1684366, EPI_ISL_1684367, EPI_ISL_1684368, EPI_ISL_1684369, EPI_ISL_1684370, EPI_ISL_1684371, EPI_ISL_1684372, EPI_ISL_1684373, EPI_ISL_1684374, EPI_ISL_1684375, EPI_ISL_1684376, EPI_ISL_1684377, EPI_ISL_1684378, EPI_ISL_1684379, EPI_ISL_1684380, EPI_ISL_1684382, EPI_ISL_1684383, EPI_ISL_1684384, EPI_ISL_1684401, EPI_ISL_1684402, EPI_ISL_1684403, EPI_ISL_1684404, EPI_ISL_1684405, EPI_ISL_1684406, EPI_ISL_1684407, EPI_ISL_1684408, EPI_ISL_1684409, EPI_ISL_1684410, EPI_ISL_1684411, EPI_ISL_1684412, EPI_ISL_1684413, EPI_ISL_1684414, EPI_ISL_1684415, EPI_ISL_1684454, EPI_ISL_1684455, EPI_ISL_1684456, EPI_ISL_1684457, EPI_ISL_1684458, EPI_ISL_1684459, EPI_ISL_1684460, EPI_ISL_1684461, EPI_ISL_1684462, EPI_ISL_1684500, EPI_ISL_1684632, EPI_ISL_1684635, EPI_ISL_1684758, EPI_ISL_1684786, EPI_ISL_1684833, EPI_ISL_1684834, EPI_ISL_1684836, EPI_ISL_1684848, EPI_ISL_1684853, EPI_ISL_1684854, EPI_ISL_1684855, EPI_ISL_1684883, EPI_ISL_1684894, EPI_ISL_1684895, EPI_ISL_1684896, EPI_ISL_1684897, EPI_ISL_1684898, EPI_ISL_1684971, EPI_ISL_1684997, EPI_ISL_1695023, EPI_ISL_1798209, EPI_ISL_1798210, EPI_ISL_1798211, EPI_ISL_1798258, EPI_ISL_1798259, EPI_ISL_1798260, EPI_ISL_1798292, EPI_ISL_1798297, EPI_ISL_1798298, EPI_ISL_1798299, EPI_ISL_1798300, EPI_ISL_1798301, EPI_ISL_1798302, EPI_ISL_1798308, EPI_ISL_1798309, EPI_ISL_1798310, EPI_ISL_1798313, EPI_ISL_1798317, EPI_ISL_1798318, EPI_ISL_1798321, EPI_ISL_1798322, EPI_ISL_1798326, EPI_ISL_1798328, EPI_ISL_1798330, EPI_ISL_1798435, EPI_ISL_1798436, EPI_ISL_1798438, EPI_ISL_1798440, EPI_ISL_1798471, EPI_ISL_1798474, EPI_ISL_1798475, EPI_ISL_1798477, EPI_ISL_1798479, EPI_ISL_1798493, EPI_ISL_1798495, EPI_ISL_1798529, EPI_ISL_1798531, EPI_ISL_1798537, EPI_ISL_1798539, EPI_ISL_1798541, EPI_ISL_1798544, EPI_ISL_1798546, EPI_ISL_1798548, EPI_ISL_1798550, EPI_ISL_1798552, EPI_ISL_1798556, EPI_ISL_1798558, EPI_ISL_1798560, EPI_ISL_1798562, EPI_ISL_1798564, EPI_ISL_1798567, EPI_ISL_1798569, EPI_ISL_1798570, EPI_ISL_1798572, EPI_ISL_1798574, EPI_ISL_1798587, EPI_ISL_1798588, EPI_ISL_1798590, EPI_ISL_1798592, EPI_ISL_1798594, EPI_ISL_1798596, EPI_ISL_1798598, EPI_ISL_1798601, EPI_ISL_1798602, EPI_ISL_1798629, EPI_ISL_1798865, EPI_ISL_1798897, EPI_ISL_1798949, EPI_ISL_1798950, EPI_ISL_1798951, EPI_ISL_1798952, EPI_ISL_1798953, EPI_ISL_1798954, EPI_ISL_1798959, | | | |

| | | | |
|---|---|---|---|
| EPI_ISL_1798960, EPI_ISL_1798976, EPI_ISL_1798978, EPI_ISL_1798979, EPI_ISL_1798980, EPI_ISL_1798981, EPI_ISL_1798982, EPI_ISL_1798983, EPI_ISL_1798984, EPI_ISL_1798985, EPI_ISL_1798986, EPI_ISL_1798987, EPI_ISL_1798988, EPI_ISL_1798989, EPI_ISL_1798990, EPI_ISL_1798991, EPI_ISL_1798992, EPI_ISL_1798993, EPI_ISL_1798994, EPI_ISL_1798995, EPI_ISL_1798996, EPI_ISL_1798997, EPI_ISL_1798998, EPI_ISL_1798999, EPI_ISL_1799000, EPI_ISL_1799001, EPI_ISL_1799002, EPI_ISL_1799003, EPI_ISL_1799004, EPI_ISL_1799006, EPI_ISL_1799007, EPI_ISL_1799008, EPI_ISL_1799009, EPI_ISL_1799142, EPI_ISL_1799143, EPI_ISL_1799153, EPI_ISL_1799167, EPI_ISL_1799168, EPI_ISL_1799169, EPI_ISL_1799259, EPI_ISL_1799277, EPI_ISL_1799290, EPI_ISL_1799291, EPI_ISL_1799292, EPI_ISL_1799335, EPI_ISL_1799336, EPI_ISL_1799337, EPI_ISL_1799338, EPI_ISL_1799399, EPI_ISL_1799400, EPI_ISL_1799401, EPI_ISL_1799425, EPI_ISL_1799426, EPI_ISL_1799427, EPI_ISL_1799428, EPI_ISL_1799447, EPI_ISL_1799448, EPI_ISL_1799449, EPI_ISL_1799454, EPI_ISL_1799455, EPI_ISL_1799456, EPI_ISL_1799457, EPI_ISL_1799471, EPI_ISL_1799472, EPI_ISL_1799473, EPI_ISL_1799474, EPI_ISL_1799484, EPI_ISL_1799485, EPI_ISL_1799551, EPI_ISL_1799552, EPI_ISL_1925654, EPI_ISL_1930643, EPI_ISL_1930646, EPI_ISL_1930650, EPI_ISL_1930653, EPI_ISL_1930656, EPI_ISL_1930659, EPI_ISL_1930674, EPI_ISL_1930675, EPI_ISL_1930676 | | | |
| see above | Laboratory Corporation of America | Centers for Disease Control and Prevention Division of Viral Diseases, Pathogen Discovery | Adrian Paskey; Amanda Douglas; Amanda Suchanek; Andrea Throop; Ayla Burns; Benjamin Rambo-Martin; Bobbi Croy; Brian Krueger; Brian Norvell; Christopher Gulvick; Christos Petropoulos; Clinton R. Paden; Craig Lukasik; Dakota Howard; Darlene Wagner; Debbie Boles; Dhwani Batra; Duncan MacCannell; Eyad Almasri; Goran Stevovic; Howard Engler; Hrushikesh Deshmukh; Jake Humphrey; Jana Schroth; Jason Caravas; Joe Voshell; John Pruitt; Jonathan Meltzer; Jonathan Williams; Kara Moser; Kimberly Wagner; Lax Iyer; Lyndon Tilson; Manoj Jain; Marcia Eisenberg; Mary Ann Cristobal; Mary Williamson; Matthew Schmerer; Michael Levandoski; Mike Sapeta; Mindy Nye; Minoo Agarwal; Mohan Kolli; Nuthawin Charoensri; Oren Cohen; Peter W. Cook; Prashant Gupta; Qian Zeng; Rama Ghatti; Scott Parker; Scott Ryan; Scott Sammons; Shatavia Morrison; Stanley Letovsky; Steven Ragan; Suresh Babu Selvaraju; Susan Countryman; Susan Hicks; Suzanne Dale; Thomas Urban; Tim Kuphal; Tricia Zwiefelhofer; Vincent Drouillon; Yvette Unoarumhi |
| EPI_ISL_1707686, EPI_ISL_1707687 | Los Angeles County PHL | Los Angeles County PHL | P. Hemarajata et al. |
| EPI_ISL_2009392 | Lurie Children's Hospital of Chicago | Northwestern University - Ozer Lab | Egon A. Ozer; Judd F. Hultquist; Lacy M. Simons; Larry K. Kociolek; Michael G. Ison; Ramon Lorenzo-Redondo; Taylor J. Dean; William J. Muller; Xiaotian; Zheng |
| EPI_ISL_1716896, EPI_ISL_1716899 | Martin Luther King Jr. Community Hospital | Los Angeles County PHL | P. Hemarajata et al. |
| EPI_ISL_1677791, EPI_ISL_1700688, EPI_ISL_1700689, EPI_ISL_1700690, EPI_ISL_1700691, EPI_ISL_1700692, EPI_ISL_1700693, EPI_ISL_1700694, EPI_ISL_2001029, EPI_ISL_2001032, EPI_ISL_2001034, EPI_ISL_2001036 | | | |
| see above | Monterey County Public Health Laboratory | Monterey County Public Health Laboratory | Monterey County Public Health Laboratory |
| EPI_ISL_2103197 | Orange County Public Health Lab | Chan-Zuckerberg Biohub | CZB Cliahub Consortium |
| EPI_ISL_1824248 | Pathology and Laboratory Medicine Institute, Cleveland Clinic, Ohio, USA | Pathology and Laboratory Medicine Institute, Cleveland Clinic, Ohio, USA | Daniel H. Farkas; Daniel Rhoads; David Bosler; David Plunkett; Gary Procop; Jay Brock; Jessica Spildener; Joy Nakitandwe; Kristen McDonnell; Yu-Wei Cheng; Zheng Jin Tu |
| EPI_ISL_1669340 | Providence Little Company of Mary Medical Center | Los Angeles County PHL | P. Hemarajata et al. |
| EPI_ISL_1525956, EPI_ISL_1526012, EPI_ISL_1526029, EPI_ISL_1526059, EPI_ISL_1552703, EPI_ISL_1552752, EPI_ISL_1552761, EPI_ISL_1552792, EPI_ISL_1552793, EPI_ISL_1552794, EPI_ISL_1552855, EPI_ISL_1559458, EPI_ISL_1559476, EPI_ISL_1559481, EPI_ISL_1559514, EPI_ISL_1559560, EPI_ISL_1581859, EPI_ISL_1581889, EPI_ISL_1581943, EPI_ISL_1581945, EPI_ISL_1581947, EPI_ISL_1581958, EPI_ISL_1581978, EPI_ISL_1582001, EPI_ISL_1582032, EPI_ISL_1582080, EPI_ISL_1622104, EPI_ISL_1648141, EPI_ISL_1648142, EPI_ISL_1648143, EPI_ISL_1648154, EPI_ISL_1648155, EPI_ISL_1648156, EPI_ISL_1648157, EPI_ISL_1648160, EPI_ISL_1648166, EPI_ISL_1648172, EPI_ISL_1648175, EPI_ISL_1648180, EPI_ISL_1648182, EPI_ISL_1648185, EPI_ISL_1648186, EPI_ISL_1648188, EPI_ISL_1648190, EPI_ISL_1648191, EPI_ISL_1648192, EPI_ISL_1648196, EPI_ISL_1648197, EPI_ISL_1648198, EPI_ISL_1648199, EPI_ISL_1648201, EPI_ISL_1648203, EPI_ISL_1648205, EPI_ISL_1648210, EPI_ISL_1648214, EPI_ISL_1648217, EPI_ISL_1648221, EPI_ISL_1648222, EPI_ISL_1648224, EPI_ISL_1648228, EPI_ISL_1648234, EPI_ISL_1648235, EPI_ISL_1648239, EPI_ISL_1648242, EPI_ISL_1648243, EPI_ISL_1648247, EPI_ISL_1648254, EPI_ISL_1648258, EPI_ISL_1648261, EPI_ISL_1648262, EPI_ISL_1648266, EPI_ISL_1648268, EPI_ISL_1648269, EPI_ISL_1648270, EPI_ISL_1648271, EPI_ISL_1648273, EPI_ISL_1648275, EPI_ISL_1648277, EPI_ISL_1648278, EPI_ISL_1648281, EPI_ISL_1648282, EPI_ISL_1648283, EPI_ISL_1648290, EPI_ISL_1648291, EPI_ISL_1648292, EPI_ISL_1648298, EPI_ISL_1648299, EPI_ISL_1648306, EPI_ISL_1648311, EPI_ISL_1648312, EPI_ISL_1648313, EPI_ISL_1648319, EPI_ISL_1648321, EPI_ISL_1648323, EPI_ISL_1648324, EPI_ISL_1648326, EPI_ISL_1648330, EPI_ISL_1648331, EPI_ISL_1648333, EPI_ISL_1648334, EPI_ISL_1648335, EPI_ISL_1648337, EPI_ISL_1648343, EPI_ISL_1648345, EPI_ISL_1648346, EPI_ISL_1648347, EPI_ISL_1648349, EPI_ISL_1648351, EPI_ISL_1648352, EPI_ISL_1648355, EPI_ISL_1648364, EPI_ISL_1648366, EPI_ISL_1648369, EPI_ISL_1648376, EPI_ISL_1648380, EPI_ISL_1648381, EPI_ISL_1648385, EPI_ISL_1648391, EPI_ISL_1648393, EPI_ISL_1648394, EPI_ISL_1648395, EPI_ISL_1648398, EPI_ISL_1648400, EPI_ISL_1648403, EPI_ISL_1648406, EPI_ISL_1648407, EPI_ISL_1648416, EPI_ISL_1648419, EPI_ISL_1648420, EPI_ISL_1648421, EPI_ISL_1648422, EPI_ISL_1648423, EPI_ISL_1648424, EPI_ISL_1648427, EPI_ISL_1648431, EPI_ISL_1648432, EPI_ISL_1648435, EPI_ISL_1648436, EPI_ISL_1648438, EPI_ISL_1648441, EPI_ISL_1648444, EPI_ISL_1648447, EPI_ISL_1648448, EPI_ISL_1648449, EPI_ISL_1648452, EPI_ISL_1648453, EPI_ISL_1648454, EPI_ISL_1648618, EPI_ISL_1648629, EPI_ISL_1648631, EPI_ISL_1648639, EPI_ISL_1648643, EPI_ISL_1648645, EPI_ISL_1648654, EPI_ISL_1648666, EPI_ISL_1648667, EPI_ISL_1648670, EPI_ISL_1648679, EPI_ISL_1648690, EPI_ISL_1648693, EPI_ISL_1648699, EPI_ISL_1648702, EPI_ISL_1648714, EPI_ISL_1648723, EPI_ISL_1648725, EPI_ISL_1648728, EPI_ISL_1648742, EPI_ISL_1648754, EPI_ISL_1648766, EPI_ISL_1648770, EPI_ISL_1648833, EPI_ISL_1648893, EPI_ISL_1668548, EPI_ISL_1668614, EPI_ISL_1668634, EPI_ISL_1668635, EPI_ISL_1668641, EPI_ISL_1668657, EPI_ISL_1668659, EPI_ISL_1668661, EPI_ISL_1668667, EPI_ISL_1668668, EPI_ISL_1668669, EPI_ISL_1668681, EPI_ISL_1668687, EPI_ISL_1668691, EPI_ISL_1668694, EPI_ISL_1668713, EPI_ISL_1668728, EPI_ISL_1668729, EPI_ISL_1668733, EPI_ISL_1668737, EPI_ISL_1668753, EPI_ISL_1668760, EPI_ISL_1668776, EPI_ISL_1668777, EPI_ISL_1668781, EPI_ISL_1668789, EPI_ISL_1668790, EPI_ISL_1668798, EPI_ISL_1668803, EPI_ISL_1668843, EPI_ISL_1668845, EPI_ISL_1668871, EPI_ISL_1668889, EPI_ISL_1668944, EPI_ISL_1668991, EPI_ISL_1669000, EPI_ISL_1669004, EPI_ISL_1669009, EPI_ISL_1669021, EPI_ISL_1669033, EPI_ISL_1669046, EPI_ISL_1669061, EPI_ISL_1669064, EPI_ISL_1669074, EPI_ISL_1669080, EPI_ISL_1669092, EPI_ISL_1669102, EPI_ISL_1669111, EPI_ISL_1669129, EPI_ISL_1669148, EPI_ISL_1669190, EPI_ISL_1669204, EPI_ISL_1669224, EPI_ISL_1669260, EPI_ISL_1669264, EPI_ISL_1669304, EPI_ISL_1691398, EPI_ISL_1691472, EPI_ISL_1691476, EPI_ISL_1691482, EPI_ISL_1691488, EPI_ISL_1694695, EPI_ISL_1695040, EPI_ISL_1695057, EPI_ISL_1695073, EPI_ISL_1695076, EPI_ISL_1695077, EPI_ISL_1695078, EPI_ISL_1695080, EPI_ISL_1695082, EPI_ISL_1695084, EPI_ISL_1695085, EPI_ISL_1695087, EPI_ISL_1695091, EPI_ISL_1695674, EPI_ISL_1695736, EPI_ISL_1702691, EPI_ISL_1738064, EPI_ISL_1738069, EPI_ISL_1738087, EPI_ISL_1738101, EPI_ISL_1738123, EPI_ISL_1738178, EPI_ISL_1738195, EPI_ISL_1738204, EPI_ISL_1738207, EPI_ISL_1738210, EPI_ISL_1738222, EPI_ISL_1738231, EPI_ISL_1738238, EPI_ISL_1738246, EPI_ISL_1738250, EPI_ISL_1738263, EPI_ISL_1738280, EPI_ISL_1738292, EPI_ISL_1738296, EPI_ISL_1738329, EPI_ISL_1738383, EPI_ISL_1738401, EPI_ISL_1738467, EPI_ISL_1738474, EPI_ISL_1738477, EPI_ISL_1738481, EPI_ISL_1738482, EPI_ISL_1738487, EPI_ISL_1738491, EPI_ISL_1738492, EPI_ISL_1738494, EPI_ISL_1738497, EPI_ISL_1738506, EPI_ISL_1738507, EPI_ISL_1738508, EPI_ISL_1738511, EPI_ISL_1738515, EPI_ISL_1738518, EPI_ISL_1738523, EPI_ISL_1738524, EPI_ISL_1738526, EPI_ISL_1738531, EPI_ISL_1738532, EPI_ISL_1738536, EPI_ISL_1738544, EPI_ISL_1738547, EPI_ISL_1738550, EPI_ISL_1738562, EPI_ISL_1738565, EPI_ISL_1738583, EPI_ISL_1738597, EPI_ISL_1738598, EPI_ISL_1738599, EPI_ISL_1738600, EPI_ISL_1738606, EPI_ISL_1738620, EPI_ISL_1738622, EPI_ISL_1738630, EPI_ISL_1738641, EPI_ISL_1738652, EPI_ISL_1738662, EPI_ISL_1738663, EPI_ISL_1738684, EPI_ISL_1738688, EPI_ISL_1738690, EPI_ISL_1738713, EPI_ISL_1738721, EPI_ISL_1738736, EPI_ISL_1738739, EPI_ISL_1738747, EPI_ISL_1753033, EPI_ISL_1753034, EPI_ISL_1753035, EPI_ISL_1753038, EPI_ISL_1753041, EPI_ISL_1753045, EPI_ISL_1753062, EPI_ISL_1753063, EPI_ISL_1753068, EPI_ISL_1753070, EPI_ISL_1753084, EPI_ISL_1753086, EPI_ISL_1753091, EPI_ISL_1753105, EPI_ISL_1753108, EPI_ISL_1753111, EPI_ISL_1753113, EPI_ISL_1753126, EPI_ISL_1753139, EPI_ISL_1753143, EPI_ISL_1753145, EPI_ISL_1753149, EPI_ISL_1753154, EPI_ISL_1753156, EPI_ISL_1753161, EPI_ISL_1753163, EPI_ISL_1753165, EPI_ISL_1753170, EPI_ISL_1753172, EPI_ISL_1753175, EPI_ISL_1753177, EPI_ISL_1753178, EPI_ISL_1753179, EPI_ISL_1753182, EPI_ISL_1753183, EPI_ISL_1753184, EPI_ISL_1753185, EPI_ISL_1753186, EPI_ISL_1753187, EPI_ISL_1753188, EPI_ISL_1753190, EPI_ISL_1753193, EPI_ISL_1753196, EPI_ISL_1753197, EPI_ISL_1753200, EPI_ISL_1753203, EPI_ISL_1753205, EPI_ISL_1753207, EPI_ISL_1753211, EPI_ISL_1753212, EPI_ISL_1753213, EPI_ISL_1753219, EPI_ISL_1753220, EPI_ISL_1753221, EPI_ISL_1753222, EPI_ISL_1753223, EPI_ISL_1753226, EPI_ISL_1753228, EPI_ISL_1753233, EPI_ISL_1753238, EPI_ISL_1753240, EPI_ISL_1753242, EPI_ISL_1753243, EPI_ISL_1753248, EPI_ISL_1753250, EPI_ISL_1753252, EPI_ISL_1753255, EPI_ISL_1753257, EPI_ISL_1753260, EPI_ISL_1753261, EPI_ISL_1753262, EPI_ISL_1753264, EPI_ISL_1753265, EPI_ISL_1753266, EPI_ISL_1753267, EPI_ISL_1753269, EPI_ISL_1753274, EPI_ISL_1753276, EPI_ISL_1753279, EPI_ISL_1753280, EPI_ISL_1753284, EPI_ISL_1753287, EPI_ISL_1753300, EPI_ISL_1753306, EPI_ISL_1753307, EPI_ISL_1753316, EPI_ISL_1753326, EPI_ISL_1753332, EPI_ISL_1753344, EPI_ISL_1753347, EPI_ISL_1753349, EPI_ISL_1753352, EPI_ISL_1753356, EPI_ISL_1753361, EPI_ISL_1753365, EPI_ISL_1753372, EPI_ISL_1753375, EPI_ISL_1753379, EPI_ISL_1753380, EPI_ISL_1753382, EPI_ISL_1753383, EPI_ISL_1753389, EPI_ISL_1753393, EPI_ISL_1753396, EPI_ISL_1753404, EPI_ISL_1753416, EPI_ISL_1753621, EPI_ISL_1753645, EPI_ISL_1753646, EPI_ISL_1753647, EPI_ISL_1753653, EPI_ISL_1753655, EPI_ISL_1753656, EPI_ISL_1753657, EPI_ISL_1761871, EPI_ISL_1761872, EPI_ISL_1761873, EPI_ISL_1761877, EPI_ISL_1761879, EPI_ISL_1761880, EPI_ISL_1761881, EPI_ISL_1761885, EPI_ISL_1761891, EPI_ISL_1761902, EPI_ISL_1761904, EPI_ISL_1761909, EPI_ISL_1761911, EPI_ISL_1761912, EPI_ISL_1761916, EPI_ISL_1761917, EPI_ISL_1761930, EPI_ISL_1761934, EPI_ISL_1761935, EPI_ISL_1761938, EPI_ISL_1761943, EPI_ISL_1761945, EPI_ISL_1761948, EPI_ISL_1761950, EPI_ISL_1761956, EPI_ISL_1761958, EPI_ISL_1761959, EPI_ISL_1761960, EPI_ISL_1761964, EPI_ISL_1761966, EPI_ISL_1761969, EPI_ISL_1761973, EPI_ISL_1761979, EPI_ISL_1761982, EPI_ISL_1761987, EPI_ISL_1761991, EPI_ISL_1761998, EPI_ISL_1762007, EPI_ISL_1762009, EPI_ISL_1762011, EPI_ISL_1762022, EPI_ISL_1762025, EPI_ISL_1762026, EPI_ISL_1797926, EPI_ISL_1798036, EPI_ISL_1798071, EPI_ISL_1798086, EPI_ISL_1798096, EPI_ISL_1798104, EPI_ISL_1798118, EPI_ISL_1798129, EPI_ISL_1798151, EPI_ISL_1798161, EPI_ISL_1798183, EPI_ISL_1804812, EPI_ISL_1804815, EPI_ISL_1804816, EPI_ISL_1804819, EPI_ISL_1818064, EPI_ISL_1818069, EPI_ISL_1818309, EPI_ISL_1818410, EPI_ISL_1834201, EPI_ISL_1834227, EPI_ISL_1834293, EPI_ISL_1834301, EPI_ISL_1834310, EPI_ISL_1834311, EPI_ISL_1834402, EPI_ISL_1840260, EPI_ISL_1840275, EPI_ISL_1840279, EPI_ISL_1840284, EPI_ISL_1840286, EPI_ISL_1840294, EPI_ISL_1840304, EPI_ISL_1840315, EPI_ISL_1840336, EPI_ISL_1840339, EPI_ISL_1840346, EPI_ISL_1840375, EPI_ISL_1840380, EPI_ISL_1840385, EPI_ISL_1840398, EPI_ISL_1840406, EPI_ISL_1900132, EPI_ISL_1900147, EPI_ISL_1900442, EPI_ISL_1900445, EPI_ISL_1900449, EPI_ISL_1900451, EPI_ISL_1905848, EPI_ISL_1905852, EPI_ISL_1905859, EPI_ISL_1905866, EPI_ISL_1905869, EPI_ISL_1905901, EPI_ISL_1905905, EPI_ISL_1905925, EPI_ISL_1905927, EPI_ISL_1906009, EPI_ISL_1906030, EPI_ISL_1906036, EPI_ISL_1906074, EPI_ISL_1906102, EPI_ISL_1906126, EPI_ISL_1906143, EPI_ISL_1906145, EPI_ISL_1906168, EPI_ISL_1906450, EPI_ISL_1906571, EPI_ISL_1906584, EPI_ISL_1906586, EPI_ISL_1906589, EPI_ISL_1906591, EPI_ISL_1906613, EPI_ISL_1906617, EPI_ISL_1906621, EPI_ISL_1906625, EPI_ISL_1906641, EPI_ISL_1906650, EPI_ISL_1906654, EPI_ISL_1906663, EPI_ISL_1906672, EPI_ISL_1906684, EPI_ISL_1906736, EPI_ISL_1906742, EPI_ISL_1906744, EPI_ISL_1906749, EPI_ISL_1906751, EPI_ISL_1906753, EPI_ISL_1906757, EPI_ISL_1906763, EPI_ISL_1906768, EPI_ISL_1906772, EPI_ISL_1906778, EPI_ISL_1906790, EPI_ISL_1906804, EPI_ISL_1906808, EPI_ISL_1906819, EPI_ISL_1906825, EPI_ISL_1906830, EPI_ISL_1906846, EPI_ISL_1906847, EPI_ISL_1906848, EPI_ISL_1906856, EPI_ISL_1906863, EPI_ISL_1906864, EPI_ISL_1906867, EPI_ISL_1906871, EPI_ISL_1906874, EPI_ISL_1906882, EPI_ISL_1935558, EPI_ISL_1935563, EPI_ISL_1935571, EPI_ISL_1935576, EPI_ISL_1935582, EPI_ISL_2133447, EPI_ISL_2133463, EPI_ISL_2133511, EPI_ISL_2133515, EPI_ISL_2133568, EPI_ISL_2133569, EPI_ISL_2133676, EPI_ISL_2133753, EPI_ISL_2133778, EPI_ISL_2140761 | | | |
| see above | Quest Diagnostics Incorporated | Centers for Disease Control and Prevention Division of Viral Diseases, Pathogen Discovery | A. Gerasimova; A. Perez; Adrian Paskey; B. Anderson; Benjamin Rambo-Martin; Christopher Gulvick; Clinton R. Paden; Dakota Howard; Darlene Wagner; Dhwani Batra; Duncan MacCannell; F. Lacbawan; I. A. Shlyakhter; Jason Caravas; K.E. Livingston; Kara Moser; L.E. Bernstein; M. Hua; Matthew Schmerer; P. Tanpaiboon; Peter W. Cook; R. M. Kagan; R. Owen; R. V. Rolando; S. H. Rosenthal; Scott Sammons; Shatavia Morrison; Y. Liu; Yvette Unoarumhi |
| EPI_ISL_1794768 | Rady Children's Hospital - San Diego | Andersen lab at Scripps Research | Christina Clarke; David Dimmock; Denise Malicki; Kathryn Bouic; Linda Luo; SEARCH Alliance San Diego with Nanda Radamchar; Teresa Mueller |
| EPI_ISL_2254425 | Respiratory Viruses Branch, Centers for Disease Control and Prevention | Respiratory Viruses Branch, Centers for Disease Control and Prevention | Batra, D.; C.R.; Campbell, P.; Caravas, J.; Case, R.; Clark, C.; Cook; Ghorpade, V.; Gulvick, C.; Hardison, M.; Houdeshell, H.; Howard, D.; Kvalvaag, O.; MacCannell, D.; Morrison, S.; Moser, K.; Nall, D.; P.W.; Paden; Paskey, A.; Rambo-Martin, B.; Sammons, S.; Sanders, E.; Schmerer, M.; Unoarumhi, Y.; Vest, A.; Wagner, D.; Westlund, S. |
| EPI_ISL_1791429, EPI_ISL_1791430, EPI_ISL_1791431, EPI_ISL_1791432, EPI_ISL_1791433, EPI_ISL_1791434, EPI_ISL_1791435, EPI_ISL_1791436, EPI_ISL_1791437, EPI_ISL_1791438, EPI_ISL_1791439, EPI_ISL_1791440, EPI_ISL_1791441, EPI_ISL_1791442, EPI_ISL_1791443, EPI_ISL_1791444, EPI_ISL_1791445, EPI_ISL_1791446, EPI_ISL_1791447, EPI_ISL_1791448, EPI_ISL_1791449, EPI_ISL_1791450, EPI_ISL_1791451, EPI_ISL_1791452, EPI_ISL_1791453, EPI_ISL_1791454, EPI_ISL_1791455, EPI_ISL_1791456, EPI_ISL_1794616, EPI_ISL_1794617, EPI_ISL_1794622, EPI_ISL_1794623, EPI_ISL_1794624, EPI_ISL_1794625, EPI_ISL_1794626, EPI_ISL_1794630, EPI_ISL_1794631, EPI_ISL_1794632, EPI_ISL_1794633, EPI_ISL_1794637, EPI_ISL_1794640, EPI_ISL_1794642, EPI_ISL_1794644, EPI_ISL_1794646, EPI_ISL_1794647, EPI_ISL_1794651, EPI_ISL_1794652, EPI_ISL_1794937, EPI_ISL_1794938, EPI_ISL_1794942, EPI_ISL_1794944, EPI_ISL_1794948, EPI_ISL_1794949, EPI_ISL_1794950, EPI_ISL_1794952, EPI_ISL_1794953, EPI_ISL_1794954, EPI_ISL_1794957, EPI_ISL_1794958, EPI_ISL_1794959, EPI_ISL_1794961, EPI_ISL_1794965, EPI_ISL_1794966, EPI_ISL_1794969, EPI_ISL_1794970, EPI_ISL_1794973, EPI_ISL_1794974, EPI_ISL_1794975, EPI_ISL_1794979, EPI_ISL_1794987, EPI_ISL_1794996, EPI_ISL_1795001, EPI_ISL_1795002, EPI_ISL_1795006, EPI_ISL_1795007, EPI_ISL_1795011, EPI_ISL_1795017, EPI_ISL_1795022, EPI_ISL_1795025, EPI_ISL_1795026, EPI_ISL_1795032, EPI_ISL_1795033, EPI_ISL_1795038, EPI_ISL_1969283, EPI_ISL_1969373, EPI_ISL_1969374, EPI_ISL_1969375, EPI_ISL_1969376, EPI_ISL_1969380, EPI_ISL_1969381, EPI_ISL_1969382, EPI_ISL_1969386, EPI_ISL_1969387, EPI_ISL_1969388, EPI_ISL_1969389, EPI_ISL_1969393, EPI_ISL_1969394, EPI_ISL_1969395, EPI_ISL_1969399, EPI_ISL_1969400, EPI_ISL_1969405, EPI_ISL_1969406, EPI_ISL_1969410, EPI_ISL_1969411, EPI_ISL_1969416, EPI_ISL_1969417, EPI_ISL_1969418, EPI_ISL_1969423, EPI_ISL_1969424, EPI_ISL_1969429, EPI_ISL_1969430, EPI_ISL_1969434, EPI_ISL_1969437, EPI_ISL_1969438, EPI_ISL_1969439, EPI_ISL_1969440, EPI_ISL_1969441, EPI_ISL_1969442, EPI_ISL_1969443, EPI_ISL_1969444, EPI_ISL_1969445, EPI_ISL_1969446, EPI_ISL_1969447, EPI_ISL_1969448, EPI_ISL_1969449, EPI_ISL_1969450, EPI_ISL_1969499, EPI_ISL_1969500 | | | |
| see above | San Diego County Public Health Laboratory | Andersen lab at Scripps Research | Brett Austin; Jovan Shephard; SEARCH Alliance San Diego with Ashleigh Murphy; SEARCH Alliance San Diego with Tracy Basler |
| EPI_ISL_2102995, EPI_ISL_2103023, EPI_ISL_2103033, EPI_ISL_2103058, EPI_ISL_2103109, EPI_ISL_2103121, EPI_ISL_2103146, EPI_ISL_2103177 | | | |
| see above | Santa Clara County Public Health Laboratory | Chan-Zuckerberg Biohub | CZB Cliahub Consortium |
| EPI_ISL_1508964, EPI_ISL_1508965, EPI_ISL_1532139, EPI_ISL_1532140, EPI_ISL_1532144, EPI_ISL_1620461 | Santa Clara County Public Health Laboratory | Santa Clara County Public Health Laboratory | Santa Clara County Public Health Department |
| EPI_ISL_1664263, EPI_ISL_1664264, EPI_ISL_1664265, EPI_ISL_1664266, EPI_ISL_1664275, EPI_ISL_1913216 | Santa Clara County Public Health Laboratory | Santa Clara County public Health Laboratory | Santa Clara County Public Health Department |
| EPI_ISL_1532127, EPI_ISL_1532128, EPI_ISL_1532129, EPI_ISL_1532130, EPI_ISL_1532132, EPI_ISL_1620456 | Santa Clara Valley Medical Center | Santa Clara County Public Health Laboratory | Santa Clara County Public Health Department |
| EPI_ISL_1664276, EPI_ISL_1664277, EPI_ISL_1664278, EPI_ISL_1664279, EPI_ISL_1664280, EPI_ISL_1664281, EPI_ISL_1664282, EPI_ISL_1664283, EPI_ISL_1664284, EPI_ISL_1664285, EPI_ISL_1664286, EPI_ISL_1664287, EPI_ISL_1664289, EPI_ISL_1664290, EPI_ISL_1664291, EPI_ISL_1664292, EPI_ISL_1664293, EPI_ISL_1664294, EPI_ISL_1664295, EPI_ISL_1664296, EPI_ISL_1664297, EPI_ISL_1664298, EPI_ISL_1664299, EPI_ISL_1913215, EPI_ISL_1913218, EPI_ISL_1913226, EPI_ISL_1913227, EPI_ISL_1913228, EPI_ISL_1913229, EPI_ISL_1913230, EPI_ISL_1913231, EPI_ISL_1913232, EPI_ISL_1913233, EPI_ISL_1913234, EPI_ISL_1913235, EPI_ISL_1913236, EPI_ISL_1913237, EPI_ISL_1913238, EPI_ISL_1913239, EPI_ISL_1913240, EPI_ISL_1913241, EPI_ISL_1913242, EPI_ISL_1913243, EPI_ISL_1913244, EPI_ISL_1913245, EPI_ISL_1913246, EPI_ISL_1913247, EPI_ISL_1913248, EPI_ISL_1913249, EPI_ISL_1913250 | | | |
| see above | Santa Clara Valley Medical Center | Santa Clara County public Health Laboratory | Santa Clara County Public Health Department |

| | | | |
|---|---|---|---|
| EPI_ISL_1794721, EPI_ISL_1794722, EPI_ISL_1794723, EPI_ISL_1794724, EPI_ISL_1794725, EPI_ISL_1794726, EPI_ISL_1794727, EPI_ISL_1794728, EPI_ISL_1794729, EPI_ISL_1794730, EPI_ISL_1794731, EPI_ISL_1794732, EPI_ISL_1794733, EPI_ISL_1794734, EPI_ISL_1794735, EPI_ISL_1794736, EPI_ISL_1794737, EPI_ISL_1794738, EPI_ISL_1794739, EPI_ISL_1794740, EPI_ISL_1794741, EPI_ISL_1794839, EPI_ISL_1794840, EPI_ISL_1794841, EPI_ISL_1794842, EPI_ISL_1794843, EPI_ISL_1794844, EPI_ISL_1794845, EPI_ISL_1794846, EPI_ISL_1794847, EPI_ISL_1794848, EPI_ISL_1794849, EPI_ISL_1794850, EPI_ISL_1794851, EPI_ISL_1794852, EPI_ISL_1794853, EPI_ISL_1794934, EPI_ISL_1794935, EPI_ISL_1969534, EPI_ISL_1969535, EPI_ISL_1969536, EPI_ISL_1969537, EPI_ISL_1969538, EPI_ISL_1969539, EPI_ISL_1969540, EPI_ISL_1969541, EPI_ISL_1969542, EPI_ISL_1969543, EPI_ISL_1969544, EPI_ISL_1969545, EPI_ISL_1969550, EPI_ISL_1969551, EPI_ISL_1969552, EPI_ISL_1969553 | | | |
| see above | Scripps Medical Laboratory | Andersen lab at Scripps Research | Ellen Stefanski; Ian Mchardy; SEARCH Alliance San Diego with Michael Quigley |
| EPI_ISL_1794700, EPI_ISL_1794701, EPI_ISL_1794702, EPI_ISL_1794705, EPI_ISL_1794706, EPI_ISL_1794707, EPI_ISL_1794708, EPI_ISL_1794709, EPI_ISL_1794710, EPI_ISL_1794711, EPI_ISL_1794712, EPI_ISL_1794742, EPI_ISL_1794743, EPI_ISL_1794744, EPI_ISL_1794745, EPI_ISL_1794746, EPI_ISL_1794747, EPI_ISL_1794748, EPI_ISL_1794749, EPI_ISL_1794750, EPI_ISL_1794751, EPI_ISL_1794752, EPI_ISL_1794753, EPI_ISL_1794754, EPI_ISL_1794755, EPI_ISL_1794756, EPI_ISL_1794757, EPI_ISL_1794758, EPI_ISL_1794759, EPI_ISL_1794760, EPI_ISL_1794761, EPI_ISL_1794762, EPI_ISL_1794763, EPI_ISL_1794764, EPI_ISL_1794765, EPI_ISL_1794766, EPI_ISL_1794767, EPI_ISL_1794782, EPI_ISL_1794783, EPI_ISL_1794784, EPI_ISL_1794785, EPI_ISL_1794786, EPI_ISL_1794885, EPI_ISL_1794886, EPI_ISL_1794887, EPI_ISL_1794888, EPI_ISL_1794889, EPI_ISL_1794890, EPI_ISL_1794891, EPI_ISL_1794892, EPI_ISL_1794893, EPI_ISL_1794894, EPI_ISL_1794895, EPI_ISL_1794896, EPI_ISL_1794897, EPI_ISL_1794898, EPI_ISL_1794899, EPI_ISL_1794900, EPI_ISL_1794901, EPI_ISL_1794902, EPI_ISL_1794903, EPI_ISL_1794904, EPI_ISL_1794905, EPI_ISL_1794906, EPI_ISL_1794907, EPI_ISL_1794908, EPI_ISL_1794909, EPI_ISL_1794910, EPI_ISL_1794911, EPI_ISL_1794912, EPI_ISL_1794913, EPI_ISL_1794914, EPI_ISL_1794915, EPI_ISL_1794916, EPI_ISL_1794917, EPI_ISL_1794918, EPI_ISL_1969563, EPI_ISL_1969564, EPI_ISL_1969565, EPI_ISL_1969566, EPI_ISL_1969567, EPI_ISL_1969568, EPI_ISL_1969569, EPI_ISL_1969574, EPI_ISL_1969576, EPI_ISL_1969577, EPI_ISL_1969578, EPI_ISL_1969579, EPI_ISL_1969580, EPI_ISL_1969581, EPI_ISL_1969582, EPI_ISL_1969583, EPI_ISL_1969584, EPI_ISL_1969585, EPI_ISL_1969586, EPI_ISL_1969587, EPI_ISL_1969588, EPI_ISL_1969590, EPI_ISL_1969595, EPI_ISL_1969596, EPI_ISL_1969597, EPI_ISL_1969598, EPI_ISL_1969599, EPI_ISL_1969601, EPI_ISL_1969606, EPI_ISL_1969609 | | | |
| see above | Sharp HealthCare Laboratory | Andersen lab at Scripps Research | Art Mendoza; Cathy Woerle; Jacquelyn Berumen; Liam McGinnis; Omid Bakhtar; SEARCH Alliance San Diego with Aaron Harding |
| EPI_ISL_1620455, EPI_ISL_1620457, EPI_ISL_1620458, EPI_ISL_1620459, EPI_ISL_1620460, EPI_ISL_1620462, EPI_ISL_1620463, EPI_ISL_1620464, EPI_ISL_1620465 | | | |
| see above | Stanford Clinical Virology Laboratory | Santa Clara County Public Health Laboratory | Santa Clara County Public Health Department |
| EPI_ISL_1924633, EPI_ISL_1924634, EPI_ISL_1924635, EPI_ISL_1924636, EPI_ISL_1924637, EPI_ISL_1924638, EPI_ISL_1924639, EPI_ISL_1924640, EPI_ISL_1924641, EPI_ISL_1924642, EPI_ISL_1924643, EPI_ISL_1924644, EPI_ISL_1924645, EPI_ISL_1924646, EPI_ISL_1924647, EPI_ISL_1924648 | | | |
| see above | Stanford Clinical Virology Laboratory | Santa Clara County public Health Laboratory | Santa Clara County Public Health Department |
| EPI_ISL_1675226, EPI_ISL_1675227, EPI_ISL_1701680, EPI_ISL_1701681 | Stanford Health Care | Stanford University School of Medicine, Clinical Virology Laboratory | Becky Jiang; Bernadette Truong; Daniel Solis; Fumiko Yamamoto; Hannah Wang; Jacob Miller; James Zehnder; Kenji O. Mfuh; Malaya K. Sahoo; Mamdouh Sibai; Marilyn Mar; Michelle Verghese; Naomi Iwai; and Benjamin A. Pinsky |
| EPI_ISL_1540751 | The Jackson Laboratory | The Jackson Laboratory | Adams M; Bergeron D; Kelly K; Li L; Long J; Omerza G; Renzette N |
| EPI_ISL_1707042, EPI_ISL_1707044, EPI_ISL_1707045, EPI_ISL_1707046, EPI_ISL_1707047, EPI_ISL_1707048, EPI_ISL_1707050, EPI_ISL_1707051, EPI_ISL_1707052, EPI_ISL_1707053, EPI_ISL_1707054, EPI_ISL_1707055, EPI_ISL_1707056, EPI_ISL_1707057, EPI_ISL_1707058, EPI_ISL_1707059, EPI_ISL_1707060, EPI_ISL_1707061, EPI_ISL_1716873, EPI_ISL_1716874, EPI_ISL_1716875, EPI_ISL_1716876 | | | |
| see above | UCLA Clinical Micro Lab | Los Angeles County PHL | P. Hemarajata et al. |
| EPI_ISL_1469852, EPI_ISL_1469875, EPI_ISL_1469876, EPI_ISL_1469878, EPI_ISL_1469879, EPI_ISL_1469880, EPI_ISL_1469881, EPI_ISL_1469883, EPI_ISL_1921080, EPI_ISL_1921081, EPI_ISL_1921082, EPI_ISL_1921083, EPI_ISL_1921084, EPI_ISL_1921112, EPI_ISL_1921113, EPI_ISL_1921114, EPI_ISL_1921115, EPI_ISL_1921128, EPI_ISL_1921146, EPI_ISL_2031772, EPI_ISL_2031773, EPI_ISL_2031774, EPI_ISL_2031775, EPI_ISL_2031776, EPI_ISL_2031777, EPI_ISL_2031778, EPI_ISL_2031779, EPI_ISL_2031780, EPI_ISL_2031781, EPI_ISL_2031782, EPI_ISL_2031783, EPI_ISL_2031784, EPI_ISL_2031785, EPI_ISL_2031786 | | | |
| see above | UCSC Genomics Institute | UCSC Genomics Institute | A. Marm Kilpatrick; Angie Hinrichs; Beth Shapiro; Bryan Thornlow; Ciara Wanket; David Haussler; Eric Beraut; Hugh Olsen; Ikenna Anigbogu; Isabel Bjork; Jakob McBroome; Jeremy Sanford; Joshua Kapp; Mark Akeson; Maximilian Haeussler; Michael Stone; Miten Jain; Molly Cassatt-Johnstone; Namrita Dhillon; Russell Corbett-Detig; Terren Chang; Yatish Turakhia |
| EPI_ISL_1508921, EPI_ISL_1508922, EPI_ISL_1508923, EPI_ISL_1508924, EPI_ISL_1508925, EPI_ISL_1508926, EPI_ISL_1508927, EPI_ISL_1508928, EPI_ISL_1669348, EPI_ISL_1706609, EPI_ISL_1707078, EPI_ISL_1707079, EPI_ISL_1707080, EPI_ISL_1707081, EPI_ISL_1707082, EPI_ISL_1707083, EPI_ISL_1707084, EPI_ISL_1707085, EPI_ISL_1707086, EPI_ISL_1707087, EPI_ISL_1707088, EPI_ISL_1707089, EPI_ISL_1707090, EPI_ISL_1707091, EPI_ISL_1707092, EPI_ISL_1707093, EPI_ISL_1707094, EPI_ISL_1707095, EPI_ISL_1707096, EPI_ISL_1707097, EPI_ISL_1707098, EPI_ISL_1716881, EPI_ISL_1716882, EPI_ISL_1716883, EPI_ISL_1716884, EPI_ISL_1716885, EPI_ISL_1716886, EPI_ISL_1716887, EPI_ISL_1716888, EPI_ISL_1716889, EPI_ISL_1716890, EPI_ISL_1716891, EPI_ISL_1716892, EPI_ISL_1716893, EPI_ISL_1716894, EPI_ISL_1716895, EPI_ISL_1716897, EPI_ISL_1716898 | | | |
| see above | USC Clinical Lab | Los Angeles County PHL | P. Hemarajata et al. |
| EPI_ISL_2031858 | VA Medical Center | Los Angeles County PHL | P. Hemarajata et al. |

We gratefully acknowledge the following Authors from the Originating laboratories responsible for obtaining the specimens, as well as the Submitting laboratories where the genome data were generated and shared via GISAID, on which this research is based.

All Submitters of data may be contacted directly via www.gisaid.org

Authors are sorted alphabetically.

| Accession ID | Originating Laboratory | Submitting Laboratory | Authors |
|---|---|---|---|
| EPI_ISL_1167668, EPI_ISL_1167669, EPI_ISL_1167670, EPI_ISL_1167671, EPI_ISL_1167672, EPI_ISL_1167673, EPI_ISL_1167674, EPI_ISL_1167675, EPI_ISL_1167676, EPI_ISL_1167677, EPI_ISL_1167678, EPI_ISL_1167679, EPI_ISL_1167680, EPI_ISL_1167681, EPI_ISL_1167682, EPI_ISL_1167683, EPI_ISL_1167684, EPI_ISL_1167685, EPI_ISL_1167686, EPI_ISL_1167687, EPI_ISL_1167688, EPI_ISL_1167689, EPI_ISL_1167691, EPI_ISL_1167692, EPI_ISL_1167693, EPI_ISL_1167694, EPI_ISL_1167695, EPI_ISL_1167698, EPI_ISL_1167699, EPI_ISL_1167700, EPI_ISL_1167701, EPI_ISL_1167702, EPI_ISL_1167703, EPI_ISL_1167704, EPI_ISL_1167705, EPI_ISL_1167706, EPI_ISL_1167707, EPI_ISL_1167708, EPI_ISL_1167709, EPI_ISL_1167710, EPI_ISL_1167715, EPI_ISL_1167716, EPI_ISL_1167717, EPI_ISL_1167718, EPI_ISL_1167719, EPI_ISL_1167720, EPI_ISL_1167721, EPI_ISL_1167722, EPI_ISL_1167723, EPI_ISL_1167724, EPI_ISL_1167725, EPI_ISL_1167726, EPI_ISL_1167727, EPI_ISL_1167728, EPI_ISL_1167729, EPI_ISL_1167730, EPI_ISL_1167731, EPI_ISL_1167732, EPI_ISL_1167733, EPI_ISL_1167734, EPI_ISL_1167735, EPI_ISL_1167736, EPI_ISL_1167737, EPI_ISL_1167738, EPI_ISL_1167739, EPI_ISL_1167740, EPI_ISL_1167741, EPI_ISL_1167742, EPI_ISL_1167743, EPI_ISL_1167744, EPI_ISL_1167745, EPI_ISL_1167746, EPI_ISL_1167747, EPI_ISL_1167748, EPI_ISL_1167749, EPI_ISL_1167750, EPI_ISL_1167751, EPI_ISL_1167752, EPI_ISL_1167753, EPI_ISL_1167754, EPI_ISL_1167756, EPI_ISL_1167757, EPI_ISL_1167758, EPI_ISL_1167759, EPI_ISL_1167760, EPI_ISL_1167761, EPI_ISL_1167762, EPI_ISL_1167763, EPI_ISL_1167764, EPI_ISL_1167765, EPI_ISL_1167766, EPI_ISL_1167767, EPI_ISL_1167768, EPI_ISL_1167769, EPI_ISL_1167770, EPI_ISL_1167771, EPI_ISL_1167772, EPI_ISL_1167773, EPI_ISL_1167774, EPI_ISL_1167775, EPI_ISL_1167776, EPI_ISL_1167777, EPI_ISL_1167778, EPI_ISL_1167780, EPI_ISL_1167790, EPI_ISL_1167794, EPI_ISL_1167796, EPI_ISL_1167803, EPI_ISL_1167804, EPI_ISL_1167805, EPI_ISL_1167806, EPI_ISL_1167810, EPI_ISL_1167812, EPI_ISL_1167819, EPI_ISL_1167822, EPI_ISL_1167823, EPI_ISL_1167825, EPI_ISL_1167827, EPI_ISL_1167829, EPI_ISL_1167835, EPI_ISL_1167836, EPI_ISL_1167837, EPI_ISL_1167839, EPI_ISL_1167843, EPI_ISL_1167844, EPI_ISL_1167847, EPI_ISL_1167848, EPI_ISL_1167849, EPI_ISL_1167851, EPI_ISL_1167852, EPI_ISL_1167854, EPI_ISL_1167857, EPI_ISL_1167858, EPI_ISL_1167860, EPI_ISL_1167861, EPI_ISL_1167863, EPI_ISL_1167864, EPI_ISL_1167865, EPI_ISL_1167866, EPI_ISL_1167867, EPI_ISL_1167871, EPI_ISL_1167872, EPI_ISL_1167874, EPI_ISL_1167875, EPI_ISL_1167876, EPI_ISL_1167877, EPI_ISL_1167878, EPI_ISL_1167879, EPI_ISL_1167881, EPI_ISL_1167883, EPI_ISL_1167884, EPI_ISL_1167885, EPI_ISL_1167886, EPI_ISL_1167887, EPI_ISL_1167888, EPI_ISL_1167890, EPI_ISL_1167891, EPI_ISL_1167892, EPI_ISL_1167893, EPI_ISL_1167895, EPI_ISL_1167896, EPI_ISL_1167897, EPI_ISL_1167899, EPI_ISL_1167901, EPI_ISL_1167902, EPI_ISL_1167903, EPI_ISL_1167904, EPI_ISL_1167905, EPI_ISL_1167906, EPI_ISL_1167907, EPI_ISL_1167909, EPI_ISL_1167911, EPI_ISL_1167912, EPI_ISL_1167914, EPI_ISL_1167915, EPI_ISL_1167918, EPI_ISL_1167921, EPI_ISL_1167922, EPI_ISL_1167923, EPI_ISL_1167924, EPI_ISL_1167925, EPI_ISL_1167927, EPI_ISL_1167928, EPI_ISL_1167929, EPI_ISL_1167930, EPI_ISL_1167931, EPI_ISL_1300449, EPI_ISL_1300452, EPI_ISL_1300454, EPI_ISL_1300455, EPI_ISL_1300456, EPI_ISL_1300457, EPI_ISL_1300458, EPI_ISL_1300459, EPI_ISL_1300460, EPI_ISL_1300461, EPI_ISL_1300462, EPI_ISL_1300463, EPI_ISL_1300464, EPI_ISL_1300465, EPI_ISL_1300466, EPI_ISL_1300468, EPI_ISL_1300469, EPI_ISL_1300470, EPI_ISL_1300471, EPI_ISL_1300472, EPI_ISL_1300473, EPI_ISL_1300474, EPI_ISL_1300475, EPI_ISL_1300476, EPI_ISL_1300477, EPI_ISL_1300478, EPI_ISL_1300479, EPI_ISL_1300480, EPI_ISL_1300481, EPI_ISL_1300482, EPI_ISL_1300483, EPI_ISL_1300484, EPI_ISL_1300485, EPI_ISL_1300486, EPI_ISL_1300487, EPI_ISL_1300488, EPI_ISL_1300489, EPI_ISL_1300490, EPI_ISL_1300491, EPI_ISL_1300492, EPI_ISL_1300493, EPI_ISL_1300495, EPI_ISL_1300497, EPI_ISL_1300498, EPI_ISL_1300499, EPI_ISL_1300500, EPI_ISL_1300501, EPI_ISL_1300502, EPI_ISL_1300503, EPI_ISL_1300504, EPI_ISL_1300505, EPI_ISL_1300506, EPI_ISL_1300507, EPI_ISL_1300508, EPI_ISL_1300509, EPI_ISL_1300510, EPI_ISL_1300511, EPI_ISL_1300512, EPI_ISL_1300513, EPI_ISL_1300514, EPI_ISL_1300515, EPI_ISL_1300516, EPI_ISL_1300517, EPI_ISL_1300518, EPI_ISL_1300519, EPI_ISL_1300520, EPI_ISL_1321447, EPI_ISL_1321448, EPI_ISL_1321449, EPI_ISL_1321450, EPI_ISL_1321451, EPI_ISL_1321452, EPI_ISL_1321453, EPI_ISL_1321454, EPI_ISL_1321455, EPI_ISL_1321456, EPI_ISL_1321457, EPI_ISL_1321458, EPI_ISL_1321459, EPI_ISL_1321460, EPI_ISL_1321461, EPI_ISL_1321462, EPI_ISL_1321463, EPI_ISL_1321464, EPI_ISL_1321465, EPI_ISL_1321466, EPI_ISL_1321467, EPI_ISL_1321468, EPI_ISL_1321469, EPI_ISL_1321470, EPI_ISL_1321471, EPI_ISL_1321472, EPI_ISL_1321473, EPI_ISL_1321474, EPI_ISL_1321475, EPI_ISL_1321476, EPI_ISL_1321477, EPI_ISL_1321478, EPI_ISL_1321479, EPI_ISL_1321480, EPI_ISL_1321481, EPI_ISL_1321482, EPI_ISL_1321483, EPI_ISL_1321484, EPI_ISL_1321485, EPI_ISL_1321486, EPI_ISL_1321487, EPI_ISL_1321488, EPI_ISL_1321489, EPI_ISL_1321490, EPI_ISL_1321491, EPI_ISL_1321492, EPI_ISL_1321493, EPI_ISL_1321494, EPI_ISL_1321495, EPI_ISL_1321496, EPI_ISL_1321497, EPI_ISL_1321498, EPI_ISL_1321499, EPI_ISL_1321500, EPI_ISL_1321501, EPI_ISL_1321502, EPI_ISL_1321503, EPI_ISL_1321504, EPI_ISL_1321505, EPI_ISL_1321506, EPI_ISL_1321507, EPI_ISL_1321508, EPI_ISL_1321509, EPI_ISL_1321510, EPI_ISL_1321511, EPI_ISL_1321512, EPI_ISL_1321513, EPI_ISL_1321514, EPI_ISL_1321515, EPI_ISL_1321516, EPI_ISL_1321517, EPI_ISL_1321518, EPI_ISL_1321519, EPI_ISL_1321520, EPI_ISL_1321521, EPI_ISL_1321522, EPI_ISL_1321523, EPI_ISL_1321524, EPI_ISL_1321525, EPI_ISL_1321526, EPI_ISL_1321527, EPI_ISL_1321528, EPI_ISL_1321529, EPI_ISL_1321530, EPI_ISL_1321531, EPI_ISL_1321532, EPI_ISL_1321533, EPI_ISL_1321534, EPI_ISL_1321535, EPI_ISL_1321536, EPI_ISL_1321537, EPI_ISL_1321538, EPI_ISL_1321539, EPI_ISL_1321540, EPI_ISL_1321541, EPI_ISL_1321542, EPI_ISL_1321543, EPI_ISL_1321544, EPI_ISL_1321545, EPI_ISL_1321546, EPI_ISL_1321547, EPI_ISL_1321548, EPI_ISL_1321549, EPI_ISL_1321550, EPI_ISL_1321551, EPI_ISL_1321552, EPI_ISL_1321553, EPI_ISL_1321554, EPI_ISL_1321555, EPI_ISL_1321556, EPI_ISL_1321557, EPI_ISL_1321558, EPI_ISL_1321559, EPI_ISL_1321560, EPI_ISL_1321561, EPI_ISL_1321562, EPI_ISL_1321563, EPI_ISL_1321564, EPI_ISL_1321565, EPI_ISL_1321566, EPI_ISL_1321567, EPI_ISL_1321568, EPI_ISL_1321569, EPI_ISL_1321570, EPI_ISL_1321571, EPI_ISL_1321572, EPI_ISL_1321573, EPI_ISL_1321574, EPI_ISL_1321575, EPI_ISL_1321576, EPI_ISL_1321577, EPI_ISL_1321578, EPI_ISL_1321579, EPI_ISL_1321580, EPI_ISL_1321581, EPI_ISL_1321582, EPI_ISL_1321583, EPI_ISL_1321584, EPI_ISL_1321585, EPI_ISL_1321586, EPI_ISL_1321587, EPI_ISL_1321588, EPI_ISL_1321589, EPI_ISL_1321590, EPI_ISL_1321591, EPI_ISL_1321592, EPI_ISL_1321593, EPI_ISL_1321594, EPI_ISL_1321595, EPI_ISL_1321596, EPI_ISL_1321597, EPI_ISL_1321598, EPI_ISL_1321599, EPI_ISL_1321600, EPI_ISL_1470415, EPI_ISL_1470416, EPI_ISL_1470417, EPI_ISL_1470418, EPI_ISL_1470419, EPI_ISL_1470420, EPI_ISL_1470421, EPI_ISL_1470422, EPI_ISL_1470423, EPI_ISL_1470424, EPI_ISL_1470425, EPI_ISL_1470426, EPI_ISL_1470427, EPI_ISL_1470428, EPI_ISL_1470429, EPI_ISL_1470430, EPI_ISL_1470431, EPI_ISL_1470432, EPI_ISL_1470433, EPI_ISL_1470434, EPI_ISL_1470435, EPI_ISL_1470436, EPI_ISL_1470437, EPI_ISL_1470438, EPI_ISL_1470439, EPI_ISL_1470440, EPI_ISL_1470441, EPI_ISL_1470442, EPI_ISL_1470443, EPI_ISL_1470444, EPI_ISL_1470445, EPI_ISL_1470446, EPI_ISL_1470447, EPI_ISL_1470448, EPI_ISL_1470449, EPI_ISL_1470450, EPI_ISL_1470451, EPI_ISL_1470452, EPI_ISL_1470453, EPI_ISL_1470454, EPI_ISL_1470455, EPI_ISL_1470456, EPI_ISL_1470457, EPI_ISL_1470458, EPI_ISL_1470459, EPI_ISL_1470460, EPI_ISL_1470461, EPI_ISL_1470462, EPI_ISL_1470463, EPI_ISL_1470464, EPI_ISL_1470465, EPI_ISL_1470466, EPI_ISL_1470467, EPI_ISL_1470468, EPI_ISL_1470469, EPI_ISL_1470470, EPI_ISL_1470471, EPI_ISL_1470472, EPI_ISL_1470473, EPI_ISL_1470474, EPI_ISL_1470475, EPI_ISL_1470476, EPI_ISL_1470477, EPI_ISL_1470478, EPI_ISL_1470479, EPI_ISL_1470480, EPI_ISL_1470481, EPI_ISL_1470482, EPI_ISL_1470483, EPI_ISL_1470484, EPI_ISL_1470485, EPI_ISL_1470486, EPI_ISL_1470487, EPI_ISL_1470488, EPI_ISL_1470489, EPI_ISL_1470490, EPI_ISL_1470491, EPI_ISL_1470492, EPI_ISL_1470493, EPI_ISL_1470494, EPI_ISL_1470495, EPI_ISL_1470496, EPI_ISL_1470497, EPI_ISL_1470498, EPI_ISL_1470499, EPI_ISL_1470500, EPI_ISL_1470501, EPI_ISL_1470502, EPI_ISL_1470503, EPI_ISL_1470504, EPI_ISL_1470505, EPI_ISL_1470506, EPI_ISL_1470507, EPI_ISL_1470508, EPI_ISL_1470509, EPI_ISL_1470510, EPI_ISL_1470511, EPI_ISL_1470512, EPI_ISL_1470513, EPI_ISL_1470514, EPI_ISL_1470515, EPI_ISL_1470516, EPI_ISL_1470517, EPI_ISL_1470518, EPI_ISL_1470519, EPI_ISL_1470520, EPI_ISL_1470521, EPI_ISL_1470522, EPI_ISL_1470523, EPI_ISL_1470524, EPI_ISL_1470525, EPI_ISL_1470526, EPI_ISL_1470527, EPI_ISL_1470528, EPI_ISL_1470529, EPI_ISL_1470530, EPI_ISL_1470531, EPI_ISL_1470532, EPI_ISL_1470533, EPI_ISL_1470534, EPI_ISL_1470535, EPI_ISL_1470536, EPI_ISL_1470537, EPI_ISL_1470538, EPI_ISL_1470539, EPI_ISL_1470540, EPI_ISL_1470541, EPI_ISL_1470542, EPI_ISL_1470543, EPI_ISL_1470544, EPI_ISL_1470545, EPI_ISL_1470546, EPI_ISL_1470547, EPI_ISL_1470548, EPI_ISL_1470549, EPI_ISL_1470550, EPI_ISL_1470551, EPI_ISL_1470552, EPI_ISL_1470553, EPI_ISL_1470554, EPI_ISL_1470555, EPI_ISL_1470556, EPI_ISL_1470557, EPI_ISL_1470558, EPI_ISL_1470559, EPI_ISL_1534544, EPI_ISL_1534545, EPI_ISL_1534546, EPI_ISL_1534547, EPI_ISL_1534548, EPI_ISL_1534549, EPI_ISL_1534550, EPI_ISL_1534551, EPI_ISL_1534552, EPI_ISL_1534553, EPI_ISL_1534554, EPI_ISL_1534555, EPI_ISL_1534556, EPI_ISL_1534557, EPI_ISL_1534558, EPI_ISL_1534559, EPI_ISL_1534560, EPI_ISL_1534561, EPI_ISL_1534562, EPI_ISL_1534563, EPI_ISL_1534564, EPI_ISL_1534565, EPI_ISL_1534566, EPI_ISL_1534567, EPI_ISL_1534568, EPI_ISL_1534569, EPI_ISL_1534570, EPI_ISL_1534571, EPI_ISL_1534572, EPI_ISL_1534573, EPI_ISL_1534574, EPI_ISL_1534575, EPI_ISL_1534576, EPI_ISL_1534577, EPI_ISL_1534578, EPI_ISL_1534579, EPI_ISL_1534580, EPI_ISL_1534581, EPI_ISL_1534582, EPI_ISL_1534583, EPI_ISL_1534584, EPI_ISL_1534585, EPI_ISL_1534586, EPI_ISL_1534587, EPI_ISL_1534588, EPI_ISL_1534589, EPI_ISL_1534590, EPI_ISL_1534591, EPI_ISL_1534592, EPI_ISL_1534593, EPI_ISL_1534594, EPI_ISL_1534595, EPI_ISL_1534596, EPI_ISL_1534597, EPI_ISL_1534598, EPI_ISL_1534599, EPI_ISL_1534600, EPI_ISL_1534601, EPI_ISL_1534602, EPI_ISL_1534603, EPI_ISL_1534604, EPI_ISL_1534605, EPI_ISL_1534606, EPI_ISL_1534607, EPI_ISL_1534608, EPI_ISL_1534609, EPI_ISL_1540996, EPI_ISL_1540997, EPI_ISL_1540998, EPI_ISL_1540999, EPI_ISL_1541000, EPI_ISL_1541001, EPI_ISL_1541002, EPI_ISL_1541003, EPI_ISL_1541004, EPI_ISL_1541005, EPI_ISL_1541006, EPI_ISL_1541007, EPI_ISL_1541008, EPI_ISL_1541009, EPI_ISL_1541010, EPI_ISL_1541011, EPI_ISL_1541012, EPI_ISL_1541015, EPI_ISL_1541016, EPI_ISL_1541017, EPI_ISL_1541018, EPI_ISL_1541019, EPI_ISL_1541020, EPI_ISL_1541021, EPI_ISL_1541022, EPI_ISL_1541023, EPI_ISL_1541024, EPI_ISL_1541035, EPI_ISL_1541036, EPI_ISL_1633370, EPI_ISL_1633371, EPI_ISL_1633372, EPI_ISL_1633373, EPI_ISL_1633374, EPI_ISL_1633375, EPI_ISL_1633376, EPI_ISL_1633377, EPI_ISL_1633411, EPI_ISL_1633412, EPI_ISL_1633413, EPI_ISL_1633473, EPI_ISL_1633474, EPI_ISL_1633475, EPI_ISL_1633476, EPI_ISL_1633477, EPI_ISL_1633478, EPI_ISL_1633479, EPI_ISL_1633480, EPI_ISL_1633481, EPI_ISL_1633482, EPI_ISL_1633483, EPI_ISL_1633484, EPI_ISL_1633485, EPI_ISL_1633486, EPI_ISL_1633487, EPI_ISL_1633488, EPI_ISL_1633489, EPI_ISL_1633490, EPI_ISL_1633491, EPI_ISL_1633492, EPI_ISL_1633493, EPI_ISL_1633494, EPI_ISL_1633495, EPI_ISL_1633496, EPI_ISL_1633497, EPI_ISL_1633498, EPI_ISL_1633499, EPI_ISL_1633500, EPI_ISL_1633501, EPI_ISL_1633502, EPI_ISL_1633503, EPI_ISL_1633504, EPI_ISL_1633505, EPI_ISL_1633506, EPI_ISL_1633507, EPI_ISL_1633508 | | | |
| see above | Genetica Molecular and Subdepartamento de Virologia ISP Chile | Instituto de Salud Publica de Chile | Javier Tognarelli, Karen Orostica, Barbara Parra, Loredana Arata, Jaime Lagos, Gisselle Barra, Patricia Bustos, Rodrigo Fasce, Andres Castillo, Jorge Fernandez |
| EPI_ISL_1712336, EPI_ISL_1712337, EPI_ISL_1712338 | Genetica Molecular and Subdepartamento de Virologia ISP Chile | Instituto de Salud Publica de Chile | Javier Tognarelli, Karen Orostica, Barbara Parra, Loredana Arata, Gisselle Barra, Patricia Bustos, Rodrigo Fasce, Andres Castillo, Soledad Ulloa, Jorge Fernandez |
| EPI_ISL_1827525 | AREA DE SALUD LOS CHILES | Inciensa, Instituto Costarricense de Investigación y Enseñanza en Nutrición y Salud | Francisco Duarte, Hebleen Porras, Claudio Soto-Garita, Estela Cordero, Adriana Godínez, Melany Calderón, José Luis Vargas, Mariela Gutiérrez, Joselyn Prado & Teresita Somogyi |
| EPI_ISL_2009220, EPI_ISL_2009225, EPI_ISL_2009226, EPI_ISL_2009227, EPI_ISL_2009228, EPI_ISL_2009229, EPI_ISL_2009230, EPI_ISL_2009615 | Genetica Molecular and Subdepartamento de Virologia ISP Chile | Instituto de Salud Publica de Chile | Javier Tognarelli, Karen Orostica, Barbara Parra, Loredana Arata, Gisselle Barra, Patricia Bustos, Rodrigo Fasce, Andres Castillo, Soledad Ulloa, Jorge Fernandez |
| EPI_ISL_625673, EPI_ISL_625674, EPI_ISL_625676, EPI_ISL_625677, EPI_ISL_625678, EPI_ISL_625679 | Laboratory of Molecular Medicine, University of Magallanes | Centro Asistencial Docente y de Investigacion, Universidad de Magallanes | Jorge Gonzalez, Jacqueline Aldridge, Diego Alvarez, Marcelo Navarrete |
| EPI_ISL_746479, EPI_ISL_746480, EPI_ISL_746481, EPI_ISL_746483, EPI_ISL_746490, EPI_ISL_746491, EPI_ISL_746492, EPI_ISL_746498, EPI_ISL_746499, EPI_ISL_746500, EPI_ISL_746501, EPI_ISL_746502, EPI_ISL_746503, EPI_ISL_746504, EPI_ISL_746505, EPI_ISL_746509, EPI_ISL_746510, EPI_ISL_746511, EPI_ISL_746512, EPI_ISL_746513, EPI_ISL_746514, EPI_ISL_746515, EPI_ISL_746516, EPI_ISL_746517, EPI_ISL_746518, EPI_ISL_746519, EPI_ISL_746521, EPI_ISL_746522, EPI_ISL_746523, EPI_ISL_746524, EPI_ISL_746525, EPI_ISL_746526, EPI_ISL_746527, EPI_ISL_746528, EPI_ISL_746529, EPI_ISL_746530, EPI_ISL_746531, EPI_ISL_746532, EPI_ISL_746533, EPI_ISL_746534, EPI_ISL_746535, EPI_ISL_746536, EPI_ISL_746537, EPI_ISL_746538, EPI_ISL_746539, EPI_ISL_746540, EPI_ISL_746541, EPI_ISL_746542, EPI_ISL_746543, EPI_ISL_746544, EPI_ISL_746724, EPI_ISL_746726, EPI_ISL_746728, EPI_ISL_746729, EPI_ISL_746730, EPI_ISL_746731, EPI_ISL_746732, EPI_ISL_746733, EPI_ISL_746734, EPI_ISL_746735, EPI_ISL_746736, EPI_ISL_746737, EPI_ISL_746738, EPI_ISL_746739, EPI_ISL_746740, EPI_ISL_746741, EPI_ISL_746742, EPI_ISL_746743, EPI_ISL_746744, EPI_ISL_746745, EPI_ISL_746746, EPI_ISL_746747, EPI_ISL_746748, EPI_ISL_746749, EPI_ISL_746750, EPI_ISL_746751, EPI_ISL_746752, EPI_ISL_746753, EPI_ISL_746754, EPI_ISL_746755, EPI_ISL_746756, EPI_ISL_746757, EPI_ISL_746758, EPI_ISL_746759, EPI_ISL_746760, EPI_ISL_746761, EPI_ISL_746762, EPI_ISL_746763, EPI_ISL_746764, EPI_ISL_746765, EPI_ISL_746766, EPI_ISL_746767, EPI_ISL_746768, EPI_ISL_746769, EPI_ISL_746770, EPI_ISL_746771, EPI_ISL_746772, EPI_ISL_746773, EPI_ISL_754393 | | | |
| see above | Genetica Molecular and Subdepartamento de Virologia ISP Chile | Instituto de Salud Publica de Chile | Javier Tognarelli, Barbara Parra, Loredana Arata, Jaime Lagos, Gisselle Barra, Patricia Bustos, Rodrigo Fasce, Andres Castillo, Jorge Fernandez |
| EPI_ISL_802545, EPI_ISL_802546, EPI_ISL_802547, EPI_ISL_802548, EPI_ISL_802549, EPI_ISL_802550, EPI_ISL_802551, EPI_ISL_802552, EPI_ISL_802553, EPI_ISL_802554, EPI_ISL_802555, EPI_ISL_802556, EPI_ISL_802557, EPI_ISL_802558, EPI_ISL_802559, EPI_ISL_802560, EPI_ISL_802561, EPI_ISL_802562, EPI_ISL_802563, EPI_ISL_802564, EPI_ISL_802565, EPI_ISL_802566 | | | |
| see above | Molecular Microbiology and Food Research Laboratory (MMFRLAB) - Universidad San Sebastián | Facultad de Ciencias de la Vida, UNAB | Dayán Sanhueza, Macarena Bastías, César Echeverría, Sebastián Wolter, Waldo Díaz, Jorge Olivares, Eduardo Castro, Claudio Meneses |

We gratefully acknowledge the following Authors from the Originating laboratories responsible for obtaining the specimens, as well as the Submitting laboratories where the genome data were generated and shared via GISAID, on which this research is based.

All Submitters of data may be contacted directly via www.gisaid.org

Authors are sorted alphabetically.

| Accession ID | Originating Laboratory | Submitting Laboratory | Authors |
|---|---|---|---|
| EPI_ISL_1015353, EPI_ISL_1015354, EPI_ISL_1015355, EPI_ISL_1015356, EPI_ISL_1015357 | UNIGEM | Universidad Nacional de Colombia - Laboratorio Genómico One Health | Andres F. Cardona-Rios, Daniel O. Maldonado-Perez, Laura Silvana Perez, Karl A Ciuoderis, Idabely Betancur Ortiz, Sandra Ines Cano, Diego A. Álvarez-Díaz, Carlos Franco-Muñoz, Marcela Mercado-Reyes, Jorge E. Osorio, Juan P. Hernandez-Ortiz |
| EPI_ISL_1015358, EPI_ISL_1015359, EPI_ISL_1015360 | Hospital Pablo Tobón Uribe | Universidad Nacional de Colombia - Laboratorio Genómico One Health | Andres F. Cardona-Rios, Daniel O. Maldonado-Perez, Laura Silvana Perez, Karl A Ciuoderis, Idabely Betancur Ortiz, Sandra Ines Cano, Diego A. Álvarez-Díaz, Carlos Franco-Muñoz, Marcela Mercado-Reyes, Jorge E. Osorio, Juan P. Hernandez-Ortiz |
| EPI_ISL_1017701 | Laboratorio de Salud Pública de Boyacá | Instituto Nacional de Salud- Dirección de Investigación en Salud Pública | Katherine Laiton-Donato, Diego A. Álvarez-Díaz, Carlos Franco-Muñoz, Hector Alejandro Ruiz-Moreno, Maria T. Herrera-Sepúlveda, Diego Andrés Prada, Jhonnatan Reales-González, Sheryll Corchuelo, Julian Naizaque, Gerardo Santamaría, Magdalena Wiesner, Martha Lucia Ospina Martinez, Marcela Mercado-Reyes |
| EPI_ISL_1017702 | Laboratorio Docente Asistencial-Universidad de Antioquia | Instituto Nacional de Salud- Dirección de Investigación en Salud Pública | Katherine Laiton-Donato, Diego A. Álvarez-Díaz, Carlos Franco-Muñoz, Hector Alejandro Ruiz-Moreno, Maria T. Herrera-Sepúlveda, Diego Andrés Prada, Jhonnatan Reales-González, Sheryll Corchuelo, Julian Naizaque, Gerardo Santamaría, Magdalena Wiesner, Martha Lucia Ospina Martinez, Marcela Mercado-Reyes |
| EPI_ISL_1017706, EPI_ISL_1017707, EPI_ISL_1017708, EPI_ISL_1017709, EPI_ISL_1017710 | Hospital Universitario Hernando Moncaleano Perdomo | Instituto Nacional de Salud- Dirección de Investigación en Salud Pública | Katherine Laiton-Donato, Diego A. Álvarez-Díaz, Carlos Franco-Muñoz, Hector Alejandro Ruiz-Moreno, Maria T. Herrera-Sepúlveda, Diego Andrés Prada, Jhonnatan Reales-González, Sheryll Corchuelo, Julian Naizaque, Gerardo Santamaría, Magdalena Wiesner, Martha Lucia Ospina Martinez, Marcela Mercado-Reyes |
| EPI_ISL_1017712 | Hemato Oncologos S.A | Instituto Nacional de Salud- Dirección de Investigación en Salud Pública | Katherine Laiton-Donato, Diego A. Álvarez-Díaz, Carlos Franco-Muñoz, Hector Alejandro Ruiz-Moreno, Maria T. Herrera-Sepúlveda, Diego Andrés Prada, Jhonnatan Reales-González, Sheryll Corchuelo, Julian Naizaque, Gerardo Santamaría, Magdalena Wiesner, Martha Lucia Ospina Martinez, Marcela Mercado-Reyes |
| EPI_ISL_1091779, EPI_ISL_1091780, EPI_ISL_1091781 | ICMT | Universidad Nacional de Colombia - Laboratorio Genómico One Health | Andres F. Cardona-Rios, Daniel O. Maldonado-Perez, Laura Silvana Perez, Karl A Ciuoderis, Idabely Betancur Ortiz, Sandra Ines Cano, Diego A. Álvarez-Díaz, Carlos Franco-Muñoz, Marcela Mercado-Reyes, Jorge E. Osorio, Juan P. Hernandez-Ortiz |
| EPI_ISL_1091782, EPI_ISL_1091783 | PECET | Universidad Nacional de Colombia - Laboratorio Genómico One Health | Andres F. Cardona-Rios, Daniel O. Maldonado-Perez, Laura Silvana Perez, Karl A Ciuoderis, Idabely Betancur Ortiz, Sandra Ines Cano, Diego A. Álvarez-Díaz, Carlos Franco-Muñoz, Marcela Mercado-Reyes, Jorge E. Osorio, Juan P. Hernandez-Ortiz |
| EPI_ISL_1091784, EPI_ISL_1091785 | LDSP | Universidad Nacional de Colombia - Laboratorio Genómico One Health | Andres F. Cardona-Rios, Daniel O. Maldonado-Perez, Laura Silvana Perez, Karl A Ciuoderis, Maria Angélica Maya, Idabely Betancur Ortiz, Sandra Ines Cano, Diego A. Álvarez-Díaz, Carlos Franco-Muñoz, Marcela Mercado-Reyes, Jorge E. Osorio, Juan P. Hernandez-Ortiz |
| EPI_ISL_1091919 | Hospital Departamental María Inmaculada | Instituto Nacional de Salud- Dirección de Investigación en Salud Pública | Katherine Laiton-Donato, Diego A. Álvarez-Díaz, Carlos Franco-Muñoz, Hector Alejandro Ruiz-Moreno, Maria T. Herrera-Sepúlveda, Diego Andrés Prada, Jhonnatan Reales-González, Sheryll Corchuelo, Julian Naizaque, Gerardo Santamaría, Magdalena Wiesner, Martha Lucia Ospina Martinez, Marcela Mercado-Reyes |
| EPI_ISL_1091922, EPI_ISL_1091923 | Laboratorio de salud público de Atlantico | Instituto Nacional de Salud- Dirección de Investigación en Salud Pública | Katherine Laiton-Donato, Diego A. Álvarez-Díaz, Carlos Franco-Muñoz, Hector Alejandro Ruiz-Moreno, Maria T. Herrera-Sepúlveda, Diego Andrés Prada, Jhonnatan Reales-González, Sheryll Corchuelo, Julian Naizaque, Gerardo Santamaría, Magdalena Wiesner, Martha Lucia Ospina Martinez, Marcela Mercado-Reyes |
| EPI_ISL_1092005 | Laboratorio de Salud Publica de Cesar | Instituto Nacional de Salud- Dirección de Investigación en Salud Pública | Katherine Laiton-Donato, Diego A. Álvarez-Díaz, Carlos Franco-Muñoz, Hector Alejandro Ruiz-Moreno, Maria T. Herrera-Sepúlveda, Diego Andrés Prada, Jhonnatan Reales-González, Sheryll Corchuelo, Julian Naizaque, Gerardo Santamaría, Magdalena Wiesner, Martha Lucia Ospina Martinez, Marcela Mercado-Reyes |
| EPI_ISL_1092006 | Laboratorio IMAT | Instituto Nacional de Salud- Dirección de Investigación en Salud Pública | Katherine Laiton-Donato, Diego A. Álvarez-Díaz, Carlos Franco-Muñoz, Hector Alejandro Ruiz-Moreno, Maria T. Herrera-Sepúlveda, Diego Andrés Prada, Jhonnatan Reales-González, Sheryll Corchuelo, Julian Naizaque, Gerardo Santamaría, Magdalena Wiesner, Martha Lucia Ospina Martinez, Marcela Mercado-Reyes |
| EPI_ISL_1092007 | Laboratorio Continental | Instituto Nacional de Salud- Dirección de Investigación en Salud Pública | Katherine Laiton-Donato, Diego A. Álvarez-Díaz, Carlos Franco-Muñoz, Hector Alejandro Ruiz-Moreno, Maria T. Herrera-Sepúlveda, Diego Andrés Prada, Jhonnatan Reales-González, Sheryll Corchuelo, Julian Naizaque, Gerardo Santamaría, Magdalena Wiesner, Martha Lucia Ospina Martinez, Marcela Mercado-Reyes |
| EPI_ISL_1092008 | Laboratorio de Salud Publica de Bogota | Instituto Nacional de Salud- Dirección de Investigación en Salud Pública | Katherine Laiton-Donato, Diego A. Álvarez-Díaz, Carlos Franco-Muñoz, Hector Alejandro Ruiz-Moreno, Maria T. Herrera-Sepúlveda, Diego Andrés Prada, Jhonnatan Reales-González, Sheryll Corchuelo, Julian Naizaque, Gerardo Santamaría, Magdalena Wiesner, Martha Lucia Ospina Martinez, Marcela Mercado-Reyes |
| EPI_ISL_1220045 | Universidad de Magdalena | Instituto Nacional de Salud- Dirección de Investigación en Salud Pública | Katherine Laiton-Donato, Carlos Franco-Muñoz, Diego A. Álvarez-Díaz, Hector Alejandro Ruiz-Moreno, Jhonnatan Reales-González, Diego Andrés Prada, Sheryll Corchuelo, Maria T. Herrera-Sepúlveda, Julian Naizaque, Gerardo Santamaría, Magdalena Wiesner, Martha Lucia Ospina Martínez, Marcela Mercado-Reyes. |
| EPI_ISL_1220051 | ANALICEMOS LABORATORIO CLINICO ESPECIALIZADO | Instituto Nacional de Salud- Dirección de Investigación en Salud Pública | Katherine Laiton-Donato, Carlos Franco-Muñoz, Diego A. Álvarez-Díaz, Hector Alejandro Ruiz-Moreno, Jhonnatan Reales-González, Diego Andrés Prada, Sheryll Corchuelo, Maria T. Herrera-Sepúlveda, Julian Naizaque, Gerardo Santamaría, Magdalena Wiesner, Martha Lucia Ospina Martínez, Marcela Mercado-Reyes. |
| EPI_ISL_1220055, EPI_ISL_1220057 | LABORATORIO BIOLOGIA MOLECULAR IMAT SAS | Instituto Nacional de Salud- Dirección de Investigación en Salud Pública | Katherine Laiton-Donato, Carlos Franco-Muñoz, Diego A. Álvarez-Díaz, Hector Alejandro Ruiz-Moreno, Jhonnatan Reales-González, Diego Andrés Prada, Sheryll Corchuelo, Maria T. Herrera-Sepúlveda, Julian Naizaque, Gerardo Santamaría, Magdalena Wiesner, Martha Lucia Ospina Martínez, Marcela Mercado-Reyes. |
| EPI_ISL_1220058, EPI_ISL_1220059, EPI_ISL_1220060 | FUNDACION VALLE DEL LILI | Instituto Nacional de Salud- Dirección de Investigación en Salud Pública | Katherine Laiton-Donato, Carlos Franco-Muñoz, Diego A. Álvarez-Díaz, Hector Alejandro Ruiz-Moreno, Jhonnatan Reales-González, Diego Andrés Prada, Sheryll Corchuelo, Maria T. Herrera-Sepúlveda, Julian Naizaque, Gerardo Santamaría, Magdalena Wiesner, Martha Lucia Ospina Martínez, Marcela Mercado-Reyes. |
| EPI_ISL_1220061, EPI_ISL_1220062, EPI_ISL_1220065 | Investigaciones Biomédicas - Universidad de Sucre | Instituto Nacional de Salud- Dirección de Investigación en Salud Pública | Katherine Laiton-Donato, Carlos Franco-Muñoz, Diego A. Álvarez-Díaz, Hector Alejandro Ruiz-Moreno, Jhonnatan Reales-González, Diego Andrés Prada, Sheryll Corchuelo, Maria T. Herrera-Sepúlveda, Julian Naizaque, Gerardo Santamaría, Magdalena Wiesner, Martha Lucia Ospina Martínez, Marcela Mercado-Reyes. |
| EPI_ISL_1303368 | Clinica de la Costa | Instituto Nacional de Salud- Dirección de Investigación en Salud Pública | Katherine Laiton-Donato, Diego A. Álvarez-Díaz, Carlos Franco-Muñoz, Hector Alejandro Ruiz-Moreno, Maria T. Herrera-Sepúlveda, Diego Andrés Prada, Jhonnatan Reales-González, Sheryll Corchuelo, Julian Naizaque, Gerardo Santamaría, Sergio Gomez, Lisseth Pardo, Juan Camilo Martinez, Marta Lopez Blanco, Ángela Alarcon Cruz, Diana Malo, Carmen Osorio, Magdalena Wiesner, Martha Lucia Ospina Martinez, Marcela Mercado-Reyes |
| EPI_ISL_1303369, EPI_ISL_1303370, EPI_ISL_1303375 | UNIVERSIDAD DE CORDOBA - INSTITUTO DE INVESTIGACIONES BIOLÓGICAS DEL TRÓPICO IIBT | Instituto Nacional de Salud- Dirección de Investigación en Salud Pública | Katherine Laiton-Donato, Diego A. Álvarez-Díaz, Carlos Franco-Muñoz, Hector Alejandro Ruiz-Moreno, Maria T. Herrera-Sepúlveda, Diego Andrés Prada, Jhonnatan Reales-González, Sheryll Corchuelo, Julian Naizaque, Gerardo Santamaría, Sergio Gomez, Lisseth Pardo, Juan Camilo Martinez, Marta Lopez Blanco, Ángela Alarcon Cruz, Diana Malo, Carmen Osorio, Magdalena Wiesner, Martha Lucia Ospina Martinez, Marcela Mercado-Reyes |
| EPI_ISL_1324134, EPI_ISL_1324135, EPI_ISL_1324136, EPI_ISL_1324138, | UW Virology Lab | UW Virology Lab | Pavitra Roychoudhury, Hong Xie, Lasata Shrestha, Shah Mohamed Bakhash, Michelle Lin, Margaret Mills, Noah Baker, Sean Ellis, Saraswathi Sathees, Meei-Li Huang, Keith R Jerome, Alexander Greninger |

| | | | |
|---|---|---|---|
| EPI_ISL_1324141, EPI_ISL_1324143, EPI_ISL_1324144, EPI_ISL_1324148 | | | |
| EPI_ISL_1557223 | Instituto Nacional de Salud | Corporacion Corpogen Universidad de los Andes Universidad Central | Chaib De Mares, Maryam; Zambrano, María Mercedes; Del Portillo, Patricia; Romero, Christian; Díaz, Nelly; Anzola, Juan Manuel; González, Laura; Duitama, Jorge; Restrepo, Silvia |
| EPI_ISL_1576835, EPI_ISL_1577026, EPI_ISL_1577182, EPI_ISL_1577390, EPI_ISL_1582589, EPI_ISL_1582590, EPI_ISL_1628474 | Universidad Industrial de Santander (Laboratorio Central de Investigaciones - Clínica Chicamocha). | Universidad Industrial de Santander. | Francisco Martinez-Perez, Cristian E. Cadena-Caballero, Diego Rueda-Plata, Carolina S. Torres-Jiménez, Lizeth J. Forero-Buitrago, Erika Lizarazo-Gutiérrez, Carlos Barrios-Hernández, Lina M. Vera-Cala. |
| EPI_ISL_1628499 | Universidad Industrial de Santander (Laboratorio Central de Investigaciones - Clínica Chicamocha). | Universidad Industrial de Santander | Francisco Martinez-Perez, Cristian E. Cadena-Caballero, Diego Rueda-Plata, Carolina S. Torres-Jiménez, Lizeth J. Forero-Buitrago, Erika Lizarazo-Gutiérrez, Carlos Barrios-Hernández, Lina M. Vera-Cala. |
| EPI_ISL_1629710 | Universidad Industrial de Santander (Laboratorio Central de Investigaciones - Clínica Chicamocha). | Universidad Industrial de Santander. | Francisco Martinez-Perez, Cristian E. Cadena-Caballero, Diego Rueda-Plata, Carolina S. Torres-Jiménez, Lizeth J. Forero-Buitrago, Erika Lizarazo-Gutiérrez, Carlos Barrios-Hernández, Lina M. Vera-Cala. |
| EPI_ISL_1629716 | Hospital Pablo Tobón Uribe | Universidad Nacional de Colombia - Laboratorio Genómico One Health | Maria Stella López, Andres F. Cardona-Rios, Daniel O. Maldonado-Perez, Simón Villegas Velásquez, Laura Silvana Perez, Karl A Ciuoderis, Idabely Betancur Ortiz, Sandra Ines Cano, Eliana Patricia Calvo Tapiero, Diego A. Álvarez-Díaz, Carlos Franco-Muñoz, Hector Alejandro Ruiz-Moreno, Marcela Mercado-Reyes, Jorge E. Osorio, Juan P. Hernandez-Ortiz |
| EPI_ISL_1629717, EPI_ISL_1629720 | ICMT | Universidad Nacional de Colombia - Laboratorio Genómico One Health | Maria Stella López, Andres F. Cardona-Rios, Daniel O. Maldonado-Perez, Simón Villegas Velásquez, Laura Silvana Perez, Karl A Ciuoderis, Idabely Betancur Ortiz, Sandra Ines Cano, Eliana Patricia Calvo Tapiero, Diego A. Álvarez-Díaz, Carlos Franco-Muñoz, Hector Alejandro Ruiz-Moreno, Marcela Mercado-Reyes, Jorge E. Osorio, Juan P. Hernandez-Ortiz |
| EPI_ISL_1629721, EPI_ISL_1629722, EPI_ISL_1629723, EPI_ISL_1629724, EPI_ISL_1629725 | SOMER | Universidad Nacional de Colombia - Laboratorio Genómico One Health | Simón Villegas Velásquez, Andres F. Cardona-Rios, Daniel O. Maldonado-Perez, Maria Stella López, Laura Silvana Perez, Karl A Ciuoderis, Idabely Betancur Ortiz, Sandra Ines Cano, Eliana Patricia Calvo Tapiero, Diego A. Álvarez-Díaz, Carlos Franco-Muñoz, Hector Alejandro Ruiz-Moreno, Marcela Mercado-Reyes, Jorge E. Osorio, Juan P. Hernandez-Ortiz |
| EPI_ISL_1629728, EPI_ISL_1629729 | UDEA | Universidad Nacional de Colombia - Laboratorio Genómico One Health | Daniel O. Maldonado-Perez, Andres F. Cardona-Rios, Maria Stella López, Simón Villegas Velásquez, Laura Silvana Perez, Karl A Ciuoderis, Idabely Betancur Ortiz, Sandra Ines Cano, Eliana Patricia Calvo Tapiero, Diego A. Álvarez-Díaz, Carlos Franco-Muñoz, Hector Alejandro Ruiz-Moreno, Marcela Mercado-Reyes, Jorge E. Osorio, Juan P. Hernandez-Ortiz |
| EPI_ISL_1629733, EPI_ISL_1629734, EPI_ISL_1629735 | SURA | Universidad Nacional de Colombia - Laboratorio Genómico One Health | Laura Silvana Perez, Andres F. Cardona-Rios, Daniel O. Maldonado-Perez, Maria Stella López, Simón Villegas Velásquez, Karl A Ciuoderis, Idabely Betancur Ortiz, Sandra Ines Cano, Eliana Patricia Calvo Tapiero, Diego A. Álvarez-Díaz, Carlos Franco-Muñoz, Hector Alejandro Ruiz-Moreno, Marcela Mercado-Reyes, Jorge E. Osorio, Juan P. Hernandez-Ortiz |
| EPI_ISL_1629738, EPI_ISL_1629739, EPI_ISL_1629740 | CLÍNICA U BOLIVARIANA | Universidad Nacional de Colombia - Laboratorio Genómico One Health | Karl A Ciuoderis, Andres F. Cardona-Rios, Daniel O. Maldonado-Perez, Maria Stella López, Simón Villegas Velásquez, Laura Silvana Perez, Idabely Betancur Ortiz, Sandra Ines Cano, Eliana Patricia Calvo Tapiero, Diego A. Álvarez-Díaz, Carlos Franco-Muñoz, Hector Alejandro Ruiz-Moreno, Marcela Mercado-Reyes, Jorge E. Osorio, Juan P. Hernandez-Ortiz |
| EPI_ISL_1629742 | CLÍNICA U BOLIVARIANA | Universidad Nacional de Colombia - Laboratorio Genómico One Health | Andres F. Cardona-Rios, Daniel O. Maldonado-Perez, Maria Stella López, Simón Villegas Velásquez, Laura Silvana Perez, Karl A Ciuoderis, Idabely Betancur Ortiz, Sandra Ines Cano, Eliana Patricia Calvo Tapiero, Diego A. Álvarez-Díaz, Carlos Franco-Muñoz, Hector Alejandro Ruiz-Moreno, Marcela Mercado-Reyes, Jorge E. Osorio, Juan P. Hernandez-Ortiz |
| EPI_ISL_1629743, EPI_ISL_1629744, EPI_ISL_1629745, EPI_ISL_1629746, EPI_ISL_1629747 | LABORATORIO ECHAVARRIA | Universidad Nacional de Colombia - Laboratorio Genómico One Health | Andres F. Cardona-Rios, Daniel O. Maldonado-Perez, Maria Stella López, Simón Villegas Velásquez, Laura Silvana Perez, Karl A Ciuoderis, Idabely Betancur Ortiz, Sandra Ines Cano, Eliana Patricia Calvo Tapiero, Diego A. Álvarez-Díaz, Carlos Franco-Muñoz, Hector Alejandro Ruiz-Moreno, Marcela Mercado-Reyes, Jorge E. Osorio, Juan P. Hernandez-Ortiz |
| EPI_ISL_1629750, EPI_ISL_1629751, EPI_ISL_1629752 | SYNLAB | Universidad Nacional de Colombia - Laboratorio Genómico One Health | Andres F. Cardona-Rios, Daniel O. Maldonado-Perez, Maria Stella López, Simón Villegas Velásquez, Laura Silvana Perez, Karl A Ciuoderis, Idabely Betancur Ortiz, Sandra Ines Cano, Eliana Patricia Calvo Tapiero, Diego A. Álvarez-Díaz, Carlos Franco-Muñoz, Hector Alejandro Ruiz-Moreno, Marcela Mercado-Reyes, Jorge E. Osorio, Juan P. Hernandez-Ortiz |
| EPI_ISL_1629753, EPI_ISL_1629754, EPI_ISL_1629755, EPI_ISL_1629756, EPI_ISL_1629757, EPI_ISL_1629758, EPI_ISL_1629759 | H.U SAN VICENTE FUNDACION | Universidad Nacional de Colombia - Laboratorio Genómico One Health | Andres F. Cardona-Rios, Daniel O. Maldonado-Perez, Maria Stella López, Simón Villegas Velásquez, Laura Silvana Perez, Karl A Ciuoderis, Idabely Betancur Ortiz, Sandra Ines Cano, Eliana Patricia Calvo Tapiero, Diego A. Álvarez-Díaz, Carlos Franco-Muñoz, Hector Alejandro Ruiz-Moreno, Marcela Mercado-Reyes, Jorge E. Osorio, Juan P. Hernandez-Ortiz |
| EPI_ISL_1629760, EPI_ISL_1629761 | LABMÉDICO | Universidad Nacional de Colombia - Laboratorio Genómico One Health | Andres F. Cardona-Rios, Daniel O. Maldonado-Perez, Maria Stella López, Simón Villegas Velásquez, Laura Silvana Perez, Karl A Ciuoderis, Idabely Betancur Ortiz, Sandra Ines Cano, Eliana Patricia Calvo Tapiero, Diego A. Álvarez-Díaz, Carlos Franco-Muñoz, Hector Alejandro Ruiz-Moreno, Marcela Mercado-Reyes, Jorge E. Osorio, Juan P. Hernandez-Ortiz |
| EPI_ISL_1673261, EPI_ISL_1673262 | LABORATORIO LAS AMERICAS | Universidad Nacional de Colombia - Laboratorio Genómico One Health | Maria Stella López, Andres F. Cardona-Rios, Daniel O. Maldonado-Perez, Simón Villegas Velásquez, Lina M. Hurtado, Laura Silvana Perez, Karl A Ciuoderis, Idabely Betancur Ortiz, Sandra Ines Cano, Eliana Patricia Calvo Tapiero, Diego A. Álvarez-Díaz, Carlos Franco-Muñoz, Hector Alejandro Ruiz-Moreno, Marcela Mercado-Reyes, Jorge E. Osorio, Juan P. Hernandez-Ortiz |
| EPI_ISL_1820901 | SURA | Universidad Nacional de Colombia - Laboratorio Genómico One Health | Andres F. Cardona-Rios, Maria Stella López, Daniel O. Maldonado-Perez, Simón Villegas Velásquez, Lina M. Hurtado, Laura Silvana Perez, Karl A Ciuoderis, Maria Angélica Maya, Rita Almanza Payares, Idabely Betancur Ortiz, Sandra Ines Cano, Diego A. Álvarez-Díaz, Carlos Franco-Muñoz, Katherine Laiton-Donato, Hector Alejandro Ruiz-Moreno, Marcela Mercado-Reyes, Jorge E. Osorio, Juan P. Hernandez-Ortiz |
| EPI_ISL_739663, EPI_ISL_739664, EPI_ISL_739665, EPI_ISL_739666, EPI_ISL_739667, EPI_ISL_739668, EPI_ISL_739669, EPI_ISL_739670, EPI_ISL_739671, EPI_ISL_739672, EPI_ISL_739673, EPI_ISL_739674, EPI_ISL_739675, EPI_ISL_739676, EPI_ISL_739677, EPI_ISL_739678, EPI_ISL_739679, EPI_ISL_739680, EPI_ISL_739681, EPI_ISL_739682, EPI_ISL_739683 | | | |
| see above | Instituto Nacional de Salud, Bogotá, Colombia | Instituto Nacional de Salud, Bogotá, Colombia | Katherine Laiton-Donato, Diego A. Álvarez-Díaz, Carlos Franco-Muñoz, Mauricio Pacheco-Montealegre, Jonathan Reales, Diego Andrés Prada, Sheryl Corchuelo, Magdalena Weisner, Martha Lucia Ospina Martinez, Marcela Mercado-Reyes |
| EPI_ISL_791084 | Instituto Nacional de Salud - Unidad de Secuenciación y Análisis Genómico | Instituto Nacional de Salud - Dirección de Investigación en Salud Pública | Katherine Laiton-Donato, Diego A. Álvarez-Díaz, Carlos Franco-Muñoz, Mauricio Pacheco-Montealegre, Jonathan Reales, Sheryl Corchuelo, Maria T. Herrera, Julian Naizaque, Gerardo Santamaría, Paola Muñoz-Laiton, Diego Andrés Prada, Magdalena Wiesner, Martha Lucia Ospina Martinez, Marcela Mercado-Reyes |
| EPI_ISL_791085 | Instituto Nacional de Salud - Unidad de Secuenciación y Análisis Genómico | Instituto Nacional de Salud - Dirección de Investigación en Salud Pública | Katherine Laiton-Donato, Diego A. Álvarez-Díaz, Carlos Franco-Muñoz, Mauricio Pacheco-Montealegre, Jeadran Malagon-Rojas, Jesith Toloza, Julia Almentero, Ronald Lopez, Jonathan Reales, Diego Andrés Prada, Magdalena Wiesner, Martha Lucia Ospina Martinez, Marcela Mercado-Reyes |
| EPI_ISL_791086, EPI_ISL_791087, EPI_ISL_791088, EPI_ISL_791089, EPI_ISL_791090, EPI_ISL_791091, EPI_ISL_791092, EPI_ISL_791093, EPI_ISL_791094, EPI_ISL_791095, EPI_ISL_791096 | | | |
| see above | Instituto Nacional de Salud - Unidad de Secuenciación y Análisis Genómico | Instituto Nacional de Salud - Dirección de Investigación en Salud Pública | Katherine Laiton-Donato, Diego A. Álvarez-Díaz, Carlos Franco-Muñoz, Mauricio Pacheco-Montealegre, Jonathan Reales, Sheryl Corchuelo, Maria T. Herrera, Julian Naizaque, Gerardo Santamaría, Paola Muñoz-Laiton, Diego Andrés Prada, Magdalena Wiesner, Martha Lucia Ospina Martinez, Marcela Mercado-Reyes |
| EPI_ISL_794649, EPI_ISL_794650, EPI_ISL_794651 | Fundación Cardio Infantil | Instituto Nacional de Salud - Dirección de Investigación en Salud Pública | Katherine Laiton-Donato, Diego A. Álvarez-Díaz, Carlos Franco-Muñoz, Mauricio Pacheco-Montealegre, Jonathan Reales, Sheryl Corchuelo, Maria T. Herrera, Julian Naizaque, Gerardo Santamaría, Paola Muñoz-Laiton, Diego Andrés Prada, Magdalena Wiesner, Martha Lucia Ospina Martinez, Marcela Mercado-Reyes |
| EPI_ISL_794652, EPI_ISL_794653 | LSP DEL TOLIMA | Instituto Nacional de Salud - Dirección de Investigación en Salud Pública | Katherine Laiton-Donato, Diego A. Álvarez-Díaz, Carlos Franco-Muñoz, Mauricio Pacheco-Montealegre, Jonathan Reales, Sheryl Corchuelo, Maria T. Herrera, Julian Naizaque, Gerardo Santamaría, Paola Muñoz-Laiton, Diego Andrés Prada, Magdalena Wiesner, Martha Lucia Ospina Martinez, Marcela Mercado-Reyes |
| EPI_ISL_794654, EPI_ISL_794655 | Fundación Valle del Lili | Instituto Nacional de Salud - Dirección de Investigación en Salud Pública | Katherine Laiton-Donato, Diego A. Álvarez-Díaz, Carlos Franco-Muñoz, Mauricio Pacheco-Montealegre, Jonathan Reales, Sheryl Corchuelo, Maria T. Herrera, Julian Naizaque, Gerardo Santamaría, Paola Muñoz-Laiton, Diego Andrés Prada, Magdalena Wiesner, Martha Lucia Ospina Martinez, Marcela Mercado-Reyes |

| | | | |
|---|---|---|---|
| EPI_ISL_794656, EPI_ISL_794657 | UNIDAD HEMATOLOGICA ESPECIALIZADA | Instituto Nacional de Salud - Dirección de Investigación en Salud Pública | Katherine Laiton-Donato, Diego A. Álvarez-Díaz, Carlos Franco-Muñoz, Mauricio Pacheco-Montealegre, Jonathan Reales, Sheryl Corchuelo, Maria T. Herrera, Julian Naizaque, Gerardo Santamaría, Paola Muñoz-Laiton, Diego Andrés Prada, Magdalena Wiesner, Martha Lucia Ospina Martinez, Marcela Mercado-Reyes |
| EPI_ISL_794658 | DIRECCION DE SANIDAD POLICIA NACIONAL | Instituto Nacional de Salud - Dirección de Investigación en Salud Pública | Katherine Laiton-Donato, Diego A. Álvarez-Díaz, Carlos Franco-Muñoz, Mauricio Pacheco-Montealegre, Jonathan Reales, Sheryl Corchuelo, Maria T. Herrera, Julian Naizaque, Gerardo Santamaría, Paola Muñoz-Laiton, Diego Andrés Prada, Magdalena Wiesner, Martha Lucia Ospina Martinez, Marcela Mercado-Reyes |
| EPI_ISL_794659, EPI_ISL_794660 | HOSPITAL UNIVERSITARIO SAN IGNACIO | Instituto Nacional de Salud - Dirección de Investigación en Salud Pública | Katherine Laiton-Donato, Diego A. Álvarez-Díaz, Carlos Franco-Muñoz, Mauricio Pacheco-Montealegre, Jonathan Reales, Sheryl Corchuelo, Maria T. Herrera, Julian Naizaque, Gerardo Santamaría, Paola Muñoz-Laiton, Diego Andrés Prada, Magdalena Wiesner, Martha Lucia Ospina Martinez, Marcela Mercado-Reyes |
| EPI_ISL_794661 | DRECCION DE SANIDAD POLICIA NACIONAL | Instituto Nacional de Salud - Dirección de Investigación en Salud Pública | Katherine Laiton-Donato, Diego A. Álvarez-Díaz, Carlos Franco-Muñoz, Mauricio Pacheco-Montealegre, Jonathan Reales, Sheryl Corchuelo, Maria T. Herrera, Julian Naizaque, Gerardo Santamaría, Paola Muñoz-Laiton, Diego Andrés Prada, Magdalena Wiesner, Martha Lucia Ospina Martinez, Marcela Mercado-Reyes |
| EPI_ISL_794662 | Fundación Cardio Infantil | Instituto Nacional de Salud - Dirección de Investigación en Salud Pública | Katherine Laiton-Donato, Diego A. Álvarez-Díaz, Carlos Franco-Muñoz, Mauricio Pacheco-Montealegre, Jonathan Reales, Sheryl Corchuelo, Maria T. Herrera, Julian Naizaque, Gerardo Santamaría, Paola Muñoz-Laiton, Diego Andrés Prada, Magdalena Wiesner, Martha Lucia Ospina Martinez, Marcela Mercado-Reyes |
| EPI_ISL_794663 | Carvajal Laboratorios | Instituto Nacional de Salud - Dirección de Investigación en Salud Pública | Katherine Laiton-Donato, Diego A. Álvarez-Díaz, Carlos Franco-Muñoz, Mauricio Pacheco-Montealegre, Jonathan Reales, Sheryl Corchuelo, Maria T. Herrera, Julian Naizaque, Gerardo Santamaría, Paola Muñoz-Laiton, Diego Andrés Prada, Magdalena Wiesner, Martha Lucia Ospina Martinez, Marcela Mercado-Reyes |
| EPI_ISL_794664, EPI_ISL_794665 | Fundación Valle del Lili | Instituto Nacional de Salud - Dirección de Investigación en Salud Pública | Katherine Laiton-Donato, Diego A. Álvarez-Díaz, Carlos Franco-Muñoz, Mauricio Pacheco-Montealegre, Jonathan Reales, Sheryl Corchuelo, Maria T. Herrera, Julian Naizaque, Gerardo Santamaría, Paola Muñoz-Laiton, Diego Andrés Prada, Magdalena Wiesner, Martha Lucia Ospina Martinez, Marcela Mercado-Reyes |
| EPI_ISL_794666, EPI_ISL_794667 | SYNLAB REGIONAL NOROCCIDENTE | Instituto Nacional de Salud - Dirección de Investigación en Salud Pública | Katherine Laiton-Donato, Diego A. Álvarez-Díaz, Carlos Franco-Muñoz, Mauricio Pacheco-Montealegre, Jonathan Reales, Sheryl Corchuelo, Maria T. Herrera, Julian Naizaque, Gerardo Santamaría, Paola Muñoz-Laiton, Diego Andrés Prada, Magdalena Wiesner, Martha Lucia Ospina Martinez, Marcela Mercado-Reyes |
| EPI_ISL_845618, EPI_ISL_845619 | Ayudas diagnósticas SURA | Instituto Nacional de Salud - Dirección de Investigación en Salud Pública | Katherine Laiton-Donato, Diego A. Álvarez-Díaz, Carlos Franco-Muñoz, Mauricio Pacheco-Montealegre, Maria T. Herrera-Sepúlveda, Jonathan Reales, Sheryll Corchuelo, Julian Naizaque, Gerardo Santamaría, Paola Muñoz-Laiton, Diego Andrés Prada, Magdalena Wiesner, Martha Lucia Ospina Martinez, Marcela Mercado-Reyes |
| EPI_ISL_845620 | compensar calle 63 | Instituto Nacional de Salud - Dirección de Investigación en Salud Pública | Katherine Laiton-Donato, Diego A. Álvarez-Díaz, Carlos Franco-Muñoz, Mauricio Pacheco-Montealegre, Maria T. Herrera-Sepúlveda, Jonathan Reales, Sheryll Corchuelo, Julian Naizaque, Gerardo Santamaría, Paola Muñoz-Laiton, Diego Andrés Prada, Magdalena Wiesner, Martha Lucia Ospina Martinez, Marcela Mercado-Reyes |
| EPI_ISL_845621, EPI_ISL_845622, EPI_ISL_845623, EPI_ISL_845624, EPI_ISL_845625 | Dirección de Sanidad Ejército | Instituto Nacional de Salud - Dirección de Investigación en Salud Pública | Katherine Laiton-Donato, Diego A. Álvarez-Díaz, Carlos Franco-Muñoz, Mauricio Pacheco-Montealegre, Maria T. Herrera-Sepúlveda, Jonathan Reales, Sheryll Corchuelo, Julian Naizaque, Gerardo Santamaría, Paola Muñoz-Laiton, Diego Andrés Prada, Magdalena Wiesner, Martha Lucia Ospina Martinez, Marcela Mercado-Reyes |
| EPI_ISL_845626 | FUNDACION CARDIOINFANTIL | Instituto Nacional de Salud - Dirección de Investigación en Salud Pública | Katherine Laiton-Donato, Diego A. Álvarez-Díaz, Carlos Franco-Muñoz, Mauricio Pacheco-Montealegre, Maria T. Herrera-Sepúlveda, Jonathan Reales, Sheryll Corchuelo, Julian Naizaque, Gerardo Santamaría, Paola Muñoz-Laiton, Diego Andrés Prada, Magdalena Wiesner, Martha Lucia Ospina Martinez, Marcela Mercado-Reyes |
| EPI_ISL_845627 | SYNLAB COLOMBIA S.A.S | Instituto Nacional de Salud - Dirección de Investigación en Salud Pública | Katherine Laiton-Donato, Diego A. Álvarez-Díaz, Carlos Franco-Muñoz, Mauricio Pacheco-Montealegre, Maria T. Herrera-Sepúlveda, Jonathan Reales, Sheryll Corchuelo, Julian Naizaque, Gerardo Santamaría, Paola Muñoz-Laiton, Diego Andrés Prada, Magdalena Wiesner, Martha Lucia Ospina Martinez, Marcela Mercado-Reyes |
| EPI_ISL_845628, EPI_ISL_845629 | Laboratorio Médico Echavarria | Instituto Nacional de Salud - Dirección de Investigación en Salud Pública | Katherine Laiton-Donato, Diego A. Álvarez-Díaz, Carlos Franco-Muñoz, Mauricio Pacheco-Montealegre, Maria T. Herrera-Sepúlveda, Jonathan Reales, Sheryll Corchuelo, Julian Naizaque, Gerardo Santamaría, Paola Muñoz-Laiton, Diego Andrés Prada, Magdalena Wiesner, Martha Lucia Ospina Martinez, Marcela Mercado-Reyes |
| EPI_ISL_845630, EPI_ISL_845631, EPI_ISL_845632, EPI_ISL_845633 | IDIME S.A | Instituto Nacional de Salud - Dirección de Investigación en Salud Pública | Katherine Laiton-Donato, Diego A. Álvarez-Díaz, Carlos Franco-Muñoz, Mauricio Pacheco-Montealegre, Maria T. Herrera-Sepúlveda, Jonathan Reales, Sheryll Corchuelo, Julian Naizaque, Gerardo Santamaría, Paola Muñoz-Laiton, Diego Andrés Prada, Magdalena Wiesner, Martha Lucia Ospina Martinez, Marcela Mercado-Reyes |
| EPI_ISL_845634 | Instituto Nacional de Cancerologia | Instituto Nacional de Salud - Dirección de Investigación en Salud Pública | Katherine Laiton-Donato, Diego A. Álvarez-Díaz, Carlos Franco-Muñoz, Mauricio Pacheco-Montealegre, Maria T. Herrera-Sepúlveda, Jonathan Reales, Sheryll Corchuelo, Julian Naizaque, Gerardo Santamaría, Paola Muñoz-Laiton, Diego Andrés Prada, Magdalena Wiesner, Martha Lucia Ospina Martinez, Marcela Mercado-Reyes |
| EPI_ISL_845635, EPI_ISL_845636, EPI_ISL_845637, EPI_ISL_845638, EPI_ISL_845639, EPI_ISL_845640, EPI_ISL_845641, EPI_ISL_845642, EPI_ISL_845643, EPI_ISL_845644, EPI_ISL_845645, EPI_ISL_845646, EPI_ISL_845647, EPI_ISL_845648, EPI_ISL_845649, EPI_ISL_845650, EPI_ISL_845651, EPI_ISL_845652, EPI_ISL_845653, EPI_ISL_845654, EPI_ISL_845655 | | | |
| see above | Laboratorio de Salud Pública - Secretaría Distrital de Salud | Instituto Nacional de Salud - Dirección de Investigación en Salud Pública | Katherine Laiton-Donato, Diego A. Álvarez-Díaz, Carlos Franco-Muñoz, Mauricio Pacheco-Montealegre, Maria T. Herrera-Sepúlveda, Jonathan Reales, Sheryll Corchuelo, Julian Naizaque, Gerardo Santamaría, Paola Muñoz-Laiton, Diego Andrés Prada, Magdalena Wiesner, Martha Lucia Ospina Martinez, Marcela Mercado-Reyes |
| EPI_ISL_845656 | UNIDAD HEMATOLÓGICA ESPECIALIZADA | Instituto Nacional de Salud - Dirección de Investigación en Salud Pública | Katherine Laiton-Donato, Diego A. Álvarez-Díaz, Carlos Franco-Muñoz, Mauricio Pacheco-Montealegre, Maria T. Herrera-Sepúlveda, Jonathan Reales, Sheryll Corchuelo, Julian Naizaque, Gerardo Santamaría, Paola Muñoz-Laiton, Diego Andrés Prada, Magdalena Wiesner, Martha Lucia Ospina Martinez, Marcela Mercado-Reyes |
| EPI_ISL_902736 | Dirección Departamental de Salud de Pública Leticia | Instituto Nacional de Salud- Dirección de Investigación en Salud Pública, Universidad de los Andes- Applied genomics research group, Vicerrectoria de Investigación y Creación, Universidad de los Andes- Systems and Computing Engineering Department | Katherine Laiton-Donato, Diego A. Álvarez-Díaz, Carlos Franco-Muñoz, Mauricio Pacheco-Montealegre, Maria T. Herrera-Sepúlveda, Diego Andrés Prada, Jorge Duitama, Laura Natalia Gonzalez, Jorge Ivan Diaz, Silvia Restrepo-Restrepo, Magdalena Wiesner, Martha Lucia Ospina Martinez, Marcela Mercado-Reyes |
| EPI_ISL_906144, EPI_ISL_906145 | Laboratorio de Salud Publica de Amazonas | Instituto Nacional de Salud- Dirección de Investigación en Salud Pública, Universidad de los Andes- Applied genomics research group, Vicerrectoria de Investigación y Creación, Universidad de los Andes- Systems and Computing Engineering Department | Katherine Laiton-Donato, Diego A. Álvarez-Díaz, Carlos Franco-Muñoz, Mauricio Pacheco-Montealegre, Héctor Alejandro Ruiz-Moreno, Maria T. Herrera-Sepúlveda, Diego Andrés Prada, Jhonnatan Reales-González, Sheryll Corchuelo, Julian Naizaque, Gerardo Santamaría Jorge Duitama, Laura Natalia Gonzalez, Jorge Ivan Diaz, Silvia Restrepo-Restrepo, Magdalena Wiesner, Martha Lucia Ospina Martinez, Marcela Mercado-Reyes |
| EPI_ISL_906529 | Direccion de partamental de salud del Amazonas | Instituto Nacional de Salud- Dirección de Investigación en Salud Pública, Universidad de los Andes- Applied genomics research group, Vicerrectoria de Investigación y Creación, Universidad de los Andes- Systems and Computing Engineering Department | Katherine Laiton-Donato, Diego A. Álvarez-Díaz, Carlos Franco-Muñoz, Mauricio Pacheco-Montealegre, Héctor Alejandro Ruiz-Moreno, Maria T. Herrera-Sepúlveda, Diego Andrés Prada, Jhonnatan Reales-González, Sheryll Corchuelo, Julian Naizaque, Gerardo Santamaría Jorge Duitama, Laura Natalia Gonzalez, Jorge Ivan Diaz, Silvia Restrepo-Restrepo, Magdalena Wiesner, Martha Lucia Ospina Martinez, Marcela Mercado-Reyes |
| EPI_ISL_906530 | Laboratorio de salud publica de Bogota | Instituto Nacional de Salud- Dirección de Investigación en Salud Pública, Universidad de los Andes- Applied genomics research group, Vicerrectoria de Investigación y Creación, Universidad de los Andes- Systems and Computing Engineering Department | Katherine Laiton-Donato, Diego A. Álvarez-Díaz, Carlos Franco-Muñoz, Mauricio Pacheco-Montealegre, Héctor Alejandro Ruiz-Moreno, Maria T. Herrera-Sepúlveda, Diego Andrés Prada, Jhonnatan Reales-González, Sheryll Corchuelo, Julian Naizaque, Gerardo Santamaría Jorge Duitama, Laura Natalia Gonzalez, Jorge Ivan Diaz, Silvia Restrepo-Restrepo, Magdalena Wiesner, Martha Lucia Ospina Martinez, Marcela Mercado-Reyes |

| | | | |
|---|---|---|---|
| EPI_ISL_906531 | Laboratorio de salud publica del Valle del Cauca | Instituto Nacional de Salud- Dirección de Investigación en Salud Pública, Universidad de los Andes- Applied genomics research group, Vicerrectoria de Investigación y Creación, Universidad de los Andes- Systems and Computing Engineering Department | Katherine Laiton-Donato, Diego A. Álvarez-Díaz, Carlos Franco-Muñoz, Mauricio Pacheco-Montealegre, Héctor Alejandro Ruiz-Moreno, Maria T. Herrera-Sepúlveda, Diego Andrés Prada, Jhonnatan Reales-González, Sheryll Corchuelo, Julian Naizaque, Gerardo Santamaría Jorge Duitama, Laura Natalia Gonzalez, Jorge Ivan Diaz, Silvia Restrepo-Restrepo, Magdalena Wiesner, Martha Lucia Ospina Martinez, Marcela Mercado-Reyes |
| EPI_ISL_906532 | Hospital San Jose de Maicao | Instituto Nacional de Salud- Dirección de Investigación en Salud Pública, Universidad de los Andes- Applied genomics research group, Vicerrectoria de Investigación y Creación, Universidad de los Andes- Systems and Computing Engineering Department | Katherine Laiton-Donato, Diego A. Álvarez-Díaz, Carlos Franco-Muñoz, Mauricio Pacheco-Montealegre, Héctor Alejandro Ruiz-Moreno, Maria T. Herrera-Sepúlveda, Diego Andrés Prada, Jhonnatan Reales-González, Sheryll Corchuelo, Julian Naizaque, Gerardo Santamaría Jorge Duitama, Laura Natalia Gonzalez, Jorge Ivan Diaz, Silvia Restrepo-Restrepo, Magdalena Wiesner, Martha Lucia Ospina Martinez, Marcela Mercado-Reyes |
| EPI_ISL_906533 | Laboratorio de Virologia-Instituto Nacional de Salud | Instituto Nacional de Salud- Dirección de Investigación en Salud Pública, Universidad de los Andes- Applied genomics research group, Vicerrectoria de Investigación y Creación, Universidad de los Andes- Systems and Computing Engineering Department | Katherine Laiton-Donato, Diego A. Álvarez-Díaz, Carlos Franco-Muñoz, Mauricio Pacheco-Montealegre, Héctor Alejandro Ruiz-Moreno, Maria T. Herrera-Sepúlveda, Diego Andrés Prada, Jhonnatan Reales-González, Sheryll Corchuelo, Julian Naizaque, Gerardo Santamaría Jorge Duitama, Laura Natalia Gonzalez, Jorge Ivan Diaz, Silvia Restrepo-Restrepo, Magdalena Wiesner, Martha Lucia Ospina Martinez, Marcela Mercado-Reyes |
| EPI_ISL_906534 | Laboratorio de Salud Publica de Amazonas | Instituto Nacional de Salud- Dirección de Investigación en Salud Pública, Universidad de los Andes- Applied genomics research group, Vicerrectoria de Investigación y Creación, Universidad de los Andes- Systems and Computing Engineering Department | Katherine Laiton-Donato, Diego A. Álvarez-Díaz, Carlos Franco-Muñoz, Mauricio Pacheco-Montealegre, Héctor Alejandro Ruiz-Moreno, Maria T. Herrera-Sepúlveda, Diego Andrés Prada, Jhonnatan Reales-González, Sheryll Corchuelo, Julian Naizaque, Gerardo Santamaría Jorge Duitama, Laura Natalia Gonzalez, Jorge Ivan Diaz, Silvia Restrepo-Restrepo, Magdalena Wiesner, Martha Lucia Ospina Martinez, Marcela Mercado-Reyes |
| EPI_ISL_906535 | HOSPITAL DEPARTAMENTAL DE VILLAVICENCIO E.S.E. | Instituto Nacional de Salud- Dirección de Investigación en Salud Pública, Universidad de los Andes- Applied genomics research group, Vicerrectoria de Investigación y Creación, Universidad de los Andes- Systems and Computing Engineering Department | Katherine Laiton-Donato, Diego A. Álvarez-Díaz, Carlos Franco-Muñoz, Mauricio Pacheco-Montealegre, Héctor Alejandro Ruiz-Moreno, Maria T. Herrera-Sepúlveda, Diego Andrés Prada, Jhonnatan Reales-González, Sheryll Corchuelo, Julian Naizaque, Gerardo Santamaría Jorge Duitama, Laura Natalia Gonzalez, Jorge Ivan Diaz, Silvia Restrepo-Restrepo, Magdalena Wiesner, Martha Lucia Ospina Martinez, Marcela Mercado-Reyes |
| EPI_ISL_906536 | CLINICA OCCIDENTE | Instituto Nacional de Salud- Dirección de Investigación en Salud Pública, Universidad de los Andes- Applied genomics research group, Vicerrectoria de Investigación y Creación, Universidad de los Andes- Systems and Computing Engineering Department | Katherine Laiton-Donato, Diego A. Álvarez-Díaz, Carlos Franco-Muñoz, Mauricio Pacheco-Montealegre, Héctor Alejandro Ruiz-Moreno, Maria T. Herrera-Sepúlveda, Diego Andrés Prada, Jhonnatan Reales-González, Sheryll Corchuelo, Julian Naizaque, Gerardo Santamaría Jorge Duitama, Laura Natalia Gonzalez, Jorge Ivan Diaz, Silvia Restrepo-Restrepo, Magdalena Wiesner, Martha Lucia Ospina Martinez, Marcela Mercado-Reyes |
| EPI_ISL_906537 | HOSPITAL DEPARTAMENTAL DE VILLAVICENCIO E.S.E. | Instituto Nacional de Salud- Dirección de Investigación en Salud Pública, Universidad de los Andes- Applied genomics research group, Vicerrectoria de Investigación y Creación, Universidad de los Andes- Systems and Computing Engineering Department | Katherine Laiton-Donato, Diego A. Álvarez-Díaz, Carlos Franco-Muñoz, Mauricio Pacheco-Montealegre, Héctor Alejandro Ruiz-Moreno, Maria T. Herrera-Sepúlveda, Diego Andrés Prada, Jhonnatan Reales-González, Sheryll Corchuelo, Julian Naizaque, Gerardo Santamaría Jorge Duitama, Laura Natalia Gonzalez, Jorge Ivan Diaz, Silvia Restrepo-Restrepo, Magdalena Wiesner, Martha Lucia Ospina Martinez, Marcela Mercado-Reyes |
| EPI_ISL_906539 | HOSPITAL DEPARTAMENTAL DE VILLAVICENCIO | Instituto Nacional de Salud- Dirección de Investigación en Salud Pública, Universidad de los Andes- Applied genomics research group, Vicerrectoria de Investigación y Creación, Universidad de los Andes- Systems and Computing Engineering Department | Katherine Laiton-Donato, Diego A. Álvarez-Díaz, Carlos Franco-Muñoz, Mauricio Pacheco-Montealegre, Héctor Alejandro Ruiz-Moreno, Maria T. Herrera-Sepúlveda, Diego Andrés Prada, Jhonnatan Reales-González, Sheryll Corchuelo, Julian Naizaque, Gerardo Santamaría Jorge Duitama, Laura Natalia Gonzalez, Jorge Ivan Diaz, Silvia Restrepo-Restrepo, Magdalena Wiesner, Martha Lucia Ospina Martinez, Marcela Mercado-Reyes |
| EPI_ISL_906540 | CLINICA DE OCCIDENTE | Instituto Nacional de Salud- Dirección de Investigación en Salud Pública, Universidad de los Andes- Applied genomics research group, Vicerrectoria de Investigación y Creación, Universidad de los Andes- Systems and Computing Engineering Department | Katherine Laiton-Donato, Diego A. Álvarez-Díaz, Carlos Franco-Muñoz, Mauricio Pacheco-Montealegre, Héctor Alejandro Ruiz-Moreno, Maria T. Herrera-Sepúlveda, Diego Andrés Prada, Jhonnatan Reales-González, Sheryll Corchuelo, Julian Naizaque, Gerardo Santamaría Jorge Duitama, Laura Natalia Gonzalez, Jorge Ivan Diaz, Silvia Restrepo-Restrepo, Magdalena Wiesner, Martha Lucia Ospina Martinez, Marcela Mercado-Reyes |
| EPI_ISL_906541 | HOSPITAL SAN JOSE DE MAICAO | Instituto Nacional de Salud- Dirección de Investigación en Salud Pública, Universidad de los Andes- Applied genomics research group, Vicerrectoria de Investigación y Creación, Universidad de los Andes- Systems and Computing Engineering Department | Katherine Laiton-Donato, Diego A. Álvarez-Díaz, Carlos Franco-Muñoz, Mauricio Pacheco-Montealegre, Héctor Alejandro Ruiz-Moreno, Maria T. Herrera-Sepúlveda, Diego Andrés Prada, Jhonnatan Reales-González, Sheryll Corchuelo, Julian Naizaque, Gerardo Santamaría Jorge Duitama, Laura Natalia Gonzalez, Jorge Ivan Diaz, Silvia Restrepo-Restrepo, Magdalena Wiesner, Martha Lucia Ospina Martinez, Marcela Mercado-Reyes |
| EPI_ISL_906542 | Laboratorio de Virologia-Instituto Nacional de Salud | Instituto Nacional de Salud- Dirección de Investigación en Salud Pública, Universidad de los Andes- Applied genomics research group, Vicerrectoria de Investigación y Creación, Universidad de los Andes- Systems and Computing Engineering Department | Katherine Laiton-Donato, Diego A. Álvarez-Díaz, Carlos Franco-Muñoz, Mauricio Pacheco-Montealegre, Héctor Alejandro Ruiz-Moreno, Maria T. Herrera-Sepúlveda, Diego Andrés Prada, Jhonnatan Reales-González, Sheryll Corchuelo, Julian Naizaque, Gerardo Santamaría Jorge Duitama, Laura Natalia Gonzalez, Jorge Ivan Diaz, Silvia Restrepo-Restrepo, Magdalena Wiesner, Martha Lucia Ospina Martinez, Marcela Mercado-Reyes |
| EPI_ISL_906543 | FUNDACION HOSPITAL SAN PEDRO | Instituto Nacional de Salud- Dirección de Investigación en Salud Pública, Universidad de los Andes- Applied genomics research group, Vicerrectoria de Investigación y Creación, Universidad de los Andes- Systems and Computing Engineering Department | Katherine Laiton-Donato, Diego A. Álvarez-Díaz, Carlos Franco-Muñoz, Mauricio Pacheco-Montealegre, Héctor Alejandro Ruiz-Moreno, Maria T. Herrera-Sepúlveda, Diego Andrés Prada, Jhonnatan Reales-González, Sheryll Corchuelo, Julian Naizaque, Gerardo Santamaría Jorge Duitama, Laura Natalia Gonzalez, Jorge Ivan Diaz, Silvia Restrepo-Restrepo, Magdalena Wiesner, Martha Lucia Ospina Martinez, Marcela Mercado-Reyes |
| EPI_ISL_906544 | CLINICA DE OCCIDENTE | Instituto Nacional de Salud- Dirección de Investigación en Salud Pública, Universidad de los Andes- Applied genomics research group, Vicerrectoria de Investigación y Creación, Universidad de los Andes- Systems and Computing Engineering Department | Katherine Laiton-Donato, Diego A. Álvarez-Díaz, Carlos Franco-Muñoz, Mauricio Pacheco-Montealegre, Héctor Alejandro Ruiz-Moreno, Maria T. Herrera-Sepúlveda, Diego Andrés Prada, Jhonnatan Reales-González, Sheryll Corchuelo, Julian Naizaque, Gerardo Santamaría Jorge Duitama, Laura Natalia Gonzalez, Jorge Ivan Diaz, Silvia Restrepo-Restrepo, Magdalena Wiesner, Martha Lucia Ospina Martinez, Marcela Mercado-Reyes |
| EPI_ISL_906545 | E.S.E. HOSPITAL SAN JOSE DE MAICAO | Instituto Nacional de Salud- Dirección de Investigación en Salud Pública, Universidad de los Andes- Applied genomics research group, Vicerrectoria de Investigación y Creación, Universidad de los Andes- Systems and Computing Engineering Department | Katherine Laiton-Donato, Diego A. Álvarez-Díaz, Carlos Franco-Muñoz, Mauricio Pacheco-Montealegre, Héctor Alejandro Ruiz-Moreno, Maria T. Herrera-Sepúlveda, Diego Andrés Prada, Jhonnatan Reales-González, Sheryll Corchuelo, Julian Naizaque, Gerardo Santamaría Jorge Duitama, Laura Natalia Gonzalez, Jorge Ivan Diaz, Silvia Restrepo-Restrepo, Magdalena Wiesner, Martha Lucia Ospina Martinez, Marcela Mercado-Reyes |
| EPI_ISL_906547 | HOSPITAL DEPARTAMENTAL DE VILLAVICENCIO E.S.E. | Instituto Nacional de Salud- Dirección de Investigación en Salud Pública, Universidad de los Andes- Applied genomics research group, Vicerrectoria de Investigación y Creación, Universidad de los Andes- Systems and Computing Engineering Department | Katherine Laiton-Donato, Diego A. Álvarez-Díaz, Carlos Franco-Muñoz, Mauricio Pacheco-Montealegre, Héctor Alejandro Ruiz-Moreno, Maria T. Herrera-Sepúlveda, Diego Andrés Prada, Jhonnatan Reales-González, Sheryll Corchuelo, Julian Naizaque, Gerardo Santamaría Jorge Duitama, Laura Natalia Gonzalez, Jorge Ivan Diaz, Silvia Restrepo-Restrepo, Magdalena Wiesner, Martha Lucia Ospina Martinez, Marcela Mercado-Reyes |
| EPI_ISL_906548 | Laboratorio de Salud Pública Fronterizo de Arauca | Instituto Nacional de Salud- Dirección de Investigación en Salud Pública, Universidad de los Andes- Applied genomics research group, Vicerrectoria de Investigación y Creación, Universidad de los Andes- Systems and Computing Engineering Department | Katherine Laiton-Donato, Diego A. Álvarez-Díaz, Carlos Franco-Muñoz, Mauricio Pacheco-Montealegre, Héctor Alejandro Ruiz-Moreno, Maria T. Herrera-Sepúlveda, Diego Andrés Prada, Jhonnatan Reales-González, Sheryll Corchuelo, Julian Naizaque, Gerardo Santamaría Jorge Duitama, Laura Natalia Gonzalez, Jorge Ivan Diaz, Silvia Restrepo-Restrepo, Magdalena Wiesner, Martha Lucia Ospina Martinez, Marcela Mercado-Reyes |

| | | | |
|---|---|---|---|
| EPI_ISL_906549 | Laboratorio de Virologia-Instituto Nacional de Salud | Instituto Nacional de Salud- Dirección de Investigación en Salud Pública, Universidad de los Andes- Applied genomics research group, Vicerrectoria de Investigación y Creación, Universidad de los Andes- Systems and Computing Engineering Department | Katherine Laiton-Donato, Diego A. Álvarez-Díaz, Carlos Franco-Muñoz, Mauricio Pacheco-Montealegre, Héctor Alejandro Ruiz-Moreno, Maria T. Herrera-Sepúlveda, Diego Andrés Prada, Jhonnatan Reales-González, Sheryll Corchuelo, Julian Naizaque, Gerardo Santamaría Jorge Duitama, Laura Natalia Gonzalez, Jorge Ivan Diaz, Silvia Restrepo-Restrepo, Magdalena Wiesner, Martha Lucia Ospina Martinez, Marcela Mercado-Reyes |
| EPI_ISL_906550, EPI_ISL_906551 | DIRECCION DEPARTAMENTAL DE SALUD DE AMAZONAS | Instituto Nacional de Salud- Dirección de Investigación en Salud Pública, Universidad de los Andes- Applied genomics research group, Vicerrectoria de Investigación y Creación, Universidad de los Andes- Systems and Computing Engineering Department | Katherine Laiton-Donato, Diego A. Álvarez-Díaz, Carlos Franco-Muñoz, Mauricio Pacheco-Montealegre, Héctor Alejandro Ruiz-Moreno, Maria T. Herrera-Sepúlveda, Diego Andrés Prada, Jhonnatan Reales-González, Sheryll Corchuelo, Julian Naizaque, Gerardo Santamaría Jorge Duitama, Laura Natalia Gonzalez, Jorge Ivan Diaz, Silvia Restrepo-Restrepo, Magdalena Wiesner, Martha Lucia Ospina Martinez, Marcela Mercado-Reyes |
| EPI_ISL_906552 | Laboratorio Angel Diagnostica | Instituto Nacional de Salud- Dirección de Investigación en Salud Pública, Universidad de los Andes- Applied genomics research group, Vicerrectoria de Investigación y Creación, Universidad de los Andes- Systems and Computing Engineering Department | Katherine Laiton-Donato, Diego A. Álvarez-Díaz, Carlos Franco-Muñoz, Mauricio Pacheco-Montealegre, Héctor Alejandro Ruiz-Moreno, Maria T. Herrera-Sepúlveda, Diego Andrés Prada, Jhonnatan Reales-González, Sheryll Corchuelo, Julian Naizaque, Gerardo Santamaría Jorge Duitama, Laura Natalia Gonzalez, Jorge Ivan Diaz, Silvia Restrepo-Restrepo, Magdalena Wiesner, Martha Lucia Ospina Martinez, Marcela Mercado-Reyes |
| EPI_ISL_906553, EPI_ISL_906554, EPI_ISL_906555 | Laboratorio Bienestar | Instituto Nacional de Salud- Dirección de Investigación en Salud Pública, Universidad de los Andes- Applied genomics research group, Vicerrectoria de Investigación y Creación, Universidad de los Andes- Systems and Computing Engineering Department | Katherine Laiton-Donato, Diego A. Álvarez-Díaz, Carlos Franco-Muñoz, Mauricio Pacheco-Montealegre, Héctor Alejandro Ruiz-Moreno, Maria T. Herrera-Sepúlveda, Diego Andrés Prada, Jhonnatan Reales-González, Sheryll Corchuelo, Julian Naizaque, Gerardo Santamaría Jorge Duitama, Laura Natalia Gonzalez, Jorge Ivan Diaz, Silvia Restrepo-Restrepo, Magdalena Wiesner, Martha Lucia Ospina Martinez, Marcela Mercado-Reyes |
| EPI_ISL_906556 | DIRECCION DEPARTAMENTAL DE SALUD DE GUAJIRA | Instituto Nacional de Salud- Dirección de Investigación en Salud Pública, Universidad de los Andes- Applied genomics research group, Vicerrectoria de Investigación y Creación, Universidad de los Andes- Systems and Computing Engineering Department | Katherine Laiton-Donato, Diego A. Álvarez-Díaz, Carlos Franco-Muñoz, Mauricio Pacheco-Montealegre, Héctor Alejandro Ruiz-Moreno, Maria T. Herrera-Sepúlveda, Diego Andrés Prada, Jhonnatan Reales-González, Sheryll Corchuelo, Julian Naizaque, Gerardo Santamaría Jorge Duitama, Laura Natalia Gonzalez, Jorge Ivan Diaz, Silvia Restrepo-Restrepo, Magdalena Wiesner, Martha Lucia Ospina Martinez, Marcela Mercado-Reyes |
| EPI_ISL_906557, EPI_ISL_906558, EPI_ISL_906559, EPI_ISL_906560, EPI_ISL_906561 | Laboratorio de Salud Publica de Amazonas | Instituto Nacional de Salud- Dirección de Investigación en Salud Pública, Universidad de los Andes- Applied genomics research group, Vicerrectoria de Investigación y Creación, Universidad de los Andes- Systems and Computing Engineering Department | Katherine Laiton-Donato, Diego A. Álvarez-Díaz, Carlos Franco-Muñoz, Mauricio Pacheco-Montealegre, Héctor Alejandro Ruiz-Moreno, Maria T. Herrera-Sepúlveda, Diego Andrés Prada, Jhonnatan Reales-González, Sheryll Corchuelo, Julian Naizaque, Gerardo Santamaría Jorge Duitama, Laura Natalia Gonzalez, Jorge Ivan Diaz, Silvia Restrepo-Restrepo, Magdalena Wiesner, Martha Lucia Ospina Martinez, Marcela Mercado-Reyes |
| EPI_ISL_956282, EPI_ISL_956288, EPI_ISL_956289, EPI_ISL_956290, EPI_ISL_956297 | Instituto Nacional de Salud- Dirección de Redes de Laboratorios de Salud Pública | Instituto Nacional de Salud- Dirección de Investigación en Salud Pública | Katherine Laiton-Donato, Diego A. Álvarez-Díaz, Carlos Franco-Muñoz, Mauricio Pacheco-Montealegre, Hector Alejandro Ruiz-Moreno, Maria T. Herrera-Sepúlveda, Diego Andrés Prada, Jhonnatan Reales-González, Sheryll Corchuelo, Julian Naizaque, Gerardo Santamaría, Magdalena Wiesner, Martha Lucia Ospina Martinez, Marcela Mercado-Reyes |
| EPI_ISL_956298 | Laboratorio de salud publica del Valle del Cauca | Instituto Nacional de Salud- Dirección de Investigación en Salud Pública | Katherine Laiton-Donato, Diego A. Álvarez-Díaz, Carlos Franco-Muñoz, Mauricio Pacheco-Montealegre, Hector Alejandro Ruiz-Moreno, Maria T. Herrera-Sepúlveda, Diego Andrés Prada, Jhonnatan Reales-González, Sheryll Corchuelo, Julian Naizaque, Gerardo Santamaría, Magdalena Wiesner, Martha Lucia Ospina Martinez, Marcela Mercado-Reyes |
| EPI_ISL_956299, EPI_ISL_956300 | Laboratorio de salud publica Arauca | Instituto Nacional de Salud- Dirección de Investigación en Salud Pública | Katherine Laiton-Donato, Diego A. Álvarez-Díaz, Carlos Franco-Muñoz, Mauricio Pacheco-Montealegre, Hector Alejandro Ruiz-Moreno, Maria T. Herrera-Sepúlveda, Diego Andrés Prada, Jhonnatan Reales-González, Sheryll Corchuelo, Julian Naizaque, Gerardo Santamaría, Magdalena Wiesner, Martha Lucia Ospina Martinez, Marcela Mercado-Reyes |
| EPI_ISL_956301 | Laboratorio Gencore- Universidad de los Andes | Instituto Nacional de Salud- Dirección de Investigación en Salud Pública, Universidad de los Andes- Gencore | Katherine Laiton-Donato, Diego A. Álvarez-Díaz, Carlos Franco-Muñoz, Mauricio Pacheco-Montealegre, Hector Alejandro Ruiz-Moreno, Maria T. Herrera-Sepúlveda, Diego Andrés Prada, Jhonnatan Reales-González, Sheryll Corchuelo, Julian Naizaque, Gerardo Santamaría, Magdalena Wiesner, Martha Lucia Ospina Martinez, Marcela Mercado-Reyes |
| EPI_ISL_956304, EPI_ISL_956305 | ESE INSTITUTO NACIONAL DE CANCEROLOGIA | Instituto Nacional de Salud- Dirección de Investigación en Salud Pública | Katherine Laiton-Donato, Diego A. Álvarez-Díaz, Carlos Franco-Muñoz, Mauricio Pacheco-Montealegre, Hector Alejandro Ruiz-Moreno, Maria T. Herrera-Sepúlveda, Diego Andrés Prada, Jhonnatan Reales-González, Sheryll Corchuelo, Julian Naizaque, Gerardo Santamaría, Magdalena Wiesner, Martha Lucia Ospina Martinez, Marcela Mercado-Reyes |
| EPI_ISL_956306 | Laboratorio Gencore- Universidad de los Andes | Instituto Nacional de Salud- Dirección de Investigación en Salud Pública, Universidad de los Andes- Gencore | Katherine Laiton-Donato, Diego A. Álvarez-Díaz, Carlos Franco-Muñoz, Mauricio Pacheco-Montealegre, Hector Alejandro Ruiz-Moreno, Maria T. Herrera-Sepúlveda, Diego Andrés Prada, Jhonnatan Reales-González, Sheryll Corchuelo, Julian Naizaque, Gerardo Santamaría, Magdalena Wiesner, Martha Lucia Ospina Martinez, Marcela Mercado-Reyes |

We gratefully acknowledge the following Authors from the Originating laboratories responsible for obtaining the specimens, as well as the Submitting laboratories where the genome data were generated and shared via GISAID, on which this research is based.

All Submitters of data may be contacted directly via www.gisaid.org

Authors are sorted alphabetically.

| Accession ID | Originating Laboratory | Submitting Laboratory | Authors |
|---|---|---|---|
| EPI_ISL_1939975, EPI_ISL_1939976, EPI_ISL_1939977, EPI_ISL_1939978, EPI_ISL_1939979, EPI_ISL_1940022, EPI_ISL_1940023, EPI_ISL_1940024, EPI_ISL_1940025, EPI_ISL_1940026, EPI_ISL_1940027, EPI_ISL_1940028, EPI_ISL_1940036, EPI_ISL_2015537, EPI_ISL_2015598 | | | |
| see above | A D HALLI | inStem NCBS - INSACOG | Uma Ramakrishnan Dasaradhi Palakodeti Aswin SaiNarain |
| EPI_ISL_1940091, EPI_ISL_1940092, EPI_ISL_1940130, EPI_ISL_1940131, EPI_ISL_1940132, EPI_ISL_2015553 | AD HALLI | inStem NCBS - INSACOG | Uma Ramakrishnan Dasaradhi Palakodeti Aswin SaiNarain |
| EPI_ISL_2015498 | AGAR PHC | inStem NCBS - INSACOG | Uma Ramakrishnan Dasaradhi Palakodeti Aswin SaiNarain |
| EPI_ISL_2015501 | AGARA UPHC | inStem NCBS - INSACOG | Uma Ramakrishnan Dasaradhi Palakodeti Aswin SaiNarain |
| EPI_ISL_2015473, EPI_ISL_2015474, EPI_ISL_2015476, EPI_ISL_2015477, EPI_ISL_2015484, EPI_ISL_2015488, EPI_ISL_2015491, EPI_ISL_2015496, EPI_ISL_2015535 | | | |
| see above | AGRAHARA PHC | inStem NCBS - INSACOG | Uma Ramakrishnan Dasaradhi Palakodeti Aswin SaiNarain |
| EPI_ISL_1663504, EPI_ISL_1663505, EPI_ISL_1663506, EPI_ISL_1663507, EPI_ISL_1663508, EPI_ISL_1663509, EPI_ISL_1663510, EPI_ISL_1663511, EPI_ISL_1663512, EPI_ISL_1663513, EPI_ISL_1663514, EPI_ISL_1663515, EPI_ISL_1663516, EPI_ISL_1663517, EPI_ISL_1663518, EPI_ISL_1663519, EPI_ISL_1663520, EPI_ISL_1663521, EPI_ISL_1663522, EPI_ISL_1663523 | | | |
| see above | AIIMS, Patna | Institute of Life Sciences - INSACOG | Ajay Parida; Amol M. Kanampalliwar; Arup Ghosh; Atimukta Jha; INSACOG Consortium; Punit Prasad; Rajeeb Swain; Rupesh Dash; Safal Walia; Shifu Aggarwal; Sunil K. Raghav |
| EPI_ISL_1372268, EPI_ISL_1372271, EPI_ISL_1372273, EPI_ISL_1372275, EPI_ISL_1372277, EPI_ISL_1547799, EPI_ISL_1547800 | | | |
| see above | AP SSO | CSIR-Centre for Cellular and Molecular Biology - INSACOG | Amareshwar Vodapalli; Ara Sreenivas; B Himasri; Divya Tej Sowpati; Karthik Bharadwaj Tallapaka; Lamuk Zaveri; Onkar Kulkarni; Rakesh K Mishra; Sharath Chandra Thota; Shreekant Verma; Sofia Banu; Viswagithe S L |
| EPI_ISL_1940093, EPI_ISL_1940094 | APOLLO | inStem NCBS - INSACOG | Uma Ramakrishnan Dasaradhi Palakodeti Aswin SaiNarain |
| EPI_ISL_1972130, EPI_ISL_1972131, EPI_ISL_1972132, EPI_ISL_1972133, EPI_ISL_1972134, EPI_ISL_1972135, EPI_ISL_1972136, EPI_ISL_1972137, EPI_ISL_1972138, EPI_ISL_1972139, EPI_ISL_1972140, EPI_ISL_1972141, EPI_ISL_1972142, EPI_ISL_1972143, EPI_ISL_1972144, EPI_ISL_1972145, EPI_ISL_1972146, EPI_ISL_1972147 | | | |
| see above | All India Institute of Medical Sciences, Bhopal | All India Institute of Medical Sciences, Bhopal | Anvita Gupta Malhotra; Arun Raghuvanshi; Ashvini Kumar Yadav; Debasis Biswas and Sarman Singh; Dipesh Kale; Jitendra Singh; Prem Shankar; Shashwati Nema |
| EPI_ISL_909958, EPI_ISL_909962 | Apollo Hospitals | CSIR-Centre for Cellular and Molecular Biology | Divya Tej Sowpati; Irawathy Goud; Karthik Bharadwaj Tallapaka; Lamuk Zaveri; Onkar Kulkarni; Payel Mukherjee; Sofia Banu; Suneetha Narreddy |
| EPI_ISL_1708325, EPI_ISL_1708436, EPI_ISL_1708441, EPI_ISL_1708444 | Atturu PHC | inStem NCBS - INSACOG | Uma Ramakrishnan Dasaradhi Palakodeti Aswin SaiNarain |
| EPI_ISL_825034, EPI_ISL_825035, EPI_ISL_825036, EPI_ISL_825042, EPI_ISL_1544110, EPI_ISL_1544111, EPI_ISL_1544112, EPI_ISL_1544113, EPI_ISL_1544114, EPI_ISL_1544115, EPI_ISL_1544116, EPI_ISL_1544117 | | | |
| see above | B.J. Medical College and Civil hospital, Ahmedabad | Gujarat Biotechnology Research Centre | Afzal Ansari; Apurvasinh Puvar; Chaitanya Joshi; Dinesh Kumar; Dipa Kinariwala; Janvi Raval; Kamlesh J Upadhyay; Labdhi Pandya; Madhvi Joshi; Nikha Trivedi; Nilima Shah; Nitesh Shah; Nitin Savaliya; Pranay Shah; Ramesh Pandit; Sanjay Kapadia; Sonal Sharma; Twinkle Soni; Umang Mishra; Zarna Patel; Zuber Saiyed |
| EPI_ISL_995696, EPI_ISL_995708, EPI_ISL_995709, EPI_ISL_995710, EPI_ISL_995711, EPI_ISL_995712, EPI_ISL_995724, EPI_ISL_995748, EPI_ISL_995763 | | | |
| see above | BANGALORE MEDICAL COLLEGE AND RESEARCH INSTITUTE | Department of Neurovirology, National Institute of Mental Health and Neurosciences (NIMHANS) | Anita S Desai; Anson Kunjumon George; Chitra Pattabiraman; Darshan Sreenivas; Harsha.P.K; Nakka Vijay Kiran Reddy; Pramada Prasad; Risha Rasheed; V Ravi |
| EPI_ISL_1385820, EPI_ISL_1595865, EPI_ISL_1595866 | BANGALORE MEDICAL COLLEGE AND RESEARCH INSTITUTE | INSACOG-KA, NIMHANS | Ananthapadmanabha Kotambail; Anita S Desai; Anson Kunjumon George; Chetan G K; Chitra Pattabiraman; Darshan Sreenivas; Gautham Arunachal Udupi; Nakka Vijay Kiran Reddy; Pramada Prasad; V Ravi |
| EPI_ISL_747244, EPI_ISL_803901, EPI_ISL_803902, EPI_ISL_803904, EPI_ISL_803908, EPI_ISL_803911, EPI_ISL_803912, EPI_ISL_807158, EPI_ISL_825141, EPI_ISL_825142, EPI_ISL_825143, EPI_ISL_825148, EPI_ISL_845874, EPI_ISL_845875, EPI_ISL_845876, EPI_ISL_845878, EPI_ISL_845879, EPI_ISL_845880, EPI_ISL_845883, EPI_ISL_845884, EPI_ISL_845887, EPI_ISL_845888, EPI_ISL_845889, EPI_ISL_845890, EPI_ISL_860189, EPI_ISL_860194, EPI_ISL_860195, EPI_ISL_860197, EPI_ISL_860199, EPI_ISL_860200, EPI_ISL_860201, EPI_ISL_995173, EPI_ISL_995174, EPI_ISL_995175, EPI_ISL_995176, EPI_ISL_995177, EPI_ISL_995178, EPI_ISL_995179, EPI_ISL_995180, EPI_ISL_995181, EPI_ISL_995182, EPI_ISL_995183, EPI_ISL_995184, EPI_ISL_995704, EPI_ISL_995705, EPI_ISL_995715, EPI_ISL_995716, EPI_ISL_995717, EPI_ISL_995718, EPI_ISL_995719, EPI_ISL_995725, EPI_ISL_995728, EPI_ISL_995729, EPI_ISL_995732, EPI_ISL_995735, EPI_ISL_995736, EPI_ISL_995739, EPI_ISL_995740, EPI_ISL_995741, EPI_ISL_995746, EPI_ISL_995750, EPI_ISL_995751, EPI_ISL_995752, EPI_ISL_995754, EPI_ISL_995755, EPI_ISL_995756, EPI_ISL_995757, EPI_ISL_1007660, EPI_ISL_1007661, EPI_ISL_1250853, EPI_ISL_1250854, EPI_ISL_1250855, EPI_ISL_1250856, EPI_ISL_1250857, EPI_ISL_1250858 | | | |
| see above | BBMP Urban PHC | Department of Neurovirology, National Institute of Mental Health and Neurosciences (NIMHANS) | Anita S Desai; Anson Kunjumon George; Chitra Pattabiraman; Darshan Sreenivas; Harsha.P.K; Nakka Vijay Kiran Reddy; Pramada Prasad; Risha Rasheed; V Ravi |
| EPI_ISL_1385817, EPI_ISL_1385818, EPI_ISL_1385819, EPI_ISL_1385821, EPI_ISL_1385822, EPI_ISL_1385823, EPI_ISL_1385824, EPI_ISL_1385826, EPI_ISL_1385827, EPI_ISL_1385828, EPI_ISL_1385837, EPI_ISL_1385838, EPI_ISL_1396654, EPI_ISL_1595854, EPI_ISL_1595855, EPI_ISL_1595860, EPI_ISL_1595861, EPI_ISL_1595862, EPI_ISL_1595863, EPI_ISL_1595894, EPI_ISL_1595901, EPI_ISL_1595902, EPI_ISL_1595903, EPI_ISL_1595904, EPI_ISL_1595905, EPI_ISL_1595906, EPI_ISL_1595907, EPI_ISL_1595908, EPI_ISL_1595909, EPI_ISL_1595910, EPI_ISL_1595911, EPI_ISL_1595912, EPI_ISL_1595915, EPI_ISL_1595916, EPI_ISL_1818595, EPI_ISL_1818603, EPI_ISL_1818604, EPI_ISL_1818605, EPI_ISL_1818609, EPI_ISL_1818610, EPI_ISL_1818611, EPI_ISL_1818612, EPI_ISL_1818613, EPI_ISL_1818624, EPI_ISL_1818625, EPI_ISL_1818626, EPI_ISL_1818627, EPI_ISL_1818628, EPI_ISL_1818631, EPI_ISL_1818632, EPI_ISL_1818633, EPI_ISL_1818634, EPI_ISL_1818635, EPI_ISL_1818636, EPI_ISL_1818637, EPI_ISL_1818638, EPI_ISL_1818651, EPI_ISL_1818652 | | | |
| see above | BBMP Urban PHC | INSACOG-KA, NIMHANS | Ananthapadmanabha Kotambail; Anita S Desai; Anson Kunjumon George; Chetan G K; Chitra Pattabiraman; Darshan Sreenivas; Gautham Arunachal Udupi; Nakka Vijay Kiran Reddy; Pramada Prasad; V Ravi |
| EPI_ISL_2017765, EPI_ISL_2017766, EPI_ISL_2017767, EPI_ISL_2017768, EPI_ISL_2017769, EPI_ISL_2017770, EPI_ISL_2017771, EPI_ISL_2017772, EPI_ISL_2017773, EPI_ISL_2017774, EPI_ISL_2017775, EPI_ISL_2017776, EPI_ISL_2017777, EPI_ISL_2017778, EPI_ISL_2017779, EPI_ISL_2017780, EPI_ISL_2017781, EPI_ISL_2017782 | | | |
| see above | BJ Medical College and Civil Hospital, Ahmedabad | Gujarat Biotechnology Research Centre | Chaitanya Joshi; Dinesh Kumar; Janvi Raval; Madhvi Joshi; Nitesh Shah; Nitin Savaliya; Pranay Shah; Ramesh Pandit; Sonal Sharma; Twinkle Soni; Umang Mishra; Zarna Patel; Zuber Saiyed |
| EPI_ISL_1662384, EPI_ISL_1662385, EPI_ISL_1662386, EPI_ISL_1662387, EPI_ISL_1662388, EPI_ISL_1662389, EPI_ISL_1662390, EPI_ISL_1662391, EPI_ISL_1662392, EPI_ISL_1662393, EPI_ISL_1662394, EPI_ISL_1662395, EPI_ISL_1662396, EPI_ISL_1662397, EPI_ISL_1662398, EPI_ISL_1662399, EPI_ISL_1662400, EPI_ISL_1662401, EPI_ISL_1662402, EPI_ISL_1662403, EPI_ISL_1662404, EPI_ISL_1662405, EPI_ISL_1662406, EPI_ISL_1662407, EPI_ISL_1662408, EPI_ISL_1662409, EPI_ISL_1662410, EPI_ISL_1662411, EPI_ISL_1662412, EPI_ISL_1662413, EPI_ISL_1662414, EPI_ISL_1662415, EPI_ISL_1662416, EPI_ISL_1662417, EPI_ISL_1662418, EPI_ISL_1662419, EPI_ISL_1662420, EPI_ISL_1662421, EPI_ISL_1663235, EPI_ISL_1663236, EPI_ISL_1663237, EPI_ISL_1663238, EPI_ISL_1663239, EPI_ISL_1663240, EPI_ISL_1663241, EPI_ISL_1663242, EPI_ISL_1663243, EPI_ISL_1663244, EPI_ISL_1663245, EPI_ISL_1663246, EPI_ISL_1663247, EPI_ISL_1663248, EPI_ISL_1663249, EPI_ISL_1663250, EPI_ISL_1663251, EPI_ISL_1663252, EPI_ISL_1663253, EPI_ISL_1663254, EPI_ISL_1663255, EPI_ISL_1663256, EPI_ISL_1663257, EPI_ISL_1663258, EPI_ISL_1663259, EPI_ISL_1663260, EPI_ISL_1663261, EPI_ISL_1663262 | | | |
| see above | BRLSABVM Government Medical College, Rajnandgaon, Chhattisgarh | Institute of Life Sciences - INSACOG | Ajay Parida; Amol M. Kanampalliwar; Arup Ghosh; Atimukta Jha; INSACOG Consortium; Punit Prasad; Rajeeb Swain; Rupesh Dash; Safal Walia; Shifu Aggarwal; Sunil K. Raghav |
| EPI_ISL_825144, EPI_ISL_860182, EPI_ISL_860183, EPI_ISL_860184, EPI_ISL_860185, EPI_ISL_860186 | Bangalore Medical College and Research Institute | Department of Neurovirology, National Institute of Mental Health and Neurosciences (NIMHANS) | Anita S Desai; Anson Kunjumon George; Chitra Pattabiraman; Darshan Sreenivas; Nakka Vijay Kiran Reddy; Pramada Prasad; Risha Rasheed; V Ravi |
| EPI_ISL_1908685, EPI_ISL_1908686, EPI_ISL_1908687, EPI_ISL_1908688, EPI_ISL_1908689, EPI_ISL_1908690, EPI_ISL_1908691, EPI_ISL_1908692, EPI_ISL_1908693, EPI_ISL_1908694, EPI_ISL_1908695, EPI_ISL_1908696, EPI_ISL_1908697, EPI_ISL_1908698, EPI_ISL_1908699, EPI_ISL_1908700, EPI_ISL_1908701, EPI_ISL_1908702, EPI_ISL_1908703, EPI_ISL_1908704, EPI_ISL_1908705, EPI_ISL_1908706, EPI_ISL_1908707, EPI_ISL_1908708, EPI_ISL_1908709, EPI_ISL_1908710, EPI_ISL_1908711, EPI_ISL_1908712, EPI_ISL_1908713, EPI_ISL_1908714, EPI_ISL_1908715, EPI_ISL_1908716, EPI_ISL_1908717, EPI_ISL_1908718, EPI_ISL_1908719, EPI_ISL_1908720, EPI_ISL_1908721, EPI_ISL_1908722, EPI_ISL_1908723, EPI_ISL_1908724, EPI_ISL_1908725, EPI_ISL_1908726, EPI_ISL_1908727, EPI_ISL_1908728, EPI_ISL_1908729, EPI_ISL_1908730, EPI_ISL_1908731, EPI_ISL_1908732, EPI_ISL_1908733, EPI_ISL_1908734, EPI_ISL_1908736, EPI_ISL_1908737, EPI_ISL_1908738, EPI_ISL_1908739, EPI_ISL_1908740, EPI_ISL_1908741, EPI_ISL_1908742, EPI_ISL_1908743, EPI_ISL_1908744, EPI_ISL_1908745, EPI_ISL_1908746, EPI_ISL_1908747, EPI_ISL_1908748, EPI_ISL_1908749, EPI_ISL_1908750, EPI_ISL_1908751, EPI_ISL_1908752, EPI_ISL_1908753, EPI_ISL_1908754, EPI_ISL_1908755, EPI_ISL_1908756, EPI_ISL_1908757, EPI_ISL_1908758, EPI_ISL_1908759, EPI_ISL_1908760, EPI_ISL_1908761, EPI_ISL_1908762, EPI_ISL_1908763, EPI_ISL_1908764, EPI_ISL_1908765, EPI_ISL_1908766, EPI_ISL_1908767, EPI_ISL_1908768, EPI_ISL_1908769, EPI_ISL_1908770, EPI_ISL_1908771, EPI_ISL_1908772, EPI_ISL_1908773, EPI_ISL_1908774, EPI_ISL_1908775, EPI_ISL_1908776, EPI_ISL_1908777, EPI_ISL_1908778, EPI_ISL_1908779, EPI_ISL_1908780, EPI_ISL_1908781, EPI_ISL_1908782, EPI_ISL_1908783, EPI_ISL_1908784, EPI_ISL_1908785, EPI_ISL_1908786, EPI_ISL_1970088, EPI_ISL_1970089, EPI_ISL_1970090, EPI_ISL_1970091, EPI_ISL_1970092, EPI_ISL_1970093, EPI_ISL_1970094, EPI_ISL_1970095, EPI_ISL_1970096, EPI_ISL_1970097, EPI_ISL_1970098, EPI_ISL_1970099, EPI_ISL_1970100, EPI_ISL_1970101, EPI_ISL_1970102, EPI_ISL_1970103, EPI_ISL_1970104, EPI_ISL_1970105, EPI_ISL_1970106, EPI_ISL_1970107, EPI_ISL_1970108, EPI_ISL_1970109, EPI_ISL_1970110, EPI_ISL_1970111, EPI_ISL_1970112, EPI_ISL_1970113, EPI_ISL_1970114, EPI_ISL_1970115, EPI_ISL_1970116, EPI_ISL_1970117, EPI_ISL_1970118, EPI_ISL_1970119, EPI_ISL_1970120, EPI_ISL_1970121, EPI_ISL_1970122, EPI_ISL_1970123, EPI_ISL_1970124, EPI_ISL_1970125, EPI_ISL_1970126, EPI_ISL_1970127, EPI_ISL_1970128, EPI_ISL_1970129, EPI_ISL_1970130, EPI_ISL_1970131, EPI_ISL_1970132, EPI_ISL_1970133, EPI_ISL_1970134, EPI_ISL_1970135, EPI_ISL_1970136, EPI_ISL_1970137, EPI_ISL_1970138, EPI_ISL_1970139, EPI_ISL_1970140, EPI_ISL_1970141, EPI_ISL_1970142, EPI_ISL_1970143, EPI_ISL_1970144, EPI_ISL_1970145, EPI_ISL_1970146, EPI_ISL_1970147, EPI_ISL_1970148, EPI_ISL_1970149, EPI_ISL_1970150, EPI_ISL_1970151, EPI_ISL_1970152, EPI_ISL_1970153, EPI_ISL_1970154, EPI_ISL_1970155, EPI_ISL_1970156, EPI_ISL_1970157, EPI_ISL_1970158, EPI_ISL_1970159, EPI_ISL_1970160, EPI_ISL_1970161, EPI_ISL_1970162, EPI_ISL_1970163, EPI_ISL_1970164, EPI_ISL_1970165, EPI_ISL_1970166, EPI_ISL_1970167, EPI_ISL_1970168, EPI_ISL_1970169, EPI_ISL_1970170, EPI_ISL_1970171, EPI_ISL_1970172, EPI_ISL_1970173, EPI_ISL_1970174, EPI_ISL_1970175, EPI_ISL_1970176, EPI_ISL_1970177, EPI_ISL_1970178, EPI_ISL_1970179, EPI_ISL_1970180, EPI_ISL_1970181, EPI_ISL_1970182, EPI_ISL_1970183, EPI_ISL_1970184, EPI_ISL_1970185, EPI_ISL_1970186, EPI_ISL_1970187, EPI_ISL_1970188, EPI_ISL_1970189, EPI_ISL_1970190, EPI_ISL_1970191, EPI_ISL_1970192, EPI_ISL_1970193, EPI_ISL_1970194, EPI_ISL_1970195, EPI_ISL_1970196, EPI_ISL_1970197, EPI_ISL_1970198, EPI_ISL_1970199, EPI_ISL_1970200, EPI_ISL_1970201, EPI_ISL_1970202, EPI_ISL_1970203, EPI_ISL_1970204, EPI_ISL_1970205, EPI_ISL_1970206, EPI_ISL_1970207, EPI_ISL_1970208, EPI_ISL_1970209, EPI_ISL_1970210, EPI_ISL_1970211, EPI_ISL_1970212, EPI_ISL_1970213, EPI_ISL_1970214, EPI_ISL_1970215, EPI_ISL_1970216, EPI_ISL_1970217, EPI_ISL_1970218, EPI_ISL_1970219, EPI_ISL_1970220, EPI_ISL_1970221, EPI_ISL_1970222, EPI_ISL_1970223, EPI_ISL_1970224, EPI_ISL_1970225, EPI_ISL_1970226, EPI_ISL_1970227, EPI_ISL_1970228, EPI_ISL_1970229, EPI_ISL_1970230, EPI_ISL_1970231, EPI_ISL_1970232, EPI_ISL_1970233, EPI_ISL_1970234, EPI_ISL_1970235, EPI_ISL_1970236, EPI_ISL_1970237, EPI_ISL_1970238, EPI_ISL_1970239, EPI_ISL_1970240, EPI_ISL_1970241, EPI_ISL_1970242, EPI_ISL_1970243, EPI_ISL_1970244, EPI_ISL_1970245, EPI_ISL_1970246, EPI_ISL_1970247, EPI_ISL_1970248, EPI_ISL_1970249, EPI_ISL_1970250, EPI_ISL_1970251, EPI_ISL_1970252, EPI_ISL_1970253, EPI_ISL_1970254, EPI_ISL_1970255, EPI_ISL_1970256, EPI_ISL_1970257, EPI_ISL_1970258, EPI_ISL_1970259, EPI_ISL_1970260, EPI_ISL_1970261, EPI_ISL_1970262, EPI_ISL_1970263, EPI_ISL_1970264, EPI_ISL_1970265, EPI_ISL_1970266, EPI_ISL_1970267, EPI_ISL_1970268, EPI_ISL_1970269, EPI_ISL_1970270, EPI_ISL_1970271, EPI_ISL_1970272, EPI_ISL_1970273, EPI_ISL_1970274, EPI_ISL_1970275, EPI_ISL_1970276, EPI_ISL_1970277, EPI_ISL_1970278, EPI_ISL_1970279, EPI_ISL_1970280, EPI_ISL_1970281, EPI_ISL_1970282, EPI_ISL_1970283, EPI_ISL_1970284, EPI_ISL_1970285, EPI_ISL_1970286, EPI_ISL_1970287, EPI_ISL_1970288, EPI_ISL_1970289, EPI_ISL_1970290, EPI_ISL_1970291, EPI_ISL_1970292, EPI_ISL_1970293, EPI_ISL_1970294, EPI_ISL_1970295, EPI_ISL_1970296, EPI_ISL_1970297, EPI_ISL_1970298, EPI_ISL_1970299, EPI_ISL_1970300, EPI_ISL_1970301, EPI_ISL_1970302, EPI_ISL_1970303, EPI_ISL_1970304, EPI_ISL_1970305, EPI_ISL_1970306, EPI_ISL_1970307, EPI_ISL_1970308, EPI_ISL_1970309, EPI_ISL_1970310, EPI_ISL_1970311, EPI_ISL_1970312, EPI_ISL_1970313, EPI_ISL_1970314, EPI_ISL_1970315, EPI_ISL_1970316, EPI_ISL_1970317, EPI_ISL_1970318, EPI_ISL_1970319, EPI_ISL_1970320, EPI_ISL_1970321, EPI_ISL_1970322, EPI_ISL_1970323, EPI_ISL_1970324, EPI_ISL_1970325, EPI_ISL_1970326, EPI_ISL_1970327, EPI_ISL_1970328, EPI_ISL_1970329, EPI_ISL_1970330, EPI_ISL_1970331, EPI_ISL_1970332, EPI_ISL_1970333, EPI_ISL_1970334, EPI_ISL_1970335, EPI_ISL_1970336, EPI_ISL_1970337, EPI_ISL_1970338, EPI_ISL_1970339, EPI_ISL_1970340, EPI_ISL_1970341, EPI_ISL_1970342, EPI_ISL_1970343 | | | |
| see above | Biotechnology Division, NCDC Delhi | NCDC Delhi, Biotechnology Division | Hema Gogia; Hemlata Lall; Kalaiarasan Ponnusamy; Mahesh S Dhar; Manoj K Singh; Meena Datta; Partha Rakshit; Preeti Madan; Priyanka Singh; Radhakrishnan V. S; Robin Marwal; Sandhya Kabra; Sujeet K Singh; Uma Sharma |
| EPI_ISL_1708429, EPI_ISL_1708430, EPI_ISL_1708438, EPI_ISL_1708442, EPI_ISL_1708443 | Byatarayanapura PHC | inStem NCBS - INSACOG | Uma Ramakrishnan Dasaradhi Palakodeti Aswin SaiNarain |
| EPI_ISL_1663263, EPI_ISL_1663264, EPI_ISL_1663265, EPI_ISL_1663266, EPI_ISL_1663267, EPI_ISL_1663268, EPI_ISL_1663269, EPI_ISL_1663270, EPI_ISL_1663271, EPI_ISL_1663272, EPI_ISL_1663273, EPI_ISL_1663274, EPI_ISL_1663275, EPI_ISL_1663276, EPI_ISL_1663277, EPI_ISL_1663278, EPI_ISL_1663279, EPI_ISL_1663280, EPI_ISL_1663281, EPI_ISL_1663282, EPI_ISL_1663283, EPI_ISL_1663284, EPI_ISL_1663285, EPI_ISL_1663286, EPI_ISL_1663287, EPI_ISL_1663288, EPI_ISL_1663289, EPI_ISL_1663290, EPI_ISL_1663291, EPI_ISL_1663292, EPI_ISL_1663293, EPI_ISL_1663294, EPI_ISL_1663295, EPI_ISL_1663296, EPI_ISL_1663297, EPI_ISL_1663298, EPI_ISL_1663299, EPI_ISL_1663300, EPI_ISL_1663301, EPI_ISL_1663302, EPI_ISL_1663303, EPI_ISL_1663304, EPI_ISL_1663305, EPI_ISL_1663306, EPI_ISL_1663307, EPI_ISL_1663308, EPI_ISL_1663309, EPI_ISL_1663310, EPI_ISL_1663311, EPI_ISL_1663312, EPI_ISL_1663313, EPI_ISL_1663314, EPI_ISL_1663315, EPI_ISL_1663316, EPI_ISL_1663317, EPI_ISL_1663318, EPI_ISL_1663319, EPI_ISL_1663320, EPI_ISL_1663321, EPI_ISL_1663322 | | | |
| see above | CIIMS, Bilaspur, Chhattisgarh | Institute of Life Sciences - INSACOG | Ajay Parida; Amol M. Kanampalliwar; Arup Ghosh; Atimukta Jha; INSACOG Consortium; Punit Prasad; Rajeeb Swain; Rupesh Dash; Safal Walia; Shifu Aggarwal; Sunil K. Raghav |
| EPI_ISL_2015468, EPI_ISL_2015502, EPI_ISL_2015503, EPI_ISL_2015504, EPI_ISL_2015505, EPI_ISL_2015507, EPI_ISL_2015508, EPI_ISL_2015509, EPI_ISL_2015510, EPI_ISL_2015513, EPI_ISL_2015517, EPI_ISL_2015519, EPI_ISL_2015521, EPI_ISL_2015524, EPI_ISL_2015525, EPI_ISL_2015526, EPI_ISL_2015531, EPI_ISL_2015533, EPI_ISL_2015536, EPI_ISL_2015539, EPI_ISL_2015540, EPI_ISL_2015552, EPI_ISL_2015557, EPI_ISL_2015559, EPI_ISL_2015560, EPI_ISL_2015561, EPI_ISL_2015562, EPI_ISL_2015563, EPI_ISL_2015566, EPI_ISL_2015568, EPI_ISL_2015569, EPI_ISL_2015570, EPI_ISL_2015572, EPI_ISL_2015574, EPI_ISL_2015579, EPI_ISL_2015581, EPI_ISL_2015582, EPI_ISL_2015583, EPI_ISL_2015584, EPI_ISL_2015585, EPI_ISL_2015586, EPI_ISL_2015589, EPI_ISL_2015590, EPI_ISL_2015592, EPI_ISL_2015593, EPI_ISL_2015595, EPI_ISL_2015596, EPI_ISL_2015597, EPI_ISL_2015600, EPI_ISL_2015601, EPI_ISL_2015602, EPI_ISL_2015606, EPI_ISL_2015607, EPI_ISL_2015608, EPI_ISL_2015610, EPI_ISL_2015611, EPI_ISL_2015612, EPI_ISL_2015613, EPI_ISL_2015617, EPI_ISL_2015618 | | | |
| see above | CMC Vellore | inStem NCBS - INSACOG | Uma Ramakrishnan Dasaradhi Palakodeti Aswin SaiNarain |
| EPI_ISL_1818586 | COMMAND HOSPITAL | INSACOG-KA, NIMHANS | Ananthapadmanabha Kotambail; Anita S Desai; Anson Kunjumon George; Chetan G K; Chitra Pattabiraman; Darshan Sreenivas; Gautham Arunachal Udupi; Pramada Prasad; V Ravi |
| EPI_ISL_1672391 | CSIR-Centre for Cellular and Molecular Biology | CSIR-Centre for Cellular and Molecular Biology | Amareshwar Vodapalli; Ara Sreenivas; B Himasri; Dhiviya Vedagiri; Divya Gupta; Divya Tej Sowpati; Karthik Bharadwaj Tallapaka; Krishnan Harinivas Harshan; Lamuk Zaveri; Onkar Kulkarni; Rakesh K Mishra; Sharath Chandra Thota; Shreekant Verma; Sofia Banu; Vishal Sah; Viswagithe S L |
| EPI_ISL_1380285, EPI_ISL_1380286, EPI_ISL_1380287, EPI_ISL_1380288, EPI_ISL_1380289, EPI_ISL_1380290, EPI_ISL_1380291, EPI_ISL_1380292, EPI_ISL_1380293, EPI_ISL_1380294, EPI_ISL_1380295, EPI_ISL_1380296, EPI_ISL_1380297, EPI_ISL_1380298, EPI_ISL_1380299, EPI_ISL_1380300, EPI_ISL_1380301, EPI_ISL_1380302, EPI_ISL_1380303, EPI_ISL_1380304, EPI_ISL_1380305, EPI_ISL_1380306, EPI_ISL_1380307, EPI_ISL_1380308, EPI_ISL_1380309, EPI_ISL_1380310, EPI_ISL_1380311, EPI_ISL_1380312, EPI_ISL_1380313, EPI_ISL_1380314, EPI_ISL_1380315, EPI_ISL_1380316, EPI_ISL_1380317, EPI_ISL_1380318, EPI_ISL_1380319, EPI_ISL_1380320, EPI_ISL_1380321, EPI_ISL_1380322, EPI_ISL_1380323, EPI_ISL_1380324, EPI_ISL_1380325, EPI_ISL_1380326, EPI_ISL_1380327, EPI_ISL_1380328, EPI_ISL_1380329, EPI_ISL_1380330, EPI_ISL_1380331, EPI_ISL_1380332, EPI_ISL_1380333, EPI_ISL_1380334, EPI_ISL_1380335, EPI_ISL_1380336, EPI_ISL_1380337, EPI_ISL_1380338, EPI_ISL_1380339, EPI_ISL_1380340, EPI_ISL_1380341, EPI_ISL_1380342, EPI_ISL_1380343, EPI_ISL_1380344, EPI_ISL_1380345, EPI_ISL_1380346, EPI_ISL_1380347, EPI_ISL_1380348, EPI_ISL_1380349, EPI_ISL_1380350, | | | |

| | | | |
|---|---|---|---|
| EPI_ISL_1380351, EPI_ISL_1380352, EPI_ISL_1380353, EPI_ISL_1380354, EPI_ISL_1380355, EPI_ISL_1380356, EPI_ISL_1380357, EPI_ISL_1380358, EPI_ISL_1380359, EPI_ISL_1380360, EPI_ISL_1380361, EPI_ISL_1380362, EPI_ISL_1380363, EPI_ISL_1380364, EPI_ISL_1380365, EPI_ISL_1380366, EPI_ISL_1380367, EPI_ISL_1380368, EPI_ISL_1380369, EPI_ISL_1380370, EPI_ISL_1380371, EPI_ISL_1380372, EPI_ISL_1380373, EPI_ISL_1380374, EPI_ISL_1380375, EPI_ISL_1380376, EPI_ISL_1380377, EPI_ISL_1380378, EPI_ISL_1380379, EPI_ISL_1380380, EPI_ISL_1380381, EPI_ISL_1380382, EPI_ISL_1380383, EPI_ISL_1380384, EPI_ISL_1380385, EPI_ISL_1380386, EPI_ISL_1380387, EPI_ISL_1380388, EPI_ISL_1380389, EPI_ISL_1380390, EPI_ISL_1380391, EPI_ISL_1380392, EPI_ISL_1380393, EPI_ISL_1380394, EPI_ISL_1380395, EPI_ISL_1380396, EPI_ISL_1380397, EPI_ISL_1380398, EPI_ISL_1380399, EPI_ISL_1380400, EPI_ISL_1380401, EPI_ISL_1380402, EPI_ISL_1380403, EPI_ISL_1380404, EPI_ISL_1380405, EPI_ISL_1380406, EPI_ISL_1380407, EPI_ISL_1380408, EPI_ISL_1380409, EPI_ISL_1380410, EPI_ISL_1380411, EPI_ISL_1380412, EPI_ISL_1380413, EPI_ISL_1380414, EPI_ISL_1380415, EPI_ISL_1380416, EPI_ISL_1380417, EPI_ISL_1380418, EPI_ISL_1380419, EPI_ISL_1380420, EPI_ISL_1380421, EPI_ISL_1380422, EPI_ISL_1380423, EPI_ISL_1380424, EPI_ISL_1380425, EPI_ISL_1380426, EPI_ISL_1380427, EPI_ISL_1380428, EPI_ISL_1380429, EPI_ISL_1380430, EPI_ISL_1380431, EPI_ISL_1380432, EPI_ISL_1380433, EPI_ISL_1380434, EPI_ISL_1380435, EPI_ISL_1380436, EPI_ISL_1380437, EPI_ISL_1380438, EPI_ISL_1380439, EPI_ISL_1380440, EPI_ISL_1380441, EPI_ISL_1380442, EPI_ISL_1380443, EPI_ISL_1380444, EPI_ISL_1380445, EPI_ISL_1380446, EPI_ISL_1380447, EPI_ISL_1380448, EPI_ISL_1380449, EPI_ISL_1380450, EPI_ISL_1380451, EPI_ISL_1380452, EPI_ISL_1380453, EPI_ISL_1380454, EPI_ISL_1380455, EPI_ISL_1380456, EPI_ISL_1380457, EPI_ISL_1380458, EPI_ISL_1380459, EPI_ISL_1380460, EPI_ISL_1380461, EPI_ISL_1380462, EPI_ISL_1380463, EPI_ISL_1380464, EPI_ISL_1380465, EPI_ISL_1380466, EPI_ISL_1380467, EPI_ISL_1380468, EPI_ISL_1380469, EPI_ISL_1380470, EPI_ISL_1380471, EPI_ISL_1380472, EPI_ISL_1380473, EPI_ISL_1380474, EPI_ISL_1380475, EPI_ISL_1380476, EPI_ISL_1380477, EPI_ISL_1380478, EPI_ISL_1380479, EPI_ISL_1380480, EPI_ISL_1380481, EPI_ISL_1380482, EPI_ISL_1380483, EPI_ISL_1380484, EPI_ISL_1380485, EPI_ISL_1380486, EPI_ISL_1380487, EPI_ISL_1380488, EPI_ISL_1380489, EPI_ISL_1380490, EPI_ISL_1380491, EPI_ISL_1380492, EPI_ISL_1380493, EPI_ISL_1380494, EPI_ISL_1380495, EPI_ISL_1380496, EPI_ISL_1380497, EPI_ISL_1380498, EPI_ISL_1398950, EPI_ISL_1398951, EPI_ISL_1398952, EPI_ISL_1398953, EPI_ISL_1398954, EPI_ISL_1838348, EPI_ISL_1838350, EPI_ISL_1838352, EPI_ISL_1838356, EPI_ISL_1838359, EPI_ISL_1838361, EPI_ISL_1838362, EPI_ISL_1838364, EPI_ISL_1838366, EPI_ISL_1838367, EPI_ISL_1838369, EPI_ISL_1838370, EPI_ISL_1838372, EPI_ISL_1838374, EPI_ISL_1838377, EPI_ISL_1838380, EPI_ISL_1838384, EPI_ISL_1838385, EPI_ISL_1838387, EPI_ISL_1838389, EPI_ISL_1838390, EPI_ISL_1838392, EPI_ISL_1838393, EPI_ISL_1838395, EPI_ISL_1838397, EPI_ISL_1838398, EPI_ISL_1838400, EPI_ISL_1838402, EPI_ISL_1838403, EPI_ISL_1838405, EPI_ISL_1838406, EPI_ISL_1838408, EPI_ISL_1838410, EPI_ISL_1838411, EPI_ISL_1838413, EPI_ISL_1838414, EPI_ISL_1838418, EPI_ISL_1838422, EPI_ISL_1838426, EPI_ISL_1838430, EPI_ISL_1838435, EPI_ISL_1838439, EPI_ISL_1838443, EPI_ISL_1838445, EPI_ISL_1838446, EPI_ISL_1838448, EPI_ISL_1838450, EPI_ISL_1838452, EPI_ISL_1838453, EPI_ISL_1838454, EPI_ISL_1838455, EPI_ISL_1838456, EPI_ISL_1838457, EPI_ISL_1838458, EPI_ISL_1838459, EPI_ISL_1838460, EPI_ISL_1838461, EPI_ISL_1838462, EPI_ISL_1838463, EPI_ISL_1838464, EPI_ISL_1838465, EPI_ISL_1838466, EPI_ISL_1838467, EPI_ISL_1838468, EPI_ISL_1838469, EPI_ISL_1838470, EPI_ISL_1838471, EPI_ISL_1838472, EPI_ISL_1838473, EPI_ISL_1838474, EPI_ISL_1838475, EPI_ISL_1838476, EPI_ISL_1838477, EPI_ISL_1838478, EPI_ISL_1838479, EPI_ISL_1838480, EPI_ISL_1838481, EPI_ISL_1838482, EPI_ISL_1838483, EPI_ISL_1838484, EPI_ISL_1838485, EPI_ISL_1838486, EPI_ISL_1838487, EPI_ISL_1838488, EPI_ISL_1838489, EPI_ISL_1838490, EPI_ISL_1838491, EPI_ISL_1838492, EPI_ISL_1838493, EPI_ISL_1838494, EPI_ISL_1838495, EPI_ISL_1838496, EPI_ISL_1838497, EPI_ISL_1838498, EPI_ISL_1838499, EPI_ISL_1838500, EPI_ISL_1838501, EPI_ISL_1838502, EPI_ISL_1838503, EPI_ISL_1838504, EPI_ISL_1838505, EPI_ISL_1838506, EPI_ISL_1838507, EPI_ISL_1838508, EPI_ISL_1838509, EPI_ISL_1838510, EPI_ISL_1838511, EPI_ISL_1838512, EPI_ISL_1838513, EPI_ISL_1838514, EPI_ISL_1838515, EPI_ISL_1838516, EPI_ISL_1838517, EPI_ISL_1838518, EPI_ISL_1838519, EPI_ISL_1838520, EPI_ISL_1838521, EPI_ISL_1838522, EPI_ISL_1838523, EPI_ISL_1838524, EPI_ISL_1838525, EPI_ISL_1838526, EPI_ISL_1838527, EPI_ISL_1838528, EPI_ISL_1838529, EPI_ISL_1838530, EPI_ISL_1838531, EPI_ISL_1838532, EPI_ISL_1838533, EPI_ISL_1838534, EPI_ISL_1838535, EPI_ISL_1838536, EPI_ISL_1838537, EPI_ISL_1838538, EPI_ISL_1838539, EPI_ISL_1838541, EPI_ISL_1838542, EPI_ISL_1838543, EPI_ISL_1838544, EPI_ISL_1838545, EPI_ISL_1838546, EPI_ISL_1838547, EPI_ISL_1838548, EPI_ISL_1838549, EPI_ISL_1838550, EPI_ISL_1838551, EPI_ISL_1838552, EPI_ISL_1838553, EPI_ISL_1838554, EPI_ISL_1838555, EPI_ISL_1838556, EPI_ISL_1838557, EPI_ISL_1838558, EPI_ISL_1838559, EPI_ISL_1838560, EPI_ISL_1838561, EPI_ISL_1838562, EPI_ISL_1838563, EPI_ISL_1838564, EPI_ISL_1838565, EPI_ISL_1838566, EPI_ISL_1838567, EPI_ISL_1838568, EPI_ISL_1838569, EPI_ISL_1838570, EPI_ISL_1838571, EPI_ISL_1838572, EPI_ISL_1838573, EPI_ISL_1838574, EPI_ISL_1838575, EPI_ISL_1838576, EPI_ISL_1838577, EPI_ISL_1838578, EPI_ISL_1838579, EPI_ISL_1838580, EPI_ISL_1838581, EPI_ISL_1838582, EPI_ISL_1838583, EPI_ISL_1838584, EPI_ISL_1838585, EPI_ISL_1838586, EPI_ISL_1838587, EPI_ISL_1838588, EPI_ISL_1838589, EPI_ISL_1838590, EPI_ISL_1838591, EPI_ISL_1838592, EPI_ISL_1838593, EPI_ISL_1838594, EPI_ISL_1838595, EPI_ISL_1838596, EPI_ISL_1838597, EPI_ISL_1838598, EPI_ISL_1838599, EPI_ISL_1838600, EPI_ISL_1838601, EPI_ISL_1838602, EPI_ISL_1838603, EPI_ISL_1838604, EPI_ISL_1838605, EPI_ISL_1838606, EPI_ISL_1838607, EPI_ISL_1838608, EPI_ISL_1838609, EPI_ISL_1838610, EPI_ISL_1838611, EPI_ISL_1838612, EPI_ISL_1838613, EPI_ISL_1838614, EPI_ISL_1838615, EPI_ISL_1838616, EPI_ISL_1838617, EPI_ISL_1838618, EPI_ISL_1838619, EPI_ISL_1838620, EPI_ISL_1838621, EPI_ISL_1838622, EPI_ISL_1838623, EPI_ISL_1838624, EPI_ISL_1838625, EPI_ISL_1838626, EPI_ISL_1838627, EPI_ISL_1838628, EPI_ISL_1838629, EPI_ISL_1838630, EPI_ISL_1838631, EPI_ISL_1838632, EPI_ISL_1838633, EPI_ISL_1838634, EPI_ISL_1838635, EPI_ISL_1838636, EPI_ISL_1838637, EPI_ISL_1838638, EPI_ISL_1838639, EPI_ISL_1838640, EPI_ISL_1838641, EPI_ISL_1838642, EPI_ISL_1838643, EPI_ISL_1838644, EPI_ISL_1838645, EPI_ISL_1838646, EPI_ISL_1838647, EPI_ISL_1838648, EPI_ISL_1838649, EPI_ISL_1838650, EPI_ISL_1838651, EPI_ISL_1838652, EPI_ISL_1838653, EPI_ISL_1838654, EPI_ISL_1838655, EPI_ISL_1838656, EPI_ISL_1838657, EPI_ISL_1838658, EPI_ISL_1838659, EPI_ISL_1838660, EPI_ISL_1838661, EPI_ISL_1838662, EPI_ISL_1838663, EPI_ISL_1838664, EPI_ISL_1838665, EPI_ISL_1838666, EPI_ISL_1838667, EPI_ISL_1838668, EPI_ISL_1838669, EPI_ISL_1838670, EPI_ISL_1838671, EPI_ISL_1838672, EPI_ISL_1838673, EPI_ISL_1838674, EPI_ISL_1838675, EPI_ISL_1838676, EPI_ISL_1838677, EPI_ISL_1838678, EPI_ISL_1838679, EPI_ISL_1838680, EPI_ISL_1838681, EPI_ISL_1838682, EPI_ISL_1838683, EPI_ISL_1838684, EPI_ISL_1838685, EPI_ISL_1838686, EPI_ISL_1838687, EPI_ISL_1838688, EPI_ISL_1838689, EPI_ISL_1838690, EPI_ISL_1838691, EPI_ISL_1838692, EPI_ISL_1838693, EPI_ISL_1838694, EPI_ISL_1838695, EPI_ISL_1838696, EPI_ISL_1838697, EPI_ISL_1838698, EPI_ISL_1838699, EPI_ISL_1838700, EPI_ISL_1838701, EPI_ISL_1838702, EPI_ISL_1838703, EPI_ISL_1838704, EPI_ISL_1838705, EPI_ISL_1838706, EPI_ISL_1838707, EPI_ISL_1838708, EPI_ISL_1838709, EPI_ISL_1838710, EPI_ISL_1838711, EPI_ISL_1838712, EPI_ISL_1838713, EPI_ISL_1838714, EPI_ISL_1838715, EPI_ISL_1838716, EPI_ISL_1838717, EPI_ISL_1838718, EPI_ISL_1838719, EPI_ISL_1838720, EPI_ISL_1838721, EPI_ISL_1838722, EPI_ISL_1838723, EPI_ISL_1838724, EPI_ISL_1838725, EPI_ISL_1838726, EPI_ISL_1838727, EPI_ISL_1838728, EPI_ISL_1838729, EPI_ISL_1838730, EPI_ISL_1838731, EPI_ISL_1838732, EPI_ISL_1838733, EPI_ISL_1838734, EPI_ISL_1838735, EPI_ISL_1838736, EPI_ISL_1838737, EPI_ISL_1838738, EPI_ISL_1838739, EPI_ISL_1838740, EPI_ISL_1838741, EPI_ISL_1838742, EPI_ISL_1838743, EPI_ISL_1838744, EPI_ISL_1838745, EPI_ISL_1838746, EPI_ISL_1838747, EPI_ISL_1838748, EPI_ISL_1838749, EPI_ISL_1838750, EPI_ISL_1838751, EPI_ISL_1838752, EPI_ISL_1838753, EPI_ISL_1838754, EPI_ISL_1838755, EPI_ISL_1838756, EPI_ISL_1838757, EPI_ISL_1838758, EPI_ISL_1838759, EPI_ISL_1838760, EPI_ISL_1838761, EPI_ISL_1838762, EPI_ISL_1838763, EPI_ISL_1838764, EPI_ISL_1838765, EPI_ISL_1838766, EPI_ISL_1838767, EPI_ISL_1838768, EPI_ISL_1838769, EPI_ISL_1838770, EPI_ISL_1838771, EPI_ISL_1838772, EPI_ISL_1838773, EPI_ISL_1838774, EPI_ISL_1838775, EPI_ISL_1838776, EPI_ISL_1838777, EPI_ISL_1838778, EPI_ISL_1838779, EPI_ISL_1838780, EPI_ISL_1838781, EPI_ISL_1838782, EPI_ISL_1838783, EPI_ISL_1838784, EPI_ISL_1838785, EPI_ISL_1838786, EPI_ISL_1838787, EPI_ISL_1838788, EPI_ISL_1838789, EPI_ISL_1838790, EPI_ISL_1838792, EPI_ISL_1838793, EPI_ISL_1838794, EPI_ISL_1838795, EPI_ISL_1838796, EPI_ISL_1838797, EPI_ISL_1838798, EPI_ISL_1838799, EPI_ISL_1838800, EPI_ISL_1838801, EPI_ISL_1838802, EPI_ISL_1838803, EPI_ISL_1838804, EPI_ISL_1838805, EPI_ISL_1838806, EPI_ISL_1838807, EPI_ISL_1838808, EPI_ISL_1838809, EPI_ISL_1838810, EPI_ISL_1838811, EPI_ISL_1838812, EPI_ISL_1838813, EPI_ISL_1838814, EPI_ISL_1838815, EPI_ISL_1838816, EPI_ISL_1838817, EPI_ISL_1838818, EPI_ISL_1838819, EPI_ISL_1838820, EPI_ISL_1838821, EPI_ISL_1838822, EPI_ISL_1838823, EPI_ISL_1838824, EPI_ISL_1838825, EPI_ISL_1838826, EPI_ISL_1838827, EPI_ISL_1838828, EPI_ISL_1838829, EPI_ISL_1838830, EPI_ISL_1838831, EPI_ISL_1838832, EPI_ISL_1838833, EPI_ISL_1838834, EPI_ISL_1838835, EPI_ISL_1838836, EPI_ISL_1838837, EPI_ISL_1838838, EPI_ISL_1838839, EPI_ISL_1838840, EPI_ISL_1838841, EPI_ISL_1838842, EPI_ISL_1838843, EPI_ISL_1838844, EPI_ISL_1838845, EPI_ISL_1838846, EPI_ISL_1838847, EPI_ISL_1838848, EPI_ISL_1838849, EPI_ISL_1838850, EPI_ISL_1838851, EPI_ISL_1838852, EPI_ISL_1838853, EPI_ISL_1838854, EPI_ISL_1838855, EPI_ISL_1838856, EPI_ISL_1838857, EPI_ISL_1838858, EPI_ISL_1838859, EPI_ISL_1838860, EPI_ISL_1838861, EPI_ISL_1838862, EPI_ISL_1838863, EPI_ISL_1838864, EPI_ISL_1838865, EPI_ISL_1838866, EPI_ISL_1838867, EPI_ISL_1838868, EPI_ISL_1838869, EPI_ISL_1838870, EPI_ISL_1838871, EPI_ISL_1838872, EPI_ISL_1838873, EPI_ISL_1838874, EPI_ISL_1838875, EPI_ISL_1838876, EPI_ISL_1838877, EPI_ISL_1838878, EPI_ISL_1838879, EPI_ISL_1838880, EPI_ISL_1838881, EPI_ISL_1838882, EPI_ISL_1838883, EPI_ISL_1838884, EPI_ISL_1838885, EPI_ISL_1838886, EPI_ISL_1838887, EPI_ISL_1838888, EPI_ISL_1838889, EPI_ISL_1838890, EPI_ISL_1838891, EPI_ISL_1838892, EPI_ISL_1838893, EPI_ISL_1838894, EPI_ISL_1838895, EPI_ISL_1838896, EPI_ISL_1838897, EPI_ISL_1838898, EPI_ISL_1838899, EPI_ISL_1838900, EPI_ISL_1838901, EPI_ISL_1838902, EPI_ISL_1838903, EPI_ISL_1838904, EPI_ISL_1838905, EPI_ISL_1838906, EPI_ISL_1838907, EPI_ISL_1838908, EPI_ISL_1838909, EPI_ISL_1838910, EPI_ISL_1838911, EPI_ISL_1838912, EPI_ISL_1838913, EPI_ISL_1838914, EPI_ISL_1838915, EPI_ISL_1838916, EPI_ISL_1838917, EPI_ISL_1838918, EPI_ISL_1838919, EPI_ISL_1972343, EPI_ISL_1972344, EPI_ISL_1972345, EPI_ISL_1972347, EPI_ISL_1972349, EPI_ISL_1972351, EPI_ISL_1972353, EPI_ISL_1972354, EPI_ISL_1972355 | | | |
| see above | CSIR-Centre for Cellular and Molecular Biology | CSIR-Centre for Cellular and Molecular Biology-INSACOG | Amareshwar Vodapalli; Ara Sreenivas; Archana Bharadwaj Siva; B Himasri; Blessy B John; Divya Tej Sowpati; Karthik Bharadwaj Tallapaka; Lamuk Zaveri; Namami Gaur; Onkar Kulkarni; Payel Mukherjee; Pratheusa Maccha; Priya Singh; Purushotham Vodnala; Rakesh K Mishra; Sharath Chandra Thota; Shreekant Verma; Sofia Banu; Tulasi Nagabandi; Valli Nagalakshmi Undamatla; Viswagithe S L |
| EPI_ISL_911532, EPI_ISL_911533, EPI_ISL_911534, EPI_ISL_911535, EPI_ISL_911536, EPI_ISL_911537, EPI_ISL_911538, EPI_ISL_911539, EPI_ISL_911540, EPI_ISL_911541, EPI_ISL_911542, EPI_ISL_911543, EPI_ISL_911544, EPI_ISL_911545, EPI_ISL_911546, EPI_ISL_911547, EPI_ISL_1716738, EPI_ISL_1716739, EPI_ISL_1716740, EPI_ISL_1716741, EPI_ISL_1716742, EPI_ISL_1716743, EPI_ISL_1716744, EPI_ISL_1716745, EPI_ISL_1716746, EPI_ISL_1716747, EPI_ISL_1716748, EPI_ISL_1716749, EPI_ISL_1716750, EPI_ISL_1716751, EPI_ISL_1716752, EPI_ISL_1716753, EPI_ISL_1716754, EPI_ISL_1716755, EPI_ISL_1716756, EPI_ISL_1716757, EPI_ISL_1716758, EPI_ISL_1716759, EPI_ISL_1716760, EPI_ISL_1716761, EPI_ISL_1716762, EPI_ISL_1716763, EPI_ISL_1716764, EPI_ISL_1716765, EPI_ISL_1716766, EPI_ISL_1716767, EPI_ISL_1716768, EPI_ISL_1716769, EPI_ISL_1716770, EPI_ISL_1716771, EPI_ISL_1716772, EPI_ISL_1716773, EPI_ISL_1716774, EPI_ISL_1716775, EPI_ISL_1716776, EPI_ISL_1716777, EPI_ISL_1716778, EPI_ISL_1716779, EPI_ISL_1716780, EPI_ISL_1716781, EPI_ISL_1716782, EPI_ISL_1716783, EPI_ISL_1716784, EPI_ISL_1716785, EPI_ISL_1716786, EPI_ISL_1716787, EPI_ISL_1716788, EPI_ISL_1716789, EPI_ISL_1716790, EPI_ISL_1716791, EPI_ISL_1716792, EPI_ISL_1716793, EPI_ISL_1716794, EPI_ISL_1716795, EPI_ISL_1716796, EPI_ISL_1716797, EPI_ISL_1716798, EPI_ISL_1716799, EPI_ISL_1716800, EPI_ISL_1716801, EPI_ISL_1716802, EPI_ISL_1716803, EPI_ISL_1716804, EPI_ISL_1716805, EPI_ISL_1716806, EPI_ISL_1716807, EPI_ISL_1716808, EPI_ISL_1716809, EPI_ISL_1716810, EPI_ISL_1716811, EPI_ISL_1716812, EPI_ISL_1716813, EPI_ISL_1716814, EPI_ISL_1716815, EPI_ISL_1716816, EPI_ISL_1716817, EPI_ISL_1716818, EPI_ISL_1716819, EPI_ISL_1716820 | | | |
| see above | CSIR-IGIB | CSIR-IGIB | Akshay Kanakan; Anil Kumar; Anish Maitra; Aparna S Murali; Azka Khan; Janani SV; Neha Jha; Nisha Rawat; Partha Chattopadhyay; Priti Devi; Priyanka Mehta; Rajesh Pandey; Ranjeet Maurya; Shweta Sahni |
| EPI_ISL_1360295, EPI_ISL_1360296, EPI_ISL_1360297, EPI_ISL_1360298, EPI_ISL_1360299, EPI_ISL_1360300, EPI_ISL_1360301, EPI_ISL_1360302, EPI_ISL_1360303, EPI_ISL_1360304, EPI_ISL_1360305, EPI_ISL_1360306, EPI_ISL_1360307, EPI_ISL_1360308, EPI_ISL_1360309, EPI_ISL_1360310, EPI_ISL_1360311, EPI_ISL_1360312, EPI_ISL_1360313, EPI_ISL_1360314, EPI_ISL_1360315, EPI_ISL_1360316, EPI_ISL_1360317, EPI_ISL_1360318, EPI_ISL_1360319, EPI_ISL_1360320, EPI_ISL_1360321, EPI_ISL_1360322, EPI_ISL_1360323, EPI_ISL_1360324, EPI_ISL_1360325, EPI_ISL_1360326, EPI_ISL_1360327, EPI_ISL_1360328, EPI_ISL_1360329, EPI_ISL_1360330, EPI_ISL_1360331, EPI_ISL_1360332, EPI_ISL_1360333, EPI_ISL_1360334, EPI_ISL_1360335, EPI_ISL_1360336, EPI_ISL_1360337, EPI_ISL_1360338, EPI_ISL_1360339, EPI_ISL_1360340, EPI_ISL_1360341, EPI_ISL_1360342, EPI_ISL_1360343, EPI_ISL_1360344, EPI_ISL_1360345, EPI_ISL_1360346, EPI_ISL_1360347, EPI_ISL_1360348, EPI_ISL_1360349, EPI_ISL_1360350, EPI_ISL_1360351, EPI_ISL_1360352, EPI_ISL_1360353, EPI_ISL_1360354, EPI_ISL_1360355, EPI_ISL_1360356, EPI_ISL_1360357, EPI_ISL_1360358, EPI_ISL_1360359, EPI_ISL_1360360, EPI_ISL_1360361, EPI_ISL_1360362, EPI_ISL_1360363, EPI_ISL_1360364, EPI_ISL_1360365, EPI_ISL_1360366, EPI_ISL_1360367, EPI_ISL_1360368, EPI_ISL_1360369, EPI_ISL_1360370, EPI_ISL_1360371, EPI_ISL_1360372, EPI_ISL_1360373, EPI_ISL_1360374, EPI_ISL_1360375, EPI_ISL_1360376, EPI_ISL_1360377, EPI_ISL_1360378, EPI_ISL_1360379, EPI_ISL_1360380, EPI_ISL_1360381, EPI_ISL_1360382, EPI_ISL_1360383, EPI_ISL_1360384, EPI_ISL_1360385, EPI_ISL_1360386, EPI_ISL_1360387, EPI_ISL_1360388, EPI_ISL_1372279, EPI_ISL_1547802, EPI_ISL_1547803, EPI_ISL_1547804, EPI_ISL_1547805, EPI_ISL_1718261, EPI_ISL_1916266, EPI_ISL_1916267, EPI_ISL_1916268, EPI_ISL_1916269, EPI_ISL_1916270, EPI_ISL_1916271, EPI_ISL_1916272, EPI_ISL_1916273, EPI_ISL_1916274, EPI_ISL_1916275, EPI_ISL_1916276, EPI_ISL_1916277, EPI_ISL_1916278, EPI_ISL_1916279, EPI_ISL_1916280, EPI_ISL_1916281, EPI_ISL_1916282, EPI_ISL_1916283, EPI_ISL_1916284, EPI_ISL_1916285, EPI_ISL_1916286, EPI_ISL_1916287, EPI_ISL_1916288, EPI_ISL_1916289, EPI_ISL_1916290, EPI_ISL_1916291, EPI_ISL_1916292, EPI_ISL_1916293, EPI_ISL_1916294, EPI_ISL_1916295, EPI_ISL_1916296, EPI_ISL_1916297, EPI_ISL_1916298, EPI_ISL_1916299, EPI_ISL_1916300, EPI_ISL_1916301, EPI_ISL_1916302, EPI_ISL_1916303, EPI_ISL_1916304, EPI_ISL_1916305, EPI_ISL_1916306, EPI_ISL_1916307, EPI_ISL_1916308, EPI_ISL_1916309, EPI_ISL_1916310, EPI_ISL_1916311, EPI_ISL_1916312, EPI_ISL_1916313, EPI_ISL_1916361, EPI_ISL_1916362, EPI_ISL_1916363, EPI_ISL_1916364, EPI_ISL_1916365, EPI_ISL_1916366, EPI_ISL_1916367, EPI_ISL_1916368, EPI_ISL_1916369, EPI_ISL_1916370, EPI_ISL_1916371, EPI_ISL_1916372, EPI_ISL_1916373, EPI_ISL_1916374, EPI_ISL_1916375, EPI_ISL_1916376, EPI_ISL_1916377, EPI_ISL_1916378, EPI_ISL_1916379, EPI_ISL_1916380, EPI_ISL_1916381, EPI_ISL_1916382, EPI_ISL_1916383, EPI_ISL_1916384, EPI_ISL_1916385, EPI_ISL_1916386, EPI_ISL_1916387, EPI_ISL_1916388, EPI_ISL_1916389, EPI_ISL_1916390, EPI_ISL_1916391, EPI_ISL_1916392, EPI_ISL_1916393, EPI_ISL_1916394, EPI_ISL_1916395, EPI_ISL_1916396, EPI_ISL_1916397, EPI_ISL_1916398, EPI_ISL_1916399, EPI_ISL_1916400, EPI_ISL_1916401, EPI_ISL_1916402, EPI_ISL_1916403, EPI_ISL_1916404, EPI_ISL_1916405, EPI_ISL_1916406, EPI_ISL_1916407, EPI_ISL_1916408, EPI_ISL_1916409, EPI_ISL_1916410, EPI_ISL_1916411, EPI_ISL_1916412, EPI_ISL_1916413, EPI_ISL_1916414, EPI_ISL_1916415, EPI_ISL_1916416, EPI_ISL_1916417, EPI_ISL_1916418, EPI_ISL_1916419, EPI_ISL_1916420, EPI_ISL_1916421, EPI_ISL_1916422, EPI_ISL_1916423, EPI_ISL_1916424, EPI_ISL_1916425, EPI_ISL_1916426, EPI_ISL_1916427, EPI_ISL_1916428, EPI_ISL_1916429, EPI_ISL_1916430, EPI_ISL_1916431, EPI_ISL_1916432, EPI_ISL_1916433, EPI_ISL_1916434, EPI_ISL_1916435, EPI_ISL_1916436, EPI_ISL_1916437, EPI_ISL_1916438, EPI_ISL_1916439, EPI_ISL_1916440, EPI_ISL_1916441, EPI_ISL_1916442, EPI_ISL_1916443, EPI_ISL_1916444, EPI_ISL_1916445, EPI_ISL_1916446, EPI_ISL_1916447, EPI_ISL_1916448, EPI_ISL_1916449, EPI_ISL_1916450, EPI_ISL_1916451, EPI_ISL_1916452, EPI_ISL_1916453, EPI_ISL_1916454, EPI_ISL_1916455, EPI_ISL_1916456, EPI_ISL_1916457, EPI_ISL_1916458, EPI_ISL_1916459, EPI_ISL_1916460, EPI_ISL_1916461, EPI_ISL_1916462, EPI_ISL_1916463, EPI_ISL_1916464, EPI_ISL_1916465, EPI_ISL_1916466, EPI_ISL_1916467, EPI_ISL_1916468, EPI_ISL_1916469, EPI_ISL_1916470, EPI_ISL_1916471, EPI_ISL_1916472, EPI_ISL_1916473, EPI_ISL_1916474, EPI_ISL_1916475, EPI_ISL_1916476, EPI_ISL_1916477, EPI_ISL_1916478, EPI_ISL_1916479, EPI_ISL_1916480, EPI_ISL_1916481, EPI_ISL_1916482, EPI_ISL_1916483, EPI_ISL_1916484, EPI_ISL_1916485, EPI_ISL_1916486, EPI_ISL_1916487, EPI_ISL_1916488, EPI_ISL_1916489, EPI_ISL_1916490, EPI_ISL_1916491, EPI_ISL_1916492, EPI_ISL_1916493, EPI_ISL_1916494, EPI_ISL_1916495, EPI_ISL_1916496, EPI_ISL_1916497, EPI_ISL_1916498, EPI_ISL_1970625, EPI_ISL_1970626, EPI_ISL_1970627, EPI_ISL_1970628, EPI_ISL_1970629 | | | |
| see above | CSIR-National Environmental Engineering Research Institute | CSIR-Centre for Cellular and Molecular Biology - INSACOG | Amareshwar Vodapalli; Ara Sreenivas; B Himasri; Divya Tej Sowpati; Karthik Bharadwaj Tallapaka; Krishna Khairnar; Lamuk Zaveri; Onkar Kulkarni; Rakesh K Mishra; Sharath Chandra Thota; Shreekant Verma; Sofia Banu; Viswagithe S L |
| EPI_ISL_1372011, EPI_ISL_1662424, EPI_ISL_1662425, EPI_ISL_1662426, EPI_ISL_1662427, EPI_ISL_1662428, EPI_ISL_1662429, EPI_ISL_1662430, EPI_ISL_1662431, EPI_ISL_1662432, EPI_ISL_1662433, EPI_ISL_1662434, EPI_ISL_1662435, EPI_ISL_1662436, EPI_ISL_1662437, EPI_ISL_1662438, EPI_ISL_1662439, EPI_ISL_1662440, EPI_ISL_1662441, EPI_ISL_1662442, EPI_ISL_1662443, EPI_ISL_1662444, EPI_ISL_1663323, EPI_ISL_1663324, EPI_ISL_1663356, EPI_ISL_1663357, EPI_ISL_1663488, EPI_ISL_1663489, EPI_ISL_1663490, EPI_ISL_1663491, EPI_ISL_1663492, EPI_ISL_1663493, EPI_ISL_1663494, EPI_ISL_1663495, EPI_ISL_1663496 | | | |
| see above | Capital hospital, Bhubaneswar | Institute of Life Sciences - INSACOG | Ajay Parida; Amol M. Kanampalliwar; Arup Ghosh; Atimukta Jha; INSACOG Consortium; Punit Prasad; Rajeeb Swain; Rupesh Dash; Safal Walia; Shifu Aggarwal; Sunil K. Raghav |
| EPI_ISL_2106937, EPI_ISL_2106938, EPI_ISL_2106939, EPI_ISL_2106940, EPI_ISL_2106941, EPI_ISL_2106942, EPI_ISL_2106943, EPI_ISL_2106944, EPI_ISL_2106945, EPI_ISL_2106946, EPI_ISL_2106947, EPI_ISL_2106948, EPI_ISL_2106949, EPI_ISL_2106950, EPI_ISL_2106951, EPI_ISL_2106952, EPI_ISL_2106953, EPI_ISL_2106954, EPI_ISL_2106955, EPI_ISL_2106956, EPI_ISL_2106957, EPI_ISL_2106958, EPI_ISL_2106959, EPI_ISL_2106960, EPI_ISL_2106961, EPI_ISL_2106962, EPI_ISL_2106963, EPI_ISL_2106964, EPI_ISL_2106965, EPI_ISL_2106966, EPI_ISL_2106967, EPI_ISL_2106968, EPI_ISL_2106969, EPI_ISL_2106970, EPI_ISL_2106971, EPI_ISL_2106972, EPI_ISL_2106973, EPI_ISL_2106974 | | | |
| see above | Centre for DNA Fingerprinting and Diagnostics | CDFD -INSACOG | Ashwin Dalal; Asmita Gupta; Divya Vashisht; Murali Bashyam; Pratyusha Bala; Vinay Donipadi |
| EPI_ISL_1334325 | Clinical Specimen Feces | Enteric Viruses Group, ICMR-National Institute of Virology | Dr. Mallika Lavania; Dr. Varsha Potdar |
| EPI_ISL_2714644, EPI_ISL_2714645, EPI_ISL_2714646, EPI_ISL_2714647, EPI_ISL_2714648, EPI_ISL_2714649, EPI_ISL_2714650, EPI_ISL_2714651, EPI_ISL_2714652, EPI_ISL_2714653, EPI_ISL_2714654, EPI_ISL_2714655, EPI_ISL_2714656, EPI_ISL_2714657, EPI_ISL_2714658, EPI_ISL_2714659, EPI_ISL_2714660, EPI_ISL_2714661, EPI_ISL_2714662, EPI_ISL_2714663, EPI_ISL_2714664, EPI_ISL_2714665, EPI_ISL_2714666, EPI_ISL_2714667, EPI_ISL_2714668, EPI_ISL_2714669, EPI_ISL_2714670, EPI_ISL_2714671, EPI_ISL_2714672, EPI_ISL_2714673, EPI_ISL_2714674, EPI_ISL_2714675, EPI_ISL_2714676, EPI_ISL_2714677, EPI_ISL_2714678, EPI_ISL_2714679, EPI_ISL_2714680, EPI_ISL_2714681, EPI_ISL_2714682, EPI_ISL_2714683, EPI_ISL_2714684, EPI_ISL_2714685, EPI_ISL_2714686, EPI_ISL_2714687, EPI_ISL_2714688, EPI_ISL_2714689, EPI_ISL_2714690, EPI_ISL_2714691, EPI_ISL_2714692, EPI_ISL_2714693, EPI_ISL_2714694, EPI_ISL_2714695, EPI_ISL_2714696, EPI_ISL_2714697, EPI_ISL_2714698 | | | |
| see above | Communicable Diseases, Interactive Research School for Health Affairs (IRSHA) | Communicable Diseases, Interactive Research School for Health Affairs (IRSHA) | A.C.; Arankalle; Mishra; Modak, M.; Pisal; S.R.; Shrivastava, S.; V.A.; Virkar, R. |
| EPI_ISL_1710598 | Communicable Diseases, Interactive Research School for Health Affairs | Communicable Diseases, Interactive Research School for Health Affairs | Akhilesh Chandra Mishra; Meera Modak; Rashmi Virkar; Shamburaje S Pisal; Shubham Shrivastava; Vidya A Arankalle |
| EPI_ISL_1595867, EPI_ISL_1595868, EPI_ISL_1595869, EPI_ISL_1595870, EPI_ISL_1595871, EPI_ISL_1595872, EPI_ISL_1595873 | | | |
| see above | DSU KALABURAGI | INSACOG-KA, NIMHANS | Ananthapadmanabha Kotambail; Anita S Desai; Anson Kunjumon George; Chetan G K; Chitra Pattabiraman; Darshan Sreenivas; Gautham Arunachal Udupi; Pramada Prasad; V Ravi |
| EPI_ISL_2100635, EPI_ISL_2100673, EPI_ISL_2100674, EPI_ISL_2100675 | Delhi North District | CSIR-Institute of Genomics and Integrative Biology | Animesh Ray; Asangala Kamai; Ashwin Varadarajan; Ayush Goel; Bharathram Uppilli*; Devasish Desai; Manish Kumar; Mohammed Ahmed; Mohammed Faruq; Naveet Wig; Pooja Sharma*; Rajesh Pandey; Sarafaraz Alam; Saruchi Wadhwa; Satish Swain; Sheeba Saifi; Sushma Rajpoot; Umang Arora |
| EPI_ISL_1844693, EPI_ISL_1844698, EPI_ISL_1844703, EPI_ISL_1844707, EPI_ISL_1844711 | Department of Microbiology, Dr YSPGMC | Biotechnology Division, NCDC | Hema Gogia; Hemlata Lall; Kalaiarasan Ponnusamy; Mahesh S Dhar; Manoj K Singh; Meena Datta; Partha Rakshit; Preeti Madan; Priyanka Singh; Radhakrishnan V. S; Robin Marwal; Sandhya Kabra; Sujeet K Singh; Uma Sharma |
| EPI_ISL_1662287, EPI_ISL_1662288, EPI_ISL_1662289, EPI_ISL_1662290, EPI_ISL_1662291 | Dept. Of Microbiology, Lt.. Baliram Kashyap Memorial Govt. Medical college, Dimrapal, Jagdalpur | Institute of Life Sciences - INSACOG | Ajay Parida; Amol M. Kanampalliwar; Arup Ghosh; Atimukta Jha; INSACOG Consortium; Punit Prasad; Rajeeb Swain; Rupesh Dash; Safal Walia; Shifu Aggarwal; Sunil K. Raghav |
| EPI_ISL_1708463 | Deputy Director of health survices erode | inStem NCBS - INSACOG | Uma Ramakrishnan Dasaradhi Palakodeti Aswin SaiNarain |
| EPI_ISL_751270 | Devki Devi Foundation, a unit of Max Healthcare | CSIR-IGIB/Max | Akansha Tyagi; Akshay Kanakan; Anurag Agrawal; Bansidhar Tarai; Janani Srinivasa Vasudevan; Mitali Mukerji; Mohammed Faruq; Poonam Das; Rajesh Pandey#; Ranjeet Maurya; Samreen Siddiqui; Sujeet Jha; Uzma Shamim |
| EPI_ISL_1939849, EPI_ISL_1939850, EPI_ISL_1939851, EPI_ISL_1939852, EPI_ISL_1939853, EPI_ISL_1939854, EPI_ISL_1939855, EPI_ISL_1939856, EPI_ISL_1939857, EPI_ISL_1939858, EPI_ISL_1939859, EPI_ISL_1939860, EPI_ISL_1939861, EPI_ISL_1939862 | | | |
| see above | Dr S raju, Director of public heath and preventive medicine | inStem NCBS - INSACOG | Uma Ramakrishnan Dasaradhi Palakodeti Aswin SaiNarain |
| EPI_ISL_1708474, EPI_ISL_1708475, EPI_ISL_1708476, EPI_ISL_1708477, EPI_ISL_1708478, EPI_ISL_1708479, EPI_ISL_1708480, EPI_ISL_1708481, EPI_ISL_1708482, EPI_ISL_1708483, EPI_ISL_1708484, EPI_ISL_1708485, EPI_ISL_1708486, EPI_ISL_1708487, EPI_ISL_1708488, EPI_ISL_1708489, EPI_ISL_1708490, EPI_ISL_1708491, EPI_ISL_1708492, EPI_ISL_1708493, EPI_ISL_1708494, EPI_ISL_1708495, EPI_ISL_1708496, EPI_ISL_1708497 | | | |
| see above | Dr.Madhusudhan MBBD MD ICMR-VRDL | inStem NCBS - INSACOG | Uma Ramakrishnan Dasaradhi Palakodeti Aswin SaiNarain |
| EPI_ISL_1708445, EPI_ISL_1708446 | Dr.P.venkateshwarlu nodal officer VRDL lab Microbiology Dept | inStem NCBS - INSACOG | Uma Ramakrishnan Dasaradhi Palakodeti Aswin SaiNarain |
| EPI_ISL_1708449, EPI_ISL_1708450, EPI_ISL_1708451, EPI_ISL_1708452, EPI_ISL_1708453, EPI_ISL_1708454, EPI_ISL_1708455, EPI_ISL_1708456, EPI_ISL_1708457, EPI_ISL_1708458, EPI_ISL_1708459, EPI_ISL_1708460, EPI_ISL_1708461, EPI_ISL_1708464, EPI_ISL_1708465, EPI_ISL_1708466, EPI_ISL_1708467, EPI_ISL_1708468, EPI_ISL_1708469, EPI_ISL_1708470, EPI_ISL_1708471, EPI_ISL_1708472, EPI_ISL_1708473, EPI_ISL_1708498, EPI_ISL_1708499, EPI_ISL_1708500, EPI_ISL_1708501, EPI_ISL_1708502, EPI_ISL_1708503, EPI_ISL_1708504 | | | |
| see above | Dr.S.raju Director of public heath and preventive medicine | inStem NCBS - INSACOG | Aswin Seshasayee"; Uma Ramakrishnan Dasaradhi Palakodeti Aswin SaiNarain |
| EPI_ISL_745198 | ESI Hospital | CSIR-Centre for Cellular and Molecular Biology | Archana Bharadwaj Siva; B Himasri; Blessy B John; Divya Tej Sowpati; Dr.P.Shashikala Reddy; Dr.S.Pavani; Dr.Satyaprasad; Dr.V.Sudha Rani; Karthik Bharadwaj Tallapaka; Lamuk Zaveri; M Soujanya Reddy; Namami Gaur; Nikhil Hajirnis; Onkar Kulkarni; Payel Mukherjee; Pratheusa Maccha; Priya Singh; Purushotham Vodnala; Rakesh K Mishra; Sakshi Shambhavi; Shagufta Khan; Sofia Banu; Tulasi Nagabandi; Viswagithe S L |
| EPI_ISL_1940051, EPI_ISL_1940052 | GANGONAHALLI | inStem NCBS - INSACOG | Uma Ramakrishnan Dasaradhi Palakodeti Aswin SaiNarain |
| EPI_ISL_1940077 | GANGONDAHALLI | inStem NCBS - INSACOG | Uma Ramakrishnan Dasaradhi Palakodeti Aswin SaiNarain |

| | | | |
|---|---|---|---|
| EPI_ISL_2015532 | GANGONDNAHALLI | inStem NCBS - INSACOG | Uma Ramakrishnan Dasaradhi Palakodeti Aswin SaiNarain |
| EPI_ISL_1677770, EPI_ISL_1677771, EPI_ISL_1677772, EPI_ISL_1677773, EPI_ISL_1677774, EPI_ISL_1677775, EPI_ISL_1677776, EPI_ISL_1677777, EPI_ISL_1677778, EPI_ISL_1677779, EPI_ISL_1677780, EPI_ISL_1677781, EPI_ISL_1677782, EPI_ISL_1677783, EPI_ISL_1677784, EPI_ISL_1677785, EPI_ISL_1677786, EPI_ISL_1677787, EPI_ISL_1677788, EPI_ISL_1677789, EPI_ISL_1677790, EPI_ISL_1677792, EPI_ISL_1677793, EPI_ISL_1677794, EPI_ISL_1677795, EPI_ISL_1677796, EPI_ISL_1677797, EPI_ISL_1677798 | | | |
| see above | GMERS Medical Collage, GMERS Medical College,Civil Hospital, Sola, Ahmedabad | Gujarat Biotechnology Research Centre | Chaitanya Joshi; Dinesh Kumar; Janvi Raval; Madhvi Joshi; Nidhi Sood; Nitesh Shah; Nitin Savaliya; Ramesh Pandit; Sonal Sharma; Twinkle Soni; Umang Mishra; Zarna Patel; Zuber Saiyed |
| EPI_ISL_1706595, EPI_ISL_1706596, EPI_ISL_1706597 | GMERS Medical Collage, GMERS Medical College,Civil Hospital, Sola, Ahmedabad | Gujarat Biotechnology Research Centre | Chaitanya Joshi; Dinesh Kumar; Janvi Raval; Madhvi Joshi; Nidhi Sood; Nitesh Shah; Nitin Savaliya; Ramesh Pandit; Sonal Sharma; Twinkle Soni; Umang Mishra; Zarna Patel; Zuber Saiyed |
| EPI_ISL_1677741, EPI_ISL_1677742, EPI_ISL_1677743, EPI_ISL_1677744, EPI_ISL_1677745, EPI_ISL_1677746, EPI_ISL_1677747, EPI_ISL_1677748, EPI_ISL_1677749, EPI_ISL_1677750, EPI_ISL_1677751, EPI_ISL_1677752, EPI_ISL_1677753, EPI_ISL_1677754, EPI_ISL_1677755, EPI_ISL_1677756, EPI_ISL_1677758, EPI_ISL_1677759, EPI_ISL_1677760, EPI_ISL_1677761, EPI_ISL_1677762, EPI_ISL_1677763, EPI_ISL_1677764, EPI_ISL_1677765, EPI_ISL_1677766, EPI_ISL_1706593, EPI_ISL_1706594 | | | |
| see above | GMERS Medical College and Hospital, Gotri | Gujarat Biotechnology Research Centre | Bithika Duttaroy; Chaitanya Joshi; Dinesh Kumar; Janvi Raval; Madhvi Joshi; Nitesh Shah; Nitin Savaliya; Ramesh Pandit; Sonal Sharma; Twinkle Soni; Umang Mishra; Zarna Patel; Zuber Saiyed |
| EPI_ISL_825048, EPI_ISL_825049, EPI_ISL_825054, EPI_ISL_825055, EPI_ISL_825056, EPI_ISL_825057, EPI_ISL_825058, EPI_ISL_825061, EPI_ISL_825062 | | | |
| see above | GMERS Medical College and Hospital, Gotri, Vadodara | Gujarat Biotechnology Research Centre | Afzal Ansari; Apurvasinh Puvar; Bithika Duttaroy; Chaitanya Joshi; Dinesh Kumar; Janvi Raval; Kalpesh Mistry; Labdhi Pandya; Madhvi Joshi; Mitesh Kamothi; Nikha Trivedi; Nitin Savaliya; Ramesh Pandit; Zarna Patel; Zuber Saiyed |
| EPI_ISL_2001178, EPI_ISL_2001179, EPI_ISL_2001180, EPI_ISL_2001181, EPI_ISL_2001182, EPI_ISL_2001183, EPI_ISL_2001184, EPI_ISL_2001185, EPI_ISL_2001186, EPI_ISL_2017752, EPI_ISL_2017753, EPI_ISL_2017754, EPI_ISL_2017755, EPI_ISL_2017756, EPI_ISL_2017757, EPI_ISL_2017758 | | | |
| see above | GMERS, Government Medical College (GMC), Gotri | Gujarat Biotechnology Research Centre | Bithika Duttaroy; Chaitanya Joshi; Dinesh Kumar; Janvi Raval; Madhvi Joshi; Nitesh Shah; Nitin Savaliya; Ramesh Pandit; Sonal Sharma; Twinkle Soni; Umang Mishra; Zarna Patel; Zuber Saiyed |
| EPI_ISL_2001121, EPI_ISL_2001122, EPI_ISL_2001123, EPI_ISL_2001124, EPI_ISL_2001125, EPI_ISL_2001126, EPI_ISL_2001127, EPI_ISL_2001128, EPI_ISL_2001129, EPI_ISL_2001130, EPI_ISL_2001131, EPI_ISL_2001132, EPI_ISL_2001133, EPI_ISL_2001134, EPI_ISL_2001135, EPI_ISL_2001136, EPI_ISL_2001137, EPI_ISL_2001138, EPI_ISL_2001139, EPI_ISL_2001140, EPI_ISL_2001141, EPI_ISL_2001142, EPI_ISL_2001143, EPI_ISL_2001144, EPI_ISL_2001145, EPI_ISL_2001146, EPI_ISL_2001147, EPI_ISL_2001148 | | | |
| see above | GMERS, Government Medical College, Gandhinagar | Gujarat Biotechnology Research Centre | Chaitanya Joshi; Dinesh Kumar; Gaurishankar Shrimali; Janvi Raval; Madhvi Joshi; Nitesh Shah; Nitin Savaliya; Ramesh Pandit; Sonal Sharma; Twinkle Soni; Umang Mishra; Zarna Patel; Zuber Saiyed |
| EPI_ISL_1544133, EPI_ISL_1544134 | GMERS, Medical College, Gandhinagar | Gujarat Biotechnology Research Centre | Chaitanya Joshi; Dinesh Kumar; Gaurishankar Shrimali; Janvi Raval; Madhvi Joshi; Nitesh Shah; Nitin Savaliya; Ramesh Pandit; Sonal Sharma; Twinkle Soni; Umang Mishra; Zarna Patel; Zuber Saiyed |
| EPI_ISL_2015549, EPI_ISL_2015639 | GOVIDRAJNAGAR | inStem NCBS - INSACOG | Uma Ramakrishnan Dasaradhi Palakodeti Aswin SaiNarain |
| EPI_ISL_1940017, EPI_ISL_1940018, EPI_ISL_1940019, EPI_ISL_1940020, EPI_ISL_1940021 | GOVIMDRAJ NAGAR | inStem NCBS - INSACOG | Uma Ramakrishnan Dasaradhi Palakodeti Aswin SaiNarain |
| EPI_ISL_1940071, EPI_ISL_1940072, EPI_ISL_1940073, EPI_ISL_1940074, EPI_ISL_1940075, EPI_ISL_1940076 | GOVINDRAJ NAGAR | inStem NCBS - INSACOG | Uma Ramakrishnan Dasaradhi Palakodeti Aswin SaiNarain |
| EPI_ISL_2151313 | GOVINDRAJ NAGAR UPHC | inStem NCBS - INSACOG | Uma Ramakrishnan Dasaradhi Palakodeti Aswin SaiNarain |
| EPI_ISL_1940049, EPI_ISL_1940050, EPI_ISL_1940109, EPI_ISL_1940110, EPI_ISL_1940111, EPI_ISL_2015470, EPI_ISL_2015500, EPI_ISL_2015514, EPI_ISL_2015538, EPI_ISL_2015619 | | | |
| see above | GOVINDRAJNAGAR | inStem NCBS - INSACOG | Uma Ramakrishnan Dasaradhi Palakodeti Aswin SaiNarain |
| EPI_ISL_1544179, EPI_ISL_1544180, EPI_ISL_1544181, EPI_ISL_1544182, EPI_ISL_1544183, EPI_ISL_1544184, EPI_ISL_1544185, EPI_ISL_1544186, EPI_ISL_1544187, EPI_ISL_1544188, EPI_ISL_1544189, EPI_ISL_1544190, EPI_ISL_1544191, EPI_ISL_1544192, EPI_ISL_1544193, EPI_ISL_1544194, EPI_ISL_1544195, EPI_ISL_1677767 | | | |
| see above | Government Medical College (GMC), Surat | Gujarat Biotechnology Research Centre | Chaitanya Joshi; Dinesh Kumar; Janvi Raval; Madhvi Joshi; Neeta Khandelwal; Nitesh Shah; Nitin Savaliya; Ramesh Pandit; Sonal Sharma; Twinkle Soni; Umang Mishra; Zarna Patel; Zuber Saiyed |
| EPI_ISL_2000678, EPI_ISL_2000679 | Government Medical College Ernakulam | CSIR Institute of Genomics and Integrative Biology | Abhinav Jain; Afra Shamnath; Anjali Bajaj; Anoop Joseph; Arvinden V.R.; Bani Jolly; Beena Philomina J; Bindhu Vasudevan; Chandni Radhakrishnan; Fairoz C. P; Mercy Rophina; Mohamed Imran; Mohit Kumar Divakar; Neethu John; Nisha Majeed; P. Geetha; P. Jayesh Kumar; Prasanth Viswanathan; Priyanka R Nair; Rahul C. Bhoyar; Samatha Mathew; Srashti Jyoti Agrawal; Sridhar Sivasubbu; Vigneshwar Senthivel; Vinod Scaria; Vishu Gupta |
| EPI_ISL_2000674 | Government Medical College Idukki | CSIR Institute of Genomics and Integrative Biology | Abhinav Jain; Afra Shamnath; Anjali Bajaj; Anoop Joseph; Arvinden V.R.; Bani Jolly; Beena Philomina J; Bindhu Vasudevan; Chandni Radhakrishnan; Fairoz C. P; Mercy Rophina; Mohamed Imran; Mohit Kumar Divakar; Neethu John; Nisha Majeed; P. Geetha; P. Jayesh Kumar; Prasanth Viswanathan; Priyanka R Nair; Rahul C. Bhoyar; Samatha Mathew; Srashti Jyoti Agrawal; Sridhar Sivasubbu; Vigneshwar Senthivel; Vinod Scaria; Vishu Gupta |
| EPI_ISL_2000675, EPI_ISL_2000676, EPI_ISL_2000677 | Government Medical College Kozhikode | CSIR Institute of Genomics and Integrative Biology | Abhinav Jain; Afra Shamnath; Anjali Bajaj; Anoop Joseph; Arvinden V.R.; Bani Jolly; Beena Philomina J; Bindhu Vasudevan; Chandni Radhakrishnan; Fairoz C. P; Mercy Rophina; Mohamed Imran; Mohit Kumar Divakar; Neethu John; Nisha Majeed; P. Geetha; P. Jayesh Kumar; Prasanth Viswanathan; Priyanka R Nair; Rahul C. Bhoyar; Samatha Mathew; Srashti Jyoti Agrawal; Sridhar Sivasubbu; Vigneshwar Senthivel; Vinod Scaria; Vishu Gupta |
| EPI_ISL_1662345, EPI_ISL_1662346, EPI_ISL_1662347, EPI_ISL_1662348, EPI_ISL_1662349, EPI_ISL_1662350, EPI_ISL_1662351, EPI_ISL_1662352, EPI_ISL_1662353, EPI_ISL_1662354, EPI_ISL_1662355, EPI_ISL_1662357, EPI_ISL_1662358, EPI_ISL_1662359, EPI_ISL_1662360, EPI_ISL_1662361, EPI_ISL_1662362, EPI_ISL_1662363, EPI_ISL_1662364, EPI_ISL_1662365, EPI_ISL_1662366, EPI_ISL_1662367, EPI_ISL_1662368, EPI_ISL_1662369, EPI_ISL_1662370, EPI_ISL_1662371, EPI_ISL_1662372, EPI_ISL_1662373, EPI_ISL_1662374, EPI_ISL_1662375, EPI_ISL_1662376, EPI_ISL_1662377, EPI_ISL_1662378, EPI_ISL_1662379, EPI_ISL_1662380, EPI_ISL_1662381, EPI_ISL_1662382, EPI_ISL_1662383 | | | |
| see above | Govt. Medical College, Ambikapur, Surguja | Institute of Life Sciences - INSACOG | Ajay Parida; Amol M. Kanampalliwar; Arup Ghosh; Atimukta Jha; INSACOG Consortium; Punit Prasad; Rajeeb Swain; Rupesh Dash; Safal Walia; Shifu Aggarwal; Sunil K. Raghav |
| EPI_ISL_1677768, EPI_ISL_1677769 | Gujarat Biotechnology Research Centre (GBRC), Gandhinagar | Gujarat Biotechnology Research Centre | Chaitanya Joshi; Dinesh Kumar; Janvi Raval; Madhvi Joshi; Nitesh Shah; Nitin Savaliya; Ramesh Pandit; Sonal Sharma; Twinkle Soni; Umang Mishra; Zarna Patel; Zuber Saiyed |
| EPI_ISL_745201 | Guntur General Hospital | CSIR-Centre for Cellular and Molecular Biology | Archana Bharadwaj Siva; B Himasri; Blessy B John; Divya Tej Sowpati; Dr.P.Shashikala Reddy; Dr.S.Pavani; Dr.Satyaprasad; Dr.V.Sudha Rani; Karthik Bharadwaj Tallapaka; Lamuk Zaveri; M Soujanya Reddy; Namami Gaur; Nikhil Hajirnis; Onkar Kulkarni; Payel Mukherjee; Pratheusa Maccha; Priya Singh; Purushotham Vodnala; Rakesh K Mishra; Sakshi Shambhavi; Shagufta Khan; Sofia Banu; Tulasi Nagabandi; Viswagithe S L |
| EPI_ISL_2015471, EPI_ISL_2015499, EPI_ISL_2015506, EPI_ISL_2015520, EPI_ISL_2015523, EPI_ISL_2015545, EPI_ISL_2015573, EPI_ISL_2015614, EPI_ISL_2015634 | | | |
| see above | HOSA HALLI | inStem NCBS - INSACOG | Uma Ramakrishnan Dasaradhi Palakodeti Aswin SaiNarain |
| EPI_ISL_1939980, EPI_ISL_1939981, EPI_ISL_1939982, EPI_ISL_1939983, EPI_ISL_1939984, EPI_ISL_1940000, EPI_ISL_1940001, EPI_ISL_1940002, EPI_ISL_1940003, EPI_ISL_1940004, EPI_ISL_1940005, EPI_ISL_1940006, EPI_ISL_1940007, EPI_ISL_1940008, EPI_ISL_1940009, EPI_ISL_1940010, EPI_ISL_1940011, EPI_ISL_1940012, EPI_ISL_1940013, EPI_ISL_1940014, EPI_ISL_1940015, EPI_ISL_1940016, EPI_ISL_1940037, EPI_ISL_1940038, EPI_ISL_1940039, EPI_ISL_1940040, EPI_ISL_1940041, EPI_ISL_1940042, EPI_ISL_1940043, EPI_ISL_1940044, EPI_ISL_1940045, EPI_ISL_1940046, EPI_ISL_1940047, EPI_ISL_1940048, EPI_ISL_1940062, EPI_ISL_1940063, EPI_ISL_1940064, EPI_ISL_1940065, EPI_ISL_1940066, EPI_ISL_1940067, EPI_ISL_1940068, EPI_ISL_1940078, EPI_ISL_1940079, EPI_ISL_1940080, EPI_ISL_1940081, EPI_ISL_1940082, EPI_ISL_1940083, EPI_ISL_1940084, EPI_ISL_1940085, EPI_ISL_1940086, EPI_ISL_1940087, EPI_ISL_1940088, EPI_ISL_1940089, EPI_ISL_1940090, EPI_ISL_1940112, EPI_ISL_1940113, EPI_ISL_1940114, EPI_ISL_1940115, EPI_ISL_1940116, EPI_ISL_1940117, EPI_ISL_1940118, EPI_ISL_1940119, EPI_ISL_1940120, EPI_ISL_1940121, EPI_ISL_1940122, EPI_ISL_1940123, EPI_ISL_1940124, EPI_ISL_1940125, EPI_ISL_1940126, EPI_ISL_1940127, EPI_ISL_1940128, EPI_ISL_1940129, EPI_ISL_1940142, EPI_ISL_2015493, EPI_ISL_2015516, EPI_ISL_2015527, EPI_ISL_2015547, EPI_ISL_2015554, EPI_ISL_2015565, EPI_ISL_2015567, EPI_ISL_2015575, EPI_ISL_2015616 | | | |
| see above | HOSAHALLI | inStem NCBS - INSACOG | Uma Ramakrishnan Dasaradhi Palakodeti Aswin SaiNarain |
| EPI_ISL_2100187, EPI_ISL_2100188, EPI_ISL_2100190, EPI_ISL_2100192, EPI_ISL_2100193, EPI_ISL_2100195, EPI_ISL_2100197, EPI_ISL_2100198, EPI_ISL_2100202, EPI_ISL_2100205, EPI_ISL_2100208, EPI_ISL_2100212, EPI_ISL_2100216 | | | |
| see above | ICMR-National Institute for Research In Tuberculosis | NIV Influenza | Dr Luke Elizabeth Hanna; Dr Padmapriyadarsini.C; Dr Radha Gopalaswamy; Dr S Siva Kumar; Mr Ramesh K |
| EPI_ISL_1703635, EPI_ISL_1703636, EPI_ISL_1703637, EPI_ISL_1703638, EPI_ISL_1703639, EPI_ISL_1703640, EPI_ISL_1703641, EPI_ISL_1703642, EPI_ISL_1703643, EPI_ISL_1703644, EPI_ISL_1703645, EPI_ISL_1703646, EPI_ISL_1703647, EPI_ISL_1703648, EPI_ISL_1703649, EPI_ISL_1703650, EPI_ISL_1703651, EPI_ISL_1703652, EPI_ISL_1703653, EPI_ISL_1703654, EPI_ISL_1703655, EPI_ISL_1703656, EPI_ISL_1703657, EPI_ISL_1703658, EPI_ISL_1703659, EPI_ISL_1703660, EPI_ISL_1703661, EPI_ISL_1703662, EPI_ISL_1703663, EPI_ISL_1703664, EPI_ISL_1703665, EPI_ISL_1703666, EPI_ISL_1703667, EPI_ISL_1703668, EPI_ISL_1703669, EPI_ISL_1703670, EPI_ISL_1703671, EPI_ISL_1703672, EPI_ISL_1703673, EPI_ISL_1703674, EPI_ISL_1703675, EPI_ISL_1703676, EPI_ISL_1703677, EPI_ISL_1703678, EPI_ISL_1703679, EPI_ISL_1703680, EPI_ISL_1703681, EPI_ISL_1703682, EPI_ISL_1703683, EPI_ISL_1703684, EPI_ISL_1703685, EPI_ISL_1703686, EPI_ISL_1703687, EPI_ISL_1703688, EPI_ISL_1703689, EPI_ISL_1703690, EPI_ISL_1703691, EPI_ISL_1703692, EPI_ISL_1703693, EPI_ISL_1703694, EPI_ISL_1703695, EPI_ISL_1703696, EPI_ISL_1703697, EPI_ISL_1703698, EPI_ISL_1703699, EPI_ISL_1703700, EPI_ISL_1703701, EPI_ISL_1703702, EPI_ISL_1703703, EPI_ISL_1703704, EPI_ISL_1703705, EPI_ISL_1703706, EPI_ISL_1703707, EPI_ISL_1703708, EPI_ISL_1703709, EPI_ISL_1703710, EPI_ISL_1703711, EPI_ISL_1703712, EPI_ISL_1703713, EPI_ISL_1703714, EPI_ISL_1703715, EPI_ISL_1703716, EPI_ISL_1703717, EPI_ISL_1703718, EPI_ISL_1703719, EPI_ISL_1703720, EPI_ISL_1703721, EPI_ISL_1703722, EPI_ISL_1703723, EPI_ISL_1703724, EPI_ISL_1703725, EPI_ISL_1703726, EPI_ISL_1703727, EPI_ISL_1703728, EPI_ISL_1703729, EPI_ISL_1703730, EPI_ISL_1703731, EPI_ISL_1703732, EPI_ISL_1703733, EPI_ISL_1703734, EPI_ISL_1703735, EPI_ISL_1703736, EPI_ISL_1703737, EPI_ISL_1703738, EPI_ISL_1703739, EPI_ISL_1703740, EPI_ISL_1703741, EPI_ISL_1703742, EPI_ISL_1703743, EPI_ISL_1703744, EPI_ISL_1703745, EPI_ISL_1703746, EPI_ISL_1703747, EPI_ISL_1703748, EPI_ISL_1703749, EPI_ISL_1703750, EPI_ISL_1703751, EPI_ISL_1703752, EPI_ISL_1703753, EPI_ISL_1703754, EPI_ISL_1703755, EPI_ISL_1703756, EPI_ISL_1703757, EPI_ISL_1703758, EPI_ISL_1703759, EPI_ISL_1703760, EPI_ISL_1703761, EPI_ISL_1703762, EPI_ISL_1703763, EPI_ISL_1703764, EPI_ISL_1703765, EPI_ISL_1703766, EPI_ISL_1703767, EPI_ISL_1703768, EPI_ISL_1703769, EPI_ISL_1703770, EPI_ISL_1703771, EPI_ISL_1703772, EPI_ISL_1703773, EPI_ISL_1703774, EPI_ISL_1703775, EPI_ISL_1703776, EPI_ISL_1703777, EPI_ISL_1703778, EPI_ISL_1703779, EPI_ISL_1703780, EPI_ISL_1703781, EPI_ISL_1703782, EPI_ISL_1703783, EPI_ISL_1703784, EPI_ISL_1703785, EPI_ISL_1703786, EPI_ISL_1703787, EPI_ISL_1703788, EPI_ISL_1703789, EPI_ISL_1703790, EPI_ISL_1703791, EPI_ISL_1703792, EPI_ISL_1703793, EPI_ISL_1703794, EPI_ISL_1703795, EPI_ISL_1703796, EPI_ISL_1703797, EPI_ISL_1703798, EPI_ISL_1703799, EPI_ISL_1703800, EPI_ISL_1703801, EPI_ISL_1703802, EPI_ISL_1703803, EPI_ISL_1703804, EPI_ISL_1703805, EPI_ISL_1703806, EPI_ISL_1703808, EPI_ISL_1703809, EPI_ISL_1703810, EPI_ISL_1703811, EPI_ISL_1703812, EPI_ISL_1703813, EPI_ISL_1703814, EPI_ISL_1703815, EPI_ISL_1703816, EPI_ISL_1703817, EPI_ISL_1703818, EPI_ISL_1703819, EPI_ISL_1703820, EPI_ISL_1703821, EPI_ISL_1703822, EPI_ISL_1703823, EPI_ISL_1703824, EPI_ISL_1703825, EPI_ISL_1703826, EPI_ISL_1703827, EPI_ISL_1703828, EPI_ISL_1703829, EPI_ISL_1703830, EPI_ISL_1703831, EPI_ISL_1703832, EPI_ISL_1703833, EPI_ISL_1703834, EPI_ISL_1703835, EPI_ISL_1703836, EPI_ISL_1703837, EPI_ISL_1703838, EPI_ISL_1703839, EPI_ISL_1703841, EPI_ISL_1703842, EPI_ISL_1703843, EPI_ISL_1703844, EPI_ISL_1703845, EPI_ISL_1703846, EPI_ISL_1703847, EPI_ISL_1703848, EPI_ISL_1703850, EPI_ISL_1703851, EPI_ISL_1703852, EPI_ISL_1703853, EPI_ISL_1703854, EPI_ISL_1703855, EPI_ISL_1703856, EPI_ISL_1703858, EPI_ISL_1703859, EPI_ISL_1703860, EPI_ISL_1703861, EPI_ISL_1703862, EPI_ISL_1703863, EPI_ISL_1703864, EPI_ISL_1703865, EPI_ISL_1703866, EPI_ISL_1703868, EPI_ISL_1703869, EPI_ISL_1703870, EPI_ISL_1703871, EPI_ISL_1703872, EPI_ISL_1703873, EPI_ISL_1703875, EPI_ISL_1703876, EPI_ISL_1703877, EPI_ISL_1703878, EPI_ISL_1703879, EPI_ISL_1703880, EPI_ISL_1703881, EPI_ISL_1703882, EPI_ISL_1703883, EPI_ISL_1703884, EPI_ISL_1703886, EPI_ISL_1703887, EPI_ISL_1703888, EPI_ISL_1703889, EPI_ISL_1703890, EPI_ISL_1703891, EPI_ISL_1703892, EPI_ISL_1703893, EPI_ISL_1703894, EPI_ISL_1703895, EPI_ISL_1703896, EPI_ISL_1703898, EPI_ISL_1703899, EPI_ISL_1703900, EPI_ISL_1703901, EPI_ISL_1703902, EPI_ISL_1703903, EPI_ISL_1703904, EPI_ISL_1703906, EPI_ISL_1703907, EPI_ISL_1703908, EPI_ISL_1703909, EPI_ISL_1703910, EPI_ISL_1703911, EPI_ISL_1703912, EPI_ISL_1703913, EPI_ISL_1703914, EPI_ISL_1703915, EPI_ISL_1703916, EPI_ISL_1703917, EPI_ISL_1703918, EPI_ISL_1703920, EPI_ISL_1703921, EPI_ISL_1703922, EPI_ISL_1703923, EPI_ISL_1703924, EPI_ISL_1703925, EPI_ISL_1703927, EPI_ISL_1703928, EPI_ISL_1703929, EPI_ISL_1703930, EPI_ISL_1703931, EPI_ISL_1703932, EPI_ISL_1703933, EPI_ISL_1703934, EPI_ISL_1703935, EPI_ISL_1703937, EPI_ISL_1703938, EPI_ISL_1703939, EPI_ISL_1703940, EPI_ISL_1703941, EPI_ISL_1703942, EPI_ISL_1703943, EPI_ISL_1703944, EPI_ISL_1703945, EPI_ISL_1703946, EPI_ISL_1703948, EPI_ISL_1703949, EPI_ISL_1703950, EPI_ISL_1703951, EPI_ISL_1703952, EPI_ISL_1703953, EPI_ISL_1703954, EPI_ISL_1703955, EPI_ISL_1703956, EPI_ISL_1703957, EPI_ISL_1703959, EPI_ISL_1703960, EPI_ISL_1703961, EPI_ISL_1703962, EPI_ISL_1703963, EPI_ISL_1703964, EPI_ISL_1703965, EPI_ISL_1703966, EPI_ISL_1703967, EPI_ISL_1703968, EPI_ISL_1703969, EPI_ISL_1703971, EPI_ISL_1703972, EPI_ISL_1703973, EPI_ISL_1703974, EPI_ISL_1703975, EPI_ISL_1703976, EPI_ISL_1703977, EPI_ISL_1703978, EPI_ISL_1703979, EPI_ISL_1703981, EPI_ISL_1703982, EPI_ISL_1703983, EPI_ISL_1703984, EPI_ISL_1703985, EPI_ISL_1703986, EPI_ISL_1703987, EPI_ISL_1703988, EPI_ISL_1703989, EPI_ISL_1703990, EPI_ISL_1703992, EPI_ISL_1703993, EPI_ISL_1703994, EPI_ISL_1703995, EPI_ISL_1703996, EPI_ISL_1703997, EPI_ISL_1703999, EPI_ISL_1704000, EPI_ISL_1704001, EPI_ISL_1704002, EPI_ISL_1704003, EPI_ISL_1704004, EPI_ISL_1704005, EPI_ISL_1704006, EPI_ISL_1704007, EPI_ISL_1704008, EPI_ISL_1704009, EPI_ISL_1704011, EPI_ISL_1704012, EPI_ISL_1704013, EPI_ISL_1704014, EPI_ISL_1704015, EPI_ISL_1704016, EPI_ISL_1704017, EPI_ISL_1704018, EPI_ISL_1704019, EPI_ISL_1704020, EPI_ISL_1704021, EPI_ISL_1704022, EPI_ISL_1704024, EPI_ISL_1704025, EPI_ISL_1704026, EPI_ISL_1704027, EPI_ISL_1704028, EPI_ISL_1704029, EPI_ISL_1704030, EPI_ISL_1704032, EPI_ISL_1704033, EPI_ISL_1704034, EPI_ISL_1704035, EPI_ISL_1704036, EPI_ISL_1704037, EPI_ISL_1704038, EPI_ISL_1704039, EPI_ISL_1704040, EPI_ISL_1704042, EPI_ISL_1704043, EPI_ISL_1704044, EPI_ISL_1704045, EPI_ISL_1704046, EPI_ISL_1704047, EPI_ISL_1704048, EPI_ISL_1704049, EPI_ISL_1704050, EPI_ISL_1704052, EPI_ISL_1704053, EPI_ISL_1704055, EPI_ISL_1704056, EPI_ISL_1704057, EPI_ISL_1704058, EPI_ISL_1704060, EPI_ISL_1704061, EPI_ISL_1704062, EPI_ISL_1704063, EPI_ISL_1704064, EPI_ISL_1704065, EPI_ISL_1704066, EPI_ISL_1704067, EPI_ISL_1704069, EPI_ISL_1704070, EPI_ISL_1704071, EPI_ISL_1704072, EPI_ISL_1704073, EPI_ISL_1704074, EPI_ISL_1704076, EPI_ISL_1704077, EPI_ISL_1704078, EPI_ISL_1704079, EPI_ISL_1704080, EPI_ISL_1704081, EPI_ISL_1704082, EPI_ISL_1704083, EPI_ISL_1704084, EPI_ISL_1704085, EPI_ISL_1704086, EPI_ISL_1704088, EPI_ISL_1704089, EPI_ISL_1704090, EPI_ISL_1704091, EPI_ISL_1704092, EPI_ISL_1704093, EPI_ISL_1704094, EPI_ISL_1704095, EPI_ISL_1704096, EPI_ISL_1704097, EPI_ISL_1704099, EPI_ISL_1704100, EPI_ISL_1704101, EPI_ISL_1704102, EPI_ISL_1704103, EPI_ISL_1704104, EPI_ISL_1704105, EPI_ISL_1704106, EPI_ISL_1704107, EPI_ISL_1704108, EPI_ISL_1704109, EPI_ISL_1704110, EPI_ISL_1704111, EPI_ISL_1704113, EPI_ISL_1704114, EPI_ISL_1704115, EPI_ISL_1704116, EPI_ISL_1704117, EPI_ISL_1704118, EPI_ISL_1704119, EPI_ISL_1704120, EPI_ISL_1704121, EPI_ISL_1704122, EPI_ISL_1704123, EPI_ISL_1704125, EPI_ISL_1704126, EPI_ISL_1704127, EPI_ISL_1704128, EPI_ISL_1704129, EPI_ISL_1704130, EPI_ISL_1704131, EPI_ISL_1704132, EPI_ISL_1704133, EPI_ISL_1704134, EPI_ISL_1704135, EPI_ISL_1704137, EPI_ISL_1704138, EPI_ISL_1704139, EPI_ISL_1704140, EPI_ISL_1704141, EPI_ISL_1704142, EPI_ISL_1704143, EPI_ISL_1704144, EPI_ISL_1704145, EPI_ISL_1704146, EPI_ISL_1704148, EPI_ISL_1704149, EPI_ISL_1704150, EPI_ISL_1704151, EPI_ISL_1704152, EPI_ISL_1704153, EPI_ISL_1704154, EPI_ISL_1704155, EPI_ISL_1704156, EPI_ISL_1704157, EPI_ISL_1704158, EPI_ISL_1704159, EPI_ISL_1704161, EPI_ISL_1704162, EPI_ISL_1704163, EPI_ISL_1704164, EPI_ISL_1704165, EPI_ISL_1704166, EPI_ISL_1704167, EPI_ISL_1704168, EPI_ISL_1704169, EPI_ISL_1704170, EPI_ISL_1704172, EPI_ISL_1704173, EPI_ISL_1704174, EPI_ISL_1704175, EPI_ISL_1704176, EPI_ISL_1704177, EPI_ISL_1704178, EPI_ISL_1704179, EPI_ISL_1704180, EPI_ISL_1704181, EPI_ISL_1704182, EPI_ISL_1704183, EPI_ISL_1704185, EPI_ISL_1704186, EPI_ISL_1704187, EPI_ISL_1704188, EPI_ISL_1704189, EPI_ISL_1704190, EPI_ISL_1704191, EPI_ISL_1704193, EPI_ISL_1704194, EPI_ISL_1704195, EPI_ISL_1704196, EPI_ISL_1704197, EPI_ISL_1704198, EPI_ISL_1704199, EPI_ISL_1704200, EPI_ISL_1704201, EPI_ISL_1704202, EPI_ISL_1704203, EPI_ISL_1704205, EPI_ISL_1704206, EPI_ISL_1704207, EPI_ISL_1704208, EPI_ISL_1704209, EPI_ISL_1704210, EPI_ISL_1704211, EPI_ISL_1704212, EPI_ISL_1704213, EPI_ISL_1704214, EPI_ISL_1704215, EPI_ISL_1704217, EPI_ISL_1704218, EPI_ISL_1704219, EPI_ISL_1704220, EPI_ISL_1704221, EPI_ISL_1704222, EPI_ISL_1704223, EPI_ISL_1704224, EPI_ISL_1704225, EPI_ISL_1704226, EPI_ISL_1704227, EPI_ISL_1704229, EPI_ISL_1704230, EPI_ISL_1704231, EPI_ISL_1704232, EPI_ISL_1704233, EPI_ISL_1704234, EPI_ISL_1704235, EPI_ISL_1704236, EPI_ISL_1704237, EPI_ISL_1704238, EPI_ISL_1704240, EPI_ISL_1704241, EPI_ISL_1704242, EPI_ISL_1704243, EPI_ISL_1704244, EPI_ISL_1704245, EPI_ISL_1704246, EPI_ISL_1704247, EPI_ISL_1704249, EPI_ISL_1704250, EPI_ISL_1704251, EPI_ISL_1704252, EPI_ISL_1704253, EPI_ISL_1704254, EPI_ISL_1704255, EPI_ISL_1704256, EPI_ISL_1704257, EPI_ISL_1704258, EPI_ISL_1704259, EPI_ISL_1704261, EPI_ISL_1704262, EPI_ISL_1704263, EPI_ISL_1704264, EPI_ISL_1704265, EPI_ISL_1704266, EPI_ISL_1704267, EPI_ISL_1704268, EPI_ISL_1704269, EPI_ISL_1704270, EPI_ISL_1704271, EPI_ISL_1704272, EPI_ISL_1704274, EPI_ISL_1704275, EPI_ISL_1704276, EPI_ISL_1704277, EPI_ISL_1704278, EPI_ISL_1704279, EPI_ISL_1704280, EPI_ISL_1704281, EPI_ISL_1704282, EPI_ISL_1704283, EPI_ISL_1704284, EPI_ISL_1704285, EPI_ISL_1704287, EPI_ISL_1704288, EPI_ISL_1704289, EPI_ISL_1704290, EPI_ISL_1704291, EPI_ISL_1704292, EPI_ISL_1704293, EPI_ISL_1704294, EPI_ISL_1704295, EPI_ISL_1704297, EPI_ISL_1704298, EPI_ISL_1704299, EPI_ISL_1704300, EPI_ISL_1704301, EPI_ISL_1704302, EPI_ISL_1704303, EPI_ISL_1704304, EPI_ISL_1704306, EPI_ISL_1704307, EPI_ISL_1704308, EPI_ISL_1704309, EPI_ISL_1704310, EPI_ISL_1704311, EPI_ISL_1704312, EPI_ISL_1704313, EPI_ISL_1704315, EPI_ISL_1704316, EPI_ISL_1704318, EPI_ISL_1704319, EPI_ISL_1704320, EPI_ISL_1704321, EPI_ISL_1704322, EPI_ISL_1704323, EPI_ISL_1704324, EPI_ISL_1704325, EPI_ISL_1704326, EPI_ISL_1704327, EPI_ISL_1704328, EPI_ISL_1704329, EPI_ISL_1704330, EPI_ISL_1704331, EPI_ISL_1704332, EPI_ISL_1704333, EPI_ISL_1704334, EPI_ISL_1704335, EPI_ISL_1704336, EPI_ISL_1704337, EPI_ISL_1704338, EPI_ISL_1704339, EPI_ISL_1704340, EPI_ISL_1704341, EPI_ISL_1704342, EPI_ISL_1704343, EPI_ISL_1704344, EPI_ISL_1704345, EPI_ISL_1704346, EPI_ISL_1704347, EPI_ISL_1704348, EPI_ISL_1704349, EPI_ISL_1704350, EPI_ISL_1704351, EPI_ISL_1704352, EPI_ISL_1704353, EPI_ISL_1704354, EPI_ISL_1704355, EPI_ISL_1704356, EPI_ISL_1704357, EPI_ISL_1704358, EPI_ISL_1704359, EPI_ISL_1704360, EPI_ISL_1704361, EPI_ISL_1704362, EPI_ISL_1704363, EPI_ISL_1704364, EPI_ISL_1704365, EPI_ISL_1704366, EPI_ISL_1704367, EPI_ISL_1704368, EPI_ISL_1704369, EPI_ISL_1704370, EPI_ISL_1704371, EPI_ISL_1704372, EPI_ISL_1704373, EPI_ISL_1704374, EPI_ISL_1704375, EPI_ISL_1704376, EPI_ISL_1704377, EPI_ISL_1704378, EPI_ISL_1704379, EPI_ISL_1704380, EPI_ISL_1704381, EPI_ISL_1704382, EPI_ISL_1704383, EPI_ISL_1704384, EPI_ISL_1704385, EPI_ISL_1704386, EPI_ISL_1704387, EPI_ISL_1704388, EPI_ISL_1704389, EPI_ISL_1704390, EPI_ISL_1704391, EPI_ISL_1704392, EPI_ISL_1704393, EPI_ISL_1704394, EPI_ISL_1704395, EPI_ISL_1704396, EPI_ISL_1704397, EPI_ISL_1704398, EPI_ISL_1704399, EPI_ISL_1704400, EPI_ISL_1704401, EPI_ISL_1704402, EPI_ISL_1704403, EPI_ISL_1704404, EPI_ISL_1704405, EPI_ISL_1704406, EPI_ISL_1704407, EPI_ISL_1704408, EPI_ISL_1704410, EPI_ISL_1704411, EPI_ISL_1704412, EPI_ISL_1704413, EPI_ISL_1704414, EPI_ISL_1704415, EPI_ISL_1704416, EPI_ISL_1704417, EPI_ISL_1704418, EPI_ISL_1704419, EPI_ISL_1704420, EPI_ISL_1704421, EPI_ISL_1704422, EPI_ISL_1704423, EPI_ISL_1704424, EPI_ISL_1704425, EPI_ISL_1704426, EPI_ISL_1704427, EPI_ISL_1704428, EPI_ISL_1704429, EPI_ISL_1704430, EPI_ISL_1704431, EPI_ISL_1704432, EPI_ISL_1704433, EPI_ISL_1704434, EPI_ISL_1704435, EPI_ISL_1704436, EPI_ISL_1704437, EPI_ISL_1704438, EPI_ISL_1704439, EPI_ISL_1704440, EPI_ISL_1704441, EPI_ISL_1704442, EPI_ISL_1704443, EPI_ISL_1704444, EPI_ISL_1704445, EPI_ISL_1704446, EPI_ISL_1704447, EPI_ISL_1704448, EPI_ISL_1704449, EPI_ISL_1704450, EPI_ISL_1704451, EPI_ISL_1704452, EPI_ISL_1704453, EPI_ISL_1704454, EPI_ISL_1704455, EPI_ISL_1704456, EPI_ISL_1704457, EPI_ISL_1704458, EPI_ISL_1704459, EPI_ISL_1704460, EPI_ISL_1704461, EPI_ISL_1704462, EPI_ISL_1704463, EPI_ISL_1704464, EPI_ISL_1704465, EPI_ISL_1704466, EPI_ISL_1704467, EPI_ISL_1704468, EPI_ISL_1704469, EPI_ISL_1704470, EPI_ISL_1704471, EPI_ISL_1704472, EPI_ISL_1704473, EPI_ISL_1704474, EPI_ISL_1704475, EPI_ISL_1704476, EPI_ISL_1704477, EPI_ISL_1704478, EPI_ISL_1704479, EPI_ISL_1704480, EPI_ISL_1704481, EPI_ISL_1704482, EPI_ISL_1704483, EPI_ISL_1704484, EPI_ISL_1704485, EPI_ISL_1704486, EPI_ISL_1704487, EPI_ISL_1704488, EPI_ISL_1704489, EPI_ISL_1704490, EPI_ISL_1704491, EPI_ISL_1704492, EPI_ISL_1704494, EPI_ISL_1704495, EPI_ISL_1704496, EPI_ISL_1704497, EPI_ISL_1704498, EPI_ISL_1704499, EPI_ISL_1704500, EPI_ISL_1704501, EPI_ISL_1704502, EPI_ISL_1704503, EPI_ISL_1704504, EPI_ISL_1704505, EPI_ISL_1704506, EPI_ISL_1704507, EPI_ISL_1704508, EPI_ISL_1704509, EPI_ISL_1704510, EPI_ISL_1704511, EPI_ISL_1704512, EPI_ISL_1704513, EPI_ISL_1704514, EPI_ISL_1704515, EPI_ISL_1704516, EPI_ISL_1704517, EPI_ISL_1704518, EPI_ISL_1704519, EPI_ISL_1704520, EPI_ISL_1704521, EPI_ISL_1704522, EPI_ISL_1704523, EPI_ISL_1704524, EPI_ISL_1704525, EPI_ISL_1704526, EPI_ISL_1704527, EPI_ISL_1704528, EPI_ISL_1704529, EPI_ISL_1704530, EPI_ISL_1704531, EPI_ISL_1704532, EPI_ISL_1704533, EPI_ISL_1704534, EPI_ISL_1704535, EPI_ISL_1704536, EPI_ISL_1704537, EPI_ISL_1704538, EPI_ISL_1704539, EPI_ISL_1704540, EPI_ISL_1704541, EPI_ISL_1704542, EPI_ISL_1704543, EPI_ISL_1704544, EPI_ISL_1704545, EPI_ISL_1704546, EPI_ISL_1704547, EPI_ISL_1704548, EPI_ISL_1704549, EPI_ISL_1704550, EPI_ISL_1704551, EPI_ISL_1704552, EPI_ISL_1704553, EPI_ISL_1704554, EPI_ISL_1704555, EPI_ISL_1704556, EPI_ISL_1704557, EPI_ISL_1704558, EPI_ISL_1704559, EPI_ISL_1704560, EPI_ISL_1704561, EPI_ISL_1704562, EPI_ISL_1704563, EPI_ISL_1704564, EPI_ISL_1704565, EPI_ISL_1704566, EPI_ISL_1704567, EPI_ISL_1704568, EPI_ISL_1704569, EPI_ISL_1704570, EPI_ISL_1704571, EPI_ISL_1704572, EPI_ISL_1704573, EPI_ISL_1704574, EPI_ISL_1704575, EPI_ISL_1704576, EPI_ISL_1704577, EPI_ISL_1704578, EPI_ISL_1704579, EPI_ISL_1704580, EPI_ISL_1704581, EPI_ISL_1704582, EPI_ISL_1704583, EPI_ISL_1704584, EPI_ISL_1704585, EPI_ISL_1704586, EPI_ISL_1704587, EPI_ISL_1704588, EPI_ISL_1704589, EPI_ISL_1704590, EPI_ISL_1704591, EPI_ISL_1704592, EPI_ISL_1704593, EPI_ISL_1704594, EPI_ISL_1704595, EPI_ISL_1704596, EPI_ISL_1704597, EPI_ISL_1704598, EPI_ISL_1704599, EPI_ISL_1704600, EPI_ISL_1704601, EPI_ISL_1704602, EPI_ISL_1704603, EPI_ISL_1704604, EPI_ISL_1704605, EPI_ISL_1704606, EPI_ISL_1704607, EPI_ISL_1704608, EPI_ISL_1704609, EPI_ISL_1704610, EPI_ISL_1704611, EPI_ISL_1704612, EPI_ISL_1704613, EPI_ISL_1704614, EPI_ISL_1704615, EPI_ISL_1704616, EPI_ISL_1704617, EPI_ISL_1704618, EPI_ISL_1704619, EPI_ISL_1704620, EPI_ISL_1704621, EPI_ISL_1704622, EPI_ISL_1704623, EPI_ISL_1704624, EPI_ISL_1704625, EPI_ISL_1704626, EPI_ISL_1704627, EPI_ISL_1704628, EPI_ISL_1704629, EPI_ISL_1704630, EPI_ISL_1704631, | | | |

| | | | |
|---|---|---|---|
| EPI_ISL_1704632, EPI_ISL_1704633, EPI_ISL_1704634, EPI_ISL_1704635, EPI_ISL_1704636, EPI_ISL_1704637, EPI_ISL_1704638, EPI_ISL_1704639, EPI_ISL_1704640, EPI_ISL_1704641, EPI_ISL_1704642, EPI_ISL_1704643, EPI_ISL_1704644, EPI_ISL_1704645, EPI_ISL_1704646, EPI_ISL_1704647, EPI_ISL_1704648, EPI_ISL_1704649, EPI_ISL_1704650, EPI_ISL_1704651, EPI_ISL_1704652, EPI_ISL_1704653, EPI_ISL_1704654, EPI_ISL_1704655, EPI_ISL_1704656, EPI_ISL_1704657, EPI_ISL_1704658, EPI_ISL_1704659, EPI_ISL_1704660, EPI_ISL_1704661, EPI_ISL_1704662, EPI_ISL_1704663, EPI_ISL_1704664, EPI_ISL_1704665, EPI_ISL_1704666, EPI_ISL_1704667, EPI_ISL_1704668, EPI_ISL_1704669, EPI_ISL_1704670, EPI_ISL_1704671, EPI_ISL_1704672, EPI_ISL_1841236, EPI_ISL_1841237, EPI_ISL_1841238, EPI_ISL_1841239, EPI_ISL_1841240, EPI_ISL_1841241, EPI_ISL_1841242, EPI_ISL_1841243, EPI_ISL_1841244, EPI_ISL_1841245, EPI_ISL_1841246, EPI_ISL_1841247, EPI_ISL_1841248, EPI_ISL_1841249, EPI_ISL_1841250, EPI_ISL_1841251, EPI_ISL_1841252, EPI_ISL_1841253, EPI_ISL_1841254, EPI_ISL_1841255, EPI_ISL_1841256, EPI_ISL_1841257, EPI_ISL_1841258, EPI_ISL_1841259, EPI_ISL_1841260, EPI_ISL_1841261, EPI_ISL_1841262, EPI_ISL_1841263, EPI_ISL_1841264, EPI_ISL_1841265, EPI_ISL_1841266, EPI_ISL_1841267, EPI_ISL_1841268, EPI_ISL_1841269, EPI_ISL_1841270, EPI_ISL_1841271, EPI_ISL_1841272, EPI_ISL_1841273, EPI_ISL_1841274, EPI_ISL_1841275, EPI_ISL_1841276, EPI_ISL_1841277, EPI_ISL_1841278, EPI_ISL_1841279, EPI_ISL_1841280, EPI_ISL_1841281, EPI_ISL_1841282, EPI_ISL_1841283, EPI_ISL_1841286, EPI_ISL_1841287, EPI_ISL_1841288, EPI_ISL_1841289, EPI_ISL_1841290, EPI_ISL_1841291, EPI_ISL_1841292, EPI_ISL_1841293, EPI_ISL_1841294, EPI_ISL_1841295, EPI_ISL_1841296, EPI_ISL_1841297, EPI_ISL_1841298, EPI_ISL_1841299, EPI_ISL_1841300, EPI_ISL_1841301, EPI_ISL_1841302, EPI_ISL_1841303, EPI_ISL_1841304, EPI_ISL_1841305, EPI_ISL_1841306, EPI_ISL_1841307, EPI_ISL_1841308, EPI_ISL_1841309, EPI_ISL_1841310, EPI_ISL_1841311, EPI_ISL_1841312, EPI_ISL_1841313, EPI_ISL_1841314, EPI_ISL_1841315, EPI_ISL_1841316, EPI_ISL_1841317, EPI_ISL_1841318, EPI_ISL_1841319, EPI_ISL_1841320, EPI_ISL_1841321, EPI_ISL_1841322, EPI_ISL_1841323, EPI_ISL_1841324, EPI_ISL_1841325, EPI_ISL_1841326, EPI_ISL_1841327, EPI_ISL_1841328, EPI_ISL_1841329, EPI_ISL_1841330, EPI_ISL_1841331, EPI_ISL_1841332, EPI_ISL_1841333, EPI_ISL_1841334, EPI_ISL_1841335, EPI_ISL_1841336, EPI_ISL_1841337, EPI_ISL_1841338, EPI_ISL_1841339, EPI_ISL_1841340, EPI_ISL_1841341, EPI_ISL_1841342, EPI_ISL_1841343, EPI_ISL_1841344, EPI_ISL_1841345, EPI_ISL_1841346, EPI_ISL_1841347, EPI_ISL_1841348, EPI_ISL_1841349, EPI_ISL_1841350, EPI_ISL_1841351, EPI_ISL_1841352, EPI_ISL_1841353, EPI_ISL_1841354, EPI_ISL_1841355, EPI_ISL_1841356, EPI_ISL_1841357, EPI_ISL_1841358, EPI_ISL_1841359, EPI_ISL_1841360, EPI_ISL_1841361, EPI_ISL_1841362, EPI_ISL_1841363, EPI_ISL_1841364, EPI_ISL_1841365, EPI_ISL_1841366, EPI_ISL_1841367, EPI_ISL_1841368, EPI_ISL_1841369, EPI_ISL_1841370, EPI_ISL_1841371, EPI_ISL_1841376, EPI_ISL_1841377, EPI_ISL_1841378, EPI_ISL_1841379, EPI_ISL_1841380, EPI_ISL_1841381, EPI_ISL_1841382, EPI_ISL_1841383, EPI_ISL_1928387, EPI_ISL_1928388, EPI_ISL_1928389, EPI_ISL_1928391, EPI_ISL_1928392, EPI_ISL_1928394, EPI_ISL_1928395, EPI_ISL_1928396, EPI_ISL_1928398, EPI_ISL_1928399, EPI_ISL_1928401, EPI_ISL_1928402, EPI_ISL_1928404, EPI_ISL_1928405, EPI_ISL_1928407, EPI_ISL_1928408, EPI_ISL_1928410, EPI_ISL_1928411, EPI_ISL_1928413, EPI_ISL_1928414, EPI_ISL_1928416, EPI_ISL_1928417, EPI_ISL_1928419, EPI_ISL_1928420, EPI_ISL_1928422, EPI_ISL_1928423, EPI_ISL_1928425, EPI_ISL_1928426, EPI_ISL_1928427, EPI_ISL_1928429, EPI_ISL_1928430, EPI_ISL_1928432, EPI_ISL_1928433, EPI_ISL_1928435, EPI_ISL_1928436, EPI_ISL_1928438, EPI_ISL_1928439, EPI_ISL_1928441, EPI_ISL_1928442, EPI_ISL_1928444, EPI_ISL_1928445, EPI_ISL_1928447, EPI_ISL_1928448, EPI_ISL_1928450, EPI_ISL_1928451, EPI_ISL_1928453, EPI_ISL_1928454, EPI_ISL_1928456, EPI_ISL_1928457, EPI_ISL_1928459, EPI_ISL_1928460, EPI_ISL_1928462, EPI_ISL_1928463, EPI_ISL_1928464, EPI_ISL_1928465, EPI_ISL_1928466, EPI_ISL_1928467, EPI_ISL_1928468, EPI_ISL_1928469, EPI_ISL_1928471, EPI_ISL_1928472, EPI_ISL_1928474, EPI_ISL_1928475, EPI_ISL_1928477, EPI_ISL_1928478, EPI_ISL_1928480, EPI_ISL_1928481, EPI_ISL_1928483, EPI_ISL_1928484, EPI_ISL_1928485, EPI_ISL_1928487, EPI_ISL_1928488, EPI_ISL_1928490, EPI_ISL_1928491, EPI_ISL_1928493, EPI_ISL_1928494, EPI_ISL_1928496, EPI_ISL_1928497, EPI_ISL_1928499, EPI_ISL_1928500, EPI_ISL_1928502, EPI_ISL_1928503, EPI_ISL_1928505, EPI_ISL_1928506, EPI_ISL_1928507, EPI_ISL_1928509, EPI_ISL_1928510, EPI_ISL_1928512, EPI_ISL_1928513, EPI_ISL_1928515, EPI_ISL_1928517, EPI_ISL_1928518, EPI_ISL_1928520, EPI_ISL_1928521, EPI_ISL_1928522, EPI_ISL_1928524, EPI_ISL_1928525, EPI_ISL_1928527, EPI_ISL_1928528, EPI_ISL_1928530, EPI_ISL_1928531, EPI_ISL_1928533, EPI_ISL_1928534, EPI_ISL_1928536, EPI_ISL_1928537, EPI_ISL_1928539, EPI_ISL_1928540, EPI_ISL_1928541, EPI_ISL_1928543, EPI_ISL_1928544, EPI_ISL_1928546, EPI_ISL_1928547, EPI_ISL_1928549, EPI_ISL_1928550, EPI_ISL_1928552, EPI_ISL_1928553, EPI_ISL_1928555, EPI_ISL_1928556, EPI_ISL_1928558, EPI_ISL_1928559, EPI_ISL_1928561, EPI_ISL_1928562, EPI_ISL_1928563, EPI_ISL_1928565, EPI_ISL_1928566, EPI_ISL_1928568, EPI_ISL_1928569, EPI_ISL_1928571, EPI_ISL_1928572, EPI_ISL_1928573, EPI_ISL_1928575, EPI_ISL_1928576, EPI_ISL_1928578, EPI_ISL_1928579, EPI_ISL_1928581, EPI_ISL_1928583, EPI_ISL_1928584, EPI_ISL_1928586, EPI_ISL_1928587, EPI_ISL_1928588, EPI_ISL_1928590, EPI_ISL_1928592, EPI_ISL_1928593, EPI_ISL_1928595, EPI_ISL_1928596, EPI_ISL_1928598, EPI_ISL_1928599, EPI_ISL_1928601, EPI_ISL_1928602, EPI_ISL_1928603, EPI_ISL_1928605, EPI_ISL_1928607, EPI_ISL_1928608, EPI_ISL_1928610, EPI_ISL_1928611, EPI_ISL_1928613, EPI_ISL_1928614, EPI_ISL_1970419, EPI_ISL_1970420, EPI_ISL_1970421, EPI_ISL_1970422, EPI_ISL_1970423, EPI_ISL_1970424, EPI_ISL_1970425, EPI_ISL_1970426, EPI_ISL_1970427, EPI_ISL_1970428, EPI_ISL_1970429, EPI_ISL_1970430, EPI_ISL_1970431, EPI_ISL_1970432, EPI_ISL_1970433, EPI_ISL_1970434, EPI_ISL_1970435, EPI_ISL_1970436, EPI_ISL_1970437, EPI_ISL_1970438, EPI_ISL_1970439, EPI_ISL_1970440, EPI_ISL_1970441, EPI_ISL_1970442, EPI_ISL_1970443, EPI_ISL_1970445, EPI_ISL_1970446, EPI_ISL_1970447, EPI_ISL_1970448, EPI_ISL_1970449, EPI_ISL_1970450, EPI_ISL_1970451, EPI_ISL_1970452, EPI_ISL_1970453, EPI_ISL_1970455, EPI_ISL_1970456, EPI_ISL_1970458, EPI_ISL_1970460, EPI_ISL_1970461, EPI_ISL_1970463, EPI_ISL_1970464, EPI_ISL_1970465, EPI_ISL_1970466, EPI_ISL_1970467, EPI_ISL_1970468, EPI_ISL_1970470, EPI_ISL_1970471, EPI_ISL_1970472, EPI_ISL_1970473, EPI_ISL_1970474, EPI_ISL_1970475, EPI_ISL_1970476, EPI_ISL_1970477, EPI_ISL_1970478, EPI_ISL_1970479, EPI_ISL_1970480, EPI_ISL_1970481, EPI_ISL_1970482, EPI_ISL_1970483, EPI_ISL_1970484, EPI_ISL_1970486, EPI_ISL_1970487, EPI_ISL_1970488, EPI_ISL_1970489, EPI_ISL_1970490, EPI_ISL_1970491, EPI_ISL_1970492, EPI_ISL_1970493, EPI_ISL_1970494, EPI_ISL_1970495, EPI_ISL_1970496, EPI_ISL_1970497, EPI_ISL_1970499, EPI_ISL_1970500, EPI_ISL_1970501, EPI_ISL_1970502, EPI_ISL_1970503, EPI_ISL_1970504, EPI_ISL_1970505, EPI_ISL_1970506, EPI_ISL_1970507, EPI_ISL_1970513, EPI_ISL_1970536, EPI_ISL_1970537, EPI_ISL_1970538, EPI_ISL_1970539, EPI_ISL_1970540, EPI_ISL_1970541, EPI_ISL_1970542, EPI_ISL_2131509, EPI_ISL_2131510, EPI_ISL_2131511, EPI_ISL_2131512, EPI_ISL_2131513, EPI_ISL_2131535, EPI_ISL_2131536, EPI_ISL_2131537, EPI_ISL_2131538, EPI_ISL_2131539, EPI_ISL_2131540, EPI_ISL_2131541, EPI_ISL_2131542, EPI_ISL_2131543, EPI_ISL_2131544, EPI_ISL_2131545, EPI_ISL_2131546 | | | |
| see above | ICMR-National Institute of Virology - INSACOG | NIV Influenza | Dr. Varsha Potdar |
| EPI_ISL_1672644 | ID and BG Hospital / CSIR - Indian Institute of Chemical Biology (CSIR-IICB) | CSIR Institute of Genomics and Integrative Biology (CSIR-IGIB) | Akshay Kanakan; Azka Khan; Debaleena Bhowmik; Dipyaman Ganguly; Janani SV; Neha Jha; Partha Chattopadhyay; Priti Devi; Priyanka Mehta; Rajesh Pandey; Ranjeet Maurya; Sachin Sharma; Sandip Paul; Shekhar Ranjan Paul; Shweta Sahni; Yogiraj Ray |
| EPI_ISL_1419053, EPI_ISL_1419054, EPI_ISL_1419055, EPI_ISL_1419056, EPI_ISL_1419057, EPI_ISL_1419058, EPI_ISL_1419059, EPI_ISL_1419060, EPI_ISL_1419061, EPI_ISL_1419062, EPI_ISL_1419063, EPI_ISL_1419064, EPI_ISL_1419065, EPI_ISL_1419066, EPI_ISL_1419067, EPI_ISL_1419068, EPI_ISL_1419069, EPI_ISL_1419070, EPI_ISL_1419071, EPI_ISL_1419072, EPI_ISL_1419073, EPI_ISL_1419074, EPI_ISL_1419075, EPI_ISL_1419076, EPI_ISL_1419078, EPI_ISL_1419079, EPI_ISL_1419080, EPI_ISL_1419084, EPI_ISL_1419095, EPI_ISL_1419097, EPI_ISL_1419098, EPI_ISL_1419099, EPI_ISL_1419100, EPI_ISL_1419101, EPI_ISL_1419102, EPI_ISL_1419108, EPI_ISL_1419109, EPI_ISL_1419110, EPI_ISL_1419112, EPI_ISL_1419113, EPI_ISL_2132678 | | | |
| see above | INSACOG-Mizoram | National Institute of Biomedical Genomics – INSACOG | Arindam Maitra; Gracy Laldinmawii; N Senthil Kumar; Nidhan Kumar Biswas; Saumitra Das; Sreedhar Chinnaswamy; Swagnik Roy |
| EPI_ISL_1419116, EPI_ISL_1419117, EPI_ISL_1419118, EPI_ISL_1419119, EPI_ISL_1419120, EPI_ISL_1419121, EPI_ISL_1419122, EPI_ISL_1419123, EPI_ISL_1419124, EPI_ISL_1419125, EPI_ISL_1419126, EPI_ISL_1419127, EPI_ISL_1419128, EPI_ISL_1419129, EPI_ISL_1419130, EPI_ISL_1419131, EPI_ISL_1419132, EPI_ISL_1419133, EPI_ISL_1419134, EPI_ISL_1419135, EPI_ISL_1419136, EPI_ISL_1419137, EPI_ISL_1419138, EPI_ISL_1419139, EPI_ISL_1419140, EPI_ISL_1419141, EPI_ISL_1419142, EPI_ISL_2151073, EPI_ISL_2151074, EPI_ISL_2151075, EPI_ISL_2151076, EPI_ISL_2151077, EPI_ISL_2151078, EPI_ISL_2151079, EPI_ISL_2151080, EPI_ISL_2151081, EPI_ISL_2151082, EPI_ISL_2151083, EPI_ISL_2151084, EPI_ISL_2151085, EPI_ISL_2151086, EPI_ISL_2151087, EPI_ISL_2151088, EPI_ISL_2151089, EPI_ISL_2151090, EPI_ISL_2151091, EPI_ISL_2151092, EPI_ISL_2151093, EPI_ISL_2151094, EPI_ISL_2151095, EPI_ISL_2151096, EPI_ISL_2151097, EPI_ISL_2151098, EPI_ISL_2151099, EPI_ISL_2151100, EPI_ISL_2151101, EPI_ISL_2151102, EPI_ISL_2151103, EPI_ISL_2151104 | | | |
| see above | INSACOG-Sikkim | National Institute of Biomedical Genomics – INSACOG | Arindam Maitra; Kaden Zangmu Bhutia; Nidhan Kumar Biswas; Saumitra Das; Sreedhar Chinnaswamy Tshering Ongmu Bhutia; Tara D Sharma |
| EPI_ISL_1357691, EPI_ISL_1357692, EPI_ISL_1357693, EPI_ISL_1357694, EPI_ISL_1357695, EPI_ISL_1357696, EPI_ISL_1357697, EPI_ISL_1357698, EPI_ISL_1357699, EPI_ISL_1357700, EPI_ISL_1357701, EPI_ISL_1357702, EPI_ISL_1357703, EPI_ISL_1357704, EPI_ISL_1357705, EPI_ISL_1357706 | | | |
| see above | INSACOG-WB | National Institute of Biomedical Genomics | Ajay Chakraborti; Arindam Maitra; Bhaswati Bandyopadhyay; Nidhan Kumar Biswas; Saumitra Das; Sreedhar Chinnaswamy; Tamal Ghosh |
| EPI_ISL_1419048, EPI_ISL_1419049, EPI_ISL_1419050, EPI_ISL_1419051, EPI_ISL_1419052, EPI_ISL_1419143, EPI_ISL_1419147, EPI_ISL_1419148, EPI_ISL_1419149, EPI_ISL_1419150, EPI_ISL_1419151, EPI_ISL_1419152, EPI_ISL_1419153, EPI_ISL_1419154, EPI_ISL_1419155, EPI_ISL_1419156, EPI_ISL_1419157, EPI_ISL_1419158, EPI_ISL_1419159, EPI_ISL_1419160, EPI_ISL_1419161, EPI_ISL_1419162, EPI_ISL_1419163, EPI_ISL_1419164, EPI_ISL_1419165, EPI_ISL_1419166, EPI_ISL_1419167, EPI_ISL_1419168, EPI_ISL_1419169, EPI_ISL_1419170, EPI_ISL_1419171, EPI_ISL_1419172, EPI_ISL_1419173, EPI_ISL_1419174, EPI_ISL_1419175, EPI_ISL_1419176, EPI_ISL_1419177, EPI_ISL_1419178, EPI_ISL_1419179, EPI_ISL_1419180, EPI_ISL_1419181, EPI_ISL_1419182, EPI_ISL_1419183, EPI_ISL_1419184, EPI_ISL_1419185, EPI_ISL_1419186, EPI_ISL_1419187, EPI_ISL_1419188, EPI_ISL_1419189, EPI_ISL_1419190, EPI_ISL_1419191, EPI_ISL_1419192, EPI_ISL_1419193, EPI_ISL_1419194, EPI_ISL_1419195, EPI_ISL_1419196, EPI_ISL_1419197, EPI_ISL_1419204, EPI_ISL_1419205, EPI_ISL_1419206, EPI_ISL_1419207, EPI_ISL_1419208, EPI_ISL_1419209, EPI_ISL_1419210, EPI_ISL_1419211, EPI_ISL_1419212, EPI_ISL_1419213, EPI_ISL_1419214, EPI_ISL_1419215, EPI_ISL_1419216, EPI_ISL_1419217, EPI_ISL_1419218, EPI_ISL_1419219, EPI_ISL_1419220, EPI_ISL_1419221, EPI_ISL_1419222, EPI_ISL_1419223, EPI_ISL_1419224, EPI_ISL_1419225, EPI_ISL_1419226, EPI_ISL_1419227, EPI_ISL_1419228, EPI_ISL_1419229, EPI_ISL_1419230, EPI_ISL_1419231, EPI_ISL_1419232, EPI_ISL_1419233, EPI_ISL_1419234, EPI_ISL_1419235, EPI_ISL_1419236, EPI_ISL_1419237, EPI_ISL_1419238, EPI_ISL_1419239, EPI_ISL_1419240, EPI_ISL_1419241, EPI_ISL_1419242, EPI_ISL_1419243, EPI_ISL_1419244, EPI_ISL_1419245, EPI_ISL_1419246, EPI_ISL_1419247, EPI_ISL_1419248, EPI_ISL_1419249, EPI_ISL_1419250, EPI_ISL_1419251, EPI_ISL_1419291, EPI_ISL_1419292, EPI_ISL_1419293, EPI_ISL_1419295, EPI_ISL_1419296, EPI_ISL_1419297, EPI_ISL_1419298, EPI_ISL_1419347, EPI_ISL_1419348, EPI_ISL_1419349, EPI_ISL_1419350, EPI_ISL_1419351, EPI_ISL_1419352, EPI_ISL_1419353, EPI_ISL_1419354, EPI_ISL_1419355, EPI_ISL_1419356, EPI_ISL_1419357, EPI_ISL_1419404, EPI_ISL_1419405, EPI_ISL_1419406, EPI_ISL_1419407, EPI_ISL_1419408, EPI_ISL_1419409, EPI_ISL_1419410, EPI_ISL_1419411, EPI_ISL_1419412, EPI_ISL_1419413, EPI_ISL_1419414, EPI_ISL_1419415, EPI_ISL_1419416, EPI_ISL_1419417, EPI_ISL_1419418, EPI_ISL_1419419, EPI_ISL_1419420, EPI_ISL_1419421, EPI_ISL_1419422, EPI_ISL_1419423, EPI_ISL_1419424, EPI_ISL_1419425, EPI_ISL_1419426, EPI_ISL_1419427, EPI_ISL_1419428, EPI_ISL_1419429, EPI_ISL_1419430, EPI_ISL_1419431, EPI_ISL_1419432, EPI_ISL_1419433, EPI_ISL_1419434, EPI_ISL_1419435, EPI_ISL_1419436, EPI_ISL_1419437, EPI_ISL_1419438, EPI_ISL_1419439, EPI_ISL_1419440, EPI_ISL_1419441, EPI_ISL_1419442, EPI_ISL_1419443, EPI_ISL_1419444, EPI_ISL_1419445, EPI_ISL_1419446, EPI_ISL_1419447, EPI_ISL_1419448, EPI_ISL_1419449, EPI_ISL_1419450, EPI_ISL_1419451, EPI_ISL_1419452, EPI_ISL_1419453, EPI_ISL_1419454, EPI_ISL_1419455, EPI_ISL_1419456, EPI_ISL_1419457, EPI_ISL_1419458, EPI_ISL_1419459, EPI_ISL_1419460, EPI_ISL_1419461, EPI_ISL_1419462, EPI_ISL_1419463, EPI_ISL_1419464, EPI_ISL_1419465, EPI_ISL_1419466, EPI_ISL_1419467, EPI_ISL_1419468, EPI_ISL_1419469, EPI_ISL_1419470, EPI_ISL_1419471, EPI_ISL_1419472, EPI_ISL_1419473, EPI_ISL_1419474, EPI_ISL_1419475, EPI_ISL_1419476, EPI_ISL_1419477, EPI_ISL_1419478, EPI_ISL_1419479, EPI_ISL_1419480, EPI_ISL_1419481, EPI_ISL_1419482, EPI_ISL_1419483, EPI_ISL_1419484, EPI_ISL_1419485, EPI_ISL_1419494, EPI_ISL_1419496, EPI_ISL_1419497, EPI_ISL_1419498, EPI_ISL_1419499, EPI_ISL_1419500, EPI_ISL_1419501, EPI_ISL_1419510, EPI_ISL_1419511, EPI_ISL_1419512, EPI_ISL_1419513, EPI_ISL_1419514, EPI_ISL_1419515, EPI_ISL_1419516, EPI_ISL_1419517, EPI_ISL_1419518, EPI_ISL_1419519, EPI_ISL_1419520, EPI_ISL_1419521, EPI_ISL_1419522, EPI_ISL_1419523, EPI_ISL_1419524, EPI_ISL_1419525, EPI_ISL_1419526, EPI_ISL_1419527, EPI_ISL_1419528, EPI_ISL_1419529, EPI_ISL_1419530, EPI_ISL_1419531, EPI_ISL_1419532, EPI_ISL_1419533, EPI_ISL_1419534, EPI_ISL_1419535, EPI_ISL_1419536, EPI_ISL_1419537, EPI_ISL_1419538, EPI_ISL_1419539, EPI_ISL_1419540, EPI_ISL_1419541, EPI_ISL_1419542, EPI_ISL_1419543, EPI_ISL_1419544, EPI_ISL_1419545, EPI_ISL_1419546, EPI_ISL_1419547, EPI_ISL_1419548, EPI_ISL_1419549, EPI_ISL_1419550, EPI_ISL_1419551, EPI_ISL_1419552, EPI_ISL_1419553, EPI_ISL_1419554, EPI_ISL_1419555, EPI_ISL_1419556, EPI_ISL_1419557, EPI_ISL_1419558, EPI_ISL_1419559, EPI_ISL_1419560, EPI_ISL_1419561, EPI_ISL_1419562, EPI_ISL_1419563, EPI_ISL_1419564, EPI_ISL_1419565, EPI_ISL_1419566, EPI_ISL_1419567, EPI_ISL_1419568, EPI_ISL_1419569, EPI_ISL_1419570, EPI_ISL_1419571, EPI_ISL_1419572, EPI_ISL_1419573, EPI_ISL_1419574, EPI_ISL_1419575, EPI_ISL_1419576, EPI_ISL_1419577, EPI_ISL_1419578, EPI_ISL_1419582, EPI_ISL_1419583, EPI_ISL_1419584, EPI_ISL_1419585, EPI_ISL_1419596, EPI_ISL_1419597, EPI_ISL_1419598, EPI_ISL_1419599, EPI_ISL_1419600, EPI_ISL_1419601, EPI_ISL_1419602, EPI_ISL_1419603, EPI_ISL_1419604, EPI_ISL_1419605, EPI_ISL_1419606, EPI_ISL_1419607, EPI_ISL_1419608, EPI_ISL_1419609, EPI_ISL_1419610, EPI_ISL_1419611, EPI_ISL_1419612, EPI_ISL_1419613, EPI_ISL_1419614, EPI_ISL_1419615, EPI_ISL_1419616, EPI_ISL_1419617, EPI_ISL_1419618, EPI_ISL_1419619, EPI_ISL_1419620, EPI_ISL_1419621, EPI_ISL_1419622, EPI_ISL_1419623, EPI_ISL_1419624, EPI_ISL_1419625, EPI_ISL_1419626, EPI_ISL_1419627, EPI_ISL_1419628, EPI_ISL_1419629, EPI_ISL_1419630, EPI_ISL_1419631, EPI_ISL_1419632, EPI_ISL_1419633, EPI_ISL_1419634, EPI_ISL_1419635, EPI_ISL_1419636, EPI_ISL_1419637, EPI_ISL_1419638, EPI_ISL_1419639, EPI_ISL_1419640, EPI_ISL_1419641, EPI_ISL_1419642, EPI_ISL_1419643, EPI_ISL_1419644, EPI_ISL_1419645, EPI_ISL_1419646, EPI_ISL_1419647, EPI_ISL_1419648, EPI_ISL_1419649, EPI_ISL_1419650, EPI_ISL_1419653, EPI_ISL_1419654, EPI_ISL_1419655, EPI_ISL_1419656, EPI_ISL_1419657, EPI_ISL_1419658, EPI_ISL_1419659, EPI_ISL_1419660, EPI_ISL_1419661, EPI_ISL_1419662, EPI_ISL_1419663, EPI_ISL_1419664, EPI_ISL_1419665, EPI_ISL_1419666, EPI_ISL_1419667, EPI_ISL_1419668, EPI_ISL_1419669, EPI_ISL_1419670, EPI_ISL_1419671, EPI_ISL_1419672, EPI_ISL_1419673, EPI_ISL_1419674, EPI_ISL_1419675, EPI_ISL_1419676, EPI_ISL_1419677, EPI_ISL_1419678, EPI_ISL_1419679, EPI_ISL_1419681, EPI_ISL_1419682, EPI_ISL_1419683, EPI_ISL_1419684, EPI_ISL_1419685, EPI_ISL_1419686, EPI_ISL_1419687, EPI_ISL_1419688, EPI_ISL_1419689, EPI_ISL_1419690, EPI_ISL_1419691, EPI_ISL_1419692, EPI_ISL_1419693, EPI_ISL_1419694, EPI_ISL_1419695, EPI_ISL_1419696, EPI_ISL_1419697, EPI_ISL_1419698, EPI_ISL_1419699, EPI_ISL_1419700, EPI_ISL_1419701, EPI_ISL_1419702, EPI_ISL_1419703, EPI_ISL_1419704, EPI_ISL_1419705, EPI_ISL_1419706, EPI_ISL_1419707, EPI_ISL_1419708, EPI_ISL_1419709, EPI_ISL_1419710, EPI_ISL_1419711, EPI_ISL_1419712, EPI_ISL_1419713, EPI_ISL_1419714, EPI_ISL_1419715, EPI_ISL_1419716, EPI_ISL_1419717, EPI_ISL_1419718, EPI_ISL_1419719, EPI_ISL_1419720, EPI_ISL_1419721, EPI_ISL_1419722, EPI_ISL_1419723, EPI_ISL_1419724, EPI_ISL_1419725, EPI_ISL_1419726, EPI_ISL_1419727, EPI_ISL_1419728, EPI_ISL_1419729, EPI_ISL_1419730, EPI_ISL_1419731, EPI_ISL_1419732, EPI_ISL_1419733, EPI_ISL_1419734, EPI_ISL_1419735, EPI_ISL_1419736, EPI_ISL_1419737, EPI_ISL_1419738, EPI_ISL_1419739, EPI_ISL_1419740, EPI_ISL_1419741, EPI_ISL_1419742, EPI_ISL_1419743, EPI_ISL_1419744, EPI_ISL_1419745, EPI_ISL_1419746, EPI_ISL_1419747, EPI_ISL_1419748, EPI_ISL_1419749, EPI_ISL_1419750, EPI_ISL_1419751, EPI_ISL_1419752, EPI_ISL_1419753, EPI_ISL_1419754, EPI_ISL_1419755, EPI_ISL_1419756, EPI_ISL_1419757, EPI_ISL_1419758, EPI_ISL_1419759, EPI_ISL_1419760, EPI_ISL_1419764, EPI_ISL_1419765, EPI_ISL_1419766, EPI_ISL_1419767, EPI_ISL_1419768, EPI_ISL_1419769, EPI_ISL_1419770, EPI_ISL_1419771, EPI_ISL_1419772, EPI_ISL_1419773, EPI_ISL_1419774, EPI_ISL_1419775, EPI_ISL_1419776, EPI_ISL_1419777, EPI_ISL_1419778, EPI_ISL_1419779, EPI_ISL_1419780, EPI_ISL_1419781, EPI_ISL_1419782, EPI_ISL_1419783, EPI_ISL_1419784, EPI_ISL_1419785, EPI_ISL_1419786, EPI_ISL_1419787, EPI_ISL_1419788, EPI_ISL_1419789, EPI_ISL_1419790, EPI_ISL_1419791, EPI_ISL_1419792, EPI_ISL_1419793, EPI_ISL_1419794, EPI_ISL_1419795, EPI_ISL_1419796, EPI_ISL_1419797, EPI_ISL_1419798, EPI_ISL_1419799, EPI_ISL_1419800, EPI_ISL_1419801, EPI_ISL_1419802, EPI_ISL_1419803, EPI_ISL_1419804, EPI_ISL_1419805, EPI_ISL_1419806, EPI_ISL_1419807, EPI_ISL_1419808, EPI_ISL_1419809, EPI_ISL_1419810, EPI_ISL_1419811, EPI_ISL_1419812, EPI_ISL_1419813, EPI_ISL_1419814, EPI_ISL_1419847, EPI_ISL_1419848, EPI_ISL_1419849, EPI_ISL_1419850, EPI_ISL_1419851, EPI_ISL_1419852, EPI_ISL_1419853, EPI_ISL_1419854, EPI_ISL_1419855, EPI_ISL_1419862, EPI_ISL_1419873, EPI_ISL_1419874, EPI_ISL_1419875, EPI_ISL_1419876, EPI_ISL_1419877, EPI_ISL_1419878, EPI_ISL_1419879, EPI_ISL_1419880, EPI_ISL_1419881, EPI_ISL_1419882, EPI_ISL_1419883, EPI_ISL_1419884, EPI_ISL_1419886, EPI_ISL_1419887, EPI_ISL_1419888, EPI_ISL_1419889, EPI_ISL_1419890, EPI_ISL_1419891, EPI_ISL_1419892, EPI_ISL_1419893, EPI_ISL_1419894, EPI_ISL_1419895, EPI_ISL_1419896, EPI_ISL_1419897, EPI_ISL_1419898, EPI_ISL_1419899, EPI_ISL_1419900, EPI_ISL_1419901, EPI_ISL_1419902, EPI_ISL_1419903, EPI_ISL_1419904, EPI_ISL_1419905, EPI_ISL_1419906, EPI_ISL_1419908, EPI_ISL_1419909, EPI_ISL_1419910, EPI_ISL_1419911, EPI_ISL_1419912, EPI_ISL_1419913, EPI_ISL_1419914, EPI_ISL_1419917, EPI_ISL_1419918, EPI_ISL_1419919, EPI_ISL_1419920, EPI_ISL_1419921, EPI_ISL_1419922, EPI_ISL_1419923, EPI_ISL_1419924, EPI_ISL_1589655, EPI_ISL_1589656, EPI_ISL_1589657, EPI_ISL_1589658, EPI_ISL_1589659, EPI_ISL_1589660, EPI_ISL_1589661, EPI_ISL_1589662, EPI_ISL_1589663, EPI_ISL_1589664, EPI_ISL_1589665, EPI_ISL_1589666, EPI_ISL_1589667, EPI_ISL_1589668, EPI_ISL_1589669, EPI_ISL_1589670, EPI_ISL_1589671, EPI_ISL_1589672, EPI_ISL_1589673, EPI_ISL_1589674, EPI_ISL_1589675, EPI_ISL_1589676, EPI_ISL_1589677, EPI_ISL_1589678, EPI_ISL_1589679, EPI_ISL_1589680, EPI_ISL_1589681, EPI_ISL_1589682, EPI_ISL_1589683, EPI_ISL_1589684, EPI_ISL_1589685, EPI_ISL_1589686, EPI_ISL_1589687, EPI_ISL_1589688, EPI_ISL_1589689, EPI_ISL_1589690, EPI_ISL_1589691, EPI_ISL_1589692, EPI_ISL_1589693, EPI_ISL_1589694, EPI_ISL_1589695, EPI_ISL_1589696, EPI_ISL_1589697, EPI_ISL_1589698, EPI_ISL_1589699, EPI_ISL_1589700, EPI_ISL_1589701, EPI_ISL_1589702, EPI_ISL_1589703, EPI_ISL_1589704, EPI_ISL_1589705, EPI_ISL_1589706, EPI_ISL_1589707, EPI_ISL_1589708, EPI_ISL_1589709, EPI_ISL_1589710, EPI_ISL_1589711, EPI_ISL_1589712, EPI_ISL_1589713, EPI_ISL_1589714, EPI_ISL_1589715, EPI_ISL_1589716, EPI_ISL_1589717, EPI_ISL_1589718, EPI_ISL_1589719, EPI_ISL_1589720, EPI_ISL_1589721, EPI_ISL_1589722, EPI_ISL_1589723, EPI_ISL_1589724, EPI_ISL_1589725, EPI_ISL_1589726, EPI_ISL_1589727, EPI_ISL_1589728, EPI_ISL_1589729, EPI_ISL_1589730, EPI_ISL_1589731, EPI_ISL_1589732, EPI_ISL_1589733, EPI_ISL_1589734, EPI_ISL_1589735, EPI_ISL_1589736, EPI_ISL_1589737, EPI_ISL_1589738, EPI_ISL_1589739, EPI_ISL_1589740, EPI_ISL_1589741, EPI_ISL_1589742, EPI_ISL_1589743, EPI_ISL_1589744, EPI_ISL_1589745, EPI_ISL_1589746, EPI_ISL_1589747, EPI_ISL_1589748, EPI_ISL_1589749, EPI_ISL_1589750, EPI_ISL_1589751, EPI_ISL_1589752, EPI_ISL_1589753, EPI_ISL_1589754, EPI_ISL_1589755, EPI_ISL_1589756, EPI_ISL_1589757, EPI_ISL_1589758, EPI_ISL_1589759, EPI_ISL_1589760, EPI_ISL_1589761, EPI_ISL_1589762, EPI_ISL_1589763, EPI_ISL_1589764, EPI_ISL_1589765, EPI_ISL_1589766, EPI_ISL_1589767, EPI_ISL_1589768, EPI_ISL_1589769, EPI_ISL_1589770, EPI_ISL_1589771, EPI_ISL_1589772, EPI_ISL_1589773, EPI_ISL_1589774, EPI_ISL_1589775, EPI_ISL_1589776, EPI_ISL_1589777, EPI_ISL_1589778, EPI_ISL_1589779, EPI_ISL_1589780, EPI_ISL_1589781, EPI_ISL_1589782, EPI_ISL_1589783, EPI_ISL_1589784, EPI_ISL_1589785, EPI_ISL_1589786, EPI_ISL_1589787, EPI_ISL_1589788, EPI_ISL_1589789, EPI_ISL_1589790, EPI_ISL_1589791, EPI_ISL_1589792, EPI_ISL_1589793, EPI_ISL_1589794, EPI_ISL_1589795, EPI_ISL_1589796, EPI_ISL_1589797, EPI_ISL_1589798, EPI_ISL_1589799, EPI_ISL_1589800, EPI_ISL_1589801, EPI_ISL_1589802, EPI_ISL_1589803, EPI_ISL_1589804, EPI_ISL_1589805, EPI_ISL_1589806, EPI_ISL_1589807, EPI_ISL_1589808, EPI_ISL_1589809, EPI_ISL_1589810, EPI_ISL_1589811, EPI_ISL_1589812, EPI_ISL_1589813, EPI_ISL_1589814, EPI_ISL_1589815, EPI_ISL_1589816, EPI_ISL_1589817, EPI_ISL_1589818, EPI_ISL_1589819, EPI_ISL_1589820, EPI_ISL_1589821, EPI_ISL_1589822, EPI_ISL_1589823, EPI_ISL_1589824, EPI_ISL_1589825, EPI_ISL_1589826, EPI_ISL_1589827, EPI_ISL_1589828, EPI_ISL_1589829, EPI_ISL_1589830, EPI_ISL_1589831, EPI_ISL_1589832, EPI_ISL_1589833, EPI_ISL_1589834, EPI_ISL_1589835, EPI_ISL_1589836, EPI_ISL_1589837, EPI_ISL_1589838, EPI_ISL_1589839, EPI_ISL_1589840, EPI_ISL_1589841, EPI_ISL_1589842, EPI_ISL_1589843, EPI_ISL_1589844, EPI_ISL_1589845, EPI_ISL_1589846, EPI_ISL_1589847, EPI_ISL_1589848, EPI_ISL_1589849, EPI_ISL_1589850, EPI_ISL_1589851, EPI_ISL_1589852, EPI_ISL_1589853, EPI_ISL_1589854, EPI_ISL_1589855, EPI_ISL_1589856, EPI_ISL_1589857, EPI_ISL_1589858, EPI_ISL_1589859, EPI_ISL_1589860, EPI_ISL_1589861, EPI_ISL_1589862, EPI_ISL_1589863, EPI_ISL_1589864, EPI_ISL_1589865, EPI_ISL_1589866, EPI_ISL_1589867, EPI_ISL_1589868, EPI_ISL_1589869, EPI_ISL_1589870, EPI_ISL_1589871, EPI_ISL_1589872, EPI_ISL_1589873, EPI_ISL_1589874, EPI_ISL_1589875, EPI_ISL_1589876, EPI_ISL_1589877, EPI_ISL_1589878, EPI_ISL_1589879, EPI_ISL_1589880, EPI_ISL_1589881, EPI_ISL_1589882, EPI_ISL_1589883, EPI_ISL_1589884, EPI_ISL_1589885, EPI_ISL_1589886, EPI_ISL_1589887, EPI_ISL_1589888, EPI_ISL_1589889, EPI_ISL_1589890, EPI_ISL_1589891, EPI_ISL_1589892, EPI_ISL_1589893, EPI_ISL_1589894, EPI_ISL_1589895, EPI_ISL_1589896, EPI_ISL_1589897, EPI_ISL_1589898, EPI_ISL_1589899, EPI_ISL_1589900, EPI_ISL_1589901, EPI_ISL_1589902, EPI_ISL_1589903, EPI_ISL_1589904, EPI_ISL_1589905, EPI_ISL_1589906, EPI_ISL_1589907, EPI_ISL_1589908, EPI_ISL_1589909, EPI_ISL_1589910, EPI_ISL_1589911, EPI_ISL_1589912, EPI_ISL_1589913, EPI_ISL_1589914, EPI_ISL_1589915, EPI_ISL_1589916, EPI_ISL_1589917, EPI_ISL_1589918, EPI_ISL_1589919, EPI_ISL_1589920, EPI_ISL_1589921, EPI_ISL_1589922, EPI_ISL_1589923, EPI_ISL_1589924, EPI_ISL_1589925, EPI_ISL_1589926, EPI_ISL_1589927, EPI_ISL_1589928, EPI_ISL_1589929, EPI_ISL_1589930, EPI_ISL_1589931, EPI_ISL_1589932, EPI_ISL_1589933, EPI_ISL_1589934, EPI_ISL_1589935, EPI_ISL_1589936, EPI_ISL_1589937, EPI_ISL_1589938, EPI_ISL_1589939, EPI_ISL_1589940, EPI_ISL_1589941, EPI_ISL_1589942, EPI_ISL_1589943, EPI_ISL_1589944, EPI_ISL_1589945, EPI_ISL_1589946, EPI_ISL_1589947, EPI_ISL_1589948, EPI_ISL_1589949, EPI_ISL_1589950, EPI_ISL_1589951, EPI_ISL_1589952, EPI_ISL_1589953, EPI_ISL_1589954, EPI_ISL_1589955, EPI_ISL_1589956, EPI_ISL_1589957, EPI_ISL_1589958, EPI_ISL_1589959, EPI_ISL_1589960, EPI_ISL_1589961, EPI_ISL_1589962, EPI_ISL_1589963, EPI_ISL_1589964, EPI_ISL_1589965, EPI_ISL_1589966, EPI_ISL_1589967, EPI_ISL_1589968, EPI_ISL_1589969, EPI_ISL_1589970, EPI_ISL_1589971, EPI_ISL_1589972, EPI_ISL_1589973, EPI_ISL_1589974, EPI_ISL_1589975, EPI_ISL_1589976, EPI_ISL_2132672, EPI_ISL_2132673, EPI_ISL_2132674, EPI_ISL_2132675, EPI_ISL_2132676, EPI_ISL_2132677, EPI_ISL_2132679, EPI_ISL_2132680, EPI_ISL_2132681, EPI_ISL_2132682, EPI_ISL_2132683, EPI_ISL_2132684, EPI_ISL_2132685, EPI_ISL_2132686, EPI_ISL_2132687, EPI_ISL_2132688, EPI_ISL_2132689, EPI_ISL_2132692, EPI_ISL_2132693, EPI_ISL_2132694, EPI_ISL_2132695, EPI_ISL_2132696, EPI_ISL_2132697, EPI_ISL_2132698, EPI_ISL_2132699, EPI_ISL_2132700, EPI_ISL_2132701, EPI_ISL_2132706, EPI_ISL_2132707, EPI_ISL_2132708, EPI_ISL_2132709, EPI_ISL_2132710, EPI_ISL_2132711, EPI_ISL_2132712, EPI_ISL_2132713, EPI_ISL_2132714, EPI_ISL_2132715, EPI_ISL_2132716, EPI_ISL_2132717, EPI_ISL_2132718, EPI_ISL_2132719, EPI_ISL_2132720, EPI_ISL_2132721 | | | |
| see above | INSACOG-WB | National Institute of Biomedical Genomics – INSACOG | Ajay Chakraborti; Arindam Maitra; Bhaswati Bandyopadhyay; Nidhan Kumar Biswas; Saumitra Das; Sreedhar Chinnaswamy; Tamal Ghosh |
| EPI_ISL_1663354, EPI_ISL_1663355 | ITKI Arogyashala | Institute of Life Sciences - INSACOG | Ajay Parida; Amol M. Kanampalliwar; Arup Ghosh; Atimukta Jha; INSACOG Consortium; Punit Prasad; Rajeeb Swain; Rupesh Dash; Safal Walia; Shifu Aggarwal; Sunil K. Raghav |
| EPI_ISL_1063810, EPI_ISL_1063811, EPI_ISL_1063812, EPI_ISL_1063814, EPI_ISL_1063815, EPI_ISL_1063816, EPI_ISL_1063873, EPI_ISL_1063874, EPI_ISL_1063875, EPI_ISL_1063876, EPI_ISL_1063877, EPI_ISL_1063891, EPI_ISL_1063892 | | | |
| see above | Immunogenomics lab, Institute of Life Sciences, Bhubaneswar | Immunogenomics lab, Institute of Life Sciences, Bhubaneswar | Ajay Parida; Amol M. Kanampalliwar; Arup Ghosh; Atimukta Jha; INSACOG Consortium; Kirtal Hansdah; Punit Prasad; Rajeeb Swain; Rupesh Dash; Safal Walia; Shifu Aggarwal; Sunil K. Raghav |
| EPI_ISL_1372010, EPI_ISL_1372086, EPI_ISL_1372087, EPI_ISL_1372088, EPI_ISL_1372090, EPI_ISL_1372092, EPI_ISL_1372093, EPI_ISL_1662445, EPI_ISL_1662446, EPI_ISL_1662447, EPI_ISL_1662448, EPI_ISL_1662449, EPI_ISL_1662450, EPI_ISL_1662451, EPI_ISL_1662452, EPI_ISL_1662453, EPI_ISL_1662454, EPI_ISL_1662455, EPI_ISL_1662456, EPI_ISL_1662457, EPI_ISL_1662458, EPI_ISL_1662459, EPI_ISL_1662460, EPI_ISL_1662461, EPI_ISL_1662462, EPI_ISL_1662463, EPI_ISL_1663552, EPI_ISL_1663553, EPI_ISL_1663554, EPI_ISL_1663555, EPI_ISL_1663557, EPI_ISL_1663558, EPI_ISL_1663559, EPI_ISL_1663560, EPI_ISL_1663561, EPI_ISL_1663562, EPI_ISL_1663563, EPI_ISL_1663564 | | | |
| see above | Immunogenomics lab, Institute of Life Sciences, Bhubaneswar | Institute of Life Sciences - INSACOG | Ajay Parida; Amol M. Kanampalliwar; Arup Ghosh; Atimukta Jha; INSACOG Consortium; Punit Prasad; Rajeeb Swain; Rupesh Dash; Safal Walia; Shifu Aggarwal; Sunil K. Raghav |
| EPI_ISL_1647423, EPI_ISL_1647424, EPI_ISL_1647425, EPI_ISL_1647426, EPI_ISL_1647427, EPI_ISL_1647428, EPI_ISL_1647429, EPI_ISL_1647430, EPI_ISL_1647431, EPI_ISL_1647432, EPI_ISL_1647433, EPI_ISL_1647434, EPI_ISL_1647435, EPI_ISL_1647436, EPI_ISL_1647437, EPI_ISL_1647438, EPI_ISL_1647439, EPI_ISL_1647440, EPI_ISL_1647441, EPI_ISL_1647442, EPI_ISL_1647443, EPI_ISL_1647444, EPI_ISL_1647445, EPI_ISL_1647446, EPI_ISL_1647447, EPI_ISL_1647448, EPI_ISL_1647449, EPI_ISL_1647450, EPI_ISL_1647451, EPI_ISL_1647452, EPI_ISL_1647453, EPI_ISL_1647454, EPI_ISL_1647455, EPI_ISL_1669766, EPI_ISL_1669767, EPI_ISL_1669768, EPI_ISL_1669769, EPI_ISL_1669770, EPI_ISL_1669771, EPI_ISL_1669772, EPI_ISL_1669773, EPI_ISL_1669774, EPI_ISL_1669775, EPI_ISL_1669776, EPI_ISL_1669777, EPI_ISL_1669778, EPI_ISL_2099769, EPI_ISL_2099770 | | | |
| see above | Indian Council of Medical Research-National Institute of Virology, Microbial Containment Complex | Indian Council of Medical Research-National Institute of Virology, Microbial Containment Complex | Pragya D Yadav |
| EPI_ISL_825084, EPI_ISL_825086, EPI_ISL_825087, EPI_ISL_825088, EPI_ISL_825089 | Indian Council of Medical Research-National Institute of Virology,Maximum Containment Laboratory | Indian Council of Medical Research-National Institute of Virology,Maximum Containment Laboratory | Deepak Y Patil; Dimpal A Nyayanit; Jayshri Pethani; Pragya D Yadav; Prasad Sarkale; Rima R Sahay; Savita Patil; Shrikant Baradkar; Varsha Potdar |
| EPI_ISL_1278285, EPI_ISL_1278286, EPI_ISL_1312365, EPI_ISL_1312366, EPI_ISL_2013028, EPI_ISL_2013029, EPI_ISL_2013030, EPI_ISL_2013031, EPI_ISL_2013032, EPI_ISL_2013033, EPI_ISL_2013034, EPI_ISL_2014131, EPI_ISL_2014132, EPI_ISL_2014133, EPI_ISL_2014135, EPI_ISL_2032336, EPI_ISL_2032338, EPI_ISL_2036277, EPI_ISL_2036278, EPI_ISL_2036279, EPI_ISL_2036280, EPI_ISL_2036281, EPI_ISL_2036282, EPI_ISL_2036283, EPI_ISL_2036284, EPI_ISL_2036285, EPI_ISL_2036286, EPI_ISL_2036287, EPI_ISL_2036288, EPI_ISL_2036289, EPI_ISL_2036290, EPI_ISL_2036291, EPI_ISL_2036292, EPI_ISL_2036293, EPI_ISL_2036294, EPI_ISL_2036295, EPI_ISL_2036296, EPI_ISL_2036297, EPI_ISL_2036298, EPI_ISL_2036299, EPI_ISL_2036300, EPI_ISL_2036301, EPI_ISL_2036302, EPI_ISL_2036303, EPI_ISL_2036304 | | | |
| see above | Indian Council of Medical Research-National Institute of Virology, Microbial Containment | Indian Council of Medical Research-National Institute of Virology, Microbial Containment | Pragya D Yadav; Pragya D. Yadav |

| | | | |
|---|---|---|---|
| | Complex | Complex | |
| EPI_ISL_1372094 | Ispat General Hospital (IGH), Rourkela | Institute of Life Sciences - INSACOG | Ajay Parida; Amol M. Kanampalliwar; Arup Ghosh; Atimukta Jha; INSACOG Consortium; Punit Prasad; Rajeeb Swain; Rupesh Dash; Safal Walia; Shifu Aggarwal; Sunil K. Raghav |
| EPI_ISL_995700, EPI_ISL_995701, EPI_ISL_995720, EPI_ISL_995721 | JAYANAGAR GENERAL HOSPITAL | Department of Neurovirology, National Institute of Mental Health and Neurosciences (NIMHANS) | Anita S Desai; Anson Kunjumon George; Chitra Pattabiraman; Darshan Sreenivas; Nakka Vijay Kiran Reddy; Pramada Prasad; Risha Rasheed; V Ravi |
| EPI_ISL_1939839, EPI_ISL_1939840, EPI_ISL_1939841, EPI_ISL_1939842, EPI_ISL_1939843, EPI_ISL_1939844, EPI_ISL_1939845, EPI_ISL_1939846, EPI_ISL_1939847, EPI_ISL_1939848, EPI_ISL_1939863, EPI_ISL_1939864, EPI_ISL_1939865, EPI_ISL_1939866, EPI_ISL_1939867, EPI_ISL_1939868, EPI_ISL_1939869, EPI_ISL_1939870, EPI_ISL_1939871, EPI_ISL_1939872, EPI_ISL_1939873, EPI_ISL_1939874, EPI_ISL_1939929, EPI_ISL_1939930, EPI_ISL_1939931, EPI_ISL_1939932, EPI_ISL_1939933, EPI_ISL_1939934, EPI_ISL_1939935, EPI_ISL_1939936, EPI_ISL_1939937, EPI_ISL_1939938, EPI_ISL_1939939, EPI_ISL_1939940, EPI_ISL_1939941, EPI_ISL_1939942, EPI_ISL_1939943, EPI_ISL_1939944, EPI_ISL_1939945, EPI_ISL_1939946, EPI_ISL_1939947, EPI_ISL_1939948, EPI_ISL_1939949, EPI_ISL_1939950, EPI_ISL_1939951, EPI_ISL_1939952, EPI_ISL_1939953, EPI_ISL_1939954, EPI_ISL_1939955, EPI_ISL_1939956, EPI_ISL_1939957, EPI_ISL_1939958, EPI_ISL_1939959, EPI_ISL_1939960, EPI_ISL_1939961, EPI_ISL_1939962, EPI_ISL_1939963, EPI_ISL_1939964, EPI_ISL_1939965, EPI_ISL_1939966, EPI_ISL_1939967, EPI_ISL_1939968 | | | |
| see above | JIPMER Puducherry | inStem NCBS - INSACOG | Uma Ramakrishnan Dasaradhi Palakodeti Aswin SaiNarain |
| EPI_ISL_2151274, EPI_ISL_2151275, EPI_ISL_2151276 | JJR NAGAR UPHC | inStem NCBS - INSACOG | Uma Ramakrishnan Dasaradhi Palakodeti Aswin SaiNarain |
| EPI_ISL_860193, EPI_ISL_995731, EPI_ISL_995743, EPI_ISL_995744, EPI_ISL_995745 | Jayanagar General Hospital | Department of Neurovirology, National Institute of Mental Health and Neurosciences (NIMHANS) | Anita S Desai; Anson Kunjumon George; Chitra Pattabiraman; Darshan Sreenivas; Harsha.P.K; Nakka Vijay Kiran Reddy; Pramada Prasad; Risha Rasheed; V Ravi |
| EPI_ISL_1818630 | Jayanagar General Hospital | INSACOG-KA, NIMHANS | Ananthapadmanabha Kotambail; Anita S Desai; Anson Kunjumon George; Chetan G K; Chitra Pattabiraman; Darshan Sreenivas; Gautham Arunachal Udupi; Pramada Prasad; V Ravi |
| EPI_ISL_1939969, EPI_ISL_1939970, EPI_ISL_1939971, EPI_ISL_1939972 | KAMAKSHIPALYA | inStem NCBS - INSACOG | Uma Ramakrishnan Dasaradhi Palakodeti Aswin SaiNarain |
| EPI_ISL_995185, EPI_ISL_995186, EPI_ISL_995187, EPI_ISL_995188 | KARWAR INSTITUTE OF MEDICAL SCIENCES | Department of Neurovirology, National Institute of Mental Health and Neurosciences (NIMHANS) | Anita S Desai; Anson Kunjumon George; Chitra Pattabiraman; Darshan Sreenivas; Nakka Vijay Kiran Reddy; Pramada Prasad; Risha Rasheed; V Ravi |
| EPI_ISL_995695 | KASTURBA MEDICAL COLLEGE, MANGALORE | Department of Neurovirology, National Institute of Mental Health and Neurosciences (NIMHANS) | Anita S Desai; Anson Kunjumon George; Chitra Pattabiraman; Darshan Sreenivas; Nakka Vijay Kiran Reddy; Pramada Prasad; Risha Rasheed; V Ravi |
| EPI_ISL_995189, EPI_ISL_995190, EPI_ISL_995694, EPI_ISL_995697, EPI_ISL_995698, EPI_ISL_995699, EPI_ISL_995713, EPI_ISL_995714, EPI_ISL_995759, EPI_ISL_995760, EPI_ISL_995761, EPI_ISL_995762, EPI_ISL_995764 | | | |
| see above | KEMPEGOWDA INTERNATIONAL AIRPORT | Department of Neurovirology, National Institute of Mental Health and Neurosciences (NIMHANS) | Anita S Desai; Anson Kunjumon George; Chitra Pattabiraman; Darshan Sreenivas; Harsha.P.K; Nakka Vijay Kiran Reddy; Pramada Prasad; Risha Rasheed; V Ravi |
| EPI_ISL_1385814, EPI_ISL_1385815, EPI_ISL_1385816, EPI_ISL_1385825, EPI_ISL_1385829, EPI_ISL_1385830, EPI_ISL_1385831, EPI_ISL_1385832, EPI_ISL_1385839, EPI_ISL_1385840, EPI_ISL_1396655, EPI_ISL_1595857, EPI_ISL_1595858, EPI_ISL_1595859, EPI_ISL_1595891, EPI_ISL_1595892, EPI_ISL_1595893, EPI_ISL_1595895, EPI_ISL_1595896, EPI_ISL_1595897, EPI_ISL_1595898, EPI_ISL_1595899, EPI_ISL_1595900, EPI_ISL_1818583, EPI_ISL_1818584, EPI_ISL_1818585, EPI_ISL_1818587, EPI_ISL_1818588, EPI_ISL_1818589, EPI_ISL_1818614, EPI_ISL_1818615, EPI_ISL_1818616, EPI_ISL_1818617, EPI_ISL_1818618, EPI_ISL_1818622 | | | |
| see above | KEMPEGOWDA INTERNATIONAL AIRPORT | INSACOG-KA, NIMHANS | Ananthapadmanabha Kotambail; Anita S Desai; Anson Kunjumon George; Chetan G K; Chitra Pattabiraman; Darshan Sreenivas; Gautham Arunachal Udupi; Nakka Vijay Kiran Reddy; Pramada Prasad; V Ravi |
| EPI_ISL_1940100, EPI_ISL_2015555, EPI_ISL_2015633 | KEMPEGOWDA INTERNATIONAL AIRPORT | inStem NCBS - INSACOG | Uma Ramakrishnan Dasaradhi Palakodeti Aswin SaiNarain |
| EPI_ISL_1595914 | KIMS, KARWAR | INSACOG-KA, NIMHANS | Ananthapadmanabha Kotambail; Anita S Desai; Anson Kunjumon George; Chetan G K; Chitra Pattabiraman; Darshan Sreenivas; Gautham Arunachal Udupi; Pramada Prasad; V Ravi |
| EPI_ISL_2015467, EPI_ISL_2015483, EPI_ISL_2015485, EPI_ISL_2015489, EPI_ISL_2015494, EPI_ISL_2015534, EPI_ISL_2015541, EPI_ISL_2015543, EPI_ISL_2015551 | | | |
| see above | KMC MANIPAL | inStem NCBS - INSACOG | Uma Ramakrishnan Dasaradhi Palakodeti Aswin SaiNarain |
| EPI_ISL_745197 | Kakatiya Medical College | CSIR-Centre for Cellular and Molecular Biology | Archana Bharadwaj Siva; B Himasri; Blessy B John; Divya Tej Sowpati; Dr.P.Shashikala Reddy; Dr.S.Pavani; Dr.Satyaprasad; Dr.V.Sudha Rani; Karthik Bharadwaj Tallapaka; Lamuk Zaveri; M Soujanya Reddy; Namami Gaur; Nikhil Hajirnis; Onkar Kulkarni; Payel Mukherjee; Pratheusa Maccha; Priya Singh; Purushotham Vodnala; Rakesh K Mishra; Sakshi Shambhavi; Shagufta Khan; Sofia Banu; Tulasi Nagabandi; Viswagithe S L |
| EPI_ISL_803906, EPI_ISL_803907, EPI_ISL_803910, EPI_ISL_803914, EPI_ISL_803915, EPI_ISL_825146, EPI_ISL_825150, EPI_ISL_825151, EPI_ISL_845881 | | | |
| see above | Kidwai Memorial Institute of Oncology | Department of Neurovirology, National Institute of Mental Health and Neurosciences (NIMHANS) | Anita S Desai; Chitra Pattabiraman; Darshan Sreenivas; Nakka Vijay Kiran Reddy; Pramada Prasad; Risha Rasheed; V Ravi |
| EPI_ISL_1708322, EPI_ISL_1708328, EPI_ISL_1708433, EPI_ISL_1708437, EPI_ISL_1708439, EPI_ISL_1708440, EPI_ISL_2151156, EPI_ISL_2151157, EPI_ISL_2151158, EPI_ISL_2151159, EPI_ISL_2151160, EPI_ISL_2151161, EPI_ISL_2151163, EPI_ISL_2151165, EPI_ISL_2151166, EPI_ISL_2151167, EPI_ISL_2151168, EPI_ISL_2151169, EPI_ISL_2151170, EPI_ISL_2151171, EPI_ISL_2151172, EPI_ISL_2151174, EPI_ISL_2151176, EPI_ISL_2151177, EPI_ISL_2151178, EPI_ISL_2151179, EPI_ISL_2151180, EPI_ISL_2151183, EPI_ISL_2151184, EPI_ISL_2151185, EPI_ISL_2151190, EPI_ISL_2151191, EPI_ISL_2151192, EPI_ISL_2151193, EPI_ISL_2151194, EPI_ISL_2151195, EPI_ISL_2151196, EPI_ISL_2151197, EPI_ISL_2151198, EPI_ISL_2151199, EPI_ISL_2151200, EPI_ISL_2151201, EPI_ISL_2151202, EPI_ISL_2151203, EPI_ISL_2151204, EPI_ISL_2151205, EPI_ISL_2151206, EPI_ISL_2151207, EPI_ISL_2151208, EPI_ISL_2151209, EPI_ISL_2151210, EPI_ISL_2151211, EPI_ISL_2151212, EPI_ISL_2151213, EPI_ISL_2151214, EPI_ISL_2151215, EPI_ISL_2151216, EPI_ISL_2151217, EPI_ISL_2151218, EPI_ISL_2151219, EPI_ISL_2151220, EPI_ISL_2151221, EPI_ISL_2151222, EPI_ISL_2151223, EPI_ISL_2151224, EPI_ISL_2151225, EPI_ISL_2151226, EPI_ISL_2151227, EPI_ISL_2151228, EPI_ISL_2151229, EPI_ISL_2151230, EPI_ISL_2151231, EPI_ISL_2151232, EPI_ISL_2151233, EPI_ISL_2151236, EPI_ISL_2151237, EPI_ISL_2151238, EPI_ISL_2151239, EPI_ISL_2151240, EPI_ISL_2151241, EPI_ISL_2151242, EPI_ISL_2151243, EPI_ISL_2151244 | | | |
| see above | Kodigehalli PHC | inStem NCBS - INSACOG | Uma Ramakrishnan Dasaradhi Palakodeti Aswin SaiNarain; Uma Ramakrishnan Dasaradhi Palakodeti Aswin SaiNarain |
| EPI_ISL_1663538, EPI_ISL_1663539, EPI_ISL_1663540, EPI_ISL_1663541, EPI_ISL_1663542, EPI_ISL_1663543, EPI_ISL_1663544, EPI_ISL_1663545, EPI_ISL_1663546, EPI_ISL_1663547, EPI_ISL_1663548, EPI_ISL_1663549, EPI_ISL_1663550, EPI_ISL_1663551 | | | |
| see above | MGM Medical College, Jamshedpur | Institute of Life Sciences - INSACOG | Ajay Parida; Amol M. Kanampalliwar; Arup Ghosh; Atimukta Jha; INSACOG Consortium; Punit Prasad; Rajeeb Swain; Rupesh Dash; Safal Walia; Shifu Aggarwal; Sunil K. Raghav |
| EPI_ISL_2015469, EPI_ISL_2015472, EPI_ISL_2015479, EPI_ISL_2015480, EPI_ISL_2015482, EPI_ISL_2015487, EPI_ISL_2015528, EPI_ISL_2015556, EPI_ISL_2015577 | | | |
| see above | MIV, MAHE | inStem NCBS - INSACOG | Uma Ramakrishnan Dasaradhi Palakodeti Aswin SaiNarain |
| EPI_ISL_1708323, EPI_ISL_1708324, EPI_ISL_1708326, EPI_ISL_1708327, EPI_ISL_1708431, EPI_ISL_1708432, EPI_ISL_1708435, EPI_ISL_1708447, EPI_ISL_1708448, EPI_ISL_2151162, EPI_ISL_2151164 | | | |
| see above | MS Palya PHC | inStem NCBS - INSACOG | Uma Ramakrishnan Dasaradhi Palakodeti Aswin SaiNarain; Uma Ramakrishnan Dasaradhi Palakodeti Aswin SaiNarain |
| EPI_ISL_1385833, EPI_ISL_1385834, EPI_ISL_1385835 | Mandya Institute of Medical Sciences | INSACOG-KA, NIMHANS | Anita S Desai; Anson Kunjumon George; Chitra Pattabiraman; Darshan Sreenivas; Nakka Vijay Kiran Reddy; Pramada Prasad; V Ravi |
| EPI_ISL_1372269, EPI_ISL_1372270, EPI_ISL_1372272, EPI_ISL_1372274, EPI_ISL_1372276, EPI_ISL_1372278, EPI_ISL_1547801 | | | |
| see above | Mapmygenome | CSIR-Centre for Cellular and Molecular Biology - INSACOG | Amareshwar Vodapalli; Ara Sreenivas; B Himasri; Divya Tej Sowpati; Karthik Bharadwaj Tallapaka; Lamuk Zaveri; Onkar Kulkarni; Rakesh K Mishra; Sharath Chandra Thota; Shreekant Verma; Sofia Banu; Viswagithe S L |
| EPI_ISL_1844778, EPI_ISL_1844782 | Maulana Azad Medical College | Biotechnology Division, NCDC | Hema Gogia; Hemlata Lall; Kalaiarasan Ponnusamy; Mahesh S Dhar; Manoj K Singh; Meena Datta; Partha Rakshit; Preeti Madan; Priyanka Singh; Radhakrishnan V. S; Robin Marwal; Sandhya Kabra; Sujeet K Singh; Uma Sharma |
| EPI_ISL_1663536, EPI_ISL_1663537 | Medini Rai Medical College,Palamu, Jharkhand | Institute of Life Sciences - INSACOG | Ajay Parida; Amol M. Kanampalliwar; Arup Ghosh; Atimukta Jha; INSACOG Consortium; Punit Prasad; Rajeeb Swain; Rupesh Dash; Safal Walia; Shifu Aggarwal; Sunil K. Raghav |
| EPI_ISL_1372012, EPI_ISL_1372013, EPI_ISL_1372014, EPI_ISL_1372015, EPI_ISL_1372016, EPI_ISL_1372017, EPI_ISL_1372018, EPI_ISL_1372019, EPI_ISL_1372020, EPI_ISL_1372021, EPI_ISL_1372022, EPI_ISL_1372023, EPI_ISL_1372024, EPI_ISL_1372025, EPI_ISL_1372026, EPI_ISL_1372027, EPI_ISL_1372028, EPI_ISL_1372029, EPI_ISL_1372030, EPI_ISL_1372031, EPI_ISL_1372032, EPI_ISL_1372033, EPI_ISL_1372034, EPI_ISL_1372035, EPI_ISL_1372036, EPI_ISL_1372037, EPI_ISL_1372038, EPI_ISL_1372039, EPI_ISL_1372040, EPI_ISL_1372041, EPI_ISL_1372042, EPI_ISL_1372043, EPI_ISL_1372044, EPI_ISL_1372045, EPI_ISL_1372046, EPI_ISL_1372047, EPI_ISL_1372048, EPI_ISL_1372049, EPI_ISL_1372050, EPI_ISL_1372051, EPI_ISL_1372052, EPI_ISL_1372053, EPI_ISL_1372054, EPI_ISL_1372055, EPI_ISL_1372056, EPI_ISL_1372057, EPI_ISL_1372058, EPI_ISL_1372059, EPI_ISL_1372060, EPI_ISL_1372061, EPI_ISL_1372062, EPI_ISL_1372063, EPI_ISL_1372064, EPI_ISL_1372065, EPI_ISL_1372066, EPI_ISL_1372067, EPI_ISL_1372068, EPI_ISL_1372069, EPI_ISL_1372070, EPI_ISL_1372071, EPI_ISL_1372072, EPI_ISL_1372073, EPI_ISL_1372074, EPI_ISL_1372075, EPI_ISL_1372076, EPI_ISL_1372077, EPI_ISL_1372078, EPI_ISL_1372079, EPI_ISL_1372080, EPI_ISL_1372081, EPI_ISL_1372082, EPI_ISL_1372083, EPI_ISL_1372084, EPI_ISL_1372085, EPI_ISL_1372096 | | | |
| see above | Microbiology Department , Late Shri Lakhiram Agrawal Memory Hospital & College, Raigarh, Chhattisgarh | Institute of Life Sciences - INSACOG | Ajay Parida; Amol M. Kanampalliwar; Arup Ghosh; Atimukta Jha; INSACOG Consortium; Punit Prasad; Rajeeb Swain; Rupesh Dash; Safal Walia; Shifu Aggarwal; Sunil K. Raghav |
| EPI_ISL_2001112, EPI_ISL_2001113 | Microbiology, Indian Council Of Medical Research-Regional Medical Research center (ICMR-RMRC) | Microbiology, Indian Council Of Medical Research-Regional Medical Research center (ICMR-RMRC) | Bali; Bhattacharya; Biswas, D.; Borkakoty, B.; C.K.; Hazarika, R.; M.D.; N.K.; Sarmah |
| EPI_ISL_1063806, EPI_ISL_1063855, EPI_ISL_1063856, EPI_ISL_1063857, EPI_ISL_1063858, EPI_ISL_1063859, EPI_ISL_1063860, EPI_ISL_1063861 | | | |
| see above | Molecular Laboratory, Vikash Multispeciality Hospital, Bargarh | Immunogenomics lab, Institute of Life Sciences, Bhubaneswar | Ajay Parida; Amol M. Kanampalliwar; Arup Ghosh; Atimukta Jha; INSACOG Consortium; Kirtal Hansdah; Punit Prasad; Rajeeb Swain; Rupesh Dash; Safal Walia; Shifu Aggarwal; Sunil K. Raghav |
| EPI_ISL_1662270, EPI_ISL_1662271 | Molecular Laboratory, Vikash Multispeciality Hospital, Bargarh | Institute of Life Sciences - INSACOG | Ajay Parida; Amol M. Kanampalliwar; Arup Ghosh; Atimukta Jha; INSACOG Consortium; Punit Prasad; Rajeeb Swain; Rupesh Dash; Safal Walia; Shifu Aggarwal; Sunil K. Raghav |
| EPI_ISL_862051 | Multidisciplinary Research Unit, DHR-ICMR, Institute of Medical Sciences, Banaras Hindu University | Multidisciplinary Research Unit, DHR-ICMR, Institute of Medical Sciences, Banaras Hindu University | Abhay Kumar Yadav; Arup Acharjee; Ashish; Chetan Sahni; Deepa Devadas; Jay Prakash Maurya; Manpreet Kaur; Nitish Kumar Singh; Prashant Singh; Priyoneel Basu; Richa Arya; Royana Singh; Sanjay Kumar; Surendra Pratap Mishra; Tribhuwan Mohan Mahapatra |
| EPI_ISL_2017759, EPI_ISL_2017760, EPI_ISL_2017761, EPI_ISL_2017762 | N.H.L Municipal Medical College, Ahmedabad | Gujarat Biotechnology Research Centre | Chaitanya Joshi; Dinesh Kumar; Janvi Raval; Jayshri Pethani; Madhvi Joshi; Nitesh Shah; Nitin Savaliya; Ramesh Pandit; Sonal Sharma; Twinkle Soni; Umang Mishra; Zarna Patel; Zuber Saiyed |
| EPI_ISL_1939875, EPI_ISL_1939876, EPI_ISL_1939877, EPI_ISL_1939878, EPI_ISL_1939879, EPI_ISL_1939880, EPI_ISL_1939881, EPI_ISL_1939882, EPI_ISL_1939883, EPI_ISL_1939884, EPI_ISL_1939885, EPI_ISL_1939886, EPI_ISL_1939887, EPI_ISL_1939888, EPI_ISL_1939889, EPI_ISL_1939890, EPI_ISL_1939891, EPI_ISL_1939892, EPI_ISL_1939893, EPI_ISL_1939894, EPI_ISL_1939895, EPI_ISL_1939896, EPI_ISL_1939897, EPI_ISL_1939898, EPI_ISL_1939899, EPI_ISL_1939900, EPI_ISL_1939901, EPI_ISL_1939902, EPI_ISL_1939903, EPI_ISL_1939904, EPI_ISL_1939905, EPI_ISL_1939906, EPI_ISL_1939907, EPI_ISL_1939908, EPI_ISL_1939909, EPI_ISL_1939910, EPI_ISL_1939911, EPI_ISL_1939912, EPI_ISL_1939913, EPI_ISL_1939914, EPI_ISL_1939915, EPI_ISL_1939916, EPI_ISL_1939917, EPI_ISL_1939918, EPI_ISL_1939919, EPI_ISL_1939920, EPI_ISL_1939921, EPI_ISL_1939922, EPI_ISL_1939923, EPI_ISL_1939924, EPI_ISL_1939925, EPI_ISL_1939926, EPI_ISL_1939927, EPI_ISL_1939928 | | | |
| see above | NCCS | inStem NCBS - INSACOG | Uma Ramakrishnan Dasaradhi Palakodeti Aswin SaiNarain |
| EPI_ISL_1663363, EPI_ISL_1663364, EPI_ISL_1663365, EPI_ISL_1663366, EPI_ISL_1663367, EPI_ISL_1663368, EPI_ISL_1663369, EPI_ISL_1663370, EPI_ISL_1663371, EPI_ISL_1663372, EPI_ISL_1663373, EPI_ISL_1663374, EPI_ISL_1663375, EPI_ISL_1663376, EPI_ISL_1663377, EPI_ISL_1663378, EPI_ISL_1663379, EPI_ISL_1663380, EPI_ISL_1663381, EPI_ISL_1663382, EPI_ISL_1663383, EPI_ISL_1663384, EPI_ISL_1663385, EPI_ISL_1663386, EPI_ISL_1663387, EPI_ISL_1663388, EPI_ISL_1663389, EPI_ISL_1663390, EPI_ISL_1663391, EPI_ISL_1663392, EPI_ISL_1663393, EPI_ISL_1663394, EPI_ISL_1663395, EPI_ISL_1663396, EPI_ISL_1663397, EPI_ISL_1663398, EPI_ISL_1663399, EPI_ISL_1663400, EPI_ISL_1663401, EPI_ISL_1663402, EPI_ISL_1663403, EPI_ISL_1663405, EPI_ISL_1663406, EPI_ISL_1663407, EPI_ISL_1663408, EPI_ISL_1663409, EPI_ISL_1663410, EPI_ISL_1663411, EPI_ISL_1663412, EPI_ISL_1663413, EPI_ISL_1663414, EPI_ISL_1663415, EPI_ISL_1663416, EPI_ISL_1663417, EPI_ISL_1663418, EPI_ISL_1663419, EPI_ISL_1663420, EPI_ISL_1663421, EPI_ISL_1663422, EPI_ISL_1663423, EPI_ISL_1663424, EPI_ISL_1663425, EPI_ISL_1663426, EPI_ISL_1663427, EPI_ISL_1663428, EPI_ISL_1663429, EPI_ISL_1663430, EPI_ISL_1663431, EPI_ISL_1663432, EPI_ISL_1663433, EPI_ISL_1663434, EPI_ISL_1663435, EPI_ISL_1663436, EPI_ISL_1663437, EPI_ISL_1663438, EPI_ISL_1663439, EPI_ISL_1663440, EPI_ISL_1663441, EPI_ISL_1663442, EPI_ISL_1663443, EPI_ISL_1663444, EPI_ISL_1663445, EPI_ISL_1663446, EPI_ISL_1663447, EPI_ISL_1663448, EPI_ISL_1663449, EPI_ISL_1663450, EPI_ISL_1663451, EPI_ISL_1663452, EPI_ISL_1663453, EPI_ISL_1663454, EPI_ISL_1663455, EPI_ISL_1663456, EPI_ISL_1663457, EPI_ISL_1663458, EPI_ISL_1663459, EPI_ISL_1663460, EPI_ISL_1663461, EPI_ISL_1663462, EPI_ISL_1663463, EPI_ISL_1663464, EPI_ISL_1663465, EPI_ISL_1663466, EPI_ISL_1663467, EPI_ISL_1663468, EPI_ISL_1663469, EPI_ISL_1663470, EPI_ISL_1663471, EPI_ISL_1663472, EPI_ISL_1663473, EPI_ISL_1663474, EPI_ISL_1663475, EPI_ISL_1663476, EPI_ISL_1663477, EPI_ISL_1663478, EPI_ISL_1663479, EPI_ISL_1663480, EPI_ISL_1663481, EPI_ISL_1663482, EPI_ISL_1663483, EPI_ISL_1663484, EPI_ISL_1663485, EPI_ISL_1663486, EPI_ISL_1663487 | | | |
| see above | NCCS, Pune | Institute of Life Sciences - INSACOG | Ajay Parida; Amol M. Kanampalliwar; Arup Ghosh; Atimukta Jha; INSACOG Consortium; Punit Prasad; Rajeeb Swain; Rupesh Dash; Safal Walia; Shifu Aggarwal; Sunil K. Raghav |
| EPI_ISL_825043, EPI_ISL_825044, EPI_ISL_825045, EPI_ISL_825046, EPI_ISL_825047, EPI_ISL_825050, EPI_ISL_825051, EPI_ISL_825052, EPI_ISL_825053, EPI_ISL_825059, EPI_ISL_825060, EPI_ISL_825063 | | | |
| see above | NHL Municipal Medical College, Ahmedbad | Gujarat Biotechnology Research Centre | Afzal Ansari; Apurvasinh Puvar; Atit Shah; Bhavin Prajapati; Bimal Chauhan; Chaitanya Joshi; Dinesh Kumar; Janvi Raval; Jayshri Pethani; Labdhi Pandya; Madhvi Joshi; Monila Patel; NM Shaikh; Nikha Trivedi; Nitin Savaliya; Ramesh Pandit; Tanmay Mehta; Zarna Patel; Zuber Saiyed |
| EPI_ISL_1939985, EPI_ISL_1939986, EPI_ISL_1939987, EPI_ISL_1939988, EPI_ISL_1939989, EPI_ISL_1939999, EPI_ISL_1940029, EPI_ISL_1940035, EPI_ISL_1940053, EPI_ISL_1940054, EPI_ISL_1940055, EPI_ISL_1940056, EPI_ISL_1940057, EPI_ISL_1940058, EPI_ISL_1940069, EPI_ISL_1940070, EPI_ISL_1940095, EPI_ISL_1940096, EPI_ISL_1940097, EPI_ISL_1940098, EPI_ISL_1940099, EPI_ISL_1940107, EPI_ISL_1940108, EPI_ISL_1940133, EPI_ISL_1940134, EPI_ISL_1940135, EPI_ISL_1940136, EPI_ISL_1940137, EPI_ISL_1940138, EPI_ISL_1940139, EPI_ISL_1940140, EPI_ISL_1940141, EPI_ISL_2015475, EPI_ISL_2015478, EPI_ISL_2015486, EPI_ISL_2015492, EPI_ISL_2015497, EPI_ISL_2015542, EPI_ISL_2015546, EPI_ISL_2015564, EPI_ISL_2015576, EPI_ISL_2015578, EPI_ISL_2015580, EPI_ISL_2015588, EPI_ISL_2015594, EPI_ISL_2015603, EPI_ISL_2015604, EPI_ISL_2015621, EPI_ISL_2015628, EPI_ISL_2015631, EPI_ISL_2151247, EPI_ISL_2151249, EPI_ISL_2151250, EPI_ISL_2151253, EPI_ISL_2151254, EPI_ISL_2151255, EPI_ISL_2151256, EPI_ISL_2151257, EPI_ISL_2151268, EPI_ISL_2151269, EPI_ISL_2151270, EPI_ISL_2151271, EPI_ISL_2151277, EPI_ISL_2151278, EPI_ISL_2151279, EPI_ISL_2151284, EPI_ISL_2151288, EPI_ISL_2151289, EPI_ISL_2151290, EPI_ISL_2151291, EPI_ISL_2151292, EPI_ISL_2151293, EPI_ISL_2151294, EPI_ISL_2151300, EPI_ISL_2151302, EPI_ISL_2151303, EPI_ISL_2151304, EPI_ISL_2151305, EPI_ISL_2151306, EPI_ISL_2151307, EPI_ISL_2151308, EPI_ISL_2151309, EPI_ISL_2151310, EPI_ISL_2151311, EPI_ISL_2151312, EPI_ISL_2151323, EPI_ISL_2151329, EPI_ISL_2151330, EPI_ISL_2151331, EPI_ISL_2151334 | | | |

| | | | |
|---|---|---|---|
| see above | NIMHANS | inStem NCBS - INSACOG | Uma Ramakrishnan Dasaradhi Palakodeti Aswin SaiNarain |
| EPI_ISL_1034154, EPI_ISL_1034155, EPI_ISL_1034156, EPI_ISL_1034161, EPI_ISL_1034162, EPI_ISL_1034163, EPI_ISL_1034164, EPI_ISL_1034165, EPI_ISL_1034166, EPI_ISL_1034168, EPI_ISL_1034169, EPI_ISL_1034170, EPI_ISL_1034171, EPI_ISL_1034172, EPI_ISL_1034173, EPI_ISL_1034174, EPI_ISL_1034175, EPI_ISL_1034176, EPI_ISL_1034177, EPI_ISL_1034178, EPI_ISL_1034179, EPI_ISL_1034180, EPI_ISL_1034181, EPI_ISL_1034182, EPI_ISL_1034183, EPI_ISL_1034184, EPI_ISL_1034185, EPI_ISL_1034186, EPI_ISL_1034188, EPI_ISL_1034189 | | | |
| see above | NIV Influenza | NIV Influenza | Potdar V |
| EPI_ISL_1844765, EPI_ISL_1844767, EPI_ISL_1844769, EPI_ISL_1844771, EPI_ISL_1844773 | NORTHERN RAILWAY CENTRAL HOSPITAL | Biotechnology Division, NCDC | Hema Gogia; Hemlata Lall; Kalaiarasan Ponnusamy; Mahesh S Dhar; Manoj K Singh; Meena Datta; Partha Rakshit; Preeti Madan; Priyanka Singh; Radhakrishnan V. S; Robin Marwal; Sandhya Kabra; Sujeet K Singh; Uma Sharma |
| EPI_ISL_1662277, EPI_ISL_1662278, EPI_ISL_1662279, EPI_ISL_1662280, EPI_ISL_1662281 | Nalanda Medical College & Hospital, Patna | Institute of Life Sciences - INSACOG | Ajay Parida; Amol M. Kanampalliwar; Arup Ghosh; Atimukta Jha; INSACOG Consortium; Punit Prasad; Rajeeb Swain; Rupesh Dash; Safal Walia; Shifu Aggarwal; Sunil K. Raghav |
| EPI_ISL_1372095 | Nalanda Medical College Hospital, Patna | Institute of Life Sciences - INSACOG | Ajay Parida; Amol M. Kanampalliwar; Arup Ghosh; Atimukta Jha; INSACOG Consortium; Punit Prasad; Rajeeb Swain; Rupesh Dash; Safal Walia; Shifu Aggarwal; Sunil K. Raghav |
| EPI_ISL_825147 | Narayana Nethralaya lab | Department of Neurovirology, National Institute of Mental Health and Neurosciences (NIMHANS) | Anita S Desai; Chitra Pattabiraman; Pramada Prasad; V Ravi |
| EPI_ISL_1384818 | National Centre For Cell Science | National Centre For Cell Science | Ajay Pillai; Dhiraj Paul; Manoj Kumar Bhat; Mitali Inamdar; Mohak P Gujare; Shivang P. Bhanushali; Sonal Manik Chavan; Yogesh Shouche |
| EPI_ISL_1384815, EPI_ISL_1384816, EPI_ISL_1384817, EPI_ISL_1384819, EPI_ISL_1384820, EPI_ISL_1384821, EPI_ISL_1384822, EPI_ISL_1384823, EPI_ISL_1384824, EPI_ISL_1384825, EPI_ISL_1384826, EPI_ISL_1384827, EPI_ISL_1384828, EPI_ISL_1384829, EPI_ISL_1384830, EPI_ISL_1384831, EPI_ISL_1384832, EPI_ISL_1384833, EPI_ISL_1384834, EPI_ISL_1384835, EPI_ISL_1384843, EPI_ISL_1384844, EPI_ISL_1384845, EPI_ISL_1384846, EPI_ISL_1384847, EPI_ISL_1384850, EPI_ISL_1384851, EPI_ISL_1384852, EPI_ISL_1384853, EPI_ISL_1384854, EPI_ISL_1384855, EPI_ISL_1384856, EPI_ISL_1384857, EPI_ISL_1384858, EPI_ISL_1384860, EPI_ISL_1384861, EPI_ISL_1384862, EPI_ISL_1384864, EPI_ISL_1384865, EPI_ISL_1384866, EPI_ISL_1384867, EPI_ISL_1384868, EPI_ISL_1384869, EPI_ISL_1384870, EPI_ISL_1384871, EPI_ISL_1384872, EPI_ISL_1384873, EPI_ISL_1384874, EPI_ISL_1384875, EPI_ISL_1384876, EPI_ISL_1384877, EPI_ISL_1384878, EPI_ISL_1384879, EPI_ISL_1384880, EPI_ISL_1384881, EPI_ISL_1384882, EPI_ISL_1384883, EPI_ISL_1384884, EPI_ISL_1415086, EPI_ISL_1415087, EPI_ISL_1415088, EPI_ISL_1415090, EPI_ISL_1415091, EPI_ISL_1415093, EPI_ISL_1415094, EPI_ISL_1415096, EPI_ISL_1415097, EPI_ISL_1415099, EPI_ISL_1415100, EPI_ISL_1415101, EPI_ISL_1415102, EPI_ISL_1415103, EPI_ISL_1415104, EPI_ISL_1415105, EPI_ISL_1415108, EPI_ISL_1415110, EPI_ISL_1415111, EPI_ISL_1415112, EPI_ISL_1415113, EPI_ISL_1415114, EPI_ISL_1415116, EPI_ISL_1415117, EPI_ISL_1415118, EPI_ISL_1415119, EPI_ISL_1415120, EPI_ISL_1415121, EPI_ISL_1415122, EPI_ISL_1415123, EPI_ISL_1415124, EPI_ISL_1415125, EPI_ISL_1415126, EPI_ISL_1415127, EPI_ISL_1415128, EPI_ISL_1415132, EPI_ISL_1415133, EPI_ISL_1415137, EPI_ISL_1415138, EPI_ISL_1415139, EPI_ISL_1415140, EPI_ISL_1415141, EPI_ISL_1415143, EPI_ISL_1415144, EPI_ISL_1415147, EPI_ISL_1415148, EPI_ISL_1415149, EPI_ISL_1415152, EPI_ISL_1415153, EPI_ISL_1415155, EPI_ISL_1415156, EPI_ISL_1415157, EPI_ISL_1415158, EPI_ISL_1415159, EPI_ISL_1415160, EPI_ISL_1415161, EPI_ISL_1415162, EPI_ISL_1415163, EPI_ISL_1415164, EPI_ISL_1415165, EPI_ISL_1415166, EPI_ISL_1415167, EPI_ISL_1415169, EPI_ISL_1415170, EPI_ISL_1415171, EPI_ISL_1415172, EPI_ISL_1415173, EPI_ISL_1415174, EPI_ISL_1415175, EPI_ISL_1415176, EPI_ISL_1415177, EPI_ISL_1415178, EPI_ISL_1415179, EPI_ISL_1415180, EPI_ISL_1415181, EPI_ISL_1415182, EPI_ISL_1415183, EPI_ISL_1415184, EPI_ISL_1415186, EPI_ISL_1415188, EPI_ISL_1415189, EPI_ISL_1415190, EPI_ISL_1415191, EPI_ISL_1415192, EPI_ISL_1415193, EPI_ISL_1415194, EPI_ISL_1415195, EPI_ISL_1415196, EPI_ISL_1415197, EPI_ISL_1415198, EPI_ISL_1415199, EPI_ISL_1415200, EPI_ISL_1415201, EPI_ISL_1415202, EPI_ISL_1415203, EPI_ISL_1415204, EPI_ISL_1415205, EPI_ISL_1415206, EPI_ISL_1415207, EPI_ISL_1415208, EPI_ISL_1415209, EPI_ISL_1415210, EPI_ISL_1415211, EPI_ISL_1415212, EPI_ISL_1415213, EPI_ISL_1415214, EPI_ISL_1415215, EPI_ISL_1415216, EPI_ISL_1415217, EPI_ISL_1415218, EPI_ISL_1415219, EPI_ISL_1415220, EPI_ISL_1415221, EPI_ISL_1415222, EPI_ISL_1415223, EPI_ISL_1415224, EPI_ISL_1415225, EPI_ISL_1415226, EPI_ISL_1415227, EPI_ISL_1415228, EPI_ISL_1415229, EPI_ISL_1415231, EPI_ISL_1415232, EPI_ISL_1415233, EPI_ISL_1415235, EPI_ISL_1415237, EPI_ISL_1415238, EPI_ISL_1415239, EPI_ISL_1415240, EPI_ISL_1415241, EPI_ISL_1415242, EPI_ISL_1415243, EPI_ISL_1415244, EPI_ISL_1415245, EPI_ISL_1415246, EPI_ISL_1415247, EPI_ISL_1415248, EPI_ISL_1415249, EPI_ISL_1415250, EPI_ISL_1415251, EPI_ISL_1415252, EPI_ISL_1415253, EPI_ISL_1415254, EPI_ISL_1415255, EPI_ISL_1415256, EPI_ISL_1415257, EPI_ISL_1415258, EPI_ISL_1415259, EPI_ISL_1415260, EPI_ISL_1415261, EPI_ISL_1415262, EPI_ISL_1415263, EPI_ISL_1415264, EPI_ISL_1415265, EPI_ISL_1415266, EPI_ISL_1415267, EPI_ISL_1415268, EPI_ISL_1415269, EPI_ISL_1415270, EPI_ISL_1415271, EPI_ISL_1415272, EPI_ISL_1415273, EPI_ISL_1415274, EPI_ISL_1415275, EPI_ISL_1415276, EPI_ISL_1415277, EPI_ISL_1415278, EPI_ISL_1415279, EPI_ISL_1415280, EPI_ISL_1415281, EPI_ISL_1415282, EPI_ISL_1415283, EPI_ISL_1415284, EPI_ISL_1415285, EPI_ISL_1415286, EPI_ISL_1415287, EPI_ISL_1415288, EPI_ISL_1415289, EPI_ISL_1415290, EPI_ISL_1415291, EPI_ISL_1415292, EPI_ISL_1415294, EPI_ISL_1415295, EPI_ISL_1415296, EPI_ISL_1415297, EPI_ISL_1415298, EPI_ISL_1415299, EPI_ISL_1415300, EPI_ISL_1415301, EPI_ISL_1415302, EPI_ISL_1415303, EPI_ISL_1415304, EPI_ISL_1415305, EPI_ISL_1415306, EPI_ISL_1415307, EPI_ISL_1415308, EPI_ISL_1415309, EPI_ISL_1415310, EPI_ISL_1415311, EPI_ISL_1415312, EPI_ISL_1415313, EPI_ISL_1415314, EPI_ISL_1415315, EPI_ISL_1415316, EPI_ISL_1415317, EPI_ISL_1415318, EPI_ISL_1415319, EPI_ISL_1415320, EPI_ISL_1415321, EPI_ISL_1415322, EPI_ISL_1415323, EPI_ISL_1415324, EPI_ISL_1415326, EPI_ISL_1415327, EPI_ISL_1415328, EPI_ISL_1415329, EPI_ISL_1415331, EPI_ISL_1415332, EPI_ISL_1415333, EPI_ISL_1415334, EPI_ISL_1415335, EPI_ISL_1415336, EPI_ISL_1415337, EPI_ISL_1415338, EPI_ISL_1415339, EPI_ISL_1415340, EPI_ISL_1415341, EPI_ISL_1415342, EPI_ISL_1415343, EPI_ISL_1415344, EPI_ISL_1415345, EPI_ISL_1415346, EPI_ISL_1415348, EPI_ISL_1415349, EPI_ISL_1415350, EPI_ISL_1415351, EPI_ISL_1415353, EPI_ISL_1415354, EPI_ISL_1415355, EPI_ISL_1415356, EPI_ISL_1415357, EPI_ISL_1415358, EPI_ISL_1415359, EPI_ISL_1415360, EPI_ISL_1415361, EPI_ISL_1415362, EPI_ISL_1415363, EPI_ISL_1415364, EPI_ISL_1415365, EPI_ISL_1415366, EPI_ISL_1415367, EPI_ISL_1415368, EPI_ISL_1415369, EPI_ISL_1415370, EPI_ISL_1415371, EPI_ISL_1415372, EPI_ISL_1415374, EPI_ISL_1415376, EPI_ISL_1415377, EPI_ISL_1415378, EPI_ISL_1415379, EPI_ISL_1415380, EPI_ISL_1415381, EPI_ISL_1415383, EPI_ISL_1415385, EPI_ISL_1415386, EPI_ISL_1415387, EPI_ISL_1415388, EPI_ISL_1415389, EPI_ISL_1415390, EPI_ISL_1415391, EPI_ISL_1511198, EPI_ISL_1511199, EPI_ISL_1511200, EPI_ISL_1511201, EPI_ISL_1511202, EPI_ISL_1533729, EPI_ISL_1533730, EPI_ISL_1533731, EPI_ISL_1533732, EPI_ISL_1533733, EPI_ISL_1533734, EPI_ISL_1533735, EPI_ISL_1533736, EPI_ISL_1533737, EPI_ISL_1533738, EPI_ISL_1533739, EPI_ISL_1533740, EPI_ISL_1533741, EPI_ISL_1533742, EPI_ISL_1533743, EPI_ISL_1533744, EPI_ISL_1533745, EPI_ISL_1533746, EPI_ISL_1533747, EPI_ISL_1533748, EPI_ISL_1533749, EPI_ISL_1533750, EPI_ISL_1533751, EPI_ISL_1533752, EPI_ISL_1533753, EPI_ISL_1533754, EPI_ISL_1533755, EPI_ISL_1533756, EPI_ISL_1533757, EPI_ISL_1533758, EPI_ISL_1533759, EPI_ISL_1533760, EPI_ISL_1533761, EPI_ISL_1533762, EPI_ISL_1533763, EPI_ISL_1533764, EPI_ISL_1533765, EPI_ISL_1533766, EPI_ISL_1533767, EPI_ISL_1533768, EPI_ISL_1533769, EPI_ISL_1533770, EPI_ISL_1533771, EPI_ISL_1533772, EPI_ISL_1533773, EPI_ISL_1533774, EPI_ISL_1533775, EPI_ISL_1533776, EPI_ISL_1533777, EPI_ISL_1533778, EPI_ISL_1533779, EPI_ISL_1533780, EPI_ISL_1533781, EPI_ISL_1533782, EPI_ISL_1533783, EPI_ISL_1533784, EPI_ISL_1533785, EPI_ISL_1533786, EPI_ISL_1533787, EPI_ISL_1533788, EPI_ISL_1533789, EPI_ISL_1533790, EPI_ISL_1533791, EPI_ISL_1533792, EPI_ISL_1533793, EPI_ISL_1533794, EPI_ISL_1533795, EPI_ISL_1533796, EPI_ISL_1533797, EPI_ISL_1533798, EPI_ISL_1533799, EPI_ISL_1533800, EPI_ISL_1533801, EPI_ISL_1540451, EPI_ISL_1540452, EPI_ISL_1540453, EPI_ISL_1540454, EPI_ISL_1540455, EPI_ISL_1540456, EPI_ISL_1540457, EPI_ISL_1543988, EPI_ISL_1543989, EPI_ISL_1543990, EPI_ISL_1543991, EPI_ISL_1543992, EPI_ISL_1543993, EPI_ISL_1543994, EPI_ISL_1543995, EPI_ISL_1543996, EPI_ISL_1543997, EPI_ISL_1543998, EPI_ISL_1543999, EPI_ISL_1544000, EPI_ISL_1544001, EPI_ISL_1544002, EPI_ISL_1544003, EPI_ISL_1544004, EPI_ISL_1544005, EPI_ISL_1544006, EPI_ISL_1544007, EPI_ISL_1544008, EPI_ISL_1544009, EPI_ISL_1544010, EPI_ISL_1544011, EPI_ISL_1544012, EPI_ISL_1544013, EPI_ISL_1544014, EPI_ISL_1544015, EPI_ISL_1544016, EPI_ISL_1544017, EPI_ISL_1544018, EPI_ISL_1544019, EPI_ISL_1544020, EPI_ISL_1544021, EPI_ISL_1544022, EPI_ISL_1544023, EPI_ISL_1544024, EPI_ISL_1544025, EPI_ISL_1544026, EPI_ISL_1544027, EPI_ISL_1544028, EPI_ISL_1544029, EPI_ISL_1544030, EPI_ISL_1544031, EPI_ISL_1544032, EPI_ISL_1544033, EPI_ISL_1544034, EPI_ISL_1544035, EPI_ISL_1544036, EPI_ISL_1544037, EPI_ISL_1544038, EPI_ISL_1544039, EPI_ISL_1544040, EPI_ISL_1544041, EPI_ISL_1544042, EPI_ISL_1544043, EPI_ISL_1544044, EPI_ISL_1544045, EPI_ISL_1544046, EPI_ISL_1544047, EPI_ISL_1544048, EPI_ISL_1544049, EPI_ISL_1544050, EPI_ISL_1544051, EPI_ISL_1544052, EPI_ISL_1544053, EPI_ISL_1544054, EPI_ISL_1544055, EPI_ISL_1544056, EPI_ISL_1544057, EPI_ISL_1544058, EPI_ISL_1544059, EPI_ISL_1544060, EPI_ISL_1544061, EPI_ISL_1544062, EPI_ISL_1544063, EPI_ISL_1544064, EPI_ISL_1544065, EPI_ISL_1544066, EPI_ISL_1544067, EPI_ISL_1544068, EPI_ISL_1544069, EPI_ISL_1544070, EPI_ISL_1544071, EPI_ISL_1544072, EPI_ISL_1544073, EPI_ISL_1544074 | | | |
| see above | National Centre For Cell Science | National Centre For Cell Science – INSACOG | Ajay Pillai; Dhiraj Paul; INSACOG Consortium team; Manoj Kumar Bhat; Mitali Inamdar; Mohak P Gujare; Shivang P. Bhanushali; Sonal Manik Chavan; Yogesh Shouche; Yogesh Shouche. |
| EPI_ISL_1384859, EPI_ISL_1384863 | National Centre For Cell Science | National Institute of Virology, Pune | Ajay Pillai; Dhiraj Paul; Manoj Kumar Bhat; Mitali Inamdar; Mohak P Gujare; Shivang P. Bhanushali; Sonal Manik Chavan; Yogesh Shouche |
| EPI_ISL_2107008, EPI_ISL_2107009, EPI_ISL_2107010, EPI_ISL_2107011, EPI_ISL_2107012, EPI_ISL_2107013, EPI_ISL_2107014, EPI_ISL_2107015, EPI_ISL_2107016, EPI_ISL_2107017, EPI_ISL_2107018, EPI_ISL_2107019, EPI_ISL_2107020, EPI_ISL_2107021, EPI_ISL_2107022, EPI_ISL_2107023, EPI_ISL_2107024, EPI_ISL_2107025, EPI_ISL_2107026, EPI_ISL_2107027, EPI_ISL_2107028, EPI_ISL_2107029, EPI_ISL_2107030, EPI_ISL_2107031, EPI_ISL_2107032, EPI_ISL_2107033, EPI_ISL_2107034, EPI_ISL_2107035, EPI_ISL_2107036, EPI_ISL_2107037, EPI_ISL_2107038, EPI_ISL_2107039, EPI_ISL_2107040, EPI_ISL_2107041, EPI_ISL_2107042, EPI_ISL_2107043, EPI_ISL_2107044, EPI_ISL_2107045, EPI_ISL_2107046, EPI_ISL_2107047, EPI_ISL_2107048, EPI_ISL_2107049, EPI_ISL_2107050, EPI_ISL_2107051, EPI_ISL_2107052, EPI_ISL_2107053, EPI_ISL_2107054, EPI_ISL_2107055, EPI_ISL_2107056, EPI_ISL_2107057, EPI_ISL_2107058, EPI_ISL_2107059, EPI_ISL_2107060, EPI_ISL_2107061, EPI_ISL_2107062, EPI_ISL_2107063, EPI_ISL_2107064, EPI_ISL_2107065, EPI_ISL_2107066, EPI_ISL_2107067, EPI_ISL_2107068, EPI_ISL_2107069, EPI_ISL_2107070, EPI_ISL_2107071, EPI_ISL_2107072, EPI_ISL_2107073, EPI_ISL_2107074, EPI_ISL_2107075, EPI_ISL_2107076, EPI_ISL_2107077, EPI_ISL_2107078, EPI_ISL_2107079, EPI_ISL_2107080, EPI_ISL_2107081, EPI_ISL_2107082, EPI_ISL_2107083, EPI_ISL_2107084, EPI_ISL_2107085, EPI_ISL_2107086, EPI_ISL_2107087, EPI_ISL_2107088, EPI_ISL_2107089, EPI_ISL_2107090, EPI_ISL_2107091, EPI_ISL_2107092 | | | |
| see above | National Centre For Cell Science – INSACOG | National Centre For Cell Science | Ajay Pillai; Dhiraj Paul; INSACOG Consortium team; Manoj Kumar Bhat; Mitali Inamdar; Mohak P Gujare; Shivang P. Bhanushali; Yogesh Shouche |
| EPI_ISL_2099534, EPI_ISL_2099535, EPI_ISL_2099536, EPI_ISL_2099537, EPI_ISL_2099538, EPI_ISL_2099539, EPI_ISL_2099540, EPI_ISL_2099541, EPI_ISL_2099542, EPI_ISL_2099543, EPI_ISL_2099544, EPI_ISL_2099545, EPI_ISL_2099546, EPI_ISL_2099547, EPI_ISL_2099548, EPI_ISL_2099549, EPI_ISL_2099550, EPI_ISL_2099551, EPI_ISL_2099552, EPI_ISL_2099553, EPI_ISL_2099554, EPI_ISL_2099555, EPI_ISL_2099556, EPI_ISL_2099557, EPI_ISL_2099558, EPI_ISL_2099559, EPI_ISL_2099560, EPI_ISL_2099561, EPI_ISL_2099562, EPI_ISL_2099563, EPI_ISL_2099564, EPI_ISL_2099565, EPI_ISL_2099566, EPI_ISL_2099567, EPI_ISL_2099568, EPI_ISL_2099569, EPI_ISL_2099570, EPI_ISL_2099571, EPI_ISL_2099572, EPI_ISL_2099573, EPI_ISL_2099574, EPI_ISL_2099575, EPI_ISL_2099576, EPI_ISL_2099577, EPI_ISL_2099578, EPI_ISL_2099579, EPI_ISL_2099580, EPI_ISL_2099581, EPI_ISL_2099582, EPI_ISL_2099583, EPI_ISL_2099584, EPI_ISL_2099585, EPI_ISL_2099586, EPI_ISL_2099587, EPI_ISL_2099588, EPI_ISL_2099589, EPI_ISL_2099590, EPI_ISL_2099591, EPI_ISL_2099592, EPI_ISL_2099593, EPI_ISL_2099594, EPI_ISL_2099595, EPI_ISL_2099596, EPI_ISL_2099597, EPI_ISL_2099598, EPI_ISL_2099599, EPI_ISL_2099600, EPI_ISL_2099601, EPI_ISL_2099602, EPI_ISL_2099603, EPI_ISL_2099604, EPI_ISL_2099605, EPI_ISL_2099606, EPI_ISL_2099607, EPI_ISL_2099608, EPI_ISL_2099609, EPI_ISL_2099610, EPI_ISL_2099611, EPI_ISL_2099612, EPI_ISL_2099613, EPI_ISL_2099614, EPI_ISL_2099615, EPI_ISL_2099616, EPI_ISL_2099617, EPI_ISL_2099618, EPI_ISL_2099619, EPI_ISL_2099620, EPI_ISL_2099621, EPI_ISL_2099622, EPI_ISL_2099623, EPI_ISL_2099624, EPI_ISL_2099625, EPI_ISL_2099626, EPI_ISL_2099627, EPI_ISL_2099628, EPI_ISL_2099629, EPI_ISL_2099630, EPI_ISL_2099631, EPI_ISL_2099632, EPI_ISL_2099633, EPI_ISL_2099634, EPI_ISL_2099635, EPI_ISL_2099636, EPI_ISL_2099637, EPI_ISL_2099638, EPI_ISL_2099639, EPI_ISL_2099640, EPI_ISL_2099641, EPI_ISL_2099642, EPI_ISL_2099643, EPI_ISL_2099644, EPI_ISL_2099645, EPI_ISL_2099646, EPI_ISL_2099647, EPI_ISL_2099648, EPI_ISL_2099649, EPI_ISL_2099650, EPI_ISL_2099651, EPI_ISL_2099652, EPI_ISL_2099653, EPI_ISL_2099654, EPI_ISL_2099655, EPI_ISL_2099656, EPI_ISL_2099657, EPI_ISL_2099658, EPI_ISL_2099659, EPI_ISL_2099660, EPI_ISL_2099661, EPI_ISL_2099662, EPI_ISL_2099663, EPI_ISL_2099664, EPI_ISL_2099665, EPI_ISL_2099666, EPI_ISL_2099667, EPI_ISL_2099668, EPI_ISL_2099669, EPI_ISL_2099670, EPI_ISL_2099671, EPI_ISL_2099672, EPI_ISL_2099673, EPI_ISL_2099674, EPI_ISL_2099675, EPI_ISL_2099676, EPI_ISL_2099677, EPI_ISL_2099678, EPI_ISL_2099679, EPI_ISL_2099680, EPI_ISL_2099681, EPI_ISL_2099682, EPI_ISL_2099683, EPI_ISL_2099684, EPI_ISL_2099685, EPI_ISL_2099686, EPI_ISL_2099687, EPI_ISL_2099689, EPI_ISL_2099690, EPI_ISL_2099691, EPI_ISL_2099692, EPI_ISL_2099693, EPI_ISL_2099694, EPI_ISL_2099695, EPI_ISL_2099696, EPI_ISL_2099697, EPI_ISL_2099698, EPI_ISL_2099699, EPI_ISL_2099700, EPI_ISL_2099701, EPI_ISL_2099702, EPI_ISL_2099703, EPI_ISL_2099704, EPI_ISL_2099705, EPI_ISL_2099706, EPI_ISL_2099707, EPI_ISL_2099708, EPI_ISL_2099709, EPI_ISL_2099710, EPI_ISL_2099711, EPI_ISL_2099713, EPI_ISL_2099714, EPI_ISL_2099715, EPI_ISL_2099716, EPI_ISL_2099717, EPI_ISL_2099718, EPI_ISL_2099719, EPI_ISL_2099720, EPI_ISL_2099721, EPI_ISL_2099722, EPI_ISL_2099723, EPI_ISL_2099724, EPI_ISL_2099725, EPI_ISL_2099726, EPI_ISL_2099727, EPI_ISL_2099728, EPI_ISL_2099729, EPI_ISL_2099733, EPI_ISL_2099734, EPI_ISL_2099735, EPI_ISL_2099737, EPI_ISL_2099738, EPI_ISL_2099739, EPI_ISL_2099740, EPI_ISL_2099741, EPI_ISL_2099742, EPI_ISL_2099743, EPI_ISL_2099744, EPI_ISL_2099745, EPI_ISL_2099746, EPI_ISL_2099747, EPI_ISL_2099748, EPI_ISL_2099749, EPI_ISL_2099752, EPI_ISL_2099757, EPI_ISL_2099758, EPI_ISL_2099760, EPI_ISL_2099761, EPI_ISL_2099762, EPI_ISL_2099764, EPI_ISL_2099765, EPI_ISL_2099766, EPI_ISL_2099767, EPI_ISL_2099768 | | | |
| see above | National Centre for Disease Control | CDFD -INSACOG | To be addded later |
| EPI_ISL_803905, EPI_ISL_825152, EPI_ISL_845877, EPI_ISL_860191, EPI_ISL_860196, EPI_ISL_995707, EPI_ISL_995734, EPI_ISL_995737, EPI_ISL_995738 | | | |
| see above | National Institute of Mental Health and Neurosciences (NIMHANS) | Department of Neurovirology, National Institute of Mental Health and Neurosciences (NIMHANS) | Anita S Desai; Anson Kunjumon George; Chitra Pattabiraman; Darshan Sreenivas; Harsha.P.K; Nakka Vijay Kiran Reddy; Pramada Prasad; Risha Rasheed; V Ravi |
| EPI_ISL_1595856, EPI_ISL_1595889, EPI_ISL_1595917, EPI_ISL_1818590, EPI_ISL_1818596, EPI_ISL_1818597, EPI_ISL_1818598, EPI_ISL_1818599, EPI_ISL_1818600, EPI_ISL_1818601, EPI_ISL_1818602, EPI_ISL_1818607, EPI_ISL_1818608, EPI_ISL_1818629, EPI_ISL_1818639, EPI_ISL_1818640, EPI_ISL_1818641, EPI_ISL_1818642, EPI_ISL_1818643, EPI_ISL_1818644 | | | |
| see above | National Institute of Mental Health and Neurosciences (NIMHANS) | INSACOG-KA, NIMHANS | Ananthapadmanabha Kotambail; Anita S Desai; Anson Kunjumon George; Chetan G K; Chitra Pattabiraman; Darshan Sreenivas; Gautham Arunachal Udupi; Pramada Prasad; V Ravi |
| EPI_ISL_825149 | National Institute of Virology Bangalore Unit (NIV BU) | Department of Neurovirology, National Institute of Mental Health and Neurosciences (NIMHANS) | Anita S Desai; Chitra Pattabiraman; Pramada Prasad; V Ravi |
| EPI_ISL_745200 | Nizams Institute of Medical Sciences | CSIR-Centre for Cellular and Molecular Biology | Archana Bharadwaj Siva; B Himasri; Blessy B John; Divya Tej Sowpati; Dr.P.Shashikala Reddy; Dr.S.Pavani; Dr.Satyaprasad; Dr.V.Sudha Rani; Karthik Bharadwaj Tallapaka; Lamuk Zaveri; M Soujanya Reddy; Namami Gaur; Nikhil Hajirnis; Onkar Kulkarni; Payel Mukherjee; Pratheusa Maccha; Priya Singh; Purushotham Vodnala; Rakesh K Mishra; Sakshi Shambhavi; Shagufta Khan; Sofia Banu; Tulasi Nagabandi; Viswagithe S L |
| EPI_ISL_745194, EPI_ISL_745195, EPI_ISL_745196 | Osmania Medical College | CSIR-Centre for Cellular and Molecular Biology | Archana Bharadwaj Siva; B Himasri; Blessy B John; Divya Tej Sowpati; Dr.P.Shashikala Reddy; Dr.S.Pavani; Dr.Satyaprasad; Dr.V.Sudha Rani; Karthik Bharadwaj Tallapaka; Lamuk Zaveri; M Soujanya Reddy; Namami Gaur; Nikhil Hajirnis; Onkar Kulkarni; Payel Mukherjee; Pratheusa Maccha; Priya Singh; Purushotham Vodnala; Rakesh K Mishra; Sakshi Shambhavi; Shagufta Khan; Sofia Banu; Tulasi Nagabandi; Viswagithe S L |
| EPI_ISL_2151283, EPI_ISL_2151295 | PHC AGARA | inStem NCBS - INSACOG | Uma Ramakrishnan Dasaradhi Palakodeti Aswin SaiNarain |
| EPI_ISL_1663333, EPI_ISL_1663334, EPI_ISL_1663335, EPI_ISL_1663336, EPI_ISL_1663337 | PRMMCCH, Baripada | Institute of Life Sciences - INSACOG | Ajay Parida; Amol M. Kanampalliwar; Arup Ghosh; Atimukta Jha; INSACOG Consortium; Punit Prasad; Rajeeb Swain; Rupesh Dash; Safal Walia; Shifu Aggarwal; Sunil K. Raghav |
| EPI_ISL_2001153, EPI_ISL_2001154, EPI_ISL_2001155, EPI_ISL_2001156, EPI_ISL_2001157, EPI_ISL_2001158, EPI_ISL_2001159, EPI_ISL_2001160, EPI_ISL_2001161, EPI_ISL_2001162, EPI_ISL_2001163, EPI_ISL_2001164, EPI_ISL_2001165, EPI_ISL_2001166, EPI_ISL_2001167, EPI_ISL_2001168, EPI_ISL_2001169, EPI_ISL_2001170, EPI_ISL_2001171, EPI_ISL_2001172, EPI_ISL_2001173, EPI_ISL_2001174, EPI_ISL_2001175, EPI_ISL_2001176, EPI_ISL_2001177, EPI_ISL_2013598 | | | |
| see above | PanGenomics International Pvt. Ltd. | Gujarat Biotechnology Research Centre | Chaitanya Joshi; Dinesh Kumar; Dipali Dhawan; Janvi Raval; Madhvi Joshi; Nitesh Shah; Nitin Savaliya; Ramesh Pandit; Sonal Sharma; Twinkle Soni; Umang Mishra; Zarna Patel; Zuber Saiyed |
| EPI_ISL_1372009 | Patna Medical College, Patna | Institute of Life Sciences - INSACOG | Ajay Parida; Amol M. Kanampalliwar; Arup Ghosh; Atimukta Jha; INSACOG Consortium; Punit Prasad; Rajeeb Swain; Rupesh Dash; Safal Walia; Shifu Aggarwal; Sunil K. Raghav |
| EPI_ISL_1662292, EPI_ISL_1662293, EPI_ISL_1662294, EPI_ISL_1662295, EPI_ISL_1662296, EPI_ISL_1662297, EPI_ISL_1662298, EPI_ISL_1662299, EPI_ISL_1662300, EPI_ISL_1662301, EPI_ISL_1662302, EPI_ISL_1662303, EPI_ISL_1662304, EPI_ISL_1662305, EPI_ISL_1662306, EPI_ISL_1662307, EPI_ISL_1662308, EPI_ISL_1662309, EPI_ISL_1662310, EPI_ISL_1662311, EPI_ISL_1662312, EPI_ISL_1662313, EPI_ISL_1662314, EPI_ISL_1662315, EPI_ISL_1662316, EPI_ISL_1662317, EPI_ISL_1662318, EPI_ISL_1662319, EPI_ISL_1662320, EPI_ISL_1662321, EPI_ISL_1662322, EPI_ISL_1662323, EPI_ISL_1662324, EPI_ISL_1662325, EPI_ISL_1662326, EPI_ISL_1662327, EPI_ISL_1662328, EPI_ISL_1662329, EPI_ISL_1662330, EPI_ISL_1662331, EPI_ISL_1662332, EPI_ISL_1662333, EPI_ISL_1662334, EPI_ISL_1662335, EPI_ISL_1662336, EPI_ISL_1662337, EPI_ISL_1662338, EPI_ISL_1662339, EPI_ISL_1662340, EPI_ISL_1662341, EPI_ISL_1662342, EPI_ISL_1662343, EPI_ISL_1662344 | | | |
| see above | Pt. Jawahar Lal Nehru Memorial Medical College, Raipur, Chhattisgarh | Institute of Life Sciences - INSACOG | Ajay Parida; Amol M. Kanampalliwar; Arup Ghosh; Atimukta Jha; INSACOG Consortium; Punit Prasad; Rajeeb Swain; Rupesh Dash; Safal Walia; Shifu Aggarwal; Sunil K. Raghav |
| EPI_ISL_1939992, EPI_ISL_1939993, EPI_ISL_1939994, EPI_ISL_1939995, EPI_ISL_1939996, EPI_ISL_1939997, EPI_ISL_1939998, EPI_ISL_1940030, EPI_ISL_1940031, EPI_ISL_1940032, EPI_ISL_1940033, EPI_ISL_1940034, EPI_ISL_1940102, EPI_ISL_1940103, EPI_ISL_1940104, EPI_ISL_1940105, EPI_ISL_1940106, EPI_ISL_2015511, EPI_ISL_2015548, EPI_ISL_2015558, EPI_ISL_2015605, EPI_ISL_2015620, EPI_ISL_2015622, EPI_ISL_2015623, EPI_ISL_2015624, EPI_ISL_2015625, EPI_ISL_2015626, EPI_ISL_2015629, EPI_ISL_2015630, EPI_ISL_2015632, EPI_ISL_2015635, EPI_ISL_2015638, EPI_ISL_2151297, EPI_ISL_2151298, EPI_ISL_2151320, EPI_ISL_2151321, EPI_ISL_2151322, EPI_ISL_2151328, EPI_ISL_2151332, EPI_ISL_2151333 | | | |
| see above | RAILWAY | inStem NCBS - INSACOG | Uma Ramakrishnan Dasaradhi Palakodeti Aswin SaiNarain |
| EPI_ISL_1818591, EPI_ISL_1818592, EPI_ISL_1818593, EPI_ISL_1818594, EPI_ISL_1818606, EPI_ISL_1818619, EPI_ISL_1818620, EPI_ISL_1818621, EPI_ISL_1818623, EPI_ISL_1818645, EPI_ISL_1818646, EPI_ISL_1818647, EPI_ISL_1818648, EPI_ISL_1818649 | | | |
| see above | RAILWAY H | INSACOG-KA, NIMHANS | Ananthapadmanabha Kotambail; Anita S Desai; Anson Kunjumon George; Chetan G K; Chitra Pattabiraman; Darshan Sreenivas; Gautham Arunachal Udupi; Pramada Prasad; V Ravi |
| EPI_ISL_1940059, EPI_ISL_1940060, EPI_ISL_1940061, EPI_ISL_2015490, EPI_ISL_2015512, EPI_ISL_2015522, EPI_ISL_2015529, EPI_ISL_2015530, EPI_ISL_2015550, EPI_ISL_2015571, EPI_ISL_2015587, EPI_ISL_2015591, EPI_ISL_2015599, EPI_ISL_2015609 | | | |
| see above | RAILWAY H | inStem NCBS - INSACOG | Uma Ramakrishnan Dasaradhi Palakodeti Aswin SaiNarain |
| EPI_ISL_995191, EPI_ISL_995702, EPI_ISL_995703, EPI_ISL_995706, EPI_ISL_995722, EPI_ISL_995723, EPI_ISL_995749, EPI_ISL_995753 | | | |
| see above | RAILWAY HOSPITAL | Department of Neurovirology, National Institute of Mental Health and Neurosciences (NIMHANS) | Anita S Desai; Anson Kunjumon George; Chitra Pattabiraman; Darshan Sreenivas; Harsha.P.K; Nakka Vijay Kiran Reddy; Pramada Prasad; Risha Rasheed; V Ravi |
| EPI_ISL_2015515, EPI_ISL_2015518, EPI_ISL_2015615, EPI_ISL_2151245, EPI_ISL_2151248, EPI_ISL_2151251, EPI_ISL_2151252, EPI_ISL_2151259, EPI_ISL_2151260, EPI_ISL_2151261, EPI_ISL_2151262, EPI_ISL_2151263, EPI_ISL_2151272, EPI_ISL_2151273, EPI_ISL_2151281, EPI_ISL_2151282, EPI_ISL_2151285, EPI_ISL_2151286, EPI_ISL_2151287, EPI_ISL_2151296, EPI_ISL_2151299, EPI_ISL_2151301, EPI_ISL_2151315, EPI_ISL_2151316, EPI_ISL_2151317, EPI_ISL_2151318, EPI_ISL_2151319, EPI_ISL_2151326, EPI_ISL_2151327 | | | |
| see above | RAILWAY HOSPITAL | inStem NCBS - INSACOG | Uma Ramakrishnan Dasaradhi Palakodeti Aswin SaiNarain |
| EPI_ISL_1063807, EPI_ISL_1063809, EPI_ISL_1063840, EPI_ISL_1063841, EPI_ISL_1063842, EPI_ISL_1063843, EPI_ISL_1063844, EPI_ISL_1063845, EPI_ISL_1063846, EPI_ISL_1063847, EPI_ISL_1063848, EPI_ISL_1063849, EPI_ISL_1063850, EPI_ISL_1063851, EPI_ISL_1063897, EPI_ISL_1063898 | | | |

| | | | |
|---|---|---|---|
| see above | REGIONAL VRDL, ICMR-RMRC BBSR | Immunogenomics lab, Institute of Life Sciences, Bhubaneswar | Ajay Parida; Amol M. Kanampalliwar; Arup Ghosh; Atimukta Jha; INSACOG Consortium; Kirtal Hansdah; Punit Prasad; Rajeeb Swain; Rupesh Dash; Safal Walia; Shifu Aggarwal; Sunil K. Raghav |
| EPI_ISL_1662265, EPI_ISL_1662266, EPI_ISL_1662267, EPI_ISL_1662268, EPI_ISL_1662269, EPI_ISL_1663524, EPI_ISL_1663525, EPI_ISL_1663526, EPI_ISL_1663527, EPI_ISL_1663528, EPI_ISL_1663529, EPI_ISL_1663530, EPI_ISL_1663531, EPI_ISL_1663532, EPI_ISL_1663533, EPI_ISL_1663534, EPI_ISL_1663535 | | | |
| see above | REGIONAL VRDL, ICMR-RMRC BBSR | Institute of Life Sciences - INSACOG | Ajay Parida; Amol M. Kanampalliwar; Arup Ghosh; Atimukta Jha; INSACOG Consortium; Punit Prasad; Rajeeb Swain; Rupesh Dash; Safal Walia; Shifu Aggarwal; Sunil K. Raghav |
| EPI_ISL_2151264, EPI_ISL_2151265, EPI_ISL_2151266, EPI_ISL_2151314, EPI_ISL_2151325 | RH HOSAHALLI | inStem NCBS - INSACOG | Uma Ramakrishnan Dasaradhi Palakodeti Aswin SaiNarain |
| EPI_ISL_1663338, EPI_ISL_1663339, EPI_ISL_1663340, EPI_ISL_1663341, EPI_ISL_1663342, EPI_ISL_1663343, EPI_ISL_1663344, EPI_ISL_1663345, EPI_ISL_1663346, EPI_ISL_1663347, EPI_ISL_1663348, EPI_ISL_1663349, EPI_ISL_1663350, EPI_ISL_1663351, EPI_ISL_1663352, EPI_ISL_1663353 | | | |
| see above | RIMS, Ranchi | Institute of Life Sciences - INSACOG | Ajay Parida; Amol M. Kanampalliwar; Arup Ghosh; Atimukta Jha; INSACOG Consortium; Punit Prasad; Rajeeb Swain; Rupesh Dash; Safal Walia; Shifu Aggarwal; Sunil K. Raghav |
| EPI_ISL_1662260, EPI_ISL_1662261, EPI_ISL_1662262, EPI_ISL_1662263, EPI_ISL_1662264, EPI_ISL_1663358, EPI_ISL_1663359, EPI_ISL_1663360, EPI_ISL_1663361, EPI_ISL_1663362 | | | |
| see above | RMRI, Patna | Institute of Life Sciences - INSACOG | Ajay Parida; Amol M. Kanampalliwar; Arup Ghosh; Atimukta Jha; INSACOG Consortium; Punit Prasad; Rajeeb Swain; Rupesh Dash; Safal Walia; Shifu Aggarwal; Sunil K. Raghav |
| EPI_ISL_2015627 | Railway | inStem NCBS - INSACOG | Uma Ramakrishnan Dasaradhi Palakodeti Aswin SaiNarain |
| EPI_ISL_845885, EPI_ISL_845886, EPI_ISL_860190, EPI_ISL_860192, EPI_ISL_860198, EPI_ISL_995726, EPI_ISL_995727, EPI_ISL_995733, EPI_ISL_995742, EPI_ISL_995747 | | | |
| see above | Railway Hospital | Department of Neurovirology, National Institute of Mental Health and Neurosciences (NIMHANS) | Anita S Desai; Anson Kunjumon George; Chitra Pattabiraman; Darshan Sreenivas; Harsha.P.K; Nakka Vijay Kiran Reddy; Pramada Prasad; Risha Rasheed; V Ravi |
| EPI_ISL_745199 | Rangaraya Medical College | CSIR-Centre for Cellular and Molecular Biology | Archana Bharadwaj Siva; B Himasri; Blessy B John; Divya Tej Sowpati; Dr.P.Shashikala Reddy; Dr.S.Pavani; Dr.Satyaprasad; Dr.V.Sudha Rani; Karthik Bharadwaj Tallapaka; Lamuk Zaveri; M Soujanya Reddy; Namami Gaur; Nikhil Hajirnis; Onkar Kulkarni; Payel Mukherjee; Pratheusa Maccha; Priya Singh; Purushotham Vodnala; Rakesh K Mishra; Sakshi Shambhavi; Shagufta Khan; Sofia Banu; Tulasi Nagabandi; Viswagithe S L |
| EPI_ISL_1662275, EPI_ISL_1662276, EPI_ISL_1662422, EPI_ISL_1662423 | S.C.B. Medical College and Hospital, Cuttack | Institute of Life Sciences - INSACOG | Ajay Parida; Amol M. Kanampalliwar; Arup Ghosh; Atimukta Jha; INSACOG Consortium; Punit Prasad; Rajeeb Swain; Rupesh Dash; Safal Walia; Shifu Aggarwal; Sunil K. Raghav |
| EPI_ISL_1372008 | S.L.N MEDICAL COLLEGE & HOSPITAL, KORAPUT | Institute of Life Sciences - INSACOG | Ajay Parida; Amol M. Kanampalliwar; Arup Ghosh; Atimukta Jha; INSACOG Consortium; Punit Prasad; Rajeeb Swain; Rupesh Dash; Safal Walia; Shifu Aggarwal; Sunil K. Raghav |
| EPI_ISL_857395 | S.M.S.Medical College, Jaipur, Rajasthan | S.M.S.Medical College, Jaipur, Rajasthan | Dr. Bharti Malhotra; Dr. Swati Gautam |
| EPI_ISL_1708462 | S.Roghayya Quality manager Path Gene Healthcare Private Limited | inStem NCBS - INSACOG | Uma Ramakrishnan Dasaradhi Palakodeti Aswin SaiNarain |
| EPI_ISL_1939990, EPI_ISL_1939991, EPI_ISL_1940101, EPI_ISL_2015640 | SANJAY GANDHI | inStem NCBS - INSACOG | Uma Ramakrishnan Dasaradhi Palakodeti Aswin SaiNarain |
| EPI_ISL_2151246, EPI_ISL_2151258, EPI_ISL_2151280 | SANJAY GANDHI INSTITUTE OF TRAUMA AND ORTHOPAEDICS | inStem NCBS - INSACOG | Uma Ramakrishnan Dasaradhi Palakodeti Aswin SaiNarain |
| EPI_ISL_1844689 | SAROJ HOSPITAL | Biotechnology Division, NCDC | Hema Gogia; Hemlata Lall; Kalaiarasan Ponnusamy; Mahesh S Dhar; Manoj K Singh; Meena Datta; Partha Rakshit; Preeti Madan; Priyanka Singh; Radhakrishnan V. S; Robin Marwal; Sandhya Kabra; Sujeet K Singh; Uma Sharma |
| EPI_ISL_1662272, EPI_ISL_1662273, EPI_ISL_1662274 | SK Medical College, Muzaffarpur | Institute of Life Sciences - INSACOG | Ajay Parida; Amol M. Kanampalliwar; Arup Ghosh; Atimukta Jha; INSACOG Consortium; Punit Prasad; Rajeeb Swain; Rupesh Dash; Safal Walia; Shifu Aggarwal; Sunil K. Raghav |
| EPI_ISL_891152, EPI_ISL_891213, EPI_ISL_891214, EPI_ISL_891219 | SMS Medical College Jaipur | SMS Medical College Jaipur | Bharti Malhotra; Himanshu Sharma; Neha Bhomia; Nivedita Gupta; Pragya Yadav; Pratibha Sharma; Swati Gautam; Varsha Potdar |
| EPI_ISL_891207, EPI_ISL_891211 | SMS Medical College Jaipur | SMS Medical College jaipur | Bharti Malhotra; Himanshu Sharma; Neha Bhomia; Nivedita Gupta; Pragya Yadav; Pratibha Sharma; Swati Gautam; Varsha Potdar |
| EPI_ISL_891210 | SMS Medical College jaipur | SMS Medical College Jaipur | Bharti Malhotra; Himanshu Sharma; Neha Bhomia; Nivedita Gupta; Pragya Yadav; Pratibha Sharma; Swati Gautam; Varsha Potdar |
| EPI_ISL_1580503 | SMS Medical College, Jaipur | S.M.S. Medical College | ; Bharti Malhotra; Himanshu Sharma; Pratibha Mishra; Swati Gautam |
| EPI_ISL_2151267 | SRI JAYADEVA INSTITUTE OF CARDIOLOGY | inStem NCBS - INSACOG | Uma Ramakrishnan Dasaradhi Palakodeti Aswin SaiNarain |
| EPI_ISL_995758 | SRI JAYADEVA INSTITUTE OF CARDIOVASCULAR SCIENCES AND RESEARCH | Department of Neurovirology, National Institute of Mental Health and Neurosciences (NIMHANS) | Anita S Desai; Anson Kunjumon George; Chitra Pattabiraman; Darshan Sreenivas; Harsha.P.K; Nakka Vijay Kiran Reddy; Pramada Prasad; Risha Rasheed; V Ravi |
| EPI_ISL_1544142, EPI_ISL_1544143, EPI_ISL_1544144, EPI_ISL_1544145, EPI_ISL_1544146, EPI_ISL_1544147, EPI_ISL_1544148, EPI_ISL_1544149, EPI_ISL_1544150, EPI_ISL_1544151, EPI_ISL_1544152, EPI_ISL_1544153, EPI_ISL_1544154, EPI_ISL_1544155, EPI_ISL_1544156, EPI_ISL_1544157, EPI_ISL_1544158, EPI_ISL_1544159, EPI_ISL_1544160, EPI_ISL_1544161, EPI_ISL_1544162, EPI_ISL_1544163, EPI_ISL_1544164, EPI_ISL_1544165, EPI_ISL_1544170, EPI_ISL_1544171, EPI_ISL_1544172, EPI_ISL_1544173, EPI_ISL_1544174, EPI_ISL_1544175, EPI_ISL_1544176, EPI_ISL_1544177, EPI_ISL_1544178 | | | |
| see above | SSG Medical College, Baroda | Gujarat Biotechnology Research Centre | Chaitanya Joshi; Dinesh Kumar; Janvi Raval; Madhvi Joshi; Nitesh Shah; Nitin Savaliya; Ramesh Pandit; Sonal Sharma; Tanuja B. Javadekar; Twinkle Soni; Umang Mishra; Zarna Patel; Zuber Saiyed |
| EPI_ISL_1939973 | SUBRAMANYA NAGAR | inStem NCBS - INSACOG | Uma Ramakrishnan Dasaradhi Palakodeti Aswin SaiNarain |
| EPI_ISL_1939974 | SUBRAMNAYA NAGAR | inStem NCBS - INSACOG | Uma Ramakrishnan Dasaradhi Palakodeti Aswin SaiNarain |
| EPI_ISL_2151173, EPI_ISL_2151175, EPI_ISL_2151181, EPI_ISL_2151182, EPI_ISL_2151186, EPI_ISL_2151187, EPI_ISL_2151188, EPI_ISL_2151189, EPI_ISL_2151234, EPI_ISL_2151235 | | | |
| see above | Sahakara Nagar PHC | inStem NCBS - INSACOG | Uma Ramakrishnan Dasaradhi Palakodeti Aswin SaiNarain |
| EPI_ISL_825140 | Sanjay Gandhi Institute of Trauma & Orthopaedics | Department of Neurovirology, National Institute of Mental Health and Neurosciences (NIMHANS) | Anita S Desai; Chitra Pattabiraman; Pramada Prasad; V Ravi |
| EPI_ISL_995730 | Sanjay Gandhi Institute of Trauma and Orthopaedics | Department of Neurovirology, National Institute of Mental Health and Neurosciences (NIMHANS) | Anita S Desai; Anson Kunjumon George; Chitra Pattabiraman; Darshan Sreenivas; Harsha.P.K; Nakka Vijay Kiran Reddy; Pramada Prasad; Risha Rasheed; V Ravi |
| EPI_ISL_803903, EPI_ISL_803909, EPI_ISL_803913, EPI_ISL_803916, EPI_ISL_860187, EPI_ISL_860188 | Shimoga Institute of Medical Sciences | Department of Neurovirology, National Institute of Mental Health and Neurosciences (NIMHANS) | Anita S Desai; Anson Kunjumon George; Chitra Pattabiraman; Darshan Sreenivas; Nakka Vijay Kiran Reddy; Pramada Prasad; Risha Rasheed; V Ravi |
| EPI_ISL_1595864, EPI_ISL_1595913, EPI_ISL_1818650 | Sri Jayadeva Institute of Cardiovascular Sciences and Research | INSACOG-KA, NIMHANS | Ananthapadmanabha Kotambail; Anita S Desai; Anson Kunjumon George; Chetan G K; Chitra Pattabiraman; Darshan Sreenivas; Gautham Arunachal Udupi; Pramada Prasad; V Ravi |
| EPI_ISL_944749, EPI_ISL_944750, EPI_ISL_944751, EPI_ISL_944752, EPI_ISL_944753, EPI_ISL_944754, EPI_ISL_944755, EPI_ISL_1731751, EPI_ISL_1731752, EPI_ISL_1731753, EPI_ISL_1731754, EPI_ISL_1731755, EPI_ISL_1731756, EPI_ISL_1731757, EPI_ISL_1914579, EPI_ISL_1914580, EPI_ISL_1914581, EPI_ISL_1914582, EPI_ISL_1914583, EPI_ISL_1914584, EPI_ISL_1914585, EPI_ISL_1914586, EPI_ISL_1914587, EPI_ISL_1914588, EPI_ISL_1914589, EPI_ISL_1914590, EPI_ISL_1914591, EPI_ISL_1914592, EPI_ISL_1914593, EPI_ISL_1914594, EPI_ISL_1914595, EPI_ISL_1914596, EPI_ISL_1914597, EPI_ISL_1914598, EPI_ISL_1969752, EPI_ISL_1969753, EPI_ISL_2098711, EPI_ISL_2098712, EPI_ISL_2098713, EPI_ISL_2098714, EPI_ISL_2098715, EPI_ISL_2098716, EPI_ISL_2098717, EPI_ISL_2098718, EPI_ISL_2098719, EPI_ISL_2098720, EPI_ISL_2098721, EPI_ISL_2098722 | | | |
| see above | State Virus Research and Diagnostic Laboratory (VRDL), AIIMS Raipur | State Virus Research and Diagnostic Laboratory (VRDL), AIIMS Raipur | Anudita Bhargava; Jiyan Chandrawanshi; Kuldeep Sharma; MD Rafiiqullah Khan; Pragya Agarwala; Priyanka Singh; Pushpendra Singh; Sanjay Singh Negi; Somya Sharma |
| EPI_ISL_826275, EPI_ISL_826276 | Sterling Hospital, Memnagar, Ahmedabad | Gujarat Biotechnology Research Centre | Afzal Ansari; Apurvasinh Puvar; Atul K Patel; Chaitanya Joshi; Dinesh Kumar; Janvi Raval; Labdhi Pandya; Madhvi Joshi; Naman Shashtri; Nikha Trivedi; Nitin Savaliya; Ramesh Pandit; Zarna Patel; Zuber Saiyed |
| EPI_ISL_1544118, EPI_ISL_1544119, EPI_ISL_1544120, EPI_ISL_1544121, EPI_ISL_1544122, EPI_ISL_1544123, EPI_ISL_1544124, EPI_ISL_1544125, EPI_ISL_1544126, EPI_ISL_1544127, EPI_ISL_1544128, EPI_ISL_1544129, EPI_ISL_1544130, EPI_ISL_1544131, EPI_ISL_1544132, EPI_ISL_1544135, EPI_ISL_1544136, EPI_ISL_1544137, EPI_ISL_1544138, EPI_ISL_1544139, EPI_ISL_1544140, EPI_ISL_1544141, EPI_ISL_1544166, EPI_ISL_1544167, EPI_ISL_1544168, EPI_ISL_1544169 | | | |
| see above | Supratech Micropath Laboratory Research Institute Pvt Ltd, Ahmedabad | Gujarat Biotechnology Research Centre | Chaitanya Joshi; Dinesh Kumar; Janvi Raval; Madhvi Joshi; Mahendra Parikh; Nitesh Shah; Nitin Savaliya; Ramesh Pandit; Sonal Sharma; Twinkle Soni; Umang Mishra; Zarna Patel; Zuber Saiyed |
| EPI_ISL_1385836 | Syngene | INSACOG-KA, NIMHANS | Anita S Desai; Anson Kunjumon George; Chitra Pattabiraman; Darshan Sreenivas; Nakka Vijay Kiran Reddy; Pramada Prasad; V Ravi |
| EPI_ISL_2023383 | The Foundation for Medical Research | The Foundation for Medical Research | Ambreen Shaikh; Ayan Mandal; Grishma Patel; Kayzad Nilgiriwala; Nerges Mistry; Smriti Vaswani; Swapneil Parikh |
| EPI_ISL_1837988, EPI_ISL_1837991, EPI_ISL_1837993, EPI_ISL_1837996, EPI_ISL_1837998, EPI_ISL_1838001, EPI_ISL_1838004, EPI_ISL_1838007, EPI_ISL_1838010, EPI_ISL_1838012, EPI_ISL_1838015, EPI_ISL_1838018, EPI_ISL_1838021, EPI_ISL_1838023, EPI_ISL_1838026, EPI_ISL_1838029, EPI_ISL_1838032, EPI_ISL_1838035, EPI_ISL_1838037, EPI_ISL_1838040, EPI_ISL_1838042, EPI_ISL_1838045, EPI_ISL_1838047, EPI_ISL_1838050, EPI_ISL_1838052, EPI_ISL_1838055, EPI_ISL_1838057, EPI_ISL_1838060, EPI_ISL_1838063, EPI_ISL_1838065, EPI_ISL_1838068, EPI_ISL_1838070, EPI_ISL_1838073, EPI_ISL_1838075, EPI_ISL_1838078, EPI_ISL_1838080, EPI_ISL_1838083, EPI_ISL_1838086, EPI_ISL_1838088, EPI_ISL_1838091, EPI_ISL_1838094, EPI_ISL_1838097, EPI_ISL_1838100, EPI_ISL_1838102, EPI_ISL_1838106, EPI_ISL_1838109, EPI_ISL_1838111, EPI_ISL_1838114, EPI_ISL_1838117, EPI_ISL_1838119, EPI_ISL_1838122, EPI_ISL_1838123, EPI_ISL_1838125, EPI_ISL_1838126, EPI_ISL_1838128, EPI_ISL_1838129, EPI_ISL_1838131, EPI_ISL_1838132, EPI_ISL_1838134, EPI_ISL_1838136, EPI_ISL_1838139, EPI_ISL_1838140, EPI_ISL_1838142, EPI_ISL_1838143, EPI_ISL_1838145, EPI_ISL_1838148, EPI_ISL_1838149, EPI_ISL_1838151, EPI_ISL_1838152, EPI_ISL_1838153, EPI_ISL_1838155, EPI_ISL_1838156, EPI_ISL_1838158, EPI_ISL_1838159, EPI_ISL_1838161, EPI_ISL_1838162, EPI_ISL_1838164, EPI_ISL_1838165, EPI_ISL_1838166, EPI_ISL_1838168, EPI_ISL_1838169, EPI_ISL_1838171, EPI_ISL_1838172, EPI_ISL_1838173, EPI_ISL_1838175, EPI_ISL_1838176, EPI_ISL_1838178, EPI_ISL_1838179, EPI_ISL_1838182, EPI_ISL_1838185, EPI_ISL_1838186, EPI_ISL_1838188, EPI_ISL_1838189, EPI_ISL_1838191, EPI_ISL_1838194, EPI_ISL_1838195, EPI_ISL_1838196, EPI_ISL_1838198, EPI_ISL_1838200, EPI_ISL_1838201, EPI_ISL_1838203, EPI_ISL_1838205, EPI_ISL_1838207, EPI_ISL_1838209, EPI_ISL_1838210, EPI_ISL_1838212, EPI_ISL_1838214, EPI_ISL_1838215, EPI_ISL_1838217, EPI_ISL_1838219, EPI_ISL_1838220, EPI_ISL_1838222, EPI_ISL_1838224, EPI_ISL_1838225, EPI_ISL_1838227, EPI_ISL_1838229, EPI_ISL_1838230, EPI_ISL_1838232, EPI_ISL_1838234, EPI_ISL_1838235, EPI_ISL_1838237, EPI_ISL_1838239, EPI_ISL_1838241, EPI_ISL_1838242, EPI_ISL_1838244, EPI_ISL_1838246, EPI_ISL_1838248, EPI_ISL_1838250, EPI_ISL_1838251, EPI_ISL_1838253, EPI_ISL_1838254, EPI_ISL_1838256, EPI_ISL_1838257, EPI_ISL_1838259, EPI_ISL_1838260, EPI_ISL_1838262, EPI_ISL_1838264, EPI_ISL_1838265, EPI_ISL_1838267, EPI_ISL_1838269, EPI_ISL_1838271, EPI_ISL_1838272, EPI_ISL_1838274, EPI_ISL_1838275, EPI_ISL_1838277, EPI_ISL_1838278, EPI_ISL_1838280, EPI_ISL_1838282, EPI_ISL_1838284, EPI_ISL_1838285, EPI_ISL_1838287, EPI_ISL_1838288, EPI_ISL_1838290, EPI_ISL_1838292, EPI_ISL_1838293, EPI_ISL_1838295, EPI_ISL_1838296, EPI_ISL_1838298, EPI_ISL_1838299, EPI_ISL_1838301, EPI_ISL_1838303, EPI_ISL_1838304, EPI_ISL_1838306, EPI_ISL_1838307, EPI_ISL_1838309, EPI_ISL_1838311, EPI_ISL_1838312, EPI_ISL_1838314, EPI_ISL_1838315, EPI_ISL_1838317, EPI_ISL_1838318, EPI_ISL_1838320, EPI_ISL_1838322, EPI_ISL_1838323, EPI_ISL_1838325, EPI_ISL_1838327, EPI_ISL_1838328, EPI_ISL_1838330, EPI_ISL_1838332, EPI_ISL_1838333, EPI_ISL_1838335, EPI_ISL_1838337, EPI_ISL_1838338, EPI_ISL_1838340, EPI_ISL_1838342, EPI_ISL_1838343, EPI_ISL_1838345, EPI_ISL_1838346, EPI_ISL_1972346, EPI_ISL_1972348, EPI_ISL_1972350 | | | |
| see above | The National Centre for Cell Science | CSIR-Centre for Cellular and Molecular Biology-INSACOG | Ajay Pillai; Amareshwar Vodapalli; Ara Sreenivas; Archana Bharadwaj Siva; B Himasri; Blessy B John; Dhiraj Paul; Divya Tej Sowpati; INSACOG Consortium team; Karthik Bharadwaj Tallapaka; Lamuk Zaveri; Manoj Kumar Bhat; Mitali Inamdar; Mohak P Gujare; Onkar Kulkarni; Payel Mukherjee; Rakesh K Mishra; Sharath Chandra Thota; Shivang P. Bhanushali; Shreekant Verma; Sofia Banu; Sonal Manik Chavan; Tulasi Nagabandi; Valli Nagalakshmi Undamatla; Viswagithe S L; Yogesh Shouche |
| EPI_ISL_2033743, EPI_ISL_2033745, EPI_ISL_2033747 | Translational Health Science and Technology Institute -ESIC medical college and hospital, Faridabad | THSTI Bioassay laboratory | Akshay Kanakan; Anil Pandey; Guruprasad R Medigeshi; Heena Shaman; Janani Srinivasa Vasudevan; Jigme Wangchuk; Naseem Ahmed Khan; Neha Jha; Priyanka Mehta; Rajesh Pandey; Ranjeet Maurya |
| EPI_ISL_2151324 | UPHC PANTHARPALYA | inStem NCBS - INSACOG | Uma Ramakrishnan Dasaradhi Palakodeti Aswin SaiNarain |
| EPI_ISL_2020930, EPI_ISL_2020931, EPI_ISL_2020932, EPI_ISL_2020933, EPI_ISL_2020934, EPI_ISL_2020935, EPI_ISL_2020936, EPI_ISL_2020937, EPI_ISL_2020938, EPI_ISL_2020939, EPI_ISL_2020940, EPI_ISL_2020941, EPI_ISL_2020942, EPI_ISL_2020943, EPI_ISL_2020944, EPI_ISL_2020945, EPI_ISL_2020946, EPI_ISL_2020947, EPI_ISL_2020948, EPI_ISL_2020949, EPI_ISL_2020950, EPI_ISL_2020951, EPI_ISL_2020952, EPI_ISL_2020953, EPI_ISL_2020954, EPI_ISL_2020955, EPI_ISL_2020956, EPI_ISL_2020957, EPI_ISL_2020958, EPI_ISL_2020959, EPI_ISL_2020960 | | | |
| see above | Urban Public Health Centre (UPHC), SECTOR-2, Gandhinagar | Gujarat Biotechnology Research Centre | Amit Kanani; Amrutlal Patel; Bhargav Waghela; Bhumika Prajapati; Chaitanya Joshi; Dinesh Kumar; Janvi Raval; Kalpesh Goswami; Madhvi Joshi; Nilen Patel; Nitesh Shah; Nitin Savaliya; Ramesh Pandit; Sonal Sharma; Tejendra Purohit; Twinkle Soni; Umang Mishra; Zarna Patel; Zuber Saiyed |
| EPI_ISL_1844738, EPI_ISL_1844743, EPI_ISL_1844747, EPI_ISL_1844752, EPI_ISL_1844756, EPI_ISL_1844761 | VIRAL RESEARCH & DIAGNOSTIC LAB, GMC | Biotechnology Division, NCDC | Hema Gogia; Hemlata Lall; Kalaiarasan Ponnusamy; Mahesh S Dhar; Manoj K Singh; Meena Datta; Partha Rakshit; Preeti Madan; Priyanka Singh; Radhakrishnan V. S; Robin Marwal; Sandhya Kabra; Sujeet K Singh; Uma Sharma |
| EPI_ISL_2017746, EPI_ISL_2017747, | VRDL, Goverment Medical College (GMC) and | Gujarat Biotechnology Research Centre | Chaitanya Joshi; Dinesh Kumar; Janvi Raval; Madhvi Joshi; Nitesh Shah; Nitin Savaliya; Ramesh Pandit; Sonal Sharma; Twinkle Soni; Umang Mishra; Vicky Gandhi; Zarna Patel; Zuber Saiyed |

| | | | |
|---|---|---|---|
| EPI_ISL_2017748, EPI_ISL_2017749, EPI_ISL_2017750, EPI_ISL_2017751 | Hospital, Valsad | | |
| EPI_ISL_2001116, EPI_ISL_2001117, EPI_ISL_2001118, EPI_ISL_2001119, EPI_ISL_2001120, EPI_ISL_2001149, EPI_ISL_2001150, EPI_ISL_2001151, EPI_ISL_2001152 | | | |
| see above | VRDL, Goverment Medical College (GMC), Bhavnagar | Gujarat Biotechnology Research Centre | Chaitanya Joshi; Dinesh Kumar; Janvi Raval; Kairavi Jwalantbhai Desai; Madhvi Joshi; Nitesh Shah; Nitin Savaliya; Ramesh Pandit; Sonal Sharma; Twinkle Soni; Umang Mishra; Zarna Patel; Zuber Saiyed |
| EPI_ISL_2001187, EPI_ISL_2001188, EPI_ISL_2001189, EPI_ISL_2001190, EPI_ISL_2001191, EPI_ISL_2001192, EPI_ISL_2001193, EPI_ISL_2001194, EPI_ISL_2001195, EPI_ISL_2001196, EPI_ISL_2001197, EPI_ISL_2001198, EPI_ISL_2001199, EPI_ISL_2001200, EPI_ISL_2001201, EPI_ISL_2001202, EPI_ISL_2001203, EPI_ISL_2001204, EPI_ISL_2001205, EPI_ISL_2001206, EPI_ISL_2001207, EPI_ISL_2001208, EPI_ISL_2001209, EPI_ISL_2001210, EPI_ISL_2001211, EPI_ISL_2001212, EPI_ISL_2017728, EPI_ISL_2017729, EPI_ISL_2017730, EPI_ISL_2017731, EPI_ISL_2017732, EPI_ISL_2017733, EPI_ISL_2017734, EPI_ISL_2017735, EPI_ISL_2017736, EPI_ISL_2017737, EPI_ISL_2017738, EPI_ISL_2017739, EPI_ISL_2017740, EPI_ISL_2017741, EPI_ISL_2017742, EPI_ISL_2017743, EPI_ISL_2017744, EPI_ISL_2017745, EPI_ISL_2017763, EPI_ISL_2017764 | | | |
| see above | VRDL, Goverment Medical College (GMC), Surat | Gujarat Biotechnology Research Centre | Chaitanya Joshi; Dinesh Kumar; Janvi Raval; Madhvi Joshi; Neeta Khandelwal; Nitesh Shah; Nitin Savaliya; Ramesh Pandit; Sonal Sharma; Twinkle Soni; Umang Mishra; Zarna Patel; Zuber Saiyed |
| EPI_ISL_845882 | VRDL, Mysore Medical College | Department of Neurovirology, National Institute of Mental Health and Neurosciences (NIMHANS) | Anita S Desai; Chitra Pattabiraman; Darshan Sreenivas; Nakka Vijay Kiran Reddy; Pramada Prasad; Risha Rasheed; V Ravi |
| EPI_ISL_1662282, EPI_ISL_1662283, EPI_ISL_1662284, EPI_ISL_1662285, EPI_ISL_1662286, EPI_ISL_1663325, EPI_ISL_1663326, EPI_ISL_1663327, EPI_ISL_1663328, EPI_ISL_1663329, EPI_ISL_1663330, EPI_ISL_1663331, EPI_ISL_1663332, EPI_ISL_1663497, EPI_ISL_1663498, EPI_ISL_1663499, EPI_ISL_1663500, EPI_ISL_1663501, EPI_ISL_1663502, EPI_ISL_1663503 | | | |
| see above | Veer Surendra Sai Institute of Medical Sciences and Research, Burla, Sambalpur | Institute of Life Sciences - INSACOG | Ajay Parida; Amol M. Kanampalliwar; Arup Ghosh; Atimukta Jha; INSACOG Consortium; Punit Prasad; Rajeeb Swain; Rupesh Dash; Safal Walia; Shifu Aggarwal; Sunil K. Raghav |
| EPI_ISL_1844715, EPI_ISL_1844720, EPI_ISL_1844724, EPI_ISL_1844729, EPI_ISL_1844733 | Viral Testing Laboratory, GADVASU | Biotechnology Division, NCDC | Hema Gogia; Hemlata Lall; Kalaiarasan Ponnusamy; Mahesh S Dhar; Manoj K Singh; Meena Datta; Partha Rakshit; Preeti Madan; Priyanka Singh; Radhakrishnan V. S; Robin Marwal; Sandhya Kabra; Sujeet K Singh; Uma Sharma |
| EPI_ISL_1662241, EPI_ISL_1662242, EPI_ISL_1662243, EPI_ISL_1662244, EPI_ISL_1662245, EPI_ISL_1662246, EPI_ISL_1662247, EPI_ISL_1662248, EPI_ISL_1662249, EPI_ISL_1662250, EPI_ISL_1662251, EPI_ISL_1662252, EPI_ISL_1662253, EPI_ISL_1662254, EPI_ISL_1662255, EPI_ISL_1662256, EPI_ISL_1662257, EPI_ISL_1662258, EPI_ISL_1662259 | | | |
| see above | Virology Lab, Ispat General Hospital, Rourkela | Institute of Life Sciences - INSACOG | Ajay Parida; Amol M. Kanampalliwar; Arup Ghosh; Atimukta Jha; INSACOG Consortium; Punit Prasad; Rajeeb Swain; Rupesh Dash; Safal Walia; Shifu Aggarwal; Sunil K. Raghav |
| EPI_ISL_825145, EPI_ISL_825153, EPI_ISL_825154 | Virus Research and Diagnostic Laboratory (DH-VRDL) | Department of Neurovirology, National Institute of Mental Health and Neurosciences (NIMHANS) | Anita S Desai; Chitra Pattabiraman; Pramada Prasad; V Ravi |
| EPI_ISL_1595874, EPI_ISL_1595875, EPI_ISL_1595876, EPI_ISL_1595877, EPI_ISL_1595878, EPI_ISL_1595879, EPI_ISL_1595880, EPI_ISL_1595881, EPI_ISL_1595882, EPI_ISL_1595883, EPI_ISL_1595884, EPI_ISL_1595885, EPI_ISL_1595886, EPI_ISL_1595887, EPI_ISL_1595888, EPI_ISL_1595890 | | | |
| see above | WENLOCK HOSPITAL, MANGALORE | INSACOG-KA, NIMHANS | Ananthapadmanabha Kotambail; Anita S Desai; Anson Kunjumon George; Chetan G K; Chitra Pattabiraman; Darshan Sreenivas; Gautham Arunachal Udupi; Pramada Prasad; V Ravi |
| EPI_ISL_1708434 | Yelahanka APTS PHC | inStem NCBS - INSACOG | Uma Ramakrishnan Dasaradhi Palakodeti Aswin SaiNarain |
| EPI_ISL_1372007 | ZEUSNJOVE DIAGNOSTICS LLP, C/O, Kalinga Hospital,Chandrasekharpur,Bhubaneswar | Institute of Life Sciences - INSACOG | Ajay Parida; Amol M. Kanampalliwar; Arup Ghosh; Atimukta Jha; INSACOG Consortium; Punit Prasad; Rajeeb Swain; Rupesh Dash; Safal Walia; Shifu Aggarwal; Sunil K. Raghav |
| EPI_ISL_1063852, EPI_ISL_1063853, EPI_ISL_1063854 | inDNA Life Sciences Private Limited, Bhubaneswar | Immunogenomics lab, Institute of Life Sciences, Bhubaneswar | Ajay Parida; Amol M. Kanampalliwar; Arup Ghosh; Atimukta Jha; INSACOG Consortium; Kirtal Hansdah; Punit Prasad; Rajeeb Swain; Rupesh Dash; Safal Walia; Shifu Aggarwal; Sunil K. Raghav |

We gratefully acknowledge the following Authors from the Originating laboratories responsible for obtaining the specimens, as well as the Submitting laboratories where the genome data were generated and shared via GISAID, on which this research is based.

All Submitters of data may be contacted directly via www.gisaid.org

Authors are sorted alphabetically.

| Accession ID | Originating Laboratory | Submitting Laboratory | Authors |
|---|---|---|---|
| EPI_ISL_1005695 | RSUD Dr. Soedirman | Genetics Working Group (Pokja Genetik) Faculty of Medicine, Public Health and Nursing Universitas Gadjah Mada (FK-KMK UGM); Disease Investigation Center Wates Ministry of Agriculture Indonesia; Department of Microbiology FK-KMK UGM; Laboratorium Diagnostik Yayasan Tahija World Mosquito Program (WMP) Yogyakarta Center for Tropical Medicine FK-KMK UGM; Integrated Research Center FK-KMK UGM; Department of Computer Science and Electronics FMIPA UGM; Balai Besar Teknik Kesehatan Lingkungan dan Pengendalian Penyakit (BBTKLPP) Yogyakarta | Gunadi, Hendra Wibawa, Marcellus, Mohamad S. Hakim, Edwin W. Daniwijaya, Ludhang P. Rizki, Endah Supriyati, Eggi Arguni, Titik Nuryastuti, Tri Wibawa, Dwi AA Nugrahaningsih, Afiahayati, Siswanto, Kristy Iskandar, Nungki Anggorowati, Irene, Indaryati, Havid Setyawan, Arry Setyowati, Chusni Mubarakh, Try Nirmala Sari, Audric Kenny Tedja, Maria Patricia Inggriani |
| EPI_ISL_1005696 | RSUD Dr. Tjitrowardojo | Genetics Working Group (Pokja Genetik) Faculty of Medicine, Public Health and Nursing Universitas Gadjah Mada (FK-KMK UGM); Disease Investigation Center Wates Ministry of Agriculture Indonesia; Department of Microbiology FK-KMK UGM; Laboratorium Diagnostik Yayasan Tahija World Mosquito Program (WMP) Yogyakarta Center for Tropical Medicine FK-KMK UGM; Integrated Research Center FK-KMK UGM; Department of Computer Science and Electronics FMIPA UGM; Balai Besar Teknik Kesehatan Lingkungan dan Pengendalian Penyakit (BBTKLPP) Yogyakarta | Gunadi, Hendra Wibawa, Marcellus, Mohamad S. Hakim, Edwin W. Daniwijaya, Ludhang P. Rizki, Endah Supriyati, Eggi Arguni, Titik Nuryastuti, Tri Wibawa, Dwi AA Nugrahaningsih, Afiahayati, Siswanto, Kristy Iskandar, Nungki Anggorowati, Irene, Indaryati, Havid Setyawan, Arry Setyowati, Chusni Mubarakh, Try Nirmala Sari, Susan Simanjaya, Alvin Santoso Kalim |
| EPI_ISL_1005697 | RSUD Dr. Tjitrowardojo | Genetics Working Group (Pokja Genetik) Faculty of Medicine, Public Health and Nursing Universitas Gadjah Mada (FK-KMK UGM); Disease Investigation Center Wates Ministry of Agriculture Indonesia; Department of Microbiology FK-KMK UGM; Laboratorium Diagnostik Yayasan Tahija World Mosquito Program (WMP) Yogyakarta Center for Tropical Medicine FK-KMK UGM; Integrated Research Center FK-KMK UGM; Department of Computer Science and Electronics FMIPA UGM; Balai Besar Teknik Kesehatan Lingkungan dan Pengendalian Penyakit (BBTKLPP) Yogyakarta | Gunadi, Hendra Wibawa, Marcellus, Mohamad S. Hakim, Edwin W. Daniwijaya, Ludhang P. Rizki, Endah Supriyati, Eggi Arguni, Titik Nuryastuti, Tri Wibawa, Dwi AA Nugrahaningsih, Afiahayati, Siswanto, Kristy Iskandar, Nungki Anggorowati, Irene, Indaryati, Havid Setyawan, Arry Setyowati, Chusni Mubarakh, Try Nirmala Sari, Kemala Athollah, Dyah Ayu Puspitarani |
| EPI_ISL_1005698 | RSUD Dr. Tjitrowardojo | Genetics Working Group (Pokja Genetik) Faculty of | Gunadi, Hendra Wibawa, Marcellus, Mohamad S. Hakim, Edwin W. Daniwijaya, Ludhang P. Rizki, Endah Supriyati, Eggi Arguni, Titik Nuryastuti, Tri Wibawa, Dwi AA Nugrahaningsih, Afiahayati, Siswanto, Kristy Iskandar, Nungki Anggorowati, Irene, Indaryati, Havid Setyawan, Arry Setyowati, Chusni Mubarakh, Try Nirmala Sari, Aditya Rifqi Fauzi, Fadil Fahri |

| | | | |
|---|---|---|---|
| | | Medicine, Public Health and Nursing Universitas Gadjah Mada (FK-KMK UGM); Disease Investigation Center Wates Ministry of Agriculture Indonesia; Department of Microbiology FK-KMK UGM; Laboratorium Diagnostik Yayasan Tahija World Mosquito Program (WMP) Yogyakarta Center for Tropical Medicine FK-KMK UGM; Integrated Research Center FK-KMK UGM; Department of Computer Science and Electronics FMIPA UGM; Balai Besar Teknik Kesehatan Lingkungan dan Pengendalian Penyakit (BBTKLPP) Yogyakarta | |
| EPI_ISL_1020186 | BSL3 Laboratory, Bogor, West Java | Biosafety Level-3 Laboratory, Indonesian Institute of Sciences (LIPI) | Anggia Prasetyoputri, Isa Nuryana, Sri Swasthikawati, Sugiyono Saputra, Syam Budi Iryanto, Gita Syahputra, Ratih Asmana Ningrum |
| EPI_ISL_1020192 | Melania Hospital, Bagor , west Java | Biosafety Level-3 Laboratory, Indonesian Institute of Sciences (LIPI) | Anik Budhi Dharmayanthi, Ade Andriani, Hariyatun, Asep M. Ridwanuloh, Syam Budi Iryanto, Eko Wahyu Putro, Gita Syahputra, Susanti, Inswasti Cahyani, Matthew W. Loose, Mohammad Ilyas, Wien Kusharyoto |
| EPI_ISL_1020193 | RS PMI, Bogor, West Java | Biosafety Level-3 Laboratory, Indonesian Institute of Sciences (LIPI) | Anggia Prasetyoputri, Isa Nuryana, Sri Swasthikawati, Sugiyono Saputra, Syam Budi Iryanto, Eko Wahyu Putro, Fahrurrozi, Ratih Asmana Ningrum |
| EPI_ISL_1020194 | UMMI Hospital, Bogor, West Java | Biosafety Level-3 Laboratory, Indonesian Institute of Sciences (LIPI) | Anggia Prasetyoputri, Isa Nuryana, Sri Swasthikawati, Sugiyono Saputra, Syam Budi Iryanto, Gita Syahputra, Ratih Asmana Ningrum |
| EPI_ISL_1020195 | Melania Hospital, Bogor , West Java | Biosafety Level-3 Laboratory, Indonesian Institute of Sciences (LIPI) | Anggia Prasetyoputri, Erwin Hasrianda, Herjuno A. Nugroho, Nova Dhilla Yanthi, Fahrurrozi, Syam Budi Iryanto, Gita Syahputra, Ratih Asmana Ningrum |
| EPI_ISL_1020196 | RS PMI, Bogor, West Java | Biosafety Level-3 Laboratory, Indonesian Institute of Sciences (LIPI) | Anggia Prasetyoputri, Isa Nuryana, Sri Swasthikawati, Sugiyono Saputra, Syam Budi Iryanto, Eko Wahyu Putro, Ratih Asmana Ningrum |
| EPI_ISL_1020197 | Dinkes, Bogor, West Java | Biosafety Level-3 Laboratory, Indonesian Institute of Sciences (LIPI) | Anik Budhi Dharmayanthi, Ade Andriani, Hariyatun, Asep M. Ridwanuloh, Syam Budi Iryanto, Eko Wahyu Putro, Gita Syahputra, Susanti, Inswasti Cahyani, Matthew W. Loose, Mohammad Ilyas, Wien Kusharyoto |
| EPI_ISL_1020203 | Klinik Apotik Dien, Jakarta | Biosafety Level-3 Laboratory, Indonesian Institute of Sciences (LIPI) | Anik Budhi Dharmayanthi, Ade Andriani, Hariyatun, Asep M. Ridwanuloh, Syam Budi Iryanto, Eko Wahyu Putro, Gita Syahputra, Susanti, Inswasti Cahyani, Matthew W. Loose, Mohammad Ilyas, Wien Kusharyoto |
| EPI_ISL_1020204 | DD Klinik Ciputat, South Tangerang, Banten | Biosafety Level-3 Laboratory, Indonesian Institute of Sciences (LIPI) | Anggia Prasetyoputri, Isa Nuryana, Sri Swasthikawati, Sugiyono Saputra, Syam Budi Iryanto, Eko Wahyu Putro, Fahrurrozi, Ratih Asmana Ningrum |
| EPI_ISL_1020205 | DMC Dompet Dhuafa, South Tangerang, Banten | Biosafety Level-3 Laboratory, Indonesian Institute of Sciences (LIPI) | Anggia Prasetyoputri, Isa Nuryana, Sri Swasthikawati, Sugiyono Saputra, Syam Budi Iryanto, Anik B. Dharmayanthi, Nuruliawaty Utami, Ratih Asmana Ningrum |
| EPI_ISL_1020206 | Qadr Hospital, Tangerang, Banten | Biosafety Level-3 Laboratory, Indonesian Institute of Sciences (LIPI) | Anggia Prasetyoputri, Isa Nuryana, Sri Swasthikawati, Sugiyono Saputra, Syam Budi Iryanto, Eko Wahyu Putro, Gita Syahputra, Ari Sulistyo Wulandari, Ratih Asmana Ningrum |
| EPI_ISL_1041030 | Dr. Cipto Mangunkusumo (RSCM) Hospital | Faculty of Medicine, Universitas Indonesia | Rina La Distia Nora, Mei Riasanti, Rani Dwi Ningtias, Ari Fahrial Syam |
| EPI_ISL_1048372 | Farmalab Indoutama Jakarta | National Institute of Health Research and Development | Hana Apsari Pawestri, Subangkit, Kartika Dewi Puspa, Arie Ardiansyah Nugraha, Hartanti Dian Ikawati, Krisna Nur Andriana Pangesti, Nelly Puspandari, Vivi Setiawaty. |
| EPI_ISL_1048375 | Farmalab Indoutama Jakarta | National Institute of Health Research and Development | Hana Apsari Pawestri, Subangkit, Kartika Dewi Puspa, Arie Ardiansyah Nugraha, Hartanti Dian Ikawati, Krisna Nur Andriana Pangesti, Nelly Puspandari, Vivi Setiawaty. |
| EPI_ISL_1048406 | Balai Penelitian dan Pengembangan Biomedis Papua | National Institute of Health Research and Development | Hana Apsari Pawestri, Subangkit, Kartika Dewi Puspa, Arie Ardiansyah Nugraha, Hartanti Dian Ikawati, Krisna Nur Andriana Pangesti, Nelly Puspandari, Anton Oktavianus, Hotma Sitompul, Vivi Setiawaty. |
| EPI_ISL_1069153 | Binjai Health Office | Institute of Tropical Disease, Universitas Airlangga; Fakultas Kedokteran, Universitas Sumatra Utara | Krisnoadi Rahardjo, Aldise M Nastri, Jezzy R Dewantari, Rima R Prasetya, Inke N D Lubis, R Lia Kusumawati, Muhammad Ichwan, Franciscus Ginting, Meliani, Ramadhan Bestari, Mirzan Hasibuan, Irbah R Nainggolan, R Andika D Cahyadi, Yasuko Mori, Soetjipto, Kazufumi Shimizu, Maria I Lusida |
| EPI_ISL_1072962 | Kota Nopan Primary Health Centre, Mandailing Natal District | Institute of Tropical Disease, Universitas Airlangga; Faculty of Medicine, Universitas Sumatera Utara | Aldise M Nastri, Jezzy R Dewantari, Rima R Prasetya, Krisnoadi Rahardjo, Inke N D Lubis, R Lia Kusumawati, Muhammad Ichwan, Franciscus Ginting, Meliani, Ramadhan Bestari, Mirzan Hasibuan, Irbah R Nainggolan, R Andika D Cahyadi, Yasuko Mori, Soetjipto, Kazufumi Shimizu, Maria I Lusida |
| EPI_ISL_1072963 | North Sumatera Health Office | Institute of Tropical Disease, Universitas | Jezzy R Dewantari, Rima R Prasetya, Krisnoadi Rahardjo, Aldise M Nastri, Inke N D Lubis, R Lia Kusumawati, Muhammad Ichwan, Franciscus Ginting, Meliani, Ramadhan Bestari, Mirzan Hasibuan, Irbah R Nainggolan, R Andika D Cahyadi, Yasuko Mori, Soetjipto, Kazufumi Shimizu, Maria I Lusida |

| | | | |
|---|---|---|---|
| | | Airlangga; Faculty of Medicine, Universitas Sumatera Utara | |
| EPI_ISL_1072964 | Bhayangkara Hospital Medan | Institute of Tropical Disease, Universitas Airlangga; Faculty of Medicine, Universitas Sumatera Utara | Rima R Prasetya, Krisnoadi Rahardjo, Aldise M Nastri, Jezzy R Dewantari, Inke N D Lubis, R Lia Kusumawati, Muhammad Ichwan, Franciscus Ginting, Meliani, Ramadhan Bestari, Mirzan Hasibuan, Irbah R Nainggolan, R Andika D Cahyadi, Yasuko Mori, Soetjipto, Kazufumi Shimizu, Maria I Lusida |
| EPI_ISL_1072965 | Department of Microbiology, Faculty of Medicine, Universitas Sumatera Utara | Institute of Tropical Disease, Universitas Airlangga; Faculty of Medicine, Universitas Sumatera Utara | Kazufumi Shimizu, Krisnoadi Rahardjo, Aldise M Nastri, Jezzy R Dewantari, Rima R Prasetya, Inke N D Lubis, R Lia Kusumawati, Muhammad Ichwan, Franciscus Ginting, Meliani, Ramadhan Bestari, Mirzan Hasibuan, Irbah R Nainggolan, R Andika D Cahyadi, Yasuko Mori, Soetjipto, Maria I Lusida |
| EPI_ISL_1072986 | Department of Microbiology, Faculty of Medicine, Universitas Sumatera Utara | Institute of Tropical Disease, Universitas Airlangga; Faculty of Medicine, Universitas Sumatera Utara | Krisnoadi Rahardjo, Aldise M Nastri, Jezzy R Dewantari, Rima R Prasetya, Inke N D Lubis, R Lia Kusumawati, Muhammad Ichwan, Franciscus Ginting, Meliani, Ramadhan Bestari, Mirzan Hasibuan, Irbah R Nainggolan, R Andika D Cahyadi, Yasuko Mori, Soetjipto, Kazufumi Shimizu, Maria I Lusida |
| EPI_ISL_1117450 | Melania Hospital, Bagor , west Java | Biosafety Level-3 Laboratory, Indonesian Institute of Sciences (LIPI) | Anggia Prasetyoputri, Isa Nuryana, Sri Swasthikawati, Sugiyono Saputra, Syam Budi Iryanto, Eko Wahyu Putro, Gita Syahputra, Ari Sulistyo Wulandari, Ratih Asmana Ningrum |
| EPI_ISL_1117451 | RS PMI, Bogor, West Java | Biosafety Level-3 Laboratory, Indonesian Institute of Sciences (LIPI) | Anik Budhi Dharmayanthi, Ade Andriani, Hariyatun, Asep M. Ridwanuloh, Syam Budi Iryanto, Eko Wahyu Putro, Gita Syahputra, Susanti, Inswasti Cahyani, Matthew W. Loose, Mohammad Ilyas, Wien Kusharyoto |
| EPI_ISL_1117456 | Murni Teguh Hospital, Jakarta | Biosafety Level-3 Laboratory, Indonesian Institute of Sciences (LIPI) | Anik Budhi Dharmayanthi, Ade Andriani, Hariyatun, Asep M. Ridwanuloh, Syam Budi Iryanto, Eko Wahyu Putro, Gita Syahputra, Susanti, Inswasti Cahyani, Matthew W. Loose, Mohammad Ilyas, Wien Kusharyoto |
| EPI_ISL_1117457 | Klinik Apotik Dien, Jakarta | Biosafety Level-3 Laboratory, Indonesian Institute of Sciences (LIPI) | Anik Budhi Dharmayanthi, Ade Andriani, Hariyatun, Asep M. Ridwanuloh, Syam Budi Iryanto, Eko Wahyu Putro, Gita Syahputra, Susanti, Inswasti Cahyani, Matthew W. Loose, Mohammad Ilyas, Wien Kusharyoto |
| EPI_ISL_1117589 | RS PMI, Bogor, West Java | Biosafety Level-3 Laboratory, Indonesian Institute of Sciences (LIPI) | Anggia Prasetyoputri, Isa Nuryana, Sri Swasthikawati, Sugiyono Saputra, Syam Budi Iryanto, Gita Syahputra, Ratih Asmana Ningrum |
| EPI_ISL_1117590 | RS Trimitra, Bogor, West java | Biosafety Level-3 Laboratory, Indonesian Institute of Sciences (LIPI) | Anik Budhi Dharmayanthi, Ade Andriani, Hariyatun, Asep M. Ridwanuloh, Syam Budi Iryanto, Eko Wahyu Putro, Gita Syahputra, Susanti, Inswasti Cahyani, Matthew W. Loose, Mohammad Ilyas, Wien Kusharyoto |
| EPI_ISL_1117591 | Dinkes, Bogor, West Java | Biosafety Level-3 Laboratory, Indonesian Institute of Sciences (LIPI) | Anik Budhi Dharmayanthi, Ade Andriani, Hariyatun, Asep M. Ridwanuloh, Syam Budi Iryanto, Eko Wahyu Putro, Gita Syahputra, Susanti, Inswasti Cahyani, Matthew W. Loose, Mohammad Ilyas, Wien Kusharyoto |
| EPI_ISL_1117592 | Melania Hospital, Bagor , West Java | Biosafety Level-3 Laboratory, Indonesian Institute of Sciences (LIPI) | Anik Budhi Dharmayanthi, Ade Andriani, Hariyatun, Asep M. Ridwanuloh, Syam Budi Iryanto, Eko Wahyu Putro, Gita Syahputra, Susanti, Inswasti Cahyani, Matthew W. Loose, Mohammad Ilyas, Wien Kusharyoto |
| EPI_ISL_1118206 | National Institute of Health Research and Development | National Institute of Health Research and Development | Hana Apsari Pawestri, Subangkit, Kartika Dewi Puspa, Arie Ardiansyah Nugraha, Hartanti Dian Ikawati, Krisna Nur Andriana Pangesti, Yuni Rukminiati, Ririn Ramadhany, Agustiningsih, Kindi Adam, Holy Arif Wibowo, Triyani Soekarso, Ni Ketut Susilarini, Nurika Hariastuti, Ully Alfi Nikmah, Reni Herman, Nike Susanti, Herna, Tati Febriyanti, Natalie Laurencia Kipuw, Fauzul Muna, Irene Lorinda Indalao, Nelly Puspandari, Vivi Setiawaty. |
| EPI_ISL_1118280 | Merial Health Hospital, Jakarta | Biosafety Level-3 Laboratory, Indonesian Institute of Sciences (LIPI) | Anik Budhi Dharmayanthi, Ade Andriani, Hariyatun, Asep M. Ridwanuloh, Syam Budi Iryanto, Eko Wahyu Putro, Gita Syahputra, Susanti, Inswasti Cahyani, Matthew W. Loose, Mohammad Ilyas, Wien Kusharyoto |
| EPI_ISL_1122424 | Clinical Microbiology Laboratory, Faculty of Medicine, Universitas Indonesia | Clinical Microbiology Laboratory, Faculty of Medicine, Universitas Indonesia | Fera Ibrahim, Augustine Natasha, Andi Yasmon, Fithriyah , Yulia Rosa Saharman, Erni J. Nelwan, Anis Karuniawati, Pratiwi Sudarmono |
| EPI_ISL_1137600 | BSL3 Laboratory, Bogor, West Java | Biosafety Level-3 Laboratory, Indonesian Institute of Sciences (LIPI) | Anggia Prasetyoputri, Isa Nuryana, Sri Swasthikawati, Sugiyono Saputra, Syam Budi Iryanto, Eko Wahyu Putro, Ratih Asmana Ningrum |
| EPI_ISL_1137601 | RS PMI, Bogor, West Java | Biosafety Level-3 Laboratory, Indonesian Institute of Sciences (LIPI) | Anik Budhi Dharmayanthi, Ade Andriani, Hariyatun, Asep M. Ridwanuloh, Syam Budi Iryanto, Eko Wahyu Putro, Gita Syahputra, Susanti, Inswasti Cahyani, Matthew W. Loose, Mohammad Ilyas, Wien Kusharyoto |
| EPI_ISL_1159381 | Surabaya Public Health Office | Institute of Tropical Disease, Universitas Airlangga | Aldise M Nastri, Jezzy R Dewantari, Rima R Prasetya, Krisnoadi Rahardjo, Febria Rachmanita, Gatot Soegiarto, Laksmi Wulandari, Resti Yudhawati, Yasuko Mori, Soetjipto, Kazufumi Shimizu, Maria I Lusida |
| EPI_ISL_1159382 | Adi Husada Kapasari Hospital | Institute of Tropical Disease, Universitas Airlangga | Maria I Lusida, Krisnoadi Rahardjo, Aldise M Nastri, Jezzy R Dewantari, Rima R Prasetya, Hermanto Wijaya, Gatot Soegiarto, Laksmi Wulandari, Resti Yudhawati, Soetjipto, Yasuko Mori, Kazufumi Shimizu |
| EPI_ISL_1159383 | Siti Khodijah Hospital | Institute of Tropical Disease, Universitas Airlangga | Aldise M Nastri, Jezzy R Dewantari, Rima R Prasetya, Krisnoadi Rahardjo, Muhammad Hamdan, Gatot Soegiarto, Laksmi Wulandari, Resti Yudhawati, Soetjipto, Yasuko Mori, Maria I Lusida, Kazufumi Shimizu |
| EPI_ISL_1159384 | Institute of Tropical Disease | Institute of Tropical Disease, Universitas Airlangga | Jezzy R Dewantari, Rima R Prasetya, Krisnoadi Rahardjo, Aldise M Nastri, Gatot Soegiarto, Laksmi Wulandari, Resti Yudhawati, Soetjipto, Yasuko Mori, Maria I Lusida, Kazufumi Shimizu |
| EPI_ISL_1159385 | Graha Medika Maternity Hospital | Institute of Tropical Disease, Universitas Airlangga | Rima R Prasetya, Krisnoadi Rahardjo, Aldise M Nastri, Jezzy R Dewantari, Diah R Kusumawati, Gatot Soegiarto, Laksmi Wulandari, Resti Yudhawati, Soetjipto, Yasuko Mori, Maria I Lusida, Kazufumi Shimizu |
| EPI_ISL_1159386 | Siti Khodijah Hospital | Institute of Tropical Disease, Universitas Airlangga | Krisnoadi Rahardjo, Aldise M Nastri, Jezzy R Dewantari, Rima R Prasetya, Muhammad Hamdan, Gatot Soegiarto, Laksmi Wulandari, Resti Yudhawati, Yasuko Mori, Soetjipto, Kazufumi Shimizu, Maria I Lusida |

| | | | |
|---|---|---|---|
| EPI_ISL_1159387 | Adi Husada Kapasari Hospital | Institute of Tropical Disease, Universitas Airlangga | Maria I Lusida, Krisnoadi Rahardjo, Aldise M Nastri, Jezzy R Dewantari, Rima R Prasetya, Hermanto Wijaya, Gatot Soegiarto, Laksmi Wulandari, Resti Yudhawati, Soetjipto, Yasuko Mori, Kazufumi Shimizu |
| EPI_ISL_1159388 | Paru Hospital | Institute of Tropical Disease, Universitas Airlangga | Aldise M Nastri, Jezzy R Dewantari, Rima R Prasetya, Krisnoadi Rahardjo, Dyah Retno, Gatot Soegiarto, Laksmi Wulandari, Resti Yudhawati, Soetjipto, Yasuko Mori, Maria I Lusida, Kazufumi Shimizu |
| EPI_ISL_1159389 | Darmo Hospital | Institute of Tropical Disease, Universitas Airlangga | Jezzy R Dewantari, Rima R Prasetya, Krisnoadi Rahardjo, Aldise M Nastri, Sulung Budianto, Gatot Soegiarto, Laksmi Wulandari, Resti Yudhawati, Soetjipto, Yasuko Mori, Maria I Lusida, Kazufumi Shimizu |
| EPI_ISL_1165068 | Anwar Medika General Hospital | Institute of Tropical Disease, Universitas Airlangga | Jezzy R Dewantari, Rima R Prasetya, Krisnoadi Rahardjo, Aldise M Nastri, Nungky Taniasari ,Gatot Soegiarto, Laksmi Wulandari, Resti Yudhawati, Yasuko Mori, Soetjipto, Kazufumi Shimizu, Maria I Lusida |
| EPI_ISL_1165069, EPI_ISL_1165070 | Siti Khodijah Hospital | Institute of Tropical Disease, Universitas Airlangga | Rima R Prasetya, Krisnoadi Rahardjo, Aldise M Nastri, Jezzy R Dewantari, Muhammad Hamdan, Gatot Soegiarto, Laksmi Wulandari, Resti Yudhawati, Yasuko Mori, Soetjipto, Kazufumi Shimizu, Maria I Lusida |
| EPI_ISL_1165071 | Anwar Medika General Hospital | Institute of Tropical Disease, Universitas Airlangga | Kazufumi Shimizu, Krisnoadi Rahardjo, Aldise M Nastri, Jezzy R Dewantari, Rima R Prasetya, Nungky Taniasari, Gatot Soegiarto, Laksmi Wulandari, Resti Yudhawati, Yasuko Mori, Soetjipto, Maria I Lusida |
| EPI_ISL_1165076 | Rahman Rahim Hospital | Institute of Tropical Disease, Universitas Airlangga | Jezzy R Dewantari, Rima R Prasetya, Krisnoadi Rahardjo, Aldise M Nastri, Rendra E Febrianto, Gatot Soegiarto, Laksmi Wulandari, Resti Yudhawati, Yasuko Mori, Soetjipto, Kazufumi Shimizu, Maria I Lusida |
| EPI_ISL_1169047 | RSUP Dr. Mohammad Hoesin Palembang Sumatera Selatan | National Institute of Health Research and Development | Vivi Setiawaty, Hana Apsari Pawestri, Subangkit, Kartika Dewi Puspa, Arie Ardiansyah Nugraha, Hartanti Dian Ikawati, Nelly Puspandari |
| EPI_ISL_1169048 | BBTKLPP Banjarbaru, Kalimantan Selatan | National Institute of Health Research and Development | Vivi Setiawaty, Hana Apsari Pawestri, Subangkit, Kartika Dewi Puspa, Arie Ardiansyah Nugraha, Hartanti Dian Ikawati, Nelly Puspandari |
| EPI_ISL_1247019 | Clinical microbiology lab, RSPTN Universitas Hasanuddin | Clinical microbiology lab, RSPTN Universitas Hasanuddin | Muhammad Nasrum Massi, Rizalinda Sjahril, Gita Vita Soraya, Triwibawa, Handayani, Muhammad Yusuf Usman, Rufika Shari |
| EPI_ISL_1257818, EPI_ISL_1257819, EPI_ISL_1257820, EPI_ISL_1257822 | Balai Litbangkes Papua | National Institute of Health Research and Development | Hana Apsari Pawestri, Subangkit, Kartika Dewi Puspa, Arie Ardiansyah Nugraha, Hartanti Dian Ikawati, Krisna Pangesti, Nelly Puspandari, Hotma Sitompul, Antonius Octavian, Vivi Setiawaty |
| EPI_ISL_1257823, EPI_ISL_1257824, EPI_ISL_1257825, EPI_ISL_1257826 | RSUP DR Mohammad Hoesin | National Institute of Health Research and Development | Hana Apsari Pawestri, Subangkit, Kartika Dewi Puspa, Arie Ardiansyah Nugraha, Hartanti Dian Ikawati, Krisna Pangesti, Nelly Puspandari, Eny Rahmawati, Rogaya, Vivi Setiawaty |
| EPI_ISL_1257832 | BTKL-PP Ambon | National Institute of Health Research and Development | Vivi Setiawaty, Hana Apsari Pawestri, Subangkit, Kartika Dewi Puspa, Arie Ardiansyah Nugraha, Hartanti Dian Ikawati, Nelly Puspandari, Krisna Pangesti |
| EPI_ISL_1257851, EPI_ISL_1257852 | BBTKLPP Banjarbaru | National Institute of Health Research and Development | Hartanti Dian Ikawati, Hana Apsari Pawestri, Subangkit, Kartika Dewi Puspa, Arie Ardiansyah Nugraha, Krisna Pangesti, Nelly Puspandari, Dewi Hermawati, Mutia Raudah, Vivi Setiawaty |
| EPI_ISL_1257863 | RSUP Dr. Sardjito | National Institute of Health Research and Development | Kartika Dewi Puspa, Hana Apsari Pawestri, Subangkit, Arie Ardiansyah Nugraha, Hartanti Dian Ikawati, Nelly Puspandari, Krisna Pangesti, Vivi Setiawaty |
| EPI_ISL_1257864 | BTKL-PP Ambon | National Institute of Health Research and Development | Kartika Dewi Puspa, Hana Apsari Pawestri, Subangkit, Arie Ardiansyah Nugraha, Hartanti Dian Ikawati, Nelly Puspandari, Krisna Pangesti, Vivi Setiawaty |
| EPI_ISL_1283923, EPI_ISL_1283924 | RSIA Sayyidah | Eijkman Institute for Molecular Biology, Ministry of Research and Technology/National Agency for Research and Innovation | Edison Johar, Frilasita A Yudhaputri, Hidayat Trimarsanto, Iskandar Adnan, Lydia V. Panggalo, Sukma Oktavianthi, Willy Agustine, Safarina G Malik, Khin Saw Myint, Amin Soebandrio |
| EPI_ISL_1283927 | RSU Harapan Bunda | Eijkman Institute for Molecular Biology, Ministry of Research and Technology/National Agency for Research and Innovation | Edison Johar, Frilasita A Yudhaputri, Hidayat Trimarsanto, Iskandar Adnan, Lydia V. Panggalo, Sukma Oktavianthi, Willy Agustine, Safarina G Malik, Khin Saw Myint, Amin Soebandrio |
| EPI_ISL_1283929 | RS Mitra Keluarga Kelapa Gading | Eijkman Institute for Molecular Biology, Ministry of Research and Technology/National Agency for Research and Innovation | Edison Johar, Frilasita A Yudhaputri, Hidayat Trimarsanto, Iskandar Adnan, Lydia V. Panggalo, Sukma Oktavianthi, Willy Agustine, Safarina G Malik, Khin Saw Myint, Amin Soebandrio |
| EPI_ISL_1283932 | RS Mulya Tangerang | Eijkman Institute for Molecular Biology, Ministry of Research and Technology/National Agency for Research and Innovation | Edison Johar, Frilasita A Yudhaputri, Hidayat Trimarsanto, Iskandar Adnan, Lydia V. Panggalo, Sukma Oktavianthi, Willy Agustine, Safarina G Malik, Khin Saw Myint, Amin Soebandrio |
| EPI_ISL_1283933 | RS Mitra Keluarga Gading Serpong | Eijkman Institute for Molecular Biology, Ministry of Research and | Edison Johar, Frilasita A Yudhaputri, Hidayat Trimarsanto, Iskandar Adnan, Lydia V. Panggalo, Sukma Oktavianthi, Willy Agustine, Safarina G Malik, Khin Saw Myint, Amin Soebandrio |

| | | | |
|---|---|---|---|
| | | Technology/National Agency for Research and Innovation | |
| EPI_ISL_1283936 | RS Sentra Medika Cikarang | Eijkman Institute for Molecular Biology, Ministry of Research and Technology/National Agency for Research and Innovation | Edison Johar, Frilasita A Yudhaputri, Hidayat Trimarsanto, Iskandar Adnan, Lydia V. Panggalo, Sukma Oktavianthi, Willy Agustine, Safarina G Malik, Khin Saw Myint, Amin Soebandrio |
| EPI_ISL_1283938 | RSUD Cileungsi | Eijkman Institute for Molecular Biology, Ministry of Research and Technology/National Agency for Research and Innovation | Frilasita A Yudhaputri, Hidayat Trimarsanto, Iskandar Adnan, Lydia V. Panggalo, Sukma Oktavianthi, Willy Agustine, Edison Johar, Safarina G Malik, Khin Saw Myint, Amin Soebandrio |
| EPI_ISL_1283941, EPI_ISL_1283945, EPI_ISL_1283949, EPI_ISL_1283953 | RS Mitra Keluarga Gading Serpong | Eijkman Institute for Molecular Biology, Ministry of Research and Technology/National Agency for Research and Innovation | Frilasita A Yudhaputri, Hidayat Trimarsanto, Iskandar Adnan, Lydia V. Panggalo, Sukma Oktavianthi, Willy Agustine, Edison Johar, Safarina G Malik, Khin Saw Myint, Amin Soebandrio |
| EPI_ISL_1283957 | RSIA Selasih Medika | Eijkman Institute for Molecular Biology, Ministry of Research and Technology/National Agency for Research and Innovation | Frilasita A Yudhaputri, Hidayat Trimarsanto, Iskandar Adnan, Lydia V. Panggalo, Sukma Oktavianthi, Willy Agustine, Edison Johar, Safarina G Malik, Khin Saw Myint, Amin Soebandrio |
| EPI_ISL_1283960 | RS Asy-Syifaa Leuwiliang | Eijkman Institute for Molecular Biology, Ministry of Research and Technology/National Agency for Research and Innovation | Hidayat Trimarsanto, Iskandar Adnan, Lydia V. Panggalo, Sukma Oktavianthi, Willy Agustine, Edison Johar, Frilasita A Yudhaputri, Safarina G Malik, Khin Saw Myint, Amin Soebandrio |
| EPI_ISL_1283964 | RS Mitra Keluarga Gading Serpong | Eijkman Institute for Molecular Biology, Ministry of Research and Technology/National Agency for Research and Innovation | Hidayat Trimarsanto, Iskandar Adnan, Lydia V. Panggalo, Sukma Oktavianthi, Willy Agustine, Edison Johar, Frilasita A Yudhaputri, Safarina G Malik, Khin Saw Myint, Amin Soebandrio |
| EPI_ISL_1283967 | RS Bina Husada Bogor | Eijkman Institute for Molecular Biology, Ministry of Research and Technology/National Agency for Research and Innovation | Hidayat Trimarsanto, Iskandar Adnan, Lydia V. Panggalo, Sukma Oktavianthi, Willy Agustine, Edison Johar, Frilasita A Yudhaputri, Safarina G Malik, Khin Saw Myint, Amin Soebandrio |
| EPI_ISL_1283970 | RSIA Selasih Medika | Eijkman Institute for Molecular Biology, Ministry of Research and Technology/National Agency for Research and Innovation | Hidayat Trimarsanto, Iskandar Adnan, Lydia V. Panggalo, Sukma Oktavianthi, Willy Agustine, Edison Johar, Frilasita A Yudhaputri, Safarina G Malik, Khin Saw Myint, Amin Soebandrio |
| EPI_ISL_1283973 | RSU Bunda Mulia Cikarang | Eijkman Institute for Molecular Biology, Ministry of Research and Technology/National Agency for Research and Innovation | Hidayat Trimarsanto, Iskandar Adnan, Lydia V. Panggalo, Sukma Oktavianthi, Willy Agustine, Edison Johar, Frilasita A Yudhaputri, Safarina G Malik, Khin Saw Myint, Amin Soebandrio |
| EPI_ISL_1283979 | RS Cendana | Eijkman Institute for Molecular Biology, Ministry of Research and Technology/National Agency for Research and Innovation | Iskandar Adnan, Lydia V. Panggalo, Sukma Oktavianthi, Willy Agustine, Edison Johar, Hidayat Trimarsanto, Frilasita A Yudhaputri, Safarina G Malik, Khin Saw Myint, Amin Soebandrio |
| EPI_ISL_1283982, EPI_ISL_1283984 | RS Mitra Keluarga Gading Serpong | Eijkman Institute for Molecular Biology, Ministry of Research and Technology/National Agency for Research and Innovation | Iskandar Adnan, Lydia V. Panggalo, Sukma Oktavianthi, Willy Agustine, Edison Johar, Hidayat Trimarsanto, Frilasita A Yudhaputri, Safarina G Malik, Khin Saw Myint, Amin Soebandrio |
| EPI_ISL_1297194 | RSUD Pasarminggu | Stem Cell Lab. Universitas Pembangunan Nasional Veteran Jakarta (UPNVJ) | Andri Pramesyanti Pramono, Yudhi Nugraha, Ria Maria Theresa, Yudi Amiarno, Megadianty Mokoginta |
| EPI_ISL_1300496 | RSUD Pasarminggu | Stem Cell Lab, Universitas Pembangunan Nasional Veteran Jakarta (UPNVJ) | Andri Pramesyanti Pramono, Yudhi Nugraha, Ria Maria Theresa, Yudi Amiarno, Megadianty Mokoginta |
| EPI_ISL_1300521 | RSUD Pasarminggu | Stem Cell Lab, Universitas Pembangunan Nasional Veteran Jakarta | Andri Pramesyanti Pramono, Yudhi Nugraha, Ria Maria Theresa, Yudi Amiarno, Megadianty Mokoginta |
| EPI_ISL_1300522, EPI_ISL_1300535, | RSUD Pasarminggu | Stem Cell Lab, Universitas Pembangunan Nasional | Andri Pramesyanti Pramono, Yudhi Nugraha, Ria Maria Theresa, Yudi Amiarno, Megadianty Mokoginta |

| | | | |
|---|---|---|---|
| EPI_ISL_1300540, EPI_ISL_1300644, EPI_ISL_1300660, EPI_ISL_1300661 | | Veteran Jakarta (UPNVJ) | |
| EPI_ISL_1335883 | North Sumatera Health Office | Faculty of Medicine, Universitas Sumatera Utara; Institute of Tropical Disease, Universitas Airlangga | Aldise M Nastri, Jezzy R Dewantari, Rima R Prasetya, Krisnoadi Rahardjo, Yasuko Mori, Soetjipto, Kazufumi Shimizu, Maria I Lusida, Meliani, R Andika D Cahyadi, Mirzan Hasibuan, Irbah R Nainggolan, Ramadhan Bestari, Franciscus Ginting, Muhammad Ichwan, R Lia Kusumawati |
| EPI_ISL_1335884 | Department of Microbiology, Faculty of Medicine, Universitas Sumatera Utara | Faculty of Medicine, Universitas Sumatera Utara; Institute of Tropical Disease, Universitas Airlangga | Jezzy R Dewantari, Rima R Prasetya, Krisnoadi Rahardjo, Aldise M Nastri, Yasuko Mori, Soetjipto, Kazufumi Shimizu, Maria I Lusida, Meliani, R Andika D Cahyadi, Mirzan Hasibuan, Irbah R Nainggolan, Ramadhan Bestari, Franciscus Ginting, Muhammad Ichwan, R Lia Kusumawati |
| EPI_ISL_1335885 | North Sumatera Health Office | Faculty of Medicine, Universitas Sumatera Utara; Institute of Tropical Disease, Universitas Airlangga | Rima R Prasetya, Krisnoadi Rahardjo, Aldise M Nastri, Jezzy R Dewantari, Yasuko Mori, Soetjipto, Kazufumi Shimizu, Maria I Lusida, Meliani, R Andika D Cahyadi, Mirzan Hasibuan, Irbah R Nainggolan, Ramadhan Bestari, Franciscus Ginting, Muhammad Ichwan, R Lia Kusumawati, Inke N D Lubis |
| EPI_ISL_1335886 | Sundari Hospital Medan | Faculty of Medicine, Universitas Sumatera Utara; Institute of Tropical Disease, Universitas Airlangga | Kazufumi Shimizu, Krisnoadi Rahardjo, Aldise M Nastri, Jezzy R Dewantari, Rima R Prasetya, Yasuko Mori, Soetjipto, Maria I Lusida, Meliani, R Andika D Cahyadi, Mirzan Hasibuan, Irbah R Nainggolan, Ramadhan Bestari, Franciscus Ginting, Muhammad Ichwan, R Lia Kusumawati, Inke N D Lubis |
| EPI_ISL_1335887 | North Sumatera Health Office | Faculty of Medicine, Universitas Sumatera Utara; Institute of Tropical Disease, Universitas Airlangga | Maria I Lusida, Krisnoadi Rahardjo, Aldise M Nastri, Jezzy R Dewantari, Rima R Prasetya, Yasuko Mori, Soetjipto, Kazufumi Shimizu, Meliani, R Andika D Cahyadi, Mirzan Hasibuan, Irbah R Nainggolan, Ramadhan Bestari, Franciscus Ginting, Muhammad Ichwan, R Lia Kusumawati, Inke N D Lubis |
| EPI_ISL_1335888 | Martha Friska Multatuli Hospital Medan | Faculty of Medicine, Universitas Sumatera Utara; Institute of Tropical Disease, Universitas Airlangga | Krisnoadi Rahardjo, Aldise M Nastri, Jezzy R Dewantari, Rima R Prasetya, Yasuko Mori, Soetjipto, Kazufumi Shimizu, Maria I Lusida, Meliani, R Andika D Cahyadi, Mirzan Hasibuan, Irbah R Nainggolan, Ramadhan Bestari, Franciscus Ginting, Muhammad Ichwan, R Lia Kusumawati, Inke N D Lubis |
| EPI_ISL_1335889 | Advent Hospital Medan | Faculty of Medicine, Universitas Sumatera Utara; Institute of Tropical Disease, Universitas Airlangga | Krisnoadi Rahardjo, Aldise M Nastri, Jezzy R Dewantari, Rima R Prasetya, Yasuko Mori, Soetjipto, Kazufumi Shimizu, Maria I Lusida, Meliani, R Andika D Cahyadi, Mirzan Hasibuan, Irbah R Nainggolan, Ramadhan Bestari, Franciscus Ginting, Muhammad Ichwan, R Lia Kusumawati, Inke N D Lubis |
| EPI_ISL_1335890 | Elisabeth Hospital Medan | Faculty of Medicine, Universitas Sumatera Utara; Institute of Tropical Disease, Universitas Airlangga | Aldise M Nastri, Jezzy R Dewantari, Rima R Prasetya, Krisnoadi Rahardjo, Yasuko Mori, Soetjipto, Kazufumi Shimizu, Maria I Lusida, Meliani, R Andika D Cahyadi, Mirzan Hasibuan, Irbah R Nainggolan, Ramadhan Bestari, Franciscus Ginting, Muhammad Ichwan, R Lia Kusumawati, Inke N D Lubis |
| EPI_ISL_1335891 | Advent Hospital Medan | Faculty of Medicine, Universitas Sumatera Utara; Institute of Tropical Disease, Universitas Airlangga | Jezzy R Dewantari, Rima R Prasetya, Krisnoadi Rahardjo, Aldise M Nastri, Yasuko Mori, Soetjipto, Kazufumi Shimizu, Maria I Lusida, Meliani, R Andika D Cahyadi, Mirzan Hasibuan, Irbah R Nainggolan, Ramadhan Bestari, Franciscus Ginting, Muhammad Ichwan, R Lia Kusumawati, Inke N D Lubis |
| EPI_ISL_1335892 | Department of Microbiology, Faculty of Medicine, Universitas Sumatera Utara | Faculty of Medicine, Universitas Sumatera Utara; Institute of Tropical Disease, Universitas Airlangga | Rima R Prasetya, Krisnoadi Rahardjo, Aldise M Nastri, Jezzy R Dewantari, Yasuko Mori, Soetjipto, Kazufumi Shimizu, Maria I Lusida, Meliani, R Andika D Cahyadi, Mirzan Hasibuan, Irbah R Nainggolan, Ramadhan Bestari, Franciscus Ginting, Muhammad Ichwan, R Lia Kusumawati, Inke N D Lubis |
| EPI_ISL_1335893 | Department of Microbiology, Faculty of Medicine, Universitas Sumatera Utara | Faculty of Medicine, Universitas Sumatera Utara; Institute of Tropical Disease, Universitas Airlangga | Kazufumi Shimizu, Krisnoadi Rahardjo, Aldise M Nastri, Jezzy R Dewantari, Rima R Prasetya, Yasuko Mori, Soetjipto, Maria I Lusida, Meliani, R Andika D Cahyadi, Mirzan Hasibuan, Irbah R Nainggolan, Ramadhan Bestari, Franciscus Ginting, Muhammad Ichwan, R Lia Kusumawati, Inke N D Lubis |
| EPI_ISL_1335894 | Department of Microbiology, Faculty of Medicine, Universitas Sumatera Utara | Faculty of Medicine, Universitas Sumatera Utara; Institute of Tropical Disease, Universitas Airlangga | Maria I Lusida, Krisnoadi Rahardjo, Aldise M Nastri, Jezzy R Dewantari, Rima R Prasetya, Yasuko Mori, Soetjipto, Kazufumi Shimizu, Meliani, R Andika D Cahyadi, Mirzan Hasibuan, Irbah R Nainggolan, Ramadhan Bestari, Franciscus Ginting, Muhammad Ichwan, R Lia Kusumawati, Inke N D Lubis |
| EPI_ISL_1335895 | Martha Friska Multatuli Hospital Medan | Faculty of Medicine, Universitas Sumatera Utara; Institute of Tropical Disease, Universitas Airlangga | Krisnoadi Rahardjo, Aldise M Nastri, Jezzy R Dewantari, Rima R Prasetya, Yasuko Mori, Soetjipto, Kazufumi Shimizu, Maria I Lusida, Meliani, R Andika D Cahyadi, Mirzan Hasibuan, Irbah R Nainggolan, Ramadhan Bestari, Franciscus Ginting, Muhammad Ichwan, R Lia Kusumawati, Inke N D Lubis |
| EPI_ISL_1335897 | North Sumatera Health Office | Faculty of Medicine, Universitas Sumatera Utara; Institute of Tropical Disease, Universitas Airlangga | Jezzy R Dewantari, Rima R Prasetya, Krisnoadi Rahardjo, Aldise M Nastri, Yasuko Mori, Soetjipto, Kazufumi Shimizu, Maria I Lusida, Meliani, R Andika D Cahyadi, Mirzan Hasibuan, Irbah R Nainggolan, Ramadhan Bestari, Franciscus Ginting, Muhammad Ichwan, R Lia Kusumawati, Inke N D Lubis |
| EPI_ISL_1335898 | Department of Microbiology, Faculty of Medicine, Universitas Sumatera Utara | Faculty of Medicine, Universitas Sumatera Utara; Institute of Tropical Disease, Universitas Airlangga | Rima R Prasetya, Krisnoadi Rahardjo, Aldise M Nastri, Jezzy R Dewantari, Yasuko Mori, Soetjipto, Kazufumi Shimizu, Maria I Lusida, Meliani, R Andika D Cahyadi, Mirzan Hasibuan, Irbah R Nainggolan, Ramadhan Bestari, Franciscus Ginting, Muhammad Ichwan, R Lia Kusumawati, Inke N D Lubis |

| | | | |
|---|---|---|---|
| EPI_ISL_1335899 | Department of Microbiology, Faculty of Medicine, Universitas Sumatera Utara | Faculty of Medicine, Universitas Sumatera Utara; Institute of Tropical Disease, Universitas Airlangga | Kazufumi Shimizu, Krisnoadi Rahardjo, Aldise M Nastri, Jezzy R Dewantari, Rima R Prasetya, Yasuko Mori, Soetjipto, Maria I Lusida, Meliani, R Andika D Cahyadi, Mirzan Hasibuan, Irbah R Nainggolan, Ramadhan Bestari, Franciscus Ginting, Muhammad Ichwan, R Lia Kusumawati, Inke N D Lubis |
| EPI_ISL_1340862, EPI_ISL_1340863, EPI_ISL_1340864, EPI_ISL_1340865, EPI_ISL_1340866, EPI_ISL_1340867, EPI_ISL_1345232 | West Bandung Public Health | West Java Health Laboratory; School of Life Sciences and Technology, Institut Teknologi Bandung | Azzania Fibriani, Ema Rahmawati, Ryan Bayusantika Ristandi, Rifky Waluyajati Rachman, Cut Nur Cinthia Alamanda, Isak Solihin, Rini Robiani, Miftahul Faridl, Karimatu Khoirunnisa |
| EPI_ISL_1346047 | West Java Health Laboratory | West Java Health Laboratory; School of Life Sciences and Technology, Institut Teknologi Bandung | Azzania Fibriani, Ema Rahmawati, Ryan Bayusantika Ristandi, Rifky Waluyajati Rachman, Cut Nur Cinthia Alamanda, Isak Solihin, Rini Robiani, Miftahul Faridl, Karimatu Khoirunnisa |
| EPI_ISL_1346484 | West Bandung Public Health | West Java Health Laboratory; School of Life Sciences and Technology, Institut Teknologi Bandung | Azzania Fibriani, Ema Rahmawati, Ryan Bayusantika Ristandi, Rifky Waluyajati Rachman, Cut Nur Cinthia Alamanda, Isak Solihin, Rini Robiani, Miftahul Faridl, Karimatu Khoirunnisa |
| EPI_ISL_1346485 | Cianjur Public Health | West Java Health Laboratory; School of Life Sciences and Technology, Institut Teknologi Bandung | Azzania Fibriani, Ema Rahmawati, Ryan Bayusantika Ristandi, Rifky Waluyajati Rachman, Cut Nur Cinthia Alamanda, Isak Solihin, Rini Robiani, Miftahul Faridl, Karimatu Khoirunnisa |
| EPI_ISL_1346627 | Hasan Sadikin Hospital | West Java Health Laboratory; School of Life Sciences and Technology, Institut Teknologi Bandung | Azzania Fibriani, Ema Rahmawati, Ryan Bayusantika Ristandi, Rifky Waluyajati Rachman, Cut Nur Cinthia Alamanda, Isak Solihin, Rini Robiani, Miftahul Faridl, Karimatu Khoirunnisa, Nina Susana Dewi, Anggraini Alam, Basti Andriyoko, Djatnika Setiabudi, Ida Parwati |
| EPI_ISL_1447331, EPI_ISL_1447332, EPI_ISL_1447333, EPI_ISL_1447334, EPI_ISL_1447335, EPI_ISL_1447337, EPI_ISL_1447338, EPI_ISL_1447339, EPI_ISL_1447340, EPI_ISL_1447341, EPI_ISL_1447343, EPI_ISL_1447345, EPI_ISL_1447346, EPI_ISL_1447347, EPI_ISL_1447348, EPI_ISL_1447349, EPI_ISL_1447350, EPI_ISL_1447353, EPI_ISL_1447354, EPI_ISL_1447355, EPI_ISL_1447356, EPI_ISL_1447357, EPI_ISL_1447358, EPI_ISL_1447359, EPI_ISL_1447360, EPI_ISL_1447361, EPI_ISL_1447362, EPI_ISL_1469232, EPI_ISL_1469233, EPI_ISL_1469235, EPI_ISL_1469237 | | | |
| see above | Diagnostic and Research Center of Infectious Diseases, Medical Faculty, Andalas University | Diagnostic and Research Center of Infectious Diseases, Medical Faculty, Andalas University | Andani Eka Putra, Syafrizayanti, Ikhwan R. Sudji, Linosefa, Dede Rahman Agustian, Ayu Novita Trisnawati, Siskalil Fahma, Nia Ayuni Putri, Sekar Asri Tresnaningtyas, SM Rezvi, Fauzul Azhim, Dessy Arisanty, Gestina Aliska, Desmawati, Nita Afriani ,Syandrez Prima Putra, Juane Plantika Menra, Yolani Syaputri, Mutia Lailani |
| EPI_ISL_1469290, EPI_ISL_1469291 | National Institute Health Research and Development | National Institute of Health Research and Development | Nur Ika Hariastuti, Nike Susanti, Hana Apsari Pawestri, Subangkit, Kartika Dewi Puspa, Arie Ardiansyah Nugraha, Hartanti Dian Ikawati, Nelly Puspandari, Vivi Setiawaty |
| EPI_ISL_1533034, EPI_ISL_1533035, EPI_ISL_1533038, EPI_ISL_1533039, EPI_ISL_1533041, EPI_ISL_1533042, EPI_ISL_1533043, EPI_ISL_1533045, EPI_ISL_1533047, EPI_ISL_1533048, EPI_ISL_1533049, EPI_ISL_1533050, EPI_ISL_1533051, EPI_ISL_1533052, EPI_ISL_1533053, EPI_ISL_1533054, EPI_ISL_1533055, EPI_ISL_1533056, EPI_ISL_1533057, EPI_ISL_1533059, EPI_ISL_1533060, EPI_ISL_1533063, EPI_ISL_1533064, EPI_ISL_1533067, EPI_ISL_1533068, EPI_ISL_1533070, EPI_ISL_1533071, EPI_ISL_1533072, EPI_ISL_1533073, EPI_ISL_1533074, EPI_ISL_1533075, EPI_ISL_1533076, EPI_ISL_1533078, EPI_ISL_1533079, EPI_ISL_1533080, EPI_ISL_1533081, EPI_ISL_1533083, EPI_ISL_1533084, EPI_ISL_1533085, EPI_ISL_1533086, EPI_ISL_1533087, EPI_ISL_1533088, EPI_ISL_1533090, EPI_ISL_1533091, EPI_ISL_1533092, EPI_ISL_1533093, EPI_ISL_1533095, EPI_ISL_1533096, EPI_ISL_1533097, EPI_ISL_1533098, EPI_ISL_1533100, EPI_ISL_1533101 | | | |
| see above | Diagnostic and Research Center of Infectious Diseases, Medical Faculty, Andalas University | Diagnostic and Research Center of Infectious Diseases, Medical Faculty, Andalas University | Andani Eka Putra, Syafrizayanti, Ikhwan R. Sudji, Linosefa, Dede Rahman Agustian, Ayu Novita Trisnawati, Siskalil Fahma, Nia Ayuni Putri, Sekar Asri Tresnaningtyas, SM Rezvi, Fauzul Azhim, Dessy Arisanty, Gestina Aliska, Desmawati, Nita Afriani ,Syandrez Prima Putra, Juane Plantika Menra, Yolani Syaputri, Mutia Lailani |
| EPI_ISL_1622360, EPI_ISL_1622371 | RS Mitra Keluarga Gading Serpong | Eijkman Institute for Molecular Biology, Ministry of Research and Technology/National Agency for Research and Innovation | Willy Agustine, Hidayat Trimarsanto, Lydia V. Panggalo, Iskandar Adnan, Sukma Oktavianthi, Edison Johar, Frilasita A Yudhaputri, Muhammad Rezki Rasyak, Safarina G Malik, Khin Saw Myint, Amin Soebandrio |
| EPI_ISL_1622372, EPI_ISL_1622379, EPI_ISL_1622380 | RS Mitra Keluarga Gading Serpong | Eijkman Institute for Molecular Biology, Ministry of Research and Technology/National Agency for Research and Innovation | Hidayat Trimarsanto, Lydia V. Panggalo, Iskandar Adnan, Sukma Oktavianthi, Edison Johar, Frilasita A Yudhaputri, Muhammad Rezki Rasyak, Willy Agustine, Safarina G Malik, Khin Saw Myint, Amin Soebandrio |
| EPI_ISL_1630280 | Clinical Microbiology Lab RSPTN Universitas Hasanuddin | Clinical Microbiology Lab RSPTN Universitas Hasanuddin | Muhammad Nasrum Massi, Rizalinda Sjahril, Gita Vita Soraya, Triwibawa, Handayani Halik, Muhammad Yusuf Usman, Rufika Shari |
| EPI_ISL_1789113 | Siti Khodijah Hospital | Institute of Tropical Disease, Universitas Airlangga | Aldise M Nastri, Jezzy R Dewantari, Rima R Prasetya, Krisnoadi Rahardjo, Muhammad Hamdan, Gatot Soegiarto, Laksmi Wulandari, Resti Yudhawati, Soetjipto, Yasuko Mori, Maria I Lusida, Kazufumi Shimizu |
| EPI_ISL_1789114 | Siti Khodijah Hospital | Institute of Tropical Disease, Universitas Airlangga | Jezzy R Dewantari, Rima R Prasetya, Krisnoadi Rahardjo, Aldise M Nastri, Muhammad Hamdan, Gatot Soegiarto, Laksmi Wulandari, Resti Yudhawati, Soetjipto, Yasuko Mori, Maria I Lusida, Kazufumi Shimizu |
| EPI_ISL_1789115 | Siti Khodijah Hospital | Institute of Tropical Disease, Universitas Airlangga | Rima R Prasetya, Krisnoadi Rahardjo, Aldise M Nastri, Jezzy R Dewantari, Muhammad Hamdan, Gatot Soegiarto, Laksmi Wulandari, Resti Yudhawati, Soetjipto, Yasuko Mori, Maria I Lusida, Kazufumi Shimizu |
| EPI_ISL_1789116 | Anwar Medika General Hospital | Institute of Tropical Disease, Universitas Airlangga | Maria I Lusida, Krisnoadi Rahardjo, Aldise M Nastri, Jezzy R Dewantari, Rima R Prasetya, Nungky Taniasari, Gatot Soegiarto, Laksmi Wulandari, Resti Yudhawati, Soetjipto, Yasuko Mori, Kazufumi Shimizu |
| EPI_ISL_1789117 | Anwar Medika General Hospital | Institute of Tropical Disease, Universitas Airlangga | Krisnoadi Rahardjo, Aldise M Nastri, Jezzy R Dewantari, Rima R Prasetya, Muhammad Hamdan, Gatot Soegiarto, Laksmi Wulandari, Resti Yudhawati, Yasuko Mori, Soetjipto, Kazufumi Shimizu, Maria I Lusida |
| EPI_ISL_1795053 | Anwar Medika General Hospital | Institute of Tropical Disease, Universitas Airlangga | Aldise M Nastri, Jezzy R Dewantari, Rima R Prasetya, Krisnoadi Rahardjo, Nungky Taniasari, Gatot Soegiarto, Laksmi Wulandari, Resti Yudhawati, Yasuko Mori, Soetjipto, Kazufumi Shimizu, Maria I Lusida |

| | | | |
|---|---|---|---|
| EPI_ISL_1795054 | Darus Syifa' Islamic Hospital | Institute of Tropical Disease, Universitas Airlangga | Krisnoadi Rahardjo, Aldise M Nastri, Jezzy R Dewantari, Rima R Prasetya, HM Faiz, Gatot Soegiarto, Laksmi Wulandari, Resti Yudhawati, Yasuko Mori, Soetjipto, Kazufumi Shimizu, Maria I Lusida |
| EPI_ISL_1795055 | Muhammadiyah Hospital | Institute of Tropical Disease, Universitas Airlangga | Rima R Prasetya, Krisnoadi Rahardjo, Aldise M Nastri, Jezzy R Dewantari, Imam Suyuthi, Gatot Soegiarto, Laksmi Wulandari, Resti Yudhawati, Yasuko Mori, Soetjipto, Kazufumi Shimizu, Maria I Lusida |
| EPI_ISL_1795056 | Siloam Hospital | Institute of Tropical Disease, Universitas Airlangga | Kazufumi Shimizu, Krisnoadi Rahardjo, Aldise M Nastri, Jezzy R Dewantari, Rima R Prasetya, Maria M Padmidewi, Gatot Soegiarto, Laksmi Wulandari, Retno A Setyoningrum, Resti Yudhawati, Yohko K Shimizu, Mitsuhiro Nishimura, Yasuko Mori, Soetjipto, Maria I Lusida |
| EPI_ISL_1795057 | Muhammadiyah Hospital | Institute of Tropical Disease, Universitas Airlangga | Jezzy R Dewantari, Rima R Prasetya, Krisnoadi Rahardjo, Aldise M Nastri, Imam Suyuthi, Gatot Soegiarto, Laksmi Wulandari, Resti Yudhawati, Yasuko Mori, Soetjipto, Kazufumi Shimizu, Maria I Lusida |
| EPI_ISL_1795058 | Institute of Tropical Disease, Universitas Airlangga | Institute of Tropical Disease, Universitas Airlangga | Krisnoadi Rahardjo, Aldise M Nastri, Jezzy R Dewantari, Rima R Prasetya, Gatot Soegiarto, Laksmi Wulandari, Resti Yudhawati, Yasuko Mori, Soetjipto, Kazufumi Shimizu, Maria I Lusida |
| EPI_ISL_1795059 | Citra Medika Hospital | Institute of Tropical Disease, Universitas Airlangga | Aldise M Nastri, Jezzy R Dewantari, Rima R Prasetya, Krisnoadi Rahardjo, Wahidah Rachmaniyah, Gatot Soegiarto, Laksmi Wulandari, Resti Yudhawati, Yasuko Mori, Soetjipto, Kazufumi Shimizu, Maria I Lusida |
| EPI_ISL_1795060 | Institute of Tropical Disease, Universitas Airlangga | Institute of Tropical Disease, Universitas Airlangga | Kazufumi Shimizu, Krisnoadi Rahardjo, Aldise M Nastri, Jezzy R Dewantari, Rima R Prasetya, Gatot Soegiarto, Laksmi Wulandari, Retno A Setyoningrum, Resti Yudhawati, Yohko K Shimizu, Mitsuhiro Nishimura, Yasuko Mori, Soetjipto, Maria I Lusida |
| EPI_ISL_1824616, EPI_ISL_1824617 | RSUD Bahteramas Kendari | National Institute of Health Research and Development | Hana Apsari Pawestri, Subangkit, Kartika Dewi Puspa, Arie Ardiansyah Nugraha, Hartanti Dian Ikawati, Krisna Pangesti, Nelly Puspandari, Vivi Setiawaty |
| EPI_ISL_1969239, EPI_ISL_1969240 | Hotel Grand Sahid Jaya | National Institute of Health Research and Development | Vivi Setiawaty, Hana Apsari Pawestri, Subangkit, Kartika Dewi Puspa, Arie Ardiansyah Nugraha, Hartanti Dian Ikawati, Krisna Pangesti, Triyani Soekarso, Nelly Puspandari |
| EPI_ISL_1969241 | Hotel Grand Tjokro | National Institute of Health Research and Development | Vivi Setiawaty, Hana Apsari Pawestri, Subangkit, Kartika Dewi Puspa, Arie Ardiansyah Nugraha, Hartanti Dian Ikawati, Krisna Pangesti, Triyani Soekarso, Nelly Puspandari |
| EPI_ISL_1969242, EPI_ISL_1969243 | Swissbel Hotel Airport | National Institute of Health Research and Development | Vivi Setiawaty, Hana Apsari Pawestri, Subangkit, Kartika Dewi Puspa, Arie Ardiansyah Nugraha, Hartanti Dian Ikawati, Krisna Pangesti, Triyani Soekarso, Nelly Puspandari |
| EPI_ISL_1969244 | Lab RSUP DR Mohammad Hoesin Palembang | National Institute of Health Research and Development | Vivi Setiawaty, Hana Apsari Pawestri, Subangkit, Kartika Dewi Puspa, Arie Ardiansyah Nugraha, Hartanti Dian Ikawati, Krisna Pangesti, Triyani Soekarso, Nelly Puspandari |
| EPI_ISL_2013043 | RS Sentra Medika Cikarang | Eijkman Institute for Molecular Biology, National Agency for Research and Innovation | Edison Johar, Frilasita A Yudhaputri, Muhammad Rezki Rasyak, Willy Agustine, Hidayat Trimarsanto, Lydia V. Panggalo, Iskandar Adnan, Sukma Oktavianthi, Safarina G Malik, Khin Saw Myint, Amin Soebandrio |
| EPI_ISL_2013044 | RS Sentra Medika Cikarang | Eijkman Institute for Molecular Biology, National Agency for Research and Innovation | Willy Agustine, Hidayat Trimarsanto, Lydia V. Panggalo, Iskandar Adnan, Sukma Oktavianthi, Edison Johar, Frilasita A Yudhaputri, Muhammad Rezki Rasyak, Safarina G Malik, Khin Saw Myint, Amin Soebandrio |
| EPI_ISL_2013045 | RS Mitra Keluarga Cikarang | Eijkman Institute for Molecular Biology, National Agency for Research and Innovation | Edison Johar, Frilasita A Yudhaputri, Muhammad Rezki Rasyak, Willy Agustine, Hidayat Trimarsanto, Lydia V. Panggalo, Iskandar Adnan, Sukma Oktavianthi, Safarina G Malik, Khin Saw Myint, Amin Soebandrio |
| EPI_ISL_2013046 | RSUD Cileungsi | Eijkman Institute for Molecular Biology, National Agency for Research and Innovation | Lydia V. Panggalo, Iskandar Adnan, Sukma Oktavianthi, Edison Johar, Frilasita A Yudhaputri, Muhammad Rezki Rasyak, Willy Agustine, Hidayat Trimarsanto, Safarina G Malik, Khin Saw Myint, Amin Soebandrio |
| EPI_ISL_2013047 | RS Mitra Keluarga Kelapa Gading | Eijkman Institute for Molecular Biology, National Agency for Research and Innovation | Hidayat Trimarsanto, Lydia V. Panggalo, Iskandar Adnan, Sukma Oktavianthi, Edison Johar, Frilasita A Yudhaputri, Muhammad Rezki Rasyak, Willy Agustine, Safarina G Malik, Khin Saw Myint, Amin Soebandrio |
| EPI_ISL_2013509 | RS Harapan Bunda | Eijkman Institute for Molecular Biology, National Agency for Research and Innovation | Lydia V. Panggalo, Iskandar Adnan, Sukma Oktavianthi, Edison Johar, Frilasita A Yudhaputri, Muhammad Rezki Rasyak, Willy Agustine, Hidayat Trimarsanto, Safarina G Malik, Khin Saw Myint, Amin Soebandrio |
| EPI_ISL_2047507, EPI_ISL_2047508, EPI_ISL_2047509, EPI_ISL_2047510, EPI_ISL_2047511, EPI_ISL_2047512 | Lab RSUP DR Mohammad Hoesin Palembang | National Institute of Health Research and Development | Arie Ardiansyah Nugraha, Kartika Dewi Puspa, Hana Apsari Pawestri, Hartanti Dian Ikawati, Krisna Pangesti, Triyani Soekarso, Nelly Puspandari, Subangkit, Eny Rahmawati, Rogaya, Vivi Setiawaty |
| EPI_ISL_2101022 | Molecular Biology Laboratory, Faculty Medicine and Health Sciences, Warmadewa University | Eijkman Institute for Molecular Biology, National Agency for Research and Innovation; Molecular Biology Laboratory, Faculty Medicine and Health Sciences, Warmadewa University | Frilasita A Yudhaputri, Muhammad Rezki Rasyak, Willy Agustine, Hidayat Trimarsanto, Lydia V. Panggalo, Iskandar Adnan, Sukma Oktavianthi, Lidwina Priliani, Edison Johar, Sri Masyeni, Erly Sintya, Safarina G Malik, Khin Saw Myint, Amin Soebandrio |
| EPI_ISL_2101023 | Molecular Biology Laboratory, Faculty Medicine and Health | Eijkman Institute for Molecular Biology, National Agency for Research and | Muhammad Rezki Rasyak, Willy Agustine, Hidayat Trimarsanto, Lydia V. Panggalo, Iskandar Adnan, Sukma Oktavianthi, Lidwina Priliani, Edison Johar, Frilasita A Yudhaputri, Sri Masyeni, Erly Sintya, Safarina G Malik, Khin Saw Myint, Amin Soebandrio |

| | | | |
|---|---|---|---|
| | Sciences, Warmadewa University | Innovation; Molecular Biology Laboratory, Faculty Medicine and Health Sciences, Warmadewa University | |
| EPI_ISL_2101024 | Molecular Biology Laboratory, Faculty Medicine and Health Sciences, Warmadewa University | Eijkman Institute for Molecular Biology, National Agency for Research and Innovation; Molecular Biology Laboratory, Faculty Medicine and Health Sciences, Warmadewa University | Willy Agustine, Hidayat Trimarsanto, Lydia V. Panggalo, Iskandar Adnan, Sukma Oktavianthi, Lidwina Priliani, Edison Johar, Frilasita A Yudhaputri, Muhammad Rezki Rasyak, Sri Masyeni, Erly Sintya, Safarina G Malik, Khin Saw Myint, Amin Soebandrio |
| EPI_ISL_2101025 | Molecular Biology Laboratory, Faculty Medicine and Health Sciences, Warmadewa University | Eijkman Institute for Molecular Biology, National Agency for Research and Innovation; Molecular Biology Laboratory, Faculty Medicine and Health Sciences, Warmadewa University | Hidayat Trimarsanto, Lydia V. Panggalo, Iskandar Adnan, Sukma Oktavianthi, Lidwina Priliani, Edison Johar, Frilasita A Yudhaputri, Muhammad Rezki Rasyak, Willy Agustine, Sri Masyeni, Erly Sintya, Safarina G Malik, Khin Saw Myint, Amin Soebandrio |
| EPI_ISL_2101026 | Molecular Biology Laboratory, Faculty Medicine and Health Sciences, Warmadewa University | Eijkman Institute for Molecular Biology, National Agency for Research and Innovation; Molecular Biology Laboratory, Faculty Medicine and Health Sciences, Warmadewa University | Willy Agustine, Hidayat Trimarsanto, Lydia V. Panggalo, Iskandar Adnan, Sukma Oktavianthi, Lidwina Priliani, Edison Johar, Frilasita A Yudhaputri, Muhammad Rezki Rasyak, Sri Masyeni, Erly Sintya, Safarina G Malik, Khin Saw Myint, Amin Soebandrio |
| EPI_ISL_2101027 | Molecular Biology Laboratory, Faculty Medicine and Health Sciences, Warmadewa University | Eijkman Institute for Molecular Biology, National Agency for Research and Innovation; Molecular Biology Laboratory, Faculty Medicine and Health Sciences, Warmadewa University | Iskandar Adnan, Sukma Oktavianthi, Lidwina Priliani, Edison Johar, Frilasita A Yudhaputri, Muhammad Rezki Rasyak, Willy Agustine, Hidayat Trimarsanto, Lydia V. Panggalo, Sri Masyeni, Erly Sintya, Safarina G Malik, Khin Saw Myint, Amin Soebandrio |
| EPI_ISL_2101028 | Molecular Biology Laboratory, Faculty Medicine and Health Sciences, Warmadewa University | Eijkman Institute for Molecular Biology, National Agency for Research and Innovation; Molecular Biology Laboratory, Faculty Medicine and Health Sciences, Warmadewa University | Edison Johar, Frilasita A Yudhaputri, Muhammad Rezki Rasyak, Willy Agustine, Hidayat Trimarsanto, Lydia V. Panggalo, Iskandar Adnan, Sukma Oktavianthi, Lidwina Priliani, Sri Masyeni, Erly Sintya, Safarina G Malik, Khin Saw Myint, Amin Soebandrio |
| EPI_ISL_2101029 | Molecular Biology Laboratory, Faculty Medicine and Health Sciences, Warmadewa University | Eijkman Institute for Molecular Biology, National Agency for Research and Innovation; Molecular Biology Laboratory, Faculty Medicine and Health Sciences, Warmadewa University | Hidayat Trimarsanto, Lydia V. Panggalo, Iskandar Adnan, Sukma Oktavianthi, Lidwina Priliani, Edison Johar, Frilasita A Yudhaputri, Muhammad Rezki Rasyak, Willy Agustine, Sri Masyeni, Erly Sintya, Safarina G Malik, Khin Saw Myint, Amin Soebandrio |
| EPI_ISL_2101030 | Molecular Biology Laboratory, Faculty Medicine and Health Sciences, Warmadewa University | Eijkman Institute for Molecular Biology, National Agency for Research and Innovation; Molecular Biology Laboratory, Faculty Medicine and Health Sciences, Warmadewa University | Sukma Oktavianthi, Lidwina Priliani, Edison Johar, Frilasita A Yudhaputri, Muhammad Rezki Rasyak, Willy Agustine, Hidayat Trimarsanto, Lydia V. Panggalo, Iskandar Adnan, Sri Masyeni, Erly Sintya, Safarina G Malik, Khin Saw Myint, Amin Soebandrio |
| EPI_ISL_2101031 | Molecular Biology Laboratory, Faculty Medicine and Health Sciences, Warmadewa University | Eijkman Institute for Molecular Biology, National Agency for Research and Innovation; Molecular Biology Laboratory, Faculty Medicine and Health Sciences, Warmadewa University | Frilasita A Yudhaputri, Muhammad Rezki Rasyak, Willy Agustine, Hidayat Trimarsanto, Lydia V. Panggalo, Iskandar Adnan, Sukma Oktavianthi, Lidwina Priliani, Edison Johar, Sri Masyeni, Erly Sintya, Safarina G Malik, Khin Saw Myint, Amin Soebandrio |
| EPI_ISL_2101032 | Molecular Biology Laboratory, Faculty Medicine and Health Sciences, Warmadewa University | Eijkman Institute for Molecular Biology, National Agency for Research and Innovation; Molecular Biology Laboratory, Faculty Medicine and Health Sciences, Warmadewa University | Lydia V. Panggalo, Iskandar Adnan, Sukma Oktavianthi, Lidwina Priliani, Edison Johar, Frilasita A Yudhaputri, Muhammad Rezki Rasyak, Willy Agustine, Hidayat Trimarsanto, Sri Masyeni, Erly Sintya, Safarina G Malik, Khin Saw Myint, Amin Soebandrio |
| EPI_ISL_2101034 | Molecular Biology Laboratory, Faculty Medicine and Health Sciences, Warmadewa University | Eijkman Institute for Molecular Biology, National Agency for Research and Innovation; Molecular Biology Laboratory, Faculty | Willy Agustine, Hidayat Trimarsanto, Lydia V. Panggalo, Iskandar Adnan, Sukma Oktavianthi, Lidwina Priliani, Edison Johar, Frilasita A Yudhaputri, Muhammad Rezki Rasyak, Sri Masyeni, Erly Sintya, Safarina G Malik, Khin Saw Myint, Amin Soebandrio |

| | | | |
|---|---|---|---|
| | | Medicine and Health Sciences, Warmadewa University | |
| EPI_ISL_2101035, EPI_ISL_2101036, EPI_ISL_2101037 | Molecular Biology Laboratory, Faculty Medicine and Health Sciences, Warmadewa University | Eijkman Institute for Molecular Biology, National Agency for Research and Innovation; Molecular Biology Laboratory, Faculty Medicine and Health Sciences, Warmadewa University | Lydia V. Panggalo, Iskandar Adnan, Sukma Oktavianthi, Lidwina Priliani, Edison Johar, Filasita A Yudhaputri, Muhammad Rezki Rasyak, Willy Agustine, Hidayat Trimarsanto, Sri Masyeni, Erly Sintya, Safarina G Malik, Khin Saw Myint, Amin Soebandrio |
| EPI_ISL_2101038 | Molecular Biology Laboratory, Faculty Medicine and Health Sciences, Warmadewa University | Eijkman Institute for Molecular Biology, National Agency for Research and Innovation; Molecular Biology Laboratory, Faculty Medicine and Health Sciences, Warmadewa University | Iskandar Adnan, Sukma Oktavianthi, Lidwina Priliani, Edison Johar, Filasita A Yudhaputri, Muhammad Rezki Rasyak, Willy Agustine, Hidayat Trimarsanto, Lydia V. Panggalo, Sri Masyeni, Erly Sintya, Safarina G Malik, Khin Saw Myint, Amin Soebandrio |
| EPI_ISL_2101039 | Molecular Biology Laboratory, Faculty Medicine and Health Sciences, Warmadewa University | Eijkman Institute for Molecular Biology, National Agency for Research and Innovation; Molecular Biology Laboratory, Faculty Medicine and Health Sciences, Warmadewa University | Edison Johar, Filasita A Yudhaputri, Muhammad Rezki Rasyak, Willy Agustine, Hidayat Trimarsanto, Lydia V. Panggalo, Iskandar Adnan, Sukma Oktavianthi, Lidwina Priliani, Sri Masyeni, Erly Sintya, Safarina G Malik, Khin Saw Myint, Amin Soebandrio |
| EPI_ISL_2101040 | Molecular Biology Laboratory, Faculty Medicine and Health Sciences, Warmadewa University | Eijkman Institute for Molecular Biology, National Agency for Research and Innovation; Molecular Biology Laboratory, Faculty Medicine and Health Sciences, Warmadewa University | Hidayat Trimarsanto, Lydia V. Panggalo, Iskandar Adnan, Sukma Oktavianthi, Lidwina Priliani, Edison Johar, Filasita A Yudhaputri, Muhammad Rezki Rasyak, Willy Agustine, Sri Masyeni, Erly Sintya, Safarina G Malik, Khin Saw Myint, Amin Soebandrio |
| EPI_ISL_2101041 | Molecular Biology Laboratory, Faculty Medicine and Health Sciences, Warmadewa University | Eijkman Institute for Molecular Biology, National Agency for Research and Innovation; Molecular Biology Laboratory, Faculty Medicine and Health Sciences, Warmadewa University | Sukma Oktavianthi, Lidwina Priliani, Edison Johar, Filasita A Yudhaputri, Muhammad Rezki Rasyak, Willy Agustine, Hidayat Trimarsanto, Lydia V. Panggalo, Iskandar Adnan, Sri Masyeni, Erly Sintya, Safarina G Malik, Khin Saw Myint, Amin Soebandrio |
| EPI_ISL_2101042 | Molecular Biology Laboratory, Faculty Medicine and Health Sciences, Warmadewa University | Eijkman Institute for Molecular Biology, National Agency for Research and Innovation; Molecular Biology Laboratory, Faculty Medicine and Health Sciences, Warmadewa University | Willy Agustine, Hidayat Trimarsanto, Lydia V. Panggalo, Iskandar Adnan, Sukma Oktavianthi, Lidwina Priliani, Edison Johar, Filasita A Yudhaputri, Muhammad Rezki Rasyak, Sri Masyeni, Erly Sintya, Safarina G Malik, Khin Saw Myint, Amin Soebandrio |
| EPI_ISL_2101043 | Molecular Biology Laboratory, Faculty Medicine and Health Sciences, Warmadewa University | Eijkman Institute for Molecular Biology, National Agency for Research and Innovation; Molecular Biology Laboratory, Faculty Medicine and Health Sciences, Warmadewa University | Hidayat Trimarsanto, Lydia V. Panggalo, Iskandar Adnan, Sukma Oktavianthi, Lidwina Priliani, Edison Johar, Filasita A Yudhaputri, Muhammad Rezki Rasyak, Willy Agustine, Sri Masyeni, Erly Sintya, Safarina G Malik, Khin Saw Myint, Amin Soebandrio |
| EPI_ISL_2101044 | Molecular Biology Laboratory, Faculty Medicine and Health Sciences, Warmadewa University | Eijkman Institute for Molecular Biology, National Agency for Research and Innovation; Molecular Biology Laboratory, Faculty Medicine and Health Sciences, Warmadewa University | Lydia V. Panggalo, Iskandar Adnan, Sukma Oktavianthi, Lidwina Priliani, Edison Johar, Filasita A Yudhaputri, Muhammad Rezki Rasyak, Willy Agustine, Hidayat Trimarsanto, Sri Masyeni, Erly Sintya, Safarina G Malik, Khin Saw Myint, Amin Soebandrio |
| EPI_ISL_2101045 | Molecular Biology Laboratory, Faculty Medicine and Health Sciences, Warmadewa University | Eijkman Institute for Molecular Biology, National Agency for Research and Innovation; Molecular Biology Laboratory, Faculty Medicine and Health Sciences, Warmadewa University | Iskandar Adnan, Sukma Oktavianthi, Lidwina Priliani, Edison Johar, Filasita A Yudhaputri, Muhammad Rezki Rasyak, Willy Agustine, Hidayat Trimarsanto, Lydia V. Panggalo, Sri Masyeni, Erly Sintya, Safarina G Malik, Khin Saw Myint, Amin Soebandrio |
| EPI_ISL_2101046 | Molecular Biology Laboratory, Faculty Medicine and Health Sciences, Warmadewa University | Eijkman Institute for Molecular Biology, National Agency for Research and Innovation; Molecular Biology Laboratory, Faculty | Sukma Oktavianthi, Lidwina Priliani, Edison Johar, Filasita A Yudhaputri, Muhammad Rezki Rasyak, Willy Agustine, Hidayat Trimarsanto, Lydia V. Panggalo, Iskandar Adnan, Sri Masyeni, Erly Sintya, Safarina G Malik, Khin Saw Myint, Amin Soebandrio |

| | | | |
|---|---|---|---|
| | | Medicine and Health Sciences, Warmadewa University | |
| EPI_ISL_2101047 | Molecular Biology Laboratory, Faculty Medicine and Health Sciences, Warmadewa University | Eijkman Institute for Molecular Biology, National Agency for Research and Innovation; Molecular Biology Laboratory, Faculty Medicine and Health Sciences, Warmadewa University | Lidwina Priliani, Edison Johar, Frilasita A Yudhaputri, Muhammad Rezki Rasyak, Willy Agustine, Hidayat Trimarsanto, Lydia V. Panggalo, Iskandar Adnan, Sukma Oktavianthi, Sri Masyeni, Erly Sintya, Safarina G Malik, Khin Saw Myint, Amin Soebandrio |
| EPI_ISL_2101048 | Molecular Biology Laboratory, Faculty Medicine and Health Sciences, Warmadewa University | Eijkman Institute for Molecular Biology, National Agency for Research and Innovation; Molecular Biology Laboratory, Faculty Medicine and Health Sciences, Warmadewa University | Edison Johar, Frilasita A Yudhaputri, Muhammad Rezki Rasyak, Willy Agustine, Hidayat Trimarsanto, Lydia V. Panggalo, Iskandar Adnan, Sukma Oktavianthi, Lidwina Priliani, Sri Masyeni, Erly Sintya, Safarina G Malik, Khin Saw Myint, Amin Soebandrio |
| EPI_ISL_2101049 | Molecular Biology Laboratory, Faculty Medicine and Health Sciences, Warmadewa University | Eijkman Institute for Molecular Biology, National Agency for Research and Innovation; Molecular Biology Laboratory, Faculty Medicine and Health Sciences, Warmadewa University | Frilasita A Yudhaputri, Muhammad Rezki Rasyak, Willy Agustine, Hidayat Trimarsanto, Lydia V. Panggalo, Iskandar Adnan, Sukma Oktavianthi, Lidwina Priliani, Edison Johar, Sri Masyeni, Erly Sintya, Safarina G Malik, Khin Saw Myint, Amin Soebandrio |
| EPI_ISL_2101050 | Molecular Biology Laboratory, Faculty Medicine and Health Sciences, Warmadewa University | Eijkman Institute for Molecular Biology, National Agency for Research and Innovation; Molecular Biology Laboratory, Faculty Medicine and Health Sciences, Warmadewa University | Willy Agustine, Hidayat Trimarsanto, Lydia V. Panggalo, Iskandar Adnan, Sukma Oktavianthi, Lidwina Priliani, Edison Johar, Frilasita A Yudhaputri, Muhammad Rezki Rasyak, Sri Masyeni, Erly Sintya, Safarina G Malik, Khin Saw Myint, Amin Soebandrio |
| EPI_ISL_2101051 | Molecular Biology Laboratory, Faculty Medicine and Health Sciences, Warmadewa University | Eijkman Institute for Molecular Biology, National Agency for Research and Innovation; Molecular Biology Laboratory, Faculty Medicine and Health Sciences, Warmadewa University | Hidayat Trimarsanto, Lydia V. Panggalo, Iskandar Adnan, Sukma Oktavianthi, Lidwina Priliani, Edison Johar, Frilasita A Yudhaputri, Muhammad Rezki Rasyak, Willy Agustine, Sri Masyeni, Erly Sintya, Safarina G Malik, Khin Saw Myint, Amin Soebandrio |
| EPI_ISL_2101052 | Molecular Biology Laboratory, Faculty Medicine and Health Sciences, Warmadewa University | Eijkman Institute for Molecular Biology, National Agency for Research and Innovation; Molecular Biology Laboratory, Faculty Medicine and Health Sciences, Warmadewa University | Lydia V. Panggalo, Iskandar Adnan, Sukma Oktavianthi, Lidwina Priliani, Edison Johar, Frilasita A Yudhaputri, Muhammad Rezki Rasyak, Willy Agustine, Hidayat Trimarsanto, Sri Masyeni, Erly Sintya, Safarina G Malik, Khin Saw Myint, Amin Soebandrio |
| EPI_ISL_2101053 | Molecular Biology Laboratory, Faculty Medicine and Health Sciences, Warmadewa University | Eijkman Institute for Molecular Biology, National Agency for Research and Innovation; Molecular Biology Laboratory, Faculty Medicine and Health Sciences, Warmadewa University | Iskandar Adnan, Sukma Oktavianthi, Lidwina Priliani, Edison Johar, Frilasita A Yudhaputri, Muhammad Rezki Rasyak, Willy Agustine, Hidayat Trimarsanto, Lydia V. Panggalo, Sri Masyeni, Erly Sintya, Safarina G Malik, Khin Saw Myint, Amin Soebandrio |
| EPI_ISL_2101054 | Molecular Biology Laboratory, Faculty Medicine and Health Sciences, Warmadewa University | Eijkman Institute for Molecular Biology, National Agency for Research and Innovation; Molecular Biology Laboratory, Faculty Medicine and Health Sciences, Warmadewa University | Sukma Oktavianthi, Lidwina Priliani, Edison Johar, Frilasita A Yudhaputri, Muhammad Rezki Rasyak, Willy Agustine, Hidayat Trimarsanto, Lydia V. Panggalo, Iskandar Adnan, Sri Masyeni, Erly Sintya, Safarina G Malik, Khin Saw Myint, Amin Soebandrio |
| EPI_ISL_2101060 | Molecular Biology Laboratory, Faculty Medicine and Health Sciences, Warmadewa University | Eijkman Institute for Molecular Biology, National Agency for Research and Innovation; Molecular Biology Laboratory, Faculty Medicine and Health Sciences, Warmadewa University | Lydia V. Panggalo, Iskandar Adnan, Sukma Oktavianthi, Lidwina Priliani, Edison Johar, Frilasita A Yudhaputri, Muhammad Rezki Rasyak, Willy Agustine, Hidayat Trimarsanto, Sri Masyeni, Erly Sintya, Safarina G Malik, Khin Saw Myint, Amin Soebandrio |
| EPI_ISL_2101061 | Molecular Biology Laboratory, Faculty Medicine and Health Sciences, Warmadewa University | Eijkman Institute for Molecular Biology, National Agency for Research and Innovation; Molecular Biology Laboratory, Faculty | Iskandar Adnan, Sukma Oktavianthi, Lidwina Priliani, Edison Johar, Frilasita A Yudhaputri, Muhammad Rezki Rasyak, Willy Agustine, Hidayat Trimarsanto, Lydia V. Panggalo, Sri Masyeni, Erly Sintya, Safarina G Malik, Khin Saw Myint, Amin Soebandrio |

| | | | |
|---|---|---|---|
| | | Medicine and Health Sciences, Warmadewa University | |
| EPI_ISL_2101170 | Molecular Biology Laboratory, Faculty Medicine and Health Sciences, Warmadewa University | Eijkman Institute for Molecular Biology, National Agency for Research and Innovation; Molecular Biology Laboratory, Faculty Medicine and Health Sciences, Warmadewa University | Lidwina Priliani, Edison Johar, Frilasita A Yudhaputri, Muhammad Rezki Rasyak, Willy Agustine, Hidayat Trimarsanto, Lydia V. Panggalo, Iskandar Adnan, Sukma Oktavianthi, Sri Masyeni, Erly Sintya, Safarina G Malik, Khin Saw Myint, Amin Soebandrio |
| EPI_ISL_2101171 | Molecular Biology Laboratory, Faculty Medicine and Health Sciences, Warmadewa University | Eijkman Institute for Molecular Biology, National Agency for Research and Innovation; Molecular Biology Laboratory, Faculty Medicine and Health Sciences, Warmadewa University | Edison Johar, Frilasita A Yudhaputri, Muhammad Rezki Rasyak, Willy Agustine, Hidayat Trimarsanto, Lydia V. Panggalo, Iskandar Adnan, Sukma Oktavianthi, Lidwina Priliani, Sri Masyeni, Erly Sintya, Safarina G Malik, Khin Saw Myint, Amin Soebandrio |
| EPI_ISL_2101172 | Molecular Biology Laboratory, Faculty Medicine and Health Sciences, Warmadewa University | Eijkman Institute for Molecular Biology, National Agency for Research and Innovation; Molecular Biology Laboratory, Faculty Medicine and Health Sciences, Warmadewa University | Iskandar Adnan, Sukma Oktavianthi, Lidwina Priliani, Edison Johar, Frilasita A Yudhaputri, Muhammad Rezki Rasyak, Willy Agustine, Hidayat Trimarsanto, Lydia V. Panggalo, Sri Masyeni, Erly Sintya, Safarina G Malik, Khin Saw Myint, Amin Soebandrio |
| EPI_ISL_2101173 | Molecular Biology Laboratory, Faculty Medicine and Health Sciences, Warmadewa University | Eijkman Institute for Molecular Biology, National Agency for Research and Innovation; Molecular Biology Laboratory, Faculty Medicine and Health Sciences, Warmadewa University | Sukma Oktavianthi, Lidwina Priliani, Edison Johar, Frilasita A Yudhaputri, Muhammad Rezki Rasyak, Willy Agustine, Hidayat Trimarsanto, Lydia V. Panggalo, Iskandar Adnan, Sri Masyeni, Erly Sintya, Safarina G Malik, Khin Saw Myint, Amin Soebandrio |
| EPI_ISL_2101174 | Molecular Biology Laboratory, Faculty Medicine and Health Sciences, Warmadewa University | Eijkman Institute for Molecular Biology, National Agency for Research and Innovation; Molecular Biology Laboratory, Faculty Medicine and Health Sciences, Warmadewa University | Lidwina Priliani, Edison Johar, Frilasita A Yudhaputri, Muhammad Rezki Rasyak, Willy Agustine, Hidayat Trimarsanto, Lydia V. Panggalo, Iskandar Adnan, Sukma Oktavianthi, Sri Masyeni, Erly Sintya, Safarina G Malik, Khin Saw Myint, Amin Soebandrio |
| EPI_ISL_2101175 | Molecular Biology Laboratory, Faculty Medicine and Health Sciences, Warmadewa University | Eijkman Institute for Molecular Biology, National Agency for Research and Innovation; Molecular Biology Laboratory, Faculty Medicine and Health Sciences, Warmadewa University | Edison Johar, Frilasita A Yudhaputri, Muhammad Rezki Rasyak, Willy Agustine, Hidayat Trimarsanto, Lydia V. Panggalo, Iskandar Adnan, Sukma Oktavianthi, Lidwina Priliani, Sri Masyeni, Erly Sintya, Safarina G Malik, Khin Saw Myint, Amin Soebandrio |
| EPI_ISL_2101176 | Molecular Biology Laboratory, Faculty Medicine and Health Sciences, Warmadewa University | Eijkman Institute for Molecular Biology, National Agency for Research and Innovation; Molecular Biology Laboratory, Faculty Medicine and Health Sciences, Warmadewa University | Frilasita A Yudhaputri, Muhammad Rezki Rasyak, Willy Agustine, Hidayat Trimarsanto, Lydia V. Panggalo, Iskandar Adnan, Sukma Oktavianthi, Lidwina Priliani, Edison Johar, Sri Masyeni, Erly Sintya, Safarina G Malik, Khin Saw Myint, Amin Soebandrio |
| EPI_ISL_2101177 | Molecular Biology Laboratory, Faculty Medicine and Health Sciences, Warmadewa University | Eijkman Institute for Molecular Biology, National Agency for Research and Innovation; Molecular Biology Laboratory, Faculty Medicine and Health Sciences, Warmadewa University | Willy Agustine, Hidayat Trimarsanto, Lydia V. Panggalo, Iskandar Adnan, Sukma Oktavianthi, Lidwina Priliani, Edison Johar, Frilasita A Yudhaputri, Muhammad Rezki Rasyak, Sri Masyeni, Erly Sintya, Safarina G Malik, Khin Saw Myint, Amin Soebandrio |
| EPI_ISL_2101178 | Molecular Biology Laboratory, Faculty Medicine and Health Sciences, Warmadewa University | Eijkman Institute for Molecular Biology, National Agency for Research and Innovation; Molecular Biology Laboratory, Faculty Medicine and Health Sciences, Warmadewa University | Hidayat Trimarsanto, Lydia V. Panggalo, Iskandar Adnan, Sukma Oktavianthi, Lidwina Priliani, Edison Johar, Frilasita A Yudhaputri, Muhammad Rezki Rasyak, Willy Agustine, Sri Masyeni, Erly Sintya, Safarina G Malik, Khin Saw Myint, Amin Soebandrio |
| EPI_ISL_2101179 | Molecular Biology Laboratory, Faculty Medicine and Health Sciences, Warmadewa University | Eijkman Institute for Molecular Biology, National Agency for Research and Innovation; Molecular Biology Laboratory, Faculty | Lydia V. Panggalo, Iskandar Adnan, Sukma Oktavianthi, Lidwina Priliani, Edison Johar, Frilasita A Yudhaputri, Muhammad Rezki Rasyak, Willy Agustine, Hidayat Trimarsanto, Sri Masyeni, Erly Sintya, Safarina G Malik, Khin Saw Myint, Amin Soebandrio |

| | | | |
|---|---|---|---|
| | | Medicine and Health Sciences, Warmadewa University | |
| EPI_ISL_2101180 | Molecular Biology Laboratory, Faculty Medicine and Health Sciences, Warmadewa University | Eijkman Institute for Molecular Biology, National Agency for Research and Innovation; Molecular Biology Laboratory, Faculty Medicine and Health Sciences, Warmadewa University | Sukma Oktavianthi, Lidwina Priliani, Edison Johar, Frilasita A Yudhaputri, Muhammad Rezki Rasyak, Willy Agustine, Hidayat Trimarsanto, Lydia V. Panggalo, Iskandar Adnan, Sri Masyeni, Erly Sintya, Safarina G Malik, Khin Saw Myint, Amin Soebandrio |
| EPI_ISL_2101181 | Molecular Biology Laboratory, Faculty Medicine and Health Sciences, Warmadewa University | Eijkman Institute for Molecular Biology, National Agency for Research and Innovation; Molecular Biology Laboratory, Faculty Medicine and Health Sciences, Warmadewa University | Lidwina Priliani, Edison Johar, Frilasita A Yudhaputri, Muhammad Rezki Rasyak, Willy Agustine, Hidayat Trimarsanto, Lydia V. Panggalo, Iskandar Adnan, Sukma Oktavianthi, Sri Masyeni, Erly Sintya, Safarina G Malik, Khin Saw Myint, Amin Soebandrio |
| EPI_ISL_2101182 | Molecular Biology Laboratory, Faculty Medicine and Health Sciences, Warmadewa University | Eijkman Institute for Molecular Biology, National Agency for Research and Innovation; Molecular Biology Laboratory, Faculty Medicine and Health Sciences, Warmadewa University | Edison Johar, Frilasita A Yudhaputri, Muhammad Rezki Rasyak, Willy Agustine, Hidayat Trimarsanto, Lydia V. Panggalo, Iskandar Adnan, Sukma Oktavianthi, Lidwina Priliani, Sri Masyeni, Erly Sintya, Safarina G Malik, Khin Saw Myint, Amin Soebandrio |
| EPI_ISL_2101183 | Molecular Biology Laboratory, Faculty Medicine and Health Sciences, Warmadewa University | Eijkman Institute for Molecular Biology, National Agency for Research and Innovation; Molecular Biology Laboratory, Faculty Medicine and Health Sciences, Warmadewa University | Frilasita A Yudhaputri, Muhammad Rezki Rasyak, Willy Agustine, Hidayat Trimarsanto, Lydia V. Panggalo, Iskandar Adnan, Sukma Oktavianthi, Lidwina Priliani, Edison Johar, Sri Masyeni, Erly Sintya, Safarina G Malik, Khin Saw Myint, Amin Soebandrio |
| EPI_ISL_2101184 | Molecular Biology Laboratory, Faculty Medicine and Health Sciences, Warmadewa University | Eijkman Institute for Molecular Biology, National Agency for Research and Innovation; Molecular Biology Laboratory, Faculty Medicine and Health Sciences, Warmadewa University | Willy Agustine, Hidayat Trimarsanto, Lydia V. Panggalo, Iskandar Adnan, Sukma Oktavianthi, Lidwina Priliani, Edison Johar, Frilasita A Yudhaputri, Muhammad Rezki Rasyak, Sri Masyeni, Erly Sintya, Safarina G Malik, Khin Saw Myint, Amin Soebandrio |
| EPI_ISL_2101185 | Molecular Biology Laboratory, Faculty Medicine and Health Sciences, Warmadewa University | Eijkman Institute for Molecular Biology, National Agency for Research and Innovation; Molecular Biology Laboratory, Faculty Medicine and Health Sciences, Warmadewa University | Hidayat Trimarsanto, Lydia V. Panggalo, Iskandar Adnan, Sukma Oktavianthi, Lidwina Priliani, Edison Johar, Frilasita A Yudhaputri, Muhammad Rezki Rasyak, Willy Agustine, Sri Masyeni, Erly Sintya, Safarina G Malik, Khin Saw Myint, Amin Soebandrio |
| EPI_ISL_2101186 | Molecular Biology Laboratory, Faculty Medicine and Health Sciences, Warmadewa University | Eijkman Institute for Molecular Biology, National Agency for Research and Innovation; Molecular Biology Laboratory, Faculty Medicine and Health Sciences, Warmadewa University | Lydia V. Panggalo, Iskandar Adnan, Sukma Oktavianthi, Lidwina Priliani, Edison Johar, Frilasita A Yudhaputri, Muhammad Rezki Rasyak, Willy Agustine, Hidayat Trimarsanto, Sri Masyeni, Erly Sintya, Safarina G Malik, Khin Saw Myint, Amin Soebandrio |
| EPI_ISL_2101187 | Molecular Biology Laboratory, Faculty Medicine and Health Sciences, Warmadewa University | Eijkman Institute for Molecular Biology, National Agency for Research and Innovation; Molecular Biology Laboratory, Faculty Medicine and Health Sciences, Warmadewa University | Iskandar Adnan, Sukma Oktavianthi, Lidwina Priliani, Edison Johar, Frilasita A Yudhaputri, Muhammad Rezki Rasyak, Willy Agustine, Hidayat Trimarsanto, Lydia V. Panggalo, Sri Masyeni, Erly Sintya, Safarina G Malik, Khin Saw Myint, Amin Soebandrio |
| EPI_ISL_2101188 | Molecular Biology Laboratory, Faculty Medicine and Health Sciences, Warmadewa University | Eijkman Institute for Molecular Biology, National Agency for Research and Innovation; Molecular Biology Laboratory, Faculty Medicine and Health Sciences, Warmadewa University | Sukma Oktavianthi, Lidwina Priliani, Edison Johar, Frilasita A Yudhaputri, Muhammad Rezki Rasyak, Willy Agustine, Hidayat Trimarsanto, Lydia V. Panggalo, Iskandar Adnan, Sri Masyeni, Erly Sintya, Safarina G Malik, Khin Saw Myint, Amin Soebandrio |
| EPI_ISL_2101189 | Molecular Biology Laboratory, Faculty Medicine and Health Sciences, Warmadewa University | Eijkman Institute for Molecular Biology, National Agency for Research and Innovation; Molecular Biology Laboratory, Faculty | Lidwina Priliani, Edison Johar, Frilasita A Yudhaputri, Muhammad Rezki Rasyak, Willy Agustine, Hidayat Trimarsanto, Lydia V. Panggalo, Iskandar Adnan, Sukma Oktavianthi, Sri Masyeni, Erly Sintya, Safarina G Malik, Khin Saw Myint, Amin Soebandrio |

| | | | |
|---|---|---|---|
| | | Medicine and Health Sciences, Warmadewa University | |
| EPI_ISL_2101190 | Molecular Biology Laboratory, Faculty Medicine and Health Sciences, Warmadewa University | Eijkman Institute for Molecular Biology, National Agency for Research and Innovation; Molecular Biology Laboratory, Faculty Medicine and Health Sciences, Warmadewa University | Frilasita A Yudhaputri, Muhammad Rezki Rasyak, Willy Agustine, Hidayat Trimarsanto, Lydia V. Panggalo, Iskandar Adnan, Sukma Oktavianthi, Lidwina Priliani, Edison Johar, Sri Masyeni, Erly Sintya, Safarina G Malik, Khin Saw Myint, Amin Soebandrio |
| EPI_ISL_2101191 | Molecular Biology Laboratory, Faculty Medicine and Health Sciences, Warmadewa University | Eijkman Institute for Molecular Biology, National Agency for Research and Innovation; Molecular Biology Laboratory, Faculty Medicine and Health Sciences, Warmadewa University | Willy Agustine, Hidayat Trimarsanto, Lydia V. Panggalo, Iskandar Adnan, Sukma Oktavianthi, Lidwina Priliani, Edison Johar, Frilasita A Yudhaputri, Muhammad Rezki Rasyak, Sri Masyeni, Erly Sintya, Safarina G Malik, Khin Saw Myint, Amin Soebandrio |
| EPI_ISL_2101192 | Molecular Biology Laboratory, Faculty Medicine and Health Sciences, Warmadewa University | Eijkman Institute for Molecular Biology, National Agency for Research and Innovation; Molecular Biology Laboratory, Faculty Medicine and Health Sciences, Warmadewa University | Hidayat Trimarsanto, Lydia V. Panggalo, Iskandar Adnan, Sukma Oktavianthi, Lidwina Priliani, Edison Johar, Frilasita A Yudhaputri, Muhammad Rezki Rasyak, Willy Agustine, Sri Masyeni, Erly Sintya, Safarina G Malik, Khin Saw Myint, Amin Soebandrio |
| EPI_ISL_2101193 | Molecular Biology Laboratory, Faculty Medicine and Health Sciences, Warmadewa University | Eijkman Institute for Molecular Biology, National Agency for Research and Innovation; Molecular Biology Laboratory, Faculty Medicine and Health Sciences, Warmadewa University | Lydia V. Panggalo, Iskandar Adnan, Sukma Oktavianthi, Lidwina Priliani, Edison Johar, Frilasita A Yudhaputri, Muhammad Rezki Rasyak, Willy Agustine, Hidayat Trimarsanto, Sri Masyeni, Erly Sintya, Safarina G Malik, Khin Saw Myint, Amin Soebandrio |
| EPI_ISL_2101194 | Molecular Biology Laboratory, Faculty Medicine and Health Sciences, Warmadewa University | Eijkman Institute for Molecular Biology, National Agency for Research and Innovation; Molecular Biology Laboratory, Faculty Medicine and Health Sciences, Warmadewa University | Iskandar Adnan, Sukma Oktavianthi, Lidwina Priliani, Edison Johar, Frilasita A Yudhaputri, Muhammad Rezki Rasyak, Willy Agustine, Hidayat Trimarsanto, Lydia V. Panggalo, Sri Masyeni, Erly Sintya, Safarina G Malik, Khin Saw Myint, Amin Soebandrio |
| EPI_ISL_2101195 | Molecular Biology Laboratory, Faculty Medicine and Health Sciences, Warmadewa University | Eijkman Institute for Molecular Biology, National Agency for Research and Innovation; Molecular Biology Laboratory, Faculty Medicine and Health Sciences, Warmadewa University | Sukma Oktavianthi, Lidwina Priliani, Edison Johar, Frilasita A Yudhaputri, Muhammad Rezki Rasyak, Willy Agustine, Hidayat Trimarsanto, Lydia V. Panggalo, Iskandar Adnan, Sri Masyeni, Erly Sintya, Safarina G Malik, Khin Saw Myint, Amin Soebandrio |
| EPI_ISL_2101196 | Molecular Biology Laboratory, Faculty Medicine and Health Sciences, Warmadewa University | Eijkman Institute for Molecular Biology, National Agency for Research and Innovation; Molecular Biology Laboratory, Faculty Medicine and Health Sciences, Warmadewa University | Lidwina Priliani, Edison Johar, Frilasita A Yudhaputri, Muhammad Rezki Rasyak, Willy Agustine, Hidayat Trimarsanto, Lydia V. Panggalo, Iskandar Adnan, Sukma Oktavianthi, Sri Masyeni, Erly Sintya, Safarina G Malik, Khin Saw Myint, Amin Soebandrio |
| EPI_ISL_2101197 | Molecular Biology Laboratory, Faculty Medicine and Health Sciences, Warmadewa University | Eijkman Institute for Molecular Biology, National Agency for Research and Innovation; Molecular Biology Laboratory, Faculty Medicine and Health Sciences, Warmadewa University | Edison Johar, Frilasita A Yudhaputri, Muhammad Rezki Rasyak, Willy Agustine, Hidayat Trimarsanto, Lydia V. Panggalo, Iskandar Adnan, Sukma Oktavianthi, Lidwina Priliani, Sri Masyeni, Erly Sintya, Safarina G Malik, Khin Saw Myint, Amin Soebandrio |
| EPI_ISL_2101198 | Molecular Biology Laboratory, Faculty Medicine and Health Sciences, Warmadewa University | Eijkman Institute for Molecular Biology, National Agency for Research and Innovation; Molecular Biology Laboratory, Faculty Medicine and Health Sciences, Warmadewa University | Frilasita A Yudhaputri, Muhammad Rezki Rasyak, Willy Agustine, Hidayat Trimarsanto, Lydia V. Panggalo, Iskandar Adnan, Sukma Oktavianthi, Lidwina Priliani, Edison Johar, Sri Masyeni, Erly Sintya, Safarina G Malik, Khin Saw Myint, Amin Soebandrio |
| EPI_ISL_2101199 | Molecular Biology Laboratory, Faculty Medicine and Health Sciences, Warmadewa University | Eijkman Institute for Molecular Biology, National Agency for Research and Innovation; Molecular Biology Laboratory, Faculty | Willy Agustine, Hidayat Trimarsanto, Lydia V. Panggalo, Iskandar Adnan, Sukma Oktavianthi, Lidwina Priliani, Edison Johar, Frilasita A Yudhaputri, Muhammad Rezki Rasyak, Sri Masyeni, Erly Sintya, Safarina G Malik, Khin Saw Myint, Amin Soebandrio |

| | | | |
|---|---|---|---|
| | | Medicine and Health Sciences, Warmadewa University | |
| EPI_ISL_2101200 | Molecular Biology Laboratory, Faculty Medicine and Health Sciences, Warmadewa University | Eijkman Institute for Molecular Biology, National Agency for Research and Innovation; Molecular Biology Laboratory, Faculty Medicine and Health Sciences, Warmadewa University | Lydia V. Panggalo, Iskandar Adnan, Sukma Oktavianthi, Lidwina Priliani, Edison Johar, Frilasita A Yudhaputri, Muhammad Rezki Rasyak, Willy Agustine, Hidayat Trimarsanto, Sri Masyeni, Erly Sintya, Safarina G Malik, Khin Saw Myint, Amin Soebandrio |
| EPI_ISL_2101202 | Molecular Biology Laboratory, Faculty Medicine and Health Sciences, Warmadewa University | Eijkman Institute for Molecular Biology, National Agency for Research and Innovation; Molecular Biology Laboratory, Faculty Medicine and Health Sciences, Warmadewa University | Iskandar Adnan, Sukma Oktavianthi, Lidwina Priliani, Edison Johar, Frilasita A Yudhaputri, Muhammad Rezki Rasyak, Willy Agustine, Hidayat Trimarsanto, Lydia V. Panggalo, Sri Masyeni, Erly Sintya, Safarina G Malik, Khin Saw Myint, Amin Soebandrio |
| EPI_ISL_2101203 | Molecular Biology Laboratory, Faculty Medicine and Health Sciences, Warmadewa University | Eijkman Institute for Molecular Biology, National Agency for Research and Innovation; Molecular Biology Laboratory, Faculty Medicine and Health Sciences, Warmadewa University | Sukma Oktavianthi, Lidwina Priliani, Edison Johar, Frilasita A Yudhaputri, Muhammad Rezki Rasyak, Willy Agustine, Hidayat Trimarsanto, Lydia V. Panggalo, Iskandar Adnan, Sri Masyeni, Erly Sintya, Safarina G Malik, Khin Saw Myint, Amin Soebandrio |
| EPI_ISL_2101204 | Molecular Biology Laboratory, Faculty Medicine and Health Sciences, Warmadewa University | Eijkman Institute for Molecular Biology, National Agency for Research and Innovation; Molecular Biology Laboratory, Faculty Medicine and Health Sciences, Warmadewa University | Lidwina Priliani, Edison Johar, Frilasita A Yudhaputri, Muhammad Rezki Rasyak, Willy Agustine, Hidayat Trimarsanto, Lydia V. Panggalo, Iskandar Adnan, Sukma Oktavianthi, Sri Masyeni, Erly Sintya, Safarina G Malik, Khin Saw Myint, Amin Soebandrio |
| EPI_ISL_2101205 | Molecular Biology Laboratory, Faculty Medicine and Health Sciences, Warmadewa University | Eijkman Institute for Molecular Biology, National Agency for Research and Innovation; Molecular Biology Laboratory, Faculty Medicine and Health Sciences, Warmadewa University | Edison Johar, Frilasita A Yudhaputri, Muhammad Rezki Rasyak, Willy Agustine, Hidayat Trimarsanto, Lydia V. Panggalo, Iskandar Adnan, Sukma Oktavianthi, Lidwina Priliani, Sri Masyeni, Erly Sintya, Safarina G Malik, Khin Saw Myint, Amin Soebandrio |
| EPI_ISL_2101206 | Molecular Biology Laboratory, Faculty Medicine and Health Sciences, Warmadewa University | Eijkman Institute for Molecular Biology, National Agency for Research and Innovation; Molecular Biology Laboratory, Faculty Medicine and Health Sciences, Warmadewa University | Frilasita A Yudhaputri, Muhammad Rezki Rasyak, Willy Agustine, Hidayat Trimarsanto, Lydia V. Panggalo, Iskandar Adnan, Sukma Oktavianthi, Lidwina Priliani, Edison Johar, Sri Masyeni, Erly Sintya, Safarina G Malik, Khin Saw Myint, Amin Soebandrio |
| EPI_ISL_2101207 | Molecular Biology Laboratory, Faculty Medicine and Health Sciences, Warmadewa University | Eijkman Institute for Molecular Biology, National Agency for Research and Innovation; Molecular Biology Laboratory, Faculty Medicine and Health Sciences, Warmadewa University | Willy Agustine, Hidayat Trimarsanto, Lydia V. Panggalo, Iskandar Adnan, Sukma Oktavianthi, Lidwina Priliani, Edison Johar, Frilasita A Yudhaputri, Muhammad Rezki Rasyak, Sri Masyeni, Erly Sintya, Safarina G Malik, Khin Saw Myint, Amin Soebandrio |
| EPI_ISL_2101208 | Molecular Biology Laboratory, Faculty Medicine and Health Sciences, Warmadewa University | Eijkman Institute for Molecular Biology, National Agency for Research and Innovation; Molecular Biology Laboratory, Faculty Medicine and Health Sciences, Warmadewa University | Hidayat Trimarsanto, Lydia V. Panggalo, Iskandar Adnan, Sukma Oktavianthi, Lidwina Priliani, Edison Johar, Frilasita A Yudhaputri, Muhammad Rezki Rasyak, Willy Agustine, Sri Masyeni, Erly Sintya, Safarina G Malik, Khin Saw Myint, Amin Soebandrio |
| EPI_ISL_2101209 | Molecular Biology Laboratory, Faculty Medicine and Health Sciences, Warmadewa University | Eijkman Institute for Molecular Biology, National Agency for Research and Innovation; Molecular Biology Laboratory, Faculty Medicine and Health Sciences, Warmadewa University | Lydia V. Panggalo, Iskandar Adnan, Sukma Oktavianthi, Lidwina Priliani, Edison Johar, Frilasita A Yudhaputri, Muhammad Rezki Rasyak, Willy Agustine, Hidayat Trimarsanto, Sri Masyeni, Erly Sintya, Safarina G Malik, Khin Saw Myint, Amin Soebandrio |
| EPI_ISL_2101210 | Molecular Biology Laboratory, Faculty Medicine and Health Sciences, Warmadewa University | Eijkman Institute for Molecular Biology, National Agency for Research and Innovation; Molecular Biology Laboratory, Faculty | Iskandar Adnan, Sukma Oktavianthi, Lidwina Priliani, Edison Johar, Frilasita A Yudhaputri, Muhammad Rezki Rasyak, Willy Agustine, Hidayat Trimarsanto, Lydia V. Panggalo, Sri Masyeni, Erly Sintya, Safarina G Malik, Khin Saw Myint, Amin Soebandrio |

| | | | |
|---|---|---|---|
| | | Medicine and Health Sciences, Warmadewa University | |
| EPI_ISL_2101211 | Molecular Biology Laboratory, Faculty Medicine and Health Sciences, Warmadewa University | Eijkman Institute for Molecular Biology, National Agency for Research and Innovation; Molecular Biology Laboratory, Faculty Medicine and Health Sciences, Warmadewa University | Lidwina Priliani, Edison Johar, Frilasita A Yudhaputri, Muhammad Rezki Rasyak, Willy Agustine, Hidayat Trimarsanto, Lydia V. Panggalo, Iskandar Adnan, Sukma Oktavianthi, Sri Masyeni, Erly Sintya, Safarina G Malik, Khin Saw Myint, Amin Soebandrio |
| EPI_ISL_2101212 | Molecular Biology Laboratory, Faculty Medicine and Health Sciences, Warmadewa University | Eijkman Institute for Molecular Biology, National Agency for Research and Innovation; Molecular Biology Laboratory, Faculty Medicine and Health Sciences, Warmadewa University | Edison Johar, Frilasita A Yudhaputri, Muhammad Rezki Rasyak, Willy Agustine, Hidayat Trimarsanto, Lydia V. Panggalo, Iskandar Adnan, Sukma Oktavianthi, Lidwina Priliani, Sri Masyeni, Erly Sintya, Safarina G Malik, Khin Saw Myint, Amin Soebandrio |
| EPI_ISL_2101213 | Molecular Biology Laboratory, Faculty Medicine and Health Sciences, Warmadewa University | Eijkman Institute for Molecular Biology, National Agency for Research and Innovation; Molecular Biology Laboratory, Faculty Medicine and Health Sciences, Warmadewa University | Frilasita A Yudhaputri, Muhammad Rezki Rasyak, Willy Agustine, Hidayat Trimarsanto, Lydia V. Panggalo, Iskandar Adnan, Sukma Oktavianthi, Lidwina Priliani, Edison Johar, Sri Masyeni, Erly Sintya, Safarina G Malik, Khin Saw Myint, Amin Soebandrio |
| EPI_ISL_2101214 | Molecular Biology Laboratory, Faculty Medicine and Health Sciences, Warmadewa University | Eijkman Institute for Molecular Biology, National Agency for Research and Innovation; Molecular Biology Laboratory, Faculty Medicine and Health Sciences, Warmadewa University | Willy Agustine, Hidayat Trimarsanto, Lydia V. Panggalo, Iskandar Adnan, Sukma Oktavianthi, Lidwina Priliani, Edison Johar, Frilasita A Yudhaputri, Muhammad Rezki Rasyak, Sri Masyeni, Erly Sintya, Safarina G Malik, Khin Saw Myint, Amin Soebandrio |
| EPI_ISL_2101215 | Molecular Biology Laboratory, Faculty Medicine and Health Sciences, Warmadewa University | Eijkman Institute for Molecular Biology, National Agency for Research and Innovation; Molecular Biology Laboratory, Faculty Medicine and Health Sciences, Warmadewa University | Hidayat Trimarsanto, Lydia V. Panggalo, Iskandar Adnan, Sukma Oktavianthi, Lidwina Priliani, Edison Johar, Frilasita A Yudhaputri, Muhammad Rezki Rasyak, Willy Agustine, Sri Masyeni, Erly Sintya, Safarina G Malik, Khin Saw Myint, Amin Soebandrio |
| EPI_ISL_2101236 | RSUD Klungkung | Eijkman Institute for Molecular Biology, National Agency for Research and Innovation; Molecular Biology Laboratory, Faculty Medicine and Health Sciences, Warmadewa University | Frilasita A Yudhaputri, Muhammad Rezki Rasyak, Willy Agustine, Hidayat Trimarsanto, Lydia V. Panggalo, Iskandar Adnan, Sukma Oktavianthi, Lidwina Priliani, Edison Johar, Sri Masyeni, Erly Sintya, I Komang Parwata, Safarina G Malik, Khin Saw Myint, Amin Soebandrio |
| EPI_ISL_2101237 | RSUD Klungkung | Eijkman Institute for Molecular Biology, National Agency for Research and Innovation; Molecular Biology Laboratory, Faculty Medicine and Health Sciences, Warmadewa University | Willy Agustine, Hidayat Trimarsanto, Lydia V. Panggalo, Iskandar Adnan, Sukma Oktavianthi, Lidwina Priliani, Edison Johar, Frilasita A Yudhaputri, Muhammad Rezki Rasyak, Sri Masyeni, Erly Sintya, I Komang Parwata, Safarina G Malik, Khin Saw Myint, Amin Soebandrio |
| EPI_ISL_2101238 | RSUD Klungkung | Eijkman Institute for Molecular Biology, National Agency for Research and Innovation; Molecular Biology Laboratory, Faculty Medicine and Health Sciences, Warmadewa University | Iskandar Adnan, Sukma Oktavianthi, Lidwina Priliani, Edison Johar, Frilasita A Yudhaputri, Muhammad Rezki Rasyak, Willy Agustine, Hidayat Trimarsanto, Lydia V. Panggalo, Sri Masyeni, Erly Sintya, I Komang Parwata, Safarina G Malik, Khin Saw Myint, Amin Soebandrio |
| EPI_ISL_2101239 | RSUD Klungkung | Eijkman Institute for Molecular Biology, National Agency for Research and Innovation; Molecular Biology Laboratory, Faculty Medicine and Health Sciences, Warmadewa University | Lidwina Priliani, Edison Johar, Frilasita A Yudhaputri, Muhammad Rezki Rasyak, Willy Agustine, Hidayat Trimarsanto, Lydia V. Panggalo, Iskandar Adnan, Sukma Oktavianthi, Sri Masyeni, Erly Sintya, I Komang Parwata, Safarina G Malik, Khin Saw Myint, Amin Soebandrio |
| EPI_ISL_2101240 | RSUD Klungkung | Eijkman Institute for Molecular Biology, National Agency for Research and Innovation; Molecular Biology Laboratory, Faculty | Edison Johar, Frilasita A Yudhaputri, Muhammad Rezki Rasyak, Willy Agustine, Hidayat Trimarsanto, Lydia V. Panggalo, Iskandar Adnan, Sukma Oktavianthi, Lidwina Priliani, Sri Masyeni, Erly Sintya, I Komang Parwata, Safarina G Malik, Khin Saw Myint, Amin Soebandrio |

| | | | |
|---|---|---|---|
| | | Medicine and Health Sciences, Warmadewa University | |
| EPI_ISL_2101241 | RSUD Klungkung | Eijkman Institute for Molecular Biology, National Agency for Research and Innovation; Molecular Biology Laboratory, Faculty Medicine and Health Sciences, Warmadewa University | Frilasita A Yudhaputri, Muhammad Rezki Rasyak, Willy Agustine, Hidayat Trimarsanto, Lydia V. Panggalo, Iskandar Adnan, Sukma Oktavianthi, Lidwina Priliani, Edison Johar, Sri Masyeni, Erly Sintya, I Komang Parwata, Safarina G Malik, Khin Saw Myint, Amin Soebandrio |
| EPI_ISL_2101242 | RSUD Klungkung | Eijkman Institute for Molecular Biology, National Agency for Research and Innovation; Molecular Biology Laboratory, Faculty Medicine and Health Sciences, Warmadewa University | Muhammad Rezki Rasyak, Willy Agustine, Hidayat Trimarsanto, Lydia V. Panggalo, Iskandar Adnan, Sukma Oktavianthi, Lidwina Priliani, Edison Johar, Frilasita A Yudhaputri, Sri Masyeni, Erly Sintya, I Komang Parwata, Safarina G Malik, Khin Saw Myint, Amin Soebandrio |
| EPI_ISL_2101243 | RSUD Klungkung | Eijkman Institute for Molecular Biology, National Agency for Research and Innovation; Molecular Biology Laboratory, Faculty Medicine and Health Sciences, Warmadewa University | Willy Agustine, Hidayat Trimarsanto, Lydia V. Panggalo, Iskandar Adnan, Sukma Oktavianthi, Lidwina Priliani, Edison Johar, Frilasita A Yudhaputri, Muhammad Rezki Rasyak, Sri Masyeni, Erly Sintya, I Komang Parwata, Safarina G Malik, Khin Saw Myint, Amin Soebandrio |
| EPI_ISL_2101244 | RSUD Klungkung | Eijkman Institute for Molecular Biology, National Agency for Research and Innovation; Molecular Biology Laboratory, Faculty Medicine and Health Sciences, Warmadewa University | Lydia V. Panggalo, Iskandar Adnan, Sukma Oktavianthi, Lidwina Priliani, Edison Johar, Frilasita A Yudhaputri, Muhammad Rezki Rasyak, Willy Agustine, Hidayat Trimarsanto, Sri Masyeni, Erly Sintya, I Komang Parwata, Safarina G Malik, Khin Saw Myint, Amin Soebandrio |
| EPI_ISL_2101245 | RSUD Klungkung | Eijkman Institute for Molecular Biology, National Agency for Research and Innovation; Molecular Biology Laboratory, Faculty Medicine and Health Sciences, Warmadewa University | Iskandar Adnan, Sukma Oktavianthi, Lidwina Priliani, Edison Johar, Frilasita A Yudhaputri, Muhammad Rezki Rasyak, Willy Agustine, Hidayat Trimarsanto, Lydia V. Panggalo, Sri Masyeni, Erly Sintya, I Komang Parwata, Safarina G Malik, Khin Saw Myint, Amin Soebandrio |
| EPI_ISL_2101256 | RSUD Mangusada | Eijkman Institute for Molecular Biology, National Agency for Research and Innovation; Molecular Biology Laboratory, Faculty Medicine and Health Sciences, Warmadewa University | Edison Johar, Frilasita A Yudhaputri, Muhammad Rezki Rasyak, Willy Agustine, Hidayat Trimarsanto, Lydia V. Panggalo, Iskandar Adnan, Sukma Oktavianthi, Lidwina Priliani, Sri Masyeni, Erly Sintya, Ida Ayu Wayan Mahayani, Safarina G Malik, Khin Saw Myint, Amin Soebandrio |
| EPI_ISL_2101257 | RSUD Mangusada | Eijkman Institute for Molecular Biology, National Agency for Research and Innovation; Molecular Biology Laboratory, Faculty Medicine and Health Sciences, Warmadewa University | Frilasita A Yudhaputri, Muhammad Rezki Rasyak, Willy Agustine, Hidayat Trimarsanto, Lydia V. Panggalo, Iskandar Adnan, Sukma Oktavianthi, Lidwina Priliani, Edison Johar, Sri Masyeni, Erly Sintya, Ida Ayu Wayan Mahayani, Safarina G Malik, Khin Saw Myint, Amin Soebandrio |
| EPI_ISL_2101258 | RSUD Mangusada | Eijkman Institute for Molecular Biology, National Agency for Research and Innovation; Molecular Biology Laboratory, Faculty Medicine and Health Sciences, Warmadewa University | Muhammad Rezki Rasyak, Willy Agustine, Hidayat Trimarsanto, Lydia V. Panggalo, Iskandar Adnan, Sukma Oktavianthi, Lidwina Priliani, Edison Johar, Frilasita A Yudhaputri, Sri Masyeni, Erly Sintya, Ida Ayu Wayan Mahayani, Safarina G Malik, Khin Saw Myint, Amin Soebandrio |
| EPI_ISL_2101259 | RSUD Mangusada | Eijkman Institute for Molecular Biology, National Agency for Research and Innovation; Molecular Biology Laboratory, Faculty Medicine and Health Sciences, Warmadewa University | Willy Agustine, Hidayat Trimarsanto, Lydia V. Panggalo, Iskandar Adnan, Sukma Oktavianthi, Lidwina Priliani, Edison Johar, Frilasita A Yudhaputri, Muhammad Rezki Rasyak, Sri Masyeni, Erly Sintya, Ida Ayu Wayan Mahayani, Safarina G Malik, Khin Saw Myint, Amin Soebandrio |
| EPI_ISL_2101260 | RSUD Mangusada | Eijkman Institute for Molecular Biology, National Agency for Research and Innovation; Molecular Biology Laboratory, Faculty | Hidayat Trimarsanto, Lydia V. Panggalo, Iskandar Adnan, Sukma Oktavianthi, Lidwina Priliani, Edison Johar, Frilasita A Yudhaputri, Muhammad Rezki Rasyak, Willy Agustine, Sri Masyeni, Erly Sintya, Ida Ayu Wayan Mahayani, Safarina G Malik, Khin Saw Myint, Amin Soebandrio |

| | | | |
|---|---|---|---|
| | | Medicine and Health Sciences, Warmadewa University | |
| EPI_ISL_2101261 | RSUD Mangusada | Eijkman Institute for Molecular Biology, National Agency for Research and Innovation; Molecular Biology Laboratory, Faculty Medicine and Health Sciences, Warmadewa University | Lydia V. Panggalo, Iskandar Adnan, Sukma Oktavianthi, Lidwina Priliani, Edison Johar, Frilasita A Yudhaputri, Muhammad Rezki Rasyak, Willy Agustine, Hidayat Trimarsanto, Sri Masyeni, Erly Sintya, Ida Ayu Wayan Mahayani, Safarina G Malik, Khin Saw Myint, Amin Soebandrio |
| EPI_ISL_2101262 | RSUD Mangusada | Eijkman Institute for Molecular Biology, National Agency for Research and Innovation; Molecular Biology Laboratory, Faculty Medicine and Health Sciences, Warmadewa University | Iskandar Adnan, Sukma Oktavianthi, Lidwina Priliani, Edison Johar, Frilasita A Yudhaputri, Muhammad Rezki Rasyak, Willy Agustine, Hidayat Trimarsanto, Lydia V. Panggalo, Sri Masyeni, Erly Sintya, Ida Ayu Wayan Mahayani, Safarina G Malik, Khin Saw Myint, Amin Soebandrio |
| EPI_ISL_2101263 | RSUD Mangusada | Eijkman Institute for Molecular Biology, National Agency for Research and Innovation; Molecular Biology Laboratory, Faculty Medicine and Health Sciences, Warmadewa University | Sukma Oktavianthi, Lidwina Priliani, Edison Johar, Frilasita A Yudhaputri, Muhammad Rezki Rasyak, Willy Agustine, Hidayat Trimarsanto, Lydia V. Panggalo, Iskandar Adnan, Sri Masyeni, Erly Sintya, Ida Ayu Wayan Mahayani, Safarina G Malik, Khin Saw Myint, Amin Soebandrio |
| EPI_ISL_2101264 | RSUD Mangusada | Eijkman Institute for Molecular Biology, National Agency for Research and Innovation; Molecular Biology Laboratory, Faculty Medicine and Health Sciences, Warmadewa University | Lidwina Priliani, Edison Johar, Frilasita A Yudhaputri, Muhammad Rezki Rasyak, Willy Agustine, Hidayat Trimarsanto, Lydia V. Panggalo, Iskandar Adnan, Sukma Oktavianthi, Sri Masyeni, Erly Sintya, Ida Ayu Wayan Mahayani, Safarina G Malik, Khin Saw Myint, Amin Soebandrio |
| EPI_ISL_2101265 | RSUD Mangusada | Eijkman Institute for Molecular Biology, National Agency for Research and Innovation; Molecular Biology Laboratory, Faculty Medicine and Health Sciences, Warmadewa University | Edison Johar, Frilasita A Yudhaputri, Muhammad Rezki Rasyak, Willy Agustine, Hidayat Trimarsanto, Lydia V. Panggalo, Iskandar Adnan, Sukma Oktavianthi, Lidwina Priliani, Sri Masyeni, Erly Sintya, Ida Ayu Wayan Mahayani, Safarina G Malik, Khin Saw Myint, Amin Soebandrio |
| EPI_ISL_2101266 | RSUD Mangusada | Eijkman Institute for Molecular Biology, National Agency for Research and Innovation; Molecular Biology Laboratory, Faculty Medicine and Health Sciences, Warmadewa University | Frilasita A Yudhaputri, Muhammad Rezki Rasyak, Willy Agustine, Hidayat Trimarsanto, Lydia V. Panggalo, Iskandar Adnan, Sukma Oktavianthi, Lidwina Priliani, Edison Johar, Sri Masyeni, Erly Sintya, Ida Ayu Wayan Mahayani, Safarina G Malik, Khin Saw Myint, Amin Soebandrio |
| EPI_ISL_2101267 | RSUD Mangusada | Eijkman Institute for Molecular Biology, National Agency for Research and Innovation; Molecular Biology Laboratory, Faculty Medicine and Health Sciences, Warmadewa University | Muhammad Rezki Rasyak, Willy Agustine, Hidayat Trimarsanto, Lydia V. Panggalo, Iskandar Adnan, Sukma Oktavianthi, Lidwina Priliani, Edison Johar, Frilasita A Yudhaputri, Sri Masyeni, Erly Sintya, Ida Ayu Wayan Mahayani, Safarina G Malik, Khin Saw Myint, Amin Soebandrio |
| EPI_ISL_2101268 | RSUD Mangusada | Eijkman Institute for Molecular Biology, National Agency for Research and Innovation; Molecular Biology Laboratory, Faculty Medicine and Health Sciences, Warmadewa University | Willy Agustine, Hidayat Trimarsanto, Lydia V. Panggalo, Iskandar Adnan, Sukma Oktavianthi, Lidwina Priliani, Edison Johar, Frilasita A Yudhaputri, Muhammad Rezki Rasyak, Sri Masyeni, Erly Sintya, Ida Ayu Wayan Mahayani, Safarina G Malik, Khin Saw Myint, Amin Soebandrio |
| EPI_ISL_2101269 | RSUD Mangusada | Eijkman Institute for Molecular Biology, National Agency for Research and Innovation; Molecular Biology Laboratory, Faculty Medicine and Health Sciences, Warmadewa University | Hidayat Trimarsanto, Lydia V. Panggalo, Iskandar Adnan, Sukma Oktavianthi, Lidwina Priliani, Edison Johar, Frilasita A Yudhaputri, Muhammad Rezki Rasyak, Willy Agustine, Sri Masyeni, Erly Sintya, Ida Ayu Wayan Mahayani, Safarina G Malik, Khin Saw Myint, Amin Soebandrio |
| EPI_ISL_2106976 | RSUP Sanglah | National Institute of Health Research and Development | Vivi Setiawaty, Hana Apsari Pawestri, Subangkit, Kartika Dewi Puspa, Arie Ardiansyah Nugraha, Hartanti Dian Ikawati, Krisna Pangesti, Triyani Soekarso, Nelly Puspandari, I Nengah Tony Rustawan, I Gede Ketut Sajinadiyasa |
| EPI_ISL_516829 | RSUD Nyi Ageng Serang | Genetics Working Group (Pokja Genetik) Faculty of Medicine, Public Health and Nursing Universitas | Gunadi, Hendra Wibawa, . Marcellus, Mohamad S. Hakim, Edwin W. Daniwijaya, Ludhang P. Rizki, Endah Supriyati, Eggi Arguni, Titik Nuryastuti, Tri Wibawa, Dwi AA Nugrahaningsih, Afiahayati , . Siswanto, Beby Dewi Sartika, Dyah Ayu Puspitarani |

| | | | |
|---|---|---|---|
| | | Gadjah Mada (FK-KMK UGM), Disease Investigation Center Wates Ministry of Agriculture Indonesia, Department of Microbiology FK-KMK UGM, Laboratorium Diagnostik Yayasan Tahija World Mosquito Program (WMP) Yogyakarta Center for Tropical Medicine FK-KMK UGM, Integrated Research Center FK-KMK UGM, Department of Computer Science and Electronics FMIPA UGM | |
| EPI_ISL_518819 | Qadr Hospital, Tangerang, Banten | Biosafety Level-3 Laboratory, Indonesian Institute of Sciences (LIPI) | Anik Budhi Dharmayanthi, Syam Budi Iryanto, Andri Wardiana, Anggia Prasetyoputri, Isa Nuryana, Ade Andriani, Ario Betha Juanssilfero, Asep M Ridwanuloh, Ahmad Fathoni, Rifki Sadikin, Ratih Asmana Ningrum, Wien Kusharyoto, Puspita Lisdiyanti |
| EPI_ISL_518820 | Klinik Apotek Dein, Jakarta, Indonesia | Biosafety Level-3 Laboratory, Indonesian Institute of Sciences (LIPI) | Anik Budhi Dharmayanthi, Syam Budi Iryanto, Andri Wardiana, Anggia Prasetyoputri, Isa Nuryana, Ade Andriani, Ario Betha Juanssilfero, Asep M Ridwanuloh, Ahmad Fathoni, Rifki Sadikin, Ratih Asmana Ningrum, Wien Kusharyoto, Puspita Lisdiyanti |
| EPI_ISL_538505, EPI_ISL_538509 | National Institute of Health Research and Development | National Institute of Health Research and Development | Pawestri, HA; Subangkit; Puspa, KD; Nugraha, AA; Ikawati, HD; Pangesti, KNA; Soekarso, T; Susilarini, NK; Hariastuti, NI; Nikmah, UA; Mursinah; Febriyani, A; Herman, R; Susanti, N; Herna; Febriyanti, T; Nurhadi, M; Paisal; Ramadhany, R; Agustiningsih; Kurniawati, J; Kipuw, NL; Muna, F; Indalau, IL; Adam, K; Wibowo, HA; Rizki, A; Puspandary, N; Setiawaty,V. |
| EPI_ISL_576113 | RSUP Dr. Sardjito | Genetics Working Group (Pokja Genetik) Faculty of Medicine, Public Health and Nursing Universitas Gadjah Mada (FK-KMK UGM); Disease Investigation Center Wates Ministry of Agriculture Indonesia; Department of Microbiology FK-KMK UGM; Laboratorium Diagnostik Yayasan Tahija World Mosquito Program (WMP) Yogyakarta Center for Tropical Medicine FK-KMK UGM; Integrated Research Center FK-KMK UGM; Department of Computer Science and Electronics FMIPA UGM | Gunadi, Hendra Wibawa, . Marcellus, Mohamad S. Hakim, Edwin W. Daniwijaya, Ludhang P. Rizki, Endah Supriyati, Eggi Arguni, Titik Nuryastuti, Tri Wibawa, Dwi AA Nugrahaningsih, Afiahayati, Siswanto, Kristy Iskandar, Nungki Anggorowati, Ika Trisnawati, Riat El Khair, William Widitjiarso, Fadil Fahri |
| EPI_ISL_576114 | RSUP Dr. Sardjito | Genetics Working Group (Pokja Genetik) Faculty of Medicine, Public Health and Nursing Universitas Gadjah Mada (FK-KMK UGM); Disease Investigation Center Wates Ministry of Agriculture Indonesia; Department of Microbiology FK-KMK UGM; Laboratorium Diagnostik Yayasan Tahija World Mosquito Program (WMP) Yogyakarta Center for Tropical Medicine FK-KMK UGM; Integrated Research Center FK-KMK UGM; Department of Computer Science and Electronics FMIPA UGM | Gunadi, Hendra Wibawa, . Marcellus, Mohamad S. Hakim, Edwin W. Daniwijaya, Ludhang P. Rizki, Endah Supriyati, Eggi Arguni, Titik Nuryastuti, Tri Wibawa, Dwi AA Nugrahaningsih, Afiahayati, Siswanto, Kristy Iskandar, Nungki Anggorowati, Ika Trisnawati, Riat El Khair, Audric Kenny Tedja, Aditya Rifqi Fauzi |
| EPI_ISL_576115 | RSUP Dr Sardjito | Genetics Working Group (Pokja Genetik) Faculty of Medicine, Public Health and Nursing Universitas Gadjah Mada (FK-KMK UGM); Disease Investigation Center Wates Ministry of Agriculture Indonesia; Department of Microbiology FK-KMK UGM; Laboratorium Diagnostik Yayasan Tahija World Mosquito Program (WMP) Yogyakarta Center for Tropical Medicine FK-KMK UGM; Integrated Research Center FK-KMK | Gunadi, Hendra Wibawa, . Marcellus, Mohamad S. Hakim, Edwin W. Daniwijaya, Ludhang P. Rizki, Endah Supriyati, Eggi Arguni, Titik Nuryastuti, Tri Wibawa, Dwi AA Nugrahaningsih, Afiahayati, Siswanto, Kristy Iskandar, Nungki Anggorowati, Ika Trisnawati, Riat El Khair, Yunika Puspadewi, Osman Sianipar, Umi Solekhah Intansari, Elizabeth Henny Herningtiyas, Ira Puspitawati, Nur Imma Fatimah Harahap, Untung Wirawan, Maria Patricia Inggriani |

| | | | |
|---|---|---|---|
| | | UGM; Department of Computer Science and Electronics FMIPA UGM | |
| EPI_ISL_576116 | RSUP Dr. Sardjito | Genetics Working Group (Pokja Genetik) Faculty of Medicine, Public Health and Nursing Universitas Gadjah Mada (FK-KMK UGM); Disease Investigation Center Wates Ministry of Agriculture Indonesia; Department of Microbiology FK-KMK UGM; Laboratorium Diagnostik Yayasan Tahija World Mosquito Program (WMP) Yogyakarta Center for Tropical Medicine FK-KMK UGM; Integrated Research Center FK-KMK UGM; Department of Computer Science and Electronics FMIPA UGM | Gunadi, Hendra Wibawa, . Marcellus, Mohamad S. Hakim, Edwin W. Daniwijaya, Ludhang P. Rizki, Endah Supriyati, Eggi Arguni, Titik Nuryastuti, Tri Wibawa, Dwi AA Nugrahaningsih, Afiahayati, Siswanto, Kristy Iskandar, Nungki Anggorowati, Ika Trisnawati, Sumardi, Eko Budiono, Bambang Sigit Riyanto, Heni Retnowulan, Munawar Gani, Satria Maulana, Nur Rahmi Ananda, Riat El Khair, Yunika Puspadewi, Osman Sianipar, Umi Solekhah Intansari, Elizabeth Henny Herningtiyas, Ira Puspitawati, Nur Imma Fatimah Harahap, Ririn Enggy Yuliyanti, Alvin Santoso Kalim |
| EPI_ISL_576128 | RSUP Dr. Sardjito | Genetics Working Group (Pokja Genetik) Faculty of Medicine, Public Health and Nursing Universitas Gadjah Mada (FK-KMK UGM); Disease Investigation Center Wates Ministry of Agriculture Indonesia; Department of Microbiology FK-KMK UGM; Laboratorium Diagnostik Yayasan Tahija World Mosquito Program (WMP) Yogyakarta Center for Tropical Medicine FK-KMK UGM; Integrated Research Center FK-KMK UGM; Department of Computer Science and Electronics FMIPA UGM | Gunadi, Hendra Wibawa, Marcellus, Mohamad S. Hakim, Edwin W. Daniwijaya, Ludhang P. Rizki, Endah Supriyati, Eggi Arguni, Titik Nuryastuti, Tri Wibawa, Dwi AA Nugrahaningsih, Afiahayati, Siswanto, Kristy Iskandar, Nungki Anggorowati, Ika Trisnawati, Sumardi, Eko Budiono, Bambang Sigit Riyanto, Heni Retnowulan, Munawar Gani, Satria Maulana, Nur Rahmi Ananda, Riat El Khair, Yunika Puspadewi, Osman Sianipar, Umi Solekhah Intansari, Elizabeth Henny Herningtiyas, Ira Puspitawati, Nur Imma Fatimah Harahap, Dyah Ayu Puspitarani, Kemala Athollah |
| EPI_ISL_576130 | RSUP Dr. Sardjito | Genetics Working Group (Pokja Genetik) Faculty of Medicine, Public Health and Nursing Universitas Gadjah Mada (FK-KMK UGM); Disease Investigation Center Wates Ministry of Agriculture Indonesia; Department of Microbiology FK-KMK UGM; Laboratorium Diagnostik Yayasan Tahija World Mosquito Program (WMP) Yogyakarta Center for Tropical Medicine FK-KMK UGM; Integrated Research Center FK-KMK UGM; Department of Computer Science and Electronics FMIPA UGM | Gunadi, Hendra Wibawa, Marcellus, Mohamad S. Hakim, Edwin W. Daniwijaya, Ludhang P. Rizki, Endah Supriyati, Eggi Arguni, Titik Nuryastuti, Tri Wibawa, Dwi AA Nugrahaningsih, Afiahayati, Siswanto, Kristy Iskandar, Nungki Anggorowati, Rina Triasih, Amalia Setyati, Dwikisworo Setyowireni, Ida Safitri Laksanawati, Cahya Dewi Satria, Titis Widowati, Elisabeth Siti Herini, Riat El Khair, Yunika Puspadewi, Osman Sianipar, Umi Solekhah Intansari, Elizabeth Henny Herningtiyas, Ira Puspitawati, Nur Imma Fatimah Harahap, Alvin S. Kalim, Susan Simanjaya, Desyifa Mursalin |
| EPI_ISL_576383 | RSUD Budi Rahayu Kota Magelang | Genetics Working Group (Pokja Genetik) Faculty of Medicine, Public Health and Nursing Universitas Gadjah Mada (FK-KMK UGM); Disease Investigation Center Wates Ministry of Agriculture Indonesia; Department of Microbiology FK-KMK UGM; Laboratorium Diagnostik Yayasan Tahija World Mosquito Program (WMP) Yogyakarta Center for Tropical Medicine FK-KMK UGM; Integrated Research Center FK-KMK UGM; Department of Computer Science and Electronics FMIPA UGM; Balai Besar Teknik Kesehatan Lingkungan dan | Gunadi, Hendra Wibawa, Marcellus, Mohamad S. Hakim, Edwin W. Daniwijaya, Ludhang P. Rizki, Endah Supriyati, Eggi Arguni, Titik Nuryastuti, Tri Wibawa, Dwi AA Nugrahaningsih, Afiahayati, Siswanto, Kristy Iskandar, Nungki Anggorowati, Irene, Indaryati, Havid Setyawan, Ari Meliyanti, Merliana Sari Situmeang, Audric Kenny Tedja, Aditya Rifqi Fauzi |

| | | | |
|---|---|---|---|
| | | Pengendalian Penyakit (BBTKLPP) Yogyakarta | |
| EPI_ISL_576387 | National Institute of Health Research and Development | National Institute of Health Research and Development | Pawestri,HA;Subangkit;Puspa,KD;Nugraha,AA;Ikawati,HD;Pangesti, KNA;Soekarso,T;Susilarini,NK;Hariastuti,NI;Nikmah,UA;Mursinah;Febriyani,A;Herman,R;Susanti,N;Herna;Febriyanti,T; Nurhadi,M; Paisal;Ramadhany,R;Agustiningsih;Kurniawati,J;Kipuw,NL;Muna,F;Indalau,IL;Adam,K;Wibowo,HA;Rizki,A;Puspandari,N;Setiawaty,V |
| EPI_ISL_610155 | Puskesmas Cangkrep | Genetics Working Group (Pokja Genetik) Faculty of Medicine, Public Health and Nursing Universitas Gadjah Mada (FK-KMK UGM); Disease Investigation Center Wates Ministry of Agriculture Indonesia; Department of Microbiology FK-KMK UGM; Laboratorium Diagnostik Yayasan Tahija World Mosquito Program (WMP) Yogyakarta Center for Tropical Medicine FK-KMK UGM; Integrated Research Center FK-KMK UGM; Department of Computer Science and Electronics FMIPA UGM; Balai Besar Teknik Kesehatan Lingkungan dan Pengendalian Penyakit (BBTKLPP) Yogyakarta | Gunadi, Hendra Wibawa, Marcellus, Mohamad S. Hakim, Edwin W. Daniwijaya, Ludhang P. Rizki, Endah Supriyati, Eggi Arguni, Titik Nuryastuti, Tri Wibawa, Dwi AA Nugrahaningsih, Afiahayati, Siswanto, Kristy Iskandar, Nungki Anggorowati, Irene, Indaryati, Havid Setyawan, Nawang Sukestiningsih, Fadil Fahri, Ririn Enggy Yuliyanti |
| EPI_ISL_610158 | RS Panti Rini | Genetics Working Group (Pokja Genetik) Faculty of Medicine, Public Health and Nursing Universitas Gadjah Mada (FK-KMK UGM); Disease Investigation Center Wates Ministry of Agriculture Indonesia; Department of Microbiology FK-KMK UGM; Laboratorium Diagnostik Yayasan Tahija World Mosquito Program (WMP) Yogyakarta Center for Tropical Medicine FK-KMK UGM; Integrated Research Center FK-KMK UGM; Department of Computer Science and Electronics FMIPA UGM | Gunadi, Hendra Wibawa, Marcellus, Mohamad S. Hakim, Edwin W. Daniwijaya, Ludhang P. Rizki, Endah Supriyati, Eggi Arguni, Titik Nuryastuti, Tri Wibawa, Dwi AA Nugrahaningsih, Afiahayati, Siswanto, Kristy Iskandar, Nungki Anggorowati, V. Noegroho Isti Donodjati, Wawan Triharjanto, Dwiki Afandy, Dyah Ayu Puspitarani |
| EPI_ISL_610161 | RSUD Dr. Tjitrowardojo | Genetics Working Group (Pokja Genetik) Faculty of Medicine, Public Health and Nursing Universitas Gadjah Mada (FK-KMK UGM); Disease Investigation Center Wates Ministry of Agriculture Indonesia; Department of Microbiology FK-KMK UGM; Laboratorium Diagnostik Yayasan Tahija World Mosquito Program (WMP) Yogyakarta Center for Tropical Medicine FK-KMK UGM; Integrated Research Center FK-KMK UGM; Department of Computer Science and Electronics FMIPA UGM; Balai Besar Teknik Kesehatan Lingkungan dan Pengendalian Penyakit (BBTKLPP) Yogyakarta | Gunadi, Hendra Wibawa, Marcellus, Mohamad S. Hakim, Edwin W. Daniwijaya, Ludhang P. Rizki, Endah Supriyati, Eggi Arguni, Titik Nuryastuti, Tri Wibawa, Dwi AA Nugrahaningsih, Afiahayati, Siswanto, Kristy Iskandar, Nungki Anggorowati, Irene, Indaryati, Havid Setyawan, Wuryanto, Kemala Athollah, Desyifa Mursalin |
| EPI_ISL_610162 | RSUD Dr. Tjitrowardojo | Genetics Working Group (Pokja Genetik) Faculty of Medicine, Public Health and Nursing Universitas Gadjah Mada (FK-KMK UGM); Disease Investigation Center Wates Ministry of Agriculture Indonesia; Department of Microbiology FK-KMK UGM; Laboratorium Diagnostik Yayasan Tahija | Gunadi, Hendra Wibawa, Marcellus, Mohamad S. Hakim, Edwin W. Daniwijaya, Ludhang P. Rizki, Endah Supriyati, Eggi Arguni, Titik Nuryastuti, Tri Wibawa, Dwi AA Nugrahaningsih, Afiahayati, Siswanto, Kristy Iskandar, Nungki Anggorowati, Irene, Indaryati, Havid Setyawan, Wuryanto, Susan Simanjaya, Alvin Santoso Kalim |

| | | | |
|---|---|---|---|
| | | World Mosquito Program (WMP) Yogyakarta Center for Tropical Medicine FK-KMK UGM; Integrated Research Center FK-KMK UGM; Department of Computer Science and Electronics FMIPA UGM; Balai Besar Teknik Kesehatan Lingkungan dan Pengendalian Penyakit (BBTKLPP) Yogyakarta | |
| EPI_ISL_632936 | Puskesmas Mlati 1 Sleman | Genetics Working Group (Pokja Genetik) Faculty of Medicine, Public Health and Nursing Universitas Gadjah Mada (FK-KMK UGM); Disease Investigation Center Wates Ministry of Agriculture Indonesia; Department of Microbiology FK-KMK UGM; Laboratorium Diagnostik Yayasan Tahija World Mosquito Program (WMP) Yogyakarta Center for Tropical Medicine FK-KMK UGM; Integrated Research Center FK-KMK UGM; Department of Computer Science and Electronics FMIPA UGM; RSUP Dr. Sardjito | Gunadi, Hendra Wibawa, Marcellus, Mohamad S. Hakim, Edwin W. Daniwijaya, Ludhang P. Rizki, Endah Supriyati, Eggi Arguni, Titik Nuryastuti, Tri Wibawa, Dwi AA Nugrahaningsih, Afiahayati, Siswanto, Kristy Iskandar, Nungki Anggorowati, Ika Trisnawati, Sumardi, Eko Budiono, Bambang Sigit Riyanto, Heni Retnowulan, Munawar Gani, Satria Maulana, Nur Rahmi Ananda, Riat El Khair, Yunika Puspadewi, Osman Sianipar, Umi Solekhah Intansari, Elizabeth Henny Herningtiyas, Ira Puspitawati, Nur Imma Fatimah Harahap, Ernawati, Untung Wirawan , William Widitjiarso |
| EPI_ISL_632937 | RSUD Saptosari Gunung Kidul | Genetics Working Group (Pokja Genetik) Faculty of Medicine, Public Health and Nursing Universitas Gadjah Mada (FK-KMK UGM); Disease Investigation Center Wates Ministry of Agriculture Indonesia; Department of Microbiology FK-KMK UGM; Laboratorium Diagnostik Yayasan Tahija World Mosquito Program (WMP) Yogyakarta Center for Tropical Medicine FK-KMK UGM; Integrated Research Center FK-KMK UGM; Department of Computer Science and Electronics FMIPA UGM; Balai Besar Teknik Kesehatan Lingkungan dan Pengendalian Penyakit (BBTKLPP) Yogyakarta | Gunadi, Hendra Wibawa, Marcellus, Mohamad S. Hakim, Edwin W. Daniwijaya, Ludhang P. Rizki, Endah Supriyati, Eggi Arguni, Titik Nuryastuti, Tri Wibawa, Dwi AA Nugrahaningsih, Afiahayati, Siswanto, Kristy Iskandar, Nungki Anggorowati, Irene, Indaryati, Havid Setyawan, Eko Darmawan, Maria Patricia Inggriani, Audric Kenny Tedja |
| EPI_ISL_707692 | Medical Research Center, Faculty of Medicine, Syarif Hidayatullah State Islamic University Jakarta | Medical Research Center, Faculty of Medicine, Syarif Hidayatullah State Islamic University Jakarta | Chris Adhiyanto, Laifa Hendarmin, Erike Suwarsono, Zeti Harriyati, Rini Puspitaningrum, Ferania Mela, Dennis Nurjadi |
| EPI_ISL_707696 | Medical Research Center, Faculty of Medicine, Syarif Hidayatullah State Islamic University Jakarta | Medical Research Center, Faculty of Medicine, Syarif Hidayatullah State Islamic University Jakarta | Laifa Hendarmin, Chris Adhiyanto, Erike Suwarsono, Zeti Harriyati, Rini Puspitaningrum, Ferania Mela, Dennis Nurjadi |
| EPI_ISL_707778 | Medical Research Center, Faculty of Medicine, Syarif Hidayatullah State Islamic University Jakarta | Medical Research Center, Faculty of Medicine, Syarif Hidayatullah State Islamic University Jakarta | Erike A Suwarsono, Chris Adhiyanto, Laifa Hendarmin, Zeti Harriyati, Rini Puspitaningrum, Ferania Mela, Dennis Nurjadi |
| EPI_ISL_707890 | Medical Research Center, Faculty of Medicine, Syarif Hidayatullah State Islamic University Jakarta | Medical Research Center, Faculty of Medicine, Syarif Hidayatullah State Islamic University Jakarta | Zeti Harriyati, Chris Adhiyanto, Laifa Hendarmin, Erike A Suwarsono, Dennis Nurjadi |
| EPI_ISL_708652 | Medical Research Center, Faculty of Medicine, Syarif Hidayatullah State Islamic University Jakarta | Medical Research Center, Faculty of Medicine, Syarif Hidayatullah State Islamic University Jakarta | Chris Adhiyanto, Laifa Hendarmin, Erike A Suwarsono, Zeti Harriyati, Endah Wulandari, Flori R Sari, Hari Hendarto |
| EPI_ISL_745033 | West Java Health Laboratory | West Java Health Laboratory | Azzania Fibriani, Ema Rahmawati, Ryan Bayusantika Ristandi, Rifky Waluyajati Rachman, Cut Nur Cinthia Alamanda, Isak Solihin, Rini Robiani, Miftahul Faridl, Karimatu Khoirunnisa |
| EPI_ISL_747194 | Clinical Microbiology Laboratory, Faculty of Medicine, Universitas Indonesia | Clinical Microbiology Laboratory, Faculty of Medicine, Universitas Indonesia | Fera Ibrahim, Augustine Natasha, Andi Yasmon, Pratiwi Sudarmono, Yulia Rosa Saharman, Anis Karuniawati, Fithriyah |

| | | | |
|---|---|---|---|
| EPI_ISL_747233 | National Institute of Health Research and Development | National Institute of Health Research and Development | Subangkit; Indrasari,ND; Wulandari,D; Monika,M; Pawestri,HA; Puspa,KD; Nugraha,AA; Ikawati,HD; Pangesti,KNA; Soekarso,T; Paisal; Setiawaty,Vivi |
| EPI_ISL_747235 | Sukabumi Health Laboratory | West Java Health Laboratory; School of Life Sciences and Technology, Institut Teknologi Bandung | Azzania Fibriani, Ema Rahmawati, Ryan Bayusantika Ristandi, Rifky Waluyajati Rachman, Cut Nur Cinthia Alamanda, Isak Solihin, Rini Robiani, Miftahul Faridl, Karimatu Khoirunnisa |
| EPI_ISL_747236 | Sukabumi Public Health | West Java Health Laboratory; School of Life Sciences and Technology, Institut Teknologi Bandung | Azzania Fibriani, Ema Rahmawati, Ryan Bayusantika Ristandi, Rifky Waluyajati Rachman, Cut Nur Cinthia Alamanda, Isak Solihin, Rini Robiani, Miftahul Faridl, Karimatu Khoirunnisa |
| EPI_ISL_747237 | National Institute of Health Research and Development | National Institute of Health Research and Development | Subangkit,Nafiandi,Estaria,V;Pawestri,HA;Puspa,KD;Nugraha,AA;Ikawati,HD;Pangesti,KNA;Soekarso,T;Paisal;Puspandari,N;Setiawaty,V |
| EPI_ISL_747238 | Bogor Public Health | West Java Health Laboratory; School of Life Sciences and Technology, Institut Teknologi Bandung | Azzania Fibriani, Ema Rahmawati, Ryan Bayusantika Ristandi, Rifky Waluyajati Rachman, Cut Nur Cinthia Alamanda, Isak Solihin, Rini Robiani, Miftahul Faridl, Karimatu Khoirunnisa |
| EPI_ISL_747239 | Kuningan Publlic Health Office | West Java Health Laboratory; School of Life Sciences and Technology, Institut Teknologi Bandung | Azzania Fibriani, Ema Rahmawati, Ryan Bayusantika Ristandi, Rifky Waluyajati Rachman, Cut Nur Cinthia Alamanda, Isak Solihin, Rini Robiani, Miftahul Faridl, Karimatu Khoirunnisa |
| EPI_ISL_747240 | Al Islam Hospital | West Java Health Laboratory; School of Life Sciences and Technology, Institut Teknologi Bandung | Azzania Fibriani, Ema Rahmawati, Ryan Bayusantika Ristandi, Rifky Waluyajati Rachman, Cut Nur Cinthia Alamanda, Isak Solihin, Rini Robiani, Miftahul Faridl, Karimatu Khoirunnisa |
| EPI_ISL_747241 | Subang Public Health Office | West Java Health Laboratory; School of Life Sciences and Technology, Institut Teknologi Bandung | Azzania Fibriani, Ema Rahmawati, Ryan Bayusantika Ristandi, Rifky Waluyajati Rachman, Cut Nur Cinthia Alamanda, Isak Solihin, Rini Robiani, Miftahul Faridl, Karimatu Khoirunnisa |
| EPI_ISL_756376 | National Institute of Health Research and Development | National Institute of Health Research and Development | Ramadhany,R; Rukminiati,Y; Agustiningsih; Adam,K; Wibowo,HA; Pawestri,HA; Subangkit;Puspa,KD; Nugraha,AA; Ikawati,HD; Pangesti,KNA; Soekarso,T; Susilarini,NK; Hariastuti,NI; Nikmah,UA;Mursinah; Febriyani,A; Herman,R; Susanti,N;Herna; Febriyanti,T; Nurhadi,M; Kurniawati,J; Kipuw,NL; Muna,F; Indalao,IL; Rizki,A; Puspandari,N; Setiawaty,Vivi |
| EPI_ISL_756401 | National Institute of Health Research and Development | National Institute of Health Research and Development | Adam,K; Wibowo,HA; Ramadhany,R; Rukminiati,Y; Agustiningsih; Pawestri,HA; Subangkit; Puspa,KD; Nugraha,AA; Ikawati,HD; Pangesti,KNA; Soekarso,T; Susilarini,NK; Hariastuti,NI; Nikmah,UA; Mursinah; Febriyani,A; Herman,R; Susanti,N; Herna; Febriyanti,T; Nurhadi,M; Kurniawati,J; Kipuw,NL; Muna,F; Indalao,IL; Rizki,A; Puspandari,N; Setiawaty,Vivi |
| EPI_ISL_757287 | National Institute of Health Research and Development | National Institute of Health Research and Development | Rukminiati,Y;Agustiningsih;Adam,K;Wibowo,HA;Ramadhany,R;Pawestri,HA;Subangkit;Puspa,KD;Nugraha,AA;Ikawati,HD;Pangesti,KNA;Soekarso,T;Susilarini,NK;Hariastuti,NI;Nikmah,UA;Mursinah;Febriyani,A;Herman,R;Susanti,N;Herna;Febriyanti,T; Nurhadi,M;Kurniawati,J;Kipuw,NL;Muna,F;Indalao,IL;Rizki,A;Puspandari,N;Setiawaty,V |
| EPI_ISL_759701 | RS UNS, Surakarta | UNS/RS-UNS, Faculty of Medicine | Betty Suryawati, Yulia Sari, Hartono, Maryani, Afif Avicenna Gufron, HanaHana Apsari Pawestri, Kartika Dewi Puspa, Hartanti Dian Ika, Arie Ardiansyah Nugraha, Vivi Setiawaty. |
| EPI_ISL_759702 | RS UNS Surakarta | Universitas Sebelas Maret (UNS), Rumah sakit Universitas Sebelas Maret (RS-UNS) Surakarta, National Institute of Health Research and Development, Indonesian Ministry of Health | Betty Suryawati, Yulia sari, hartono, maryani, Afif Avicena Gufron, Hana Apsari Pawestri, Kartika, Dewi Puspa, Hartanti Dian Ika, Arie Ardiansyah Nugraha, Vivi Setiawaty |
| EPI_ISL_759703 | RSUD Bung Karno | Universitas Sebelas Maret (UNS), Rumah sakit Universitas Sebelas Maret (RS-UNS) Surakarta, National Institute of Health Research and Development, Indonesian Ministry of Health | Betty Suryawati, Yulia sari, hartono, maryani, Afif Avicena Gufron, Hana Apsari Pawestri, Kartika, Dewi Puspa, Hartanti Dian Ika, Arie Ardiansyah Nugraha, Vivi Setiawaty |
| EPI_ISL_759705 | RSUD Bung Karno, Surakarta | Universitas Sebelas Maret (UNS); Rumah Sakit Universitas Sebelas Maret (RS-UNS), Surakarta; National Institute of Health Research and Development, Indonesian Ministry of Health , Jakarta). | Betty Suryawati, Yulia Sari, Hartono, Maryani, Afif Avicena Gufron, Hana Apsari Pawestri, Kartika, Dewi Puspa, Hartanti Dian Ika, Arie Ardiansyah Nugraha, Vivi Setiawaty. |
| EPI_ISL_759842 | Rumah Sakit Universitas Sebelas Maret (RS-UNS) | Universitas Sebelas Maret (UNS); Rumah Sakit Universitas Sebelas Maret (RS-UNS), Surakarta; National Institute of Health Research and Development, Indonesian Ministry of Health, Jakarta. | Betty Suryawati, Yulia Sari, Hartono, Maryani, Afif Avicena Ghufron, Hana Apsari Pawestri, Kartika, Dewi Puspa, Hartanti Dian Ika, Arie Ardiansyah Nugraha, Vivi Setiawaty. |
| EPI_ISL_759968 | RSI Darus Syifa' | Institute of Tropical Disease, Universitas Airlangga | Maria I Lusida, Krisnoadi Rahardjo, Aldise M Nastri, Jezzy R Dewantari, Rima R Prasetya, HM Faiz, Gatot Soegiarto, Laksmi Wulandari, Resti Yudhawati, Soetjipto, Yasuko Mori, Kazufumi Shimizu |
| EPI_ISL_763063 | RS Panti Waluyo, Surakarta | Universitas Sebelas Maret (UNS); Rumah Sakit UNS | Betty Suryawati, Yulia Sari, Hartono, Maryani, Afif A Ghufron, Revi G H Novika, Hana A Pawestri, Kartika, Dewi Puspa, Hartanti Dian Ika, Arie A Nugraha, Vivi Setiawaty. |

| | | | |
|---|---|---|---|
| | | (RS-UNS); National Institute of Health Research and Development, Indonesian Ministry of Health. | |
| EPI_ISL_763064 | RS Kasih Ibu, Surakarta | Universitas Sebelas Maret (UNS); Rumah Sakit UNS (RS-UNS); National Institute of Health Research and Development, Indonesian Ministry of Health. | Betty Suryawati, Yulia Sari, Hartono, Maryani, Afif A Ghufron, Revi G H Novika, Hana A Pawestri, Kartika, Dewi Puspa, Hartanti Dian Ika, Arie A Nugraha, Vivi Setiawaty. |
| EPI_ISL_763066 | Puskesmas Grogol, Sukoharjo | Universitas Sebelas Maret (UNS); Rumah Sakit UNS (RS-UNS); National Institute of Health Research and Development, Indonesian Ministry of Health. | Yulia Sari, Hartono, Maryani, Betty Suryawati, Afif A Ghufron, Revi G H Novika, Hana A Pawestri, Kartika, Dewi Puspa, Hartanti Dian Ika, Arie A Nugraha, Vivi Setiawaty. |
| EPI_ISL_763068 | RSUD Kota Surakarta | Universitas Sebelas Maret (UNS); Rumah Sakit UNS (RS-UNS); National Institute of Health Research and Development, Indonesian Ministry of Health. | Maryani, Yulia Sari, Hartono, Afif A Ghufron, Revi G H Novika, Hana A Pawestri, Kartika, Dewi Puspa, Hartanti Dian Ika, Arie A Nugraha, Vivi Setiawaty, Betty Suryawati. |
| EPI_ISL_763070 | Puskesmas Jambu Kulon, Klaten | Universitas Sebelas Maret (UNS); Rumah Sakit UNS (RS-UNS); National Institute of Health Research and Development, Indonesian Ministry of Health. | Yulia Sari, Hartono, Maryani, Afif A Ghufron, Hana A Pawestri, Kartika, Dewi Puspa, Hartanti Dian Ika, Arie A Nugraha, Vivi Setiawaty, Betty Suryawati. |
| EPI_ISL_775594 | Balitvet Lampung | National Institute of Health Research and Development | Nugraha,AA;Subangkit;Pawestri,HA;Ikawati,HD;Puspa,KD;Saswiyanti,E;Srihanto,EA;Pangesti,KNA;Soekarso,T;Puspandari,N;Setiawaty,V |
| EPI_ISL_775597 | RS Pertamina Sorong | National Institute of Health Research and Development | Puspa,KD;Subangkit;Pawestri,HA;Ikawati,HD;Nugraha,AA;Yosepin;Pangesti,KNA;Soekarso,T;Puspandari,N;Setiawaty,V |
| EPI_ISL_791977 | Laboratorium Mikrobiologi Samarinda | National Institute of Health Research and Development | Nugraha,AA;Subangkit;Pawestri,HA;Ikawati,HD;Puspa,KD;Tandirogang,N;Yadi;Pangesti,KNA;Soekarso,T;Puspandari,N;Setiawaty,V |
| EPI_ISL_791979 | RSUD Curup Bengkulu | National Institute of Health Research and Development | Nugraha,AA;Subangkit;Pawestri,HA;Ikawati,HD;Puspa,KD;Mardiani;Pangesti,KNA;Soekarso,T;Puspandari,N;Setiawaty,V |
| EPI_ISL_791980 | RSUD Blambangan Banyuwangi | National Institute of Health Research and Development | Ikawati,HD;Subangkit;Pawestri,HA;Nugraha,AA;Puspa,KD;Noor,RI;Pangesti,KNA;Soekarso,T;Puspandari,N;Setiawaty,V |
| EPI_ISL_791983 | RSUD dr. Kanujoso Djatiwibowo Balikpapan | National Institute of Health Research and Development | Subangkit;Pawestri,HA;Ikawati,HD;Nugraha,AA;Puspa,KD;Ivanna;Pangesti,KNA;Soekarso,T;Puspandari,N;Setiawaty,V |
| EPI_ISL_791985 | RS Santa Maria Pekanbaru | National Institute of Health Research and Development | Puspa,KD;Subangkit;Pawestri,HA;Ikawati,HD;Nugraha,AA;Fridayenti;Pangesti,KNA;Soekarso,T;Puspandari,N;Setiawaty,V |
| EPI_ISL_791986 | RSUD Morotai Maluku Utara | National Institute of Health Research and Development | Subangkit;Pawestri,HA;Ikawati,HD;Nugraha,AA;Puspa,KD;Denggo,SD;Pangesti,KNA;Soekarso,T;Puspandari,N;Setiawaty,V |
| EPI_ISL_791988 | Balai Besar POM Semarang | National Institute of Health Research and Development | Subangkit;Pawestri,HA;Ikawati,HD;Nugraha,AA;Puspa,KD;Aryanti;Pangesti,KNA;Soekarso,T;Puspandari,N;Setiawaty,V |
| EPI_ISL_791989 | RSUD Waluyo Jati Kota Kraksaan, Jawa Timur | National Institute of Health Research and Development | Subangkit;Pawestri,HA;Ikawati,HD;Nugraha,AA;Puspa,KD;Muhashonah,I;Pangesti,KNA;Soekarso,T;Puspandari,N;Setiawaty,V |
| EPI_ISL_803876 | RSUD Tarakan Kalimantan Utara | National Institute of Health Research and Development | Subangkit;Pawestri,HA;Ikawati,HD;Nugraha,AA;Puspa,KD;Sriwedari,K;Sindunata,R;Pangesti,KNA;Soekarso,T;Puspandari,N;Setiawaty,V |
| EPI_ISL_833039 | RS TK II 02.05.01 dr. AK Gani Palembang | National Institute of Health Research and Development | Subangkit;Pawestri,HA;Ikawati,HD;Nugraha,AA;Puspa,KD;Pangesti,KNA;Soekarso,T;Tantula,A;Wiradharma,RP;Puspandari,N;Setiawaty,V |
| EPI_ISL_833040 | RSUD Dr. Abdul Aziz Singkawang | National Institute of Health Research and Development | Subangkit;Pawestri,HA;Ikawati,HD;Nugraha,AA;Puspa,KD;Pangesti,KNA;Soekarso,T;Junus,HN;Puspandari,N;Setiawaty,V |
| EPI_ISL_833388 | Klinik Apotek Dein, Jakarta, Indonesia | Biosafety Level-3 Laboratory, Indonesian Institute of Sciences (LIPI) | Syam Budi Iryanto, Andri Wardiana, Anggia Prasetyoputri, Isa Nuryana, Ade Andriani, Anik Budhi Dharmayanthi, Ahmad Fathoni, Dian Fitria Agustiyanti, Asrul M. Fuad, Ratih Asmana Ningrum |
| EPI_ISL_833389 | Klinik Apotek Dein, Jakarta, Indonesia | Biosafety Level-3 Laboratory, Indonesian Institute of Sciences (LIPI) | Syam Budi Iryanto, Andri Wardiana, Anggia Prasetyoputri, Isa Nuryana, Ade Andriani, Anik Budhi Dharmayanthi, Ahmad Fathoni, Popi Hadi Wisnuwardhani, Sri Swasthikawati, Ratih Asmana Ningrum |

| | | | |
|---|---|---|---|
| EPI_ISL_833390 | Klinik Apotek Dein, Jakarta, Indonesia | Biosafety Level-3 Laboratory, Indonesian Institute of Sciences (LIPI) | Andri Wardiana, Anggia Prasetyoputri, Isa Nuryana, Ade Andriani, Anik Budhi Dharmayanthi, Syam Budi Iryanto,Ahmad Fathoni, Fahrurrozy, Alfi Taufik Fathurahman, Ratih Asmana Ningrum |
| EPI_ISL_833495 | Klinik Apotek Dein, Jakarta, Indonesia | Biosafety Level-3 Laboratory, Indonesian Institute of Sciences (LIPI) | Ade Andriani, Anik Budhi Dharmayanthi, Syam Budi Iryanto, Andri Wardiana, Anggia Prasetyoputri, Isa Nuryana, Ahmad Fathoni, Hariyatun, Akhirta Atikana, Ratih Asmana Ningrum |
| EPI_ISL_833496 | RS PMI Bogor, Indonesia | Biosafety Level-3 Laboratory, Indonesian Institute of Sciences (LIPI) | Anik Budhi Dharmayanthi, Syam Budi Iryanto, Andri Wardiana, Anggia Prasetyoputri, Isa Nuryana, Ade Andriani, Ahmad Fathoni, Arie Wulandari, Yana Rubiyana, Ratih Asmana Ningrum |
| EPI_ISL_833497 | Klinik Apotek Dein, Jakarta, Indonesia | Biosafety Level-3 Laboratory, Indonesian Institute of Sciences (LIPI) | Syam Budi Iryanto, Andri Wardiana, Anggia Prasetyoputri, Anik Budhi Dharmayanthi, Isa Nuryana, Ade Andriani, Ahmad Fathoni, Saiful anwar, Pamella Apriliana, Ratih Asmana Ningrum |
| EPI_ISL_833498 | Kartika Pulomas Hospital, Jakarta, Indonesia | Biosafety Level-3 Laboratory, Indonesian Institute of Sciences (LIPI) | Anggia Prasetyoputri, Isa Nuryana, Ade Andriani, Anik Budhi Dharmayanthi, Syam Budi Iryanto, Andri Wardiana, Ahmad Fathoni, Winda Tasia, Pinka Alisa Diena Shafa, Ratih Asmana Ningrum |
| EPI_ISL_833499 | RS Qadr, Tangerang, Indonesia | Biosafety Level-3 Laboratory, Indonesian Institute of Sciences (LIPI) | Isa Nuryana, Ade Andriani, Ahmad Fathoni, Anik Budhi Dharmayanthi, Syam Budi Iryanto, Andri Wardiana, Anggia Prasetyoputri, Ratna Dwi Ramadani, Sugiyono Saputra, Ratih Asmana Ningrum |
| EPI_ISL_833509 | RS Melania, Bogor, Indonesia | Biosafety Level-3 Laboratory, Indonesian Institute of Sciences (LIPI) | Syam Budi Iryanto, Andri Wardiana, Anik Budhi Dharmayanthi, Anggia Prasetyoputri, Isa Nuryana, Ade Andriani, Ahmad Fathoni, Ratih Asmana Ningrum |
| EPI_ISL_833510 | RS Melania, Bogor, Indonesia | Biosafety Level-3 Laboratory, Indonesian Institute of Sciences (LIPI) | Anik Budhi Dharmayanthi, Syam Budi Iryanto, Andri Wardiana, Anggia Prasetyoputri, Isa Nuryana, Ade Andriani, Ahmad Fathoni, Ratih Asmana Ningrum |
| EPI_ISL_833571 | RS Qadr, Tangerang, Indonesia | Biosafety Level-3 Laboratory, Indonesian Institute of Sciences (LIPI) | Ade Andriani, Ahmad Fathoni, Anik Budhi Dharmayanthi, Syam Budi Iryanto, Andri Wardiana, Anggia Prasetyoputri, Isa Nuryana, Erwin Fajar Hasrianda, Ryan Haryo Setiawan, Ratih Asmana Ningrum |
| EPI_ISL_862039 | RSUP Dr. Sardjito | Genetics Working Group (Pokja Genetik) Faculty of Medicine, Public Health and Nursing Universitas Gadjah Mada (FK-KMK UGM); Disease Investigation Center Wates Ministry of Agriculture Indonesia; Department of Microbiology FK-KMK UGM; Laboratorium Diagnostik Yayasan Tahija World Mosquito Program (WMP) Yogyakarta Center for Tropical Medicine FK-KMK UGM; Integrated Research Center FK-KMK UGM; Department of Computer Science and Electronics FMIPA UGM; RSUP Dr. Sardjito | Gunadi, Hendra Wibawa, . Marcellus, Mohamad S. Hakim, Edwin W. Daniwijaya, Ludhang P. Rizki, Endah Supriyati, Eggi Arguni, Titik Nuryastuti, Tri Wibawa, Dwi AA Nugrahaningsih, Afiahayati, Siswanto, Kristy Iskandar, Nungki Anggorowati, Ika Trisnawati, Sumardi, Eko Budiono, Bambang Sigit Riyanto, Heni Retnowulan, Munawar Gani, Satria Maulana, Nur Rahmi Ananda, Riat El Khair, Yunika Puspadewi, Osman Sianipar, Umi Solekhah Intansari, Elizabeth Henny Herningtiyas, Ira Puspitawati, Nur Imma Fatimah Harahap,William Widitjiarso, Maria Patricia Inggriani |
| EPI_ISL_862040 | RSUP Dr. Sardjito | Genetics Working Group (Pokja Genetik) Faculty of Medicine, Public Health and Nursing Universitas Gadjah Mada (FK-KMK UGM); Disease Investigation Center Wates Ministry of Agriculture Indonesia; Department of Microbiology FK-KMK UGM; Laboratorium Diagnostik Yayasan Tahija World Mosquito Program (WMP) Yogyakarta Center for Tropical Medicine FK-KMK UGM; Integrated Research Center FK-KMK UGM; Department of Computer Science and Electronics FMIPA UGM; RSUP Dr. Sardjito | Gunadi, Hendra Wibawa, . Marcellus, Mohamad S. Hakim, Edwin W. Daniwijaya, Ludhang P. Rizki, Endah Supriyati, Eggi Arguni, Titik Nuryastuti, Tri Wibawa, Dwi AA Nugrahaningsih, Afiahayati, Siswanto, Kristy Iskandar, Nungki Anggorowati, Ika Trisnawati, Sumardi, Eko Budiono, Bambang Sigit Riyanto, Heni Retnowulan, Munawar Gani, Satria Maulana, Nur Rahmi Ananda, Riat El Khair, Yunika Puspadewi, Osman Sianipar, Umi Solekhah Intansari, Elizabeth Henny Herningtiyas, Ira Puspitawati, Nur Imma Fatimah Harahap,Alvin Santoso Kalim, Untung Riawan |
| EPI_ISL_862041 | RSUP Dr. Sardjito | Genetics Working Group (Pokja Genetik) Faculty of Medicine, Public Health and Nursing Universitas Gadjah Mada (FK-KMK UGM); Disease Investigation Center Wates Ministry of Agriculture Indonesia; Department of Microbiology FK-KMK UGM; Laboratorium Diagnostik Yayasan Tahija World Mosquito Program | Gunadi, Hendra Wibawa, . Marcellus, Mohamad S. Hakim, Edwin W. Daniwijaya, Ludhang P. Rizki, Endah Supriyati, Eggi Arguni, Titik Nuryastuti, Tri Wibawa, Dwi AA Nugrahaningsih, Afiahayati, Siswanto, Kristy Iskandar, Nungki Anggorowati, Ika Trisnawati, Sumardi, Eko Budiono, Bambang Sigit Riyanto, Heni Retnowulan, Munawar Gani, Satria Maulana, Nur Rahmi Ananda, Riat El Khair, Yunika Puspadewi, Osman Sianipar, Umi Solekhah Intansari, Elizabeth Henny Herningtiyas, Ira Puspitawati, Nur Imma Fatimah Harahap, Dyah Ayu Puspitarani, Audric Kenny Tedja |

| | | | |
|---|---|---|---|
| | | (WMP) Yogyakarta Center for Tropical Medicine FK-KMK UGM; Integrated Research Center FK-KMK UGM; Department of Computer Science and Electronics FMIPA UGM; RSUP Dr. Sardjito | |
| EPI_ISL_872188 | RSUP Dr. Sardjito | Genetics Working Group (Pokja Genetik) Faculty of Medicine, Public Health and Nursing Universitas Gadjah Mada (FK-KMK UGM); Disease Investigation Center Wates Ministry of Agriculture Indonesia; Department of Microbiology FK-KMK UGM; Laboratorium Diagnostik Yayasan Tahija World Mosquito Program (WMP) Yogyakarta Center for Tropical Medicine FK-KMK UGM; Integrated Research Center FK-KMK UGM; Department of Computer Science and Electronics FMIPA UGM; RSUP Dr. Sardjito | Gunadi, Hendra Wibawa, . Marcellus, Mohamad S. Hakim, Edwin W. Daniwijaya, Ludhang P. Rizki, Endah Supriyati, Eggi Arguni, Titik Nuryastuti, Tri Wibawa, Dwi AA Nugrahaningsih, Afiahayati, Siswanto, Kristy Iskandar, Nungki Anggorowati, Ika Trisnawati, Sumardi, Eko Budiono, Bambang Sigit Riyanto, Heni Retnowulan, Munawar Gani, Satria Maulana, Nur Rahmi Ananda, Riat El Khair, Yunika Puspadewi, Osman Sianipar, Umi Solekhah Intansari, Elizabeth Henny Herningtiyas, Ira Puspitawati, Nur Imma Fatimah Harahap, Aditya Rifqi Fauzi, Fadil Fahri |
| EPI_ISL_872189 | RSUP Dr. Sardjito | Genetics Working Group (Pokja Genetik) Faculty of Medicine, Public Health and Nursing Universitas Gadjah Mada (FK-KMK UGM); Disease Investigation Center Wates Ministry of Agriculture Indonesia; Department of Microbiology FK-KMK UGM; Laboratorium Diagnostik Yayasan Tahija World Mosquito Program (WMP) Yogyakarta Center for Tropical Medicine FK-KMK UGM; Integrated Research Center FK-KMK UGM; Department of Computer Science and Electronics FMIPA UGM; RSUP Dr. Sardjito | Gunadi, Hendra Wibawa, . Marcellus, Mohamad S. Hakim, Edwin W. Daniwijaya, Ludhang P. Rizki, Endah Supriyati, Eggi Arguni, Titik Nuryastuti, Tri Wibawa, Dwi AA Nugrahaningsih, Afiahayati, Siswanto, Kristy Iskandar, Nungki Anggorowati, Ika Trisnawati, Sumardi, Eko Budiono, Bambang Sigit Riyanto, Heni Retnowulan, Munawar Gani, Satria Maulana, Nur Rahmi Ananda, Riat El Khair, Yunika Puspadewi, Osman Sianipar, Umi Solekhah Intansari, Elizabeth Henny Herningtiyas, Ira Puspitawati, Nur Imma Fatimah Harahap, Alvin S. Kalim, Dwiki afandy |
| EPI_ISL_872190 | RSUP Dr. Sardjito | Genetics Working Group (Pokja Genetik) Faculty of Medicine, Public Health and Nursing Universitas Gadjah Mada (FK-KMK UGM); Disease Investigation Center Wates Ministry of Agriculture Indonesia; Department of Microbiology FK-KMK UGM; Laboratorium Diagnostik Yayasan Tahija World Mosquito Program (WMP) Yogyakarta Center for Tropical Medicine FK-KMK UGM; Integrated Research Center FK-KMK UGM; Department of Computer Science and Electronics FMIPA UGM; RSUP Dr. Sardjito | Gunadi, Hendra Wibawa, . Marcellus, Mohamad S. Hakim, Edwin W. Daniwijaya, Ludhang P. Rizki, Endah Supriyati, Eggi Arguni, Titik Nuryastuti, Tri Wibawa, Dwi AA Nugrahaningsih, Afiahayati, Siswanto, Kristy Iskandar, Nungki Anggorowati, Ika Trisnawati, Sumardi, Eko Budiono, Bambang Sigit Riyanto, Heni Retnowulan, Munawar Gani, Satria Maulana, Nur Rahmi Ananda, Riat El Khair, Yunika Puspadewi, Osman Sianipar, Umi Solekhah Intansari, Elizabeth Henny Herningtiyas, Ira Puspitawati, Nur Imma Fatimah Harahap, Dyah Ayu Puspitarani, Kemala Athollah |
| EPI_ISL_877126 | Academic Hospital of Gajdah Mada University (RSA UGM) | Genetics Working Group (Pokja Genetik) Faculty of Medicine, Public Health and Nursing Universitas Gadjah Mada (FK-KMK UGM); Disease Investigation Center Wates Ministry of Agriculture Indonesia; Department of Microbiology FK-KMK UGM; Laboratorium Diagnostik Yayasan Tahija World Mosquito Program (WMP) Yogyakarta Center | Gunadi, Hendra Wibawa, Marcellus, Mohamad S. Hakim, Edwin W. Daniwijaya, Ludhang P. Rizki, Endah Supriyati, Eggi Arguni, Titik Nuryastuti, Tri Wibawa, Dwi AA Nugrahaningsih, Afiahayati, Siswanto,Untung Riawan, William Widitjiarso |

| | | | |
|---|---|---|---|
| | | for Tropical Medicine FK-KMK UGM; Integrated Research center FK-KMK UGM; Department of Computer Science and Electronics FMIPA UGM | |
| EPI_ISL_877127 | Academic Hospital of Gajdah Mada University (RSA UGM) | Genetics Working Group (Pokja Genetik) Faculty of Medicine, Public Health and Nursing Universitas Gadjah Mada (FK-KMK UGM); Disease Investigation Center Wates Ministry of Agriculture Indonesia; Department of Microbiology FK-KMK UGM; Laboratorium Diagnostik Yayasan Tahija World Mosquito Program (WMP) Yogyakarta Center for Tropical Medicine FK-KMK UGM; Integrated Research center FK-KMK UGM; Department of Computer Science and Electronics FMIPA UGM | Gunadi, Hendra Wibawa, Marcellus, Mohamad S. Hakim, Edwin W. Daniwijaya, Ludhang P. Rizki, Endah Supriyati, Eggi Arguni, Titik Nuryastuti, Tri Wibawa, Dwi AA Nugrahaningsih, Afiahayati, Siswanto,Audric Kenny Tedja, Maria Patricia Inggriani |
| EPI_ISL_877128 | Academic Hospital of Gajdah Mada University (RSA UGM) | Genetics Working Group (Pokja Genetik) Faculty of Medicine, Public Health and Nursing Universitas Gadjah Mada (FK-KMK UGM); Disease Investigation Center Wates Ministry of Agriculture Indonesia; Department of Microbiology FK-KMK UGM; Laboratorium Diagnostik Yayasan Tahija World Mosquito Program (WMP) Yogyakarta Center for Tropical Medicine FK-KMK UGM; Integrated Research center FK-KMK UGM; Department of Computer Science and Electronics FMIPA UGM | Gunadi, Hendra Wibawa, Marcellus, Mohamad S. Hakim, Edwin W. Daniwijaya, Ludhang P. Rizki, Endah Supriyati, Eggi Arguni, Titik Nuryastuti, Tri Wibawa, Dwi AA Nugrahaningsih, Afiahayati, Siswanto, Kristy Iskandar, Nungki Anggorowati, Aditya Rifqi Fauzi, Fadil Fahri |
| EPI_ISL_877129 | Academic Hospital of Gajdah Mada University (RSA UGM) | Genetics Working Group (Pokja Genetik) Faculty of Medicine, Public Health and Nursing Universitas Gadjah Mada (FK-KMK UGM); Disease Investigation Center Wates Ministry of Agriculture Indonesia; Department of Microbiology FK-KMK UGM; Laboratorium Diagnostik Yayasan Tahija World Mosquito Program (WMP) Yogyakarta Center for Tropical Medicine FK-KMK UGM; Integrated Research Center FK-KMK UGM; Department of Computer Science and Electronics FMIPA UGM | Gunadi, Hendra Wibawa, Marcellus, Mohamad S. Hakim, Edwin W. Daniwijaya, Ludhang P. Rizki, Endah Supriyati, Eggi Arguni, Titik Nuryastuti, Tri Wibawa, Dwi AA Nugrahaningsih, Afiahayati, Siswanto, Kristy Iskandar, Nungki Anggorowati, Dwiki afandy, Dyah Ayu Puspitarani |
| EPI_ISL_877130 | Academic Hospital of Gajdah Mada University (RSA UGM) | Genetics Working Group (Pokja Genetik) Faculty of Medicine, Public Health and Nursing Universitas Gadjah Mada (FK-KMK UGM); Disease Investigation Center Wates Ministry of Agriculture Indonesia; Department of Microbiology FK-KMK UGM; Laboratorium Diagnostik Yayasan Tahija World Mosquito Program (WMP) Yogyakarta Center for Tropical Medicine FK-KMK UGM; Integrated Research center FK-KMK | Gunadi, Hendra Wibawa, Marcellus, Mohamad S. Hakim, Edwin W. Daniwijaya, Ludhang P. Rizki, Endah Supriyati, Eggi Arguni, Titik Nuryastuti, Tri Wibawa, Dwi AA Nugrahaningsih, Afiahayati, Siswanto, Kristy Iskandar, Nungki Anggorowati, Kemala Athollah, Desyifa Mursalin |

| | | | |
|---|---|---|---|
| | | UGM; Department of Computer Science and Electronics FMIPA UGM | |
| EPI_ISL_877131 | Academic Hospital of Gajdah Mada University (RSA UGM) | Genetics Working Group (Pokja Genetik) Faculty of Medicine, Public Health and Nursing Universitas Gadjah Mada (FK-KMK UGM); Disease Investigation Center Wates Ministry of Agriculture Indonesia; Department of Microbiology FK-KMK UGM; Laboratorium Diagnostik Yayasan Tahija World Mosquito Program (WMP) Yogyakarta Center for Tropical Medicine FK-KMK UGM; Integrated Research center FK-KMK UGM; Department of Computer Science and Electronics FMIPA UGM | Gunadi, Hendra Wibawa, Marcellus, Mohamad S. Hakim, Edwin W. Daniwijaya, Ludhang P. Rizki, Endah Supriyati, Eggi Arguni, Titik Nuryastuti, Tri Wibawa, Dwi AA Nugrahaningsih, Afiahayati, Siswanto, Kristy Iskandar, Nungki Anggorowati, Susan Simanjaya, Alvin Santoso Kalim |
| EPI_ISL_877419 | Siti Khodijah Hospital | Institute of Tropical Disease, Universitas Airlangga | Krisnoadi Rahardjo, Aldise M Nastri, Jezzy R Dewantari, Rima R Prasetya, Muhammad Hamdan, Gatot Soegiarto, Laksmi Wulandari, Resti Yudhawati, Yasuko Mori, Soetjipto, Kazufumi Shimizu, Maria I Lusida |
| EPI_ISL_877420 | Darus Syifa' Islamic Hospital | Institute of Tropical Disease, Universitas Airlangga | Aldise M Nastri, Jezzy R Dewantari, Rima R Prasetya, Krisnoadi Rahardjo, HM Faiz, Gatot Soegiarto, Laksmi Wulandari, Resti Yudhawati, Soetjipto, Yasuko Mori, Maria I Lusida, Kazufumi Shimizu |
| EPI_ISL_877421 | Institute of Tropical Disease | Institute of Tropical Disease, Universitas Airlangga | Jezzy R Dewantari, Rima R Prasetya, Krisnoadi Rahardjo, Aldise M Nastri, Gatot Soegiarto, Laksmi Wulandari, Resti Yudhawati, Soetjipto, Yasuko Mori, Maria I Lusida, Kazufumi Shimizu |
| EPI_ISL_877422 | Citra Medika Hospital | Institute of Tropical Disease, Universitas Airlangga | Rima R Prasetya, Krisnoadi Rahardjo, Aldise M Nastri, Jezzy R Dewantari, Wahidah Rachmaniyah, Gatot Soegiarto, Laksmi Wulandari, Resti Yudhawati, Soetjipto, Yasuko Mori, Maria I Lusida, Kazufumi Shimizu |
| EPI_ISL_877423 | Primasatya Husada Citra Hospital | Institute of Tropical Disease, Universitas Airlangga | Maria I Lusida, Krisnoadi Rahardjo, Aldise M Nastri, Jezzy R Dewantari, Rima R Prasetya, Pudji Djanuartono, Gatot Soegiarto, Laksmi Wulandari, Resti Yudhawati, Soetjipto, Yasuko Mori, Kazufumi Shimizu |
| EPI_ISL_877456 | Primasatya Husada Citra Hospital | Institute of Tropical Disease, Universitas Airlangga | Krisnoadi Rahardjo, Aldise M Nastri, Jezzy R Dewantari, Rima R Prasetya, Pudji Djanuartono, Gatot Soegiarto, Laksmi Wulandari, Resti Yudhawati, Soetjipto, Yasuko Mori, Maria I Lusida, Kazufumi Shimizu |
| EPI_ISL_877457 | Primasatya Husada Citra Hospital | Institute of Tropical Disease, Universitas Airlangga | Aldise M Nastri, Jezzy R Dewantari, Rima R Prasetya, Krisnoadi Rahardjo, Pudji Djanuartono, Gatot Soegiarto, Laksmi Wulandari, Resti Yudhawati, Soetjipto, Yasuko Mori, Maria I Lusida, Kazufumi Shimizu |
| EPI_ISL_877458 | Anwar Medika General Hospital | Institute of Tropical Disease, Universitas Airlangga | Jezzy R Dewantari, Rima R Prasetya, Krisnoadi Rahardjo, Aldise M Nastri, Nungky Taniasari, Gatot Soegiarto, Laksmi Wulandari, Resti Yudhawati, Soetjipto, Yasuko Mori, Maria I Lusida, Kazufumi Shimizu |
| EPI_ISL_877459 | Darmo Hospital | Institute of Tropical Disease, Universitas Airlangga | Rima R Prasetya, Krisnoadi Rahardjo, Aldise M Nastri, Jezzy R Dewantari, Sulung Budianto, Gatot Soegiarto, Laksmi Wulandari, Resti Yudhawati, Soetjipto, Yasuko Mori, Maria I Lusida, Kazufumi Shimizu |
| EPI_ISL_877460 | Siti Khodijah Hospital | Institute of Tropical Disease, Universitas Airlangga | Maria I Lusida, Krisnoadi Rahardjo, Aldise M Nastri, Jezzy R Dewantari, Rima R Prasetya, Muhammad Hamdan, Gatot Soegiarto, Laksmi Wulandari, Resti Yudhawati, Soetjipto, Yasuko Mori, Kazufumi Shimizu |
| EPI_ISL_877538 | Siti Khodijah Hospital | Institute of Tropical Disease, Universitas Airlangga | Krisnoadi Rahardjo, Aldise M Nastri, Jezzy R Dewantari, Rima R Prasetya, Muhammad Hamdan, Gatot Soegiarto, Laksmi Wulandari, Resti Yudhawati, Soetjipto, Yasuko Mori, Maria I Lusida, Kazufumi Shimizu |
| EPI_ISL_882645 | Medical Research Center, Faculty of Medicine, Syarif Hidayatullah State Islamic University Jakarta | Medical Research Center, Faculty of Medicine, Syarif Hidayatullah State Islamic University Jakarta | Chris Adhiyanto, Erike A Suwarsono, Laifa Hendarmin, Zeti Harriyati, Endah Wulandari, Flori Ratna Sari, Fika Ekayanti, Hari Hendarto |
| EPI_ISL_882646 | Medical Research Center, Faculty of Medicine, Syarif Hidayatullah State Islamic University Jakarta | Medical Research Center, Faculty of Medicine, Syarif Hidayatullah State Islamic University Jakarta | Chris Adhiyanto, Laifa Hendarmin, Zeti Harriyati, Endah Wulandari, Flori Ratna Sari, Fika Ekayanti, Hari Hendarto |
| EPI_ISL_882656 | Medical Research Center, Faculty of Medicine, Syarif Hidayatullah State Islamic University Jakarta | Medical Research Center, Faculty of Medicine, Syarif Hidayatullah State Islamic University Jakarta | Erike A Suwarsono, Chris Adhiyanto, Laifa Hendarmin, Zeti Harriyati, Flori Ratna Sari, Endah Wulandari, Fika Ekayanti |
| EPI_ISL_882769 | Institute of Tropical Disease | Institute of Tropical Disease, Universitas Airlangga | Kazufumi Shimizu, Krisnoadi Rahardjo, Aldise M Nastri, Jezzy R Dewantari, Rima R Prasetya, Gatot Soegiarto, Laksmi Wulandari, Resti Yudhawati, Yasuko Mori, Soetjipto, Maria I Lusida |
| EPI_ISL_882770 | Darus Syifa' Islamic Hospital | Institute of Tropical Disease, Universitas Airlangga | Aldise M Nastri, Jezzy R Dewantari, Rima R Prasetya, Krisnoadi Rahardjo, HM Faiz, Gatot Soegiarto, Laksmi Wulandari, Resti Yudhawati, Soetjipto, Yasuko Mori, Kazufumi Shimizu, Maria I Lusida |
| EPI_ISL_882771 | Siti Khodijah Hospital | Institute of Tropical Disease, Universitas Airlangga | Kazufumi Shimizu, Krisnoadi Rahardjo, Aldise M Nastri, Jezzy R Dewantari, Rima R Prasetya, Muhammad Hamdan, Gatot Soegiarto, Laksmi Wulandari, Resti Yudhawati, Yasuko Mori, Soetjipto, Maria I Lusida |
| EPI_ISL_883957 | Medical Research Center, | Medical Research Center, | Erike A Suwarsono, Chris Adhiyanto, Laifa Hendarmin, Zeti Harriyati, Endah Wulandari, Flori Ratna Sari, Fika Ekayanti, Hari Hendarto |

| | | | |
|---|---|---|---|
| | Faculty of Medicine, Syarif Hidayatullah State Islamic University Jakarta | Faculty of Medicine, Syarif Hidayatullah State Islamic University Jakarta | |
| EPI_ISL_885142 | RSUP Dr. Sardjito | Genetics Working Group (Pokja Genetik) Faculty of Medicine, Public Health and Nursing Universitas Gadjah Mada (FK-KMK UGM); Disease Investigation Center Wates Ministry of Agriculture Indonesia; Department of Microbiology FK-KMK UGM; Laboratorium Diagnostik Yayasan Tahija World Mosquito Program (WMP) Yogyakarta Center for Tropical Medicine FK-KMK UGM; Integrated Research Center FK-KMK UGM; Department of Computer Science and Electronics FMIPA UGM; RSUP Dr. Sardjito | Gunadi, Hendra Wibawa, . Marcellus, Mohamad S. Hakim, Edwin W. Daniwijaya, Ludhang P. Rizki, Endah Supriyati, Eggi Arguni, Titik Nuryastuti, Tri Wibawa, Dwi AA Nugrahaningsih, Afiahayati, Siswanto, Kristy Iskandar, Nungki Anggorowati, Ika Trisnawati, Sumardi, Eko Budiono, Bambang Sigit Riyanto, Heni Retnowulan, Munawar Gani, Satria Maulana, Nur Rahmi Ananda, Riat El Khair, Yunika Puspadewi, Osman Sianipar, Umi Solekhah Intansari, Elizabeth Henny Herningtiyas, Ira Puspitawati, Nur Imma Fatimah Harahap, Alvin Santoso Kalim, Susan Simanjaya |
| EPI_ISL_888591 | Siti Khodijah Hospital | Institute of Tropical Disease, Universitas Airlangga | Krisnoadi Rahardjo, Aldise M Nastri, Jezzy R Dewantari, Rima R Prasetya, Muhammad Hamdan, Gatot Soegiarto, Laksmi Wulandari, Resti Yudhawati, Yasuko Mori, Soetjipto, Kazufumi Shimizu, Maria I Lusida |
| EPI_ISL_888593 | Siti Khodijah Hospital | Institute of Tropical Disease, Universitas Airlangga | Rima R Prasetya, Krisnoadi Rahardjo, Aldise M Nastri, Jezzy R Dewantari, Muhammad Hamdan, Gatot Soegiarto, Laksmi Wulandari, Resti Yudhawati, Soetjipto, Yasuko Mori, Kazufumi Shimizu, Maria I Lusida |
| EPI_ISL_888979 | RS Bhakti Mulia | Eijkman Institute for Molecular Biology, Ministry of Research and Technology/National Agency for Research and Innovation | Willy Agustine, Edison Johar, Hidayat Trimarsanto, Iskandar Adnan, Lydia V. Panggalo, Sukma Oktavianthi, Frilasita A Yudhaputri, Safarina G Malik, Khin Saw Myint, Amin Soebandrio |
| EPI_ISL_888980 | RS Anna | Eijkman Institute for Molecular Biology, Ministry of Research and Technology/National Agency for Research and Innovation | Hidayat Trimarsanto, Iskandar Adnan, Lydia V. Panggalo, Sukma Oktavianthi, Willy Agustine, Edison Johar, Frilasita A Yudhaputri, Safarina G Malik, Khin Saw Myint, Amin Soebandrio |
| EPI_ISL_888981 | RS Omni Pulomas | Eijkman Institute for Molecular Biology, Ministry of Research and Technology/National Agency for Research and Innovation | Iskandar Adnan, Lydia V. Panggalo, Sukma Oktavianthi, Willy Agustine, Edison Johar, Hidayat Trimarsanto, Frilasita A Yudhaputri, Safarina G Malik, Khin Saw Myint, Amin Soebandrio |
| EPI_ISL_888983 | RS Hermina Daan Mogot | Eijkman Institute for Molecular Biology, Ministry of Research and Technology/National Agency for Research and Innovation | Edison Johar, Frilasita A Yudhaputri, Hidayat Trimarsanto, Iskandar Adnan, Lydia V. Panggalo, Sukma Oktavianthi, Willy Agustine, Safarina G Malik, Khin Saw Myint, Amin Soebandrio |
| EPI_ISL_888984 | RS Hermina Depok | Eijkman Institute for Molecular Biology, Ministry of Research and Technology/National Agency for Research and Innovation | Edison Johar, Frilasita A Yudhaputri, Hidayat Trimarsanto, Iskandar Adnan, Lydia V. Panggalo, Sukma Oktavianthi, Willy Agustine, Safarina G Malik, Khin Saw Myint, Amin Soebandrio |
| EPI_ISL_888985 | RS Hermina Serpong | Eijkman Institute for Molecular Biology, Ministry of Research and Technology/National Agency for Research and Innovation | Edison Johar, Frilasita A Yudhaputri, Hidayat Trimarsanto, Iskandar Adnan, Lydia V. Panggalo, Sukma Oktavianthi, Willy Agustine, Safarina G Malik, Khin Saw Myint, Amin Soebandrio |
| EPI_ISL_888986 | RSU Bhakti Kartini | Eijkman Institute for Molecular Biology, Ministry of Research and Technology/National Agency for Research and Innovation | Edison Johar, Frilasita A Yudhaputri, Hidayat Trimarsanto, Iskandar Adnan, Lydia V. Panggalo, Sukma Oktavianthi, Willy Agustine, Safarina G Malik, Khin Saw Myint, Amin Soebandrio |
| EPI_ISL_888987 | RS Citra Medika Depok | Eijkman Institute for Molecular Biology, Ministry of Research and Technology/National Agency for Research and Innovation | Frilasita A Yudhaputri, Hidayat Trimarsanto, Iskandar Adnan, Lydia V. Panggalo, Sukma Oktavianthi, Willy Agustine, Edison Johar, Safarina G Malik, Khin Saw Myint, Amin Soebandrio |
| EPI_ISL_888988 | RS Qadr | Eijkman Institute for Molecular Biology, Ministry of Research and | Frilasita A Yudhaputri, Hidayat Trimarsanto, Iskandar Adnan, Lydia V. Panggalo, Sukma Oktavianthi, Willy Agustine, Edison Johar, Safarina G Malik, Khin Saw Myint, Amin Soebandrio |

| | | | |
|---|---|---|---|
| | | Technology/National Agency for Research and Innovation | |
| EPI_ISL_888989, EPI_ISL_888990 | RS Pondok Indah Bintaro Jaya | Eijkman Institute for Molecular Biology, Ministry of Research and Technology/National Agency for Research and Innovation | Frilasita A Yudhaputri, Hidayat Trimarsanto, Iskandar Adnan, Lydia V. Panggalo, Sukma Oktavianthi, Willy Agustine, Edison Johar, Safarina G Malik, Khin Saw Myint, Amin Soebandrio |
| EPI_ISL_888991 | RS Mitra Keluarga Gading Serpong | Eijkman Institute for Molecular Biology, Ministry of Research and Technology/National Agency for Research and Innovation | Frilasita A Yudhaputri, Hidayat Trimarsanto, Iskandar Adnan, Lydia V. Panggalo, Sukma Oktavianthi, Willy Agustine, Edison Johar, Safarina G Malik, Khin Saw Myint, Amin Soebandrio |
| EPI_ISL_888992 | RS Mitra Keluarga Gading Serpong | Eijkman Institute for Molecular Biology, Ministry of Research and Technology/National Agency for Research and Innovation | Hidayat Trimarsanto, Iskandar Adnan, Lydia V. Panggalo, Sukma Oktavianthi, Willy Agustine, Edison Johar, Frilasita A Yudhaputri, Safarina G Malik, Khin Saw Myint, Amin Soebandrio |
| EPI_ISL_888993 | RSU Medirossa | Eijkman Institute for Molecular Biology, Ministry of Research and Technology/National Agency for Research and Innovation | Iskandar Adnan, Lydia V. Panggalo, Sukma Oktavianthi, Willy Agustine, Edison Johar, Hidayat Trimarsanto, Frilasita A Yudhaputri, Safarina G Malik, Khin Saw Myint, Amin Soebandrio |
| EPI_ISL_888994 | RSU MMC | Eijkman Institute for Molecular Biology, Ministry of Research and Technology/National Agency for Research and Innovation | Lydia V. Panggalo, Sukma Oktavianthi, Willy Agustine, Edison Johar, Hidayat Trimarsanto, Iskandar Adnan, Frilasita A Yudhaputri, Safarina G Malik, Khin Saw Myint, Amin Soebandrio |
| EPI_ISL_888995 | RSU Sumber Waras | Eijkman Institute for Molecular Biology, Ministry of Research and Technology/National Agency for Research and Innovation | Lydia V. Panggalo, Sukma Oktavianthi, Willy Agustine, Edison Johar, Hidayat Trimarsanto, Iskandar Adnan, Frilasita A Yudhaputri, Safarina G Malik, Khin Saw Myint, Amin Soebandrio |
| EPI_ISL_888996 | RSU Graha Juanda | Eijkman Institute for Molecular Biology, Ministry of Research and Technology/National Agency for Research and Innovation | Lydia V. Panggalo, Sukma Oktavianthi, Willy Agustine, Edison Johar, Hidayat Trimarsanto, Iskandar Adnan, Frilasita A Yudhaputri, Safarina G Malik, Khin Saw Myint, Amin Soebandrio |
| EPI_ISL_888997 | RSU Asysyifaa | Eijkman Institute for Molecular Biology, Ministry of Research and Technology/National Agency for Research and Innovation | Sukma Oktavianthi, Willy Agustine, Edison Johar, Hidayat Trimarsanto, Iskandar Adnan, Lydia V. Panggalo, Frilasita A Yudhaputri, Safarina G Malik, Khin Saw Myint, Amin Soebandrio |
| EPI_ISL_888998 | RSUD Cileungsi | Eijkman Institute for Molecular Biology, Ministry of Research and Technology/National Agency for Research and Innovation | Sukma Oktavianthi, Willy Agustine, Edison Johar, Hidayat Trimarsanto, Iskandar Adnan, Lydia V. Panggalo, Frilasita A Yudhaputri, Safarina G Malik, Khin Saw Myint, Amin Soebandrio |
| EPI_ISL_888999 | RS Mitra Keluarga Cibubur | Eijkman Institute for Molecular Biology, Ministry of Research and Technology/National Agency for Research and Innovation | Sukma Oktavianthi, Willy Agustine, Edison Johar, Hidayat Trimarsanto, Iskandar Adnan, Lydia V. Panggalo, Frilasita A Yudhaputri, Safarina G Malik, Khin Saw Myint, Amin Soebandrio |
| EPI_ISL_889000 | RSUD Cileungsi | Eijkman Institute for Molecular Biology, Ministry of Research and Technology/National Agency for Research and Innovation | Sukma Oktavianthi, Willy Agustine, Edison Johar, Hidayat Trimarsanto, Iskandar Adnan, Lydia V. Panggalo, Frilasita A Yudhaputri, Safarina G Malik, Khin Saw Myint, Amin Soebandrio |
| EPI_ISL_889001 | RSU Azra | Eijkman Institute for Molecular Biology, Ministry of Research and Technology/National Agency for Research and Innovation | Sukma Oktavianthi, Willy Agustine, Edison Johar, Hidayat Trimarsanto, Iskandar Adnan, Lydia V. Panggalo, Frilasita A Yudhaputri, Safarina G Malik, Khin Saw Myint, Amin Soebandrio |
| EPI_ISL_889002 | RSU Sumber Waras | Eijkman Institute for Molecular Biology, Ministry of Research and Technology/National Agency for Research and Innovation | Sukma Oktavianthi, Willy Agustine, Edison Johar, Hidayat Trimarsanto, Iskandar Adnan, Lydia V. Panggalo, Frilasita A Yudhaputri, Safarina G Malik, Khin Saw Myint, Amin Soebandrio |

| | | | |
|---|---|---|---|
| EPI_ISL_889003, EPI_ISL_889004 | RS Mitra Keluarga Gading Serpong | Eijkman Institute for Molecular Biology, Ministry of Research and Technology/National Agency for Research and Innovation | Willy Agustine, Edison Johar, Hidayat Trimarsanto, Iskandar Adnan, Lydia V. Panggalo, Sukma Oktavianthi, Frilasita A Yudhaputri, Safarina G Malik, Khin Saw Myint, Amin Soebandrio |
| EPI_ISL_889009 | RS Mitra Keluarga Bintaro | Eijkman Institute for Molecular Biology, Ministry of Research and Technology/National Agency for Research and Innovation | Edison Johar, Frilasita A Yudhaputri, Hidayat Trimarsanto, Iskandar Adnan, Lydia V. Panggalo, Sukma Oktavianthi, Willy Agustine, Safarina G Malik, Khin Saw Myint, Amin Soebandrio |
| EPI_ISL_889010 | RSU Medika Dramaga | Eijkman Institute for Molecular Biology, Ministry of Research and Technology/National Agency for Research and Innovation | Frilasita A Yudhaputri, Hidayat Trimarsanto, Iskandar Adnan, Lydia V. Panggalo, Sukma Oktavianthi, Willy Agustine, Edison Johar, Safarina G Malik, Khin Saw Myint, Amin Soebandrio |
| EPI_ISL_889011 | RSU Permata Cibubur | Eijkman Institute for Molecular Biology, Ministry of Research and Technology/National Agency for Research and Innovation | Hidayat Trimarsanto, Iskandar Adnan, Lydia V. Panggalo, Sukma Oktavianthi, Willy Agustine, Edison Johar, Frilasita A Yudhaputri, Safarina G Malik, Khin Saw Myint, Amin Soebandrio |
| EPI_ISL_889012 | RS Mitra Keluarga Pratama Jatiasih | Eijkman Institute for Molecular Biology, Ministry of Research and Technology/National Agency for Research and Innovation | Hidayat Trimarsanto, Iskandar Adnan, Lydia V. Panggalo, Sukma Oktavianthi, Willy Agustine, Edison Johar, Frilasita A Yudhaputri, Safarina G Malik, Khin Saw Myint, Amin Soebandrio |
| EPI_ISL_889013 | RS Rawa Lumbu | Eijkman Institute for Molecular Biology, Ministry of Research and Technology/National Agency for Research and Innovation | Iskandar Adnan, Lydia V. Panggalo, Sukma Oktavianthi, Willy Agustine, Edison Johar, Hidayat Trimarsanto, Frilasita A Yudhaputri, Safarina G Malik, Khin Saw Myint, Amin Soebandrio |
| EPI_ISL_889014 | RSU Medika Dramaga | Eijkman Institute for Molecular Biology, Ministry of Research and Technology/National Agency for Research and Innovation | Iskandar Adnan, Lydia V. Panggalo, Sukma Oktavianthi, Willy Agustine, Edison Johar, Hidayat Trimarsanto, Frilasita A Yudhaputri, Safarina G Malik, Khin Saw Myint, Amin Soebandrio |
| EPI_ISL_889015 | RS Anna | Eijkman Institute for Molecular Biology, Ministry of Research and Technology/National Agency for Research and Innovation | Iskandar Adnan, Lydia V. Panggalo, Sukma Oktavianthi, Willy Agustine, Edison Johar, Hidayat Trimarsanto, Frilasita A Yudhaputri, Safarina G Malik, Khin Saw Myint, Amin Soebandrio |
| EPI_ISL_889016 | RS Mitra Husada | Eijkman Institute for Molecular Biology, Ministry of Research and Technology/National Agency for Research and Innovation | Iskandar Adnan, Lydia V. Panggalo, Sukma Oktavianthi, Willy Agustine, Edison Johar, Hidayat Trimarsanto, Frilasita A Yudhaputri, Safarina G Malik, Khin Saw Myint, Amin Soebandrio |
| EPI_ISL_889017 | RS Hermina Tangerang | Eijkman Institute for Molecular Biology, Ministry of Research and Technology/National Agency for Research and Innovation | Lydia V. Panggalo, Sukma Oktavianthi, Willy Agustine, Edison Johar, Hidayat Trimarsanto, Iskandar Adnan, Frilasita A Yudhaputri, Safarina G Malik, Khin Saw Myint, Amin Soebandrio |
| EPI_ISL_889018 | RSU Medika Dramaga | Eijkman Institute for Molecular Biology, Ministry of Research and Technology/National Agency for Research and Innovation | Lydia V. Panggalo, Sukma Oktavianthi, Willy Agustine, Edison Johar, Hidayat Trimarsanto, Iskandar Adnan, Frilasita A Yudhaputri, Safarina G Malik, Khin Saw Myint, Amin Soebandrio |
| EPI_ISL_889019 | RSIA Sayyidah | Eijkman Institute for Molecular Biology, Ministry of Research and Technology/National Agency for Research and Innovation | Lydia V. Panggalo, Sukma Oktavianthi, Willy Agustine, Edison Johar, Hidayat Trimarsanto, Iskandar Adnan, Frilasita A Yudhaputri, Safarina G Malik, Khin Saw Myint, Amin Soebandrio |
| EPI_ISL_889020 | RS Omni Cikarang | Eijkman Institute for Molecular Biology, Ministry of Research and Technology/National Agency for Research and Innovation | Edison Johar, Frilasita A Yudhaputri, Hidayat Trimarsanto, Iskandar Adnan, Lydia V. Panggalo, Sukma Oktavianthi, Willy Agustine, Safarina G Malik, Khin Saw Myint, Amin Soebandrio |
| EPI_ISL_889021 | RSIA Kenari Graha Medika | Eijkman Institute for Molecular Biology, Ministry of Research and | Hidayat Trimarsanto, Iskandar Adnan, Lydia V. Panggalo, Sukma Oktavianthi, Willy Agustine, Edison Johar, Frilasita A Yudhaputri, Safarina G Malik, Khin Saw Myint, Amin Soebandrio |

| | | | |
|---|---|---|---|
| | | Technology/National Agency for Research and Innovation | |
| EPI_ISL_889022 | RS Anna | Eijkman Institute for Molecular Biology, Ministry of Research and Technology/National Agency for Research and Innovation | Hidayat Trimarsanto, Iskandar Adnan, Lydia V. Panggalo, Sukma Oktavianthi, Willy Agustine, Edison Johar, Frilasita A Yudhaputri, Safarina G Malik, Khin Saw Myint, Amin Soebandrio |
| EPI_ISL_890185 | Academic Hospital of Gadjah Mada University (RSA UGM) | Genetics Working Group (Pokja Genetik) Faculty of Medicine, Public Health and Nursing Universitas Gadjah Mada (FK-KMK UGM); Disease Investigation Center Wates Ministry of Agriculture Indonesia; Department of Microbiology FK-KMK UGM; Laboratorium Diagnostik Yayasan Tahija World Mosquito Program (WMP) Yogyakarta Center for Tropical Medicine FK-KMK UGM; Integrated Research center FK-KMK UGM; Department of Computer Science and Electronics FMIPA UGM | Gunadi, Hendra Wibawa, Marcellus, Mohamad S. Hakim, Edwin W. Daniwijaya, Ludhang P. Rizki, Endah Supriyati, Eggi Arguni, Titik Nuryastuti, Tri Wibawa, Dwi AA Nugrahaningsih, Afiahayati, Siswanto, Kristy Iskandar, Nungki Anggorowati, Aditya Rifqi Fauzi, Fadil Fahri |
| EPI_ISL_890186 | Academic Hospital of Gadjah Mada University (RSA UGM) | Genetics Working Group (Pokja Genetik) Faculty of Medicine, Public Health and Nursing Universitas Gadjah Mada (FK-KMK UGM); Disease Investigation Center Wates Ministry of Agriculture Indonesia; Department of Microbiology FK-KMK UGM; Laboratorium Diagnostik Yayasan Tahija World Mosquito Program (WMP) Yogyakarta Center for Tropical Medicine FK-KMK UGM; Integrated Research center FK-KMK UGM; Department of Computer Science and Electronics FMIPA UGM | Gunadi, Hendra Wibawa, Marcellus, Mohamad S. Hakim, Edwin W. Daniwijaya, Ludhang P. Rizki, Endah Supriyati, Eggi Arguni, Titik Nuryastuti, Tri Wibawa, Dwi AA Nugrahaningsih, Afiahayati, Siswanto, Kristy Iskandar, Nungki Anggorowati, Audric Kenny Tedja, Maria Patricia Inggriani |
| EPI_ISL_890187 | Academic Hospital of Gadjah Mada University (RSA UGM) | Genetics Working Group (Pokja Genetik) Faculty of Medicine, Public Health and Nursing Universitas Gadjah Mada (FK-KMK UGM); Disease Investigation Center Wates Ministry of Agriculture Indonesia; Department of Microbiology FK-KMK UGM; Laboratorium Diagnostik Yayasan Tahija World Mosquito Program (WMP) Yogyakarta Center for Tropical Medicine FK-KMK UGM; Integrated Research center FK-KMK UGM; Department of Computer Science and Electronics FMIPA UGM | Gunadi, Hendra Wibawa, Marcellus, Mohamad S. Hakim, Edwin W. Daniwijaya, Ludhang P. Rizki, Endah Supriyati, Eggi Arguni, Titik Nuryastuti, Tri Wibawa, Dwi AA Nugrahaningsih, Afiahayati, Siswanto, Kristy Iskandar, Nungki Anggorowati, Untung Riawan, William Widitjiarso |
| EPI_ISL_891151 | Academic Hospital of Gadjah Mada University (RSA UGM) | Genetics Working Group (Pokja Genetik) Faculty of Medicine, Public Health and Nursing Universitas Gadjah Mada (FK-KMK UGM); Disease Investigation Center Wates Ministry of Agriculture Indonesia; Department of Microbiology FK-KMK UGM; Laboratorium Diagnostik Yayasan Tahija World Mosquito Program (WMP) Yogyakarta Center for Tropical Medicine FK-KMK UGM; Integrated | Gunadi, Hendra Wibawa, Marcellus, Mohamad S. Hakim, Edwin W. Daniwijaya, Ludhang P. Rizki, Endah Supriyati, Eggi Arguni, Titik Nuryastuti, Tri Wibawa, Dwi AA Nugrahaningsih, Afiahayati, Siswanto, Kristy Iskandar, Nungki Anggorowati, Alvin Santoso Kalim, Susan Simanjaya |

| | | | |
|---|---|---|---|
| | | Research center FK-KMK UGM; Department of Computer Science and Electronics FMIPA UGM | |
| EPI_ISL_902737 | RSA UGM | Genetics Working Group (Pokja Genetik) Faculty of Medicine, Public Health and Nursing Universitas Gadjah Mada (FK-KMK UGM); Disease Investigation Center Wates Ministry of Agriculture Indonesia; Department of Microbiology FK-KMK UGM; Laboratorium Diagnostik Yayasan Tahija World Mosquito Program (WMP) Yogyakarta Center for Tropical Medicine FK-KMK UGM; Integrated Research center FK-KMK UGM; Department of Computer Science and Electronics FMIPA UGM | Gunadi, Hendra Wibawa, Marcellus, Mohamad S. Hakim, Edwin W. Daniwijaya, Ludhang P. Rizki, Endah Supriyati, Eggi Arguni, Titik Nuryastuti, Tri Wibawa, Dwi AA Nugrahaningsih, Afiahayati, Siswanto, Kristy Iskandar, Nungki Anggorowati, Aditya Rifqi Fauzi, Fadil Fahri |
| EPI_ISL_902749 | RSA UGM | Genetics Working Group (Pokja Genetik) Faculty of Medicine, Public Health and Nursing Universitas Gadjah Mada (FK-KMK UGM); Disease Investigation Center Wates Ministry of Agriculture Indonesia; Department of Microbiology FK-KMK UGM; Laboratorium Diagnostik Yayasan Tahija World Mosquito Program (WMP) Yogyakarta Center for Tropical Medicine FK-KMK UGM; Integrated Research center FK-KMK UGM; Department of Computer Science and Electronics FMIPA UGM | Gunadi, Hendra Wibawa, Marcellus, Mohamad S. Hakim, Edwin W. Daniwijaya, Ludhang P. Rizki, Endah Supriyati, Eggi Arguni, Titik Nuryastuti, Tri Wibawa, Dwi AA Nugrahaningsih, Afiahayati, Siswanto, Kristy Iskandar, Nungki Anggorowati, Dyah Ayu Puspitarani, Dwiki afandy |
| EPI_ISL_902917 | Tanjungpura University Hospital | Tanjungpura University Hospital | Andriani; Mahyarudin; Novianry V; Liana DF; Shofiyah SS Astuti P; Kahtan MI; Rialita A; Putri EA; Windarti W; Sastriawan W |
| EPI_ISL_902918 | Tanjungpura University Hospital | Tanjungpura University Hospital | Andriani; Mahyarudin; Virhan Novianry ; Delima Fajar Liana; Sofi Siti Shofiyah; Puji Astuti P; Muhammad Ibnu Kahtan; Ambar Rialita; Eka Ardiani Putri EA; Wiwik Windarti ; Helmi Sastriawan; Willy Handoko |
| EPI_ISL_902919 | Tanjungpura University Hospital | Tanjungpura University Hospital | Andriani; Mahyarudin; Virhan Novianry ; Delima Fajar Liana; Sofi Siti Shofiyah; Puji Astuti P; Muhammad Ibnu Kahtan; Ambar Rialita; Eka Ardiani Putri EA; Wiwik Windarti ; Helmi Sastriawan; Arif Wicaksono |
| EPI_ISL_902921 | Tanjungpura University Hospital | Tanjungpura University Hospital | Puji Astuti; Andriani; Mahyarudin; Delima Fajar Liana; Virhan Novianry ; Sofi Siti Shofiyah |
| EPI_ISL_902922 | Tanjungpura University Hospital | Tanjungpura University Hospital | Andriani; Mahyarudin; Virhan Novianry ; Delima Fajar Liana; Sofi Siti Shofiyah; Puji Astuti; Muhammad Ibnu Kahtan; Ambar Rialita; Eka Ardiani Putri; Wiwik Windarti ; Helmi Sastriawan |
| EPI_ISL_902924 | Tanjungpura University Hospital | Tanjungpura University Hospital | Andriani; Mahyarudin; Virhan Novianry; Delima Fajar Liana; Sofi Siti Shofiyah; Puji Astuti ; Muhammad Ibnu Kahtan; Ambar Rialita; Eka Ardiani Putri; Wiwik Windarti; Helmi Sastriawan |
| EPI_ISL_905731 | Academic Hospital of Gadjah Mada University (RSA UGM) | Genetics Working Group (Pokja Genetik) Faculty of Medicine, Public Health and Nursing Universitas Gadjah Mada (FK-KMK UGM); Disease Investigation Center Wates Ministry of Agriculture Indonesia; Department of Microbiology FK-KMK UGM; Laboratorium Diagnostik Yayasan Tahija World Mosquito Program (WMP) Yogyakarta Center for Tropical Medicine FK-KMK UGM; Integrated Research center FK-KMK UGM; Department of Computer Science and Electronics FMIPA UGM | Gunadi, Hendra Wibawa, Marcellus, Mohamad S. Hakim, Edwin W. Daniwijaya, Ludhang P. Rizki, Endah Supriyati, Eggi Arguni, Titik Nuryastuti, Tri Wibawa, Dwi AA Nugrahaningsih, Afiahayati, Siswanto, Kristy Iskandar,Nungki Anggorowati, Dyah Ayu Puspitarani, Dwiki afandy |
| EPI_ISL_906050 | RSA UGM | Genetics Working Group (Pokja Genetik) Faculty of Medicine, Public Health and Nursing Universitas Gadjah Mada (FK-KMK UGM); Disease | Gunadi, Hendra Wibawa, Marcellus, Mohamad S. Hakim, Edwin W. Daniwijaya, Ludhang P. Rizki, Endah Supriyati, Eggi Arguni, Titik Nuryastuti, Tri Wibawa, Dwi AA Nugrahaningsih, Afiahayati, Siswanto,Kristy Iskandar,Nungki Anggorowati, Susan Simanjaya, Kemala Athollah |

| | | | |
|---|---|---|---|
| | | Investigation Center Wates Ministry of Agriculture Indonesia; Department of Microbiology FK-KMK UGM; Laboratorium Diagnostik Yayasan Tahija World Mosquito Program (WMP) Yogyakarta Center for Tropical Medicine FK-KMK UGM; Integrated Research center FK-KMK UGM; Department of Computer Science and Electronics FMIPA UGM | |
| EPI_ISL_906051 | RSA UGM | Genetics Working Group (Pokja Genetik) Faculty of Medicine, Public Health and Nursing Universitas Gadjah Mada (FK-KMK UGM); Disease Investigation Center Wates Ministry of Agriculture Indonesia; Department of Microbiology FK-KMK UGM; Laboratorium Diagnostik Yayasan Tahija World Mosquito Program (WMP) Yogyakarta Center for Tropical Medicine FK-KMK UGM; Integrated Research center FK-KMK UGM; Department of Computer Science and Electronics FMIPA UGM | Gunadi, Hendra Wibawa, Marcellus, Mohamad S. Hakim, Edwin W. Daniwijaya, Ludhang P. Rizki, Endah Supriyati, Eggi Arguni, Titik Nuryastuti, Tri Wibawa, Dwi AA Nugrahaningsih, Afiahayati, Siswanto, Kristy Iskandar, Nungki Anggorowati, Alvin Santoso Kalim, Dwiki Afandy |
| EPI_ISL_906052 | RSA UGM | Genetics Working Group (Pokja Genetik) Faculty of Medicine, Public Health and Nursing Universitas Gadjah Mada (FK-KMK UGM); Disease Investigation Center Wates Ministry of Agriculture Indonesia; Department of Microbiology FK-KMK UGM; Laboratorium Diagnostik Yayasan Tahija World Mosquito Program (WMP) Yogyakarta Center for Tropical Medicine FK-KMK UGM; Integrated Research center FK-KMK UGM; Department of Computer Science and Electronics FMIPA UGM | Gunadi, Hendra Wibawa, Marcellus, Mohamad S. Hakim, Edwin W. Daniwijaya, Ludhang P. Rizki, Endah Supriyati, Eggi Arguni, Titik Nuryastuti, Tri Wibawa, Dwi AA Nugrahaningsih, Afiahayati, Siswanto, Kristy Iskandar, Nungki Anggorowati, William Widitjiarso, Untung Riawan |
| EPI_ISL_909772 | National Institute of Health Research and Development | National Institute of Health Research and Development | Holy Arif Wibowo; Ririn Ramadhany; Yuni Rukminiati; Agustiningsih; Kindi Adam; Hana Apsari Pawestri; Subangkit; Kartika Dewi Puspa; Arie Ardiansyah Nugraha; Hartanti Dian Ikawati; Krisna Nur Andriana Pangesti; Triyani Soekarso; Ni Ketut Susilarini; Nur Ika Hariastuti; Uli Alfi Nikmah; Mursinah; Asri Febriyani; Reni Herman; Nike Susanti; Herna; Tati Febriyanti; Juwita Kurniawati; Natalie Laurencia Kipuw; Fauzul Muna; Irene Lorinda Indalao; Aulia Rizki; Nelly Puspandari; vivi Setiawaty |
| EPI_ISL_911675 | Tanjungpura University Hospital | Tanjungpura University Hospital | Mahyarudin; Andriani; Virhan Novianry; Delima Fajar Liana ; Sofi Siti Shofiyah; Puji Astuti ; Muhammad Ibnu Kahtan ; Ambar Rialita; Eka Ardiani Putri; Wiwik Windarti; Helmi Sastriawan |
| EPI_ISL_911707 | Academic Hospital of Gadjah Mada University (RSA UGM) | Genetics Working Group (Pokja Genetik) Faculty of Medicine, Public Health and Nursing Universitas Gadjah Mada (FK-KMK UGM); Disease Investigation Center Wates Ministry of Agriculture Indonesia; Department of Microbiology FK-KMK UGM; Laboratorium Diagnostik Yayasan Tahija World Mosquito Program (WMP) Yogyakarta Center for Tropical Medicine FK-KMK UGM; Integrated Research center FK-KMK UGM; Department of Computer Science and Electronics FMIPA UGM | Gunadi, Hendra Wibawa, Marcellus, Mohamad S. Hakim, Edwin W. Daniwijaya, Ludhang P. Rizki, Endah Supriyati, Eggi Arguni, Titik Nuryastuti, Tri Wibawa, Dwi AA Nugrahaningsih, Afiahayati, Siswanto, Kristy Iskandar, Nungki Anggorowati, Untung Riawan, Kemala Athollah |
| EPI_ISL_911708 | Tanjungpura University Hospital | Tanjungpura University Hospital | Puji Astuti; Andriani; Mahyarudin; Delima Fajar Liana; Virhan Novianry ; Sofi Siti Shofiyah |
| EPI_ISL_911709 | Academic Hospital of Gadjah Mada University (RSA UGM) | Genetics Working Group (Pokja Genetik) Faculty of Medicine, Public Health | Gunadi, Hendra Wibawa, Marcellus, Mohamad S. Hakim, Edwin W. Daniwijaya, Ludhang P. Rizki, Endah Supriyati, Eggi Arguni, Titik Nuryastuti, Tri Wibawa, Dwi AA Nugrahaningsih, Afiahayati, Siswanto, Kristy Iskandar, Nungki Anggorowati, Dyah Ayu Puspitarani, Dwiki afandy |

| | | | |
|---|---|---|---|
| | | and Nursing Universitas Gadjah Mada (FK-KMK UGM); Disease Investigation Center Wates Ministry of Agriculture Indonesia; Department of Microbiology FK-KMK UGM; Laboratorium Diagnostik Yayasan Tahija World Mosquito Program (WMP) Yogyakarta Center for Tropical Medicine FK-KMK UGM; Integrated Research center FK-KMK UGM; Department of Computer Science and Electronics FMIPA UGM | |
| EPI_ISL_911732 | Tanjungpura University Hospital | Tanjungpura University Hospital | Delima Fajar Liana; Mahyarudin; Andriani; Virhan Novianry; Sofi Siti Shofiyah; Puji Astuti; Muhammad Ibnu Kahtan; Ambar Rialita; Eka Ardiani Putri; Wiwik Windarti; Helmi Sastriawan |
| EPI_ISL_911750 | Tanjungpura University Hospital | Tanjungpura University Hospital | Virhan Novianry; Delima Fajar Liana; Mahyarudin; Andriani; Sofi Siti Shofiyah; Puji Astuti; Muhammad Kahtan; Ambar Rialita; Eka Ardiani Putri; Wiwik Windarti; Helmi Sastriawan, Rini Andriani |
| EPI_ISL_933575, EPI_ISL_933576 | Medical Research Center, Faculty of Medicine, Syarif Hidayatullah State Islamic University Jakarta | Medical Research Center, Faculty of Medicine, Syarif Hidayatullah State Islamic University Jakarta | Erike A Suwarsono, Chris Adhiyanto, Laifa Hendarmin, Zeti Harriyati, Endah Wulandari, Flori Ratna Sari, Fika Ekayanti, Hari Hendarto |
| EPI_ISL_942960, EPI_ISL_942971 | National Institute of Health Research and Development | National Institute of Health Research and Development | Subangkit, Hana Apsari Pawestri, Kartika Dewi Puspa, Arie Ardiansyah Nugraha, Hartanti Dian Ikawati, Krisna Nur Andriana Pangesti, Yuni Rukminiati, Ririn Ramadhany, Agustiningsih, Kindi Adam, Holy Arif Wibowo, Triyani Soekarso, Ni Ketut Susilarini, Nurika Hariastuti, Ully Alfi Nikmah, Reni Herman, Nike Susanti, Herna, Tati Febriyanti, Natalie Laurencia Kipuw, Fauzul Muna, Irene Lorinda Indalao, Nelly Puspandari, Vivi Setiawaty. |
| EPI_ISL_943548 | National Institute of Health Research and Development | National Institute of Health Research and Development | Subangkit, Hana Apsari Pawestri, Kartika Dewi Puspa, Arie Ardiansyah Nugraha, Hartanti Dian Ikawati, Krisna Nur Andriana Pangesti, Yuni Rukminiati, Ririn Ramadhany, Agustiningsih, Kindi Adam, Holy Arif Wibowo, Triyani Soekarso, Ni Ketut Susilarini, Nurika Hariastuti, Ully Alfi Nikmah, Reni Herman, Nike Susanti, Herna, Tati Febriyanti, Natalie Laurencia Kipuw, Fauzul Muna, Irene Lorinda Indalao, Nelly Puspandari, Vivi Setiawaty |
| EPI_ISL_947247 | RSU Bunda Mulia | Eijkman Institute for Molecular Biology, Ministry of Research and Technology/National Agency for Research and Innovation | Edison Johar, Frilasita A Yudhaputri, Hidayat Trimarsanto, Iskandar Adnan, Lydia V. Panggalo, Sukma Oktavianthi, Willy Agustine, Safarina G Malik, Khin Saw Myint, Amin Soebandrio |
| EPI_ISL_947249, EPI_ISL_947252 | RS Bina Husada Cibinong | Eijkman Institute for Molecular Biology, Ministry of Research and Technology/National Agency for Research and Innovation | Edison Johar, Frilasita A Yudhaputri, Hidayat Trimarsanto, Iskandar Adnan, Lydia V. Panggalo, Sukma Oktavianthi, Willy Agustine, Safarina G Malik, Khin Saw Myint, Amin Soebandrio |
| EPI_ISL_947255 | RSU Meilia | Eijkman Institute for Molecular Biology, Ministry of Research and Technology/National Agency for Research and Innovation | Edison Johar, Frilasita A Yudhaputri, Hidayat Trimarsanto, Iskandar Adnan, Lydia V. Panggalo, Sukma Oktavianthi, Willy Agustine, Safarina G Malik, Khin Saw Myint, Amin Soebandrio |
| EPI_ISL_947257 | RS Bina Husada Cibinong | Eijkman Institute for Molecular Biology, Ministry of Research and Technology/National Agency for Research and Innovation | Edison Johar, Frilasita A Yudhaputri, Hidayat Trimarsanto, Iskandar Adnan, Lydia V. Panggalo, Sukma Oktavianthi, Willy Agustine, Safarina G Malik, Khin Saw Myint, Amin Soebandrio |
| EPI_ISL_947260 | RSU Bunda Mulia | Eijkman Institute for Molecular Biology, Ministry of Research and Technology/National Agency for Research and Innovation | Iskandar Adnan, Lydia V. Panggalo, Sukma Oktavianthi, Willy Agustine, Edison Johar, Hidayat Trimarsanto, Frilasita A Yudhaputri, Safarina G Malik, Khin Saw Myint, Amin Soebandrio |
| EPI_ISL_947262 | RS Qadr | Eijkman Institute for Molecular Biology, Ministry of Research and Technology/National Agency for Research and Innovation | Frilasita A Yudhaputri, Hidayat Trimarsanto, Iskandar Adnan, Lydia V. Panggalo, Sukma Oktavianthi, Willy Agustine, Edison Johar, Safarina G Malik, Khin Saw Myint, Amin Soebandrio |
| EPI_ISL_947265 | RS Sari Asih Ciledug | Eijkman Institute for Molecular Biology, Ministry of Research and Technology/National Agency for Research and Innovation | Frilasita A Yudhaputri, Hidayat Trimarsanto, Iskandar Adnan, Lydia V. Panggalo, Sukma Oktavianthi, Willy Agustine, Edison Johar, Safarina G Malik, Khin Saw Myint, Amin Soebandrio |
| EPI_ISL_947268 | RSU Anggrek Mas | Eijkman Institute for Molecular Biology, Ministry of Research and Technology/National Agency for Research and Innovation | Frilasita A Yudhaputri, Hidayat Trimarsanto, Iskandar Adnan, Lydia V. Panggalo, Sukma Oktavianthi, Willy Agustine, Edison Johar, Safarina G Malik, Khin Saw Myint, Amin Soebandrio |
| EPI_ISL_947270 | RS Mitra Keluarga Gading | Eijkman Institute for | Frilasita A Yudhaputri, Hidayat Trimarsanto, Iskandar Adnan, Lydia V. Panggalo, Sukma Oktavianthi, Willy Agustine, Edison Johar, Safarina G Malik, Khin Saw Myint, Amin Soebandrio |

| | | | |
|---|---|---|---|
| | Serpong | Molecular Biology, Ministry of Research and Technology/National Agency for Research and Innovation | |
| EPI_ISL_947273 | RSIA Sayyidah | Eijkman Institute for Molecular Biology, Ministry of Research and Technology/National Agency for Research and Innovation | Frilasita A Yudhaputri, Hidayat Trimarsanto, Iskandar Adnan, Lydia V. Panggalo, Sukma Oktavianthi, Willy Agustine, Edison Johar, Safarina G Malik, Khin Saw Myint, Amin Soebandrio |
| EPI_ISL_947275 | RS Umum Mulia Pajajaran | Eijkman Institute for Molecular Biology, Ministry of Research and Technology/National Agency for Research and Innovation | Frilasita A Yudhaputri, Hidayat Trimarsanto, Iskandar Adnan, Lydia V. Panggalo, Sukma Oktavianthi, Willy Agustine, Edison Johar, Safarina G Malik, Khin Saw Myint, Amin Soebandrio |
| EPI_ISL_947278 | RS Umum Ciputra Hospital Citragarden City | Eijkman Institute for Molecular Biology, Ministry of Research and Technology/National Agency for Research and Innovation | Hidayat Trimarsanto, Iskandar Adnan, Lydia V. Panggalo, Sukma Oktavianthi, Willy Agustine, Edison Johar, Frilasita A Yudhaputri, Safarina G Malik, Khin Saw Myint, Amin Soebandrio |
| EPI_ISL_947281 | RSU Medika Dramaga | Eijkman Institute for Molecular Biology, Ministry of Research and Technology/National Agency for Research and Innovation | Hidayat Trimarsanto, Iskandar Adnan, Lydia V. Panggalo, Sukma Oktavianthi, Willy Agustine, Edison Johar, Frilasita A Yudhaputri, Safarina G Malik, Khin Saw Myint, Amin Soebandrio |
| EPI_ISL_947284 | RSIA Bunda Sejati | Eijkman Institute for Molecular Biology, Ministry of Research and Technology/National Agency for Research and Innovation | Hidayat Trimarsanto, Iskandar Adnan, Lydia V. Panggalo, Sukma Oktavianthi, Willy Agustine, Edison Johar, Frilasita A Yudhaputri, Safarina G Malik, Khin Saw Myint, Amin Soebandrio |
| EPI_ISL_947287 | RSIA PKU Muhammadiyah | Eijkman Institute for Molecular Biology, Ministry of Research and Technology/National Agency for Research and Innovation | Hidayat Trimarsanto, Iskandar Adnan, Lydia V. Panggalo, Sukma Oktavianthi, Willy Agustine, Edison Johar, Frilasita A Yudhaputri, Safarina G Malik, Khin Saw Myint, Amin Soebandrio |
| EPI_ISL_947290 | RS Hermina Bitung | Eijkman Institute for Molecular Biology, Ministry of Research and Technology/National Agency for Research and Innovation | Hidayat Trimarsanto, Iskandar Adnan, Lydia V. Panggalo, Sukma Oktavianthi, Willy Agustine, Edison Johar, Frilasita A Yudhaputri, Safarina G Malik, Khin Saw Myint, Amin Soebandrio |
| EPI_ISL_947293 | Eijkman Institute for Molecular Biology, Ministry of Research and Technology/National Agency for Research and Innovation | Eijkman Institute for Molecular Biology, Ministry of Research and Technology/National Agency for Research and Innovation | Edison Johar, Frilasita A Yudhaputri, Hidayat Trimarsanto, Iskandar Adnan, Lydia V. Panggalo, Sukma Oktavianthi, Willy Agustine, Safarina G Malik, Khin Saw Myint, Amin Soebandrio |
| EPI_ISL_947296 | RSU Kartika Husada | Eijkman Institute for Molecular Biology, Ministry of Research and Technology/National Agency for Research and Innovation | Iskandar Adnan, Lydia V. Panggalo, Sukma Oktavianthi, Willy Agustine, Edison Johar, Hidayat Trimarsanto, Frilasita A Yudhaputri, Safarina G Malik, Khin Saw Myint, Amin Soebandrio |
| EPI_ISL_947298 | RSU Medirossa | Eijkman Institute for Molecular Biology, Ministry of Research and Technology/National Agency for Research and Innovation | Iskandar Adnan, Lydia V. Panggalo, Sukma Oktavianthi, Willy Agustine, Edison Johar, Hidayat Trimarsanto, Frilasita A Yudhaputri, Safarina G Malik, Khin Saw Myint, Amin Soebandrio |
| EPI_ISL_947301 | RS Mitra Keluarga Cibubur | Eijkman Institute for Molecular Biology, Ministry of Research and Technology/National Agency for Research and Innovation | Iskandar Adnan, Lydia V. Panggalo, Sukma Oktavianthi, Willy Agustine, Edison Johar, Hidayat Trimarsanto, Frilasita A Yudhaputri, Safarina G Malik, Khin Saw Myint, Amin Soebandrio |
| EPI_ISL_947304 | RSU Harapan Bunda | Eijkman Institute for Molecular Biology, Ministry of Research and Technology/National Agency for Research and Innovation | Iskandar Adnan, Lydia V. Panggalo, Sukma Oktavianthi, Willy Agustine, Edison Johar, Hidayat Trimarsanto, Frilasita A Yudhaputri, Safarina G Malik, Khin Saw Myint, Amin Soebandrio |
| EPI_ISL_947307, EPI_ISL_947309 | RS Sentra Medika Cikarang | Eijkman Institute for Molecular Biology, Ministry of Research and Technology/National | Lydia V. Panggalo, Sukma Oktavianthi, Willy Agustine, Edison Johar, Hidayat Trimarsanto, Iskandar Adnan, Frilasita A Yudhaputri, Safarina G Malik, Khin Saw Myint, Amin Soebandrio |

| | | | |
|---|---|---|---|
| | | Agency for Research and Innovation | |
| EPI_ISL_947312 | RSU Bunda Mulia | Eijkman Institute for Molecular Biology, Ministry of Research and Technology/National Agency for Research and Innovation | Lydia V. Panggalo, Sukma Oktavianthi, Willy Agustine, Edison Johar, Hidayat Trimarsanto, Iskandar Adnan, Frilasita A Yudhaputri, Safarina G Malik, Khin Saw Myint, Amin Soebandrio |
| EPI_ISL_947315 | RSU Harapan Bunda | Eijkman Institute for Molecular Biology, Ministry of Research and Technology/National Agency for Research and Innovation | Lydia V. Panggalo, Sukma Oktavianthi, Willy Agustine, Edison Johar, Hidayat Trimarsanto, Iskandar Adnan, Frilasita A Yudhaputri, Safarina G Malik, Khin Saw Myint, Amin Soebandrio |
| EPI_ISL_947317 | RSU Medirossa | Eijkman Institute for Molecular Biology, Ministry of Research and Technology/National Agency for Research and Innovation | Lydia V. Panggalo, Sukma Oktavianthi, Willy Agustine, Edison Johar, Hidayat Trimarsanto, Iskandar Adnan, Frilasita A Yudhaputri, Safarina G Malik, Khin Saw Myint, Amin Soebandrio |
| EPI_ISL_947320 | RS Sari Asih Ciledug | Eijkman Institute for Molecular Biology, Ministry of Research and Technology/National Agency for Research and Innovation | Sukma Oktavianthi, Willy Agustine, Edison Johar, Hidayat Trimarsanto, Iskandar Adnan, Lydia V. Panggalo, Frilasita A Yudhaputri, Safarina G Malik, Khin Saw Myint, Amin Soebandrio |
| EPI_ISL_947322 | RS Mitra Keluarga Kelapa Gading | Eijkman Institute for Molecular Biology, Ministry of Research and Technology/National Agency for Research and Innovation | Sukma Oktavianthi, Willy Agustine, Edison Johar, Hidayat Trimarsanto, Iskandar Adnan, Lydia V. Panggalo, Frilasita A Yudhaputri, Safarina G Malik, Khin Saw Myint, Amin Soebandrio |
| EPI_ISL_947323 | RS Siloam Kebon Jeruk | Eijkman Institute for Molecular Biology, Ministry of Research and Technology/National Agency for Research and Innovation | Sukma Oktavianthi, Willy Agustine, Edison Johar, Hidayat Trimarsanto, Iskandar Adnan, Lydia V. Panggalo, Frilasita A Yudhaputri, Safarina G Malik, Khin Saw Myint, Amin Soebandrio |
| EPI_ISL_947324 | RSU Medirossa | Eijkman Institute for Molecular Biology, Ministry of Research and Technology/National Agency for Research and Innovation | Sukma Oktavianthi, Willy Agustine, Edison Johar, Hidayat Trimarsanto, Iskandar Adnan, Lydia V. Panggalo, Frilasita A Yudhaputri, Safarina G Malik, Khin Saw Myint, Amin Soebandrio |
| EPI_ISL_947325 | RSIA PKU Muhammadiyah | Eijkman Institute for Molecular Biology, Ministry of Research and Technology/National Agency for Research and Innovation | Sukma Oktavianthi, Willy Agustine, Edison Johar, Hidayat Trimarsanto, Iskandar Adnan, Lydia V. Panggalo, Frilasita A Yudhaputri, Safarina G Malik, Khin Saw Myint, Amin Soebandrio |
| EPI_ISL_947326, EPI_ISL_947327 | RS Mitra Keluarga Gading Serpong | Eijkman Institute for Molecular Biology, Ministry of Research and Technology/National Agency for Research and Innovation | Willy Agustine, Edison Johar, Hidayat Trimarsanto, Iskandar Adnan, Lydia V. Panggalo, Sukma Oktavianthi, Frilasita A Yudhaputri, Safarina G Malik, Khin Saw Myint, Amin Soebandrio |
| EPI_ISL_947328 | RS Mitra Keluarga Depok | Eijkman Institute for Molecular Biology, Ministry of Research and Technology/National Agency for Research and Innovation | Willy Agustine, Edison Johar, Hidayat Trimarsanto, Iskandar Adnan, Lydia V. Panggalo, Sukma Oktavianthi, Frilasita A Yudhaputri, Safarina G Malik, Khin Saw Myint, Amin Soebandrio |
| EPI_ISL_947329, EPI_ISL_947330 | RSU Bunda Mulia | Eijkman Institute for Molecular Biology, Ministry of Research and Technology/National Agency for Research and Innovation | Willy Agustine, Edison Johar, Hidayat Trimarsanto, Iskandar Adnan, Lydia V. Panggalo, Sukma Oktavianthi, Frilasita A Yudhaputri, Safarina G Malik, Khin Saw Myint, Amin Soebandrio |
| EPI_ISL_949183 | National Institute of Health Research and Development | National Institute of Health Research and Development | Kindi Adam; Holy Arif Wibowo; Ririn Ramadhany; Yuni Rukminiati; Agustiningsih; Hana Apsari Pawestri; Subangkit; Kartika Dewi Puspa; Arie Ardiansyah Nugraha; Hartanti Dian Ikawati; Krisna Nur Andriana Pangesti; Triyani Soekarso; Ni Ketut Susilarini; Nur Ika Hariastuti; Uly Alfi Nikmah; Mursinah; Asri Febriyani; Reni Herman; Nike Susanti; Herna; Tati Febriyanti; Natalie Laurencia Kipuw; Fauzul Muna; Irene Lorinda Indalao; Aulia Rizki; Nelly Puspandari; Vivi Setiawaty |
| EPI_ISL_949192 | National Institute of Health Research and Development | National Institute of Health Research and Development | Ririn Ramadhany; Yuni Rukminiati; Agustiningsih; Kindi Adam; Holy Arif Wibowo; Hana Apsari Pawestri; Subangkit; Kartika Dewi Puspa; Arie Ardiansyah Nugraha; Hartanti Dian Ikawati; Krisna Nur Andriana Pangesti; Ni Ketut Susilarini; Nur Ika Hariastuti; Uly Alfi Nikmah; Mursinah; Asri Febriyani; Reni Herman; Nike Susanti; Herna; Tati Febriyanti; Natalie Laurentia Kipuw; Fauzul Muna; Irene Lorinda Indalao; Aulia Rizki; Nelly Puspandari; Vivi Setiawaty |
| EPI_ISL_953427 | National Institute of Health Research and Development | National Institute of Health Research and Development | Subangkit, Hana Apsari Pawestri, Kartika Dewi Puspa, Arie Ardiansyah Nugraha, Hartanti Dian Ikawati, Krisna Nur Andriana Pangesti, Yuni Rukminiati, Ririn Ramadhany, Agustiningsih, Kindi Adam, Holy Arif Wibowo, Triyani Soekarso, Ni Ketut Susilarini, Nurika Hariastuti, Ully Alfi Nikmah, Reni Herman, Nike Susanti, Herna, Tati Febriyanti, Natalie Laurencia Kipuw, Fauzul Muna, Irene Lorinda Indalao, Nelly Puspandari, Vivi Setiawaty |
| EPI_ISL_956279 | Siti Khodijah Hospital | Institute of Tropical Disease, Universitas Airlangga | Jezzy R Dewantari, Rima R Prasetya, Krisnoadi Rahardjo, Aldise M Nastri, Muhammad Hamdan, Gatot Soegiarto, Laksmi Wulandari, Resti Yudhawati, Soetjipto, Yasuko Mori, Maria I Lusida, Kazufumi Shimizu |

| | | | |
|---|---|---|---|
| EPI_ISL_956280 | Institute of Tropical Disease | Institute of Tropical Disease, Universitas Airlangga | Rima R Prasetya, Krisnoadi Rahardjo, Aldise M Nastri, Jezzy R Dewantari, Gatot Soegiarto, Laksmi Wulandari, Resti Yudhawati, Soetjipto, Yasuko Mori, Maria I Lusida, Kazufumi Shimizu |
| EPI_ISL_956281 | Siti Khodijah Hospital | Institute of Tropical Disease, Universitas Airlangga | Maria I Lusida, Krisnoadi Rahardjo, Aldise M Nastri, Jezzy R Dewantari, Rima R Prasetya, Muhammad Hamdan, Gatot Soegiarto, Laksmi Wulandari, Resti Yudhawati, Soetjipto, Yasuko Mori, Kazufumi Shimizu |
| EPI_ISL_956312 | Institute of Tropical Disease | Institute of Tropical Disease, Universitas Airlangga | Krisnoadi Rahardjo, Aldise M Nastri, Jezzy R Dewantari, Rima R Prasetya, Gatot Soegiarto, Laksmi Wulandari, Resti Yudhawati, Yasuko Mori, Soetjipto, Kazufumi Shimizu, Maria I Lusida |
| EPI_ISL_956313 | Siti Khodijah Hospital | Institute of Tropical Disease, Universitas Airlangga | Jezzy R Dewantari, Rima R Prasetya, Krisnoadi Rahardjo, Aldise M Nastri, Muhammad Hamdan, Gatot Soegiarto, Laksmi Wulandari, Resti Yudhawati, Yasuko Mori, Soetjipto, Kazufumi Shimizu, Maria I Lusida |
| EPI_ISL_956315 | Siti Khodijah Hospital | Institute of Tropical Disease, Universitas Airlangga | Kazufumi Shimizu, Krisnoadi Rahardjo, Aldise M Nastri, Jezzy R Dewantari, Rima R Prasetya, Muhammad Hamdan, Gatot Soegiarto, Laksmi Wulandari, Resti Yudhawati, Yasuko Mori, Soetjipto, Maria I Lusida |
| EPI_ISL_956316 | Institute of Tropical Disease | Institute of Tropical Disease, Universitas Airlangga | Krisnoadi Rahardjo, Aldise M Nastri, Jezzy R Dewantari, Rima R Prasetya, Gatot Soegiarto, Laksmi Wulandari, Resti Yudhawati, Yasuko Mori, Soetjipto, Kazufumi Shimizu, Maria I Lusida |
| EPI_ISL_984276 | National Institute of Health Research and Development | National Institute of Health Research and Development | Holy Arif Wibowo, Ririn Ramadhany, Yuni Rukminiati, Agustiningsih, Kindi Adam, Hana Apsari Pawestri, Subangkit, Kartika Dewi Puspa, Arie Ardiansyah Nugraha, Hartanti Dian Ikawati, Krisna Nur Andriana Pangesti, Ni Ketut Susilarini, Nur Ika Hariastuti, Uly Alfi Nikmah, Mursinah, Asri Febriyani, Reni Herman, Nike Susanti, Herna, Tati Febriyanti, Natalie Laurencia Kipuw, Fauzul Muna, Irene Lorinda Indalao, Aulia Rizki, Nelly Puspandari, Vivi Setiawaty |
| EPI_ISL_984309 | National Institute of Health Research and Development | National Institute of Health Research and Development | Agustiningsih, Kindi Adam, Holy Arif Wibowo, Ririn Ramadhany, Yuni Rukminiati, Hana Apsari Pawestri, Subangkit, Kartika Dewi Puspa, Arie Ardiansyah Nugraha, Hartanti Dian Ikawati, Krisna Nur Andriana Pangesti, Ni Ketut Susilarini, Nur Ika Hariastuti, Uly Alfi Nikmah, Mursinah, Asri Febriyani, Reni Herman, Nike Susanti, Herna, Tati Febriyanti, Natalie Laurencia Kipuw, Fauzul Muna, Irene Lorinda Indalao, Aulia Rizki, Nelly Puspandari, Vivi Setiawaty |
| EPI_ISL_984313 | National Institute of Health Research and Development | National Institute of Health Research and Development | Yuni Rukminiati, Agustiningsih, Kindi Adam, Holy Arif Wibowo, Ririn Ramadhany, Hana Apsari Pawestri, Subangkit, Kartika Dewi Puspa, Arie Ardiansyah Nugraha, Hartanti Dian Ikawati, Krisna Nur Andriana Pangesti, Ni Ketut Susilarini, Nur Ika Hariastuti, Uly Alfi Nikmah, Mursinah, Asri Febriyani, Reni Herman, Nike Susanti, Herna, Tati Febriyanti, Natalie Laurencia Kipuw, Fauzul Muna, Irene Lorinda Indalao, Aulia Rizki, Nelly Puspandari, Vivi Setiawaty |
| EPI_ISL_985396 | RSUD Dr. Tjitrowardojo | Genetics Working Group (Pokja Genetik) Faculty of Medicine, Public Health and Nursing Universitas Gadjah Mada (FK-KMK UGM); Disease Investigation Center Wates Ministry of Agriculture Indonesia; Department of Microbiology FK-KMK UGM; Laboratorium Diagnostik Yayasan Tahija World Mosquito Program (WMP) Yogyakarta Center for Tropical Medicine FK-KMK UGM; Integrated Research Center FK-KMK UGM; Department of Computer Science and Electronics FMIPA UGM; Balai Besar Teknik Kesehatan Lingkungan dan Pengendalian Penyakit (BBTKLPP) Yogyakarta | Gunadi, Hendra Wibawa, Marcellus, Mohamad S. Hakim, Edwin W. Daniwijaya, Ludhang P. Rizki, Endah Supriyati, Eggi Arguni, Titik Nuryastuti, Tri Wibawa, Dwi AA Nugrahaningsih, Afiahayati, Siswanto, Kristy Iskandar, Nungki Anggorowati, Irene, Indaryati, Havid Setyawan, Arry Setyowati, Chusni Mubarakh, Try Nirmala Sari, Dwiki afandy, Dyah Ayu Puspitarani |
| EPI_ISL_985397 | RSUD Dr. Tjitrowardojo | Genetics Working Group (Pokja Genetik) Faculty of Medicine, Public Health and Nursing Universitas Gadjah Mada (FK-KMK UGM); Disease Investigation Center Wates Ministry of Agriculture Indonesia; Department of Microbiology FK-KMK UGM; Laboratorium Diagnostik Yayasan Tahija World Mosquito Program (WMP) Yogyakarta Center for Tropical Medicine FK-KMK UGM; Integrated Research Center FK-KMK UGM; Department of Computer Science and Electronics FMIPA UGM; Balai Besar Teknik Kesehatan Lingkungan dan Pengendalian Penyakit (BBTKLPP) Yogyakarta | Gunadi, Hendra Wibawa, Marcellus, Mohamad S. Hakim, Edwin W. Daniwijaya, Ludhang P. Rizki, Endah Supriyati, Eggi Arguni, Titik Nuryastuti, Tri Wibawa, Dwi AA Nugrahaningsih, Afiahayati, Siswanto, Kristy Iskandar, Nungki Anggorowati, Irene, Indaryati, Havid Setyawan, Arry Setyowati, Chusni Mubarakh, Try Nirmala Sari, Untung Riawan, William Widitjiarso |
| EPI_ISL_985398 | RSA UGM | Genetics Working Group (Pokja Genetik) Faculty of Medicine, Public Health and Nursing Universitas Gadjah Mada (FK-KMK UGM); Disease Investigation Center Wates | Gunadi, Hendra Wibawa, Marcellus, Mohamad S. Hakim, Edwin W. Daniwijaya, Ludhang P. Rizki, Endah Supriyati, Eggi Arguni, Titik Nuryastuti, Tri Wibawa, Dwi AA Nugrahaningsih, Afiahayati, Siswanto, Kristy Iskandar, Nungki Anggorowati, Susan Simanjaya, Maria Patricia Inggriani |

Ministry of Agriculture Indonesia; Department of Microbiology FK-KMK UGM; Laboratorium Diagnostik Yayasan Tahija World Mosquito Program (WMP) Yogyakarta Center for Tropical Medicine FK-KMK UGM; Integrated Research center FK-KMK UGM; Department of Computer Science and Electronics FMIPA UGM

We gratefully acknowledge the following Authors from the Originating laboratories responsible for obtaining the specimens, as well as the Submitting laboratories where the genome data were generated and shared via GISAID, on which this research is based.

All Submitters of data may be contacted directly via www.gisaid.org

Authors are sorted alphabetically.

| Accession ID | Originating Laboratory | Submitting Laboratory | Authors |
|---|---|---|---|

EPI_ISL_1073142, EPI_ISL_1073143, EPI_ISL_1073144, EPI_ISL_1073145, EPI_ISL_1073146, EPI_ISL_1073147, EPI_ISL_1073148, EPI_ISL_1073149, EPI_ISL_1073150, EPI_ISL_1073151, EPI_ISL_1073152, EPI_ISL_1073153, EPI_ISL_1073154, EPI_ISL_1073155, EPI_ISL_1073156, EPI_ISL_1073157, EPI_ISL_1073158,
EPI_ISL_1073159, EPI_ISL_1073160, EPI_ISL_1073161, EPI_ISL_1073162, EPI_ISL_1073163, EPI_ISL_1073164, EPI_ISL_1073165, EPI_ISL_1073166, EPI_ISL_1073167, EPI_ISL_1073168, EPI_ISL_1073169, EPI_ISL_1073170, EPI_ISL_1073171, EPI_ISL_1073172, EPI_ISL_1073173, EPI_ISL_1073174, EPI_ISL_1073175,
EPI_ISL_1073176, EPI_ISL_1073177, EPI_ISL_1073178, EPI_ISL_1073179, EPI_ISL_1073180, EPI_ISL_1073181, EPI_ISL_1073182, EPI_ISL_1073183, EPI_ISL_1073184, EPI_ISL_1073185, EPI_ISL_1073186, EPI_ISL_1073187, EPI_ISL_1073188, EPI_ISL_1073189, EPI_ISL_1073190, EPI_ISL_1073191, EPI_ISL_1073192,
EPI_ISL_1073193, EPI_ISL_1073194, EPI_ISL_1073195, EPI_ISL_1073196, EPI_ISL_1073197, EPI_ISL_1073198, EPI_ISL_1073199, EPI_ISL_1073200, EPI_ISL_1073201, EPI_ISL_1073202, EPI_ISL_1073203, EPI_ISL_1073204, EPI_ISL_1073205, EPI_ISL_1073206, EPI_ISL_1073207, EPI_ISL_1073208, EPI_ISL_1073209,
EPI_ISL_1073210, EPI_ISL_1073211, EPI_ISL_1073212, EPI_ISL_1073213, EPI_ISL_1073214, EPI_ISL_1073215, EPI_ISL_1073216, EPI_ISL_1073217, EPI_ISL_1073218, EPI_ISL_1073219, EPI_ISL_1073220, EPI_ISL_1073221, EPI_ISL_1073222, EPI_ISL_1073223, EPI_ISL_1073224, EPI_ISL_1073225, EPI_ISL_1073226,
EPI_ISL_1073227, EPI_ISL_1073228, EPI_ISL_1073229, EPI_ISL_1073230, EPI_ISL_1073231, EPI_ISL_1073232, EPI_ISL_1073233, EPI_ISL_1073234, EPI_ISL_1073235, EPI_ISL_1073236, EPI_ISL_1073237, EPI_ISL_1073238, EPI_ISL_1073239, EPI_ISL_1073240, EPI_ISL_1073241, EPI_ISL_1073242, EPI_ISL_1073243,
EPI_ISL_1073244, EPI_ISL_1073245, EPI_ISL_1073246, EPI_ISL_1073247, EPI_ISL_1073248, EPI_ISL_1073249, EPI_ISL_1073250, EPI_ISL_1073251, EPI_ISL_1073252, EPI_ISL_1073253, EPI_ISL_1073254, EPI_ISL_1073255, EPI_ISL_1073256, EPI_ISL_1073257, EPI_ISL_1073258, EPI_ISL_1073259, EPI_ISL_1073260,
EPI_ISL_1073261, EPI_ISL_1073262, EPI_ISL_1073263, EPI_ISL_1073264, EPI_ISL_1073265, EPI_ISL_1073266, EPI_ISL_1073267, EPI_ISL_1073268, EPI_ISL_1073269, EPI_ISL_1073270, EPI_ISL_1073271, EPI_ISL_1073272, EPI_ISL_1073273, EPI_ISL_1073274, EPI_ISL_1073275, EPI_ISL_1073276, EPI_ISL_1073277,
EPI_ISL_1073278, EPI_ISL_1073279, EPI_ISL_1073280, EPI_ISL_1073281, EPI_ISL_1073282, EPI_ISL_1073283, EPI_ISL_1073284, EPI_ISL_1073285, EPI_ISL_1073286, EPI_ISL_1073287, EPI_ISL_1073288, EPI_ISL_1073289, EPI_ISL_1073290, EPI_ISL_1073291, EPI_ISL_1073292, EPI_ISL_1073293, EPI_ISL_1073294,
EPI_ISL_1073295, EPI_ISL_1073296, EPI_ISL_1073297, EPI_ISL_1073298, EPI_ISL_1073299, EPI_ISL_1073300, EPI_ISL_1073301, EPI_ISL_1073302, EPI_ISL_1073303, EPI_ISL_1073304, EPI_ISL_1073305, EPI_ISL_1073306, EPI_ISL_1073307, EPI_ISL_1073308, EPI_ISL_1073309, EPI_ISL_1073310, EPI_ISL_1073311,
EPI_ISL_1073312, EPI_ISL_1073313, EPI_ISL_1073314, EPI_ISL_1073315, EPI_ISL_1073316, EPI_ISL_1073317, EPI_ISL_1073318, EPI_ISL_1073319, EPI_ISL_1073320, EPI_ISL_1073321, EPI_ISL_1073322, EPI_ISL_1073323, EPI_ISL_1073324, EPI_ISL_1073325, EPI_ISL_1073326, EPI_ISL_1073327, EPI_ISL_1073328,
EPI_ISL_1073329, EPI_ISL_1073330, EPI_ISL_1073331, EPI_ISL_1073332, EPI_ISL_1073333, EPI_ISL_1073334, EPI_ISL_1073335, EPI_ISL_1073336, EPI_ISL_1073337, EPI_ISL_1073338, EPI_ISL_1073339, EPI_ISL_1073340, EPI_ISL_1073341, EPI_ISL_1073342, EPI_ISL_1073343, EPI_ISL_1073344, EPI_ISL_1073345,
EPI_ISL_1073346, EPI_ISL_1073347, EPI_ISL_1073348, EPI_ISL_1073349, EPI_ISL_1073350, EPI_ISL_1073351, EPI_ISL_1073352, EPI_ISL_1073353, EPI_ISL_1073354, EPI_ISL_1073355, EPI_ISL_1073356, EPI_ISL_1073357, EPI_ISL_1073358, EPI_ISL_1073359, EPI_ISL_1073360, EPI_ISL_1073361, EPI_ISL_1073362,
EPI_ISL_1073363, EPI_ISL_1073364, EPI_ISL_1073365, EPI_ISL_1073366, EPI_ISL_1073367, EPI_ISL_1073368, EPI_ISL_1073369, EPI_ISL_1073370, EPI_ISL_1073371, EPI_ISL_1073372, EPI_ISL_1073373, EPI_ISL_1073374, EPI_ISL_1073375, EPI_ISL_1073376, EPI_ISL_1073377, EPI_ISL_1073378, EPI_ISL_1073379,
EPI_ISL_1073380, EPI_ISL_1073381, EPI_ISL_1073382, EPI_ISL_1073383, EPI_ISL_1073384, EPI_ISL_1073385, EPI_ISL_1073387, EPI_ISL_1073388, EPI_ISL_1073389, EPI_ISL_1073390, EPI_ISL_1073391, EPI_ISL_1073392, EPI_ISL_1073393, EPI_ISL_1073394, EPI_ISL_1073395, EPI_ISL_1073396, EPI_ISL_1073397,
EPI_ISL_1073398, EPI_ISL_1073399, EPI_ISL_1073400, EPI_ISL_1073401, EPI_ISL_1073402, EPI_ISL_1073403, EPI_ISL_1073404, EPI_ISL_1073405, EPI_ISL_1073406, EPI_ISL_1073407, EPI_ISL_1073408, EPI_ISL_1073409, EPI_ISL_1073410, EPI_ISL_1073411, EPI_ISL_1073412, EPI_ISL_1073413, EPI_ISL_1073414,
EPI_ISL_1073415, EPI_ISL_1073416, EPI_ISL_1073417, EPI_ISL_1073418, EPI_ISL_1073419, EPI_ISL_1073420, EPI_ISL_1073421, EPI_ISL_1073422, EPI_ISL_1073423, EPI_ISL_1073424, EPI_ISL_1073425, EPI_ISL_1073426, EPI_ISL_1073427, EPI_ISL_1073428, EPI_ISL_1073429, EPI_ISL_1073430, EPI_ISL_1073431,
EPI_ISL_1073432, EPI_ISL_1073433, EPI_ISL_1073434, EPI_ISL_1073435, EPI_ISL_1073436, EPI_ISL_1073437, EPI_ISL_1073438, EPI_ISL_1073439, EPI_ISL_1073440, EPI_ISL_1073441, EPI_ISL_1073442, EPI_ISL_1073443, EPI_ISL_1073444, EPI_ISL_1073445, EPI_ISL_1073446, EPI_ISL_1073447, EPI_ISL_1073448,
EPI_ISL_1073449, EPI_ISL_1073450, EPI_ISL_1073451, EPI_ISL_1073452, EPI_ISL_1073453, EPI_ISL_1073454, EPI_ISL_1073455, EPI_ISL_1073456, EPI_ISL_1073457, EPI_ISL_1073458, EPI_ISL_1073459, EPI_ISL_1073460, EPI_ISL_1073461, EPI_ISL_1073462, EPI_ISL_1073463, EPI_ISL_1073464, EPI_ISL_1073465,
EPI_ISL_1073466, EPI_ISL_1073467, EPI_ISL_1073468, EPI_ISL_1073469, EPI_ISL_1073470, EPI_ISL_1073471, EPI_ISL_1073472, EPI_ISL_1073473, EPI_ISL_1073474, EPI_ISL_1073475, EPI_ISL_1073476, EPI_ISL_1073477, EPI_ISL_1073478, EPI_ISL_1073479, EPI_ISL_1073480, EPI_ISL_1073481, EPI_ISL_1073482,
EPI_ISL_1073483, EPI_ISL_1073484, EPI_ISL_1073485, EPI_ISL_1073486, EPI_ISL_1073487, EPI_ISL_1073488, EPI_ISL_1073489, EPI_ISL_1073490, EPI_ISL_1073491, EPI_ISL_1073492, EPI_ISL_1073493, EPI_ISL_1073494, EPI_ISL_1073495, EPI_ISL_1073496, EPI_ISL_1073497, EPI_ISL_1073498, EPI_ISL_1073499,
EPI_ISL_1073500, EPI_ISL_1073501, EPI_ISL_1073502, EPI_ISL_1073503, EPI_ISL_1073504, EPI_ISL_1073505, EPI_ISL_1073506, EPI_ISL_1073507, EPI_ISL_1073508, EPI_ISL_1073509, EPI_ISL_1073510, EPI_ISL_1073511, EPI_ISL_1073512, EPI_ISL_1073513, EPI_ISL_1073514, EPI_ISL_1073515, EPI_ISL_1073516,
EPI_ISL_1073517, EPI_ISL_1073518, EPI_ISL_1073519, EPI_ISL_1073520, EPI_ISL_1073521, EPI_ISL_1073522, EPI_ISL_1073523, EPI_ISL_1073524, EPI_ISL_1073525, EPI_ISL_1073526, EPI_ISL_1073527, EPI_ISL_1073528, EPI_ISL_1073529, EPI_ISL_1073530, EPI_ISL_1073531, EPI_ISL_1073532, EPI_ISL_1073533,
EPI_ISL_1073534, EPI_ISL_1073535, EPI_ISL_1073536, EPI_ISL_1073537, EPI_ISL_1073538, EPI_ISL_1073539, EPI_ISL_1073540, EPI_ISL_1073541, EPI_ISL_1073542, EPI_ISL_1073543, EPI_ISL_1073544, EPI_ISL_1073545, EPI_ISL_1073546, EPI_ISL_1073548, EPI_ISL_1073551, EPI_ISL_1073552, EPI_ISL_1073553,
EPI_ISL_1073555, EPI_ISL_1073556, EPI_ISL_1073559, EPI_ISL_1073560, EPI_ISL_1073561, EPI_ISL_1073562, EPI_ISL_1073563, EPI_ISL_1073564, EPI_ISL_1073565, EPI_ISL_1073566, EPI_ISL_1073567, EPI_ISL_1073568, EPI_ISL_1073569, EPI_ISL_1073570, EPI_ISL_1073571, EPI_ISL_1073572, EPI_ISL_1073573,
EPI_ISL_1073574, EPI_ISL_1073575, EPI_ISL_1073576, EPI_ISL_1073577, EPI_ISL_1073578, EPI_ISL_1073579, EPI_ISL_1073580, EPI_ISL_1073581, EPI_ISL_1073582, EPI_ISL_1073583, EPI_ISL_1073584, EPI_ISL_1073585, EPI_ISL_1073586, EPI_ISL_1073587, EPI_ISL_1073588, EPI_ISL_1073589, EPI_ISL_1073590,
EPI_ISL_1073591, EPI_ISL_1073592, EPI_ISL_1073593, EPI_ISL_1073594, EPI_ISL_1073595, EPI_ISL_1073596, EPI_ISL_1073597, EPI_ISL_1073598, EPI_ISL_1073599, EPI_ISL_1073600, EPI_ISL_1073601, EPI_ISL_1073602, EPI_ISL_1073603, EPI_ISL_1073604, EPI_ISL_1073605, EPI_ISL_1073606, EPI_ISL_1073607,
EPI_ISL_1073608, EPI_ISL_1073609, EPI_ISL_1073610, EPI_ISL_1073611, EPI_ISL_1073612, EPI_ISL_1073613, EPI_ISL_1073614, EPI_ISL_1073615, EPI_ISL_1073616, EPI_ISL_1073617, EPI_ISL_1073618, EPI_ISL_1073619, EPI_ISL_1073620, EPI_ISL_1073621, EPI_ISL_1073622, EPI_ISL_1073623, EPI_ISL_1073624,
EPI_ISL_1073625, EPI_ISL_1073626, EPI_ISL_1073627, EPI_ISL_1073628, EPI_ISL_1073629, EPI_ISL_1073630, EPI_ISL_1073631, EPI_ISL_1073632, EPI_ISL_1073633, EPI_ISL_1073634, EPI_ISL_1073635, EPI_ISL_1073636, EPI_ISL_1073637, EPI_ISL_1209469, EPI_ISL_1209470, EPI_ISL_1209471, EPI_ISL_1209472,
EPI_ISL_1209473, EPI_ISL_1209474, EPI_ISL_1209475, EPI_ISL_1209476, EPI_ISL_1209477, EPI_ISL_1209478, EPI_ISL_1209479, EPI_ISL_1209480, EPI_ISL_1209481, EPI_ISL_1209482, EPI_ISL_1209483, EPI_ISL_1209484, EPI_ISL_1209485, EPI_ISL_1209486, EPI_ISL_1209487, EPI_ISL_1209488, EPI_ISL_1209489,
EPI_ISL_1209490, EPI_ISL_1209491, EPI_ISL_1209492, EPI_ISL_1209493, EPI_ISL_1209494, EPI_ISL_1209495, EPI_ISL_1209496, EPI_ISL_1209497, EPI_ISL_1209498, EPI_ISL_1209499, EPI_ISL_1209500, EPI_ISL_1209501, EPI_ISL_1209502, EPI_ISL_1209503, EPI_ISL_1209504, EPI_ISL_1209505, EPI_ISL_1209506,
EPI_ISL_1209507, EPI_ISL_1209508, EPI_ISL_1209509, EPI_ISL_1209510, EPI_ISL_1209511, EPI_ISL_1209512, EPI_ISL_1209513, EPI_ISL_1209514, EPI_ISL_1209515, EPI_ISL_1209516, EPI_ISL_1209517, EPI_ISL_1209518, EPI_ISL_1209519, EPI_ISL_1209520, EPI_ISL_1209521, EPI_ISL_1209522, EPI_ISL_1209523,
EPI_ISL_1209524, EPI_ISL_1209525, EPI_ISL_1209526, EPI_ISL_1209527, EPI_ISL_1209528, EPI_ISL_1209529, EPI_ISL_1209530, EPI_ISL_1209531, EPI_ISL_1209532, EPI_ISL_1209533, EPI_ISL_1209534, EPI_ISL_1209535, EPI_ISL_1209536, EPI_ISL_1209537, EPI_ISL_1209538, EPI_ISL_1209539, EPI_ISL_1209540,
EPI_ISL_1209541, EPI_ISL_1209542, EPI_ISL_1209543, EPI_ISL_1209544, EPI_ISL_1209545, EPI_ISL_1209546, EPI_ISL_1209547, EPI_ISL_1209548, EPI_ISL_1209549, EPI_ISL_1209550, EPI_ISL_1209551, EPI_ISL_1209552, EPI_ISL_1209553, EPI_ISL_1209554, EPI_ISL_1209555, EPI_ISL_1209556, EPI_ISL_1209557,
EPI_ISL_1209558, EPI_ISL_1209559, EPI_ISL_1209560, EPI_ISL_1209561, EPI_ISL_1209562, EPI_ISL_1209563, EPI_ISL_1209564, EPI_ISL_1209565, EPI_ISL_1209566, EPI_ISL_1209567, EPI_ISL_1209568, EPI_ISL_1209569, EPI_ISL_1209570, EPI_ISL_1209571, EPI_ISL_1209572, EPI_ISL_1209573, EPI_ISL_1209574,
EPI_ISL_1209575, EPI_ISL_1209576, EPI_ISL_1209577, EPI_ISL_1209578, EPI_ISL_1209579, EPI_ISL_1209580, EPI_ISL_1209581, EPI_ISL_1209582, EPI_ISL_1209583, EPI_ISL_1209584, EPI_ISL_1209585, EPI_ISL_1209586, EPI_ISL_1209587, EPI_ISL_1209588, EPI_ISL_1209589, EPI_ISL_1209590, EPI_ISL_1209591,
EPI_ISL_1209592, EPI_ISL_1209593, EPI_ISL_1209594, EPI_ISL_1209595, EPI_ISL_1209596, EPI_ISL_1209597, EPI_ISL_1209598, EPI_ISL_1209599, EPI_ISL_1209600, EPI_ISL_1209601, EPI_ISL_1209602, EPI_ISL_1209603, EPI_ISL_1209604, EPI_ISL_1209605, EPI_ISL_1209606, EPI_ISL_1209607, EPI_ISL_1209608,
EPI_ISL_1209609, EPI_ISL_1209610, EPI_ISL_1209611, EPI_ISL_1209612, EPI_ISL_1209613, EPI_ISL_1209614, EPI_ISL_1209615, EPI_ISL_1209616, EPI_ISL_1209617, EPI_ISL_1209618, EPI_ISL_1209619, EPI_ISL_1209620, EPI_ISL_1209621, EPI_ISL_1209622, EPI_ISL_1209623, EPI_ISL_1209624, EPI_ISL_1209625,
EPI_ISL_1209626, EPI_ISL_1209627, EPI_ISL_1209628, EPI_ISL_1209629, EPI_ISL_1209630, EPI_ISL_1209631, EPI_ISL_1209632, EPI_ISL_1209633, EPI_ISL_1209634, EPI_ISL_1209635, EPI_ISL_1209636, EPI_ISL_1209637, EPI_ISL_1209638, EPI_ISL_1209639, EPI_ISL_1209640, EPI_ISL_1209641, EPI_ISL_1209642,
EPI_ISL_1209643, EPI_ISL_1209644, EPI_ISL_1209645, EPI_ISL_1209646, EPI_ISL_1209647, EPI_ISL_1209648, EPI_ISL_1209649, EPI_ISL_1209650, EPI_ISL_1209651, EPI_ISL_1209652, EPI_ISL_1209653, EPI_ISL_1209654, EPI_ISL_1209655, EPI_ISL_1209656, EPI_ISL_1209657, EPI_ISL_1209658, EPI_ISL_1209659,
EPI_ISL_1209660, EPI_ISL_1209661, EPI_ISL_1209662, EPI_ISL_1209663, EPI_ISL_1209664, EPI_ISL_1209665, EPI_ISL_1209666, EPI_ISL_1209667, EPI_ISL_1209668, EPI_ISL_1209669, EPI_ISL_1209670, EPI_ISL_1209671, EPI_ISL_1209672, EPI_ISL_1209673, EPI_ISL_1209674, EPI_ISL_1209675, EPI_ISL_1209676,
EPI_ISL_1209677, EPI_ISL_1209678, EPI_ISL_1209679, EPI_ISL_1209680, EPI_ISL_1209681, EPI_ISL_1209682, EPI_ISL_1209683, EPI_ISL_1209684, EPI_ISL_1209685, EPI_ISL_1209686, EPI_ISL_1209687, EPI_ISL_1209688, EPI_ISL_1209689, EPI_ISL_1209690, EPI_ISL_1209691, EPI_ISL_1209692, EPI_ISL_1209693,
EPI_ISL_1209694, EPI_ISL_1209695, EPI_ISL_1209696, EPI_ISL_1209697, EPI_ISL_1209698, EPI_ISL_1209699, EPI_ISL_1209700, EPI_ISL_1209701, EPI_ISL_1209702, EPI_ISL_1209703, EPI_ISL_1209704, EPI_ISL_1209705, EPI_ISL_1209706, EPI_ISL_1209707, EPI_ISL_1209708, EPI_ISL_1209709, EPI_ISL_1209710,
EPI_ISL_1209711, EPI_ISL_1209712, EPI_ISL_1209713, EPI_ISL_1209714, EPI_ISL_1209715, EPI_ISL_1209716, EPI_ISL_1209717, EPI_ISL_1209718, EPI_ISL_1209719, EPI_ISL_1209720, EPI_ISL_1209721, EPI_ISL_1209722, EPI_ISL_1209723, EPI_ISL_1209724, EPI_ISL_1209725, EPI_ISL_1209726, EPI_ISL_1209727,
EPI_ISL_1209728, EPI_ISL_1209729, EPI_ISL_1209730, EPI_ISL_1209731, EPI_ISL_1209732, EPI_ISL_1209733, EPI_ISL_1209734, EPI_ISL_1209735, EPI_ISL_1209736, EPI_ISL_1209737, EPI_ISL_1209738, EPI_ISL_1209739, EPI_ISL_1209740, EPI_ISL_1209741, EPI_ISL_1209742, EPI_ISL_1209743, EPI_ISL_1209744,
EPI_ISL_1209745, EPI_ISL_1209746, EPI_ISL_1209747, EPI_ISL_1209748, EPI_ISL_1209749, EPI_ISL_1209750, EPI_ISL_1209751, EPI_ISL_1209752, EPI_ISL_1209753, EPI_ISL_1209754, EPI_ISL_1209755, EPI_ISL_1209756, EPI_ISL_1209757, EPI_ISL_1209758, EPI_ISL_1209759, EPI_ISL_1209760, EPI_ISL_1209761,
EPI_ISL_1209762, EPI_ISL_1209763, EPI_ISL_1209764, EPI_ISL_1209765, EPI_ISL_1209766, EPI_ISL_1209767, EPI_ISL_1209768, EPI_ISL_1209769, EPI_ISL_1209770, EPI_ISL_1209771, EPI_ISL_1209772, EPI_ISL_1209773, EPI_ISL_1209774, EPI_ISL_1209775, EPI_ISL_1209776, EPI_ISL_1209777, EPI_ISL_1209778,
EPI_ISL_1209779, EPI_ISL_1209780, EPI_ISL_1209781, EPI_ISL_1209782, EPI_ISL_1209783, EPI_ISL_1209784, EPI_ISL_1209785, EPI_ISL_1209786, EPI_ISL_1209787, EPI_ISL_1209788, EPI_ISL_1209789, EPI_ISL_1209790, EPI_ISL_1209791, EPI_ISL_1209792, EPI_ISL_1209793, EPI_ISL_1209794, EPI_ISL_1209795,
EPI_ISL_1209796, EPI_ISL_1209797, EPI_ISL_1209798, EPI_ISL_1209799, EPI_ISL_1209800, EPI_ISL_1209801, EPI_ISL_1209802, EPI_ISL_1209803, EPI_ISL_1209804, EPI_ISL_1209805, EPI_ISL_1209806, EPI_ISL_1209807, EPI_ISL_1209808, EPI_ISL_1209809, EPI_ISL_1209810, EPI_ISL_1209811, EPI_ISL_1209812,
EPI_ISL_1209813, EPI_ISL_1209814, EPI_ISL_1209815, EPI_ISL_1209816, EPI_ISL_1209817, EPI_ISL_1209818, EPI_ISL_1209819, EPI_ISL_1209820, EPI_ISL_1209821, EPI_ISL_1209822, EPI_ISL_1209823, EPI_ISL_1209824, EPI_ISL_1209825, EPI_ISL_1209826, EPI_ISL_1209827, EPI_ISL_1209828, EPI_ISL_1209829,
EPI_ISL_1209830, EPI_ISL_1209831, EPI_ISL_1209832, EPI_ISL_1209833, EPI_ISL_1209834, EPI_ISL_1209835, EPI_ISL_1209836, EPI_ISL_1209837, EPI_ISL_1209838, EPI_ISL_1209839, EPI_ISL_1209840, EPI_ISL_1209841, EPI_ISL_1209842, EPI_ISL_1209843, EPI_ISL_1209844, EPI_ISL_1209845, EPI_ISL_1209846,
EPI_ISL_1209847, EPI_ISL_1209848, EPI_ISL_1209849, EPI_ISL_1209850, EPI_ISL_1209851, EPI_ISL_1209852, EPI_ISL_1209853, EPI_ISL_1209854, EPI_ISL_1209855, EPI_ISL_1209856, EPI_ISL_1209857, EPI_ISL_1209858, EPI_ISL_1209859, EPI_ISL_1209860, EPI_ISL_1209861, EPI_ISL_1209862, EPI_ISL_1209863,
EPI_ISL_1209864, EPI_ISL_1209865, EPI_ISL_1209866, EPI_ISL_1209867, EPI_ISL_1209868, EPI_ISL_1209869, EPI_ISL_1209870, EPI_ISL_1209871, EPI_ISL_1209872, EPI_ISL_1209873, EPI_ISL_1209874, EPI_ISL_1209875, EPI_ISL_1209876, EPI_ISL_1209877, EPI_ISL_1209878, EPI_ISL_1209879, EPI_ISL_1209880,
EPI_ISL_1209881, EPI_ISL_1209882, EPI_ISL_1209883, EPI_ISL_1209884, EPI_ISL_1209885, EPI_ISL_1209886, EPI_ISL_1209887, EPI_ISL_1209888, EPI_ISL_1209889, EPI_ISL_1209890, EPI_ISL_1209891, EPI_ISL_1209892, EPI_ISL_1209893, EPI_ISL_1209894, EPI_ISL_1209895, EPI_ISL_1209896, EPI_ISL_1209897,
EPI_ISL_1209898, EPI_ISL_1209899, EPI_ISL_1209900, EPI_ISL_1209901, EPI_ISL_1209902, EPI_ISL_1209903, EPI_ISL_1209904, EPI_ISL_1209905, EPI_ISL_1209906, EPI_ISL_1209907, EPI_ISL_1209908, EPI_ISL_1209909, EPI_ISL_1209910, EPI_ISL_1209911, EPI_ISL_1209912, EPI_ISL_1209913, EPI_ISL_1209914,
EPI_ISL_1209915, EPI_ISL_1209916, EPI_ISL_1209917, EPI_ISL_1209918, EPI_ISL_1209919, EPI_ISL_1209920, EPI_ISL_1209921, EPI_ISL_1209922, EPI_ISL_1209923, EPI_ISL_1209924, EPI_ISL_1209925, EPI_ISL_1209926, EPI_ISL_1209927, EPI_ISL_1209928, EPI_ISL_1209929, EPI_ISL_1209930, EPI_ISL_1209931,
EPI_ISL_1209932, EPI_ISL_1209933, EPI_ISL_1209934, EPI_ISL_1209935, EPI_ISL_1209936, EPI_ISL_1209937, EPI_ISL_1209938, EPI_ISL_1209939, EPI_ISL_1209940, EPI_ISL_1209941, EPI_ISL_1209942, EPI_ISL_1209943, EPI_ISL_1209944, EPI_ISL_1209945, EPI_ISL_1209946, EPI_ISL_1209947, EPI_ISL_1209948,
EPI_ISL_1209949, EPI_ISL_1209950, EPI_ISL_1209951, EPI_ISL_1209952, EPI_ISL_1209953, EPI_ISL_1209954, EPI_ISL_1209955, EPI_ISL_1209956, EPI_ISL_1209957, EPI_ISL_1209958, EPI_ISL_1209959, EPI_ISL_1209960, EPI_ISL_1209961, EPI_ISL_1209962, EPI_ISL_1209963, EPI_ISL_1209964, EPI_ISL_1209965,
EPI_ISL_1209966, EPI_ISL_1209967, EPI_ISL_1209968, EPI_ISL_1209969, EPI_ISL_1209970, EPI_ISL_1209971, EPI_ISL_1209972, EPI_ISL_1209973, EPI_ISL_1209974, EPI_ISL_1209975, EPI_ISL_1209976, EPI_ISL_1209977, EPI_ISL_1209978, EPI_ISL_1209979, EPI_ISL_1209980, EPI_ISL_1209981, EPI_ISL_1209982,
EPI_ISL_1209983, EPI_ISL_1209984, EPI_ISL_1209985, EPI_ISL_1209986, EPI_ISL_1209987, EPI_ISL_1209988, EPI_ISL_1209989, EPI_ISL_1209990, EPI_ISL_1209991, EPI_ISL_1209992, EPI_ISL_1209993, EPI_ISL_1209994, EPI_ISL_1209995, EPI_ISL_1209996, EPI_ISL_1209997, EPI_ISL_1209998, EPI_ISL_1209999,
EPI_ISL_1210000, EPI_ISL_1210001, EPI_ISL_1210002, EPI_ISL_1210003, EPI_ISL_1210004, EPI_ISL_1210005, EPI_ISL_1210006, EPI_ISL_1210007, EPI_ISL_1210008, EPI_ISL_1210009, EPI_ISL_1210010, EPI_ISL_1210011, EPI_ISL_1210012, EPI_ISL_1210013, EPI_ISL_1210014, EPI_ISL_1210015, EPI_ISL_1210016,
EPI_ISL_1210017, EPI_ISL_1210018, EPI_ISL_1210019, EPI_ISL_1210020, EPI_ISL_1210021, EPI_ISL_1210022, EPI_ISL_1210023, EPI_ISL_1210024, EPI_ISL_1210025, EPI_ISL_1210026, EPI_ISL_1210027, EPI_ISL_1210028, EPI_ISL_1210029, EPI_ISL_1210030, EPI_ISL_1210031, EPI_ISL_1210032, EPI_ISL_1210033,
EPI_ISL_1210034, EPI_ISL_1210035, EPI_ISL_1210036, EPI_ISL_1210037, EPI_ISL_1210038, EPI_ISL_1210039, EPI_ISL_1210040, EPI_ISL_1210041, EPI_ISL_1210042, EPI_ISL_1210043, EPI_ISL_1210044, EPI_ISL_1210045, EPI_ISL_1210046, EPI_ISL_1210047, EPI_ISL_1210048, EPI_ISL_1210049, EPI_ISL_1210050,
EPI_ISL_1210051, EPI_ISL_1210052, EPI_ISL_1210053, EPI_ISL_1210054, EPI_ISL_1210055, EPI_ISL_1210056, EPI_ISL_1210057, EPI_ISL_1210058, EPI_ISL_1210059, EPI_ISL_1210060, EPI_ISL_1210061, EPI_ISL_1210062, EPI_ISL_1210063, EPI_ISL_1210064, EPI_ISL_1210065, EPI_ISL_1210066, EPI_ISL_1210067,
EPI_ISL_1210068, EPI_ISL_1210069, EPI_ISL_1210070, EPI_ISL_1210071, EPI_ISL_1210072, EPI_ISL_1210073, EPI_ISL_1210074, EPI_ISL_1210075, EPI_ISL_1210076, EPI_ISL_1210077, EPI_ISL_1210078, EPI_ISL_1210079, EPI_ISL_1210080, EPI_ISL_1210081, EPI_ISL_1210082, EPI_ISL_1210083, EPI_ISL_1210084,
EPI_ISL_1210085, EPI_ISL_1210086, EPI_ISL_1210087, EPI_ISL_1210088, EPI_ISL_1210089, EPI_ISL_1210090, EPI_ISL_1210091, EPI_ISL_1210092, EPI_ISL_1210093, EPI_ISL_1210094, EPI_ISL_1210095, EPI_ISL_1210096, EPI_ISL_1210097, EPI_ISL_1210098, EPI_ISL_1210099, EPI_ISL_1210100, EPI_ISL_1210101,
EPI_ISL_1210102, EPI_ISL_1210103, EPI_ISL_1210104, EPI_ISL_1210105, EPI_ISL_1210106, EPI_ISL_1210107, EPI_ISL_1210108, EPI_ISL_1210109, EPI_ISL_1210110, EPI_ISL_1210111, EPI_ISL_1210112, EPI_ISL_1210113, EPI_ISL_1210114, EPI_ISL_1210115, EPI_ISL_1210116, EPI_ISL_1210117, EPI_ISL_1210118,
EPI_ISL_1210119, EPI_ISL_1210120, EPI_ISL_1210121, EPI_ISL_1210122, EPI_ISL_1210123, EPI_ISL_1210124, EPI_ISL_1210125, EPI_ISL_1210126, EPI_ISL_1210127, EPI_ISL_1210128, EPI_ISL_1210129, EPI_ISL_1210130, EPI_ISL_1210131, EPI_ISL_1210132, EPI_ISL_1210133, EPI_ISL_1210134, EPI_ISL_1210135,
EPI_ISL_1210136, EPI_ISL_1210137, EPI_ISL_1210138, EPI_ISL_1210139, EPI_ISL_1210140, EPI_ISL_1210141, EPI_ISL_1210142, EPI_ISL_1210143, EPI_ISL_1210144, EPI_ISL_1210145, EPI_ISL_1210146, EPI_ISL_1210147, EPI_ISL_1210148, EPI_ISL_1210149, EPI_ISL_1210150, EPI_ISL_1210151, EPI_ISL_1210152,
EPI_ISL_1210153, EPI_ISL_1210154, EPI_ISL_1210155, EPI_ISL_1210156, EPI_ISL_1210157, EPI_ISL_1210158, EPI_ISL_1210159, EPI_ISL_1210160, EPI_ISL_1210161, EPI_ISL_1210162, EPI_ISL_1210163, EPI_ISL_1210164, EPI_ISL_1210165, EPI_ISL_1210166, EPI_ISL_1210167, EPI_ISL_1210168, EPI_ISL_1210169,
EPI_ISL_1210170, EPI_ISL_1210171, EPI_ISL_1210172, EPI_ISL_1210173, EPI_ISL_1210174, EPI_ISL_1210175, EPI_ISL_1210176, EPI_ISL_1210177, EPI_ISL_1210178, EPI_ISL_1210179, EPI_ISL_1210180, EPI_ISL_1210181, EPI_ISL_1210182, EPI_ISL_1210183, EPI_ISL_1210184, EPI_ISL_1210185, EPI_ISL_1210186,
EPI_ISL_1210187, EPI_ISL_1210188, EPI_ISL_1210189, EPI_ISL_1210190, EPI_ISL_1210191, EPI_ISL_1210192, EPI_ISL_1210193, EPI_ISL_1210194, EPI_ISL_1210195, EPI_ISL_1210196, EPI_ISL_1210197, EPI_ISL_1210198, EPI_ISL_1210199, EPI_ISL_1210200, EPI_ISL_1210201, EPI_ISL_1210202, EPI_ISL_1210203,
EPI_ISL_1210204, EPI_ISL_1210205, EPI_ISL_1210206, EPI_ISL_1210207, EPI_ISL_1210208, EPI_ISL_1210209, EPI_ISL_1210210, EPI_ISL_1210211, EPI_ISL_1210212, EPI_ISL_1210213, EPI_ISL_1210214, EPI_ISL_1210215, EPI_ISL_1210216, EPI_ISL_1210218, EPI_ISL_1210219, EPI_ISL_1210220, EPI_ISL_1210221,
EPI_ISL_1210222, EPI_ISL_1210223, EPI_ISL_1210224, EPI_ISL_1210225, EPI_ISL_1210226, EPI_ISL_1210227, EPI_ISL_1210228, EPI_ISL_1210229, EPI_ISL_1210230, EPI_ISL_1210231, EPI_ISL_1210232, EPI_ISL_1210233, EPI_ISL_1210234, EPI_ISL_1210235, EPI_ISL_1210236, EPI_ISL_1210237, EPI_ISL_1210238,
EPI_ISL_1210239, EPI_ISL_1210240, EPI_ISL_1210241, EPI_ISL_1210242, EPI_ISL_1210243, EPI_ISL_1210244, EPI_ISL_1210245, EPI_ISL_1210246, EPI_ISL_1210247, EPI_ISL_1210248, EPI_ISL_1210249, EPI_ISL_1210250, EPI_ISL_1210251, EPI_ISL_1210252, EPI_ISL_1210253, EPI_ISL_1210254, EPI_ISL_1210255,

EPI_ISL_1210256, EPI_ISL_1210257, EPI_ISL_1210258, EPI_ISL_1210259, EPI_ISL_1210260, EPI_ISL_1210261, EPI_ISL_1210262, EPI_ISL_1210263, EPI_ISL_1210264, EPI_ISL_1210265, EPI_ISL_1210266, EPI_ISL_1210267, EPI_ISL_1210268, EPI_ISL_1210269, EPI_ISL_1210270, EPI_ISL_1210271, EPI_ISL_1210272, EPI_ISL_1210273, EPI_ISL_1210274, EPI_ISL_1210275, EPI_ISL_1210276, EPI_ISL_1210277, EPI_ISL_1210278, EPI_ISL_1210279, EPI_ISL_1210280, EPI_ISL_1210281, EPI_ISL_1210282, EPI_ISL_1210283, EPI_ISL_1210284, EPI_ISL_1210285, EPI_ISL_1210286, EPI_ISL_1210287, EPI_ISL_1210288, EPI_ISL_1210289, EPI_ISL_1210290, EPI_ISL_1210291, EPI_ISL_1210292, EPI_ISL_1210293, EPI_ISL_1210294, EPI_ISL_1210295, EPI_ISL_1210296, EPI_ISL_1210297, EPI_ISL_1210299, EPI_ISL_1210300, EPI_ISL_1210301, EPI_ISL_1210302, EPI_ISL_1210303, EPI_ISL_1210304, EPI_ISL_1210305, EPI_ISL_1210306, EPI_ISL_1210307, EPI_ISL_1210308, EPI_ISL_1210309, EPI_ISL_1210310, EPI_ISL_1210311, EPI_ISL_1210312, EPI_ISL_1210313, EPI_ISL_1210314, EPI_ISL_1210315, EPI_ISL_1210316, EPI_ISL_1210317, EPI_ISL_1210318, EPI_ISL_1210319, EPI_ISL_1210320, EPI_ISL_1210321, EPI_ISL_1210322, EPI_ISL_1210323, EPI_ISL_1210324, EPI_ISL_1210325, EPI_ISL_1210326, EPI_ISL_1210327, EPI_ISL_1210328, EPI_ISL_1210329, EPI_ISL_1210330, EPI_ISL_1210331, EPI_ISL_1210332, EPI_ISL_1210333, EPI_ISL_1210334, EPI_ISL_1210335, EPI_ISL_1210336, EPI_ISL_1210337, EPI_ISL_1210338, EPI_ISL_1210339, EPI_ISL_1210340, EPI_ISL_1210341, EPI_ISL_1210342, EPI_ISL_1210343, EPI_ISL_1210344, EPI_ISL_1210345, EPI_ISL_1210346, EPI_ISL_1210347, EPI_ISL_1210348, EPI_ISL_1210349, EPI_ISL_1210350, EPI_ISL_1240652, EPI_ISL_1240653, EPI_ISL_1240654, EPI_ISL_1240656, EPI_ISL_1240657, EPI_ISL_1240678, EPI_ISL_1240679

| | | | |
|---|---|---|---|
| see above | Israel Central Virology laboratory | Israel National Consortium for SARS-CoV-2 sequencing | Neta Zuckerman, Efrat Dahan Bucris, Michal Mandelboim, Dana Bar-Ilan, Oran Erster, Tzvia Mann, Omer Murik, David A. Zeevi, Assaf Rokney, Joseph Jaffe, Eva Nachum, Maya Davidovich Cohen, Ephraim Fass, Gal Zizelski Valenci, Mor Rubinstein, Efrat Rorman, Israel Nissan, Efrat Glick-Saar, Omri Nayshool, Gideon Rechavi, Ella Mendelson, Orna Mor |
| EPI_ISL_1259181 | Israel Central Virology laboratory | Israel National Consortium for SARS-CoV-2 sequencing | "Neta Zuckerman, Efrat Dahan Bucris, Michal Mandelboim, Dana Bar-Ilan, Oran Erster, Tzvia Mann, Omer Murik, David A. Zeevi, Assaf Rokney, Joseph Jaffe, Eva Nachum, Maya Davidovich Cohen, Ephraim Fass, Gal Zizelski Valenci, Mor Rubinstein, Efrat Rorman, Israel Nissan, Efrat Glick-Saar, Omri Nayshool, Gideon Rechavi, Ella Mendelson, Orna Mor" |
| EPI_ISL_1259208 | Israel Central Virology laboratory | Israel National Consortium for SARS-CoV-2 sequencing | Neta Zuckerman, Efrat Dahan Bucris, Michal Mandelboim, Dana Bar-Ilan, Oran Erster, Tzvia Mann, Omer Murik, David A. Zeevi, Assaf Rokney, Joseph Jaffe, Eva Nachum, Maya Davidovich Cohen, Ephraim Fass, Gal Zizelski Valenci, Mor Rubinstein, Efrat Rorman, Israel Nissan, Efrat Glick-Saar, Omri Nayshool, Gideon Rechavi, Ella Mendelson, Orna Mor |

EPI_ISL_1259209, EPI_ISL_1259210, EPI_ISL_1259211, EPI_ISL_1259212, EPI_ISL_1259213, EPI_ISL_1259263, EPI_ISL_1259264, EPI_ISL_1259265, EPI_ISL_1259278, EPI_ISL_1259279, EPI_ISL_1259280, EPI_ISL_1259281

| | | | |
|---|---|---|---|
| see above | Israel Central Virology laboratory | Israel National Consortium for SARS-CoV-2 sequencing | "Neta Zuckerman, Efrat Dahan Bucris, Michal Mandelboim, Dana Bar-Ilan, Oran Erster, Tzvia Mann, Omer Murik, David A. Zeevi, Assaf Rokney, Joseph Jaffe, Eva Nachum, Maya Davidovich Cohen, Ephraim Fass, Gal Zizelski Valenci, Mor Rubinstein, Efrat Rorman, Israel Nissan, Efrat Glick-Saar, Omri Nayshool, Gideon Rechavi, Ella Mendelson, Orna Mor" |

EPI_ISL_1278373, EPI_ISL_1278374, EPI_ISL_1278375, EPI_ISL_1278376, EPI_ISL_1278377, EPI_ISL_1278378, EPI_ISL_1278379, EPI_ISL_1278380, EPI_ISL_1278381, EPI_ISL_1278382, EPI_ISL_1278383, EPI_ISL_1278384, EPI_ISL_1278385, EPI_ISL_1278386, EPI_ISL_1278387, EPI_ISL_1278388, EPI_ISL_1278389, EPI_ISL_1278390, EPI_ISL_1278391, EPI_ISL_1278392, EPI_ISL_1278393, EPI_ISL_1278394, EPI_ISL_1278395, EPI_ISL_1278396, EPI_ISL_1278397, EPI_ISL_1278398, EPI_ISL_1278399, EPI_ISL_1278400, EPI_ISL_1278401, EPI_ISL_1278402, EPI_ISL_1278403, EPI_ISL_1278404, EPI_ISL_1278405, EPI_ISL_1278406, EPI_ISL_1278407, EPI_ISL_1278408, EPI_ISL_1278409, EPI_ISL_1278410, EPI_ISL_1278411, EPI_ISL_1278412, EPI_ISL_1278413, EPI_ISL_1278414, EPI_ISL_1278415, EPI_ISL_1278416, EPI_ISL_1278417, EPI_ISL_1278418, EPI_ISL_1278419, EPI_ISL_1278420, EPI_ISL_1278421, EPI_ISL_1278422, EPI_ISL_1278423, EPI_ISL_1278424, EPI_ISL_1278425, EPI_ISL_1278426, EPI_ISL_1278427, EPI_ISL_1278428, EPI_ISL_1278429, EPI_ISL_1278430, EPI_ISL_1278431, EPI_ISL_1278432, EPI_ISL_1278433, EPI_ISL_1278434, EPI_ISL_1278435, EPI_ISL_1278436, EPI_ISL_1278437, EPI_ISL_1278438, EPI_ISL_1278439, EPI_ISL_1278440, EPI_ISL_1278441, EPI_ISL_1278442, EPI_ISL_1278443, EPI_ISL_1278444, EPI_ISL_1278445, EPI_ISL_1278446, EPI_ISL_1278447, EPI_ISL_1278448, EPI_ISL_1278449, EPI_ISL_1278450, EPI_ISL_1278451, EPI_ISL_1278452, EPI_ISL_1278453, EPI_ISL_1278454, EPI_ISL_1278455, EPI_ISL_1278456, EPI_ISL_1278457, EPI_ISL_1278458, EPI_ISL_1278459, EPI_ISL_1278460, EPI_ISL_1278461, EPI_ISL_1278462, EPI_ISL_1278463, EPI_ISL_1278464, EPI_ISL_1278465, EPI_ISL_1278466, EPI_ISL_1278467, EPI_ISL_1278468, EPI_ISL_1278469, EPI_ISL_1278470, EPI_ISL_1278471, EPI_ISL_1278472, EPI_ISL_1278473, EPI_ISL_1278474, EPI_ISL_1278475, EPI_ISL_1278476, EPI_ISL_1278477, EPI_ISL_1278478, EPI_ISL_1278479, EPI_ISL_1278480, EPI_ISL_1278481, EPI_ISL_1278482, EPI_ISL_1278483, EPI_ISL_1278484, EPI_ISL_1278485, EPI_ISL_1278486, EPI_ISL_1278487, EPI_ISL_1278488, EPI_ISL_1278489, EPI_ISL_1278490, EPI_ISL_1278491, EPI_ISL_1278492, EPI_ISL_1278493, EPI_ISL_1278494, EPI_ISL_1278495, EPI_ISL_1278496, EPI_ISL_1278497, EPI_ISL_1278498, EPI_ISL_1278499, EPI_ISL_1278500, EPI_ISL_1278501, EPI_ISL_1278502, EPI_ISL_1278503, EPI_ISL_1278504, EPI_ISL_1278505, EPI_ISL_1278506, EPI_ISL_1278507, EPI_ISL_1278508, EPI_ISL_1278509, EPI_ISL_1278510, EPI_ISL_1278511, EPI_ISL_1278512, EPI_ISL_1278513, EPI_ISL_1278514, EPI_ISL_1278515, EPI_ISL_1278516, EPI_ISL_1278517, EPI_ISL_1278518, EPI_ISL_1278519, EPI_ISL_1278520, EPI_ISL_1278521, EPI_ISL_1278522, EPI_ISL_1278523, EPI_ISL_1278524, EPI_ISL_1278525, EPI_ISL_1278526, EPI_ISL_1278527, EPI_ISL_1278528, EPI_ISL_1278529, EPI_ISL_1278530, EPI_ISL_1278531, EPI_ISL_1278532, EPI_ISL_1278533, EPI_ISL_1278534, EPI_ISL_1278535, EPI_ISL_1278536, EPI_ISL_1278537, EPI_ISL_1278538, EPI_ISL_1278539, EPI_ISL_1278540, EPI_ISL_1278541, EPI_ISL_1278542, EPI_ISL_1278543, EPI_ISL_1278544, EPI_ISL_1278545, EPI_ISL_1278546, EPI_ISL_1278547, EPI_ISL_1278548, EPI_ISL_1278549, EPI_ISL_1278550, EPI_ISL_1278551, EPI_ISL_1278552, EPI_ISL_1278553, EPI_ISL_1278554, EPI_ISL_1278555, EPI_ISL_1278556, EPI_ISL_1278557, EPI_ISL_1278558, EPI_ISL_1278559, EPI_ISL_1278560, EPI_ISL_1278561, EPI_ISL_1278562, EPI_ISL_1278563, EPI_ISL_1278564, EPI_ISL_1278565, EPI_ISL_1278566, EPI_ISL_1278567, EPI_ISL_1278568, EPI_ISL_1278569, EPI_ISL_1278570, EPI_ISL_1278571, EPI_ISL_1278572, EPI_ISL_1278573, EPI_ISL_1278574, EPI_ISL_1278575, EPI_ISL_1278576, EPI_ISL_1278577, EPI_ISL_1278578, EPI_ISL_1278579, EPI_ISL_1278580, EPI_ISL_1278581, EPI_ISL_1278582, EPI_ISL_1278583, EPI_ISL_1278584, EPI_ISL_1278585, EPI_ISL_1278586, EPI_ISL_1278587, EPI_ISL_1278588, EPI_ISL_1278589, EPI_ISL_1278590, EPI_ISL_1278591, EPI_ISL_1278592, EPI_ISL_1278593, EPI_ISL_1278594, EPI_ISL_1278595, EPI_ISL_1278596, EPI_ISL_1278597, EPI_ISL_1278598, EPI_ISL_1278599, EPI_ISL_1278600, EPI_ISL_1278601, EPI_ISL_1278602, EPI_ISL_1278603, EPI_ISL_1278604, EPI_ISL_1278605, EPI_ISL_1278606, EPI_ISL_1278607, EPI_ISL_1278608, EPI_ISL_1278609, EPI_ISL_1278610, EPI_ISL_1278611, EPI_ISL_1278612, EPI_ISL_1278613, EPI_ISL_1278614, EPI_ISL_1278615, EPI_ISL_1278616, EPI_ISL_1278617, EPI_ISL_1278618, EPI_ISL_1278619, EPI_ISL_1278620, EPI_ISL_1278621, EPI_ISL_1278622, EPI_ISL_1278623, EPI_ISL_1278624, EPI_ISL_1278625, EPI_ISL_1278626, EPI_ISL_1278627, EPI_ISL_1278628, EPI_ISL_1278629, EPI_ISL_1278630, EPI_ISL_1278631, EPI_ISL_1278632, EPI_ISL_1278633, EPI_ISL_1278634, EPI_ISL_1278635, EPI_ISL_1278636, EPI_ISL_1278637, EPI_ISL_1278638, EPI_ISL_1278639, EPI_ISL_1278640, EPI_ISL_1278641, EPI_ISL_1278642, EPI_ISL_1278643, EPI_ISL_1278644, EPI_ISL_1278645, EPI_ISL_1278646, EPI_ISL_1278647, EPI_ISL_1278648, EPI_ISL_1278649, EPI_ISL_1278650, EPI_ISL_1278651, EPI_ISL_1278652, EPI_ISL_1278653, EPI_ISL_1278654, EPI_ISL_1278655, EPI_ISL_1278656, EPI_ISL_1278657, EPI_ISL_1278658, EPI_ISL_1278659, EPI_ISL_1278660, EPI_ISL_1278661, EPI_ISL_1278662, EPI_ISL_1278663, EPI_ISL_1278664, EPI_ISL_1278665, EPI_ISL_1278666, EPI_ISL_1278667, EPI_ISL_1278668, EPI_ISL_1278669, EPI_ISL_1278670, EPI_ISL_1278671, EPI_ISL_1278672, EPI_ISL_1278673, EPI_ISL_1278674, EPI_ISL_1278675, EPI_ISL_1278676, EPI_ISL_1278677, EPI_ISL_1278678, EPI_ISL_1278679, EPI_ISL_1278680, EPI_ISL_1278681, EPI_ISL_1278682, EPI_ISL_1278683, EPI_ISL_1278684, EPI_ISL_1278685, EPI_ISL_1278686, EPI_ISL_1278687, EPI_ISL_1278688, EPI_ISL_1278689, EPI_ISL_1278690, EPI_ISL_1278691, EPI_ISL_1278692, EPI_ISL_1278693, EPI_ISL_1278694, EPI_ISL_1278695, EPI_ISL_1278696, EPI_ISL_1278697, EPI_ISL_1278698, EPI_ISL_1278699, EPI_ISL_1278700, EPI_ISL_1278701, EPI_ISL_1278702, EPI_ISL_1278703, EPI_ISL_1278704, EPI_ISL_1278705, EPI_ISL_1278706, EPI_ISL_1278707, EPI_ISL_1278708, EPI_ISL_1278709, EPI_ISL_1278710, EPI_ISL_1278711, EPI_ISL_1278712, EPI_ISL_1278713, EPI_ISL_1278714, EPI_ISL_1278715, EPI_ISL_1278716, EPI_ISL_1278717, EPI_ISL_1278718, EPI_ISL_1278719, EPI_ISL_1278720, EPI_ISL_1278721, EPI_ISL_1278722, EPI_ISL_1278723, EPI_ISL_1278724, EPI_ISL_1278725, EPI_ISL_1278726, EPI_ISL_1278727, EPI_ISL_1278728, EPI_ISL_1278729, EPI_ISL_1278730, EPI_ISL_1278731, EPI_ISL_1278732, EPI_ISL_1278733, EPI_ISL_1278734, EPI_ISL_1278735, EPI_ISL_1278736, EPI_ISL_1278737, EPI_ISL_1278738, EPI_ISL_1278739, EPI_ISL_1278740, EPI_ISL_1278741, EPI_ISL_1278742, EPI_ISL_1278743, EPI_ISL_1278744, EPI_ISL_1278745, EPI_ISL_1278746, EPI_ISL_1278747, EPI_ISL_1278748, EPI_ISL_1278749, EPI_ISL_1278750, EPI_ISL_1278751, EPI_ISL_1278752, EPI_ISL_1278753, EPI_ISL_1278754, EPI_ISL_1278755, EPI_ISL_1278756, EPI_ISL_1278757, EPI_ISL_1278758, EPI_ISL_1278759, EPI_ISL_1278760, EPI_ISL_1278761, EPI_ISL_1278762, EPI_ISL_1278763, EPI_ISL_1278764, EPI_ISL_1278765, EPI_ISL_1278766, EPI_ISL_1278767, EPI_ISL_1278768, EPI_ISL_1278769, EPI_ISL_1278770, EPI_ISL_1278771, EPI_ISL_1278772, EPI_ISL_1278773, EPI_ISL_1278774, EPI_ISL_1278775, EPI_ISL_1278776, EPI_ISL_1278777, EPI_ISL_1278778, EPI_ISL_1278779, EPI_ISL_1278780, EPI_ISL_1278781, EPI_ISL_1278782, EPI_ISL_1278783, EPI_ISL_1278784, EPI_ISL_1278785, EPI_ISL_1278786, EPI_ISL_1278787, EPI_ISL_1278788, EPI_ISL_1278789, EPI_ISL_1278790, EPI_ISL_1278791, EPI_ISL_1278792, EPI_ISL_1278793, EPI_ISL_1278794, EPI_ISL_1278795, EPI_ISL_1278796, EPI_ISL_1278797, EPI_ISL_1278798, EPI_ISL_1278799, EPI_ISL_1278800, EPI_ISL_1278801, EPI_ISL_1278802, EPI_ISL_1278803, EPI_ISL_1278804, EPI_ISL_1278805, EPI_ISL_1278806, EPI_ISL_1278807, EPI_ISL_1278808, EPI_ISL_1278809, EPI_ISL_1278810, EPI_ISL_1278811, EPI_ISL_1278812, EPI_ISL_1278813, EPI_ISL_1278814, EPI_ISL_1278815, EPI_ISL_1278816, EPI_ISL_1278817, EPI_ISL_1278818, EPI_ISL_1278819, EPI_ISL_1278820, EPI_ISL_1278821, EPI_ISL_1278822, EPI_ISL_1278823, EPI_ISL_1278824, EPI_ISL_1278825, EPI_ISL_1278826, EPI_ISL_1278827, EPI_ISL_1278828, EPI_ISL_1278829, EPI_ISL_1278830, EPI_ISL_1278831, EPI_ISL_1278832, EPI_ISL_1278833, EPI_ISL_1278834, EPI_ISL_1278835, EPI_ISL_1278836, EPI_ISL_1278837, EPI_ISL_1278838, EPI_ISL_1278839, EPI_ISL_1278840, EPI_ISL_1278841, EPI_ISL_1278842, EPI_ISL_1278843, EPI_ISL_1278844, EPI_ISL_1278845, EPI_ISL_1278846, EPI_ISL_1278847, EPI_ISL_1278848, EPI_ISL_1278849, EPI_ISL_1278850, EPI_ISL_1278851, EPI_ISL_1278852, EPI_ISL_1278853, EPI_ISL_1278854, EPI_ISL_1278855, EPI_ISL_1278856, EPI_ISL_1278857, EPI_ISL_1278858, EPI_ISL_1278859, EPI_ISL_1278860, EPI_ISL_1278861, EPI_ISL_1278862, EPI_ISL_1278863, EPI_ISL_1278864, EPI_ISL_1278865, EPI_ISL_1278866, EPI_ISL_1278867, EPI_ISL_1278868, EPI_ISL_1278869, EPI_ISL_1278870, EPI_ISL_1278871, EPI_ISL_1278872, EPI_ISL_1278873, EPI_ISL_1278874, EPI_ISL_1278875, EPI_ISL_1278877, EPI_ISL_1278878, EPI_ISL_1278879, EPI_ISL_1278880, EPI_ISL_1278881, EPI_ISL_1278882, EPI_ISL_1278884, EPI_ISL_1278885, EPI_ISL_1278887, EPI_ISL_1278888, EPI_ISL_1278889, EPI_ISL_1278890, EPI_ISL_1278891, EPI_ISL_1278892, EPI_ISL_1278893, EPI_ISL_1278894, EPI_ISL_1278895, EPI_ISL_1278896, EPI_ISL_1278897, EPI_ISL_1278898, EPI_ISL_1278899, EPI_ISL_1278900, EPI_ISL_1278901, EPI_ISL_1278902, EPI_ISL_1278903, EPI_ISL_1278904, EPI_ISL_1278905, EPI_ISL_1278906, EPI_ISL_1278907, EPI_ISL_1278908, EPI_ISL_1278909, EPI_ISL_1278910, EPI_ISL_1278911, EPI_ISL_1278912, EPI_ISL_1278913, EPI_ISL_1278914, EPI_ISL_1278915, EPI_ISL_1278916, EPI_ISL_1278917, EPI_ISL_1278918, EPI_ISL_1278919, EPI_ISL_1278920, EPI_ISL_1278921, EPI_ISL_1278922, EPI_ISL_1278923, EPI_ISL_1278924, EPI_ISL_1278925, EPI_ISL_1278926, EPI_ISL_1278927, EPI_ISL_1278928, EPI_ISL_1278929, EPI_ISL_1278930, EPI_ISL_1278931, EPI_ISL_1278932, EPI_ISL_1278933, EPI_ISL_1278934, EPI_ISL_1278935, EPI_ISL_1278936, EPI_ISL_1278937, EPI_ISL_1278938, EPI_ISL_1278939, EPI_ISL_1278940, EPI_ISL_1278941, EPI_ISL_1278942

| | | | |
|---|---|---|---|
| see above | Clalit Health Services Laboratories, Israel | Stern Lab | Stern Lab |

EPI_ISL_1357777, EPI_ISL_1357778, EPI_ISL_1357779, EPI_ISL_1357780, EPI_ISL_1357781, EPI_ISL_1357782, EPI_ISL_1357783, EPI_ISL_1357784, EPI_ISL_1357785, EPI_ISL_1357786, EPI_ISL_1357787, EPI_ISL_1357788, EPI_ISL_1357789, EPI_ISL_1357790, EPI_ISL_1357791, EPI_ISL_1357792, EPI_ISL_1357793, EPI_ISL_1357794, EPI_ISL_1357795, EPI_ISL_1357796, EPI_ISL_1357797, EPI_ISL_1357798, EPI_ISL_1357799, EPI_ISL_1357800, EPI_ISL_1357801, EPI_ISL_1357802, EPI_ISL_1357803, EPI_ISL_1357804, EPI_ISL_1357805, EPI_ISL_1357806, EPI_ISL_1357807, EPI_ISL_1357808, EPI_ISL_1357809, EPI_ISL_1357810, EPI_ISL_1357811, EPI_ISL_1357812, EPI_ISL_1357813, EPI_ISL_1357814, EPI_ISL_1357815, EPI_ISL_1357816, EPI_ISL_1357817, EPI_ISL_1357818, EPI_ISL_1357819, EPI_ISL_1357820, EPI_ISL_1357821, EPI_ISL_1357822, EPI_ISL_1357823, EPI_ISL_1357824, EPI_ISL_1357825, EPI_ISL_1357826, EPI_ISL_1357827, EPI_ISL_1357828, EPI_ISL_1357829, EPI_ISL_1357830, EPI_ISL_1357831, EPI_ISL_1357832, EPI_ISL_1357833, EPI_ISL_1357834, EPI_ISL_1357835, EPI_ISL_1357836, EPI_ISL_1357837, EPI_ISL_1357838, EPI_ISL_1357839, EPI_ISL_1357840, EPI_ISL_1357841, EPI_ISL_1358415, EPI_ISL_1358416, EPI_ISL_1358417, EPI_ISL_1358418, EPI_ISL_1358419, EPI_ISL_1358420, EPI_ISL_1358421, EPI_ISL_1358422, EPI_ISL_1358423, EPI_ISL_1358424, EPI_ISL_1358425, EPI_ISL_1358426, EPI_ISL_1358427, EPI_ISL_1358428, EPI_ISL_1358429, EPI_ISL_1358430, EPI_ISL_1358431, EPI_ISL_1358432, EPI_ISL_1358433, EPI_ISL_1358434, EPI_ISL_1358435, EPI_ISL_1358436, EPI_ISL_1358437, EPI_ISL_1358438, EPI_ISL_1358439, EPI_ISL_1358440, EPI_ISL_1358441, EPI_ISL_1358442, EPI_ISL_1358443, EPI_ISL_1358444, EPI_ISL_1358445, EPI_ISL_1358446, EPI_ISL_1358447, EPI_ISL_1358448, EPI_ISL_1358449, EPI_ISL_1358450, EPI_ISL_1358451, EPI_ISL_1358452, EPI_ISL_1358453, EPI_ISL_1358454, EPI_ISL_1358455, EPI_ISL_1358456, EPI_ISL_1358457, EPI_ISL_1358458, EPI_ISL_1358459, EPI_ISL_1358460, EPI_ISL_1358461, EPI_ISL_1358462, EPI_ISL_1358463, EPI_ISL_1358464, EPI_ISL_1358465, EPI_ISL_1358466, EPI_ISL_1358467, EPI_ISL_1358468, EPI_ISL_1358469, EPI_ISL_1358470, EPI_ISL_1358471, EPI_ISL_1358472, EPI_ISL_1358473, EPI_ISL_1358474, EPI_ISL_1358475, EPI_ISL_1358476, EPI_ISL_1358477, EPI_ISL_1358478, EPI_ISL_1358479, EPI_ISL_1358480, EPI_ISL_1358481, EPI_ISL_1358482, EPI_ISL_1358483, EPI_ISL_1358484, EPI_ISL_1358485, EPI_ISL_1358486, EPI_ISL_1358487, EPI_ISL_1358488, EPI_ISL_1358489, EPI_ISL_1358490, EPI_ISL_1358491, EPI_ISL_1358492, EPI_ISL_1358493, EPI_ISL_1358494, EPI_ISL_1358495, EPI_ISL_1358496, EPI_ISL_1358497, EPI_ISL_1358498, EPI_ISL_1358499, EPI_ISL_1358500, EPI_ISL_1358501, EPI_ISL_1358502, EPI_ISL_1358503, EPI_ISL_1358504, EPI_ISL_1358505, EPI_ISL_1358506, EPI_ISL_1358507, EPI_ISL_1358508, EPI_ISL_1358509, EPI_ISL_1358510, EPI_ISL_1358511, EPI_ISL_1358512, EPI_ISL_1358513, EPI_ISL_1358514, EPI_ISL_1358515, EPI_ISL_1358516, EPI_ISL_1358517, EPI_ISL_1358518, EPI_ISL_1358519, EPI_ISL_1358520, EPI_ISL_1358521, EPI_ISL_1358522, EPI_ISL_1358523, EPI_ISL_1358524, EPI_ISL_1358525, EPI_ISL_1358526, EPI_ISL_1358527, EPI_ISL_1358528, EPI_ISL_1358529, EPI_ISL_1358530, EPI_ISL_1358531, EPI_ISL_1358532, EPI_ISL_1358533, EPI_ISL_1358534, EPI_ISL_1358535, EPI_ISL_1358536, EPI_ISL_1358537, EPI_ISL_1358538, EPI_ISL_1358539, EPI_ISL_1358540, EPI_ISL_1358541, EPI_ISL_1358542, EPI_ISL_1358543, EPI_ISL_1358544, EPI_ISL_1358545, EPI_ISL_1358546, EPI_ISL_1358547, EPI_ISL_1358548, EPI_ISL_1358549, EPI_ISL_1358550, EPI_ISL_1358551, EPI_ISL_1358552, EPI_ISL_1358553, EPI_ISL_1358554, EPI_ISL_1358555, EPI_ISL_1358556, EPI_ISL_1358557, EPI_ISL_1358558, EPI_ISL_1358559, EPI_ISL_1358560, EPI_ISL_1358561, EPI_ISL_1358562, EPI_ISL_1358563, EPI_ISL_1358564, EPI_ISL_1358565, EPI_ISL_1358566, EPI_ISL_1358567, EPI_ISL_1358568, EPI_ISL_1358569, EPI_ISL_1358570, EPI_ISL_1358571, EPI_ISL_1358572, EPI_ISL_1358573, EPI_ISL_1358574, EPI_ISL_1358575, EPI_ISL_1358576, EPI_ISL_1358577, EPI_ISL_1358578, EPI_ISL_1358579, EPI_ISL_1358580, EPI_ISL_1358581, EPI_ISL_1358582, EPI_ISL_1358583, EPI_ISL_1358584, EPI_ISL_1358585, EPI_ISL_1358586, EPI_ISL_1358587, EPI_ISL_1358588, EPI_ISL_1358589, EPI_ISL_1358590, EPI_ISL_1358591, EPI_ISL_1358592, EPI_ISL_1358593, EPI_ISL_1358594, EPI_ISL_1358595, EPI_ISL_1358596, EPI_ISL_1358597, EPI_ISL_1358598, EPI_ISL_1358599, EPI_ISL_1358600, EPI_ISL_1358601, EPI_ISL_1358602, EPI_ISL_1358603, EPI_ISL_1358604, EPI_ISL_1358605, EPI_ISL_1358606, EPI_ISL_1358607, EPI_ISL_1358608, EPI_ISL_1358609, EPI_ISL_1358610, EPI_ISL_1358611, EPI_ISL_1358612, EPI_ISL_1358613, EPI_ISL_1358614, EPI_ISL_1358615, EPI_ISL_1358616, EPI_ISL_1358617, EPI_ISL_1358618, EPI_ISL_1358619, EPI_ISL_1358620, EPI_ISL_1358621, EPI_ISL_1358622, EPI_ISL_1358623, EPI_ISL_1358624, EPI_ISL_1358625, EPI_ISL_1358626, EPI_ISL_1358627, EPI_ISL_1358628, EPI_ISL_1358629, EPI_ISL_1358630, EPI_ISL_1358631, EPI_ISL_1358632, EPI_ISL_1358633, EPI_ISL_1358634, EPI_ISL_1358635, EPI_ISL_1358636, EPI_ISL_1358637, EPI_ISL_1358638, EPI_ISL_1358639, EPI_ISL_1358640, EPI_ISL_1358641, EPI_ISL_1358642, EPI_ISL_1358643, EPI_ISL_1358644, EPI_ISL_1358645, EPI_ISL_1358646, EPI_ISL_1358647, EPI_ISL_1358648, EPI_ISL_1358649, EPI_ISL_1358650, EPI_ISL_1358651, EPI_ISL_1358652, EPI_ISL_1358653, EPI_ISL_1358654, EPI_ISL_1358655, EPI_ISL_1358656, EPI_ISL_1358657, EPI_ISL_1358658, EPI_ISL_1358659, EPI_ISL_1358660, EPI_ISL_1358661, EPI_ISL_1358662, EPI_ISL_1358663, EPI_ISL_1358664, EPI_ISL_1358665, EPI_ISL_1358666, EPI_ISL_1358667, EPI_ISL_1358668, EPI_ISL_1358669, EPI_ISL_1358670, EPI_ISL_1358671, EPI_ISL_1358672, EPI_ISL_1358673, EPI_ISL_1358674, EPI_ISL_1358675, EPI_ISL_1358676, EPI_ISL_1358677, EPI_ISL_1358678, EPI_ISL_1358679, EPI_ISL_1358680, EPI_ISL_1358681, EPI_ISL_1358682, EPI_ISL_1358683, EPI_ISL_1358684, EPI_ISL_1358685, EPI_ISL_1358686, EPI_ISL_1358687, EPI_ISL_1358688, EPI_ISL_1358689, EPI_ISL_1358690, EPI_ISL_1358691, EPI_ISL_1358692, EPI_ISL_1358693, EPI_ISL_1358694, EPI_ISL_1358695, EPI_ISL_1358696, EPI_ISL_1358697, EPI_ISL_1358698, EPI_ISL_1358699, EPI_ISL_1358700, EPI_ISL_1358701, EPI_ISL_1358702, EPI_ISL_1358703, EPI_ISL_1358704, EPI_ISL_1358705, EPI_ISL_1358706, EPI_ISL_1358707, EPI_ISL_1358708, EPI_ISL_1358709, EPI_ISL_1358710, EPI_ISL_1358711, EPI_ISL_1358712, EPI_ISL_1358713, EPI_ISL_1358714, EPI_ISL_1358715, EPI_ISL_1358716, EPI_ISL_1358717, EPI_ISL_1358718, EPI_ISL_1358719, EPI_ISL_1358720, EPI_ISL_1358721, EPI_ISL_1358722, EPI_ISL_1358723, EPI_ISL_1358724, EPI_ISL_1358725, EPI_ISL_1358726, EPI_ISL_1358727, EPI_ISL_1358728, EPI_ISL_1358729, EPI_ISL_1358730, EPI_ISL_1358731, EPI_ISL_1358732, EPI_ISL_1358733, EPI_ISL_1358734, EPI_ISL_1358735, EPI_ISL_1358736, EPI_ISL_1358737, EPI_ISL_1358738, EPI_ISL_1358739, EPI_ISL_1358740, EPI_ISL_1358741, EPI_ISL_1358742, EPI_ISL_1358743, EPI_ISL_1358744, EPI_ISL_1358745, EPI_ISL_1358746, EPI_ISL_1358747, EPI_ISL_1358748, EPI_ISL_1358749, EPI_ISL_1358750, EPI_ISL_1358751, EPI_ISL_1358752, EPI_ISL_1358753, EPI_ISL_1358754, EPI_ISL_1358755, EPI_ISL_1358756, EPI_ISL_1358757, EPI_ISL_1358758, EPI_ISL_1358759, EPI_ISL_1358760, EPI_ISL_1358761, EPI_ISL_1358762, EPI_ISL_1358763, EPI_ISL_1358764, EPI_ISL_1358765, EPI_ISL_1358766, EPI_ISL_1358767, EPI_ISL_1358768, EPI_ISL_1358769, EPI_ISL_1358770, EPI_ISL_1358771, EPI_ISL_1358772, EPI_ISL_1358773, EPI_ISL_1358774, EPI_ISL_1358775, EPI_ISL_1358776, EPI_ISL_1358777, EPI_ISL_1358778, EPI_ISL_1358779, EPI_ISL_1358780, EPI_ISL_1358781, EPI_ISL_1358782, EPI_ISL_1358783, EPI_ISL_1358784, EPI_ISL_1358785, EPI_ISL_1358786, EPI_ISL_1358787, EPI_ISL_1358788, EPI_ISL_1358789, EPI_ISL_1358790, EPI_ISL_1358791, EPI_ISL_1358792, EPI_ISL_1358793, EPI_ISL_1358794, EPI_ISL_1358795, EPI_ISL_1358796, EPI_ISL_1358797, EPI_ISL_1358798, EPI_ISL_1358799, EPI_ISL_1358800, EPI_ISL_1358801, EPI_ISL_1358802, EPI_ISL_1358803, EPI_ISL_1358804, EPI_ISL_1358805, EPI_ISL_1358806, EPI_ISL_1358807, EPI_ISL_1358808, EPI_ISL_1358809, EPI_ISL_1358810, EPI_ISL_1358811, EPI_ISL_1358812, EPI_ISL_1358813, EPI_ISL_1358814, EPI_ISL_1358815, EPI_ISL_1358816, EPI_ISL_1358817, EPI_ISL_1358818, EPI_ISL_1358819, EPI_ISL_1358820, EPI_ISL_1358821, EPI_ISL_1358822, EPI_ISL_1358823, EPI_ISL_1358824, EPI_ISL_1358825, EPI_ISL_1358826, EPI_ISL_1358827, EPI_ISL_1358828, EPI_ISL_1358829, EPI_ISL_1358830, EPI_ISL_1358831, EPI_ISL_1358832, EPI_ISL_1358833, EPI_ISL_1358834, EPI_ISL_1358835, EPI_ISL_1358836, EPI_ISL_1358837, EPI_ISL_1358838, EPI_ISL_1358839, EPI_ISL_1358840, EPI_ISL_1358841, EPI_ISL_1358842,

EPI_ISL_1358843, EPI_ISL_1358844, EPI_ISL_1358845, EPI_ISL_1358846, EPI_ISL_1358847, EPI_ISL_1358848, EPI_ISL_1358849, EPI_ISL_1358850, EPI_ISL_1358851, EPI_ISL_1358852, EPI_ISL_1358853, EPI_ISL_1358854, EPI_ISL_1358855, EPI_ISL_1358856, EPI_ISL_1358857, EPI_ISL_1358858, EPI_ISL_1358859, EPI_ISL_1358860, EPI_ISL_1358861, EPI_ISL_1358862, EPI_ISL_1358863, EPI_ISL_1358864, EPI_ISL_1358865, EPI_ISL_1358866, EPI_ISL_1358867, EPI_ISL_1358868, EPI_ISL_1358869, EPI_ISL_1358870, EPI_ISL_1358871, EPI_ISL_1358872, EPI_ISL_1358873, EPI_ISL_1358874, EPI_ISL_1358875, EPI_ISL_1358876, EPI_ISL_1358877, EPI_ISL_1358878, EPI_ISL_1358879, EPI_ISL_1358880, EPI_ISL_1358881, EPI_ISL_1358882, EPI_ISL_1358883, EPI_ISL_1358884, EPI_ISL_1358885, EPI_ISL_1358886, EPI_ISL_1358887, EPI_ISL_1358888, EPI_ISL_1358889, EPI_ISL_1358890, EPI_ISL_1358891, EPI_ISL_1358892, EPI_ISL_1358893, EPI_ISL_1358894, EPI_ISL_1358895, EPI_ISL_1358896, EPI_ISL_1358897, EPI_ISL_1358898, EPI_ISL_1358899, EPI_ISL_1358900, EPI_ISL_1358901, EPI_ISL_1358902, EPI_ISL_1358903

| | | | |
|---|---|---|---|
| see above | Israel Central Virology laboratory | Israel National Consortium for SARS-CoV-2 sequencing | Neta Zuckerman, Efrat Dahan Bucris, Michal Mandelboim, Dana Bar-Ilan, Oran Erster, Tzvia Mann, Omer Murik, David A. Zeevi, Assaf Rokney, Joseph Jaffe, Eva Nachum, Maya Davidovich Cohen, Ephraim Fass, Gal Zizelski Valenci, Mor Rubinstein, Efrat Rorman, Israel Nissan, Efrat Glick-Saar, Omri Nayshool, Gideon Rechavi, Ella Mendelson, Orna Mor |

EPI_ISL_1424882, EPI_ISL_1424883, EPI_ISL_1424884, EPI_ISL_1424885, EPI_ISL_1424886, EPI_ISL_1424887, EPI_ISL_1424888, EPI_ISL_1424889, EPI_ISL_1424890, EPI_ISL_1424891, EPI_ISL_1424892, EPI_ISL_1424893, EPI_ISL_1424894, EPI_ISL_1424895, EPI_ISL_1424896, EPI_ISL_1424897, EPI_ISL_1424898, EPI_ISL_1424899, EPI_ISL_1424900, EPI_ISL_1424901, EPI_ISL_1424902, EPI_ISL_1424903, EPI_ISL_1424904, EPI_ISL_1424905, EPI_ISL_1424906, EPI_ISL_1424907, EPI_ISL_1424908, EPI_ISL_1424909, EPI_ISL_1424910, EPI_ISL_1424911, EPI_ISL_1424912, EPI_ISL_1424913, EPI_ISL_1424914, EPI_ISL_1424915, EPI_ISL_1424916, EPI_ISL_1424917, EPI_ISL_1424918, EPI_ISL_1424919, EPI_ISL_1424920, EPI_ISL_1424921, EPI_ISL_1424922, EPI_ISL_1424923, EPI_ISL_1424924, EPI_ISL_1424925, EPI_ISL_1424926, EPI_ISL_1424927, EPI_ISL_1424928, EPI_ISL_1424929, EPI_ISL_1424930, EPI_ISL_1424931, EPI_ISL_1424932, EPI_ISL_1424933, EPI_ISL_1424934, EPI_ISL_1424935, EPI_ISL_1424936, EPI_ISL_1424937, EPI_ISL_1424938, EPI_ISL_1424939, EPI_ISL_1424940, EPI_ISL_1424941, EPI_ISL_1424942, EPI_ISL_1424943, EPI_ISL_1424944, EPI_ISL_1424945, EPI_ISL_1424946, EPI_ISL_1424947, EPI_ISL_1424948, EPI_ISL_1424949, EPI_ISL_1424950, EPI_ISL_1424951, EPI_ISL_1424952, EPI_ISL_1424953, EPI_ISL_1424954, EPI_ISL_1424955, EPI_ISL_1424956, EPI_ISL_1424957, EPI_ISL_1424958, EPI_ISL_1424959, EPI_ISL_1424960, EPI_ISL_1424961, EPI_ISL_1424962, EPI_ISL_1424963, EPI_ISL_1424964, EPI_ISL_1424965, EPI_ISL_1424966, EPI_ISL_1424967, EPI_ISL_1424968, EPI_ISL_1424969, EPI_ISL_1424970, EPI_ISL_1424971, EPI_ISL_1424972, EPI_ISL_1424973, EPI_ISL_1424974, EPI_ISL_1424975, EPI_ISL_1424976, EPI_ISL_1424977, EPI_ISL_1424978, EPI_ISL_1424979, EPI_ISL_1424980, EPI_ISL_1424981, EPI_ISL_1424982, EPI_ISL_1424983, EPI_ISL_1424984, EPI_ISL_1424985, EPI_ISL_1424986, EPI_ISL_1424987, EPI_ISL_1424988, EPI_ISL_1424989, EPI_ISL_1424990, EPI_ISL_1424991, EPI_ISL_1424992, EPI_ISL_1424993, EPI_ISL_1424994, EPI_ISL_1424995, EPI_ISL_1424996, EPI_ISL_1424997, EPI_ISL_1424998, EPI_ISL_1424999, EPI_ISL_1425000, EPI_ISL_1425001, EPI_ISL_1425002, EPI_ISL_1425003, EPI_ISL_1425004, EPI_ISL_1425005, EPI_ISL_1425006, EPI_ISL_1425007, EPI_ISL_1425008, EPI_ISL_1425009, EPI_ISL_1425010, EPI_ISL_1425011, EPI_ISL_1425012, EPI_ISL_1425013, EPI_ISL_1425014, EPI_ISL_1425015, EPI_ISL_1425016, EPI_ISL_1425017, EPI_ISL_1425018, EPI_ISL_1425019, EPI_ISL_1425020, EPI_ISL_1425021, EPI_ISL_1425022, EPI_ISL_1425023, EPI_ISL_1425024, EPI_ISL_1425025, EPI_ISL_1425026, EPI_ISL_1425027, EPI_ISL_1425028, EPI_ISL_1425029, EPI_ISL_1425030, EPI_ISL_1425031, EPI_ISL_1425032, EPI_ISL_1425033, EPI_ISL_1425034, EPI_ISL_1425035, EPI_ISL_1425036, EPI_ISL_1425037, EPI_ISL_1425038, EPI_ISL_1425039, EPI_ISL_1425040, EPI_ISL_1425041, EPI_ISL_1425042, EPI_ISL_1425043, EPI_ISL_1425044, EPI_ISL_1425045, EPI_ISL_1425046, EPI_ISL_1425047, EPI_ISL_1425048, EPI_ISL_1425049, EPI_ISL_1425050, EPI_ISL_1425051, EPI_ISL_1425052, EPI_ISL_1425053, EPI_ISL_1425054, EPI_ISL_1425055, EPI_ISL_1425056, EPI_ISL_1425057, EPI_ISL_1425058, EPI_ISL_1425059, EPI_ISL_1425060, EPI_ISL_1425061, EPI_ISL_1425062, EPI_ISL_1425063, EPI_ISL_1425064, EPI_ISL_1425065, EPI_ISL_1425066, EPI_ISL_1425067, EPI_ISL_1425068, EPI_ISL_1425069, EPI_ISL_1425070, EPI_ISL_1425071, EPI_ISL_1425072, EPI_ISL_1425073, EPI_ISL_1425074, EPI_ISL_1425075, EPI_ISL_1435646, EPI_ISL_1435649, EPI_ISL_1435651, EPI_ISL_1435653, EPI_ISL_1435655, EPI_ISL_1435657, EPI_ISL_1435660, EPI_ISL_1435662, EPI_ISL_1656477, EPI_ISL_1656478, EPI_ISL_1656479, EPI_ISL_1656480, EPI_ISL_1656481, EPI_ISL_1656482, EPI_ISL_1656483, EPI_ISL_1656484, EPI_ISL_1656485, EPI_ISL_1656486, EPI_ISL_1656487, EPI_ISL_1656488, EPI_ISL_1656489, EPI_ISL_1656490, EPI_ISL_1656491, EPI_ISL_1656492, EPI_ISL_1656493, EPI_ISL_1656494, EPI_ISL_1656495, EPI_ISL_1656496, EPI_ISL_1656497, EPI_ISL_1656499, EPI_ISL_1656500, EPI_ISL_1656501, EPI_ISL_1656502, EPI_ISL_1656504, EPI_ISL_1656505, EPI_ISL_1656507, EPI_ISL_1656508, EPI_ISL_1656509, EPI_ISL_1656510, EPI_ISL_1656511, EPI_ISL_1656512, EPI_ISL_1656513, EPI_ISL_1656514, EPI_ISL_1656515, EPI_ISL_1656516, EPI_ISL_1656517, EPI_ISL_1656518, EPI_ISL_1656519

| | | | |
|---|---|---|---|
| see above | Clalit Health Services Laboratories, Israel | Stern Lab | Stern Lab |

EPI_ISL_1673763, EPI_ISL_1673764, EPI_ISL_1673765, EPI_ISL_1673766, EPI_ISL_1673767, EPI_ISL_1673768, EPI_ISL_1673769, EPI_ISL_1673770, EPI_ISL_1673771, EPI_ISL_1673772, EPI_ISL_1673773, EPI_ISL_1673774, EPI_ISL_1673775, EPI_ISL_1673776, EPI_ISL_1673777, EPI_ISL_1673778, EPI_ISL_1673779, EPI_ISL_1673780, EPI_ISL_1673781, EPI_ISL_1673782, EPI_ISL_1673783, EPI_ISL_1673784, EPI_ISL_1673785, EPI_ISL_1673786, EPI_ISL_1673787, EPI_ISL_1673788, EPI_ISL_1673789, EPI_ISL_1673790, EPI_ISL_1673791, EPI_ISL_1673792, EPI_ISL_1673793, EPI_ISL_1673794, EPI_ISL_1673795, EPI_ISL_1673796, EPI_ISL_1673797, EPI_ISL_1673798, EPI_ISL_1673799, EPI_ISL_1673800, EPI_ISL_1673801, EPI_ISL_1673802, EPI_ISL_1673803, EPI_ISL_1673804, EPI_ISL_1673805, EPI_ISL_1673806, EPI_ISL_1673807, EPI_ISL_1673808, EPI_ISL_1673809, EPI_ISL_1673810, EPI_ISL_1673811, EPI_ISL_1673812, EPI_ISL_1673813, EPI_ISL_1673814, EPI_ISL_1673815, EPI_ISL_1673816, EPI_ISL_1673817, EPI_ISL_1673818, EPI_ISL_1673819, EPI_ISL_1673820, EPI_ISL_1673821, EPI_ISL_1673822, EPI_ISL_1673823, EPI_ISL_1673824, EPI_ISL_1673825, EPI_ISL_1673826, EPI_ISL_1673827, EPI_ISL_1673828, EPI_ISL_1673829, EPI_ISL_1673830, EPI_ISL_1673831, EPI_ISL_1673832, EPI_ISL_1673833, EPI_ISL_1673834, EPI_ISL_1673835, EPI_ISL_1673836, EPI_ISL_1673837, EPI_ISL_1673838, EPI_ISL_1673839, EPI_ISL_1673840, EPI_ISL_1673841, EPI_ISL_1673842, EPI_ISL_1673843, EPI_ISL_1673844, EPI_ISL_1673845, EPI_ISL_1673846, EPI_ISL_1673847, EPI_ISL_1673848, EPI_ISL_1673849, EPI_ISL_1673850, EPI_ISL_1673851, EPI_ISL_1673852, EPI_ISL_1673853, EPI_ISL_1673854, EPI_ISL_1673855, EPI_ISL_1673856, EPI_ISL_1673857, EPI_ISL_1673858, EPI_ISL_1673859, EPI_ISL_1673860, EPI_ISL_1673861, EPI_ISL_1673862, EPI_ISL_1673863, EPI_ISL_1673864, EPI_ISL_1673865, EPI_ISL_1673866, EPI_ISL_1673867, EPI_ISL_1673868, EPI_ISL_1673869, EPI_ISL_1673870, EPI_ISL_1673871, EPI_ISL_1673872, EPI_ISL_1673873, EPI_ISL_1673874, EPI_ISL_1673875, EPI_ISL_1673876, EPI_ISL_1673877, EPI_ISL_1673878, EPI_ISL_1673879, EPI_ISL_1673880, EPI_ISL_1673881, EPI_ISL_1673882, EPI_ISL_1673883, EPI_ISL_1673884, EPI_ISL_1673885, EPI_ISL_1673886, EPI_ISL_1673887, EPI_ISL_1673888, EPI_ISL_1673889, EPI_ISL_1673890, EPI_ISL_1673891, EPI_ISL_1673892, EPI_ISL_1673893, EPI_ISL_1673894, EPI_ISL_1673895, EPI_ISL_1673896, EPI_ISL_1673897, EPI_ISL_1673898, EPI_ISL_1673899, EPI_ISL_1673900, EPI_ISL_1673901, EPI_ISL_1673902, EPI_ISL_1673903, EPI_ISL_1673904, EPI_ISL_1673905, EPI_ISL_1673906, EPI_ISL_1673907, EPI_ISL_1673908, EPI_ISL_1673909, EPI_ISL_1673910, EPI_ISL_1673911, EPI_ISL_1673912, EPI_ISL_1673913, EPI_ISL_1673914, EPI_ISL_1673915, EPI_ISL_1673916, EPI_ISL_1673917, EPI_ISL_1673918, EPI_ISL_1673919, EPI_ISL_1673920, EPI_ISL_1673921, EPI_ISL_1673922, EPI_ISL_1673923, EPI_ISL_1673924, EPI_ISL_1673925, EPI_ISL_1673926, EPI_ISL_1673927, EPI_ISL_1673928, EPI_ISL_1673929, EPI_ISL_1673930, EPI_ISL_1673931, EPI_ISL_1673932, EPI_ISL_1673933, EPI_ISL_1673934, EPI_ISL_1673935, EPI_ISL_1673936, EPI_ISL_1673937, EPI_ISL_1673938, EPI_ISL_1673939, EPI_ISL_1673940, EPI_ISL_1673941, EPI_ISL_1673942, EPI_ISL_1673943, EPI_ISL_1673944, EPI_ISL_1673945, EPI_ISL_1673946, EPI_ISL_1673947, EPI_ISL_1673948, EPI_ISL_1673949, EPI_ISL_1673950, EPI_ISL_1673951, EPI_ISL_1673952, EPI_ISL_1673953, EPI_ISL_1673954, EPI_ISL_1673955, EPI_ISL_1673956, EPI_ISL_1673957, EPI_ISL_1673958, EPI_ISL_1673959, EPI_ISL_1673960, EPI_ISL_1673961, EPI_ISL_1673962, EPI_ISL_1673963, EPI_ISL_1673964, EPI_ISL_1673965, EPI_ISL_1673966, EPI_ISL_1673967, EPI_ISL_1673968, EPI_ISL_1673969, EPI_ISL_1673970, EPI_ISL_1673971, EPI_ISL_1673972, EPI_ISL_1673973, EPI_ISL_1673974, EPI_ISL_1673975, EPI_ISL_1673976, EPI_ISL_1673977, EPI_ISL_1673978, EPI_ISL_1673979, EPI_ISL_1673980, EPI_ISL_1673981, EPI_ISL_1673982, EPI_ISL_1673983, EPI_ISL_1673984, EPI_ISL_1673985, EPI_ISL_1673986, EPI_ISL_1673987, EPI_ISL_1673988, EPI_ISL_1673989, EPI_ISL_1673990, EPI_ISL_1673991, EPI_ISL_1673992, EPI_ISL_1673993, EPI_ISL_1673994, EPI_ISL_1673995, EPI_ISL_1673996, EPI_ISL_1673997, EPI_ISL_1673998, EPI_ISL_1673999, EPI_ISL_1674000, EPI_ISL_1674001, EPI_ISL_1674002, EPI_ISL_1674003, EPI_ISL_1674004, EPI_ISL_1674005, EPI_ISL_1674006, EPI_ISL_1674007, EPI_ISL_1674008, EPI_ISL_1674009, EPI_ISL_1674011, EPI_ISL_1674012, EPI_ISL_1674013, EPI_ISL_1674014, EPI_ISL_1674015, EPI_ISL_1674016, EPI_ISL_1674017, EPI_ISL_1674018, EPI_ISL_1674019, EPI_ISL_1674020, EPI_ISL_1674021, EPI_ISL_1674022, EPI_ISL_1674023, EPI_ISL_1674024, EPI_ISL_1674025, EPI_ISL_1674026, EPI_ISL_1674027, EPI_ISL_1674028, EPI_ISL_1674029, EPI_ISL_1674030, EPI_ISL_1674031, EPI_ISL_1674032, EPI_ISL_1674033, EPI_ISL_1674034, EPI_ISL_1674035, EPI_ISL_1674036, EPI_ISL_1674037, EPI_ISL_1674038, EPI_ISL_1674039, EPI_ISL_1674040, EPI_ISL_1674041, EPI_ISL_1674042, EPI_ISL_1674043, EPI_ISL_1674044, EPI_ISL_1674045, EPI_ISL_1674046, EPI_ISL_1674047, EPI_ISL_1674048, EPI_ISL_1674049, EPI_ISL_1674050, EPI_ISL_1674051, EPI_ISL_1674052, EPI_ISL_1674053, EPI_ISL_1674054, EPI_ISL_1674055, EPI_ISL_1674056, EPI_ISL_1674057, EPI_ISL_1674058, EPI_ISL_1674059, EPI_ISL_1674060, EPI_ISL_1674061, EPI_ISL_1674062, EPI_ISL_1674063, EPI_ISL_1674064, EPI_ISL_1674065, EPI_ISL_1674066, EPI_ISL_1674067, EPI_ISL_1674068, EPI_ISL_1674069, EPI_ISL_1674070, EPI_ISL_1674071, EPI_ISL_1674072, EPI_ISL_1674073, EPI_ISL_1674074, EPI_ISL_1674075, EPI_ISL_1674076, EPI_ISL_1674077, EPI_ISL_1674078, EPI_ISL_1674079, EPI_ISL_1674080, EPI_ISL_1674081, EPI_ISL_1674082, EPI_ISL_1674083, EPI_ISL_1674084, EPI_ISL_1674085, EPI_ISL_1674086, EPI_ISL_1674087, EPI_ISL_1674088, EPI_ISL_1674089, EPI_ISL_1674090, EPI_ISL_1674091, EPI_ISL_1674092, EPI_ISL_1674093, EPI_ISL_1674094, EPI_ISL_1674095, EPI_ISL_1674096, EPI_ISL_1674097, EPI_ISL_1674098, EPI_ISL_1674099, EPI_ISL_1674100, EPI_ISL_1674101, EPI_ISL_1674102, EPI_ISL_1674103, EPI_ISL_1674104, EPI_ISL_1674105, EPI_ISL_1674106, EPI_ISL_1674107, EPI_ISL_1674108, EPI_ISL_1674109, EPI_ISL_1674110, EPI_ISL_1674111, EPI_ISL_1674112, EPI_ISL_1674113, EPI_ISL_1674114, EPI_ISL_1674115, EPI_ISL_1674116, EPI_ISL_1674117, EPI_ISL_1674118, EPI_ISL_1674119, EPI_ISL_1674120, EPI_ISL_1674121, EPI_ISL_1674122, EPI_ISL_1674123, EPI_ISL_1674124, EPI_ISL_1674125, EPI_ISL_1674126, EPI_ISL_1674127, EPI_ISL_1674128, EPI_ISL_1674129, EPI_ISL_1674130, EPI_ISL_1674131, EPI_ISL_1674132, EPI_ISL_1674133, EPI_ISL_1674134, EPI_ISL_1674135, EPI_ISL_1674136, EPI_ISL_1674137, EPI_ISL_1674138, EPI_ISL_1674139, EPI_ISL_1674140, EPI_ISL_1674141, EPI_ISL_1674142, EPI_ISL_1674143, EPI_ISL_1674144, EPI_ISL_1674145, EPI_ISL_1674146, EPI_ISL_1674147, EPI_ISL_1674148, EPI_ISL_1674149, EPI_ISL_1674150, EPI_ISL_1674151, EPI_ISL_1674152, EPI_ISL_1674153, EPI_ISL_1674154, EPI_ISL_1674155, EPI_ISL_1674156, EPI_ISL_1674157, EPI_ISL_1674158, EPI_ISL_1674159, EPI_ISL_1674160, EPI_ISL_1674161, EPI_ISL_1674162, EPI_ISL_1674163, EPI_ISL_1674164, EPI_ISL_1674165, EPI_ISL_1674166, EPI_ISL_1674167, EPI_ISL_1674168, EPI_ISL_1674169, EPI_ISL_1674170, EPI_ISL_1674171, EPI_ISL_1674172, EPI_ISL_1674173, EPI_ISL_1674174, EPI_ISL_1674175, EPI_ISL_1674176, EPI_ISL_1674177, EPI_ISL_1674178, EPI_ISL_1674179, EPI_ISL_1674180, EPI_ISL_1674181, EPI_ISL_1674182, EPI_ISL_1674183, EPI_ISL_1674184, EPI_ISL_1674185, EPI_ISL_1674186, EPI_ISL_1674187, EPI_ISL_1674188, EPI_ISL_1674189, EPI_ISL_1674190, EPI_ISL_1674191, EPI_ISL_1674192, EPI_ISL_1674193, EPI_ISL_1674194, EPI_ISL_1674195, EPI_ISL_1674196, EPI_ISL_1674197, EPI_ISL_1674198, EPI_ISL_1674199, EPI_ISL_1674200, EPI_ISL_1674201, EPI_ISL_1674202, EPI_ISL_1674203, EPI_ISL_1674204, EPI_ISL_1674205, EPI_ISL_1674206, EPI_ISL_1674207, EPI_ISL_1674208, EPI_ISL_1674209, EPI_ISL_1674210, EPI_ISL_1674211, EPI_ISL_1674212, EPI_ISL_1674213, EPI_ISL_1674214, EPI_ISL_1674215, EPI_ISL_1674216, EPI_ISL_1674217, EPI_ISL_1674218, EPI_ISL_1674219, EPI_ISL_1674220, EPI_ISL_1674221, EPI_ISL_1674222, EPI_ISL_1674223, EPI_ISL_1674224, EPI_ISL_1674225, EPI_ISL_1674226, EPI_ISL_1674227, EPI_ISL_1674228, EPI_ISL_1674229, EPI_ISL_1674230, EPI_ISL_1674231, EPI_ISL_1674232, EPI_ISL_1674233, EPI_ISL_1674234, EPI_ISL_1674235, EPI_ISL_1674236, EPI_ISL_1674237, EPI_ISL_1674238, EPI_ISL_1674239, EPI_ISL_1674240, EPI_ISL_1674241, EPI_ISL_1674242, EPI_ISL_1674243, EPI_ISL_1674244, EPI_ISL_1674245, EPI_ISL_1674246, EPI_ISL_1674247, EPI_ISL_1674248, EPI_ISL_1674249, EPI_ISL_1674250, EPI_ISL_1674251, EPI_ISL_1674252, EPI_ISL_1674253, EPI_ISL_1674254, EPI_ISL_1674255, EPI_ISL_1674256, EPI_ISL_1674257, EPI_ISL_1674258, EPI_ISL_1674259, EPI_ISL_1674260, EPI_ISL_1674261, EPI_ISL_1674262, EPI_ISL_1674263, EPI_ISL_1674264, EPI_ISL_1674265, EPI_ISL_1674266, EPI_ISL_1674267, EPI_ISL_1674268, EPI_ISL_1674269, EPI_ISL_1674270, EPI_ISL_1674271, EPI_ISL_1674272, EPI_ISL_1674273, EPI_ISL_1674274, EPI_ISL_1674275, EPI_ISL_1674276, EPI_ISL_1674277, EPI_ISL_1674278, EPI_ISL_1674279, EPI_ISL_1674280, EPI_ISL_1674281, EPI_ISL_1674282, EPI_ISL_1674283, EPI_ISL_1674284, EPI_ISL_1674285, EPI_ISL_1674286, EPI_ISL_1674287, EPI_ISL_1674288, EPI_ISL_1674289, EPI_ISL_1674290, EPI_ISL_1674291, EPI_ISL_1674292, EPI_ISL_1674293, EPI_ISL_1674294, EPI_ISL_1674295, EPI_ISL_1674296, EPI_ISL_1674297, EPI_ISL_1674298, EPI_ISL_1674299, EPI_ISL_1674300, EPI_ISL_1674301, EPI_ISL_1674302, EPI_ISL_1674303, EPI_ISL_1674304, EPI_ISL_1674305, EPI_ISL_1674306, EPI_ISL_1674307, EPI_ISL_1674308, EPI_ISL_1674309, EPI_ISL_1674310, EPI_ISL_1674311, EPI_ISL_1674312, EPI_ISL_1674313, EPI_ISL_1674314, EPI_ISL_1674315, EPI_ISL_1674316, EPI_ISL_1674317, EPI_ISL_1674318, EPI_ISL_1674319, EPI_ISL_1674320, EPI_ISL_1674321, EPI_ISL_1674322, EPI_ISL_1674323, EPI_ISL_1674324, EPI_ISL_1674325, EPI_ISL_1674326, EPI_ISL_1674327, EPI_ISL_1674328, EPI_ISL_1674329, EPI_ISL_1674330, EPI_ISL_1674331, EPI_ISL_1674332, EPI_ISL_1674333, EPI_ISL_1674334, EPI_ISL_1674335, EPI_ISL_1674336, EPI_ISL_1674337, EPI_ISL_1674338, EPI_ISL_1674339, EPI_ISL_1674340, EPI_ISL_1674341, EPI_ISL_1674342, EPI_ISL_1674343, EPI_ISL_1674344, EPI_ISL_1674345, EPI_ISL_1674346, EPI_ISL_1674347, EPI_ISL_1674348, EPI_ISL_1674349, EPI_ISL_1674350, EPI_ISL_1674351, EPI_ISL_1674352, EPI_ISL_1674353, EPI_ISL_1674354, EPI_ISL_1674355, EPI_ISL_1674356, EPI_ISL_1674357, EPI_ISL_1674358, EPI_ISL_1674359, EPI_ISL_1674360, EPI_ISL_1674361, EPI_ISL_1674362, EPI_ISL_1674363, EPI_ISL_1674364, EPI_ISL_1674365, EPI_ISL_1674366, EPI_ISL_1674367, EPI_ISL_1674368, EPI_ISL_1674369, EPI_ISL_1674370, EPI_ISL_1674371, EPI_ISL_1674372, EPI_ISL_1674373, EPI_ISL_1674374, EPI_ISL_1674375, EPI_ISL_1674376, EPI_ISL_1674377, EPI_ISL_1674378, EPI_ISL_1674379, EPI_ISL_1674380, EPI_ISL_1674381, EPI_ISL_1674382, EPI_ISL_1674383, EPI_ISL_1674384, EPI_ISL_1674385, EPI_ISL_1674386, EPI_ISL_1674387, EPI_ISL_1674388, EPI_ISL_1674389, EPI_ISL_1674390, EPI_ISL_1674391, EPI_ISL_1674392, EPI_ISL_1674393, EPI_ISL_1674394, EPI_ISL_1674395, EPI_ISL_1674396, EPI_ISL_1674397, EPI_ISL_1674398, EPI_ISL_1674399, EPI_ISL_1674400, EPI_ISL_1674401, EPI_ISL_1674402, EPI_ISL_1674403, EPI_ISL_1674404, EPI_ISL_1674405, EPI_ISL_1674406, EPI_ISL_1674407, EPI_ISL_1674408, EPI_ISL_1674409, EPI_ISL_1674410, EPI_ISL_1674411, EPI_ISL_1674412, EPI_ISL_1674413, EPI_ISL_1674414, EPI_ISL_1674415, EPI_ISL_1674416, EPI_ISL_1674417, EPI_ISL_1674418, EPI_ISL_1674419, EPI_ISL_1674420, EPI_ISL_1674421, EPI_ISL_1674422, EPI_ISL_1674423, EPI_ISL_1674424, EPI_ISL_1674425, EPI_ISL_1674426, EPI_ISL_1674427, EPI_ISL_1674428, EPI_ISL_1674429, EPI_ISL_1674430, EPI_ISL_1674431, EPI_ISL_1674432, EPI_ISL_1674433, EPI_ISL_1674434, EPI_ISL_1674435, EPI_ISL_1674436, EPI_ISL_1674437, EPI_ISL_1674438, EPI_ISL_1674439, EPI_ISL_1674440, EPI_ISL_1674441, EPI_ISL_1674442, EPI_ISL_1674443, EPI_ISL_1674444, EPI_ISL_1674445, EPI_ISL_1674446, EPI_ISL_1674447, EPI_ISL_1674448, EPI_ISL_1674449, EPI_ISL_1674450, EPI_ISL_1674451, EPI_ISL_1674452, EPI_ISL_1674453, EPI_ISL_1674454, EPI_ISL_1674455, EPI_ISL_1674456, EPI_ISL_1674457, EPI_ISL_1674458, EPI_ISL_1674459, EPI_ISL_1674460, EPI_ISL_1674461, EPI_ISL_1674462, EPI_ISL_1674463, EPI_ISL_1674464, EPI_ISL_1674465, EPI_ISL_1674466, EPI_ISL_1674467, EPI_ISL_1674468, EPI_ISL_1674469, EPI_ISL_1674470, EPI_ISL_1674471, EPI_ISL_1674472, EPI_ISL_1674473, EPI_ISL_1674474, EPI_ISL_1674475, EPI_ISL_1674476, EPI_ISL_1674477, EPI_ISL_1674478, EPI_ISL_1674479, EPI_ISL_1674480, EPI_ISL_1674481, EPI_ISL_1674482, EPI_ISL_1674483, EPI_ISL_1674484, EPI_ISL_1674485, EPI_ISL_1674486, EPI_ISL_1674487, EPI_ISL_1674488, EPI_ISL_1674489, EPI_ISL_1674490, EPI_ISL_1674491, EPI_ISL_1674492, EPI_ISL_1674493, EPI_ISL_1674494, EPI_ISL_1674495, EPI_ISL_1674496, EPI_ISL_1674497, EPI_ISL_1674498, EPI_ISL_1674499, EPI_ISL_1674500, EPI_ISL_1674501, EPI_ISL_1674502, EPI_ISL_1674503, EPI_ISL_1674504, EPI_ISL_1674505, EPI_ISL_1674506, EPI_ISL_1674507, EPI_ISL_1674508, EPI_ISL_1674509, EPI_ISL_1674510, EPI_ISL_1674511, EPI_ISL_1674512, EPI_ISL_1674513, EPI_ISL_1674514, EPI_ISL_1674515, EPI_ISL_1674516, EPI_ISL_1674517, EPI_ISL_1674518, EPI_ISL_1674519, EPI_ISL_1674520, EPI_ISL_1674521, EPI_ISL_1674522, EPI_ISL_1674523, EPI_ISL_1674524, EPI_ISL_1674525, EPI_ISL_1674526, EPI_ISL_1674527, EPI_ISL_1674528, EPI_ISL_1674529, EPI_ISL_1674530

| | | | |
|---|---|---|---|
| see above | Israel Central Virology laboratory | Israel National Consortium for SARS-CoV-2 sequencing | Neta Zuckerman, Efrat Dahan Bucris, Michal Mandelboim, Dana Bar-Ilan, Oran Erster, Tzvia Mann, Omer Murik, David A. Zeevi, Assaf Rokney, Joseph Jaffe, Eva Nachum, Maya Davidovich Cohen, Ephraim Fass, Gal Zizelski Valenci, Mor Rubinstein, Efrat Rorman, Israel Nissan, Efrat Glick-Saar, Omri Nayshool, Gideon Rechavi, Ella Mendelson, Orna Mor |

EPI_ISL_1762089, EPI_ISL_1762090, EPI_ISL_1762091, EPI_ISL_1762092, EPI_ISL_1762093, EPI_ISL_1762094, EPI_ISL_1762095, EPI_ISL_1762096, EPI_ISL_1762097, EPI_ISL_1762098, EPI_ISL_1762099, EPI_ISL_1762100, EPI_ISL_1762101, EPI_ISL_1762102, EPI_ISL_1762103, EPI_ISL_1762104, EPI_ISL_1762105, EPI_ISL_1762106, EPI_ISL_1762107, EPI_ISL_1762108, EPI_ISL_1762109, EPI_ISL_1762110, EPI_ISL_1762111, EPI_ISL_1762112, EPI_ISL_1762113, EPI_ISL_1762114, EPI_ISL_1762115, EPI_ISL_1762116, EPI_ISL_1762117, EPI_ISL_1762118, EPI_ISL_1762119, EPI_ISL_1762120, EPI_ISL_1762121, EPI_ISL_1762122, EPI_ISL_1762123, EPI_ISL_1762124, EPI_ISL_1762125, EPI_ISL_1762126, EPI_ISL_1762127, EPI_ISL_1762128, EPI_ISL_1762129, EPI_ISL_1762130, EPI_ISL_1762131, EPI_ISL_1762132, EPI_ISL_1762133, EPI_ISL_1762134, EPI_ISL_1762135, EPI_ISL_1762136, EPI_ISL_1762137, EPI_ISL_1762138, EPI_ISL_1762139, EPI_ISL_1762140, EPI_ISL_1762141, EPI_ISL_1762142, EPI_ISL_1762143, EPI_ISL_1762144, EPI_ISL_1762145, EPI_ISL_1762146, EPI_ISL_1762147, EPI_ISL_1762148, EPI_ISL_1762149, EPI_ISL_1762150, EPI_ISL_1762151, EPI_ISL_1762152, EPI_ISL_1762153, EPI_ISL_1762154, EPI_ISL_1762155, EPI_ISL_1762156, EPI_ISL_1762157, EPI_ISL_1762158, EPI_ISL_1762159, EPI_ISL_1762160, EPI_ISL_1762161, EPI_ISL_1762162, EPI_ISL_1762163, EPI_ISL_1762164, EPI_ISL_1762165, EPI_ISL_1762166, EPI_ISL_1762167, EPI_ISL_1762168, EPI_ISL_1762169, EPI_ISL_1762170, EPI_ISL_1762171, EPI_ISL_1762172, EPI_ISL_1762173, EPI_ISL_1762174, EPI_ISL_1762175, EPI_ISL_1762176, EPI_ISL_1762177, EPI_ISL_1762178, EPI_ISL_1762179, EPI_ISL_1762180, EPI_ISL_1762181, EPI_ISL_1762182, EPI_ISL_1762183, EPI_ISL_1762184, EPI_ISL_1762185, EPI_ISL_1762186, EPI_ISL_1762187, EPI_ISL_1762188, EPI_ISL_1762189, EPI_ISL_1762190, EPI_ISL_1762191, EPI_ISL_1762192, EPI_ISL_1762193, EPI_ISL_1762194, EPI_ISL_1762195, EPI_ISL_1762196, EPI_ISL_1762197, EPI_ISL_1762198, EPI_ISL_1762199, EPI_ISL_1762200, EPI_ISL_1762201, EPI_ISL_1762202, EPI_ISL_1762203, EPI_ISL_1762204, EPI_ISL_1762205, EPI_ISL_1762206, EPI_ISL_1762207, EPI_ISL_1762208, EPI_ISL_1762209, EPI_ISL_1762210, EPI_ISL_1762211, EPI_ISL_1762212, EPI_ISL_1762213, EPI_ISL_1762214, EPI_ISL_1762215, EPI_ISL_1762216, EPI_ISL_1762217, EPI_ISL_1762218, EPI_ISL_1762219, EPI_ISL_1762220, EPI_ISL_1762221, EPI_ISL_1762222, EPI_ISL_1762223, EPI_ISL_1762224, EPI_ISL_1762225, EPI_ISL_1762226, EPI_ISL_1762227, EPI_ISL_1762228, EPI_ISL_1762229, EPI_ISL_1762230, EPI_ISL_1762231, EPI_ISL_1762232, EPI_ISL_1762233, EPI_ISL_1762234, EPI_ISL_1762235, EPI_ISL_1762236, EPI_ISL_1762237, EPI_ISL_1762238, EPI_ISL_1762239, EPI_ISL_1762240, EPI_ISL_1762241, EPI_ISL_1762242, EPI_ISL_1762243, EPI_ISL_1762244, EPI_ISL_1762245, EPI_ISL_1762246, EPI_ISL_1762247, EPI_ISL_1762248, EPI_ISL_1762249, EPI_ISL_1762250, EPI_ISL_1762251, EPI_ISL_1762252, EPI_ISL_1762253, EPI_ISL_1762254, EPI_ISL_1762255, EPI_ISL_1762256, EPI_ISL_1762257, EPI_ISL_1762258, EPI_ISL_1762259, EPI_ISL_1762260, EPI_ISL_1762261, EPI_ISL_1762262, EPI_ISL_1762263, EPI_ISL_1762264, EPI_ISL_1762265, EPI_ISL_1762266, EPI_ISL_1762267, EPI_ISL_1762268, EPI_ISL_1762269, EPI_ISL_1762270, EPI_ISL_1762271, EPI_ISL_1762272, EPI_ISL_1762273, EPI_ISL_1762274, EPI_ISL_1762275, EPI_ISL_1762276, EPI_ISL_1762277, EPI_ISL_1762278, EPI_ISL_1762279, EPI_ISL_1762280, EPI_ISL_1762281, EPI_ISL_1762282, EPI_ISL_1762283, EPI_ISL_1762284, EPI_ISL_1762285, EPI_ISL_1762286, EPI_ISL_1762287, EPI_ISL_1762288, EPI_ISL_1762289, EPI_ISL_1762290, EPI_ISL_1762291, EPI_ISL_1762292,

EPI_ISL_1762293, EPI_ISL_1762294, EPI_ISL_1762295, EPI_ISL_1762296, EPI_ISL_1762297, EPI_ISL_1762298, EPI_ISL_1762299, EPI_ISL_1762300, EPI_ISL_1762301, EPI_ISL_1762302, EPI_ISL_1762303, EPI_ISL_1762304, EPI_ISL_1762305, EPI_ISL_1762306, EPI_ISL_1762307, EPI_ISL_1762308, EPI_ISL_1762309,
EPI_ISL_1762310, EPI_ISL_1762311, EPI_ISL_1762312, EPI_ISL_1762313, EPI_ISL_1762314, EPI_ISL_1762315, EPI_ISL_1762316, EPI_ISL_1762317, EPI_ISL_1762318, EPI_ISL_1762319, EPI_ISL_1762320, EPI_ISL_1762321, EPI_ISL_1762322, EPI_ISL_1762323, EPI_ISL_1762324, EPI_ISL_1762325, EPI_ISL_1762326,
EPI_ISL_1762327, EPI_ISL_1762328, EPI_ISL_1762329, EPI_ISL_1762330, EPI_ISL_1762331, EPI_ISL_1762332, EPI_ISL_1762333, EPI_ISL_1762334, EPI_ISL_1762335, EPI_ISL_1762336, EPI_ISL_1762337, EPI_ISL_1762338, EPI_ISL_1762339, EPI_ISL_1762340, EPI_ISL_1762341, EPI_ISL_1762342, EPI_ISL_1762343,
EPI_ISL_1762344, EPI_ISL_1762345, EPI_ISL_1762346, EPI_ISL_1762347, EPI_ISL_1762348, EPI_ISL_1762349, EPI_ISL_1762350, EPI_ISL_1762351, EPI_ISL_1762352, EPI_ISL_1762353, EPI_ISL_1762354, EPI_ISL_1762355, EPI_ISL_1762356, EPI_ISL_1762357, EPI_ISL_1762358, EPI_ISL_1762359, EPI_ISL_1762360,
EPI_ISL_1762361, EPI_ISL_1762362, EPI_ISL_1762363, EPI_ISL_1762364, EPI_ISL_1762365, EPI_ISL_1762366, EPI_ISL_1762367, EPI_ISL_1762368, EPI_ISL_1762369, EPI_ISL_1762370, EPI_ISL_1762371, EPI_ISL_1762372, EPI_ISL_1762373, EPI_ISL_1762374, EPI_ISL_1762375, EPI_ISL_1762376, EPI_ISL_1762377,
EPI_ISL_1762378, EPI_ISL_1762379, EPI_ISL_1762380, EPI_ISL_1762381, EPI_ISL_1762382, EPI_ISL_1762383, EPI_ISL_1762384, EPI_ISL_1762385, EPI_ISL_1762386, EPI_ISL_1762387, EPI_ISL_1762388, EPI_ISL_1762389, EPI_ISL_1762390, EPI_ISL_1762391, EPI_ISL_1762392, EPI_ISL_1762393, EPI_ISL_1762394,
EPI_ISL_1762395, EPI_ISL_1762396, EPI_ISL_1762397, EPI_ISL_1762398, EPI_ISL_1762399, EPI_ISL_1762400, EPI_ISL_1762401, EPI_ISL_1762402, EPI_ISL_1762403, EPI_ISL_1762404, EPI_ISL_1762405, EPI_ISL_1762406, EPI_ISL_1762407, EPI_ISL_1762408, EPI_ISL_1762409, EPI_ISL_1762410, EPI_ISL_1762411,
EPI_ISL_1762412, EPI_ISL_1762413, EPI_ISL_1762414, EPI_ISL_1762415, EPI_ISL_1762416, EPI_ISL_1762417, EPI_ISL_1762418, EPI_ISL_1762419, EPI_ISL_1762420, EPI_ISL_1762421, EPI_ISL_1762422, EPI_ISL_1762423, EPI_ISL_1762424, EPI_ISL_1762425, EPI_ISL_1762426, EPI_ISL_1762427, EPI_ISL_1762428,
EPI_ISL_1762429, EPI_ISL_1762430, EPI_ISL_1762431, EPI_ISL_1762432, EPI_ISL_1762433, EPI_ISL_1762434, EPI_ISL_1762435, EPI_ISL_1762436, EPI_ISL_1762437, EPI_ISL_1762438, EPI_ISL_1762439, EPI_ISL_1762440, EPI_ISL_1762441, EPI_ISL_1762442, EPI_ISL_1762443, EPI_ISL_1762444, EPI_ISL_1762445,
EPI_ISL_1762446, EPI_ISL_1762447, EPI_ISL_1762448, EPI_ISL_1762449, EPI_ISL_1762450, EPI_ISL_1762451, EPI_ISL_1762452, EPI_ISL_1762453, EPI_ISL_1762454, EPI_ISL_1762455, EPI_ISL_1762456, EPI_ISL_1762457, EPI_ISL_1762458, EPI_ISL_1762459, EPI_ISL_1762460, EPI_ISL_1762461, EPI_ISL_1762462,
EPI_ISL_1762463, EPI_ISL_1762464, EPI_ISL_1762465, EPI_ISL_1762466, EPI_ISL_1762467, EPI_ISL_1762468, EPI_ISL_1762469, EPI_ISL_1762470, EPI_ISL_1762471, EPI_ISL_1762472, EPI_ISL_1762473, EPI_ISL_1762474, EPI_ISL_1762475, EPI_ISL_1762476, EPI_ISL_1762477, EPI_ISL_1762478, EPI_ISL_1762479,
EPI_ISL_1762480, EPI_ISL_1762481, EPI_ISL_1762482, EPI_ISL_1762483, EPI_ISL_1762484, EPI_ISL_1762485, EPI_ISL_1762486, EPI_ISL_1762487, EPI_ISL_1762488, EPI_ISL_1762489, EPI_ISL_1762490, EPI_ISL_1762491, EPI_ISL_1762492, EPI_ISL_1762493, EPI_ISL_1762494, EPI_ISL_1762495, EPI_ISL_1762496,
EPI_ISL_1762497, EPI_ISL_1762498, EPI_ISL_1762499, EPI_ISL_1762500, EPI_ISL_1762501, EPI_ISL_1762502, EPI_ISL_1762503, EPI_ISL_1762504, EPI_ISL_1762505, EPI_ISL_1762506, EPI_ISL_1762507, EPI_ISL_1762508, EPI_ISL_1762509, EPI_ISL_1762510, EPI_ISL_1762511, EPI_ISL_1762512, EPI_ISL_1762513,
EPI_ISL_1762514, EPI_ISL_1762515, EPI_ISL_1762516, EPI_ISL_1762517, EPI_ISL_1762518, EPI_ISL_1762519, EPI_ISL_1762520, EPI_ISL_1762521, EPI_ISL_1762522, EPI_ISL_1762523, EPI_ISL_1762524, EPI_ISL_1762525, EPI_ISL_1762526, EPI_ISL_1762527, EPI_ISL_1762528, EPI_ISL_1762529, EPI_ISL_1762530,
EPI_ISL_1762531, EPI_ISL_1762532, EPI_ISL_1762533, EPI_ISL_1762534, EPI_ISL_1762535, EPI_ISL_1762536, EPI_ISL_1762537, EPI_ISL_1762538, EPI_ISL_1762539, EPI_ISL_1762540, EPI_ISL_1762541, EPI_ISL_1762542, EPI_ISL_1762543, EPI_ISL_1762544, EPI_ISL_1762545, EPI_ISL_1762546, EPI_ISL_1762547,
EPI_ISL_1762548, EPI_ISL_1762549, EPI_ISL_1762550, EPI_ISL_1762551, EPI_ISL_1762552, EPI_ISL_1762553, EPI_ISL_1762554, EPI_ISL_1762555, EPI_ISL_1762556, EPI_ISL_1762557, EPI_ISL_1762558, EPI_ISL_1762559, EPI_ISL_1762560, EPI_ISL_1762561, EPI_ISL_1762562, EPI_ISL_1762563, EPI_ISL_1762564,
EPI_ISL_1762565, EPI_ISL_1762566, EPI_ISL_1762567, EPI_ISL_1762568, EPI_ISL_1762569, EPI_ISL_1762570, EPI_ISL_1762571, EPI_ISL_1762572, EPI_ISL_1762573, EPI_ISL_1762574, EPI_ISL_1762575, EPI_ISL_1762576, EPI_ISL_1762577, EPI_ISL_1762578, EPI_ISL_1762579, EPI_ISL_1762580, EPI_ISL_1762581,
EPI_ISL_1762582, EPI_ISL_1762583, EPI_ISL_1762584, EPI_ISL_1762585, EPI_ISL_1762586, EPI_ISL_1762587, EPI_ISL_1762588, EPI_ISL_1762589, EPI_ISL_1762590, EPI_ISL_1762591, EPI_ISL_1762592, EPI_ISL_1762593, EPI_ISL_1762594, EPI_ISL_1762595, EPI_ISL_1762596, EPI_ISL_1762597, EPI_ISL_1762598,
EPI_ISL_1762599, EPI_ISL_1762600, EPI_ISL_1762601, EPI_ISL_1762602, EPI_ISL_1762603, EPI_ISL_1762604, EPI_ISL_1762605, EPI_ISL_1762606, EPI_ISL_1762607, EPI_ISL_1762608, EPI_ISL_1762609, EPI_ISL_1762610, EPI_ISL_1762611, EPI_ISL_1762612, EPI_ISL_1762613, EPI_ISL_1762614, EPI_ISL_1762615,
EPI_ISL_1762616, EPI_ISL_1762617, EPI_ISL_1762618, EPI_ISL_1762619, EPI_ISL_1762620, EPI_ISL_1762621, EPI_ISL_1762622, EPI_ISL_1762623, EPI_ISL_1762624, EPI_ISL_1762625, EPI_ISL_1762626, EPI_ISL_1762627, EPI_ISL_1762628, EPI_ISL_1762629, EPI_ISL_1762630, EPI_ISL_1762631, EPI_ISL_1762632,
EPI_ISL_1762633, EPI_ISL_1762634, EPI_ISL_1762635, EPI_ISL_1762636, EPI_ISL_1762637, EPI_ISL_1762638, EPI_ISL_1762639, EPI_ISL_1762640, EPI_ISL_1762641, EPI_ISL_1762642, EPI_ISL_1762643, EPI_ISL_1762644, EPI_ISL_1762645, EPI_ISL_1762646, EPI_ISL_1762647, EPI_ISL_1762648, EPI_ISL_1762649,
EPI_ISL_1762650, EPI_ISL_1762651, EPI_ISL_1762652, EPI_ISL_1762653, EPI_ISL_1762654, EPI_ISL_1762655, EPI_ISL_1762656, EPI_ISL_1762657, EPI_ISL_1762658, EPI_ISL_1762659, EPI_ISL_1762660, EPI_ISL_1762661, EPI_ISL_1762662, EPI_ISL_1762663, EPI_ISL_1762664, EPI_ISL_1762665, EPI_ISL_1762666,
EPI_ISL_1762667, EPI_ISL_1762668, EPI_ISL_1762669, EPI_ISL_1762670, EPI_ISL_1762671, EPI_ISL_1762672, EPI_ISL_1762673, EPI_ISL_1762674, EPI_ISL_1762675, EPI_ISL_1762676, EPI_ISL_1762677, EPI_ISL_1762678, EPI_ISL_1762679, EPI_ISL_1762680, EPI_ISL_1762681, EPI_ISL_1762682, EPI_ISL_1762683,
EPI_ISL_1762684, EPI_ISL_1762685, EPI_ISL_1762686, EPI_ISL_1762687, EPI_ISL_1762688, EPI_ISL_1762689, EPI_ISL_1762690, EPI_ISL_1762691, EPI_ISL_1762692, EPI_ISL_1762693, EPI_ISL_1762694, EPI_ISL_1762695, EPI_ISL_1762696, EPI_ISL_1762697, EPI_ISL_1762698, EPI_ISL_1762699, EPI_ISL_1762700,
EPI_ISL_1762701, EPI_ISL_1762702, EPI_ISL_1762703, EPI_ISL_1762704, EPI_ISL_1762705, EPI_ISL_1762706, EPI_ISL_1762707, EPI_ISL_1762708, EPI_ISL_1762709, EPI_ISL_1762710, EPI_ISL_1762711, EPI_ISL_1762712, EPI_ISL_1762713, EPI_ISL_1762714, EPI_ISL_1762715, EPI_ISL_1762716, EPI_ISL_1762717,
EPI_ISL_1762718, EPI_ISL_1762719, EPI_ISL_1762720, EPI_ISL_1762721, EPI_ISL_1762722, EPI_ISL_1762723, EPI_ISL_1762724, EPI_ISL_1762725, EPI_ISL_1762726, EPI_ISL_1762727, EPI_ISL_1762728, EPI_ISL_1762729, EPI_ISL_1762730, EPI_ISL_1762731, EPI_ISL_1762732, EPI_ISL_1762733, EPI_ISL_1762734,
EPI_ISL_1762735, EPI_ISL_1762736, EPI_ISL_1762737, EPI_ISL_1762738, EPI_ISL_1762739, EPI_ISL_1762740, EPI_ISL_1762741, EPI_ISL_1762742, EPI_ISL_1762743, EPI_ISL_1762744, EPI_ISL_1762745, EPI_ISL_1762746, EPI_ISL_1762747, EPI_ISL_1762748, EPI_ISL_1762749, EPI_ISL_1762750, EPI_ISL_1762751,
EPI_ISL_1762752, EPI_ISL_1762753, EPI_ISL_1762754, EPI_ISL_1762755, EPI_ISL_1762756, EPI_ISL_1762757, EPI_ISL_1762758, EPI_ISL_1762759, EPI_ISL_1762760, EPI_ISL_1762761, EPI_ISL_1762762, EPI_ISL_1762763, EPI_ISL_1762764, EPI_ISL_1762765, EPI_ISL_1762766, EPI_ISL_1762767, EPI_ISL_1762768,
EPI_ISL_1762769, EPI_ISL_1762770, EPI_ISL_1762771, EPI_ISL_1762772, EPI_ISL_1762773, EPI_ISL_1762774, EPI_ISL_1762775, EPI_ISL_1762776, EPI_ISL_1762777, EPI_ISL_1762778, EPI_ISL_1762779, EPI_ISL_1762780, EPI_ISL_1762781, EPI_ISL_1762782, EPI_ISL_1762783, EPI_ISL_1762784, EPI_ISL_1762785,
EPI_ISL_1762786, EPI_ISL_1762787, EPI_ISL_1762788, EPI_ISL_1762789, EPI_ISL_1762790, EPI_ISL_1762791, EPI_ISL_1762792, EPI_ISL_1762793, EPI_ISL_1762794, EPI_ISL_1762795, EPI_ISL_1762796, EPI_ISL_1762797, EPI_ISL_1762798, EPI_ISL_1762799, EPI_ISL_1762800, EPI_ISL_1762801, EPI_ISL_1762802,
EPI_ISL_1762803, EPI_ISL_1762804, EPI_ISL_1762805, EPI_ISL_1762806, EPI_ISL_1762807, EPI_ISL_1762808, EPI_ISL_1762809, EPI_ISL_1762810, EPI_ISL_1762811, EPI_ISL_1762812, EPI_ISL_1762813, EPI_ISL_1762814, EPI_ISL_1762815, EPI_ISL_1762816, EPI_ISL_1762817, EPI_ISL_1762818, EPI_ISL_1762819,
EPI_ISL_1762820, EPI_ISL_1762821, EPI_ISL_1762822, EPI_ISL_1762823, EPI_ISL_1762824, EPI_ISL_1762825, EPI_ISL_1762826, EPI_ISL_1762827, EPI_ISL_1762828, EPI_ISL_1762829, EPI_ISL_1762830, EPI_ISL_1762831, EPI_ISL_1762832, EPI_ISL_1762833, EPI_ISL_1762834, EPI_ISL_1762835, EPI_ISL_1762836,
EPI_ISL_1762837, EPI_ISL_1762838, EPI_ISL_1762839, EPI_ISL_1762840, EPI_ISL_1762841, EPI_ISL_1762842, EPI_ISL_1762843, EPI_ISL_1762844, EPI_ISL_1762845, EPI_ISL_1762846, EPI_ISL_1762847, EPI_ISL_1762848, EPI_ISL_1762849, EPI_ISL_1762850, EPI_ISL_1762851, EPI_ISL_1762852, EPI_ISL_1762853,
EPI_ISL_1762854, EPI_ISL_1762855, EPI_ISL_1762856, EPI_ISL_1762857, EPI_ISL_1762858, EPI_ISL_1762859, EPI_ISL_1762860, EPI_ISL_1762861, EPI_ISL_1762862, EPI_ISL_1762863, EPI_ISL_1762864, EPI_ISL_1762865, EPI_ISL_1762866, EPI_ISL_1762867, EPI_ISL_1762868, EPI_ISL_1762869, EPI_ISL_1762870,
EPI_ISL_1762871, EPI_ISL_1762872, EPI_ISL_1762873, EPI_ISL_1762874, EPI_ISL_1762875, EPI_ISL_1762876, EPI_ISL_1762877, EPI_ISL_1762878, EPI_ISL_1762879, EPI_ISL_1762880, EPI_ISL_1762881, EPI_ISL_1762882, EPI_ISL_1762883, EPI_ISL_1762884, EPI_ISL_1762885, EPI_ISL_1762886, EPI_ISL_1762887,
EPI_ISL_1762888, EPI_ISL_1762889, EPI_ISL_1762890, EPI_ISL_1762891, EPI_ISL_1762892, EPI_ISL_1762893, EPI_ISL_1762894, EPI_ISL_1762895, EPI_ISL_1762896, EPI_ISL_1762897, EPI_ISL_1762898, EPI_ISL_1762899, EPI_ISL_1762900, EPI_ISL_1762901, EPI_ISL_1762902, EPI_ISL_1762903, EPI_ISL_1762904,
EPI_ISL_1762905, EPI_ISL_1762906, EPI_ISL_1762907, EPI_ISL_1762908, EPI_ISL_1762909, EPI_ISL_1762910, EPI_ISL_1762911, EPI_ISL_1762912, EPI_ISL_1762913, EPI_ISL_1762914, EPI_ISL_1762915, EPI_ISL_1762916, EPI_ISL_1762917, EPI_ISL_1762918, EPI_ISL_1762919, EPI_ISL_1762920, EPI_ISL_1762921,
EPI_ISL_1762922, EPI_ISL_1762923, EPI_ISL_1762924, EPI_ISL_1762925, EPI_ISL_1762926, EPI_ISL_1762927, EPI_ISL_1762928, EPI_ISL_1762929, EPI_ISL_1762930, EPI_ISL_1762931, EPI_ISL_1762932, EPI_ISL_1762933, EPI_ISL_1762934, EPI_ISL_1762935, EPI_ISL_1762936, EPI_ISL_1762937, EPI_ISL_1762938,
EPI_ISL_1762939, EPI_ISL_1762940, EPI_ISL_1762941, EPI_ISL_1762942, EPI_ISL_1762943, EPI_ISL_1762944, EPI_ISL_1762945, EPI_ISL_1762946, EPI_ISL_1762947, EPI_ISL_1762948, EPI_ISL_1762949, EPI_ISL_1762950, EPI_ISL_1762951, EPI_ISL_1762952, EPI_ISL_1762953, EPI_ISL_1762954, EPI_ISL_1762955,
EPI_ISL_1762956, EPI_ISL_1762957, EPI_ISL_1762958, EPI_ISL_1762959, EPI_ISL_1762960, EPI_ISL_1762961, EPI_ISL_1762962, EPI_ISL_1762963, EPI_ISL_1762964, EPI_ISL_1762965, EPI_ISL_1762966, EPI_ISL_1762967, EPI_ISL_1762968, EPI_ISL_1762969, EPI_ISL_1762970, EPI_ISL_1762971, EPI_ISL_1762972,
EPI_ISL_1762973, EPI_ISL_1762974, EPI_ISL_1762975, EPI_ISL_1762976, EPI_ISL_1762977, EPI_ISL_1762978, EPI_ISL_1762979, EPI_ISL_1762980, EPI_ISL_1762981, EPI_ISL_1762982, EPI_ISL_1762983, EPI_ISL_1762984, EPI_ISL_1762985, EPI_ISL_1762986, EPI_ISL_1762987, EPI_ISL_1762988, EPI_ISL_1762989,
EPI_ISL_1762990, EPI_ISL_1762991, EPI_ISL_1762992, EPI_ISL_1762993, EPI_ISL_1762994, EPI_ISL_1762995, EPI_ISL_1762996, EPI_ISL_1762997, EPI_ISL_1762998, EPI_ISL_1762999, EPI_ISL_1763000, EPI_ISL_1763001, EPI_ISL_1763002, EPI_ISL_1763003, EPI_ISL_1763004, EPI_ISL_1763005, EPI_ISL_1763006,
EPI_ISL_1763007, EPI_ISL_1763008, EPI_ISL_1763009, EPI_ISL_1763010, EPI_ISL_1763011, EPI_ISL_1763012, EPI_ISL_1763013, EPI_ISL_1763014, EPI_ISL_1763015, EPI_ISL_1763016, EPI_ISL_1763017, EPI_ISL_1763018, EPI_ISL_1763019, EPI_ISL_1763020, EPI_ISL_1763021, EPI_ISL_1763022, EPI_ISL_1763023,
EPI_ISL_1763024, EPI_ISL_1763025, EPI_ISL_1763026, EPI_ISL_1763027, EPI_ISL_1763028, EPI_ISL_1763029, EPI_ISL_1763030, EPI_ISL_1763031, EPI_ISL_1763032, EPI_ISL_1763033, EPI_ISL_1763034, EPI_ISL_1763035, EPI_ISL_1763036, EPI_ISL_1763037, EPI_ISL_1763038, EPI_ISL_1763039, EPI_ISL_1763040,
EPI_ISL_1763041, EPI_ISL_1763042, EPI_ISL_1763043, EPI_ISL_1763044, EPI_ISL_1763045, EPI_ISL_1763046, EPI_ISL_1763047, EPI_ISL_1763048, EPI_ISL_1763049, EPI_ISL_1763050, EPI_ISL_1763051, EPI_ISL_1763052, EPI_ISL_1763053, EPI_ISL_1763054, EPI_ISL_1763055, EPI_ISL_1763056, EPI_ISL_1763057,
EPI_ISL_1763058, EPI_ISL_1763059, EPI_ISL_1763060, EPI_ISL_1763061, EPI_ISL_1763062, EPI_ISL_1763063, EPI_ISL_1763064, EPI_ISL_1763065, EPI_ISL_1763066, EPI_ISL_1763067, EPI_ISL_1763068, EPI_ISL_1763069, EPI_ISL_1763070, EPI_ISL_1763071, EPI_ISL_1763072, EPI_ISL_1763073, EPI_ISL_1763074,
EPI_ISL_1763075, EPI_ISL_1763076, EPI_ISL_1763077, EPI_ISL_1763078, EPI_ISL_1763079, EPI_ISL_1763080, EPI_ISL_1763081, EPI_ISL_1763082, EPI_ISL_1763083, EPI_ISL_1763084, EPI_ISL_1763085, EPI_ISL_1763086, EPI_ISL_1763087, EPI_ISL_1763088, EPI_ISL_1763089, EPI_ISL_1763090, EPI_ISL_1763091,
EPI_ISL_1763092, EPI_ISL_1763093, EPI_ISL_1763094, EPI_ISL_1763095, EPI_ISL_1763096, EPI_ISL_1763097, EPI_ISL_1763098, EPI_ISL_1763099, EPI_ISL_1763100, EPI_ISL_1763101, EPI_ISL_1763102, EPI_ISL_1763103, EPI_ISL_1763104, EPI_ISL_1763105, EPI_ISL_1763106, EPI_ISL_1763107, EPI_ISL_1763108,
EPI_ISL_1763109, EPI_ISL_1763110, EPI_ISL_1763111, EPI_ISL_1763112, EPI_ISL_1763113, EPI_ISL_1763114, EPI_ISL_1763115, EPI_ISL_1763116, EPI_ISL_1763117, EPI_ISL_1763118, EPI_ISL_1763119, EPI_ISL_1763120, EPI_ISL_1763121, EPI_ISL_1763122, EPI_ISL_1763123, EPI_ISL_1763124, EPI_ISL_1763125,
EPI_ISL_1763126, EPI_ISL_1763127, EPI_ISL_1763128, EPI_ISL_1763129, EPI_ISL_1763130, EPI_ISL_1763131, EPI_ISL_1763132, EPI_ISL_1763133, EPI_ISL_1763134, EPI_ISL_1763135, EPI_ISL_1763136, EPI_ISL_1763137, EPI_ISL_1763138, EPI_ISL_1763139, EPI_ISL_1763140, EPI_ISL_1763141, EPI_ISL_1763142,
EPI_ISL_1763143, EPI_ISL_1763144, EPI_ISL_1763145, EPI_ISL_1763146, EPI_ISL_1763147, EPI_ISL_1763148, EPI_ISL_1763149, EPI_ISL_1763150, EPI_ISL_1763151, EPI_ISL_1763152, EPI_ISL_1763153, EPI_ISL_1763154, EPI_ISL_1763155, EPI_ISL_1763156, EPI_ISL_1763157, EPI_ISL_1763158, EPI_ISL_1763159,
EPI_ISL_1763160, EPI_ISL_1763161, EPI_ISL_1763162, EPI_ISL_1763163, EPI_ISL_1763164, EPI_ISL_1763165, EPI_ISL_1763166, EPI_ISL_1763167, EPI_ISL_1763168, EPI_ISL_1763169, EPI_ISL_1763170, EPI_ISL_1763171, EPI_ISL_1763172, EPI_ISL_1763173, EPI_ISL_1763174, EPI_ISL_1763175, EPI_ISL_1763176,
EPI_ISL_1763177, EPI_ISL_1763178, EPI_ISL_1763179, EPI_ISL_1763180, EPI_ISL_1763181, EPI_ISL_1763182, EPI_ISL_1763183, EPI_ISL_1763184, EPI_ISL_1763185, EPI_ISL_1763186, EPI_ISL_1763187, EPI_ISL_1763188, EPI_ISL_1763189, EPI_ISL_1763190, EPI_ISL_1763191, EPI_ISL_1763192, EPI_ISL_1763193,
EPI_ISL_1763194, EPI_ISL_1763195, EPI_ISL_1763196, EPI_ISL_1763197, EPI_ISL_1763198, EPI_ISL_1763199, EPI_ISL_1763200, EPI_ISL_1763201, EPI_ISL_1763202, EPI_ISL_1763203, EPI_ISL_1763204, EPI_ISL_1763205, EPI_ISL_1763206, EPI_ISL_1763207, EPI_ISL_1763208, EPI_ISL_1763209, EPI_ISL_1763210,
EPI_ISL_1763211, EPI_ISL_1763212, EPI_ISL_1763213, EPI_ISL_1763214, EPI_ISL_1763215, EPI_ISL_1763216, EPI_ISL_1763217, EPI_ISL_1763218, EPI_ISL_1763219, EPI_ISL_1763220, EPI_ISL_1763221, EPI_ISL_1763222, EPI_ISL_1763223, EPI_ISL_1763224, EPI_ISL_1763225, EPI_ISL_1763226, EPI_ISL_1763227,
EPI_ISL_1763228, EPI_ISL_1763229, EPI_ISL_1763230, EPI_ISL_1763231, EPI_ISL_1763232, EPI_ISL_1763233, EPI_ISL_1763234, EPI_ISL_1763235, EPI_ISL_1763236, EPI_ISL_1763237, EPI_ISL_1763238, EPI_ISL_1763239, EPI_ISL_1763240, EPI_ISL_1763241, EPI_ISL_1763242, EPI_ISL_1763243, EPI_ISL_1763244,
EPI_ISL_1763245, EPI_ISL_1763246, EPI_ISL_1763247, EPI_ISL_1763248, EPI_ISL_1763249, EPI_ISL_1763250, EPI_ISL_1763251, EPI_ISL_1763252, EPI_ISL_1763253, EPI_ISL_1763254, EPI_ISL_1763255, EPI_ISL_1763256, EPI_ISL_1763257, EPI_ISL_1763258, EPI_ISL_1763259, EPI_ISL_1763260, EPI_ISL_1763261,
EPI_ISL_1763262, EPI_ISL_1763263, EPI_ISL_1763264, EPI_ISL_1763265, EPI_ISL_1763266, EPI_ISL_1763267, EPI_ISL_1763268, EPI_ISL_1763269, EPI_ISL_1763270, EPI_ISL_1763271, EPI_ISL_1763272, EPI_ISL_1763273, EPI_ISL_1763274, EPI_ISL_1763275, EPI_ISL_1763276, EPI_ISL_1763277, EPI_ISL_1763278,
EPI_ISL_1763279, EPI_ISL_1763280, EPI_ISL_1763281, EPI_ISL_1763282, EPI_ISL_1763283, EPI_ISL_1763284, EPI_ISL_1763285, EPI_ISL_1763286, EPI_ISL_1763287, EPI_ISL_1763288, EPI_ISL_1763289, EPI_ISL_1763290, EPI_ISL_1763291, EPI_ISL_1763292, EPI_ISL_1763293, EPI_ISL_1763294, EPI_ISL_1763295,
EPI_ISL_1763296, EPI_ISL_1763297, EPI_ISL_1763298, EPI_ISL_1763299, EPI_ISL_1763300, EPI_ISL_1763301, EPI_ISL_1763302, EPI_ISL_1763303, EPI_ISL_1763304, EPI_ISL_1763305, EPI_ISL_1763306, EPI_ISL_1763307, EPI_ISL_1763308, EPI_ISL_1763309, EPI_ISL_1763310, EPI_ISL_1763311, EPI_ISL_1763312,
EPI_ISL_1763313, EPI_ISL_1763314, EPI_ISL_1763315, EPI_ISL_1763316, EPI_ISL_1763317, EPI_ISL_1763318, EPI_ISL_1763319, EPI_ISL_1763320, EPI_ISL_1763321, EPI_ISL_1763322, EPI_ISL_1763323, EPI_ISL_1763324, EPI_ISL_1763325, EPI_ISL_1763326, EPI_ISL_1763327, EPI_ISL_1763328, EPI_ISL_1763329,
EPI_ISL_1763330, EPI_ISL_1763331, EPI_ISL_1763332, EPI_ISL_1763333, EPI_ISL_1763334, EPI_ISL_1763335, EPI_ISL_1763336, EPI_ISL_1763337, EPI_ISL_1763338, EPI_ISL_1763339, EPI_ISL_1763340, EPI_ISL_1763341, EPI_ISL_1763342, EPI_ISL_1763343, EPI_ISL_1763344, EPI_ISL_1763345, EPI_ISL_1763346,
EPI_ISL_1763347, EPI_ISL_1763348, EPI_ISL_1763349, EPI_ISL_1763350, EPI_ISL_1763351, EPI_ISL_1763352, EPI_ISL_1763353, EPI_ISL_1763354, EPI_ISL_1763355, EPI_ISL_1763356, EPI_ISL_1763357, EPI_ISL_1763358, EPI_ISL_1763359, EPI_ISL_1763360, EPI_ISL_1763361, EPI_ISL_1763362, EPI_ISL_1763363,
EPI_ISL_1763364, EPI_ISL_1763365, EPI_ISL_1763366, EPI_ISL_1763367, EPI_ISL_1763368, EPI_ISL_1763369, EPI_ISL_1763370, EPI_ISL_1763371, EPI_ISL_1763372, EPI_ISL_1763373, EPI_ISL_1763374, EPI_ISL_1763375, EPI_ISL_1763376, EPI_ISL_1763377, EPI_ISL_1763378, EPI_ISL_1763379, EPI_ISL_1763380,
EPI_ISL_1763381, EPI_ISL_1763382, EPI_ISL_1763383, EPI_ISL_1763384, EPI_ISL_1763385, EPI_ISL_1763386, EPI_ISL_1763387, EPI_ISL_1763388, EPI_ISL_1763389, EPI_ISL_1763390, EPI_ISL_1763391, EPI_ISL_1763392, EPI_ISL_1763393, EPI_ISL_1763394, EPI_ISL_1763395, EPI_ISL_1763396, EPI_ISL_1763397,
EPI_ISL_1763398, EPI_ISL_1763399, EPI_ISL_1763400, EPI_ISL_1763401, EPI_ISL_1763402, EPI_ISL_1763403, EPI_ISL_1763404, EPI_ISL_1763405, EPI_ISL_1763406, EPI_ISL_1763407, EPI_ISL_1763408, EPI_ISL_1763409, EPI_ISL_1763410, EPI_ISL_1763411, EPI_ISL_1763412, EPI_ISL_1763413, EPI_ISL_1763414,
EPI_ISL_1763415, EPI_ISL_1763416, EPI_ISL_1763417, EPI_ISL_1763418, EPI_ISL_1763419, EPI_ISL_1763420, EPI_ISL_1763421, EPI_ISL_1763422, EPI_ISL_1763423, EPI_ISL_1763424, EPI_ISL_1763425, EPI_ISL_1763426, EPI_ISL_1763427, EPI_ISL_1763428, EPI_ISL_1763429, EPI_ISL_1763430, EPI_ISL_1763431,
EPI_ISL_1763432, EPI_ISL_1763433, EPI_ISL_1763434, EPI_ISL_1763435, EPI_ISL_1763436, EPI_ISL_1763437, EPI_ISL_1763438, EPI_ISL_1763439, EPI_ISL_1763440, EPI_ISL_1763441, EPI_ISL_1763442, EPI_ISL_1763443, EPI_ISL_1763444, EPI_ISL_1763445, EPI_ISL_1763446, EPI_ISL_1763447, EPI_ISL_1763448,
EPI_ISL_1763449, EPI_ISL_1763450, EPI_ISL_1763451, EPI_ISL_1763452, EPI_ISL_1763453, EPI_ISL_1763454, EPI_ISL_1763455, EPI_ISL_1763456, EPI_ISL_1763457, EPI_ISL_1763458, EPI_ISL_1763459, EPI_ISL_1763460, EPI_ISL_1763461, EPI_ISL_1763462, EPI_ISL_1763463, EPI_ISL_1763464, EPI_ISL_1763465,
EPI_ISL_1763466, EPI_ISL_1763467, EPI_ISL_1763468, EPI_ISL_1763469, EPI_ISL_1763470, EPI_ISL_1763471, EPI_ISL_1763472, EPI_ISL_1763473, EPI_ISL_1763474, EPI_ISL_1763475, EPI_ISL_1763476, EPI_ISL_1763477, EPI_ISL_1763478, EPI_ISL_1763479, EPI_ISL_1763480, EPI_ISL_1763481, EPI_ISL_1763482,
EPI_ISL_1763483, EPI_ISL_1763484, EPI_ISL_1763485, EPI_ISL_1763486, EPI_ISL_1763487, EPI_ISL_1763488, EPI_ISL_1763489, EPI_ISL_1763490, EPI_ISL_1763491, EPI_ISL_1763492, EPI_ISL_1763493, EPI_ISL_1763494, EPI_ISL_1763495, EPI_ISL_1763496, EPI_ISL_1763497, EPI_ISL_1763498, EPI_ISL_1763499,
EPI_ISL_1763500, EPI_ISL_1763501, EPI_ISL_1763502, EPI_ISL_1763503, EPI_ISL_1763504, EPI_ISL_1763505, EPI_ISL_1763506, EPI_ISL_1763507, EPI_ISL_1763508, EPI_ISL_1763509, EPI_ISL_1763510, EPI_ISL_1763511, EPI_ISL_1763512, EPI_ISL_1763513, EPI_ISL_1763514, EPI_ISL_1763515, EPI_ISL_1763516,
EPI_ISL_1763517, EPI_ISL_1763518, EPI_ISL_1763519, EPI_ISL_1763520, EPI_ISL_1763521, EPI_ISL_1763522, EPI_ISL_1763523, EPI_ISL_1763524, EPI_ISL_1763525, EPI_ISL_1763526, EPI_ISL_1763527, EPI_ISL_1763528, EPI_ISL_1763529, EPI_ISL_1763530, EPI_ISL_1763531, EPI_ISL_1763532, EPI_ISL_1763533,
EPI_ISL_1763534, EPI_ISL_1763535, EPI_ISL_1763536, EPI_ISL_1763537, EPI_ISL_1763538, EPI_ISL_1763539, EPI_ISL_1763540, EPI_ISL_1763541, EPI_ISL_1763542, EPI_ISL_1763543, EPI_ISL_1763544, EPI_ISL_1763545, EPI_ISL_1763546, EPI_ISL_1763547, EPI_ISL_1763548, EPI_ISL_1763549, EPI_ISL_1763550,
EPI_ISL_1763551, EPI_ISL_1763552, EPI_ISL_1763553, EPI_ISL_1763554, EPI_ISL_1763555, EPI_ISL_1763556, EPI_ISL_1763557, EPI_ISL_1763558, EPI_ISL_1763559, EPI_ISL_1763560, EPI_ISL_1763561, EPI_ISL_1763562, EPI_ISL_1763563, EPI_ISL_1763564, EPI_ISL_1763565, EPI_ISL_1763566, EPI_ISL_1763567,
EPI_ISL_1763568, EPI_ISL_1763569, EPI_ISL_1763570, EPI_ISL_1763571, EPI_ISL_1763572, EPI_ISL_1763573, EPI_ISL_1763574, EPI_ISL_1763575, EPI_ISL_1763576, EPI_ISL_1763577, EPI_ISL_1763578, EPI_ISL_1763579, EPI_ISL_1763580, EPI_ISL_1763581, EPI_ISL_1763582, EPI_ISL_1763583, EPI_ISL_1763584,
EPI_ISL_1763585, EPI_ISL_1763586, EPI_ISL_1763587, EPI_ISL_1763588, EPI_ISL_1763589, EPI_ISL_1763590, EPI_ISL_1763591, EPI_ISL_1763592, EPI_ISL_1763593, EPI_ISL_1763594, EPI_ISL_1763595, EPI_ISL_1763596, EPI_ISL_1763597, EPI_ISL_1763598, EPI_ISL_1763599, EPI_ISL_1763600, EPI_ISL_1763601,
EPI_ISL_1763602, EPI_ISL_1763603, EPI_ISL_1763604, EPI_ISL_1763605, EPI_ISL_1763606, EPI_ISL_1763607, EPI_ISL_1763608, EPI_ISL_1763609, EPI_ISL_1763610, EPI_ISL_1763611, EPI_ISL_1763613, EPI_ISL_1763614, EPI_ISL_1763615, EPI_ISL_1763616, EPI_ISL_1763617, EPI_ISL_1763618, EPI_ISL_1763619,
EPI_ISL_1763620, EPI_ISL_1763621, EPI_ISL_1763622, EPI_ISL_1763623, EPI_ISL_1763624, EPI_ISL_1763625, EPI_ISL_1763626, EPI_ISL_1763627, EPI_ISL_1763628, EPI_ISL_1763629, EPI_ISL_1763630, EPI_ISL_1763631, EPI_ISL_1763632, EPI_ISL_1763633, EPI_ISL_1763634, EPI_ISL_1763635, EPI_ISL_1763636,
EPI_ISL_1763637, EPI_ISL_1763638, EPI_ISL_1763639, EPI_ISL_1763640, EPI_ISL_1763641, EPI_ISL_1763642, EPI_ISL_1763643, EPI_ISL_1763644, EPI_ISL_1763645, EPI_ISL_1763646, EPI_ISL_1763647, EPI_ISL_1763648, EPI_ISL_1763649, EPI_ISL_1763650, EPI_ISL_1763651, EPI_ISL_1763652, EPI_ISL_1763653,
EPI_ISL_1763654, EPI_ISL_1763655, EPI_ISL_1763656, EPI_ISL_1763657, EPI_ISL_1763658, EPI_ISL_1763659, EPI_ISL_1763660, EPI_ISL_1763661, EPI_ISL_1763662, EPI_ISL_1763663, EPI_ISL_1763664, EPI_ISL_1763665, EPI_ISL_1763666, EPI_ISL_1763667, EPI_ISL_1763668, EPI_ISL_1763669, EPI_ISL_1763670,
EPI_ISL_1763671, EPI_ISL_1763672, EPI_ISL_1763673, EPI_ISL_1763674, EPI_ISL_1763675, EPI_ISL_1763676, EPI_ISL_1763677, EPI_ISL_1763678, EPI_ISL_1763679, EPI_ISL_1763680, EPI_ISL_1763681, EPI_ISL_1763682, EPI_ISL_1763683, EPI_ISL_1763684, EPI_ISL_1763685, EPI_ISL_1763686, EPI_ISL_1763687,
EPI_ISL_1763688, EPI_ISL_1763689, EPI_ISL_1763690, EPI_ISL_1763691, EPI_ISL_1763692, EPI_ISL_1763693, EPI_ISL_1763694, EPI_ISL_1763695, EPI_ISL_1763696, EPI_ISL_1763697, EPI_ISL_1763698, EPI_ISL_1763699, EPI_ISL_1763700, EPI_ISL_1763701, EPI_ISL_1763702, EPI_ISL_1763703, EPI_ISL_1763704,
EPI_ISL_1763705, EPI_ISL_1763706, EPI_ISL_1763707, EPI_ISL_1763708, EPI_ISL_1763709, EPI_ISL_1763710, EPI_ISL_1763711, EPI_ISL_1763712, EPI_ISL_1763713, EPI_ISL_1763714, EPI_ISL_1763715, EPI_ISL_1763716, EPI_ISL_1763717, EPI_ISL_1763718, EPI_ISL_1763719, EPI_ISL_1763720, EPI_ISL_1763721,
EPI_ISL_1763722, EPI_ISL_1763723, EPI_ISL_1763724, EPI_ISL_1763725, EPI_ISL_1763726, EPI_ISL_1763727, EPI_ISL_1763728, EPI_ISL_1763729, EPI_ISL_1763730, EPI_ISL_1763731, EPI_ISL_1763732, EPI_ISL_1763733, EPI_ISL_1763734, EPI_ISL_1763735, EPI_ISL_1763736, EPI_ISL_1763737, EPI_ISL_1763738,
EPI_ISL_1763739, EPI_ISL_1763740, EPI_ISL_1763741, EPI_ISL_1763742, EPI_ISL_1763743, EPI_ISL_1763744, EPI_ISL_1763745, EPI_ISL_1763746, EPI_ISL_1763747, EPI_ISL_1763748, EPI_ISL_1763749, EPI_ISL_1763750, EPI_ISL_1763751, EPI_ISL_1763752, EPI_ISL_1763753, EPI_ISL_1763754, EPI_ISL_1763755,
EPI_ISL_1763756, EPI_ISL_1763757, EPI_ISL_1763758, EPI_ISL_1763759, EPI_ISL_1763760, EPI_ISL_1763761, EPI_ISL_1763762, EPI_ISL_1763763, EPI_ISL_1763764, EPI_ISL_1763765, EPI_ISL_1763766, EPI_ISL_1763767, EPI_ISL_1763768, EPI_ISL_1763769, EPI_ISL_1763770, EPI_ISL_1763771, EPI_ISL_1763772,

EPI_ISL_1763773, EPI_ISL_1763774, EPI_ISL_1763775, EPI_ISL_1763776, EPI_ISL_1763777, EPI_ISL_1763778, EPI_ISL_1763779, EPI_ISL_1763780, EPI_ISL_1763781, EPI_ISL_1763782, EPI_ISL_1763783, EPI_ISL_1763784, EPI_ISL_1763785, EPI_ISL_1763786, EPI_ISL_1763787, EPI_ISL_1763788, EPI_ISL_1763789,
EPI_ISL_1763790, EPI_ISL_1763791, EPI_ISL_1763792, EPI_ISL_1763793, EPI_ISL_1763794, EPI_ISL_1763795, EPI_ISL_1763796, EPI_ISL_1763797, EPI_ISL_1763798, EPI_ISL_1763799, EPI_ISL_1763800, EPI_ISL_1763801, EPI_ISL_1763802, EPI_ISL_1763803, EPI_ISL_1763804, EPI_ISL_1763805, EPI_ISL_1763806,
EPI_ISL_1763807, EPI_ISL_1763808, EPI_ISL_1763809, EPI_ISL_1763810, EPI_ISL_1763811, EPI_ISL_1763812, EPI_ISL_1763813, EPI_ISL_1763814, EPI_ISL_1763815, EPI_ISL_1763816, EPI_ISL_1763817, EPI_ISL_1763818, EPI_ISL_1763819, EPI_ISL_1763820, EPI_ISL_1763821, EPI_ISL_1763822, EPI_ISL_1763823,
EPI_ISL_1763824, EPI_ISL_1763825, EPI_ISL_1763826, EPI_ISL_1763827, EPI_ISL_1763828, EPI_ISL_1763829, EPI_ISL_1763830, EPI_ISL_1763831, EPI_ISL_1763832, EPI_ISL_1763833, EPI_ISL_1763834, EPI_ISL_1763835, EPI_ISL_1763836, EPI_ISL_1763837, EPI_ISL_1763838, EPI_ISL_1763839, EPI_ISL_1763840,
EPI_ISL_1763841, EPI_ISL_1763842, EPI_ISL_1763843, EPI_ISL_1763844, EPI_ISL_1763845, EPI_ISL_1763846, EPI_ISL_1763847, EPI_ISL_1763848, EPI_ISL_1763849, EPI_ISL_1763850, EPI_ISL_1763851, EPI_ISL_1763852, EPI_ISL_1763853, EPI_ISL_1763854, EPI_ISL_1763855, EPI_ISL_1763856, EPI_ISL_1763857,
EPI_ISL_1763858, EPI_ISL_1763859, EPI_ISL_1763860, EPI_ISL_1763861, EPI_ISL_1763862, EPI_ISL_1763863, EPI_ISL_1763864, EPI_ISL_1763865, EPI_ISL_1763866, EPI_ISL_1763867, EPI_ISL_1763868, EPI_ISL_1763869, EPI_ISL_1763870, EPI_ISL_1763871, EPI_ISL_1763872, EPI_ISL_1763873, EPI_ISL_1763874,
EPI_ISL_1763875, EPI_ISL_1763876, EPI_ISL_1763877, EPI_ISL_1763878, EPI_ISL_1763879, EPI_ISL_1763880, EPI_ISL_1763881, EPI_ISL_1763882, EPI_ISL_1763883, EPI_ISL_1763884, EPI_ISL_1763885, EPI_ISL_1763886, EPI_ISL_1763887, EPI_ISL_1763888, EPI_ISL_1763889, EPI_ISL_1763890, EPI_ISL_1763891,
EPI_ISL_1763892, EPI_ISL_1763893, EPI_ISL_1763894, EPI_ISL_1763895, EPI_ISL_1763896, EPI_ISL_1763897, EPI_ISL_1763898, EPI_ISL_1763899, EPI_ISL_1763900, EPI_ISL_1763901, EPI_ISL_1763902, EPI_ISL_1763903, EPI_ISL_1763904, EPI_ISL_1763905, EPI_ISL_1763906, EPI_ISL_1763907, EPI_ISL_1763908,
EPI_ISL_1763909, EPI_ISL_1763910, EPI_ISL_1763911, EPI_ISL_1763912, EPI_ISL_1763913, EPI_ISL_1763914, EPI_ISL_1763915, EPI_ISL_1763916, EPI_ISL_1763917, EPI_ISL_1763918, EPI_ISL_1763919, EPI_ISL_1763920, EPI_ISL_1763921, EPI_ISL_1763922, EPI_ISL_1763923, EPI_ISL_1763924, EPI_ISL_1763925,
EPI_ISL_1763926, EPI_ISL_1763927, EPI_ISL_1763928, EPI_ISL_1763929, EPI_ISL_1763930, EPI_ISL_1763931, EPI_ISL_1763932, EPI_ISL_1763933, EPI_ISL_1763934, EPI_ISL_1763935, EPI_ISL_1763936, EPI_ISL_1763937, EPI_ISL_1763938, EPI_ISL_1763939, EPI_ISL_1763940, EPI_ISL_1763941, EPI_ISL_1763942,
EPI_ISL_1763943, EPI_ISL_1763944, EPI_ISL_1763945, EPI_ISL_1763946, EPI_ISL_1763947, EPI_ISL_1763948, EPI_ISL_1763949, EPI_ISL_1763950, EPI_ISL_1763951, EPI_ISL_1763952, EPI_ISL_1763953, EPI_ISL_1763954, EPI_ISL_1763955, EPI_ISL_1763956, EPI_ISL_1763957, EPI_ISL_1763958, EPI_ISL_1763959,
EPI_ISL_1763960, EPI_ISL_1763961, EPI_ISL_1763962, EPI_ISL_1763963, EPI_ISL_1763964, EPI_ISL_1763965, EPI_ISL_1763966, EPI_ISL_1763967, EPI_ISL_1763968, EPI_ISL_1763969, EPI_ISL_1763970, EPI_ISL_1763971, EPI_ISL_1763972, EPI_ISL_1763973, EPI_ISL_1763974, EPI_ISL_1763975, EPI_ISL_1763976,
EPI_ISL_1763977, EPI_ISL_1763978, EPI_ISL_1763979, EPI_ISL_1763980, EPI_ISL_1763981, EPI_ISL_1763982, EPI_ISL_1763983, EPI_ISL_1763984, EPI_ISL_1763985, EPI_ISL_1763986, EPI_ISL_1763987, EPI_ISL_1763988, EPI_ISL_1763989, EPI_ISL_1763990, EPI_ISL_1763991, EPI_ISL_1763992, EPI_ISL_1763993,
EPI_ISL_1763994, EPI_ISL_1763995, EPI_ISL_1763996, EPI_ISL_1763997, EPI_ISL_1763998, EPI_ISL_1763999, EPI_ISL_1764000, EPI_ISL_1764001, EPI_ISL_1764002, EPI_ISL_1764003, EPI_ISL_1764004, EPI_ISL_1764005, EPI_ISL_1764006, EPI_ISL_1764007, EPI_ISL_1764008, EPI_ISL_1764009, EPI_ISL_1764010,
EPI_ISL_1764011, EPI_ISL_1764012, EPI_ISL_1764013, EPI_ISL_1764014, EPI_ISL_1764015, EPI_ISL_1764016, EPI_ISL_1764017, EPI_ISL_1764018, EPI_ISL_1764019, EPI_ISL_1764020, EPI_ISL_1764021, EPI_ISL_1764022, EPI_ISL_1764023, EPI_ISL_1764024, EPI_ISL_1764025, EPI_ISL_1764026, EPI_ISL_1764027,
EPI_ISL_1764028, EPI_ISL_1764029, EPI_ISL_1764030, EPI_ISL_1764031, EPI_ISL_1764032, EPI_ISL_1764033, EPI_ISL_1764034, EPI_ISL_1764035, EPI_ISL_1764036, EPI_ISL_1764037, EPI_ISL_1764038, EPI_ISL_1764039, EPI_ISL_1764040, EPI_ISL_1764041, EPI_ISL_1764042, EPI_ISL_1764043, EPI_ISL_1764044,
EPI_ISL_1764045, EPI_ISL_1764046, EPI_ISL_1764047, EPI_ISL_1764048, EPI_ISL_1764049, EPI_ISL_1764050, EPI_ISL_1764051, EPI_ISL_1764052, EPI_ISL_1764053, EPI_ISL_1764054, EPI_ISL_1764055, EPI_ISL_1764056, EPI_ISL_1764057, EPI_ISL_1764058, EPI_ISL_1764059, EPI_ISL_1764060, EPI_ISL_1764061,
EPI_ISL_1764062, EPI_ISL_1764063, EPI_ISL_1764064, EPI_ISL_1764065, EPI_ISL_1764066, EPI_ISL_1764067, EPI_ISL_1764068, EPI_ISL_1764069, EPI_ISL_1764070, EPI_ISL_1764071, EPI_ISL_1764072, EPI_ISL_1764073, EPI_ISL_1764074, EPI_ISL_1764075, EPI_ISL_1764076, EPI_ISL_1764077, EPI_ISL_1764078,
EPI_ISL_1764079, EPI_ISL_1764080, EPI_ISL_1764081, EPI_ISL_1764082, EPI_ISL_1764083, EPI_ISL_1764084, EPI_ISL_1764085, EPI_ISL_1764086, EPI_ISL_1764087, EPI_ISL_1764088, EPI_ISL_1764089, EPI_ISL_1764090, EPI_ISL_1764091, EPI_ISL_1764092, EPI_ISL_1764093, EPI_ISL_1764094, EPI_ISL_1764095,
EPI_ISL_1764096, EPI_ISL_1764097, EPI_ISL_1764098, EPI_ISL_1764099, EPI_ISL_1764100, EPI_ISL_1764101, EPI_ISL_1764102, EPI_ISL_1764103, EPI_ISL_1764104, EPI_ISL_1764105, EPI_ISL_1764106, EPI_ISL_1764107, EPI_ISL_1764108, EPI_ISL_1764109, EPI_ISL_1764110, EPI_ISL_1764111, EPI_ISL_1764112,
EPI_ISL_1764113, EPI_ISL_1764114, EPI_ISL_1764115, EPI_ISL_1764116, EPI_ISL_1764117, EPI_ISL_1764118, EPI_ISL_1764119, EPI_ISL_1764120, EPI_ISL_1764121, EPI_ISL_1764122, EPI_ISL_1764123, EPI_ISL_1764124, EPI_ISL_1764125, EPI_ISL_1764126, EPI_ISL_1764127, EPI_ISL_1764128, EPI_ISL_1764129,
EPI_ISL_1764130, EPI_ISL_1764131, EPI_ISL_1764132, EPI_ISL_1764133, EPI_ISL_1764134, EPI_ISL_1764135, EPI_ISL_1764136, EPI_ISL_1764137, EPI_ISL_1764138, EPI_ISL_1764139, EPI_ISL_1764140, EPI_ISL_1764141, EPI_ISL_1764142, EPI_ISL_1764143, EPI_ISL_2083087, EPI_ISL_2083088, EPI_ISL_2083089,
EPI_ISL_2083090, EPI_ISL_2083091, EPI_ISL_2083092, EPI_ISL_2083093, EPI_ISL_2083094, EPI_ISL_2083095, EPI_ISL_2083096, EPI_ISL_2083097, EPI_ISL_2083098, EPI_ISL_2083099, EPI_ISL_2083100, EPI_ISL_2083101, EPI_ISL_2083102, EPI_ISL_2083103, EPI_ISL_2083104, EPI_ISL_2083105, EPI_ISL_2083106,
EPI_ISL_2083107, EPI_ISL_2083108, EPI_ISL_2083109, EPI_ISL_2083110, EPI_ISL_2083111, EPI_ISL_2083112, EPI_ISL_2083113, EPI_ISL_2083114, EPI_ISL_2083115, EPI_ISL_2083116, EPI_ISL_2083117, EPI_ISL_2083118, EPI_ISL_2083119, EPI_ISL_2083120, EPI_ISL_2083121, EPI_ISL_2083122, EPI_ISL_2083123,
EPI_ISL_2083124, EPI_ISL_2083125, EPI_ISL_2083126, EPI_ISL_2083127, EPI_ISL_2083128, EPI_ISL_2083129, EPI_ISL_2083130, EPI_ISL_2083131, EPI_ISL_2083132, EPI_ISL_2083133, EPI_ISL_2083134, EPI_ISL_2083135, EPI_ISL_2083136, EPI_ISL_2083137, EPI_ISL_2083138, EPI_ISL_2083139, EPI_ISL_2083140,
EPI_ISL_2083141, EPI_ISL_2083142, EPI_ISL_2083143, EPI_ISL_2083144, EPI_ISL_2083145, EPI_ISL_2083146, EPI_ISL_2083147, EPI_ISL_2083148, EPI_ISL_2083149, EPI_ISL_2083150, EPI_ISL_2083151, EPI_ISL_2083152, EPI_ISL_2083153, EPI_ISL_2083154, EPI_ISL_2083155, EPI_ISL_2083156, EPI_ISL_2083157,
EPI_ISL_2083158, EPI_ISL_2083159, EPI_ISL_2083160, EPI_ISL_2083161, EPI_ISL_2083162, EPI_ISL_2083163, EPI_ISL_2083164, EPI_ISL_2083165, EPI_ISL_2083166, EPI_ISL_2083167, EPI_ISL_2083168, EPI_ISL_2083169, EPI_ISL_2083170, EPI_ISL_2083171, EPI_ISL_2083172, EPI_ISL_2083173, EPI_ISL_2083174,
EPI_ISL_2083175, EPI_ISL_2083176, EPI_ISL_2083177, EPI_ISL_2083178, EPI_ISL_2083179, EPI_ISL_2083180, EPI_ISL_2083181, EPI_ISL_2083182, EPI_ISL_2083183, EPI_ISL_2083184, EPI_ISL_2083185, EPI_ISL_2083186, EPI_ISL_2083187, EPI_ISL_2083188, EPI_ISL_2083189, EPI_ISL_2083190, EPI_ISL_2083191,
EPI_ISL_2083192, EPI_ISL_2083193, EPI_ISL_2083194, EPI_ISL_2083195, EPI_ISL_2083196, EPI_ISL_2083197, EPI_ISL_2083198, EPI_ISL_2083199, EPI_ISL_2083200, EPI_ISL_2083201, EPI_ISL_2083202, EPI_ISL_2083203, EPI_ISL_2083204, EPI_ISL_2083205, EPI_ISL_2083206, EPI_ISL_2083207, EPI_ISL_2083208,
EPI_ISL_2083209, EPI_ISL_2083210, EPI_ISL_2083211, EPI_ISL_2083212, EPI_ISL_2083213, EPI_ISL_2083214, EPI_ISL_2083215, EPI_ISL_2083216, EPI_ISL_2083217, EPI_ISL_2083218, EPI_ISL_2083219, EPI_ISL_2083220, EPI_ISL_2083221, EPI_ISL_2083222, EPI_ISL_2083223, EPI_ISL_2083224, EPI_ISL_2083225,
EPI_ISL_2083226, EPI_ISL_2083227, EPI_ISL_2083228, EPI_ISL_2083229, EPI_ISL_2083230, EPI_ISL_2083231, EPI_ISL_2083232, EPI_ISL_2083233, EPI_ISL_2083234, EPI_ISL_2083235, EPI_ISL_2083236, EPI_ISL_2083237, EPI_ISL_2083238, EPI_ISL_2083239, EPI_ISL_2083240, EPI_ISL_2083241, EPI_ISL_2083242,
EPI_ISL_2083243, EPI_ISL_2083244, EPI_ISL_2083245, EPI_ISL_2083246, EPI_ISL_2083247, EPI_ISL_2083248, EPI_ISL_2083249, EPI_ISL_2083250, EPI_ISL_2083251, EPI_ISL_2083252, EPI_ISL_2083253, EPI_ISL_2083254, EPI_ISL_2083255, EPI_ISL_2083256, EPI_ISL_2083257, EPI_ISL_2083258, EPI_ISL_2083259,
EPI_ISL_2083260, EPI_ISL_2083261, EPI_ISL_2083262, EPI_ISL_2083263, EPI_ISL_2083264, EPI_ISL_2083265, EPI_ISL_2083266, EPI_ISL_2083267, EPI_ISL_2083268, EPI_ISL_2083269, EPI_ISL_2083270, EPI_ISL_2083271, EPI_ISL_2083272, EPI_ISL_2083273, EPI_ISL_2083274, EPI_ISL_2083275, EPI_ISL_2083276,
EPI_ISL_2083277, EPI_ISL_2083278, EPI_ISL_2083279, EPI_ISL_2083280, EPI_ISL_2083281, EPI_ISL_2083282, EPI_ISL_2083283, EPI_ISL_2083284, EPI_ISL_2083285, EPI_ISL_2083286, EPI_ISL_2083287, EPI_ISL_2083288, EPI_ISL_2083289, EPI_ISL_2083290, EPI_ISL_2083291, EPI_ISL_2083292, EPI_ISL_2083293,
EPI_ISL_2083294, EPI_ISL_2083295, EPI_ISL_2083296, EPI_ISL_2083297, EPI_ISL_2083298, EPI_ISL_2083299, EPI_ISL_2083300, EPI_ISL_2083301, EPI_ISL_2083302, EPI_ISL_2083303, EPI_ISL_2083304, EPI_ISL_2083305, EPI_ISL_2083306, EPI_ISL_2083307, EPI_ISL_2083308, EPI_ISL_2083309, EPI_ISL_2083310,
EPI_ISL_2083311, EPI_ISL_2083312, EPI_ISL_2083313, EPI_ISL_2083314, EPI_ISL_2083315, EPI_ISL_2083316, EPI_ISL_2083317, EPI_ISL_2083318, EPI_ISL_2083319, EPI_ISL_2083320, EPI_ISL_2083321, EPI_ISL_2083322, EPI_ISL_2083323, EPI_ISL_2083324, EPI_ISL_2083325, EPI_ISL_2083326, EPI_ISL_2083327,
EPI_ISL_2083328, EPI_ISL_2083329, EPI_ISL_2083330, EPI_ISL_2083331, EPI_ISL_2083332, EPI_ISL_2083333, EPI_ISL_2083334, EPI_ISL_2083335, EPI_ISL_2083336, EPI_ISL_2083337, EPI_ISL_2083338, EPI_ISL_2083339, EPI_ISL_2083340, EPI_ISL_2083341, EPI_ISL_2083342, EPI_ISL_2083343, EPI_ISL_2083344,
EPI_ISL_2083345, EPI_ISL_2083346, EPI_ISL_2083347, EPI_ISL_2083348, EPI_ISL_2083349, EPI_ISL_2083350, EPI_ISL_2083351, EPI_ISL_2083352, EPI_ISL_2083353, EPI_ISL_2083354, EPI_ISL_2083355, EPI_ISL_2083356, EPI_ISL_2083357, EPI_ISL_2083358, EPI_ISL_2083359, EPI_ISL_2083360, EPI_ISL_2083361,
EPI_ISL_2083362, EPI_ISL_2083363, EPI_ISL_2083364, EPI_ISL_2083365, EPI_ISL_2083366, EPI_ISL_2083367, EPI_ISL_2083368, EPI_ISL_2083369, EPI_ISL_2083370, EPI_ISL_2083371, EPI_ISL_2083372, EPI_ISL_2083373, EPI_ISL_2083374, EPI_ISL_2083375, EPI_ISL_2083376, EPI_ISL_2083377, EPI_ISL_2083378,
EPI_ISL_2083379, EPI_ISL_2083380, EPI_ISL_2083381, EPI_ISL_2083382, EPI_ISL_2083383, EPI_ISL_2083384, EPI_ISL_2083385, EPI_ISL_2083386, EPI_ISL_2083387, EPI_ISL_2083388, EPI_ISL_2083389, EPI_ISL_2083390, EPI_ISL_2083391, EPI_ISL_2083392, EPI_ISL_2083393, EPI_ISL_2083394, EPI_ISL_2083395,
EPI_ISL_2083396, EPI_ISL_2083397, EPI_ISL_2083398, EPI_ISL_2083399, EPI_ISL_2083400, EPI_ISL_2083401, EPI_ISL_2083402, EPI_ISL_2083403, EPI_ISL_2083404, EPI_ISL_2083405, EPI_ISL_2083406, EPI_ISL_2083407, EPI_ISL_2083408, EPI_ISL_2083409, EPI_ISL_2083410, EPI_ISL_2083411, EPI_ISL_2083412,
EPI_ISL_2083413, EPI_ISL_2083414, EPI_ISL_2083415, EPI_ISL_2083416, EPI_ISL_2083417, EPI_ISL_2083418, EPI_ISL_2083419, EPI_ISL_2083420, EPI_ISL_2083421, EPI_ISL_2083422, EPI_ISL_2083423, EPI_ISL_2083424, EPI_ISL_2083425, EPI_ISL_2083426, EPI_ISL_2083427, EPI_ISL_2083428, EPI_ISL_2083429,
EPI_ISL_2083430, EPI_ISL_2083431, EPI_ISL_2083432, EPI_ISL_2083433, EPI_ISL_2083434, EPI_ISL_2083435, EPI_ISL_2083436, EPI_ISL_2083437, EPI_ISL_2083438, EPI_ISL_2083439, EPI_ISL_2083440, EPI_ISL_2083441, EPI_ISL_2083442, EPI_ISL_2083443, EPI_ISL_2083444, EPI_ISL_2083445, EPI_ISL_2083446,
EPI_ISL_2083447, EPI_ISL_2083448, EPI_ISL_2083449, EPI_ISL_2083450, EPI_ISL_2083451, EPI_ISL_2083452, EPI_ISL_2083453, EPI_ISL_2083454, EPI_ISL_2083455, EPI_ISL_2083456, EPI_ISL_2083457, EPI_ISL_2083458, EPI_ISL_2083459, EPI_ISL_2083460, EPI_ISL_2083461, EPI_ISL_2083462, EPI_ISL_2083463,
EPI_ISL_2083464, EPI_ISL_2083465, EPI_ISL_2083466, EPI_ISL_2083467, EPI_ISL_2083468, EPI_ISL_2083469, EPI_ISL_2083470, EPI_ISL_2083471, EPI_ISL_2083472, EPI_ISL_2083473, EPI_ISL_2083474, EPI_ISL_2083475, EPI_ISL_2083476, EPI_ISL_2083477, EPI_ISL_2083478, EPI_ISL_2083479, EPI_ISL_2083480,
EPI_ISL_2083481, EPI_ISL_2083482, EPI_ISL_2083483, EPI_ISL_2083484, EPI_ISL_2083485, EPI_ISL_2083486, EPI_ISL_2083487, EPI_ISL_2083488, EPI_ISL_2083489, EPI_ISL_2083490, EPI_ISL_2083491, EPI_ISL_2083492, EPI_ISL_2083493, EPI_ISL_2083494, EPI_ISL_2083495, EPI_ISL_2083496, EPI_ISL_2083497,
EPI_ISL_2083498, EPI_ISL_2083499, EPI_ISL_2083500, EPI_ISL_2083501, EPI_ISL_2083502, EPI_ISL_2083503, EPI_ISL_2083504, EPI_ISL_2083505, EPI_ISL_2083507, EPI_ISL_2083508, EPI_ISL_2083509, EPI_ISL_2083510, EPI_ISL_2083511, EPI_ISL_2083512, EPI_ISL_2083513, EPI_ISL_2083514, EPI_ISL_2083515,
EPI_ISL_2083516, EPI_ISL_2083517, EPI_ISL_2083518, EPI_ISL_2083519, EPI_ISL_2083520, EPI_ISL_2083521, EPI_ISL_2083522, EPI_ISL_2083523, EPI_ISL_2083524, EPI_ISL_2083525, EPI_ISL_2083526, EPI_ISL_2083527, EPI_ISL_2083528, EPI_ISL_2083529, EPI_ISL_2083530, EPI_ISL_2083531, EPI_ISL_2083532,
EPI_ISL_2083533, EPI_ISL_2083534, EPI_ISL_2083535, EPI_ISL_2083536, EPI_ISL_2083537, EPI_ISL_2083538, EPI_ISL_2083539, EPI_ISL_2083540, EPI_ISL_2083541, EPI_ISL_2083542, EPI_ISL_2083543, EPI_ISL_2083544, EPI_ISL_2083545, EPI_ISL_2083546, EPI_ISL_2083547, EPI_ISL_2083548, EPI_ISL_2083549,
EPI_ISL_2083550, EPI_ISL_2083551, EPI_ISL_2083552, EPI_ISL_2083553, EPI_ISL_2083554, EPI_ISL_2083555, EPI_ISL_2083556, EPI_ISL_2083557, EPI_ISL_2083558, EPI_ISL_2083559, EPI_ISL_2083560, EPI_ISL_2083561, EPI_ISL_2083562, EPI_ISL_2083563, EPI_ISL_2083564, EPI_ISL_2083565, EPI_ISL_2083566,
EPI_ISL_2083567, EPI_ISL_2083568, EPI_ISL_2083569, EPI_ISL_2083570, EPI_ISL_2083571, EPI_ISL_2083572, EPI_ISL_2083573, EPI_ISL_2083574, EPI_ISL_2083575, EPI_ISL_2083576, EPI_ISL_2083577, EPI_ISL_2083578, EPI_ISL_2083579, EPI_ISL_2083580, EPI_ISL_2083581, EPI_ISL_2083582, EPI_ISL_2083583,
EPI_ISL_2083584, EPI_ISL_2083585, EPI_ISL_2083586, EPI_ISL_2083587, EPI_ISL_2083588, EPI_ISL_2083589, EPI_ISL_2083590, EPI_ISL_2083591, EPI_ISL_2083592, EPI_ISL_2083593, EPI_ISL_2083594, EPI_ISL_2083595, EPI_ISL_2083596, EPI_ISL_2083597, EPI_ISL_2083598, EPI_ISL_2083599, EPI_ISL_2083600,
EPI_ISL_2083601, EPI_ISL_2083602, EPI_ISL_2083603, EPI_ISL_2083604, EPI_ISL_2083605, EPI_ISL_2083606, EPI_ISL_2083607, EPI_ISL_2083608, EPI_ISL_2083609, EPI_ISL_2083610, EPI_ISL_2083611, EPI_ISL_2083612, EPI_ISL_2083613, EPI_ISL_2083614, EPI_ISL_2083615, EPI_ISL_2083616, EPI_ISL_2083617,
EPI_ISL_2083618, EPI_ISL_2083619, EPI_ISL_2083620, EPI_ISL_2083621, EPI_ISL_2083622, EPI_ISL_2083623, EPI_ISL_2083624, EPI_ISL_2083625, EPI_ISL_2083626, EPI_ISL_2083627, EPI_ISL_2083628, EPI_ISL_2083629, EPI_ISL_2083630, EPI_ISL_2083631, EPI_ISL_2083632, EPI_ISL_2083633, EPI_ISL_2083634,
EPI_ISL_2083635, EPI_ISL_2083636, EPI_ISL_2083637, EPI_ISL_2083638, EPI_ISL_2083639, EPI_ISL_2083640, EPI_ISL_2083641, EPI_ISL_2083642, EPI_ISL_2083643, EPI_ISL_2083644, EPI_ISL_2083645, EPI_ISL_2083646, EPI_ISL_2083647, EPI_ISL_2083648, EPI_ISL_2083649, EPI_ISL_2083650, EPI_ISL_2083651,
EPI_ISL_2083652, EPI_ISL_2083653, EPI_ISL_2083654, EPI_ISL_2083655, EPI_ISL_2083656, EPI_ISL_2083657, EPI_ISL_2083658, EPI_ISL_2083659, EPI_ISL_2083660, EPI_ISL_2083661, EPI_ISL_2083662, EPI_ISL_2083663, EPI_ISL_2083664, EPI_ISL_2083665, EPI_ISL_2083666, EPI_ISL_2083667, EPI_ISL_2083668,
EPI_ISL_2083669, EPI_ISL_2083670, EPI_ISL_2083671, EPI_ISL_2083672, EPI_ISL_2083673, EPI_ISL_2083674, EPI_ISL_2083675, EPI_ISL_2083676, EPI_ISL_2083677, EPI_ISL_2083678, EPI_ISL_2083679, EPI_ISL_2083680, EPI_ISL_2083681, EPI_ISL_2083682, EPI_ISL_2083683, EPI_ISL_2083684, EPI_ISL_2083685,
EPI_ISL_2083686, EPI_ISL_2083687, EPI_ISL_2083688, EPI_ISL_2083689, EPI_ISL_2083690, EPI_ISL_2083691, EPI_ISL_2083692, EPI_ISL_2083693, EPI_ISL_2083694, EPI_ISL_2083695, EPI_ISL_2083696, EPI_ISL_2083697, EPI_ISL_2083698, EPI_ISL_2083699, EPI_ISL_2083700, EPI_ISL_2083701, EPI_ISL_2083702,
EPI_ISL_2083703, EPI_ISL_2083704, EPI_ISL_2083705, EPI_ISL_2083706, EPI_ISL_2083707, EPI_ISL_2083708, EPI_ISL_2083709, EPI_ISL_2083710, EPI_ISL_2083711, EPI_ISL_2083712, EPI_ISL_2083713, EPI_ISL_2083714, EPI_ISL_2083715, EPI_ISL_2083716, EPI_ISL_2083717, EPI_ISL_2083718, EPI_ISL_2083719,
EPI_ISL_2083720, EPI_ISL_2083721, EPI_ISL_2083722, EPI_ISL_2083723, EPI_ISL_2083724, EPI_ISL_2083725, EPI_ISL_2083726, EPI_ISL_2083727, EPI_ISL_2083728, EPI_ISL_2083729, EPI_ISL_2083730, EPI_ISL_2083731, EPI_ISL_2083732, EPI_ISL_2083733, EPI_ISL_2083734, EPI_ISL_2083735, EPI_ISL_2083736,
EPI_ISL_2083737, EPI_ISL_2083738, EPI_ISL_2083739, EPI_ISL_2083740, EPI_ISL_2083741, EPI_ISL_2083742, EPI_ISL_2083743, EPI_ISL_2083744, EPI_ISL_2083745, EPI_ISL_2083746, EPI_ISL_2083747, EPI_ISL_2083748, EPI_ISL_2083749, EPI_ISL_2083750, EPI_ISL_2083751, EPI_ISL_2083752, EPI_ISL_2083753,
EPI_ISL_2083754, EPI_ISL_2083755, EPI_ISL_2083756, EPI_ISL_2083757, EPI_ISL_2083758, EPI_ISL_2083759, EPI_ISL_2083760, EPI_ISL_2083761, EPI_ISL_2083762, EPI_ISL_2083763, EPI_ISL_2083764, EPI_ISL_2083765, EPI_ISL_2083766, EPI_ISL_2083767, EPI_ISL_2083768, EPI_ISL_2083769, EPI_ISL_2083770,
EPI_ISL_2083771, EPI_ISL_2083772, EPI_ISL_2083773, EPI_ISL_2083774, EPI_ISL_2083775, EPI_ISL_2083776, EPI_ISL_2083777, EPI_ISL_2083778, EPI_ISL_2083779, EPI_ISL_2083780, EPI_ISL_2083781, EPI_ISL_2083782, EPI_ISL_2083783, EPI_ISL_2083784, EPI_ISL_2083785, EPI_ISL_2083786, EPI_ISL_2083787,
EPI_ISL_2083788, EPI_ISL_2083789, EPI_ISL_2083790, EPI_ISL_2083791, EPI_ISL_2083792, EPI_ISL_2083793, EPI_ISL_2083794, EPI_ISL_2083795, EPI_ISL_2083796, EPI_ISL_2083797, EPI_ISL_2083798, EPI_ISL_2083799, EPI_ISL_2083800, EPI_ISL_2083801, EPI_ISL_2083802, EPI_ISL_2083803, EPI_ISL_2083804,
EPI_ISL_2083805, EPI_ISL_2083806, EPI_ISL_2083807, EPI_ISL_2083808, EPI_ISL_2083809, EPI_ISL_2083810, EPI_ISL_2083811, EPI_ISL_2083812, EPI_ISL_2083813, EPI_ISL_2083814, EPI_ISL_2083815, EPI_ISL_2083816, EPI_ISL_2083817, EPI_ISL_2083818, EPI_ISL_2083819, EPI_ISL_2083820, EPI_ISL_2083821,
EPI_ISL_2083822, EPI_ISL_2083823, EPI_ISL_2083824, EPI_ISL_2083825, EPI_ISL_2083826, EPI_ISL_2083827, EPI_ISL_2083828, EPI_ISL_2083829, EPI_ISL_2083830, EPI_ISL_2083831, EPI_ISL_2083832, EPI_ISL_2083833, EPI_ISL_2083834, EPI_ISL_2083835, EPI_ISL_2083836, EPI_ISL_2083837, EPI_ISL_2083838,
EPI_ISL_2083839, EPI_ISL_2083840, EPI_ISL_2083841, EPI_ISL_2083842, EPI_ISL_2083843, EPI_ISL_2083844, EPI_ISL_2083845, EPI_ISL_2083846, EPI_ISL_2083847, EPI_ISL_2083848, EPI_ISL_2083849, EPI_ISL_2083850, EPI_ISL_2083851, EPI_ISL_2083852, EPI_ISL_2083853, EPI_ISL_2083854, EPI_ISL_2083855,
EPI_ISL_2083856, EPI_ISL_2083857, EPI_ISL_2083858, EPI_ISL_2083859, EPI_ISL_2083860, EPI_ISL_2083861, EPI_ISL_2083862, EPI_ISL_2083863, EPI_ISL_2083864, EPI_ISL_2083865, EPI_ISL_2083866, EPI_ISL_2083867, EPI_ISL_2083868, EPI_ISL_2083869, EPI_ISL_2083870, EPI_ISL_2083871, EPI_ISL_2083872,
EPI_ISL_2083873, EPI_ISL_2083874, EPI_ISL_2083875, EPI_ISL_2083876, EPI_ISL_2083877, EPI_ISL_2083878, EPI_ISL_2083879, EPI_ISL_2083880, EPI_ISL_2083881, EPI_ISL_2083882, EPI_ISL_2083883, EPI_ISL_2083884, EPI_ISL_2083885, EPI_ISL_2083886, EPI_ISL_2083887, EPI_ISL_2083888, EPI_ISL_2083889,
EPI_ISL_2083890, EPI_ISL_2083891, EPI_ISL_2083892, EPI_ISL_2083894, EPI_ISL_2083895, EPI_ISL_2083896, EPI_ISL_2083897, EPI_ISL_2083898, EPI_ISL_2083899, EPI_ISL_2083900, EPI_ISL_2083901, EPI_ISL_2083902, EPI_ISL_2083903, EPI_ISL_2083904, EPI_ISL_2083905, EPI_ISL_2083906, EPI_ISL_2083907,
EPI_ISL_2083908, EPI_ISL_2083909, EPI_ISL_2083910, EPI_ISL_2083911, EPI_ISL_2083912, EPI_ISL_2083913, EPI_ISL_2083914, EPI_ISL_2083915, EPI_ISL_2083916, EPI_ISL_2083917, EPI_ISL_2083918, EPI_ISL_2083919, EPI_ISL_2083920, EPI_ISL_2083921, EPI_ISL_2083922, EPI_ISL_2083923, EPI_ISL_2083924,
EPI_ISL_2083925, EPI_ISL_2083926, EPI_ISL_2083927, EPI_ISL_2083928, EPI_ISL_2083929, EPI_ISL_2083930, EPI_ISL_2083931, EPI_ISL_2083932, EPI_ISL_2083933, EPI_ISL_2083934, EPI_ISL_2083935, EPI_ISL_2083936, EPI_ISL_2083937, EPI_ISL_2083938, EPI_ISL_2083939, EPI_ISL_2083940, EPI_ISL_2083941,
EPI_ISL_2083942, EPI_ISL_2083943, EPI_ISL_2083944, EPI_ISL_2083945, EPI_ISL_2083946, EPI_ISL_2083947, EPI_ISL_2083948, EPI_ISL_2083949, EPI_ISL_2083950, EPI_ISL_2083951, EPI_ISL_2083952, EPI_ISL_2083953, EPI_ISL_2083954, EPI_ISL_2083955, EPI_ISL_2083956, EPI_ISL_2083957, EPI_ISL_2083958,
EPI_ISL_2083959, EPI_ISL_2083960, EPI_ISL_2083961, EPI_ISL_2083962, EPI_ISL_2083963, EPI_ISL_2083964, EPI_ISL_2083965, EPI_ISL_2083966, EPI_ISL_2083967, EPI_ISL_2083968, EPI_ISL_2083969, EPI_ISL_2083970, EPI_ISL_2083971, EPI_ISL_2083972, EPI_ISL_2083973, EPI_ISL_2083974, EPI_ISL_2083975,
EPI_ISL_2083976, EPI_ISL_2083977, EPI_ISL_2083978, EPI_ISL_2083979, EPI_ISL_2083980, EPI_ISL_2083981, EPI_ISL_2083982, EPI_ISL_2083983, EPI_ISL_2083984, EPI_ISL_2083985, EPI_ISL_2083986, EPI_ISL_2083987, EPI_ISL_2083988, EPI_ISL_2083989, EPI_ISL_2083990, EPI_ISL_2083991, EPI_ISL_2083992,
EPI_ISL_2083993, EPI_ISL_2083994, EPI_ISL_2083995, EPI_ISL_2083996, EPI_ISL_2083997, EPI_ISL_2083998, EPI_ISL_2083999, EPI_ISL_2084000, EPI_ISL_2084001, EPI_ISL_2084002, EPI_ISL_2084003, EPI_ISL_2084004, EPI_ISL_2084005, EPI_ISL_2084006, EPI_ISL_2084007, EPI_ISL_2084008, EPI_ISL_2084009,
EPI_ISL_2084010, EPI_ISL_2084011, EPI_ISL_2084012, EPI_ISL_2084013, EPI_ISL_2084014, EPI_ISL_2084015, EPI_ISL_2084016, EPI_ISL_2084017, EPI_ISL_2084018, EPI_ISL_2084019, EPI_ISL_2084020, EPI_ISL_2084021, EPI_ISL_2084022, EPI_ISL_2084023, EPI_ISL_2084024, EPI_ISL_2084025, EPI_ISL_2084026,
EPI_ISL_2084027, EPI_ISL_2084028, EPI_ISL_2084029, EPI_ISL_2084030, EPI_ISL_2084031, EPI_ISL_2084032, EPI_ISL_2084033, EPI_ISL_2084034, EPI_ISL_2084035, EPI_ISL_2084036, EPI_ISL_2084037, EPI_ISL_2084038, EPI_ISL_2084039, EPI_ISL_2084040, EPI_ISL_2084041, EPI_ISL_2084042, EPI_ISL_2084043,
EPI_ISL_2084044, EPI_ISL_2084045, EPI_ISL_2084046, EPI_ISL_2084047, EPI_ISL_2084048, EPI_ISL_2084049, EPI_ISL_2084050, EPI_ISL_2084051, EPI_ISL_2084052, EPI_ISL_2084053, EPI_ISL_2084054, EPI_ISL_2084055, EPI_ISL_2084056, EPI_ISL_2084057, EPI_ISL_2084058, EPI_ISL_2084059, EPI_ISL_2084060,
EPI_ISL_2084061, EPI_ISL_2084062, EPI_ISL_2084063, EPI_ISL_2084064, EPI_ISL_2084065, EPI_ISL_2084066, EPI_ISL_2084067, EPI_ISL_2084068, EPI_ISL_2084069, EPI_ISL_2084070, EPI_ISL_2084071, EPI_ISL_2084072, EPI_ISL_2084073, EPI_ISL_2084074, EPI_ISL_2084075, EPI_ISL_2084076, EPI_ISL_2084077,
EPI_ISL_2084078, EPI_ISL_2084079, EPI_ISL_2084080, EPI_ISL_2084081, EPI_ISL_2084082, EPI_ISL_2084083, EPI_ISL_2084084, EPI_ISL_2084085, EPI_ISL_2084086, EPI_ISL_2084087, EPI_ISL_2084088, EPI_ISL_2084089, EPI_ISL_2084090, EPI_ISL_2084091, EPI_ISL_2084092, EPI_ISL_2084093, EPI_ISL_2084094,
EPI_ISL_2084095, EPI_ISL_2084096, EPI_ISL_2084097, EPI_ISL_2084098, EPI_ISL_2084099, EPI_ISL_2084100, EPI_ISL_2084101, EPI_ISL_2084102, EPI_ISL_2084103, EPI_ISL_2084104, EPI_ISL_2084105, EPI_ISL_2084106, EPI_ISL_2084107, EPI_ISL_2084108, EPI_ISL_2084109, EPI_ISL_2084110, EPI_ISL_2084111,
EPI_ISL_2084112, EPI_ISL_2084113, EPI_ISL_2084114, EPI_ISL_2084115, EPI_ISL_2084116, EPI_ISL_2084117, EPI_ISL_2084118, EPI_ISL_2084119, EPI_ISL_2084120, EPI_ISL_2084121, EPI_ISL_2084122, EPI_ISL_2084123, EPI_ISL_2084124, EPI_ISL_2084125, EPI_ISL_2084126, EPI_ISL_2084127, EPI_ISL_2084128,
EPI_ISL_2084129, EPI_ISL_2084130, EPI_ISL_2084131, EPI_ISL_2084132, EPI_ISL_2084133, EPI_ISL_2084134, EPI_ISL_2084135, EPI_ISL_2084136, EPI_ISL_2084137, EPI_ISL_2084138, EPI_ISL_2084139, EPI_ISL_2084140, EPI_ISL_2084141, EPI_ISL_2084142, EPI_ISL_2084143, EPI_ISL_2084144, EPI_ISL_2084145,
EPI_ISL_2084146, EPI_ISL_2084147, EPI_ISL_2084148, EPI_ISL_2084149, EPI_ISL_2084150, EPI_ISL_2084151, EPI_ISL_2084152, EPI_ISL_2084153, EPI_ISL_2084154, EPI_ISL_2084155, EPI_ISL_2084156, EPI_ISL_2084157, EPI_ISL_2084158, EPI_ISL_2084159, EPI_ISL_2084160, EPI_ISL_2084161, EPI_ISL_2084162,
EPI_ISL_2084163, EPI_ISL_2084164, EPI_ISL_2084165, EPI_ISL_2084166, EPI_ISL_2084167, EPI_ISL_2084168, EPI_ISL_2084169, EPI_ISL_2084170, EPI_ISL_2084171, EPI_ISL_2084172, EPI_ISL_2084173, EPI_ISL_2084174, EPI_ISL_2084175, EPI_ISL_2084176, EPI_ISL_2084177, EPI_ISL_2084178, EPI_ISL_2084179,
EPI_ISL_2084180, EPI_ISL_2084181, EPI_ISL_2084182, EPI_ISL_2084183, EPI_ISL_2084184, EPI_ISL_2084185, EPI_ISL_2084186, EPI_ISL_2084187, EPI_ISL_2084188, EPI_ISL_2084189, EPI_ISL_2084190, EPI_ISL_2084191, EPI_ISL_2084192, EPI_ISL_2084193, EPI_ISL_2084194, EPI_ISL_2084195, EPI_ISL_2084196,

EPI_ISL_2084197, EPI_ISL_2084198, EPI_ISL_2084199, EPI_ISL_2084200, EPI_ISL_2084201, EPI_ISL_2084202, EPI_ISL_2084203, EPI_ISL_2084204, EPI_ISL_2084205, EPI_ISL_2084206, EPI_ISL_2084207, EPI_ISL_2084208, EPI_ISL_2084209, EPI_ISL_2084210, EPI_ISL_2084211, EPI_ISL_2084212, EPI_ISL_2084213, EPI_ISL_2084214, EPI_ISL_2084215, EPI_ISL_2084216, EPI_ISL_2084217, EPI_ISL_2084218, EPI_ISL_2084219, EPI_ISL_2084220, EPI_ISL_2084221, EPI_ISL_2084222, EPI_ISL_2084223, EPI_ISL_2084224, EPI_ISL_2084225, EPI_ISL_2084226, EPI_ISL_2084227, EPI_ISL_2084228, EPI_ISL_2084229, EPI_ISL_2084230, EPI_ISL_2084231, EPI_ISL_2084232, EPI_ISL_2084233, EPI_ISL_2084234, EPI_ISL_2084235, EPI_ISL_2084236, EPI_ISL_2084237, EPI_ISL_2084238, EPI_ISL_2084239, EPI_ISL_2084240, EPI_ISL_2084241, EPI_ISL_2084242, EPI_ISL_2084243, EPI_ISL_2084244, EPI_ISL_2084245, EPI_ISL_2084246, EPI_ISL_2084247, EPI_ISL_2084248, EPI_ISL_2084249, EPI_ISL_2084250, EPI_ISL_2084251, EPI_ISL_2084252, EPI_ISL_2084253, EPI_ISL_2084254, EPI_ISL_2084255, EPI_ISL_2084256, EPI_ISL_2084257, EPI_ISL_2084258, EPI_ISL_2084259, EPI_ISL_2084260, EPI_ISL_2084261, EPI_ISL_2084262, EPI_ISL_2084263, EPI_ISL_2084264, EPI_ISL_2084265, EPI_ISL_2084266, EPI_ISL_2084267, EPI_ISL_2084268, EPI_ISL_2084269, EPI_ISL_2084270, EPI_ISL_2084271, EPI_ISL_2084272, EPI_ISL_2084273, EPI_ISL_2084274, EPI_ISL_2084275, EPI_ISL_2084276, EPI_ISL_2084277, EPI_ISL_2084278, EPI_ISL_2084279, EPI_ISL_2084280, EPI_ISL_2084281, EPI_ISL_2084282, EPI_ISL_2084283, EPI_ISL_2084284, EPI_ISL_2084285, EPI_ISL_2084286, EPI_ISL_2084287, EPI_ISL_2084288, EPI_ISL_2084289, EPI_ISL_2084290, EPI_ISL_2084291, EPI_ISL_2084292, EPI_ISL_2084293, EPI_ISL_2084294, EPI_ISL_2084295, EPI_ISL_2084296, EPI_ISL_2084297, EPI_ISL_2084298, EPI_ISL_2084299, EPI_ISL_2084300, EPI_ISL_2084301, EPI_ISL_2084302, EPI_ISL_2084303, EPI_ISL_2084304, EPI_ISL_2084305, EPI_ISL_2084306, EPI_ISL_2084307, EPI_ISL_2084308, EPI_ISL_2084309, EPI_ISL_2084310, EPI_ISL_2084311, EPI_ISL_2084312, EPI_ISL_2084313, EPI_ISL_2084314, EPI_ISL_2084315, EPI_ISL_2084316, EPI_ISL_2084317, EPI_ISL_2084318, EPI_ISL_2084319, EPI_ISL_2084320, EPI_ISL_2084321, EPI_ISL_2084322, EPI_ISL_2084323, EPI_ISL_2084324, EPI_ISL_2084325, EPI_ISL_2084326, EPI_ISL_2084327, EPI_ISL_2084328, EPI_ISL_2084329, EPI_ISL_2084330, EPI_ISL_2084331, EPI_ISL_2084332, EPI_ISL_2084333, EPI_ISL_2084334, EPI_ISL_2084335, EPI_ISL_2084336, EPI_ISL_2084337, EPI_ISL_2084338, EPI_ISL_2084339, EPI_ISL_2084340, EPI_ISL_2084341, EPI_ISL_2084342, EPI_ISL_2084343, EPI_ISL_2084344, EPI_ISL_2084345, EPI_ISL_2084346, EPI_ISL_2084347, EPI_ISL_2084348, EPI_ISL_2084349, EPI_ISL_2084350, EPI_ISL_2084351, EPI_ISL_2084352, EPI_ISL_2084353, EPI_ISL_2084354, EPI_ISL_2084355, EPI_ISL_2084356, EPI_ISL_2084357, EPI_ISL_2084358, EPI_ISL_2084359, EPI_ISL_2084360, EPI_ISL_2084361, EPI_ISL_2084362, EPI_ISL_2084363, EPI_ISL_2084364, EPI_ISL_2084365, EPI_ISL_2084366, EPI_ISL_2084367, EPI_ISL_2084368, EPI_ISL_2084369, EPI_ISL_2084370, EPI_ISL_2084371, EPI_ISL_2084372, EPI_ISL_2084373, EPI_ISL_2084374, EPI_ISL_2084375, EPI_ISL_2084376, EPI_ISL_2084377, EPI_ISL_2084378, EPI_ISL_2084379, EPI_ISL_2084380, EPI_ISL_2084381, EPI_ISL_2084382, EPI_ISL_2084383, EPI_ISL_2084384, EPI_ISL_2084385, EPI_ISL_2084386, EPI_ISL_2084387, EPI_ISL_2084388, EPI_ISL_2084389, EPI_ISL_2084390, EPI_ISL_2084391, EPI_ISL_2084392, EPI_ISL_2084393, EPI_ISL_2084394, EPI_ISL_2084395, EPI_ISL_2084396, EPI_ISL_2084397, EPI_ISL_2084398, EPI_ISL_2084399, EPI_ISL_2084400, EPI_ISL_2084401, EPI_ISL_2084402, EPI_ISL_2084403, EPI_ISL_2084404, EPI_ISL_2084405, EPI_ISL_2084406, EPI_ISL_2084407, EPI_ISL_2084408, EPI_ISL_2084409, EPI_ISL_2084410, EPI_ISL_2084411, EPI_ISL_2084412, EPI_ISL_2084413, EPI_ISL_2084414, EPI_ISL_2084415, EPI_ISL_2084416, EPI_ISL_2084417, EPI_ISL_2084418, EPI_ISL_2084419, EPI_ISL_2084420, EPI_ISL_2084421, EPI_ISL_2084422, EPI_ISL_2084423, EPI_ISL_2084424, EPI_ISL_2084425, EPI_ISL_2084426, EPI_ISL_2084427, EPI_ISL_2084428, EPI_ISL_2084429, EPI_ISL_2084430, EPI_ISL_2084431, EPI_ISL_2084432, EPI_ISL_2084433, EPI_ISL_2084434, EPI_ISL_2084435, EPI_ISL_2084436, EPI_ISL_2084437, EPI_ISL_2084438, EPI_ISL_2084439, EPI_ISL_2084440, EPI_ISL_2084441, EPI_ISL_2084442, EPI_ISL_2084443, EPI_ISL_2084444, EPI_ISL_2084445, EPI_ISL_2084446, EPI_ISL_2084447, EPI_ISL_2084448, EPI_ISL_2084449, EPI_ISL_2084450, EPI_ISL_2084451, EPI_ISL_2084452, EPI_ISL_2084453, EPI_ISL_2084454, EPI_ISL_2084455, EPI_ISL_2084456, EPI_ISL_2084457, EPI_ISL_2084458, EPI_ISL_2084459, EPI_ISL_2084460, EPI_ISL_2084461, EPI_ISL_2084462, EPI_ISL_2084463, EPI_ISL_2084464, EPI_ISL_2084465, EPI_ISL_2084466, EPI_ISL_2084467, EPI_ISL_2084468, EPI_ISL_2084469, EPI_ISL_2084470, EPI_ISL_2084471, EPI_ISL_2084472, EPI_ISL_2084473, EPI_ISL_2084474, EPI_ISL_2084475, EPI_ISL_2084476, EPI_ISL_2084477, EPI_ISL_2084478, EPI_ISL_2084479, EPI_ISL_2084480, EPI_ISL_2084481, EPI_ISL_2084482, EPI_ISL_2084483, EPI_ISL_2084484, EPI_ISL_2084485, EPI_ISL_2084486, EPI_ISL_2084487, EPI_ISL_2084488, EPI_ISL_2084489, EPI_ISL_2084490, EPI_ISL_2084491, EPI_ISL_2084492, EPI_ISL_2084493, EPI_ISL_2084494, EPI_ISL_2084495, EPI_ISL_2084496, EPI_ISL_2084497, EPI_ISL_2084498, EPI_ISL_2084499, EPI_ISL_2084500, EPI_ISL_2084501, EPI_ISL_2084502, EPI_ISL_2084503, EPI_ISL_2084504, EPI_ISL_2084505, EPI_ISL_2084506, EPI_ISL_2084507, EPI_ISL_2084508, EPI_ISL_2084509, EPI_ISL_2084510, EPI_ISL_2084511, EPI_ISL_2084512, EPI_ISL_2084513, EPI_ISL_2084514, EPI_ISL_2084515, EPI_ISL_2084516, EPI_ISL_2084517, EPI_ISL_2084518, EPI_ISL_2084519, EPI_ISL_2084520, EPI_ISL_2084521, EPI_ISL_2084522, EPI_ISL_2084523, EPI_ISL_2084524, EPI_ISL_2084525, EPI_ISL_2084526, EPI_ISL_2084527, EPI_ISL_2084528, EPI_ISL_2084529, EPI_ISL_2084530, EPI_ISL_2084531, EPI_ISL_2084532, EPI_ISL_2084533, EPI_ISL_2084534, EPI_ISL_2084535, EPI_ISL_2084536, EPI_ISL_2084537, EPI_ISL_2084538, EPI_ISL_2084539, EPI_ISL_2084540, EPI_ISL_2084541, EPI_ISL_2084542, EPI_ISL_2084543, EPI_ISL_2084544, EPI_ISL_2084545, EPI_ISL_2084546, EPI_ISL_2084547, EPI_ISL_2084548, EPI_ISL_2084549, EPI_ISL_2084550, EPI_ISL_2084551, EPI_ISL_2084552, EPI_ISL_2084553, EPI_ISL_2084554, EPI_ISL_2084555, EPI_ISL_2084556, EPI_ISL_2084557, EPI_ISL_2084558, EPI_ISL_2084559, EPI_ISL_2084560, EPI_ISL_2084561, EPI_ISL_2084562, EPI_ISL_2084563, EPI_ISL_2084564, EPI_ISL_2084565, EPI_ISL_2084566, EPI_ISL_2084567, EPI_ISL_2084568, EPI_ISL_2084569, EPI_ISL_2084570, EPI_ISL_2084571, EPI_ISL_2084572, EPI_ISL_2084573, EPI_ISL_2084574, EPI_ISL_2084575, EPI_ISL_2084576, EPI_ISL_2084577, EPI_ISL_2084578, EPI_ISL_2084579, EPI_ISL_2084580, EPI_ISL_2084581, EPI_ISL_2084582, EPI_ISL_2084583, EPI_ISL_2084584, EPI_ISL_2084585, EPI_ISL_2084586, EPI_ISL_2084587, EPI_ISL_2084588, EPI_ISL_2084589, EPI_ISL_2084590, EPI_ISL_2084591, EPI_ISL_2084592, EPI_ISL_2084593, EPI_ISL_2084594, EPI_ISL_2084595, EPI_ISL_2084596, EPI_ISL_2084597, EPI_ISL_2084598, EPI_ISL_2084599, EPI_ISL_2084600, EPI_ISL_2084601, EPI_ISL_2084602, EPI_ISL_2084603, EPI_ISL_2084604, EPI_ISL_2084605, EPI_ISL_2084606, EPI_ISL_2084607, EPI_ISL_2084608, EPI_ISL_2084609, EPI_ISL_2084610, EPI_ISL_2084611, EPI_ISL_2084612, EPI_ISL_2084613, EPI_ISL_2084614, EPI_ISL_2084615, EPI_ISL_2084616, EPI_ISL_2084617, EPI_ISL_2084618, EPI_ISL_2084619, EPI_ISL_2084620, EPI_ISL_2084621, EPI_ISL_2084622, EPI_ISL_2084623, EPI_ISL_2084624, EPI_ISL_2084625, EPI_ISL_2084626, EPI_ISL_2084627, EPI_ISL_2084628, EPI_ISL_2084629, EPI_ISL_2084630, EPI_ISL_2084631, EPI_ISL_2084632, EPI_ISL_2084633, EPI_ISL_2084634, EPI_ISL_2084635, EPI_ISL_2084636, EPI_ISL_2084637, EPI_ISL_2084638, EPI_ISL_2084639, EPI_ISL_2084640, EPI_ISL_2084641, EPI_ISL_2084642, EPI_ISL_2084643, EPI_ISL_2084644, EPI_ISL_2084645, EPI_ISL_2084646, EPI_ISL_2084647, EPI_ISL_2084648, EPI_ISL_2084649, EPI_ISL_2084650, EPI_ISL_2084651, EPI_ISL_2084652, EPI_ISL_2084653, EPI_ISL_2084654, EPI_ISL_2084655, EPI_ISL_2084656, EPI_ISL_2084657, EPI_ISL_2084658, EPI_ISL_2084659, EPI_ISL_2084660, EPI_ISL_2084661, EPI_ISL_2084662, EPI_ISL_2084663, EPI_ISL_2084664, EPI_ISL_2084665, EPI_ISL_2084666, EPI_ISL_2084667, EPI_ISL_2084668, EPI_ISL_2084669, EPI_ISL_2084670, EPI_ISL_2084671, EPI_ISL_2084672, EPI_ISL_2084673, EPI_ISL_2084674, EPI_ISL_2084675, EPI_ISL_2084676, EPI_ISL_2084677, EPI_ISL_2084678, EPI_ISL_2084679, EPI_ISL_2084680, EPI_ISL_2084681, EPI_ISL_2084682, EPI_ISL_2084683, EPI_ISL_2084684, EPI_ISL_2084685, EPI_ISL_2084686, EPI_ISL_2084687, EPI_ISL_2084688, EPI_ISL_2084689, EPI_ISL_2084690, EPI_ISL_2084691, EPI_ISL_2084692, EPI_ISL_2084693, EPI_ISL_2084694, EPI_ISL_2084695, EPI_ISL_2084696, EPI_ISL_2084697, EPI_ISL_2084698, EPI_ISL_2084699, EPI_ISL_2084700, EPI_ISL_2084701, EPI_ISL_2084702, EPI_ISL_2084703, EPI_ISL_2084704, EPI_ISL_2084705, EPI_ISL_2084706, EPI_ISL_2084707, EPI_ISL_2084708, EPI_ISL_2084709, EPI_ISL_2084710, EPI_ISL_2084711, EPI_ISL_2084712, EPI_ISL_2084713, EPI_ISL_2084714, EPI_ISL_2084715, EPI_ISL_2084716, EPI_ISL_2084717, EPI_ISL_2084718, EPI_ISL_2084719, EPI_ISL_2084720, EPI_ISL_2084721, EPI_ISL_2084722, EPI_ISL_2084723, EPI_ISL_2084724, EPI_ISL_2084725, EPI_ISL_2084726, EPI_ISL_2084727, EPI_ISL_2084728, EPI_ISL_2084729, EPI_ISL_2084730, EPI_ISL_2084731, EPI_ISL_2084732

| | | | |
|---|---|---|---|
| see above | Israel Central Virology laboratory | Israel National Consortium for SARS-CoV-2 sequencing | Neta Zuckerman, Efrat Dahan Bucris, Michal Mandelboim, Dana Bar-Ilan, Oran Erster, Tzvia Mann, Omer Murik, David A. Zeevi, Assaf Rokney, Joseph Jaffe, Eva Nachum, Maya Davidovich Cohen, Ephraim Fass, Gal Zizelski Valenci, Mor Rubinstein, Efrat Rorman, Israel Nissan, Efrat Glick-Saar, Omri Nayshool, Gideon Rechavi, Ella Mendelson, Orna Mor |

EPI_ISL_2084733, EPI_ISL_2084734, EPI_ISL_2084735, EPI_ISL_2084736, EPI_ISL_2084737, EPI_ISL_2084738, EPI_ISL_2084739, EPI_ISL_2084740, EPI_ISL_2084741, EPI_ISL_2084742, EPI_ISL_2084743, EPI_ISL_2084744, EPI_ISL_2084745, EPI_ISL_2084746, EPI_ISL_2084747, EPI_ISL_2084748, EPI_ISL_2084749, EPI_ISL_2084750, EPI_ISL_2084751, EPI_ISL_2084752, EPI_ISL_2084753, EPI_ISL_2084754, EPI_ISL_2084755, EPI_ISL_2084756, EPI_ISL_2084757, EPI_ISL_2084758, EPI_ISL_2084759, EPI_ISL_2084760, EPI_ISL_2084761, EPI_ISL_2084762, EPI_ISL_2084763, EPI_ISL_2084764, EPI_ISL_2084765, EPI_ISL_2084766, EPI_ISL_2084767, EPI_ISL_2084768, EPI_ISL_2084769, EPI_ISL_2084770, EPI_ISL_2084771, EPI_ISL_2084772, EPI_ISL_2084773, EPI_ISL_2084774, EPI_ISL_2084775, EPI_ISL_2084776, EPI_ISL_2084777, EPI_ISL_2084778, EPI_ISL_2084779, EPI_ISL_2084780, EPI_ISL_2084781, EPI_ISL_2084782, EPI_ISL_2084783, EPI_ISL_2084784, EPI_ISL_2084785, EPI_ISL_2084786, EPI_ISL_2084787, EPI_ISL_2084788, EPI_ISL_2084789, EPI_ISL_2084790, EPI_ISL_2084791, EPI_ISL_2084792, EPI_ISL_2084793, EPI_ISL_2084794, EPI_ISL_2084795, EPI_ISL_2084796, EPI_ISL_2084797, EPI_ISL_2084798, EPI_ISL_2084799, EPI_ISL_2084800, EPI_ISL_2084801, EPI_ISL_2084802, EPI_ISL_2084803, EPI_ISL_2084804, EPI_ISL_2084805, EPI_ISL_2084806, EPI_ISL_2084807, EPI_ISL_2084808, EPI_ISL_2084809, EPI_ISL_2084810, EPI_ISL_2084811, EPI_ISL_2084812, EPI_ISL_2084813, EPI_ISL_2084814, EPI_ISL_2084815, EPI_ISL_2084816, EPI_ISL_2084817, EPI_ISL_2084818, EPI_ISL_2084819, EPI_ISL_2084820, EPI_ISL_2084821, EPI_ISL_2084822, EPI_ISL_2084823, EPI_ISL_2084824, EPI_ISL_2084825, EPI_ISL_2084826, EPI_ISL_2084827, EPI_ISL_2084828, EPI_ISL_2084829, EPI_ISL_2084830, EPI_ISL_2084831, EPI_ISL_2084832, EPI_ISL_2084833, EPI_ISL_2084834, EPI_ISL_2084835, EPI_ISL_2084836, EPI_ISL_2084837, EPI_ISL_2084838, EPI_ISL_2084839, EPI_ISL_2084840, EPI_ISL_2084841, EPI_ISL_2084842, EPI_ISL_2084843, EPI_ISL_2084844, EPI_ISL_2084845, EPI_ISL_2084846, EPI_ISL_2084847, EPI_ISL_2084848, EPI_ISL_2084849, EPI_ISL_2084850, EPI_ISL_2084851, EPI_ISL_2084852, EPI_ISL_2084853, EPI_ISL_2084854, EPI_ISL_2084855, EPI_ISL_2084856, EPI_ISL_2084857, EPI_ISL_2084858, EPI_ISL_2084859, EPI_ISL_2084860, EPI_ISL_2084861, EPI_ISL_2084862, EPI_ISL_2084863, EPI_ISL_2084864, EPI_ISL_2084865, EPI_ISL_2084866, EPI_ISL_2084867, EPI_ISL_2084868, EPI_ISL_2084869, EPI_ISL_2084870, EPI_ISL_2084871, EPI_ISL_2084872, EPI_ISL_2084873, EPI_ISL_2084874, EPI_ISL_2084875, EPI_ISL_2084876, EPI_ISL_2084877, EPI_ISL_2084878, EPI_ISL_2084879, EPI_ISL_2084880, EPI_ISL_2084881, EPI_ISL_2084882, EPI_ISL_2084883, EPI_ISL_2084884, EPI_ISL_2084885, EPI_ISL_2084886, EPI_ISL_2084887, EPI_ISL_2084888, EPI_ISL_2084889, EPI_ISL_2084890, EPI_ISL_2084891, EPI_ISL_2084892, EPI_ISL_2084893, EPI_ISL_2084894, EPI_ISL_2084895, EPI_ISL_2084896, EPI_ISL_2084897, EPI_ISL_2084898, EPI_ISL_2084899, EPI_ISL_2084900, EPI_ISL_2084901, EPI_ISL_2084902, EPI_ISL_2084903, EPI_ISL_2084904, EPI_ISL_2084905, EPI_ISL_2084906, EPI_ISL_2084907, EPI_ISL_2084908, EPI_ISL_2084909, EPI_ISL_2084910, EPI_ISL_2084911, EPI_ISL_2084912, EPI_ISL_2084913, EPI_ISL_2084914, EPI_ISL_2084915, EPI_ISL_2084916, EPI_ISL_2084917, EPI_ISL_2084918, EPI_ISL_2084919, EPI_ISL_2084920, EPI_ISL_2084921, EPI_ISL_2084922, EPI_ISL_2084923, EPI_ISL_2084924, EPI_ISL_2084925, EPI_ISL_2084926, EPI_ISL_2084927, EPI_ISL_2084928, EPI_ISL_2084929, EPI_ISL_2084930, EPI_ISL_2084931, EPI_ISL_2084932, EPI_ISL_2084933, EPI_ISL_2084934, EPI_ISL_2084935, EPI_ISL_2084936, EPI_ISL_2084937, EPI_ISL_2084938, EPI_ISL_2084939, EPI_ISL_2084940, EPI_ISL_2084941, EPI_ISL_2084942, EPI_ISL_2084943, EPI_ISL_2084944, EPI_ISL_2084945, EPI_ISL_2084946, EPI_ISL_2084947, EPI_ISL_2084948, EPI_ISL_2084949, EPI_ISL_2084950, EPI_ISL_2084951, EPI_ISL_2084952, EPI_ISL_2084953, EPI_ISL_2084954, EPI_ISL_2084955, EPI_ISL_2084956, EPI_ISL_2084957, EPI_ISL_2084958, EPI_ISL_2084959, EPI_ISL_2084960, EPI_ISL_2084961, EPI_ISL_2084962, EPI_ISL_2084963, EPI_ISL_2084964, EPI_ISL_2084965, EPI_ISL_2084966, EPI_ISL_2084967, EPI_ISL_2084968, EPI_ISL_2084969, EPI_ISL_2084970, EPI_ISL_2084971, EPI_ISL_2084972, EPI_ISL_2084973, EPI_ISL_2084974, EPI_ISL_2084975, EPI_ISL_2084976, EPI_ISL_2084977, EPI_ISL_2084978, EPI_ISL_2084979, EPI_ISL_2084980, EPI_ISL_2084981, EPI_ISL_2084982, EPI_ISL_2084983, EPI_ISL_2084984, EPI_ISL_2084985, EPI_ISL_2084986, EPI_ISL_2084987, EPI_ISL_2084988, EPI_ISL_2084989, EPI_ISL_2084990, EPI_ISL_2084991, EPI_ISL_2084992, EPI_ISL_2084993, EPI_ISL_2084994, EPI_ISL_2084995, EPI_ISL_2084996, EPI_ISL_2084997, EPI_ISL_2084998, EPI_ISL_2084999, EPI_ISL_2085000, EPI_ISL_2085001, EPI_ISL_2085002, EPI_ISL_2085003, EPI_ISL_2085004, EPI_ISL_2085005, EPI_ISL_2085006, EPI_ISL_2085007, EPI_ISL_2085008, EPI_ISL_2085009, EPI_ISL_2085010, EPI_ISL_2085011, EPI_ISL_2085012, EPI_ISL_2085013, EPI_ISL_2085014, EPI_ISL_2085015, EPI_ISL_2085016, EPI_ISL_2085017, EPI_ISL_2085018, EPI_ISL_2085019, EPI_ISL_2085020, EPI_ISL_2085021, EPI_ISL_2085022, EPI_ISL_2085023, EPI_ISL_2085024, EPI_ISL_2085025, EPI_ISL_2085026, EPI_ISL_2085027, EPI_ISL_2085028, EPI_ISL_2085029, EPI_ISL_2085030, EPI_ISL_2085031, EPI_ISL_2085032, EPI_ISL_2085033, EPI_ISL_2085034, EPI_ISL_2085035, EPI_ISL_2085036, EPI_ISL_2085037, EPI_ISL_2085038, EPI_ISL_2085039, EPI_ISL_2085040, EPI_ISL_2085041, EPI_ISL_2085042, EPI_ISL_2085043, EPI_ISL_2085044, EPI_ISL_2085045, EPI_ISL_2085046, EPI_ISL_2085047, EPI_ISL_2085048, EPI_ISL_2085049, EPI_ISL_2085050, EPI_ISL_2085051, EPI_ISL_2085052, EPI_ISL_2085053, EPI_ISL_2085054, EPI_ISL_2085055, EPI_ISL_2085056, EPI_ISL_2085057, EPI_ISL_2085058, EPI_ISL_2085059, EPI_ISL_2085060, EPI_ISL_2085061, EPI_ISL_2085062, EPI_ISL_2085063, EPI_ISL_2085064, EPI_ISL_2085065, EPI_ISL_2085066, EPI_ISL_2085067, EPI_ISL_2085068, EPI_ISL_2085069, EPI_ISL_2085070, EPI_ISL_2085071, EPI_ISL_2085072, EPI_ISL_2085073, EPI_ISL_2085074, EPI_ISL_2085075, EPI_ISL_2085076, EPI_ISL_2085077, EPI_ISL_2085078, EPI_ISL_2085079, EPI_ISL_2085080, EPI_ISL_2085081, EPI_ISL_2085082, EPI_ISL_2085083, EPI_ISL_2085084, EPI_ISL_2085085, EPI_ISL_2085086, EPI_ISL_2085087, EPI_ISL_2085088, EPI_ISL_2085089, EPI_ISL_2085090, EPI_ISL_2085091, EPI_ISL_2085092, EPI_ISL_2085093, EPI_ISL_2085094, EPI_ISL_2085095, EPI_ISL_2085096, EPI_ISL_2085097, EPI_ISL_2085098, EPI_ISL_2085099, EPI_ISL_2085100, EPI_ISL_2085101, EPI_ISL_2085102, EPI_ISL_2085103, EPI_ISL_2085104, EPI_ISL_2085105, EPI_ISL_2085106, EPI_ISL_2085107, EPI_ISL_2085108, EPI_ISL_2085109, EPI_ISL_2085110, EPI_ISL_2085111, EPI_ISL_2085112, EPI_ISL_2085113, EPI_ISL_2085114, EPI_ISL_2085115, EPI_ISL_2085116, EPI_ISL_2085117, EPI_ISL_2085118, EPI_ISL_2085119, EPI_ISL_2085120, EPI_ISL_2085121, EPI_ISL_2085122, EPI_ISL_2085123, EPI_ISL_2085124, EPI_ISL_2085125, EPI_ISL_2085126, EPI_ISL_2085127, EPI_ISL_2085128, EPI_ISL_2085129, EPI_ISL_2085130, EPI_ISL_2085131, EPI_ISL_2085132, EPI_ISL_2085133, EPI_ISL_2085134, EPI_ISL_2085135, EPI_ISL_2085136, EPI_ISL_2085137, EPI_ISL_2085138, EPI_ISL_2085139, EPI_ISL_2085140, EPI_ISL_2085141, EPI_ISL_2085142, EPI_ISL_2085143, EPI_ISL_2085144, EPI_ISL_2085145, EPI_ISL_2085146, EPI_ISL_2085147, EPI_ISL_2085148, EPI_ISL_2085149, EPI_ISL_2085150, EPI_ISL_2085151, EPI_ISL_2085152, EPI_ISL_2085153, EPI_ISL_2085154, EPI_ISL_2085155, EPI_ISL_2085156, EPI_ISL_2085157, EPI_ISL_2085158, EPI_ISL_2085159, EPI_ISL_2085160, EPI_ISL_2085161, EPI_ISL_2085162, EPI_ISL_2085163, EPI_ISL_2085164, EPI_ISL_2085165, EPI_ISL_2085166, EPI_ISL_2085167, EPI_ISL_2085168, EPI_ISL_2085169, EPI_ISL_2085170, EPI_ISL_2085171, EPI_ISL_2085172, EPI_ISL_2085173, EPI_ISL_2085174, EPI_ISL_2085175, EPI_ISL_2085176, EPI_ISL_2085177, EPI_ISL_2085178, EPI_ISL_2085179, EPI_ISL_2085180, EPI_ISL_2085181, EPI_ISL_2085182, EPI_ISL_2085183, EPI_ISL_2085184, EPI_ISL_2085185, EPI_ISL_2085186, EPI_ISL_2085187, EPI_ISL_2085188, EPI_ISL_2085189, EPI_ISL_2085190, EPI_ISL_2085191, EPI_ISL_2085192, EPI_ISL_2085193, EPI_ISL_2085194, EPI_ISL_2085195, EPI_ISL_2085196, EPI_ISL_2085197, EPI_ISL_2085198, EPI_ISL_2085199, EPI_ISL_2085200, EPI_ISL_2085201, EPI_ISL_2085202, EPI_ISL_2085203, EPI_ISL_2085204, EPI_ISL_2085205, EPI_ISL_2085206, EPI_ISL_2085207, EPI_ISL_2085208, EPI_ISL_2085209, EPI_ISL_2085210, EPI_ISL_2085211, EPI_ISL_2085212, EPI_ISL_2085213, EPI_ISL_2085214, EPI_ISL_2085215, EPI_ISL_2085216, EPI_ISL_2085217, EPI_ISL_2085218, EPI_ISL_2085219, EPI_ISL_2085220, EPI_ISL_2085221, EPI_ISL_2085222, EPI_ISL_2085223, EPI_ISL_2085224, EPI_ISL_2085225, EPI_ISL_2085226, EPI_ISL_2085227, EPI_ISL_2085228, EPI_ISL_2085229, EPI_ISL_2085230, EPI_ISL_2085231, EPI_ISL_2085232, EPI_ISL_2085233, EPI_ISL_2085234, EPI_ISL_2085235, EPI_ISL_2085236, EPI_ISL_2085237, EPI_ISL_2085238, EPI_ISL_2085239, EPI_ISL_2085240, EPI_ISL_2085241, EPI_ISL_2085242, EPI_ISL_2085243, EPI_ISL_2085244, EPI_ISL_2085245, EPI_ISL_2085246, EPI_ISL_2085247, EPI_ISL_2085248, EPI_ISL_2085249, EPI_ISL_2085250, EPI_ISL_2085251, EPI_ISL_2085252, EPI_ISL_2085253, EPI_ISL_2085254, EPI_ISL_2085255, EPI_ISL_2085256, EPI_ISL_2085257, EPI_ISL_2085258, EPI_ISL_2085259, EPI_ISL_2085260, EPI_ISL_2085261, EPI_ISL_2085262, EPI_ISL_2085263, EPI_ISL_2085264, EPI_ISL_2085265, EPI_ISL_2085266, EPI_ISL_2085267, EPI_ISL_2085268, EPI_ISL_2085269, EPI_ISL_2085270, EPI_ISL_2085271, EPI_ISL_2085272, EPI_ISL_2085273, EPI_ISL_2085274, EPI_ISL_2085275, EPI_ISL_2085276, EPI_ISL_2085277, EPI_ISL_2085278, EPI_ISL_2085279, EPI_ISL_2085280, EPI_ISL_2085281, EPI_ISL_2085282, EPI_ISL_2085283, EPI_ISL_2085284, EPI_ISL_2085285, EPI_ISL_2085286, EPI_ISL_2085287, EPI_ISL_2085288, EPI_ISL_2085289, EPI_ISL_2085290, EPI_ISL_2085291, EPI_ISL_2085292, EPI_ISL_2085293, EPI_ISL_2085294, EPI_ISL_2085295, EPI_ISL_2085296, EPI_ISL_2085297, EPI_ISL_2085298, EPI_ISL_2085299, EPI_ISL_2085300, EPI_ISL_2085301, EPI_ISL_2085302, EPI_ISL_2085303, EPI_ISL_2085304, EPI_ISL_2085305, EPI_ISL_2085306, EPI_ISL_2085307, EPI_ISL_2085308, EPI_ISL_2085309, EPI_ISL_2085310, EPI_ISL_2085311, EPI_ISL_2085312, EPI_ISL_2085313, EPI_ISL_2085314, EPI_ISL_2085315, EPI_ISL_2085316, EPI_ISL_2085317, EPI_ISL_2085318, EPI_ISL_2085319, EPI_ISL_2085320, EPI_ISL_2085321, EPI_ISL_2085322, EPI_ISL_2085323, EPI_ISL_2085324, EPI_ISL_2085325, EPI_ISL_2085326, EPI_ISL_2085327, EPI_ISL_2085328, EPI_ISL_2085329, EPI_ISL_2085330, EPI_ISL_2085331, EPI_ISL_2085332, EPI_ISL_2085333, EPI_ISL_2085334, EPI_ISL_2085335, EPI_ISL_2085336, EPI_ISL_2085337, EPI_ISL_2085338, EPI_ISL_2085339, EPI_ISL_2085340, EPI_ISL_2085341, EPI_ISL_2085342, EPI_ISL_2085343, EPI_ISL_2085344, EPI_ISL_2085345, EPI_ISL_2085346, EPI_ISL_2085347, EPI_ISL_2085348, EPI_ISL_2085349, EPI_ISL_2085350, EPI_ISL_2085351, EPI_ISL_2085352, EPI_ISL_2085353, EPI_ISL_2085354, EPI_ISL_2085355, EPI_ISL_2085356, EPI_ISL_2085357, EPI_ISL_2085358, EPI_ISL_2085359, EPI_ISL_2085360, EPI_ISL_2085361, EPI_ISL_2085362, EPI_ISL_2085363, EPI_ISL_2085364, EPI_ISL_2085365, EPI_ISL_2085366, EPI_ISL_2085367, EPI_ISL_2085368, EPI_ISL_2085369, EPI_ISL_2085370, EPI_ISL_2085371, EPI_ISL_2085372, EPI_ISL_2085373, EPI_ISL_2085374, EPI_ISL_2085375, EPI_ISL_2085376, EPI_ISL_2085377, EPI_ISL_2085378, EPI_ISL_2085379, EPI_ISL_2085380, EPI_ISL_2085381, EPI_ISL_2085382, EPI_ISL_2085383, EPI_ISL_2085384, EPI_ISL_2085385, EPI_ISL_2085386, EPI_ISL_2085387, EPI_ISL_2085388, EPI_ISL_2085389, EPI_ISL_2085390, EPI_ISL_2085391, EPI_ISL_2085392, EPI_ISL_2085393, EPI_ISL_2085394, EPI_ISL_2085395, EPI_ISL_2085396, EPI_ISL_2085397, EPI_ISL_2085398, EPI_ISL_2085399, EPI_ISL_2085400, EPI_ISL_2085401, EPI_ISL_2085402, EPI_ISL_2085403, EPI_ISL_2085404, EPI_ISL_2085405, EPI_ISL_2085406, EPI_ISL_2085407, EPI_ISL_2085408, EPI_ISL_2085409, EPI_ISL_2085410, EPI_ISL_2085411, EPI_ISL_2085412, EPI_ISL_2085413, EPI_ISL_2085414, EPI_ISL_2085415, EPI_ISL_2085416, EPI_ISL_2085417, EPI_ISL_2085418, EPI_ISL_2085419, EPI_ISL_2085420, EPI_ISL_2085421, EPI_ISL_2085422, EPI_ISL_2085423, EPI_ISL_2085424, EPI_ISL_2085425, EPI_ISL_2085426, EPI_ISL_2085427, EPI_ISL_2085428, EPI_ISL_2085429, EPI_ISL_2085430, EPI_ISL_2085431, EPI_ISL_2085432, EPI_ISL_2085433, EPI_ISL_2085434, EPI_ISL_2085435, EPI_ISL_2085436, EPI_ISL_2085437, EPI_ISL_2085438, EPI_ISL_2085439, EPI_ISL_2085440, EPI_ISL_2085441, EPI_ISL_2085442, EPI_ISL_2085443, EPI_ISL_2085444, EPI_ISL_2085445, EPI_ISL_2085446, EPI_ISL_2085447, EPI_ISL_2085448, EPI_ISL_2085449, EPI_ISL_2085450, EPI_ISL_2085451, EPI_ISL_2085452, EPI_ISL_2085453, EPI_ISL_2085454, EPI_ISL_2085455, EPI_ISL_2085456, EPI_ISL_2085457, EPI_ISL_2085458, EPI_ISL_2085459, EPI_ISL_2085460, EPI_ISL_2085461, EPI_ISL_2085462, EPI_ISL_2085463, EPI_ISL_2085464, EPI_ISL_2085465, EPI_ISL_2085466, EPI_ISL_2085467, EPI_ISL_2085468, EPI_ISL_2085469, EPI_ISL_2085470, EPI_ISL_2085471, EPI_ISL_2085472, EPI_ISL_2085473, EPI_ISL_2085474, EPI_ISL_2085475, EPI_ISL_2085476, EPI_ISL_2085477, EPI_ISL_2085478, EPI_ISL_2085479, EPI_ISL_2085480, EPI_ISL_2085481, EPI_ISL_2085482, EPI_ISL_2085483, EPI_ISL_2085484, EPI_ISL_2085485, EPI_ISL_2085486, EPI_ISL_2085487, EPI_ISL_2085488, EPI_ISL_2085489, EPI_ISL_2085490, EPI_ISL_2085491, EPI_ISL_2085492, EPI_ISL_2085493, EPI_ISL_2085494, EPI_ISL_2085495, EPI_ISL_2085496, EPI_ISL_2085497, EPI_ISL_2085498, EPI_ISL_2085499, EPI_ISL_2085500, EPI_ISL_2085501, EPI_ISL_2085502, EPI_ISL_2085503, EPI_ISL_2085504, EPI_ISL_2085505, EPI_ISL_2085506, EPI_ISL_2085507, EPI_ISL_2085508, EPI_ISL_2085509, EPI_ISL_2085510, EPI_ISL_2085511, EPI_ISL_2085512, EPI_ISL_2085513, EPI_ISL_2085514, EPI_ISL_2085515, EPI_ISL_2085516, EPI_ISL_2085517, EPI_ISL_2085518, EPI_ISL_2085519, EPI_ISL_2085520, EPI_ISL_2085521, EPI_ISL_2085522, EPI_ISL_2085523, EPI_ISL_2085524, EPI_ISL_2085525, EPI_ISL_2085526, EPI_ISL_2085527, EPI_ISL_2085528, EPI_ISL_2085529, EPI_ISL_2085530, EPI_ISL_2085531, EPI_ISL_2085532, EPI_ISL_2085533, EPI_ISL_2085534, EPI_ISL_2085535, EPI_ISL_2085536, EPI_ISL_2085537, EPI_ISL_2085538, EPI_ISL_2085539, EPI_ISL_2085540, EPI_ISL_2085541, EPI_ISL_2085542, EPI_ISL_2085543, EPI_ISL_2085544, EPI_ISL_2085545, EPI_ISL_2085546, EPI_ISL_2085547, EPI_ISL_2085548, EPI_ISL_2085549, EPI_ISL_2085550, EPI_ISL_2085551, EPI_ISL_2085552, EPI_ISL_2085553, EPI_ISL_2085554, EPI_ISL_2085555, EPI_ISL_2085556, EPI_ISL_2085557, EPI_ISL_2085558, EPI_ISL_2085559, EPI_ISL_2085560, EPI_ISL_2085561, EPI_ISL_2085562, EPI_ISL_2085563, EPI_ISL_2085564, EPI_ISL_2085565, EPI_ISL_2085566, EPI_ISL_2085567, EPI_ISL_2085568, EPI_ISL_2085569, EPI_ISL_2085570, EPI_ISL_2085571, EPI_ISL_2085572, EPI_ISL_2085573, EPI_ISL_2085574, EPI_ISL_2085575, EPI_ISL_2085576, EPI_ISL_2085577, EPI_ISL_2085578, EPI_ISL_2085579, EPI_ISL_2085580, EPI_ISL_2085581, EPI_ISL_2085582, EPI_ISL_2085583, EPI_ISL_2085584, EPI_ISL_2085585, EPI_ISL_2085586, EPI_ISL_2085587, EPI_ISL_2085588, EPI_ISL_2085589, EPI_ISL_2085590, EPI_ISL_2085591, EPI_ISL_2085592, EPI_ISL_2085593, EPI_ISL_2085594, EPI_ISL_2085595, EPI_ISL_2085596, EPI_ISL_2085597, EPI_ISL_2085598, EPI_ISL_2085599,

| | | | |
|---|---|---|---|
| EPI_ISL_2085600, EPI_ISL_2085601, EPI_ISL_2085602, EPI_ISL_2085603, EPI_ISL_2085604, EPI_ISL_2085605, EPI_ISL_2085606, EPI_ISL_2085607, EPI_ISL_2085608, EPI_ISL_2085609, EPI_ISL_2085610, EPI_ISL_2085611, EPI_ISL_2085612, EPI_ISL_2085613, EPI_ISL_2085614, EPI_ISL_2085615, EPI_ISL_2085616, EPI_ISL_2085617, EPI_ISL_2085618, EPI_ISL_2085619, EPI_ISL_2085620, EPI_ISL_2085621, EPI_ISL_2085622, EPI_ISL_2085623, EPI_ISL_2085624, EPI_ISL_2085625, EPI_ISL_2085626, EPI_ISL_2085627, EPI_ISL_2085628, EPI_ISL_2085629, EPI_ISL_2085630, EPI_ISL_2085631, EPI_ISL_2085632, EPI_ISL_2085633, EPI_ISL_2085634, EPI_ISL_2085635, EPI_ISL_2085636, EPI_ISL_2085637, EPI_ISL_2085638, EPI_ISL_2085639, EPI_ISL_2085640, EPI_ISL_2085641, EPI_ISL_2085642, EPI_ISL_2085643, EPI_ISL_2085644, EPI_ISL_2085645, EPI_ISL_2085646, EPI_ISL_2085647, EPI_ISL_2085648, EPI_ISL_2085649, EPI_ISL_2085650, EPI_ISL_2085651, EPI_ISL_2085652, EPI_ISL_2085653, EPI_ISL_2085654, EPI_ISL_2085655, EPI_ISL_2085656, EPI_ISL_2085657, EPI_ISL_2085658, EPI_ISL_2085659, EPI_ISL_2085660, EPI_ISL_2085661, EPI_ISL_2085662, EPI_ISL_2085663, EPI_ISL_2085664, EPI_ISL_2085665, EPI_ISL_2085666, EPI_ISL_2085667, EPI_ISL_2085668, EPI_ISL_2085669, EPI_ISL_2085670, EPI_ISL_2085671, EPI_ISL_2085672, EPI_ISL_2085673, EPI_ISL_2085674, EPI_ISL_2085675, EPI_ISL_2085676, EPI_ISL_2085677, EPI_ISL_2085678, EPI_ISL_2085679, EPI_ISL_2085680, EPI_ISL_2085681, EPI_ISL_2085682, EPI_ISL_2085683, EPI_ISL_2085684, EPI_ISL_2085685, EPI_ISL_2085686, EPI_ISL_2085687, EPI_ISL_2085688, EPI_ISL_2085689, EPI_ISL_2085690, EPI_ISL_2085691, EPI_ISL_2085692, EPI_ISL_2085693, EPI_ISL_2085694, EPI_ISL_2085695, EPI_ISL_2085696, EPI_ISL_2085697, EPI_ISL_2085698, EPI_ISL_2085699, EPI_ISL_2085700, EPI_ISL_2085701, EPI_ISL_2085702, EPI_ISL_2085703, EPI_ISL_2085704, EPI_ISL_2085705, EPI_ISL_2085706, EPI_ISL_2085707, EPI_ISL_2085708, EPI_ISL_2085709, EPI_ISL_2085710, EPI_ISL_2085711, EPI_ISL_2085712, EPI_ISL_2085713, EPI_ISL_2085714, EPI_ISL_2085715, EPI_ISL_2085716, EPI_ISL_2085717, EPI_ISL_2085718, EPI_ISL_2085719, EPI_ISL_2085720, EPI_ISL_2085721, EPI_ISL_2085722, EPI_ISL_2085723, EPI_ISL_2085724, EPI_ISL_2085725, EPI_ISL_2085726, EPI_ISL_2085727, EPI_ISL_2085728, EPI_ISL_2085729, EPI_ISL_2085730, EPI_ISL_2085731, EPI_ISL_2085732, EPI_ISL_2085733, EPI_ISL_2085734, EPI_ISL_2085735, EPI_ISL_2085736, EPI_ISL_2085737, EPI_ISL_2085738, EPI_ISL_2085739, EPI_ISL_2085740, EPI_ISL_2085741, EPI_ISL_2085742, EPI_ISL_2085743, EPI_ISL_2085744, EPI_ISL_2085745, EPI_ISL_2085746, EPI_ISL_2085747, EPI_ISL_2085748, EPI_ISL_2085749, EPI_ISL_2085750, EPI_ISL_2085751, EPI_ISL_2085752, EPI_ISL_2085753, EPI_ISL_2085754, EPI_ISL_2085755, EPI_ISL_2085756, EPI_ISL_2085757, EPI_ISL_2085758, EPI_ISL_2085759, EPI_ISL_2085760, EPI_ISL_2085761, EPI_ISL_2085762, EPI_ISL_2085763, EPI_ISL_2085764, EPI_ISL_2085765, EPI_ISL_2085766, EPI_ISL_2085767, EPI_ISL_2085768, EPI_ISL_2085769, EPI_ISL_2085770, EPI_ISL_2085771, EPI_ISL_2085772, EPI_ISL_2085773, EPI_ISL_2085774, EPI_ISL_2085775, EPI_ISL_2085776, EPI_ISL_2085777, EPI_ISL_2085815, EPI_ISL_2085816, EPI_ISL_2085817, EPI_ISL_2085818, EPI_ISL_2085982, EPI_ISL_2085983, EPI_ISL_2085984, EPI_ISL_2085985, EPI_ISL_2085986, EPI_ISL_2085987, EPI_ISL_2085988, EPI_ISL_2085989, EPI_ISL_2085993, EPI_ISL_2085994 | | | |
| see above | Israel Central Virology laboratory | Israel National Consortium for SARS-CoV-2 sequencing | Neta Zuckerman, Efrat Dahan Bucris, Michal Mandelboim, Dana Bar-Ilan, Miranda Geva, Netanel Abu, Oran Erster, Efrat Glick-Saar, Omri Nayshool, Gideon Rechavi, Ella Mendelson, Orna Mor |
| EPI_ISL_2096781, EPI_ISL_2096782, EPI_ISL_2096783, EPI_ISL_2096784, EPI_ISL_2096785, EPI_ISL_2096786, EPI_ISL_2096787, EPI_ISL_2096788, EPI_ISL_2096789, EPI_ISL_2096790, EPI_ISL_2096791, EPI_ISL_2096792, EPI_ISL_2096793, EPI_ISL_2096794, EPI_ISL_2096795, EPI_ISL_2096796, EPI_ISL_2096797, EPI_ISL_2096798, EPI_ISL_2096799, EPI_ISL_2096800, EPI_ISL_2096801, EPI_ISL_2096802, EPI_ISL_2096803, EPI_ISL_2096804, EPI_ISL_2096805, EPI_ISL_2096806, EPI_ISL_2096807, EPI_ISL_2096808, EPI_ISL_2096809, EPI_ISL_2096810, EPI_ISL_2096811, EPI_ISL_2096812, EPI_ISL_2096813, EPI_ISL_2096814, EPI_ISL_2096815, EPI_ISL_2096816, EPI_ISL_2096817, EPI_ISL_2096818, EPI_ISL_2096819, EPI_ISL_2096820, EPI_ISL_2096821, EPI_ISL_2096822, EPI_ISL_2096823, EPI_ISL_2096824, EPI_ISL_2096825, EPI_ISL_2096826, EPI_ISL_2096827, EPI_ISL_2096828, EPI_ISL_2096829, EPI_ISL_2096830, EPI_ISL_2096831, EPI_ISL_2096832, EPI_ISL_2096833, EPI_ISL_2096834, EPI_ISL_2096835, EPI_ISL_2096836, EPI_ISL_2096837, EPI_ISL_2096838, EPI_ISL_2096839, EPI_ISL_2096840, EPI_ISL_2096841, EPI_ISL_2096842, EPI_ISL_2096843, EPI_ISL_2096844, EPI_ISL_2096845, EPI_ISL_2096846, EPI_ISL_2096847, EPI_ISL_2096848, EPI_ISL_2096849, EPI_ISL_2096850, EPI_ISL_2096851, EPI_ISL_2096852, EPI_ISL_2096853, EPI_ISL_2096854, EPI_ISL_2096855, EPI_ISL_2096856, EPI_ISL_2096857, EPI_ISL_2096858, EPI_ISL_2096859, EPI_ISL_2096860, EPI_ISL_2096861, EPI_ISL_2096862, EPI_ISL_2096863, EPI_ISL_2096864, EPI_ISL_2096865, EPI_ISL_2096866, EPI_ISL_2096867, EPI_ISL_2096868, EPI_ISL_2096869, EPI_ISL_2096870, EPI_ISL_2096871, EPI_ISL_2096872, EPI_ISL_2096873, EPI_ISL_2096874, EPI_ISL_2096875, EPI_ISL_2096876, EPI_ISL_2096877, EPI_ISL_2096878, EPI_ISL_2096880, EPI_ISL_2096881, EPI_ISL_2096882, EPI_ISL_2096883, EPI_ISL_2096884, EPI_ISL_2096885, EPI_ISL_2096886, EPI_ISL_2096887, EPI_ISL_2096888, EPI_ISL_2096889, EPI_ISL_2096890, EPI_ISL_2096891, EPI_ISL_2096892, EPI_ISL_2096893, EPI_ISL_2096894, EPI_ISL_2096895, EPI_ISL_2096896, EPI_ISL_2096897, EPI_ISL_2096898, EPI_ISL_2096899, EPI_ISL_2096900, EPI_ISL_2096901, EPI_ISL_2096902, EPI_ISL_2096903, EPI_ISL_2096904, EPI_ISL_2096905, EPI_ISL_2096906, EPI_ISL_2096907, EPI_ISL_2096908, EPI_ISL_2096909, EPI_ISL_2096910, EPI_ISL_2096911, EPI_ISL_2096917, EPI_ISL_2096918, EPI_ISL_2096919, EPI_ISL_2096920, EPI_ISL_2096921, EPI_ISL_2096922, EPI_ISL_2096923, EPI_ISL_2096924, EPI_ISL_2096925, EPI_ISL_2096926, EPI_ISL_2096949 | | | |
| see above | Hadassah Medical Center Clinical Virology Laboratory, Hadassah Ein Kerem | Hadassah Hebrew University Viral Sequencing Group, Hadassah Hebrew University Medical Center | Hadar Golan Berman, Esther Oiknine-Djian, Mila Rivkin, Sheera Adar, Dana G. Wolf |
| EPI_ISL_2096951, EPI_ISL_2096952, EPI_ISL_2096953, EPI_ISL_2096954, EPI_ISL_2096955, EPI_ISL_2096956, EPI_ISL_2096957, EPI_ISL_2096958, EPI_ISL_2096959, EPI_ISL_2096960, EPI_ISL_2096961, EPI_ISL_2096962, EPI_ISL_2096963, EPI_ISL_2096964, EPI_ISL_2096965, EPI_ISL_2096966, EPI_ISL_2096967, EPI_ISL_2096968, EPI_ISL_2096969, EPI_ISL_2096970, EPI_ISL_2096971, EPI_ISL_2096972, EPI_ISL_2096973, EPI_ISL_2096974, EPI_ISL_2096975, EPI_ISL_2096976, EPI_ISL_2096977, EPI_ISL_2096978, EPI_ISL_2096979, EPI_ISL_2096980, EPI_ISL_2096981 | | | |
| see above | Kaplan Medical Center Infectious Diseases Unit, Rehovot, Israel | Hadassah Hebrew University Viral Sequencing Group, Hadassah Hebrew University Medical Center | Hadar Golan Berman, Esther Oiknine-Djian, Mila Rivkin, Sheera Adar, Dana G. Wolf |
| EPI_ISL_2096982, EPI_ISL_2096983, EPI_ISL_2096984, EPI_ISL_2096985, EPI_ISL_2096986, EPI_ISL_2096987, EPI_ISL_2096988, EPI_ISL_2096989, EPI_ISL_2096990, EPI_ISL_2096991 | Hadassah Medical Center Clinical Virology Laboratory, Hadassah Ein Kerem | Hadassah Hebrew University Viral Sequencing Group, Hadassah Hebrew University Medical Center | Hadar Golan Berman, Esther Oiknine-Djian, Mila Rivkin, Sheera Adar, Dana G. Wolf |
| EPI_ISL_737202, EPI_ISL_737203, EPI_ISL_737204 | Central Virology Laboratory, Israel Ministry of Health | Central Virology Laboratory, Israel Ministry of Health | Neta S. Zuckerman, Efrat Dahan Bucris, Oran Erster, Michal Mandelboim, Orna Mor, Ella Mendelson |
| EPI_ISL_776569, EPI_ISL_776570, EPI_ISL_776571, EPI_ISL_776572, EPI_ISL_776573, EPI_ISL_776574, EPI_ISL_776575, EPI_ISL_776576, EPI_ISL_776577, EPI_ISL_776578, EPI_ISL_776579, EPI_ISL_776580, EPI_ISL_776581, EPI_ISL_776582, EPI_ISL_776583, EPI_ISL_776584, EPI_ISL_776585, EPI_ISL_776586, EPI_ISL_776587, EPI_ISL_776588, EPI_ISL_776589, EPI_ISL_776590, EPI_ISL_776591, EPI_ISL_776592, EPI_ISL_776593, EPI_ISL_776594, EPI_ISL_776595, EPI_ISL_776596, EPI_ISL_776597, EPI_ISL_776598, EPI_ISL_776599, EPI_ISL_776600, EPI_ISL_776601, EPI_ISL_776602, EPI_ISL_776603, EPI_ISL_776604, EPI_ISL_776605, EPI_ISL_776606, EPI_ISL_776607, EPI_ISL_776608, EPI_ISL_776609, EPI_ISL_776610, EPI_ISL_776611, EPI_ISL_776612, EPI_ISL_776613, EPI_ISL_776614, EPI_ISL_776615, EPI_ISL_776616, EPI_ISL_776617, EPI_ISL_776618, EPI_ISL_776619, EPI_ISL_776620, EPI_ISL_776621, EPI_ISL_776622, EPI_ISL_776623, EPI_ISL_776624, EPI_ISL_776625, EPI_ISL_776626, EPI_ISL_776627, EPI_ISL_776628, EPI_ISL_776629, EPI_ISL_776630, EPI_ISL_776631, EPI_ISL_776632, EPI_ISL_776633, EPI_ISL_776634, EPI_ISL_776635, EPI_ISL_776636, EPI_ISL_776637, EPI_ISL_776638, EPI_ISL_776639, EPI_ISL_776640, EPI_ISL_776641, EPI_ISL_776642, EPI_ISL_776643, EPI_ISL_776644, EPI_ISL_776645, EPI_ISL_776646, EPI_ISL_776647, EPI_ISL_776648, EPI_ISL_776649, EPI_ISL_776650, EPI_ISL_776651, EPI_ISL_776652, EPI_ISL_776653, EPI_ISL_776654, EPI_ISL_776655, EPI_ISL_776656, EPI_ISL_776775, EPI_ISL_776776, EPI_ISL_776777, EPI_ISL_776778, EPI_ISL_776779, EPI_ISL_776780, EPI_ISL_776781, EPI_ISL_776782, EPI_ISL_776783, EPI_ISL_776784, EPI_ISL_776785, EPI_ISL_776786, EPI_ISL_776787, EPI_ISL_776788, EPI_ISL_776789, EPI_ISL_776790, EPI_ISL_776791, EPI_ISL_776792, EPI_ISL_776793, EPI_ISL_776794, EPI_ISL_776795, EPI_ISL_776796, EPI_ISL_776797, EPI_ISL_776798, EPI_ISL_776799, EPI_ISL_776800, EPI_ISL_776801, EPI_ISL_776802, EPI_ISL_776803, EPI_ISL_776804, EPI_ISL_776805, EPI_ISL_776806, EPI_ISL_776807, EPI_ISL_776808, EPI_ISL_776809, EPI_ISL_776810, EPI_ISL_776811, EPI_ISL_776812, EPI_ISL_776813, EPI_ISL_776814, EPI_ISL_776815, EPI_ISL_776816, EPI_ISL_776817, EPI_ISL_776818, EPI_ISL_776819, EPI_ISL_776820, EPI_ISL_776821, EPI_ISL_776822, EPI_ISL_776823, EPI_ISL_776824, EPI_ISL_776825, EPI_ISL_776826, EPI_ISL_776827, EPI_ISL_776828, EPI_ISL_776829, EPI_ISL_776830, EPI_ISL_776831, EPI_ISL_776832, EPI_ISL_776833, EPI_ISL_776834, EPI_ISL_776835, EPI_ISL_776836, EPI_ISL_776837, EPI_ISL_776838, EPI_ISL_776839, EPI_ISL_776840, EPI_ISL_776841, EPI_ISL_776842, EPI_ISL_776843, EPI_ISL_776844, EPI_ISL_776845, EPI_ISL_776846, EPI_ISL_776847, EPI_ISL_776848, EPI_ISL_776849, EPI_ISL_776850, EPI_ISL_776851, EPI_ISL_776852, EPI_ISL_776853, EPI_ISL_776854, EPI_ISL_776855, EPI_ISL_776856, EPI_ISL_776857, EPI_ISL_776858, EPI_ISL_776859, EPI_ISL_776860, EPI_ISL_776861, EPI_ISL_776862, EPI_ISL_776863, EPI_ISL_776864, EPI_ISL_776865, EPI_ISL_776866, EPI_ISL_776867, EPI_ISL_776868 | | | |
| see above | Israel Central Virology laboratory | Israel Central Virology laboratory | Neta Zuckerman, Efrat Dahan Bucris, Oran Erster, Michal Mandelboim, Orna Mor, Ella Mendelson |
| EPI_ISL_804056, EPI_ISL_804057, EPI_ISL_804058, EPI_ISL_804059, EPI_ISL_804060, EPI_ISL_804061, EPI_ISL_804062, EPI_ISL_804063, EPI_ISL_804064, EPI_ISL_804065, EPI_ISL_804066, EPI_ISL_804067, EPI_ISL_804068, EPI_ISL_804069, EPI_ISL_804070, EPI_ISL_804071, EPI_ISL_804072, EPI_ISL_804073, EPI_ISL_804074, EPI_ISL_804075, EPI_ISL_804076, EPI_ISL_804077, EPI_ISL_804078, EPI_ISL_804079, EPI_ISL_804080, EPI_ISL_804081, EPI_ISL_804082, EPI_ISL_804083, EPI_ISL_804085, EPI_ISL_804086, EPI_ISL_804087, EPI_ISL_804088, EPI_ISL_804089, EPI_ISL_804090, EPI_ISL_804091, EPI_ISL_804092, EPI_ISL_804093, EPI_ISL_804094, EPI_ISL_804095, EPI_ISL_804096, EPI_ISL_804097, EPI_ISL_804098, EPI_ISL_804099, EPI_ISL_804100, EPI_ISL_804101, EPI_ISL_804102, EPI_ISL_804103, EPI_ISL_804104, EPI_ISL_804105, EPI_ISL_804106, EPI_ISL_804107, EPI_ISL_804108, EPI_ISL_804109, EPI_ISL_804110, EPI_ISL_804111, EPI_ISL_804112, EPI_ISL_804113, EPI_ISL_804114, EPI_ISL_804115, EPI_ISL_804116, EPI_ISL_804117, EPI_ISL_804118, EPI_ISL_804119, EPI_ISL_804120, EPI_ISL_804121, EPI_ISL_804122, EPI_ISL_804123, EPI_ISL_804124, EPI_ISL_804125, EPI_ISL_804126, EPI_ISL_804127, EPI_ISL_804128, EPI_ISL_804129, EPI_ISL_804130, EPI_ISL_804131, EPI_ISL_804132, EPI_ISL_804133, EPI_ISL_804134, EPI_ISL_804135, EPI_ISL_804136, EPI_ISL_804137, EPI_ISL_804138, EPI_ISL_804139, EPI_ISL_804140, EPI_ISL_804141, EPI_ISL_804142, EPI_ISL_804143, EPI_ISL_804144, EPI_ISL_804145, EPI_ISL_804146, EPI_ISL_804147, EPI_ISL_804148, EPI_ISL_804149, EPI_ISL_804150, EPI_ISL_804151, EPI_ISL_804152, EPI_ISL_804153, EPI_ISL_804154, EPI_ISL_804155, EPI_ISL_804156, EPI_ISL_804157, EPI_ISL_804158, EPI_ISL_804159, EPI_ISL_804160, EPI_ISL_804161, EPI_ISL_804162, EPI_ISL_804163, EPI_ISL_804164, EPI_ISL_804165, EPI_ISL_804166, EPI_ISL_804167, EPI_ISL_804168, EPI_ISL_804169, EPI_ISL_804170, EPI_ISL_804171, EPI_ISL_804172, EPI_ISL_804173, EPI_ISL_804174, EPI_ISL_804175, EPI_ISL_804176, EPI_ISL_804177, EPI_ISL_804178, EPI_ISL_804179, EPI_ISL_804180, EPI_ISL_804181, EPI_ISL_804182, EPI_ISL_804183, EPI_ISL_804184, EPI_ISL_804185, EPI_ISL_804186, EPI_ISL_804187, EPI_ISL_804188, EPI_ISL_804189, EPI_ISL_804190, EPI_ISL_804191, EPI_ISL_804192, EPI_ISL_804193, EPI_ISL_804194, EPI_ISL_804195, EPI_ISL_804196, EPI_ISL_804197, EPI_ISL_804198, EPI_ISL_804199, EPI_ISL_804200, EPI_ISL_804201, EPI_ISL_804202, EPI_ISL_804203, EPI_ISL_804204, EPI_ISL_804205, EPI_ISL_804206, EPI_ISL_804207, EPI_ISL_804208, EPI_ISL_804209, EPI_ISL_804210, EPI_ISL_804211, EPI_ISL_804212, EPI_ISL_804213, EPI_ISL_814092, EPI_ISL_814093, EPI_ISL_814094, EPI_ISL_814095, EPI_ISL_814096, EPI_ISL_814097, EPI_ISL_814098, EPI_ISL_814099, EPI_ISL_814100, EPI_ISL_814101, EPI_ISL_814102, EPI_ISL_814103, EPI_ISL_814104, EPI_ISL_814105, EPI_ISL_814106, EPI_ISL_814107, EPI_ISL_814108, EPI_ISL_814109, EPI_ISL_814110, EPI_ISL_814111, EPI_ISL_814112, EPI_ISL_814113, EPI_ISL_814114, EPI_ISL_814115, EPI_ISL_814116, EPI_ISL_814117, EPI_ISL_814118, EPI_ISL_814119, EPI_ISL_814120, EPI_ISL_814121, EPI_ISL_814122, EPI_ISL_814123, EPI_ISL_814124, EPI_ISL_814125, EPI_ISL_814126, EPI_ISL_814127, EPI_ISL_814128, EPI_ISL_814129, EPI_ISL_814130, EPI_ISL_814131, EPI_ISL_814132, EPI_ISL_814133, EPI_ISL_814134, EPI_ISL_814135, EPI_ISL_814136, EPI_ISL_814137, EPI_ISL_814138, EPI_ISL_814139, EPI_ISL_814140, EPI_ISL_814141, EPI_ISL_814142, EPI_ISL_814143, EPI_ISL_814144, EPI_ISL_814145, EPI_ISL_814146, EPI_ISL_814147, EPI_ISL_814148, EPI_ISL_814149, EPI_ISL_814150, EPI_ISL_814151, EPI_ISL_814152, EPI_ISL_814153, EPI_ISL_814154, EPI_ISL_814155, EPI_ISL_814156, EPI_ISL_814157, EPI_ISL_814158, EPI_ISL_814159, EPI_ISL_814160, EPI_ISL_814161, EPI_ISL_814162, EPI_ISL_814163, EPI_ISL_814164, EPI_ISL_814165, EPI_ISL_814166, EPI_ISL_814167, EPI_ISL_814168, EPI_ISL_814169, EPI_ISL_814170, EPI_ISL_814171, EPI_ISL_814172, EPI_ISL_814173, EPI_ISL_814174, EPI_ISL_814175, EPI_ISL_814176, EPI_ISL_814177, EPI_ISL_814178, EPI_ISL_814179, EPI_ISL_814180, EPI_ISL_814181, EPI_ISL_814182, EPI_ISL_814183, EPI_ISL_814184, EPI_ISL_814185, EPI_ISL_814186, EPI_ISL_814187, EPI_ISL_814188, EPI_ISL_814189, EPI_ISL_814190, EPI_ISL_814191, EPI_ISL_814192, EPI_ISL_814193, EPI_ISL_814194, EPI_ISL_814195, EPI_ISL_814196, EPI_ISL_814197, EPI_ISL_814198, EPI_ISL_814199, EPI_ISL_814200, EPI_ISL_814201, EPI_ISL_814202, EPI_ISL_814203, EPI_ISL_814204, EPI_ISL_814205, EPI_ISL_814206, EPI_ISL_814207, EPI_ISL_814208, EPI_ISL_814209, EPI_ISL_814210, EPI_ISL_814211, EPI_ISL_814212, EPI_ISL_814213, EPI_ISL_814214, EPI_ISL_814215, EPI_ISL_814216, EPI_ISL_814217, EPI_ISL_814218, EPI_ISL_814219, EPI_ISL_814220, EPI_ISL_814221, EPI_ISL_814222, EPI_ISL_814223, EPI_ISL_814224, EPI_ISL_814225, EPI_ISL_814226, EPI_ISL_814227, EPI_ISL_814228, EPI_ISL_814229, EPI_ISL_814230, EPI_ISL_814231, EPI_ISL_814232, EPI_ISL_814233, EPI_ISL_814234, EPI_ISL_814235, EPI_ISL_814236, EPI_ISL_814237, EPI_ISL_814238, EPI_ISL_814239, EPI_ISL_814240, EPI_ISL_814241, EPI_ISL_814242, EPI_ISL_814243, EPI_ISL_814244, EPI_ISL_814245, EPI_ISL_814246, EPI_ISL_814247, EPI_ISL_814248, EPI_ISL_814249, EPI_ISL_814250, EPI_ISL_814251, EPI_ISL_814252, EPI_ISL_814253, EPI_ISL_814254, EPI_ISL_814255, EPI_ISL_814256, EPI_ISL_814257, EPI_ISL_814258, EPI_ISL_814259, EPI_ISL_814260, EPI_ISL_814261, EPI_ISL_814262, EPI_ISL_814263, EPI_ISL_814264, EPI_ISL_814265, EPI_ISL_814266, EPI_ISL_814267, EPI_ISL_814268, EPI_ISL_814269, EPI_ISL_814270, EPI_ISL_814271, EPI_ISL_814272, EPI_ISL_814273, EPI_ISL_889023, EPI_ISL_889024, EPI_ISL_889025, EPI_ISL_889026, EPI_ISL_889027, EPI_ISL_889028, EPI_ISL_889029, EPI_ISL_889030, EPI_ISL_889031, EPI_ISL_889032, EPI_ISL_889033, EPI_ISL_889034, EPI_ISL_889035, EPI_ISL_889036, EPI_ISL_889037, EPI_ISL_889038, EPI_ISL_889039, EPI_ISL_889040, EPI_ISL_889041, EPI_ISL_889042, EPI_ISL_889043, EPI_ISL_889044, EPI_ISL_889045, EPI_ISL_889046, EPI_ISL_889047, EPI_ISL_889048, EPI_ISL_889049, EPI_ISL_889050, EPI_ISL_889051, EPI_ISL_889052, EPI_ISL_889053, EPI_ISL_889054, EPI_ISL_889055, EPI_ISL_889056, EPI_ISL_889057, EPI_ISL_889058, EPI_ISL_889059, EPI_ISL_889060, EPI_ISL_889061, EPI_ISL_889062, EPI_ISL_889063, EPI_ISL_889064, EPI_ISL_889065, EPI_ISL_889066, EPI_ISL_889067, EPI_ISL_889068, EPI_ISL_889069, EPI_ISL_889070, EPI_ISL_889071, EPI_ISL_889072, EPI_ISL_889073, EPI_ISL_889074, EPI_ISL_889075, EPI_ISL_889076, EPI_ISL_889077, EPI_ISL_889078, EPI_ISL_889079, EPI_ISL_889080, EPI_ISL_889081, EPI_ISL_889082, EPI_ISL_889083, EPI_ISL_889084, EPI_ISL_889085, EPI_ISL_889086, EPI_ISL_889087, EPI_ISL_889088, EPI_ISL_889089, EPI_ISL_889090, EPI_ISL_889091, EPI_ISL_889092, EPI_ISL_889093, EPI_ISL_889094, EPI_ISL_889095, EPI_ISL_889096, EPI_ISL_889097, EPI_ISL_889098, EPI_ISL_889099, EPI_ISL_889100, EPI_ISL_889101, EPI_ISL_889102, EPI_ISL_889103, EPI_ISL_889104, EPI_ISL_889105, EPI_ISL_889106, EPI_ISL_889107, EPI_ISL_889108, EPI_ISL_889109, EPI_ISL_889110, EPI_ISL_889111, EPI_ISL_889112, EPI_ISL_889113, EPI_ISL_889114, EPI_ISL_889115, EPI_ISL_889116, EPI_ISL_889117, EPI_ISL_889118, EPI_ISL_889119, EPI_ISL_889120, EPI_ISL_889121, EPI_ISL_889122, EPI_ISL_889123, EPI_ISL_889124, EPI_ISL_889125, EPI_ISL_889126, EPI_ISL_889127, EPI_ISL_889128, EPI_ISL_889129, EPI_ISL_889130, EPI_ISL_889131, EPI_ISL_889132, EPI_ISL_889133, EPI_ISL_889134, EPI_ISL_889135, EPI_ISL_889136, EPI_ISL_889137, EPI_ISL_889138, EPI_ISL_889139, EPI_ISL_889140, EPI_ISL_889141, EPI_ISL_889142, EPI_ISL_889143, EPI_ISL_889144, EPI_ISL_889145, EPI_ISL_889146, EPI_ISL_889147, EPI_ISL_889148, EPI_ISL_889149, EPI_ISL_889150, EPI_ISL_889151, EPI_ISL_889152, EPI_ISL_889153, EPI_ISL_889154, EPI_ISL_889155, EPI_ISL_889156, EPI_ISL_889157, EPI_ISL_889158, EPI_ISL_889159, EPI_ISL_889160, EPI_ISL_889161, EPI_ISL_889162, EPI_ISL_889163, EPI_ISL_889164, EPI_ISL_889165, EPI_ISL_889166, EPI_ISL_889167, EPI_ISL_889168, EPI_ISL_889169, EPI_ISL_889170, EPI_ISL_889171, EPI_ISL_889172, EPI_ISL_889173, EPI_ISL_889174, EPI_ISL_889175, EPI_ISL_889176, EPI_ISL_889177, EPI_ISL_889178, EPI_ISL_889179, EPI_ISL_889180, EPI_ISL_889181, EPI_ISL_889182, EPI_ISL_889183, EPI_ISL_889184, EPI_ISL_889185, EPI_ISL_889186, EPI_ISL_889187, EPI_ISL_889188, EPI_ISL_889189, EPI_ISL_889190, EPI_ISL_889191, EPI_ISL_889192, EPI_ISL_889193, EPI_ISL_889194, EPI_ISL_889195, EPI_ISL_889196, EPI_ISL_889197, EPI_ISL_889198, EPI_ISL_889199, EPI_ISL_889200, EPI_ISL_889201, EPI_ISL_889202, EPI_ISL_889203, EPI_ISL_889204, EPI_ISL_889205, EPI_ISL_889206, EPI_ISL_889207, EPI_ISL_889208, EPI_ISL_889209, EPI_ISL_889210, EPI_ISL_889211, EPI_ISL_889212, EPI_ISL_889213, EPI_ISL_889214, EPI_ISL_889215, EPI_ISL_889216, EPI_ISL_889217, EPI_ISL_889218, EPI_ISL_889219, EPI_ISL_889220, EPI_ISL_889221, EPI_ISL_889222, EPI_ISL_889223, EPI_ISL_889224, EPI_ISL_889225, EPI_ISL_889226, EPI_ISL_889227, EPI_ISL_889228, EPI_ISL_889229, EPI_ISL_889230, EPI_ISL_889231, EPI_ISL_889232, EPI_ISL_889233, EPI_ISL_889234, EPI_ISL_889235, EPI_ISL_889236, EPI_ISL_889237, EPI_ISL_889238, EPI_ISL_889239, EPI_ISL_889240, EPI_ISL_889241, EPI_ISL_889242, EPI_ISL_889243, EPI_ISL_889244, EPI_ISL_889245, EPI_ISL_889246, EPI_ISL_889247, EPI_ISL_889248, EPI_ISL_889249, EPI_ISL_889250, EPI_ISL_889251, EPI_ISL_889252, EPI_ISL_889253, EPI_ISL_889254, EPI_ISL_889255, EPI_ISL_889256, EPI_ISL_889257, EPI_ISL_889258, EPI_ISL_889259, EPI_ISL_889260, EPI_ISL_889261, EPI_ISL_889262, EPI_ISL_889263, EPI_ISL_889264, EPI_ISL_889265, EPI_ISL_889266, EPI_ISL_889267, EPI_ISL_889268, EPI_ISL_889269, EPI_ISL_889270, EPI_ISL_889271, EPI_ISL_889272, EPI_ISL_889273, EPI_ISL_889274, EPI_ISL_889275, EPI_ISL_889276, EPI_ISL_889277, EPI_ISL_889278, EPI_ISL_889279, EPI_ISL_889280, EPI_ISL_889281, EPI_ISL_889282, EPI_ISL_889283, EPI_ISL_889284, EPI_ISL_889285, EPI_ISL_889286, EPI_ISL_889287, EPI_ISL_889288, EPI_ISL_889289, EPI_ISL_889290, EPI_ISL_889291, EPI_ISL_889292, EPI_ISL_889293, EPI_ISL_889294, EPI_ISL_889295, | | | |

| | | | |
|---|---|---|---|
| EPI_ISL_889296, EPI_ISL_889297, EPI_ISL_889298, EPI_ISL_889299, EPI_ISL_889300, EPI_ISL_889301, EPI_ISL_889302, EPI_ISL_889303, EPI_ISL_889304, EPI_ISL_889305, EPI_ISL_889306, EPI_ISL_944188, EPI_ISL_944189, EPI_ISL_944190, EPI_ISL_944191, EPI_ISL_944192, EPI_ISL_944193, EPI_ISL_944194, EPI_ISL_944195, EPI_ISL_944196, EPI_ISL_944197, EPI_ISL_944198, EPI_ISL_944199, EPI_ISL_944200, EPI_ISL_944201, EPI_ISL_944202, EPI_ISL_944203, EPI_ISL_944204, EPI_ISL_944205, EPI_ISL_944206, EPI_ISL_944207, EPI_ISL_944208, EPI_ISL_944209, EPI_ISL_944210, EPI_ISL_944211, EPI_ISL_944212, EPI_ISL_944213, EPI_ISL_944214, EPI_ISL_944215, EPI_ISL_944216, EPI_ISL_944217, EPI_ISL_944218, EPI_ISL_944219, EPI_ISL_944220, EPI_ISL_944221, EPI_ISL_944222, EPI_ISL_944223, EPI_ISL_944224, EPI_ISL_944225, EPI_ISL_944226, EPI_ISL_944227, EPI_ISL_944228, EPI_ISL_944229, EPI_ISL_944230, EPI_ISL_944231, EPI_ISL_944232, EPI_ISL_944233, EPI_ISL_944234, EPI_ISL_944235, EPI_ISL_944236, EPI_ISL_944237, EPI_ISL_944238, EPI_ISL_944239, EPI_ISL_944240, EPI_ISL_944241, EPI_ISL_944242, EPI_ISL_944243, EPI_ISL_944244, EPI_ISL_944245, EPI_ISL_944246, EPI_ISL_944247, EPI_ISL_944248, EPI_ISL_944249, EPI_ISL_944250, EPI_ISL_944251, EPI_ISL_944252, EPI_ISL_944253, EPI_ISL_944254, EPI_ISL_944255, EPI_ISL_944256, EPI_ISL_944257, EPI_ISL_944258, EPI_ISL_944259, EPI_ISL_944260, EPI_ISL_944261, EPI_ISL_944262, EPI_ISL_944263, EPI_ISL_944264, EPI_ISL_944265, EPI_ISL_944266, EPI_ISL_944267, EPI_ISL_944268, EPI_ISL_944269, EPI_ISL_944270, EPI_ISL_944271, EPI_ISL_944272, EPI_ISL_944273, EPI_ISL_944274, EPI_ISL_944275, EPI_ISL_944276, EPI_ISL_944277, EPI_ISL_944278, EPI_ISL_944279, EPI_ISL_944280, EPI_ISL_944281, EPI_ISL_944282, EPI_ISL_944283, EPI_ISL_944284, EPI_ISL_944285, EPI_ISL_944286, EPI_ISL_944287, EPI_ISL_944288, EPI_ISL_944289, EPI_ISL_944290, EPI_ISL_944291, EPI_ISL_944292, EPI_ISL_944293, EPI_ISL_944294, EPI_ISL_944295, EPI_ISL_944296, EPI_ISL_944297, EPI_ISL_944298, EPI_ISL_944299, EPI_ISL_944300, EPI_ISL_944301, EPI_ISL_944302, EPI_ISL_944303, EPI_ISL_944304, EPI_ISL_944305, EPI_ISL_944306, EPI_ISL_944307, EPI_ISL_944308, EPI_ISL_944309, EPI_ISL_944310, EPI_ISL_944311, EPI_ISL_944312, EPI_ISL_944313, EPI_ISL_944314, EPI_ISL_944315, EPI_ISL_944316, EPI_ISL_944317, EPI_ISL_944318, EPI_ISL_944319, EPI_ISL_944320, EPI_ISL_944321, EPI_ISL_944322, EPI_ISL_944323, EPI_ISL_944324, EPI_ISL_944325, EPI_ISL_944326, EPI_ISL_944327, EPI_ISL_944328, EPI_ISL_944329, EPI_ISL_944330, EPI_ISL_944331, EPI_ISL_944332, EPI_ISL_944333, EPI_ISL_944334, EPI_ISL_944335, EPI_ISL_944336, EPI_ISL_944337, EPI_ISL_944338, EPI_ISL_944339, EPI_ISL_944340, EPI_ISL_944341, EPI_ISL_944342, EPI_ISL_944343, EPI_ISL_944344, EPI_ISL_944345, EPI_ISL_944346, EPI_ISL_944347, EPI_ISL_944348, EPI_ISL_944349, EPI_ISL_944350, EPI_ISL_944351, EPI_ISL_944352, EPI_ISL_944353, EPI_ISL_944354, EPI_ISL_944355, EPI_ISL_944356, EPI_ISL_944357, EPI_ISL_944358, EPI_ISL_944359, EPI_ISL_944360, EPI_ISL_944361, EPI_ISL_944362, EPI_ISL_944363, EPI_ISL_944364, EPI_ISL_944365, EPI_ISL_944366, EPI_ISL_944367, EPI_ISL_944368, EPI_ISL_944369, EPI_ISL_944370, EPI_ISL_944371, EPI_ISL_944372, EPI_ISL_944373, EPI_ISL_944374, EPI_ISL_944375, EPI_ISL_944376, EPI_ISL_944377, EPI_ISL_944378, EPI_ISL_944379, EPI_ISL_944381, EPI_ISL_944382, EPI_ISL_944383, EPI_ISL_944384, EPI_ISL_944385, EPI_ISL_944386, EPI_ISL_944387, EPI_ISL_944388, EPI_ISL_944389, EPI_ISL_944390, EPI_ISL_944391, EPI_ISL_944392, EPI_ISL_944393, EPI_ISL_944394, EPI_ISL_944395, EPI_ISL_944396, EPI_ISL_944397, EPI_ISL_944398, EPI_ISL_944399, EPI_ISL_944400, EPI_ISL_944401, EPI_ISL_944402, EPI_ISL_944403, EPI_ISL_944404, EPI_ISL_944405, EPI_ISL_944406, EPI_ISL_944407, EPI_ISL_944408, EPI_ISL_944409, EPI_ISL_944410, EPI_ISL_944411, EPI_ISL_944412, EPI_ISL_944413, EPI_ISL_944414, EPI_ISL_944415, EPI_ISL_944416, EPI_ISL_944417, EPI_ISL_944418, EPI_ISL_944419, EPI_ISL_944420, EPI_ISL_944421, EPI_ISL_944422, EPI_ISL_944423, EPI_ISL_944424, EPI_ISL_944425, EPI_ISL_944426, EPI_ISL_944427, EPI_ISL_944428, EPI_ISL_944429, EPI_ISL_944430, EPI_ISL_944431, EPI_ISL_944432, EPI_ISL_944433, EPI_ISL_944434, EPI_ISL_944435, EPI_ISL_944436, EPI_ISL_944437, EPI_ISL_944438, EPI_ISL_944439, EPI_ISL_944440, EPI_ISL_944441, EPI_ISL_944442, EPI_ISL_944443, EPI_ISL_944444, EPI_ISL_944445, EPI_ISL_944446, EPI_ISL_944447, EPI_ISL_944448, EPI_ISL_944449, EPI_ISL_944450, EPI_ISL_944451, EPI_ISL_944452, EPI_ISL_944453, EPI_ISL_944454, EPI_ISL_944455, EPI_ISL_944456, EPI_ISL_944457, EPI_ISL_944458, EPI_ISL_944459, EPI_ISL_944460, EPI_ISL_944461, EPI_ISL_944462, EPI_ISL_944463, EPI_ISL_944464, EPI_ISL_944465, EPI_ISL_944466, EPI_ISL_944467, EPI_ISL_944468, EPI_ISL_944469, EPI_ISL_944470, EPI_ISL_944471, EPI_ISL_944472, EPI_ISL_944473, EPI_ISL_944474, EPI_ISL_944475, EPI_ISL_944476, EPI_ISL_944477, EPI_ISL_944478, EPI_ISL_944479, EPI_ISL_944480, EPI_ISL_944481, EPI_ISL_944482, EPI_ISL_944483, EPI_ISL_944484, EPI_ISL_944485, EPI_ISL_944486, EPI_ISL_944487, EPI_ISL_944488, EPI_ISL_944489, EPI_ISL_944490, EPI_ISL_944491, EPI_ISL_944492, EPI_ISL_944493, EPI_ISL_944494, EPI_ISL_944495, EPI_ISL_944496, EPI_ISL_944497, EPI_ISL_944498, EPI_ISL_944499, EPI_ISL_944500, EPI_ISL_944501, EPI_ISL_944502, EPI_ISL_944503, EPI_ISL_944504, EPI_ISL_944505, EPI_ISL_944506, EPI_ISL_944507, EPI_ISL_944508, EPI_ISL_944509, EPI_ISL_944510, EPI_ISL_944511, EPI_ISL_944512, EPI_ISL_944513, EPI_ISL_944514, EPI_ISL_944515, EPI_ISL_944516, EPI_ISL_944517, EPI_ISL_944518, EPI_ISL_944519, EPI_ISL_944520, EPI_ISL_944521, EPI_ISL_944522, EPI_ISL_944523, EPI_ISL_944524, EPI_ISL_944525, EPI_ISL_944526, EPI_ISL_944527, EPI_ISL_944528, EPI_ISL_944529, EPI_ISL_944530, EPI_ISL_944531, EPI_ISL_944532, EPI_ISL_944533, EPI_ISL_944534, EPI_ISL_944535, EPI_ISL_944536, EPI_ISL_944537, EPI_ISL_944538, EPI_ISL_944539, EPI_ISL_944540, EPI_ISL_944541, EPI_ISL_944542, EPI_ISL_944543, EPI_ISL_944544, EPI_ISL_944545, EPI_ISL_944546, EPI_ISL_944547, EPI_ISL_944548, EPI_ISL_944549, EPI_ISL_944550, EPI_ISL_944551, EPI_ISL_944552, EPI_ISL_944553, EPI_ISL_944554, EPI_ISL_944555, EPI_ISL_944556, EPI_ISL_944557, EPI_ISL_944558, EPI_ISL_944559, EPI_ISL_944560, EPI_ISL_944561, EPI_ISL_944562, EPI_ISL_944563, EPI_ISL_944564, EPI_ISL_944565, EPI_ISL_944566, EPI_ISL_944567, EPI_ISL_944568, EPI_ISL_944569, EPI_ISL_944570 | | | |
| see above | Israel Central Virology laboratory | Israel National Consortium for SARS-CoV-2 sequencing | Neta Zuckerman, Efrat Dahan Bucris, Michal Mandelboim, Dana Bar-Ilan, Oran Erster, Tzvia Mann, Omer Murik, David A. Zeevi, Assaf Rokney, Joseph Jaffe, Eva Nachum, Maya Davidovich Cohen, Ephraim Fass, Gal Zizelski Valenci, Mor Rubinstein, Efrat Rorman, Israel Nissan, Efrat Glick-Saar, Omri Nayshool, Gideon Rechavi, Ella Mendelson, Orna Mor |

We gratefully acknowledge the following Authors from the Originating laboratories responsible for obtaining the specimens, as well as the Submitting laboratories where the genome data were generated and shared via GISAID, on which this research is based.

All Submitters of data may be contacted directly via www.gisaid.org

Authors are sorted alphabetically.

| Accession ID | Originating Laboratory | Submitting Laboratory | Authors |
|---|---|---|---|
| EPI_ISL_1000969, EPI_ISL_1000970 | Presidio di Brindisi Di Summa - Perrino | Istituto Zooprofilattico Sperimentale della Puglia e della Basilicata | Parisi A., Bianco A., Capozzi L., Del Sambro L., Simone D., Manzulli V, Rondinone V., Pace L., Cipolletta D., Galante D. |
| EPI_ISL_1007642 | University of Bari Biomedical Sciences and Human Oncology | University of Bari Biomedical Sciences and Human Oncology | Chironna M., Sallustio A., Loconsole D., Accogli M. |
| EPI_ISL_1008663 | Istituto Zooprofilattico Sperimentale della Puglia e della Basilicata | Istituto Zooprofilattico Sperimentale della Puglia e della Basilicata | Parisi A., Bianco A., Capozzi L., Del Sambro L., Simone D., Chironna M., Loconsole D., Sallustio A., Giannico A. |
| EPI_ISL_1008664 | Ospedale Santa Caterina Novella | Istituto Zooprofilattico Sperimentale della Puglia e della Basilicata | Parisi A., Bianco A., Capozzi L., Del Sambro L., Simone D., Chironna M., Loconsole D., Sallustio A., Ridolfi D. |
| EPI_ISL_1008665, EPI_ISL_1008666, EPI_ISL_1008667, EPI_ISL_1008668 | Istituto Zooprofilattico Sperimentale della Puglia e della Basilicata | Istituto Zooprofilattico Sperimentale della Puglia e della Basilicata | Parisi A., Bianco A., Capozzi L., Del Sambro L., Simone D., Chironna M., Loconsole D., Sallustio A., Ridolfi D. |
| EPI_ISL_1008669, EPI_ISL_1008670, EPI_ISL_1008671, EPI_ISL_1008672, EPI_ISL_1008673, EPI_ISL_1008674, EPI_ISL_1008675, EPI_ISL_1008676, EPI_ISL_1008677 | Istituto Zooprofilattico Sperimentale della Puglia e della Basilicata | Istituto Zooprofilattico Sperimentale della Puglia e della Basilicata | Parisi A., Bianco A., Capozzi L., Del Sambro L., Simone D., Chironna M., Loconsole D., Sallustio A., Giannico A. |
| EPI_ISL_1008678 | Presidio di Brindisi Di Summa - Perrino | Istituto Zooprofilattico Sperimentale della Puglia e della Basilicata | Parisi A., Bianco A., Capozzi L., Del Sambro L., Simone D., Chironna M., Loconsole D., Sallustio A., Ridolfi D. |
| EPI_ISL_1008679, EPI_ISL_1008680, EPI_ISL_1008681, EPI_ISL_1008682, EPI_ISL_1008683, EPI_ISL_1008684, EPI_ISL_1008685, EPI_ISL_1008686, EPI_ISL_1008687 | Istituto Zooprofilattico Sperimentale della Puglia e della Basilicata | Istituto Zooprofilattico Sperimentale della Puglia e della Basilicata | Parisi A., Bianco A., Capozzi L., Del Sambro L., Simone D., Chironna M., Loconsole D., Sallustio A., Ridolfi D. |
| EPI_ISL_1008688, EPI_ISL_1008689, EPI_ISL_1008690, EPI_ISL_1008691, EPI_ISL_1008692, EPI_ISL_1008693, EPI_ISL_1008694, EPI_ISL_1008695, EPI_ISL_1008696, EPI_ISL_1008697, EPI_ISL_1008698, EPI_ISL_1008699 | | | |
| see above | Istituto Zooprofilattico Sperimentale della Puglia e della Basilicata | Istituto Zooprofilattico Sperimentale della Puglia e della Basilicata | Parisi A., Bianco A., Capozzi L., Del Sambro L., Simone D., Chironna M., Loconsole D., Sallustio A., Giannico A. |
| EPI_ISL_1008700 | Ospedale Santa Caterina Novella | Istituto Zooprofilattico Sperimentale della Puglia e della Basilicata | Parisi A., Bianco A., Capozzi L., Del Sambro L., Simone D., Chironna M., Loconsole D., Sallustio A., Ridolfi D. |
| EPI_ISL_1008701, EPI_ISL_1008702 | Istituto Zooprofilattico Sperimentale della Puglia e della Basilicata | Istituto Zooprofilattico Sperimentale della Puglia e della Basilicata | Parisi A., Bianco A., Capozzi L., Del Sambro L., Simone D., Chironna M., Loconsole D., Sallustio A., Giannico A. |
| EPI_ISL_1008703, EPI_ISL_1008704, EPI_ISL_1008705 | Presidio di Brindisi Di Summa - Perrino | Istituto Zooprofilattico Sperimentale della Puglia e della Basilicata | Parisi A., Bianco A., Capozzi L., Del Sambro L., Simone D., Chironna M., Loconsole D., Sallustio A., Ridolfi D. |
| EPI_ISL_1008706 | Ospedale Santissima Annunziata | Istituto Zooprofilattico Sperimentale della Puglia e della Basilicata | Parisi A., Bianco A., Capozzi L., Del Sambro L., Simone D., Chironna M., Loconsole D., Sallustio A., Ridolfi D. |
| EPI_ISL_1008707, EPI_ISL_1008708, EPI_ISL_1008709, EPI_ISL_1008710, EPI_ISL_1008711, EPI_ISL_1008712 | Presidio di Brindisi Di Summa - Perrino | Istituto Zooprofilattico Sperimentale della Puglia e della Basilicata | Parisi A., Bianco A., Capozzi L., Del Sambro L., Simone D., Chironna M., Loconsole D., Sallustio A., Ridolfi D. |
| EPI_ISL_1009018 | Istituto Zooprofilattico Sperimentale Lazio e Toscana "M. Aleandri" | INMI Lazzaro Spallanzani IRCCS | CEM Gruber, B Bartolini, E Giombini, M Rueca, O Butera, F Messina, MT Scicluna, G Manna, A Cersini, A Di Caro, MR Capobianchi |
| EPI_ISL_1009019 | Azienda Ospedaliera San Camillo Forlanini | INMI Lazzaro Spallanzani IRCCS | E Giombini, M. Rueca, B Bartolini, O Butera, C.E.M Gruber, F Messina, G Parisi, M Irno Consalvo, A Di Caro, MR Capobianchi |
| EPI_ISL_1009020 | Azienda Ospedaliera San Camillo Forlanini | INMI Lazzaro Spallanzani IRCCS | M. Rueca, O Butera, B Bartolini, E Giombini,C.E.M Gruber, F Messina, G Parisi, ML Guarino, A Di Caro, MR Capobianchi |
| EPI_ISL_1009021 | San Gallicano Dermatological Institute I.F.O. | INMI Lazzaro Spallanzani IRCCS | B Bartolini, O Butera, C.E.M Gruber, M Rueca, F Messina, E Giombini, F Pimpinelli, F Ensoli, EG Di Domenico, MR Capobianchi, A Di Caro |
| EPI_ISL_1009022 | San Gallicano Dermatological Institute I.F.O. | INMI Lazzaro Spallanzani IRCCS | F Messina, C.E.M Gruber, B Bartolini, E Giombini, M Rueca, O Butera, F Pimpinelli, F Ensoli, M pontone, MR Capobianchi, A Di Caro |
| EPI_ISL_1009023 | San Gallicano Dermatological Institute I.F.O. | INMI Lazzaro Spallanzani IRCCS | O Butera, F Messina, CEM Gruber, B Bartolini, E Giombini, M Rueca, F Pimpinelli, F Ensoli, G D'Agosto, MR Capobianchi, A Di Caro |
| EPI_ISL_1009024 | Fondazione Policlinico Universitario "A. Gemelli" IRCCS | INMI Lazzaro Spallanzani IRCCS | E Giombini, M. Rueca, B Bartolini, O Butera, C.E.M Gruber, F Messina, P Cattani,M Sanguinetti, A Di Caro, MR Capobianchi |
| EPI_ISL_1009025 | Fondazione Policlinico Universitario "A. Gemelli" IRCCS | INMI Lazzaro Spallanzani IRCCS | B Bartolini, O Butera, C.E.M Gruber, M Rueca, F Messina, E Giombini, P Cattani,M Sanguinetti, MR Capobianchi, A Di Caro |
| EPI_ISL_1009026 | Fondazione Policlinico Universitario "A. Gemelli" IRCCS | INMI Lazzaro Spallanzani IRCCS | F Messina, C.E.M Gruber, B Bartolini, E Giombini, M Rueca, O Butera, P Cattani,M Sanguinetti, A Di Caro, MR Capobianchi |
| EPI_ISL_1009027 | Fondazione Policlinico Universitario "A. Gemelli" IRCCS | INMI Lazzaro Spallanzani IRCCS | M Rueca, O Butera, F Messina, CEM Gruber, B Bartolini, E Giombini, P Cattani,M Sanguinetti, A Di Caro, MR Capobianchi |
| EPI_ISL_1009028 | AOU Policlinico Umberto I; Sapienza Università di Roma | INMI Lazzaro Spallanzani IRCCS | CEM Gruber, B Bartolini, E Giombini, M Rueca, O Butera, F Messina, G Antonelli, O Turriziani, A Di Caro, MR Capobianchi |
| EPI_ISL_1009029 | AOU Policlinico Umberto I; Sapienza Università di Roma | INMI Lazzaro Spallanzani IRCCS | B Bartolini, O Butera, C.E.M Gruber, M Rueca, F Messina, E Giombini, G Antonelli, O Turriziani, MR Capobianchi, A Di Caro |
| EPI_ISL_1009030 | AOU Policlinico Umberto I; Sapienza Università di Roma | INMI Lazzaro Spallanzani IRCCS | M Rueca, O Butera, F Messina, CEM Gruber, B Bartolini, E Giombini, G Antonelli, O Turriziani, A Di Caro, MR Capobianchi |
| EPI_ISL_1009031 | AOU Policlinico Umberto I; Sapienza Università di Roma | INMI Lazzaro Spallanzani IRCCS | E Giombini, M. Rueca, B Bartolini, O Butera, C.E.M Gruber, F Messina, G Antonelli, O Turriziani, MR Capobianchi, A Di Caro |
| EPI_ISL_1012923, EPI_ISL_1012924 | Department of Infectious Diseases, Istituto Superiore di Sanità, Rome, Italy; ASST Sette Laghi, Varese, Italy | Istituto Superiore di Sanità (ISS) | Paola Stefanelli, Angela Di Martino, Alessandra Lo Presti, Stefano Fiore, Fabrizio Maggi, Federica Novazzi, Andreina Baj, Angelo Genoni, Manuela Marra, Maria Carollo, Marco Crescenzi |
| EPI_ISL_1013095 | Azienda Sanitaria dell'Alto Adige Laboratorio Aziendale di Microbiologia e Virologia | Istituto di Genomica Applicata | Elisabetta Pagani, Irene Bianconi, Elisabetta Giacobazzi, Elisa Masi, Stefanie Wieser, Irena Jurman, Vera Vendramin, Eleonora Paparelli, Davide Scaglione, Michele Morgante |
| EPI_ISL_1013523, EPI_ISL_1013524, EPI_ISL_1013525, EPI_ISL_1013526, EPI_ISL_1013527, EPI_ISL_1013528, EPI_ISL_1013529, EPI_ISL_1013530, EPI_ISL_1013531, EPI_ISL_1013532, EPI_ISL_1013533, EPI_ISL_1013534, EPI_ISL_1013535, EPI_ISL_1013536, EPI_ISL_1013537, EPI_ISL_1013538, EPI_ISL_1013539, EPI_ISL_1013540, EPI_ISL_1013541, EPI_ISL_1013542, EPI_ISL_1013543, EPI_ISL_1013544, EPI_ISL_1013545, EPI_ISL_1013546, EPI_ISL_1013547, EPI_ISL_1013548, EPI_ISL_1013549, EPI_ISL_1013550, EPI_ISL_1013551, EPI_ISL_1013552, EPI_ISL_1013553, EPI_ISL_1013554, EPI_ISL_1013555, EPI_ISL_1013556, EPI_ISL_1013557, EPI_ISL_1013558, EPI_ISL_1013559, EPI_ISL_1013560, EPI_ISL_1013561, EPI_ISL_1013562, EPI_ISL_1013563, EPI_ISL_1013564, EPI_ISL_1013565, EPI_ISL_1013566, EPI_ISL_1013567, EPI_ISL_1013568, EPI_ISL_1013569, EPI_ISL_1013570, EPI_ISL_1013571, EPI_ISL_1013572, EPI_ISL_1013573, EPI_ISL_1013574, EPI_ISL_1013575, EPI_ISL_1013576, EPI_ISL_1013577, EPI_ISL_1013578, EPI_ISL_1013579, EPI_ISL_1013580, EPI_ISL_1013581, EPI_ISL_1013582 | | | |
| see above | 1.AO Universitaria 'S. Giovanni di Dio e Ruggi D'Aragona, | 1. Genome Research Center for Health (CRGS) / 2. | Giorgio Giurato, Francesca Rizzo, Alessandro Weisz, Gianluigi Franci, Giovanni Nassa, Pasquale Pagliano, Roberta Tarallo, Elena Alexandrova, Ylenia |

| | | | |
|---|---|---|---|
| | Scuola Medica Salernitana' Hospital / 2.UOC di Virologia e Microbiologia, Università della Campania 'L. Vanvitelli' / 3.AO Universitaria 'Federico II' Napoli Hospital / 4.AORN 'San Giuseppe Moscati' Avellino Hospital / 5.AO 'San Pio - presidio G. Rummo' Benevento Hospital / 6.AO 'Sant'Anna e San Sebastiano' Caserta Hospital / 7.PO 'Maria Santissima Addolorata' Eboli Hospital / 8.Biogem Istituto di Ricerche Genetiche | Laboratory of Molecular Medicine and Genomics(LMMGe) / 3. Center for Research in Pure and Applied Mathematics (CRMPA) | D'Agostino, Carlo Ferravante, Jessica Lamberti, Viola Melone, Domenico Memoli, Valeria Mirici Cappa, Domenico Palumbo, Giovanni Pecoraro, Assunta Sellitto, Oriana Strianese, Ilaria Terenzi, Giuseppe Fenza, Aniello Gentile, Antonello Saccomanno, Sonia Amabile, Teresa Rocco, Annamaria Salvati, Emilia Vaccaro, Massimiliano Galdiero, Michele Cennamo, Giuseppe Portella, Maria Grazia Foti, Mariarosaria Ingino, Maria Landi, Maurizio Fumi, Vincenzo Rocco, Rita Greco, Vittoria Letizia, Arnolfo Petruzziello, Maddalena Schioppa, Gregorio Goffredi, Francesca Marciano, Michele Caraglia, Alessia Cossu, Marianna Scrima, Edmondo Adorisio, Morena D'Avenia, Michela Iacobellis, Rosanna Piluscio, Giorgio Dirani, Vittorio Sambri, Simona Semprini, Silvia Zanoli, Francesco Curcio, Stefania Marzinotto, Andreina Baj, Fausto Sessa. |
| EPI_ISL_1014237 | University of Bari Biomedical Sciences and Human Oncology | University of Bari Biomedical Sciences and Human Oncology | Chironna M., Sallustio A., Loconsole D., Accogli M. |
| EPI_ISL_1014545 | Department of Infectious Diseases, Istituto Superiore di Sanità, Rome, Italy; Università degli Studi di Perugia, Perugia, Italy | Istituto Superiore di Sanità (ISS) | Paola Stefanelli, Alessandra Lo Presti, Angela Di Martino, Stefano Fiore, Antonella Mencacci, Barbara Camilloni, Manuela Marra, Maria Carollo, Marco Crescenzi |
| EPI_ISL_1014675 | Department of Infectious Diseases, Istituto Superiore di Sanità, Rome, Italy; Università degli Studi di Perugia, Perugia, Italy | Istituto Superiore di Sanità (ISS) | Paola Stefanelli, Alessandra Lo Presti, Angela Di Martino, Stefano Fiore, Antonella Mencacci, Barbara Camilloni, Manuela Marra, Maria Carollo, Marco Crescenzi, Luca De Sabato |
| EPI_ISL_1014690 | University of Bari Biomedical Sciences and Human Oncology | University of Bari Biomedical Sciences and Human Oncology | Chironna M., Sallustio A., Loconsole D., Accogli M. |
| EPI_ISL_1015515, EPI_ISL_1015516, EPI_ISL_1015517, EPI_ISL_1015518, EPI_ISL_1015519, EPI_ISL_1015520, EPI_ISL_1015521, EPI_ISL_1015532, EPI_ISL_1015533, EPI_ISL_1015534, EPI_ISL_1015535, EPI_ISL_1015536, EPI_ISL_1015537, EPI_ISL_1015538, EPI_ISL_1015539, EPI_ISL_1015540, EPI_ISL_1015541, EPI_ISL_1015542, EPI_ISL_1015543, EPI_ISL_1015544, EPI_ISL_1015545, EPI_ISL_1015546, EPI_ISL_1015547, EPI_ISL_1015548, EPI_ISL_1015549, EPI_ISL_1015550, EPI_ISL_1015551, EPI_ISL_1015552, EPI_ISL_1015553, EPI_ISL_1015554, EPI_ISL_1015555, EPI_ISL_1015556, EPI_ISL_1015557, EPI_ISL_1015558, EPI_ISL_1015559, EPI_ISL_1015560 | | | |
| see above | Azienda Sanitaria dell'Alto Adige Laboratorio Aziendale di Microbiologia e Virologia | Istituto di Genomica Applicata | Elisabetta Pagani, Irene Bianconi, Elisabetta Giacobazzi, Elisa Masi, Stefanie Wieser, Irena Jurman, Vera Vendramin, Gabriele Magris, Eleonora Paparelli, Davide Scaglione, Michele Morgante |
| EPI_ISL_1020306 | Istituto Zooprofilattico Sperimentale dell' Umbria e delle Marche -Togo Rosati Università degli Studi di Perugia | Department of Food safety, Nutrition and Veterinary public health - Istituto Superiore di Sanità | Massimo Biagetti , Monica Giammarioli, Luca De Sabato, Gabriele Vaccari, Ilaria Di Bartolo, Giovanni Ianiro, Antonella Mencacci, Barbara Camilloni |
| EPI_ISL_1020310 | Laboratorio di Riferimento Regionale della Sicilia Occidentale per l'Emergenza COVID-19 | Laboratorio di Riferimento Regionale della Sicilia Occidentale per l'Emergenza COVID-19 | Fabio Tramuto, Carmelo Massimo Maida, Daniela Di Naro, Giulia Randazzo, Walter Mazzucco, Giorgio Graziano, Vincenzo Restivo, Claudio Costantino, Francesco Vitale |
| EPI_ISL_1020311, EPI_ISL_1020313 | Istituto Zooprofilattico Sperimentale dell' Umbria e delle Marche -Togo Rosati Università degli Studi di Perugia | Department of Food safety, Nutrition and Veterinary public health - Istituto Superiore di Sanità | Massimo Biagetti , Monica Giammarioli, Luca De Sabato, Gabriele Vaccari, Ilaria Di Bartolo, Giovanni Ianiro, Antonella Mencacci, Barbara Camilloni |
| EPI_ISL_1020314 | Laboratorio di Riferimento Regionale della Sicilia Occidentale per l'Emergenza COVID-19 | Laboratorio di Riferimento Regionale della Sicilia Occidentale per l'Emergenza COVID-19 | Fabio Tramuto, Carmelo Massimo Maida, Daniela Di Naro, Giulia Randazzo, Walter Mazzucco, Giorgio Graziano, Vincenzo Restivo, Claudio Costantino, Francesco Vitale |
| EPI_ISL_1020328 | Istituto Zooprofilattico Sperimentale dell' Umbria e delle Marche -Togo Rosati Università degli Studi di Perugia | Department of Food safety, Nutrition and Veterinary public health - Istituto Superiore di Sanità | Massimo Biagetti , Monica Giammarioli, Luca De Sabato, Gabriele Vaccari, Ilaria Di Bartolo, Giovanni Ianiro, Antonella Mencacci, Barbara Camilloni |
| EPI_ISL_1020329 | Laboratorio di Riferimento Regionale della Sicilia Occidentale per l'Emergenza COVID-19 | Laboratorio di Riferimento Regionale della Sicilia Occidentale per l'Emergenza COVID-19 | Fabio Tramuto, Carmelo Massimo Maida, Daniela Di Naro, Giulia Randazzo, Walter Mazzucco, Giorgio Graziano, Vincenzo Restivo, Claudio Costantino, Francesco Vitale |
| EPI_ISL_1020330 | Istituto Zooprofilattico Sperimentale dell' Umbria e delle Marche -Togo Rosati Università degli Studi di Perugia | Department of Food safety, Nutrition and Veterinary public health - Istituto Superiore di Sanità | Massimo Biagetti , Monica Giammarioli, Luca De Sabato, Gabriele Vaccari, Ilaria Di Bartolo, Giovanni Ianiro, Antonella Mencacci, Barbara Camilloni |
| EPI_ISL_1020331, EPI_ISL_1020333, EPI_ISL_1020334, EPI_ISL_1020335, EPI_ISL_1020336, EPI_ISL_1020337, EPI_ISL_1020339 | Laboratorio di Riferimento Regionale della Sicilia Occidentale per l'Emergenza COVID-19 | Laboratorio di Riferimento Regionale della Sicilia Occidentale per l'Emergenza COVID-19 | Fabio Tramuto, Carmelo Massimo Maida, Daniela Di Naro, Giulia Randazzo, Walter Mazzucco, Giorgio Graziano, Vincenzo Restivo, Claudio Costantino, Francesco Vitale |
| EPI_ISL_1023524 | Ospedale San Camillo De Lellis di Rieti | INMI Lazzaro Spallanzani IRCCS | Cesare E.M. Gruber, Barbara Bartolini, Emanuela Giombini, Francesco Messina, Martina Rueca, Ornella Butera, Stefano Venarubea, Luca Casertano, Luisa Marchioni, Antonino Di Caro, Maria R. Capobianchi |
| EPI_ISL_1034144 | MEDICO COMPETENTE P.O. L'AQUILA | Istituto Zooprofilattico Sperimentale dell'Abruzzo e Molise "G. Caporale" | Lorusso A, Marcacci M, Di Domenico M, Ancora M, Curini V, Mangone I, Rinaldi A, Scialabba S, Di Pasquale A, Cammà C, Puglia I, Calistri P, Savini G |
| EPI_ISL_1034280 | INMI Lazzaro Spallanzani IRCCS | INMI Lazzaro Spallanzani IRCCS | Cesare E.M. Gruber, Barbara Bartolini, Emanuela Giombini, Francesco Messina, Martina Rueca, Ornella Butera, Emanuele Nicastri, Alessandra D'Abramo, Antonino Di Caro, Maria R. Capobianchi |
| EPI_ISL_1034740, EPI_ISL_1034741, EPI_ISL_1034742, EPI_ISL_1034743, EPI_ISL_1034744, EPI_ISL_1034745, EPI_ISL_1034746, EPI_ISL_1034747, EPI_ISL_1034748, EPI_ISL_1034749, EPI_ISL_1034750, EPI_ISL_1034751, EPI_ISL_1034752, EPI_ISL_1034753 | | | |
| see above | Center of Advanced Studies and Technology, Molecular Genetics Laboratory | Center of Advanced Studies and Technology, Molecular Genetics Laboratory | Ferrante Rossella, Mandatori Domitilla, De Fabritiis Simone, Damiani Verena, Anaclerio Federico |
| EPI_ISL_1034793 | Virologia Dipartimento di Scienze Biomediche Università di Sassari Viale San Pietro, 43/B - Sassari | Laboratorio Specialistico di Ematologia, Ospedale "San Francesco", via Manníroni 1, 08100 Nuoro | Giovanna Piras, Tatiana Fancello, Maria Monne, Rosanna Asproni, Caterina Serra, Elena Rimini, Salvatore Rubino |
| EPI_ISL_1034916 | Laboratorio Biologia Molecolare Sars Cov2 - UOC Laboratorio Analisi - Servizio Medicina di Laboratorio, Ospedale "San Francesco" - ATS-ASSL Nuoro | Laboratorio specialistico UOC Ematologia - Ospedale "San Francesco" - ATS-ASSL Nuoro | Piras Giovanna, Asproni Rosanna, Malune Paolo, Fiamma Maura, Monne Maria Itria, Palmas Angelo Domenico, Lo Maglio Iana, Mameli Giuseppe |
| EPI_ISL_1035871, EPI_ISL_1035900, EPI_ISL_1035901, EPI_ISL_1035966 | Department of Infectious Diseases, Istituto Superiore di Sanità, Rome, Italy; Università degli Studi di Siena, Siena, Italy | Istituto Superiore di Sanità (ISS) | Paola Stefanelli, Angela Di Martino, Alessandra Lo Presti, Stefano Fiore, Maria Grazia Cusi, Gabriele Anichini, Claudia Gandolfo, Gianni Gori Savellini, Manuela Marra, Maria Carollo, Marco Crescenzi |
| EPI_ISL_1036140 | SIESP TERAMO | Istituto Zooprofilattico Sperimentale dell'Abruzzo e Molise "G. Caporale" | Lorusso A, Marcacci M, Di Domenico M, Ancora M, Curini V, Mangone I, Rinaldi A, Scialabba S, Di Pasquale A, Cammà C, Puglia I, Calistri P, Savini G |
| EPI_ISL_1036141 | SIESP SULM | Istituto Zooprofilattico Sperimentale dell'Abruzzo e Molise "G. Caporale" | Lorusso A, Marcacci M, Di Domenico M, Ancora M, Curini V, Mangone I, Rinaldi A, Scialabba S, Di Pasquale A, Cammà C, Puglia I, Calistri P, Savini G |
| EPI_ISL_1036142 | OSP CIV ATRI MEDICINA INT | Istituto Zooprofilattico Sperimentale dell'Abruzzo e Molise "G. Caporale" | Lorusso A, Marcacci M, Di Domenico M, Ancora M, Curini V, Mangone I, Rinaldi A, Scialabba S, Di Pasquale A, Cammà C, Puglia I, Calistri P, Savini G |
| EPI_ISL_1036143 | OSP SAN SALVATORE MED INT | Istituto Zooprofilattico Sperimentale dell'Abruzzo e Molise "G. Caporale" | Lorusso A, Marcacci M, Di Domenico M, Ancora M, Curini V, Mangone I, Rinaldi A, Scialabba S, Di Pasquale A, Cammà C, Puglia I, Calistri P, Savini G |
| EPI_ISL_1036144, EPI_ISL_1036145 | SIESP SULM | Istituto Zooprofilattico Sperimentale dell'Abruzzo e Molise "G. Caporale" | Lorusso A, Marcacci M, Di Domenico M, Ancora M, Curini V, Mangone I, Rinaldi A, Scialabba S, Di Pasquale A, Cammà C, Puglia I, Calistri P, Savini G |
| EPI_ISL_1036146 | SIESP CH | Istituto Zooprofilattico Sperimentale dell'Abruzzo e Molise "G. Caporale" | Lorusso A, Marcacci M, Di Domenico M, Ancora M, Curini V, Mangone I, Rinaldi A, Scialabba S, Di Pasquale A, Cammà C, Puglia I, Calistri P, Savini G |
| EPI_ISL_1036147 | SIESP DIP PREV CHIETI | Istituto Zooprofilattico Sperimentale dell'Abruzzo e Molise "G. Caporale" | Lorusso A, Marcacci M, Di Domenico M, Ancora M, Curini V, Mangone I, Rinaldi A, Scialabba S, Di Pasquale A, Cammà C, Puglia I, Calistri P, Savini G |
| EPI_ISL_1036148 | USCA AVEZZANO | Istituto Zooprofilattico Sperimentale dell'Abruzzo e Molise "G. Caporale" | Lorusso A, Marcacci M, Di Domenico M, Ancora M, Curini V, Mangone I, Rinaldi A, Scialabba S, Di Pasquale A, Cammà C, Puglia I, Calistri P, Savini G |
| EPI_ISL_1036149 | OSP SAN SALVATORE MED INT | Istituto Zooprofilattico Sperimentale dell'Abruzzo e Molise "G. Caporale" | Lorusso A, Marcacci M, Di Domenico M, Ancora M, Curini V, Mangone I, Rinaldi A, Scialabba S, Di Pasquale A, Cammà C, Puglia I, Calistri P, Savini G |
| EPI_ISL_1036150 | SIESP CH | Istituto Zooprofilattico Sperimentale dell'Abruzzo e Molise "G. | Lorusso A, Marcacci M, Di Domenico M, Ancora M, Curini V, Mangone I, Rinaldi A, Scialabba S, Di Pasquale A, Cammà C, Puglia I, Calistri P, Savini G |

| | | | |
|---|---|---|---|
| | | Caporale" | |
| EPI_ISL_1036151 | PRES OSP TAGLIACOZZO | Istituto Zooprofilattico Sperimentale dell'Abruzzo e Molise "G. Caporale" | Lorusso A, Marcacci M, Di Domenico M, Ancora M, Curini V, Mangone I, Rinaldi A, Scialabba S, Di Pasquale A, Cammà C, Puglia I, Calistri P, Savini G |
| EPI_ISL_1036152 | OSP CIV GIULIANOVA PRONTO SOCC | Istituto Zooprofilattico Sperimentale dell'Abruzzo e Molise "G. Caporale" | Lorusso A, Marcacci M, Di Domenico M, Ancora M, Curini V, Mangone I, Rinaldi A, Scialabba S, Di Pasquale A, Cammà C, Puglia I, Calistri P, Savini G |
| EPI_ISL_1036153 | SIESP AQ | Istituto Zooprofilattico Sperimentale dell'Abruzzo e Molise "G. Caporale" | Lorusso A, Marcacci M, Di Domenico M, Ancora M, Curini V, Mangone I, Rinaldi A, Scialabba S, Di Pasquale A, Cammà C, Puglia I, Calistri P, Savini G |
| EPI_ISL_1036154, EPI_ISL_1036155 | SIESP TERAMO | Istituto Zooprofilattico Sperimentale dell'Abruzzo e Molise "G. Caporale" | Lorusso A, Marcacci M, Di Domenico M, Ancora M, Curini V, Mangone I, Rinaldi A, Scialabba S, Di Pasquale A, Cammà C, Puglia I, Calistri P, Savini G |
| EPI_ISL_1036156 | SIESP DIP PREV CHIETI | Istituto Zooprofilattico Sperimentale dell'Abruzzo e Molise "G. Caporale" | Lorusso A, Marcacci M, Di Domenico M, Ancora M, Curini V, Mangone I, Rinaldi A, Scialabba S, Di Pasquale A, Cammà C, Puglia I, Calistri P, Savini G |
| EPI_ISL_1036157 | SIESP TERAMO | Istituto Zooprofilattico Sperimentale dell'Abruzzo e Molise "G. Caporale" | Lorusso A, Marcacci M, Di Domenico M, Ancora M, Curini V, Mangone I, Rinaldi A, Scialabba S, Di Pasquale A, Cammà C, Puglia I, Calistri P, Savini G |
| EPI_ISL_1036158 | OSP SAN SALVATORE MED INT | Istituto Zooprofilattico Sperimentale dell'Abruzzo e Molise "G. Caporale" | Lorusso A, Marcacci M, Di Domenico M, Ancora M, Curini V, Mangone I, Rinaldi A, Scialabba S, Di Pasquale A, Cammà C, Puglia I, Calistri P, Savini G |
| EPI_ISL_1036159 | OSP CIV GIULIANOVA PRONTO SOCC | Istituto Zooprofilattico Sperimentale dell'Abruzzo e Molise "G. Caporale" | Lorusso A, Marcacci M, Di Domenico M, Ancora M, Curini V, Mangone I, Rinaldi A, Scialabba S, Di Pasquale A, Cammà C, Puglia I, Calistri P, Savini G |
| EPI_ISL_1036160 | SIESP SULM | Istituto Zooprofilattico Sperimentale dell'Abruzzo e Molise "G. Caporale" | Lorusso A, Marcacci M, Di Domenico M, Ancora M, Curini V, Mangone I, Rinaldi A, Scialabba S, Di Pasquale A, Cammà C, Puglia I, Calistri P, Savini G |
| EPI_ISL_1036161 | OSP CIV TERAMO RIANIMAZIONE CARDIOCHIR | Istituto Zooprofilattico Sperimentale dell'Abruzzo e Molise "G. Caporale" | Lorusso A, Marcacci M, Di Domenico M, Ancora M, Curini V, Mangone I, Rinaldi A, Scialabba S, Di Pasquale A, Cammà C, Puglia I, Calistri P, Savini G |
| EPI_ISL_1036162 | SIESP TERAMO | Istituto Zooprofilattico Sperimentale dell'Abruzzo e Molise "G. Caporale" | Lorusso A, Marcacci M, Di Domenico M, Ancora M, Curini V, Mangone I, Rinaldi A, Scialabba S, Di Pasquale A, Cammà C, Puglia I, Calistri P, Savini G |
| EPI_ISL_1036163, EPI_ISL_1036164 | SIESP CH | Istituto Zooprofilattico Sperimentale dell'Abruzzo e Molise "G. Caporale" | Lorusso A, Marcacci M, Di Domenico M, Ancora M, Curini V, Mangone I, Rinaldi A, Scialabba S, Di Pasquale A, Cammà C, Puglia I, Calistri P, Savini G |
| EPI_ISL_1036165 | SIESP TERAMO | Istituto Zooprofilattico Sperimentale dell'Abruzzo e Molise "G. Caporale" | Lorusso A, Marcacci M, Di Domenico M, Ancora M, Curini V, Mangone I, Rinaldi A, Scialabba S, Di Pasquale A, Cammà C, Puglia I, Calistri P, Savini G |
| EPI_ISL_1036166, EPI_ISL_1036167 | SIESP CH | Istituto Zooprofilattico Sperimentale dell'Abruzzo e Molise "G. Caporale" | Lorusso A, Marcacci M, Di Domenico M, Ancora M, Curini V, Mangone I, Rinaldi A, Scialabba S, Di Pasquale A, Cammà C, Puglia I, Calistri P, Savini G |
| EPI_ISL_1036168, EPI_ISL_1036169 | SIESP TERAMO | Istituto Zooprofilattico Sperimentale dell'Abruzzo e Molise "G. Caporale" | Lorusso A, Marcacci M, Di Domenico M, Ancora M, Curini V, Mangone I, Rinaldi A, Scialabba S, Di Pasquale A, Cammà C, Puglia I, Calistri P, Savini G |
| EPI_ISL_1036170, EPI_ISL_1036171 | SIESP CH | Istituto Zooprofilattico Sperimentale dell'Abruzzo e Molise "G. Caporale" | Lorusso A, Marcacci M, Di Domenico M, Ancora M, Curini V, Mangone I, Rinaldi A, Scialabba S, Di Pasquale A, Cammà C, Puglia I, Calistri P, Savini G |
| EPI_ISL_1036172 | SIESP TERAMO | Istituto Zooprofilattico Sperimentale dell'Abruzzo e Molise "G. Caporale" | Lorusso A, Marcacci M, Di Domenico M, Ancora M, Curini V, Mangone I, Rinaldi A, Scialabba S, Di Pasquale A, Cammà C, Puglia I, Calistri P, Savini G |
| EPI_ISL_1036173, EPI_ISL_1036174, EPI_ISL_1036175, EPI_ISL_1036176 | SIESP CH | Istituto Zooprofilattico Sperimentale dell'Abruzzo e Molise "G. Caporale" | Lorusso A, Marcacci M, Di Domenico M, Ancora M, Curini V, Mangone I, Rinaldi A, Scialabba S, Di Pasquale A, Cammà C, Puglia I, Calistri P, Savini G |
| EPI_ISL_1036177 | SIESP DIP PREV CHIETI | Istituto Zooprofilattico Sperimentale dell'Abruzzo e Molise "G. Caporale" | Lorusso A, Marcacci M, Di Domenico M, Ancora M, Curini V, Mangone I, Rinaldi A, Scialabba S, Di Pasquale A, Cammà C, Puglia I, Calistri P, Savini G |
| EPI_ISL_1036178 | SIESP SULM | Istituto Zooprofilattico Sperimentale dell'Abruzzo e Molise "G. Caporale" | Lorusso A, Marcacci M, Di Domenico M, Ancora M, Curini V, Mangone I, Rinaldi A, Scialabba S, Di Pasquale A, Cammà C, Puglia I, Calistri P, Savini G |
| EPI_ISL_1036179 | SIESP CH DISTRETTO SANITARIO CHIETI | Istituto Zooprofilattico Sperimentale dell'Abruzzo e Molise "G. Caporale" | Lorusso A, Marcacci M, Di Domenico M, Ancora M, Curini V, Mangone I, Rinaldi A, Scialabba S, Di Pasquale A, Cammà C, Puglia I, Calistri P, Savini G |
| EPI_ISL_1036180, EPI_ISL_1036181 | SIESP TERAMO | Istituto Zooprofilattico Sperimentale dell'Abruzzo e Molise "G. Caporale" | Lorusso A, Marcacci M, Di Domenico M, Ancora M, Curini V, Mangone I, Rinaldi A, Scialabba S, Di Pasquale A, Cammà C, Puglia I, Calistri P, Savini G |
| EPI_ISL_1036182 | SIESP CH | Istituto Zooprofilattico Sperimentale dell'Abruzzo e Molise "G. Caporale" | Lorusso A, Marcacci M, Di Domenico M, Ancora M, Curini V, Mangone I, Rinaldi A, Scialabba S, Di Pasquale A, Cammà C, Puglia I, Calistri P, Savini G |
| EPI_ISL_1036183, EPI_ISL_1036184 | SIESP SULM | Istituto Zooprofilattico Sperimentale dell'Abruzzo e Molise "G. Caporale" | Lorusso A, Marcacci M, Di Domenico M, Ancora M, Curini V, Mangone I, Rinaldi A, Scialabba S, Di Pasquale A, Cammà C, Puglia I, Calistri P, Savini G |
| EPI_ISL_1036185 | SIESP DIP PREV CHIETI | Istituto Zooprofilattico Sperimentale dell'Abruzzo e Molise "G. Caporale" | Lorusso A, Marcacci M, Di Domenico M, Ancora M, Curini V, Mangone I, Rinaldi A, Scialabba S, Di Pasquale A, Cammà C, Puglia I, Calistri P, Savini G |
| EPI_ISL_1036186 | OSP SAN SALVATORE MED INT | Istituto Zooprofilattico Sperimentale dell'Abruzzo e Molise "G. Caporale" | Lorusso A, Marcacci M, Di Domenico M, Ancora M, Curini V, Mangone I, Rinaldi A, Scialabba S, Di Pasquale A, Cammà C, Puglia I, Calistri P, Savini G |
| EPI_ISL_1036187 | SIESP TERAMO | Istituto Zooprofilattico Sperimentale dell'Abruzzo e Molise "G. Caporale" | Lorusso A, Marcacci M, Di Domenico M, Ancora M, Curini V, Mangone I, Rinaldi A, Scialabba S, Di Pasquale A, Cammà C, Puglia I, Calistri P, Savini G |
| EPI_ISL_1036188 | OSP SAN SALVATORE UOC MAL INF | Istituto Zooprofilattico Sperimentale dell'Abruzzo e Molise "G. Caporale" | Lorusso A, Marcacci M, Di Domenico M, Ancora M, Curini V, Mangone I, Rinaldi A, Scialabba S, Di Pasquale A, Cammà C, Puglia I, Calistri P, Savini G |
| EPI_ISL_1036189 | OSP CIV TERAMO PNEUMOLOGIA | Istituto Zooprofilattico Sperimentale dell'Abruzzo e Molise "G. Caporale" | Lorusso A, Marcacci M, Di Domenico M, Ancora M, Curini V, Mangone I, Rinaldi A, Scialabba S, Di Pasquale A, Cammà C, Puglia I, Calistri P, Savini G |
| EPI_ISL_1036190 | SIESP CH | Istituto Zooprofilattico Sperimentale dell'Abruzzo e Molise "G. Caporale" | Lorusso A, Marcacci M, Di Domenico M, Ancora M, Curini V, Mangone I, Rinaldi A, Scialabba S, Di Pasquale A, Cammà C, Puglia I, Calistri P, Savini G |
| EPI_ISL_1036191 | SIESP SULM | Istituto Zooprofilattico Sperimentale dell'Abruzzo e Molise "G. Caporale" | Lorusso A, Marcacci M, Di Domenico M, Ancora M, Curini V, Mangone I, Rinaldi A, Scialabba S, Di Pasquale A, Cammà C, Puglia I, Calistri P, Savini G |
| EPI_ISL_1036192 | SIESP TERAMO | Istituto Zooprofilattico Sperimentale dell'Abruzzo e Molise "G. Caporale" | Lorusso A, Marcacci M, Di Domenico M, Ancora M, Curini V, Mangone I, Rinaldi A, Scialabba S, Di Pasquale A, Cammà C, Puglia I, Calistri P, Savini G |
| EPI_ISL_1036193, EPI_ISL_1036194, EPI_ISL_1036195, EPI_ISL_1036196, EPI_ISL_1036197 | SIESP CH | Istituto Zooprofilattico Sperimentale dell'Abruzzo e Molise "G. Caporale" | Lorusso A, Marcacci M, Di Domenico M, Ancora M, Curini V, Mangone I, Rinaldi A, Scialabba S, Di Pasquale A, Cammà C, Puglia I, Calistri P, Savini G |
| EPI_ISL_1036198 | ASL AVEZZANO-SULMONA-L'AQUILA DIP PREV SERV IGIENE E SAN PUBBLICA | Istituto Zooprofilattico Sperimentale dell'Abruzzo e Molise "G. Caporale" | Lorusso A, Marcacci M, Di Domenico M, Ancora M, Curini V, Mangone I, Rinaldi A, Scialabba S, Di Pasquale A, Cammà C, Puglia I, Calistri P, Savini G |

| | | | |
|---|---|---|---|
| EPI_ISL_1036199, EPI_ISL_1036200, EPI_ISL_1036201 | SIESP CH | Istituto Zooprofilattico Sperimentale dell'Abruzzo e Molise "G. Caporale" | Lorusso A, Marcacci M, Di Domenico M, Ancora M, Curini V, Mangone I, Rinaldi A, Scialabba S, Di Pasquale A, Cammà C, Puglia I, Calistri P, Savini G |
| EPI_ISL_1036202 | SIESP TERAMO | Istituto Zooprofilattico Sperimentale dell'Abruzzo e Molise "G. Caporale" | Lorusso A, Marcacci M, Di Domenico M, Ancora M, Curini V, Mangone I, Rinaldi A, Scialabba S, Di Pasquale A, Cammà C, Puglia I, Calistri P, Savini G |
| EPI_ISL_1036203, EPI_ISL_1036204 | SIESP CH | Istituto Zooprofilattico Sperimentale dell'Abruzzo e Molise "G. Caporale" | Lorusso A, Marcacci M, Di Domenico M, Ancora M, Curini V, Mangone I, Rinaldi A, Scialabba S, Di Pasquale A, Cammà C, Puglia I, Calistri P, Savini G |
| EPI_ISL_1036205 | PRES TERR CASOLI | Istituto Zooprofilattico Sperimentale dell'Abruzzo e Molise "G. Caporale" | Lorusso A, Marcacci M, Di Domenico M, Ancora M, Curini V, Mangone I, Rinaldi A, Scialabba S, Di Pasquale A, Cammà C, Puglia I, Calistri P, Savini G |
| EPI_ISL_1036206 | SIESP AQ | Istituto Zooprofilattico Sperimentale dell'Abruzzo e Molise "G. Caporale" | Lorusso A, Marcacci M, Di Domenico M, Ancora M, Curini V, Mangone I, Rinaldi A, Scialabba S, Di Pasquale A, Cammà C, Puglia I, Calistri P, Savini G |
| EPI_ISL_1036207, EPI_ISL_1036208 | SIESP TERAMO | Istituto Zooprofilattico Sperimentale dell'Abruzzo e Molise "G. Caporale" | Lorusso A, Marcacci M, Di Domenico M, Ancora M, Curini V, Mangone I, Rinaldi A, Scialabba S, Di Pasquale A, Cammà C, Puglia I, Calistri P, Savini G |
| EPI_ISL_1036209 | SIESP DIP PREV CHIETI | Istituto Zooprofilattico Sperimentale dell'Abruzzo e Molise "G. Caporale" | Lorusso A, Marcacci M, Di Domenico M, Ancora M, Curini V, Mangone I, Rinaldi A, Scialabba S, Di Pasquale A, Cammà C, Puglia I, Calistri P, Savini G |
| EPI_ISL_1036210 | SIESP TERAMO | Istituto Zooprofilattico Sperimentale dell'Abruzzo e Molise "G. Caporale" | Lorusso A, Marcacci M, Di Domenico M, Ancora M, Curini V, Mangone I, Rinaldi A, Scialabba S, Di Pasquale A, Cammà C, Puglia I, Calistri P, Savini G |
| EPI_ISL_1036211 | ASL AVEZZANO-SULMONA-L'AQUILA | Istituto Zooprofilattico Sperimentale dell'Abruzzo e Molise "G. Caporale" | Lorusso A, Marcacci M, Di Domenico M, Ancora M, Curini V, Mangone I, Rinaldi A, Scialabba S, Di Pasquale A, Cammà C, Puglia I, Calistri P, Savini G |
| EPI_ISL_1036212, EPI_ISL_1036213, EPI_ISL_1036214, EPI_ISL_1036215, EPI_ISL_1036216 | SIESP TERAMO | Istituto Zooprofilattico Sperimentale dell'Abruzzo e Molise "G. Caporale" | Lorusso A, Marcacci M, Di Domenico M, Ancora M, Curini V, Mangone I, Rinaldi A, Scialabba S, Di Pasquale A, Cammà C, Puglia I, Calistri P, Savini G |
| EPI_ISL_1036217 | FONDAZIONE PICCOLA OPERA CARITAS | Istituto Zooprofilattico Sperimentale dell'Abruzzo e Molise "G. Caporale" | Lorusso A, Marcacci M, Di Domenico M, Ancora M, Curini V, Mangone I, Rinaldi A, Scialabba S, Di Pasquale A, Cammà C, Puglia I, Calistri P, Savini G |
| EPI_ISL_1036218, EPI_ISL_1036219, EPI_ISL_1036220 | SIESP TERAMO | Istituto Zooprofilattico Sperimentale dell'Abruzzo e Molise "G. Caporale" | Lorusso A, Marcacci M, Di Domenico M, Ancora M, Curini V, Mangone I, Rinaldi A, Scialabba S, Di Pasquale A, Cammà C, Puglia I, Calistri P, Savini G |
| EPI_ISL_1036221 | OSP SAN SALVATORE UOC MAL INF | Istituto Zooprofilattico Sperimentale dell'Abruzzo e Molise "G. Caporale" | Lorusso A, Marcacci M, Di Domenico M, Ancora M, Curini V, Mangone I, Rinaldi A, Scialabba S, Di Pasquale A, Cammà C, Puglia I, Calistri P, Savini G |
| EPI_ISL_1036222 | SIESP TERAMO | Istituto Zooprofilattico Sperimentale dell'Abruzzo e Molise "G. Caporale" | Lorusso A, Marcacci M, Di Domenico M, Ancora M, Curini V, Mangone I, Rinaldi A, Scialabba S, Di Pasquale A, Cammà C, Puglia I, Calistri P, Savini G |
| EPI_ISL_1036223 | OSP SAN SALVATORE MED INT | Istituto Zooprofilattico Sperimentale dell'Abruzzo e Molise "G. Caporale" | Lorusso A, Marcacci M, Di Domenico M, Ancora M, Curini V, Mangone I, Rinaldi A, Scialabba S, Di Pasquale A, Cammà C, Puglia I, Calistri P, Savini G |
| EPI_ISL_1036224, EPI_ISL_1036225, EPI_ISL_1036226, EPI_ISL_1036227, EPI_ISL_1036228, EPI_ISL_1036229 | P.O.CARDARELLI | Istituto Zooprofilattico Sperimentale dell'Abruzzo e Molise "G. Caporale" | Scutellà M, Niro G, Lorusso A, Marcacci M, Di Domenico M, Ancora M, Curini V, Mangone I, Rinaldi A, Scialabba S, Di Pasquale A, Cammà C, Puglia I, Calistri P, Savini G |
| EPI_ISL_1036238, EPI_ISL_1036239 | Center of Advanced Studies and Technology, Molecular Genetics Laboratory | Center of Advanced Studies and Technology, Molecular Genetics Laboratory | Ferrante Rossella, Mandatori Domitilla, De Fabritiis Simone, Damiani Verena, Anaclerio Federico |
| EPI_ISL_1036755 | Unità Operativa di Microbiologia, IRCCS Policlinico di Sant'Orsola, Azienda Ospedaliero-Universitaria di Bologna | Unità di Analisi del Rischio ed Epidemiologia Genomica, Istituto Zooprofilattico Sperimentale dell'Emilia Romagna e della Lombardia (IZSLER) | Giada Rossini, Giuliano Furlini, Tiziana Lazzarotto, Marina Morganti, Ilaria Menozzi, Erika Scaltriti, Stefano Pongolini |
| EPI_ISL_1039256, EPI_ISL_1039257, EPI_ISL_1039258, EPI_ISL_1039259, EPI_ISL_1039260, EPI_ISL_1039261, EPI_ISL_1039262, EPI_ISL_1039263, EPI_ISL_1039264, EPI_ISL_1039265, EPI_ISL_1039266, EPI_ISL_1039267, EPI_ISL_1039268, EPI_ISL_1039269, EPI_ISL_1039270, EPI_ISL_1039271, EPI_ISL_1039272, EPI_ISL_1039273, EPI_ISL_1039274, EPI_ISL_1039275, EPI_ISL_1039276, EPI_ISL_1039277, EPI_ISL_1039278, EPI_ISL_1039279, EPI_ISL_1039280, EPI_ISL_1039281, EPI_ISL_1039282, EPI_ISL_1039283, EPI_ISL_1039284, EPI_ISL_1039285, EPI_ISL_1039286, EPI_ISL_1039287, EPI_ISL_1039288, EPI_ISL_1039289, EPI_ISL_1039290, EPI_ISL_1039291, EPI_ISL_1039292, EPI_ISL_1039293, EPI_ISL_1039294, EPI_ISL_1039295, EPI_ISL_1039296, EPI_ISL_1039297, EPI_ISL_1039298, EPI_ISL_1039299, EPI_ISL_1039300, EPI_ISL_1039301, EPI_ISL_1039302, EPI_ISL_1039303, EPI_ISL_1039304, EPI_ISL_1039305, EPI_ISL_1039306, EPI_ISL_1039307, EPI_ISL_1039308, EPI_ISL_1039309, EPI_ISL_1039310, EPI_ISL_1039311, EPI_ISL_1039312, EPI_ISL_1039313, EPI_ISL_1039314, EPI_ISL_1039315, EPI_ISL_1039316, EPI_ISL_1039317, EPI_ISL_1039318, EPI_ISL_1039319, EPI_ISL_1039320, EPI_ISL_1039321, EPI_ISL_1039322, EPI_ISL_1039323, EPI_ISL_1039324, EPI_ISL_1039325, EPI_ISL_1039326, EPI_ISL_1039327, EPI_ISL_1039328, EPI_ISL_1039329, EPI_ISL_1039330, EPI_ISL_1039331, EPI_ISL_1039332, EPI_ISL_1039333, EPI_ISL_1039334, EPI_ISL_1039335, EPI_ISL_1039336, EPI_ISL_1039337, EPI_ISL_1039338, EPI_ISL_1039339, EPI_ISL_1039340, EPI_ISL_1039341, EPI_ISL_1039342, EPI_ISL_1039343, EPI_ISL_1039344, EPI_ISL_1039345, EPI_ISL_1039346, EPI_ISL_1039347, EPI_ISL_1039348, EPI_ISL_1039349, EPI_ISL_1039350, EPI_ISL_1039351, EPI_ISL_1039352, EPI_ISL_1039353, EPI_ISL_1039354, EPI_ISL_1039355, EPI_ISL_1039356, EPI_ISL_1039357, EPI_ISL_1039358, EPI_ISL_1039359, EPI_ISL_1039360, EPI_ISL_1039361, EPI_ISL_1039362, EPI_ISL_1039363, EPI_ISL_1039364, EPI_ISL_1039365, EPI_ISL_1039366, EPI_ISL_1039367, EPI_ISL_1039368, EPI_ISL_1039369, EPI_ISL_1039370, EPI_ISL_1039371, EPI_ISL_1039372, EPI_ISL_1039373, EPI_ISL_1039374, EPI_ISL_1039375, EPI_ISL_1039376, EPI_ISL_1039377, EPI_ISL_1039378, EPI_ISL_1039379, EPI_ISL_1039380, EPI_ISL_1039381, EPI_ISL_1039382, EPI_ISL_1039383, EPI_ISL_1039384, EPI_ISL_1039385, EPI_ISL_1039386, EPI_ISL_1039387, EPI_ISL_1039388, EPI_ISL_1039389, EPI_ISL_1039390, EPI_ISL_1039391, EPI_ISL_1039392, EPI_ISL_1039393, EPI_ISL_1039394, EPI_ISL_1039395, EPI_ISL_1039396, EPI_ISL_1039397, EPI_ISL_1039398, EPI_ISL_1039399, EPI_ISL_1039400, EPI_ISL_1039401, EPI_ISL_1039402, EPI_ISL_1039403, EPI_ISL_1039404, EPI_ISL_1039405, EPI_ISL_1039406, EPI_ISL_1039407, EPI_ISL_1039408, EPI_ISL_1039409, EPI_ISL_1039410, EPI_ISL_1039411, EPI_ISL_1039412, EPI_ISL_1039413, EPI_ISL_1039414, EPI_ISL_1039415, EPI_ISL_1039416, EPI_ISL_1039417, EPI_ISL_1039418, EPI_ISL_1039419, EPI_ISL_1039420, EPI_ISL_1039421, EPI_ISL_1039422, EPI_ISL_1039423, EPI_ISL_1039424, EPI_ISL_1039425, EPI_ISL_1039426, EPI_ISL_1039427, EPI_ISL_1039428, EPI_ISL_1039429, EPI_ISL_1039430, EPI_ISL_1039431, EPI_ISL_1039432, EPI_ISL_1039433, EPI_ISL_1039434, EPI_ISL_1039435, EPI_ISL_1039436, EPI_ISL_1039437, EPI_ISL_1039438, EPI_ISL_1039439, EPI_ISL_1039440, EPI_ISL_1039441, EPI_ISL_1039442, EPI_ISL_1039443, EPI_ISL_1039444, EPI_ISL_1039445, EPI_ISL_1039446, EPI_ISL_1039447, EPI_ISL_1039448, EPI_ISL_1039449, EPI_ISL_1039450, EPI_ISL_1039451, EPI_ISL_1039452, EPI_ISL_1039453, EPI_ISL_1039454, EPI_ISL_1039455, EPI_ISL_1039456, EPI_ISL_1039457, EPI_ISL_1039458, EPI_ISL_1039459, EPI_ISL_1039460, EPI_ISL_1039461, EPI_ISL_1039462, EPI_ISL_1039463, EPI_ISL_1039464, EPI_ISL_1039465, EPI_ISL_1039466, EPI_ISL_1039467, EPI_ISL_1039468, EPI_ISL_1039469, EPI_ISL_1039470, EPI_ISL_1039471, EPI_ISL_1039472, EPI_ISL_1039473, EPI_ISL_1039474, EPI_ISL_1039475, EPI_ISL_1039476, EPI_ISL_1039477, EPI_ISL_1039478, EPI_ISL_1039479, EPI_ISL_1039480, EPI_ISL_1039481, EPI_ISL_1039482, EPI_ISL_1039483, EPI_ISL_1039484, EPI_ISL_1039485, EPI_ISL_1039486, EPI_ISL_1039487, EPI_ISL_1039488, EPI_ISL_1039489, EPI_ISL_1039490, EPI_ISL_1039491, EPI_ISL_1039492, EPI_ISL_1039493, EPI_ISL_1039494, EPI_ISL_1039495, EPI_ISL_1039496, EPI_ISL_1039497, EPI_ISL_1039498, EPI_ISL_1039499, EPI_ISL_1039500, EPI_ISL_1039501, EPI_ISL_1039502, EPI_ISL_1039503, EPI_ISL_1039504, EPI_ISL_1039505, EPI_ISL_1039506, EPI_ISL_1039507, EPI_ISL_1039508, EPI_ISL_1039509, EPI_ISL_1039510, EPI_ISL_1039511, EPI_ISL_1039512, EPI_ISL_1039513, EPI_ISL_1039514, EPI_ISL_1039515, EPI_ISL_1039516, EPI_ISL_1039517 | | | |
| see above | Istituto Zooprofilattico Sperimentale del Mezzogiorno | TIGEM | Antonio Grimaldi, Patrizia Annunziata, Francesco Panariello, Biancamaria Pierri, Valentina Bouche, Chiara Colantuono, Maria Concetta Cuomo, Denise Di Concilio, Lucio Di Filippo, Anna Manfredi, Marcello Salvi, Antonio Limone, Pellegrino Cerino, Andrea Ballabio, Davide Cacchiarelli. |
| EPI_ISL_1039782 | INMI Lazzaro Spallanzani IRCCS | INMI Lazzaro Spallanzani IRCCS | CEM Gruber, B Bartolini, E Giombini, F Messina, M Rueca, O Butera, A Di Caro, MR Capobianchi |
| EPI_ISL_1039783 | INMI Lazzaro Spallanzani IRCCS | INMI Lazzaro Spallanzani IRCCS | B Bartolini, M. Rueca, E Giombini, O Butera, C.E.M Gruber, F Messina, MR Capobianchi, A Di Caro |
| EPI_ISL_1039784 | Azienda Ospedaliera San Camillo Forlanini | INMI Lazzaro Spallanzani IRCCS | CEM Gruber, B Bartolini, E Giombini, M Rueca, O Butera, F Messina, D.Gallone, G Parisi, ML Guarino, A Di Caro, MR Capobianchi |
| EPI_ISL_1039785 | Ospedale "F. Spaziani" Frosinone | INMI Lazzaro Spallanzani IRCCS | B Bartolini, E Giombini, C.E.M Gruber, F Messina, M Rueca, O Butera, C. Gargiulo, C Sias, R Pulselli, A Di Caro, MR Capobianchi |
| EPI_ISL_1048362, EPI_ISL_1048363, EPI_ISL_1048364, EPI_ISL_1048365, EPI_ISL_1048366, EPI_ISL_1048368, EPI_ISL_1048369 | INT Fondazione Pascale | INT Fondazione Pascale | INT Fondazione Pascale |
| EPI_ISL_1048798, EPI_ISL_1048799, EPI_ISL_1048800, EPI_ISL_1048802 | U.O. Genomics, S.S. Genetics and Advanced Omics Techniques, Istituto Zooprofilattico Sperimentale del Piemonte Liguria e Valle d'Aosta | U.O. Genomics, S.S. Genetics and Advanced Omics Techniques, Istituto Zooprofilattico Sperimentale del Piemonte Liguria e Valle d'Aosta | Chiara Beltramo, Elena Bozzetta, Pier Luigi Acutis, Simone Peletto |
| EPI_ISL_1048805, EPI_ISL_1048807, EPI_ISL_1048808, EPI_ISL_1048809, EPI_ISL_1048813 | Laboratory of Clinical Microbiology, Virology and Bioemergencies - Fatebenefratelli Sacco Hospital | Virology Laboratory, Scientific Department, Army Medical Center | Valeria Micheli, Alessandro Mancon, Alberto Rizzo, Maria Rita Gismondo, Silvia Fillo, Anella Monte, Anna Anselmo, Vanessa Vera Fain, Giovanni Faggioni, Riccardo De Sanctis, Florigio Lista |
| EPI_ISL_1048822, EPI_ISL_1048823, EPI_ISL_1048824, EPI_ISL_1048825, EPI_ISL_1048826, EPI_ISL_1048828 | Azienda Sanitaria dell'Alto Adige Laboratorio Aziendale di Microbiologia e Virologia | Istituto di Genomica Applicata | Elisabetta Pagani, Irene Bianconi, Elisabetta Giacobazzi, Elisa Masi, Stefanie Wieser, Irena Jurman, Vera Vendramin, Gabriele Magris, Eleonora Paparelli, Davide Scaglione, Michele Morgante |

| | | | |
|---|---|---|---|
| EPI_ISL_1049260 | Struttura Semplice Dipartimentale di Virologia e Microbiologia molecolare, Azienda Ospedaliero-Universitaria, Policlinico di Modena | U.O. Microbiologia, Laboratorio Unico Centro Servizi - AUSL della Romagna | Pecorari Monica, Gennari William, Fregni Serpini Giulia, Giorgio Dirani, Silvia Zannoli, Vittorio Sambri |
| EPI_ISL_1049261 | U.O. Microbiologia Laboratorio Unico Centro Servizi AUSL della Romagna | U.O. Microbiologia Laboratorio Unico Centro Servizi AUSL della Romagna | Silvia Zannoli, Giorgio Dirani, Vittorio Sambri |
| EPI_ISL_1049340 | Istituto Zooprofilattico Sperimentale del Mezzogiorno - Istituto Zooprofilattico Sperimentale del Mezzogiorno - IZSM | Telethon Institute of Genetics and Medicine - Telethon Institute of Genetics and Medicine - TIGEM | Antonio Grimaldi, Patrizia Annunziata, Francesco Panariello, Biancamaria Pierri, Valentina Bouche, Chiara Colantuono, Maria Concetta Cuomo, Denise Di Concilio, Lucio Di Filippo, Anna Manfredi, Marcello Salvi, Antonio Limone, Pellegrino Cerino, Andrea Ballabio, Davide Cacchiarelli. |
| EPI_ISL_1049341, EPI_ISL_1049342, EPI_ISL_1049343, EPI_ISL_1049344, EPI_ISL_1049345, EPI_ISL_1049346, EPI_ISL_1049347, EPI_ISL_1049348, EPI_ISL_1049349, EPI_ISL_1049350, EPI_ISL_1049351, EPI_ISL_1049352, EPI_ISL_1049353, EPI_ISL_1049354, EPI_ISL_1049355, EPI_ISL_1049356, EPI_ISL_1049357, EPI_ISL_1049358, EPI_ISL_1049359, EPI_ISL_1049360, EPI_ISL_1049361, EPI_ISL_1049362, EPI_ISL_1049363, EPI_ISL_1049364, EPI_ISL_1049365, EPI_ISL_1049366, EPI_ISL_1049367, EPI_ISL_1049368, EPI_ISL_1049369, EPI_ISL_1049370, EPI_ISL_1049371, EPI_ISL_1049372, EPI_ISL_1049373, EPI_ISL_1049374, EPI_ISL_1049375, EPI_ISL_1049376, EPI_ISL_1049377, EPI_ISL_1049378, EPI_ISL_1049379, EPI_ISL_1049380, EPI_ISL_1049381, EPI_ISL_1049382, EPI_ISL_1049383, EPI_ISL_1049384, EPI_ISL_1049385, EPI_ISL_1049386, EPI_ISL_1049387, EPI_ISL_1049388, EPI_ISL_1049389, EPI_ISL_1049390, EPI_ISL_1049391, EPI_ISL_1049392, EPI_ISL_1049393, EPI_ISL_1049394, EPI_ISL_1049395, EPI_ISL_1049396, EPI_ISL_1049397, EPI_ISL_1049398, EPI_ISL_1049399, EPI_ISL_1049400, EPI_ISL_1049401, EPI_ISL_1049402, EPI_ISL_1049403, EPI_ISL_1049404, EPI_ISL_1049405, EPI_ISL_1049406, EPI_ISL_1049407, EPI_ISL_1049408, EPI_ISL_1049409, EPI_ISL_1049410, EPI_ISL_1049411, EPI_ISL_1049412, EPI_ISL_1049413, EPI_ISL_1049414, EPI_ISL_1049415, EPI_ISL_1049416, EPI_ISL_1049417, EPI_ISL_1049418, EPI_ISL_1049419, EPI_ISL_1049420, EPI_ISL_1049421, EPI_ISL_1049422, EPI_ISL_1049423, EPI_ISL_1049424, EPI_ISL_1049425, EPI_ISL_1049426, EPI_ISL_1049427, EPI_ISL_1049428, EPI_ISL_1049429, EPI_ISL_1049430, EPI_ISL_1049431, EPI_ISL_1049432, EPI_ISL_1049433, EPI_ISL_1049434, EPI_ISL_1049435, EPI_ISL_1049436, EPI_ISL_1049437, EPI_ISL_1049438, EPI_ISL_1049439, EPI_ISL_1049440, EPI_ISL_1049441, EPI_ISL_1049442, EPI_ISL_1049443, EPI_ISL_1049444, EPI_ISL_1049445, EPI_ISL_1049446, EPI_ISL_1049447, EPI_ISL_1049448, EPI_ISL_1049449, EPI_ISL_1049450, EPI_ISL_1049451, EPI_ISL_1049452, EPI_ISL_1049453, EPI_ISL_1049454, EPI_ISL_1049455, EPI_ISL_1049456, EPI_ISL_1049457, EPI_ISL_1049458, EPI_ISL_1049459, EPI_ISL_1049460, EPI_ISL_1049461, EPI_ISL_1049462, EPI_ISL_1049463, EPI_ISL_1049464, EPI_ISL_1049465, EPI_ISL_1049466, EPI_ISL_1049467, EPI_ISL_1049468, EPI_ISL_1049469, EPI_ISL_1049470, EPI_ISL_1049471, EPI_ISL_1049472, EPI_ISL_1049473, EPI_ISL_1049474, EPI_ISL_1049475, EPI_ISL_1049476, EPI_ISL_1049477, EPI_ISL_1049478, EPI_ISL_1049479, EPI_ISL_1049480, EPI_ISL_1049481 | | | |
| see above | Istituto Zooprofilattico Sperimentale del Mezzogiorno - IZSM | Telethon Institute of Genetics and Medicine - TIGEM | Antonio Grimaldi, Patrizia Annunziata, Francesco Panariello, Biancamaria Pierri, Valentina Bouche, Chiara Colantuono, Maria Concetta Cuomo, Denise Di Concilio, Lucio Di Filippo, Anna Manfredi, Marcello Salvi, Antonio Limone, Pellegrino Cerino, Andrea Ballabio, Davide Cacchiarelli. |
| EPI_ISL_1055772, EPI_ISL_1055774, EPI_ISL_1055781 | U.O. Genomics, S.S. Genetics and Advanced Omics Techniques, Istituto Zooprofilattico Sperimentale del Piemonte Liguria e Valle d'Aosta | U.O. Genomics, S.S. Genetics and Advanced Omics Techniques, Istituto Zooprofilattico Sperimentale del Piemonte Liguria e Valle d'Aosta | Chiara Beltramo, Elena Bozzetta, Pier Luigi Acutis, Simone Peletto |
| EPI_ISL_1055796 | Ospedale A.S.O. Alessandria | U.O. Genomics, S.S. Genetics and Advanced Omics Techniques, Istituto Zooprofilattico Sperimentale del Piemonte Liguria e Valle d'Aosta | Chiara Beltramo, Elena Bozzetta, Pier Luigi Acutis, Simone Peletto |
| EPI_ISL_1055797, EPI_ISL_1055798, EPI_ISL_1055803, EPI_ISL_1055804, EPI_ISL_1055805, EPI_ISL_1055806, EPI_ISL_1055807 | A.O.U. San Luigi Gonzaga, S.S. Unità di Prevenzione Rischio Infettivo | U.O. Genomics, S.S. Genetics and Advanced Omics Techniques, Istituto Zooprofilattico Sperimentale del Piemonte Liguria e Valle d'Aosta | Chiara Beltramo, Elena Bozzetta, Pier Luigi Acutis, Simone Peletto |
| EPI_ISL_1055808 | Laboratory of Microbiology and Virology, Ospedale Amedeo di Savoia, ASL "Città di Torino" | U.O. Genomics, S.S. Genetics and Advanced Omics Techniques, Istituto Zooprofilattico Sperimentale del Piemonte Liguria e Valle d'Aosta | Chiara Beltramo, Francesco Cerutti, Valeria Ghisetti, Elena Bozzetta, Pier Luigi Acutis, Simone Peletto |
| EPI_ISL_1055817 | SIESP DIPARTIMENTO DI PREVENZIONE CHIETI | Istituto Zooprofilattico Sperimentale dell'Abruzzo e Molise "G. Caporale" | Lorusso A, Marcacci M, Di Domenico M, Ancora M, Curini V, Mangone I, Rinaldi A, Delli Compagni E, Scialabba S, Di Pasquale A, Cammà C, Puglia I, Calistri P, Savini G |
| EPI_ISL_1056655, EPI_ISL_1056656, EPI_ISL_1056657, EPI_ISL_1056658, EPI_ISL_1056659, EPI_ISL_1056660, EPI_ISL_1056661, EPI_ISL_1056662, EPI_ISL_1056663, EPI_ISL_1056664, EPI_ISL_1056665, EPI_ISL_1056666, EPI_ISL_1056667, EPI_ISL_1056668, EPI_ISL_1056669, EPI_ISL_1056670, EPI_ISL_1056671, EPI_ISL_1056672, EPI_ISL_1056673, EPI_ISL_1056674, EPI_ISL_1056675, EPI_ISL_1056676, EPI_ISL_1056677, EPI_ISL_1056678, EPI_ISL_1056679, EPI_ISL_1056680, EPI_ISL_1056681, EPI_ISL_1056682, EPI_ISL_1056683, EPI_ISL_1056684, EPI_ISL_1056685, EPI_ISL_1056686, EPI_ISL_1056687, EPI_ISL_1056688, EPI_ISL_1056689, EPI_ISL_1056690, EPI_ISL_1056691, EPI_ISL_1056692, EPI_ISL_1056693, EPI_ISL_1056694, EPI_ISL_1056695, EPI_ISL_1056696, EPI_ISL_1056697, EPI_ISL_1056698, EPI_ISL_1056699, EPI_ISL_1056700, EPI_ISL_1056701, EPI_ISL_1056702, EPI_ISL_1056703, EPI_ISL_1056704, EPI_ISL_1056705, EPI_ISL_1056706, EPI_ISL_1056707, EPI_ISL_1056708, EPI_ISL_1056709, EPI_ISL_1056710, EPI_ISL_1056711, EPI_ISL_1056712, EPI_ISL_1056713, EPI_ISL_1056714, EPI_ISL_1056715, EPI_ISL_1056716, EPI_ISL_1056717, EPI_ISL_1056718, EPI_ISL_1056719, EPI_ISL_1056720, EPI_ISL_1056721, EPI_ISL_1056722, EPI_ISL_1056723, EPI_ISL_1056724, EPI_ISL_1056725, EPI_ISL_1056726, EPI_ISL_1056727, EPI_ISL_1056728, EPI_ISL_1056729, EPI_ISL_1056730, EPI_ISL_1056731, EPI_ISL_1056732, EPI_ISL_1056733, EPI_ISL_1056734, EPI_ISL_1056735, EPI_ISL_1056736, EPI_ISL_1056737, EPI_ISL_1056738, EPI_ISL_1056739, EPI_ISL_1056740, EPI_ISL_1056741, EPI_ISL_1056742, EPI_ISL_1056743, EPI_ISL_1056744, EPI_ISL_1056745, EPI_ISL_1056746, EPI_ISL_1056747, EPI_ISL_1056748, EPI_ISL_1056749, EPI_ISL_1056750, EPI_ISL_1056751, EPI_ISL_1056752, EPI_ISL_1056753, EPI_ISL_1056754, EPI_ISL_1056755, EPI_ISL_1056756, EPI_ISL_1056757, EPI_ISL_1056758, EPI_ISL_1056759, EPI_ISL_1056760, EPI_ISL_1056761, EPI_ISL_1056762, EPI_ISL_1056763, EPI_ISL_1056764, EPI_ISL_1056765, EPI_ISL_1056766, EPI_ISL_1056767, EPI_ISL_1056768, EPI_ISL_1056769, EPI_ISL_1056770, EPI_ISL_1056771, EPI_ISL_1056772, EPI_ISL_1056773, EPI_ISL_1056774, EPI_ISL_1056775, EPI_ISL_1056776, EPI_ISL_1056777, EPI_ISL_1056778, EPI_ISL_1056779, EPI_ISL_1056780, EPI_ISL_1056781, EPI_ISL_1056782, EPI_ISL_1056783, EPI_ISL_1056784, EPI_ISL_1056785, EPI_ISL_1056786, EPI_ISL_1056787, EPI_ISL_1056788, EPI_ISL_1056789, EPI_ISL_1056790, EPI_ISL_1056791, EPI_ISL_1056792, EPI_ISL_1056793, EPI_ISL_1056794, EPI_ISL_1056795, EPI_ISL_1056796, EPI_ISL_1056797, EPI_ISL_1056798, EPI_ISL_1056799, EPI_ISL_1056800, EPI_ISL_1056801, EPI_ISL_1056802, EPI_ISL_1056803, EPI_ISL_1056804, EPI_ISL_1056805, EPI_ISL_1056806, EPI_ISL_1056807, EPI_ISL_1056808, EPI_ISL_1056809, EPI_ISL_1056810, EPI_ISL_1056811, EPI_ISL_1056812, EPI_ISL_1056813, EPI_ISL_1056814, EPI_ISL_1056815, EPI_ISL_1056816, EPI_ISL_1056817, EPI_ISL_1056818, EPI_ISL_1056819, EPI_ISL_1056820, EPI_ISL_1056821, EPI_ISL_1056822, EPI_ISL_1056823, EPI_ISL_1056824, EPI_ISL_1056825, EPI_ISL_1056826, EPI_ISL_1056827, EPI_ISL_1056828, EPI_ISL_1056829, EPI_ISL_1056830, EPI_ISL_1056831, EPI_ISL_1056832, EPI_ISL_1056833, EPI_ISL_1056834, EPI_ISL_1056835, EPI_ISL_1056836, EPI_ISL_1056837, EPI_ISL_1056838, EPI_ISL_1056839, EPI_ISL_1056840, EPI_ISL_1056841, EPI_ISL_1056842, EPI_ISL_1056843, EPI_ISL_1056844, EPI_ISL_1056845, EPI_ISL_1056846, EPI_ISL_1056847, EPI_ISL_1056848, EPI_ISL_1056849, EPI_ISL_1056850, EPI_ISL_1056851, EPI_ISL_1056852, EPI_ISL_1056853, EPI_ISL_1056854, EPI_ISL_1056855, EPI_ISL_1056856, EPI_ISL_1056857, EPI_ISL_1056858, EPI_ISL_1056859, EPI_ISL_1056860, EPI_ISL_1056861, EPI_ISL_1056862, EPI_ISL_1056863, EPI_ISL_1056864, EPI_ISL_1056865, EPI_ISL_1056866, EPI_ISL_1056867, EPI_ISL_1056868, EPI_ISL_1056869, EPI_ISL_1056870, EPI_ISL_1056871, EPI_ISL_1056872, EPI_ISL_1056873, EPI_ISL_1056874, EPI_ISL_1056875, EPI_ISL_1056876, EPI_ISL_1056877, EPI_ISL_1056878, EPI_ISL_1056879, EPI_ISL_1056880, EPI_ISL_1056881, EPI_ISL_1056882, EPI_ISL_1056883, EPI_ISL_1056884, EPI_ISL_1056885, EPI_ISL_1056886, EPI_ISL_1056887, EPI_ISL_1056888, EPI_ISL_1056889, EPI_ISL_1056890, EPI_ISL_1056891, EPI_ISL_1056892, EPI_ISL_1056893, EPI_ISL_1056894, EPI_ISL_1056895, EPI_ISL_1056896, EPI_ISL_1056897, EPI_ISL_1056898, EPI_ISL_1056899, EPI_ISL_1056900, EPI_ISL_1056901, EPI_ISL_1056902, EPI_ISL_1056903, EPI_ISL_1056904, EPI_ISL_1056905, EPI_ISL_1056906, EPI_ISL_1056907, EPI_ISL_1056908, EPI_ISL_1056909, EPI_ISL_1056910, EPI_ISL_1056911, EPI_ISL_1056912, EPI_ISL_1056913, EPI_ISL_1056914, EPI_ISL_1056915, EPI_ISL_1056916, EPI_ISL_1056917, EPI_ISL_1056918, EPI_ISL_1056919, EPI_ISL_1056920, EPI_ISL_1056921, EPI_ISL_1056922, EPI_ISL_1056923, EPI_ISL_1056924, EPI_ISL_1056925, EPI_ISL_1056926, EPI_ISL_1056927, EPI_ISL_1056928, EPI_ISL_1056929, EPI_ISL_1056930, EPI_ISL_1056931, EPI_ISL_1056932, EPI_ISL_1056933, EPI_ISL_1056934, EPI_ISL_1056935, EPI_ISL_1056936, EPI_ISL_1056937, EPI_ISL_1056938, EPI_ISL_1056939, EPI_ISL_1056940, EPI_ISL_1056941, EPI_ISL_1056942, EPI_ISL_1056943, EPI_ISL_1056944, EPI_ISL_1056945, EPI_ISL_1056946, EPI_ISL_1056947, EPI_ISL_1056948, EPI_ISL_1056949, EPI_ISL_1056950, EPI_ISL_1056951, EPI_ISL_1056952, EPI_ISL_1057099, EPI_ISL_1057100, EPI_ISL_1057101 | | | |
| see above | Istituto Zooprofilattico Sperimentale del Mezzogiorno - IZSM | Telethon Institute of Genetics and Medicine - TIGEM | Antonio Grimaldi, Patrizia Annunziata, Francesco Panariello, Biancamaria Pierri, Valentina Bouche, Chiara Colantuono, Maria Concetta Cuomo, Denise Di Concilio, Lucio Di Filippo, Anna Manfredi, Marcello Salvi, Antonio Limone, Pellegrino Cerino, Andrea Ballabio, Davide Cacchiarelli. |
| EPI_ISL_1058030 | SIESP CHIETI - DRIVE IN CHIETI | Istituto Zooprofilattico Sperimentale dell'Abruzzo e Molise "G. Caporale" | Lorusso A, Marcacci M, Di Domenico M, Ancora M, Curini V, Mangone I, Rinaldi A, Delli Compagni E, Scialabba S, Di Pasquale A, Cammà C, Puglia I, Calistri P, Savini G |
| EPI_ISL_1058031, EPI_ISL_1058032, EPI_ISL_1058033 | SIESP CHIETI - DRIVE IN ORTONA | Istituto Zooprofilattico Sperimentale dell'Abruzzo e Molise "G. Caporale" | Lorusso A, Marcacci M, Di Domenico M, Ancora M, Curini V, Mangone I, Rinaldi A, Delli Compagni E, Scialabba S, Di Pasquale A, Cammà C, Puglia I, Calistri P, Savini G |
| EPI_ISL_1058040 | SIESP CHIETI DRIVE IN LANCIANO | Istituto Zooprofilattico Sperimentale dell'Abruzzo e Molise "G. Caporale" | Lorusso A, Marcacci M, Di Domenico M, Ancora M, Curini V, Mangone I, Rinaldi A, Scialabba S, Di Pasquale A, Cammà C, Puglia I, Calistri P, Savini G |
| EPI_ISL_1058041 | SIESP CHIETI | Istituto Zooprofilattico Sperimentale dell'Abruzzo e Molise "G. Caporale" | Lorusso A, Marcacci M, Di Domenico M, Ancora M, Curini V, Mangone I, Rinaldi A, Scialabba S, Di Pasquale A, Cammà C, Puglia I, Calistri P, Savini G |
| EPI_ISL_1058042 | SIESP CHIETI - DRIVE IN ORTONA | Istituto Zooprofilattico Sperimentale dell'Abruzzo e Molise "G. Caporale" | Lorusso A, Marcacci M, Di Domenico M, Ancora M, Curini V, Mangone I, Rinaldi A, Scialabba S, Di Pasquale A, Cammà C, Puglia I, Calistri P, Savini G |
| EPI_ISL_1058043 | SIESP CHIETI - DRIVE IN CHIETI | Istituto Zooprofilattico Sperimentale dell'Abruzzo e Molise "G. Caporale" | Lorusso A, Marcacci M, Di Domenico M, Ancora M, Curini V, Mangone I, Rinaldi A, Scialabba S, Di Pasquale A, Cammà C, Puglia I, Calistri P, Savini G |
| EPI_ISL_1058044, EPI_ISL_1058045 | SIESP CHIETI - DRIVE IN ORTONA | Istituto Zooprofilattico Sperimentale dell'Abruzzo e Molise "G. Caporale" | Lorusso A, Marcacci M, Di Domenico M, Ancora M, Curini V, Mangone I, Rinaldi A, Scialabba S, Di Pasquale A, Cammà C, Puglia I, Calistri P, Savini G |
| EPI_ISL_1061038 | Istituto Zooprofilattico Sperimentale del Mezzogiorno (Istituto Zooprofilattico Sperimentale del Mezzogiorno (IZSM)) | Telethon Institute of Genetics and Medicine (TIGEM) | Antonio Grimaldi, Patrizia Annunziata, Francesco Panariello, Biancamaria Pierri, Valentina Bouche, Chiara Colantuono, Maria Concetta Cuomo, Denise Di Concilio, Lucio Di Filippo, Anna Manfredi, Marcello Salvi, Antonio Limone, Pellegrino Cerino, Andrea Ballabio, Davide Cacchiarelli. |
| EPI_ISL_1061039, EPI_ISL_1061040, EPI_ISL_1061041, EPI_ISL_1061042, EPI_ISL_1061043, EPI_ISL_1061044, EPI_ISL_1061045, EPI_ISL_1061046, EPI_ISL_1061047, EPI_ISL_1061048, EPI_ISL_1061049, EPI_ISL_1061050, EPI_ISL_1061051, EPI_ISL_1061052, EPI_ISL_1061053, EPI_ISL_1061054, EPI_ISL_1061055, EPI_ISL_1061056, EPI_ISL_1061057, EPI_ISL_1061058, EPI_ISL_1061059, EPI_ISL_1061060, EPI_ISL_1061061, EPI_ISL_1061062, EPI_ISL_1061063, EPI_ISL_1061064, EPI_ISL_1061065, EPI_ISL_1061066, EPI_ISL_1061067, EPI_ISL_1061068, EPI_ISL_1061069, EPI_ISL_1061070, EPI_ISL_1061071, EPI_ISL_1061072, EPI_ISL_1061073, EPI_ISL_1061074, EPI_ISL_1061075, EPI_ISL_1061076, EPI_ISL_1061077, EPI_ISL_1061078, EPI_ISL_1061079, EPI_ISL_1061080, EPI_ISL_1061081, EPI_ISL_1061082, EPI_ISL_1061083, EPI_ISL_1061084, EPI_ISL_1061085, EPI_ISL_1061086, EPI_ISL_1061087, EPI_ISL_1061088, EPI_ISL_1061089, EPI_ISL_1061090, EPI_ISL_1061091, EPI_ISL_1061092, EPI_ISL_1061093, EPI_ISL_1061094, EPI_ISL_1061095, EPI_ISL_1061096, EPI_ISL_1061097, EPI_ISL_1061098, EPI_ISL_1061099, EPI_ISL_1061100, EPI_ISL_1061101, EPI_ISL_1061102, EPI_ISL_1061103, EPI_ISL_1061104, EPI_ISL_1061105, EPI_ISL_1061106, EPI_ISL_1061107, EPI_ISL_1061108, EPI_ISL_1061109, EPI_ISL_1061110, EPI_ISL_1061111, EPI_ISL_1061112, EPI_ISL_1061113, EPI_ISL_1061114, EPI_ISL_1061115, EPI_ISL_1061116, EPI_ISL_1061117, EPI_ISL_1061118, EPI_ISL_1061119, EPI_ISL_1061120, EPI_ISL_1061121, EPI_ISL_1061122, EPI_ISL_1061123, EPI_ISL_1061124, EPI_ISL_1061125, EPI_ISL_1061126, EPI_ISL_1061127, EPI_ISL_1061128, EPI_ISL_1061129, EPI_ISL_1061130, EPI_ISL_1061131, EPI_ISL_1061132, EPI_ISL_1061133, EPI_ISL_1061134, EPI_ISL_1061135, EPI_ISL_1061136, EPI_ISL_1061137, EPI_ISL_1061138, EPI_ISL_1061139, EPI_ISL_1061140, EPI_ISL_1061141, EPI_ISL_1061142, EPI_ISL_1061143, EPI_ISL_1061144, EPI_ISL_1061145, EPI_ISL_1061146, EPI_ISL_1061147, EPI_ISL_1061148, EPI_ISL_1061149, EPI_ISL_1061150, EPI_ISL_1061151, EPI_ISL_1061152, EPI_ISL_1061153, EPI_ISL_1061154, EPI_ISL_1061155, EPI_ISL_1061156, EPI_ISL_1061157, EPI_ISL_1061158, EPI_ISL_1061159, EPI_ISL_1061160, EPI_ISL_1061161, EPI_ISL_1061162, EPI_ISL_1061163, EPI_ISL_1061164, EPI_ISL_1061165, EPI_ISL_1061166, EPI_ISL_1061167, EPI_ISL_1061168, EPI_ISL_1061169, EPI_ISL_1061170, EPI_ISL_1061171, EPI_ISL_1061172, EPI_ISL_1061173, EPI_ISL_1061174, | | | |

| | | | |
|---|---|---|---|
| EPI_ISL_1061175, EPI_ISL_1061176, EPI_ISL_1061177, EPI_ISL_1061178, EPI_ISL_1061179, EPI_ISL_1061180, EPI_ISL_1061181, EPI_ISL_1061182, EPI_ISL_1061183, EPI_ISL_1061184, EPI_ISL_1061185, EPI_ISL_1061186, EPI_ISL_1061187, EPI_ISL_1061188, EPI_ISL_1061189, EPI_ISL_1061190, EPI_ISL_1061191, EPI_ISL_1061192, EPI_ISL_1061193, EPI_ISL_1061194, EPI_ISL_1061195, EPI_ISL_1061196, EPI_ISL_1061197, EPI_ISL_1061198, EPI_ISL_1061199, EPI_ISL_1061200, EPI_ISL_1061201, EPI_ISL_1061202, EPI_ISL_1061203, EPI_ISL_1061204, EPI_ISL_1061205, EPI_ISL_1061206, EPI_ISL_1061207, EPI_ISL_1061208, EPI_ISL_1061209, EPI_ISL_1061210, EPI_ISL_1061211, EPI_ISL_1061212, EPI_ISL_1061213, EPI_ISL_1061214, EPI_ISL_1061215, EPI_ISL_1061216, EPI_ISL_1061217, EPI_ISL_1061218, EPI_ISL_1061219, EPI_ISL_1061220, EPI_ISL_1061221, EPI_ISL_1061222, EPI_ISL_1061223, EPI_ISL_1061224, EPI_ISL_1061225, EPI_ISL_1061226, EPI_ISL_1061227, EPI_ISL_1061228, EPI_ISL_1061229, EPI_ISL_1061230, EPI_ISL_1061231, EPI_ISL_1061232, EPI_ISL_1061233, EPI_ISL_1061234, EPI_ISL_1061235, EPI_ISL_1061236, EPI_ISL_1061237, EPI_ISL_1061238, EPI_ISL_1061239, EPI_ISL_1061240, EPI_ISL_1061241, EPI_ISL_1061242, EPI_ISL_1061243, EPI_ISL_1061244, EPI_ISL_1061245, EPI_ISL_1061246, EPI_ISL_1061247, EPI_ISL_1061248, EPI_ISL_1061249, EPI_ISL_1061250, EPI_ISL_1061251, EPI_ISL_1061252, EPI_ISL_1061253, EPI_ISL_1061254, EPI_ISL_1061255, EPI_ISL_1061256, EPI_ISL_1061257, EPI_ISL_1061258, EPI_ISL_1061259, EPI_ISL_1061260, EPI_ISL_1061261, EPI_ISL_1061262, EPI_ISL_1061263, EPI_ISL_1061264, EPI_ISL_1061265, EPI_ISL_1061266, EPI_ISL_1061267, EPI_ISL_1061268, EPI_ISL_1061269, EPI_ISL_1061270, EPI_ISL_1061271, EPI_ISL_1061272, EPI_ISL_1061273, EPI_ISL_1061274, EPI_ISL_1061275, EPI_ISL_1061276, EPI_ISL_1061311, EPI_ISL_1061312, EPI_ISL_1061313, EPI_ISL_1061314, EPI_ISL_1061315, EPI_ISL_1061316, EPI_ISL_1061317, EPI_ISL_1061318, EPI_ISL_1061319, EPI_ISL_1061320, EPI_ISL_1061321, EPI_ISL_1061322, EPI_ISL_1061323, EPI_ISL_1061324, EPI_ISL_1061325, EPI_ISL_1061326, EPI_ISL_1061327, EPI_ISL_1061328, EPI_ISL_1061329, EPI_ISL_1061330, EPI_ISL_1061331, EPI_ISL_1061332, EPI_ISL_1061333, EPI_ISL_1061334, EPI_ISL_1061335, EPI_ISL_1061336, EPI_ISL_1061337, EPI_ISL_1061338, EPI_ISL_1061339, EPI_ISL_1061340, EPI_ISL_1061341, EPI_ISL_1061342, EPI_ISL_1061343, EPI_ISL_1061344, EPI_ISL_1061345, EPI_ISL_1061346, EPI_ISL_1061347, EPI_ISL_1061348, EPI_ISL_1061349, EPI_ISL_1061350, EPI_ISL_1061351, EPI_ISL_1061352, EPI_ISL_1061353, EPI_ISL_1061354, EPI_ISL_1061355, EPI_ISL_1061356, EPI_ISL_1061357, EPI_ISL_1061358, EPI_ISL_1061359, EPI_ISL_1061360, EPI_ISL_1061361, EPI_ISL_1061362, EPI_ISL_1061363, EPI_ISL_1061364, EPI_ISL_1061365, EPI_ISL_1061366, EPI_ISL_1061367, EPI_ISL_1061368, EPI_ISL_1061369, EPI_ISL_1061370, EPI_ISL_1061371, EPI_ISL_1061372, EPI_ISL_1061373, EPI_ISL_1061374, EPI_ISL_1061375, EPI_ISL_1061376, EPI_ISL_1061377, EPI_ISL_1061378, EPI_ISL_1061379, EPI_ISL_1061380, EPI_ISL_1061381, EPI_ISL_1061382, EPI_ISL_1061383, EPI_ISL_1061384, EPI_ISL_1061385, EPI_ISL_1061386, EPI_ISL_1061387, EPI_ISL_1061388, EPI_ISL_1061389, EPI_ISL_1061390, EPI_ISL_1061391, EPI_ISL_1061392, EPI_ISL_1061393, EPI_ISL_1061394, EPI_ISL_1061395, EPI_ISL_1061396, EPI_ISL_1061397, EPI_ISL_1061398 | | | |
| see above | Istituto Zooprofilattico Sperimentale del Mezzogiorno (IZSM) | Telethon Institute of Genetics and Medicine (TIGEM) | Antonio Grimaldi, Patrizia Annunziata, Francesco Panariello, Biancamaria Pierri, Valentina Bouche, Chiara Colantuono, Maria Concetta Cuomo, Denise Di Concilio, Lucio Di Filippo, Anna Manfredi, Marcello Salvi, Antonio Limone, Pellegrino Cerino, Andrea Ballabio, Davide Cacchiarelli. |
| EPI_ISL_1063404, EPI_ISL_1063405, EPI_ISL_1063406, EPI_ISL_1063407, EPI_ISL_1063408, EPI_ISL_1063409, EPI_ISL_1063410, EPI_ISL_1063411, EPI_ISL_1063412, EPI_ISL_1063413, EPI_ISL_1063414, EPI_ISL_1063415, EPI_ISL_1063416, EPI_ISL_1063417, EPI_ISL_1063418, EPI_ISL_1063419, EPI_ISL_1063420, EPI_ISL_1063421, EPI_ISL_1063422, EPI_ISL_1063423, EPI_ISL_1063424, EPI_ISL_1063425, EPI_ISL_1063426, EPI_ISL_1063427, EPI_ISL_1063428, EPI_ISL_1063429, EPI_ISL_1063430, EPI_ISL_1063431, EPI_ISL_1063432, EPI_ISL_1063433, EPI_ISL_1063434, EPI_ISL_1063435, EPI_ISL_1063436, EPI_ISL_1063437, EPI_ISL_1063438, EPI_ISL_1063439, EPI_ISL_1063440, EPI_ISL_1063441, EPI_ISL_1063442, EPI_ISL_1063443, EPI_ISL_1063444, EPI_ISL_1063445, EPI_ISL_1063446, EPI_ISL_1063447, EPI_ISL_1063448, EPI_ISL_1063449, EPI_ISL_1063450, EPI_ISL_1063451, EPI_ISL_1063452, EPI_ISL_1063453, EPI_ISL_1063454, EPI_ISL_1063455, EPI_ISL_1063456, EPI_ISL_1063457, EPI_ISL_1063458, EPI_ISL_1063459, EPI_ISL_1063460, EPI_ISL_1063461, EPI_ISL_1063462, EPI_ISL_1063463, EPI_ISL_1063464, EPI_ISL_1063465, EPI_ISL_1063466, EPI_ISL_1063467, EPI_ISL_1063468, EPI_ISL_1063469, EPI_ISL_1063470, EPI_ISL_1063471, EPI_ISL_1063472, EPI_ISL_1063473, EPI_ISL_1063474, EPI_ISL_1063475, EPI_ISL_1063476, EPI_ISL_1063477, EPI_ISL_1063478, EPI_ISL_1063479, EPI_ISL_1063480, EPI_ISL_1063481, EPI_ISL_1063482, EPI_ISL_1063483, EPI_ISL_1063484, EPI_ISL_1063485, EPI_ISL_1063486, EPI_ISL_1063487, EPI_ISL_1063488, EPI_ISL_1063489, EPI_ISL_1063490, EPI_ISL_1063491, EPI_ISL_1063492, EPI_ISL_1063493 | | | |
| see above | Azienda Sanitaria dell'Alto Adige Laboratorio Aziendale di Microbiologia e Virologia | Istituto di Genomica Applicata | Elisabetta Pagani, Irene Bianconi, Elisabetta Giacobazzi, Elisa Masi, Stefanie Wieser, Irena Jurman, Vera Vendramin, Gabriele Magris, Eleonora Paparelli, Davide Scaglione, Michele Morgante |
| EPI_ISL_1063784 | OSP SS ANNIUNZIATA CHIETI, PRONTO SOCCORSO | Istituto Zooprofilattico Sperimentale dell'Abruzzo e Molise "G. Caporale" | Lorusso A, Marcacci M, Di Domenico M, Ancora M, Curini V, Mangone I, Rinaldi A, Scialabba S, Di Pasquale A, Cammà C, Puglia I, Calistri P, Savini G |
| EPI_ISL_1063785, EPI_ISL_1063786, EPI_ISL_1063787 | SIESP CHIETI - DRIVE IN ORTONA | Istituto Zooprofilattico Sperimentale dell'Abruzzo e Molise "G. Caporale" | Lorusso A, Marcacci M, Di Domenico M, Ancora M, Curini V, Mangone I, Rinaldi A, Scialabba S, Di Pasquale A, Cammà C, Puglia I, Calistri P, Savini G |
| EPI_ISL_1063788 | P.O.CARDARELLI | Istituto Zooprofilattico Sperimentale dell'Abruzzo e Molise "G. Caporale" | Scutellà M, Niro G, Lorusso A, Marcacci M, Di Domenico M, Ancora M, Curini V, Mangone I, Rinaldi A, Scialabba S, Di Pasquale A, Cammà C, Puglia I, Calistri P, Savini G |
| EPI_ISL_1063909 | Laboratorio Genzano - ASL RM 6 | INMI Lazzaro Spallanzani IRCCS | B Bartolini, O Butera, C.E.M Gruber, M Rueca, F Messina, E Giombini, G Tramini, E Conti, MR Capobianchi, A Di Caro |
| EPI_ISL_1063910 | Ospedale San Filippo Neri | INMI Lazzaro Spallanzani IRCCS | M Rueca, O Butera, F Messina, CEM Gruber, B Bartolini, E Giombini, M Melandri, ML Schiavone, A Di Caro, MR Capobianchi |
| EPI_ISL_1063911 | INMI Lazzaro Spallanzani IRCCS | INMI Lazzaro Spallanzani IRCCS | E Giombini, M. Rueca, B Bartolini, O Butera, CEM Gruber, F Messina, A Di Caro, MR Capobianchi |
| EPI_ISL_1063912 | Laboratorio Biologia Molecolare Sars Cov2 - UOC Laboratorio Analisi - Servizio Medicina di Laboratorio, Ospedale "San Francesco" - ATS-ASSL Nuoro | Laboratorio specialistico UOC Ematologia - Ospedale "San Francesco" - ATS-ASSL Nuoro | Piras Giovanna, Asproni Rosanna, Malune Paolo, Fiamma Maura, Monne Maria Itria, Palmas Angelo Domenico, Lo Maglio Iana, Mameli Giuseppe |
| EPI_ISL_1069204 | Azienda Sanitaria Locale Napoli 1 | Centro Polidiagnostico strumentale AMES | Sirica Roberto |
| EPI_ISL_1073034 | ASL Napoli 1 Centro | AMES Centro Polidiagnostico Strumentale S.r.l. | Giovanni Savarese, Raffaella Ruggiero, Eloisa Evangelista, Antonella Di Carlo, Luisa Circelli, Luigi D'Amore, Roberto Sirica, Antonio Fico |
| EPI_ISL_1074622, EPI_ISL_1074623 | ASL Napoli 1 Centro | AMES Centro Polidiagnostico Strumentale S.r.l. | "Giovanni Savarese, Raffaella Ruggiero, Eloisa Evangelista, Antonella Di Carlo, Luisa Circelli, Luigi D'Amore, Roberto Sirica, Antonio Fico" |
| EPI_ISL_1079352 | Genomic Medicine Laboratory, IRCCS Santa Lucia Foundation | INMI Lazzaro Spallanzani IRCCS | CEM Gruber, B Bartolini, E Giombini, F Messina, M Rueca, O Butera, E Giardina, A Di Caro, MR Capobianchi |
| EPI_ISL_1079353 | IRCCS San Raffaele | INMI Lazzaro Spallanzani IRCCS | B Bartolini, E Giombini, F Messina, M Rueca, O Butera, CEM Gruber, D Russo, D Limongi, MR Capobianchi, A Di Caro |
| EPI_ISL_1079355 | Genomic Medicine Laboratory, IRCCS Santa Lucia Foundation | INMI Lazzaro Spallanzani IRCCS | B Bartolini, E Giombini, F Messina, M Rueca, O Butera, CEM Gruber, E Giardina, MR Capobianchi, A Di Caro |
| EPI_ISL_1079356 | Ospedale S.M. Goretti di Latina - ASL Latina presidio ospedaliero Nord | INMI Lazzaro Spallanzani IRCCS | F Messina, M Rueca, O Butera, CEM Gruber, B Bartolini, E Giombini, L Corso, L Di Biase, MR Capobianchi, A Di Caro |
| EPI_ISL_1079358 | Ospedale S.M. Goretti di Latina - ASL Latina presidio ospedaliero Nord | INMI Lazzaro Spallanzani IRCCS | CEM Gruber, B Bartolini, E Giombini, F Messina, O Butera, M Rueca, L Corso, A Lucci, MR Capobianchi, A Di Caro |
| EPI_ISL_1079359 | Fondazione Policlinico Tor Vergata | INMI Lazzaro Spallanzani IRCCS | CEM Gruber, B Bartolini, E Giombini, F Messina, M Rueca, O Butera, S Grelli, M Ciotti, C D'Agostini, A Di Caro, MR Capobianchi |
| EPI_ISL_1079360 | Ospedale "F. Spaziani" Frosinone | INMI Lazzaro Spallanzani IRCCS | F Messina, M Rueca, O Butera, CEM Gruber, B Bartolini, E Giombini, C. Sias, R Pulselli, MR Capobianchi, A Di Caro |
| EPI_ISL_1079362 | Azienda Ospedaliera San Camillo Forlanini | INMI Lazzaro Spallanzani IRCCS | M Rueca, O Butera, CEM Gruber, B Bartolini, E Giombini, F Messina, D Gallone, A D'Agostino, A Di Caro, MR Capobianchi |
| EPI_ISL_1079363 | Azienda Ospedaliera San Camillo Forlanini | INMI Lazzaro Spallanzani IRCCS | B Bartolini, E Giombini, M Rueca, O Butera, CEM Gruber,F Messina, D Gallone, G Meoni, A Di Caro, MR Capobianchi |
| EPI_ISL_1079364 | Ospedale San Camillo De Lellis di Rieti | INMI Lazzaro Spallanzani IRCCS | B Bartolini, E Giombini, F Messina, M Rueca, O Butera, CEM Gruber, S Venarubea, A De Luca, MR Capobianchi, A Di Caro |
| EPI_ISL_1079366 | Ospedale San Camillo De Lellis di Rieti | INMI Lazzaro Spallanzani IRCCS | E Giombini, F Messina, M Rueca, O Butera, CEM Gruber, B Bartolini, A De Luca, L Casertano, A Di Caro, MR Capobianchi |
| EPI_ISL_1079368 | Ospedale Sandro Pertini ASL Roma2 | INMI Lazzaro Spallanzani IRCCS | F Messina, M Rueca, O Butera, CEM Gruber, B Bartolini, E Giombini, MC Cava, G Cappiello, MR Capobianchi, A Di Caro |
| EPI_ISL_1079370 | Ospedale Sandro Pertini ASL Roma2 | INMI Lazzaro Spallanzani IRCCS | M Rueca, O Butera, CEM Gruber, B Bartolini, E Giombini, F Messina, MC Cava, S Romano, A Di Caro, MR Capobianchi |
| EPI_ISL_1079371 | Fondazione Policlinico Tor Vergata | INMI Lazzaro Spallanzani IRCCS | CEM Gruber, B Bartolini, E Giombini, F Messina, O Butera, M Rueca, S Grelli, P Paba, D Ombres, MR Capobianchi, A Di Caro |
| EPI_ISL_1079373 | Fondazione Policlinico Tor Vergata | INMI Lazzaro Spallanzani IRCCS | E Giombini, F Messina, M Rueca, O Butera, CEM Gruber, B Bartolini, S Grelli, C Fontana, D Di Cave, A Di Caro, MR Capobianchi |
| EPI_ISL_1079376 | San Gallicano Dermatological Institute I.F.O. | INMI Lazzaro Spallanzani IRCCS | B Bartolini, E Giombini, F Messina, M Rueca, O Butera, CEM Gruber, F Pimpinelli, F Ensoli, G D'Agosto, MR Capobianchi, A Di Caro |
| EPI_ISL_1079378 | San Gallicano Dermatological Institute I.F.O. | INMI Lazzaro Spallanzani IRCCS | E Giombini, F Messina, M Rueca, O Butera, CEM Gruber, B Bartolini, F Ensoli, F Pimpinelli, E Trento, A Di Caro, MR Capobianchi |
| EPI_ISL_1079380 | Sant'Eugenio/CTO ASL Roma 2 | INMI Lazzaro Spallanzani IRCCS | F Messina, M Rueca, O Butera, CEM Gruber, B Bartolini, E Giombini, F Bondanini, GC Cocciolillo, MR Capobianchi, A Di Caro |
| EPI_ISL_1079382 | Sant'Eugenio/CTO ASL Roma 2 | INMI Lazzaro Spallanzani IRCCS | M Rueca, O Butera, CEM Gruber, B Bartolini, E Giombini, F Messina, F Bondanini, C Disegni, A Di Caro, MR Capobianchi |
| EPI_ISL_1079383 | Azienda Ospedaliera San Camillo Forlanini | INMI Lazzaro Spallanzani IRCCS | CEM Gruber, B Bartolini, O Butera, E Giombini, F Messina, M Rueca, R.Sperning, D Gallone, MR Capobianchi, A Di Caro |
| EPI_ISL_1079386 | Azienda Ospedaliera San Camillo Forlanini | INMI Lazzaro Spallanzani IRCCS | E Giombini, M Rueca, O Butera, CEM Gruber, B Bartolini,F Messina, D Gallone, M Irno Consalvo, A Di Caro, MR Capobianchi |
| EPI_ISL_1079388 | University Hospital Sant'Andrea-Sapienza | INMI Lazzaro Spallanzani IRCCS | E Giombini, F Messina, M Rueca, O Butera, CEM Gruber, B Bartolini, I Santino, M Simmaco, A Di Caro, MR Capobianchi |
| EPI_ISL_1079390 | Laboratorio Genzano - ASL RM 6 | INMI Lazzaro Spallanzani IRCCS | F Messina, M Rueca, O Butera, CEM Gruber, B Bartolini, E Giombini, G Tramini, E Conti, MR Capobianchi, A Di Caro |
| EPI_ISL_1079391 | Laboratorio Genzano - ASL RM 6 | INMI Lazzaro Spallanzani IRCCS | M Rueca, O Butera, CEM Gruber, B Bartolini, E Giombini, F Messina, E Conti, G Tramini, A Di Caro, MR Capobianchi |
| EPI_ISL_1079392 | Policlinico Universitario Campus Bio-Medico | INMI Lazzaro Spallanzani IRCCS | O Butera, CEM Gruber, B Bartolini, E Giombini, F Messina, M Rueca, E Riva, S Angeletti, MR Capobianchi, A Di Caro |
| EPI_ISL_1079394 | Policlinico Universitario Campus Bio-Medico | INMI Lazzaro Spallanzani IRCCS | CEM Gruber, B Bartolini, E Giombini, F Messina, M Rueca, O Butera, M Fogolari, L De Florio, E Riva, A Di Caro, MR Capobianchi |
| EPI_ISL_1079395 | IRCCS San Raffaele | INMI Lazzaro Spallanzani IRCCS | E Giombini, F Messina, M Rueca, O Butera, CEM Gruber, B Bartolini, D Limongi, D Russo, A Di Caro, MR Capobianchi |
| EPI_ISL_1079398 | Ospedale "F. Spaziani" Frosinone | INMI Lazzaro Spallanzani IRCCS | F Messina, M Rueca, O Butera, CEM Gruber, B Bartolini, E Giombini, R Pulselli, G Brocco, MR Capobianchi, A Di Caro |

| | | | |
|---|---|---|---|
| EPI_ISL_1079400 | Ospedale San Filippo Neri | INMI Lazzaro Spallanzani IRCCS | M Rueca, O Butera, F Messina, CEM Gruber, B Bartolini, E Giombini, M Melandri, ML Schiavone, A Tamburro, A Di Caro, MR Capobianchi |
| EPI_ISL_1079992 | Ospedale S.M. Goretti di Latina - ASL Latina presidio ospedaliero Nord | INMI Lazzaro Spallanzani IRCCS | CEM Gruber, B Bartolini, E Giombini, F Messina, O Butera, M Rueca, S Pignalosa, L Corso, MR Capobianchi, A Di Caro |
| EPI_ISL_1079993 | Ospedale S.M. Goretti di Latina - ASL Latina presidio ospedaliero Nord | INMI Lazzaro Spallanzani IRCCS | B Bartolini, E Giombini, F Messina, O Butera, M Rueca, CEM Gruber, L Di Biase, A Lucci, A Di Caro, MR Capobianchi |
| EPI_ISL_1079994 | Azienda Ospedaliera San Camillo Forlanini | INMI Lazzaro Spallanzani IRCCS | O Butera, E Giombini, F Messina, M Rueca, CEM Gruber, B Bartolini, G Parisi, MI Consalvo, MR Capobianchi, A Di Caro |
| EPI_ISL_1079995 | INMI Lazzaro Spallanzani IRCCS | INMI Lazzaro Spallanzani IRCCS | E Giombini, F Messina, M Rueca, O Butera, CEM Gruber, B Bartolini, A Di Caro, MR Capobianchi |
| EPI_ISL_1079996 | Ospedale Sandro Pertini ASL Roma2 | INMI Lazzaro Spallanzani IRCCS | F Messina, M Rueca, O Butera, CEM Gruber, B Bartolini, E Giombini, MC Cava, G Cappiello, MR Capobianchi, A Di Caro |
| EPI_ISL_1079998 | Ospedale San Filippo Neri | INMI Lazzaro Spallanzani IRCCS | M Rueca, O Butera, F Messina, CEM Gruber, B Bartolini, E Giombini, M Meledandri, ML Schiavone, MR Capobianchi |
| EPI_ISL_1079999 | Ospedale San Filippo Neri | INMI Lazzaro Spallanzani IRCCS | CEM Gruber, B Bartolini, E Giombini, F Messina, O Butera, M Rueca, ML Schiavone, A Tamburro, A Di Caro, MR Capobianchi |
| EPI_ISL_1080001 | Ospedale Sandro Pertini ASL Roma2 | INMI Lazzaro Spallanzani IRCCS | B Bartolini, E Giombini, F Messina, O Butera, M Rueca, CEM Gruber, R Longo, V Michela, MR Capobianchi, A Di Caro |
| EPI_ISL_1080605, EPI_ISL_1080606, EPI_ISL_1080607, EPI_ISL_1080608, EPI_ISL_1080609, EPI_ISL_1080610, EPI_ISL_1080611, EPI_ISL_1080612, EPI_ISL_1080613, EPI_ISL_1080614, EPI_ISL_1080615, EPI_ISL_1080616, EPI_ISL_1080617, EPI_ISL_1080618, EPI_ISL_1080619, EPI_ISL_1080620, EPI_ISL_1080621, EPI_ISL_1080622, EPI_ISL_1080623, EPI_ISL_1080624, EPI_ISL_1080625, EPI_ISL_1080626, EPI_ISL_1080627, EPI_ISL_1080628, EPI_ISL_1080629, EPI_ISL_1080630, EPI_ISL_1080631, EPI_ISL_1080632, EPI_ISL_1080633, EPI_ISL_1080635, EPI_ISL_1080636, EPI_ISL_1080637, EPI_ISL_1080639, EPI_ISL_1080640, EPI_ISL_1080641, EPI_ISL_1080642, EPI_ISL_1080643, EPI_ISL_1080644, EPI_ISL_1080645, EPI_ISL_1080646, EPI_ISL_1080647, EPI_ISL_1080648, EPI_ISL_1080649, EPI_ISL_1080650, EPI_ISL_1080651, EPI_ISL_1080652, EPI_ISL_1080653, EPI_ISL_1080654, EPI_ISL_1080655, EPI_ISL_1080656, EPI_ISL_1080657, EPI_ISL_1080658, EPI_ISL_1080659, EPI_ISL_1080660, EPI_ISL_1080661, EPI_ISL_1080662, EPI_ISL_1080663, EPI_ISL_1080664, EPI_ISL_1080665, EPI_ISL_1080666, EPI_ISL_1080667, EPI_ISL_1080669, EPI_ISL_1080670, EPI_ISL_1080671, EPI_ISL_1080672, EPI_ISL_1080673, EPI_ISL_1080674, EPI_ISL_1080675, EPI_ISL_1080676, EPI_ISL_1080677, EPI_ISL_1080678, EPI_ISL_1080679, EPI_ISL_1080681, EPI_ISL_1080682, EPI_ISL_1080683, EPI_ISL_1080684, EPI_ISL_1080685, EPI_ISL_1080686, EPI_ISL_1080687, EPI_ISL_1080688, EPI_ISL_1080689, EPI_ISL_1080690, EPI_ISL_1080691, EPI_ISL_1080692, EPI_ISL_1080693, EPI_ISL_1080694, EPI_ISL_1080695, EPI_ISL_1080696, EPI_ISL_1080697, EPI_ISL_1080698, EPI_ISL_1080699, EPI_ISL_1080700, EPI_ISL_1080701, EPI_ISL_1080702, EPI_ISL_1080703, EPI_ISL_1080704, EPI_ISL_1080705, EPI_ISL_1080706, EPI_ISL_1080707, EPI_ISL_1080708, EPI_ISL_1080709, EPI_ISL_1080710, EPI_ISL_1080711, EPI_ISL_1080712, EPI_ISL_1080713, EPI_ISL_1080714, EPI_ISL_1080715, EPI_ISL_1080716, EPI_ISL_1080717, EPI_ISL_1080718, EPI_ISL_1080719, EPI_ISL_1080720, EPI_ISL_1080721, EPI_ISL_1080722, EPI_ISL_1080723, EPI_ISL_1080724, EPI_ISL_1080725, EPI_ISL_1080726, EPI_ISL_1080727, EPI_ISL_1080728, EPI_ISL_1080729, EPI_ISL_1080730, EPI_ISL_1080731, EPI_ISL_1080732, EPI_ISL_1080733, EPI_ISL_1080734, EPI_ISL_1080735, EPI_ISL_1080736, EPI_ISL_1080737, EPI_ISL_1080738, EPI_ISL_1080739, EPI_ISL_1080740, EPI_ISL_1080741, EPI_ISL_1080742, EPI_ISL_1080743, EPI_ISL_1080744, EPI_ISL_1080745, EPI_ISL_1080748 | | | |
| see above | ASL Napoli 1 Centro | AMES Centro Polidiagnostico Strumentale S.r.l. | "Giovanni Savarese, Raffaella Ruggiero, Eloisa Evangelista, Antonella Di Carlo, Luisa Circelli, Luigi D'Amore, Roberto Sirica, Antonio Fico" |
| EPI_ISL_1081953 | Laboratorio di Riferimento Regionale della Sicilia Occidentale per l'Emergenza COVID-19 | Laboratorio di Riferimento Regionale della Sicilia Occidentale per l'Emergenza COVID-19 | Fabio Tramuto, Carmelo Massimo Maida, Daniela Di Naro, Giulia Randazzo, Walter Mazzucco, Giorgio Graziano, Vincenzo Restivo, Claudio Costantino, Stefano Giaramida, Francesco Vitale |
| EPI_ISL_1082253 | Laboratorio Biologia Molecolare Sars Cov2 - UOC Laboratorio Analisi - Servizio Medicina di Laboratorio, Ospedale "San Francesco" - ATS-ASSL Nuoro | Laboratorio specialistico UOC Ematologia - Ospedale "San Francesco" - ATS-ASSL Nuoro | Giovanna Piras, Rosanna Asproni, Paolo Malune, Maura Fiamma, Maria Itria Monne, Angelo Domenico Palmas, Iana Lo Maglio, Giuseppe Mameli |
| EPI_ISL_1082254 | Virologia, Dipartimento di Scienze Biomediche, Università di Sassari, Viale San Pietro, 43/B - Sassari | Laboratorio specialistico UOC Ematologia - Ospedale "San Francesco" - ATS-ASSL Nuoro | Giovanna Piras, Asproni Rosanna, Paolo Malune, Maria Itria Monne, Angelo Domenico Palmas, Caterina Serra, Elena Rimini, Salvatore Rubino |
| EPI_ISL_1082292, EPI_ISL_1082293, EPI_ISL_1082295 | INT Fondazione Pascale | INT Fondazione Pascale | INT Fondazione Pascale |
| EPI_ISL_1082454, EPI_ISL_1082455, EPI_ISL_1082456, EPI_ISL_1082457, EPI_ISL_1082458, EPI_ISL_1082459, EPI_ISL_1082460, EPI_ISL_1082461, EPI_ISL_1082462, EPI_ISL_1082463, EPI_ISL_1082464, EPI_ISL_1082465, EPI_ISL_1082466, EPI_ISL_1082467, EPI_ISL_1082468, EPI_ISL_1082469, EPI_ISL_1082470, EPI_ISL_1082471, EPI_ISL_1082472, EPI_ISL_1082473, EPI_ISL_1082474, EPI_ISL_1082475, EPI_ISL_1082476, EPI_ISL_1082477, EPI_ISL_1082478, EPI_ISL_1082479, EPI_ISL_1082480, EPI_ISL_1082481, EPI_ISL_1082482, EPI_ISL_1082483, EPI_ISL_1082484, EPI_ISL_1082485, EPI_ISL_1082486, EPI_ISL_1082487, EPI_ISL_1082488, EPI_ISL_1082489, EPI_ISL_1082490, EPI_ISL_1082491, EPI_ISL_1082492, EPI_ISL_1082493, EPI_ISL_1082494, EPI_ISL_1082495, EPI_ISL_1082496, EPI_ISL_1082497, EPI_ISL_1082499, EPI_ISL_1082500, EPI_ISL_1082502, EPI_ISL_1082503, EPI_ISL_1082504, EPI_ISL_1082505, EPI_ISL_1082506, EPI_ISL_1082507, EPI_ISL_1082508, EPI_ISL_1082509, EPI_ISL_1082510, EPI_ISL_1082511, EPI_ISL_1082512, EPI_ISL_1082513, EPI_ISL_1082514, EPI_ISL_1082515, EPI_ISL_1082516, EPI_ISL_1082517, EPI_ISL_1082518, EPI_ISL_1082519, EPI_ISL_1082520, EPI_ISL_1082521, EPI_ISL_1082522, EPI_ISL_1082523, EPI_ISL_1082524, EPI_ISL_1082525 | | | |
| see above | ASL Napoli 1 Centro | AMES Centro Polidiagnostico Strumentale S.r.l. | "Giovanni Savarese, Raffaella Ruggiero, Eloisa Evangelista, Antonella Di Carlo, Luisa Circelli, Luigi D'Amore, Roberto Sirica, Nadia Petrillo, Monica Ianniello, Antonio Fico" |
| EPI_ISL_1085009, EPI_ISL_1085010, EPI_ISL_1085011, EPI_ISL_1085012 | Azienda USL Valle d'Aosta | U.O. Genomics, S.S. Genetics and Advanced Omics Techniques, Istituto Zooprofilattico Sperimentale del Piemonte Liguria e Valle d'Aosta | Chiara Beltramo, Elena Bozzetta, Pier Luigi Acutis, Simone Peletto |
| EPI_ISL_1085015, EPI_ISL_1085016, EPI_ISL_1085017 | ASL Verbano Cusio Ossola - Laboratorio analisi | U.O. Genomics, S.S. Genetics and Advanced Omics Techniques, Istituto Zooprofilattico Sperimentale del Piemonte Liguria e Valle d'Aosta | Chiara Beltramo, Elena Bozzetta, Pier Luigi Acutis, Simone Peletto |
| EPI_ISL_1085025, EPI_ISL_1085026, EPI_ISL_1085027, EPI_ISL_1085028, EPI_ISL_1085029, EPI_ISL_1085030, EPI_ISL_1085032, EPI_ISL_1085033, EPI_ISL_1085034, EPI_ISL_1085035, EPI_ISL_1085036, EPI_ISL_1085037, EPI_ISL_1085038, EPI_ISL_1085039, EPI_ISL_1085040, EPI_ISL_1085041, EPI_ISL_1085042, EPI_ISL_1085043, EPI_ISL_1085044, EPI_ISL_1085045, EPI_ISL_1085046, EPI_ISL_1085047, EPI_ISL_1085048, EPI_ISL_1085049, EPI_ISL_1085050, EPI_ISL_1085051, EPI_ISL_1085052, EPI_ISL_1085053, EPI_ISL_1085054, EPI_ISL_1085055, EPI_ISL_1085056, EPI_ISL_1085057, EPI_ISL_1085058, EPI_ISL_1085059, EPI_ISL_1085060, EPI_ISL_1085061, EPI_ISL_1085062, EPI_ISL_1085063, EPI_ISL_1085064, EPI_ISL_1085065, EPI_ISL_1085066, EPI_ISL_1085067, EPI_ISL_1085068, EPI_ISL_1085069, EPI_ISL_1085070, EPI_ISL_1085071, EPI_ISL_1085072, EPI_ISL_1085073, EPI_ISL_1085074, EPI_ISL_1085075, EPI_ISL_1085076, EPI_ISL_1085077, EPI_ISL_1085078, EPI_ISL_1085079, EPI_ISL_1085080, EPI_ISL_1085081, EPI_ISL_1085082, EPI_ISL_1085083, EPI_ISL_1085084, EPI_ISL_1085085, EPI_ISL_1085086, EPI_ISL_1085087, EPI_ISL_1085088, EPI_ISL_1085089, EPI_ISL_1085090 | | | |
| see above | ASL Napoli 1 Centro | AMES Centro Polidiagnostico Strumentale S.r.l. | "Giovanni Savarese, Raffaella Ruggiero, Eloisa Evangelista, Antonella Di Carlo, Luisa Circelli, Luigi D'Amore, Roberto Sirica, Nadia Petrillo, Monica Ianniello, Antonio Fico" |
| EPI_ISL_1085094 | Laboratory of Infectious Diseases, Department of Biomedical and Clinical Sciences L. Sacco, University of Milan | Laboratory of Infectious Diseases, Department of Biomedical and Clinical Sciences L. Sacco, University of Milan | Alessia Lai, Annalisa Bergna, Carla Della Ventura, Claudia Balotta, Massimo Galli, Gianguglielmo Zehender on behalf of SARS-CoV-2 ITALIAN RESEARCH ENTERPRISE-(SCIRE) Collaborative Group |
| EPI_ISL_1085096, EPI_ISL_1085097, EPI_ISL_1085098, EPI_ISL_1085099, EPI_ISL_1085100, EPI_ISL_1085101, EPI_ISL_1085102, EPI_ISL_1085103, EPI_ISL_1085104, EPI_ISL_1085105, EPI_ISL_1085106, EPI_ISL_1085107, EPI_ISL_1085108, EPI_ISL_1085109, EPI_ISL_1085110, EPI_ISL_1085111, EPI_ISL_1085112, EPI_ISL_1085113, EPI_ISL_1085114, EPI_ISL_1085115, EPI_ISL_1085116, EPI_ISL_1085117, EPI_ISL_1085118, EPI_ISL_1085119, EPI_ISL_1085120, EPI_ISL_1085121, EPI_ISL_1085122, EPI_ISL_1085123, EPI_ISL_1085124, EPI_ISL_1085125, EPI_ISL_1085126, EPI_ISL_1085127, EPI_ISL_1085128, EPI_ISL_1085129, EPI_ISL_1085130, EPI_ISL_1085131, EPI_ISL_1085132, EPI_ISL_1085133, EPI_ISL_1085134, EPI_ISL_1085135, EPI_ISL_1085136, EPI_ISL_1085137, EPI_ISL_1085138, EPI_ISL_1085139, EPI_ISL_1085140, EPI_ISL_1085141, EPI_ISL_1085142, EPI_ISL_1085143, EPI_ISL_1085144, EPI_ISL_1085145, EPI_ISL_1085146, EPI_ISL_1085147, EPI_ISL_1085148, EPI_ISL_1085149, EPI_ISL_1085150, EPI_ISL_1085151, EPI_ISL_1085152, EPI_ISL_1085153, EPI_ISL_1085154, EPI_ISL_1085155, EPI_ISL_1085156, EPI_ISL_1085157, EPI_ISL_1085158, EPI_ISL_1085159, EPI_ISL_1085160 | | | |
| see above | ASL Napoli 1 Centro | AMES Centro Polidiagnostico Strumentale S.r.l. | "Giovanni Savarese, Raffaella Ruggiero, Eloisa Evangelista, Antonella Di Carlo, Luisa Circelli, Luigi D'Amore, Roberto Sirica, Nadia Petrillo, Monica Ianniello, Antonio Fico" |
| EPI_ISL_1085162, EPI_ISL_1085164, EPI_ISL_1085165, EPI_ISL_1085167, EPI_ISL_1085169, EPI_ISL_1085170, EPI_ISL_1085173, EPI_ISL_1085174 | Laboratory of Infectious Diseases, Department of Biomedical and Clinical Sciences L. Sacco, University of Milan | Laboratory of Infectious Diseases, Department of Biomedical and Clinical Sciences L. Sacco, University of Milan | Alessia Lai, Annalisa Bergna, Carla Della Ventura, Claudia Balotta, Massimo Galli, Gianguglielmo Zehender on behalf of SARS-CoV-2 ITALIAN RESEARCH ENTERPRISE-(SCIRE) Collaborative Group |
| EPI_ISL_1085176, EPI_ISL_1085177, EPI_ISL_1085178, EPI_ISL_1085179, EPI_ISL_1085180, EPI_ISL_1085181, EPI_ISL_1085182, EPI_ISL_1085183, EPI_ISL_1085184, EPI_ISL_1085185, EPI_ISL_1085186, EPI_ISL_1085187, EPI_ISL_1085188, EPI_ISL_1085189, EPI_ISL_1085190, EPI_ISL_1085191, EPI_ISL_1085192, EPI_ISL_1085193, EPI_ISL_1085194, EPI_ISL_1085195, EPI_ISL_1085196, EPI_ISL_1085197, EPI_ISL_1085198, EPI_ISL_1085199, EPI_ISL_1085200, EPI_ISL_1085201, EPI_ISL_1085202, EPI_ISL_1085203, EPI_ISL_1085204, EPI_ISL_1085205, EPI_ISL_1085206, EPI_ISL_1085207, EPI_ISL_1085208, EPI_ISL_1085209, EPI_ISL_1085210, EPI_ISL_1085211, EPI_ISL_1085212, EPI_ISL_1085213, EPI_ISL_1085214, EPI_ISL_1085215, EPI_ISL_1085216, EPI_ISL_1085217, EPI_ISL_1085218, EPI_ISL_1085219, EPI_ISL_1085220, EPI_ISL_1085221, EPI_ISL_1085222, EPI_ISL_1085223, EPI_ISL_1085224, EPI_ISL_1085225, EPI_ISL_1085226, EPI_ISL_1085227, EPI_ISL_1085228, EPI_ISL_1085229 | | | |
| see above | ASL Napoli 1 Centro | AMES Centro Polidiagnostico Strumentale S.r.l. | "Giovanni Savarese, Raffaella Ruggiero, Eloisa Evangelista, Antonella Di Carlo, Luisa Circelli, Luigi D'Amore, Roberto Sirica, Nadia Petrillo, Monica Ianniello, Antonio Fico" |
| EPI_ISL_1085230 | Laboratory of Infectious Diseases, Department of Biomedical and Clinical Sciences L. Sacco, University of Milan | Laboratory of Infectious Diseases, Department of Biomedical and Clinical Sciences L. Sacco, University of Milan | Alessia Lai, Annalisa Bergna, Carla Della Ventura, Claudia Balotta, Massimo Galli, Gianguglielmo Zehender on behalf of SARS-CoV-2 ITALIAN RESEARCH ENTERPRISE-(SCIRE) Collaborative Group |
| EPI_ISL_1086068, EPI_ISL_1086073, EPI_ISL_1086076, EPI_ISL_1086082, EPI_ISL_1086083, EPI_ISL_1086084, EPI_ISL_1086092, EPI_ISL_1086095, EPI_ISL_1086097, EPI_ISL_1086099, EPI_ISL_1086100, EPI_ISL_1086103, EPI_ISL_1086107, EPI_ISL_1086110, EPI_ISL_1086113, EPI_ISL_1086115, EPI_ISL_1086119, EPI_ISL_1086120, EPI_ISL_1086123, EPI_ISL_1086124, EPI_ISL_1086125, EPI_ISL_1086126, EPI_ISL_1086127, EPI_ISL_1086128, EPI_ISL_1086129, EPI_ISL_1086130, EPI_ISL_1086132, EPI_ISL_1086143, EPI_ISL_1086147, EPI_ISL_1086148, EPI_ISL_1086151, EPI_ISL_1086152, EPI_ISL_1086156, EPI_ISL_1086161, EPI_ISL_1086162, EPI_ISL_1086167, EPI_ISL_1086174, EPI_ISL_1086176, EPI_ISL_1086178, EPI_ISL_1086189, EPI_ISL_1086196, EPI_ISL_1086197, EPI_ISL_1086215, EPI_ISL_1086216 | | | |
| see above | IZSM | TIGEM | Antonio Grimaldi, Patrizia Annunziata, Francesco Panariello, Biancamaria Pierri, Valentina Bouche, Chiara Colantuono, Maria Concetta Cuomo, Denise Di |

Concilio, Lucio Di Filippo, Anna Manfredi, Marcello Salvi, Antonio Limone, Pellegrino Cerino, Andrea Ballabio, Davide Cacchiarelli.

EPI_ISL_1087353, EPI_ISL_1087354, EPI_ISL_1087355, EPI_ISL_1087356, EPI_ISL_1087357, EPI_ISL_1087358, EPI_ISL_1087359, EPI_ISL_1087360, EPI_ISL_1087361, EPI_ISL_1087362, EPI_ISL_1087363, EPI_ISL_1087364, EPI_ISL_1087365, EPI_ISL_1087366, EPI_ISL_1087367, EPI_ISL_1087368, EPI_ISL_1087369, EPI_ISL_1087370, EPI_ISL_1087371, EPI_ISL_1087372, EPI_ISL_1087373, EPI_ISL_1087374, EPI_ISL_1087375, EPI_ISL_1087376, EPI_ISL_1087377, EPI_ISL_1087378, EPI_ISL_1087379, EPI_ISL_1087380, EPI_ISL_1087381, EPI_ISL_1087383, EPI_ISL_1087384, EPI_ISL_1087385, EPI_ISL_1087386, EPI_ISL_1087387, EPI_ISL_1087388, EPI_ISL_1087389, EPI_ISL_1087390, EPI_ISL_1087391, EPI_ISL_1087392, EPI_ISL_1087393, EPI_ISL_1087394, EPI_ISL_1087395, EPI_ISL_1087396, EPI_ISL_1087397, EPI_ISL_1087398, EPI_ISL_1087399, EPI_ISL_1087400, EPI_ISL_1087401, EPI_ISL_1087402, EPI_ISL_1087403, EPI_ISL_1087404, EPI_ISL_1087405, EPI_ISL_1087406, EPI_ISL_1087407, EPI_ISL_1087408, EPI_ISL_1087409, EPI_ISL_1087410, EPI_ISL_1087411, EPI_ISL_1087412, EPI_ISL_1087413, EPI_ISL_1088008, EPI_ISL_1088009, EPI_ISL_1088010, EPI_ISL_1088011, EPI_ISL_1088012, EPI_ISL_1088013, EPI_ISL_1088014, EPI_ISL_1088015, EPI_ISL_1088016, EPI_ISL_1088017, EPI_ISL_1088018, EPI_ISL_1088019, EPI_ISL_1088020, EPI_ISL_1088021, EPI_ISL_1088022, EPI_ISL_1088023, EPI_ISL_1088024, EPI_ISL_1088025, EPI_ISL_1088026, EPI_ISL_1088027, EPI_ISL_1088028, EPI_ISL_1088029, EPI_ISL_1088030, EPI_ISL_1088031, EPI_ISL_1088032, EPI_ISL_1088033, EPI_ISL_1088034, EPI_ISL_1088035, EPI_ISL_1088036, EPI_ISL_1088037, EPI_ISL_1088038, EPI_ISL_1088039, EPI_ISL_1088040, EPI_ISL_1088041, EPI_ISL_1088042, EPI_ISL_1088043, EPI_ISL_1088044, EPI_ISL_1088045, EPI_ISL_1088046, EPI_ISL_1088047, EPI_ISL_1088048, EPI_ISL_1088049, EPI_ISL_1088050, EPI_ISL_1088051, EPI_ISL_1088052, EPI_ISL_1088053, EPI_ISL_1088054, EPI_ISL_1088055, EPI_ISL_1088056, EPI_ISL_1088057, EPI_ISL_1088058, EPI_ISL_1088059, EPI_ISL_1088060, EPI_ISL_1088061, EPI_ISL_1088062, EPI_ISL_1088063, EPI_ISL_1088064, EPI_ISL_1088065, EPI_ISL_1088066, EPI_ISL_1088067, EPI_ISL_1088068, EPI_ISL_1088069, EPI_ISL_1088070, EPI_ISL_1088071, EPI_ISL_1088072, EPI_ISL_1088073, EPI_ISL_1088074, EPI_ISL_1088075, EPI_ISL_1088076

| | | | |
|---|---|---|---|
| see above | ASL Napoli 1 Centro | AMES Centro Polidiagnostico Strumentale S.r.l. | "Giovanni Savarese, Raffaella Ruggiero, Eloisa Evangelista, Antonella Di Carlo, Luisa Circelli, Luigi D'Amore, Roberto Sirica, Nadia Petrillo, Monica Ianniello, Antonio Fico" |
| EPI_ISL_1091275 | Ospedale S.M. Goretti di Latina - ASL Latina presidio ospedaliero Nord | INMI Lazzaro Spallanzani IRCCS | CEM Gruber, B Bartolini, E Giombini, F Messina, O Butera, M Rueca, S Pignalosa, L Corso, MR Capobianchi, A Di Caro |
| EPI_ISL_1091276 | INMI Lazzaro Spallanzani IRCCS | INMI Lazzaro Spallanzani IRCCS | O Butera, E Giombini, F Messina, M Rueca, CEM Gruber, B Bartolini, MR Capobianchi, A Di Caro |
| EPI_ISL_1091277 | INMI Lazzaro Spallanzani IRCCS | INMI Lazzaro Spallanzani IRCCS | CEM Gruber, B Bartolini, O Butera, E Giombini, F Messina, M Rueca, A Di Caro, MR Capobianchi |
| EPI_ISL_1091278 | INMI Lazzaro Spallanzani IRCCS | INMI Lazzaro Spallanzani IRCCS | B Bartolini, E Giombini, F Messina, O Butera, M Rueca, CEM Gruber, A Di Caro, MR Capobianchi |
| EPI_ISL_1091279 | INMI Lazzaro Spallanzani IRCCS | INMI Lazzaro Spallanzani IRCCS | M Rueca, CEM Gruber, B Bartolini, O Butera, E Giombini, F Messina, A Di Caro, MR Capobianchi |
| EPI_ISL_1091280 | Azienda Ospedaliera San Camillo Forlanini | INMI Lazzaro Spallanzani IRCCS | F Messina, M Rueca, O Butera, CEM Gruber, B Bartolini, E Giombini, G Parisi, D Gallone, MR Capobianchi, A Di Caro |
| EPI_ISL_1091281 | Azienda Ospedaliera San Camillo Forlanini | INMI Lazzaro Spallanzani IRCCS | M Rueca, CEM Gruber, B Bartolini, O Butera, E Giombini, F Messina, G Parisi, A D'Agostino, A Di Caro, MR Capobianchi |
| EPI_ISL_1091282 | Azienda Ospedaliera San Camillo Forlanini | INMI Lazzaro Spallanzani IRCCS | O Butera, E Giombini, F Messina, M Rueca, CEM Gruber, B Bartolini, G Parisi, ML Guarino, MR Capobianchi, A Di Caro |
| EPI_ISL_1091283 | Azienda Ospedaliera San Camillo Forlanini | INMI Lazzaro Spallanzani IRCCS | E Giombini, F Messina, M Rueca, O Butera, CEM Gruber, B Bartolini, G Parisi, MI Consalvo, A Di Caro, MR Capobianchi |
| EPI_ISL_1091284 | Azienda Ospedaliera San Camillo Forlanini | INMI Lazzaro Spallanzani IRCCS | B Bartolini, E Giombini, F Messina, O Butera, M Rueca, CEM Gruber, G Parisi, F Basile, MR Capobianchi, A Di Caro |
| EPI_ISL_1091285 | Azienda Ospedaliera San Camillo Forlanini | INMI Lazzaro Spallanzani IRCCS | F Messina, M Rueca, O Butera, CEM Gruber, B Bartolini, E Giombini, G Parisi, G Meoni, A Di Caro, MR Capobianchi |
| EPI_ISL_1091286 | Azienda Ospedaliera San Camillo Forlanini | INMI Lazzaro Spallanzani IRCCS | CEM Gruber, B Bartolini, E Giombini, F Messina, O Butera, M Rueca, G Parisi, ML Guarino, A Di Caro, MR Capobianchi |
| EPI_ISL_1091287 | Ospedale Sandro Pertini ASL Roma2 | INMI Lazzaro Spallanzani IRCCS | CEM Gruber, B Bartolini, O Butera, E Giombini, F Messina, M Rueca, MC Cava, R Longo, A Di Caro, MR Capobianchi |
| EPI_ISL_1091288 | Ospedale Sandro Pertini ASL Roma3 | INMI Lazzaro Spallanzani IRCCS | B Bartolini, E Giombini, F Messina, O Butera, M Rueca, CEM Gruber, G Cappiello, V Michela, MR Capobianchi, A Di Caro |
| EPI_ISL_1091289 | Ospedale Sandro Pertini ASL Roma4 | INMI Lazzaro Spallanzani IRCCS | O Butera, E Giombini, F Messina, M Rueca, CEM Gruber, B Bartolini, MC Cava, S Romano, MR Capobianchi, A Di Caro |
| EPI_ISL_1091290 | Ospedale San Filippo Neri | INMI Lazzaro Spallanzani IRCCS | M Rueca, CEM Gruber, B Bartolini, O Butera, E Giombini, F Messina, ML Schiavone, A Tamburro, MR Capobianchi, A Di Caro |
| EPI_ISL_1091291 | Ospedale San Filippo Neri | INMI Lazzaro Spallanzani IRCCS | F Messina, M Rueca, O Butera, CEM Gruber, B Bartolini, E Giombini, A Tamburro, M Meledandri, MR Capobianchi, A Di Caro |
| EPI_ISL_1091787 | San Gallicano Dermatological Institute I.F.O. | INMI Lazzaro Spallanzani IRCCS | B Bartolini, E Giombini, F Messina, M Rueca, O Butera, CEM Gruber, F Pimpinelli, F Ensoli, G Prignano, A Di Caro, MR Capobianchi |
| EPI_ISL_1091788 | INMI Lazzaro Spallanzani IRCCS | INMI Lazzaro Spallanzani IRCCS | E Giombini, F Messina, M Rueca, O Butera, CEM Gruber, B Bartolini, A Di Caro, MR Capobianchi |
| EPI_ISL_1091789 | INMI Lazzaro Spallanzani IRCCS | INMI Lazzaro Spallanzani IRCCS | O Butera, CEM Gruber, B Bartolini, E Giombini, F Messina, M Rueca, MR Capobianchi, A Di Caro |
| EPI_ISL_1091790 | San Gallicano Dermatological Institute I.F.O. | INMI Lazzaro Spallanzani IRCCS | M Rueca, O Butera, CEM Gruber, B Bartolini, E Giombini, F Messina, F Pimpinelli, F Ensoli, G D'Agosto, A Di Caro, MR Capobianchi |
| EPI_ISL_1091791 | Laboratorio Genzano - ASL RM 6 | INMI Lazzaro Spallanzani IRCCS | O Butera, CEM Gruber, B Bartolini, E Giombini, F Messina, M Rueca, G Tramini, E Conti, MR Capobianchi, A Di Caro |
| EPI_ISL_1091792 | Laboratorio Genzano - ASL RM 6 | INMI Lazzaro Spallanzani IRCCS | B Bartolini, E Giombini, F Messina, M Rueca, O Butera, CEM Gruber, G Tramini, E Conti, A Di Caro, MR Capobianchi |
| EPI_ISL_1091793 | Laboratorio Genzano - ASL RM 6 | INMI Lazzaro Spallanzani IRCCS | CEM Gruber, B Bartolini, E Giombini, F Messina, M Rueca, O Butera, G Tramini, E Conti, MR Capobianchi, A Di Caro |
| EPI_ISL_1091794 | Ospedale "F. Spaziani" Frosinone | INMI Lazzaro Spallanzani IRCCS | F Messina, M Rueca, O Butera, CEM Gruber, B Bartolini, E Giombini, R Pulselli, C Sias, G Brocco, MR Capobianchi, A Di Caro |
| EPI_ISL_1091795 | Laboratorio Genzano - ASL RM 6 | INMI Lazzaro Spallanzani IRCCS | M Rueca, O Butera, CEM Gruber, B Bartolini, E Giombini, F Messina, G Tramini, E Conti, A Di Caro, MR Capobianchi |
| EPI_ISL_1091796 | Ospedale S.M. Goretti di Latina - ASL Latina presidio ospedaliero Nord | INMI Lazzaro Spallanzani IRCCS | B Bartolini, E Giombini, F Messina, M Rueca, O Butera, CEM Gruber, A Lucci, S Pignalosa, L Corso, A Di Caro, MR Capobianchi |
| EPI_ISL_1091797 | Azienda Ospedaliera San Camillo Forlanini | INMI Lazzaro Spallanzani IRCCS | M Rueca, O Butera, CEM Gruber, B Bartolini, E Giombini, F Messina, G Parisi, D Gallone, F Basile, A Di Caro, MR Capobianchi |
| EPI_ISL_1091798 | Synlab Lazio S.r.l. | INMI Lazzaro Spallanzani IRCCS | E Giombini, F Messina, M Rueca, O Butera, CEM Gruber, B Bartolini, E Trappolini, SA Santini, A Di Caro, MR Capobianchi |

EPI_ISL_1093477, EPI_ISL_1093478, EPI_ISL_1093479, EPI_ISL_1093480, EPI_ISL_1093481, EPI_ISL_1093482, EPI_ISL_1093483, EPI_ISL_1093484, EPI_ISL_1093485, EPI_ISL_1093486, EPI_ISL_1093487, EPI_ISL_1093488, EPI_ISL_1093489, EPI_ISL_1093490, EPI_ISL_1093491

| | | | |
|---|---|---|---|
| see above | Center of Advanced Studies and Technology, Molecular Genetics Laboratory | Center of Advanced Studies and Technology, Molecular Genetics Laboratory | Ferrante Rossella, Mandatori Domitilla, De Fabritiis Simone, Damiani Verena, Anaclerio Federico |
| EPI_ISL_1095364 | Istituto Zooprofilattico Sperimentale del Mezzogiorno - Azienda Ospedaliera "Pugliese Ciaccio" di Catanzaro | Lab. Microbiologia e Virologia, Cotugno, A.O. dei Colli | Luigi Atripaldi, Claudia Tiberio, Anna Perfetti, Pellegrino Cerino, Biancamaria Pierri, Maria Concetta Cuomo, Pasquale Minchella |

EPI_ISL_1095366, EPI_ISL_1095367, EPI_ISL_1095368, EPI_ISL_1095369, EPI_ISL_1095370, EPI_ISL_1095371, EPI_ISL_1095372, EPI_ISL_1095373, EPI_ISL_1095374, EPI_ISL_1095375, EPI_ISL_1095376, EPI_ISL_1095377, EPI_ISL_1095378, EPI_ISL_1095379, EPI_ISL_1095380, EPI_ISL_1095381, EPI_ISL_1095382, EPI_ISL_1095383

| | | | |
|---|---|---|---|
| see above | Istituto Zooprofilattico Sperimentale del Mezzogiorno | Lab. Microbiologia e Virologia, Cotugno, A.O. dei Colli | Luigi Atripaldi, Claudia Tiberio, Anna Perfetti, Pellegrino Cerino, Biancamaria Pierri, Maria Concetta Cuomo |
| EPI_ISL_1095384 | Laboratorio Microbiologia e Virologia P.O. Cotugno A.O. dei Colli | Laboratorio Microbiologia e Virologia P.O. Cotugno A.O. dei Colli | Luigi Atripaldi, Claudia Tiberio, Anna Perfetti, Pellegrino Cerino, Biancamaria Pierri, Maria Concetta Cuomo |

EPI_ISL_1095385, EPI_ISL_1095386, EPI_ISL_1095387, EPI_ISL_1095388, EPI_ISL_1095389, EPI_ISL_1095390, EPI_ISL_1095391, EPI_ISL_1095392, EPI_ISL_1095393, EPI_ISL_1095394, EPI_ISL_1095395, EPI_ISL_1095396, EPI_ISL_1095397, EPI_ISL_1095398, EPI_ISL_1095399, EPI_ISL_1095400, EPI_ISL_1095401, EPI_ISL_1095402, EPI_ISL_1095403, EPI_ISL_1095404, EPI_ISL_1095405, EPI_ISL_1095406, EPI_ISL_1095407

| | | | |
|---|---|---|---|
| see above | Istituto Zooprofilattico Sperimentale del Mezzogiorno | Lab. Microbiologia e Virologia, Cotugno, A.O. dei Colli | Luigi Atripaldi, Claudia Tiberio, Anna Perfetti, Pellegrino Cerino, Biancamaria Pierri, Maria Concetta Cuomo |
| EPI_ISL_1095449, EPI_ISL_1096228, EPI_ISL_1096229, EPI_ISL_1096230, EPI_ISL_1096231, EPI_ISL_1096232, EPI_ISL_1096233, EPI_ISL_1096342 | Laboratorio Microbiologia e Virologia P.O. Cotugno A.O. dei Colli | Laboratorio Microbiologia e Virologia P.O. Cotugno A.O. dei Colli | Luigi Atripaldi, Claudia Tiberio, Anna Perfetti, Pellegrino Cerino, Biancamaria Pierri, Maria Concetta Cuomo |

EPI_ISL_1096349, EPI_ISL_1096350, EPI_ISL_1096351, EPI_ISL_1096352, EPI_ISL_1096353, EPI_ISL_1096354, EPI_ISL_1096355, EPI_ISL_1096356, EPI_ISL_1096357, EPI_ISL_1096358, EPI_ISL_1096359, EPI_ISL_1096360, EPI_ISL_1096361, EPI_ISL_1096362, EPI_ISL_1096363, EPI_ISL_1096364, EPI_ISL_1096365, EPI_ISL_1096366, EPI_ISL_1096367, EPI_ISL_1096368, EPI_ISL_1096369, EPI_ISL_1096370, EPI_ISL_1096371, EPI_ISL_1096372, EPI_ISL_1096373, EPI_ISL_1096374, EPI_ISL_1096375, EPI_ISL_1096376, EPI_ISL_1096377, EPI_ISL_1096378, EPI_ISL_1096379, EPI_ISL_1096380, EPI_ISL_1096381, EPI_ISL_1096382, EPI_ISL_1096383, EPI_ISL_1096384, EPI_ISL_1096385, EPI_ISL_1096386, EPI_ISL_1096387, EPI_ISL_1096388, EPI_ISL_1096389, EPI_ISL_1096390, EPI_ISL_1096391, EPI_ISL_1096392, EPI_ISL_1096393, EPI_ISL_1096394, EPI_ISL_1096395, EPI_ISL_1096396, EPI_ISL_1096397, EPI_ISL_1096398, EPI_ISL_1096399, EPI_ISL_1096400, EPI_ISL_1096401, EPI_ISL_1096402, EPI_ISL_1096403, EPI_ISL_1096404, EPI_ISL_1096405, EPI_ISL_1096406, EPI_ISL_1096407, EPI_ISL_1096408, EPI_ISL_1096409, EPI_ISL_1096410, EPI_ISL_1096411, EPI_ISL_1096412, EPI_ISL_1096413, EPI_ISL_1096414, EPI_ISL_1096415, EPI_ISL_1096416, EPI_ISL_1096417, EPI_ISL_1096418, EPI_ISL_1096419, EPI_ISL_1096420, EPI_ISL_1096421, EPI_ISL_1096422, EPI_ISL_1096423, EPI_ISL_1096424, EPI_ISL_1096425, EPI_ISL_1096426, EPI_ISL_1096427, EPI_ISL_1096428, EPI_ISL_1096429, EPI_ISL_1096430, EPI_ISL_1096431, EPI_ISL_1096432, EPI_ISL_1096433, EPI_ISL_1096434, EPI_ISL_1096435, EPI_ISL_1096436, EPI_ISL_1096437, EPI_ISL_1096438, EPI_ISL_1096439, EPI_ISL_1096440, EPI_ISL_1096441, EPI_ISL_1096442, EPI_ISL_1096443, EPI_ISL_1096444, EPI_ISL_1096445, EPI_ISL_1096446, EPI_ISL_1096447, EPI_ISL_1096448, EPI_ISL_1096449, EPI_ISL_1096450, EPI_ISL_1096451, EPI_ISL_1096452, EPI_ISL_1096453, EPI_ISL_1096454, EPI_ISL_1096455, EPI_ISL_1096456, EPI_ISL_1096457, EPI_ISL_1096458, EPI_ISL_1096459, EPI_ISL_1096460, EPI_ISL_1096461, EPI_ISL_1096462, EPI_ISL_1096463, EPI_ISL_1096464, EPI_ISL_1096465, EPI_ISL_1096466, EPI_ISL_1096467, EPI_ISL_1096468, EPI_ISL_1096469, EPI_ISL_1096470, EPI_ISL_1096471, EPI_ISL_1096472, EPI_ISL_1096473, EPI_ISL_1096474, EPI_ISL_1096475, EPI_ISL_1096476, EPI_ISL_1096477, EPI_ISL_1096478, EPI_ISL_1096479, EPI_ISL_1096480, EPI_ISL_1096481, EPI_ISL_1096482, EPI_ISL_1096483, EPI_ISL_1096484,

EPI_ISL_1096485, EPI_ISL_1096486, EPI_ISL_1096487, EPI_ISL_1096488, EPI_ISL_1096489, EPI_ISL_1096490, EPI_ISL_1096491, EPI_ISL_1096492, EPI_ISL_1096493, EPI_ISL_1096494, EPI_ISL_1096495, EPI_ISL_1096496, EPI_ISL_1096497, EPI_ISL_1096498, EPI_ISL_1096499, EPI_ISL_1096500, EPI_ISL_1096501, EPI_ISL_1096502, EPI_ISL_1096503, EPI_ISL_1096504, EPI_ISL_1096505, EPI_ISL_1096506, EPI_ISL_1096507, EPI_ISL_1096508, EPI_ISL_1096509, EPI_ISL_1096510, EPI_ISL_1096511, EPI_ISL_1096512, EPI_ISL_1096513, EPI_ISL_1096514, EPI_ISL_1096515, EPI_ISL_1096516, EPI_ISL_1096517, EPI_ISL_1096518, EPI_ISL_1096519, EPI_ISL_1096520, EPI_ISL_1096521, EPI_ISL_1096522, EPI_ISL_1096523, EPI_ISL_1096524, EPI_ISL_1096525, EPI_ISL_1096526, EPI_ISL_1096527, EPI_ISL_1096528, EPI_ISL_1096529, EPI_ISL_1096530, EPI_ISL_1096531, EPI_ISL_1096532, EPI_ISL_1096533, EPI_ISL_1096534, EPI_ISL_1096535, EPI_ISL_1096536, EPI_ISL_1096537, EPI_ISL_1096538, EPI_ISL_1096539, EPI_ISL_1096540, EPI_ISL_1096541, EPI_ISL_1096542, EPI_ISL_1096543, EPI_ISL_1096544, EPI_ISL_1096545, EPI_ISL_1096546, EPI_ISL_1096547, EPI_ISL_1096548, EPI_ISL_1096549, EPI_ISL_1096550, EPI_ISL_1096551, EPI_ISL_1096552, EPI_ISL_1096553, EPI_ISL_1096554, EPI_ISL_1096555, EPI_ISL_1096556, EPI_ISL_1096557, EPI_ISL_1096558, EPI_ISL_1096559, EPI_ISL_1096560, EPI_ISL_1096561, EPI_ISL_1096562, EPI_ISL_1096563, EPI_ISL_1096564, EPI_ISL_1096565, EPI_ISL_1096566, EPI_ISL_1096567, EPI_ISL_1096568, EPI_ISL_1096569, EPI_ISL_1096570, EPI_ISL_1096571, EPI_ISL_1096572, EPI_ISL_1096573, EPI_ISL_1096574, EPI_ISL_1096575, EPI_ISL_1096576, EPI_ISL_1096577, EPI_ISL_1096578, EPI_ISL_1096579, EPI_ISL_1096580, EPI_ISL_1096581, EPI_ISL_1096582, EPI_ISL_1096583, EPI_ISL_1096584, EPI_ISL_1096585, EPI_ISL_1096586, EPI_ISL_1096587, EPI_ISL_1096588, EPI_ISL_1096589, EPI_ISL_1096590, EPI_ISL_1096591, EPI_ISL_1096592, EPI_ISL_1096593, EPI_ISL_1096594, EPI_ISL_1096595, EPI_ISL_1096596, EPI_ISL_1096597, EPI_ISL_1096598, EPI_ISL_1096599, EPI_ISL_1096600, EPI_ISL_1096601, EPI_ISL_1096602, EPI_ISL_1096603, EPI_ISL_1096604, EPI_ISL_1096605, EPI_ISL_1096606, EPI_ISL_1096607, EPI_ISL_1096608, EPI_ISL_1096609, EPI_ISL_1096610, EPI_ISL_1096611, EPI_ISL_1096612, EPI_ISL_1096613, EPI_ISL_1096614, EPI_ISL_1096615, EPI_ISL_1096616, EPI_ISL_1096617, EPI_ISL_1096618, EPI_ISL_1096619, EPI_ISL_1096620, EPI_ISL_1096621, EPI_ISL_1096622, EPI_ISL_1096623, EPI_ISL_1096624, EPI_ISL_1096625, EPI_ISL_1096626, EPI_ISL_1096627, EPI_ISL_1096628, EPI_ISL_1096629, EPI_ISL_1096630, EPI_ISL_1096631, EPI_ISL_1096632, EPI_ISL_1096633, EPI_ISL_1096634, EPI_ISL_1096635, EPI_ISL_1096636, EPI_ISL_1096637, EPI_ISL_1096638, EPI_ISL_1096639, EPI_ISL_1096640, EPI_ISL_1096641, EPI_ISL_1096642, EPI_ISL_1096643, EPI_ISL_1096644, EPI_ISL_1096645, EPI_ISL_1096646, EPI_ISL_1096647, EPI_ISL_1096648, EPI_ISL_1096649, EPI_ISL_1096650, EPI_ISL_1096651, EPI_ISL_1096652, EPI_ISL_1096653, EPI_ISL_1096654, EPI_ISL_1096655, EPI_ISL_1096656, EPI_ISL_1096657, EPI_ISL_1096658, EPI_ISL_1096659, EPI_ISL_1096660, EPI_ISL_1096661, EPI_ISL_1096662, EPI_ISL_1096663, EPI_ISL_1096664, EPI_ISL_1096665, EPI_ISL_1096666, EPI_ISL_1096667, EPI_ISL_1096668, EPI_ISL_1096669, EPI_ISL_1096670, EPI_ISL_1096671, EPI_ISL_1096672, EPI_ISL_1096673, EPI_ISL_1096674, EPI_ISL_1096675, EPI_ISL_1096676, EPI_ISL_1096677, EPI_ISL_1096678, EPI_ISL_1096679, EPI_ISL_1096680, EPI_ISL_1096681, EPI_ISL_1096682, EPI_ISL_1096683, EPI_ISL_1096684, EPI_ISL_1096685, EPI_ISL_1096686, EPI_ISL_1096687, EPI_ISL_1096688, EPI_ISL_1096689, EPI_ISL_1096690, EPI_ISL_1096691, EPI_ISL_1096692, EPI_ISL_1096693, EPI_ISL_1096694, EPI_ISL_1096695, EPI_ISL_1096696, EPI_ISL_1096697, EPI_ISL_1096698, EPI_ISL_1096699, EPI_ISL_1096700, EPI_ISL_1096701, EPI_ISL_1096702, EPI_ISL_1096703, EPI_ISL_1096704, EPI_ISL_1096705, EPI_ISL_1096706, EPI_ISL_1096707, EPI_ISL_1096708, EPI_ISL_1096709, EPI_ISL_1096710, EPI_ISL_1096711, EPI_ISL_1096712, EPI_ISL_1096713, EPI_ISL_1096714, EPI_ISL_1096715, EPI_ISL_1096716, EPI_ISL_1096717, EPI_ISL_1096718, EPI_ISL_1096719, EPI_ISL_1096720, EPI_ISL_1096721, EPI_ISL_1096722, EPI_ISL_1096723, EPI_ISL_1096724, EPI_ISL_1096725, EPI_ISL_1096726, EPI_ISL_1096727, EPI_ISL_1096728, EPI_ISL_1096729, EPI_ISL_1096730, EPI_ISL_1096731, EPI_ISL_1096732, EPI_ISL_1096733, EPI_ISL_1096734, EPI_ISL_1096735, EPI_ISL_1096736, EPI_ISL_1096737, EPI_ISL_1096738, EPI_ISL_1096739, EPI_ISL_1096740, EPI_ISL_1096741, EPI_ISL_1096742, EPI_ISL_1096743, EPI_ISL_1096744, EPI_ISL_1096745, EPI_ISL_1096746, EPI_ISL_1096747, EPI_ISL_1096748, EPI_ISL_1096749, EPI_ISL_1096750, EPI_ISL_1096751, EPI_ISL_1096752, EPI_ISL_1096753, EPI_ISL_1096754, EPI_ISL_1096755, EPI_ISL_1096756, EPI_ISL_1096757, EPI_ISL_1096758, EPI_ISL_1096759, EPI_ISL_1096760, EPI_ISL_1096761, EPI_ISL_1096762, EPI_ISL_1096763, EPI_ISL_1096764, EPI_ISL_1096765, EPI_ISL_1096766, EPI_ISL_1096767, EPI_ISL_1096768, EPI_ISL_1096769, EPI_ISL_1096770, EPI_ISL_1096771, EPI_ISL_1096772, EPI_ISL_1096773, EPI_ISL_1096774, EPI_ISL_1096775, EPI_ISL_1096776, EPI_ISL_1096777, EPI_ISL_1096778, EPI_ISL_1096779, EPI_ISL_1096780, EPI_ISL_1096781, EPI_ISL_1096782, EPI_ISL_1096783, EPI_ISL_1096784, EPI_ISL_1096785, EPI_ISL_1096786, EPI_ISL_1096787, EPI_ISL_1096788, EPI_ISL_1096789, EPI_ISL_1096790, EPI_ISL_1096791, EPI_ISL_1096792, EPI_ISL_1096793, EPI_ISL_1096794, EPI_ISL_1096795, EPI_ISL_1096796, EPI_ISL_1096797, EPI_ISL_1096798, EPI_ISL_1096799, EPI_ISL_1096800, EPI_ISL_1096801, EPI_ISL_1096802, EPI_ISL_1096803, EPI_ISL_1096804, EPI_ISL_1096805, EPI_ISL_1096806, EPI_ISL_1096807, EPI_ISL_1096808, EPI_ISL_1096809, EPI_ISL_1096810, EPI_ISL_1096811, EPI_ISL_1096812, EPI_ISL_1096813, EPI_ISL_1096814, EPI_ISL_1096815, EPI_ISL_1096816, EPI_ISL_1096817, EPI_ISL_1096818, EPI_ISL_1096819, EPI_ISL_1096820, EPI_ISL_1096821, EPI_ISL_1096822, EPI_ISL_1096823, EPI_ISL_1096824, EPI_ISL_1096825, EPI_ISL_1096826, EPI_ISL_1096827, EPI_ISL_1096828, EPI_ISL_1096829, EPI_ISL_1096830, EPI_ISL_1096831, EPI_ISL_1096832, EPI_ISL_1096833, EPI_ISL_1096834, EPI_ISL_1096835, EPI_ISL_1096836, EPI_ISL_1096837, EPI_ISL_1096838, EPI_ISL_1096839, EPI_ISL_1096840, EPI_ISL_1096841, EPI_ISL_1096842, EPI_ISL_1096843, EPI_ISL_1096844, EPI_ISL_1096845, EPI_ISL_1096846, EPI_ISL_1096847, EPI_ISL_1096848, EPI_ISL_1096849, EPI_ISL_1096850, EPI_ISL_1096851, EPI_ISL_1096852, EPI_ISL_1096853, EPI_ISL_1096854, EPI_ISL_1096855, EPI_ISL_1096856, EPI_ISL_1096857, EPI_ISL_1096858, EPI_ISL_1096859, EPI_ISL_1096860, EPI_ISL_1096861, EPI_ISL_1096862, EPI_ISL_1096863, EPI_ISL_1096864, EPI_ISL_1096865, EPI_ISL_1096866, EPI_ISL_1096867, EPI_ISL_1096868, EPI_ISL_1096869, EPI_ISL_1096870, EPI_ISL_1096871, EPI_ISL_1096872, EPI_ISL_1096873, EPI_ISL_1096874, EPI_ISL_1096875, EPI_ISL_1096876, EPI_ISL_1096877, EPI_ISL_1096878, EPI_ISL_1096879, EPI_ISL_1096880, EPI_ISL_1096881, EPI_ISL_1096882, EPI_ISL_1096883, EPI_ISL_1096884, EPI_ISL_1096885, EPI_ISL_1096886, EPI_ISL_1096887, EPI_ISL_1096888, EPI_ISL_1096889, EPI_ISL_1096890, EPI_ISL_1096891, EPI_ISL_1096892, EPI_ISL_1096893, EPI_ISL_1096894, EPI_ISL_1096895, EPI_ISL_1096896, EPI_ISL_1096897, EPI_ISL_1096898, EPI_ISL_1096899, EPI_ISL_1096900, EPI_ISL_1096901, EPI_ISL_1096902, EPI_ISL_1096903, EPI_ISL_1096904, EPI_ISL_1096905, EPI_ISL_1096906, EPI_ISL_1096907, EPI_ISL_1096908, EPI_ISL_1096909, EPI_ISL_1096910, EPI_ISL_1096911, EPI_ISL_1096912, EPI_ISL_1096913, EPI_ISL_1096914, EPI_ISL_1096915, EPI_ISL_1096916, EPI_ISL_1096917, EPI_ISL_1096918, EPI_ISL_1096919, EPI_ISL_1096920, EPI_ISL_1096921, EPI_ISL_1096922, EPI_ISL_1096923, EPI_ISL_1096924, EPI_ISL_1096925, EPI_ISL_1096926, EPI_ISL_1096927, EPI_ISL_1096928, EPI_ISL_1096929, EPI_ISL_1096930, EPI_ISL_1096931, EPI_ISL_1096932, EPI_ISL_1096933, EPI_ISL_1096934, EPI_ISL_1096935, EPI_ISL_1096936, EPI_ISL_1096937, EPI_ISL_1096938, EPI_ISL_1096939, EPI_ISL_1096940, EPI_ISL_1096941, EPI_ISL_1096942, EPI_ISL_1096943, EPI_ISL_1096944, EPI_ISL_1096945, EPI_ISL_1096946, EPI_ISL_1096947, EPI_ISL_1096948, EPI_ISL_1096949, EPI_ISL_1096950, EPI_ISL_1096951, EPI_ISL_1096952, EPI_ISL_1096953, EPI_ISL_1096954, EPI_ISL_1096955, EPI_ISL_1096956, EPI_ISL_1096957, EPI_ISL_1096958, EPI_ISL_1096959, EPI_ISL_1096960, EPI_ISL_1096961, EPI_ISL_1096962, EPI_ISL_1096963, EPI_ISL_1096964, EPI_ISL_1096965, EPI_ISL_1096966, EPI_ISL_1096967, EPI_ISL_1096968, EPI_ISL_1096969, EPI_ISL_1096970, EPI_ISL_1096971, EPI_ISL_1096972, EPI_ISL_1096973, EPI_ISL_1096974, EPI_ISL_1096975, EPI_ISL_1096976, EPI_ISL_1096977, EPI_ISL_1096978, EPI_ISL_1096979, EPI_ISL_1096980, EPI_ISL_1096981

| | | | |
|---|---|---|---|
| see above | ASL Napoli 1 Centro | AMES Centro Polidiagnostico Strumentale S.r.l. | "Giovanni Savarese, Raffaella Ruggiero, Eloisa Evangelista, Antonella Di Carlo, Luisa Circelli, Luigi D'Amore, Roberto Sirica, Antonio Fico" |
| EPI_ISL_1096982, EPI_ISL_1096984, EPI_ISL_1096986, EPI_ISL_1096989, EPI_ISL_1096991, EPI_ISL_1096993, EPI_ISL_1096994, EPI_ISL_1096996, EPI_ISL_1096998, EPI_ISL_1097000 | Laboratorio Microbiologia e Virologia P.O. Cotugno A.O. dei Colli | Laboratorio Microbiologia e Virologia P.O. Cotugno A.O. dei Colli | Luigi Atripaldi, Claudia Tiberio, Anna Perfetti, Pellegrino Cerino, Biancamaria Pierri, Maria Concetta Cuomo |
| EPI_ISL_1097002 | Center of Advanced Studies and Technology, Molecular Genetics Laboratory | Center of Advanced Studies and Technology, Molecular Genetics Laboratory | Ferrante Rossella, Mandatori Domitilla, De Fabritiis Simone, Damiani Verena, Anaclerio Federico |

EPI_ISL_1097004, EPI_ISL_1097005, EPI_ISL_1097007, EPI_ISL_1097008, EPI_ISL_1097009, EPI_ISL_1097010, EPI_ISL_1097011, EPI_ISL_1097012, EPI_ISL_1097013, EPI_ISL_1097014, EPI_ISL_1097015

| | | | |
|---|---|---|---|
| see above | Laboratorio Microbiologia e Virologia P.O. Cotugno A.O. dei Colli | Laboratorio Microbiologia e Virologia P.O. Cotugno A.O. dei Colli | Luigi Atripaldi, Claudia Tiberio, Anna Perfetti, Pellegrino Cerino, Biancamaria Pierri, Maria Concetta Cuomo |
| EPI_ISL_1097016 | Laboratorio Microbiologia e Virologia P.O. Cotugno A.O. dei Col | Laboratorio Microbiologia e Virologia P.O. Cotugno A.O. dei Colli | Luigi Atripaldi, Claudia Tiberio, Anna Perfetti, Pellegrino Cerino, Biancamaria Pierri, Maria Concetta Cuomo |
| EPI_ISL_1097017, EPI_ISL_1097019, EPI_ISL_1097020, EPI_ISL_1097021, EPI_ISL_1097022 | Laboratorio Microbiologia e Virologia P.O. Cotugno A.O. dei Colli | Laboratorio Microbiologia e Virologia P.O. Cotugno A.O. dei Colli | Luigi Atripaldi, Claudia Tiberio, Anna Perfetti, Pellegrino Cerino, Biancamaria Pierri, Maria Concetta Cuomo |
| EPI_ISL_1104645, EPI_ISL_1104647, EPI_ISL_1104649 | Virologia, Dipartimento di Scienze Biomediche, Università di Sassari, Viale San Pietro, 43/B - Sassari | Laboratorio specialistico UOC Ematologia - Ospedale "San Francesco" - ATS-ASSL Nuoro | Giovanna Piras, Asproni Rosanna, Paolo Malune, Maria Itria Monne, Angelo Domenico Palmas, Caterina Serra, Elena Rimini, Salvatore Rubino |
| EPI_ISL_1104650 | Laboratorio Biologia Molecolare Sars Cov2 - UOC Laboratorio Analisi - Servizio Medicina di Laboratorio, Ospedale "San Francesco" - ATS-ASSL Nuoro | Laboratorio specialistico UOC Ematologia - Ospedale "San Francesco" - ATS-ASSL Nuoro | Giovanna Piras, Rosanna Asproni, Paolo Malune, Maura Fiamma, Maria Itria Monne, Angelo Domenico Palmas, Iana Lo Maglio, Giuseppe Mameli |
| EPI_ISL_1104652, EPI_ISL_1104654, EPI_ISL_1104656 | Virologia, Dipartimento di Scienze Biomediche, Università di Sassari, Viale San Pietro, 43/B - Sassari | Laboratorio specialistico UOC Ematologia - Ospedale "San Francesco" - ATS-ASSL Nuoro | Giovanna Piras, Asproni Rosanna, Paolo Malune, Maria Itria Monne, Angelo Domenico Palmas, Caterina Serra, Elena Rimini, Salvatore Rubino |
| EPI_ISL_1109574 | SC (UCO) Igiene e Sanità Pubblica (funzione integrata con SC Microbiologia e Virologia) | ARGO Laboratorio Genomica ed Epigenomica | Licastro D, Dal Monego S, Degasperi M, Marcello A, D'Agaro P, Lombardo F |
| EPI_ISL_1109575 | SC (UCO) Igiene e Sanità Pubblica (funzione integrata con SC Microbiologia e Virologia) e Azienda Ospedaliera Pordenone | ARGO Laboratorio Genomica ed Epigenomica | Licastro D, Dal Monego S, Degasperi M, Marcello A, D'Agaro P, De Rosa R |
| EPI_ISL_1109576 | SC (UCO) Igiene e Sanità Pubblica (funzione integrata con SC Microbiologia e Virologia) | ARGO Laboratorio Genomica ed Epigenomica | Licastro D, Dal Monego S, Degasperi M, Marcello A, Segat L, Piscianz E, D'Agaro P |
| EPI_ISL_1109577 | SC (UCO) Igiene e Sanità Pubblica (funzione integrata con SC Microbiologia e Virologia) | ARGO Laboratorio Genomica ed Epigenomica | Licastro D, Dal Monego S, Degasperi M, Marcello A, D'Agaro P, Lombardo F |
| EPI_ISL_1109578, EPI_ISL_1109579, EPI_ISL_1109580, EPI_ISL_1109581 | SC (UCO) Igiene e Sanità Pubblica (funzione integrata con SC Microbiologia e Virologia) e Az. Ospedaliero-Universitaria Udine | ARGO Laboratorio Genomica ed Epigenomica | Licastro D, Dal Monego S, Degasperi M, Marcello A, D'Agaro P, Pipan C |
| EPI_ISL_1109582 | SC (UCO) Igiene e Sanità Pubblica (funzione integrata con SC Microbiologia e Virologia) | ARGO Laboratorio Genomica ed Epigenomica | Licastro D, Dal Monego S, Degasperi M, Marcello A, D'Agaro P, Lombardo F |
| EPI_ISL_1109583, EPI_ISL_1109584, EPI_ISL_1109585, EPI_ISL_1109586 | SC (UCO) Igiene e Sanità Pubblica (funzione integrata con SC Microbiologia e Virologia) e Az. Ospedaliero-Universitaria Udine | ARGO Laboratorio Genomica ed Epigenomica | Licastro D, Dal Monego S, Degasperi M, Marcello A, D'Agaro P, Pipan C |
| EPI_ISL_1109587 | SC (UCO) Igiene e Sanità Pubblica (funzione integrata con SC Microbiologia e Virologia) e Azienda Ospedaliera Pordenone | ARGO Laboratorio Genomica ed Epigenomica | Licastro D, Dal Monego S, Degasperi M, Marcello A, D'Agaro P, De Rosa R |
| EPI_ISL_1109588 | SC (UCO) Igiene e Sanità Pubblica (funzione integrata con SC Microbiologia e Virologia) | ARGO Laboratorio Genomica ed Epigenomica | Licastro D, Dal Monego S, Degasperi M, Marcello A, D'Agaro P, Lombardo F |
| EPI_ISL_1109589, EPI_ISL_1109590 | SC (UCO) Igiene e Sanità Pubblica (funzione integrata con SC Microbiologia e Virologia) e Az. Ospedaliero-Universitaria Udine | ARGO Laboratorio Genomica ed Epigenomica | Licastro D, Dal Monego S, Degasperi M, Marcello A, D'Agaro P, Pipan C |
| EPI_ISL_1109591 | SC (UCO) Igiene e Sanità Pubblica (funzione integrata con SC | ARGO Laboratorio Genomica ed Epigenomica | Licastro D, Dal Monego S, Degasperi M, Marcello A, D'Agaro P, Lombardo F |

| | | | |
|---|---|---|---|
| | Microbiologia e Virologia) | | |
| EPI_ISL_1109592, EPI_ISL_1109593 | SC (UCO) Igiene e Sanità Pubblica (funzione integrata con SC Microbiologia e Virologia) | ARGO Laboratorio Genomica ed Epigenomica | Licastro D, Dal Monego S, Degasperi M, Marcello A, Segat L, Piscianz E, D'Agaro P |
| EPI_ISL_1109594 | SC (UCO) Igiene e Sanità Pubblica (funzione integrata con SC Microbiologia e Virologia) e Azienda Ospedaliera Pordenone | ARGO Laboratorio Genomica ed Epigenomica | Licastro D, Dal Monego S, Degasperi M, Marcello A, D'Agaro P, De Rosa R |
| EPI_ISL_1109595, EPI_ISL_1109596, EPI_ISL_1109597, EPI_ISL_1109598, EPI_ISL_1109599 | SC (UCO) Igiene e Sanità Pubblica (funzione integrata con SC Microbiologia e Virologia) e Az. Ospedaliero-Universitaria Udine | ARGO Laboratorio Genomica ed Epigenomica | Licastro D, Dal Monego S, Degasperi M, Marcello A, D'Agaro P, Pipan C |
| EPI_ISL_1109600 | SC (UCO) Igiene e Sanità Pubblica (funzione integrata con SC Microbiologia e Virologia) | ARGO Laboratorio Genomica ed Epigenomica | Licastro D, Dal Monego S, Degasperi M, Marcello A, D'Agaro P, Lombardo F |
| EPI_ISL_1109601, EPI_ISL_1109602, EPI_ISL_1109603, EPI_ISL_1109604, EPI_ISL_1109605, EPI_ISL_1109606, EPI_ISL_1109607 | SC (UCO) Igiene e Sanità Pubblica (funzione integrata con SC Microbiologia e Virologia) e Az. Ospedaliero-Universitaria Udine | ARGO Laboratorio Genomica ed Epigenomica | Licastro D, Dal Monego S, Degasperi M, Marcello A, D'Agaro P, Pipan C |
| EPI_ISL_1109608, EPI_ISL_1109609 | SC (UCO) Igiene e Sanità Pubblica (funzione integrata con SC Microbiologia e Virologia) | ARGO Laboratorio Genomica ed Epigenomica | Licastro D, Dal Monego S, Degasperi M, Marcello A, Segat L, Piscianz E, D'Agaro P |
| EPI_ISL_1109610, EPI_ISL_1109611, EPI_ISL_1109612 | SC (UCO) Igiene e Sanità Pubblica (funzione integrata con SC Microbiologia e Virologia) e Azienda Ospedaliera Pordenone | ARGO Laboratorio Genomica ed Epigenomica | Licastro D, Dal Monego S, Degasperi M, Marcello A, D'Agaro P, De Rosa R |
| EPI_ISL_1109613 | SC (UCO) Igiene e Sanità Pubblica (funzione integrata con SC Microbiologia e Virologia) e Az. Ospedaliero-Universitaria Udine | ARGO Laboratorio Genomica ed Epigenomica | Licastro D, Dal Monego S, Degasperi M, Marcello A, D'Agaro P, Pipan C |
| EPI_ISL_1109614, EPI_ISL_1109615 | SC (UCO) Igiene e Sanità Pubblica (funzione integrata con SC Microbiologia e Virologia) e Azienda Ospedaliera Pordenone | ARGO Laboratorio Genomica ed Epigenomica | Licastro D, Dal Monego S, Degasperi M, Marcello A, D'Agaro P, De Rosa R |
| EPI_ISL_1109616, EPI_ISL_1109617, EPI_ISL_1109618, EPI_ISL_1109619 | SC (UCO) Igiene e Sanità Pubblica (funzione integrata con SC Microbiologia e Virologia) e Az. Ospedaliero-Universitaria Udine | ARGO Laboratorio Genomica ed Epigenomica | Licastro D, Dal Monego S, Degasperi M, Marcello A, D'Agaro P, Pipan C |
| EPI_ISL_1109620 | SC (UCO) Igiene e Sanità Pubblica (funzione integrata con SC Microbiologia e Virologia) | ARGO Laboratorio Genomica ed Epigenomica | Licastro D, Dal Monego S, Degasperi M, Marcello A, Segat L, Piscianz E, D'Agaro P |
| EPI_ISL_1109621 | SC (UCO) Igiene e Sanità Pubblica (funzione integrata con SC Microbiologia e Virologia) e Az. Ospedaliero-Universitaria Udine | ARGO Laboratorio Genomica ed Epigenomica | Licastro D, Dal Monego S, Degasperi M, Marcello A, D'Agaro P, Pipan C |
| EPI_ISL_1110295, EPI_ISL_1110296, EPI_ISL_1110297, EPI_ISL_1110298, EPI_ISL_1110299, EPI_ISL_1110300, EPI_ISL_1110301, EPI_ISL_1110302, EPI_ISL_1110303, EPI_ISL_1110304, EPI_ISL_1110305, EPI_ISL_1110306, EPI_ISL_1110307, EPI_ISL_1110308, EPI_ISL_1110309, EPI_ISL_1110310, EPI_ISL_1110311, EPI_ISL_1110312, EPI_ISL_1110313, EPI_ISL_1110314, EPI_ISL_1110315, EPI_ISL_1110316, EPI_ISL_1110317, EPI_ISL_1110318, EPI_ISL_1110319, EPI_ISL_1110320, EPI_ISL_1110321, EPI_ISL_1110322, EPI_ISL_1110323, EPI_ISL_1110324, EPI_ISL_1110325, EPI_ISL_1110326, EPI_ISL_1110327, EPI_ISL_1110328, EPI_ISL_1110329, EPI_ISL_1110330, EPI_ISL_1110331, EPI_ISL_1110332, EPI_ISL_1110333, EPI_ISL_1110334, EPI_ISL_1110335, EPI_ISL_1110336, EPI_ISL_1110337, EPI_ISL_1110338, EPI_ISL_1110339, EPI_ISL_1110340, EPI_ISL_1110341, EPI_ISL_1110342, EPI_ISL_1110343, EPI_ISL_1110344, EPI_ISL_1110345, EPI_ISL_1110346, EPI_ISL_1110347, EPI_ISL_1110348, EPI_ISL_1110349, EPI_ISL_1110350, EPI_ISL_1110351, EPI_ISL_1110352, EPI_ISL_1110353, EPI_ISL_1110354, EPI_ISL_1110355, EPI_ISL_1110356, EPI_ISL_1110357, EPI_ISL_1110358, EPI_ISL_1110359, EPI_ISL_1110360, EPI_ISL_1110361, EPI_ISL_1110362, EPI_ISL_1110363, EPI_ISL_1110364, EPI_ISL_1110365, EPI_ISL_1110366, EPI_ISL_1110367, EPI_ISL_1110368, EPI_ISL_1110369, EPI_ISL_1110370, EPI_ISL_1110371, EPI_ISL_1110372, EPI_ISL_1110373, EPI_ISL_1110374, EPI_ISL_1110375, EPI_ISL_1110376, EPI_ISL_1110377, EPI_ISL_1110378, EPI_ISL_1110379, EPI_ISL_1110380, EPI_ISL_1110381, EPI_ISL_1110382, EPI_ISL_1110383, EPI_ISL_1110384, EPI_ISL_1110385, EPI_ISL_1110386, EPI_ISL_1110387, EPI_ISL_1110388 | | | |
| see above | Azienda Sanitaria dell'Alto Adige Laboratorio Aziendale di Microbiologia e Virologia | Istituto di Genomica Applicata | Elisabetta Pagani, Irene Bianconi, Elisabetta Giacobazzi, Elisa Masi, Stefanie Wieser, Irena Jurman, Vera Vendramin, Gabriele Magris, Eleonora Paparelli, Davide Scaglione, Michele Morgante |
| EPI_ISL_1110581, EPI_ISL_1110582, EPI_ISL_1110583, EPI_ISL_1110584, EPI_ISL_1110585, EPI_ISL_1110586, EPI_ISL_1110587, EPI_ISL_1110588, EPI_ISL_1110589, EPI_ISL_1110590, EPI_ISL_1110591, EPI_ISL_1110592, EPI_ISL_1110593, EPI_ISL_1110594, EPI_ISL_1110595 | | | |
| see above | LAB ANALISI PO CARDARELLI CB | Istituto Zooprofilattico Sperimentale dell'Abruzzo e Molise "G. Caporale" | Scutellà M, Niro G, Lorusso A, Marcacci M, Di Domenico M, Ancora M, Curini V, Mangone I, Rinaldi A, Scialabba S, Di Pasquale A, Cammà C, Puglia I, Calistri P, Savini G |
| EPI_ISL_1110596 | SIESP L'Aquila | Istituto Zooprofilattico Sperimentale dell'Abruzzo e Molise "G. Caporale" | Lorusso A, Marcacci M, Di Domenico M, Ancora M, Curini V, Mangone I, Rinaldi A, Scialabba S, Di Pasquale A, Cammà C, Puglia I, Calistri P, Savini G |
| EPI_ISL_1110597 | SIESP CHIETI-Drive in Chieti | Istituto Zooprofilattico Sperimentale dell'Abruzzo e Molise "G. Caporale" | Lorusso A, Marcacci M, Di Domenico M, Ancora M, Curini V, Mangone I, Rinaldi A, Scialabba S, Di Pasquale A, Cammà C, Puglia I, Calistri P, Savini G |
| EPI_ISL_1110598, EPI_ISL_1110599, EPI_ISL_1110600 | SIESP CH-Drive in VASTO | Istituto Zooprofilattico Sperimentale dell'Abruzzo e Molise "G. Caporale" | Lorusso A, Marcacci M, Di Domenico M, Ancora M, Curini V, Mangone I, Rinaldi A, Scialabba S, Di Pasquale A, Cammà C, Puglia I, Calistri P, Savini G |
| EPI_ISL_1110601 | Ospedale Civile Teramo | Istituto Zooprofilattico Sperimentale dell'Abruzzo e Molise "G. Caporale" | Lorusso A, Marcacci M, Di Domenico M, Ancora M, Curini V, Mangone I, Rinaldi A, Scialabba S, Di Pasquale A, Cammà C, Puglia I, Calistri P, Savini G |
| EPI_ISL_1110602 | SIESP Dip. Prev Teramo | Istituto Zooprofilattico Sperimentale dell'Abruzzo e Molise "G. Caporale" | Lorusso A, Marcacci M, Di Domenico M, Ancora M, Curini V, Mangone I, Rinaldi A, Scialabba S, Di Pasquale A, Cammà C, Puglia I, Calistri P, Savini G |
| EPI_ISL_1112221, EPI_ISL_1112222 | Ospedale Santissima Annunziata | Istituto Zooprofilattico Sperimentale della Puglia e della Basilicata | Parisi A., Bianco A., Capozzi L., Del Sambro L., Simone D., Giannico A., Ridolfi D. |
| EPI_ISL_1112223, EPI_ISL_1112224, EPI_ISL_1112225, EPI_ISL_1112226 | Ospedale Di Venere - Carbonara | Istituto Zooprofilattico Sperimentale della Puglia e della Basilicata | Parisi A., Bianco A., Capozzi L., Del Sambro L., Simone D., Giannico A., Ridolfi D. |
| EPI_ISL_1112227, EPI_ISL_1112228, EPI_ISL_1112229, EPI_ISL_1112230, EPI_ISL_1112231, EPI_ISL_1112232, EPI_ISL_1112233, EPI_ISL_1112234 | Istituto Zooprofilattico Sperimentale della Puglia e della Basilicata | Istituto Zooprofilattico Sperimentale della Puglia e della Basilicata | Parisi A., Bianco A., Capozzi L., Del Sambro L., Simone D., Giannico A., Ridolfi D. |
| EPI_ISL_1112235 | Ospedali Riuniti Azienda Ospedaliera Universitaria - Foggia | Istituto Zooprofilattico Sperimentale della Puglia e della Basilicata | Parisi A., Bianco A., Capozzi L., Del Sambro L., Simone D., Giannico A., Ridolfi D. |
| EPI_ISL_1112236 | Dipartimento di Scienze Biomediche e Oncologia Umana - Azienda Ospedaliero Universitaria Consorziale Policlinico | Istituto Zooprofilattico Sperimentale della Puglia e della Basilicata | Parisi A., Bianco A., Capozzi L., Del Sambro L., Simone D., Giannico A., Ridolfi D. |
| EPI_ISL_1112237, EPI_ISL_1112238, EPI_ISL_1112239, EPI_ISL_1112240, EPI_ISL_1112241, EPI_ISL_1112242, EPI_ISL_1112243, EPI_ISL_1112244, EPI_ISL_1112245, EPI_ISL_1112246, EPI_ISL_1112247, EPI_ISL_1112248, EPI_ISL_1112249, EPI_ISL_1112250, EPI_ISL_1112251, EPI_ISL_1112252, EPI_ISL_1112253, EPI_ISL_1112254, EPI_ISL_1112255, EPI_ISL_1112256, EPI_ISL_1112257, EPI_ISL_1112258, EPI_ISL_1112259, EPI_ISL_1112260, EPI_ISL_1112261, EPI_ISL_1112262, EPI_ISL_1112263, EPI_ISL_1112264, EPI_ISL_1112265, EPI_ISL_1112266, EPI_ISL_1112267, EPI_ISL_1112268, EPI_ISL_1112269, EPI_ISL_1112270, EPI_ISL_1112271, EPI_ISL_1112272, EPI_ISL_1112273, EPI_ISL_1112274, EPI_ISL_1112275, EPI_ISL_1112276, EPI_ISL_1112277, EPI_ISL_1112278, EPI_ISL_1112279, EPI_ISL_1112280, EPI_ISL_1112281, EPI_ISL_1112282, EPI_ISL_1112283, EPI_ISL_1112284, EPI_ISL_1112285, EPI_ISL_1112286, EPI_ISL_1112287, EPI_ISL_1112288, EPI_ISL_1112289, EPI_ISL_1112290, EPI_ISL_1112291 | | | |
| see above | Dipartimento di Scienze Biomediche e Oncologia Umana - Azienda Ospedaliero Universitaria Consorziale Policlinico | Istituto Zooprofilattico Sperimentale della Puglia e della Basilicata | Parisi A., Bianco A., Capozzi L., Del Sambro L., Simone D., Chironna M., Loconsole D., Sallustio A. |
| EPI_ISL_1114766 | SIESP L'Aquila | Istituto Zooprofilattico Sperimentale dell'Abruzzo e Molise "G. Caporale" | Lorusso A, Marcacci M, Di Domenico M, Ancora M, Curini V, Mangone I, Rinaldi A, Scialabba S, Di Pasquale A, Cammà C, Puglia I, Calistri P, Savini G |
| EPI_ISL_1116471, EPI_ISL_1116472 | Dipartimento di Scienze Biomediche e Oncologia Umana - | Istituto Zooprofilattico Sperimentale della Puglia e della | Parisi A., Bianco A., Capozzi L., Del Sambro L., Simone D., Chironna M., Loconsole D., Sallustio A. |

| | | | |
|---|---|---|---|
| | Azienda Ospedaliero Universitaria Consorziale Policlinico | Basilicata | |
| EPI_ISL_1116473 | Istituto Zooprofilattico Sperimentale della Puglia e della Basilicata | Istituto Zooprofilattico Sperimentale della Puglia e della Basilicata | Parisi A., Bianco A., Capozzi L., Del Sambro L., Simone D., Chironna M., Loconsole D., Sallustio A., Ridolfi D. |
| EPI_ISL_1116474 | Istituto Zooprofilattico Sperimentale della Puglia e della Basilicata | Istituto Zooprofilattico Sperimentale della Puglia e della Basilicata | Parisi A., Bianco A., Capozzi L., Del Sambro L., Simone D., Chironna M., Loconsole D., Sallustio A., Giannico A. |
| EPI_ISL_1116475 | Dipartimento di Scienze Biomediche e Oncologia Umana - Azienda Ospedaliero Universitaria Consorziale Policlinico | Istituto Zooprofilattico Sperimentale della Puglia e della Basilicata | Parisi A., Bianco A., Capozzi L., Del Sambro L., Simone D., Chironna M., Loconsole D., Sallustio A. |
| EPI_ISL_1117458 | OSP SAN SALVATORE, MEDICINA INTERNA | Istituto Zooprofilattico Sperimentale dell'Abruzzo e Molise "G. Caporale" | Lorusso A, Marcacci M, Di Domenico M, Ancora M, Curini V, Mangone I, Rinaldi A, Scialabba S, Di Pasquale A, Cammà C, Scialabba S, Puglia I, Calistri P, Savini G |
| EPI_ISL_1117459 | SIESP L'AQUILA, | Istituto Zooprofilattico Sperimentale dell'Abruzzo e Molise "G. Caporale" | Lorusso A, Marcacci M, Di Domenico M, Ancora M, Curini V, Mangone I, Rinaldi A, Scialabba S, Di Pasquale A, Cammà C, Scialabba S, Puglia I, Calistri P, Savini G |
| EPI_ISL_1117460 | SIESP DIP PREV CHIETI | Istituto Zooprofilattico Sperimentale dell'Abruzzo e Molise "G. Caporale" | Lorusso A, Marcacci M, Di Domenico M, Ancora M, Curini V, Mangone I, Rinaldi A, Scialabba S, Di Pasquale A, Cammà C, Scialabba S, Puglia I, Calistri P, Savini G |
| EPI_ISL_1117461 | SIESP CHIETI,DRIVE IN CHIETI | Istituto Zooprofilattico Sperimentale dell'Abruzzo e Molise "G. Caporale" | Lorusso A, Marcacci M, Di Domenico M, Ancora M, Curini V, Mangone I, Rinaldi A, Scialabba S, Di Pasquale A, Cammà C, Scialabba S, Puglia I, Calistri P, Savini G |
| EPI_ISL_1117462 | SIESP CHIETI, DRIVE IN CHIETI | Istituto Zooprofilattico Sperimentale dell'Abruzzo e Molise "G. Caporale" | Lorusso A, Marcacci M, Di Domenico M, Ancora M, Curini V, Mangone I, Rinaldi A, Scialabba S, Di Pasquale A, Cammà C, Scialabba S, Puglia I, Calistri P, Savini G |
| EPI_ISL_1117463 | SIESP CHIETI, DRIVE IN LANCIANO | Istituto Zooprofilattico Sperimentale dell'Abruzzo e Molise "G. Caporale" | Lorusso A, Marcacci M, Di Domenico M, Ancora M, Curini V, Mangone I, Rinaldi A, Scialabba S, Di Pasquale A, Cammà C, Scialabba S, Puglia I, Calistri P, Savini G |
| EPI_ISL_1117464 | SIESP CHIETI, DRIVE IN CHIETI | Istituto Zooprofilattico Sperimentale dell'Abruzzo e Molise "G. Caporale" | Lorusso A, Marcacci M, Di Domenico M, Ancora M, Curini V, Mangone I, Rinaldi A, Scialabba S, Di Pasquale A, Cammà C, Scialabba S, Puglia I, Calistri P, Savini G |
| EPI_ISL_1117465 | SIESP DIP PREV CHIETI | Istituto Zooprofilattico Sperimentale dell'Abruzzo e Molise "G. Caporale" | Lorusso A, Marcacci M, Di Domenico M, Ancora M, Curini V, Mangone I, Rinaldi A, Scialabba S, Di Pasquale A, Cammà C, Scialabba S, Puglia I, Calistri P, Savini G |
| EPI_ISL_1117466, EPI_ISL_1117467 | SIESP DIP PREV TERAMO | Istituto Zooprofilattico Sperimentale dell'Abruzzo e Molise "G. Caporale" | Lorusso A, Marcacci M, Di Domenico M, Ancora M, Curini V, Mangone I, Rinaldi A, Scialabba S, Di Pasquale A, Cammà C, Scialabba S, Puglia I, Calistri P, Savini G |
| EPI_ISL_1117468 | OSP SAN SALVATORE, MEDICINA INTERNA | Istituto Zooprofilattico Sperimentale dell'Abruzzo e Molise "G. Caporale" | Lorusso A, Marcacci M, Di Domenico M, Ancora M, Curini V, Mangone I, Rinaldi A, Scialabba S, Di Pasquale A, Cammà C, Scialabba S, Puglia I, Calistri P, Savini G |
| EPI_ISL_1117469, EPI_ISL_1117470, EPI_ISL_1117471, EPI_ISL_1117472 | SIESP DIP PREV TERAMO | Istituto Zooprofilattico Sperimentale dell'Abruzzo e Molise "G. Caporale" | Lorusso A, Marcacci M, Di Domenico M, Ancora M, Curini V, Mangone I, Rinaldi A, Scialabba S, Di Pasquale A, Cammà C, Scialabba S, Puglia I, Calistri P, Savini G |
| EPI_ISL_1117473 | OSP SAN SALVATORE, MEDICINA INTERNA | Istituto Zooprofilattico Sperimentale dell'Abruzzo e Molise "G. Caporale" | Lorusso A, Marcacci M, Di Domenico M, Ancora M, Curini V, Mangone I, Rinaldi A, Scialabba S, Di Pasquale A, Cammà C, Scialabba S, Puglia I, Calistri P, Savini G |
| EPI_ISL_1117474 | SIESP DIP PREV CHIETI | Istituto Zooprofilattico Sperimentale dell'Abruzzo e Molise "G. Caporale" | Lorusso A, Marcacci M, Di Domenico M, Ancora M, Curini V, Mangone I, Rinaldi A, Scialabba S, Di Pasquale A, Cammà C, Scialabba S, Puglia I, Calistri P, Savini G |
| EPI_ISL_1117475 | SIESP SULMONA | Istituto Zooprofilattico Sperimentale dell'Abruzzo e Molise "G. Caporale" | Lorusso A, Marcacci M, Di Domenico M, Ancora M, Curini V, Mangone I, Rinaldi A, Scialabba S, Di Pasquale A, Cammà C, Scialabba S, Puglia I, Calistri P, Savini G |
| EPI_ISL_1117476, EPI_ISL_1117477 | SIESP CHIETI, DRIVE IN CHIETI | Istituto Zooprofilattico Sperimentale dell'Abruzzo e Molise "G. Caporale" | Lorusso A, Marcacci M, Di Domenico M, Ancora M, Curini V, Mangone I, Rinaldi A, Scialabba S, Di Pasquale A, Cammà C, Scialabba S, Puglia I, Calistri P, Savini G |
| EPI_ISL_1117478 | SIESP CHIETI, DRIVE IN ORTONA | Istituto Zooprofilattico Sperimentale dell'Abruzzo e Molise "G. Caporale" | Lorusso A, Marcacci M, Di Domenico M, Ancora M, Curini V, Mangone I, Rinaldi A, Scialabba S, Di Pasquale A, Cammà C, Scialabba S, Puglia I, Calistri P, Savini G |
| EPI_ISL_1117479 | SIESP CHIETI, DRIVE IN LANCIANO | Istituto Zooprofilattico Sperimentale dell'Abruzzo e Molise "G. Caporale" | Lorusso A, Marcacci M, Di Domenico M, Ancora M, Curini V, Mangone I, Rinaldi A, Scialabba S, Di Pasquale A, Cammà C, Scialabba S, Puglia I, Calistri P, Savini G |
| EPI_ISL_1117480, EPI_ISL_1117481, EPI_ISL_1117482 | SIESP DIP PREV TERAMO | Istituto Zooprofilattico Sperimentale dell'Abruzzo e Molise "G. Caporale" | Lorusso A, Marcacci M, Di Domenico M, Ancora M, Curini V, Mangone I, Rinaldi A, Scialabba S, Di Pasquale A, Cammà C, Scialabba S, Puglia I, Calistri P, Savini G |
| EPI_ISL_1117483, EPI_ISL_1117484 | OSP SAN SALVATORE, MEDICINA INTERNA | Istituto Zooprofilattico Sperimentale dell'Abruzzo e Molise "G. Caporale" | Lorusso A, Marcacci M, Di Domenico M, Ancora M, Curini V, Mangone I, Rinaldi A, Scialabba S, Di Pasquale A, Cammà C, Scialabba S, Puglia I, Calistri P, Savini G |
| EPI_ISL_1117485 | SIESP DIP PREV TERAMO | Istituto Zooprofilattico Sperimentale dell'Abruzzo e Molise "G. Caporale" | Lorusso A, Marcacci M, Di Domenico M, Ancora M, Curini V, Mangone I, Rinaldi A, Scialabba S, Di Pasquale A, Cammà C, Scialabba S, Puglia I, Calistri P, Savini G |
| EPI_ISL_1117486, EPI_ISL_1117487 | SIESP DIP PREV CHIETI | Istituto Zooprofilattico Sperimentale dell'Abruzzo e Molise "G. Caporale" | Lorusso A, Marcacci M, Di Domenico M, Ancora M, Curini V, Mangone I, Rinaldi A, Scialabba S, Di Pasquale A, Cammà C, Scialabba S, Puglia I, Calistri P, Savini G |
| EPI_ISL_1117488 | DIP PREV AVEZZANO, SERVIZIO DI IGIENE EPIDEMIOLOGIA E SANITÀ PUBBLICA | Istituto Zooprofilattico Sperimentale dell'Abruzzo e Molise "G. Caporale" | Lorusso A, Marcacci M, Di Domenico M, Ancora M, Curini V, Mangone I, Rinaldi A, Scialabba S, Di Pasquale A, Cammà C, Scialabba S, Puglia I, Calistri P, Savini G |
| EPI_ISL_1117489 | SIESP DIP PREV TERAMO | Istituto Zooprofilattico Sperimentale dell'Abruzzo e Molise "G. Caporale" | Lorusso A, Marcacci M, Di Domenico M, Ancora M, Curini V, Mangone I, Rinaldi A, Scialabba S, Di Pasquale A, Cammà C, Scialabba S, Puglia I, Calistri P, Savini G |
| EPI_ISL_1117490 | SIESP CHIETI, DRIVE IN VASTO | Istituto Zooprofilattico Sperimentale dell'Abruzzo e Molise "G. Caporale" | Lorusso A, Marcacci M, Di Domenico M, Ancora M, Curini V, Mangone I, Rinaldi A, Scialabba S, Di Pasquale A, Cammà C, Scialabba S, Puglia I, Calistri P, Savini G |
| EPI_ISL_1117491, EPI_ISL_1117492 | SIESP DIP PREV CHIETI | Istituto Zooprofilattico Sperimentale dell'Abruzzo e Molise "G. Caporale" | Lorusso A, Marcacci M, Di Domenico M, Ancora M, Curini V, Mangone I, Rinaldi A, Scialabba S, Di Pasquale A, Cammà C, Scialabba S, Puglia I, Calistri P, Savini G |
| EPI_ISL_1117493 | OSP SAN SALVATORE, MEDICINA INTERNA | Istituto Zooprofilattico Sperimentale dell'Abruzzo e Molise "G. Caporale" | Lorusso A, Marcacci M, Di Domenico M, Ancora M, Curini V, Mangone I, Rinaldi A, Scialabba S, Di Pasquale A, Cammà C, Scialabba S, Puglia I, Calistri P, Savini G |
| EPI_ISL_1117494 | SIESP DIP PREV CHIETI | Istituto Zooprofilattico Sperimentale dell'Abruzzo e Molise "G. Caporale" | Lorusso A, Marcacci M, Di Domenico M, Ancora M, Curini V, Mangone I, Rinaldi A, Scialabba S, Di Pasquale A, Cammà C, Scialabba S, Puglia I, Calistri P, Savini G |
| EPI_ISL_1117495 | SIESP DIP PREV TERAMO | Istituto Zooprofilattico Sperimentale dell'Abruzzo e Molise "G. Caporale" | Lorusso A, Marcacci M, Di Domenico M, Ancora M, Curini V, Mangone I, Rinaldi A, Scialabba S, Di Pasquale A, Cammà C, Scialabba S, Puglia I, Calistri P, Savini G |
| EPI_ISL_1117496 | USCA PESCINA | Istituto Zooprofilattico Sperimentale dell'Abruzzo e Molise "G. Caporale" | Lorusso A, Marcacci M, Di Domenico M, Ancora M, Curini V, Mangone I, Rinaldi A, Scialabba S, Di Pasquale A, Cammà C, Scialabba S, Puglia I, Calistri P, Savini G |
| EPI_ISL_1117497 | SIESP CHIETI, DRIVE IN ORTONA | Istituto Zooprofilattico Sperimentale dell'Abruzzo e Molise "G. Caporale" | Lorusso A, Marcacci M, Di Domenico M, Ancora M, Curini V, Mangone I, Rinaldi A, Scialabba S, Di Pasquale A, Cammà C, Scialabba S, Puglia I, Calistri P, Savini G |
| EPI_ISL_1117498 | PRESIDIO TERRITORIALE CASOLI | Istituto Zooprofilattico Sperimentale dell'Abruzzo e Molise "G. Caporale" | Lorusso A, Marcacci M, Di Domenico M, Ancora M, Curini V, Mangone I, Rinaldi A, Scialabba S, Di Pasquale A, Cammà C, Scialabba S, Puglia I, Calistri P, Savini G |
| EPI_ISL_1117499 | SIESP CHIETI, DRIVE IN VASTO | Istituto Zooprofilattico Sperimentale dell'Abruzzo e Molise "G. | Lorusso A, Marcacci M, Di Domenico M, Ancora M, Curini V, Mangone I, Rinaldi A, Scialabba S, Di Pasquale A, Cammà C, Scialabba S, Puglia I, Calistri |

| | | | |
|---|---|---|---|
| | | Caporale" | P, Savini G |
| EPI_ISL_1117500 | SIESP CHIETI, DRIVE IN ORTONA | Istituto Zooprofilattico Sperimentale dell'Abruzzo e Molise "G. Caporale" | Lorusso A, Marcacci M, Di Domenico M, Ancora M, Curini V, Mangone I, Rinaldi A, Scialabba S, Di Pasquale A, Cammà C, Scialabba S, Puglia I, Calistri P, Savini G |
| EPI_ISL_1117501 | OSP SAN SALVATORE, MEDICINA INTERNA | Istituto Zooprofilattico Sperimentale dell'Abruzzo e Molise "G. Caporale" | Lorusso A, Marcacci M, Di Domenico M, Ancora M, Curini V, Mangone I, Rinaldi A, Scialabba S, Di Pasquale A, Cammà C, Scialabba S, Puglia I, Calistri P, Savini G |
| EPI_ISL_1117502 | SIESP DIP PREV CHIETI | Istituto Zooprofilattico Sperimentale dell'Abruzzo e Molise "G. Caporale" | Lorusso A, Marcacci M, Di Domenico M, Ancora M, Curini V, Mangone I, Rinaldi A, Scialabba S, Di Pasquale A, Cammà C, Scialabba S, Puglia I, Calistri P, Savini G |
| EPI_ISL_1117503 | SIESP DIP PREV TERAMO | Istituto Zooprofilattico Sperimentale dell'Abruzzo e Molise "G. Caporale" | Lorusso A, Marcacci M, Di Domenico M, Ancora M, Curini V, Mangone I, Rinaldi A, Scialabba S, Di Pasquale A, Cammà C, Scialabba S, Puglia I, Calistri P, Savini G |
| EPI_ISL_1117504, EPI_ISL_1117505 | OSP SAN SALVATORE, MEDICINA INTERNA | Istituto Zooprofilattico Sperimentale dell'Abruzzo e Molise "G. Caporale" | Lorusso A, Marcacci M, Di Domenico M, Ancora M, Curini V, Mangone I, Rinaldi A, Scialabba S, Di Pasquale A, Cammà C, Scialabba S, Puglia I, Calistri P, Savini G |
| EPI_ISL_1117506 | SIESP DIP PREV CHIETI | Istituto Zooprofilattico Sperimentale dell'Abruzzo e Molise "G. Caporale" | Lorusso A, Marcacci M, Di Domenico M, Ancora M, Curini V, Mangone I, Rinaldi A, Scialabba S, Di Pasquale A, Cammà C, Scialabba S, Puglia I, Calistri P, Savini G |
| EPI_ISL_1117507, EPI_ISL_1117508 | SIESP CHIETI, DRIVE IN LANCIANO | Istituto Zooprofilattico Sperimentale dell'Abruzzo e Molise "G. Caporale" | Lorusso A, Marcacci M, Di Domenico M, Ancora M, Curini V, Mangone I, Rinaldi A, Scialabba S, Di Pasquale A, Cammà C, Scialabba S, Puglia I, Calistri P, Savini G |
| EPI_ISL_1117510, EPI_ISL_1117511, EPI_ISL_1117512, EPI_ISL_1117513 | SIESP DIP PREV CHIETI | Istituto Zooprofilattico Sperimentale dell'Abruzzo e Molise "G. Caporale" | Lorusso A, Marcacci M, Di Domenico M, Ancora M, Curini V, Mangone I, Rinaldi A, Scialabba S, Di Pasquale A, Cammà C, Scialabba S, Puglia I, Calistri P, Savini G |
| EPI_ISL_1117514 | PRES OSP SAN LIBERATORE | Istituto Zooprofilattico Sperimentale dell'Abruzzo e Molise "G. Caporale" | Lorusso A, Marcacci M, Di Domenico M, Ancora M, Curini V, Mangone I, Rinaldi A, Scialabba S, Di Pasquale A, Cammà C, Scialabba S, Puglia I, Calistri P, Savini G |
| EPI_ISL_1117515 | SIESP DIP PREV TERAMO | Istituto Zooprofilattico Sperimentale dell'Abruzzo e Molise "G. Caporale" | Lorusso A, Marcacci M, Di Domenico M, Ancora M, Curini V, Mangone I, Rinaldi A, Scialabba S, Di Pasquale A, Cammà C, Scialabba S, Puglia I, Calistri P, Savini G |
| EPI_ISL_1117516 | SIESP CHIETI, DRIVE IN LANCIANO | Istituto Zooprofilattico Sperimentale dell'Abruzzo e Molise "G. Caporale" | Lorusso A, Marcacci M, Di Domenico M, Ancora M, Curini V, Mangone I, Rinaldi A, Scialabba S, Di Pasquale A, Cammà C, Scialabba S, Puglia I, Calistri P, Savini G |
| EPI_ISL_1117517 | SIESP CHIETI, DRIVE IN ORTONA | Istituto Zooprofilattico Sperimentale dell'Abruzzo e Molise "G. Caporale" | Lorusso A, Marcacci M, Di Domenico M, Ancora M, Curini V, Mangone I, Rinaldi A, Scialabba S, Di Pasquale A, Cammà C, Scialabba S, Puglia I, Calistri P, Savini G |
| EPI_ISL_1117518 | SIESP DIP PREV TERAMO | Istituto Zooprofilattico Sperimentale dell'Abruzzo e Molise "G. Caporale" | Lorusso A, Marcacci M, Di Domenico M, Ancora M, Curini V, Mangone I, Rinaldi A, Scialabba S, Di Pasquale A, Cammà C, Scialabba S, Puglia I, Calistri P, Savini G |
| EPI_ISL_1117519 | SIESP CHIETI, DRIVE IN CHIETI | Istituto Zooprofilattico Sperimentale dell'Abruzzo e Molise "G. Caporale" | Lorusso A, Marcacci M, Di Domenico M, Ancora M, Curini V, Mangone I, Rinaldi A, Scialabba S, Di Pasquale A, Cammà C, Scialabba S, Puglia I, Calistri P, Savini G |
| EPI_ISL_1117520 | USCA AVEZZANO | Istituto Zooprofilattico Sperimentale dell'Abruzzo e Molise "G. Caporale" | Lorusso A, Marcacci M, Di Domenico M, Ancora M, Curini V, Mangone I, Rinaldi A, Scialabba S, Di Pasquale A, Cammà C, Scialabba S, Puglia I, Calistri P, Savini G |
| EPI_ISL_1117521 | DIP PREV AVEZZANO, SERVIZIO DI IGIENE EPIDEMIOLOGIA E SANITÀ PUBBLICA | Istituto Zooprofilattico Sperimentale dell'Abruzzo e Molise "G. Caporale" | Lorusso A, Marcacci M, Di Domenico M, Ancora M, Curini V, Mangone I, Rinaldi A, Scialabba S, Di Pasquale A, Cammà C, Scialabba S, Puglia I, Calistri P, Savini G |
| EPI_ISL_1117522, EPI_ISL_1117523, EPI_ISL_1117524 | SIESP DIP PREV TERAMO | Istituto Zooprofilattico Sperimentale dell'Abruzzo e Molise "G. Caporale" | Lorusso A, Marcacci M, Di Domenico M, Ancora M, Curini V, Mangone I, Rinaldi A, Scialabba S, Di Pasquale A, Cammà C, Scialabba S, Puglia I, Calistri P, Savini G |
| EPI_ISL_1117525 | OSP SAN SALVATORE, MEDICINA INTERNA | Istituto Zooprofilattico Sperimentale dell'Abruzzo e Molise "G. Caporale" | Lorusso A, Marcacci M, Di Domenico M, Ancora M, Curini V, Mangone I, Rinaldi A, Scialabba S, Di Pasquale A, Cammà C, Scialabba S, Puglia I, Calistri P, Savini G |
| EPI_ISL_1117526 | IZS TERAMO | Istituto Zooprofilattico Sperimentale dell'Abruzzo e Molise "G. Caporale" | Lorusso A, Marcacci M, Di Domenico M, Ancora M, Curini V, Mangone I, Rinaldi A, Scialabba S, Di Pasquale A, Cammà C, Scialabba S, Puglia I, Calistri P, Savini G |
| EPI_ISL_1117661, EPI_ISL_1117662 | SIESP DIP PREV TERAMO | Istituto Zooprofilattico Sperimentale dell'Abruzzo e Molise "G. Caporale" | Lorusso A, Marcacci M, Di Domenico M, Ancora M, Curini V, Mangone I, Rinaldi A, Scialabba S, Di Pasquale A, Cammà C, Puglia I, Calistri P, Savini G |
| EPI_ISL_1117663 | SIESP CHIETI, DRIVE IN ORTONA | Istituto Zooprofilattico Sperimentale dell'Abruzzo e Molise "G. Caporale" | Lorusso A, Marcacci M, Di Domenico M, Ancora M, Curini V, Mangone I, Rinaldi A, Scialabba S, Di Pasquale A, Cammà C, Puglia I, Calistri P, Savini G |
| EPI_ISL_1117664 | SIESP DIP PREV CHIETI | Istituto Zooprofilattico Sperimentale dell'Abruzzo e Molise "G. Caporale" | Lorusso A, Marcacci M, Di Domenico M, Ancora M, Curini V, Mangone I, Rinaldi A, Scialabba S, Di Pasquale A, Cammà C, Puglia I, Calistri P, Savini G |
| EPI_ISL_1117665, EPI_ISL_1117666 | SIESP DIP PREV TERAMO | Istituto Zooprofilattico Sperimentale dell'Abruzzo e Molise "G. Caporale" | Lorusso A, Marcacci M, Di Domenico M, Ancora M, Curini V, Mangone I, Rinaldi A, Scialabba S, Di Pasquale A, Cammà C, Puglia I, Calistri P, Savini G |
| EPI_ISL_1117667 | SIESP CHIETI, DRIVE IN ORTONA | Istituto Zooprofilattico Sperimentale dell'Abruzzo e Molise "G. Caporale" | Lorusso A, Marcacci M, Di Domenico M, Ancora M, Curini V, Mangone I, Rinaldi A, Scialabba S, Di Pasquale A, Cammà C, Puglia I, Calistri P, Savini G |
| EPI_ISL_1117668 | SIESP DIP PREV TERAMO | Istituto Zooprofilattico Sperimentale dell'Abruzzo e Molise "G. Caporale" | Lorusso A, Marcacci M, Di Domenico M, Ancora M, Curini V, Mangone I, Rinaldi A, Scialabba S, Di Pasquale A, Cammà C, Puglia I, Calistri P, Savini G |
| EPI_ISL_1117669 | SIESP CHIETI, DRIVE IN CHIETI | Istituto Zooprofilattico Sperimentale dell'Abruzzo e Molise "G. Caporale" | Lorusso A, Marcacci M, Di Domenico M, Ancora M, Curini V, Mangone I, Rinaldi A, Scialabba S, Di Pasquale A, Cammà C, Puglia I, Calistri P, Savini G |
| EPI_ISL_1117670, EPI_ISL_1117671 | SIESP DIP PREV TERAMO | Istituto Zooprofilattico Sperimentale dell'Abruzzo e Molise "G. Caporale" | Lorusso A, Marcacci M, Di Domenico M, Ancora M, Curini V, Mangone I, Rinaldi A, Scialabba S, Di Pasquale A, Cammà C, Puglia I, Calistri P, Savini G |
| EPI_ISL_1117672 | SIESP DIP PREV CHIETI | Istituto Zooprofilattico Sperimentale dell'Abruzzo e Molise "G. Caporale" | Lorusso A, Marcacci M, Di Domenico M, Ancora M, Curini V, Mangone I, Rinaldi A, Scialabba S, Di Pasquale A, Cammà C, Puglia I, Calistri P, Savini G |
| EPI_ISL_1117673 | SIESP DIP PREV TERAMO | Istituto Zooprofilattico Sperimentale dell'Abruzzo e Molise "G. Caporale" | Lorusso A, Marcacci M, Di Domenico M, Ancora M, Curini V, Mangone I, Rinaldi A, Scialabba S, Di Pasquale A, Cammà C, Puglia I, Calistri P, Savini G |
| EPI_ISL_1117674 | SIESP CHIETI, DRIVE IN CHIETI | Istituto Zooprofilattico Sperimentale dell'Abruzzo e Molise "G. Caporale" | Lorusso A, Marcacci M, Di Domenico M, Ancora M, Curini V, Mangone I, Rinaldi A, Scialabba S, Di Pasquale A, Cammà C, Puglia I, Calistri P, Savini G |
| EPI_ISL_1117675 | DIP PREV AVEZZANO-SERVIZIO DI IGIENE EPIDEMIOLOGIA E SANITÀ PUBBLICA | Istituto Zooprofilattico Sperimentale dell'Abruzzo e Molise "G. Caporale" | Lorusso A, Marcacci M, Di Domenico M, Ancora M, Curini V, Mangone I, Rinaldi A, Scialabba S, Di Pasquale A, Cammà C, Puglia I, Calistri P, Savini G |
| EPI_ISL_1117676 | SIESP CHIETI, DRIVE IN LANCIANO | Istituto Zooprofilattico Sperimentale dell'Abruzzo e Molise "G. Caporale" | Lorusso A, Marcacci M, Di Domenico M, Ancora M, Curini V, Mangone I, Rinaldi A, Scialabba S, Di Pasquale A, Cammà C, Puglia I, Calistri P, Savini G |
| EPI_ISL_1117677 | USCA CASTEL DI SANGRO | Istituto Zooprofilattico Sperimentale dell'Abruzzo e Molise "G. Caporale" | Lorusso A, Marcacci M, Di Domenico M, Ancora M, Curini V, Mangone I, Rinaldi A, Scialabba S, Di Pasquale A, Cammà C, Puglia I, Calistri P, Savini G |
| EPI_ISL_1117678, EPI_ISL_1117679 | SIESP CHIETI, DRIVE IN VASTO | Istituto Zooprofilattico Sperimentale dell'Abruzzo e Molise "G. Caporale" | Lorusso A, Marcacci M, Di Domenico M, Ancora M, Curini V, Mangone I, Rinaldi A, Scialabba S, Di Pasquale A, Cammà C, Puglia I, Calistri P, Savini G |
| EPI_ISL_1117680 | SIESP DIP PREV TERAMO | Istituto Zooprofilattico Sperimentale dell'Abruzzo e Molise "G. | Lorusso A, Marcacci M, Di Domenico M, Ancora M, Curini V, Mangone I, Rinaldi A, Scialabba S, Di Pasquale A, Cammà C, Puglia I, Calistri P, Savini G |

| | | | |
|---|---|---|---|
| | | Caporale" | |
| EPI_ISL_1117681 | SIESP CHIETI, DRIVE IN CHIETI | Istituto Zooprofilattico Sperimentale dell'Abruzzo e Molise "G. Caporale" | Lorusso A, Marcacci M, Di Domenico M, Ancora M, Curini V, Mangone I, Rinaldi A, Scialabba S, Di Pasquale A, Cammà C, Puglia I, Calistri P, Savini G |
| EPI_ISL_1117682 | SIESP CHIETI, DRIVE IN LANCIANO | Istituto Zooprofilattico Sperimentale dell'Abruzzo e Molise "G. Caporale" | Lorusso A, Marcacci M, Di Domenico M, Ancora M, Curini V, Mangone I, Rinaldi A, Scialabba S, Di Pasquale A, Cammà C, Puglia I, Calistri P, Savini G |
| EPI_ISL_1117683, EPI_ISL_1117684 | USCA CASTEL DI SANGRO | Istituto Zooprofilattico Sperimentale dell'Abruzzo e Molise "G. Caporale" | Lorusso A, Marcacci M, Di Domenico M, Ancora M, Curini V, Mangone I, Rinaldi A, Scialabba S, Di Pasquale A, Cammà C, Puglia I, Calistri P, Savini G |
| EPI_ISL_1117685 | SIESP DIP PREV TERAMO | Istituto Zooprofilattico Sperimentale dell'Abruzzo e Molise "G. Caporale" | Lorusso A, Marcacci M, Di Domenico M, Ancora M, Curini V, Mangone I, Rinaldi A, Scialabba S, Di Pasquale A, Cammà C, Puglia I, Calistri P, Savini G |
| EPI_ISL_1117686 | DIP PREV AVEZZANO-SERVIZIO DI IGIENE EPIDEMIOLOGIA E SANITÀ PUBBLICA | Istituto Zooprofilattico Sperimentale dell'Abruzzo e Molise "G. Caporale" | Lorusso A, Marcacci M, Di Domenico M, Ancora M, Curini V, Mangone I, Rinaldi A, Scialabba S, Di Pasquale A, Cammà C, Puglia I, Calistri P, Savini G |
| EPI_ISL_1117687 | SIESP CHIETI, DRIVE IN ORTONA | Istituto Zooprofilattico Sperimentale dell'Abruzzo e Molise "G. Caporale" | Lorusso A, Marcacci M, Di Domenico M, Ancora M, Curini V, Mangone I, Rinaldi A, Scialabba S, Di Pasquale A, Cammà C, Puglia I, Calistri P, Savini G |
| EPI_ISL_1117688 | SIESP DIP PREV CHIETI | Istituto Zooprofilattico Sperimentale dell'Abruzzo e Molise "G. Caporale" | Lorusso A, Marcacci M, Di Domenico M, Ancora M, Curini V, Mangone I, Rinaldi A, Scialabba S, Di Pasquale A, Cammà C, Puglia I, Calistri P, Savini G |
| EPI_ISL_1117689 | SIESP CHIETI, DRIVE IN LANCIANO | Istituto Zooprofilattico Sperimentale dell'Abruzzo e Molise "G. Caporale" | Lorusso A, Marcacci M, Di Domenico M, Ancora M, Curini V, Mangone I, Rinaldi A, Scialabba S, Di Pasquale A, Cammà C, Puglia I, Calistri P, Savini G |
| EPI_ISL_1117691 | SIESP DIP PREV TERAMO | Istituto Zooprofilattico Sperimentale dell'Abruzzo e Molise "G. Caporale" | Lorusso A, Marcacci M, Di Domenico M, Ancora M, Curini V, Mangone I, Rinaldi A, Scialabba S, Di Pasquale A, Cammà C, Puglia I, Calistri P, Savini G |
| EPI_ISL_1117692 | PRESIDIO OSP SAN LIBERATORE | Istituto Zooprofilattico Sperimentale dell'Abruzzo e Molise "G. Caporale" | Lorusso A, Marcacci M, Di Domenico M, Ancora M, Curini V, Mangone I, Rinaldi A, Scialabba S, Di Pasquale A, Cammà C, Puglia I, Calistri P, Savini G |
| EPI_ISL_1117693 | SIESP DIP PREV TERAMO | Istituto Zooprofilattico Sperimentale dell'Abruzzo e Molise "G. Caporale" | Lorusso A, Marcacci M, Di Domenico M, Ancora M, Curini V, Mangone I, Rinaldi A, Scialabba S, Di Pasquale A, Cammà C, Puglia I, Calistri P, Savini G |
| EPI_ISL_1117694, EPI_ISL_1117695 | SIESP CHIETI, DRIVE IN LANCIANO | Istituto Zooprofilattico Sperimentale dell'Abruzzo e Molise "G. Caporale" | Lorusso A, Marcacci M, Di Domenico M, Ancora M, Curini V, Mangone I, Rinaldi A, Scialabba S, Di Pasquale A, Cammà C, Puglia I, Calistri P, Savini G |
| EPI_ISL_1117696 | SIESP CHIETI, DRIVE IN VASTO | Istituto Zooprofilattico Sperimentale dell'Abruzzo e Molise "G. Caporale" | Lorusso A, Marcacci M, Di Domenico M, Ancora M, Curini V, Mangone I, Rinaldi A, Scialabba S, Di Pasquale A, Cammà C, Puglia I, Calistri P, Savini G |
| EPI_ISL_1117697 | DIP PREV AVEZZANO-SERVIZIO DI IGIENE EPIDEMIOLOGIA E SANITÀ PUBBLICA | Istituto Zooprofilattico Sperimentale dell'Abruzzo e Molise "G. Caporale" | Lorusso A, Marcacci M, Di Domenico M, Ancora M, Curini V, Mangone I, Rinaldi A, Scialabba S, Di Pasquale A, Cammà C, Puglia I, Calistri P, Savini G |
| EPI_ISL_1117698 | SIESP CHIETI, DRIVE IN LANCIANO | Istituto Zooprofilattico Sperimentale dell'Abruzzo e Molise "G. Caporale" | Lorusso A, Marcacci M, Di Domenico M, Ancora M, Curini V, Mangone I, Rinaldi A, Scialabba S, Di Pasquale A, Cammà C, Puglia I, Calistri P, Savini G |
| EPI_ISL_1117699 | SIESP CHIETI, DRIVE IN VASTO | Istituto Zooprofilattico Sperimentale dell'Abruzzo e Molise "G. Caporale" | Lorusso A, Marcacci M, Di Domenico M, Ancora M, Curini V, Mangone I, Rinaldi A, Scialabba S, Di Pasquale A, Cammà C, Puglia I, Calistri P, Savini G |
| EPI_ISL_1118159, EPI_ISL_1118160, EPI_ISL_1118161, EPI_ISL_1118162, EPI_ISL_1118163, EPI_ISL_1118164, EPI_ISL_1118165, EPI_ISL_1118166, EPI_ISL_1118167, EPI_ISL_1118168, EPI_ISL_1118169, EPI_ISL_1118170, EPI_ISL_1118171, EPI_ISL_1118172, EPI_ISL_1118173, EPI_ISL_1118174, EPI_ISL_1118175, EPI_ISL_1118176, EPI_ISL_1118177, EPI_ISL_1118178, EPI_ISL_1118179, EPI_ISL_1118180, EPI_ISL_1118181, EPI_ISL_1118182, EPI_ISL_1118183, EPI_ISL_1118184, EPI_ISL_1118185, EPI_ISL_1118186, EPI_ISL_1118187, EPI_ISL_1118188, EPI_ISL_1118189, EPI_ISL_1118190, EPI_ISL_1118191, EPI_ISL_1118192, EPI_ISL_1118193, EPI_ISL_1118194, EPI_ISL_1118195, EPI_ISL_1118196, EPI_ISL_1118197, EPI_ISL_1118198, EPI_ISL_1118199, EPI_ISL_1118200, EPI_ISL_1118201, EPI_ISL_1118202, EPI_ISL_1118203, EPI_ISL_1118204 | | | |
| see above | LAB ANALISI PO CARDARELLI CAMPOBASSO | Istituto Zooprofilattico Sperimentale dell'Abruzzo e Molise "G. Caporale" | Scutellà M, Niro G, Felice V. Lorusso A, Marcacci M, Di Domenico M, Ancora M, Curini V, Mangone I, Rinaldi A, Scialabba S, Di Pasquale A, Cammà C, Puglia I, Calistri P, Savini G |
| EPI_ISL_1118930, EPI_ISL_1118932, EPI_ISL_1118934, EPI_ISL_1119567 | Laboratorio Microbiologia e Virologia P.O. Cotugno A.O. dei Colli | Laboratorio Microbiologia e Virologia P.O. Cotugno A.O. dei Colli | Luigi Atripaldi, Claudia Tiberio, Anna Perfetti, Pellegrino Cerino, Biancamaria Pierri, Maria Concetta Cuomo |
| EPI_ISL_1120120 | Area Biologia Molecolare - Istituto Zooprofilattico Sperimentale della Sicilia | Area Biologia Molecolare - Istituto Zooprofilattico Sperimentale della Sicilia | REALE Stefano, SCIBETTA Silvia, PIAZZA Angela, BRUNO Gabriella, CASTELLI Germano, BRUNO Federica, VITALE Francesco, TRAMUTO Fabio, MAIDA Carmelo Massimo, MAZZUCCO Walter, DI NARO Daniela, RANDAZZO Giulia, VITALE Fabrizio |
| EPI_ISL_1121032 | SIESP CHIETI, DRIVE IN LANCIANO | Istituto Zooprofilattico Sperimentale dell'Abruzzo e Molise "G. Caporale" | Lorusso A, Marcacci M, Di Domenico M, Ancora M, Curini V, Mangone I, Rinaldi A, Scialabba S, Di Pasquale A, Cammà C, Puglia I, Calistri P, Savini G |
| EPI_ISL_1121033 | USCA CASTEL DI SANGRO | Istituto Zooprofilattico Sperimentale dell'Abruzzo e Molise "G. Caporale" | Lorusso A, Marcacci M, Di Domenico M, Ancora M, Curini V, Mangone I, Rinaldi A, Scialabba S, Di Pasquale A, Cammà C, Puglia I, Calistri P, Savini G |
| EPI_ISL_1121034, EPI_ISL_1121035, EPI_ISL_1121036 | LAB ANALISI PO CARDARELLI CAMPOBASSO | Istituto Zooprofilattico Sperimentale dell'Abruzzo e Molise "G. Caporale" | Scutellà M, Niro G, Felice V. Lorusso A, Marcacci M, Di Domenico M, Ancora M, Curini V, Mangone I, Rinaldi A, Scialabba S, Di Pasquale A, Cammà C, Puglia I, Calistri P, Savini G |
| EPI_ISL_1121037 | SIESP DIP PREV TERAMO | Istituto Zooprofilattico Sperimentale dell'Abruzzo e Molise "G. Caporale" | Lorusso A, Marcacci M, Di Domenico M, Ancora M, Curini V, Mangone I, Rinaldi A, Scialabba S, Di Pasquale A, Cammà C, Scialabba S, Puglia I, Calistri P, Savini G |
| EPI_ISL_1121038 | OSP SAN SALVATORE, MEDICINA INTERNA | Istituto Zooprofilattico Sperimentale dell'Abruzzo e Molise "G. Caporale" | Lorusso A, Marcacci M, Di Domenico M, Ancora M, Curini V, Mangone I, Rinaldi A, Scialabba S, Di Pasquale A, Cammà C, Scialabba S, Puglia I, Calistri P, Savini G |
| EPI_ISL_1129215 | Istituto Zooprofilattico Sperimentale del Mezzogiorno | Lab. Microbiologia e Virologia, Cotugno, A.O. dei Colli | Luigi Atripaldi, Claudia Tiberio, Anna Perfetti, Pellegrino Cerino, Biancamaria Pierri, Maria Concetta Cuomo |
| EPI_ISL_1132668 | Azienda Ospedaliero - Universitaria di Modena Policlinico - Virologia e Microbiologia Molecolare | Zooprofilattico Sperimentale dell'Emilia Romagna e della Lombardia (IZSLER), Risk Analysis and Genomic Epidemiology Unit | Monica Pecorari, William Gennari, Giulia Fregni Serpini, Marina Morganti, Ilaria Menozzi, Erika Scaltriti, Stefano Pongolini |
| EPI_ISL_1140102, EPI_ISL_1140103, EPI_ISL_1140104, EPI_ISL_1140105, EPI_ISL_1140106, EPI_ISL_1140107, EPI_ISL_1140108, EPI_ISL_1140109, EPI_ISL_1140110, EPI_ISL_1140111, EPI_ISL_1140112, EPI_ISL_1140113, EPI_ISL_1140114, EPI_ISL_1140115, EPI_ISL_1140116, EPI_ISL_1140117, EPI_ISL_1140118, EPI_ISL_1140119 | | | |
| see above | Università di Parma, Laboratorio di Igiene e Sanità Pubblica | Istituto Zooprofilattico Sperimentale della Lombardia e dell'Emilia Romagna (IZSLER), Risk Analysis and Genomic Epidemiology Unit | Maria Eugenia Colucci, Licia Veronesi, Paola Affanni, Marina Morganti, Ilaria Menozzi, Erika Scaltriti, Stefano Pongolini |
| EPI_ISL_1154361, EPI_ISL_1154890 | Area Biologia Molecolare - Istituto Zooprofilattico Sperimentale della Sicilia | Area Biologia Molecolare - Istituto Zooprofilattico Sperimentale della Sicilia | REALE Stefano, SCIBETTA Silvia, PIAZZA Angela, BRUNO Gabriella, CASTELLI Germano, BRUNO Federica, VITALE Francesco, TRAMUTO Fabio, MAIDA Carmelo Massimo, MAZZUCCO Walter, DI NARO Daniela, RANDAZZO Giulia, VITALE Fabrizio |
| EPI_ISL_1157894, EPI_ISL_1157895, EPI_ISL_1157896, EPI_ISL_1157897, EPI_ISL_1157898, EPI_ISL_1157899, EPI_ISL_1157900, EPI_ISL_1157901, EPI_ISL_1157902, EPI_ISL_1157903, EPI_ISL_1157904, EPI_ISL_1157905, EPI_ISL_1157906, EPI_ISL_1157907, EPI_ISL_1157908, EPI_ISL_1157909, EPI_ISL_1157910, EPI_ISL_1157911, EPI_ISL_1157912, EPI_ISL_1157913, EPI_ISL_1157914, EPI_ISL_1157915, EPI_ISL_1157916, EPI_ISL_1157917, EPI_ISL_1157918, EPI_ISL_1157919, EPI_ISL_1157920, EPI_ISL_1157921, EPI_ISL_1157922, EPI_ISL_1157923, EPI_ISL_1157924, EPI_ISL_1157925, EPI_ISL_1157926, EPI_ISL_1157927, EPI_ISL_1157928, EPI_ISL_1157929, EPI_ISL_1157930, EPI_ISL_1157931, EPI_ISL_1157932, EPI_ISL_1157933, EPI_ISL_1157934, EPI_ISL_1157935, EPI_ISL_1157936, EPI_ISL_1157937, EPI_ISL_1157938, EPI_ISL_1157939, EPI_ISL_1157940, EPI_ISL_1157941, EPI_ISL_1157942, EPI_ISL_1157943, EPI_ISL_1157944, EPI_ISL_1157945, EPI_ISL_1157946, EPI_ISL_1157947, EPI_ISL_1157948, EPI_ISL_1157949, EPI_ISL_1157950, EPI_ISL_1157951, EPI_ISL_1157952, EPI_ISL_1157953, EPI_ISL_1157954, EPI_ISL_1157955, EPI_ISL_1157956, EPI_ISL_1157957, EPI_ISL_1157958, EPI_ISL_1157959, EPI_ISL_1157960, EPI_ISL_1157961, EPI_ISL_1157962, EPI_ISL_1157963, EPI_ISL_1157964, EPI_ISL_1157965, EPI_ISL_1157966, EPI_ISL_1157967, EPI_ISL_1157968, EPI_ISL_1157969, EPI_ISL_1157970, EPI_ISL_1157971, EPI_ISL_1157972, EPI_ISL_1157973, EPI_ISL_1157974, EPI_ISL_1157975, EPI_ISL_1157976, EPI_ISL_1157977, EPI_ISL_1157978, EPI_ISL_1157979, EPI_ISL_1157980, EPI_ISL_1157981, EPI_ISL_1157982, EPI_ISL_1157983, EPI_ISL_1157984, EPI_ISL_1157985, EPI_ISL_1157986, EPI_ISL_1157987, EPI_ISL_1157988, EPI_ISL_1157989, EPI_ISL_1157990, EPI_ISL_1157991, EPI_ISL_1157992, EPI_ISL_1157993, EPI_ISL_1157994, EPI_ISL_1157995, EPI_ISL_1157996, EPI_ISL_1157997, EPI_ISL_1157998, EPI_ISL_1157999, EPI_ISL_1158000, EPI_ISL_1158001, EPI_ISL_1158002, EPI_ISL_1158003, EPI_ISL_1158004, EPI_ISL_1158005, EPI_ISL_1158006, EPI_ISL_1158007, EPI_ISL_1158008, EPI_ISL_1158009, EPI_ISL_1158010, EPI_ISL_1158011, EPI_ISL_1158012, EPI_ISL_1158013, EPI_ISL_1158014, EPI_ISL_1158015, EPI_ISL_1158016, EPI_ISL_1158017, EPI_ISL_1158018, EPI_ISL_1158019, EPI_ISL_1158020, EPI_ISL_1158021, EPI_ISL_1158022, EPI_ISL_1158023, EPI_ISL_1158024, EPI_ISL_1158025, EPI_ISL_1158026, EPI_ISL_1158027, EPI_ISL_1158028, EPI_ISL_1158029, EPI_ISL_1158030 | | | |

| | | | |
|---|---|---|---|
| see above | ASL Napoli 1 Centro | AMES Centro Polidiagnostico Strumentale S.r.l. | "Giovanni Savarese, Raffaella Ruggiero, Eloisa Evangelista, Antonella Di Carlo, Luisa Circelli, Luigi D'Amore, Roberto Sirica, Nadia Petrillo, Monica Ianniello, Antonio Fico" |
| EPI_ISL_1158846 | Unità Operativa di Microbiologia, IRCCS Policlinico di Sant'Orsola, Azienda Ospedaliero-Universitaria di Bologna | U.O. Microbiologia Laboratorio Unico Centro Servizi AUSL della Romagna | Giada Rossini, Giuliano Furlini, Tiziana Lazzarotto, Giorgio Dirani, Silvia Zannoli, Vittorio Sambri |
| EPI_ISL_1163688, EPI_ISL_1163689, EPI_ISL_1163690, EPI_ISL_1163691, EPI_ISL_1163692 | UOC Microbiologia e Virologia, Azienda Ospedaliera Universitaria Senese, Siena, Italy | Dipartimento di Biotecnologie Mediche | Maria Grazia Cusi, David Pinzauti, Claudia Gandolfo, Gabriele Anichini, Gianni Pozzi, Gianni Gori Savellini, Francesco Santoro |
| EPI_ISL_1163795, EPI_ISL_1163796, EPI_ISL_1163797 | Azienda USL Umbria 2 | Istituto Zooprofilattico Sperimentale dell'Abruzzo e Molise "G. Caporale" | Proietti A., Pistoni E., Lorusso A, Marcacci M, Di Domenico M, Ancora M, Curini V, Mangone I, Rinaldi A, Scialabba S, Di Pasquale A, Cammà C, Puglia I, Calistri P, Savini G |
| EPI_ISL_1163798, EPI_ISL_1163799, EPI_ISL_1163800, EPI_ISL_1163801, EPI_ISL_1163802, EPI_ISL_1163803, EPI_ISL_1163804, EPI_ISL_1163805, EPI_ISL_1163806, EPI_ISL_1163807, EPI_ISL_1163808, EPI_ISL_1163809, EPI_ISL_1163810, EPI_ISL_1163811, EPI_ISL_1163812, EPI_ISL_1163813, EPI_ISL_1163814, EPI_ISL_1163815, EPI_ISL_1163816, EPI_ISL_1163817, EPI_ISL_1163818, EPI_ISL_1163819, EPI_ISL_1163820, EPI_ISL_1163821, EPI_ISL_1163822, EPI_ISL_1163823, EPI_ISL_1163824, EPI_ISL_1163825, EPI_ISL_1163826, EPI_ISL_1163827, EPI_ISL_1163828 | | | |
| see above | Università degli Studi di Perugia | Istituto Zooprofilattico Sperimentale dell'Abruzzo e Molise "G. Caporale" | Mencacci A., Camilloni B., Lorusso A, Marcacci M, Di Domenico M, Ancora M, Curini V, Mangone I, Rinaldi A, Scialabba S, Di Pasquale A, Cammà C, Puglia I, Calistri P, Savini G |
| EPI_ISL_1163829, EPI_ISL_1163830 | Istituto Zooprofilattico Sperimentale Umbria e Marche "Togo Rosati" | Istituto Zooprofilattico Sperimentale dell'Abruzzo e Molise "G. Caporale" | Biagetti M., Giammarioli M., Lorusso A, Marcacci M, Di Domenico M, Ancora M, Curini V, Mangone I, Rinaldi A, Scialabba S, Di Pasquale A, Cammà C, Puglia I, Calistri P, Savini G |
| EPI_ISL_1164754, EPI_ISL_1164755, EPI_ISL_1164756 | Azienda USL Umbria 2 | Istituto Zooprofilattico Sperimentale dell'Abruzzo e Molise "G. Caporale" | Proietti A, Pistoni E, Lorusso A, Marcacci M, Di Domenico M, Ancora M, Curini V, Mangone I, Rinaldi A, Scialabba S, Di Pasquale A, Cammà C, Puglia I, Calistri P, Savini G |
| EPI_ISL_1164816, EPI_ISL_1164853, EPI_ISL_1164854, EPI_ISL_1164930, EPI_ISL_1164969, EPI_ISL_1165000, EPI_ISL_1165066, EPI_ISL_1165077, EPI_ISL_1165138, EPI_ISL_1165143, EPI_ISL_1165899, EPI_ISL_1166083 | | | |
| see above | Struttura Semplice Dipartimentale di Virologia e Microbiologia molecolare, Azienda Ospedaliero-Universitaria, Policlinico di Modena | U.O. Microbiologia, Laboratorio Unico Centro Servizi - AUSL della Romagna | Pecorari Monica, Gennari William, Fregni Serpini Giulia, Giorgio Dirani, Silvia Zannoli, Vittorio Sambri |
| EPI_ISL_1166147, EPI_ISL_1166148 | Laboratorio Analisi Osp. Città di Castello - Azienda USL Umbria1 | Istituto Zooprofilattico Sperimentale dell'Abruzzo e Molise "G. Caporale" | Malagigi V, Tacconi P, Lorusso A, Marcacci M, Di Domenico M, Ancora M, Curini V, Mangone I, Rinaldi A, Scialabba S, Di Pasquale A, Cammà C, Puglia I, Calistri P, Savini G |
| EPI_ISL_1166149, EPI_ISL_1166150, EPI_ISL_1166151, EPI_ISL_1166152, EPI_ISL_1166153, EPI_ISL_1166154, EPI_ISL_1166155, EPI_ISL_1166156, EPI_ISL_1166157, EPI_ISL_1166158, EPI_ISL_1166159, EPI_ISL_1166160, EPI_ISL_1166161, EPI_ISL_1166162, EPI_ISL_1166163, EPI_ISL_1166164, EPI_ISL_1166165, EPI_ISL_1166166, EPI_ISL_1166167, EPI_ISL_1166168, EPI_ISL_1166169, EPI_ISL_1166170, EPI_ISL_1166171 | | | |
| see above | Università degli Studi di Perugia | Istituto Zooprofilattico Sperimentale dell'Abruzzo e Molise "G. Caporale" | Mencacci A, Camilloni B, Lorusso A, Marcacci M, Di Domenico M, Ancora M, Curini V, Mangone I, Rinaldi A, Scialabba S, Di Pasquale A, Cammà C, Puglia I, Calistri P, Savini G |
| EPI_ISL_1166172, EPI_ISL_1166173, EPI_ISL_1166174, EPI_ISL_1166175, EPI_ISL_1166176, EPI_ISL_1166177, EPI_ISL_1166178, EPI_ISL_1166179, EPI_ISL_1166180, EPI_ISL_1166181, EPI_ISL_1166182, EPI_ISL_1166183, EPI_ISL_1166184, EPI_ISL_1166185, EPI_ISL_1166186, EPI_ISL_1166187, EPI_ISL_1166188 | | | |
| see above | Istituto Zooprofilattico Sperimentale Umbria e Marche "Togo Rosati" | Istituto Zooprofilattico Sperimentale dell'Abruzzo e Molise "G. Caporale" | Biagetti M, Giammarioli M, Lorusso A, Marcacci M, Di Domenico M, Ancora M, Curini V, Mangone I, Rinaldi A, Scialabba S, Di Pasquale A, Cammà C, Puglia I, Calistri P, Savini G |
| EPI_ISL_1166221, EPI_ISL_1166222, EPI_ISL_1166223, EPI_ISL_1166225, EPI_ISL_1166226, EPI_ISL_1166227, EPI_ISL_1166228, EPI_ISL_1166229, EPI_ISL_1166231, EPI_ISL_1166232, EPI_ISL_1166233, EPI_ISL_1166238, EPI_ISL_1166239, EPI_ISL_1166240, EPI_ISL_1166241, EPI_ISL_1166242, EPI_ISL_1166245, EPI_ISL_1166247, EPI_ISL_1166249, EPI_ISL_1166251, EPI_ISL_1166252, EPI_ISL_1166253, EPI_ISL_1166255, EPI_ISL_1166256, EPI_ISL_1166261, EPI_ISL_1166263, EPI_ISL_1166264, EPI_ISL_1166265, EPI_ISL_1166267, EPI_ISL_1166268, EPI_ISL_1166269, EPI_ISL_1166271, EPI_ISL_1166272, EPI_ISL_1166273, EPI_ISL_1166275, EPI_ISL_1166276, EPI_ISL_1166277, EPI_ISL_1166279, EPI_ISL_1166280, EPI_ISL_1166281, EPI_ISL_1166282, EPI_ISL_1166283, EPI_ISL_1166284, EPI_ISL_1166285, EPI_ISL_1166286, EPI_ISL_1166287, EPI_ISL_1166289, EPI_ISL_1166290, EPI_ISL_1166291, EPI_ISL_1166293, EPI_ISL_1166294, EPI_ISL_1166296, EPI_ISL_1166297, EPI_ISL_1166298, EPI_ISL_1166299, EPI_ISL_1166300, EPI_ISL_1166301, EPI_ISL_1166302, EPI_ISL_1166303, EPI_ISL_1166304, EPI_ISL_1166305, EPI_ISL_1166306, EPI_ISL_1166307, EPI_ISL_1166308, EPI_ISL_1166311, EPI_ISL_1166312, EPI_ISL_1166313, EPI_ISL_1166314, EPI_ISL_1166316, EPI_ISL_1166318, EPI_ISL_1166319, EPI_ISL_1166320, EPI_ISL_1166322, EPI_ISL_1166323, EPI_ISL_1166324, EPI_ISL_1166325, EPI_ISL_1166326, EPI_ISL_1166327, EPI_ISL_1166328, EPI_ISL_1166329, EPI_ISL_1166330, EPI_ISL_1166331, EPI_ISL_1166332, EPI_ISL_1166333, EPI_ISL_1166334, EPI_ISL_1166335, EPI_ISL_1166336, EPI_ISL_1166338, EPI_ISL_1166339, EPI_ISL_1166340, EPI_ISL_1166341, EPI_ISL_1166342, EPI_ISL_1166343, EPI_ISL_1166344, EPI_ISL_1166345, EPI_ISL_1166346, EPI_ISL_1166347, EPI_ISL_1166348, EPI_ISL_1166349, EPI_ISL_1166350, EPI_ISL_1166351, EPI_ISL_1166353, EPI_ISL_1166354, EPI_ISL_1166356, EPI_ISL_1166358, EPI_ISL_1166359, EPI_ISL_1166361, EPI_ISL_1166362, EPI_ISL_1166363, EPI_ISL_1166364, EPI_ISL_1166366, EPI_ISL_1166367, EPI_ISL_1166368, EPI_ISL_1166371, EPI_ISL_1166373, EPI_ISL_1166374, EPI_ISL_1166375, EPI_ISL_1166376, EPI_ISL_1166377, EPI_ISL_1166379, EPI_ISL_1166380, EPI_ISL_1166382, EPI_ISL_1166383, EPI_ISL_1166384, EPI_ISL_1166386, EPI_ISL_1166389, EPI_ISL_1166390, EPI_ISL_1166391, EPI_ISL_1166394, EPI_ISL_1166397, EPI_ISL_1166399, EPI_ISL_1166400, EPI_ISL_1166401, EPI_ISL_1166402, EPI_ISL_1166403, EPI_ISL_1166404, EPI_ISL_1166405, EPI_ISL_1166406, EPI_ISL_1166407, EPI_ISL_1166408, EPI_ISL_1166409, EPI_ISL_1166410, EPI_ISL_1166411, EPI_ISL_1166412, EPI_ISL_1166413, EPI_ISL_1166415, EPI_ISL_1166416, EPI_ISL_1166417, EPI_ISL_1166418, EPI_ISL_1166419, EPI_ISL_1166420, EPI_ISL_1166421, EPI_ISL_1166422, EPI_ISL_1166423, EPI_ISL_1166424, EPI_ISL_1166425, EPI_ISL_1166426, EPI_ISL_1166427, EPI_ISL_1166428, EPI_ISL_1166429, EPI_ISL_1166430, EPI_ISL_1166431, EPI_ISL_1166432, EPI_ISL_1166433, EPI_ISL_1166434, EPI_ISL_1166435, EPI_ISL_1166436, EPI_ISL_1166437, EPI_ISL_1166438, EPI_ISL_1166440, EPI_ISL_1166443, EPI_ISL_1166444, EPI_ISL_1166446, EPI_ISL_1166447, EPI_ISL_1166449, EPI_ISL_1166450, EPI_ISL_1166451, EPI_ISL_1166453, EPI_ISL_1166454, EPI_ISL_1166455, EPI_ISL_1166456, EPI_ISL_1166457, EPI_ISL_1166458, EPI_ISL_1166459, EPI_ISL_1166460, EPI_ISL_1166461, EPI_ISL_1166462, EPI_ISL_1166463, EPI_ISL_1166464, EPI_ISL_1166465, EPI_ISL_1166466, EPI_ISL_1166467, EPI_ISL_1166468, EPI_ISL_1166469, EPI_ISL_1166470, EPI_ISL_1166471, EPI_ISL_1166472, EPI_ISL_1166474, EPI_ISL_1166475, EPI_ISL_1166476, EPI_ISL_1166477, EPI_ISL_1166478, EPI_ISL_1166479, EPI_ISL_1166480, EPI_ISL_1166481, EPI_ISL_1166482, EPI_ISL_1166483, EPI_ISL_1166484, EPI_ISL_1166485, EPI_ISL_1166486, EPI_ISL_1166487, EPI_ISL_1166488, EPI_ISL_1166489, EPI_ISL_1166490, EPI_ISL_1166491, EPI_ISL_1166492, EPI_ISL_1166493, EPI_ISL_1166494, EPI_ISL_1166495, EPI_ISL_1166496, EPI_ISL_1166497, EPI_ISL_1166498, EPI_ISL_1166499, EPI_ISL_1166500, EPI_ISL_1166501, EPI_ISL_1166502, EPI_ISL_1166503, EPI_ISL_1166504, EPI_ISL_1166505, EPI_ISL_1166506, EPI_ISL_1166507, EPI_ISL_1166509, EPI_ISL_1166510, EPI_ISL_1166511, EPI_ISL_1166512, EPI_ISL_1166513, EPI_ISL_1166514, EPI_ISL_1166515, EPI_ISL_1166516, EPI_ISL_1166517, EPI_ISL_1166518, EPI_ISL_1166519, EPI_ISL_1166520, EPI_ISL_1166521, EPI_ISL_1166522, EPI_ISL_1166523, EPI_ISL_1166524, EPI_ISL_1166525, EPI_ISL_1166526, EPI_ISL_1166527, EPI_ISL_1166528, EPI_ISL_1166529, EPI_ISL_1166530, EPI_ISL_1166531, EPI_ISL_1166532, EPI_ISL_1166533, EPI_ISL_1167309, EPI_ISL_1167310, EPI_ISL_1167312, EPI_ISL_1167313, EPI_ISL_1167314, EPI_ISL_1167316, EPI_ISL_1167317, EPI_ISL_1167318, EPI_ISL_1167320, EPI_ISL_1167323, EPI_ISL_1167325, EPI_ISL_1167328, EPI_ISL_1167335, EPI_ISL_1167337, EPI_ISL_1167342, EPI_ISL_1167343, EPI_ISL_1167344, EPI_ISL_1167346, EPI_ISL_1167347, EPI_ISL_1167352, EPI_ISL_1167353, EPI_ISL_1167354, EPI_ISL_1167355, EPI_ISL_1167357, EPI_ISL_1167359, EPI_ISL_1167360, EPI_ISL_1167362, EPI_ISL_1167363, EPI_ISL_1167366, EPI_ISL_1167368, EPI_ISL_1167372, EPI_ISL_1167373, EPI_ISL_1167374, EPI_ISL_1167375, EPI_ISL_1167376, EPI_ISL_1167377, EPI_ISL_1167378, EPI_ISL_1167379, EPI_ISL_1167380, EPI_ISL_1167381, EPI_ISL_1167382, EPI_ISL_1167383, EPI_ISL_1167385, EPI_ISL_1167388, EPI_ISL_1167389, EPI_ISL_1167390, EPI_ISL_1167391, EPI_ISL_1167392, EPI_ISL_1167394, EPI_ISL_1167397, EPI_ISL_1167398, EPI_ISL_1167399, EPI_ISL_1167400, EPI_ISL_1167401, EPI_ISL_1167402, EPI_ISL_1167403, EPI_ISL_1167408, EPI_ISL_1167409, EPI_ISL_1167410, EPI_ISL_1167411, EPI_ISL_1167412, EPI_ISL_1167413, EPI_ISL_1167414, EPI_ISL_1167415, EPI_ISL_1167416, EPI_ISL_1167420, EPI_ISL_1167421, EPI_ISL_1167422, EPI_ISL_1167423, EPI_ISL_1167424, EPI_ISL_1167426, EPI_ISL_1167427, EPI_ISL_1167428, EPI_ISL_1167430, EPI_ISL_1167431, EPI_ISL_1167432, EPI_ISL_1167433, EPI_ISL_1167434, EPI_ISL_1167435, EPI_ISL_1167437, EPI_ISL_1167441, EPI_ISL_1167442, EPI_ISL_1167443, EPI_ISL_1167444, EPI_ISL_1167446, EPI_ISL_1167451, EPI_ISL_1167455, EPI_ISL_1167458, EPI_ISL_1167459, EPI_ISL_1167460, EPI_ISL_1167461, EPI_ISL_1167465, EPI_ISL_1167466, EPI_ISL_1167467, EPI_ISL_1167468, EPI_ISL_1167469, EPI_ISL_1167471, EPI_ISL_1167472, EPI_ISL_1167475, EPI_ISL_1167476, EPI_ISL_1167478, EPI_ISL_1167479, EPI_ISL_1167480, EPI_ISL_1167482, EPI_ISL_1167484, EPI_ISL_1167486, EPI_ISL_1167487, EPI_ISL_1167488, EPI_ISL_1167489, EPI_ISL_1167490, EPI_ISL_1167491, EPI_ISL_1167494, EPI_ISL_1167496, EPI_ISL_1167497, EPI_ISL_1167498, EPI_ISL_1167499, EPI_ISL_1167501, EPI_ISL_1167502, EPI_ISL_1167505, EPI_ISL_1167507, EPI_ISL_1167508, EPI_ISL_1167511, EPI_ISL_1167512, EPI_ISL_1167513, EPI_ISL_1167516, EPI_ISL_1167518, EPI_ISL_1167519, EPI_ISL_1167520, EPI_ISL_1167522, EPI_ISL_1167523, EPI_ISL_1167525, EPI_ISL_1167526, EPI_ISL_1167529, EPI_ISL_1167531, EPI_ISL_1167532, EPI_ISL_1167534, EPI_ISL_1167535, EPI_ISL_1167536, EPI_ISL_1167537, EPI_ISL_1167538, EPI_ISL_1167539, EPI_ISL_1167540, EPI_ISL_1167543, EPI_ISL_1167544 | | | |
| see above | ASL Napoli 1 Centro | AMES Centro Polidiagnostico Strumentale S.r.l. | "Giovanni Savarese, Raffaella Ruggiero, Eloisa Evangelista, Antonella Di Carlo, Luisa Circelli, Luigi D'Amore, Nadia Petrillo, Monica Ianniello, Roberto Sirica,Maurizio D'Amora, Antonio Fico" |
| EPI_ISL_1169883 | INMI Lazzaro Spallanzani IRCCS | INMI Lazzaro Spallanzani IRCCS | CEM Gruber, B Bartolini, E Giombini, F Messina, M Rueca, O Butera, MR Capobianchi, A Di Caro |
| EPI_ISL_1169884 | INMI Lazzaro Spallanzani IRCCS | INMI Lazzaro Spallanzani IRCCS | F Messina, M Rueca, O Butera, CEM Gruber, B Bartolini, E Giombini, MR Capobianchi, A Di Caro |
| EPI_ISL_1169885 | INMI Lazzaro Spallanzani IRCCS | INMI Lazzaro Spallanzani IRCCS | M Rueca, O Butera, CEM Gruber, B Bartolini, E Giombini, F Messina, A Di Caro, MR Capobianchi |
| EPI_ISL_1169886 | INMI Lazzaro Spallanzani IRCCS | INMI Lazzaro Spallanzani IRCCS | B Bartolini, E Giombini, F Messina, M Rueca, O Butera, CEM Gruber, A Di Caro, MR Capobianchi |
| EPI_ISL_1169887 | San Gallicano Dermatological Institute I.F.O. | INMI Lazzaro Spallanzani IRCCS | E Giombini, F Messina, M Rueca, O Butera, CEM Gruber, B Bartolini, F Pimpinelli, F Ensoli, EG Di Domenico, A Di Caro, MR Capobianchi |
| EPI_ISL_1169889 | San Gallicano Dermatological Institute I.F.O. | INMI Lazzaro Spallanzani IRCCS | CEM Gruber, B Bartolini, E Giombini, F Messina, M Rueca, O Butera, F Pimpinelli, F Ensoli, G D'Agosto, MR Capobianchi, A Di Caro |
| EPI_ISL_1169890 | Laboratorio Genzano - ASL RM 6 | INMI Lazzaro Spallanzani IRCCS | O Butera, CEM Gruber, B Bartolini, E Giombini, F Messina, M Rueca, G Tramini, E Conti, MR Capobianchi, A Di Caro |
| EPI_ISL_1169891 | Synlab Lazio S.r.l. | INMI Lazzaro Spallanzani IRCCS | F Messina, M Rueca, O Butera, CEM Gruber, B Bartolini, E Giombini, E Trappolini, SA Santini, MR Capobianchi, A Di Caro |
| EPI_ISL_1169892, EPI_ISL_1169893 | Università degli Studi di Perugia | Istituto Zooprofilattico Sperimentale dell'Abruzzo e Molise "G. Caporale" | Mencacci A., Camilloni B., Lorusso A, Marcacci M, Di Domenico M, Ancora M, Curini V, Mangone I, Rinaldi A, Scialabba S, Di Pasquale A, Cammà C, Puglia I, Calistri P, Savini G |
| EPI_ISL_1169894, EPI_ISL_1169895 | Istituto Zooprofilattico Sperimentale Umbria e Marche "Togo Rosati" | Istituto Zooprofilattico Sperimentale dell'Abruzzo e Molise "G. Caporale" | Biagetti M., Giammarioli M., Lorusso A, Marcacci M, Di Domenico M, Ancora M, Curini V, Mangone I, Rinaldi A, Scialabba S, Di Pasquale A, Cammà C, Puglia I, Calistri P, Savini G |
| EPI_ISL_1169905, EPI_ISL_1169906, EPI_ISL_1169907, EPI_ISL_1169908, EPI_ISL_1169909, EPI_ISL_1169910, EPI_ISL_1169911 | UOC Microbiologia e Virologia, Azienda Ospedaliera Universitaria Senese, Siena, Italy | Dipartimento di Biotecnologie Mediche | Maria Grazia Cusi, David Pinzauti, Claudia Gandolfo, Gabriele Anichini, Gianni Pozzi, Gianni Gori Savellini, Francesco Santoro |
| EPI_ISL_1171675, EPI_ISL_1171676, EPI_ISL_1171677, EPI_ISL_1171678, EPI_ISL_1171679, EPI_ISL_1171680, EPI_ISL_1171681, EPI_ISL_1171682, EPI_ISL_1171683, EPI_ISL_1171684, EPI_ISL_1171685, EPI_ISL_1171686, EPI_ISL_1171687, EPI_ISL_1171688, EPI_ISL_1171689, EPI_ISL_1171690, EPI_ISL_1171691, | | | |

| | | | |
|---|---|---|---|
| EPI_ISL_1171692, EPI_ISL_1171693, EPI_ISL_1171694, EPI_ISL_1171695, EPI_ISL_1171696, EPI_ISL_1171697, EPI_ISL_1171698, EPI_ISL_1171699, EPI_ISL_1171700, EPI_ISL_1171701, EPI_ISL_1171702, EPI_ISL_1171703, EPI_ISL_1171704, EPI_ISL_1171705 | | | |
| see above | UOC LAB ANALISI PO CARDARELLI CAMPOBASSO | Istituto Zooprofilattico Sperimentale dell'Abruzzo e Molise "G. Caporale" | Scutellà M, Niro G, Felice V. Lorusso A, Marcacci M, Di Domenico M, Ancora M, Curini V, Mangone I, Rinaldi A, Scialabba S, Di Pasquale A, Cammà C, Puglia I, Calistri P, Savini G |
| EPI_ISL_1173202 | Laboratorio Biologia Molecolare Sars Cov2 - UOC Laboratorio Analisi - Servizio Medicina di Laboratorio, Ospedale "San Francesco" - ATS-ASSL Nuoro | Laboratorio Specialistico Ematologia, UOC Ematologia, Ospedale "San Francesco" - ATS-ASSL Nuoro | Pira Giovanna, Rosu Valentina, Fancello Patrizia, Asproni Rosanna, Carta Franco, Serra Silvia, Piras Giovanna, Monne Maria Itria, Malune Paolo, Fiamma Maura, Palmas Angelo Domenico, Lo Maglio Iana, Mameli Giuseppe |
| EPI_ISL_1173258, EPI_ISL_1173259, EPI_ISL_1173260, EPI_ISL_1173261, EPI_ISL_1173262, EPI_ISL_1173263, EPI_ISL_1173264, EPI_ISL_1173265, EPI_ISL_1173266, EPI_ISL_1173267, EPI_ISL_1173268, EPI_ISL_1173269, EPI_ISL_1173270, EPI_ISL_1173271, EPI_ISL_1173272, EPI_ISL_1173273, EPI_ISL_1173274, EPI_ISL_1173275, EPI_ISL_1173276, EPI_ISL_1173277, EPI_ISL_1173278, EPI_ISL_1173279, EPI_ISL_1173280, EPI_ISL_1173281, EPI_ISL_1173282, EPI_ISL_1173283, EPI_ISL_1173284, EPI_ISL_1173285, EPI_ISL_1173286, EPI_ISL_1173287, EPI_ISL_1173288, EPI_ISL_1173289, EPI_ISL_1173290, EPI_ISL_1173291, EPI_ISL_1173292, EPI_ISL_1173293, EPI_ISL_1173294, EPI_ISL_1173295, EPI_ISL_1173296, EPI_ISL_1173297, EPI_ISL_1173298, EPI_ISL_1173299, EPI_ISL_1173300, EPI_ISL_1173301, EPI_ISL_1173302, EPI_ISL_1173303, EPI_ISL_1173304, EPI_ISL_1173305, EPI_ISL_1173306, EPI_ISL_1173307, EPI_ISL_1173308, EPI_ISL_1173309, EPI_ISL_1173310, EPI_ISL_1173311, EPI_ISL_1173312, EPI_ISL_1173313, EPI_ISL_1173314, EPI_ISL_1173315, EPI_ISL_1173316, EPI_ISL_1173317, EPI_ISL_1173318, EPI_ISL_1173319, EPI_ISL_1173320, EPI_ISL_1173321, EPI_ISL_1173322, EPI_ISL_1173323, EPI_ISL_1173324, EPI_ISL_1173325, EPI_ISL_1173326, EPI_ISL_1173327, EPI_ISL_1173328, EPI_ISL_1173329, EPI_ISL_1173330, EPI_ISL_1173331, EPI_ISL_1173332, EPI_ISL_1173333, EPI_ISL_1173334, EPI_ISL_1173335, EPI_ISL_1173336, EPI_ISL_1173337, EPI_ISL_1173338, EPI_ISL_1173339, EPI_ISL_1173340, EPI_ISL_1173341, EPI_ISL_1173342, EPI_ISL_1173343, EPI_ISL_1173344, EPI_ISL_1173345, EPI_ISL_1173346, EPI_ISL_1173347, EPI_ISL_1173348, EPI_ISL_1173349, EPI_ISL_1173350, EPI_ISL_1173351, EPI_ISL_1173352, EPI_ISL_1173353, EPI_ISL_1173354, EPI_ISL_1173356, EPI_ISL_1173357, EPI_ISL_1173358, EPI_ISL_1173359, EPI_ISL_1173360, EPI_ISL_1173361, EPI_ISL_1173362, EPI_ISL_1173363, EPI_ISL_1173364, EPI_ISL_1173365, EPI_ISL_1173366, EPI_ISL_1173367, EPI_ISL_1173368, EPI_ISL_1173369, EPI_ISL_1173370, EPI_ISL_1173371, EPI_ISL_1173372, EPI_ISL_1173373, EPI_ISL_1173374, EPI_ISL_1173375, EPI_ISL_1173376, EPI_ISL_1173377, EPI_ISL_1173378, EPI_ISL_1173379, EPI_ISL_1173380, EPI_ISL_1173381, EPI_ISL_1173382, EPI_ISL_1173383, EPI_ISL_1173384, EPI_ISL_1173385, EPI_ISL_1173386, EPI_ISL_1173387, EPI_ISL_1173388, EPI_ISL_1173389, EPI_ISL_1173390, EPI_ISL_1173391, EPI_ISL_1173392, EPI_ISL_1173393, EPI_ISL_1173394, EPI_ISL_1173395, EPI_ISL_1173396, EPI_ISL_1173397, EPI_ISL_1173398, EPI_ISL_1173399, EPI_ISL_1173400, EPI_ISL_1173401, EPI_ISL_1173402, EPI_ISL_1173403, EPI_ISL_1173404, EPI_ISL_1173405, EPI_ISL_1173406, EPI_ISL_1173407, EPI_ISL_1173408, EPI_ISL_1173409, EPI_ISL_1173410, EPI_ISL_1173411, EPI_ISL_1173412, EPI_ISL_1173413, EPI_ISL_1173414, EPI_ISL_1173415, EPI_ISL_1173416, EPI_ISL_1173417, EPI_ISL_1173418, EPI_ISL_1173419, EPI_ISL_1173420, EPI_ISL_1173421, EPI_ISL_1173422, EPI_ISL_1173423, EPI_ISL_1173424, EPI_ISL_1173425, EPI_ISL_1173426, EPI_ISL_1173427, EPI_ISL_1173428, EPI_ISL_1173429, EPI_ISL_1173430, EPI_ISL_1173431, EPI_ISL_1173432, EPI_ISL_1173433, EPI_ISL_1173434, EPI_ISL_1173435, EPI_ISL_1173436, EPI_ISL_1173437, EPI_ISL_1173438, EPI_ISL_1173439, EPI_ISL_1173440, EPI_ISL_1173441, EPI_ISL_1173442, EPI_ISL_1173443, EPI_ISL_1173444, EPI_ISL_1173445, EPI_ISL_1173446, EPI_ISL_1173447, EPI_ISL_1173448, EPI_ISL_1173449, EPI_ISL_1173450, EPI_ISL_1173451, EPI_ISL_1173452, EPI_ISL_1173453, EPI_ISL_1173454, EPI_ISL_1173455, EPI_ISL_1173456, EPI_ISL_1173457, EPI_ISL_1173458, EPI_ISL_1173459, EPI_ISL_1173460, EPI_ISL_1173461, EPI_ISL_1173462, EPI_ISL_1173463, EPI_ISL_1173464, EPI_ISL_1173465, EPI_ISL_1173466, EPI_ISL_1173467, EPI_ISL_1173468, EPI_ISL_1173469, EPI_ISL_1173470, EPI_ISL_1173471, EPI_ISL_1173472, EPI_ISL_1173473, EPI_ISL_1173474, EPI_ISL_1173475, EPI_ISL_1173476, EPI_ISL_1173477, EPI_ISL_1173478, EPI_ISL_1173479, EPI_ISL_1173480, EPI_ISL_1173481, EPI_ISL_1173482, EPI_ISL_1173483, EPI_ISL_1173484, EPI_ISL_1173485, EPI_ISL_1173486, EPI_ISL_1173487, EPI_ISL_1173488, EPI_ISL_1173489, EPI_ISL_1173490, EPI_ISL_1173491, EPI_ISL_1173492, EPI_ISL_1173493, EPI_ISL_1173494, EPI_ISL_1173495, EPI_ISL_1173496, EPI_ISL_1173497, EPI_ISL_1173498, EPI_ISL_1173499, EPI_ISL_1173500, EPI_ISL_1173501, EPI_ISL_1173502, EPI_ISL_1173503, EPI_ISL_1173504 | | | |
| see above | Istituto Zooprofilattico Sperimentale del Mezzogiorno (IZSM) | Telethon Institute of Genetics and Medicine (TIGEM) | Antonio Grimaldi Patrizia Annunziata Francesco Panariello Biancamaria Pierri Valentina Bouche Chiara Colantuono Maria Concetta Cuomo Denise Di Concilio Lucio Di Filippo Anna Manfredi Marcello Salvi Antonio Limone Pellegrino Cerino Andrea Ballabio Davide Cacchiarelli |
| EPI_ISL_1173519, EPI_ISL_1173520, EPI_ISL_1173521, EPI_ISL_1173522, EPI_ISL_1173523, EPI_ISL_1173524, EPI_ISL_1173525, EPI_ISL_1173526, EPI_ISL_1173527, EPI_ISL_1173528, EPI_ISL_1173529, EPI_ISL_1173530, EPI_ISL_1173531, EPI_ISL_1173532, EPI_ISL_1173533, EPI_ISL_1173534, EPI_ISL_1173535, EPI_ISL_1173536, EPI_ISL_1173537, EPI_ISL_1173538, EPI_ISL_1173539, EPI_ISL_1173540, EPI_ISL_1173541, EPI_ISL_1173542, EPI_ISL_1173543, EPI_ISL_1173544, EPI_ISL_1173545, EPI_ISL_1173546, EPI_ISL_1173547, EPI_ISL_1173548, EPI_ISL_1173549, EPI_ISL_1173550, EPI_ISL_1173551, EPI_ISL_1173552, EPI_ISL_1173553, EPI_ISL_1173554, EPI_ISL_1173555, EPI_ISL_1173556, EPI_ISL_1173557, EPI_ISL_1173558, EPI_ISL_1173559, EPI_ISL_1173560, EPI_ISL_1173561, EPI_ISL_1173562, EPI_ISL_1173563, EPI_ISL_1173564, EPI_ISL_1173565, EPI_ISL_1173566, EPI_ISL_1173567, EPI_ISL_1173568, EPI_ISL_1173569, EPI_ISL_1173570, EPI_ISL_1173571, EPI_ISL_1173572, EPI_ISL_1173573, EPI_ISL_1173574, EPI_ISL_1173575, EPI_ISL_1173576, EPI_ISL_1173577, EPI_ISL_1173578, EPI_ISL_1173579, EPI_ISL_1173580, EPI_ISL_1173581, EPI_ISL_1173582, EPI_ISL_1173583, EPI_ISL_1173584, EPI_ISL_1173585, EPI_ISL_1173586, EPI_ISL_1173587, EPI_ISL_1173588, EPI_ISL_1173589, EPI_ISL_1173590, EPI_ISL_1173591, EPI_ISL_1173592, EPI_ISL_1173593, EPI_ISL_1173594, EPI_ISL_1173595, EPI_ISL_1173596, EPI_ISL_1173597, EPI_ISL_1173598, EPI_ISL_1173599, EPI_ISL_1173600, EPI_ISL_1173601, EPI_ISL_1173602, EPI_ISL_1173603, EPI_ISL_1173604, EPI_ISL_1173605, EPI_ISL_1173606, EPI_ISL_1173607, EPI_ISL_1173608, EPI_ISL_1173609, EPI_ISL_1173610, EPI_ISL_1173611, EPI_ISL_1173612, EPI_ISL_1173613, EPI_ISL_1173614, EPI_ISL_1173615, EPI_ISL_1173616, EPI_ISL_1173617, EPI_ISL_1173618, EPI_ISL_1173619, EPI_ISL_1173620, EPI_ISL_1173621, EPI_ISL_1173622, EPI_ISL_1173623, EPI_ISL_1173624, EPI_ISL_1173625, EPI_ISL_1173626, EPI_ISL_1173627, EPI_ISL_1173628, EPI_ISL_1173629, EPI_ISL_1173630, EPI_ISL_1173631, EPI_ISL_1173632, EPI_ISL_1173633, EPI_ISL_1173634, EPI_ISL_1173635, EPI_ISL_1173636, EPI_ISL_1173637, EPI_ISL_1173638, EPI_ISL_1173639, EPI_ISL_1173640, EPI_ISL_1173641, EPI_ISL_1173642, EPI_ISL_1173643, EPI_ISL_1173644, EPI_ISL_1173645, EPI_ISL_1173646, EPI_ISL_1173647, EPI_ISL_1173648, EPI_ISL_1173649, EPI_ISL_1173650, EPI_ISL_1173651, EPI_ISL_1173652, EPI_ISL_1173653, EPI_ISL_1173654, EPI_ISL_1173655, EPI_ISL_1173656, EPI_ISL_1173657, EPI_ISL_1173658, EPI_ISL_1173659, EPI_ISL_1173660, EPI_ISL_1173661, EPI_ISL_1173662, EPI_ISL_1173663, EPI_ISL_1173664, EPI_ISL_1173665, EPI_ISL_1173666, EPI_ISL_1173667, EPI_ISL_1173668, EPI_ISL_1173669, EPI_ISL_1173670, EPI_ISL_1173671, EPI_ISL_1173672, EPI_ISL_1173673, EPI_ISL_1173674, EPI_ISL_1173675, EPI_ISL_1173676, EPI_ISL_1173677, EPI_ISL_1173678, EPI_ISL_1173679, EPI_ISL_1173680, EPI_ISL_1173681, EPI_ISL_1173682, EPI_ISL_1173683, EPI_ISL_1173684, EPI_ISL_1173685, EPI_ISL_1173686, EPI_ISL_1173687, EPI_ISL_1173688, EPI_ISL_1173689, EPI_ISL_1173690, EPI_ISL_1173691, EPI_ISL_1173692, EPI_ISL_1173693, EPI_ISL_1173694, EPI_ISL_1173695, EPI_ISL_1173696, EPI_ISL_1173697, EPI_ISL_1173698, EPI_ISL_1173699, EPI_ISL_1173700, EPI_ISL_1173701, EPI_ISL_1173702, EPI_ISL_1173703, EPI_ISL_1173704, EPI_ISL_1173705, EPI_ISL_1173706, EPI_ISL_1173707, EPI_ISL_1173708, EPI_ISL_1173709, EPI_ISL_1173710, EPI_ISL_1173711, EPI_ISL_1173712, EPI_ISL_1173713, EPI_ISL_1173714, EPI_ISL_1173715, EPI_ISL_1173716, EPI_ISL_1173717, EPI_ISL_1173718, EPI_ISL_1173719, EPI_ISL_1173720, EPI_ISL_1173721, EPI_ISL_1173722, EPI_ISL_1173723, EPI_ISL_1173724, EPI_ISL_1173725, EPI_ISL_1173726, EPI_ISL_1173727, EPI_ISL_1173728, EPI_ISL_1173729, EPI_ISL_1173730, EPI_ISL_1173731, EPI_ISL_1173732, EPI_ISL_1173733, EPI_ISL_1173734, EPI_ISL_1173735, EPI_ISL_1173736, EPI_ISL_1173737, EPI_ISL_1173738, EPI_ISL_1173739, EPI_ISL_1173740, EPI_ISL_1173741, EPI_ISL_1173742, EPI_ISL_1173743, EPI_ISL_1173744, EPI_ISL_1173745, EPI_ISL_1173746, EPI_ISL_1173747, EPI_ISL_1173748, EPI_ISL_1173749, EPI_ISL_1173750, EPI_ISL_1173751, EPI_ISL_1173752, EPI_ISL_1173753, EPI_ISL_1173754, EPI_ISL_1173755, EPI_ISL_1173756, EPI_ISL_1173757, EPI_ISL_1173758, EPI_ISL_1173759, EPI_ISL_1173760, EPI_ISL_1173761, EPI_ISL_1173762, EPI_ISL_1173763, EPI_ISL_1173764 | | | |
| see above | IZSM | TIGEM | Antonio Grimaldi Patrizia Annunziata Francesco Panariello Biancamaria Pierri Valentina Bouche Chiara Colantuono Maria Concetta Cuomo Denise Di Concilio Lucio Di Filippo Anna Manfredi Marcello Salvi Antonio Limone Pellegrino Cerino Andrea Ballabio Davide Cacchiarelli |
| EPI_ISL_1173769, EPI_ISL_1173770, EPI_ISL_1173771, EPI_ISL_1173772, EPI_ISL_1173773, EPI_ISL_1173774, EPI_ISL_1173775, EPI_ISL_1173776, EPI_ISL_1173777, EPI_ISL_1173778, EPI_ISL_1173779, EPI_ISL_1173780, EPI_ISL_1173781, EPI_ISL_1173784, EPI_ISL_1173787, EPI_ISL_1173789, EPI_ISL_1173792, EPI_ISL_1173795, EPI_ISL_1173798, EPI_ISL_1173801, EPI_ISL_1173804, EPI_ISL_1173808, EPI_ISL_1173811, EPI_ISL_1173814, EPI_ISL_1173817, EPI_ISL_1173820, EPI_ISL_1173822, EPI_ISL_1173825, EPI_ISL_1173828, EPI_ISL_1173832, EPI_ISL_1173835, EPI_ISL_1173838, EPI_ISL_1173840, EPI_ISL_1173843, EPI_ISL_1173847, EPI_ISL_1173850, EPI_ISL_1173853, EPI_ISL_1173856, EPI_ISL_1173859, EPI_ISL_1173862, EPI_ISL_1173865, EPI_ISL_1173868, EPI_ISL_1173871, EPI_ISL_1173874, EPI_ISL_1173876, EPI_ISL_1173879, EPI_ISL_1173882, EPI_ISL_1173886, EPI_ISL_1173889, EPI_ISL_1173891, EPI_ISL_1173895, EPI_ISL_1173897, EPI_ISL_1173901, EPI_ISL_1173904, EPI_ISL_1173907, EPI_ISL_1173910, EPI_ISL_1173914, EPI_ISL_1173916, EPI_ISL_1173919, EPI_ISL_1173922, EPI_ISL_1173925, EPI_ISL_1173928, EPI_ISL_1173931, EPI_ISL_1173935, EPI_ISL_1173938, EPI_ISL_1173940, EPI_ISL_1173943, EPI_ISL_1173946, EPI_ISL_1173950, EPI_ISL_1173953, EPI_ISL_1173956, EPI_ISL_1173959, EPI_ISL_1173962, EPI_ISL_1173965, EPI_ISL_1173968, EPI_ISL_1173974, EPI_ISL_1173985, EPI_ISL_1173988, EPI_ISL_1173991, EPI_ISL_1174004, EPI_ISL_1174007 | | | |
| see above | Università Federico II - Dipartimento di scienze mediche traslazionali - Napoli | TIGEM | Antonio Grimaldi Patrizia Annunziata Francesco Panariello Michele Cennamo Valentina Bouche Chiara Colantuono Lucio Di Filippo Mariano Fiorenza Anna Manfredi Marcello Salvi Giuseppe Portella Andrea Ballabio Davide Cacchiarelli |
| EPI_ISL_1175969, EPI_ISL_1175970, EPI_ISL_1175971, EPI_ISL_1175972, EPI_ISL_1175973, EPI_ISL_1175974, EPI_ISL_1175975, EPI_ISL_1175976, EPI_ISL_1175977, EPI_ISL_1175978, EPI_ISL_1175979, EPI_ISL_1175980, EPI_ISL_1175981, EPI_ISL_1175982, EPI_ISL_1175983, EPI_ISL_1175984, EPI_ISL_1175985, EPI_ISL_1175986, EPI_ISL_1175987, EPI_ISL_1175988, EPI_ISL_1175989, EPI_ISL_1175990, EPI_ISL_1175991, EPI_ISL_1175992, EPI_ISL_1175993, EPI_ISL_1175994, EPI_ISL_1175995, EPI_ISL_1175996, EPI_ISL_1175997, EPI_ISL_1175998, EPI_ISL_1175999, EPI_ISL_1176000, EPI_ISL_1176001, EPI_ISL_1176002, EPI_ISL_1176003, EPI_ISL_1176004, EPI_ISL_1176005, EPI_ISL_1176006, EPI_ISL_1176007, EPI_ISL_1176008, EPI_ISL_1176009, EPI_ISL_1176010, EPI_ISL_1176011, EPI_ISL_1176012, EPI_ISL_1176013, EPI_ISL_1176014, EPI_ISL_1176015, EPI_ISL_1176016, EPI_ISL_1176017, EPI_ISL_1176018, EPI_ISL_1176019, EPI_ISL_1176020, EPI_ISL_1176021, EPI_ISL_1176022, EPI_ISL_1176023, EPI_ISL_1176024, EPI_ISL_1176025, EPI_ISL_1176026, EPI_ISL_1176027, EPI_ISL_1176028, EPI_ISL_1176029, EPI_ISL_1176030, EPI_ISL_1176031, EPI_ISL_1176032, EPI_ISL_1176033, EPI_ISL_1176034, EPI_ISL_1176035, EPI_ISL_1176036, EPI_ISL_1176037, EPI_ISL_1176038, EPI_ISL_1176039, EPI_ISL_1176040, EPI_ISL_1176041, EPI_ISL_1176042, EPI_ISL_1176043, EPI_ISL_1176044, EPI_ISL_1176045, EPI_ISL_1176046, EPI_ISL_1176047, EPI_ISL_1176048, EPI_ISL_1176049, EPI_ISL_1176050, EPI_ISL_1176051, EPI_ISL_1176052, EPI_ISL_1176053, EPI_ISL_1176054, EPI_ISL_1176055, EPI_ISL_1176056, EPI_ISL_1176057, EPI_ISL_1176058, EPI_ISL_1176059, EPI_ISL_1176060, EPI_ISL_1176061, EPI_ISL_1176062, EPI_ISL_1176063, EPI_ISL_1176064, EPI_ISL_1176065, EPI_ISL_1176066, EPI_ISL_1176067, EPI_ISL_1176068, EPI_ISL_1176069, EPI_ISL_1176070, EPI_ISL_1176071, EPI_ISL_1176072, EPI_ISL_1176073, EPI_ISL_1176074, EPI_ISL_1176075, EPI_ISL_1176076, EPI_ISL_1176077, EPI_ISL_1176078, EPI_ISL_1176079, EPI_ISL_1176080, EPI_ISL_1176081, EPI_ISL_1176082, EPI_ISL_1176083, EPI_ISL_1176084, EPI_ISL_1176085, EPI_ISL_1176086, EPI_ISL_1176087, EPI_ISL_1176088, EPI_ISL_1176089, EPI_ISL_1176090, EPI_ISL_1176091, EPI_ISL_1176092, EPI_ISL_1176093, EPI_ISL_1176094, EPI_ISL_1176095, EPI_ISL_1176096, EPI_ISL_1176097, EPI_ISL_1176098, EPI_ISL_1176099, EPI_ISL_1176100, EPI_ISL_1176101, EPI_ISL_1176102, EPI_ISL_1176103, EPI_ISL_1176104, EPI_ISL_1176105, EPI_ISL_1176106, EPI_ISL_1176107, EPI_ISL_1176108, EPI_ISL_1176109, EPI_ISL_1176110, EPI_ISL_1176111, EPI_ISL_1176112, EPI_ISL_1176113, EPI_ISL_1176114, EPI_ISL_1176115, EPI_ISL_1176116, EPI_ISL_1176117, EPI_ISL_1176118, EPI_ISL_1176119, EPI_ISL_1176120, EPI_ISL_1176121, EPI_ISL_1176122, EPI_ISL_1176123, EPI_ISL_1176124, EPI_ISL_1176125, EPI_ISL_1176126, EPI_ISL_1176127, EPI_ISL_1176128, EPI_ISL_1176129, EPI_ISL_1176130, EPI_ISL_1176131, EPI_ISL_1176132, EPI_ISL_1176133, EPI_ISL_1176134, EPI_ISL_1176135, EPI_ISL_1176136, EPI_ISL_1176137, EPI_ISL_1176138, EPI_ISL_1176139, EPI_ISL_1176140, EPI_ISL_1176141, EPI_ISL_1176142, EPI_ISL_1176143, EPI_ISL_1176144, EPI_ISL_1176145, EPI_ISL_1176146, EPI_ISL_1176147, EPI_ISL_1176148, EPI_ISL_1176149, EPI_ISL_1176150, EPI_ISL_1176151, EPI_ISL_1176152, EPI_ISL_1176153, EPI_ISL_1176154, EPI_ISL_1176155, EPI_ISL_1176156, EPI_ISL_1176157, EPI_ISL_1176158, EPI_ISL_1176159, EPI_ISL_1176160, EPI_ISL_1176161, EPI_ISL_1176162, EPI_ISL_1176163, EPI_ISL_1176164, EPI_ISL_1176165, EPI_ISL_1176166, EPI_ISL_1176167, EPI_ISL_1176168, EPI_ISL_1176169, EPI_ISL_1176170, EPI_ISL_1176171, EPI_ISL_1176172, EPI_ISL_1176173, EPI_ISL_1176174, EPI_ISL_1176175, EPI_ISL_1176176, EPI_ISL_1176177, EPI_ISL_1176178, EPI_ISL_1176179, EPI_ISL_1176180, EPI_ISL_1176181, EPI_ISL_1176182, EPI_ISL_1176183, EPI_ISL_1176184, EPI_ISL_1176185, EPI_ISL_1176186, EPI_ISL_1176187, EPI_ISL_1176188, EPI_ISL_1176189, EPI_ISL_1176190, EPI_ISL_1176191, EPI_ISL_1176192, EPI_ISL_1176193, EPI_ISL_1176194, EPI_ISL_1176195, EPI_ISL_1176196, EPI_ISL_1176197, EPI_ISL_1176198, EPI_ISL_1176199, EPI_ISL_1176200, EPI_ISL_1176201, EPI_ISL_1176202, EPI_ISL_1176203, EPI_ISL_1176204, EPI_ISL_1176205, EPI_ISL_1176206, EPI_ISL_1176207, EPI_ISL_1176208, EPI_ISL_1176209, EPI_ISL_1176210, EPI_ISL_1176211, EPI_ISL_1176212, EPI_ISL_1176213, EPI_ISL_1176214, EPI_ISL_1176215, EPI_ISL_1176216, EPI_ISL_1176217, EPI_ISL_1176218, EPI_ISL_1176219, EPI_ISL_1176220, EPI_ISL_1176221, EPI_ISL_1176222, EPI_ISL_1176223, EPI_ISL_1176224, EPI_ISL_1176225, EPI_ISL_1176226, EPI_ISL_1176227, EPI_ISL_1176228, EPI_ISL_1176229, EPI_ISL_1176230, EPI_ISL_1176231, EPI_ISL_1176232, EPI_ISL_1176233, EPI_ISL_1176234, EPI_ISL_1176235, EPI_ISL_1176236, EPI_ISL_1176237, EPI_ISL_1176238, EPI_ISL_1176239, EPI_ISL_1176240, EPI_ISL_1176241, EPI_ISL_1176242, EPI_ISL_1176243, EPI_ISL_1176244, EPI_ISL_1176245, EPI_ISL_1176246, EPI_ISL_1176247, EPI_ISL_1176248, EPI_ISL_1176249, EPI_ISL_1176250, EPI_ISL_1176251, EPI_ISL_1176252, EPI_ISL_1176253, EPI_ISL_1176254, EPI_ISL_1176255, EPI_ISL_1176256, EPI_ISL_1176257, EPI_ISL_1176258, EPI_ISL_1176259, EPI_ISL_1176260, EPI_ISL_1176261, EPI_ISL_1176262, EPI_ISL_1176263, EPI_ISL_1176264, EPI_ISL_1176265, EPI_ISL_1176266, EPI_ISL_1176267, EPI_ISL_1176268, EPI_ISL_1176269, EPI_ISL_1176270, EPI_ISL_1176271, EPI_ISL_1176272, EPI_ISL_1176273, EPI_ISL_1176274, EPI_ISL_1176275, EPI_ISL_1176276, EPI_ISL_1176277, EPI_ISL_1176278, EPI_ISL_1176279, EPI_ISL_1176280, EPI_ISL_1176281, EPI_ISL_1176282, EPI_ISL_1176283, EPI_ISL_1176284, EPI_ISL_1176285, EPI_ISL_1176286, EPI_ISL_1176287, EPI_ISL_1176288, EPI_ISL_1176289, EPI_ISL_1176290, EPI_ISL_1176291, EPI_ISL_1176292, EPI_ISL_1176293, EPI_ISL_1176294, EPI_ISL_1176295, EPI_ISL_1176296 | | | |
| see above | IZSM | TIGEM | Antonio Grimaldi Patrizia Annunziata Francesco Panariello Biancamaria Pierri Valentina Bouche Chiara Colantuono Maria Concetta Cuomo Denise Di Concilio Lucio Di Filippo Anna Manfredi Marcello Salvi Antonio Limone Pellegrino Cerino Andrea Ballabio Davide Cacchiarelli |
| EPI_ISL_1178582 | Department of Infectious Diseases, Istituto Superiore di Sanità, Rome, Italy; Università degli Studi di Perugia, Perugia, Italy | Istituto Superiore di Sanità (ISS) | Paola Stefanelli, Alessandra Lo Presti, Angela Di Martino, Stefano Fiore, Antonella Mencacci, Barbara Camilloni, Manuela Marra, Maria Carollo, Marco Crescenzi |
| EPI_ISL_1180141, EPI_ISL_1180142 | Laboratorio Biologia Molecolare Sars Cov2 - UOC Laboratorio Analisi - Servizio Medicina di Laboratorio, Ospedale "San Francesco" - ATS-ASSL Nuoro | Laboratorio specialistico UOC Ematologia - Ospedale "San Francesco" - ATS-ASSL Nuoro | Giovanna Piras, Rosanna Asproni, Paolo Malune, Maria Itria Monne, Maura Fiamma, Angelo Domenico Palmas, Iana Lo Maglio, Giuseppe Mameli |
| EPI_ISL_1181341, EPI_ISL_1181342, EPI_ISL_1181343 | U.O. Genomics, S.S. Genetics and Advanced Omics Techniques, Istituto Zooprofilattico Sperimentale del Piemonte Liguria e Valle d'Aosta | U.O. Genomics, S.S. Genetics and Advanced Omics Techniques, Istituto Zooprofilattico Sperimentale del Piemonte Liguria e Valle d'Aosta | Chiara Beltramo, Elena Bozzetta, Pier Luigi Acutis, Simone Peletto |
| EPI_ISL_1181344 | Department of Infectious Diseases, Istituto Superiore di Sanità, Rome, Italy; AOR San Carlo, Potenza, Italy | Istituto Superiore di Sanità (ISS) | Paola Stefanelli, Angela Di Martino, Alessandra Lo Presti, Stefano Fiore, Antonio Picerno, Lopizzo Teresa, Anna Curci, Angela Menchise, Manuela Marra, Maria Carollo, Marco Crescenzi |
| EPI_ISL_1181346 | U.O. Genomics, S.S. Genetics and Advanced Omics | U.O. Genomics, S.S. Genetics and Advanced Omics | Chiara Beltramo, Elena Bozzetta, Pier Luigi Acutis, Simone Peletto |

| | | | |
|---|---|---|---|
| | Techniques, Istituto Zooprofilattico Sperimentale del Piemonte Liguria e Valle d'Aosta | Techniques, Istituto Zooprofilattico Sperimentale del Piemonte Liguria e Valle d'Aosta | |
| EPI_ISL_1181347, EPI_ISL_1181349 | Department of Infectious Diseases, Istituto Superiore di Sanità, Rome, Italy; U.O.S.D. Laboratorio Analisi Distretto 4 Ovest AULSS 9 Scaligera, Italy | Istituto Superiore di Sanità (ISS) | Paola Stefanelli, Angela Di Martino, Alessandra Lo Presti, Stefano Fiore, Maria Pompilia Visconti, Rosa Sforza, Manuela Marra, Maria Carollo, Marco Crescenzi |
| EPI_ISL_1181350 | U.O. Genomics, S.S. Genetics and Advanced Omics Techniques, Istituto Zooprofilattico Sperimentale del Piemonte Liguria e Valle d'Aosta | U.O. Genomics, S.S. Genetics and Advanced Omics Techniques, Istituto Zooprofilattico Sperimentale del Piemonte Liguria e Valle d'Aosta | Chiara Beltramo, Elena Bozzetta, Pier Luigi Acutis, Simone Peletto |
| EPI_ISL_1181352 | Department of Infectious Diseases, Istituto Superiore di Sanità, Rome, Italy; Azienda Ospedaliera Santa Maria di Terni, Terni, Italy | Istituto Superiore di Sanità (ISS) | Paola Stefanelli, Angela Di Martino, Alessandra Lo Presti, Stefano Fiore, Augusto Scaccetti, Michele Palumbo, Cinzia Di Giuli, Manuela Marra, Maria Carollo, Marco Crescenzi |
| EPI_ISL_1181417, EPI_ISL_1181625 | U.O. Genomics, S.S. Genetics and Advanced Omics Techniques, Istituto Zooprofilattico Sperimentale del Piemonte Liguria e Valle d'Aosta | U.O. Genomics, S.S. Genetics and Advanced Omics Techniques, Istituto Zooprofilattico Sperimentale del Piemonte Liguria e Valle d'Aosta | Chiara Beltramo, Elena Bozzetta, Pier Luigi Acutis, Simone Peletto |
| EPI_ISL_1181627 | Department of Infectious Diseases, Istituto Superiore di Sanità, Rome, Italy; Azienda Usl Toscana sud est, Arezzo, Italy | Istituto Superiore di Sanità (ISS) | Paola Stefanelli, Angela Di Martino, Alessandra Lo Presti, Stefano Fiore, Agostino Ognibene, Manuela Marra, Maria Carollo, Marco Crescenzi |
| EPI_ISL_1181933, EPI_ISL_1181934, EPI_ISL_1181935, EPI_ISL_1181936, EPI_ISL_1181937, EPI_ISL_1181938, EPI_ISL_1181939, EPI_ISL_1181940, EPI_ISL_1181941, EPI_ISL_1181942, EPI_ISL_1181943, EPI_ISL_1181944, EPI_ISL_1181945, EPI_ISL_1181946, EPI_ISL_1181947 | | | |
| see above | Azienda Ospedaliero - Universitaria di Modena Policlinico - Virologia e Microbiologia Molecolare | Istituto Zooprofilattico Sperimentale della Lombardia e dell'Emilia Romagna (IZSLER), Risk Analysis and Genomic Epidemiology Unit | Monica Pecorari, William Gennari, Giulia Fregni Serpini, Marina Morganti, Ilaria Menozzi, Erika Scaltriti, Stefano Pongolini |
| EPI_ISL_1185536, EPI_ISL_1185537, EPI_ISL_1185538, EPI_ISL_1185539, EPI_ISL_1185540, EPI_ISL_1185541, EPI_ISL_1185542, EPI_ISL_1185543, EPI_ISL_1185544, EPI_ISL_1185545, EPI_ISL_1185546, EPI_ISL_1185547, EPI_ISL_1185548, EPI_ISL_1185549, EPI_ISL_1185550, EPI_ISL_1185551, EPI_ISL_1185552, EPI_ISL_1185553, EPI_ISL_1185554, EPI_ISL_1185555, EPI_ISL_1185556, EPI_ISL_1185557, EPI_ISL_1185558, EPI_ISL_1185559, EPI_ISL_1185560, EPI_ISL_1185561, EPI_ISL_1185562, EPI_ISL_1185563, EPI_ISL_1185564, EPI_ISL_1185565, EPI_ISL_1185566, EPI_ISL_1185567, EPI_ISL_1185568, EPI_ISL_1185569, EPI_ISL_1185570, EPI_ISL_1185571, EPI_ISL_1185572, EPI_ISL_1185573, EPI_ISL_1185574, EPI_ISL_1185575, EPI_ISL_1185576, EPI_ISL_1185577, EPI_ISL_1185578, EPI_ISL_1185579, EPI_ISL_1185580, EPI_ISL_1185581, EPI_ISL_1185582, EPI_ISL_1185583, EPI_ISL_1185584, EPI_ISL_1185585, EPI_ISL_1185586, EPI_ISL_1185587, EPI_ISL_1185588, EPI_ISL_1185589, EPI_ISL_1185590, EPI_ISL_1185591, EPI_ISL_1185592, EPI_ISL_1185593, EPI_ISL_1185594, EPI_ISL_1185595, EPI_ISL_1185596, EPI_ISL_1185597, EPI_ISL_1185598, EPI_ISL_1185599, EPI_ISL_1185600, EPI_ISL_1185601, EPI_ISL_1185602, EPI_ISL_1185603, EPI_ISL_1185604, EPI_ISL_1185605, EPI_ISL_1185606, EPI_ISL_1185607, EPI_ISL_1185608, EPI_ISL_1185609, EPI_ISL_1185610, EPI_ISL_1185611, EPI_ISL_1185612, EPI_ISL_1185613, EPI_ISL_1185614, EPI_ISL_1185615, EPI_ISL_1185616, EPI_ISL_1185617, EPI_ISL_1185618, EPI_ISL_1185619, EPI_ISL_1185620, EPI_ISL_1185621, EPI_ISL_1185622, EPI_ISL_1185623, EPI_ISL_1185624, EPI_ISL_1185625, EPI_ISL_1185626, EPI_ISL_1185627, EPI_ISL_1185628, EPI_ISL_1185629, EPI_ISL_1185630, EPI_ISL_1185631, EPI_ISL_1185632, EPI_ISL_1185633, EPI_ISL_1185634, EPI_ISL_1185635, EPI_ISL_1185636, EPI_ISL_1185637, EPI_ISL_1185638, EPI_ISL_1185639, EPI_ISL_1185640, EPI_ISL_1185641, EPI_ISL_1185642, EPI_ISL_1185643, EPI_ISL_1185644, EPI_ISL_1185645, EPI_ISL_1185646, EPI_ISL_1185647, EPI_ISL_1185648, EPI_ISL_1185649, EPI_ISL_1185650, EPI_ISL_1185651, EPI_ISL_1185652, EPI_ISL_1185653, EPI_ISL_1185654, EPI_ISL_1185655, EPI_ISL_1185656, EPI_ISL_1185657, EPI_ISL_1185658, EPI_ISL_1185659, EPI_ISL_1185660, EPI_ISL_1185661, EPI_ISL_1185662, EPI_ISL_1185663, EPI_ISL_1185664, EPI_ISL_1185665, EPI_ISL_1185666, EPI_ISL_1185667, EPI_ISL_1185668, EPI_ISL_1185669, EPI_ISL_1185670, EPI_ISL_1185671, EPI_ISL_1185672, EPI_ISL_1185673, EPI_ISL_1185674, EPI_ISL_1185675, EPI_ISL_1185676, EPI_ISL_1185677, EPI_ISL_1185678, EPI_ISL_1185679, EPI_ISL_1185680, EPI_ISL_1185681, EPI_ISL_1185682, EPI_ISL_1185683, EPI_ISL_1185684, EPI_ISL_1185685, EPI_ISL_1185686, EPI_ISL_1185687, EPI_ISL_1185688, EPI_ISL_1185689, EPI_ISL_1185690, EPI_ISL_1185691, EPI_ISL_1185692, EPI_ISL_1185693, EPI_ISL_1185694, EPI_ISL_1185695, EPI_ISL_1185696, EPI_ISL_1185697, EPI_ISL_1185698, EPI_ISL_1185699, EPI_ISL_1185700, EPI_ISL_1185701, EPI_ISL_1185702, EPI_ISL_1185703, EPI_ISL_1185704, EPI_ISL_1185705, EPI_ISL_1185706, EPI_ISL_1185707, EPI_ISL_1185708, EPI_ISL_1185709, EPI_ISL_1185710, EPI_ISL_1185711, EPI_ISL_1185712, EPI_ISL_1185713, EPI_ISL_1185714, EPI_ISL_1185715, EPI_ISL_1185716, EPI_ISL_1185717, EPI_ISL_1185718, EPI_ISL_1185719, EPI_ISL_1185720, EPI_ISL_1185721, EPI_ISL_1185722, EPI_ISL_1185723, EPI_ISL_1185724, EPI_ISL_1185725, EPI_ISL_1185726, EPI_ISL_1185727, EPI_ISL_1185728, EPI_ISL_1185729, EPI_ISL_1185730, EPI_ISL_1185731, EPI_ISL_1185732, EPI_ISL_1185733, EPI_ISL_1185734, EPI_ISL_1185735, EPI_ISL_1185736, EPI_ISL_1185737, EPI_ISL_1185738, EPI_ISL_1185739, EPI_ISL_1185740, EPI_ISL_1185741, EPI_ISL_1185742, EPI_ISL_1185743, EPI_ISL_1185744, EPI_ISL_1185745, EPI_ISL_1185746, EPI_ISL_1185747, EPI_ISL_1185748, EPI_ISL_1185749, EPI_ISL_1185750, EPI_ISL_1185751, EPI_ISL_1185752, EPI_ISL_1185753, EPI_ISL_1185754, EPI_ISL_1185755, EPI_ISL_1185756, EPI_ISL_1185757, EPI_ISL_1185758, EPI_ISL_1185759, EPI_ISL_1185760, EPI_ISL_1185761, EPI_ISL_1185762, EPI_ISL_1185763, EPI_ISL_1185764, EPI_ISL_1185765, EPI_ISL_1185766, EPI_ISL_1185767, EPI_ISL_1185768, EPI_ISL_1185769, EPI_ISL_1185770, EPI_ISL_1185771, EPI_ISL_1185772 | | | |
| see above | IZSM | TIGEM | Antonio Grimaldi Patrizia Annunziata Francesco Panariello Biancamaria Pierri Valentina Bouche Chiara Colantuono Maria Concetta Cuomo Denise Di Concilio Lucio Di Filippo Anna Manfredi Marcello Salvi Antonio Limone Pellegrino Cerino Andrea Ballabio Davide Cacchiarelli |
| EPI_ISL_1185777, EPI_ISL_1185778, EPI_ISL_1185779, EPI_ISL_1185780, EPI_ISL_1185781, EPI_ISL_1185782, EPI_ISL_1185783, EPI_ISL_1185784, EPI_ISL_1185785, EPI_ISL_1185786, EPI_ISL_1185787, EPI_ISL_1185788, EPI_ISL_1185789, EPI_ISL_1185790, EPI_ISL_1185791, EPI_ISL_1185792, EPI_ISL_1185793, EPI_ISL_1185794, EPI_ISL_1185795, EPI_ISL_1185796, EPI_ISL_1185797, EPI_ISL_1185798, EPI_ISL_1185799, EPI_ISL_1185800, EPI_ISL_1185801, EPI_ISL_1185802, EPI_ISL_1185803, EPI_ISL_1185804, EPI_ISL_1185805, EPI_ISL_1185806, EPI_ISL_1185807, EPI_ISL_1185808, EPI_ISL_1185809, EPI_ISL_1185810, EPI_ISL_1185811, EPI_ISL_1185812, EPI_ISL_1185813, EPI_ISL_1185814, EPI_ISL_1185815, EPI_ISL_1185816, EPI_ISL_1185817, EPI_ISL_1185818, EPI_ISL_1185819, EPI_ISL_1185820, EPI_ISL_1185821, EPI_ISL_1185822, EPI_ISL_1185823, EPI_ISL_1185824, EPI_ISL_1185825, EPI_ISL_1185826, EPI_ISL_1185827, EPI_ISL_1185829, EPI_ISL_1185830, EPI_ISL_1185831, EPI_ISL_1185832, EPI_ISL_1185833, EPI_ISL_1185834, EPI_ISL_1185835, EPI_ISL_1185836, EPI_ISL_1185837, EPI_ISL_1185838, EPI_ISL_1185839, EPI_ISL_1185840, EPI_ISL_1185841, EPI_ISL_1185842, EPI_ISL_1185843, EPI_ISL_1185844, EPI_ISL_1185845, EPI_ISL_1185846 | | | |
| see above | Azienda Sanitaria dell'Alto Adige Laboratorio Aziendale di Microbiologia e Virologia | Istituto di Genomica Applicata | Elisabetta Pagani, Irene Bianconi, Elisabetta Giacobazzi, Elisa Masi, Stefanie Wieser, Irena Jurman, Vera Vendramin, Gabriele Magris, Eleonora Paparelli, Davide Scaglione, Michele Morgante |
| EPI_ISL_1186218, EPI_ISL_1186219, EPI_ISL_1186220, EPI_ISL_1186221, EPI_ISL_1186222, EPI_ISL_1186223, EPI_ISL_1186224, EPI_ISL_1186225, EPI_ISL_1186226, EPI_ISL_1186227, EPI_ISL_1186228, EPI_ISL_1186229, EPI_ISL_1186230, EPI_ISL_1186231, EPI_ISL_1186232, EPI_ISL_1186233, EPI_ISL_1186234, EPI_ISL_1186235, EPI_ISL_1186236, EPI_ISL_1186237, EPI_ISL_1186238, EPI_ISL_1186239, EPI_ISL_1186240, EPI_ISL_1186241, EPI_ISL_1186242, EPI_ISL_1186243, EPI_ISL_1186244, EPI_ISL_1186245, EPI_ISL_1186246, EPI_ISL_1186247 | | | |
| see above | IZSM | TIGEM | Antonio Grimaldi Patrizia Annunziata Francesco Panariello Biancamaria Pierri Valentina Bouche Chiara Colantuono Maria Concetta Cuomo Denise Di Concilio Lucio Di Filippo Anna Manfredi Marcello Salvi Antonio Limone Pellegrino Cerino Andrea Ballabio Davide Cacchiarelli |
| EPI_ISL_1191142, EPI_ISL_1191143, EPI_ISL_1191144, EPI_ISL_1191145 | Laboratorio Biologia Molecolare Sars Cov2 - UOC Laboratorio Analisi - Servizio Medicina di Laboratorio, Ospedale "San Francesco" - ATS-ASSL Nuoro | Laboratorio specialistico UOC Ematologia - Ospedale "San Francesco" - ATS-ASSL Nuoro | Piras Giovanna, Asproni Rosanna,,Fiamma Maura, Monne Maria Itria, Palmas Angelo Domenico, Lo Maglio Iana, Mameli Giuseppe |
| EPI_ISL_1195427, EPI_ISL_1195428, EPI_ISL_1195429, EPI_ISL_1195430, EPI_ISL_1195431, EPI_ISL_1195432, EPI_ISL_1195433, EPI_ISL_1195434, EPI_ISL_1195435, EPI_ISL_1195436, EPI_ISL_1195437, EPI_ISL_1195438, EPI_ISL_1195439, EPI_ISL_1195440, EPI_ISL_1195441, EPI_ISL_1195442, EPI_ISL_1195443, EPI_ISL_1195444, EPI_ISL_1195445, EPI_ISL_1195446, EPI_ISL_1195447, EPI_ISL_1195448, EPI_ISL_1195449, EPI_ISL_1195450, EPI_ISL_1195451, EPI_ISL_1195452, EPI_ISL_1195453, EPI_ISL_1195454, EPI_ISL_1195455, EPI_ISL_1195456, EPI_ISL_1195457, EPI_ISL_1195458, EPI_ISL_1195459, EPI_ISL_1195460, EPI_ISL_1195461, EPI_ISL_1195462, EPI_ISL_1195463, EPI_ISL_1195464, EPI_ISL_1195465, EPI_ISL_1195466, EPI_ISL_1195467, EPI_ISL_1195468, EPI_ISL_1195469, EPI_ISL_1195470, EPI_ISL_1195471, EPI_ISL_1195472, EPI_ISL_1195473, EPI_ISL_1195474, EPI_ISL_1195475, EPI_ISL_1195476 | | | |
| see above | SC Microbiologia e Virologia AOUSS | AMES Centro Polidiagnostico Strumentale S.r.l. | Gabriele Ibba, Rosangela Sau, Flavia Angioj, Marcello Abbondio, Salvatore Rubino, Sergio Uzzau. |
| EPI_ISL_1195950 | INMI Lazzaro Spallanzani IRCCS | INMI Lazzaro Spallanzani IRCCS | F Messina, M Rueca, O Butera, CEM Gruber, B Bartolini, E Giombini, MR Capobianchi, A Di Caro |
| EPI_ISL_1195951 | Laboratorio Genzano - ASL RM 9 | INMI Lazzaro Spallanzani IRCCS | F Messina, M Rueca, O Butera, CEM Gruber, B Bartolini, E Giombini, G Tramini, E Conti, MR Capobianchi, A Di Caro |
| EPI_ISL_1195952 | Laboratorio Genzano - ASL RM 10 | INMI Lazzaro Spallanzani IRCCS | M Rueca, O Butera, CEM Gruber, B Bartolini, E Giombini, F Messina, G Tramini, E Conti, A Di Caro, MR Capobianchi |
| EPI_ISL_1195953 | Ospedale "F. Spaziani" Frosinone | INMI Lazzaro Spallanzani IRCCS | O Butera, CEM Gruber, B Bartolini, E Giombini, F Messina, M Rueca, R Pulselli, C Gargiulo, C Sias, MR Capobianchi, A Di Caro |
| EPI_ISL_1195954 | Ospedale "F. Spaziani" Frosinone | INMI Lazzaro Spallanzani IRCCS | CEM Gruber, B Bartolini, E Giombini, F Messina, M Rueca, O Butera, R Pulselli, G Brocco, C Gargiulo, MR Capobianchi, A Di Caro |
| EPI_ISL_1195955 | San Gallicano Dermatological Institute I.F.O. | INMI Lazzaro Spallanzani IRCCS | M Rueca, O Butera, CEM Gruber, B Bartolini, E Giombini, F Messina, F Pimpinelli, F Ensoli, G Prignano, A Di Caro, MR Capobianchi |
| EPI_ISL_1195956 | San Gallicano Dermatological Institute I.F.O. | INMI Lazzaro Spallanzani IRCCS | O Butera, CEM Gruber, B Bartolini, E Giombini, F Messina, M Rueca, F Pimpinelli, F Ensoli, A Mastrofrancesco, MR Capobianchi, A Di Caro |
| EPI_ISL_1195957 | Laboratorio Genzano - ASL RM 6 | INMI Lazzaro Spallanzani IRCCS | B Bartolini, E Giombini, F Messina, M Rueca, O Butera, CEM Gruber, G Tramini, E Conti, A Di Caro, MR Capobianchi |
| EPI_ISL_1195958 | Laboratorio Genzano - ASL RM 8 | INMI Lazzaro Spallanzani IRCCS | E Giombini, F Messina, M Rueca, O Butera, CEM Gruber, B Bartolini, G Tramini, E Conti, A Di Caro, MR Capobianchi |
| EPI_ISL_1195959 | Ospedale "F. Spaziani" Frosinone | INMI Lazzaro Spallanzani IRCCS | E Giombini, F Messina, M Rueca, O Butera, CEM Gruber, B Bartolini, R Pulselli, C Gargiulo, C Sias, A Di Caro, MR Capobianchi |
| EPI_ISL_1195960 | Ospedale "F. Spaziani" Frosinone | INMI Lazzaro Spallanzani IRCCS | F Messina, M Rueca, O Butera, CEM Gruber, B Bartolini, E Giombini, R Pulselli, C Sias, G Brocco, MR Capobianchi, A Di Caro |
| EPI_ISL_1195961, EPI_ISL_1195962, EPI_ISL_1195963 | Microbiology and Virology Unit, Florence Careggi University Hospital | Microbiology and Virology Unit, Florence Careggi University Hospital | Vincenzo Di Pilato, Marco Coppi, Fabio Morecchiato, Noemi Aiezza, Ilaria Baccani, Nicla Giovacchini, Alberto Antonelli, Emanuele Gori, Gian Maria Rossolini |
| EPI_ISL_1196931, EPI_ISL_1196932, EPI_ISL_1196933, EPI_ISL_1196934, EPI_ISL_1196935, EPI_ISL_1196936, EPI_ISL_1196937, EPI_ISL_1196938, EPI_ISL_1196939, EPI_ISL_1196940, EPI_ISL_1196941, EPI_ISL_1196942, EPI_ISL_1196943, EPI_ISL_1196944, EPI_ISL_1196945, EPI_ISL_1196946, EPI_ISL_1196947, EPI_ISL_1196948, EPI_ISL_1196949, EPI_ISL_1196950, EPI_ISL_1196951, EPI_ISL_1196952, EPI_ISL_1196953, EPI_ISL_1196954, EPI_ISL_1196955, EPI_ISL_1196956, EPI_ISL_1196957, EPI_ISL_1196958, EPI_ISL_1196959, EPI_ISL_1196960, EPI_ISL_1196961, EPI_ISL_1196962, EPI_ISL_1196963, EPI_ISL_1196964, EPI_ISL_1196965, EPI_ISL_1196966, EPI_ISL_1196967, EPI_ISL_1196968, EPI_ISL_1196969, EPI_ISL_1196970, EPI_ISL_1196971, EPI_ISL_1196972, EPI_ISL_1196973, EPI_ISL_1196974, EPI_ISL_1196975, EPI_ISL_1196976, EPI_ISL_1196977, EPI_ISL_1196978, EPI_ISL_1196979, EPI_ISL_1196980, EPI_ISL_1196981, EPI_ISL_1196982, EPI_ISL_1196983, EPI_ISL_1196984, EPI_ISL_1196985, EPI_ISL_1196986, EPI_ISL_1196987, EPI_ISL_1196988, EPI_ISL_1196989, EPI_ISL_1196990, EPI_ISL_1196991, EPI_ISL_1196992, EPI_ISL_1196993, EPI_ISL_1196994, EPI_ISL_1196995, EPI_ISL_1196996, EPI_ISL_1196997, EPI_ISL_1196998, | | | |

| | | | |
|---|---|---|---|
| EPI_ISL_1196999, EPI_ISL_1197000, EPI_ISL_1197001, EPI_ISL_1197002, EPI_ISL_1197003, EPI_ISL_1197004, EPI_ISL_1197005, EPI_ISL_1197006, EPI_ISL_1197007, EPI_ISL_1197008, EPI_ISL_1197009, EPI_ISL_1197010, EPI_ISL_1197011, EPI_ISL_1197012, EPI_ISL_1197013, EPI_ISL_1197014, EPI_ISL_1197015, EPI_ISL_1197016, EPI_ISL_1197017, EPI_ISL_1197018, EPI_ISL_1197019, EPI_ISL_1197020, EPI_ISL_1197021, EPI_ISL_1197022, EPI_ISL_1197023, EPI_ISL_1197024, EPI_ISL_1197025, EPI_ISL_1197026, EPI_ISL_1197027, EPI_ISL_1197028, EPI_ISL_1197029, EPI_ISL_1197030, EPI_ISL_1197031, EPI_ISL_1197032, EPI_ISL_1197033, EPI_ISL_1197034, EPI_ISL_1197035, EPI_ISL_1197036 | | | |
| see above | IZSM | TIGEM | Antonio Grimaldi Patrizia Annunziata Francesco Panariello Biancamaria Pierri Valentina Bouche Chiara Colantuono Maria Concetta Cuomo Denise Di Concilio Lucio Di Filippo Anna Manfredi Marcello Salvi Antonio Limone Pellegrino Cerino Andrea Ballabio Davide Cacchiarelli |
| EPI_ISL_1197038, EPI_ISL_1197040 | Department of Infectious Diseases, Istituto Superiore di Sanità, Rome, Italy; Azienda Sanitaria Locale TO5, Torino, Italy | Istituto Superiore di Sanità (ISS) | Paola Stefanelli, Angela Di Martino, Alessandra Lo Presti, Stefano Fiore, Alessandra Allocco, Cagnoli Claudia, Manuela Marra, Maria Carollo, Marco Crescenzi |
| EPI_ISL_1198007 | Department of Infectious Diseases, Istituto Superiore di Sanità, Rome, Italy; Azienda Ospedaliera Santa Maria di Terni, Terni, Italy | Istituto Superiore di Sanità (ISS) | Paola Stefanelli, Angela Di Martino, Alessandra Lo Presti, Stefano Fiore, Augusto Scaccetti, Michele Palumbo, Cinzia Di Giuli, Manuela Marra, Maria Carollo, Marco Crescenzi |
| EPI_ISL_1198698 | INMI Lazzaro Spallanzani IRCCS | INMI Lazzaro Spallanzani IRCCS | E Giombini, F Messina, M Rueca, O Butera, CEM Gruber, B Bartolini, A Di Caro, MR Capobianchi |
| EPI_ISL_1198794, EPI_ISL_1198795, EPI_ISL_1198796 | Department of Infectious Diseases, Istituto Superiore di Sanità, Rome, Italy; U.O.S.D. Laboratorio Analisi Distretto 4 Ovest AULSS 9 Scaligera, Italy | Istituto Superiore di Sanità (ISS) | Paola Stefanelli, Angela Di Martino, Alessandra Lo Presti, Stefano Fiore, Maria Pompilia Visconti, Rosa Sforza, Manuela Marra, Maria Carollo, Marco Crescenzi |
| EPI_ISL_1198918, EPI_ISL_1199386, EPI_ISL_1199633 | Department of Infectious Diseases, Istituto Superiore di Sanità, Rome, Italy; L.C. Laboratori Campisi s.r.l., Siracusa, Sicily, Italy | Istituto Superiore di Sanità (ISS) | Paola Stefanelli, Angela Di Martino, Alessandra Lo Presti, Stefano Fiore, Campisi Corrado, Iemmolo Rosario, Manuela Marra, Maria Carollo, Marco Crescenzi |
| EPI_ISL_1199634, EPI_ISL_1199635 | Department of Infectious Diseases, Istituto Superiore di Sanità, Rome, Italy; Università degli Studi di Perugia, Perugia, Italy | Istituto Superiore di Sanità (ISS) | Paola Stefanelli, Alessandra Lo Presti, Angela Di Martino, Stefano Fiore, Antonella Mencacci, Barbara Camilloni, Manuela Marra, Maria Carollo, Marco Crescenzi |
| EPI_ISL_1199636 | Unità Operativa di Microbiologia e Virologia, Ospedale Civico e Di Cristina, ARNAS, Palermo, Italy | Tumor Immunology Unit, Department of Health Sciences, University of Palermo School of Medicine; Section of Microbiology, University of Palermo School of Medicine; National Research Council of Italy - High Performance Computing and Networking Institute (CNR-ICAR) of Palermo | Vacca Davide, Fiannaca Antonino, Bonura Celestino, Cancila Valeria, La Paglia Laura, Bonura Floriana, Morello Gaia, Massimo La Rosa, Di Bernardo Francesca, Alfonso Maurizio Urso, Tripodo Claudio, Craxi Antonio, Vitale Francesco, Giammanco Maurizio Giovanni |
| EPI_ISL_1199707, EPI_ISL_1200004 | Department of Infectious Diseases, Istituto Superiore di Sanità, Rome, Italy; Università degli Studi di Perugia, Perugia, Italy | Istituto Superiore di Sanità (ISS) | Paola Stefanelli, Alessandra Lo Presti, Angela Di Martino, Stefano Fiore, Antonella Mencacci, Barbara Camilloni, Manuela Marra, Maria Carollo, Marco Crescenzi |
| EPI_ISL_1200086 | Unità Operativa di Microbiologia e Virologia, Ospedale Civico e Di Cristina, ARNAS, Palermo, Italy | Tumor Immunology Unit, Department of Health Sciences, University of Palermo School of Medicine; Section of Microbiology, University of Palermo School of Medicine; National Research Council of Italy - High Performance Computing and Networking Institute (CNR-ICAR) of Palermo | Vacca Davide, Fiannaca Antonino, Bonura Celestino, Cancila Valeria, La Paglia Laura, Bonura Floriana, Morello Gaia, Massimo La Rosa, Di Bernardo Francesca, Alfonso Maurizio Urso, Tripodo Claudio, Craxi Antonio, Vitale Francesco, Giammanco Maurizio Giovanni |
| EPI_ISL_1200515, EPI_ISL_1200516, EPI_ISL_1200517 | Department of Infectious Diseases, Istituto Superiore di Sanità, Rome, Italy; Università degli Studi di Perugia, Perugia, Italy | Istituto Superiore di Sanità (ISS) | Paola Stefanelli, Alessandra Lo Presti, Angela Di Martino, Stefano Fiore, Antonella Mencacci, Barbara Camilloni, Manuela Marra, Maria Carollo, Marco Crescenzi |
| EPI_ISL_1200635, EPI_ISL_1200636, EPI_ISL_1200637 | Department of Infectious Diseases, Istituto Superiore di Sanità, Rome, Italy; AOR San Carlo, Potenza, Italy | Department of Infectious Diseases, Istituto Superiore di Sanità | Paola Stefanelli, Angela Di Martino, Alessandra Lo Presti, Stefano Fiore, Antonio Picerno, Lopizzo Teresa, Anna Curci, Angela Menchise, Manuela Marra, Maria Carollo, Marco Crescenzi |
| EPI_ISL_1201280, EPI_ISL_1201282, EPI_ISL_1201283, EPI_ISL_1201284 | Center of Advanced Studies and Technology, Molecular Genetics Laboratory | Center of Advanced Studies and Technology, Molecular Genetics Laboratory | Ferrante Rossella, Mandatori Domitilla, De Fabritiis Simone, Damiani Verena, Anaclerio Federico |
| EPI_ISL_1209466, EPI_ISL_1209467, EPI_ISL_1209468 | Unità Operativa di Microbiologia e Virologia, Ospedale Civico e Di Cristina, ARNAS, Palermo, Italy | Tumor Immunology Unit, Department of Health Sciences, University of Palermo School of Medicine; Section of Microbiology, University of Palermo School of Medicine; National Research Council of Italy - High Performance Computing and Networking Institute (CNR-ICAR) of Palermo | Vacca, Davide; Fiannaca, Antonino; Bonura, Celestino; Cancila, Valeria; La Paglia, Laura; Bonura, Floriana; Morello, Gaia; La Rosa, Massimo; Di Bernardo, Francesca; Urso, Alfonso Maurizio; Tripodo, Claudio; Craxi, Antonio; Vitale, Francesco; Giammanco, Maurizio Giovanni. |
| EPI_ISL_1210494 | INMI Lazzaro Spallanzani IRCCS | INMI Lazzaro Spallanzani IRCCS | B Bartolini, E Giombini, F Messina, M Rueca, O Butera, CEM Gruber, A Di Caro, MR Capobianchi |
| EPI_ISL_1210495 | INMI Lazzaro Spallanzani IRCCS | INMI Lazzaro Spallanzani IRCCS | CEM Gruber, B Bartolini, E Giombini, F Messina, M Rueca, O Butera, MR Capobianchi, A Di Caro |
| EPI_ISL_1210496 | INMI Lazzaro Spallanzani IRCCS | INMI Lazzaro Spallanzani IRCCS | E Giombini, F Messina, M Rueca, O Butera, CEM Gruber, B Bartolini, A Di Caro, MR Capobianchi |
| EPI_ISL_1210497 | San Gallicano Dermatological Institute I.F.O. | INMI Lazzaro Spallanzani IRCCS | B Bartolini, E Giombini, F Messina, M Rueca, O Butera, CEM Gruber, F Pimpinelli, F Ensoli, E Trento, A Di Caro, MR Capobianchi |
| EPI_ISL_1210498 | Laboratorio Genzano - ASL RM 7 | INMI Lazzaro Spallanzani IRCCS | CEM Gruber, B Bartolini, E Giombini, F Messina, M Rueca, O Butera, G Tramini, E Conti, MR Capobianchi, A Di Caro |
| EPI_ISL_1210499 | Ospedale "F. Spaziani" Frosinone | INMI Lazzaro Spallanzani IRCCS | B Bartolini, E Giombini, F Messina, M Rueca, O Butera, CEM Gruber, R Pulselli, C Sias, G Brocco, A Di Caro, MR Capobianchi |
| EPI_ISL_1210500 | Ospedale "F. Spaziani" Frosinone | INMI Lazzaro Spallanzani IRCCS | M Rueca, O Butera, CEM Gruber, B Bartolini, E Giombini, F Messina, R Pulselli, G Brocco, C Gargiulo, A Di Caro, MR Capobianchi |
| EPI_ISL_1212619, EPI_ISL_1212621, EPI_ISL_1212623, EPI_ISL_1212626, EPI_ISL_1212628, EPI_ISL_1212630, EPI_ISL_1212632, EPI_ISL_1212634, EPI_ISL_1212636, EPI_ISL_1212638, EPI_ISL_1212641, EPI_ISL_1212643, EPI_ISL_1212645, EPI_ISL_1212647, EPI_ISL_1212650, EPI_ISL_1212652, EPI_ISL_1212654, EPI_ISL_1212656, EPI_ISL_1212659, EPI_ISL_1212661, EPI_ISL_1212664, EPI_ISL_1212666, EPI_ISL_1212668, EPI_ISL_1212670 | | | |
| see above | Università degli Studi di Perugia | Istituto Zooprofilattico Sperimentale dell'Abruzzo e Molise "G. Caporale" | Mencacci A., Camilloni B., Lorusso A, Marcacci M, Di Domenico M, Ancora M, Curini V, Mangone I, Rinaldi A, Scialabba S, Di Pasquale A, Cammà C, Puglia I, Calistri P, Savini G |
| EPI_ISL_1212673, EPI_ISL_1212675, EPI_ISL_1212677, EPI_ISL_1212679, EPI_ISL_1212681, EPI_ISL_1212684, EPI_ISL_1212686, EPI_ISL_1212689, EPI_ISL_1212691, EPI_ISL_1212693, EPI_ISL_1212696, EPI_ISL_1212698, EPI_ISL_1212700 | | | |
| see above | Istituto Zooprofilattico Sperimentale Umbria e Marche "Togo Rosati" | Istituto Zooprofilattico Sperimentale dell'Abruzzo e Molise "G. Caporale" | Biagetti M., Giammarioli M., Lorusso A, Marcacci M, Di Domenico M, Ancora M, Curini V, Mangone I, Rinaldi A, Scialabba S, Di Pasquale A, Cammà C, Puglia I, Calistri P, Savini G |
| EPI_ISL_1215947, EPI_ISL_1216923, EPI_ISL_1217356 | Department of Sciences for Health Promotion and Mother-Child Care "G. D'Alessandro", University of Palermo | Tumor Immunology Unit, Department of Health Sciences, University of Palermo School of Medicine; Section of Microbiology, University of Palermo School of Medicine; National Research Council of Italy - High Performance Computing and Networking Institute (CNR-ICAR) of Palermo | Vacca, Davide; Fiannaca, Antonino; Bonura, Celestino; Cancila, Valeria; La Paglia, Laura; Bonura, Floriana; Morello, Gaia; La Rosa, Massimo; Di Bernardo, Francesca; Urso, Alfonso Maurizio; Tripodo, Claudio; Craxi, Antonio; Vitale, Francesco; Giammanco, Maurizio Giovanni. |
| EPI_ISL_1217377 | San Gallicano Dermatological Institute I.F.O. | INMI Lazzaro Spallanzani IRCCS | E Giombini, F Messina, M Rueca, O Butera, CEM Gruber, B Bartolini, F Pimpinelli, F Ensoli, M Pontone, A Di Caro, MR Capobianchi |
| EPI_ISL_1217380 | Laboratorio Genzano - ASL RM 8 | INMI Lazzaro Spallanzani IRCCS | O Butera, CEM Gruber, B Bartolini, E Giombini, F Messina, M Rueca, G Tramini, E Conti, MR Capobianchi, A Di Caro |
| EPI_ISL_1217383 | Laboratorio Genzano - ASL RM 10 | INMI Lazzaro Spallanzani IRCCS | CEM Gruber, B Bartolini, E Giombini, F Messina, M Rueca, O Butera, G Tramini, E Conti, MR Capobianchi, A Di Caro |
| EPI_ISL_1217386 | Ospedale "F. Spaziani" Frosinone | INMI Lazzaro Spallanzani IRCCS | M Rueca, O Butera, CEM Gruber, B Bartolini, E Giombini, F Messina, R Pulselli, C Sias, G Brocco, A Di Caro, MR Capobianchi |
| EPI_ISL_1217388 | Laboratorio Genzano - ASL RM 6 | INMI Lazzaro Spallanzani IRCCS | CEM Gruber, B Bartolini, E Giombini, F Messina, M Rueca, O Butera, G Tramini, E Conti, MR Capobianchi, A Di Caro |
| EPI_ISL_1217709 | Department of Sciences for Health Promotion and Mother-Child Care "G. D'Alessandro", University of Palermo | Tumor Immunology Unit, Department of Health Sciences, University of Palermo School of Medicine; Section of Microbiology, University of Palermo School of Medicine; National Research Council of Italy - High Performance Computing and Networking Institute (CNR-ICAR) of Palermo | Vacca, Davide; Fiannaca, Antonino; Bonura, Celestino; Cancila, Valeria; La Paglia, Laura; Bonura, Floriana; Morello, Gaia; La Rosa, Massimo; Di Bernardo, Francesca; Urso, Alfonso Maurizio; Tripodo, Claudio; Craxi, Antonio; Vitale, Francesco; Giammanco, Maurizio Giovanni. |

| | | | |
|---|---|---|---|
| EPI_ISL_1219707, EPI_ISL_1219708, EPI_ISL_1219709, EPI_ISL_1219710 | Laboratorio Biologia Molecolare Sars Cov2 - UOC Laboratorio Analisi - Servizio Medicina di Laboratorio, Ospedale "San Francesco" - ATS-ASSL Nuoro | Laboratorio specialistico UOC Ematologia - Ospedale "San Francesco" - ATS-ASSL Nuoro | Piras Giovanna, Asproni Rosanna,,Fiamma Maura, Monne Maria Itria, Palmas Angelo Domenico, Lo Maglio Iana, Mameli Giuseppe |
| EPI_ISL_1222711 | Laboratorio Genzano - ASL RM 6 | INMI Lazzaro Spallanzani IRCCS | F Messina, M Rueca, O Butera, CEM Gruber, B Bartolini, E Giombini, G Tramini, E Conti, MR Capobianchi, A Di Caro |
| EPI_ISL_1222712 | Laboratorio Genzano - ASL RM 7 | INMI Lazzaro Spallanzani IRCCS | M Rueca, O Butera, CEM Gruber, B Bartolini, E Giombini, F Messina, G Tramini, E Conti, A Di Caro, MR Capobianchi |
| EPI_ISL_1222713 | Laboratorio Genzano - ASL RM 9 | INMI Lazzaro Spallanzani IRCCS | B Bartolini, E Giombini, F Messina, M Rueca, O Butera, CEM Gruber, G Tramini, E Conti, A Di Caro, MR Capobianchi |
| EPI_ISL_1222714 | Laboratorio Genzano - ASL RM 11 | INMI Lazzaro Spallanzani IRCCS | E Giombini, F Messina, M Rueca, O Butera, CEM Gruber, B Bartolini, G Tramini, E Conti, A Di Caro, MR Capobianchi |
| EPI_ISL_1222715 | Ospedale "F. Spaziani" Frosinone | INMI Lazzaro Spallanzani IRCCS | O Butera, CEM Gruber, B Bartolini, E Giombini, F Messina, M Rueca, R Pulselli, G Brocco, C Gargiulo, MR Capobianchi, A Di Caro |
| EPI_ISL_1222716 | Ospedale "F. Spaziani" Frosinone | INMI Lazzaro Spallanzani IRCCS | B Bartolini, E Giombini, F Messina, M Rueca, O Butera, CEM Gruber, R Pulselli, C Gargiulo, C Sias, A Di Caro, MR Capobianchi |
| EPI_ISL_1222717 | Ospedale "F. Spaziani" Frosinone | INMI Lazzaro Spallanzani IRCCS | E Giombini, F Messina, M Rueca, O Butera, CEM Gruber, B Bartolini, R Pulselli, G Brocco, C Gargiulo, A Di Caro, MR Capobianchi |
| EPI_ISL_1222718 | Ospedale "F. Spaziani" Frosinone | INMI Lazzaro Spallanzani IRCCS | F Messina, M Rueca, O Butera, CEM Gruber, B Bartolini, E Giombini, R Pulselli, C Gargiulo, C Sias, MR Capobianchi, A Di Caro |
| EPI_ISL_1222719 | Ospedale "F. Spaziani" Frosinone | INMI Lazzaro Spallanzani IRCCS | B Bartolini, E Giombini, F Messina, M Rueca, O Butera, CEM Gruber, R Pulselli, C Gargiulo, C Sias, A Di Caro, MR Capobianchi |
| EPI_ISL_1222783, EPI_ISL_1222784, EPI_ISL_1222785, EPI_ISL_1222787, EPI_ISL_1222788, EPI_ISL_1222789, EPI_ISL_1222790, EPI_ISL_1222791, EPI_ISL_1222792, EPI_ISL_1222793, EPI_ISL_1222794, EPI_ISL_1222795, EPI_ISL_1222796, EPI_ISL_1222797, EPI_ISL_1222799, EPI_ISL_1222800, EPI_ISL_1222801, EPI_ISL_1222802, EPI_ISL_1222803, EPI_ISL_1222804, EPI_ISL_1222805, EPI_ISL_1222806, EPI_ISL_1222807, EPI_ISL_1222808, EPI_ISL_1222809, EPI_ISL_1222810, EPI_ISL_1222811, EPI_ISL_1222812, EPI_ISL_1222813, EPI_ISL_1222814, EPI_ISL_1222815, EPI_ISL_1222816, EPI_ISL_1222817, EPI_ISL_1222818, EPI_ISL_1222819, EPI_ISL_1222820, EPI_ISL_1222822, EPI_ISL_1222823, EPI_ISL_1222824, EPI_ISL_1222825, EPI_ISL_1222826, EPI_ISL_1222827, EPI_ISL_1222828, EPI_ISL_1222829, EPI_ISL_1222831, EPI_ISL_1222832, EPI_ISL_1222833, EPI_ISL_1222834, EPI_ISL_1222835, EPI_ISL_1222836, EPI_ISL_1222837, EPI_ISL_1222838, EPI_ISL_1222839, EPI_ISL_1222840, EPI_ISL_1222841, EPI_ISL_1222842, EPI_ISL_1222843, EPI_ISL_1222845, EPI_ISL_1222846, EPI_ISL_1222847, EPI_ISL_1222848, EPI_ISL_1222849, EPI_ISL_1222850, EPI_ISL_1222851, EPI_ISL_1222852, EPI_ISL_1222853, EPI_ISL_1222854, EPI_ISL_1222855, EPI_ISL_1222856, EPI_ISL_1222857, EPI_ISL_1222858, EPI_ISL_1222859, EPI_ISL_1222860, EPI_ISL_1222861, EPI_ISL_1222863, EPI_ISL_1222864, EPI_ISL_1222865, EPI_ISL_1222866, EPI_ISL_1222868, EPI_ISL_1222869, EPI_ISL_1222871, EPI_ISL_1222872, EPI_ISL_1222873, EPI_ISL_1222874, EPI_ISL_1222875, EPI_ISL_1222876, EPI_ISL_1222877, EPI_ISL_1222878, EPI_ISL_1222879, EPI_ISL_1222880, EPI_ISL_1222881, EPI_ISL_1222882, EPI_ISL_1222883, EPI_ISL_1222884, EPI_ISL_1222885, EPI_ISL_1222886, EPI_ISL_1222888, EPI_ISL_1222889, EPI_ISL_1222890, EPI_ISL_1222891, EPI_ISL_1222893, EPI_ISL_1222894, EPI_ISL_1222895, EPI_ISL_1222896, EPI_ISL_1222897, EPI_ISL_1222898, EPI_ISL_1222899, EPI_ISL_1222900, EPI_ISL_1222901, EPI_ISL_1222902, EPI_ISL_1222903, EPI_ISL_1222904, EPI_ISL_1222905, EPI_ISL_1222906, EPI_ISL_1222907, EPI_ISL_1222908, EPI_ISL_1222909, EPI_ISL_1222910, EPI_ISL_1222911, EPI_ISL_1222912, EPI_ISL_1222913, EPI_ISL_1222914, EPI_ISL_1222916, EPI_ISL_1222917, EPI_ISL_1222918, EPI_ISL_1222919, EPI_ISL_1222924, EPI_ISL_1222925, EPI_ISL_1222926 | | | |
| see above | Azienda Sanitaria dell'Alto Adige Laboratorio Aziendale di Microbiologia e Virologia | Istituto di Genomica Applicata | Elisabetta Pagani, Irene Bianconi, Elisabetta Giacobazzi, Elisa Masi, Stefanie Wieser, Irena Jurman, Vera Vendramin, Gabriele Magris, Eleonora Paparelli, Davide Scaglione, Michele Morgante |
| EPI_ISL_1224943 | Department of Infectious Diseases, Istituto Superiore di Sanità, Rome, Italy; Azienda Ospedaliera Universitaria Pisana, Pisa, Italy | Istituto Superiore di Sanità (ISS) | Paola Stefanelli, Angela Di Martino, Alessandra Lo Presti, Stefano Fiore, Paola Mazzetti, Maria Linda Vatteroni, Anna Lisa Capria, Flavia Favilli, Sabrina Gabbriellini, Lisa Macera, Giovanni Segatori, Mauro Pistello, Manuela Marra, Maria Carollo, Marco Crescenzi |
| EPI_ISL_1224990 | Virology Unit, AOUP | Department of Infectious Diseases, Istituto Superiore di Sanità | Paola Stefanelli, Angela Di Martino, Alessandra Lo Presti, Stefano Fiore, Marialinda Vatteroni, Susi Frateschi, Mauro Pistello, Manuela Marra, Maria Carollo, Marco Crescenzi |
| EPI_ISL_1229130 | IRCCS San Raffaele | INMI Lazzaro Spallanzani IRCCS | B Bartolini, E Giombini, F Messina, M Rueca, O Butera, CEM Gruber, F Ciatti, D Russo, D Limongi, A Di Caro, MR Capobianchi |
| EPI_ISL_1229134 | Infectious Diseases Clinic, Udine University Hospital | INMI Lazzaro Spallanzani IRCCS | M Rueca, O Butera, CEM Gruber, B Bartolini, E Giombini, F Messina, C Tascini, D Pecori, A Di Caro, MR Capobianchi |
| EPI_ISL_1229135 | Infectious Diseases Clinic, Udine University Hospital | INMI Lazzaro Spallanzani IRCCS | O Butera, CEM Gruber, B Bartolini, E Giombini, F Messina, M Rueca, C Tascini, D Pecori, MR Capobianchi, A Di Caro |
| EPI_ISL_1229136 | Ospedale "F. Spaziani" Frosinone | INMI Lazzaro Spallanzani IRCCS | F Messina, M Rueca, O Butera, CEM Gruber, B Bartolini, E Giombini, R Pulselli, C Sias, G Brocco, MR Capobianchi, A Di Caro |
| EPI_ISL_1229137 | Ospedale "F. Spaziani" Frosinone | INMI Lazzaro Spallanzani IRCCS | M Rueca, O Butera, CEM Gruber, B Bartolini, E Giombini, F Messina, R Pulselli, G Brocco, C Gargiulo, A Di Caro, MR Capobianchi |
| EPI_ISL_1229138 | University Hospital Sant'Andrea-Sapienza | INMI Lazzaro Spallanzani IRCCS | O Butera, CEM Gruber, B Bartolini, E Giombini, F Messina, M Rueca, M Simmaco, I Santino, MR Capobianchi, A Di Caro |
| EPI_ISL_1229139 | Laboratorio Genzano - ASL RM 6 | INMI Lazzaro Spallanzani IRCCS | B Bartolini, E Giombini, F Messina, M Rueca, O Butera, CEM Gruber, G Tramini, E Conti, A Di Caro, MR Capobianchi |
| EPI_ISL_1234383 | San Gallicano Dermatological Institute I.F.O. | INMI Lazzaro Spallanzani IRCCS | B Bartolini, E Giombini, F Messina, M Rueca, O Butera, CEM Gruber, F Pimpinelli, I Cavallo, MR Capobianchi, A Di Caro |
| EPI_ISL_1250945, EPI_ISL_1250956, EPI_ISL_1250966, EPI_ISL_1250975, EPI_ISL_1250976, EPI_ISL_1250977, EPI_ISL_1250978, EPI_ISL_1250979, EPI_ISL_1250980 | Siesp dipartimento di prevenzione | Istituto Zooprofilattico Sperimentale dell'Abruzzo e Molise "G. Caporale" | Lorusso A, Marcacci M, Di Domenico M, Ancora M, Curini V, Mangone I, Rinaldi A, Scialabba S, Di Pasquale A, Cammà C, Puglia I, Calistri P, Savini G |
| EPI_ISL_1250981, EPI_ISL_1250982 | DIP. PREV. AVEZZANO | Istituto Zooprofilattico Sperimentale dell'Abruzzo e Molise "G. Caporale" | Lorusso A, Marcacci M, Di Domenico M, Ancora M, Curini V, Mangone I, Rinaldi A, Scialabba S, Di Pasquale A, Cammà C, Puglia I, Calistri P, Savini G |
| EPI_ISL_1250983 | Siesp dipartimento di prevenzione | Istituto Zooprofilattico Sperimentale dell'Abruzzo e Molise "G. Caporale" | Lorusso A, Marcacci M, Di Domenico M, Ancora M, Curini V, Mangone I, Rinaldi A, Scialabba S, Di Pasquale A, Cammà C, Puglia I, Calistri P, Savini G |
| EPI_ISL_1250984 | SIESP CHIETI - LANCIANO | Istituto Zooprofilattico Sperimentale dell'Abruzzo e Molise "G. Caporale" | Lorusso A, Marcacci M, Di Domenico M, Ancora M, Curini V, Mangone I, Rinaldi A, Scialabba S, Di Pasquale A, Cammà C, Puglia I, Calistri P, Savini G |
| EPI_ISL_1250985 | Usca castel di sangro | Istituto Zooprofilattico Sperimentale dell'Abruzzo e Molise "G. Caporale" | Lorusso A, Marcacci M, Di Domenico M, Ancora M, Curini V, Mangone I, Rinaldi A, Scialabba S, Di Pasquale A, Cammà C, Puglia I, Calistri P, Savini G |
| EPI_ISL_1250986 | USCA Pescina | Istituto Zooprofilattico Sperimentale dell'Abruzzo e Molise "G. Caporale" | Lorusso A, Marcacci M, Di Domenico M, Ancora M, Curini V, Mangone I, Rinaldi A, Scialabba S, Di Pasquale A, Cammà C, Puglia I, Calistri P, Savini G |
| EPI_ISL_1250987 | siesp Chieti | Istituto Zooprofilattico Sperimentale dell'Abruzzo e Molise "G. Caporale" | Lorusso A, Marcacci M, Di Domenico M, Ancora M, Curini V, Mangone I, Rinaldi A, Scialabba S, Di Pasquale A, Cammà C, Puglia I, Calistri P, Savini G |
| EPI_ISL_1250988 | Siesp dipartimento di prevenzione | Istituto Zooprofilattico Sperimentale dell'Abruzzo e Molise "G. Caporale" | Lorusso A, Marcacci M, Di Domenico M, Ancora M, Curini V, Mangone I, Rinaldi A, Scialabba S, Di Pasquale A, Cammà C, Puglia I, Calistri P, Savini G |
| EPI_ISL_1250989 | SIESP CHIETI - DRIVE IN ORTONA | Istituto Zooprofilattico Sperimentale dell'Abruzzo e Molise "G. Caporale" | Lorusso A, Marcacci M, Di Domenico M, Ancora M, Curini V, Mangone I, Rinaldi A, Scialabba S, Di Pasquale A, Cammà C, Puglia I, Calistri P, Savini G |
| EPI_ISL_1250990 | SIESP CHIETI - LANCIANO | Istituto Zooprofilattico Sperimentale dell'Abruzzo e Molise "G. Caporale" | Lorusso A, Marcacci M, Di Domenico M, Ancora M, Curini V, Mangone I, Rinaldi A, Scialabba S, Di Pasquale A, Cammà C, Puglia I, Calistri P, Savini G |
| EPI_ISL_1250991, EPI_ISL_1250992 | Siesp dipartimento di prevenzione | Istituto Zooprofilattico Sperimentale dell'Abruzzo e Molise "G. Caporale" | Lorusso A, Marcacci M, Di Domenico M, Ancora M, Curini V, Mangone I, Rinaldi A, Scialabba S, Di Pasquale A, Cammà C, Puglia I, Calistri P, Savini G |
| EPI_ISL_1250993 | SIESP CHIETI - DRIVE IN ORTONA | Istituto Zooprofilattico Sperimentale dell'Abruzzo e Molise "G. Caporale" | Lorusso A, Marcacci M, Di Domenico M, Ancora M, Curini V, Mangone I, Rinaldi A, Scialabba S, Di Pasquale A, Cammà C, Puglia I, Calistri P, Savini G |
| EPI_ISL_1250994, EPI_ISL_1250995, EPI_ISL_1250996, EPI_ISL_1250997, EPI_ISL_1250998 | Siesp dipartimento di prevenzione | Istituto Zooprofilattico Sperimentale dell'Abruzzo e Molise "G. Caporale" | Lorusso A, Marcacci M, Di Domenico M, Ancora M, Curini V, Mangone I, Rinaldi A, Scialabba S, Di Pasquale A, Cammà C, Puglia I, Calistri P, Savini G |
| EPI_ISL_1250999, EPI_ISL_1251000 | SIESP CHIETI - LANCIANO | Istituto Zooprofilattico Sperimentale dell'Abruzzo e Molise "G. Caporale" | Lorusso A, Marcacci M, Di Domenico M, Ancora M, Curini V, Mangone I, Rinaldi A, Scialabba S, Di Pasquale A, Cammà C, Puglia I, Calistri P, Savini G |
| EPI_ISL_1251001 | SIESP CHIETI - DRIVE IN ORTONA | Istituto Zooprofilattico Sperimentale dell'Abruzzo e Molise "G. Caporale" | Lorusso A, Marcacci M, Di Domenico M, Ancora M, Curini V, Mangone I, Rinaldi A, Scialabba S, Di Pasquale A, Cammà C, Puglia I, Calistri P, Savini G |

| | | | |
|---|---|---|---|
| EPI_ISL_1251002 | OSPEDALE SAN SALVATORE L'AQUILA UOC PNEUMOLOGIA | Istituto Zooprofilattico Sperimentale dell'Abruzzo e Molise "G. Caporale" | Lorusso A, Marcacci M, Di Domenico M, Ancora M, Curini V, Mangone I, Rinaldi A, Scialabba S, Di Pasquale A, Cammà C, Puglia I, Calistri P, Savini G |
| EPI_ISL_1251003 | SIESP CHIETI - DRIVE IN CHIET | Istituto Zooprofilattico Sperimentale dell'Abruzzo e Molise "G. Caporale" | Lorusso A, Marcacci M, Di Domenico M, Ancora M, Curini V, Mangone I, Rinaldi A, Scialabba S, Di Pasquale A, Cammà C, Puglia I, Calistri P, Savini G |
| EPI_ISL_1251004 | siesp Chieti | Istituto Zooprofilattico Sperimentale dell'Abruzzo e Molise "G. Caporale" | Lorusso A, Marcacci M, Di Domenico M, Ancora M, Curini V, Mangone I, Rinaldi A, Scialabba S, Di Pasquale A, Cammà C, Puglia I, Calistri P, Savini G |
| EPI_ISL_1251005, EPI_ISL_1251006, EPI_ISL_1251007 | OSPEDALE SAN SALVATORE | Istituto Zooprofilattico Sperimentale dell'Abruzzo e Molise "G. Caporale" | Lorusso A, Marcacci M, Di Domenico M, Ancora M, Curini V, Mangone I, Rinaldi A, Scialabba S, Di Pasquale A, Cammà C, Puglia I, Calistri P, Savini G |
| EPI_ISL_1251008, EPI_ISL_1251009, EPI_ISL_1251010, EPI_ISL_1251011, EPI_ISL_1251012 | Siesp dipartimento di prevenzione | Istituto Zooprofilattico Sperimentale dell'Abruzzo e Molise "G. Caporale" | Lorusso A, Marcacci M, Di Domenico M, Ancora M, Curini V, Mangone I, Rinaldi A, Scialabba S, Di Pasquale A, Cammà C, Puglia I, Calistri P, Savini G |
| EPI_ISL_1251013 | OSPEDALE CIVILE GIULIANOVA | Istituto Zooprofilattico Sperimentale dell'Abruzzo e Molise "G. Caporale" | Lorusso A, Marcacci M, Di Domenico M, Ancora M, Curini V, Mangone I, Rinaldi A, Scialabba S, Di Pasquale A, Cammà C, Puglia I, Calistri P, Savini G |
| EPI_ISL_1251014, EPI_ISL_1251015, EPI_ISL_1251016, EPI_ISL_1251017 | Siesp dipartimento di prevenzione | Istituto Zooprofilattico Sperimentale dell'Abruzzo e Molise "G. Caporale" | Lorusso A, Marcacci M, Di Domenico M, Ancora M, Curini V, Mangone I, Rinaldi A, Scialabba S, Di Pasquale A, Cammà C, Puglia I, Calistri P, Savini G |
| EPI_ISL_1251018 | "DIP. PREV. AVEZZANO SERVIZIO DI IGIENE EPIDEMIOLOGIA E SANITA' PUBBLICA | Istituto Zooprofilattico Sperimentale dell'Abruzzo e Molise "G. Caporale" | Lorusso A, Marcacci M, Di Domenico M, Ancora M, Curini V, Mangone I, Rinaldi A, Scialabba S, Di Pasquale A, Cammà C, Puglia I, Calistri P, Savini G |
| EPI_ISL_1251019 | Usca castel di sangro | Istituto Zooprofilattico Sperimentale dell'Abruzzo e Molise "G. Caporale" | Lorusso A, Marcacci M, Di Domenico M, Ancora M, Curini V, Mangone I, Rinaldi A, Scialabba S, Di Pasquale A, Cammà C, Puglia I, Calistri P, Savini G |
| EPI_ISL_1251020 | Siesp dipartimento di prevenzione | Istituto Zooprofilattico Sperimentale dell'Abruzzo e Molise "G. Caporale" | Lorusso A, Marcacci M, Di Domenico M, Ancora M, Curini V, Mangone I, Rinaldi A, Scialabba S, Di Pasquale A, Cammà C, Puglia I, Calistri P, Savini G |
| EPI_ISL_1251021, EPI_ISL_1251022 | Usca castel di sangro | Istituto Zooprofilattico Sperimentale dell'Abruzzo e Molise "G. Caporale" | Lorusso A, Marcacci M, Di Domenico M, Ancora M, Curini V, Mangone I, Rinaldi A, Scialabba S, Di Pasquale A, Cammà C, Puglia I, Calistri P, Savini G |
| EPI_ISL_1251023 | USCA Pescina | Istituto Zooprofilattico Sperimentale dell'Abruzzo e Molise "G. Caporale" | Lorusso A, Marcacci M, Di Domenico M, Ancora M, Curini V, Mangone I, Rinaldi A, Scialabba S, Di Pasquale A, Cammà C, Puglia I, Calistri P, Savini G |
| EPI_ISL_1251024 | OSPEDALE CIVILE GIULIANOVA | Istituto Zooprofilattico Sperimentale dell'Abruzzo e Molise "G. Caporale" | Lorusso A, Marcacci M, Di Domenico M, Ancora M, Curini V, Mangone I, Rinaldi A, Scialabba S, Di Pasquale A, Cammà C, Puglia I, Calistri P, Savini G |
| EPI_ISL_1251025 | PRESIDIO TERRITORIALE CASOLI | Istituto Zooprofilattico Sperimentale dell'Abruzzo e Molise "G. Caporale" | Lorusso A, Marcacci M, Di Domenico M, Ancora M, Curini V, Mangone I, Rinaldi A, Scialabba S, Di Pasquale A, Cammà C, Puglia I, Calistri P, Savini G |
| EPI_ISL_1251026 | siesp Chieti | Istituto Zooprofilattico Sperimentale dell'Abruzzo e Molise "G. Caporale" | Lorusso A, Marcacci M, Di Domenico M, Ancora M, Curini V, Mangone I, Rinaldi A, Scialabba S, Di Pasquale A, Cammà C, Puglia I, Calistri P, Savini G |
| EPI_ISL_1251027 | USCA Tagliacozzo | Istituto Zooprofilattico Sperimentale dell'Abruzzo e Molise "G. Caporale" | Lorusso A, Marcacci M, Di Domenico M, Ancora M, Curini V, Mangone I, Rinaldi A, Scialabba S, Di Pasquale A, Cammà C, Puglia I, Calistri P, Savini G |
| EPI_ISL_1251028 | siesp Chieti | Istituto Zooprofilattico Sperimentale dell'Abruzzo e Molise "G. Caporale" | Lorusso A, Marcacci M, Di Domenico M, Ancora M, Curini V, Mangone I, Rinaldi A, Scialabba S, Di Pasquale A, Cammà C, Puglia I, Calistri P, Savini G |
| EPI_ISL_1251029, EPI_ISL_1251030 | SIESP CHIETI - DRIVE IN VASTO | Istituto Zooprofilattico Sperimentale dell'Abruzzo e Molise "G. Caporale" | Lorusso A, Marcacci M, Di Domenico M, Ancora M, Curini V, Mangone I, Rinaldi A, Scialabba S, Di Pasquale A, Cammà C, Puglia I, Calistri P, Savini G |
| EPI_ISL_1251031 | Siesp dipartimento di prevenzione | Istituto Zooprofilattico Sperimentale dell'Abruzzo e Molise "G. Caporale" | Lorusso A, Marcacci M, Di Domenico M, Ancora M, Curini V, Mangone I, Rinaldi A, Scialabba S, Di Pasquale A, Cammà C, Puglia I, Calistri P, Savini G |
| EPI_ISL_1251032 | SIESP CHIETI - DRIVE IN VASTO | Istituto Zooprofilattico Sperimentale dell'Abruzzo e Molise "G. Caporale" | Lorusso A, Marcacci M, Di Domenico M, Ancora M, Curini V, Mangone I, Rinaldi A, Scialabba S, Di Pasquale A, Cammà C, Puglia I, Calistri P, Savini G |
| EPI_ISL_1251033, EPI_ISL_1251034, EPI_ISL_1251035, EPI_ISL_1251036 | Siesp dipartimento di prevenzione | Istituto Zooprofilattico Sperimentale dell'Abruzzo e Molise "G. Caporale" | Lorusso A, Marcacci M, Di Domenico M, Ancora M, Curini V, Mangone I, Rinaldi A, Scialabba S, Di Pasquale A, Cammà C, Puglia I, Calistri P, Savini G |
| EPI_ISL_1251037 | SIESP SULMONA | Istituto Zooprofilattico Sperimentale dell'Abruzzo e Molise "G. Caporale" | Lorusso A, Marcacci M, Di Domenico M, Ancora M, Curini V, Mangone I, Rinaldi A, Scialabba S, Di Pasquale A, Cammà C, Puglia I, Calistri P, Savini G |
| EPI_ISL_1251038 | SIESP CHIETI - DRIVE IN ORTONA | Istituto Zooprofilattico Sperimentale dell'Abruzzo e Molise "G. Caporale" | Lorusso A, Marcacci M, Di Domenico M, Ancora M, Curini V, Mangone I, Rinaldi A, Scialabba S, Di Pasquale A, Cammà C, Puglia I, Calistri P, Savini G |
| EPI_ISL_1251039 | DIP. PREV. AVEZZANO SERVIZIO DI IGIENE EPIDEMIOLOGIA E SANITA' PUBBLICA | Istituto Zooprofilattico Sperimentale dell'Abruzzo e Molise "G. Caporale" | Lorusso A, Marcacci M, Di Domenico M, Ancora M, Curini V, Mangone I, Rinaldi A, Scialabba S, Di Pasquale A, Cammà C, Puglia I, Calistri P, Savini G |
| EPI_ISL_1251040 | SIESP L'AQUILA | Istituto Zooprofilattico Sperimentale dell'Abruzzo e Molise "G. Caporale" | Lorusso A, Marcacci M, Di Domenico M, Ancora M, Curini V, Mangone I, Rinaldi A, Scialabba S, Di Pasquale A, Cammà C, Puglia I, Calistri P, Savini G |
| EPI_ISL_1251041, EPI_ISL_1251042 | Siesp dipartimento di prevenzione | Istituto Zooprofilattico Sperimentale dell'Abruzzo e Molise "G. Caporale" | Lorusso A, Marcacci M, Di Domenico M, Ancora M, Curini V, Mangone I, Rinaldi A, Scialabba S, Di Pasquale A, Cammà C, Puglia I, Calistri P, Savini G |
| EPI_ISL_1251043 | SIESP CHIETI - LANCIANO | Istituto Zooprofilattico Sperimentale dell'Abruzzo e Molise "G. Caporale" | Lorusso A, Marcacci M, Di Domenico M, Ancora M, Curini V, Mangone I, Rinaldi A, Scialabba S, Di Pasquale A, Cammà C, Puglia I, Calistri P, Savini G |
| EPI_ISL_1251044 | USCA Avezzano | Istituto Zooprofilattico Sperimentale dell'Abruzzo e Molise "G. Caporale" | Lorusso A, Marcacci M, Di Domenico M, Ancora M, Curini V, Mangone I, Rinaldi A, Scialabba S, Di Pasquale A, Cammà C, Puglia I, Calistri P, Savini G |
| EPI_ISL_1251045 | OSPEDALE CIVILE GIULIANOVA | Istituto Zooprofilattico Sperimentale dell'Abruzzo e Molise "G. Caporale" | Lorusso A, Marcacci M, Di Domenico M, Ancora M, Curini V, Mangone I, Rinaldi A, Scialabba S, Di Pasquale A, Cammà C, Puglia I, Calistri P, Savini G |
| EPI_ISL_1251046, EPI_ISL_1251047, EPI_ISL_1251048, EPI_ISL_1251049, EPI_ISL_1251050, EPI_ISL_1251051 | Siesp dipartimento di prevenzione | Istituto Zooprofilattico Sperimentale dell'Abruzzo e Molise "G. Caporale" | Lorusso A, Marcacci M, Di Domenico M, Ancora M, Curini V, Mangone I, Rinaldi A, Scialabba S, Di Pasquale A, Cammà C, Puglia I, Calistri P, Savini G |
| EPI_ISL_1251052 | SIESP SULMONA | Istituto Zooprofilattico Sperimentale dell'Abruzzo e Molise "G. Caporale" | Lorusso A, Marcacci M, Di Domenico M, Ancora M, Curini V, Mangone I, Rinaldi A, Scialabba S, Di Pasquale A, Cammà C, Puglia I, Calistri P, Savini G |
| EPI_ISL_1251053 | Siesp dipartimento di prevenzione | Istituto Zooprofilattico Sperimentale dell'Abruzzo e Molise "G. Caporale" | Lorusso A, Marcacci M, Di Domenico M, Ancora M, Curini V, Mangone I, Rinaldi A, Scialabba S, Di Pasquale A, Cammà C, Puglia I, Calistri P, Savini G |
| EPI_ISL_1251054 | MEDICO COMPETENTE P.O. L'AQUILA | Istituto Zooprofilattico Sperimentale dell'Abruzzo e Molise "G. Caporale" | Lorusso A, Marcacci M, Di Domenico M, Ancora M, Curini V, Mangone I, Rinaldi A, Scialabba S, Di Pasquale A, Cammà C, Puglia I, Calistri P, Savini G |
| EPI_ISL_1251062 | USCA Pescina | Istituto Zooprofilattico Sperimentale dell'Abruzzo e Molise "G. Caporale" | Lorusso A, Marcacci M, Di Domenico M, Ancora M, Curini V, Mangone I, Rinaldi A, Scialabba S, Di Pasquale A, Cammà C, Puglia I, Calistri P, Savini G |
| EPI_ISL_1251239 | SC (UCO) Igiene e Sanità Pubblica | ARGO Laboratorio Genomica ed Epigenomica | Licastro D, Dal Monego S, Degasperi M, D'Agaro P, Lombardo F |

| | | | |
|---|---|---|---|
| EPI_ISL_1251240, EPI_ISL_1251241, EPI_ISL_1251242, EPI_ISL_1251243, EPI_ISL_1251244, EPI_ISL_1251246 | SC (UCO) Igiene e Sanità Pubblica | ARGO Laboratorio Genomica ed Epigenomica | Licastro D, Dal Monego S, Degasperi M, Segat L, Piscianz E, D'Agaro P |
| EPI_ISL_1251247, EPI_ISL_1251248, EPI_ISL_1251249 | SC (UCO) Igiene e Sanità Pubblica | ARGO Laboratorio Genomica ed Epigenomica | Licastro D, Dal Monego S, Degasperi M, Marcello A, D'Agaro P, Pipan C |
| EPI_ISL_1251250, EPI_ISL_1251252, EPI_ISL_1251253, EPI_ISL_1251254, EPI_ISL_1251255, EPI_ISL_1251256, EPI_ISL_1251257, EPI_ISL_1251258, EPI_ISL_1251259 | SC (UCO) Igiene e Sanità Pubblica | ARGO Laboratorio Genomica ed Epigenomica | Licastro D, Dal Monego S, Degasperi M, Segat L, Piscianz E, D'Agaro P |
| EPI_ISL_1252892, EPI_ISL_1252904, EPI_ISL_1253399 | Struttura Semplice Dipartimentale di Virologia e Microbiologia molecolare, Azienda Ospedaliero-Universitaria, Policlinico di Modena | U.O. Microbiologia, Laboratorio Unico Centro Servizi - AUSL della Romagna | Pecorari Monica, Gennari William, Fregni Serpini Giulia, Giorgio Dirani, Silvia Zannoli, Vittorio Sambri |
| EPI_ISL_1253705 | Laboratorio Unico Provinciale Azienda Ospedaliero-Universitaria di Ferrara | U.O. Microbiologia, Laboratorio Unico Centro Servizi - AUSL della Romagna | Michela Boni, Sara Ghisellini, Giorgio Dirani, Silvia Zannoli, Vittorio Sambri |
| EPI_ISL_1254647, EPI_ISL_1254648, EPI_ISL_1254649, EPI_ISL_1254650, EPI_ISL_1254651 | Ospedale di Venere | Istituto Zooprofilattico Sperimentale della Puglia e della Basilicata | Parisi A., Bianco A., Capozzi L., Del Sambro L., Simone D., Iacobellis M., Ridolfi D., Giannico A. |
| EPI_ISL_1254652, EPI_ISL_1254653, EPI_ISL_1254654, EPI_ISL_1254655, EPI_ISL_1254656, EPI_ISL_1254657 | Istituto Zooprofilattico Sperimentale della Puglia e della Basilicata | Istituto Zooprofilattico Sperimentale della Puglia e della Basilicata | Parisi A., Bianco A., Capozzi L., Del Sambro L., Simone D., Manzulli V, Rondinone V., Pace L., Cipolletta D., Galante D. |
| EPI_ISL_1254658, EPI_ISL_1254659, EPI_ISL_1254660, EPI_ISL_1254661, EPI_ISL_1254662, EPI_ISL_1254663, EPI_ISL_1254664 | Presidio di Brindisi Di Summa - Perrino | Istituto Zooprofilattico Sperimentale della Puglia e della Basilicata | Parisi A., Bianco A., Capozzi L., Del Sambro L., Simone D., Difato L., Ridolfi D., Giannico A. |
| EPI_ISL_1254665 | Ospedale Santa Caterina Novella | Istituto Zooprofilattico Sperimentale della Puglia e della Basilicata | Parisi A., Bianco A., Capozzi L., Del Sambro L., Simone D., Difato L., Ridolfi D., Giannico A. |
| EPI_ISL_1254666, EPI_ISL_1254667, EPI_ISL_1254668, EPI_ISL_1254669, EPI_ISL_1254670 | Ospedale di Venere | Istituto Zooprofilattico Sperimentale della Puglia e della Basilicata | Parisi A., Bianco A., Capozzi L., Del Sambro L., Simone D., Iacobellis M., Ridolfi D., Giannico A. |
| EPI_ISL_1254671, EPI_ISL_1254672, EPI_ISL_1254673, EPI_ISL_1254674 | Istituto Zooprofilattico Sperimentale della Puglia e della Basilicata | Istituto Zooprofilattico Sperimentale della Puglia e della Basilicata | Parisi A., Bianco A., Capozzi L., Del Sambro L., Simone D., Manzulli V, Rondinone V., Pace L., Cipolletta D., Galante D. |
| EPI_ISL_1254675 | Presidio di Brindisi Di Summa - Perrino | Istituto Zooprofilattico Sperimentale della Puglia e della Basilicata | Parisi A., Bianco A., Capozzi L., Del Sambro L., Simone D., Difato L., Ridolfi D., Giannico A. |
| EPI_ISL_1254682 | Ospedale Generale Regionale F. Miulli | Istituto Zooprofilattico Sperimentale della Puglia e della Basilicata | Parisi A., Bianco A., Capozzi L., Del Sambro L., Simone D., Difato L., Ridolfi D., Giannico A. |
| EPI_ISL_1254688, EPI_ISL_1254689, EPI_ISL_1254690, EPI_ISL_1254691, EPI_ISL_1254695 | Istituto Zooprofilattico Sperimentale della Puglia e della Basilicata | Istituto Zooprofilattico Sperimentale della Puglia e della Basilicata | Parisi A., Bianco A., Capozzi L., Del Sambro L., Simone D., Manzulli V, Rondinone V., Pace L., Cipolletta D., Galante D. |
| EPI_ISL_1254715, EPI_ISL_1254716, EPI_ISL_1254717, EPI_ISL_1254718, EPI_ISL_1254719, EPI_ISL_1254720, EPI_ISL_1254721, EPI_ISL_1254722, EPI_ISL_1254723, EPI_ISL_1254724, EPI_ISL_1254725, EPI_ISL_1254726, EPI_ISL_1254727, EPI_ISL_1254728, EPI_ISL_1254729, EPI_ISL_1254730, EPI_ISL_1254731, EPI_ISL_1254732, EPI_ISL_1254733, EPI_ISL_1254734, EPI_ISL_1254735, EPI_ISL_1254736, EPI_ISL_1254737, EPI_ISL_1254738, EPI_ISL_1254747, EPI_ISL_1254749, EPI_ISL_1254753, EPI_ISL_1254754, EPI_ISL_1254755, EPI_ISL_1254756, EPI_ISL_1254757, EPI_ISL_1254758, EPI_ISL_1254759, EPI_ISL_1254760, EPI_ISL_1254761, EPI_ISL_1254762, EPI_ISL_1254763, EPI_ISL_1254764, EPI_ISL_1254765, EPI_ISL_1254766, EPI_ISL_1254767, EPI_ISL_1254768, EPI_ISL_1254769, EPI_ISL_1254770, EPI_ISL_1254771, EPI_ISL_1254772, EPI_ISL_1254773, EPI_ISL_1254774, EPI_ISL_1254775, EPI_ISL_1254776, EPI_ISL_1254777, EPI_ISL_1254778, EPI_ISL_1254779, EPI_ISL_1254780, EPI_ISL_1254781, EPI_ISL_1254783, EPI_ISL_1254784, EPI_ISL_1254785, EPI_ISL_1254786, EPI_ISL_1254787, EPI_ISL_1254788, EPI_ISL_1254789, EPI_ISL_1254790, EPI_ISL_1254791, EPI_ISL_1254792, EPI_ISL_1254793, EPI_ISL_1254794, EPI_ISL_1254795, EPI_ISL_1254796, EPI_ISL_1254797, EPI_ISL_1254798, EPI_ISL_1254799, EPI_ISL_1254800, EPI_ISL_1254801, EPI_ISL_1254802, EPI_ISL_1254803, EPI_ISL_1254804, EPI_ISL_1254805, EPI_ISL_1254806, EPI_ISL_1254807, EPI_ISL_1254808, EPI_ISL_1254809, EPI_ISL_1254810, EPI_ISL_1254811, EPI_ISL_1254812, EPI_ISL_1254813, EPI_ISL_1254814, EPI_ISL_1254815, EPI_ISL_1254816, EPI_ISL_1254817, EPI_ISL_1254818, EPI_ISL_1254819, EPI_ISL_1254820, EPI_ISL_1254821, EPI_ISL_1254822, EPI_ISL_1254823, EPI_ISL_1254824, EPI_ISL_1254825, EPI_ISL_1254826, EPI_ISL_1254827, EPI_ISL_1254828, EPI_ISL_1254829, EPI_ISL_1254830, EPI_ISL_1254831, EPI_ISL_1254832, EPI_ISL_1254833, EPI_ISL_1254834, EPI_ISL_1254835, EPI_ISL_1254836, EPI_ISL_1254837, EPI_ISL_1254838, EPI_ISL_1254839, EPI_ISL_1254840, EPI_ISL_1254841, EPI_ISL_1254842, EPI_ISL_1254843, EPI_ISL_1254844, EPI_ISL_1254845, EPI_ISL_1254846, EPI_ISL_1254847, EPI_ISL_1254848, EPI_ISL_1254849, EPI_ISL_1254850, EPI_ISL_1254851, EPI_ISL_1254852, EPI_ISL_1254853, EPI_ISL_1254854, EPI_ISL_1254855, EPI_ISL_1254856, EPI_ISL_1254857, EPI_ISL_1254858, EPI_ISL_1254859, EPI_ISL_1254860, EPI_ISL_1254861, EPI_ISL_1254862, EPI_ISL_1254863, EPI_ISL_1254864, EPI_ISL_1254865, EPI_ISL_1254866, EPI_ISL_1254867, EPI_ISL_1254868, EPI_ISL_1254869, EPI_ISL_1254870, EPI_ISL_1254871, EPI_ISL_1254872, EPI_ISL_1254873, EPI_ISL_1254874, EPI_ISL_1254875, EPI_ISL_1254876, EPI_ISL_1254877, EPI_ISL_1254878, EPI_ISL_1254879, EPI_ISL_1254880, EPI_ISL_1254881, EPI_ISL_1254882, EPI_ISL_1254883, EPI_ISL_1254884, EPI_ISL_1254885, EPI_ISL_1254886, EPI_ISL_1254887, EPI_ISL_1254888, EPI_ISL_1254889, EPI_ISL_1254890, EPI_ISL_1254891, EPI_ISL_1254892, EPI_ISL_1254893, EPI_ISL_1254894, EPI_ISL_1254895, EPI_ISL_1254896, EPI_ISL_1254897, EPI_ISL_1254898, EPI_ISL_1254899, EPI_ISL_1254900, EPI_ISL_1254901, EPI_ISL_1254902, EPI_ISL_1254903, EPI_ISL_1254904, EPI_ISL_1254905, EPI_ISL_1254906, EPI_ISL_1254907, EPI_ISL_1254908, EPI_ISL_1254909, EPI_ISL_1254910, EPI_ISL_1254911, EPI_ISL_1254912, EPI_ISL_1254913, EPI_ISL_1254914, EPI_ISL_1254915, EPI_ISL_1254916, EPI_ISL_1254917, EPI_ISL_1254918, EPI_ISL_1254919, EPI_ISL_1254920, EPI_ISL_1254921, EPI_ISL_1254922, EPI_ISL_1254923, EPI_ISL_1254924, EPI_ISL_1254925, EPI_ISL_1254926, EPI_ISL_1254927, EPI_ISL_1254928, EPI_ISL_1254929, EPI_ISL_1254930, EPI_ISL_1254931, EPI_ISL_1254932, EPI_ISL_1254933, EPI_ISL_1254934, EPI_ISL_1254935, EPI_ISL_1254936, EPI_ISL_1254937, EPI_ISL_1254938, EPI_ISL_1254939, EPI_ISL_1254940, EPI_ISL_1254941, EPI_ISL_1254942, EPI_ISL_1254943, EPI_ISL_1254944, EPI_ISL_1254945, EPI_ISL_1254946, EPI_ISL_1254947, EPI_ISL_1254948, EPI_ISL_1254949, EPI_ISL_1254950, EPI_ISL_1254951, EPI_ISL_1254952, EPI_ISL_1254953, EPI_ISL_1254954, EPI_ISL_1254955, EPI_ISL_1254956, EPI_ISL_1254957, EPI_ISL_1254958, EPI_ISL_1254959, EPI_ISL_1254960, EPI_ISL_1254961, EPI_ISL_1254962, EPI_ISL_1254963, EPI_ISL_1254964, EPI_ISL_1254965, EPI_ISL_1254966, EPI_ISL_1254967, EPI_ISL_1254968, EPI_ISL_1254969, EPI_ISL_1254970, EPI_ISL_1254971, EPI_ISL_1254972, EPI_ISL_1254973, EPI_ISL_1254974, EPI_ISL_1254975, EPI_ISL_1254976, EPI_ISL_1254977, EPI_ISL_1254978, EPI_ISL_1254979, EPI_ISL_1254980, EPI_ISL_1254981, EPI_ISL_1254982, EPI_ISL_1254983, EPI_ISL_1254984, EPI_ISL_1254985, EPI_ISL_1254986, EPI_ISL_1254987, EPI_ISL_1254988, EPI_ISL_1254989, EPI_ISL_1254990, EPI_ISL_1254991, EPI_ISL_1254992, EPI_ISL_1254993, EPI_ISL_1254994, EPI_ISL_1254995, EPI_ISL_1254996, EPI_ISL_1254997, EPI_ISL_1254998, EPI_ISL_1254999, EPI_ISL_1255000, EPI_ISL_1255001, EPI_ISL_1255002, EPI_ISL_1255003, EPI_ISL_1255004, EPI_ISL_1255005, EPI_ISL_1255006, EPI_ISL_1255007, EPI_ISL_1255008, EPI_ISL_1255009, EPI_ISL_1255010, EPI_ISL_1255011, EPI_ISL_1255012, EPI_ISL_1255013, EPI_ISL_1255014, EPI_ISL_1255015, EPI_ISL_1255016, EPI_ISL_1255017, EPI_ISL_1255018, EPI_ISL_1255019, EPI_ISL_1255020, EPI_ISL_1255021, EPI_ISL_1255022, EPI_ISL_1255023, EPI_ISL_1255024, EPI_ISL_1255025, EPI_ISL_1255026, EPI_ISL_1255027, EPI_ISL_1255028, EPI_ISL_1255029, EPI_ISL_1255030, EPI_ISL_1255031, EPI_ISL_1255032, EPI_ISL_1255033, EPI_ISL_1255034, EPI_ISL_1255035, EPI_ISL_1255036, EPI_ISL_1255037, EPI_ISL_1255038, EPI_ISL_1255039, EPI_ISL_1255040, EPI_ISL_1255041, EPI_ISL_1255042, EPI_ISL_1255043, EPI_ISL_1255044, EPI_ISL_1255045, EPI_ISL_1255046, EPI_ISL_1255047, EPI_ISL_1255048, EPI_ISL_1255049, EPI_ISL_1255050 | | | |
| see above | Istituto Zooprofilattico Sperimentale del Mezzogiorno | Telethon Institute of Genetics and Medicine (TIGEM) | Antonio Grimaldi Patrizia Annunziata Francesco Panariello Biancamaria Pierri Claudia Tiberio Valentina Bouche Chiara Colantuono Maria Concetta Cuomo Denise Di Concilio Lucio Di Filippo Anna Manfredi Marcello Salvi Antonio Limone Luigi Atripaldi Pellegrino Cerino Andrea Ballabio Davide Cacchiarelli |
| EPI_ISL_1258988, EPI_ISL_1258989, EPI_ISL_1258990, EPI_ISL_1258991, EPI_ISL_1258992, EPI_ISL_1258993, EPI_ISL_1258994, EPI_ISL_1258995, EPI_ISL_1258996, EPI_ISL_1258997, EPI_ISL_1258998, EPI_ISL_1258999, EPI_ISL_1259000, EPI_ISL_1259001, EPI_ISL_1259002, EPI_ISL_1259003, EPI_ISL_1259004, EPI_ISL_1259005, EPI_ISL_1259006, EPI_ISL_1259007, EPI_ISL_1259008, EPI_ISL_1259009, EPI_ISL_1259010, EPI_ISL_1259011, EPI_ISL_1259012, EPI_ISL_1259013, EPI_ISL_1259014, EPI_ISL_1259015, EPI_ISL_1259016, EPI_ISL_1259017, EPI_ISL_1259018, EPI_ISL_1259019 | | | |
| see above | Azienda Ospedaliero - Universitaria di Bologna IRCCS Policlinico di Sant'Orsola - Unità di Microbiologia | Istituto Zooprofilattico Sperimentale della Lombardia e dell'Emilia Romagna (IZSLER), Risk Analysis and Genomic Epidemiology Unit | Giada Rossini, Giuliano Furlini, Tiziana Lazzarotto, Marina Morganti, Ilaria Menozzi, Erika Scaltriti, Stefano Pongolini |
| EPI_ISL_1259182, EPI_ISL_1259183, EPI_ISL_1259184, EPI_ISL_1259185, EPI_ISL_1259186, EPI_ISL_1259187, EPI_ISL_1259188, EPI_ISL_1259189, EPI_ISL_1259190, EPI_ISL_1259191, EPI_ISL_1259192, EPI_ISL_1259193 | | | |
| see above | Department of Virology, Istituto Zooprofilattico Sperimentale del Lazio e della Toscana (IZSLT) | Department of General Diagnostics; Department of Virology; Istituto Zooprofilattico Sperimentale del Lazio e della Toscana (IZSLT) | Patricia Alba, Giuseppe Manna, Elena L. Diaconu, Fabiola Feltrin, Raffaela Conti, Teresa Scicluna, Virginia Carfora, Antonella Cersini, Alessia Franco, Antonio Battisti. |
| EPI_ISL_1260662, EPI_ISL_1260663, EPI_ISL_1260664, EPI_ISL_1260665, EPI_ISL_1260666, EPI_ISL_1260667, EPI_ISL_1260668, EPI_ISL_1260669, EPI_ISL_1260670, EPI_ISL_1260671, EPI_ISL_1260672, EPI_ISL_1260673, EPI_ISL_1260674, EPI_ISL_1260675, EPI_ISL_1260676, EPI_ISL_1260677, EPI_ISL_1260678, EPI_ISL_1260679, EPI_ISL_1260680, EPI_ISL_1260681, EPI_ISL_1260682, EPI_ISL_1260683, EPI_ISL_1260684, EPI_ISL_1260685, EPI_ISL_1260686 | | | |
| see above | Azienda Sanitaria Locale di Piacenza - Presidio Ospedaliero - Laboratorio di Microbiologia | Istituto Zooprofilattico Sperimentale della Lombardia e dell'Emilia Romagna (IZSLER), Risk Analysis and Genomic Epidemiology Unit | Lo Cascio Giuliana, Schiavo Roberta, Peroni Irene, Marina Morganti, Ilaria Menozzi, Erika Scaltriti, Stefano Pongolini |
| EPI_ISL_1260840, EPI_ISL_1260841, EPI_ISL_1260842, EPI_ISL_1260843, EPI_ISL_1260844, EPI_ISL_1260845, EPI_ISL_1260846, EPI_ISL_1260847, EPI_ISL_1260848, EPI_ISL_1260849, EPI_ISL_1260850, EPI_ISL_1260851, EPI_ISL_1260852, EPI_ISL_1260853, EPI_ISL_1260854 | | | |

| | | | |
|---|---|---|---|
| see above | Center of Advanced Studies and Technology, Molecular Genetics Laboratory | Center of Advanced Studies and Technology, Molecular Genetics Laboratory | Ferrante Rossella, Mandatori Domitilla, De Fabritiis Simone, Damiani Verena, Anaclerio Federico |
| EPI_ISL_1260895, EPI_ISL_1260896 | ULSS 7 Pedemontana - Distretto 1 | Istituto Zooprofilattico Sperimentale delle Venezie | Adelaide Milani, Alessia Schivo, Annalisa Salviato, Erika Giorgia Quaranta, Luca Tassoni, Ambra Pastori, Edoardo Giussani, Alice Fusaro, Isabella Monne, Calogero Terregino, Antonia Ricci |
| EPI_ISL_1260897, EPI_ISL_1260898, EPI_ISL_1260899, EPI_ISL_1260900 | ULSS 7 Pedemontana - Distretto 2 | Istituto Zooprofilattico Sperimentale delle Venezie | Adelaide Milani, Alessia Schivo, Annalisa Salviato, Erika Giorgia Quaranta, Luca Tassoni, Ambra Pastori, Edoardo Giussani, Alice Fusaro, Isabella Monne, Calogero Terregino, Antonia Ricci |
| EPI_ISL_1260901 | Microbiologia e Virologia | Istituto Zooprofilattico Sperimentale delle Venezie | Adelaide Milani, Alessia Schivo, Annalisa Salviato, Erika Giorgia Quaranta, Luca Tassoni, Ambra Pastori, Edoardo Giussani, Alice Fusaro, Isabella Monne, Calogero Terregino, Antonia Ricci |
| EPI_ISL_1260902 | ULSS 03 Venezia | Istituto Zooprofilattico Sperimentale delle Venezie | Adelaide Milani, Alessia Schivo, Annalisa Salviato, Erika Giorgia Quaranta, Luca Tassoni, Ambra Pastori, Edoardo Giussani, Alice Fusaro, Isabella Monne, Calogero Terregino, Antonia Ricci |
| EPI_ISL_1260904, EPI_ISL_1260905, EPI_ISL_1260906, EPI_ISL_1260907, EPI_ISL_1260908, EPI_ISL_1260909, EPI_ISL_1260910, EPI_ISL_1260911, EPI_ISL_1260912 | ULSS 6 Euganea | Istituto Zooprofilattico Sperimentale delle Venezie | Adelaide Milani, Alessia Schivo, Annalisa Salviato, Erika Giorgia Quaranta, Luca Tassoni, Ambra Pastori, Edoardo Giussani, Alice Fusaro, Isabella Monne, Calogero Terregino, Antonia Ricci |
| EPI_ISL_1260917, EPI_ISL_1260919, EPI_ISL_1260920, EPI_ISL_1260921, EPI_ISL_1260922, EPI_ISL_1260923, EPI_ISL_1260924, EPI_ISL_1260925, EPI_ISL_1260926, EPI_ISL_1260927, EPI_ISL_1260928 | | | |
| see above | ULSS 03 Venezia | Istituto Zooprofilattico Sperimentale delle Venezie | Adelaide Milani, Alessia Schivo, Annalisa Salviato, Erika Giorgia Quaranta, Luca Tassoni, Ambra Pastori, Edoardo Giussani, Alice Fusaro, Isabella Monne, Calogero Terregino, Antonia Ricci |
| EPI_ISL_1260929 | ULSS 1 Dolomiti | Istituto Zooprofilattico Sperimentale delle Venezie | Adelaide Milani, Alessia Schivo, Annalisa Salviato, Erika Giorgia Quaranta, Luca Tassoni, Ambra Pastori, Edoardo Giussani, Alice Fusaro, Isabella Monne, Calogero Terregino, Antonia Ricci |
| EPI_ISL_1260930 | Microbiologia e Virologia | Istituto Zooprofilattico Sperimentale delle Venezie | Adelaide Milani, Alessia Schivo, Annalisa Salviato, Erika Giorgia Quaranta, Luca Tassoni, Ambra Pastori, Edoardo Giussani, Alice Fusaro, Isabella Monne, Calogero Terregino, Antonia Ricci |
| EPI_ISL_1260931 | ULSS 6 Euganea | Istituto Zooprofilattico Sperimentale delle Venezie | Adelaide Milani, Alessia Schivo, Annalisa Salviato, Erika Giorgia Quaranta, Luca Tassoni, Ambra Pastori, Edoardo Giussani, Alice Fusaro, Isabella Monne, Calogero Terregino, Antonia Ricci |
| EPI_ISL_1260932, EPI_ISL_1260933, EPI_ISL_1260934, EPI_ISL_1260935, EPI_ISL_1260936 | ULSS 5 Polesana | Istituto Zooprofilattico Sperimentale delle Venezie | Adelaide Milani, Alessia Schivo, Annalisa Salviato, Erika Giorgia Quaranta, Luca Tassoni, Ambra Pastori, Edoardo Giussani, Alice Fusaro, Isabella Monne, Calogero Terregino, Antonia Ricci |
| EPI_ISL_1260938 | ULSS 1 Dolomiti | Istituto Zooprofilattico Sperimentale delle Venezie | Adelaide Milani, Alessia Schivo, Annalisa Salviato, Erika Giorgia Quaranta, Luca Tassoni, Ambra Pastori, Edoardo Giussani, Alice Fusaro, Isabella Monne, Calogero Terregino, Antonia Ricci |
| EPI_ISL_1260939, EPI_ISL_1260940 | Microbiologia e Virologia | Istituto Zooprofilattico Sperimentale delle Venezie | Adelaide Milani, Alessia Schivo, Annalisa Salviato, Erika Giorgia Quaranta, Luca Tassoni, Ambra Pastori, Edoardo Giussani, Alice Fusaro, Isabella Monne, Calogero Terregino, Antonia Ricci |
| EPI_ISL_1260942 | ULSS 8 Berica | Istituto Zooprofilattico Sperimentale delle Venezie | Adelaide Milani, Alessia Schivo, Annalisa Salviato, Erika Giorgia Quaranta, Luca Tassoni, Ambra Pastori, Edoardo Giussani, Alice Fusaro, Isabella Monne, Calogero Terregino, Antonia Ricci |
| EPI_ISL_1260943 | ULSS 5 Polesana | Istituto Zooprofilattico Sperimentale delle Venezie | Adelaide Milani, Alessia Schivo, Annalisa Salviato, Erika Giorgia Quaranta, Luca Tassoni, Ambra Pastori, Edoardo Giussani, Alice Fusaro, Isabella Monne, Calogero Terregino, Antonia Ricci |
| EPI_ISL_1260944, EPI_ISL_1260945 | ULSS 7 Pedemontana - Distretto 1 | Istituto Zooprofilattico Sperimentale delle Venezie | Adelaide Milani, Alessia Schivo, Annalisa Salviato, Erika Giorgia Quaranta, Luca Tassoni, Ambra Pastori, Edoardo Giussani, Alice Fusaro, Isabella Monne, Calogero Terregino, Antonia Ricci |
| EPI_ISL_1260946, EPI_ISL_1260947, EPI_ISL_1260948 | ULSS 03 Venezia | Istituto Zooprofilattico Sperimentale delle Venezie | Adelaide Milani, Alessia Schivo, Annalisa Salviato, Erika Giorgia Quaranta, Luca Tassoni, Ambra Pastori, Edoardo Giussani, Alice Fusaro, Isabella Monne, Calogero Terregino, Antonia Ricci |
| EPI_ISL_1260949, EPI_ISL_1260950 | Microbiologia e Virologia | Istituto Zooprofilattico Sperimentale delle Venezie | Adelaide Milani, Alessia Schivo, Annalisa Salviato, Erika Giorgia Quaranta, Luca Tassoni, Ambra Pastori, Edoardo Giussani, Alice Fusaro, Isabella Monne, Calogero Terregino, Antonia Ricci |
| EPI_ISL_1260951, EPI_ISL_1260952, EPI_ISL_1260953 | ULSS 03 Venezia | Istituto Zooprofilattico Sperimentale delle Venezie | Adelaide Milani, Alessia Schivo, Annalisa Salviato, Erika Giorgia Quaranta, Luca Tassoni, Ambra Pastori, Edoardo Giussani, Alice Fusaro, Isabella Monne, Calogero Terregino, Antonia Ricci |
| EPI_ISL_1260954, EPI_ISL_1260955 | ULSS 1 Dolomiti | Istituto Zooprofilattico Sperimentale delle Venezie | Adelaide Milani, Alessia Schivo, Annalisa Salviato, Erika Giorgia Quaranta, Luca Tassoni, Ambra Pastori, Edoardo Giussani, Alice Fusaro, Isabella Monne, Calogero Terregino, Antonia Ricci |
| EPI_ISL_1260956 | ULSS 2 Treviso | Istituto Zooprofilattico Sperimentale delle Venezie | Adelaide Milani, Alessia Schivo, Annalisa Salviato, Erika Giorgia Quaranta, Luca Tassoni, Ambra Pastori, Edoardo Giussani, Alice Fusaro, Isabella Monne, Calogero Terregino, Antonia Ricci |
| EPI_ISL_1260957, EPI_ISL_1260958, EPI_ISL_1260959 | ULSS 04 Veneto Orientale | Istituto Zooprofilattico Sperimentale delle Venezie | Adelaide Milani, Alessia Schivo, Annalisa Salviato, Erika Giorgia Quaranta, Luca Tassoni, Ambra Pastori, Edoardo Giussani, Alice Fusaro, Isabella Monne, Calogero Terregino, Antonia Ricci |
| EPI_ISL_1260960, EPI_ISL_1260961, EPI_ISL_1260962 | ULSS 7 Pedemontana - Distretto 2 | Istituto Zooprofilattico Sperimentale delle Venezie | Adelaide Milani, Alessia Schivo, Annalisa Salviato, Erika Giorgia Quaranta, Luca Tassoni, Ambra Pastori, Edoardo Giussani, Alice Fusaro, Isabella Monne, Calogero Terregino, Antonia Ricci |
| EPI_ISL_1260963 | ULSS 04 Veneto Orientale | Istituto Zooprofilattico Sperimentale delle Venezie | Adelaide Milani, Alessia Schivo, Annalisa Salviato, Erika Giorgia Quaranta, Luca Tassoni, Ambra Pastori, Edoardo Giussani, Alice Fusaro, Isabella Monne, Calogero Terregino, Antonia Ricci |
| EPI_ISL_1260964 | Microbiologia e Virologia | Istituto Zooprofilattico Sperimentale delle Venezie | Adelaide Milani, Alessia Schivo, Annalisa Salviato, Erika Giorgia Quaranta, Luca Tassoni, Ambra Pastori, Edoardo Giussani, Alice Fusaro, Isabella Monne, Calogero Terregino, Antonia Ricci |
| EPI_ISL_1261427, EPI_ISL_1261447 | US Air Force School of Aerospace Medicine | US Air Force School of Aerospace Medicine | Anthony Fries, Jennifer Meyer, William Gruner, William Buggele, Amanda Javorina, Sarah Purves, Clarise Starr, Elizabeth Macias |
| EPI_ISL_1261868 | Ospedale di Venere | Istituto Zooprofilattico Sperimentale della Puglia e della Basilicata | Parisi A., Bianco A., Capozzi L., Del Sambro L., Simone D., Iacobellis M., Ridolfi D., Giannico A. |
| EPI_ISL_1265899, EPI_ISL_1265901 | Laboratorio Unico Provinciale Azienda Ospedaliero-Universitaria di Ferrara | U.O. Microbiologia, Laboratorio Unico Centro Servizi - AUSL della Romagna | Michela Boni, Sara Ghisellini, Giorgio Dirani, Silvia Zannoli, Vittorio Sambri |
| EPI_ISL_1290803 | INMI Lazzaro Spallanzani IRCCS | INMI Lazzaro Spallanzani IRCCS | E Giombini, F Messina, M Rueca, O Butera, CEM Gruber, F Santini, B Bartolini, G Bonfiglio, A Di Caro, MR Capobianchi |
| EPI_ISL_1295607 | Laboratorio Genzano - ASL RM 6 | INMI Lazzaro Spallanzani IRCCS | F Santini, B Bartolini, E Giombini, F Messina, M Rueca, G Bonfiglio, O Butera, CEM Gruber, G Tramini, E Conti, MR Capobianchi, A Di Caro |
| EPI_ISL_1295608 | Ospedale "F. Spaziani" Frosinone | INMI Lazzaro Spallanzani IRCCS | B Bartolini, E Giombini, F Messina, M Rueca, G Bonfiglio, O Butera, CEM Gruber, F Santini, R Pulselli, C Gargiulo, C Sias, A Di Caro, MR Capobianchi |
| EPI_ISL_1295614 | Ospedale "F. Spaziani" Frosinone | INMI Lazzaro Spallanzani IRCCS | E Giombini, F Messina, M Rueca, G Bonfiglio, O Butera, CEM Gruber, F Santini, B Bartolini, R Pulselli, C Sias, G Brocco, MR Capobianchi, A Di Caro |
| EPI_ISL_1296238, EPI_ISL_1296239, EPI_ISL_1296240, EPI_ISL_1296241, EPI_ISL_1296242, EPI_ISL_1296243, EPI_ISL_1296244, EPI_ISL_1296245, EPI_ISL_1296246, EPI_ISL_1296247, EPI_ISL_1296248, EPI_ISL_1296249, EPI_ISL_1296250, EPI_ISL_1296251, EPI_ISL_1296252 | | | |
| see above | Università degli Studi di Perugia | Istituto Zooprofilattico Sperimentale dell'Abruzzo e Molise "G. Caporale" | Mencacci A, Camilloni B,Lorusso A, Marcacci M, Di Domenico M, Ancora M, Curini V, Mangone I, Rinaldi A, Scialabba S, Di Pasquale A, Cammà C, Puglia I, Calistri P, Savini G |
| EPI_ISL_1296253, EPI_ISL_1296254, EPI_ISL_1296255, EPI_ISL_1296256, EPI_ISL_1296257 | Laboratorio Analisi Osp. Città di Castello - Azienda USL Umbria1 | Istituto Zooprofilattico Sperimentale dell'Abruzzo e Molise "G. Caporale" | Malagigi V, Tacconi P, Mencacci A, Camilloni B,Lorusso A, Marcacci M, Di Domenico M, Ancora M, Curini V, Mangone I, Rinaldi A, Scialabba S, Di Pasquale A, Cammà C, Puglia I, Calistri P, Savini G |

| | | | |
|---|---|---|---|
| EPI_ISL_1296258 | Università degli Studi di Perugia | Istituto Zooprofilattico Sperimentale dell'Abruzzo e Molise "G. Caporale" | Mencacci A, Camilloni B,Lorusso A, Marcacci M, Di Domenico M, Ancora M, Curini V, Mangone I, Rinaldi A, Scialabba S, Di Pasquale A, Cammà C, Puglia I, Calistri P, Savini G |
| EPI_ISL_1296259, EPI_ISL_1296260 | Laboratorio Analisi Osp. Città di Castello - Azienda USL Umbria1 | Istituto Zooprofilattico Sperimentale dell'Abruzzo e Molise "G. Caporale" | Malagigi V, Tacconi P, Mencacci A, Camilloni B,Lorusso A, Marcacci M, Di Domenico M, Ancora M, Curini V, Mangone I, Rinaldi A, Scialabba S, Di Pasquale A, Cammà C, Puglia I, Calistri P, Savini G |
| EPI_ISL_1296261, EPI_ISL_1296262 | Istituto Zooprofilattico Sperimentale Umbria e Marche "Togo Rosati" | Istituto Zooprofilattico Sperimentale dell'Abruzzo e Molise "G. Caporale" | Biagetti M,Giammarioli M,Lorusso A, Marcacci M, Di Domenico M, Ancora M, Curini V, Mangone I, Rinaldi A, Scialabba S, Di Pasquale A, Cammà C, Puglia I, Calistri P, Savini G |
| EPI_ISL_1296263, EPI_ISL_1296264, EPI_ISL_1296265, EPI_ISL_1296266, EPI_ISL_1296267 | Azienda Ospedaliera Terni | Istituto Zooprofilattico Sperimentale dell'Abruzzo e Molise "G. Caporale" | Palumbo M, ScaccettiA,Lorusso A, Marcacci M, Di Domenico M, Ancora M, Curini V, Mangone I, Rinaldi A, Scialabba S, Di Pasquale A, Cammà C, Puglia I, Calistri P, Savini G |
| EPI_ISL_1296268 | Università degli Studi di Perugia | Istituto Zooprofilattico Sperimentale dell'Abruzzo e Molise "G. Caporale" | Mencacci A, Camilloni B, Lorusso A, Marcacci M, Di Domenico M, Ancora M, Curini V, Mangone I, Rinaldi A, Scialabba S, Di Pasquale A, Cammà C, Puglia I, Calistri P, Savini G |
| EPI_ISL_1296269, EPI_ISL_1296270, EPI_ISL_1296271, EPI_ISL_1296272, EPI_ISL_1296273, EPI_ISL_1296274, EPI_ISL_1296275, EPI_ISL_1296276, EPI_ISL_1296277, EPI_ISL_1296278 | Azienda Ospedaliera Terni | Istituto Zooprofilattico Sperimentale dell'Abruzzo e Molise "G. Caporale" | Palumbo M, ScaccettiA, Lorusso A, Marcacci M, Di Domenico M, Ancora M, Curini V, Mangone I, Rinaldi A, Scialabba S, Di Pasquale A, Cammà C, Puglia I, Calistri P, Savini G |
| EPI_ISL_1296279 | Università degli Studi di Perugia | Istituto Zooprofilattico Sperimentale dell'Abruzzo e Molise "G. Caporale" | Mencacci A, Camilloni B, Lorusso A, Marcacci M, Di Domenico M, Ancora M, Curini V, Mangone I, Rinaldi A, Scialabba S, Di Pasquale A, Cammà C, Puglia I, Calistri P, Savini G |
| EPI_ISL_1296280 | Azienda Ospedaliera Terni | Istituto Zooprofilattico Sperimentale dell'Abruzzo e Molise "G. Caporale" | Palumbo M, Scaccetti A,,Lorusso A, Marcacci M, Di Domenico M, Ancora M, Curini V, Mangone I, Rinaldi A, Scialabba S, Di Pasquale A, Cammà C, Puglia I, Calistri P, Savini G |
| EPI_ISL_1296281, EPI_ISL_1296282, EPI_ISL_1296283, EPI_ISL_1296284, EPI_ISL_1296285, EPI_ISL_1296286, EPI_ISL_1296287 | Azienda Ospedaliera Terni | Istituto Zooprofilattico Sperimentale dell'Abruzzo e Molise "G. Caporale" | Palumbo M, Scaccetti A, Lorusso A, Marcacci M, Di Domenico M, Ancora M, Curini V, Mangone I, Rinaldi A, Scialabba S, Di Pasquale A, Cammà C, Puglia I, Calistri P, Savini G |
| EPI_ISL_1296288 | Università degli Studi di Perugia | Istituto Zooprofilattico Sperimentale dell'Abruzzo e Molise "G. Caporale" | Mencacci A,Camilloni B,Lorusso A, Marcacci M, Di Domenico M, Ancora M, Curini V, Mangone I, Rinaldi A, Scialabba S, Di Pasquale A, Cammà C, Puglia I, Calistri P, Savini G |
| EPI_ISL_1296289, EPI_ISL_1296290, EPI_ISL_1296291 | Università degli Studi di Perugia | Istituto Zooprofilattico Sperimentale dell'Abruzzo e Molise "G. Caporale" | Mencacci A, Camilloni B,Lorusso A, Marcacci M, Di Domenico M, Ancora M, Curini V, Mangone I, Rinaldi A, Scialabba S, Di Pasquale A, Cammà C, Puglia I, Calistri P, Savini G |
| EPI_ISL_1297827, EPI_ISL_1297828, EPI_ISL_1297829, EPI_ISL_1297830, EPI_ISL_1297831, EPI_ISL_1297832, EPI_ISL_1297833, EPI_ISL_1297834, EPI_ISL_1297835, EPI_ISL_1297836, EPI_ISL_1297837, EPI_ISL_1297838, EPI_ISL_1297839, EPI_ISL_1297840, EPI_ISL_1297841, EPI_ISL_1297842, EPI_ISL_1297843, EPI_ISL_1297844, EPI_ISL_1297845, EPI_ISL_1297846, EPI_ISL_1297847, EPI_ISL_1297848, EPI_ISL_1297849, EPI_ISL_1297850 | | | |
| see above | Arcispedale Santa Maria Nuova, Autoimmunità, Allergologia e Biotecnologie Innovative | Istituto Zooprofilattico Sperimentale della Lombardia e dell'Emilia Romagna (IZSLER), Risk Analysis and Genomic Epidemiology Unit | Alessandro Zerbini, Lucia Belloni, Stefania Croci, Marina Morganti, Ilaria Menozzi, Erika Scaltriti, Stefano Pongolini |
| EPI_ISL_1299500 | Azienda USL Umbria 2 | Istituto Zooprofilattico Sperimentale dell'Abruzzo e Molise "G. Caporale" | Proietti A, Pistoni E, Lorusso A, Marcacci M, Di Domenico M, Ancora M, Curini V, Mangone I, Rinaldi A, Scialabba S, Di Pasquale A, Cammà C, Puglia I, Calistri P, Savini G |
| EPI_ISL_1299506, EPI_ISL_1299507, EPI_ISL_1299508, EPI_ISL_1299509, EPI_ISL_1299510 | Azienda Ospedaliera Terni | Istituto Zooprofilattico Sperimentale dell'Abruzzo e Molise "G. Caporale" | Palumbo M, ScaccettiA, Lorusso A, Marcacci M, Di Domenico M, Ancora M, Curini V, Mangone I, Rinaldi A, Scialabba S, Di Pasquale A, Cammà C, Puglia I, Calistri P, Savini G |
| EPI_ISL_1299511, EPI_ISL_1299512, EPI_ISL_1299513, EPI_ISL_1299514, EPI_ISL_1299515, EPI_ISL_1299516, EPI_ISL_1299517, EPI_ISL_1299518, EPI_ISL_1299519, EPI_ISL_1299520 | Azienda Ospedaliera Terni | Istituto Zooprofilattico Sperimentale dell'Abruzzo e Molise "G. Caporale" | Palumbo m, Scaccetti A, Lorusso A, Marcacci M, Di Domenico M, Ancora M, Curini V, Mangone I, Rinaldi A, Scialabba S, Di Pasquale A, Cammà C, Puglia I, Calistri P, Savini G |
| EPI_ISL_1299521 | Azienda USL Umbria 2 | Istituto Zooprofilattico Sperimentale dell'Abruzzo e Molise "G. Caporale" | Proietti A, Pistoni E, Lorusso A, Marcacci M, Di Domenico M, Ancora M, Curini V, Mangone I, Rinaldi A, Scialabba S, Di Pasquale A, Cammà C, Puglia I, Calistri P, Savini G |
| EPI_ISL_1299522, EPI_ISL_1299523, EPI_ISL_1299524, EPI_ISL_1299525, EPI_ISL_1299526 | Azienda Ospedaliera Terni | Istituto Zooprofilattico Sperimentale dell'Abruzzo e Molise "G. Caporale" | Palumbo m, Scaccetti A, Lorusso A, Marcacci M, Di Domenico M, Ancora M, Curini V, Mangone I, Rinaldi A, Scialabba S, Di Pasquale A, Cammà C, Puglia I, Calistri P, Savini G |
| EPI_ISL_1299527 | Istituto Zooprofilattico Sperimentale Umbria e Marche "Togo Rosati" | Istituto Zooprofilattico Sperimentale dell'Abruzzo e Molise "G. Caporale" | Biagetti m, Giammarioli M, Lorusso A, Marcacci M, Di Domenico M, Ancora M, Curini V, Mangone I, Rinaldi A, Scialabba S, Di Pasquale A, Cammà C, Puglia I, Calistri P, Savini G |
| EPI_ISL_1299532, EPI_ISL_1299543 | Azienda USL Umbria 2 | Istituto Zooprofilattico Sperimentale dell'Abruzzo e Molise "G. Caporale" | Proietti A, Pistoni E, Lorusso A, Marcacci M, Di Domenico M, Ancora M, Curini V, Mangone I, Rinaldi A, Scialabba S, Di Pasquale A, Cammà C, Puglia I, Calistri P, Savini G |
| EPI_ISL_1299545, EPI_ISL_1299546, EPI_ISL_1299547, EPI_ISL_1299548, EPI_ISL_1299551, EPI_ISL_1299552 | Laboratorio Analisi Osp. Città di Castello - Azienda USL Umbria1 | Istituto Zooprofilattico Sperimentale dell'Abruzzo e Molise "G. Caporale" | Malagigi V, Tacconi P,Lorusso A, Marcacci M, Di Domenico M, Ancora M, Curini V, Mangone I, Rinaldi A, Scialabba S, Di Pasquale A, Cammà C, Puglia I, Calistri P, Savini G |
| EPI_ISL_1299554 | Azienda USL Umbria 2 | Istituto Zooprofilattico Sperimentale dell'Abruzzo e Molise "G. Caporale" | Proietti A, Pistoni E, Lorusso A, Marcacci M, Di Domenico M, Ancora M, Curini V, Mangone I, Rinaldi A, Scialabba S, Di Pasquale A, Cammà C, Puglia I, Calistri P, Savini G |
| EPI_ISL_1299555 | Laboratorio Analisi Osp. Città di Castello - Azienda USL Umbria1 | Istituto Zooprofilattico Sperimentale dell'Abruzzo e Molise "G. Caporale" | Malagigi V, Tacconi P,Lorusso A, Marcacci M, Di Domenico M, Ancora M, Curini V, Mangone I, Rinaldi A, Scialabba S, Di Pasquale A, Cammà C, Puglia I, Calistri P, Savini G |
| EPI_ISL_1299557, EPI_ISL_1299558, EPI_ISL_1299560, EPI_ISL_1299561 | Università degli Studi di Perugia | Istituto Zooprofilattico Sperimentale dell'Abruzzo e Molise "G. Caporale" | Mencacci A, Camilloni B,Lorusso A, Marcacci M, Di Domenico M, Ancora M, Curini V, Mangone I, Rinaldi A, Scialabba S, Di Pasquale A, Cammà C, Puglia I, Calistri P, Savini G |
| EPI_ISL_1299563 | Azienda Ospedaliera Terni | Istituto Zooprofilattico Sperimentale dell'Abruzzo e Molise "G. Caporale" | Palumbo m, Scaccetti A, Lorusso A, Marcacci M, Di Domenico M, Ancora M, Curini V, Mangone I, Rinaldi A, Scialabba S, Di Pasquale A, Cammà C, Puglia I, Calistri P, Savini G |
| EPI_ISL_1302402, EPI_ISL_1302403, EPI_ISL_1302404, EPI_ISL_1302405, EPI_ISL_1302406, EPI_ISL_1302407, EPI_ISL_1302408, EPI_ISL_1302409, EPI_ISL_1302410, EPI_ISL_1302411, EPI_ISL_1302412, EPI_ISL_1302413, EPI_ISL_1302414, EPI_ISL_1302415, EPI_ISL_1302416, EPI_ISL_1302417, EPI_ISL_1302418, EPI_ISL_1302419, EPI_ISL_1302420, EPI_ISL_1302421, EPI_ISL_1302422, EPI_ISL_1302423, EPI_ISL_1302424, EPI_ISL_1302425, EPI_ISL_1302426, EPI_ISL_1302427, EPI_ISL_1302428, EPI_ISL_1302430, EPI_ISL_1302431, EPI_ISL_1302432, EPI_ISL_1302433, EPI_ISL_1302434, EPI_ISL_1302435, EPI_ISL_1302436, EPI_ISL_1302437, EPI_ISL_1302438, EPI_ISL_1302439, EPI_ISL_1302440, EPI_ISL_1302441, EPI_ISL_1302442, EPI_ISL_1302443 | | | |
| see above | SC Microbiologia e Virologia AOUSS | AMES Centro Polidiagnostico Strumentale S.r.l. | Salvatore Rubino; Flavia Angioj; Laura Firino; Rosalba Govoni; Gabriele Ibba; Vincenzo Lai; Erica Mura; Bianca Paglietti; Claudia Piu; Anna Puggioni; Elena Rimini; Giulia Rocca; Caterina Serra; Sergio Uzzau. |
| EPI_ISL_1303427 | Microbiology Unit, ASST Monza - San Gerardo Hospital | Laboratory of Clinical Microbiology, Virology and Bioemergencies, ASST Fatebenefratelli Sacco - Sacco Hospital | Valeria Micheli, Alberto Rizzo, Alessandro Mancon, Fiorenza Bracchitta, Luca Rizzuto, Sergio Malandrin, Annalisa Cavallero, Maria Rita Gismondo |
| EPI_ISL_1321809 | Azienda Sanitaria Locale di Piacenza - Presidio Ospedaliero - Laboratorio di Microbiologia | U.O. Microbiologia, Laboratorio Unico Centro Servizi - AUSL della Romagna | Lo Cascio Giuliana, Schiavo Roberta, Giorgio Dirani, Silvia Zannoli, Vittorio Sambri |

| | | | |
|---|---|---|---|
| EPI_ISL_1336797 | DIP. PREV. AVEZZANO SERVIZIO DI IGIENE EPIDEMIOLOGIA E SANITA' PUBBLICA | Istituto Zooprofilattico Sperimentale dell'Abruzzo e Molise "G. Caporale" | Lorusso A, Marcacci M, Di Domenico M, Ancora M, Curini V, Di Lollo V, Mangone I, Rinaldi A, Scialabba S, Caporale M, Di Pasquale A, Cammà C, Puglia I, Calistri P, Savini G |
| EPI_ISL_1360389, EPI_ISL_1360390, EPI_ISL_1360391, EPI_ISL_1360392, EPI_ISL_1361510 | 1.AO Universitaria 'S. Giovanni di Dio e Ruggi D'Aragona, Scuola Medica Salernitana' Hospital / 2.UOC di Virologia e Microbiologia, Università della Campania 'L. Vanvitelli' / 3.AO Universitaria 'Federico II' Napoli Hospital / 4.AORN 'San Giuseppe Moscati' Avellino Hospital / 5.AO 'San Pio - presidio G. Rummo' Benevento Hospital / 6.AO 'Sant'Anna e San Sebastiano' Caserta Hospital / 7.PO 'Maria Santissima Addolorata' Eboli Hospital / 8.Biogem Istituto di Ricerche Genetiche / 9. U.O.C. di Genetica Medica e di Laboratorio A.O.R.N., Azienda Ospedaliera di Rilievo Nazionale Antonio Cardarelli, Napoli / 10. Centro di riferimento Oncologico della Basilicata (IRCCS-CROB), Rionero in Vulture (PZ) / 11. Presidio Ospedaliero di Agropoli, Agropoli (SA). | 1. Genome Research Center for Health (CRGS) / 2. Laboratory of Molecular Medicine and Genomics(LMMGe) / 3. Center for Research in Pure and Applied Mathematics (CRMPA) | Giorgio Giurato (Corresponding Author), Francesca Rizzo (Corresponding Author), Alessandro Weisz (Corresponding Author), Gianluigi Franci, Giovanni Nassa, Pasquale Pagliano, Roberta Tarallo, Elena Alexandrova, Ylenia D'Agostino, Carlo Ferravante, Jessica Lamberti, Viola Melone, Domenico Memoli, Valeria Mirici Cappa, Domenico Palumbo, Giovanni Pecoraro, Assunta Sellitto, Oriana Strianese, Ilaria Terenzi, Giuseppe Fenza, Aniello Gentile, Antonello Saccomanno, Sonia Amabile, Teresa Rocco, Annamaria Salvati, Emilia Vaccaro, Massimiliano Galdiero, Michele Cennamo, Giuseppe Portella, Maria Grazia Foti, Mariarosaria Ingino, Maria Landi, Maurizio Fumi, Vincenzo Rocco, Rita Greco, Vittoria Letizia, Arnolfo Petruzziello, Maddalena Schioppa, Gregorio Goffredi, Francesca Marciano, Michele Caraglia, Alessia Cossu, Marianna Scrima, Edmondo Adorisio, Morena D'Avenia, Michela Iacobellis, Rosanna Piluscio, Giorgio Dirani, Vittorio Sambri, Simona Semprini, Silvia Zanoli, Francesco Curcio, Stefania Marzinotto, Andreina Baj, Fausto Sessa, Massimiliano Chetta, Marina Tarsitano, Giusy Giordano, Marcello Ametrano, Mariano Fiorenza, Caivano Antonella, D'Auria Fiorella, De Luca Luciana. |
| EPI_ISL_1361511, EPI_ISL_1361512, EPI_ISL_1361513, EPI_ISL_1361514 | 1.AO Universitaria 'S. Giovanni di Dio e Ruggi D'Aragona, Scuola Medica Salernitana' Hospital / 2.UOC di Virologia e Microbiologia, Università della Campania 'L. Vanvitelli' / 3.AO Universitaria 'Federico II' Napoli Hospital / 4.AORN 'San Giuseppe Moscati' Avellino Hospital / 5.AO 'San Pio - presidio G. Rummo' Benevento Hospital / 6.AO 'Sant'Anna e San Sebastiano' Caserta Hospital / 7.PO 'Maria Santissima Addolorata' Eboli Hospital / 8.Biogem Istituto di Ricerche Genetiche / 9. U.O.C. di Genetica Medica e di Laboratorio A.O.R.N., Azienda Ospedaliera di Rilievo Nazionale Antonio Cardarelli, Napoli / 10. Centro di riferimento Oncologico della Basilicata (IRCCS-CROB), Rionero in Vulture (PZ) / 11. Presidio Ospedaliero di Agropoli, Agropoli (SA). | 1. Genome Research Center for Health (CRGS) / 2. Laboratory of Molecular Medicine and Genomics(LMMGe) / 3. Center for Research in Pure and Applied Mathematics (CRMPA) | Giorgio Giurato (Corresponding Author), Francesca Rizzo (Corresponding Author), Alessandro Weisz (Corresponding Author), Gianluigi Franci, Giovanni Nassa, Pasquale Pagliano, Roberta Tarallo, Elena Alexandrova, Ylenia D'Agostino, Carlo Ferravante, Jessica Lamberti, Viola Melone, Domenico Memoli, Valeria Mirici Cappa, Domenico Palumbo, Giovanni Pecoraro, Assunta Sellitto, Oriana Strianese, Ilaria Terenzi, Giuseppe Fenza, Aniello Gentile, Antonello Saccomanno, Sonia Amabile, Teresa Rocco, Annamaria Salvati, Emilia Vaccaro, Massimiliano Galdiero, Michele Cennamo, Giuseppe Portella, Maria Grazia Foti, Mariarosaria Ingino, Maria Landi, Maurizio Fumi, Vincenzo Rocco, Rita Greco, Vittoria Letizia, Arnolfo Petruzziello, Maddalena Schioppa, Gregorio Goffredi, Francesca Marciano, Michele Caraglia, Alessia Cossu, Marianna Scrima, Edmondo Adorisio, Morena D'Avenia, Michela Iacobellis, Rosanna Piluscio, Giorgio Dirani, Vittorio Sambri, Simona Semprini, Silvia Zanoli, Francesco Curcio, Stefania Marzinotto, Andreina Baj, Fausto Sessa. |
| EPI_ISL_1361515 | 1.AO Universitaria 'S. Giovanni di Dio e Ruggi D'Aragona, Scuola Medica Salernitana' Hospital / 2.UOC di Virologia e Microbiologia, Università della Campania 'L. Vanvitelli' / 3.AO Universitaria 'Federico II' Napoli Hospital / 4.AORN 'San Giuseppe Moscati' Avellino Hospital / 5.AO 'San Pio - presidio G. Rummo' Benevento Hospital / 6.AO 'Sant'Anna e San Sebastiano' Caserta Hospital / 7.PO 'Maria Santissima Addolorata' Eboli Hospital / 8.Biogem Istituto di Ricerche Genetiche / 9. U.O.C. di Genetica Medica e di Laboratorio A.O.R.N., Azienda Ospedaliera di Rilievo Nazionale Antonio Cardarelli, Napoli / 10. Centro di riferimento Oncologico della Basilicata (IRCCS-CROB), Rionero in Vulture (PZ) / 11. Presidio Ospedaliero di Agropoli, Agropoli (SA). | 1. Genome Research Center for Health (CRGS) / 2. Laboratory of Molecular Medicine and Genomics(LMMGe) / 3. Center for Research in Pure and Applied Mathematics (CRMPA) | Giorgio Giurato (Corresponding Author), Francesca Rizzo (Corresponding Author), Alessandro Weisz (Corresponding Author), Gianluigi Franci, Giovanni Nassa, Pasquale Pagliano, Roberta Tarallo, Elena Alexandrova, Ylenia D'Agostino, Carlo Ferravante, Jessica Lamberti, Viola Melone, Domenico Memoli, Valeria Mirici Cappa, Domenico Palumbo, Giovanni Pecoraro, Assunta Sellitto, Oriana Strianese, Ilaria Terenzi, Giuseppe Fenza, Aniello Gentile, Antonello Saccomanno, Sonia Amabile, Teresa Rocco, Annamaria Salvati, Emilia Vaccaro, Massimiliano Galdiero, Michele Cennamo, Giuseppe Portella, Maria Grazia Foti, Mariarosaria Ingino, Maria Landi, Maurizio Fumi, Vincenzo Rocco, Rita Greco, Vittoria Letizia, Arnolfo Petruzziello, Maddalena Schioppa, Gregorio Goffredi, Francesca Marciano, Michele Caraglia, Alessia Cossu, Marianna Scrima, Edmondo Adorisio, Morena D'Avenia, Michela Iacobellis, Rosanna Piluscio, Giorgio Dirani, Vittorio Sambri, Simona Semprini, Silvia Zanoli, Francesco Curcio, Stefania Marzinotto, Andreina Baj, Fausto Sessa, Massimiliano Chetta, Marina Tarsitano, Giusy Giordano, Marcello Ametrano, Mariano Fiorenza, Caivano Antonella, D'Auria Fiorella, De Luca Luciana. |
| EPI_ISL_1361516, EPI_ISL_1361517, EPI_ISL_1361518, EPI_ISL_1361519 | 1.AO Universitaria 'S. Giovanni di Dio e Ruggi D'Aragona, Scuola Medica Salernitana' Hospital / 2.UOC di Virologia e Microbiologia, Università della Campania 'L. Vanvitelli' / 3.AO Universitaria 'Federico II' Napoli Hospital / 4.AORN 'San Giuseppe Moscati' Avellino Hospital / 5.AO 'San Pio - presidio G. Rummo' Benevento Hospital / 6.AO 'Sant'Anna e San Sebastiano' Caserta Hospital / 7.PO 'Maria Santissima Addolorata' Eboli Hospital / 8.Biogem Istituto di Ricerche Genetiche / 9. U.O.C. di Genetica Medica e di Laboratorio A.O.R.N., Azienda Ospedaliera di Rilievo Nazionale Antonio Cardarelli, Napoli / 10. Centro di riferimento Oncologico della Basilicata (IRCCS-CROB), Rionero in Vulture (PZ) / 11. Presidio Ospedaliero di Agropoli, Agropoli (SA). | 1. Genome Research Center for Health (CRGS) / 2. Laboratory of Molecular Medicine and Genomics(LMMGe) / 3. Center for Research in Pure and Applied Mathematics (CRMPA) | Giorgio Giurato (Corresponding Author), Francesca Rizzo (Corresponding Author), Alessandro Weisz (Corresponding Author), Gianluigi Franci, Giovanni Nassa, Pasquale Pagliano, Roberta Tarallo, Elena Alexandrova, Ylenia D'Agostino, Carlo Ferravante, Jessica Lamberti, Viola Melone, Domenico Memoli, Valeria Mirici Cappa, Domenico Palumbo, Giovanni Pecoraro, Assunta Sellitto, Oriana Strianese, Ilaria Terenzi, Giuseppe Fenza, Aniello Gentile, Antonello Saccomanno, Sonia Amabile, Teresa Rocco, Annamaria Salvati, Emilia Vaccaro, Massimiliano Galdiero, Michele Cennamo, Giuseppe Portella, Maria Grazia Foti, Mariarosaria Ingino, Maria Landi, Maurizio Fumi, Vincenzo Rocco, Rita Greco, Vittoria Letizia, Arnolfo Petruzziello, Maddalena Schioppa, Gregorio Goffredi, Francesca Marciano, Michele Caraglia, Alessia Cossu, Marianna Scrima, Edmondo Adorisio, Morena D'Avenia, Michela Iacobellis, Rosanna Piluscio, Giorgio Dirani, Vittorio Sambri, Simona Semprini, Silvia Zanoli, Francesco Curcio, Stefania Marzinotto, Andreina Baj, Fausto Sessa. |
| EPI_ISL_1361520 | 1.AO Universitaria 'S. Giovanni di Dio e Ruggi D'Aragona, Scuola Medica Salernitana' Hospital / 2.UOC di Virologia e Microbiologia, Università della Campania 'L. Vanvitelli' / 3.AO Universitaria 'Federico II' Napoli Hospital / 4.AORN 'San Giuseppe Moscati' Avellino Hospital / 5.AO 'San Pio - presidio G. Rummo' Benevento Hospital / 6.AO 'Sant'Anna e San Sebastiano' Caserta Hospital / 7.PO 'Maria Santissima Addolorata' Eboli Hospital / 8.Biogem Istituto di Ricerche Genetiche / 9. U.O.C. di Genetica Medica e di Laboratorio A.O.R.N., Azienda Ospedaliera di Rilievo Nazionale Antonio Cardarelli, Napoli / 10. Centro di riferimento Oncologico della Basilicata (IRCCS-CROB), Rionero in Vulture (PZ) / 11. Presidio Ospedaliero di Agropoli, Agropoli (SA). | 1. Genome Research Center for Health (CRGS) / 2. Laboratory of Molecular Medicine and Genomics(LMMGe) / 3. Center for Research in Pure and Applied Mathematics (CRMPA) | Giorgio Giurato (Corresponding Author), Francesca Rizzo (Corresponding Author), Alessandro Weisz (Corresponding Author), Gianluigi Franci, Giovanni Nassa, Pasquale Pagliano, Roberta Tarallo, Elena Alexandrova, Ylenia D'Agostino, Carlo Ferravante, Jessica Lamberti, Viola Melone, Domenico Memoli, Valeria Mirici Cappa, Domenico Palumbo, Giovanni Pecoraro, Assunta Sellitto, Oriana Strianese, Ilaria Terenzi, Giuseppe Fenza, Aniello Gentile, Antonello Saccomanno, Sonia Amabile, Teresa Rocco, Annamaria Salvati, Emilia Vaccaro, Massimiliano Galdiero, Michele Cennamo, Giuseppe Portella, Maria Grazia Foti, Mariarosaria Ingino, Maria Landi, Maurizio Fumi, Vincenzo Rocco, Rita Greco, Vittoria Letizia, Arnolfo Petruzziello, Maddalena Schioppa, Gregorio Goffredi, Francesca Marciano, Michele Caraglia, Alessia Cossu, Marianna Scrima, Edmondo Adorisio, Morena D'Avenia, Michela Iacobellis, Rosanna Piluscio, Giorgio Dirani, Vittorio Sambri, Simona Semprini, Silvia Zanoli, Francesco Curcio, Stefania Marzinotto, Andreina Baj, Fausto Sessa, Massimiliano Chetta, Marina Tarsitano, Giusy Giordano, Marcello Ametrano, Mariano Fiorenza, Caivano Antonella, D'Auria Fiorella, De Luca Luciana. |
| EPI_ISL_1361521, EPI_ISL_1361522, EPI_ISL_1361523, EPI_ISL_1361524 | 1.AO Universitaria 'S. Giovanni di Dio e Ruggi D'Aragona, Scuola Medica Salernitana' Hospital / 2.UOC di Virologia e Microbiologia, Università della Campania 'L. Vanvitelli' / 3.AO Universitaria 'Federico II' Napoli Hospital / 4.AORN 'San Giuseppe Moscati' Avellino Hospital / 5.AO 'San Pio - presidio G. Rummo' Benevento Hospital / 6.AO 'Sant'Anna e San Sebastiano' Caserta Hospital / 7.PO 'Maria Santissima Addolorata' Eboli Hospital / 8.Biogem Istituto di Ricerche Genetiche / 9. U.O.C. di Genetica Medica e di Laboratorio A.O.R.N., Azienda Ospedaliera di Rilievo Nazionale Antonio Cardarelli, Napoli / 10. Centro di riferimento Oncologico della Basilicata (IRCCS-CROB), Rionero in Vulture (PZ) / 11. Presidio Ospedaliero di Agropoli, Agropoli (SA). | 1. Genome Research Center for Health (CRGS) / 2. Laboratory of Molecular Medicine and Genomics(LMMGe) / 3. Center for Research in Pure and Applied Mathematics (CRMPA) | Giorgio Giurato (Corresponding Author), Francesca Rizzo (Corresponding Author), Alessandro Weisz (Corresponding Author), Gianluigi Franci, Giovanni Nassa, Pasquale Pagliano, Roberta Tarallo, Elena Alexandrova, Ylenia D'Agostino, Carlo Ferravante, Jessica Lamberti, Viola Melone, Domenico Memoli, Valeria Mirici Cappa, Domenico Palumbo, Giovanni Pecoraro, Assunta Sellitto, Oriana Strianese, Ilaria Terenzi, Giuseppe Fenza, Aniello Gentile, Antonello Saccomanno, Sonia Amabile, Teresa Rocco, Annamaria Salvati, Emilia Vaccaro, Massimiliano Galdiero, Michele Cennamo, Giuseppe Portella, Maria Grazia Foti, Mariarosaria Ingino, Maria Landi, Maurizio Fumi, Vincenzo Rocco, Rita Greco, Vittoria Letizia, Arnolfo Petruzziello, Maddalena Schioppa, Gregorio Goffredi, Francesca Marciano, Michele Caraglia, Alessia Cossu, Marianna Scrima, Edmondo Adorisio, Morena D'Avenia, Michela Iacobellis, Rosanna Piluscio, Giorgio Dirani, Vittorio Sambri, Simona Semprini, Silvia Zanoli, Francesco Curcio, Stefania Marzinotto, Andreina Baj, Fausto Sessa. |
| EPI_ISL_1361525 | 1.AO Universitaria 'S. Giovanni di Dio e Ruggi D'Aragona, Scuola Medica Salernitana' Hospital / 2.UOC di Virologia e Microbiologia, Università della Campania 'L. Vanvitelli' / 3.AO Universitaria 'Federico II' Napoli Hospital / 4.AORN 'San | 1. Genome Research Center for Health (CRGS) / 2. Laboratory of Molecular Medicine and Genomics(LMMGe) / 3. Center for Research in Pure and Applied Mathematics (CRMPA) | Giorgio Giurato (Corresponding Author), Francesca Rizzo (Corresponding Author), Alessandro Weisz (Corresponding Author), Gianluigi Franci, Giovanni Nassa, Pasquale Pagliano, Roberta Tarallo, Elena Alexandrova, Ylenia D'Agostino, Carlo Ferravante, Jessica Lamberti, Viola Melone, Domenico Memoli, Valeria Mirici Cappa, Domenico Palumbo, Giovanni Pecoraro, Assunta Sellitto, Oriana Strianese, Ilaria Terenzi, Giuseppe Fenza, Aniello Gentile, Antonello Saccomanno, Sonia Amabile, Teresa Rocco, Annamaria Salvati, Emilia Vaccaro, Massimiliano Galdiero, Michele Cennamo, Giuseppe Portella, Maria |

| | | | |
|---|---|---|---|
| | Giuseppe Moscati' Avellino Hospital / 5.AO 'San Pio - presidio G. Rummo' Benevento Hospital / 6.AO 'Sant'Anna e San Sebastiano' Caserta Hospital / 7.PO 'Maria Santissima Addolorata' Eboli Hospital / 8.Biogem Istituto di Ricerche Genetiche / 9. U.O.C. di Genetica Medica e di Laboratorio A.O.R.N., Azienda Ospedaliera di Rilievo Nazionale Antonio Cardarelli, Napoli / 10. Centro di riferimento Oncologico della Basilicata (IRCCS-CROB), Rionero in Vulture (PZ) / 11. Presidio Ospedaliero di Agropoli, Agropoli (SA). | | Grazia Foti, Mariarosaria Ingino, Maria Landi, Maurizio Fumi, Vincenzo Rocco, Rita Greco, Vittoria Letizia, Arnolfo Petruzziello, Maddalena Schioppa, Gregorio Goffredi, Francesca Marciano, Michele Caraglia, Alessia Cossu, Marianna Scrima, Edmondo Adorisio, Morena D'Avenia, Michela Iacobellis, Rosanna Piluscio, Giorgio Dirani, Vittorio Sambri, Simona Semprini, Silvia Zanoli, Francesco Curcio, Stefania Marzinotto, Andreina Baj, Fausto Sessa, Massimiliano Chetta, Marina Tarsitano, Giusy Giordano, Marcello Ametrano, Mariano Fiorenza, Caivano Antonella, D'Auria Fiorella, De Luca Luciana. |
| EPI_ISL_1361526, EPI_ISL_1361527, EPI_ISL_1361528, EPI_ISL_1361529 | 1.AO Universitaria 'S. Giovanni di Dio e Ruggi D'Aragona, Scuola Medica Salernitana' Hospital / 2.UOC di Virologia e Microbiologia, Università della Campania 'L. Vanvitelli' / 3.AO Universitaria 'Federico II' Napoli Hospital / 4.AORN 'San Giuseppe Moscati' Avellino Hospital / 5.AO 'San Pio - presidio G. Rummo' Benevento Hospital / 6.AO 'Sant'Anna e San Sebastiano' Caserta Hospital / 7.PO 'Maria Santissima Addolorata' Eboli Hospital / 8.Biogem Istituto di Ricerche Genetiche / 9. U.O.C. di Genetica Medica e di Laboratorio A.O.R.N., Azienda Ospedaliera di Rilievo Nazionale Antonio Cardarelli, Napoli / 10. Centro di riferimento Oncologico della Basilicata (IRCCS-CROB), Rionero in Vulture (PZ) / 11. Presidio Ospedaliero di Agropoli, Agropoli (SA). | 1. Genome Research Center for Health (CRGS) / 2. Laboratory of Molecular Medicine and Genomics(LMMGe) / 3. Center for Research in Pure and Applied Mathematics (CRMPA) | Giorgio Giurato (Corresponding Author), Francesca Rizzo (Corresponding Author), Alessandro Weisz (Corresponding Author), Gianluigi Franci, Giovanni Nassa, Pasquale Pagliano, Roberta Tarallo, Elena Alexandrova, Ylenia D'Agostino, Carlo Ferravante, Jessica Lamberti, Viola Melone, Domenico Memoli, Valeria Mirici Cappa, Domenico Palumbo, Giovanni Pecoraro, Assunta Sellitto, Oriana Strianese, Ilaria Terenzi, Giuseppe Fenza, Aniello Gentile, Antonello Saccomanno, Sonia Amabile, Teresa Rocco, Annamaria Salvati, Emilia Vaccaro, Massimiliano Galdiero, Michele Cennamo, Giuseppe Portella, Maria Grazia Foti, Mariarosaria Ingino, Maria Landi, Maurizio Fumi, Vincenzo Rocco, Rita Greco, Vittoria Letizia, Arnolfo Petruzziello, Maddalena Schioppa, Gregorio Goffredi, Francesca Marciano, Michele Caraglia, Alessia Cossu, Marianna Scrima, Edmondo Adorisio, Morena D'Avenia, Michela Iacobellis, Rosanna Piluscio, Giorgio Dirani, Vittorio Sambri, Simona Semprini, Silvia Zanoli, Francesco Curcio, Stefania Marzinotto, Andreina Baj, Fausto Sessa. |
| EPI_ISL_1361530 | 1.AO Universitaria 'S. Giovanni di Dio e Ruggi D'Aragona, Scuola Medica Salernitana' Hospital / 2.UOC di Virologia e Microbiologia, Università della Campania 'L. Vanvitelli' / 3.AO Universitaria 'Federico II' Napoli Hospital / 4.AORN 'San Giuseppe Moscati' Avellino Hospital / 5.AO 'San Pio - presidio G. Rummo' Benevento Hospital / 6.AO 'Sant'Anna e San Sebastiano' Caserta Hospital / 7.PO 'Maria Santissima Addolorata' Eboli Hospital / 8.Biogem Istituto di Ricerche Genetiche / 9. U.O.C. di Genetica Medica e di Laboratorio A.O.R.N., Azienda Ospedaliera di Rilievo Nazionale Antonio Cardarelli, Napoli / 10. Centro di riferimento Oncologico della Basilicata (IRCCS-CROB), Rionero in Vulture (PZ) / 11. Presidio Ospedaliero di Agropoli, Agropoli (SA). | 1. Genome Research Center for Health (CRGS) / 2. Laboratory of Molecular Medicine and Genomics(LMMGe) / 3. Center for Research in Pure and Applied Mathematics (CRMPA) | Giorgio Giurato (Corresponding Author), Francesca Rizzo (Corresponding Author), Alessandro Weisz (Corresponding Author), Gianluigi Franci, Giovanni Nassa, Pasquale Pagliano, Roberta Tarallo, Elena Alexandrova, Ylenia D'Agostino, Carlo Ferravante, Jessica Lamberti, Viola Melone, Domenico Memoli, Valeria Mirici Cappa, Domenico Palumbo, Giovanni Pecoraro, Assunta Sellitto, Oriana Strianese, Ilaria Terenzi, Giuseppe Fenza, Aniello Gentile, Antonello Saccomanno, Sonia Amabile, Teresa Rocco, Annamaria Salvati, Emilia Vaccaro, Massimiliano Galdiero, Michele Cennamo, Giuseppe Portella, Maria Grazia Foti, Mariarosaria Ingino, Maria Landi, Maurizio Fumi, Vincenzo Rocco, Rita Greco, Vittoria Letizia, Arnolfo Petruzziello, Maddalena Schioppa, Gregorio Goffredi, Francesca Marciano, Michele Caraglia, Alessia Cossu, Marianna Scrima, Edmondo Adorisio, Morena D'Avenia, Michela Iacobellis, Rosanna Piluscio, Giorgio Dirani, Vittorio Sambri, Simona Semprini, Silvia Zanoli, Francesco Curcio, Stefania Marzinotto, Andreina Baj, Fausto Sessa, Massimiliano Chetta, Marina Tarsitano, Giusy Giordano, Marcello Ametrano, Mariano Fiorenza, Caivano Antonella, D'Auria Fiorella, De Luca Luciana. |
| EPI_ISL_1361531, EPI_ISL_1361532, EPI_ISL_1361533, EPI_ISL_1361534 | 1.AO Universitaria 'S. Giovanni di Dio e Ruggi D'Aragona, Scuola Medica Salernitana' Hospital / 2.UOC di Virologia e Microbiologia, Università della Campania 'L. Vanvitelli' / 3.AO Universitaria 'Federico II' Napoli Hospital / 4.AORN 'San Giuseppe Moscati' Avellino Hospital / 5.AO 'San Pio - presidio G. Rummo' Benevento Hospital / 6.AO 'Sant'Anna e San Sebastiano' Caserta Hospital / 7.PO 'Maria Santissima Addolorata' Eboli Hospital / 8.Biogem Istituto di Ricerche Genetiche / 9. U.O.C. di Genetica Medica e di Laboratorio A.O.R.N., Azienda Ospedaliera di Rilievo Nazionale Antonio Cardarelli, Napoli / 10. Centro di riferimento Oncologico della Basilicata (IRCCS-CROB), Rionero in Vulture (PZ) / 11. Presidio Ospedaliero di Agropoli, Agropoli (SA). | 1. Genome Research Center for Health (CRGS) / 2. Laboratory of Molecular Medicine and Genomics(LMMGe) / 3. Center for Research in Pure and Applied Mathematics (CRMPA) | Giorgio Giurato (Corresponding Author), Francesca Rizzo (Corresponding Author), Alessandro Weisz (Corresponding Author), Gianluigi Franci, Giovanni Nassa, Pasquale Pagliano, Roberta Tarallo, Elena Alexandrova, Ylenia D'Agostino, Carlo Ferravante, Jessica Lamberti, Viola Melone, Domenico Memoli, Valeria Mirici Cappa, Domenico Palumbo, Giovanni Pecoraro, Assunta Sellitto, Oriana Strianese, Ilaria Terenzi, Giuseppe Fenza, Aniello Gentile, Antonello Saccomanno, Sonia Amabile, Teresa Rocco, Annamaria Salvati, Emilia Vaccaro, Massimiliano Galdiero, Michele Cennamo, Giuseppe Portella, Maria Grazia Foti, Mariarosaria Ingino, Maria Landi, Maurizio Fumi, Vincenzo Rocco, Rita Greco, Vittoria Letizia, Arnolfo Petruzziello, Maddalena Schioppa, Gregorio Goffredi, Francesca Marciano, Michele Caraglia, Alessia Cossu, Marianna Scrima, Edmondo Adorisio, Morena D'Avenia, Michela Iacobellis, Rosanna Piluscio, Giorgio Dirani, Vittorio Sambri, Simona Semprini, Silvia Zanoli, Francesco Curcio, Stefania Marzinotto, Andreina Baj, Fausto Sessa. |
| EPI_ISL_1361535 | 1.AO Universitaria 'S. Giovanni di Dio e Ruggi D'Aragona, Scuola Medica Salernitana' Hospital / 2.UOC di Virologia e Microbiologia, Università della Campania 'L. Vanvitelli' / 3.AO Universitaria 'Federico II' Napoli Hospital / 4.AORN 'San Giuseppe Moscati' Avellino Hospital / 5.AO 'San Pio - presidio G. Rummo' Benevento Hospital / 6.AO 'Sant'Anna e San Sebastiano' Caserta Hospital / 7.PO 'Maria Santissima Addolorata' Eboli Hospital / 8.Biogem Istituto di Ricerche Genetiche / 9. U.O.C. di Genetica Medica e di Laboratorio A.O.R.N., Azienda Ospedaliera di Rilievo Nazionale Antonio Cardarelli, Napoli / 10. Centro di riferimento Oncologico della Basilicata (IRCCS-CROB), Rionero in Vulture (PZ) / 11. Presidio Ospedaliero di Agropoli, Agropoli (SA). | 1. Genome Research Center for Health (CRGS) / 2. Laboratory of Molecular Medicine and Genomics(LMMGe) / 3. Center for Research in Pure and Applied Mathematics (CRMPA) | Giorgio Giurato (Corresponding Author), Francesca Rizzo (Corresponding Author), Alessandro Weisz (Corresponding Author), Gianluigi Franci, Giovanni Nassa, Pasquale Pagliano, Roberta Tarallo, Elena Alexandrova, Ylenia D'Agostino, Carlo Ferravante, Jessica Lamberti, Viola Melone, Domenico Memoli, Valeria Mirici Cappa, Domenico Palumbo, Giovanni Pecoraro, Assunta Sellitto, Oriana Strianese, Ilaria Terenzi, Giuseppe Fenza, Aniello Gentile, Antonello Saccomanno, Sonia Amabile, Teresa Rocco, Annamaria Salvati, Emilia Vaccaro, Massimiliano Galdiero, Michele Cennamo, Giuseppe Portella, Maria Grazia Foti, Mariarosaria Ingino, Maria Landi, Maurizio Fumi, Vincenzo Rocco, Rita Greco, Vittoria Letizia, Arnolfo Petruzziello, Maddalena Schioppa, Gregorio Goffredi, Francesca Marciano, Michele Caraglia, Alessia Cossu, Marianna Scrima, Edmondo Adorisio, Morena D'Avenia, Michela Iacobellis, Rosanna Piluscio, Giorgio Dirani, Vittorio Sambri, Simona Semprini, Silvia Zanoli, Francesco Curcio, Stefania Marzinotto, Andreina Baj, Fausto Sessa, Massimiliano Chetta, Marina Tarsitano, Giusy Giordano, Marcello Ametrano, Mariano Fiorenza, Caivano Antonella, D'Auria Fiorella, De Luca Luciana. |
| EPI_ISL_1361536, EPI_ISL_1361537, EPI_ISL_1361538, EPI_ISL_1361539 | 1.AO Universitaria 'S. Giovanni di Dio e Ruggi D'Aragona, Scuola Medica Salernitana' Hospital / 2.UOC di Virologia e Microbiologia, Università della Campania 'L. Vanvitelli' / 3.AO Universitaria 'Federico II' Napoli Hospital / 4.AORN 'San Giuseppe Moscati' Avellino Hospital / 5.AO 'San Pio - presidio G. Rummo' Benevento Hospital / 6.AO 'Sant'Anna e San Sebastiano' Caserta Hospital / 7.PO 'Maria Santissima Addolorata' Eboli Hospital / 8.Biogem Istituto di Ricerche Genetiche / 9. U.O.C. di Genetica Medica e di Laboratorio A.O.R.N., Azienda Ospedaliera di Rilievo Nazionale Antonio Cardarelli, Napoli / 10. Centro di riferimento Oncologico della Basilicata (IRCCS-CROB), Rionero in Vulture (PZ) / 11. Presidio Ospedaliero di Agropoli, Agropoli (SA). | 1. Genome Research Center for Health (CRGS) / 2. Laboratory of Molecular Medicine and Genomics(LMMGe) / 3. Center for Research in Pure and Applied Mathematics (CRMPA) | Giorgio Giurato (Corresponding Author), Francesca Rizzo (Corresponding Author), Alessandro Weisz (Corresponding Author), Gianluigi Franci, Giovanni Nassa, Pasquale Pagliano, Roberta Tarallo, Elena Alexandrova, Ylenia D'Agostino, Carlo Ferravante, Jessica Lamberti, Viola Melone, Domenico Memoli, Valeria Mirici Cappa, Domenico Palumbo, Giovanni Pecoraro, Assunta Sellitto, Oriana Strianese, Ilaria Terenzi, Giuseppe Fenza, Aniello Gentile, Antonello Saccomanno, Sonia Amabile, Teresa Rocco, Annamaria Salvati, Emilia Vaccaro, Massimiliano Galdiero, Michele Cennamo, Giuseppe Portella, Maria Grazia Foti, Mariarosaria Ingino, Maria Landi, Maurizio Fumi, Vincenzo Rocco, Rita Greco, Vittoria Letizia, Arnolfo Petruzziello, Maddalena Schioppa, Gregorio Goffredi, Francesca Marciano, Michele Caraglia, Alessia Cossu, Marianna Scrima, Edmondo Adorisio, Morena D'Avenia, Michela Iacobellis, Rosanna Piluscio, Giorgio Dirani, Vittorio Sambri, Simona Semprini, Silvia Zanoli, Francesco Curcio, Stefania Marzinotto, Andreina Baj, Fausto Sessa. |
| EPI_ISL_1361540 | 1.AO Universitaria 'S. Giovanni di Dio e Ruggi D'Aragona, Scuola Medica Salernitana' Hospital / 2.UOC di Virologia e Microbiologia, Università della Campania 'L. Vanvitelli' / 3.AO Universitaria 'Federico II' Napoli Hospital / 4.AORN 'San Giuseppe Moscati' Avellino Hospital / 5.AO 'San Pio - presidio G. Rummo' Benevento Hospital / 6.AO 'Sant'Anna e San Sebastiano' Caserta Hospital / 7.PO 'Maria Santissima Addolorata' Eboli Hospital / 8.Biogem Istituto di Ricerche Genetiche / 9. U.O.C. di Genetica Medica e di Laboratorio A.O.R.N., Azienda Ospedaliera di Rilievo Nazionale Antonio | 1. Genome Research Center for Health (CRGS) / 2. Laboratory of Molecular Medicine and Genomics(LMMGe) / 3. Center for Research in Pure and Applied Mathematics (CRMPA) | Giorgio Giurato (Corresponding Author), Francesca Rizzo (Corresponding Author), Alessandro Weisz (Corresponding Author), Gianluigi Franci, Giovanni Nassa, Pasquale Pagliano, Roberta Tarallo, Elena Alexandrova, Ylenia D'Agostino, Carlo Ferravante, Jessica Lamberti, Viola Melone, Domenico Memoli, Valeria Mirici Cappa, Domenico Palumbo, Giovanni Pecoraro, Assunta Sellitto, Oriana Strianese, Ilaria Terenzi, Giuseppe Fenza, Aniello Gentile, Antonello Saccomanno, Sonia Amabile, Teresa Rocco, Annamaria Salvati, Emilia Vaccaro, Massimiliano Galdiero, Michele Cennamo, Giuseppe Portella, Maria Grazia Foti, Mariarosaria Ingino, Maria Landi, Maurizio Fumi, Vincenzo Rocco, Rita Greco, Vittoria Letizia, Arnolfo Petruzziello, Maddalena Schioppa, Gregorio Goffredi, Francesca Marciano, Michele Caraglia, Alessia Cossu, Marianna Scrima, Edmondo Adorisio, Morena D'Avenia, Michela Iacobellis, Rosanna Piluscio, Giorgio Dirani, Vittorio Sambri, Simona Semprini, Silvia Zanoli, Francesco Curcio, Stefania Marzinotto, Andreina Baj, Fausto Sessa, Massimiliano Chetta, Marina Tarsitano, Giusy Giordano, Marcello Ametrano, Mariano Fiorenza, Caivano Antonella, D'Auria Fiorella, De Luca Luciana. |

| | | | |
|---|---|---|---|
| | Cardarelli, Napoli / 10. Centro di riferimento Oncologico della Basilicata (IRCCS-CROB), Rionero in Vulture (PZ) / 11. Presidio Ospedaliero di Agropoli, Agropoli (SA). | | |
| EPI_ISL_1361541, EPI_ISL_1361542, EPI_ISL_1361543, EPI_ISL_1361544 | 1.AO Universitaria 'S. Giovanni di Dio e Ruggi D'Aragona, Scuola Medica Salernitana' Hospital / 2.UOC di Virologia e Microbiologia, Università della Campania 'L. Vanvitelli' / 3.AO Universitaria 'Federico II' Napoli Hospital / 4.AORN 'San Giuseppe Moscati' Avellino Hospital / 5.AO 'San Pio - presidio G. Rummo' Benevento Hospital / 6.AO 'Sant'Anna e San Sebastiano' Caserta Hospital / 7.PO 'Maria Santissima Addolorata' Eboli Hospital / 8.Biogem Istituto di Ricerche Genetiche / 9. U.O.C. di Genetica Medica e di Laboratorio A.O.R.N., Azienda Ospedaliera di Rilievo Nazionale Antonio Cardarelli, Napoli / 10. Centro di riferimento Oncologico della Basilicata (IRCCS-CROB), Rionero in Vulture (PZ) / 11. Presidio Ospedaliero di Agropoli, Agropoli (SA). | 1. Genome Research Center for Health (CRGS) / 2. Laboratory of Molecular Medicine and Genomics(LMMGe) / 3. Center for Research in Pure and Applied Mathematics (CRMPA) | Giorgio Giurato (Corresponding Author), Francesca Rizzo (Corresponding Author), Alessandro Weisz (Corresponding Author), Gianluigi Franci, Giovanni Nassa, Pasquale Pagliano, Roberta Tarallo, Elena Alexandrova, Ylenia D'Agostino, Carlo Ferravante, Jessica Lamberti, Viola Melone, Domenico Memoli, Valeria Mirici Cappa, Domenico Palumbo, Giovanni Pecoraro, Assunta Sellitto, Oriana Strianese, Ilaria Terenzi, Giuseppe Fenza, Aniello Gentile, Antonello Saccomanno, Sonia Amabile, Teresa Rocco, Annamaria Salvati, Emilia Vaccaro, Massimiliano Galdiero, Michele Cennamo, Giuseppe Portella, Maria Grazia Foti, Mariarosaria Ingino, Maria Landi, Maurizio Fumi, Vincenzo Rocco, Rita Greco, Vittoria Letizia, Arnolfo Petruzziello, Maddalena Schioppa, Gregorio Goffredi, Francesca Marciano, Michele Caraglia, Alessia Cossu, Marianna Scrima, Edmondo Adorisio, Morena D'Avenia, Michela Iacobellis, Rosanna Piluscio, Giorgio Dirani, Vittorio Sambri, Simona Semprini, Silvia Zanoli, Francesco Curcio, Stefania Marzinotto, Andreina Baj, Fausto Sessa. |
| EPI_ISL_1361545 | 1.AO Universitaria 'S. Giovanni di Dio e Ruggi D'Aragona, Scuola Medica Salernitana' Hospital / 2.UOC di Virologia e Microbiologia, Università della Campania 'L. Vanvitelli' / 3.AO Universitaria 'Federico II' Napoli Hospital / 4.AORN 'San Giuseppe Moscati' Avellino Hospital / 5.AO 'San Pio - presidio G. Rummo' Benevento Hospital / 6.AO 'Sant'Anna e San Sebastiano' Caserta Hospital / 7.PO 'Maria Santissima Addolorata' Eboli Hospital / 8.Biogem Istituto di Ricerche Genetiche / 9. U.O.C. di Genetica Medica e di Laboratorio A.O.R.N., Azienda Ospedaliera di Rilievo Nazionale Antonio Cardarelli, Napoli / 10. Centro di riferimento Oncologico della Basilicata (IRCCS-CROB), Rionero in Vulture (PZ) / 11. Presidio Ospedaliero di Agropoli, Agropoli (SA). | 1. Genome Research Center for Health (CRGS) / 2. Laboratory of Molecular Medicine and Genomics(LMMGe) / 3. Center for Research in Pure and Applied Mathematics (CRMPA) | Giorgio Giurato (Corresponding Author), Francesca Rizzo (Corresponding Author), Alessandro Weisz (Corresponding Author), Gianluigi Franci, Giovanni Nassa, Pasquale Pagliano, Roberta Tarallo, Elena Alexandrova, Ylenia D'Agostino, Carlo Ferravante, Jessica Lamberti, Viola Melone, Domenico Memoli, Valeria Mirici Cappa, Domenico Palumbo, Giovanni Pecoraro, Assunta Sellitto, Oriana Strianese, Ilaria Terenzi, Giuseppe Fenza, Aniello Gentile, Antonello Saccomanno, Sonia Amabile, Teresa Rocco, Annamaria Salvati, Emilia Vaccaro, Massimiliano Galdiero, Michele Cennamo, Giuseppe Portella, Maria Grazia Foti, Mariarosaria Ingino, Maria Landi, Maurizio Fumi, Vincenzo Rocco, Rita Greco, Vittoria Letizia, Arnolfo Petruzziello, Maddalena Schioppa, Gregorio Goffredi, Francesca Marciano, Michele Caraglia, Alessia Cossu, Marianna Scrima, Edmondo Adorisio, Morena D'Avenia, Michela Iacobellis, Rosanna Piluscio, Giorgio Dirani, Vittorio Sambri, Simona Semprini, Silvia Zanoli, Francesco Curcio, Stefania Marzinotto, Andreina Baj, Fausto Sessa, Massimiliano Chetta, Marina Tarsitano, Giusy Giordano, Marcello Ametrano, Mariano Fiorenza, Caivano Antonella, D'Auria Fiorella, De Luca Luciana. |
| EPI_ISL_1361546, EPI_ISL_1361547, EPI_ISL_1361548, EPI_ISL_1361549 | 1.AO Universitaria 'S. Giovanni di Dio e Ruggi D'Aragona, Scuola Medica Salernitana' Hospital / 2.UOC di Virologia e Microbiologia, Università della Campania 'L. Vanvitelli' / 3.AO Universitaria 'Federico II' Napoli Hospital / 4.AORN 'San Giuseppe Moscati' Avellino Hospital / 5.AO 'San Pio - presidio G. Rummo' Benevento Hospital / 6.AO 'Sant'Anna e San Sebastiano' Caserta Hospital / 7.PO 'Maria Santissima Addolorata' Eboli Hospital / 8.Biogem Istituto di Ricerche Genetiche / 9. U.O.C. di Genetica Medica e di Laboratorio A.O.R.N., Azienda Ospedaliera di Rilievo Nazionale Antonio Cardarelli, Napoli / 10. Centro di riferimento Oncologico della Basilicata (IRCCS-CROB), Rionero in Vulture (PZ) / 11. Presidio Ospedaliero di Agropoli, Agropoli (SA). | 1. Genome Research Center for Health (CRGS) / 2. Laboratory of Molecular Medicine and Genomics(LMMGe) / 3. Center for Research in Pure and Applied Mathematics (CRMPA) | Giorgio Giurato (Corresponding Author), Francesca Rizzo (Corresponding Author), Alessandro Weisz (Corresponding Author), Gianluigi Franci, Giovanni Nassa, Pasquale Pagliano, Roberta Tarallo, Elena Alexandrova, Ylenia D'Agostino, Carlo Ferravante, Jessica Lamberti, Viola Melone, Domenico Memoli, Valeria Mirici Cappa, Domenico Palumbo, Giovanni Pecoraro, Assunta Sellitto, Oriana Strianese, Ilaria Terenzi, Giuseppe Fenza, Aniello Gentile, Antonello Saccomanno, Sonia Amabile, Teresa Rocco, Annamaria Salvati, Emilia Vaccaro, Massimiliano Galdiero, Michele Cennamo, Giuseppe Portella, Maria Grazia Foti, Mariarosaria Ingino, Maria Landi, Maurizio Fumi, Vincenzo Rocco, Rita Greco, Vittoria Letizia, Arnolfo Petruzziello, Maddalena Schioppa, Gregorio Goffredi, Francesca Marciano, Michele Caraglia, Alessia Cossu, Marianna Scrima, Edmondo Adorisio, Morena D'Avenia, Michela Iacobellis, Rosanna Piluscio, Giorgio Dirani, Vittorio Sambri, Simona Semprini, Silvia Zanoli, Francesco Curcio, Stefania Marzinotto, Andreina Baj, Fausto Sessa. |
| EPI_ISL_1361550 | 1.AO Universitaria 'S. Giovanni di Dio e Ruggi D'Aragona, Scuola Medica Salernitana' Hospital / 2.UOC di Virologia e Microbiologia, Università della Campania 'L. Vanvitelli' / 3.AO Universitaria 'Federico II' Napoli Hospital / 4.AORN 'San Giuseppe Moscati' Avellino Hospital / 5.AO 'San Pio - presidio G. Rummo' Benevento Hospital / 6.AO 'Sant'Anna e San Sebastiano' Caserta Hospital / 7.PO 'Maria Santissima Addolorata' Eboli Hospital / 8.Biogem Istituto di Ricerche Genetiche / 9. U.O.C. di Genetica Medica e di Laboratorio A.O.R.N., Azienda Ospedaliera di Rilievo Nazionale Antonio Cardarelli, Napoli / 10. Centro di riferimento Oncologico della Basilicata (IRCCS-CROB), Rionero in Vulture (PZ) / 11. Presidio Ospedaliero di Agropoli, Agropoli (SA). | 1. Genome Research Center for Health (CRGS) / 2. Laboratory of Molecular Medicine and Genomics(LMMGe) / 3. Center for Research in Pure and Applied Mathematics (CRMPA) | Giorgio Giurato (Corresponding Author), Francesca Rizzo (Corresponding Author), Alessandro Weisz (Corresponding Author), Gianluigi Franci, Giovanni Nassa, Pasquale Pagliano, Roberta Tarallo, Elena Alexandrova, Ylenia D'Agostino, Carlo Ferravante, Jessica Lamberti, Viola Melone, Domenico Memoli, Valeria Mirici Cappa, Domenico Palumbo, Giovanni Pecoraro, Assunta Sellitto, Oriana Strianese, Ilaria Terenzi, Giuseppe Fenza, Aniello Gentile, Antonello Saccomanno, Sonia Amabile, Teresa Rocco, Annamaria Salvati, Emilia Vaccaro, Massimiliano Galdiero, Michele Cennamo, Giuseppe Portella, Maria Grazia Foti, Mariarosaria Ingino, Maria Landi, Maurizio Fumi, Vincenzo Rocco, Rita Greco, Vittoria Letizia, Arnolfo Petruzziello, Maddalena Schioppa, Gregorio Goffredi, Francesca Marciano, Michele Caraglia, Alessia Cossu, Marianna Scrima, Edmondo Adorisio, Morena D'Avenia, Michela Iacobellis, Rosanna Piluscio, Giorgio Dirani, Vittorio Sambri, Simona Semprini, Silvia Zanoli, Francesco Curcio, Stefania Marzinotto, Andreina Baj, Fausto Sessa, Massimiliano Chetta, Marina Tarsitano, Giusy Giordano, Marcello Ametrano, Mariano Fiorenza, Caivano Antonella, D'Auria Fiorella, De Luca Luciana. |
| EPI_ISL_1361551, EPI_ISL_1361552, EPI_ISL_1361553, EPI_ISL_1361554 | 1.AO Universitaria 'S. Giovanni di Dio e Ruggi D'Aragona, Scuola Medica Salernitana' Hospital / 2.UOC di Virologia e Microbiologia, Università della Campania 'L. Vanvitelli' / 3.AO Universitaria 'Federico II' Napoli Hospital / 4.AORN 'San Giuseppe Moscati' Avellino Hospital / 5.AO 'San Pio - presidio G. Rummo' Benevento Hospital / 6.AO 'Sant'Anna e San Sebastiano' Caserta Hospital / 7.PO 'Maria Santissima Addolorata' Eboli Hospital / 8.Biogem Istituto di Ricerche Genetiche / 9. U.O.C. di Genetica Medica e di Laboratorio A.O.R.N., Azienda Ospedaliera di Rilievo Nazionale Antonio Cardarelli, Napoli / 10. Centro di riferimento Oncologico della Basilicata (IRCCS-CROB), Rionero in Vulture (PZ) / 11. Presidio Ospedaliero di Agropoli, Agropoli (SA). | 1. Genome Research Center for Health (CRGS) / 2. Laboratory of Molecular Medicine and Genomics(LMMGe) / 3. Center for Research in Pure and Applied Mathematics (CRMPA) | Giorgio Giurato (Corresponding Author), Francesca Rizzo (Corresponding Author), Alessandro Weisz (Corresponding Author), Gianluigi Franci, Giovanni Nassa, Pasquale Pagliano, Roberta Tarallo, Elena Alexandrova, Ylenia D'Agostino, Carlo Ferravante, Jessica Lamberti, Viola Melone, Domenico Memoli, Valeria Mirici Cappa, Domenico Palumbo, Giovanni Pecoraro, Assunta Sellitto, Oriana Strianese, Ilaria Terenzi, Giuseppe Fenza, Aniello Gentile, Antonello Saccomanno, Sonia Amabile, Teresa Rocco, Annamaria Salvati, Emilia Vaccaro, Massimiliano Galdiero, Michele Cennamo, Giuseppe Portella, Maria Grazia Foti, Mariarosaria Ingino, Maria Landi, Maurizio Fumi, Vincenzo Rocco, Rita Greco, Vittoria Letizia, Arnolfo Petruzziello, Maddalena Schioppa, Gregorio Goffredi, Francesca Marciano, Michele Caraglia, Alessia Cossu, Marianna Scrima, Edmondo Adorisio, Morena D'Avenia, Michela Iacobellis, Rosanna Piluscio, Giorgio Dirani, Vittorio Sambri, Simona Semprini, Silvia Zanoli, Francesco Curcio, Stefania Marzinotto, Andreina Baj, Fausto Sessa. |
| EPI_ISL_1361555 | 1.AO Universitaria 'S. Giovanni di Dio e Ruggi D'Aragona, Scuola Medica Salernitana' Hospital / 2.UOC di Virologia e Microbiologia, Università della Campania 'L. Vanvitelli' / 3.AO Universitaria 'Federico II' Napoli Hospital / 4.AORN 'San Giuseppe Moscati' Avellino Hospital / 5.AO 'San Pio - presidio G. Rummo' Benevento Hospital / 6.AO 'Sant'Anna e San Sebastiano' Caserta Hospital / 7.PO 'Maria Santissima Addolorata' Eboli Hospital / 8.Biogem Istituto di Ricerche Genetiche / 9. U.O.C. di Genetica Medica e di Laboratorio A.O.R.N., Azienda Ospedaliera di Rilievo Nazionale Antonio Cardarelli, Napoli / 10. Centro di riferimento Oncologico della Basilicata (IRCCS-CROB), Rionero in Vulture (PZ) / 11. Presidio Ospedaliero di Agropoli, Agropoli (SA). | 1. Genome Research Center for Health (CRGS) / 2. Laboratory of Molecular Medicine and Genomics(LMMGe) / 3. Center for Research in Pure and Applied Mathematics (CRMPA) | Giorgio Giurato (Corresponding Author), Francesca Rizzo (Corresponding Author), Alessandro Weisz (Corresponding Author), Gianluigi Franci, Giovanni Nassa, Pasquale Pagliano, Roberta Tarallo, Elena Alexandrova, Ylenia D'Agostino, Carlo Ferravante, Jessica Lamberti, Viola Melone, Domenico Memoli, Valeria Mirici Cappa, Domenico Palumbo, Giovanni Pecoraro, Assunta Sellitto, Oriana Strianese, Ilaria Terenzi, Giuseppe Fenza, Aniello Gentile, Antonello Saccomanno, Sonia Amabile, Teresa Rocco, Annamaria Salvati, Emilia Vaccaro, Massimiliano Galdiero, Michele Cennamo, Giuseppe Portella, Maria Grazia Foti, Mariarosaria Ingino, Maria Landi, Maurizio Fumi, Vincenzo Rocco, Rita Greco, Vittoria Letizia, Arnolfo Petruzziello, Maddalena Schioppa, Gregorio Goffredi, Francesca Marciano, Michele Caraglia, Alessia Cossu, Marianna Scrima, Edmondo Adorisio, Morena D'Avenia, Michela Iacobellis, Rosanna Piluscio, Giorgio Dirani, Vittorio Sambri, Simona Semprini, Silvia Zanoli, Francesco Curcio, Stefania Marzinotto, Andreina Baj, Fausto Sessa, Massimiliano Chetta, Marina Tarsitano, Giusy Giordano, Marcello Ametrano, Mariano Fiorenza, Caivano Antonella, D'Auria Fiorella, De Luca Luciana. |
| EPI_ISL_1361556, EPI_ISL_1361557, EPI_ISL_1361558, EPI_ISL_1361559 | 1.AO Universitaria 'S. Giovanni di Dio e Ruggi D'Aragona, Scuola Medica Salernitana' Hospital / 2.UOC di Virologia e Microbiologia, Università della Campania 'L. Vanvitelli' / 3.AO | 1. Genome Research Center for Health (CRGS) / 2. Laboratory of Molecular Medicine and Genomics(LMMGe) / 3. Center for Research in Pure and Applied Mathematics | Giorgio Giurato (Corresponding Author), Francesca Rizzo (Corresponding Author), Alessandro Weisz (Corresponding Author), Gianluigi Franci, Giovanni Nassa, Pasquale Pagliano, Roberta Tarallo, Elena Alexandrova, Ylenia D'Agostino, Carlo Ferravante, Jessica Lamberti, Viola Melone, Domenico Memoli, Valeria Mirici Cappa, Domenico Palumbo, Giovanni Pecoraro, Assunta Sellitto, Oriana Strianese, Ilaria Terenzi, Giuseppe Fenza, Aniello Gentile, Antonello |

| | | | |
|---|---|---|---|
| | Universitaria 'Federico II' Napoli Hospital / 4.AORN 'San Giuseppe Moscati' Avellino Hospital / 5.AO 'San Pio - presidio G. Rummo' Benevento Hospital / 6.AO 'Sant'Anna e San Sebastiano' Caserta Hospital / 7.PO 'Maria Santissima Addolorata' Eboli Hospital / 8.Biogem Istituto di Ricerche Genetiche / 9. U.O.C. di Genetica Medica e di Laboratorio A.O.R.N., Azienda Ospedaliera di Rilievo Nazionale Antonio Cardarelli, Napoli / 10. Centro di riferimento Oncologico della Basilicata (IRCCS-CROB), Rionero in Vulture (PZ) / 11. Presidio Ospedaliero di Agropoli, Agropoli (SA). | (CRMPA) | Saccomanno, Sonia Amabile, Teresa Rocco, Annamaria Salvati, Emilia Vaccaro, Massimiliano Galdiero, Michele Cennamo, Giuseppe Portella, Maria Grazia Foti, Mariarosaria Ingino, Maria Landi, Maurizio Fumi, Vincenzo Rocco, Rita Greco, Vittoria Letizia, Arnolfo Petruzziello, Maddalena Schioppa, Gregorio Goffredi, Francesca Marciano, Michele Caraglia, Alessia Cossu, Marianna Scrima, Edmondo Adorisio, Morena D'Avenia, Michela Iacobellis, Rosanna Piluscio, Giorgio Dirani, Vittorio Sambri, Simona Semprini, Silvia Zanoli, Francesco Curcio, Stefania Marzinotto, Andreina Baj, Fausto Sessa. |
| EPI_ISL_1361560 | 1.AO Universitaria 'S. Giovanni di Dio e Ruggi D'Aragona, Scuola Medica Salernitana' Hospital / 2.UOC di Virologia e Microbiologia, Università della Campania 'L. Vanvitelli' / 3.AO Universitaria 'Federico II' Napoli Hospital / 4.AORN 'San Giuseppe Moscati' Avellino Hospital / 5.AO 'San Pio - presidio G. Rummo' Benevento Hospital / 6.AO 'Sant'Anna e San Sebastiano' Caserta Hospital / 7.PO 'Maria Santissima Addolorata' Eboli Hospital / 8.Biogem Istituto di Ricerche Genetiche / 9. U.O.C. di Genetica Medica e di Laboratorio A.O.R.N., Azienda Ospedaliera di Rilievo Nazionale Antonio Cardarelli, Napoli / 10. Centro di riferimento Oncologico della Basilicata (IRCCS-CROB), Rionero in Vulture (PZ) / 11. Presidio Ospedaliero di Agropoli, Agropoli (SA). | 1. Genome Research Center for Health (CRGS) / 2. Laboratory of Molecular Medicine and Genomics(LMMGe) / 3. Center for Research in Pure and Applied Mathematics (CRMPA) | Giorgio Giurato (Corresponding Author), Francesca Rizzo (Corresponding Author), Alessandro Weisz (Corresponding Author), Gianluigi Franci, Giovanni Nassa, Pasquale Pagliano, Roberta Tarallo, Elena Alexandrova, Ylenia D'Agostino, Carlo Ferravante, Jessica Lamberti, Viola Melone, Domenico Memoli, Valeria Mirici Cappa, Domenico Palumbo, Giovanni Pecoraro, Assunta Sellitto, Oriana Strianese, Ilaria Terenzi, Giuseppe Fenza, Aniello Gentile, Antonello Saccomanno, Sonia Amabile, Teresa Rocco, Annamaria Salvati, Emilia Vaccaro, Massimiliano Galdiero, Michele Cennamo, Giuseppe Portella, Maria Grazia Foti, Mariarosaria Ingino, Maria Landi, Maurizio Fumi, Vincenzo Rocco, Rita Greco, Vittoria Letizia, Arnolfo Petruzziello, Maddalena Schioppa, Gregorio Goffredi, Francesca Marciano, Michele Caraglia, Alessia Cossu, Marianna Scrima, Edmondo Adorisio, Morena D'Avenia, Michela Iacobellis, Rosanna Piluscio, Giorgio Dirani, Vittorio Sambri, Simona Semprini, Silvia Zanoli, Francesco Curcio, Stefania Marzinotto, Andreina Baj, Fausto Sessa, Massimiliano Chetta, Marina Tarsitano, Giusy Giordano, Marcello Ametrano, Mariano Fiorenza, Caivano Antonella, D'Auria Fiorella, De Luca Luciana. |
| EPI_ISL_1361561, EPI_ISL_1361562, EPI_ISL_1361563, EPI_ISL_1361564 | 1.AO Universitaria 'S. Giovanni di Dio e Ruggi D'Aragona, Scuola Medica Salernitana' Hospital / 2.UOC di Virologia e Microbiologia, Università della Campania 'L. Vanvitelli' / 3.AO Universitaria 'Federico II' Napoli Hospital / 4.AORN 'San Giuseppe Moscati' Avellino Hospital / 5.AO 'San Pio - presidio G. Rummo' Benevento Hospital / 6.AO 'Sant'Anna e San Sebastiano' Caserta Hospital / 7.PO 'Maria Santissima Addolorata' Eboli Hospital / 8.Biogem Istituto di Ricerche Genetiche / 9. U.O.C. di Genetica Medica e di Laboratorio A.O.R.N., Azienda Ospedaliera di Rilievo Nazionale Antonio Cardarelli, Napoli / 10. Centro di riferimento Oncologico della Basilicata (IRCCS-CROB), Rionero in Vulture (PZ) / 11. Presidio Ospedaliero di Agropoli, Agropoli (SA). | 1. Genome Research Center for Health (CRGS) / 2. Laboratory of Molecular Medicine and Genomics(LMMGe) / 3. Center for Research in Pure and Applied Mathematics (CRMPA) | Giorgio Giurato (Corresponding Author), Francesca Rizzo (Corresponding Author), Alessandro Weisz (Corresponding Author), Gianluigi Franci, Giovanni Nassa, Pasquale Pagliano, Roberta Tarallo, Elena Alexandrova, Ylenia D'Agostino, Carlo Ferravante, Jessica Lamberti, Viola Melone, Domenico Memoli, Valeria Mirici Cappa, Domenico Palumbo, Giovanni Pecoraro, Assunta Sellitto, Oriana Strianese, Ilaria Terenzi, Giuseppe Fenza, Aniello Gentile, Antonello Saccomanno, Sonia Amabile, Teresa Rocco, Annamaria Salvati, Emilia Vaccaro, Massimiliano Galdiero, Michele Cennamo, Giuseppe Portella, Maria Grazia Foti, Mariarosaria Ingino, Maria Landi, Maurizio Fumi, Vincenzo Rocco, Rita Greco, Vittoria Letizia, Arnolfo Petruzziello, Maddalena Schioppa, Gregorio Goffredi, Francesca Marciano, Michele Caraglia, Alessia Cossu, Marianna Scrima, Edmondo Adorisio, Morena D'Avenia, Michela Iacobellis, Rosanna Piluscio, Giorgio Dirani, Vittorio Sambri, Simona Semprini, Silvia Zanoli, Francesco Curcio, Stefania Marzinotto, Andreina Baj, Fausto Sessa. |
| EPI_ISL_1361565 | 1.AO Universitaria 'S. Giovanni di Dio e Ruggi D'Aragona, Scuola Medica Salernitana' Hospital / 2.UOC di Virologia e Microbiologia, Università della Campania 'L. Vanvitelli' / 3.AO Universitaria 'Federico II' Napoli Hospital / 4.AORN 'San Giuseppe Moscati' Avellino Hospital / 5.AO 'San Pio - presidio G. Rummo' Benevento Hospital / 6.AO 'Sant'Anna e San Sebastiano' Caserta Hospital / 7.PO 'Maria Santissima Addolorata' Eboli Hospital / 8.Biogem Istituto di Ricerche Genetiche / 9. U.O.C. di Genetica Medica e di Laboratorio A.O.R.N., Azienda Ospedaliera di Rilievo Nazionale Antonio Cardarelli, Napoli / 10. Centro di riferimento Oncologico della Basilicata (IRCCS-CROB), Rionero in Vulture (PZ) / 11. Presidio Ospedaliero di Agropoli, Agropoli (SA). | 1. Genome Research Center for Health (CRGS) / 2. Laboratory of Molecular Medicine and Genomics(LMMGe) / 3. Center for Research in Pure and Applied Mathematics (CRMPA) | Giorgio Giurato (Corresponding Author), Francesca Rizzo (Corresponding Author), Alessandro Weisz (Corresponding Author), Gianluigi Franci, Giovanni Nassa, Pasquale Pagliano, Roberta Tarallo, Elena Alexandrova, Ylenia D'Agostino, Carlo Ferravante, Jessica Lamberti, Viola Melone, Domenico Memoli, Valeria Mirici Cappa, Domenico Palumbo, Giovanni Pecoraro, Assunta Sellitto, Oriana Strianese, Ilaria Terenzi, Giuseppe Fenza, Aniello Gentile, Antonello Saccomanno, Sonia Amabile, Teresa Rocco, Annamaria Salvati, Emilia Vaccaro, Massimiliano Galdiero, Michele Cennamo, Giuseppe Portella, Maria Grazia Foti, Mariarosaria Ingino, Maria Landi, Maurizio Fumi, Vincenzo Rocco, Rita Greco, Vittoria Letizia, Arnolfo Petruzziello, Maddalena Schioppa, Gregorio Goffredi, Francesca Marciano, Michele Caraglia, Alessia Cossu, Marianna Scrima, Edmondo Adorisio, Morena D'Avenia, Michela Iacobellis, Rosanna Piluscio, Giorgio Dirani, Vittorio Sambri, Simona Semprini, Silvia Zanoli, Francesco Curcio, Stefania Marzinotto, Andreina Baj, Fausto Sessa, Massimiliano Chetta, Marina Tarsitano, Giusy Giordano, Marcello Ametrano, Mariano Fiorenza, Caivano Antonella, D'Auria Fiorella, De Luca Luciana. |
| EPI_ISL_1361566, EPI_ISL_1361567, EPI_ISL_1361568, EPI_ISL_1361569 | 1.AO Universitaria 'S. Giovanni di Dio e Ruggi D'Aragona, Scuola Medica Salernitana' Hospital / 2.UOC di Virologia e Microbiologia, Università della Campania 'L. Vanvitelli' / 3.AO Universitaria 'Federico II' Napoli Hospital / 4.AORN 'San Giuseppe Moscati' Avellino Hospital / 5.AO 'San Pio - presidio G. Rummo' Benevento Hospital / 6.AO 'Sant'Anna e San Sebastiano' Caserta Hospital / 7.PO 'Maria Santissima Addolorata' Eboli Hospital / 8.Biogem Istituto di Ricerche Genetiche / 9. U.O.C. di Genetica Medica e di Laboratorio A.O.R.N., Azienda Ospedaliera di Rilievo Nazionale Antonio Cardarelli, Napoli / 10. Centro di riferimento Oncologico della Basilicata (IRCCS-CROB), Rionero in Vulture (PZ) / 11. Presidio Ospedaliero di Agropoli, Agropoli (SA). | 1. Genome Research Center for Health (CRGS) / 2. Laboratory of Molecular Medicine and Genomics(LMMGe) / 3. Center for Research in Pure and Applied Mathematics (CRMPA) | Giorgio Giurato (Corresponding Author), Francesca Rizzo (Corresponding Author), Alessandro Weisz (Corresponding Author), Gianluigi Franci, Giovanni Nassa, Pasquale Pagliano, Roberta Tarallo, Elena Alexandrova, Ylenia D'Agostino, Carlo Ferravante, Jessica Lamberti, Viola Melone, Domenico Memoli, Valeria Mirici Cappa, Domenico Palumbo, Giovanni Pecoraro, Assunta Sellitto, Oriana Strianese, Ilaria Terenzi, Giuseppe Fenza, Aniello Gentile, Antonello Saccomanno, Sonia Amabile, Teresa Rocco, Annamaria Salvati, Emilia Vaccaro, Massimiliano Galdiero, Michele Cennamo, Giuseppe Portella, Maria Grazia Foti, Mariarosaria Ingino, Maria Landi, Maurizio Fumi, Vincenzo Rocco, Rita Greco, Vittoria Letizia, Arnolfo Petruzziello, Maddalena Schioppa, Gregorio Goffredi, Francesca Marciano, Michele Caraglia, Alessia Cossu, Marianna Scrima, Edmondo Adorisio, Morena D'Avenia, Michela Iacobellis, Rosanna Piluscio, Giorgio Dirani, Vittorio Sambri, Simona Semprini, Silvia Zanoli, Francesco Curcio, Stefania Marzinotto, Andreina Baj, Fausto Sessa. |
| EPI_ISL_1361570 | 1.AO Universitaria 'S. Giovanni di Dio e Ruggi D'Aragona, Scuola Medica Salernitana' Hospital / 2.UOC di Virologia e Microbiologia, Università della Campania 'L. Vanvitelli' / 3.AO Universitaria 'Federico II' Napoli Hospital / 4.AORN 'San Giuseppe Moscati' Avellino Hospital / 5.AO 'San Pio - presidio G. Rummo' Benevento Hospital / 6.AO 'Sant'Anna e San Sebastiano' Caserta Hospital / 7.PO 'Maria Santissima Addolorata' Eboli Hospital / 8.Biogem Istituto di Ricerche Genetiche / 9. U.O.C. di Genetica Medica e di Laboratorio A.O.R.N., Azienda Ospedaliera di Rilievo Nazionale Antonio Cardarelli, Napoli / 10. Centro di riferimento Oncologico della Basilicata (IRCCS-CROB), Rionero in Vulture (PZ) / 11. Presidio Ospedaliero di Agropoli, Agropoli (SA). | 1. Genome Research Center for Health (CRGS) / 2. Laboratory of Molecular Medicine and Genomics(LMMGe) / 3. Center for Research in Pure and Applied Mathematics (CRMPA) | Giorgio Giurato (Corresponding Author), Francesca Rizzo (Corresponding Author), Alessandro Weisz (Corresponding Author), Gianluigi Franci, Giovanni Nassa, Pasquale Pagliano, Roberta Tarallo, Elena Alexandrova, Ylenia D'Agostino, Carlo Ferravante, Jessica Lamberti, Viola Melone, Domenico Memoli, Valeria Mirici Cappa, Domenico Palumbo, Giovanni Pecoraro, Assunta Sellitto, Oriana Strianese, Ilaria Terenzi, Giuseppe Fenza, Aniello Gentile, Antonello Saccomanno, Sonia Amabile, Teresa Rocco, Annamaria Salvati, Emilia Vaccaro, Massimiliano Galdiero, Michele Cennamo, Giuseppe Portella, Maria Grazia Foti, Mariarosaria Ingino, Maria Landi, Maurizio Fumi, Vincenzo Rocco, Rita Greco, Vittoria Letizia, Arnolfo Petruzziello, Maddalena Schioppa, Gregorio Goffredi, Francesca Marciano, Michele Caraglia, Alessia Cossu, Marianna Scrima, Edmondo Adorisio, Morena D'Avenia, Michela Iacobellis, Rosanna Piluscio, Giorgio Dirani, Vittorio Sambri, Simona Semprini, Silvia Zanoli, Francesco Curcio, Stefania Marzinotto, Andreina Baj, Fausto Sessa, Massimiliano Chetta, Marina Tarsitano, Giusy Giordano, Marcello Ametrano, Mariano Fiorenza, Caivano Antonella, D'Auria Fiorella, De Luca Luciana. |
| EPI_ISL_1361571, EPI_ISL_1361572, EPI_ISL_1361573, EPI_ISL_1361574 | 1.AO Universitaria 'S. Giovanni di Dio e Ruggi D'Aragona, Scuola Medica Salernitana' Hospital / 2.UOC di Virologia e Microbiologia, Università della Campania 'L. Vanvitelli' / 3.AO Universitaria 'Federico II' Napoli Hospital / 4.AORN 'San Giuseppe Moscati' Avellino Hospital / 5.AO 'San Pio - presidio G. Rummo' Benevento Hospital / 6.AO 'Sant'Anna e San Sebastiano' Caserta Hospital / 7.PO 'Maria Santissima Addolorata' Eboli Hospital / 8.Biogem Istituto di Ricerche Genetiche / 9. U.O.C. di Genetica Medica e di Laboratorio | 1. Genome Research Center for Health (CRGS) / 2. Laboratory of Molecular Medicine and Genomics(LMMGe) / 3. Center for Research in Pure and Applied Mathematics (CRMPA) | Giorgio Giurato (Corresponding Author), Francesca Rizzo (Corresponding Author), Alessandro Weisz (Corresponding Author), Gianluigi Franci, Giovanni Nassa, Pasquale Pagliano, Roberta Tarallo, Elena Alexandrova, Ylenia D'Agostino, Carlo Ferravante, Jessica Lamberti, Viola Melone, Domenico Memoli, Valeria Mirici Cappa, Domenico Palumbo, Giovanni Pecoraro, Assunta Sellitto, Oriana Strianese, Ilaria Terenzi, Giuseppe Fenza, Aniello Gentile, Antonello Saccomanno, Sonia Amabile, Teresa Rocco, Annamaria Salvati, Emilia Vaccaro, Massimiliano Galdiero, Michele Cennamo, Giuseppe Portella, Maria Grazia Foti, Mariarosaria Ingino, Maria Landi, Maurizio Fumi, Vincenzo Rocco, Rita Greco, Vittoria Letizia, Arnolfo Petruzziello, Maddalena Schioppa, Gregorio Goffredi, Francesca Marciano, Michele Caraglia, Alessia Cossu, Marianna Scrima, Edmondo Adorisio, Morena D'Avenia, Michela Iacobellis, Rosanna Piluscio, Giorgio Dirani, Vittorio Sambri, Simona Semprini, Silvia Zanoli, Francesco Curcio, Stefania Marzinotto, Andreina Baj, Fausto Sessa. |

| | | | |
|---|---|---|---|
| | A.O.R.N., Azienda Ospedaliera di Rilievo Nazionale Antonio Cardarelli, Napoli / 10. Centro di riferimento Oncologico della Basilicata (IRCCS-CROB), Rionero in Vulture (PZ) / 11. Presidio Ospedaliero di Agropoli, Agropoli (SA). | | |
| EPI_ISL_1361575 | 1.AO Universitaria 'S. Giovanni di Dio e Ruggi D'Aragona, Scuola Medica Salernitana' Hospital / 2.UOC di Virologia e Microbiologia, Università della Campania 'L. Vanvitelli' / 3.AO Universitaria 'Federico II' Napoli Hospital / 4.AORN 'San Giuseppe Moscati' Avellino Hospital / 5.AO 'San Pio - presidio G. Rummo' Benevento Hospital / 6.AO 'Sant'Anna e San Sebastiano' Caserta Hospital / 7.PO 'Maria Santissima Addolorata' Eboli Hospital / 8.Biogem Istituto di Ricerche Genetiche / 9. U.O.C. di Genetica Medica e di Laboratorio A.O.R.N., Azienda Ospedaliera di Rilievo Nazionale Antonio Cardarelli, Napoli / 10. Centro di riferimento Oncologico della Basilicata (IRCCS-CROB), Rionero in Vulture (PZ) / 11. Presidio Ospedaliero di Agropoli, Agropoli (SA). | 1. Genome Research Center for Health (CRGS) / 2. Laboratory of Molecular Medicine and Genomics(LMMGe) / 3. Center for Research in Pure and Applied Mathematics (CRMPA) | Giorgio Giurato (Corresponding Author), Francesca Rizzo (Corresponding Author), Alessandro Weisz (Corresponding Author), Gianluigi Franci, Giovanni Nassa, Pasquale Pagliano, Roberta Tarallo, Elena Alexandrova, Ylenia D'Agostino, Carlo Ferravante, Jessica Lamberti, Viola Melone, Domenico Memoli, Valeria Mirici Cappa, Domenico Palumbo, Giovanni Pecoraro, Assunta Sellitto, Oriana Strianese, Ilaria Terenzi, Giuseppe Fenza, Aniello Gentile, Antonello Saccomanno, Sonia Amabile, Teresa Rocco, Annamaria Salvati, Emilia Vaccaro, Massimiliano Galdiero, Michele Cennamo, Giuseppe Portella, Maria Grazia Foti, Mariarosaria Ingino, Maria Landi, Maurizio Fumi, Vincenzo Rocco, Rita Greco, Vittoria Letizia, Arnolfo Petruzziello, Maddalena Schioppa, Gregorio Goffredi, Francesca Marciano, Michele Caraglia, Alessia Cossu, Marianna Scrima, Edmondo Adorisio, Morena D'Avenia, Michela Iacobellis, Rosanna Piluscio, Giorgio Dirani, Vittorio Sambri, Simona Semprini, Silvia Zanoli, Francesco Curcio, Stefania Marzinotto, Andreina Baj, Fausto Sessa, Massimiliano Chetta, Marina Tarsitano, Giusy Giordano, Marcello Ametrano, Mariano Fiorenza, Caivano Antonella, D'Auria Fiorella, De Luca Luciana. |
| EPI_ISL_1361576, EPI_ISL_1361577, EPI_ISL_1361578, EPI_ISL_1361579 | 1.AO Universitaria 'S. Giovanni di Dio e Ruggi D'Aragona, Scuola Medica Salernitana' Hospital / 2.UOC di Virologia e Microbiologia, Università della Campania 'L. Vanvitelli' / 3.AO Universitaria 'Federico II' Napoli Hospital / 4.AORN 'San Giuseppe Moscati' Avellino Hospital / 5.AO 'San Pio - presidio G. Rummo' Benevento Hospital / 6.AO 'Sant'Anna e San Sebastiano' Caserta Hospital / 7.PO 'Maria Santissima Addolorata' Eboli Hospital / 8.Biogem Istituto di Ricerche Genetiche / 9. U.O.C. di Genetica Medica e di Laboratorio A.O.R.N., Azienda Ospedaliera di Rilievo Nazionale Antonio Cardarelli, Napoli / 10. Centro di riferimento Oncologico della Basilicata (IRCCS-CROB), Rionero in Vulture (PZ) / 11. Presidio Ospedaliero di Agropoli, Agropoli (SA). | 1. Genome Research Center for Health (CRGS) / 2. Laboratory of Molecular Medicine and Genomics(LMMGe) / 3. Center for Research in Pure and Applied Mathematics (CRMPA) | Giorgio Giurato (Corresponding Author), Francesca Rizzo (Corresponding Author), Alessandro Weisz (Corresponding Author), Gianluigi Franci, Giovanni Nassa, Pasquale Pagliano, Roberta Tarallo, Elena Alexandrova, Ylenia D'Agostino, Carlo Ferravante, Jessica Lamberti, Viola Melone, Domenico Memoli, Valeria Mirici Cappa, Domenico Palumbo, Giovanni Pecoraro, Assunta Sellitto, Oriana Strianese, Ilaria Terenzi, Giuseppe Fenza, Aniello Gentile, Antonello Saccomanno, Sonia Amabile, Teresa Rocco, Annamaria Salvati, Emilia Vaccaro, Massimiliano Galdiero, Michele Cennamo, Giuseppe Portella, Maria Grazia Foti, Mariarosaria Ingino, Maria Landi, Maurizio Fumi, Vincenzo Rocco, Rita Greco, Vittoria Letizia, Arnolfo Petruzziello, Maddalena Schioppa, Gregorio Goffredi, Francesca Marciano, Michele Caraglia, Alessia Cossu, Marianna Scrima, Edmondo Adorisio, Morena D'Avenia, Michela Iacobellis, Rosanna Piluscio, Giorgio Dirani, Vittorio Sambri, Simona Semprini, Silvia Zanoli, Francesco Curcio, Stefania Marzinotto, Andreina Baj, Fausto Sessa. |
| EPI_ISL_1361580 | 1.AO Universitaria 'S. Giovanni di Dio e Ruggi D'Aragona, Scuola Medica Salernitana' Hospital / 2.UOC di Virologia e Microbiologia, Università della Campania 'L. Vanvitelli' / 3.AO Universitaria 'Federico II' Napoli Hospital / 4.AORN 'San Giuseppe Moscati' Avellino Hospital / 5.AO 'San Pio - presidio G. Rummo' Benevento Hospital / 6.AO 'Sant'Anna e San Sebastiano' Caserta Hospital / 7.PO 'Maria Santissima Addolorata' Eboli Hospital / 8.Biogem Istituto di Ricerche Genetiche / 9. U.O.C. di Genetica Medica e di Laboratorio A.O.R.N., Azienda Ospedaliera di Rilievo Nazionale Antonio Cardarelli, Napoli / 10. Centro di riferimento Oncologico della Basilicata (IRCCS-CROB), Rionero in Vulture (PZ) / 11. Presidio Ospedaliero di Agropoli, Agropoli (SA). | 1. Genome Research Center for Health (CRGS) / 2. Laboratory of Molecular Medicine and Genomics(LMMGe) / 3. Center for Research in Pure and Applied Mathematics (CRMPA) | Giorgio Giurato (Corresponding Author), Francesca Rizzo (Corresponding Author), Alessandro Weisz (Corresponding Author), Gianluigi Franci, Giovanni Nassa, Pasquale Pagliano, Roberta Tarallo, Elena Alexandrova, Ylenia D'Agostino, Carlo Ferravante, Jessica Lamberti, Viola Melone, Domenico Memoli, Valeria Mirici Cappa, Domenico Palumbo, Giovanni Pecoraro, Assunta Sellitto, Oriana Strianese, Ilaria Terenzi, Giuseppe Fenza, Aniello Gentile, Antonello Saccomanno, Sonia Amabile, Teresa Rocco, Annamaria Salvati, Emilia Vaccaro, Massimiliano Galdiero, Michele Cennamo, Giuseppe Portella, Maria Grazia Foti, Mariarosaria Ingino, Maria Landi, Maurizio Fumi, Vincenzo Rocco, Rita Greco, Vittoria Letizia, Arnolfo Petruzziello, Maddalena Schioppa, Gregorio Goffredi, Francesca Marciano, Michele Caraglia, Alessia Cossu, Marianna Scrima, Edmondo Adorisio, Morena D'Avenia, Michela Iacobellis, Rosanna Piluscio, Giorgio Dirani, Vittorio Sambri, Simona Semprini, Silvia Zanoli, Francesco Curcio, Stefania Marzinotto, Andreina Baj, Fausto Sessa, Massimiliano Chetta, Marina Tarsitano, Giusy Giordano, Marcello Ametrano, Mariano Fiorenza, Caivano Antonella, D'Auria Fiorella, De Luca Luciana. |
| EPI_ISL_1361581, EPI_ISL_1361582, EPI_ISL_1361583, EPI_ISL_1361584 | 1.AO Universitaria 'S. Giovanni di Dio e Ruggi D'Aragona, Scuola Medica Salernitana' Hospital / 2.UOC di Virologia e Microbiologia, Università della Campania 'L. Vanvitelli' / 3.AO Universitaria 'Federico II' Napoli Hospital / 4.AORN 'San Giuseppe Moscati' Avellino Hospital / 5.AO 'San Pio - presidio G. Rummo' Benevento Hospital / 6.AO 'Sant'Anna e San Sebastiano' Caserta Hospital / 7.PO 'Maria Santissima Addolorata' Eboli Hospital / 8.Biogem Istituto di Ricerche Genetiche / 9. U.O.C. di Genetica Medica e di Laboratorio A.O.R.N., Azienda Ospedaliera di Rilievo Nazionale Antonio Cardarelli, Napoli / 10. Centro di riferimento Oncologico della Basilicata (IRCCS-CROB), Rionero in Vulture (PZ) / 11. Presidio Ospedaliero di Agropoli, Agropoli (SA). | 1. Genome Research Center for Health (CRGS) / 2. Laboratory of Molecular Medicine and Genomics(LMMGe) / 3. Center for Research in Pure and Applied Mathematics (CRMPA) | Giorgio Giurato (Corresponding Author), Francesca Rizzo (Corresponding Author), Alessandro Weisz (Corresponding Author), Gianluigi Franci, Giovanni Nassa, Pasquale Pagliano, Roberta Tarallo, Elena Alexandrova, Ylenia D'Agostino, Carlo Ferravante, Jessica Lamberti, Viola Melone, Domenico Memoli, Valeria Mirici Cappa, Domenico Palumbo, Giovanni Pecoraro, Assunta Sellitto, Oriana Strianese, Ilaria Terenzi, Giuseppe Fenza, Aniello Gentile, Antonello Saccomanno, Sonia Amabile, Teresa Rocco, Annamaria Salvati, Emilia Vaccaro, Massimiliano Galdiero, Michele Cennamo, Giuseppe Portella, Maria Grazia Foti, Mariarosaria Ingino, Maria Landi, Maurizio Fumi, Vincenzo Rocco, Rita Greco, Vittoria Letizia, Arnolfo Petruzziello, Maddalena Schioppa, Gregorio Goffredi, Francesca Marciano, Michele Caraglia, Alessia Cossu, Marianna Scrima, Edmondo Adorisio, Morena D'Avenia, Michela Iacobellis, Rosanna Piluscio, Giorgio Dirani, Vittorio Sambri, Simona Semprini, Silvia Zanoli, Francesco Curcio, Stefania Marzinotto, Andreina Baj, Fausto Sessa. |
| EPI_ISL_1361585 | 1.AO Universitaria 'S. Giovanni di Dio e Ruggi D'Aragona, Scuola Medica Salernitana' Hospital / 2.UOC di Virologia e Microbiologia, Università della Campania 'L. Vanvitelli' / 3.AO Universitaria 'Federico II' Napoli Hospital / 4.AORN 'San Giuseppe Moscati' Avellino Hospital / 5.AO 'San Pio - presidio G. Rummo' Benevento Hospital / 6.AO 'Sant'Anna e San Sebastiano' Caserta Hospital / 7.PO 'Maria Santissima Addolorata' Eboli Hospital / 8.Biogem Istituto di Ricerche Genetiche / 9. U.O.C. di Genetica Medica e di Laboratorio A.O.R.N., Azienda Ospedaliera di Rilievo Nazionale Antonio Cardarelli, Napoli / 10. Centro di riferimento Oncologico della Basilicata (IRCCS-CROB), Rionero in Vulture (PZ) / 11. Presidio Ospedaliero di Agropoli, Agropoli (SA). | 1. Genome Research Center for Health (CRGS) / 2. Laboratory of Molecular Medicine and Genomics(LMMGe) / 3. Center for Research in Pure and Applied Mathematics (CRMPA) | Giorgio Giurato (Corresponding Author), Francesca Rizzo (Corresponding Author), Alessandro Weisz (Corresponding Author), Gianluigi Franci, Giovanni Nassa, Pasquale Pagliano, Roberta Tarallo, Elena Alexandrova, Ylenia D'Agostino, Carlo Ferravante, Jessica Lamberti, Viola Melone, Domenico Memoli, Valeria Mirici Cappa, Domenico Palumbo, Giovanni Pecoraro, Assunta Sellitto, Oriana Strianese, Ilaria Terenzi, Giuseppe Fenza, Aniello Gentile, Antonello Saccomanno, Sonia Amabile, Teresa Rocco, Annamaria Salvati, Emilia Vaccaro, Massimiliano Galdiero, Michele Cennamo, Giuseppe Portella, Maria Grazia Foti, Mariarosaria Ingino, Maria Landi, Maurizio Fumi, Vincenzo Rocco, Rita Greco, Vittoria Letizia, Arnolfo Petruzziello, Maddalena Schioppa, Gregorio Goffredi, Francesca Marciano, Michele Caraglia, Alessia Cossu, Marianna Scrima, Edmondo Adorisio, Morena D'Avenia, Michela Iacobellis, Rosanna Piluscio, Giorgio Dirani, Vittorio Sambri, Simona Semprini, Silvia Zanoli, Francesco Curcio, Stefania Marzinotto, Andreina Baj, Fausto Sessa, Massimiliano Chetta, Marina Tarsitano, Giusy Giordano, Marcello Ametrano, Mariano Fiorenza, Caivano Antonella, D'Auria Fiorella, De Luca Luciana. |
| EPI_ISL_1361586, EPI_ISL_1361587, EPI_ISL_1361588, EPI_ISL_1361589 | 1.AO Universitaria 'S. Giovanni di Dio e Ruggi D'Aragona, Scuola Medica Salernitana' Hospital / 2.UOC di Virologia e Microbiologia, Università della Campania 'L. Vanvitelli' / 3.AO Universitaria 'Federico II' Napoli Hospital / 4.AORN 'San Giuseppe Moscati' Avellino Hospital / 5.AO 'San Pio - presidio G. Rummo' Benevento Hospital / 6.AO 'Sant'Anna e San Sebastiano' Caserta Hospital / 7.PO 'Maria Santissima Addolorata' Eboli Hospital / 8.Biogem Istituto di Ricerche Genetiche / 9. U.O.C. di Genetica Medica e di Laboratorio A.O.R.N., Azienda Ospedaliera di Rilievo Nazionale Antonio Cardarelli, Napoli / 10. Centro di riferimento Oncologico della Basilicata (IRCCS-CROB), Rionero in Vulture (PZ) / 11. Presidio Ospedaliero di Agropoli, Agropoli (SA). | 1. Genome Research Center for Health (CRGS) / 2. Laboratory of Molecular Medicine and Genomics(LMMGe) / 3. Center for Research in Pure and Applied Mathematics (CRMPA) | Giorgio Giurato (Corresponding Author), Francesca Rizzo (Corresponding Author), Alessandro Weisz (Corresponding Author), Gianluigi Franci, Giovanni Nassa, Pasquale Pagliano, Roberta Tarallo, Elena Alexandrova, Ylenia D'Agostino, Carlo Ferravante, Jessica Lamberti, Viola Melone, Domenico Memoli, Valeria Mirici Cappa, Domenico Palumbo, Giovanni Pecoraro, Assunta Sellitto, Oriana Strianese, Ilaria Terenzi, Giuseppe Fenza, Aniello Gentile, Antonello Saccomanno, Sonia Amabile, Teresa Rocco, Annamaria Salvati, Emilia Vaccaro, Massimiliano Galdiero, Michele Cennamo, Giuseppe Portella, Maria Grazia Foti, Mariarosaria Ingino, Maria Landi, Maurizio Fumi, Vincenzo Rocco, Rita Greco, Vittoria Letizia, Arnolfo Petruzziello, Maddalena Schioppa, Gregorio Goffredi, Francesca Marciano, Michele Caraglia, Alessia Cossu, Marianna Scrima, Edmondo Adorisio, Morena D'Avenia, Michela Iacobellis, Rosanna Piluscio, Giorgio Dirani, Vittorio Sambri, Simona Semprini, Silvia Zanoli, Francesco Curcio, Stefania Marzinotto, Andreina Baj, Fausto Sessa. |
| EPI_ISL_1361590 | 1.AO Universitaria 'S. Giovanni di Dio e Ruggi D'Aragona, Scuola Medica Salernitana' Hospital / 2.UOC di Virologia e | 1. Genome Research Center for Health (CRGS) / 2. Laboratory of Molecular Medicine and Genomics(LMMGe) / 3. | Giorgio Giurato (Corresponding Author), Francesca Rizzo (Corresponding Author), Alessandro Weisz (Corresponding Author), Gianluigi Franci, Giovanni Nassa, Pasquale Pagliano, Roberta Tarallo, Elena Alexandrova, Ylenia D'Agostino, Carlo Ferravante, Jessica Lamberti, Viola Melone, Domenico Memoli, |

| | | | |
|---|---|---|---|
| | Microbiologia, Università della Campania 'L. Vanvitelli' / 3.AO Universitaria 'Federico II' Napoli Hospital / 4.AORN 'San Giuseppe Moscati' Avellino Hospital / 5.AO 'San Pio - presidio G. Rummo' Benevento Hospital / 6.AO 'Sant'Anna e San Sebastiano' Caserta Hospital / 7.PO 'Maria Santissima Addolorata' Eboli Hospital / 8.Biogem Istituto di Ricerche Genetiche / 9. U.O.C. di Genetica Medica e di Laboratorio A.O.R.N., Azienda Ospedaliera di Rilievo Nazionale Antonio Cardarelli, Napoli / 10. Centro di riferimento Oncologico della Basilicata (IRCCS-CROB), Rionero in Vulture (PZ) / 11. Presidio Ospedaliero di Agropoli, Agropoli (SA). | Center for Research in Pure and Applied Mathematics (CRMPA) | Valeria Mirici Cappa, Domenico Palumbo, Giovanni Pecoraro, Assunta Sellitto, Oriana Strianese, Ilaria Terenzi, Giuseppe Fenza, Aniello Gentile, Antonello Saccomanno, Sonia Amabile, Teresa Rocco, Annamaria Salvati, Emilia Vaccaro, Massimiliano Galdiero, Michele Cennamo, Giuseppe Portella, Maria Grazia Foti, Mariarosaria Ingino, Maria Landi, Maurizio Fumi, Vincenzo Rocco, Rita Greco, Vittoria Letizia, Arnolfo Petruzziello, Maddalena Schioppa, Gregorio Goffredi, Francesca Marciano, Michele Caraglia, Alessia Cossu, Marianna Scrima, Edmondo Adorisio, Morena D'Avenia, Michela Iacobellis, Rosanna Piluscio, Giorgio Dirani, Vittorio Sambri, Simona Semprini, Silvia Zanoli, Francesco Curcio, Stefania Marzinotto, Andreina Baj, Fausto Sessa, Massimiliano Chetta, Marina Tarsitano, Giusy Giordano, Marcello Ametrano, Mariano Fiorenza, Caivano Antonella, D'Auria Fiorella, De Luca Luciana. |
| EPI_ISL_1361591, EPI_ISL_1361592, EPI_ISL_1361594, EPI_ISL_1361595, EPI_ISL_1361597, EPI_ISL_1361598, EPI_ISL_1361599, EPI_ISL_1361600 | 1.AO Universitaria 'S. Giovanni di Dio e Ruggi D'Aragona, Scuola Medica Salernitana' Hospital / 2.UOC di Virologia e Microbiologia, Università della Campania 'L. Vanvitelli' / 3.AO Universitaria 'Federico II' Napoli Hospital / 4.AORN 'San Giuseppe Moscati' Avellino Hospital / 5.AO 'San Pio - presidio G. Rummo' Benevento Hospital / 6.AO 'Sant'Anna e San Sebastiano' Caserta Hospital / 7.PO 'Maria Santissima Addolorata' Eboli Hospital / 8.Biogem Istituto di Ricerche Genetiche / 9. U.O.C. di Genetica Medica e di Laboratorio A.O.R.N., Azienda Ospedaliera di Rilievo Nazionale Antonio Cardarelli, Napoli / 10. Centro di riferimento Oncologico della Basilicata (IRCCS-CROB), Rionero in Vulture (PZ) / 11. Presidio Ospedaliero di Agropoli, Agropoli (SA). | 1. Genome Research Center for Health (CRGS) / 2. Laboratory of Molecular Medicine and Genomics(LMMGe) / 3. Center for Research in Pure and Applied Mathematics (CRMPA) | Giorgio Giurato (Corresponding Author), Francesca Rizzo (Corresponding Author), Alessandro Weisz (Corresponding Author), Gianluigi Franci, Giovanni Nassa, Pasquale Pagliano, Roberta Tarallo, Elena Alexandrova, Ylenia D'Agostino, Carlo Ferravante, Jessica Lamberti, Viola Melone, Domenico Memoli, Valeria Mirici Cappa, Domenico Palumbo, Giovanni Pecoraro, Assunta Sellitto, Oriana Strianese, Ilaria Terenzi, Giuseppe Fenza, Aniello Gentile, Antonello Saccomanno, Sonia Amabile, Teresa Rocco, Annamaria Salvati, Emilia Vaccaro, Massimiliano Galdiero, Michele Cennamo, Giuseppe Portella, Maria Grazia Foti, Mariarosaria Ingino, Maria Landi, Maurizio Fumi, Vincenzo Rocco, Rita Greco, Vittoria Letizia, Arnolfo Petruzziello, Maddalena Schioppa, Gregorio Goffredi, Francesca Marciano, Michele Caraglia, Alessia Cossu, Marianna Scrima, Edmondo Adorisio, Morena D'Avenia, Michela Iacobellis, Rosanna Piluscio, Giorgio Dirani, Vittorio Sambri, Simona Semprini, Silvia Zanoli, Francesco Curcio, Stefania Marzinotto, Andreina Baj, Fausto Sessa. |
| EPI_ISL_1361601 | 1.AO Universitaria 'S. Giovanni di Dio e Ruggi D'Aragona, Scuola Medica Salernitana' Hospital / 2.UOC di Virologia e Microbiologia, Università della Campania 'L. Vanvitelli' / 3.AO Universitaria 'Federico II' Napoli Hospital / 4.AORN 'San Giuseppe Moscati' Avellino Hospital / 5.AO 'San Pio - presidio G. Rummo' Benevento Hospital / 6.AO 'Sant'Anna e San Sebastiano' Caserta Hospital / 7.PO 'Maria Santissima Addolorata' Eboli Hospital / 8.Biogem Istituto di Ricerche Genetiche / 9. U.O.C. di Genetica Medica e di Laboratorio A.O.R.N., Azienda Ospedaliera di Rilievo Nazionale Antonio Cardarelli, Napoli / 10. Centro di riferimento Oncologico della Basilicata (IRCCS-CROB), Rionero in Vulture (PZ) / 11. Presidio Ospedaliero di Agropoli, Agropoli (SA). | 1. Genome Research Center for Health (CRGS) / 2. Laboratory of Molecular Medicine and Genomics(LMMGe) / 3. Center for Research in Pure and Applied Mathematics (CRMPA) | Giorgio Giurato (Corresponding Author), Francesca Rizzo (Corresponding Author), Alessandro Weisz (Corresponding Author), Gianluigi Franci, Giovanni Nassa, Pasquale Pagliano, Roberta Tarallo, Elena Alexandrova, Ylenia D'Agostino, Carlo Ferravante, Jessica Lamberti, Viola Melone, Domenico Memoli, Valeria Mirici Cappa, Domenico Palumbo, Giovanni Pecoraro, Assunta Sellitto, Oriana Strianese, Ilaria Terenzi, Giuseppe Fenza, Aniello Gentile, Antonello Saccomanno, Sonia Amabile, Teresa Rocco, Annamaria Salvati, Emilia Vaccaro, Massimiliano Galdiero, Michele Cennamo, Giuseppe Portella, Maria Grazia Foti, Mariarosaria Ingino, Maria Landi, Maurizio Fumi, Vincenzo Rocco, Rita Greco, Vittoria Letizia, Arnolfo Petruzziello, Maddalena Schioppa, Gregorio Goffredi, Francesca Marciano, Michele Caraglia, Alessia Cossu, Marianna Scrima, Edmondo Adorisio, Morena D'Avenia, Michela Iacobellis, Rosanna Piluscio, Giorgio Dirani, Vittorio Sambri, Simona Semprini, Silvia Zanoli, Francesco Curcio, Stefania Marzinotto, Andreina Baj, Fausto Sessa, Massimiliano Chetta, Marina Tarsitano, Giusy Giordano, Marcello Ametrano, Mariano Fiorenza, Caivano Antonella, D'Auria Fiorella, De Luca Luciana. |
| EPI_ISL_1361602, EPI_ISL_1361603, EPI_ISL_1361604, EPI_ISL_1361605 | 1.AO Universitaria 'S. Giovanni di Dio e Ruggi D'Aragona, Scuola Medica Salernitana' Hospital / 2.UOC di Virologia e Microbiologia, Università della Campania 'L. Vanvitelli' / 3.AO Universitaria 'Federico II' Napoli Hospital / 4.AORN 'San Giuseppe Moscati' Avellino Hospital / 5.AO 'San Pio - presidio G. Rummo' Benevento Hospital / 6.AO 'Sant'Anna e San Sebastiano' Caserta Hospital / 7.PO 'Maria Santissima Addolorata' Eboli Hospital / 8.Biogem Istituto di Ricerche Genetiche / 9. U.O.C. di Genetica Medica e di Laboratorio A.O.R.N., Azienda Ospedaliera di Rilievo Nazionale Antonio Cardarelli, Napoli / 10. Centro di riferimento Oncologico della Basilicata (IRCCS-CROB), Rionero in Vulture (PZ) / 11. Presidio Ospedaliero di Agropoli, Agropoli (SA). | 1. Genome Research Center for Health (CRGS) / 2. Laboratory of Molecular Medicine and Genomics(LMMGe) / 3. Center for Research in Pure and Applied Mathematics (CRMPA) | Giorgio Giurato (Corresponding Author), Francesca Rizzo (Corresponding Author), Alessandro Weisz (Corresponding Author), Gianluigi Franci, Giovanni Nassa, Pasquale Pagliano, Roberta Tarallo, Elena Alexandrova, Ylenia D'Agostino, Carlo Ferravante, Jessica Lamberti, Viola Melone, Domenico Memoli, Valeria Mirici Cappa, Domenico Palumbo, Giovanni Pecoraro, Assunta Sellitto, Oriana Strianese, Ilaria Terenzi, Giuseppe Fenza, Aniello Gentile, Antonello Saccomanno, Sonia Amabile, Teresa Rocco, Annamaria Salvati, Emilia Vaccaro, Massimiliano Galdiero, Michele Cennamo, Giuseppe Portella, Maria Grazia Foti, Mariarosaria Ingino, Maria Landi, Maurizio Fumi, Vincenzo Rocco, Rita Greco, Vittoria Letizia, Arnolfo Petruzziello, Maddalena Schioppa, Gregorio Goffredi, Francesca Marciano, Michele Caraglia, Alessia Cossu, Marianna Scrima, Edmondo Adorisio, Morena D'Avenia, Michela Iacobellis, Rosanna Piluscio, Giorgio Dirani, Vittorio Sambri, Simona Semprini, Silvia Zanoli, Francesco Curcio, Stefania Marzinotto, Andreina Baj, Fausto Sessa. |
| EPI_ISL_1361606 | 1.AO Universitaria 'S. Giovanni di Dio e Ruggi D'Aragona, Scuola Medica Salernitana' Hospital / 2.UOC di Virologia e Microbiologia, Università della Campania 'L. Vanvitelli' / 3.AO Universitaria 'Federico II' Napoli Hospital / 4.AORN 'San Giuseppe Moscati' Avellino Hospital / 5.AO 'San Pio - presidio G. Rummo' Benevento Hospital / 6.AO 'Sant'Anna e San Sebastiano' Caserta Hospital / 7.PO 'Maria Santissima Addolorata' Eboli Hospital / 8.Biogem Istituto di Ricerche Genetiche / 9. U.O.C. di Genetica Medica e di Laboratorio A.O.R.N., Azienda Ospedaliera di Rilievo Nazionale Antonio Cardarelli, Napoli / 10. Centro di riferimento Oncologico della Basilicata (IRCCS-CROB), Rionero in Vulture (PZ) / 11. Presidio Ospedaliero di Agropoli, Agropoli (SA). | 1. Genome Research Center for Health (CRGS) / 2. Laboratory of Molecular Medicine and Genomics(LMMGe) / 3. Center for Research in Pure and Applied Mathematics (CRMPA) | Giorgio Giurato (Corresponding Author), Francesca Rizzo (Corresponding Author), Alessandro Weisz (Corresponding Author), Gianluigi Franci, Giovanni Nassa, Pasquale Pagliano, Roberta Tarallo, Elena Alexandrova, Ylenia D'Agostino, Carlo Ferravante, Jessica Lamberti, Viola Melone, Domenico Memoli, Valeria Mirici Cappa, Domenico Palumbo, Giovanni Pecoraro, Assunta Sellitto, Oriana Strianese, Ilaria Terenzi, Giuseppe Fenza, Aniello Gentile, Antonello Saccomanno, Sonia Amabile, Teresa Rocco, Annamaria Salvati, Emilia Vaccaro, Massimiliano Galdiero, Michele Cennamo, Giuseppe Portella, Maria Grazia Foti, Mariarosaria Ingino, Maria Landi, Maurizio Fumi, Vincenzo Rocco, Rita Greco, Vittoria Letizia, Arnolfo Petruzziello, Maddalena Schioppa, Gregorio Goffredi, Francesca Marciano, Michele Caraglia, Alessia Cossu, Marianna Scrima, Edmondo Adorisio, Morena D'Avenia, Michela Iacobellis, Rosanna Piluscio, Giorgio Dirani, Vittorio Sambri, Simona Semprini, Silvia Zanoli, Francesco Curcio, Stefania Marzinotto, Andreina Baj, Fausto Sessa, Massimiliano Chetta, Marina Tarsitano, Giusy Giordano, Marcello Ametrano, Mariano Fiorenza, Caivano Antonella, D'Auria Fiorella, De Luca Luciana. |
| EPI_ISL_1361607, EPI_ISL_1361608, EPI_ISL_1361609, EPI_ISL_1361610 | 1.AO Universitaria 'S. Giovanni di Dio e Ruggi D'Aragona, Scuola Medica Salernitana' Hospital / 2.UOC di Virologia e Microbiologia, Università della Campania 'L. Vanvitelli' / 3.AO Universitaria 'Federico II' Napoli Hospital / 4.AORN 'San Giuseppe Moscati' Avellino Hospital / 5.AO 'San Pio - presidio G. Rummo' Benevento Hospital / 6.AO 'Sant'Anna e San Sebastiano' Caserta Hospital / 7.PO 'Maria Santissima Addolorata' Eboli Hospital / 8.Biogem Istituto di Ricerche Genetiche / 9. U.O.C. di Genetica Medica e di Laboratorio A.O.R.N., Azienda Ospedaliera di Rilievo Nazionale Antonio Cardarelli, Napoli / 10. Centro di riferimento Oncologico della Basilicata (IRCCS-CROB), Rionero in Vulture (PZ) / 11. Presidio Ospedaliero di Agropoli, Agropoli (SA). | 1. Genome Research Center for Health (CRGS) / 2. Laboratory of Molecular Medicine and Genomics(LMMGe) / 3. Center for Research in Pure and Applied Mathematics (CRMPA) | Giorgio Giurato (Corresponding Author), Francesca Rizzo (Corresponding Author), Alessandro Weisz (Corresponding Author), Gianluigi Franci, Giovanni Nassa, Pasquale Pagliano, Roberta Tarallo, Elena Alexandrova, Ylenia D'Agostino, Carlo Ferravante, Jessica Lamberti, Viola Melone, Domenico Memoli, Valeria Mirici Cappa, Domenico Palumbo, Giovanni Pecoraro, Assunta Sellitto, Oriana Strianese, Ilaria Terenzi, Giuseppe Fenza, Aniello Gentile, Antonello Saccomanno, Sonia Amabile, Teresa Rocco, Annamaria Salvati, Emilia Vaccaro, Massimiliano Galdiero, Michele Cennamo, Giuseppe Portella, Maria Grazia Foti, Mariarosaria Ingino, Maria Landi, Maurizio Fumi, Vincenzo Rocco, Rita Greco, Vittoria Letizia, Arnolfo Petruzziello, Maddalena Schioppa, Gregorio Goffredi, Francesca Marciano, Michele Caraglia, Alessia Cossu, Marianna Scrima, Edmondo Adorisio, Morena D'Avenia, Michela Iacobellis, Rosanna Piluscio, Giorgio Dirani, Vittorio Sambri, Simona Semprini, Silvia Zanoli, Francesco Curcio, Stefania Marzinotto, Andreina Baj, Fausto Sessa. |
| EPI_ISL_1361611 | 1.AO Universitaria 'S. Giovanni di Dio e Ruggi D'Aragona, Scuola Medica Salernitana' Hospital / 2.UOC di Virologia e Microbiologia, Università della Campania 'L. Vanvitelli' / 3.AO Universitaria 'Federico II' Napoli Hospital / 4.AORN 'San Giuseppe Moscati' Avellino Hospital / 5.AO 'San Pio - presidio G. Rummo' Benevento Hospital / 6.AO 'Sant'Anna e San Sebastiano' Caserta Hospital / 7.PO 'Maria Santissima Addolorata' Eboli Hospital / 8.Biogem Istituto di Ricerche | 1. Genome Research Center for Health (CRGS) / 2. Laboratory of Molecular Medicine and Genomics(LMMGe) / 3. Center for Research in Pure and Applied Mathematics (CRMPA) | Giorgio Giurato (Corresponding Author), Francesca Rizzo (Corresponding Author), Alessandro Weisz (Corresponding Author), Gianluigi Franci, Giovanni Nassa, Pasquale Pagliano, Roberta Tarallo, Elena Alexandrova, Ylenia D'Agostino, Carlo Ferravante, Jessica Lamberti, Viola Melone, Domenico Memoli, Valeria Mirici Cappa, Domenico Palumbo, Giovanni Pecoraro, Assunta Sellitto, Oriana Strianese, Ilaria Terenzi, Giuseppe Fenza, Aniello Gentile, Antonello Saccomanno, Sonia Amabile, Teresa Rocco, Annamaria Salvati, Emilia Vaccaro, Massimiliano Galdiero, Michele Cennamo, Giuseppe Portella, Maria Grazia Foti, Mariarosaria Ingino, Maria Landi, Maurizio Fumi, Vincenzo Rocco, Rita Greco, Vittoria Letizia, Arnolfo Petruzziello, Maddalena Schioppa, Gregorio Goffredi, Francesca Marciano, Michele Caraglia, Alessia Cossu, Marianna Scrima, Edmondo Adorisio, Morena D'Avenia, Michela Iacobellis, Rosanna Piluscio, Giorgio Dirani, Vittorio Sambri, Simona Semprini, Silvia Zanoli, Francesco Curcio, Stefania Marzinotto, Andreina Baj, Fausto Sessa, Massimiliano Chetta, Marina Tarsitano, Giusy Giordano, Marcello Ametrano, Mariano Fiorenza, Caivano Antonella, D'Auria Fiorella, De Luca Luciana. |

| | | | |
|---|---|---|---|
| | Genetiche / 9. U.O.C. di Genetica Medica e di Laboratorio A.O.R.N., Azienda Ospedaliera di Rilievo Nazionale Antonio Cardarelli, Napoli / 10. Centro di riferimento Oncologico della Basilicata (IRCCS-CROB), Rionero in Vulture (PZ) / 11. Presidio Ospedaliero di Agropoli, Agropoli (SA). | | |
| EPI_ISL_1361612, EPI_ISL_1361613, EPI_ISL_1361614, EPI_ISL_1361615 | 1.AO Universitaria 'S. Giovanni di Dio e Ruggi D'Aragona, Scuola Medica Salernitana' Hospital / 2.UOC di Virologia e Microbiologia, Università della Campania 'L. Vanvitelli' / 3.AO Universitaria 'Federico II' Napoli Hospital / 4.AORN 'San Giuseppe Moscati' Avellino Hospital / 5.AO 'San Pio - presidio G. Rummo' Benevento Hospital / 6.AO 'Sant'Anna e San Sebastiano' Caserta Hospital / 7.PO 'Maria Santissima Addolorata' Eboli Hospital / 8.Biogem Istituto di Ricerche Genetiche / 9. U.O.C. di Genetica Medica e di Laboratorio A.O.R.N., Azienda Ospedaliera di Rilievo Nazionale Antonio Cardarelli, Napoli / 10. Centro di riferimento Oncologico della Basilicata (IRCCS-CROB), Rionero in Vulture (PZ) / 11. Presidio Ospedaliero di Agropoli, Agropoli (SA). | 1. Genome Research Center for Health (CRGS) / 2. Laboratory of Molecular Medicine and Genomics(LMMGe) / 3. Center for Research in Pure and Applied Mathematics (CRMPA) | Giorgio Giurato (Corresponding Author), Francesca Rizzo (Corresponding Author), Alessandro Weisz (Corresponding Author), Gianluigi Franci, Giovanni Nassa, Pasquale Pagliano, Roberta Tarallo, Elena Alexandrova, Ylenia D'Agostino, Carlo Ferravante, Jessica Lamberti, Viola Melone, Domenico Memoli, Valeria Mirici Cappa, Domenico Palumbo, Giovanni Pecoraro, Assunta Sellitto, Oriana Strianese, Ilaria Terenzi, Giuseppe Fenza, Aniello Gentile, Antonello Saccomanno, Sonia Amabile, Teresa Rocco, Annamaria Salvati, Emilia Vaccaro, Massimiliano Galdiero, Michele Cennamo, Giuseppe Portella, Maria Grazia Foti, Mariarosaria Ingino, Maria Landi, Maurizio Fumi, Vincenzo Rocco, Rita Greco, Vittoria Letizia, Arnolfo Petruzziello, Maddalena Schioppa, Gregorio Goffredi, Francesca Marciano, Michele Caraglia, Alessia Cossu, Marianna Scrima, Edmondo Adorisio, Morena D'Avenia, Michela Iacobellis, Rosanna Piluscio, Giorgio Dirani, Vittorio Sambri, Simona Semprini, Silvia Zanoli, Francesco Curcio, Stefania Marzinotto, Andreina Baj, Fausto Sessa. |
| EPI_ISL_1361616 | 1.AO Universitaria 'S. Giovanni di Dio e Ruggi D'Aragona, Scuola Medica Salernitana' Hospital / 2.UOC di Virologia e Microbiologia, Università della Campania 'L. Vanvitelli' / 3.AO Universitaria 'Federico II' Napoli Hospital / 4.AORN 'San Giuseppe Moscati' Avellino Hospital / 5.AO 'San Pio - presidio G. Rummo' Benevento Hospital / 6.AO 'Sant'Anna e San Sebastiano' Caserta Hospital / 7.PO 'Maria Santissima Addolorata' Eboli Hospital / 8.Biogem Istituto di Ricerche Genetiche / 9. U.O.C. di Genetica Medica e di Laboratorio A.O.R.N., Azienda Ospedaliera di Rilievo Nazionale Antonio Cardarelli, Napoli / 10. Centro di riferimento Oncologico della Basilicata (IRCCS-CROB), Rionero in Vulture (PZ) / 11. Presidio Ospedaliero di Agropoli, Agropoli (SA). | 1. Genome Research Center for Health (CRGS) / 2. Laboratory of Molecular Medicine and Genomics(LMMGe) / 3. Center for Research in Pure and Applied Mathematics (CRMPA) | Giorgio Giurato (Corresponding Author), Francesca Rizzo (Corresponding Author), Alessandro Weisz (Corresponding Author), Gianluigi Franci, Giovanni Nassa, Pasquale Pagliano, Roberta Tarallo, Elena Alexandrova, Ylenia D'Agostino, Carlo Ferravante, Jessica Lamberti, Viola Melone, Domenico Memoli, Valeria Mirici Cappa, Domenico Palumbo, Giovanni Pecoraro, Assunta Sellitto, Oriana Strianese, Ilaria Terenzi, Giuseppe Fenza, Aniello Gentile, Antonello Saccomanno, Sonia Amabile, Teresa Rocco, Annamaria Salvati, Emilia Vaccaro, Massimiliano Galdiero, Michele Cennamo, Giuseppe Portella, Maria Grazia Foti, Mariarosaria Ingino, Maria Landi, Maurizio Fumi, Vincenzo Rocco, Rita Greco, Vittoria Letizia, Arnolfo Petruzziello, Maddalena Schioppa, Gregorio Goffredi, Francesca Marciano, Michele Caraglia, Alessia Cossu, Marianna Scrima, Edmondo Adorisio, Morena D'Avenia, Michela Iacobellis, Rosanna Piluscio, Giorgio Dirani, Vittorio Sambri, Simona Semprini, Silvia Zanoli, Francesco Curcio, Stefania Marzinotto, Andreina Baj, Fausto Sessa, Massimiliano Chetta, Marina Tarsitano, Giusy Giordano, Marcello Ametrano, Mariano Fiorenza, Caivano Antonella, D'Auria Fiorella, De Luca Luciana. |
| EPI_ISL_1361617, EPI_ISL_1361618, EPI_ISL_1361619, EPI_ISL_1361620 | 1.AO Universitaria 'S. Giovanni di Dio e Ruggi D'Aragona, Scuola Medica Salernitana' Hospital / 2.UOC di Virologia e Microbiologia, Università della Campania 'L. Vanvitelli' / 3.AO Universitaria 'Federico II' Napoli Hospital / 4.AORN 'San Giuseppe Moscati' Avellino Hospital / 5.AO 'San Pio - presidio G. Rummo' Benevento Hospital / 6.AO 'Sant'Anna e San Sebastiano' Caserta Hospital / 7.PO 'Maria Santissima Addolorata' Eboli Hospital / 8.Biogem Istituto di Ricerche Genetiche / 9. U.O.C. di Genetica Medica e di Laboratorio A.O.R.N., Azienda Ospedaliera di Rilievo Nazionale Antonio Cardarelli, Napoli / 10. Centro di riferimento Oncologico della Basilicata (IRCCS-CROB), Rionero in Vulture (PZ) / 11. Presidio Ospedaliero di Agropoli, Agropoli (SA). | 1. Genome Research Center for Health (CRGS) / 2. Laboratory of Molecular Medicine and Genomics(LMMGe) / 3. Center for Research in Pure and Applied Mathematics (CRMPA) | Giorgio Giurato (Corresponding Author), Francesca Rizzo (Corresponding Author), Alessandro Weisz (Corresponding Author), Gianluigi Franci, Giovanni Nassa, Pasquale Pagliano, Roberta Tarallo, Elena Alexandrova, Ylenia D'Agostino, Carlo Ferravante, Jessica Lamberti, Viola Melone, Domenico Memoli, Valeria Mirici Cappa, Domenico Palumbo, Giovanni Pecoraro, Assunta Sellitto, Oriana Strianese, Ilaria Terenzi, Giuseppe Fenza, Aniello Gentile, Antonello Saccomanno, Sonia Amabile, Teresa Rocco, Annamaria Salvati, Emilia Vaccaro, Massimiliano Galdiero, Michele Cennamo, Giuseppe Portella, Maria Grazia Foti, Mariarosaria Ingino, Maria Landi, Maurizio Fumi, Vincenzo Rocco, Rita Greco, Vittoria Letizia, Arnolfo Petruzziello, Maddalena Schioppa, Gregorio Goffredi, Francesca Marciano, Michele Caraglia, Alessia Cossu, Marianna Scrima, Edmondo Adorisio, Morena D'Avenia, Michela Iacobellis, Rosanna Piluscio, Giorgio Dirani, Vittorio Sambri, Simona Semprini, Silvia Zanoli, Francesco Curcio, Stefania Marzinotto, Andreina Baj, Fausto Sessa. |
| EPI_ISL_1361621 | 1.AO Universitaria 'S. Giovanni di Dio e Ruggi D'Aragona, Scuola Medica Salernitana' Hospital / 2.UOC di Virologia e Microbiologia, Università della Campania 'L. Vanvitelli' / 3.AO Universitaria 'Federico II' Napoli Hospital / 4.AORN 'San Giuseppe Moscati' Avellino Hospital / 5.AO 'San Pio - presidio G. Rummo' Benevento Hospital / 6.AO 'Sant'Anna e San Sebastiano' Caserta Hospital / 7.PO 'Maria Santissima Addolorata' Eboli Hospital / 8.Biogem Istituto di Ricerche Genetiche / 9. U.O.C. di Genetica Medica e di Laboratorio A.O.R.N., Azienda Ospedaliera di Rilievo Nazionale Antonio Cardarelli, Napoli / 10. Centro di riferimento Oncologico della Basilicata (IRCCS-CROB), Rionero in Vulture (PZ) / 11. Presidio Ospedaliero di Agropoli, Agropoli (SA). | 1. Genome Research Center for Health (CRGS) / 2. Laboratory of Molecular Medicine and Genomics(LMMGe) / 3. Center for Research in Pure and Applied Mathematics (CRMPA) | Giorgio Giurato (Corresponding Author), Francesca Rizzo (Corresponding Author), Alessandro Weisz (Corresponding Author), Gianluigi Franci, Giovanni Nassa, Pasquale Pagliano, Roberta Tarallo, Elena Alexandrova, Ylenia D'Agostino, Carlo Ferravante, Jessica Lamberti, Viola Melone, Domenico Memoli, Valeria Mirici Cappa, Domenico Palumbo, Giovanni Pecoraro, Assunta Sellitto, Oriana Strianese, Ilaria Terenzi, Giuseppe Fenza, Aniello Gentile, Antonello Saccomanno, Sonia Amabile, Teresa Rocco, Annamaria Salvati, Emilia Vaccaro, Massimiliano Galdiero, Michele Cennamo, Giuseppe Portella, Maria Grazia Foti, Mariarosaria Ingino, Maria Landi, Maurizio Fumi, Vincenzo Rocco, Rita Greco, Vittoria Letizia, Arnolfo Petruzziello, Maddalena Schioppa, Gregorio Goffredi, Francesca Marciano, Michele Caraglia, Alessia Cossu, Marianna Scrima, Edmondo Adorisio, Morena D'Avenia, Michela Iacobellis, Rosanna Piluscio, Giorgio Dirani, Vittorio Sambri, Simona Semprini, Silvia Zanoli, Francesco Curcio, Stefania Marzinotto, Andreina Baj, Fausto Sessa, Massimiliano Chetta, Marina Tarsitano, Giusy Giordano, Marcello Ametrano, Mariano Fiorenza, Caivano Antonella, D'Auria Fiorella, De Luca Luciana. |
| EPI_ISL_1361622, EPI_ISL_1361623, EPI_ISL_1361624, EPI_ISL_1361625 | 1.AO Universitaria 'S. Giovanni di Dio e Ruggi D'Aragona, Scuola Medica Salernitana' Hospital / 2.UOC di Virologia e Microbiologia, Università della Campania 'L. Vanvitelli' / 3.AO Universitaria 'Federico II' Napoli Hospital / 4.AORN 'San Giuseppe Moscati' Avellino Hospital / 5.AO 'San Pio - presidio G. Rummo' Benevento Hospital / 6.AO 'Sant'Anna e San Sebastiano' Caserta Hospital / 7.PO 'Maria Santissima Addolorata' Eboli Hospital / 8.Biogem Istituto di Ricerche Genetiche / 9. U.O.C. di Genetica Medica e di Laboratorio A.O.R.N., Azienda Ospedaliera di Rilievo Nazionale Antonio Cardarelli, Napoli / 10. Centro di riferimento Oncologico della Basilicata (IRCCS-CROB), Rionero in Vulture (PZ) / 11. Presidio Ospedaliero di Agropoli, Agropoli (SA). | 1. Genome Research Center for Health (CRGS) / 2. Laboratory of Molecular Medicine and Genomics(LMMGe) / 3. Center for Research in Pure and Applied Mathematics (CRMPA) | Giorgio Giurato (Corresponding Author), Francesca Rizzo (Corresponding Author), Alessandro Weisz (Corresponding Author), Gianluigi Franci, Giovanni Nassa, Pasquale Pagliano, Roberta Tarallo, Elena Alexandrova, Ylenia D'Agostino, Carlo Ferravante, Jessica Lamberti, Viola Melone, Domenico Memoli, Valeria Mirici Cappa, Domenico Palumbo, Giovanni Pecoraro, Assunta Sellitto, Oriana Strianese, Ilaria Terenzi, Giuseppe Fenza, Aniello Gentile, Antonello Saccomanno, Sonia Amabile, Teresa Rocco, Annamaria Salvati, Emilia Vaccaro, Massimiliano Galdiero, Michele Cennamo, Giuseppe Portella, Maria Grazia Foti, Mariarosaria Ingino, Maria Landi, Maurizio Fumi, Vincenzo Rocco, Rita Greco, Vittoria Letizia, Arnolfo Petruzziello, Maddalena Schioppa, Gregorio Goffredi, Francesca Marciano, Michele Caraglia, Alessia Cossu, Marianna Scrima, Edmondo Adorisio, Morena D'Avenia, Michela Iacobellis, Rosanna Piluscio, Giorgio Dirani, Vittorio Sambri, Simona Semprini, Silvia Zanoli, Francesco Curcio, Stefania Marzinotto, Andreina Baj, Fausto Sessa. |
| EPI_ISL_1361626 | 1.AO Universitaria 'S. Giovanni di Dio e Ruggi D'Aragona, Scuola Medica Salernitana' Hospital / 2.UOC di Virologia e Microbiologia, Università della Campania 'L. Vanvitelli' / 3.AO Universitaria 'Federico II' Napoli Hospital / 4.AORN 'San Giuseppe Moscati' Avellino Hospital / 5.AO 'San Pio - presidio G. Rummo' Benevento Hospital / 6.AO 'Sant'Anna e San Sebastiano' Caserta Hospital / 7.PO 'Maria Santissima Addolorata' Eboli Hospital / 8.Biogem Istituto di Ricerche Genetiche / 9. U.O.C. di Genetica Medica e di Laboratorio A.O.R.N., Azienda Ospedaliera di Rilievo Nazionale Antonio Cardarelli, Napoli / 10. Centro di riferimento Oncologico della Basilicata (IRCCS-CROB), Rionero in Vulture (PZ) / 11. Presidio Ospedaliero di Agropoli, Agropoli (SA). | 1. Genome Research Center for Health (CRGS) / 2. Laboratory of Molecular Medicine and Genomics(LMMGe) / 3. Center for Research in Pure and Applied Mathematics (CRMPA) | Giorgio Giurato (Corresponding Author), Francesca Rizzo (Corresponding Author), Alessandro Weisz (Corresponding Author), Gianluigi Franci, Giovanni Nassa, Pasquale Pagliano, Roberta Tarallo, Elena Alexandrova, Ylenia D'Agostino, Carlo Ferravante, Jessica Lamberti, Viola Melone, Domenico Memoli, Valeria Mirici Cappa, Domenico Palumbo, Giovanni Pecoraro, Assunta Sellitto, Oriana Strianese, Ilaria Terenzi, Giuseppe Fenza, Aniello Gentile, Antonello Saccomanno, Sonia Amabile, Teresa Rocco, Annamaria Salvati, Emilia Vaccaro, Massimiliano Galdiero, Michele Cennamo, Giuseppe Portella, Maria Grazia Foti, Mariarosaria Ingino, Maria Landi, Maurizio Fumi, Vincenzo Rocco, Rita Greco, Vittoria Letizia, Arnolfo Petruzziello, Maddalena Schioppa, Gregorio Goffredi, Francesca Marciano, Michele Caraglia, Alessia Cossu, Marianna Scrima, Edmondo Adorisio, Morena D'Avenia, Michela Iacobellis, Rosanna Piluscio, Giorgio Dirani, Vittorio Sambri, Simona Semprini, Silvia Zanoli, Francesco Curcio, Stefania Marzinotto, Andreina Baj, Fausto Sessa, Massimiliano Chetta, Marina Tarsitano, Giusy Giordano, Marcello Ametrano, Mariano Fiorenza, Caivano Antonella, D'Auria Fiorella, De Luca Luciana. |
| EPI_ISL_1361627, EPI_ISL_1361628, | 1.AO Universitaria 'S. Giovanni di Dio e Ruggi D'Aragona, | 1. Genome Research Center for Health (CRGS) / 2. | Giorgio Giurato (Corresponding Author), Francesca Rizzo (Corresponding Author), Alessandro Weisz (Corresponding Author), Gianluigi Franci, Giovanni |

| | | | |
|---|---|---|---|
| EPI_ISL_1361629, EPI_ISL_1361630 | Scuola Medica Salernitana' Hospital / 2.UOC di Virologia e Microbiologia, Università della Campania 'L. Vanvitelli' / 3.AO Universitaria 'Federico II' Napoli Hospital / 4.AORN 'San Giuseppe Moscati' Avellino Hospital / 5.AO 'San Pio - presidio G. Rummo' Benevento Hospital / 6.AO 'Sant'Anna e San Sebastiano' Caserta Hospital / 7.PO 'Maria Santissima Addolorata' Eboli Hospital / 8.Biogem Istituto di Ricerche Genetiche / 9. U.O.C. di Genetica Medica e di Laboratorio A.O.R.N., Azienda Ospedaliera di Rilievo Nazionale Antonio Cardarelli, Napoli / 10. Centro di riferimento Oncologico della Basilicata (IRCCS-CROB), Rionero in Vulture (PZ) / 11. Presidio Ospedaliero di Agropoli, Agropoli (SA). | Laboratory of Molecular Medicine and Genomics(LMMGe) / 3. Center for Research in Pure and Applied Mathematics (CRMPA) | Nassa, Pasquale Pagliano, Roberta Tarallo, Elena Alexandrova, Ylenia D'Agostino, Carlo Ferravante, Jessica Lamberti, Viola Melone, Domenico Memoli, Valeria Mirici Cappa, Domenico Palumbo, Giovanni Pecoraro, Assunta Sellitto, Oriana Strianese, Ilaria Terenzi, Giuseppe Fenza, Aniello Gentile, Antonello Saccomanno, Sonia Amabile, Teresa Rocco, Annamaria Salvati, Emilia Vaccaro, Massimiliano Galdiero, Michele Cennamo, Giuseppe Portella, Maria Grazia Foti, Mariarosaria Ingino, Maria Landi, Maurizio Fumi, Vincenzo Rocco, Rita Greco, Vittoria Letizia, Arnolfo Petruzziello, Maddalena Schioppa, Gregorio Goffredi, Francesca Marciano, Michele Caraglia, Alessia Cossu, Marianna Scrima, Edmondo Adorisio, Morena D'Avenia, Michela Iacobellis, Rosanna Piluscio, Giorgio Dirani, Vittorio Sambri, Simona Semprini, Silvia Zanoli, Francesco Curcio, Stefania Marzinotto, Andreina Baj, Fausto Sessa. |
| EPI_ISL_1366498, EPI_ISL_1366499, EPI_ISL_1366500, EPI_ISL_1366501 | 1.AO Universitaria 'S. Giovanni di Dio e Ruggi D'Aragona, Scuola Medica Salernitana' Hospital / 2.UOC di Virologia e Microbiologia, Università della Campania 'L. Vanvitelli' / 3.AO Universitaria 'Federico II' Napoli Hospital / 4.AORN 'San Giuseppe Moscati' Avellino Hospital / 5.AO 'San Pio - presidio G. Rummo' Benevento Hospital / 6.AO 'Sant'Anna e San Sebastiano' Caserta Hospital / 7.PO 'Maria Santissima Addolorata' Eboli Hospital / 8.Biogem Istituto di Ricerche Genetiche / 9. U.O.C. di Genetica Medica e di Laboratorio A.O.R.N., Azienda Ospedaliera di Rilievo Nazionale Antonio Cardarelli, Napoli / 10. Centro di riferimento Oncologico della Basilicata (IRCCS-CROB), Rionero in Vulture (PZ) / 11. Presidio Ospedaliero di Agropoli, Agropoli (SA). | 1. Genome Research Center for Health (CRGS) / 2. Laboratory of Molecular Medicine and Genomics(LMMGe) / 3. Center for Research in Pure and Applied Mathematics (CRMPA) | Giorgio Giurato, Francesca Rizzo, Alessandro Weisz, Gianluigi Franci, Giovanni Nassa, Pasquale Pagliano, Roberta Tarallo, Elena Alexandrova, Ylenia D'Agostino, Carlo Ferravante, Jessica Lamberti, Viola Melone, Domenico Memoli, Valeria Mirici Cappa, Domenico Palumbo, Giovanni Pecoraro, Assunta Sellitto, Oriana Strianese, Ilaria Terenzi, Giuseppe Fenza, Aniello Gentile, Antonello Saccomanno, Sonia Amabile, Teresa Rocco, Annamaria Salvati, Emilia Vaccaro, Massimiliano Galdiero, Michele Cennamo, Giuseppe Portella, Maria Grazia Foti, Mariarosaria Ingino, Maria Landi, Maurizio Fumi, Vincenzo Rocco, Rita Greco, Vittoria Letizia, Arnolfo Petruzziello, Maddalena Schioppa, Gregorio Goffredi, Francesca Marciano, Michele Caraglia, Alessia Cossu, Marianna Scrima, Edmondo Adorisio, Morena D'Avenia, Michela Iacobellis, Rosanna Piluscio, Giorgio Dirani, Vittorio Sambri, Simona Semprini, Silvia Zanoli, Francesco Curcio, Stefania Marzinotto, Andreina Baj, Fausto Sessa, Massimiliano Chetta, Marina Tarsitano, Giusy Giordano, Marcello Ametrano, Mariano Fiorenza, Caivano Antonella, D'Auria Fiorella, De Luca Luciana. |
| EPI_ISL_1366890, EPI_ISL_1366891, EPI_ISL_1366892, EPI_ISL_1366893, EPI_ISL_1366894, EPI_ISL_1366895, EPI_ISL_1366896, EPI_ISL_1366897, EPI_ISL_1366898, EPI_ISL_1366899, EPI_ISL_1366900, EPI_ISL_1366901, EPI_ISL_1366902, EPI_ISL_1366903, EPI_ISL_1366904, EPI_ISL_1366905, EPI_ISL_1366906, EPI_ISL_1366907, EPI_ISL_1366908, EPI_ISL_1366909, EPI_ISL_1366910, EPI_ISL_1366911, EPI_ISL_1366912, EPI_ISL_1366913, EPI_ISL_1366914, EPI_ISL_1366915, EPI_ISL_1366916, EPI_ISL_1366917, EPI_ISL_1366918, EPI_ISL_1366919, EPI_ISL_1366920 | | | |
| see above | 1.AO Universitaria 'S. Giovanni di Dio e Ruggi D'Aragona, Scuola Medica Salernitana' Hospital / 2.UOC di Virologia e Microbiologia, Università della Campania 'L. Vanvitelli' / 3.AO Universitaria 'Federico II' Napoli Hospital / 4.AORN 'San Giuseppe Moscati' Avellino Hospital / 5.AO 'San Pio - presidio G. Rummo' Benevento Hospital / 6.AO 'Sant'Anna e San Sebastiano' Caserta Hospital / 7.PO 'Maria Santissima Addolorata' Eboli Hospital / 8.Biogem Istituto di Ricerche Genetiche | 1. Genome Research Center for Health (CRGS) / 2. Laboratory of Molecular Medicine and Genomics(LMMGe) / 3. Center for Research in Pure and Applied Mathematics (CRMPA) | Giorgio Giurato (Corresponding Author), Francesca Rizzo (Corresponding Author), Alessandro Weisz (Corresponding Author), Gianluigi Franci, Giovanni Nassa, Pasquale Pagliano, Roberta Tarallo, Elena Alexandrova, Ylenia D'Agostino, Carlo Ferravante, Jessica Lamberti, Viola Melone, Domenico Memoli, Valeria Mirici Cappa, Domenico Palumbo, Giovanni Pecoraro, Assunta Sellitto, Oriana Strianese, Ilaria Terenzi, Giuseppe Fenza, Aniello Gentile, Antonello Saccomanno, Sonia Amabile, Teresa Rocco, Annamaria Salvati, Emilia Vaccaro, Massimiliano Galdiero, Michele Cennamo, Giuseppe Portella, Maria Grazia Foti, Mariarosaria Ingino, Maria Landi, Maurizio Fumi, Vincenzo Rocco, Rita Greco, Vittoria Letizia, Arnolfo Petruzziello, Maddalena Schioppa, Gregorio Goffredi, Francesca Marciano, Michele Caraglia, Alessia Cossu, Marianna Scrima, Edmondo Adorisio, Morena D'Avenia, Michela Iacobellis, Rosanna Piluscio, Giorgio Dirani, Vittorio Sambri, Simona Semprini, Silvia Zanoli, Francesco Curcio, Stefania Marzinotto, Andreina Baj, Fausto Sessa. |
| EPI_ISL_1372414 | 1.AO Universitaria 'S. Giovanni di Dio e Ruggi D'Aragona, Scuola Medica Salernitana' Hospital / 2.UOC di Virologia e Microbiologia, Università della Campania 'L. Vanvitelli' / 3.AO Universitaria 'Federico II' Napoli Hospital / 4.AORN 'San Giuseppe Moscati' Avellino Hospital / 5.AO 'San Pio - presidio G. Rummo' Benevento Hospital / 6.AO 'Sant'Anna e San Sebastiano' Caserta Hospital / 7.PO 'Maria Santissima Addolorata' Eboli Hospital / 8.Biogem Istituto di Ricerche Genetiche / 9. U.O.C. di Genetica Medica e di Laboratorio A.O.R.N., Azienda Ospedaliera di Rilievo Nazionale Antonio Cardarelli, Napoli / 10. Centro di riferimento Oncologico della Basilicata (IRCCS-CROB), Rionero in Vulture (PZ) / 11. Presidio Ospedaliero di Agropoli, Agropoli (SA). | 1. Genome Research Center for Health (CRGS) / 2. Laboratory of Molecular Medicine and Genomics(LMMGe) / 3. Center for Research in Pure and Applied Mathematics (CRMPA) | Giorgio Giurato (Corresponding Author), Francesca Rizzo (Corresponding Author), Alessandro Weisz (Corresponding Author), Gianluigi Franci, Giovanni Nassa, Pasquale Pagliano, Roberta Tarallo, Elena Alexandrova, Ylenia D'Agostino, Carlo Ferravante, Jessica Lamberti, Viola Melone, Domenico Memoli, Valeria Mirici Cappa, Domenico Palumbo, Giovanni Pecoraro, Assunta Sellitto, Oriana Strianese, Ilaria Terenzi, Giuseppe Fenza, Aniello Gentile, Antonello Saccomanno, Sonia Amabile, Teresa Rocco, Annamaria Salvati, Emilia Vaccaro, Massimiliano Galdiero, Michele Cennamo, Giuseppe Portella, Maria Grazia Foti, Mariarosaria Ingino, Maria Landi, Maurizio Fumi, Vincenzo Rocco, Rita Greco, Vittoria Letizia, Arnolfo Petruzziello, Maddalena Schioppa, Gregorio Goffredi, Francesca Marciano, Michele Caraglia, Alessia Cossu, Marianna Scrima, Edmondo Adorisio, Morena D'Avenia, Michela Iacobellis, Rosanna Piluscio, Giorgio Dirani, Vittorio Sambri, Simona Semprini, Silvia Zanoli, Francesco Curcio, Stefania Marzinotto, Andreina Baj, Fausto Sessa, Massimiliano Chetta, Marina Tarsitano, Giusy Giordano, Marcello Ametrano, Mariano Fiorenza, Caivano Antonella, D'Auria Fiorella, De Luca Luciana. |
| EPI_ISL_1379436, EPI_ISL_1379437, EPI_ISL_1379439, EPI_ISL_1379440, EPI_ISL_1379441, EPI_ISL_1379442 | Department of Molecular and Translational Medicine, Section of Microbiology, University of Brescia, ASST Spedali Civili, Brescia | Department of Molecular and Translational Medicine, Section of Microbiology, University of Brescia, ASST Spedali Civili, Brescia | Francesca Caccuri, Alberto Zani, Serena Messali, Anna Bertelli, Simona Fiorentini, Arnaldo Caruso |
| EPI_ISL_1390395, EPI_ISL_1390681 | IZSM | TIGEM | Antonio Grimaldi,Patrizia Annunziata,Francesco Panariello,Biancamaria Pierri Claudia Tiberio Valentina Bouche,Chiara Colantuono,Maria Concetta Cuomo,Denise Di Concilio,Lucio Di Filippo,Anna Manfredi,Marcello Salvi,Antonio Limone Luigi Atripaldi Pellegrino Cerino,Andrea Ballabio,Davide Cacchiarelli |
| EPI_ISL_1392923, EPI_ISL_1392924, EPI_ISL_1392925, EPI_ISL_1392926, EPI_ISL_1392927, EPI_ISL_1392928, EPI_ISL_1392929, EPI_ISL_1392930, EPI_ISL_1392931, EPI_ISL_1392932, EPI_ISL_1392933, EPI_ISL_1392934, EPI_ISL_1392935, EPI_ISL_1392936, EPI_ISL_1392937, EPI_ISL_1392938, EPI_ISL_1392939, EPI_ISL_1392940, EPI_ISL_1392941, EPI_ISL_1392942, EPI_ISL_1392943 | | | |
| see above | SC Microbiologia e Virologia AOUSS | AMES Centro Polidiagnostico Strumentale S.r.l. | Salvatore Rubino; Flavia Angioj; Laura Firino; Rosalba Govoni; Gabriele Ibba; Vincenzo Lai; Erica Mura; Bianca Paglietti; Claudia Piu; Anna Puggioni; Elena Rimini; Giulia Rocca; Caterina Serra; Sergio Uzzau. |
| EPI_ISL_1403609, EPI_ISL_1403610, EPI_ISL_1403611 | Molecular Virology Unit, Microbiology and Virology Depertment, Fondazione IRCCS Policlinico San Matteo, Pavia | Molecular Virology Unit, Microbiology and Virology Depertment, Fondazione IRCCS Policlinico San Matteo, Pavia | Federica Giardina, Guglielmo Ferrari, Stefano Gaiarsa, Gherard Batisti Biffignandi, Stefania Paolucci, Antonio Piralla, Fausto Baldanti |
| EPI_ISL_1416329, EPI_ISL_1416330, EPI_ISL_1416331 | Microbiology and Virology Laboratory, "Policlinico Riuniti, Azienda Ospedaliero Universitaria, Foggia" | Microbiology and Virology Laboratory, "Policlinico Riuniti, Azienda Ospedaliero Universitaria, Foggia" | Fabio Arena, Settimia Altamura, Donatella Cedola, Valeria Delli Carri, Annamaria D'Aprile, Maria Rosaria Lipsi, Loris Micelli, Maria Pia Patrizio, Daniela Pisanelli, Tiziana Rollo, Maurizio Margaglione and Rosella De Nittis. |
| EPI_ISL_1417439 | Microbiology and Virology Laboratory, "Policlinico Riuniti, Azienda Ospedaliero Universitaria, Foggia" | Microbiology and Virology Laboratory, "Policlinico Riuniti, Azienda Ospedaliero Universitaria, Foggia" | Fabio Arena, Settimia Altamura, Donatella Cedola, Valeria Delli Carri, Annamaria D'Aprile, Maria Rosaria Lipsi, Loris Micelli, Maria Pia Patrizio, Daniela Pisanelli, Tiziana Rollo, Maurizio Margaglione and Rosella De Nittis. |
| EPI_ISL_1424144, EPI_ISL_1424145, EPI_ISL_1424146, EPI_ISL_1424147, EPI_ISL_1424148, EPI_ISL_1424149, EPI_ISL_1424150, EPI_ISL_1424151, EPI_ISL_1424152, EPI_ISL_1424153, EPI_ISL_1424154, EPI_ISL_1424155, EPI_ISL_1424156, EPI_ISL_1424157, EPI_ISL_1424158, EPI_ISL_1424159, EPI_ISL_1424160, EPI_ISL_1424161, EPI_ISL_1424162, EPI_ISL_1424163, EPI_ISL_1424164, EPI_ISL_1424165, EPI_ISL_1424166, EPI_ISL_1424167, EPI_ISL_1424168, EPI_ISL_1424169, EPI_ISL_1424170, EPI_ISL_1424171, EPI_ISL_1424172, EPI_ISL_1424173, EPI_ISL_1424174, EPI_ISL_1424175, EPI_ISL_1424176, EPI_ISL_1424177, EPI_ISL_1424178, EPI_ISL_1424179, EPI_ISL_1424185, EPI_ISL_1424186, EPI_ISL_1424187, EPI_ISL_1424188, EPI_ISL_1424189, EPI_ISL_1424190, EPI_ISL_1424192, EPI_ISL_1424193, EPI_ISL_1424194, EPI_ISL_1424195, EPI_ISL_1424196, EPI_ISL_1424197, EPI_ISL_1424198, EPI_ISL_1424199, EPI_ISL_1424200, EPI_ISL_1424201, EPI_ISL_1424202, EPI_ISL_1424203, EPI_ISL_1424204, EPI_ISL_1424205, EPI_ISL_1424207, EPI_ISL_1424208, EPI_ISL_1424209, EPI_ISL_1424210, EPI_ISL_1424212, EPI_ISL_1424214, EPI_ISL_1424215, EPI_ISL_1424216, EPI_ISL_1424217, EPI_ISL_1424219, EPI_ISL_1424227, EPI_ISL_1424228, EPI_ISL_1424231, EPI_ISL_1424232, EPI_ISL_1424233, EPI_ISL_1424234, EPI_ISL_1424235, EPI_ISL_1424236, EPI_ISL_1424237, EPI_ISL_1424238, EPI_ISL_1424239, EPI_ISL_1424240, EPI_ISL_1424241, EPI_ISL_1424242, EPI_ISL_1424243, EPI_ISL_1424244, EPI_ISL_1424245, EPI_ISL_1424246, EPI_ISL_1424247, EPI_ISL_1424248, EPI_ISL_1424249, EPI_ISL_1424250, EPI_ISL_1424251, EPI_ISL_1424252, EPI_ISL_1424253, EPI_ISL_1424254, EPI_ISL_1424255, EPI_ISL_1424256, EPI_ISL_1424257, EPI_ISL_1424258, EPI_ISL_1424259, EPI_ISL_1424260, EPI_ISL_1424261, EPI_ISL_1424262, EPI_ISL_1424263, EPI_ISL_1424264, EPI_ISL_1424265, EPI_ISL_1424266, EPI_ISL_1424267, EPI_ISL_1424268, EPI_ISL_1424269, EPI_ISL_1424270, EPI_ISL_1424271, EPI_ISL_1424272, EPI_ISL_1424273, EPI_ISL_1424274, EPI_ISL_1424275, EPI_ISL_1424276, EPI_ISL_1424277, EPI_ISL_1424278, EPI_ISL_1424279, EPI_ISL_1424280, EPI_ISL_1424281, EPI_ISL_1424282, EPI_ISL_1424283, EPI_ISL_1424284, EPI_ISL_1424285, EPI_ISL_1424286, EPI_ISL_1424287, EPI_ISL_1424288, EPI_ISL_1424289, EPI_ISL_1424290, EPI_ISL_1424291, EPI_ISL_1424292, EPI_ISL_1424293, EPI_ISL_1424294, EPI_ISL_1424295, EPI_ISL_1424296, EPI_ISL_1424297, EPI_ISL_1424298, EPI_ISL_1424299, EPI_ISL_1424300, EPI_ISL_1424301, EPI_ISL_1424302, EPI_ISL_1424303, EPI_ISL_1424304, EPI_ISL_1424305, EPI_ISL_1424306, EPI_ISL_1424307, EPI_ISL_1424308, EPI_ISL_1424309, EPI_ISL_1424310, EPI_ISL_1424311, EPI_ISL_1424312, EPI_ISL_1424313, EPI_ISL_1424314, EPI_ISL_1424315, EPI_ISL_1424316, EPI_ISL_1424317, EPI_ISL_1424318, EPI_ISL_1424319, EPI_ISL_1424320, EPI_ISL_1424321, EPI_ISL_1424322, EPI_ISL_1424323, EPI_ISL_1424324, EPI_ISL_1424325, EPI_ISL_1424326, EPI_ISL_1424327, EPI_ISL_1424328, EPI_ISL_1424329, EPI_ISL_1424330, EPI_ISL_1424331, EPI_ISL_1424332, EPI_ISL_1424333, EPI_ISL_1424334, EPI_ISL_1424335, EPI_ISL_1424336, EPI_ISL_1424337, EPI_ISL_1424338, EPI_ISL_1424339, EPI_ISL_1424340, EPI_ISL_1424341, EPI_ISL_1424342, EPI_ISL_1424343, EPI_ISL_1424344, EPI_ISL_1424345, EPI_ISL_1424346, EPI_ISL_1424347, EPI_ISL_1424348, EPI_ISL_1424349, EPI_ISL_1424350, EPI_ISL_1424351, EPI_ISL_1424352, EPI_ISL_1424353, EPI_ISL_1424354, EPI_ISL_1424355, EPI_ISL_1424356, EPI_ISL_1424357, EPI_ISL_1424358, EPI_ISL_1424359, EPI_ISL_1424360, EPI_ISL_1424361, EPI_ISL_1424362, EPI_ISL_1424363, EPI_ISL_1424364, EPI_ISL_1424365, EPI_ISL_1424366, EPI_ISL_1424367, EPI_ISL_1424368, EPI_ISL_1424369, EPI_ISL_1424370, EPI_ISL_1424371, EPI_ISL_1424372, EPI_ISL_1424373, EPI_ISL_1424374, EPI_ISL_1424375, EPI_ISL_1424376 | | | |
| see above | Istituto Zooprofilattico Sperimentale del Mezzogiorno | TIGEM | Antonio Grimaldi,Patrizia Annunziata,Francesco Panariello,Biancamaria Pierri Claudia Tiberio Valentina Bouche,Chiara Colantuono,Maria Concetta Cuomo,Denise Di Concilio,Lucio Di Filippo,Anna Manfredi,Marcello Salvi,Antonio Limone Luigi Atripaldi Pellegrino Cerino,Andrea Ballabio,Davide Cacchiarelli |

| | | | |
|---|---|---|---|
| EPI_ISL_1424397, EPI_ISL_1424398, EPI_ISL_1424399, EPI_ISL_1424400, EPI_ISL_1424403, EPI_ISL_1424414, EPI_ISL_1424425, EPI_ISL_1424431, EPI_ISL_1424432 | Università Federico II - Dipartimento di scienze mediche traslazionali - Napoli | TIGEM | Antonio Grimaldi Patrizia Annunziata Francesco Panariello Michele Cennamo Valentina Bouche Chiara Colantuono Lucio Di Filippo Mariano Fiorenza Anna Manfredi Marcello Salvi Giuseppe Portella Andrea Ballabio Davide Cacchiarelli |
| EPI_ISL_1442121, EPI_ISL_1442128, EPI_ISL_1442134, EPI_ISL_1442138, EPI_ISL_1442141, EPI_ISL_1442143, EPI_ISL_1442145 | Azienda Ospedaliero - Universitaria di Modena Policlinico - Virologia e Microbiologia Molecolare | Istituto Zooprofilattico Sperimentale della Lombardia e dell'Emilia Romagna (IZSLER), Risk Analysis and Genomic Epidemiology Unit | Monica Pecorari, William Gennari, Giulia Fregni Serpini, Marina Morganti, Ilaria Menozzi, Erika Scaltriti, Stefano Pongolini |
| EPI_ISL_1443157, EPI_ISL_1443158, EPI_ISL_1443159, EPI_ISL_1443160, EPI_ISL_1443161, EPI_ISL_1443162, EPI_ISL_1443163, EPI_ISL_1443164, EPI_ISL_1443165, EPI_ISL_1443166, EPI_ISL_1443167, EPI_ISL_1443168, EPI_ISL_1443169 | | | |
| see above | Istituto Zooprofilattico Sperimentale della Lombardia e dell'Emilia Romagna (IZSLER), Sede Territoriale di Modena | Istituto Zooprofilattico Sperimentale della Lombardia e dell'Emilia Romagna (IZSLER), Risk Analysis and Genomic Epidemiology Unit | Carra Elena, Gianluca Rugna, Marina Morganti, Ilaria Menozzi, Erika Scaltriti, Stefano Pongolini |
| EPI_ISL_1447950 | 20210213554 | TIGEM | Antonio Grimaldi Patrizia Annunziata Francesco Panariello Biancamaria Pierri Claudia Tiberio Valentina Bouche Chiara Colantuono Maria Concetta Cuomo Denise Di Concilio Lucio Di Filippo Anna Manfredi Marcello Salvi Antonio Limone Luigi Atripaldi Pellegrino Cerino Andrea Ballabio Davide Cacchiarelli |
| EPI_ISL_1447951 | 20210213565 | TIGEM | Antonio Grimaldi Patrizia Annunziata Francesco Panariello Biancamaria Pierri Claudia Tiberio Valentina Bouche Chiara Colantuono Maria Concetta Cuomo Denise Di Concilio Lucio Di Filippo Anna Manfredi Marcello Salvi Antonio Limone Luigi Atripaldi Pellegrino Cerino Andrea Ballabio Davide Cacchiarelli |
| EPI_ISL_1447953 | 20210213607 | TIGEM | Antonio Grimaldi Patrizia Annunziata Francesco Panariello Biancamaria Pierri Claudia Tiberio Valentina Bouche Chiara Colantuono Maria Concetta Cuomo Denise Di Concilio Lucio Di Filippo Anna Manfredi Marcello Salvi Antonio Limone Luigi Atripaldi Pellegrino Cerino Andrea Ballabio Davide Cacchiarelli |
| EPI_ISL_1447957 | 20210213570 | TIGEM | Antonio Grimaldi Patrizia Annunziata Francesco Panariello Biancamaria Pierri Claudia Tiberio Valentina Bouche Chiara Colantuono Maria Concetta Cuomo Denise Di Concilio Lucio Di Filippo Anna Manfredi Marcello Salvi Antonio Limone Luigi Atripaldi Pellegrino Cerino Andrea Ballabio Davide Cacchiarelli |
| EPI_ISL_1447958 | 20210213566 | TIGEM | Antonio Grimaldi Patrizia Annunziata Francesco Panariello Biancamaria Pierri Claudia Tiberio Valentina Bouche Chiara Colantuono Maria Concetta Cuomo Denise Di Concilio Lucio Di Filippo Anna Manfredi Marcello Salvi Antonio Limone Luigi Atripaldi Pellegrino Cerino Andrea Ballabio Davide Cacchiarelli |
| EPI_ISL_1447961 | 20210213546 | TIGEM | Antonio Grimaldi Patrizia Annunziata Francesco Panariello Biancamaria Pierri Claudia Tiberio Valentina Bouche Chiara Colantuono Maria Concetta Cuomo Denise Di Concilio Lucio Di Filippo Anna Manfredi Marcello Salvi Antonio Limone Luigi Atripaldi Pellegrino Cerino Andrea Ballabio Davide Cacchiarelli |
| EPI_ISL_1500436, EPI_ISL_1500437, EPI_ISL_1500438, EPI_ISL_1500439 | Laboratory of Clinical Microbiology, Virology and Bioemergencies, ASST Fatebenefratelli Sacco - Sacco Hospital | Laboratory of Clinical Microbiology, Virology and Bioemergencies, ASST Fatebenefratelli Sacco - Sacco Hospital | Valeria Micheli, Alessandro Mancon, Alberto Rizzo, Fiorenza Bracchitta, Luca Rizzuto, Maria Rita Gismondo |
| EPI_ISL_1500446, EPI_ISL_1500447 | ASST della Brianza | Laboratory of Clinical Microbiology, Virology and Bioemergencies, ASST Fatebenefratelli Sacco - Sacco Hospital | Valeria Micheli, Alessandro Mancon, Alberto Rizzo, Fiorenza Bracchitta, Luca Rizzuto, Maria Rita Gismondo |
| EPI_ISL_1509319 | IZSM | TIGEM | Antonio Grimaldi Patrizia Annunziata Francesco Panariello Biancamaria Pierri Claudia Tiberio Valentina Bouche Chiara Colantuono Maria Concetta Cuomo Denise Di Concilio Lucio Di Filippo Anna Manfredi Marcello Salvi Antonio Limone Luigi Atripaldi Pellegrino Cerino Andrea Ballabio Davide Cacchiarelli |
| EPI_ISL_1509757, EPI_ISL_1509758 | SC (UCO) Igiene e Sanità Pubblica (funzione integrata con SC Microbiologia e Virologia) e Az. Ospedaliero-Universitaria Udine | ARGO Laboratorio Genomica ed Epigenomica | Licastro D, Dal Monego S, Degasperi M, Marcello A, Segat L, Piscianz E, D'Agaro P |
| EPI_ISL_1509923 | Dipartimento di Medicina e Chirurgia Universita' degli Studi dell'Insubria Laboratorio di Microbiologia Medica Varese Italia | Laboratorio di Microbiologia e Virologia, Università Vita-Salute San Raffaele, Milano | Fabrizio Maggi, Matteo Castelli, Roberta Antonia Diotti, Michela Sampaolo, Enzo Boeri, Elena Criscuolo, Roberto Ferrarese, Massimo Clementi, Nicasio Mancini and Nicola Clementi |
| EPI_ISL_1510620, EPI_ISL_1510621, EPI_ISL_1510622 | Azienda Sanitaria Locale di Piacenza - Presidio Ospedaliero - Laboratorio di Microbiologia | Istituto Zooprofilattico Sperimentale della Lombardia e dell'Emilia Romagna (IZSLER), Risk Analysis and Genomic Epidemiology Unit | Lo Cascio Giuliana, Schiavo Roberta, Peroni Irene, Marina Morganti, Ilaria Menozzi, Erika Scaltriti, Stefano Pongolini |
| EPI_ISL_1524386 | ULSS 1 Dolomiti | Istituto Zooprofilattico Sperimentale delle Venezie | Adelaide Milani, Alessia Schivo, Annalisa Salviato, Elisa Palumbo, Erika Giorgia Quaranta, Luca Tassoni, Ambra Pastori, Edoardo Giussani, Alice Fusaro, Isabella Monne, Calogero Terregino, Antonia Ricci |
| EPI_ISL_1524392 | ULSS 6 Euganea | Istituto Zooprofilattico Sperimentale delle Venezie | Adelaide Milani, Alessia Schivo, Annalisa Salviato, Elisa Palumbo, Erika Giorgia Quaranta, Luca Tassoni, Ambra Pastori, Edoardo Giussani, Alice Fusaro, Isabella Monne, Calogero Terregino, Antonia Ricci |
| EPI_ISL_1524393, EPI_ISL_1524394, EPI_ISL_1524398 | Microbiologia e Virologia | Istituto Zooprofilattico Sperimentale delle Venezie | Adelaide Milani, Alessia Schivo, Annalisa Salviato, Elisa Palumbo, Erika Giorgia Quaranta, Luca Tassoni, Ambra Pastori, Edoardo Giussani, Alice Fusaro, Isabella Monne, Calogero Terregino, Antonia Ricci |
| EPI_ISL_1524400, EPI_ISL_1524401, EPI_ISL_1524402, EPI_ISL_1524403 | ULSS 8 Berica | Istituto Zooprofilattico Sperimentale delle Venezie | Adelaide Milani, Alessia Schivo, Annalisa Salviato, Elisa Palumbo, Erika Giorgia Quaranta, Luca Tassoni, Ambra Pastori, Edoardo Giussani, Alice Fusaro, Isabella Monne, Calogero Terregino, Antonia Ricci |
| EPI_ISL_1524404, EPI_ISL_1524405 | ULSS 5 Polesana | Istituto Zooprofilattico Sperimentale delle Venezie | Adelaide Milani, Alessia Schivo, Annalisa Salviato, Elisa Palumbo, Erika Giorgia Quaranta, Luca Tassoni, Ambra Pastori, Edoardo Giussani, Alice Fusaro, Isabella Monne, Calogero Terregino, Antonia Ricci |
| EPI_ISL_1524406 | Ospedale San Bonifacio | Istituto Zooprofilattico Sperimentale delle Venezie | Adelaide Milani, Alessia Schivo, Annalisa Salviato, Elisa Palumbo, Erika Giorgia Quaranta, Luca Tassoni, Ambra Pastori, Edoardo Giussani, Alice Fusaro, Isabella Monne, Calogero Terregino, Antonia Ricci |
| EPI_ISL_1524409, EPI_ISL_1524410 | Ospedale Magalini | Istituto Zooprofilattico Sperimentale delle Venezie | Adelaide Milani, Alessia Schivo, Annalisa Salviato, Elisa Palumbo, Erika Giorgia Quaranta, Luca Tassoni, Ambra Pastori, Edoardo Giussani, Alice Fusaro, Isabella Monne, Calogero Terregino, Antonia Ricci |
| EPI_ISL_1524412, EPI_ISL_1524413, EPI_ISL_1524414, EPI_ISL_1524415, EPI_ISL_1524417 | ULSS 6 Euganea | Istituto Zooprofilattico Sperimentale delle Venezie | Adelaide Milani, Alessia Schivo, Annalisa Salviato, Elisa Palumbo, Erika Giorgia Quaranta, Luca Tassoni, Ambra Pastori, Edoardo Giussani, Alice Fusaro, Isabella Monne, Calogero Terregino, Antonia Ricci |
| EPI_ISL_1524419, EPI_ISL_1524420, EPI_ISL_1524421, EPI_ISL_1524423 | ULSS 7 Pedemontana - Distretto 2 | Istituto Zooprofilattico Sperimentale delle Venezie | Adelaide Milani, Alessia Schivo, Annalisa Salviato, Elisa Palumbo, Erika Giorgia Quaranta, Luca Tassoni, Ambra Pastori, Edoardo Giussani, Alice Fusaro, Isabella Monne, Calogero Terregino, Antonia Ricci |
| EPI_ISL_1524424, EPI_ISL_1524425, EPI_ISL_1524426, EPI_ISL_1524427, EPI_ISL_1524428, EPI_ISL_1524429 | ULSS 6 Euganea | Istituto Zooprofilattico Sperimentale delle Venezie | Adelaide Milani, Alessia Schivo, Annalisa Salviato, Elisa Palumbo, Erika Giorgia Quaranta, Luca Tassoni, Ambra Pastori, Edoardo Giussani, Alice Fusaro, Isabella Monne, Calogero Terregino, Antonia Ricci |
| EPI_ISL_1524430, EPI_ISL_1524431, EPI_ISL_1524432 | ULSS 1 Dolomiti | Istituto Zooprofilattico Sperimentale delle Venezie | Adelaide Milani, Alessia Schivo, Annalisa Salviato, Elisa Palumbo, Erika Giorgia Quaranta, Luca Tassoni, Ambra Pastori, Edoardo Giussani, Alice Fusaro, Isabella Monne, Calogero Terregino, Antonia Ricci |
| EPI_ISL_1524435 | ULSS 8 Berica | Istituto Zooprofilattico Sperimentale delle Venezie | Adelaide Milani, Alessia Schivo, Annalisa Salviato, Elisa Palumbo, Erika Giorgia Quaranta, Luca Tassoni, Ambra Pastori, Edoardo Giussani, Alice Fusaro, Isabella Monne, Calogero Terregino, Antonia Ricci |

| | | | |
|---|---|---|---|
| EPI_ISL_1524436 | ULSS 5 Polesana | Istituto Zooprofilattico Sperimentale delle Venezie | Adelaide Milani, Alessia Schivo, Annalisa Salviato, Elisa Palumbo, Erika Giorgia Quaranta, Luca Tassoni, Ambra Pastori, Edoardo Giussani, Alice Fusaro, Isabella Monne, Calogero Terregino, Antonia Ricci |
| EPI_ISL_1524437, EPI_ISL_1524438, EPI_ISL_1524439, EPI_ISL_1524440, EPI_ISL_1524441, EPI_ISL_1524442, EPI_ISL_1524443, EPI_ISL_1524444, EPI_ISL_1524445 | ULSS 3 Venezia | Istituto Zooprofilattico Sperimentale delle Venezie | Adelaide Milani, Alessia Schivo, Annalisa Salviato, Elisa Palumbo, Erika Giorgia Quaranta, Luca Tassoni, Ambra Pastori, Edoardo Giussani, Alice Fusaro, Isabella Monne, Calogero Terregino, Antonia Ricci |
| EPI_ISL_1524446, EPI_ISL_1524447 | ULSS 6 Euganea | Istituto Zooprofilattico Sperimentale delle Venezie | Adelaide Milani, Alessia Schivo, Annalisa Salviato, Elisa Palumbo, Erika Giorgia Quaranta, Luca Tassoni, Ambra Pastori, Edoardo Giussani, Alice Fusaro, Isabella Monne, Calogero Terregino, Antonia Ricci |
| EPI_ISL_1524448, EPI_ISL_1524449 | ULSS 7 Pedemontana - Distretto 1 | Istituto Zooprofilattico Sperimentale delle Venezie | Adelaide Milani, Alessia Schivo, Annalisa Salviato, Elisa Palumbo, Erika Giorgia Quaranta, Luca Tassoni, Ambra Pastori, Edoardo Giussani, Alice Fusaro, Isabella Monne, Calogero Terregino, Antonia Ricci |
| EPI_ISL_1524450 | ULSS 8 Berica | Istituto Zooprofilattico Sperimentale delle Venezie | Adelaide Milani, Alessia Schivo, Annalisa Salviato, Elisa Palumbo, Erika Giorgia Quaranta, Luca Tassoni, Ambra Pastori, Edoardo Giussani, Alice Fusaro, Isabella Monne, Calogero Terregino, Antonia Ricci |
| EPI_ISL_1524451 | ULSS 3 Venezia | Istituto Zooprofilattico Sperimentale delle Venezie | Adelaide Milani, Alessia Schivo, Annalisa Salviato, Elisa Palumbo, Erika Giorgia Quaranta, Luca Tassoni, Ambra Pastori, Edoardo Giussani, Alice Fusaro, Isabella Monne, Calogero Terregino, Antonia Ricci |
| EPI_ISL_1524452 | ULSS 1 Dolomiti | Istituto Zooprofilattico Sperimentale delle Venezie | Adelaide Milani, Alessia Schivo, Annalisa Salviato, Elisa Palumbo, Erika Giorgia Quaranta, Luca Tassoni, Ambra Pastori, Edoardo Giussani, Alice Fusaro, Isabella Monne, Calogero Terregino, Antonia Ricci |
| EPI_ISL_1524453 | ULSS 4 Veneto Orientale | Istituto Zooprofilattico Sperimentale delle Venezie | Adelaide Milani, Alessia Schivo, Annalisa Salviato, Elisa Palumbo, Erika Giorgia Quaranta, Luca Tassoni, Ambra Pastori, Edoardo Giussani, Alice Fusaro, Isabella Monne, Calogero Terregino, Antonia Ricci |
| EPI_ISL_1524454 | ULSS 5 Polesana | Istituto Zooprofilattico Sperimentale delle Venezie | Adelaide Milani, Alessia Schivo, Annalisa Salviato, Elisa Palumbo, Erika Giorgia Quaranta, Luca Tassoni, Ambra Pastori, Edoardo Giussani, Alice Fusaro, Isabella Monne, Calogero Terregino, Antonia Ricci |
| EPI_ISL_1524455 | Microbiologia e Virologia | Istituto Zooprofilattico Sperimentale delle Venezie | Adelaide Milani, Alessia Schivo, Annalisa Salviato, Elisa Palumbo, Erika Giorgia Quaranta, Luca Tassoni, Ambra Pastori, Edoardo Giussani, Alice Fusaro, Isabella Monne, Calogero Terregino, Antonia Ricci |
| EPI_ISL_1547388, EPI_ISL_1547389, EPI_ISL_1547390, EPI_ISL_1547391 | 1.AO Universitaria 'S. Giovanni di Dio e Ruggi D'Aragona, Scuola Medica Salernitana' Hospital / 2.UOC di Virologia e Microbiologia, Università della Campania 'L. Vanvitelli' / 3.AO Universitaria 'Federico II' Napoli Hospital / 4.AORN 'San Giuseppe Moscati' Avellino Hospital / 5.AO 'San Pio - presidio G. Rummo' Benevento Hospital / 6.AO 'Sant'Anna e San Sebastiano' Caserta Hospital / 7.PO 'Maria Santissima Addolorata' Eboli Hospital / 8.Biogem Istituto di Ricerche Genetiche / 9. U.O.C. di Genetica Medica e di Laboratorio A.O.R.N., Azienda Ospedaliera di Rilievo Nazionale Antonio Cardarelli, Napoli / 10. Centro di riferimento Oncologico della Basilicata (IRCCS-CROB), Rionero in Vulture (PZ) / 11. Presidio Ospedaliero di Agropoli, Agropoli (SA). | 1. Genome Research Center for Health (CRGS) / 2. Laboratory of Molecular Medicine and Genomics(LMMGe) / 3. Center for Research in Pure and Applied Mathematics (CRMPA) | Giorgio Giurato, Francesca Rizzo, Alessandro Weisz, Gianluigi Franci, Giovanni Nassa, Pasquale Pagliano, Roberta Tarallo, Elena Alexandrova, Ylenia D'Agostino, Carlo Ferravante, Jessica Lamberti, Viola Melone, Domenico Memoli, Valeria Mirici Cappa, Domenico Palumbo, Giovanni Pecoraro, Assunta Sellitto, Oriana Strianese, Ilaria Terenzi, Giuseppe Fenza, Aniello Gentile, Antonello Saccomanno, Sonia Amabile, Teresa Rocco, Annamaria Salvati, Emilia Vaccaro, Massimiliano Galdiero, Michele Cennamo, Giuseppe Portella, Maria Grazia Foti, Mariarosaria Ingino, Maria Landi, Maurizio Fumi, Vincenzo Rocco, Rita Greco, Vittoria Letizia, Arnolfo Petruzziello, Maddalena Schioppa, Gregorio Goffredi, Francesca Marciano, Michele Caraglia, Alessia Cossu, Marianna Scrima, Edmondo Adorisio, Morena D'Avenia, Michela Iacobellis, Rosanna Piluscio, Giorgio Dirani, Vittorio Sambri, Simona Semprini, Silvia Zanoli, Francesco Curcio, Stefania Marzinotto, Andreina Baj, Fausto Sessa, Massimiliano Chetta, Marina Tarsitano, Giusy Giordano, Marcello Ametrano, Mariano Fiorenza, Caivano Antonella, D'Auria Fiorella, De Luca Luciana. |
| EPI_ISL_1577214 | 1.AO Universitaria 'S. Giovanni di Dio e Ruggi D'Aragona, Scuola Medica Salernitana' Hospital / 2.UOC di Virologia e Microbiologia, Università della Campania 'L. Vanvitelli' / 3.AO Universitaria 'Federico II' Napoli Hospital / 4.AORN 'San Giuseppe Moscati' Avellino Hospital / 5.AO 'San Pio - presidio G. Rummo' Benevento Hospital / 6.AO 'Sant'Anna e San Sebastiano' Caserta Hospital / 7.PO 'Maria Santissima Addolorata' Eboli Hospital / 8.Biogem Istituto di Ricerche Genetiche / 9. U.O.C. di Genetica Medica e di Laboratorio A.O.R.N., Azienda Ospedaliera di Rilievo Nazionale Antonio Cardarelli, Napoli / 10. Centro di riferimento Oncologico della Basilicata (IRCCS-CROB), Rionero in Vulture (PZ) / 11. Presidio Ospedaliero di Agropoli, Agropoli (SA). | 1. Genome Research Center for Health (CRGS) / 2. Laboratory of Molecular Medicine and Genomics(LMMGe) / 3. Center for Research in Pure and Applied Mathematics (CRMPA) | Giorgio Giurato (Corresponding Author), Francesca Rizzo (Corresponding Author), Alessandro Weisz (Corresponding Author), Gianluigi Franci, Giovanni Nassa, Pasquale Pagliano, Roberta Tarallo, Elena Alexandrova, Ylenia D'Agostino, Carlo Ferravante, Jessica Lamberti, Viola Melone, Domenico Memoli, Valeria Mirici Cappa, Domenico Palumbo, Giovanni Pecoraro, Assunta Sellitto, Oriana Strianese, Ilaria Terenzi, Giuseppe Fenza, Aniello Gentile, Antonello Saccomanno, Sonia Amabile, Teresa Rocco, Annamaria Salvati, Emilia Vaccaro, Massimiliano Galdiero, Michele Cennamo, Giuseppe Portella, Maria Grazia Foti, Mariarosaria Ingino, Maria Landi, Maurizio Fumi, Vincenzo Rocco, Rita Greco, Vittoria Letizia, Arnolfo Petruzziello, Maddalena Schioppa, Gregorio Goffredi, Francesca Marciano, Michele Caraglia, Alessia Cossu, Marianna Scrima, Edmondo Adorisio, Morena D'Avenia, Michela Iacobellis, Rosanna Piluscio, Giorgio Dirani, Vittorio Sambri, Simona Semprini, Silvia Zanoli, Francesco Curcio, Stefania Marzinotto, Andreina Baj, Fausto Sessa, Massimiliano Chetta, Marina Tarsitano, Giusy Giordano, Marcello Ametrano, Mariano Fiorenza, Caivano Antonella, D'Auria Fiorella, De Luca Luciana. |
| EPI_ISL_1577215, EPI_ISL_1577216, EPI_ISL_1577217, EPI_ISL_1577218 | 1.AO Universitaria 'S. Giovanni di Dio e Ruggi D'Aragona, Scuola Medica Salernitana' Hospital / 2.UOC di Virologia e Microbiologia, Università della Campania 'L. Vanvitelli' / 3.AO Universitaria 'Federico II' Napoli Hospital / 4.AORN 'San Giuseppe Moscati' Avellino Hospital / 5.AO 'San Pio - presidio G. Rummo' Benevento Hospital / 6.AO 'Sant'Anna e San Sebastiano' Caserta Hospital / 7.PO 'Maria Santissima Addolorata' Eboli Hospital / 8.Biogem Istituto di Ricerche Genetiche / 9. U.O.C. di Genetica Medica e di Laboratorio A.O.R.N., Azienda Ospedaliera di Rilievo Nazionale Antonio Cardarelli, Napoli / 10. Centro di riferimento Oncologico della Basilicata (IRCCS-CROB), Rionero in Vulture (PZ) / 11. Presidio Ospedaliero di Agropoli, Agropoli (SA). | 1. Genome Research Center for Health (CRGS) / 2. Laboratory of Molecular Medicine and Genomics(LMMGe) / 3. Center for Research in Pure and Applied Mathematics (CRMPA) | Giorgio Giurato (Corresponding Author), Francesca Rizzo (Corresponding Author), Alessandro Weisz (Corresponding Author), Gianluigi Franci, Giovanni Nassa, Pasquale Pagliano, Roberta Tarallo, Elena Alexandrova, Ylenia D'Agostino, Carlo Ferravante, Jessica Lamberti, Viola Melone, Domenico Memoli, Valeria Mirici Cappa, Domenico Palumbo, Giovanni Pecoraro, Assunta Sellitto, Oriana Strianese, Ilaria Terenzi, Giuseppe Fenza, Aniello Gentile, Antonello Saccomanno, Sonia Amabile, Teresa Rocco, Annamaria Salvati, Emilia Vaccaro, Massimiliano Galdiero, Michele Cennamo, Giuseppe Portella, Maria Grazia Foti, Mariarosaria Ingino, Maria Landi, Maurizio Fumi, Vincenzo Rocco, Rita Greco, Vittoria Letizia, Arnolfo Petruzziello, Maddalena Schioppa, Gregorio Goffredi, Francesca Marciano, Michele Caraglia, Alessia Cossu, Marianna Scrima, Edmondo Adorisio, Morena D'Avenia, Michela Iacobellis, Rosanna Piluscio, Giorgio Dirani, Vittorio Sambri, Simona Semprini, Silvia Zanoli, Francesco Curcio, Stefania Marzinotto, Andreina Baj, Fausto Sessa. |
| EPI_ISL_1577219 | 1.AO Universitaria 'S. Giovanni di Dio e Ruggi D'Aragona, Scuola Medica Salernitana' Hospital / 2.UOC di Virologia e Microbiologia, Università della Campania 'L. Vanvitelli' / 3.AO Universitaria 'Federico II' Napoli Hospital / 4.AORN 'San Giuseppe Moscati' Avellino Hospital / 5.AO 'San Pio - presidio G. Rummo' Benevento Hospital / 6.AO 'Sant'Anna e San Sebastiano' Caserta Hospital / 7.PO 'Maria Santissima Addolorata' Eboli Hospital / 8.Biogem Istituto di Ricerche Genetiche / 9. U.O.C. di Genetica Medica e di Laboratorio A.O.R.N., Azienda Ospedaliera di Rilievo Nazionale Antonio Cardarelli, Napoli / 10. Centro di riferimento Oncologico della Basilicata (IRCCS-CROB), Rionero in Vulture (PZ) / 11. Presidio Ospedaliero di Agropoli, Agropoli (SA). | 1. Genome Research Center for Health (CRGS) / 2. Laboratory of Molecular Medicine and Genomics(LMMGe) / 3. Center for Research in Pure and Applied Mathematics (CRMPA) | Giorgio Giurato (Corresponding Author), Francesca Rizzo (Corresponding Author), Alessandro Weisz (Corresponding Author), Gianluigi Franci, Giovanni Nassa, Pasquale Pagliano, Roberta Tarallo, Elena Alexandrova, Ylenia D'Agostino, Carlo Ferravante, Jessica Lamberti, Viola Melone, Domenico Memoli, Valeria Mirici Cappa, Domenico Palumbo, Giovanni Pecoraro, Assunta Sellitto, Oriana Strianese, Ilaria Terenzi, Giuseppe Fenza, Aniello Gentile, Antonello Saccomanno, Sonia Amabile, Teresa Rocco, Annamaria Salvati, Emilia Vaccaro, Massimiliano Galdiero, Michele Cennamo, Giuseppe Portella, Maria Grazia Foti, Mariarosaria Ingino, Maria Landi, Maurizio Fumi, Vincenzo Rocco, Rita Greco, Vittoria Letizia, Arnolfo Petruzziello, Maddalena Schioppa, Gregorio Goffredi, Francesca Marciano, Michele Caraglia, Alessia Cossu, Marianna Scrima, Edmondo Adorisio, Morena D'Avenia, Michela Iacobellis, Rosanna Piluscio, Giorgio Dirani, Vittorio Sambri, Simona Semprini, Silvia Zanoli, Francesco Curcio, Stefania Marzinotto, Andreina Baj, Fausto Sessa, Massimiliano Chetta, Marina Tarsitano, Giusy Giordano, Marcello Ametrano, Mariano Fiorenza, Caivano Antonella, D'Auria Fiorella, De Luca Luciana. |
| EPI_ISL_1577222, EPI_ISL_1577223 | 1.AO Universitaria 'S. Giovanni di Dio e Ruggi D'Aragona, Scuola Medica Salernitana' Hospital / 2.UOC di Virologia e Microbiologia, Università della Campania 'L. Vanvitelli' / 3.AO Universitaria 'Federico II' Napoli Hospital / 4.AORN 'San Giuseppe Moscati' Avellino Hospital / 5.AO 'San Pio - presidio | 1. Genome Research Center for Health (CRGS) / 2. Laboratory of Molecular Medicine and Genomics(LMMGe) / 3. Center for Research in Pure and Applied Mathematics (CRMPA) | Giorgio Giurato (Corresponding Author), Francesca Rizzo (Corresponding Author), Alessandro Weisz (Corresponding Author), Gianluigi Franci, Giovanni Nassa, Pasquale Pagliano, Roberta Tarallo, Elena Alexandrova, Ylenia D'Agostino, Carlo Ferravante, Jessica Lamberti, Viola Melone, Domenico Memoli, Valeria Mirici Cappa, Domenico Palumbo, Giovanni Pecoraro, Assunta Sellitto, Oriana Strianese, Ilaria Terenzi, Giuseppe Fenza, Aniello Gentile, Antonello Saccomanno, Sonia Amabile, Teresa Rocco, Annamaria Salvati, Emilia Vaccaro, Massimiliano Galdiero, Michele Cennamo, Giuseppe Portella, Maria Grazia Foti, Mariarosaria Ingino, Maria Landi, Maurizio Fumi, Vincenzo Rocco, Rita Greco, Vittoria Letizia, Arnolfo Petruzziello, Maddalena Schioppa, |

| | | | |
|---|---|---|---|
| | G. Rummo' Benevento Hospital / 6.AO 'Sant'Anna e San Sebastiano' Caserta Hospital / 7.PO 'Maria Santissima Addolorata' Eboli Hospital / 8.Biogem Istituto di Ricerche Genetiche / 9. U.O.C. di Genetica Medica e di Laboratorio A.O.R.N., Azienda Ospedaliera di Rilievo Nazionale Antonio Cardarelli, Napoli / 10. Centro di riferimento Oncologico della Basilicata (IRCCS-CROB), Rionero in Vulture (PZ) / 11. Presidio Ospedaliero di Agropoli, Agropoli (SA). | | Gregorio Goffredi, Francesca Marciano, Michele Caraglia, Alessia Cossu, Marianna Scrima, Edmondo Adorisio, Morena D'Avenia, Michela Iacobellis, Rosanna Piluscio, Giorgio Dirani, Vittorio Sambri, Simona Semprini, Silvia Zanoli, Francesco Curcio, Stefania Marzinotto, Andreina Baj, Fausto Sessa. |
| EPI_ISL_1577224 | 1.AO Universitaria 'S. Giovanni di Dio e Ruggi D'Aragona, Scuola Medica Salernitana' Hospital / 2.UOC di Virologia e Microbiologia, Università della Campania 'L. Vanvitelli' / 3.AO Universitaria 'Federico II' Napoli Hospital / 4.AORN 'San Giuseppe Moscati' Avellino Hospital / 5.AO 'San Pio - presidio G. Rummo' Benevento Hospital / 6.AO 'Sant'Anna e San Sebastiano' Caserta Hospital / 7.PO 'Maria Santissima Addolorata' Eboli Hospital / 8.Biogem Istituto di Ricerche Genetiche / 9. U.O.C. di Genetica Medica e di Laboratorio A.O.R.N., Azienda Ospedaliera di Rilievo Nazionale Antonio Cardarelli, Napoli / 10. Centro di riferimento Oncologico della Basilicata (IRCCS-CROB), Rionero in Vulture (PZ) / 11. Presidio Ospedaliero di Agropoli, Agropoli (SA). | 1. Genome Research Center for Health (CRGS) / 2. Laboratory of Molecular Medicine and Genomics(LMMGe) / 3. Center for Research in Pure and Applied Mathematics (CRMPA) | Giorgio Giurato (Corresponding Author), Francesca Rizzo (Corresponding Author), Alessandro Weisz (Corresponding Author), Gianluigi Franci, Giovanni Nassa, Pasquale Pagliano, Roberta Tarallo, Elena Alexandrova, Ylenia D'Agostino, Carlo Ferravante, Jessica Lamberti, Viola Melone, Domenico Memoli, Valeria Mirici Cappa, Domenico Palumbo, Giovanni Pecoraro, Assunta Sellitto, Oriana Strianese, Ilaria Terenzi, Giuseppe Fenza, Aniello Gentile, Antonello Saccomanno, Sonia Amabile, Teresa Rocco, Annamaria Salvati, Emilia Vaccaro, Massimiliano Galdiero, Michele Cennamo, Giuseppe Portella, Maria Grazia Foti, Mariarosaria Ingino, Maria Landi, Maurizio Fumi, Vincenzo Rocco, Rita Greco, Vittoria Letizia, Arnolfo Petruzziello, Maddalena Schioppa, Gregorio Goffredi, Francesca Marciano, Michele Caraglia, Alessia Cossu, Marianna Scrima, Edmondo Adorisio, Morena D'Avenia, Michela Iacobellis, Rosanna Piluscio, Giorgio Dirani, Vittorio Sambri, Simona Semprini, Silvia Zanoli, Francesco Curcio, Stefania Marzinotto, Andreina Baj, Fausto Sessa, Massimiliano Chetta, Marina Tarsitano, Giusy Giordano, Marcello Ametrano, Mariano Fiorenza, Caivano Antonella, D'Auria Fiorella, De Luca Luciana. |
| EPI_ISL_1577225, EPI_ISL_1577226, EPI_ISL_1577227, EPI_ISL_1577228 | 1.AO Universitaria 'S. Giovanni di Dio e Ruggi D'Aragona, Scuola Medica Salernitana' Hospital / 2.UOC di Virologia e Microbiologia, Università della Campania 'L. Vanvitelli' / 3.AO Universitaria 'Federico II' Napoli Hospital / 4.AORN 'San Giuseppe Moscati' Avellino Hospital / 5.AO 'San Pio - presidio G. Rummo' Benevento Hospital / 6.AO 'Sant'Anna e San Sebastiano' Caserta Hospital / 7.PO 'Maria Santissima Addolorata' Eboli Hospital / 8.Biogem Istituto di Ricerche Genetiche / 9. U.O.C. di Genetica Medica e di Laboratorio A.O.R.N., Azienda Ospedaliera di Rilievo Nazionale Antonio Cardarelli, Napoli / 10. Centro di riferimento Oncologico della Basilicata (IRCCS-CROB), Rionero in Vulture (PZ) / 11. Presidio Ospedaliero di Agropoli, Agropoli (SA). | 1. Genome Research Center for Health (CRGS) / 2. Laboratory of Molecular Medicine and Genomics(LMMGe) / 3. Center for Research in Pure and Applied Mathematics (CRMPA) | Giorgio Giurato (Corresponding Author), Francesca Rizzo (Corresponding Author), Alessandro Weisz (Corresponding Author), Gianluigi Franci, Giovanni Nassa, Pasquale Pagliano, Roberta Tarallo, Elena Alexandrova, Ylenia D'Agostino, Carlo Ferravante, Jessica Lamberti, Viola Melone, Domenico Memoli, Valeria Mirici Cappa, Domenico Palumbo, Giovanni Pecoraro, Assunta Sellitto, Oriana Strianese, Ilaria Terenzi, Giuseppe Fenza, Aniello Gentile, Antonello Saccomanno, Sonia Amabile, Teresa Rocco, Annamaria Salvati, Emilia Vaccaro, Massimiliano Galdiero, Michele Cennamo, Giuseppe Portella, Maria Grazia Foti, Mariarosaria Ingino, Maria Landi, Maurizio Fumi, Vincenzo Rocco, Rita Greco, Vittoria Letizia, Arnolfo Petruzziello, Maddalena Schioppa, Gregorio Goffredi, Francesca Marciano, Michele Caraglia, Alessia Cossu, Marianna Scrima, Edmondo Adorisio, Morena D'Avenia, Michela Iacobellis, Rosanna Piluscio, Giorgio Dirani, Vittorio Sambri, Simona Semprini, Silvia Zanoli, Francesco Curcio, Stefania Marzinotto, Andreina Baj, Fausto Sessa. |
| EPI_ISL_1577229 | 1.AO Universitaria 'S. Giovanni di Dio e Ruggi D'Aragona, Scuola Medica Salernitana' Hospital / 2.UOC di Virologia e Microbiologia, Università della Campania 'L. Vanvitelli' / 3.AO Universitaria 'Federico II' Napoli Hospital / 4.AORN 'San Giuseppe Moscati' Avellino Hospital / 5.AO 'San Pio - presidio G. Rummo' Benevento Hospital / 6.AO 'Sant'Anna e San Sebastiano' Caserta Hospital / 7.PO 'Maria Santissima Addolorata' Eboli Hospital / 8.Biogem Istituto di Ricerche Genetiche / 9. U.O.C. di Genetica Medica e di Laboratorio A.O.R.N., Azienda Ospedaliera di Rilievo Nazionale Antonio Cardarelli, Napoli / 10. Centro di riferimento Oncologico della Basilicata (IRCCS-CROB), Rionero in Vulture (PZ) / 11. Presidio Ospedaliero di Agropoli, Agropoli (SA). | 1. Genome Research Center for Health (CRGS) / 2. Laboratory of Molecular Medicine and Genomics(LMMGe) / 3. Center for Research in Pure and Applied Mathematics (CRMPA) | Giorgio Giurato (Corresponding Author), Francesca Rizzo (Corresponding Author), Alessandro Weisz (Corresponding Author), Gianluigi Franci, Giovanni Nassa, Pasquale Pagliano, Roberta Tarallo, Elena Alexandrova, Ylenia D'Agostino, Carlo Ferravante, Jessica Lamberti, Viola Melone, Domenico Memoli, Valeria Mirici Cappa, Domenico Palumbo, Giovanni Pecoraro, Assunta Sellitto, Oriana Strianese, Ilaria Terenzi, Giuseppe Fenza, Aniello Gentile, Antonello Saccomanno, Sonia Amabile, Teresa Rocco, Annamaria Salvati, Emilia Vaccaro, Massimiliano Galdiero, Michele Cennamo, Giuseppe Portella, Maria Grazia Foti, Mariarosaria Ingino, Maria Landi, Maurizio Fumi, Vincenzo Rocco, Rita Greco, Vittoria Letizia, Arnolfo Petruzziello, Maddalena Schioppa, Gregorio Goffredi, Francesca Marciano, Michele Caraglia, Alessia Cossu, Marianna Scrima, Edmondo Adorisio, Morena D'Avenia, Michela Iacobellis, Rosanna Piluscio, Giorgio Dirani, Vittorio Sambri, Simona Semprini, Silvia Zanoli, Francesco Curcio, Stefania Marzinotto, Andreina Baj, Fausto Sessa, Massimiliano Chetta, Marina Tarsitano, Giusy Giordano, Marcello Ametrano, Mariano Fiorenza, Caivano Antonella, D'Auria Fiorella, De Luca Luciana. |
| EPI_ISL_1577230, EPI_ISL_1577231, EPI_ISL_1577232, EPI_ISL_1577233 | 1.AO Universitaria 'S. Giovanni di Dio e Ruggi D'Aragona, Scuola Medica Salernitana' Hospital / 2.UOC di Virologia e Microbiologia, Università della Campania 'L. Vanvitelli' / 3.AO Universitaria 'Federico II' Napoli Hospital / 4.AORN 'San Giuseppe Moscati' Avellino Hospital / 5.AO 'San Pio - presidio G. Rummo' Benevento Hospital / 6.AO 'Sant'Anna e San Sebastiano' Caserta Hospital / 7.PO 'Maria Santissima Addolorata' Eboli Hospital / 8.Biogem Istituto di Ricerche Genetiche / 9. U.O.C. di Genetica Medica e di Laboratorio A.O.R.N., Azienda Ospedaliera di Rilievo Nazionale Antonio Cardarelli, Napoli / 10. Centro di riferimento Oncologico della Basilicata (IRCCS-CROB), Rionero in Vulture (PZ) / 11. Presidio Ospedaliero di Agropoli, Agropoli (SA). | 1. Genome Research Center for Health (CRGS) / 2. Laboratory of Molecular Medicine and Genomics(LMMGe) / 3. Center for Research in Pure and Applied Mathematics (CRMPA) | Giorgio Giurato (Corresponding Author), Francesca Rizzo (Corresponding Author), Alessandro Weisz (Corresponding Author), Gianluigi Franci, Giovanni Nassa, Pasquale Pagliano, Roberta Tarallo, Elena Alexandrova, Ylenia D'Agostino, Carlo Ferravante, Jessica Lamberti, Viola Melone, Domenico Memoli, Valeria Mirici Cappa, Domenico Palumbo, Giovanni Pecoraro, Assunta Sellitto, Oriana Strianese, Ilaria Terenzi, Giuseppe Fenza, Aniello Gentile, Antonello Saccomanno, Sonia Amabile, Teresa Rocco, Annamaria Salvati, Emilia Vaccaro, Massimiliano Galdiero, Michele Cennamo, Giuseppe Portella, Maria Grazia Foti, Mariarosaria Ingino, Maria Landi, Maurizio Fumi, Vincenzo Rocco, Rita Greco, Vittoria Letizia, Arnolfo Petruzziello, Maddalena Schioppa, Gregorio Goffredi, Francesca Marciano, Michele Caraglia, Alessia Cossu, Marianna Scrima, Edmondo Adorisio, Morena D'Avenia, Michela Iacobellis, Rosanna Piluscio, Giorgio Dirani, Vittorio Sambri, Simona Semprini, Silvia Zanoli, Francesco Curcio, Stefania Marzinotto, Andreina Baj, Fausto Sessa. |
| EPI_ISL_1577234 | 1.AO Universitaria 'S. Giovanni di Dio e Ruggi D'Aragona, Scuola Medica Salernitana' Hospital / 2.UOC di Virologia e Microbiologia, Università della Campania 'L. Vanvitelli' / 3.AO Universitaria 'Federico II' Napoli Hospital / 4.AORN 'San Giuseppe Moscati' Avellino Hospital / 5.AO 'San Pio - presidio G. Rummo' Benevento Hospital / 6.AO 'Sant'Anna e San Sebastiano' Caserta Hospital / 7.PO 'Maria Santissima Addolorata' Eboli Hospital / 8.Biogem Istituto di Ricerche Genetiche / 9. U.O.C. di Genetica Medica e di Laboratorio A.O.R.N., Azienda Ospedaliera di Rilievo Nazionale Antonio Cardarelli, Napoli / 10. Centro di riferimento Oncologico della Basilicata (IRCCS-CROB), Rionero in Vulture (PZ) / 11. Presidio Ospedaliero di Agropoli, Agropoli (SA). | 1. Genome Research Center for Health (CRGS) / 2. Laboratory of Molecular Medicine and Genomics(LMMGe) / 3. Center for Research in Pure and Applied Mathematics (CRMPA) | Giorgio Giurato (Corresponding Author), Francesca Rizzo (Corresponding Author), Alessandro Weisz (Corresponding Author), Gianluigi Franci, Giovanni Nassa, Pasquale Pagliano, Roberta Tarallo, Elena Alexandrova, Ylenia D'Agostino, Carlo Ferravante, Jessica Lamberti, Viola Melone, Domenico Memoli, Valeria Mirici Cappa, Domenico Palumbo, Giovanni Pecoraro, Assunta Sellitto, Oriana Strianese, Ilaria Terenzi, Giuseppe Fenza, Aniello Gentile, Antonello Saccomanno, Sonia Amabile, Teresa Rocco, Annamaria Salvati, Emilia Vaccaro, Massimiliano Galdiero, Michele Cennamo, Giuseppe Portella, Maria Grazia Foti, Mariarosaria Ingino, Maria Landi, Maurizio Fumi, Vincenzo Rocco, Rita Greco, Vittoria Letizia, Arnolfo Petruzziello, Maddalena Schioppa, Gregorio Goffredi, Francesca Marciano, Michele Caraglia, Alessia Cossu, Marianna Scrima, Edmondo Adorisio, Morena D'Avenia, Michela Iacobellis, Rosanna Piluscio, Giorgio Dirani, Vittorio Sambri, Simona Semprini, Silvia Zanoli, Francesco Curcio, Stefania Marzinotto, Andreina Baj, Fausto Sessa, Massimiliano Chetta, Marina Tarsitano, Giusy Giordano, Marcello Ametrano, Mariano Fiorenza, Caivano Antonella, D'Auria Fiorella, De Luca Luciana. |
| EPI_ISL_1577235, EPI_ISL_1577236, EPI_ISL_1577237, EPI_ISL_1577238 | 1.AO Universitaria 'S. Giovanni di Dio e Ruggi D'Aragona, Scuola Medica Salernitana' Hospital / 2.UOC di Virologia e Microbiologia, Università della Campania 'L. Vanvitelli' / 3.AO Universitaria 'Federico II' Napoli Hospital / 4.AORN 'San Giuseppe Moscati' Avellino Hospital / 5.AO 'San Pio - presidio G. Rummo' Benevento Hospital / 6.AO 'Sant'Anna e San Sebastiano' Caserta Hospital / 7.PO 'Maria Santissima Addolorata' Eboli Hospital / 8.Biogem Istituto di Ricerche Genetiche / 9. U.O.C. di Genetica Medica e di Laboratorio A.O.R.N., Azienda Ospedaliera di Rilievo Nazionale Antonio Cardarelli, Napoli / 10. Centro di riferimento Oncologico della | 1. Genome Research Center for Health (CRGS) / 2. Laboratory of Molecular Medicine and Genomics(LMMGe) / 3. Center for Research in Pure and Applied Mathematics (CRMPA) | Giorgio Giurato (Corresponding Author), Francesca Rizzo (Corresponding Author), Alessandro Weisz (Corresponding Author), Gianluigi Franci, Giovanni Nassa, Pasquale Pagliano, Roberta Tarallo, Elena Alexandrova, Ylenia D'Agostino, Carlo Ferravante, Jessica Lamberti, Viola Melone, Domenico Memoli, Valeria Mirici Cappa, Domenico Palumbo, Giovanni Pecoraro, Assunta Sellitto, Oriana Strianese, Ilaria Terenzi, Giuseppe Fenza, Aniello Gentile, Antonello Saccomanno, Sonia Amabile, Teresa Rocco, Annamaria Salvati, Emilia Vaccaro, Massimiliano Galdiero, Michele Cennamo, Giuseppe Portella, Maria Grazia Foti, Mariarosaria Ingino, Maria Landi, Maurizio Fumi, Vincenzo Rocco, Rita Greco, Vittoria Letizia, Arnolfo Petruzziello, Maddalena Schioppa, Gregorio Goffredi, Francesca Marciano, Michele Caraglia, Alessia Cossu, Marianna Scrima, Edmondo Adorisio, Morena D'Avenia, Michela Iacobellis, Rosanna Piluscio, Giorgio Dirani, Vittorio Sambri, Simona Semprini, Silvia Zanoli, Francesco Curcio, Stefania Marzinotto, Andreina Baj, Fausto Sessa. |

| | | | |
|---|---|---|---|
| | Basilicata (IRCCS-CROB), Rionero in Vulture (PZ) / 11. Presidio Ospedaliero di Agropoli, Agropoli (SA). | | |
| EPI_ISL_1577239 | 1.AO Universitaria 'S. Giovanni di Dio e Ruggi D'Aragona, Scuola Medica Salernitana' Hospital / 2.UOC di Virologia e Microbiologia, Università della Campania 'L. Vanvitelli' / 3.AO Universitaria 'Federico II' Napoli Hospital / 4.AORN 'San Giuseppe Moscati' Avellino Hospital / 5.AO 'San Pio - presidio G. Rummo' Benevento Hospital / 6.AO 'Sant'Anna e San Sebastiano' Caserta Hospital / 7.PO 'Maria Santissima Addolorata' Eboli Hospital / 8.Biogem Istituto di Ricerche Genetiche / 9. U.O.C. di Genetica Medica e di Laboratorio A.O.R.N., Azienda Ospedaliera di Rilievo Nazionale Antonio Cardarelli, Napoli / 10. Centro di riferimento Oncologico della Basilicata (IRCCS-CROB), Rionero in Vulture (PZ) / 11. Presidio Ospedaliero di Agropoli, Agropoli (SA). | 1. Genome Research Center for Health (CRGS) / 2. Laboratory of Molecular Medicine and Genomics(LMMGe) / 3. Center for Research in Pure and Applied Mathematics (CRMPA) | Giorgio Giurato (Corresponding Author), Francesca Rizzo (Corresponding Author), Alessandro Weisz (Corresponding Author), Gianluigi Franci, Giovanni Nassa, Pasquale Pagliano, Roberta Tarallo, Elena Alexandrova, Ylenia D'Agostino, Carlo Ferravante, Jessica Lamberti, Viola Melone, Domenico Memoli, Valeria Mirici Cappa, Domenico Palumbo, Giovanni Pecoraro, Assunta Sellitto, Oriana Strianese, Ilaria Terenzi, Giuseppe Fenza, Aniello Gentile, Antonello Saccomanno, Sonia Amabile, Teresa Rocco, Annamaria Salvati, Emilia Vaccaro, Massimiliano Galdiero, Michele Cennamo, Giuseppe Portella, Maria Grazia Foti, Mariarosaria Ingino, Maria Landi, Maurizio Fumi, Vincenzo Rocco, Rita Greco, Vittoria Letizia, Arnolfo Petruzziello, Maddalena Schioppa, Gregorio Goffredi, Francesca Marciano, Michele Caraglia, Alessia Cossu, Marianna Scrima, Edmondo Adorisio, Morena D'Avenia, Michela Iacobellis, Rosanna Piluscio, Giorgio Dirani, Vittorio Sambri, Simona Semprini, Silvia Zanoli, Francesco Curcio, Stefania Marzinotto, Andreina Baj, Fausto Sessa, Massimiliano Chetta, Marina Tarsitano, Giusy Giordano, Marcello Ametrano, Mariano Fiorenza, Caivano Antonella, D'Auria Fiorella, De Luca Luciana. |
| EPI_ISL_1577240, EPI_ISL_1577241, EPI_ISL_1577242, EPI_ISL_1577243 | 1.AO Universitaria 'S. Giovanni di Dio e Ruggi D'Aragona, Scuola Medica Salernitana' Hospital / 2.UOC di Virologia e Microbiologia, Università della Campania 'L. Vanvitelli' / 3.AO Universitaria 'Federico II' Napoli Hospital / 4.AORN 'San Giuseppe Moscati' Avellino Hospital / 5.AO 'San Pio - presidio G. Rummo' Benevento Hospital / 6.AO 'Sant'Anna e San Sebastiano' Caserta Hospital / 7.PO 'Maria Santissima Addolorata' Eboli Hospital / 8.Biogem Istituto di Ricerche Genetiche / 9. U.O.C. di Genetica Medica e di Laboratorio A.O.R.N., Azienda Ospedaliera di Rilievo Nazionale Antonio Cardarelli, Napoli / 10. Centro di riferimento Oncologico della Basilicata (IRCCS-CROB), Rionero in Vulture (PZ) / 11. Presidio Ospedaliero di Agropoli, Agropoli (SA). | 1. Genome Research Center for Health (CRGS) / 2. Laboratory of Molecular Medicine and Genomics(LMMGe) / 3. Center for Research in Pure and Applied Mathematics (CRMPA) | Giorgio Giurato (Corresponding Author), Francesca Rizzo (Corresponding Author), Alessandro Weisz (Corresponding Author), Gianluigi Franci, Giovanni Nassa, Pasquale Pagliano, Roberta Tarallo, Elena Alexandrova, Ylenia D'Agostino, Carlo Ferravante, Jessica Lamberti, Viola Melone, Domenico Memoli, Valeria Mirici Cappa, Domenico Palumbo, Giovanni Pecoraro, Assunta Sellitto, Oriana Strianese, Ilaria Terenzi, Giuseppe Fenza, Aniello Gentile, Antonello Saccomanno, Sonia Amabile, Teresa Rocco, Annamaria Salvati, Emilia Vaccaro, Massimiliano Galdiero, Michele Cennamo, Giuseppe Portella, Maria Grazia Foti, Mariarosaria Ingino, Maria Landi, Maurizio Fumi, Vincenzo Rocco, Rita Greco, Vittoria Letizia, Arnolfo Petruzziello, Maddalena Schioppa, Gregorio Goffredi, Francesca Marciano, Michele Caraglia, Alessia Cossu, Marianna Scrima, Edmondo Adorisio, Morena D'Avenia, Michela Iacobellis, Rosanna Piluscio, Giorgio Dirani, Vittorio Sambri, Simona Semprini, Silvia Zanoli, Francesco Curcio, Stefania Marzinotto, Andreina Baj, Fausto Sessa. |
| EPI_ISL_1577244 | 1.AO Universitaria 'S. Giovanni di Dio e Ruggi D'Aragona, Scuola Medica Salernitana' Hospital / 2.UOC di Virologia e Microbiologia, Università della Campania 'L. Vanvitelli' / 3.AO Universitaria 'Federico II' Napoli Hospital / 4.AORN 'San Giuseppe Moscati' Avellino Hospital / 5.AO 'San Pio - presidio G. Rummo' Benevento Hospital / 6.AO 'Sant'Anna e San Sebastiano' Caserta Hospital / 7.PO 'Maria Santissima Addolorata' Eboli Hospital / 8.Biogem Istituto di Ricerche Genetiche / 9. U.O.C. di Genetica Medica e di Laboratorio A.O.R.N., Azienda Ospedaliera di Rilievo Nazionale Antonio Cardarelli, Napoli / 10. Centro di riferimento Oncologico della Basilicata (IRCCS-CROB), Rionero in Vulture (PZ) / 11. Presidio Ospedaliero di Agropoli, Agropoli (SA). | 1. Genome Research Center for Health (CRGS) / 2. Laboratory of Molecular Medicine and Genomics(LMMGe) / 3. Center for Research in Pure and Applied Mathematics (CRMPA) | Giorgio Giurato (Corresponding Author), Francesca Rizzo (Corresponding Author), Alessandro Weisz (Corresponding Author), Gianluigi Franci, Giovanni Nassa, Pasquale Pagliano, Roberta Tarallo, Elena Alexandrova, Ylenia D'Agostino, Carlo Ferravante, Jessica Lamberti, Viola Melone, Domenico Memoli, Valeria Mirici Cappa, Domenico Palumbo, Giovanni Pecoraro, Assunta Sellitto, Oriana Strianese, Ilaria Terenzi, Giuseppe Fenza, Aniello Gentile, Antonello Saccomanno, Sonia Amabile, Teresa Rocco, Annamaria Salvati, Emilia Vaccaro, Massimiliano Galdiero, Michele Cennamo, Giuseppe Portella, Maria Grazia Foti, Mariarosaria Ingino, Maria Landi, Maurizio Fumi, Vincenzo Rocco, Rita Greco, Vittoria Letizia, Arnolfo Petruzziello, Maddalena Schioppa, Gregorio Goffredi, Francesca Marciano, Michele Caraglia, Alessia Cossu, Marianna Scrima, Edmondo Adorisio, Morena D'Avenia, Michela Iacobellis, Rosanna Piluscio, Giorgio Dirani, Vittorio Sambri, Simona Semprini, Silvia Zanoli, Francesco Curcio, Stefania Marzinotto, Andreina Baj, Fausto Sessa, Massimiliano Chetta, Marina Tarsitano, Giusy Giordano, Marcello Ametrano, Mariano Fiorenza, Caivano Antonella, D'Auria Fiorella, De Luca Luciana. |
| EPI_ISL_1577245, EPI_ISL_1577246, EPI_ISL_1577247, EPI_ISL_1577248 | 1.AO Universitaria 'S. Giovanni di Dio e Ruggi D'Aragona, Scuola Medica Salernitana' Hospital / 2.UOC di Virologia e Microbiologia, Università della Campania 'L. Vanvitelli' / 3.AO Universitaria 'Federico II' Napoli Hospital / 4.AORN 'San Giuseppe Moscati' Avellino Hospital / 5.AO 'San Pio - presidio G. Rummo' Benevento Hospital / 6.AO 'Sant'Anna e San Sebastiano' Caserta Hospital / 7.PO 'Maria Santissima Addolorata' Eboli Hospital / 8.Biogem Istituto di Ricerche Genetiche / 9. U.O.C. di Genetica Medica e di Laboratorio A.O.R.N., Azienda Ospedaliera di Rilievo Nazionale Antonio Cardarelli, Napoli / 10. Centro di riferimento Oncologico della Basilicata (IRCCS-CROB), Rionero in Vulture (PZ) / 11. Presidio Ospedaliero di Agropoli, Agropoli (SA). | 1. Genome Research Center for Health (CRGS) / 2. Laboratory of Molecular Medicine and Genomics(LMMGe) / 3. Center for Research in Pure and Applied Mathematics (CRMPA) | Giorgio Giurato (Corresponding Author), Francesca Rizzo (Corresponding Author), Alessandro Weisz (Corresponding Author), Gianluigi Franci, Giovanni Nassa, Pasquale Pagliano, Roberta Tarallo, Elena Alexandrova, Ylenia D'Agostino, Carlo Ferravante, Jessica Lamberti, Viola Melone, Domenico Memoli, Valeria Mirici Cappa, Domenico Palumbo, Giovanni Pecoraro, Assunta Sellitto, Oriana Strianese, Ilaria Terenzi, Giuseppe Fenza, Aniello Gentile, Antonello Saccomanno, Sonia Amabile, Teresa Rocco, Annamaria Salvati, Emilia Vaccaro, Massimiliano Galdiero, Michele Cennamo, Giuseppe Portella, Maria Grazia Foti, Mariarosaria Ingino, Maria Landi, Maurizio Fumi, Vincenzo Rocco, Rita Greco, Vittoria Letizia, Arnolfo Petruzziello, Maddalena Schioppa, Gregorio Goffredi, Francesca Marciano, Michele Caraglia, Alessia Cossu, Marianna Scrima, Edmondo Adorisio, Morena D'Avenia, Michela Iacobellis, Rosanna Piluscio, Giorgio Dirani, Vittorio Sambri, Simona Semprini, Silvia Zanoli, Francesco Curcio, Stefania Marzinotto, Andreina Baj, Fausto Sessa. |
| EPI_ISL_1577249 | 1.AO Universitaria 'S. Giovanni di Dio e Ruggi D'Aragona, Scuola Medica Salernitana' Hospital / 2.UOC di Virologia e Microbiologia, Università della Campania 'L. Vanvitelli' / 3.AO Universitaria 'Federico II' Napoli Hospital / 4.AORN 'San Giuseppe Moscati' Avellino Hospital / 5.AO 'San Pio - presidio G. Rummo' Benevento Hospital / 6.AO 'Sant'Anna e San Sebastiano' Caserta Hospital / 7.PO 'Maria Santissima Addolorata' Eboli Hospital / 8.Biogem Istituto di Ricerche Genetiche / 9. U.O.C. di Genetica Medica e di Laboratorio A.O.R.N., Azienda Ospedaliera di Rilievo Nazionale Antonio Cardarelli, Napoli / 10. Centro di riferimento Oncologico della Basilicata (IRCCS-CROB), Rionero in Vulture (PZ) / 11. Presidio Ospedaliero di Agropoli, Agropoli (SA). | 1. Genome Research Center for Health (CRGS) / 2. Laboratory of Molecular Medicine and Genomics(LMMGe) / 3. Center for Research in Pure and Applied Mathematics (CRMPA) | Giorgio Giurato (Corresponding Author), Francesca Rizzo (Corresponding Author), Alessandro Weisz (Corresponding Author), Gianluigi Franci, Giovanni Nassa, Pasquale Pagliano, Roberta Tarallo, Elena Alexandrova, Ylenia D'Agostino, Carlo Ferravante, Jessica Lamberti, Viola Melone, Domenico Memoli, Valeria Mirici Cappa, Domenico Palumbo, Giovanni Pecoraro, Assunta Sellitto, Oriana Strianese, Ilaria Terenzi, Giuseppe Fenza, Aniello Gentile, Antonello Saccomanno, Sonia Amabile, Teresa Rocco, Annamaria Salvati, Emilia Vaccaro, Massimiliano Galdiero, Michele Cennamo, Giuseppe Portella, Maria Grazia Foti, Mariarosaria Ingino, Maria Landi, Maurizio Fumi, Vincenzo Rocco, Rita Greco, Vittoria Letizia, Arnolfo Petruzziello, Maddalena Schioppa, Gregorio Goffredi, Francesca Marciano, Michele Caraglia, Alessia Cossu, Marianna Scrima, Edmondo Adorisio, Morena D'Avenia, Michela Iacobellis, Rosanna Piluscio, Giorgio Dirani, Vittorio Sambri, Simona Semprini, Silvia Zanoli, Francesco Curcio, Stefania Marzinotto, Andreina Baj, Fausto Sessa, Massimiliano Chetta, Marina Tarsitano, Giusy Giordano, Marcello Ametrano, Mariano Fiorenza, Caivano Antonella, D'Auria Fiorella, De Luca Luciana. |
| EPI_ISL_1577250, EPI_ISL_1577251, EPI_ISL_1577252, EPI_ISL_1577253 | 1.AO Universitaria 'S. Giovanni di Dio e Ruggi D'Aragona, Scuola Medica Salernitana' Hospital / 2.UOC di Virologia e Microbiologia, Università della Campania 'L. Vanvitelli' / 3.AO Universitaria 'Federico II' Napoli Hospital / 4.AORN 'San Giuseppe Moscati' Avellino Hospital / 5.AO 'San Pio - presidio G. Rummo' Benevento Hospital / 6.AO 'Sant'Anna e San Sebastiano' Caserta Hospital / 7.PO 'Maria Santissima Addolorata' Eboli Hospital / 8.Biogem Istituto di Ricerche Genetiche / 9. U.O.C. di Genetica Medica e di Laboratorio A.O.R.N., Azienda Ospedaliera di Rilievo Nazionale Antonio Cardarelli, Napoli / 10. Centro di riferimento Oncologico della Basilicata (IRCCS-CROB), Rionero in Vulture (PZ) / 11. Presidio Ospedaliero di Agropoli, Agropoli (SA). | 1. Genome Research Center for Health (CRGS) / 2. Laboratory of Molecular Medicine and Genomics(LMMGe) / 3. Center for Research in Pure and Applied Mathematics (CRMPA) | Giorgio Giurato (Corresponding Author), Francesca Rizzo (Corresponding Author), Alessandro Weisz (Corresponding Author), Gianluigi Franci, Giovanni Nassa, Pasquale Pagliano, Roberta Tarallo, Elena Alexandrova, Ylenia D'Agostino, Carlo Ferravante, Jessica Lamberti, Viola Melone, Domenico Memoli, Valeria Mirici Cappa, Domenico Palumbo, Giovanni Pecoraro, Assunta Sellitto, Oriana Strianese, Ilaria Terenzi, Giuseppe Fenza, Aniello Gentile, Antonello Saccomanno, Sonia Amabile, Teresa Rocco, Annamaria Salvati, Emilia Vaccaro, Massimiliano Galdiero, Michele Cennamo, Giuseppe Portella, Maria Grazia Foti, Mariarosaria Ingino, Maria Landi, Maurizio Fumi, Vincenzo Rocco, Rita Greco, Vittoria Letizia, Arnolfo Petruzziello, Maddalena Schioppa, Gregorio Goffredi, Francesca Marciano, Michele Caraglia, Alessia Cossu, Marianna Scrima, Edmondo Adorisio, Morena D'Avenia, Michela Iacobellis, Rosanna Piluscio, Giorgio Dirani, Vittorio Sambri, Simona Semprini, Silvia Zanoli, Francesco Curcio, Stefania Marzinotto, Andreina Baj, Fausto Sessa. |
| EPI_ISL_1577254 | 1.AO Universitaria 'S. Giovanni di Dio e Ruggi D'Aragona, Scuola Medica Salernitana' Hospital / 2.UOC di Virologia e Microbiologia, Università della Campania 'L. Vanvitelli' / 3.AO Universitaria 'Federico II' Napoli Hospital / 4.AORN 'San | 1. Genome Research Center for Health (CRGS) / 2. Laboratory of Molecular Medicine and Genomics(LMMGe) / 3. Center for Research in Pure and Applied Mathematics (CRMPA) | Giorgio Giurato (Corresponding Author), Francesca Rizzo (Corresponding Author), Alessandro Weisz (Corresponding Author), Gianluigi Franci, Giovanni Nassa, Pasquale Pagliano, Roberta Tarallo, Elena Alexandrova, Ylenia D'Agostino, Carlo Ferravante, Jessica Lamberti, Viola Melone, Domenico Memoli, Valeria Mirici Cappa, Domenico Palumbo, Giovanni Pecoraro, Assunta Sellitto, Oriana Strianese, Ilaria Terenzi, Giuseppe Fenza, Aniello Gentile, Antonello Saccomanno, Sonia Amabile, Teresa Rocco, Annamaria Salvati, Emilia Vaccaro, Massimiliano Galdiero, Michele Cennamo, Giuseppe Portella, Maria |

| | | | |
|---|---|---|---|
| | Giuseppe Moscati' Avellino Hospital / 5.AO 'San Pio - presidio G. Rummo' Benevento Hospital / 6.AO 'Sant'Anna e San Sebastiano' Caserta Hospital / 7.PO 'Maria Santissima Addolorata' Eboli Hospital / 8.Biogem Istituto di Ricerche Genetiche / 9. U.O.C. di Genetica Medica e di Laboratorio A.O.R.N., Azienda Ospedaliera di Rilievo Nazionale Antonio Cardarelli, Napoli / 10. Centro di riferimento Oncologico della Basilicata (IRCCS-CROB), Rionero in Vulture (PZ) / 11. Presidio Ospedaliero di Agropoli, Agropoli (SA). | | Grazia Foti, Mariarosaria Ingino, Maria Landi, Maurizio Fumi, Vincenzo Rocco, Rita Greco, Vittoria Letizia, Arnolfo Petruzziello, Maddalena Schioppa, Gregorio Goffredi, Francesca Marciano, Michele Caraglia, Alessia Cossu, Marianna Scrima, Edmondo Adorisio, Morena D'Avenia, Michela Iacobellis, Rosanna Piluscio, Giorgio Dirani, Vittorio Sambri, Simona Semprini, Silvia Zanoli, Francesco Curcio, Stefania Marzinotto, Andreina Baj, Fausto Sessa, Massimiliano Chetta, Marina Tarsitano, Giusy Giordano, Marcello Ametrano, Mariano Fiorenza, Caivano Antonella, D'Auria Fiorella, De Luca Luciana. |
| EPI_ISL_1577255, EPI_ISL_1577256, EPI_ISL_1577278 | 1.AO Universitaria 'S. Giovanni di Dio e Ruggi D'Aragona, Scuola Medica Salernitana' Hospital / 2.UOC di Virologia e Microbiologia, Università della Campania 'L. Vanvitelli' / 3.AO Universitaria 'Federico II' Napoli Hospital / 4.AORN 'San Giuseppe Moscati' Avellino Hospital / 5.AO 'San Pio - presidio G. Rummo' Benevento Hospital / 6.AO 'Sant'Anna e San Sebastiano' Caserta Hospital / 7.PO 'Maria Santissima Addolorata' Eboli Hospital / 8.Biogem Istituto di Ricerche Genetiche / 9. U.O.C. di Genetica Medica e di Laboratorio A.O.R.N., Azienda Ospedaliera di Rilievo Nazionale Antonio Cardarelli, Napoli / 10. Centro di riferimento Oncologico della Basilicata (IRCCS-CROB), Rionero in Vulture (PZ) / 11. Presidio Ospedaliero di Agropoli, Agropoli (SA). | 1. Genome Research Center for Health (CRGS) / 2. Laboratory of Molecular Medicine and Genomics(LMMGe) / 3. Center for Research in Pure and Applied Mathematics (CRMPA) | Giorgio Giurato (Corresponding Author), Francesca Rizzo (Corresponding Author), Alessandro Weisz (Corresponding Author), Gianluigi Franci, Giovanni Nassa, Pasquale Pagliano, Roberta Tarallo, Elena Alexandrova, Ylenia D'Agostino, Carlo Ferravante, Jessica Lamberti, Viola Melone, Domenico Memoli, Valeria Mirici Cappa, Domenico Palumbo, Giovanni Pecoraro, Assunta Sellitto, Oriana Strianese, Ilaria Terenzi, Giuseppe Fenza, Aniello Gentile, Antonello Saccomanno, Sonia Amabile, Teresa Rocco, Annamaria Salvati, Emilia Vaccaro, Massimiliano Galdiero, Michele Cennamo, Giuseppe Portella, Maria Grazia Foti, Mariarosaria Ingino, Maria Landi, Maurizio Fumi, Vincenzo Rocco, Rita Greco, Vittoria Letizia, Arnolfo Petruzziello, Maddalena Schioppa, Gregorio Goffredi, Francesca Marciano, Michele Caraglia, Alessia Cossu, Marianna Scrima, Edmondo Adorisio, Morena D'Avenia, Michela Iacobellis, Rosanna Piluscio, Giorgio Dirani, Vittorio Sambri, Simona Semprini, Silvia Zanoli, Francesco Curcio, Stefania Marzinotto, Andreina Baj, Fausto Sessa. |
| EPI_ISL_1577279 | 1.AO Universitaria 'S. Giovanni di Dio e Ruggi D'Aragona, Scuola Medica Salernitana' Hospital / 2.UOC di Virologia e Microbiologia, Università della Campania 'L. Vanvitelli' / 3.AO Universitaria 'Federico II' Napoli Hospital / 4.AORN 'San Giuseppe Moscati' Avellino Hospital / 5.AO 'San Pio - presidio G. Rummo' Benevento Hospital / 6.AO 'Sant'Anna e San Sebastiano' Caserta Hospital / 7.PO 'Maria Santissima Addolorata' Eboli Hospital / 8.Biogem Istituto di Ricerche Genetiche / 9. U.O.C. di Genetica Medica e di Laboratorio A.O.R.N., Azienda Ospedaliera di Rilievo Nazionale Antonio Cardarelli, Napoli / 10. Centro di riferimento Oncologico della Basilicata (IRCCS-CROB), Rionero in Vulture (PZ) / 11. Presidio Ospedaliero di Agropoli, Agropoli (SA). | 1. Genome Research Center for Health (CRGS) / 2. Laboratory of Molecular Medicine and Genomics(LMMGe) / 3. Center for Research in Pure and Applied Mathematics (CRMPA) | Giorgio Giurato (Corresponding Author), Francesca Rizzo (Corresponding Author), Alessandro Weisz (Corresponding Author), Gianluigi Franci, Giovanni Nassa, Pasquale Pagliano, Roberta Tarallo, Elena Alexandrova, Ylenia D'Agostino, Carlo Ferravante, Jessica Lamberti, Viola Melone, Domenico Memoli, Valeria Mirici Cappa, Domenico Palumbo, Giovanni Pecoraro, Assunta Sellitto, Oriana Strianese, Ilaria Terenzi, Giuseppe Fenza, Aniello Gentile, Antonello Saccomanno, Sonia Amabile, Teresa Rocco, Annamaria Salvati, Emilia Vaccaro, Massimiliano Galdiero, Michele Cennamo, Giuseppe Portella, Maria Grazia Foti, Mariarosaria Ingino, Maria Landi, Maurizio Fumi, Vincenzo Rocco, Rita Greco, Vittoria Letizia, Arnolfo Petruzziello, Maddalena Schioppa, Gregorio Goffredi, Francesca Marciano, Michele Caraglia, Alessia Cossu, Marianna Scrima, Edmondo Adorisio, Morena D'Avenia, Michela Iacobellis, Rosanna Piluscio, Giorgio Dirani, Vittorio Sambri, Simona Semprini, Silvia Zanoli, Francesco Curcio, Stefania Marzinotto, Andreina Baj, Fausto Sessa, Massimiliano Chetta, Marina Tarsitano, Giusy Giordano, Marcello Ametrano, Mariano Fiorenza, Caivano Antonella, D'Auria Fiorella, De Luca Luciana. |
| EPI_ISL_1577280, EPI_ISL_1577281, EPI_ISL_1577282, EPI_ISL_1577283, EPI_ISL_1577284, EPI_ISL_1577285, EPI_ISL_1577286, EPI_ISL_1577287 | 1.AO Universitaria 'S. Giovanni di Dio e Ruggi D'Aragona, Scuola Medica Salernitana' Hospital / 2.UOC di Virologia e Microbiologia, Università della Campania 'L. Vanvitelli' / 3.AO Universitaria 'Federico II' Napoli Hospital / 4.AORN 'San Giuseppe Moscati' Avellino Hospital / 5.AO 'San Pio - presidio G. Rummo' Benevento Hospital / 6.AO 'Sant'Anna e San Sebastiano' Caserta Hospital / 7.PO 'Maria Santissima Addolorata' Eboli Hospital / 8.Biogem Istituto di Ricerche Genetiche / 9. U.O.C. di Genetica Medica e di Laboratorio A.O.R.N., Azienda Ospedaliera di Rilievo Nazionale Antonio Cardarelli, Napoli / 10. Centro di riferimento Oncologico della Basilicata (IRCCS-CROB), Rionero in Vulture (PZ) / 11. Presidio Ospedaliero di Agropoli, Agropoli (SA). | 1. Genome Research Center for Health (CRGS) / 2. Laboratory of Molecular Medicine and Genomics(LMMGe) / 3. Center for Research in Pure and Applied Mathematics (CRMPA) | Giorgio Giurato (Corresponding Author), Francesca Rizzo (Corresponding Author), Alessandro Weisz (Corresponding Author), Gianluigi Franci, Giovanni Nassa, Pasquale Pagliano, Roberta Tarallo, Elena Alexandrova, Ylenia D'Agostino, Carlo Ferravante, Jessica Lamberti, Viola Melone, Domenico Memoli, Valeria Mirici Cappa, Domenico Palumbo, Giovanni Pecoraro, Assunta Sellitto, Oriana Strianese, Ilaria Terenzi, Giuseppe Fenza, Aniello Gentile, Antonello Saccomanno, Sonia Amabile, Teresa Rocco, Annamaria Salvati, Emilia Vaccaro, Massimiliano Galdiero, Michele Cennamo, Giuseppe Portella, Maria Grazia Foti, Mariarosaria Ingino, Maria Landi, Maurizio Fumi, Vincenzo Rocco, Rita Greco, Vittoria Letizia, Arnolfo Petruzziello, Maddalena Schioppa, Gregorio Goffredi, Francesca Marciano, Michele Caraglia, Alessia Cossu, Marianna Scrima, Edmondo Adorisio, Morena D'Avenia, Michela Iacobellis, Rosanna Piluscio, Giorgio Dirani, Vittorio Sambri, Simona Semprini, Silvia Zanoli, Francesco Curcio, Stefania Marzinotto, Andreina Baj, Fausto Sessa. |
| EPI_ISL_1577288 | 1.AO Universitaria 'S. Giovanni di Dio e Ruggi D'Aragona, Scuola Medica Salernitana' Hospital / 2.UOC di Virologia e Microbiologia, Università della Campania 'L. Vanvitelli' / 3.AO Universitaria 'Federico II' Napoli Hospital / 4.AORN 'San Giuseppe Moscati' Avellino Hospital / 5.AO 'San Pio - presidio G. Rummo' Benevento Hospital / 6.AO 'Sant'Anna e San Sebastiano' Caserta Hospital / 7.PO 'Maria Santissima Addolorata' Eboli Hospital / 8.Biogem Istituto di Ricerche Genetiche / 9. U.O.C. di Genetica Medica e di Laboratorio A.O.R.N., Azienda Ospedaliera di Rilievo Nazionale Antonio Cardarelli, Napoli / 10. Centro di riferimento Oncologico della Basilicata (IRCCS-CROB), Rionero in Vulture (PZ) / 11. Presidio Ospedaliero di Agropoli, Agropoli (SA). | 1. Genome Research Center for Health (CRGS) / 2. Laboratory of Molecular Medicine and Genomics(LMMGe) / 3. Center for Research in Pure and Applied Mathematics (CRMPA) | Giorgio Giurato (Corresponding Author), Francesca Rizzo (Corresponding Author), Alessandro Weisz (Corresponding Author), Gianluigi Franci, Giovanni Nassa, Pasquale Pagliano, Roberta Tarallo, Elena Alexandrova, Ylenia D'Agostino, Carlo Ferravante, Jessica Lamberti, Viola Melone, Domenico Memoli, Valeria Mirici Cappa, Domenico Palumbo, Giovanni Pecoraro, Assunta Sellitto, Oriana Strianese, Ilaria Terenzi, Giuseppe Fenza, Aniello Gentile, Antonello Saccomanno, Sonia Amabile, Teresa Rocco, Annamaria Salvati, Emilia Vaccaro, Massimiliano Galdiero, Michele Cennamo, Giuseppe Portella, Maria Grazia Foti, Mariarosaria Ingino, Maria Landi, Maurizio Fumi, Vincenzo Rocco, Rita Greco, Vittoria Letizia, Arnolfo Petruzziello, Maddalena Schioppa, Gregorio Goffredi, Francesca Marciano, Michele Caraglia, Alessia Cossu, Marianna Scrima, Edmondo Adorisio, Morena D'Avenia, Michela Iacobellis, Rosanna Piluscio, Giorgio Dirani, Vittorio Sambri, Simona Semprini, Silvia Zanoli, Francesco Curcio, Stefania Marzinotto, Andreina Baj, Fausto Sessa, Massimiliano Chetta, Marina Tarsitano, Giusy Giordano, Marcello Ametrano, Mariano Fiorenza, Caivano Antonella, D'Auria Fiorella, De Luca Luciana. |
| EPI_ISL_1577289, EPI_ISL_1577290, EPI_ISL_1577291, EPI_ISL_1577292 | 1.AO Universitaria 'S. Giovanni di Dio e Ruggi D'Aragona, Scuola Medica Salernitana' Hospital / 2.UOC di Virologia e Microbiologia, Università della Campania 'L. Vanvitelli' / 3.AO Universitaria 'Federico II' Napoli Hospital / 4.AORN 'San Giuseppe Moscati' Avellino Hospital / 5.AO 'San Pio - presidio G. Rummo' Benevento Hospital / 6.AO 'Sant'Anna e San Sebastiano' Caserta Hospital / 7.PO 'Maria Santissima Addolorata' Eboli Hospital / 8.Biogem Istituto di Ricerche Genetiche / 9. U.O.C. di Genetica Medica e di Laboratorio A.O.R.N., Azienda Ospedaliera di Rilievo Nazionale Antonio Cardarelli, Napoli / 10. Centro di riferimento Oncologico della Basilicata (IRCCS-CROB), Rionero in Vulture (PZ) / 11. Presidio Ospedaliero di Agropoli, Agropoli (SA). | 1. Genome Research Center for Health (CRGS) / 2. Laboratory of Molecular Medicine and Genomics(LMMGe) / 3. Center for Research in Pure and Applied Mathematics (CRMPA) | Giorgio Giurato (Corresponding Author), Francesca Rizzo (Corresponding Author), Alessandro Weisz (Corresponding Author), Gianluigi Franci, Giovanni Nassa, Pasquale Pagliano, Roberta Tarallo, Elena Alexandrova, Ylenia D'Agostino, Carlo Ferravante, Jessica Lamberti, Viola Melone, Domenico Memoli, Valeria Mirici Cappa, Domenico Palumbo, Giovanni Pecoraro, Assunta Sellitto, Oriana Strianese, Ilaria Terenzi, Giuseppe Fenza, Aniello Gentile, Antonello Saccomanno, Sonia Amabile, Teresa Rocco, Annamaria Salvati, Emilia Vaccaro, Massimiliano Galdiero, Michele Cennamo, Giuseppe Portella, Maria Grazia Foti, Mariarosaria Ingino, Maria Landi, Maurizio Fumi, Vincenzo Rocco, Rita Greco, Vittoria Letizia, Arnolfo Petruzziello, Maddalena Schioppa, Gregorio Goffredi, Francesca Marciano, Michele Caraglia, Alessia Cossu, Marianna Scrima, Edmondo Adorisio, Morena D'Avenia, Michela Iacobellis, Rosanna Piluscio, Giorgio Dirani, Vittorio Sambri, Simona Semprini, Silvia Zanoli, Francesco Curcio, Stefania Marzinotto, Andreina Baj, Fausto Sessa. |
| EPI_ISL_1577293 | 1.AO Universitaria 'S. Giovanni di Dio e Ruggi D'Aragona, Scuola Medica Salernitana' Hospital / 2.UOC di Virologia e Microbiologia, Università della Campania 'L. Vanvitelli' / 3.AO Universitaria 'Federico II' Napoli Hospital / 4.AORN 'San Giuseppe Moscati' Avellino Hospital / 5.AO 'San Pio - presidio G. Rummo' Benevento Hospital / 6.AO 'Sant'Anna e San Sebastiano' Caserta Hospital / 7.PO 'Maria Santissima Addolorata' Eboli Hospital / 8.Biogem Istituto di Ricerche Genetiche / 9. U.O.C. di Genetica Medica e di Laboratorio A.O.R.N., Azienda Ospedaliera di Rilievo Nazionale Antonio | 1. Genome Research Center for Health (CRGS) / 2. Laboratory of Molecular Medicine and Genomics(LMMGe) / 3. Center for Research in Pure and Applied Mathematics (CRMPA) | Giorgio Giurato (Corresponding Author), Francesca Rizzo (Corresponding Author), Alessandro Weisz (Corresponding Author), Gianluigi Franci, Giovanni Nassa, Pasquale Pagliano, Roberta Tarallo, Elena Alexandrova, Ylenia D'Agostino, Carlo Ferravante, Jessica Lamberti, Viola Melone, Domenico Memoli, Valeria Mirici Cappa, Domenico Palumbo, Giovanni Pecoraro, Assunta Sellitto, Oriana Strianese, Ilaria Terenzi, Giuseppe Fenza, Aniello Gentile, Antonello Saccomanno, Sonia Amabile, Teresa Rocco, Annamaria Salvati, Emilia Vaccaro, Massimiliano Galdiero, Michele Cennamo, Giuseppe Portella, Maria Grazia Foti, Mariarosaria Ingino, Maria Landi, Maurizio Fumi, Vincenzo Rocco, Rita Greco, Vittoria Letizia, Arnolfo Petruzziello, Maddalena Schioppa, Gregorio Goffredi, Francesca Marciano, Michele Caraglia, Alessia Cossu, Marianna Scrima, Edmondo Adorisio, Morena D'Avenia, Michela Iacobellis, Rosanna Piluscio, Giorgio Dirani, Vittorio Sambri, Simona Semprini, Silvia Zanoli, Francesco Curcio, Stefania Marzinotto, Andreina Baj, Fausto Sessa, Massimiliano Chetta, Marina Tarsitano, Giusy Giordano, Marcello Ametrano, Mariano Fiorenza, Caivano Antonella, D'Auria Fiorella, De Luca Luciana. |

| | | | |
|---|---|---|---|
| | Cardarelli, Napoli / 10. Centro di riferimento Oncologico della Basilicata (IRCCS-CROB), Rionero in Vulture (PZ) / 11. Presidio Ospedaliero di Agropoli, Agropoli (SA). | | |
| EPI_ISL_1577294, EPI_ISL_1577295, EPI_ISL_1577296 | 1.AO Universitaria 'S. Giovanni di Dio e Ruggi D'Aragona, Scuola Medica Salernitana' Hospital / 2.UOC di Virologia e Microbiologia, Università della Campania 'L. Vanvitelli' / 3.AO Universitaria 'Federico II' Napoli Hospital / 4.AORN 'San Giuseppe Moscati' Avellino Hospital / 5.AO 'San Pio - presidio G. Rummo' Benevento Hospital / 6.AO 'Sant'Anna e San Sebastiano' Caserta Hospital / 7.PO 'Maria Santissima Addolorata' Eboli Hospital / 8.Biogem Istituto di Ricerche Genetiche / 9. U.O.C. di Genetica Medica e di Laboratorio A.O.R.N., Azienda Ospedaliera di Rilievo Nazionale Antonio Cardarelli, Napoli / 10. Centro di riferimento Oncologico della Basilicata (IRCCS-CROB), Rionero in Vulture (PZ) / 11. Presidio Ospedaliero di Agropoli, Agropoli (SA). | 1. Genome Research Center for Health (CRGS) / 2. Laboratory of Molecular Medicine and Genomics(LMMGe) / 3. Center for Research in Pure and Applied Mathematics (CRMPA) | Giorgio Giurato (Corresponding Author), Francesca Rizzo (Corresponding Author), Alessandro Weisz (Corresponding Author), Gianluigi Franci, Giovanni Nassa, Pasquale Pagliano, Roberta Tarallo, Elena Alexandrova, Ylenia D'Agostino, Carlo Ferravante, Jessica Lamberti, Viola Melone, Domenico Memoli, Valeria Mirici Cappa, Domenico Palumbo, Giovanni Pecoraro, Assunta Sellitto, Oriana Strianese, Ilaria Terenzi, Giuseppe Fenza, Aniello Gentile, Antonello Saccomanno, Sonia Amabile, Teresa Rocco, Annamaria Salvati, Emilia Vaccaro, Massimiliano Galdiero, Michele Cennamo, Giuseppe Portella, Maria Grazia Foti, Mariarosaria Ingino, Maria Landi, Maurizio Fumi, Vincenzo Rocco, Rita Greco, Vittoria Letizia, Arnolfo Petruzziello, Maddalena Schioppa, Gregorio Goffredi, Francesca Marciano, Michele Caraglia, Alessia Cossu, Marianna Scrima, Edmondo Adorisio, Morena D'Avenia, Michela Iacobellis, Rosanna Piluscio, Giorgio Dirani, Vittorio Sambri, Simona Semprini, Silvia Zanoli, Francesco Curcio, Stefania Marzinotto, Andreina Baj, Fausto Sessa. |
| EPI_ISL_1577297 | 1.AO Universitaria 'S. Giovanni di Dio e Ruggi D'Aragona, Scuola Medica Salernitana' Hospital / 2.UOC di Virologia e Microbiologia, Università della Campania 'L. Vanvitelli' / 3.AO Universitaria 'Federico II' Napoli Hospital / 4.AORN 'San Giuseppe Moscati' Avellino Hospital / 5.AO 'San Pio - presidio G. Rummo' Benevento Hospital / 6.AO 'Sant'Anna e San Sebastiano' Caserta Hospital / 7.PO 'Maria Santissima Addolorata' Eboli Hospital / 8.Biogem Istituto di Ricerche Genetiche / 9. U.O.C. di Genetica Medica e di Laboratorio A.O.R.N., Azienda Ospedaliera di Rilievo Nazionale Antonio Cardarelli, Napoli / 10. Centro di riferimento Oncologico della Basilicata (IRCCS-CROB), Rionero in Vulture (PZ) / 11. Presidio Ospedaliero di Agropoli, Agropoli (SA). | 1. Genome Research Center for Health (CRGS) / 2. Laboratory of Molecular Medicine and Genomics(LMMGe) / 3. Center for Research in Pure and Applied Mathematics (CRMPA) | Giorgio Giurato (Corresponding Author), Francesca Rizzo (Corresponding Author), Alessandro Weisz (Corresponding Author), Gianluigi Franci, Giovanni Nassa, Pasquale Pagliano, Roberta Tarallo, Elena Alexandrova, Ylenia D'Agostino, Carlo Ferravante, Jessica Lamberti, Viola Melone, Domenico Memoli, Valeria Mirici Cappa, Domenico Palumbo, Giovanni Pecoraro, Assunta Sellitto, Oriana Strianese, Ilaria Terenzi, Giuseppe Fenza, Aniello Gentile, Antonello Saccomanno, Sonia Amabile, Teresa Rocco, Annamaria Salvati, Emilia Vaccaro, Massimiliano Galdiero, Michele Cennamo, Giuseppe Portella, Maria Grazia Foti, Mariarosaria Ingino, Maria Landi, Maurizio Fumi, Vincenzo Rocco, Rita Greco, Vittoria Letizia, Arnolfo Petruzziello, Maddalena Schioppa, Gregorio Goffredi, Francesca Marciano, Michele Caraglia, Alessia Cossu, Marianna Scrima, Edmondo Adorisio, Morena D'Avenia, Michela Iacobellis, Rosanna Piluscio, Giorgio Dirani, Vittorio Sambri, Simona Semprini, Silvia Zanoli, Francesco Curcio, Stefania Marzinotto, Andreina Baj, Fausto Sessa, Massimiliano Chetta, Marina Tarsitano, Giusy Giordano, Marcello Ametrano, Mariano Fiorenza, Caivano Antonella, D'Auria Fiorella, De Luca Luciana. |
| EPI_ISL_1577298, EPI_ISL_1577299, EPI_ISL_1577300, EPI_ISL_1577301 | 1.AO Universitaria 'S. Giovanni di Dio e Ruggi D'Aragona, Scuola Medica Salernitana' Hospital / 2.UOC di Virologia e Microbiologia, Università della Campania 'L. Vanvitelli' / 3.AO Universitaria 'Federico II' Napoli Hospital / 4.AORN 'San Giuseppe Moscati' Avellino Hospital / 5.AO 'San Pio - presidio G. Rummo' Benevento Hospital / 6.AO 'Sant'Anna e San Sebastiano' Caserta Hospital / 7.PO 'Maria Santissima Addolorata' Eboli Hospital / 8.Biogem Istituto di Ricerche Genetiche / 9. U.O.C. di Genetica Medica e di Laboratorio A.O.R.N., Azienda Ospedaliera di Rilievo Nazionale Antonio Cardarelli, Napoli / 10. Centro di riferimento Oncologico della Basilicata (IRCCS-CROB), Rionero in Vulture (PZ) / 11. Presidio Ospedaliero di Agropoli, Agropoli (SA). | 1. Genome Research Center for Health (CRGS) / 2. Laboratory of Molecular Medicine and Genomics(LMMGe) / 3. Center for Research in Pure and Applied Mathematics (CRMPA) | Giorgio Giurato (Corresponding Author), Francesca Rizzo (Corresponding Author), Alessandro Weisz (Corresponding Author), Gianluigi Franci, Giovanni Nassa, Pasquale Pagliano, Roberta Tarallo, Elena Alexandrova, Ylenia D'Agostino, Carlo Ferravante, Jessica Lamberti, Viola Melone, Domenico Memoli, Valeria Mirici Cappa, Domenico Palumbo, Giovanni Pecoraro, Assunta Sellitto, Oriana Strianese, Ilaria Terenzi, Giuseppe Fenza, Aniello Gentile, Antonello Saccomanno, Sonia Amabile, Teresa Rocco, Annamaria Salvati, Emilia Vaccaro, Massimiliano Galdiero, Michele Cennamo, Giuseppe Portella, Maria Grazia Foti, Mariarosaria Ingino, Maria Landi, Maurizio Fumi, Vincenzo Rocco, Rita Greco, Vittoria Letizia, Arnolfo Petruzziello, Maddalena Schioppa, Gregorio Goffredi, Francesca Marciano, Michele Caraglia, Alessia Cossu, Marianna Scrima, Edmondo Adorisio, Morena D'Avenia, Michela Iacobellis, Rosanna Piluscio, Giorgio Dirani, Vittorio Sambri, Simona Semprini, Silvia Zanoli, Francesco Curcio, Stefania Marzinotto, Andreina Baj, Fausto Sessa. |
| EPI_ISL_1577302 | 1.AO Universitaria 'S. Giovanni di Dio e Ruggi D'Aragona, Scuola Medica Salernitana' Hospital / 2.UOC di Virologia e Microbiologia, Università della Campania 'L. Vanvitelli' / 3.AO Universitaria 'Federico II' Napoli Hospital / 4.AORN 'San Giuseppe Moscati' Avellino Hospital / 5.AO 'San Pio - presidio G. Rummo' Benevento Hospital / 6.AO 'Sant'Anna e San Sebastiano' Caserta Hospital / 7.PO 'Maria Santissima Addolorata' Eboli Hospital / 8.Biogem Istituto di Ricerche Genetiche / 9. U.O.C. di Genetica Medica e di Laboratorio A.O.R.N., Azienda Ospedaliera di Rilievo Nazionale Antonio Cardarelli, Napoli / 10. Centro di riferimento Oncologico della Basilicata (IRCCS-CROB), Rionero in Vulture (PZ) / 11. Presidio Ospedaliero di Agropoli, Agropoli (SA). | 1. Genome Research Center for Health (CRGS) / 2. Laboratory of Molecular Medicine and Genomics(LMMGe) / 3. Center for Research in Pure and Applied Mathematics (CRMPA) | Giorgio Giurato (Corresponding Author), Francesca Rizzo (Corresponding Author), Alessandro Weisz (Corresponding Author), Gianluigi Franci, Giovanni Nassa, Pasquale Pagliano, Roberta Tarallo, Elena Alexandrova, Ylenia D'Agostino, Carlo Ferravante, Jessica Lamberti, Viola Melone, Domenico Memoli, Valeria Mirici Cappa, Domenico Palumbo, Giovanni Pecoraro, Assunta Sellitto, Oriana Strianese, Ilaria Terenzi, Giuseppe Fenza, Aniello Gentile, Antonello Saccomanno, Sonia Amabile, Teresa Rocco, Annamaria Salvati, Emilia Vaccaro, Massimiliano Galdiero, Michele Cennamo, Giuseppe Portella, Maria Grazia Foti, Mariarosaria Ingino, Maria Landi, Maurizio Fumi, Vincenzo Rocco, Rita Greco, Vittoria Letizia, Arnolfo Petruzziello, Maddalena Schioppa, Gregorio Goffredi, Francesca Marciano, Michele Caraglia, Alessia Cossu, Marianna Scrima, Edmondo Adorisio, Morena D'Avenia, Michela Iacobellis, Rosanna Piluscio, Giorgio Dirani, Vittorio Sambri, Simona Semprini, Silvia Zanoli, Francesco Curcio, Stefania Marzinotto, Andreina Baj, Fausto Sessa, Massimiliano Chetta, Marina Tarsitano, Giusy Giordano, Marcello Ametrano, Mariano Fiorenza, Caivano Antonella, D'Auria Fiorella, De Luca Luciana. |
| EPI_ISL_1577303, EPI_ISL_1577305, EPI_ISL_1577306, EPI_ISL_1577313, EPI_ISL_1577314, EPI_ISL_1577315, EPI_ISL_1577316 | 1.AO Universitaria 'S. Giovanni di Dio e Ruggi D'Aragona, Scuola Medica Salernitana' Hospital / 2.UOC di Virologia e Microbiologia, Università della Campania 'L. Vanvitelli' / 3.AO Universitaria 'Federico II' Napoli Hospital / 4.AORN 'San Giuseppe Moscati' Avellino Hospital / 5.AO 'San Pio - presidio G. Rummo' Benevento Hospital / 6.AO 'Sant'Anna e San Sebastiano' Caserta Hospital / 7.PO 'Maria Santissima Addolorata' Eboli Hospital / 8.Biogem Istituto di Ricerche Genetiche / 9. U.O.C. di Genetica Medica e di Laboratorio A.O.R.N., Azienda Ospedaliera di Rilievo Nazionale Antonio Cardarelli, Napoli / 10. Centro di riferimento Oncologico della Basilicata (IRCCS-CROB), Rionero in Vulture (PZ) / 11. Presidio Ospedaliero di Agropoli, Agropoli (SA). | 1. Genome Research Center for Health (CRGS) / 2. Laboratory of Molecular Medicine and Genomics(LMMGe) / 3. Center for Research in Pure and Applied Mathematics (CRMPA) | Giorgio Giurato (Corresponding Author), Francesca Rizzo (Corresponding Author), Alessandro Weisz (Corresponding Author), Gianluigi Franci, Giovanni Nassa, Pasquale Pagliano, Roberta Tarallo, Elena Alexandrova, Ylenia D'Agostino, Carlo Ferravante, Jessica Lamberti, Viola Melone, Domenico Memoli, Valeria Mirici Cappa, Domenico Palumbo, Giovanni Pecoraro, Assunta Sellitto, Oriana Strianese, Ilaria Terenzi, Giuseppe Fenza, Aniello Gentile, Antonello Saccomanno, Sonia Amabile, Teresa Rocco, Annamaria Salvati, Emilia Vaccaro, Massimiliano Galdiero, Michele Cennamo, Giuseppe Portella, Maria Grazia Foti, Mariarosaria Ingino, Maria Landi, Maurizio Fumi, Vincenzo Rocco, Rita Greco, Vittoria Letizia, Arnolfo Petruzziello, Maddalena Schioppa, Gregorio Goffredi, Francesca Marciano, Michele Caraglia, Alessia Cossu, Marianna Scrima, Edmondo Adorisio, Morena D'Avenia, Michela Iacobellis, Rosanna Piluscio, Giorgio Dirani, Vittorio Sambri, Simona Semprini, Silvia Zanoli, Francesco Curcio, Stefania Marzinotto, Andreina Baj, Fausto Sessa. |
| EPI_ISL_1577317 | 1.AO Universitaria 'S. Giovanni di Dio e Ruggi D'Aragona, Scuola Medica Salernitana' Hospital / 2.UOC di Virologia e Microbiologia, Università della Campania 'L. Vanvitelli' / 3.AO Universitaria 'Federico II' Napoli Hospital / 4.AORN 'San Giuseppe Moscati' Avellino Hospital / 5.AO 'San Pio - presidio G. Rummo' Benevento Hospital / 6.AO 'Sant'Anna e San Sebastiano' Caserta Hospital / 7.PO 'Maria Santissima Addolorata' Eboli Hospital / 8.Biogem Istituto di Ricerche Genetiche / 9. U.O.C. di Genetica Medica e di Laboratorio A.O.R.N., Azienda Ospedaliera di Rilievo Nazionale Antonio Cardarelli, Napoli / 10. Centro di riferimento Oncologico della Basilicata (IRCCS-CROB), Rionero in Vulture (PZ) / 11. Presidio Ospedaliero di Agropoli, Agropoli (SA). | 1. Genome Research Center for Health (CRGS) / 2. Laboratory of Molecular Medicine and Genomics(LMMGe) / 3. Center for Research in Pure and Applied Mathematics (CRMPA) | Giorgio Giurato (Corresponding Author), Francesca Rizzo (Corresponding Author), Alessandro Weisz (Corresponding Author), Gianluigi Franci, Giovanni Nassa, Pasquale Pagliano, Roberta Tarallo, Elena Alexandrova, Ylenia D'Agostino, Carlo Ferravante, Jessica Lamberti, Viola Melone, Domenico Memoli, Valeria Mirici Cappa, Domenico Palumbo, Giovanni Pecoraro, Assunta Sellitto, Oriana Strianese, Ilaria Terenzi, Giuseppe Fenza, Aniello Gentile, Antonello Saccomanno, Sonia Amabile, Teresa Rocco, Annamaria Salvati, Emilia Vaccaro, Massimiliano Galdiero, Michele Cennamo, Giuseppe Portella, Maria Grazia Foti, Mariarosaria Ingino, Maria Landi, Maurizio Fumi, Vincenzo Rocco, Rita Greco, Vittoria Letizia, Arnolfo Petruzziello, Maddalena Schioppa, Gregorio Goffredi, Francesca Marciano, Michele Caraglia, Alessia Cossu, Marianna Scrima, Edmondo Adorisio, Morena D'Avenia, Michela Iacobellis, Rosanna Piluscio, Giorgio Dirani, Vittorio Sambri, Simona Semprini, Silvia Zanoli, Francesco Curcio, Stefania Marzinotto, Andreina Baj, Fausto Sessa, Massimiliano Chetta, Marina Tarsitano, Giusy Giordano, Marcello Ametrano, Mariano Fiorenza, Caivano Antonella, D'Auria Fiorella, De Luca Luciana. |
| EPI_ISL_1577318, EPI_ISL_1577319, EPI_ISL_1577320, EPI_ISL_1577321 | 1.AO Universitaria 'S. Giovanni di Dio e Ruggi D'Aragona, Scuola Medica Salernitana' Hospital / 2.UOC di Virologia e Microbiologia, Università della Campania 'L. Vanvitelli' / 3.AO | 1. Genome Research Center for Health (CRGS) / 2. Laboratory of Molecular Medicine and Genomics(LMMGe) / 3. Center for Research in Pure and Applied Mathematics | Giorgio Giurato (Corresponding Author), Francesca Rizzo (Corresponding Author), Alessandro Weisz (Corresponding Author), Gianluigi Franci, Giovanni Nassa, Pasquale Pagliano, Roberta Tarallo, Elena Alexandrova, Ylenia D'Agostino, Carlo Ferravante, Jessica Lamberti, Viola Melone, Domenico Memoli, Valeria Mirici Cappa, Domenico Palumbo, Giovanni Pecoraro, Assunta Sellitto, Oriana Strianese, Ilaria Terenzi, Giuseppe Fenza, Aniello Gentile, Antonello |

| | | | |
|---|---|---|---|
| | Universitaria 'Federico II' Napoli Hospital / 4.AORN 'San Giuseppe Moscati' Avellino Hospital / 5.AO 'San Pio - presidio G. Rummo' Benevento Hospital / 6.AO 'Sant'Anna e San Sebastiano' Caserta Hospital / 7.PO 'Maria Santissima Addolorata' Eboli Hospital / 8.Biogem Istituto di Ricerche Genetiche / 9. U.O.C. di Genetica Medica e di Laboratorio A.O.R.N., Azienda Ospedaliera di Rilievo Nazionale Antonio Cardarelli, Napoli / 10. Centro di riferimento Oncologico della Basilicata (IRCCS-CROB), Rionero in Vulture (PZ) / 11. Presidio Ospedaliero di Agropoli, Agropoli (SA). | (CRMPA) | Saccomanno, Sonia Amabile, Teresa Rocco, Annamaria Salvati, Emilia Vaccaro, Massimiliano Galdiero, Michele Cennamo, Giuseppe Portella, Maria Grazia Foti, Mariarosaria Ingino, Maria Landi, Maurizio Fumi, Vincenzo Rocco, Rita Greco, Vittoria Letizia, Arnolfo Petruzziello, Maddalena Schioppa, Gregorio Goffredi, Francesca Marciano, Michele Caraglia, Alessia Cossu, Marianna Scrima, Edmondo Adorisio, Morena D'Avenia, Michela Iacobellis, Rosanna Piluscio, Giorgio Dirani, Vittorio Sambri, Simona Semprini, Silvia Zanoli, Francesco Curcio, Stefania Marzinotto, Andreina Baj, Fausto Sessa. |
| EPI_ISL_1577322 | 1.AO Universitaria 'S. Giovanni di Dio e Ruggi D'Aragona, Scuola Medica Salernitana' Hospital / 2.UOC di Virologia e Microbiologia, Università della Campania 'L. Vanvitelli' / 3.AO Universitaria 'Federico II' Napoli Hospital / 4.AORN 'San Giuseppe Moscati' Avellino Hospital / 5.AO 'San Pio - presidio G. Rummo' Benevento Hospital / 6.AO 'Sant'Anna e San Sebastiano' Caserta Hospital / 7.PO 'Maria Santissima Addolorata' Eboli Hospital / 8.Biogem Istituto di Ricerche Genetiche / 9. U.O.C. di Genetica Medica e di Laboratorio A.O.R.N., Azienda Ospedaliera di Rilievo Nazionale Antonio Cardarelli, Napoli / 10. Centro di riferimento Oncologico della Basilicata (IRCCS-CROB), Rionero in Vulture (PZ) / 11. Presidio Ospedaliero di Agropoli, Agropoli (SA). | 1. Genome Research Center for Health (CRGS) / 2. Laboratory of Molecular Medicine and Genomics(LMMGe) / 3. Center for Research in Pure and Applied Mathematics (CRMPA) | Giorgio Giurato (Corresponding Author), Francesca Rizzo (Corresponding Author), Alessandro Weisz (Corresponding Author), Gianluigi Franci, Giovanni Nassa, Pasquale Pagliano, Roberta Tarallo, Elena Alexandrova, Ylenia D'Agostino, Carlo Ferravante, Jessica Lamberti, Viola Melone, Domenico Memoli, Valeria Mirici Cappa, Domenico Palumbo, Giovanni Pecoraro, Assunta Sellitto, Oriana Strianese, Ilaria Terenzi, Giuseppe Fenza, Aniello Gentile, Antonello Saccomanno, Sonia Amabile, Teresa Rocco, Annamaria Salvati, Emilia Vaccaro, Massimiliano Galdiero, Michele Cennamo, Giuseppe Portella, Maria Grazia Foti, Mariarosaria Ingino, Maria Landi, Maurizio Fumi, Vincenzo Rocco, Rita Greco, Vittoria Letizia, Arnolfo Petruzziello, Maddalena Schioppa, Gregorio Goffredi, Francesca Marciano, Michele Caraglia, Alessia Cossu, Marianna Scrima, Edmondo Adorisio, Morena D'Avenia, Michela Iacobellis, Rosanna Piluscio, Giorgio Dirani, Vittorio Sambri, Simona Semprini, Silvia Zanoli, Francesco Curcio, Stefania Marzinotto, Andreina Baj, Fausto Sessa, Massimiliano Chetta, Marina Tarsitano, Giusy Giordano, Marcello Ametrano, Mariano Fiorenza, Caivano Antonella, D'Auria Fiorella, De Luca Luciana. |
| EPI_ISL_1577323, EPI_ISL_1577324, EPI_ISL_1577325, EPI_ISL_1577326 | 1.AO Universitaria 'S. Giovanni di Dio e Ruggi D'Aragona, Scuola Medica Salernitana' Hospital / 2.UOC di Virologia e Microbiologia, Università della Campania 'L. Vanvitelli' / 3.AO Universitaria 'Federico II' Napoli Hospital / 4.AORN 'San Giuseppe Moscati' Avellino Hospital / 5.AO 'San Pio - presidio G. Rummo' Benevento Hospital / 6.AO 'Sant'Anna e San Sebastiano' Caserta Hospital / 7.PO 'Maria Santissima Addolorata' Eboli Hospital / 8.Biogem Istituto di Ricerche Genetiche / 9. U.O.C. di Genetica Medica e di Laboratorio A.O.R.N., Azienda Ospedaliera di Rilievo Nazionale Antonio Cardarelli, Napoli / 10. Centro di riferimento Oncologico della Basilicata (IRCCS-CROB), Rionero in Vulture (PZ) / 11. Presidio Ospedaliero di Agropoli, Agropoli (SA). | 1. Genome Research Center for Health (CRGS) / 2. Laboratory of Molecular Medicine and Genomics(LMMGe) / 3. Center for Research in Pure and Applied Mathematics (CRMPA) | Giorgio Giurato (Corresponding Author), Francesca Rizzo (Corresponding Author), Alessandro Weisz (Corresponding Author), Gianluigi Franci, Giovanni Nassa, Pasquale Pagliano, Roberta Tarallo, Elena Alexandrova, Ylenia D'Agostino, Carlo Ferravante, Jessica Lamberti, Viola Melone, Domenico Memoli, Valeria Mirici Cappa, Domenico Palumbo, Giovanni Pecoraro, Assunta Sellitto, Oriana Strianese, Ilaria Terenzi, Giuseppe Fenza, Aniello Gentile, Antonello Saccomanno, Sonia Amabile, Teresa Rocco, Annamaria Salvati, Emilia Vaccaro, Massimiliano Galdiero, Michele Cennamo, Giuseppe Portella, Maria Grazia Foti, Mariarosaria Ingino, Maria Landi, Maurizio Fumi, Vincenzo Rocco, Rita Greco, Vittoria Letizia, Arnolfo Petruzziello, Maddalena Schioppa, Gregorio Goffredi, Francesca Marciano, Michele Caraglia, Alessia Cossu, Marianna Scrima, Edmondo Adorisio, Morena D'Avenia, Michela Iacobellis, Rosanna Piluscio, Giorgio Dirani, Vittorio Sambri, Simona Semprini, Silvia Zanoli, Francesco Curcio, Stefania Marzinotto, Andreina Baj, Fausto Sessa. |
| EPI_ISL_1577327 | 1.AO Universitaria 'S. Giovanni di Dio e Ruggi D'Aragona, Scuola Medica Salernitana' Hospital / 2.UOC di Virologia e Microbiologia, Università della Campania 'L. Vanvitelli' / 3.AO Universitaria 'Federico II' Napoli Hospital / 4.AORN 'San Giuseppe Moscati' Avellino Hospital / 5.AO 'San Pio - presidio G. Rummo' Benevento Hospital / 6.AO 'Sant'Anna e San Sebastiano' Caserta Hospital / 7.PO 'Maria Santissima Addolorata' Eboli Hospital / 8.Biogem Istituto di Ricerche Genetiche / 9. U.O.C. di Genetica Medica e di Laboratorio A.O.R.N., Azienda Ospedaliera di Rilievo Nazionale Antonio Cardarelli, Napoli / 10. Centro di riferimento Oncologico della Basilicata (IRCCS-CROB), Rionero in Vulture (PZ) / 11. Presidio Ospedaliero di Agropoli, Agropoli (SA). | 1. Genome Research Center for Health (CRGS) / 2. Laboratory of Molecular Medicine and Genomics(LMMGe) / 3. Center for Research in Pure and Applied Mathematics (CRMPA) | Giorgio Giurato (Corresponding Author), Francesca Rizzo (Corresponding Author), Alessandro Weisz (Corresponding Author), Gianluigi Franci, Giovanni Nassa, Pasquale Pagliano, Roberta Tarallo, Elena Alexandrova, Ylenia D'Agostino, Carlo Ferravante, Jessica Lamberti, Viola Melone, Domenico Memoli, Valeria Mirici Cappa, Domenico Palumbo, Giovanni Pecoraro, Assunta Sellitto, Oriana Strianese, Ilaria Terenzi, Giuseppe Fenza, Aniello Gentile, Antonello Saccomanno, Sonia Amabile, Teresa Rocco, Annamaria Salvati, Emilia Vaccaro, Massimiliano Galdiero, Michele Cennamo, Giuseppe Portella, Maria Grazia Foti, Mariarosaria Ingino, Maria Landi, Maurizio Fumi, Vincenzo Rocco, Rita Greco, Vittoria Letizia, Arnolfo Petruzziello, Maddalena Schioppa, Gregorio Goffredi, Francesca Marciano, Michele Caraglia, Alessia Cossu, Marianna Scrima, Edmondo Adorisio, Morena D'Avenia, Michela Iacobellis, Rosanna Piluscio, Giorgio Dirani, Vittorio Sambri, Simona Semprini, Silvia Zanoli, Francesco Curcio, Stefania Marzinotto, Andreina Baj, Fausto Sessa, Massimiliano Chetta, Marina Tarsitano, Giusy Giordano, Marcello Ametrano, Mariano Fiorenza, Caivano Antonella, D'Auria Fiorella, De Luca Luciana. |
| EPI_ISL_1577328 | 1.AO Universitaria 'S. Giovanni di Dio e Ruggi D'Aragona, Scuola Medica Salernitana' Hospital / 2.UOC di Virologia e Microbiologia, Università della Campania 'L. Vanvitelli' / 3.AO Universitaria 'Federico II' Napoli Hospital / 4.AORN 'San Giuseppe Moscati' Avellino Hospital / 5.AO 'San Pio - presidio G. Rummo' Benevento Hospital / 6.AO 'Sant'Anna e San Sebastiano' Caserta Hospital / 7.PO 'Maria Santissima Addolorata' Eboli Hospital / 8.Biogem Istituto di Ricerche Genetiche / 9. U.O.C. di Genetica Medica e di Laboratorio A.O.R.N., Azienda Ospedaliera di Rilievo Nazionale Antonio Cardarelli, Napoli / 10. Centro di riferimento Oncologico della Basilicata (IRCCS-CROB), Rionero in Vulture (PZ) / 11. Presidio Ospedaliero di Agropoli, Agropoli (SA). | 1. Genome Research Center for Health (CRGS) / 2. Laboratory of Molecular Medicine and Genomics(LMMGe) / 3. Center for Research in Pure and Applied Mathematics (CRMPA) | Giorgio Giurato (Corresponding Author), Francesca Rizzo (Corresponding Author), Alessandro Weisz (Corresponding Author), Gianluigi Franci, Giovanni Nassa, Pasquale Pagliano, Roberta Tarallo, Elena Alexandrova, Ylenia D'Agostino, Carlo Ferravante, Jessica Lamberti, Viola Melone, Domenico Memoli, Valeria Mirici Cappa, Domenico Palumbo, Giovanni Pecoraro, Assunta Sellitto, Oriana Strianese, Ilaria Terenzi, Giuseppe Fenza, Aniello Gentile, Antonello Saccomanno, Sonia Amabile, Teresa Rocco, Annamaria Salvati, Emilia Vaccaro, Massimiliano Galdiero, Michele Cennamo, Giuseppe Portella, Maria Grazia Foti, Mariarosaria Ingino, Maria Landi, Maurizio Fumi, Vincenzo Rocco, Rita Greco, Vittoria Letizia, Arnolfo Petruzziello, Maddalena Schioppa, Gregorio Goffredi, Francesca Marciano, Michele Caraglia, Alessia Cossu, Marianna Scrima, Edmondo Adorisio, Morena D'Avenia, Michela Iacobellis, Rosanna Piluscio, Giorgio Dirani, Vittorio Sambri, Simona Semprini, Silvia Zanoli, Francesco Curcio, Stefania Marzinotto, Andreina Baj, Fausto Sessa. |
| EPI_ISL_1585281, EPI_ISL_1585282 | Laboratorio di Riferimento Regionale della Sicilia Occidentale per l'Emergenza COVID-19 | Laboratorio di Riferimento Regionale della Sicilia Occidentale per l'Emergenza COVID-19 | Fabio Tramuto, Carmelo Massimo Maida, Daniela Di Naro, Giulia Randazzo, Walter Mazzucco, Giorgio Graziano, Vincenzo Restivo, Claudio Costantino, Francesco Vitale |
| EPI_ISL_1599180 | Laboratorio di Microbiologia e Virologia, Università Vita-Salute San Raffaele, Milano | Laboratorio di Microbiologia e Virologia, Università Vita-Salute San Raffaele, Milano | Nicola Clementi, Roberto Ferrarese, Elena Criscuolo, Michela Sampaolo, Matteo Castelli, Roberta Antonia Diotti, Enzo Boeri, Massimo Clementi, Nicasio Mancini |
| EPI_ISL_1627321, EPI_ISL_1627322, EPI_ISL_1627323 | Istituto Zooprofilattico Sperimentale della Puglia e della Basilicata | Istituto Zooprofilattico Sperimentale della Puglia e della Basilicata | Parisi A., Bianco A., Capozzi L., Del Sambro L., Simone D., Galante D., Pace L. |
| EPI_ISL_1627344 | Ospedale Vito Fazzi | Istituto Zooprofilattico Sperimentale della Puglia e della Basilicata | Parisi A., Bianco A., Capozzi L., Del Sambro L., Simone D., Difato L., Ridolfi D., Giannico A. |
| EPI_ISL_1627345, EPI_ISL_1627346 | Presidio di Brindisi Di Summa - Perrino | Istituto Zooprofilattico Sperimentale della Puglia e della Basilicata | Parisi A., Bianco A., Capozzi L., Del Sambro L., Simone D., Difato L., Santoro A., Ridolfi D., Giannico A. |
| EPI_ISL_1627347 | Ospedale di Venere | Istituto Zooprofilattico Sperimentale della Puglia e della Basilicata | Parisi A., Bianco A., Capozzi L., Del Sambro L., Simone D., Iacobellis M., Ridolfi D., Giannico A. |
| EPI_ISL_1627348, EPI_ISL_1627349, EPI_ISL_1627350, EPI_ISL_1627351 | Presidio di Brindisi Di Summa - Perrino | Istituto Zooprofilattico Sperimentale della Puglia e della Basilicata | Parisi A., Bianco A., Capozzi L., Del Sambro L., Simone D., Difato L., Santoro A., Ridolfi D., Giannico A. |
| EPI_ISL_1627352, EPI_ISL_1627353, EPI_ISL_1627354, EPI_ISL_1627355, EPI_ISL_1627356, EPI_ISL_1627357, EPI_ISL_1627358, EPI_ISL_1627359, EPI_ISL_1627360 | Istituto Zooprofilattico Sperimentale della Puglia e della Basilicata | Istituto Zooprofilattico Sperimentale della Puglia e della Basilicata | Parisi A., Bianco A., Capozzi L., Del Sambro L., Simone D., Galante D., Pace L. |

| | | | |
|---|---|---|---|
| EPI_ISL_1632706 | Presidio di Brindisi Di Summa - Perrino | Istituto Zooprofilattico Sperimentale della Puglia e della Basilicata | Parisi A., Bianco A., Capozzi L., Del Sambro L., Simone D., Difato L., Santoro A., Ridolfi D., Giannico A. |
| EPI_ISL_1632774, EPI_ISL_1632775, EPI_ISL_1632776, EPI_ISL_1632777, EPI_ISL_1632778, EPI_ISL_1632779, EPI_ISL_1632780, EPI_ISL_1632781, EPI_ISL_1632782, EPI_ISL_1632783, EPI_ISL_1632784, EPI_ISL_1632785, EPI_ISL_1632786, EPI_ISL_1632787, EPI_ISL_1632788, EPI_ISL_1632789, EPI_ISL_1632790, EPI_ISL_1632791, EPI_ISL_1632792, EPI_ISL_1632793, EPI_ISL_1632794, EPI_ISL_1632795, EPI_ISL_1632796, EPI_ISL_1632797, EPI_ISL_1632798, EPI_ISL_1632799, EPI_ISL_1632800, EPI_ISL_1632801, EPI_ISL_1632802, EPI_ISL_1632803, EPI_ISL_1632805, EPI_ISL_1632806, EPI_ISL_1632807, EPI_ISL_1632808, EPI_ISL_1632809, EPI_ISL_1632810, EPI_ISL_1632811, EPI_ISL_1632812, EPI_ISL_1632813, EPI_ISL_1632814, EPI_ISL_1632815, EPI_ISL_1632816, EPI_ISL_1632817, EPI_ISL_1632818, EPI_ISL_1632819, EPI_ISL_1632820, EPI_ISL_1632821, EPI_ISL_1632822 | | | |
| see above | Istituto Zooprofilattico Sperimentale del Mezzogiorno | Telethon Institute of Genetics and Medicine (TIGEM) | Antonio Grimaldi Patrizia Annunziata Francesco Panariello Biancamaria Pierri Claudia Tiberio Teresa Giuliano Valentina Bouche Chiara Colantuono Maria Concetta Cuomo Denise Di Concilio Lucio Di Filippo Anna Manfredi Marcello Salvi Antonio Limone Luigi Atripaldi Pellegrino Cerino Andrea Ballabio Davide Cacchiarelli |
| EPI_ISL_1633029, EPI_ISL_1633030, EPI_ISL_1633031, EPI_ISL_1633032, EPI_ISL_1633033, EPI_ISL_1633034, EPI_ISL_1633035, EPI_ISL_1633036, EPI_ISL_1633037, EPI_ISL_1633038, EPI_ISL_1633039, EPI_ISL_1633040, EPI_ISL_1633041, EPI_ISL_1633042, EPI_ISL_1633043, EPI_ISL_1633044, EPI_ISL_1633045, EPI_ISL_1633046, EPI_ISL_1633047, EPI_ISL_1633048, EPI_ISL_1633049, EPI_ISL_1633050, EPI_ISL_1633051, EPI_ISL_1633052, EPI_ISL_1633053, EPI_ISL_1633054, EPI_ISL_1633055, EPI_ISL_1633056, EPI_ISL_1633057, EPI_ISL_1633058, EPI_ISL_1633059, EPI_ISL_1633060, EPI_ISL_1633061, EPI_ISL_1633062, EPI_ISL_1633063, EPI_ISL_1633064, EPI_ISL_1633065, EPI_ISL_1633066, EPI_ISL_1633067, EPI_ISL_1633068, EPI_ISL_1633069, EPI_ISL_1633070, EPI_ISL_1633071, EPI_ISL_1633072, EPI_ISL_1633073, EPI_ISL_1633074 | | | |
| see above | Università Federico II - Dipartimento di scienze mediche traslazionali - Napoli | Telethon Institute of Genetics and Medicine (TIGEM) | Antonio Grimaldi Patrizia Annunziata Francesco Panariello Teresa Giuliano Michele Cennamo Valentina Bouche Chiara Colantuono Lucio Di Filippo Mariano Fiorenza Anna Manfredi Marcello Salvi Giuseppe Portella Andrea Ballabio Davide Cacchiarelli |
| EPI_ISL_1633121, EPI_ISL_1633122, EPI_ISL_1633123 | Istituto Zooprofilattico Sperimentale del Mezzogiorno | Telethon Institute of Genetics and Medicine (TIGEM) | Antonio Grimaldi Patrizia Annunziata Francesco Panariello Biancamaria Pierri Claudia Tiberio Teresa Giuliano Valentina Bouche Chiara Colantuono Maria Concetta Cuomo Denise Di Concilio Lucio Di Filippo Anna Manfredi Marcello Salvi Antonio Limone Luigi Atripaldi Pellegrino Cerino Andrea Ballabio Davide Cacchiarelli |
| EPI_ISL_1649779, EPI_ISL_1649780, EPI_ISL_1649782, EPI_ISL_1649783, EPI_ISL_1649785, EPI_ISL_1649786, EPI_ISL_1649788, EPI_ISL_1649789, EPI_ISL_1649791, EPI_ISL_1649793, EPI_ISL_1649794, EPI_ISL_1649795, EPI_ISL_1649797, EPI_ISL_1649798, EPI_ISL_1649800, EPI_ISL_1649802, EPI_ISL_1649803, EPI_ISL_1649805, EPI_ISL_1649806, EPI_ISL_1649808, EPI_ISL_1649809, EPI_ISL_1649811, EPI_ISL_1649812, EPI_ISL_1649814 | | | |
| see above | Laboratory of Clinical Microbiology, Virology and Bioemergencies, ASST Fatebenefratelli Sacco - Sacco Hospital | Laboratory of Clinical Microbiology, Virology and Bioemergencies, ASST Fatebenefratelli Sacco - Sacco Hospital | Valeria Micheli, Alessandro Mancon, Alberto Rizzo, Fiorenza Bracchitta, Luca Rizzuto, Maria Rita Gismondo |
| EPI_ISL_1654586 | Laboratorio HUB -Azienda Ospedaliero Universitaria - AOU - Cagliari | Laboratorio SPOKE Biologia Molecolare -Azienda Ospedaliero Universitaria - AOU - Cagliari | Germano Orrù, Sara Fais, Alessandra Scano, Miriam Loddo, Riccardo Cappai, Ferdinando Coghe |
| EPI_ISL_1660341, EPI_ISL_1660342, EPI_ISL_1660343, EPI_ISL_1660344, EPI_ISL_1660345, EPI_ISL_1660346, EPI_ISL_1660347, EPI_ISL_1660348, EPI_ISL_1660349, EPI_ISL_1660350, EPI_ISL_1660351, EPI_ISL_1660352, EPI_ISL_1660353, EPI_ISL_1660354, EPI_ISL_1660355, EPI_ISL_1660356, EPI_ISL_1660357, EPI_ISL_1660358, EPI_ISL_1660359, EPI_ISL_1660360, EPI_ISL_1660361, EPI_ISL_1660362, EPI_ISL_1660363, EPI_ISL_1660364, EPI_ISL_1660365, EPI_ISL_1660366, EPI_ISL_1660367, EPI_ISL_1660368, EPI_ISL_1660369, EPI_ISL_1660370, EPI_ISL_1660371, EPI_ISL_1660372, EPI_ISL_1660373, EPI_ISL_1660374, EPI_ISL_1660375 | | | |
| see above | Laboratory of Clinical Microbiology, Virology and Bioemergencies, ASST Fatebenefratelli Sacco - Sacco Hospital | Laboratory of Clinical Microbiology, Virology and Bioemergencies, ASST Fatebenefratelli Sacco - Sacco Hospital | Valeria Micheli, Alessandro Mancon, Alberto Rizzo, Fiorenza Bracchitta, Luca Rizzuto, Maria Rita Gismondo |
| EPI_ISL_1670883 | Laboratorio Analisi Osp. Città di Castello - Azienda USL Umbria1 | Istituto Zooprofilattico Sperimentale dell'Abruzzo e Molise "G. Caporale" | Malagigi V., Tacconi P., Lorusso A, Marcacci M, Di Domenico M, Ancora M, Curini V, Mangone I, Rinaldi A, Delli Compagni E, Scialabba S, Di Pasquale A, Cammà C, Puglia I, Calistri P, Savini G |
| EPI_ISL_1670913, EPI_ISL_1670914, EPI_ISL_1670915, EPI_ISL_1670916, EPI_ISL_1670917, EPI_ISL_1670918, EPI_ISL_1670919, EPI_ISL_1670920, EPI_ISL_1670921, EPI_ISL_1670922, EPI_ISL_1670923, EPI_ISL_1670924, EPI_ISL_1670925, EPI_ISL_1670926, EPI_ISL_1670927, EPI_ISL_1670931, EPI_ISL_1670934, EPI_ISL_1670936, EPI_ISL_1670937, EPI_ISL_1670938 | | | |
| see above | IRCCS San Gallicano Dermatological Institute | IRCCS Regina Elena National Cancer Institute | Francesca De Nicola, Sara Donzelli, Grazia Prignano, Matteo Pallocca, Sabrina Strano, Fabrizio Ensoli, Fulvia Pimpinelli, Maurizio Fanciulli, Giovanni Blandino, Aldo Morrone, Gennaro Ciliberto |
| EPI_ISL_1692726, EPI_ISL_1692733, EPI_ISL_1692736, EPI_ISL_1692751, EPI_ISL_1692754, EPI_ISL_1692785, EPI_ISL_1692808 | Università Federico II - Dipartimento di scienze mediche traslazionali - Napoli | Telethon Institute of Genetics and Medicine (TIGEM) | Antonio Grimaldi Patrizia Annunziata Francesco Panariello Teresa Giuliano Michele Cennamo Valentina Bouche Chiara Colantuono Lucio Di Filippo Mariano Fiorenza Anna Manfredi Marcello Salvi Giuseppe Portella Andrea Ballabio Davide Cacchiarelli |
| EPI_ISL_1715059, EPI_ISL_1715060, EPI_ISL_1715061, EPI_ISL_1715062, EPI_ISL_1715064, EPI_ISL_1715065, EPI_ISL_1715066, EPI_ISL_1715067, EPI_ISL_1715068, EPI_ISL_1715069, EPI_ISL_1715070, EPI_ISL_1715071, EPI_ISL_1715072, EPI_ISL_1715073, EPI_ISL_1715074, EPI_ISL_1715075, EPI_ISL_1715076, EPI_ISL_1715077, EPI_ISL_1715078, EPI_ISL_1715080, EPI_ISL_1715081, EPI_ISL_1715082, EPI_ISL_1715083, EPI_ISL_1715084, EPI_ISL_1715086, EPI_ISL_1715087, EPI_ISL_1715088, EPI_ISL_1715089, EPI_ISL_1715090, EPI_ISL_1715091, EPI_ISL_1715092, EPI_ISL_1715093, EPI_ISL_1715094, EPI_ISL_1715095, EPI_ISL_1715096, EPI_ISL_1715097, EPI_ISL_1715098, EPI_ISL_1715099, EPI_ISL_1715101, EPI_ISL_1715102, EPI_ISL_1715103, EPI_ISL_1715104, EPI_ISL_1715105, EPI_ISL_1715107, EPI_ISL_1715108, EPI_ISL_1715109, EPI_ISL_1715110, EPI_ISL_1715111, EPI_ISL_1715112, EPI_ISL_1715115, EPI_ISL_1715116, EPI_ISL_1715117, EPI_ISL_1715118, EPI_ISL_1715119, EPI_ISL_1715121, EPI_ISL_1715122, EPI_ISL_1715123, EPI_ISL_1715124, EPI_ISL_1715125, EPI_ISL_1715126, EPI_ISL_1715128, EPI_ISL_1715129, EPI_ISL_1715130, EPI_ISL_1715131, EPI_ISL_1715132 | | | |
| see above | UOC laboratorio di analisi Istituto Giannina Gaslini | TIGEM | Antonio Grimaldi Patrizia Annunziata Francesco Panariello Teresa Giuliano Valentina Bouche Chiara Colantuono Lucio Di Filippo Anna Manfredi Marcello Salvi Andrea Ballabio Davide Cacchiarelli |
| EPI_ISL_1761103 | Medicine and Surgery, University of Insubria | Department of Molecular and Translational Medicine, Section of Microbiology, University of Brescia, ASST Spedali Civili, Brescia | Alberto Zani, Francesca Caccuri, Anna Bertelli, Serena Messali, Carlo Bonfanti, Arnaldo Caruso |
| EPI_ISL_1789729, EPI_ISL_1789730, EPI_ISL_1789731, EPI_ISL_1789732, EPI_ISL_1789734, EPI_ISL_1789767 | SC Microbiologia e Virologia AOUSS | AMES Centro Polidiagnostico Strumentale S.r.l. | Salvatore Rubino; Flavia Angioj; Laura Firino; Rosalba Govoni; Gabriele Ibba; Vincenzo Lai; Erica Mura; Bianca Paglietti; Claudia Piu; Anna Puggioni; Elena Rimini; Giulia Rocca; Caterina Serra; Sergio Uzzau. |
| EPI_ISL_1791141, EPI_ISL_1791142, EPI_ISL_1791314, EPI_ISL_1791315, EPI_ISL_1791457 | U.O. Microbiologia, Laboratorio Unico Centro Servizi - AUSL della Romagna | U.O. Microbiologia, Laboratorio Unico Centro Servizi - AUSL della Romagna | Giorgio Dirani, Silvia Zannoli, Vittorio Sambri |
| EPI_ISL_1818938 | Carate Brianza Hospital | Laboratory of Clinical Microbiology, Virology and Bioemergencies, ASST Fatebenefratelli Sacco - Sacco Hospital | Valeria Micheli, Alessandro Mancon, Alberto Rizzo, Fiorenza Bracchitta, Luca Rizzuto, Maria Rita Gismondo |
| EPI_ISL_1818939 | Laboratory of Clinical Microbiology, Virology and Bioemergencies, ASST Fatebenefratelli Sacco - Sacco Hospital | Laboratory of Clinical Microbiology, Virology and Bioemergencies, ASST Fatebenefratelli Sacco - Sacco Hospital | Valeria Micheli, Alessandro Mancon, Alberto Rizzo, Fiorenza Bracchitta, Luca Rizzuto, Maria Rita Gismondo |
| EPI_ISL_1841230, EPI_ISL_1841231, EPI_ISL_1841233, EPI_ISL_1841234, EPI_ISL_1841235 | Laboratorio Biologia Molecolare Sars Cov2 - UOC Laboratorio Analisi - Servizio Medicina di Laboratorio, Ospedale "San Francesco" - ATS-ASSL Nuoro | Laboratorio specialistico UOC Ematologia - Ospedale "San Francesco" - ATS-ASSL Nuoro | Giovanna Piras, Rosanna Asproni, Paolo Malune, Maura Fiamma, Maria Itria Monne, Giuseppe Mameli |
| EPI_ISL_1855099, EPI_ISL_1855100, EPI_ISL_1855101, EPI_ISL_1855102 | Virology Laboratory, Scientific Department, Army Medical Center | Virology Laboratory, Scientific Department, Army Medical Center | Silvia Fillo, Riccardo De Sanctis, Antonella Fortunato, Anella Monte, Anna Anselmo, Vanessa Vera Fain, Francesco Giordani, Giandomenico Cerreto, Filippo Molinari, Giancarlo Petralito, Florigio Lista |
| EPI_ISL_1857287, EPI_ISL_1857288, EPI_ISL_1857289, EPI_ISL_1857290, EPI_ISL_1857291, EPI_ISL_1857292, EPI_ISL_1857294 | Microbiology and Virology Laboratory, "Policlinico Riuniti, Azienda Ospedaliero Universitaria, Foggia" | Microbiology and Virology Laboratory, "Policlinico Riuniti, Azienda Ospedaliero Universitaria, Foggia" | Fabio Arena, Settimia Altamura, Donatella Cedola, Valeria Delli Carri, Annamaria D'Aprile, Maria Rosaria Lipsi, Loris Micelli, Maria Pia Patrizio, Daniela Pisanelli, Tiziana Rollo, Maurizio Margaglione and Rosella De Nittis. |
| EPI_ISL_1908157 | U.O. Microbiologia, Laboratorio Unico Centro Servizi - AUSL della Romagna | U.O. Microbiologia, Laboratorio Unico Centro Servizi - AUSL della Romagna | Giorgio Dirani, Silvia Zannoli, Vittorio Sambri |
| EPI_ISL_1924880 | Laboratorio di Microbiologia e Virologia, Università Vita-Salute San Raffaele, Milano | Laboratorio di Microbiologia e Virologia, Università Vita-Salute San Raffaele, Milano | Roberto Ferrarese, Elena Criscuolo, Michela Sampaolo, Matteo Castelli, Roberta Antonia Diotti, Enzo Boeri, Massimo Clementi, Nicasio Mancini, Nicola Clementi |
| EPI_ISL_2007588 | Dipartimento di Medicina e Chirurgia Universita' degli Studi | Laboratorio di Microbiologia e Virologia, Università Vita-Salute | Matteo Castelli, Roberto Ferrarese, Elena Criscuolo, Roberta Antonia Diotti, Massimo Clementi, Nicasio Mancini, Nicola Clementi |

| | | | |
|---|---|---|---|
| | dell'Insubria Laboratorio di Microbiologia Medica Varese Italia | San Raffaele, Milano | |
| EPI_ISL_2020643, EPI_ISL_2020644, EPI_ISL_2020645, EPI_ISL_2020646, EPI_ISL_2020647, EPI_ISL_2020648, EPI_ISL_2020649, EPI_ISL_2020650, EPI_ISL_2020651, EPI_ISL_2020652, EPI_ISL_2020653, EPI_ISL_2020654, EPI_ISL_2020655, EPI_ISL_2020656, EPI_ISL_2020657, EPI_ISL_2020658, EPI_ISL_2020659, EPI_ISL_2020660, EPI_ISL_2020661, EPI_ISL_2020662, EPI_ISL_2020663, EPI_ISL_2020664, EPI_ISL_2020665, EPI_ISL_2020666, EPI_ISL_2020667, EPI_ISL_2020668, EPI_ISL_2020669, EPI_ISL_2020670, EPI_ISL_2020671, EPI_ISL_2020672, EPI_ISL_2020673, EPI_ISL_2020677, EPI_ISL_2020678, EPI_ISL_2020680, EPI_ISL_2020681, EPI_ISL_2020682, EPI_ISL_2020683 | | | |
| see above | Università Federico II - Dipartimento di scienze mediche traslazionali - Napoli | TIGEM | Antonio Grimaldi Patrizia Annunziata Francesco Panariello Teresa Giuliano Michele Cennamo Valentina Bouche Chiara Colantuono Lucio Di Filippo Mariano Fiorenza Anna Manfredi Marcello Salvi Giuseppe Portella Andrea Ballabio Davide Cacchiarelli |
| EPI_ISL_2036762, EPI_ISL_2036763, EPI_ISL_2036764, EPI_ISL_2036765, EPI_ISL_2036766, EPI_ISL_2036767, EPI_ISL_2036768, EPI_ISL_2036769, EPI_ISL_2036770, EPI_ISL_2036771, EPI_ISL_2036772, EPI_ISL_2036773, EPI_ISL_2036774, EPI_ISL_2036775, EPI_ISL_2036776, EPI_ISL_2036777, EPI_ISL_2036778, EPI_ISL_2036779, EPI_ISL_2036780, EPI_ISL_2036781, EPI_ISL_2036782, EPI_ISL_2036783, EPI_ISL_2036784, EPI_ISL_2036785, EPI_ISL_2036786, EPI_ISL_2036787, EPI_ISL_2036788, EPI_ISL_2036789, EPI_ISL_2036790, EPI_ISL_2036791, EPI_ISL_2036792, EPI_ISL_2036793, EPI_ISL_2036794, EPI_ISL_2036795, EPI_ISL_2036796, EPI_ISL_2036797, EPI_ISL_2036798, EPI_ISL_2036799, EPI_ISL_2036800, EPI_ISL_2036801, EPI_ISL_2036802, EPI_ISL_2036803, EPI_ISL_2036804, EPI_ISL_2036805, EPI_ISL_2036806, EPI_ISL_2036807, EPI_ISL_2036808, EPI_ISL_2036809, EPI_ISL_2036810, EPI_ISL_2036811, EPI_ISL_2036812, EPI_ISL_2036813, EPI_ISL_2036814, EPI_ISL_2036815, EPI_ISL_2036816, EPI_ISL_2036817, EPI_ISL_2036818, EPI_ISL_2036819, EPI_ISL_2036820, EPI_ISL_2036821, EPI_ISL_2036822, EPI_ISL_2036823, EPI_ISL_2036824, EPI_ISL_2036825, EPI_ISL_2036826, EPI_ISL_2036827, EPI_ISL_2036828, EPI_ISL_2036829, EPI_ISL_2036830, EPI_ISL_2036831, EPI_ISL_2036832, EPI_ISL_2036833 | | | |
| see above | Virology Laboratory, Scientific Department, Army Medical Center | Virology Laboratory, Scientific Department, Army Medical Center | Silvia Fillo, Riccardo De Sanctis, Antonella Fortunato, Anella Monte, Anna Anselmo, Vanessa Vera Fain, Francesco Giordani, Giandomenico Cerreto, Filippo Molinari, Giancarlo Petralito, Florigio Lista |
| EPI_ISL_2036834, EPI_ISL_2036835, EPI_ISL_2036836, EPI_ISL_2036837, EPI_ISL_2036838, EPI_ISL_2036839, EPI_ISL_2036840, EPI_ISL_2036841, EPI_ISL_2036842, EPI_ISL_2036843, EPI_ISL_2036844, EPI_ISL_2036845, EPI_ISL_2036846, EPI_ISL_2036847, EPI_ISL_2036848, EPI_ISL_2036849, EPI_ISL_2036850, EPI_ISL_2036851, EPI_ISL_2036852, EPI_ISL_2036853, EPI_ISL_2036854, EPI_ISL_2036855, EPI_ISL_2036856, EPI_ISL_2036857, EPI_ISL_2036858, EPI_ISL_2036859, EPI_ISL_2036860, EPI_ISL_2036861, EPI_ISL_2036862, EPI_ISL_2036863, EPI_ISL_2036864, EPI_ISL_2036865, EPI_ISL_2036866, EPI_ISL_2036867, EPI_ISL_2036868, EPI_ISL_2036869, EPI_ISL_2036870, EPI_ISL_2036871, EPI_ISL_2036898, EPI_ISL_2036902, EPI_ISL_2036904, EPI_ISL_2036905, EPI_ISL_2036906, EPI_ISL_2036907, EPI_ISL_2036908, EPI_ISL_2036909, EPI_ISL_2036910, EPI_ISL_2036911, EPI_ISL_2036914 | | | |
| see above | Virology Laboratory, Scientific Department, Army Medical Center | Virology Laboratory, Scientific Department, Army Medical Center | Silvia Fillo, Riccardo De Sanctis, Antonella Fortunato, Anella Monte, Rossella Brandi, Giulia Campoli, Marzia Cavalli, Lucia Nicosia, Anna Anselmo, Vanessa Vera Fain, Francesco Giordani, Giandomenico Cerreto, Filippo Molinari, Giancarlo Petralito, Florigio Lista |
| EPI_ISL_2047852 | Università degli Studi di Perugia | Istituto Zooprofilattico Sperimentale dell'Abruzzo e Molise "G. Caporale" | Mencacci A., Camilloni B., Lorusso A, Marcacci M, Di Domenico M, Ancora M, Curini V, Mangone I, Rinaldi A, Delli Compagni E, Scialabba S, Di Pasquale A, Cammà C, Puglia I, Calistri P, Savini G |
| EPI_ISL_2047853 | Istituto Zooprofilattico Sperimentale Umbria e Marche "Togo Rosati" | Istituto Zooprofilattico Sperimentale dell'Abruzzo e Molise "G. Caporale" | Biagetti M., Giammarioli M., Lorusso A, Marcacci M, Di Domenico M, Ancora M, Curini V, Mangone I, Rinaldi A, Delli Compagni E, Scialabba S, Di Pasquale A, Cammà C, Puglia I, Calistri P, Savini G |
| EPI_ISL_2047854 | Istituto Zooprofilattico Sperimentale Umbria e Marche "Togo Rosati" | Istituto Zooprofilattico Sperimentale dell'Abruzzo e Molise "G. Caporale" | Biagetti M, Giammarioli M, Lorusso A, Marcacci M, Di Domenico M, Ancora M, Curini V, Mangone I, Rinaldi A, Delli Compagni E, Scialabba S, Di Pasquale A, Cammà C, Puglia I, Calistri P, Savini G |
| EPI_ISL_2047855, EPI_ISL_2047856 | Azienda Ospedaliera Terni | Istituto Zooprofilattico Sperimentale dell'Abruzzo e Molise "G. Caporale" | Palumbo M, Scaccetti A, Lorusso A, Marcacci M, Di Domenico M, Ancora M, Curini V, Mangone I, Rinaldi A, Delli Compagni E, Scialabba S, Di Pasquale A, Cammà C, Puglia I, Calistri P, Savini G |
| EPI_ISL_2047857 | Azienda Ospedaliera Terni | Istituto Zooprofilattico Sperimentale dell'Abruzzo e Molise "G. Caporale" | Palumbo A, Scaccetti A, Lorusso A, Marcacci M, Di Domenico M, Ancora M, Curini V, Mangone I, Rinaldi A, Delli Compagni E, Scialabba S, Di Pasquale A, Cammà C, Puglia I, Calistri P, Savini G |
| EPI_ISL_2047858 | Azienda USL Umbria 2 | Istituto Zooprofilattico Sperimentale dell'Abruzzo e Molise "G. Caporale" | Proietti A, PistoniE, Lorusso A, Marcacci M, Di Domenico M, Ancora M, Curini V, Mangone I, Rinaldi A, Delli Compagni E, Scialabba S, Di Pasquale A, Cammà C, Puglia I, Calistri P, Savini G |
| EPI_ISL_2047873 | Azienda USL Umbria 2 | Istituto Zooprofilattico Sperimentale dell'Abruzzo e Molise "G. Caporale" | Proietti A, Pistoni E, Lorusso A, Marcacci M, Di Domenico M, Ancora M, Curini V, Mangone I, Rinaldi A, Delli Compagni E, Scialabba S, Di Pasquale A, Cammà C, Puglia I, Calistri P, Savini G |
| EPI_ISL_2080867, EPI_ISL_2080868, EPI_ISL_2080869, EPI_ISL_2080870, EPI_ISL_2080871, EPI_ISL_2080872 | Center of Advanced Studies and Technology, Molecular Genetics Laboratory | Center of Advanced Studies and Technology, Molecular Genetics Laboratory | Anaclerio Federico, Ferrante Rossella, De Fabritiis Simone, Damiani Verena, Mandatori Domitilla |
| EPI_ISL_2086187, EPI_ISL_2086188, EPI_ISL_2086189, EPI_ISL_2086190, EPI_ISL_2086191, EPI_ISL_2086192, EPI_ISL_2086193, EPI_ISL_2086194, EPI_ISL_2086195, EPI_ISL_2086196 | CQRC_QUALITY CONTROL CHEMICAL BIOLOGICAL RISK_AOOR Villa Sofia Cervello Palermo | CQRC_QUALITY CONTROL CHEMICAL BIOLOGICAL RISK_AOOR Villa Sofia Cervello Palermo | Di Gaudio, F., Contino, F., Brunacci, G.. |
| EPI_ISL_2086197, EPI_ISL_2086198, EPI_ISL_2086199, EPI_ISL_2086200, EPI_ISL_2086201, EPI_ISL_2086202, EPI_ISL_2086203, EPI_ISL_2086204, EPI_ISL_2086205, EPI_ISL_2086206, EPI_ISL_2086207, EPI_ISL_2086208, EPI_ISL_2086209, EPI_ISL_2086210 | | | |
| see above | CQRC_QUALITY CONTROL CHEMICAL BIOLOGICAL RISK_AOOR Villa Sofia Cervello Palermo | CQRC_QUALITY CONTROL CHEMICAL BIOLOGICAL RISK_AOOR Villa Sofia Cervello Palermo | Di Gaudio, F., Brunacci, G., Contino, F. |
| EPI_ISL_2098847, EPI_ISL_2098848, EPI_ISL_2098849, EPI_ISL_2098850, EPI_ISL_2098851, EPI_ISL_2098852, EPI_ISL_2098853, EPI_ISL_2098854, EPI_ISL_2098855, EPI_ISL_2098856, EPI_ISL_2098857, EPI_ISL_2098858, EPI_ISL_2098859 | | | |
| see above | CQRC_QUALITY CONTROL CHEMICAL BIOLOGICAL RISK_AOOR Villa Sofia Cervello Palermo | CQRC_QUALITY CONTROL CHEMICAL BIOLOGICAL RISK_AOOR Villa Sofia Cervello Palermo | Di Gaudio, F., Contino, F., Brunacci, G. |
| EPI_ISL_2098860, EPI_ISL_2098861, EPI_ISL_2098862, EPI_ISL_2098863, EPI_ISL_2098864, EPI_ISL_2098865, EPI_ISL_2098866, EPI_ISL_2098867, EPI_ISL_2098868, EPI_ISL_2098869, EPI_ISL_2098870, EPI_ISL_2098871 | | | |
| see above | CQRC_QUALITY CONTROL CHEMICAL BIOLOGICAL RISK_AOOR Villa Sofia Cervello Palermo | CQRC_QUALITY CONTROL CHEMICAL BIOLOGICAL RISK_AOOR Villa Sofia Cervello Palermo | Di Gaudio, F., Brunacci G., Contino, F. |
| EPI_ISL_2099949, EPI_ISL_2099950, EPI_ISL_2099951, EPI_ISL_2099952, EPI_ISL_2099953, EPI_ISL_2099954 | CQRC_QUALITY CONTROL CHEMICAL BIOLOGICAL RISK_AOOR Villa Sofia Cervello Palermo | CQRC_QUALITY CONTROL CHEMICAL BIOLOGICAL RISK_AOOR Villa Sofia Cervello Palermo | Di Gaudio, F., Contino, F., Brunacci, G. |
| EPI_ISL_2099955 | CQRC_QUALITY CONTROL CHEMICAL BIOLOGICAL RISK_AOOR Villa Sofia Cervello Palermo | CQRC_QUALITY CONTROL CHEMICAL BIOLOGICAL RISK_AOOR Villa Sofia Cervello Palermo | Di Gaudio, F., Contino, F., Gasparro G., Rinaudo G., Brunacci, G. |
| EPI_ISL_2099956, EPI_ISL_2099957, EPI_ISL_2099958, EPI_ISL_2099959, EPI_ISL_2099960, EPI_ISL_2099961, EPI_ISL_2099962, EPI_ISL_2099963, EPI_ISL_2099964 | CQRC_QUALITY CONTROL CHEMICAL BIOLOGICAL RISK_AOOR Villa Sofia Cervello Palermo | CQRC_QUALITY CONTROL CHEMICAL BIOLOGICAL RISK_AOOR Villa Sofia Cervello Palermo | Di Gaudio, Brunacci, G., F. Contino, F. |
| EPI_ISL_2099965 | CQRC_QUALITY CONTROL CHEMICAL BIOLOGICAL RISK_AOOR Villa Sofia Cervello Palermo | CQRC_QUALITY CONTROL CHEMICAL BIOLOGICAL RISK_AOOR Villa Sofia Cervello Palermo | Di Gaudio, F., Contino, F., Gasparro G., Rinaudo G., Brunacci, G. |
| EPI_ISL_2099966 | CQRC_QUALITY CONTROL CHEMICAL BIOLOGICAL RISK_AOOR Villa Sofia Cervello Palermo | CQRC_QUALITY CONTROL CHEMICAL BIOLOGICAL RISK_AOOR Villa Sofia Cervello Palermo | Di Gaudio, Brunacci, G., F. Contino, F. |
| EPI_ISL_2137620, EPI_ISL_2137621, EPI_ISL_2137622, EPI_ISL_2137623, EPI_ISL_2137624, EPI_ISL_2137625, EPI_ISL_2137626, EPI_ISL_2137627, EPI_ISL_2137628, EPI_ISL_2137629, EPI_ISL_2137630, EPI_ISL_2137631, EPI_ISL_2137632, EPI_ISL_2137633, EPI_ISL_2137634, EPI_ISL_2137635, EPI_ISL_2137636, EPI_ISL_2137637, EPI_ISL_2137638, EPI_ISL_2137639, EPI_ISL_2137640, EPI_ISL_2137641 | | | |
| see above | Istituto Zooprofilattico Sperimentale del Mezzogiorno - Azienda Ospedaliera "Pugliese Ciaccio" di Catanzaro | TIGEM | Antonio Grimaldi Patrizia Annunziata Francesco Panariello Biancamaria Pierri Claudia Tiberio Teresa Giuliano Valentina Bouche Chiara Colantuono Maria Concetta Cuomo Denise Di Concilio Lucio Di Filippo Anna Manfredi Pasquale Minchella Marcello Salvi Antonio Limone Luigi Atripaldi Pellegrino Cerino Andrea Ballabio Davide Cacchiarelli |
| EPI_ISL_2143846 | Istituto Zooprofilattico Sperimentale del Mezzogiorno | Telethon Institute of Genetics and Medicine (TIGEM) | Antonio Grimaldi Patrizia Annunziata Francesco Panariello Biancamaria Pierri Claudia Tiberio Teresa Giuliano Valentina Bouche Chiara Colantuono Maria Concetta Cuomo Denise Di Concilio Lucio Di Filippo Anna Manfredi Marcello Salvi Antonio Limone Luigi Atripaldi Pellegrino Cerino Andrea Ballabio Davide Cacchiarelli |
| EPI_ISL_755574 | Center of Advanced Studies and Technology, CAST | Center of Advanced Studies and Technology, CAST | Ferrante,R., Mandatori,D., De Fabritiis,S. |

| | | | |
|---|---|---|---|
| EPI_ISL_765571 | National Institute for Infectious Diseases, INMI, "L. Spallanzani" IRCCS | National Institute for Infectious Diseases, INMI, "L. Spallanzani" IRCCS | E Giombini, B Bartolini, O Butera, C.E.M Gruber, M Rueca, F Messina, MR Capobianchi, A Di Caro |
| EPI_ISL_767013 | University of Bari Biomedical Sciences and Human Oncology | University of Bari Biomedical Sciences and Human Oncology | Maria Chironna, Anna Sallustio, Daniela Loconsole, Marisa Accogli |
| EPI_ISL_803900 | National Institute for Infectious Diseases, INMI, "L. Spallanzani" IRCCS | National Institute for Infectious Diseases, INMI, "L. Spallanzani" IRCCS | B Bartolini, O Butera, C.E.M Gruber, M Rueca, F Messina, E Giombini, MR Capobianchi, A Di Caro |
| EPI_ISL_804040 | SC (UCO) Igiene e Sanità Pubblica (funzione integrata con SC Microbiologia e Virologia) e Laboratory of Molecular Virology of the International Centre for Genetic Engineering and Biotechnology (ICGEB) | ARGO Laboratorio Genomica ed Epigenomica | Licastro D, Dal Monego S, Degasperi M, Marcello A, D'Agaro P |
| EPI_ISL_825156 | Microbiology and Virology Unit, Florence Careggi University Hospital | Microbiology and Virology Unit, Florence Careggi University Hospital | Vincenzo Di Pilato, Marco Coppi, Fabio Morecchiato, Alberto Antonelli, Emanuele Gori, Gian Maria Rossolini |
| EPI_ISL_826458 | University of Bari Biomedical Sciences and Human Oncology | University of Bari Biomedical Sciences and Human Oncology | Chironna Maria, Sallustio Anna, Loconsole Daniela, Accogli Marisa |
| EPI_ISL_833228, EPI_ISL_833229 | R.P. GUARDIAGRELE Ospedale di Comunità | Istitituto Zooprofilattico Sperimentale dell'Abruzzo e Molise "G.Caporale" | Lorusso A, Marcacci M, Di Domenico M, Curini V, Ancora M, Cammà C, Rinaldi A, Mangone I, Di Pasquale A, Puglia I, Calistri P, Savini G. |
| EPI_ISL_833230 | OSPEDALE S.S. ANNUNZIATA CHIETI - CLINICA MEDICA (MEDICINA GENERALE 1) | Istitituto Zooprofilattico Sperimentale dell'Abruzzo e Molise "G.Caporale" | Lorusso A, Marcacci M, Di Domenico M, Curini V, Ancora M, Cammà C, Rinaldi A, Mangone I, Di Pasquale A, Puglia I, Calistri P, Savini G. |
| EPI_ISL_833231 | SIESP CHIETI - DRIVE IN LANCIANO | Istitituto Zooprofilattico Sperimentale dell'Abruzzo e Molise "G.Caporale" | Lorusso A, Marcacci M, Di Domenico M, Curini V, Ancora M, Cammà C, Rinaldi A, Mangone I, Di Pasquale A, Puglia I, Calistri P, Savini G. |
| EPI_ISL_833232 | SIESP CHIETI - DRIVE IN CHIETI | Istitituto Zooprofilattico Sperimentale dell'Abruzzo e Molise "G.Caporale" | Lorusso A, Marcacci M, Di Domenico M, Curini V, Ancora M, Cammà C, Rinaldi A, Mangone I, Di Pasquale A, Puglia I, Calistri P, Savini G. |
| EPI_ISL_833233, EPI_ISL_833234, EPI_ISL_833235, EPI_ISL_833236, EPI_ISL_833237, EPI_ISL_833238 | SIESP DIPARTIMENTO DI PREVENZIONE CHIETI | Istitituto Zooprofilattico Sperimentale dell'Abruzzo e Molise "G.Caporale" | Lorusso A, Marcacci M, Di Domenico M, Curini V, Ancora M, Cammà C, Rinaldi A, Mangone I, Di Pasquale A, Puglia I, Calistri P, Savini G. |
| EPI_ISL_833239, EPI_ISL_833240 | SIESP DIPARTIMENTO DI PREVENZIONE TERAMO | Istitituto Zooprofilattico Sperimentale dell'Abruzzo e Molise "G.Caporale" | Lorusso A, Marcacci M, Di Domenico M, Curini V, Ancora M, Cammà C, Rinaldi A, Mangone I, Di Pasquale A, Puglia I, Calistri P, Savini G. |
| EPI_ISL_833241 | SIESP DIPARTIMENTO DI PREVENZIONE CHIETI | Istitituto Zooprofilattico Sperimentale dell'Abruzzo e Molise "G.Caporale" | Lorusso A, Marcacci M, Di Domenico M, Curini V, Ancora M, Cammà C, Rinaldi A, Mangone I, Di Pasquale A, Puglia I, Calistri P, Savini G. |
| EPI_ISL_833242 | SIESP CHIETI - DRIVE IN LANCIANO | Istitituto Zooprofilattico Sperimentale dell'Abruzzo e Molise "G.Caporale" | Lorusso A, Marcacci M, Di Domenico M, Curini V, Ancora M, Cammà C, Rinaldi A, Mangone I, Di Pasquale A, Puglia I, Calistri P, Savini G. |
| EPI_ISL_833243, EPI_ISL_833244, EPI_ISL_833245, EPI_ISL_833246, EPI_ISL_833247 | SIESP DIPARTIMENTO DI PREVENZIONE CHIETI | Istitituto Zooprofilattico Sperimentale dell'Abruzzo e Molise "G.Caporale" | Lorusso A, Marcacci M, Di Domenico M, Curini V, Ancora M, Cammà C, Rinaldi A, Mangone I, Di Pasquale A, Puglia I, Calistri P, Savini G. |
| EPI_ISL_833251 | Ospedale Civile Atri-Medicina Interna | Istituto Zooprofilattico Sperimentale dell'Abruzzo e Molise "G. Caporale" | Lorusso A, Marcacci M, Di Domenico M, Ancora M, Curini V, Mangone I, Rinaldi A, Di Pasquale A, Cammà C, Puglia I, Calistri P, Savini G |
| EPI_ISL_833252 | Ospedale Civile Giulianova-Pronto Soccorso | Istituto Zooprofilattico Sperimentale dell'Abruzzo e Molise "G. Caporale" | Lorusso A, Marcacci M, Di Domenico M, Ancora M, Curini V, Mangone I, Rinaldi A, Di Pasquale A, Cammà C, Puglia I, Calistri P, Savini G |
| EPI_ISL_833253 | USCA-Avezzano | Istituto Zooprofilattico Sperimentale dell'Abruzzo e Molise "G. Caporale" | Lorusso A, Marcacci M, Di Domenico M, Ancora M, Curini V, Mangone I, Rinaldi A, Di Pasquale A, Cammà C, Puglia I, Calistri P, Savini G |
| EPI_ISL_833254 | USCA-Pescina | Istituto Zooprofilattico Sperimentale dell'Abruzzo e Molise "G. Caporale" | Lorusso A, Marcacci M, Di Domenico M, Ancora M, Curini V, Mangone I, Rinaldi A, Di Pasquale A, Cammà C, Puglia I, Calistri P, Savini G |
| EPI_ISL_833255, EPI_ISL_833256, EPI_ISL_833257 | Ospedale Civile Atri | Istituto Zooprofilattico Sperimentale dell'Abruzzo e Molise "G. Caporale" | Lorusso A, Marcacci M, Di Domenico M, Ancora M, Curini V, Mangone I, Rinaldi A, Di Pasquale A, Cammà C, Puglia I, Calistri P, Savini G |
| EPI_ISL_833258, EPI_ISL_833259 | SIESP DIPARTIMENTO DI PREVENZIONE TERAMO | Istituto Zooprofilattico Sperimentale dell'Abruzzo e Molise "G. Caporale" | Lorusso A, Marcacci M, Di Domenico M, Ancora M, Curini V, Mangone I, Rinaldi A, Di Pasquale A, Cammà C, Puglia I, Calistri P, Savini G |
| EPI_ISL_833260, EPI_ISL_833261 | Giulianova | Istituto Zooprofilattico Sperimentale dell'Abruzzo e Molise "G. Caporale" | Lorusso A, Marcacci M, Di Domenico M, Ancora M, Curini V, Mangone I, Rinaldi A, Di Pasquale A, Cammà C, Puglia I, Calistri P, Savini G |
| EPI_ISL_833262, EPI_ISL_833263, EPI_ISL_833264 | SIESP DIPARTIMENTO DI PREVENZIONE CHIETI | Istituto Zooprofilattico Sperimentale dell'Abruzzo e Molise "G. Caporale" | Lorusso A, Marcacci M, Di Domenico M, Ancora M, Curini V, Mangone I, Rinaldi A, Di Pasquale A, Cammà C, Puglia I, Calistri P, Savini G |
| EPI_ISL_833265, EPI_ISL_833266, EPI_ISL_833267, EPI_ISL_833268, EPI_ISL_833269 | Ospedale SS Annunziata Chieti | Istituto Zooprofilattico Sperimentale dell'Abruzzo e Molise "G. Caporale" | Lorusso A, Marcacci M, Di Domenico M, Ancora M, Curini V, Mangone I, Rinaldi A, Di Pasquale A, Cammà C, Puglia I, Calistri P, Savini G |
| EPI_ISL_833270, EPI_ISL_833272, EPI_ISL_833273, EPI_ISL_833274, EPI_ISL_833275 | SIESP CHIETI - DRIVE IN ORTONA | Istituto Zooprofilattico Sperimentale dell'Abruzzo e Molise "G. Caporale" | Lorusso A, Marcacci M, Di Domenico M, Ancora M, Curini V, Mangone I, Rinaldi A, Di Pasquale A, Cammà C, Puglia I, Calistri P, Savini G |
| EPI_ISL_833276 | Ospedale Civile G.Mazzini-Teramo | Istituto Zooprofilattico Sperimentale dell'Abruzzo e Molise "G. Caporale" | Lorusso A, Marcacci M, Di Domenico M, Ancora M, Curini V, Mangone I, Rinaldi A, Di Pasquale A, Cammà C, Puglia I, Calistri P, Savini G |
| EPI_ISL_833278, EPI_ISL_833279, EPI_ISL_833280 | SIESP CHIETI- DRIVE IN CHIETI | Istituto Zooprofilattico Sperimentale dell'Abruzzo e Molise "G. Caporale" | Lorusso A, Marcacci M, Di Domenico M, Ancora M, Curini V, Mangone I, Rinaldi A, Di Pasquale A, Cammà C, Puglia I, Calistri P, Savini G |
| EPI_ISL_833281, EPI_ISL_833282 | SIESP DIPARTIMENTO DI PREVENZIONE TERAMO | Istituto Zooprofilattico Sperimentale dell'Abruzzo e Molise "G. Caporale" | Lorusso A, Marcacci M, Di Domenico M, Ancora M, Curini V, Mangone I, Rinaldi A, Di Pasquale A, Cammà C, Puglia I, Calistri P, Savini G |
| EPI_ISL_833283 | DIPARTIMENTO PREVENZIONE AVEZZANO-SERVIZIO DI IGIENE EPIDEMIOLOGIA E SANITA' PUBBLICA | Istituto Zooprofilattico Sperimentale dell'Abruzzo e Molise "G. Caporale" | Lorusso A, Marcacci M, Di Domenico M, Ancora M, Curini V, Mangone I, Rinaldi A, Di Pasquale A, Cammà C, Puglia I, Calistri P, Savini G |
| EPI_ISL_833284, EPI_ISL_833285, EPI_ISL_833286, EPI_ISL_833287 | SIESP CHIETI - DRIVE IN ORTONA | Istituto Zooprofilattico Sperimentale dell'Abruzzo e Molise "G. Caporale" | Lorusso A, Marcacci M, Di Domenico M, Ancora M, Curini V, Mangone I, Rinaldi A, Di Pasquale A, Cammà C, Puglia I, Calistri P, Savini G |
| EPI_ISL_833288, EPI_ISL_833290 | PRESIDIO TERRITORIALE CASOLI CH | Istituto Zooprofilattico Sperimentale dell'Abruzzo e Molise "G. Caporale" | Lorusso A, Marcacci M, Di Domenico M, Ancora M, Curini V, Mangone I, Rinaldi A, Di Pasquale A, Cammà C, Puglia I, Calistri P, Savini G |
| EPI_ISL_833291 | SIESP DIPARTIMENTO DI PREVENZIONE DELL'AQUILA | Istituto Zooprofilattico Sperimentale dell'Abruzzo e Molise "G. Caporale" | Lorusso A, Marcacci M, Di Domenico M, Ancora M, Curini V, Mangone I, Rinaldi A, Di Pasquale A, Cammà C, Puglia I, Calistri P, Savini G |
| EPI_ISL_833292, EPI_ISL_833293, EPI_ISL_833294 | SIESP CHIETI - DRIVE IN LANCIANO | Istituto Zooprofilattico Sperimentale dell'Abruzzo e Molise "G. Caporale" | Lorusso A, Marcacci M, Di Domenico M, Ancora M, Curini V, Mangone I, Rinaldi A, Di Pasquale A, Cammà C, Puglia I, Calistri P, Savini G |
| EPI_ISL_833295, EPI_ISL_833296 | SIESP DIPARTIMENTO DI PREVENZIONE TERAMO | Istituto Zooprofilattico Sperimentale dell'Abruzzo e Molise "G. | Lorusso A, Marcacci M, Di Domenico M, Ancora M, Curini V, Mangone I, Rinaldi A, Di Pasquale A, Cammà C, Puglia I, Calistri P, Savini G |

| | | | |
|---|---|---|---|
| | | Caporale" | |
| EPI_ISL_833297 | Ospedale Civile Giulianova | Istituto Zooprofilattico Sperimentale dell'Abruzzo e Molise "G. Caporale" | Lorusso A, Marcacci M, Di Domenico M, Ancora M, Curini V, Mangone I, Rinaldi A, Di Pasquale A, Cammà C, Puglia I, Calistri P, Savini G |
| EPI_ISL_833298, EPI_ISL_833300 | SIESP CHIETI - DRIVE IN LANCIANO | Istituto Zooprofilattico Sperimentale dell'Abruzzo e Molise "G. Caporale" | Lorusso A, Marcacci M, Di Domenico M, Ancora M, Curini V, Mangone I, Rinaldi A, Di Pasquale A, Cammà C, Puglia I, Calistri P, Savini G |
| EPI_ISL_833301 | DIPARTIMENTO PREVENZIONE AVEZZANO-SERVIZIO DI IGIENE EPIDEMIOLOGIA E SANITA' PUBBLICA | Istituto Zooprofilattico Sperimentale dell'Abruzzo e Molise "G. Caporale" | Lorusso A, Marcacci M, Di Domenico M, Ancora M, Curini V, Mangone I, Rinaldi A, Di Pasquale A, Cammà C, Puglia I, Calistri P, Savini G |
| EPI_ISL_833302, EPI_ISL_833303, EPI_ISL_833304, EPI_ISL_833305 | SIESP DIPARTIMENTO DI PREVENZIONE TERAMO | Istituto Zooprofilattico Sperimentale dell'Abruzzo e Molise "G. Caporale" | Lorusso A, Marcacci M, Di Domenico M, Ancora M, Curini V, Mangone I, Rinaldi A, Di Pasquale A, Cammà C, Puglia I, Calistri P, Savini G |
| EPI_ISL_833306, EPI_ISL_833307, EPI_ISL_833308, EPI_ISL_833309 | SIESP CHIETI - DRIVE IN ORTONA | Istituto Zooprofilattico Sperimentale dell'Abruzzo e Molise "G. Caporale" | Lorusso A, Marcacci M, Di Domenico M, Ancora M, Curini V, Mangone I, Rinaldi A, Di Pasquale A, Cammà C, Puglia I, Calistri P, Savini G |
| EPI_ISL_833311, EPI_ISL_833312 | SIESP CHIETI- DRIVE IN CHIETI | Istituto Zooprofilattico Sperimentale dell'Abruzzo e Molise "G. Caporale" | Lorusso A, Marcacci M, Di Domenico M, Ancora M, Curini V, Mangone I, Rinaldi A, Di Pasquale A, Cammà C, Puglia I, Calistri P, Savini G |
| EPI_ISL_833313, EPI_ISL_833314, EPI_ISL_833315 | SIESP CHIETI - DRIVE IN LANCIANO | Istituto Zooprofilattico Sperimentale dell'Abruzzo e Molise "G. Caporale" | Lorusso A, Marcacci M, Di Domenico M, Ancora M, Curini V, Mangone I, Rinaldi A, Di Pasquale A, Cammà C, Puglia I, Calistri P, Savini G |
| EPI_ISL_833316 | SIESP CHIETI - DRIVE IN GISSI | Istituto Zooprofilattico Sperimentale dell'Abruzzo e Molise "G. Caporale" | Lorusso A, Marcacci M, Di Domenico M, Ancora M, Curini V, Mangone I, Rinaldi A, Di Pasquale A, Cammà C, Puglia I, Calistri P, Savini G |
| EPI_ISL_833317, EPI_ISL_833318 | SIESP CHIETI - DRIVE IN CHIETI | Istituto Zooprofilattico Sperimentale dell'Abruzzo e Molise "G. Caporale" | Lorusso A, Marcacci M, Di Domenico M, Ancora M, Curini V, Mangone I, Rinaldi A, Di Pasquale A, Cammà C, Puglia I, Calistri P, Savini G |
| EPI_ISL_833319, EPI_ISL_833320, EPI_ISL_833322 | SIESP DIPARTIMENTO DI PREVENZIONE SULMONA | Istituto Zooprofilattico Sperimentale dell'Abruzzo e Molise "G. Caporale" | Lorusso A, Marcacci M, Di Domenico M, Ancora M, Curini V, Mangone I, Rinaldi A, Di Pasquale A, Cammà C, Puglia I, Calistri P, Savini G |
| EPI_ISL_833323 | SIESP CHIETI - DRIVE IN CHIETI | Istituto Zooprofilattico Sperimentale dell'Abruzzo e Molise "G. Caporale" | Lorusso A, Marcacci M, Di Domenico M, Ancora M, Curini V, Mangone I, Rinaldi A, Di Pasquale A, Cammà C, Puglia I, Calistri P, Savini G |
| EPI_ISL_833324 | SIESP DIPARTIMENTO DI PREVENZIONE CHIETI | Istituto Zooprofilattico Sperimentale dell'Abruzzo e Molise "G. Caporale" | Lorusso A, Marcacci M, Di Domenico M, Ancora M, Curini V, Mangone I, Rinaldi A, Di Pasquale A, Cammà C, Puglia I, Calistri P, Savini G |
| EPI_ISL_833325 | Ospedale SS Annunziata Chieti | Istituto Zooprofilattico Sperimentale dell'Abruzzo e Molise "G. Caporale" | Lorusso A, Marcacci M, Di Domenico M, Ancora M, Curini V, Mangone I, Rinaldi A, Di Pasquale A, Cammà C, Puglia I, Calistri P, Savini G |
| EPI_ISL_833326 | SIESP DIPARTIMENTO DI PREVENZIONE CHIETI | Istituto Zooprofilattico Sperimentale dell'Abruzzo e Molise "G. Caporale" | Lorusso A, Marcacci M, Di Domenico M, Ancora M, Curini V, Mangone I, Rinaldi A, Di Pasquale A, Cammà C, Puglia I, Calistri P, Savini G |
| EPI_ISL_833327 | Ospedale SS Annunziata Chieti | Istituto Zooprofilattico Sperimentale dell'Abruzzo e Molise "G. Caporale" | Lorusso A, Marcacci M, Di Domenico M, Ancora M, Curini V, Mangone I, Rinaldi A, Di Pasquale A, Cammà C, Puglia I, Calistri P, Savini G |
| EPI_ISL_833328, EPI_ISL_833329, EPI_ISL_833330 | SIESP CHIETI - DRIVE IN ORTONA | Istituto Zooprofilattico Sperimentale dell'Abruzzo e Molise "G. Caporale" | Lorusso A, Marcacci M, Di Domenico M, Ancora M, Curini V, Mangone I, Rinaldi A, Di Pasquale A, Cammà C, Puglia I, Calistri P, Savini G |
| EPI_ISL_833331 | Ospedale Civile Atri | Istituto Zooprofilattico Sperimentale dell'Abruzzo e Molise "G. Caporale" | Lorusso A, Marcacci M, Di Domenico M, Ancora M, Curini V, Mangone I, Rinaldi A, Di Pasquale A, Cammà C, Puglia I, Calistri P, Savini G |
| EPI_ISL_833435, EPI_ISL_833436, EPI_ISL_833437, EPI_ISL_833438, EPI_ISL_833439, EPI_ISL_833440, EPI_ISL_833441, EPI_ISL_833442, EPI_ISL_833443, EPI_ISL_833444, EPI_ISL_833445, EPI_ISL_833446 | | | |
| see above | SC (UCO) Igiene e Sanità Pubblica (funzione integrata con SC Microbiologia e Virologia) e Laboratory of Molecular Virology of the International Centre for Genetic Engineering and Biotechnology (ICGEB) | ARGO Laboratorio Genomica ed Epigenomica | Licastro D, Dal Monego S, Degasperi M, Marcello A, D'Agaro P |
| EPI_ISL_837254, EPI_ISL_837326, EPI_ISL_837327, EPI_ISL_837328, EPI_ISL_837329, EPI_ISL_837330, EPI_ISL_837448, EPI_ISL_837483, EPI_ISL_837484, EPI_ISL_837485 | Istituto Zooprofilattico Sperimentale del Mezzogiorno | TIGEM | Antonio Grimaldi, Patrizia Annunziata, Francesco Panariello, Biancamaria Pierri, Valentina Bouche, Chiara Colantuono, Maria Concetta Cuomo, Denise Di Concilio, Lucio Di Filippo, Anna Manfredi, Marcello Salvi, Antonio Limone, Pellegrino Cerino, Andrea Ballabio, Davide Cacchiarelli. |
| EPI_ISL_847825 | Lab. Microbiologia e Virologia, Cotugno, A.O. dei Colli | Lab. Microbiologia e Virologia, Cotugno, A.O. dei Colli | Luigi Atripaldi, Claudia Tiberio, Anna Perfetti |
| EPI_ISL_849634 | SIESP DIPARTIMENTO DI PREVENZIONE CHIETI | Istitituto Zooprofilattico Sperimentale dell'Abruzzo e Molise "G.Caporale" | Lorusso A, Marcacci M, Di Domenico M, Curini V, Ancora M, Cammà C, Rinaldi A, Mangone I, Di Pasquale A, Puglia I, Savini G. |
| EPI_ISL_849635, EPI_ISL_849636 | DIPARTIMENTO PREVENZIONE AVEZZANO-SERVIZIO DI IGIENE EPIDEMIOLOGIA E SANITA' PUBBLICA | Istitituto Zooprofilattico Sperimentale dell'Abruzzo e Molise "G.Caporale" | Lorusso A, Marcacci M, Di Domenico M, Curini V, Ancora M, Cammà C, Rinaldi A, Mangone I, Di Pasquale A, Puglia I, Savini G. |
| EPI_ISL_849637, EPI_ISL_849638, EPI_ISL_849639, EPI_ISL_849640, EPI_ISL_849641, EPI_ISL_849642, EPI_ISL_849643, EPI_ISL_849644, EPI_ISL_849645, EPI_ISL_849646 | SIESP DIPARTIMENTO DI PREVENZIONE CHIETI | Istitituto Zooprofilattico Sperimentale dell'Abruzzo e Molise "G.Caporale" | Lorusso A, Marcacci M, Di Domenico M, Curini V, Ancora M, Cammà C, Rinaldi A, Mangone I, Di Pasquale A, Puglia I, Savini G. |
| EPI_ISL_849647, EPI_ISL_849648, EPI_ISL_849649 | SIESP- DRIVE IN CHIETI | Istitituto Zooprofilattico Sperimentale dell'Abruzzo e Molise "G.Caporale" | Lorusso A, Marcacci M, Di Domenico M, Curini V, Ancora M, Cammà C, Rinaldi A, Mangone I, Di Pasquale A, Puglia I, Savini G. |
| EPI_ISL_849763 | Molecular Biology, IZS Sicilia | Molecular Biology, IZS Sicilia | Reale,S., Seidita,G., Di Gaudio,F., Scibetta,S. and Vitale,F. |
| EPI_ISL_852619, EPI_ISL_852622, EPI_ISL_852628, EPI_ISL_852629, EPI_ISL_852630, EPI_ISL_852631, EPI_ISL_852632, EPI_ISL_852813, EPI_ISL_852819, EPI_ISL_852821, EPI_ISL_852823, EPI_ISL_852825, EPI_ISL_852828, EPI_ISL_855556, EPI_ISL_860203, EPI_ISL_860204, EPI_ISL_860205, EPI_ISL_860257 | | | |
| see above | Lab. Microbiologia e Virologia, Cotugno, A.O. dei Colli | Lab. Microbiologia e Virologia, Cotugno, A.O. dei Colli | Luigi Atripaldi, Claudia Tiberio, Anna Perfetti |
| EPI_ISL_875557 | ULSS 2 Treviso | Istituto Zooprofilattico Sperimentale delle Venezie | Adelaide Milani, Alessia Schivo, Annalisa Salviato, Erika Giorgia Quaranta, Ambra Pastori, Bianca Zecchin, Alice Fusaro, Isabella Monne, Calogero Terregino, Antonia Ricci |
| EPI_ISL_875558 | Microbiologia e Virologia | Istituto Zooprofilattico Sperimentale delle Venezie | Adelaide Milani, Alessia Schivo, Annalisa Salviato, Erika Giorgia Quaranta, Ambra Pastori, Bianca Zecchin, Alice Fusaro, Isabella Monne, Calogero Terregino, Antonia Ricci |
| EPI_ISL_875566, EPI_ISL_875567, EPI_ISL_875568 | SIESP L'AQUILA | Istituto Zooprofilattico Sperimentale dell'Abruzzo e Molise "G. Caporale" | Lorusso A, Marcacci M, Di Domenico M, Ancora M, Curini V, Mangone I, Rinaldi A, Scialabba S, Di Pasquale A, Cammà C, Puglia I, Calistri P, Savini G |
| EPI_ISL_876738, EPI_ISL_876741 | Istituto Zooprofilattico Sperimentale della Puglia e della Basilicata | Istituto Zooprofilattico Sperimentale della Puglia e della Basilicata | Parisi A., Bianco A., Capozzi L., Del Sambro L., Manzulli V, Rondinone V., Pace L., Cipolletta D., Galante D. |
| EPI_ISL_876742 | Ospedale Santa Caterina Novella | Istituto Zooprofilattico Sperimentale della Puglia e della Basilicata | Parisi A., Bianco A., Capozzi L., Del Sambro L., Manzulli V, Rondinone V., Pace L., Cipolletta D., Galante D. |
| EPI_ISL_876743 | Presidio di Brindisi Di Summa - Perrino | Istituto Zooprofilattico Sperimentale della Puglia e della | Parisi A., Bianco A., Capozzi L., Del Sambro L., Manzulli V, Rondinone V., Pace L., Cipolletta D., Galante D. |

| | | | |
|---|---|---|---|
| | | Basilicata | |
| EPI_ISL_876744, EPI_ISL_876745, EPI_ISL_876746, EPI_ISL_876747 | Istituto Zooprofilattico Sperimentale della Puglia e della Basilicata | Istituto Zooprofilattico Sperimentale della Puglia e della Basilicata | Parisi A., Bianco A., Capozzi L., Del Sambro L., Manzulli V, Rondinone V., Pace L., Cipolletta D., Galante D. |
| EPI_ISL_882635 | SC (UCO) Igiene e Sanità Pubblica (funzione integrata con SC Microbiologia e Virologia) e Laboratory of Molecular Virology of the International Centre for Genetic Engineering and Biotechnology (ICGEB) | ARGO Laboratorio Genomica ed EpigenomicA | Licastro D, Dal Monego S, Degasperi M, Marcello A, D'Agaro P |
| EPI_ISL_882636 | SC (UCO) Igiene e Sanità Pubblica (funzione integrata con SC Microbiologia e Virologia) e Laboratory of Molecular Virology of the International Centre for Genetic Engineering and Biotechnology (ICGEB) | ARGO Laboratorio Genomica ed Epigenomica | Licastro D, Dal Monego S, Degasperi M, Marcello A, D'Agaro P |
| EPI_ISL_882652 | Laboratory of Microbiology and Virology, Ospedale Amedeo di Savoia, ASL "Città di Torino" | U.O. Genomics, S.S. Genetics and Advanced Omics Techniques, Istituto Zooprofilattico Sperimentale del Piemonte, Liguria e Valle d'Aosta | Chiara Beltramo, Francesco Cerutti, Valeria Ghisetti, Elena Bozzetta, Pier Luigi Acutis, Simone Peletto |
| EPI_ISL_882654 | Laboratory of Microbiology and Virology, Ospedale Amedeo di Savoia, ASL "Città di Torino" | U.O. Genomics, S.S. Genetics and Advanced Omics Techniques Istituto Zooprofilattico Sperimentale del Piemonte, Liguria e Valle d'Aosta | Chiara Beltramo, Francesco Cerutti, Valeria Ghisetti, Elena Bozzetta, Pier Luigi Acutis, Simone Peletto |
| EPI_ISL_882918 | INMI Lazzaro Spallanzani IRCCS | INMI Lazzaro Spallanzani IRCCS | E Giombini, M. Rueca, B Bartolini, O Butera, C.E.M Gruber, F Messina, MR Capobianchi, A Di Caro |
| EPI_ISL_882919 | INMI Lazzaro Spallanzani IRCCS | INMI Lazzaro Spallanzani IRCCS | C.E.M Gruber, B Bartolini, E Giombini, M Rueca, O Butera, F Messina, A Di Caro, MR Capobianchi |
| EPI_ISL_882920 | INMI Lazzaro Spallanzani IRCCS | INMI Lazzaro Spallanzani IRCCS | M Rueca, E Giombini, C.E.M Gruber, B Bartolini, O Butera, F Messina, A Di Caro, MR Capobianchi |
| EPI_ISL_882921 | Genomic Medicine Laboratory, IRCCS Santa Lucia Foundation | INMI Lazzaro Spallanzani IRCCS | M Rueca, E Giombini, C.E.M Gruber, B Bartolini, O Butera, F Messina, E Giardina, A Di Caro, MR Capobianchi |
| EPI_ISL_882922 | Sant'Eugenio/CTO ASL Roma 2 | INMI Lazzaro Spallanzani IRCCS | E Giombini, M Rueca, B Bartolini, O Butera, C.E.M Gruber, F Messina, F Bondanini, GC Cocciolillo, C Disegni, MR Capobianchi, A Di Caro |
| EPI_ISL_882924 | INMI Lazzaro Spallanzani IRCCS | INMI Lazzaro Spallanzani IRCCS | F Messina, O Butera, E Giombini, M Rueca, B Bartolini, C.E.M Gruber, MR Capobianchi, A Di Caro |
| EPI_ISL_882925 | INMI Lazzaro Spallanzani IRCCS | INMI Lazzaro Spallanzani IRCCS | C.E.M Gruber, B Bartolini, E Giombini, M Rueca, O Butera, F Messina, A Di Caro, MR Capobianchi |
| EPI_ISL_882926 | INMI Lazzaro Spallanzani IRCCS | INMI Lazzaro Spallanzani IRCCS | B Bartolini, O Butera, C.E.M Gruber, M Rueca, F Messina, E Giombini, MR Capobianchi, A Di Caro |
| EPI_ISL_882927 | INMI Lazzaro Spallanzani IRCCS | INMI Lazzaro Spallanzani IRCCS | M Rueca, E Giombini, C.E.M Gruber, B Bartolini, O Butera, F Messina, A Di Caro, MR Capobianchi |
| EPI_ISL_882928 | INMI Lazzaro Spallanzani IRCCS | INMI Lazzaro Spallanzani IRCCS | F Messina, O Butera, E Giombini, M Rueca, B Bartolini, C.E.M Gruber, MR Capobianchi, A Di Caro |
| EPI_ISL_882929 | INMI Lazzaro Spallanzani IRCCS | INMI Lazzaro Spallanzani IRCCS | C.E.M Gruber, B Bartolini, E Giombini, M Rueca, O Butera, F Messina, A Di Caro, MR Capobianchi |
| EPI_ISL_882930 | INMI Lazzaro Spallanzani IRCCS | INMI Lazzaro Spallanzani IRCCS | E Giombini, M. Rueca, B Bartolini, O Butera, C.E.M Gruber, F Messina, MR Capobianchi, A Di Caro |
| EPI_ISL_882931 | INMI Lazzaro Spallanzani IRCCS | INMI Lazzaro Spallanzani IRCCS | B Bartolini, O Butera, C.E.M Gruber, M Rueca, F Messina, E Giombini, MR Capobianchi, A Di Caro |
| EPI_ISL_882932 | INMI Lazzaro Spallanzani IRCCS | INMI Lazzaro Spallanzani IRCCS | F Messina, O Butera, E Giombini, M Rueca, B Bartolini, C.E.M Gruber, MR Capobianchi, A Di Caro |
| EPI_ISL_882933 | Sant'Eugenio/CTO ASL Roma 2 | INMI Lazzaro Spallanzani IRCCS | E Giombini, M. Rueca, B Bartolini, O Butera, C.E.M Gruber, F Bondanini, GC Cocciolillo, C Disegni, F Messina, MR Capobianchi, A Di Caro |
| EPI_ISL_882934 | Sant'Eugenio/CTO ASL Roma 2 | INMI Lazzaro Spallanzani IRCCS | B Bartolini, O Butera, C.E.M Gruber, M Rueca, F Messina, E Giombini, F Bondanini, GC Cocciolillo, C Disegni, MR Capobianchi, A Di Caro |
| EPI_ISL_882935 | Sant'Eugenio/CTO ASL Roma 2 | INMI Lazzaro Spallanzani IRCCS | F Messina, O Butera, E Giombini, M Rueca, B Bartolini, C.E.M Gruber, F Bondanini, C Disegni, GC Cocciolillo, MR Capobianchi, A Di Caro |
| EPI_ISL_882936 | Sant'Eugenio/CTO ASL Roma 2 | INMI Lazzaro Spallanzani IRCCS | E Giombini, M Rueca, B Bartolini, O Butera, C.E.M Gruber, F Messina, F Bondanini, GC Cocciolillo, C Disegni, MR Capobianchi, A Di Caro |
| EPI_ISL_882937 | Azienda Ospedaliera San Giovanni Addolorata | INMI Lazzaro Spallanzani IRCCS | F Messina, O Butera, E Giombini, M Rueca, B Bartolini, C.E.M Gruber, M Gaudio, PM Placanica, MR Capobianchi, A Di Caro |
| EPI_ISL_882938 | Azienda Ospedaliera San Giovanni Addolorata | INMI Lazzaro Spallanzani IRCCS | C.E.M Gruber, B Bartolini, E Giombini, M Rueca, O Butera, F Messina, PM Placanica, M Gaudio, A Di Caro, MR Capobianchi |
| EPI_ISL_882939 | Azienda Ospedaliera San Giovanni Addolorata | INMI Lazzaro Spallanzani IRCCS | B Bartolini, O Butera, C.E.M Gruber, M Rueca, F Messina, E Giombini, M Gaudio, PM Placanica, MR Capobianchi, A Di Caro |
| EPI_ISL_882940 | Ospedale San Camillo De Lellis Rieti | INMI Lazzaro Spallanzani IRCCS | M Rueca, E Giombini, C.E.M Gruber, B Bartolini, O Butera, S Venarubea, A De Luca, L Casertano, F Messina, A Di Caro, MR Capobianchi |
| EPI_ISL_882941 | Ospedale San Camillo De Lellis Rieti | INMI Lazzaro Spallanzani IRCCS | C.E.M Gruber, B Bartolini, E Giombini, M Rueca, O Butera, F Messina, S Venarubea, L Casertano, A De Luca, A Di Caro, MR Capobianchi |
| EPI_ISL_882942 | Ospedale San Camillo De Lellis Rieti | INMI Lazzaro Spallanzani IRCCS | M Rueca, E Giombini, C.E.M Gruber, B Bartolini, O Butera, F Messina, S Venarubea, A De Luca, L Casertano, A Di Caro, MR Capobianchi |
| EPI_ISL_883286, EPI_ISL_883287 | SIESP DIPARTIMENTO DI PREVENZIONE CHIE | Istituto Zooprofilattico Sperimentale dell'Abruzzo e Molise "G. Caporale" | Lorusso A, Marcacci M, Di Domenico M, Ancora M, Curini V, Mangone I, Rinaldi A, Scialabba S, Di Pasquale A, Cammà C, Puglia I, Calistri P, Savini G |
| EPI_ISL_883288, EPI_ISL_883289 | SIESP CHIETI-DRIVE IN ORTONA | Istituto Zooprofilattico Sperimentale dell'Abruzzo e Molise "G. Caporale" | Lorusso A, Marcacci M, Di Domenico M, Ancora M, Curini V, Mangone I, Rinaldi A, Scialabba S, Di Pasquale A, Cammà C, Puglia I, Calistri P, Savini G |
| EPI_ISL_883290 | SIESP CHIETI-DRIVE IN CHIETI | Istituto Zooprofilattico Sperimentale dell'Abruzzo e Molise "G. Caporale" | Lorusso A, Marcacci M, Di Domenico M, Ancora M, Curini V, Mangone I, Rinaldi A, Scialabba S, Di Pasquale A, Cammà C, Puglia I, Calistri P, Savini G |
| EPI_ISL_883291 | RP Guardiagrele-Ospedale di Comunità | Istituto Zooprofilattico Sperimentale dell'Abruzzo e Molise "G. Caporale" | Lorusso A, Marcacci M, Di Domenico M, Ancora M, Curini V, Mangone I, Rinaldi A, Scialabba S, Di Pasquale A, Cammà C, Puglia I, Calistri P, Savini G |
| EPI_ISL_883292 | SIESP CHIETI-DRIVE IN ORTONA | Istituto Zooprofilattico Sperimentale dell'Abruzzo e Molise "G. Caporale" | Lorusso A, Marcacci M, Di Domenico M, Ancora M, Curini V, Mangone I, Rinaldi A, Scialabba S, Di Pasquale A, Cammà C, Puglia I, Calistri P, Savini G |
| EPI_ISL_883293 | SIESP CHIETI-DRIVE IN LANCIANO | Istituto Zooprofilattico Sperimentale dell'Abruzzo e Molise "G. Caporale" | Lorusso A, Marcacci M, Di Domenico M, Ancora M, Curini V, Mangone I, Rinaldi A, Scialabba S, Di Pasquale A, Cammà C, Puglia I, Calistri P, Savini G |
| EPI_ISL_883294, EPI_ISL_883295 | RP Guardiagrele-Ospedale di Comunità | Istituto Zooprofilattico Sperimentale dell'Abruzzo e Molise "G. Caporale" | Lorusso A, Marcacci M, Di Domenico M, Ancora M, Curini V, Mangone I, Rinaldi A, Scialabba S, Di Pasquale A, Cammà C, Puglia I, Calistri P, Savini G |
| EPI_ISL_883296 | SIESP CHIETI-DRIVE IN ORTONA | Istituto Zooprofilattico Sperimentale dell'Abruzzo e Molise "G. Caporale" | Lorusso A, Marcacci M, Di Domenico M, Ancora M, Curini V, Mangone I, Rinaldi A, Scialabba S, Di Pasquale A, Cammà C, Puglia I, Calistri P, Savini G |
| EPI_ISL_883297 | RP Guardiagrele-Ospedale di Comunità | Istituto Zooprofilattico Sperimentale dell'Abruzzo e Molise "G. Caporale" | Lorusso A, Marcacci M, Di Domenico M, Ancora M, Curini V, Mangone I, Rinaldi A, Scialabba S, Di Pasquale A, Cammà C, Puglia I, Calistri P, Savini G |
| EPI_ISL_883298, EPI_ISL_883299 | SIESP DIPARTIMENTO DI PREVENZIONE CHIE | Istituto Zooprofilattico Sperimentale dell'Abruzzo e Molise "G. Caporale" | Lorusso A, Marcacci M, Di Domenico M, Ancora M, Curini V, Mangone I, Rinaldi A, Scialabba S, Di Pasquale A, Cammà C, Puglia I, Calistri P, Savini G |
| EPI_ISL_883300 | SIESP CHIETI-DRIVE IN CHIETI | Istituto Zooprofilattico Sperimentale dell'Abruzzo e Molise "G. Caporale" | Lorusso A, Marcacci M, Di Domenico M, Ancora M, Curini V, Mangone I, Rinaldi A, Scialabba S, Di Pasquale A, Cammà C, Puglia I, Calistri P, Savini G |
| EPI_ISL_883301, EPI_ISL_883302 | RP Guardiagrele-Ospedale di Comunità | Istituto Zooprofilattico Sperimentale dell'Abruzzo e Molise "G. Caporale" | Lorusso A, Marcacci M, Di Domenico M, Ancora M, Curini V, Mangone I, Rinaldi A, Scialabba S, Di Pasquale A, Cammà C, Puglia I, Calistri P, Savini G |
| EPI_ISL_883303, EPI_ISL_883304, EPI_ISL_883305 | SIESP DIPARTIMENTO DI PREVENZIONE CHIE | Istituto Zooprofilattico Sperimentale dell'Abruzzo e Molise "G. Caporale" | Lorusso A, Marcacci M, Di Domenico M, Ancora M, Curini V, Mangone I, Rinaldi A, Scialabba S, Di Pasquale A, Cammà C, Puglia I, Calistri P, Savini G |

| | | | |
|---|---|---|---|
| EPI_ISL_888596 | Istituto Zooprofilattico Sperimentale del Piemonte, Liguria e Valle d'Aosta | U.O. Genomics, S.S. Genetics and Advanced Omics Techniques Istituto Zooprofilattico Sperimentale del Piemonte, Liguria e Valle d'Aosta | Chiara Beltramo, Pier Luigi Acutis, Maria Goria, Elena Bozzetta, Simone Peletto |
| EPI_ISL_888676 | Center of Advanced Studies and Technology, Molecular Genetics Laboratory | Center of Advanced Studies and Technology, Molecular Genetics Laboratory | De Fabritiis Simone, Mandatori Domitilla, Ferrante Rossella |
| EPI_ISL_900583, EPI_ISL_900633, EPI_ISL_900634, EPI_ISL_900635, EPI_ISL_900636, EPI_ISL_900637, EPI_ISL_900638, EPI_ISL_900639, EPI_ISL_900640, EPI_ISL_900641, EPI_ISL_900642, EPI_ISL_900643, EPI_ISL_900644, EPI_ISL_900645, EPI_ISL_900646, EPI_ISL_900648, EPI_ISL_900649, EPI_ISL_900650, EPI_ISL_900685, EPI_ISL_900686, EPI_ISL_900687, EPI_ISL_900688, EPI_ISL_900689, EPI_ISL_900690 | | | |
| see above | IZSM | TIGEM | Antonio Grimaldi, Patrizia Annunziata, Francesco Panariello, Biancamaria Pierri, Valentina Bouche, Chiara Colantuono, Maria Concetta Cuomo, Denise Di Concilio, Lucio Di Filippo, Anna Manfredi, Marcello Salvi, Antonio Limone, Pellegrino Cerino, Andrea Ballabio, Davide Cacchiarelli. |
| EPI_ISL_902750 | Ospedale "F. Spaziani" Frosinone | INMI Lazzaro Spallanzani IRCCS | F Messina, C.E.M Gruber, B Bartolini, E Giombini, M Rueca, O Butera, C. Gargiulo, C Sias, A Di Caro, MR Capobianchi |
| EPI_ISL_902751 | Ospedale Sandro Pertini ASL Roma2 | INMI Lazzaro Spallanzani IRCCS | B Bartolini, O Butera, C.E.M Gruber, M Rueca, F Messina, E Giombini, G Cappiello, MC Cava, MR Capobianchi, A Di Caro |
| EPI_ISL_902752 | Ospedale Sandro Pertini ASL Roma2 | INMI Lazzaro Spallanzani IRCCS | O Butera, F Messina, C.E.M Gruber, B Bartolini, E Giombini, M Rueca, S Romano, MC Cava, A Di Caro, MR Capobianchi |
| EPI_ISL_902753 | Azienda Ospedaliera San Camillo Forlanini | INMI Lazzaro Spallanzani IRCCS | C.E.M Gruber, B Bartolini, E Giombini, M Rueca, O Butera, F Messina, D.Gallone, G Parisi, A Di Caro, MR Capobianchi |
| EPI_ISL_902754 | University Hospital Sant'Andrea-Sapienza | INMI Lazzaro Spallanzani IRCCS | B Bartolini, E Giombini, M Rueca, O Butera, F Messina, C.E.M Gruber, M Simmaco, I Santino, A Di Caro, MR Capobianchi |
| EPI_ISL_902755 | IRCCS San Raffaele | INMI Lazzaro Spallanzani IRCCS | E Giombini, M Rueca, O Butera, F Messina, C.E.M Gruber, B Bartolini, D Russo, D Limongi, MR Capobianchi, A Di Caro |
| EPI_ISL_902756 | Fondazione Policlinico Universitario "A. Gemelli" IRCCS | INMI Lazzaro Spallanzani IRCCS | M Rueca, O Butera, F Messina, C.E.M Gruber, B Bartolini, E Giombini, P Cattani, M Sanguinetti, MR Capobianchi, A Di Caro |
| EPI_ISL_902757 | San Camillo De Lellis Rieti Hospital | INMI Lazzaro Spallanzani IRCCS | C.E.M Gruber, B Bartolini, E Giombini, M Rueca, O Butera, F Messina, S Venarubea, A De Luca, A Di Caro, MR Capobianchi |
| EPI_ISL_902926 | SC (UCO) Igiene e Sanità Pubblica (funzione integrata con SC Microbiologia e Virologia) e Laboratory of Molecular Virology of the International Centre for Genetic Engineering and Biotechnology (ICGEB) | ARGO Laboratorio Genomica ed Epigenomica | Licastro D, Dal Monego S, Degasperi M, Marcello A, D'Agaro P |
| EPI_ISL_911524, EPI_ISL_911525, EPI_ISL_911526, EPI_ISL_911527 | Microbiology and Virology Unit, Florence Careggi University Hospital | Microbiology and Virology Unit, Florence Careggi University Hospital | Vincenzo Di Pilato, Marco Coppi, Fabio Morecchiato, Noemi Aiezza, Ilaria Baccani, Alberto Antonelli, Emanuele Gori, Gian Maria Rossolini |
| EPI_ISL_913446 | Laboratorio Genzano - ASL RM 6 | INMI Lazzaro Spallanzani IRCCS | Emanuela Giombini, Martina Rueca, Barbara Bartolini, Ornella Butera, Cesare E.M. Gruber, Francesco Messina, Grazia Tramini, Emanuela Conti, Antonino Di Caro, Maria R. Capobianchi |
| EPI_ISL_913447 | Laboratorio Genzano - ASL RM 6 | INMI Lazzaro Spallanzani IRCCS | Francesco Messina, Emanuela Giombini, Ornella Butera, Cesare EM Gruber, Martina Rueca, Barbara Bartolini, Grazia Tramini, Emanuela Conti, Maria R Capobianchi, Antonino Di Caro |
| EPI_ISL_918263 | SIESP DIPARTIMENTO DI PREVENZIONE CHIE | Istituto Zooprofilattico Sperimentale dell'Abruzzo e Molise "G. Caporale" | Lorusso A, Marcacci M, Di Domenico M, Ancora M, Curini V, Mangone I, Rinaldi A, Scialabba S, Di Pasquale A, Cammà C, Puglia I, Calistri P, Savini G |
| EPI_ISL_918264 | SIESP CHIETI-DRIVE IN CHIETI | Istituto Zooprofilattico Sperimentale dell'Abruzzo e Molise "G. Caporale" | Lorusso A, Marcacci M, Di Domenico M, Ancora M, Curini V, Mangone I, Rinaldi A, Scialabba S, Di Pasquale A, Cammà C, Puglia I, Calistri P, Savini G |
| EPI_ISL_918265 | OSPEDALE ATRI | Istituto Zooprofilattico Sperimentale dell'Abruzzo e Molise "G. Caporale" | Lorusso A, Marcacci M, Di Domenico M, Ancora M, Curini V, Mangone I, Rinaldi A, Scialabba S, Di Pasquale A, Cammà C, Puglia I, Calistri P, Savini G |
| EPI_ISL_918266 | SIESP CHIETI-Drive in Lanciano | Istituto Zooprofilattico Sperimentale dell'Abruzzo e Molise "G. Caporale" | Lorusso A, Marcacci M, Di Domenico M, Ancora M, Curini V, Mangone I, Rinaldi A, Scialabba S, Di Pasquale A, Cammà C, Puglia I, Calistri P, Savini G |
| EPI_ISL_918267 | SIESP CHIETI - DRIVE IN CHIETI | Istituto Zooprofilattico Sperimentale dell'Abruzzo e Molise "G. Caporale" | Lorusso A, Marcacci M, Di Domenico M, Ancora M, Curini V, Mangone I, Rinaldi A, Scialabba S, Di Pasquale A, Cammà C, Puglia I, Calistri P, Savini G |
| EPI_ISL_918268 | SIESP CHIETI - DRIVE IN ORTONA | Istituto Zooprofilattico Sperimentale dell'Abruzzo e Molise "G. Caporale" | Lorusso A, Marcacci M, Di Domenico M, Ancora M, Curini V, Mangone I, Rinaldi A, Scialabba S, Di Pasquale A, Cammà C, Puglia I, Calistri P, Savini G |
| EPI_ISL_918269 | SIESP DIPARTIMENTO DI PREVENZIONE TERAMO | Istituto Zooprofilattico Sperimentale dell'Abruzzo e Molise "G. Caporale" | Lorusso A, Marcacci M, Di Domenico M, Ancora M, Curini V, Mangone I, Rinaldi A, Scialabba S, Di Pasquale A, Cammà C, Puglia I, Calistri P, Savini G |
| EPI_ISL_918270 | SIESP CHIETI - DRIVE IN LANCIANO | Istituto Zooprofilattico Sperimentale dell'Abruzzo e Molise "G. Caporale" | Lorusso A, Marcacci M, Di Domenico M, Ancora M, Curini V, Mangone I, Rinaldi A, Scialabba S, Di Pasquale A, Cammà C, Puglia I, Calistri P, Savini G |
| EPI_ISL_918271 | SIESP DIPARTIMENTO DI PREVENZIONE CHIETI | Istituto Zooprofilattico Sperimentale dell'Abruzzo e Molise "G. Caporale" | Lorusso A, Marcacci M, Di Domenico M, Ancora M, Curini V, Mangone I, Rinaldi A, Scialabba S, Di Pasquale A, Cammà C, Puglia I, Calistri P, Savini G |
| EPI_ISL_918408, EPI_ISL_918409 | Ospedale Santa Caterina Novella | Istituto Zooprofilattico Sperimentale della Puglia e della Basilicata | Parisi A., Bianco A., Capozzi L., Del Sambro L., Simone D., Manzulli V, Rondinone V., Pace L., Cipolletta D., Galante D. |
| EPI_ISL_918410 | Ospedale Di Venere - Carbonara | Istituto Zooprofilattico Sperimentale della Puglia e della Basilicata | Parisi A., Bianco A., Capozzi L., Del Sambro L., Simone D., Manzulli V, Rondinone V., Pace L., Cipolletta D., Galante D. |
| EPI_ISL_925502, EPI_ISL_925503, EPI_ISL_925504, EPI_ISL_925505, EPI_ISL_925506, EPI_ISL_925507, EPI_ISL_925508 | Microbiology and Virology Laboratory, "Policlinico Riuniti, Azienda Ospedaliero Universitaria, Foggia" | Microbiology and Virology Laboratory, "Policlinico Riuniti, Azienda Ospedaliero Universitaria, Foggia" | Fabio Arena, Carmela Paolillo, Settimia Altamura, Donatella Cedola, Valeria Delli Carri, Annamaria D'Aprile, Maria Rosaria Lipsi, Loris Micelli, Maria Pia Patrizio, Daniela Pisanelli, Tiziana Rollo, Iole Natalicchio, Gaetano Corso, Maurizio Margaglione and Rosella De Nittis. |
| EPI_ISL_935156 | SIESP CHIETI | Istituto Zooprofilattico Sperimentale dell'Abruzzo e Molise "G. Caporale" | Lorusso A, Marcacci M, Di Domenico M, Ancora M, Curini V, Mangone I, Rinaldi A, Scialabba S, Di Pasquale A, Cammà C, Puglia I, Calistri P, Savini G |
| EPI_ISL_935157 | SIESP CHIETI-DRIVE IN ORTONA | Istituto Zooprofilattico Sperimentale dell'Abruzzo e Molise "G. Caporale" | Lorusso A, Marcacci M, Di Domenico M, Ancora M, Curini V, Mangone I, Rinaldi A, Scialabba S, Di Pasquale A, Cammà C, Puglia I, Calistri P, Savini G |
| EPI_ISL_935158, EPI_ISL_935159 | SIESP CHIETI | Istituto Zooprofilattico Sperimentale dell'Abruzzo e Molise "G. Caporale" | Lorusso A, Marcacci M, Di Domenico M, Ancora M, Curini V, Mangone I, Rinaldi A, Scialabba S, Di Pasquale A, Cammà C, Puglia I, Calistri P, Savini G |
| EPI_ISL_935160 | SIESP CHIETI-DRIVE IN ORTONA | Istituto Zooprofilattico Sperimentale dell'Abruzzo e Molise "G. Caporale" | Lorusso A, Marcacci M, Di Domenico M, Ancora M, Curini V, Mangone I, Rinaldi A, Scialabba S, Di Pasquale A, Cammà C, Puglia I, Calistri P, Savini G |
| EPI_ISL_935161, EPI_ISL_935162 | SIESP CHIETI | Istituto Zooprofilattico Sperimentale dell'Abruzzo e Molise "G. Caporale" | Lorusso A, Marcacci M, Di Domenico M, Ancora M, Curini V, Mangone I, Rinaldi A, Scialabba S, Di Pasquale A, Cammà C, Puglia I, Calistri P, Savini G |
| EPI_ISL_935163, EPI_ISL_935164 | SIESP CHIETI-DRIVE IN ORTONA | Istituto Zooprofilattico Sperimentale dell'Abruzzo e Molise "G. Caporale" | Lorusso A, Marcacci M, Di Domenico M, Ancora M, Curini V, Mangone I, Rinaldi A, Scialabba S, Di Pasquale A, Cammà C, Puglia I, Calistri P, Savini G |
| EPI_ISL_935165, EPI_ISL_935166, EPI_ISL_935167, EPI_ISL_935168, EPI_ISL_935169, EPI_ISL_935971, EPI_ISL_935972 | SIESP CHIETI | Istituto Zooprofilattico Sperimentale dell'Abruzzo e Molise "G. Caporale" | Lorusso A, Marcacci M, Di Domenico M, Ancora M, Curini V, Mangone I, Rinaldi A, Scialabba S, Di Pasquale A, Cammà C, Puglia I, Calistri P, Savini G |
| EPI_ISL_940565 | University of Bari Biomedical Sciences and Human Oncology | University of Bari Biomedical Sciences and Human Oncology | Chironna M., Sallustio A., Loconsole D., Accogli M. |

| | | | |
|---|---|---|---|
| EPI_ISL_940607 | U.O. Genomics, S.S. Genetics and Advanced Omics Techniques, Istituto Zooprofilattico Sperimentale del Piemonte Liguria e Valle d'Aosta | U.O. Genomics, S.S. Genetics and Advanced Omics Techniques, Istituto Zooprofilattico Sperimentale del Piemonte Liguria e Valle d'Aosta | Chiara Beltramo, Francesco Cerutti, Valeria Ghisetti, Elena Bozzetta, Pier Luigi Acutis, Simone Peletto |
| EPI_ISL_940631, EPI_ISL_940632, EPI_ISL_940739, EPI_ISL_940749 | University of Bari Biomedical Sciences and Human Oncology | University of Bari Biomedical Sciences and Human Oncology | Chironna M., Sallustio A., Loconsole D., Accogli M. |
| EPI_ISL_941336 | Ospedale San Camillo De Lellis di Rieti | INMI Lazzaro Spallanzani IRCCS | Cesare E.M. Gruber, Francesco Messina, Martina Rueca, Emanuela Giombini, Barbara Bartolini, Ornella Butera, Stefano Venarubea, Luca Casertano, Antonino Di Caro, Maria R. Capobianchi |
| EPI_ISL_941337 | Ospedale San Camillo De Lellis di Rieti | INMI Lazzaro Spallanzani IRCCS | Ornella Butera, Barbara Bartolini, Cesare E.M. Gruber, Martina Rueca, Francesco Messina, Emanuela Giombini, Stefano Venarubea, Assunta De Luca, Maria R. Capobianchi, Antonino Di Caro |
| EPI_ISL_941338 | INMI Lazzaro Spallanzani IRCCS | INMI Lazzaro Spallanzani IRCCS | Emanuela Giombini, Ornella Butera, Cesare E.M. Gruber, Martina Rueca, Barbara Bartolini, Francesco Messina, Giandomenico Russo, Daniele Castiglia , Stefania Madonna, Antonino Di Caro, Maria R. Capobianchi |
| EPI_ISL_944785 | Laboratory of Microbiology and Virology, Ospedale Amedeo di Savoia, ASL "Città di Torino" | U.O. Genomics, S.S. Genetics and Advanced Omics Techniques, Istituto Zooprofilattico Sperimentale del Piemonte Liguria e Valle d'Aosta | Chiara Beltramo, Francesco Cerutti, Valeria Ghisetti, Elena Bozzetta, Pier Luigi Acutis, Simone Peletto |
| EPI_ISL_949124 | SC (UCO) Igiene e Sanità Pubblica (funzione integrata con SC Microbiologia e Virologia) e Laboratory of Molecular Virology of the International Centre for Genetic Engineering and Biotechnology (ICGEB) | ARGO Laboratorio Genomica ed Epigenomica | Licastro D, Dal Monego S, Degasperi M, Marcello A, D'Agaro P |
| EPI_ISL_949182 | University of Bari Biomedical Sciences and Human Oncology | University of Bari Biomedical Sciences and Human Oncology | Chironna M., Sallustio A., Loconsole D., Accogli M. |
| EPI_ISL_949184 | University of Bari Biomedical Sciences and Human Oncology | University of Bari Biomedical Sciences and Human Oncology | Chironna M., Sallustio A., Loconsole D., Accogli A. |
| EPI_ISL_949185 | University of Bari Biomedical Sciences and Human Oncology | University of Bari Biomedical Sciences and Human Oncology | Chironna M., Sallustio A., Loconsole D., Accogli M. |
| EPI_ISL_949188 | University of Bari Biomedical Sciences and Human Oncology | University of Bari Biomedical Sciences and Human Oncology | Chironna A., Sallustio A., Loconsole D., Accogli M. |
| EPI_ISL_949191 | University of Bari Biomedical Sciences and Human Oncology | University of Bari Biomedical Sciences and Human Oncology | Chironna M., Sallustio A., Loconsole D., Accogli M. |
| EPI_ISL_954210, EPI_ISL_954211, EPI_ISL_954212, EPI_ISL_954213, EPI_ISL_954214, EPI_ISL_954215, EPI_ISL_954218, EPI_ISL_954219, EPI_ISL_954220, EPI_ISL_954221, EPI_ISL_954222, EPI_ISL_954223, EPI_ISL_954224, EPI_ISL_954225 | | | |
| see above | 1.AO Universitaria 'S. Giovanni di Dio e Ruggi D'Aragona, Scuola Medica Salernitana' Hospital / 2.UOC di Virologia e Microbiologia, Università della Campania 'L. Vanvitelli' / 3.AO Universitaria 'Federico II' Napoli Hospital / 4.AORN 'San Giuseppe Moscati' Avellino Hospital / 5.AO 'San Pio - presidio G. Rummo' Benevento Hospital / 6.AO 'Sant'Anna e San Sebastiano' Caserta Hospital / 7.PO 'Maria Santissima Addolorata' Eboli Hospital / 8.Biogem Istituto di Ricerche Genetiche | 1. Genome Research Center for Health (CRGS) / 2. Laboratory of Molecular Medicine and Genomics(LMMGe) / 3. Center for Research in Pure and Applied Mathematics (CRMPA) | Giorgio Giurato, Francesca Rizzo, Alessandro Weisz, Gianluigi Franci, Giovanni Nassa, Pasquale Pagliano, Roberta Tarallo, Elena Alexandrova, Ylenia D'Agostino, Carlo Ferravante, Jessica Lamberti, Viola Melone, Domenico Memoli, Valeria Mirici Cappa, Domenico Palumbo, Giovanni Pecoraro, Assunta Sellitto, Oriana Strianese, Ilaria Terenzi, Giuseppe Fenza, Aniello Gentile, Antonello Saccomanno, Sonia Amabile, Teresa Rocco, Annamaria Salvati, Emilia Vaccaro, Massimiliano Galdiero, Michele Cennamo, Giuseppe Portella, Maria Grazia Foti, Mariarosaria Ingino, Maria Landi, Maurizio Fumi, Vincenzo Rocco, Rita Greco, Vittoria Letizia, Arnolfo Petruzziello, Maddalena Schioppa, Gregorio Goffredi, Francesca Marciano, Michele Caraglia, Alessia Cossu, Marianna Scrima, Edmondo Adorisio, Morena D'Avenia, Michela Iacobellis, Rosanna Piluscio, Giorgio Dirani, Vittorio Sambri, Simona Semprini, Silvia Zanoli, Francesco Curcio, Stefania Marzinotto, Andreina Baj, Fausto Sessa. |
| EPI_ISL_959384 | INT Fondazione Pascale | INT Fondazione Pascale | INT Fondazione Pascale |
| EPI_ISL_960459, EPI_ISL_960460, EPI_ISL_960461, EPI_ISL_960462, EPI_ISL_960463, EPI_ISL_960464, EPI_ISL_960465, EPI_ISL_960466, EPI_ISL_960467, EPI_ISL_960470, EPI_ISL_960471, EPI_ISL_960479, EPI_ISL_960560, EPI_ISL_960561, EPI_ISL_960562, EPI_ISL_960563, EPI_ISL_960564, EPI_ISL_960565, EPI_ISL_960566, EPI_ISL_960567, EPI_ISL_960571, EPI_ISL_960572, EPI_ISL_960573, EPI_ISL_960649, EPI_ISL_960650, EPI_ISL_960651, EPI_ISL_960652, EPI_ISL_960653, EPI_ISL_960654, EPI_ISL_960655, EPI_ISL_960656, EPI_ISL_960657, EPI_ISL_960659 | | | |
| see above | Istituto Zooprofilattico Sperimentale del Mezzogiorno | TIGEM | Antonio Grimaldi, Patrizia Annunziata, Francesco Panariello, Biancamaria Pierri, Valentina Bouche, Chiara Colantuono, Maria Concetta Cuomo, Denise Di Concilio, Lucio Di Filippo, Anna Manfredi, Marcello Salvi, Antonio Limone, Pellegrino Cerino, Andrea Ballabio, Davide Cacchiarelli. |
| EPI_ISL_961018 | SIESP CHIETI - DRIVE IN CHIETI | Istituto Zooprofilattico Sperimentale dell'Abruzzo e Molise "G. Caporale" | Lorusso A, Marcacci M, Di Domenico M, Ancora M, Curini V, Mangone I, Rinaldi A, Scialabba S, Di Pasquale A, Cammà C, Puglia I, Calistri P, Savini G |
| EPI_ISL_961019, EPI_ISL_961020 | SIESP CHIETI - DRIVE IN ORTONA | Istituto Zooprofilattico Sperimentale dell'Abruzzo e Molise "G. Caporale" | Lorusso A, Marcacci M, Di Domenico M, Ancora M, Curini V, Mangone I, Rinaldi A, Scialabba S, Di Pasquale A, Cammà C, Puglia I, Calistri P, Savini G |
| EPI_ISL_961021, EPI_ISL_961022, EPI_ISL_961023, EPI_ISL_961024, EPI_ISL_961025, EPI_ISL_961026 | SIESP CHIETI - DRIVE IN CHIETI | Istituto Zooprofilattico Sperimentale dell'Abruzzo e Molise "G. Caporale" | Lorusso A, Marcacci M, Di Domenico M, Ancora M, Curini V, Mangone I, Rinaldi A, Scialabba S, Di Pasquale A, Cammà C, Puglia I, Calistri P, Savini G |
| EPI_ISL_961027 | SIESP CHIETI-DRIVE IN VASTO | Istituto Zooprofilattico Sperimentale dell'Abruzzo e Molise "G. Caporale" | Lorusso A, Marcacci M, Di Domenico M, Ancora M, Curini V, Mangone I, Rinaldi A, Scialabba S, Di Pasquale A, Cammà C, Puglia I, Calistri P, Savini G |
| EPI_ISL_961028, EPI_ISL_961029, EPI_ISL_961030, EPI_ISL_961031 | SIESP CHIETI - DRIVE IN CHIETI | Istituto Zooprofilattico Sperimentale dell'Abruzzo e Molise "G. Caporale" | Lorusso A, Marcacci M, Di Domenico M, Ancora M, Curini V, Mangone I, Rinaldi A, Scialabba S, Di Pasquale A, Cammà C, Puglia I, Calistri P, Savini G |
| EPI_ISL_961032 | SIESP DIPARTIMENTO DI PREVENZIONE CHIETI | Istituto Zooprofilattico Sperimentale dell'Abruzzo e Molise "G. Caporale" | Lorusso A, Marcacci M, Di Domenico M, Ancora M, Curini V, Mangone I, Rinaldi A, Scialabba S, Di Pasquale A, Cammà C, Puglia I, Calistri P, Savini G |
| EPI_ISL_961033 | SIESP CHIETI - DRIVE IN ORTONA | Istituto Zooprofilattico Sperimentale dell'Abruzzo e Molise "G. Caporale" | Lorusso A, Marcacci M, Di Domenico M, Ancora M, Curini V, Mangone I, Rinaldi A, Scialabba S, Di Pasquale A, Cammà C, Puglia I, Calistri P, Savini G |
| EPI_ISL_961034, EPI_ISL_961035 | SIESP CHIETI - DRIVE IN CHIETI | Istituto Zooprofilattico Sperimentale dell'Abruzzo e Molise "G. Caporale" | Lorusso A, Marcacci M, Di Domenico M, Ancora M, Curini V, Mangone I, Rinaldi A, Scialabba S, Di Pasquale A, Cammà C, Puglia I, Calistri P, Savini G |
| EPI_ISL_961036 | SIESP CHIETI - DRIVE IN ORTONA | Istituto Zooprofilattico Sperimentale dell'Abruzzo e Molise "G. Caporale" | Lorusso A, Marcacci M, Di Domenico M, Ancora M, Curini V, Mangone I, Rinaldi A, Scialabba S, Di Pasquale A, Cammà C, Puglia I, Calistri P, Savini G |
| EPI_ISL_961037, EPI_ISL_961038, EPI_ISL_961039, EPI_ISL_961040, EPI_ISL_961041 | SIESP CHIETI - DRIVE IN CHIETI | Istituto Zooprofilattico Sperimentale dell'Abruzzo e Molise "G. Caporale" | Lorusso A, Marcacci M, Di Domenico M, Ancora M, Curini V, Mangone I, Rinaldi A, Scialabba S, Di Pasquale A, Cammà C, Puglia I, Calistri P, Savini G |
| EPI_ISL_961042 | SIESP TERAMO | Istituto Zooprofilattico Sperimentale dell'Abruzzo e Molise "G. Caporale" | Lorusso A, Marcacci M, Di Domenico M, Ancora M, Curini V, Mangone I, Rinaldi A, Scialabba S, Di Pasquale A, Cammà C, Puglia I, Calistri P, Savini G |
| EPI_ISL_961043 | SIESP CHIETI - DRIVE IN CHIETI | Istituto Zooprofilattico Sperimentale dell'Abruzzo e Molise "G. Caporale" | Lorusso A, Marcacci M, Di Domenico M, Ancora M, Curini V, Mangone I, Rinaldi A, Scialabba S, Di Pasquale A, Cammà C, Puglia I, Calistri P, Savini G |
| EPI_ISL_961044, EPI_ISL_961045, EPI_ISL_961046 | SIESP CHIETI - DRIVE IN ORTONA | Istituto Zooprofilattico Sperimentale dell'Abruzzo e Molise "G. Caporale" | Lorusso A, Marcacci M, Di Domenico M, Ancora M, Curini V, Mangone I, Rinaldi A, Scialabba S, Di Pasquale A, Cammà C, Puglia I, Calistri P, Savini G |
| EPI_ISL_961047 | SIESP CHIETI - DRIVE IN CHIETI | Istituto Zooprofilattico Sperimentale dell'Abruzzo e Molise "G. Caporale" | Lorusso A, Marcacci M, Di Domenico M, Ancora M, Curini V, Mangone I, Rinaldi A, Scialabba S, Di Pasquale A, Cammà C, Puglia I, Calistri P, Savini G |
| EPI_ISL_961048, EPI_ISL_961049 | SIESP CHIETI - DRIVE IN ORTONA | Istituto Zooprofilattico Sperimentale dell'Abruzzo e Molise "G. Caporale" | Lorusso A, Marcacci M, Di Domenico M, Ancora M, Curini V, Mangone I, Rinaldi A, Scialabba S, Di Pasquale A, Cammà C, Puglia I, Calistri P, Savini G |
| EPI_ISL_961050, EPI_ISL_961051, | SIESP DIPARTIMENTO DI PREVENZIONE CHIETI | Istituto Zooprofilattico Sperimentale dell'Abruzzo e Molise "G. | Lorusso A, Marcacci M, Di Domenico M, Ancora M, Curini V, Mangone I, Rinaldi A, Scialabba S, Di Pasquale A, Cammà C, Puglia I, Calistri P, Savini G |

| | | | |
|---|---|---|---|
| EPI_ISL_961052, EPI_ISL_961053, EPI_ISL_961054 | | Caporale" | |
| EPI_ISL_961055, EPI_ISL_961056 | SIESP CHIETI - DRIVE IN ORTONA | Istituto Zooprofilattico Sperimentale dell'Abruzzo e Molise "G. Caporale" | Lorusso A, Marcacci M, Di Domenico M, Ancora M, Curini V, Mangone I, Rinaldi A, Scialabba S, Di Pasquale A, Cammà C, Puglia I, Calistri P, Savini G |
| EPI_ISL_961057, EPI_ISL_961058, EPI_ISL_961059, EPI_ISL_961060 | SIESP CHIETI - DRIVE IN CHIETI | Istituto Zooprofilattico Sperimentale dell'Abruzzo e Molise "G. Caporale" | Lorusso A, Marcacci M, Di Domenico M, Ancora M, Curini V, Mangone I, Rinaldi A, Scialabba S, Di Pasquale A, Cammà C, Puglia I, Calistri P, Savini G |
| EPI_ISL_961061 | SIESP CHIETI - DRIVE IN ORTONA | Istituto Zooprofilattico Sperimentale dell'Abruzzo e Molise "G. Caporale" | Lorusso A, Marcacci M, Di Domenico M, Ancora M, Curini V, Mangone I, Rinaldi A, Scialabba S, Di Pasquale A, Cammà C, Puglia I, Calistri P, Savini G |
| EPI_ISL_961062 | SIESP CHIETI - DRIVE IN CHIETI | Istituto Zooprofilattico Sperimentale dell'Abruzzo e Molise "G. Caporale" | Lorusso A, Marcacci M, Di Domenico M, Ancora M, Curini V, Mangone I, Rinaldi A, Scialabba S, Di Pasquale A, Cammà C, Puglia I, Calistri P, Savini G |
| EPI_ISL_961063 | SIESP DIPARTIMENTO DI PREVENZIONE CHIETI | Istituto Zooprofilattico Sperimentale dell'Abruzzo e Molise "G. Caporale" | Lorusso A, Marcacci M, Di Domenico M, Ancora M, Curini V, Mangone I, Rinaldi A, Scialabba S, Di Pasquale A, Cammà C, Puglia I, Calistri P, Savini G |
| EPI_ISL_961064, EPI_ISL_961065 | SIESP CHIETI - DRIVE IN ORTONA | Istituto Zooprofilattico Sperimentale dell'Abruzzo e Molise "G. Caporale" | Lorusso A, Marcacci M, Di Domenico M, Ancora M, Curini V, Mangone I, Rinaldi A, Scialabba S, Di Pasquale A, Cammà C, Puglia I, Calistri P, Savini G |
| EPI_ISL_961066 | SIESP CHIETI - DRIVE IN LANCIANO | Istituto Zooprofilattico Sperimentale dell'Abruzzo e Molise "G. Caporale" | Lorusso A, Marcacci M, Di Domenico M, Ancora M, Curini V, Mangone I, Rinaldi A, Scialabba S, Di Pasquale A, Cammà C, Puglia I, Calistri P, Savini G |
| EPI_ISL_961067, EPI_ISL_961068 | SIESP DIPARTIMENTO DI PREVENZIONE CHIETI | Istituto Zooprofilattico Sperimentale dell'Abruzzo e Molise "G. Caporale" | Lorusso A, Marcacci M, Di Domenico M, Ancora M, Curini V, Mangone I, Rinaldi A, Scialabba S, Di Pasquale A, Cammà C, Puglia I, Calistri P, Savini G |
| EPI_ISL_961069 | SIESP CHIETI - DRIVE IN CHIETI | Istituto Zooprofilattico Sperimentale dell'Abruzzo e Molise "G. Caporale" | Lorusso A, Marcacci M, Di Domenico M, Ancora M, Curini V, Mangone I, Rinaldi A, Scialabba S, Di Pasquale A, Cammà C, Puglia I, Calistri P, Savini G |
| EPI_ISL_961070 | SIESP DIPARTIMENTO DI PREVENZIONE CHIETI | Istituto Zooprofilattico Sperimentale dell'Abruzzo e Molise "G. Caporale" | Lorusso A, Marcacci M, Di Domenico M, Ancora M, Curini V, Mangone I, Rinaldi A, Scialabba S, Di Pasquale A, Cammà C, Puglia I, Calistri P, Savini G |
| EPI_ISL_961071, EPI_ISL_961072, EPI_ISL_961073 | SIESP CHIETI - DRIVE IN CHIETI | Istituto Zooprofilattico Sperimentale dell'Abruzzo e Molise "G. Caporale" | Lorusso A, Marcacci M, Di Domenico M, Ancora M, Curini V, Mangone I, Rinaldi A, Scialabba S, Di Pasquale A, Cammà C, Puglia I, Calistri P, Savini G |
| EPI_ISL_961074 | SIESP CHIETI - DRIVE IN ORTONA | Istituto Zooprofilattico Sperimentale dell'Abruzzo e Molise "G. Caporale" | Lorusso A, Marcacci M, Di Domenico M, Ancora M, Curini V, Mangone I, Rinaldi A, Scialabba S, Di Pasquale A, Cammà C, Puglia I, Calistri P, Savini G |
| EPI_ISL_961075 | SIESP DIPARTIMENTO DI PREVENZIONE CHIETI | Istituto Zooprofilattico Sperimentale dell'Abruzzo e Molise "G. Caporale" | Lorusso A, Marcacci M, Di Domenico M, Ancora M, Curini V, Mangone I, Rinaldi A, Scialabba S, Di Pasquale A, Cammà C, Puglia I, Calistri P, Savini G |
| EPI_ISL_961076 | SIESP CHIETI - DRIVE IN ORTONA | Istituto Zooprofilattico Sperimentale dell'Abruzzo e Molise "G. Caporale" | Lorusso A, Marcacci M, Di Domenico M, Ancora M, Curini V, Mangone I, Rinaldi A, Scialabba S, Di Pasquale A, Cammà C, Puglia I, Calistri P, Savini G |
| EPI_ISL_961077 | SIESP TERAMO | Istituto Zooprofilattico Sperimentale dell'Abruzzo e Molise "G. Caporale" | Lorusso A, Marcacci M, Di Domenico M, Ancora M, Curini V, Mangone I, Rinaldi A, Scialabba S, Di Pasquale A, Cammà C, Puglia I, Calistri P, Savini G |
| EPI_ISL_961078, EPI_ISL_961079, EPI_ISL_961080 | DIP. PREV. AVEZZANO SERVIZIO DI IGIENE EPIDEMIOLOGIAE SANITA' PUBBLICA | Istituto Zooprofilattico Sperimentale dell'Abruzzo e Molise "G. Caporale" | Lorusso A, Marcacci M, Di Domenico M, Ancora M, Curini V, Mangone I, Rinaldi A, Scialabba S, Di Pasquale A, Cammà C, Puglia I, Calistri P, Savini G |
| EPI_ISL_961081 | SIESP CHIETI - DRIVE IN CHIETI | Istituto Zooprofilattico Sperimentale dell'Abruzzo e Molise "G. Caporale" | Lorusso A, Marcacci M, Di Domenico M, Ancora M, Curini V, Mangone I, Rinaldi A, Scialabba S, Di Pasquale A, Cammà C, Puglia I, Calistri P, Savini G |
| EPI_ISL_961082 | SIESP DIPARTIMENTO DI PREVENZIONE CHIETI | Istituto Zooprofilattico Sperimentale dell'Abruzzo e Molise "G. Caporale" | Lorusso A, Marcacci M, Di Domenico M, Ancora M, Curini V, Mangone I, Rinaldi A, Scialabba S, Di Pasquale A, Cammà C, Puglia I, Calistri P, Savini G |
| EPI_ISL_961083 | DIP. PREV. AVEZZANO SERVIZIO DI IGIENE EPIDEMIOLOGIAE SANITA' PUBBLICA | Istituto Zooprofilattico Sperimentale dell'Abruzzo e Molise "G. Caporale" | Lorusso A, Marcacci M, Di Domenico M, Ancora M, Curini V, Mangone I, Rinaldi A, Scialabba S, Di Pasquale A, Cammà C, Puglia I, Calistri P, Savini G |
| EPI_ISL_961084 | SIESP TERAMO | Istituto Zooprofilattico Sperimentale dell'Abruzzo e Molise "G. Caporale" | Lorusso A, Marcacci M, Di Domenico M, Ancora M, Curini V, Mangone I, Rinaldi A, Scialabba S, Di Pasquale A, Cammà C, Puglia I, Calistri P, Savini G |
| EPI_ISL_961085 | SIESP CHIETI - DRIVE IN ORTONA | Istituto Zooprofilattico Sperimentale dell'Abruzzo e Molise "G. Caporale" | Lorusso A, Marcacci M, Di Domenico M, Ancora M, Curini V, Mangone I, Rinaldi A, Scialabba S, Di Pasquale A, Cammà C, Puglia I, Calistri P, Savini G |
| EPI_ISL_961086 | SIESP CHIETI - DRIVE IN CHIETI | Istituto Zooprofilattico Sperimentale dell'Abruzzo e Molise "G. Caporale" | Lorusso A, Marcacci M, Di Domenico M, Ancora M, Curini V, Mangone I, Rinaldi A, Scialabba S, Di Pasquale A, Cammà C, Puglia I, Calistri P, Savini G |
| EPI_ISL_961087, EPI_ISL_961088, EPI_ISL_961089 | SIESP DIPARTIMENTO DI PREVENZIONE CHIETI | Istituto Zooprofilattico Sperimentale dell'Abruzzo e Molise "G. Caporale" | Lorusso A, Marcacci M, Di Domenico M, Ancora M, Curini V, Mangone I, Rinaldi A, Scialabba S, Di Pasquale A, Cammà C, Puglia I, Calistri P, Savini G |
| EPI_ISL_961090, EPI_ISL_961091, EPI_ISL_961092 | SIESP CHIETI - DRIVE IN CHIETI | Istituto Zooprofilattico Sperimentale dell'Abruzzo e Molise "G. Caporale" | Lorusso A, Marcacci M, Di Domenico M, Ancora M, Curini V, Mangone I, Rinaldi A, Scialabba S, Di Pasquale A, Cammà C, Puglia I, Calistri P, Savini G |
| EPI_ISL_961093 | Ospedale SS Annunziata | Istituto Zooprofilattico Sperimentale dell'Abruzzo e Molise "G. Caporale" | Lorusso A, Marcacci M, Di Domenico M, Ancora M, Curini V, Mangone I, Rinaldi A, Scialabba S, Di Pasquale A, Cammà C, Puglia I, Calistri P, Savini G |
| EPI_ISL_961094, EPI_ISL_961095 | DIP. PREV. AVEZZANO SERVIZIO DI IGIENE EPIDEMIOLOGIAE SANITA' PUBBLICA | Istituto Zooprofilattico Sperimentale dell'Abruzzo e Molise "G. Caporale" | Lorusso A, Marcacci M, Di Domenico M, Ancora M, Curini V, Mangone I, Rinaldi A, Scialabba S, Di Pasquale A, Cammà C, Puglia I, Calistri P, Savini G |
| EPI_ISL_961096 | SIESP L'AQUILA | Istituto Zooprofilattico Sperimentale dell'Abruzzo e Molise "G. Caporale" | Lorusso A, Marcacci M, Di Domenico M, Ancora M, Curini V, Mangone I, Rinaldi A, Scialabba S, Di Pasquale A, Cammà C, Puglia I, Calistri P, Savini G |
| EPI_ISL_961097, EPI_ISL_961098, EPI_ISL_961099, EPI_ISL_961100, EPI_ISL_961101, EPI_ISL_961102, EPI_ISL_961103 | SIESP TERAMO | Istituto Zooprofilattico Sperimentale dell'Abruzzo e Molise "G. Caporale" | Lorusso A, Marcacci M, Di Domenico M, Ancora M, Curini V, Mangone I, Rinaldi A, Scialabba S, Di Pasquale A, Cammà C, Puglia I, Calistri P, Savini G |
| EPI_ISL_961673 | SIESP CHIETI DRIVE IN LANCIANO | Istituto Zooprofilattico Sperimentale dell'Abruzzo e Molise "G. Caporale" | Lorusso A, Marcacci M, Di Domenico M, Ancora M, Curini V, Mangone I, Rinaldi A, Scialabba S, Di Pasquale A, Cammà C, Puglia I, Calistri P, Savini G |
| EPI_ISL_961674, EPI_ISL_961675, EPI_ISL_961676, EPI_ISL_961677 | SIESP CHIETI - DRIVE IN CHIETI | Istituto Zooprofilattico Sperimentale dell'Abruzzo e Molise "G. Caporale" | Lorusso A, Marcacci M, Di Domenico M, Ancora M, Curini V, Mangone I, Rinaldi A, Scialabba S, Di Pasquale A, Cammà C, Puglia I, Calistri P, Savini G |
| EPI_ISL_961678 | SIESP TERAMO | Istituto Zooprofilattico Sperimentale dell'Abruzzo e Molise "G. Caporale" | Lorusso A, Marcacci M, Di Domenico M, Ancora M, Curini V, Mangone I, Rinaldi A, Scialabba S, Di Pasquale A, Cammà C, Puglia I, Calistri P, Savini G |
| EPI_ISL_961679, EPI_ISL_961680, EPI_ISL_961681, EPI_ISL_961682 | SIESP CHIETI - DRIVE IN CHIETI | Istituto Zooprofilattico Sperimentale dell'Abruzzo e Molise "G. Caporale" | Lorusso A, Marcacci M, Di Domenico M, Ancora M, Curini V, Mangone I, Rinaldi A, Scialabba S, Di Pasquale A, Cammà C, Puglia I, Calistri P, Savini G |
| EPI_ISL_961683 | SIESP TERAMO | Istituto Zooprofilattico Sperimentale dell'Abruzzo e Molise "G. Caporale" | Lorusso A, Marcacci M, Di Domenico M, Ancora M, Curini V, Mangone I, Rinaldi A, Scialabba S, Di Pasquale A, Cammà C, Puglia I, Calistri P, Savini G |
| EPI_ISL_961684 | SIESP CHIETI - DRIVE IN CHIETI | Istituto Zooprofilattico Sperimentale dell'Abruzzo e Molise "G. | Lorusso A, Marcacci M, Di Domenico M, Ancora M, Curini V, Mangone I, Rinaldi A, Scialabba S, Di Pasquale A, Cammà C, Puglia I, Calistri P, Savini G |

| | | | |
|---|---|---|---|
| | | Caporale" | |
| EPI_ISL_961685 | SIESP DIPARTIMENTO DI PREVENZIONE CHIETI | Istituto Zooprofilattico Sperimentale dell'Abruzzo e Molise "G. Caporale" | Lorusso A, Marcacci M, Di Domenico M, Ancora M, Curini V, Mangone I, Rinaldi A, Scialabba S, Di Pasquale A, Cammà C, Puglia I, Calistri P, Savini G |
| EPI_ISL_961686, EPI_ISL_961687 | SIESP CHIETI - DRIVE IN ORTONA | Istituto Zooprofilattico Sperimentale dell'Abruzzo e Molise "G. Caporale" | Lorusso A, Marcacci M, Di Domenico M, Ancora M, Curini V, Mangone I, Rinaldi A, Scialabba S, Di Pasquale A, Cammà C, Puglia I, Calistri P, Savini G |
| EPI_ISL_961688 | SIESP TERAMO | Istituto Zooprofilattico Sperimentale dell'Abruzzo e Molise "G. Caporale" | Lorusso A, Marcacci M, Di Domenico M, Ancora M, Curini V, Mangone I, Rinaldi A, Scialabba S, Di Pasquale A, Cammà C, Puglia I, Calistri P, Savini G |
| EPI_ISL_961689, EPI_ISL_961690 | SIESP CHIETI - DRIVE IN CHIETI | Istituto Zooprofilattico Sperimentale dell'Abruzzo e Molise "G. Caporale" | Lorusso A, Marcacci M, Di Domenico M, Ancora M, Curini V, Mangone I, Rinaldi A, Scialabba S, Di Pasquale A, Cammà C, Puglia I, Calistri P, Savini G |
| EPI_ISL_961691, EPI_ISL_961692, EPI_ISL_961693 | SIESP CHIETI - DRIVE IN ORTONA | Istituto Zooprofilattico Sperimentale dell'Abruzzo e Molise "G. Caporale" | Lorusso A, Marcacci M, Di Domenico M, Ancora M, Curini V, Mangone I, Rinaldi A, Scialabba S, Di Pasquale A, Cammà C, Puglia I, Calistri P, Savini G |
| EPI_ISL_961694 | SIESP CHIETI - DRIVE IN CHIETI | Istituto Zooprofilattico Sperimentale dell'Abruzzo e Molise "G. Caporale" | Lorusso A, Marcacci M, Di Domenico M, Ancora M, Curini V, Mangone I, Rinaldi A, Scialabba S, Di Pasquale A, Cammà C, Puglia I, Calistri P, Savini G |
| EPI_ISL_961695 | SIESP DIPARTIMENTO DI PREVENZIONE CHIETI | Istituto Zooprofilattico Sperimentale dell'Abruzzo e Molise "G. Caporale" | Lorusso A, Marcacci M, Di Domenico M, Ancora M, Curini V, Mangone I, Rinaldi A, Scialabba S, Di Pasquale A, Cammà C, Puglia I, Calistri P, Savini G |
| EPI_ISL_961696 | SIESP SULMONA | Istituto Zooprofilattico Sperimentale dell'Abruzzo e Molise "G. Caporale" | Lorusso A, Marcacci M, Di Domenico M, Ancora M, Curini V, Mangone I, Rinaldi A, Scialabba S, Di Pasquale A, Cammà C, Puglia I, Calistri P, Savini G |
| EPI_ISL_961697, EPI_ISL_961698 | SIESP CHIETI - DRIVE IN CHIETI | Istituto Zooprofilattico Sperimentale dell'Abruzzo e Molise "G. Caporale" | Lorusso A, Marcacci M, Di Domenico M, Ancora M, Curini V, Mangone I, Rinaldi A, Scialabba S, Di Pasquale A, Cammà C, Puglia I, Calistri P, Savini G |
| EPI_ISL_961699 | SIESP CHIETI - DRIVE IN ORTONA | Istituto Zooprofilattico Sperimentale dell'Abruzzo e Molise "G. Caporale" | Lorusso A, Marcacci M, Di Domenico M, Ancora M, Curini V, Mangone I, Rinaldi A, Scialabba S, Di Pasquale A, Cammà C, Puglia I, Calistri P, Savini G |
| EPI_ISL_961700 | SIESP CHIETI - DRIVE IN CHIETI | Istituto Zooprofilattico Sperimentale dell'Abruzzo e Molise "G. Caporale" | Lorusso A, Marcacci M, Di Domenico M, Ancora M, Curini V, Mangone I, Rinaldi A, Scialabba S, Di Pasquale A, Cammà C, Puglia I, Calistri P, Savini G |
| EPI_ISL_961701 | Ospedale Civile Teramo | Istituto Zooprofilattico Sperimentale dell'Abruzzo e Molise "G. Caporale" | Lorusso A, Marcacci M, Di Domenico M, Ancora M, Curini V, Mangone I, Rinaldi A, Scialabba S, Di Pasquale A, Cammà C, Puglia I, Calistri P, Savini G |
| EPI_ISL_961702 | SIESP CHIETI - DRIVE IN CHIETI | Istituto Zooprofilattico Sperimentale dell'Abruzzo e Molise "G. Caporale" | Lorusso A, Marcacci M, Di Domenico M, Ancora M, Curini V, Mangone I, Rinaldi A, Scialabba S, Di Pasquale A, Cammà C, Puglia I, Calistri P, Savini G |
| EPI_ISL_961703 | SIESP DIPARTIMENTO DI PREVENZIONE CHIETI | Istituto Zooprofilattico Sperimentale dell'Abruzzo e Molise "G. Caporale" | Lorusso A, Marcacci M, Di Domenico M, Ancora M, Curini V, Mangone I, Rinaldi A, Scialabba S, Di Pasquale A, Cammà C, Puglia I, Calistri P, Savini G |
| EPI_ISL_961704 | SIESP SULMONA | Istituto Zooprofilattico Sperimentale dell'Abruzzo e Molise "G. Caporale" | Lorusso A, Marcacci M, Di Domenico M, Ancora M, Curini V, Mangone I, Rinaldi A, Scialabba S, Di Pasquale A, Cammà C, Puglia I, Calistri P, Savini G |
| EPI_ISL_961705 | Ospedale San Salvatore UOC Malattie Infettive | Istituto Zooprofilattico Sperimentale dell'Abruzzo e Molise "G. Caporale" | Lorusso A, Marcacci M, Di Domenico M, Ancora M, Curini V, Mangone I, Rinaldi A, Scialabba S, Di Pasquale A, Cammà C, Puglia I, Calistri P, Savini G |
| EPI_ISL_961706 | SIESP CHIETI - DRIVE IN CHIETI | Istituto Zooprofilattico Sperimentale dell'Abruzzo e Molise "G. Caporale" | Lorusso A, Marcacci M, Di Domenico M, Ancora M, Curini V, Mangone I, Rinaldi A, Scialabba S, Di Pasquale A, Cammà C, Puglia I, Calistri P, Savini G |
| EPI_ISL_961707, EPI_ISL_961708 | SIESP DIPARTIMENTO DI PREVENZIONE CHIETI | Istituto Zooprofilattico Sperimentale dell'Abruzzo e Molise "G. Caporale" | Lorusso A, Marcacci M, Di Domenico M, Ancora M, Curini V, Mangone I, Rinaldi A, Scialabba S, Di Pasquale A, Cammà C, Puglia I, Calistri P, Savini G |
| EPI_ISL_961709, EPI_ISL_961710, EPI_ISL_961711 | SIESP CHIETI - DRIVE IN CHIETI | Istituto Zooprofilattico Sperimentale dell'Abruzzo e Molise "G. Caporale" | Lorusso A, Marcacci M, Di Domenico M, Ancora M, Curini V, Mangone I, Rinaldi A, Scialabba S, Di Pasquale A, Cammà C, Puglia I, Calistri P, Savini G |
| EPI_ISL_961712, EPI_ISL_961713 | SIESP CHIETI DRIVE IN LANCIANO | Istituto Zooprofilattico Sperimentale dell'Abruzzo e Molise "G. Caporale" | Lorusso A, Marcacci M, Di Domenico M, Ancora M, Curini V, Mangone I, Rinaldi A, Scialabba S, Di Pasquale A, Cammà C, Puglia I, Calistri P, Savini G |
| EPI_ISL_961714 | SIESP CHIETI - DRIVE IN CHIETI | Istituto Zooprofilattico Sperimentale dell'Abruzzo e Molise "G. Caporale" | Lorusso A, Marcacci M, Di Domenico M, Ancora M, Curini V, Mangone I, Rinaldi A, Scialabba S, Di Pasquale A, Cammà C, Puglia I, Calistri P, Savini G |
| EPI_ISL_961715, EPI_ISL_961716 | SIESP CHIETI - DRIVE IN ORTONA | Istituto Zooprofilattico Sperimentale dell'Abruzzo e Molise "G. Caporale" | Lorusso A, Marcacci M, Di Domenico M, Ancora M, Curini V, Mangone I, Rinaldi A, Scialabba S, Di Pasquale A, Cammà C, Puglia I, Calistri P, Savini G |
| EPI_ISL_961717 | SIESP CHIETI - DRIVE IN CHIETI | Istituto Zooprofilattico Sperimentale dell'Abruzzo e Molise "G. Caporale" | Lorusso A, Marcacci M, Di Domenico M, Ancora M, Curini V, Mangone I, Rinaldi A, Scialabba S, Di Pasquale A, Cammà C, Puglia I, Calistri P, Savini G |
| EPI_ISL_961718, EPI_ISL_961719, EPI_ISL_961720, EPI_ISL_961721, EPI_ISL_961722 | SIESP CHIETI - DRIVE IN ORTONA | Istituto Zooprofilattico Sperimentale dell'Abruzzo e Molise "G. Caporale" | Lorusso A, Marcacci M, Di Domenico M, Ancora M, Curini V, Mangone I, Rinaldi A, Scialabba S, Di Pasquale A, Cammà C, Puglia I, Calistri P, Savini G |
| EPI_ISL_961723 | SIESP CHIETI DRIVE IN LANCIANO | Istituto Zooprofilattico Sperimentale dell'Abruzzo e Molise "G. Caporale" | Lorusso A, Marcacci M, Di Domenico M, Ancora M, Curini V, Mangone I, Rinaldi A, Scialabba S, Di Pasquale A, Cammà C, Puglia I, Calistri P, Savini G |
| EPI_ISL_961724 | SIESP CHIETI - DRIVE IN ORTONA | Istituto Zooprofilattico Sperimentale dell'Abruzzo e Molise "G. Caporale" | Lorusso A, Marcacci M, Di Domenico M, Ancora M, Curini V, Mangone I, Rinaldi A, Scialabba S, Di Pasquale A, Cammà C, Puglia I, Calistri P, Savini G |
| EPI_ISL_961725 | SIESP CHIETI DRIVE IN GISSI | Istituto Zooprofilattico Sperimentale dell'Abruzzo e Molise "G. Caporale" | Lorusso A, Marcacci M, Di Domenico M, Ancora M, Curini V, Mangone I, Rinaldi A, Scialabba S, Di Pasquale A, Cammà C, Puglia I, Calistri P, Savini G |
| EPI_ISL_961726, EPI_ISL_961727 | SIESP CHIETI - DRIVE IN CHIETI | Istituto Zooprofilattico Sperimentale dell'Abruzzo e Molise "G. Caporale" | Lorusso A, Marcacci M, Di Domenico M, Ancora M, Curini V, Mangone I, Rinaldi A, Scialabba S, Di Pasquale A, Cammà C, Puglia I, Calistri P, Savini G |
| EPI_ISL_961728 | SIESP DIPARTIMENTO DI PREVENZIONE CHIETI | Istituto Zooprofilattico Sperimentale dell'Abruzzo e Molise "G. Caporale" | Lorusso A, Marcacci M, Di Domenico M, Ancora M, Curini V, Mangone I, Rinaldi A, Scialabba S, Di Pasquale A, Cammà C, Puglia I, Calistri P, Savini G |
| EPI_ISL_961729 | SIESP CHIETI - DRIVE IN CHIETI | Istituto Zooprofilattico Sperimentale dell'Abruzzo e Molise "G. Caporale" | Lorusso A, Marcacci M, Di Domenico M, Ancora M, Curini V, Mangone I, Rinaldi A, Scialabba S, Di Pasquale A, Cammà C, Puglia I, Calistri P, Savini G |
| EPI_ISL_961730 | Dipartimento Prevenzione Avezzano-Servizio Igiene epidemiologia Sanità Pubblica | Istituto Zooprofilattico Sperimentale dell'Abruzzo e Molise "G. Caporale" | Lorusso A, Marcacci M, Di Domenico M, Ancora M, Curini V, Mangone I, Rinaldi A, Scialabba S, Di Pasquale A, Cammà C, Puglia I, Calistri P, Savini G |
| EPI_ISL_961731, EPI_ISL_961732 | SIESP CHIETI - DRIVE IN CHIETI | Istituto Zooprofilattico Sperimentale dell'Abruzzo e Molise "G. Caporale" | Lorusso A, Marcacci M, Di Domenico M, Ancora M, Curini V, Mangone I, Rinaldi A, Scialabba S, Di Pasquale A, Cammà C, Puglia I, Calistri P, Savini G |
| EPI_ISL_961733 | SIESP CHIETI - DRIVE IN ORTONA | Istituto Zooprofilattico Sperimentale dell'Abruzzo e Molise "G. Caporale" | Lorusso A, Marcacci M, Di Domenico M, Ancora M, Curini V, Mangone I, Rinaldi A, Scialabba S, Di Pasquale A, Cammà C, Puglia I, Calistri P, Savini G |
| EPI_ISL_961734 | Dipartimento Prevenzione Avezzano-Servizio Igiene epidemiologia Sanità Pubblica | Istituto Zooprofilattico Sperimentale dell'Abruzzo e Molise "G. Caporale" | Lorusso A, Marcacci M, Di Domenico M, Ancora M, Curini V, Mangone I, Rinaldi A, Scialabba S, Di Pasquale A, Cammà C, Puglia I, Calistri P, Savini G |

| | | | |
|---|---|---|---|
| EPI_ISL_961735, EPI_ISL_961736 | SIESP CHIETI - DRIVE IN CHIETI | Istituto Zooprofilattico Sperimentale dell'Abruzzo e Molise "G. Caporale" | Lorusso A, Marcacci M, Di Domenico M, Ancora M, Curini V, Mangone I, Rinaldi A, Scialabba S, Di Pasquale A, Cammà C, Puglia I, Calistri P, Savini G |
| EPI_ISL_961737 | SIESP CHIETI- DRIVE IN VASTO | Istituto Zooprofilattico Sperimentale dell'Abruzzo e Molise "G. Caporale" | Lorusso A, Marcacci M, Di Domenico M, Ancora M, Curini V, Mangone I, Rinaldi A, Scialabba S, Di Pasquale A, Cammà C, Puglia I, Calistri P, Savini G |
| EPI_ISL_961738, EPI_ISL_961739 | SIESP CHIETI DRIVE IN LANCIANO | Istituto Zooprofilattico Sperimentale dell'Abruzzo e Molise "G. Caporale" | Lorusso A, Marcacci M, Di Domenico M, Ancora M, Curini V, Mangone I, Rinaldi A, Scialabba S, Di Pasquale A, Cammà C, Puglia I, Calistri P, Savini G |
| EPI_ISL_961740 | SIESP CHIETI - DRIVE IN CHIETI | Istituto Zooprofilattico Sperimentale dell'Abruzzo e Molise "G. Caporale" | Lorusso A, Marcacci M, Di Domenico M, Ancora M, Curini V, Mangone I, Rinaldi A, Scialabba S, Di Pasquale A, Cammà C, Puglia I, Calistri P, Savini G |
| EPI_ISL_961741 | SIESP CHIETI- DRIVE IN VASTO | Istituto Zooprofilattico Sperimentale dell'Abruzzo e Molise "G. Caporale" | Lorusso A, Marcacci M, Di Domenico M, Ancora M, Curini V, Mangone I, Rinaldi A, Scialabba S, Di Pasquale A, Cammà C, Puglia I, Calistri P, Savini G |
| EPI_ISL_961742 | SIESP CHIETI - DRIVE IN CHIETI | Istituto Zooprofilattico Sperimentale dell'Abruzzo e Molise "G. Caporale" | Lorusso A, Marcacci M, Di Domenico M, Ancora M, Curini V, Mangone I, Rinaldi A, Scialabba S, Di Pasquale A, Cammà C, Puglia I, Calistri P, Savini G |
| EPI_ISL_961743 | SIESP CHIETI DRIVE IN GISSI | Istituto Zooprofilattico Sperimentale dell'Abruzzo e Molise "G. Caporale" | Lorusso A, Marcacci M, Di Domenico M, Ancora M, Curini V, Mangone I, Rinaldi A, Scialabba S, Di Pasquale A, Cammà C, Puglia I, Calistri P, Savini G |
| EPI_ISL_961744 | SIESP CHIETI - DRIVE IN ORTONA | Istituto Zooprofilattico Sperimentale dell'Abruzzo e Molise "G. Caporale" | Lorusso A, Marcacci M, Di Domenico M, Ancora M, Curini V, Mangone I, Rinaldi A, Scialabba S, Di Pasquale A, Cammà C, Puglia I, Calistri P, Savini G |
| EPI_ISL_961745 | SIESP CHIETI DRIVE IN GISSI | Istituto Zooprofilattico Sperimentale dell'Abruzzo e Molise "G. Caporale" | Lorusso A, Marcacci M, Di Domenico M, Ancora M, Curini V, Mangone I, Rinaldi A, Scialabba S, Di Pasquale A, Cammà C, Puglia I, Calistri P, Savini G |
| EPI_ISL_961746 | SIESP CHIETI - DRIVE IN CHIETI | Istituto Zooprofilattico Sperimentale dell'Abruzzo e Molise "G. Caporale" | Lorusso A, Marcacci M, Di Domenico M, Ancora M, Curini V, Mangone I, Rinaldi A, Scialabba S, Di Pasquale A, Cammà C, Puglia I, Calistri P, Savini G |
| EPI_ISL_961747 | Dipartimento Prevenzione Avezzano-Servizio Igiene epidemiologia Sanità Pubblica | Istituto Zooprofilattico Sperimentale dell'Abruzzo e Molise "G. Caporale" | Lorusso A, Marcacci M, Di Domenico M, Ancora M, Curini V, Mangone I, Rinaldi A, Scialabba S, Di Pasquale A, Cammà C, Puglia I, Calistri P, Savini G |
| EPI_ISL_961748 | SIESP CHIETI DRIVE IN LANCIANO | Istituto Zooprofilattico Sperimentale dell'Abruzzo e Molise "G. Caporale" | Lorusso A, Marcacci M, Di Domenico M, Ancora M, Curini V, Mangone I, Rinaldi A, Scialabba S, Di Pasquale A, Cammà C, Puglia I, Calistri P, Savini G |
| EPI_ISL_961749 | SIESP DIPARTIMENTO DI PREVENZIONE CHIETI | Istituto Zooprofilattico Sperimentale dell'Abruzzo e Molise "G. Caporale" | Lorusso A, Marcacci M, Di Domenico M, Ancora M, Curini V, Mangone I, Rinaldi A, Scialabba S, Di Pasquale A, Cammà C, Puglia I, Calistri P, Savini G |
| EPI_ISL_961750 | Ospedale Cvile Atri Med. Interna | Istituto Zooprofilattico Sperimentale dell'Abruzzo e Molise "G. Caporale" | Lorusso A, Marcacci M, Di Domenico M, Ancora M, Curini V, Mangone I, Rinaldi A, Scialabba S, Di Pasquale A, Cammà C, Puglia I, Calistri P, Savini G |
| EPI_ISL_961751 | SIESP DIPARTIMENTO DI PREVENZIONE CHIETI | Istituto Zooprofilattico Sperimentale dell'Abruzzo e Molise "G. Caporale" | Lorusso A, Marcacci M, Di Domenico M, Ancora M, Curini V, Mangone I, Rinaldi A, Scialabba S, Di Pasquale A, Cammà C, Puglia I, Calistri P, Savini G |
| EPI_ISL_961752, EPI_ISL_961753 | SIESP SULMONA | Istituto Zooprofilattico Sperimentale dell'Abruzzo e Molise "G. Caporale" | Lorusso A, Marcacci M, Di Domenico M, Ancora M, Curini V, Mangone I, Rinaldi A, Scialabba S, Di Pasquale A, Cammà C, Puglia I, Calistri P, Savini G |
| EPI_ISL_961754 | Presidio Ospedaliero Sulmona | Istituto Zooprofilattico Sperimentale dell'Abruzzo e Molise "G. Caporale" | Lorusso A, Marcacci M, Di Domenico M, Ancora M, Curini V, Mangone I, Rinaldi A, Scialabba S, Di Pasquale A, Cammà C, Puglia I, Calistri P, Savini G |
| EPI_ISL_961755, EPI_ISL_961756, EPI_ISL_961757, EPI_ISL_961758 | SIESP SULMONA | Istituto Zooprofilattico Sperimentale dell'Abruzzo e Molise "G. Caporale" | Lorusso A, Marcacci M, Di Domenico M, Ancora M, Curini V, Mangone I, Rinaldi A, Scialabba S, Di Pasquale A, Cammà C, Puglia I, Calistri P, Savini G |
| EPI_ISL_965025 | S.C. Microbiologia e Virologia Laboratorio Virologia -Speciale Centro Influenza - AOU di Sassari - Viale san Pietro 43/B Palazzo Infettivologia | Laboratorio specialistico UOC Ematologia - Ospedale "San Francesco" - ATS-ASSL Nuoro | Piras Giovanna, Malune Paolo, Asproni Rosanna, Monne Maria Itria, Palmas Angelo Domenico Serra Caterina, Rimini Elena, Rubino Salvatore |
| EPI_ISL_965028, EPI_ISL_965031 | Laboratorio Biologia Molecolare Sars Cov2 - UOC Laboratorio Analisi - Servizio Medicina di Laboratorio, Ospedale "San Francesco" - ATS-ASSL Nuoro | Laboratorio specialistico UOC Ematologia - Ospedale "San Francesco" - ATS-ASSL Nuoro | Piras Giovanna, Asproni Rosanna, Malune Paolo, Fiamma Maura, Monne Maria Itria, Palmas Angelo Domenico, Lo Maglio Iana, Mameli Giuseppe |
| EPI_ISL_965126 | San Gallicano Dermatological Institute I.F.O. | INMI Lazzaro Spallanzani IRCCS | E Giombini, M. Rueca, B Bartolini, O Butera, C.E.M Gruber, F Messina, F Pimpinelli, F Ensoli, G Prignano, , A Di Caro, MR Capobianchi |
| EPI_ISL_965127 | INMI Lazzaro Spallanzani IRCCS | INMI Lazzaro Spallanzani IRCCS | CEM Gruber, B Bartolini, E Giombini, M Rueca, O Butera, F Messina, A Di Caro, MR Capobianchi |
| EPI_ISL_965128 | INMI Lazzaro Spallanzani IRCCS | INMI Lazzaro Spallanzani IRCCS | M. Rueca, E Giombini, B Bartolini, O Butera, C.E.M Gruber, F Messina, A Di Caro, MR Capobianchi |
| EPI_ISL_965129 | Sant'Eugenio/CTO ASL Roma 2. | INMI Lazzaro Spallanzani IRCCS | F Messina, C.E.M Gruber, B Bartolini, E Giombini, M Rueca, O Butera, F Bondanini, GC Cocciolillo, C Disegni, A Di Caro, MR Capobianchi |
| EPI_ISL_965130 | Ospedale Sandro Pertini ASL Roma2 | INMI Lazzaro Spallanzani IRCCS | B Bartolini, O Butera, C.E.M Gruber, M Rueca, F Messina, E Giombini, R Longo, V Michela, MR Capobianchi, A Di Caro |
| EPI_ISL_965131 | Azienda Ospedaliera San Giovanni Addolorata | INMI Lazzaro Spallanzani IRCCS | O Butera, F Messina, CEM Gruber, B Bartolini, E Giombini, M Rueca, M Gaudio, PM Placanica, MR Capobianchi, A Di Caro |
| EPI_ISL_965132 | Azienda Ospedaliera San Giovanni Addolorata | INMI Lazzaro Spallanzani IRCCS | CEM Gruber, B Bartolini, E Giombini, M Rueca, O Butera, F Messina, M Gaudio, PM Placanica, A Di Caro, MR Capobianchi |
| EPI_ISL_965133 | Azienda Ospedaliera San Camillo Forlanini | INMI Lazzaro Spallanzani IRCCS | B Bartolini, O Butera, C.E.M Gruber, M Rueca, F Messina, E Giombini, F Basile, A D'Agostino, G Parisi, MR Capobianchi, A Di Caro |
| EPI_ISL_965134 | San Gallicano Dermatological Institute I.F.O. | INMI Lazzaro Spallanzani IRCCS | M Rueca, O Butera, F Messina, CEM Gruber, B Bartolini, E Giombini, F Pimpinelli, F Ensoli, A Mastrofrancesco, A Di Caro, MR Capobianchi |
| EPI_ISL_965135 | Presidio Ospedaliero G. B. Grassi ASL RM3 | INMI Lazzaro Spallanzani IRCCS | F Messina, C.E.M Gruber, B Bartolini, E Giombini, M Rueca, O Butera, F Tabacco, E Ristori, A Di Caro, MR Capobianchi |
| EPI_ISL_965136 | Azienda Sanitaria Locale Roma 5 | INMI Lazzaro Spallanzani IRCCS | O Butera, F Messina, CEM Gruber, B Bartolini, E Giombini, M Rueca, D Di Fusco, D Cerini, MR Capobianchi, A Di Caro |
| EPI_ISL_965137 | Fondazione Policlinico Universitario "A. Gemelli" IRCCS | INMI Lazzaro Spallanzani IRCCS | E Giombini, M. Rueca, B Bartolini, O Butera, C.E.M Gruber, F Messina, P Cattani,M Sanguinetti, A Di Caro, MR Capobianchi |
| EPI_ISL_965139 | Fondazione Policlinico Universitario "A. Gemelli" IRCCS | INMI Lazzaro Spallanzani IRCCS | B Bartolini, O Butera, C.E.M Gruber, M Rueca, F Messina, E Giombini, P Cattani,M Sanguinetti, MR Capobianchi, A Di Caro |
| EPI_ISL_965140 | UOC "Microbiologia e Virologia" AOU "Policlinico Umberto I", Roma; Dipartimento di Medicina Molecolare Sapienza Università di Roma | INMI Lazzaro Spallanzani IRCCS | F Messina, C.E.M Gruber, B Bartolini, E Giombini, M Rueca, O Butera, G Antonelli, O Turriziani, MR Capobianchi, A Di Caro |
| EPI_ISL_965223 | SIESP SULMONA | Istituto Zooprofilattico Sperimentale dell'Abruzzo e Molise "G. Caporale" | Lorusso A, Marcacci M, Di Domenico M, Ancora M, Curini V, Mangone I, Rinaldi A, Scialabba S, Di Pasquale A, Cammà C, Puglia I, Calistri P, Savini G |
| EPI_ISL_965224 | OSPEDALE SS. ANNUNZIATA CHIETI | Istituto Zooprofilattico Sperimentale dell'Abruzzo e Molise "G. Caporale" | Lorusso A, Marcacci M, Di Domenico M, Ancora M, Curini V, Mangone I, Rinaldi A, Scialabba S, Di Pasquale A, Cammà C, Puglia I, Calistri P, Savini G |
| EPI_ISL_965225, EPI_ISL_965226 | USCA SULMONA | Istituto Zooprofilattico Sperimentale dell'Abruzzo e Molise "G. Caporale" | Lorusso A, Marcacci M, Di Domenico M, Ancora M, Curini V, Mangone I, Rinaldi A, Scialabba S, Di Pasquale A, Cammà C, Puglia I, Calistri P, Savini G |
| EPI_ISL_965227, EPI_ISL_965228, EPI_ISL_965229 | SIESP CHIETI - DRIVE IN CHIETI | Istituto Zooprofilattico Sperimentale dell'Abruzzo e Molise "G. Caporale" | Lorusso A, Marcacci M, Di Domenico M, Ancora M, Curini V, Mangone I, Rinaldi A, Scialabba S, Di Pasquale A, Cammà C, Puglia I, Calistri P, Savini G |
| EPI_ISL_965230 | SIESP TERAMO DIP PREV | Istituto Zooprofilattico Sperimentale dell'Abruzzo e Molise "G. Caporale" | Lorusso A, Marcacci M, Di Domenico M, Ancora M, Curini V, Mangone I, Rinaldi A, Scialabba S, Di Pasquale A, Cammà C, Puglia I, Calistri P, Savini G |

| | | | |
|---|---|---|---|
| EPI_ISL_965231 | Ospedale San Salvatore UOC MED INT. | Istituto Zooprofilattico Sperimentale dell'Abruzzo e Molise "G. Caporale" | Lorusso A, Marcacci M, Di Domenico M, Ancora M, Curini V, Mangone I, Rinaldi A, Scialabba S, Di Pasquale A, Cammà C, Puglia I, Calistri P, Savini G |
| EPI_ISL_965232, EPI_ISL_965233 | SIESP CHIETI - DRIVE IN CHIETI | Istituto Zooprofilattico Sperimentale dell'Abruzzo e Molise "G. Caporale" | Lorusso A, Marcacci M, Di Domenico M, Ancora M, Curini V, Mangone I, Rinaldi A, Scialabba S, Di Pasquale A, Cammà C, Puglia I, Calistri P, Savini G |
| EPI_ISL_965234 | SIESP SULMONA | Istituto Zooprofilattico Sperimentale dell'Abruzzo e Molise "G. Caporale" | Lorusso A, Marcacci M, Di Domenico M, Ancora M, Curini V, Mangone I, Rinaldi A, Scialabba S, Di Pasquale A, Cammà C, Puglia I, Calistri P, Savini G |
| EPI_ISL_965235 | SIESP TERAMO DIP PREV | Istituto Zooprofilattico Sperimentale dell'Abruzzo e Molise "G. Caporale" | Lorusso A, Marcacci M, Di Domenico M, Ancora M, Curini V, Mangone I, Rinaldi A, Scialabba S, Di Pasquale A, Cammà C, Puglia I, Calistri P, Savini G |
| EPI_ISL_965236, EPI_ISL_965237, EPI_ISL_965238, EPI_ISL_965239 | SIESP CHIETI - DRIVE IN CHIETI | Istituto Zooprofilattico Sperimentale dell'Abruzzo e Molise "G. Caporale" | Lorusso A, Marcacci M, Di Domenico M, Ancora M, Curini V, Mangone I, Rinaldi A, Scialabba S, Di Pasquale A, Cammà C, Puglia I, Calistri P, Savini G |
| EPI_ISL_965240 | SIESP CHIETI | Istituto Zooprofilattico Sperimentale dell'Abruzzo e Molise "G. Caporale" | Lorusso A, Marcacci M, Di Domenico M, Ancora M, Curini V, Mangone I, Rinaldi A, Scialabba S, Di Pasquale A, Cammà C, Puglia I, Calistri P, Savini G |
| EPI_ISL_965241, EPI_ISL_965242 | USCA SULMONA | Istituto Zooprofilattico Sperimentale dell'Abruzzo e Molise "G. Caporale" | Lorusso A, Marcacci M, Di Domenico M, Ancora M, Curini V, Mangone I, Rinaldi A, Scialabba S, Di Pasquale A, Cammà C, Puglia I, Calistri P, Savini G |
| EPI_ISL_965243, EPI_ISL_965244 | SIESP CHIETI-DRIVE IN ORTONA | Istituto Zooprofilattico Sperimentale dell'Abruzzo e Molise "G. Caporale" | Lorusso A, Marcacci M, Di Domenico M, Ancora M, Curini V, Mangone I, Rinaldi A, Scialabba S, Di Pasquale A, Cammà C, Puglia I, Calistri P, Savini G |
| EPI_ISL_965245 | OSPEDALE CIVILE TERAMO, DIV. PNEUMOLOGIA, PIA | Istituto Zooprofilattico Sperimentale dell'Abruzzo e Molise "G. Caporale" | Lorusso A, Marcacci M, Di Domenico M, Ancora M, Curini V, Mangone I, Rinaldi A, Scialabba S, Di Pasquale A, Cammà C, Puglia I, Calistri P, Savini G |
| EPI_ISL_965246 | OSP CIV GIULIANOVA, MED INT | Istituto Zooprofilattico Sperimentale dell'Abruzzo e Molise "G. Caporale" | Lorusso A, Marcacci M, Di Domenico M, Ancora M, Curini V, Mangone I, Rinaldi A, Scialabba S, Di Pasquale A, Cammà C, Puglia I, Calistri P, Savini G |
| EPI_ISL_965247, EPI_ISL_965248, EPI_ISL_965249, EPI_ISL_965250, EPI_ISL_965251, EPI_ISL_965252, EPI_ISL_965253 | SIESP CHIETI | Istituto Zooprofilattico Sperimentale dell'Abruzzo e Molise "G. Caporale" | Lorusso A, Marcacci M, Di Domenico M, Ancora M, Curini V, Mangone I, Rinaldi A, Scialabba S, Di Pasquale A, Cammà C, Puglia I, Calistri P, Savini G |
| EPI_ISL_965254, EPI_ISL_965255 | SIESP CHIETI - DRIVE IN CHIETI | Istituto Zooprofilattico Sperimentale dell'Abruzzo e Molise "G. Caporale" | Lorusso A, Marcacci M, Di Domenico M, Ancora M, Curini V, Mangone I, Rinaldi A, Scialabba S, Di Pasquale A, Cammà C, Puglia I, Calistri P, Savini G |
| EPI_ISL_965256 | SIESP CHIETI-DRIVE IN VASTO | Istituto Zooprofilattico Sperimentale dell'Abruzzo e Molise "G. Caporale" | Lorusso A, Marcacci M, Di Domenico M, Ancora M, Curini V, Mangone I, Rinaldi A, Scialabba S, Di Pasquale A, Cammà C, Puglia I, Calistri P, Savini G |
| EPI_ISL_965257 | SIESP CHIETI-DRIVE IN GISSI | Istituto Zooprofilattico Sperimentale dell'Abruzzo e Molise "G. Caporale" | Lorusso A, Marcacci M, Di Domenico M, Ancora M, Curini V, Mangone I, Rinaldi A, Scialabba S, Di Pasquale A, Cammà C, Puglia I, Calistri P, Savini G |
| EPI_ISL_965258, EPI_ISL_965259 | SIESP CHIETI-DRIVE IN VASTO | Istituto Zooprofilattico Sperimentale dell'Abruzzo e Molise "G. Caporale" | Lorusso A, Marcacci M, Di Domenico M, Ancora M, Curini V, Mangone I, Rinaldi A, Scialabba S, Di Pasquale A, Cammà C, Puglia I, Calistri P, Savini G |
| EPI_ISL_965260 | SIESP CHIETI-DRIVE IN GISSI | Istituto Zooprofilattico Sperimentale dell'Abruzzo e Molise "G. Caporale" | Lorusso A, Marcacci M, Di Domenico M, Ancora M, Curini V, Mangone I, Rinaldi A, Scialabba S, Di Pasquale A, Cammà C, Puglia I, Calistri P, Savini G |
| EPI_ISL_965261 | SIESP CHIETI | Istituto Zooprofilattico Sperimentale dell'Abruzzo e Molise "G. Caporale" | Lorusso A, Marcacci M, Di Domenico M, Ancora M, Curini V, Mangone I, Rinaldi A, Scialabba S, Di Pasquale A, Cammà C, Puglia I, Calistri P, Savini G |
| EPI_ISL_965262 | SIESP CHIETI, DISTRETTO SAN CHIETI | Istituto Zooprofilattico Sperimentale dell'Abruzzo e Molise "G. Caporale" | Lorusso A, Marcacci M, Di Domenico M, Ancora M, Curini V, Mangone I, Rinaldi A, Scialabba S, Di Pasquale A, Cammà C, Puglia I, Calistri P, Savini G |
| EPI_ISL_965263 | SIESP CHIETI-DRIVE IN ORTONA | Istituto Zooprofilattico Sperimentale dell'Abruzzo e Molise "G. Caporale" | Lorusso A, Marcacci M, Di Domenico M, Ancora M, Curini V, Mangone I, Rinaldi A, Scialabba S, Di Pasquale A, Cammà C, Puglia I, Calistri P, Savini G |
| EPI_ISL_965265 | SIESP CHIETI - DRIVE IN CHIETI | Istituto Zooprofilattico Sperimentale dell'Abruzzo e Molise "G. Caporale" | Lorusso A, Marcacci M, Di Domenico M, Ancora M, Curini V, Mangone I, Rinaldi A, Scialabba S, Di Pasquale A, Cammà C, Puglia I, Calistri P, Savini G |
| EPI_ISL_965266 | SIESP CHIETI | Istituto Zooprofilattico Sperimentale dell'Abruzzo e Molise "G. Caporale" | Lorusso A, Marcacci M, Di Domenico M, Ancora M, Curini V, Mangone I, Rinaldi A, Scialabba S, Di Pasquale A, Cammà C, Puglia I, Calistri P, Savini G |
| EPI_ISL_965267 | SIESP CHIETI, DISTRETTON SAN CHIETI | Istituto Zooprofilattico Sperimentale dell'Abruzzo e Molise "G. Caporale" | Lorusso A, Marcacci M, Di Domenico M, Ancora M, Curini V, Mangone I, Rinaldi A, Scialabba S, Di Pasquale A, Cammà C, Puglia I, Calistri P, Savini G |
| EPI_ISL_965268, EPI_ISL_965269 | SIESP CHIETI, DISTRETTO SAN CHIETI | Istituto Zooprofilattico Sperimentale dell'Abruzzo e Molise "G. Caporale" | Lorusso A, Marcacci M, Di Domenico M, Ancora M, Curini V, Mangone I, Rinaldi A, Scialabba S, Di Pasquale A, Cammà C, Puglia I, Calistri P, Savini G |
| EPI_ISL_965270 | SIESP CHIETI | Istituto Zooprofilattico Sperimentale dell'Abruzzo e Molise "G. Caporale" | Lorusso A, Marcacci M, Di Domenico M, Ancora M, Curini V, Mangone I, Rinaldi A, Scialabba S, Di Pasquale A, Cammà C, Puglia I, Calistri P, Savini G |
| EPI_ISL_965271 | SIESP CHIETI, DISTRETTO SAN CHIETI | Istituto Zooprofilattico Sperimentale dell'Abruzzo e Molise "G. Caporale" | Lorusso A, Marcacci M, Di Domenico M, Ancora M, Curini V, Mangone I, Rinaldi A, Scialabba S, Di Pasquale A, Cammà C, Puglia I, Calistri P, Savini G |
| EPI_ISL_965272 | SIESP CHIETI | Istituto Zooprofilattico Sperimentale dell'Abruzzo e Molise "G. Caporale" | Lorusso A, Marcacci M, Di Domenico M, Ancora M, Curini V, Mangone I, Rinaldi A, Scialabba S, Di Pasquale A, Cammà C, Puglia I, Calistri P, Savini G |
| EPI_ISL_965273, EPI_ISL_965274, EPI_ISL_965275 | SIESP CHIETI, DISTRETTO SAN CHIETI | Istituto Zooprofilattico Sperimentale dell'Abruzzo e Molise "G. Caporale" | Lorusso A, Marcacci M, Di Domenico M, Ancora M, Curini V, Mangone I, Rinaldi A, Scialabba S, Di Pasquale A, Cammà C, Puglia I, Calistri P, Savini G |
| EPI_ISL_965276 | OSP CIV ATRI | Istituto Zooprofilattico Sperimentale dell'Abruzzo e Molise "G. Caporale" | Lorusso A, Marcacci M, Di Domenico M, Ancora M, Curini V, Mangone I, Rinaldi A, Scialabba S, Di Pasquale A, Cammà C, Puglia I, Calistri P, Savini G |
| EPI_ISL_965278, EPI_ISL_965279, EPI_ISL_965280, EPI_ISL_965281, EPI_ISL_965282, EPI_ISL_965283, EPI_ISL_965284, EPI_ISL_965285, EPI_ISL_965286, EPI_ISL_965287, EPI_ISL_965288, EPI_ISL_965289, EPI_ISL_965290, EPI_ISL_965291, EPI_ISL_965292, EPI_ISL_965293, EPI_ISL_965294, EPI_ISL_965295, EPI_ISL_965296, EPI_ISL_965297, EPI_ISL_965298, EPI_ISL_965299, EPI_ISL_965300 | | | |
| see above | P.O.CARDARELLI | Istituto Zooprofilattico Sperimentale dell'Abruzzo e Molise "G. Caporale" | Scutellà M, Niro G, Lorusso A, Marcacci M, Di Domenico M, Ancora M, Curini V, Mangone I, Rinaldi A, Scialabba S, Di Pasquale A, Cammà C, Puglia I, Calistri P, Savini G |
| EPI_ISL_969131 | Laboratorio Microbiologia e Virologia P.O. Cotugno A.O. dei Colli | Laboratorio Microbiologia e Virologia P.O. Cotugno A.O. dei Colli | Luigi Atripaldi, Claudia Tiberio, Anna Perfetti, |
| EPI_ISL_969227 | Laboratorio Microbiologia e Virologia P.O. Cotugno A.O. dei Colli | Laboratorio Microbiologia e Virologia P.O. Cotugno A.O. dei Colli | Luigi Atripaldi, Claudia Tiberio, Anna Perfetti |
| EPI_ISL_969297 | Laboratorio Microbiologia e Virologia, P.O. Cotugno, A.O. dei Colli | Laboratorio Microbiologia e Virologia, P.O. Cotugno, A.O. dei Colli | Luigi Atripaldi, Claudia Tiberio, Anna Perfetti |
| EPI_ISL_969884 | Laboratorio Microbiologia e Virologia P.O. Cotugno A.O. dei Colli | Laboratorio Microbiologia e Virologia P.O. Cotugno A.O. dei Colli | Luigi Atripaldi,Claudia Tiberio, Anna Perfetti |
| EPI_ISL_970647 | Laboratorio Microbiologia e Virologia P.O. Cotugno A.O. dei | Laboratorio Microbiologia e Virologia P.O. Cotugno A.O. dei | Luigi Atripaldi, Claudia Tiberio,Anna Perfetti |

| | | | |
|---|---|---|---|
| | Colli | Colli | |
| EPI_ISL_974745 | Laboratorio Microbiologia e Virologia P.O. Cotugno A.O. dei Colli | Laboratorio Microbiologia e Virologia P.O. Cotugno A.O. dei Colli | Luigi Atripaldi, Claudia Tiberio, Anna Perfetti, |
| EPI_ISL_977155, EPI_ISL_977156, EPI_ISL_977157, EPI_ISL_977158 | ULSS 03 Venezia | Istituto Zooprofilattico Sperimentale delle Venezie | Adelaide Milani, Alessia Schivo, Annalisa Salviato, Erika Giorgia Quaranta, Ambra Pastori, Bianca Zecchin, Alice Fusaro, Isabella Monne, Calogero Terregino, Antonia Ricci |
| EPI_ISL_977160, EPI_ISL_977210, EPI_ISL_977211 | Microbiologia e Virologia | Istituto Zooprofilattico Sperimentale delle Venezie | Adelaide Milani, Alessia Schivo, Annalisa Salviato, Erika Giorgia Quaranta, Ambra Pastori, Bianca Zecchin, Alice Fusaro, Isabella Monne, Calogero Terregino, Antonia Ricci |
| EPI_ISL_977240, EPI_ISL_977241 | ULSS 5 Polesana | Istituto Zooprofilattico Sperimentale delle Venezie | Adelaide Milani, Alessia Schivo, Annalisa Salviato, Erika Giorgia Quaranta, Ambra Pastori, Bianca Zecchin, Alice Fusaro, Isabella Monne, Calogero Terregino, Antonia Ricci |
| EPI_ISL_977246 | Microbiologia e Virologia | Istituto Zooprofilattico Sperimentale delle Venezie | Adelaide Milani, Alessia Schivo, Annalisa Salviato, Erika Giorgia Quaranta, Ambra Pastori, Bianca Zecchin, Alice Fusaro, Isabella Monne, Calogero Terregino, Antonia Ricci |
| EPI_ISL_977249, EPI_ISL_977250 | ULSS 03 Venezia | Istituto Zooprofilattico Sperimentale delle Venezie | Adelaide Milani, Alessia Schivo, Annalisa Salviato, Erika Giorgia Quaranta, Ambra Pastori, Bianca Zecchin, Alice Fusaro, Isabella Monne, Calogero Terregino, Antonia Ricci |
| EPI_ISL_977495, EPI_ISL_977496, EPI_ISL_977497, EPI_ISL_977498 | University of Bari Biomedical Sciences and Human Oncology | University of Bari Biomedical Sciences and Human Oncology | Chironna M., Sallustio A., Loconsole D., Accogli M. |
| EPI_ISL_977598 | Laboratorio Microbiologia e Virologia P.O. Cotugno A.O. dei Colli | Laboratorio Microbiologia e Virologia P.O. Cotugno A.O. dei Colli | Luigi Atripaldi, Claudia Tiberio, Anna Perfetti |
| EPI_ISL_977601, EPI_ISL_977603 | Laboratorio Microbiologia e Virologia P.O. Cotugno A.O. dei Colli | Laboratorio Microbiologia e Virologia P.O. Cotugno A.O. dei Colli | Luigi Atripaldi, Claudia Tiberio, Anna Perfetti, |
| EPI_ISL_977604, EPI_ISL_977605, EPI_ISL_977606, EPI_ISL_977607, EPI_ISL_977608, EPI_ISL_977609, EPI_ISL_977610, EPI_ISL_977611 | SC (UCO) Igiene e Sanità Pubblica (funzione integrata con SC Microbiologia e Virologia) e Laboratory of Molecular Virology of the International Centre for Genetic Engineering and Biotechnology (ICGEB) | ARGO Laboratorio Genomica ed Epigenomica | Licastro D, Dal Monego S, Degasperi M, Marcello A, D'Agaro P, De Rosa R |
| EPI_ISL_977612, EPI_ISL_977613, EPI_ISL_977614, EPI_ISL_977615, EPI_ISL_977616, EPI_ISL_977617, EPI_ISL_977618, EPI_ISL_977619, EPI_ISL_977620 | SC (UCO) Igiene e Sanità Pubblica (funzione integrata con SC Microbiologia e Virologia) e Laboratory of Molecular Virology of the International Centre for Genetic Engineering and Biotechnology (ICGEB) | ARGO Laboratorio Genomica ed Epigenomica | Licastro D, Dal Monego S, Degasperi M, Marcello A, D'Agaro P, Pipan C |
| EPI_ISL_977621 | SC (UCO) Igiene e Sanità Pubblica (funzione integrata con SC Microbiologia e Virologia) e Laboratory of Molecular Virology of the International Centre for Genetic Engineering and Biotechnology (ICGEB) | ARGO Laboratorio Genomica ed Epigenomica | Licastro D, Dal Monego S, Degasperi M, Marcello A, D'Agaro P |
| EPI_ISL_977622, EPI_ISL_977623, EPI_ISL_977624, EPI_ISL_977625, EPI_ISL_977626, EPI_ISL_977627, EPI_ISL_977628, EPI_ISL_977629, EPI_ISL_977630, EPI_ISL_977631, EPI_ISL_977632, EPI_ISL_977633, EPI_ISL_977634, EPI_ISL_977635, EPI_ISL_977636, EPI_ISL_977637, EPI_ISL_977638, EPI_ISL_977639, EPI_ISL_977640 | | | |
| see above | SC (UCO) Igiene e Sanità Pubblica (funzione integrata con SC Microbiologia e Virologia) e Laboratory of Molecular Virology of the International Centre for Genetic Engineering and Biotechnology (ICGEB) | ARGO Laboratorio Genomica ed Epigenomica | Licastro D, Dal Monego S, Degasperi M, Marcello A, Segat L, Piscianz E, D'Agaro P |
| EPI_ISL_977641, EPI_ISL_977642, EPI_ISL_977643, EPI_ISL_977644, EPI_ISL_977645, EPI_ISL_977646, EPI_ISL_977647, EPI_ISL_977648, EPI_ISL_977649, EPI_ISL_977650 | SC (UCO) Igiene e Sanità Pubblica (funzione integrata con SC Microbiologia e Virologia) e Laboratory of Molecular Virology of the International Centre for Genetic Engineering and Biotechnology (ICGEB) | ARGO Laboratorio Genomica ed Epigenomica | Licastro D, Dal Monego S, Degasperi M, Marcello A, D'Agaro P, Lombardo F |
| EPI_ISL_977651 | Laboratorio Microbiologia e Virologia P.O. Cotugno A.O. dei Colli | Laboratorio Microbiologia e Virologia P.O. Cotugno A.O. dei Colli | Luigi Atripaldi, Claudia Tiberio, Anna Perfetti, |
| EPI_ISL_983096, EPI_ISL_983097, EPI_ISL_983098 | Microbiology and Virology Unit, Florence Careggi University Hospital | Microbiology and Virology Unit, Florence Careggi University Hospital | Vincenzo Di Pilato, Marco Coppi, Fabio Morecchiato, Noemi Aiezza, Ilaria Baccani, Alberto Antonelli, Emanuele Gori, Gian Maria Rossolini |
| EPI_ISL_983321 | Eurofins Genoma Group s.r.l. | INMI Lazzaro Spallanzani IRCCS | O Butera, F Messina, CEM Gruber, B Bartolini, E Giombini, M Rueca, F Spinella, F Fiorentino, MR Capobianchi, A Di Caro |
| EPI_ISL_983322 | ASL LATINA PRESIDIO OSPEDALIERO NORD | INMI Lazzaro Spallanzani IRCCS | E Giombini, M. Rueca, B Bartolini, O Butera, C.E.M Gruber, F Messina, A Lucci, S Pignalosa, A Di Caro, MR Capobianchi |
| EPI_ISL_983323 | INMI Lazzaro Spallanzani IRCCS | INMI Lazzaro Spallanzani IRCCS | CEM Gruber, B Bartolini, E Giombini, M Rueca, O Butera, F Messina, A Di Caro, MR Capobianchi |
| EPI_ISL_983324 | INMI Lazzaro Spallanzani IRCCS | INMI Lazzaro Spallanzani IRCCS | B Bartolini, O Butera, C.E.M Gruber, M Rueca, F Messina, E Giombini, MR Capobianchi, A Di Caro |
| EPI_ISL_983325 | INMI Lazzaro Spallanzani IRCCS | INMI Lazzaro Spallanzani IRCCS | M Rueca, O Butera, F Messina, CEM Gruber, B Bartolini, E Giombini, A Di Caro, MR Capobianchi |
| EPI_ISL_983326 | Laboratorio di Genetica Medica Ospedale Belcolle | INMI Lazzaro Spallanzani IRCCS | F Messina, C.E.M Gruber, B Bartolini, E Giombini, M Rueca, O Butera, F Natoni, G Pessina, A Di Caro, MR Capobianchi |
| EPI_ISL_983327 | Dipartimento di Prevenzione ASL Roma 4 | INMI Lazzaro Spallanzani IRCCS | O Butera, F Messina, CEM Gruber, B Bartolini, E Giombini, M Rueca, S Ursino, MR Capobianchi, A Di Caro |
| EPI_ISL_983328 | Azienda Ospedaliera San Camillo Forlanini | INMI Lazzaro Spallanzani IRCCS | E Giombini, M. Rueca, B Bartolini, O Butera, C.E.M Gruber, F Messina, G Parisi, ML Guarino, A Di Caro, MR Capobianchi |
| EPI_ISL_983329 | Istituto Zooprofilattico Sperimentale Lazio e Toscana "M. Aleandri" | INMI Lazzaro Spallanzani IRCCS | CEM Gruber, B Bartolini, E Giombini, M Rueca, O Butera, F Messina, MT Scicluna, G Manna, A Cersini, A Di Caro, MR Capobianchi |
| EPI_ISL_984995, EPI_ISL_984996 | U.O. Igiene, Ospedale Policlinico San Martino | U.O. Igiene, Ospedale Policlinico San Martino | Bruzzone Bianca, Caligiuri Patrizia, De Pace Vanessa, Domnich Alexander, Orsi Andrea, Ricucci Valentina, Icardi Giancarlo |
| EPI_ISL_984997, EPI_ISL_984998, EPI_ISL_984999 | S.C. Laboratorio Analisi, ASL 3 Liguria | U.O. Igiene, Ospedale Policlinico San Martino | Bruzzone Bianca, Caligiuri Patrizia, De Pace Vanessa, Domnich Alexander, Orsi Andrea, Ricucci Valentina, Spitaleri Antonino, Icardi Giancarlo |
| EPI_ISL_985000 | U.O. Igiene, Ospedale Policlinico San Martino | U.O. Igiene, Ospedale Policlinico San Martino | Bruzzone Bianca, Caligiuri Patrizia, De Pace Vanessa, Domnich Alexander, Orsi Andrea, Ricucci Valentina, Spitaleri Antonino, Icardi Giancarlo |
| EPI_ISL_985001, EPI_ISL_985002, EPI_ISL_985003, EPI_ISL_985004, EPI_ISL_985005, EPI_ISL_985006, EPI_ISL_985007, EPI_ISL_985008, EPI_ISL_985009, EPI_ISL_985010, EPI_ISL_985011, EPI_ISL_985012, EPI_ISL_985013, EPI_ISL_985014, EPI_ISL_985015, EPI_ISL_985016 | | | |
| see above | S.C. Laboratorio Analisi, ASL 3 Liguria | U.O. Igiene, Ospedale Policlinico San Martino | Bruzzone Bianca, Caligiuri Patrizia, De Pace Vanessa, Domnich Alexander, Orsi Andrea, Ricucci Valentina, Spitaleri Antonino, Icardi Giancarlo |
| EPI_ISL_985017 | S.C. Patologia Clinica, Ospedale Sant'Andrea, ASL 5 | U.O. Igiene, Ospedale Policlinico San Martino | Bruzzone Bianca, Battolla Enrico, Caligiuri Patrizia, De Pace Vanessa, Domnich Alexander, Orsi Andrea, Ricucci Valentina, Icardi Giancarlo |
| EPI_ISL_985018, EPI_ISL_985019, EPI_ISL_985020, EPI_ISL_985021, EPI_ISL_985022, EPI_ISL_985023, EPI_ISL_985024 | S.S.D. Microbiologia, Stabilimento ospedaliero di Sanremo, ASL 1 Liguria | U.O. Igiene, Ospedale Policlinico San Martino | Bruzzone Bianca, Caligiuri Patrizia, De Pace Vanessa, Domnich Alexander, Dusi Pier Andrea, Orsi Andrea, Ricucci Valentina, Icardi Giancarlo |
| EPI_ISL_985025, EPI_ISL_985026 | S.S.D. Microbiologia, Ospedale Santa Corona di Pietra Ligure, ASL 2 Liguria | U.O. Igiene, Ospedale Policlinico San Martino | Bruzzone Bianca, Caligiuri Patrizia, De Pace Vanessa, Domnich Alexander, Orsi Andrea, Ricucci Valentina, Valle Caterina, Icardi Giancarlo |

| | | | |
|---|---|---|---|
| EPI_ISL_985027, EPI_ISL_985028, EPI_ISL_985029, EPI_ISL_985030, EPI_ISL_985031 | Laboratorio di Patologia Clinica, Ospedale San Paolo in Valloria, ASL 2 Liguria | U.O. Igiene, Ospedale Policlinico San Martino | Bruzzone Bianca, Caligiuri Patrizia, De Pace Vanessa, Domnich Alexander, Lillo Flavia, Orsi Andrea, Ricucci Valentina, Icardi Giancarlo |
| EPI_ISL_985032 | U.O. Igiene, Ospedale Policlinico San Martino | U.O. Igiene, Ospedale Policlinico San Martino | Bruzzone Bianca, Caligiuri Patrizia, De Pace Vanessa, Domnich Alexander, Orsi Andrea, Ricucci Valentina, Icardi Giancarlo |

We gratefully acknowledge the following Authors from the Originating laboratories responsible for obtaining the specimens, as well as the Submitting laboratories where the genome data were generated and shared via GISAID, on which this research is based.

All Submitters of data may be contacted directly via www.gisaid.org

Authors are sorted alphabetically.

| Accession ID | Originating Laboratory | Submitting Laboratory | Authors |
|---|---|---|---|
| EPI_ISL_1167308, EPI_ISL_1167311, EPI_ISL_1167315, EPI_ISL_1167319, EPI_ISL_1167321, EPI_ISL_1167322, EPI_ISL_1167324, EPI_ISL_1167326, EPI_ISL_1167327, EPI_ISL_1167329, EPI_ISL_1167330, EPI_ISL_1167331, EPI_ISL_1167332, EPI_ISL_1167333, EPI_ISL_1167334, EPI_ISL_1167336, EPI_ISL_1167338, EPI_ISL_1167339, EPI_ISL_1167340, EPI_ISL_1167341, EPI_ISL_1167345, EPI_ISL_1167348, EPI_ISL_1167349, EPI_ISL_1167350, EPI_ISL_1167351, EPI_ISL_1167356, EPI_ISL_1167358, EPI_ISL_1167361, EPI_ISL_1167364, EPI_ISL_1167365, EPI_ISL_1167367, EPI_ISL_1167369, EPI_ISL_1167370, EPI_ISL_1167371, EPI_ISL_1167384, EPI_ISL_1167386, EPI_ISL_1167387, EPI_ISL_1167393, EPI_ISL_1167395, EPI_ISL_1167396, EPI_ISL_1167404, EPI_ISL_1167405, EPI_ISL_1167406, EPI_ISL_1167407, EPI_ISL_1167417, EPI_ISL_1167418, EPI_ISL_1167419, EPI_ISL_1167425, EPI_ISL_1167429, EPI_ISL_1167436, EPI_ISL_1167438, EPI_ISL_1167439, EPI_ISL_1167440, EPI_ISL_1167445, EPI_ISL_1167450, EPI_ISL_1167452, EPI_ISL_1167456, EPI_ISL_1167464, EPI_ISL_1167470, EPI_ISL_1167473, EPI_ISL_1167474, EPI_ISL_1167477, EPI_ISL_1167481, EPI_ISL_1167483, EPI_ISL_1167485, EPI_ISL_1167492, EPI_ISL_1167493, EPI_ISL_1167495, EPI_ISL_1167500, EPI_ISL_1167503, EPI_ISL_1167504, EPI_ISL_1167506, EPI_ISL_1167509, EPI_ISL_1167510, EPI_ISL_1167514, EPI_ISL_1167515, EPI_ISL_1167517, EPI_ISL_1167521, EPI_ISL_1167524, EPI_ISL_1167527, EPI_ISL_1167528, EPI_ISL_1167530, EPI_ISL_1167533, EPI_ISL_1167541, EPI_ISL_1167542, EPI_ISL_1167545, EPI_ISL_1167546, EPI_ISL_1169061, EPI_ISL_1169062, EPI_ISL_1169063, EPI_ISL_1169064, EPI_ISL_1169065, EPI_ISL_1169066, EPI_ISL_1169067, EPI_ISL_1169068, EPI_ISL_1169069, EPI_ISL_1169070, EPI_ISL_1169071, EPI_ISL_1169072, EPI_ISL_1169073, EPI_ISL_1169074, EPI_ISL_1169075, EPI_ISL_1169076, EPI_ISL_1169077, EPI_ISL_1169078, EPI_ISL_1169079, EPI_ISL_1169080, EPI_ISL_1169081, EPI_ISL_1169082, EPI_ISL_1169083, EPI_ISL_1169084, EPI_ISL_1169085, EPI_ISL_1169086, EPI_ISL_1169087, EPI_ISL_1169088, EPI_ISL_1169089, EPI_ISL_1169090, EPI_ISL_1169091, EPI_ISL_1169092, EPI_ISL_1169093, EPI_ISL_1169094, EPI_ISL_1169095, EPI_ISL_1169096, EPI_ISL_1169097, EPI_ISL_1169098, EPI_ISL_1169099, EPI_ISL_1169100, EPI_ISL_1169101, EPI_ISL_1169102, EPI_ISL_1169103, EPI_ISL_1169104, EPI_ISL_1169105, EPI_ISL_1169106, EPI_ISL_1169107, EPI_ISL_1169108, EPI_ISL_1169109, EPI_ISL_1169110, EPI_ISL_1169111, EPI_ISL_1169112, EPI_ISL_1169113, EPI_ISL_1169114, EPI_ISL_1169115, EPI_ISL_1169116, EPI_ISL_1169117, EPI_ISL_1169118, EPI_ISL_1169119, EPI_ISL_1169120, EPI_ISL_1169121, EPI_ISL_1169122, EPI_ISL_1169123, EPI_ISL_1169124, EPI_ISL_1169125, EPI_ISL_1169126, EPI_ISL_1169127, EPI_ISL_1169128, EPI_ISL_1169129, EPI_ISL_1169130, EPI_ISL_1169131, EPI_ISL_1169132, EPI_ISL_1169133, EPI_ISL_1169134, EPI_ISL_1169135, EPI_ISL_1169136, EPI_ISL_1169137, EPI_ISL_1169138, EPI_ISL_1169139, EPI_ISL_1169140, EPI_ISL_1169141, EPI_ISL_1169142, EPI_ISL_1169143, EPI_ISL_1169144, EPI_ISL_1169145, EPI_ISL_1169146, EPI_ISL_1169147, EPI_ISL_1169148, EPI_ISL_1169149, EPI_ISL_1169150, EPI_ISL_1169151, EPI_ISL_1169152, EPI_ISL_1169153, EPI_ISL_1169154, EPI_ISL_1169155, EPI_ISL_1169156, EPI_ISL_1169157, EPI_ISL_1169158, EPI_ISL_1169159, EPI_ISL_1169160, EPI_ISL_1169161, EPI_ISL_1169162, EPI_ISL_1169163, EPI_ISL_1169164, EPI_ISL_1169165, EPI_ISL_1169166, EPI_ISL_1169167, EPI_ISL_1169168, EPI_ISL_1169169, EPI_ISL_1169170, EPI_ISL_1169171, EPI_ISL_1169172, EPI_ISL_1169173, EPI_ISL_1169174, EPI_ISL_1169175, EPI_ISL_1169176, EPI_ISL_1169177, EPI_ISL_1169178, EPI_ISL_1169179, EPI_ISL_1169180, EPI_ISL_1169181, EPI_ISL_1169182, EPI_ISL_1169183, EPI_ISL_1169184, EPI_ISL_1169185, EPI_ISL_1169186, EPI_ISL_1169187, EPI_ISL_1169188, EPI_ISL_1169189, EPI_ISL_1169190, EPI_ISL_1169191, EPI_ISL_1169192, EPI_ISL_1169193, EPI_ISL_1169194, EPI_ISL_1169195, EPI_ISL_1169196, EPI_ISL_1169197, EPI_ISL_1169198, EPI_ISL_1169199, EPI_ISL_1169200, EPI_ISL_1169201, EPI_ISL_1169202, EPI_ISL_1169203, EPI_ISL_1169204, EPI_ISL_1169205, EPI_ISL_1169206, EPI_ISL_1169207, EPI_ISL_1169208, EPI_ISL_1169209, EPI_ISL_1169210, EPI_ISL_1169211, EPI_ISL_1169212, EPI_ISL_1169213, EPI_ISL_1169214, EPI_ISL_1169215, EPI_ISL_1169216, EPI_ISL_1169217, EPI_ISL_1169218, EPI_ISL_1169219, EPI_ISL_1169220, EPI_ISL_1169221, EPI_ISL_1169222, EPI_ISL_1169223, EPI_ISL_1169224, EPI_ISL_1169225, EPI_ISL_1169226, EPI_ISL_1169227, EPI_ISL_1169228, EPI_ISL_1169229, EPI_ISL_1169230, EPI_ISL_1169231, EPI_ISL_1169232, EPI_ISL_1169233, EPI_ISL_1169234, EPI_ISL_1169235, EPI_ISL_1169236, EPI_ISL_1169237, EPI_ISL_1169238, EPI_ISL_1169239, EPI_ISL_1169240, EPI_ISL_1169241, EPI_ISL_1169242, EPI_ISL_1169243, EPI_ISL_1169244, EPI_ISL_1169245, EPI_ISL_1169246, EPI_ISL_1169247, EPI_ISL_1169248, EPI_ISL_1169249, EPI_ISL_1169250, EPI_ISL_1169251, EPI_ISL_1169252, EPI_ISL_1169253, EPI_ISL_1169254, EPI_ISL_1169255, EPI_ISL_1169256, EPI_ISL_1169257, EPI_ISL_1169258, EPI_ISL_1169259, EPI_ISL_1169260, EPI_ISL_1169261, EPI_ISL_1169262, EPI_ISL_1169263, EPI_ISL_1169264, EPI_ISL_1169265, EPI_ISL_1169266, EPI_ISL_1169267, EPI_ISL_1169268, EPI_ISL_1169269, EPI_ISL_1169270, EPI_ISL_1169271, EPI_ISL_1169272, EPI_ISL_1169273, EPI_ISL_1169274, EPI_ISL_1169275, EPI_ISL_1169276, EPI_ISL_1169277, EPI_ISL_1169278, EPI_ISL_1169279, EPI_ISL_1169280, EPI_ISL_1169281, EPI_ISL_1169282, EPI_ISL_1169283, EPI_ISL_1169284, EPI_ISL_1169285, EPI_ISL_1169286, EPI_ISL_1169287, EPI_ISL_1169288, EPI_ISL_1169289, EPI_ISL_1169290, EPI_ISL_1169291, EPI_ISL_1169292, EPI_ISL_1169293, EPI_ISL_1169294, EPI_ISL_1169295, EPI_ISL_1169296, EPI_ISL_1169297, EPI_ISL_1169298, EPI_ISL_1169299, EPI_ISL_1169300, EPI_ISL_1169301, EPI_ISL_1169302, EPI_ISL_1169303, EPI_ISL_1169304, EPI_ISL_1169305, EPI_ISL_1169306, EPI_ISL_1169307, EPI_ISL_1169308, EPI_ISL_1169309, EPI_ISL_1169310, EPI_ISL_1169311, EPI_ISL_1169312, EPI_ISL_1169313, EPI_ISL_1169314, EPI_ISL_1169315, EPI_ISL_1169316, EPI_ISL_1169317, EPI_ISL_1169318, EPI_ISL_1169319, EPI_ISL_1169320, EPI_ISL_1169321, EPI_ISL_1169322, EPI_ISL_1169323, EPI_ISL_1169324, EPI_ISL_1169325, EPI_ISL_1169326, EPI_ISL_1169327, EPI_ISL_1169328, EPI_ISL_1169329, EPI_ISL_1169330, EPI_ISL_1169331, EPI_ISL_1169332, EPI_ISL_1169333, EPI_ISL_1169334, EPI_ISL_1169335, EPI_ISL_1169336, EPI_ISL_1169337, EPI_ISL_1169338, EPI_ISL_1169339, EPI_ISL_1169340, EPI_ISL_1169341, EPI_ISL_1169342, EPI_ISL_1169343, EPI_ISL_1169344, EPI_ISL_1169345, EPI_ISL_1169346, EPI_ISL_1169347, EPI_ISL_1169348, EPI_ISL_1169349, EPI_ISL_1169350, EPI_ISL_1169351, EPI_ISL_1169352, EPI_ISL_1169353, EPI_ISL_1169354, EPI_ISL_1169355, EPI_ISL_1169356, EPI_ISL_1169357, EPI_ISL_1169358, EPI_ISL_1169359, EPI_ISL_1169360, EPI_ISL_1169361, EPI_ISL_1169362, EPI_ISL_1169363, EPI_ISL_1169364, EPI_ISL_1169365, EPI_ISL_1169366, EPI_ISL_1169367, EPI_ISL_1169368, EPI_ISL_1169369, EPI_ISL_1169370, EPI_ISL_1169371, EPI_ISL_1169372, EPI_ISL_1169373, EPI_ISL_1169374, EPI_ISL_1169376, EPI_ISL_1169377, EPI_ISL_1169378, EPI_ISL_1169379, EPI_ISL_1169380, EPI_ISL_1169381, EPI_ISL_1169382, EPI_ISL_1169383, EPI_ISL_1169384, EPI_ISL_1169385, EPI_ISL_1169386, EPI_ISL_1169387, EPI_ISL_1169388, EPI_ISL_1169389, EPI_ISL_1169390, EPI_ISL_1169391, EPI_ISL_1169392, EPI_ISL_1169393, EPI_ISL_1169394, EPI_ISL_1169395, EPI_ISL_1169396, EPI_ISL_1169397, EPI_ISL_1169398, EPI_ISL_1169399, EPI_ISL_1169400, EPI_ISL_1169401, EPI_ISL_1169402, EPI_ISL_1169403, EPI_ISL_1169404, EPI_ISL_1169405, EPI_ISL_1169406, EPI_ISL_1169407, EPI_ISL_1169408, EPI_ISL_1169409, EPI_ISL_1169410, EPI_ISL_1169411, EPI_ISL_1169412, EPI_ISL_1169413, EPI_ISL_1169414, EPI_ISL_1169415, EPI_ISL_1169416, EPI_ISL_1169417, EPI_ISL_1169418, EPI_ISL_1169419, EPI_ISL_1169420, EPI_ISL_1169421, EPI_ISL_1169422, EPI_ISL_1169423, EPI_ISL_1169424, EPI_ISL_1169425, EPI_ISL_1169426, EPI_ISL_1169427, EPI_ISL_1169428, EPI_ISL_1169429, EPI_ISL_1169430, EPI_ISL_1169431, EPI_ISL_1169432, EPI_ISL_1169433, EPI_ISL_1169434, EPI_ISL_1169435, EPI_ISL_1169436, EPI_ISL_1169437, EPI_ISL_1169438, EPI_ISL_1169439, EPI_ISL_1169440, EPI_ISL_1169441, EPI_ISL_1169442, EPI_ISL_1169443, EPI_ISL_1169444, EPI_ISL_1169445, EPI_ISL_1169446, EPI_ISL_1169447, EPI_ISL_1169448, EPI_ISL_1169449, EPI_ISL_1169450, EPI_ISL_1169451, EPI_ISL_1169452, EPI_ISL_1169453, EPI_ISL_1169454, EPI_ISL_1169455, EPI_ISL_1169456, EPI_ISL_1169457, EPI_ISL_1169458, EPI_ISL_1169459, EPI_ISL_1169460, EPI_ISL_1169461, EPI_ISL_1169462, EPI_ISL_1169463, EPI_ISL_1169464, EPI_ISL_1169465, EPI_ISL_1169466, EPI_ISL_1169467, EPI_ISL_1169468, EPI_ISL_1169469, EPI_ISL_1169470, EPI_ISL_1169471, EPI_ISL_1169472, EPI_ISL_1169473, EPI_ISL_1169474, EPI_ISL_1169475, EPI_ISL_1169476, EPI_ISL_1169477, EPI_ISL_1169478, EPI_ISL_1169479, EPI_ISL_1169480, EPI_ISL_1169481, EPI_ISL_1169482 | | | |
| see above | ASL Napoli 1 Centro | AMES Centro Polidiagnostico Strumentale S.r.l. | "Giovanni Savarese, Raffaella Ruggiero, Eloisa Evangelista, Antonella Di Carlo, Luisa Circelli, Luigi D'Amore, Nadia Petrillo, Monica Ianniello, Roberto Sirica,Maurizio D'Amora, Antonio Fico" |
| EPI_ISL_1169888 | Ospedale "F. Spaziani" Frosinone | INMI Lazzaro Spallanzani IRCCS | E Giombini, F Messina, M Rueca, O Butera, CEM Gruber, B Bartolini, R Pulselli, C Gargiulo, C Sias, A Di Caro, MR Capobianchi |
| EPI_ISL_1176355 | Laboratorio di Riferimento Regionale della Sicilia Occidentale per l'Emergenza COVID-19 | Laboratorio di Riferimento Regionale della Sicilia Occidentale per l'Emergenza COVID-19 | Fabio Tramuto, Carmelo Massimo Maida, Daniela Di Naro, Giulia Randazzo, Walter Mazzucco, Giorgio Graziano, Vincenzo Restivo, Claudio Costantino, Francesco Vitale |
| EPI_ISL_1180143, EPI_ISL_1180144 | Laboratorio Biologia Molecolare Sars Cov2 - UOC Laboratorio Analisi - Servizio Medicina di Laboratorio, Ospedale "San Francesco" - ATS-ASSL Nuoro | Laboratorio specialistico UOC Ematologia - Ospedale "San Francesco" - ATS-ASSL Nuoro | Giovanna Piras, Rosanna Asproni, Paolo Malune, Maria Itria Monne, Maura Fiamma, Angelo Domenico Palmas, Iana Lo Maglio, Giuseppe Mameli |
| EPI_ISL_1201281, EPI_ISL_1201285, EPI_ISL_1201286 | Center of Advanced Studies and Technology, Molecular Genetics Laboratory | Center of Advanced Studies and Technology, Molecular Genetics Laboratory | Ferrante Rossella, Mandatori Domitilla, De Fabritiis Simone, Damiani Verena, Anaclerio Federico |
| EPI_ISL_1209014, EPI_ISL_1209020, EPI_ISL_1209025 | University of Bari Biomedical Sciences and Human Oncology | University of Bari Biomedical Sciences and Human Oncology | Chironna M., Sallustio A., Loconsole D., Accogli M., Parisi A. |
| EPI_ISL_1219040, EPI_ISL_1219041, EPI_ISL_1219042, EPI_ISL_1219043, EPI_ISL_1219044, EPI_ISL_1219045, EPI_ISL_1219046, EPI_ISL_1219047, EPI_ISL_1219048, EPI_ISL_1219049, EPI_ISL_1219050, EPI_ISL_1219051, EPI_ISL_1219052, EPI_ISL_1219053, EPI_ISL_1219054, EPI_ISL_1219055, EPI_ISL_1219056, EPI_ISL_1219057, EPI_ISL_1219058, EPI_ISL_1219059, EPI_ISL_1219060, EPI_ISL_1219061, EPI_ISL_1219062, EPI_ISL_1219063, EPI_ISL_1219064, EPI_ISL_1219065, EPI_ISL_1219066, EPI_ISL_1219067, EPI_ISL_1219068, EPI_ISL_1219069, EPI_ISL_1219070, EPI_ISL_1219071, EPI_ISL_1219072, EPI_ISL_1219073, EPI_ISL_1219074, EPI_ISL_1219075, EPI_ISL_1219076, EPI_ISL_1219077, EPI_ISL_1219078, EPI_ISL_1219079, EPI_ISL_1219080, EPI_ISL_1219081, EPI_ISL_1219082, EPI_ISL_1219083, EPI_ISL_1219084, EPI_ISL_1219085, EPI_ISL_1219086, EPI_ISL_1219087, EPI_ISL_1219088, EPI_ISL_1219089, EPI_ISL_1219090, EPI_ISL_1219091, EPI_ISL_1219092, EPI_ISL_1219093, EPI_ISL_1219094, EPI_ISL_1219095, EPI_ISL_1219096, EPI_ISL_1219097, EPI_ISL_1219098, EPI_ISL_1219099, EPI_ISL_1219100, EPI_ISL_1219101, EPI_ISL_1219102, EPI_ISL_1219103, EPI_ISL_1219104, EPI_ISL_1219105, EPI_ISL_1219106, EPI_ISL_1219107, EPI_ISL_1219108, EPI_ISL_1219109, EPI_ISL_1219110, EPI_ISL_1219111, EPI_ISL_1219112, EPI_ISL_1219113, EPI_ISL_1219114, EPI_ISL_1219115, EPI_ISL_1219116, EPI_ISL_1219117, EPI_ISL_1219118, EPI_ISL_1219119, EPI_ISL_1219120, EPI_ISL_1219121, EPI_ISL_1219122, EPI_ISL_1219123, EPI_ISL_1219124, EPI_ISL_1219125, EPI_ISL_1219126, EPI_ISL_1219127, EPI_ISL_1219128, EPI_ISL_1219129, EPI_ISL_1219130, EPI_ISL_1219131 | | | |
| see above | ASL Napoli 1 Centro | AMES Centro Polidiagnostico Strumentale S.r.l. | "Giovanni Savarese, Raffaella Ruggiero, Eloisa Evangelista, Antonella Di Carlo, Luisa Circelli, Luigi D'Amore, Nadia Petrillo, Monica Ianniello, Roberto Sirica,Maurizio D'Amora, Antonio Fico" |
| EPI_ISL_1222920, EPI_ISL_1222921, EPI_ISL_1222922 | Azienda Sanitaria dell'Alto Adige Laboratorio Aziendale di Microbiologia e Virologia | Istituto di Genomica Applicata | Elisabetta Pagani, Irene Bianconi, Elisabetta Giacobazzi, Elisa Masi, Stefanie Wieser, Irena Jurman, Vera Vendramin, Gabriele Magris, Eleonora Paparelli, Davide Scaglione, Michele Morgante |
| EPI_ISL_1229131 | INMI Lazzaro Spallanzani IRCCS | INMI Lazzaro Spallanzani IRCCS | CEM Gruber, B Bartolini, E Giombini, F Messina, M Rueca, O Butera, MR Capobianchi, A Di Caro |
| EPI_ISL_1229132 | Ospedale San Camillo De Lellis di Rieti | INMI Lazzaro Spallanzani IRCCS | E Giombini, F Messina, M Rueca, O Butera, CEM Gruber, B Bartolini, S Venarubea, A De Luca , L Casertano, A Di Caro, MR Capobianchi |
| EPI_ISL_1229133 | San Gallicano Dermatological Institute I.F.O. | INMI Lazzaro Spallanzani IRCCS | F Messina, M Rueca, O Butera, CEM Gruber, B Bartolini, E Giombini, F Pimpinelli, F Ensoli, G D'Agosto, MR Capobianchi, A Di Caro |
| EPI_ISL_1229140 | Presidio Ospedaliero G. B. Grassi ASL RM3 | INMI Lazzaro Spallanzani IRCCS | E Giombini, F Messina, M Rueca, O Butera, CEM Gruber, B Bartolini, F Tabacco, E Ristori, A Di Caro, MR Capobianchi |
| EPI_ISL_1229141 | Ospedale "F. Spaziani" Frosinone | INMI Lazzaro Spallanzani IRCCS | F Messina, M Rueca, O Butera, CEM Gruber, B Bartolini, E Giombini, R Pulselli, C Gargiulo, C Sias, MR Capobianchi, A Di Caro |
| EPI_ISL_1229142 | IDI-IRCCS | INMI Lazzaro Spallanzani IRCCS | M Rueca, O Butera, CEM Gruber, B Bartolini, E Giombini, F Messina, G Russo, D Castiglia, S Madonna, A Di Caro, MR Capobianchi |
| EPI_ISL_1229143 | IDI-IRCCS | INMI Lazzaro Spallanzani IRCCS | O Butera, CEM Gruber, B Bartolini, E Giombini, F Messina, M Rueca, G Russo, D Castiglia, S Madonna, MR Capobianchi, A Di Caro |
| EPI_ISL_1229179, EPI_ISL_1229180, EPI_ISL_1229181, EPI_ISL_1229182, EPI_ISL_1229183, EPI_ISL_1229184, EPI_ISL_1229185, EPI_ISL_1229186, EPI_ISL_1229187, EPI_ISL_1229188, EPI_ISL_1229189, EPI_ISL_1229190, EPI_ISL_1229191, EPI_ISL_1229192, EPI_ISL_1229193, EPI_ISL_1229194, EPI_ISL_1229195, EPI_ISL_1229196, EPI_ISL_1229197, EPI_ISL_1229198, EPI_ISL_1229199, EPI_ISL_1229200, EPI_ISL_1229201, EPI_ISL_1229202, EPI_ISL_1229203, EPI_ISL_1229204, EPI_ISL_1229205, EPI_ISL_1229206, EPI_ISL_1229207, EPI_ISL_1229208, EPI_ISL_1229209, EPI_ISL_1229210, EPI_ISL_1229211, EPI_ISL_1229212, EPI_ISL_1229213, EPI_ISL_1229214, EPI_ISL_1229215, EPI_ISL_1229216, EPI_ISL_1229217, EPI_ISL_1229218, EPI_ISL_1229219, EPI_ISL_1229220, EPI_ISL_1229221, EPI_ISL_1229222, EPI_ISL_1229223, EPI_ISL_1229224, EPI_ISL_1229225, EPI_ISL_1229226, EPI_ISL_1229227, EPI_ISL_1229228, EPI_ISL_1229229, EPI_ISL_1229230, EPI_ISL_1229231, EPI_ISL_1229232, EPI_ISL_1229233, EPI_ISL_1229234, EPI_ISL_1229235, EPI_ISL_1229236, EPI_ISL_1229237, EPI_ISL_1229238, EPI_ISL_1229239, EPI_ISL_1229240, EPI_ISL_1229241, EPI_ISL_1229242, EPI_ISL_1229243, EPI_ISL_1229244, EPI_ISL_1229245, EPI_ISL_1229246, EPI_ISL_1229247, EPI_ISL_1229248, EPI_ISL_1229249, EPI_ISL_1229250, EPI_ISL_1229251, EPI_ISL_1229252, EPI_ISL_1229253, EPI_ISL_1229254, EPI_ISL_1229255, EPI_ISL_1229256, EPI_ISL_1229257, EPI_ISL_1229258, EPI_ISL_1229259, EPI_ISL_1229260, EPI_ISL_1229261, EPI_ISL_1229262, EPI_ISL_1229263, EPI_ISL_1229264, EPI_ISL_1229265, EPI_ISL_1229266, EPI_ISL_1229267, EPI_ISL_1229268, EPI_ISL_1229269, EPI_ISL_1229270, EPI_ISL_1229271, EPI_ISL_1229272, EPI_ISL_1229273, EPI_ISL_1229274, EPI_ISL_1229275, EPI_ISL_1229276, EPI_ISL_1229277, EPI_ISL_1229278, EPI_ISL_1229279, EPI_ISL_1229280, EPI_ISL_1229281, EPI_ISL_1229282, EPI_ISL_1229283, EPI_ISL_1229284, EPI_ISL_1229285, EPI_ISL_1229286, EPI_ISL_1229287, EPI_ISL_1229288, EPI_ISL_1229289, EPI_ISL_1229290, EPI_ISL_1229291, EPI_ISL_1229292, EPI_ISL_1229293, EPI_ISL_1229294, EPI_ISL_1229295, EPI_ISL_1229296, EPI_ISL_1229297, EPI_ISL_1229298, EPI_ISL_1229299, EPI_ISL_1229300, EPI_ISL_1229301, EPI_ISL_1229302, EPI_ISL_1229303, EPI_ISL_1229304, EPI_ISL_1229305, EPI_ISL_1229306, EPI_ISL_1229307, EPI_ISL_1229308, EPI_ISL_1229309, EPI_ISL_1229310, EPI_ISL_1229311, EPI_ISL_1229312, EPI_ISL_1229313, EPI_ISL_1229314, | | | |

| | | | |
|---|---|---|---|
| EPI_ISL_1229315, EPI_ISL_1229316, EPI_ISL_1229317, EPI_ISL_1229318, EPI_ISL_1229319, EPI_ISL_1229320, EPI_ISL_1229321, EPI_ISL_1229322, EPI_ISL_1229323, EPI_ISL_1229324, EPI_ISL_1229325, EPI_ISL_1229326, EPI_ISL_1229327, EPI_ISL_1229328, EPI_ISL_1229329, EPI_ISL_1229330, EPI_ISL_1229331, EPI_ISL_1229332, EPI_ISL_1229333, EPI_ISL_1229334, EPI_ISL_1229335, EPI_ISL_1229336, EPI_ISL_1229337, EPI_ISL_1229338, EPI_ISL_1229339, EPI_ISL_1229340, EPI_ISL_1229341, EPI_ISL_1229342, EPI_ISL_1229343, EPI_ISL_1229344, EPI_ISL_1229345, EPI_ISL_1229346, EPI_ISL_1229347, EPI_ISL_1229348, EPI_ISL_1229349, EPI_ISL_1229350, EPI_ISL_1229351, EPI_ISL_1229352, EPI_ISL_1229353, EPI_ISL_1229354, EPI_ISL_1229355, EPI_ISL_1229356, EPI_ISL_1229357, EPI_ISL_1229358, EPI_ISL_1229359, EPI_ISL_1229360, EPI_ISL_1229361, EPI_ISL_1229362, EPI_ISL_1229363, EPI_ISL_1229364, EPI_ISL_1229365, EPI_ISL_1229366 | | | |
| see above | ASL Napoli 1 Centro | AMES Centro Polidiagnostico Strumentale S.r.l. | "Giovanni Savarese, Raffaella Ruggiero, Eloisa Evangelista, Antonella Di Carlo, Luisa Circelli, Luigi D'Amore, Roberto Sirica,Nadia Petrillo, Monica Ianniello, Maurizio D'Amora, Antonio Fico" |
| EPI_ISL_1229535, EPI_ISL_1229536, EPI_ISL_1229537, EPI_ISL_1229538, EPI_ISL_1229539, EPI_ISL_1229540, EPI_ISL_1229541, EPI_ISL_1229542, EPI_ISL_1229543, EPI_ISL_1229544, EPI_ISL_1229545, EPI_ISL_1229546, EPI_ISL_1229547, EPI_ISL_1229548, EPI_ISL_1229549, EPI_ISL_1229550, EPI_ISL_1229551, EPI_ISL_1229552, EPI_ISL_1229553, EPI_ISL_1229554, EPI_ISL_1229555, EPI_ISL_1229556, EPI_ISL_1229557, EPI_ISL_1229558, EPI_ISL_1229559, EPI_ISL_1229560, EPI_ISL_1229561, EPI_ISL_1229562, EPI_ISL_1229563, EPI_ISL_1229564, EPI_ISL_1229565, EPI_ISL_1229566, EPI_ISL_1229567, EPI_ISL_1229568, EPI_ISL_1229569, EPI_ISL_1229570, EPI_ISL_1229571, EPI_ISL_1229572, EPI_ISL_1229573, EPI_ISL_1229574, EPI_ISL_1229575, EPI_ISL_1229576, EPI_ISL_1229577, EPI_ISL_1229578, EPI_ISL_1229579, EPI_ISL_1229580, EPI_ISL_1229581, EPI_ISL_1229582, EPI_ISL_1229583, EPI_ISL_1229584, EPI_ISL_1229585, EPI_ISL_1229586, EPI_ISL_1229587, EPI_ISL_1229588, EPI_ISL_1229589, EPI_ISL_1229590, EPI_ISL_1229591, EPI_ISL_1229592, EPI_ISL_1229593, EPI_ISL_1229594, EPI_ISL_1229595, EPI_ISL_1229596, EPI_ISL_1229597, EPI_ISL_1229598, EPI_ISL_1229599, EPI_ISL_1229600, EPI_ISL_1229601, EPI_ISL_1229602, EPI_ISL_1229603, EPI_ISL_1229604, EPI_ISL_1229605, EPI_ISL_1229606, EPI_ISL_1229607, EPI_ISL_1229608, EPI_ISL_1229609, EPI_ISL_1229610, EPI_ISL_1229611, EPI_ISL_1229612, EPI_ISL_1229613, EPI_ISL_1229614, EPI_ISL_1229615, EPI_ISL_1229616, EPI_ISL_1229617, EPI_ISL_1229618, EPI_ISL_1229619, EPI_ISL_1229620, EPI_ISL_1229621, EPI_ISL_1229622, EPI_ISL_1229623, EPI_ISL_1229624, EPI_ISL_1229625, EPI_ISL_1229626, EPI_ISL_1229627, EPI_ISL_1229628, EPI_ISL_1229629, EPI_ISL_1229630, EPI_ISL_1229631, EPI_ISL_1229632, EPI_ISL_1229633, EPI_ISL_1229634, EPI_ISL_1229635, EPI_ISL_1229636, EPI_ISL_1229637, EPI_ISL_1229638, EPI_ISL_1229639, EPI_ISL_1229640, EPI_ISL_1229641, EPI_ISL_1229642, EPI_ISL_1229643, EPI_ISL_1229644, EPI_ISL_1229645, EPI_ISL_1229646, EPI_ISL_1229647, EPI_ISL_1229648, EPI_ISL_1229649, EPI_ISL_1229650, EPI_ISL_1229651, EPI_ISL_1229652, EPI_ISL_1229653, EPI_ISL_1229654, EPI_ISL_1229655, EPI_ISL_1229656, EPI_ISL_1229657, EPI_ISL_1229658, EPI_ISL_1229659, EPI_ISL_1229660, EPI_ISL_1229661, EPI_ISL_1229662, EPI_ISL_1229663, EPI_ISL_1229664, EPI_ISL_1229665, EPI_ISL_1229666, EPI_ISL_1229667, EPI_ISL_1229668, EPI_ISL_1229669, EPI_ISL_1229670, EPI_ISL_1229671, EPI_ISL_1229672, EPI_ISL_1229673, EPI_ISL_1229674, EPI_ISL_1229675, EPI_ISL_1229676, EPI_ISL_1229677, EPI_ISL_1229678, EPI_ISL_1229679, EPI_ISL_1229680, EPI_ISL_1229681, EPI_ISL_1229682, EPI_ISL_1229683, EPI_ISL_1229684, EPI_ISL_1229685, EPI_ISL_1229686, EPI_ISL_1229687, EPI_ISL_1229688, EPI_ISL_1229689, EPI_ISL_1229690, EPI_ISL_1229691, EPI_ISL_1229692, EPI_ISL_1229693, EPI_ISL_1229694, EPI_ISL_1229695, EPI_ISL_1229696, EPI_ISL_1229697, EPI_ISL_1229698, EPI_ISL_1229699, EPI_ISL_1229700, EPI_ISL_1229701, EPI_ISL_1229702, EPI_ISL_1229703, EPI_ISL_1229704, EPI_ISL_1229705, EPI_ISL_1229706, EPI_ISL_1229707, EPI_ISL_1229708, EPI_ISL_1229709, EPI_ISL_1229710, EPI_ISL_1229711, EPI_ISL_1229712, EPI_ISL_1229713, EPI_ISL_1229714, EPI_ISL_1229715, EPI_ISL_1229716, EPI_ISL_1229717, EPI_ISL_1229718, EPI_ISL_1229719, EPI_ISL_1229720, EPI_ISL_1229721, EPI_ISL_1229722, EPI_ISL_1229723, EPI_ISL_1229724, EPI_ISL_1229725, EPI_ISL_1229726, EPI_ISL_1229727, EPI_ISL_1229728, EPI_ISL_1229729, EPI_ISL_1229730, EPI_ISL_1229731, EPI_ISL_1229732, EPI_ISL_1229733, EPI_ISL_1229734, EPI_ISL_1229735, EPI_ISL_1229736, EPI_ISL_1229737, EPI_ISL_1229738, EPI_ISL_1229739, EPI_ISL_1229740, EPI_ISL_1229741, EPI_ISL_1229742, EPI_ISL_1229743, EPI_ISL_1229744, EPI_ISL_1229745, EPI_ISL_1229746, EPI_ISL_1229747, EPI_ISL_1229748, EPI_ISL_1229749, EPI_ISL_1229750, EPI_ISL_1229751, EPI_ISL_1229752, EPI_ISL_1229753, EPI_ISL_1229754, EPI_ISL_1229755, EPI_ISL_1229756, EPI_ISL_1229757, EPI_ISL_1229758, EPI_ISL_1229759, EPI_ISL_1229760, EPI_ISL_1229761, EPI_ISL_1229762, EPI_ISL_1229763, EPI_ISL_1229764, EPI_ISL_1229765, EPI_ISL_1229766, EPI_ISL_1229767, EPI_ISL_1229768, EPI_ISL_1229769, EPI_ISL_1229770, EPI_ISL_1229771, EPI_ISL_1229772, EPI_ISL_1229773, EPI_ISL_1229774, EPI_ISL_1229775, EPI_ISL_1229776, EPI_ISL_1229777, EPI_ISL_1229778, EPI_ISL_1229779, EPI_ISL_1229780, EPI_ISL_1229781, EPI_ISL_1229782, EPI_ISL_1229783, EPI_ISL_1229784, EPI_ISL_1229785, EPI_ISL_1229786, EPI_ISL_1229787, EPI_ISL_1229788, EPI_ISL_1229789, EPI_ISL_1229790, EPI_ISL_1229791, EPI_ISL_1229792, EPI_ISL_1229793, EPI_ISL_1229794, EPI_ISL_1229795, EPI_ISL_1229796, EPI_ISL_1229797, EPI_ISL_1229798, EPI_ISL_1229799, EPI_ISL_1229800, EPI_ISL_1229801, EPI_ISL_1229802, EPI_ISL_1229803, EPI_ISL_1229804, EPI_ISL_1229805, EPI_ISL_1229806, EPI_ISL_1229807, EPI_ISL_1229808, EPI_ISL_1229809, EPI_ISL_1229810, EPI_ISL_1229811, EPI_ISL_1229812, EPI_ISL_1229813, EPI_ISL_1229814, EPI_ISL_1229815, EPI_ISL_1229816, EPI_ISL_1229817, EPI_ISL_1229818, EPI_ISL_1229819, EPI_ISL_1229820, EPI_ISL_1229821, EPI_ISL_1229822, EPI_ISL_1229823, EPI_ISL_1229824, EPI_ISL_1229825, EPI_ISL_1229826, EPI_ISL_1229827, EPI_ISL_1229828, EPI_ISL_1229829, EPI_ISL_1229830, EPI_ISL_1229831, EPI_ISL_1229832, EPI_ISL_1229833, EPI_ISL_1229834, EPI_ISL_1229835, EPI_ISL_1229836, EPI_ISL_1229837, EPI_ISL_1229838, EPI_ISL_1229839, EPI_ISL_1229840, EPI_ISL_1229841, EPI_ISL_1229842, EPI_ISL_1229843, EPI_ISL_1229844, EPI_ISL_1229845, EPI_ISL_1229846, EPI_ISL_1229847, EPI_ISL_1229848, EPI_ISL_1229849, EPI_ISL_1229850, EPI_ISL_1229851, EPI_ISL_1229852, EPI_ISL_1229853, EPI_ISL_1229854, EPI_ISL_1229855, EPI_ISL_1229856, EPI_ISL_1229857, EPI_ISL_1229858, EPI_ISL_1229859, EPI_ISL_1229860, EPI_ISL_1229861, EPI_ISL_1229862, EPI_ISL_1229863, EPI_ISL_1229864, EPI_ISL_1229865, EPI_ISL_1229866, EPI_ISL_1229867, EPI_ISL_1229868, EPI_ISL_1229869, EPI_ISL_1229870, EPI_ISL_1229871, EPI_ISL_1229872, EPI_ISL_1229873, EPI_ISL_1229874, EPI_ISL_1229875, EPI_ISL_1229876, EPI_ISL_1229877, EPI_ISL_1229878, EPI_ISL_1229879, EPI_ISL_1229880, EPI_ISL_1229881, EPI_ISL_1229882, EPI_ISL_1229883, EPI_ISL_1229884, EPI_ISL_1229885, EPI_ISL_1229886, EPI_ISL_1229887, EPI_ISL_1229888, EPI_ISL_1229889, EPI_ISL_1229890, EPI_ISL_1229891, EPI_ISL_1229892, EPI_ISL_1229893, EPI_ISL_1229894, EPI_ISL_1229895, EPI_ISL_1229896, EPI_ISL_1229897, EPI_ISL_1229898, EPI_ISL_1229899, EPI_ISL_1229900, EPI_ISL_1229901, EPI_ISL_1229902, EPI_ISL_1229903, EPI_ISL_1229904, EPI_ISL_1229905, EPI_ISL_1229906, EPI_ISL_1229907, EPI_ISL_1229908, EPI_ISL_1229909, EPI_ISL_1229910, EPI_ISL_1229911, EPI_ISL_1229912, EPI_ISL_1229913, EPI_ISL_1229914, EPI_ISL_1229915, EPI_ISL_1229916, EPI_ISL_1229917, EPI_ISL_1229918, EPI_ISL_1229919, EPI_ISL_1229920, EPI_ISL_1229921, EPI_ISL_1229922, EPI_ISL_1229923, EPI_ISL_1229924, EPI_ISL_1229925, EPI_ISL_1229926, EPI_ISL_1229927, EPI_ISL_1229928, EPI_ISL_1229929, EPI_ISL_1229930, EPI_ISL_1229931, EPI_ISL_1229932, EPI_ISL_1229933, EPI_ISL_1229934, EPI_ISL_1229935, EPI_ISL_1229936, EPI_ISL_1229937, EPI_ISL_1229938 | | | |
| see above | ASL Napoli 1 Centro | AMES Centro Polidiagnostico Strumentale S.r.l. | "Giovanni Savarese, Raffaella Ruggiero, Eloisa Evangelista, Antonella Di Carlo, Luisa Circelli, Luigi D'Amore, Roberto Sirica, Nadia Petrillo, Monica Ianniello, Maurizio D'Amora, Antonio Fico" |
| EPI_ISL_1241860 | Laboratorio di Riferimento Regionale della Sicilia Occidentale per l'Emergenza COVID-19 | Laboratorio di Riferimento Regionale della Sicilia Occidentale per l'Emergenza COVID-19 | Fabio Tramuto, Carmelo Massimo Maida, Daniela Di Naro, Giulia Randazzo, Walter Mazzucco, Giorgio Graziano, Vincenzo Restivo, Claudio Costantino, Francesco Vitale |
| EPI_ISL_1250946 | Usca castel di sangro | Istituto Zooprofilattico Sperimentale dell'Abruzzo e Molise "G. Caporale" | Lorusso A, Marcacci M, Di Domenico M, Ancora M, Curini V, Mangone I, Rinaldi A, Scialabba S, Di Pasquale A, Cammà C, Puglia I, Calistri P, Savini G |
| EPI_ISL_1250947 | siesp Chieti | Istituto Zooprofilattico Sperimentale dell'Abruzzo e Molise "G. Caporale" | Lorusso A, Marcacci M, Di Domenico M, Ancora M, Curini V, Mangone I, Rinaldi A, Scialabba S, Di Pasquale A, Cammà C, Puglia I, Calistri P, Savini G |
| EPI_ISL_1250948 | Usca castel di sangro | Istituto Zooprofilattico Sperimentale dell'Abruzzo e Molise "G. Caporale" | Lorusso A, Marcacci M, Di Domenico M, Ancora M, Curini V, Mangone I, Rinaldi A, Scialabba S, Di Pasquale A, Cammà C, Puglia I, Calistri P, Savini G |
| EPI_ISL_1250949 | Siesp dipartimento di prevenzione | Istituto Zooprofilattico Sperimentale dell'Abruzzo e Molise "G. Caporale" | Lorusso A, Marcacci M, Di Domenico M, Ancora M, Curini V, Mangone I, Rinaldi A, Scialabba S, Di Pasquale A, Cammà C, Puglia I, Calistri P, Savini G |
| EPI_ISL_1250950, EPI_ISL_1250951 | siesp dipartimento di prevenzione | Istituto Zooprofilattico Sperimentale dell'Abruzzo e Molise "G. Caporale" | Lorusso A, Marcacci M, Di Domenico M, Ancora M, Curini V, Mangone I, Rinaldi A, Scialabba S, Di Pasquale A, Cammà C, Puglia I, Calistri P, Savini G |
| EPI_ISL_1250952 | ospedale civile teramo | Istituto Zooprofilattico Sperimentale dell'Abruzzo e Molise "G. Caporale" | Lorusso A, Marcacci M, Di Domenico M, Ancora M, Curini V, Mangone I, Rinaldi A, Scialabba S, Di Pasquale A, Cammà C, Puglia I, Calistri P, Savini G |
| EPI_ISL_1250953 | dip prev avezzano | Istituto Zooprofilattico Sperimentale dell'Abruzzo e Molise "G. Caporale" | Lorusso A, Marcacci M, Di Domenico M, Ancora M, Curini V, Mangone I, Rinaldi A, Scialabba S, Di Pasquale A, Cammà C, Puglia I, Calistri P, Savini G |
| EPI_ISL_1250954 | siesp Chieti | Istituto Zooprofilattico Sperimentale dell'Abruzzo e Molise "G. Caporale" | Lorusso A, Marcacci M, Di Domenico M, Ancora M, Curini V, Mangone I, Rinaldi A, Scialabba S, Di Pasquale A, Cammà C, Puglia I, Calistri P, Savini G |
| EPI_ISL_1250955 | OSPEDALE CIVILE GIULIANOVA | Istituto Zooprofilattico Sperimentale dell'Abruzzo e Molise "G. Caporale" | Lorusso A, Marcacci M, Di Domenico M, Ancora M, Curini V, Mangone I, Rinaldi A, Scialabba S, Di Pasquale A, Cammà C, Puglia I, Calistri P, Savini G |
| EPI_ISL_1250957 | Siesp dipartimento di prevenzione | Istituto Zooprofilattico Sperimentale dell'Abruzzo e Molise "G. Caporale" | Lorusso A, Marcacci M, Di Domenico M, Ancora M, Curini V, Mangone I, Rinaldi A, Scialabba S, Di Pasquale A, Cammà C, Puglia I, Calistri P, Savini G |
| EPI_ISL_1250958 | dip prev avezzano | Istituto Zooprofilattico Sperimentale dell'Abruzzo e Molise "G. Caporale" | Lorusso A, Marcacci M, Di Domenico M, Ancora M, Curini V, Mangone I, Rinaldi A, Scialabba S, Di Pasquale A, Cammà C, Puglia I, Calistri P, Savini G |
| EPI_ISL_1250959 | siesp Chieti | Istituto Zooprofilattico Sperimentale dell'Abruzzo e Molise "G. Caporale" | Lorusso A, Marcacci M, Di Domenico M, Ancora M, Curini V, Mangone I, Rinaldi A, Scialabba S, Di Pasquale A, Cammà C, Puglia I, Calistri P, Savini G |
| EPI_ISL_1250960 | SIESP SULMONA | Istituto Zooprofilattico Sperimentale dell'Abruzzo e Molise "G. Caporale" | Lorusso A, Marcacci M, Di Domenico M, Ancora M, Curini V, Mangone I, Rinaldi A, Scialabba S, Di Pasquale A, Cammà C, Puglia I, Calistri P, Savini G |
| EPI_ISL_1250961 | siesp Chieti | Istituto Zooprofilattico Sperimentale dell'Abruzzo e Molise "G. Caporale" | Lorusso A, Marcacci M, Di Domenico M, Ancora M, Curini V, Mangone I, Rinaldi A, Scialabba S, Di Pasquale A, Cammà C, Puglia I, Calistri P, Savini G |
| EPI_ISL_1250962, EPI_ISL_1250963, EPI_ISL_1250964, EPI_ISL_1250965 | Siesp dipartimento di prevenzione | Istituto Zooprofilattico Sperimentale dell'Abruzzo e Molise "G. Caporale" | Lorusso A, Marcacci M, Di Domenico M, Ancora M, Curini V, Mangone I, Rinaldi A, Scialabba S, Di Pasquale A, Cammà C, Puglia I, Calistri P, Savini G |
| EPI_ISL_1250967, EPI_ISL_1250968 | SIESP CHIETI - LANCIANO | Istituto Zooprofilattico Sperimentale dell'Abruzzo e Molise "G. Caporale" | Lorusso A, Marcacci M, Di Domenico M, Ancora M, Curini V, Mangone I, Rinaldi A, Scialabba S, Di Pasquale A, Cammà C, Puglia I, Calistri P, Savini G |
| EPI_ISL_1250969 | Siesp dipartimento di prevenzione | Istituto Zooprofilattico Sperimentale dell'Abruzzo e Molise "G. Caporale" | Lorusso A, Marcacci M, Di Domenico M, Ancora M, Curini V, Mangone I, Rinaldi A, Scialabba S, Di Pasquale A, Cammà C, Puglia I, Calistri P, Savini G |
| EPI_ISL_1250970, EPI_ISL_1250971 | SIESP CHIETI - LANCIANO | Istituto Zooprofilattico Sperimentale dell'Abruzzo e Molise "G. Caporale" | Lorusso A, Marcacci M, Di Domenico M, Ancora M, Curini V, Mangone I, Rinaldi A, Scialabba S, Di Pasquale A, Cammà C, Puglia I, Calistri P, Savini G |
| EPI_ISL_1250972 | OSPEDALE SANT'OMERO | Istituto Zooprofilattico Sperimentale dell'Abruzzo e Molise "G. Caporale" | Lorusso A, Marcacci M, Di Domenico M, Ancora M, Curini V, Mangone I, Rinaldi A, Scialabba S, Di Pasquale A, Cammà C, Puglia I, Calistri P, Savini G |
| EPI_ISL_1250973 | siesp Chieti | Istituto Zooprofilattico Sperimentale dell'Abruzzo e Molise "G. Caporale" | Lorusso A, Marcacci M, Di Domenico M, Ancora M, Curini V, Mangone I, Rinaldi A, Scialabba S, Di Pasquale A, Cammà C, Puglia I, Calistri P, Savini G |
| EPI_ISL_1250974 | SIESP CHIETI - LANCIANO | Istituto Zooprofilattico Sperimentale dell'Abruzzo e Molise "G. | Lorusso A, Marcacci M, Di Domenico M, Ancora M, Curini V, Mangone I, Rinaldi A, Scialabba S, Di Pasquale A, Cammà C, Puglia I, Calistri P, Savini G |

| | | | |
|---|---|---|---|
| | | Caporale" | |
| EPI_ISL_1251055 | OSPEDALE CIVILE GIULIANOVA - MEDICINA INTERNA | Istituto Zooprofilattico Sperimentale dell'Abruzzo e Molise "G. Caporale" | Lorusso A, Marcacci M, Di Domenico M, Ancora M, Curini V, Mangone I, Rinaldi A, Scialabba S, Di Pasquale A, Cammà C, Puglia I, Calistri P, Savini G |
| EPI_ISL_1251056 | SIESP CHIETI - DRIVE IN CHIETI | Istituto Zooprofilattico Sperimentale dell'Abruzzo e Molise "G. Caporale" | Lorusso A, Marcacci M, Di Domenico M, Ancora M, Curini V, Mangone I, Rinaldi A, Scialabba S, Di Pasquale A, Cammà C, Puglia I, Calistri P, Savini G |
| EPI_ISL_1251057, EPI_ISL_1251058, EPI_ISL_1251059, EPI_ISL_1251060 | Siesp dipartimento di prevenzione | Istituto Zooprofilattico Sperimentale dell'Abruzzo e Molise "G. Caporale" | Lorusso A, Marcacci M, Di Domenico M, Ancora M, Curini V, Mangone I, Rinaldi A, Scialabba S, Di Pasquale A, Cammà C, Puglia I, Calistri P, Savini G |
| EPI_ISL_1251061 | DIP. PREV. AVEZZANO SERVIZIO DI IGIENE EPIDEMIOLOGIA E SANITA' PUBBLICA | Istituto Zooprofilattico Sperimentale dell'Abruzzo e Molise "G. Caporale" | Lorusso A, Marcacci M, Di Domenico M, Ancora M, Curini V, Mangone I, Rinaldi A, Scialabba S, Di Pasquale A, Cammà C, Puglia I, Calistri P, Savini G |
| EPI_ISL_1251063, EPI_ISL_1251064, EPI_ISL_1251065 | Siesp dipartimento di prevenzione | Istituto Zooprofilattico Sperimentale dell'Abruzzo e Molise "G. Caporale" | Lorusso A, Marcacci M, Di Domenico M, Ancora M, Curini V, Mangone I, Rinaldi A, Scialabba S, Di Pasquale A, Cammà C, Puglia I, Calistri P, Savini G |
| EPI_ISL_1251066, EPI_ISL_1251067 | USCA Castel di Sangro | Istituto Zooprofilattico Sperimentale dell'Abruzzo e Molise "G. Caporale" | Lorusso A, Marcacci M, Di Domenico M, Ancora M, Curini V, Mangone I, Rinaldi A, Scialabba S, Di Pasquale A, Cammà C, Puglia I, Calistri P, Savini G |
| EPI_ISL_1251068, EPI_ISL_1251069, EPI_ISL_1251070, EPI_ISL_1251071 | SIESP CHIETI - DRIVE IN ORTONA | Istituto Zooprofilattico Sperimentale dell'Abruzzo e Molise "G. Caporale" | Lorusso A, Marcacci M, Di Domenico M, Ancora M, Curini V, Mangone I, Rinaldi A, Scialabba S, Di Pasquale A, Cammà C, Puglia I, Calistri P, Savini G |
| EPI_ISL_1251072 | USCA Castel di Sangro | Istituto Zooprofilattico Sperimentale dell'Abruzzo e Molise "G. Caporale" | Lorusso A, Marcacci M, Di Domenico M, Ancora M, Curini V, Mangone I, Rinaldi A, Scialabba S, Di Pasquale A, Cammà C, Puglia I, Calistri P, Savini G |
| EPI_ISL_1251073, EPI_ISL_1251074, EPI_ISL_1251075 | SIESP L'AQUILA | Istituto Zooprofilattico Sperimentale dell'Abruzzo e Molise "G. Caporale" | Lorusso A, Marcacci M, Di Domenico M, Ancora M, Curini V, Mangone I, Rinaldi A, Scialabba S, Di Pasquale A, Cammà C, Puglia I, Calistri P, Savini G |
| EPI_ISL_1251076 | SIESP SULMONA | Istituto Zooprofilattico Sperimentale dell'Abruzzo e Molise "G. Caporale" | Lorusso A, Marcacci M, Di Domenico M, Ancora M, Curini V, Mangone I, Rinaldi A, Scialabba S, Di Pasquale A, Cammà C, Puglia I, Calistri P, Savini G |
| EPI_ISL_1251077 | siesp Chieti | Istituto Zooprofilattico Sperimentale dell'Abruzzo e Molise "G. Caporale" | Lorusso A, Marcacci M, Di Domenico M, Ancora M, Curini V, Mangone I, Rinaldi A, Scialabba S, Di Pasquale A, Cammà C, Puglia I, Calistri P, Savini G |
| EPI_ISL_1251078 | "Presidio Ospedaliero "San Liberatore" Atri | Istituto Zooprofilattico Sperimentale dell'Abruzzo e Molise "G. Caporale" | Lorusso A, Marcacci M, Di Domenico M, Ancora M, Curini V, Mangone I, Rinaldi A, Scialabba S, Di Pasquale A, Cammà C, Puglia I, Calistri P, Savini G |
| EPI_ISL_1251079 | RSA GIULIANOVA | Istituto Zooprofilattico Sperimentale dell'Abruzzo e Molise "G. Caporale" | Lorusso A, Marcacci M, Di Domenico M, Ancora M, Curini V, Mangone I, Rinaldi A, Scialabba S, Di Pasquale A, Cammà C, Puglia I, Calistri P, Savini G |
| EPI_ISL_1251080 | Siesp dipartimento di prevenzione | Istituto Zooprofilattico Sperimentale dell'Abruzzo e Molise "G. Caporale" | Lorusso A, Marcacci M, Di Domenico M, Ancora M, Curini V, Mangone I, Rinaldi A, Scialabba S, Di Pasquale A, Cammà C, Puglia I, Calistri P, Savini G |
| EPI_ISL_1251081 | SIESP CHIETI - DRIVE IN CHIETI | Istituto Zooprofilattico Sperimentale dell'Abruzzo e Molise "G. Caporale" | Lorusso A, Marcacci M, Di Domenico M, Ancora M, Curini V, Mangone I, Rinaldi A, Scialabba S, Di Pasquale A, Cammà C, Puglia I, Calistri P, Savini G |
| EPI_ISL_1251082 | Siesp dipartimento di prevenzione | Istituto Zooprofilattico Sperimentale dell'Abruzzo e Molise "G. Caporale" | Lorusso A, Marcacci M, Di Domenico M, Ancora M, Curini V, Mangone I, Rinaldi A, Scialabba S, Di Pasquale A, Cammà C, Puglia I, Calistri P, Savini G |
| EPI_ISL_1251083 | SIESP CHIETI - DRIVE IN CHIETI | Istituto Zooprofilattico Sperimentale dell'Abruzzo e Molise "G. Caporale" | Lorusso A, Marcacci M, Di Domenico M, Ancora M, Curini V, Mangone I, Rinaldi A, Scialabba S, Di Pasquale A, Cammà C, Puglia I, Calistri P, Savini G |
| EPI_ISL_1251084, EPI_ISL_1251085, EPI_ISL_1251086, EPI_ISL_1251087, EPI_ISL_1251088, EPI_ISL_1251089, EPI_ISL_1251090, EPI_ISL_1251091, EPI_ISL_1251092 | Siesp dipartimento di prevenzione | Istituto Zooprofilattico Sperimentale dell'Abruzzo e Molise "G. Caporale" | Lorusso A, Marcacci M, Di Domenico M, Ancora M, Curini V, Mangone I, Rinaldi A, Scialabba S, Di Pasquale A, Cammà C, Puglia I, Calistri P, Savini G |
| EPI_ISL_1251093 | SIESP L'AQUILA | Istituto Zooprofilattico Sperimentale dell'Abruzzo e Molise "G. Caporale" | Lorusso A, Marcacci M, Di Domenico M, Ancora M, Curini V, Mangone I, Rinaldi A, Scialabba S, Di Pasquale A, Cammà C, Puglia I, Calistri P, Savini G |
| EPI_ISL_1251094 | SIESP CHIETI - LANCIANO | Istituto Zooprofilattico Sperimentale dell'Abruzzo e Molise "G. Caporale" | Lorusso A, Marcacci M, Di Domenico M, Ancora M, Curini V, Mangone I, Rinaldi A, Scialabba S, Di Pasquale A, Cammà C, Puglia I, Calistri P, Savini G |
| EPI_ISL_1251095 | SIESP CHIETI - DRIVE IN CHIETI | Istituto Zooprofilattico Sperimentale dell'Abruzzo e Molise "G. Caporale" | Lorusso A, Marcacci M, Di Domenico M, Ancora M, Curini V, Mangone I, Rinaldi A, Scialabba S, Di Pasquale A, Cammà C, Puglia I, Calistri P, Savini G |
| EPI_ISL_1251096, EPI_ISL_1251097 | Presidio Ospedaliero "San Liberatore" Atri | Istituto Zooprofilattico Sperimentale dell'Abruzzo e Molise "G. Caporale" | Lorusso A, Marcacci M, Di Domenico M, Ancora M, Curini V, Mangone I, Rinaldi A, Scialabba S, Di Pasquale A, Cammà C, Puglia I, Calistri P, Savini G |
| EPI_ISL_1251098, EPI_ISL_1251099, EPI_ISL_1251100 | Siesp dipartimento di prevenzione | Istituto Zooprofilattico Sperimentale dell'Abruzzo e Molise "G. Caporale" | Lorusso A, Marcacci M, Di Domenico M, Ancora M, Curini V, Mangone I, Rinaldi A, Scialabba S, Di Pasquale A, Cammà C, Puglia I, Calistri P, Savini G |
| EPI_ISL_1251101 | SIESP CHIETI - LANCIANO | Istituto Zooprofilattico Sperimentale dell'Abruzzo e Molise "G. Caporale" | Lorusso A, Marcacci M, Di Domenico M, Ancora M, Curini V, Mangone I, Rinaldi A, Scialabba S, Di Pasquale A, Cammà C, Puglia I, Calistri P, Savini G |
| EPI_ISL_1251102 | Siesp dipartimento di prevenzione | Istituto Zooprofilattico Sperimentale dell'Abruzzo e Molise "G. Caporale" | Lorusso A, Marcacci M, Di Domenico M, Ancora M, Curini V, Mangone I, Rinaldi A, Scialabba S, Di Pasquale A, Cammà C, Puglia I, Calistri P, Savini G |
| EPI_ISL_1251103 | SIESP L'AQUILA | Istituto Zooprofilattico Sperimentale dell'Abruzzo e Molise "G. Caporale" | Lorusso A, Marcacci M, Di Domenico M, Ancora M, Curini V, Mangone I, Rinaldi A, Scialabba S, Di Pasquale A, Cammà C, Puglia I, Calistri P, Savini G |
| EPI_ISL_1251104, EPI_ISL_1251105, EPI_ISL_1251106 | Siesp dipartimento di prevenzione | Istituto Zooprofilattico Sperimentale dell'Abruzzo e Molise "G. Caporale" | Lorusso A, Marcacci M, Di Domenico M, Ancora M, Curini V, Mangone I, Rinaldi A, Scialabba S, Di Pasquale A, Cammà C, Puglia I, Calistri P, Savini G |
| EPI_ISL_1251107, EPI_ISL_1251108 | SIESP CHIETI - DRIVE IN ORTONA | Istituto Zooprofilattico Sperimentale dell'Abruzzo e Molise "G. Caporale" | Lorusso A, Marcacci M, Di Domenico M, Ancora M, Curini V, Mangone I, Rinaldi A, Scialabba S, Di Pasquale A, Cammà C, Puglia I, Calistri P, Savini G |
| EPI_ISL_1251109, EPI_ISL_1251110, EPI_ISL_1251111, EPI_ISL_1251112 | SIESP CHIETI - LANCIANO | Istituto Zooprofilattico Sperimentale dell'Abruzzo e Molise "G. Caporale" | Lorusso A, Marcacci M, Di Domenico M, Ancora M, Curini V, Mangone I, Rinaldi A, Scialabba S, Di Pasquale A, Cammà C, Puglia I, Calistri P, Savini G |
| EPI_ISL_1251113 | SIESP CHIETI - DRIVE IN GISSI | Istituto Zooprofilattico Sperimentale dell'Abruzzo e Molise "G. Caporale" | Lorusso A, Marcacci M, Di Domenico M, Ancora M, Curini V, Mangone I, Rinaldi A, Scialabba S, Di Pasquale A, Cammà C, Puglia I, Calistri P, Savini G |
| EPI_ISL_1251114 | RSA GIULIANOVA | Istituto Zooprofilattico Sperimentale dell'Abruzzo e Molise "G. Caporale" | Lorusso A, Marcacci M, Di Domenico M, Ancora M, Curini V, Mangone I, Rinaldi A, Scialabba S, Di Pasquale A, Cammà C, Puglia I, Calistri P, Savini G |
| EPI_ISL_1251115, EPI_ISL_1251116, EPI_ISL_1251117 | SIESP L'AQUILA | Istituto Zooprofilattico Sperimentale dell'Abruzzo e Molise "G. Caporale" | Lorusso A, Marcacci M, Di Domenico M, Ancora M, Curini V, Mangone I, Rinaldi A, Scialabba S, Di Pasquale A, Cammà C, Puglia I, Calistri P, Savini G |
| EPI_ISL_1251245, EPI_ISL_1251251 | SC (UCO) Igiene e Sanità Pubblica | ARGO Laboratorio Genomica ed Epigenomica | Licastro D, Dal Monego S, Degasperi M, Segat L, Piscianz E, D'Agaro P |
| EPI_ISL_1253609 | Laboratorio Unico Provinciale Azienda Ospedaliero-Universitaria di Ferrara | U.O. Microbiologia, Laboratorio Unico Centro Servizi - AUSL della Romagna | Michela Boni, Sara Ghisellini, Giorgio Dirani, Silvia Zannoli, Vittorio Sambri |

| | | | |
|---|---|---|---|
| EPI_ISL_1254676, EPI_ISL_1254677, EPI_ISL_1254678, EPI_ISL_1254679, EPI_ISL_1254680 | Presidio di Brindisi Di Summa - Perrino | Istituto Zooprofilattico Sperimentale della Puglia e della Basilicata | Parisi A., Bianco A., Capozzi L., Del Sambro L., Simone D., Difato L., Ridolfi D., Giannico A. |
| EPI_ISL_1254681, EPI_ISL_1254683, EPI_ISL_1254684, EPI_ISL_1254685, EPI_ISL_1254686, EPI_ISL_1254687 | Ospedale Generale Regionale F. Miulli | Istituto Zooprofilattico Sperimentale della Puglia e della Basilicata | Parisi A., Bianco A., Capozzi L., Del Sambro L., Simone D., Difato L., Ridolfi D., Giannico A. |
| EPI_ISL_1254692, EPI_ISL_1254693, EPI_ISL_1254694 | Istituto Zooprofilattico Sperimentale della Puglia e della Basilicata | Istituto Zooprofilattico Sperimentale della Puglia e della Basilicata | Parisi A., Bianco A., Capozzi L., Del Sambro L., Simone D., Manzulli V, Rondinone V., Pace L., Cipolletta D., Galante D. |
| EPI_ISL_1254696, EPI_ISL_1254697, EPI_ISL_1254698, EPI_ISL_1254699, EPI_ISL_1254700 | Ospedale Santissima Annunziata | Istituto Zooprofilattico Sperimentale della Puglia e della Basilicata | Parisi A., Bianco A., Capozzi L., Del Sambro L., Simone D., Tampoia M., Difato L., Ridolfi D., Giannico A. |
| EPI_ISL_1254701 | Ospedale Santa Caterina Novella | Istituto Zooprofilattico Sperimentale della Puglia e della Basilicata | Parisi A., Bianco A., Capozzi L., Del Sambro L., Simone D., Difato L., Ridolfi D., Giannico A. |
| EPI_ISL_1254702, EPI_ISL_1254703 | Ospedale Casa Sollievo della Sofferenza | Istituto Zooprofilattico Sperimentale della Puglia e della Basilicata | Parisi A., Bianco A., Capozzi L., Del Sambro L., Simone D., Difato L., Ridolfi D., Giannico A. |
| EPI_ISL_1257900 | INT Fondazione Pascale | INT Fondazione Pascale | INT Fondazione Pascale |
| EPI_ISL_1259020, EPI_ISL_1259021, EPI_ISL_1259022, EPI_ISL_1259023, EPI_ISL_1259024 | Azienda Ospedaliero - Universitaria di Bologna IRCCS Policlinico di Sant'Orsola - Unità di Microbiologia | Istituto Zooprofilattico Sperimentale della Lombardia e dell'Emilia Romagna (IZSLER), Risk Analysis and Genomic Epidemiology Unit | Giada Rossini, Giuliano Furlini, Tiziana Lazzarotto, Marina Morganti, Ilaria Menozzi, Erika Scaltriti, Stefano Pongolini |
| EPI_ISL_1259194, EPI_ISL_1259195, EPI_ISL_1259196, EPI_ISL_1259197, EPI_ISL_1259198, EPI_ISL_1259199, EPI_ISL_1259200, EPI_ISL_1259201, EPI_ISL_1259202, EPI_ISL_1259203, EPI_ISL_1259204, EPI_ISL_1259205, EPI_ISL_1259206, EPI_ISL_1259207 | | | |
| see above | Department of Virology, Istituto Zooprofilattico Sperimentale del Lazio e della Toscana (IZSLT) | Department of General Diagnostics; Department of Virology; Istituto Zooprofilattico Sperimentale del Lazio e della Toscana (IZSLT) | Patricia Alba, Giuseppe Manna, Elena L. Diaconu, Fabiola Feltrin, Raffaela Conti, Teresa Scicluna, Virginia Carfora, Antonella Cersini, Alessia Franco, Antonio Battisti. |
| EPI_ISL_1259405 | INMI Lazzaro Spallanzani IRCCS | INMI Lazzaro Spallanzani IRCCS | O Butera, CEM Gruber, B Bartolini, E Giombini, F Santini, F Messina, M Rueca, G Bonfiglio, MR Capobianchi, A Di Caro |
| EPI_ISL_1259406 | INMI Lazzaro Spallanzani IRCCS | INMI Lazzaro Spallanzani IRCCS | CEM Gruber, B Bartolini, E Giombini, F Santini, F Messina, M Rueca, O Butera, G Bonfiglio, A Di Caro, MR Capobianchi |
| EPI_ISL_1259407 | INMI Lazzaro Spallanzani IRCCS | INMI Lazzaro Spallanzani IRCCS | M Rueca, O Butera, CEM Gruber, B Bartolini, F Santini, E Giombini, G Bonfiglio, F Messina, MR Capobianchi, A Di Caro |
| EPI_ISL_1259408 | INMI Lazzaro Spallanzani IRCCS | INMI Lazzaro Spallanzani IRCCS | CEM Gruber, B Bartolini, E Giombini, F Messina, F Santini, M Rueca0, G Bonfiglio, O Butera, MR Capobianchi, A Di Caro |
| EPI_ISL_1259409 | Ospedale Sandro Pertini ASL Roma2 | INMI Lazzaro Spallanzani IRCCS | B Bartolini, E Giombini, F Messina, M Rueca, O Butera, F Santini, CEM Gruber, G Bonfiglio, MC Cava, G Cappiello, S Romano, A Di Caro, MR Capobianchi |
| EPI_ISL_1259410 | INMI Lazzaro Spallanzani IRCCS | INMI Lazzaro Spallanzani IRCCS | M Rueca, O Butera, CEM Gruber, B Bartolini, E Giombini, F Santini, G Bonfiglio, F Messina, A Di Caro, MR Capobianchi |
| EPI_ISL_1259411 | INMI Lazzaro Spallanzani IRCCS | INMI Lazzaro Spallanzani IRCCS | F Messina, M Rueca, O Butera, CEM Gruber, B Bartolini, F Santini, G Bonfiglio, E Giombini, MR Capobianchi, A Di Caro |
| EPI_ISL_1259412 | Sant'Eugenio/CTO ASL Roma 2 | INMI Lazzaro Spallanzani IRCCS | M Rueca, O Butera, CEM Gruber, B Bartolini, E Giombini, F Messina, F Santini, G Bonfiglio, F Bondanini, GC Cocciolillo, C Disegni, A Di Caro, MR Capobianchi |
| EPI_ISL_1259413 | INMI Lazzaro Spallanzani IRCCS | INMI Lazzaro Spallanzani IRCCS | CEM Gruber, B Bartolini, E Giombini, F Messina, M Rueca, O Butera, F Santini, G Bonfiglio, A Di Caro, MR Capobianchi |
| EPI_ISL_1259414 | INMI Lazzaro Spallanzani IRCCS | INMI Lazzaro Spallanzani IRCCS | F Messina, M Rueca, O Butera, CEM Gruber, B Bartolini, G Bonfiglio, E Giombini, F Santini, A Di Caro, MR Capobianchi |
| EPI_ISL_1259415 | INMI Lazzaro Spallanzani IRCCS | INMI Lazzaro Spallanzani IRCCS | O Butera, CEM Gruber, B Bartolini, E Giombini, G Bonfiglio, F Messina, M Rueca, F Santini, A Di Caro, MR Capobianchi |
| EPI_ISL_1260687, EPI_ISL_1260688, EPI_ISL_1260689, EPI_ISL_1260690, EPI_ISL_1260691, EPI_ISL_1260692, EPI_ISL_1260693 | Azienda Sanitaria Locale di Piacenza - Presidio Ospedaliero - Laboratorio di Microbiologia | Istituto Zooprofilattico Sperimentale della Lombardia e dell'Emilia Romagna (IZSLER), Risk Analysis and Genomic Epidemiology Unit | Lo Cascio Giuliana, Schiavo Roberta, Peroni Irene, Marina Morganti, Ilaria Menozzi, Erika Scaltriti, Stefano Pongolini |
| EPI_ISL_1261869 | Ospedale Generale Regionale F. Miulli | Istituto Zooprofilattico Sperimentale della Puglia e della Basilicata | Parisi A., Bianco A., Capozzi L., Del Sambro L., Simone D., Difato L., Ridolfi D., Giannico A. |
| EPI_ISL_1261870 | Istituto Zooprofilattico Sperimentale della Puglia e della Basilicata | Istituto Zooprofilattico Sperimentale della Puglia e della Basilicata | Parisi A., Bianco A., Capozzi L., Del Sambro L., Simone D., Manzulli V, Rondinone V., Pace L., Cipolletta D., Galante D. |
| EPI_ISL_1287411, EPI_ISL_1287412, EPI_ISL_1287413, EPI_ISL_1287414, EPI_ISL_1287415, EPI_ISL_1287416, EPI_ISL_1287417, EPI_ISL_1287418, EPI_ISL_1287419, EPI_ISL_1287420, EPI_ISL_1287421, EPI_ISL_1287422, EPI_ISL_1287423, EPI_ISL_1287424, EPI_ISL_1287425, EPI_ISL_1287426, EPI_ISL_1287427, EPI_ISL_1287428, EPI_ISL_1287429, EPI_ISL_1287430, EPI_ISL_1287431, EPI_ISL_1287432, EPI_ISL_1287433, EPI_ISL_1287434, EPI_ISL_1287435, EPI_ISL_1287436, EPI_ISL_1287437, EPI_ISL_1287438, EPI_ISL_1287439, EPI_ISL_1287440, EPI_ISL_1287441, EPI_ISL_1287442, EPI_ISL_1287443, EPI_ISL_1287444, EPI_ISL_1287445, EPI_ISL_1287446, EPI_ISL_1287447, EPI_ISL_1287448, EPI_ISL_1287449, EPI_ISL_1287450, EPI_ISL_1287451, EPI_ISL_1287452, EPI_ISL_1287453, EPI_ISL_1287454, EPI_ISL_1287455, EPI_ISL_1287456, EPI_ISL_1287457, EPI_ISL_1287458, EPI_ISL_1287459, EPI_ISL_1287460, EPI_ISL_1287461, EPI_ISL_1287462, EPI_ISL_1287463, EPI_ISL_1287464, EPI_ISL_1287465, EPI_ISL_1287466, EPI_ISL_1287467, EPI_ISL_1287468, EPI_ISL_1287469, EPI_ISL_1287470, EPI_ISL_1287471, EPI_ISL_1287472, EPI_ISL_1287473, EPI_ISL_1287474, EPI_ISL_1287475, EPI_ISL_1287476, EPI_ISL_1287477, EPI_ISL_1287478, EPI_ISL_1287479, EPI_ISL_1287480, EPI_ISL_1287481, EPI_ISL_1287482, EPI_ISL_1287483, EPI_ISL_1287484, EPI_ISL_1287485, EPI_ISL_1287486, EPI_ISL_1287487, EPI_ISL_1287488, EPI_ISL_1287489, EPI_ISL_1287490, EPI_ISL_1287491, EPI_ISL_1287492, EPI_ISL_1287493, EPI_ISL_1287494, EPI_ISL_1287495, EPI_ISL_1287496, EPI_ISL_1287497, EPI_ISL_1287498, EPI_ISL_1287499, EPI_ISL_1287500, EPI_ISL_1287501, EPI_ISL_1287502, EPI_ISL_1287503, EPI_ISL_1287504, EPI_ISL_1287505, EPI_ISL_1287506, EPI_ISL_1287507, EPI_ISL_1287508, EPI_ISL_1287509, EPI_ISL_1287510, EPI_ISL_1287511, EPI_ISL_1287512, EPI_ISL_1287513, EPI_ISL_1287514, EPI_ISL_1287515, EPI_ISL_1287516, EPI_ISL_1287517, EPI_ISL_1287518, EPI_ISL_1287519, EPI_ISL_1287520, EPI_ISL_1287521, EPI_ISL_1287522, EPI_ISL_1287523, EPI_ISL_1287524, EPI_ISL_1287525, EPI_ISL_1287526, EPI_ISL_1287527, EPI_ISL_1287528, EPI_ISL_1287529, EPI_ISL_1287530, EPI_ISL_1287531, EPI_ISL_1287532, EPI_ISL_1287533, EPI_ISL_1287534, EPI_ISL_1287535, EPI_ISL_1287536, EPI_ISL_1287537, EPI_ISL_1287538, EPI_ISL_1287539, EPI_ISL_1287540, EPI_ISL_1287541, EPI_ISL_1287542, EPI_ISL_1287543, EPI_ISL_1287544, EPI_ISL_1287545, EPI_ISL_1287546, EPI_ISL_1287547, EPI_ISL_1287548, EPI_ISL_1287549, EPI_ISL_1287550, EPI_ISL_1287551, EPI_ISL_1287552, EPI_ISL_1287553, EPI_ISL_1287554, EPI_ISL_1287555, EPI_ISL_1287556, EPI_ISL_1287557, EPI_ISL_1287558, EPI_ISL_1287559, EPI_ISL_1287560, EPI_ISL_1287561, EPI_ISL_1287562, EPI_ISL_1287563, EPI_ISL_1287564, EPI_ISL_1287565, EPI_ISL_1287566, EPI_ISL_1287567, EPI_ISL_1287568, EPI_ISL_1287569, EPI_ISL_1287570, EPI_ISL_1287571, EPI_ISL_1287572, EPI_ISL_1287573, EPI_ISL_1287574, EPI_ISL_1287575, EPI_ISL_1287576, EPI_ISL_1287577, EPI_ISL_1287578, EPI_ISL_1287579, EPI_ISL_1287580, EPI_ISL_1287581, EPI_ISL_1287582, EPI_ISL_1287583, EPI_ISL_1287584, EPI_ISL_1287585, EPI_ISL_1287586, EPI_ISL_1287587, EPI_ISL_1287588, EPI_ISL_1287589, EPI_ISL_1287590, EPI_ISL_1287591, EPI_ISL_1287592, EPI_ISL_1287593, EPI_ISL_1287594, EPI_ISL_1287595, EPI_ISL_1287596, EPI_ISL_1287597, EPI_ISL_1287598, EPI_ISL_1287599, EPI_ISL_1287600, EPI_ISL_1287601, EPI_ISL_1287602, EPI_ISL_1287603, EPI_ISL_1287604, EPI_ISL_1287605, EPI_ISL_1287606, EPI_ISL_1287607, EPI_ISL_1287608, EPI_ISL_1287609, EPI_ISL_1287610, EPI_ISL_1287611, EPI_ISL_1287612, EPI_ISL_1287613, EPI_ISL_1287614, EPI_ISL_1287615, EPI_ISL_1287616, EPI_ISL_1287617, EPI_ISL_1287618, EPI_ISL_1287619, EPI_ISL_1287620, EPI_ISL_1287621, EPI_ISL_1287622, EPI_ISL_1287623, EPI_ISL_1287624, EPI_ISL_1287625, EPI_ISL_1287626, EPI_ISL_1287627, EPI_ISL_1287628, EPI_ISL_1287629, EPI_ISL_1287630, EPI_ISL_1287631, EPI_ISL_1287632, EPI_ISL_1287633, EPI_ISL_1287634, EPI_ISL_1287635, EPI_ISL_1287636, EPI_ISL_1287637, EPI_ISL_1287638, EPI_ISL_1287639, EPI_ISL_1287640, EPI_ISL_1287641, EPI_ISL_1287642, EPI_ISL_1287643, EPI_ISL_1287644, EPI_ISL_1287645, EPI_ISL_1287646, EPI_ISL_1287647, EPI_ISL_1287648, EPI_ISL_1287649, EPI_ISL_1287650, EPI_ISL_1287651, EPI_ISL_1287652, EPI_ISL_1287653, EPI_ISL_1287654, EPI_ISL_1287655, EPI_ISL_1287656, EPI_ISL_1287657, EPI_ISL_1287658, EPI_ISL_1287659, EPI_ISL_1287660, EPI_ISL_1287661, EPI_ISL_1287662, EPI_ISL_1287663, EPI_ISL_1287664, EPI_ISL_1287665, EPI_ISL_1287666, EPI_ISL_1287667, EPI_ISL_1287668, EPI_ISL_1287669, EPI_ISL_1287670, EPI_ISL_1287671, EPI_ISL_1287672, EPI_ISL_1287673, EPI_ISL_1287674, EPI_ISL_1287675, EPI_ISL_1287676, EPI_ISL_1287677, EPI_ISL_1287678, EPI_ISL_1287679, EPI_ISL_1287680, EPI_ISL_1287681, EPI_ISL_1287682, EPI_ISL_1287683, EPI_ISL_1287684, EPI_ISL_1287685, EPI_ISL_1287686, EPI_ISL_1287687, EPI_ISL_1287688, EPI_ISL_1287689, EPI_ISL_1287690, EPI_ISL_1287691, EPI_ISL_1287692, EPI_ISL_1287693, EPI_ISL_1287694, EPI_ISL_1287695, EPI_ISL_1287696, EPI_ISL_1287697, EPI_ISL_1287698, EPI_ISL_1287699, EPI_ISL_1287700, EPI_ISL_1287701, EPI_ISL_1287702, EPI_ISL_1287703, EPI_ISL_1287704, EPI_ISL_1287705, EPI_ISL_1287706, EPI_ISL_1287707, EPI_ISL_1287708, EPI_ISL_1287709, EPI_ISL_1287710, EPI_ISL_1287711, EPI_ISL_1287712, EPI_ISL_1287713, EPI_ISL_1287714, EPI_ISL_1287715, EPI_ISL_1287716, EPI_ISL_1287717, EPI_ISL_1287718, EPI_ISL_1287719, EPI_ISL_1287720, EPI_ISL_1287721, EPI_ISL_1287722, EPI_ISL_1287723, EPI_ISL_1287724, EPI_ISL_1287725, EPI_ISL_1287726, EPI_ISL_1287727, EPI_ISL_1287728, EPI_ISL_1287729, EPI_ISL_1287730, EPI_ISL_1287731, EPI_ISL_1287732, EPI_ISL_1287733, EPI_ISL_1287734, EPI_ISL_1287735, EPI_ISL_1287736 | | | |
| see above | Istituto Zooprofilattico Sperimentale del Mezzogiorno | TIGEM | Antonio Grimaldi Patrizia Annunziata Francesco Panariello Biancamaria Pierri Claudia Tiberio Valentina Bouche Chiara Colantuono Maria Concetta Cuomo Denise Di Concilio Lucio Di Filippo Anna Manfredi Marcello Salvi Antonio Limone Luigi Atripaldi Pellegrino Cerino Andrea Ballabio Davide Cacchiarelli |
| EPI_ISL_1290805 | DIP. PREV. AVEZZANO SERVIZIO DI IGIENE EPIDEMIOLOGIA E SANITA' PUBBLICA | Istituto Zooprofilattico Sperimentale dell'Abruzzo e Molise "G. Caporale" | Lorusso A, Marcacci M, Di Domenico M, Ancora M, Curini V, Mangone I, Rinaldi A, Scialabba S, Di Pasquale A, Cammà C, Puglia I, Calistri P, Savini G |
| EPI_ISL_1290806 | siesp Chieti | Istituto Zooprofilattico Sperimentale dell'Abruzzo e Molise "G. Caporale" | Lorusso A, Marcacci M, Di Domenico M, Ancora M, Curini V, Mangone I, Rinaldi A, Scialabba S, Di Pasquale A, Cammà C, Puglia I, Calistri P, Savini G |
| EPI_ISL_1290807 | Siesp dipartimento di prevenzione | Istituto Zooprofilattico Sperimentale dell'Abruzzo e Molise "G. Caporale" | Lorusso A, Marcacci M, Di Domenico M, Ancora M, Curini V, Mangone I, Rinaldi A, Scialabba S, Di Pasquale A, Cammà C, Puglia I, Calistri P, Savini G |

| | | | |
|---|---|---|---|
| EPI_ISL_1290808 | SIESP L'AQUILA | Istituto Zooprofilattico Sperimentale dell'Abruzzo e Molise "G. Caporale" | Lorusso A, Marcacci M, Di Domenico M, Ancora M, Curini V, Mangone I, Rinaldi A, Scialabba S, Di Pasquale A, Cammà C, Puglia I, Calistri P, Savini G |
| EPI_ISL_1295606 | INMI Lazzaro Spallanzani IRCCS | INMI Lazzaro Spallanzani IRCCS | M Rueca, G Bonfiglio, O Butera, CEM Gruber, F Santini, B Bartolini, E Giombini, F Messina, MR Capobianchi, A Di Caro |
| EPI_ISL_1295609 | INMI Lazzaro Spallanzani IRCCS | INMI Lazzaro Spallanzani IRCCS | F Messina, M Rueca, G Bonfiglio, O Butera, CEM Gruber, F Santini, B Bartolini, E Giombini, A Di Caro, MR Capobianchi |
| EPI_ISL_1295610 | Presidio Ospedaliero G. B. Grassi ASL RM3 | INMI Lazzaro Spallanzani IRCCS | O Butera, CEM Gruber, F Santini, B Bartolini, E Giombini, F Messina, M Rueca, G Bonfiglio, F Tabacco, E Ristori, MR Capobianchi, A Di Caro |
| EPI_ISL_1295611 | Ospedale "F. Spaziani" Frosinone | INMI Lazzaro Spallanzani IRCCS | M Rueca, G Bonfiglio, O Butera, CEM Gruber, F Santini, B Bartolini, E Giombini, F Messina, R Pulselli, G Brocco, C Gargiulo, MR Capobianchi, A Di Caro |
| EPI_ISL_1295612 | Ospedale "F. Spaziani" Frosinone | INMI Lazzaro Spallanzani IRCCS | F Messina, M Rueca, G Bonfiglio, O Butera, CEM Gruber, F Santini, B Bartolini, E Giombini, R Pulselli, C Sias, G Brocco, A Di Caro, MR Capobianchi |
| EPI_ISL_1295613 | Fondazione Policlinico Universitario "A. Gemelli" IRCCS | INMI Lazzaro Spallanzani IRCCS | B Bartolini, E Giombini, F Messina, M Rueca, G Bonfiglio, O Butera, CEM Gruber, F Santini, P Cattani, M Sanguinetti, A Di Caro, MR Capobianchi |
| EPI_ISL_1295615 | Azienda Sanitaria Locale Roma 5 | INMI Lazzaro Spallanzani IRCCS | E Giombini, F Messina, M Rueca, G Bonfiglio, O Butera, CEM Gruber, F Santini, B Bartolini, D Di Fusco, D Cerini, MR Capobianchi, A Di Caro |
| EPI_ISL_1295616 | INMI Lazzaro Spallanzani IRCCS | INMI Lazzaro Spallanzani IRCCS | E Giombini, F Messina, M Rueca, G Bonfiglio, O Butera, CEM Gruber, F Santini, B Bartolini, MR Capobianchi, A Di Caro |
| EPI_ISL_1295617 | INMI Lazzaro Spallanzani IRCCS | INMI Lazzaro Spallanzani IRCCS | F Santini, B Bartolini, E Giombini, F Messina, M Rueca, G Bonfiglio, O Butera, CEM Gruber, MR Capobianchi, A Di Caro |
| EPI_ISL_1295642 | INMI Lazzaro Spallanzani IRCCS | INMI Lazzaro Spallanzani IRCCS | G Bonfiglio, O Butera, CEM Gruber, F Santini,B Bartolini, E Giombini, F Messina, M Rueca, A Di Caro, MR Capobianchi |
| EPI_ISL_1296237 | INMI Lazzaro Spallanzani IRCCS | INMI Lazzaro Spallanzani IRCCS | CEM Gruber, F Santini, B Bartolini, E Giombini, F Messina, M Rueca, G Bonfiglio, O Butera, A Di Caro, MR Capobianchi |
| EPI_ISL_1296292 | INMI Lazzaro Spallanzani IRCCS | INMI Lazzaro Spallanzani IRCCS | B Bartolini, E Giombini, F Messina, M Rueca, G Bonfiglio, O Butera, CEM Gruber, F Santini, A Di Caro, MR Capobianchi |
| EPI_ISL_1298493, EPI_ISL_1298494, EPI_ISL_1298495, EPI_ISL_1298496, EPI_ISL_1298497, EPI_ISL_1298498, EPI_ISL_1298499, EPI_ISL_1298500, EPI_ISL_1298501, EPI_ISL_1298502, EPI_ISL_1298503, EPI_ISL_1298504, EPI_ISL_1298505, EPI_ISL_1298506, EPI_ISL_1298507, EPI_ISL_1298508, EPI_ISL_1298509, EPI_ISL_1298510, EPI_ISL_1298511, EPI_ISL_1298512, EPI_ISL_1298513, EPI_ISL_1298514, EPI_ISL_1298515, EPI_ISL_1298516, EPI_ISL_1298517, EPI_ISL_1298518, EPI_ISL_1298519, EPI_ISL_1298520, EPI_ISL_1298521, EPI_ISL_1298522, EPI_ISL_1298523, EPI_ISL_1298524, EPI_ISL_1298525, EPI_ISL_1298526, EPI_ISL_1298527, EPI_ISL_1298528, EPI_ISL_1298529, EPI_ISL_1298530, EPI_ISL_1298531, EPI_ISL_1298532, EPI_ISL_1298533, EPI_ISL_1298534, EPI_ISL_1298535, EPI_ISL_1298536, EPI_ISL_1298537, EPI_ISL_1298538, EPI_ISL_1298539, EPI_ISL_1298540, EPI_ISL_1298541, EPI_ISL_1298542, EPI_ISL_1298543, EPI_ISL_1298544, EPI_ISL_1298545, EPI_ISL_1298546, EPI_ISL_1298547, EPI_ISL_1298548, EPI_ISL_1298549, EPI_ISL_1298550, EPI_ISL_1298551, EPI_ISL_1298552, EPI_ISL_1298553, EPI_ISL_1298554, EPI_ISL_1298555, EPI_ISL_1298556, EPI_ISL_1298557, EPI_ISL_1298558, EPI_ISL_1298559, EPI_ISL_1298560, EPI_ISL_1298561, EPI_ISL_1298562, EPI_ISL_1298563, EPI_ISL_1298564, EPI_ISL_1298565, EPI_ISL_1298566, EPI_ISL_1298567, EPI_ISL_1298568, EPI_ISL_1298569, EPI_ISL_1298570, EPI_ISL_1298571, EPI_ISL_1298572, EPI_ISL_1298573, EPI_ISL_1298574, EPI_ISL_1298575, EPI_ISL_1298576, EPI_ISL_1298577, EPI_ISL_1298578, EPI_ISL_1298579, EPI_ISL_1298580, EPI_ISL_1298581, EPI_ISL_1298582, EPI_ISL_1298583, EPI_ISL_1298584, EPI_ISL_1298585, EPI_ISL_1298586, EPI_ISL_1298587, EPI_ISL_1298588, EPI_ISL_1298589, EPI_ISL_1298590, EPI_ISL_1298591, EPI_ISL_1298592, EPI_ISL_1298593, EPI_ISL_1298594, EPI_ISL_1298595, EPI_ISL_1298596, EPI_ISL_1298597, EPI_ISL_1298598, EPI_ISL_1298599, EPI_ISL_1298600, EPI_ISL_1298601, EPI_ISL_1298602, EPI_ISL_1298603, EPI_ISL_1298604, EPI_ISL_1298605, EPI_ISL_1298606, EPI_ISL_1298607, EPI_ISL_1298608, EPI_ISL_1298609, EPI_ISL_1298610, EPI_ISL_1298611, EPI_ISL_1298612, EPI_ISL_1298613, EPI_ISL_1298614, EPI_ISL_1298615, EPI_ISL_1298616, EPI_ISL_1298617, EPI_ISL_1298618, EPI_ISL_1298619, EPI_ISL_1298620, EPI_ISL_1298621, EPI_ISL_1298622, EPI_ISL_1298623, EPI_ISL_1298624, EPI_ISL_1298625, EPI_ISL_1298626, EPI_ISL_1298627, EPI_ISL_1298628, EPI_ISL_1298629, EPI_ISL_1298630, EPI_ISL_1298631, EPI_ISL_1298632, EPI_ISL_1298633, EPI_ISL_1298634, EPI_ISL_1298635, EPI_ISL_1298636, EPI_ISL_1298637, EPI_ISL_1298638, EPI_ISL_1298639, EPI_ISL_1298640, EPI_ISL_1298641, EPI_ISL_1298642, EPI_ISL_1298643, EPI_ISL_1298644, EPI_ISL_1298645, EPI_ISL_1298646, EPI_ISL_1298647, EPI_ISL_1298648, EPI_ISL_1298649, EPI_ISL_1298650, EPI_ISL_1298651, EPI_ISL_1298652, EPI_ISL_1298653, EPI_ISL_1298654, EPI_ISL_1298655, EPI_ISL_1298656, EPI_ISL_1298657, EPI_ISL_1298658, EPI_ISL_1298659, EPI_ISL_1298660, EPI_ISL_1298661, EPI_ISL_1298662, EPI_ISL_1298663, EPI_ISL_1298664, EPI_ISL_1298665, EPI_ISL_1298666, EPI_ISL_1298667, EPI_ISL_1298668, EPI_ISL_1298669, EPI_ISL_1298670, EPI_ISL_1298671, EPI_ISL_1298672, EPI_ISL_1298673, EPI_ISL_1298674, EPI_ISL_1298675, EPI_ISL_1298676, EPI_ISL_1298677, EPI_ISL_1298678, EPI_ISL_1298679, EPI_ISL_1298680, EPI_ISL_1298681, EPI_ISL_1298682, EPI_ISL_1298683, EPI_ISL_1298684, EPI_ISL_1298685, EPI_ISL_1298686, EPI_ISL_1298687, EPI_ISL_1298688, EPI_ISL_1298689, EPI_ISL_1298690, EPI_ISL_1298691, EPI_ISL_1298692, EPI_ISL_1298693, EPI_ISL_1298694, EPI_ISL_1298695, EPI_ISL_1298696, EPI_ISL_1298697, EPI_ISL_1298698, EPI_ISL_1298699, EPI_ISL_1298700, EPI_ISL_1298701, EPI_ISL_1298702, EPI_ISL_1298703, EPI_ISL_1298704, EPI_ISL_1298705, EPI_ISL_1298706, EPI_ISL_1298707, EPI_ISL_1298708, EPI_ISL_1298709, EPI_ISL_1298710, EPI_ISL_1298711, EPI_ISL_1298712, EPI_ISL_1298713, EPI_ISL_1298714, EPI_ISL_1298715, EPI_ISL_1298716, EPI_ISL_1298717, EPI_ISL_1298718, EPI_ISL_1298719, EPI_ISL_1298720, EPI_ISL_1298721, EPI_ISL_1298722, EPI_ISL_1298723, EPI_ISL_1298724, EPI_ISL_1298725, EPI_ISL_1298726, EPI_ISL_1298727, EPI_ISL_1298728, EPI_ISL_1298729, EPI_ISL_1298730, EPI_ISL_1298731, EPI_ISL_1298732, EPI_ISL_1298733, EPI_ISL_1298734, EPI_ISL_1298735, EPI_ISL_1298736, EPI_ISL_1298737, EPI_ISL_1298738, EPI_ISL_1298739, EPI_ISL_1298740, EPI_ISL_1298741, EPI_ISL_1298742, EPI_ISL_1298743, EPI_ISL_1298744, EPI_ISL_1298745, EPI_ISL_1298746, EPI_ISL_1298747, EPI_ISL_1298748, EPI_ISL_1298749, EPI_ISL_1298750, EPI_ISL_1298751, EPI_ISL_1298752, EPI_ISL_1298753, EPI_ISL_1298754, EPI_ISL_1298755, EPI_ISL_1298756, EPI_ISL_1298757, EPI_ISL_1298758, EPI_ISL_1298759, EPI_ISL_1298760, EPI_ISL_1298761, EPI_ISL_1298762, EPI_ISL_1298763, EPI_ISL_1298764, EPI_ISL_1298765, EPI_ISL_1298766, EPI_ISL_1298767, EPI_ISL_1298768, EPI_ISL_1298769, EPI_ISL_1298770, EPI_ISL_1298771, EPI_ISL_1298772, EPI_ISL_1298773, EPI_ISL_1298774, EPI_ISL_1298775, EPI_ISL_1298776, EPI_ISL_1298777, EPI_ISL_1298778, EPI_ISL_1298779, EPI_ISL_1298780, EPI_ISL_1298781, EPI_ISL_1298782, EPI_ISL_1298783, EPI_ISL_1298784, EPI_ISL_1298785, EPI_ISL_1298786, EPI_ISL_1298787, EPI_ISL_1298788, EPI_ISL_1298789, EPI_ISL_1298790, EPI_ISL_1298791, EPI_ISL_1298792, EPI_ISL_1298793, EPI_ISL_1298794, EPI_ISL_1298795, EPI_ISL_1298796, EPI_ISL_1298797, EPI_ISL_1298798, EPI_ISL_1298799, EPI_ISL_1298800, EPI_ISL_1298801, EPI_ISL_1298802, EPI_ISL_1298803, EPI_ISL_1298804, EPI_ISL_1298805, EPI_ISL_1298806, EPI_ISL_1298807, EPI_ISL_1298808, EPI_ISL_1298809, EPI_ISL_1298810, EPI_ISL_1298811, EPI_ISL_1298812, EPI_ISL_1298813, EPI_ISL_1298814, EPI_ISL_1298815, EPI_ISL_1298816, EPI_ISL_1298817, EPI_ISL_1298818, EPI_ISL_1298819, EPI_ISL_1298820, EPI_ISL_1298821, EPI_ISL_1298822, EPI_ISL_1298823, EPI_ISL_1298824, EPI_ISL_1298825, EPI_ISL_1298826, EPI_ISL_1298827, EPI_ISL_1298828, EPI_ISL_1298829, EPI_ISL_1298830, EPI_ISL_1298831, EPI_ISL_1298832, EPI_ISL_1298833, EPI_ISL_1298834, EPI_ISL_1298835, EPI_ISL_1298836, EPI_ISL_1298837, EPI_ISL_1298838, EPI_ISL_1298839, EPI_ISL_1298840, EPI_ISL_1298841, EPI_ISL_1298842, EPI_ISL_1298843, EPI_ISL_1298844, EPI_ISL_1298845, EPI_ISL_1298846, EPI_ISL_1298847, EPI_ISL_1298848, EPI_ISL_1298849, EPI_ISL_1298850, EPI_ISL_1298851, EPI_ISL_1298852, EPI_ISL_1298853, EPI_ISL_1298854, EPI_ISL_1298855, EPI_ISL_1298856, EPI_ISL_1298857, EPI_ISL_1298858, EPI_ISL_1298859, EPI_ISL_1298860, EPI_ISL_1298861, EPI_ISL_1298862, EPI_ISL_1298863, EPI_ISL_1298864, EPI_ISL_1298865, EPI_ISL_1298866, EPI_ISL_1298867, EPI_ISL_1298868, EPI_ISL_1298869, EPI_ISL_1298870, EPI_ISL_1298871, EPI_ISL_1298872, EPI_ISL_1298873, EPI_ISL_1298874, EPI_ISL_1298875, EPI_ISL_1298876, EPI_ISL_1298877, EPI_ISL_1298878, EPI_ISL_1298879, EPI_ISL_1298880, EPI_ISL_1298881, EPI_ISL_1298882, EPI_ISL_1298883, EPI_ISL_1298884, EPI_ISL_1298885, EPI_ISL_1298886, EPI_ISL_1298887, EPI_ISL_1298888, EPI_ISL_1298889, EPI_ISL_1298890, EPI_ISL_1298891, EPI_ISL_1298892, EPI_ISL_1298893, EPI_ISL_1298894, EPI_ISL_1298895, EPI_ISL_1298896, EPI_ISL_1298897, EPI_ISL_1298898, EPI_ISL_1298899, EPI_ISL_1298900, EPI_ISL_1298901, EPI_ISL_1298902, EPI_ISL_1298903, EPI_ISL_1298904, EPI_ISL_1298905, EPI_ISL_1298906, EPI_ISL_1298907, EPI_ISL_1298908, EPI_ISL_1298909, EPI_ISL_1298910, EPI_ISL_1298911, EPI_ISL_1298912, EPI_ISL_1298913, EPI_ISL_1298914, EPI_ISL_1298915, EPI_ISL_1298916, EPI_ISL_1298917, EPI_ISL_1298918, EPI_ISL_1298919, EPI_ISL_1298920, EPI_ISL_1298921, EPI_ISL_1298922, EPI_ISL_1298923, EPI_ISL_1298924, EPI_ISL_1298925, EPI_ISL_1298926, EPI_ISL_1298927, EPI_ISL_1298928, EPI_ISL_1298929, EPI_ISL_1298930, EPI_ISL_1298931, EPI_ISL_1298932, EPI_ISL_1298933, EPI_ISL_1298934, EPI_ISL_1298935, EPI_ISL_1298936, EPI_ISL_1298937, EPI_ISL_1298938, EPI_ISL_1298939, EPI_ISL_1298940, EPI_ISL_1298941, EPI_ISL_1298942, EPI_ISL_1298943, EPI_ISL_1298944, EPI_ISL_1298945, EPI_ISL_1298946, EPI_ISL_1298947, EPI_ISL_1298948, EPI_ISL_1298949, EPI_ISL_1298950, EPI_ISL_1298951, EPI_ISL_1298952, EPI_ISL_1298953, EPI_ISL_1298954, EPI_ISL_1298955, EPI_ISL_1298956, EPI_ISL_1298957, EPI_ISL_1298958, EPI_ISL_1298959, EPI_ISL_1298960, EPI_ISL_1298961, EPI_ISL_1298962, EPI_ISL_1298963, EPI_ISL_1298964, EPI_ISL_1298965, EPI_ISL_1298966, EPI_ISL_1298967, EPI_ISL_1298968, EPI_ISL_1298969, EPI_ISL_1298970, EPI_ISL_1298971, EPI_ISL_1298972, EPI_ISL_1298973, EPI_ISL_1298974, EPI_ISL_1298975, EPI_ISL_1298976, EPI_ISL_1298977, EPI_ISL_1298978, EPI_ISL_1298979, EPI_ISL_1298980, EPI_ISL_1298981, EPI_ISL_1298982, EPI_ISL_1298983, EPI_ISL_1298984, EPI_ISL_1298985, EPI_ISL_1298986, EPI_ISL_1298987, EPI_ISL_1298988, EPI_ISL_1298989, EPI_ISL_1298990, EPI_ISL_1298991, EPI_ISL_1298992, EPI_ISL_1298993, EPI_ISL_1298994, EPI_ISL_1298995, EPI_ISL_1298996, EPI_ISL_1298997, EPI_ISL_1298998, EPI_ISL_1298999, EPI_ISL_1299000, EPI_ISL_1299001, EPI_ISL_1299002, EPI_ISL_1299003, EPI_ISL_1299004, EPI_ISL_1299005, EPI_ISL_1299006, EPI_ISL_1299007, EPI_ISL_1299008, EPI_ISL_1299009, EPI_ISL_1299010, EPI_ISL_1299011, EPI_ISL_1299012, EPI_ISL_1299013, EPI_ISL_1299014, EPI_ISL_1299015, EPI_ISL_1299016, EPI_ISL_1299017, EPI_ISL_1299018, EPI_ISL_1299019, EPI_ISL_1299020, EPI_ISL_1299021, EPI_ISL_1299022, EPI_ISL_1299023, EPI_ISL_1299024, EPI_ISL_1299025, EPI_ISL_1299026, EPI_ISL_1299027, EPI_ISL_1299028, EPI_ISL_1299029, EPI_ISL_1299030, EPI_ISL_1299031, EPI_ISL_1299032, EPI_ISL_1299033, EPI_ISL_1299034, EPI_ISL_1299035, EPI_ISL_1299036, EPI_ISL_1299037, EPI_ISL_1299038, EPI_ISL_1299039, EPI_ISL_1299040, EPI_ISL_1299041, EPI_ISL_1299042, EPI_ISL_1299043, EPI_ISL_1299044, EPI_ISL_1299045, EPI_ISL_1299046, EPI_ISL_1299047, EPI_ISL_1299048, EPI_ISL_1299049, EPI_ISL_1299050, EPI_ISL_1299051, EPI_ISL_1299052, EPI_ISL_1299053, EPI_ISL_1299054, EPI_ISL_1299055, EPI_ISL_1299056, EPI_ISL_1299057, EPI_ISL_1299058, EPI_ISL_1299059, EPI_ISL_1299060, EPI_ISL_1299061, EPI_ISL_1299062, EPI_ISL_1299063, EPI_ISL_1299064, EPI_ISL_1299065, EPI_ISL_1299066, EPI_ISL_1299067, EPI_ISL_1299068, EPI_ISL_1299069, EPI_ISL_1299070, EPI_ISL_1299071, EPI_ISL_1299072, EPI_ISL_1299073, EPI_ISL_1299074, EPI_ISL_1299075, EPI_ISL_1299076, EPI_ISL_1299077, EPI_ISL_1299078, EPI_ISL_1299079, EPI_ISL_1299080, EPI_ISL_1299081, EPI_ISL_1299082, EPI_ISL_1299083, EPI_ISL_1299084, EPI_ISL_1299085, EPI_ISL_1299086, EPI_ISL_1299087, EPI_ISL_1299088, EPI_ISL_1299089, EPI_ISL_1299090, EPI_ISL_1299091, EPI_ISL_1299092, EPI_ISL_1299093, EPI_ISL_1299094, EPI_ISL_1299095, EPI_ISL_1299096, EPI_ISL_1299097, EPI_ISL_1299098, EPI_ISL_1299099, EPI_ISL_1299100, EPI_ISL_1299101, EPI_ISL_1299102, EPI_ISL_1299103, EPI_ISL_1299104, EPI_ISL_1299105, EPI_ISL_1299106, EPI_ISL_1299107, EPI_ISL_1299108, EPI_ISL_1299109, EPI_ISL_1299110, EPI_ISL_1299111, EPI_ISL_1299112, EPI_ISL_1299113, EPI_ISL_1299114, EPI_ISL_1299115, EPI_ISL_1299116, EPI_ISL_1299117, EPI_ISL_1299118, EPI_ISL_1299119, EPI_ISL_1299120, EPI_ISL_1299121, EPI_ISL_1299122, EPI_ISL_1299123, EPI_ISL_1299124, EPI_ISL_1299125, EPI_ISL_1299126, EPI_ISL_1299127, EPI_ISL_1299128, EPI_ISL_1299129, EPI_ISL_1299130, EPI_ISL_1299131, EPI_ISL_1299132, EPI_ISL_1299133, EPI_ISL_1299134, EPI_ISL_1299135, EPI_ISL_1299136, EPI_ISL_1299137, EPI_ISL_1299138, EPI_ISL_1299139, EPI_ISL_1299140, EPI_ISL_1299141, EPI_ISL_1299142, EPI_ISL_1299143, EPI_ISL_1299144, EPI_ISL_1299145, EPI_ISL_1299146, EPI_ISL_1299147, EPI_ISL_1299148, EPI_ISL_1299149, EPI_ISL_1299150, EPI_ISL_1299151, EPI_ISL_1299152, EPI_ISL_1299153, EPI_ISL_1299154, EPI_ISL_1299155, EPI_ISL_1299156, EPI_ISL_1299157, EPI_ISL_1299158, EPI_ISL_1299159, EPI_ISL_1299160, EPI_ISL_1299161, EPI_ISL_1299162, EPI_ISL_1299163, EPI_ISL_1299164, EPI_ISL_1299165, EPI_ISL_1299166, EPI_ISL_1299167, EPI_ISL_1299168, EPI_ISL_1299169, EPI_ISL_1299170, EPI_ISL_1299171, EPI_ISL_1299172, EPI_ISL_1299173, EPI_ISL_1299174, EPI_ISL_1299175, EPI_ISL_1299176, EPI_ISL_1299177, EPI_ISL_1299178, EPI_ISL_1299179, EPI_ISL_1299180, EPI_ISL_1299181, EPI_ISL_1299182, EPI_ISL_1299183, EPI_ISL_1299184, EPI_ISL_1299185, EPI_ISL_1299186, EPI_ISL_1299187, EPI_ISL_1299188, EPI_ISL_1299189, EPI_ISL_1299190, EPI_ISL_1299191, EPI_ISL_1299192, EPI_ISL_1299193, EPI_ISL_1299194 | | | |
| see above | ASL Napoli 1 Centro | AMES Centro Polidiagnostico Strumentale S.r.l. | "Giovanni Savarese, Raffaella Ruggiero, Eloisa Evangelista, Antonella Di Carlo, Luisa Circelli, Luigi D'Amore, Nadia Petrillo, Monica Ianniello, Roberto Sirica, Maurizio D'Amora, Antonio Fico" |
| EPI_ISL_1299217 | IDI-IRCCS | INMI Lazzaro Spallanzani IRCCS | G Bonfiglio, O Butera, CEM Gruber, F Santini, B Bartolini, E Giombini, F Messina, M Rueca, G Russo, D Castiglia, S Madonna, A Di Caro, MR Capobianchi |
| EPI_ISL_1299218 | INMI Lazzaro Spallanzani IRCCS | INMI Lazzaro Spallanzani IRCCS | CEM Gruber, F Santini, B Bartolini, E Giombini, F Messina, M Rueca, G Bonfiglio, O Butera, A Di Caro, MR Capobianchi |
| EPI_ISL_1299219 | Ospedale S.M. Goretti di Latina - ASL Latina presidio ospedaliero Nord | INMI Lazzaro Spallanzani IRCCS | O Butera, CEM Gruber, F Santini, B Bartolini, E Giombini, F Messina, M Rueca, G Bonfiglio, A Lucci, S Pignalosa, L Corso, MR Capobianchi, A Di Caro |
| EPI_ISL_1299501, EPI_ISL_1299502, EPI_ISL_1299503, EPI_ISL_1299504, EPI_ISL_1299505 | Azienda Ospedaliera Terni | Istituto Zooprofilattico Sperimentale dell'Abruzzo e Molise "G. Caporale" | Palumbo M, ScaccettiA, Lorusso A, Marcacci M, Di Domenico M, Ancora M, Curini V, Mangone I, Rinaldi A, Scialabba S, Di Pasquale A, Cammà C, Puglia I, Calistri P, Savini G |
| EPI_ISL_1299528, EPI_ISL_1299529, EPI_ISL_1299530, EPI_ISL_1299531 | Istituto Zooprofilattico Sperimentale Umbria e Marche "Togo Rosati" | Istituto Zooprofilattico Sperimentale dell'Abruzzo e Molise "G. Caporale" | Biagetti m, Giammarioli M, Lorusso A, Marcacci M, Di Domenico M, Ancora M, Curini V, Mangone I, Rinaldi A, Scialabba S, Di Pasquale A, Cammà C, Puglia I, Calistri P, Savini G |
| EPI_ISL_1299533, EPI_ISL_1299534, EPI_ISL_1299535, EPI_ISL_1299536, EPI_ISL_1299537, EPI_ISL_1299538, EPI_ISL_1299539, EPI_ISL_1299540, EPI_ISL_1299541, EPI_ISL_1299542 | Istituto Zooprofilattico Sperimentale Umbria e Marche "Togo Rosati" | Istituto Zooprofilattico Sperimentale dell'Abruzzo e Molise "G. Caporale" | Biagetti M, Giammarioli M, Lorusso A, Marcacci M, Di Domenico M, Ancora M, Curini V, Mangone I, Rinaldi A, Scialabba S, Di Pasquale A, Cammà C, Puglia I, Calistri P, Savini G |
| EPI_ISL_1299544 | Istituto Zooprofilattico Sperimentale Umbria e Marche "Togo Rosati" | Istituto Zooprofilattico Sperimentale dell'Abruzzo e Molise "G. Caporale" | Biagetti M, Giammarioli M,Lorusso A, Marcacci M, Di Domenico M, Ancora M, Curini V, Mangone I, Rinaldi A, Scialabba S, Di Pasquale A, Cammà C, Puglia I, Calistri P, Savini G |
| EPI_ISL_1299549, EPI_ISL_1299550, | Laboratorio Analisi Osp. Città di Castello - Azienda USL | Istituto Zooprofilattico Sperimentale dell'Abruzzo e Molise "G. | Malagigi V, Tacconi P,Lorusso A, Marcacci M, Di Domenico M, Ancora M, Curini V, Mangone I, Rinaldi A, Scialabba S, Di Pasquale A, Cammà C, Puglia I, |

| | | | |
|---|---|---|---|
| EPI_ISL_1299553 | Umbria1 | Caporale" | Calistri P, Savini G |
| EPI_ISL_1299556, EPI_ISL_1299559, EPI_ISL_1299562, EPI_ISL_1299564, EPI_ISL_1299565, EPI_ISL_1299566, EPI_ISL_1299567, EPI_ISL_1299568, EPI_ISL_1299569 | Università degli Studi di Perugia | Istituto Zooprofilattico Sperimentale dell'Abruzzo e Molise "G. Caporale" | Mencacci A, Camilloni B,Lorusso A, Marcacci M, Di Domenico M, Ancora M, Curini V, Mangone I, Rinaldi A, Scialabba S, Di Pasquale A, Cammà C, Puglia I, Calistri P, Savini G |
| EPI_ISL_1299570 | Azienda Ospedaliera Terni | Istituto Zooprofilattico Sperimentale dell'Abruzzo e Molise "G. Caporale" | Palumbo M, Scaccetti A, Lorusso A, Marcacci M, Di Domenico M, Ancora M, Curini V, Mangone I, Rinaldi A, Scialabba S, Di Pasquale A, Cammà C, Puglia I, Calistri P, Savini G |
| EPI_ISL_1305774 | QUALITY CONTROL CHEMICAL BIOLOGICAL RISK, AOOR Villa Sofia Cervello Palermo | QUALITY CONTROL CHEMICAL BIOLOGICAL RISK, AOOR Villa Sofia Cervello Palermo | Di Gaudio,F., Seidita,G., Contino,F., and Brunacci,G. |
| EPI_ISL_1307722, EPI_ISL_1307723, EPI_ISL_1307724, EPI_ISL_1307725, EPI_ISL_1307726, EPI_ISL_1307727, EPI_ISL_1307728, EPI_ISL_1307729, EPI_ISL_1307730, EPI_ISL_1307731, EPI_ISL_1307732, EPI_ISL_1307733, EPI_ISL_1307734, EPI_ISL_1307735, EPI_ISL_1307736, EPI_ISL_1307737, EPI_ISL_1307738, EPI_ISL_1307739, EPI_ISL_1307740, EPI_ISL_1307741, EPI_ISL_1307742, EPI_ISL_1307743, EPI_ISL_1307744, EPI_ISL_1307745, EPI_ISL_1307746, EPI_ISL_1307747, EPI_ISL_1307748, EPI_ISL_1307749, EPI_ISL_1307750, EPI_ISL_1307751, EPI_ISL_1307752, EPI_ISL_1307753, EPI_ISL_1307754, EPI_ISL_1307755, EPI_ISL_1307756, EPI_ISL_1307757, EPI_ISL_1307758, EPI_ISL_1307759, EPI_ISL_1307760, EPI_ISL_1307761, EPI_ISL_1307762, EPI_ISL_1307763, EPI_ISL_1307764, EPI_ISL_1307765, EPI_ISL_1307766, EPI_ISL_1307767, EPI_ISL_1307768, EPI_ISL_1307769, EPI_ISL_1307770, EPI_ISL_1307771, EPI_ISL_1307772, EPI_ISL_1307773, EPI_ISL_1307774, EPI_ISL_1307775, EPI_ISL_1307776, EPI_ISL_1307777, EPI_ISL_1307778, EPI_ISL_1307779, EPI_ISL_1307780, EPI_ISL_1307781, EPI_ISL_1307782, EPI_ISL_1307783, EPI_ISL_1307784, EPI_ISL_1307785, EPI_ISL_1307786, EPI_ISL_1307787, EPI_ISL_1307788, EPI_ISL_1307789, EPI_ISL_1307790, EPI_ISL_1307791, EPI_ISL_1307792, EPI_ISL_1307793, EPI_ISL_1307794, EPI_ISL_1307795, EPI_ISL_1307796, EPI_ISL_1307797, EPI_ISL_1307798, EPI_ISL_1307799 | | | |
| see above | Istituto Zooprofilattico Sperimentale del Mezzogiorno | TIGEM | Antonio Grimaldi Patrizia Annunziata Francesco Panariello Biancamaria Pierri Claudia Tiberio Valentina Bouche Chiara Colantuono Maria Concetta Cuomo Denise Di Concilio Lucio Di Filippo Anna Manfredi Marcello Salvi Antonio Limone Luigi Atripaldi Pellegrino Cerino Andrea Ballabio Davide Cacchiarelli |
| EPI_ISL_1307800, EPI_ISL_1307801, EPI_ISL_1307802, EPI_ISL_1307803, EPI_ISL_1307804, EPI_ISL_1307805, EPI_ISL_1307806, EPI_ISL_1307807, EPI_ISL_1307808, EPI_ISL_1307809, EPI_ISL_1307810, EPI_ISL_1307811, EPI_ISL_1307812, EPI_ISL_1307813, EPI_ISL_1307814, EPI_ISL_1307815, EPI_ISL_1307816, EPI_ISL_1307817, EPI_ISL_1307818, EPI_ISL_1307819, EPI_ISL_1307820, EPI_ISL_1307821, EPI_ISL_1307822, EPI_ISL_1307823, EPI_ISL_1307824, EPI_ISL_1307825, EPI_ISL_1307826, EPI_ISL_1307827, EPI_ISL_1307828, EPI_ISL_1307829, EPI_ISL_1307830, EPI_ISL_1307831, EPI_ISL_1307832, EPI_ISL_1307833, EPI_ISL_1307834, EPI_ISL_1307835, EPI_ISL_1307836, EPI_ISL_1307837, EPI_ISL_1307838, EPI_ISL_1307839, EPI_ISL_1307840, EPI_ISL_1307841 | | | |
| see above | Lab. Microbiologia e Virologia - Cotugno - A.O. dei Colli | TIGEM | Antonio Grimaldi Patrizia Annunziata Francesco Panariello Biancamaria Pierri Claudia Tiberio Valentina Bouche Chiara Colantuono Maria Concetta Cuomo Denise Di Concilio Lucio Di Filippo Anna Manfredi Marcello Salvi Antonio Limone Luigi Atripaldi Pellegrino Cerino Andrea Ballabio Davide Cacchiarelli |
| EPI_ISL_1307842, EPI_ISL_1307843, EPI_ISL_1307844, EPI_ISL_1307845, EPI_ISL_1307846, EPI_ISL_1307847, EPI_ISL_1307848, EPI_ISL_1307849, EPI_ISL_1307850, EPI_ISL_1307851, EPI_ISL_1307852, EPI_ISL_1307853, EPI_ISL_1307854, EPI_ISL_1307855, EPI_ISL_1307856, EPI_ISL_1307857, EPI_ISL_1307858, EPI_ISL_1307859, EPI_ISL_1307860, EPI_ISL_1307861, EPI_ISL_1307862, EPI_ISL_1307863, EPI_ISL_1307864, EPI_ISL_1307865, EPI_ISL_1307866, EPI_ISL_1307867, EPI_ISL_1307868, EPI_ISL_1307869, EPI_ISL_1307870, EPI_ISL_1307871, EPI_ISL_1307872, EPI_ISL_1307873, EPI_ISL_1307874, EPI_ISL_1307875, EPI_ISL_1307876, EPI_ISL_1307877, EPI_ISL_1307878, EPI_ISL_1307879, EPI_ISL_1307880, EPI_ISL_1307881, EPI_ISL_1307882, EPI_ISL_1307883, EPI_ISL_1307884, EPI_ISL_1307885, EPI_ISL_1307886, EPI_ISL_1307887, EPI_ISL_1307888, EPI_ISL_1307889, EPI_ISL_1307890, EPI_ISL_1307891, EPI_ISL_1307892, EPI_ISL_1307893, EPI_ISL_1307894, EPI_ISL_1307895, EPI_ISL_1307896, EPI_ISL_1307897, EPI_ISL_1307898, EPI_ISL_1307899, EPI_ISL_1307900, EPI_ISL_1307901, EPI_ISL_1307902, EPI_ISL_1307903, EPI_ISL_1307904, EPI_ISL_1307905, EPI_ISL_1307906, EPI_ISL_1307907, EPI_ISL_1307908, EPI_ISL_1307909, EPI_ISL_1307910, EPI_ISL_1307911, EPI_ISL_1307912, EPI_ISL_1307913, EPI_ISL_1307914, EPI_ISL_1307915, EPI_ISL_1307916, EPI_ISL_1307917, EPI_ISL_1307918, EPI_ISL_1307919, EPI_ISL_1307920, EPI_ISL_1307921, EPI_ISL_1307922, EPI_ISL_1307923, EPI_ISL_1307924, EPI_ISL_1307925, EPI_ISL_1307926, EPI_ISL_1307927, EPI_ISL_1307928, EPI_ISL_1307929, EPI_ISL_1307930, EPI_ISL_1307931, EPI_ISL_1307932, EPI_ISL_1307933, EPI_ISL_1307934, EPI_ISL_1307935, EPI_ISL_1307936, EPI_ISL_1307937, EPI_ISL_1307938, EPI_ISL_1307939, EPI_ISL_1307940, EPI_ISL_1307941 | | | |
| see above | Istituto Zooprofilattico Sperimentale del Mezzogiorno | TIGEM | Antonio Grimaldi Patrizia Annunziata Francesco Panariello Biancamaria Pierri Claudia Tiberio Valentina Bouche Chiara Colantuono Maria Concetta Cuomo Denise Di Concilio Lucio Di Filippo Anna Manfredi Marcello Salvi Antonio Limone Luigi Atripaldi Pellegrino Cerino Andrea Ballabio Davide Cacchiarelli |
| EPI_ISL_1307942, EPI_ISL_1307943, EPI_ISL_1307944, EPI_ISL_1307945, EPI_ISL_1307946, EPI_ISL_1307947, EPI_ISL_1307948, EPI_ISL_1307949, EPI_ISL_1307950, EPI_ISL_1307951, EPI_ISL_1307952, EPI_ISL_1307953, EPI_ISL_1307954, EPI_ISL_1307955, EPI_ISL_1307956, EPI_ISL_1307957, EPI_ISL_1307958, EPI_ISL_1307959, EPI_ISL_1307960, EPI_ISL_1307961, EPI_ISL_1307962, EPI_ISL_1307963, EPI_ISL_1307964, EPI_ISL_1307965, EPI_ISL_1307966, EPI_ISL_1307967, EPI_ISL_1307968, EPI_ISL_1307969, EPI_ISL_1307970, EPI_ISL_1307971, EPI_ISL_1307972, EPI_ISL_1307973, EPI_ISL_1307974, EPI_ISL_1307975, EPI_ISL_1307976, EPI_ISL_1307977, EPI_ISL_1307978, EPI_ISL_1307979, EPI_ISL_1307980, EPI_ISL_1307981, EPI_ISL_1307982, EPI_ISL_1307983, EPI_ISL_1307984, EPI_ISL_1307985, EPI_ISL_1307986, EPI_ISL_1307987, EPI_ISL_1307988, EPI_ISL_1307989, EPI_ISL_1307990, EPI_ISL_1307991, EPI_ISL_1307992, EPI_ISL_1307993, EPI_ISL_1307994, EPI_ISL_1307995, EPI_ISL_1307996, EPI_ISL_1307997, EPI_ISL_1307998, EPI_ISL_1307999, EPI_ISL_1308000, EPI_ISL_1308001, EPI_ISL_1308002, EPI_ISL_1308003, EPI_ISL_1308004, EPI_ISL_1308005, EPI_ISL_1308006, EPI_ISL_1308007, EPI_ISL_1308008, EPI_ISL_1308009 | | | |
| see above | Lab. Microbiologia e Virologia Cotugno A.O. dei Colli | TIGEM | Antonio Grimaldi Patrizia Annunziata Francesco Panariello Biancamaria Pierri Claudia Tiberio Valentina Bouche Chiara Colantuono Maria Concetta Cuomo Denise Di Concilio Lucio Di Filippo Anna Manfredi Marcello Salvi Antonio Limone Luigi Atripaldi Pellegrino Cerino Andrea Ballabio Davide Cacchiarelli |
| EPI_ISL_1310791 | QUALITY CONTROL CHEMICAL BIOLOGICAL RISK, AOOR Villa Sofia Cervello Palermo | QUALITY CONTROL CHEMICAL BIOLOGICAL RISK, AOOR Villa Sofia Cervello Palermo | Di Gaudio,F., Contino,F., Brunacci,G., Orlando, V., Buffa, V., Di Chiara, V. and Seidita,G. |
| EPI_ISL_1310795, EPI_ISL_1310799 | Istituto Zooprofilattico Sperimentale del Mezzogiorno | TIGEM | Antonio Grimaldi Patrizia Annunziata Francesco Panariello Biancamaria Pierri Claudia Tiberio Valentina Bouche Chiara Colantuono Maria Concetta Cuomo Denise Di Concilio Lucio Di Filippo Anna Manfredi Marcello Salvi Antonio Limone Luigi Atripaldi Pellegrino Cerino Andrea Ballabio Davide Cacchiarelli |
| EPI_ISL_1311860, EPI_ISL_1311861, EPI_ISL_1311862 | Laboratorio Biologia Molecolare Sars Cov2 - UOC Laboratorio Analisi - Servizio Medicina di Laboratorio, Ospedale "San Francesco" - ATS-ASSL Nuoro | Laboratorio specialistico UOC Ematologia - Ospedale "San Francesco" - ATS-ASSL Nuoro | Piras Giovanna, Asproni Rosanna,Fiamma Maura, Monne Maria Itria, Malune Paolo, Palmas Angelo Domenico, Lo Maglio Iana, Mameli Giuseppe |
| EPI_ISL_1311964 | QUALITY CONTROL CHEMICAL BIOLOGICAL RISK, AOOR Villa Sofia Cervello Palermo | QUALITY CONTROL CHEMICAL BIOLOGICAL RISK, AOOR Villa Sofia Cervello Palermo | Di Gaudio,F., Contino,F. and Brunacci,G. |
| EPI_ISL_1311973 | QUALITY CONTROL CHEMICAL BIOLOGICAL RISK, AOOR Villa Sofia Cervello Palermo | QUALITY CONTROL CHEMICAL BIOLOGICAL RISK, AOOR Villa Sofia Cervello Palermo | Di Gaudio,F., Brunacci, G.Seidita,G., and Contino,F. |
| EPI_ISL_1312063 | QUALITY CONTROL CHEMICAL BIOLOGICAL RISK, AOOR Villa Sofia Cervello Palermo | QUALITY CONTROL CHEMICAL BIOLOGICAL RISK, AOOR Villa Sofia Cervello Palermo | Di Gaudio,F., Seidita,G., Contino,F., Orlando, V., Di Chiara, V., Buffa, V., Todaro, I., Vasto, S., Pace, E., Cangemi, G. and Brunacci,G. |
| EPI_ISL_1312389 | QUALITY CONTROL CHEMICAL BIOLOGICAL RISK, AOOR Villa Sofia Cervello Palermo | QUALITY CONTROL CHEMICAL BIOLOGICAL RISK, AOOR Villa Sofia Cervello Palermo | Di Gaudio,F., Contino,F., Seidita,G. and Brunacci,G. |
| EPI_ISL_1312772 | QUALITY CONTROL CHEMICAL BIOLOGICAL RISK, AOOR Villa Sofia Cervello Palermo | QUALITY CONTROL CHEMICAL BIOLOGICAL RISK, AOOR Villa Sofia Cervello Palermo | Di Gaudio,F., Brunacci,G., Contino,F., Orlando, V., Buffa, V., Di Chiara, V., Todaro, I. and Seidita,G. |
| EPI_ISL_1315620 | Laboratorio di Riferimento Regionale della Sicilia Occidentale per l'Emergenza COVID-19 | Laboratorio di Riferimento Regionale della Sicilia Occidentale per l'Emergenza COVID-19 | Fabio Tramuto, Carmelo Massimo Maida, Daniela Di Naro, Giulia Randazzo, Walter Mazzucco, Giorgio Graziano, Vincenzo Restivo, Claudio Costantino, Francesco Vitale |
| EPI_ISL_1320193, EPI_ISL_1320198 | IZSM | IZSM-U.O.C. Virologia | Maurizio Viscardi, Lorena Cardillo, Claudio de Martinis, Giovanna Fusco, Esterina De Carlo, Antonio Limone |
| EPI_ISL_1321734, EPI_ISL_1321735, EPI_ISL_1321736, EPI_ISL_1321737, EPI_ISL_1321738, EPI_ISL_1321739, EPI_ISL_1321740, EPI_ISL_1321741, EPI_ISL_1321742, EPI_ISL_1321743, EPI_ISL_1321744, EPI_ISL_1321745, EPI_ISL_1321746, EPI_ISL_1321747, EPI_ISL_1321748, EPI_ISL_1321749, EPI_ISL_1321750, EPI_ISL_1321751, EPI_ISL_1321752, EPI_ISL_1321753, EPI_ISL_1321754, EPI_ISL_1321755 | | | |
| see above | Università degli Studi di Perugia | Istituto Zooprofilattico Sperimentale dell'Abruzzo e Molise "G. Caporale" | Mencacci A, Camilloni B, Lorusso A, Marcacci M, Di Domenico M, Ancora M, Curini V, Mangone I, Rinaldi A, Scialabba S, Di Pasquale A, Cammà C, Puglia I, Calistri P, Savini G |
| EPI_ISL_1321756, EPI_ISL_1321757, EPI_ISL_1321758, EPI_ISL_1321759, EPI_ISL_1321760, EPI_ISL_1321761, EPI_ISL_1321762, EPI_ISL_1321763 | Azienda Ospedaliera Terni | Istituto Zooprofilattico Sperimentale dell'Abruzzo e Molise "G. Caporale" | Palumbo M, Scaccetti A, Lorusso A, Marcacci M, Di Domenico M, Ancora M, Curini V, Mangone I, Rinaldi A, Scialabba S, Di Pasquale A, Cammà C, Puglia I, Calistri P, Savini G |
| EPI_ISL_1321764 | Università degli Studi di Perugia | Istituto Zooprofilattico Sperimentale dell'Abruzzo e Molise "G. Caporale" | Mencacci A, Camilloni B, Lorusso A, Marcacci M, Di Domenico M, Ancora M, Curini V, Mangone I, Rinaldi A, Scialabba S, Di Pasquale A, Cammà C, Puglia I, Calistri P, Savini G |
| EPI_ISL_1321765, EPI_ISL_1321766, EPI_ISL_1321767, EPI_ISL_1321768, EPI_ISL_1321769, EPI_ISL_1321770, | Azienda Ospedaliera Terni | Istituto Zooprofilattico Sperimentale dell'Abruzzo e Molise "G. Caporale" | Palumbo M, Scaccetti A, Lorusso A, Marcacci M, Di Domenico M, Ancora M, Curini V, Mangone I, Rinaldi A, Scialabba S, Di Pasquale A, Cammà C, Puglia I, Calistri P, Savini G |

| | | | |
|---|---|---|---|
| EPI_ISL_1321771, EPI_ISL_1321772, EPI_ISL_1321773 | | | |
| EPI_ISL_1321774 | Università degli Studi di Perugia | Istituto Zooprofilattico Sperimentale dell'Abruzzo e Molise "G. Caporale" | Mencacci A, Camilloni B, Lorusso A, Marcacci M, Di Domenico M, Ancora M, Curini V, Mangone I, Rinaldi A, Scialabba S, Di Pasquale A, Cammà C, Puglia I, Calistri P, Savini G |
| EPI_ISL_1321775, EPI_ISL_1321776, EPI_ISL_1321777, EPI_ISL_1321778, EPI_ISL_1321779, EPI_ISL_1321780, EPI_ISL_1321781, EPI_ISL_1321782, EPI_ISL_1321783, EPI_ISL_1321784 | Azienda Ospedaliera Terni | Istituto Zooprofilattico Sperimentale dell'Abruzzo e Molise "G. Caporale" | Palumbo M, Scaccetti A, Lorusso A, Marcacci M, Di Domenico M, Ancora M, Curini V, Mangone I, Rinaldi A, Scialabba S, Di Pasquale A, Cammà C, Puglia I, Calistri P, Savini G |
| EPI_ISL_1321785 | Università degli Studi di Perugia | Istituto Zooprofilattico Sperimentale dell'Abruzzo e Molise "G. Caporale" | Mencacci A, Camilloni B, Lorusso A, Marcacci M, Di Domenico M, Ancora M, Curini V, Mangone I, Rinaldi A, Scialabba S, Di Pasquale A, Cammà C, Puglia I, Calistri P, Savini G |
| EPI_ISL_1321786, EPI_ISL_1321787, EPI_ISL_1321788, EPI_ISL_1321789, EPI_ISL_1321790, EPI_ISL_1321791, EPI_ISL_1321792, EPI_ISL_1321793, EPI_ISL_1321794 | Azienda Ospedaliera Terni | Istituto Zooprofilattico Sperimentale dell'Abruzzo e Molise "G. Caporale" | Palumbo M, Scaccetti A, Lorusso A, Marcacci M, Di Domenico M, Ancora M, Curini V, Mangone I, Rinaldi A, Scialabba S, Di Pasquale A, Cammà C, Puglia I, Calistri P, Savini G |
| EPI_ISL_1321795 | Istituto Zooprofilattico Sperimentale Umbria e Marche "Togo Rosati" | Istituto Zooprofilattico Sperimentale dell'Abruzzo e Molise "G. Caporale" | Biagetti M, Giammarioli M,Lorusso A, Marcacci M, Di Domenico M, Ancora M, Curini V, Mangone I, Rinaldi A, Scialabba S, Di Pasquale A, Cammà C, Puglia I, Calistri P, Savini G |
| EPI_ISL_1321796 | Università degli Studi di Perugia | Istituto Zooprofilattico Sperimentale dell'Abruzzo e Molise "G. Caporale" | Mencacci A, Camilloni B, Lorusso A, Marcacci M, Di Domenico M, Ancora M, Curini V, Mangone I, Rinaldi A, Scialabba S, Di Pasquale A, Cammà C, Puglia I, Calistri P, Savini G |
| EPI_ISL_1321797, EPI_ISL_1321798, EPI_ISL_1321799, EPI_ISL_1321800, EPI_ISL_1321801, EPI_ISL_1321802, EPI_ISL_1321803 | Istituto Zooprofilattico Sperimentale Umbria e Marche "Togo Rosati" | Istituto Zooprofilattico Sperimentale dell'Abruzzo e Molise "G. Caporale" | Biagetti M, Giammarioli M,Lorusso A, Marcacci M, Di Domenico M, Ancora M, Curini V, Mangone I, Rinaldi A, Scialabba S, Di Pasquale A, Cammà C, Puglia I, Calistri P, Savini G |
| EPI_ISL_1321804, EPI_ISL_1321805, EPI_ISL_1321806 | Università degli Studi di Perugia | Istituto Zooprofilattico Sperimentale dell'Abruzzo e Molise "G. Caporale" | Mencacci A, Camilloni B, Lorusso A, Marcacci M, Di Domenico M, Ancora M, Curini V, Mangone I, Rinaldi A, Scialabba S, Di Pasquale A, Cammà C, Puglia I, Calistri P, Savini G |
| EPI_ISL_1323980 | Istituto Zooprofilattico Sperimentale del Mezzogiorno (IZSM) | IZSM-U.O.C. Virologia | Maurizio Viscardi, Lorena Cardillo, Claudio de Martinis, Giovanna Fusco, Esterina De Carlo, Antonio Limone |
| EPI_ISL_1324019, EPI_ISL_1324020, EPI_ISL_1324021, EPI_ISL_1324022, EPI_ISL_1324023, EPI_ISL_1324024, EPI_ISL_1324025, EPI_ISL_1324026, EPI_ISL_1324027, EPI_ISL_1324028 | SC Microbiologia e Virologia AOUSS | SC Microbiologia e Virologia AOUSS | Salvatore Rubino; Marcello Abbondio; Flavia Angioj; Laura Firino; Rosalba Govoni; Gabriele Ibba; Vincenzo Lai; Erica Mura; Bianca Paglietti; Claudia Piu; Anna Puggioni; Elena Rimini; Giulia Rocca; Rosangela Sau; Caterina Serra; Sergio Uzzau. |
| EPI_ISL_1336083, EPI_ISL_1336084 | SIESP DIPARTIMENTO DI PREVENZIONE TERAMO | Istituto Zooprofilattico Sperimentale dell'Abruzzo e Molise "G. Caporale" | Lorusso A, Marcacci M, Di Domenico M, Ancora M, Curini V, Mangone I, Rinaldi A, Scialabba S, Di Pasquale A, Cammà C, Puglia I, Calistri P, Savini G |
| EPI_ISL_1336178 | Azienda Ospedaliera Terni | Istituto Zooprofilattico Sperimentale dell'Abruzzo e Molise "G. Caporale" | Palumbo M, Scaccetti A, Lorusso A, Marcacci M, Di Domenico M, Ancora M, Curini V, Mangone I, Rinaldi A, Scialabba S, Di Pasquale A, Cammà C, Puglia I, Calistri P, Savini G |
| EPI_ISL_1336179 | Istituto Zooprofilattico Sperimentale Umbria e Marche "Togo Rosati" | Istituto Zooprofilattico Sperimentale dell'Abruzzo e Molise "G. Caporale" | Biagetti M, Giammarioli M,Lorusso A, Marcacci M, Di Domenico M, Ancora M, Curini V, Mangone I, Rinaldi A, Scialabba S, Di Pasquale A, Cammà C, Puglia I, Calistri P, Savini G |
| EPI_ISL_1336650 | Università degli Studi di Perugia | Istituto Zooprofilattico Sperimentale dell'Abruzzo e Molise "G. Caporale" | Mencacci A, Camilloni B, Lorusso A, Marcacci M, Di Domenico M, Ancora M, Curini V, Mangone I, Rinaldi A, Scialabba S, Di Pasquale A, Cammà C, Puglia I, Calistri P, Savini G |
| EPI_ISL_1336651 | Azienda Ospedaliera Terni | Istituto Zooprofilattico Sperimentale dell'Abruzzo e Molise "G. Caporale" | Palumbo M, Scaccetti A, Lorusso A, Marcacci M, Di Domenico M, Ancora M, Curini V, Mangone I, Rinaldi A, Scialabba S, Di Pasquale A, Cammà C, Puglia I, Calistri P, Savini G |
| EPI_ISL_1336653, EPI_ISL_1336654, EPI_ISL_1336655, EPI_ISL_1336656, EPI_ISL_1336657, EPI_ISL_1336658 | Center of Advanced Studies and Technology, Molecular Genetics Laboratory | Center of Advanced Studies and Technology, Molecular Genetics Laboratory | Anaclerio Federico, Ferrante Rossella, Mandatori Domitilla, De Fabritiis Simone, Damiani Verena |
| EPI_ISL_1336659 | RSA GIULIANOVA | Istituto Zooprofilattico Sperimentale dell'Abruzzo e Molise "G. Caporale" | Lorusso A, Marcacci M, Di Domenico M, Ancora M, Curini V, Di Lollo V, Mangone I, Rinaldi A, Scialabba S, Caporale M, Di Pasquale A, Cammà C, Puglia I, Calistri P, Savini G |
| EPI_ISL_1336660 | OSPEDALE SAN SALVATORE - MEDICINA DI LABORATORIO | Istituto Zooprofilattico Sperimentale dell'Abruzzo e Molise "G. Caporale" | Lorusso A, Marcacci M, Di Domenico M, Ancora M, Curini V, Di Lollo V, Mangone I, Rinaldi A, Scialabba S, Caporale M, Di Pasquale A, Cammà C, Puglia I, Calistri P, Savini G |
| EPI_ISL_1336661 | USCA Tagliacozzo | Istituto Zooprofilattico Sperimentale dell'Abruzzo e Molise "G. Caporale" | Lorusso A, Marcacci M, Di Domenico M, Ancora M, Curini V, Di Lollo V, Mangone I, Rinaldi A, Scialabba S, Caporale M, Di Pasquale A, Cammà C, Puglia I, Calistri P, Savini G |
| EPI_ISL_1336662 | SIESP DIPARTIMENTO DI PREVENZIONE CHIETI | Istituto Zooprofilattico Sperimentale dell'Abruzzo e Molise "G. Caporale" | Lorusso A, Marcacci M, Di Domenico M, Ancora M, Curini V, Di Lollo V, Mangone I, Rinaldi A, Scialabba S, Caporale M, Di Pasquale A, Cammà C, Puglia I, Calistri P, Savini G |
| EPI_ISL_1336663 | USCA Sulmona | Istituto Zooprofilattico Sperimentale dell'Abruzzo e Molise "G. Caporale" | Lorusso A, Marcacci M, Di Domenico M, Ancora M, Curini V, Di Lollo V, Mangone I, Rinaldi A, Scialabba S, Caporale M, Di Pasquale A, Cammà C, Puglia I, Calistri P, Savini G |
| EPI_ISL_1336664, EPI_ISL_1336665 | SIESP DIPARTIMENTO DI PREVENZIONE TERAMO | Istituto Zooprofilattico Sperimentale dell'Abruzzo e Molise "G. Caporale" | Lorusso A, Marcacci M, Di Domenico M, Ancora M, Curini V, Di Lollo V, Mangone I, Rinaldi A, Scialabba S, Caporale M, Di Pasquale A, Cammà C, Puglia I, Calistri P, Savini G |
| EPI_ISL_1336666 | SIESP DIPARTIMENTO DI PREVENZIONE CHIETI | Istituto Zooprofilattico Sperimentale dell'Abruzzo e Molise "G. Caporale" | Lorusso A, Marcacci M, Di Domenico M, Ancora M, Curini V, Di Lollo V, Mangone I, Rinaldi A, Scialabba S, Caporale M, Di Pasquale A, Cammà C, Puglia I, Calistri P, Savini G |
| EPI_ISL_1336667 | SIESP L'AQUILA | Istituto Zooprofilattico Sperimentale dell'Abruzzo e Molise "G. Caporale" | Lorusso A, Marcacci M, Di Domenico M, Ancora M, Curini V, Di Lollo V, Mangone I, Rinaldi A, Scialabba S, Caporale M, Di Pasquale A, Cammà C, Puglia I, Calistri P, Savini G |
| EPI_ISL_1336668 | SIESP DIPARTIMENTO DI PREVENZIONE CHIETI | Istituto Zooprofilattico Sperimentale dell'Abruzzo e Molise "G. Caporale" | Lorusso A, Marcacci M, Di Domenico M, Ancora M, Curini V, Di Lollo V, Mangone I, Rinaldi A, Scialabba S, Caporale M, Di Pasquale A, Cammà C, Puglia I, Calistri P, Savini G |
| EPI_ISL_1336669 | SIESP CHIETI - DRIVE IN ORTONA | Istituto Zooprofilattico Sperimentale dell'Abruzzo e Molise "G. Caporale" | Lorusso A, Marcacci M, Di Domenico M, Ancora M, Curini V, Di Lollo V, Mangone I, Rinaldi A, Scialabba S, Caporale M, Di Pasquale A, Cammà C, Puglia I, Calistri P, Savini G |
| EPI_ISL_1336670 | SIESP CHIETI - DRIVE IN LANCIANO | Istituto Zooprofilattico Sperimentale dell'Abruzzo e Molise "G. Caporale" | Lorusso A, Marcacci M, Di Domenico M, Ancora M, Curini V, Di Lollo V, Mangone I, Rinaldi A, Scialabba S, Caporale M, Di Pasquale A, Cammà C, Puglia I, Calistri P, Savini G |
| EPI_ISL_1336671 | USCA Sulmona | Istituto Zooprofilattico Sperimentale dell'Abruzzo e Molise "G. Caporale" | Lorusso A, Marcacci M, Di Domenico M, Ancora M, Curini V, Di Lollo V, Mangone I, Rinaldi A, Scialabba S, Caporale M, Di Pasquale A, Cammà C, Puglia I, Calistri P, Savini G |
| EPI_ISL_1336672 | SIESP CHIETI - DRIVE IN LANCIANO | Istituto Zooprofilattico Sperimentale dell'Abruzzo e Molise "G. Caporale" | Lorusso A, Marcacci M, Di Domenico M, Ancora M, Curini V, Di Lollo V, Mangone I, Rinaldi A, Scialabba S, Caporale M, Di Pasquale A, Cammà C, Puglia I, Calistri P, Savini G |
| EPI_ISL_1336673 | USCA Sulmona | Istituto Zooprofilattico Sperimentale dell'Abruzzo e Molise "G. Caporale" | Lorusso A, Marcacci M, Di Domenico M, Ancora M, Curini V, Di Lollo V, Mangone I, Rinaldi A, Scialabba S, Caporale M, Di Pasquale A, Cammà C, Puglia I, Calistri P, Savini G |

| | | | |
|---|---|---|---|
| EPI_ISL_1336674, EPI_ISL_1336675 | SIESP CHIETI - DRIVE IN CHIETI | Istituto Zooprofilattico Sperimentale dell'Abruzzo e Molise "G. Caporale" | Lorusso A, Marcacci M, Di Domenico M, Ancora M, Curini V, Di Lollo V, Mangone I, Rinaldi A, Scialabba S, Caporale M, Di Pasquale A, Cammà C, Puglia I, Calistri P, Savini G |
| EPI_ISL_1336676 | DIP. PREV. AVEZZANO SERVIZIO DI IGIENE EPIDEMIOLOGIA E SANITA' PUBBLICA | Istituto Zooprofilattico Sperimentale dell'Abruzzo e Molise "G. Caporale" | Lorusso A, Marcacci M, Di Domenico M, Ancora M, Curini V, Di Lollo V, Mangone I, Rinaldi A, Scialabba S, Caporale M, Di Pasquale A, Cammà C, Puglia I, Calistri P, Savini G |
| EPI_ISL_1336677 | SIESP SULMONA | Istituto Zooprofilattico Sperimentale dell'Abruzzo e Molise "G. Caporale" | Lorusso A, Marcacci M, Di Domenico M, Ancora M, Curini V, Di Lollo V, Mangone I, Rinaldi A, Scialabba S, Caporale M, Di Pasquale A, Cammà C, Puglia I, Calistri P, Savini G |
| EPI_ISL_1336678 | SIESP DIPARTIMENTO DI PREVENZIONE CHIETI | Istituto Zooprofilattico Sperimentale dell'Abruzzo e Molise "G. Caporale" | Lorusso A, Marcacci M, Di Domenico M, Ancora M, Curini V, Di Lollo V, Mangone I, Rinaldi A, Scialabba S, Caporale M, Di Pasquale A, Cammà C, Puglia I, Calistri P, Savini G |
| EPI_ISL_1336679 | RSA GIULIANOVA | Istituto Zooprofilattico Sperimentale dell'Abruzzo e Molise "G. Caporale" | Lorusso A, Marcacci M, Di Domenico M, Ancora M, Curini V, Di Lollo V, Mangone I, Rinaldi A, Scialabba S, Caporale M, Di Pasquale A, Cammà C, Puglia I, Calistri P, Savini G |
| EPI_ISL_1336680 | Presidio Ospedaliero "San Liberatore" Atri | Istituto Zooprofilattico Sperimentale dell'Abruzzo e Molise "G. Caporale" | Lorusso A, Marcacci M, Di Domenico M, Ancora M, Curini V, Di Lollo V, Mangone I, Rinaldi A, Scialabba S, Caporale M, Di Pasquale A, Cammà C, Puglia I, Calistri P, Savini G |
| EPI_ISL_1336681 | SIESP SULMONA | Istituto Zooprofilattico Sperimentale dell'Abruzzo e Molise "G. Caporale" | Lorusso A, Marcacci M, Di Domenico M, Ancora M, Curini V, Di Lollo V, Mangone I, Rinaldi A, Scialabba S, Caporale M, Di Pasquale A, Cammà C, Puglia I, Calistri P, Savini G |
| EPI_ISL_1336682 | USCA Sulmona | Istituto Zooprofilattico Sperimentale dell'Abruzzo e Molise "G. Caporale" | Lorusso A, Marcacci M, Di Domenico M, Ancora M, Curini V, Di Lollo V, Mangone I, Rinaldi A, Scialabba S, Caporale M, Di Pasquale A, Cammà C, Puglia I, Calistri P, Savini G |
| EPI_ISL_1336683 | SIESP CHIETI - DRIVE IN CHIETI | Istituto Zooprofilattico Sperimentale dell'Abruzzo e Molise "G. Caporale" | Lorusso A, Marcacci M, Di Domenico M, Ancora M, Curini V, Di Lollo V, Mangone I, Rinaldi A, Scialabba S, Caporale M, Di Pasquale A, Cammà C, Puglia I, Calistri P, Savini G |
| EPI_ISL_1336684, EPI_ISL_1336685 | SIESP DIPARTIMENTO DI PREVENZIONE TERAMO | Istituto Zooprofilattico Sperimentale dell'Abruzzo e Molise "G. Caporale" | Lorusso A, Marcacci M, Di Domenico M, Ancora M, Curini V, Di Lollo V, Mangone I, Rinaldi A, Scialabba S, Caporale M, Di Pasquale A, Cammà C, Puglia I, Calistri P, Savini G |
| EPI_ISL_1336686, EPI_ISL_1336687, EPI_ISL_1336688, EPI_ISL_1336689 | SIESP DIPARTIMENTO DI PREVENZIONE CHIETI | Istituto Zooprofilattico Sperimentale dell'Abruzzo e Molise "G. Caporale" | Lorusso A, Marcacci M, Di Domenico M, Ancora M, Curini V, Di Lollo V, Mangone I, Rinaldi A, Scialabba S, Caporale M, Di Pasquale A, Cammà C, Puglia I, Calistri P, Savini G |
| EPI_ISL_1336690 | Presidio Ospedaliero "San Liberatore" Atri | Istituto Zooprofilattico Sperimentale dell'Abruzzo e Molise "G. Caporale" | Lorusso A, Marcacci M, Di Domenico M, Ancora M, Curini V, Di Lollo V, Mangone I, Rinaldi A, Scialabba S, Caporale M, Di Pasquale A, Cammà C, Puglia I, Calistri P, Savini G |
| EPI_ISL_1336691 | SIESP DIPARTIMENTO DI PREVENZIONE CHIETI | Istituto Zooprofilattico Sperimentale dell'Abruzzo e Molise "G. Caporale" | Lorusso A, Marcacci M, Di Domenico M, Ancora M, Curini V, Di Lollo V, Mangone I, Rinaldi A, Scialabba S, Caporale M, Di Pasquale A, Cammà C, Puglia I, Calistri P, Savini G |
| EPI_ISL_1336692, EPI_ISL_1336693 | SIESP CHIETI - DRIVE IN CHIETI | Istituto Zooprofilattico Sperimentale dell'Abruzzo e Molise "G. Caporale" | Lorusso A, Marcacci M, Di Domenico M, Ancora M, Curini V, Di Lollo V, Mangone I, Rinaldi A, Scialabba S, Caporale M, Di Pasquale A, Cammà C, Puglia I, Calistri P, Savini G |
| EPI_ISL_1336694 | SIESP DIPARTIMENTO DI PREVENZIONE CHIETI | Istituto Zooprofilattico Sperimentale dell'Abruzzo e Molise "G. Caporale" | Lorusso A, Marcacci M, Di Domenico M, Ancora M, Curini V, Di Lollo V, Mangone I, Rinaldi A, Scialabba S, Caporale M, Di Pasquale A, Cammà C, Puglia I, Calistri P, Savini G |
| EPI_ISL_1336695 | SIESP DIPARTIMENTO DI PREVENZIONE TERAMO | Istituto Zooprofilattico Sperimentale dell'Abruzzo e Molise "G. Caporale" | Lorusso A, Marcacci M, Di Domenico M, Ancora M, Curini V, Di Lollo V, Mangone I, Rinaldi A, Scialabba S, Caporale M, Di Pasquale A, Cammà C, Puglia I, Calistri P, Savini G |
| EPI_ISL_1336696 | SIESP CHIETI - DRIVE IN CHIETI | Istituto Zooprofilattico Sperimentale dell'Abruzzo e Molise "G. Caporale" | Lorusso A, Marcacci M, Di Domenico M, Ancora M, Curini V, Di Lollo V, Mangone I, Rinaldi A, Scialabba S, Caporale M, Di Pasquale A, Cammà C, Puglia I, Calistri P, Savini G |
| EPI_ISL_1336697 | OSPEDALE "F. RENZETTI" LANCIANO - ORTOPEDIA E TRAUMATOLOGIA | Istituto Zooprofilattico Sperimentale dell'Abruzzo e Molise "G. Caporale" | Lorusso A, Marcacci M, Di Domenico M, Ancora M, Curini V, Di Lollo V, Mangone I, Rinaldi A, Scialabba S, Caporale M, Di Pasquale A, Cammà C, Puglia I, Calistri P, Savini G |
| EPI_ISL_1336698 | USCA Sulmona | Istituto Zooprofilattico Sperimentale dell'Abruzzo e Molise "G. Caporale" | Lorusso A, Marcacci M, Di Domenico M, Ancora M, Curini V, Di Lollo V, Mangone I, Rinaldi A, Scialabba S, Caporale M, Di Pasquale A, Cammà C, Puglia I, Calistri P, Savini G |
| EPI_ISL_1336699 | SIESP DIPARTIMENTO DI PREVENZIONE CHIETI | Istituto Zooprofilattico Sperimentale dell'Abruzzo e Molise "G. Caporale" | Lorusso A, Marcacci M, Di Domenico M, Ancora M, Curini V, Di Lollo V, Mangone I, Rinaldi A, Scialabba S, Caporale M, Di Pasquale A, Cammà C, Puglia I, Calistri P, Savini G |
| EPI_ISL_1336700 | SIESP CHIETI - DRIVE IN GISSI | Istituto Zooprofilattico Sperimentale dell'Abruzzo e Molise "G. Caporale" | Lorusso A, Marcacci M, Di Domenico M, Ancora M, Curini V, Di Lollo V, Mangone I, Rinaldi A, Scialabba S, Caporale M, Di Pasquale A, Cammà C, Puglia I, Calistri P, Savini G |
| EPI_ISL_1336701 | SIESP CHIETI - DRIVE IN VASTO | Istituto Zooprofilattico Sperimentale dell'Abruzzo e Molise "G. Caporale" | Lorusso A, Marcacci M, Di Domenico M, Ancora M, Curini V, Di Lollo V, Mangone I, Rinaldi A, Scialabba S, Caporale M, Di Pasquale A, Cammà C, Puglia I, Calistri P, Savini G |
| EPI_ISL_1336702 | USCA Sulmona | Istituto Zooprofilattico Sperimentale dell'Abruzzo e Molise "G. Caporale" | Lorusso A, Marcacci M, Di Domenico M, Ancora M, Curini V, Di Lollo V, Mangone I, Rinaldi A, Scialabba S, Caporale M, Di Pasquale A, Cammà C, Puglia I, Calistri P, Savini G |
| EPI_ISL_1336703 | SIESP CHIETI - DRIVE IN ORTONA | Istituto Zooprofilattico Sperimentale dell'Abruzzo e Molise "G. Caporale" | Lorusso A, Marcacci M, Di Domenico M, Ancora M, Curini V, Di Lollo V, Mangone I, Rinaldi A, Scialabba S, Caporale M, Di Pasquale A, Cammà C, Puglia I, Calistri P, Savini G |
| EPI_ISL_1336704 | OSPEDALE CIVILE TERAMO - PRONTO SOCCORSO | Istituto Zooprofilattico Sperimentale dell'Abruzzo e Molise "G. Caporale" | Lorusso A, Marcacci M, Di Domenico M, Ancora M, Curini V, Di Lollo V, Mangone I, Rinaldi A, Scialabba S, Caporale M, Di Pasquale A, Cammà C, Puglia I, Calistri P, Savini G |
| EPI_ISL_1336705 | SIESP DIPARTIMENTO DI PREVENZIONE TERAMO | Istituto Zooprofilattico Sperimentale dell'Abruzzo e Molise "G. Caporale" | Lorusso A, Marcacci M, Di Domenico M, Ancora M, Curini V, Di Lollo V, Mangone I, Rinaldi A, Scialabba S, Caporale M, Di Pasquale A, Cammà C, Puglia I, Calistri P, Savini G |
| EPI_ISL_1336706 | DIP. PREV. AVEZZANO SERVIZIO DI IGIENE EPIDEMIOLOGIA E SANITA' PUBBLICA | Istituto Zooprofilattico Sperimentale dell'Abruzzo e Molise "G. Caporale" | Lorusso A, Marcacci M, Di Domenico M, Ancora M, Curini V, Di Lollo V, Mangone I, Rinaldi A, Scialabba S, Caporale M, Di Pasquale A, Cammà C, Puglia I, Calistri P, Savini G |
| EPI_ISL_1336707 | SIESP CHIETI - DRIVE IN ORTONA | Istituto Zooprofilattico Sperimentale dell'Abruzzo e Molise "G. Caporale" | Lorusso A, Marcacci M, Di Domenico M, Ancora M, Curini V, Di Lollo V, Mangone I, Rinaldi A, Scialabba S, Caporale M, Di Pasquale A, Cammà C, Puglia I, Calistri P, Savini G |
| EPI_ISL_1336708 | USCA Pescina | Istituto Zooprofilattico Sperimentale dell'Abruzzo e Molise "G. Caporale" | Lorusso A, Marcacci M, Di Domenico M, Ancora M, Curini V, Di Lollo V, Mangone I, Rinaldi A, Scialabba S, Caporale M, Di Pasquale A, Cammà C, Puglia I, Calistri P, Savini G |
| EPI_ISL_1336709, EPI_ISL_1336710, EPI_ISL_1336711 | SIESP DIPARTIMENTO DI PREVENZIONE TERAMO | Istituto Zooprofilattico Sperimentale dell'Abruzzo e Molise "G. Caporale" | Lorusso A, Marcacci M, Di Domenico M, Ancora M, Curini V, Di Lollo V, Mangone I, Rinaldi A, Scialabba S, Caporale M, Di Pasquale A, Cammà C, Puglia I, Calistri P, Savini G |
| EPI_ISL_1336712 | SIESP CHIETI - DRIVE IN CHIETI | Istituto Zooprofilattico Sperimentale dell'Abruzzo e Molise "G. Caporale" | Lorusso A, Marcacci M, Di Domenico M, Ancora M, Curini V, Di Lollo V, Mangone I, Rinaldi A, Scialabba S, Caporale M, Di Pasquale A, Cammà C, Puglia I, Calistri P, Savini G |
| EPI_ISL_1336713 | SIESP DIPARTIMENTO DI PREVENZIONE TERAMO | Istituto Zooprofilattico Sperimentale dell'Abruzzo e Molise "G. Caporale" | Lorusso A, Marcacci M, Di Domenico M, Ancora M, Curini V, Di Lollo V, Mangone I, Rinaldi A, Scialabba S, Caporale M, Di Pasquale A, Cammà C, Puglia I, Calistri P, Savini G |
| EPI_ISL_1336714 | SIESP CHIETI - DRIVE IN CHIETI | Istituto Zooprofilattico Sperimentale dell'Abruzzo e Molise "G. Caporale" | Lorusso A, Marcacci M, Di Domenico M, Ancora M, Curini V, Di Lollo V, Mangone I, Rinaldi A, Scialabba S, Caporale M, Di Pasquale A, Cammà C, Puglia I, Calistri P, Savini G |
| EPI_ISL_1336715, EPI_ISL_1336716, EPI_ISL_1336717 | USCA Castel di Sangro | Istituto Zooprofilattico Sperimentale dell'Abruzzo e Molise "G. Caporale" | Lorusso A, Marcacci M, Di Domenico M, Ancora M, Curini V, Di Lollo V, Mangone I, Rinaldi A, Scialabba S, Caporale M, Di Pasquale A, Cammà C, Puglia I, Calistri P, Savini G |
| EPI_ISL_1336718 | SIESP DIPARTIMENTO DI PREVENZIONE TERAMO | Istituto Zooprofilattico Sperimentale dell'Abruzzo e Molise "G. Caporale" | Lorusso A, Marcacci M, Di Domenico M, Ancora M, Curini V, Di Lollo V, Mangone I, Rinaldi A, Scialabba S, Caporale M, Di Pasquale A, Cammà C, Puglia I, Calistri P, Savini G |

| | | | |
|---|---|---|---|
| EPI_ISL_1336719, EPI_ISL_1336720 | SIESP CHIETI - DRIVE IN ORTONA | Istituto Zooprofilattico Sperimentale dell'Abruzzo e Molise "G. Caporale" | Lorusso A, Marcacci M, Di Domenico M, Ancora M, Curini V, Di Lollo V, Mangone I, Rinaldi A, Scialabba S, Caporale M, Di Pasquale A, Cammà C, Puglia I, Calistri P, Savini G |
| EPI_ISL_1336721 | USCA Pescina | Istituto Zooprofilattico Sperimentale dell'Abruzzo e Molise "G. Caporale" | Lorusso A, Marcacci M, Di Domenico M, Ancora M, Curini V, Di Lollo V, Mangone I, Rinaldi A, Scialabba S, Caporale M, Di Pasquale A, Cammà C, Puglia I, Calistri P, Savini G |
| EPI_ISL_1336722 | USCA Castel di Sangro | Istituto Zooprofilattico Sperimentale dell'Abruzzo e Molise "G. Caporale" | Lorusso A, Marcacci M, Di Domenico M, Ancora M, Curini V, Di Lollo V, Mangone I, Rinaldi A, Scialabba S, Caporale M, Di Pasquale A, Cammà C, Puglia I, Calistri P, Savini G |
| EPI_ISL_1336723 | SIESP DIPARTIMENTO DI PREVENZIONE TERAMO | Istituto Zooprofilattico Sperimentale dell'Abruzzo e Molise "G. Caporale" | Lorusso A, Marcacci M, Di Domenico M, Ancora M, Curini V, Di Lollo V, Mangone I, Rinaldi A, Scialabba S, Caporale M, Di Pasquale A, Cammà C, Puglia I, Calistri P, Savini G |
| EPI_ISL_1336724 | USCA Castel di Sangro | Istituto Zooprofilattico Sperimentale dell'Abruzzo e Molise "G. Caporale" | Lorusso A, Marcacci M, Di Domenico M, Ancora M, Curini V, Di Lollo V, Mangone I, Rinaldi A, Scialabba S, Caporale M, Di Pasquale A, Cammà C, Puglia I, Calistri P, Savini G |
| EPI_ISL_1336725 | SIESP CHIETI - DRIVE IN VASTO | Istituto Zooprofilattico Sperimentale dell'Abruzzo e Molise "G. Caporale" | Lorusso A, Marcacci M, Di Domenico M, Ancora M, Curini V, Di Lollo V, Mangone I, Rinaldi A, Scialabba S, Caporale M, Di Pasquale A, Cammà C, Puglia I, Calistri P, Savini G |
| EPI_ISL_1336726 | DIP. PREV. AVEZZANO SERVIZIO DI IGIENE EPIDEMIOLOGIA E SANITA' PUBBLICA | Istituto Zooprofilattico Sperimentale dell'Abruzzo e Molise "G. Caporale" | Lorusso A, Marcacci M, Di Domenico M, Ancora M, Curini V, Di Lollo V, Mangone I, Rinaldi A, Scialabba S, Caporale M, Di Pasquale A, Cammà C, Puglia I, Calistri P, Savini G |
| EPI_ISL_1336727 | USCA Tagliacozzo | Istituto Zooprofilattico Sperimentale dell'Abruzzo e Molise "G. Caporale" | Lorusso A, Marcacci M, Di Domenico M, Ancora M, Curini V, Di Lollo V, Mangone I, Rinaldi A, Scialabba S, Caporale M, Di Pasquale A, Cammà C, Puglia I, Calistri P, Savini G |
| EPI_ISL_1336728, EPI_ISL_1336729 | SIESP DIPARTIMENTO DI PREVENZIONE TERAMO | Istituto Zooprofilattico Sperimentale dell'Abruzzo e Molise "G. Caporale" | Lorusso A, Marcacci M, Di Domenico M, Ancora M, Curini V, Di Lollo V, Mangone I, Rinaldi A, Scialabba S, Caporale M, Di Pasquale A, Cammà C, Puglia I, Calistri P, Savini G |
| EPI_ISL_1336730, EPI_ISL_1336731 | Simeoni - La Casa del Sole | Istituto Zooprofilattico Sperimentale dell'Abruzzo e Molise "G. Caporale" | Lorusso A, Marcacci M, Di Domenico M, Ancora M, Curini V, Di Lollo V, Mangone I, Rinaldi A, Scialabba S, Caporale M, Di Pasquale A, Cammà C, Puglia I, Calistri P, Savini G |
| EPI_ISL_1336732 | OSPEDALE CIVILE GIULIANOVA - PRONTO SOCCORSO | Istituto Zooprofilattico Sperimentale dell'Abruzzo e Molise "G. Caporale" | Lorusso A, Marcacci M, Di Domenico M, Ancora M, Curini V, Di Lollo V, Mangone I, Rinaldi A, Scialabba S, Caporale M, Di Pasquale A, Cammà C, Puglia I, Calistri P, Savini G |
| EPI_ISL_1336733, EPI_ISL_1336734 | SIESP DIPARTIMENTO DI PREVENZIONE TERAMO | Istituto Zooprofilattico Sperimentale dell'Abruzzo e Molise "G. Caporale" | Lorusso A, Marcacci M, Di Domenico M, Ancora M, Curini V, Di Lollo V, Mangone I, Rinaldi A, Scialabba S, Caporale M, Di Pasquale A, Cammà C, Puglia I, Calistri P, Savini G |
| EPI_ISL_1336735 | DIP. PREV. AVEZZANO SERVIZIO DI IGIENE EPIDEMIOLOGIA E SANITA' PUBBLICA | Istituto Zooprofilattico Sperimentale dell'Abruzzo e Molise "G. Caporale" | Lorusso A, Marcacci M, Di Domenico M, Ancora M, Curini V, Di Lollo V, Mangone I, Rinaldi A, Scialabba S, Caporale M, Di Pasquale A, Cammà C, Puglia I, Calistri P, Savini G |
| EPI_ISL_1336736 | SIESP CHIETI - DRIVE IN CHIETI | Istituto Zooprofilattico Sperimentale dell'Abruzzo e Molise "G. Caporale" | Lorusso A, Marcacci M, Di Domenico M, Ancora M, Curini V, Di Lollo V, Mangone I, Rinaldi A, Scialabba S, Caporale M, Di Pasquale A, Cammà C, Puglia I, Calistri P, Savini G |
| EPI_ISL_1336737 | USCA Tagliacozzo | Istituto Zooprofilattico Sperimentale dell'Abruzzo e Molise "G. Caporale" | Lorusso A, Marcacci M, Di Domenico M, Ancora M, Curini V, Di Lollo V, Mangone I, Rinaldi A, Scialabba S, Caporale M, Di Pasquale A, Cammà C, Puglia I, Calistri P, Savini G |
| EPI_ISL_1336738, EPI_ISL_1336739 | SIESP DIPARTIMENTO DI PREVENZIONE TERAMO | Istituto Zooprofilattico Sperimentale dell'Abruzzo e Molise "G. Caporale" | Lorusso A, Marcacci M, Di Domenico M, Ancora M, Curini V, Di Lollo V, Mangone I, Rinaldi A, Scialabba S, Caporale M, Di Pasquale A, Cammà C, Puglia I, Calistri P, Savini G |
| EPI_ISL_1336740, EPI_ISL_1336741 | SIESP CHIETI - DRIVE IN CHIETI | Istituto Zooprofilattico Sperimentale dell'Abruzzo e Molise "G. Caporale" | Lorusso A, Marcacci M, Di Domenico M, Ancora M, Curini V, Di Lollo V, Mangone I, Rinaldi A, Scialabba S, Caporale M, Di Pasquale A, Cammà C, Puglia I, Calistri P, Savini G |
| EPI_ISL_1336742 | USCA Castel di Sangro | Istituto Zooprofilattico Sperimentale dell'Abruzzo e Molise "G. Caporale" | Lorusso A, Marcacci M, Di Domenico M, Ancora M, Curini V, Di Lollo V, Mangone I, Rinaldi A, Scialabba S, Caporale M, Di Pasquale A, Cammà C, Puglia I, Calistri P, Savini G |
| EPI_ISL_1336743 | RSA GIULIANOVA | Istituto Zooprofilattico Sperimentale dell'Abruzzo e Molise "G. Caporale" | Lorusso A, Marcacci M, Di Domenico M, Ancora M, Curini V, Di Lollo V, Mangone I, Rinaldi A, Scialabba S, Caporale M, Di Pasquale A, Cammà C, Puglia I, Calistri P, Savini G |
| EPI_ISL_1336744, EPI_ISL_1336745 | USCA Castel di Sangro | Istituto Zooprofilattico Sperimentale dell'Abruzzo e Molise "G. Caporale" | Lorusso A, Marcacci M, Di Domenico M, Ancora M, Curini V, Di Lollo V, Mangone I, Rinaldi A, Scialabba S, Caporale M, Di Pasquale A, Cammà C, Puglia I, Calistri P, Savini G |
| EPI_ISL_1336746 | SIESP DIPARTIMENTO DI PREVENZIONE TERAMO | Istituto Zooprofilattico Sperimentale dell'Abruzzo e Molise "G. Caporale" | Lorusso A, Marcacci M, Di Domenico M, Ancora M, Curini V, Di Lollo V, Mangone I, Rinaldi A, Scialabba S, Caporale M, Di Pasquale A, Cammà C, Puglia I, Calistri P, Savini G |
| EPI_ISL_1336747, EPI_ISL_1336748 | SIESP CHIETI - DRIVE IN ORTONA | Istituto Zooprofilattico Sperimentale dell'Abruzzo e Molise "G. Caporale" | Lorusso A, Marcacci M, Di Domenico M, Ancora M, Curini V, Di Lollo V, Mangone I, Rinaldi A, Scialabba S, Caporale M, Di Pasquale A, Cammà C, Puglia I, Calistri P, Savini G |
| EPI_ISL_1336749 | DIP. PREV. AVEZZANO SERVIZIO DI IGIENE EPIDEMIOLOGIA E SANITA' PUBBLICA | Istituto Zooprofilattico Sperimentale dell'Abruzzo e Molise "G. Caporale" | Lorusso A, Marcacci M, Di Domenico M, Ancora M, Curini V, Di Lollo V, Mangone I, Rinaldi A, Scialabba S, Caporale M, Di Pasquale A, Cammà C, Puglia I, Calistri P, Savini G |
| EPI_ISL_1336750 | USCA Pescina | Istituto Zooprofilattico Sperimentale dell'Abruzzo e Molise "G. Caporale" | Lorusso A, Marcacci M, Di Domenico M, Ancora M, Curini V, Di Lollo V, Mangone I, Rinaldi A, Scialabba S, Caporale M, Di Pasquale A, Cammà C, Puglia I, Calistri P, Savini G |
| EPI_ISL_1336751 | SIESP CHIETI - DRIVE IN CHIETI | Istituto Zooprofilattico Sperimentale dell'Abruzzo e Molise "G. Caporale" | Lorusso A, Marcacci M, Di Domenico M, Ancora M, Curini V, Di Lollo V, Mangone I, Rinaldi A, Scialabba S, Caporale M, Di Pasquale A, Cammà C, Puglia I, Calistri P, Savini G |
| EPI_ISL_1336752, EPI_ISL_1336753 | USCA Castel di Sangro | Istituto Zooprofilattico Sperimentale dell'Abruzzo e Molise "G. Caporale" | Lorusso A, Marcacci M, Di Domenico M, Ancora M, Curini V, Di Lollo V, Mangone I, Rinaldi A, Scialabba S, Caporale M, Di Pasquale A, Cammà C, Puglia I, Calistri P, Savini G |
| EPI_ISL_1336754 | SIESP CHIETI - DRIVE IN VASTO | Istituto Zooprofilattico Sperimentale dell'Abruzzo e Molise "G. Caporale" | Lorusso A, Marcacci M, Di Domenico M, Ancora M, Curini V, Di Lollo V, Mangone I, Rinaldi A, Scialabba S, Caporale M, Di Pasquale A, Cammà C, Puglia I, Calistri P, Savini G |
| EPI_ISL_1336755 | SIESP DIPARTIMENTO DI PREVENZIONE TERAMO | Istituto Zooprofilattico Sperimentale dell'Abruzzo e Molise "G. Caporale" | Lorusso A, Marcacci M, Di Domenico M, Ancora M, Curini V, Di Lollo V, Mangone I, Rinaldi A, Scialabba S, Caporale M, Di Pasquale A, Cammà C, Puglia I, Calistri P, Savini G |
| EPI_ISL_1336756 | Simeoni - La Casa del Sole | Istituto Zooprofilattico Sperimentale dell'Abruzzo e Molise "G. Caporale" | Lorusso A, Marcacci M, Di Domenico M, Ancora M, Curini V, Di Lollo V, Mangone I, Rinaldi A, Scialabba S, Caporale M, Di Pasquale A, Cammà C, Puglia I, Calistri P, Savini G |
| EPI_ISL_1336757, EPI_ISL_1336758 | SIESP DIPARTIMENTO DI PREVENZIONE TERAMO | Istituto Zooprofilattico Sperimentale dell'Abruzzo e Molise "G. Caporale" | Lorusso A, Marcacci M, Di Domenico M, Ancora M, Curini V, Di Lollo V, Mangone I, Rinaldi A, Scialabba S, Caporale M, Di Pasquale A, Cammà C, Puglia I, Calistri P, Savini G |
| EPI_ISL_1336759 | SIESP CHIETI - DRIVE IN LANCIANO | Istituto Zooprofilattico Sperimentale dell'Abruzzo e Molise "G. Caporale" | Lorusso A, Marcacci M, Di Domenico M, Ancora M, Curini V, Di Lollo V, Mangone I, Rinaldi A, Scialabba S, Caporale M, Di Pasquale A, Cammà C, Puglia I, Calistri P, Savini G |
| EPI_ISL_1336760 | SIESP DIPARTIMENTO DI PREVENZIONE TERAMO | Istituto Zooprofilattico Sperimentale dell'Abruzzo e Molise "G. Caporale" | Lorusso A, Marcacci M, Di Domenico M, Ancora M, Curini V, Di Lollo V, Mangone I, Rinaldi A, Scialabba S, Caporale M, Di Pasquale A, Cammà C, Puglia I, Calistri P, Savini G |
| EPI_ISL_1336761 | DIP. PREV. AVEZZANO SERVIZIO DI IGIENE EPIDEMIOLOGIA E SANITA' PUBBLICA | Istituto Zooprofilattico Sperimentale dell'Abruzzo e Molise "G. Caporale" | Lorusso A, Marcacci M, Di Domenico M, Ancora M, Curini V, Di Lollo V, Mangone I, Rinaldi A, Scialabba S, Caporale M, Di Pasquale A, Cammà C, Puglia I, Calistri P, Savini G |
| EPI_ISL_1336762, EPI_ISL_1336763, EPI_ISL_1336764, EPI_ISL_1336765 | SIESP CHIETI - DRIVE IN LANCIANO | Istituto Zooprofilattico Sperimentale dell'Abruzzo e Molise "G. Caporale" | Lorusso A, Marcacci M, Di Domenico M, Ancora M, Curini V, Di Lollo V, Mangone I, Rinaldi A, Scialabba S, Caporale M, Di Pasquale A, Cammà C, Puglia I, Calistri P, Savini G |
| EPI_ISL_1336766 | SIESP DIPARTIMENTO DI PREVENZIONE TERAMO | Istituto Zooprofilattico Sperimentale dell'Abruzzo e Molise "G. Caporale" | Lorusso A, Marcacci M, Di Domenico M, Ancora M, Curini V, Di Lollo V, Mangone I, Rinaldi A, Scialabba S, Caporale M, Di Pasquale A, Cammà C, Puglia I, Calistri P, Savini G |

| | | | |
|---|---|---|---|
| EPI_ISL_1336767 | PRESIDIO TERRITORIALE CASOLI | Istituto Zooprofilattico Sperimentale dell'Abruzzo e Molise "G. Caporale" | Lorusso A, Marcacci M, Di Domenico M, Ancora M, Curini V, Di Lollo V, Mangone I, Rinaldi A, Scialabba S, Caporale M, Di Pasquale A, Cammà C, Puglia I, Calistri P, Savini G |
| EPI_ISL_1336768, EPI_ISL_1336769, EPI_ISL_1336770, EPI_ISL_1336771 | SIESP DIPARTIMENTO DI PREVENZIONE TERAMO | Istituto Zooprofilattico Sperimentale dell'Abruzzo e Molise "G. Caporale" | Lorusso A, Marcacci M, Di Domenico M, Ancora M, Curini V, Di Lollo V, Mangone I, Rinaldi A, Scialabba S, Caporale M, Di Pasquale A, Cammà C, Puglia I, Calistri P, Savini G |
| EPI_ISL_1336772, EPI_ISL_1336773 | SIESP CHIETI - DRIVE IN LANCIANO | Istituto Zooprofilattico Sperimentale dell'Abruzzo e Molise "G. Caporale" | Lorusso A, Marcacci M, Di Domenico M, Ancora M, Curini V, Di Lollo V, Mangone I, Rinaldi A, Scialabba S, Caporale M, Di Pasquale A, Cammà C, Puglia I, Calistri P, Savini G |
| EPI_ISL_1336774, EPI_ISL_1336775 | SIESP DIPARTIMENTO DI PREVENZIONE TERAMO | Istituto Zooprofilattico Sperimentale dell'Abruzzo e Molise "G. Caporale" | Lorusso A, Marcacci M, Di Domenico M, Ancora M, Curini V, Di Lollo V, Mangone I, Rinaldi A, Scialabba S, Caporale M, Di Pasquale A, Cammà C, Puglia I, Calistri P, Savini G |
| EPI_ISL_1336776, EPI_ISL_1336777 | RSA GIULIANOVA | Istituto Zooprofilattico Sperimentale dell'Abruzzo e Molise "G. Caporale" | Lorusso A, Marcacci M, Di Domenico M, Ancora M, Curini V, Di Lollo V, Mangone I, Rinaldi A, Scialabba S, Caporale M, Di Pasquale A, Cammà C, Puglia I, Calistri P, Savini G |
| EPI_ISL_1336778, EPI_ISL_1336779 | SIESP DIPARTIMENTO DI PREVENZIONE TERAMO | Istituto Zooprofilattico Sperimentale dell'Abruzzo e Molise "G. Caporale" | Lorusso A, Marcacci M, Di Domenico M, Ancora M, Curini V, Di Lollo V, Mangone I, Rinaldi A, Scialabba S, Caporale M, Di Pasquale A, Cammà C, Puglia I, Calistri P, Savini G |
| EPI_ISL_1336780 | SIESP CHIETI - DRIVE IN LANCIANO | Istituto Zooprofilattico Sperimentale dell'Abruzzo e Molise "G. Caporale" | Lorusso A, Marcacci M, Di Domenico M, Ancora M, Curini V, Di Lollo V, Mangone I, Rinaldi A, Scialabba S, Caporale M, Di Pasquale A, Cammà C, Puglia I, Calistri P, Savini G |
| EPI_ISL_1336781 | SIESP DIPARTIMENTO DI PREVENZIONE TERAMO | Istituto Zooprofilattico Sperimentale dell'Abruzzo e Molise "G. Caporale" | Lorusso A, Marcacci M, Di Domenico M, Ancora M, Curini V, Di Lollo V, Mangone I, Rinaldi A, Scialabba S, Caporale M, Di Pasquale A, Cammà C, Puglia I, Calistri P, Savini G |
| EPI_ISL_1336782 | SIESP DIPARTIMENTO DI PREVENZIONE CHIETI | Istituto Zooprofilattico Sperimentale dell'Abruzzo e Molise "G. Caporale" | Lorusso A, Marcacci M, Di Domenico M, Ancora M, Curini V, Di Lollo V, Mangone I, Rinaldi A, Scialabba S, Caporale M, Di Pasquale A, Cammà C, Puglia I, Calistri P, Savini G |
| EPI_ISL_1336783, EPI_ISL_1336784 | SIESP DIPARTIMENTO DI PREVENZIONE TERAMO | Istituto Zooprofilattico Sperimentale dell'Abruzzo e Molise "G. Caporale" | Lorusso A, Marcacci M, Di Domenico M, Ancora M, Curini V, Di Lollo V, Mangone I, Rinaldi A, Scialabba S, Caporale M, Di Pasquale A, Cammà C, Puglia I, Calistri P, Savini G |
| EPI_ISL_1336785 | DIP. PREV. AVEZZANO SERVIZIO DI IGIENE EPIDEMIOLOGIA E SANITA' PUBBLICA | Istituto Zooprofilattico Sperimentale dell'Abruzzo e Molise "G. Caporale" | Lorusso A, Marcacci M, Di Domenico M, Ancora M, Curini V, Di Lollo V, Mangone I, Rinaldi A, Scialabba S, Caporale M, Di Pasquale A, Cammà C, Puglia I, Calistri P, Savini G |
| EPI_ISL_1336786 | USCA Avezzano | Istituto Zooprofilattico Sperimentale dell'Abruzzo e Molise "G. Caporale" | Lorusso A, Marcacci M, Di Domenico M, Ancora M, Curini V, Di Lollo V, Mangone I, Rinaldi A, Scialabba S, Caporale M, Di Pasquale A, Cammà C, Puglia I, Calistri P, Savini G |
| EPI_ISL_1336787 | SIESP CHIETI - DRIVE IN ORTONA | Istituto Zooprofilattico Sperimentale dell'Abruzzo e Molise "G. Caporale" | Lorusso A, Marcacci M, Di Domenico M, Ancora M, Curini V, Di Lollo V, Mangone I, Rinaldi A, Scialabba S, Caporale M, Di Pasquale A, Cammà C, Puglia I, Calistri P, Savini G |
| EPI_ISL_1336788 | OSPEDALE CIVILE SANT'OMERO - PRONTO SOCCORSO | Istituto Zooprofilattico Sperimentale dell'Abruzzo e Molise "G. Caporale" | Lorusso A, Marcacci M, Di Domenico M, Ancora M, Curini V, Di Lollo V, Mangone I, Rinaldi A, Scialabba S, Caporale M, Di Pasquale A, Cammà C, Puglia I, Calistri P, Savini G |
| EPI_ISL_1336789, EPI_ISL_1336790, EPI_ISL_1336791 | SIESP DIPARTIMENTO DI PREVENZIONE TERAMO | Istituto Zooprofilattico Sperimentale dell'Abruzzo e Molise "G. Caporale" | Lorusso A, Marcacci M, Di Domenico M, Ancora M, Curini V, Di Lollo V, Mangone I, Rinaldi A, Scialabba S, Caporale M, Di Pasquale A, Cammà C, Puglia I, Calistri P, Savini G |
| EPI_ISL_1336792 | SIESP CHIETI - DRIVE IN LANCIANO | Istituto Zooprofilattico Sperimentale dell'Abruzzo e Molise "G. Caporale" | Lorusso A, Marcacci M, Di Domenico M, Ancora M, Curini V, Di Lollo V, Mangone I, Rinaldi A, Scialabba S, Caporale M, Di Pasquale A, Cammà C, Puglia I, Calistri P, Savini G |
| EPI_ISL_1336793 | SIESP CHIETI - DRIVE IN VASTO | Istituto Zooprofilattico Sperimentale dell'Abruzzo e Molise "G. Caporale" | Lorusso A, Marcacci M, Di Domenico M, Ancora M, Curini V, Di Lollo V, Mangone I, Rinaldi A, Scialabba S, Caporale M, Di Pasquale A, Cammà C, Puglia I, Calistri P, Savini G |
| EPI_ISL_1336794, EPI_ISL_1336795 | SIESP CHIETI - DRIVE IN LANCIANO | Istituto Zooprofilattico Sperimentale dell'Abruzzo e Molise "G. Caporale" | Lorusso A, Marcacci M, Di Domenico M, Ancora M, Curini V, Di Lollo V, Mangone I, Rinaldi A, Scialabba S, Caporale M, Di Pasquale A, Cammà C, Puglia I, Calistri P, Savini G |
| EPI_ISL_1336796 | SIESP DIPARTIMENTO DI PREVENZIONE TERAMO | Istituto Zooprofilattico Sperimentale dell'Abruzzo e Molise "G. Caporale" | Lorusso A, Marcacci M, Di Domenico M, Ancora M, Curini V, Di Lollo V, Mangone I, Rinaldi A, Scialabba S, Caporale M, Di Pasquale A, Cammà C, Puglia I, Calistri P, Savini G |
| EPI_ISL_1336800, EPI_ISL_1336801, EPI_ISL_1336802, EPI_ISL_1336803, EPI_ISL_1336804, EPI_ISL_1336805, EPI_ISL_1336806, EPI_ISL_1336807, EPI_ISL_1336808, EPI_ISL_1336809, EPI_ISL_1336810, EPI_ISL_1336811, EPI_ISL_1336812, EPI_ISL_1336813, EPI_ISL_1336814, EPI_ISL_1336815, EPI_ISL_1336816, EPI_ISL_1336817, EPI_ISL_1336818, EPI_ISL_1336819, EPI_ISL_1336820 | | | |
| see above | Azienda Ospedaliera Terni | Istituto Zooprofilattico Sperimentale dell'Abruzzo e Molise "G. Caporale" | Palumbo M, Scaccetti A, Lorusso A, Marcacci M, Di Domenico M, Ancora M, Curini V, Mangone I, Rinaldi A, Scialabba S, Caporale M, Di Pasquale A, Cammà C, Puglia I, Calistri P, Savini G |
| EPI_ISL_1336821 | Istituto Zooprofilattico Sperimentale Umbria e Marche "Togo Rosati" | Istituto Zooprofilattico Sperimentale dell'Abruzzo e Molise "G. Caporale" | Biagetti M, Giammarioli M, Lorusso A, Marcacci M, Di Domenico M, Ancora M, Curini V, Mangone I, Rinaldi A, Scialabba S, Caporale M, Di Pasquale A, Cammà C, Puglia I, Calistri P, Savini G |
| EPI_ISL_1336822, EPI_ISL_1336823, EPI_ISL_1336824, EPI_ISL_1336825, EPI_ISL_1336826, EPI_ISL_1336827, EPI_ISL_1336828, EPI_ISL_1336829, EPI_ISL_1336830, EPI_ISL_1336831 | Azienda Ospedaliera Terni | Istituto Zooprofilattico Sperimentale dell'Abruzzo e Molise "G. Caporale" | Palumbo M, Scaccetti A, Lorusso A, Marcacci M, Di Domenico M, Ancora M, Curini V, Mangone I, Rinaldi A, Scialabba S, Caporale M, Di Pasquale A, Cammà C, Puglia I, Calistri P, Savini G |
| EPI_ISL_1336832 | Istituto Zooprofilattico Sperimentale Umbria e Marche "Togo Rosati" | Istituto Zooprofilattico Sperimentale dell'Abruzzo e Molise "G. Caporale" | Biagetti M, Giammarioli M, Lorusso A, Marcacci M, Di Domenico M, Ancora M, Curini V, Mangone I, Rinaldi A, Scialabba S, Caporale M, Di Pasquale A, Cammà C, Puglia I, Calistri P, Savini G |
| EPI_ISL_1336833, EPI_ISL_1336834, EPI_ISL_1336835, EPI_ISL_1336836, EPI_ISL_1336837, EPI_ISL_1336838, EPI_ISL_1336839, EPI_ISL_1336840, EPI_ISL_1336841 | Azienda Ospedaliera Terni | Istituto Zooprofilattico Sperimentale dell'Abruzzo e Molise "G. Caporale" | Palumbo M, Scaccetti A, Lorusso A, Marcacci M, Di Domenico M, Ancora M, Curini V, Mangone I, Rinaldi A, Scialabba S, Caporale M, Di Pasquale A, Cammà C, Puglia I, Calistri P, Savini G |
| EPI_ISL_1336842 | Istituto Zooprofilattico Sperimentale Umbria e Marche "Togo Rosati" | Istituto Zooprofilattico Sperimentale dell'Abruzzo e Molise "G. Caporale" | Biagetti M, Giammarioli M, Lorusso A, Marcacci M, Di Domenico M, Ancora M, Curini V, Mangone I, Rinaldi A, Scialabba S, Caporale M, Di Pasquale A, Cammà C, Puglia I, Calistri P, Savini G |
| EPI_ISL_1336843, EPI_ISL_1336844, EPI_ISL_1336845, EPI_ISL_1336846, EPI_ISL_1336847, EPI_ISL_1336848, EPI_ISL_1336849, EPI_ISL_1336850, EPI_ISL_1336851, EPI_ISL_1336852 | Azienda Ospedaliera Terni | Istituto Zooprofilattico Sperimentale dell'Abruzzo e Molise "G. Caporale" | Palumbo M, Scaccetti A, Lorusso A, Marcacci M, Di Domenico M, Ancora M, Curini V, Mangone I, Rinaldi A, Scialabba S, Caporale M, Di Pasquale A, Cammà C, Puglia I, Calistri P, Savini G |
| EPI_ISL_1336853 | Istituto Zooprofilattico Sperimentale Umbria e Marche "Togo Rosati" | Istituto Zooprofilattico Sperimentale dell'Abruzzo e Molise "G. Caporale" | Biagetti M, Giammarioli M, Lorusso A, Marcacci M, Di Domenico M, Ancora M, Curini V, Mangone I, Rinaldi A, Scialabba S, Caporale M, Di Pasquale A, Cammà C, Puglia I, Calistri P, Savini G |
| EPI_ISL_1336854, EPI_ISL_1336855, EPI_ISL_1336856, EPI_ISL_1336857, EPI_ISL_1336858, EPI_ISL_1336859 | Azienda Ospedaliera Terni | Istituto Zooprofilattico Sperimentale dell'Abruzzo e Molise "G. Caporale" | Palumbo M, Scaccetti A, Lorusso A, Marcacci M, Di Domenico M, Ancora M, Curini V, Mangone I, Rinaldi A, Scialabba S, Caporale M, Di Pasquale A, Cammà C, Puglia I, Calistri P, Savini G |
| EPI_ISL_1336860 | Azienda USL Umbria 2 | Istituto Zooprofilattico Sperimentale dell'Abruzzo e Molise "G. Caporale" | Proietti A, Pistoni E, Lorusso A, Marcacci M, Di Domenico M, Ancora M, Curini V, Mangone I, Rinaldi A, Scialabba S, Caporale M, Di Pasquale A, Cammà C, Puglia I, Calistri P, Savini G |
| EPI_ISL_1336861 | Azienda USL Umbria 3 | Istituto Zooprofilattico Sperimentale dell'Abruzzo e Molise "G. Caporale" | Proietti A, Pistoni E, Lorusso A, Marcacci M, Di Domenico M, Ancora M, Curini V, Mangone I, Rinaldi A, Scialabba S, Caporale M, Di Pasquale A, Cammà C, Puglia I, Calistri P, Savini G |

| | | | |
|---|---|---|---|
| EPI_ISL_1336862 | Azienda Ospedaliera Terni | Istituto Zooprofilattico Sperimentale dell'Abruzzo e Molise "G. Caporale" | Palumbo M, Scaccetti A, Lorusso A, Marcacci M, Di Domenico M, Ancora M, Curini V, Mangone I, Rinaldi A, Scialabba S, Caporale M, Di Pasquale A, Cammà C, Puglia I, Calistri P, Savini G |
| EPI_ISL_1336863 | Laboratorio Analisi Osp. Città di Castello - Azienda USL Umbria1 | Istituto Zooprofilattico Sperimentale dell'Abruzzo e Molise "G. Caporale" | Malagigi V, Tacconi P, Lorusso A, Marcacci M, Di Domenico M, Ancora M, Curini V, Mangone I, Rinaldi A, Scialabba S, Caporale M, Di Pasquale A, Cammà C, Puglia I, Calistri P, Savini G |
| EPI_ISL_1336864 | Laboratorio Analisi Osp. Città di Castello - Azienda USL Umbria2 | Istituto Zooprofilattico Sperimentale dell'Abruzzo e Molise "G. Caporale" | Malagigi V, Tacconi P, Lorusso A, Marcacci M, Di Domenico M, Ancora M, Curini V, Mangone I, Rinaldi A, Scialabba S, Caporale M, Di Pasquale A, Cammà C, Puglia I, Calistri P, Savini G |
| EPI_ISL_1336865 | Laboratorio Analisi Osp. Città di Castello - Azienda USL Umbria3 | Istituto Zooprofilattico Sperimentale dell'Abruzzo e Molise "G. Caporale" | Malagigi V, Tacconi P, Lorusso A, Marcacci M, Di Domenico M, Ancora M, Curini V, Mangone I, Rinaldi A, Scialabba S, Caporale M, Di Pasquale A, Cammà C, Puglia I, Calistri P, Savini G |
| EPI_ISL_1336866 | Laboratorio Analisi Osp. Città di Castello - Azienda USL Umbria4 | Istituto Zooprofilattico Sperimentale dell'Abruzzo e Molise "G. Caporale" | Malagigi V, Tacconi P, Lorusso A, Marcacci M, Di Domenico M, Ancora M, Curini V, Mangone I, Rinaldi A, Scialabba S, Caporale M, Di Pasquale A, Cammà C, Puglia I, Calistri P, Savini G |
| EPI_ISL_1336867 | Laboratorio Analisi Osp. Città di Castello - Azienda USL Umbria5 | Istituto Zooprofilattico Sperimentale dell'Abruzzo e Molise "G. Caporale" | Malagigi V, Tacconi P, Lorusso A, Marcacci M, Di Domenico M, Ancora M, Curini V, Mangone I, Rinaldi A, Scialabba S, Caporale M, Di Pasquale A, Cammà C, Puglia I, Calistri P, Savini G |
| EPI_ISL_1336868 | Laboratorio Analisi Osp. Città di Castello - Azienda USL Umbria6 | Istituto Zooprofilattico Sperimentale dell'Abruzzo e Molise "G. Caporale" | Malagigi V, Tacconi P, Lorusso A, Marcacci M, Di Domenico M, Ancora M, Curini V, Mangone I, Rinaldi A, Scialabba S, Caporale M, Di Pasquale A, Cammà C, Puglia I, Calistri P, Savini G |
| EPI_ISL_1336869 | Laboratorio Analisi Osp. Città di Castello - Azienda USL Umbria7 | Istituto Zooprofilattico Sperimentale dell'Abruzzo e Molise "G. Caporale" | Malagigi V, Tacconi P, Lorusso A, Marcacci M, Di Domenico M, Ancora M, Curini V, Mangone I, Rinaldi A, Scialabba S, Caporale M, Di Pasquale A, Cammà C, Puglia I, Calistri P, Savini G |
| EPI_ISL_1336870 | Laboratorio Analisi Osp. Città di Castello - Azienda USL Umbria8 | Istituto Zooprofilattico Sperimentale dell'Abruzzo e Molise "G. Caporale" | Malagigi V, Tacconi P, Lorusso A, Marcacci M, Di Domenico M, Ancora M, Curini V, Mangone I, Rinaldi A, Scialabba S, Caporale M, Di Pasquale A, Cammà C, Puglia I, Calistri P, Savini G |
| EPI_ISL_1336871, EPI_ISL_1336872 | Università degli Studi di Perugia | Istituto Zooprofilattico Sperimentale dell'Abruzzo e Molise "G. Caporale" | Mencacci A, Camilloni B, Lorusso A, Marcacci M, Di Domenico M, Ancora M, Curini V, Mangone I, Rinaldi A, Scialabba S, Caporale M, Di Pasquale A, Cammà C, Puglia I, Calistri P, Savini G |
| EPI_ISL_1336873 | Istituto Zooprofilattico Sperimentale Umbria e Marche "Togo Rosati" | Istituto Zooprofilattico Sperimentale dell'Abruzzo e Molise "G. Caporale" | Biagetti M, Giammarioli M, Lorusso A, Marcacci M, Di Domenico M, Ancora M, Curini V, Mangone I, Rinaldi A, Scialabba S, Caporale M, Di Pasquale A, Cammà C, Puglia I, Calistri P, Savini G |
| EPI_ISL_1336874, EPI_ISL_1336875, EPI_ISL_1336876, EPI_ISL_1336877, EPI_ISL_1336878, EPI_ISL_1336879 | Università degli Studi di Perugia | Istituto Zooprofilattico Sperimentale dell'Abruzzo e Molise "G. Caporale" | Mencacci A, Camilloni B, Lorusso A, Marcacci M, Di Domenico M, Ancora M, Curini V, Mangone I, Rinaldi A, Scialabba S, Caporale M, Di Pasquale A, Cammà C, Puglia I, Calistri P, Savini G |
| EPI_ISL_1336880 | Istituto Zooprofilattico Sperimentale Umbria e Marche "Togo Rosati" | Istituto Zooprofilattico Sperimentale dell'Abruzzo e Molise "G. Caporale" | Biagetti M, Giammarioli M, Lorusso A, Marcacci M, Di Domenico M, Ancora M, Curini V, Mangone I, Rinaldi A, Scialabba S, Caporale M, Di Pasquale A, Cammà C, Puglia I, Calistri P, Savini G |
| EPI_ISL_1336881 | SIESP DIPARTIMENTO DI PREVENZIONE TERAMO | Istituto Zooprofilattico Sperimentale dell'Abruzzo e Molise "G. Caporale" | Lorusso A, Marcacci M, Di Domenico M, Ancora M, Curini V, Di Lollo Valeria, Mangone I, Rinaldi A, Scialabba S, Caporale M, Di Pasquale A, Cammà C, Puglia I, Calistri P, Savini G |
| EPI_ISL_1336882 | USCA Castel di Sangro | Istituto Zooprofilattico Sperimentale dell'Abruzzo e Molise "G. Caporale" | Lorusso A, Marcacci M, Di Domenico M, Ancora M, Curini V, Di Lollo Valeria, Mangone I, Rinaldi A, Scialabba S, Caporale M, Di Pasquale A, Cammà C, Puglia I, Calistri P, Savini G |
| EPI_ISL_1336883 | SIESP CHIETI - DRIVE IN GISSI | Istituto Zooprofilattico Sperimentale dell'Abruzzo e Molise "G. Caporale" | Lorusso A, Marcacci M, Di Domenico M, Ancora M, Curini V, Di Lollo Valeria, Mangone I, Rinaldi A, Scialabba S, Caporale M, Di Pasquale A, Cammà C, Puglia I, Calistri P, Savini G |
| EPI_ISL_1336884 | SIESP CHIETI - DRIVE IN LANCIANO | Istituto Zooprofilattico Sperimentale dell'Abruzzo e Molise "G. Caporale" | Lorusso A, Marcacci M, Di Domenico M, Ancora M, Curini V, Di Lollo Valeria, Mangone I, Rinaldi A, Scialabba S, Caporale M, Di Pasquale A, Cammà C, Puglia I, Calistri P, Savini G |
| EPI_ISL_1336885 | SIESP DIPARTIMENTO DI PREVENZIONE TERAMO | Istituto Zooprofilattico Sperimentale dell'Abruzzo e Molise "G. Caporale" | Lorusso A, Marcacci M, Di Domenico M, Ancora M, Curini V, Di Lollo Valeria, Mangone I, Rinaldi A, Scialabba S, Caporale M, Di Pasquale A, Cammà C, Puglia I, Calistri P, Savini G |
| EPI_ISL_1336886 | SIESP SULMONA | Istituto Zooprofilattico Sperimentale dell'Abruzzo e Molise "G. Caporale" | Lorusso A, Marcacci M, Di Domenico M, Ancora M, Curini V, Di Lollo Valeria, Mangone I, Rinaldi A, Scialabba S, Caporale M, Di Pasquale A, Cammà C, Puglia I, Calistri P, Savini G |
| EPI_ISL_1336887 | SIESP CHIETI - DRIVE IN LANCIANO | Istituto Zooprofilattico Sperimentale dell'Abruzzo e Molise "G. Caporale" | Lorusso A, Marcacci M, Di Domenico M, Ancora M, Curini V, Di Lollo Valeria, Mangone I, Rinaldi A, Scialabba S, Caporale M, Di Pasquale A, Cammà C, Puglia I, Calistri P, Savini G |
| EPI_ISL_1336888 | DIP. PREV. AVEZZANO SERVIZIO DI IGIENE EPIDEMIOLOGIA E SANITA' PUBBLICA | Istituto Zooprofilattico Sperimentale dell'Abruzzo e Molise "G. Caporale" | Lorusso A, Marcacci M, Di Domenico M, Ancora M, Curini V, Di Lollo Valeria, Mangone I, Rinaldi A, Scialabba S, Caporale M, Di Pasquale A, Cammà C, Puglia I, Calistri P, Savini G |
| EPI_ISL_1336889, EPI_ISL_1336890, EPI_ISL_1336891 | SIESP DIPARTIMENTO DI PREVENZIONE TERAMO | Istituto Zooprofilattico Sperimentale dell'Abruzzo e Molise "G. Caporale" | Lorusso A, Marcacci M, Di Domenico M, Ancora M, Curini V, Di Lollo Valeria, Mangone I, Rinaldi A, Scialabba S, Caporale M, Di Pasquale A, Cammà C, Puglia I, Calistri P, Savini G |
| EPI_ISL_1336892 | SIESP CHIETI - DRIVE IN ATESSA | Istituto Zooprofilattico Sperimentale dell'Abruzzo e Molise "G. Caporale" | Lorusso A, Marcacci M, Di Domenico M, Ancora M, Curini V, Di Lollo Valeria, Mangone I, Rinaldi A, Scialabba S, Caporale M, Di Pasquale A, Cammà C, Puglia I, Calistri P, Savini G |
| EPI_ISL_1336893, EPI_ISL_1336894 | SIESP DIPARTIMENTO DI PREVENZIONE TERAMO | Istituto Zooprofilattico Sperimentale dell'Abruzzo e Molise "G. Caporale" | Lorusso A, Marcacci M, Di Domenico M, Ancora M, Curini V, Di Lollo Valeria, Mangone I, Rinaldi A, Scialabba S, Caporale M, Di Pasquale A, Cammà C, Puglia I, Calistri P, Savini G |
| EPI_ISL_1336895 | USCA Pescina | Istituto Zooprofilattico Sperimentale dell'Abruzzo e Molise "G. Caporale" | Lorusso A, Marcacci M, Di Domenico M, Ancora M, Curini V, Di Lollo Valeria, Mangone I, Rinaldi A, Scialabba S, Caporale M, Di Pasquale A, Cammà C, Puglia I, Calistri P, Savini G |
| EPI_ISL_1336896 | USCA Castel di Sangro | Istituto Zooprofilattico Sperimentale dell'Abruzzo e Molise "G. Caporale" | Lorusso A, Marcacci M, Di Domenico M, Ancora M, Curini V, Di Lollo Valeria, Mangone I, Rinaldi A, Scialabba S, Caporale M, Di Pasquale A, Cammà C, Puglia I, Calistri P, Savini G |
| EPI_ISL_1336897 | SIESP CHIETI - DRIVE IN GISSI | Istituto Zooprofilattico Sperimentale dell'Abruzzo e Molise "G. Caporale" | Lorusso A, Marcacci M, Di Domenico M, Ancora M, Curini V, Di Lollo Valeria, Mangone I, Rinaldi A, Scialabba S, Caporale M, Di Pasquale A, Cammà C, Puglia I, Calistri P, Savini G |
| EPI_ISL_1336898 | USCA Sulmona | Istituto Zooprofilattico Sperimentale dell'Abruzzo e Molise "G. Caporale" | Lorusso A, Marcacci M, Di Domenico M, Ancora M, Curini V, Di Lollo Valeria, Mangone I, Rinaldi A, Scialabba S, Caporale M, Di Pasquale A, Cammà C, Puglia I, Calistri P, Savini G |
| EPI_ISL_1336899 | SIESP DIPARTIMENTO DI PREVENZIONE TERAMO | Istituto Zooprofilattico Sperimentale dell'Abruzzo e Molise "G. Caporale" | Lorusso A, Marcacci M, Di Domenico M, Ancora M, Curini V, Di Lollo Valeria, Mangone I, Rinaldi A, Scialabba S, Caporale M, Di Pasquale A, Cammà C, Puglia I, Calistri P, Savini G |
| EPI_ISL_1336900 | OSPEDALE "SAN CAMILLO" ATESSA - MEDICINA E LUNGODEGENZA | Istituto Zooprofilattico Sperimentale dell'Abruzzo e Molise "G. Caporale" | Lorusso A, Marcacci M, Di Domenico M, Ancora M, Curini V, Di Lollo Valeria, Mangone I, Rinaldi A, Scialabba S, Caporale M, Di Pasquale A, Cammà C, Puglia I, Calistri P, Savini G |
| EPI_ISL_1336901 | DIP. PREV. AVEZZANO SERVIZIO DI IGIENE EPIDEMIOLOGIA E SANITA' PUBBLICA | Istituto Zooprofilattico Sperimentale dell'Abruzzo e Molise "G. Caporale" | Lorusso A, Marcacci M, Di Domenico M, Ancora M, Curini V, Di Lollo Valeria, Mangone I, Rinaldi A, Scialabba S, Caporale M, Di Pasquale A, Cammà C, Puglia I, Calistri P, Savini G |
| EPI_ISL_1336902 | SIESP CHIETI - DRIVE IN GISSI | Istituto Zooprofilattico Sperimentale dell'Abruzzo e Molise "G. Caporale" | Lorusso A, Marcacci M, Di Domenico M, Ancora M, Curini V, Di Lollo Valeria, Mangone I, Rinaldi A, Scialabba S, Caporale M, Di Pasquale A, Cammà C, Puglia I, Calistri P, Savini G |
| EPI_ISL_1336903 | SIESP CHIETI - DRIVE IN LANCIANO | Istituto Zooprofilattico Sperimentale dell'Abruzzo e Molise "G. Caporale" | Lorusso A, Marcacci M, Di Domenico M, Ancora M, Curini V, Di Lollo Valeria, Mangone I, Rinaldi A, Scialabba S, Caporale M, Di Pasquale A, Cammà C, Puglia I, Calistri P, Savini G |
| EPI_ISL_1336904 | SIESP CHIETI - DRIVE IN ORTONA | Istituto Zooprofilattico Sperimentale dell'Abruzzo e Molise "G. Caporale" | Lorusso A, Marcacci M, Di Domenico M, Ancora M, Curini V, Di Lollo Valeria, Mangone I, Rinaldi A, Scialabba S, Caporale M, Di Pasquale A, Cammà C, Puglia I, Calistri P, Savini G |
| EPI_ISL_1336905 | SIESP DIPARTIMENTO DI PREVENZIONE TERAMO | Istituto Zooprofilattico Sperimentale dell'Abruzzo e Molise "G. Caporale" | Lorusso A, Marcacci M, Di Domenico M, Ancora M, Curini V, Di Lollo Valeria, Mangone I, Rinaldi A, Scialabba S, Caporale M, Di Pasquale A, Cammà C, Puglia I, Calistri P, Savini G |

| | | | |
|---|---|---|---|
| EPI_ISL_1336906 | SIESP DIPARTIMENTO DI PREVENZIONE CHIETI | Istituto Zooprofilattico Sperimentale dell'Abruzzo e Molise "G. Caporale" | Lorusso A, Marcacci M, Di Domenico M, Ancora M, Curini V, Di Lollo Valeria, Mangone I, Rinaldi A, Scialabba S, Caporale M, Di Pasquale A, Cammà C, Puglia I, Calistri P, Savini G |
| EPI_ISL_1336907, EPI_ISL_1336908, EPI_ISL_1336909 | SIESP SULMONA | Istituto Zooprofilattico Sperimentale dell'Abruzzo e Molise "G. Caporale" | Lorusso A, Marcacci M, Di Domenico M, Ancora M, Curini V, Di Lollo Valeria, Mangone I, Rinaldi A, Scialabba S, Caporale M, Di Pasquale A, Cammà C, Puglia I, Calistri P, Savini G |
| EPI_ISL_1336910 | SIESP CHIETI - DRIVE IN CHIETI | Istituto Zooprofilattico Sperimentale dell'Abruzzo e Molise "G. Caporale" | Lorusso A, Marcacci M, Di Domenico M, Ancora M, Curini V, Di Lollo Valeria, Mangone I, Rinaldi A, Scialabba S, Caporale M, Di Pasquale A, Cammà C, Puglia I, Calistri P, Savini G |
| EPI_ISL_1336911 | L'AQUILONE | Istituto Zooprofilattico Sperimentale dell'Abruzzo e Molise "G. Caporale" | Lorusso A, Marcacci M, Di Domenico M, Ancora M, Curini V, Di Lollo Valeria, Mangone I, Rinaldi A, Scialabba S, Caporale M, Di Pasquale A, Cammà C, Puglia I, Calistri P, Savini G |
| EPI_ISL_1336912 | SIESP DIPARTIMENTO DI PREVENZIONE TERAMO | Istituto Zooprofilattico Sperimentale dell'Abruzzo e Molise "G. Caporale" | Lorusso A, Marcacci M, Di Domenico M, Ancora M, Curini V, Di Lollo Valeria, Mangone I, Rinaldi A, Scialabba S, Caporale M, Di Pasquale A, Cammà C, Puglia I, Calistri P, Savini G |
| EPI_ISL_1336913 | OSPEDALE CIVILE SANT'OMERO - PRONTO SOCCORSO | Istituto Zooprofilattico Sperimentale dell'Abruzzo e Molise "G. Caporale" | Lorusso A, Marcacci M, Di Domenico M, Ancora M, Curini V, Di Lollo Valeria, Mangone I, Rinaldi A, Scialabba S, Caporale M, Di Pasquale A, Cammà C, Puglia I, Calistri P, Savini G |
| EPI_ISL_1336914 | SIESP DIPARTIMENTO DI PREVENZIONE TERAMO | Istituto Zooprofilattico Sperimentale dell'Abruzzo e Molise "G. Caporale" | Lorusso A, Marcacci M, Di Domenico M, Ancora M, Curini V, Di Lollo Valeria, Mangone I, Rinaldi A, Scialabba S, Caporale M, Di Pasquale A, Cammà C, Puglia I, Calistri P, Savini G |
| EPI_ISL_1336915 | SIESP CHIETI - DRIVE IN LANCIANO | Istituto Zooprofilattico Sperimentale dell'Abruzzo e Molise "G. Caporale" | Lorusso A, Marcacci M, Di Domenico M, Ancora M, Curini V, Di Lollo Valeria, Mangone I, Rinaldi A, Scialabba S, Caporale M, Di Pasquale A, Cammà C, Puglia I, Calistri P, Savini G |
| EPI_ISL_1336916, EPI_ISL_1336917, EPI_ISL_1336918, EPI_ISL_1336919 | SIESP DIPARTIMENTO DI PREVENZIONE TERAMO | Istituto Zooprofilattico Sperimentale dell'Abruzzo e Molise "G. Caporale" | Lorusso A, Marcacci M, Di Domenico M, Ancora M, Curini V, Di Lollo Valeria, Mangone I, Rinaldi A, Scialabba S, Caporale M, Di Pasquale A, Cammà C, Puglia I, Calistri P, Savini G |
| EPI_ISL_1336920 | SIESP CHIETI - DRIVE IN VASTO | Istituto Zooprofilattico Sperimentale dell'Abruzzo e Molise "G. Caporale" | Lorusso A, Marcacci M, Di Domenico M, Ancora M, Curini V, Di Lollo Valeria, Mangone I, Rinaldi A, Scialabba S, Caporale M, Di Pasquale A, Cammà C, Puglia I, Calistri P, Savini G |
| EPI_ISL_1336921 | SIESP DIPARTIMENTO DI PREVENZIONE TERAMO | Istituto Zooprofilattico Sperimentale dell'Abruzzo e Molise "G. Caporale" | Lorusso A, Marcacci M, Di Domenico M, Ancora M, Curini V, Di Lollo Valeria, Mangone I, Rinaldi A, Scialabba S, Caporale M, Di Pasquale A, Cammà C, Puglia I, Calistri P, Savini G |
| EPI_ISL_1336922 | DIP. PREV. AVEZZANO SERVIZIO DI IGIENE EPIDEMIOLOGIA E SANITA' PUBBLICA | Istituto Zooprofilattico Sperimentale dell'Abruzzo e Molise "G. Caporale" | Lorusso A, Marcacci M, Di Domenico M, Ancora M, Curini V, Di Lollo Valeria, Mangone I, Rinaldi A, Scialabba S, Caporale M, Di Pasquale A, Cammà C, Puglia I, Calistri P, Savini G |
| EPI_ISL_1336923 | Casa di Cura Di Lorenzo | Istituto Zooprofilattico Sperimentale dell'Abruzzo e Molise "G. Caporale" | Lorusso A, Marcacci M, Di Domenico M, Ancora M, Curini V, Di Lollo Valeria, Mangone I, Rinaldi A, Scialabba S, Caporale M, Di Pasquale A, Cammà C, Puglia I, Calistri P, Savini G |
| EPI_ISL_1336924 | SIESP SULMONA | Istituto Zooprofilattico Sperimentale dell'Abruzzo e Molise "G. Caporale" | Lorusso A, Marcacci M, Di Domenico M, Ancora M, Curini V, Di Lollo Valeria, Mangone I, Rinaldi A, Scialabba S, Caporale M, Di Pasquale A, Cammà C, Puglia I, Calistri P, Savini G |
| EPI_ISL_1336925 | DIP. PREV. AVEZZANO SERVIZIO DI IGIENE EPIDEMIOLOGIA E SANITA' PUBBLICA | Istituto Zooprofilattico Sperimentale dell'Abruzzo e Molise "G. Caporale" | Lorusso A, Marcacci M, Di Domenico M, Ancora M, Curini V, Di Lollo Valeria, Mangone I, Rinaldi A, Scialabba S, Caporale M, Di Pasquale A, Cammà C, Puglia I, Calistri P, Savini G |
| EPI_ISL_1336926 | SIESP CHIETI - DRIVE IN CHIETI | Istituto Zooprofilattico Sperimentale dell'Abruzzo e Molise "G. Caporale" | Lorusso A, Marcacci M, Di Domenico M, Ancora M, Curini V, Di Lollo Valeria, Mangone I, Rinaldi A, Scialabba S, Caporale M, Di Pasquale A, Cammà C, Puglia I, Calistri P, Savini G |
| EPI_ISL_1336927 | USCA Castel di Sangro | Istituto Zooprofilattico Sperimentale dell'Abruzzo e Molise "G. Caporale" | Lorusso A, Marcacci M, Di Domenico M, Ancora M, Curini V, Di Lollo Valeria, Mangone I, Rinaldi A, Scialabba S, Caporale M, Di Pasquale A, Cammà C, Puglia I, Calistri P, Savini G |
| EPI_ISL_1336928 | SIESP CHIETI - DRIVE IN VASTO | Istituto Zooprofilattico Sperimentale dell'Abruzzo e Molise "G. Caporale" | Lorusso A, Marcacci M, Di Domenico M, Ancora M, Curini V, Di Lollo Valeria, Mangone I, Rinaldi A, Scialabba S, Caporale M, Di Pasquale A, Cammà C, Puglia I, Calistri P, Savini G |
| EPI_ISL_1336929, EPI_ISL_1336930 | SIESP DIPARTIMENTO DI PREVENZIONE CHIETI | Istituto Zooprofilattico Sperimentale dell'Abruzzo e Molise "G. Caporale" | Lorusso A, Marcacci M, Di Domenico M, Ancora M, Curini V, Di Lollo Valeria, Mangone I, Rinaldi A, Scialabba S, Caporale M, Di Pasquale A, Cammà C, Puglia I, Calistri P, Savini G |
| EPI_ISL_1336931 | USCA Sulmona | Istituto Zooprofilattico Sperimentale dell'Abruzzo e Molise "G. Caporale" | Lorusso A, Marcacci M, Di Domenico M, Ancora M, Curini V, Di Lollo Valeria, Mangone I, Rinaldi A, Scialabba S, Caporale M, Di Pasquale A, Cammà C, Puglia I, Calistri P, Savini G |
| EPI_ISL_1336932 | DIP. PREV. AVEZZANO SERVIZIO DI IGIENE EPIDEMIOLOGIA E SANITA' PUBBLICA | Istituto Zooprofilattico Sperimentale dell'Abruzzo e Molise "G. Caporale" | Lorusso A, Marcacci M, Di Domenico M, Ancora M, Curini V, Di Lollo Valeria, Mangone I, Rinaldi A, Scialabba S, Caporale M, Di Pasquale A, Cammà C, Puglia I, Calistri P, Savini G |
| EPI_ISL_1336933 | SIESP CHIETI - DRIVE IN CHIETI | Istituto Zooprofilattico Sperimentale dell'Abruzzo e Molise "G. Caporale" | Lorusso A, Marcacci M, Di Domenico M, Ancora M, Curini V, Di Lollo Valeria, Mangone I, Rinaldi A, Scialabba S, Caporale M, Di Pasquale A, Cammà C, Puglia I, Calistri P, Savini G |
| EPI_ISL_1336934 | SIESP CHIETI - DRIVE IN VASTO | Istituto Zooprofilattico Sperimentale dell'Abruzzo e Molise "G. Caporale" | Lorusso A, Marcacci M, Di Domenico M, Ancora M, Curini V, Di Lollo Valeria, Mangone I, Rinaldi A, Scialabba S, Caporale M, Di Pasquale A, Cammà C, Puglia I, Calistri P, Savini G |
| EPI_ISL_1336935 | SIESP CHIETI - DRIVE IN LANCIANO | Istituto Zooprofilattico Sperimentale dell'Abruzzo e Molise "G. Caporale" | Lorusso A, Marcacci M, Di Domenico M, Ancora M, Curini V, Di Lollo Valeria, Mangone I, Rinaldi A, Scialabba S, Caporale M, Di Pasquale A, Cammà C, Puglia I, Calistri P, Savini G |
| EPI_ISL_1336936 | SIESP CHIETI - DRIVE IN ORTONA | Istituto Zooprofilattico Sperimentale dell'Abruzzo e Molise "G. Caporale" | Lorusso A, Marcacci M, Di Domenico M, Ancora M, Curini V, Di Lollo Valeria, Mangone I, Rinaldi A, Scialabba S, Caporale M, Di Pasquale A, Cammà C, Puglia I, Calistri P, Savini G |
| EPI_ISL_1336937 | SIESP CHIETI - DRIVE IN GISSI | Istituto Zooprofilattico Sperimentale dell'Abruzzo e Molise "G. Caporale" | Lorusso A, Marcacci M, Di Domenico M, Ancora M, Curini V, Di Lollo Valeria, Mangone I, Rinaldi A, Scialabba S, Caporale M, Di Pasquale A, Cammà C, Puglia I, Calistri P, Savini G |
| EPI_ISL_1336938 | SIESP CHIETI - DRIVE IN LANCIANO | Istituto Zooprofilattico Sperimentale dell'Abruzzo e Molise "G. Caporale" | Lorusso A, Marcacci M, Di Domenico M, Ancora M, Curini V, Di Lollo Valeria, Mangone I, Rinaldi A, Scialabba S, Caporale M, Di Pasquale A, Cammà C, Puglia I, Calistri P, Savini G |
| EPI_ISL_1336939, EPI_ISL_1336940 | USCA Castel di Sangro | Istituto Zooprofilattico Sperimentale dell'Abruzzo e Molise "G. Caporale" | Lorusso A, Marcacci M, Di Domenico M, Ancora M, Curini V, Di Lollo Valeria, Mangone I, Rinaldi A, Scialabba S, Caporale M, Di Pasquale A, Cammà C, Puglia I, Calistri P, Savini G |
| EPI_ISL_1336941 | USCA Avezzano | Istituto Zooprofilattico Sperimentale dell'Abruzzo e Molise "G. Caporale" | Lorusso A, Marcacci M, Di Domenico M, Ancora M, Curini V, Di Lollo Valeria, Mangone I, Rinaldi A, Scialabba S, Caporale M, Di Pasquale A, Cammà C, Puglia I, Calistri P, Savini G |
| EPI_ISL_1336942 | SIESP DIPARTIMENTO DI PREVENZIONE TERAMO | Istituto Zooprofilattico Sperimentale dell'Abruzzo e Molise "G. Caporale" | Lorusso A, Marcacci M, Di Domenico M, Ancora M, Curini V, Di Lollo Valeria, Mangone I, Rinaldi A, Scialabba S, Caporale M, Di Pasquale A, Cammà C, Puglia I, Calistri P, Savini G |
| EPI_ISL_1336943 | SIESP CHIETI - DRIVE IN VASTO | Istituto Zooprofilattico Sperimentale dell'Abruzzo e Molise "G. Caporale" | Lorusso A, Marcacci M, Di Domenico M, Ancora M, Curini V, Di Lollo Valeria, Mangone I, Rinaldi A, Scialabba S, Caporale M, Di Pasquale A, Cammà C, Puglia I, Calistri P, Savini G |
| EPI_ISL_1336944 | USCA Tagliacozzo | Istituto Zooprofilattico Sperimentale dell'Abruzzo e Molise "G. Caporale" | Lorusso A, Marcacci M, Di Domenico M, Ancora M, Curini V, Di Lollo Valeria, Mangone I, Rinaldi A, Scialabba S, Caporale M, Di Pasquale A, Cammà C, Puglia I, Calistri P, Savini G |
| EPI_ISL_1336945 | SIESP DIPARTIMENTO DI PREVENZIONE CHIETI | Istituto Zooprofilattico Sperimentale dell'Abruzzo e Molise "G. Caporale" | Lorusso A, Marcacci M, Di Domenico M, Ancora M, Curini V, Di Lollo Valeria, Mangone I, Rinaldi A, Scialabba S, Caporale M, Di Pasquale A, Cammà C, Puglia I, Calistri P, Savini G |
| EPI_ISL_1336946 | SIESP DIPARTIMENTO DI PREVENZIONE TERAMO | Istituto Zooprofilattico Sperimentale dell'Abruzzo e Molise "G. Caporale" | Lorusso A, Marcacci M, Di Domenico M, Ancora M, Curini V, Di Lollo Valeria, Mangone I, Rinaldi A, Scialabba S, Caporale M, Di Pasquale A, Cammà C, Puglia I, Calistri P, Savini G |
| EPI_ISL_1336947 | USCA Pescina | Istituto Zooprofilattico Sperimentale dell'Abruzzo e Molise "G. Caporale" | Lorusso A, Marcacci M, Di Domenico M, Ancora M, Curini V, Di Lollo Valeria, Mangone I, Rinaldi A, Scialabba S, Caporale M, Di Pasquale A, Cammà C, Puglia I, Calistri P, Savini G |

| | | | |
|---|---|---|---|
| EPI_ISL_1336948, EPI_ISL_1336949 | SIESP DIPARTIMENTO DI PREVENZIONE TERAMO | Istituto Zooprofilattico Sperimentale dell'Abruzzo e Molise "G. Caporale" | Lorusso A, Marcacci M, Di Domenico M, Ancora M, Curini V, Di Lollo Valeria, Mangone I, Rinaldi A, Scialabba S, Caporale M, Di Pasquale A, Cammà C, Puglia I, Calistri P, Savini G |
| EPI_ISL_1336950 | SIESP CHIETI - DRIVE IN LANCIANO | Istituto Zooprofilattico Sperimentale dell'Abruzzo e Molise "G. Caporale" | Lorusso A, Marcacci M, Di Domenico M, Ancora M, Curini V, Di Lollo Valeria, Mangone I, Rinaldi A, Scialabba S, Caporale M, Di Pasquale A, Cammà C, Puglia I, Calistri P, Savini G |
| EPI_ISL_1336951 | OSPEDALE CIVILE TERAMO - CENTRO TRASFUSIONALE | Istituto Zooprofilattico Sperimentale dell'Abruzzo e Molise "G. Caporale" | Lorusso A, Marcacci M, Di Domenico M, Ancora M, Curini V, Di Lollo Valeria, Mangone I, Rinaldi A, Scialabba S, Caporale M, Di Pasquale A, Cammà C, Puglia I, Calistri P, Savini G |
| EPI_ISL_1336952, EPI_ISL_1336953, EPI_ISL_1336954 | SIESP DIPARTIMENTO DI PREVENZIONE TERAMO | Istituto Zooprofilattico Sperimentale dell'Abruzzo e Molise "G. Caporale" | Lorusso A, Marcacci M, Di Domenico M, Ancora M, Curini V, Di Lollo Valeria, Mangone I, Rinaldi A, Scialabba S, Caporale M, Di Pasquale A, Cammà C, Puglia I, Calistri P, Savini G |
| EPI_ISL_1336955 | SIESP CHIETI - DRIVE IN CHIETI | Istituto Zooprofilattico Sperimentale dell'Abruzzo e Molise "G. Caporale" | Lorusso A, Marcacci M, Di Domenico M, Ancora M, Curini V, Di Lollo Valeria, Mangone I, Rinaldi A, Scialabba S, Caporale M, Di Pasquale A, Cammà C, Puglia I, Calistri P, Savini G |
| EPI_ISL_1336956 | SIESP DIPARTIMENTO DI PREVENZIONE TERAMO | Istituto Zooprofilattico Sperimentale dell'Abruzzo e Molise "G. Caporale" | Lorusso A, Marcacci M, Di Domenico M, Ancora M, Curini V, Di Lollo Valeria, Mangone I, Rinaldi A, Scialabba S, Caporale M, Di Pasquale A, Cammà C, Puglia I, Calistri P, Savini G |
| EPI_ISL_1336957 | SIESP CHIETI - DRIVE IN VASTO | Istituto Zooprofilattico Sperimentale dell'Abruzzo e Molise "G. Caporale" | Lorusso A, Marcacci M, Di Domenico M, Ancora M, Curini V, Di Lollo Valeria, Mangone I, Rinaldi A, Scialabba S, Caporale M, Di Pasquale A, Cammà C, Puglia I, Calistri P, Savini G |
| EPI_ISL_1336958 | SIESP CHIETI - DRIVE IN ORTONA | Istituto Zooprofilattico Sperimentale dell'Abruzzo e Molise "G. Caporale" | Lorusso A, Marcacci M, Di Domenico M, Ancora M, Curini V, Di Lollo Valeria, Mangone I, Rinaldi A, Scialabba S, Caporale M, Di Pasquale A, Cammà C, Puglia I, Calistri P, Savini G |
| EPI_ISL_1336959 | SIESP CHIETI - DRIVE IN VASTO | Istituto Zooprofilattico Sperimentale dell'Abruzzo e Molise "G. Caporale" | Lorusso A, Marcacci M, Di Domenico M, Ancora M, Curini V, Di Lollo Valeria, Mangone I, Rinaldi A, Scialabba S, Caporale M, Di Pasquale A, Cammà C, Puglia I, Calistri P, Savini G |
| EPI_ISL_1336960 | SIESP DIPARTIMENTO DI PREVENZIONE TERAMO | Istituto Zooprofilattico Sperimentale dell'Abruzzo e Molise "G. Caporale" | Lorusso A, Marcacci M, Di Domenico M, Ancora M, Curini V, Di Lollo Valeria, Mangone I, Rinaldi A, Scialabba S, Caporale M, Di Pasquale A, Cammà C, Puglia I, Calistri P, Savini G |
| EPI_ISL_1336961 | SIESP CHIETI - DRIVE IN CHIETI | Istituto Zooprofilattico Sperimentale dell'Abruzzo e Molise "G. Caporale" | Lorusso A, Marcacci M, Di Domenico M, Ancora M, Curini V, Di Lollo Valeria, Mangone I, Rinaldi A, Scialabba S, Caporale M, Di Pasquale A, Cammà C, Puglia I, Calistri P, Savini G |
| EPI_ISL_1336962 | OSPEDALE CIVILE GIULIANOVA - PRONTO SOCCORSO | Istituto Zooprofilattico Sperimentale dell'Abruzzo e Molise "G. Caporale" | Lorusso A, Marcacci M, Di Domenico M, Ancora M, Curini V, Di Lollo Valeria, Mangone I, Rinaldi A, Scialabba S, Caporale M, Di Pasquale A, Cammà C, Puglia I, Calistri P, Savini G |
| EPI_ISL_1336963 | SIESP CHIETI - DRIVE IN ORTONA | Istituto Zooprofilattico Sperimentale dell'Abruzzo e Molise "G. Caporale" | Lorusso A, Marcacci M, Di Domenico M, Ancora M, Curini V, Di Lollo Valeria, Mangone I, Rinaldi A, Scialabba S, Caporale M, Di Pasquale A, Cammà C, Puglia I, Calistri P, Savini G |
| EPI_ISL_1347245 | INT Fondazione Pascale | INT Fondazione Pascale | INT Fondazione Pascale |
| EPI_ISL_1347505 | S.C. ANALISI CLINICHE - Settore Biologia Molecolare AUSL Valle d'Aosta | S.S. Genetica e Tecniche Omiche Avanzate Istituto Zooprofilattico Sperimentale del Piemonte, Liguria e Valle d'Aosta | Chiara Beltramo, Elena Bozzetta, Maria Goria, Pier Luigi Acutis, Simone Peletto |
| EPI_ISL_1347549 | S.C. ANALISI CLINICHE - Settore Biologia Molecolare | S.S. Genetica e Tecniche Omiche Avanzate Istituto Zooprofilattico Sperimentale del Piemonte, Liguria e Valle d'Aosta | Chiara Beltramo, Elena Bozzetta, Maria Goria, Pier Luigi Acutis, Simone Peletto |
| EPI_ISL_1359088, EPI_ISL_1359089, EPI_ISL_1359090, EPI_ISL_1359091, EPI_ISL_1359092, EPI_ISL_1359093, EPI_ISL_1359094, EPI_ISL_1359095, EPI_ISL_1359096, EPI_ISL_1359097, EPI_ISL_1359098, EPI_ISL_1359099, EPI_ISL_1359100, EPI_ISL_1359101, EPI_ISL_1359102, EPI_ISL_1359103, EPI_ISL_1359104, EPI_ISL_1359105, EPI_ISL_1359106, EPI_ISL_1359107, EPI_ISL_1359108, EPI_ISL_1359109, EPI_ISL_1359110, EPI_ISL_1359111, EPI_ISL_1359112, EPI_ISL_1359113, EPI_ISL_1359114, EPI_ISL_1359115, EPI_ISL_1359116, EPI_ISL_1359117, EPI_ISL_1359118, EPI_ISL_1359119, EPI_ISL_1359120, EPI_ISL_1359121, EPI_ISL_1359122, EPI_ISL_1359123, EPI_ISL_1359124, EPI_ISL_1359125, EPI_ISL_1359126, EPI_ISL_1359127, EPI_ISL_1359128, EPI_ISL_1359129, EPI_ISL_1359130, EPI_ISL_1359131, EPI_ISL_1359132, EPI_ISL_1359133, EPI_ISL_1359135, EPI_ISL_1359136, EPI_ISL_1359137, EPI_ISL_1359138, EPI_ISL_1359139, EPI_ISL_1359140, EPI_ISL_1359141, EPI_ISL_1359142, EPI_ISL_1359143, EPI_ISL_1359144, EPI_ISL_1359145, EPI_ISL_1359146, EPI_ISL_1359147, EPI_ISL_1359148, EPI_ISL_1359149, EPI_ISL_1359150, EPI_ISL_1359151, EPI_ISL_1359152, EPI_ISL_1359153, EPI_ISL_1359154, EPI_ISL_1359155, EPI_ISL_1359156, EPI_ISL_1359157, EPI_ISL_1359158, EPI_ISL_1359159, EPI_ISL_1359160, EPI_ISL_1359161, EPI_ISL_1359162, EPI_ISL_1359163, EPI_ISL_1359164, EPI_ISL_1359165, EPI_ISL_1359166, EPI_ISL_1359167, EPI_ISL_1359168, EPI_ISL_1359169, EPI_ISL_1359170, EPI_ISL_1359171, EPI_ISL_1359172, EPI_ISL_1359173, EPI_ISL_1359174, EPI_ISL_1359175, EPI_ISL_1359176, EPI_ISL_1359177, EPI_ISL_1359178, EPI_ISL_1359179, EPI_ISL_1359180, EPI_ISL_1359181, EPI_ISL_1359182, EPI_ISL_1359183, EPI_ISL_1359184, EPI_ISL_1359185, EPI_ISL_1359186, EPI_ISL_1359187, EPI_ISL_1359188, EPI_ISL_1359189, EPI_ISL_1359190, EPI_ISL_1359191, EPI_ISL_1359192, EPI_ISL_1359193, EPI_ISL_1359194, EPI_ISL_1359195, EPI_ISL_1359196, EPI_ISL_1359197, EPI_ISL_1359198, EPI_ISL_1359199, EPI_ISL_1359200, EPI_ISL_1359202, EPI_ISL_1359203, EPI_ISL_1359204, EPI_ISL_1359205, EPI_ISL_1359206, EPI_ISL_1359207, EPI_ISL_1359208, EPI_ISL_1359209, EPI_ISL_1359210, EPI_ISL_1359211, EPI_ISL_1359212, EPI_ISL_1359213, EPI_ISL_1359215, EPI_ISL_1359216, EPI_ISL_1359217, EPI_ISL_1359218, EPI_ISL_1359219, EPI_ISL_1359220, EPI_ISL_1359221, EPI_ISL_1359222, EPI_ISL_1359223, EPI_ISL_1359224, EPI_ISL_1359225, EPI_ISL_1359226, EPI_ISL_1359227, EPI_ISL_1359228, EPI_ISL_1359229, EPI_ISL_1359231, EPI_ISL_1359232, EPI_ISL_1359233, EPI_ISL_1359234, EPI_ISL_1359235, EPI_ISL_1359236, EPI_ISL_1359237, EPI_ISL_1359238, EPI_ISL_1359239, EPI_ISL_1359240, EPI_ISL_1359241, EPI_ISL_1359242, EPI_ISL_1359243, EPI_ISL_1359244, EPI_ISL_1359245, EPI_ISL_1359246, EPI_ISL_1359247, EPI_ISL_1359248, EPI_ISL_1359249, EPI_ISL_1359250, EPI_ISL_1359251, EPI_ISL_1359252, EPI_ISL_1359253, EPI_ISL_1359254, EPI_ISL_1359255, EPI_ISL_1359256, EPI_ISL_1359257, EPI_ISL_1359258, EPI_ISL_1359259, EPI_ISL_1359260, EPI_ISL_1359261, EPI_ISL_1359262, EPI_ISL_1359263, EPI_ISL_1359264, EPI_ISL_1359265, EPI_ISL_1359266, EPI_ISL_1359267, EPI_ISL_1359268, EPI_ISL_1359269, EPI_ISL_1359270, EPI_ISL_1359271, EPI_ISL_1359272, EPI_ISL_1359273, EPI_ISL_1359274, EPI_ISL_1359275, EPI_ISL_1359276, EPI_ISL_1359277, EPI_ISL_1359278, EPI_ISL_1359279, EPI_ISL_1359280, EPI_ISL_1359281, EPI_ISL_1359282, EPI_ISL_1359283, EPI_ISL_1359284, EPI_ISL_1359285, EPI_ISL_1359286, EPI_ISL_1359287, EPI_ISL_1359288, EPI_ISL_1359289, EPI_ISL_1359290, EPI_ISL_1359291, EPI_ISL_1359292, EPI_ISL_1359293, EPI_ISL_1359294, EPI_ISL_1359295, EPI_ISL_1359296, EPI_ISL_1359297, EPI_ISL_1359298, EPI_ISL_1359299, EPI_ISL_1359300, EPI_ISL_1359301, EPI_ISL_1359302, EPI_ISL_1359303, EPI_ISL_1359304, EPI_ISL_1359305, EPI_ISL_1359306, EPI_ISL_1359307, EPI_ISL_1359308, EPI_ISL_1359309, EPI_ISL_1359310, EPI_ISL_1359311, EPI_ISL_1359312, EPI_ISL_1359313, EPI_ISL_1359314, EPI_ISL_1359315, EPI_ISL_1359316, EPI_ISL_1359317, EPI_ISL_1359318, EPI_ISL_1359319, EPI_ISL_1359320, EPI_ISL_1359321, EPI_ISL_1359322, EPI_ISL_1359323, EPI_ISL_1359324, EPI_ISL_1359325, EPI_ISL_1359326, EPI_ISL_1359327, EPI_ISL_1359328, EPI_ISL_1359329, EPI_ISL_1359330, EPI_ISL_1359331, EPI_ISL_1359332, EPI_ISL_1359333, EPI_ISL_1359334, EPI_ISL_1359335, EPI_ISL_1359336, EPI_ISL_1359337, EPI_ISL_1359338, EPI_ISL_1359339, EPI_ISL_1359340, EPI_ISL_1359341, EPI_ISL_1359342, EPI_ISL_1359343, EPI_ISL_1359344, EPI_ISL_1359345, EPI_ISL_1359346, EPI_ISL_1359347, EPI_ISL_1359348, EPI_ISL_1359349, EPI_ISL_1359350, EPI_ISL_1359351, EPI_ISL_1359352, EPI_ISL_1359353, EPI_ISL_1359354, EPI_ISL_1359355, EPI_ISL_1359356, EPI_ISL_1359357, EPI_ISL_1359358, EPI_ISL_1359359, EPI_ISL_1359360, EPI_ISL_1359361, EPI_ISL_1359362, EPI_ISL_1359363, EPI_ISL_1359364, EPI_ISL_1359365, EPI_ISL_1359366, EPI_ISL_1359367, EPI_ISL_1359368, EPI_ISL_1359369, EPI_ISL_1359370, EPI_ISL_1359371, EPI_ISL_1359372, EPI_ISL_1359373, EPI_ISL_1359374, EPI_ISL_1359375, EPI_ISL_1359376, EPI_ISL_1359377, EPI_ISL_1359378, EPI_ISL_1359379, EPI_ISL_1359380, EPI_ISL_1359381, EPI_ISL_1359382, EPI_ISL_1359383, EPI_ISL_1359384, EPI_ISL_1359385, EPI_ISL_1359386, EPI_ISL_1359387, EPI_ISL_1359388, EPI_ISL_1359389, EPI_ISL_1359390, EPI_ISL_1359391, EPI_ISL_1359392, EPI_ISL_1359393, EPI_ISL_1359394, EPI_ISL_1359395, EPI_ISL_1359396, EPI_ISL_1359397, EPI_ISL_1359398, EPI_ISL_1359399, EPI_ISL_1359400, EPI_ISL_1359401, EPI_ISL_1359402, EPI_ISL_1359403, EPI_ISL_1359404, EPI_ISL_1359405, EPI_ISL_1359406, EPI_ISL_1359407, EPI_ISL_1359408, EPI_ISL_1359409, EPI_ISL_1359410, EPI_ISL_1359411, EPI_ISL_1359412, EPI_ISL_1359413, EPI_ISL_1359414, EPI_ISL_1359415, EPI_ISL_1359416, EPI_ISL_1359417, EPI_ISL_1359418, EPI_ISL_1359419, EPI_ISL_1359420, EPI_ISL_1359421, EPI_ISL_1359422, EPI_ISL_1359423, EPI_ISL_1359424, EPI_ISL_1359425, EPI_ISL_1359426, EPI_ISL_1359427, EPI_ISL_1359428, EPI_ISL_1359429, EPI_ISL_1359430, EPI_ISL_1359431, EPI_ISL_1359432, EPI_ISL_1359433, EPI_ISL_1359434, EPI_ISL_1359435, EPI_ISL_1359436, EPI_ISL_1359437, EPI_ISL_1359438, EPI_ISL_1359439, EPI_ISL_1359440, EPI_ISL_1359441, EPI_ISL_1359442, EPI_ISL_1359443, EPI_ISL_1359444, EPI_ISL_1359445, EPI_ISL_1359446, EPI_ISL_1359447, EPI_ISL_1359448, EPI_ISL_1359449, EPI_ISL_1359450, EPI_ISL_1359451, EPI_ISL_1359452, EPI_ISL_1359453, EPI_ISL_1359454, EPI_ISL_1359455, EPI_ISL_1359456, EPI_ISL_1359457, EPI_ISL_1359458, EPI_ISL_1359459, EPI_ISL_1359460, EPI_ISL_1359461, EPI_ISL_1359462, EPI_ISL_1359463, EPI_ISL_1359464, EPI_ISL_1359465, EPI_ISL_1359466, EPI_ISL_1359467, EPI_ISL_1359468, EPI_ISL_1359469, EPI_ISL_1359470, EPI_ISL_1359471, EPI_ISL_1359472, EPI_ISL_1359473, EPI_ISL_1359474, EPI_ISL_1359475, EPI_ISL_1359476, EPI_ISL_1359477, EPI_ISL_1359478, EPI_ISL_1359479, EPI_ISL_1359480, EPI_ISL_1359481, EPI_ISL_1359482, EPI_ISL_1359483, EPI_ISL_1359484, EPI_ISL_1359485, EPI_ISL_1359486, EPI_ISL_1359487, EPI_ISL_1359489, EPI_ISL_1359490, EPI_ISL_1359491, EPI_ISL_1359492, EPI_ISL_1359493, EPI_ISL_1359494, EPI_ISL_1359495, EPI_ISL_1359496, EPI_ISL_1359497, EPI_ISL_1359498, EPI_ISL_1359499, EPI_ISL_1359500, EPI_ISL_1359501, EPI_ISL_1359502, EPI_ISL_1359503, EPI_ISL_1359504, EPI_ISL_1359505, EPI_ISL_1359506, EPI_ISL_1359507, EPI_ISL_1359508, EPI_ISL_1359509, EPI_ISL_1359510, EPI_ISL_1359511, EPI_ISL_1359512, EPI_ISL_1359513, EPI_ISL_1359514, EPI_ISL_1359515, EPI_ISL_1359516, EPI_ISL_1359517, EPI_ISL_1359518, EPI_ISL_1359519, EPI_ISL_1359520, EPI_ISL_1359521, EPI_ISL_1359522, EPI_ISL_1359523, EPI_ISL_1359524, EPI_ISL_1359525, EPI_ISL_1359526, EPI_ISL_1359527, EPI_ISL_1359528, EPI_ISL_1359529, EPI_ISL_1359530, EPI_ISL_1359531, EPI_ISL_1359532, EPI_ISL_1359533, EPI_ISL_1359534, EPI_ISL_1359535, EPI_ISL_1359536, EPI_ISL_1359537, EPI_ISL_1359538, EPI_ISL_1359539, EPI_ISL_1359540, EPI_ISL_1359541, EPI_ISL_1359542, EPI_ISL_1359543, EPI_ISL_1359544, EPI_ISL_1359545, EPI_ISL_1359546, EPI_ISL_1359547, EPI_ISL_1359548, EPI_ISL_1359549, EPI_ISL_1359550, EPI_ISL_1359551, EPI_ISL_1359552, EPI_ISL_1359553, EPI_ISL_1359554, EPI_ISL_1359555, EPI_ISL_1359556, EPI_ISL_1359557, EPI_ISL_1359558, EPI_ISL_1359559, EPI_ISL_1359560, EPI_ISL_1359561, EPI_ISL_1359562, EPI_ISL_1359563, EPI_ISL_1359564, EPI_ISL_1359565, EPI_ISL_1359566, EPI_ISL_1359567, EPI_ISL_1359568, EPI_ISL_1359569, EPI_ISL_1359570, EPI_ISL_1359571, EPI_ISL_1359572, EPI_ISL_1359573, EPI_ISL_1359574, EPI_ISL_1359575, EPI_ISL_1359576, EPI_ISL_1359577, EPI_ISL_1359578, EPI_ISL_1359579, EPI_ISL_1359580, EPI_ISL_1359581, EPI_ISL_1359582, EPI_ISL_1359583, EPI_ISL_1359584, EPI_ISL_1359585, EPI_ISL_1359586, EPI_ISL_1359587, EPI_ISL_1359588, EPI_ISL_1359589, EPI_ISL_1359590, EPI_ISL_1359591, EPI_ISL_1359592, EPI_ISL_1359593, EPI_ISL_1359594, EPI_ISL_1359595, EPI_ISL_1359596, EPI_ISL_1359597, EPI_ISL_1359598, EPI_ISL_1359599, EPI_ISL_1359600, EPI_ISL_1359601, EPI_ISL_1359602, EPI_ISL_1359603, EPI_ISL_1359604, EPI_ISL_1359605, EPI_ISL_1359606, EPI_ISL_1359607, EPI_ISL_1359608, EPI_ISL_1359609, EPI_ISL_1359610, EPI_ISL_1359611, EPI_ISL_1359612, EPI_ISL_1359613, EPI_ISL_1359614, EPI_ISL_1359615, EPI_ISL_1359616, EPI_ISL_1359617, EPI_ISL_1359618, EPI_ISL_1359619, EPI_ISL_1359620, EPI_ISL_1359621, EPI_ISL_1359622, EPI_ISL_1359623, EPI_ISL_1359624, EPI_ISL_1359625, EPI_ISL_1359626, EPI_ISL_1359627, EPI_ISL_1359628, EPI_ISL_1359629, EPI_ISL_1359630, EPI_ISL_1359631, EPI_ISL_1359632, EPI_ISL_1359633, EPI_ISL_1359634, EPI_ISL_1359635, EPI_ISL_1359636, EPI_ISL_1359637, EPI_ISL_1359638, EPI_ISL_1359639, EPI_ISL_1359640, EPI_ISL_1359641, EPI_ISL_1359642, EPI_ISL_1359643, EPI_ISL_1359644, EPI_ISL_1359645, EPI_ISL_1359646, EPI_ISL_1359647, EPI_ISL_1359648, EPI_ISL_1359649, EPI_ISL_1359650, EPI_ISL_1359651, EPI_ISL_1359652, EPI_ISL_1359653, EPI_ISL_1359654, EPI_ISL_1359655, EPI_ISL_1359656, EPI_ISL_1359657, EPI_ISL_1359658, EPI_ISL_1359659, EPI_ISL_1359660, EPI_ISL_1359661, EPI_ISL_1359662, EPI_ISL_1359663, EPI_ISL_1359664, EPI_ISL_1359665, EPI_ISL_1359666, EPI_ISL_1359667, EPI_ISL_1359668, EPI_ISL_1359669, EPI_ISL_1359670, EPI_ISL_1359671, EPI_ISL_1359672, EPI_ISL_1359673, EPI_ISL_1359674, EPI_ISL_1359675, EPI_ISL_1359676, EPI_ISL_1359677, EPI_ISL_1359678, EPI_ISL_1359679, EPI_ISL_1359680, EPI_ISL_1359681, EPI_ISL_1359682, EPI_ISL_1359683, EPI_ISL_1359684, EPI_ISL_1359685, EPI_ISL_1359686, EPI_ISL_1359687, EPI_ISL_1359688, EPI_ISL_1359689, EPI_ISL_1359690, EPI_ISL_1359691, EPI_ISL_1359692, EPI_ISL_1359693, EPI_ISL_1359694, EPI_ISL_1359695, EPI_ISL_1359696, EPI_ISL_1359697, EPI_ISL_1359698, EPI_ISL_1359699, EPI_ISL_1359700, EPI_ISL_1359701, EPI_ISL_1359702, EPI_ISL_1359703, EPI_ISL_1359704, EPI_ISL_1359705, EPI_ISL_1359706, EPI_ISL_1359707, EPI_ISL_1359708, EPI_ISL_1359709, EPI_ISL_1359710, EPI_ISL_1359711, EPI_ISL_1359712, EPI_ISL_1359713, EPI_ISL_1359714, EPI_ISL_1359715, EPI_ISL_1359716, EPI_ISL_1359717, EPI_ISL_1359718, EPI_ISL_1359719, EPI_ISL_1359720, EPI_ISL_1359721, EPI_ISL_1359722, EPI_ISL_1359723, EPI_ISL_1359724, EPI_ISL_1359725, EPI_ISL_1359726, EPI_ISL_1359727, EPI_ISL_1359728, EPI_ISL_1359729, EPI_ISL_1359730, EPI_ISL_1359731, EPI_ISL_1359732, EPI_ISL_1359733, EPI_ISL_1359734, EPI_ISL_1359735, EPI_ISL_1359736, EPI_ISL_1359737, EPI_ISL_1359738, EPI_ISL_1359740, EPI_ISL_1359741, EPI_ISL_1359742 | | | |
| see above | ASL Napoli 1 Centro | AMES Centro Polidiagnostico Strumentale S.r.l. | "Giovanni Savarese, Raffaella Ruggiero, Eloisa Evangelista, Antonella Di Carlo, Luisa Circelli, Luigi D'Amore, Nadia Petrillo, Monica Ianniello, Roberto Sirica,Maurizio D'Amora, Antonio Fico" |
| EPI_ISL_1364620 | Villa Tiberia Hospital | INMI Lazzaro Spallanzani IRCCS | F Messina, M Rueca, O Butera, G Bonfiglio, F Santini, CEM Gruber, B Bartolini, E Giombini, A Di Caro, MR Capobianchi |
| EPI_ISL_1365746 | Laboratorio di Riferimento Regionale della Sicilia Occidentale per l'Emergenza COVID-19 | Laboratorio di Riferimento Regionale della Sicilia Occidentale per l'Emergenza COVID-19 | Fabio Tramuto, Carmelo Massimo Maida, Daniela Di Naro, Giulia Randazzo, Walter Mazzucco, Giorgio Graziano, Vincenzo Restivo, Claudio Costantino, Francesco Vitale |

| | | | |
|---|---|---|---|
| EPI_ISL_1372540 | Laboratorio Biologia Molecolare Sars Cov2 - UOC Laboratorio Analisi - Servizio Medicina di Laboratorio, Ospedale "San Francesco" - ATS-ASSL Nuoro | Laboratorio specialistico UOC Ematologia - Ospedale "San Francesco" - ATS-ASSL Nuoro | Piras Giovanna, Asproni Rosanna,Fiamma Maura, Monne Maria Itria, Malune Paolo, Palmas Angelo Domenico, Lo Maglio Iana, Mameli Giuseppe |
| EPI_ISL_1372541, EPI_ISL_1372542, EPI_ISL_1372543, EPI_ISL_1372544, EPI_ISL_1372545 | SC Microbiologia e Virologia AOUSS | Laboratorio specialistico UOC Ematologia - Ospedale "San Francesco" - ATS-ASSL Nuoro | Piras Giovanna, Asproni Rosanna,Fiamma Maura, Monne Maria Itria, Malune Paolo, Palmas Angelo Domenico, Lo Maglio Iana, Mameli Giuseppe Salvatore Rubino; Marcello Abbondio; Flavia Angioj; Laura Firino; Rosalba Govoni; Gabriele Ibba; Vincenzo Lai; Erica Mura; Bianca Paglietti; Claudia Piu; Anna Puggioni; Elena Rimini; Giulia Rocca; Rosangela Sau; Caterina Serra; Sergio Uzzau. |
| EPI_ISL_1372546 | Laboratorio Biologia Molecolare Sars Cov2 - UOC Laboratorio Analisi - Servizio Medicina di Laboratorio, Ospedale "San Francesco" - ATS-ASSL Nuoro | Laboratorio specialistico UOC Ematologia - Ospedale "San Francesco" - ATS-ASSL Nuoro | Piras Giovanna, Asproni Rosanna,Fiamma Maura, Monne Maria Itria, Malune Paolo, Palmas Angelo Domenico, Lo Maglio Iana, Mameli Giuseppe |
| EPI_ISL_1372547, EPI_ISL_1372548, EPI_ISL_1372549 | SC Microbiologia e Virologia AOUSS | Laboratorio specialistico UOC Ematologia - Ospedale "San Francesco" - ATS-ASSL Nuoro | Piras Giovanna, Asproni Rosanna,Fiamma Maura, Monne Maria Itria, Malune Paolo, Palmas Angelo Domenico, Lo Maglio Iana, Mameli Giuseppe Salvatore Rubino; Marcello Abbondio; Flavia Angioj; Laura Firino; Rosalba Govoni; Gabriele Ibba; Vincenzo Lai; Erica Mura; Bianca Paglietti; Claudia Piu; Anna Puggioni; Elena Rimini; Giulia Rocca; Rosangela Sau; Caterina Serra; Sergio Uzzau. |
| EPI_ISL_1372550, EPI_ISL_1372551, EPI_ISL_1372552, EPI_ISL_1372553, EPI_ISL_1372554, EPI_ISL_1372555, EPI_ISL_1372556, EPI_ISL_1372557, EPI_ISL_1372558, EPI_ISL_1372559, EPI_ISL_1372560, EPI_ISL_1372561 | | | |
| see above | Department of Infectious Diseases, Istituto Superiore di Sanità, Rome, Italy; AOR San Carlo, Potenza, Italy | Department of Infectious Diseases, Istituto Superiore di Sanità | Paola Stefanelli, Angela Di Martino, Alessandra Lo Presti, Stefano Fiore, Antonio Picerno, Lopizzo Teresa, Anna Curci, Angela Menchise, Manuela Marra, Maria Carollo, Marco Crescenzi |
| EPI_ISL_1379979, EPI_ISL_1379980, EPI_ISL_1379981, EPI_ISL_1379982, EPI_ISL_1379983, EPI_ISL_1379984, EPI_ISL_1379985, EPI_ISL_1379986, EPI_ISL_1379987, EPI_ISL_1379988, EPI_ISL_1379989, EPI_ISL_1379990, EPI_ISL_1379991, EPI_ISL_1379992, EPI_ISL_1379993, EPI_ISL_1379994, EPI_ISL_1379995, EPI_ISL_1379996, EPI_ISL_1379997, EPI_ISL_1379998, EPI_ISL_1379999, EPI_ISL_1380000, EPI_ISL_1380001, EPI_ISL_1380002, EPI_ISL_1380003, EPI_ISL_1380004, EPI_ISL_1380005, EPI_ISL_1380006, EPI_ISL_1380007, EPI_ISL_1380008, EPI_ISL_1380009, EPI_ISL_1380010, EPI_ISL_1380011, EPI_ISL_1380012, EPI_ISL_1380013, EPI_ISL_1380014, EPI_ISL_1380015, EPI_ISL_1380016, EPI_ISL_1380017, EPI_ISL_1380018, EPI_ISL_1380019, EPI_ISL_1380020, EPI_ISL_1380021, EPI_ISL_1380022, EPI_ISL_1380023, EPI_ISL_1380024, EPI_ISL_1380025, EPI_ISL_1380026, EPI_ISL_1380027, EPI_ISL_1380028, EPI_ISL_1380029, EPI_ISL_1380030, EPI_ISL_1380031, EPI_ISL_1380032, EPI_ISL_1380033, EPI_ISL_1380034, EPI_ISL_1380035, EPI_ISL_1380036, EPI_ISL_1380037, EPI_ISL_1380038, EPI_ISL_1380039, EPI_ISL_1380040, EPI_ISL_1380041, EPI_ISL_1380042, EPI_ISL_1380043, EPI_ISL_1380044, EPI_ISL_1380045 | | | |
| see above | SC Microbiologia e Virologia AOUSS | AMES Centro Polidiagnostico Strumentale S.r.l. | Salvatore Rubino; Flavia Angioj; Laura Firino; Rosalba Govoni; Gabriele Ibba; Vincenzo Lai; Erica Mura; Bianca Paglietti; Claudia Piu; Anna Puggioni; Elena Rimini; Giulia Rocca; Caterina Serra; Sergio Uzzau. |
| EPI_ISL_1385803 | Laboratorio Microbiologia P.O.Cardarelli | Laboratorio Microbiologia P.O. Cardarelli | Valentina Felice, Giovanna Niro, Scutellà Massimiliano |
| EPI_ISL_1385805 | Laboratorio Microbiologia P.O. Cardarelli | Laboratorio Microbiologia P.O. Cardarelli | Valentina Felice, Giovanna Niro, Massimiliano Scutellà |
| EPI_ISL_1386123, EPI_ISL_1386124, EPI_ISL_1386125, EPI_ISL_1386126, EPI_ISL_1386129, EPI_ISL_1386130, EPI_ISL_1386131, EPI_ISL_1386132, EPI_ISL_1386133, EPI_ISL_1386224, EPI_ISL_1386225, EPI_ISL_1386226, EPI_ISL_1386227, EPI_ISL_1386228, EPI_ISL_1386229, EPI_ISL_1386230, EPI_ISL_1386231, EPI_ISL_1386232, EPI_ISL_1386233, EPI_ISL_1386234 | | | |
| see above | Area Biologia Molecolare Istituto Zooprofilattico Sperimentale della Sicilia | Area Biologia Molecolare Istituto Zooprofilattico Sperimentale della Sicilia | REALE Stefano, SCIBETTA Silvia, PIAZZA Angela, COLLURA Rosaria, BRUNO Gabriella, CASTELLI Germano, BRUNO Federica, VITALE Fabrizio |
| EPI_ISL_1390377, EPI_ISL_1390378, EPI_ISL_1390379, EPI_ISL_1390380, EPI_ISL_1390381, EPI_ISL_1390382, EPI_ISL_1390383, EPI_ISL_1390384, EPI_ISL_1390385, EPI_ISL_1390386, EPI_ISL_1390387, EPI_ISL_1390388, EPI_ISL_1390389, EPI_ISL_1390390, EPI_ISL_1390391, EPI_ISL_1390392, EPI_ISL_1390393, EPI_ISL_1390394, EPI_ISL_1390396, EPI_ISL_1390397, EPI_ISL_1390398, EPI_ISL_1390399, EPI_ISL_1390400, EPI_ISL_1390401, EPI_ISL_1390402, EPI_ISL_1390403, EPI_ISL_1390404, EPI_ISL_1390405, EPI_ISL_1390406, EPI_ISL_1390407, EPI_ISL_1390408, EPI_ISL_1390409, EPI_ISL_1390410, EPI_ISL_1390411, EPI_ISL_1390412, EPI_ISL_1390413, EPI_ISL_1390414, EPI_ISL_1390415, EPI_ISL_1390416, EPI_ISL_1390417, EPI_ISL_1390418, EPI_ISL_1390419, EPI_ISL_1390420, EPI_ISL_1390421, EPI_ISL_1390422, EPI_ISL_1390423, EPI_ISL_1390424, EPI_ISL_1390425, EPI_ISL_1390426, EPI_ISL_1390427, EPI_ISL_1390428, EPI_ISL_1390429, EPI_ISL_1390430, EPI_ISL_1390431, EPI_ISL_1390432, EPI_ISL_1390433, EPI_ISL_1390434, EPI_ISL_1390435, EPI_ISL_1390436, EPI_ISL_1390437, EPI_ISL_1390438, EPI_ISL_1390439, EPI_ISL_1390440, EPI_ISL_1390441, EPI_ISL_1390442, EPI_ISL_1390443, EPI_ISL_1390444, EPI_ISL_1390445, EPI_ISL_1390446, EPI_ISL_1390447, EPI_ISL_1390448, EPI_ISL_1390449, EPI_ISL_1390450, EPI_ISL_1390451, EPI_ISL_1390452, EPI_ISL_1390453, EPI_ISL_1390454, EPI_ISL_1390455, EPI_ISL_1390456, EPI_ISL_1390457, EPI_ISL_1390458, EPI_ISL_1390459, EPI_ISL_1390460, EPI_ISL_1390461, EPI_ISL_1390462, EPI_ISL_1390463, EPI_ISL_1390464, EPI_ISL_1390465, EPI_ISL_1390466, EPI_ISL_1390467, EPI_ISL_1390468, EPI_ISL_1390469, EPI_ISL_1390470, EPI_ISL_1390471, EPI_ISL_1390472, EPI_ISL_1390473, EPI_ISL_1390474, EPI_ISL_1390475, EPI_ISL_1390476, EPI_ISL_1390477, EPI_ISL_1390478, EPI_ISL_1390479, EPI_ISL_1390480, EPI_ISL_1390481, EPI_ISL_1390482, EPI_ISL_1390483, EPI_ISL_1390484, EPI_ISL_1390485, EPI_ISL_1390486, EPI_ISL_1390487, EPI_ISL_1390488, EPI_ISL_1390489, EPI_ISL_1390490, EPI_ISL_1390491, EPI_ISL_1390492, EPI_ISL_1390493, EPI_ISL_1390494, EPI_ISL_1390495, EPI_ISL_1390496, EPI_ISL_1390497, EPI_ISL_1390498, EPI_ISL_1390499, EPI_ISL_1390500, EPI_ISL_1390501, EPI_ISL_1390502, EPI_ISL_1390503, EPI_ISL_1390504, EPI_ISL_1390505, EPI_ISL_1390506, EPI_ISL_1390507, EPI_ISL_1390508, EPI_ISL_1390509, EPI_ISL_1390510, EPI_ISL_1390511, EPI_ISL_1390512, EPI_ISL_1390513, EPI_ISL_1390514, EPI_ISL_1390515, EPI_ISL_1390516, EPI_ISL_1390517, EPI_ISL_1390518, EPI_ISL_1390519, EPI_ISL_1390520, EPI_ISL_1390521, EPI_ISL_1390522, EPI_ISL_1390523, EPI_ISL_1390524, EPI_ISL_1390525, EPI_ISL_1390526, EPI_ISL_1390527, EPI_ISL_1390528, EPI_ISL_1390529, EPI_ISL_1390530, EPI_ISL_1390531, EPI_ISL_1390532, EPI_ISL_1390533, EPI_ISL_1390534, EPI_ISL_1390535, EPI_ISL_1390536, EPI_ISL_1390537, EPI_ISL_1390538, EPI_ISL_1390539, EPI_ISL_1390540, EPI_ISL_1390541, EPI_ISL_1390542, EPI_ISL_1390543, EPI_ISL_1390544, EPI_ISL_1390545, EPI_ISL_1390547, EPI_ISL_1390548, EPI_ISL_1390549, EPI_ISL_1390550, EPI_ISL_1390551, EPI_ISL_1390552, EPI_ISL_1390553, EPI_ISL_1390554, EPI_ISL_1390555, EPI_ISL_1390556, EPI_ISL_1390557, EPI_ISL_1390558, EPI_ISL_1390559, EPI_ISL_1390560, EPI_ISL_1390561, EPI_ISL_1390562, EPI_ISL_1390563, EPI_ISL_1390564, EPI_ISL_1390565, EPI_ISL_1390566, EPI_ISL_1390567, EPI_ISL_1390568, EPI_ISL_1390569, EPI_ISL_1390570, EPI_ISL_1390571, EPI_ISL_1390572, EPI_ISL_1390573, EPI_ISL_1390574, EPI_ISL_1390575, EPI_ISL_1390576, EPI_ISL_1390577, EPI_ISL_1390578, EPI_ISL_1390579, EPI_ISL_1390580, EPI_ISL_1390581, EPI_ISL_1390582, EPI_ISL_1390583, EPI_ISL_1390584, EPI_ISL_1390585, EPI_ISL_1390586, EPI_ISL_1390587, EPI_ISL_1390588, EPI_ISL_1390589, EPI_ISL_1390590, EPI_ISL_1390591, EPI_ISL_1390592, EPI_ISL_1390593, EPI_ISL_1390594, EPI_ISL_1390595, EPI_ISL_1390596, EPI_ISL_1390597, EPI_ISL_1390598, EPI_ISL_1390599, EPI_ISL_1390600, EPI_ISL_1390601, EPI_ISL_1390602, EPI_ISL_1390603, EPI_ISL_1390604, EPI_ISL_1390605, EPI_ISL_1390606, EPI_ISL_1390607, EPI_ISL_1390608, EPI_ISL_1390609, EPI_ISL_1390610, EPI_ISL_1390611, EPI_ISL_1390612, EPI_ISL_1390613, EPI_ISL_1390614, EPI_ISL_1390615, EPI_ISL_1390616, EPI_ISL_1390617, EPI_ISL_1390618, EPI_ISL_1390619, EPI_ISL_1390620, EPI_ISL_1390621, EPI_ISL_1390622, EPI_ISL_1390623, EPI_ISL_1390624, EPI_ISL_1390625, EPI_ISL_1390626, EPI_ISL_1390627, EPI_ISL_1390628, EPI_ISL_1390629, EPI_ISL_1390630, EPI_ISL_1390631, EPI_ISL_1390632, EPI_ISL_1390633, EPI_ISL_1390634, EPI_ISL_1390635, EPI_ISL_1390636, EPI_ISL_1390637, EPI_ISL_1390638, EPI_ISL_1390639, EPI_ISL_1390640, EPI_ISL_1390641, EPI_ISL_1390642, EPI_ISL_1390643, EPI_ISL_1390644, EPI_ISL_1390645, EPI_ISL_1390646, EPI_ISL_1390647, EPI_ISL_1390648, EPI_ISL_1390649, EPI_ISL_1390650, EPI_ISL_1390651, EPI_ISL_1390652, EPI_ISL_1390653, EPI_ISL_1390654, EPI_ISL_1390655, EPI_ISL_1390656, EPI_ISL_1390657, EPI_ISL_1390658, EPI_ISL_1390659, EPI_ISL_1390660, EPI_ISL_1390661, EPI_ISL_1390662, EPI_ISL_1390663, EPI_ISL_1390664, EPI_ISL_1390665, EPI_ISL_1390666, EPI_ISL_1390667, EPI_ISL_1390668, EPI_ISL_1390669, EPI_ISL_1390670, EPI_ISL_1390671, EPI_ISL_1390672, EPI_ISL_1390673, EPI_ISL_1390674, EPI_ISL_1390675, EPI_ISL_1390676, EPI_ISL_1390677, EPI_ISL_1390678, EPI_ISL_1390679, EPI_ISL_1390680, EPI_ISL_1390682, EPI_ISL_1390683, EPI_ISL_1390684, EPI_ISL_1390685, EPI_ISL_1390686, EPI_ISL_1390687, EPI_ISL_1390688, EPI_ISL_1390689 | | | |
| see above | IZSM | TIGEM | Antonio Grimaldi,Patrizia Annunziata,Francesco Panariello,Biancamaria Pierri Claudia Tiberio Valentina Bouche,Chiara Colantuono,Maria Concetta Cuomo,Denise Di Concilio,Lucio Di Filippo,Anna Manfredi,Marcello Salvi,Antonio Limone Luigi Atripaldi Pellegrino Cerino,Andrea Ballabio,Davide Cacchiarelli |
| EPI_ISL_1390690, EPI_ISL_1390691, EPI_ISL_1390692, EPI_ISL_1390693, EPI_ISL_1390694, EPI_ISL_1390695, EPI_ISL_1390696, EPI_ISL_1390697, EPI_ISL_1390698, EPI_ISL_1390699, EPI_ISL_1390700, EPI_ISL_1390701, EPI_ISL_1390702, EPI_ISL_1390703, EPI_ISL_1390704, EPI_ISL_1390705, EPI_ISL_1390706, EPI_ISL_1390707, EPI_ISL_1390708, EPI_ISL_1390709, EPI_ISL_1390710, EPI_ISL_1390711, EPI_ISL_1390712, EPI_ISL_1390713, EPI_ISL_1390714 | | | |
| see above | Istituto Zooprofilattico Sperimentale del Mezzogiorno - Azienda Ospedaliera "Pugliese Ciaccio" di Catanzaro | TIGEM | Antonio Grimaldi,Patrizia Annunziata,Francesco Panariello,Biancamaria Pierri Claudia Tiberio Valentina Bouche,Chiara Colantuono,Maria Concetta Cuomo,Denise Di Concilio,Lucio Di Filippo,Anna Manfredi,Marcello Salvi,Antonio Limone Pasquale Minchella Luigi Atripaldi Pellegrino Cerino,Andrea Ballabio,Davide Cacchiarelli |
| EPI_ISL_1390715 | Istituto Zooprofilattico Sperimentale del Mezzogiorno - Azienda Ospedaliera "Pugliese Ciaccio" di Catanzaro | TIGEM | Antonio Grimaldi,Patrizia Annunziata,Francesco Panariello,Biancamaria Pierri Claudia Tiberio Valentina Bouche,Chiara Colantuono,Maria Concetta Cuomo,Denise Di Concilio,Lucio Di Filippo,Anna Manfredi,Marcello Salvi,Antonio Limone Luigi Atripaldi Pellegrino Cerino,Andrea Ballabio,Davide Cacchiarelli |
| EPI_ISL_1390716, EPI_ISL_1390717, EPI_ISL_1390718, EPI_ISL_1390719, EPI_ISL_1390720, EPI_ISL_1390721, EPI_ISL_1390722, EPI_ISL_1390723, EPI_ISL_1390724, EPI_ISL_1390725, EPI_ISL_1390726, EPI_ISL_1390727, EPI_ISL_1390728, EPI_ISL_1390729, EPI_ISL_1390730 | | | |
| see above | IZSM | TIGEM | Antonio Grimaldi,Patrizia Annunziata,Francesco Panariello,Biancamaria Pierri Claudia Tiberio Valentina Bouche,Chiara Colantuono,Maria Concetta Cuomo,Denise Di Concilio,Lucio Di Filippo,Anna Manfredi,Marcello Salvi,Antonio Limone Luigi Atripaldi Pellegrino Cerino,Andrea Ballabio,Davide Cacchiarelli |
| EPI_ISL_1390769 | Azienda Ospedaliera Terni | Istituto Zooprofilattico Sperimentale dell'Abruzzo e Molise "G. Caporale" | Palumbo M, Scaccetti A, Lorusso A, Marcacci M, Di Domenico M, Ancora M, Curini V, Mangone I, Rinaldi A, Scialabba S, Caporale M, Di Pasquale A, Cammà C, Puglia I, Calistri P, Savini G |
| EPI_ISL_1391020, EPI_ISL_1391021, EPI_ISL_1391022, EPI_ISL_1391023 | Ospedale Generale Regionale F. Miulli | Istituto Zooprofilattico Sperimentale della Puglia e della Basilicata | Parisi A., Bianco A., Capozzi L., Del Sambro L., Simone D., Ceci E. |
| EPI_ISL_1391024, EPI_ISL_1391025, EPI_ISL_1391026, EPI_ISL_1391027, EPI_ISL_1391028 | I.R.C.C.S. "S. De Bellis" - Ente Ospedaliero | Istituto Zooprofilattico Sperimentale della Puglia e della Basilicata | Parisi A., Bianco A., Capozzi L., Del Sambro L., Simone D., Lippolis A., Notarnicola M. |
| EPI_ISL_1391029, EPI_ISL_1391030, EPI_ISL_1391031, EPI_ISL_1391032, EPI_ISL_1391033, EPI_ISL_1391034, EPI_ISL_1391035, EPI_ISL_1391036, EPI_ISL_1391037 | Ospedale Generale Regionale F. Miulli | Istituto Zooprofilattico Sperimentale della Puglia e della Basilicata | Parisi A., Bianco A., Capozzi L., Del Sambro L., Simone D., Ceci E. |
| EPI_ISL_1391038, EPI_ISL_1391039, EPI_ISL_1391040, EPI_ISL_1391041, EPI_ISL_1391042, EPI_ISL_1391043, EPI_ISL_1391044, EPI_ISL_1391045, EPI_ISL_1391046, EPI_ISL_1391047, EPI_ISL_1391048, EPI_ISL_1391049, EPI_ISL_1391050, EPI_ISL_1391051, EPI_ISL_1391052 | | | |
| see above | Ospedale di Venere | Istituto Zooprofilattico Sperimentale della Puglia e della | Parisi A., Bianco A., Capozzi L., Del Sambro L., Simone D., Iacobellis M., Ridolfi D., Giannico A. |

| | | | |
|---|---|---|---|
| | | Basilicata | |
| EPI_ISL_1391053, EPI_ISL_1391054, EPI_ISL_1391055, EPI_ISL_1391056, EPI_ISL_1391057 | Presidio di Brindisi Di Summa - Perrino | Istituto Zooprofilattico Sperimentale della Puglia e della Basilicata | Parisi A., Bianco A., Capozzi L., Del Sambro L., Simone D., Difato L., Santoro A., Ridolfi D., Giannico A. |
| EPI_ISL_1391058, EPI_ISL_1391059, EPI_ISL_1391060, EPI_ISL_1391061, EPI_ISL_1391062, EPI_ISL_1391063, EPI_ISL_1391064 | Ospedale Vito Fazzi | Istituto Zooprofilattico Sperimentale della Puglia e della Basilicata | Parisi A., Bianco A., Capozzi L., Del Sambro L., Simone D., Difato L., Lobreglio G. |
| EPI_ISL_1391065, EPI_ISL_1391066, EPI_ISL_1391067, EPI_ISL_1391068, EPI_ISL_1391069, EPI_ISL_1391070, EPI_ISL_1391071, EPI_ISL_1391072 | Ospedale Santa Caterina Novella | Istituto Zooprofilattico Sperimentale della Puglia e della Basilicata | Parisi A., Bianco A., Capozzi L., Del Sambro L., Simone D., Difato L., Bruno A. R. |
| EPI_ISL_1391073, EPI_ISL_1391074, EPI_ISL_1391075, EPI_ISL_1391076 | Ospedale Casa Sollievo della Sofferenza | Istituto Zooprofilattico Sperimentale della Puglia e della Basilicata | Parisi A., Bianco A., Capozzi L., Del Sambro L., Simone D., Difato L., Ridolfi D., Giannico A, Labonia M. |
| EPI_ISL_1391077, EPI_ISL_1391078, EPI_ISL_1391079, EPI_ISL_1391080, EPI_ISL_1391081, EPI_ISL_1391082, EPI_ISL_1391083, EPI_ISL_1391084, EPI_ISL_1391085, EPI_ISL_1391086, EPI_ISL_1391087, EPI_ISL_1391088, EPI_ISL_1391089, EPI_ISL_1391090, EPI_ISL_1391091, EPI_ISL_1391092, EPI_ISL_1391093, EPI_ISL_1391094, EPI_ISL_1391095, EPI_ISL_1391096, EPI_ISL_1391097 | | | |
| see above | Ospedale Santissima Annunziata | Istituto Zooprofilattico Sperimentale della Puglia e della Basilicata | Parisi A., Bianco A., Capozzi L., Del Sambro L., Simone D., Difato L., Ridolfi D., Giannico A, Tampoia M. |
| EPI_ISL_1391204, EPI_ISL_1391205, EPI_ISL_1391206, EPI_ISL_1391207, EPI_ISL_1391208, EPI_ISL_1391209, EPI_ISL_1391210, EPI_ISL_1391211, EPI_ISL_1391212, EPI_ISL_1391213, EPI_ISL_1391214, EPI_ISL_1391215, EPI_ISL_1391216, EPI_ISL_1391217, EPI_ISL_1391218, EPI_ISL_1391219, EPI_ISL_1391220, EPI_ISL_1391221, EPI_ISL_1391222, EPI_ISL_1391223, EPI_ISL_1391224, EPI_ISL_1391225, EPI_ISL_1391226, EPI_ISL_1391227, EPI_ISL_1391228, EPI_ISL_1391229 | | | |
| see above | SIESP DIPARTIMENTO DI PREVENZIONE TERAMO TERAMO(TERAMO) | Istituto Zooprofilattico Sperimentale dell'Abruzzo e Molise "G. Caporale" | Lorusso A, Marcacci M, Di Domenico M, Ancora M, Curini V, Di Lollo Valeria, Mangone I, Rinaldi A, Delli Compagni E, Scialabba S, Caporale M, Di Pasquale A, Cammà C, Puglia I, Calistri P, Savini G |
| EPI_ISL_1391230, EPI_ISL_1391231, EPI_ISL_1391232, EPI_ISL_1391233 | SIESP CHIETI - DRIVE IN LANCIANO LANCIANO(CHIETI) | Istituto Zooprofilattico Sperimentale dell'Abruzzo e Molise "G. Caporale" | Lorusso A, Marcacci M, Di Domenico M, Ancora M, Curini V, Di Lollo Valeria, Mangone I, Rinaldi A, Delli Compagni E, Scialabba S, Caporale M, Di Pasquale A, Cammà C, Puglia I, Calistri P, Savini G |
| EPI_ISL_1391234, EPI_ISL_1391235, EPI_ISL_1391236, EPI_ISL_1391237 | SIESP SULMONA SULMONA(L'AQUILA) | Istituto Zooprofilattico Sperimentale dell'Abruzzo e Molise "G. Caporale" | Lorusso A, Marcacci M, Di Domenico M, Ancora M, Curini V, Di Lollo Valeria, Mangone I, Rinaldi A, Delli Compagni E, Scialabba S, Caporale M, Di Pasquale A, Cammà C, Puglia I, Calistri P, Savini G |
| EPI_ISL_1391238 | DIP. PREV. AVEZZANO SERVIZIO DI IGIENE EPIDEMIOLOGIA E SANITA' PUBBLICA AVEZZANO(L'AQUILA) | Istituto Zooprofilattico Sperimentale dell'Abruzzo e Molise "G. Caporale" | Lorusso A, Marcacci M, Di Domenico M, Ancora M, Curini V, Di Lollo Valeria, Mangone I, Rinaldi A, Delli Compagni E, Scialabba S, Caporale M, Di Pasquale A, Cammà C, Puglia I, Calistri P, Savini G |
| EPI_ISL_1391239, EPI_ISL_1391240 | SIESP SULMONA SULMONA(L'AQUILA) | Istituto Zooprofilattico Sperimentale dell'Abruzzo e Molise "G. Caporale" | Lorusso A, Marcacci M, Di Domenico M, Ancora M, Curini V, Di Lollo Valeria, Mangone I, Rinaldi A, Delli Compagni E, Scialabba S, Caporale M, Di Pasquale A, Cammà C, Puglia I, Calistri P, Savini G |
| EPI_ISL_1391241, EPI_ISL_1391242, EPI_ISL_1391243, EPI_ISL_1391244, EPI_ISL_1391245, EPI_ISL_1391246, EPI_ISL_1391247 | DIP. PREV. AVEZZANO SERVIZIO DI IGIENE EPIDEMIOLOGIA E SANITA' PUBBLICA AVEZZANO(L'AQUILA) | Istituto Zooprofilattico Sperimentale dell'Abruzzo e Molise "G. Caporale" | Lorusso A, Marcacci M, Di Domenico M, Ancora M, Curini V, Di Lollo Valeria, Mangone I, Rinaldi A, Delli Compagni E, Scialabba S, Caporale M, Di Pasquale A, Cammà C, Puglia I, Calistri P, Savini G |
| EPI_ISL_1391248 | SIESP SULMONA SULMONA(L'AQUILA) | Istituto Zooprofilattico Sperimentale dell'Abruzzo e Molise "G. Caporale" | Lorusso A, Marcacci M, Di Domenico M, Ancora M, Curini V, Di Lollo Valeria, Mangone I, Rinaldi A, Delli Compagni E, Scialabba S, Caporale M, Di Pasquale A, Cammà C, Puglia I, Calistri P, Savini G |
| EPI_ISL_1391249, EPI_ISL_1391250, EPI_ISL_1391251 | SIESP DIPARTIMENTO DI PREVENZIONE CHIETI CHIETI(CHIETI) | Istituto Zooprofilattico Sperimentale dell'Abruzzo e Molise "G. Caporale" | Lorusso A, Marcacci M, Di Domenico M, Ancora M, Curini V, Di Lollo Valeria, Mangone I, Rinaldi A, Delli Compagni E, Scialabba S, Caporale M, Di Pasquale A, Cammà C, Puglia I, Calistri P, Savini G |
| EPI_ISL_1391252 | SIESP CHIETI - DRIVE IN LANCIANO LANCIANO(CHIETI) | Istituto Zooprofilattico Sperimentale dell'Abruzzo e Molise "G. Caporale" | Lorusso A, Marcacci M, Di Domenico M, Ancora M, Curini V, Di Lollo Valeria, Mangone I, Rinaldi A, Delli Compagni E, Scialabba S, Caporale M, Di Pasquale A, Cammà C, Puglia I, Calistri P, Savini G |
| EPI_ISL_1391253, EPI_ISL_1391254, EPI_ISL_1391255, EPI_ISL_1391256, EPI_ISL_1391257, EPI_ISL_1391258, EPI_ISL_1391259 | SIESP DIPARTIMENTO DI PREVENZIONE CHIETI CHIETI(CHIETI) | Istituto Zooprofilattico Sperimentale dell'Abruzzo e Molise "G. Caporale" | Lorusso A, Marcacci M, Di Domenico M, Ancora M, Curini V, Di Lollo Valeria, Mangone I, Rinaldi A, Delli Compagni E, Scialabba S, Caporale M, Di Pasquale A, Cammà C, Puglia I, Calistri P, Savini G |
| EPI_ISL_1391260, EPI_ISL_1391261, EPI_ISL_1391262 | SIESP CHIETI - DRIVE IN LANCIANO LANCIANO(CHIETI) | Istituto Zooprofilattico Sperimentale dell'Abruzzo e Molise "G. Caporale" | Lorusso A, Marcacci M, Di Domenico M, Ancora M, Curini V, Di Lollo Valeria, Mangone I, Rinaldi A, Delli Compagni E, Scialabba S, Caporale M, Di Pasquale A, Cammà C, Puglia I, Calistri P, Savini G |
| EPI_ISL_1391263, EPI_ISL_1391264 | SIESP DIPARTIMENTO DI PREVENZIONE CHIETI CHIETI(CHIETI) | Istituto Zooprofilattico Sperimentale dell'Abruzzo e Molise "G. Caporale" | Lorusso A, Marcacci M, Di Domenico M, Ancora M, Curini V, Di Lollo Valeria, Mangone I, Rinaldi A, Delli Compagni E, Scialabba S, Caporale M, Di Pasquale A, Cammà C, Puglia I, Calistri P, Savini G |
| EPI_ISL_1391265 | SIESP CHIETI - DRIVE IN LANCIANO LANCIANO(CHIETI) | Istituto Zooprofilattico Sperimentale dell'Abruzzo e Molise "G. Caporale" | Lorusso A, Marcacci M, Di Domenico M, Ancora M, Curini V, Di Lollo Valeria, Mangone I, Rinaldi A, Delli Compagni E, Scialabba S, Caporale M, Di Pasquale A, Cammà C, Puglia I, Calistri P, Savini G |
| EPI_ISL_1391266, EPI_ISL_1391267, EPI_ISL_1391268, EPI_ISL_1391269, EPI_ISL_1391270 | SIESP CHIETI - DRIVE IN CHIETI CHIETI(CHIETI) | Istituto Zooprofilattico Sperimentale dell'Abruzzo e Molise "G. Caporale" | Lorusso A, Marcacci M, Di Domenico M, Ancora M, Curini V, Di Lollo Valeria, Mangone I, Rinaldi A, Delli Compagni E, Scialabba S, Caporale M, Di Pasquale A, Cammà C, Puglia I, Calistri P, Savini G |
| EPI_ISL_1391271, EPI_ISL_1391272, EPI_ISL_1391273 | SIESP CHIETI - DRIVE IN ORTONA ORTONA(CHIETI) | Istituto Zooprofilattico Sperimentale dell'Abruzzo e Molise "G. Caporale" | Lorusso A, Marcacci M, Di Domenico M, Ancora M, Curini V, Di Lollo Valeria, Mangone I, Rinaldi A, Delli Compagni E, Scialabba S, Caporale M, Di Pasquale A, Cammà C, Puglia I, Calistri P, Savini G |
| EPI_ISL_1391274 | SIESP DIPARTIMENTO DI PREVENZIONE TERAMO TERAMO(TERAMO) | Istituto Zooprofilattico Sperimentale dell'Abruzzo e Molise "G. Caporale" | Lorusso A, Marcacci M, Di Domenico M, Ancora M, Curini V, Di Lollo Valeria, Mangone I, Rinaldi A, Delli Compagni E, Scialabba S, Caporale M, Di Pasquale A, Cammà C, Puglia I, Calistri P, Savini G |
| EPI_ISL_1391275 | SIESP CHIETI - DRIVE IN GISSI GISSI(CHIETI) | Istituto Zooprofilattico Sperimentale dell'Abruzzo e Molise "G. Caporale" | Lorusso A, Marcacci M, Di Domenico M, Ancora M, Curini V, Di Lollo Valeria, Mangone I, Rinaldi A, Delli Compagni E, Scialabba S, Caporale M, Di Pasquale A, Cammà C, Puglia I, Calistri P, Savini G |
| EPI_ISL_1391393 | SIESP DIPARTIMENTO DI PREVENZIONE CHIETI | Istituto Zooprofilattico Sperimentale dell'Abruzzo e Molise "G. Caporale" | Lorusso A, Marcacci M, Di Domenico M, Ancora M, Curini V, Di Lollo Valeria, Mangone I, Rinaldi A, Scialabba S, Caporale M, Di Pasquale A, Cammà C, Puglia I, Calistri P, Savini G |
| EPI_ISL_1391394 | SIESP DIPARTIMENTO DI PREVENZIONE TERAMO | Istituto Zooprofilattico Sperimentale dell'Abruzzo e Molise "G. Caporale" | Lorusso A, Marcacci M, Di Domenico M, Ancora M, Curini V, Di Lollo Valeria, Mangone I, Rinaldi A, Scialabba S, Caporale M, Di Pasquale A, Cammà C, Puglia I, Calistri P, Savini G |
| EPI_ISL_1391395 | OSPEDALE S.S. ANNUNZIATA CHIETI | Istituto Zooprofilattico Sperimentale dell'Abruzzo e Molise "G. Caporale" | Lorusso A, Marcacci M, Di Domenico M, Ancora M, Curini V, Di Lollo Valeria, Mangone I, Rinaldi A, Scialabba S, Caporale M, Di Pasquale A, Cammà C, Puglia I, Calistri P, Savini G |
| EPI_ISL_1391396 | SIESP CHIETI - DRIVE IN CHIETI | Istituto Zooprofilattico Sperimentale dell'Abruzzo e Molise "G. Caporale" | Lorusso A, Marcacci M, Di Domenico M, Ancora M, Curini V, Di Lollo Valeria, Mangone I, Rinaldi A, Scialabba S, Caporale M, Di Pasquale A, Cammà C, Puglia I, Calistri P, Savini G |
| EPI_ISL_1391397 | SIESP CHIETI - DRIVE IN ORTONA | Istituto Zooprofilattico Sperimentale dell'Abruzzo e Molise "G. Caporale" | Lorusso A, Marcacci M, Di Domenico M, Ancora M, Curini V, Di Lollo Valeria, Mangone I, Rinaldi A, Scialabba S, Caporale M, Di Pasquale A, Cammà C, Puglia I, Calistri P, Savini G |
| EPI_ISL_1391398 | SIESP SULMONA | Istituto Zooprofilattico Sperimentale dell'Abruzzo e Molise "G. Caporale" | Lorusso A, Marcacci M, Di Domenico M, Ancora M, Curini V, Di Lollo Valeria, Mangone I, Rinaldi A, Scialabba S, Caporale M, Di Pasquale A, Cammà C, Puglia I, Calistri P, Savini G |

| | | | |
|---|---|---|---|
| EPI_ISL_1391399, EPI_ISL_1391400 | SIESP CHIETI - DRIVE IN ORTONA | Istituto Zooprofilattico Sperimentale dell'Abruzzo e Molise "G. Caporale" | Lorusso A, Marcacci M, Di Domenico M, Ancora M, Curini V, Di Lollo Valeria, Mangone I, Rinaldi A, Scialabba S, Caporale M, Di Pasquale A, Cammà C, Puglia I, Calistri P, Savini G |
| EPI_ISL_1391401, EPI_ISL_1391402 | SIESP DIPARTIMENTO DI PREVENZIONE TERAMO | Istituto Zooprofilattico Sperimentale dell'Abruzzo e Molise "G. Caporale" | Lorusso A, Marcacci M, Di Domenico M, Ancora M, Curini V, Di Lollo Valeria, Mangone I, Rinaldi A, Scialabba S, Caporale M, Di Pasquale A, Cammà C, Puglia I, Calistri P, Savini G |
| EPI_ISL_1391403 | DIP. PREV. AVEZZANO SERVIZIO DI IGIENE EPIDEMIOLOGIA E SANITA' PUBBLICA | Istituto Zooprofilattico Sperimentale dell'Abruzzo e Molise "G. Caporale" | Lorusso A, Marcacci M, Di Domenico M, Ancora M, Curini V, Di Lollo Valeria, Mangone I, Rinaldi A, Scialabba S, Caporale M, Di Pasquale A, Cammà C, Puglia I, Calistri P, Savini G |
| EPI_ISL_1391404 | SIESP DIPARTIMENTO DI PREVENZIONE CHIETI | Istituto Zooprofilattico Sperimentale dell'Abruzzo e Molise "G. Caporale" | Lorusso A, Marcacci M, Di Domenico M, Ancora M, Curini V, Di Lollo Valeria, Mangone I, Rinaldi A, Scialabba S, Caporale M, Di Pasquale A, Cammà C, Puglia I, Calistri P, Savini G |
| EPI_ISL_1391405 | SIESP CHIETI - DRIVE IN CHIETI | Istituto Zooprofilattico Sperimentale dell'Abruzzo e Molise "G. Caporale" | Lorusso A, Marcacci M, Di Domenico M, Ancora M, Curini V, Di Lollo Valeria, Mangone I, Rinaldi A, Scialabba S, Caporale M, Di Pasquale A, Cammà C, Puglia I, Calistri P, Savini G |
| EPI_ISL_1391406 | SIESP SULMONA | Istituto Zooprofilattico Sperimentale dell'Abruzzo e Molise "G. Caporale" | Lorusso A, Marcacci M, Di Domenico M, Ancora M, Curini V, Di Lollo Valeria, Mangone I, Rinaldi A, Scialabba S, Caporale M, Di Pasquale A, Cammà C, Puglia I, Calistri P, Savini G |
| EPI_ISL_1391407 | SIESP DIPARTIMENTO DI PREVENZIONE TERAMO | Istituto Zooprofilattico Sperimentale dell'Abruzzo e Molise "G. Caporale" | Lorusso A, Marcacci M, Di Domenico M, Ancora M, Curini V, Di Lollo Valeria, Mangone I, Rinaldi A, Scialabba S, Caporale M, Di Pasquale A, Cammà C, Puglia I, Calistri P, Savini G |
| EPI_ISL_1391408 | SIESP CHIETI - DRIVE IN ORTONA | Istituto Zooprofilattico Sperimentale dell'Abruzzo e Molise "G. Caporale" | Lorusso A, Marcacci M, Di Domenico M, Ancora M, Curini V, Di Lollo Valeria, Mangone I, Rinaldi A, Scialabba S, Caporale M, Di Pasquale A, Cammà C, Puglia I, Calistri P, Savini G |
| EPI_ISL_1392650, EPI_ISL_1392651 | OSPEDALE CIVILE GIULIANOVA - PRONTO SOCCORSO GIULIANOVA(TERAMO) | Istituto Zooprofilattico Sperimentale dell'Abruzzo e Molise "G. Caporale" | Lorusso A, Marcacci M, Di Domenico M, Ancora M, Curini V, Di Lollo Valeria, Mangone I, Rinaldi A, Delli Compagni E, Scialabba S, Caporale M, Di Pasquale A, Cammà C, Puglia I, Calistri P, Savini G |
| EPI_ISL_1392652, EPI_ISL_1392653, EPI_ISL_1392654, EPI_ISL_1392655, EPI_ISL_1392656, EPI_ISL_1392657, EPI_ISL_1392658, EPI_ISL_1392659, EPI_ISL_1392660, EPI_ISL_1392661, EPI_ISL_1392662 | | | |
| see above | SIESP DIPARTIMENTO DI PREVENZIONE TERAMO TERAMO(TERAMO) | Istituto Zooprofilattico Sperimentale dell'Abruzzo e Molise "G. Caporale" | Lorusso A, Marcacci M, Di Domenico M, Ancora M, Curini V, Di Lollo Valeria, Mangone I, Rinaldi A, Delli Compagni E, Scialabba S, Caporale M, Di Pasquale A, Cammà C, Puglia I, Calistri P, Savini G |
| EPI_ISL_1392663, EPI_ISL_1392664, EPI_ISL_1392665 | RSA GIULIANOVA GIULIANOVA(TERAMO) | Istituto Zooprofilattico Sperimentale dell'Abruzzo e Molise "G. Caporale" | Lorusso A, Marcacci M, Di Domenico M, Ancora M, Curini V, Di Lollo Valeria, Mangone I, Rinaldi A, Delli Compagni E, Scialabba S, Caporale M, Di Pasquale A, Cammà C, Puglia I, Calistri P, Savini G |
| EPI_ISL_1392666 | SIESP DIPARTIMENTO DI PREVENZIONE CHIETI CHIETI(CHIETI) | Istituto Zooprofilattico Sperimentale dell'Abruzzo e Molise "G. Caporale" | Lorusso A, Marcacci M, Di Domenico M, Ancora M, Curini V, Di Lollo Valeria, Mangone I, Rinaldi A, Delli Compagni E, Scialabba S, Caporale M, Di Pasquale A, Cammà C, Puglia I, Calistri P, Savini G |
| EPI_ISL_1392667, EPI_ISL_1392668, EPI_ISL_1392669 | DIP. PREV. AVEZZANO SERVIZIO DI IGIENE EPIDEMIOLOGIA E SANITA' PUBBLICA AVEZZANO(L'AQUILA) | Istituto Zooprofilattico Sperimentale dell'Abruzzo e Molise "G. Caporale" | Lorusso A, Marcacci M, Di Domenico M, Ancora M, Curini V, Di Lollo Valeria, Mangone I, Rinaldi A, Delli Compagni E, Scialabba S, Caporale M, Di Pasquale A, Cammà C, Puglia I, Calistri P, Savini G |
| EPI_ISL_1392670, EPI_ISL_1392671 | SIESP SULMONA SULMONA(L'AQUILA) | Istituto Zooprofilattico Sperimentale dell'Abruzzo e Molise "G. Caporale" | Lorusso A, Marcacci M, Di Domenico M, Ancora M, Curini V, Di Lollo Valeria, Mangone I, Rinaldi A, Delli Compagni E, Scialabba S, Caporale M, Di Pasquale A, Cammà C, Puglia I, Calistri P, Savini G |
| EPI_ISL_1392672, EPI_ISL_1392673 | DIP. PREV. AVEZZANO SERVIZIO DI IGIENE EPIDEMIOLOGIA E SANITA' PUBBLICA AVEZZANO(L'AQUILA) | Istituto Zooprofilattico Sperimentale dell'Abruzzo e Molise "G. Caporale" | Lorusso A, Marcacci M, Di Domenico M, Ancora M, Curini V, Di Lollo Valeria, Mangone I, Rinaldi A, Delli Compagni E, Scialabba S, Caporale M, Di Pasquale A, Cammà C, Puglia I, Calistri P, Savini G |
| EPI_ISL_1392674 | SIESP SULMONA SULMONA(L'AQUILA) | Istituto Zooprofilattico Sperimentale dell'Abruzzo e Molise "G. Caporale" | Lorusso A, Marcacci M, Di Domenico M, Ancora M, Curini V, Di Lollo Valeria, Mangone I, Rinaldi A, Delli Compagni E, Scialabba S, Caporale M, Di Pasquale A, Cammà C, Puglia I, Calistri P, Savini G |
| EPI_ISL_1392675, EPI_ISL_1392676, EPI_ISL_1392677, EPI_ISL_1392678, EPI_ISL_1392679 | DIP. PREV. AVEZZANO SERVIZIO DI IGIENE EPIDEMIOLOGIA E SANITA' PUBBLICA AVEZZANO(L'AQUILA) | Istituto Zooprofilattico Sperimentale dell'Abruzzo e Molise "G. Caporale" | Lorusso A, Marcacci M, Di Domenico M, Ancora M, Curini V, Di Lollo Valeria, Mangone I, Rinaldi A, Delli Compagni E, Scialabba S, Caporale M, Di Pasquale A, Cammà C, Puglia I, Calistri P, Savini G |
| EPI_ISL_1392680 | SIESP CHIETI - DRIVE IN CHIETI CHIETI(CHIETI) | Istituto Zooprofilattico Sperimentale dell'Abruzzo e Molise "G. Caporale" | Lorusso A, Marcacci M, Di Domenico M, Ancora M, Curini V, Di Lollo Valeria, Mangone I, Rinaldi A, Delli Compagni E, Scialabba S, Caporale M, Di Pasquale A, Cammà C, Puglia I, Calistri P, Savini G |
| EPI_ISL_1392681 | SIESP CHIETI - DRIVE IN LANCIANO LANCIANO(CHIETI) | Istituto Zooprofilattico Sperimentale dell'Abruzzo e Molise "G. Caporale" | Lorusso A, Marcacci M, Di Domenico M, Ancora M, Curini V, Di Lollo Valeria, Mangone I, Rinaldi A, Delli Compagni E, Scialabba S, Caporale M, Di Pasquale A, Cammà C, Puglia I, Calistri P, Savini G |
| EPI_ISL_1392682 | SIESP DIPARTIMENTO DI PREVENZIONE TERAMO TERAMO(TERAMO) | Istituto Zooprofilattico Sperimentale dell'Abruzzo e Molise "G. Caporale" | Lorusso A, Marcacci M, Di Domenico M, Ancora M, Curini V, Di Lollo Valeria, Mangone I, Rinaldi A, Delli Compagni E, Scialabba S, Caporale M, Di Pasquale A, Cammà C, Puglia I, Calistri P, Savini G |
| EPI_ISL_1392683, EPI_ISL_1392684 | USCA Tagliacozzo TAGLIACOZZO(L'AQUILA) | Istituto Zooprofilattico Sperimentale dell'Abruzzo e Molise "G. Caporale" | Lorusso A, Marcacci M, Di Domenico M, Ancora M, Curini V, Di Lollo Valeria, Mangone I, Rinaldi A, Delli Compagni E, Scialabba S, Caporale M, Di Pasquale A, Cammà C, Puglia I, Calistri P, Savini G |
| EPI_ISL_1392685, EPI_ISL_1392686, EPI_ISL_1392687 | USCA Pescina PESCINA(L'AQUILA) | Istituto Zooprofilattico Sperimentale dell'Abruzzo e Molise "G. Caporale" | Lorusso A, Marcacci M, Di Domenico M, Ancora M, Curini V, Di Lollo Valeria, Mangone I, Rinaldi A, Delli Compagni E, Scialabba S, Caporale M, Di Pasquale A, Cammà C, Puglia I, Calistri P, Savini G |
| EPI_ISL_1392688, EPI_ISL_1392689 | SIESP CHIETI - DRIVE IN VASTO VASTO(CHIETI) | Istituto Zooprofilattico Sperimentale dell'Abruzzo e Molise "G. Caporale" | Lorusso A, Marcacci M, Di Domenico M, Ancora M, Curini V, Di Lollo Valeria, Mangone I, Rinaldi A, Delli Compagni E, Scialabba S, Caporale M, Di Pasquale A, Cammà C, Puglia I, Calistri P, Savini G |
| EPI_ISL_1392690 | SIESP DIPARTIMENTO DI PREVENZIONE CHIETI CHIETI(CHIETI) | Istituto Zooprofilattico Sperimentale dell'Abruzzo e Molise "G. Caporale" | Lorusso A, Marcacci M, Di Domenico M, Ancora M, Curini V, Di Lollo Valeria, Mangone I, Rinaldi A, Delli Compagni E, Scialabba S, Caporale M, Di Pasquale A, Cammà C, Puglia I, Calistri P, Savini G |
| EPI_ISL_1392691, EPI_ISL_1392692, EPI_ISL_1392693 | SIESP CHIETI - DRIVE IN GISSI GISSI(CHIETI) | Istituto Zooprofilattico Sperimentale dell'Abruzzo e Molise "G. Caporale" | Lorusso A, Marcacci M, Di Domenico M, Ancora M, Curini V, Di Lollo Valeria, Mangone I, Rinaldi A, Delli Compagni E, Scialabba S, Caporale M, Di Pasquale A, Cammà C, Puglia I, Calistri P, Savini G |
| EPI_ISL_1392694 | SIESP CHIETI - DRIVE IN VASTO VASTO(CHIETI) | Istituto Zooprofilattico Sperimentale dell'Abruzzo e Molise "G. Caporale" | Lorusso A, Marcacci M, Di Domenico M, Ancora M, Curini V, Di Lollo Valeria, Mangone I, Rinaldi A, Delli Compagni E, Scialabba S, Caporale M, Di Pasquale A, Cammà C, Puglia I, Calistri P, Savini G |
| EPI_ISL_1392695, EPI_ISL_1392696, EPI_ISL_1392697 | SIESP CHIETI - DRIVE IN LANCIANO LANCIANO(CHIETI) | Istituto Zooprofilattico Sperimentale dell'Abruzzo e Molise "G. Caporale" | Lorusso A, Marcacci M, Di Domenico M, Ancora M, Curini V, Di Lollo Valeria, Mangone I, Rinaldi A, Delli Compagni E, Scialabba S, Caporale M, Di Pasquale A, Cammà C, Puglia I, Calistri P, Savini G |
| EPI_ISL_1392698, EPI_ISL_1392699, EPI_ISL_1392700, EPI_ISL_1392701 | SIESP CHIETI - DRIVE IN CHIETI CHIETI(CHIETI) | Istituto Zooprofilattico Sperimentale dell'Abruzzo e Molise "G. Caporale" | Lorusso A, Marcacci M, Di Domenico M, Ancora M, Curini V, Di Lollo Valeria, Mangone I, Rinaldi A, Delli Compagni E, Scialabba S, Caporale M, Di Pasquale A, Cammà C, Puglia I, Calistri P, Savini G |
| EPI_ISL_1392702 | SIESP CHIETI - DRIVE IN ORTONA ORTONA(CHIETI) | Istituto Zooprofilattico Sperimentale dell'Abruzzo e Molise "G. Caporale" | Lorusso A, Marcacci M, Di Domenico M, Ancora M, Curini V, Di Lollo Valeria, Mangone I, Rinaldi A, Delli Compagni E, Scialabba S, Caporale M, Di Pasquale A, Cammà C, Puglia I, Calistri P, Savini G |
| EPI_ISL_1392703, EPI_ISL_1392704, EPI_ISL_1392705, EPI_ISL_1392706, EPI_ISL_1392707, EPI_ISL_1392708, EPI_ISL_1392709 | SIESP DIPARTIMENTO DI PREVENZIONE TERAMO TERAMO(TERAMO) | Istituto Zooprofilattico Sperimentale dell'Abruzzo e Molise "G. Caporale" | Lorusso A, Marcacci M, Di Domenico M, Ancora M, Curini V, Di Lollo Valeria, Mangone I, Rinaldi A, Delli Compagni E, Scialabba S, Caporale M, Di Pasquale A, Cammà C, Puglia I, Calistri P, Savini G |
| EPI_ISL_1392710 | SIESP CHIETI - DRIVE IN CHIETI CHIETI(CHIETI) | Istituto Zooprofilattico Sperimentale dell'Abruzzo e Molise "G. Caporale" | Lorusso A, Marcacci M, Di Domenico M, Ancora M, Curini V, Di Lollo Valeria, Mangone I, Rinaldi A, Delli Compagni E, Scialabba S, Caporale M, Di Pasquale A, Cammà C, Puglia I, Calistri P, Savini G |
| EPI_ISL_1392711, EPI_ISL_1392712 | DIP. PREV. AVEZZANO SERVIZIO DI IGIENE EPIDEMIOLOGIA E SANITA' PUBBLICA AVEZZANO(L'AQUILA) | Istituto Zooprofilattico Sperimentale dell'Abruzzo e Molise "G. Caporale" | Lorusso A, Marcacci M, Di Domenico M, Ancora M, Curini V, Di Lollo Valeria, Mangone I, Rinaldi A, Delli Compagni E, Scialabba S, Caporale M, Di Pasquale A, Cammà C, Puglia I, Calistri P, Savini G |

| | | | |
|---|---|---|---|
| EPI_ISL_1392713 | SIESP DIPARTIMENTO DI PREVENZIONE TERAMO TERAMO(TERAMO) | Istituto Zooprofilattico Sperimentale dell'Abruzzo e Molise "G. Caporale" | Lorusso A, Marcacci M, Di Domenico M, Ancora M, Curini V, Di Lollo Valeria, Mangone I, Rinaldi A, Delli Compagni E, Scialabba S, Caporale M, Di Pasquale A, Cammà C, Puglia I, Calistri P, Savini G |
| EPI_ISL_1394738 | Laboratorio Analisi Osp. Città di Castello - Azienda USL Umbria1 | Istituto Zooprofilattico Sperimentale dell'Abruzzo e Molise "G. Caporale" | Malagigi V, Tacconi P, Lorusso A, Marcacci M, Di Domenico M, Ancora M, Curini V, Di Lollo Valeria, Mangone I, Rinaldi A, Scialabba S, Caporale M, Di Pasquale A, Cammà C, Puglia I, Calistri P, Savini G |
| EPI_ISL_1394739, EPI_ISL_1394740, EPI_ISL_1394741, EPI_ISL_1394742, EPI_ISL_1394743 | Azienda USL Umbria 2 | Istituto Zooprofilattico Sperimentale dell'Abruzzo e Molise "G. Caporale" | Proietti A, Pistoni E, Lorusso A, Marcacci M, Di Domenico M, Ancora M, Curini V, Di Lollo Valeria, Mangone I, Rinaldi A, Scialabba S, Caporale M, Di Pasquale A, Cammà C, Puglia I, Calistri P, Savini G |
| EPI_ISL_1394744, EPI_ISL_1394745, EPI_ISL_1394746, EPI_ISL_1394747, EPI_ISL_1394748 | Azienda Ospedaliera Terni | Istituto Zooprofilattico Sperimentale dell'Abruzzo e Molise "G. Caporale" | Palumbo M, Scaccetti A, Lorusso A, Marcacci M, Di Domenico M, Ancora M, Curini V, Di Lollo Valeria, Mangone I, Rinaldi A, Scialabba S, Caporale M, Di Pasquale A, Cammà C, Puglia I, Calistri P, Savini G |
| EPI_ISL_1394749, EPI_ISL_1394750, EPI_ISL_1394751, EPI_ISL_1394752 | Istituto Zooprofilattico Sperimentale Umbria e Marche "Togo Rosati" | Istituto Zooprofilattico Sperimentale dell'Abruzzo e Molise "G. Caporale" | Biagetti M, Giammarioli M, Lorusso A, Marcacci M, Di Domenico M, Ancora M, Curini V, Di Lollo Valeria, Mangone I, Rinaldi A, Scialabba S, Caporale M, Di Pasquale A, Cammà C, Puglia I, Calistri P, Savini G |
| EPI_ISL_1394753, EPI_ISL_1394754, EPI_ISL_1394755 | Università degli Studi di Perugia | Istituto Zooprofilattico Sperimentale dell'Abruzzo e Molise "G. Caporale" | Mencacci A, Camilloni B, Lorusso A, Marcacci M, Di Domenico M, Ancora M, Curini V, Di Lollo Valeria, Mangone I, Rinaldi A, Scialabba S, Caporale M, Di Pasquale A, Cammà C, Puglia I, Calistri P, Savini G |
| EPI_ISL_1394756, EPI_ISL_1394757 | Laboratorio Analisi Osp. Città di Castello - Azienda USL Umbria1 | Istituto Zooprofilattico Sperimentale dell'Abruzzo e Molise "G. Caporale" | Malagigi V, Tacconi P, Lorusso A, Marcacci M, Di Domenico M, Ancora M, Curini V, Di Lollo Valeria, Mangone I, Rinaldi A, Scialabba S, Caporale M, Di Pasquale A, Cammà C, Puglia I, Calistri P, Savini G |
| EPI_ISL_1394758 | Università degli Studi di Perugia | Istituto Zooprofilattico Sperimentale dell'Abruzzo e Molise "G. Caporale" | Mencacci A, Camilloni B, Lorusso A, Marcacci M, Di Domenico M, Ancora M, Curini V, Di Lollo Valeria, Mangone I, Rinaldi A, Scialabba S, Caporale M, Di Pasquale A, Cammà C, Puglia I, Calistri P, Savini G |
| EPI_ISL_1396408 | IZSM | TIGEM | Antonio Grimaldi,Patrizia Annunziata,Francesco Panariello,Biancamaria Pierri Claudia Tiberio Valentina Bouche,Chiara Colantuono,Maria Concetta Cuomo,Denise Di Concilio,Lucio Di Filippo,Anna Manfredi,Marcello Salvi,Antonio Limone Luigi Atripaldi Pellegrino Cerino,Andrea Ballabio,Davide Cacchiarelli |
| EPI_ISL_1396556, EPI_ISL_1396557 | SC Microbiologia e Virologia AOUSS | AMES Centro Polidiagnostico Strumentale S.r.l. | Salvatore Rubino; Flavia Angioj; Laura Firino; Rosalba Govoni; Gabriele Ibba; Vincenzo Lai; Erica Mura; Bianca Paglietti; Claudia Piu; Anna Puggioni; Elena Rimini; Giulia Rocca; Caterina Serra; Sergio Uzzau. |
| EPI_ISL_1396650 | SIESP CHIETI - DRIVE IN ORTONA | Istituto Zooprofilattico Sperimentale dell'Abruzzo e Molise "G. Caporale" | Lorusso A, Marcacci M, Di Domenico M, Ancora M, Curini V, Di Lollo Valeria, Mangone I, Rinaldi A, Scialabba S, Caporale M, Di Pasquale A, Cammà C, Puglia I, Calistri P, Savini G |
| EPI_ISL_1396651, EPI_ISL_1396652 | SIESP DIPARTIMENTO DI PREVENZIONE TERAMO | Istituto Zooprofilattico Sperimentale dell'Abruzzo e Molise "G. Caporale" | Lorusso A, Marcacci M, Di Domenico M, Ancora M, Curini V, Di Lollo Valeria, Mangone I, Rinaldi A, Scialabba S, Caporale M, Di Pasquale A, Cammà C, Puglia I, Calistri P, Savini G |
| EPI_ISL_1396653 | SIESP DIPARTIMENTO DI PREVENZIONE CHIETI | Istituto Zooprofilattico Sperimentale dell'Abruzzo e Molise "G. Caporale" | Lorusso A, Marcacci M, Di Domenico M, Ancora M, Curini V, Di Lollo Valeria, Mangone I, Rinaldi A, Scialabba S, Caporale M, Di Pasquale A, Cammà C, Puglia I, Calistri P, Savini G |
| EPI_ISL_1399230, EPI_ISL_1399246 | INT Fondazione Pascale | INT Fondazione Pascale | INT Fondazione Pascale |
| EPI_ISL_1403500, EPI_ISL_1403501 | PRESIDIO TERRITORIALE CASOLI CASOLI(CHIETI) | Istituto Zooprofilattico Sperimentale dell'Abruzzo e Molise "G. Caporale" | Lorusso A, Marcacci M, Di Domenico M, Ancora M, Curini V, Mangone I, Delli Compagni E, Rinaldi A, Scialabba S, Di Pasquale A, Cammà C, Puglia I, Calistri P, Savini G |
| EPI_ISL_1403502 | SIESP DIPARTIMENTO DI PREVENZIONE TERAMO TERAMO(TERAMO) | Istituto Zooprofilattico Sperimentale dell'Abruzzo e Molise "G. Caporale" | Lorusso A, Marcacci M, Di Domenico M, Ancora M, Curini V, Mangone I, Delli Compagni E, Rinaldi A, Scialabba S, Di Pasquale A, Cammà C, Puglia I, Calistri P, Savini G |
| EPI_ISL_1403503 | SIESP CHIETI - DRIVE IN GISSI GISSI(CHIETI) | Istituto Zooprofilattico Sperimentale dell'Abruzzo e Molise "G. Caporale" | Lorusso A, Marcacci M, Di Domenico M, Ancora M, Curini V, Mangone I, Delli Compagni E, Rinaldi A, Scialabba S, Di Pasquale A, Cammà C, Puglia I, Calistri P, Savini G |
| EPI_ISL_1403504 | USCA Tagliacozzo TAGLIACOZZO(L'AQUILA) | Istituto Zooprofilattico Sperimentale dell'Abruzzo e Molise "G. Caporale" | Lorusso A, Marcacci M, Di Domenico M, Ancora M, Curini V, Mangone I, Delli Compagni E, Rinaldi A, Scialabba S, Di Pasquale A, Cammà C, Puglia I, Calistri P, Savini G |
| EPI_ISL_1403505, EPI_ISL_1403506, EPI_ISL_1403507 | SIESP CHIETI - DRIVE IN GISSI GISSI(CHIETI) | Istituto Zooprofilattico Sperimentale dell'Abruzzo e Molise "G. Caporale" | Lorusso A, Marcacci M, Di Domenico M, Ancora M, Curini V, Mangone I, Delli Compagni E, Rinaldi A, Scialabba S, Di Pasquale A, Cammà C, Puglia I, Calistri P, Savini G |
| EPI_ISL_1403508 | OSPEDALE CIVILE GIULIANOVA - PRONTO SOCCORSO GIULIANOVA(TERAMO) | Istituto Zooprofilattico Sperimentale dell'Abruzzo e Molise "G. Caporale" | Lorusso A, Marcacci M, Di Domenico M, Ancora M, Curini V, Mangone I, Delli Compagni E, Rinaldi A, Scialabba S, Di Pasquale A, Cammà C, Puglia I, Calistri P, Savini G |
| EPI_ISL_1403509 | SIESP CHIETI - DRIVE IN ORTONA ORTONA(CHIETI) | Istituto Zooprofilattico Sperimentale dell'Abruzzo e Molise "G. Caporale" | Lorusso A, Marcacci M, Di Domenico M, Ancora M, Curini V, Mangone I, Delli Compagni E, Rinaldi A, Scialabba S, Di Pasquale A, Cammà C, Puglia I, Calistri P, Savini G |
| EPI_ISL_1403510 | MEDICO COMPETENTE P.O. SULMONA - CASTEL DI SANGRO SULMONA(L'AQUILA) | Istituto Zooprofilattico Sperimentale dell'Abruzzo e Molise "G. Caporale" | Lorusso A, Marcacci M, Di Domenico M, Ancora M, Curini V, Mangone I, Delli Compagni E, Rinaldi A, Scialabba S, Di Pasquale A, Cammà C, Puglia I, Calistri P, Savini G |
| EPI_ISL_1403511 | SIESP SULMONA SULMONA(L'AQUILA) | Istituto Zooprofilattico Sperimentale dell'Abruzzo e Molise "G. Caporale" | Lorusso A, Marcacci M, Di Domenico M, Ancora M, Curini V, Mangone I, Delli Compagni E, Rinaldi A, Scialabba S, Di Pasquale A, Cammà C, Puglia I, Calistri P, Savini G |
| EPI_ISL_1403512, EPI_ISL_1403513 | DIP. PREV. AVEZZANO SERVIZIO DI IGIENE EPIDEMIOLOGIA E SANITA' PUBBLICA AVEZZANO(L'AQUILA) | Istituto Zooprofilattico Sperimentale dell'Abruzzo e Molise "G. Caporale" | Lorusso A, Marcacci M, Di Domenico M, Ancora M, Curini V, Mangone I, Delli Compagni E, Rinaldi A, Scialabba S, Di Pasquale A, Cammà C, Puglia I, Calistri P, Savini G |
| EPI_ISL_1403514 | SIESP SULMONA SULMONA(L'AQUILA) | Istituto Zooprofilattico Sperimentale dell'Abruzzo e Molise "G. Caporale" | Lorusso A, Marcacci M, Di Domenico M, Ancora M, Curini V, Mangone I, Delli Compagni E, Rinaldi A, Scialabba S, Di Pasquale A, Cammà C, Puglia I, Calistri P, Savini G |
| EPI_ISL_1403515, EPI_ISL_1403516, EPI_ISL_1403517 | USCA Castel di Sangro CASTEL DI SANGRO(L'AQUILA) | Istituto Zooprofilattico Sperimentale dell'Abruzzo e Molise "G. Caporale" | Lorusso A, Marcacci M, Di Domenico M, Ancora M, Curini V, Mangone I, Delli Compagni E, Rinaldi A, Scialabba S, Di Pasquale A, Cammà C, Puglia I, Calistri P, Savini G |
| EPI_ISL_1403518 | SIESP DIPARTIMENTO DI PREVENZIONE TERAMO TERAMO(TERAMO) | Istituto Zooprofilattico Sperimentale dell'Abruzzo e Molise "G. Caporale" | Lorusso A, Marcacci M, Di Domenico M, Ancora M, Curini V, Mangone I, Delli Compagni E, Rinaldi A, Scialabba S, Di Pasquale A, Cammà C, Puglia I, Calistri P, Savini G |
| EPI_ISL_1403519 | SIESP CHIETI - DRIVE IN CHIETI CHIETI(CHIETI) | Istituto Zooprofilattico Sperimentale dell'Abruzzo e Molise "G. Caporale" | Lorusso A, Marcacci M, Di Domenico M, Ancora M, Curini V, Mangone I, Delli Compagni E, Rinaldi A, Scialabba S, Di Pasquale A, Cammà C, Puglia I, Calistri P, Savini G |
| EPI_ISL_1403520 | SIESP DIPARTIMENTO DI PREVENZIONE TERAMO TERAMO(TERAMO) | Istituto Zooprofilattico Sperimentale dell'Abruzzo e Molise "G. Caporale" | Lorusso A, Marcacci M, Di Domenico M, Ancora M, Curini V, Mangone I, Delli Compagni E, Rinaldi A, Scialabba S, Di Pasquale A, Cammà C, Puglia I, Calistri P, Savini G |
| EPI_ISL_1403521 | SIESP CHIETI - DRIVE IN ORTONA ORTONA(CHIETI) | Istituto Zooprofilattico Sperimentale dell'Abruzzo e Molise "G. Caporale" | Lorusso A, Marcacci M, Di Domenico M, Ancora M, Curini V, Mangone I, Delli Compagni E, Rinaldi A, Scialabba S, Di Pasquale A, Cammà C, Puglia I, Calistri P, Savini G |
| EPI_ISL_1403522 | SIESP DIPARTIMENTO DI PREVENZIONE TERAMO TERAMO(TERAMO) | Istituto Zooprofilattico Sperimentale dell'Abruzzo e Molise "G. Caporale" | Lorusso A, Marcacci M, Di Domenico M, Ancora M, Curini V, Mangone I, Delli Compagni E, Rinaldi A, Scialabba S, Di Pasquale A, Cammà C, Puglia I, Calistri P, Savini G |
| EPI_ISL_1403523, EPI_ISL_1403524 | SIESP SULMONA SULMONA(L'AQUILA) | Istituto Zooprofilattico Sperimentale dell'Abruzzo e Molise "G. Caporale" | Lorusso A, Marcacci M, Di Domenico M, Ancora M, Curini V, Mangone I, Delli Compagni E, Rinaldi A, Scialabba S, Di Pasquale A, Cammà C, Puglia I, Calistri P, Savini G |
| EPI_ISL_1403525 | SIESP CHIETI - DRIVE IN CHIETI CHIETI(CHIETI) | Istituto Zooprofilattico Sperimentale dell'Abruzzo e Molise "G. Caporale" | Lorusso A, Marcacci M, Di Domenico M, Ancora M, Curini V, Mangone I, Delli Compagni E, Rinaldi A, Scialabba S, Di Pasquale A, Cammà C, Puglia I, Calistri P, Savini G |
| EPI_ISL_1403526 | USCA Pescina PESCINA(L'AQUILA) | Istituto Zooprofilattico Sperimentale dell'Abruzzo e Molise "G. Caporale" | Lorusso A, Marcacci M, Di Domenico M, Ancora M, Curini V, Mangone I, Delli Compagni E, Rinaldi A, Scialabba S, Di Pasquale A, Cammà C, Puglia I, Calistri P, Savini G |

| | | | |
|---|---|---|---|
| EPI_ISL_1403527 | SIESP SULMONA SULMONA(L'AQUILA) | Istituto Zooprofilattico Sperimentale dell'Abruzzo e Molise "G. Caporale" | Lorusso A, Marcacci M, Di Domenico M, Ancora M, Curini V, Mangone I, Delli Compagni E, Rinaldi A, Scialabba S, Di Pasquale A, Cammà C, Puglia I, Calistri P, Savini G |
| EPI_ISL_1403528 | SIESP CHIETI - DRIVE IN CHIETI CHIETI(CHIETI) | Istituto Zooprofilattico Sperimentale dell'Abruzzo e Molise "G. Caporale" | Lorusso A, Marcacci M, Di Domenico M, Ancora M, Curini V, Mangone I, Delli Compagni E, Rinaldi A, Scialabba S, Di Pasquale A, Cammà C, Puglia I, Calistri P, Savini G |
| EPI_ISL_1403529 | USCA Sulmona SULMONA(L'AQUILA) | Istituto Zooprofilattico Sperimentale dell'Abruzzo e Molise "G. Caporale" | Lorusso A, Marcacci M, Di Domenico M, Ancora M, Curini V, Mangone I, Delli Compagni E, Rinaldi A, Scialabba S, Di Pasquale A, Cammà C, Puglia I, Calistri P, Savini G |
| EPI_ISL_1403530, EPI_ISL_1403531 | MEDICO COMPETENTE P.O. SULMONA - CASTEL DI SANGRO SULMONA(L'AQUILA) | Istituto Zooprofilattico Sperimentale dell'Abruzzo e Molise "G. Caporale" | Lorusso A, Marcacci M, Di Domenico M, Ancora M, Curini V, Mangone I, Delli Compagni E, Rinaldi A, Scialabba S, Di Pasquale A, Cammà C, Puglia I, Calistri P, Savini G |
| EPI_ISL_1403532 | SIESP CHIETI - DRIVE IN LANCIANO LANCIANO(CHIETI) | Istituto Zooprofilattico Sperimentale dell'Abruzzo e Molise "G. Caporale" | Lorusso A, Marcacci M, Di Domenico M, Ancora M, Curini V, Mangone I, Delli Compagni E, Rinaldi A, Scialabba S, Di Pasquale A, Cammà C, Puglia I, Calistri P, Savini G |
| EPI_ISL_1403533 | SIESP DIPARTIMENTO DI PREVENZIONE CHIETI CHIETI(CHIETI) | Istituto Zooprofilattico Sperimentale dell'Abruzzo e Molise "G. Caporale" | Lorusso A, Marcacci M, Di Domenico M, Ancora M, Curini V, Mangone I, Delli Compagni E, Rinaldi A, Scialabba S, Di Pasquale A, Cammà C, Puglia I, Calistri P, Savini G |
| EPI_ISL_1403534 | SIESP CHIETI - DRIVE IN ATESSA ATESSA(CHIETI) | Istituto Zooprofilattico Sperimentale dell'Abruzzo e Molise "G. Caporale" | Lorusso A, Marcacci M, Di Domenico M, Ancora M, Curini V, Mangone I, Delli Compagni E, Rinaldi A, Scialabba S, Di Pasquale A, Cammà C, Puglia I, Calistri P, Savini G |
| EPI_ISL_1403535 | SIESP CHIETI - DRIVE IN VASTO VASTO(CHIETI) | Istituto Zooprofilattico Sperimentale dell'Abruzzo e Molise "G. Caporale" | Lorusso A, Marcacci M, Di Domenico M, Ancora M, Curini V, Mangone I, Delli Compagni E, Rinaldi A, Scialabba S, Di Pasquale A, Cammà C, Puglia I, Calistri P, Savini G |
| EPI_ISL_1403536 | SIESP SULMONA SULMONA(L'AQUILA) | Istituto Zooprofilattico Sperimentale dell'Abruzzo e Molise "G. Caporale" | Lorusso A, Marcacci M, Di Domenico M, Ancora M, Curini V, Mangone I, Delli Compagni E, Rinaldi A, Scialabba S, Di Pasquale A, Cammà C, Puglia I, Calistri P, Savini G |
| EPI_ISL_1403537 | USCA Sulmona SULMONA(L'AQUILA) | Istituto Zooprofilattico Sperimentale dell'Abruzzo e Molise "G. Caporale" | Lorusso A, Marcacci M, Di Domenico M, Ancora M, Curini V, Mangone I, Delli Compagni E, Rinaldi A, Scialabba S, Di Pasquale A, Cammà C, Puglia I, Calistri P, Savini G |
| EPI_ISL_1403538, EPI_ISL_1403539 | SIESP SULMONA SULMONA(L'AQUILA) | Istituto Zooprofilattico Sperimentale dell'Abruzzo e Molise "G. Caporale" | Lorusso A, Marcacci M, Di Domenico M, Ancora M, Curini V, Mangone I, Delli Compagni E, Rinaldi A, Scialabba S, Di Pasquale A, Cammà C, Puglia I, Calistri P, Savini G |
| EPI_ISL_1403540 | SIESP DIPARTIMENTO DI PREVENZIONE TERAMO TERAMO(TERAMO) | Istituto Zooprofilattico Sperimentale dell'Abruzzo e Molise "G. Caporale" | Lorusso A, Marcacci M, Di Domenico M, Ancora M, Curini V, Mangone I, Delli Compagni E, Rinaldi A, Scialabba S, Di Pasquale A, Cammà C, Puglia I, Calistri P, Savini G |
| EPI_ISL_1403541 | DIP. PREV. AVEZZANO SERVIZIO DI IGIENE EPIDEMIOLOGIA E SANITA' PUBBLICA AVEZZANO(L'AQUILA) | Istituto Zooprofilattico Sperimentale dell'Abruzzo e Molise "G. Caporale" | Lorusso A, Marcacci M, Di Domenico M, Ancora M, Curini V, Mangone I, Delli Compagni E, Rinaldi A, Scialabba S, Di Pasquale A, Cammà C, Puglia I, Calistri P, Savini G |
| EPI_ISL_1403542 | USCA Pescina PESCINA(L'AQUILA) | Istituto Zooprofilattico Sperimentale dell'Abruzzo e Molise "G. Caporale" | Lorusso A, Marcacci M, Di Domenico M, Ancora M, Curini V, Mangone I, Delli Compagni E, Rinaldi A, Scialabba S, Di Pasquale A, Cammà C, Puglia I, Calistri P, Savini G |
| EPI_ISL_1403543 | SIESP CHIETI - DRIVE IN ORTONA ORTONA(CHIETI) | Istituto Zooprofilattico Sperimentale dell'Abruzzo e Molise "G. Caporale" | Lorusso A, Marcacci M, Di Domenico M, Ancora M, Curini V, Mangone I, Delli Compagni E, Rinaldi A, Scialabba S, Di Pasquale A, Cammà C, Puglia I, Calistri P, Savini G |
| EPI_ISL_1403544 | DIP. PREV. AVEZZANO SERVIZIO DI IGIENE EPIDEMIOLOGIA E SANITA' PUBBLICA AVEZZANO(L'AQUILA) | Istituto Zooprofilattico Sperimentale dell'Abruzzo e Molise "G. Caporale" | Lorusso A, Marcacci M, Di Domenico M, Ancora M, Curini V, Mangone I, Delli Compagni E, Rinaldi A, Scialabba S, Di Pasquale A, Cammà C, Puglia I, Calistri P, Savini G |
| EPI_ISL_1403545 | USCA Pescina PESCINA(L'AQUILA) | Istituto Zooprofilattico Sperimentale dell'Abruzzo e Molise "G. Caporale" | Lorusso A, Marcacci M, Di Domenico M, Ancora M, Curini V, Mangone I, Delli Compagni E, Rinaldi A, Scialabba S, Di Pasquale A, Cammà C, Puglia I, Calistri P, Savini G |
| EPI_ISL_1403546 | DIP. PREV. AVEZZANO SERVIZIO DI IGIENE EPIDEMIOLOGIA E SANITA' PUBBLICA AVEZZANO(L'AQUILA) | Istituto Zooprofilattico Sperimentale dell'Abruzzo e Molise "G. Caporale" | Lorusso A, Marcacci M, Di Domenico M, Ancora M, Curini V, Mangone I, Delli Compagni E, Rinaldi A, Scialabba S, Di Pasquale A, Cammà C, Puglia I, Calistri P, Savini G |
| EPI_ISL_1403547 | USCA Pescina PESCINA(L'AQUILA) | Istituto Zooprofilattico Sperimentale dell'Abruzzo e Molise "G. Caporale" | Lorusso A, Marcacci M, Di Domenico M, Ancora M, Curini V, Mangone I, Delli Compagni E, Rinaldi A, Scialabba S, Di Pasquale A, Cammà C, Puglia I, Calistri P, Savini G |
| EPI_ISL_1403548 | OSPEDALE CIVILE SANT'OMERO - PRONTO SOCCORSO SANT'OMERO(TERAMO) | Istituto Zooprofilattico Sperimentale dell'Abruzzo e Molise "G. Caporale" | Lorusso A, Marcacci M, Di Domenico M, Ancora M, Curini V, Mangone I, Delli Compagni E, Rinaldi A, Scialabba S, Di Pasquale A, Cammà C, Puglia I, Calistri P, Savini G |
| EPI_ISL_1403549, EPI_ISL_1403550, EPI_ISL_1403551 | SIESP DIPARTIMENTO DI PREVENZIONE TERAMO TERAMO(TERAMO) | Istituto Zooprofilattico Sperimentale dell'Abruzzo e Molise "G. Caporale" | Lorusso A, Marcacci M, Di Domenico M, Ancora M, Curini V, Mangone I, Delli Compagni E, Rinaldi A, Scialabba S, Di Pasquale A, Cammà C, Puglia I, Calistri P, Savini G |
| EPI_ISL_1403552 | OSPEDALE CIVILE GIULIANOVA - MEDICINA INTERNA GIULIANOVA(TERAMO) | Istituto Zooprofilattico Sperimentale dell'Abruzzo e Molise "G. Caporale" | Lorusso A, Marcacci M, Di Domenico M, Ancora M, Curini V, Mangone I, Delli Compagni E, Rinaldi A, Scialabba S, Di Pasquale A, Cammà C, Puglia I, Calistri P, Savini G |
| EPI_ISL_1403553, EPI_ISL_1403554 | SIESP DIPARTIMENTO DI PREVENZIONE TERAMO TERAMO(TERAMO) | Istituto Zooprofilattico Sperimentale dell'Abruzzo e Molise "G. Caporale" | Lorusso A, Marcacci M, Di Domenico M, Ancora M, Curini V, Mangone I, Delli Compagni E, Rinaldi A, Scialabba S, Di Pasquale A, Cammà C, Puglia I, Calistri P, Savini G |
| EPI_ISL_1403555 | OSPEDALE CIVILE SANT'OMERO - PRONTO SOCCORSO SANT'OMERO(TERAMO) | Istituto Zooprofilattico Sperimentale dell'Abruzzo e Molise "G. Caporale" | Lorusso A, Marcacci M, Di Domenico M, Ancora M, Curini V, Mangone I, Delli Compagni E, Rinaldi A, Scialabba S, Di Pasquale A, Cammà C, Puglia I, Calistri P, Savini G |
| EPI_ISL_1403556, EPI_ISL_1403557, EPI_ISL_1403558 | SIESP DIPARTIMENTO DI PREVENZIONE TERAMO TERAMO(TERAMO) | Istituto Zooprofilattico Sperimentale dell'Abruzzo e Molise "G. Caporale" | Lorusso A, Marcacci M, Di Domenico M, Ancora M, Curini V, Mangone I, Delli Compagni E, Rinaldi A, Scialabba S, Di Pasquale A, Cammà C, Puglia I, Calistri P, Savini G |
| EPI_ISL_1403559 | SIESP CHIETI - DRIVE IN VASTO VASTO(CHIETI) | Istituto Zooprofilattico Sperimentale dell'Abruzzo e Molise "G. Caporale" | Lorusso A, Marcacci M, Di Domenico M, Ancora M, Curini V, Mangone I, Delli Compagni E, Rinaldi A, Scialabba S, Di Pasquale A, Cammà C, Puglia I, Calistri P, Savini G |
| EPI_ISL_1403560 | SIESP CHIETI - DRIVE IN LANCIANO LANCIANO(CHIETI) | Istituto Zooprofilattico Sperimentale dell'Abruzzo e Molise "G. Caporale" | Lorusso A, Marcacci M, Di Domenico M, Ancora M, Curini V, Mangone I, Delli Compagni E, Rinaldi A, Scialabba S, Di Pasquale A, Cammà C, Puglia I, Calistri P, Savini G |
| EPI_ISL_1403561, EPI_ISL_1403562, EPI_ISL_1403563, EPI_ISL_1403564, EPI_ISL_1403565 | SIESP DIPARTIMENTO DI PREVENZIONE TERAMO TERAMO(TERAMO) | Istituto Zooprofilattico Sperimentale dell'Abruzzo e Molise "G. Caporale" | Lorusso A, Marcacci M, Di Domenico M, Ancora M, Curini V, Mangone I, Delli Compagni E, Rinaldi A, Scialabba S, Di Pasquale A, Cammà C, Puglia I, Calistri P, Savini G |
| EPI_ISL_1403566 | SIESP CHIETI - DRIVE IN CHIETI CHIETI(CHIETI) | Istituto Zooprofilattico Sperimentale dell'Abruzzo e Molise "G. Caporale" | Lorusso A, Marcacci M, Di Domenico M, Ancora M, Curini V, Mangone I, Delli Compagni E, Rinaldi A, Scialabba S, Di Pasquale A, Cammà C, Puglia I, Calistri P, Savini G |
| EPI_ISL_1403567 | SIESP DIPARTIMENTO DI PREVENZIONE TERAMO TERAMO(TERAMO) | Istituto Zooprofilattico Sperimentale dell'Abruzzo e Molise "G. Caporale" | Lorusso A, Marcacci M, Di Domenico M, Ancora M, Curini V, Mangone I, Delli Compagni E, Rinaldi A, Scialabba S, Di Pasquale A, Cammà C, Puglia I, Calistri P, Savini G |
| EPI_ISL_1403568 | SIESP CHIETI - DRIVE IN ORTONA ORTONA(CHIETI) | Istituto Zooprofilattico Sperimentale dell'Abruzzo e Molise "G. Caporale" | Lorusso A, Marcacci M, Di Domenico M, Ancora M, Curini V, Mangone I, Delli Compagni E, Rinaldi A, Scialabba S, Di Pasquale A, Cammà C, Puglia I, Calistri P, Savini G |
| EPI_ISL_1403569, EPI_ISL_1403570, EPI_ISL_1403571, EPI_ISL_1403572, EPI_ISL_1403573, EPI_ISL_1403574, EPI_ISL_1403575 | SIESP DIPARTIMENTO DI PREVENZIONE TERAMO TERAMO(TERAMO) | Istituto Zooprofilattico Sperimentale dell'Abruzzo e Molise "G. Caporale" | Lorusso A, Marcacci M, Di Domenico M, Ancora M, Curini V, Mangone I, Delli Compagni E, Rinaldi A, Scialabba S, Di Pasquale A, Cammà C, Puglia I, Calistri P, Savini G |
| EPI_ISL_1403576 | SIESP SULMONA SULMONA(L'AQUILA) | Istituto Zooprofilattico Sperimentale dell'Abruzzo e Molise "G. Caporale" | Lorusso A, Marcacci M, Di Domenico M, Ancora M, Curini V, Mangone I, Delli Compagni E, Rinaldi A, Scialabba S, Di Pasquale A, Cammà C, Puglia I, Calistri P, Savini G |

| | | | |
|---|---|---|---|
| EPI_ISL_1403577 | SIESP DIPARTIMENTO DI PREVENZIONE CHIETI CHIETI(CHIETI) | Istituto Zooprofilattico Sperimentale dell'Abruzzo e Molise "G. Caporale" | Lorusso A, Marcacci M, Di Domenico M, Ancora M, Curini V, Mangone I, Delli Compagni E, Rinaldi A, Scialabba S, Di Pasquale A, Cammà C, Puglia I, Calistri P, Savini G |
| EPI_ISL_1403578, EPI_ISL_1403579, EPI_ISL_1403580 | SIESP DIPARTIMENTO DI PREVENZIONE TERAMO TERAMO(TERAMO) | Istituto Zooprofilattico Sperimentale dell'Abruzzo e Molise "G. Caporale" | Lorusso A, Marcacci M, Di Domenico M, Ancora M, Curini V, Mangone I, Delli Compagni E, Rinaldi A, Scialabba S, Di Pasquale A, Cammà C, Puglia I, Calistri P, Savini G |
| EPI_ISL_1403581 | SIESP CHIETI - DRIVE IN ORTONA ORTONA(CHIETI) | Istituto Zooprofilattico Sperimentale dell'Abruzzo e Molise "G. Caporale" | Lorusso A, Marcacci M, Di Domenico M, Ancora M, Curini V, Mangone I, Delli Compagni E, Rinaldi A, Scialabba S, Di Pasquale A, Cammà C, Puglia I, Calistri P, Savini G |
| EPI_ISL_1403582, EPI_ISL_1403583 | SIESP DIPARTIMENTO DI PREVENZIONE TERAMO TERAMO(TERAMO) | Istituto Zooprofilattico Sperimentale dell'Abruzzo e Molise "G. Caporale" | Lorusso A, Marcacci M, Di Domenico M, Ancora M, Curini V, Mangone I, Delli Compagni E, Rinaldi A, Scialabba S, Di Pasquale A, Cammà C, Puglia I, Calistri P, Savini G |
| EPI_ISL_1403584 | DIP. PREV. AVEZZANO SERVIZIO DI IGIENE EPIDEMIOLOGIA E SANITA' PUBBLICA AVEZZANO(L'AQUILA) | Istituto Zooprofilattico Sperimentale dell'Abruzzo e Molise "G. Caporale" | Lorusso A, Marcacci M, Di Domenico M, Ancora M, Curini V, Mangone I, Delli Compagni E, Rinaldi A, Scialabba S, Di Pasquale A, Cammà C, Puglia I, Calistri P, Savini G |
| EPI_ISL_1403585, EPI_ISL_1403586, EPI_ISL_1403587, EPI_ISL_1403588, EPI_ISL_1403589, EPI_ISL_1403590, EPI_ISL_1403591 | PRESIDIO TERRITORIALE CASOLI CASOLI(CHIETI) | Istituto Zooprofilattico Sperimentale dell'Abruzzo e Molise "G. Caporale" | Lorusso A, Marcacci M, Di Domenico M, Ancora M, Curini V, Mangone I, Delli Compagni E, Rinaldi A, Scialabba S, Di Pasquale A, Cammà C, Puglia I, Calistri P, Savini G |
| EPI_ISL_1403592 | SIESP DIPARTIMENTO DI PREVENZIONE TERAMO TERAMO(TERAMO) | Istituto Zooprofilattico Sperimentale dell'Abruzzo e Molise "G. Caporale" | Lorusso A, Marcacci M, Di Domenico M, Ancora M, Curini V, Mangone I, Delli Compagni E, Rinaldi A, Scialabba S, Di Pasquale A, Cammà C, Puglia I, Calistri P, Savini G |
| EPI_ISL_1403593, EPI_ISL_1403594, EPI_ISL_1403595, EPI_ISL_1403596, EPI_ISL_1403597, EPI_ISL_1403598, EPI_ISL_1403599, EPI_ISL_1403600 | PRESIDIO TERRITORIALE CASOLI CASOLI(CHIETI) | Istituto Zooprofilattico Sperimentale dell'Abruzzo e Molise "G. Caporale" | Lorusso A, Marcacci M, Di Domenico M, Ancora M, Curini V, Mangone I, Delli Compagni E, Rinaldi A, Scialabba S, Di Pasquale A, Cammà C, Puglia I, Calistri P, Savini G |
| EPI_ISL_1403601 | DIP. PREV. AVEZZANO SERVIZIO DI IGIENE EPIDEMIOLOGIA E SANITA' PUBBLICA AVEZZANO(L'AQUILA) | Istituto Zooprofilattico Sperimentale dell'Abruzzo e Molise "G. Caporale" | Lorusso A, Marcacci M, Di Domenico M, Ancora M, Curini V, Mangone I, Delli Compagni E, Rinaldi A, Scialabba S, Di Pasquale A, Cammà C, Puglia I, Calistri P, Savini G |
| EPI_ISL_1403602, EPI_ISL_1403603 | SIESP DIPARTIMENTO DI PREVENZIONE TERAMO TERAMO(TERAMO) | Istituto Zooprofilattico Sperimentale dell'Abruzzo e Molise "G. Caporale" | Lorusso A, Marcacci M, Di Domenico M, Ancora M, Curini V, Mangone I, Delli Compagni E, Rinaldi A, Scialabba S, Di Pasquale A, Cammà C, Puglia I, Calistri P, Savini G |
| EPI_ISL_1403604 | SIESP CHIETI - DRIVE IN LANCIANO LANCIANO(CHIETI) | Istituto Zooprofilattico Sperimentale dell'Abruzzo e Molise "G. Caporale" | Lorusso A, Marcacci M, Di Domenico M, Ancora M, Curini V, Mangone I, Delli Compagni E, Rinaldi A, Scialabba S, Di Pasquale A, Cammà C, Puglia I, Calistri P, Savini G |
| EPI_ISL_1403605 | SIESP DIPARTIMENTO DI PREVENZIONE TERAMO TERAMO(TERAMO) | Istituto Zooprofilattico Sperimentale dell'Abruzzo e Molise "G. Caporale" | Lorusso A, Marcacci M, Di Domenico M, Ancora M, Curini V, Mangone I, Delli Compagni E, Rinaldi A, Scialabba S, Di Pasquale A, Cammà C, Puglia I, Calistri P, Savini G |
| EPI_ISL_1403606 | SIESP CHIETI - DRIVE IN LANCIANO LANCIANO(CHIETI) | Istituto Zooprofilattico Sperimentale dell'Abruzzo e Molise "G. Caporale" | Lorusso A, Marcacci M, Di Domenico M, Ancora M, Curini V, Mangone I, Delli Compagni E, Rinaldi A, Scialabba S, Di Pasquale A, Cammà C, Puglia I, Calistri P, Savini G |
| EPI_ISL_1404435 | Università di Parma, Laboratorio di Igiene e Sanità Pubblica | Istituto Zooprofilattico Sperimentale della Lombardia e dell'Emilia Romagna (IZSLER), Risk Analysis and Genomic Epidemiology Unit | Maria Eugenia Colucci, Licia Veronesi, Paola Affanni, Marina Morganti, Ilaria Menozzi, Erika Scaltriti, Stefano Pongolini |
| EPI_ISL_1406249 | INT Fondazione Pascale | INT Fondazione Pascale | INT Fondazione Pascale |
| EPI_ISL_1406296, EPI_ISL_1406297, EPI_ISL_1406298, EPI_ISL_1406299, EPI_ISL_1406300, EPI_ISL_1406301, EPI_ISL_1406302, EPI_ISL_1406303, EPI_ISL_1406304, EPI_ISL_1406305, EPI_ISL_1406306, EPI_ISL_1406307, EPI_ISL_1406308, EPI_ISL_1406309 | | | |
| see above | Laboratorio Aziendale di Microbiologia e Virologia, Azienda Sanitaria dell'Alto Adige | Laboratorio Aziendale di Microbiologia e Virologia, Azienda Sanitaria dell'Alto Adige | Irene Bianconi, Bartolomeo Mobilio Rodriguez, Elisabetta Giacobazzi, Elisa Masi, Stefanie Wieser, Anne Picard, Claudia Volpato, Chiara Cantaloni, Elisabetta Pagani |
| EPI_ISL_1406778 | Laboratorio Microbiologia PO Cardarelli | laboratorio Microbiologia PO Cardarelli | Felice Valentina,Giovanna Niro, Massimiliano Scutellà |
| EPI_ISL_1406779 | Laboratorio microbiologia PO Cardarelli | laboratorio microbiologia PO Cardarelli | Valentina Felice, Giovanna Niro, Massimiliano Scutellà |
| EPI_ISL_1406780 | Laboratorio microbiologia PO Cardarelli | Laboratorio microbiologia PO Cardarelli | Valentina Felice, Giovanna Niro, Massimiliano Scutellà |
| EPI_ISL_1406781 | Laboratorio microbiologia PO Cardarelli | laboratorio microbiologia PO Cardarelli | Valentina Felice, Giovanna Niro, Massimiliano Scutellà |
| EPI_ISL_1408885, EPI_ISL_1408886 | UOC Microbiologia e Virologia, Azienda Ospedaliera Universitaria Senese, Siena, Italy | Dipartimento di Biotecnologie Mediche | Maria Grazia Cusi, David Pinzauti, Claudia Gandolfo, Gabriele Anichini, Gianni Pozzi, Gianni Gori Savellini, Francesco Santoro |
| EPI_ISL_1412929 | SIESP CHIETI - DRIVE IN ORTONA ORTONA(CHIETI) | Istituto Zooprofilattico Sperimentale dell'Abruzzo e Molise "G. Caporale" | Lorusso A, Marcacci M, Di Domenico M, Ancora M, Curini V, Di Lollo Valeria, Mangone I, Rinaldi A, Delli Compagni E, Scialabba S, Caporale M, Di Pasquale A, Cammà C, Puglia I, Calistri P, Savini G |
| EPI_ISL_1412932 | SIESP CHIETI - DRIVE IN LANCIANO LANCIANO(CHIETI) | Istituto Zooprofilattico Sperimentale dell'Abruzzo e Molise "G. Caporale" | Lorusso A, Marcacci M, Di Domenico M, Ancora M, Curini V, Di Lollo Valeria, Mangone I, Rinaldi A, Delli Compagni E, Scialabba S, Caporale M, Di Pasquale A, Cammà C, Puglia I, Calistri P, Savini G |
| EPI_ISL_1412935 | SIESP CHIETI - DRIVE IN CHIETI CHIETI(CHIETI) | Istituto Zooprofilattico Sperimentale dell'Abruzzo e Molise "G. Caporale" | Lorusso A, Marcacci M, Di Domenico M, Ancora M, Curini V, Di Lollo Valeria, Mangone I, Rinaldi A, Delli Compagni E, Scialabba S, Caporale M, Di Pasquale A, Cammà C, Puglia I, Calistri P, Savini G |
| EPI_ISL_1415420 | SIESP DIPARTIMENTO DI PREVENZIONE TERAMO TERAMO(TERAMO) | Istituto Zooprofilattico Sperimentale dell'Abruzzo e Molise "G. Caporale" | Lorusso A, Marcacci M, Di Domenico M, Ancora M, Curini V, Di Lollo Valeria, Mangone I, Rinaldi A, Delli Compagni E, Scialabba S, Caporale M, Di Pasquale A, Cammà C, Puglia I, Calistri P, Savini G |
| EPI_ISL_1415421 | OSPEDALE CIVILE SANT'OMERO - PRONTO SOCCORSO SANT'OMERO(TERAMO) | Istituto Zooprofilattico Sperimentale dell'Abruzzo e Molise "G. Caporale" | Lorusso A, Marcacci M, Di Domenico M, Ancora M, Curini V, Di Lollo Valeria, Mangone I, Rinaldi A, Delli Compagni E, Scialabba S, Caporale M, Di Pasquale A, Cammà C, Puglia I, Calistri P, Savini G |
| EPI_ISL_1415422 | USCA Avezzano AVEZZANO(L'AQUILA) | Istituto Zooprofilattico Sperimentale dell'Abruzzo e Molise "G. Caporale" | Lorusso A, Marcacci M, Di Domenico M, Ancora M, Curini V, Di Lollo Valeria, Mangone I, Rinaldi A, Delli Compagni E, Scialabba S, Caporale M, Di Pasquale A, Cammà C, Puglia I, Calistri P, Savini G |
| EPI_ISL_1415423 | SIESP CHIETI - DRIVE IN ORTONA ORTONA(CHIETI) | Istituto Zooprofilattico Sperimentale dell'Abruzzo e Molise "G. Caporale" | Lorusso A, Marcacci M, Di Domenico M, Ancora M, Curini V, Di Lollo Valeria, Mangone I, Rinaldi A, Delli Compagni E, Scialabba S, Caporale M, Di Pasquale A, Cammà C, Puglia I, Calistri P, Savini G |
| EPI_ISL_1415424 | SIESP DIPARTIMENTO DI PREVENZIONE TERAMO TERAMO(TERAMO) | Istituto Zooprofilattico Sperimentale dell'Abruzzo e Molise "G. Caporale" | Lorusso A, Marcacci M, Di Domenico M, Ancora M, Curini V, Di Lollo Valeria, Mangone I, Rinaldi A, Delli Compagni E, Scialabba S, Caporale M, Di Pasquale A, Cammà C, Puglia I, Calistri P, Savini G |
| EPI_ISL_1415426 | SIESP CHIETI - DRIVE IN LANCIANO | Istituto Zooprofilattico Sperimentale dell'Abruzzo e Molise "G. Caporale" | Lorusso A, Marcacci M, Di Domenico M, Ancora M, Curini V, Di Lollo Valeria, Mangone I, Rinaldi A, Delli Compagni E, Scialabba S, Caporale M, Di Pasquale A, Cammà C, Puglia I, Calistri P, Savini G |
| EPI_ISL_1416312, EPI_ISL_1416313, EPI_ISL_1416314, EPI_ISL_1416315, EPI_ISL_1416316, EPI_ISL_1416317, EPI_ISL_1416318, EPI_ISL_1416319, EPI_ISL_1416320 | UOC Microbiologia e Virologia, Azienda Ospedaliera Universitaria Senese, Siena, Italy | Dipartimento di Biotecnologie Mediche | Maria Grazia Cusi, David Pinzauti, Claudia Gandolfo, Gabriele Anichini, Gianni Pozzi, Gianni Gori Savellini, Francesco Santoro |
| EPI_ISL_1417440, EPI_ISL_1417441 | Microbiology and Virology Laboratory, "Policlinico Riuniti, Azienda Ospedaliero Universitaria, Foggia" | Microbiology and Virology Laboratory, "Policlinico Riuniti, Azienda Ospedaliero Universitaria, Foggia" | Fabio Arena, Settimia Altamura, Donatella Cedola, Valeria Delli Carri, Annamaria D'Aprile, Maria Rosaria Lipsi, Loris Micelli, Maria Pia Patrizio, Daniela Pisanelli, Tiziana Rollo, Maurizio Margaglione and Rosella De Nittis. |
| EPI_ISL_1420829, EPI_ISL_1420830 | SIESP CHIETI - DRIVE IN ORTONA ORTONA(CHIETI) | Istituto Zooprofilattico Sperimentale dell'Abruzzo e Molise "G. | Lorusso A, Marcacci M, Di Domenico M, Ancora M, Curini V, Mangone I, Delli Compagni E, Rinaldi A, Scialabba S, Di Pasquale A, Cammà C, Puglia I, |

| | | | |
|---|---|---|---|
| | | Caporale" | Calistri P, Savini G |
| EPI_ISL_1420831 | PRESIDIO TERRITORIALE CASOLI CASOLI(CHIETI) | Istituto Zooprofilattico Sperimentale dell'Abruzzo e Molise "G. Caporale" | Lorusso A, Marcacci M, Di Domenico M, Ancora M, Curini V, Mangone I, Delli Compagni E, Rinaldi A, Scialabba S, Di Pasquale A, Cammà C, Puglia I, Calistri P, Savini G |
| EPI_ISL_1420832 | USCA Tagliacozzo TAGLIACOZZO(L'AQUILA) | Istituto Zooprofilattico Sperimentale dell'Abruzzo e Molise "G. Caporale" | Lorusso A, Marcacci M, Di Domenico M, Ancora M, Curini V, Mangone I, Delli Compagni E, Rinaldi A, Scialabba S, Di Pasquale A, Cammà C, Puglia I, Calistri P, Savini G |
| EPI_ISL_1424377, EPI_ISL_1424378, EPI_ISL_1424379, EPI_ISL_1424380, EPI_ISL_1424381, EPI_ISL_1424382, EPI_ISL_1424383, EPI_ISL_1424384, EPI_ISL_1424385, EPI_ISL_1424386, EPI_ISL_1424387, EPI_ISL_1424388, EPI_ISL_1424389, EPI_ISL_1424390, EPI_ISL_1424391, EPI_ISL_1424392, EPI_ISL_1424393, EPI_ISL_1424394, EPI_ISL_1424395, EPI_ISL_1424396 | | | |
| see above | Istituto Zooprofilattico Sperimentale del Mezzogiorno | TIGEM | Antonio Grimaldi,Patrizia Annunziata,Francesco Panariello,Biancamaria Pierri Claudia Tiberio Valentina Bouche,Chiara Colantuono,Maria Concetta Cuomo,Denise Di Concilio,Lucio Di Filippo,Anna Manfredi,Marcello Salvi,Antonio Limone Luigi Atripaldi Pellegrino Cerino,Andrea Ballabio,Davide Cacchiarelli |
| EPI_ISL_1424401, EPI_ISL_1424402, EPI_ISL_1424404, EPI_ISL_1424405, EPI_ISL_1424406, EPI_ISL_1424407, EPI_ISL_1424408, EPI_ISL_1424409, EPI_ISL_1424410, EPI_ISL_1424411, EPI_ISL_1424412, EPI_ISL_1424413, EPI_ISL_1424415, EPI_ISL_1424416, EPI_ISL_1424417, EPI_ISL_1424418, EPI_ISL_1424419, EPI_ISL_1424420, EPI_ISL_1424421, EPI_ISL_1424422, EPI_ISL_1424423, EPI_ISL_1424424, EPI_ISL_1424426, EPI_ISL_1424427, EPI_ISL_1424428, EPI_ISL_1424429, EPI_ISL_1424430, EPI_ISL_1424433, EPI_ISL_1424434, EPI_ISL_1424435, EPI_ISL_1424436, EPI_ISL_1424437, EPI_ISL_1424438, EPI_ISL_1424439, EPI_ISL_1424440, EPI_ISL_1424441, EPI_ISL_1424442, EPI_ISL_1424443, EPI_ISL_1424444, EPI_ISL_1424445, EPI_ISL_1424446, EPI_ISL_1424447, EPI_ISL_1424448, EPI_ISL_1424449, EPI_ISL_1424450, EPI_ISL_1424451, EPI_ISL_1424452, EPI_ISL_1424453, EPI_ISL_1424454, EPI_ISL_1424455, EPI_ISL_1424456, EPI_ISL_1424457, EPI_ISL_1424458, EPI_ISL_1424459, EPI_ISL_1424460, EPI_ISL_1424461, EPI_ISL_1424462, EPI_ISL_1424463, EPI_ISL_1424464, EPI_ISL_1424465 | | | |
| see above | Università Federico II - Dipartimento di scienze mediche traslazionali - Napoli | TIGEM | Antonio Grimaldi Patrizia Annunziata Francesco Panariello Michele Cennamo Valentina Bouche Chiara Colantuono Lucio Di Filippo Mariano Fiorenza Anna Manfredi Marcello Salvi Giuseppe Portella Andrea Ballabio Davide Cacchiarelli |
| EPI_ISL_1436338, EPI_ISL_1436340, EPI_ISL_1436342, EPI_ISL_1436343, EPI_ISL_1436345, EPI_ISL_1436347, EPI_ISL_1436349, EPI_ISL_1436351, EPI_ISL_1436353, EPI_ISL_1436355, EPI_ISL_1436357, EPI_ISL_1436358, EPI_ISL_1436360 | | | |
| see above | Università di Parma, Laboratorio di Igiene e Sanità Pubblica | Istituto Zooprofilattico Sperimentale della Lombardia e dell'Emilia Romagna (IZSLER), Risk Analysis and Genomic Epidemiology Unit | Maria Eugenia Colucci, Licia Veronesi, Paola Affanni, Marina Morganti, Ilaria Menozzi, Erika Scaltriti, Stefano Pongolini |
| EPI_ISL_1442147, EPI_ISL_1442149, EPI_ISL_1442151, EPI_ISL_1442153, EPI_ISL_1442155, EPI_ISL_1442162, EPI_ISL_1442166, EPI_ISL_1442169, EPI_ISL_1442174, EPI_ISL_1442179, EPI_ISL_1442185, EPI_ISL_1442187, EPI_ISL_1442190, EPI_ISL_1442192, EPI_ISL_1442194, EPI_ISL_1442196, EPI_ISL_1442198, EPI_ISL_1442200, EPI_ISL_1442202, EPI_ISL_1442207, EPI_ISL_1442216, EPI_ISL_1442218, EPI_ISL_1442220, EPI_ISL_1442222, EPI_ISL_1442224, EPI_ISL_1442227, EPI_ISL_1442229, EPI_ISL_1442231, EPI_ISL_1442233, EPI_ISL_1442235, EPI_ISL_1442237, EPI_ISL_1442238, EPI_ISL_1442244, EPI_ISL_1442249, EPI_ISL_1442254 | | | |
| see above | Azienda Ospedaliero - Universitaria di Modena Policlinico - Virologia e Microbiologia Molecolare | Istituto Zooprofilattico Sperimentale della Lombardia e dell'Emilia Romagna (IZSLER), Risk Analysis and Genomic Epidemiology Unit | Monica Pecorari, William Gennari, Giulia Fregni Serpini, Marina Morganti, Ilaria Menozzi, Erika Scaltriti, Stefano Pongolini |
| EPI_ISL_1442426, EPI_ISL_1442433, EPI_ISL_1442438, EPI_ISL_1442440, EPI_ISL_1442442, EPI_ISL_1442445, EPI_ISL_1442447, EPI_ISL_1442448, EPI_ISL_1442451, EPI_ISL_1442453, EPI_ISL_1442455, EPI_ISL_1442462, EPI_ISL_1442464, EPI_ISL_1442466, EPI_ISL_1442469, EPI_ISL_1442471, EPI_ISL_1442473, EPI_ISL_1442475, EPI_ISL_1442476, EPI_ISL_1442479, EPI_ISL_1442481, EPI_ISL_1442483, EPI_ISL_1442485, EPI_ISL_1442487, EPI_ISL_1442489 | | | |
| see above | Azienda Sanitaria Locale di Piacenza - Presidio Ospedaliero - Laboratorio di Microbiologia | Istituto Zooprofilattico Sperimentale della Lombardia e dell'Emilia Romagna (IZSLER), Risk Analysis and Genomic Epidemiology Unit | Lo Cascio Giuliana, Schiavo Roberta, Peroni Irene, Marina Morganti, Ilaria Menozzi, Erika Scaltriti, Stefano Pongolini |
| EPI_ISL_1442600, EPI_ISL_1442606, EPI_ISL_1442612, EPI_ISL_1442617, EPI_ISL_1442622, EPI_ISL_1442626, EPI_ISL_1442631, EPI_ISL_1442636, EPI_ISL_1442639, EPI_ISL_1442644, EPI_ISL_1442650, EPI_ISL_1442654, EPI_ISL_1442656, EPI_ISL_1442658, EPI_ISL_1442660, EPI_ISL_1442662, EPI_ISL_1442664, EPI_ISL_1442666, EPI_ISL_1442667, EPI_ISL_1442671, EPI_ISL_1442676, EPI_ISL_1442678, EPI_ISL_1442680, EPI_ISL_1442682 | | | |
| see above | SC Microbiologia e Virologia AOUSS | AMES Centro Polidiagnostico Strumentale S.r.l. | Salvatore Rubino; Flavia Angioj; Laura Firino; Rosalba Govoni; Gabriele Ibba; Vincenzo Lai; Erica Mura; Bianca Paglietti; Claudia Piu; Anna Puggioni; Elena Rimini; Giulia Rocca; Caterina Serra; Sergio Uzzau. |
| EPI_ISL_1442994, EPI_ISL_1442995, EPI_ISL_1442996, EPI_ISL_1442997, EPI_ISL_1442998, EPI_ISL_1442999 | Laboratorio di Genetica Medica Matera, Italy | Laboratorio di Genetica Medica Matera, Italy | Domenico Dell'Edera, Arianna Allegretti, Angela Mitidieri, Simone Francesca |
| EPI_ISL_1443004, EPI_ISL_1443005, EPI_ISL_1443006, EPI_ISL_1443007, EPI_ISL_1443008, EPI_ISL_1443009, EPI_ISL_1443010, EPI_ISL_1443011, EPI_ISL_1443012, EPI_ISL_1443013, EPI_ISL_1443014, EPI_ISL_1443015, EPI_ISL_1443016, EPI_ISL_1443017, EPI_ISL_1443018, EPI_ISL_1443019, EPI_ISL_1443020, EPI_ISL_1443021, EPI_ISL_1443022, EPI_ISL_1443023, EPI_ISL_1443024, EPI_ISL_1443025, EPI_ISL_1443026, EPI_ISL_1443027, EPI_ISL_1443028, EPI_ISL_1443029 | | | |
| see above | Arcispedale Santa Maria Nuova, Autoimmunità, Allergologia e Biotecnologie Innovative | Istituto Zooprofilattico Sperimentale della Lombardia e dell'Emilia Romagna (IZSLER), Risk Analysis and Genomic Epidemiology Unit | Alessandro Zerbini, Lucia Belloni, Stefania Croci, Marina Morganti, Ilaria Menozzi, Erika Scaltriti, Stefano Pongolini |
| EPI_ISL_1444732, EPI_ISL_1444733, EPI_ISL_1444734, EPI_ISL_1444735, EPI_ISL_1444736, EPI_ISL_1444737, EPI_ISL_1444738, EPI_ISL_1444739, EPI_ISL_1444740, EPI_ISL_1444741, EPI_ISL_1444742, EPI_ISL_1444743, EPI_ISL_1444744, EPI_ISL_1444745, EPI_ISL_1444746, EPI_ISL_1444747, EPI_ISL_1444748, EPI_ISL_1444749, EPI_ISL_1444750, EPI_ISL_1444751, EPI_ISL_1444752, EPI_ISL_1444753, EPI_ISL_1444754, EPI_ISL_1444755, EPI_ISL_1444756, EPI_ISL_1444757, EPI_ISL_1444758, EPI_ISL_1444759, EPI_ISL_1444760, EPI_ISL_1444761, EPI_ISL_1444762, EPI_ISL_1444763, EPI_ISL_1444764, EPI_ISL_1444765, EPI_ISL_1444766, EPI_ISL_1444767, EPI_ISL_1444768, EPI_ISL_1444769, EPI_ISL_1444770, EPI_ISL_1444771, EPI_ISL_1444772, EPI_ISL_1444773, EPI_ISL_1444774, EPI_ISL_1444775, EPI_ISL_1444776 | | | |
| see above | Laboratorio Aziendale di Microbiologia e Virologia, Azienda Sanitaria dell'Alto Adige | Laboratorio Aziendale di Microbiologia e Virologia, Azienda Sanitaria dell'Alto Adige | Irene Bianconi, Bartolomeo Mobilio Rodriguez, Elisabetta Giacobazzi, Elisa Masi, Stefanie Wieser, Anne Picard, Claudia Volpato, Chiara Cantaloni, Elisabetta Pagani |
| EPI_ISL_1447677, EPI_ISL_1447678, EPI_ISL_1447679, EPI_ISL_1447680, EPI_ISL_1447681, EPI_ISL_1447682, EPI_ISL_1447683, EPI_ISL_1447684, EPI_ISL_1447685, EPI_ISL_1447686, EPI_ISL_1447687, EPI_ISL_1447688, EPI_ISL_1447689, EPI_ISL_1447690, EPI_ISL_1447691, EPI_ISL_1447692, EPI_ISL_1447693, EPI_ISL_1447694, EPI_ISL_1447695, EPI_ISL_1447696, EPI_ISL_1447697, EPI_ISL_1447698, EPI_ISL_1447699, EPI_ISL_1447700, EPI_ISL_1447701, EPI_ISL_1447702, EPI_ISL_1447703, EPI_ISL_1447704, EPI_ISL_1447705, EPI_ISL_1447706, EPI_ISL_1447707, EPI_ISL_1447708, EPI_ISL_1447709, EPI_ISL_1447710, EPI_ISL_1447711, EPI_ISL_1447712, EPI_ISL_1447713, EPI_ISL_1447714, EPI_ISL_1447715, EPI_ISL_1447716, EPI_ISL_1447717, EPI_ISL_1447718, EPI_ISL_1447719, EPI_ISL_1447720, EPI_ISL_1447721, EPI_ISL_1447722, EPI_ISL_1447723, EPI_ISL_1447724, EPI_ISL_1447725, EPI_ISL_1447726, EPI_ISL_1447727, EPI_ISL_1447728, EPI_ISL_1447729, EPI_ISL_1447730, EPI_ISL_1447731, EPI_ISL_1447732, EPI_ISL_1447733, EPI_ISL_1447734, EPI_ISL_1447735, EPI_ISL_1447736, EPI_ISL_1447737, EPI_ISL_1447738, EPI_ISL_1447739, EPI_ISL_1447740, EPI_ISL_1447741, EPI_ISL_1447742, EPI_ISL_1447743, EPI_ISL_1447744, EPI_ISL_1447745, EPI_ISL_1447746, EPI_ISL_1447747, EPI_ISL_1447748, EPI_ISL_1447749, EPI_ISL_1447750, EPI_ISL_1447751, EPI_ISL_1447752, EPI_ISL_1447753, EPI_ISL_1447754, EPI_ISL_1447755, EPI_ISL_1447756, EPI_ISL_1447757, EPI_ISL_1447758, EPI_ISL_1447759, EPI_ISL_1447760, EPI_ISL_1447761, EPI_ISL_1447762, EPI_ISL_1447763, EPI_ISL_1447764, EPI_ISL_1447765, EPI_ISL_1447766, EPI_ISL_1447767, EPI_ISL_1447768, EPI_ISL_1447769, EPI_ISL_1447770, EPI_ISL_1447771, EPI_ISL_1447772, EPI_ISL_1447773, EPI_ISL_1447774, EPI_ISL_1447775, EPI_ISL_1447776, EPI_ISL_1447777, EPI_ISL_1447778, EPI_ISL_1447779, EPI_ISL_1447780, EPI_ISL_1447781, EPI_ISL_1447782, EPI_ISL_1447783, EPI_ISL_1447784, EPI_ISL_1447785, EPI_ISL_1447786, EPI_ISL_1447787, EPI_ISL_1447788, EPI_ISL_1447789, EPI_ISL_1447790, EPI_ISL_1447791, EPI_ISL_1447792, EPI_ISL_1447793, EPI_ISL_1447794, EPI_ISL_1447795, EPI_ISL_1447796, EPI_ISL_1447797, EPI_ISL_1447798, EPI_ISL_1447799, EPI_ISL_1447800, EPI_ISL_1447801, EPI_ISL_1447802, EPI_ISL_1447803, EPI_ISL_1447804, EPI_ISL_1447805, EPI_ISL_1447806, EPI_ISL_1447807, EPI_ISL_1447808, EPI_ISL_1447809, EPI_ISL_1447810, EPI_ISL_1447811, EPI_ISL_1447812, EPI_ISL_1447813, EPI_ISL_1447814, EPI_ISL_1447815, EPI_ISL_1447816, EPI_ISL_1447817, EPI_ISL_1447818, EPI_ISL_1447819, EPI_ISL_1447820, EPI_ISL_1447821, EPI_ISL_1447822, EPI_ISL_1447823, EPI_ISL_1447824, EPI_ISL_1447825, EPI_ISL_1447826, EPI_ISL_1447827, EPI_ISL_1447828, EPI_ISL_1447829, EPI_ISL_1447830, EPI_ISL_1447831, EPI_ISL_1447832, EPI_ISL_1447833, EPI_ISL_1447834, EPI_ISL_1447835, EPI_ISL_1447836, EPI_ISL_1447837, EPI_ISL_1447838, EPI_ISL_1447839, EPI_ISL_1447840, EPI_ISL_1447841, EPI_ISL_1447842, EPI_ISL_1447843, EPI_ISL_1447844, EPI_ISL_1447845, EPI_ISL_1447846, EPI_ISL_1447847, EPI_ISL_1447848, EPI_ISL_1447849, EPI_ISL_1447850, EPI_ISL_1447851, EPI_ISL_1447852, EPI_ISL_1447853, EPI_ISL_1447854, EPI_ISL_1447855, EPI_ISL_1447856, EPI_ISL_1447857, EPI_ISL_1447858, EPI_ISL_1447859, EPI_ISL_1447860, EPI_ISL_1447861, EPI_ISL_1447862, EPI_ISL_1447863, EPI_ISL_1447864, EPI_ISL_1447865, EPI_ISL_1447866, EPI_ISL_1447867, EPI_ISL_1447868, EPI_ISL_1447869, EPI_ISL_1447870, EPI_ISL_1447871, EPI_ISL_1447872, EPI_ISL_1447873, EPI_ISL_1447874, EPI_ISL_1447875, EPI_ISL_1447876, EPI_ISL_1447877, EPI_ISL_1447878, EPI_ISL_1447879, EPI_ISL_1447880, EPI_ISL_1447881, EPI_ISL_1447882, EPI_ISL_1447883, EPI_ISL_1447884, EPI_ISL_1447885, EPI_ISL_1447886, EPI_ISL_1447887, EPI_ISL_1447888, EPI_ISL_1447889, EPI_ISL_1447890, EPI_ISL_1447891, EPI_ISL_1447892, EPI_ISL_1447893, EPI_ISL_1447894, EPI_ISL_1447895, EPI_ISL_1447896, EPI_ISL_1447897, EPI_ISL_1447898, EPI_ISL_1447899, EPI_ISL_1447900, EPI_ISL_1447901, EPI_ISL_1447902, EPI_ISL_1447903, EPI_ISL_1447904, EPI_ISL_1447905, EPI_ISL_1447906, EPI_ISL_1447907, EPI_ISL_1447908, EPI_ISL_1447909, EPI_ISL_1447910, EPI_ISL_1447911, EPI_ISL_1447912, EPI_ISL_1447913, EPI_ISL_1447914, EPI_ISL_1447915, EPI_ISL_1447916, EPI_ISL_1447917, EPI_ISL_1447918, EPI_ISL_1447919, EPI_ISL_1447920, EPI_ISL_1447921, EPI_ISL_1447922, EPI_ISL_1447923, EPI_ISL_1447924, EPI_ISL_1447925 | | | |
| see above | IZSM | TIGEM | Antonio Grimaldi Patrizia Annunziata Francesco Panariello Biancamaria Pierri Claudia Tiberio Valentina Bouche Chiara Colantuono Maria Concetta Cuomo Denise Di Concilio Lucio Di Filippo Anna Manfredi Marcello Salvi Antonio Limone Luigi Atripaldi Pellegrino Cerino Andrea Ballabio Davide Cacchiarelli |
| EPI_ISL_1447926 | 20210303049 | TIGEM | Antonio Grimaldi Patrizia Annunziata Francesco Panariello Biancamaria Pierri Claudia Tiberio Valentina Bouche Chiara Colantuono Maria Concetta Cuomo Denise Di Concilio Lucio Di Filippo Anna Manfredi Marcello Salvi Antonio Limone Luigi Atripaldi Pellegrino Cerino Andrea Ballabio Davide Cacchiarelli |
| EPI_ISL_1447927 | 20210303149 | TIGEM | Antonio Grimaldi Patrizia Annunziata Francesco Panariello Biancamaria Pierri Claudia Tiberio Valentina Bouche Chiara Colantuono Maria Concetta Cuomo Denise Di Concilio Lucio Di Filippo Anna Manfredi Marcello Salvi Antonio Limone Luigi Atripaldi Pellegrino Cerino Andrea Ballabio Davide Cacchiarelli |
| EPI_ISL_1447928 | 20210303033 | TIGEM | Antonio Grimaldi Patrizia Annunziata Francesco Panariello Biancamaria Pierri Claudia Tiberio Valentina Bouche Chiara Colantuono Maria Concetta Cuomo Denise Di Concilio Lucio Di Filippo Anna Manfredi Marcello Salvi Antonio Limone Luigi Atripaldi Pellegrino Cerino Andrea Ballabio Davide Cacchiarelli |

| | | | |
|---|---|---|---|
| EPI_ISL_1447929 | 20210302964 | TIGEM | Antonio Grimaldi Patrizia Annunziata Francesco Panariello Biancamaria Pierri Claudia Tiberio Valentina Bouche Chiara Colantuono Maria Concetta Cuomo Denise Di Concilio Lucio Di Filippo Anna Manfredi Marcello Salvi Antonio Limone Luigi Atripaldi Pellegrino Cerino Andrea Ballabio Davide Cacchiarelli |
| EPI_ISL_1447930 | 20210302931 | TIGEM | Antonio Grimaldi Patrizia Annunziata Francesco Panariello Biancamaria Pierri Claudia Tiberio Valentina Bouche Chiara Colantuono Maria Concetta Cuomo Denise Di Concilio Lucio Di Filippo Anna Manfredi Marcello Salvi Antonio Limone Luigi Atripaldi Pellegrino Cerino Andrea Ballabio Davide Cacchiarelli |
| EPI_ISL_1447931 | 20210302924 | TIGEM | Antonio Grimaldi Patrizia Annunziata Francesco Panariello Biancamaria Pierri Claudia Tiberio Valentina Bouche Chiara Colantuono Maria Concetta Cuomo Denise Di Concilio Lucio Di Filippo Anna Manfredi Marcello Salvi Antonio Limone Luigi Atripaldi Pellegrino Cerino Andrea Ballabio Davide Cacchiarelli |
| EPI_ISL_1447932 | 20210302944 | TIGEM | Antonio Grimaldi Patrizia Annunziata Francesco Panariello Biancamaria Pierri Claudia Tiberio Valentina Bouche Chiara Colantuono Maria Concetta Cuomo Denise Di Concilio Lucio Di Filippo Anna Manfredi Marcello Salvi Antonio Limone Luigi Atripaldi Pellegrino Cerino Andrea Ballabio Davide Cacchiarelli |
| EPI_ISL_1447933 | 20210302988 | TIGEM | Antonio Grimaldi Patrizia Annunziata Francesco Panariello Biancamaria Pierri Claudia Tiberio Valentina Bouche Chiara Colantuono Maria Concetta Cuomo Denise Di Concilio Lucio Di Filippo Anna Manfredi Marcello Salvi Antonio Limone Luigi Atripaldi Pellegrino Cerino Andrea Ballabio Davide Cacchiarelli |
| EPI_ISL_1447934 | 20210302925 | TIGEM | Antonio Grimaldi Patrizia Annunziata Francesco Panariello Biancamaria Pierri Claudia Tiberio Valentina Bouche Chiara Colantuono Maria Concetta Cuomo Denise Di Concilio Lucio Di Filippo Anna Manfredi Marcello Salvi Antonio Limone Luigi Atripaldi Pellegrino Cerino Andrea Ballabio Davide Cacchiarelli |
| EPI_ISL_1447935 | 20210303140 | TIGEM | Antonio Grimaldi Patrizia Annunziata Francesco Panariello Biancamaria Pierri Claudia Tiberio Valentina Bouche Chiara Colantuono Maria Concetta Cuomo Denise Di Concilio Lucio Di Filippo Anna Manfredi Marcello Salvi Antonio Limone Luigi Atripaldi Pellegrino Cerino Andrea Ballabio Davide Cacchiarelli |
| EPI_ISL_1447936 | 20210302233 | TIGEM | Antonio Grimaldi Patrizia Annunziata Francesco Panariello Biancamaria Pierri Claudia Tiberio Valentina Bouche Chiara Colantuono Maria Concetta Cuomo Denise Di Concilio Lucio Di Filippo Anna Manfredi Marcello Salvi Antonio Limone Luigi Atripaldi Pellegrino Cerino Andrea Ballabio Davide Cacchiarelli |
| EPI_ISL_1447937 | 20210302215 | TIGEM | Antonio Grimaldi Patrizia Annunziata Francesco Panariello Biancamaria Pierri Claudia Tiberio Valentina Bouche Chiara Colantuono Maria Concetta Cuomo Denise Di Concilio Lucio Di Filippo Anna Manfredi Marcello Salvi Antonio Limone Luigi Atripaldi Pellegrino Cerino Andrea Ballabio Davide Cacchiarelli |
| EPI_ISL_1447938 | 20210302281 | TIGEM | Antonio Grimaldi Patrizia Annunziata Francesco Panariello Biancamaria Pierri Claudia Tiberio Valentina Bouche Chiara Colantuono Maria Concetta Cuomo Denise Di Concilio Lucio Di Filippo Anna Manfredi Marcello Salvi Antonio Limone Luigi Atripaldi Pellegrino Cerino Andrea Ballabio Davide Cacchiarelli |
| EPI_ISL_1447939 | 20210302302 | TIGEM | Antonio Grimaldi Patrizia Annunziata Francesco Panariello Biancamaria Pierri Claudia Tiberio Valentina Bouche Chiara Colantuono Maria Concetta Cuomo Denise Di Concilio Lucio Di Filippo Anna Manfredi Marcello Salvi Antonio Limone Luigi Atripaldi Pellegrino Cerino Andrea Ballabio Davide Cacchiarelli |
| EPI_ISL_1447940 | 20210302217 | TIGEM | Antonio Grimaldi Patrizia Annunziata Francesco Panariello Biancamaria Pierri Claudia Tiberio Valentina Bouche Chiara Colantuono Maria Concetta Cuomo Denise Di Concilio Lucio Di Filippo Anna Manfredi Marcello Salvi Antonio Limone Luigi Atripaldi Pellegrino Cerino Andrea Ballabio Davide Cacchiarelli |
| EPI_ISL_1447941 | 20210302039 | TIGEM | Antonio Grimaldi Patrizia Annunziata Francesco Panariello Biancamaria Pierri Claudia Tiberio Valentina Bouche Chiara Colantuono Maria Concetta Cuomo Denise Di Concilio Lucio Di Filippo Anna Manfredi Marcello Salvi Antonio Limone Luigi Atripaldi Pellegrino Cerino Andrea Ballabio Davide Cacchiarelli |
| EPI_ISL_1447942 | 20210302244 | TIGEM | Antonio Grimaldi Patrizia Annunziata Francesco Panariello Biancamaria Pierri Claudia Tiberio Valentina Bouche Chiara Colantuono Maria Concetta Cuomo Denise Di Concilio Lucio Di Filippo Anna Manfredi Marcello Salvi Antonio Limone Luigi Atripaldi Pellegrino Cerino Andrea Ballabio Davide Cacchiarelli |
| EPI_ISL_1447943 | 20210218154 | TIGEM | Antonio Grimaldi Patrizia Annunziata Francesco Panariello Biancamaria Pierri Claudia Tiberio Valentina Bouche Chiara Colantuono Maria Concetta Cuomo Denise Di Concilio Lucio Di Filippo Anna Manfredi Marcello Salvi Antonio Limone Luigi Atripaldi Pellegrino Cerino Andrea Ballabio Davide Cacchiarelli |
| EPI_ISL_1447944 | 20210302922 | TIGEM | Antonio Grimaldi Patrizia Annunziata Francesco Panariello Biancamaria Pierri Claudia Tiberio Valentina Bouche Chiara Colantuono Maria Concetta Cuomo Denise Di Concilio Lucio Di Filippo Anna Manfredi Marcello Salvi Antonio Limone Luigi Atripaldi Pellegrino Cerino Andrea Ballabio Davide Cacchiarelli |
| EPI_ISL_1447945 | 20210303025 | TIGEM | Antonio Grimaldi Patrizia Annunziata Francesco Panariello Biancamaria Pierri Claudia Tiberio Valentina Bouche Chiara Colantuono Maria Concetta Cuomo Denise Di Concilio Lucio Di Filippo Anna Manfredi Marcello Salvi Antonio Limone Luigi Atripaldi Pellegrino Cerino Andrea Ballabio Davide Cacchiarelli |
| EPI_ISL_1447946 | 20210302266 | TIGEM | Antonio Grimaldi Patrizia Annunziata Francesco Panariello Biancamaria Pierri Claudia Tiberio Valentina Bouche Chiara Colantuono Maria Concetta Cuomo Denise Di Concilio Lucio Di Filippo Anna Manfredi Marcello Salvi Antonio Limone Luigi Atripaldi Pellegrino Cerino Andrea Ballabio Davide Cacchiarelli |
| EPI_ISL_1447947 | 20210302250 | TIGEM | Antonio Grimaldi Patrizia Annunziata Francesco Panariello Biancamaria Pierri Claudia Tiberio Valentina Bouche Chiara Colantuono Maria Concetta Cuomo Denise Di Concilio Lucio Di Filippo Anna Manfredi Marcello Salvi Antonio Limone Luigi Atripaldi Pellegrino Cerino Andrea Ballabio Davide Cacchiarelli |
| EPI_ISL_1447948 | 20210218178 | TIGEM | Antonio Grimaldi Patrizia Annunziata Francesco Panariello Biancamaria Pierri Claudia Tiberio Valentina Bouche Chiara Colantuono Maria Concetta Cuomo Denise Di Concilio Lucio Di Filippo Anna Manfredi Marcello Salvi Antonio Limone Luigi Atripaldi Pellegrino Cerino Andrea Ballabio Davide Cacchiarelli |
| EPI_ISL_1447949 | 20210302239 | TIGEM | Antonio Grimaldi Patrizia Annunziata Francesco Panariello Biancamaria Pierri Claudia Tiberio Valentina Bouche Chiara Colantuono Maria Concetta Cuomo Denise Di Concilio Lucio Di Filippo Anna Manfredi Marcello Salvi Antonio Limone Luigi Atripaldi Pellegrino Cerino Andrea Ballabio Davide Cacchiarelli |
| EPI_ISL_1447952 | 20210217579 | TIGEM | Antonio Grimaldi Patrizia Annunziata Francesco Panariello Biancamaria Pierri Claudia Tiberio Valentina Bouche Chiara Colantuono Maria Concetta Cuomo Denise Di Concilio Lucio Di Filippo Anna Manfredi Marcello Salvi Antonio Limone Luigi Atripaldi Pellegrino Cerino Andrea Ballabio Davide Cacchiarelli |
| EPI_ISL_1447954 | 20210218337 | TIGEM | Antonio Grimaldi Patrizia Annunziata Francesco Panariello Biancamaria Pierri Claudia Tiberio Valentina Bouche Chiara Colantuono Maria Concetta Cuomo Denise Di Concilio Lucio Di Filippo Anna Manfredi Marcello Salvi Antonio Limone Luigi Atripaldi Pellegrino Cerino Andrea Ballabio Davide Cacchiarelli |
| EPI_ISL_1447955 | 20210218343 | TIGEM | Antonio Grimaldi Patrizia Annunziata Francesco Panariello Biancamaria Pierri Claudia Tiberio Valentina Bouche Chiara Colantuono Maria Concetta Cuomo Denise Di Concilio Lucio Di Filippo Anna Manfredi Marcello Salvi Antonio Limone Luigi Atripaldi Pellegrino Cerino Andrea Ballabio Davide Cacchiarelli |
| EPI_ISL_1447956 | 20210218316 | TIGEM | Antonio Grimaldi Patrizia Annunziata Francesco Panariello Biancamaria Pierri Claudia Tiberio Valentina Bouche Chiara Colantuono Maria Concetta Cuomo Denise Di Concilio Lucio Di Filippo Anna Manfredi Marcello Salvi Antonio Limone Luigi Atripaldi Pellegrino Cerino Andrea Ballabio Davide Cacchiarelli |

| | | | |
|---|---|---|---|
| EPI_ISL_1447959 | 20210218322 | TIGEM | Antonio Grimaldi Patrizia Annunziata Francesco Panariello Biancamaria Pierri Claudia Tiberio Valentina Bouche Chiara Colantuono Maria Concetta Cuomo Denise Di Concilio Lucio Di Filippo Anna Manfredi Marcello Salvi Antonio Limone Luigi Atripaldi Pellegrino Cerino Andrea Ballabio Davide Cacchiarelli |
| EPI_ISL_1447960 | 20210218311 | TIGEM | Antonio Grimaldi Patrizia Annunziata Francesco Panariello Biancamaria Pierri Claudia Tiberio Valentina Bouche Chiara Colantuono Maria Concetta Cuomo Denise Di Concilio Lucio Di Filippo Anna Manfredi Marcello Salvi Antonio Limone Luigi Atripaldi Pellegrino Cerino Andrea Ballabio Davide Cacchiarelli |
| EPI_ISL_1447962 | 20210300850 | TIGEM | Antonio Grimaldi Patrizia Annunziata Francesco Panariello Biancamaria Pierri Claudia Tiberio Valentina Bouche Chiara Colantuono Maria Concetta Cuomo Denise Di Concilio Lucio Di Filippo Anna Manfredi Marcello Salvi Antonio Limone Luigi Atripaldi Pellegrino Cerino Andrea Ballabio Davide Cacchiarelli |
| EPI_ISL_1447963 | 20210301640 | TIGEM | Antonio Grimaldi Patrizia Annunziata Francesco Panariello Biancamaria Pierri Claudia Tiberio Valentina Bouche Chiara Colantuono Maria Concetta Cuomo Denise Di Concilio Lucio Di Filippo Anna Manfredi Marcello Salvi Antonio Limone Luigi Atripaldi Pellegrino Cerino Andrea Ballabio Davide Cacchiarelli |
| EPI_ISL_1447964 | 20210301624 | TIGEM | Antonio Grimaldi Patrizia Annunziata Francesco Panariello Biancamaria Pierri Claudia Tiberio Valentina Bouche Chiara Colantuono Maria Concetta Cuomo Denise Di Concilio Lucio Di Filippo Anna Manfredi Marcello Salvi Antonio Limone Luigi Atripaldi Pellegrino Cerino Andrea Ballabio Davide Cacchiarelli |
| EPI_ISL_1447965 | 20210300911 | TIGEM | Antonio Grimaldi Patrizia Annunziata Francesco Panariello Biancamaria Pierri Claudia Tiberio Valentina Bouche Chiara Colantuono Maria Concetta Cuomo Denise Di Concilio Lucio Di Filippo Anna Manfredi Marcello Salvi Antonio Limone Luigi Atripaldi Pellegrino Cerino Andrea Ballabio Davide Cacchiarelli |
| EPI_ISL_1447966 | 20210300066 | TIGEM | Antonio Grimaldi Patrizia Annunziata Francesco Panariello Biancamaria Pierri Claudia Tiberio Valentina Bouche Chiara Colantuono Maria Concetta Cuomo Denise Di Concilio Lucio Di Filippo Anna Manfredi Marcello Salvi Antonio Limone Luigi Atripaldi Pellegrino Cerino Andrea Ballabio Davide Cacchiarelli |
| EPI_ISL_1447967 | 20210218375 | TIGEM | Antonio Grimaldi Patrizia Annunziata Francesco Panariello Biancamaria Pierri Claudia Tiberio Valentina Bouche Chiara Colantuono Maria Concetta Cuomo Denise Di Concilio Lucio Di Filippo Anna Manfredi Marcello Salvi Antonio Limone Luigi Atripaldi Pellegrino Cerino Andrea Ballabio Davide Cacchiarelli |
| EPI_ISL_1447968 | 20210300057 | TIGEM | Antonio Grimaldi Patrizia Annunziata Francesco Panariello Biancamaria Pierri Claudia Tiberio Valentina Bouche Chiara Colantuono Maria Concetta Cuomo Denise Di Concilio Lucio Di Filippo Anna Manfredi Marcello Salvi Antonio Limone Luigi Atripaldi Pellegrino Cerino Andrea Ballabio Davide Cacchiarelli |
| EPI_ISL_1447969 | 20210300061 | TIGEM | Antonio Grimaldi Patrizia Annunziata Francesco Panariello Biancamaria Pierri Claudia Tiberio Valentina Bouche Chiara Colantuono Maria Concetta Cuomo Denise Di Concilio Lucio Di Filippo Anna Manfredi Marcello Salvi Antonio Limone Luigi Atripaldi Pellegrino Cerino Andrea Ballabio Davide Cacchiarelli |
| EPI_ISL_1447970 | 20210218285 | TIGEM | Antonio Grimaldi Patrizia Annunziata Francesco Panariello Biancamaria Pierri Claudia Tiberio Valentina Bouche Chiara Colantuono Maria Concetta Cuomo Denise Di Concilio Lucio Di Filippo Anna Manfredi Marcello Salvi Antonio Limone Luigi Atripaldi Pellegrino Cerino Andrea Ballabio Davide Cacchiarelli |
| EPI_ISL_1447971 | 20210218329 | TIGEM | Antonio Grimaldi Patrizia Annunziata Francesco Panariello Biancamaria Pierri Claudia Tiberio Valentina Bouche Chiara Colantuono Maria Concetta Cuomo Denise Di Concilio Lucio Di Filippo Anna Manfredi Marcello Salvi Antonio Limone Luigi Atripaldi Pellegrino Cerino Andrea Ballabio Davide Cacchiarelli |
| EPI_ISL_1447972 | 20210300761 | TIGEM | Antonio Grimaldi Patrizia Annunziata Francesco Panariello Biancamaria Pierri Claudia Tiberio Valentina Bouche Chiara Colantuono Maria Concetta Cuomo Denise Di Concilio Lucio Di Filippo Anna Manfredi Marcello Salvi Antonio Limone Luigi Atripaldi Pellegrino Cerino Andrea Ballabio Davide Cacchiarelli |
| EPI_ISL_1447973 | 20210300708 | TIGEM | Antonio Grimaldi Patrizia Annunziata Francesco Panariello Biancamaria Pierri Claudia Tiberio Valentina Bouche Chiara Colantuono Maria Concetta Cuomo Denise Di Concilio Lucio Di Filippo Anna Manfredi Marcello Salvi Antonio Limone Luigi Atripaldi Pellegrino Cerino Andrea Ballabio Davide Cacchiarelli |
| EPI_ISL_1447974 | 20210301612 | TIGEM | Antonio Grimaldi Patrizia Annunziata Francesco Panariello Biancamaria Pierri Claudia Tiberio Valentina Bouche Chiara Colantuono Maria Concetta Cuomo Denise Di Concilio Lucio Di Filippo Anna Manfredi Marcello Salvi Antonio Limone Luigi Atripaldi Pellegrino Cerino Andrea Ballabio Davide Cacchiarelli |
| EPI_ISL_1447975 | 20210305370 | TIGEM | Antonio Grimaldi Patrizia Annunziata Francesco Panariello Biancamaria Pierri Claudia Tiberio Valentina Bouche Chiara Colantuono Maria Concetta Cuomo Denise Di Concilio Lucio Di Filippo Anna Manfredi Marcello Salvi Antonio Limone Luigi Atripaldi Pellegrino Cerino Andrea Ballabio Davide Cacchiarelli |
| EPI_ISL_1447976 | 20210305554 | TIGEM | Antonio Grimaldi Patrizia Annunziata Francesco Panariello Biancamaria Pierri Claudia Tiberio Valentina Bouche Chiara Colantuono Maria Concetta Cuomo Denise Di Concilio Lucio Di Filippo Anna Manfredi Marcello Salvi Antonio Limone Luigi Atripaldi Pellegrino Cerino Andrea Ballabio Davide Cacchiarelli |
| EPI_ISL_1447977 | 20210305752 | TIGEM | Antonio Grimaldi Patrizia Annunziata Francesco Panariello Biancamaria Pierri Claudia Tiberio Valentina Bouche Chiara Colantuono Maria Concetta Cuomo Denise Di Concilio Lucio Di Filippo Anna Manfredi Marcello Salvi Antonio Limone Luigi Atripaldi Pellegrino Cerino Andrea Ballabio Davide Cacchiarelli |
| EPI_ISL_1447978 | 20210300851 | TIGEM | Antonio Grimaldi Patrizia Annunziata Francesco Panariello Biancamaria Pierri Claudia Tiberio Valentina Bouche Chiara Colantuono Maria Concetta Cuomo Denise Di Concilio Lucio Di Filippo Anna Manfredi Marcello Salvi Antonio Limone Luigi Atripaldi Pellegrino Cerino Andrea Ballabio Davide Cacchiarelli |
| EPI_ISL_1447979 | 20210300858 | TIGEM | Antonio Grimaldi Patrizia Annunziata Francesco Panariello Biancamaria Pierri Claudia Tiberio Valentina Bouche Chiara Colantuono Maria Concetta Cuomo Denise Di Concilio Lucio Di Filippo Anna Manfredi Marcello Salvi Antonio Limone Luigi Atripaldi Pellegrino Cerino Andrea Ballabio Davide Cacchiarelli |
| EPI_ISL_1447980 | 20210301675 | TIGEM | Antonio Grimaldi Patrizia Annunziata Francesco Panariello Biancamaria Pierri Claudia Tiberio Valentina Bouche Chiara Colantuono Maria Concetta Cuomo Denise Di Concilio Lucio Di Filippo Anna Manfredi Marcello Salvi Antonio Limone Luigi Atripaldi Pellegrino Cerino Andrea Ballabio Davide Cacchiarelli |
| EPI_ISL_1447981 | 20210300406 | TIGEM | Antonio Grimaldi Patrizia Annunziata Francesco Panariello Biancamaria Pierri Claudia Tiberio Valentina Bouche Chiara Colantuono Maria Concetta Cuomo Denise Di Concilio Lucio Di Filippo Anna Manfredi Marcello Salvi Antonio Limone Luigi Atripaldi Pellegrino Cerino Andrea Ballabio Davide Cacchiarelli |
| EPI_ISL_1447982 | IZSM | TIGEM | Antonio Grimaldi Patrizia Annunziata Francesco Panariello Biancamaria Pierri Claudia Tiberio Valentina Bouche Chiara Colantuono Maria Concetta Cuomo Denise Di Concilio Lucio Di Filippo Anna Manfredi Marcello Salvi Antonio Limone Luigi Atripaldi Pellegrino Cerino Andrea Ballabio Davide Cacchiarelli |
| EPI_ISL_1447983 | 20210300801 | TIGEM | Antonio Grimaldi Patrizia Annunziata Francesco Panariello Biancamaria Pierri Claudia Tiberio Valentina Bouche Chiara Colantuono Maria Concetta Cuomo Denise Di Concilio Lucio Di Filippo Anna Manfredi Marcello Salvi Antonio Limone Luigi Atripaldi Pellegrino Cerino Andrea Ballabio Davide Cacchiarelli |
| EPI_ISL_1447984 | 20210300812 | TIGEM | Antonio Grimaldi Patrizia Annunziata Francesco Panariello Biancamaria Pierri Claudia Tiberio Valentina Bouche Chiara Colantuono Maria Concetta Cuomo Denise Di Concilio Lucio Di Filippo Anna Manfredi Marcello Salvi Antonio Limone Luigi Atripaldi Pellegrino Cerino Andrea Ballabio Davide Cacchiarelli |

| | | | |
|---|---|---|---|
| EPI_ISL_1447985 | 20210300868 | TIGEM | Antonio Grimaldi Patrizia Annunziata Francesco Panariello Biancamaria Pierri Claudia Tiberio Valentina Bouche Chiara Colantuono Maria Concetta Cuomo Denise Di Concilio Lucio Di Filippo Anna Manfredi Marcello Salvi Antonio Limone Luigi Atripaldi Pellegrino Cerino Andrea Ballabio Davide Cacchiarelli |
| EPI_ISL_1447986 | 20210301662 | TIGEM | Antonio Grimaldi Patrizia Annunziata Francesco Panariello Biancamaria Pierri Claudia Tiberio Valentina Bouche Chiara Colantuono Maria Concetta Cuomo Denise Di Concilio Lucio Di Filippo Anna Manfredi Marcello Salvi Antonio Limone Luigi Atripaldi Pellegrino Cerino Andrea Ballabio Davide Cacchiarelli |
| EPI_ISL_1447987 | 20210300728 | TIGEM | Antonio Grimaldi Patrizia Annunziata Francesco Panariello Biancamaria Pierri Claudia Tiberio Valentina Bouche Chiara Colantuono Maria Concetta Cuomo Denise Di Concilio Lucio Di Filippo Anna Manfredi Marcello Salvi Antonio Limone Luigi Atripaldi Pellegrino Cerino Andrea Ballabio Davide Cacchiarelli |
| EPI_ISL_1447988 | 20210300533 | TIGEM | Antonio Grimaldi Patrizia Annunziata Francesco Panariello Biancamaria Pierri Claudia Tiberio Valentina Bouche Chiara Colantuono Maria Concetta Cuomo Denise Di Concilio Lucio Di Filippo Anna Manfredi Marcello Salvi Antonio Limone Luigi Atripaldi Pellegrino Cerino Andrea Ballabio Davide Cacchiarelli |
| EPI_ISL_1447989 | 20210300725 | TIGEM | Antonio Grimaldi Patrizia Annunziata Francesco Panariello Biancamaria Pierri Claudia Tiberio Valentina Bouche Chiara Colantuono Maria Concetta Cuomo Denise Di Concilio Lucio Di Filippo Anna Manfredi Marcello Salvi Antonio Limone Luigi Atripaldi Pellegrino Cerino Andrea Ballabio Davide Cacchiarelli |
| EPI_ISL_1447990 | 20210303795 | TIGEM | Antonio Grimaldi Patrizia Annunziata Francesco Panariello Biancamaria Pierri Claudia Tiberio Valentina Bouche Chiara Colantuono Maria Concetta Cuomo Denise Di Concilio Lucio Di Filippo Anna Manfredi Marcello Salvi Antonio Limone Luigi Atripaldi Pellegrino Cerino Andrea Ballabio Davide Cacchiarelli |
| EPI_ISL_1447991 | 20210303889 | TIGEM | Antonio Grimaldi Patrizia Annunziata Francesco Panariello Biancamaria Pierri Claudia Tiberio Valentina Bouche Chiara Colantuono Maria Concetta Cuomo Denise Di Concilio Lucio Di Filippo Anna Manfredi Marcello Salvi Antonio Limone Luigi Atripaldi Pellegrino Cerino Andrea Ballabio Davide Cacchiarelli |
| EPI_ISL_1447992 | 20210304674 | TIGEM | Antonio Grimaldi Patrizia Annunziata Francesco Panariello Biancamaria Pierri Claudia Tiberio Valentina Bouche Chiara Colantuono Maria Concetta Cuomo Denise Di Concilio Lucio Di Filippo Anna Manfredi Marcello Salvi Antonio Limone Luigi Atripaldi Pellegrino Cerino Andrea Ballabio Davide Cacchiarelli |
| EPI_ISL_1447993 | 20210304470 | TIGEM | Antonio Grimaldi Patrizia Annunziata Francesco Panariello Biancamaria Pierri Claudia Tiberio Valentina Bouche Chiara Colantuono Maria Concetta Cuomo Denise Di Concilio Lucio Di Filippo Anna Manfredi Marcello Salvi Antonio Limone Luigi Atripaldi Pellegrino Cerino Andrea Ballabio Davide Cacchiarelli |
| EPI_ISL_1447994 | 20210303793 | TIGEM | Antonio Grimaldi Patrizia Annunziata Francesco Panariello Biancamaria Pierri Claudia Tiberio Valentina Bouche Chiara Colantuono Maria Concetta Cuomo Denise Di Concilio Lucio Di Filippo Anna Manfredi Marcello Salvi Antonio Limone Luigi Atripaldi Pellegrino Cerino Andrea Ballabio Davide Cacchiarelli |
| EPI_ISL_1447995 | 20210303977 | TIGEM | Antonio Grimaldi Patrizia Annunziata Francesco Panariello Biancamaria Pierri Claudia Tiberio Valentina Bouche Chiara Colantuono Maria Concetta Cuomo Denise Di Concilio Lucio Di Filippo Anna Manfredi Marcello Salvi Antonio Limone Luigi Atripaldi Pellegrino Cerino Andrea Ballabio Davide Cacchiarelli |
| EPI_ISL_1447996 | 20210303844 | TIGEM | Antonio Grimaldi Patrizia Annunziata Francesco Panariello Biancamaria Pierri Claudia Tiberio Valentina Bouche Chiara Colantuono Maria Concetta Cuomo Denise Di Concilio Lucio Di Filippo Anna Manfredi Marcello Salvi Antonio Limone Luigi Atripaldi Pellegrino Cerino Andrea Ballabio Davide Cacchiarelli |
| EPI_ISL_1447997 | 20210303976 | TIGEM | Antonio Grimaldi Patrizia Annunziata Francesco Panariello Biancamaria Pierri Claudia Tiberio Valentina Bouche Chiara Colantuono Maria Concetta Cuomo Denise Di Concilio Lucio Di Filippo Anna Manfredi Marcello Salvi Antonio Limone Luigi Atripaldi Pellegrino Cerino Andrea Ballabio Davide Cacchiarelli |
| EPI_ISL_1447998 | 20210303978 | TIGEM | Antonio Grimaldi Patrizia Annunziata Francesco Panariello Biancamaria Pierri Claudia Tiberio Valentina Bouche Chiara Colantuono Maria Concetta Cuomo Denise Di Concilio Lucio Di Filippo Anna Manfredi Marcello Salvi Antonio Limone Luigi Atripaldi Pellegrino Cerino Andrea Ballabio Davide Cacchiarelli |
| EPI_ISL_1447999 | 20210303971 | TIGEM | Antonio Grimaldi Patrizia Annunziata Francesco Panariello Biancamaria Pierri Claudia Tiberio Valentina Bouche Chiara Colantuono Maria Concetta Cuomo Denise Di Concilio Lucio Di Filippo Anna Manfredi Marcello Salvi Antonio Limone Luigi Atripaldi Pellegrino Cerino Andrea Ballabio Davide Cacchiarelli |
| EPI_ISL_1448000 | 20210304422 | TIGEM | Antonio Grimaldi Patrizia Annunziata Francesco Panariello Biancamaria Pierri Claudia Tiberio Valentina Bouche Chiara Colantuono Maria Concetta Cuomo Denise Di Concilio Lucio Di Filippo Anna Manfredi Marcello Salvi Antonio Limone Luigi Atripaldi Pellegrino Cerino Andrea Ballabio Davide Cacchiarelli |
| EPI_ISL_1448001 | 20210304568 | TIGEM | Antonio Grimaldi Patrizia Annunziata Francesco Panariello Biancamaria Pierri Claudia Tiberio Valentina Bouche Chiara Colantuono Maria Concetta Cuomo Denise Di Concilio Lucio Di Filippo Anna Manfredi Marcello Salvi Antonio Limone Luigi Atripaldi Pellegrino Cerino Andrea Ballabio Davide Cacchiarelli |
| EPI_ISL_1448002 | 20210304543 | TIGEM | Antonio Grimaldi Patrizia Annunziata Francesco Panariello Biancamaria Pierri Claudia Tiberio Valentina Bouche Chiara Colantuono Maria Concetta Cuomo Denise Di Concilio Lucio Di Filippo Anna Manfredi Marcello Salvi Antonio Limone Luigi Atripaldi Pellegrino Cerino Andrea Ballabio Davide Cacchiarelli |
| EPI_ISL_1448003 | 20210303842 | TIGEM | Antonio Grimaldi Patrizia Annunziata Francesco Panariello Biancamaria Pierri Claudia Tiberio Valentina Bouche Chiara Colantuono Maria Concetta Cuomo Denise Di Concilio Lucio Di Filippo Anna Manfredi Marcello Salvi Antonio Limone Luigi Atripaldi Pellegrino Cerino Andrea Ballabio Davide Cacchiarelli |
| EPI_ISL_1448004 | 20210303790 | TIGEM | Antonio Grimaldi Patrizia Annunziata Francesco Panariello Biancamaria Pierri Claudia Tiberio Valentina Bouche Chiara Colantuono Maria Concetta Cuomo Denise Di Concilio Lucio Di Filippo Anna Manfredi Marcello Salvi Antonio Limone Luigi Atripaldi Pellegrino Cerino Andrea Ballabio Davide Cacchiarelli |
| EPI_ISL_1448005 | 20210304373 | TIGEM | Antonio Grimaldi Patrizia Annunziata Francesco Panariello Biancamaria Pierri Claudia Tiberio Valentina Bouche Chiara Colantuono Maria Concetta Cuomo Denise Di Concilio Lucio Di Filippo Anna Manfredi Marcello Salvi Antonio Limone Luigi Atripaldi Pellegrino Cerino Andrea Ballabio Davide Cacchiarelli |
| EPI_ISL_1448006 | 20210304339 | TIGEM | Antonio Grimaldi Patrizia Annunziata Francesco Panariello Biancamaria Pierri Claudia Tiberio Valentina Bouche Chiara Colantuono Maria Concetta Cuomo Denise Di Concilio Lucio Di Filippo Anna Manfredi Marcello Salvi Antonio Limone Luigi Atripaldi Pellegrino Cerino Andrea Ballabio Davide Cacchiarelli |
| EPI_ISL_1448007 | 20210303845 | TIGEM | Antonio Grimaldi Patrizia Annunziata Francesco Panariello Biancamaria Pierri Claudia Tiberio Valentina Bouche Chiara Colantuono Maria Concetta Cuomo Denise Di Concilio Lucio Di Filippo Anna Manfredi Marcello Salvi Antonio Limone Luigi Atripaldi Pellegrino Cerino Andrea Ballabio Davide Cacchiarelli |
| EPI_ISL_1448008 | 20210303778 | TIGEM | Antonio Grimaldi Patrizia Annunziata Francesco Panariello Biancamaria Pierri Claudia Tiberio Valentina Bouche Chiara Colantuono Maria Concetta Cuomo Denise Di Concilio Lucio Di Filippo Anna Manfredi Marcello Salvi Antonio Limone Luigi Atripaldi Pellegrino Cerino Andrea Ballabio Davide Cacchiarelli |
| EPI_ISL_1448009 | 20210303943 | TIGEM | Antonio Grimaldi Patrizia Annunziata Francesco Panariello Biancamaria Pierri Claudia Tiberio Valentina Bouche Chiara Colantuono Maria Concetta Cuomo Denise Di Concilio Lucio Di Filippo Anna Manfredi Marcello Salvi Antonio Limone Luigi Atripaldi Pellegrino Cerino Andrea Ballabio Davide Cacchiarelli |

| | | | |
|---|---|---|---|
| EPI_ISL_1448010 | 20210302199 | TIGEM | Antonio Grimaldi Patrizia Annunziata Francesco Panariello Biancamaria Pierri Claudia Tiberio Valentina Bouche Chiara Colantuono Maria Concetta Cuomo Denise Di Concilio Lucio Di Filippo Anna Manfredi Marcello Salvi Antonio Limone Luigi Atripaldi Pellegrino Cerino Andrea Ballabio Davide Cacchiarelli |
| EPI_ISL_1448011 | 20210304393 | TIGEM | Antonio Grimaldi Patrizia Annunziata Francesco Panariello Biancamaria Pierri Claudia Tiberio Valentina Bouche Chiara Colantuono Maria Concetta Cuomo Denise Di Concilio Lucio Di Filippo Anna Manfredi Marcello Salvi Antonio Limone Luigi Atripaldi Pellegrino Cerino Andrea Ballabio Davide Cacchiarelli |
| EPI_ISL_1448012 | 20210304041 | TIGEM | Antonio Grimaldi Patrizia Annunziata Francesco Panariello Biancamaria Pierri Claudia Tiberio Valentina Bouche Chiara Colantuono Maria Concetta Cuomo Denise Di Concilio Lucio Di Filippo Anna Manfredi Marcello Salvi Antonio Limone Luigi Atripaldi Pellegrino Cerino Andrea Ballabio Davide Cacchiarelli |
| EPI_ISL_1448013 | 20210303909 | TIGEM | Antonio Grimaldi Patrizia Annunziata Francesco Panariello Biancamaria Pierri Claudia Tiberio Valentina Bouche Chiara Colantuono Maria Concetta Cuomo Denise Di Concilio Lucio Di Filippo Anna Manfredi Marcello Salvi Antonio Limone Luigi Atripaldi Pellegrino Cerino Andrea Ballabio Davide Cacchiarelli |
| EPI_ISL_1448014 | 20210304324 | TIGEM | Antonio Grimaldi Patrizia Annunziata Francesco Panariello Biancamaria Pierri Claudia Tiberio Valentina Bouche Chiara Colantuono Maria Concetta Cuomo Denise Di Concilio Lucio Di Filippo Anna Manfredi Marcello Salvi Antonio Limone Luigi Atripaldi Pellegrino Cerino Andrea Ballabio Davide Cacchiarelli |
| EPI_ISL_1448015 | 20210304551 | TIGEM | Antonio Grimaldi Patrizia Annunziata Francesco Panariello Biancamaria Pierri Claudia Tiberio Valentina Bouche Chiara Colantuono Maria Concetta Cuomo Denise Di Concilio Lucio Di Filippo Anna Manfredi Marcello Salvi Antonio Limone Luigi Atripaldi Pellegrino Cerino Andrea Ballabio Davide Cacchiarelli |
| EPI_ISL_1448509 | 1) Laboratorio Cerba 2) Università Milano-Bicocca 3) TaskForce Covid19-Ceinge Regione Campania | TaskForce Covid19-Ceinge Regione Campania | Francesco Broccolo (1,2) Ettore Capoluongo (3) Alessio Colombo (2) Adelaide Bussini (1) Nicola Perrelli (2) Silvia Cavalli (2) Alice Urbani (2) Giovanni Paolella (3) Veronica Ferrucci (3) Rossella Tufano (3) Angelo Boccia (3) Massimo Zollo (3) 1) Università Milano-Bicocca 2) Laboratorio Cerba 3) TaskForce Covid19-Ceinge Regione Campania |
| EPI_ISL_1455669 | INMI Lazzaro Spallanzani IRCCS | INMI Lazzaro Spallanzani IRCCS | M Rueca, G Bonfiglio, O Butera, CEM Gruber, F Santini, B Bartolini, E Giombini, F Messina, A Di Caro, MR Capobianchi |
| EPI_ISL_1455673 | TaskForce Covid19-Ceinge Regione Campania | TaskForce Covid19-Ceinge Regione Campania | Massimo Zollo (1) Ettore Capoluongo (1) Veronica Ferrucci (1) Rossella Tufano (1) Angelo Boccia (1) Giovanni Paolella (1) 1) TaskForce Covid19-Ceinge Regione Campania |
| EPI_ISL_1461253, EPI_ISL_1461254, EPI_ISL_1461255, EPI_ISL_1461256, EPI_ISL_1461257, EPI_ISL_1461258, EPI_ISL_1461259, EPI_ISL_1461260, EPI_ISL_1461261, EPI_ISL_1461262, EPI_ISL_1461263, EPI_ISL_1461264, EPI_ISL_1461265, EPI_ISL_1461266, EPI_ISL_1461267, EPI_ISL_1461268, EPI_ISL_1461269, EPI_ISL_1461270, EPI_ISL_1461271, EPI_ISL_1461272, EPI_ISL_1461273, EPI_ISL_1461274, EPI_ISL_1461275, EPI_ISL_1461276, EPI_ISL_1461277 | | | |
| see above | Laboratorio Aziendale di Microbiologia e Virologia, Azienda Sanitaria dell'Alto Adige | Laboratorio Aziendale di Microbiologia e Virologia, Azienda Sanitaria dell'Alto Adige | Irene Bianconi, Bartolomeo Mobilio Rodriguez, Elisabetta Giacobazzi, Elisa Masi, Stefanie Wieser, Anne Picard, Claudia Volpato, Chiara Cantaloni, Elisabetta Pagani |
| EPI_ISL_1464620 | INMI Lazzaro Spallanzani IRCCS | INMI Lazzaro Spallanzani IRCCS | B Bartolini, E Giombini, F Messina, M Rueca, G Bonfiglio, O Butera, CEM Gruber, F Santini, A Di Caro, MR Capobianchi |
| EPI_ISL_1464621 | Ospedale Cristo Re | INMI Lazzaro Spallanzani IRCCS | E Giombini, F Messina, M Rueca, G Bonfiglio, O Butera, CEM Gruber, F Santini, B Bartolini, MR Capobianchi, A Di Caro |
| EPI_ISL_1464622 | Ospedale S.M. Goretti | INMI Lazzaro Spallanzani IRCCS | F Messina, M Rueca, G Bonfiglio, O Butera, CEM Gruber, F Santini, B Bartolini, E Giombini, A Lucci, L Corso, L Di Biase, A Di Caro, MR Capobianchi |
| EPI_ISL_1464623 | Ospedale San Camillo De Lellis | INMI Lazzaro Spallanzani IRCCS | O Butera, CEM Gruber, F Santini, B Bartolini, E Giombini, F Messina, M Rueca, G Bonfiglio, S Venarubea, A De Luca , L Casertano, MR Capobianchi, A Di Caro |
| EPI_ISL_1464624 | Ospedale di Genzano - ASL RM 6 | INMI Lazzaro Spallanzani IRCCS | CEM Gruber, F Santini, B Bartolini, E Giombini, F Messina, M Rueca, G Bonfiglio, O Butera, G Tramini, E Conti, MR Capobianchi, A Di Caro |
| EPI_ISL_1464625 | Ospedale di Genzano - ASL RM 6 | INMI Lazzaro Spallanzani IRCCS | F Santini, B Bartolini, E Giombini, F Messina, M Rueca, G Bonfiglio, O Butera, CEM Gruber, G Tramini, E Conti, MR Capobianchi, A Di Caro |
| EPI_ISL_1469945, EPI_ISL_1469946, EPI_ISL_1469947, EPI_ISL_1469948, EPI_ISL_1469949, EPI_ISL_1469950, EPI_ISL_1469951, EPI_ISL_1469952, EPI_ISL_1469953, EPI_ISL_1469954, EPI_ISL_1469955, EPI_ISL_1469956, EPI_ISL_1469957, EPI_ISL_1469960, EPI_ISL_1469962, EPI_ISL_1469963 | | | |
| see above | University of Bari Biomedical Sciences and Human Oncology | University of Bari Biomedical Sciences and Human Oncology | Chironna M., Sallustio A., Loconsole D., Accogli M. |
| EPI_ISL_1483701, EPI_ISL_1483702 | laboratorio microbiologia PO Cardarelli | laboratorio microbiologia PO Cardarelli | Valentina Felice, Massimiliano Scutella. Giovanna Niro |
| EPI_ISL_1483704 | laboratorio micorbiologia PO Cardarelli | laboratorio micorbiologia PO Cardarelli | Valentina Felice, Massimiliano Scutella, Giovanna Niro |
| EPI_ISL_1483705, EPI_ISL_1483706 | laboratorio microbiologia PO Cardarelli | laboratorio microbiologia PO Cardarelli | Valentina Felice, Massimiliano Scutella, Giovanna Niro |
| EPI_ISL_1483788 | laboratorio microbiologia PO Cardarelli | laboratorio microbiologia PO Cardarelli | Valentina Felice, Giovanna Niro, Massimiliano Scutella |
| EPI_ISL_1489740, EPI_ISL_1489741, EPI_ISL_1489743, EPI_ISL_1489744, EPI_ISL_1489745, EPI_ISL_1489746, EPI_ISL_1489747, EPI_ISL_1489748, EPI_ISL_1489749, EPI_ISL_1489750, EPI_ISL_1489751, EPI_ISL_1489752, EPI_ISL_1489753, EPI_ISL_1489754, EPI_ISL_1489755, EPI_ISL_1489756, EPI_ISL_1489757, EPI_ISL_1489759, EPI_ISL_1489760, EPI_ISL_1489761, EPI_ISL_1489762, EPI_ISL_1489763, EPI_ISL_1489764, EPI_ISL_1489765, EPI_ISL_1489766, EPI_ISL_1489767, EPI_ISL_1489768, EPI_ISL_1489769, EPI_ISL_1489770, EPI_ISL_1489771, EPI_ISL_1489772, EPI_ISL_1489773, EPI_ISL_1489774, EPI_ISL_1489775, EPI_ISL_1489776, EPI_ISL_1489777, EPI_ISL_1489778, EPI_ISL_1489779, EPI_ISL_1489780, EPI_ISL_1489781, EPI_ISL_1489782, EPI_ISL_1489783, EPI_ISL_1489784, EPI_ISL_1489785, EPI_ISL_1489786, EPI_ISL_1489787, EPI_ISL_1489788, EPI_ISL_1489789, EPI_ISL_1489790, EPI_ISL_1489791, EPI_ISL_1489792, EPI_ISL_1489793, EPI_ISL_1489794, EPI_ISL_1489795, EPI_ISL_1489796, EPI_ISL_1489797, EPI_ISL_1489798, EPI_ISL_1489799, EPI_ISL_1489800, EPI_ISL_1489801, EPI_ISL_1489802, EPI_ISL_1489803, EPI_ISL_1489804, EPI_ISL_1489805, EPI_ISL_1489806, EPI_ISL_1489807, EPI_ISL_1489808, EPI_ISL_1489809, EPI_ISL_1489810, EPI_ISL_1489811, EPI_ISL_1489812, EPI_ISL_1489813, EPI_ISL_1489814, EPI_ISL_1489815, EPI_ISL_1489816, EPI_ISL_1489817, EPI_ISL_1489818, EPI_ISL_1489819, EPI_ISL_1489820, EPI_ISL_1489821, EPI_ISL_1489822, EPI_ISL_1489823, EPI_ISL_1489824, EPI_ISL_1489825, EPI_ISL_1489826, EPI_ISL_1489827, EPI_ISL_1489828, EPI_ISL_1489829, EPI_ISL_1489830, EPI_ISL_1489831, EPI_ISL_1489832, EPI_ISL_1489833, EPI_ISL_1489834 | | | |
| see above | Azienda Sanitaria dell'Alto Adige Laboratorio Aziendale di Microbiologia e Virologia | Istituto di Genomica Applicata | Elisabetta Pagani, Irene Bianconi, Elisabetta Giacobazzi, Elisa Masi, Stefanie Wieser, Irena Jurman, Vera Vendramin, Gabriele Magris, Eleonora Paparelli, Davide Scaglione, Michele Morgante |
| EPI_ISL_1492564 | Ospedale F.Spaziani | INMI Lazzaro Spallanzani IRCCS | G Bonfiglio, O Butera, CEM Gruber, F Santini, B Bartolini, E Giombini, F Messina, M Rueca, R Pulselli, G Brocco, C Gargiulo, A Di Caro, MR Capobianchi |
| EPI_ISL_1492565 | Ospedale di Genzano - ASL RM 6 | INMI Lazzaro Spallanzani IRCCS | B Bartolini, E Giombini, F Messina, M Rueca, G Bonfiglio, O Butera, CEM Gruber, F Santini, G Tramini, E Conti, A Di Caro, MR Capobianchi |
| EPI_ISL_1492566 | San Gallicano Dermatological Institute I.F.O. | INMI Lazzaro Spallanzani IRCCS | E Giombini, F Messina, M Rueca, G Bonfiglio, O Butera, CEM Gruber, F Santini, B Bartolini, F Pimpinelli, F Ensoli, G Prignano, MR Capobianchi, A Di Caro |
| EPI_ISL_1492567 | Ospedale San Filippo Neri | INMI Lazzaro Spallanzani IRCCS | CEM Gruber, F Santini, B Bartolini, E Giombini, F Messina, M Rueca, G Bonfiglio, O Butera, M Meledandri, ML Schiavone, A Tamburro, A Di Caro, MR Capobianchi |
| EPI_ISL_1492568 | Policlinico Casilino | INMI Lazzaro Spallanzani IRCCS | F Messina, M Rueca, G Bonfiglio, O Butera, CEM Gruber, F Santini, B Bartolini, E Giombini, A Di Caro, MR Capobianchi |
| EPI_ISL_1492569 | Ospedale dei Castelli | INMI Lazzaro Spallanzani IRCCS | E Giombini, F Messina, M Rueca, G Bonfiglio, O Butera, CEM Gruber, F Santini, B Bartolini, MR Capobianchi, A Di Caro |
| EPI_ISL_1492570 | San Gallicano Dermatological Institute I.F.O. | INMI Lazzaro Spallanzani IRCCS | M Rueca, G Bonfiglio, O Butera, CEM Gruber, F Santini, B Bartolini, E Giombini, F Messina, F Pimpinelli, F Ensoli, A Mastrofrancesco, MR Capobianchi, A Di Caro |
| EPI_ISL_1492571 | Ospedale San Filippo Neri | INMI Lazzaro Spallanzani IRCCS | G Bonfiglio, O Butera, CEM Gruber, F Santini, B Bartolini, E Giombini, F Messina, M Rueca, M Meledandri, ML Schiavone, A Tamburro, A Di Caro, MR Capobianchi |
| EPI_ISL_1492572 | INMI Lazzaro Spallanzani IRCCS | INMI Lazzaro Spallanzani IRCCS | O Butera, CEM Gruber, F Santini, B Bartolini, E Giombini, F Messina, M Rueca, G Bonfiglio, MR Capobianchi, A Di Caro |
| EPI_ISL_1492573 | Azienda Ospedaliera San Giovanni Addolorata | INMI Lazzaro Spallanzani IRCCS | G Bonfiglio, O Butera, CEM Gruber, F Santini, B Bartolini, E Giombini, F Messina, M Rueca, M Gaudio, PM Placanica, A Di Caro, MR Capobianchi |
| EPI_ISL_1492574 | San Gallicano Dermatological Institute I.F.O. | INMI Lazzaro Spallanzani IRCCS | O Butera, CEM Gruber, F Santini, B Bartolini, E Giombini, F Messina, M Rueca, G Bonfiglio, F Pimpinelli, F Ensoli, EG Di Domenico, MR Capobianchi, A Di Caro |
| EPI_ISL_1492578 | ACISMOM | INMI Lazzaro Spallanzani IRCCS | F Messina, M Rueca, G Bonfiglio, O Butera, CEM Gruber, F Santini, B Bartolini, E Giombini, MM Todini, I Giambini, A Di Caro, MR Capobianchi |
| EPI_ISL_1494675, EPI_ISL_1494676, EPI_ISL_1494677, EPI_ISL_1494678, EPI_ISL_1494679, EPI_ISL_1494680, EPI_ISL_1494681, EPI_ISL_1494682, EPI_ISL_1494683, EPI_ISL_1494684, EPI_ISL_1494685, EPI_ISL_1494686, EPI_ISL_1494687, EPI_ISL_1494688, EPI_ISL_1494689, EPI_ISL_1494690, EPI_ISL_1494691, EPI_ISL_1494692, EPI_ISL_1494693, EPI_ISL_1494694, EPI_ISL_1494695, EPI_ISL_1494696, EPI_ISL_1494697, EPI_ISL_1494698, EPI_ISL_1494699, EPI_ISL_1494700, EPI_ISL_1494701, EPI_ISL_1494702, EPI_ISL_1494703, EPI_ISL_1494704, EPI_ISL_1494705, EPI_ISL_1494706, EPI_ISL_1494707, EPI_ISL_1494708, EPI_ISL_1494709, EPI_ISL_1494710, EPI_ISL_1494711, EPI_ISL_1494712, EPI_ISL_1494713 | | | |

| | | | |
|---|---|---|---|
| see above | Laboratorio Aziendale di Microbiologia e Virologia, Azienda Sanitaria dell'Alto Adige | Laboratorio Aziendale di Microbiologia e Virologia, Azienda Sanitaria dell'Alto Adige | Irene Bianconi, Bartolomeo Mobilio Rodriguez, Elisabetta Giacobazzi, Elisa Masi, Stefanie Wieser, Anne Picard, Claudia Volpato, Chiara Cantaloni, Elisabetta Pagani |
| EPI_ISL_1495144 | Laboratorio microbiologia PO Cardarelli | Laboratorio microbiologia PO Cardarelli | Valentina Felice, Massimiliano Scutella, Giovanna Niro |
| EPI_ISL_1495145 | Laboratorio microbiologia PO Cardarelli | Laboratorio microbiologia PO Cardarelli | Valentna Felice, Massimiliano Scutella, Giovanna niro |
| EPI_ISL_1495146 | Laboratorio microbiologia PO Cardarelli | Laboratorio microbiologia PO Cardarelli | Valentina Felice, Massimiliano Scutella, Giovanna Niro |
| EPI_ISL_1495147 | Laboratorio microbiologia PO Cardarelli | Laboratorio microbiologia PO Cardarelli | Valentina felice, Massimiliano scutella, Giovanna Niro |
| EPI_ISL_1495148 | Laboratorio microbiologia PO Cardarelli | Laboratorio microbiologia PO Cardarelli | Valentina Felice, Massimiliano Scutella,Giovanna Niro |
| EPI_ISL_1495149 | Laboratorio microbiologia PO Cardarelli | Laboratorio microbiologia PO Cardarelli | Valentina Felice, Massimiliano Scutella, giovanna Niro |
| EPI_ISL_1495151 | Laboratorio microbiologia PO Cardarelli | Laboratorio microbiologia PO Cardarelli | Valentina Felice, Massimiliano Scutella, Giovanna Niro |
| EPI_ISL_1495972 | laboratorio microbiologia PO Cardarelli | laboratorio microbiologia PO Cardarelli | Valentina Felice, Massimiliano Scutella, Giovanna Niro |
| EPI_ISL_1495999 | Ospedale S.M. Goretti | INMI Lazzaro Spallanzani IRCCS | CEM Gruber, F Santini, B Bartolini, E Giombini, F Messina, M Rueca, G Bonfiglio, O Butera, A Lucci, L Corso, L Di Biase, A Di Caro, MR Capobianchi |
| EPI_ISL_1496000 | IRCCS San Raffaele | INMI Lazzaro Spallanzani IRCCS | F Santini, B Bartolini, E Giombini, F Messina, M Rueca, G Bonfiglio, O Butera, CEM Gruber, F Ciatti, D Russo, D Limongi, MR Capobianchi, A Di Caro |
| EPI_ISL_1496001 | Azienda Ospedaliera San Giovanni Addolorata | INMI Lazzaro Spallanzani IRCCS | M Rueca, G Bonfiglio, O Butera, CEM Gruber, F Santini, B Bartolini, E Giombini, F Messina, M Gaudio, PM Placanica, MR Capobianchi, A Di Caro |
| EPI_ISL_1496002 | INMI Lazzaro Spallanzani IRCCS | INMI Lazzaro Spallanzani IRCCS | F Messina, M Rueca, G Bonfiglio, O Butera, CEM Gruber, F Santini, B Bartolini, E Giombini, A Di Caro, MR Capobianchi |
| EPI_ISL_1497610 | Ospedale di Venere | Istituto Zooprofilattico Sperimentale della Puglia e della Basilicata | Parisi A., Bianco A., Capozzi L., Del Sambro L., Simone D., Iacobellis M., Ridolfi D., Giannico A. |
| EPI_ISL_1497612 | INT Fondazione Pascale | INT Fondazione Pascale | INT Fondazione Pascale |
| EPI_ISL_1499474, EPI_ISL_1499475, EPI_ISL_1499476, EPI_ISL_1499477, EPI_ISL_1499478, EPI_ISL_1499479, EPI_ISL_1499480, EPI_ISL_1499481, EPI_ISL_1499482, EPI_ISL_1499483, EPI_ISL_1499484, EPI_ISL_1499485, EPI_ISL_1499486, EPI_ISL_1499487, EPI_ISL_1499488, EPI_ISL_1499489, EPI_ISL_1499490, EPI_ISL_1499491, EPI_ISL_1499492, EPI_ISL_1499493, EPI_ISL_1499494, EPI_ISL_1499495, EPI_ISL_1499496, EPI_ISL_1499497, EPI_ISL_1499498, EPI_ISL_1499499, EPI_ISL_1499500, EPI_ISL_1499501, EPI_ISL_1499502, EPI_ISL_1499503 | | | |
| see above | Azienda Ospedaliera Terni | Istituto Zooprofilattico Sperimentale dell'Abruzzo e Molise "G. Caporale" | Palumbo M, Scaccetti A, Lorusso A, Marcacci M, Di Domenico M, Ancora M, Curini V, Di Lollo V, Delli Compagni E, Mangone I, Rinaldi A, Scialabba S, Caporale M, Di Pasquale A, Cammà C, Puglia I, Calistri P, Savini G |
| EPI_ISL_1499621, EPI_ISL_1499622, EPI_ISL_1499623, EPI_ISL_1499624, EPI_ISL_1499625, EPI_ISL_1499626, EPI_ISL_1499627 | Azienda Ospedaliera Terni | Istituto Zooprofilattico Sperimentale dell'Abruzzo e Molise "G. Caporale" | Palumbo M, Scaccetti A, Lorusso A, Marcacci M, Di Domenico M, Ancora M, Curini V, Di Lollo Valeria, Mangone I, Rinaldi A, Delli Compagni E, Scialabba S, Caporale M, Di Pasquale A, Cammà C, Puglia I, Calistri P, Savini G |
| EPI_ISL_1499628, EPI_ISL_1499629, EPI_ISL_1499630, EPI_ISL_1499631, EPI_ISL_1499632, EPI_ISL_1499633, EPI_ISL_1499634, EPI_ISL_1499635, EPI_ISL_1499636, EPI_ISL_1499637, EPI_ISL_1499638, EPI_ISL_1499639, EPI_ISL_1499640, EPI_ISL_1499641, EPI_ISL_1499642, EPI_ISL_1499643, EPI_ISL_1499644 | | | |
| see above | Università degli Studi di Perugia | Istituto Zooprofilattico Sperimentale dell'Abruzzo e Molise "G. Caporale" | Mencacci A, Camilloni B, Lorusso A, Marcacci M, Di Domenico M, Ancora M, Curini V, Di Lollo Valeria, Mangone I, Rinaldi A, Delli Compagni E, Scialabba S, Caporale M, Di Pasquale A, Cammà C, Puglia I, Calistri P, Savini G |
| EPI_ISL_1499645, EPI_ISL_1499646, EPI_ISL_1499647, EPI_ISL_1499648, EPI_ISL_1499649, EPI_ISL_1499650, EPI_ISL_1499651 | Laboratorio Analisi Osp. Città di Castello - Azienda USL Umbria1 | Istituto Zooprofilattico Sperimentale dell'Abruzzo e Molise "G. Caporale" | Malagigi V, Tacconi P,Lorusso A, Marcacci M, Di Domenico M, Ancora M, Curini V, Di Lollo Valeria, Mangone I, Rinaldi A, Delli Compagni E, Scialabba S, Caporale M, Di Pasquale A, Cammà C, Puglia I, Calistri P, Savini G |
| EPI_ISL_1500426, EPI_ISL_1500427, EPI_ISL_1500428, EPI_ISL_1500429, EPI_ISL_1500430, EPI_ISL_1500431, EPI_ISL_1500432 | UOC Microbiologia e Virologia, Azienda Ospedaliera Universitaria Senese, Siena, Italy | Dipartimento di Biotecnologie Mediche | Maria Grazia Cusi, David Pinzauti, Claudia Gandolfo, Gabriele Anichini, Gianni Pozzi, Gianni Gori Savellini, Francesco Santoro |
| EPI_ISL_1502435, EPI_ISL_1502436, EPI_ISL_1502437, EPI_ISL_1502438 | SIESP DIPARTIMENTO DI PREVENZIONE TERAMO TERAMO(TERAMO) | Istituto Zooprofilattico Sperimentale dell'Abruzzo e Molise "G. Caporale" | Lorusso A, Marcacci M, Di Domenico M, Ancora M, Curini V, Di Lollo Valeria, Mangone I, Rinaldi A, Delli Compagni E, Scialabba S, Caporale M, Di Pasquale A, Cammà C, Puglia I, Calistri P, Savini G |
| EPI_ISL_1502439 | SIESP DIPARTIMENTO DI PREVENZIONE CHIETI CHIETI(CHIETI) | Istituto Zooprofilattico Sperimentale dell'Abruzzo e Molise "G. Caporale" | Lorusso A, Marcacci M, Di Domenico M, Ancora M, Curini V, Di Lollo Valeria, Mangone I, Rinaldi A, Delli Compagni E, Scialabba S, Caporale M, Di Pasquale A, Cammà C, Puglia I, Calistri P, Savini G |
| EPI_ISL_1502440 | SIESP CHIETI - DRIVE IN LANCIANO LANCIANO(CHIETI) | Istituto Zooprofilattico Sperimentale dell'Abruzzo e Molise "G. Caporale" | Lorusso A, Marcacci M, Di Domenico M, Ancora M, Curini V, Di Lollo Valeria, Mangone I, Rinaldi A, Delli Compagni E, Scialabba S, Caporale M, Di Pasquale A, Cammà C, Puglia I, Calistri P, Savini G |
| EPI_ISL_1502441 | SIESP CHIETI - DRIVE IN ATESSA ATESSA(CHIETI) | Istituto Zooprofilattico Sperimentale dell'Abruzzo e Molise "G. Caporale" | Lorusso A, Marcacci M, Di Domenico M, Ancora M, Curini V, Di Lollo Valeria, Mangone I, Rinaldi A, Delli Compagni E, Scialabba S, Caporale M, Di Pasquale A, Cammà C, Puglia I, Calistri P, Savini G |
| EPI_ISL_1502442, EPI_ISL_1502443, EPI_ISL_1502444 | SIESP CHIETI - DRIVE IN LANCIANO LANCIANO(CHIETI) | Istituto Zooprofilattico Sperimentale dell'Abruzzo e Molise "G. Caporale" | Lorusso A, Marcacci M, Di Domenico M, Ancora M, Curini V, Di Lollo Valeria, Mangone I, Rinaldi A, Delli Compagni E, Scialabba S, Caporale M, Di Pasquale A, Cammà C, Puglia I, Calistri P, Savini G |
| EPI_ISL_1502445, EPI_ISL_1502446 | SIESP CHIETI - DRIVE IN VASTO VASTO(CHIETI) | Istituto Zooprofilattico Sperimentale dell'Abruzzo e Molise "G. Caporale" | Lorusso A, Marcacci M, Di Domenico M, Ancora M, Curini V, Di Lollo Valeria, Mangone I, Rinaldi A, Delli Compagni E, Scialabba S, Caporale M, Di Pasquale A, Cammà C, Puglia I, Calistri P, Savini G |
| EPI_ISL_1502447, EPI_ISL_1502448 | SIESP CHIETI - DRIVE IN ORTONA ORTONA(CHIETI) | Istituto Zooprofilattico Sperimentale dell'Abruzzo e Molise "G. Caporale" | Lorusso A, Marcacci M, Di Domenico M, Ancora M, Curini V, Di Lollo Valeria, Mangone I, Rinaldi A, Delli Compagni E, Scialabba S, Caporale M, Di Pasquale A, Cammà C, Puglia I, Calistri P, Savini G |
| EPI_ISL_1502449 | OSPEDALE CIVILE GIULIANOVA - PRONTO SOCCORSO GIULIANOVA(TERAMO) | Istituto Zooprofilattico Sperimentale dell'Abruzzo e Molise "G. Caporale" | Lorusso A, Marcacci M, Di Domenico M, Ancora M, Curini V, Di Lollo Valeria, Mangone I, Rinaldi A, Delli Compagni E, Scialabba S, Caporale M, Di Pasquale A, Cammà C, Puglia I, Calistri P, Savini G |
| EPI_ISL_1502450, EPI_ISL_1502451 | SIESP CHIETI - DRIVE IN GISSI GISSI(CHIETI) | Istituto Zooprofilattico Sperimentale dell'Abruzzo e Molise "G. Caporale" | Lorusso A, Marcacci M, Di Domenico M, Ancora M, Curini V, Di Lollo Valeria, Mangone I, Rinaldi A, Delli Compagni E, Scialabba S, Caporale M, Di Pasquale A, Cammà C, Puglia I, Calistri P, Savini G |
| EPI_ISL_1502452, EPI_ISL_1502453, EPI_ISL_1502454, EPI_ISL_1502455, EPI_ISL_1502456, EPI_ISL_1502457, EPI_ISL_1502458, EPI_ISL_1502459 | SIESP DIPARTIMENTO DI PREVENZIONE TERAMO TERAMO(TERAMO) | Istituto Zooprofilattico Sperimentale dell'Abruzzo e Molise "G. Caporale" | Lorusso A, Marcacci M, Di Domenico M, Ancora M, Curini V, Di Lollo Valeria, Mangone I, Rinaldi A, Delli Compagni E, Scialabba S, Caporale M, Di Pasquale A, Cammà C, Puglia I, Calistri P, Savini G |
| EPI_ISL_1502460 | RSA GIULIANOVA GIULIANOVA(TERAMO) | Istituto Zooprofilattico Sperimentale dell'Abruzzo e Molise "G. Caporale" | Lorusso A, Marcacci M, Di Domenico M, Ancora M, Curini V, Di Lollo Valeria, Mangone I, Rinaldi A, Delli Compagni E, Scialabba S, Caporale M, Di Pasquale A, Cammà C, Puglia I, Calistri P, Savini G |
| EPI_ISL_1502461, EPI_ISL_1502462, EPI_ISL_1502463, EPI_ISL_1502464, EPI_ISL_1502465 | SIESP DIPARTIMENTO DI PREVENZIONE TERAMO TERAMO(TERAMO) | Istituto Zooprofilattico Sperimentale dell'Abruzzo e Molise "G. Caporale" | Lorusso A, Marcacci M, Di Domenico M, Ancora M, Curini V, Di Lollo Valeria, Mangone I, Rinaldi A, Delli Compagni E, Scialabba S, Caporale M, Di Pasquale A, Cammà C, Puglia I, Calistri P, Savini G |
| EPI_ISL_1502466 | SIESP CHIETI - DRIVE IN CHIETI CHIETI(CHIETI) | Istituto Zooprofilattico Sperimentale dell'Abruzzo e Molise "G. Caporale" | Lorusso A, Marcacci M, Di Domenico M, Ancora M, Curini V, Di Lollo Valeria, Mangone I, Rinaldi A, Delli Compagni E, Scialabba S, Caporale M, Di Pasquale A, Cammà C, Puglia I, Calistri P, Savini G |
| EPI_ISL_1502467, EPI_ISL_1502468, EPI_ISL_1502469, EPI_ISL_1502470 | SIESP DIPARTIMENTO DI PREVENZIONE TERAMO TERAMO(TERAMO) | Istituto Zooprofilattico Sperimentale dell'Abruzzo e Molise "G. Caporale" | Lorusso A, Marcacci M, Di Domenico M, Ancora M, Curini V, Di Lollo Valeria, Mangone I, Rinaldi A, Delli Compagni E, Scialabba S, Caporale M, Di Pasquale A, Cammà C, Puglia I, Calistri P, Savini G |
| EPI_ISL_1502471 | SIESP CHIETI - DRIVE IN ORTONA ORTONA(CHIETI) | Istituto Zooprofilattico Sperimentale dell'Abruzzo e Molise "G. Caporale" | Lorusso A, Marcacci M, Di Domenico M, Ancora M, Curini V, Di Lollo Valeria, Mangone I, Rinaldi A, Delli Compagni E, Scialabba S, Caporale M, Di Pasquale A, Cammà C, Puglia I, Calistri P, Savini G |

| | | | |
|---|---|---|---|
| EPI_ISL_1502472 | OSPEDALE "F. RENZETTI" LANCIANO - MEDICINA INTERNA LANCIANO(CHIETI) | Istituto Zooprofilattico Sperimentale dell'Abruzzo e Molise "G. Caporale" | Lorusso A, Marcacci M, Di Domenico M, Ancora M, Curini V, Di Lollo Valeria, Mangone I, Rinaldi A, Delli Compagni E, Scialabba S, Caporale M, Di Pasquale A, Cammà C, Puglia I, Calistri P, Savini G |
| EPI_ISL_1502473 | SIESP CHIETI - DRIVE IN LANCIANO LANCIANO(CHIETI) | Istituto Zooprofilattico Sperimentale dell'Abruzzo e Molise "G. Caporale" | Lorusso A, Marcacci M, Di Domenico M, Ancora M, Curini V, Di Lollo Valeria, Mangone I, Rinaldi A, Delli Compagni E, Scialabba S, Caporale M, Di Pasquale A, Cammà C, Puglia I, Calistri P, Savini G |
| EPI_ISL_1502474 | SIESP SULMONA SULMONA(L'AQUILA) | Istituto Zooprofilattico Sperimentale dell'Abruzzo e Molise "G. Caporale" | Lorusso A, Marcacci M, Di Domenico M, Ancora M, Curini V, Di Lollo Valeria, Mangone I, Rinaldi A, Delli Compagni E, Scialabba S, Caporale M, Di Pasquale A, Cammà C, Puglia I, Calistri P, Savini G |
| EPI_ISL_1502475, EPI_ISL_1502476, EPI_ISL_1502477, EPI_ISL_1502478, EPI_ISL_1502479 | DIP. PREV. AVEZZANO SERVIZIO DI IGIENE EPIDEMIOLOGIA E SANITA' PUBBLICA AVEZZANO(L'AQUILA) | Istituto Zooprofilattico Sperimentale dell'Abruzzo e Molise "G. Caporale" | Lorusso A, Marcacci M, Di Domenico M, Ancora M, Curini V, Di Lollo Valeria, Mangone I, Rinaldi A, Delli Compagni E, Scialabba S, Caporale M, Di Pasquale A, Cammà C, Puglia I, Calistri P, Savini G |
| EPI_ISL_1502480 | SIESP DIPARTIMENTO DI PREVENZIONE TERAMO TERAMO(TERAMO) | Istituto Zooprofilattico Sperimentale dell'Abruzzo e Molise "G. Caporale" | Lorusso A, Marcacci M, Di Domenico M, Ancora M, Curini V, Di Lollo Valeria, Mangone I, Rinaldi A, Delli Compagni E, Scialabba S, Caporale M, Di Pasquale A, Cammà C, Puglia I, Calistri P, Savini G |
| EPI_ISL_1502481 | DIP. PREV. AVEZZANO SERVIZIO DI IGIENE EPIDEMIOLOGIA E SANITA' PUBBLICA AVEZZANO(L'AQUILA) | Istituto Zooprofilattico Sperimentale dell'Abruzzo e Molise "G. Caporale" | Lorusso A, Marcacci M, Di Domenico M, Ancora M, Curini V, Di Lollo Valeria, Mangone I, Rinaldi A, Delli Compagni E, Scialabba S, Caporale M, Di Pasquale A, Cammà C, Puglia I, Calistri P, Savini G |
| EPI_ISL_1502482, EPI_ISL_1502483 | SIESP CHIETI - DRIVE IN LANCIANO LANCIANO(CHIETI) | Istituto Zooprofilattico Sperimentale dell'Abruzzo e Molise "G. Caporale" | Lorusso A, Marcacci M, Di Domenico M, Ancora M, Curini V, Di Lollo Valeria, Mangone I, Rinaldi A, Delli Compagni E, Scialabba S, Caporale M, Di Pasquale A, Cammà C, Puglia I, Calistri P, Savini G |
| EPI_ISL_1502484 | SIESP CHIETI - DRIVE IN VASTO VASTO(CHIETI) | Istituto Zooprofilattico Sperimentale dell'Abruzzo e Molise "G. Caporale" | Lorusso A, Marcacci M, Di Domenico M, Ancora M, Curini V, Di Lollo Valeria, Mangone I, Rinaldi A, Delli Compagni E, Scialabba S, Caporale M, Di Pasquale A, Cammà C, Puglia I, Calistri P, Savini G |
| EPI_ISL_1502485, EPI_ISL_1502486 | SIESP CHIETI - DRIVE IN ORTONA ORTONA(CHIETI) | Istituto Zooprofilattico Sperimentale dell'Abruzzo e Molise "G. Caporale" | Lorusso A, Marcacci M, Di Domenico M, Ancora M, Curini V, Di Lollo Valeria, Mangone I, Rinaldi A, Delli Compagni E, Scialabba S, Caporale M, Di Pasquale A, Cammà C, Puglia I, Calistri P, Savini G |
| EPI_ISL_1502487, EPI_ISL_1502488 | SIESP CHIETI - DRIVE IN GISSI GISSI(CHIETI) | Istituto Zooprofilattico Sperimentale dell'Abruzzo e Molise "G. Caporale" | Lorusso A, Marcacci M, Di Domenico M, Ancora M, Curini V, Di Lollo Valeria, Mangone I, Rinaldi A, Delli Compagni E, Scialabba S, Caporale M, Di Pasquale A, Cammà C, Puglia I, Calistri P, Savini G |
| EPI_ISL_1502489, EPI_ISL_1502490 | SIESP CHIETI - DRIVE IN CHIETI CHIETI(CHIETI) | Istituto Zooprofilattico Sperimentale dell'Abruzzo e Molise "G. Caporale" | Lorusso A, Marcacci M, Di Domenico M, Ancora M, Curini V, Di Lollo Valeria, Mangone I, Rinaldi A, Delli Compagni E, Scialabba S, Caporale M, Di Pasquale A, Cammà C, Puglia I, Calistri P, Savini G |
| EPI_ISL_1502491 | SIESP DIPARTIMENTO DI PREVENZIONE CHIETI CHIETI(CHIETI) | Istituto Zooprofilattico Sperimentale dell'Abruzzo e Molise "G. Caporale" | Lorusso A, Marcacci M, Di Domenico M, Ancora M, Curini V, Di Lollo Valeria, Mangone I, Rinaldi A, Delli Compagni E, Scialabba S, Caporale M, Di Pasquale A, Cammà C, Puglia I, Calistri P, Savini G |
| EPI_ISL_1502492, EPI_ISL_1502493, EPI_ISL_1502494, EPI_ISL_1502495, EPI_ISL_1502496, EPI_ISL_1502497 | SIESP DIPARTIMENTO DI PREVENZIONE TERAMO TERAMO(TERAMO) | Istituto Zooprofilattico Sperimentale dell'Abruzzo e Molise "G. Caporale" | Lorusso A, Marcacci M, Di Domenico M, Ancora M, Curini V, Di Lollo Valeria, Mangone I, Rinaldi A, Delli Compagni E, Scialabba S, Caporale M, Di Pasquale A, Cammà C, Puglia I, Calistri P, Savini G |
| EPI_ISL_1502498 | RSA GIULIANOVA GIULIANOVA(TERAMO) | Istituto Zooprofilattico Sperimentale dell'Abruzzo e Molise "G. Caporale" | Lorusso A, Marcacci M, Di Domenico M, Ancora M, Curini V, Di Lollo Valeria, Mangone I, Rinaldi A, Delli Compagni E, Scialabba S, Caporale M, Di Pasquale A, Cammà C, Puglia I, Calistri P, Savini G |
| EPI_ISL_1502499 | SIESP DIPARTIMENTO DI PREVENZIONE TERAMO TERAMO(TERAMO) | Istituto Zooprofilattico Sperimentale dell'Abruzzo e Molise "G. Caporale" | Lorusso A, Marcacci M, Di Domenico M, Ancora M, Curini V, Di Lollo Valeria, Mangone I, Rinaldi A, Delli Compagni E, Scialabba S, Caporale M, Di Pasquale A, Cammà C, Puglia I, Calistri P, Savini G |
| EPI_ISL_1502500, EPI_ISL_1502501, EPI_ISL_1502502 | SIESP DIPARTIMENTO DI PREVENZIONE CHIETI CHIETI(CHIETI) | Istituto Zooprofilattico Sperimentale dell'Abruzzo e Molise "G. Caporale" | Lorusso A, Marcacci M, Di Domenico M, Ancora M, Curini V, Di Lollo Valeria, Mangone I, Rinaldi A, Delli Compagni E, Scialabba S, Caporale M, Di Pasquale A, Cammà C, Puglia I, Calistri P, Savini G |
| EPI_ISL_1502503, EPI_ISL_1502504, EPI_ISL_1502505 | SIESP DIPARTIMENTO DI PREVENZIONE TERAMO TERAMO(TERAMO) | Istituto Zooprofilattico Sperimentale dell'Abruzzo e Molise "G. Caporale" | Lorusso A, Marcacci M, Di Domenico M, Ancora M, Curini V, Di Lollo Valeria, Mangone I, Rinaldi A, Delli Compagni E, Scialabba S, Caporale M, Di Pasquale A, Cammà C, Puglia I, Calistri P, Savini G |
| EPI_ISL_1502506 | SIESP CHIETI - DRIVE IN CHIETI CHIETI(CHIETI) | Istituto Zooprofilattico Sperimentale dell'Abruzzo e Molise "G. Caporale" | Lorusso A, Marcacci M, Di Domenico M, Ancora M, Curini V, Di Lollo Valeria, Mangone I, Rinaldi A, Delli Compagni E, Scialabba S, Caporale M, Di Pasquale A, Cammà C, Puglia I, Calistri P, Savini G |
| EPI_ISL_1502507 | SIESP CHIETI - DRIVE IN ORTONA ORTONA(CHIETI) | Istituto Zooprofilattico Sperimentale dell'Abruzzo e Molise "G. Caporale" | Lorusso A, Marcacci M, Di Domenico M, Ancora M, Curini V, Di Lollo Valeria, Mangone I, Rinaldi A, Delli Compagni E, Scialabba S, Caporale M, Di Pasquale A, Cammà C, Puglia I, Calistri P, Savini G |
| EPI_ISL_1502508, EPI_ISL_1502509, EPI_ISL_1502510, EPI_ISL_1502511 | SIESP SULMONA SULMONA(L'AQUILA) | Istituto Zooprofilattico Sperimentale dell'Abruzzo e Molise "G. Caporale" | Lorusso A, Marcacci M, Di Domenico M, Ancora M, Curini V, Di Lollo Valeria, Mangone I, Rinaldi A, Delli Compagni E, Scialabba S, Caporale M, Di Pasquale A, Cammà C, Puglia I, Calistri P, Savini G |
| EPI_ISL_1502512 | DIP. PREV. AVEZZANO SERVIZIO DI IGIENE EPIDEMIOLOGIA E SANITA' PUBBLICA AVEZZANO(L'AQUILA) | Istituto Zooprofilattico Sperimentale dell'Abruzzo e Molise "G. Caporale" | Lorusso A, Marcacci M, Di Domenico M, Ancora M, Curini V, Di Lollo Valeria, Mangone I, Rinaldi A, Delli Compagni E, Scialabba S, Caporale M, Di Pasquale A, Cammà C, Puglia I, Calistri P, Savini G |
| EPI_ISL_1502513 | SIESP SULMONA SULMONA(L'AQUILA) | Istituto Zooprofilattico Sperimentale dell'Abruzzo e Molise "G. Caporale" | Lorusso A, Marcacci M, Di Domenico M, Ancora M, Curini V, Di Lollo Valeria, Mangone I, Rinaldi A, Delli Compagni E, Scialabba S, Caporale M, Di Pasquale A, Cammà C, Puglia I, Calistri P, Savini G |
| EPI_ISL_1502514, EPI_ISL_1502515, EPI_ISL_1502516 | DIP. PREV. AVEZZANO SERVIZIO DI IGIENE EPIDEMIOLOGIA E SANITA' PUBBLICA AVEZZANO(L'AQUILA) | Istituto Zooprofilattico Sperimentale dell'Abruzzo e Molise "G. Caporale" | Lorusso A, Marcacci M, Di Domenico M, Ancora M, Curini V, Di Lollo Valeria, Mangone I, Rinaldi A, Delli Compagni E, Scialabba S, Caporale M, Di Pasquale A, Cammà C, Puglia I, Calistri P, Savini G |
| EPI_ISL_1502517 | OSPEDALE CIVILE GIULIANOVA - PRONTO SOCCORSO GIULIANOVA(TERAMO) | Istituto Zooprofilattico Sperimentale dell'Abruzzo e Molise "G. Caporale" | Lorusso A, Marcacci M, Di Domenico M, Ancora M, Curini V, Di Lollo Valeria, Mangone I, Rinaldi A, Delli Compagni E, Scialabba S, Caporale M, Di Pasquale A, Cammà C, Puglia I, Calistri P, Savini G |
| EPI_ISL_1502518 | SIESP DIPARTIMENTO DI PREVENZIONE TERAMO TERAMO(TERAMO) | Istituto Zooprofilattico Sperimentale dell'Abruzzo e Molise "G. Caporale" | Lorusso A, Marcacci M, Di Domenico M, Ancora M, Curini V, Di Lollo Valeria, Mangone I, Rinaldi A, Delli Compagni E, Scialabba S, Caporale M, Di Pasquale A, Cammà C, Puglia I, Calistri P, Savini G |
| EPI_ISL_1502519, EPI_ISL_1502520, EPI_ISL_1502521 | DIP. PREV. AVEZZANO SERVIZIO DI IGIENE EPIDEMIOLOGIA E SANITA' PUBBLICA AVEZZANO(L'AQUILA) | Istituto Zooprofilattico Sperimentale dell'Abruzzo e Molise "G. Caporale" | Lorusso A, Marcacci M, Di Domenico M, Ancora M, Curini V, Di Lollo Valeria, Mangone I, Rinaldi A, Delli Compagni E, Scialabba S, Caporale M, Di Pasquale A, Cammà C, Puglia I, Calistri P, Savini G |
| EPI_ISL_1502522 | OSPEDALE "F. RENZETTI" LANCIANO - MEDICINA INTERNA LANCIANO(CHIETI) | Istituto Zooprofilattico Sperimentale dell'Abruzzo e Molise "G. Caporale" | Lorusso A, Marcacci M, Di Domenico M, Ancora M, Curini V, Di Lollo Valeria, Mangone I, Rinaldi A, Delli Compagni E, Scialabba S, Caporale M, Di Pasquale A, Cammà C, Puglia I, Calistri P, Savini G |
| EPI_ISL_1502523 | PRESIDIO TERRITORIALE CASOLI CASOLI(CHIETI) | Istituto Zooprofilattico Sperimentale dell'Abruzzo e Molise "G. Caporale" | Lorusso A, Marcacci M, Di Domenico M, Ancora M, Curini V, Di Lollo Valeria, Mangone I, Rinaldi A, Delli Compagni E, Scialabba S, Caporale M, Di Pasquale A, Cammà C, Puglia I, Calistri P, Savini G |
| EPI_ISL_1502524, EPI_ISL_1502525 | OSPEDALE "F. RENZETTI" LANCIANO - MEDICINA INTERNA LANCIANO(CHIETI) | Istituto Zooprofilattico Sperimentale dell'Abruzzo e Molise "G. Caporale" | Lorusso A, Marcacci M, Di Domenico M, Ancora M, Curini V, Di Lollo Valeria, Mangone I, Rinaldi A, Delli Compagni E, Scialabba S, Caporale M, Di Pasquale A, Cammà C, Puglia I, Calistri P, Savini G |
| EPI_ISL_1502526, EPI_ISL_1502527 | PRESIDIO TERRITORIALE CASOLI CASOLI(CHIETI) | Istituto Zooprofilattico Sperimentale dell'Abruzzo e Molise "G. Caporale" | Lorusso A, Marcacci M, Di Domenico M, Ancora M, Curini V, Di Lollo Valeria, Mangone I, Rinaldi A, Delli Compagni E, Scialabba S, Caporale M, Di Pasquale A, Cammà C, Puglia I, Calistri P, Savini G |
| EPI_ISL_1502528, EPI_ISL_1502529 | USCA Pescina PESCINA(L'AQUILA) | Istituto Zooprofilattico Sperimentale dell'Abruzzo e Molise "G. Caporale" | Lorusso A, Marcacci M, Di Domenico M, Ancora M, Curini V, Di Lollo Valeria, Mangone I, Rinaldi A, Delli Compagni E, Scialabba S, Caporale M, Di Pasquale A, Cammà C, Puglia I, Calistri P, Savini G |
| EPI_ISL_1502530 | SIESP DIPARTIMENTO DI PREVENZIONE CHIETI CHIETI(CHIETI) | Istituto Zooprofilattico Sperimentale dell'Abruzzo e Molise "G. Caporale" | Lorusso A, Marcacci M, Di Domenico M, Ancora M, Curini V, Di Lollo Valeria, Mangone I, Rinaldi A, Delli Compagni E, Scialabba S, Caporale M, Di Pasquale A, Cammà C, Puglia I, Calistri P, Savini G |
| EPI_ISL_1502531 | SIESP CHIETI - DRIVE IN GISSI GISSI(CHIETI) | Istituto Zooprofilattico Sperimentale dell'Abruzzo e Molise "G. Caporale" | Lorusso A, Marcacci M, Di Domenico M, Ancora M, Curini V, Di Lollo Valeria, Mangone I, Rinaldi A, Delli Compagni E, Scialabba S, Caporale M, Di Pasquale A, Cammà C, Puglia I, Calistri P, Savini G |

| | | | |
|---|---|---|---|
| EPI_ISL_1502534, EPI_ISL_1502535, EPI_ISL_1502536, EPI_ISL_1502537 | VILLA NOVECENTO S.R.L LANCIANO(CHIETI) | Istituto Zooprofilattico Sperimentale dell'Abruzzo e Molise "G. Caporale" | Lorusso A, Marcacci M, Di Domenico M, Ancora M, Curini V, Di Lollo Valeria, Mangone I, Rinaldi A, Delli Compagni E, Scialabba S, Caporale M, Di Pasquale A, Cammà C, Puglia I, Calistri P, Savini G |
| EPI_ISL_1502538 | SIESP CHIETI - DRIVE IN ORTONA ORTONA(CHIETI) | Istituto Zooprofilattico Sperimentale dell'Abruzzo e Molise "G. Caporale" | Lorusso A, Marcacci M, Di Domenico M, Ancora M, Curini V, Di Lollo Valeria, Mangone I, Rinaldi A, Delli Compagni E, Scialabba S, Caporale M, Di Pasquale A, Cammà C, Puglia I, Calistri P, Savini G |
| EPI_ISL_1502539 | SIESP CHIETI - DRIVE IN CHIETI CHIETI(CHIETI) | Istituto Zooprofilattico Sperimentale dell'Abruzzo e Molise "G. Caporale" | Lorusso A, Marcacci M, Di Domenico M, Ancora M, Curini V, Di Lollo Valeria, Mangone I, Rinaldi A, Delli Compagni E, Scialabba S, Caporale M, Di Pasquale A, Cammà C, Puglia I, Calistri P, Savini G |
| EPI_ISL_1502540, EPI_ISL_1502541, EPI_ISL_1502542, EPI_ISL_1502543, EPI_ISL_1502544, EPI_ISL_1502545, EPI_ISL_1502546, EPI_ISL_1502547, EPI_ISL_1502548, EPI_ISL_1502549, EPI_ISL_1502550, EPI_ISL_1502551, EPI_ISL_1502552, EPI_ISL_1502553, EPI_ISL_1502554, EPI_ISL_1502555, EPI_ISL_1502556, EPI_ISL_1502557, EPI_ISL_1502558, EPI_ISL_1502559, EPI_ISL_1502560, EPI_ISL_1502561, EPI_ISL_1502562, EPI_ISL_1502563, EPI_ISL_1502564, EPI_ISL_1502565, EPI_ISL_1502566 | | | |
| see above | OSPEDALE SAN SALVATORE - MEDICINA DI LABORATORIO L'AQUILA(L'AQUILA) | Istituto Zooprofilattico Sperimentale dell'Abruzzo e Molise "G. Caporale" | Lorusso A, Marcacci M, Di Domenico M, Ancora M, Curini V, Di Lollo Valeria, Mangone I, Rinaldi A, Delli Compagni E, Scialabba S, Caporale M, Di Pasquale A, Cammà C, Puglia I, Calistri P, Savini G |
| EPI_ISL_1509341, EPI_ISL_1509342, EPI_ISL_1509343, EPI_ISL_1509344, EPI_ISL_1509345, EPI_ISL_1509346, EPI_ISL_1509347, EPI_ISL_1509348, EPI_ISL_1509349, EPI_ISL_1509350, EPI_ISL_1509351, EPI_ISL_1509352, EPI_ISL_1509353, EPI_ISL_1509354, EPI_ISL_1509355, EPI_ISL_1509356, EPI_ISL_1509357, EPI_ISL_1509358, EPI_ISL_1509359, EPI_ISL_1509360, EPI_ISL_1509361, EPI_ISL_1509362, EPI_ISL_1509363, EPI_ISL_1509364, EPI_ISL_1509365, EPI_ISL_1509366, EPI_ISL_1509367, EPI_ISL_1509368, EPI_ISL_1509369, EPI_ISL_1509370, EPI_ISL_1509371, EPI_ISL_1509372, EPI_ISL_1509373, EPI_ISL_1509374, EPI_ISL_1509375, EPI_ISL_1509376, EPI_ISL_1509377, EPI_ISL_1509378, EPI_ISL_1509379, EPI_ISL_1509380, EPI_ISL_1509381, EPI_ISL_1509382, EPI_ISL_1509383, EPI_ISL_1509384, EPI_ISL_1509385, EPI_ISL_1509386, EPI_ISL_1509387, EPI_ISL_1509388, EPI_ISL_1509389, EPI_ISL_1509390, EPI_ISL_1509391, EPI_ISL_1509392, EPI_ISL_1509393, EPI_ISL_1509394, EPI_ISL_1509395, EPI_ISL_1509396, EPI_ISL_1509397, EPI_ISL_1509398, EPI_ISL_1509399, EPI_ISL_1509400, EPI_ISL_1509401, EPI_ISL_1509402, EPI_ISL_1509403, EPI_ISL_1509404, EPI_ISL_1509405, EPI_ISL_1509406, EPI_ISL_1509407, EPI_ISL_1509408, EPI_ISL_1509409, EPI_ISL_1509410, EPI_ISL_1509411, EPI_ISL_1509412, EPI_ISL_1509413, EPI_ISL_1509414, EPI_ISL_1509415, EPI_ISL_1509416, EPI_ISL_1509417, EPI_ISL_1509418, EPI_ISL_1509419, EPI_ISL_1509420, EPI_ISL_1509421, EPI_ISL_1509422, EPI_ISL_1509423, EPI_ISL_1509424, EPI_ISL_1509425, EPI_ISL_1509426, EPI_ISL_1509427, EPI_ISL_1509428, EPI_ISL_1509429, EPI_ISL_1509430, EPI_ISL_1509431, EPI_ISL_1509432, EPI_ISL_1509433, EPI_ISL_1509434, EPI_ISL_1509435, EPI_ISL_1509436, EPI_ISL_1509437, EPI_ISL_1509438, EPI_ISL_1509439, EPI_ISL_1509440, EPI_ISL_1509441, EPI_ISL_1509442, EPI_ISL_1509443, EPI_ISL_1509444, EPI_ISL_1509445, EPI_ISL_1509446, EPI_ISL_1509447, EPI_ISL_1509448, EPI_ISL_1509449, EPI_ISL_1509450, EPI_ISL_1509451, EPI_ISL_1509452, EPI_ISL_1509453, EPI_ISL_1509454, EPI_ISL_1509455, EPI_ISL_1509456, EPI_ISL_1509457, EPI_ISL_1509458, EPI_ISL_1509459, EPI_ISL_1509460, EPI_ISL_1509461, EPI_ISL_1509462, EPI_ISL_1509549, EPI_ISL_1509550, EPI_ISL_1509551, EPI_ISL_1509552, EPI_ISL_1509553, EPI_ISL_1509554, EPI_ISL_1509555, EPI_ISL_1509556, EPI_ISL_1509557, EPI_ISL_1509558, EPI_ISL_1509559, EPI_ISL_1509560, EPI_ISL_1509561, EPI_ISL_1509562, EPI_ISL_1509563, EPI_ISL_1509564, EPI_ISL_1509565, EPI_ISL_1509566, EPI_ISL_1509567, EPI_ISL_1509568, EPI_ISL_1509569, EPI_ISL_1509570, EPI_ISL_1509571, EPI_ISL_1509572, EPI_ISL_1509573, EPI_ISL_1509574, EPI_ISL_1509575, EPI_ISL_1509576, EPI_ISL_1509577, EPI_ISL_1509578, EPI_ISL_1509579, EPI_ISL_1509580, EPI_ISL_1509581, EPI_ISL_1509582, EPI_ISL_1509583, EPI_ISL_1509584, EPI_ISL_1509585, EPI_ISL_1509586, EPI_ISL_1509587, EPI_ISL_1509588, EPI_ISL_1509589, EPI_ISL_1509590, EPI_ISL_1509591, EPI_ISL_1509592, EPI_ISL_1509593, EPI_ISL_1509594, EPI_ISL_1509595, EPI_ISL_1509596, EPI_ISL_1509597, EPI_ISL_1509598, EPI_ISL_1509599, EPI_ISL_1509600, EPI_ISL_1509601, EPI_ISL_1509602, EPI_ISL_1509603, EPI_ISL_1509604, EPI_ISL_1509605, EPI_ISL_1509606, EPI_ISL_1509607, EPI_ISL_1509608, EPI_ISL_1509609, EPI_ISL_1509610, EPI_ISL_1509611, EPI_ISL_1509612, EPI_ISL_1509613, EPI_ISL_1509614, EPI_ISL_1509615, EPI_ISL_1509616, EPI_ISL_1509617, EPI_ISL_1509618, EPI_ISL_1509619, EPI_ISL_1509620, EPI_ISL_1509621, EPI_ISL_1509622, EPI_ISL_1509623, EPI_ISL_1509624, EPI_ISL_1509625, EPI_ISL_1509626, EPI_ISL_1509627, EPI_ISL_1509628, EPI_ISL_1509629, EPI_ISL_1509630, EPI_ISL_1509631, EPI_ISL_1509632, EPI_ISL_1509633, EPI_ISL_1509634, EPI_ISL_1509635, EPI_ISL_1509636 | | | |
| see above | IZSM | TIGEM | Antonio Grimaldi Patrizia Annunziata Francesco Panariello Biancamaria Pierri Claudia Tiberio Valentina Bouche Chiara Colantuono Maria Concetta Cuomo Denise Di Concilio Lucio Di Filippo Anna Manfredi Marcello Salvi Antonio Limone Luigi Atripaldi Pellegrino Cerino Andrea Ballabio Davide Cacchiarelli |
| EPI_ISL_1509721, EPI_ISL_1509722, EPI_ISL_1509723, EPI_ISL_1509724, EPI_ISL_1509725, EPI_ISL_1509726, EPI_ISL_1509727, EPI_ISL_1509728, EPI_ISL_1509729, EPI_ISL_1509730 | SC (UCO) Igiene e Sanità Pubblica (funzione integrata con SC Microbiologia e Virologia) e Az. Ospedaliero-Universitaria Udine | ARGO Laboratorio Genomica ed Epigenomica | Licastro D, Dal Monego S, Degasperi M, Marcello A, Segat L, Piscianz E, D'Agaro P |
| EPI_ISL_1509734, EPI_ISL_1509735, EPI_ISL_1509736 | SC (UCO) Igiene e Sanità Pubblica (funzione integrata con SC Microbiologia e Virologia) | ARGO Laboratorio Genomica ed Epigenomica | Licastro D, Dal Monego S, Degasperi M, Marcello A, Segat L, Piscianz E, D'Agaro P |
| EPI_ISL_1509737, EPI_ISL_1509738, EPI_ISL_1509739, EPI_ISL_1509740, EPI_ISL_1509741 | SC (UCO) Igiene e Sanità Pubblica (funzione integrata con SC Microbiologia e Virologia) e Azienda Ospedaliera Pordenone | ARGO Laboratorio Genomica ed Epigenomica | Licastro D, Dal Monego S, Degasperi M, Marcello A, Segat L, Piscianz E, D'Agaro P |
| EPI_ISL_1509742, EPI_ISL_1509743, EPI_ISL_1509744, EPI_ISL_1509745 | SC (UCO) Igiene e Sanità Pubblica (funzione integrata con SC Microbiologia e Virologia) | ARGO Laboratorio Genomica ed Epigenomica | Licastro D, Dal Monego S, Degasperi M, Marcello A, Segat L, Piscianz E, D'Agaro P |
| EPI_ISL_1509746 | SC (UCO) Igiene e Sanità Pubblica (funzione integrata con SC Microbiologia e Virologia) e Az. Ospedaliero-Universitaria Udine | ARGO Laboratorio Genomica ed Epigenomica | Licastro D, Dal Monego S, Degasperi M, Marcello A, Segat L, Piscianz E, D'Agaro P |
| EPI_ISL_1509747 | SC (UCO) Igiene e Sanità Pubblica (funzione integrata con SC Microbiologia e Virologia) e Azienda Ospedaliera Pordenone | ARGO Laboratorio Genomica ed Epigenomica | Licastro D, Dal Monego S, Degasperi M, Marcello A, Segat L, Piscianz E, D'Agaro P |
| EPI_ISL_1509748, EPI_ISL_1509749, EPI_ISL_1509750 | SC (UCO) Igiene e Sanità Pubblica (funzione integrata con SC Microbiologia e Virologia) e Az. Ospedaliero-Universitaria Udine | ARGO Laboratorio Genomica ed Epigenomica | Licastro D, Dal Monego S, Degasperi M, Marcello A, Segat L, Piscianz E, D'Agaro P |
| EPI_ISL_1509751 | SC (UCO) Igiene e Sanità Pubblica (funzione integrata con SC Microbiologia e Virologia) | ARGO Laboratorio Genomica ed Epigenomica | Licastro D, Dal Monego S, Degasperi M, Marcello A, Segat L, Piscianz E, D'Agaro P |
| EPI_ISL_1509752, EPI_ISL_1509753 | SC (UCO) Igiene e Sanità Pubblica (funzione integrata con SC Microbiologia e Virologia) e Az. Ospedaliero-Universitaria Udine | ARGO Laboratorio Genomica ed Epigenomica | Licastro D, Dal Monego S, Degasperi M, Marcello A, Segat L, Piscianz E, D'Agaro P |
| EPI_ISL_1509754 | SC (UCO) Igiene e Sanità Pubblica (funzione integrata con SC Microbiologia e Virologia) e Azienda Ospedaliera Pordenone | ARGO Laboratorio Genomica ed Epigenomica | Licastro D, Dal Monego S, Degasperi M, Marcello A, Segat L, Piscianz E, D'Agaro P |
| EPI_ISL_1509756 | SC (UCO) Igiene e Sanità Pubblica (funzione integrata con SC Microbiologia e Virologia) e Az. Ospedaliero-Universitaria Udine | ARGO Laboratorio Genomica ed Epigenomica | Licastro D, Dal Monego S, Degasperi M, Marcello A, Segat L, Piscianz E, D'Agaro P |
| EPI_ISL_1510420, EPI_ISL_1510421, EPI_ISL_1510422, EPI_ISL_1510423, EPI_ISL_1510424, EPI_ISL_1510425, EPI_ISL_1510426, EPI_ISL_1510427, EPI_ISL_1510428, EPI_ISL_1510429, EPI_ISL_1510430 | | | |
| see above | SC (UCO) Igiene e Sanità Pubblica (funzione integrata con SC Microbiologia e Virologia) | ARGO Laboratorio Genomica ed Epigenomica | Licastro D, Dal Monego S, Degasperi M, Marcello A, Segat L, Piscianz E, D'Agaro P |
| EPI_ISL_1510431, EPI_ISL_1510432, EPI_ISL_1510433, EPI_ISL_1510434, EPI_ISL_1510435, EPI_ISL_1510436, EPI_ISL_1510437, EPI_ISL_1510438, EPI_ISL_1510439, EPI_ISL_1510440, EPI_ISL_1510441, EPI_ISL_1510442, EPI_ISL_1510443, EPI_ISL_1510444, EPI_ISL_1510445, EPI_ISL_1510446 | | | |
| see above | SC (UCO) Igiene e Sanità Pubblica (funzione integrata con SC Microbiologia e Virologia) | ARGO Laboratorio Genomica ed Epigenomica | Licastro D, Dal Monego S, Degasperi M, Marcello A, D'Agaro P, De Rosa R |
| EPI_ISL_1510447, EPI_ISL_1510448, EPI_ISL_1510449, EPI_ISL_1510450, EPI_ISL_1510451, EPI_ISL_1510452, EPI_ISL_1510453, EPI_ISL_1510454, EPI_ISL_1510455, EPI_ISL_1510456, EPI_ISL_1510457, EPI_ISL_1510458, EPI_ISL_1510459, EPI_ISL_1510460, EPI_ISL_1510461 | | | |
| see above | SC (UCO) Igiene e Sanità Pubblica (funzione integrata con SC Microbiologia e Virologia) | ARGO Laboratorio Genomica ed Epigenomica | Licastro D, Dal Monego S, Degasperi M, D'Agaro P, Lombardo F |
| EPI_ISL_1510462, EPI_ISL_1510463, EPI_ISL_1510464, EPI_ISL_1510465, EPI_ISL_1510466, EPI_ISL_1510467, EPI_ISL_1510468, EPI_ISL_1510469, EPI_ISL_1510470, EPI_ISL_1510471, EPI_ISL_1510472, EPI_ISL_1510473, EPI_ISL_1510474 | | | |
| see above | SC (UCO) Igiene e Sanità Pubblica (funzione integrata con SC Microbiologia e Virologia) | ARGO Laboratorio Genomica ed Epigenomica | Licastro D, Dal Monego S, Degasperi M, Marcello A, D'Agaro P, Pipan C |
| EPI_ISL_1510475, EPI_ISL_1510476, EPI_ISL_1510477, EPI_ISL_1510478 | SC (UCO) Igiene e Sanità Pubblica (funzione integrata con SC Microbiologia e Virologia) | ARGO Laboratorio Genomica ed Epigenomica | Licastro D, Dal Monego S, Degasperi M, Marcello A, D'Agaro P, De Rosa R |
| EPI_ISL_1510479 | SC (UCO) Igiene e Sanità Pubblica (funzione integrata con SC Microbiologia e Virologia) | ARGO Laboratorio Genomica ed Epigenomica | Licastro D, Dal Monego S, Degasperi M, Marcello A, Segat L, Piscianz E, D'Agaro P |
| EPI_ISL_1510563, EPI_ISL_1510564, EPI_ISL_1510565, EPI_ISL_1510566, EPI_ISL_1510567, EPI_ISL_1510568, EPI_ISL_1510569, EPI_ISL_1510570, EPI_ISL_1510571, EPI_ISL_1510572, EPI_ISL_1510573, EPI_ISL_1510574, EPI_ISL_1510575, EPI_ISL_1510576, EPI_ISL_1510577, EPI_ISL_1510578, EPI_ISL_1510579, | | | |

| | | | |
|---|---|---|---|
| EPI_ISL_1510580, EPI_ISL_1510581, EPI_ISL_1510582, EPI_ISL_1510583, EPI_ISL_1510584, EPI_ISL_1510585, EPI_ISL_1510586, EPI_ISL_1510587, EPI_ISL_1510588, EPI_ISL_1510589, EPI_ISL_1510590, EPI_ISL_1510591, EPI_ISL_1510592, EPI_ISL_1510593, EPI_ISL_1510594, EPI_ISL_1510595, EPI_ISL_1510596, EPI_ISL_1510597, EPI_ISL_1510598, EPI_ISL_1510599, EPI_ISL_1510600, EPI_ISL_1510601, EPI_ISL_1510602, EPI_ISL_1510603, EPI_ISL_1510604, EPI_ISL_1510605 | | | |
| see above | Università di Parma, Laboratorio di Igiene e Sanità Pubblica | Istituto Zooprofilattico Sperimentale della Lombardia e dell'Emilia Romagna (IZSLER), Risk Analysis and Genomic Epidemiology Unit | Maria Eugenia Colucci, Licia Veronesi, Paola Affanni, Marina Morganti, Ilaria Menozzi, Erika Scaltriti, Stefano Pongolini |
| EPI_ISL_1510623, EPI_ISL_1510624, EPI_ISL_1510625, EPI_ISL_1510626, EPI_ISL_1510627, EPI_ISL_1510628, EPI_ISL_1510629, EPI_ISL_1510630, EPI_ISL_1510631, EPI_ISL_1510632, EPI_ISL_1510633, EPI_ISL_1510634, EPI_ISL_1510635 | | | |
| see above | Azienda Sanitaria Locale di Piacenza - Presidio Ospedaliero - Laboratorio di Microbiologia | Istituto Zooprofilattico Sperimentale della Lombardia e dell'Emilia Romagna (IZSLER), Risk Analysis and Genomic Epidemiology Unit | Lo Cascio Giuliana, Schiavo Roberta, Peroni Irene, Marina Morganti, Ilaria Menozzi, Erika Scaltriti, Stefano Pongolini |
| EPI_ISL_1510651, EPI_ISL_1510652, EPI_ISL_1510653, EPI_ISL_1510654, EPI_ISL_1510655, EPI_ISL_1510656, EPI_ISL_1510657, EPI_ISL_1510658, EPI_ISL_1510659, EPI_ISL_1510660, EPI_ISL_1510661, EPI_ISL_1510662, EPI_ISL_1510663, EPI_ISL_1510664, EPI_ISL_1510665 | | | |
| see above | Arcispedale Santa Maria Nuova, Autoimmunità, Allergologia e Biotecnologie Innovative | Istituto Zooprofilattico Sperimentale della Lombardia e dell'Emilia Romagna (IZSLER), Risk Analysis and Genomic Epidemiology Unit | Alessandro Zerbini, Lucia Belloni, Stefania Croci, Marina Morganti, Ilaria Menozzi, Erika Scaltriti, Stefano Pongolini |
| EPI_ISL_1520098 | Università degli Studi di Perugia | Istituto Zooprofilattico Sperimentale dell'Abruzzo e Molise "G. Caporale" | Mencacci A, Camilloni B, Lorusso A, Marcacci M, Di Domenico M, Ancora M, Curini V, Di Lollo Valeria, Mangone I, Rinaldi A, Delli Compagni E, Scialabba S, Caporale M, Di Pasquale A, Cammà C, Puglia I, Calistri P, Savini G |
| EPI_ISL_1520140, EPI_ISL_1520169, EPI_ISL_1520172, EPI_ISL_1520174, EPI_ISL_1520177, EPI_ISL_1520180, EPI_ISL_1520366, EPI_ISL_1520368, EPI_ISL_1520370, EPI_ISL_1520371, EPI_ISL_1520373, EPI_ISL_1520375 | | | |
| see above | University of Bari Biomedical Sciences and Human Oncology | University of Bari Biomedical Sciences and Human Oncology | Chironna M., Sallustio A., Loconsole D., Accogli M. |
| EPI_ISL_1520377 | University of Bari Biomedical Sciences and Human Oncology | University of Bari Biomedical Sciences and Human Oncology | Chironna M., sallustio A., Loconsole D., Accogli M. |
| EPI_ISL_1520378, EPI_ISL_1520379, EPI_ISL_1520380 | University of Bari Biomedical Sciences and Human Oncology | University of Bari Biomedical Sciences and Human Oncology | Chironna M., Sallustio A., Loconsole D., Accogli M. |
| EPI_ISL_1523544, EPI_ISL_1523545, EPI_ISL_1523546, EPI_ISL_1523547, EPI_ISL_1523548, EPI_ISL_1523549, EPI_ISL_1523550, EPI_ISL_1523551, EPI_ISL_1523552, EPI_ISL_1523553, EPI_ISL_1523554, EPI_ISL_1523555, EPI_ISL_1523556, EPI_ISL_1523557, EPI_ISL_1523558, EPI_ISL_1523559, EPI_ISL_1523560, EPI_ISL_1523561, EPI_ISL_1523562, EPI_ISL_1523563, EPI_ISL_1523564, EPI_ISL_1523565, EPI_ISL_1523566, EPI_ISL_1523567, EPI_ISL_1523568, EPI_ISL_1523569, EPI_ISL_1523570, EPI_ISL_1523571, EPI_ISL_1523572, EPI_ISL_1523573, EPI_ISL_1523574, EPI_ISL_1523575, EPI_ISL_1523576, EPI_ISL_1523577, EPI_ISL_1523578, EPI_ISL_1523579, EPI_ISL_1523580, EPI_ISL_1523581, EPI_ISL_1523582, EPI_ISL_1523583, EPI_ISL_1523585, EPI_ISL_1523586, EPI_ISL_1523587, EPI_ISL_1523588, EPI_ISL_1523589, EPI_ISL_1523590, EPI_ISL_1523591, EPI_ISL_1523592, EPI_ISL_1523593, EPI_ISL_1523594, EPI_ISL_1523595, EPI_ISL_1523596, EPI_ISL_1523597, EPI_ISL_1523598, EPI_ISL_1523599, EPI_ISL_1523600, EPI_ISL_1523601, EPI_ISL_1523602, EPI_ISL_1523603, EPI_ISL_1523604, EPI_ISL_1523605, EPI_ISL_1523606, EPI_ISL_1523607, EPI_ISL_1523608, EPI_ISL_1523609, EPI_ISL_1523610, EPI_ISL_1523611, EPI_ISL_1523612, EPI_ISL_1523613, EPI_ISL_1523614, EPI_ISL_1523615, EPI_ISL_1523616, EPI_ISL_1523617, EPI_ISL_1523618, EPI_ISL_1523619, EPI_ISL_1523620, EPI_ISL_1523621, EPI_ISL_1523622, EPI_ISL_1523623, EPI_ISL_1523624, EPI_ISL_1523625, EPI_ISL_1523626, EPI_ISL_1523627, EPI_ISL_1523628, EPI_ISL_1523629, EPI_ISL_1523630, EPI_ISL_1523631, EPI_ISL_1523632, EPI_ISL_1523633, EPI_ISL_1523634, EPI_ISL_1523635, EPI_ISL_1523636, EPI_ISL_1523637, EPI_ISL_1523638, EPI_ISL_1523639 | | | |
| see above | Azienda Sanitaria dell'Alto Adige Laboratorio Aziendale di Microbiologia e Virologia | Istituto di Genomica Applicata | Elisabetta Pagani, Irene Bianconi, Elisabetta Giacobazzi, Elisa Masi, Stefanie Wieser, Irena Jurman, Vera Vendramin, Gabriele Magris, Eleonora Paparelli, Davide Scaglione, Michele Morgante |
| EPI_ISL_1524385 | ULSS 6 Euganea | Istituto Zooprofilattico Sperimentale delle Venezie | Adelaide Milani, Alessia Schivo, Annalisa Salviato, Elisa Palumbo, Erika Giorgia Quaranta, Luca Tassoni, Ambra Pastori, Edoardo Giussani, Alice Fusaro, Isabella Monne, Calogero Terregino, Antonia Ricci |
| EPI_ISL_1524387 | ULSS 1 Dolomiti | Istituto Zooprofilattico Sperimentale delle Venezie | Adelaide Milani, Alessia Schivo, Annalisa Salviato, Elisa Palumbo, Erika Giorgia Quaranta, Luca Tassoni, Ambra Pastori, Edoardo Giussani, Alice Fusaro, Isabella Monne, Calogero Terregino, Antonia Ricci |
| EPI_ISL_1524388, EPI_ISL_1524389, EPI_ISL_1524390, EPI_ISL_1524391 | ULSS 6 Euganea | Istituto Zooprofilattico Sperimentale delle Venezie | Adelaide Milani, Alessia Schivo, Annalisa Salviato, Elisa Palumbo, Erika Giorgia Quaranta, Luca Tassoni, Ambra Pastori, Edoardo Giussani, Alice Fusaro, Isabella Monne, Calogero Terregino, Antonia Ricci |
| EPI_ISL_1524395, EPI_ISL_1524396, EPI_ISL_1524397, EPI_ISL_1524399 | Microbiologia e Virologia | Istituto Zooprofilattico Sperimentale delle Venezie | Adelaide Milani, Alessia Schivo, Annalisa Salviato, Elisa Palumbo, Erika Giorgia Quaranta, Luca Tassoni, Ambra Pastori, Edoardo Giussani, Alice Fusaro, Isabella Monne, Calogero Terregino, Antonia Ricci |
| EPI_ISL_1524407, EPI_ISL_1524408 | Ospedale San Bonifacio | Istituto Zooprofilattico Sperimentale delle Venezie | Adelaide Milani, Alessia Schivo, Annalisa Salviato, Elisa Palumbo, Erika Giorgia Quaranta, Luca Tassoni, Ambra Pastori, Edoardo Giussani, Alice Fusaro, Isabella Monne, Calogero Terregino, Antonia Ricci |
| EPI_ISL_1524411 | Ospedale Magalini | Istituto Zooprofilattico Sperimentale delle Venezie | Adelaide Milani, Alessia Schivo, Annalisa Salviato, Elisa Palumbo, Erika Giorgia Quaranta, Luca Tassoni, Ambra Pastori, Edoardo Giussani, Alice Fusaro, Isabella Monne, Calogero Terregino, Antonia Ricci |
| EPI_ISL_1524416, EPI_ISL_1524418 | ULSS 6 Euganea | Istituto Zooprofilattico Sperimentale delle Venezie | Adelaide Milani, Alessia Schivo, Annalisa Salviato, Elisa Palumbo, Erika Giorgia Quaranta, Luca Tassoni, Ambra Pastori, Edoardo Giussani, Alice Fusaro, Isabella Monne, Calogero Terregino, Antonia Ricci |
| EPI_ISL_1524422 | ULSS 7 Pedemontana - Distretto 2 | Istituto Zooprofilattico Sperimentale delle Venezie | Adelaide Milani, Alessia Schivo, Annalisa Salviato, Elisa Palumbo, Erika Giorgia Quaranta, Luca Tassoni, Ambra Pastori, Edoardo Giussani, Alice Fusaro, Isabella Monne, Calogero Terregino, Antonia Ricci |
| EPI_ISL_1524433, EPI_ISL_1524434 | ULSS 8 Berica | Istituto Zooprofilattico Sperimentale delle Venezie | Adelaide Milani, Alessia Schivo, Annalisa Salviato, Elisa Palumbo, Erika Giorgia Quaranta, Luca Tassoni, Ambra Pastori, Edoardo Giussani, Alice Fusaro, Isabella Monne, Calogero Terregino, Antonia Ricci |
| EPI_ISL_1524456 | Microbiologia e Virologia | Istituto Zooprofilattico Sperimentale delle Venezie | Adelaide Milani, Alessia Schivo, Annalisa Salviato, Elisa Palumbo, Erika Giorgia Quaranta, Luca Tassoni, Ambra Pastori, Edoardo Giussani, Alice Fusaro, Isabella Monne, Calogero Terregino, Antonia Ricci |
| EPI_ISL_1524457, EPI_ISL_1524458, EPI_ISL_1524459, EPI_ISL_1524460, EPI_ISL_1524461 | ULSS 3 Venezia | Istituto Zooprofilattico Sperimentale delle Venezie | Adelaide Milani, Alessia Schivo, Annalisa Salviato, Elisa Palumbo, Erika Giorgia Quaranta, Luca Tassoni, Ambra Pastori, Edoardo Giussani, Alice Fusaro, Isabella Monne, Calogero Terregino, Antonia Ricci |
| EPI_ISL_1524462, EPI_ISL_1524463 | ULSS 7 Pedemontana - Distretto 1 | Istituto Zooprofilattico Sperimentale delle Venezie | Adelaide Milani, Alessia Schivo, Annalisa Salviato, Elisa Palumbo, Erika Giorgia Quaranta, Luca Tassoni, Ambra Pastori, Edoardo Giussani, Alice Fusaro, Isabella Monne, Calogero Terregino, Antonia Ricci |
| EPI_ISL_1524464 | ULSS 4 Veneto Orientale | Istituto di Genomica Applicata; Istituto Zooprofilattico Sperimentale delle Venezie | Irena Jurman, Vera Vendramin, Gabriele Magris, Eleonora Paparelli, Davide Scaglione, Michele Morgante, Silvia Ormelli, Maria Varotto, Valeria D'Amico, Francesca Bruno, Alessia Schivo, Annalisa Salviato, Erika Giorgia Quaranta, Luca Tassoni, Ambra Pastori, Edoardo Giussani, Alice Fusaro, Isabella Monne, Calogero Terregino, Antonia Ricci |
| EPI_ISL_1524465 | ULSS 7 Pedemontana - Distretto 2 | Istituto di Genomica Applicata; Istituto Zooprofilattico Sperimentale delle Venezie | Irena Jurman, Vera Vendramin, Gabriele Magris, Eleonora Paparelli, Davide Scaglione, Michele Morgante, Silvia Ormelli, Maria Varotto, Valeria D'Amico, Francesca Bruno, Alessia Schivo, Annalisa Salviato, Erika Giorgia Quaranta, Luca Tassoni, Ambra Pastori, Edoardo Giussani, Alice Fusaro, Isabella Monne, Calogero Terregino, Antonia Ricci |
| EPI_ISL_1524466 | Ospedale Magalini | Istituto di Genomica Applicata; Istituto Zooprofilattico Sperimentale delle Venezie | Irena Jurman, Vera Vendramin, Gabriele Magris, Eleonora Paparelli, Davide Scaglione, Michele Morgante, Silvia Ormelli, Maria Varotto, Valeria D'Amico, Francesca Bruno, Alessia Schivo, Annalisa Salviato, Erika Giorgia Quaranta, Luca Tassoni, Ambra Pastori, Edoardo Giussani, Alice Fusaro, Isabella Monne, Calogero Terregino, Antonia Ricci |
| EPI_ISL_1524467, EPI_ISL_1524468 | Ospedale Mater Salutis | Istituto di Genomica Applicata; Istituto Zooprofilattico Sperimentale delle Venezie | Irena Jurman, Vera Vendramin, Gabriele Magris, Eleonora Paparelli, Davide Scaglione, Michele Morgante, Silvia Ormelli, Maria Varotto, Valeria D'Amico, Francesca Bruno, Alessia Schivo, Annalisa Salviato, Erika Giorgia Quaranta, Luca Tassoni, Ambra Pastori, Edoardo Giussani, Alice Fusaro, Isabella Monne, Calogero Terregino, Antonia Ricci |
| EPI_ISL_1524469 | Microbiologia e Virologia | Istituto di Genomica Applicata; Istituto Zooprofilattico Sperimentale delle Venezie | Irena Jurman, Vera Vendramin, Gabriele Magris, Eleonora Paparelli, Davide Scaglione, Michele Morgante, Silvia Ormelli, Maria Varotto, Valeria D'Amico, Francesca Bruno, Alessia Schivo, Annalisa Salviato, Erika Giorgia Quaranta, Luca Tassoni, Ambra Pastori, Edoardo Giussani, Alice Fusaro, Isabella Monne, Calogero Terregino, Antonia Ricci |
| EPI_ISL_1524470, EPI_ISL_1524471, EPI_ISL_1524472, EPI_ISL_1524473, EPI_ISL_1524474, EPI_ISL_1524475, EPI_ISL_1524476, EPI_ISL_1524477, EPI_ISL_1524478, EPI_ISL_1524479, EPI_ISL_1524480, EPI_ISL_1524481, EPI_ISL_1524482, EPI_ISL_1524483 | | | |
| see above | ULSS 1 Dolomiti | Istituto di Genomica Applicata; Istituto Zooprofilattico Sperimentale delle Venezie | Irena Jurman, Vera Vendramin, Gabriele Magris, Eleonora Paparelli, Davide Scaglione, Michele Morgante, Silvia Ormelli, Maria Varotto, Valeria D'Amico, Francesca Bruno, Alessia Schivo, Annalisa Salviato, Erika Giorgia Quaranta, Luca Tassoni, Ambra Pastori, Edoardo Giussani, Alice Fusaro, Isabella Monne, Calogero Terregino, Antonia Ricci |

| | | | |
|---|---|---|---|
| EPI_ISL_1524484, EPI_ISL_1524485, EPI_ISL_1524486, EPI_ISL_1524487, EPI_ISL_1524488, EPI_ISL_1524489, EPI_ISL_1524490, EPI_ISL_1524491, EPI_ISL_1524492, EPI_ISL_1524493, EPI_ISL_1524494, EPI_ISL_1524495, EPI_ISL_1524496, EPI_ISL_1524497 | | | |
| see above | ULSS 5 Polesana | Istituto di Genomica Applicata; Istituto Zooprofilattico Sperimentale delle Venezie | Irena Jurman, Vera Vendramin, Gabriele Magris, Eleonora Paparelli, Davide Scaglione, Michele Morgante, Silvia Ormelli, Maria Varotto, Valeria D'Amico, Francesca Bruno, Alessia Schivo, Annalisa Salviato, Erika Giorgia Quaranta, Luca Tassoni, Ambra Pastori, Edoardo Giussani, Alice Fusaro, Isabella Monne, Calogero Terregino, Antonia Ricci |
| EPI_ISL_1524498, EPI_ISL_1524499, EPI_ISL_1524500, EPI_ISL_1524501, EPI_ISL_1524502, EPI_ISL_1524503, EPI_ISL_1524504, EPI_ISL_1524505, EPI_ISL_1524506 | Azienda Ospedaliera Universitaria Integrata Verona | Istituto di Genomica Applicata; Istituto Zooprofilattico Sperimentale delle Venezie | Irena Jurman, Vera Vendramin, Gabriele Magris, Eleonora Paparelli, Davide Scaglione, Michele Morgante, Silvia Ormelli, Maria Varotto, Valeria D'Amico, Francesca Bruno, Alessia Schivo, Annalisa Salviato, Erika Giorgia Quaranta, Luca Tassoni, Ambra Pastori, Edoardo Giussani, Alice Fusaro, Isabella Monne, Calogero Terregino, Antonia Ricci |
| EPI_ISL_1524507, EPI_ISL_1524508, EPI_ISL_1524509, EPI_ISL_1524510, EPI_ISL_1524511, EPI_ISL_1524512, EPI_ISL_1524513 | ULSS 4 Veneto Orientale | Istituto di Genomica Applicata; Istituto Zooprofilattico Sperimentale delle Venezie | Irena Jurman, Vera Vendramin, Gabriele Magris, Eleonora Paparelli, Davide Scaglione, Michele Morgante, Silvia Ormelli, Maria Varotto, Valeria D'Amico, Francesca Bruno, Alessia Schivo, Annalisa Salviato, Erika Giorgia Quaranta, Luca Tassoni, Ambra Pastori, Edoardo Giussani, Alice Fusaro, Isabella Monne, Calogero Terregino, Antonia Ricci |
| EPI_ISL_1524514, EPI_ISL_1524515, EPI_ISL_1524516, EPI_ISL_1524517, EPI_ISL_1524518, EPI_ISL_1524519 | ULSS 7 Pedemontana - Distretto 1 | Istituto di Genomica Applicata; Istituto Zooprofilattico Sperimentale delle Venezie | Irena Jurman, Vera Vendramin, Gabriele Magris, Eleonora Paparelli, Davide Scaglione, Michele Morgante, Silvia Ormelli, Maria Varotto, Valeria D'Amico, Francesca Bruno, Alessia Schivo, Annalisa Salviato, Erika Giorgia Quaranta, Luca Tassoni, Ambra Pastori, Edoardo Giussani, Alice Fusaro, Isabella Monne, Calogero Terregino, Antonia Ricci |
| EPI_ISL_1524520, EPI_ISL_1524521, EPI_ISL_1524522, EPI_ISL_1524523, EPI_ISL_1524524 | ULSS 7 Pedemontana - Distretto 2 | Istituto di Genomica Applicata; Istituto Zooprofilattico Sperimentale delle Venezie | Irena Jurman, Vera Vendramin, Gabriele Magris, Eleonora Paparelli, Davide Scaglione, Michele Morgante, Silvia Ormelli, Maria Varotto, Valeria D'Amico, Francesca Bruno, Alessia Schivo, Annalisa Salviato, Erika Giorgia Quaranta, Luca Tassoni, Ambra Pastori, Edoardo Giussani, Alice Fusaro, Isabella Monne, Calogero Terregino, Antonia Ricci |
| EPI_ISL_1524525, EPI_ISL_1524526, EPI_ISL_1524527, EPI_ISL_1524528, EPI_ISL_1524529, EPI_ISL_1524530, EPI_ISL_1524531, EPI_ISL_1524532, EPI_ISL_1524533, EPI_ISL_1524534, EPI_ISL_1524535, EPI_ISL_1524536, EPI_ISL_1524537, EPI_ISL_1524538 | | | |
| see above | ULSS 8 Berica | Istituto di Genomica Applicata; Istituto Zooprofilattico Sperimentale delle Venezie | Irena Jurman, Vera Vendramin, Gabriele Magris, Eleonora Paparelli, Davide Scaglione, Michele Morgante, Silvia Ormelli, Maria Varotto, Valeria D'Amico, Francesca Bruno, Alessia Schivo, Annalisa Salviato, Erika Giorgia Quaranta, Luca Tassoni, Ambra Pastori, Edoardo Giussani, Alice Fusaro, Isabella Monne, Calogero Terregino, Antonia Ricci |
| EPI_ISL_1524539, EPI_ISL_1524540, EPI_ISL_1524541, EPI_ISL_1524542, EPI_ISL_1524543, EPI_ISL_1524544, EPI_ISL_1524545, EPI_ISL_1524546, EPI_ISL_1524547 | Ospedale Magalini | Istituto di Genomica Applicata; Istituto Zooprofilattico Sperimentale delle Venezie | Irena Jurman, Vera Vendramin, Gabriele Magris, Eleonora Paparelli, Davide Scaglione, Michele Morgante, Silvia Ormelli, Maria Varotto, Valeria D'Amico, Francesca Bruno, Alessia Schivo, Annalisa Salviato, Erika Giorgia Quaranta, Luca Tassoni, Ambra Pastori, Edoardo Giussani, Alice Fusaro, Isabella Monne, Calogero Terregino, Antonia Ricci |
| EPI_ISL_1524548, EPI_ISL_1524549, EPI_ISL_1524550, EPI_ISL_1524551, EPI_ISL_1524552, EPI_ISL_1524553, EPI_ISL_1524554, EPI_ISL_1524555, EPI_ISL_1524556, EPI_ISL_1524557, EPI_ISL_1524558, EPI_ISL_1524559, EPI_ISL_1524560 | | | |
| see above | ULSS 6 Euganea | Istituto di Genomica Applicata; Istituto Zooprofilattico Sperimentale delle Venezie | Irena Jurman, Vera Vendramin, Gabriele Magris, Eleonora Paparelli, Davide Scaglione, Michele Morgante, Silvia Ormelli, Maria Varotto, Valeria D'Amico, Francesca Bruno, Alessia Schivo, Annalisa Salviato, Erika Giorgia Quaranta, Luca Tassoni, Ambra Pastori, Edoardo Giussani, Alice Fusaro, Isabella Monne, Calogero Terregino, Antonia Ricci |
| EPI_ISL_1524561, EPI_ISL_1524562, EPI_ISL_1524563, EPI_ISL_1524564, EPI_ISL_1524565, EPI_ISL_1524566, EPI_ISL_1524567, EPI_ISL_1524568 | Ospedale Mater Salutis | Istituto di Genomica Applicata; Istituto Zooprofilattico Sperimentale delle Venezie | Irena Jurman, Vera Vendramin, Gabriele Magris, Eleonora Paparelli, Davide Scaglione, Michele Morgante, Silvia Ormelli, Maria Varotto, Valeria D'Amico, Francesca Bruno, Alessia Schivo, Annalisa Salviato, Erika Giorgia Quaranta, Luca Tassoni, Ambra Pastori, Edoardo Giussani, Alice Fusaro, Isabella Monne, Calogero Terregino, Antonia Ricci |
| EPI_ISL_1524569, EPI_ISL_1524570, EPI_ISL_1524571, EPI_ISL_1524572, EPI_ISL_1524573, EPI_ISL_1524574, EPI_ISL_1524575, EPI_ISL_1524576, EPI_ISL_1524577, EPI_ISL_1524578, EPI_ISL_1524579, EPI_ISL_1524580, EPI_ISL_1524581, EPI_ISL_1524582 | | | |
| see above | Microbiologia e Virologia | Istituto di Genomica Applicata; Istituto Zooprofilattico Sperimentale delle Venezie | Irena Jurman, Vera Vendramin, Gabriele Magris, Eleonora Paparelli, Davide Scaglione, Michele Morgante, Silvia Ormelli, Maria Varotto, Valeria D'Amico, Francesca Bruno, Alessia Schivo, Annalisa Salviato, Erika Giorgia Quaranta, Luca Tassoni, Ambra Pastori, Edoardo Giussani, Alice Fusaro, Isabella Monne, Calogero Terregino, Antonia Ricci |
| EPI_ISL_1524583, EPI_ISL_1524584, EPI_ISL_1524585, EPI_ISL_1524586, EPI_ISL_1524587, EPI_ISL_1524588, EPI_ISL_1524589, EPI_ISL_1524590, EPI_ISL_1524591, EPI_ISL_1524592, EPI_ISL_1524593, EPI_ISL_1524594, EPI_ISL_1524595, EPI_ISL_1524596, EPI_ISL_1524597, EPI_ISL_1524598, EPI_ISL_1524599, EPI_ISL_1524600 | | | |
| see above | ULSS 2 Marca Trevigiana | Istituto di Genomica Applicata; Istituto Zooprofilattico Sperimentale delle Venezie | Irena Jurman, Vera Vendramin, Gabriele Magris, Eleonora Paparelli, Davide Scaglione, Michele Morgante, Silvia Ormelli, Maria Varotto, Valeria D'Amico, Francesca Bruno, Alessia Schivo, Annalisa Salviato, Erika Giorgia Quaranta, Luca Tassoni, Ambra Pastori, Edoardo Giussani, Alice Fusaro, Isabella Monne, Calogero Terregino, Antonia Ricci |
| EPI_ISL_1524601, EPI_ISL_1524602, EPI_ISL_1524603, EPI_ISL_1524604, EPI_ISL_1524605, EPI_ISL_1524606, EPI_ISL_1524607, EPI_ISL_1524608, EPI_ISL_1524609, EPI_ISL_1524610, EPI_ISL_1524611, EPI_ISL_1524612, EPI_ISL_1524613, EPI_ISL_1524614, EPI_ISL_1524615, EPI_ISL_1524616, EPI_ISL_1524617, EPI_ISL_1524618, EPI_ISL_1524619 | | | |
| see above | ULSS 3 Venezia | Istituto di Genomica Applicata; Istituto Zooprofilattico Sperimentale delle Venezie | Irena Jurman, Vera Vendramin, Gabriele Magris, Eleonora Paparelli, Davide Scaglione, Michele Morgante, Silvia Ormelli, Maria Varotto, Valeria D'Amico, Francesca Bruno, Alessia Schivo, Annalisa Salviato, Erika Giorgia Quaranta, Luca Tassoni, Ambra Pastori, Edoardo Giussani, Alice Fusaro, Isabella Monne, Calogero Terregino, Antonia Ricci |
| EPI_ISL_1524620, EPI_ISL_1524621, EPI_ISL_1524622, EPI_ISL_1524623 | Ospedale Magalini | Istituto Zooprofilattico Sperimentale delle Venezie | Adelaide Milani, Alessia Schivo, Annalisa Salviato, Elisa Palumbo, Erika Giorgia Quaranta, Luca Tassoni, Ambra Pastori, Edoardo Giussani, Alice Fusaro, Isabella Monne, Calogero Terregino, Antonia Ricci |
| EPI_ISL_1524624 | Ospedale San Bonifacio | Istituto Zooprofilattico Sperimentale delle Venezie | Adelaide Milani, Alessia Schivo, Annalisa Salviato, Elisa Palumbo, Erika Giorgia Quaranta, Luca Tassoni, Ambra Pastori, Edoardo Giussani, Alice Fusaro, Isabella Monne, Calogero Terregino, Antonia Ricci |
| EPI_ISL_1524625, EPI_ISL_1524626, EPI_ISL_1524627, EPI_ISL_1524628, EPI_ISL_1524629, EPI_ISL_1524630 | Ospedale Mater Salutis | Istituto Zooprofilattico Sperimentale delle Venezie | Adelaide Milani, Alessia Schivo, Annalisa Salviato, Elisa Palumbo, Erika Giorgia Quaranta, Luca Tassoni, Ambra Pastori, Edoardo Giussani, Alice Fusaro, Isabella Monne, Calogero Terregino, Antonia Ricci |
| EPI_ISL_1524631, EPI_ISL_1524632, EPI_ISL_1524633, EPI_ISL_1524634, EPI_ISL_1524635, EPI_ISL_1524636, EPI_ISL_1524637, EPI_ISL_1524638, EPI_ISL_1524639, EPI_ISL_1524640, EPI_ISL_1524641, EPI_ISL_1524642, EPI_ISL_1524643, EPI_ISL_1524644, EPI_ISL_1524645, EPI_ISL_1524646 | | | |
| see above | ULSS 6 Euganea | Istituto Zooprofilattico Sperimentale delle Venezie | Adelaide Milani, Alessia Schivo, Annalisa Salviato, Elisa Palumbo, Erika Giorgia Quaranta, Luca Tassoni, Ambra Pastori, Edoardo Giussani, Alice Fusaro, Isabella Monne, Calogero Terregino, Antonia Ricci |
| EPI_ISL_1524648, EPI_ISL_1524649, EPI_ISL_1524651, EPI_ISL_1524652, EPI_ISL_1524653 | ULSS 1 Dolomiti | Istituto Zooprofilattico Sperimentale delle Venezie | Adelaide Milani, Alessia Schivo, Annalisa Salviato, Elisa Palumbo, Erika Giorgia Quaranta, Luca Tassoni, Ambra Pastori, Edoardo Giussani, Alice Fusaro, Isabella Monne, Calogero Terregino, Antonia Ricci |
| EPI_ISL_1524654, EPI_ISL_1524655, EPI_ISL_1524656, EPI_ISL_1524657, EPI_ISL_1524658, EPI_ISL_1524659 | ULSS 5 Polesana | Istituto Zooprofilattico Sperimentale delle Venezie | Adelaide Milani, Alessia Schivo, Annalisa Salviato, Elisa Palumbo, Erika Giorgia Quaranta, Luca Tassoni, Ambra Pastori, Edoardo Giussani, Alice Fusaro, Isabella Monne, Calogero Terregino, Antonia Ricci |
| EPI_ISL_1524660, EPI_ISL_1524661, EPI_ISL_1524662, EPI_ISL_1524663 | Azienda Ospedaliera Universitaria Integrata Verona | Istituto Zooprofilattico Sperimentale delle Venezie | Adelaide Milani, Alessia Schivo, Annalisa Salviato, Elisa Palumbo, Erika Giorgia Quaranta, Luca Tassoni, Ambra Pastori, Edoardo Giussani, Alice Fusaro, Isabella Monne, Calogero Terregino, Antonia Ricci |
| EPI_ISL_1524664, EPI_ISL_1524665, EPI_ISL_1524666, EPI_ISL_1524667, EPI_ISL_1524668, EPI_ISL_1524669 | ULSS 4 Veneto Orientale | Istituto Zooprofilattico Sperimentale delle Venezie | Adelaide Milani, Alessia Schivo, Annalisa Salviato, Elisa Palumbo, Erika Giorgia Quaranta, Luca Tassoni, Ambra Pastori, Edoardo Giussani, Alice Fusaro, Isabella Monne, Calogero Terregino, Antonia Ricci |
| EPI_ISL_1524670, EPI_ISL_1524671, EPI_ISL_1524672 | ULSS 7 Pedemontana - Distretto 1 | Istituto Zooprofilattico Sperimentale delle Venezie | Adelaide Milani, Alessia Schivo, Annalisa Salviato, Elisa Palumbo, Erika Giorgia Quaranta, Luca Tassoni, Ambra Pastori, Edoardo Giussani, Alice Fusaro, Isabella Monne, Calogero Terregino, Antonia Ricci |

| | | | |
|---|---|---|---|
| EPI_ISL_1524673, EPI_ISL_1524674, EPI_ISL_1524675 | ULSS 7 Pedemontana - Distretto 2 | Istituto Zooprofilattico Sperimentale delle Venezie | Adelaide Milani, Alessia Schivo, Annalisa Salviato, Elisa Palumbo, Erika Giorgia Quaranta, Luca Tassoni, Ambra Pastori, Edoardo Giussani, Alice Fusaro, Isabella Monne, Calogero Terregino, Antonia Ricci |
| EPI_ISL_1524676, EPI_ISL_1524677, EPI_ISL_1524678 | ULSS 3 Venezia | Istituto Zooprofilattico Sperimentale delle Venezie | Adelaide Milani, Alessia Schivo, Annalisa Salviato, Elisa Palumbo, Erika Giorgia Quaranta, Luca Tassoni, Ambra Pastori, Edoardo Giussani, Alice Fusaro, Isabella Monne, Calogero Terregino, Antonia Ricci |
| EPI_ISL_1524679, EPI_ISL_1524680, EPI_ISL_1524681, EPI_ISL_1524682, EPI_ISL_1524683, EPI_ISL_1524684, EPI_ISL_1524685, EPI_ISL_1524686, EPI_ISL_1524687, EPI_ISL_1524689, EPI_ISL_1524690, EPI_ISL_1524691 | | | |
| see above | Microbiologia e Virologia | Istituto Zooprofilattico Sperimentale delle Venezie | Adelaide Milani, Alessia Schivo, Annalisa Salviato, Elisa Palumbo, Erika Giorgia Quaranta, Luca Tassoni, Ambra Pastori, Edoardo Giussani, Alice Fusaro, Isabella Monne, Calogero Terregino, Antonia Ricci |
| EPI_ISL_1524692, EPI_ISL_1524693, EPI_ISL_1524694, EPI_ISL_1524695 | ULSS 8 Berica | Istituto Zooprofilattico Sperimentale delle Venezie | Adelaide Milani, Alessia Schivo, Annalisa Salviato, Elisa Palumbo, Erika Giorgia Quaranta, Luca Tassoni, Ambra Pastori, Edoardo Giussani, Alice Fusaro, Isabella Monne, Calogero Terregino, Antonia Ricci |
| EPI_ISL_1524696, EPI_ISL_1524697 | ULSS 2 Marca Trevigiana | Istituto Zooprofilattico Sperimentale delle Venezie | Adelaide Milani, Alessia Schivo, Annalisa Salviato, Elisa Palumbo, Erika Giorgia Quaranta, Luca Tassoni, Ambra Pastori, Edoardo Giussani, Alice Fusaro, Isabella Monne, Calogero Terregino, Antonia Ricci |
| EPI_ISL_1524709, EPI_ISL_1524710 | Azienda Ospedaliera Terni | Istituto Zooprofilattico Sperimentale dell'Abruzzo e Molise "G. Caporale" | Palumbo M, Scaccetti A, Lorusso A, Marcacci M, Di Domenico M, Ancora M, Curini V, Di Lollo V, Delli Compagni E, Mangone I, Rinaldi A, Scialabba S, Caporale M, Di Pasquale A, Cammà C, Puglia I, Calistri P, Savini G |
| EPI_ISL_1524739 | Azienda Ospedaliera San Camillo Forlanini | INMI Lazzaro Spallanzani IRCCS | G Bonfiglio, O Butera, CEM Gruber, F Santini, B Bartolini, E Giombini, F Messina, M Rueca, G Parisi, F Basile, A D'Agostino, A Di Caro, MR Capobianchi |
| EPI_ISL_1524740 | Ospedale San Giovanni Evangelista | INMI Lazzaro Spallanzani IRCCS | F Messina, M Rueca, G Bonfiglio, O Butera, CEM Gruber, F Santini, B Bartolini, E Giombini, D Di Fusco, D Cerini, A Di Caro, MR Capobianchi |
| EPI_ISL_1524741 | Ospedale San Giovanni Evangelista | INMI Lazzaro Spallanzani IRCCS | E Giombini, F Messina, M Rueca, G Bonfiglio, O Butera, CEM Gruber, F Santini, B Bartolini, D Di Fusco, D Cerini, MR Capobianchi, A Di Caro |
| EPI_ISL_1524742 | Ospedale San Filippo Neri | INMI Lazzaro Spallanzani IRCCS | M Rueca, G Bonfiglio, O Butera, CEM Gruber, F Santini, B Bartolini, E Giombini, F Messina, M Meledandri, ML Schiavone, A Tamburro, MR Capobianchi, A Di Caro |
| EPI_ISL_1524851 | USCA Avezzano AVEZZANO(L'AQUILA) | Istituto Zooprofilattico Sperimentale dell'Abruzzo e Molise "G. Caporale" | Lorusso A, Marcacci M, Di Domenico M, Ancora M, Curini V, Di Lollo Valeria, Mangone I, Rinaldi A, Delli Compagni E, Scialabba S, Caporale M, Di Pasquale A, Cammà C, Puglia I, Calistri P, Savini G |
| EPI_ISL_1524852, EPI_ISL_1524853, EPI_ISL_1524854, EPI_ISL_1524855, EPI_ISL_1524856 | SIESP DIPARTIMENTO DI PREVENZIONE TERAMO TERAMO(TERAMO) | Istituto Zooprofilattico Sperimentale dell'Abruzzo e Molise "G. Caporale" | Lorusso A, Marcacci M, Di Domenico M, Ancora M, Curini V, Di Lollo Valeria, Mangone I, Rinaldi A, Delli Compagni E, Scialabba S, Caporale M, Di Pasquale A, Cammà C, Puglia I, Calistri P, Savini G |
| EPI_ISL_1524857 | SIESP CHIETI - DRIVE IN CHIETI CHIETI(CHIETI) | Istituto Zooprofilattico Sperimentale dell'Abruzzo e Molise "G. Caporale" | Lorusso A, Marcacci M, Di Domenico M, Ancora M, Curini V, Di Lollo Valeria, Mangone I, Rinaldi A, Delli Compagni E, Scialabba S, Caporale M, Di Pasquale A, Cammà C, Puglia I, Calistri P, Savini G |
| EPI_ISL_1524858 | SIESP CHIETI - DRIVE IN VASTO VASTO(CHIETI) | Istituto Zooprofilattico Sperimentale dell'Abruzzo e Molise "G. Caporale" | Lorusso A, Marcacci M, Di Domenico M, Ancora M, Curini V, Di Lollo Valeria, Mangone I, Rinaldi A, Delli Compagni E, Scialabba S, Caporale M, Di Pasquale A, Cammà C, Puglia I, Calistri P, Savini G |
| EPI_ISL_1524859 | SIESP CHIETI - DRIVE IN GISSI GISSI(CHIETI) | Istituto Zooprofilattico Sperimentale dell'Abruzzo e Molise "G. Caporale" | Lorusso A, Marcacci M, Di Domenico M, Ancora M, Curini V, Di Lollo Valeria, Mangone I, Rinaldi A, Delli Compagni E, Scialabba S, Caporale M, Di Pasquale A, Cammà C, Puglia I, Calistri P, Savini G |
| EPI_ISL_1524860 | SIESP CHIETI - DRIVE IN CHIETI CHIETI(CHIETI) | Istituto Zooprofilattico Sperimentale dell'Abruzzo e Molise "G. Caporale" | Lorusso A, Marcacci M, Di Domenico M, Ancora M, Curini V, Di Lollo Valeria, Mangone I, Rinaldi A, Delli Compagni E, Scialabba S, Caporale M, Di Pasquale A, Cammà C, Puglia I, Calistri P, Savini G |
| EPI_ISL_1524861, EPI_ISL_1524862 | SIESP DIPARTIMENTO DI PREVENZIONE TERAMO TERAMO(TERAMO) | Istituto Zooprofilattico Sperimentale dell'Abruzzo e Molise "G. Caporale" | Lorusso A, Marcacci M, Di Domenico M, Ancora M, Curini V, Di Lollo Valeria, Mangone I, Rinaldi A, Delli Compagni E, Scialabba S, Caporale M, Di Pasquale A, Cammà C, Puglia I, Calistri P, Savini G |
| EPI_ISL_1524863, EPI_ISL_1524864 | SIESP CHIETI - DRIVE IN LANCIANO LANCIANO(CHIETI) | Istituto Zooprofilattico Sperimentale dell'Abruzzo e Molise "G. Caporale" | Lorusso A, Marcacci M, Di Domenico M, Ancora M, Curini V, Di Lollo Valeria, Mangone I, Rinaldi A, Delli Compagni E, Scialabba S, Caporale M, Di Pasquale A, Cammà C, Puglia I, Calistri P, Savini G |
| EPI_ISL_1524865, EPI_ISL_1524866 | SIESP SULMONA SULMONA(L'AQUILA) | Istituto Zooprofilattico Sperimentale dell'Abruzzo e Molise "G. Caporale" | Lorusso A, Marcacci M, Di Domenico M, Ancora M, Curini V, Di Lollo Valeria, Mangone I, Rinaldi A, Delli Compagni E, Scialabba S, Caporale M, Di Pasquale A, Cammà C, Puglia I, Calistri P, Savini G |
| EPI_ISL_1524867 | SIESP CHIETI - DRIVE IN LANCIANO LANCIANO(CHIETI) | Istituto Zooprofilattico Sperimentale dell'Abruzzo e Molise "G. Caporale" | Lorusso A, Marcacci M, Di Domenico M, Ancora M, Curini V, Di Lollo Valeria, Mangone I, Rinaldi A, Delli Compagni E, Scialabba S, Caporale M, Di Pasquale A, Cammà C, Puglia I, Calistri P, Savini G |
| EPI_ISL_1524868 | Assistenza Domiciliare Integrata (ADI) TERAMO TERAMO(TERAMO) | Istituto Zooprofilattico Sperimentale dell'Abruzzo e Molise "G. Caporale" | Lorusso A, Marcacci M, Di Domenico M, Ancora M, Curini V, Di Lollo Valeria, Mangone I, Rinaldi A, Delli Compagni E, Scialabba S, Caporale M, Di Pasquale A, Cammà C, Puglia I, Calistri P, Savini G |
| EPI_ISL_1524869, EPI_ISL_1524870 | SIESP DIPARTIMENTO DI PREVENZIONE TERAMO TERAMO(TERAMO) | Istituto Zooprofilattico Sperimentale dell'Abruzzo e Molise "G. Caporale" | Lorusso A, Marcacci M, Di Domenico M, Ancora M, Curini V, Di Lollo Valeria, Mangone I, Rinaldi A, Delli Compagni E, Scialabba S, Caporale M, Di Pasquale A, Cammà C, Puglia I, Calistri P, Savini G |
| EPI_ISL_1524871 | SIESP SULMONA SULMONA(L'AQUILA) | Istituto Zooprofilattico Sperimentale dell'Abruzzo e Molise "G. Caporale" | Lorusso A, Marcacci M, Di Domenico M, Ancora M, Curini V, Di Lollo Valeria, Mangone I, Rinaldi A, Delli Compagni E, Scialabba S, Caporale M, Di Pasquale A, Cammà C, Puglia I, Calistri P, Savini G |
| EPI_ISL_1525249 | Laboratorio HUB -Azienda Ospedaliero Universitaria - AOU - Cagliari | Laboratorio SPOKE Biologia Molecolare -Azienda Ospedaliero Universitaria - AOU - Cagliari | Germano Orrù, Sara Fais, Alessandra Scano, Riccardo Cappai, Ferdinando Coghe |
| EPI_ISL_1532816, EPI_ISL_1532817, EPI_ISL_1532818, EPI_ISL_1532819, EPI_ISL_1532820, EPI_ISL_1532821, EPI_ISL_1532822, EPI_ISL_1532823, EPI_ISL_1532824, EPI_ISL_1532825, EPI_ISL_1532826, EPI_ISL_1532827, EPI_ISL_1532828, EPI_ISL_1532829, EPI_ISL_1532830, EPI_ISL_1532831, EPI_ISL_1532832, EPI_ISL_1532833, EPI_ISL_1532834, EPI_ISL_1532835, EPI_ISL_1532836, EPI_ISL_1532837, EPI_ISL_1532838, EPI_ISL_1532839, EPI_ISL_1532840 | | | |
| see above | SC Microbiologia e Virologia AOUSS | AMES Centro Polidiagnostico Strumentale S.r.l. | Salvatore Rubino; Flavia Angioj; Laura Firino; Rosalba Govoni; Gabriele Ibba; Vincenzo Lai; Erica Mura; Bianca Paglietti; Claudia Piu; Anna Puggioni; Elena Rimini; Giulia Rocca; Caterina Serra; Sergio Uzzau. |
| EPI_ISL_1544075, EPI_ISL_1544076, EPI_ISL_1544077, EPI_ISL_1544078, EPI_ISL_1544079, EPI_ISL_1544080, EPI_ISL_1544081, EPI_ISL_1544082, EPI_ISL_1544083, EPI_ISL_1544084, EPI_ISL_1544085, EPI_ISL_1544086, EPI_ISL_1544087, EPI_ISL_1544088, EPI_ISL_1544089, EPI_ISL_1544090, EPI_ISL_1544091, EPI_ISL_1544092, EPI_ISL_1544093, EPI_ISL_1544094, EPI_ISL_1544095, EPI_ISL_1544096, EPI_ISL_1544097, EPI_ISL_1544098, EPI_ISL_1544099, EPI_ISL_1544100, EPI_ISL_1544101, EPI_ISL_1544102, EPI_ISL_1544103, EPI_ISL_1544104, EPI_ISL_1544105, EPI_ISL_1544106, EPI_ISL_1544107 | | | |
| see above | SOC Igiene ed Epidemiologia Clinica, ASUFC, Udine and SC (UCO) Igiene e Sanità Pubblica, ASUGI, Trieste | Istituto di Genomica Applicata | Pierlanfranco D'Agaro, Corrado Pipan, Irena Jurman, Vera Vendramin, Gabriele Magris, Eleonora Paparelli, Davide Scaglione, Michele Morgante |
| EPI_ISL_1545451, EPI_ISL_1545452, EPI_ISL_1545453, EPI_ISL_1545454, EPI_ISL_1545455, EPI_ISL_1545456, EPI_ISL_1545457, EPI_ISL_1545458, EPI_ISL_1545459, EPI_ISL_1545460, EPI_ISL_1545461, EPI_ISL_1545462, EPI_ISL_1545463, EPI_ISL_1545464, EPI_ISL_1545465, EPI_ISL_1545466, EPI_ISL_1545467, EPI_ISL_1545468, EPI_ISL_1545469, EPI_ISL_1545470, EPI_ISL_1545471, EPI_ISL_1545472, EPI_ISL_1545473, EPI_ISL_1545474, EPI_ISL_1545475, EPI_ISL_1545476, EPI_ISL_1545477, EPI_ISL_1545478, EPI_ISL_1545479, EPI_ISL_1545480, EPI_ISL_1545481, EPI_ISL_1545482, EPI_ISL_1545483, EPI_ISL_1545484, EPI_ISL_1545485, EPI_ISL_1545486, EPI_ISL_1545487, EPI_ISL_1545488, EPI_ISL_1545489, EPI_ISL_1545490, EPI_ISL_1545491, EPI_ISL_1545492, EPI_ISL_1545493, EPI_ISL_1545494, EPI_ISL_1545495, EPI_ISL_1545496, EPI_ISL_1545497, EPI_ISL_1545498, EPI_ISL_1545499, EPI_ISL_1545500, EPI_ISL_1545501, EPI_ISL_1545502, EPI_ISL_1545503, EPI_ISL_1545504, EPI_ISL_1545505, EPI_ISL_1545506, EPI_ISL_1545507, EPI_ISL_1545508, EPI_ISL_1545509, EPI_ISL_1545510, EPI_ISL_1545511, EPI_ISL_1545512, EPI_ISL_1545513, EPI_ISL_1545514, EPI_ISL_1545515, EPI_ISL_1545516, EPI_ISL_1545517, EPI_ISL_1545518, EPI_ISL_1545519, EPI_ISL_1545520, EPI_ISL_1545521, EPI_ISL_1545522, EPI_ISL_1545523, EPI_ISL_1545524, EPI_ISL_1545525, EPI_ISL_1545526, EPI_ISL_1545527, EPI_ISL_1545528, EPI_ISL_1545529, EPI_ISL_1545530, EPI_ISL_1545531, EPI_ISL_1545532, EPI_ISL_1545533, EPI_ISL_1545534, EPI_ISL_1545535, EPI_ISL_1545536, EPI_ISL_1545537, EPI_ISL_1545538, EPI_ISL_1545539, EPI_ISL_1545540, EPI_ISL_1545541, EPI_ISL_1545542, EPI_ISL_1545543, EPI_ISL_1545544, EPI_ISL_1545545, EPI_ISL_1545546, EPI_ISL_1545547, EPI_ISL_1545548, EPI_ISL_1545549, EPI_ISL_1545550, EPI_ISL_1545551, EPI_ISL_1545552, EPI_ISL_1545553, EPI_ISL_1545554, EPI_ISL_1545555, EPI_ISL_1545556, EPI_ISL_1545557, EPI_ISL_1545558, EPI_ISL_1545559, EPI_ISL_1545560, EPI_ISL_1545561, EPI_ISL_1545562, EPI_ISL_1545563, EPI_ISL_1545564, EPI_ISL_1545565, EPI_ISL_1545566, EPI_ISL_1545567, EPI_ISL_1545568, EPI_ISL_1545569, EPI_ISL_1545570, EPI_ISL_1545571, EPI_ISL_1545572, EPI_ISL_1545573, EPI_ISL_1545574, EPI_ISL_1545575, EPI_ISL_1545576, EPI_ISL_1545577, EPI_ISL_1545578, EPI_ISL_1545579, EPI_ISL_1545580, EPI_ISL_1545581, EPI_ISL_1545582, EPI_ISL_1545583, EPI_ISL_1545584, EPI_ISL_1545585, EPI_ISL_1545586, EPI_ISL_1545587, EPI_ISL_1545588, EPI_ISL_1545589, EPI_ISL_1545590, EPI_ISL_1545591, EPI_ISL_1545592, EPI_ISL_1545593, EPI_ISL_1545594, EPI_ISL_1545595, EPI_ISL_1545596, EPI_ISL_1545597, EPI_ISL_1545598, EPI_ISL_1545599, EPI_ISL_1545600, EPI_ISL_1545601, EPI_ISL_1545602, EPI_ISL_1545603, EPI_ISL_1545604, EPI_ISL_1545605, EPI_ISL_1545606, EPI_ISL_1545607, EPI_ISL_1545608, EPI_ISL_1545609, EPI_ISL_1545610, EPI_ISL_1545611, EPI_ISL_1545612, EPI_ISL_1545613, EPI_ISL_1545614, EPI_ISL_1545615, EPI_ISL_1545616, EPI_ISL_1545617, EPI_ISL_1545618, EPI_ISL_1545619, EPI_ISL_1545620, EPI_ISL_1545621, EPI_ISL_1545622, EPI_ISL_1545623, EPI_ISL_1545624, EPI_ISL_1545625, EPI_ISL_1545626, EPI_ISL_1545627, EPI_ISL_1545628, EPI_ISL_1545629 | | | |
| see above | Istituto Zooprofilattico Sperimentale del Mezzogiorno - Azienda Ospedaliera Pugliese Ciaccio di Catanzaro | TIGEM | Antonio Grimaldi Patrizia Annunziata Francesco Panariello Biancamaria Pierri Claudia Tiberio Teresa Giuliano Valentina Bouche Chiara Colantuono Maria Concetta Cuomo Denise Di Concilio Lucio Di Filippo Anna Manfredi Marcello Salvi Pasquale Minchella Antonio Limone Luigi Atripaldi Pellegrino Cerino Andrea Ballabio Davide Cacchiarelli |
| EPI_ISL_1545630, EPI_ISL_1545631 | Istituto Zooprofilattico Sperimentale del Mezzogiorno | TIGEM | Antonio Grimaldi Patrizia Annunziata Francesco Panariello Biancamaria Pierri Claudia Tiberio Teresa Giuliano Valentina Bouche Chiara Colantuono Maria Concetta Cuomo Denise Di Concilio Lucio Di Filippo Anna Manfredi Marcello Salvi Antonio Limone Luigi Atripaldi Pellegrino Cerino Andrea Ballabio Davide Cacchiarelli |

| | | | |
|---|---|---|---|
| EPI_ISL_1545669, EPI_ISL_1545670, EPI_ISL_1545671, EPI_ISL_1545672, EPI_ISL_1545673, EPI_ISL_1545675, EPI_ISL_1545676, EPI_ISL_1545677, EPI_ISL_1545678, EPI_ISL_1545679, EPI_ISL_1545680, EPI_ISL_1545681, EPI_ISL_1545682, EPI_ISL_1545683, EPI_ISL_1545684, EPI_ISL_1545685, EPI_ISL_1545686, EPI_ISL_1545687, EPI_ISL_1545688, EPI_ISL_1545689, EPI_ISL_1545690, EPI_ISL_1545691, EPI_ISL_1545692, EPI_ISL_1545693, EPI_ISL_1545694, EPI_ISL_1545695, EPI_ISL_1545696, EPI_ISL_1545697, EPI_ISL_1545698, EPI_ISL_1545699, EPI_ISL_1545700, EPI_ISL_1545701, EPI_ISL_1545702, EPI_ISL_1545703, EPI_ISL_1545705, EPI_ISL_1545706, EPI_ISL_1545707, EPI_ISL_1545708, EPI_ISL_1545709, EPI_ISL_1545710, EPI_ISL_1545711, EPI_ISL_1545712, EPI_ISL_1545713, EPI_ISL_1545714, EPI_ISL_1545715, EPI_ISL_1545716, EPI_ISL_1545731, EPI_ISL_1545761, EPI_ISL_1545762, EPI_ISL_1545763 | | | |
| see above | Azienda Sanitaria dell'Alto Adige Laboratorio Aziendale di Microbiologia e Virologia | Istituto di Genomica Applicata | Elisabetta Pagani, Irene Bianconi, Elisabetta Giacobazzi, Elisa Masi, Stefanie Wieser, Irena Jurman, Vera Vendramin, Gabriele Magris, Eleonora Paparelli, Davide Scaglione, Michele Morgante |
| EPI_ISL_1547375 | 1.AO Universitaria 'S. Giovanni di Dio e Ruggi D'Aragona, Scuola Medica Salernitana' Hospital / 2.UOC di Virologia e Microbiologia, Università della Campania 'L. Vanvitelli' / 3.AO Universitaria 'Federico II' Napoli Hospital / 4.AORN 'San Giuseppe Moscati' Avellino Hospital / 5.AO 'San Pio - presidio G. Rummo' Benevento Hospital / 6.AO 'Sant'Anna e San Sebastiano' Caserta Hospital / 7.PO 'Maria Santissima Addolorata' Eboli Hospital / 8.Biogem Istituto di Ricerche Genetiche / 9. U.O.C. di Genetica Medica e di Laboratorio A.O.R.N., Azienda Ospedaliera di Rilievo Nazionale Antonio Cardarelli, Napoli / 10. Centro di riferimento Oncologico della Basilicata (IRCCS-CROB), Rionero in Vulture (PZ) / 11. Presidio Ospedaliero di Agropoli, Agropoli (SA). | 1. Genome Research Center for Health (CRGS) / 2. Laboratory of Molecular Medicine and Genomics(LMMGe) / 3. Center for Research in Pure and Applied Mathematics (CRMPA) | Giorgio Giurato (Corresponding Author), Francesca Rizzo (Corresponding Author), Alessandro Weisz (Corresponding Author), Gianluigi Franci, Giovanni Nassa, Pasquale Pagliano, Roberta Tarallo, Elena Alexandrova, Ylenia D'Agostino, Carlo Ferravante, Jessica Lamberti, Viola Melone, Domenico Memoli, Valeria Mirici Cappa, Domenico Palumbo, Giovanni Pecoraro, Assunta Sellitto, Oriana Strianese, Ilaria Terenzi, Giuseppe Fenza, Aniello Gentile, Antonello Saccomanno, Sonia Amabile, Teresa Rocco, Annamaria Salvati, Emilia Vaccaro, Massimiliano Galdiero, Michele Cennamo, Giuseppe Portella, Maria Grazia Foti, Mariarosaria Ingino, Maria Landi, Maurizio Fumi, Vincenzo Rocco, Rita Greco, Vittoria Letizia, Arnolfo Petruzziello, Maddalena Schioppa, Gregorio Goffredi, Francesca Marciano, Michele Caraglia, Alessia Cossu, Marianna Scrima, Edmondo Adorisio, Morena D'Avenia, Michela Iacobellis, Rosanna Piluscio, Giorgio Dirani, Vittorio Sambri, Simona Semprini, Silvia Zanoli, Francesco Curcio, Stefania Marzinotto, Andreina Baj, Fau |
| EPI_ISL_1547392 | 1.AO Universitaria 'S. Giovanni di Dio e Ruggi D'Aragona, Scuola Medica Salernitana' Hospital / 2.UOC di Virologia e Microbiologia, Università della Campania 'L. Vanvitelli' / 3.AO Universitaria 'Federico II' Napoli Hospital / 4.AORN 'San Giuseppe Moscati' Avellino Hospital / 5.AO 'San Pio - presidio G. Rummo' Benevento Hospital / 6.AO 'Sant'Anna e San Sebastiano' Caserta Hospital / 7.PO 'Maria Santissima Addolorata' Eboli Hospital / 8.Biogem Istituto di Ricerche Genetiche / 9. U.O.C. di Genetica Medica e di Laboratorio A.O.R.N., Azienda Ospedaliera di Rilievo Nazionale Antonio Cardarelli, Napoli / 10. Centro di riferimento Oncologico della Basilicata (IRCCS-CROB), Rionero in Vulture (PZ) / 11. Presidio Ospedaliero di Agropoli, Agropoli (SA). | 1. Genome Research Center for Health (CRGS) / 2. Laboratory of Molecular Medicine and Genomics(LMMGe) / 3. Center for Research in Pure and Applied Mathematics (CRMPA) | Giorgio Giurato, Francesca Rizzo, Alessandro Weisz, Gianluigi Franci, Giovanni Nassa, Pasquale Pagliano, Roberta Tarallo, Elena Alexandrova, Ylenia D'Agostino, Carlo Ferravante, Jessica Lamberti, Viola Melone, Domenico Memoli, Valeria Mirici Cappa, Domenico Palumbo, Giovanni Pecoraro, Assunta Sellitto, Oriana Strianese, Ilaria Terenzi, Giuseppe Fenza, Aniello Gentile, Antonello Saccomanno, Sonia Amabile, Teresa Rocco, Annamaria Salvati, Emilia Vaccaro, Massimiliano Galdiero, Michele Cennamo, Giuseppe Portella, Maria Grazia Foti, Mariarosaria Ingino, Maria Landi, Maurizio Fumi, Vincenzo Rocco, Rita Greco, Vittoria Letizia, Arnolfo Petruzziello, Maddalena Schioppa, Gregorio Goffredi, Francesca Marciano, Michele Caraglia, Alessia Cossu, Marianna Scrima, Edmondo Adorisio, Morena D'Avenia, Michela Iacobellis, Rosanna Piluscio, Giorgio Dirani, Vittorio Sambri, Simona Semprini, Silvia Zanoli, Francesco Curcio, Stefania Marzinotto, Andreina Baj, Fausto Sessa, Massimiliano Chetta, Marina Tarsitano, Giusy Giordano, Marcello Ametrano, Mariano Fiorenza, Caivano Antonella, D'Auria Fiorella, De Luca Luciana. |
| EPI_ISL_1547806, EPI_ISL_1547807, EPI_ISL_1547808, EPI_ISL_1547809, EPI_ISL_1547810, EPI_ISL_1547811, EPI_ISL_1547812, EPI_ISL_1547813, EPI_ISL_1547814, EPI_ISL_1547815, EPI_ISL_1547816, EPI_ISL_1547817, EPI_ISL_1547818, EPI_ISL_1547819, EPI_ISL_1547820, EPI_ISL_1547821, EPI_ISL_1547822, EPI_ISL_1547823, EPI_ISL_1547824, EPI_ISL_1547825, EPI_ISL_1547826 | | | |
| see above | Laboratory of Clinical Microbiology, Virology and Bioemergencies, ASST Fatebenefratelli Sacco - Sacco Hospital | Laboratory of Clinical Microbiology, Virology and Bioemergencies, ASST Fatebenefratelli Sacco - Sacco Hospital | Valeria Micheli, Alessandro Mancon, Alberto Rizzo, Fiorenza Bracchitta, Luca Rizzuto, Maria Rita Gismondo |
| EPI_ISL_1557254, EPI_ISL_1557256, EPI_ISL_1557257, EPI_ISL_1557258, EPI_ISL_1557259, EPI_ISL_1557260, EPI_ISL_1557261, EPI_ISL_1557262, EPI_ISL_1557264, EPI_ISL_1557265, EPI_ISL_1557266, EPI_ISL_1557267, EPI_ISL_1557268, EPI_ISL_1557269, EPI_ISL_1557270, EPI_ISL_1557271, EPI_ISL_1557272, EPI_ISL_1557273, EPI_ISL_1557274, EPI_ISL_1557275, EPI_ISL_1557276, EPI_ISL_1557277, EPI_ISL_1557278, EPI_ISL_1557279, EPI_ISL_1557280, EPI_ISL_1557282, EPI_ISL_1557283, EPI_ISL_1557284, EPI_ISL_1557285, EPI_ISL_1557286, EPI_ISL_1557287, EPI_ISL_1557288, EPI_ISL_1557289, EPI_ISL_1557290, EPI_ISL_1557291, EPI_ISL_1557292, EPI_ISL_1557293, EPI_ISL_1557294, EPI_ISL_1557295, EPI_ISL_1557296, EPI_ISL_1557297, EPI_ISL_1557298, EPI_ISL_1557299, EPI_ISL_1557300, EPI_ISL_1557301, EPI_ISL_1557302, EPI_ISL_1557303, EPI_ISL_1557304, EPI_ISL_1557305, EPI_ISL_1557306, EPI_ISL_1557309, EPI_ISL_1557310, EPI_ISL_1557311, EPI_ISL_1557312, EPI_ISL_1557313, EPI_ISL_1557314, EPI_ISL_1557315, EPI_ISL_1557316, EPI_ISL_1557317, EPI_ISL_1557318, EPI_ISL_1557319, EPI_ISL_1557320, EPI_ISL_1557321, EPI_ISL_1557322, EPI_ISL_1557323, EPI_ISL_1557324, EPI_ISL_1557325, EPI_ISL_1557326, EPI_ISL_1557327, EPI_ISL_1557328, EPI_ISL_1557329, EPI_ISL_1557330, EPI_ISL_1557331, EPI_ISL_1557333, EPI_ISL_1557335, EPI_ISL_1557336, EPI_ISL_1557337, EPI_ISL_1557338, EPI_ISL_1557339, EPI_ISL_1557340, EPI_ISL_1557341, EPI_ISL_1557342, EPI_ISL_1557343, EPI_ISL_1557344, EPI_ISL_1557345, EPI_ISL_1557346, EPI_ISL_1557347, EPI_ISL_1557348, EPI_ISL_1557349, EPI_ISL_1557350, EPI_ISL_1557351, EPI_ISL_1557352, EPI_ISL_1557353, EPI_ISL_1557354, EPI_ISL_1557355, EPI_ISL_1557356, EPI_ISL_1557358, EPI_ISL_1557359, EPI_ISL_1557361, EPI_ISL_1557362, EPI_ISL_1557363, EPI_ISL_1557364, EPI_ISL_1557365, EPI_ISL_1557366, EPI_ISL_1557367, EPI_ISL_1557368, EPI_ISL_1557369, EPI_ISL_1557370, EPI_ISL_1557371, EPI_ISL_1557373, EPI_ISL_1557375, EPI_ISL_1557376, EPI_ISL_1557377, EPI_ISL_1557378, EPI_ISL_1557379, EPI_ISL_1557380, EPI_ISL_1557381, EPI_ISL_1557382, EPI_ISL_1557383, EPI_ISL_1557384, EPI_ISL_1557385, EPI_ISL_1557386, EPI_ISL_1557390, EPI_ISL_1557391, EPI_ISL_1557392, EPI_ISL_1557393, EPI_ISL_1557394, EPI_ISL_1557395, EPI_ISL_1557396, EPI_ISL_1557397, EPI_ISL_1557398, EPI_ISL_1557399, EPI_ISL_1557401, EPI_ISL_1557403, EPI_ISL_1557404, EPI_ISL_1557405, EPI_ISL_1557406, EPI_ISL_1557407, EPI_ISL_1557408, EPI_ISL_1557409, EPI_ISL_1557410, EPI_ISL_1557411, EPI_ISL_1557412, EPI_ISL_1557413, EPI_ISL_1557414, EPI_ISL_1557417, EPI_ISL_1557418, EPI_ISL_1557419, EPI_ISL_1557420, EPI_ISL_1557421, EPI_ISL_1557422, EPI_ISL_1557423, EPI_ISL_1557424, EPI_ISL_1557425, EPI_ISL_1557426, EPI_ISL_1557427, EPI_ISL_1557430, EPI_ISL_1557431, EPI_ISL_1557432, EPI_ISL_1557433, EPI_ISL_1557434, EPI_ISL_1557435, EPI_ISL_1557436, EPI_ISL_1557437, EPI_ISL_1557438, EPI_ISL_1557439, EPI_ISL_1557440, EPI_ISL_1557442, EPI_ISL_1557443, EPI_ISL_1557445, EPI_ISL_1557446, EPI_ISL_1557447, EPI_ISL_1557448, EPI_ISL_1557449, EPI_ISL_1557450, EPI_ISL_1557451, EPI_ISL_1557452, EPI_ISL_1557453, EPI_ISL_1557454, EPI_ISL_1557455, EPI_ISL_1557457, EPI_ISL_1557458, EPI_ISL_1557459, EPI_ISL_1557460, EPI_ISL_1557461, EPI_ISL_1557462, EPI_ISL_1557463, EPI_ISL_1557464, EPI_ISL_1557465, EPI_ISL_1557466, EPI_ISL_1557467, EPI_ISL_1557469, EPI_ISL_1557470, EPI_ISL_1557471, EPI_ISL_1557473, EPI_ISL_1557474, EPI_ISL_1557475, EPI_ISL_1557476, EPI_ISL_1557477, EPI_ISL_1557478, EPI_ISL_1557479, EPI_ISL_1557480, EPI_ISL_1557481, EPI_ISL_1557482, EPI_ISL_1557483, EPI_ISL_1557484, EPI_ISL_1557485, EPI_ISL_1557486, EPI_ISL_1557487, EPI_ISL_1557488, EPI_ISL_1557489, EPI_ISL_1557490, EPI_ISL_1557491, EPI_ISL_1557492, EPI_ISL_1557493, EPI_ISL_1557494, EPI_ISL_1557495, EPI_ISL_1557497, EPI_ISL_1557498, EPI_ISL_1557499, EPI_ISL_1557500, EPI_ISL_1557501, EPI_ISL_1557502, EPI_ISL_1557503, EPI_ISL_1557504, EPI_ISL_1557505, EPI_ISL_1557506, EPI_ISL_1557507, EPI_ISL_1557508, EPI_ISL_1557509, EPI_ISL_1557510, EPI_ISL_1557511, EPI_ISL_1557512, EPI_ISL_1557513, EPI_ISL_1557514, EPI_ISL_1557515, EPI_ISL_1557516, EPI_ISL_1557517, EPI_ISL_1557518, EPI_ISL_1557519, EPI_ISL_1557520, EPI_ISL_1557521, EPI_ISL_1557523, EPI_ISL_1557524, EPI_ISL_1557525, EPI_ISL_1557526, EPI_ISL_1557528, EPI_ISL_1557529, EPI_ISL_1557530, EPI_ISL_1557531, EPI_ISL_1557532, EPI_ISL_1557533, EPI_ISL_1557534, EPI_ISL_1557535, EPI_ISL_1557536, EPI_ISL_1557537, EPI_ISL_1557538, EPI_ISL_1557539, EPI_ISL_1557540, EPI_ISL_1557541, EPI_ISL_1557542, EPI_ISL_1557543, EPI_ISL_1557544, EPI_ISL_1557545, EPI_ISL_1557547, EPI_ISL_1557548, EPI_ISL_1557549, EPI_ISL_1557550, EPI_ISL_1557552, EPI_ISL_1557553, EPI_ISL_1557554, EPI_ISL_1557555, EPI_ISL_1557556, EPI_ISL_1557557, EPI_ISL_1557558, EPI_ISL_1557559, EPI_ISL_1557560, EPI_ISL_1557561, EPI_ISL_1557562, EPI_ISL_1557563, EPI_ISL_1557564, EPI_ISL_1557565, EPI_ISL_1557566, EPI_ISL_1557567, EPI_ISL_1557568, EPI_ISL_1557569, EPI_ISL_1557570, EPI_ISL_1557571, EPI_ISL_1557572, EPI_ISL_1557573, EPI_ISL_1557574, EPI_ISL_1557577, EPI_ISL_1557578, EPI_ISL_1557579, EPI_ISL_1557580, EPI_ISL_1557581, EPI_ISL_1557582, EPI_ISL_1557583, EPI_ISL_1557584, EPI_ISL_1557585, EPI_ISL_1557586, EPI_ISL_1557587, EPI_ISL_1557588, EPI_ISL_1557589, EPI_ISL_1557590, EPI_ISL_1557591, EPI_ISL_1557592, EPI_ISL_1557593, EPI_ISL_1557594, EPI_ISL_1557595, EPI_ISL_1557596, EPI_ISL_1557597, EPI_ISL_1557598, EPI_ISL_1557599, EPI_ISL_1557600, EPI_ISL_1557603, EPI_ISL_1557604, EPI_ISL_1557605, EPI_ISL_1557606, EPI_ISL_1557607, EPI_ISL_1557608, EPI_ISL_1557609, EPI_ISL_1557610, EPI_ISL_1557611, EPI_ISL_1557612, EPI_ISL_1557613, EPI_ISL_1557614, EPI_ISL_1557615, EPI_ISL_1557616, EPI_ISL_1557617, EPI_ISL_1557618, EPI_ISL_1557619, EPI_ISL_1557620, EPI_ISL_1557621, EPI_ISL_1557622, EPI_ISL_1557623, EPI_ISL_1557624, EPI_ISL_1557625, EPI_ISL_1557626, EPI_ISL_1557627, EPI_ISL_1557629, EPI_ISL_1557630, EPI_ISL_1557631, EPI_ISL_1557632, EPI_ISL_1557633, EPI_ISL_1557634, EPI_ISL_1557635, EPI_ISL_1557636, EPI_ISL_1557637, EPI_ISL_1557638, EPI_ISL_1557639, EPI_ISL_1557640, EPI_ISL_1557641, EPI_ISL_1557642, EPI_ISL_1557643, EPI_ISL_1557644, EPI_ISL_1557645, EPI_ISL_1557646, EPI_ISL_1557647, EPI_ISL_1557648, EPI_ISL_1557649, EPI_ISL_1557650, EPI_ISL_1557651, EPI_ISL_1557652, EPI_ISL_1557653, EPI_ISL_1557654, EPI_ISL_1557657, EPI_ISL_1557658, EPI_ISL_1557659, EPI_ISL_1557660, EPI_ISL_1557661, EPI_ISL_1557662, EPI_ISL_1557663, EPI_ISL_1557664, EPI_ISL_1557665, EPI_ISL_1557667, EPI_ISL_1557668, EPI_ISL_1557669, EPI_ISL_1557670, EPI_ISL_1557671, EPI_ISL_1557672, EPI_ISL_1557673, EPI_ISL_1557675, EPI_ISL_1557676, EPI_ISL_1557677, EPI_ISL_1557678, EPI_ISL_1557679, EPI_ISL_1557680, EPI_ISL_1557681, EPI_ISL_1557682, EPI_ISL_1557685, EPI_ISL_1557686, EPI_ISL_1557687, EPI_ISL_1557688, EPI_ISL_1557690, EPI_ISL_1557691, EPI_ISL_1557692, EPI_ISL_1557693, EPI_ISL_1557694, EPI_ISL_1557695, EPI_ISL_1557696, EPI_ISL_1557697, EPI_ISL_1557698, EPI_ISL_1557699, EPI_ISL_1557700, EPI_ISL_1557701, EPI_ISL_1557702, EPI_ISL_1557703, EPI_ISL_1557704, EPI_ISL_1557705, EPI_ISL_1557706, EPI_ISL_1557707, EPI_ISL_1557708, EPI_ISL_1557711, EPI_ISL_1557712, EPI_ISL_1557713, EPI_ISL_1557714, EPI_ISL_1557715, EPI_ISL_1557716, EPI_ISL_1557717, EPI_ISL_1557718, EPI_ISL_1557719, EPI_ISL_1557720, EPI_ISL_1557721, EPI_ISL_1557722, EPI_ISL_1557723, EPI_ISL_1557724, EPI_ISL_1557725, EPI_ISL_1557726, EPI_ISL_1557727, EPI_ISL_1557728, EPI_ISL_1557729, EPI_ISL_1557730, EPI_ISL_1557731, EPI_ISL_1557732, EPI_ISL_1557733, EPI_ISL_1557734, EPI_ISL_1557735, EPI_ISL_1557738, EPI_ISL_1557739, EPI_ISL_1557740, EPI_ISL_1557741, EPI_ISL_1557742, EPI_ISL_1557743, EPI_ISL_1557744, EPI_ISL_1557745, EPI_ISL_1557746, EPI_ISL_1557747, EPI_ISL_1557748, EPI_ISL_1557749, EPI_ISL_1557750, EPI_ISL_1557751, EPI_ISL_1557752, EPI_ISL_1557753, EPI_ISL_1557754, EPI_ISL_1557755, EPI_ISL_1557756, EPI_ISL_1557757, EPI_ISL_1557758, EPI_ISL_1557759, EPI_ISL_1557760, EPI_ISL_1557761, EPI_ISL_1557762, EPI_ISL_1557765, EPI_ISL_1557766, EPI_ISL_1557767, EPI_ISL_1557768, EPI_ISL_1557769, EPI_ISL_1557770, EPI_ISL_1557771, EPI_ISL_1557772, EPI_ISL_1557773, EPI_ISL_1557774, EPI_ISL_1557775, EPI_ISL_1557776, EPI_ISL_1557777, EPI_ISL_1557778, EPI_ISL_1557779, EPI_ISL_1557780, EPI_ISL_1557781, EPI_ISL_1557782, EPI_ISL_1557783, EPI_ISL_1557784, EPI_ISL_1557785, EPI_ISL_1557786, EPI_ISL_1557787, EPI_ISL_1557788, EPI_ISL_1557790, EPI_ISL_1557791, EPI_ISL_1557792, EPI_ISL_1557793, EPI_ISL_1557794, EPI_ISL_1557795, EPI_ISL_1557796, EPI_ISL_1557797, EPI_ISL_1557798, EPI_ISL_1557799, EPI_ISL_1557800, EPI_ISL_1557801, EPI_ISL_1557802, EPI_ISL_1557803, EPI_ISL_1557804, EPI_ISL_1557805, EPI_ISL_1557806, EPI_ISL_1557807, EPI_ISL_1557808, EPI_ISL_1557809, EPI_ISL_1557810, EPI_ISL_1557811, EPI_ISL_1557812, EPI_ISL_1557813, EPI_ISL_1557814, EPI_ISL_1557815, EPI_ISL_1557818, EPI_ISL_1557819, EPI_ISL_1557820, EPI_ISL_1557821, EPI_ISL_1557822, EPI_ISL_1557823, EPI_ISL_1557824, EPI_ISL_1557825, EPI_ISL_1557826, EPI_ISL_1557827, EPI_ISL_1557828, EPI_ISL_1557829, EPI_ISL_1557830, EPI_ISL_1557831, EPI_ISL_1557832, EPI_ISL_1557833, EPI_ISL_1557834, EPI_ISL_1557835, EPI_ISL_1557836, EPI_ISL_1557837, EPI_ISL_1557838, EPI_ISL_1557839, EPI_ISL_1557840, EPI_ISL_1557841, EPI_ISL_1557842, EPI_ISL_1557843, EPI_ISL_1557846, EPI_ISL_1557847, EPI_ISL_1557848, EPI_ISL_1557849, EPI_ISL_1557850, EPI_ISL_1557851, EPI_ISL_1557852, EPI_ISL_1557853, EPI_ISL_1557854, EPI_ISL_1557855, EPI_ISL_1557856, EPI_ISL_1557859, EPI_ISL_1557860, EPI_ISL_1557861, EPI_ISL_1557862, EPI_ISL_1557863, EPI_ISL_1557864, EPI_ISL_1557865, EPI_ISL_1557866, EPI_ISL_1557867, EPI_ISL_1557868, EPI_ISL_1557869, EPI_ISL_1557870, EPI_ISL_1557873, EPI_ISL_1557874, EPI_ISL_1557875, EPI_ISL_1557876, EPI_ISL_1557877, EPI_ISL_1557878, EPI_ISL_1557879, EPI_ISL_1557880, EPI_ISL_1557881, EPI_ISL_1557882, EPI_ISL_1557883, EPI_ISL_1557884, EPI_ISL_1557885, EPI_ISL_1557886, EPI_ISL_1557887, EPI_ISL_1557888, EPI_ISL_1557889, EPI_ISL_1557890, EPI_ISL_1557891, EPI_ISL_1557892, EPI_ISL_1557893, EPI_ISL_1557894, EPI_ISL_1557895, EPI_ISL_1557896, EPI_ISL_1557898, EPI_ISL_1557899, EPI_ISL_1557900, EPI_ISL_1557902, EPI_ISL_1557903, EPI_ISL_1557904, EPI_ISL_1557905, EPI_ISL_1557906, EPI_ISL_1557907, EPI_ISL_1557908, EPI_ISL_1557909, EPI_ISL_1557910, EPI_ISL_1557911, EPI_ISL_1557912, EPI_ISL_1557913, EPI_ISL_1557914, EPI_ISL_1557915, EPI_ISL_1557916, EPI_ISL_1557917, EPI_ISL_1557918, EPI_ISL_1557919, EPI_ISL_1557920, EPI_ISL_1557921, EPI_ISL_1557922, EPI_ISL_1557923, EPI_ISL_1557924, EPI_ISL_1557926, EPI_ISL_1557927, EPI_ISL_1557928, EPI_ISL_1557929, EPI_ISL_1557931, EPI_ISL_1557932, EPI_ISL_1557933, EPI_ISL_1557934, EPI_ISL_1557935, EPI_ISL_1557936, EPI_ISL_1557937, EPI_ISL_1557938, EPI_ISL_1557941, EPI_ISL_1557942, EPI_ISL_1557943, EPI_ISL_1557944, EPI_ISL_1557945, EPI_ISL_1557946, EPI_ISL_1557947, EPI_ISL_1557948, EPI_ISL_1557950, EPI_ISL_1557951, EPI_ISL_1557952, EPI_ISL_1557953, EPI_ISL_1557954, EPI_ISL_1557956, EPI_ISL_1557957, EPI_ISL_1557958, EPI_ISL_1557959, EPI_ISL_1557960, EPI_ISL_1557961, EPI_ISL_1557962, EPI_ISL_1557963, EPI_ISL_1557964, EPI_ISL_1557965, EPI_ISL_1557966, EPI_ISL_1557967, EPI_ISL_1557968, EPI_ISL_1557969, EPI_ISL_1557970, EPI_ISL_1557971, EPI_ISL_1557972, EPI_ISL_1557973, EPI_ISL_1557974, EPI_ISL_1557975, EPI_ISL_1557976, EPI_ISL_1557977, EPI_ISL_1557978, EPI_ISL_1557979, EPI_ISL_1557980, EPI_ISL_1557981, EPI_ISL_1557983, EPI_ISL_1557984, EPI_ISL_1557985, EPI_ISL_1557986, EPI_ISL_1557987, EPI_ISL_1557988, EPI_ISL_1557989, EPI_ISL_1557990, EPI_ISL_1557991, EPI_ISL_1557992, EPI_ISL_1557993, EPI_ISL_1557994, EPI_ISL_1557995, EPI_ISL_1557996, EPI_ISL_1557997, EPI_ISL_1557998, EPI_ISL_1557999, EPI_ISL_1558000, EPI_ISL_1558001, EPI_ISL_1558003, EPI_ISL_1558004, EPI_ISL_1558005, EPI_ISL_1558006, EPI_ISL_1558007, EPI_ISL_1558008, EPI_ISL_1558009, EPI_ISL_1558011, EPI_ISL_1558012, EPI_ISL_1558013, EPI_ISL_1558014, EPI_ISL_1558015, EPI_ISL_1558016, EPI_ISL_1558017, EPI_ISL_1558018, EPI_ISL_1558019, EPI_ISL_1558020, EPI_ISL_1558021, EPI_ISL_1558022, EPI_ISL_1558023, EPI_ISL_1558024, EPI_ISL_1558025, EPI_ISL_1558026, EPI_ISL_1558027, EPI_ISL_1558028, EPI_ISL_1558029, EPI_ISL_1558031, EPI_ISL_1558032, EPI_ISL_1558033, EPI_ISL_1558035, EPI_ISL_1558036, EPI_ISL_1558037, EPI_ISL_1558038, EPI_ISL_1558039, EPI_ISL_1558040, EPI_ISL_1558041, EPI_ISL_1558042, EPI_ISL_1558043, EPI_ISL_1558044, EPI_ISL_1558045, EPI_ISL_1558046, EPI_ISL_1558047, EPI_ISL_1558048, EPI_ISL_1558049, EPI_ISL_1558050, EPI_ISL_1558051, EPI_ISL_1558052, EPI_ISL_1558053, EPI_ISL_1558054, EPI_ISL_1558055, EPI_ISL_1558056, EPI_ISL_1558057, EPI_ISL_1558058, EPI_ISL_1558061, EPI_ISL_1558062, EPI_ISL_1558063, EPI_ISL_1558064, EPI_ISL_1558065, EPI_ISL_1558066, EPI_ISL_1558067, EPI_ISL_1558068, EPI_ISL_1558069, EPI_ISL_1558070, EPI_ISL_1558071, EPI_ISL_1558072, EPI_ISL_1558073, EPI_ISL_1558075, EPI_ISL_1558076, EPI_ISL_1558077, EPI_ISL_1558078, EPI_ISL_1558079, EPI_ISL_1558080, EPI_ISL_1558081, EPI_ISL_1558082, EPI_ISL_1558083, EPI_ISL_1558085, EPI_ISL_1558086, EPI_ISL_1558087, EPI_ISL_1558088, EPI_ISL_1558090, EPI_ISL_1558091, EPI_ISL_1558092, EPI_ISL_1558093, EPI_ISL_1558094, EPI_ISL_1558095, EPI_ISL_1558096, EPI_ISL_1558097, EPI_ISL_1558098, EPI_ISL_1558099, EPI_ISL_1558100, EPI_ISL_1558101, EPI_ISL_1558102, EPI_ISL_1558103, EPI_ISL_1558104, EPI_ISL_1558105, EPI_ISL_1558106, EPI_ISL_1558107, EPI_ISL_1558108, EPI_ISL_1558110, EPI_ISL_1558111, EPI_ISL_1558112, EPI_ISL_1558113, EPI_ISL_1558115, EPI_ISL_1558116, EPI_ISL_1558117, EPI_ISL_1558118, EPI_ISL_1558119, EPI_ISL_1558120, EPI_ISL_1558121, EPI_ISL_1558122, EPI_ISL_1558123, EPI_ISL_1558124, EPI_ISL_1558125, EPI_ISL_1558126, EPI_ISL_1558127, EPI_ISL_1558128, EPI_ISL_1558129, EPI_ISL_1558130, EPI_ISL_1558131, EPI_ISL_1558132, EPI_ISL_1558133, EPI_ISL_1558134, EPI_ISL_1558136, EPI_ISL_1558137, EPI_ISL_1558138, EPI_ISL_1558139, EPI_ISL_1558140, EPI_ISL_1558141, EPI_ISL_1558142, EPI_ISL_1558143, EPI_ISL_1558144, EPI_ISL_1558146, EPI_ISL_1558147, EPI_ISL_1558148, | | | |

| | | | |
|---|---|---|---|
| EPI_ISL_1558149, EPI_ISL_1558150, EPI_ISL_1558151, EPI_ISL_1558152, EPI_ISL_1558153, EPI_ISL_1558154, EPI_ISL_1558155, EPI_ISL_1558156, EPI_ISL_1558157, EPI_ISL_1558158, EPI_ISL_1558160, EPI_ISL_1558161, EPI_ISL_1558162, EPI_ISL_1558163, EPI_ISL_1558164, EPI_ISL_1558165, EPI_ISL_1558166, EPI_ISL_1558167, EPI_ISL_1558168, EPI_ISL_1558169, EPI_ISL_1558170, EPI_ISL_1558171, EPI_ISL_1558173, EPI_ISL_1558174, EPI_ISL_1558175, EPI_ISL_1558176, EPI_ISL_1558177, EPI_ISL_1558178, EPI_ISL_1558179, EPI_ISL_1558180, EPI_ISL_1558181, EPI_ISL_1558182, EPI_ISL_1558183, EPI_ISL_1558184, EPI_ISL_1558185, EPI_ISL_1558186, EPI_ISL_1558187, EPI_ISL_1558189, EPI_ISL_1558191, EPI_ISL_1558192, EPI_ISL_1558193, EPI_ISL_1558194, EPI_ISL_1558195, EPI_ISL_1558196, EPI_ISL_1558198, EPI_ISL_1558199, EPI_ISL_1558200, EPI_ISL_1558201, EPI_ISL_1558202, EPI_ISL_1558203, EPI_ISL_1558204, EPI_ISL_1558205, EPI_ISL_1558206, EPI_ISL_1558207, EPI_ISL_1558208, EPI_ISL_1558209, EPI_ISL_1558210, EPI_ISL_1558211, EPI_ISL_1558212, EPI_ISL_1558213, EPI_ISL_1558214, EPI_ISL_1558216, EPI_ISL_1558217, EPI_ISL_1558218, EPI_ISL_1558219, EPI_ISL_1558220, EPI_ISL_1558221, EPI_ISL_1558222, EPI_ISL_1558223, EPI_ISL_1558225, EPI_ISL_1558226, EPI_ISL_1558227, EPI_ISL_1558228, EPI_ISL_1558229, EPI_ISL_1558230, EPI_ISL_1558231, EPI_ISL_1558232, EPI_ISL_1558233, EPI_ISL_1558234, EPI_ISL_1558235, EPI_ISL_1558236, EPI_ISL_1558237, EPI_ISL_1558238, EPI_ISL_1558239, EPI_ISL_1558240, EPI_ISL_1558241, EPI_ISL_1558242, EPI_ISL_1558243, EPI_ISL_1558244, EPI_ISL_1558246, EPI_ISL_1558247, EPI_ISL_1558248, EPI_ISL_1558249, EPI_ISL_1558250, EPI_ISL_1558251, EPI_ISL_1558252, EPI_ISL_1558254, EPI_ISL_1558255, EPI_ISL_1558256, EPI_ISL_1558257, EPI_ISL_1558258, EPI_ISL_1558259, EPI_ISL_1558260, EPI_ISL_1558261, EPI_ISL_1558262, EPI_ISL_1558263, EPI_ISL_1558264, EPI_ISL_1558265, EPI_ISL_1558266, EPI_ISL_1558267, EPI_ISL_1558268, EPI_ISL_1558269, EPI_ISL_1558270, EPI_ISL_1558272, EPI_ISL_1558273, EPI_ISL_1558274, EPI_ISL_1558275, EPI_ISL_1558276, EPI_ISL_1558277, EPI_ISL_1558278, EPI_ISL_1558279, EPI_ISL_1558280, EPI_ISL_1558281, EPI_ISL_1558282, EPI_ISL_1558283, EPI_ISL_1558284, EPI_ISL_1558285, EPI_ISL_1558286, EPI_ISL_1558287, EPI_ISL_1558288, EPI_ISL_1558289, EPI_ISL_1558290, EPI_ISL_1558291, EPI_ISL_1558292, EPI_ISL_1558293, EPI_ISL_1558294, EPI_ISL_1558295, EPI_ISL_1558296, EPI_ISL_1558297, EPI_ISL_1558298, EPI_ISL_1558299, EPI_ISL_1558301, EPI_ISL_1558302, EPI_ISL_1558303, EPI_ISL_1558304, EPI_ISL_1558305, EPI_ISL_1558306, EPI_ISL_1558307, EPI_ISL_1558308, EPI_ISL_1558309, EPI_ISL_1558310, EPI_ISL_1558311, EPI_ISL_1558312, EPI_ISL_1558313, EPI_ISL_1558314, EPI_ISL_1558315, EPI_ISL_1558316, EPI_ISL_1558317, EPI_ISL_1558318, EPI_ISL_1558319, EPI_ISL_1558320, EPI_ISL_1558321, EPI_ISL_1558322, EPI_ISL_1558324, EPI_ISL_1558325, EPI_ISL_1558327, EPI_ISL_1558328, EPI_ISL_1558329, EPI_ISL_1558330, EPI_ISL_1558331, EPI_ISL_1558332, EPI_ISL_1558333, EPI_ISL_1558334, EPI_ISL_1558335, EPI_ISL_1558336, EPI_ISL_1558337, EPI_ISL_1558338, EPI_ISL_1558339, EPI_ISL_1558340, EPI_ISL_1558341, EPI_ISL_1558342, EPI_ISL_1558343, EPI_ISL_1558344, EPI_ISL_1558345, EPI_ISL_1558346, EPI_ISL_1558347, EPI_ISL_1558348, EPI_ISL_1558349, EPI_ISL_1558350, EPI_ISL_1558351, EPI_ISL_1558352, EPI_ISL_1558353, EPI_ISL_1558354, EPI_ISL_1558355, EPI_ISL_1558356, EPI_ISL_1558357, EPI_ISL_1558358, EPI_ISL_1558359, EPI_ISL_1558360, EPI_ISL_1558361, EPI_ISL_1558362, EPI_ISL_1558363, EPI_ISL_1558364, EPI_ISL_1558365, EPI_ISL_1558366, EPI_ISL_1558367, EPI_ISL_1558368, EPI_ISL_1558369, EPI_ISL_1558370, EPI_ISL_1558371, EPI_ISL_1558372, EPI_ISL_1558373, EPI_ISL_1558374, EPI_ISL_1558375, EPI_ISL_1558376, EPI_ISL_1558377, EPI_ISL_1558378, EPI_ISL_1558379, EPI_ISL_1558380, EPI_ISL_1558381, EPI_ISL_1558382, EPI_ISL_1558383, EPI_ISL_1558384, EPI_ISL_1558385, EPI_ISL_1558386, EPI_ISL_1558387, EPI_ISL_1558388, EPI_ISL_1558389, EPI_ISL_1558390, EPI_ISL_1558391, EPI_ISL_1558392, EPI_ISL_1558393, EPI_ISL_1558394, EPI_ISL_1558395, EPI_ISL_1558396, EPI_ISL_1558397, EPI_ISL_1558398, EPI_ISL_1558399, EPI_ISL_1558400, EPI_ISL_1558401, EPI_ISL_1558402, EPI_ISL_1558404, EPI_ISL_1558405, EPI_ISL_1558406, EPI_ISL_1558407, EPI_ISL_1558409, EPI_ISL_1558410, EPI_ISL_1558411, EPI_ISL_1558412, EPI_ISL_1558413, EPI_ISL_1558414, EPI_ISL_1558415, EPI_ISL_1558416, EPI_ISL_1558417, EPI_ISL_1558418, EPI_ISL_1558419, EPI_ISL_1558420, EPI_ISL_1558421, EPI_ISL_1558422, EPI_ISL_1558423, EPI_ISL_1558424, EPI_ISL_1558425, EPI_ISL_1558426, EPI_ISL_1558427, EPI_ISL_1558428, EPI_ISL_1558429, EPI_ISL_1558430, EPI_ISL_1558431, EPI_ISL_1558432, EPI_ISL_1558433, EPI_ISL_1558434, EPI_ISL_1558435, EPI_ISL_1558436, EPI_ISL_1558437, EPI_ISL_1558438, EPI_ISL_1558439, EPI_ISL_1558440, EPI_ISL_1558441, EPI_ISL_1558442, EPI_ISL_1558443, EPI_ISL_1558444, EPI_ISL_1558445, EPI_ISL_1558446, EPI_ISL_1558447, EPI_ISL_1558448, EPI_ISL_1558449, EPI_ISL_1558450, EPI_ISL_1558451, EPI_ISL_1558452, EPI_ISL_1558453, EPI_ISL_1558454, EPI_ISL_1558455, EPI_ISL_1558456, EPI_ISL_1558457, EPI_ISL_1558459, EPI_ISL_1558460, EPI_ISL_1558461, EPI_ISL_1558462, EPI_ISL_1558463, EPI_ISL_1558464, EPI_ISL_1558465, EPI_ISL_1558466, EPI_ISL_1558467, EPI_ISL_1558468, EPI_ISL_1558469, EPI_ISL_1558470, EPI_ISL_1558471, EPI_ISL_1558472, EPI_ISL_1558473, EPI_ISL_1558474, EPI_ISL_1558475, EPI_ISL_1558476, EPI_ISL_1558477, EPI_ISL_1558478, EPI_ISL_1558479, EPI_ISL_1558480, EPI_ISL_1558481, EPI_ISL_1558482, EPI_ISL_1558483, EPI_ISL_1558484, EPI_ISL_1558485, EPI_ISL_1558486, EPI_ISL_1558488, EPI_ISL_1558489, EPI_ISL_1558490, EPI_ISL_1558491, EPI_ISL_1558492, EPI_ISL_1558493, EPI_ISL_1558494, EPI_ISL_1558495, EPI_ISL_1558496, EPI_ISL_1558497, EPI_ISL_1558498, EPI_ISL_1558499, EPI_ISL_1558500, EPI_ISL_1558501, EPI_ISL_1558502, EPI_ISL_1558503, EPI_ISL_1558504, EPI_ISL_1558505, EPI_ISL_1558506, EPI_ISL_1558507, EPI_ISL_1558508, EPI_ISL_1558509, EPI_ISL_1558510, EPI_ISL_1558511, EPI_ISL_1558512, EPI_ISL_1558513, EPI_ISL_1558515, EPI_ISL_1558516, EPI_ISL_1558517, EPI_ISL_1558518, EPI_ISL_1558520 | | | |
| see above | ASL Napoli 1 Centro | AMES Centro Polidiagnostico Strumentale S.r.l. | "Giovanni Savarese, Raffaella Ruggiero, Eloisa Evangelista, Antonella Di Carlo, Luisa Circelli, Luigi D'Amore, Nadia Petrillo, Monica Ianniello, Roberto Sirica,Maurizio D'Amora, Antonio Fico" |
| EPI_ISL_1558595, EPI_ISL_1558596, EPI_ISL_1558597, EPI_ISL_1558598, EPI_ISL_1558599, EPI_ISL_1558600, EPI_ISL_1558601, EPI_ISL_1558602, EPI_ISL_1558603, EPI_ISL_1558604, EPI_ISL_1558605, EPI_ISL_1558606, EPI_ISL_1558607, EPI_ISL_1558608, EPI_ISL_1558609, EPI_ISL_1558610, EPI_ISL_1558611, EPI_ISL_1558612, EPI_ISL_1558613, EPI_ISL_1558614 | | | |
| see above | Department of Virology, Istituto Zooprofilattico Sperimentale del Lazio e della Toscana (IZSLT) | Department of General Diagnostics; Department of Virology; Istituto Zooprofilattico Sperimentale del Lazio e della Toscana (IZSLT) | Patricia Alba, Giuseppe Manna, Elena L. Diaconu, Fabiola Feltrin, Raffaela Conti, Teresa Scicluna, Virginia Carfora, Antonella Cersini, Alessia Franco, Antonio Battisti. |
| EPI_ISL_1577221, EPI_ISL_1577257, EPI_ISL_1577258 | 1.AO Universitaria 'S. Giovanni di Dio e Ruggi D'Aragona, Scuola Medica Salernitana' Hospital / 2.UOC di Virologia e Microbiologia, Università della Campania 'L. Vanvitelli' / 3.AO Universitaria 'Federico II' Napoli Hospital / 4.AORN 'San Giuseppe Moscati' Avellino Hospital / 5.AO 'San Pio - presidio G. Rummo' Benevento Hospital / 6.AO 'Sant'Anna e San Sebastiano' Caserta Hospital / 7.PO 'Maria Santissima Addolorata' Eboli Hospital / 8.Biogem Istituto di Ricerche Genetiche / 9. U.O.C. di Genetica Medica e di Laboratorio A.O.R.N., Azienda Ospedaliera di Rilievo Nazionale Antonio Cardarelli, Napoli / 10. Centro di riferimento Oncologico della Basilicata (IRCCS-CROB), Rionero in Vulture (PZ) / 11. Presidio Ospedaliero di Agropoli, Agropoli (SA). | 1. Genome Research Center for Health (CRGS) / 2. Laboratory of Molecular Medicine and Genomics(LMMGe) / 3. Center for Research in Pure and Applied Mathematics (CRMPA) | Giorgio Giurato (Corresponding Author), Francesca Rizzo (Corresponding Author), Alessandro Weisz (Corresponding Author), Gianluigi Franci, Giovanni Nassa, Pasquale Pagliano, Roberta Tarallo, Elena Alexandrova, Ylenia D'Agostino, Carlo Ferravante, Jessica Lamberti, Viola Melone, Domenico Memoli, Valeria Mirici Cappa, Domenico Palumbo, Giovanni Pecoraro, Assunta Sellitto, Oriana Strianese, Ilaria Terenzi, Giuseppe Fenza, Aniello Gentile, Antonello Saccomanno, Sonia Amabile, Teresa Rocco, Annamaria Salvati, Emilia Vaccaro, Massimiliano Galdiero, Michele Cennamo, Giuseppe Portella, Maria Grazia Foti, Mariarosaria Ingino, Maria Landi, Maurizio Fumi, Vincenzo Rocco, Rita Greco, Vittoria Letizia, Arnolfo Petruzziello, Maddalena Schioppa, Gregorio Goffredi, Francesca Marciano, Michele Caraglia, Alessia Cossu, Marianna Scrima, Edmondo Adorisio, Morena D'Avenia, Michela Iacobellis, Rosanna Piluscio, Giorgio Dirani, Vittorio Sambri, Simona Semprini, Silvia Zanoli, Francesco Curcio, Stefania Marzinotto, Andreina Baj, Fausto Sessa. |
| EPI_ISL_1577259 | 1.AO Universitaria 'S. Giovanni di Dio e Ruggi D'Aragona, Scuola Medica Salernitana' Hospital / 2.UOC di Virologia e Microbiologia, Università della Campania 'L. Vanvitelli' / 3.AO Universitaria 'Federico II' Napoli Hospital / 4.AORN 'San Giuseppe Moscati' Avellino Hospital / 5.AO 'San Pio - presidio G. Rummo' Benevento Hospital / 6.AO 'Sant'Anna e San Sebastiano' Caserta Hospital / 7.PO 'Maria Santissima Addolorata' Eboli Hospital / 8.Biogem Istituto di Ricerche Genetiche / 9. U.O.C. di Genetica Medica e di Laboratorio A.O.R.N., Azienda Ospedaliera di Rilievo Nazionale Antonio Cardarelli, Napoli / 10. Centro di riferimento Oncologico della Basilicata (IRCCS-CROB), Rionero in Vulture (PZ) / 11. Presidio Ospedaliero di Agropoli, Agropoli (SA). | 1. Genome Research Center for Health (CRGS) / 2. Laboratory of Molecular Medicine and Genomics(LMMGe) / 3. Center for Research in Pure and Applied Mathematics (CRMPA) | Giorgio Giurato (Corresponding Author), Francesca Rizzo (Corresponding Author), Alessandro Weisz (Corresponding Author), Gianluigi Franci, Giovanni Nassa, Pasquale Pagliano, Roberta Tarallo, Elena Alexandrova, Ylenia D'Agostino, Carlo Ferravante, Jessica Lamberti, Viola Melone, Domenico Memoli, Valeria Mirici Cappa, Domenico Palumbo, Giovanni Pecoraro, Assunta Sellitto, Oriana Strianese, Ilaria Terenzi, Giuseppe Fenza, Aniello Gentile, Antonello Saccomanno, Sonia Amabile, Teresa Rocco, Annamaria Salvati, Emilia Vaccaro, Massimiliano Galdiero, Michele Cennamo, Giuseppe Portella, Maria Grazia Foti, Mariarosaria Ingino, Maria Landi, Maurizio Fumi, Vincenzo Rocco, Rita Greco, Vittoria Letizia, Arnolfo Petruzziello, Maddalena Schioppa, Gregorio Goffredi, Francesca Marciano, Michele Caraglia, Alessia Cossu, Marianna Scrima, Edmondo Adorisio, Morena D'Avenia, Michela Iacobellis, Rosanna Piluscio, Giorgio Dirani, Vittorio Sambri, Simona Semprini, Silvia Zanoli, Francesco Curcio, Stefania Marzinotto, Andreina Baj, Fausto Sessa, Massimiliano Chetta, Marina Tarsitano, Giusy Giordano, Marcello Ametrano, Mariano Fiorenza, Caivano Antonella, D'Auria Fiorella, De Luca Luciana. |
| EPI_ISL_1577260, EPI_ISL_1577261, EPI_ISL_1577262, EPI_ISL_1577263 | 1.AO Universitaria 'S. Giovanni di Dio e Ruggi D'Aragona, Scuola Medica Salernitana' Hospital / 2.UOC di Virologia e Microbiologia, Università della Campania 'L. Vanvitelli' / 3.AO Universitaria 'Federico II' Napoli Hospital / 4.AORN 'San Giuseppe Moscati' Avellino Hospital / 5.AO 'San Pio - presidio G. Rummo' Benevento Hospital / 6.AO 'Sant'Anna e San Sebastiano' Caserta Hospital / 7.PO 'Maria Santissima Addolorata' Eboli Hospital / 8.Biogem Istituto di Ricerche Genetiche / 9. U.O.C. di Genetica Medica e di Laboratorio A.O.R.N., Azienda Ospedaliera di Rilievo Nazionale Antonio Cardarelli, Napoli / 10. Centro di riferimento Oncologico della Basilicata (IRCCS-CROB), Rionero in Vulture (PZ) / 11. Presidio Ospedaliero di Agropoli, Agropoli (SA). | 1. Genome Research Center for Health (CRGS) / 2. Laboratory of Molecular Medicine and Genomics(LMMGe) / 3. Center for Research in Pure and Applied Mathematics (CRMPA) | Giorgio Giurato (Corresponding Author), Francesca Rizzo (Corresponding Author), Alessandro Weisz (Corresponding Author), Gianluigi Franci, Giovanni Nassa, Pasquale Pagliano, Roberta Tarallo, Elena Alexandrova, Ylenia D'Agostino, Carlo Ferravante, Jessica Lamberti, Viola Melone, Domenico Memoli, Valeria Mirici Cappa, Domenico Palumbo, Giovanni Pecoraro, Assunta Sellitto, Oriana Strianese, Ilaria Terenzi, Giuseppe Fenza, Aniello Gentile, Antonello Saccomanno, Sonia Amabile, Teresa Rocco, Annamaria Salvati, Emilia Vaccaro, Massimiliano Galdiero, Michele Cennamo, Giuseppe Portella, Maria Grazia Foti, Mariarosaria Ingino, Maria Landi, Maurizio Fumi, Vincenzo Rocco, Rita Greco, Vittoria Letizia, Arnolfo Petruzziello, Maddalena Schioppa, Gregorio Goffredi, Francesca Marciano, Michele Caraglia, Alessia Cossu, Marianna Scrima, Edmondo Adorisio, Morena D'Avenia, Michela Iacobellis, Rosanna Piluscio, Giorgio Dirani, Vittorio Sambri, Simona Semprini, Silvia Zanoli, Francesco Curcio, Stefania Marzinotto, Andreina Baj, Fausto Sessa. |
| EPI_ISL_1577264 | 1.AO Universitaria 'S. Giovanni di Dio e Ruggi D'Aragona, Scuola Medica Salernitana' Hospital / 2.UOC di Virologia e Microbiologia, Università della Campania 'L. Vanvitelli' / 3.AO Universitaria 'Federico II' Napoli Hospital / 4.AORN 'San Giuseppe Moscati' Avellino Hospital / 5.AO 'San Pio - presidio G. Rummo' Benevento Hospital / 6.AO 'Sant'Anna e San Sebastiano' Caserta Hospital / 7.PO 'Maria Santissima Addolorata' Eboli Hospital / 8.Biogem Istituto di Ricerche Genetiche / 9. U.O.C. di Genetica Medica e di Laboratorio A.O.R.N., Azienda Ospedaliera di Rilievo Nazionale Antonio Cardarelli, Napoli / 10. Centro di riferimento Oncologico della Basilicata (IRCCS-CROB), Rionero in Vulture (PZ) / 11. Presidio Ospedaliero di Agropoli, Agropoli (SA). | 1. Genome Research Center for Health (CRGS) / 2. Laboratory of Molecular Medicine and Genomics(LMMGe) / 3. Center for Research in Pure and Applied Mathematics (CRMPA) | Giorgio Giurato (Corresponding Author), Francesca Rizzo (Corresponding Author), Alessandro Weisz (Corresponding Author), Gianluigi Franci, Giovanni Nassa, Pasquale Pagliano, Roberta Tarallo, Elena Alexandrova, Ylenia D'Agostino, Carlo Ferravante, Jessica Lamberti, Viola Melone, Domenico Memoli, Valeria Mirici Cappa, Domenico Palumbo, Giovanni Pecoraro, Assunta Sellitto, Oriana Strianese, Ilaria Terenzi, Giuseppe Fenza, Aniello Gentile, Antonello Saccomanno, Sonia Amabile, Teresa Rocco, Annamaria Salvati, Emilia Vaccaro, Massimiliano Galdiero, Michele Cennamo, Giuseppe Portella, Maria Grazia Foti, Mariarosaria Ingino, Maria Landi, Maurizio Fumi, Vincenzo Rocco, Rita Greco, Vittoria Letizia, Arnolfo Petruzziello, Maddalena Schioppa, Gregorio Goffredi, Francesca Marciano, Michele Caraglia, Alessia Cossu, Marianna Scrima, Edmondo Adorisio, Morena D'Avenia, Michela Iacobellis, Rosanna Piluscio, Giorgio Dirani, Vittorio Sambri, Simona Semprini, Silvia Zanoli, Francesco Curcio, Stefania Marzinotto, Andreina Baj, Fausto Sessa, Massimiliano Chetta, Marina Tarsitano, Giusy Giordano, Marcello Ametrano, Mariano Fiorenza, Caivano Antonella, D'Auria Fiorella, De Luca Luciana. |
| EPI_ISL_1577265, EPI_ISL_1577266, EPI_ISL_1577267, EPI_ISL_1577268 | 1.AO Universitaria 'S. Giovanni di Dio e Ruggi D'Aragona, Scuola Medica Salernitana' Hospital / 2.UOC di Virologia e Microbiologia, Università della Campania 'L. Vanvitelli' / 3.AO | 1. Genome Research Center for Health (CRGS) / 2. Laboratory of Molecular Medicine and Genomics(LMMGe) / 3. Center for Research in Pure and Applied Mathematics | Giorgio Giurato (Corresponding Author), Francesca Rizzo (Corresponding Author), Alessandro Weisz (Corresponding Author), Gianluigi Franci, Giovanni Nassa, Pasquale Pagliano, Roberta Tarallo, Elena Alexandrova, Ylenia D'Agostino, Carlo Ferravante, Jessica Lamberti, Viola Melone, Domenico Memoli, Valeria Mirici Cappa, Domenico Palumbo, Giovanni Pecoraro, Assunta Sellitto, Oriana Strianese, Ilaria Terenzi, Giuseppe Fenza, Aniello Gentile, Antonello |

| | | | |
|---|---|---|---|
| | Universitaria 'Federico II' Napoli Hospital / 4.AORN 'San Giuseppe Moscati' Avellino Hospital / 5.AO 'San Pio - presidio G. Rummo' Benevento Hospital / 6.AO 'Sant'Anna e San Sebastiano' Caserta Hospital / 7.PO 'Maria Santissima Addolorata' Eboli Hospital / 8.Biogem Istituto di Ricerche Genetiche / 9. U.O.C. di Genetica Medica e di Laboratorio A.O.R.N., Azienda Ospedaliera di Rilievo Nazionale Antonio Cardarelli, Napoli / 10. Centro di riferimento Oncologico della Basilicata (IRCCS-CROB), Rionero in Vulture (PZ) / 11. Presidio Ospedaliero di Agropoli, Agropoli (SA). | (CRMPA) | Saccomanno, Sonia Amabile, Teresa Rocco, Annamaria Salvati, Emilia Vaccaro, Massimiliano Galdiero, Michele Cennamo, Giuseppe Portella, Maria Grazia Foti, Mariarosaria Ingino, Maria Landi, Maurizio Fumi, Vincenzo Rocco, Rita Greco, Vittoria Letizia, Arnolfo Petruzziello, Maddalena Schioppa, Gregorio Goffredi, Francesca Marciano, Michele Caraglia, Alessia Cossu, Marianna Scrima, Edmondo Adorisio, Morena D'Avenia, Michela Iacobellis, Rosanna Piluscio, Giorgio Dirani, Vittorio Sambri, Simona Semprini, Silvia Zanoli, Francesco Curcio, Stefania Marzinotto, Andreina Baj, Fausto Sessa. |
| EPI_ISL_1577269 | 1.AO Universitaria 'S. Giovanni di Dio e Ruggi D'Aragona, Scuola Medica Salernitana' Hospital / 2.UOC di Virologia e Microbiologia, Università della Campania 'L. Vanvitelli' / 3.AO Universitaria 'Federico II' Napoli Hospital / 4.AORN 'San Giuseppe Moscati' Avellino Hospital / 5.AO 'San Pio - presidio G. Rummo' Benevento Hospital / 6.AO 'Sant'Anna e San Sebastiano' Caserta Hospital / 7.PO 'Maria Santissima Addolorata' Eboli Hospital / 8.Biogem Istituto di Ricerche Genetiche / 9. U.O.C. di Genetica Medica e di Laboratorio A.O.R.N., Azienda Ospedaliera di Rilievo Nazionale Antonio Cardarelli, Napoli / 10. Centro di riferimento Oncologico della Basilicata (IRCCS-CROB), Rionero in Vulture (PZ) / 11. Presidio Ospedaliero di Agropoli, Agropoli (SA). | 1. Genome Research Center for Health (CRGS) / 2. Laboratory of Molecular Medicine and Genomics(LMMGe) / 3. Center for Research in Pure and Applied Mathematics (CRMPA) | Giorgio Giurato (Corresponding Author), Francesca Rizzo (Corresponding Author), Alessandro Weisz (Corresponding Author), Gianluigi Franci, Giovanni Nassa, Pasquale Pagliano, Roberta Tarallo, Elena Alexandrova, Ylenia D'Agostino, Carlo Ferravante, Jessica Lamberti, Viola Melone, Domenico Memoli, Valeria Mirici Cappa, Domenico Palumbo, Giovanni Pecoraro, Assunta Sellitto, Oriana Strianese, Ilaria Terenzi, Giuseppe Fenza, Aniello Gentile, Antonello Saccomanno, Sonia Amabile, Teresa Rocco, Annamaria Salvati, Emilia Vaccaro, Massimiliano Galdiero, Michele Cennamo, Giuseppe Portella, Maria Grazia Foti, Mariarosaria Ingino, Maria Landi, Maurizio Fumi, Vincenzo Rocco, Rita Greco, Vittoria Letizia, Arnolfo Petruzziello, Maddalena Schioppa, Gregorio Goffredi, Francesca Marciano, Michele Caraglia, Alessia Cossu, Marianna Scrima, Edmondo Adorisio, Morena D'Avenia, Michela Iacobellis, Rosanna Piluscio, Giorgio Dirani, Vittorio Sambri, Simona Semprini, Silvia Zanoli, Francesco Curcio, Stefania Marzinotto, Andreina Baj, Fausto Sessa, Massimiliano Chetta, Marina Tarsitano, Giusy Giordano, Marcello Ametrano, Mariano Fiorenza, Caivano Antonella, D'Auria Fiorella, De Luca Luciana. |
| EPI_ISL_1577270, EPI_ISL_1577271, EPI_ISL_1577272, EPI_ISL_1577273 | 1.AO Universitaria 'S. Giovanni di Dio e Ruggi D'Aragona, Scuola Medica Salernitana' Hospital / 2.UOC di Virologia e Microbiologia, Università della Campania 'L. Vanvitelli' / 3.AO Universitaria 'Federico II' Napoli Hospital / 4.AORN 'San Giuseppe Moscati' Avellino Hospital / 5.AO 'San Pio - presidio G. Rummo' Benevento Hospital / 6.AO 'Sant'Anna e San Sebastiano' Caserta Hospital / 7.PO 'Maria Santissima Addolorata' Eboli Hospital / 8.Biogem Istituto di Ricerche Genetiche / 9. U.O.C. di Genetica Medica e di Laboratorio A.O.R.N., Azienda Ospedaliera di Rilievo Nazionale Antonio Cardarelli, Napoli / 10. Centro di riferimento Oncologico della Basilicata (IRCCS-CROB), Rionero in Vulture (PZ) / 11. Presidio Ospedaliero di Agropoli, Agropoli (SA). | 1. Genome Research Center for Health (CRGS) / 2. Laboratory of Molecular Medicine and Genomics(LMMGe) / 3. Center for Research in Pure and Applied Mathematics (CRMPA) | Giorgio Giurato (Corresponding Author), Francesca Rizzo (Corresponding Author), Alessandro Weisz (Corresponding Author), Gianluigi Franci, Giovanni Nassa, Pasquale Pagliano, Roberta Tarallo, Elena Alexandrova, Ylenia D'Agostino, Carlo Ferravante, Jessica Lamberti, Viola Melone, Domenico Memoli, Valeria Mirici Cappa, Domenico Palumbo, Giovanni Pecoraro, Assunta Sellitto, Oriana Strianese, Ilaria Terenzi, Giuseppe Fenza, Aniello Gentile, Antonello Saccomanno, Sonia Amabile, Teresa Rocco, Annamaria Salvati, Emilia Vaccaro, Massimiliano Galdiero, Michele Cennamo, Giuseppe Portella, Maria Grazia Foti, Mariarosaria Ingino, Maria Landi, Maurizio Fumi, Vincenzo Rocco, Rita Greco, Vittoria Letizia, Arnolfo Petruzziello, Maddalena Schioppa, Gregorio Goffredi, Francesca Marciano, Michele Caraglia, Alessia Cossu, Marianna Scrima, Edmondo Adorisio, Morena D'Avenia, Michela Iacobellis, Rosanna Piluscio, Giorgio Dirani, Vittorio Sambri, Simona Semprini, Silvia Zanoli, Francesco Curcio, Stefania Marzinotto, Andreina Baj, Fausto Sessa. |
| EPI_ISL_1577274 | 1.AO Universitaria 'S. Giovanni di Dio e Ruggi D'Aragona, Scuola Medica Salernitana' Hospital / 2.UOC di Virologia e Microbiologia, Università della Campania 'L. Vanvitelli' / 3.AO Universitaria 'Federico II' Napoli Hospital / 4.AORN 'San Giuseppe Moscati' Avellino Hospital / 5.AO 'San Pio - presidio G. Rummo' Benevento Hospital / 6.AO 'Sant'Anna e San Sebastiano' Caserta Hospital / 7.PO 'Maria Santissima Addolorata' Eboli Hospital / 8.Biogem Istituto di Ricerche Genetiche / 9. U.O.C. di Genetica Medica e di Laboratorio A.O.R.N., Azienda Ospedaliera di Rilievo Nazionale Antonio Cardarelli, Napoli / 10. Centro di riferimento Oncologico della Basilicata (IRCCS-CROB), Rionero in Vulture (PZ) / 11. Presidio Ospedaliero di Agropoli, Agropoli (SA). | 1. Genome Research Center for Health (CRGS) / 2. Laboratory of Molecular Medicine and Genomics(LMMGe) / 3. Center for Research in Pure and Applied Mathematics (CRMPA) | Giorgio Giurato (Corresponding Author), Francesca Rizzo (Corresponding Author), Alessandro Weisz (Corresponding Author), Gianluigi Franci, Giovanni Nassa, Pasquale Pagliano, Roberta Tarallo, Elena Alexandrova, Ylenia D'Agostino, Carlo Ferravante, Jessica Lamberti, Viola Melone, Domenico Memoli, Valeria Mirici Cappa, Domenico Palumbo, Giovanni Pecoraro, Assunta Sellitto, Oriana Strianese, Ilaria Terenzi, Giuseppe Fenza, Aniello Gentile, Antonello Saccomanno, Sonia Amabile, Teresa Rocco, Annamaria Salvati, Emilia Vaccaro, Massimiliano Galdiero, Michele Cennamo, Giuseppe Portella, Maria Grazia Foti, Mariarosaria Ingino, Maria Landi, Maurizio Fumi, Vincenzo Rocco, Rita Greco, Vittoria Letizia, Arnolfo Petruzziello, Maddalena Schioppa, Gregorio Goffredi, Francesca Marciano, Michele Caraglia, Alessia Cossu, Marianna Scrima, Edmondo Adorisio, Morena D'Avenia, Michela Iacobellis, Rosanna Piluscio, Giorgio Dirani, Vittorio Sambri, Simona Semprini, Silvia Zanoli, Francesco Curcio, Stefania Marzinotto, Andreina Baj, Fausto Sessa, Massimiliano Chetta, Marina Tarsitano, Giusy Giordano, Marcello Ametrano, Mariano Fiorenza, Caivano Antonella, D'Auria Fiorella, De Luca Luciana. |
| EPI_ISL_1577275, EPI_ISL_1577276, EPI_ISL_1577277, EPI_ISL_1577304 | 1.AO Universitaria 'S. Giovanni di Dio e Ruggi D'Aragona, Scuola Medica Salernitana' Hospital / 2.UOC di Virologia e Microbiologia, Università della Campania 'L. Vanvitelli' / 3.AO Universitaria 'Federico II' Napoli Hospital / 4.AORN 'San Giuseppe Moscati' Avellino Hospital / 5.AO 'San Pio - presidio G. Rummo' Benevento Hospital / 6.AO 'Sant'Anna e San Sebastiano' Caserta Hospital / 7.PO 'Maria Santissima Addolorata' Eboli Hospital / 8.Biogem Istituto di Ricerche Genetiche / 9. U.O.C. di Genetica Medica e di Laboratorio A.O.R.N., Azienda Ospedaliera di Rilievo Nazionale Antonio Cardarelli, Napoli / 10. Centro di riferimento Oncologico della Basilicata (IRCCS-CROB), Rionero in Vulture (PZ) / 11. Presidio Ospedaliero di Agropoli, Agropoli (SA). | 1. Genome Research Center for Health (CRGS) / 2. Laboratory of Molecular Medicine and Genomics(LMMGe) / 3. Center for Research in Pure and Applied Mathematics (CRMPA) | Giorgio Giurato (Corresponding Author), Francesca Rizzo (Corresponding Author), Alessandro Weisz (Corresponding Author), Gianluigi Franci, Giovanni Nassa, Pasquale Pagliano, Roberta Tarallo, Elena Alexandrova, Ylenia D'Agostino, Carlo Ferravante, Jessica Lamberti, Viola Melone, Domenico Memoli, Valeria Mirici Cappa, Domenico Palumbo, Giovanni Pecoraro, Assunta Sellitto, Oriana Strianese, Ilaria Terenzi, Giuseppe Fenza, Aniello Gentile, Antonello Saccomanno, Sonia Amabile, Teresa Rocco, Annamaria Salvati, Emilia Vaccaro, Massimiliano Galdiero, Michele Cennamo, Giuseppe Portella, Maria Grazia Foti, Mariarosaria Ingino, Maria Landi, Maurizio Fumi, Vincenzo Rocco, Rita Greco, Vittoria Letizia, Arnolfo Petruzziello, Maddalena Schioppa, Gregorio Goffredi, Francesca Marciano, Michele Caraglia, Alessia Cossu, Marianna Scrima, Edmondo Adorisio, Morena D'Avenia, Michela Iacobellis, Rosanna Piluscio, Giorgio Dirani, Vittorio Sambri, Simona Semprini, Silvia Zanoli, Francesco Curcio, Stefania Marzinotto, Andreina Baj, Fausto Sessa. |
| EPI_ISL_1577307 | 1.AO Universitaria 'S. Giovanni di Dio e Ruggi D'Aragona, Scuola Medica Salernitana' Hospital / 2.UOC di Virologia e Microbiologia, Università della Campania 'L. Vanvitelli' / 3.AO Universitaria 'Federico II' Napoli Hospital / 4.AORN 'San Giuseppe Moscati' Avellino Hospital / 5.AO 'San Pio - presidio G. Rummo' Benevento Hospital / 6.AO 'Sant'Anna e San Sebastiano' Caserta Hospital / 7.PO 'Maria Santissima Addolorata' Eboli Hospital / 8.Biogem Istituto di Ricerche Genetiche / 9. U.O.C. di Genetica Medica e di Laboratorio A.O.R.N., Azienda Ospedaliera di Rilievo Nazionale Antonio Cardarelli, Napoli / 10. Centro di riferimento Oncologico della Basilicata (IRCCS-CROB), Rionero in Vulture (PZ) / 11. Presidio Ospedaliero di Agropoli, Agropoli (SA). | 1. Genome Research Center for Health (CRGS) / 2. Laboratory of Molecular Medicine and Genomics(LMMGe) / 3. Center for Research in Pure and Applied Mathematics (CRMPA) | Giorgio Giurato (Corresponding Author), Francesca Rizzo (Corresponding Author), Alessandro Weisz (Corresponding Author), Gianluigi Franci, Giovanni Nassa, Pasquale Pagliano, Roberta Tarallo, Elena Alexandrova, Ylenia D'Agostino, Carlo Ferravante, Jessica Lamberti, Viola Melone, Domenico Memoli, Valeria Mirici Cappa, Domenico Palumbo, Giovanni Pecoraro, Assunta Sellitto, Oriana Strianese, Ilaria Terenzi, Giuseppe Fenza, Aniello Gentile, Antonello Saccomanno, Sonia Amabile, Teresa Rocco, Annamaria Salvati, Emilia Vaccaro, Massimiliano Galdiero, Michele Cennamo, Giuseppe Portella, Maria Grazia Foti, Mariarosaria Ingino, Maria Landi, Maurizio Fumi, Vincenzo Rocco, Rita Greco, Vittoria Letizia, Arnolfo Petruzziello, Maddalena Schioppa, Gregorio Goffredi, Francesca Marciano, Michele Caraglia, Alessia Cossu, Marianna Scrima, Edmondo Adorisio, Morena D'Avenia, Michela Iacobellis, Rosanna Piluscio, Giorgio Dirani, Vittorio Sambri, Simona Semprini, Silvia Zanoli, Francesco Curcio, Stefania Marzinotto, Andreina Baj, Fausto Sessa, Massimiliano Chetta, Marina Tarsitano, Giusy Giordano, Marcello Ametrano, Mariano Fiorenza, Caivano Antonella, D'Auria Fiorella, De Luca Luciana. |
| EPI_ISL_1577308, EPI_ISL_1577309, EPI_ISL_1577310, EPI_ISL_1577311 | 1.AO Universitaria 'S. Giovanni di Dio e Ruggi D'Aragona, Scuola Medica Salernitana' Hospital / 2.UOC di Virologia e Microbiologia, Università della Campania 'L. Vanvitelli' / 3.AO Universitaria 'Federico II' Napoli Hospital / 4.AORN 'San Giuseppe Moscati' Avellino Hospital / 5.AO 'San Pio - presidio G. Rummo' Benevento Hospital / 6.AO 'Sant'Anna e San Sebastiano' Caserta Hospital / 7.PO 'Maria Santissima Addolorata' Eboli Hospital / 8.Biogem Istituto di Ricerche Genetiche / 9. U.O.C. di Genetica Medica e di Laboratorio | 1. Genome Research Center for Health (CRGS) / 2. Laboratory of Molecular Medicine and Genomics(LMMGe) / 3. Center for Research in Pure and Applied Mathematics (CRMPA) | Giorgio Giurato (Corresponding Author), Francesca Rizzo (Corresponding Author), Alessandro Weisz (Corresponding Author), Gianluigi Franci, Giovanni Nassa, Pasquale Pagliano, Roberta Tarallo, Elena Alexandrova, Ylenia D'Agostino, Carlo Ferravante, Jessica Lamberti, Viola Melone, Domenico Memoli, Valeria Mirici Cappa, Domenico Palumbo, Giovanni Pecoraro, Assunta Sellitto, Oriana Strianese, Ilaria Terenzi, Giuseppe Fenza, Aniello Gentile, Antonello Saccomanno, Sonia Amabile, Teresa Rocco, Annamaria Salvati, Emilia Vaccaro, Massimiliano Galdiero, Michele Cennamo, Giuseppe Portella, Maria Grazia Foti, Mariarosaria Ingino, Maria Landi, Maurizio Fumi, Vincenzo Rocco, Rita Greco, Vittoria Letizia, Arnolfo Petruzziello, Maddalena Schioppa, Gregorio Goffredi, Francesca Marciano, Michele Caraglia, Alessia Cossu, Marianna Scrima, Edmondo Adorisio, Morena D'Avenia, Michela Iacobellis, Rosanna Piluscio, Giorgio Dirani, Vittorio Sambri, Simona Semprini, Silvia Zanoli, Francesco Curcio, Stefania Marzinotto, Andreina Baj, Fausto Sessa. |

| | | | |
|---|---|---|---|
| | A.O.R.N., Azienda Ospedaliera di Rilievo Nazionale Antonio Cardarelli, Napoli / 10. Centro di riferimento Oncologico della Basilicata (IRCCS-CROB), Rionero in Vulture (PZ) / 11. Presidio Ospedaliero di Agropoli, Agropoli (SA). | | |
| EPI_ISL_1577312 | 1.AO Universitaria 'S. Giovanni di Dio e Ruggi D'Aragona, Scuola Medica Salernitana' Hospital / 2.UOC di Virologia e Microbiologia, Università della Campania 'L. Vanvitelli' / 3.AO Universitaria 'Federico II' Napoli Hospital / 4.AORN 'San Giuseppe Moscati' Avellino Hospital / 5.AO 'San Pio - presidio G. Rummo' Benevento Hospital / 6.AO 'Sant'Anna e San Sebastiano' Caserta Hospital / 7.PO 'Maria Santissima Addolorata' Eboli Hospital / 8.Biogem Istituto di Ricerche Genetiche / 9. U.O.C. di Genetica Medica e di Laboratorio A.O.R.N., Azienda Ospedaliera di Rilievo Nazionale Antonio Cardarelli, Napoli / 10. Centro di riferimento Oncologico della Basilicata (IRCCS-CROB), Rionero in Vulture (PZ) / 11. Presidio Ospedaliero di Agropoli, Agropoli (SA). | 1. Genome Research Center for Health (CRGS) / 2. Laboratory of Molecular Medicine and Genomics(LMMGe) / 3. Center for Research in Pure and Applied Mathematics (CRMPA) | Giorgio Giurato (Corresponding Author), Francesca Rizzo (Corresponding Author), Alessandro Weisz (Corresponding Author), Gianluigi Franci, Giovanni Nassa, Pasquale Pagliano, Roberta Tarallo, Elena Alexandrova, Ylenia D'Agostino, Carlo Ferravante, Jessica Lamberti, Viola Melone, Domenico Memoli, Valeria Mirici Cappa, Domenico Palumbo, Giovanni Pecoraro, Assunta Sellitto, Oriana Strianese, Ilaria Terenzi, Giuseppe Fenza, Aniello Gentile, Antonello Saccomanno, Sonia Amabile, Teresa Rocco, Annamaria Salvati, Emilia Vaccaro, Massimiliano Galdiero, Michele Cennamo, Giuseppe Portella, Maria Grazia Foti, Mariarosaria Ingino, Maria Landi, Maurizio Fumi, Vincenzo Rocco, Rita Greco, Vittoria Letizia, Arnolfo Petruzziello, Maddalena Schioppa, Gregorio Goffredi, Francesca Marciano, Michele Caraglia, Alessia Cossu, Marianna Scrima, Edmondo Adorisio, Morena D'Avenia, Michela Iacobellis, Rosanna Piluscio, Giorgio Dirani, Vittorio Sambri, Simona Semprini, Silvia Zanoli, Francesco Curcio, Stefania Marzinotto, Andreina Baj, Fausto Sessa, Massimiliano Chetta, Marina Tarsitano, Giusy Giordano, Marcello Ametrano, Mariano Fiorenza, Caivano Antonella, D'Auria Fiorella, De Luca Luciana. |
| EPI_ISL_1577915 | PRESIDIO TERRITORIALE CASOLI CASOLI(CHIETI) | Istituto Zooprofilattico Sperimentale dell'Abruzzo e Molise "G. Caporale" | Lorusso A, Marcacci M, Di Domenico M, Ancora M, Curini V, Mangone I, Rinaldi A, Scialabba S, Di Pasquale A, Cammà C, Puglia I, Calistri P, Savini G |
| EPI_ISL_1577916 | SIESP DIPARTIMENTO DI PREVENZIONE CHIETI CHIETI(CHIETI) | Istituto Zooprofilattico Sperimentale dell'Abruzzo e Molise "G. Caporale" | Lorusso A, Marcacci M, Di Domenico M, Ancora M, Curini V, Mangone I, Rinaldi A, Scialabba S, Di Pasquale A, Cammà C, Puglia I, Calistri P, Savini G |
| EPI_ISL_1577927 | USCA Tagliacozzo TAGLIACOZZO(L'AQUILA) | Istituto Zooprofilattico Sperimentale dell'Abruzzo e Molise "G. Caporale" | Lorusso A, Marcacci M, Di Domenico M, Ancora M, Curini V, Mangone I, Rinaldi A, Scialabba S, Di Pasquale A, Cammà C, Puglia I, Calistri P, Savini G |
| EPI_ISL_1578454 | Laboratory of Clinical Microbiology, Virology and Bioemergencies, ASST Fatebenefratelli Sacco - Sacco Hospital | Laboratory of Clinical Microbiology, Virology and Bioemergencies, ASST Fatebenefratelli Sacco - Sacco Hospital | Valeria Micheli, Alessandro Mancon, Alberto Rizzo, Fiorenza Bracchitta, Luca Rizzuto, Maria Rita Gismondo |
| EPI_ISL_1578455 | Laboratory of Clinical Microbiology, Virology and Bioemergencies, ASST Fatebenefratelli Sacco - Sacco Hospital | Laboratory of Clinical Microbiology, Virology and Bioemergencies, ASST Fatebenefratelli Sacco - Sacco Hospital | Alberto Rizzo, Valeria Micheli, Alessandro Mancon, Fiorenza Bracchitta, Luca Rizzuto, Maria Rita Gismondo |
| EPI_ISL_1578456 | Laboratory of Clinical Microbiology, Virology and Bioemergencies, ASST Fatebenefratelli Sacco - Sacco Hospital | Laboratory of Clinical Microbiology, Virology and Bioemergencies, ASST Fatebenefratelli Sacco - Sacco Hospital | Valeria Micheli, Alessandro Mancon, Alberto Rizzo, Fiorenza Bracchitta, Luca Rizzuto, Maria Rita Gismondo |
| EPI_ISL_1578457, EPI_ISL_1578458, EPI_ISL_1578459, EPI_ISL_1578460 | Laboratory of Clinical Microbiology, Virology and Bioemergencies, ASST Fatebenefratelli Sacco - Sacco Hospital | Laboratory of Clinical Microbiology, Virology and Bioemergencies, ASST Fatebenefratelli Sacco - Sacco Hospital | Alessandro Mancon, Valeria Micheli, Alberto Rizzo, Fiorenza Bracchitta, Luca Rizzuto, Maria Rita Gismondo |
| EPI_ISL_1578461, EPI_ISL_1578462, EPI_ISL_1578463 | Laboratory of Clinical Microbiology, Virology and Bioemergencies, ASST Fatebenefratelli Sacco - Sacco Hospital | Laboratory of Clinical Microbiology, Virology and Bioemergencies, ASST Fatebenefratelli Sacco - Sacco Hospital | Alberto Rizzo, Valeria Micheli, Alessandro Mancon, Fiorenza Bracchitta, Luca Rizzuto, Maria Rita Gismondo |
| EPI_ISL_1578464 | Laboratory of Clinical Microbiology, Virology and Bioemergencies, ASST Fatebenefratelli Sacco - Sacco Hospital | Laboratory of Clinical Microbiology, Virology and Bioemergencies, ASST Fatebenefratelli Sacco - Sacco Hospital | Valeria Micheli, Alessandro Mancon, Alberto Rizzo, Fiorenza Bracchitta, Luca Rizzuto, Maria Rita Gismondo |
| EPI_ISL_1580533 | OSPEDALE CIVILE TERAMO - CENTRO TRASFUSIONALE TERAMO(TERAMO) | Istituto Zooprofilattico Sperimentale dell'Abruzzo e Molise "G. Caporale" | Lorusso A, Marcacci M, Di Domenico M, Ancora M, Curini V, Di Lollo Valeria, Mangone I, Rinaldi A, Delli Compagni E, Scialabba S, Caporale M, Di Pasquale A, Cammà C, Puglia I, Calistri P, Savini G |
| EPI_ISL_1585690 | Azienda USL Valle d'Aosta | U.O. Genomics, S.S. Genetics and Advanced Omics Techniques, Istituto Zooprofilattico Sperimentale del Piemonte Liguria e Valle d'Aosta | Chiara Beltramo, Elena Bozzetta, Pier Luigi Acutis, Simone Peletto |
| EPI_ISL_1591251, EPI_ISL_1591252, EPI_ISL_1591253, EPI_ISL_1591254, EPI_ISL_1591255, EPI_ISL_1591256, EPI_ISL_1591257, EPI_ISL_1591258, EPI_ISL_1591259, EPI_ISL_1591260, EPI_ISL_1591261, EPI_ISL_1591262, EPI_ISL_1591263, EPI_ISL_1591264, EPI_ISL_1607869 | | | |
| see above | Department of Virology, Istituto Zooprofilattico Sperimentale del Lazio e della Toscana (IZSLT) | Department of General Diagnostics; Department of Virology; Istituto Zooprofilattico Sperimentale del Lazio e della Toscana (IZSLT) | Patricia Alba, Giuseppe Manna, Elena L. Diaconu, Fabiola Feltrin, Raffaela Conti, Teresa Scicluna, Virginia Carfora, Antonella Cersini, Alessia Franco, Antonio Battisti. |
| EPI_ISL_1607968 | SIESP CHIETI - DRIVE IN LANCIANO LANCIANO(CHIETI) | Istituto Zooprofilattico Sperimentale dell'Abruzzo e Molise "G. Caporale" | Lorusso A, Marcacci M, Di Domenico M, Ancora M, Curini V, Mangone I, Rinaldi A, Scialabba S, Di Pasquale A, Cammà C, Puglia I, Calistri P, Savini G |
| EPI_ISL_1616837 | U.O. Microbiologia, Laboratorio Unico Centro Servizi - AUSL della Romagna | U.O. Microbiologia, Laboratorio Unico Centro Servizi - AUSL della Romagna | Giorgio Dirani, Silvia Zannoli, Vittorio Sambri, Giulia Gatti, Francesca Taddei, Ilaria Poggianti |
| EPI_ISL_1619453 | University of Bari Biomedical Sciences and Human Oncology | University of Bari Biomedical Sciences and Human Oncology | Chironna Maria., Sallustio A., Loconsole Daniela., Accogli M. |
| EPI_ISL_1620238 | University of Bari Biomedical Sciences and Human Oncology | University of Bari Biomedical Sciences and Human Oncology | Chironna M., Sallustio A., loconsole D., Accogli M. |
| EPI_ISL_1620663 | Ospedale S.M. Goretti di Latina - ASL Latina presidio ospedaliero Nord | INMI Lazzaro Spallanzani IRCCS | F Santini, B Bartolini, E Giombini, F Messina, M Rueca, G Bonfiglio, O Butera, CEM Gruber, A Lucci, S Pignalosa, L Corso, MR Capobianchi, A Di Caro |
| EPI_ISL_1626572 | INMI Lazzaro Spallanzani IRCCS | INMI Lazzaro Spallanzani IRCCS | G Bonfiglio, O Butera, CEM Gruber, F Santini, B Bartolini, E Giombini, F Messina, M Rueca, A Di Caro, MR Capobianchi |
| EPI_ISL_1627320 | Dipartimento di prevenzione Bari | Istituto Zooprofilattico Sperimentale della Puglia e della Basilicata | Parisi A., Bianco A., Capozzi L., Del Sambro L., Simone D., Difato L., Ridolfi D., Giannico A. |
| EPI_ISL_1627326, EPI_ISL_1627327, EPI_ISL_1627328, EPI_ISL_1627329, EPI_ISL_1627330, EPI_ISL_1627331, EPI_ISL_1627332 | Presidio di Brindisi Di Summa - Perrino | Istituto Zooprofilattico Sperimentale della Puglia e della Basilicata | Parisi A., Bianco A., Capozzi L., Del Sambro L., Simone D., Difato L., Santoro A., Ridolfi D., Giannico A. |
| EPI_ISL_1627333, EPI_ISL_1627334 | Dipartimento di prevenzione Bari | Istituto Zooprofilattico Sperimentale della Puglia e della Basilicata | Parisi A., Bianco A., Capozzi L., Del Sambro L., Simone D., Difato L., Ridolfi D., Giannico A. |
| EPI_ISL_1627342, EPI_ISL_1627343 | Presidio di Brindisi Di Summa - Perrino | Istituto Zooprofilattico Sperimentale della Puglia e della Basilicata | Parisi A., Bianco A., Capozzi L., Del Sambro L., Simone D., Difato L., Santoro A., Ridolfi D., Giannico A. |
| EPI_ISL_1627361 | Istituto Zooprofilattico Sperimentale della Puglia e della Basilicata | Istituto Zooprofilattico Sperimentale della Puglia e della Basilicata | Parisi A., Bianco A., Capozzi L., Del Sambro L., Simone D., Galante D., Pace L. |
| EPI_ISL_1627362 | Presidio di Brindisi Di Summa - Perrino | Istituto Zooprofilattico Sperimentale della Puglia e della Basilicata | Parisi A., Bianco A., Capozzi L., Del Sambro L., Simone D., Difato L., Santoro A., Ridolfi D., Giannico A. |
| EPI_ISL_1627363 | Ospedale Santissima Annunziata | Istituto Zooprofilattico Sperimentale della Puglia e della | Parisi A., Bianco A., Capozzi L., Del Sambro L., Simone D., Difato L., Ridolfi D., Giannico A, Tampoia M. |

Basilicata

| | | | |
|---|---|---|---|
| EPI_ISL_1632708 | Laboratorio HUB -Azienda Ospedaliero Universitaria - AOU - Cagliari | Laboratorio SPOKE Biologia Molecolare -Azienda Ospedaliero Universitaria - AOU - Cagliari | Germano Orrù, Sara Fais, Alessandra Scano, Miriam Loddo, Riccardo Cappai, Ferdinando Coghe |
| EPI_ISL_1632773 | Laboratorio HUB -Azienda Ospedaliero Universitaria - AOU - Cagliari | Laboratorio SPOKE Biologia Molecolare -Azienda Ospedaliero Universitaria - AOU - Cagliari | Germano Orrù, Alessandra Scano, Sara Fais, Miriam Loddo, Riccardo Cappai, Ferdinando Coghe |
| EPI_ISL_1632823, EPI_ISL_1632824, EPI_ISL_1632825, EPI_ISL_1632826, EPI_ISL_1632827 | Lab. Microbiologia e Virologia - Cotugno - A.O. dei Colli | Telethon Institute of Genetics and Medicine (TIGEM) | Antonio Grimaldi Patrizia Annunziata Francesco Panariello Biancamaria Pierri Claudia Tiberio Teresa Giuliano Valentina Bouche Chiara Colantuono Maria Concetta Cuomo Denise Di Concilio Lucio Di Filippo Anna Manfredi Marcello Salvi Antonio Limone Luigi Atripaldi Pellegrino Cerino Andrea Ballabio Davide Cacchiarelli |
| EPI_ISL_1633075, EPI_ISL_1633076, EPI_ISL_1633077, EPI_ISL_1633078, EPI_ISL_1633079, EPI_ISL_1633080, EPI_ISL_1633081, EPI_ISL_1633082, EPI_ISL_1633083, EPI_ISL_1633084, EPI_ISL_1633085, EPI_ISL_1633086, EPI_ISL_1633087, EPI_ISL_1633088, EPI_ISL_1633089, EPI_ISL_1633090, EPI_ISL_1633091, EPI_ISL_1633092, EPI_ISL_1633093, EPI_ISL_1633094, EPI_ISL_1633095, EPI_ISL_1633096, EPI_ISL_1633097, EPI_ISL_1633098, EPI_ISL_1633099, EPI_ISL_1633100, EPI_ISL_1633101, EPI_ISL_1633102, EPI_ISL_1633103, EPI_ISL_1633104, EPI_ISL_1633105, EPI_ISL_1633106, EPI_ISL_1633107, EPI_ISL_1633108, EPI_ISL_1633109, EPI_ISL_1633110, EPI_ISL_1633111, EPI_ISL_1633112, EPI_ISL_1633113, EPI_ISL_1633114, EPI_ISL_1633115, EPI_ISL_1633116, EPI_ISL_1633117, EPI_ISL_1633118, EPI_ISL_1633119 | | | |
| see above | Università Federico II - Dipartimento di scienze mediche traslazionali - Napoli | Telethon Institute of Genetics and Medicine (TIGEM) | Antonio Grimaldi Patrizia Annunziata Francesco Panariello Teresa Giuliano Michele Cennamo Valentina Bouche Chiara Colantuono Lucio Di Filippo Mariano Fiorenza Anna Manfredi Marcello Salvi Giuseppe Portella Andrea Ballabio Davide Cacchiarelli |
| EPI_ISL_1636399 | QUALITY CONTROL CHEMICAL BIOLOGICAL RISK, AOOR Villa Sofia Cervello Palermo | QUALITY CONTROL CHEMICAL BIOLOGICAL RISK, AOOR Villa Sofia Cervello Palermo | Di Gaudio, F., Contino, F., Brunacci, G. |
| EPI_ISL_1647032 | SC (UCO) Igiene e Sanità Pubblica (funzione integrata con SC Microbiologia e Virologia) | ARGO Laboratorio Genomica ed Epigenomica | Licastro D, Dal Monego S, Degasperi M, Marcello A, Segat L, Piscianz E, D'Agaro P |
| EPI_ISL_1647191 | Laboratorio HUB -Azienda Ospedaliero Universitaria - AOU - Cagliari | Laboratorio SPOKE Biologia Molecolare -Azienda Ospedaliero Universitaria - AOU - Cagliari | Germano Orrù, Sara Fais, Alessandra Scano,Miriam Loddo, Riccardo Cappai, Ferdinando Coghe |
| EPI_ISL_1647346 | Laboratorio HUB -Azienda Ospedaliero Universitaria - AOU - Cagliari | Laboratorio SPOKE Biologia Molecolare -Azienda Ospedaliero Universitaria - AOU - Cagliari | Germano Orrù, Sara Fais, Alessandra Scano, Miriam Loddo, Riccardo Cappai, Ferdinando Coghe |
| EPI_ISL_1653934, EPI_ISL_1653936 | University of Bari Biomedical Sciences and Human Oncology | University of Bari Biomedical Sciences and Human Oncology | Chironna M., Sallustio A., Loconsole D., Accogli M. |
| EPI_ISL_1654209, EPI_ISL_1660397, EPI_ISL_1663140 | Laboratorio HUB -Azienda Ospedaliero Universitaria - AOU - Cagliari | Laboratorio SPOKE Biologia Molecolare -Azienda Ospedaliero Universitaria - AOU - Cagliari | Germano Orrù, Sara Fais, Alessandra Scano, Miriam Loddo, Riccardo Cappai, Ferdinando Coghe |
| EPI_ISL_1663141 | Laboratorio HUB -Azienda Ospedaliero Universitaria - AOU - Cagliari | Submitting lab: Laboratorio SPOKE Biologia Molecolare -Azienda Ospedaliero Universitaria - AOU - Cagliari | Germano Orrù, Sara Fais, Alessandra Scano, Miriam Loddo, Riccardo Cappai, Ferdinando Coghe |
| EPI_ISL_1663232, EPI_ISL_1663404 | Laboratorio HUB -Azienda Ospedaliero Universitaria - AOU - Cagliari | Laboratorio SPOKE Biologia Molecolare -Azienda Ospedaliero Universitaria - AOU - Cagliari | Germano Orrù, Sara Fais, Alessandra Scano, Miriam Loddo, Riccardo Cappai, Ferdinando Coghe |
| EPI_ISL_1669779, EPI_ISL_1669780, EPI_ISL_1669781, EPI_ISL_1669782, EPI_ISL_1669783, EPI_ISL_1669784, EPI_ISL_1669785, EPI_ISL_1669786, EPI_ISL_1669787, EPI_ISL_1669788, EPI_ISL_1669789 | | | |
| see above | Istituto Zooprofilattico Sperimentale della Lombardia e dell'Emilia Romagna (IZSLER), Sede Territoriale di Modena | Istituto Zooprofilattico Sperimentale della Lombardia e dell'Emilia Romagna (IZSLER), Risk Analysis and Genomic Epidemiology Unit | Carra Elena, Gianluca Rugna, Marina Morganti, Ilaria Menozzi, Erika Scaltriti, Stefano Pongolini |
| EPI_ISL_1669839, EPI_ISL_1669840, EPI_ISL_1669841, EPI_ISL_1669842, EPI_ISL_1669843, EPI_ISL_1669844, EPI_ISL_1669845, EPI_ISL_1669846, EPI_ISL_1669847, EPI_ISL_1669848, EPI_ISL_1669849, EPI_ISL_1669850, EPI_ISL_1669851, EPI_ISL_1669852, EPI_ISL_1669853, EPI_ISL_1669854, EPI_ISL_1669855, EPI_ISL_1669856, EPI_ISL_1669857, EPI_ISL_1669858, EPI_ISL_1669859, EPI_ISL_1669860 | | | |
| see above | Azienda Ospedaliero - Universitaria di Modena Policlinico - Virologia e Microbiologia Molecolare | Istituto Zooprofilattico Sperimentale della Lombardia e dell'Emilia Romagna (IZSLER), Risk Analysis and Genomic Epidemiology Unit | Monica Pecorari, William Gennari, Giulia Fregni Serpini, Marina Morganti, Ilaria Menozzi, Erika Scaltriti, Stefano Pongolini |
| EPI_ISL_1669919, EPI_ISL_1669929 | Azienda Ospedaliera Terni | Istituto Zooprofilattico Sperimentale dell'Abruzzo e Molise "G. Caporale" | Palumbo M, Scaccetti A, Lorusso A, Marcacci M, Di Domenico M, Ancora M, Curini V, Mangone I, Rinaldi A, Scialabba S, Di Pasquale A, Cammà C, Puglia I, Calistri P, Savini G |
| EPI_ISL_1669930 | Azienda USL Umbria 2 | Istituto Zooprofilattico Sperimentale dell'Abruzzo e Molise "G. Caporale" | Proietti A, Pistoni E, Lorusso A, Marcacci M, Di Domenico M, Ancora M, Curini V, Mangone I, Rinaldi A, Scialabba S, Di Pasquale A, Cammà C, Puglia I, Calistri P, Savini G |
| EPI_ISL_1669933, EPI_ISL_1669934, EPI_ISL_1669936, EPI_ISL_1669937, EPI_ISL_1669938, EPI_ISL_1669939 | Università degli Studi di Perugia | Istituto Zooprofilattico Sperimentale dell'Abruzzo e Molise "G. Caporale" | Mencacci A, Camilloni B, Lorusso A, Marcacci M, Di Domenico M, Ancora M, Curini V, Mangone I, Rinaldi A, Scialabba S, Di Pasquale A, Cammà C, Puglia I, Calistri P, Savini G |
| EPI_ISL_1669940, EPI_ISL_1669948, EPI_ISL_1669949, EPI_ISL_1669950, EPI_ISL_1669951, EPI_ISL_1669952, EPI_ISL_1669953 | Azienda Ospedaliera Terni | Istituto Zooprofilattico Sperimentale dell'Abruzzo e Molise "G. Caporale" | Palumbo M, Scaccetti A, Lorusso A, Marcacci M, Di Domenico M, Ancora M, Curini V, Mangone I, Rinaldi A, Scialabba S, Di Pasquale A, Cammà C, Puglia I, Calistri P, Savini G |
| EPI_ISL_1670667, EPI_ISL_1670668, EPI_ISL_1670669, EPI_ISL_1670670, EPI_ISL_1670671, EPI_ISL_1670672, EPI_ISL_1670673, EPI_ISL_1670674, EPI_ISL_1670675, EPI_ISL_1670676, EPI_ISL_1670677, EPI_ISL_1670678, EPI_ISL_1670679, EPI_ISL_1670680, EPI_ISL_1670681, EPI_ISL_1670682, EPI_ISL_1670683, EPI_ISL_1670684, EPI_ISL_1670685, EPI_ISL_1670686, EPI_ISL_1670687, EPI_ISL_1670688, EPI_ISL_1670689, EPI_ISL_1670690, EPI_ISL_1670691, EPI_ISL_1670692, EPI_ISL_1670693, EPI_ISL_1670694, EPI_ISL_1670695, EPI_ISL_1670696, EPI_ISL_1670697, EPI_ISL_1670698, EPI_ISL_1670699, EPI_ISL_1670700, EPI_ISL_1670701, EPI_ISL_1670702, EPI_ISL_1670703, EPI_ISL_1670704, EPI_ISL_1670705, EPI_ISL_1670706, EPI_ISL_1670707, EPI_ISL_1670708 | | | |
| see above | Azienda Sanitaria Locale di Piacenza - Presidio Ospedaliero - Laboratorio di Microbiologia | Istituto Zooprofilattico Sperimentale della Lombardia e dell'Emilia Romagna (IZSLER), Risk Analysis and Genomic Epidemiology Unit | Lo Cascio Giuliana, Schiavo Roberta, Peroni Irene, Marina Morganti, Ilaria Menozzi, Erika Scaltriti, Stefano Pongolini |
| EPI_ISL_1670910, EPI_ISL_1670911, EPI_ISL_1670912, EPI_ISL_1670928, EPI_ISL_1670929, EPI_ISL_1670939, EPI_ISL_1670940, EPI_ISL_1670941, EPI_ISL_1670942, EPI_ISL_1670943, EPI_ISL_1670944, EPI_ISL_1670945 | | | |
| see above | IRCCS San Gallicano Dermatological Institute | IRCCS Regina Elena National Cancer Institute | Francesca De Nicola, Sara Donzelli, Grazia Prignano, Matteo Pallocca, Sabrina Strano, Fabrizio Ensoli, Fulvia Pimpinelli, Maurizio Fanciulli, Giovanni Blandino, Aldo Morrone, Gennaro Ciliberto |
| EPI_ISL_1678002 | CQRC_QUALITY CONTROL CHEMICAL BIOLOGICAL RISK_AOOR Villa Sofia Cervello Palermo | CQRC_QUALITY CONTROL CHEMICAL BIOLOGICAL RISK_AOOR Villa Sofia Cervello Palermo | Di Gaudio, F., Brunacci, G., Contino, F., |
| EPI_ISL_1678003 | CQRC_QUALITY CONTROL CHEMICAL BIOLOGICAL RISK_AOOR Villa Sofia Cervello Palermo | CQRC_QUALITY CONTROL CHEMICAL BIOLOGICAL RISK_AOOR Villa Sofia Cervello Palermo | Di Gaudio, F., Contino, F., Brunacci, G., |
| EPI_ISL_1678004, EPI_ISL_1678005, EPI_ISL_1678006 | CQRC_QUALITY CONTROL CHEMICAL BIOLOGICAL RISK_AOOR Villa Sofia Cervello Palermo | CQRC_QUALITY CONTROL CHEMICAL BIOLOGICAL RISK_AOOR Villa Sofia Cervello Palermo | Di Gaudio, F., Brunacci, G., Contino, F., |
| EPI_ISL_1678007 | CQRC_QUALITY CONTROL CHEMICAL BIOLOGICAL RISK_AOOR Villa Sofia Cervello Palermo | CQRC_QUALITY CONTROL CHEMICAL BIOLOGICAL RISK_AOOR Villa Sofia Cervello Palermo | Di Gaudio, F., Contino, F., Brunacci, G., |
| EPI_ISL_1678008 | CQRC_QUALITY CONTROL CHEMICAL BIOLOGICAL RISK_AOOR Villa Sofia Cervello Palermo | CQRC_QUALITY CONTROL CHEMICAL BIOLOGICAL RISK_AOOR Villa Sofia Cervello Palermo | Di Gaudio, F., Brunacci, G., Contino, F., |
| EPI_ISL_1678009 | CQRC_QUALITY CONTROL CHEMICAL BIOLOGICAL RISK_AOOR Villa Sofia Cervello Palermo | CQRC_QUALITY CONTROL CHEMICAL BIOLOGICAL RISK_AOOR Villa Sofia Cervello Palermo | Di Gaudio, F., Contino, F., Brunacci, G., |
| EPI_ISL_1678010 | CQRC_QUALITY CONTROL CHEMICAL BIOLOGICAL RISK_AOOR Villa Sofia Cervello Palermo | CQRC_QUALITY CONTROL CHEMICAL BIOLOGICAL RISK_AOOR Villa Sofia Cervello Palermo | Di Gaudio, F., Brunacci, G., Contino, F., |

| | | | |
|---|---|---|---|
| EPI_ISL_1678011 | CQRC_QUALITY CONTROL CHEMICAL BIOLOGICAL RISK_AOOR Villa Sofia Cervello Palermo | CQRC_QUALITY CONTROL CHEMICAL BIOLOGICAL RISK_AOOR Villa Sofia Cervello Palermo | Di Gaudio, F., Contino, F., Brunacci, G., |
| EPI_ISL_1678012 | CQRC_QUALITY CONTROL CHEMICAL BIOLOGICAL RISK_AOOR Villa Sofia Cervello Palermo | CQRC_QUALITY CONTROL CHEMICAL BIOLOGICAL RISK_AOOR Villa Sofia Cervello Palermo | Di Gaudio, F., Brunacci, G., Contino, F., |
| EPI_ISL_1678013 | CQRC_QUALITY CONTROL CHEMICAL BIOLOGICAL RISK_AOOR Villa Sofia Cervello Palermo | CQRC_QUALITY CONTROL CHEMICAL BIOLOGICAL RISK_AOOR Villa Sofia Cervello Palermo | Di Gaudio, F., Contino, F., Brunacci, G., |
| EPI_ISL_1678014 | CQRC_QUALITY CONTROL CHEMICAL BIOLOGICAL RISK_AOOR Villa Sofia Cervello Palermo | CQRC_QUALITY CONTROL CHEMICAL BIOLOGICAL RISK_AOOR Villa Sofia Cervello Palermo | Di Gaudio, F., Brunacci, G., Contino, F., |
| EPI_ISL_1678015 | CQRC_QUALITY CONTROL CHEMICAL BIOLOGICAL RISK_AOOR Villa Sofia Cervello Palermo | CQRC_QUALITY CONTROL CHEMICAL BIOLOGICAL RISK_AOOR Villa Sofia Cervello Palermo | Di Gaudio, F., Contino, F., Brunacci, G., |
| EPI_ISL_1678016 | CQRC_QUALITY CONTROL CHEMICAL BIOLOGICAL RISK_AOOR Villa Sofia Cervello Palermo | CQRC_QUALITY CONTROL CHEMICAL BIOLOGICAL RISK_AOOR Villa Sofia Cervello Palermo | Di Gaudio, F., Brunacci, G., Contino, F., |
| EPI_ISL_1678017 | CQRC_QUALITY CONTROL CHEMICAL BIOLOGICAL RISK_AOOR Villa Sofia Cervello Palermo | CQRC_QUALITY CONTROL CHEMICAL BIOLOGICAL RISK_AOOR Villa Sofia Cervello Palermo | Di Gaudio, F., Contino, F., Brunacci, G., |
| EPI_ISL_1678018 | CQRC_QUALITY CONTROL CHEMICAL BIOLOGICAL RISK_AOOR Villa Sofia Cervello Palermo | CQRC_QUALITY CONTROL CHEMICAL BIOLOGICAL RISK_AOOR Villa Sofia Cervello Palermo | Di Gaudio, F., Contino, F., Orlando, V., Buffa, V., Bonvissuto, M., Di Chiara, V., Todaro, I., Seidita, G., Di Sclafani, A., Lungari, A., Brunacci, G. |
| EPI_ISL_1678019 | CQRC_QUALITY CONTROL CHEMICAL BIOLOGICAL RISK_AOOR Villa Sofia Cervello Palermo | CQRC_QUALITY CONTROL CHEMICAL BIOLOGICAL RISK_AOOR Villa Sofia Cervello Palermo | Di Gaudio, F., Contino, F., Brunacci, G. |
| EPI_ISL_1678020 | CQRC_QUALITY CONTROL CHEMICAL BIOLOGICAL RISK_AOOR Villa Sofia Cervello Palermo | CQRC_QUALITY CONTROL CHEMICAL BIOLOGICAL RISK_AOOR Villa Sofia Cervello Palermo | Di Gaudio, F., Brunacci, G., Contino, F., |
| EPI_ISL_1678021, EPI_ISL_1678022, EPI_ISL_1678023 | CQRC_QUALITY CONTROL CHEMICAL BIOLOGICAL RISK_AOOR Villa Sofia Cervello Palermo | CQRC_QUALITY CONTROL CHEMICAL BIOLOGICAL RISK_AOOR Villa Sofia Cervello Palermo | Di Gaudio, F., Contino, F., Brunacci, G., |
| EPI_ISL_1678024 | CQRC_QUALITY CONTROL CHEMICAL BIOLOGICAL RISK_AOOR Villa Sofia Cervello Palermo | CQRC_QUALITY CONTROL CHEMICAL BIOLOGICAL RISK_AOOR Villa Sofia Cervello Palermo | Di Gaudio, F., Brunacci, G., Contino, F., |
| EPI_ISL_1706362, EPI_ISL_1706368, EPI_ISL_1706370 | Laboratorio HUB -Azienda Ospedaliero Universitaria - AOU - Cagliari | Laboratorio SPOKE Biologia Molecolare -Azienda Ospedaliero Universitaria - AOU - Cagliari | Germano Orrù, Sara Fais, Alessandra Scano, Miriam Loddo, Riccardo Cappai, Ferdinando Coghe |
| EPI_ISL_1706373, EPI_ISL_1706375 | Laboratorio HUB -Azienda Ospedaliero Universitaria - AOU - Cagliari | Laboratorio SPOKE Biologia Molecolare -Azienda Ospedaliero Universitaria - AOU - Cagliari | Germano Orrù, Sara Fais, Alessandra Scano, Valentina Medda, Riccardo Cappai, Ferdinando Coghe |
| EPI_ISL_1706549, EPI_ISL_1706550, EPI_ISL_1706580 | Laboratorio HUB -Azienda Ospedaliero Universitaria - AOU - Cagliari | Laboratorio SPOKE Biologia Molecolare -Azienda Ospedaliero Universitaria - AOU - Cagliari | Germano Orrù, Sara Fais, Valentina Medda, Alessandra Scano, Riccardo Cappai, Ferdinando Coghe |
| EPI_ISL_1711177, EPI_ISL_1711193, EPI_ISL_1711194, EPI_ISL_1711195, EPI_ISL_1711196, EPI_ISL_1711197, EPI_ISL_1711198 | Landstuhl Regional Medical Center | US Air Force School of Aerospace Medicine | Anthony Fries, Jennifer Meyer, William Gruner, William Buggele, Amanda Javorina, Sarah Purves, Fritz Castillo, Cole Anderson, Clarise Starr, Elizabeth Macias |
| EPI_ISL_1715063, EPI_ISL_1715079, EPI_ISL_1715085, EPI_ISL_1715100, EPI_ISL_1715106, EPI_ISL_1715113, EPI_ISL_1715114, EPI_ISL_1715127, EPI_ISL_1715133 | UOC laboratorio di analisi Istituto Giannina Gaslini | TIGEM | Antonio Grimaldi Patrizia Annunziata Francesco Panariello Teresa Giuliano Valentina Bouche Chiara Colantuono Lucio Di Filippo Anna Manfredi Marcello Salvi Andrea Ballabio Davide Cacchiarelli |
| EPI_ISL_1789733, EPI_ISL_1789735, EPI_ISL_1789742, EPI_ISL_1789751, EPI_ISL_1789752, EPI_ISL_1789753, EPI_ISL_1789754, EPI_ISL_1789755, EPI_ISL_1789756 | SC Microbiologia e Virologia AOUSS | AMES Centro Polidiagnostico Strumentale S.r.l. | Salvatore Rubino; Flavia Angioj; Laura Firino; Rosalba Govoni; Gabriele Ibba; Vincenzo Lai; Erica Mura; Bianca Paglietti; Claudia Piu; Anna Puggioni; Elena Rimini; Giulia Rocca; Caterina Serra; Sergio Uzzau. |
| EPI_ISL_1791316 | COVID Laboratory AOU Messina | Molecular Hepatology Lab AOU Messina | Giuseppina Raffa, Cristina Musolino, Valeria Chines, Daniele Lombardo, Domenico Giosa, Teresa Pollicino. |
| EPI_ISL_1807718 | Laboratorio Analisi Praecilia srl | Eurofins Genoma Group | Francesca Spinella, Riccardo Giannico, Valentina Andrioletti, Ettore Cotroneo (Eurofins Genoma Group) |
| EPI_ISL_1818940, EPI_ISL_1818941, EPI_ISL_1818942, EPI_ISL_1818943 | Laboratory of Clinical Microbiology, Virology and Bioemergencies, ASST Fatebenefratelli Sacco - Sacco Hospital | Laboratory of Clinical Microbiology, Virology and Bioemergencies, ASST Fatebenefratelli Sacco - Sacco Hospital | Valeria Micheli, Alessandro Mancon, Alberto Rizzo, Fiorenza Bracchitta, Luca Rizzuto, Maria Rita Gismondo |
| EPI_ISL_1818944 | Laboratory of Clinical Chemistry and Microbiology, IRCCS Orthopedic Institute Galeazzi | Laboratory of Clinical Microbiology, Virology and Bioemergencies, ASST Fatebenefratelli Sacco - Sacco Hospital | Valeria Micheli, Alessandro Mancon, Alberto Rizzo, Fiorenza Bracchitta, Luca Rizzuto, Maria Rita Gismondo |
| EPI_ISL_1819186 | Presidio di Brindisi Di Summa - Perrino | Istituto Zooprofilattico Sperimentale della Puglia e della Basilicata | Parisi A., Bianco A., Capozzi L., Del Sambro L., Simone D., Difato L., Santoro A., Ridolfi D., Giannico A. |
| EPI_ISL_1824636 | COVID Laboratory AOU Messina | Molecular Hepatology Lab AOU Messina | Giuseppina Raffa, Cristina Musolino, Valeria Chines, Daniele Lombardo, Domenico Giosa, Teresa Pollicino. |
| EPI_ISL_1841232 | Laboratorio Biologia Molecolare Sars Cov2 - UOC Laboratorio Analisi - Servizio Medicina di Laboratorio, Ospedale "San Francesco" - ATS-ASSL Nuoro | Laboratorio specialistico UOC Ematologia - Ospedale "San Francesco" - ATS-ASSL Nuoro | Giovanna Piras, Rosanna Asproni, Paolo Malune, Maura Fiamma, Maria Itria Monne, Giuseppe Mameli |
| EPI_ISL_1857293 | Microbiology and Virology Laboratory, "Policlinico Riuniti, Azienda Ospedaliero Universitaria, Foggia" | Microbiology and Virology Laboratory, "Policlinico Riuniti, Azienda Ospedaliero Universitaria, Foggia" | Fabio Arena, Settimia Altamura, Donatella Cedola, Valeria Delli Carri, Annamaria D'Aprile, Maria Rosaria Lipsi, Loris Micelli, Maria Pia Patrizio, Daniela Pisanelli, Tiziana Rollo, Maurizio Margaglione and Rosella De Nittis. |
| EPI_ISL_1909215, EPI_ISL_1909216, EPI_ISL_1909217, EPI_ISL_1909218, EPI_ISL_1909219, EPI_ISL_1909220, EPI_ISL_1909221, EPI_ISL_1909222, EPI_ISL_1909223, EPI_ISL_1909224, EPI_ISL_1909225, EPI_ISL_1909237, EPI_ISL_1909238 | | | |
| see above | Laboratory of Clinical Microbiology, Virology and Bioemergencies, ASST Fatebenefratelli Sacco - Sacco Hospital | Laboratory of Clinical Microbiology, Virology and Bioemergencies, ASST Fatebenefratelli Sacco - Sacco Hospital | Valeria Micheli, Alessandro Mancon, Alberto Rizzo, Fiorenza Bracchitta, Luca Rizzuto, Maria Rita Gismondo |
| EPI_ISL_1972898 | Department of Biomedical, Surgical and Dental Sciences and Department of Biomedical Sciences for Health, University of Milano | Department of Biomedical, Surgical and Dental Sciences and Department of Biomedical Sciences for Health, University of Milano | Serena Delbue, Elena Pariani, Cristina Galli, Lucia Signorini, Maria Dolci, Sandro Binda, Laura Bubba, Laura Pellegrinelli, Pasquale Ferrante |
| EPI_ISL_2020619 | Università Federico II - Dipartimento di scienze mediche traslazionali - Napoli | TIGEM | Antonio Grimaldi Patrizia Annunziata Francesco Panariello Teresa Giuliano Michele Cennamo Valentina Bouche Chiara Colantuono Lucio Di Filippo Mariano Fiorenza Anna Manfredi Marcello Salvi Giuseppe Portella Andrea Ballabio Davide Cacchiarelli |
| EPI_ISL_2036872, EPI_ISL_2036873, EPI_ISL_2036874, EPI_ISL_2036875, EPI_ISL_2036876, EPI_ISL_2036877, EPI_ISL_2036878, EPI_ISL_2036879, EPI_ISL_2036880, EPI_ISL_2036881, EPI_ISL_2036882, EPI_ISL_2036883, EPI_ISL_2036884, EPI_ISL_2036885, EPI_ISL_2036886, EPI_ISL_2036887, EPI_ISL_2036888, EPI_ISL_2036889, EPI_ISL_2036890, EPI_ISL_2036891, EPI_ISL_2036892, EPI_ISL_2036893, EPI_ISL_2036894, EPI_ISL_2036895, EPI_ISL_2036896, EPI_ISL_2036897, EPI_ISL_2036899, EPI_ISL_2036900, EPI_ISL_2036901, EPI_ISL_2036903, EPI_ISL_2036912, EPI_ISL_2036913, EPI_ISL_2036915, EPI_ISL_2036916, EPI_ISL_2036917, EPI_ISL_2036918, EPI_ISL_2036919, EPI_ISL_2036920, EPI_ISL_2036921, EPI_ISL_2036922, EPI_ISL_2036923, EPI_ISL_2036924, EPI_ISL_2036925, EPI_ISL_2036926, EPI_ISL_2036927, EPI_ISL_2036928, EPI_ISL_2036929, EPI_ISL_2036930, EPI_ISL_2036931, EPI_ISL_2036932, EPI_ISL_2036933, EPI_ISL_2036934, EPI_ISL_2036935 | | | |

| | | | |
|---|---|---|---|
| see above | Virology Laboratory, Scientific Department, Army Medical Center | Virology Laboratory, Scientific Department, Army Medical Center | Silvia Fillo, Riccardo De Sanctis, Antonella Fortunato, Anella Monte, Rossella Brandi, Giulia Campoli, Marzia Cavalli, Lucia Nicosia, Anna Anselmo, Vanessa Vera Fain, Francesco Giordani, Giandomenico Cerreto, Filippo Molinari, Giancarlo Petralito, Florigio Lista |
| EPI_ISL_2047859 | Azienda Ospedaliera Terni | Istituto Zooprofilattico Sperimentale dell'Abruzzo e Molise "G. Caporale" | Palumbo M, Scaccetti A, Lorusso A, Marcacci M, Di Domenico M, Ancora M, Curini V, Mangone I, Rinaldi A, Delli Compagni E, Scialabba S, Di Pasquale A, Cammà C, Puglia I, Calistri P, Savini G |
| EPI_ISL_2047860 | Azienda USL Umbria 2 | Istituto Zooprofilattico Sperimentale dell'Abruzzo e Molise "G. Caporale" | Proietti A, Pistoni E, Lorusso A, Marcacci M, Di Domenico M, Ancora M, Curini V, Di Lollo Valeria, Mangone I, Rinaldi A, Delli Compagni E, Scialabba S, Caporale M, Di Pasquale A, Cammà C, Puglia I, Calistri P, Savini G |
| EPI_ISL_2047861 | Università degli Studi di Perugia | Istituto Zooprofilattico Sperimentale dell'Abruzzo e Molise "G. Caporale" | Mencacci A, Camilloni B, Lorusso A, Marcacci M, Di Domenico M, Ancora M, Curini V, Di Lollo Valeria, Mangone I, Rinaldi A, Delli Compagni E, Scialabba S, Caporale M, Di Pasquale A, Cammà C, Puglia I, Calistri P, Savini G |
| EPI_ISL_2047862, EPI_ISL_2047863 | Laboratorio Analisi Osp. Città di Castello - Azienda USL Umbria1 | Istituto Zooprofilattico Sperimentale dell'Abruzzo e Molise "G. Caporale" | Malagigi V, Tacconi P, Lorusso A, Marcacci M, Di Domenico M, Ancora M, Curini V, Di Lollo Valeria, Mangone I, Rinaldi A, Delli Compagni E, Scialabba S, Caporale M, Di Pasquale A, Cammà C, Puglia I, Calistri P, Savini G |
| EPI_ISL_2047864 | Azienda USL Umbria 2 | Istituto Zooprofilattico Sperimentale dell'Abruzzo e Molise "G. Caporale" | Proietti A, Pistoni E, Lorusso A, Marcacci M, Di Domenico M, Ancora M, Curini V, Mangone I, Rinaldi A, Delli Compagni E, Scialabba S, Di Pasquale A, Cammà C, Puglia I, Calistri P, Savini G |
| EPI_ISL_2080863, EPI_ISL_2080864, EPI_ISL_2080865, EPI_ISL_2080866 | Center of Advanced Studies and Technology, Molecular Genetics Laboratory | Center of Advanced Studies and Technology, Molecular Genetics Laboratory | Anaclerio Federico, Ferrante Rossella, De Fabritiis Simone, Damiani Verena, Mandatori Domitilla |
| EPI_ISL_2092866, EPI_ISL_2092867, EPI_ISL_2092868 | CQRC_QUALITY CONTROL CHEMICAL BIOLOGICAL RISK_AOOR Villa Sofia Cervello Palermo | CQRC_QUALITY CONTROL CHEMICAL BIOLOGICAL RISK_AOOR Villa Sofia Cervello Palermo | Di Gaudio, F., Contino, F., Brunacci, G., |
| EPI_ISL_2092869, EPI_ISL_2092870 | CQRC_QUALITY CONTROL CHEMICAL BIOLOGICAL RISK_AOOR Villa Sofia Cervello Palermo | CQRC_QUALITY CONTROL CHEMICAL BIOLOGICAL RISK_AOOR Villa Sofia Cervello Palermo | Di Gaudio, F., Brunacci, G., Contino, F., |
| EPI_ISL_2092871 | CQRC_QUALITY CONTROL CHEMICAL BIOLOGICAL RISK_AOOR Villa Sofia Cervello Palermo | CQRC_QUALITY CONTROL CHEMICAL BIOLOGICAL RISK_AOOR Villa Sofia Cervello Palermo | Di Gaudio, F., Contino, F., Brunacci, G., |
| EPI_ISL_2092872 | CQRC_QUALITY CONTROL CHEMICAL BIOLOGICAL RISK_AOOR Villa Sofia Cervello Palermo | CQRC_QUALITY CONTROL CHEMICAL BIOLOGICAL RISK_AOOR Villa Sofia Cervello Palermo | Di Gaudio, F., Brunacci, G., Contino, F., |
| EPI_ISL_2143517 | Istituto Zooprofilattico Sperimentale del Mezzogiorno | Telethon Institute of Genetics and Medicine (TIGEM) | Antonio Grimaldi Patrizia Annunziata Francesco Panariello Biancamaria Pierri Claudia Tiberio Teresa Giuliano Valentina Bouche Chiara Colantuono Maria Concetta Cuomo Denise Di Concilio Lucio Di Filippo Anna Manfredi Marcello Salvi Antonio Limone Luigi Atripaldi Pellegrino Cerino Andrea Ballabio Davide Cacchiarelli |

We gratefully acknowledge the following Authors from the Originating laboratories responsible for obtaining the specimens, as well as the Submitting laboratories where the genome data were generated and shared via GISAID, on which this research is based.

All Submitters of data may be contacted directly via www.gisaid.org

Authors are sorted alphabetically.

| Accession ID | Originating Laboratory | Submitting Laboratory | Authors |
|---|---|---|---|
| EPI_ISL_1005559, EPI_ISL_1005560, EPI_ISL_1005561, EPI_ISL_1005562, EPI_ISL_1005563, EPI_ISL_1005564, EPI_ISL_1005565, EPI_ISL_1005566, EPI_ISL_1005567, EPI_ISL_1005568, EPI_ISL_1005569, EPI_ISL_1005570, EPI_ISL_1005571, EPI_ISL_1005572, EPI_ISL_1005573, EPI_ISL_1005574, EPI_ISL_1005575, EPI_ISL_1005576, EPI_ISL_1005577, EPI_ISL_1005578, EPI_ISL_1005579, EPI_ISL_1005580, EPI_ISL_1005581, EPI_ISL_1005582, EPI_ISL_1005656, EPI_ISL_1005657, EPI_ISL_1005658, EPI_ISL_1005659, EPI_ISL_1005660, EPI_ISL_1005661, EPI_ISL_1005662, EPI_ISL_1005663, EPI_ISL_1005664, EPI_ISL_1005665, EPI_ISL_1005666, EPI_ISL_1005667, EPI_ISL_1005668, EPI_ISL_1005669, EPI_ISL_1005670, EPI_ISL_1005671, EPI_ISL_1005672, EPI_ISL_1005673 | | | |
| see above | Instituto Nacional de Medicina Genomica | Instituto Nacional de Medicina Genomica | Hidalgo-Miranda A, Mendoza-Vargas A, Reyes-Grajeda JP, Cisneros-Villanueva M, Cedro-Tanda A,Peñaloza-Figueroa F, Herrera-Montalvo LA |
| EPI_ISL_1015335, EPI_ISL_1015336, EPI_ISL_1015337, EPI_ISL_1015338, EPI_ISL_1015339, EPI_ISL_1015340, EPI_ISL_1015341 | Univeristy of New Mexico Hospital | Center for Global Health, University of New Mexico Health Sciences Center | Daryl Domman, Kurt Schwalm, Justin Bacca, Jon Femling, Darrell Dinwiddie |
| EPI_ISL_1015342, EPI_ISL_1015343, EPI_ISL_1015344, EPI_ISL_1015345, EPI_ISL_1015346, EPI_ISL_1015347, EPI_ISL_1015348, EPI_ISL_1015349, EPI_ISL_1015350, EPI_ISL_1015351, EPI_ISL_1015352 | | | |
| see above | New Mexico Department of Health Scientific Laboratory | Center for Global Health, University of New Mexico Health Sciences Center | Daryl Domman, Kurt Schwalm, Twila Kunde, Joseph Hicks, Anastacia Griego, Michael Edwards, Darrell Dinwiddie |
| EPI_ISL_1021984, EPI_ISL_1022063, EPI_ISL_1022117, EPI_ISL_1022137, EPI_ISL_1022196, EPI_ISL_1022200, EPI_ISL_1022257, EPI_ISL_1022283, EPI_ISL_1022304, EPI_ISL_1022361, EPI_ISL_1022362, EPI_ISL_1022403, EPI_ISL_1025968, EPI_ISL_1025991, EPI_ISL_1025992, EPI_ISL_1026016, EPI_ISL_1026100, EPI_ISL_1026140, EPI_ISL_1026163, EPI_ISL_1026178, EPI_ISL_1026189, EPI_ISL_1026198, EPI_ISL_1026201, EPI_ISL_1026205, EPI_ISL_1026220, EPI_ISL_1026228, EPI_ISL_1026239, EPI_ISL_1026251, EPI_ISL_1026261, EPI_ISL_1026292, EPI_ISL_1026361, EPI_ISL_1026400, EPI_ISL_1026452, EPI_ISL_1026510, EPI_ISL_1026591, EPI_ISL_1026741, EPI_ISL_1026748, EPI_ISL_1026841, EPI_ISL_1026862, EPI_ISL_1027725, EPI_ISL_1027757, EPI_ISL_1027764, EPI_ISL_1027765, EPI_ISL_1027775, EPI_ISL_1027786, EPI_ISL_1027823, EPI_ISL_1027824, EPI_ISL_1027833, EPI_ISL_1027846, EPI_ISL_1027875, EPI_ISL_1027891, EPI_ISL_1027914, EPI_ISL_1027945, EPI_ISL_1027974, EPI_ISL_1027991, EPI_ISL_1028009, EPI_ISL_1028050, EPI_ISL_1028076, EPI_ISL_1028143, EPI_ISL_1028146, EPI_ISL_1028178, EPI_ISL_1028180, EPI_ISL_1028188, EPI_ISL_1028198, EPI_ISL_1028261, EPI_ISL_1028298, EPI_ISL_1028301, EPI_ISL_1028326, EPI_ISL_1028335, EPI_ISL_1028355, EPI_ISL_1028390, EPI_ISL_1028399, EPI_ISL_1028484, EPI_ISL_1028489, EPI_ISL_1028498, EPI_ISL_1028522, EPI_ISL_1028551, EPI_ISL_1028557, EPI_ISL_1028689, EPI_ISL_1028715, EPI_ISL_1028723, EPI_ISL_1028747, EPI_ISL_1028750, EPI_ISL_1028820, EPI_ISL_1028836, EPI_ISL_1028839, EPI_ISL_1028858, EPI_ISL_1028928, EPI_ISL_1028946, EPI_ISL_1028993, EPI_ISL_1028994, EPI_ISL_1028997, EPI_ISL_1029038, EPI_ISL_1029107, EPI_ISL_1029108, EPI_ISL_1029114, EPI_ISL_1029117, EPI_ISL_1029118, EPI_ISL_1029119, EPI_ISL_1029120, EPI_ISL_1029121, EPI_ISL_1029124, EPI_ISL_1029125, EPI_ISL_1029126, EPI_ISL_1029127, EPI_ISL_1029136, EPI_ISL_1029138, EPI_ISL_1029139, EPI_ISL_1029141, EPI_ISL_1029150, EPI_ISL_1029229, EPI_ISL_1029561, EPI_ISL_1029562, EPI_ISL_1029563, EPI_ISL_1029771, EPI_ISL_1029772, EPI_ISL_1029820, EPI_ISL_1029821, EPI_ISL_1029919, EPI_ISL_1029920, EPI_ISL_1030641, EPI_ISL_1030736, EPI_ISL_1030920, EPI_ISL_1030990, EPI_ISL_1031192, EPI_ISL_1031201, EPI_ISL_1031288, EPI_ISL_1031393, EPI_ISL_1031395, EPI_ISL_1031429, EPI_ISL_1031450, EPI_ISL_1031551, EPI_ISL_1031669, EPI_ISL_1031682, EPI_ISL_1031703, EPI_ISL_1031800, EPI_ISL_1031808, EPI_ISL_1032254, EPI_ISL_1032255, EPI_ISL_1032291, EPI_ISL_1032321, EPI_ISL_1032392, EPI_ISL_1032416, EPI_ISL_1032417, EPI_ISL_1032418, EPI_ISL_1032420, EPI_ISL_1032530, EPI_ISL_1032573 | | | |
| see above | Laboratory Corporation of America | Respiratory Viruses Branch, Division of Viral Diseases, Centers for Disease Control and Prevention | Peter W. Cook, Dakota Howard, Dhwani Batra, Ben L. Rambo-Martin, Clinton R. Paden, Suxiang Tong, Duncan MacCannell |
| EPI_ISL_1038925, EPI_ISL_1038926, EPI_ISL_1038927, EPI_ISL_1038928, EPI_ISL_1038929, EPI_ISL_1038930, EPI_ISL_1038931, EPI_ISL_1038932, EPI_ISL_1038933, EPI_ISL_1038934, EPI_ISL_1038935, EPI_ISL_1038936, EPI_ISL_1038937, EPI_ISL_1038938, EPI_ISL_1038939, EPI_ISL_1038940, EPI_ISL_1038941, EPI_ISL_1038942, EPI_ISL_1038943, EPI_ISL_1038944, EPI_ISL_1038945, EPI_ISL_1038946, EPI_ISL_1038947, EPI_ISL_1038948, EPI_ISL_1038949, EPI_ISL_1038950, EPI_ISL_1038951, EPI_ISL_1038952, EPI_ISL_1038953, EPI_ISL_1038954, EPI_ISL_1038955, EPI_ISL_1038956, EPI_ISL_1038957, EPI_ISL_1038958, EPI_ISL_1038959, EPI_ISL_1038960, EPI_ISL_1038961, EPI_ISL_1038962, EPI_ISL_1038963, EPI_ISL_1038964, EPI_ISL_1038965, EPI_ISL_1038966, EPI_ISL_1038967 | | | |
| see above | New Mexico Department of Health Scientific Laboratory | New Mexico Department of Health Scientific Laboratory | Ellie Johnson, Anastacia Griego-Fisher, D'eldra Malone, Jennifer Benoit |
| EPI_ISL_1040544, EPI_ISL_1040545, EPI_ISL_1040546, EPI_ISL_1040547, EPI_ISL_1040548, EPI_ISL_1040549, EPI_ISL_1040550, EPI_ISL_1040551, EPI_ISL_1040552, EPI_ISL_1040553, EPI_ISL_1040554, EPI_ISL_1040555, EPI_ISL_1040556, EPI_ISL_1040557, EPI_ISL_1040558, EPI_ISL_1040559, EPI_ISL_1040560, EPI_ISL_1040561, EPI_ISL_1040562, EPI_ISL_1040563, EPI_ISL_1040564, EPI_ISL_1040565, EPI_ISL_1040566, EPI_ISL_1040567, EPI_ISL_1040568, EPI_ISL_1040569, EPI_ISL_1040570, EPI_ISL_1040571, EPI_ISL_1040572, EPI_ISL_1040573, EPI_ISL_1040574, EPI_ISL_1040575, EPI_ISL_1040576, EPI_ISL_1040577, EPI_ISL_1040578, EPI_ISL_1040579, EPI_ISL_1040580, EPI_ISL_1040581, EPI_ISL_1040582, EPI_ISL_1040583, EPI_ISL_1040584, EPI_ISL_1040585, EPI_ISL_1040586, EPI_ISL_1040587, EPI_ISL_1040588, EPI_ISL_1040589, EPI_ISL_1040590, EPI_ISL_1040591, EPI_ISL_1040592, EPI_ISL_1040593, EPI_ISL_1040594, EPI_ISL_1040595, EPI_ISL_1040596, EPI_ISL_1040597, EPI_ISL_1040598, EPI_ISL_1040599, EPI_ISL_1040600, EPI_ISL_1040602, EPI_ISL_1040603, EPI_ISL_1040611, EPI_ISL_1040612, EPI_ISL_1040613, EPI_ISL_1040614, EPI_ISL_1040615, EPI_ISL_1040616, EPI_ISL_1040617, EPI_ISL_1040618, EPI_ISL_1040619, EPI_ISL_1040620, EPI_ISL_1040621, EPI_ISL_1040622 | | | |
| see above | Instituto Nacional de Medicina Genomica | Instituto Nacional de Medicina Genomica | Hidalgo-Miranda A, Mendoza-Vargas A, Reyes-Grajeda JP, Cisneros-Villanueva M, Cedro-Tanda A,Peñaloza-Figueroa F, Alcaraz-Millman M,Herrera-Montalvo LA |
| EPI_ISL_1040927 | Biologia molecular de enfermedades emergentes y EPOC, Instituto Nacional de Enfermedades Respiratorias | Biologia molecular de enfermedades emergentes y EPOC, Instituto Nacional de Enfermedades Respiratorias | Vazquez-Perez Joel Armando, Mejía-Nepomuceno Fidencio, Hernández-Teran Alejandra, Serna Ricardo, Salas-Hernández Jorge, Wong-Arambula Claudia, Rodríguez-Maldonado Abril, Perez-Padilla Rogelio, Ramirez-Gonzalez Ernesto. |
| EPI_ISL_1060612, EPI_ISL_1060613, EPI_ISL_1060614, EPI_ISL_1060615 | Instituto Nacional de Medicina Genomica | Instituto Nacional de Medicina Genomica | Hidalgo-Miranda A, Mendoza-Vargas A, Reyes-Grajeda JP, Cedro-Tanda A, Alcaraz N, Gonzalez-Barrera D, Rangel-DeLeon D, Ramirez-Vega O, Sifuentes-Rojas C, Cisneros-Villanueva M, Herrera-Montalvo LA |
| EPI_ISL_1060676, EPI_ISL_1060677, EPI_ISL_1060678, EPI_ISL_1060679, EPI_ISL_1060680, EPI_ISL_1060681, EPI_ISL_1060682, EPI_ISL_1060683, EPI_ISL_1060686, EPI_ISL_1060688, EPI_ISL_1060689, EPI_ISL_1060690, EPI_ISL_1060691, EPI_ISL_1060692, EPI_ISL_1060693, EPI_ISL_1060694, EPI_ISL_1060696, EPI_ISL_1060697, EPI_ISL_1060699, EPI_ISL_1060701, EPI_ISL_1060703, EPI_ISL_1060704, EPI_ISL_1060705, EPI_ISL_1060706, EPI_ISL_1060707, EPI_ISL_1060709, EPI_ISL_1060710, EPI_ISL_1060712, EPI_ISL_1060713, EPI_ISL_1060718, EPI_ISL_1060719, EPI_ISL_1060720, EPI_ISL_1060721, EPI_ISL_1060722, EPI_ISL_1060723, EPI_ISL_1060724, EPI_ISL_1060725, EPI_ISL_1060726, EPI_ISL_1060727, EPI_ISL_1060728, EPI_ISL_1060729, EPI_ISL_1060730 | | | |
| see above | Instituto de Diagnostico y Referencia Epidemiologicos (INDRE)_RNLSP | Instituto de Diagnostico y Referencia Epidemiologicos (INDRE) | Claudia Wong-Arambula, Abril Rodriguez-Maldonado, Fabiola Garces-Ayala, Adnan Araiza-Rodriguez, David Fragoso-Fonseca, Sergio Rangel-Guerrero, Mayra Jimenez-Morales, Nancy Munoz-Hernandez, Natividad Cruz-Ortiz, Tatiana Nunez-Garcia, Gisela Barrera-Badillo, Lucia Hernandez-Rivas, Irma Lopez-Martinez, Ernesto Ramirez-Gonzalez. |
| EPI_ISL_1064200, EPI_ISL_1064201, EPI_ISL_1064202, EPI_ISL_1064203, EPI_ISL_1064204, EPI_ISL_1064205, EPI_ISL_1064206, EPI_ISL_1064207, EPI_ISL_1064208, EPI_ISL_1064209, EPI_ISL_1064210, EPI_ISL_1064211, EPI_ISL_1064212, EPI_ISL_1064213, EPI_ISL_1064214, EPI_ISL_1064215, EPI_ISL_1064216, EPI_ISL_1064217, EPI_ISL_1064218, EPI_ISL_1064219, EPI_ISL_1064220, EPI_ISL_1064221, EPI_ISL_1064222, EPI_ISL_1064223, EPI_ISL_1064224, EPI_ISL_1064225, EPI_ISL_1064226, EPI_ISL_1064227, EPI_ISL_1064228, EPI_ISL_1064229, EPI_ISL_1064230, EPI_ISL_1064231, EPI_ISL_1064232, EPI_ISL_1064233, EPI_ISL_1064234, EPI_ISL_1064235, EPI_ISL_1064236, EPI_ISL_1064237 | | | |
| see above | New Mexico Department of Health Scientific Laboratory | Center for Global Health, University of New Mexico Health Sciences Center | Daryl Domman, Kurt Schwalm, Twila Kunde, Joseph Hicks, Anastacia Griego, Michael Edwards, Darrell Dinwiddie |
| EPI_ISL_1064916 | Univeristy of New Mexico Hospital | Center for Global Health, University of New Mexico Health Sciences Center | Daryl Domman, Kurt Schwalm, Justin Bacca, Jon Femling, Darrell Dinwiddie |
| EPI_ISL_1064918, EPI_ISL_1064919, EPI_ISL_1064920, EPI_ISL_1064921, EPI_ISL_1064923, EPI_ISL_1064924, EPI_ISL_1064925, EPI_ISL_1064928, EPI_ISL_1064929, EPI_ISL_1064930, EPI_ISL_1064931, EPI_ISL_1064932, EPI_ISL_1064933, EPI_ISL_1064934, EPI_ISL_1064935, EPI_ISL_1064936, EPI_ISL_1064937, EPI_ISL_1064938, EPI_ISL_1064939, EPI_ISL_1064940, EPI_ISL_1064941, EPI_ISL_1064942, EPI_ISL_1064943, EPI_ISL_1064944, EPI_ISL_1064945, EPI_ISL_1064946, EPI_ISL_1064947, EPI_ISL_1064948, EPI_ISL_1064949, EPI_ISL_1064950, EPI_ISL_1064951, EPI_ISL_1064952, EPI_ISL_1064953, EPI_ISL_1064954, EPI_ISL_1064955, EPI_ISL_1064956 | | | |
| see above | New Mexico Department of Health Scientific Laboratory | Center for Global Health, University of New Mexico Health Sciences Center | Daryl Domman, Kurt Schwalm, Twila Kunde, Joseph Hicks, Anastacia Griego, Michael Edwards, Darrell Dinwiddie |
| EPI_ISL_1064962, EPI_ISL_1064963, EPI_ISL_1064970, EPI_ISL_1064974, EPI_ISL_1064977, EPI_ISL_1064978, EPI_ISL_1064980, EPI_ISL_1064982 | Univeristy of New Mexico Hospital | Center for Global Health, University of New Mexico Health Sciences Center | Daryl Domman, Kurt Schwalm, Justin Bacca, Jon Femling, Darrell Dinwiddie |
| EPI_ISL_1075657 | Instituto Nacional de Medicina Genomica | Instituto Nacional de Medicina Genomica | Hidalgo-Miranda A, Mendoza-Vargas A, Reyes-Grajeda JP, Cedro-Tanda A, Alcaraz N, Gonzalez-Barrera D, Rangel-DeLeon D, Ramirez-Vega O, Sifuentes-Rojas C, Cisneros-Villanueva M, Herrera-Montalvo LA. |
| EPI_ISL_1081416 | Sharp HealthCare Laboratory | Andersen lab at Scripps Research | SEARCH Alliance San Diego with Aaron Harding, Jacquelyn Berumen, Cathy Woerle, Liam McGinnis, Art Mendoza, Omid Bakhtar |
| EPI_ISL_1081423, EPI_ISL_1081424, EPI_ISL_1081425, EPI_ISL_1081426, EPI_ISL_1081427, EPI_ISL_1081428, EPI_ISL_1081429, EPI_ISL_1081430, EPI_ISL_1081431, EPI_ISL_1081432, EPI_ISL_1081433, EPI_ISL_1081434, EPI_ISL_1081435, EPI_ISL_1081436, EPI_ISL_1081437, EPI_ISL_1081438, EPI_ISL_1081439, EPI_ISL_1081440, EPI_ISL_1081441, EPI_ISL_1081442, EPI_ISL_1081443, EPI_ISL_1081444, EPI_ISL_1081445, EPI_ISL_1081446, EPI_ISL_1081447, EPI_ISL_1081448, EPI_ISL_1081449, EPI_ISL_1081450, EPI_ISL_1081451, EPI_ISL_1081452, EPI_ISL_1081453, EPI_ISL_1081454, EPI_ISL_1081455, EPI_ISL_1081456, | | | |

| | | | |
|---|---|---|---|
| EPI_ISL_1081473, EPI_ISL_1081474, EPI_ISL_1081475, EPI_ISL_1081476, EPI_ISL_1081477, EPI_ISL_1081478, EPI_ISL_1081479, EPI_ISL_1081480, EPI_ISL_1081481, EPI_ISL_1081482, EPI_ISL_1081483, EPI_ISL_1081484, EPI_ISL_1081485, EPI_ISL_1081486, EPI_ISL_1081487, EPI_ISL_1081488, EPI_ISL_1081489, EPI_ISL_1081490, EPI_ISL_1081491, EPI_ISL_1081492, EPI_ISL_1081493, EPI_ISL_1081494, EPI_ISL_1081495 | | | |
| see above | Genomica Lab Molecular, M©xico | Andersen lab at Scripps Research | SEARCH Alliance San Diego with Jonathan Gonzalez Garcia, Jose Roman Chavez Mendez, Jose Horacio Reyna Verdugo, Martin Gonzalez Ibarra, Luis Alberto Rangel Gonzalez |
| EPI_ISL_1081506, EPI_ISL_1081544 | Sharp HealthCare Laboratory | Andersen lab at Scripps Research | SEARCH Alliance San Diego with Aaron Harding, Jacquelyn Berumen, Cathy Woerle, Liam McGinnis, Art Mendoza, Omid Bakhtar |
| EPI_ISL_1086714, EPI_ISL_1087005, EPI_ISL_1087066, EPI_ISL_1087086, EPI_ISL_1087101 | Quest Diagnostics Incorporated | Respiratory Viruses Branch, Division of Viral Diseases, Centers for Disease Control and Prevention | Peter W. Cook, Dakota Howard, Dhwani Batra, Ben L. Rambo-Martin, S. H. Rosenthal, A. Gerasimova, R. M. Kagan, B. Anderson, M. Hua, Y. Liu, L.E. Bernstein, K.E. Livingston, A. Perez, I. A. Shlyakhter, R. V. Rolando, R. Owen, P. Tanpaiboon, F. Lacbawan, Clinton R. Paden, Suxiang Tong, Duncan MacCannell |
| EPI_ISL_1091241, EPI_ISL_1091242, EPI_ISL_1091244, EPI_ISL_1091245 | Laboratorio Estatal de Salud Pública de Nuevo León | Laboratorio de Infectología Molecular, Departamento de Bioquímica y Medicina Molecular,Facultad de Medicina - Universidad Autónoma de Nuevo León | Kame A. Galán-Huerta, María F. Herrera-Saldivar, Natalia Martínez-Acuña, Sonia A. Lozano-Sepúlveda, Daniel Arellanos-Soto, Ana M. Rivas-Estilla, Samuel Buentello-Wong, Else del Carmen García-García, Gloria A. Jasso-de-la-Peña, Roberto Montes-de-Oca, Consuelo Treviño-Garza, Manuel E. de-la-O-Cavazos |
| EPI_ISL_1091247, EPI_ISL_1091248 | Laboratorio PGM | Laboratorio de Infectología Molecular, Departamento de Bioquímica y Medicina Molecular,Facultad de Medicina - Universidad Autónoma de Nuevo León | Kame A. Galán-Huerta, María F. Herrera-Saldivar, Natalia Martínez-Acuña, Sonia A. Lozano-Sepúlveda, Daniel Arellanos-Soto, Ana M. Rivas-Estilla, Javier Ramos-Jimenez, Gabriela Elizondo, Eduardo Garza-de-la-Peña |
| EPI_ISL_1091249 | Laboratorio Estatal de Salud Pública de Nuevo León | Laboratorio de Infectología Molecular, Departamento de Bioquímica y Medicina Molecular,Facultad de Medicina - Universidad Autónoma de Nuevo León | Kame A. Galán-Huerta, María F. Herrera-Saldivar, Natalia Martínez-Acuña, Sonia A. Lozano-Sepúlveda, Daniel Arellanos-Soto, Ana M. Rivas-Estilla, Samuel Buentello-Wong, Else del Carmen García-García, Gloria A. Jasso-de-la-Peña, Roberto Montes-de-Oca, Consuelo Treviño-Garza, Manuel E. de-la-O-Cavazos |
| EPI_ISL_1091250, EPI_ISL_1091251, EPI_ISL_1091252 | Laboratorio PGM | Laboratorio de Infectología Molecular, Departamento de Bioquímica y Medicina Molecular,Facultad de Medicina - Universidad Autónoma de Nuevo León | Kame A. Galán-Huerta, María F. Herrera-Saldivar, Natalia Martínez-Acuña, Sonia A. Lozano-Sepúlveda, Daniel Arellanos-Soto, Ana M. Rivas-Estilla, Javier Ramos-Jimenez, Gabriela Elizondo, Eduardo Garza-de-la-Peña |
| EPI_ISL_1091253, EPI_ISL_1091254, EPI_ISL_1091255, EPI_ISL_1091256, EPI_ISL_1091257 | Laboratorio Estatal de Salud Pública de Nuevo León | Laboratorio de Infectología Molecular, Departamento de Bioquímica y Medicina Molecular,Facultad de Medicina - Universidad Autónoma de Nuevo León | Kame A. Galán-Huerta, María F. Herrera-Saldivar, Natalia Martínez-Acuña, Sonia A. Lozano-Sepúlveda, Daniel Arellanos-Soto, Ana M. Rivas-Estilla, Samuel Buentello-Wong, Else del Carmen García-García, Gloria A. Jasso-de-la-Peña, Roberto Montes-de-Oca, Consuelo Treviño-Garza, Manuel E. de-la-O-Cavazos |
| EPI_ISL_1091258, EPI_ISL_1091259, EPI_ISL_1091260 | Laboratorio PGM | Laboratorio de Infectología Molecular, Departamento de Bioquímica y Medicina Molecular,Facultad de Medicina - Universidad Autónoma de Nuevo León | Kame A. Galán-Huerta, María F. Herrera-Saldivar, Natalia Martínez-Acuña, Sonia A. Lozano-Sepúlveda, Daniel Arellanos-Soto, Ana M. Rivas-Estilla, Javier Ramos-Jimenez, Gabriela Elizondo, Eduardo Garza-de-la-Peña |
| EPI_ISL_1091262 | Laboratorio Estatal de Salud Pública de Nuevo León | Laboratorio de Infectología Molecular, Departamento de Bioquímica y Medicina Molecular,Facultad de Medicina - Universidad Autónoma de Nuevo León | Kame A. Galán-Huerta, María F. Herrera-Saldivar, Natalia Martínez-Acuña, Sonia A. Lozano-Sepúlveda, Daniel Arellanos-Soto, Ana M. Rivas-Estilla, Samuel Buentello-Wong, Else del Carmen García-García, Gloria A. Jasso-de-la-Peña, Roberto Montes-de-Oca, Consuelo Treviño-Garza, Manuel E. de-la-O-Cavazos |
| EPI_ISL_1091263, EPI_ISL_1091264, EPI_ISL_1091265, EPI_ISL_1091266, EPI_ISL_1091267, EPI_ISL_1091268, EPI_ISL_1091269, EPI_ISL_1091270, EPI_ISL_1091271, EPI_ISL_1091273, EPI_ISL_1091274 | | | |
| see above | Laboratorio PGM | Laboratorio de Infectología Molecular, Departamento de Bioquímica y Medicina Molecular,Facultad de Medicina - Universidad Autónoma de Nuevo León | Kame A. Galán-Huerta, María F. Herrera-Saldivar, Natalia Martínez-Acuña, Sonia A. Lozano-Sepúlveda, Daniel Arellanos-Soto, Ana M. Rivas-Estilla, Javier Ramos-Jimenez, Gabriela Elizondo, Eduardo Garza-de-la-Peña |
| EPI_ISL_1092458 | Laboratorio Estatal de Salud Pública de Nuevo León | Laboratorio de Infectología Molecular, Departamento de Bioquímica y Medicina Molecular,Facultad de Medicina - Universidad Autónoma de Nuevo León | Kame A. Galán-Huerta, María F. Herrera-Saldivar, Natalia Martínez-Acuña, Sonia A. Lozano-Sepúlveda, Daniel Arellanos-Soto, Ana M. Rivas-Estilla, Samuel Buentello-Wong, Else del Carmen García-García, Gloria A. Jasso-de-la-Peña, Roberto Montes-de-Oca, Consuelo Treviño-Garza, Manuel E. de-la-O-Cavazos |
| EPI_ISL_1093145, EPI_ISL_1093146, EPI_ISL_1093147 | Laboratorio de Diagnostico de Enfermedades Emergentes y Reemergentes | Instituto de Diagnostico y Referencia Epidemiologicos (INDRE) | Claudia Wong-Arambula, Abril Rodriguez-Maldonado, Fabiola Garces-Ayala, Natividad Cruz-Ortiz, Tatiana Nunez-Garcia, Natali Vega-Magana, Gisela Barrera-Badillo, Lucia Hernandez-Rivas, Irma Lopez-Martinez, Ernesto Ramirez-Gonzalez. |
| EPI_ISL_1095516 | Instituto Nacional de Medicina Genomica | Instituto Nacional de Medicina Genomica | Hidalgo-Miranda A, Mendoza-Vargas A, Reyes-Grajeda JP, Cedro-Tanda A, Alcaraz N, Gonzalez-Barrera D, Rangel-DeLeon D, Ramirez-Vega O, Sifuentes-Rojas C, Cisneros-Villanueva M, Herrera-Montalvo LA |
| EPI_ISL_1096141 | Departamento de Cuidados Intensivos-Unidad de Investigación Médica en Inmunoquímica, UMAE Hospital de Especialidades "Bernardo Sepúlveda Gutiérrez", Centro Médico Nacional Siglo XXI, Instituto Mexicano del Seguro Social (IMSS), México. | Laboratorio de Secuenciación-Centro de Instrumentos, IMSS México /Unidad de Investigación Médica en Inmunoquímica, UMAE Hospital de Especialidades "Bernardo Sepúlveda Gutiérrez", Centro Médico Nacional Siglo XXI, Instituto Mexicano del Seguro Social (IMSS), México. | Horacio Zamudio-Meza, Javier Gaytán-Cervantes, Carolina González-Torres, Alejandro Torres Flores, Lourdes Arriaga-Pizano, Eduardo Ferat-Osorio, Arturo Cérbulo-Vázquez, Luis Sánchez Hurtado, Elizabeth Rubí Duran Castañeda, Jesús Mena López, Juan Carlos Guerrero Escobar, Zuleima Maldonado Galeana, Aura Angélica López García, José Manuel Ruiz Rincón, Joaquín González-Ibarra, César Raúl González-Bonilla, Constantino López-Macías. |
| EPI_ISL_1096142 | Departamento de Cuidados Intensivos-Unidad de Investigación Médica en Inmunoquímica, UMAE Hospital de Especialidades "Bernardo Sepúlveda Gutiérrez", Centro Médico Nacional Siglo XXI, Instituto Mexicano del Seguro Social (IMSS) México. | Laboratorio de Secuenciación-Centro de Instrumentos, IMSS México /Unidad de Investigación Médica en Inmunoquímica, UMAE Hospital de Especialidades "Bernardo Sepúlveda Gutiérrez", Centro Médico Nacional Siglo XXI, Instituto Mexicano del Seguro Social (IMSS), México. | Carolina González-Torres, Horacio Zamudio-Meza, Javier Gaytán-Cervantes, Alejandro Torres Flores, Lourdes Arriaga-Pizano, Eduardo Ferat-Osorio, Arturo Cérbulo-Vázquez, Luis Sánchez Hurtado, Elizabeth Rubí Duran Castañeda, Jesús Mena López, Juan Carlos Guerrero Escobar, Zuleima Maldonado Galeana, Aura Angélica López García, José Manuel Ruiz Rincón, Joaquín González-Ibarra, César Raúl González-Bonilla, Constantino López-Macías. |
| EPI_ISL_1096983 | Departamento de Cuidados Intensivos-Unidad de Investigación Médica en Inmunoquímica, UMAE Hospital de Especialidades "Bernardo Sepúlveda Gutiérrez", Centro Médico Nacional Siglo XXI, Instituto Mexicano del Seguro Social (IMSS) México. | Laboratorio de Secuenciación-Centro de Instrumentos, IMSS México /Unidad de Investigación Médica en Inmunoquímica, UMAE Hospital de Especialidades "Bernardo Sepúlveda Gutiérrez", Centro Médico Nacional Siglo XXI, Instituto Mexicano del Seguro Social (IMSS), México. | Carolina González-Torres, Javier Gaytán-Cervantes, Horacio Zamudio-Meza, Alejandro Torres Flores, Lourdes Arriaga-Pizano, Eduardo Ferat-Osorio, Arturo Cérbulo-Vázquez, Luis Sánchez Hurtado, Elizabeth Rubí Duran Castañeda, Jesús Mena López, Juan Carlos Guerrero Escobar, Zuleima Maldonado Galeana, Aura Angélica López García, José Manuel Ruiz Rincón, Joaquín González-Ibarra, César Raúl González-Bonilla, Constantino López-Macías. |
| EPI_ISL_1096985 | Departamento de Cuidados Intensivos-Unidad de Investigación Médica en Inmunoquímica, UMAE Hospital de Especialidades "Bernardo Sepúlveda Gutiérrez", Centro Médico Nacional Siglo XXI, Instituto Mexicano del Seguro Social (IMSS) México. | Laboratorio de Secuenciación-Centro de Instrumentos, IMSS México /Unidad de Investigación Médica en Inmunoquímica, UMAE Hospital de Especialidades "Bernardo Sepúlveda Gutiérrez", Centro Médico Nacional Siglo XXI, Instituto Mexicano del Seguro Social (IMSS), México. | Javier Gaytán-Cervantes, Carolina González-Torres, Horacio Zamudio-Meza, Alejandro Torres Flores, Lourdes Arriaga-Pizano, Eduardo Ferat-Osorio, Arturo Cérbulo-Vázquez, Luis Sánchez Hurtado, Elizabeth Rubí Duran Castañeda, Jesús Mena López, Juan Carlos Guerrero Escobar, Zuleima Maldonado Galeana, Aura Angélica López García, José Manuel Ruiz Rincón, Joaquín González-Ibarra, César Raúl González-Bonilla, Constantino López-Macías. |
| EPI_ISL_1097744, EPI_ISL_1097745, EPI_ISL_1097746, EPI_ISL_1097747, EPI_ISL_1113709, EPI_ISL_1113710, EPI_ISL_1113711, EPI_ISL_1113712, EPI_ISL_1113713, EPI_ISL_1113714, EPI_ISL_1113715, EPI_ISL_1113716, EPI_ISL_1113717, EPI_ISL_1113718, EPI_ISL_1113719, EPI_ISL_1113720, EPI_ISL_1113721, EPI_ISL_1113722, EPI_ISL_1113723, EPI_ISL_1113724, EPI_ISL_1113725, EPI_ISL_1113726, EPI_ISL_1113727, EPI_ISL_1113728, EPI_ISL_1113729, EPI_ISL_1113730, EPI_ISL_1113731, EPI_ISL_1113732, EPI_ISL_1113733, EPI_ISL_1113734, EPI_ISL_1113735, EPI_ISL_1113736, EPI_ISL_1113737, EPI_ISL_1113738, EPI_ISL_1113739, EPI_ISL_1113740, EPI_ISL_1113741, EPI_ISL_1113742, EPI_ISL_1113743, EPI_ISL_1113744, EPI_ISL_1113745, EPI_ISL_1113746, EPI_ISL_1113747, EPI_ISL_1113748, EPI_ISL_1113749, EPI_ISL_1113750, EPI_ISL_1113751, EPI_ISL_1113752, EPI_ISL_1113753 | | | |
| see above | New Mexico Department of Health Scientific Laboratory | New Mexico Department of Health Scientific Laboratory | Ellie Johnson, Anastacia Griego-Fisher, D'eldra Malone, Jennifer Benoit |
| EPI_ISL_1121488, EPI_ISL_1121515, EPI_ISL_1121518, EPI_ISL_1121519, EPI_ISL_1121520, EPI_ISL_1121521, EPI_ISL_1121522, EPI_ISL_1121523, EPI_ISL_1121524, EPI_ISL_1121525, EPI_ISL_1121526, EPI_ISL_1121527, EPI_ISL_1121528, EPI_ISL_1121529, EPI_ISL_1121530, EPI_ISL_1121531, EPI_ISL_1121532, EPI_ISL_1121533, EPI_ISL_1121534, EPI_ISL_1121535, EPI_ISL_1121536, EPI_ISL_1121538 | | | |
| see above | New Mexico Department of Health Scientific Laboratory | Center for Global Health, University of New Mexico Health Sciences Center | Daryl Domman, Kurt Schwalm, Twila Kunde, Joseph Hicks, Anastacia Griego, Michael Edwards, Darrell Dinwiddie |
| EPI_ISL_1137459, EPI_ISL_1137477 | LABDIS-UAGro | Instituto Nacional de Medicina Genomica | Hidalgo-Miranda A, Mendoza-Vargas A, Reyes-Grajeda JP, Cedro-Tanda A, Alcaraz N, Gonzalez-Barrera D, Rangel-DeLeon D, Ramirez-Vega O, Sifuentes-Rojas C, Cisneros-Villanueva M, Herrera-Montalvo LA, Berenice Illades-Aguiar, Oscar del Moral-Hernandez, Adolfo Roman-Roman, Marco Antonio-Leyva, Daniel Hernandez-Sotelo, Hugo A. Rodriguez-Ruiz, Fredy O Beltran-Anaya, Roberto Dircio-Maldonado, Julio C. Azucar-Hesiquio, Azucena D. Alvarez-Diaz, Ciresthel Bello-Rios, Gladys W. Valente-Nio, Jaqueline Loaeza-Loaeza, Madian Valadez-Quinones, Luis I. Basilio-Leyva. |
| EPI_ISL_1168452, EPI_ISL_1168453, EPI_ISL_1168454, EPI_ISL_1168455, EPI_ISL_1168456, EPI_ISL_1168457, EPI_ISL_1168458, EPI_ISL_1168459, EPI_ISL_1168460, EPI_ISL_1168461, EPI_ISL_1168462, EPI_ISL_1168463, EPI_ISL_1168464, EPI_ISL_1168466, EPI_ISL_1168467, EPI_ISL_1168468, EPI_ISL_1168469, EPI_ISL_1168470, EPI_ISL_1168471, EPI_ISL_1168472, EPI_ISL_1168473, EPI_ISL_1168474, EPI_ISL_1168475, EPI_ISL_1168476, EPI_ISL_1168477, EPI_ISL_1168478, EPI_ISL_1168479, EPI_ISL_1168480, EPI_ISL_1168481, EPI_ISL_1168482, EPI_ISL_1168483, EPI_ISL_1168484, EPI_ISL_1168485, EPI_ISL_1168486, EPI_ISL_1168487, EPI_ISL_1168488, EPI_ISL_1168489, EPI_ISL_1168490, EPI_ISL_1168491, EPI_ISL_1168492, EPI_ISL_1168493, EPI_ISL_1168494, EPI_ISL_1168495, EPI_ISL_1168496, EPI_ISL_1168497, EPI_ISL_1168498, EPI_ISL_1168499, EPI_ISL_1168500, EPI_ISL_1168501, EPI_ISL_1168502, EPI_ISL_1168503, | | | |

| | | | |
|---|---|---|---|
| EPI_ISL_1168504, EPI_ISL_1168505, EPI_ISL_1168506, EPI_ISL_1168507, EPI_ISL_1168508, EPI_ISL_1168509, EPI_ISL_1168510, EPI_ISL_1168511, EPI_ISL_1168512, EPI_ISL_1168513, EPI_ISL_1168514, EPI_ISL_1168515, EPI_ISL_1168516, EPI_ISL_1168517, EPI_ISL_1168518, EPI_ISL_1168519, EPI_ISL_1168520, EPI_ISL_1168521, EPI_ISL_1168522, EPI_ISL_1168523, EPI_ISL_1168524, EPI_ISL_1168525, EPI_ISL_1168526, EPI_ISL_1168527, EPI_ISL_1168528, EPI_ISL_1168529, EPI_ISL_1168530, EPI_ISL_1168531, EPI_ISL_1168532, EPI_ISL_1168533, EPI_ISL_1168534, EPI_ISL_1168535, EPI_ISL_1168536, EPI_ISL_1168537, EPI_ISL_1168538, EPI_ISL_1168539, EPI_ISL_1168540, EPI_ISL_1168541, EPI_ISL_1168542, EPI_ISL_1168543, EPI_ISL_1168544, EPI_ISL_1168545, EPI_ISL_1168546, EPI_ISL_1168547, EPI_ISL_1168548, EPI_ISL_1168549, EPI_ISL_1168550, EPI_ISL_1168551, EPI_ISL_1168552, EPI_ISL_1168553, EPI_ISL_1168554, EPI_ISL_1168555, EPI_ISL_1168556, EPI_ISL_1168557, EPI_ISL_1168558, EPI_ISL_1168559, EPI_ISL_1168560, EPI_ISL_1168561, EPI_ISL_1168562, EPI_ISL_1168563, EPI_ISL_1168564, EPI_ISL_1168565, EPI_ISL_1168566, EPI_ISL_1168567, EPI_ISL_1168568, EPI_ISL_1168569, EPI_ISL_1168570, EPI_ISL_1168571, EPI_ISL_1168572, EPI_ISL_1168573, EPI_ISL_1168574, EPI_ISL_1168575, EPI_ISL_1168576, EPI_ISL_1168577, EPI_ISL_1168578, EPI_ISL_1168579, EPI_ISL_1168580, EPI_ISL_1168581, EPI_ISL_1168582, EPI_ISL_1168583, EPI_ISL_1168586, EPI_ISL_1168587, EPI_ISL_1168588, EPI_ISL_1168589, EPI_ISL_1168590, EPI_ISL_1168591, EPI_ISL_1168592, EPI_ISL_1168593, EPI_ISL_1168594, EPI_ISL_1168595, EPI_ISL_1168596, EPI_ISL_1168597, EPI_ISL_1168598, EPI_ISL_1168599, EPI_ISL_1168600, EPI_ISL_1168601, EPI_ISL_1168602, EPI_ISL_1168603, EPI_ISL_1168604, EPI_ISL_1168605, EPI_ISL_1168606, EPI_ISL_1168607, EPI_ISL_1168608, EPI_ISL_1168609, EPI_ISL_1168610, EPI_ISL_1168611, EPI_ISL_1168612, EPI_ISL_1168613, EPI_ISL_1168614, EPI_ISL_1168615, EPI_ISL_1168616, EPI_ISL_1168617, EPI_ISL_1168618, EPI_ISL_1168619, EPI_ISL_1168620, EPI_ISL_1168621, EPI_ISL_1168622, EPI_ISL_1168623, EPI_ISL_1168624, EPI_ISL_1168625, EPI_ISL_1168626, EPI_ISL_1168627, EPI_ISL_1168628, EPI_ISL_1168629, EPI_ISL_1168630, EPI_ISL_1168631, EPI_ISL_1168632, EPI_ISL_1168633 | | | |
| see above | Instituto de Diagnostico y Referencia Epidemiologicos INDRE_RNLSP | Instituto de Diagnostico y Referencia Epidemiologicos (INDRE) | Claudia Wong-Arambula, Abril Rodriguez-Maldonado, Vanessa Rivero-Arredondo, Ariadna Medina-Benitez, Joaquin Quiroz-Mercado,David Fragoso-Fonseca, Sergio Rangel-Guerrero, Natividad Cruz-Ortiz, Tatiana Nunez-Garcia, Gisela Barrera-Badillo, Lucia Hernandez-Rivas, Irma Lopez-Martinez, Ernesto Ramirez-Gonzalez. |
| EPI_ISL_1185929, EPI_ISL_1185931, EPI_ISL_1185934, EPI_ISL_1185935, EPI_ISL_1185936 | Sharp HealthCare Laboratory | Andersen lab at Scripps Research | SEARCH Alliance San Diego with Aaron Harding, Jacquelyn Berumen, Cathy Woerle, Liam McGinnis, Art Mendoza, Omid Bakhtar |
| EPI_ISL_1196439, EPI_ISL_1196440, EPI_ISL_1196441, EPI_ISL_1196442, EPI_ISL_1196443, EPI_ISL_1196444, EPI_ISL_1196445, EPI_ISL_1196446, EPI_ISL_1196447, EPI_ISL_1196448, EPI_ISL_1196449, EPI_ISL_1196450, EPI_ISL_1196451, EPI_ISL_1196452, EPI_ISL_1196453, EPI_ISL_1196454, EPI_ISL_1196455, EPI_ISL_1196456, EPI_ISL_1196457, EPI_ISL_1196458, EPI_ISL_1196459, EPI_ISL_1196460, EPI_ISL_1196461, EPI_ISL_1196462, EPI_ISL_1196463, EPI_ISL_1196464, EPI_ISL_1196465, EPI_ISL_1196466, EPI_ISL_1196467, EPI_ISL_1196468, EPI_ISL_1196469, EPI_ISL_1196470, EPI_ISL_1196471, EPI_ISL_1196472, EPI_ISL_1196473, EPI_ISL_1196475, EPI_ISL_1196476, EPI_ISL_1196477, EPI_ISL_1196478, EPI_ISL_1196479, EPI_ISL_1196480, EPI_ISL_1196481, EPI_ISL_1196482, EPI_ISL_1196483, EPI_ISL_1201780 | | | |
| see above | New Mexico Department of Health Scientific Laboratory | New Mexico Department of Health Scientific Laboratory | Ellie Johnson, Anastacia Griego-Fisher, D'eldra Malone, Jennifer Benoit |
| EPI_ISL_1219711, EPI_ISL_1219712 | Instituto de Diagnostico y Referencia Epidemiologicos INDRE_RNLSP | Instituto de Diagnostico y Referencia Epidemiologicos (INDRE) | Claudia Wong-Arambula, Abril Rodriguez-Maldonado, Vanessa Rivero-Arredondo, Ariadna Medina-Benitez, Joaquin Quiroz-Mercado,David Fragoso-Fonseca, Sergio Rangel-Guerrero, Natividad Cruz-Ortiz, Tatiana Nunez-Garcia, Gisela Barrera-Badillo, Lucia Hernandez-Rivas, Irma Lopez-Martinez, Ernesto Ramirez-Gonzalez. |
| EPI_ISL_1219714 | CICSaB/UASLP/LESP SLP | Instituto de Diagnostico y Referencia Epidemiologicos (INDRE) | Claudia Wong-Arambula, Abril Rodriguez-Maldonado, Vanessa Rivero-Arredondo, Ariadna Medina-Benitez, Joaquin Quiroz-Mercado,David Fragoso-Fonseca, Sergio Rangel-Guerrero, Andreu Comas-Garcia, Natividad Cruz-Ortiz, Tatiana Nunez-Garcia, Gisela Barrera-Badillo, Lucia Hernandez-Rivas, Irma Lopez-Martinez, Ernesto Ramirez-Gonzalez. |
| EPI_ISL_1225241, EPI_ISL_1225242, EPI_ISL_1225243, EPI_ISL_1225244, EPI_ISL_1225245, EPI_ISL_1225246, EPI_ISL_1225247, EPI_ISL_1225248, EPI_ISL_1225249, EPI_ISL_1225250, EPI_ISL_1225251, EPI_ISL_1225252, EPI_ISL_1225253, EPI_ISL_1225254, EPI_ISL_1225255, EPI_ISL_1225256, EPI_ISL_1225257, EPI_ISL_1225258, EPI_ISL_1225259, EPI_ISL_1225260, EPI_ISL_1225261 | | | |
| see above | TriCore Reference Laboratories | Center for Global Health, University of New Mexico Health Sciences Center | Daryl Domman, Kurt Schwalm, Cecilia Thompson, Kendra Pesko, Karissa Culbreath, Darrell Dinwiddie |
| EPI_ISL_1225262 | TriCore Reference Laboratories | Center for Global Health, University of New Mexico Health Sciences Center | Daryl Domman, Kurt Schwalm, Cecilia Thompson, Kendra Pesko, Karissa Culbreath, Darrell Dinwiddie |
| EPI_ISL_1225263, EPI_ISL_1225264, EPI_ISL_1225265, EPI_ISL_1225266, EPI_ISL_1225267, EPI_ISL_1225268, EPI_ISL_1225269, EPI_ISL_1225270, EPI_ISL_1225271, EPI_ISL_1225272, EPI_ISL_1225273, EPI_ISL_1225274, EPI_ISL_1225275, EPI_ISL_1225276, EPI_ISL_1225277, EPI_ISL_1225278, EPI_ISL_1225279, EPI_ISL_1225280, EPI_ISL_1225281, EPI_ISL_1225282, EPI_ISL_1225283, EPI_ISL_1225284, EPI_ISL_1225285, EPI_ISL_1225286, EPI_ISL_1225287, EPI_ISL_1225288, EPI_ISL_1225289, EPI_ISL_1225290, EPI_ISL_1225291, EPI_ISL_1225292, EPI_ISL_1225293, EPI_ISL_1225294, EPI_ISL_1225295, EPI_ISL_1225296, EPI_ISL_1225297, EPI_ISL_1225298, EPI_ISL_1225299, EPI_ISL_1225300, EPI_ISL_1225301, EPI_ISL_1225302, EPI_ISL_1225303, EPI_ISL_1225304, EPI_ISL_1225305, EPI_ISL_1225306, EPI_ISL_1225307, EPI_ISL_1225308, EPI_ISL_1225309, EPI_ISL_1225310, EPI_ISL_1225311, EPI_ISL_1225312, EPI_ISL_1225313, EPI_ISL_1225314, EPI_ISL_1225315, EPI_ISL_1225316, EPI_ISL_1225317, EPI_ISL_1225318, EPI_ISL_1225319 | | | |
| see above | TriCore Reference Laboratories | Center for Global Health, University of New Mexico Health Sciences Center | Daryl Domman, Kurt Schwalm, Cecilia Thompson, Kendra Pesko, Karissa Culbreath, Darrell Dinwiddie |
| EPI_ISL_1239769, EPI_ISL_1239770, EPI_ISL_1239771, EPI_ISL_1239772, EPI_ISL_1239773, EPI_ISL_1239774, EPI_ISL_1239775, EPI_ISL_1239776, EPI_ISL_1239777, EPI_ISL_1239778, EPI_ISL_1239779, EPI_ISL_1239780, EPI_ISL_1239781, EPI_ISL_1239782, EPI_ISL_1239783, EPI_ISL_1239784, EPI_ISL_1239785, EPI_ISL_1239786, EPI_ISL_1239787, EPI_ISL_1239788, EPI_ISL_1239789, EPI_ISL_1239790, EPI_ISL_1239791, EPI_ISL_1239792, EPI_ISL_1239793, EPI_ISL_1239794, EPI_ISL_1239795, EPI_ISL_1239796, EPI_ISL_1239797, EPI_ISL_1239798, EPI_ISL_1239799, EPI_ISL_1239800, EPI_ISL_1239801, EPI_ISL_1239802, EPI_ISL_1239803, EPI_ISL_1239804, EPI_ISL_1239805, EPI_ISL_1239806, EPI_ISL_1239807, EPI_ISL_1239808, EPI_ISL_1239809, EPI_ISL_1239810, EPI_ISL_1239811, EPI_ISL_1239812, EPI_ISL_1239813, EPI_ISL_1239814, EPI_ISL_1239815, EPI_ISL_1239816, EPI_ISL_1239817, EPI_ISL_1239818, EPI_ISL_1239819, EPI_ISL_1239820, EPI_ISL_1239821, EPI_ISL_1239822, EPI_ISL_1239823, EPI_ISL_1239824, EPI_ISL_1239825, EPI_ISL_1239826, EPI_ISL_1239827, EPI_ISL_1239828, EPI_ISL_1239829, EPI_ISL_1239830, EPI_ISL_1239831, EPI_ISL_1239832, EPI_ISL_1239833, EPI_ISL_1239834, EPI_ISL_1239835, EPI_ISL_1239836, EPI_ISL_1239837, EPI_ISL_1239838, EPI_ISL_1239839, EPI_ISL_1239840, EPI_ISL_1239841, EPI_ISL_1239842, EPI_ISL_1239843, EPI_ISL_1239844, EPI_ISL_1239845, EPI_ISL_1239846 | | | |
| see above | TriCore Reference Laboratories | Center for Global Health, University of New Mexico Health Sciences Center | Daryl Domman, Kurt Schwalm, Cecilia Thompson, Kendra Pesko, Karissa Culbreath, Darrell Dinwiddie |
| EPI_ISL_1255932, EPI_ISL_1255935, EPI_ISL_1261543, EPI_ISL_1261544, EPI_ISL_1261546, EPI_ISL_1261547, EPI_ISL_1261548, EPI_ISL_1261549, EPI_ISL_1261550, EPI_ISL_1261551, EPI_ISL_1261552, EPI_ISL_1261553, EPI_ISL_1261554, EPI_ISL_1261555, EPI_ISL_1261556, EPI_ISL_1261557, EPI_ISL_1261558, EPI_ISL_1261559, EPI_ISL_1261560, EPI_ISL_1261561, EPI_ISL_1261562, EPI_ISL_1261563, EPI_ISL_1261564, EPI_ISL_1261565, EPI_ISL_1261566, EPI_ISL_1261567, EPI_ISL_1261568, EPI_ISL_1261569, EPI_ISL_1261570, EPI_ISL_1261571, EPI_ISL_1261572, EPI_ISL_1261573, EPI_ISL_1261574, EPI_ISL_1261575, EPI_ISL_1261576, EPI_ISL_1261577, EPI_ISL_1261578, EPI_ISL_1261579, EPI_ISL_1261580, EPI_ISL_1261581, EPI_ISL_1261582, EPI_ISL_1261583 | | | |
| see above | New Mexico Department of Health Scientific Laboratory | New Mexico Department of Health Scientific Laboratory | Ellie Johnson, Anastacia Griego-Fisher, D'eldra Malone, Jennifer Benoit |
| EPI_ISL_1291609, EPI_ISL_1291612, EPI_ISL_1291630, EPI_ISL_1291649 | Univeristy of New Mexico Hospital | Center for Global Health, University of New Mexico Health Sciences Center | Daryl Domman, Kurt Schwalm, Justin Bacca, Jon Femling, Darrell Dinwiddie |
| EPI_ISL_1298453 | Instituto Nacional de Medicina Genomica | Instituto Nacional de Medicina Genomica | Hidalgo-Miranda A, Mendoza-Vargas A, Reyes-Grajeda JP, Cedro-Tanda A, Alcaraz N, Gonzalez-Barrera D, Rangel-DeLeon D, Ramirez-Vega O, Herrera-Montalvo LA |
| EPI_ISL_1301453, EPI_ISL_1301539, EPI_ISL_1302152 | Instituto Nacional de Enfermedades Respiratorias (INER) | Instituto de Biotecnología de la UNAM | Authors from IBT, IMSS, InDRE and INER (in alphabetical order): Carlos F. Arias, Santiago Ávila-Ríos, Gisela Barrera-Badillo, Eduardo Becerril-Vargas, Celia Boukadida, Natividad Cruz-Ortiz, Larissa Fernandes-Matano, Ricardo Grande, Lucia Hernandez-Rivas, Alejandra Hernández-Terán, Pavel Isa, Irma Lopez-Martinez, José Arturo Martínez-Orozco, Margarita Matías-Florentino, Fidencio Mejía-Nepomuceno, Edgar Mendieta-Condado, Mario Mújica-Sánchez, José Esteban Muñoz-Medina, Tatiana Nunez-Garcia, Luis Alberto Ochoa-Carrera, Hector Esteban Paz-Juárez, Francisco Pulido, José Ernesto Ramírez-González, Alma Rincón-Rubio, Teresita Rojas-Mendoza, Jorge Salas-Hernández, Alejandro Sanchez-Flores, Clara Esperanza Santacruz-Tinoco, Andrea Santos Coy-Arechavaleta, Blanca Taboada, Gloria Vazquez, Joel Armando Vázquez-Pérez, Jerome Jean Verleyen |
| EPI_ISL_1302154 | Instituto de Diagnostico y Referencia Epidemiologicos InDRE_RNLSP | Instituto de Biotecnología de la UNAM | Authors from IBT, IMSS, InDRE and INER (in alphabetical order): Carlos F. Arias, Santiago Ávila-Ríos, Gisela Barrera-Badillo, Eduardo Becerril-Vargas, Celia Boukadida, Natividad Cruz-Ortiz, Larissa Fernandes-Matano, Ricardo Grande, Lucia Hernandez-Rivas, Alejandra Hernández-Terán, Pavel Isa, Irma Lopez-Martinez, José Arturo Martínez-Orozco, Margarita Matías-Florentino, Fidencio Mejía-Nepomuceno, Edgar Mendieta-Condado, Mario Mújica-Sánchez, José Esteban Muñoz-Medina, Tatiana Nunez-Garcia, Luis Alberto Ochoa-Carrera, Hector Esteban Paz-Juárez, Francisco Pulido, José Ernesto Ramírez-González, Alma Rincón-Rubio, Teresita Rojas-Mendoza, Jorge Salas-Hernández, Alejandro Sanchez-Flores, Clara Esperanza Santacruz-Tinoco, Andrea Santos Coy-Arechavaleta, Blanca Taboada, Gloria Vazquez, Joel Armando Vázquez-Pérez, Jerome Jean Verleyen |
| EPI_ISL_1302159 | Instituto Nacional de Enfermedades Respiratorias (INER) | Instituto de Biotecnología de la UNAM | Authors from IBT, IMSS, InDRE and INER (in alphabetical order): Carlos F. Arias, Santiago Ávila-Ríos, Gisela Barrera-Badillo, Eduardo Becerril-Vargas, Celia Boukadida, Natividad Cruz-Ortiz, Larissa Fernandes-Matano, Ricardo Grande, Lucia Hernandez-Rivas, Alejandra Hernández-Terán, Pavel Isa, Irma Lopez-Martinez, José Arturo Martínez-Orozco, Margarita Matías-Florentino, Fidencio Mejía-Nepomuceno, Edgar Mendieta-Condado, Mario Mújica-Sánchez, José Esteban Muñoz-Medina, Tatiana Nunez-Garcia, Luis Alberto Ochoa-Carrera, Hector Esteban Paz-Juárez, Francisco Pulido, José Ernesto Ramírez-González, Alma Rincón-Rubio, Teresita Rojas-Mendoza, Jorge Salas-Hernández, Alejandro Sanchez-Flores, Clara Esperanza Santacruz-Tinoco, Andrea Santos Coy-Arechavaleta, Blanca Taboada, Gloria Vazquez, Joel Armando Vázquez-Pérez, Jerome Jean Verleyen |
| EPI_ISL_1302162 | Instituto de Diagnostico y Referencia Epidemiologicos InDRE_RNLSP | Instituto de Biotecnología de la UNAM | Authors from IBT, IMSS, InDRE and INER (in alphabetical order): Carlos F. Arias, Santiago Ávila-Ríos, Gisela Barrera-Badillo, Eduardo Becerril-Vargas, Celia Boukadida, Natividad Cruz-Ortiz, Larissa Fernandes-Matano, Ricardo Grande, Lucia Hernandez-Rivas, Alejandra Hernández-Terán, Pavel Isa, Irma Lopez-Martinez, José Arturo Martínez-Orozco, Margarita Matías-Florentino, Fidencio Mejía-Nepomuceno, Edgar Mendieta-Condado, Mario Mújica-Sánchez, José Esteban Muñoz-Medina, Tatiana Nunez-Garcia, Luis Alberto Ochoa-Carrera, Hector Esteban Paz-Juárez, Francisco Pulido, José Ernesto Ramírez-González, Alma Rincón-Rubio, Teresita Rojas-Mendoza, Jorge Salas-Hernández, Alejandro Sanchez-Flores, Clara Esperanza Santacruz-Tinoco, Andrea Santos Coy-Arechavaleta, Blanca Taboada, Gloria Vazquez, Joel Armando Vázquez-Pérez, Jerome Jean Verleyen |
| EPI_ISL_1302163 | Instituto Nacional de Enfermedades Respiratorias (INER) | Instituto de Biotecnología de la UNAM | Authors from IBT, IMSS, InDRE and INER (in alphabetical order): Carlos F. Arias, Santiago Ávila-Ríos, Gisela Barrera-Badillo, Eduardo Becerril-Vargas, |

| | | | |
|---|---|---|---|
| | | | Celia Boukadida, Natividad Cruz-Ortiz, Larissa Fernandes-Matano, Ricardo Grande, Lucia Hernandez-Rivas, Alejandra Hernández-Terán, Pavel Isa, Irma Lopez-Martinez, José Arturo Martínez-Orozco, Margarita Matías-Florentino, Fidencio Mejía-Nepomuceno, Edgar Mendieta-Condado, Mario Mújica-Sánchez, José Esteban Muñoz-Medina, Tatiana Nunez-Garcia, Luis Alberto Ochoa-Carrera, Hector Esteban Paz-Juárez, Francisco Pulido, José Ernesto Ramírez-González, Alma Rincón-Rubio, Teresita Rojas-Mendoza, Jorge Salas-Hernández, Alejandro Sanchez-Flores, Clara Esperanza Santacruz-Tinoco, Andrea Santos Coy-Arechavaleta, Blanca Taboada, Gloria Vazquez, Joel Armando Vázquez-Pérez, Jerome Jean Verleyen |
| EPI_ISL_1302166, EPI_ISL_1302167, EPI_ISL_1302168 | Instituto de Diagnostico y Referencia Epidemiologicos InDRE_RNLSP | Instituto de Biotecnología de la UNAM | Authors from IBT, IMSS, InDRE and INER (in alphabetical order): Carlos F. Arias, Santiago Ávila-Ríos, Gisela Barrera-Badillo, Eduardo Becerril-Vargas, Celia Boukadida, Natividad Cruz-Ortiz, Larissa Fernandes-Matano, Ricardo Grande, Lucia Hernandez-Rivas, Alejandra Hernández-Terán, Pavel Isa, Irma Lopez-Martinez, José Arturo Martínez-Orozco, Margarita Matías-Florentino, Fidencio Mejía-Nepomuceno, Edgar Mendieta-Condado, Mario Mújica-Sánchez, José Esteban Muñoz-Medina, Tatiana Nunez-Garcia, Luis Alberto Ochoa-Carrera, Hector Esteban Paz-Juárez, Francisco Pulido, José Ernesto Ramírez-González, Alma Rincón-Rubio, Teresita Rojas-Mendoza, Jorge Salas-Hernández, Alejandro Sanchez-Flores, Clara Esperanza Santacruz-Tinoco, Andrea Santos Coy-Arechavaleta, Blanca Taboada, Gloria Vazquez, Joel Armando Vázquez-Pérez, Jerome Jean Verleyen |
| EPI_ISL_1302170, EPI_ISL_1302171 | Instituto Nacional de Enfermedades Respiratorias (INER) | Instituto de Biotecnología de la UNAM | Authors from IBT, IMSS, InDRE and INER (in alphabetical order): Carlos F. Arias, Santiago Ávila-Ríos, Gisela Barrera-Badillo, Eduardo Becerril-Vargas, Celia Boukadida, Natividad Cruz-Ortiz, Larissa Fernandes-Matano, Ricardo Grande, Lucia Hernandez-Rivas, Alejandra Hernández-Terán, Pavel Isa, Irma Lopez-Martinez, José Arturo Martínez-Orozco, Margarita Matías-Florentino, Fidencio Mejía-Nepomuceno, Edgar Mendieta-Condado, Mario Mújica-Sánchez, José Esteban Muñoz-Medina, Tatiana Nunez-Garcia, Luis Alberto Ochoa-Carrera, Hector Esteban Paz-Juárez, Francisco Pulido, José Ernesto Ramírez-González, Alma Rincón-Rubio, Teresita Rojas-Mendoza, Jorge Salas-Hernández, Alejandro Sanchez-Flores, Clara Esperanza Santacruz-Tinoco, Andrea Santos Coy-Arechavaleta, Blanca Taboada, Gloria Vazquez, Joel Armando Vázquez-Pérez, Jerome Jean Verleyen |
| EPI_ISL_1302174, EPI_ISL_1302175, EPI_ISL_1302176, EPI_ISL_1302177 | Instituto de Diagnostico y Referencia Epidemiologicos InDRE_RNLSP | Instituto de Biotecnología de la UNAM | Authors from IBT, IMSS, InDRE and INER (in alphabetical order): Carlos F. Arias, Santiago Ávila-Ríos, Gisela Barrera-Badillo, Eduardo Becerril-Vargas, Celia Boukadida, Natividad Cruz-Ortiz, Larissa Fernandes-Matano, Ricardo Grande, Lucia Hernandez-Rivas, Alejandra Hernández-Terán, Pavel Isa, Irma Lopez-Martinez, José Arturo Martínez-Orozco, Margarita Matías-Florentino, Fidencio Mejía-Nepomuceno, Edgar Mendieta-Condado, Mario Mújica-Sánchez, José Esteban Muñoz-Medina, Tatiana Nunez-Garcia, Luis Alberto Ochoa-Carrera, Hector Esteban Paz-Juárez, Francisco Pulido, José Ernesto Ramírez-González, Alma Rincón-Rubio, Teresita Rojas-Mendoza, Jorge Salas-Hernández, Alejandro Sanchez-Flores, Clara Esperanza Santacruz-Tinoco, Andrea Santos Coy-Arechavaleta, Blanca Taboada, Gloria Vazquez, Joel Armando Vázquez-Pérez, Jerome Jean Verleyen |
| EPI_ISL_1302179, EPI_ISL_1302180 | Instituto Nacional de Enfermedades Respiratorias (INER) | Instituto de Biotecnología de la UNAM | Authors from IBT, IMSS, InDRE and INER (in alphabetical order): Carlos F. Arias, Santiago Ávila-Ríos, Gisela Barrera-Badillo, Eduardo Becerril-Vargas, Celia Boukadida, Natividad Cruz-Ortiz, Larissa Fernandes-Matano, Ricardo Grande, Lucia Hernandez-Rivas, Alejandra Hernández-Terán, Pavel Isa, Irma Lopez-Martinez, José Arturo Martínez-Orozco, Margarita Matías-Florentino, Fidencio Mejía-Nepomuceno, Edgar Mendieta-Condado, Mario Mújica-Sánchez, José Esteban Muñoz-Medina, Tatiana Nunez-Garcia, Luis Alberto Ochoa-Carrera, Hector Esteban Paz-Juárez, Francisco Pulido, José Ernesto Ramírez-González, Alma Rincón-Rubio, Teresita Rojas-Mendoza, Jorge Salas-Hernández, Alejandro Sanchez-Flores, Clara Esperanza Santacruz-Tinoco, Andrea Santos Coy-Arechavaleta, Blanca Taboada, Gloria Vazquez, Joel Armando Vázquez-Pérez, Jerome Jean Verleyen |
| EPI_ISL_1302181, EPI_ISL_1302182, EPI_ISL_1302183, EPI_ISL_1302184 | Instituto de Diagnostico y Referencia Epidemiologicos InDRE_RNLSP | Instituto de Biotecnología de la UNAM | Authors from IBT, IMSS, InDRE and INER (in alphabetical order): Carlos F. Arias, Santiago Ávila-Ríos, Gisela Barrera-Badillo, Eduardo Becerril-Vargas, Celia Boukadida, Natividad Cruz-Ortiz, Larissa Fernandes-Matano, Ricardo Grande, Lucia Hernandez-Rivas, Alejandra Hernández-Terán, Pavel Isa, Irma Lopez-Martinez, José Arturo Martínez-Orozco, Margarita Matías-Florentino, Fidencio Mejía-Nepomuceno, Edgar Mendieta-Condado, Mario Mújica-Sánchez, José Esteban Muñoz-Medina, Tatiana Nunez-Garcia, Luis Alberto Ochoa-Carrera, Hector Esteban Paz-Juárez, Francisco Pulido, José Ernesto Ramírez-González, Alma Rincón-Rubio, Teresita Rojas-Mendoza, Jorge Salas-Hernández, Alejandro Sanchez-Flores, Clara Esperanza Santacruz-Tinoco, Andrea Santos Coy-Arechavaleta, Blanca Taboada, Gloria Vazquez, Joel Armando Vázquez-Pérez, Jerome Jean Verleyen |
| EPI_ISL_1302186 | Instituto Nacional de Enfermedades Respiratorias (INER) | Instituto de Biotecnología de la UNAM | Authors from IBT, IMSS, InDRE and INER (in alphabetical order): Carlos F. Arias, Santiago Ávila-Ríos, Gisela Barrera-Badillo, Eduardo Becerril-Vargas, Celia Boukadida, Natividad Cruz-Ortiz, Larissa Fernandes-Matano, Ricardo Grande, Lucia Hernandez-Rivas, Alejandra Hernández-Terán, Pavel Isa, Irma Lopez-Martinez, José Arturo Martínez-Orozco, Margarita Matías-Florentino, Fidencio Mejía-Nepomuceno, Edgar Mendieta-Condado, Mario Mújica-Sánchez, José Esteban Muñoz-Medina, Tatiana Nunez-Garcia, Luis Alberto Ochoa-Carrera, Hector Esteban Paz-Juárez, Francisco Pulido, José Ernesto Ramírez-González, Alma Rincón-Rubio, Teresita Rojas-Mendoza, Jorge Salas-Hernández, Alejandro Sanchez-Flores, Clara Esperanza Santacruz-Tinoco, Andrea Santos Coy-Arechavaleta, Blanca Taboada, Gloria Vazquez, Joel Armando Vázquez-Pérez, Jerome Jean Verleyen |
| EPI_ISL_1302187, EPI_ISL_1302188, EPI_ISL_1302189 | Instituto de Diagnostico y Referencia Epidemiologicos InDRE_RNLSP | Instituto de Biotecnología de la UNAM | Authors from IBT, IMSS, InDRE and INER (in alphabetical order): Carlos F. Arias, Santiago Ávila-Ríos, Gisela Barrera-Badillo, Eduardo Becerril-Vargas, Celia Boukadida, Natividad Cruz-Ortiz, Larissa Fernandes-Matano, Ricardo Grande, Lucia Hernandez-Rivas, Alejandra Hernández-Terán, Pavel Isa, Irma Lopez-Martinez, José Arturo Martínez-Orozco, Margarita Matías-Florentino, Fidencio Mejía-Nepomuceno, Edgar Mendieta-Condado, Mario Mújica-Sánchez, José Esteban Muñoz-Medina, Tatiana Nunez-Garcia, Luis Alberto Ochoa-Carrera, Hector Esteban Paz-Juárez, Francisco Pulido, José Ernesto Ramírez-González, Alma Rincón-Rubio, Teresita Rojas-Mendoza, Jorge Salas-Hernández, Alejandro Sanchez-Flores, Clara Esperanza Santacruz-Tinoco, Andrea Santos Coy-Arechavaleta, Blanca Taboada, Gloria Vazquez, Joel Armando Vázquez-Pérez, Jerome Jean Verleyen |
| EPI_ISL_1302191, EPI_ISL_1302192, EPI_ISL_1302193, EPI_ISL_1302194 | Instituto Nacional de Enfermedades Respiratorias (INER) | Instituto de Biotecnología de la UNAM | Authors from IBT, IMSS, InDRE and INER (in alphabetical order): Carlos F. Arias, Santiago Ávila-Ríos, Gisela Barrera-Badillo, Eduardo Becerril-Vargas, Celia Boukadida, Natividad Cruz-Ortiz, Larissa Fernandes-Matano, Ricardo Grande, Lucia Hernandez-Rivas, Alejandra Hernández-Terán, Pavel Isa, Irma Lopez-Martinez, José Arturo Martínez-Orozco, Margarita Matías-Florentino, Fidencio Mejía-Nepomuceno, Edgar Mendieta-Condado, Mario Mújica-Sánchez, José Esteban Muñoz-Medina, Tatiana Nunez-Garcia, Luis Alberto Ochoa-Carrera, Hector Esteban Paz-Juárez, Francisco Pulido, José Ernesto Ramírez-González, Alma Rincón-Rubio, Teresita Rojas-Mendoza, Jorge Salas-Hernández, Alejandro Sanchez-Flores, Clara Esperanza Santacruz-Tinoco, Andrea Santos Coy-Arechavaleta, Blanca Taboada, Gloria Vazquez, Joel Armando Vázquez-Pérez, Jerome Jean Verleyen |
| EPI_ISL_1302197 | Instituto de Diagnostico y Referencia Epidemiologicos InDRE_RNLSP | Instituto de Biotecnología de la UNAM | Authors from IBT, IMSS, InDRE and INER (in alphabetical order): Carlos F. Arias, Santiago Ávila-Ríos, Gisela Barrera-Badillo, Eduardo Becerril-Vargas, Celia Boukadida, Natividad Cruz-Ortiz, Larissa Fernandes-Matano, Ricardo Grande, Lucia Hernandez-Rivas, Alejandra Hernández-Terán, Pavel Isa, Irma Lopez-Martinez, José Arturo Martínez-Orozco, Margarita Matías-Florentino, Fidencio Mejía-Nepomuceno, Edgar Mendieta-Condado, Mario Mújica-Sánchez, José Esteban Muñoz-Medina, Tatiana Nunez-Garcia, Luis Alberto Ochoa-Carrera, Hector Esteban Paz-Juárez, Francisco Pulido, José Ernesto Ramírez-González, Alma Rincón-Rubio, Teresita Rojas-Mendoza, Jorge Salas-Hernández, Alejandro Sanchez-Flores, Clara Esperanza Santacruz-Tinoco, Andrea Santos Coy-Arechavaleta, Blanca Taboada, Gloria Vazquez, Joel Armando Vázquez-Pérez, Jerome Jean Verleyen |
| EPI_ISL_1302198 | Instituto Nacional de Enfermedades Respiratorias (INER) | Instituto de Biotecnología de la UNAM | Authors from IBT, IMSS, InDRE and INER (in alphabetical order): Carlos F. Arias, Santiago Ávila-Ríos, Gisela Barrera-Badillo, Eduardo Becerril-Vargas, Celia Boukadida, Natividad Cruz-Ortiz, Larissa Fernandes-Matano, Ricardo Grande, Lucia Hernandez-Rivas, Alejandra Hernández-Terán, Pavel Isa, Irma Lopez-Martinez, José Arturo Martínez-Orozco, Margarita Matías-Florentino, Fidencio Mejía-Nepomuceno, Edgar Mendieta-Condado, Mario Mújica-Sánchez, José Esteban Muñoz-Medina, Tatiana Nunez-Garcia, Luis Alberto Ochoa-Carrera, Hector Esteban Paz-Juárez, Francisco Pulido, José Ernesto Ramírez-González, Alma Rincón-Rubio, Teresita Rojas-Mendoza, Jorge Salas-Hernández, Alejandro Sanchez-Flores, Clara Esperanza Santacruz-Tinoco, Andrea Santos Coy-Arechavaleta, Blanca Taboada, Gloria Vazquez, Joel Armando Vázquez-Pérez, Jerome Jean Verleyen |
| EPI_ISL_1302199, EPI_ISL_1302200, EPI_ISL_1302201, EPI_ISL_1302202 | Instituto de Diagnostico y Referencia Epidemiologicos InDRE_RNLSP | Instituto de Biotecnología de la UNAM | Authors from IBT, IMSS, InDRE and INER (in alphabetical order): Carlos F. Arias, Santiago Ávila-Ríos, Gisela Barrera-Badillo, Eduardo Becerril-Vargas, Celia Boukadida, Natividad Cruz-Ortiz, Larissa Fernandes-Matano, Ricardo Grande, Lucia Hernandez-Rivas, Alejandra Hernández-Terán, Pavel Isa, Irma Lopez-Martinez, José Arturo Martínez-Orozco, Margarita Matías-Florentino, Fidencio Mejía-Nepomuceno, Edgar Mendieta-Condado, Mario Mújica-Sánchez, José Esteban Muñoz-Medina, Tatiana Nunez-Garcia, Luis Alberto Ochoa-Carrera, Hector Esteban Paz-Juárez, Francisco Pulido, José Ernesto Ramírez-González, Alma Rincón-Rubio, Teresita Rojas-Mendoza, Jorge Salas-Hernández, Alejandro Sanchez-Flores, Clara Esperanza Santacruz-Tinoco, Andrea Santos Coy-Arechavaleta, Blanca Taboada, Gloria Vazquez, Joel Armando Vázquez-Pérez, Jerome Jean Verleyen |
| EPI_ISL_1302206, EPI_ISL_1302207, EPI_ISL_1302208 | Instituto Nacional de Enfermedades Respiratorias (INER) | Instituto de Biotecnología de la UNAM | Authors from IBT, IMSS, InDRE and INER (in alphabetical order): Carlos F. Arias, Santiago Ávila-Ríos, Gisela Barrera-Badillo, Eduardo Becerril-Vargas, Celia Boukadida, Natividad Cruz-Ortiz, Larissa Fernandes-Matano, Ricardo Grande, Lucia Hernandez-Rivas, Alejandra Hernández-Terán, Pavel Isa, Irma Lopez-Martinez, José Arturo Martínez-Orozco, Margarita Matías-Florentino, Fidencio Mejía-Nepomuceno, Edgar Mendieta-Condado, Mario Mújica-Sánchez, José Esteban Muñoz-Medina, Tatiana Nunez-Garcia, Luis Alberto Ochoa-Carrera, Hector Esteban Paz-Juárez, Francisco Pulido, José Ernesto Ramírez-González, Alma Rincón-Rubio, Teresita Rojas-Mendoza, Jorge Salas-Hernández, Alejandro Sanchez-Flores, Clara Esperanza Santacruz-Tinoco, Andrea Santos Coy-Arechavaleta, Blanca Taboada, Gloria Vazquez, Joel Armando Vázquez-Pérez, Jerome Jean Verleyen |
| EPI_ISL_1302209, EPI_ISL_1302210, EPI_ISL_1302211, EPI_ISL_1302212, EPI_ISL_1302213, EPI_ISL_1302214, EPI_ISL_1302215, EPI_ISL_1302216, EPI_ISL_1302217, EPI_ISL_1302218, EPI_ISL_1302219 | | | |
| see above | Instituto de Diagnostico y Referencia Epidemiologicos InDRE_RNLSP | Instituto de Biotecnología de la UNAM | Authors from IBT, IMSS, InDRE and INER (in alphabetical order): Carlos F. Arias, Santiago Ávila-Ríos, Gisela Barrera-Badillo, Eduardo Becerril-Vargas, Celia Boukadida, Natividad Cruz-Ortiz, Larissa Fernandes-Matano, Ricardo Grande, Lucia Hernandez-Rivas, Alejandra Hernández-Terán, Pavel Isa, Irma |

| | | | |
|---|---|---|---|
| | | | Lopez-Martinez, José Arturo Martínez-Orozco, Margarita Matías-Florentino, Fidencio Mejía-Nepomuceno, Edgar Mendieta-Condado, Mario Mújica-Sánchez, José Esteban Muñoz-Medina, Tatiana Nunez-Garcia, Luis Alberto Ochoa-Carrera, Hector Esteban Paz-Juárez, Francisco Pulido, José Ernesto Ramírez-González, Alma Rincón-Rubio, Teresita Rojas-Mendoza, Jorge Salas-Hernández, Alejandro Sanchez-Flores, Clara Esperanza Santacruz-Tinoco, Andrea Santos Coy-Arechavaleta, Blanca Taboada, Gloria Vazquez, Joel Armando Vázquez-Pérez, Jerome Jean Verleyen |
| EPI_ISL_1302227, EPI_ISL_1302228, EPI_ISL_1302230, EPI_ISL_1302231, EPI_ISL_1302232, EPI_ISL_1302233, EPI_ISL_1302234 | Instituto Nacional de Enfermedades Respiratorias (INER) | Instituto de Biotecnología de la UNAM | Authors from IBT, IMSS, InDRE and INER (in alphabetical order): Carlos F. Arias, Santiago Ávila-Ríos, Gisela Barrera-Badillo, Eduardo Becerril-Vargas, Celia Boukadida, Natividad Cruz-Ortiz, Larissa Fernandes-Matano, Ricardo Grande, Lucia Hernandez-Rivas, Alejandra Hernández-Terán, Pavel Isa, Irma Lopez-Martinez, José Arturo Martínez-Orozco, Margarita Matías-Florentino, Fidencio Mejía-Nepomuceno, Edgar Mendieta-Condado, Mario Mújica-Sánchez, José Esteban Muñoz-Medina, Tatiana Nunez-Garcia, Luis Alberto Ochoa-Carrera, Hector Esteban Paz-Juárez, Francisco Pulido, José Ernesto Ramírez-González, Alma Rincón-Rubio, Teresita Rojas-Mendoza, Jorge Salas-Hernández, Alejandro Sanchez-Flores, Clara Esperanza Santacruz-Tinoco, Andrea Santos Coy-Arechavaleta, Blanca Taboada, Gloria Vazquez, Joel Armando Vázquez-Pérez, Jerome Jean Verleyen |
| EPI_ISL_1302241, EPI_ISL_1302242 | Instituto de Diagnostico y Referencia Epidemiologicos InDRE_RNLSP | Instituto de Biotecnología de la UNAM | Authors from IBT, IMSS, InDRE and INER (in alphabetical order): Carlos F. Arias, Santiago Ávila-Ríos, Gisela Barrera-Badillo, Eduardo Becerril-Vargas, Celia Boukadida, Natividad Cruz-Ortiz, Larissa Fernandes-Matano, Ricardo Grande, Lucia Hernandez-Rivas, Alejandra Hernández-Terán, Pavel Isa, Irma Lopez-Martinez, José Arturo Martínez-Orozco, Margarita Matías-Florentino, Fidencio Mejía-Nepomuceno, Edgar Mendieta-Condado, Mario Mújica-Sánchez, José Esteban Muñoz-Medina, Tatiana Nunez-Garcia, Luis Alberto Ochoa-Carrera, Hector Esteban Paz-Juárez, Francisco Pulido, José Ernesto Ramírez-González, Alma Rincón-Rubio, Teresita Rojas-Mendoza, Jorge Salas-Hernández, Alejandro Sanchez-Flores, Clara Esperanza Santacruz-Tinoco, Andrea Santos Coy-Arechavaleta, Blanca Taboada, Gloria Vazquez, Joel Armando Vázquez-Pérez, Jerome Jean Verleyen |
| EPI_ISL_1302245, EPI_ISL_1302246, EPI_ISL_1302247, EPI_ISL_1302249, EPI_ISL_1302250, EPI_ISL_1302253 | Instituto Nacional de Enfermedades Respiratorias (INER) | Instituto de Biotecnología de la UNAM | Authors from IBT, IMSS, InDRE and INER (in alphabetical order): Carlos F. Arias, Santiago Ávila-Ríos, Gisela Barrera-Badillo, Eduardo Becerril-Vargas, Celia Boukadida, Natividad Cruz-Ortiz, Larissa Fernandes-Matano, Ricardo Grande, Lucia Hernandez-Rivas, Alejandra Hernández-Terán, Pavel Isa, Irma Lopez-Martinez, José Arturo Martínez-Orozco, Margarita Matías-Florentino, Fidencio Mejía-Nepomuceno, Edgar Mendieta-Condado, Mario Mújica-Sánchez, José Esteban Muñoz-Medina, Tatiana Nunez-Garcia, Luis Alberto Ochoa-Carrera, Hector Esteban Paz-Juárez, Francisco Pulido, José Ernesto Ramírez-González, Alma Rincón-Rubio, Teresita Rojas-Mendoza, Jorge Salas-Hernández, Alejandro Sanchez-Flores, Clara Esperanza Santacruz-Tinoco, Andrea Santos Coy-Arechavaleta, Blanca Taboada, Gloria Vazquez, Joel Armando Vázquez-Pérez, Jerome Jean Verleyen |
| EPI_ISL_1302254 | Instituto de Diagnostico y Referencia Epidemiologicos InDRE_RNLSP | Instituto de Biotecnología de la UNAM | Authors from IBT, IMSS, InDRE and INER (in alphabetical order): Carlos F. Arias, Santiago Ávila-Ríos, Gisela Barrera-Badillo, Eduardo Becerril-Vargas, Celia Boukadida, Natividad Cruz-Ortiz, Larissa Fernandes-Matano, Ricardo Grande, Lucia Hernandez-Rivas, Alejandra Hernández-Terán, Pavel Isa, Irma Lopez-Martinez, José Arturo Martínez-Orozco, Margarita Matías-Florentino, Fidencio Mejía-Nepomuceno, Edgar Mendieta-Condado, Mario Mújica-Sánchez, José Esteban Muñoz-Medina, Tatiana Nunez-Garcia, Luis Alberto Ochoa-Carrera, Hector Esteban Paz-Juárez, Francisco Pulido, José Ernesto Ramírez-González, Alma Rincón-Rubio, Teresita Rojas-Mendoza, Jorge Salas-Hernández, Alejandro Sanchez-Flores, Clara Esperanza Santacruz-Tinoco, Andrea Santos Coy-Arechavaleta, Blanca Taboada, Gloria Vazquez, Joel Armando Vázquez-Pérez, Jerome Jean Verleyen |
| EPI_ISL_1302255, EPI_ISL_1302256, EPI_ISL_1302257, EPI_ISL_1302258 | Instituto Nacional de Enfermedades Respiratorias (INER) | Instituto de Biotecnología de la UNAM | Authors from IBT, IMSS, InDRE and INER (in alphabetical order): Carlos F. Arias, Santiago Ávila-Ríos, Gisela Barrera-Badillo, Eduardo Becerril-Vargas, Celia Boukadida, Natividad Cruz-Ortiz, Larissa Fernandes-Matano, Ricardo Grande, Lucia Hernandez-Rivas, Alejandra Hernández-Terán, Pavel Isa, Irma Lopez-Martinez, José Arturo Martínez-Orozco, Margarita Matías-Florentino, Fidencio Mejía-Nepomuceno, Edgar Mendieta-Condado, Mario Mújica-Sánchez, José Esteban Muñoz-Medina, Tatiana Nunez-Garcia, Luis Alberto Ochoa-Carrera, Hector Esteban Paz-Juárez, Francisco Pulido, José Ernesto Ramírez-González, Alma Rincón-Rubio, Teresita Rojas-Mendoza, Jorge Salas-Hernández, Alejandro Sanchez-Flores, Clara Esperanza Santacruz-Tinoco, Andrea Santos Coy-Arechavaleta, Blanca Taboada, Gloria Vazquez, Joel Armando Vázquez-Pérez, Jerome Jean Verleyen |
| EPI_ISL_1302259, EPI_ISL_1302262 | Instituto de Diagnostico y Referencia Epidemiologicos InDRE_RNLSP | Instituto de Biotecnología de la UNAM | Authors from IBT, IMSS, InDRE and INER (in alphabetical order): Carlos F. Arias, Santiago Ávila-Ríos, Gisela Barrera-Badillo, Eduardo Becerril-Vargas, Celia Boukadida, Natividad Cruz-Ortiz, Larissa Fernandes-Matano, Ricardo Grande, Lucia Hernandez-Rivas, Alejandra Hernández-Terán, Pavel Isa, Irma Lopez-Martinez, José Arturo Martínez-Orozco, Margarita Matías-Florentino, Fidencio Mejía-Nepomuceno, Edgar Mendieta-Condado, Mario Mújica-Sánchez, José Esteban Muñoz-Medina, Tatiana Nunez-Garcia, Luis Alberto Ochoa-Carrera, Hector Esteban Paz-Juárez, Francisco Pulido, José Ernesto Ramírez-González, Alma Rincón-Rubio, Teresita Rojas-Mendoza, Jorge Salas-Hernández, Alejandro Sanchez-Flores, Clara Esperanza Santacruz-Tinoco, Andrea Santos Coy-Arechavaleta, Blanca Taboada, Gloria Vazquez, Joel Armando Vázquez-Pérez, Jerome Jean Verleyen |
| EPI_ISL_1302263 | Instituto Nacional de Enfermedades Respiratorias (INER) | Instituto de Biotecnología de la UNAM | Authors from IBT, IMSS, InDRE and INER (in alphabetical order): Carlos F. Arias, Santiago Ávila-Ríos, Gisela Barrera-Badillo, Eduardo Becerril-Vargas, Celia Boukadida, Natividad Cruz-Ortiz, Larissa Fernandes-Matano, Ricardo Grande, Lucia Hernandez-Rivas, Alejandra Hernández-Terán, Pavel Isa, Irma Lopez-Martinez, José Arturo Martínez-Orozco, Margarita Matías-Florentino, Fidencio Mejía-Nepomuceno, Edgar Mendieta-Condado, Mario Mújica-Sánchez, José Esteban Muñoz-Medina, Tatiana Nunez-Garcia, Luis Alberto Ochoa-Carrera, Hector Esteban Paz-Juárez, Francisco Pulido, José Ernesto Ramírez-González, Alma Rincón-Rubio, Teresita Rojas-Mendoza, Jorge Salas-Hernández, Alejandro Sanchez-Flores, Clara Esperanza Santacruz-Tinoco, Andrea Santos Coy-Arechavaleta, Blanca Taboada, Gloria Vazquez, Joel Armando Vázquez-Pérez, Jerome Jean Verleyen |
| EPI_ISL_1302265 | Instituto de Diagnostico y Referencia Epidemiologicos InDRE_RNLSP | Instituto de Biotecnología de la UNAM | Authors from IBT, IMSS, InDRE and INER (in alphabetical order): Carlos F. Arias, Santiago Ávila-Ríos, Gisela Barrera-Badillo, Eduardo Becerril-Vargas, Celia Boukadida, Natividad Cruz-Ortiz, Larissa Fernandes-Matano, Ricardo Grande, Lucia Hernandez-Rivas, Alejandra Hernández-Terán, Pavel Isa, Irma Lopez-Martinez, José Arturo Martínez-Orozco, Margarita Matías-Florentino, Fidencio Mejía-Nepomuceno, Edgar Mendieta-Condado, Mario Mújica-Sánchez, José Esteban Muñoz-Medina, Tatiana Nunez-Garcia, Luis Alberto Ochoa-Carrera, Hector Esteban Paz-Juárez, Francisco Pulido, José Ernesto Ramírez-González, Alma Rincón-Rubio, Teresita Rojas-Mendoza, Jorge Salas-Hernández, Alejandro Sanchez-Flores, Clara Esperanza Santacruz-Tinoco, Andrea Santos Coy-Arechavaleta, Blanca Taboada, Gloria Vazquez, Joel Armando Vázquez-Pérez, Jerome Jean Verleyen |
| EPI_ISL_1302266, EPI_ISL_1302267, EPI_ISL_1302273 | Instituto Nacional de Enfermedades Respiratorias (INER) | Instituto de Biotecnología de la UNAM | Authors from IBT, IMSS, InDRE and INER (in alphabetical order): Carlos F. Arias, Santiago Ávila-Ríos, Gisela Barrera-Badillo, Eduardo Becerril-Vargas, Celia Boukadida, Natividad Cruz-Ortiz, Larissa Fernandes-Matano, Ricardo Grande, Lucia Hernandez-Rivas, Alejandra Hernández-Terán, Pavel Isa, Irma Lopez-Martinez, José Arturo Martínez-Orozco, Margarita Matías-Florentino, Fidencio Mejía-Nepomuceno, Edgar Mendieta-Condado, Mario Mújica-Sánchez, José Esteban Muñoz-Medina, Tatiana Nunez-Garcia, Luis Alberto Ochoa-Carrera, Hector Esteban Paz-Juárez, Francisco Pulido, José Ernesto Ramírez-González, Alma Rincón-Rubio, Teresita Rojas-Mendoza, Jorge Salas-Hernández, Alejandro Sanchez-Flores, Clara Esperanza Santacruz-Tinoco, Andrea Santos Coy-Arechavaleta, Blanca Taboada, Gloria Vazquez, Joel Armando Vázquez-Pérez, Jerome Jean Verleyen |
| EPI_ISL_1302274, EPI_ISL_1302276 | Instituto de Diagnostico y Referencia Epidemiologicos InDRE_RNLSP | Instituto de Biotecnología de la UNAM | Authors from IBT, IMSS, InDRE and INER (in alphabetical order): Carlos F. Arias, Santiago Ávila-Ríos, Gisela Barrera-Badillo, Eduardo Becerril-Vargas, Celia Boukadida, Natividad Cruz-Ortiz, Larissa Fernandes-Matano, Ricardo Grande, Lucia Hernandez-Rivas, Alejandra Hernández-Terán, Pavel Isa, Irma Lopez-Martinez, José Arturo Martínez-Orozco, Margarita Matías-Florentino, Fidencio Mejía-Nepomuceno, Edgar Mendieta-Condado, Mario Mújica-Sánchez, José Esteban Muñoz-Medina, Tatiana Nunez-Garcia, Luis Alberto Ochoa-Carrera, Hector Esteban Paz-Juárez, Francisco Pulido, José Ernesto Ramírez-González, Alma Rincón-Rubio, Teresita Rojas-Mendoza, Jorge Salas-Hernández, Alejandro Sanchez-Flores, Clara Esperanza Santacruz-Tinoco, Andrea Santos Coy-Arechavaleta, Blanca Taboada, Gloria Vazquez, Joel Armando Vázquez-Pérez, Jerome Jean Verleyen |
| EPI_ISL_1302278 | Instituto Nacional de Enfermedades Respiratorias (INER) | Instituto de Biotecnología de la UNAM | Authors from IBT, IMSS, InDRE and INER (in alphabetical order): Carlos F. Arias, Santiago Ávila-Ríos, Gisela Barrera-Badillo, Eduardo Becerril-Vargas, Celia Boukadida, Natividad Cruz-Ortiz, Larissa Fernandes-Matano, Ricardo Grande, Lucia Hernandez-Rivas, Alejandra Hernández-Terán, Pavel Isa, Irma Lopez-Martinez, José Arturo Martínez-Orozco, Margarita Matías-Florentino, Fidencio Mejía-Nepomuceno, Edgar Mendieta-Condado, Mario Mújica-Sánchez, José Esteban Muñoz-Medina, Tatiana Nunez-Garcia, Luis Alberto Ochoa-Carrera, Hector Esteban Paz-Juárez, Francisco Pulido, José Ernesto Ramírez-González, Alma Rincón-Rubio, Teresita Rojas-Mendoza, Jorge Salas-Hernández, Alejandro Sanchez-Flores, Clara Esperanza Santacruz-Tinoco, Andrea Santos Coy-Arechavaleta, Blanca Taboada, Gloria Vazquez, Joel Armando Vázquez-Pérez, Jerome Jean Verleyen |
| EPI_ISL_1302279, EPI_ISL_1302283, EPI_ISL_1302286, EPI_ISL_1302287, EPI_ISL_1302289 | Instituto de Diagnostico y Referencia Epidemiologicos InDRE_RNLSP | Instituto de Biotecnología de la UNAM | Authors from IBT, IMSS, InDRE and INER (in alphabetical order): Carlos F. Arias, Santiago Ávila-Ríos, Gisela Barrera-Badillo, Eduardo Becerril-Vargas, Celia Boukadida, Natividad Cruz-Ortiz, Larissa Fernandes-Matano, Ricardo Grande, Lucia Hernandez-Rivas, Alejandra Hernández-Terán, Pavel Isa, Irma Lopez-Martinez, José Arturo Martínez-Orozco, Margarita Matías-Florentino, Fidencio Mejía-Nepomuceno, Edgar Mendieta-Condado, Mario Mújica-Sánchez, José Esteban Muñoz-Medina, Tatiana Nunez-Garcia, Luis Alberto Ochoa-Carrera, Hector Esteban Paz-Juárez, Francisco Pulido, José Ernesto Ramírez-González, Alma Rincón-Rubio, Teresita Rojas-Mendoza, Jorge Salas-Hernández, Alejandro Sanchez-Flores, Clara Esperanza Santacruz-Tinoco, Andrea Santos Coy-Arechavaleta, Blanca Taboada, Gloria Vazquez, Joel Armando Vázquez-Pérez, Jerome Jean Verleyen |
| EPI_ISL_1302290, EPI_ISL_1302292 | Instituto Nacional de Enfermedades Respiratorias (INER) | Instituto de Biotecnología de la UNAM | Authors from IBT, IMSS, InDRE and INER (in alphabetical order): Carlos F. Arias, Santiago Ávila-Ríos, Gisela Barrera-Badillo, Eduardo Becerril-Vargas, Celia Boukadida, Natividad Cruz-Ortiz, Larissa Fernandes-Matano, Ricardo Grande, Lucia Hernandez-Rivas, Alejandra Hernández-Terán, Pavel Isa, Irma Lopez-Martinez, José Arturo Martínez-Orozco, Margarita Matías-Florentino, Fidencio Mejía-Nepomuceno, Edgar Mendieta-Condado, Mario |

| | | | |
|---|---|---|---|
| | | | Mújica-Sánchez, José Esteban Muñoz-Medina, Tatiana Nunez-Garcia, Luis Alberto Ochoa-Carrera, Hector Esteban Paz-Juárez, Francisco Pulido, José Ernesto Ramírez-González, Alma Rincón-Rubio, Teresita Rojas-Mendoza, Jorge Salas-Hernández, Alejandro Sanchez-Flores, Clara Esperanza Santacruz-Tinoco, Andrea Santos Coy-Arechavaleta, Blanca Taboada, Gloria Vazquez, Joel Armando Vázquez-Pérez, Jerome Jean Verleyen |
| EPI_ISL_1302293, EPI_ISL_1302295, EPI_ISL_1302296 | Instituto de Diagnostico y Referencia Epidemiologicos InDRE_RNLSP | Instituto de Biotecnología de la UNAM | Authors from IBT, IMSS, InDRE and INER (in alphabetical order): Carlos F. Arias, Santiago Ávila-Ríos, Gisela Barrera-Badillo, Eduardo Becerril-Vargas, Celia Boukadida, Natividad Cruz-Ortiz, Larissa Fernandes-Matano, Ricardo Grande, Lucia Hernandez-Rivas, Alejandra Hernández-Terán, Pavel Isa, Irma Lopez-Martinez, José Arturo Martínez-Orozco, Margarita Matías-Florentino, Fidencio Mejía-Nepomuceno, Edgar Mendieta-Condado, Mario Mújica-Sánchez, José Esteban Muñoz-Medina, Tatiana Nunez-Garcia, Luis Alberto Ochoa-Carrera, Hector Esteban Paz-Juárez, Francisco Pulido, José Ernesto Ramírez-González, Alma Rincón-Rubio, Teresita Rojas-Mendoza, Jorge Salas-Hernández, Alejandro Sanchez-Flores, Clara Esperanza Santacruz-Tinoco, Andrea Santos Coy-Arechavaleta, Blanca Taboada, Gloria Vazquez, Joel Armando Vázquez-Pérez, Jerome Jean Verleyen |
| EPI_ISL_1302297, EPI_ISL_1302298, EPI_ISL_1302299 | Instituto Nacional de Enfermedades Respiratorias (INER) | Instituto de Biotecnología de la UNAM | Authors from IBT, IMSS, InDRE and INER (in alphabetical order): Carlos F. Arias, Santiago Ávila-Ríos, Gisela Barrera-Badillo, Eduardo Becerril-Vargas, Celia Boukadida, Natividad Cruz-Ortiz, Larissa Fernandes-Matano, Ricardo Grande, Lucia Hernandez-Rivas, Alejandra Hernández-Terán, Pavel Isa, Irma Lopez-Martinez, José Arturo Martínez-Orozco, Margarita Matías-Florentino, Fidencio Mejía-Nepomuceno, Edgar Mendieta-Condado, Mario Mújica-Sánchez, José Esteban Muñoz-Medina, Tatiana Nunez-Garcia, Luis Alberto Ochoa-Carrera, Hector Esteban Paz-Juárez, Francisco Pulido, José Ernesto Ramírez-González, Alma Rincón-Rubio, Teresita Rojas-Mendoza, Jorge Salas-Hernández, Alejandro Sanchez-Flores, Clara Esperanza Santacruz-Tinoco, Andrea Santos Coy-Arechavaleta, Blanca Taboada, Gloria Vazquez, Joel Armando Vázquez-Pérez, Jerome Jean Verleyen |
| EPI_ISL_1302300 | Instituto de Diagnostico y Referencia Epidemiologicos InDRE_RNLSP | Instituto de Biotecnología de la UNAM | Authors from IBT, IMSS, InDRE and INER (in alphabetical order): Carlos F. Arias, Santiago Ávila-Ríos, Gisela Barrera-Badillo, Eduardo Becerril-Vargas, Celia Boukadida, Natividad Cruz-Ortiz, Larissa Fernandes-Matano, Ricardo Grande, Lucia Hernandez-Rivas, Alejandra Hernández-Terán, Pavel Isa, Irma Lopez-Martinez, José Arturo Martínez-Orozco, Margarita Matías-Florentino, Fidencio Mejía-Nepomuceno, Edgar Mendieta-Condado, Mario Mújica-Sánchez, José Esteban Muñoz-Medina, Tatiana Nunez-Garcia, Luis Alberto Ochoa-Carrera, Hector Esteban Paz-Juárez, Francisco Pulido, José Ernesto Ramírez-González, Alma Rincón-Rubio, Teresita Rojas-Mendoza, Jorge Salas-Hernández, Alejandro Sanchez-Flores, Clara Esperanza Santacruz-Tinoco, Andrea Santos Coy-Arechavaleta, Blanca Taboada, Gloria Vazquez, Joel Armando Vázquez-Pérez, Jerome Jean Verleyen |
| EPI_ISL_1302301, EPI_ISL_1302302, EPI_ISL_1302304, EPI_ISL_1302306, EPI_ISL_1302307, EPI_ISL_1302308 | Instituto Nacional de Enfermedades Respiratorias (INER) | Instituto de Biotecnología de la UNAM | Authors from IBT, IMSS, InDRE and INER (in alphabetical order): Carlos F. Arias, Santiago Ávila-Ríos, Gisela Barrera-Badillo, Eduardo Becerril-Vargas, Celia Boukadida, Natividad Cruz-Ortiz, Larissa Fernandes-Matano, Ricardo Grande, Lucia Hernandez-Rivas, Alejandra Hernández-Terán, Pavel Isa, Irma Lopez-Martinez, José Arturo Martínez-Orozco, Margarita Matías-Florentino, Fidencio Mejía-Nepomuceno, Edgar Mendieta-Condado, Mario Mújica-Sánchez, José Esteban Muñoz-Medina, Tatiana Nunez-Garcia, Luis Alberto Ochoa-Carrera, Hector Esteban Paz-Juárez, Francisco Pulido, José Ernesto Ramírez-González, Alma Rincón-Rubio, Teresita Rojas-Mendoza, Jorge Salas-Hernández, Alejandro Sanchez-Flores, Clara Esperanza Santacruz-Tinoco, Andrea Santos Coy-Arechavaleta, Blanca Taboada, Gloria Vazquez, Joel Armando Vázquez-Pérez, Jerome Jean Verleyen |
| EPI_ISL_1302309, EPI_ISL_1302310, EPI_ISL_1302313, EPI_ISL_1302314 | Instituto de Diagnostico y Referencia Epidemiologicos InDRE_RNLSP | Instituto de Biotecnología de la UNAM | Authors from IBT, IMSS, InDRE and INER (in alphabetical order): Carlos F. Arias, Santiago Ávila-Ríos, Gisela Barrera-Badillo, Eduardo Becerril-Vargas, Celia Boukadida, Natividad Cruz-Ortiz, Larissa Fernandes-Matano, Ricardo Grande, Lucia Hernandez-Rivas, Alejandra Hernández-Terán, Pavel Isa, Irma Lopez-Martinez, José Arturo Martínez-Orozco, Margarita Matías-Florentino, Fidencio Mejía-Nepomuceno, Edgar Mendieta-Condado, Mario Mújica-Sánchez, José Esteban Muñoz-Medina, Tatiana Nunez-Garcia, Luis Alberto Ochoa-Carrera, Hector Esteban Paz-Juárez, Francisco Pulido, José Ernesto Ramírez-González, Alma Rincón-Rubio, Teresita Rojas-Mendoza, Jorge Salas-Hernández, Alejandro Sanchez-Flores, Clara Esperanza Santacruz-Tinoco, Andrea Santos Coy-Arechavaleta, Blanca Taboada, Gloria Vazquez, Joel Armando Vázquez-Pérez, Jerome Jean Verleyen |
| EPI_ISL_1302316, EPI_ISL_1302317, EPI_ISL_1302319, EPI_ISL_1302320 | Instituto Nacional de Enfermedades Respiratorias (INER) | Instituto de Biotecnología de la UNAM | Authors from IBT, IMSS, InDRE and INER (in alphabetical order): Carlos F. Arias, Santiago Ávila-Ríos, Gisela Barrera-Badillo, Eduardo Becerril-Vargas, Celia Boukadida, Natividad Cruz-Ortiz, Larissa Fernandes-Matano, Ricardo Grande, Lucia Hernandez-Rivas, Alejandra Hernández-Terán, Pavel Isa, Irma Lopez-Martinez, José Arturo Martínez-Orozco, Margarita Matías-Florentino, Fidencio Mejía-Nepomuceno, Edgar Mendieta-Condado, Mario Mújica-Sánchez, José Esteban Muñoz-Medina, Tatiana Nunez-Garcia, Luis Alberto Ochoa-Carrera, Hector Esteban Paz-Juárez, Francisco Pulido, José Ernesto Ramírez-González, Alma Rincón-Rubio, Teresita Rojas-Mendoza, Jorge Salas-Hernández, Alejandro Sanchez-Flores, Clara Esperanza Santacruz-Tinoco, Andrea Santos Coy-Arechavaleta, Blanca Taboada, Gloria Vazquez, Joel Armando Vázquez-Pérez, Jerome Jean Verleyen |
| EPI_ISL_1302321, EPI_ISL_1302322, EPI_ISL_1302323 | Instituto de Diagnostico y Referencia Epidemiologicos InDRE_RNLSP | Instituto de Biotecnología de la UNAM | Authors from IBT, IMSS, InDRE and INER (in alphabetical order): Carlos F. Arias, Santiago Ávila-Ríos, Gisela Barrera-Badillo, Eduardo Becerril-Vargas, Celia Boukadida, Natividad Cruz-Ortiz, Larissa Fernandes-Matano, Ricardo Grande, Lucia Hernandez-Rivas, Alejandra Hernández-Terán, Pavel Isa, Irma Lopez-Martinez, José Arturo Martínez-Orozco, Margarita Matías-Florentino, Fidencio Mejía-Nepomuceno, Edgar Mendieta-Condado, Mario Mújica-Sánchez, José Esteban Muñoz-Medina, Tatiana Nunez-Garcia, Luis Alberto Ochoa-Carrera, Hector Esteban Paz-Juárez, Francisco Pulido, José Ernesto Ramírez-González, Alma Rincón-Rubio, Teresita Rojas-Mendoza, Jorge Salas-Hernández, Alejandro Sanchez-Flores, Clara Esperanza Santacruz-Tinoco, Andrea Santos Coy-Arechavaleta, Blanca Taboada, Gloria Vazquez, Joel Armando Vázquez-Pérez, Jerome Jean Verleyen |
| EPI_ISL_1302325, EPI_ISL_1302326, EPI_ISL_1302327, EPI_ISL_1302329, EPI_ISL_1302330 | Instituto Nacional de Enfermedades Respiratorias (INER) | Instituto de Biotecnología de la UNAM | Authors from IBT, IMSS, InDRE and INER (in alphabetical order): Carlos F. Arias, Santiago Ávila-Ríos, Gisela Barrera-Badillo, Eduardo Becerril-Vargas, Celia Boukadida, Natividad Cruz-Ortiz, Larissa Fernandes-Matano, Ricardo Grande, Lucia Hernandez-Rivas, Alejandra Hernández-Terán, Pavel Isa, Irma Lopez-Martinez, José Arturo Martínez-Orozco, Margarita Matías-Florentino, Fidencio Mejía-Nepomuceno, Edgar Mendieta-Condado, Mario Mújica-Sánchez, José Esteban Muñoz-Medina, Tatiana Nunez-Garcia, Luis Alberto Ochoa-Carrera, Hector Esteban Paz-Juárez, Francisco Pulido, José Ernesto Ramírez-González, Alma Rincón-Rubio, Teresita Rojas-Mendoza, Jorge Salas-Hernández, Alejandro Sanchez-Flores, Clara Esperanza Santacruz-Tinoco, Andrea Santos Coy-Arechavaleta, Blanca Taboada, Gloria Vazquez, Joel Armando Vázquez-Pérez, Jerome Jean Verleyen |
| EPI_ISL_1302331, EPI_ISL_1302333 | Instituto de Diagnostico y Referencia Epidemiologicos InDRE_RNLSP | Instituto de Biotecnología de la UNAM | Authors from IBT, IMSS, InDRE and INER (in alphabetical order): Carlos F. Arias, Santiago Ávila-Ríos, Gisela Barrera-Badillo, Eduardo Becerril-Vargas, Celia Boukadida, Natividad Cruz-Ortiz, Larissa Fernandes-Matano, Ricardo Grande, Lucia Hernandez-Rivas, Alejandra Hernández-Terán, Pavel Isa, Irma Lopez-Martinez, José Arturo Martínez-Orozco, Margarita Matías-Florentino, Fidencio Mejía-Nepomuceno, Edgar Mendieta-Condado, Mario Mújica-Sánchez, José Esteban Muñoz-Medina, Tatiana Nunez-Garcia, Luis Alberto Ochoa-Carrera, Hector Esteban Paz-Juárez, Francisco Pulido, José Ernesto Ramírez-González, Alma Rincón-Rubio, Teresita Rojas-Mendoza, Jorge Salas-Hernández, Alejandro Sanchez-Flores, Clara Esperanza Santacruz-Tinoco, Andrea Santos Coy-Arechavaleta, Blanca Taboada, Gloria Vazquez, Joel Armando Vázquez-Pérez, Jerome Jean Verleyen |
| EPI_ISL_1302334 | Instituto Nacional de Enfermedades Respiratorias (INER) | Instituto de Biotecnología de la UNAM | Authors from IBT, IMSS, InDRE and INER (in alphabetical order): Carlos F. Arias, Santiago Ávila-Ríos, Gisela Barrera-Badillo, Eduardo Becerril-Vargas, Celia Boukadida, Natividad Cruz-Ortiz, Larissa Fernandes-Matano, Ricardo Grande, Lucia Hernandez-Rivas, Alejandra Hernández-Terán, Pavel Isa, Irma Lopez-Martinez, José Arturo Martínez-Orozco, Margarita Matías-Florentino, Fidencio Mejía-Nepomuceno, Edgar Mendieta-Condado, Mario Mújica-Sánchez, José Esteban Muñoz-Medina, Tatiana Nunez-Garcia, Luis Alberto Ochoa-Carrera, Hector Esteban Paz-Juárez, Francisco Pulido, José Ernesto Ramírez-González, Alma Rincón-Rubio, Teresita Rojas-Mendoza, Jorge Salas-Hernández, Alejandro Sanchez-Flores, Clara Esperanza Santacruz-Tinoco, Andrea Santos Coy-Arechavaleta, Blanca Taboada, Gloria Vazquez, Joel Armando Vázquez-Pérez, Jerome Jean Verleyen |
| EPI_ISL_1302335 | Instituto de Diagnostico y Referencia Epidemiologicos InDRE_RNLSP | Instituto de Biotecnología de la UNAM | Authors from IBT, IMSS, InDRE and INER (in alphabetical order): Carlos F. Arias, Santiago Ávila-Ríos, Gisela Barrera-Badillo, Eduardo Becerril-Vargas, Celia Boukadida, Natividad Cruz-Ortiz, Larissa Fernandes-Matano, Ricardo Grande, Lucia Hernandez-Rivas, Alejandra Hernández-Terán, Pavel Isa, Irma Lopez-Martinez, José Arturo Martínez-Orozco, Margarita Matías-Florentino, Fidencio Mejía-Nepomuceno, Edgar Mendieta-Condado, Mario Mújica-Sánchez, José Esteban Muñoz-Medina, Tatiana Nunez-Garcia, Luis Alberto Ochoa-Carrera, Hector Esteban Paz-Juárez, Francisco Pulido, José Ernesto Ramírez-González, Alma Rincón-Rubio, Teresita Rojas-Mendoza, Jorge Salas-Hernández, Alejandro Sanchez-Flores, Clara Esperanza Santacruz-Tinoco, Andrea Santos Coy-Arechavaleta, Blanca Taboada, Gloria Vazquez, Joel Armando Vázquez-Pérez, Jerome Jean Verleyen |
| EPI_ISL_1302336 | Instituto Nacional de Enfermedades Respiratorias (INER) | Instituto de Biotecnología de la UNAM | Authors from IBT, IMSS, InDRE and INER (in alphabetical order): Carlos F. Arias, Santiago Ávila-Ríos, Gisela Barrera-Badillo, Eduardo Becerril-Vargas, Celia Boukadida, Natividad Cruz-Ortiz, Larissa Fernandes-Matano, Ricardo Grande, Lucia Hernandez-Rivas, Alejandra Hernández-Terán, Pavel Isa, Irma Lopez-Martinez, José Arturo Martínez-Orozco, Margarita Matías-Florentino, Fidencio Mejía-Nepomuceno, Edgar Mendieta-Condado, Mario Mújica-Sánchez, José Esteban Muñoz-Medina, Tatiana Nunez-Garcia, Luis Alberto Ochoa-Carrera, Hector Esteban Paz-Juárez, Francisco Pulido, José Ernesto Ramírez-González, Alma Rincón-Rubio, Teresita Rojas-Mendoza, Jorge Salas-Hernández, Alejandro Sanchez-Flores, Clara Esperanza Santacruz-Tinoco, Andrea Santos Coy-Arechavaleta, Blanca Taboada, Gloria Vazquez, Joel Armando Vázquez-Pérez, Jerome Jean Verleyen |
| EPI_ISL_1302337 | Instituto de Diagnostico y Referencia Epidemiologicos InDRE_RNLSP | Instituto de Biotecnología de la UNAM | Authors from IBT, IMSS, InDRE and INER (in alphabetical order): Carlos F. Arias, Santiago Ávila-Ríos, Gisela Barrera-Badillo, Eduardo Becerril-Vargas, Celia Boukadida, Natividad Cruz-Ortiz, Larissa Fernandes-Matano, Ricardo Grande, Lucia Hernandez-Rivas, Alejandra Hernández-Terán, Pavel Isa, Irma Lopez-Martinez, José Arturo Martínez-Orozco, Margarita Matías-Florentino, Fidencio Mejía-Nepomuceno, Edgar Mendieta-Condado, Mario Mújica-Sánchez, José Esteban Muñoz-Medina, Tatiana Nunez-Garcia, Luis Alberto Ochoa-Carrera, Hector Esteban Paz-Juárez, Francisco Pulido, José Ernesto Ramírez-González, Alma Rincón-Rubio, Teresita Rojas-Mendoza, Jorge Salas-Hernández, Alejandro Sanchez-Flores, Clara Esperanza Santacruz-Tinoco, Andrea Santos Coy-Arechavaleta, Blanca Taboada, Gloria Vazquez, Joel Armando Vázquez-Pérez, Jerome Jean Verleyen |

| | | | |
|---|---|---|---|
| EPI_ISL_1302339 | Instituto Nacional de Enfermedades Respiratorias (INER) | Instituto de Biotecnología de la UNAM | Authors from IBT, IMSS, InDRE and INER (in alphabetical order): Carlos F. Arias, Santiago Ávila-Ríos, Gisela Barrera-Badillo, Eduardo Becerril-Vargas, Celia Boukadida, Natividad Cruz-Ortiz, Larissa Fernandes-Matano, Ricardo Grande, Lucia Hernandez-Rivas, Alejandra Hernández-Terán, Pavel Isa, Irma Lopez-Martinez, José Arturo Martínez-Orozco, Margarita Matías-Florentino, Fidencio Mejía-Nepomuceno, Edgar Mendieta-Condado, Mario Mújica-Sánchez, José Esteban Muñoz-Medina, Tatiana Nunez-Garcia, Luis Alberto Ochoa-Carrera, Hector Esteban Paz-Juárez, Francisco Pulido, José Ernesto Ramírez-González, Alma Rincón-Rubio, Teresita Rojas-Mendoza, Jorge Salas-Hernández, Alejandro Sanchez-Flores, Clara Esperanza Santacruz-Tinoco, Andrea Santos Coy-Arechavaleta, Blanca Taboada, Gloria Vazquez, Joel Armando Vázquez-Pérez, Jerome Jean Verleyen |
| EPI_ISL_1302340, EPI_ISL_1302342, EPI_ISL_1302343, EPI_ISL_1302344 | Instituto de Diagnostico y Referencia Epidemiologicos InDRE_RNLSP | Instituto de Biotecnología de la UNAM | Authors from IBT, IMSS, InDRE and INER (in alphabetical order): Carlos F. Arias, Santiago Ávila-Ríos, Gisela Barrera-Badillo, Eduardo Becerril-Vargas, Celia Boukadida, Natividad Cruz-Ortiz, Larissa Fernandes-Matano, Ricardo Grande, Lucia Hernandez-Rivas, Alejandra Hernández-Terán, Pavel Isa, Irma Lopez-Martinez, José Arturo Martínez-Orozco, Margarita Matías-Florentino, Fidencio Mejía-Nepomuceno, Edgar Mendieta-Condado, Mario Mújica-Sánchez, José Esteban Muñoz-Medina, Tatiana Nunez-Garcia, Luis Alberto Ochoa-Carrera, Hector Esteban Paz-Juárez, Francisco Pulido, José Ernesto Ramírez-González, Alma Rincón-Rubio, Teresita Rojas-Mendoza, Jorge Salas-Hernández, Alejandro Sanchez-Flores, Clara Esperanza Santacruz-Tinoco, Andrea Santos Coy-Arechavaleta, Blanca Taboada, Gloria Vazquez, Joel Armando Vázquez-Pérez, Jerome Jean Verleyen |
| EPI_ISL_1302345, EPI_ISL_1302346, EPI_ISL_1302349, EPI_ISL_1302350, EPI_ISL_1302356, EPI_ISL_1302357 | Instituto Nacional de Enfermedades Respiratorias (INER) | Instituto de Biotecnología de la UNAM | Authors from IBT, IMSS, InDRE and INER (in alphabetical order): Carlos F. Arias, Santiago Ávila-Ríos, Gisela Barrera-Badillo, Eduardo Becerril-Vargas, Celia Boukadida, Natividad Cruz-Ortiz, Larissa Fernandes-Matano, Ricardo Grande, Lucia Hernandez-Rivas, Alejandra Hernández-Terán, Pavel Isa, Irma Lopez-Martinez, José Arturo Martínez-Orozco, Margarita Matías-Florentino, Fidencio Mejía-Nepomuceno, Edgar Mendieta-Condado, Mario Mújica-Sánchez, José Esteban Muñoz-Medina, Tatiana Nunez-Garcia, Luis Alberto Ochoa-Carrera, Hector Esteban Paz-Juárez, Francisco Pulido, José Ernesto Ramírez-González, Alma Rincón-Rubio, Teresita Rojas-Mendoza, Jorge Salas-Hernández, Alejandro Sanchez-Flores, Clara Esperanza Santacruz-Tinoco, Andrea Santos Coy-Arechavaleta, Blanca Taboada, Gloria Vazquez, Joel Armando Vázquez-Pérez, Jerome Jean Verleyen |
| EPI_ISL_1302358 | Instituto de Diagnostico y Referencia Epidemiologicos InDRE_RNLSP | Instituto de Biotecnología de la UNAM | Authors from IBT, IMSS, InDRE and INER (in alphabetical order): Carlos F. Arias, Santiago Ávila-Ríos, Gisela Barrera-Badillo, Eduardo Becerril-Vargas, Celia Boukadida, Natividad Cruz-Ortiz, Larissa Fernandes-Matano, Ricardo Grande, Lucia Hernandez-Rivas, Alejandra Hernández-Terán, Pavel Isa, Irma Lopez-Martinez, José Arturo Martínez-Orozco, Margarita Matías-Florentino, Fidencio Mejía-Nepomuceno, Edgar Mendieta-Condado, Mario Mújica-Sánchez, José Esteban Muñoz-Medina, Tatiana Nunez-Garcia, Luis Alberto Ochoa-Carrera, Hector Esteban Paz-Juárez, Francisco Pulido, José Ernesto Ramírez-González, Alma Rincón-Rubio, Teresita Rojas-Mendoza, Jorge Salas-Hernández, Alejandro Sanchez-Flores, Clara Esperanza Santacruz-Tinoco, Andrea Santos Coy-Arechavaleta, Blanca Taboada, Gloria Vazquez, Joel Armando Vázquez-Pérez, Jerome Jean Verleyen |
| EPI_ISL_1302360, EPI_ISL_1302361 | Instituto Nacional de Enfermedades Respiratorias (INER) | Instituto de Biotecnología de la UNAM | Authors from IBT, IMSS, InDRE and INER (in alphabetical order): Carlos F. Arias, Santiago Ávila-Ríos, Gisela Barrera-Badillo, Eduardo Becerril-Vargas, Celia Boukadida, Natividad Cruz-Ortiz, Larissa Fernandes-Matano, Ricardo Grande, Lucia Hernandez-Rivas, Alejandra Hernández-Terán, Pavel Isa, Irma Lopez-Martinez, José Arturo Martínez-Orozco, Margarita Matías-Florentino, Fidencio Mejía-Nepomuceno, Edgar Mendieta-Condado, Mario Mújica-Sánchez, José Esteban Muñoz-Medina, Tatiana Nunez-Garcia, Luis Alberto Ochoa-Carrera, Hector Esteban Paz-Juárez, Francisco Pulido, José Ernesto Ramírez-González, Alma Rincón-Rubio, Teresita Rojas-Mendoza, Jorge Salas-Hernández, Alejandro Sanchez-Flores, Clara Esperanza Santacruz-Tinoco, Andrea Santos Coy-Arechavaleta, Blanca Taboada, Gloria Vazquez, Joel Armando Vázquez-Pérez, Jerome Jean Verleyen |
| EPI_ISL_1302363 | Instituto de Diagnostico y Referencia Epidemiologicos InDRE_RNLSP | Instituto de Biotecnología de la UNAM | Authors from IBT, IMSS, InDRE and INER (in alphabetical order): Carlos F. Arias, Santiago Ávila-Ríos, Gisela Barrera-Badillo, Eduardo Becerril-Vargas, Celia Boukadida, Natividad Cruz-Ortiz, Larissa Fernandes-Matano, Ricardo Grande, Lucia Hernandez-Rivas, Alejandra Hernández-Terán, Pavel Isa, Irma Lopez-Martinez, José Arturo Martínez-Orozco, Margarita Matías-Florentino, Fidencio Mejía-Nepomuceno, Edgar Mendieta-Condado, Mario Mújica-Sánchez, José Esteban Muñoz-Medina, Tatiana Nunez-Garcia, Luis Alberto Ochoa-Carrera, Hector Esteban Paz-Juárez, Francisco Pulido, José Ernesto Ramírez-González, Alma Rincón-Rubio, Teresita Rojas-Mendoza, Jorge Salas-Hernández, Alejandro Sanchez-Flores, Clara Esperanza Santacruz-Tinoco, Andrea Santos Coy-Arechavaleta, Blanca Taboada, Gloria Vazquez, Joel Armando Vázquez-Pérez, Jerome Jean Verleyen |
| EPI_ISL_1302364, EPI_ISL_1302365, EPI_ISL_1302366, EPI_ISL_1302368, EPI_ISL_1302369 | Instituto Nacional de Enfermedades Respiratorias (INER) | Instituto de Biotecnología de la UNAM | Authors from IBT, IMSS, InDRE and INER (in alphabetical order): Carlos F. Arias, Santiago Ávila-Ríos, Gisela Barrera-Badillo, Eduardo Becerril-Vargas, Celia Boukadida, Natividad Cruz-Ortiz, Larissa Fernandes-Matano, Ricardo Grande, Lucia Hernandez-Rivas, Alejandra Hernández-Terán, Pavel Isa, Irma Lopez-Martinez, José Arturo Martínez-Orozco, Margarita Matías-Florentino, Fidencio Mejía-Nepomuceno, Edgar Mendieta-Condado, Mario Mújica-Sánchez, José Esteban Muñoz-Medina, Tatiana Nunez-Garcia, Luis Alberto Ochoa-Carrera, Hector Esteban Paz-Juárez, Francisco Pulido, José Ernesto Ramírez-González, Alma Rincón-Rubio, Teresita Rojas-Mendoza, Jorge Salas-Hernández, Alejandro Sanchez-Flores, Clara Esperanza Santacruz-Tinoco, Andrea Santos Coy-Arechavaleta, Blanca Taboada, Gloria Vazquez, Joel Armando Vázquez-Pérez, Jerome Jean Verleyen |
| EPI_ISL_1302371 | Instituto de Diagnostico y Referencia Epidemiologicos InDRE_RNLSP | Instituto de Biotecnología de la UNAM | Authors from IBT, IMSS, InDRE and INER (in alphabetical order): Carlos F. Arias, Santiago Ávila-Ríos, Gisela Barrera-Badillo, Eduardo Becerril-Vargas, Celia Boukadida, Natividad Cruz-Ortiz, Larissa Fernandes-Matano, Ricardo Grande, Lucia Hernandez-Rivas, Alejandra Hernández-Terán, Pavel Isa, Irma Lopez-Martinez, José Arturo Martínez-Orozco, Margarita Matías-Florentino, Fidencio Mejía-Nepomuceno, Edgar Mendieta-Condado, Mario Mújica-Sánchez, José Esteban Muñoz-Medina, Tatiana Nunez-Garcia, Luis Alberto Ochoa-Carrera, Hector Esteban Paz-Juárez, Francisco Pulido, José Ernesto Ramírez-González, Alma Rincón-Rubio, Teresita Rojas-Mendoza, Jorge Salas-Hernández, Alejandro Sanchez-Flores, Clara Esperanza Santacruz-Tinoco, Andrea Santos Coy-Arechavaleta, Blanca Taboada, Gloria Vazquez, Joel Armando Vázquez-Pérez, Jerome Jean Verleyen |
| EPI_ISL_1302373, EPI_ISL_1302374, EPI_ISL_1302375, EPI_ISL_1302376, EPI_ISL_1302377 | Instituto Nacional de Enfermedades Respiratorias (INER) | Instituto de Biotecnología de la UNAM | Authors from IBT, IMSS, InDRE and INER (in alphabetical order): Carlos F. Arias, Santiago Ávila-Ríos, Gisela Barrera-Badillo, Eduardo Becerril-Vargas, Celia Boukadida, Natividad Cruz-Ortiz, Larissa Fernandes-Matano, Ricardo Grande, Lucia Hernandez-Rivas, Alejandra Hernández-Terán, Pavel Isa, Irma Lopez-Martinez, José Arturo Martínez-Orozco, Margarita Matías-Florentino, Fidencio Mejía-Nepomuceno, Edgar Mendieta-Condado, Mario Mújica-Sánchez, José Esteban Muñoz-Medina, Tatiana Nunez-Garcia, Luis Alberto Ochoa-Carrera, Hector Esteban Paz-Juárez, Francisco Pulido, José Ernesto Ramírez-González, Alma Rincón-Rubio, Teresita Rojas-Mendoza, Jorge Salas-Hernández, Alejandro Sanchez-Flores, Clara Esperanza Santacruz-Tinoco, Andrea Santos Coy-Arechavaleta, Blanca Taboada, Gloria Vazquez, Joel Armando Vázquez-Pérez, Jerome Jean Verleyen |
| EPI_ISL_1302380, EPI_ISL_1302382 | Instituto de Diagnostico y Referencia Epidemiologicos InDRE_RNLSP | Instituto de Biotecnología de la UNAM | Authors from IBT, IMSS, InDRE and INER (in alphabetical order): Carlos F. Arias, Santiago Ávila-Ríos, Gisela Barrera-Badillo, Eduardo Becerril-Vargas, Celia Boukadida, Natividad Cruz-Ortiz, Larissa Fernandes-Matano, Ricardo Grande, Lucia Hernandez-Rivas, Alejandra Hernández-Terán, Pavel Isa, Irma Lopez-Martinez, José Arturo Martínez-Orozco, Margarita Matías-Florentino, Fidencio Mejía-Nepomuceno, Edgar Mendieta-Condado, Mario Mújica-Sánchez, José Esteban Muñoz-Medina, Tatiana Nunez-Garcia, Luis Alberto Ochoa-Carrera, Hector Esteban Paz-Juárez, Francisco Pulido, José Ernesto Ramírez-González, Alma Rincón-Rubio, Teresita Rojas-Mendoza, Jorge Salas-Hernández, Alejandro Sanchez-Flores, Clara Esperanza Santacruz-Tinoco, Andrea Santos Coy-Arechavaleta, Blanca Taboada, Gloria Vazquez, Joel Armando Vázquez-Pérez, Jerome Jean Verleyen |
| EPI_ISL_1302383, EPI_ISL_1302384, EPI_ISL_1302385, EPI_ISL_1302386, EPI_ISL_1302387 | Instituto Nacional de Enfermedades Respiratorias (INER) | Instituto de Biotecnología de la UNAM | Authors from IBT, IMSS, InDRE and INER (in alphabetical order): Carlos F. Arias, Santiago Ávila-Ríos, Gisela Barrera-Badillo, Eduardo Becerril-Vargas, Celia Boukadida, Natividad Cruz-Ortiz, Larissa Fernandes-Matano, Ricardo Grande, Lucia Hernandez-Rivas, Alejandra Hernández-Terán, Pavel Isa, Irma Lopez-Martinez, José Arturo Martínez-Orozco, Margarita Matías-Florentino, Fidencio Mejía-Nepomuceno, Edgar Mendieta-Condado, Mario Mújica-Sánchez, José Esteban Muñoz-Medina, Tatiana Nunez-Garcia, Luis Alberto Ochoa-Carrera, Hector Esteban Paz-Juárez, Francisco Pulido, José Ernesto Ramírez-González, Alma Rincón-Rubio, Teresita Rojas-Mendoza, Jorge Salas-Hernández, Alejandro Sanchez-Flores, Clara Esperanza Santacruz-Tinoco, Andrea Santos Coy-Arechavaleta, Blanca Taboada, Gloria Vazquez, Joel Armando Vázquez-Pérez, Jerome Jean Verleyen |
| EPI_ISL_1302388 | Instituto de Diagnostico y Referencia Epidemiologicos InDRE_RNLSP | Instituto de Biotecnología de la UNAM | Authors from IBT, IMSS, InDRE and INER (in alphabetical order): Carlos F. Arias, Santiago Ávila-Ríos, Gisela Barrera-Badillo, Eduardo Becerril-Vargas, Celia Boukadida, Natividad Cruz-Ortiz, Larissa Fernandes-Matano, Ricardo Grande, Lucia Hernandez-Rivas, Alejandra Hernández-Terán, Pavel Isa, Irma Lopez-Martinez, José Arturo Martínez-Orozco, Margarita Matías-Florentino, Fidencio Mejía-Nepomuceno, Edgar Mendieta-Condado, Mario Mújica-Sánchez, José Esteban Muñoz-Medina, Tatiana Nunez-Garcia, Luis Alberto Ochoa-Carrera, Hector Esteban Paz-Juárez, Francisco Pulido, José Ernesto Ramírez-González, Alma Rincón-Rubio, Teresita Rojas-Mendoza, Jorge Salas-Hernández, Alejandro Sanchez-Flores, Clara Esperanza Santacruz-Tinoco, Andrea Santos Coy-Arechavaleta, Blanca Taboada, Gloria Vazquez, Joel Armando Vázquez-Pérez, Jerome Jean Verleyen |
| EPI_ISL_1302389, EPI_ISL_1302391, EPI_ISL_1302394, EPI_ISL_1302395 | Instituto Nacional de Enfermedades Respiratorias (INER) | Instituto de Biotecnología de la UNAM | Authors from IBT, IMSS, InDRE and INER (in alphabetical order): Carlos F. Arias, Santiago Ávila-Ríos, Gisela Barrera-Badillo, Eduardo Becerril-Vargas, Celia Boukadida, Natividad Cruz-Ortiz, Larissa Fernandes-Matano, Ricardo Grande, Lucia Hernandez-Rivas, Alejandra Hernández-Terán, Pavel Isa, Irma Lopez-Martinez, José Arturo Martínez-Orozco, Margarita Matías-Florentino, Fidencio Mejía-Nepomuceno, Edgar Mendieta-Condado, Mario Mújica-Sánchez, José Esteban Muñoz-Medina, Tatiana Nunez-Garcia, Luis Alberto Ochoa-Carrera, Hector Esteban Paz-Juárez, Francisco Pulido, José Ernesto Ramírez-González, Alma Rincón-Rubio, Teresita Rojas-Mendoza, Jorge Salas-Hernández, Alejandro Sanchez-Flores, Clara Esperanza Santacruz-Tinoco, Andrea Santos Coy-Arechavaleta, Blanca Taboada, Gloria Vazquez, Joel Armando Vázquez-Pérez, Jerome Jean Verleyen |
| EPI_ISL_1302396, EPI_ISL_1302397, EPI_ISL_1302398 | Instituto de Diagnostico y Referencia Epidemiologicos InDRE_RNLSP | Instituto de Biotecnología de la UNAM | Authors from IBT, IMSS, InDRE and INER (in alphabetical order): Carlos F. Arias, Santiago Ávila-Ríos, Gisela Barrera-Badillo, Eduardo Becerril-Vargas, Celia Boukadida, Natividad Cruz-Ortiz, Larissa Fernandes-Matano, Ricardo Grande, Lucia Hernandez-Rivas, Alejandra Hernández-Terán, Pavel Isa, Irma Lopez-Martinez, José Arturo Martínez-Orozco, Margarita Matías-Florentino, Fidencio Mejía-Nepomuceno, Edgar Mendieta-Condado, Mario |

| | | | |
|---|---|---|---|
| | | | Mújica-Sánchez, José Esteban Muñoz-Medina, Tatiana Nunez-Garcia, Luis Alberto Ochoa-Carrera, Hector Esteban Paz-Juárez, Francisco Pulido, José Ernesto Ramírez-González, Alma Rincón-Rubio, Teresita Rojas-Mendoza, Jorge Salas-Hernández, Alejandro Sanchez-Flores, Clara Esperanza Santacruz-Tinoco, Andrea Santos Coy-Arechavaleta, Blanca Taboada, Gloria Vazquez, Joel Armando Vázquez-Pérez, Jerome Jean Verleyen |
| EPI_ISL_1302399, EPI_ISL_1302400 | Instituto Nacional de Enfermedades Respiratorias (INER) | Instituto de Biotecnología de la UNAM | Authors from IBT, IMSS, InDRE and INER (in alphabetical order): Carlos F. Arias, Santiago Ávila-Ríos, Gisela Barrera-Badillo, Eduardo Becerril-Vargas, Celia Boukadida, Natividad Cruz-Ortiz, Larissa Fernandes-Matano, Ricardo Grande, Lucia Hernandez-Rivas, Alejandra Hernández-Terán, Pavel Isa, Irma Lopez-Martinez, José Arturo Martínez-Orozco, Margarita Matías-Florentino, Fidencio Mejía-Nepomuceno, Edgar Mendieta-Condado, Mario Mújica-Sánchez, José Esteban Muñoz-Medina, Tatiana Nunez-Garcia, Luis Alberto Ochoa-Carrera, Hector Esteban Paz-Juárez, Francisco Pulido, José Ernesto Ramírez-González, Alma Rincón-Rubio, Teresita Rojas-Mendoza, Jorge Salas-Hernández, Alejandro Sanchez-Flores, Clara Esperanza Santacruz-Tinoco, Andrea Santos Coy-Arechavaleta, Blanca Taboada, Gloria Vazquez, Joel Armando Vázquez-Pérez, Jerome Jean Verleyen |
| EPI_ISL_1324764, EPI_ISL_1324765, EPI_ISL_1324766 | Laboratorio de Diagnostico de Enfermedades Emergentes y Reemergentes | Instituto de Diagnostico y Referencia Epidemiologicos (INDRE) | Claudia Wong-Arambula, Abril Rodriguez-Maldonado, Vanessa Rivero-Arredondo, Ariadna Medina-Benitez, Joaquin Quiroz-Mercado, Sergio Rangel-Guerrero,Natali Vega-Magana, Natividad Cruz-Ortiz, Tatiana Nunez-Garcia, Gisela Barrera-Badillo, Lucia Hernandez-Rivas, Irma Lopez-Martinez, Ernesto Ramirez-Gonzalez. |
| EPI_ISL_1334381 | LESP Guerrero | Instituto de Diagnostico y Referencia Epidemiologicos (INDRE) | Claudia Wong-Arambula, Abril Rodriguez-Maldonado, Vanessa Rivero-Arredondo, Ariadna Medina-Benitez, Joaquin Quiroz-Mercado, Sergio Rangel-Guerrero, Natividad Cruz-Ortiz, Tatiana Nunez-Garcia, Gisela Barrera-Badillo, Lucia Hernandez-Rivas, Irma Lopez-Martinez, Ernesto Ramirez-Gonzalez. |
| EPI_ISL_1337395 | LESP Colima | Instituto de Diagnostico y Referencia Epidemiologicos (INDRE) | Claudia Wong-Arambula, Abril Rodriguez-Maldonado, Vanessa Rivero-Arredondo, Ariadna Medina-Benitez, Joaquin Quiroz-Mercado, Sergio Rangel-Guerrero, Natividad Cruz-Ortiz, Tatiana Nunez-Garcia, Gisela Barrera-Badillo, Lucia Hernandez-Rivas, Irma Lopez-Martinez, Ernesto Ramirez-Gonzalez. |
| EPI_ISL_1360407, EPI_ISL_1360408, EPI_ISL_1360409, EPI_ISL_1360410, EPI_ISL_1360411, EPI_ISL_1360412 | Laboratorio de Enfermedades Emergentes y Reemergentes | Laboratorio de Enfermedades Emergentes y Reemergentes | Vega-Magaña Natali, Peña-Rodriguez Marcela, Viera-Segura Oliver, García-Chagollan Mariel, Hernández-Bello Jorge, Muñoz-Valle José Francisco |
| EPI_ISL_1366326, EPI_ISL_1366327, EPI_ISL_1366328, EPI_ISL_1366329, EPI_ISL_1366330 | Genomica Lab Molecular, Mexico | Andersen lab at Scripps Research | SEARCH Alliance San Diego with Jonathan Gonzalez Garcia, Jose Roman Chavez Mendez, Jose Horacio Reyna Verdugo, Martin Gonzalez Ibarra, Luis Alberto Rangel Gonzalez |
| EPI_ISL_1446868, EPI_ISL_1446874, EPI_ISL_1446876, EPI_ISL_1446878, EPI_ISL_1446884, EPI_ISL_1446885, EPI_ISL_1446886, EPI_ISL_1446888, EPI_ISL_1446901, EPI_ISL_1446903, EPI_ISL_1446907, EPI_ISL_1446908, EPI_ISL_1446912 | | | |
| see above | "NM Dept. Health, Scientific Laboratory Division " | Centers for Disease Control and Prevention Division of Viral Diseases, Pathogen Discovery | Mili Sheth, Sarah Nobles, Jasmine Padilla, Mark Burroughs, Shoshona Le, Katie Dillon, Peter Cook, Clinton R. Paden, Dhwani Batra, Krista Queen, Kristen Knipe, Dakota Howard, Yvette Unoarumhi, Darlene Wagner, Matthew Schmerer, Ben L. Rambo-Martin, Kristine Lacek, Sam Shepard, Alison Laufer Halpin, Dave Wentworth, Vivien Dugan, Suxiang Tong, Justin Lee |
| EPI_ISL_1469108, EPI_ISL_1469109, EPI_ISL_1469110, EPI_ISL_1469111, EPI_ISL_1469112, EPI_ISL_1469113, EPI_ISL_1469114, EPI_ISL_1469115, EPI_ISL_1469116, EPI_ISL_1469117, EPI_ISL_1469118, EPI_ISL_1469119 | | | |
| see above | Centro de Investigacion en Ciencias de la Salud y Biomedicina | CINVESTAV | Sofia Bernal, Andreu Comas |
| EPI_ISL_1493389, EPI_ISL_1493390, EPI_ISL_1493391, EPI_ISL_1493392, EPI_ISL_1493393, EPI_ISL_1493394, EPI_ISL_1493395, EPI_ISL_1493396, EPI_ISL_1493397, EPI_ISL_1493398, EPI_ISL_1493400, EPI_ISL_1493401, EPI_ISL_1493402, EPI_ISL_1493403, EPI_ISL_1493404, EPI_ISL_1493405, EPI_ISL_1493406, EPI_ISL_1493407, EPI_ISL_1493408, EPI_ISL_1493409, EPI_ISL_1493410, EPI_ISL_1493411, EPI_ISL_1493412, EPI_ISL_1493413, EPI_ISL_1493414, EPI_ISL_1493415, EPI_ISL_1493416, EPI_ISL_1493417, EPI_ISL_1493418, EPI_ISL_1493419, EPI_ISL_1493420, EPI_ISL_1493421, EPI_ISL_1493422, EPI_ISL_1493423, EPI_ISL_1493424, EPI_ISL_1493425, EPI_ISL_1493426, EPI_ISL_1493427, EPI_ISL_1493428, EPI_ISL_1493429, EPI_ISL_1493430, EPI_ISL_1493431, EPI_ISL_1493432, EPI_ISL_1493433, EPI_ISL_1493434, EPI_ISL_1493435 | | | |
| see above | New Mexico Department of Health Scientific Laboratory | New Mexico Department of Health Scientific Laboratory | Ellie Johnson, Anastacia Griego-Fisher, D'eldra Malone, Jennifer Benoit |
| EPI_ISL_1494728, EPI_ISL_1494729, EPI_ISL_1494730 | Centro de Investigación en Ciencias de la Salud y Biomedicina, U.A.S.LP. | Centro de Investigación en Ciencias de la Salud y Biomedicina, U.A.S.LP. | MD PhD Sofía Bernal Silva |
| EPI_ISL_1502814, EPI_ISL_1502816, EPI_ISL_1502817, EPI_ISL_1503143 | Infectious Diseases Department , Instituto Nacional de Ciencias Medicas y Nutrición | Instituto Nacional de Ciencias Medicas y Nutricion Infectious Diseases | Guillermo M. Ruiz-Palacios, Pilar Ramos Cervantes, Violeta Ibarra Gonzalez, Fernando Ledesma Barrientos, Miriam Arciniega Fuentes , Fernando Arteaga Cabello, Diana Paola Davalos Martinez, Luis Alberto Garcia Andrade, Luz Elena Cervantes Villar |
| EPI_ISL_1516777, EPI_ISL_1516778, EPI_ISL_1516779, EPI_ISL_1516780, EPI_ISL_1516781, EPI_ISL_1516782, EPI_ISL_1516783, EPI_ISL_1516784, EPI_ISL_1516785 | LESP Aguascalientes | Instituto de Diagnostico y Referencia Epidemiologicos (INDRE) | Claudia Wong-Arambula, Abril Rodriguez-Maldonado, Vanessa Rivero-Arredondo, Ariadna Medina-Benitez, Joaquin Quiroz-Mercado, Sergio Rangel-Guerrero, Natividad Cruz-Ortiz, Tatiana Nunez-Garcia, Gisela Barrera-Badillo, Lucia Hernandez-Rivas, Irma Lopez-Martinez, Ernesto Ramirez-Gonzalez. |
| EPI_ISL_1531892, EPI_ISL_1531893, EPI_ISL_1531894, EPI_ISL_1531895, EPI_ISL_1531896, EPI_ISL_1531897, EPI_ISL_1531898, EPI_ISL_1531899, EPI_ISL_1531900, EPI_ISL_1531901, EPI_ISL_1531902, EPI_ISL_1531903, EPI_ISL_1531904, EPI_ISL_1531905, EPI_ISL_1531906, EPI_ISL_1531907, EPI_ISL_1531908, EPI_ISL_1531909, EPI_ISL_1531910, EPI_ISL_1531911, EPI_ISL_1531912, EPI_ISL_1531913, EPI_ISL_1531914, EPI_ISL_1531915, EPI_ISL_1531916, EPI_ISL_1531917, EPI_ISL_1531918, EPI_ISL_1531919, EPI_ISL_1531920, EPI_ISL_1531921, EPI_ISL_1531922, EPI_ISL_1531923, EPI_ISL_1531924, EPI_ISL_1531925, EPI_ISL_1531926, EPI_ISL_1531927, EPI_ISL_1531928, EPI_ISL_1531929, EPI_ISL_1531930, EPI_ISL_1531931, EPI_ISL_1531932, EPI_ISL_1531933, EPI_ISL_1531934, EPI_ISL_1531935, EPI_ISL_1531936, EPI_ISL_1531937, EPI_ISL_1531938, EPI_ISL_1531939, EPI_ISL_1531940, EPI_ISL_1531941 | | | |
| see above | Centro de Diagnostico COVID-19 UABC Tijuana | Andersen lab at Scripps Research | SEARCH Alliance San Diego with Idanya Rubi Serafin Higuera, Manuel Sanchez Alavez, Jorge Luis Jimenez Niebla, German Ibarra, Jonathan Vincent Baena, Oscar Efren Zazueta Fierro |
| EPI_ISL_1591520 | Instituto Nacional de Medicina Genomica | Instituto Nacional de Medicina Genomica | Hidalgo-Miranda A, Mendoza-Vargas A, Reyes-Grajeda JP, Cedro-Tanda A, Alcaraz N, Gonzalez-Barrera D, Rangel-DeLeon D, Miranda-Ortiz H, Rosas-Escobar P, Canseco Mendez JC, Ramirez-Vega O, Munguia-Garza P, Garcia-Cardenas FJ, Gonzalez-Woge MA, Herrera-Montalvo LA |
| EPI_ISL_1626818 | LESP Ciudad de Mexico | Instituto de Diagnostico y Referencia Epidemiologicos (INDRE) | Claudia Wong-Arambula, Abril Rodriguez-Maldonado, Vanessa Rivero-Arredondo, Ariadna Medina-Benitez, Joaquin Quiroz-Mercado, Sergio Rangel-Guerrero, Natividad Cruz-Ortiz, Tatiana Nunez-Garcia, Gisela Barrera-Badillo, Lucia Hernandez-Rivas, Irma Lopez-Martinez, Ernesto Ramirez-Gonzalez. |
| EPI_ISL_1664478, EPI_ISL_1664479, EPI_ISL_1664480, EPI_ISL_1664482, EPI_ISL_1664484, EPI_ISL_1664485, EPI_ISL_1664487, EPI_ISL_1664488, EPI_ISL_1664490, EPI_ISL_1664491, EPI_ISL_1664493, EPI_ISL_1664494, EPI_ISL_1664496, EPI_ISL_1664497, EPI_ISL_1664499, EPI_ISL_1664500, EPI_ISL_1664502, EPI_ISL_1664503, EPI_ISL_1664505, EPI_ISL_1664507, EPI_ISL_1664508, EPI_ISL_1664510, EPI_ISL_1664511, EPI_ISL_1664513, EPI_ISL_1664514, EPI_ISL_1664516, EPI_ISL_1664517, EPI_ISL_1664519, EPI_ISL_1664521, EPI_ISL_1664522, EPI_ISL_1664524, EPI_ISL_1664525, EPI_ISL_1664527, EPI_ISL_1664528, EPI_ISL_1664530, EPI_ISL_1664531, EPI_ISL_1664533, EPI_ISL_1664534, EPI_ISL_1664536, EPI_ISL_1664537, EPI_ISL_1664539, EPI_ISL_1664540, EPI_ISL_1664542, EPI_ISL_1664543, EPI_ISL_1664545, EPI_ISL_1664546, EPI_ISL_1664548, EPI_ISL_1664549, EPI_ISL_1664551, EPI_ISL_1664552, EPI_ISL_1664554, EPI_ISL_1664555, EPI_ISL_1664557, EPI_ISL_1664558, EPI_ISL_1664560, EPI_ISL_1664561, EPI_ISL_1664563, EPI_ISL_1664564, EPI_ISL_1664566, EPI_ISL_1664567, EPI_ISL_1664569, EPI_ISL_1664571, EPI_ISL_1664572, EPI_ISL_1664574, EPI_ISL_1664576, EPI_ISL_1664577, EPI_ISL_1664578, EPI_ISL_1664580, EPI_ISL_1664582, EPI_ISL_1664583, EPI_ISL_1664585, EPI_ISL_1664586, EPI_ISL_1664588 | | | |
| see above | New Mexico Department of Health Scientific Laboratory | Center for Global Health, University of New Mexico Health Sciences Center | Daryl Domman, Kurt Schwalm, Twila Kunde, Joseph Hicks, Anastacia Griego, Michael Edwards, Valerie Morley, Darrell Dinwiddie |
| EPI_ISL_1673275 | LESP Ciudad de Mexico/DIAGNOMOL | Instituto de Diagnostico y Referencia Epidemiologicos (INDRE) | Claudia Wong-Arambula, Abril Rodriguez-Maldonado, Vanessa Rivero-Arredondo, Ariadna Medina-Benitez, Joaquin Quiroz-Mercado, Sergio Rangel-Guerrero, Natividad Cruz-Ortiz, Tatiana Nunez-Garcia, Gisela Barrera-Badillo, Lucia Hernandez-Rivas, Irma Lopez-Martinez, Ernesto Ramirez-Gonzalez. |
| EPI_ISL_1711720, EPI_ISL_1711721, EPI_ISL_1711722, EPI_ISL_1711724, EPI_ISL_1711725, EPI_ISL_1711726, EPI_ISL_1711727, EPI_ISL_1711728, EPI_ISL_1711730, EPI_ISL_1711731, EPI_ISL_1711732, EPI_ISL_1711733, EPI_ISL_1711734, EPI_ISL_1711735, EPI_ISL_1711736, EPI_ISL_1711738, EPI_ISL_1711739, EPI_ISL_1711740, EPI_ISL_1711742, EPI_ISL_1711744, EPI_ISL_1711745, EPI_ISL_1711747, EPI_ISL_1711750, EPI_ISL_1711751, EPI_ISL_1711752, EPI_ISL_1711753 | | | |
| see above | "NM Dept. Health, Scientific Laboratory Division " | Centers for Disease Control and Prevention Division of Viral Diseases, Pathogen Discovery | Mili Sheth, Sarah Nobles, Jasmine Padilla, Mark Burroughs, Shoshona Le, Katie Dillon, Peter Cook, Clinton R. Paden, Dhwani Batra, Krista Queen, Kristen Knipe, Dakota Howard, Yvette Unoarumhi, Darlene Wagner, Matthew Schmerer, Ben L. Rambo-Martin, Kristine Lacek, Sam Shepard, Alison Laufer Halpin, Dave Wentworth, Vivien Dugan, Suxiang Tong, Justin Lee |
| EPI_ISL_1791075, EPI_ISL_1791076, EPI_ISL_1791077, EPI_ISL_1791078, EPI_ISL_1791079, EPI_ISL_1791080, EPI_ISL_1791081, EPI_ISL_1791082, EPI_ISL_1791083, EPI_ISL_1791084, EPI_ISL_1791085, EPI_ISL_1791086, EPI_ISL_1791087, EPI_ISL_1791088, EPI_ISL_1791089, EPI_ISL_1791090, EPI_ISL_1791091, EPI_ISL_1791092, EPI_ISL_1791093, EPI_ISL_1791094, EPI_ISL_1791095, EPI_ISL_1791096, EPI_ISL_1791097, EPI_ISL_1791098, EPI_ISL_1791099, EPI_ISL_1791100, EPI_ISL_1791101, EPI_ISL_1791102, EPI_ISL_1791103, EPI_ISL_1791104, EPI_ISL_1791105, EPI_ISL_1791106, EPI_ISL_1791107, EPI_ISL_1791108, EPI_ISL_1791109, EPI_ISL_1791110, EPI_ISL_1791111, EPI_ISL_1791112, EPI_ISL_1791113, EPI_ISL_1791114, EPI_ISL_1791115, EPI_ISL_1791116, EPI_ISL_1791117, EPI_ISL_1791118, EPI_ISL_1791119, EPI_ISL_1791120, EPI_ISL_1791121, EPI_ISL_1791122, EPI_ISL_1791123, EPI_ISL_1791124, EPI_ISL_1791125, EPI_ISL_1791126, EPI_ISL_1791127, EPI_ISL_1791128, EPI_ISL_1791129, EPI_ISL_1791130, EPI_ISL_1791131, EPI_ISL_1791132, EPI_ISL_1791133, EPI_ISL_1791134, EPI_ISL_1791135, EPI_ISL_1791136, EPI_ISL_1791137, EPI_ISL_1791138, EPI_ISL_1791139, EPI_ISL_1791140 | | | |
| see above | New Mexico Department of Health Scientific Laboratory | Center for Global Health, University of New Mexico Health | Daryl Domman, Kurt Schwalm, Twila Kunde, Joseph Hicks, Anastacia Griego, Michael Edwards, Valerie Morley, Darrell Dinwiddie |

| | | | |
|---|---|---|---|
| | | Sciences Center | |
| EPI_ISL_1794925 | Sharp HealthCare Laboratory | Andersen lab at Scripps Research | SEARCH Alliance San Diego with Aaron Harding, Jacquelyn Berumen, Cathy Woerle, Liam McGinnis, Art Mendoza, Omid Bakhtar |
| EPI_ISL_1807187 | New Mexico Department of Health Scientific Laboratory | Center for Global Health, University of New Mexico Health Sciences Center | Daryl Domman, Kurt Schwalm, Twila Kunde, Joseph Hicks, Anastacia Griego, Michael Edwards, Valerie Morley, Darrell Dinwiddie |
| EPI_ISL_2105695, EPI_ISL_2105696, EPI_ISL_2105697, EPI_ISL_2105698, EPI_ISL_2105699, EPI_ISL_2105700, EPI_ISL_2105701, EPI_ISL_2105702, EPI_ISL_2105703, EPI_ISL_2105704, EPI_ISL_2105705, EPI_ISL_2105706, EPI_ISL_2105707, EPI_ISL_2105708, EPI_ISL_2105709, EPI_ISL_2105710, EPI_ISL_2105711, EPI_ISL_2105712, EPI_ISL_2105713, EPI_ISL_2105714 | | | |
| see above | LESP Jalisco | Instituto de Diagnostico y Referencia Epidemiologicos (INDRE) | Claudia Wong-Arambula, Abril Rodriguez-Maldonado, Vanessa Rivero-Arredondo, Ariadna Medina-Benitez, Joaquin Quiroz-Mercado, Sergio Rangel-Guerrero, Natividad Cruz-Ortiz, Tatiana Nunez-Garcia, Gisela Barrera-Badillo, Lucia Hernandez-Rivas, Irma Lopez-Martinez, Ernesto Ramirez-Gonzalez. |
| EPI_ISL_658863, EPI_ISL_658865, EPI_ISL_658868, EPI_ISL_658870 | Instituto de Diagnostico y Referencia Epidemiologicos (INDRE) | Instituto de Diagnostico y Referencia Epidemiologicos (INDRE) | Gisela Barrera-Badillo , Abril Rodriguez-Maldonado, Claudia Wong-Arambula , Natividad Cruz-Ortiz, Tatiana Nunez-Garcia, Dayanira Arellano-Suarez, Fabiola Garces-Ayala, Lucia Hernandez-Rivas, Irma Lopez-Martinez, Ernesto Ramirez-Gonzalez. |
| EPI_ISL_658873, EPI_ISL_658875 | Instituto de Diagnostico y Referencia Epidemiologicos (INDRE) | Instituto de Diagnostico y Referencia Epidemiologicos (INDRE) | Gisela Barrera-Badillo , Abril Rodriguez-Maldonado, Claudia Wong-Arambula , Natividad Cruz-Ortiz, Tatiana Nunez-Garcia, Dayanira Arellano-Suarez, Fabiola Garces-Ayala, Lucia Hernandez-Rivas, Irma Lopez-Martinez, Ernesto Ramirez-Gonzalez. |
| EPI_ISL_658878, EPI_ISL_658880, EPI_ISL_658883, EPI_ISL_658886, EPI_ISL_658888, EPI_ISL_658891, EPI_ISL_658893, EPI_ISL_658901 | Instituto de Diagnostico y Referencia Epidemiologicos (INDRE) | Instituto de Diagnostico y Referencia Epidemiologicos (INDRE) | Ernesto Ramirez-Gonzalez, Abril Rodriguez-Maldonado, Claudia Wong-Arambula , Natividad Cruz-Ortiz, Tatiana Nunez-Garcia, Dayanira Arellano-Suarez, Fabiola Garces-Ayala, Lucia Hernandez-Rivas, Irma Lopez-Martinez, Gisela Barrera-Badillo. |
| EPI_ISL_660068 | Instituto de Diagnostico y Referencia Epidemiologicos (INDRE) | Instituto de diagnóstico y Referencia Epidemiologicos (INDRE) Departamento de Virología | Gisela Barrera-Badillo , Abril Rodriguez-Maldonado, Claudia Wong-Arambula , Natividad Cruz-Ortiz, Tatiana Nunez-Garcia, Dayanira Arellano-Suarez, Fabiola Garces-Ayala, Lucia Hernandez-Rivas, Irma Lopez-Martinez, Ernesto Ramirez-Gonzalez. |
| EPI_ISL_747243 | General Hospital Laboratory | Pathogen Laboratory (BSL3), Biomedical Innovation Department, Experimental and Applied Biology Division, Scientific Research Center and High Education from Ensenada (CICESE) | Cervantes-Luevano K., Martinez M.A., Saavedra-Flores A., Galindo C. and Licea-Navarro A. |
| EPI_ISL_751802, EPI_ISL_751803, EPI_ISL_751804, EPI_ISL_751805, EPI_ISL_751806, EPI_ISL_751807, EPI_ISL_751808, EPI_ISL_751809, EPI_ISL_751810, EPI_ISL_751811, EPI_ISL_751812, EPI_ISL_751813, EPI_ISL_751814, EPI_ISL_751815, EPI_ISL_751816, EPI_ISL_751817, EPI_ISL_751818, EPI_ISL_751819, EPI_ISL_751820, EPI_ISL_751821, EPI_ISL_751822, EPI_ISL_751823, EPI_ISL_751824, EPI_ISL_751825, EPI_ISL_751826, EPI_ISL_751827, EPI_ISL_751828, EPI_ISL_751829, EPI_ISL_751830, EPI_ISL_751831, EPI_ISL_751832, EPI_ISL_751833, EPI_ISL_751834, EPI_ISL_751835, EPI_ISL_751836, EPI_ISL_751837, EPI_ISL_751838, EPI_ISL_751839, EPI_ISL_751840, EPI_ISL_751841, EPI_ISL_751842, EPI_ISL_751843, EPI_ISL_751844, EPI_ISL_751845, EPI_ISL_751846, EPI_ISL_766883, EPI_ISL_766884, EPI_ISL_766885, EPI_ISL_766886, EPI_ISL_766887, EPI_ISL_766888, EPI_ISL_766889, EPI_ISL_766890, EPI_ISL_766891, EPI_ISL_766892, EPI_ISL_766893, EPI_ISL_766894, EPI_ISL_766895, EPI_ISL_766896, EPI_ISL_766897, EPI_ISL_766898, EPI_ISL_766899, EPI_ISL_766900, EPI_ISL_766901, EPI_ISL_766902, EPI_ISL_766903, EPI_ISL_766904, EPI_ISL_766905, EPI_ISL_766906, EPI_ISL_766907, EPI_ISL_766908, EPI_ISL_766909, EPI_ISL_766910, EPI_ISL_766911, EPI_ISL_766912, EPI_ISL_766913, EPI_ISL_766914, EPI_ISL_766915, EPI_ISL_766916, EPI_ISL_766917, EPI_ISL_766918, EPI_ISL_766919, EPI_ISL_766920, EPI_ISL_766921, EPI_ISL_766922, EPI_ISL_766923, EPI_ISL_767044, EPI_ISL_767045, EPI_ISL_767046, EPI_ISL_767047, EPI_ISL_767048, EPI_ISL_767049, EPI_ISL_767050, EPI_ISL_767051, EPI_ISL_767052, EPI_ISL_767053, EPI_ISL_767054, EPI_ISL_767055, EPI_ISL_767056, EPI_ISL_767057, EPI_ISL_767058, EPI_ISL_767059, EPI_ISL_767060, EPI_ISL_767061, EPI_ISL_767062, EPI_ISL_767063, EPI_ISL_767064, EPI_ISL_767065, EPI_ISL_767066, EPI_ISL_767067, EPI_ISL_767068, EPI_ISL_767069, EPI_ISL_767070, EPI_ISL_767071, EPI_ISL_767072, EPI_ISL_767073, EPI_ISL_767074, EPI_ISL_767075, EPI_ISL_767076, EPI_ISL_767077, EPI_ISL_767078, EPI_ISL_767079, EPI_ISL_767080, EPI_ISL_767081, EPI_ISL_767082, EPI_ISL_767083, EPI_ISL_767084, EPI_ISL_767085, EPI_ISL_767086, EPI_ISL_767087, EPI_ISL_767088 | | | |
| see above | New Mexico Department of Health Scientific Laboratory | New Mexico Department of Health Scientific Laboratory | D'eldra Malone, Ellie Johnson, Anastacia Griego-Fisher |
| EPI_ISL_779185, EPI_ISL_779186, EPI_ISL_779187, EPI_ISL_779188, EPI_ISL_779189, EPI_ISL_779190, EPI_ISL_779191, EPI_ISL_779192, EPI_ISL_779193, EPI_ISL_779194, EPI_ISL_779195, EPI_ISL_779196, EPI_ISL_779197, EPI_ISL_779198 | | | |
| see above | Laboratorio Estatal de Salud Pública de Nuevo León | Laboratorio de Infectología Molecular, Departamento de Bioquímica y Medicina Molecular,Facultad de Medicina - Universidad Autónoma de Nuevo León | Kame A. Galán-Huerta, María F. Herrera-Saldivar, Natalia Martínez-Acuña, Sonia A. Lozano-Sepúlveda, Daniel Arellanos-Soto, Ana M. Rivas-Estilla, Samuel Buentello-Wong, Else del Carmen García-García, Gloria A. Jasso-de-la-Peña, Roberto Montes-de-Oca, Consuelo Treviño-Garza, Manuel E. de-la-O-Cavazos |
| EPI_ISL_794592 | Laboratorio Estatal de Salud Pública de Tamaulipas | Instituto de diagnóstico y Referencia Epidemiologicos (INDRE) | Abril Rodriguez-Maldonado, Claudia Wong-Arambula, Fabiola Garces-Ayala, Gisela Barrera-Badillo, Ana Maria Cortez-Calderon, Bernardita Reyes-Berrones, Hilda del Carmen-Selvera, Gloria Molina-Gamboa, Lucia Hernandez-Rivas, Irma Lopez-Martinez, Celia Alpuche-Aranda, Jose Luis Alomia-Zegarra, Hugo Lopez Gatell-Ramirez, Ernesto Ramirez-Gonzalez. |
| EPI_ISL_824513, EPI_ISL_824514, EPI_ISL_824515, EPI_ISL_824516, EPI_ISL_824517, EPI_ISL_824518, EPI_ISL_824519, EPI_ISL_824520, EPI_ISL_824521, EPI_ISL_824522, EPI_ISL_824523, EPI_ISL_824524, EPI_ISL_824525, EPI_ISL_824526, EPI_ISL_824527, EPI_ISL_824528, EPI_ISL_824529, EPI_ISL_824530, EPI_ISL_824531, EPI_ISL_824532, EPI_ISL_824533, EPI_ISL_824534, EPI_ISL_824535, EPI_ISL_824536, EPI_ISL_824537, EPI_ISL_824538, EPI_ISL_824539, EPI_ISL_824540, EPI_ISL_824541, EPI_ISL_824542, EPI_ISL_824543, EPI_ISL_824544, EPI_ISL_824545, EPI_ISL_824546, EPI_ISL_824547, EPI_ISL_824548, EPI_ISL_824549, EPI_ISL_824550, EPI_ISL_824551, EPI_ISL_824552, EPI_ISL_824553, EPI_ISL_824554 | | | |
| see above | New Mexico Department of Health Scientific Laboratory | New Mexico Department of Health Scientific Laboratory | Ellie Johnson, Anastacia Griego-Fisher, D'eldra Malone |
| EPI_ISL_831753, EPI_ISL_831769, EPI_ISL_831791 | United States Air Force School of Aerospace Medicine | United States Air Force School of Aerospace Medicine | Anthony Fries, Jennifer Meyer, William Gruner, Amanda Javorina, Sarah Purves, Clarise Starr, Elizabeth Macias |
| EPI_ISL_831900, EPI_ISL_831901, EPI_ISL_831903, EPI_ISL_831905, EPI_ISL_831906, EPI_ISL_831907, EPI_ISL_831908, EPI_ISL_831909, EPI_ISL_831910, EPI_ISL_831911, EPI_ISL_831920, EPI_ISL_831921, EPI_ISL_831922, EPI_ISL_831923, EPI_ISL_831924, EPI_ISL_831925, EPI_ISL_831926, EPI_ISL_831927, EPI_ISL_831928, EPI_ISL_831929, EPI_ISL_831930, EPI_ISL_831931, EPI_ISL_831932, EPI_ISL_831933, EPI_ISL_831934, EPI_ISL_831935, EPI_ISL_831936, EPI_ISL_831937 | | | |
| see above | New Mexico Department of Health Scientific Laboratory | New Mexico Department of Health Scientific Laboratory | Ellie Johnson, Anastacia Griego-Fisher, D'eldra Malone |
| EPI_ISL_845872 | TGen North | TGen North | Jolene Bowers, Megan Folkerts, Chris French, Hayley Yaglom, Ashlyn Pfeiffer, Darrin Lemmer, Dave Engelthaler, The Arizona COVID Genomics Union (ACGU) |
| EPI_ISL_862669, EPI_ISL_862670, EPI_ISL_862671, EPI_ISL_862672, EPI_ISL_862673, EPI_ISL_862674, EPI_ISL_862675, EPI_ISL_862676, EPI_ISL_862677, EPI_ISL_862678, EPI_ISL_862679, EPI_ISL_862680, EPI_ISL_862681, EPI_ISL_862682, EPI_ISL_862683, EPI_ISL_862684, EPI_ISL_862685, EPI_ISL_862686, EPI_ISL_862687, EPI_ISL_862688, EPI_ISL_862689, EPI_ISL_862690, EPI_ISL_862691, EPI_ISL_862692, EPI_ISL_862693, EPI_ISL_862694, EPI_ISL_862695, EPI_ISL_862696, EPI_ISL_862697, EPI_ISL_862698, EPI_ISL_862699, EPI_ISL_862700, EPI_ISL_862701, EPI_ISL_862702, EPI_ISL_862703, EPI_ISL_862704, EPI_ISL_862705, EPI_ISL_862706, EPI_ISL_862707, EPI_ISL_862708, EPI_ISL_862709, EPI_ISL_862710, EPI_ISL_862711, EPI_ISL_862712, EPI_ISL_862713 | | | |
| see above | New Mexico Department of Health Scientific Laboratory | New Mexico Department of Health Scientific Laboratory | D'eldra Malone, Ellie Johnson, Anastacia Griego-Fisher |
| EPI_ISL_869167, EPI_ISL_869168, EPI_ISL_869169, EPI_ISL_869170, EPI_ISL_869171, EPI_ISL_869172, EPI_ISL_869173, EPI_ISL_869174, EPI_ISL_869175, EPI_ISL_869176, EPI_ISL_869177, EPI_ISL_869178, EPI_ISL_869179, EPI_ISL_869180, EPI_ISL_869181, EPI_ISL_869182, EPI_ISL_869183, EPI_ISL_869184, EPI_ISL_869185, EPI_ISL_869186, EPI_ISL_869187, EPI_ISL_869188, EPI_ISL_869189, EPI_ISL_869190, EPI_ISL_869191, EPI_ISL_869192, EPI_ISL_869193, EPI_ISL_869194, EPI_ISL_869195, EPI_ISL_869196, EPI_ISL_869197, EPI_ISL_869198, EPI_ISL_869199, EPI_ISL_869200, EPI_ISL_869201, EPI_ISL_869202, EPI_ISL_869203, EPI_ISL_869204, EPI_ISL_869205, EPI_ISL_869206, EPI_ISL_869207, EPI_ISL_869208, EPI_ISL_869209, EPI_ISL_869210, EPI_ISL_869211, EPI_ISL_869212, EPI_ISL_869213, EPI_ISL_869214, EPI_ISL_869215, EPI_ISL_869216, EPI_ISL_869217, EPI_ISL_869218, EPI_ISL_869219, EPI_ISL_869220, EPI_ISL_869221, EPI_ISL_869222, EPI_ISL_869223, EPI_ISL_869224, EPI_ISL_869225, EPI_ISL_869226, EPI_ISL_869227, EPI_ISL_869228, EPI_ISL_869229, EPI_ISL_869230, EPI_ISL_869231, EPI_ISL_869232, EPI_ISL_869233, EPI_ISL_869234, EPI_ISL_869235, EPI_ISL_869236, EPI_ISL_869237, EPI_ISL_869238, EPI_ISL_869239, EPI_ISL_872479, EPI_ISL_872480, EPI_ISL_872481, EPI_ISL_872482, EPI_ISL_872483, EPI_ISL_872484, EPI_ISL_872485, EPI_ISL_872486, EPI_ISL_872487, EPI_ISL_872488, EPI_ISL_872489, EPI_ISL_872490, EPI_ISL_872491, EPI_ISL_872492, EPI_ISL_872493, EPI_ISL_872494, EPI_ISL_872495, EPI_ISL_872496, EPI_ISL_872497, EPI_ISL_872498, EPI_ISL_872499, EPI_ISL_872500, EPI_ISL_872501, EPI_ISL_872502, EPI_ISL_872503, EPI_ISL_872504, EPI_ISL_872505, EPI_ISL_872506, EPI_ISL_872507, EPI_ISL_872508, EPI_ISL_872509, EPI_ISL_872510, EPI_ISL_872511, EPI_ISL_872512, EPI_ISL_872513, EPI_ISL_872514, EPI_ISL_872515, EPI_ISL_872516, EPI_ISL_872517, EPI_ISL_872518, EPI_ISL_872519, EPI_ISL_872520, EPI_ISL_872521, EPI_ISL_872522, EPI_ISL_872523, EPI_ISL_872524, EPI_ISL_872525, EPI_ISL_872526, EPI_ISL_872527, EPI_ISL_872528, EPI_ISL_872529, EPI_ISL_872530, EPI_ISL_872531, EPI_ISL_872532, EPI_ISL_872533, EPI_ISL_872534, EPI_ISL_872535, EPI_ISL_872536, EPI_ISL_872537, EPI_ISL_872538, EPI_ISL_872539, EPI_ISL_872540, EPI_ISL_872541, EPI_ISL_872542, EPI_ISL_872543, EPI_ISL_872544, EPI_ISL_872545, EPI_ISL_872546, EPI_ISL_872547, EPI_ISL_872548, EPI_ISL_872549, EPI_ISL_872550, EPI_ISL_872551, EPI_ISL_872552, EPI_ISL_872553, EPI_ISL_872554, EPI_ISL_872555, EPI_ISL_872556, EPI_ISL_872557, EPI_ISL_872558, EPI_ISL_872559, EPI_ISL_872560, EPI_ISL_872561, EPI_ISL_872562, EPI_ISL_872563, EPI_ISL_872564, EPI_ISL_872565, EPI_ISL_872566 | | | |
| see above | New Mexico Department of Health Scientific Laboratory | Center for Global Health, University of New Mexico Health Sciences Center | Daryl Domman, Kurt Schwalm, Twila Kunde, Joseph Hicks, Anastacia Griego, Michael Edwards, Darrell Dinwiddie |
| EPI_ISL_876710 | Helix/Illumina | Genomics and Discovery, Respiratory Viruses Branch, Division of Viral Diseases, Centers for Disease Control and Prevention | Peter W. Cook,Dhwani Batra,Ben L. Rambo-Martin,Eileen de Feo,Jan Antico,Christine Tran,Matthew Tolentino,Shannon Wickline,Kim Gietzen,Brad Sickler,Jingtao Liu,Eric Allen,Phil Febbo,Summer Galloway,Nicole L. Washington,Simon White,Geraint Levan,Kelly Schiabor Barrett,Elizabeth Cirulli,Alexandre Bolze,Ary Ascencio,Charlotte Rivera-Garcia,Ryan Cho,Jason Nguyen,Sherry Wang,Jimmy Ramirez,Tyler Cassens,Efren Sandoval,Magnus Isaksson,William Lee,David Becker,Marc Laurent,James Lu,Clinton R. Paden,Suxiang Tong,Duncan MacCannell, |
| EPI_ISL_877133, EPI_ISL_877134, EPI_ISL_877135, EPI_ISL_877136, EPI_ISL_877137, EPI_ISL_877138, EPI_ISL_877139, EPI_ISL_877140, EPI_ISL_877141, EPI_ISL_877142, EPI_ISL_877143, EPI_ISL_877144, EPI_ISL_877145, EPI_ISL_877146, EPI_ISL_877147, EPI_ISL_877148, EPI_ISL_877149, EPI_ISL_877150, EPI_ISL_877151, EPI_ISL_877152, EPI_ISL_877153, EPI_ISL_877154, EPI_ISL_877155, EPI_ISL_877156, EPI_ISL_877157, EPI_ISL_877158, EPI_ISL_877159, EPI_ISL_877160, EPI_ISL_877161, EPI_ISL_877162, EPI_ISL_877163, EPI_ISL_877164, EPI_ISL_877165, EPI_ISL_877166, EPI_ISL_877167, EPI_ISL_877168, EPI_ISL_877169, EPI_ISL_877170, EPI_ISL_877171, EPI_ISL_877172, EPI_ISL_877173, EPI_ISL_877174, EPI_ISL_877175, EPI_ISL_877176, EPI_ISL_877177, EPI_ISL_877178, EPI_ISL_877179, EPI_ISL_877180, EPI_ISL_877181, EPI_ISL_877182, EPI_ISL_877183, EPI_ISL_877184, EPI_ISL_877185 | | | |
| see above | Univeristy of New Mexico Hospital | Center for Global Health, University of New Mexico Health Sciences Center | Daryl Domman, Kurt Schwalm, Justin Bacca, Jon Femling, Darrell Dinwiddie |

| | | | |
|---|---|---|---|
| EPI_ISL_878212, EPI_ISL_878226, EPI_ISL_878300, EPI_ISL_878367, EPI_ISL_878375, EPI_ISL_878390, EPI_ISL_878400, EPI_ISL_878401, EPI_ISL_878404, EPI_ISL_878408, EPI_ISL_878437, EPI_ISL_879950, EPI_ISL_879977 | | | |
| see above | San Diego County Public Health Laboratory | Andersen lab at Scripps Research | SEARCH Alliance San Diego with Tracy Basler, Jovan Shephard, Brett Austin |
| EPI_ISL_884718 | Respiratory Viruses Branch, Centers for Disease Control and Prevention | Respiratory Viruses Branch, Centers for Disease Control and Prevention | Cook,P.W., Batra,D., Rambo-Martin,B.L., de Feo,E., Antico,J., Tran,C., Tolentino,M., Wickline,S., Gietzen,K., Sickler,B., Liu,J., Allen,E., Febbo,P., Galloway,S., Washington,N.L., White,S., Levan,G., Barret,K.S., Cirulli,E., Bolze,A., Ascencio,A., Rivera-Garcia,C., Cho,R., Nguyen,J., Wang,S., Ramirez,J., Cassens,T., Sandoval,E., Isaksson,M., Lee,W., Becker,D., Laurent,M., Lu,J., Paden,C.R., Tong,S., MacCannell,D. |
| EPI_ISL_886174, EPI_ISL_886243, EPI_ISL_886406, EPI_ISL_886438, EPI_ISL_886443, EPI_ISL_886480, EPI_ISL_886481, EPI_ISL_886500, EPI_ISL_886504, EPI_ISL_886528, EPI_ISL_886548, EPI_ISL_886549, EPI_ISL_886550, EPI_ISL_886625, EPI_ISL_886638, EPI_ISL_886704, EPI_ISL_886733, EPI_ISL_886738, EPI_ISL_886739, EPI_ISL_886783, EPI_ISL_886847, EPI_ISL_886848, EPI_ISL_886861, EPI_ISL_886869, EPI_ISL_886910, EPI_ISL_886936, EPI_ISL_886941, EPI_ISL_886961, EPI_ISL_886964, EPI_ISL_886986, EPI_ISL_887052, EPI_ISL_887637, EPI_ISL_887644, EPI_ISL_887668, EPI_ISL_887687, EPI_ISL_887701, EPI_ISL_887722, EPI_ISL_887738, EPI_ISL_887760, EPI_ISL_887781, EPI_ISL_887788, EPI_ISL_887874, EPI_ISL_887951, EPI_ISL_887959, EPI_ISL_887961, EPI_ISL_888090, EPI_ISL_888154, EPI_ISL_888228, EPI_ISL_888356, EPI_ISL_888365, EPI_ISL_888409, EPI_ISL_888464, EPI_ISL_888488, EPI_ISL_888492, EPI_ISL_888496, EPI_ISL_888523, EPI_ISL_888534, EPI_ISL_888539, EPI_ISL_888552, EPI_ISL_888555, EPI_ISL_888573 | | | |
| see above | Labcorp | Genomics and Discovery, Respiratory Viruses Branch, Division of Viral Diseases, Centers for Disease Control and Prevention | Peter W. Cook,Dhwani Batra,Ben L. Rambo-Martin,Summer Galloway,Brian Krueger,Minoo Agarwal,Eyad Almasri,Debbie Boles,Ayla Burns,Nuthawin Charoensri,Oren Cohen,Susan Countryman,Mary Ann Cristobal,Bobbi Croy,Suzanne Dale,Hrushikesh Deshmukh,Amanda Douglas,Vincent Drouillon,Marcia Eisenberg,Howard Engler,Rama Ghatti,Prashant Gupta,Susan Hicks,Jake Humphrey,Lax Iyer,Manoj Jain,Mohan Kolli,Tim Kuphal,Stanley Letovsky,Michael Levandoski,Craig Lukasik,Jonathan Meltzer,Brian Norvell,Mindy Nye,Scott Parker,Christos Petropoulos,John Pruitt,Steven Ragan,Scott Ryan,Mike Sapeta,Jana Schroth,Suresh Babu Selvaraju,Goran Stevovic,Amanda Suchanek,Andrea Throop,Lyndon Tilson,Thomas Urban,Joe Voshell,Kimberly Wagner,Jonathan Williams,Mary Williamson,Qian Zeng,Tricia Zwiefelhofer,Clinton R. Paden,Suxiang Tong,Duncan MacCannell, |
| EPI_ISL_888628, EPI_ISL_888629, EPI_ISL_888630, EPI_ISL_888631, EPI_ISL_888632, EPI_ISL_888633, EPI_ISL_888634, EPI_ISL_888635, EPI_ISL_888636, EPI_ISL_888637, EPI_ISL_888638, EPI_ISL_888639, EPI_ISL_888640, EPI_ISL_888641, EPI_ISL_888642, EPI_ISL_888643, EPI_ISL_888644, EPI_ISL_888645, EPI_ISL_888646, EPI_ISL_888647, EPI_ISL_888648, EPI_ISL_888649, EPI_ISL_888650, EPI_ISL_888651, EPI_ISL_888791, EPI_ISL_888792, EPI_ISL_888793, EPI_ISL_888794, EPI_ISL_888795, EPI_ISL_888796, EPI_ISL_888797, EPI_ISL_888798, EPI_ISL_888799, EPI_ISL_888800, EPI_ISL_888801, EPI_ISL_888802, EPI_ISL_888803, EPI_ISL_888804, EPI_ISL_888805, EPI_ISL_888806, EPI_ISL_888807, EPI_ISL_888808, EPI_ISL_888809, EPI_ISL_888810, EPI_ISL_888811, EPI_ISL_888812, EPI_ISL_888813, EPI_ISL_888814, EPI_ISL_888815, EPI_ISL_888816, EPI_ISL_888817 | | | |
| see above | Univeristy of New Mexico Hospital | Center for Global Health, University of New Mexico Health Sciences Center | Daryl Domman, Kurt Schwalm, Justin Bacca, Jon Femling, Darrell Dinwiddie |
| EPI_ISL_904035, EPI_ISL_904036, EPI_ISL_904037, EPI_ISL_904038, EPI_ISL_904039, EPI_ISL_904040, EPI_ISL_904041, EPI_ISL_904042, EPI_ISL_904043, EPI_ISL_904044, EPI_ISL_904045, EPI_ISL_904046, EPI_ISL_904047, EPI_ISL_904048, EPI_ISL_904049, EPI_ISL_904050, EPI_ISL_904051, EPI_ISL_904052, EPI_ISL_904053, EPI_ISL_904054, EPI_ISL_904055, EPI_ISL_904056, EPI_ISL_904057, EPI_ISL_904058, EPI_ISL_904059, EPI_ISL_904060, EPI_ISL_904061, EPI_ISL_904062, EPI_ISL_904063, EPI_ISL_904064, EPI_ISL_904065, EPI_ISL_904066, EPI_ISL_904067, EPI_ISL_904068, EPI_ISL_904069, EPI_ISL_904070, EPI_ISL_904071, EPI_ISL_904072, EPI_ISL_904073, EPI_ISL_904074, EPI_ISL_904075, EPI_ISL_904076, EPI_ISL_904077, EPI_ISL_904078, EPI_ISL_904079, EPI_ISL_904080, EPI_ISL_904081, EPI_ISL_904082, EPI_ISL_904083, EPI_ISL_904084, EPI_ISL_904085, EPI_ISL_904086, EPI_ISL_904087, EPI_ISL_904088, EPI_ISL_904089, EPI_ISL_904090, EPI_ISL_904091, EPI_ISL_904092, EPI_ISL_904093, EPI_ISL_904094, EPI_ISL_904095, EPI_ISL_904096, EPI_ISL_904097, EPI_ISL_904098, EPI_ISL_904099, EPI_ISL_904100, EPI_ISL_904101, EPI_ISL_904102, EPI_ISL_904103, EPI_ISL_904104, EPI_ISL_904105, EPI_ISL_904106, EPI_ISL_904107, EPI_ISL_904108, EPI_ISL_904109, EPI_ISL_904110, EPI_ISL_904111, EPI_ISL_904112, EPI_ISL_904113, EPI_ISL_904114, EPI_ISL_904115, EPI_ISL_904116, EPI_ISL_904117, EPI_ISL_904118, EPI_ISL_904119, EPI_ISL_906777, EPI_ISL_906778, EPI_ISL_906779 | | | |
| see above | New Mexico Department of Health Scientific Laboratory | New Mexico Department of Health Scientific Laboratory | Ellie Johnson, Anastacia Griego-Fisher, D'eldra Malone |
| EPI_ISL_907069, EPI_ISL_907070 | Infectious Diseases, Quest Diagnostics | Infectious Diseases, Quest Diagnostics | Rosenthal,S.H., Gerasimova,A., Kagan,R.M., Anderson,B., Bernstein,L.E., Livingston,K.E., Hua,M., Liu,Y., Shalhout,D.F., Owen,R., Lacbawan,F. |
| EPI_ISL_913915, EPI_ISL_913916, EPI_ISL_913920, EPI_ISL_913921, EPI_ISL_913922, EPI_ISL_913923, EPI_ISL_913924, EPI_ISL_913925, EPI_ISL_913926, EPI_ISL_913927, EPI_ISL_913928, EPI_ISL_913929, EPI_ISL_913930, EPI_ISL_913931, EPI_ISL_913932, EPI_ISL_913933, EPI_ISL_913934, EPI_ISL_913935, EPI_ISL_913936, EPI_ISL_913937, EPI_ISL_913938, EPI_ISL_913939, EPI_ISL_913940, EPI_ISL_913941, EPI_ISL_913942, EPI_ISL_913943, EPI_ISL_913944, EPI_ISL_913945, EPI_ISL_913946, EPI_ISL_913947, EPI_ISL_913948, EPI_ISL_913949, EPI_ISL_913950, EPI_ISL_913951, EPI_ISL_913952, EPI_ISL_913953, EPI_ISL_913954, EPI_ISL_913955, EPI_ISL_913957, EPI_ISL_913958, EPI_ISL_913959, EPI_ISL_913960, EPI_ISL_913961, EPI_ISL_913962, EPI_ISL_913963, EPI_ISL_913964, EPI_ISL_913965, EPI_ISL_913966, EPI_ISL_913967, EPI_ISL_913968, EPI_ISL_913970, EPI_ISL_913971, EPI_ISL_913972, EPI_ISL_913973, EPI_ISL_913974, EPI_ISL_913975, EPI_ISL_913976, EPI_ISL_913977, EPI_ISL_913978, EPI_ISL_913979, EPI_ISL_913980, EPI_ISL_913981, EPI_ISL_913982, EPI_ISL_913983, EPI_ISL_913984, EPI_ISL_914880 | | | |
| see above | Instituto de Diagnostico y Referencia Epidemiologicos INDRE_RNLSP | Instituto de Diagnostico y Referencia Epidemiologicos (INDRE) | Claudia Wong-Arambula, Abril Rodriguez-Maldonado, Fabiola Garces-Ayala, Adnan Araiza-Rodriguez, David Fragoso-Fonseca, Sergio Rangel-Guerrero, Mayra Jimenez-Morales, Nancy Munoz-Hernandez, Natividad Cruz-Ortiz, Tatiana Nunez-Garcia, Gisela Barrera-Badillo, Lucia Hernandez-Rivas, Irma Lopez-Martinez, Ernesto Ramirez-Gonzalez. |
| EPI_ISL_915335, EPI_ISL_915336, EPI_ISL_915337, EPI_ISL_915338, EPI_ISL_915339 | Quest Diagnostics | Quest Diagnostics | Rosenthal,S.H., Gerasimova,A., Kagan,R.M., Anderson, B., Hua, M., Liu Y., Bernstein, L.E., Livingston, K.E., Perez, A., Shalhout, D.F., Shlyakhter, I.A., Owen, R., Tanpaiboon, P., Lacbawan, F. |
| EPI_ISL_925209, EPI_ISL_925210, EPI_ISL_925211, EPI_ISL_925212, EPI_ISL_925213, EPI_ISL_925214, EPI_ISL_925215, EPI_ISL_925216, EPI_ISL_925217, EPI_ISL_925218, EPI_ISL_925219, EPI_ISL_925220, EPI_ISL_925221, EPI_ISL_925222, EPI_ISL_925223, EPI_ISL_925224, EPI_ISL_925225, EPI_ISL_925226, EPI_ISL_925227, EPI_ISL_925228, EPI_ISL_925229, EPI_ISL_925230, EPI_ISL_925231, EPI_ISL_925232, EPI_ISL_925233, EPI_ISL_925234, EPI_ISL_925235, EPI_ISL_925236, EPI_ISL_925237, EPI_ISL_925238, EPI_ISL_925239, EPI_ISL_925240, EPI_ISL_925241, EPI_ISL_925242, EPI_ISL_925243, EPI_ISL_925244, EPI_ISL_925245, EPI_ISL_925246, EPI_ISL_925247, EPI_ISL_925248, EPI_ISL_925249, EPI_ISL_925250, EPI_ISL_925251, EPI_ISL_925252, EPI_ISL_925253, EPI_ISL_925254, EPI_ISL_925255, EPI_ISL_925256, EPI_ISL_925257 | | | |
| see above | New Mexico Department of Health Scientific Laboratory | New Mexico Department of Health Scientific Laboratory | Ellie Johnson, Anastacia Griego-Fisher, D'eldra Malone, Jennifer Benoit |
| EPI_ISL_930783, EPI_ISL_933512, EPI_ISL_933513, EPI_ISL_933514, EPI_ISL_933515, EPI_ISL_933516, EPI_ISL_933517, EPI_ISL_933518, EPI_ISL_933519, EPI_ISL_933520, EPI_ISL_933521, EPI_ISL_933522, EPI_ISL_933523, EPI_ISL_933524, EPI_ISL_933525, EPI_ISL_933526, EPI_ISL_933527, EPI_ISL_933528, EPI_ISL_933529, EPI_ISL_933530, EPI_ISL_933531 | | | |
| see above | Instituto Nacional de Medicina Genomica | Instituto Nacional de Medicina Genomica | Hidalgo-Miranda A, Mendoza-Vargas A, Reyes-Grajeda JP, Cisneros-Villanueva M, Cedro-Tanda A,Peñaloza-Figueroa F, Herrera-Montalvo LA |
| EPI_ISL_933661, EPI_ISL_933662, EPI_ISL_933663, EPI_ISL_933666, EPI_ISL_933667, EPI_ISL_933668, EPI_ISL_933669, EPI_ISL_933670, EPI_ISL_933671, EPI_ISL_933672, EPI_ISL_933673, EPI_ISL_933674, EPI_ISL_933675, EPI_ISL_933676, EPI_ISL_933677, EPI_ISL_933678, EPI_ISL_933679, EPI_ISL_933680, EPI_ISL_933681, EPI_ISL_933682, EPI_ISL_933683, EPI_ISL_933684, EPI_ISL_933685, EPI_ISL_933686, EPI_ISL_933687, EPI_ISL_933688, EPI_ISL_933689, EPI_ISL_933690, EPI_ISL_933691, EPI_ISL_933692, EPI_ISL_933693, EPI_ISL_933694, EPI_ISL_933695, EPI_ISL_933696, EPI_ISL_933697, EPI_ISL_933698, EPI_ISL_933699, EPI_ISL_933700, EPI_ISL_933701, EPI_ISL_933702, EPI_ISL_933703, EPI_ISL_933704, EPI_ISL_933705, EPI_ISL_933706, EPI_ISL_933707, EPI_ISL_933708, EPI_ISL_933709, EPI_ISL_933710, EPI_ISL_933711, EPI_ISL_933712 | | | |
| see above | Instituto de Diagnostico y Referencia Epidemiologicos INDRE_RNLSP | Instituto de Diagnostico y Referencia Epidemiologicos (INDRE) | Claudia Wong-Arambula, Abril Rodriguez-Maldonado, Fabiola Garces-Ayala, Adnan Araiza-Rodriguez, David Fragoso-Fonseca, Sergio Rangel-Guerrero, Mayra Jimenez-Morales, Nancy Munoz-Hernandez, Natividad Cruz-Ortiz, Tatiana Nunez-Garcia, Gisela Barrera-Badillo, Lucia Hernandez-Rivas, Irma Lopez-Martinez, Ernesto Ramirez-Gonzalez. |
| EPI_ISL_941300, EPI_ISL_941301, EPI_ISL_941303, EPI_ISL_941305, EPI_ISL_941306, EPI_ISL_941307, EPI_ISL_941308, EPI_ISL_941309, EPI_ISL_941312, EPI_ISL_941313, EPI_ISL_941314, EPI_ISL_941315, EPI_ISL_941316, EPI_ISL_941317, EPI_ISL_941318, EPI_ISL_941319, EPI_ISL_941320, EPI_ISL_941321, EPI_ISL_941322, EPI_ISL_941323, EPI_ISL_941324, EPI_ISL_941325, EPI_ISL_941326, EPI_ISL_941327, EPI_ISL_941328, EPI_ISL_941329 | | | |
| see above | New Mexico Department of Health Scientific Laboratory | New Mexico Department of Health Scientific Laboratory | Ellie Johnson, Anastacia Griego-Fisher, D'eldra Malone, Jennifer Benoit |
| EPI_ISL_942927 | LESP Nuevo Leon/Grupo de Diagnostico ARIES | Instituto de Diagnostico y Referencia Epidemiologicos (INDRE) | Claudia Wong-Arambula, Abril Rodriguez-Maldonado, Fabiola Garces-Ayala, Natividad Cruz-Ortiz, Tatiana Nunez-Garcia, Gisela Barrera-Badillo, Lucia Hernandez-Rivas, Irma Lopez-Martinez, Ernesto Ramirez-Gonzalez. |
| EPI_ISL_942928 | Instituto de Diagnostico y Referencia Epidemiologicos INDRE_RNLSP | Instituto de Diagnostico y Referencia Epidemiologicos (INDRE) | Claudia Wong-Arambula, Abril Rodriguez-Maldonado, Fabiola Garces-Ayala, Natividad Cruz-Ortiz, Tatiana Nunez-Garcia, Gisela Barrera-Badillo, Lucia Hernandez-Rivas, Irma Lopez-Martinez, Ernesto Ramirez-Gonzalez. |
| EPI_ISL_944611, EPI_ISL_944612, EPI_ISL_944613, EPI_ISL_944614, EPI_ISL_944615, EPI_ISL_944616, EPI_ISL_944617, EPI_ISL_944618, EPI_ISL_944619, EPI_ISL_944620, EPI_ISL_944621, EPI_ISL_944622, EPI_ISL_944623, EPI_ISL_944624, EPI_ISL_944625, EPI_ISL_944626, EPI_ISL_944627, EPI_ISL_944628, EPI_ISL_944629, EPI_ISL_944630, EPI_ISL_944631, EPI_ISL_944632, EPI_ISL_944633, EPI_ISL_955189, EPI_ISL_955190, EPI_ISL_955191, EPI_ISL_955192, EPI_ISL_955193, EPI_ISL_955194, EPI_ISL_955195, EPI_ISL_955196, EPI_ISL_955197, EPI_ISL_955198, EPI_ISL_955199, EPI_ISL_955200, EPI_ISL_955201, EPI_ISL_955202, EPI_ISL_955203, EPI_ISL_955204, EPI_ISL_955205, EPI_ISL_955206, EPI_ISL_955207, EPI_ISL_955208, EPI_ISL_955209 | | | |
| see above | Instituto Nacional de Medicina Genomica | Instituto Nacional de Medicina Genomica | Hidalgo-Miranda A, Mendoza-Vargas A, Reyes-Grajeda JP, Cisneros-Villanueva M, Cedro-Tanda A,Peñaloza-Figueroa F, Herrera-Montalvo LA |
| EPI_ISL_961466 | Laboratorios Lister | Instituto de Diagnostico y Referencia Epidemiologicos (INDRE) | Claudia Wong-Arambula, Abril Rodriguez-Maldonado, Fabiola Garces-Ayala, Natividad Cruz-Ortiz, Tatiana Nunez-Garcia, Gisela Barrera-Badillo, Lucia Hernandez-Rivas, Irma Lopez-Martinez, Ernesto Ramirez-Gonzalez. |
| EPI_ISL_961467, EPI_ISL_961468, EPI_ISL_961469 | LESP Nuevo Leon/Grupo de Diagnostico ARIES | Instituto de Diagnostico y Referencia Epidemiologicos (INDRE) | Claudia Wong-Arambula, Abril Rodriguez-Maldonado, Fabiola Garces-Ayala, Natividad Cruz-Ortiz, Tatiana Nunez-Garcia, Gisela Barrera-Badillo, Lucia Hernandez-Rivas, Irma Lopez-Martinez, Ernesto Ramirez-Gonzalez. |
| EPI_ISL_962662, EPI_ISL_962663 | Sharp HealthCare Laboratory | Andersen lab at Scripps Research | SEARCH Alliance San Diego with Aaron Harding, Jacquelyn Berumen, Cathy Woerle, Liam McGinnis, Art Mendoza, Omid Bakhtar |
| EPI_ISL_966621, EPI_ISL_966747, EPI_ISL_967050, EPI_ISL_967275, EPI_ISL_967879 | Helix/Illumina | Respiratory Viruses Branch, Division of Viral Diseases, Centers for Disease Control and Prevention | Peter W. Cook,Dakota Howard,Dhwani Batra,Ben L. Rambo-Martin,Eileen de Feo,Jan Antico,Christine Tran,Matthew Tolentino,Shannon Wickline,Kim Gietzen,Brad Sickler,Jingtao Liu,Eric Allen,Phil Febbo,Summer Galloway,Nicole L. Washington,Simon White,Geraint Levan,Kelly Schiabor Barrett,Elizabeth Cirulli,Alexandre Bolze,Ary Ascencio,Charlotte Rivera-Garcia,Ryan Cho,Jason Nguyen,Sherry Wang,Jimmy Ramirez,Tyler Cassens,Efren Sandoval,Magnus Isaksson,William Lee,David Becker,Marc Laurent,James Lu,Clinton R. Paden,Suxiang Tong,Duncan MacCannell, |
| EPI_ISL_979327, EPI_ISL_979328, EPI_ISL_979329, EPI_ISL_979330, EPI_ISL_979331, EPI_ISL_979332, EPI_ISL_979333, EPI_ISL_979334, EPI_ISL_979335, EPI_ISL_979336, EPI_ISL_979337, EPI_ISL_979338, EPI_ISL_979339, EPI_ISL_979340, EPI_ISL_979341, EPI_ISL_979342, EPI_ISL_979343, EPI_ISL_979344, EPI_ISL_979345, EPI_ISL_979346, EPI_ISL_979347, EPI_ISL_979348, EPI_ISL_979349, EPI_ISL_979350, EPI_ISL_979351, EPI_ISL_979352 | | | |

| | | | |
|---|---|---|---|
| see above | Laboratorio Estatal de Salud Pública de Nuevo León | Laboratorio de Infectología Molecular, Departamento de Bioquímica y Medicina Molecular,Facultad de Medicina - Universidad Autónoma de Nuevo León | Kame A. Galán-Huerta, María F. Herrera-Saldivar, Natalia Martínez-Acuña, Sonia A. Lozano-Sepúlveda, Daniel Arellanos-Soto, Ana M. Rivas-Estilla, Samuel Buentello-Wong, Else del Carmen García-García, Gloria A. Jasso-de-la-Peña, Roberto Montes-de-Oca, Consuelo Treviño-Garza, Manuel E. de-la-O-Cavazos |
| EPI_ISL_979410, EPI_ISL_979411 | New Mexico Department of Health Scientific Laboratory | New Mexico Department of Health Scientific Laboratory | Ellie Johnson, Anastacia Griego-Fisher, D'eldra Malone, Jennifer Benoit |
| EPI_ISL_985149, EPI_ISL_985150, EPI_ISL_985151, EPI_ISL_985152, EPI_ISL_985153, EPI_ISL_985154, EPI_ISL_985155, EPI_ISL_985156, EPI_ISL_985157, EPI_ISL_985158, EPI_ISL_985159, EPI_ISL_985160, EPI_ISL_985161, EPI_ISL_985162, EPI_ISL_985163, EPI_ISL_985164, EPI_ISL_985165, EPI_ISL_985166, EPI_ISL_985167 | | | |
| see above | Instituto Nacional de Medicina Genomica | Instituto Nacional de Medicina Genomica | Hidalgo-Miranda A, Mendoza-Vargas A, Reyes-Grajeda JP, Cisneros-Villanueva M, Cedro-Tanda A,Peñaloza-Figueroa F, Herrera-Montalvo LA |

We gratefully acknowledge the following Authors from the Originating laboratories responsible for obtaining the specimens, as well as the Submitting laboratories where the genome data were generated and shared via GISAID, on which this research is based.

All Submitters of data may be contacted directly via www.gisaid.org

Authors are sorted alphabetically.

| Accession ID | Originating Laboratory | Submitting Laboratory | Authors |
|---|---|---|---|
| EPI_ISL_1711717, EPI_ISL_1711718 | "NYSDOH Wadsworth Center, Virology Lab" | Centers for Disease Control and Prevention Division of Viral Diseases, Pathogen Discovery | Alison Laufer Halpin; Ben L. Rambo-Martin; Clinton R. Paden; Dakota Howard; Darlene Wagner; Dave Wentworth; Dhwani Batra; Jasmine Padilla; Justin Lee; Katie Dillon; Krista Queen; Kristen Knipe; Kristine Lacek; Mark Burroughs; Matthew Schmerer; Mili Sheth; Peter Cook; Sam Shepard; Sarah Nobles; Shoshona Le; Suxiang Tong; Vivien Dugan; Yvette Unoarumhi |
| EPI_ISL_1226003, EPI_ISL_1226004, EPI_ISL_1226005, EPI_ISL_1226006, EPI_ISL_1226048, EPI_ISL_1226049, EPI_ISL_1226050, EPI_ISL_1226051, EPI_ISL_1226052, EPI_ISL_1226053 | | | |
| see above | "NYSDOH Wadsworth Center, Virology Lab" | Genomics and Discovery, Respiratory Viruses Branch, Division of Viral Diseases, Centers for Disease Control and Prevention | Anna Montmayeur; Anna Uehara; Ben L. Rambo-Martin; Clinton R. Paden; Dhwani Batra; Haibin Wang; Jasmine Padilla; Jing Zhang; Justin Lee; Katie Dillon; Krista Queen; Kristen Knipe; Kristine Lacek; Lori Rowe; Mark Burroughs; Matthew Schmerer; Mili Sheth; Peter W. Cook; Rachel Marine; Sam Shepard; Sarah Nobles; Shoshona Le; Suxiang Tong; Yan Li; Ying Tao |
| EPI_ISL_830724, EPI_ISL_830725, EPI_ISL_853676, EPI_ISL_853677, EPI_ISL_853678, EPI_ISL_853679, EPI_ISL_853680, EPI_ISL_853681, EPI_ISL_853682 | | | |
| see above | ACUTIS DIAGNOSTICS | Wadsworth Center, New York State Department of Health | Alexis Russel; Daryl M. Lamson; Erasmus Schneider; Erica Lasek-Nesselquist; John Kelly; Jonathan Plitnick; Kirsten St. George; Matthew Shudt; Melissa A Leisner; Navjot Singh |
| EPI_ISL_861135, EPI_ISL_861136, EPI_ISL_861137, EPI_ISL_861138, EPI_ISL_861139, EPI_ISL_861140, EPI_ISL_861141, EPI_ISL_861142, EPI_ISL_861143, EPI_ISL_883436, EPI_ISL_883437, EPI_ISL_883438, EPI_ISL_883439, EPI_ISL_883440, EPI_ISL_883441, EPI_ISL_883442, EPI_ISL_883443, EPI_ISL_883444, EPI_ISL_883473, EPI_ISL_936046, EPI_ISL_936047, EPI_ISL_936048, EPI_ISL_936049, EPI_ISL_936050, EPI_ISL_936051, EPI_ISL_936052, EPI_ISL_936053, EPI_ISL_936054, EPI_ISL_936055, EPI_ISL_936056, EPI_ISL_936057, EPI_ISL_936058, EPI_ISL_936059, EPI_ISL_982499, EPI_ISL_982500, EPI_ISL_982501, EPI_ISL_982502, EPI_ISL_982503, EPI_ISL_982504, EPI_ISL_1227555, EPI_ISL_1228292, EPI_ISL_1397424, EPI_ISL_1397425, EPI_ISL_1397426, EPI_ISL_1397427, EPI_ISL_1397428, EPI_ISL_1397429, EPI_ISL_1397430, EPI_ISL_1397431, EPI_ISL_1397432, EPI_ISL_1397433, EPI_ISL_1397434, EPI_ISL_1397435, EPI_ISL_1397436, EPI_ISL_1397437, EPI_ISL_1397438, EPI_ISL_1397439, EPI_ISL_1397440, EPI_ISL_1397441, EPI_ISL_1397442, EPI_ISL_1397443, EPI_ISL_1397711, EPI_ISL_1397721, EPI_ISL_1397723, EPI_ISL_1397731, EPI_ISL_1397733, EPI_ISL_1397734, EPI_ISL_1397735, EPI_ISL_1397738, EPI_ISL_1397739, EPI_ISL_1397741, EPI_ISL_1397743, EPI_ISL_1397745, EPI_ISL_1397746, EPI_ISL_1397766 | | | |
| see above | ADIRONDACK MEDICAL CENTER | Wadsworth Center, New York State Department of Health | Alexis Russel; Daryl M. Lamson; Erasmus Schneider; Erica Lasek-Nesselquist; John Kelly; Jonathan Plitnick; Kirsten St. George; Matthew Shudt; Melissa A Leisner; Navjot Singh |
| EPI_ISL_936180, EPI_ISL_936181, EPI_ISL_1226704, EPI_ISL_1226705, EPI_ISL_1227249, EPI_ISL_1227250, EPI_ISL_1227251, EPI_ISL_1227252, EPI_ISL_1227253, EPI_ISL_1227256, EPI_ISL_1227458, EPI_ISL_1227459, EPI_ISL_1227460, EPI_ISL_1227556, EPI_ISL_1227564, EPI_ISL_1227565, EPI_ISL_1228218, EPI_ISL_1228270, EPI_ISL_1397707, EPI_ISL_1397708, EPI_ISL_1397709, EPI_ISL_1397710, EPI_ISL_1397712, EPI_ISL_1397713, EPI_ISL_1397714, EPI_ISL_1397715, EPI_ISL_1397716, EPI_ISL_1397717, EPI_ISL_1397718, EPI_ISL_1397719, EPI_ISL_1397720, EPI_ISL_1397722, EPI_ISL_1397724, EPI_ISL_1397725, EPI_ISL_1397726, EPI_ISL_1397727, EPI_ISL_1397728, EPI_ISL_1397729, EPI_ISL_1397730, EPI_ISL_1397732, EPI_ISL_1397736, EPI_ISL_1397737, EPI_ISL_1397740, EPI_ISL_1397742, EPI_ISL_1397744, EPI_ISL_1397752, EPI_ISL_1397753, EPI_ISL_1397754, EPI_ISL_1397755, EPI_ISL_1397756, EPI_ISL_1397757, EPI_ISL_1397758, EPI_ISL_1397759, EPI_ISL_1397760, EPI_ISL_1397761, EPI_ISL_1397762, EPI_ISL_1397763, EPI_ISL_1397764, EPI_ISL_1397765, EPI_ISL_1397767, EPI_ISL_1755939 | | | |
| see above | ALBANY MEDICAL CENTER | Wadsworth Center, New York State Department of Health | Alexis Russel; Alexis Russell; Catharine Prussing; Daryl M. Lamson; Erasmus Schneider; Erica Lasek-Nesselquist; John Kelly; Jonathan Plitnick; Kirsten St. George; Matthew Shudt; Melissa A Leisner; Navjot Singh |
| EPI_ISL_830683, EPI_ISL_830684, EPI_ISL_830685, EPI_ISL_830686, EPI_ISL_830687, EPI_ISL_830688, EPI_ISL_830689, EPI_ISL_830690, EPI_ISL_830691, EPI_ISL_830692, EPI_ISL_830693, EPI_ISL_830694, EPI_ISL_830695, EPI_ISL_830696, EPI_ISL_830697, EPI_ISL_830698, EPI_ISL_830699, EPI_ISL_830700, EPI_ISL_830701, EPI_ISL_830702, EPI_ISL_830703, EPI_ISL_830704, EPI_ISL_830705, EPI_ISL_830706, EPI_ISL_830707, EPI_ISL_830708, EPI_ISL_830709, EPI_ISL_830710, EPI_ISL_830711, EPI_ISL_830712, EPI_ISL_830713, EPI_ISL_830714, EPI_ISL_830715, EPI_ISL_830716, EPI_ISL_830717, EPI_ISL_830718, EPI_ISL_830719, EPI_ISL_853684, EPI_ISL_853685, EPI_ISL_853686, EPI_ISL_853687, EPI_ISL_853690, EPI_ISL_853691, EPI_ISL_853692, EPI_ISL_853693, EPI_ISL_853694, EPI_ISL_853695, EPI_ISL_853696, EPI_ISL_853697, EPI_ISL_853698, EPI_ISL_853699, EPI_ISL_853700, EPI_ISL_853701, EPI_ISL_853702, EPI_ISL_853703, EPI_ISL_853704, EPI_ISL_853709, EPI_ISL_853710, EPI_ISL_853711, EPI_ISL_884008, EPI_ISL_884009, EPI_ISL_884010, EPI_ISL_884011, EPI_ISL_884012, EPI_ISL_884017, EPI_ISL_884018, EPI_ISL_884019, EPI_ISL_884020, EPI_ISL_884021, EPI_ISL_884022, EPI_ISL_884023, EPI_ISL_884024, EPI_ISL_884025, EPI_ISL_884026, EPI_ISL_884027, EPI_ISL_884028, EPI_ISL_884029, EPI_ISL_884030, EPI_ISL_884031, EPI_ISL_884042, EPI_ISL_884043, EPI_ISL_884044, EPI_ISL_884045, EPI_ISL_884046, EPI_ISL_884048, EPI_ISL_884049, EPI_ISL_884050, EPI_ISL_884051, EPI_ISL_884052, EPI_ISL_884053, EPI_ISL_884059, EPI_ISL_884060, EPI_ISL_884061, EPI_ISL_884062, EPI_ISL_884063, EPI_ISL_884064, EPI_ISL_884065, EPI_ISL_884066, EPI_ISL_884067, EPI_ISL_884068, EPI_ISL_884069, EPI_ISL_884070, EPI_ISL_884071, EPI_ISL_884072, EPI_ISL_884073, EPI_ISL_884074, EPI_ISL_884075, EPI_ISL_884076, EPI_ISL_884077, EPI_ISL_884078, EPI_ISL_884079, EPI_ISL_884081, EPI_ISL_884082, EPI_ISL_884083, EPI_ISL_884084, EPI_ISL_884085, EPI_ISL_1016417, EPI_ISL_1016468, EPI_ISL_1016469, EPI_ISL_1016470, EPI_ISL_1016471, EPI_ISL_1016472, EPI_ISL_1016473, EPI_ISL_1016474, EPI_ISL_1016475, EPI_ISL_1499457, EPI_ISL_1499458, EPI_ISL_1499459, EPI_ISL_1499460, EPI_ISL_1499461, EPI_ISL_1499462, EPI_ISL_1499463, EPI_ISL_1499464, EPI_ISL_1499465, EPI_ISL_1499466, EPI_ISL_1499467, EPI_ISL_1499468, EPI_ISL_1499469, EPI_ISL_1499470, EPI_ISL_1499471, EPI_ISL_1499472, EPI_ISL_1499473 | | | |
| see above | ALBANY MEDICAL CENTER HOSPITAL CLINICAL LABORATORIES | Wadsworth Center, New York State Department of Health | Alexis Russel; Daryl M. Lamson; Erasmus Schneider; Erica Lasek-Nesselquist; John Kelly; Jonathan Plitnick; Kirsten St. George; Matthew Shudt; Melissa A Leisner; Navjot Singh |
| EPI_ISL_1920104, EPI_ISL_1920108, EPI_ISL_1920110, EPI_ISL_1920111 | Albany Medical Center | Wadsworth Center, New York State Department of Health | Alexis Russell; Catharine Prussing; Daryl M. Lamson; Erasmus Schneider; Erica Lasek-Nesselquist; John Kelly; Jonathan Plitnick; Kirsten St. George; Matthew Shudt; Melissa A Leisner; Navjot Singh |
| EPI_ISL_769916, EPI_ISL_769917, EPI_ISL_769918, EPI_ISL_769919, EPI_ISL_769920, EPI_ISL_769921, EPI_ISL_769922, EPI_ISL_769923, EPI_ISL_769924, EPI_ISL_769925, EPI_ISL_769926, EPI_ISL_769927, EPI_ISL_769928, EPI_ISL_769929, EPI_ISL_769930, EPI_ISL_769931, EPI_ISL_769932, EPI_ISL_769933, EPI_ISL_769934, EPI_ISL_769935, EPI_ISL_769936, EPI_ISL_769937, EPI_ISL_769938, EPI_ISL_769939, EPI_ISL_769940, EPI_ISL_769941, EPI_ISL_769942, EPI_ISL_769943, EPI_ISL_769944, EPI_ISL_769945, EPI_ISL_769946, EPI_ISL_769947, EPI_ISL_769948, EPI_ISL_769949, EPI_ISL_769950, EPI_ISL_769951, EPI_ISL_769952, EPI_ISL_769953, EPI_ISL_769954, EPI_ISL_769955, EPI_ISL_769956, EPI_ISL_769957, EPI_ISL_769958, EPI_ISL_769959, EPI_ISL_769960, EPI_ISL_769961, EPI_ISL_769962, EPI_ISL_769963, EPI_ISL_769964, EPI_ISL_769965, EPI_ISL_769966, EPI_ISL_769967, EPI_ISL_769968, EPI_ISL_769969, EPI_ISL_769970, EPI_ISL_769971, EPI_ISL_769972, EPI_ISL_769973, EPI_ISL_769974, EPI_ISL_769975, EPI_ISL_769976, EPI_ISL_769977, EPI_ISL_769978, EPI_ISL_769979, EPI_ISL_769980, EPI_ISL_769981, EPI_ISL_769982, EPI_ISL_769983, EPI_ISL_769984, EPI_ISL_769985 | | | |
| see above | Albany Medical Center Hospital Clinical Laboratories | Wadsworth Center, New York State Department.of Health | Alexis Russel; Daryl M. Lamson; Erasmus Schneider; Erica Lasek-Nesselquist; John Kelly; Jonathan Plitnick; Kirsten St. George; Matthew Shudt; Melissa A Leisner; Navjot Singh; Sara Griesemer |
| EPI_ISL_1009188, EPI_ISL_1009189, EPI_ISL_1009190, EPI_ISL_1009192, EPI_ISL_1009193 | American Esoteric Laboratories | Colleen B. Jonsson | Colleen B. Jonsson; Jyothi Parvathareddy; Mariah K. Taylor; Walter Reichard |
| EPI_ISL_955271 | American Esoteric Laboratory | Pathogen Discovery, Respiratory Viruses Branch, Division of Viral Diseases, Centers for Disease Control and Prevention | Anna Uehara; Clinton R. Paden; Haibin Wang; Jing Zhang; Krista Queen; Peter Cook; Suxiang Tong; Yan Li; Ying Tao |
| EPI_ISL_804981, EPI_ISL_804982, EPI_ISL_804983, EPI_ISL_804984, EPI_ISL_830720, EPI_ISL_830721, EPI_ISL_830722, EPI_ISL_830723, EPI_ISL_854350, EPI_ISL_854351, EPI_ISL_854352, EPI_ISL_854353, EPI_ISL_854354, EPI_ISL_854355, EPI_ISL_854356, EPI_ISL_854357, EPI_ISL_854358, EPI_ISL_854359, EPI_ISL_854360, EPI_ISL_854361, EPI_ISL_854362, EPI_ISL_854363, EPI_ISL_854364, EPI_ISL_854365, EPI_ISL_854366, EPI_ISL_854367, EPI_ISL_854369, EPI_ISL_854372, EPI_ISL_861175, EPI_ISL_861176, EPI_ISL_861177, EPI_ISL_861178, EPI_ISL_861179, EPI_ISL_861180, EPI_ISL_861181, EPI_ISL_861182, EPI_ISL_861185, EPI_ISL_861187, EPI_ISL_861210, EPI_ISL_861211, EPI_ISL_861212, EPI_ISL_861213, EPI_ISL_861214, EPI_ISL_861215, EPI_ISL_861216, EPI_ISL_861217, EPI_ISL_861218, EPI_ISL_861219, EPI_ISL_861220, EPI_ISL_861221, EPI_ISL_861222, EPI_ISL_861223, EPI_ISL_861224, EPI_ISL_861225, EPI_ISL_861226, EPI_ISL_861227, EPI_ISL_861228, EPI_ISL_861229, EPI_ISL_861230, EPI_ISL_861231, EPI_ISL_861232, EPI_ISL_896297, EPI_ISL_896300 | | | |
| see above | BIO-REFERENCE LABORATORIES | Wadsworth Center, New York State Department of Health | Alexis Russel; Daryl M. Lamson; Erasmus Schneider; Erica Lasek-Nesselquist; John Kelly; Jonathan Plitnick; Kirsten St. George; Matthew Shudt; Melissa A Leisner; Navjot Singh |
| EPI_ISL_767648, EPI_ISL_767650, EPI_ISL_767651, EPI_ISL_767652, EPI_ISL_767653, EPI_ISL_767665, EPI_ISL_767666, EPI_ISL_767667, EPI_ISL_767668, EPI_ISL_767669, EPI_ISL_767670, EPI_ISL_767671, EPI_ISL_767672, EPI_ISL_767673, EPI_ISL_767674, EPI_ISL_767675, EPI_ISL_767676, EPI_ISL_767677, EPI_ISL_767678, EPI_ISL_767679, EPI_ISL_767680, EPI_ISL_802430, EPI_ISL_802445, EPI_ISL_802446, EPI_ISL_802447, EPI_ISL_802790, EPI_ISL_802791, EPI_ISL_802792, EPI_ISL_802793, EPI_ISL_804946 | | | |
| see above | BIO-REFERENCE LABORATORIES | Wadsworth Center, New York State Department.of Health | Alexis Russel; Daryl M. Lamson; Erasmus Schneider; Erica Lasek-Nesselquist; John Kelly; Jonathan Plitnick; Kirsten St. George; Matthew Shudt; Melissa A Leisner; Navjot Singh; Sara Griesemer |
| EPI_ISL_830214, EPI_ISL_854446, EPI_ISL_883425, EPI_ISL_883429, EPI_ISL_883430, EPI_ISL_1226540 | BOSTON HEART DIAGNOSTICS CORP | Wadsworth Center, New York State Department of Health | Alexis Russel; Daryl M. Lamson; Erasmus Schneider; Erica Lasek-Nesselquist; John Kelly; Jonathan Plitnick; Kirsten St. George; Matthew Shudt; Melissa A Leisner; Navjot Singh |
| EPI_ISL_847741 | Chiu Laboratory, University of California, San Francisco | Chiu Laboratory, University of California, San Francisco | Brian Bushnell; Candace Wang; Charles Chiu; Debra Wadford; Jill Hacker; Scot Federman; Xianding (Wayne) Deng |
| EPI_ISL_1080752, EPI_ISL_1080753, EPI_ISL_1080754, EPI_ISL_1080755, EPI_ISL_1080757, EPI_ISL_1080758, EPI_ISL_1080760, EPI_ISL_1080762, EPI_ISL_1080763, EPI_ISL_1080764, EPI_ISL_1080765, EPI_ISL_1080766, EPI_ISL_1080770, EPI_ISL_1080771, EPI_ISL_1080774, EPI_ISL_1080776, EPI_ISL_1080780, EPI_ISL_1080781, EPI_ISL_1080783, EPI_ISL_1080784, EPI_ISL_1080785, EPI_ISL_1080786, EPI_ISL_1080787, EPI_ISL_1080788, EPI_ISL_1080789, EPI_ISL_1080790, EPI_ISL_1080791, EPI_ISL_1080792, EPI_ISL_1080793, EPI_ISL_1080796, EPI_ISL_1080797, EPI_ISL_1080798, EPI_ISL_1080799, EPI_ISL_1080800, EPI_ISL_1080801, EPI_ISL_1080803, EPI_ISL_1080809, EPI_ISL_1080810, EPI_ISL_1500472, EPI_ISL_1500473, EPI_ISL_1500474, EPI_ISL_1500475, EPI_ISL_1500476, EPI_ISL_1500477, EPI_ISL_1500478, EPI_ISL_1500479, EPI_ISL_1500480, EPI_ISL_1500481, EPI_ISL_1500482, EPI_ISL_1500483, EPI_ISL_1500484, EPI_ISL_1500485, EPI_ISL_1500486, EPI_ISL_1500487, EPI_ISL_1500488, EPI_ISL_1500489, EPI_ISL_1500490, EPI_ISL_1500491, EPI_ISL_1500492, EPI_ISL_1500493, EPI_ISL_1500494, EPI_ISL_1500495, EPI_ISL_1500496, EPI_ISL_1500497, EPI_ISL_1500498, EPI_ISL_1500499, EPI_ISL_1500500, EPI_ISL_1500501, EPI_ISL_1500502, EPI_ISL_1500503, EPI_ISL_1500504, EPI_ISL_1500505, EPI_ISL_1500506, EPI_ISL_1500507, EPI_ISL_1500508, EPI_ISL_1500509, EPI_ISL_1500510, EPI_ISL_1500511, EPI_ISL_1500512, EPI_ISL_1500513, EPI_ISL_1500514, EPI_ISL_1500516, EPI_ISL_1500517, EPI_ISL_1500518, EPI_ISL_1500519, EPI_ISL_1500520, EPI_ISL_1500521, EPI_ISL_1500522, EPI_ISL_1500523, EPI_ISL_1500524, EPI_ISL_1500525, EPI_ISL_1500526, EPI_ISL_1500527, EPI_ISL_1500528, EPI_ISL_1500529, EPI_ISL_1500530, EPI_ISL_1500531, EPI_ISL_1500532, EPI_ISL_1500533, EPI_ISL_1500534, EPI_ISL_1500535, EPI_ISL_1500536, EPI_ISL_1500537, EPI_ISL_1500538, EPI_ISL_1500539, EPI_ISL_1500540, EPI_ISL_1500541, EPI_ISL_1500542, EPI_ISL_1500543, EPI_ISL_1500544, EPI_ISL_1500545, EPI_ISL_1500546, EPI_ISL_1500547, EPI_ISL_1500548, EPI_ISL_1500549, EPI_ISL_1500550, EPI_ISL_1500551, EPI_ISL_1500552, EPI_ISL_1500553, EPI_ISL_1500554, EPI_ISL_1500555, EPI_ISL_1500556, EPI_ISL_1500557, EPI_ISL_1500558, EPI_ISL_1500559, EPI_ISL_1500560, EPI_ISL_1500561, EPI_ISL_1500562, EPI_ISL_1500563, EPI_ISL_1500564, EPI_ISL_1500565, EPI_ISL_1500566, EPI_ISL_1500567, EPI_ISL_1500568, EPI_ISL_1500569, EPI_ISL_1500624, EPI_ISL_1965884, EPI_ISL_1965918, EPI_ISL_1965919, EPI_ISL_1965920, EPI_ISL_1965921, EPI_ISL_1965922, EPI_ISL_1965923, EPI_ISL_1965929, EPI_ISL_1965930, EPI_ISL_1965937, EPI_ISL_1965938, EPI_ISL_1965943, EPI_ISL_1965945, EPI_ISL_1965946, EPI_ISL_1965947, EPI_ISL_1965948, EPI_ISL_1965949, EPI_ISL_1965950, EPI_ISL_1965951, EPI_ISL_1965952, EPI_ISL_1965953, EPI_ISL_1965954, EPI_ISL_1965955, EPI_ISL_1965956, EPI_ISL_1965957, EPI_ISL_1965958, EPI_ISL_1965959, EPI_ISL_1965960, EPI_ISL_1965961, EPI_ISL_1965962, EPI_ISL_1965963, EPI_ISL_1965964, EPI_ISL_1965965, EPI_ISL_1965966, EPI_ISL_1965967, EPI_ISL_1965968, EPI_ISL_1965969, EPI_ISL_1965970, EPI_ISL_1965971, EPI_ISL_1965972, EPI_ISL_1965973, EPI_ISL_1965974, EPI_ISL_1965975, EPI_ISL_1965976, EPI_ISL_1965977, EPI_ISL_1965978, EPI_ISL_1965979, EPI_ISL_1965980, EPI_ISL_1965981, EPI_ISL_1965982, EPI_ISL_1965983, EPI_ISL_1965984, EPI_ISL_1965985, EPI_ISL_1965986, EPI_ISL_1965987, EPI_ISL_1965988, EPI_ISL_1965989, EPI_ISL_1965990, EPI_ISL_1965991, EPI_ISL_1965992, EPI_ISL_1965993, EPI_ISL_1965994, EPI_ISL_1965995, EPI_ISL_1965996, EPI_ISL_1965997, EPI_ISL_1965998, EPI_ISL_1965999, EPI_ISL_1966000, EPI_ISL_1966001, EPI_ISL_1966002, EPI_ISL_1966003, EPI_ISL_1966004, EPI_ISL_1966005, EPI_ISL_1966006, EPI_ISL_1966007, EPI_ISL_1966008, EPI_ISL_1966009, EPI_ISL_1966010, EPI_ISL_1966011, EPI_ISL_1966012, EPI_ISL_1966013, EPI_ISL_1966014, EPI_ISL_1966015, EPI_ISL_1966016, EPI_ISL_1966017, EPI_ISL_1966018, EPI_ISL_1966019, EPI_ISL_1966020, EPI_ISL_1966021, EPI_ISL_1966022, EPI_ISL_1966023, EPI_ISL_1966024, EPI_ISL_1966025, EPI_ISL_1966026, EPI_ISL_1966027, EPI_ISL_1966028, EPI_ISL_1966029, EPI_ISL_1966030, EPI_ISL_1966031, EPI_ISL_1966032, EPI_ISL_1966033, EPI_ISL_1966034, EPI_ISL_1966035, EPI_ISL_1966036, EPI_ISL_1966037, EPI_ISL_1966038, EPI_ISL_1966039, EPI_ISL_1966040, EPI_ISL_1966041, EPI_ISL_1966042, EPI_ISL_1966043, EPI_ISL_1966044, EPI_ISL_1966045, EPI_ISL_1966046, EPI_ISL_1966047, EPI_ISL_1966048, EPI_ISL_1966049, EPI_ISL_1966050, EPI_ISL_1966051, EPI_ISL_1966052, EPI_ISL_1966053, EPI_ISL_1966054, EPI_ISL_1966055, EPI_ISL_1966056, EPI_ISL_1966057, EPI_ISL_1966058 | | | |
| see above | Clinical Microbiology Laboratory, NewYork Presbyterian Hospital/Columbia University Irving Medical Center | Uhlemann Laboratory, Columbia University Irving Medical Center | Anne Kelley; Anne-Catrin Uhlemann; David Ho; Hiroshi Mohri; Maya Tagliavia; Medini K. Annavajhala |
| EPI_ISL_804980, EPI_ISL_896216, EPI_ISL_896217, EPI_ISL_896227, EPI_ISL_896231, EPI_ISL_896232, EPI_ISL_896233, EPI_ISL_896234, EPI_ISL_896236, EPI_ISL_896237, EPI_ISL_896238, EPI_ISL_896239, EPI_ISL_896240, EPI_ISL_896241, EPI_ISL_896242, EPI_ISL_896243, EPI_ISL_896244, EPI_ISL_896245, EPI_ISL_896246, EPI_ISL_896382, EPI_ISL_896383, EPI_ISL_896384, EPI_ISL_896386, EPI_ISL_896389, EPI_ISL_896391, EPI_ISL_896392, EPI_ISL_896393, EPI_ISL_896394, EPI_ISL_896395, EPI_ISL_896422, EPI_ISL_896423, EPI_ISL_896424, EPI_ISL_896425, EPI_ISL_896426, EPI_ISL_896427, EPI_ISL_896428, EPI_ISL_896429, EPI_ISL_896430, EPI_ISL_896431, EPI_ISL_896432, EPI_ISL_896433, EPI_ISL_896434, EPI_ISL_896435, EPI_ISL_896436, EPI_ISL_896437, EPI_ISL_896438, EPI_ISL_896439, EPI_ISL_896440, EPI_ISL_896441, EPI_ISL_896442, EPI_ISL_896443, EPI_ISL_896444, EPI_ISL_896445, EPI_ISL_896446, EPI_ISL_936036, EPI_ISL_936037, EPI_ISL_936038, EPI_ISL_936039, EPI_ISL_936040, EPI_ISL_936041, EPI_ISL_936042, EPI_ISL_936043, EPI_ISL_936044, EPI_ISL_936045, EPI_ISL_936063, EPI_ISL_936064, EPI_ISL_936065, EPI_ISL_936066, EPI_ISL_936069, EPI_ISL_936082, EPI_ISL_936086, EPI_ISL_936089, EPI_ISL_936092, EPI_ISL_936093, EPI_ISL_1015653, EPI_ISL_1015654, EPI_ISL_1015655, EPI_ISL_1015656, EPI_ISL_1015657, EPI_ISL_1015658, EPI_ISL_1015659, EPI_ISL_1015660, EPI_ISL_1015661, EPI_ISL_1015662, EPI_ISL_1015663, EPI_ISL_1015664, EPI_ISL_1015665, EPI_ISL_1015666, EPI_ISL_1015667, EPI_ISL_1015668, EPI_ISL_1015669, EPI_ISL_1015670, EPI_ISL_1015671, EPI_ISL_1015672, EPI_ISL_1015673, EPI_ISL_1015674, EPI_ISL_1015675, EPI_ISL_1015676, EPI_ISL_1015677, EPI_ISL_1015678, EPI_ISL_1015679, EPI_ISL_1015680, EPI_ISL_1015681, EPI_ISL_1015682, EPI_ISL_1015683, EPI_ISL_1015684, EPI_ISL_1015685, EPI_ISL_1015686, EPI_ISL_1015687, EPI_ISL_1015688, EPI_ISL_1015689, EPI_ISL_1015690, EPI_ISL_1015691, EPI_ISL_1015692, EPI_ISL_1015693, EPI_ISL_1015694, | | | |

| | | | |
|---|---|---|---|
| EPI_ISL_1015695, EPI_ISL_1015696, EPI_ISL_1015697, EPI_ISL_1015698, EPI_ISL_1015699, EPI_ISL_1015700, EPI_ISL_1015701, EPI_ISL_1015702, EPI_ISL_1015703, EPI_ISL_1015704, EPI_ISL_1015705, EPI_ISL_1015706, EPI_ISL_1015707, EPI_ISL_1015708, EPI_ISL_1015709, EPI_ISL_1015710, EPI_ISL_1015711, EPI_ISL_1015712, EPI_ISL_1016111, EPI_ISL_1016112, EPI_ISL_1016113, EPI_ISL_1016114, EPI_ISL_1016115, EPI_ISL_1016116, EPI_ISL_1016117, EPI_ISL_1016260, EPI_ISL_1016261, EPI_ISL_1016267, EPI_ISL_1016268, EPI_ISL_1016270, EPI_ISL_1016281, EPI_ISL_1016282, EPI_ISL_1016283, EPI_ISL_1016284, EPI_ISL_1016287, EPI_ISL_1016288, EPI_ISL_1016289, EPI_ISL_1016290, EPI_ISL_1016291, EPI_ISL_1016292, EPI_ISL_1016293, EPI_ISL_1016294, EPI_ISL_1016295, EPI_ISL_1016296, EPI_ISL_1016297, EPI_ISL_1016323, EPI_ISL_1016324, EPI_ISL_1016325, EPI_ISL_1016326, EPI_ISL_1016328, EPI_ISL_1016329, EPI_ISL_1016330, EPI_ISL_1016331, EPI_ISL_1020487, EPI_ISL_1020488, EPI_ISL_1020489, EPI_ISL_1020490, EPI_ISL_1020491, EPI_ISL_1020492, EPI_ISL_1020493, EPI_ISL_1020494, EPI_ISL_1020495, EPI_ISL_1020496, EPI_ISL_1020497, EPI_ISL_1020498, EPI_ISL_1020499, EPI_ISL_1020500, EPI_ISL_1020501, EPI_ISL_1020502, EPI_ISL_1020503, EPI_ISL_1020504, EPI_ISL_1020505, EPI_ISL_1020506, EPI_ISL_1020507, EPI_ISL_1020508, EPI_ISL_1020509, EPI_ISL_1020510, EPI_ISL_1020511, EPI_ISL_1020512, EPI_ISL_1020513, EPI_ISL_1020514, EPI_ISL_1020515, EPI_ISL_1020516, EPI_ISL_1020517, EPI_ISL_1020518, EPI_ISL_1020519, EPI_ISL_1020520, EPI_ISL_1020521, EPI_ISL_1020522, EPI_ISL_1020523, EPI_ISL_1020524, EPI_ISL_1020525, EPI_ISL_1020526, EPI_ISL_1020527, EPI_ISL_1020528, EPI_ISL_1020529, EPI_ISL_1020530, EPI_ISL_1020531, EPI_ISL_1020532, EPI_ISL_1020533, EPI_ISL_1020534, EPI_ISL_1020535, EPI_ISL_1020536, EPI_ISL_1020537, EPI_ISL_1020538, EPI_ISL_1020539, EPI_ISL_1020540, EPI_ISL_1020541, EPI_ISL_1020542, EPI_ISL_1020543, EPI_ISL_1020544, EPI_ISL_1020545, EPI_ISL_1020546, EPI_ISL_1020547, EPI_ISL_1020548, EPI_ISL_1020549, EPI_ISL_1020550, EPI_ISL_1226456, EPI_ISL_1226463, EPI_ISL_1226466, EPI_ISL_1226502, EPI_ISL_1226503, EPI_ISL_1226512, EPI_ISL_1226596, EPI_ISL_1226599, EPI_ISL_1226671, EPI_ISL_1226674, EPI_ISL_1226743, EPI_ISL_1226800, EPI_ISL_1226801, EPI_ISL_1226802, EPI_ISL_1226803, EPI_ISL_1226804, EPI_ISL_1226823, EPI_ISL_1226824, EPI_ISL_1226874, EPI_ISL_1226875, EPI_ISL_1226901, EPI_ISL_1226902, EPI_ISL_1226903, EPI_ISL_1226904, EPI_ISL_1226905, EPI_ISL_1226906, EPI_ISL_1226926, EPI_ISL_1226988, EPI_ISL_1226989, EPI_ISL_1226990, EPI_ISL_1226991, EPI_ISL_1226992, EPI_ISL_1226993, EPI_ISL_1226994, EPI_ISL_1226995, EPI_ISL_1226996, EPI_ISL_1226997, EPI_ISL_1227163, EPI_ISL_1227237, EPI_ISL_1227283, EPI_ISL_1227289, EPI_ISL_1227290, EPI_ISL_1227293, EPI_ISL_1227295, EPI_ISL_1227296, EPI_ISL_1227304, EPI_ISL_1227305, EPI_ISL_1227314, EPI_ISL_1227320, EPI_ISL_1227326, EPI_ISL_1227357, EPI_ISL_1227360, EPI_ISL_1227361, EPI_ISL_1227362, EPI_ISL_1227363, EPI_ISL_1227675, EPI_ISL_1227895, EPI_ISL_1227896, EPI_ISL_1227897, EPI_ISL_1227898, EPI_ISL_1227899, EPI_ISL_1227900, EPI_ISL_1227901, EPI_ISL_1227902, EPI_ISL_1227904, EPI_ISL_1227905, EPI_ISL_1227906, EPI_ISL_1227907, EPI_ISL_1227908, EPI_ISL_1227910, EPI_ISL_1227913, EPI_ISL_1227914, EPI_ISL_1227915, EPI_ISL_1227916, EPI_ISL_1227917, EPI_ISL_1227918, EPI_ISL_1227919, EPI_ISL_1227921, EPI_ISL_1227922, EPI_ISL_1227923, EPI_ISL_1227924, EPI_ISL_1227925, EPI_ISL_1227926, EPI_ISL_1227927, EPI_ISL_1227928, EPI_ISL_1227929, EPI_ISL_1227930, EPI_ISL_1227931, EPI_ISL_1228158, EPI_ISL_1228253, EPI_ISL_1228509, EPI_ISL_1228514, EPI_ISL_1228529, EPI_ISL_1228543, EPI_ISL_1228544, EPI_ISL_1395726, EPI_ISL_1395727, EPI_ISL_1395728, EPI_ISL_1395729, EPI_ISL_1395730, EPI_ISL_1395731, EPI_ISL_1395732, EPI_ISL_1395733, EPI_ISL_1395734, EPI_ISL_1395735, EPI_ISL_1395736, EPI_ISL_1395737, EPI_ISL_1395738, EPI_ISL_1395739, EPI_ISL_1395740, EPI_ISL_1395741, EPI_ISL_1395742, EPI_ISL_1395743, EPI_ISL_1395744, EPI_ISL_1395745, EPI_ISL_1395746, EPI_ISL_1395747, EPI_ISL_1395748, EPI_ISL_1395749, EPI_ISL_1398044, EPI_ISL_1398045, EPI_ISL_1398046, EPI_ISL_1398047, EPI_ISL_1623952, EPI_ISL_1671162, EPI_ISL_1671211, EPI_ISL_1671212, EPI_ISL_1671217, EPI_ISL_1920142, EPI_ISL_1920145, EPI_ISL_1920154, EPI_ISL_1920160 | | | |
| see above | Columbia University Irving Medical Center | Wadsworth Center, New York State Department of Health | Alexis Russel; Alexis Russell; Catharine Prussing; Daryl M. Lamson; Erasmus Schneider; Erica Lasek-Nesselquist; John Kelly; Jonathan Plitnick; Kirsten St. George; Matthew Shudt; Melissa A Leisner; Navjot Singh |
| EPI_ISL_794283, EPI_ISL_794284, EPI_ISL_794285, EPI_ISL_794286, EPI_ISL_794287, EPI_ISL_794288, EPI_ISL_794289, EPI_ISL_794290, EPI_ISL_794291, EPI_ISL_794292, EPI_ISL_794293, EPI_ISL_794294, EPI_ISL_794295, EPI_ISL_794296, EPI_ISL_794297, EPI_ISL_794298, EPI_ISL_794299, EPI_ISL_794300, EPI_ISL_794301, EPI_ISL_794302, EPI_ISL_794303, EPI_ISL_794304, EPI_ISL_794305, EPI_ISL_794306, EPI_ISL_794307, EPI_ISL_802777, EPI_ISL_802778, EPI_ISL_802779, EPI_ISL_802780, EPI_ISL_802781, EPI_ISL_802782, EPI_ISL_802783, EPI_ISL_802784, EPI_ISL_802785, EPI_ISL_802786, EPI_ISL_802787, EPI_ISL_802788, EPI_ISL_802789 | | | |
| see above | Columbia University Irving Medical Center | Wadsworth Center, New York State Department.of Health | Alexis Russel; Daryl M. Lamson; Erasmus Schneider; Erica Lasek-Nesselquist; John Kelly; Jonathan Plitnick; Kirsten St. George; Matthew Shudt; Melissa A Leisner; Navjot Singh; Sara Griesemer |
| EPI_ISL_633009, EPI_ISL_633010, EPI_ISL_633018, EPI_ISL_633037, EPI_ISL_633056, EPI_ISL_671689, EPI_ISL_671704, EPI_ISL_683756, EPI_ISL_683795, EPI_ISL_683813, EPI_ISL_683829, EPI_ISL_683834, EPI_ISL_683842, EPI_ISL_683848, EPI_ISL_683859, EPI_ISL_683877, EPI_ISL_683882, EPI_ISL_683883, EPI_ISL_683924, EPI_ISL_683927, EPI_ISL_683928, EPI_ISL_683933, EPI_ISL_683937, EPI_ISL_683962, EPI_ISL_683964, EPI_ISL_683968, EPI_ISL_683974, EPI_ISL_683976, EPI_ISL_683977, EPI_ISL_683982, EPI_ISL_683983, EPI_ISL_683984, EPI_ISL_683985, EPI_ISL_683991, EPI_ISL_745400, EPI_ISL_745404, EPI_ISL_745413, EPI_ISL_745425, EPI_ISL_745451, EPI_ISL_745469, EPI_ISL_812729, EPI_ISL_812739, EPI_ISL_812741, EPI_ISL_812749, EPI_ISL_823832, EPI_ISL_823833, EPI_ISL_823834, EPI_ISL_823838, EPI_ISL_823839, EPI_ISL_823840, EPI_ISL_823843, EPI_ISL_823844, EPI_ISL_823865, EPI_ISL_823869, EPI_ISL_823884, EPI_ISL_823885, EPI_ISL_823886, EPI_ISL_823887, EPI_ISL_823901, EPI_ISL_823902, EPI_ISL_823912, EPI_ISL_823927, EPI_ISL_823950, EPI_ISL_824298, EPI_ISL_824317, EPI_ISL_824318, EPI_ISL_832256, EPI_ISL_832266, EPI_ISL_832267, EPI_ISL_832284, EPI_ISL_832300, EPI_ISL_832312, EPI_ISL_832336, EPI_ISL_832337, EPI_ISL_832346, EPI_ISL_832347, EPI_ISL_832348, EPI_ISL_832349, EPI_ISL_832350, EPI_ISL_857077, EPI_ISL_857078, EPI_ISL_857079, EPI_ISL_857080, EPI_ISL_857085, EPI_ISL_857086, EPI_ISL_857094, EPI_ISL_857100, EPI_ISL_857116, EPI_ISL_857118, EPI_ISL_857120, EPI_ISL_857126, EPI_ISL_857132, EPI_ISL_857133, EPI_ISL_857142, EPI_ISL_857163, EPI_ISL_857180, EPI_ISL_857181, EPI_ISL_857182, EPI_ISL_857183, EPI_ISL_857194, EPI_ISL_857200, EPI_ISL_857204, EPI_ISL_857205, EPI_ISL_857209, EPI_ISL_857231, EPI_ISL_857232, EPI_ISL_857233, EPI_ISL_857255, EPI_ISL_857258, EPI_ISL_857259, EPI_ISL_857285, EPI_ISL_857286, EPI_ISL_861332, EPI_ISL_883309, EPI_ISL_883324, EPI_ISL_883327, EPI_ISL_883329, EPI_ISL_883371, EPI_ISL_883372, EPI_ISL_883376, EPI_ISL_883377, EPI_ISL_883378, EPI_ISL_883379, EPI_ISL_883409, EPI_ISL_883413, EPI_ISL_883414, EPI_ISL_883415, EPI_ISL_937132, EPI_ISL_937133, EPI_ISL_937167, EPI_ISL_937172, EPI_ISL_937204, EPI_ISL_937205, EPI_ISL_937216, EPI_ISL_966389, EPI_ISL_966413, EPI_ISL_966416, EPI_ISL_966450, EPI_ISL_966475 | | | |
| see above | DOHMH Central Harlem | New York City Public Health Laboratory | Jade Wang; et al. |
| EPI_ISL_633014, EPI_ISL_633016, EPI_ISL_633017, EPI_ISL_633023, EPI_ISL_633025, EPI_ISL_633031, EPI_ISL_633033, EPI_ISL_633039, EPI_ISL_633040, EPI_ISL_633046, EPI_ISL_633047, EPI_ISL_633052, EPI_ISL_633066, EPI_ISL_633069, EPI_ISL_633076, EPI_ISL_633077, EPI_ISL_633078, EPI_ISL_671684, EPI_ISL_671693, EPI_ISL_671702, EPI_ISL_671713, EPI_ISL_671722, EPI_ISL_671763, EPI_ISL_683765, EPI_ISL_683780, EPI_ISL_683788, EPI_ISL_683792, EPI_ISL_683807, EPI_ISL_683808, EPI_ISL_683828, EPI_ISL_683830, EPI_ISL_683865, EPI_ISL_683923, EPI_ISL_683931, EPI_ISL_683936, EPI_ISL_683941, EPI_ISL_683957, EPI_ISL_683961, EPI_ISL_683967, EPI_ISL_683971, EPI_ISL_683972, EPI_ISL_683980, EPI_ISL_683981, EPI_ISL_683989, EPI_ISL_745410, EPI_ISL_745421, EPI_ISL_745422, EPI_ISL_745436, EPI_ISL_745453, EPI_ISL_745457, EPI_ISL_823815, EPI_ISL_823816, EPI_ISL_823864, EPI_ISL_823924, EPI_ISL_823930, EPI_ISL_823934, EPI_ISL_823948, EPI_ISL_823954, EPI_ISL_823962, EPI_ISL_824300, EPI_ISL_824319, EPI_ISL_832282, EPI_ISL_832301, EPI_ISL_832302, EPI_ISL_832313, EPI_ISL_832314, EPI_ISL_832341, EPI_ISL_832362, EPI_ISL_832363, EPI_ISL_857076, EPI_ISL_857106, EPI_ISL_857107, EPI_ISL_857108, EPI_ISL_857109, EPI_ISL_857110, EPI_ISL_857117, EPI_ISL_857130, EPI_ISL_857131, EPI_ISL_857136, EPI_ISL_857137, EPI_ISL_857138, EPI_ISL_857139, EPI_ISL_857230, EPI_ISL_857253, EPI_ISL_857254, EPI_ISL_857260, EPI_ISL_857261, EPI_ISL_857262, EPI_ISL_883335, EPI_ISL_883339, EPI_ISL_883368, EPI_ISL_883369, EPI_ISL_937136, EPI_ISL_937137, EPI_ISL_937174, EPI_ISL_937202, EPI_ISL_937203, EPI_ISL_937217, EPI_ISL_937218, EPI_ISL_966468, EPI_ISL_966483 | | | |
| see above | DOHMH Chelsea | New York City Public Health Laboratory | Jade Wang; et al. |
| EPI_ISL_632980, EPI_ISL_632981, EPI_ISL_632982, EPI_ISL_632983, EPI_ISL_632984, EPI_ISL_632985, EPI_ISL_632986, EPI_ISL_632987, EPI_ISL_632988, EPI_ISL_632989, EPI_ISL_632990, EPI_ISL_632991, EPI_ISL_632992, EPI_ISL_632993, EPI_ISL_633059, EPI_ISL_633074, EPI_ISL_671674, EPI_ISL_671677, EPI_ISL_671681, EPI_ISL_671682, EPI_ISL_671683, EPI_ISL_671685, EPI_ISL_671691, EPI_ISL_671695, EPI_ISL_671697, EPI_ISL_671699, EPI_ISL_671700, EPI_ISL_671703, EPI_ISL_671705, EPI_ISL_671710, EPI_ISL_671711, EPI_ISL_671717, EPI_ISL_671718, EPI_ISL_671719, EPI_ISL_671720, EPI_ISL_671721, EPI_ISL_671731, EPI_ISL_671732, EPI_ISL_671746, EPI_ISL_671747, EPI_ISL_671761, EPI_ISL_683753, EPI_ISL_683757, EPI_ISL_683758, EPI_ISL_683761, EPI_ISL_683764, EPI_ISL_683766, EPI_ISL_683770, EPI_ISL_683773, EPI_ISL_683777, EPI_ISL_683779, EPI_ISL_683785, EPI_ISL_683793, EPI_ISL_683794, EPI_ISL_683796, EPI_ISL_683798, EPI_ISL_683799, EPI_ISL_683800, EPI_ISL_683802, EPI_ISL_683809, EPI_ISL_683811, EPI_ISL_683812, EPI_ISL_683815, EPI_ISL_683817, EPI_ISL_683818, EPI_ISL_683824, EPI_ISL_683831, EPI_ISL_683833, EPI_ISL_683841, EPI_ISL_683847, EPI_ISL_683849, EPI_ISL_683851, EPI_ISL_683855, EPI_ISL_683857, EPI_ISL_683860, EPI_ISL_683867, EPI_ISL_683870, EPI_ISL_683872, EPI_ISL_683873, EPI_ISL_683876, EPI_ISL_683881, EPI_ISL_683893, EPI_ISL_683896, EPI_ISL_683897, EPI_ISL_683898, EPI_ISL_683899, EPI_ISL_683900, EPI_ISL_683908, EPI_ISL_683909, EPI_ISL_683911, EPI_ISL_683912, EPI_ISL_683926, EPI_ISL_683929, EPI_ISL_683932, EPI_ISL_683934, EPI_ISL_683952, EPI_ISL_683953, EPI_ISL_683960, EPI_ISL_683978, EPI_ISL_683979, EPI_ISL_745401, EPI_ISL_745403, EPI_ISL_745409, EPI_ISL_745411, EPI_ISL_745423, EPI_ISL_745424, EPI_ISL_745426, EPI_ISL_745427, EPI_ISL_745434, EPI_ISL_745435, EPI_ISL_745446, EPI_ISL_745447, EPI_ISL_745455, EPI_ISL_745458, EPI_ISL_745459, EPI_ISL_745460, EPI_ISL_745461, EPI_ISL_745462, EPI_ISL_745464, EPI_ISL_745466, EPI_ISL_745467, EPI_ISL_812726, EPI_ISL_812732, EPI_ISL_812733, EPI_ISL_812740, EPI_ISL_812742, EPI_ISL_812743, EPI_ISL_812744, EPI_ISL_812746, EPI_ISL_812747, EPI_ISL_812748, EPI_ISL_812750, EPI_ISL_812751, EPI_ISL_812760, EPI_ISL_823817, EPI_ISL_823818, EPI_ISL_823819, EPI_ISL_823826, EPI_ISL_823827, EPI_ISL_823828, EPI_ISL_823835, EPI_ISL_823836, EPI_ISL_823837, EPI_ISL_823850, EPI_ISL_823861, EPI_ISL_823862, EPI_ISL_823863, EPI_ISL_823868, EPI_ISL_823878, EPI_ISL_823879, EPI_ISL_823880, EPI_ISL_823881, EPI_ISL_823882, EPI_ISL_823898, EPI_ISL_823899, EPI_ISL_823905, EPI_ISL_823906, EPI_ISL_823918, EPI_ISL_823919, EPI_ISL_823920, EPI_ISL_823921, EPI_ISL_823922, EPI_ISL_823931, EPI_ISL_823933, EPI_ISL_823943, EPI_ISL_823944, EPI_ISL_823945, EPI_ISL_823953, EPI_ISL_823957, EPI_ISL_823963, EPI_ISL_823964, EPI_ISL_823965, EPI_ISL_823966, EPI_ISL_823967, EPI_ISL_824314, EPI_ISL_824315, EPI_ISL_824320, EPI_ISL_824328, EPI_ISL_824331, EPI_ISL_832243, EPI_ISL_832245, EPI_ISL_832251, EPI_ISL_832252, EPI_ISL_832253, EPI_ISL_832254, EPI_ISL_832260, EPI_ISL_832261, EPI_ISL_832262, EPI_ISL_832277, EPI_ISL_832278, EPI_ISL_832279, EPI_ISL_832290, EPI_ISL_832291, EPI_ISL_832303, EPI_ISL_832304, EPI_ISL_832320, EPI_ISL_832321, EPI_ISL_832322, EPI_ISL_832325, EPI_ISL_832331, EPI_ISL_832332, EPI_ISL_832333, EPI_ISL_832334, EPI_ISL_832335, EPI_ISL_832351, EPI_ISL_832355, EPI_ISL_832356, EPI_ISL_832359, EPI_ISL_832360, EPI_ISL_857067, EPI_ISL_857073, EPI_ISL_857082, EPI_ISL_857083, EPI_ISL_857084, EPI_ISL_857101, EPI_ISL_857102, EPI_ISL_857103, EPI_ISL_857104, EPI_ISL_857105, EPI_ISL_857135, EPI_ISL_857145, EPI_ISL_857146, EPI_ISL_857147, EPI_ISL_857153, EPI_ISL_857156, EPI_ISL_857158, EPI_ISL_857165, EPI_ISL_857168, EPI_ISL_857189, EPI_ISL_857218, EPI_ISL_857219, EPI_ISL_857220, EPI_ISL_857225, EPI_ISL_857238, EPI_ISL_857239, EPI_ISL_857240, EPI_ISL_857263, EPI_ISL_857264, EPI_ISL_857277, EPI_ISL_883312, EPI_ISL_883313, EPI_ISL_883348, EPI_ISL_883354, EPI_ISL_883355, EPI_ISL_883356, EPI_ISL_883410, EPI_ISL_883411, EPI_ISL_937139, EPI_ISL_937140, EPI_ISL_937141, EPI_ISL_937142, EPI_ISL_937143, EPI_ISL_937144, EPI_ISL_937164, EPI_ISL_937165, EPI_ISL_937178, EPI_ISL_937179, EPI_ISL_937180, EPI_ISL_937181, EPI_ISL_937182, EPI_ISL_937183, EPI_ISL_937184, EPI_ISL_937208, EPI_ISL_937214, EPI_ISL_937227, EPI_ISL_937228, EPI_ISL_937229, EPI_ISL_937230, EPI_ISL_937231, EPI_ISL_966386, EPI_ISL_966392, EPI_ISL_966394, EPI_ISL_966399, EPI_ISL_966409, EPI_ISL_966410, EPI_ISL_966415, EPI_ISL_966425, EPI_ISL_966445, EPI_ISL_966446, EPI_ISL_966459, EPI_ISL_966467, EPI_ISL_966474, EPI_ISL_966476, EPI_ISL_966477, EPI_ISL_966478, EPI_ISL_966480, EPI_ISL_966482, EPI_ISL_966491 | | | |
| see above | DOHMH Corona | New York City Public Health Laboratory | Jade Wang; et al. |
| EPI_ISL_632994, EPI_ISL_632995, EPI_ISL_632996, EPI_ISL_632997, EPI_ISL_632998, EPI_ISL_633063, EPI_ISL_671690, EPI_ISL_671760, EPI_ISL_683821, EPI_ISL_683827, EPI_ISL_683856, EPI_ISL_683891, EPI_ISL_683904, EPI_ISL_683910, EPI_ISL_683935, EPI_ISL_683970, EPI_ISL_745419, EPI_ISL_745420, EPI_ISL_745445, EPI_ISL_745448, EPI_ISL_745452, EPI_ISL_745454, EPI_ISL_812725, EPI_ISL_812730, EPI_ISL_812734, EPI_ISL_812738, EPI_ISL_812752, EPI_ISL_812758, EPI_ISL_823852, EPI_ISL_823853, EPI_ISL_823925, EPI_ISL_823935, EPI_ISL_823936, EPI_ISL_823942, EPI_ISL_824301, EPI_ISL_824310, EPI_ISL_824316, EPI_ISL_832244, EPI_ISL_832275, EPI_ISL_832287, EPI_ISL_832288, EPI_ISL_832292, EPI_ISL_832293, EPI_ISL_832295, EPI_ISL_832338, EPI_ISL_832340, EPI_ISL_857089, EPI_ISL_857097, EPI_ISL_857172, EPI_ISL_857173, EPI_ISL_857196, EPI_ISL_883337, EPI_ISL_883357, EPI_ISL_883358, EPI_ISL_883359, EPI_ISL_883360, EPI_ISL_883364, EPI_ISL_883370, EPI_ISL_883412, EPI_ISL_937138, EPI_ISL_937168, EPI_ISL_937173, EPI_ISL_966484 | | | |
| see above | DOHMH Crown Heights | New York City Public Health Laboratory | Jade Wang; et al. |
| EPI_ISL_633073, EPI_ISL_671678, EPI_ISL_671738, EPI_ISL_671759, EPI_ISL_683771, EPI_ISL_683816, EPI_ISL_683832, EPI_ISL_683868, EPI_ISL_683887, EPI_ISL_683888, EPI_ISL_683906, EPI_ISL_683913, EPI_ISL_683922, EPI_ISL_683963, EPI_ISL_683973, EPI_ISL_683996, EPI_ISL_745463, EPI_ISL_823849, EPI_ISL_823851, EPI_ISL_823866, EPI_ISL_823867, EPI_ISL_823904, EPI_ISL_823910, EPI_ISL_823911, EPI_ISL_823923, EPI_ISL_823938, EPI_ISL_824311, EPI_ISL_824312, EPI_ISL_824321, EPI_ISL_824322, EPI_ISL_832268, EPI_ISL_832269, EPI_ISL_832286, EPI_ISL_857095, EPI_ISL_857112, EPI_ISL_857113, EPI_ISL_857114, EPI_ISL_857115, EPI_ISL_857148, EPI_ISL_857166, EPI_ISL_857167, EPI_ISL_857221, EPI_ISL_857256, EPI_ISL_857265, EPI_ISL_857279, EPI_ISL_857280, EPI_ISL_883416, EPI_ISL_883417, EPI_ISL_937134, EPI_ISL_937135, EPI_ISL_937200, EPI_ISL_937201, EPI_ISL_966443 | | | |
| see above | DOHMH Fort Greene | New York City Public Health Laboratory | Jade Wang; et al. |
| EPI_ISL_632999, EPI_ISL_633000, EPI_ISL_633001, EPI_ISL_633002, EPI_ISL_633003, EPI_ISL_633004, EPI_ISL_633005, EPI_ISL_633006, EPI_ISL_633007, EPI_ISL_633008, EPI_ISL_633060, EPI_ISL_633061, EPI_ISL_633062, EPI_ISL_633064, EPI_ISL_671675, EPI_ISL_671692, EPI_ISL_671698, EPI_ISL_671706, EPI_ISL_671707, EPI_ISL_671712, EPI_ISL_671714, EPI_ISL_671723, EPI_ISL_671724, EPI_ISL_671730, EPI_ISL_671735, EPI_ISL_671739, EPI_ISL_671743, EPI_ISL_683775, EPI_ISL_683776, EPI_ISL_683781, EPI_ISL_683782, EPI_ISL_683786, EPI_ISL_683791, EPI_ISL_683822, EPI_ISL_683837, EPI_ISL_683838, EPI_ISL_683850, EPI_ISL_683858, EPI_ISL_683861, EPI_ISL_683862, EPI_ISL_683864, EPI_ISL_683874, EPI_ISL_683878, EPI_ISL_683885, EPI_ISL_683892, EPI_ISL_683901, EPI_ISL_683903, EPI_ISL_683914, EPI_ISL_683915, EPI_ISL_683916, EPI_ISL_683920, EPI_ISL_683921, EPI_ISL_683925, EPI_ISL_683930, EPI_ISL_683939, EPI_ISL_683944, EPI_ISL_683945, EPI_ISL_683948, EPI_ISL_683951, EPI_ISL_683956, EPI_ISL_683958, EPI_ISL_683975, EPI_ISL_683986, EPI_ISL_683990, EPI_ISL_745398, EPI_ISL_745402, EPI_ISL_745405, EPI_ISL_745407, EPI_ISL_745414, EPI_ISL_745415, EPI_ISL_745416, EPI_ISL_745417, EPI_ISL_745418, EPI_ISL_745440, EPI_ISL_745444, EPI_ISL_812727, EPI_ISL_812731, EPI_ISL_823813, EPI_ISL_823821, EPI_ISL_823822, EPI_ISL_823846, EPI_ISL_823847, EPI_ISL_823848, EPI_ISL_823870, EPI_ISL_823871, EPI_ISL_823876, EPI_ISL_823877, EPI_ISL_823894, EPI_ISL_823895, EPI_ISL_823907, EPI_ISL_823908, EPI_ISL_823915, EPI_ISL_823916, EPI_ISL_823917, EPI_ISL_823926, EPI_ISL_823928, EPI_ISL_823940, EPI_ISL_823941, EPI_ISL_823946, EPI_ISL_823949, EPI_ISL_823955, EPI_ISL_823956, EPI_ISL_823961, EPI_ISL_823968, EPI_ISL_823969, EPI_ISL_824296, EPI_ISL_824297, EPI_ISL_824302, EPI_ISL_824304, EPI_ISL_824305, EPI_ISL_824308, EPI_ISL_824309, EPI_ISL_824313, EPI_ISL_824329, EPI_ISL_824330, EPI_ISL_832246, EPI_ISL_832250, EPI_ISL_832263, EPI_ISL_832265, EPI_ISL_832272, EPI_ISL_832273, EPI_ISL_832274, EPI_ISL_832276, EPI_ISL_832283, EPI_ISL_832294, EPI_ISL_832297, EPI_ISL_832298, EPI_ISL_832306, EPI_ISL_832307, EPI_ISL_832308, EPI_ISL_832309, EPI_ISL_832310, EPI_ISL_832311, EPI_ISL_832317, EPI_ISL_832318, EPI_ISL_832319, EPI_ISL_832323, EPI_ISL_832324, EPI_ISL_832328, EPI_ISL_832329, EPI_ISL_832330, EPI_ISL_832354, EPI_ISL_832357, EPI_ISL_832358, EPI_ISL_832361, EPI_ISL_857069, EPI_ISL_857075, EPI_ISL_857091, EPI_ISL_857096, EPI_ISL_857149, EPI_ISL_857150, EPI_ISL_857151, EPI_ISL_857152, EPI_ISL_857154, EPI_ISL_857155, EPI_ISL_857174, EPI_ISL_857175, EPI_ISL_857176, EPI_ISL_857177, EPI_ISL_857179, EPI_ISL_857197, EPI_ISL_857201, EPI_ISL_857210, EPI_ISL_857211, EPI_ISL_857212, EPI_ISL_857213, EPI_ISL_857214, EPI_ISL_857215, EPI_ISL_857216, EPI_ISL_857217, EPI_ISL_857222, EPI_ISL_857223, EPI_ISL_857224, EPI_ISL_857226, EPI_ISL_857234, EPI_ISL_857235, EPI_ISL_857236, EPI_ISL_857237, EPI_ISL_857241, EPI_ISL_857242, EPI_ISL_857243, EPI_ISL_857244, EPI_ISL_857245, EPI_ISL_857246, EPI_ISL_857247, EPI_ISL_857248, EPI_ISL_857249, EPI_ISL_857250, EPI_ISL_857267, EPI_ISL_857268, EPI_ISL_857269, EPI_ISL_857270, EPI_ISL_857274, EPI_ISL_857275, EPI_ISL_857276, EPI_ISL_876519, EPI_ISL_883310, EPI_ISL_883314, EPI_ISL_883318, EPI_ISL_883319, EPI_ISL_883320, EPI_ISL_883321, EPI_ISL_883322, EPI_ISL_883323, EPI_ISL_883349, EPI_ISL_883373, EPI_ISL_883374, EPI_ISL_883375, EPI_ISL_883418, EPI_ISL_883419, EPI_ISL_883420, EPI_ISL_883421, EPI_ISL_883422, EPI_ISL_883423, EPI_ISL_937145, EPI_ISL_937146, EPI_ISL_937147, EPI_ISL_937148, EPI_ISL_937149, EPI_ISL_937150, EPI_ISL_937151, EPI_ISL_937157, EPI_ISL_937158, EPI_ISL_937159, EPI_ISL_937160, EPI_ISL_937161, EPI_ISL_937162, EPI_ISL_937163, EPI_ISL_937175, EPI_ISL_937176, EPI_ISL_937177, EPI_ISL_937213, EPI_ISL_937219, EPI_ISL_937220, EPI_ISL_937221, EPI_ISL_937222, EPI_ISL_937223, EPI_ISL_937224, EPI_ISL_937225, EPI_ISL_937226, EPI_ISL_966382, EPI_ISL_966384, EPI_ISL_966387, EPI_ISL_966388, EPI_ISL_966395, EPI_ISL_966398, EPI_ISL_966405, EPI_ISL_966412, EPI_ISL_966421, EPI_ISL_966422, EPI_ISL_966424, EPI_ISL_966432, EPI_ISL_966433, EPI_ISL_966447, EPI_ISL_966448, EPI_ISL_966449, EPI_ISL_966461, EPI_ISL_966462, EPI_ISL_966463, EPI_ISL_966479, EPI_ISL_966481 | | | |
| see above | DOHMH Jamaica | New York City Public Health Laboratory | Jade Wang; et al. |
| EPI_ISL_633015, EPI_ISL_633019, EPI_ISL_633020, EPI_ISL_633021, EPI_ISL_633022, EPI_ISL_633024, EPI_ISL_633027, EPI_ISL_633028, EPI_ISL_633029, EPI_ISL_633030, EPI_ISL_633032, EPI_ISL_633036, EPI_ISL_633044, EPI_ISL_633048, EPI_ISL_633049, EPI_ISL_633053, EPI_ISL_633057, EPI_ISL_633058, EPI_ISL_633065, EPI_ISL_633067, EPI_ISL_633068, EPI_ISL_633075, EPI_ISL_633080, EPI_ISL_633081, EPI_ISL_633082, EPI_ISL_671679, EPI_ISL_671680, EPI_ISL_671686, EPI_ISL_671687, EPI_ISL_671688, EPI_ISL_671708, EPI_ISL_671726, EPI_ISL_671727, EPI_ISL_671728, EPI_ISL_671729, EPI_ISL_671736, EPI_ISL_671740, EPI_ISL_671741, EPI_ISL_671742, EPI_ISL_671751, EPI_ISL_671752, EPI_ISL_671753, EPI_ISL_671754, EPI_ISL_671755, EPI_ISL_671756, EPI_ISL_671757, EPI_ISL_671758, EPI_ISL_683754, EPI_ISL_683760, EPI_ISL_683762, EPI_ISL_683763, EPI_ISL_683772, EPI_ISL_683778, EPI_ISL_683790, EPI_ISL_683801, EPI_ISL_683803, EPI_ISL_683804, EPI_ISL_683805, EPI_ISL_683806, EPI_ISL_683814, EPI_ISL_683823, EPI_ISL_683825, EPI_ISL_683826, EPI_ISL_683839, EPI_ISL_683840, EPI_ISL_683843, EPI_ISL_683844, EPI_ISL_683845, EPI_ISL_683853, EPI_ISL_683854, EPI_ISL_683863, EPI_ISL_683869, EPI_ISL_683879, EPI_ISL_683880, EPI_ISL_683884, EPI_ISL_683886, EPI_ISL_683889, EPI_ISL_683905, EPI_ISL_683917, EPI_ISL_683918, EPI_ISL_683919, EPI_ISL_683938, EPI_ISL_683940, EPI_ISL_683946, EPI_ISL_683947, EPI_ISL_683959, EPI_ISL_683965, EPI_ISL_683966, EPI_ISL_683969, EPI_ISL_683992, EPI_ISL_745399, EPI_ISL_745406, EPI_ISL_745412, EPI_ISL_745430, EPI_ISL_745431, EPI_ISL_745432, EPI_ISL_745433, EPI_ISL_745441, EPI_ISL_745442, EPI_ISL_745443, EPI_ISL_745449, EPI_ISL_745450, EPI_ISL_745470, EPI_ISL_812735, EPI_ISL_812736, EPI_ISL_812737, EPI_ISL_812753, EPI_ISL_812754, EPI_ISL_812755, EPI_ISL_812756, EPI_ISL_812757, EPI_ISL_823829, EPI_ISL_823830, EPI_ISL_823831, EPI_ISL_823845, EPI_ISL_823854, EPI_ISL_823855, EPI_ISL_823856, EPI_ISL_823857, EPI_ISL_823858, EPI_ISL_823859, EPI_ISL_823860, EPI_ISL_823872, EPI_ISL_823873, EPI_ISL_823874, EPI_ISL_823892, EPI_ISL_823893, EPI_ISL_823903, EPI_ISL_823929, EPI_ISL_823932, EPI_ISL_823947, EPI_ISL_823959, EPI_ISL_823960, EPI_ISL_824295, EPI_ISL_824299, EPI_ISL_824303, EPI_ISL_824323, EPI_ISL_824324, EPI_ISL_824325, EPI_ISL_824326, EPI_ISL_824327, EPI_ISL_832247, EPI_ISL_832248, EPI_ISL_832255, EPI_ISL_832257, EPI_ISL_832264, EPI_ISL_832280, EPI_ISL_832281, EPI_ISL_832285, EPI_ISL_832289, EPI_ISL_832299, EPI_ISL_832315, EPI_ISL_832316, EPI_ISL_832327, EPI_ISL_832342, EPI_ISL_832343, EPI_ISL_832344, EPI_ISL_832345, EPI_ISL_857071, EPI_ISL_857074, EPI_ISL_857081, EPI_ISL_857093, EPI_ISL_857098, EPI_ISL_857099, EPI_ISL_857119, EPI_ISL_857122, EPI_ISL_857123, EPI_ISL_857124, EPI_ISL_857125, EPI_ISL_857140, EPI_ISL_857141, EPI_ISL_857159, EPI_ISL_857160, EPI_ISL_857164, EPI_ISL_857169, EPI_ISL_857170, EPI_ISL_857171, EPI_ISL_857191, EPI_ISL_857207, EPI_ISL_857208, EPI_ISL_857229, EPI_ISL_857251, EPI_ISL_857252, EPI_ISL_857271, EPI_ISL_857272, EPI_ISL_857273, EPI_ISL_857282, EPI_ISL_857283, EPI_ISL_883331, EPI_ISL_883338, EPI_ISL_883347, EPI_ISL_883365, EPI_ISL_883366, EPI_ISL_883367, EPI_ISL_937129, EPI_ISL_937130, EPI_ISL_937131, EPI_ISL_937206, EPI_ISL_937207, EPI_ISL_937232, EPI_ISL_937233, EPI_ISL_937234, EPI_ISL_937235, EPI_ISL_937236, EPI_ISL_937238, EPI_ISL_937239, EPI_ISL_966407, EPI_ISL_966408, EPI_ISL_966414, EPI_ISL_966420, EPI_ISL_966423, EPI_ISL_966434, EPI_ISL_966435, EPI_ISL_966436, EPI_ISL_966437, EPI_ISL_966451, EPI_ISL_966460 | | | |
| see above | DOHMH Morrisania | New York City Public Health Laboratory | Jade Wang; et al. |
| EPI_ISL_633011, EPI_ISL_633013, EPI_ISL_633026, EPI_ISL_633035, EPI_ISL_633038, EPI_ISL_633045, EPI_ISL_633050, EPI_ISL_633051, EPI_ISL_633054, EPI_ISL_633055, EPI_ISL_633071, EPI_ISL_633079, EPI_ISL_671676, EPI_ISL_671696, EPI_ISL_671715, EPI_ISL_671716, EPI_ISL_671734, EPI_ISL_671737, EPI_ISL_671744, EPI_ISL_683755, EPI_ISL_683759, EPI_ISL_683774, EPI_ISL_683783, EPI_ISL_683787, EPI_ISL_683789, EPI_ISL_683797, EPI_ISL_683810, EPI_ISL_683836, EPI_ISL_683846, EPI_ISL_683852, EPI_ISL_683871, EPI_ISL_683875, EPI_ISL_683890, EPI_ISL_683943, EPI_ISL_683954, EPI_ISL_683987, EPI_ISL_683988, EPI_ISL_683993, EPI_ISL_683995, EPI_ISL_745437, EPI_ISL_745438, EPI_ISL_745456, EPI_ISL_745465, EPI_ISL_812722, EPI_ISL_812723, EPI_ISL_812724, EPI_ISL_823820, EPI_ISL_823842, EPI_ISL_823875, EPI_ISL_823889, EPI_ISL_823890, EPI_ISL_823891, EPI_ISL_823896, EPI_ISL_823897, EPI_ISL_823909, EPI_ISL_823913, EPI_ISL_823914, EPI_ISL_823937, EPI_ISL_823958, EPI_ISL_824306, EPI_ISL_824307, EPI_ISL_832249, EPI_ISL_832258, EPI_ISL_832259, EPI_ISL_832352, EPI_ISL_832353, EPI_ISL_857063, EPI_ISL_857064, EPI_ISL_857065, EPI_ISL_857068, EPI_ISL_857070, EPI_ISL_857072, EPI_ISL_857087, EPI_ISL_857088, EPI_ISL_857121, EPI_ISL_857127, EPI_ISL_857128, EPI_ISL_857129, EPI_ISL_857134, EPI_ISL_857157, EPI_ISL_857161, EPI_ISL_857162, EPI_ISL_857185, EPI_ISL_857187, EPI_ISL_857190, EPI_ISL_857192, EPI_ISL_857202, EPI_ISL_857206, EPI_ISL_857257, EPI_ISL_857266, EPI_ISL_857278, EPI_ISL_857284, EPI_ISL_883340, EPI_ISL_883341, EPI_ISL_883342, EPI_ISL_883343, EPI_ISL_883346, EPI_ISL_883361, EPI_ISL_883362, EPI_ISL_883363, EPI_ISL_883380, EPI_ISL_883381, EPI_ISL_883382, EPI_ISL_883424, EPI_ISL_937152, EPI_ISL_937153, EPI_ISL_937154, EPI_ISL_937155, EPI_ISL_937156, EPI_ISL_937169, EPI_ISL_937170, EPI_ISL_937171, EPI_ISL_937185, EPI_ISL_937209, EPI_ISL_937210, EPI_ISL_937211, EPI_ISL_937212, EPI_ISL_937215, EPI_ISL_966404, EPI_ISL_966426, EPI_ISL_966444, EPI_ISL_966464, EPI_ISL_966465, EPI_ISL_966466 | | | |
| see above | DOHMH PHL | New York City Public Health Laboratory | Jade Wang; et al. |
| EPI_ISL_633012, EPI_ISL_633034, EPI_ISL_633041, EPI_ISL_633042, EPI_ISL_633043, EPI_ISL_633072, EPI_ISL_671694, EPI_ISL_671701, EPI_ISL_671709, EPI_ISL_671725, EPI_ISL_671733, EPI_ISL_671745, EPI_ISL_671748, EPI_ISL_671749, EPI_ISL_671750, EPI_ISL_671762, EPI_ISL_683767, EPI_ISL_683768, EPI_ISL_683769, EPI_ISL_683784, EPI_ISL_683819, EPI_ISL_683820, EPI_ISL_683866, EPI_ISL_683894, EPI_ISL_683895, EPI_ISL_683902, EPI_ISL_683907, EPI_ISL_683942, EPI_ISL_683949, EPI_ISL_683950, EPI_ISL_683955, EPI_ISL_683994, EPI_ISL_745408, EPI_ISL_745428, EPI_ISL_745429, EPI_ISL_745439, EPI_ISL_745468, EPI_ISL_812728, EPI_ISL_812745, EPI_ISL_812759, EPI_ISL_823814, EPI_ISL_823823, EPI_ISL_823824, EPI_ISL_823825, EPI_ISL_823841, EPI_ISL_823883, EPI_ISL_823888, EPI_ISL_823900, EPI_ISL_823939, EPI_ISL_823951, EPI_ISL_823952, EPI_ISL_832270, EPI_ISL_832271, EPI_ISL_832296, EPI_ISL_832305, EPI_ISL_832326, EPI_ISL_832339, EPI_ISL_857090, EPI_ISL_857092, EPI_ISL_857111, EPI_ISL_857143, EPI_ISL_857144, EPI_ISL_857178, EPI_ISL_857184, EPI_ISL_857195, EPI_ISL_857203, EPI_ISL_857227, EPI_ISL_857228, EPI_ISL_857281, EPI_ISL_883333, EPI_ISL_883334, EPI_ISL_937126, EPI_ISL_937127, EPI_ISL_937128, EPI_ISL_937166, EPI_ISL_937237 | | | |
| see above | DOHMH Riverside | New York City Public Health Laboratory | Jade Wang; et al. |
| EPI_ISL_823970, EPI_ISL_823973, EPI_ISL_823975, EPI_ISL_857188, EPI_ISL_857198, EPI_ISL_857287, EPI_ISL_857288, EPI_ISL_857290, EPI_ISL_857293, EPI_ISL_857298, EPI_ISL_857301, EPI_ISL_857306, EPI_ISL_883326, EPI_ISL_883332, EPI_ISL_883395, EPI_ISL_883396, EPI_ISL_883403, EPI_ISL_937191, EPI_ISL_937192, EPI_ISL_937248, EPI_ISL_937249, EPI_ISL_937250, EPI_ISL_966385, EPI_ISL_966401, EPI_ISL_966469 | | | |
| see above | Department of Homeless Services | New York City Public Health Laboratory | Jade Wang; et al. |
| EPI_ISL_1233937 | Division of Viral Diseases, CDC Pathogen Discovery Team | Division of Viral Diseases, CDC Pathogen Discovery Team | B.L.; Batra, D.; Burroughs, M.; C.R.; Cook; J.S.; Knipe, K.; Lee; Li, Y.; Marine, R.; Montmayeur, A.; Nobles, S.; P.W.; Paden; Padilla, J.; Queen, K.; Rambo-Martin; Rowe, L.; Sheth, M.; Tao, Y.; Tong, S.; Uehara, A.; Wang, H.; Zhang, J. |
| EPI_ISL_854305 | ESOTERIX GENETIC LABORATORIES LLC | Wadsworth Center, New York State Department of Health | Alexis Russel; Daryl M. Lamson; Erasmus Schneider; Erica Lasek-Nesselquist; John Kelly; Jonathan Plitnick; Kirsten St. George; Matthew Shudt; Melissa A Leisner; Navjot Singh |
| EPI_ISL_912169 | East Gene | Grubaugh Lab - Yale School of Public Health | Anderson Brito; Anne Wyllie; Annie Watkins; Chaney Kalinich; Chantal Vogels; Isabel Ott; Joseph Fauver; Mallery Breban; Mary Petrone; Nathan Grubaugh; Tara Alpert |
| EPI_ISL_632976, EPI_ISL_632977, | Empire City Laboratories | New York City Public Health Laboratory | Jade Wang; et al. |

| | | | |
|---|---|---|---|
| EPI_ISL_632978, EPI_ISL_632979 | | | |
| EPI_ISL_1009434, EPI_ISL_1009435, EPI_ISL_1009436, EPI_ISL_1009437, EPI_ISL_1009438, EPI_ISL_1009439, EPI_ISL_1009440, EPI_ISL_1009441, EPI_ISL_1009442, EPI_ISL_1009443, EPI_ISL_1009444, EPI_ISL_1009445, EPI_ISL_1009446, EPI_ISL_1009447, EPI_ISL_1009448, EPI_ISL_1009449, EPI_ISL_1009450, EPI_ISL_1009451, EPI_ISL_1009452, EPI_ISL_1009453, EPI_ISL_1009454, EPI_ISL_1009455, EPI_ISL_1009456, EPI_ISL_1009457, EPI_ISL_1009458, EPI_ISL_1009459, EPI_ISL_1009460, EPI_ISL_1009461, EPI_ISL_1009462, EPI_ISL_1009463, EPI_ISL_1009464, EPI_ISL_1009465, EPI_ISL_1009466, EPI_ISL_1009467, EPI_ISL_1009468, EPI_ISL_1009469, EPI_ISL_1009470, EPI_ISL_1009471, EPI_ISL_1009472, EPI_ISL_1009473, EPI_ISL_1009474, EPI_ISL_1009475, EPI_ISL_1009476, EPI_ISL_1009477, EPI_ISL_1009478, EPI_ISL_1009479, EPI_ISL_1009480, EPI_ISL_1009481, EPI_ISL_1009482, EPI_ISL_1009483, EPI_ISL_1009484, EPI_ISL_1009485, EPI_ISL_1009486, EPI_ISL_1009487, EPI_ISL_1009488, EPI_ISL_1009489, EPI_ISL_1009490, EPI_ISL_1009491, EPI_ISL_1009492, EPI_ISL_1009493, EPI_ISL_1009494, EPI_ISL_1009495, EPI_ISL_1009496, EPI_ISL_1009497, EPI_ISL_1009498, EPI_ISL_1009499, EPI_ISL_1009500, EPI_ISL_1009501, EPI_ISL_1009502, EPI_ISL_1009503, EPI_ISL_1009504, EPI_ISL_1009505, EPI_ISL_1009506, EPI_ISL_1009507, EPI_ISL_1009508, EPI_ISL_1009509, EPI_ISL_1009510, EPI_ISL_1009511, EPI_ISL_1009512, EPI_ISL_1009513, EPI_ISL_1009514, EPI_ISL_1009515, EPI_ISL_1009516, EPI_ISL_1009517, EPI_ISL_1009518, EPI_ISL_1009519, EPI_ISL_1009520, EPI_ISL_1009521, EPI_ISL_1009522, EPI_ISL_1009523, EPI_ISL_1009524, EPI_ISL_1009525, EPI_ISL_1009526, EPI_ISL_1009527, EPI_ISL_1009528, EPI_ISL_1009529, EPI_ISL_1009530, EPI_ISL_1009531, EPI_ISL_1009532, EPI_ISL_1009533, EPI_ISL_1009534, EPI_ISL_1009535, EPI_ISL_1009536, EPI_ISL_1009537, EPI_ISL_1009538, EPI_ISL_1009539, EPI_ISL_1009540, EPI_ISL_1009541, EPI_ISL_1009542, EPI_ISL_1009543, EPI_ISL_1009544, EPI_ISL_1009545, EPI_ISL_1009546, EPI_ISL_1009547, EPI_ISL_1009548, EPI_ISL_1009549, EPI_ISL_1009550, EPI_ISL_1009551, EPI_ISL_1009552, EPI_ISL_1009553, EPI_ISL_1009554, EPI_ISL_1009555, EPI_ISL_1009556, EPI_ISL_1009557, EPI_ISL_1009558, EPI_ISL_1009559, EPI_ISL_1009560, EPI_ISL_1009561, EPI_ISL_1009562, EPI_ISL_1009563, EPI_ISL_1009564, EPI_ISL_1009565, EPI_ISL_1009566, EPI_ISL_1009567, EPI_ISL_1009568, EPI_ISL_1009569, EPI_ISL_1009570, EPI_ISL_1009571, EPI_ISL_1009572, EPI_ISL_1009573, EPI_ISL_1009574, EPI_ISL_1009575, EPI_ISL_1009576, EPI_ISL_1009577, EPI_ISL_1009578, EPI_ISL_1009579, EPI_ISL_1009580, EPI_ISL_1009581, EPI_ISL_1009582, EPI_ISL_1009583, EPI_ISL_1009584, EPI_ISL_1009585, EPI_ISL_1009586, EPI_ISL_1009587, EPI_ISL_1009588, EPI_ISL_1009589, EPI_ISL_1009590, EPI_ISL_1009591, EPI_ISL_1009592, EPI_ISL_1009593, EPI_ISL_1009594, EPI_ISL_1009595, EPI_ISL_1009596, EPI_ISL_1009597, EPI_ISL_1009598, EPI_ISL_1009599, EPI_ISL_1009600, EPI_ISL_1009601, EPI_ISL_1009602, EPI_ISL_1009603, EPI_ISL_1009604, EPI_ISL_1009605, EPI_ISL_1009606, EPI_ISL_1009607, EPI_ISL_1009608, EPI_ISL_1009609, EPI_ISL_1009610, EPI_ISL_1009611, EPI_ISL_1009612, EPI_ISL_1009613, EPI_ISL_1009614, EPI_ISL_1009615, EPI_ISL_1009616, EPI_ISL_1400985, EPI_ISL_1400986, EPI_ISL_1400987, EPI_ISL_1400988, EPI_ISL_1400989, EPI_ISL_1400990, EPI_ISL_1400991, EPI_ISL_1400992, EPI_ISL_1400993, EPI_ISL_1400994, EPI_ISL_1400995, EPI_ISL_1400996, EPI_ISL_1400997, EPI_ISL_1400998, EPI_ISL_1400999, EPI_ISL_1401000, EPI_ISL_1401001, EPI_ISL_1401002, EPI_ISL_1401003, EPI_ISL_1401004, EPI_ISL_1401005, EPI_ISL_1401006, EPI_ISL_1401007, EPI_ISL_1401008, EPI_ISL_1401009, EPI_ISL_1401010, EPI_ISL_1401011, EPI_ISL_1401012, EPI_ISL_1401013, EPI_ISL_1401014, EPI_ISL_1401015, EPI_ISL_1401016, EPI_ISL_1401017, EPI_ISL_1401018, EPI_ISL_1401019, EPI_ISL_1401020, EPI_ISL_1401021, EPI_ISL_1401022, EPI_ISL_1401023, EPI_ISL_1401024, EPI_ISL_1401025, EPI_ISL_1401026, EPI_ISL_1401027, EPI_ISL_1401028, EPI_ISL_1401029, EPI_ISL_1401030, EPI_ISL_1401031, EPI_ISL_1401032, EPI_ISL_1401033, EPI_ISL_1401034, EPI_ISL_1401035, EPI_ISL_1401036, EPI_ISL_1401037, EPI_ISL_1401038, EPI_ISL_1401039, EPI_ISL_1401040, EPI_ISL_1401041, EPI_ISL_1401042, EPI_ISL_1401043, EPI_ISL_1401044, EPI_ISL_1401045, EPI_ISL_1401046, EPI_ISL_1401047, EPI_ISL_1401048, EPI_ISL_1401049, EPI_ISL_1401050, EPI_ISL_1401051, EPI_ISL_1401052, EPI_ISL_1401053, EPI_ISL_1401054, EPI_ISL_1401055, EPI_ISL_1401056, EPI_ISL_1401057, EPI_ISL_1401058, EPI_ISL_1401059, EPI_ISL_1401060, EPI_ISL_1401061, EPI_ISL_1401062, EPI_ISL_1401063, EPI_ISL_1401064, EPI_ISL_1401065, EPI_ISL_1401066, EPI_ISL_1401067, EPI_ISL_1401068, EPI_ISL_1401069, EPI_ISL_1444968 | | | |
| see above | Erie County Public Health (ECPHL) | University at Buffalo Genomics and Bioinformatics Core | Alyssa Pohlman; Amanda Boccolucci; Brandon Marzullo; Donald Yergeau; Jennifer Surtees; Jonathan Bard; Natalie Lamb; Norma Nowak |
| EPI_ISL_935973, EPI_ISL_935974, EPI_ISL_935975, EPI_ISL_935976, EPI_ISL_935977, EPI_ISL_935978, EPI_ISL_935979, EPI_ISL_935980, EPI_ISL_935981, EPI_ISL_935982, EPI_ISL_935983, EPI_ISL_935984, EPI_ISL_935985, EPI_ISL_935986, EPI_ISL_935987, EPI_ISL_935988, EPI_ISL_935989, EPI_ISL_935990, EPI_ISL_935991, EPI_ISL_935992, EPI_ISL_1226418, EPI_ISL_1226452, EPI_ISL_1228268, EPI_ISL_1228269, EPI_ISL_1228272, EPI_ISL_1228434, EPI_ISL_1228435, EPI_ISL_1228436, EPI_ISL_1228437, EPI_ISL_1228438, EPI_ISL_1228439, EPI_ISL_1228440, EPI_ISL_1228441, EPI_ISL_1228442, EPI_ISL_1228443, EPI_ISL_1228447, EPI_ISL_1228448, EPI_ISL_1228449, EPI_ISL_1228455, EPI_ISL_1228456, EPI_ISL_1228457, EPI_ISL_1228458, EPI_ISL_1228459, EPI_ISL_1228463, EPI_ISL_1228464, EPI_ISL_1228467, EPI_ISL_1228468, EPI_ISL_1228470, EPI_ISL_1228471, EPI_ISL_1228472 | | | |
| see above | GLENS FALLS HOSPITAL LABORATORY | Wadsworth Center, New York State Department of Health | Alexis Russel; Daryl M. Lamson; Erasmus Schneider; Erica Lasek-Nesselquist; John Kelly; Jonathan Plitnick; Kirsten St. George; Matthew Shudt; Melissa A Leisner; Navjot Singh |
| EPI_ISL_794107, EPI_ISL_794108, EPI_ISL_794109, EPI_ISL_794110, EPI_ISL_794111, EPI_ISL_794112, EPI_ISL_794113, EPI_ISL_794114, EPI_ISL_794115, EPI_ISL_794116, EPI_ISL_794117, EPI_ISL_794118 | | | |
| see above | GLENS FALLS HOSPITAL LABORATORY | Wadsworth Center, New York State Department.of Health | Alexis Russel; Daryl M. Lamson; Erasmus Schneider; Erica Lasek-Nesselquist; John Kelly; Jonathan Plitnick; Kirsten St. George; Matthew Shudt; Melissa A Leisner; Navjot Singh; Sara Griesemer |
| EPI_ISL_850770, EPI_ISL_850874, EPI_ISL_850925, EPI_ISL_850934 | Helix / Illumina | Genomics and Discovery, Respiratory Viruses Branch, Division of Viral Diseases, Centers for Disease Control and Prevention | Alexandre Bolze; Ary Ascencio; Ben L. Rambo-Martin Eileen de Feo; Brad Sickler; Charlotte Rivera-Garcia; Christine Tran; Clinton R. Paden; David Becker; Dhwani Batra; Duncan MacCannell; Efren Sandoval; Elizabeth Cirulli; Eric Allen; Geraint Levan; James Lu; Jan Antico; Jason Nguyen; Jimmy Ramirez; Jingtao Liu; Kelly Schiabor Barrett; Kim Gietzen; Magnus Isaksson; Marc Laurent; Matthew Tolentino; Nicole L. Washington; Peter W. Cook; Phil Febbo; Ryan Cho; Shannon Wickline; Sherry Wang; Simon White; Summer Galloway; Suxiang Tong; Tyler Cassens; William Lee |
| EPI_ISL_850558, EPI_ISL_850565, EPI_ISL_850975, EPI_ISL_876711, EPI_ISL_876712 | Helix/Illumina | Genomics and Discovery, Respiratory Viruses Branch, Division of Viral Diseases, Centers for Disease Control and Prevention | ; Alexandre Bolze; Ary Ascencio; Ben L. Rambo-Martin; Ben L. Rambo-Martin Eileen de Feo; Brad Sickler; Charlotte Rivera-Garcia; Christine Tran; Clinton R. Paden; David Becker; Dhwani Batra; Duncan MacCannell; Efren Sandoval; Eileen de Feo; Elizabeth Cirulli; Eric Allen; Geraint Levan; James Lu; Jan Antico; Jason Nguyen; Jimmy Ramirez; Jingtao Liu; Kelly Schiabor Barrett; Kim Gietzen; Magnus Isaksson; Marc Laurent; Matthew Tolentino; Nicole L. Washington; Peter W. Cook; Phil Febbo; Ryan Cho; Shannon Wickline; Sherry Wang; Simon White; Summer Galloway; Suxiang Tong; Tyler Cassens; William Lee |
| EPI_ISL_966525, EPI_ISL_966706, EPI_ISL_966952, EPI_ISL_967055, EPI_ISL_967085, EPI_ISL_967137, EPI_ISL_967207, EPI_ISL_967230, EPI_ISL_967296, EPI_ISL_967332, EPI_ISL_967358, EPI_ISL_967678, EPI_ISL_967789, EPI_ISL_967880, EPI_ISL_978603, EPI_ISL_978726, EPI_ISL_978736, EPI_ISL_978741, EPI_ISL_1255415 | | | |
| see above | Helix/Illumina | Respiratory Viruses Branch, Division of Viral Diseases, Centers for Disease Control and Prevention | ; Alexandre Bolze; Ary Ascencio; Ben L. Rambo-Martin; Brad Sickler; Charlotte Rivera-Garcia; Christine Tran; Clinton R. Paden; Dakota Howard; David Becker; Dhwani Batra; Duncan MacCannell; Efren Sandoval; Eileen de Feo; Elizabeth Cirulli; Eric Allen; Geraint Levan; James Lu; Jan Antico; Jason Nguyen; Jimmy Ramirez; Jingtao Liu; Kelly Schiabor Barrett; Kim Gietzen; Magnus Isaksson; Marc Laurent; Matthew Tolentino; Nicole L. Washington; Peter W. Cook; Phil Febbo; Ryan Cho; Shannon Wickline; Sherry Wang; Simon White; Summer Galloway; Suxiang Tong; Tyler Cassens; William Lee |
| EPI_ISL_884390, EPI_ISL_884391, EPI_ISL_884392, EPI_ISL_884393 | Infectious Diseases, Quest Diagnostics | Infectious Diseases, Quest Diagnostics | Anderson, B.; Bernstein; D.F.; Gerasimova, A.; Hua, M.; K.E.; Kagan; L.E.; Lacbawan, F.; Liu, Y.; Livingston; Owen, R.; R.M.; Rosenthal; S.H.; Shalhout |
| EPI_ISL_830208, EPI_ISL_830210, EPI_ISL_830212, EPI_ISL_830223, EPI_ISL_830225, EPI_ISL_830227, EPI_ISL_830229, EPI_ISL_830230, EPI_ISL_830232, EPI_ISL_830234, EPI_ISL_830236, EPI_ISL_830238, EPI_ISL_830239, EPI_ISL_830241, EPI_ISL_830243, EPI_ISL_830571, EPI_ISL_830573, EPI_ISL_830575, EPI_ISL_830579, EPI_ISL_830580, EPI_ISL_830581, EPI_ISL_830582, EPI_ISL_830583, EPI_ISL_830584, EPI_ISL_830585, EPI_ISL_830586, EPI_ISL_830587, EPI_ISL_830588, EPI_ISL_830589, EPI_ISL_830590, EPI_ISL_830591, EPI_ISL_830592, EPI_ISL_830593, EPI_ISL_830594, EPI_ISL_830595, EPI_ISL_830596, EPI_ISL_830597, EPI_ISL_830598, EPI_ISL_830599, EPI_ISL_830600, EPI_ISL_830601, EPI_ISL_830602, EPI_ISL_830603, EPI_ISL_830604, EPI_ISL_830605, EPI_ISL_830606, EPI_ISL_830607, EPI_ISL_830608, EPI_ISL_830609, EPI_ISL_830610, EPI_ISL_830611, EPI_ISL_830612, EPI_ISL_830613, EPI_ISL_830614, EPI_ISL_830615, EPI_ISL_830616, EPI_ISL_830617, EPI_ISL_830618, EPI_ISL_830619, EPI_ISL_830620, EPI_ISL_830621, EPI_ISL_830622, EPI_ISL_830623, EPI_ISL_830624, EPI_ISL_830625, EPI_ISL_830626, EPI_ISL_830627, EPI_ISL_861106, EPI_ISL_861107, EPI_ISL_861108, EPI_ISL_861134, EPI_ISL_861144, EPI_ISL_861145, EPI_ISL_861146, EPI_ISL_861147, EPI_ISL_861148, EPI_ISL_861149, EPI_ISL_861150, EPI_ISL_861151, EPI_ISL_861152, EPI_ISL_861153, EPI_ISL_861154, EPI_ISL_861155, EPI_ISL_861156, EPI_ISL_861157, EPI_ISL_861158, EPI_ISL_861159, EPI_ISL_861160, EPI_ISL_861161, EPI_ISL_861162, EPI_ISL_861163, EPI_ISL_861164, EPI_ISL_861165, EPI_ISL_861166, EPI_ISL_861167, EPI_ISL_861168, EPI_ISL_861169, EPI_ISL_861170, EPI_ISL_861171, EPI_ISL_861172, EPI_ISL_861173, EPI_ISL_861174, EPI_ISL_936060, EPI_ISL_936061, EPI_ISL_936062, EPI_ISL_936067, EPI_ISL_936068, EPI_ISL_936070, EPI_ISL_936071, EPI_ISL_936072, EPI_ISL_936073, EPI_ISL_936074, EPI_ISL_936075, EPI_ISL_936076, EPI_ISL_936077, EPI_ISL_936078, EPI_ISL_936079, EPI_ISL_936080, EPI_ISL_936081, EPI_ISL_936083, EPI_ISL_936084, EPI_ISL_936085, EPI_ISL_936087, EPI_ISL_936088, EPI_ISL_936090, EPI_ISL_936091, EPI_ISL_936094, EPI_ISL_936095, EPI_ISL_936096, EPI_ISL_936097, EPI_ISL_936098, EPI_ISL_936099, EPI_ISL_936100, EPI_ISL_936101, EPI_ISL_936102, EPI_ISL_936144, EPI_ISL_983240, EPI_ISL_983241, EPI_ISL_983242, EPI_ISL_983244, EPI_ISL_983246, EPI_ISL_983247, EPI_ISL_983250, EPI_ISL_983251, EPI_ISL_983252, EPI_ISL_983253, EPI_ISL_983254, EPI_ISL_983255, EPI_ISL_983256, EPI_ISL_983257, EPI_ISL_983258, EPI_ISL_983259, EPI_ISL_983260, EPI_ISL_983261, EPI_ISL_983262, EPI_ISL_983263, EPI_ISL_983264, EPI_ISL_983265, EPI_ISL_983266, EPI_ISL_983267, EPI_ISL_983268, EPI_ISL_983269, EPI_ISL_983270, EPI_ISL_983271, EPI_ISL_983272, EPI_ISL_983273, EPI_ISL_983274, EPI_ISL_983275, EPI_ISL_983276, EPI_ISL_983277, EPI_ISL_983278, EPI_ISL_983279, EPI_ISL_983280, EPI_ISL_983281, EPI_ISL_983282, EPI_ISL_983284, EPI_ISL_983285, EPI_ISL_983286, EPI_ISL_983287, EPI_ISL_983288, EPI_ISL_983296, EPI_ISL_983297, EPI_ISL_983298, EPI_ISL_983299, EPI_ISL_1226436, EPI_ISL_1226846, EPI_ISL_1227546, EPI_ISL_1227965, EPI_ISL_1227966, EPI_ISL_1227967, EPI_ISL_1227968, EPI_ISL_1227969, EPI_ISL_1398209, EPI_ISL_1398210, EPI_ISL_1398211, EPI_ISL_1398212, EPI_ISL_1398213, EPI_ISL_1398214, EPI_ISL_1398215, EPI_ISL_1398216, EPI_ISL_1398217, EPI_ISL_1398218, EPI_ISL_1398219, EPI_ISL_1398220, EPI_ISL_1398221, EPI_ISL_1398222, EPI_ISL_1398223, EPI_ISL_1398224, EPI_ISL_1398225, EPI_ISL_1398226, EPI_ISL_1398227, EPI_ISL_1398228, EPI_ISL_1398229, EPI_ISL_1398230, EPI_ISL_1398231, EPI_ISL_1398232, EPI_ISL_1398233, EPI_ISL_1398234, EPI_ISL_1398235, EPI_ISL_1398236, EPI_ISL_1398237, EPI_ISL_1398238, EPI_ISL_1398239, EPI_ISL_1398240, EPI_ISL_1398241, EPI_ISL_1398242, EPI_ISL_1398243, EPI_ISL_1398244, EPI_ISL_1398245, EPI_ISL_1398246, EPI_ISL_1398247, EPI_ISL_1398248, EPI_ISL_1398249, EPI_ISL_1398250, EPI_ISL_1398251, EPI_ISL_1398252, EPI_ISL_1398253, EPI_ISL_1398254, EPI_ISL_1398255, EPI_ISL_1398256, EPI_ISL_1398257, EPI_ISL_1398258 | | | |
| see above | KALEIDA CENTER FOR LABORATORY MEDICINE | Wadsworth Center, New York State Department of Health | Alexis Russel; Daryl M. Lamson; Erasmus Schneider; Erica Lasek-Nesselquist; John Kelly; Jonathan Plitnick; Kirsten St. George; Matthew Shudt; Melissa A Leisner; Navjot Singh |
| EPI_ISL_1398204, EPI_ISL_1398205, EPI_ISL_1398206, EPI_ISL_1398207, EPI_ISL_1398208 | LENOX HILL HOSPITAL | Wadsworth Center, New York State Department of Health | Alexis Russel; Daryl M. Lamson; Erasmus Schneider; Erica Lasek-Nesselquist; John Kelly; Jonathan Plitnick; Kirsten St. George; Matthew Shudt; Melissa A Leisner; Navjot Singh |
| EPI_ISL_886231, EPI_ISL_886365, EPI_ISL_886366, EPI_ISL_886405, EPI_ISL_886508, EPI_ISL_886563, EPI_ISL_886666, EPI_ISL_886772, EPI_ISL_886830, EPI_ISL_886837, EPI_ISL_886838, EPI_ISL_886892, EPI_ISL_886918, EPI_ISL_886919, EPI_ISL_886920, EPI_ISL_886927, EPI_ISL_886991, EPI_ISL_886998, EPI_ISL_887004, EPI_ISL_887034, EPI_ISL_887036, EPI_ISL_887046, EPI_ISL_887075, EPI_ISL_887087, EPI_ISL_887102, EPI_ISL_887601, EPI_ISL_887612, EPI_ISL_887631, EPI_ISL_887660, EPI_ISL_887662, EPI_ISL_887665, EPI_ISL_887711, EPI_ISL_887770, EPI_ISL_887792, EPI_ISL_887814, EPI_ISL_887894, EPI_ISL_888114, EPI_ISL_888141, EPI_ISL_888144, EPI_ISL_888146, EPI_ISL_888172, EPI_ISL_888229, EPI_ISL_888271, EPI_ISL_888349, EPI_ISL_888367, EPI_ISL_888405, EPI_ISL_888416, EPI_ISL_888452, EPI_ISL_888456, EPI_ISL_888481, EPI_ISL_888513, EPI_ISL_888562 | | | |
| see above | Labcorp | Genomics and Discovery, Respiratory Viruses Branch, Division of Viral Diseases, Centers for Disease Control and Prevention | ; Amanda Douglas; Amanda Suchanek; Andrea Throop; Ayla Burns; Ben L. Rambo-Martin; Bobbi Croy; Brian Krueger; Brian Norvell; Christos Petropoulos; Clinton R. Paden; Craig Lukasik; Debbie Boles; Dhwani Batra; Duncan MacCannell; Eyad Almasri; Goran Stevovic; Howard Engler; Hrushikesh Deshmukh; Jake Humphrey; Jana Schroth; Joe Voshell; John Pruitt; Jonathan Meltzer; Jonathan Williams; Kimberly Wagner; Lax Iyer; Lyndon Tilson; Manoj Jain; Marcia Eisenberg; Mary Ann Cristobal; Mary Williamson; Michael Levandoski; Mike Sapeta; Mindy Nye; Minoo Agarwal; Mohan Kolli; Nuthawin Charoensri; Oren Cohen; Peter W. Cook; Prashant Gupta; Qian Zeng; Rama Ghatti; Scott Parker; Scott Ryan; Stanley Letovsky; Steven Ragan; Summer Galloway; Suresh Babu Selvaraju; Susan Countryman; Susan Hicks; Suxiang Tong; Suzanne Dale; Thomas Urban; Tim Kuphal; Tricia Zwiefelhofer; Vincent Drouillon |
| EPI_ISL_1193261 | Laboratory Corporation of America | Centers for Disease Control and Prevention Division of Viral Diseases, Pathogen Discovery | Amanda Douglas; Amanda Suchanek; Andrea Throop; Ayla Burns; Ben L. Rambo-Martin; Bobbi Croy; Brian Krueger; Brian Norvell; Christos Petropoulos; Clinton R. Paden; Craig Lukasik; Dakota Howard; Dhwani Batra; Duncan MacCannell; Eyad Almasri Debbie Boles; Goran Stevovic; Howard Engler; Hrushikesh Deshmukh; Jake Humphrey; Jana Schroth; Joe Voshell; John Pruitt; Jonathan Meltzer; Jonathan Williams; Kimberly Wagner; Lax Iyer; Lyndon Tilson; Manoj Jain; Marcia Eisenberg; Mary Ann Cristobal; Mary Williamson; Michael Levandoski; Mike Sapeta; Mindy Nye; Minoo Agarwal; Mohan Kolli; Nuthawin Charoensri; Oren Cohen; Peter W. Cook; Prashant Gupta; Qian Zeng; Rama Ghatti; Scott Parker; Scott Ryan; Stanley Letovsky; Steven Ragan; Suresh Babu Selvaraju; Susan Countryman; Susan Hicks; Suxiang Tong; Suzanne Dale; Thomas Urban; Tim Kuphal; Tricia Zwiefelhofer; Vincent Drouillon |
| EPI_ISL_1020593, EPI_ISL_1020774, EPI_ISL_1020778, EPI_ISL_1020810, EPI_ISL_1020839, EPI_ISL_1020872, EPI_ISL_1020911, EPI_ISL_1020934, EPI_ISL_1020989, EPI_ISL_1020994, EPI_ISL_1021030, EPI_ISL_1021033, EPI_ISL_1021041, EPI_ISL_1021054, EPI_ISL_1021057, EPI_ISL_1021066, EPI_ISL_1021081, EPI_ISL_1021094, EPI_ISL_1021098, EPI_ISL_1021121, EPI_ISL_1021123, EPI_ISL_1021125, EPI_ISL_1021128, EPI_ISL_1021135, EPI_ISL_1021136, EPI_ISL_1021168, EPI_ISL_1021181, EPI_ISL_1021210, EPI_ISL_1021229, EPI_ISL_1021257, EPI_ISL_1021265, EPI_ISL_1021359, EPI_ISL_1021360, EPI_ISL_1021361, EPI_ISL_1021368, EPI_ISL_1021383, EPI_ISL_1021384, EPI_ISL_1021394, EPI_ISL_1021413, EPI_ISL_1021428, EPI_ISL_1021437, EPI_ISL_1021501, EPI_ISL_1021507, EPI_ISL_1021508, EPI_ISL_1021540, EPI_ISL_1021542, EPI_ISL_1021561, EPI_ISL_1021562, EPI_ISL_1021592, EPI_ISL_1021630, EPI_ISL_1021655, EPI_ISL_1021669, EPI_ISL_1021722, EPI_ISL_1021735, EPI_ISL_1021736, EPI_ISL_1021750, EPI_ISL_1021751, EPI_ISL_1021761, EPI_ISL_1021803, EPI_ISL_1021819, EPI_ISL_1021824, EPI_ISL_1021832, EPI_ISL_1021857, EPI_ISL_1021865, EPI_ISL_1021868, EPI_ISL_1021928, EPI_ISL_1021963, EPI_ISL_1021969, EPI_ISL_1021970, EPI_ISL_1021981, EPI_ISL_1022042, EPI_ISL_1022048, EPI_ISL_1022085, EPI_ISL_1022107, EPI_ISL_1022108, EPI_ISL_1022130, EPI_ISL_1022131, EPI_ISL_1022132, EPI_ISL_1022133, EPI_ISL_1022167, EPI_ISL_1022176, EPI_ISL_1022205, EPI_ISL_1022209, EPI_ISL_1022214, EPI_ISL_1022227, EPI_ISL_1022261, EPI_ISL_1022269, EPI_ISL_1022288, EPI_ISL_1022311, EPI_ISL_1022325, EPI_ISL_1022336, EPI_ISL_1022371, EPI_ISL_1022372, EPI_ISL_1022373, EPI_ISL_1022374, EPI_ISL_1022395, EPI_ISL_1022396, EPI_ISL_1026002, EPI_ISL_1026006, EPI_ISL_1026023, EPI_ISL_1026103, EPI_ISL_1026107, EPI_ISL_1026212, EPI_ISL_1026215, EPI_ISL_1026241, EPI_ISL_1026300, EPI_ISL_1026306, EPI_ISL_1026346, EPI_ISL_1026443, EPI_ISL_1026456, EPI_ISL_1026490, EPI_ISL_1026532, EPI_ISL_1026540, EPI_ISL_1026613, EPI_ISL_1026630, EPI_ISL_1026672, EPI_ISL_1026695, EPI_ISL_1026726, EPI_ISL_1026732, EPI_ISL_1026854, EPI_ISL_1027684, EPI_ISL_1027727, EPI_ISL_1027736, EPI_ISL_1027771, EPI_ISL_1027772, EPI_ISL_1027774, EPI_ISL_1027777, EPI_ISL_1027793, EPI_ISL_1027812, EPI_ISL_1027837, EPI_ISL_1027866, EPI_ISL_1027919, EPI_ISL_1027979, EPI_ISL_1027990, EPI_ISL_1028017, EPI_ISL_1028026, EPI_ISL_1028037, EPI_ISL_1028113, EPI_ISL_1028119, EPI_ISL_1028181, EPI_ISL_1028310, EPI_ISL_1028315, EPI_ISL_1028330, EPI_ISL_1028384, EPI_ISL_1028401, EPI_ISL_1028439, EPI_ISL_1028460, EPI_ISL_1028469, EPI_ISL_1028524, EPI_ISL_1028539, EPI_ISL_1028544, EPI_ISL_1028547, EPI_ISL_1028558, EPI_ISL_1028572, EPI_ISL_1028615, EPI_ISL_1028632, EPI_ISL_1028640, EPI_ISL_1028645, EPI_ISL_1028656, EPI_ISL_1028687, EPI_ISL_1028690, EPI_ISL_1028716, EPI_ISL_1028743, EPI_ISL_1028773, EPI_ISL_1028812, EPI_ISL_1028847, EPI_ISL_1028870, EPI_ISL_1028936, EPI_ISL_1028951, EPI_ISL_1028954, EPI_ISL_1029021, EPI_ISL_1029031, EPI_ISL_1029056, EPI_ISL_1029070, EPI_ISL_1029074, EPI_ISL_1029084, EPI_ISL_1029086, EPI_ISL_1029087, EPI_ISL_1029088, EPI_ISL_1029244, EPI_ISL_1029245, EPI_ISL_1029287, EPI_ISL_1029288, EPI_ISL_1029289, EPI_ISL_1029328, EPI_ISL_1029401, EPI_ISL_1029411, EPI_ISL_1029440, EPI_ISL_1029441, EPI_ISL_1029442, EPI_ISL_1029485, EPI_ISL_1029486, EPI_ISL_1029487, EPI_ISL_1029488, EPI_ISL_1029501, EPI_ISL_1029502, EPI_ISL_1029917, EPI_ISL_1029939, EPI_ISL_1029941, EPI_ISL_1029943, EPI_ISL_1030530, EPI_ISL_1030622, EPI_ISL_1030672, EPI_ISL_1030682, EPI_ISL_1030748, EPI_ISL_1030785, EPI_ISL_1030833, EPI_ISL_1030880, EPI_ISL_1030908, EPI_ISL_1030946, EPI_ISL_1030998, EPI_ISL_1031026, EPI_ISL_1031031, EPI_ISL_1031049, EPI_ISL_1031122, EPI_ISL_1031224, EPI_ISL_1031246, EPI_ISL_1031247, EPI_ISL_1031251, EPI_ISL_1031276, EPI_ISL_1031295, EPI_ISL_1031374, EPI_ISL_1031503, EPI_ISL_1031511, EPI_ISL_1031544, EPI_ISL_1031563, EPI_ISL_1031660, EPI_ISL_1031678, EPI_ISL_1031687, EPI_ISL_1031765, EPI_ISL_1031766, EPI_ISL_1031797, EPI_ISL_1031873, EPI_ISL_1032050, EPI_ISL_1032060, EPI_ISL_1032067, EPI_ISL_1032163, EPI_ISL_1032196, EPI_ISL_1032197, EPI_ISL_1032201, EPI_ISL_1032205, EPI_ISL_1032344, EPI_ISL_1032393, EPI_ISL_1032394, EPI_ISL_1032395, EPI_ISL_1032399, EPI_ISL_1032401, EPI_ISL_1032405, EPI_ISL_1032567, EPI_ISL_1032580, EPI_ISL_1032581, EPI_ISL_1036954, EPI_ISL_1037009, EPI_ISL_1037086, EPI_ISL_1037167, EPI_ISL_1037195, EPI_ISL_1037232, EPI_ISL_1037250, EPI_ISL_1037255, EPI_ISL_1037475, EPI_ISL_1037497, EPI_ISL_1037510, EPI_ISL_1037655, EPI_ISL_1037859, EPI_ISL_1037868, EPI_ISL_1037980, EPI_ISL_1037988, EPI_ISL_1038042, EPI_ISL_1038121, EPI_ISL_1038215, EPI_ISL_1038302, EPI_ISL_1038344, EPI_ISL_1038361, EPI_ISL_1038367, EPI_ISL_1038368, EPI_ISL_1038485, EPI_ISL_1038490, EPI_ISL_1038491, EPI_ISL_1038492, EPI_ISL_1038493, EPI_ISL_1081200, EPI_ISL_1081208, EPI_ISL_1081215, EPI_ISL_1081270, EPI_ISL_1081271 | | | |
| see above | Laboratory Corporation of America | Respiratory Viruses Branch, Division of Viral Diseases, Centers for Disease Control and Prevention | Alexandre Bolze; Ary Ascencio; Ben L. Rambo-Martin; Brad Sickler; Charlotte Rivera-Garcia; Christine Tran; Clinton R. Paden; Dakota Howard; David Becker; Dhwani Batra; Duncan MacCannell; Efren Sandoval; Eileen de Feo; Elizabeth Cirulli; Eric Allen; Geraint Levan; James Lu; Jan Antico; Jason Nguyen; Jimmy Ramirez; Jingtao Liu; Kelly Schiabor Barrett; Kim Gietzen; Magnus Isaksson; Marc Laurent; Matthew Tolentino; Nicole L. Washington; Peter W. Cook; Phil Febbo; Ryan Cho; Shannon Wickline; Sherry Wang; Simon White; Summer Galloway; Suxiang Tong; Tyler Cassens; William Lee |
| EPI_ISL_830634 | MARY IMOGENE BASSETT HOSPITAL | Wadsworth Center, New York State Department of Health | Alexis Russel; Daryl M. Lamson; Erasmus Schneider; Erica Lasek-Nesselquist; John Kelly; Jonathan Plitnick; Kirsten St. George; Matthew Shudt; Melissa A Leisner; Navjot Singh |
| EPI_ISL_896295, EPI_ISL_896299, EPI_ISL_896310, EPI_ISL_896311, EPI_ISL_896312, EPI_ISL_896313, EPI_ISL_896314, EPI_ISL_896315, EPI_ISL_896316, EPI_ISL_896317, EPI_ISL_896318, EPI_ISL_896319, EPI_ISL_896320, EPI_ISL_896321, EPI_ISL_896322, EPI_ISL_896323, EPI_ISL_896324, EPI_ISL_896325, EPI_ISL_896326, EPI_ISL_896327, EPI_ISL_896328, EPI_ISL_896329, EPI_ISL_896330, EPI_ISL_896331, EPI_ISL_896332, EPI_ISL_896333, EPI_ISL_896334, EPI_ISL_896335, EPI_ISL_896336, EPI_ISL_896337, EPI_ISL_896343, EPI_ISL_896344, EPI_ISL_896345, EPI_ISL_896346, EPI_ISL_896349, EPI_ISL_896498, EPI_ISL_896499, EPI_ISL_896500, EPI_ISL_896501, EPI_ISL_896520, EPI_ISL_896521, EPI_ISL_896522, EPI_ISL_896523, EPI_ISL_896524, EPI_ISL_896530, EPI_ISL_896533, EPI_ISL_896535, EPI_ISL_896536, EPI_ISL_896541, EPI_ISL_896542, EPI_ISL_896543, EPI_ISL_896544, EPI_ISL_896545, EPI_ISL_896546, EPI_ISL_896547, EPI_ISL_896548, EPI_ISL_896552, EPI_ISL_896555, EPI_ISL_896562, EPI_ISL_896570, EPI_ISL_896571, EPI_ISL_936149, EPI_ISL_936150, EPI_ISL_936151, EPI_ISL_936152, EPI_ISL_936154, EPI_ISL_936155, EPI_ISL_936156, EPI_ISL_936157, EPI_ISL_936158, EPI_ISL_936159, EPI_ISL_936160, EPI_ISL_936161, EPI_ISL_936162, EPI_ISL_936163, EPI_ISL_936164, EPI_ISL_936165, EPI_ISL_936166, EPI_ISL_936167, EPI_ISL_936168, EPI_ISL_936169, EPI_ISL_936170, EPI_ISL_936171, EPI_ISL_936172, EPI_ISL_936174, EPI_ISL_936175, EPI_ISL_936176, EPI_ISL_936177, EPI_ISL_936178, EPI_ISL_936179, EPI_ISL_1016337, EPI_ISL_1016338, EPI_ISL_1016342, EPI_ISL_1016343, EPI_ISL_1016402, EPI_ISL_1016403, EPI_ISL_1016404, EPI_ISL_1016405, EPI_ISL_1016406, EPI_ISL_1016407, EPI_ISL_1016408, EPI_ISL_1016409, EPI_ISL_1016410, EPI_ISL_1016411, EPI_ISL_1226427, EPI_ISL_1226437, EPI_ISL_1226442, EPI_ISL_1226455, EPI_ISL_1226467, EPI_ISL_1226471, EPI_ISL_1226474, EPI_ISL_1226475, EPI_ISL_1226481, EPI_ISL_1226494, EPI_ISL_1226504, EPI_ISL_1226522, EPI_ISL_1226525, EPI_ISL_1226526, EPI_ISL_1226543, EPI_ISL_1226664, EPI_ISL_1226700, EPI_ISL_1226701, EPI_ISL_1226710, EPI_ISL_1226725, EPI_ISL_1226726, EPI_ISL_1226731, EPI_ISL_1226733, EPI_ISL_1226736, EPI_ISL_1226799, EPI_ISL_1226820, EPI_ISL_1226857, EPI_ISL_1226858, EPI_ISL_1226861, EPI_ISL_1226907, EPI_ISL_1226910, EPI_ISL_1226980, EPI_ISL_1226986, EPI_ISL_1227033, EPI_ISL_1227034, EPI_ISL_1227175, EPI_ISL_1227178, EPI_ISL_1227179, EPI_ISL_1227180, EPI_ISL_1227211, EPI_ISL_1227214, EPI_ISL_1227230, EPI_ISL_1227236, EPI_ISL_1227242, EPI_ISL_1227243, EPI_ISL_1227278, EPI_ISL_1227279, EPI_ISL_1227280, EPI_ISL_1227281, EPI_ISL_1227282, EPI_ISL_1227284, EPI_ISL_1227285, EPI_ISL_1227286, EPI_ISL_1227287, EPI_ISL_1227288, EPI_ISL_1227291, EPI_ISL_1227292, EPI_ISL_1227468, EPI_ISL_1227469, EPI_ISL_1227471, EPI_ISL_1227643, EPI_ISL_1227644, EPI_ISL_1227645, EPI_ISL_1227646, EPI_ISL_1227647, EPI_ISL_1227648, EPI_ISL_1227649, EPI_ISL_1227650, EPI_ISL_1227651, EPI_ISL_1227652, EPI_ISL_1227653, EPI_ISL_1227654, EPI_ISL_1227655, EPI_ISL_1227656, EPI_ISL_1227657, EPI_ISL_1227658, EPI_ISL_1227659, EPI_ISL_1227660, EPI_ISL_1227661, EPI_ISL_1227662, EPI_ISL_1227791, EPI_ISL_1227792, EPI_ISL_1227793, EPI_ISL_1227794, EPI_ISL_1227795, EPI_ISL_1227796, EPI_ISL_1227797, EPI_ISL_1227798, EPI_ISL_1227799, EPI_ISL_1227800, EPI_ISL_1227801, EPI_ISL_1227802, EPI_ISL_1227803, EPI_ISL_1227805, EPI_ISL_1227806, EPI_ISL_1227807, EPI_ISL_1227808, EPI_ISL_1227809, EPI_ISL_1227810, EPI_ISL_1227811, EPI_ISL_1227812, EPI_ISL_1227814, EPI_ISL_1227815, EPI_ISL_1227817, EPI_ISL_1227818, EPI_ISL_1227820, EPI_ISL_1227827, EPI_ISL_1227828, EPI_ISL_1227829, EPI_ISL_1227837, EPI_ISL_1227838, EPI_ISL_1227843, EPI_ISL_1227844, EPI_ISL_1227845, EPI_ISL_1227846, EPI_ISL_1227847, EPI_ISL_1227851, EPI_ISL_1227880, EPI_ISL_1227881, EPI_ISL_1227882, EPI_ISL_1227883, EPI_ISL_1227884, EPI_ISL_1227885, EPI_ISL_1227886, EPI_ISL_1227887, EPI_ISL_1227888, EPI_ISL_1227889, EPI_ISL_1227890, EPI_ISL_1227891, EPI_ISL_1227892, EPI_ISL_1227893, EPI_ISL_1227894, EPI_ISL_1228032, EPI_ISL_1228033, EPI_ISL_1228034, EPI_ISL_1228035, EPI_ISL_1228052, EPI_ISL_1228053, EPI_ISL_1228054, EPI_ISL_1228055, EPI_ISL_1228056, EPI_ISL_1228057, EPI_ISL_1228058, EPI_ISL_1228059, EPI_ISL_1228060, EPI_ISL_1228061, EPI_ISL_1228062, EPI_ISL_1228063, EPI_ISL_1228064, EPI_ISL_1228065, EPI_ISL_1228066, EPI_ISL_1228067, EPI_ISL_1228068, EPI_ISL_1228069, EPI_ISL_1228070, EPI_ISL_1228071, EPI_ISL_1228072, EPI_ISL_1228073, EPI_ISL_1228074, EPI_ISL_1228075, EPI_ISL_1228076, EPI_ISL_1228077, EPI_ISL_1228078, EPI_ISL_1228079, EPI_ISL_1228080, EPI_ISL_1228081, EPI_ISL_1228082, EPI_ISL_1228083, EPI_ISL_1228084, EPI_ISL_1228085, EPI_ISL_1228086, EPI_ISL_1228087, EPI_ISL_1228088, EPI_ISL_1228089, EPI_ISL_1228090, EPI_ISL_1228091, EPI_ISL_1228092, EPI_ISL_1228093, EPI_ISL_1228094, EPI_ISL_1228095, EPI_ISL_1228096, EPI_ISL_1228097, EPI_ISL_1228098, EPI_ISL_1228099, EPI_ISL_1228100, EPI_ISL_1228101, EPI_ISL_1228102, EPI_ISL_1228103, EPI_ISL_1228104, EPI_ISL_1228105, EPI_ISL_1228106, EPI_ISL_1228107, EPI_ISL_1228108, EPI_ISL_1228109, EPI_ISL_1228110, EPI_ISL_1228111, EPI_ISL_1228112, EPI_ISL_1228113, EPI_ISL_1228117, EPI_ISL_1228118, EPI_ISL_1228122, EPI_ISL_1228123, EPI_ISL_1228124, EPI_ISL_1228125, EPI_ISL_1228126, EPI_ISL_1228127, EPI_ISL_1228128, EPI_ISL_1228129, EPI_ISL_1228131, EPI_ISL_1228132, EPI_ISL_1228133, EPI_ISL_1228134, EPI_ISL_1228136, EPI_ISL_1228137, EPI_ISL_1228138, EPI_ISL_1228141, EPI_ISL_1228142, EPI_ISL_1228144, EPI_ISL_1228145, EPI_ISL_1228146, EPI_ISL_1228224, EPI_ISL_1228225, EPI_ISL_1228226, EPI_ISL_1228227, EPI_ISL_1228229, EPI_ISL_1228236, EPI_ISL_1228240, EPI_ISL_1228241, EPI_ISL_1228242, EPI_ISL_1228243, EPI_ISL_1228244, EPI_ISL_1228245, EPI_ISL_1228246, | | | |

EPI_ISL_1228247, EPI_ISL_1228248, EPI_ISL_1228249, EPI_ISL_1228250, EPI_ISL_1228251, EPI_ISL_1228254, EPI_ISL_1228307, EPI_ISL_1228308, EPI_ISL_1228309, EPI_ISL_1228310, EPI_ISL_1228311, EPI_ISL_1228312, EPI_ISL_1228313, EPI_ISL_1228314, EPI_ISL_1228315, EPI_ISL_1228316, EPI_ISL_1228317, EPI_ISL_1228318, EPI_ISL_1228319, EPI_ISL_1228320, EPI_ISL_1228321, EPI_ISL_1228322, EPI_ISL_1228323, EPI_ISL_1228324, EPI_ISL_1228325, EPI_ISL_1228326, EPI_ISL_1228327, EPI_ISL_1228328, EPI_ISL_1228329, EPI_ISL_1228330, EPI_ISL_1228331, EPI_ISL_1228333, EPI_ISL_1228334, EPI_ISL_1228335, EPI_ISL_1228336, EPI_ISL_1228337, EPI_ISL_1228338, EPI_ISL_1228339, EPI_ISL_1228340, EPI_ISL_1228341, EPI_ISL_1228346, EPI_ISL_1228347, EPI_ISL_1228349, EPI_ISL_1228350, EPI_ISL_1228351, EPI_ISL_1228352, EPI_ISL_1228353, EPI_ISL_1228354, EPI_ISL_1228355, EPI_ISL_1228360, EPI_ISL_1228362, EPI_ISL_1228363, EPI_ISL_1228364, EPI_ISL_1228365, EPI_ISL_1228366, EPI_ISL_1228367, EPI_ISL_1228368, EPI_ISL_1228484, EPI_ISL_1228485, EPI_ISL_1228486, EPI_ISL_1228487, EPI_ISL_1228488, EPI_ISL_1228489, EPI_ISL_1228490, EPI_ISL_1228491, EPI_ISL_1228492, EPI_ISL_1228493, EPI_ISL_1228494, EPI_ISL_1228495, EPI_ISL_1228496, EPI_ISL_1228497, EPI_ISL_1228498, EPI_ISL_1228499, EPI_ISL_1228500, EPI_ISL_1228502, EPI_ISL_1228503, EPI_ISL_1395710, EPI_ISL_1395711, EPI_ISL_1395712, EPI_ISL_1395713, EPI_ISL_1395714, EPI_ISL_1395715, EPI_ISL_1395716, EPI_ISL_1395717, EPI_ISL_1395718, EPI_ISL_1395719, EPI_ISL_1395720, EPI_ISL_1395721, EPI_ISL_1395722, EPI_ISL_1395723, EPI_ISL_1395724, EPI_ISL_1395725, EPI_ISL_1397383, EPI_ISL_1397384, EPI_ISL_1397385, EPI_ISL_1397386, EPI_ISL_1397387, EPI_ISL_1397388, EPI_ISL_1398040, EPI_ISL_1398041, EPI_ISL_1398043, EPI_ISL_1623919, EPI_ISL_1623920, EPI_ISL_1623921, EPI_ISL_1623922, EPI_ISL_1623923, EPI_ISL_1623924, EPI_ISL_1623925, EPI_ISL_1623926, EPI_ISL_1623927, EPI_ISL_1623928, EPI_ISL_1623929, EPI_ISL_1623930, EPI_ISL_1623931, EPI_ISL_1623932, EPI_ISL_1623933, EPI_ISL_1623934, EPI_ISL_1623935, EPI_ISL_1623936, EPI_ISL_1623937, EPI_ISL_1623938, EPI_ISL_1623939, EPI_ISL_1623940, EPI_ISL_1623941, EPI_ISL_1623942, EPI_ISL_1623943, EPI_ISL_1623944, EPI_ISL_1623945, EPI_ISL_1623946, EPI_ISL_1623947, EPI_ISL_1623948, EPI_ISL_1623949, EPI_ISL_1623950, EPI_ISL_1623951, EPI_ISL_1623990, EPI_ISL_1623991, EPI_ISL_1623992, EPI_ISL_1623993, EPI_ISL_1623994, EPI_ISL_1623995, EPI_ISL_1623996, EPI_ISL_1623997, EPI_ISL_1623998, EPI_ISL_1623999, EPI_ISL_1624000, EPI_ISL_1624001, EPI_ISL_1624002, EPI_ISL_1624005, EPI_ISL_1624006, EPI_ISL_1624007, EPI_ISL_1624008, EPI_ISL_1624009, EPI_ISL_1651216, EPI_ISL_1651217, EPI_ISL_1651226, EPI_ISL_1671137, EPI_ISL_1671147, EPI_ISL_1671165, EPI_ISL_1671193, EPI_ISL_1671201, EPI_ISL_1671202, EPI_ISL_1740502, EPI_ISL_1755940, EPI_ISL_1855210

see above | MEMORIAL SLOAN KETTERING CANCER CENTER | Wadsworth Center, New York State Department of Health | Alexis Russel; Alexis Russell; Catharine Prussing; Daryl M. Lamson; Erasmus Schneider; Erica Lasek-Nesselquist; John Kelly; Jonathan Plitnick; Kirsten St. George; Matthew Shudt; Melissa A Leisner; Navjot Singh

EPI_ISL_767583, EPI_ISL_767588, EPI_ISL_767591, EPI_ISL_767592, EPI_ISL_767593, EPI_ISL_767596, EPI_ISL_767597, EPI_ISL_767629, EPI_ISL_767630, EPI_ISL_767631, EPI_ISL_767632, EPI_ISL_767633, EPI_ISL_767634, EPI_ISL_767635, EPI_ISL_767636, EPI_ISL_767637, EPI_ISL_767638, EPI_ISL_767639, EPI_ISL_767640, EPI_ISL_767641, EPI_ISL_767642, EPI_ISL_767643, EPI_ISL_767644, EPI_ISL_767645, EPI_ISL_767646, EPI_ISL_767647, EPI_ISL_767649, EPI_ISL_767654, EPI_ISL_767655, EPI_ISL_767656, EPI_ISL_767657, EPI_ISL_767658, EPI_ISL_767659, EPI_ISL_767660, EPI_ISL_767661, EPI_ISL_767662, EPI_ISL_767663, EPI_ISL_767664, EPI_ISL_767681, EPI_ISL_767682, EPI_ISL_767683, EPI_ISL_767684, EPI_ISL_767685, EPI_ISL_767686, EPI_ISL_767687, EPI_ISL_767688, EPI_ISL_767689, EPI_ISL_767690, EPI_ISL_767691, EPI_ISL_767692, EPI_ISL_767693, EPI_ISL_767694, EPI_ISL_767695, EPI_ISL_767696, EPI_ISL_767697, EPI_ISL_767698, EPI_ISL_767699, EPI_ISL_767700, EPI_ISL_767701, EPI_ISL_767702, EPI_ISL_767703, EPI_ISL_767704, EPI_ISL_767705, EPI_ISL_767706, EPI_ISL_767707, EPI_ISL_767708, EPI_ISL_767709, EPI_ISL_767710, EPI_ISL_767711, EPI_ISL_767712, EPI_ISL_767713, EPI_ISL_767714, EPI_ISL_767715

see above | MEMORIAL SLOAN KETTERING CANCER CENTER | Wadsworth Center, New York State Department.of Health | Alexis Russel; Daryl M. Lamson; Erasmus Schneider; Erica Lasek-Nesselquist; John Kelly; Jonathan Plitnick; Kirsten St. George; Matthew Shudt; Melissa A Leisner; Navjot Singh; Sara Griesemer

EPI_ISL_854373, EPI_ISL_854405, EPI_ISL_854406, EPI_ISL_854407, EPI_ISL_854408, EPI_ISL_854409, EPI_ISL_854410, EPI_ISL_854411, EPI_ISL_854412, EPI_ISL_854413, EPI_ISL_854414, EPI_ISL_854415, EPI_ISL_854416, EPI_ISL_854417, EPI_ISL_854418, EPI_ISL_854419, EPI_ISL_854420, EPI_ISL_854422, EPI_ISL_854423, EPI_ISL_854424, EPI_ISL_854425, EPI_ISL_854426, EPI_ISL_854427, EPI_ISL_854428, EPI_ISL_854429, EPI_ISL_854430, EPI_ISL_854431, EPI_ISL_854432, EPI_ISL_854433, EPI_ISL_854434, EPI_ISL_854435, EPI_ISL_854450, EPI_ISL_854452, EPI_ISL_854453, EPI_ISL_854454, EPI_ISL_854455, EPI_ISL_854456, EPI_ISL_854457, EPI_ISL_854458, EPI_ISL_854459, EPI_ISL_861244, EPI_ISL_861245, EPI_ISL_861246, EPI_ISL_861247, EPI_ISL_861248, EPI_ISL_861249, EPI_ISL_861250, EPI_ISL_861251, EPI_ISL_861252, EPI_ISL_861253, EPI_ISL_861254, EPI_ISL_861255, EPI_ISL_861256, EPI_ISL_861257, EPI_ISL_861258, EPI_ISL_861259, EPI_ISL_861260, EPI_ISL_861261, EPI_ISL_861262, EPI_ISL_861263, EPI_ISL_861264, EPI_ISL_861265, EPI_ISL_861266, EPI_ISL_861267, EPI_ISL_861268, EPI_ISL_861269, EPI_ISL_861270, EPI_ISL_861271, EPI_ISL_861272, EPI_ISL_861273, EPI_ISL_861274, EPI_ISL_861275, EPI_ISL_861276, EPI_ISL_861277, EPI_ISL_861278, EPI_ISL_861279, EPI_ISL_861280, EPI_ISL_861281, EPI_ISL_861282, EPI_ISL_861283, EPI_ISL_861284, EPI_ISL_861285, EPI_ISL_861286, EPI_ISL_861287, EPI_ISL_861288, EPI_ISL_861289, EPI_ISL_861290, EPI_ISL_861291, EPI_ISL_861292, EPI_ISL_861293, EPI_ISL_861294, EPI_ISL_861295, EPI_ISL_861296, EPI_ISL_861297, EPI_ISL_861298, EPI_ISL_861299, EPI_ISL_861300, EPI_ISL_861301, EPI_ISL_861302, EPI_ISL_861303, EPI_ISL_861304, EPI_ISL_861305, EPI_ISL_861306, EPI_ISL_861307, EPI_ISL_861308, EPI_ISL_861309, EPI_ISL_861310, EPI_ISL_861311, EPI_ISL_861312, EPI_ISL_861313, EPI_ISL_861314, EPI_ISL_861315, EPI_ISL_861316, EPI_ISL_861317, EPI_ISL_861318, EPI_ISL_861319, EPI_ISL_861320, EPI_ISL_861321, EPI_ISL_861322, EPI_ISL_861323, EPI_ISL_861324, EPI_ISL_861325, EPI_ISL_861326, EPI_ISL_861327, EPI_ISL_861328, EPI_ISL_861329, EPI_ISL_861330, EPI_ISL_861331, EPI_ISL_936248, EPI_ISL_936249, EPI_ISL_936250, EPI_ISL_936251, EPI_ISL_936252, EPI_ISL_936253, EPI_ISL_936254, EPI_ISL_936255, EPI_ISL_936256, EPI_ISL_936257, EPI_ISL_936258, EPI_ISL_936259, EPI_ISL_936260, EPI_ISL_936261, EPI_ISL_936262, EPI_ISL_936263, EPI_ISL_936264, EPI_ISL_936265, EPI_ISL_936266, EPI_ISL_936267, EPI_ISL_936268, EPI_ISL_936269, EPI_ISL_936270, EPI_ISL_936271, EPI_ISL_936272, EPI_ISL_936273, EPI_ISL_936274, EPI_ISL_936275, EPI_ISL_936276, EPI_ISL_936277, EPI_ISL_936278, EPI_ISL_936279, EPI_ISL_936280, EPI_ISL_936281, EPI_ISL_936282, EPI_ISL_936283, EPI_ISL_936284, EPI_ISL_936285, EPI_ISL_936286, EPI_ISL_936287, EPI_ISL_936288, EPI_ISL_936289, EPI_ISL_936290, EPI_ISL_936291, EPI_ISL_936292, EPI_ISL_936293, EPI_ISL_936294, EPI_ISL_936295, EPI_ISL_936296, EPI_ISL_936297, EPI_ISL_936298, EPI_ISL_936299, EPI_ISL_982425, EPI_ISL_982426, EPI_ISL_982427, EPI_ISL_982428, EPI_ISL_982429, EPI_ISL_982430, EPI_ISL_982431, EPI_ISL_982432, EPI_ISL_982433, EPI_ISL_982434, EPI_ISL_982435, EPI_ISL_982436, EPI_ISL_982437, EPI_ISL_982438, EPI_ISL_982439, EPI_ISL_982440, EPI_ISL_982441, EPI_ISL_982442, EPI_ISL_982443, EPI_ISL_982444, EPI_ISL_982445, EPI_ISL_982446, EPI_ISL_982447, EPI_ISL_982448, EPI_ISL_982449, EPI_ISL_982450, EPI_ISL_982451, EPI_ISL_982452, EPI_ISL_982453, EPI_ISL_982454, EPI_ISL_982455, EPI_ISL_982456, EPI_ISL_982457, EPI_ISL_982458, EPI_ISL_982459, EPI_ISL_982460, EPI_ISL_982461, EPI_ISL_982462, EPI_ISL_982463, EPI_ISL_982464, EPI_ISL_982465, EPI_ISL_982466, EPI_ISL_982467, EPI_ISL_982468, EPI_ISL_982469, EPI_ISL_982470, EPI_ISL_982471, EPI_ISL_982472, EPI_ISL_982473, EPI_ISL_982474, EPI_ISL_982475, EPI_ISL_982476, EPI_ISL_982477, EPI_ISL_982478, EPI_ISL_982479, EPI_ISL_982480, EPI_ISL_982481, EPI_ISL_982482, EPI_ISL_982483, EPI_ISL_982484, EPI_ISL_982485, EPI_ISL_982486, EPI_ISL_982487, EPI_ISL_982488, EPI_ISL_982489, EPI_ISL_982490, EPI_ISL_982491, EPI_ISL_982492, EPI_ISL_982493, EPI_ISL_982494, EPI_ISL_982495, EPI_ISL_982496, EPI_ISL_982497, EPI_ISL_982498, EPI_ISL_983099, EPI_ISL_983100, EPI_ISL_983101, EPI_ISL_983102, EPI_ISL_983103, EPI_ISL_983104, EPI_ISL_983105, EPI_ISL_983106, EPI_ISL_983107, EPI_ISL_983108, EPI_ISL_983109, EPI_ISL_983110, EPI_ISL_983111, EPI_ISL_983112, EPI_ISL_983113, EPI_ISL_983114, EPI_ISL_983115, EPI_ISL_983132, EPI_ISL_983133, EPI_ISL_983134, EPI_ISL_983135, EPI_ISL_983136, EPI_ISL_983137, EPI_ISL_983138, EPI_ISL_983139, EPI_ISL_983140, EPI_ISL_983141, EPI_ISL_983142, EPI_ISL_983143, EPI_ISL_983144, EPI_ISL_983145, EPI_ISL_983146, EPI_ISL_983147, EPI_ISL_983148, EPI_ISL_983149, EPI_ISL_983150, EPI_ISL_983151, EPI_ISL_983152, EPI_ISL_983153, EPI_ISL_983154, EPI_ISL_983155, EPI_ISL_983156, EPI_ISL_983157, EPI_ISL_983158, EPI_ISL_983159, EPI_ISL_983160, EPI_ISL_983161, EPI_ISL_983162, EPI_ISL_983163, EPI_ISL_983164, EPI_ISL_983165, EPI_ISL_983166, EPI_ISL_983167, EPI_ISL_983168, EPI_ISL_983169, EPI_ISL_983170, EPI_ISL_1016171, EPI_ISL_1016181, EPI_ISL_1016183, EPI_ISL_1016184, EPI_ISL_1016187, EPI_ISL_1016189, EPI_ISL_1016190, EPI_ISL_1016191, EPI_ISL_1016201, EPI_ISL_1016202, EPI_ISL_1016203, EPI_ISL_1016204, EPI_ISL_1016205, EPI_ISL_1016207, EPI_ISL_1016208, EPI_ISL_1016210, EPI_ISL_1016212, EPI_ISL_1016213, EPI_ISL_1016215, EPI_ISL_1016216, EPI_ISL_1016217, EPI_ISL_1016218, EPI_ISL_1226431, EPI_ISL_1226433, EPI_ISL_1226450, EPI_ISL_1226464, EPI_ISL_1226492, EPI_ISL_1226493, EPI_ISL_1226529, EPI_ISL_1226530, EPI_ISL_1226597, EPI_ISL_1226601, EPI_ISL_1226604, EPI_ISL_1226605, EPI_ISL_1226680, EPI_ISL_1226689, EPI_ISL_1226714, EPI_ISL_1226722, EPI_ISL_1226724, EPI_ISL_1226737, EPI_ISL_1226740, EPI_ISL_1226741, EPI_ISL_1226742, EPI_ISL_1226744, EPI_ISL_1226745, EPI_ISL_1226746, EPI_ISL_1226747, EPI_ISL_1226748, EPI_ISL_1226749, EPI_ISL_1226750, EPI_ISL_1226842, EPI_ISL_1226864, EPI_ISL_1226871, EPI_ISL_1226876, EPI_ISL_1226877, EPI_ISL_1226878, EPI_ISL_1226879, EPI_ISL_1226880, EPI_ISL_1226881, EPI_ISL_1226882, EPI_ISL_1226909, EPI_ISL_1226934, EPI_ISL_1226939, EPI_ISL_1226940, EPI_ISL_1226941, EPI_ISL_1226942, EPI_ISL_1226943, EPI_ISL_1226944, EPI_ISL_1226945, EPI_ISL_1226946, EPI_ISL_1226947, EPI_ISL_1226948, EPI_ISL_1226949, EPI_ISL_1227016, EPI_ISL_1227017, EPI_ISL_1227035, EPI_ISL_1227036, EPI_ISL_1227037, EPI_ISL_1227038, EPI_ISL_1227039, EPI_ISL_1227041, EPI_ISL_1227042, EPI_ISL_1227043, EPI_ISL_1227044, EPI_ISL_1227045, EPI_ISL_1227046, EPI_ISL_1227047, EPI_ISL_1227048, EPI_ISL_1227049, EPI_ISL_1227297, EPI_ISL_1227298, EPI_ISL_1227299, EPI_ISL_1227300, EPI_ISL_1227301, EPI_ISL_1227306, EPI_ISL_1227307, EPI_ISL_1227308, EPI_ISL_1227309, EPI_ISL_1227310, EPI_ISL_1227311, EPI_ISL_1227312, EPI_ISL_1227313, EPI_ISL_1227315, EPI_ISL_1227316, EPI_ISL_1227317, EPI_ISL_1227318, EPI_ISL_1227319, EPI_ISL_1227321, EPI_ISL_1227322, EPI_ISL_1227323, EPI_ISL_1227324, EPI_ISL_1227325, EPI_ISL_1227327, EPI_ISL_1227328, EPI_ISL_1227329, EPI_ISL_1227330, EPI_ISL_1227331, EPI_ISL_1227332, EPI_ISL_1227333, EPI_ISL_1227334, EPI_ISL_1227335, EPI_ISL_1227336, EPI_ISL_1227338, EPI_ISL_1227339, EPI_ISL_1227340, EPI_ISL_1227341, EPI_ISL_1227342, EPI_ISL_1227343, EPI_ISL_1227344, EPI_ISL_1227345, EPI_ISL_1227346, EPI_ISL_1227347, EPI_ISL_1227348, EPI_ISL_1227349, EPI_ISL_1227350, EPI_ISL_1227351, EPI_ISL_1227352, EPI_ISL_1227353, EPI_ISL_1227354, EPI_ISL_1227355, EPI_ISL_1227356, EPI_ISL_1227364, EPI_ISL_1227365, EPI_ISL_1227366, EPI_ISL_1227367, EPI_ISL_1227368, EPI_ISL_1227369, EPI_ISL_1227370, EPI_ISL_1227371, EPI_ISL_1227372, EPI_ISL_1227373, EPI_ISL_1227374, EPI_ISL_1227375, EPI_ISL_1227376, EPI_ISL_1227377, EPI_ISL_1227378, EPI_ISL_1227380, EPI_ISL_1227381, EPI_ISL_1227382, EPI_ISL_1227383, EPI_ISL_1227384, EPI_ISL_1227385, EPI_ISL_1227386, EPI_ISL_1227503, EPI_ISL_1227665, EPI_ISL_1227909, EPI_ISL_1227911, EPI_ISL_1227912, EPI_ISL_1227932, EPI_ISL_1227933, EPI_ISL_1228114, EPI_ISL_1228173, EPI_ISL_1228256, EPI_ISL_1228262, EPI_ISL_1228520, EPI_ISL_1228530, EPI_ISL_1228531, EPI_ISL_1228536, EPI_ISL_1228541, EPI_ISL_1228542, EPI_ISL_1228550, EPI_ISL_1228553, EPI_ISL_1228558, EPI_ISL_1228559

see above | MONTEFIORE MEDICAL CENTER LABORATORIES | Wadsworth Center, New York State Department of Health | Alexis Russel; Daryl M. Lamson; Erasmus Schneider; Erica Lasek-Nesselquist; John Kelly; Jonathan Plitnick; Kirsten St. George; Matthew Shudt; Melissa A Leisner; Navjot Singh

EPI_ISL_765489, EPI_ISL_765490, EPI_ISL_765491, EPI_ISL_765492, EPI_ISL_765493, EPI_ISL_765494, EPI_ISL_765495, EPI_ISL_765530, EPI_ISL_765531, EPI_ISL_765532, EPI_ISL_765533, EPI_ISL_765534, EPI_ISL_765535, EPI_ISL_765536, EPI_ISL_765537, EPI_ISL_765538, EPI_ISL_765539, EPI_ISL_765540, EPI_ISL_765541, EPI_ISL_765542, EPI_ISL_765543, EPI_ISL_765544, EPI_ISL_765545, EPI_ISL_765546, EPI_ISL_765547, EPI_ISL_765548, EPI_ISL_765549, EPI_ISL_765550, EPI_ISL_765551, EPI_ISL_765552, EPI_ISL_765553, EPI_ISL_765554, EPI_ISL_765555, EPI_ISL_765556, EPI_ISL_765557, EPI_ISL_765558, EPI_ISL_765559, EPI_ISL_765560, EPI_ISL_765561, EPI_ISL_765562, EPI_ISL_765563, EPI_ISL_765564, EPI_ISL_765565, EPI_ISL_765566

see above | MONTEFIORE MEDICAL CENTER LABORATORIES | Wadsworth Center, New York State Department.of Health | Alexis Russel; Daryl M. Lamson; Erasmus Schneider; Erica Lasek-Nesselquist; John Kelly; Jonathan Plitnick; Kirsten St. George; Matthew Shudt; Melissa A Leisner; Navjot Singh; Sara Griesemer

EPI_ISL_801862, EPI_ISL_801864, EPI_ISL_801865, EPI_ISL_801866, EPI_ISL_801868, EPI_ISL_801869, EPI_ISL_801870, EPI_ISL_801871, EPI_ISL_801872, EPI_ISL_801873, EPI_ISL_801874, EPI_ISL_801876, EPI_ISL_801878, EPI_ISL_801880, EPI_ISL_801881, EPI_ISL_801882, EPI_ISL_801883, EPI_ISL_801884, EPI_ISL_801885, EPI_ISL_801886, EPI_ISL_801890, EPI_ISL_801892, EPI_ISL_801893, EPI_ISL_801897, EPI_ISL_801899, EPI_ISL_801900, EPI_ISL_801901, EPI_ISL_801903, EPI_ISL_801904, EPI_ISL_801905, EPI_ISL_801909, EPI_ISL_801911, EPI_ISL_801914, EPI_ISL_801915, EPI_ISL_801916, EPI_ISL_801918, EPI_ISL_801919, EPI_ISL_801920, EPI_ISL_801921, EPI_ISL_801922, EPI_ISL_801923, EPI_ISL_801924, EPI_ISL_801925, EPI_ISL_801926, EPI_ISL_801927, EPI_ISL_801930, EPI_ISL_801931, EPI_ISL_801933, EPI_ISL_801934, EPI_ISL_801936, EPI_ISL_801937, EPI_ISL_801939, EPI_ISL_801940, EPI_ISL_801941, EPI_ISL_801942, EPI_ISL_801943, EPI_ISL_801946, EPI_ISL_801947, EPI_ISL_801955, EPI_ISL_801956, EPI_ISL_801958, EPI_ISL_801959, EPI_ISL_801964, EPI_ISL_801965, EPI_ISL_801966, EPI_ISL_801967, EPI_ISL_801968, EPI_ISL_801969, EPI_ISL_801970, EPI_ISL_801971, EPI_ISL_801972, EPI_ISL_801973, EPI_ISL_801974, EPI_ISL_801975, EPI_ISL_801976, EPI_ISL_801979, EPI_ISL_802140, EPI_ISL_802141, EPI_ISL_802142, EPI_ISL_802143, EPI_ISL_802144, EPI_ISL_802145, EPI_ISL_802146, EPI_ISL_802147, EPI_ISL_802149, EPI_ISL_802150, EPI_ISL_802151, EPI_ISL_802152, EPI_ISL_802153, EPI_ISL_802154, EPI_ISL_802155, EPI_ISL_802156, EPI_ISL_802157, EPI_ISL_802158, EPI_ISL_802159, EPI_ISL_802160, EPI_ISL_802161, EPI_ISL_802162, EPI_ISL_802163, EPI_ISL_802164, EPI_ISL_802165, EPI_ISL_802166, EPI_ISL_802167, EPI_ISL_802168, EPI_ISL_802169, EPI_ISL_802170, EPI_ISL_802171, EPI_ISL_802172, EPI_ISL_802173, EPI_ISL_802174, EPI_ISL_802175, EPI_ISL_802176, EPI_ISL_802177, EPI_ISL_802178, EPI_ISL_802179, EPI_ISL_802180, EPI_ISL_802181, EPI_ISL_802182, EPI_ISL_802183, EPI_ISL_802184, EPI_ISL_802185, EPI_ISL_802186, EPI_ISL_802187, EPI_ISL_802189, EPI_ISL_802190, EPI_ISL_802191, EPI_ISL_802192, EPI_ISL_802193, EPI_ISL_802194, EPI_ISL_802195, EPI_ISL_802196, EPI_ISL_802197, EPI_ISL_802198, EPI_ISL_802199, EPI_ISL_802200, EPI_ISL_802201, EPI_ISL_802203, EPI_ISL_802204, EPI_ISL_802205, EPI_ISL_802206, EPI_ISL_802207, EPI_ISL_802208, EPI_ISL_802209, EPI_ISL_802210, EPI_ISL_802211, EPI_ISL_802212, EPI_ISL_802213, EPI_ISL_802214, EPI_ISL_802215, EPI_ISL_802216, EPI_ISL_802217, EPI_ISL_802218, EPI_ISL_802219, EPI_ISL_802220, EPI_ISL_802221, EPI_ISL_802222, EPI_ISL_802223, EPI_ISL_802224, EPI_ISL_802225, EPI_ISL_802226, EPI_ISL_802227, EPI_ISL_802228, EPI_ISL_802229, EPI_ISL_802230, EPI_ISL_802231, EPI_ISL_802232, EPI_ISL_802234, EPI_ISL_802235, EPI_ISL_802236, EPI_ISL_802237, EPI_ISL_802238, EPI_ISL_802239, EPI_ISL_802240, EPI_ISL_802241, EPI_ISL_802242, EPI_ISL_802243, EPI_ISL_802245, EPI_ISL_802246, EPI_ISL_802247, EPI_ISL_802248, EPI_ISL_802249, EPI_ISL_802250, EPI_ISL_802251, EPI_ISL_802252, EPI_ISL_802253, EPI_ISL_802254, EPI_ISL_802255, EPI_ISL_802256, EPI_ISL_802257, EPI_ISL_802259, EPI_ISL_802260, EPI_ISL_802261, EPI_ISL_802263, EPI_ISL_802264, EPI_ISL_802265, EPI_ISL_802266, EPI_ISL_802267, EPI_ISL_802268, EPI_ISL_802269, EPI_ISL_802270, EPI_ISL_802271, EPI_ISL_802272, EPI_ISL_802273, EPI_ISL_802274, EPI_ISL_802276, EPI_ISL_802277, EPI_ISL_802278, EPI_ISL_802279, EPI_ISL_802280, EPI_ISL_802281, EPI_ISL_802282, EPI_ISL_802283, EPI_ISL_802284, EPI_ISL_802285, EPI_ISL_802286, EPI_ISL_802287, EPI_ISL_802288, EPI_ISL_802289, EPI_ISL_802290, EPI_ISL_802291, EPI_ISL_802292, EPI_ISL_802293, EPI_ISL_802294, EPI_ISL_802295, EPI_ISL_802296, EPI_ISL_802297, EPI_ISL_802298, EPI_ISL_802299, EPI_ISL_802300, EPI_ISL_802301, EPI_ISL_802302, EPI_ISL_802303, EPI_ISL_802304, EPI_ISL_802305, EPI_ISL_802306, EPI_ISL_802307, EPI_ISL_802308, EPI_ISL_802309, EPI_ISL_802310, EPI_ISL_802311, EPI_ISL_802312, EPI_ISL_802313, EPI_ISL_802314, EPI_ISL_802315, EPI_ISL_802316, EPI_ISL_802317, EPI_ISL_802318, EPI_ISL_802319, EPI_ISL_802320, EPI_ISL_802321, EPI_ISL_802322, EPI_ISL_802323, EPI_ISL_802324, EPI_ISL_802325, EPI_ISL_802326, EPI_ISL_802327, EPI_ISL_802328, EPI_ISL_802329, EPI_ISL_802330, EPI_ISL_802331, EPI_ISL_802332, EPI_ISL_802333, EPI_ISL_802334, EPI_ISL_802335, EPI_ISL_802336, EPI_ISL_802337, EPI_ISL_802338, EPI_ISL_802339, EPI_ISL_802340, EPI_ISL_802342, EPI_ISL_802343, EPI_ISL_802344, EPI_ISL_802345, EPI_ISL_802346, EPI_ISL_802347, EPI_ISL_802348, EPI_ISL_802349, EPI_ISL_802350, EPI_ISL_802351, EPI_ISL_802352, EPI_ISL_802353, EPI_ISL_802354, EPI_ISL_802355, EPI_ISL_802356, EPI_ISL_802358, EPI_ISL_802359, EPI_ISL_802360, EPI_ISL_802361, EPI_ISL_802362, EPI_ISL_802363, EPI_ISL_802364, EPI_ISL_802365, EPI_ISL_802366, EPI_ISL_802367, EPI_ISL_802368, EPI_ISL_802369, EPI_ISL_802370, EPI_ISL_802371, EPI_ISL_802372, EPI_ISL_802373, EPI_ISL_802374, EPI_ISL_802375, EPI_ISL_802376, EPI_ISL_802377, EPI_ISL_802378, EPI_ISL_802379, EPI_ISL_802380, EPI_ISL_802381, EPI_ISL_802382, EPI_ISL_802383, EPI_ISL_802384, EPI_ISL_802385, EPI_ISL_802386, EPI_ISL_802387, EPI_ISL_802388, EPI_ISL_802389, EPI_ISL_802391, EPI_ISL_802394, EPI_ISL_802395, EPI_ISL_802396, EPI_ISL_802397, EPI_ISL_802398, EPI_ISL_802399, EPI_ISL_802400, EPI_ISL_802401, EPI_ISL_802402, EPI_ISL_802403, EPI_ISL_802404, EPI_ISL_802405, EPI_ISL_802406, EPI_ISL_802407, EPI_ISL_802408, EPI_ISL_802409, EPI_ISL_802410, EPI_ISL_802411, EPI_ISL_802412, EPI_ISL_802413, EPI_ISL_802414, EPI_ISL_802415, EPI_ISL_802416, EPI_ISL_802418, EPI_ISL_802419, EPI_ISL_802420, EPI_ISL_802421, EPI_ISL_802422, EPI_ISL_802423, EPI_ISL_802426, EPI_ISL_802427, EPI_ISL_1300756, EPI_ISL_1300758, EPI_ISL_1300759, EPI_ISL_1300760, EPI_ISL_1300761, EPI_ISL_1300763, EPI_ISL_1300764, EPI_ISL_1300765, EPI_ISL_1300766, EPI_ISL_1300767, EPI_ISL_1300769, EPI_ISL_1300771, EPI_ISL_1300772, EPI_ISL_1300773, EPI_ISL_1300774, EPI_ISL_1300776, EPI_ISL_1300777, EPI_ISL_1300778, EPI_ISL_1300779, EPI_ISL_1300780, EPI_ISL_1300784, EPI_ISL_1300786, EPI_ISL_1300787, EPI_ISL_1300788, EPI_ISL_1300789, EPI_ISL_1300790, EPI_ISL_1300792, EPI_ISL_1300793, EPI_ISL_1300795, EPI_ISL_1300796, EPI_ISL_1300799, EPI_ISL_1300800, EPI_ISL_1300801, EPI_ISL_1300803, EPI_ISL_1300812, EPI_ISL_1300813, EPI_ISL_1300814, EPI_ISL_1300815, EPI_ISL_1300816, EPI_ISL_1300817, EPI_ISL_1300822, EPI_ISL_1300823, EPI_ISL_1300824, EPI_ISL_1300825, EPI_ISL_1300826, EPI_ISL_1300827, EPI_ISL_1300828, EPI_ISL_1300829, EPI_ISL_1300830, EPI_ISL_1300831, EPI_ISL_1300832, EPI_ISL_1300833, EPI_ISL_1300834, EPI_ISL_1300835, EPI_ISL_1300836, EPI_ISL_1300838, EPI_ISL_1300839, EPI_ISL_1300840, EPI_ISL_1300841, EPI_ISL_1300842, EPI_ISL_1300843, EPI_ISL_1300846, EPI_ISL_1300847, EPI_ISL_1300848, EPI_ISL_1300850, EPI_ISL_1300851, EPI_ISL_1300852, EPI_ISL_1300853, EPI_ISL_1300854, EPI_ISL_1300856, EPI_ISL_1300857, EPI_ISL_1300858, EPI_ISL_1300859, EPI_ISL_1300860, EPI_ISL_1300861, EPI_ISL_1300862, EPI_ISL_1300863, EPI_ISL_1300868, EPI_ISL_1300869, EPI_ISL_1300870, EPI_ISL_1300871, EPI_ISL_1300872, EPI_ISL_1300873, EPI_ISL_1300874, EPI_ISL_1300875, EPI_ISL_1300877, EPI_ISL_1300879, EPI_ISL_1300880, EPI_ISL_1300881, EPI_ISL_1300882, EPI_ISL_1300883, EPI_ISL_1300884, EPI_ISL_1300885, EPI_ISL_1300886, EPI_ISL_1300887, EPI_ISL_1300888, EPI_ISL_1300889, EPI_ISL_1300890, EPI_ISL_1300891, EPI_ISL_1300892, EPI_ISL_1300893, EPI_ISL_1300894, EPI_ISL_1300895, EPI_ISL_1300896, EPI_ISL_1300897, EPI_ISL_1300898, EPI_ISL_1300899, EPI_ISL_1300900, EPI_ISL_1300901, EPI_ISL_1300931, EPI_ISL_1300932, EPI_ISL_1300933, EPI_ISL_1300948, EPI_ISL_1300953, EPI_ISL_1300954, EPI_ISL_1300955, EPI_ISL_1300956, EPI_ISL_1300957, EPI_ISL_1300958, EPI_ISL_1300959, EPI_ISL_1300960, EPI_ISL_1300961, EPI_ISL_1300963, EPI_ISL_1300964, EPI_ISL_1300965, EPI_ISL_1300966, EPI_ISL_1300967, EPI_ISL_1300968, EPI_ISL_1300969, EPI_ISL_1300970, EPI_ISL_1300971, EPI_ISL_1300972, EPI_ISL_1300973, EPI_ISL_1300974, EPI_ISL_1300975, EPI_ISL_1300976, EPI_ISL_1300977, EPI_ISL_1300978, EPI_ISL_1300979, EPI_ISL_1300980, EPI_ISL_1300981, EPI_ISL_1300982, EPI_ISL_1300983, EPI_ISL_1300984, EPI_ISL_1300985, EPI_ISL_1300986, EPI_ISL_1300987, EPI_ISL_1300988, EPI_ISL_1300989, EPI_ISL_1300990, EPI_ISL_1300991, EPI_ISL_1300992, EPI_ISL_1300993, EPI_ISL_1300994, EPI_ISL_1300995, EPI_ISL_1300996, EPI_ISL_1300997, EPI_ISL_1300998, EPI_ISL_1300999, EPI_ISL_1301000, EPI_ISL_1301001, EPI_ISL_1301002, EPI_ISL_1301003, EPI_ISL_1301004, EPI_ISL_1301005, EPI_ISL_1301006, EPI_ISL_1301007, EPI_ISL_1301008, EPI_ISL_1301009, EPI_ISL_1301010, EPI_ISL_1301011, EPI_ISL_1301012, EPI_ISL_1301013, EPI_ISL_1301014, EPI_ISL_1301015, EPI_ISL_1301016, EPI_ISL_1301018, EPI_ISL_1301019, EPI_ISL_1301020, EPI_ISL_1301021, EPI_ISL_1301022, EPI_ISL_1301023, EPI_ISL_1301024, EPI_ISL_1301025, EPI_ISL_1301026, EPI_ISL_1301027, EPI_ISL_1301028, EPI_ISL_1301029, EPI_ISL_1301030, EPI_ISL_1301031, EPI_ISL_1301032, EPI_ISL_1301034, EPI_ISL_1301035, EPI_ISL_1301036, EPI_ISL_1301037, EPI_ISL_1301038, EPI_ISL_1301039, EPI_ISL_1301040, EPI_ISL_1301041, EPI_ISL_1301042, EPI_ISL_1301043, EPI_ISL_1301044, EPI_ISL_1301045, EPI_ISL_1301046, EPI_ISL_1301047, EPI_ISL_1301048, EPI_ISL_1301049, EPI_ISL_1301050, EPI_ISL_1301051, EPI_ISL_1301053, EPI_ISL_1301054, EPI_ISL_1301055, EPI_ISL_1301056, EPI_ISL_1301057, EPI_ISL_1301058, EPI_ISL_1301059, EPI_ISL_1301060, EPI_ISL_1301061, EPI_ISL_1301062, EPI_ISL_1301063, EPI_ISL_1301064, EPI_ISL_1301065, EPI_ISL_1301066, EPI_ISL_1301067, EPI_ISL_1301068, EPI_ISL_1301069, EPI_ISL_1301070, EPI_ISL_1301071, EPI_ISL_1301072, EPI_ISL_1301073, EPI_ISL_1301074, EPI_ISL_1301075, EPI_ISL_1301077, EPI_ISL_1301078, EPI_ISL_1301079, EPI_ISL_1301080, EPI_ISL_1301081, EPI_ISL_1301082, EPI_ISL_1301083, EPI_ISL_1301084, EPI_ISL_1301085, EPI_ISL_1301086, EPI_ISL_1301087, EPI_ISL_1301088, EPI_ISL_1301089, EPI_ISL_1301090, EPI_ISL_1301091, EPI_ISL_1301093, EPI_ISL_1301095, EPI_ISL_1301097, EPI_ISL_1301098, EPI_ISL_1301099, EPI_ISL_1301100, EPI_ISL_1301101, EPI_ISL_1301102, EPI_ISL_1301103, EPI_ISL_1301104, EPI_ISL_1301105, EPI_ISL_1301106, EPI_ISL_1301107, EPI_ISL_1301108, EPI_ISL_1301109, EPI_ISL_1301110, EPI_ISL_1301111, EPI_ISL_1301112, EPI_ISL_1301113, EPI_ISL_1301114, EPI_ISL_1301115, EPI_ISL_1301116, EPI_ISL_1301117, EPI_ISL_1301118, EPI_ISL_1301119, EPI_ISL_1301120, EPI_ISL_1301121, EPI_ISL_1301122, EPI_ISL_1301123, EPI_ISL_1301124, EPI_ISL_1301125, EPI_ISL_1301126, EPI_ISL_1301127, EPI_ISL_1301128, EPI_ISL_1301129, EPI_ISL_1301130, EPI_ISL_1301131, EPI_ISL_1301132, EPI_ISL_1301133, EPI_ISL_1301134, EPI_ISL_1301135, EPI_ISL_1301136, EPI_ISL_1301137, EPI_ISL_1301138, EPI_ISL_1301139, EPI_ISL_1301140, EPI_ISL_1301141, EPI_ISL_1301142, EPI_ISL_1301144, EPI_ISL_1301145, EPI_ISL_1301146, EPI_ISL_1301147, EPI_ISL_1301148, EPI_ISL_1301149, EPI_ISL_1301150, EPI_ISL_1301151, EPI_ISL_1301152, EPI_ISL_1301153, EPI_ISL_1301154, EPI_ISL_1301155, EPI_ISL_1301156, EPI_ISL_1301157, EPI_ISL_1301158, EPI_ISL_1301159, EPI_ISL_1301160, EPI_ISL_1301161, EPI_ISL_1301162, EPI_ISL_1301163, EPI_ISL_1301164, EPI_ISL_1301165, EPI_ISL_1301166, EPI_ISL_1301167, EPI_ISL_1301168, EPI_ISL_1301169, EPI_ISL_1301170, EPI_ISL_1301171, EPI_ISL_1301172, EPI_ISL_1301173, EPI_ISL_1301174, EPI_ISL_1301175, EPI_ISL_1301176, EPI_ISL_1301177, EPI_ISL_1301178, EPI_ISL_1301179, EPI_ISL_1301180, EPI_ISL_1301181, EPI_ISL_1301182, EPI_ISL_1301183, EPI_ISL_1301184, EPI_ISL_1301185, EPI_ISL_1301186, EPI_ISL_1301187, EPI_ISL_1301188, EPI_ISL_1301189, EPI_ISL_1301190, EPI_ISL_1301191, EPI_ISL_1301193, EPI_ISL_1301194, EPI_ISL_1301195, EPI_ISL_1301196, EPI_ISL_1301197, EPI_ISL_1301198, EPI_ISL_1301199, EPI_ISL_1301200, EPI_ISL_1301201, EPI_ISL_1301202, EPI_ISL_1301203, EPI_ISL_1301204, EPI_ISL_1301205, EPI_ISL_1301206, EPI_ISL_1301207, EPI_ISL_1301208, EPI_ISL_1301209, EPI_ISL_1301210, EPI_ISL_1301211, EPI_ISL_1301212, EPI_ISL_1301213, EPI_ISL_1301214, EPI_ISL_1301215, EPI_ISL_1301216, EPI_ISL_1301217, EPI_ISL_1301218, EPI_ISL_1301219, EPI_ISL_1301220, EPI_ISL_1301221, EPI_ISL_1301222, EPI_ISL_1301223, EPI_ISL_1301224, EPI_ISL_1301225, EPI_ISL_1301226, EPI_ISL_1301227, EPI_ISL_1301228, EPI_ISL_1301229, EPI_ISL_1301230, EPI_ISL_1301231, EPI_ISL_1301232, EPI_ISL_1301233, EPI_ISL_1301234, EPI_ISL_1301235, EPI_ISL_1301236, EPI_ISL_1301237, EPI_ISL_1301238, EPI_ISL_1301239, EPI_ISL_1301240, EPI_ISL_1301241, EPI_ISL_1301242, EPI_ISL_1301243, EPI_ISL_1301244, EPI_ISL_1301245, EPI_ISL_1301246, EPI_ISL_1301247, EPI_ISL_1301248, EPI_ISL_1301249, EPI_ISL_1301250, EPI_ISL_1301251, EPI_ISL_1301252, EPI_ISL_1301253, EPI_ISL_1301254, EPI_ISL_1301255, EPI_ISL_1301257, EPI_ISL_1301258, EPI_ISL_1301259, EPI_ISL_1301260, EPI_ISL_1301261, EPI_ISL_1301262, EPI_ISL_1301263, EPI_ISL_1301264, EPI_ISL_1301265, EPI_ISL_1301266, EPI_ISL_1301267, EPI_ISL_1301268, EPI_ISL_1301269, EPI_ISL_1301270, EPI_ISL_1301271, EPI_ISL_1301272, EPI_ISL_1301273, EPI_ISL_1301274, EPI_ISL_1301275, EPI_ISL_1301276, EPI_ISL_1301277, EPI_ISL_1301278, EPI_ISL_1301279, EPI_ISL_1301280, EPI_ISL_1301281, EPI_ISL_1301282, EPI_ISL_1301283, EPI_ISL_1301284, EPI_ISL_1301285, EPI_ISL_1301286, EPI_ISL_1301287, EPI_ISL_1301288, EPI_ISL_1301289, EPI_ISL_1301290, EPI_ISL_1301291, EPI_ISL_1301292, EPI_ISL_1301293, EPI_ISL_1301294, EPI_ISL_1301295, EPI_ISL_1301296, EPI_ISL_1301297, EPI_ISL_1301298, EPI_ISL_1301299, EPI_ISL_1301300, EPI_ISL_1301301, EPI_ISL_1301302, EPI_ISL_1301303, EPI_ISL_1301304, EPI_ISL_1301305, EPI_ISL_1301306, EPI_ISL_1301307, EPI_ISL_1301308, EPI_ISL_1301309, EPI_ISL_1301310, EPI_ISL_1301311, EPI_ISL_1301312, EPI_ISL_1301313, EPI_ISL_1301314, EPI_ISL_1301315, EPI_ISL_1301316, EPI_ISL_1301317, EPI_ISL_1301318, EPI_ISL_1301319, EPI_ISL_1301320, EPI_ISL_1301321, EPI_ISL_1301322, EPI_ISL_1301323, EPI_ISL_1301325, EPI_ISL_1301326, EPI_ISL_1301327, EPI_ISL_1301328, EPI_ISL_1301329, EPI_ISL_1301330, EPI_ISL_1301331, EPI_ISL_1301332, EPI_ISL_1301333, EPI_ISL_1301334, EPI_ISL_1301335, EPI_ISL_1301336, EPI_ISL_1301337, EPI_ISL_1301338, EPI_ISL_1301339, EPI_ISL_1301340, EPI_ISL_1301341, EPI_ISL_1301342, EPI_ISL_1301343, EPI_ISL_1301344, EPI_ISL_1301345, EPI_ISL_1301346, EPI_ISL_1301347, EPI_ISL_1301348, EPI_ISL_1301349, EPI_ISL_1301350, EPI_ISL_1301351, EPI_ISL_1301352, EPI_ISL_1301353, EPI_ISL_1301354, EPI_ISL_1301355, EPI_ISL_1301356, EPI_ISL_1301357, EPI_ISL_1301358, EPI_ISL_1301359, EPI_ISL_1301360, EPI_ISL_1301361, EPI_ISL_1301362, EPI_ISL_1301363, EPI_ISL_1301364, EPI_ISL_1301365, EPI_ISL_1301366, EPI_ISL_1301367, EPI_ISL_1301368, EPI_ISL_1301369, EPI_ISL_1301370, EPI_ISL_1301371, EPI_ISL_1301372, EPI_ISL_1301373, EPI_ISL_1301374, EPI_ISL_1301375, EPI_ISL_1301376, EPI_ISL_1301377, EPI_ISL_1301378, EPI_ISL_1301379, EPI_ISL_1301380, EPI_ISL_1301381, EPI_ISL_1301382, EPI_ISL_1301383, EPI_ISL_1301384, EPI_ISL_1301385, EPI_ISL_1301386, EPI_ISL_1301387, EPI_ISL_1301388, EPI_ISL_1301389, EPI_ISL_1301390, EPI_ISL_1301392, EPI_ISL_1301393, EPI_ISL_1301394, EPI_ISL_1301395, EPI_ISL_1301396, EPI_ISL_1301397, EPI_ISL_1301398, EPI_ISL_1301400, EPI_ISL_1301401, EPI_ISL_1301402, EPI_ISL_1708745, EPI_ISL_1708747, EPI_ISL_1708748, EPI_ISL_1708750, EPI_ISL_1708751, EPI_ISL_1708752, EPI_ISL_1708753, EPI_ISL_1708755, EPI_ISL_1708756, EPI_ISL_1708757, EPI_ISL_1708758, EPI_ISL_1708759, EPI_ISL_1708760, EPI_ISL_1708761, EPI_ISL_1708762, EPI_ISL_1708763, EPI_ISL_1708764, EPI_ISL_1708765, EPI_ISL_1708767, EPI_ISL_1708768, EPI_ISL_1708769, EPI_ISL_1708770, EPI_ISL_1708771, EPI_ISL_1708772, EPI_ISL_1708773, EPI_ISL_1708774, EPI_ISL_1708775, EPI_ISL_1708776, EPI_ISL_1708777, EPI_ISL_1708778, EPI_ISL_1708779, EPI_ISL_1708781, EPI_ISL_1708782, EPI_ISL_1708783, EPI_ISL_1708784, EPI_ISL_1708785, EPI_ISL_1708786, EPI_ISL_1708787, EPI_ISL_1708788, EPI_ISL_1708789, EPI_ISL_1708790, EPI_ISL_1708791, EPI_ISL_1708792, EPI_ISL_1708794, EPI_ISL_1708795, EPI_ISL_1708796, EPI_ISL_1708797, EPI_ISL_1708798, EPI_ISL_1708799, EPI_ISL_1708800, EPI_ISL_1708801, EPI_ISL_1708802, EPI_ISL_1708803, EPI_ISL_1708804, EPI_ISL_1708805, EPI_ISL_1708806, EPI_ISL_1708807, EPI_ISL_1708808, EPI_ISL_1708809, EPI_ISL_1708810, EPI_ISL_1708811, EPI_ISL_1708812, EPI_ISL_1708813, EPI_ISL_1708815, EPI_ISL_1708816, EPI_ISL_1708817, EPI_ISL_1708818, EPI_ISL_1708819, EPI_ISL_1708820, EPI_ISL_1708821, EPI_ISL_1708822, EPI_ISL_1708823, EPI_ISL_1708824, EPI_ISL_1708825, EPI_ISL_1708826, EPI_ISL_1708827, EPI_ISL_1708828, EPI_ISL_1708829, EPI_ISL_1708830, EPI_ISL_1708831, EPI_ISL_1708832, EPI_ISL_1708833, EPI_ISL_1708834, EPI_ISL_1708835, EPI_ISL_1708836, EPI_ISL_1708837, EPI_ISL_1708838, EPI_ISL_1708839, EPI_ISL_1708841, EPI_ISL_1708842, EPI_ISL_1708843, EPI_ISL_1708844, EPI_ISL_1708845, EPI_ISL_1708846, EPI_ISL_1708847, EPI_ISL_1708848, EPI_ISL_1708849, EPI_ISL_1708850, EPI_ISL_1708851, EPI_ISL_1708852, EPI_ISL_1708854, EPI_ISL_1708855, EPI_ISL_1708856, EPI_ISL_1708857, EPI_ISL_1708858, EPI_ISL_1708859, EPI_ISL_1708860, EPI_ISL_1708861, EPI_ISL_1708862, EPI_ISL_1708863, EPI_ISL_1708864, EPI_ISL_1708865, EPI_ISL_1708866, EPI_ISL_1708867, EPI_ISL_1708868, EPI_ISL_1708869, EPI_ISL_1708870, EPI_ISL_1708871, EPI_ISL_1708872, EPI_ISL_1708873, EPI_ISL_1708874, EPI_ISL_1708876, EPI_ISL_1708877, EPI_ISL_1708878, EPI_ISL_1708879, EPI_ISL_1708880, EPI_ISL_1708881, EPI_ISL_1708882, EPI_ISL_1708883, EPI_ISL_1708884, EPI_ISL_1708885, EPI_ISL_1708886, EPI_ISL_1708887, EPI_ISL_1708888, EPI_ISL_1708889, EPI_ISL_1708890, EPI_ISL_1708891, EPI_ISL_1708892, EPI_ISL_1708893, EPI_ISL_1708894, EPI_ISL_1708895, EPI_ISL_1708896, EPI_ISL_1708897, EPI_ISL_1708898, EPI_ISL_1708899, EPI_ISL_1708900, EPI_ISL_1708901, EPI_ISL_1708902, EPI_ISL_1708903, EPI_ISL_1708904, EPI_ISL_1708905, EPI_ISL_1708906, EPI_ISL_1708907, EPI_ISL_1708908, EPI_ISL_1708909, EPI_ISL_1708910, EPI_ISL_1708911, EPI_ISL_1708912, EPI_ISL_1708913, EPI_ISL_1708914, EPI_ISL_1708915, EPI_ISL_1708916, EPI_ISL_1708917, EPI_ISL_1708918, EPI_ISL_1708919, EPI_ISL_1708920, EPI_ISL_1708921, EPI_ISL_1708922, EPI_ISL_1708923, EPI_ISL_1708924, EPI_ISL_1708925, EPI_ISL_1708926, EPI_ISL_1708928, EPI_ISL_1708929, EPI_ISL_1708930, EPI_ISL_1708931, EPI_ISL_1708932, EPI_ISL_1708933, EPI_ISL_1708934, EPI_ISL_1708935, EPI_ISL_1708936, EPI_ISL_1708937, EPI_ISL_1708938, EPI_ISL_1708939, EPI_ISL_1708940, EPI_ISL_1708941, EPI_ISL_1708942, EPI_ISL_1708943, EPI_ISL_1708944, EPI_ISL_1708945, EPI_ISL_1708946, EPI_ISL_1708947, EPI_ISL_1708948, EPI_ISL_1708949, EPI_ISL_1708950

see above | MSHS Clinical Microbiology Laboratories | MSHS Pathogen Surveillance Program | Adolfo García-Sastre; Adriana van de Guchte; Ajay Obla; Alberto Paniz-Mondolfi; Ana S. Gonzalez-Reiche; Andrew Kasarskis; Angela Amoako; Ashley S. Salimbangon; Ashley Salimbangon; Betsaida Salom Melo; Bremy Alburquerque; Brianne Ciferri; Charles Gleason; Daniel Floda; Deena R. Altman; Denise Jurczyszak; Elena Hirsch; Emilia Mia Sordillo; Emily Ferreri; Gintaras Deikus; Giulio Kleiner; Gopi Patel; Hala Alshammary; Harm van Bakel; Irina Oussenko; Jayeeta Dutta; Juan Soto; Julia Matthews; Katherine Beach; Kathryn Twyman; Kayla Russo; Komal Srivastava; Levy Sominsky; Mahmoud Awawda; Marta Luksza; Matthew M. Hernandez; Melissa Gitman; Michael D. Nowak; Mitchell J. Sullivan; Nancy Francoeur; Rachel Chernet; Robert Sebra; Sarah Schaefer; Shelcie Fabre; Shwetha Hara Sridhar; Viviana Simon; Ying-Chih Wang; Zenab Khan

EPI_ISL_2009118, EPI_ISL_2009119, EPI_ISL_2009120, EPI_ISL_2009121, EPI_ISL_2009124, EPI_ISL_2009128, EPI_ISL_2009129, EPI_ISL_2009130, EPI_ISL_2009135, EPI_ISL_2009136, EPI_ISL_2009145, EPI_ISL_2009146, EPI_ISL_2009148, EPI_ISL_2009149, EPI_ISL_2009151, EPI_ISL_2009153, EPI_ISL_2009155, EPI_ISL_2009156, EPI_ISL_2009161, EPI_ISL_2009162, EPI_ISL_2009163, EPI_ISL_2009174, EPI_ISL_2009175, EPI_ISL_2009178, EPI_ISL_2009179, EPI_ISL_2009180, EPI_ISL_2009181, EPI_ISL_2009182, EPI_ISL_2009183, EPI_ISL_2009184, EPI_ISL_2009185

see above | MSK Microbiology Lab | MSK Microbiology Lab | Esther Babady; Krupa Jani; Tracy McMillen

EPI_ISL_1226422, EPI_ISL_1226429, EPI_ISL_1226470, EPI_ISL_1226473, EPI_ISL_1226519, EPI_ISL_1227707, EPI_ISL_1227720, EPI_ISL_1227721, EPI_ISL_1227722, EPI_ISL_1227723, EPI_ISL_1227724, EPI_ISL_1227961, EPI_ISL_1227976, EPI_ISL_1227977, EPI_ISL_1227978, EPI_ISL_1227979, EPI_ISL_1227980, EPI_ISL_1227981, EPI_ISL_1227982, EPI_ISL_1227983, EPI_ISL_1227984, EPI_ISL_1227985, EPI_ISL_1227986, EPI_ISL_1227987, EPI_ISL_1227988, EPI_ISL_1227989, EPI_ISL_1227990, EPI_ISL_1228046, EPI_ISL_1228280

see above | Masonic Care Community of NY | Wadsworth Center, New York State Department of Health | Alexis Russel; Daryl M. Lamson; Erasmus Schneider; Erica Lasek-Nesselquist; John Kelly; Jonathan Plitnick; Kirsten St. George; Matthew Shudt; Melissa A Leisner; Navjot Singh

EPI_ISL_676652, EPI_ISL_676665, EPI_ISL_676673, EPI_ISL_677025, EPI_ISL_677111, EPI_ISL_677112, EPI_ISL_677113, EPI_ISL_677114, EPI_ISL_677115, EPI_ISL_677116, EPI_ISL_677117, EPI_ISL_677118, EPI_ISL_677119, EPI_ISL_677120, EPI_ISL_677121, EPI_ISL_677122, EPI_ISL_677123, EPI_ISL_677124, EPI_ISL_677125, EPI_ISL_677126, EPI_ISL_677127, EPI_ISL_677128, EPI_ISL_677129, EPI_ISL_677672,

| | | | |
|---|---|---|---|
| EPI_ISL_734447, EPI_ISL_734448, EPI_ISL_734449, EPI_ISL_734450, EPI_ISL_734451, EPI_ISL_734452, EPI_ISL_734453, EPI_ISL_734454, EPI_ISL_734455, EPI_ISL_734456, EPI_ISL_734457, EPI_ISL_734458, EPI_ISL_734459, EPI_ISL_734460, EPI_ISL_734461, EPI_ISL_734462, EPI_ISL_734463, EPI_ISL_734464, EPI_ISL_734465, EPI_ISL_734466, EPI_ISL_734467, EPI_ISL_734468, EPI_ISL_734469, EPI_ISL_734470, EPI_ISL_734471, EPI_ISL_734472, EPI_ISL_734473, EPI_ISL_734474, EPI_ISL_734475, EPI_ISL_734476, EPI_ISL_734477, EPI_ISL_734478, EPI_ISL_734479, EPI_ISL_734480, EPI_ISL_734481 | | | |
| see above | Masonic Medical Research Institute | Wadsworth Center, New York State Department.of Health | Alexis Russel; Daryl M. Lamson; Erasmus Schneider; Erica Lasek-Nesselquist; John Kelly; Jonathan Plitnick; Kirsten St. George; Nathan Tucker; Navjot Singh; Ryan D. Pfeiffer; Sara Griesemer |
| EPI_ISL_1920061, EPI_ISL_1920076, EPI_ISL_1920157 | Memorial Sloan Kettering Cancer Center | Wadsworth Center, New York State Department of Health | Alexis Russell; Catharine Prussing; Daryl M. Lamson; Erasmus Schneider; Erica Lasek-Nesselquist; John Kelly; Jonathan Plitnick; Kirsten St. George; Matthew Shudt; Melissa A Leisner; Navjot Singh |
| EPI_ISL_2006745, EPI_ISL_2009437, EPI_ISL_2019648, EPI_ISL_2019655, EPI_ISL_2019672, EPI_ISL_2019686, EPI_ISL_2019701, EPI_ISL_2019790, EPI_ISL_2019854, EPI_ISL_2019933, EPI_ISL_2019956, EPI_ISL_2019961, EPI_ISL_2019966, EPI_ISL_2019976, EPI_ISL_2019994, EPI_ISL_2020004, EPI_ISL_2020005 | | | |
| see above | Moderna Inc. | Moderna Inc. | Groves Dixon; Rolando Pajon; Yamuna Paila |
| EPI_ISL_1597704, EPI_ISL_1597705, EPI_ISL_1597707, EPI_ISL_1597708, EPI_ISL_1597709, EPI_ISL_1597710, EPI_ISL_1597728, EPI_ISL_1597729 | | | |
| see above | Montefiore Medical Center | Abbott | Amy Fox; Ana Olivo; Ana Vallari; Barbara Harris; Gavin Cloherty; Mary Rodgers; Todd Meyer; Yitz Goldstein |
| EPI_ISL_826588, EPI_ISL_826589, EPI_ISL_826590, EPI_ISL_826591, EPI_ISL_826593, EPI_ISL_826655, EPI_ISL_826656, EPI_ISL_826657, EPI_ISL_826658, EPI_ISL_826659, EPI_ISL_826660, EPI_ISL_826661, EPI_ISL_826662, EPI_ISL_826663, EPI_ISL_826664, EPI_ISL_826665, EPI_ISL_826667, EPI_ISL_826668, EPI_ISL_826669, EPI_ISL_826670, EPI_ISL_876036, EPI_ISL_876038 | | | |
| see above | Montefiore Medical Center | Albert Einstein College of Medicine, Dept. of Microbiology & Immunology, Chandran lab | Amy S. Fox; Ariel S. Wirchnianski; Aviv Bergman; Catalina Florez; D. Yitzchak Goldstein; David L. Goldman; Denise Haslwanter; Ethan Laudermilch; Hnin Khine; J. Maximilian Fels; Jason Barnhill; Johanna P. Daily; Karin A. Skalina; Kartik Chandran; Libusha Kelly; Lucia R. Wolgast; M. Eugenia Dieterle; Robert H. Bortz III; Rohit K. Jangra; Ryan Forster; Saad Khan; Surksha Sirichand; Wendy Szymczak; William B. Mitchell |
| EPI_ISL_861758, EPI_ISL_1017489, EPI_ISL_1017491, EPI_ISL_1017500, EPI_ISL_1017502, EPI_ISL_1017514, EPI_ISL_1017525, EPI_ISL_1017526, EPI_ISL_1017536, EPI_ISL_1017541, EPI_ISL_1017547, EPI_ISL_1017566, EPI_ISL_1017568, EPI_ISL_1017571, EPI_ISL_1017575, EPI_ISL_1017576, EPI_ISL_1017577, EPI_ISL_1017582, EPI_ISL_1017583, EPI_ISL_1017584, EPI_ISL_1038996, EPI_ISL_1067638, EPI_ISL_1067640, EPI_ISL_1067645, EPI_ISL_1067653, EPI_ISL_1067674, EPI_ISL_1067677, EPI_ISL_1067688, EPI_ISL_1080456, EPI_ISL_1091901, EPI_ISL_1091902, EPI_ISL_1091916, EPI_ISL_1091917, EPI_ISL_1091918, EPI_ISL_1112936 | | | |
| see above | Murphy Medical Associates | Grubaugh Lab - Yale School of Public Health | Anderson Brito; Anne Wyllie; Annie Watkins; Caleb Neal; Chaney Kalinich; Chantal Vogels; Isabel Ott; Joseph Fauver; Mallery Breban; Mary Petrone; Nathan Grubaugh; Steven Murphy; Tara Alpert |
| EPI_ISL_1293138 | Murphy Medical Association | Grubaugh Lab - Yale School of Public Health | Anderson Brito; Annie Watkins; Caleb Neal; Chaney Kalinich; Chantal Vogels; Eva Laszlo; Isabell Ott; Jessica Rothman; Joseph Fauver; Mallery Breban; Mary Petrone; Nathan Grubaugh; Steven Murphy; Tara Alpert |
| EPI_ISL_861186, EPI_ISL_861188, EPI_ISL_861189, EPI_ISL_861190, EPI_ISL_861191, EPI_ISL_861192, EPI_ISL_861193, EPI_ISL_861194, EPI_ISL_861195, EPI_ISL_861196, EPI_ISL_861197, EPI_ISL_861198, EPI_ISL_861413, EPI_ISL_861414, EPI_ISL_861415, EPI_ISL_861416, EPI_ISL_861417, EPI_ISL_861418, EPI_ISL_1226447, EPI_ISL_1227008, EPI_ISL_1227151, EPI_ISL_1227663, EPI_ISL_1227804, EPI_ISL_1227813, EPI_ISL_1227816, EPI_ISL_1227830, EPI_ISL_1227852, EPI_ISL_1228130, EPI_ISL_1228135, EPI_ISL_1228139, EPI_ISL_1228140, EPI_ISL_1228147, EPI_ISL_1228148, EPI_ISL_1228149, EPI_ISL_1228159, EPI_ISL_1228160, EPI_ISL_1228161, EPI_ISL_1228171, EPI_ISL_1228172, EPI_ISL_1228332, EPI_ISL_1228356, EPI_ISL_1397393, EPI_ISL_1397395, EPI_ISL_1397403, EPI_ISL_1397405, EPI_ISL_1397407, EPI_ISL_1397409 | | | |
| see above | NORTH SHORE UNIVERSITY HOSPITAL | Wadsworth Center, New York State Department of Health | Alexis Russel; Daryl M. Lamson; Erasmus Schneider; Erica Lasek-Nesselquist; John Kelly; Jonathan Plitnick; Kirsten St. George; Matthew Shudt; Melissa A Leisner; Navjot Singh |
| EPI_ISL_830572, EPI_ISL_830574, EPI_ISL_830576, EPI_ISL_830577, EPI_ISL_830578, EPI_ISL_830628, EPI_ISL_830629, EPI_ISL_830630, EPI_ISL_830631, EPI_ISL_830632, EPI_ISL_830633, EPI_ISL_830635, EPI_ISL_830636, EPI_ISL_830637, EPI_ISL_830638, EPI_ISL_830639, EPI_ISL_830640, EPI_ISL_830641, EPI_ISL_830642, EPI_ISL_853650, EPI_ISL_853651, EPI_ISL_853653, EPI_ISL_853654, EPI_ISL_853655, EPI_ISL_853656, EPI_ISL_853657, EPI_ISL_853658, EPI_ISL_853659, EPI_ISL_853660, EPI_ISL_853661, EPI_ISL_853662, EPI_ISL_853663, EPI_ISL_853664, EPI_ISL_853665, EPI_ISL_853666, EPI_ISL_853670, EPI_ISL_853671, EPI_ISL_853672, EPI_ISL_853673, EPI_ISL_853674, EPI_ISL_853675, EPI_ISL_853683, EPI_ISL_853705, EPI_ISL_853712, EPI_ISL_853713, EPI_ISL_853714, EPI_ISL_853716, EPI_ISL_853717, EPI_ISL_883426, EPI_ISL_883427, EPI_ISL_883428, EPI_ISL_883431, EPI_ISL_883432, EPI_ISL_883433, EPI_ISL_883434, EPI_ISL_883435, EPI_ISL_883445, EPI_ISL_883446, EPI_ISL_883447, EPI_ISL_883448, EPI_ISL_883449, EPI_ISL_883450, EPI_ISL_883451, EPI_ISL_883452, EPI_ISL_883453, EPI_ISL_883454, EPI_ISL_883455, EPI_ISL_883456, EPI_ISL_883457, EPI_ISL_883458, EPI_ISL_883459, EPI_ISL_883460, EPI_ISL_883461, EPI_ISL_883462, EPI_ISL_883463, EPI_ISL_883464, EPI_ISL_883465, EPI_ISL_883466, EPI_ISL_883467, EPI_ISL_883468, EPI_ISL_883469, EPI_ISL_883470, EPI_ISL_883471, EPI_ISL_883472, EPI_ISL_883474, EPI_ISL_883475, EPI_ISL_883476, EPI_ISL_883477, EPI_ISL_883478, EPI_ISL_883479, EPI_ISL_883480, EPI_ISL_883481, EPI_ISL_883482, EPI_ISL_883483, EPI_ISL_883484, EPI_ISL_883485, EPI_ISL_883486, EPI_ISL_883487, EPI_ISL_883488, EPI_ISL_883489, EPI_ISL_883490, EPI_ISL_883491, EPI_ISL_883492, EPI_ISL_883493, EPI_ISL_883494, EPI_ISL_883495, EPI_ISL_883496, EPI_ISL_883497, EPI_ISL_883498, EPI_ISL_1016138, EPI_ISL_1016140, EPI_ISL_1016143, EPI_ISL_1016144, EPI_ISL_1016145, EPI_ISL_1016146, EPI_ISL_1016147, EPI_ISL_1016148, EPI_ISL_1016149, EPI_ISL_1016150, EPI_ISL_1016151, EPI_ISL_1016152, EPI_ISL_1016153, EPI_ISL_1016156, EPI_ISL_1016157, EPI_ISL_1016158, EPI_ISL_1016159, EPI_ISL_1016160, EPI_ISL_1016161, EPI_ISL_1016162, EPI_ISL_1016163, EPI_ISL_1016164, EPI_ISL_1016165, EPI_ISL_1016166, EPI_ISL_1016167, EPI_ISL_1016168, EPI_ISL_1016413, EPI_ISL_1016458, EPI_ISL_1226425, EPI_ISL_1226444, EPI_ISL_1226472, EPI_ISL_1226506, EPI_ISL_1226553, EPI_ISL_1226672, EPI_ISL_1226707, EPI_ISL_1226717, EPI_ISL_1226720, EPI_ISL_1226849, EPI_ISL_1226853, EPI_ISL_1226856, EPI_ISL_1226859, EPI_ISL_1226865, EPI_ISL_1226898, EPI_ISL_1226899, EPI_ISL_1226911, EPI_ISL_1226912, EPI_ISL_1227153, EPI_ISL_1227154, EPI_ISL_1227181, EPI_ISL_1227664, EPI_ISL_1227677, EPI_ISL_1227678, EPI_ISL_1227819, EPI_ISL_1227821, EPI_ISL_1227822, EPI_ISL_1227823, EPI_ISL_1227824, EPI_ISL_1227825, EPI_ISL_1227826, EPI_ISL_1227831, EPI_ISL_1227832, EPI_ISL_1227833, EPI_ISL_1227834, EPI_ISL_1227835, EPI_ISL_1227836, EPI_ISL_1227839, EPI_ISL_1227840, EPI_ISL_1227841, EPI_ISL_1227842, EPI_ISL_1227848, EPI_ISL_1227849, EPI_ISL_1227850, EPI_ISL_1227853, EPI_ISL_1227854, EPI_ISL_1227855, EPI_ISL_1227856, EPI_ISL_1227857, EPI_ISL_1227858, EPI_ISL_1227859, EPI_ISL_1227903, EPI_ISL_1228150, EPI_ISL_1228151, EPI_ISL_1228152, EPI_ISL_1228153, EPI_ISL_1228155, EPI_ISL_1228156, EPI_ISL_1228157, EPI_ISL_1228170, EPI_ISL_1228235, EPI_ISL_1228342, EPI_ISL_1228343, EPI_ISL_1228344, EPI_ISL_1228345, EPI_ISL_1228348, EPI_ISL_1228357, EPI_ISL_1228358, EPI_ISL_1228359, EPI_ISL_1228361, EPI_ISL_1398042, EPI_ISL_1623918, EPI_ISL_1624004, EPI_ISL_1671209, EPI_ISL_1740504 | | | |
| see above | NORTHWELL HEALTH LABORATORIES | Wadsworth Center, New York State Department of Health | Alexis Russel; Alexis Russell; Catharine Prussing; Daryl M. Lamson; Erasmus Schneider; Erica Lasek-Nesselquist; John Kelly; Jonathan Plitnick; Kirsten St. George; Matthew Shudt; Melissa A Leisner; Navjot Singh |
| EPI_ISL_794119, EPI_ISL_794120, EPI_ISL_794121, EPI_ISL_794122, EPI_ISL_794123, EPI_ISL_794124, EPI_ISL_794125, EPI_ISL_794126, EPI_ISL_794127, EPI_ISL_794128, EPI_ISL_794129, EPI_ISL_794130, EPI_ISL_794131, EPI_ISL_794132, EPI_ISL_794133, EPI_ISL_794134, EPI_ISL_794135, EPI_ISL_794136, EPI_ISL_794137, EPI_ISL_794138, EPI_ISL_794139, EPI_ISL_794140, EPI_ISL_794141, EPI_ISL_794142, EPI_ISL_794143, EPI_ISL_794144, EPI_ISL_794145, EPI_ISL_794146, EPI_ISL_794147, EPI_ISL_794148, EPI_ISL_794149, EPI_ISL_794150, EPI_ISL_794151, EPI_ISL_794152, EPI_ISL_794153, EPI_ISL_794154, EPI_ISL_794155, EPI_ISL_794156, EPI_ISL_794157, EPI_ISL_794158 | | | |
| see above | NORTHWELL HEALTH LABORATORIES | Wadsworth Center, New York State Department.of Health | Alexis Russel; Daryl M. Lamson; Erasmus Schneider; Erica Lasek-Nesselquist; John Kelly; Jonathan Plitnick; Kirsten St. George; Matthew Shudt; Melissa A Leisner; Navjot Singh; Sara Griesemer |
| EPI_ISL_937256, EPI_ISL_937257, EPI_ISL_937258, EPI_ISL_937259, EPI_ISL_937260, EPI_ISL_937261, EPI_ISL_937262, EPI_ISL_937263, EPI_ISL_937264, EPI_ISL_937265, EPI_ISL_937266, EPI_ISL_937267, EPI_ISL_937268, EPI_ISL_937269, EPI_ISL_937270, EPI_ISL_937271, EPI_ISL_937272 | | | |
| see above | NYC HH Elmhurst Hospital Medical Center | New York City Public Health Laboratory | Jade Wang; et al. |
| EPI_ISL_1226439, EPI_ISL_1226448, EPI_ISL_1226476, EPI_ISL_1226508, EPI_ISL_1226513, EPI_ISL_1226665, EPI_ISL_1226699, EPI_ISL_1226703, EPI_ISL_1226789, EPI_ISL_1226860, EPI_ISL_1226913, EPI_ISL_1226914, EPI_ISL_1227134, EPI_ISL_1227135, EPI_ISL_1227136, EPI_ISL_1227152, EPI_ISL_1227205, EPI_ISL_1227666, EPI_ISL_1227667, EPI_ISL_1227668, EPI_ISL_1227669, EPI_ISL_1227670, EPI_ISL_1227671, EPI_ISL_1227672, EPI_ISL_1227673, EPI_ISL_1227674, EPI_ISL_1227920, EPI_ISL_1228143, EPI_ISL_1228162, EPI_ISL_1228163, EPI_ISL_1228164, EPI_ISL_1228165, EPI_ISL_1228166, EPI_ISL_1228167, EPI_ISL_1228168, EPI_ISL_1228169, EPI_ISL_1228228, EPI_ISL_1228230, EPI_ISL_1228231, EPI_ISL_1228232, EPI_ISL_1228233, EPI_ISL_1228234 | | | |
| see above | NYC Pandemic Response Lab | Wadsworth Center, New York State Department of Health | Alexis Russel; Daryl M. Lamson; Erasmus Schneider; Erica Lasek-Nesselquist; John Kelly; Jonathan Plitnick; Kirsten St. George; Matthew Shudt; Melissa A Leisner; Navjot Singh |
| EPI_ISL_751545, EPI_ISL_751548, EPI_ISL_751560, EPI_ISL_751562, EPI_ISL_751580, EPI_ISL_751581, EPI_ISL_751658, EPI_ISL_751682, EPI_ISL_751717 | | | |
| see above | NYSDOH Wadsworth Center, Virology Lab | Genomics and Discovery, Respiratory Viruses Branch, Division of Viral Diseases, Centers for Disease Control and Prevention | Anna Montmayeur; Anna Uehara; Clinton R. Paden; Haibin Wang; Jing Zhang; Justin Lee; Krista Queen; Mili Sheth; Peter W. Cook; Rachel Marine; Suxiang Tong; Yan Li; Ying Tao |
| EPI_ISL_1094217, EPI_ISL_1094219, EPI_ISL_1094227, EPI_ISL_1094285, EPI_ISL_1094286, EPI_ISL_1094307, EPI_ISL_1094322, EPI_ISL_1094359, EPI_ISL_1094368, EPI_ISL_1094370, EPI_ISL_1094378, EPI_ISL_1094433, EPI_ISL_1094434, EPI_ISL_1094435, EPI_ISL_1094436, EPI_ISL_1094437, EPI_ISL_1094438, EPI_ISL_1094439, EPI_ISL_1094440, EPI_ISL_1094441, EPI_ISL_1094443, EPI_ISL_1094444, EPI_ISL_1095131, EPI_ISL_1095132, EPI_ISL_1095134, EPI_ISL_1095135, EPI_ISL_1095136, EPI_ISL_1095138, EPI_ISL_1095139, EPI_ISL_1095140, EPI_ISL_1095141, EPI_ISL_1095143, EPI_ISL_1095144, EPI_ISL_1095145, EPI_ISL_1095146, EPI_ISL_1095147, EPI_ISL_1095148, EPI_ISL_1095150, EPI_ISL_1095151, EPI_ISL_1095152, EPI_ISL_1095153, EPI_ISL_1095154, EPI_ISL_1095155, EPI_ISL_1095156, EPI_ISL_1095157, EPI_ISL_1095158, EPI_ISL_1095159, EPI_ISL_1095160, EPI_ISL_1095161, EPI_ISL_1095162, EPI_ISL_1095163, EPI_ISL_1095164, EPI_ISL_1095165, EPI_ISL_1095166, EPI_ISL_1095167, EPI_ISL_1095168 | | | |
| see above | NYSDOH Wadsworth Center, Virology Lab | Respiratory Viruses Branch, Division of Viral Diseases, Centers for Disease Control and Prevention | Anna Montmayeur; Anna Uehara; Ben L. Rambo-Martin; Clinton R. Paden; Dhwani Batra; Haibin Wang; Jasmine Padilla; Jing Zhang; Justin Lee; Krista Queen; Lori Rowe; Mark Burroughs; Mili Sheth; Peter W. Cook; Rachel Marine; Sarah Nobles; Suxiang Tong; Yan Li; Ying Tao |
| EPI_ISL_965303, EPI_ISL_965528, EPI_ISL_965770, EPI_ISL_965812, EPI_ISL_965814, EPI_ISL_965826, EPI_ISL_965837, EPI_ISL_965838, EPI_ISL_965839, EPI_ISL_965910, EPI_ISL_965948, EPI_ISL_965962, EPI_ISL_965963, EPI_ISL_965964, EPI_ISL_965965, EPI_ISL_965967, EPI_ISL_965968, EPI_ISL_965969, EPI_ISL_965970, EPI_ISL_965971, EPI_ISL_965972, EPI_ISL_965973, EPI_ISL_965977, EPI_ISL_965980, EPI_ISL_965989, EPI_ISL_966020, EPI_ISL_966044, EPI_ISL_966062, EPI_ISL_966084, EPI_ISL_966108, EPI_ISL_966160, EPI_ISL_966184, EPI_ISL_966204, EPI_ISL_966227, EPI_ISL_966264, EPI_ISL_966289, EPI_ISL_966308, EPI_ISL_966311, EPI_ISL_966333, EPI_ISL_966357, EPI_ISL_966375, EPI_ISL_966376, EPI_ISL_966377, EPI_ISL_966378, EPI_ISL_966380, EPI_ISL_966406, EPI_ISL_966428, EPI_ISL_966452, EPI_ISL_966472, EPI_ISL_966489, EPI_ISL_966496, EPI_ISL_966549, EPI_ISL_966551, EPI_ISL_966552, EPI_ISL_966565, EPI_ISL_966591, EPI_ISL_966605, EPI_ISL_966623, EPI_ISL_966646, EPI_ISL_966663, EPI_ISL_966682, EPI_ISL_966707, EPI_ISL_977666, EPI_ISL_1040869, EPI_ISL_1040880, EPI_ISL_1040881, EPI_ISL_1040902, EPI_ISL_1040903, EPI_ISL_1050008, EPI_ISL_1050009, EPI_ISL_1050010, EPI_ISL_1050011, EPI_ISL_1050012, EPI_ISL_1050013, EPI_ISL_1050014, EPI_ISL_1050015, EPI_ISL_1050016, EPI_ISL_1050017, EPI_ISL_1050018, EPI_ISL_1050019, EPI_ISL_1050020, EPI_ISL_1050021, EPI_ISL_1050022, EPI_ISL_1050023, EPI_ISL_1050024, EPI_ISL_1050025, EPI_ISL_1050026, EPI_ISL_1050027, EPI_ISL_1050028, EPI_ISL_1050029, EPI_ISL_1050030, EPI_ISL_1050031, EPI_ISL_1050032, EPI_ISL_1050033, EPI_ISL_1050034, EPI_ISL_1050035, EPI_ISL_1050036, EPI_ISL_1050037, EPI_ISL_1050038, EPI_ISL_1050039, EPI_ISL_1050040, EPI_ISL_1050041, EPI_ISL_1050043, EPI_ISL_1050044, EPI_ISL_1050045, EPI_ISL_1050046, EPI_ISL_1050048, EPI_ISL_1050049, EPI_ISL_1050050, EPI_ISL_1050051, EPI_ISL_1050052, EPI_ISL_1050053, EPI_ISL_1050054, EPI_ISL_1050055, EPI_ISL_1050056, EPI_ISL_1050057, EPI_ISL_1050058, EPI_ISL_1050059, EPI_ISL_1050060, EPI_ISL_1050061, EPI_ISL_1050062, EPI_ISL_1050063, EPI_ISL_1050064, EPI_ISL_1060616, EPI_ISL_1060617, EPI_ISL_1060618, EPI_ISL_1060619, EPI_ISL_1060620, EPI_ISL_1060621, EPI_ISL_1060622, EPI_ISL_1060623, EPI_ISL_1060624, EPI_ISL_1060625, EPI_ISL_1060627, EPI_ISL_1060628, EPI_ISL_1060629, EPI_ISL_1060630, EPI_ISL_1060631, EPI_ISL_1060632, EPI_ISL_1060633, EPI_ISL_1060634, EPI_ISL_1060635, EPI_ISL_1060636, EPI_ISL_1060637, EPI_ISL_1060638, EPI_ISL_1060639, EPI_ISL_1060640, EPI_ISL_1060641, EPI_ISL_1060642, EPI_ISL_1060643, EPI_ISL_1060644, EPI_ISL_1060645, EPI_ISL_1060646, EPI_ISL_1060648, EPI_ISL_1060649, EPI_ISL_1060650, EPI_ISL_1060651, EPI_ISL_1060652, EPI_ISL_1060653, EPI_ISL_1060654, EPI_ISL_1060656, EPI_ISL_1060657, EPI_ISL_1060658, EPI_ISL_1060660, EPI_ISL_1060661, EPI_ISL_1060662, EPI_ISL_1060663, EPI_ISL_1060664, EPI_ISL_1060665, EPI_ISL_1060666, EPI_ISL_1060667, EPI_ISL_1060668, EPI_ISL_1060669, EPI_ISL_1060670, EPI_ISL_1060672, EPI_ISL_1060674, EPI_ISL_1060675, EPI_ISL_1087276, EPI_ISL_1087292, EPI_ISL_1087302, EPI_ISL_1087303, EPI_ISL_1087304, EPI_ISL_1087305, EPI_ISL_1087306, EPI_ISL_1087307, EPI_ISL_1087308, EPI_ISL_1087309, EPI_ISL_1087310, EPI_ISL_1087311, EPI_ISL_1087312, EPI_ISL_1789854, EPI_ISL_1789855, EPI_ISL_1789857, EPI_ISL_1789858, EPI_ISL_1789859, EPI_ISL_1789860, EPI_ISL_1789861, EPI_ISL_1789862, EPI_ISL_1789864, EPI_ISL_1789865, EPI_ISL_1789866, EPI_ISL_1789868, EPI_ISL_1789869, EPI_ISL_1789870, EPI_ISL_1789871, EPI_ISL_1789872, EPI_ISL_1789873, EPI_ISL_1789874, EPI_ISL_1789875, EPI_ISL_1789877, EPI_ISL_1789878, EPI_ISL_1789879, EPI_ISL_1789880, EPI_ISL_1789881, EPI_ISL_1789882, EPI_ISL_1789883, EPI_ISL_1789884, EPI_ISL_1789885, EPI_ISL_1789886, EPI_ISL_1789887, EPI_ISL_1789888, EPI_ISL_1789889, EPI_ISL_1789890, EPI_ISL_1789891, EPI_ISL_1789892, EPI_ISL_1789893, EPI_ISL_1789894, EPI_ISL_1789895, EPI_ISL_1789896, EPI_ISL_1789897, EPI_ISL_1789898, EPI_ISL_1789899, EPI_ISL_1789901, EPI_ISL_1789902, EPI_ISL_1922710, EPI_ISL_1922712, EPI_ISL_1922713, EPI_ISL_1922714, EPI_ISL_1922715, EPI_ISL_1922716, EPI_ISL_1922717, EPI_ISL_1922718, EPI_ISL_1922719, EPI_ISL_1922720, EPI_ISL_1922721, EPI_ISL_1922722, EPI_ISL_1922723, EPI_ISL_1922724, EPI_ISL_1922725, EPI_ISL_1922726, EPI_ISL_1922727, EPI_ISL_1922728, EPI_ISL_1922729, EPI_ISL_1922730, EPI_ISL_1922731, EPI_ISL_1922732, EPI_ISL_1922733, EPI_ISL_1922734, EPI_ISL_1922735, EPI_ISL_1922736, EPI_ISL_1922737, EPI_ISL_1922738, EPI_ISL_1922739, EPI_ISL_1922740, EPI_ISL_1922741, EPI_ISL_1922742, EPI_ISL_1922743, EPI_ISL_1922744, EPI_ISL_1922745, EPI_ISL_1922746, EPI_ISL_1922747, EPI_ISL_1922748, EPI_ISL_1922749, EPI_ISL_1922750, EPI_ISL_1922751, EPI_ISL_1922752, EPI_ISL_1922754, EPI_ISL_1922755, EPI_ISL_1922756, EPI_ISL_1922757, EPI_ISL_1922758, EPI_ISL_1922759, EPI_ISL_1922760, EPI_ISL_1922761, EPI_ISL_1922763, EPI_ISL_1922765, EPI_ISL_1922766, EPI_ISL_1922768, EPI_ISL_1922769, EPI_ISL_1922770, EPI_ISL_1922771, EPI_ISL_1922772, EPI_ISL_1922773, EPI_ISL_1922774, EPI_ISL_1922775, EPI_ISL_1922776, EPI_ISL_1922777, EPI_ISL_1922778, EPI_ISL_1922779, EPI_ISL_1922780, EPI_ISL_1922781, EPI_ISL_1922782, EPI_ISL_1922783, EPI_ISL_1922784, EPI_ISL_1922786, EPI_ISL_1922787, EPI_ISL_1922788, EPI_ISL_1922790, EPI_ISL_1922793, EPI_ISL_1922794, EPI_ISL_1922795, EPI_ISL_1922796, EPI_ISL_1922797, EPI_ISL_1922798, EPI_ISL_1922799 | | | |
| see above | NYU Langone Health | Departments of Pathology and Medicine, New York University School of Medicine | Adriana Heguy; Christian Marier; Dacia Dimartino; Emily Guzman; Gael Westby; Guiqing Wang; Paolo Cotzia; Paul Zappile; Peter Meyn; Sitharam Ramaswami; Yutong Zhang |
| EPI_ISL_1091898, EPI_ISL_1091899, EPI_ISL_1091900, EPI_ISL_1091903, EPI_ISL_1091904, EPI_ISL_1091905, EPI_ISL_1091906, EPI_ISL_1091907, EPI_ISL_1091908, EPI_ISL_1091909, EPI_ISL_1091910, EPI_ISL_1091911, EPI_ISL_1091912, EPI_ISL_1091913, EPI_ISL_1091914, EPI_ISL_1091915 | | | |
| see above | New York Presbetyrian / Weill Cornell Medicine | Grubaugh Lab - Yale School of Public Health | Anderson Brito; Anne Wyllie; Annie Watkins; Chaney Kalinich; Chantal Vogels; Isabel Ott; Joseph Fauver; Mallery Breban; Mary Petrone; Nathan Grubaugh; Tara Alpert |
| EPI_ISL_861109, EPI_ISL_861110, EPI_ISL_861111, EPI_ISL_861112, EPI_ISL_861113, EPI_ISL_861114, EPI_ISL_861118, EPI_ISL_861119, EPI_ISL_861120, EPI_ISL_861121, EPI_ISL_861122, EPI_ISL_861123, EPI_ISL_861124, EPI_ISL_861125, EPI_ISL_861126, EPI_ISL_861127, EPI_ISL_861128, EPI_ISL_861129, EPI_ISL_861130, EPI_ISL_861131, EPI_ISL_861132, EPI_ISL_861133, EPI_ISL_896294, EPI_ISL_896296, EPI_ISL_896298, EPI_ISL_896302, EPI_ISL_896303, EPI_ISL_896304, EPI_ISL_896305, EPI_ISL_896306, EPI_ISL_896338, EPI_ISL_896339, EPI_ISL_896340, EPI_ISL_896341, EPI_ISL_896342, EPI_ISL_896347, EPI_ISL_896348, EPI_ISL_896350, EPI_ISL_896351, EPI_ISL_896352, EPI_ISL_896353, EPI_ISL_896354, EPI_ISL_896355, EPI_ISL_896356, EPI_ISL_896357, EPI_ISL_896358, EPI_ISL_896359, EPI_ISL_896360, EPI_ISL_896361, EPI_ISL_896362, EPI_ISL_896363, EPI_ISL_896364, EPI_ISL_896365, EPI_ISL_896366, EPI_ISL_896367, EPI_ISL_896368, EPI_ISL_896369, EPI_ISL_896370, EPI_ISL_896371, EPI_ISL_896372, EPI_ISL_896504, EPI_ISL_896505, EPI_ISL_896506, EPI_ISL_896507, EPI_ISL_896525, EPI_ISL_896526, EPI_ISL_896527, EPI_ISL_896528, EPI_ISL_896529, EPI_ISL_896531, EPI_ISL_896532, EPI_ISL_896534, EPI_ISL_896537, EPI_ISL_896538, EPI_ISL_896539, EPI_ISL_896540, EPI_ISL_896549, EPI_ISL_896550, EPI_ISL_896551, EPI_ISL_896553, EPI_ISL_896554, EPI_ISL_896556, EPI_ISL_896557, EPI_ISL_896558, EPI_ISL_896559, EPI_ISL_896560, EPI_ISL_896561, EPI_ISL_936130, EPI_ISL_936131, EPI_ISL_936132, EPI_ISL_936133, EPI_ISL_936134, EPI_ISL_936135, EPI_ISL_936136, EPI_ISL_936137, EPI_ISL_936145, EPI_ISL_936146, EPI_ISL_936147, EPI_ISL_1016220, EPI_ISL_1016221, EPI_ISL_1016222, EPI_ISL_1016223, EPI_ISL_1016224, EPI_ISL_1016225, EPI_ISL_1016226, EPI_ISL_1016247, EPI_ISL_1016258, EPI_ISL_1016259, EPI_ISL_1016262, EPI_ISL_1016263, EPI_ISL_1016264, EPI_ISL_1016265, EPI_ISL_1016266, EPI_ISL_1016269, EPI_ISL_1016271, EPI_ISL_1016272, EPI_ISL_1016273, EPI_ISL_1016274, EPI_ISL_1016275, EPI_ISL_1016276, EPI_ISL_1016277, EPI_ISL_1016278, EPI_ISL_1016279, EPI_ISL_1016280, EPI_ISL_1016304, EPI_ISL_1016305, EPI_ISL_1016306, EPI_ISL_1016307, EPI_ISL_1016308, EPI_ISL_1016309, EPI_ISL_1016310, EPI_ISL_1016335, EPI_ISL_1016336, EPI_ISL_1016341, EPI_ISL_1016366, EPI_ISL_1016367, EPI_ISL_1016368, EPI_ISL_1016369, EPI_ISL_1016370, EPI_ISL_1016371, EPI_ISL_1016372, EPI_ISL_1016381, EPI_ISL_1016382, EPI_ISL_1016383, EPI_ISL_1016384, EPI_ISL_1016385, EPI_ISL_1016386, EPI_ISL_1016387, EPI_ISL_1016388, EPI_ISL_1016389, EPI_ISL_1016390, EPI_ISL_1016391, EPI_ISL_1016392, EPI_ISL_1016393, EPI_ISL_1016394, EPI_ISL_1016395, EPI_ISL_1016396, EPI_ISL_1016397, EPI_ISL_1016398, EPI_ISL_1016399, EPI_ISL_1016400, EPI_ISL_1016401, EPI_ISL_1226434, EPI_ISL_1394642, EPI_ISL_1394644, EPI_ISL_1394645, EPI_ISL_1394646, EPI_ISL_1394647, EPI_ISL_1394648, EPI_ISL_1394649, EPI_ISL_1394650, EPI_ISL_1394651, EPI_ISL_1394652, EPI_ISL_1394653, EPI_ISL_1394654, EPI_ISL_1394655, EPI_ISL_1394656, EPI_ISL_1394657, EPI_ISL_1394658, EPI_ISL_1394659, EPI_ISL_1394660, EPI_ISL_1394661, EPI_ISL_1394662, EPI_ISL_1394663, EPI_ISL_1394664, EPI_ISL_1397460, EPI_ISL_1397461, EPI_ISL_1397462, EPI_ISL_1397463, EPI_ISL_1397464, EPI_ISL_1397465, EPI_ISL_1397466, EPI_ISL_1397467, EPI_ISL_1397468, EPI_ISL_1397469, EPI_ISL_1397470, EPI_ISL_1397471, EPI_ISL_1397472, EPI_ISL_1397473, EPI_ISL_1397474, EPI_ISL_1397475, EPI_ISL_1397476, EPI_ISL_1397477, EPI_ISL_1397478, EPI_ISL_1397479, EPI_ISL_1397480, EPI_ISL_1397481, EPI_ISL_1397482, EPI_ISL_1397483, EPI_ISL_1397484, EPI_ISL_1397485, EPI_ISL_1397486, EPI_ISL_1397487, EPI_ISL_1397488, EPI_ISL_1397489, EPI_ISL_1397490, EPI_ISL_1397491, EPI_ISL_1397492, EPI_ISL_1397493, EPI_ISL_1397494, EPI_ISL_1397495, EPI_ISL_1397496, EPI_ISL_1397497, EPI_ISL_1397498, EPI_ISL_1397499, EPI_ISL_1397500, EPI_ISL_1397501, EPI_ISL_1397502, EPI_ISL_1397503, EPI_ISL_1397504, EPI_ISL_1397505, EPI_ISL_1397506, EPI_ISL_1397507, EPI_ISL_1397508, EPI_ISL_1397509, EPI_ISL_1397510, EPI_ISL_1397511, EPI_ISL_1397512, EPI_ISL_1397513, EPI_ISL_1397514, EPI_ISL_1397515, EPI_ISL_1397516, EPI_ISL_1397517, EPI_ISL_1397518, EPI_ISL_1397519, EPI_ISL_1397520, EPI_ISL_1397521, EPI_ISL_1397522, EPI_ISL_1397523, EPI_ISL_1397524, EPI_ISL_1397525, EPI_ISL_1397526, EPI_ISL_1397527, EPI_ISL_1397528, EPI_ISL_1397529, EPI_ISL_1397530, EPI_ISL_1397531, EPI_ISL_1397532, EPI_ISL_1397533, EPI_ISL_1397534, EPI_ISL_1397535, EPI_ISL_1397536, EPI_ISL_1397537, EPI_ISL_1397538, EPI_ISL_1397539, EPI_ISL_1397540, EPI_ISL_1397541, EPI_ISL_1624003, EPI_ISL_1624012, EPI_ISL_1624013, EPI_ISL_1624014, EPI_ISL_1624015, EPI_ISL_1624016, EPI_ISL_1624017, EPI_ISL_1624018, EPI_ISL_1624019, EPI_ISL_1624020, EPI_ISL_1624021, EPI_ISL_1624022, EPI_ISL_1624023, EPI_ISL_1624024, EPI_ISL_1624025, EPI_ISL_1624026, EPI_ISL_1624027, EPI_ISL_1624028, EPI_ISL_1624029, EPI_ISL_1624030, EPI_ISL_1624031, EPI_ISL_1624032, EPI_ISL_1624033, EPI_ISL_1624034, EPI_ISL_1624035, EPI_ISL_1624036, EPI_ISL_1624037, EPI_ISL_1624038, EPI_ISL_1624039, EPI_ISL_1624040, EPI_ISL_1624041, EPI_ISL_1624042, EPI_ISL_1624043, EPI_ISL_1624044, EPI_ISL_1624045, EPI_ISL_1624046, EPI_ISL_1624047, EPI_ISL_1624048, EPI_ISL_1624049, EPI_ISL_1624050, EPI_ISL_1624051, EPI_ISL_1624052, EPI_ISL_1624053, EPI_ISL_1624054, EPI_ISL_1624055, EPI_ISL_1624056, EPI_ISL_1624057, EPI_ISL_1624058, EPI_ISL_1624059, EPI_ISL_1624060, EPI_ISL_1624061, EPI_ISL_1624062, EPI_ISL_1624063, EPI_ISL_1624064, EPI_ISL_1624065, EPI_ISL_1624066, EPI_ISL_1624067, EPI_ISL_1624068, EPI_ISL_1624069, EPI_ISL_1624070, EPI_ISL_1624071, EPI_ISL_1624072, EPI_ISL_1624073, EPI_ISL_1624074, EPI_ISL_1624075, EPI_ISL_1624076, EPI_ISL_1624077, EPI_ISL_1624078, EPI_ISL_1624079, EPI_ISL_1624080, EPI_ISL_1624081, EPI_ISL_1624082, EPI_ISL_1651219, EPI_ISL_1651220, EPI_ISL_1651221, EPI_ISL_1671138, EPI_ISL_1671139, EPI_ISL_1671140, EPI_ISL_1671146, EPI_ISL_1671166, EPI_ISL_1671194, EPI_ISL_1671213, EPI_ISL_1671214, EPI_ISL_1671215, EPI_ISL_1671216, EPI_ISL_1789714, EPI_ISL_1920097, EPI_ISL_1920133, EPI_ISL_1920134, EPI_ISL_1920135, EPI_ISL_1920136, EPI_ISL_1920138, EPI_ISL_1920147, | | | |

| | | | |
|---|---|---|---|
| EPI_ISL_1920150, EPI_ISL_1920162, EPI_ISL_1920164, EPI_ISL_1920165, EPI_ISL_1920171 | | | |
| see above | New York Presbyterian Hospital | Wadsworth Center, New York State Department of Health | Alexis Russel; Alexis Russell; Catharine Prussing; Daryl M. Lamson; Erasmus Schneider; Erica Lasek-Nesselquist; John Kelly; Jonathan Plitnick; Kirsten St. George; Matthew Shudt; Melissa A Leisner; Navjot Singh |
| EPI_ISL_1034279 | Northwest Pathology | UW Virology Lab | Alexander Greninger; Hong Xie; Keith R Jerome; Lasata Shrestha; Meei-Li Huang; Michelle Lin; Pavitra Roychoudhury; Shah A Mohamed Bakhash |
| EPI_ISL_823971, EPI_ISL_823972, EPI_ISL_823974, EPI_ISL_857056, EPI_ISL_857057, EPI_ISL_857058, EPI_ISL_857059, EPI_ISL_857060, EPI_ISL_857061, EPI_ISL_857062, EPI_ISL_857066, EPI_ISL_857186, EPI_ISL_857193, EPI_ISL_857199, EPI_ISL_857289, EPI_ISL_857291, EPI_ISL_857292, EPI_ISL_857294, EPI_ISL_857295, EPI_ISL_857296, EPI_ISL_857297, EPI_ISL_857299, EPI_ISL_857300, EPI_ISL_857302, EPI_ISL_857303, EPI_ISL_857304, EPI_ISL_857305, EPI_ISL_857307, EPI_ISL_883315, EPI_ISL_883316, EPI_ISL_883325, EPI_ISL_883328, EPI_ISL_883330, EPI_ISL_883336, EPI_ISL_883344, EPI_ISL_883345, EPI_ISL_883350, EPI_ISL_883351, EPI_ISL_883352, EPI_ISL_883353, EPI_ISL_883390, EPI_ISL_883391, EPI_ISL_883392, EPI_ISL_883393, EPI_ISL_883394, EPI_ISL_883397, EPI_ISL_883398, EPI_ISL_883399, EPI_ISL_883400, EPI_ISL_883401, EPI_ISL_883402, EPI_ISL_883404, EPI_ISL_883405, EPI_ISL_883406, EPI_ISL_883407, EPI_ISL_883408, EPI_ISL_937186, EPI_ISL_937187, EPI_ISL_937188, EPI_ISL_937189, EPI_ISL_937190, EPI_ISL_937193, EPI_ISL_937194, EPI_ISL_937195, EPI_ISL_937196, EPI_ISL_937197, EPI_ISL_937198, EPI_ISL_937199, EPI_ISL_937240, EPI_ISL_937241, EPI_ISL_937242, EPI_ISL_937243, EPI_ISL_937244, EPI_ISL_937245, EPI_ISL_937246, EPI_ISL_937247, EPI_ISL_937251, EPI_ISL_937252, EPI_ISL_937253, EPI_ISL_937254, EPI_ISL_937255, EPI_ISL_966379, EPI_ISL_966381, EPI_ISL_966383, EPI_ISL_966390, EPI_ISL_966393, EPI_ISL_966396, EPI_ISL_966397, EPI_ISL_966403, EPI_ISL_966417, EPI_ISL_966418, EPI_ISL_966419, EPI_ISL_966427, EPI_ISL_966438, EPI_ISL_966439, EPI_ISL_966440, EPI_ISL_966441, EPI_ISL_966442, EPI_ISL_966453, EPI_ISL_966454, EPI_ISL_966455, EPI_ISL_966456, EPI_ISL_966457, EPI_ISL_966458, EPI_ISL_966470, EPI_ISL_966471, EPI_ISL_966473, EPI_ISL_966485, EPI_ISL_966486, EPI_ISL_966487, EPI_ISL_966488, EPI_ISL_966490, EPI_ISL_966492, EPI_ISL_966493, EPI_ISL_966494, EPI_ISL_966495 | | | |
| see above | OCME Office Of Chief Medical Examiner | New York City Public Health Laboratory | Jade Wang; et al. |
| EPI_ISL_984756, EPI_ISL_984757, EPI_ISL_984758, EPI_ISL_984759, EPI_ISL_984760, EPI_ISL_984761, EPI_ISL_984762, EPI_ISL_984763, EPI_ISL_984764, EPI_ISL_984765, EPI_ISL_984766, EPI_ISL_984767, EPI_ISL_984768, EPI_ISL_984769, EPI_ISL_984770, EPI_ISL_984771, EPI_ISL_984772, EPI_ISL_984773, EPI_ISL_984774, EPI_ISL_984775, EPI_ISL_984776, EPI_ISL_984777, EPI_ISL_984778, EPI_ISL_984779, EPI_ISL_984780, EPI_ISL_984781, EPI_ISL_984782, EPI_ISL_984783, EPI_ISL_984784, EPI_ISL_984785, EPI_ISL_984786, EPI_ISL_984787, EPI_ISL_984788, EPI_ISL_984789, EPI_ISL_984790, EPI_ISL_984791, EPI_ISL_984792, EPI_ISL_984793, EPI_ISL_984794, EPI_ISL_984795, EPI_ISL_984796, EPI_ISL_984797, EPI_ISL_984798, EPI_ISL_984799, EPI_ISL_984800, EPI_ISL_984801, EPI_ISL_984802, EPI_ISL_984803, EPI_ISL_984804, EPI_ISL_984805, EPI_ISL_984806, EPI_ISL_984807, EPI_ISL_984808, EPI_ISL_984809, EPI_ISL_984810, EPI_ISL_984811, EPI_ISL_984812, EPI_ISL_984813, EPI_ISL_984814, EPI_ISL_984815, EPI_ISL_984816, EPI_ISL_984817, EPI_ISL_984818, EPI_ISL_984819, EPI_ISL_984820, EPI_ISL_984821, EPI_ISL_984822, EPI_ISL_984823, EPI_ISL_984824, EPI_ISL_984825, EPI_ISL_984826, EPI_ISL_984827, EPI_ISL_984828, EPI_ISL_984829, EPI_ISL_984830, EPI_ISL_984831, EPI_ISL_984832, EPI_ISL_984833, EPI_ISL_984834, EPI_ISL_984835, EPI_ISL_984836, EPI_ISL_984837, EPI_ISL_984838, EPI_ISL_984839, EPI_ISL_984840, EPI_ISL_984841, EPI_ISL_984842, EPI_ISL_984843, EPI_ISL_984844, EPI_ISL_984845, EPI_ISL_984846, EPI_ISL_984847, EPI_ISL_984848, EPI_ISL_984849, EPI_ISL_984850, EPI_ISL_984851, EPI_ISL_984852, EPI_ISL_984853, EPI_ISL_984854, EPI_ISL_984855, EPI_ISL_984856, EPI_ISL_984857, EPI_ISL_984858, EPI_ISL_984859, EPI_ISL_984860, EPI_ISL_984861, EPI_ISL_984862, EPI_ISL_984863, EPI_ISL_984864, EPI_ISL_984865, EPI_ISL_984866, EPI_ISL_984867, EPI_ISL_984868, EPI_ISL_984869, EPI_ISL_984870, EPI_ISL_984871, EPI_ISL_984872, EPI_ISL_984873, EPI_ISL_984874, EPI_ISL_984875, EPI_ISL_984876, EPI_ISL_984877, EPI_ISL_984878, EPI_ISL_984879, EPI_ISL_984880, EPI_ISL_984881, EPI_ISL_984882, EPI_ISL_984883, EPI_ISL_984884, EPI_ISL_984885, EPI_ISL_984886, EPI_ISL_984887, EPI_ISL_984888, EPI_ISL_984889, EPI_ISL_984890, EPI_ISL_984891, EPI_ISL_984892, EPI_ISL_984893, EPI_ISL_984894, EPI_ISL_984895, EPI_ISL_984896, EPI_ISL_984897, EPI_ISL_984898, EPI_ISL_984899, EPI_ISL_984900, EPI_ISL_984901, EPI_ISL_984902, EPI_ISL_984903, EPI_ISL_984904, EPI_ISL_984905, EPI_ISL_984906, EPI_ISL_984907, EPI_ISL_984908, EPI_ISL_984909, EPI_ISL_984910, EPI_ISL_984911, EPI_ISL_984912, EPI_ISL_984913, EPI_ISL_984914, EPI_ISL_984915, EPI_ISL_984916, EPI_ISL_994802, EPI_ISL_994803, EPI_ISL_994804, EPI_ISL_994805, EPI_ISL_994806, EPI_ISL_994807, EPI_ISL_994812, EPI_ISL_994816, EPI_ISL_994817, EPI_ISL_994818, EPI_ISL_994819, EPI_ISL_994820, EPI_ISL_994822, EPI_ISL_994823, EPI_ISL_994827, EPI_ISL_994828, EPI_ISL_994836 | | | |
| see above | Pandemic Response Lab - NYC | Pandemic Response Lab, R&D | Cybill del Castillo; Haiping Hao; Henry Lee; Jon Laurent; Melissa Hopkins; Michael Hammerling; Pradeep Bugga; William Ward |
| EPI_ISL_883307, EPI_ISL_883308, EPI_ISL_883311, EPI_ISL_883317, EPI_ISL_883383, EPI_ISL_883384, EPI_ISL_883385, EPI_ISL_883386, EPI_ISL_883387, EPI_ISL_883388, EPI_ISL_883389 | | | |
| see above | Pandemic Response Laboratory | New York City Public Health Laboratory | Jade Wang; et al. |
| EPI_ISL_891212 | Population Medicine and Diagnostic Sciences, Cornell University | Population Medicine and Diagnostic Sciences, Cornell University | Cardia Caserta, L.; D.G.; E. and Diel; Mitchel; P.K.; Plachorzyk |
| EPI_ISL_855165, EPI_ISL_876925, EPI_ISL_876951, EPI_ISL_877121, EPI_ISL_915340, EPI_ISL_915341, EPI_ISL_915342, EPI_ISL_915343, EPI_ISL_915344, EPI_ISL_915345 | | | |
| see above | Quest Diagnostics | Quest Diagnostics | Anderson, B.; Bernstein; D.F.; Gerasimova, A.; Hua, M.; I.A.; K.E.; Kagan; L.E.; Lacbawan, F.; Liu Y.; Livingston; Owen, R.; Perez, A.; R.M.; Rosenthal; S.H.; Shalhout; Shlyakhter; Tanpaiboon, P. |
| EPI_ISL_1194374 | Quest Diagnostics Incorporated | Centers for Disease Control and Prevention Division of Viral Diseases, Pathogen Discovery | A. Gerasimova; A. Perez; B. Anderson; Ben L. Rambo-Martin; Clinton R. Paden; Dakota Howard; Dhwani Batra; Duncan MacCannell; F. Lacbawan; I. A. Shlyakhter; K.E. Livingston; L.E. Bernstein; M. Hua; P. Tanpaiboon; Peter W. Cook; R. M. Kagan; R. Owen; R. V. Rolando; S. H. Rosenthal; Suxiang Tong; Y. Liu |
| EPI_ISL_1086395, EPI_ISL_1086555, EPI_ISL_1086562, EPI_ISL_1086567, EPI_ISL_1086581, EPI_ISL_1086611, EPI_ISL_1086627, EPI_ISL_1086643, EPI_ISL_1086647, EPI_ISL_1086652, EPI_ISL_1086718, EPI_ISL_1086862, EPI_ISL_1086880, EPI_ISL_1087014, EPI_ISL_1087022, EPI_ISL_1087023, EPI_ISL_1087040, EPI_ISL_1087046, EPI_ISL_1087067, EPI_ISL_1087118, EPI_ISL_1087130, EPI_ISL_1087205, EPI_ISL_1087206, EPI_ISL_1087244, EPI_ISL_1087253 | | | |
| see above | Quest Diagnostics Incorporated | Respiratory Viruses Branch, Division of Viral Diseases, Centers for Disease Control and Prevention | A. Gerasimova; A. Perez; B. Anderson; Ben L. Rambo-Martin; Clinton R. Paden; Dakota Howard; Dhwani Batra; Duncan MacCannell; F. Lacbawan; I. A. Shlyakhter; K.E. Livingston; L.E. Bernstein; M. Hua; P. Tanpaiboon; Peter W. Cook; R. M. Kagan; R. Owen; R. V. Rolando; S. H. Rosenthal; Suxiang Tong; Y. Liu |
| EPI_ISL_747039, EPI_ISL_747040, EPI_ISL_747041, EPI_ISL_747042, EPI_ISL_747043, EPI_ISL_884713, EPI_ISL_884795, EPI_ISL_1233899 | | | |
| see above | Respiratory Viruses Branch, Centers for Disease Control and Prevention | Respiratory Viruses Branch, Centers for Disease Control and Prevention | Allen, E.; Antico, J.; Ascencio, A.; B.L.; Barret; Barrett; Batra, D.; Becker, D.; Bolze, A.; C.R.; Cassens, T.; Cho, R.; Cirulli, E.; Cook; D.T.; Febbo, P.; Galloway, S.; Gietzen, K.; Howard; Isaksson, M.; K.S.; Laurent, M.; Lee, J.; Lee, W.; Levan, G.; Li, Y.; Liu, J.; Lu, J.; MacCannell, D.; Marine, R.; Montmayeur, A.; N.L.; Nguyen, J.; P.W.; Paden; Queen, K.; Rambo-Martin; Ramirez, J.; Rivera-Garcia, C.; Sandoval, E.; Sheth, M.; Sickler, B.; Tao, Y.; Tolentino, M.; Tong, S.; Tran, C.; Uehara, A.; Wang, H.; Wang, S.; Washington; White, S.; Wickline, S.; de Feo, E. |
| EPI_ISL_1184113, EPI_ISL_1184114 | Rochester Regional Hospital | Multidrug-Resistant Organism Repository and Surveillance Network (MRSN) | Ana C. Ong; Brendan Corey; Brett E. Swierczewski; Edward E. Walsh; Emil Lesho; Francois Lebreton; Jason W. Bennett; and Patrick Mc Gann |
| EPI_ISL_854421, EPI_ISL_854436, EPI_ISL_854437, EPI_ISL_854438, EPI_ISL_854439, EPI_ISL_854440, EPI_ISL_854441, EPI_ISL_854442, EPI_ISL_854443, EPI_ISL_854444, EPI_ISL_854445 | | | |
| see above | SARATOGA HOSPITAL LABORATORY | Wadsworth Center, New York State Department of Health | Alexis Russel; Daryl M. Lamson; Erasmus Schneider; Erica Lasek-Nesselquist; John Kelly; Jonathan Plitnick; Kirsten St. George; Matthew Shudt; Melissa A Leisner; Navjot Singh |
| EPI_ISL_765481, EPI_ISL_765483, EPI_ISL_765517, EPI_ISL_765518, EPI_ISL_765519, EPI_ISL_765520, EPI_ISL_765522, EPI_ISL_765523, EPI_ISL_765524, EPI_ISL_765525, EPI_ISL_765526, EPI_ISL_765527, EPI_ISL_765528, EPI_ISL_765529, EPI_ISL_767589, EPI_ISL_767594, EPI_ISL_767599, EPI_ISL_767600, EPI_ISL_767601, EPI_ISL_767602, EPI_ISL_767603, EPI_ISL_767604, EPI_ISL_767605, EPI_ISL_767606, EPI_ISL_767607, EPI_ISL_767608, EPI_ISL_767609, EPI_ISL_767610, EPI_ISL_767611, EPI_ISL_767612, EPI_ISL_767613, EPI_ISL_767614, EPI_ISL_802428, EPI_ISL_802432, EPI_ISL_802437, EPI_ISL_802439, EPI_ISL_802467, EPI_ISL_802468, EPI_ISL_802491, EPI_ISL_802492, EPI_ISL_802493, EPI_ISL_802494, EPI_ISL_802495, EPI_ISL_802496, EPI_ISL_802497 | | | |
| see above | SARATOGA HOSPITAL LABORATORY | Wadsworth Center, New York State Department.of Health | Alexis Russel; Daryl M. Lamson; Erasmus Schneider; Erica Lasek-Nesselquist; John Kelly; Jonathan Plitnick; Kirsten St. George; Matthew Shudt; Melissa A Leisner; Navjot Singh; Sara Griesemer |
| EPI_ISL_896301, EPI_ISL_1016412, EPI_ISL_1016416, EPI_ISL_1016459, EPI_ISL_1016460, EPI_ISL_1016461, EPI_ISL_1016462, EPI_ISL_1016463, EPI_ISL_1016464, EPI_ISL_1016465, EPI_ISL_1016466, EPI_ISL_1016467, EPI_ISL_1016476, EPI_ISL_1016477, EPI_ISL_1016478 | | | |
| see above | STONY BROOK UNIVERSITY HOSPITAL | Wadsworth Center, New York State Department of Health | Alexis Russel; Daryl M. Lamson; Erasmus Schneider; Erica Lasek-Nesselquist; John Kelly; Jonathan Plitnick; Kirsten St. George; Matthew Shudt; Melissa A Leisner; Navjot Singh |
| EPI_ISL_802444 | SUNRISE MEDICAL LABORATORIES | Wadsworth Center, New York State Department.of Health | Alexis Russel; Daryl M. Lamson; Erasmus Schneider; Erica Lasek-Nesselquist; John Kelly; Jonathan Plitnick; Kirsten St. George; Matthew Shudt; Melissa A Leisner; Navjot Singh; Sara Griesemer |
| EPI_ISL_830650, EPI_ISL_830651, EPI_ISL_830652, EPI_ISL_830653, EPI_ISL_830654, EPI_ISL_830655, EPI_ISL_830656, EPI_ISL_830657, EPI_ISL_830658, EPI_ISL_830659, EPI_ISL_830660, EPI_ISL_830661, EPI_ISL_830662, EPI_ISL_830663, EPI_ISL_830664, EPI_ISL_830665, EPI_ISL_830666, EPI_ISL_830667, EPI_ISL_830668, EPI_ISL_830669, EPI_ISL_830670, EPI_ISL_830671, EPI_ISL_830672, EPI_ISL_830673, EPI_ISL_830674, EPI_ISL_830675, EPI_ISL_830676, EPI_ISL_830677, EPI_ISL_830678, EPI_ISL_830679, EPI_ISL_830680, EPI_ISL_830681, EPI_ISL_830682, EPI_ISL_853689, EPI_ISL_853706, EPI_ISL_853707, EPI_ISL_853708, EPI_ISL_853715, EPI_ISL_896218, EPI_ISL_896219, EPI_ISL_896220, EPI_ISL_896221, EPI_ISL_896222, EPI_ISL_896223, EPI_ISL_896224, EPI_ISL_896225, EPI_ISL_896228, EPI_ISL_896229, EPI_ISL_896230, EPI_ISL_896235, EPI_ISL_896247, EPI_ISL_896248, EPI_ISL_896249, EPI_ISL_896250, EPI_ISL_896251, EPI_ISL_896252, EPI_ISL_896253, EPI_ISL_896254, EPI_ISL_896255, EPI_ISL_896256, EPI_ISL_896257, EPI_ISL_896258, EPI_ISL_896259, EPI_ISL_896260, EPI_ISL_896261, EPI_ISL_896262, EPI_ISL_896263, EPI_ISL_896264, EPI_ISL_896265, EPI_ISL_896266, EPI_ISL_896267, EPI_ISL_896268, EPI_ISL_896269, EPI_ISL_896270, EPI_ISL_896271, EPI_ISL_896272, EPI_ISL_896273, EPI_ISL_896274, EPI_ISL_896275, EPI_ISL_896276, EPI_ISL_896277, EPI_ISL_896278, EPI_ISL_896279, EPI_ISL_896280, EPI_ISL_896281, EPI_ISL_896282, EPI_ISL_896283, EPI_ISL_896284, EPI_ISL_896285, EPI_ISL_896286, EPI_ISL_896287, EPI_ISL_896288, EPI_ISL_896289, EPI_ISL_896290, EPI_ISL_896291, EPI_ISL_896292, EPI_ISL_896293, EPI_ISL_935993, EPI_ISL_935994, EPI_ISL_935995, EPI_ISL_935996, EPI_ISL_935997, EPI_ISL_935998, EPI_ISL_935999, EPI_ISL_936000, EPI_ISL_936001, EPI_ISL_936002, EPI_ISL_936003, EPI_ISL_936004, EPI_ISL_936005, EPI_ISL_936006, EPI_ISL_936007, EPI_ISL_936008, EPI_ISL_936009, EPI_ISL_936010, EPI_ISL_936011, EPI_ISL_936012, EPI_ISL_936013, EPI_ISL_936014, EPI_ISL_936015, EPI_ISL_936016, EPI_ISL_936017, EPI_ISL_936018, EPI_ISL_936019, EPI_ISL_936020, EPI_ISL_936021, EPI_ISL_936022, EPI_ISL_936142, EPI_ISL_936143, EPI_ISL_983116, EPI_ISL_983117, EPI_ISL_983118, EPI_ISL_983243, EPI_ISL_983245, EPI_ISL_983248, EPI_ISL_983249, EPI_ISL_983283, EPI_ISL_983289, EPI_ISL_983290, EPI_ISL_983291, EPI_ISL_983292, EPI_ISL_983293, EPI_ISL_983294, EPI_ISL_983295, EPI_ISL_983300, EPI_ISL_983301, EPI_ISL_983302, EPI_ISL_983303, EPI_ISL_983304, EPI_ISL_983305, EPI_ISL_983306, EPI_ISL_983307, EPI_ISL_983308, EPI_ISL_983309, EPI_ISL_983310, EPI_ISL_983311, EPI_ISL_983312, EPI_ISL_983313, EPI_ISL_983314, EPI_ISL_983315, EPI_ISL_983316, EPI_ISL_983317, EPI_ISL_983318, EPI_ISL_983319, EPI_ISL_1016285, EPI_ISL_1016286, EPI_ISL_1016298, EPI_ISL_1016299, EPI_ISL_1016300, EPI_ISL_1016301, EPI_ISL_1016302, EPI_ISL_1016303, EPI_ISL_1016311, EPI_ISL_1016312, EPI_ISL_1016313, EPI_ISL_1016314, EPI_ISL_1016315, EPI_ISL_1016316, EPI_ISL_1016317, EPI_ISL_1016318, EPI_ISL_1016319, EPI_ISL_1016320, EPI_ISL_1016321, EPI_ISL_1016322, EPI_ISL_1016327, EPI_ISL_1227725, EPI_ISL_1227993, EPI_ISL_1227994, EPI_ISL_1227995, EPI_ISL_1227996, EPI_ISL_1227997, EPI_ISL_1227998, EPI_ISL_1227999, EPI_ISL_1228000, EPI_ISL_1228001, EPI_ISL_1228002, EPI_ISL_1228003, EPI_ISL_1228004, EPI_ISL_1228047, EPI_ISL_1228048, EPI_ISL_1397389, EPI_ISL_1397390, EPI_ISL_1397391, EPI_ISL_1397392, EPI_ISL_1397394, EPI_ISL_1397396, EPI_ISL_1397397, EPI_ISL_1397398, EPI_ISL_1397399, EPI_ISL_1397400, EPI_ISL_1397401, EPI_ISL_1397402, EPI_ISL_1397404, EPI_ISL_1397406, EPI_ISL_1397408, EPI_ISL_1397410, EPI_ISL_1397411, EPI_ISL_1397412, EPI_ISL_1397413, EPI_ISL_1397414, EPI_ISL_1397415, EPI_ISL_1397416, EPI_ISL_1397417, EPI_ISL_1397418, EPI_ISL_1397419, EPI_ISL_1397420, EPI_ISL_1397421, EPI_ISL_1397422, EPI_ISL_1397423, EPI_ISL_1397459, EPI_ISL_1499437, EPI_ISL_1499438, EPI_ISL_1499439, EPI_ISL_1499440, EPI_ISL_1499441, EPI_ISL_1499442, EPI_ISL_1499443, EPI_ISL_1499444, EPI_ISL_1499445, EPI_ISL_1499446, EPI_ISL_1499448, EPI_ISL_1499449, EPI_ISL_1499450 | | | |
| see above | SUNY UPSTATE MEDICAL UNIVERSITY | Wadsworth Center, New York State Department of Health | Alexis Russel; Daryl M. Lamson; Erasmus Schneider; Erica Lasek-Nesselquist; John Kelly; Jonathan Plitnick; Kirsten St. George; Matthew Shudt; Melissa A Leisner; Navjot Singh |
| EPI_ISL_955265, EPI_ISL_955300 | Sunrise Medical Laboratory | Pathogen Discovery, Respiratory Viruses Branch, Division of Viral Diseases, Centers for Disease Control and Prevention | Anna Uehara; Clinton R. Paden; Haibin Wang; Jing Zhang; Krista Queen; Peter Cook; Suxiang Tong; Yan Li; Ying Tao |
| EPI_ISL_854447, EPI_ISL_854448, EPI_ISL_884054, EPI_ISL_896226, EPI_ISL_936173, EPI_ISL_1016256 | TEMPUS LABS INC | Wadsworth Center, New York State Department of Health | Alexis Russel; Daryl M. Lamson; Erasmus Schneider; Erica Lasek-Nesselquist; John Kelly; Jonathan Plitnick; Kirsten St. George; Matthew Shudt; Melissa A Leisner; Navjot Singh |
| EPI_ISL_802436 | TEMPUS LABS INC | Wadsworth Center, New York State Department.of Health | Alexis Russel; Daryl M. Lamson; Erasmus Schneider; Erica Lasek-Nesselquist; John Kelly; Jonathan Plitnick; Kirsten St. George; Matthew Shudt; Melissa A Leisner; Navjot Singh; Sara Griesemer |
| EPI_ISL_853648, EPI_ISL_853649, EPI_ISL_853652, EPI_ISL_853667, EPI_ISL_853668, EPI_ISL_853669, EPI_ISL_853688, EPI_ISL_936023, EPI_ISL_936024, EPI_ISL_936025, EPI_ISL_936026, EPI_ISL_936027, EPI_ISL_936028, EPI_ISL_936029, EPI_ISL_936030, EPI_ISL_936031, EPI_ISL_936032, EPI_ISL_936033, EPI_ISL_936034, EPI_ISL_936035, EPI_ISL_936138, EPI_ISL_936139, EPI_ISL_936140, EPI_ISL_936141, EPI_ISL_936153, EPI_ISL_983119, EPI_ISL_983120, EPI_ISL_983121, EPI_ISL_983122, EPI_ISL_983123, EPI_ISL_983124, EPI_ISL_983125, EPI_ISL_983126, EPI_ISL_983127, EPI_ISL_983128, EPI_ISL_983129, EPI_ISL_983130, EPI_ISL_983131, EPI_ISL_983445, EPI_ISL_983446, EPI_ISL_983477, EPI_ISL_983485, EPI_ISL_983486, EPI_ISL_983487, EPI_ISL_983488, EPI_ISL_983489, EPI_ISL_983490, EPI_ISL_983491, EPI_ISL_983492, EPI_ISL_1016127, EPI_ISL_1016128, EPI_ISL_1016129, EPI_ISL_1016134, EPI_ISL_1016135, EPI_ISL_1016141, EPI_ISL_1016142, EPI_ISL_1016257, EPI_ISL_1226423, EPI_ISL_1226490, EPI_ISL_1226524, EPI_ISL_1226868, EPI_ISL_1227456, EPI_ISL_1227457, EPI_ISL_1227466, EPI_ISL_1227541, EPI_ISL_1227542, EPI_ISL_1227543, EPI_ISL_1227544, EPI_ISL_1227545, EPI_ISL_1227617, EPI_ISL_1227618, EPI_ISL_1227619, EPI_ISL_1227620, EPI_ISL_1227621, EPI_ISL_1227622, EPI_ISL_1227623, EPI_ISL_1227624, EPI_ISL_1227625, EPI_ISL_1227626, EPI_ISL_1227627, EPI_ISL_1227628, EPI_ISL_1227629, EPI_ISL_1227630, EPI_ISL_1227631, EPI_ISL_1227632, EPI_ISL_1227633, EPI_ISL_1227634, EPI_ISL_1227635, EPI_ISL_1227636, EPI_ISL_1227637, EPI_ISL_1227638, EPI_ISL_1227639, EPI_ISL_1227962, EPI_ISL_1227963, EPI_ISL_1227964, EPI_ISL_1227974, EPI_ISL_1227975, EPI_ISL_1228049, EPI_ISL_1228050, EPI_ISL_1228284, EPI_ISL_1228285, EPI_ISL_1228286, EPI_ISL_1228287, EPI_ISL_1228288, EPI_ISL_1228289, EPI_ISL_1228290 | | | |
| see above | THE MARY IMOGENE BASSETT HOSPITAL | Wadsworth Center, New York State Department of Health | Alexis Russel; Daryl M. Lamson; Erasmus Schneider; Erica Lasek-Nesselquist; John Kelly; Jonathan Plitnick; Kirsten St. George; Matthew Shudt; Melissa A Leisner; Navjot Singh |
| EPI_ISL_847830, EPI_ISL_1038971, EPI_ISL_1038972, EPI_ISL_1038973, EPI_ISL_1038974, EPI_ISL_1038975, EPI_ISL_1038976, EPI_ISL_1038977, EPI_ISL_1038978, EPI_ISL_1038979, EPI_ISL_1038980, EPI_ISL_1038981, EPI_ISL_1163665, EPI_ISL_1163666 | | | |
| see above | Tempus | Grubaugh Lab - Yale School of Public Health | Anderson Brito; Anne Wyllie; Annie Watkins; Chaney Kalinich; Chantal Vogels; Isabel Ott; Joseph Fauver; Mallery Breban; Mary Petrone; Nathan Grubaugh; Tara Alpert |
| EPI_ISL_854306, EPI_ISL_854307, EPI_ISL_854308, EPI_ISL_854309, EPI_ISL_854310, EPI_ISL_854311, EPI_ISL_854312, EPI_ISL_854313, EPI_ISL_854314, EPI_ISL_854315, EPI_ISL_854316, EPI_ISL_854317, EPI_ISL_854318, EPI_ISL_854319, EPI_ISL_854320, EPI_ISL_854321, EPI_ISL_854322, EPI_ISL_854323, EPI_ISL_854324, EPI_ISL_854325, EPI_ISL_854326, EPI_ISL_854327, EPI_ISL_854328, EPI_ISL_854329, EPI_ISL_854330, EPI_ISL_854331, EPI_ISL_854332, EPI_ISL_854333, EPI_ISL_854334, EPI_ISL_854335, EPI_ISL_854336, EPI_ISL_854337, EPI_ISL_854338, EPI_ISL_854339, EPI_ISL_854340, EPI_ISL_854341, EPI_ISL_854342, EPI_ISL_854343, EPI_ISL_854344, EPI_ISL_854345, EPI_ISL_854346, EPI_ISL_854347, EPI_ISL_854348, EPI_ISL_854349, EPI_ISL_854370, EPI_ISL_854371, EPI_ISL_854399, EPI_ISL_854404, EPI_ISL_861183, EPI_ISL_861184, EPI_ISL_861199, EPI_ISL_861200, EPI_ISL_861201, EPI_ISL_861202, EPI_ISL_861203, EPI_ISL_861204, EPI_ISL_861205, EPI_ISL_861206, EPI_ISL_861207, EPI_ISL_861208, EPI_ISL_861209, EPI_ISL_896385, EPI_ISL_896387, EPI_ISL_896388, EPI_ISL_896390, EPI_ISL_896396, EPI_ISL_896397, EPI_ISL_896398, EPI_ISL_896399, EPI_ISL_896400, EPI_ISL_896401, EPI_ISL_896402, EPI_ISL_896403, EPI_ISL_896404, EPI_ISL_896405, EPI_ISL_896406, EPI_ISL_896407, EPI_ISL_896408, EPI_ISL_896409, EPI_ISL_896410, EPI_ISL_896411, EPI_ISL_896412, EPI_ISL_896413, EPI_ISL_896414, EPI_ISL_896415, EPI_ISL_896416, EPI_ISL_896417, EPI_ISL_896418, EPI_ISL_896419, EPI_ISL_896420, EPI_ISL_896421, EPI_ISL_896495, EPI_ISL_896496, EPI_ISL_896497, EPI_ISL_896502, EPI_ISL_896503, EPI_ISL_896508, EPI_ISL_896509, EPI_ISL_896510, EPI_ISL_896511, EPI_ISL_896512, EPI_ISL_896513, EPI_ISL_896514, EPI_ISL_896515, EPI_ISL_896516, EPI_ISL_896517, EPI_ISL_896518, EPI_ISL_896519, EPI_ISL_896563, EPI_ISL_896564, EPI_ISL_896565, EPI_ISL_896566, EPI_ISL_896567, EPI_ISL_896568, EPI_ISL_896569, EPI_ISL_983438, EPI_ISL_983439, EPI_ISL_983440, EPI_ISL_983441, EPI_ISL_983442, EPI_ISL_983443, EPI_ISL_983444, EPI_ISL_983453, EPI_ISL_983454, EPI_ISL_983455, EPI_ISL_983456, EPI_ISL_983457, EPI_ISL_983458, EPI_ISL_983459, EPI_ISL_983460, EPI_ISL_983461, EPI_ISL_983462, EPI_ISL_983463, EPI_ISL_983464, EPI_ISL_983465, EPI_ISL_983466, EPI_ISL_983467, EPI_ISL_983468, EPI_ISL_983469, EPI_ISL_983470, EPI_ISL_983471, EPI_ISL_983472, EPI_ISL_983473, EPI_ISL_983474, | | | |

EPI_ISL_983475, EPI_ISL_983478, EPI_ISL_983479, EPI_ISL_983480, EPI_ISL_983481, EPI_ISL_983482, EPI_ISL_983483, EPI_ISL_983484, EPI_ISL_1016333, EPI_ISL_1016334, EPI_ISL_1016339, EPI_ISL_1016347, EPI_ISL_1016348, EPI_ISL_1016349, EPI_ISL_1016350, EPI_ISL_1016351, EPI_ISL_1016352, EPI_ISL_1016353, EPI_ISL_1016354, EPI_ISL_1016355, EPI_ISL_1016356, EPI_ISL_1016357,
EPI_ISL_1016358, EPI_ISL_1016359, EPI_ISL_1016360, EPI_ISL_1016361, EPI_ISL_1016362, EPI_ISL_1016363, EPI_ISL_1016364, EPI_ISL_1016414, EPI_ISL_1016415, EPI_ISL_1016418, EPI_ISL_1016419, EPI_ISL_1016420, EPI_ISL_1016421, EPI_ISL_1016422, EPI_ISL_1016423, EPI_ISL_1016424, EPI_ISL_1016425, EPI_ISL_1016426, EPI_ISL_1016427, EPI_ISL_1016428, EPI_ISL_1016429, EPI_ISL_1016430,
EPI_ISL_1016431, EPI_ISL_1016432, EPI_ISL_1016433, EPI_ISL_1016434, EPI_ISL_1016435, EPI_ISL_1016436, EPI_ISL_1016437, EPI_ISL_1016438, EPI_ISL_1016439, EPI_ISL_1016440, EPI_ISL_1016441, EPI_ISL_1016442, EPI_ISL_1016443, EPI_ISL_1016444, EPI_ISL_1016445, EPI_ISL_1016446, EPI_ISL_1016447, EPI_ISL_1016448, EPI_ISL_1016449, EPI_ISL_1016450, EPI_ISL_1016451, EPI_ISL_1016452,
EPI_ISL_1016453, EPI_ISL_1016454, EPI_ISL_1016455, EPI_ISL_1016456, EPI_ISL_1016457, EPI_ISL_1226428, EPI_ISL_1226435, EPI_ISL_1226445, EPI_ISL_1226451, EPI_ISL_1226460, EPI_ISL_1226477, EPI_ISL_1226479, EPI_ISL_1226505, EPI_ISL_1226615, EPI_ISL_1226662, EPI_ISL_1226663, EPI_ISL_1226675, EPI_ISL_1226676, EPI_ISL_1226677, EPI_ISL_1226713, EPI_ISL_1226851, EPI_ISL_1226852,
EPI_ISL_1226866, EPI_ISL_1227231, EPI_ISL_1227272, EPI_ISL_1227273, EPI_ISL_1227274, EPI_ISL_1227275, EPI_ISL_1227276, EPI_ISL_1227277, EPI_ISL_1227448, EPI_ISL_1227449, EPI_ISL_1227450, EPI_ISL_1227451, EPI_ISL_1227452, EPI_ISL_1227453, EPI_ISL_1227560, EPI_ISL_1227561, EPI_ISL_1227562, EPI_ISL_1227709, EPI_ISL_1227710, EPI_ISL_1227711, EPI_ISL_1227712, EPI_ISL_1227713,
EPI_ISL_1227714, EPI_ISL_1227715, EPI_ISL_1227728, EPI_ISL_1227729, EPI_ISL_1227730, EPI_ISL_1227731, EPI_ISL_1227732, EPI_ISL_1227733, EPI_ISL_1227734, EPI_ISL_1227735, EPI_ISL_1227736, EPI_ISL_1227737, EPI_ISL_1227738, EPI_ISL_1227739, EPI_ISL_1227740, EPI_ISL_1227741, EPI_ISL_1227742, EPI_ISL_1227743, EPI_ISL_1227744, EPI_ISL_1227745, EPI_ISL_1227746, EPI_ISL_1227747,
EPI_ISL_1227748, EPI_ISL_1227749, EPI_ISL_1227750, EPI_ISL_1227751, EPI_ISL_1227752, EPI_ISL_1227753, EPI_ISL_1227754, EPI_ISL_1227755, EPI_ISL_1227756, EPI_ISL_1227757, EPI_ISL_1227758, EPI_ISL_1228023, EPI_ISL_1228024, EPI_ISL_1228025, EPI_ISL_1228026, EPI_ISL_1228027, EPI_ISL_1228040, EPI_ISL_1228115, EPI_ISL_1228296, EPI_ISL_1228297, EPI_ISL_1228298, EPI_ISL_1228299,
EPI_ISL_1228300, EPI_ISL_1228301, EPI_ISL_1228302, EPI_ISL_1228303, EPI_ISL_1228304, EPI_ISL_1228305, EPI_ISL_1228480, EPI_ISL_1499395, EPI_ISL_1499404, EPI_ISL_1499405, EPI_ISL_1499406, EPI_ISL_1499407, EPI_ISL_1499409, EPI_ISL_1499410, EPI_ISL_1499411, EPI_ISL_1499412, EPI_ISL_1499413, EPI_ISL_1499414, EPI_ISL_1499415, EPI_ISL_1499416, EPI_ISL_1499417, EPI_ISL_1499418,
EPI_ISL_1499419, EPI_ISL_1499420, EPI_ISL_1499421, EPI_ISL_1499422, EPI_ISL_1499423, EPI_ISL_1499424, EPI_ISL_1499425, EPI_ISL_1499426, EPI_ISL_1499427, EPI_ISL_1499428, EPI_ISL_1499429, EPI_ISL_1499430, EPI_ISL_1499431, EPI_ISL_1499432, EPI_ISL_1499433, EPI_ISL_1499434, EPI_ISL_1499435, EPI_ISL_1499436

see above | URMC LABS | Wadsworth Center, New York State Department of Health | Alexis Russel; Daryl M. Lamson; Erasmus Schneider; Erica Lasek-Nesselquist; John Kelly; Jonathan Plitnick; Kirsten St. George; Matthew Shudt; Melissa A Leisner; Navjot Singh

EPI_ISL_767417, EPI_ISL_767419, EPI_ISL_767423, EPI_ISL_767424, EPI_ISL_767538, EPI_ISL_767539, EPI_ISL_767540, EPI_ISL_767541, EPI_ISL_767542, EPI_ISL_767543, EPI_ISL_767544, EPI_ISL_767545, EPI_ISL_767546, EPI_ISL_767547, EPI_ISL_767548, EPI_ISL_767549, EPI_ISL_767550, EPI_ISL_767551, EPI_ISL_767552, EPI_ISL_767553, EPI_ISL_767554, EPI_ISL_767555, EPI_ISL_767556, EPI_ISL_767557,
EPI_ISL_767558, EPI_ISL_767559, EPI_ISL_767560, EPI_ISL_767561, EPI_ISL_767562, EPI_ISL_767563, EPI_ISL_767564, EPI_ISL_767565, EPI_ISL_767566, EPI_ISL_767567, EPI_ISL_767568, EPI_ISL_767569, EPI_ISL_767570, EPI_ISL_767571, EPI_ISL_767572, EPI_ISL_767573, EPI_ISL_767574, EPI_ISL_767575, EPI_ISL_767576, EPI_ISL_767577, EPI_ISL_767579, EPI_ISL_794083, EPI_ISL_794084, EPI_ISL_794085,
EPI_ISL_794086, EPI_ISL_794087, EPI_ISL_794088, EPI_ISL_794089, EPI_ISL_794090, EPI_ISL_794091, EPI_ISL_794092, EPI_ISL_794093, EPI_ISL_794094, EPI_ISL_794095, EPI_ISL_794096, EPI_ISL_794097, EPI_ISL_794098, EPI_ISL_794099, EPI_ISL_794100, EPI_ISL_794101, EPI_ISL_794102, EPI_ISL_794103, EPI_ISL_794104, EPI_ISL_794105, EPI_ISL_794106, EPI_ISL_794182, EPI_ISL_794183, EPI_ISL_794184,
EPI_ISL_794185, EPI_ISL_794186, EPI_ISL_794187, EPI_ISL_794188, EPI_ISL_794189, EPI_ISL_794190, EPI_ISL_794191, EPI_ISL_794192, EPI_ISL_794193, EPI_ISL_794194, EPI_ISL_794195, EPI_ISL_794196, EPI_ISL_794197, EPI_ISL_794198, EPI_ISL_794199, EPI_ISL_794200, EPI_ISL_794201, EPI_ISL_794202, EPI_ISL_794203, EPI_ISL_794204, EPI_ISL_794205

see above | URMC LABS | Wadsworth Center, New York State Department.of Health | Alexis Russel; Daryl M. Lamson; Erasmus Schneider; Erica Lasek-Nesselquist; John Kelly; Jonathan Plitnick; Kirsten St. George; Matthew Shudt; Melissa A Leisner; Navjot Singh; Sara Griesemer

EPI_ISL_536661, EPI_ISL_536702, EPI_ISL_536714, EPI_ISL_536722, EPI_ISL_536752, EPI_ISL_536760, EPI_ISL_536773, EPI_ISL_1064272

see above | University of Wisconsin-Madison AIDS Vaccine Research Laboratories | University of Wisconsin-Madison AIDS Vaccine Research Laboratories | Gage Moreno; Katarina Braun; et al. AIDS Vaccine Research Laboratories

EPI_ISL_677695, EPI_ISL_677696, EPI_ISL_677699 | Vanda Pharmaceuticals Clinical Site | Vanda Pharmaceuticals | Vanda Pharmaceuticals

EPI_ISL_854374, EPI_ISL_854375, EPI_ISL_854376, EPI_ISL_854377, EPI_ISL_854378, EPI_ISL_854379, EPI_ISL_854380, EPI_ISL_854381, EPI_ISL_854382, EPI_ISL_854383, EPI_ISL_854384, EPI_ISL_854385, EPI_ISL_854386, EPI_ISL_854387, EPI_ISL_854388, EPI_ISL_854389, EPI_ISL_854390, EPI_ISL_854391, EPI_ISL_854392, EPI_ISL_854393, EPI_ISL_854394, EPI_ISL_854395, EPI_ISL_854396, EPI_ISL_854397,
EPI_ISL_854398, EPI_ISL_854400, EPI_ISL_854401, EPI_ISL_854402, EPI_ISL_854403, EPI_ISL_854449, EPI_ISL_854451, EPI_ISL_861333, EPI_ISL_861334, EPI_ISL_861335, EPI_ISL_861336, EPI_ISL_861337, EPI_ISL_861338, EPI_ISL_861339, EPI_ISL_861340, EPI_ISL_861341, EPI_ISL_861342, EPI_ISL_861343, EPI_ISL_861344, EPI_ISL_861345, EPI_ISL_861346, EPI_ISL_861347, EPI_ISL_861348, EPI_ISL_861349,
EPI_ISL_861350, EPI_ISL_861351, EPI_ISL_861352, EPI_ISL_861353, EPI_ISL_861354, EPI_ISL_861355, EPI_ISL_861356, EPI_ISL_861357, EPI_ISL_861358, EPI_ISL_861359, EPI_ISL_861360, EPI_ISL_861361, EPI_ISL_861362, EPI_ISL_861363, EPI_ISL_861364, EPI_ISL_861365, EPI_ISL_861366, EPI_ISL_861367, EPI_ISL_861368, EPI_ISL_861369, EPI_ISL_861370, EPI_ISL_861371, EPI_ISL_861372, EPI_ISL_861373,
EPI_ISL_861374, EPI_ISL_861375, EPI_ISL_861376, EPI_ISL_861377, EPI_ISL_861378, EPI_ISL_861379, EPI_ISL_861380, EPI_ISL_861381, EPI_ISL_861382, EPI_ISL_861383, EPI_ISL_861384, EPI_ISL_861385, EPI_ISL_861386, EPI_ISL_861387, EPI_ISL_861388, EPI_ISL_861389, EPI_ISL_861390, EPI_ISL_861391, EPI_ISL_861392, EPI_ISL_861393, EPI_ISL_861394, EPI_ISL_861395, EPI_ISL_861396, EPI_ISL_861397,
EPI_ISL_861398, EPI_ISL_861399, EPI_ISL_861400, EPI_ISL_861401, EPI_ISL_861402, EPI_ISL_861403, EPI_ISL_861404, EPI_ISL_861405, EPI_ISL_861406, EPI_ISL_861407, EPI_ISL_861408, EPI_ISL_861409, EPI_ISL_861410, EPI_ISL_861411, EPI_ISL_861412, EPI_ISL_936103, EPI_ISL_936104, EPI_ISL_936105, EPI_ISL_936106, EPI_ISL_936107, EPI_ISL_936108, EPI_ISL_936109, EPI_ISL_936110, EPI_ISL_936111,
EPI_ISL_936112, EPI_ISL_936113, EPI_ISL_936114, EPI_ISL_936115, EPI_ISL_936116, EPI_ISL_936117, EPI_ISL_936118, EPI_ISL_936119, EPI_ISL_936120, EPI_ISL_936121, EPI_ISL_936122, EPI_ISL_936123, EPI_ISL_936124, EPI_ISL_936125, EPI_ISL_936126, EPI_ISL_936127, EPI_ISL_936128, EPI_ISL_936129, EPI_ISL_1016332, EPI_ISL_1016340, EPI_ISL_1016344, EPI_ISL_1016345, EPI_ISL_1016346,
EPI_ISL_1016365, EPI_ISL_1016373, EPI_ISL_1016374, EPI_ISL_1016375, EPI_ISL_1016376, EPI_ISL_1016377, EPI_ISL_1016378, EPI_ISL_1016379, EPI_ISL_1016380, EPI_ISL_1226417, EPI_ISL_1226453, EPI_ISL_1226462, EPI_ISL_1226491, EPI_ISL_1226587, EPI_ISL_1226588, EPI_ISL_1226590, EPI_ISL_1226591, EPI_ISL_1226682, EPI_ISL_1226686, EPI_ISL_1226687, EPI_ISL_1226688, EPI_ISL_1226693,
EPI_ISL_1226694, EPI_ISL_1226697, EPI_ISL_1226702, EPI_ISL_1226708, EPI_ISL_1226721, EPI_ISL_1226727, EPI_ISL_1226730, EPI_ISL_1226781, EPI_ISL_1226782, EPI_ISL_1226822, EPI_ISL_1226825, EPI_ISL_1226826, EPI_ISL_1226827, EPI_ISL_1226828, EPI_ISL_1226845, EPI_ISL_1226891, EPI_ISL_1226892, EPI_ISL_1226908, EPI_ISL_1226924, EPI_ISL_1226927, EPI_ISL_1226928, EPI_ISL_1226929,
EPI_ISL_1227007, EPI_ISL_1227040, EPI_ISL_1227130, EPI_ISL_1227131, EPI_ISL_1227132, EPI_ISL_1227133, EPI_ISL_1227174, EPI_ISL_1227206, EPI_ISL_1227215, EPI_ISL_1227216, EPI_ISL_1227294, EPI_ISL_1227302, EPI_ISL_1227303, EPI_ISL_1227337, EPI_ISL_1227467, EPI_ISL_1227470, EPI_ISL_1227472, EPI_ISL_1227473, EPI_ISL_1227474, EPI_ISL_1227475, EPI_ISL_1227476, EPI_ISL_1227477,
EPI_ISL_1227478, EPI_ISL_1227479, EPI_ISL_1227480, EPI_ISL_1227481, EPI_ISL_1227482, EPI_ISL_1227483, EPI_ISL_1227484, EPI_ISL_1227485, EPI_ISL_1227486, EPI_ISL_1227487, EPI_ISL_1227488, EPI_ISL_1227489, EPI_ISL_1227490, EPI_ISL_1227491, EPI_ISL_1227492, EPI_ISL_1227493, EPI_ISL_1227494, EPI_ISL_1227495, EPI_ISL_1227496, EPI_ISL_1227497, EPI_ISL_1227498, EPI_ISL_1227499,
EPI_ISL_1227500, EPI_ISL_1227501, EPI_ISL_1227502, EPI_ISL_1227504, EPI_ISL_1227505, EPI_ISL_1227506, EPI_ISL_1227507, EPI_ISL_1227508, EPI_ISL_1227509, EPI_ISL_1227510, EPI_ISL_1227511, EPI_ISL_1227512, EPI_ISL_1227513, EPI_ISL_1227514, EPI_ISL_1227515, EPI_ISL_1227516, EPI_ISL_1227517, EPI_ISL_1227518, EPI_ISL_1227519, EPI_ISL_1227520, EPI_ISL_1227521, EPI_ISL_1227522,
EPI_ISL_1227523, EPI_ISL_1227524, EPI_ISL_1227525, EPI_ISL_1227526, EPI_ISL_1227527, EPI_ISL_1227528, EPI_ISL_1227529, EPI_ISL_1227569, EPI_ISL_1227676, EPI_ISL_1228036, EPI_ISL_1228037, EPI_ISL_1228038, EPI_ISL_1228039, EPI_ISL_1228119, EPI_ISL_1228154, EPI_ISL_1228252, EPI_ISL_1228255, EPI_ISL_1228257, EPI_ISL_1228258, EPI_ISL_1228259, EPI_ISL_1228261, EPI_ISL_1228378,
EPI_ISL_1228379, EPI_ISL_1228381, EPI_ISL_1228382, EPI_ISL_1228383, EPI_ISL_1228384, EPI_ISL_1228385, EPI_ISL_1228386, EPI_ISL_1228387, EPI_ISL_1228388, EPI_ISL_1228389, EPI_ISL_1228390, EPI_ISL_1228391, EPI_ISL_1228392, EPI_ISL_1228393, EPI_ISL_1228394, EPI_ISL_1228395, EPI_ISL_1228396, EPI_ISL_1228397, EPI_ISL_1228398, EPI_ISL_1228399, EPI_ISL_1228400, EPI_ISL_1228401,
EPI_ISL_1228402, EPI_ISL_1228403, EPI_ISL_1228404, EPI_ISL_1228405, EPI_ISL_1228406, EPI_ISL_1228407, EPI_ISL_1228408, EPI_ISL_1228409, EPI_ISL_1228410, EPI_ISL_1228411, EPI_ISL_1228412, EPI_ISL_1228413, EPI_ISL_1228414, EPI_ISL_1228415, EPI_ISL_1228416, EPI_ISL_1228417, EPI_ISL_1228418, EPI_ISL_1228419, EPI_ISL_1228420, EPI_ISL_1228421, EPI_ISL_1228501, EPI_ISL_1228504,
EPI_ISL_1228505, EPI_ISL_1228506, EPI_ISL_1228507, EPI_ISL_1228508, EPI_ISL_1228510, EPI_ISL_1228511, EPI_ISL_1228512, EPI_ISL_1228513, EPI_ISL_1228515, EPI_ISL_1228516, EPI_ISL_1228517, EPI_ISL_1228518, EPI_ISL_1228519, EPI_ISL_1228521, EPI_ISL_1228522, EPI_ISL_1228523, EPI_ISL_1228524, EPI_ISL_1228525, EPI_ISL_1228526, EPI_ISL_1228527, EPI_ISL_1228528, EPI_ISL_1228532,
EPI_ISL_1228533, EPI_ISL_1228534, EPI_ISL_1228535, EPI_ISL_1228537, EPI_ISL_1228538, EPI_ISL_1228539, EPI_ISL_1228540, EPI_ISL_1228545, EPI_ISL_1228546, EPI_ISL_1228547, EPI_ISL_1228548, EPI_ISL_1228549, EPI_ISL_1228551, EPI_ISL_1228552, EPI_ISL_1228554, EPI_ISL_1228555, EPI_ISL_1228556, EPI_ISL_1228557, EPI_ISL_1394593, EPI_ISL_1394594, EPI_ISL_1394595, EPI_ISL_1394596,
EPI_ISL_1394597, EPI_ISL_1394598, EPI_ISL_1394600, EPI_ISL_1394601, EPI_ISL_1394602, EPI_ISL_1397444, EPI_ISL_1397445, EPI_ISL_1397446, EPI_ISL_1397447, EPI_ISL_1397448, EPI_ISL_1397449, EPI_ISL_1397450, EPI_ISL_1397451, EPI_ISL_1397452, EPI_ISL_1397453, EPI_ISL_1397454, EPI_ISL_1397455, EPI_ISL_1397456, EPI_ISL_1397457, EPI_ISL_1397458, EPI_ISL_1397768, EPI_ISL_1397769,
EPI_ISL_1397770, EPI_ISL_1397771, EPI_ISL_1397772, EPI_ISL_1397773, EPI_ISL_1397774, EPI_ISL_1397775, EPI_ISL_1397776, EPI_ISL_1397777, EPI_ISL_1397778, EPI_ISL_1397779, EPI_ISL_1397780, EPI_ISL_1397781, EPI_ISL_1397782, EPI_ISL_1462553, EPI_ISL_1671143, EPI_ISL_1671161, EPI_ISL_1671218, EPI_ISL_1671219, EPI_ISL_1671220

see above | WESTCHESTER MEDICAL CENTER | Wadsworth Center, New York State Department of Health | Alexis Russel; Alexis Russell; Catharine Prussing; Daryl M. Lamson; Erasmus Schneider; Erica Lasek-Nesselquist; John Kelly; Jonathan Plitnick; Kirsten St. George; Matthew Shudt; Melissa A Leisner; Navjot Singh

EPI_ISL_794208, EPI_ISL_794209, EPI_ISL_794210, EPI_ISL_794211, EPI_ISL_794212, EPI_ISL_794213, EPI_ISL_794214, EPI_ISL_794215, EPI_ISL_794216, EPI_ISL_794217, EPI_ISL_794218, EPI_ISL_794219, EPI_ISL_794220, EPI_ISL_794221, EPI_ISL_794222, EPI_ISL_794223, EPI_ISL_794224, EPI_ISL_794225, EPI_ISL_794226, EPI_ISL_794227, EPI_ISL_794228, EPI_ISL_794229, EPI_ISL_794230, EPI_ISL_794231,
EPI_ISL_794232, EPI_ISL_794233, EPI_ISL_794234, EPI_ISL_794235, EPI_ISL_794236, EPI_ISL_794237, EPI_ISL_794238, EPI_ISL_794239, EPI_ISL_794240, EPI_ISL_794241, EPI_ISL_794242, EPI_ISL_794243, EPI_ISL_794244, EPI_ISL_794245, EPI_ISL_794246, EPI_ISL_794247, EPI_ISL_794248, EPI_ISL_794249, EPI_ISL_794250, EPI_ISL_794251, EPI_ISL_794252, EPI_ISL_794253, EPI_ISL_794254, EPI_ISL_794255,
EPI_ISL_794256, EPI_ISL_794257, EPI_ISL_794258, EPI_ISL_794259, EPI_ISL_794260, EPI_ISL_794261, EPI_ISL_794262, EPI_ISL_794263, EPI_ISL_794264, EPI_ISL_794265, EPI_ISL_794266, EPI_ISL_794267, EPI_ISL_794268, EPI_ISL_794269, EPI_ISL_794270, EPI_ISL_794271, EPI_ISL_794272, EPI_ISL_794273, EPI_ISL_794274, EPI_ISL_794275, EPI_ISL_794276, EPI_ISL_794277, EPI_ISL_794278, EPI_ISL_794279,
EPI_ISL_794280, EPI_ISL_794281, EPI_ISL_794282, EPI_ISL_802434, EPI_ISL_802435, EPI_ISL_802440, EPI_ISL_802441, EPI_ISL_802443, EPI_ISL_802448, EPI_ISL_802469, EPI_ISL_802470, EPI_ISL_802471, EPI_ISL_802472, EPI_ISL_802473, EPI_ISL_802474, EPI_ISL_802475, EPI_ISL_802476, EPI_ISL_802477, EPI_ISL_802478, EPI_ISL_802479, EPI_ISL_802480, EPI_ISL_802481, EPI_ISL_802482, EPI_ISL_802483,
EPI_ISL_802484, EPI_ISL_802485, EPI_ISL_802486, EPI_ISL_802487, EPI_ISL_802488, EPI_ISL_802489, EPI_ISL_802490

see above | WESTCHESTER MEDICAL CENTER | Wadsworth Center, New York State Department.of Health | Alexis Russel; Daryl M. Lamson; Erasmus Schneider; Erica Lasek-Nesselquist; John Kelly; Jonathan Plitnick; Kirsten St. George; Matthew Shudt; Melissa A Leisner; Navjot Singh; Sara Griesemer

EPI_ISL_767580, EPI_ISL_767581, EPI_ISL_767582, EPI_ISL_767584, EPI_ISL_767585, EPI_ISL_767586, EPI_ISL_767587, EPI_ISL_767590, EPI_ISL_767595, EPI_ISL_767598, EPI_ISL_767615, EPI_ISL_767616, EPI_ISL_767617, EPI_ISL_767618, EPI_ISL_767619, EPI_ISL_767620, EPI_ISL_767621, EPI_ISL_767622, EPI_ISL_767623, EPI_ISL_767624, EPI_ISL_767625, EPI_ISL_767626, EPI_ISL_767627

see above | WHITE PLAINS HOSPITAL CENTER LABORATORY | Wadsworth Center, New York State Department.of Health | Alexis Russel; Daryl M. Lamson; Erasmus Schneider; Erica Lasek-Nesselquist; John Kelly; Jonathan Plitnick; Kirsten St. George; Matthew Shudt; Melissa A Leisner; Navjot Singh; Sara Griesemer

EPI_ISL_830216, EPI_ISL_830218, EPI_ISL_830220, EPI_ISL_830221, EPI_ISL_830244, EPI_ISL_830246, EPI_ISL_830248, EPI_ISL_830250, EPI_ISL_830252, EPI_ISL_830253, EPI_ISL_830255, EPI_ISL_830257, EPI_ISL_830259, EPI_ISL_830261, EPI_ISL_830263, EPI_ISL_830265, EPI_ISL_830267, EPI_ISL_830269, EPI_ISL_830270, EPI_ISL_830272, EPI_ISL_830274, EPI_ISL_830276, EPI_ISL_830278, EPI_ISL_830280,
EPI_ISL_830282, EPI_ISL_830284, EPI_ISL_830286, EPI_ISL_830288, EPI_ISL_830289, EPI_ISL_830291, EPI_ISL_830293, EPI_ISL_830294, EPI_ISL_830296, EPI_ISL_830298, EPI_ISL_830300, EPI_ISL_830302, EPI_ISL_830304, EPI_ISL_830306, EPI_ISL_830308, EPI_ISL_830310, EPI_ISL_830312, EPI_ISL_830313, EPI_ISL_830315, EPI_ISL_830317, EPI_ISL_830319, EPI_ISL_830321, EPI_ISL_830323, EPI_ISL_830325,
EPI_ISL_830326, EPI_ISL_830329, EPI_ISL_830331, EPI_ISL_830333, EPI_ISL_830334, EPI_ISL_830335, EPI_ISL_830337, EPI_ISL_830338, EPI_ISL_830340, EPI_ISL_830342, EPI_ISL_830344, EPI_ISL_830346, EPI_ISL_830348, EPI_ISL_830349, EPI_ISL_830643, EPI_ISL_830644, EPI_ISL_830645, EPI_ISL_830646, EPI_ISL_830647, EPI_ISL_830726, EPI_ISL_854368, EPI_ISL_861115, EPI_ISL_861116, EPI_ISL_861117,
EPI_ISL_884013, EPI_ISL_884014, EPI_ISL_884015, EPI_ISL_884016, EPI_ISL_884032, EPI_ISL_884033, EPI_ISL_884034, EPI_ISL_884035, EPI_ISL_884036, EPI_ISL_884037, EPI_ISL_884038, EPI_ISL_884039, EPI_ISL_884040, EPI_ISL_884041, EPI_ISL_884047, EPI_ISL_884055, EPI_ISL_884056, EPI_ISL_884057, EPI_ISL_884058, EPI_ISL_884080, EPI_ISL_884086, EPI_ISL_896307, EPI_ISL_896308, EPI_ISL_896309,
EPI_ISL_936148, EPI_ISL_936212, EPI_ISL_936213, EPI_ISL_936214, EPI_ISL_936215, EPI_ISL_936216, EPI_ISL_936217, EPI_ISL_936218, EPI_ISL_936219, EPI_ISL_936220, EPI_ISL_936221, EPI_ISL_936222, EPI_ISL_936223, EPI_ISL_936224, EPI_ISL_936225, EPI_ISL_936226, EPI_ISL_936227, EPI_ISL_936228, EPI_ISL_936229, EPI_ISL_936230, EPI_ISL_936231, EPI_ISL_936232, EPI_ISL_936233, EPI_ISL_936234,
EPI_ISL_936235, EPI_ISL_936236, EPI_ISL_936237, EPI_ISL_936238, EPI_ISL_936239, EPI_ISL_936240, EPI_ISL_936241, EPI_ISL_936242, EPI_ISL_936243, EPI_ISL_936244, EPI_ISL_936245, EPI_ISL_936246, EPI_ISL_936247, EPI_ISL_936300, EPI_ISL_936301, EPI_ISL_936302, EPI_ISL_983476, EPI_ISL_1226468, EPI_ISL_1226469, EPI_ISL_1226482, EPI_ISL_1226483, EPI_ISL_1226484, EPI_ISL_1226698,
EPI_ISL_1226716, EPI_ISL_1226938, EPI_ISL_1227030, EPI_ISL_1227246, EPI_ISL_1227254, EPI_ISL_1227255, EPI_ISL_1227257, EPI_ISL_1227258, EPI_ISL_1227259, EPI_ISL_1227260, EPI_ISL_1227261, EPI_ISL_1227262, EPI_ISL_1227358, EPI_ISL_1227359, EPI_ISL_1227379, EPI_ISL_1227557, EPI_ISL_1227566, EPI_ISL_1227567, EPI_ISL_1227568, EPI_ISL_1227570, EPI_ISL_1227574, EPI_ISL_1227575,
EPI_ISL_1227576, EPI_ISL_1227577, EPI_ISL_1227578, EPI_ISL_1227579, EPI_ISL_1227580, EPI_ISL_1227581, EPI_ISL_1227582, EPI_ISL_1227583, EPI_ISL_1227584, EPI_ISL_1227585, EPI_ISL_1227586, EPI_ISL_1227587, EPI_ISL_1227588, EPI_ISL_1227589, EPI_ISL_1227590, EPI_ISL_1227591, EPI_ISL_1227592, EPI_ISL_1227960, EPI_ISL_1228260, EPI_ISL_1228271, EPI_ISL_1228294, EPI_ISL_1228295,
EPI_ISL_1228444, EPI_ISL_1228445, EPI_ISL_1228446, EPI_ISL_1228450, EPI_ISL_1228460, EPI_ISL_1228461, EPI_ISL_1228462, EPI_ISL_1228465, EPI_ISL_1228466, EPI_ISL_1228469

see above | Wadsworth Center, New York State Department of Health | Wadsworth Center, New York State Department of Health | Alexis Russel; Daryl M. Lamson; Erasmus Schneider; Erica Lasek-Nesselquist; John Kelly; Jonathan Plitnick; Kirsten St. George; Matthew Shudt; Melissa A Leisner; Navjot Singh

EPI_ISL_676651, EPI_ISL_676653, EPI_ISL_676660, EPI_ISL_676662, EPI_ISL_676666, EPI_ISL_676671, EPI_ISL_676707, EPI_ISL_676994, EPI_ISL_677026, EPI_ISL_677027, EPI_ISL_677028, EPI_ISL_677029, EPI_ISL_677030, EPI_ISL_677031, EPI_ISL_677032, EPI_ISL_677033, EPI_ISL_677034, EPI_ISL_677035, EPI_ISL_677036, EPI_ISL_677037, EPI_ISL_677038, EPI_ISL_677039, EPI_ISL_677040, EPI_ISL_677041,
EPI_ISL_677042, EPI_ISL_677043, EPI_ISL_677044, EPI_ISL_677045, EPI_ISL_677046, EPI_ISL_677047, EPI_ISL_677048, EPI_ISL_677077, EPI_ISL_677078, EPI_ISL_677079, EPI_ISL_677080, EPI_ISL_677081, EPI_ISL_677082, EPI_ISL_677083, EPI_ISL_677084, EPI_ISL_677085, EPI_ISL_677086, EPI_ISL_677087, EPI_ISL_677088, EPI_ISL_677089, EPI_ISL_677090, EPI_ISL_677091, EPI_ISL_677092, EPI_ISL_677093,
EPI_ISL_677094, EPI_ISL_677095, EPI_ISL_677096, EPI_ISL_677097, EPI_ISL_677098, EPI_ISL_677099, EPI_ISL_677100, EPI_ISL_677101, EPI_ISL_677102, EPI_ISL_677103, EPI_ISL_677104, EPI_ISL_677105, EPI_ISL_677106, EPI_ISL_677107, EPI_ISL_677108, EPI_ISL_677109, EPI_ISL_677110, EPI_ISL_677130, EPI_ISL_677131, EPI_ISL_677132, EPI_ISL_677133, EPI_ISL_677670, EPI_ISL_677671, EPI_ISL_734287,
EPI_ISL_734288, EPI_ISL_734289, EPI_ISL_734290, EPI_ISL_734291, EPI_ISL_734292, EPI_ISL_734293, EPI_ISL_734294, EPI_ISL_734295, EPI_ISL_734296, EPI_ISL_734297, EPI_ISL_734298, EPI_ISL_734299, EPI_ISL_734300, EPI_ISL_734301, EPI_ISL_734302, EPI_ISL_734303, EPI_ISL_734304, EPI_ISL_734305, EPI_ISL_734306, EPI_ISL_734307, EPI_ISL_734308, EPI_ISL_734309, EPI_ISL_734310, EPI_ISL_734311,
EPI_ISL_734312, EPI_ISL_734313, EPI_ISL_734314, EPI_ISL_734315, EPI_ISL_734316, EPI_ISL_734317, EPI_ISL_734318, EPI_ISL_734319, EPI_ISL_734320, EPI_ISL_734321, EPI_ISL_734322, EPI_ISL_734323, EPI_ISL_734324, EPI_ISL_734325, EPI_ISL_734326, EPI_ISL_734327, EPI_ISL_734328, EPI_ISL_734329, EPI_ISL_734330, EPI_ISL_734331, EPI_ISL_734332, EPI_ISL_734333, EPI_ISL_734334, EPI_ISL_734335,
EPI_ISL_734336, EPI_ISL_734337, EPI_ISL_734338, EPI_ISL_734339, EPI_ISL_734340, EPI_ISL_734341, EPI_ISL_734342, EPI_ISL_734343, EPI_ISL_734344, EPI_ISL_734345, EPI_ISL_734346, EPI_ISL_734347, EPI_ISL_734348, EPI_ISL_734349, EPI_ISL_734350, EPI_ISL_734351, EPI_ISL_734352, EPI_ISL_734353, EPI_ISL_734354, EPI_ISL_734355, EPI_ISL_734356, EPI_ISL_734357, EPI_ISL_734358, EPI_ISL_734359,
EPI_ISL_734360, EPI_ISL_734361, EPI_ISL_734362, EPI_ISL_734363, EPI_ISL_734364, EPI_ISL_734365, EPI_ISL_734366, EPI_ISL_734367, EPI_ISL_734368, EPI_ISL_734369, EPI_ISL_734370, EPI_ISL_734371, EPI_ISL_734372, EPI_ISL_734373, EPI_ISL_734374, EPI_ISL_734375, EPI_ISL_734376, EPI_ISL_734377, EPI_ISL_734378, EPI_ISL_734379, EPI_ISL_734380, EPI_ISL_734381, EPI_ISL_734382, EPI_ISL_734383,
EPI_ISL_734384, EPI_ISL_734385, EPI_ISL_734386, EPI_ISL_734387, EPI_ISL_734388, EPI_ISL_734389, EPI_ISL_734390, EPI_ISL_734391, EPI_ISL_734392, EPI_ISL_734393, EPI_ISL_734394, EPI_ISL_734395, EPI_ISL_734396, EPI_ISL_734397, EPI_ISL_734398, EPI_ISL_734399, EPI_ISL_734400, EPI_ISL_734401, EPI_ISL_734402, EPI_ISL_734403, EPI_ISL_734404, EPI_ISL_734405, EPI_ISL_734406, EPI_ISL_734407,
EPI_ISL_734408, EPI_ISL_734409, EPI_ISL_734410, EPI_ISL_734411, EPI_ISL_734412, EPI_ISL_734413, EPI_ISL_734414, EPI_ISL_734415, EPI_ISL_734416, EPI_ISL_734417, EPI_ISL_734418, EPI_ISL_734419, EPI_ISL_734420, EPI_ISL_734421, EPI_ISL_734422, EPI_ISL_734423, EPI_ISL_734424, EPI_ISL_734425, EPI_ISL_734426, EPI_ISL_734427, EPI_ISL_734428, EPI_ISL_734429, EPI_ISL_734430, EPI_ISL_734431,
EPI_ISL_734432, EPI_ISL_734433, EPI_ISL_734434, EPI_ISL_734435, EPI_ISL_734436, EPI_ISL_734437, EPI_ISL_734438, EPI_ISL_734439, EPI_ISL_734440, EPI_ISL_734441, EPI_ISL_734442, EPI_ISL_734443, EPI_ISL_734444, EPI_ISL_734445, EPI_ISL_734446, EPI_ISL_754394, EPI_ISL_754395, EPI_ISL_754396, EPI_ISL_754397, EPI_ISL_754398, EPI_ISL_754399, EPI_ISL_754400, EPI_ISL_754401, EPI_ISL_754402,
EPI_ISL_754403, EPI_ISL_754404, EPI_ISL_754405, EPI_ISL_754406, EPI_ISL_754407, EPI_ISL_754408, EPI_ISL_754409, EPI_ISL_754410, EPI_ISL_754411, EPI_ISL_754412, EPI_ISL_754413, EPI_ISL_754414, EPI_ISL_754415, EPI_ISL_754416, EPI_ISL_754417, EPI_ISL_754418, EPI_ISL_754419, EPI_ISL_754420, EPI_ISL_754421, EPI_ISL_754422, EPI_ISL_754423, EPI_ISL_754424, EPI_ISL_754425, EPI_ISL_754426,
EPI_ISL_754427, EPI_ISL_754428, EPI_ISL_754429, EPI_ISL_754430, EPI_ISL_754431, EPI_ISL_754432, EPI_ISL_754433, EPI_ISL_754434, EPI_ISL_754435, EPI_ISL_754436, EPI_ISL_754437, EPI_ISL_754438, EPI_ISL_754439, EPI_ISL_754440, EPI_ISL_754441, EPI_ISL_754442, EPI_ISL_754443, EPI_ISL_754444, EPI_ISL_754445, EPI_ISL_754446, EPI_ISL_754447, EPI_ISL_754448, EPI_ISL_754449, EPI_ISL_754450,
EPI_ISL_754451, EPI_ISL_754452, EPI_ISL_754453, EPI_ISL_754454, EPI_ISL_754455, EPI_ISL_754456, EPI_ISL_754457, EPI_ISL_754458, EPI_ISL_754459, EPI_ISL_754460, EPI_ISL_754461, EPI_ISL_754462, EPI_ISL_754463, EPI_ISL_754464, EPI_ISL_754465, EPI_ISL_754466, EPI_ISL_754467, EPI_ISL_754468, EPI_ISL_754469, EPI_ISL_754470, EPI_ISL_754471, EPI_ISL_754472, EPI_ISL_754473, EPI_ISL_754474,
EPI_ISL_754475, EPI_ISL_754476, EPI_ISL_754477, EPI_ISL_754478, EPI_ISL_754479, EPI_ISL_754480, EPI_ISL_754481, EPI_ISL_754482, EPI_ISL_754483, EPI_ISL_754484, EPI_ISL_754485, EPI_ISL_754487, EPI_ISL_754488, EPI_ISL_754489, EPI_ISL_754490, EPI_ISL_754491, EPI_ISL_754492, EPI_ISL_754493, EPI_ISL_754494, EPI_ISL_754495, EPI_ISL_754496, EPI_ISL_754497, EPI_ISL_754498, EPI_ISL_754499,
EPI_ISL_754500, EPI_ISL_754501, EPI_ISL_754502, EPI_ISL_754503, EPI_ISL_754504, EPI_ISL_754505, EPI_ISL_754506, EPI_ISL_754507, EPI_ISL_754508, EPI_ISL_754509, EPI_ISL_754510, EPI_ISL_754511, EPI_ISL_754512, EPI_ISL_754513, EPI_ISL_754514, EPI_ISL_754515, EPI_ISL_754516, EPI_ISL_754517, EPI_ISL_754518, EPI_ISL_754519, EPI_ISL_754520, EPI_ISL_754521, EPI_ISL_754522, EPI_ISL_754523,
EPI_ISL_754524, EPI_ISL_754525, EPI_ISL_754526, EPI_ISL_754527, EPI_ISL_754528, EPI_ISL_754529, EPI_ISL_754530, EPI_ISL_754531, EPI_ISL_754532, EPI_ISL_754533, EPI_ISL_754534, EPI_ISL_754535, EPI_ISL_754536, EPI_ISL_754537, EPI_ISL_754538, EPI_ISL_754539, EPI_ISL_754540, EPI_ISL_754541, EPI_ISL_754542, EPI_ISL_754543, EPI_ISL_754544, EPI_ISL_754545, EPI_ISL_754546, EPI_ISL_754547,
EPI_ISL_754548, EPI_ISL_754549, EPI_ISL_754550, EPI_ISL_754551, EPI_ISL_754552, EPI_ISL_754553, EPI_ISL_754554, EPI_ISL_754555, EPI_ISL_754556, EPI_ISL_754557, EPI_ISL_754558, EPI_ISL_754559, EPI_ISL_754560, EPI_ISL_754561, EPI_ISL_754562, EPI_ISL_754563, EPI_ISL_754564, EPI_ISL_754565, EPI_ISL_754566, EPI_ISL_754568, EPI_ISL_754569, EPI_ISL_754570, EPI_ISL_754571, EPI_ISL_754572,
EPI_ISL_754573, EPI_ISL_754574, EPI_ISL_754575, EPI_ISL_754576, EPI_ISL_754577, EPI_ISL_754578, EPI_ISL_754579, EPI_ISL_754580, EPI_ISL_754581, EPI_ISL_754582, EPI_ISL_754583, EPI_ISL_754584, EPI_ISL_754585, EPI_ISL_754586, EPI_ISL_754587, EPI_ISL_754588, EPI_ISL_754589, EPI_ISL_754590, EPI_ISL_754591, EPI_ISL_754592, EPI_ISL_754593, EPI_ISL_754594, EPI_ISL_754595, EPI_ISL_754596,
EPI_ISL_754597, EPI_ISL_754598, EPI_ISL_754599, EPI_ISL_754600, EPI_ISL_754601, EPI_ISL_754602, EPI_ISL_754603, EPI_ISL_754604, EPI_ISL_754605, EPI_ISL_754606, EPI_ISL_754607, EPI_ISL_754608, EPI_ISL_754609, EPI_ISL_754610, EPI_ISL_754611, EPI_ISL_754612, EPI_ISL_754613, EPI_ISL_754614, EPI_ISL_754615, EPI_ISL_754616, EPI_ISL_754617, EPI_ISL_754618, EPI_ISL_754677, EPI_ISL_754678,
EPI_ISL_754679, EPI_ISL_754680, EPI_ISL_754681, EPI_ISL_754682, EPI_ISL_754683, EPI_ISL_754684, EPI_ISL_754685, EPI_ISL_754686, EPI_ISL_754687, EPI_ISL_754688, EPI_ISL_754689, EPI_ISL_754690, EPI_ISL_754691, EPI_ISL_754692, EPI_ISL_754693, EPI_ISL_754694, EPI_ISL_754695, EPI_ISL_754696, EPI_ISL_754697, EPI_ISL_754698, EPI_ISL_754699, EPI_ISL_754700, EPI_ISL_754701, EPI_ISL_754702,
EPI_ISL_754703, EPI_ISL_754704, EPI_ISL_754705, EPI_ISL_754706, EPI_ISL_754707, EPI_ISL_754708, EPI_ISL_754709, EPI_ISL_754710, EPI_ISL_754711, EPI_ISL_754712, EPI_ISL_754713, EPI_ISL_754714, EPI_ISL_754715, EPI_ISL_754716, EPI_ISL_754717, EPI_ISL_754718, EPI_ISL_754719, EPI_ISL_754720, EPI_ISL_754721, EPI_ISL_754722, EPI_ISL_754723, EPI_ISL_754724, EPI_ISL_754725, EPI_ISL_754726,
EPI_ISL_754727, EPI_ISL_754728, EPI_ISL_754729, EPI_ISL_754730, EPI_ISL_754731, EPI_ISL_754732, EPI_ISL_754734, EPI_ISL_754735, EPI_ISL_754736, EPI_ISL_754737, EPI_ISL_754738, EPI_ISL_754739, EPI_ISL_754740, EPI_ISL_754741, EPI_ISL_754742, EPI_ISL_754743, EPI_ISL_754744, EPI_ISL_754745, EPI_ISL_754779, EPI_ISL_754780, EPI_ISL_754781, EPI_ISL_754782, EPI_ISL_754783, EPI_ISL_754784,
EPI_ISL_754785, EPI_ISL_754786, EPI_ISL_754787, EPI_ISL_754788, EPI_ISL_754789, EPI_ISL_754790, EPI_ISL_754791, EPI_ISL_754792, EPI_ISL_754793, EPI_ISL_754794, EPI_ISL_754795, EPI_ISL_754796, EPI_ISL_754797, EPI_ISL_754798, EPI_ISL_754799, EPI_ISL_754800, EPI_ISL_754801, EPI_ISL_754802, EPI_ISL_754803, EPI_ISL_754804, EPI_ISL_754805, EPI_ISL_754806, EPI_ISL_754807, EPI_ISL_754808,
EPI_ISL_754809, EPI_ISL_754810, EPI_ISL_754811, EPI_ISL_754812, EPI_ISL_754813, EPI_ISL_754814, EPI_ISL_754815, EPI_ISL_754816, EPI_ISL_754817, EPI_ISL_754818, EPI_ISL_754819, EPI_ISL_754820, EPI_ISL_754821, EPI_ISL_754822, EPI_ISL_754823, EPI_ISL_754824, EPI_ISL_754825, EPI_ISL_754826, EPI_ISL_754827, EPI_ISL_754828, EPI_ISL_754829, EPI_ISL_754830, EPI_ISL_754831, EPI_ISL_754832,
EPI_ISL_754833, EPI_ISL_754834, EPI_ISL_754835, EPI_ISL_754836, EPI_ISL_754837, EPI_ISL_754838, EPI_ISL_754839, EPI_ISL_754840, EPI_ISL_754841, EPI_ISL_765480, EPI_ISL_765482, EPI_ISL_765484, EPI_ISL_765485, EPI_ISL_765486, EPI_ISL_765487, EPI_ISL_765488, EPI_ISL_765496, EPI_ISL_765497, EPI_ISL_765498, EPI_ISL_765499, EPI_ISL_765500, EPI_ISL_765501, EPI_ISL_765502, EPI_ISL_765503,
EPI_ISL_765504, EPI_ISL_765505, EPI_ISL_765506, EPI_ISL_765507, EPI_ISL_765508, EPI_ISL_765509, EPI_ISL_765510, EPI_ISL_765511, EPI_ISL_765512, EPI_ISL_765513, EPI_ISL_765514, EPI_ISL_765515, EPI_ISL_765516, EPI_ISL_767418, EPI_ISL_767420, EPI_ISL_767421, EPI_ISL_767422, EPI_ISL_767425, EPI_ISL_767426, EPI_ISL_767427, EPI_ISL_767428, EPI_ISL_767429, EPI_ISL_767430, EPI_ISL_767431,
EPI_ISL_767432, EPI_ISL_767433, EPI_ISL_767434, EPI_ISL_767435, EPI_ISL_767436, EPI_ISL_767437, EPI_ISL_767438, EPI_ISL_767439, EPI_ISL_767440, EPI_ISL_767441, EPI_ISL_767442, EPI_ISL_767443, EPI_ISL_767444, EPI_ISL_767445, EPI_ISL_767446, EPI_ISL_767447, EPI_ISL_767448, EPI_ISL_767449, EPI_ISL_767450, EPI_ISL_767451, EPI_ISL_767452, EPI_ISL_767453, EPI_ISL_767454, EPI_ISL_767455,
EPI_ISL_767456, EPI_ISL_767457, EPI_ISL_767458, EPI_ISL_767459, EPI_ISL_767460, EPI_ISL_767461, EPI_ISL_767462, EPI_ISL_767463, EPI_ISL_767464, EPI_ISL_767465, EPI_ISL_767466, EPI_ISL_767467, EPI_ISL_767468, EPI_ISL_767469, EPI_ISL_767470, EPI_ISL_767471, EPI_ISL_767472, EPI_ISL_767473, EPI_ISL_767474, EPI_ISL_767475, EPI_ISL_767476, EPI_ISL_767477, EPI_ISL_767478, EPI_ISL_767479,
EPI_ISL_767480, EPI_ISL_767481, EPI_ISL_767482, EPI_ISL_767483, EPI_ISL_767484, EPI_ISL_767485, EPI_ISL_767486, EPI_ISL_767487, EPI_ISL_767488, EPI_ISL_767489, EPI_ISL_767490, EPI_ISL_767491, EPI_ISL_767492, EPI_ISL_767493, EPI_ISL_767494, EPI_ISL_767495, EPI_ISL_767496, EPI_ISL_767497, EPI_ISL_767498, EPI_ISL_767499, EPI_ISL_767500, EPI_ISL_767501, EPI_ISL_767502, EPI_ISL_767503,
EPI_ISL_767504, EPI_ISL_767505, EPI_ISL_767506, EPI_ISL_767507, EPI_ISL_767509, EPI_ISL_767510, EPI_ISL_767511, EPI_ISL_767512, EPI_ISL_767513, EPI_ISL_767514, EPI_ISL_767515, EPI_ISL_767516, EPI_ISL_767517, EPI_ISL_767518, EPI_ISL_767519, EPI_ISL_767520, EPI_ISL_767521, EPI_ISL_767522, EPI_ISL_767523, EPI_ISL_767524, EPI_ISL_767525, EPI_ISL_767526, EPI_ISL_767527, EPI_ISL_767528,
EPI_ISL_767529, EPI_ISL_767530, EPI_ISL_767531, EPI_ISL_767532, EPI_ISL_767533, EPI_ISL_767534, EPI_ISL_767535, EPI_ISL_767536, EPI_ISL_767537, EPI_ISL_767578, EPI_ISL_794014, EPI_ISL_794015, EPI_ISL_794016, EPI_ISL_794017, EPI_ISL_794018, EPI_ISL_794019, EPI_ISL_794020, EPI_ISL_794021, EPI_ISL_794022, EPI_ISL_794023, EPI_ISL_794024, EPI_ISL_794025, EPI_ISL_794026, EPI_ISL_794027,
EPI_ISL_794028, EPI_ISL_794029, EPI_ISL_794030, EPI_ISL_794031, EPI_ISL_794032, EPI_ISL_794033, EPI_ISL_794034, EPI_ISL_794035, EPI_ISL_794038, EPI_ISL_794039, EPI_ISL_794040, EPI_ISL_794041, EPI_ISL_794042, EPI_ISL_794043, EPI_ISL_794044, EPI_ISL_794045, EPI_ISL_794046, EPI_ISL_794047, EPI_ISL_794048, EPI_ISL_794049, EPI_ISL_794050, EPI_ISL_794051, EPI_ISL_794052, EPI_ISL_794053,
EPI_ISL_794054, EPI_ISL_794055, EPI_ISL_794056, EPI_ISL_794057, EPI_ISL_794058, EPI_ISL_794059, EPI_ISL_794060, EPI_ISL_794061, EPI_ISL_794062, EPI_ISL_794063, EPI_ISL_794064, EPI_ISL_794065, EPI_ISL_794066, EPI_ISL_794067, EPI_ISL_794068, EPI_ISL_794069, EPI_ISL_794070, EPI_ISL_794071, EPI_ISL_794072, EPI_ISL_794073, EPI_ISL_794074, EPI_ISL_794075, EPI_ISL_794076, EPI_ISL_794077,
EPI_ISL_794078, EPI_ISL_794079, EPI_ISL_794080, EPI_ISL_794081, EPI_ISL_794082, EPI_ISL_794159, EPI_ISL_794160, EPI_ISL_794161, EPI_ISL_794162, EPI_ISL_794163, EPI_ISL_794164, EPI_ISL_794165, EPI_ISL_794166, EPI_ISL_794167, EPI_ISL_794168, EPI_ISL_794169, EPI_ISL_794170, EPI_ISL_794171, EPI_ISL_794172, EPI_ISL_794173, EPI_ISL_794174, EPI_ISL_794175, EPI_ISL_794176, EPI_ISL_794177,
EPI_ISL_794178, EPI_ISL_794179, EPI_ISL_794180, EPI_ISL_794181, EPI_ISL_794206, EPI_ISL_794207, EPI_ISL_794308, EPI_ISL_794309, EPI_ISL_794310, EPI_ISL_794311, EPI_ISL_794312, EPI_ISL_794313, EPI_ISL_794314, EPI_ISL_794315, EPI_ISL_794316, EPI_ISL_794317, EPI_ISL_794318, EPI_ISL_794319, EPI_ISL_794320, EPI_ISL_802429, EPI_ISL_802431, EPI_ISL_802433, EPI_ISL_802438, EPI_ISL_802442,
EPI_ISL_802449, EPI_ISL_802450, EPI_ISL_802451, EPI_ISL_802452, EPI_ISL_802453, EPI_ISL_802454, EPI_ISL_802455, EPI_ISL_802456, EPI_ISL_802457, EPI_ISL_802458, EPI_ISL_802459, EPI_ISL_802460, EPI_ISL_802461, EPI_ISL_802462, EPI_ISL_802463, EPI_ISL_802464, EPI_ISL_802465, EPI_ISL_802466, EPI_ISL_802498, EPI_ISL_802499, EPI_ISL_802500, EPI_ISL_804945, EPI_ISL_804947, EPI_ISL_804948,
EPI_ISL_1049338, EPI_ISL_1049339

see above | Wadsworth Center, New York State Department.of Health | Wadsworth Center, New York State Department.of Health | Alexis Russel; Daryl M. Lamson; Erasmus Schneider; Erica Lasek-Nesselquist; John Kelly; Jonathan Plitnick; Kirsten St. George; Matthew Shudt; Melissa A Leisner; Navjot Singh; Sara Griesemer

| | | | |
|---|---|---|---|
| EPI_ISL_1968561, EPI_ISL_1968562, EPI_ISL_1968566, EPI_ISL_1968568, EPI_ISL_1968569, EPI_ISL_1968570, EPI_ISL_1968572, EPI_ISL_1968574, EPI_ISL_1968577, EPI_ISL_1968578, EPI_ISL_1968579, EPI_ISL_1968580, EPI_ISL_1968582, EPI_ISL_1968583, EPI_ISL_1968588, EPI_ISL_1968589, EPI_ISL_1968590, EPI_ISL_1968591, EPI_ISL_1968592, EPI_ISL_1968593, EPI_ISL_1968594, EPI_ISL_1968596, EPI_ISL_1968598, EPI_ISL_1968599, EPI_ISL_1968600, EPI_ISL_1968601, EPI_ISL_1968604, EPI_ISL_1968608, EPI_ISL_1968609, EPI_ISL_1968612, EPI_ISL_1968613, EPI_ISL_1968614, EPI_ISL_1968615, EPI_ISL_1968616, EPI_ISL_1968617, EPI_ISL_1968618, EPI_ISL_1968619, EPI_ISL_1968620, EPI_ISL_1968621, EPI_ISL_1968622, EPI_ISL_1968625, EPI_ISL_1968626, EPI_ISL_2031768, EPI_ISL_2031769, EPI_ISL_2031770 | | | |
| see above | Weill Cornell Medicine | New York Genome Center | Andre Corvelo; Arryn Craney; Chris Mason; Dayna M. Oschwald; Hanna Rennert; Lars F Westblade; Margaret Elizabeth Ross; Melissa Cushing; Michael Zody; Olivier Elemento; Priya Velu; Samantha Fennessey; Tom Maniatis |
| EPI_ISL_1920007, EPI_ISL_1920009, EPI_ISL_1920062, EPI_ISL_1920063, EPI_ISL_1920064, EPI_ISL_1920065, EPI_ISL_1920081, EPI_ISL_1920082, EPI_ISL_1920093 | | | |
| see above | Westchester Medical Center | Wadsworth Center, New York State Department of Health | Alexis Russell; Catharine Prussing; Daryl M. Lamson; Erasmus Schneider; Erica Lasek-Nesselquist; John Kelly; Jonathan Plitnick; Kirsten St. George; Matthew Shudt; Melissa A Leisner; Navjot Singh |

We gratefully acknowledge the following Authors from the Originating laboratories responsible for obtaining the specimens, as well as the Submitting laboratories where the genome data were generated and shared via GISAID, on which this research is based.

All Submitters of data may be contacted directly via www.gisaid.org

Authors are sorted alphabetically.

| Accession ID | Originating Laboratory | Submitting Laboratory | Authors |
|---|---|---|---|
| EPI_ISL_1446676, EPI_ISL_1446677, EPI_ISL_1447053 | "NYSDOH Wadsworth Center, Virology Lab" | Centers for Disease Control and Prevention Division of Viral Diseases, Pathogen Discovery | Alison Laufer Halpin; Ben L. Rambo-Martin; Clinton R. Paden; Dakota Howard; Darlene Wagner; Dave Wentworth; Dhwani Batra; Jasmine Padilla; Justin Lee; Katie Dillon; Krista Queen; Kristen Knipe; Kristine Lacek; Mark Burroughs; Matthew Schmerer; Mili Sheth; Peter Cook; Sam Shepard; Sarah Nobles; Shoshona Le; Suxiang Tong; Vivien Dugan; Yvette Unoarumhi |
| EPI_ISL_1226054, EPI_ISL_1226055, EPI_ISL_1226056, EPI_ISL_1226057, EPI_ISL_1226058, EPI_ISL_1226059, EPI_ISL_1226060, EPI_ISL_1226061, EPI_ISL_1226062, EPI_ISL_1226063, EPI_ISL_1226064, EPI_ISL_1226065, EPI_ISL_1226066, EPI_ISL_1226067, EPI_ISL_1226068, EPI_ISL_1226069, EPI_ISL_1226070, EPI_ISL_1226071, EPI_ISL_1226072, EPI_ISL_1226073, EPI_ISL_1226074, EPI_ISL_1226075, EPI_ISL_1226076, EPI_ISL_1226077, EPI_ISL_1226078, EPI_ISL_1226079, EPI_ISL_1226080, EPI_ISL_1226081 | | | |
| see above | "NYSDOH Wadsworth Center, Virology Lab" | Genomics and Discovery, Respiratory Viruses Branch, Division of Viral Diseases, Centers for Disease Control and Prevention | Anna Montmayeur; Anna Uehara; Ben L. Rambo-Martin; Clinton R. Paden; Dhwani Batra; Haibin Wang; Jasmine Padilla; Jing Zhang; Justin Lee; Katie Dillon; Krista Queen; Kristen Knipe; Kristine Lacek; Lori Rowe; Mark Burroughs; Matthew Schmerer; Mili Sheth; Peter W. Cook; Rachel Marine; Sam Shepard; Sarah Nobles; Shoshona Le; Suxiang Tong; Yan Li; Ying Tao |
| EPI_ISL_1397747, EPI_ISL_1397748, EPI_ISL_1397749, EPI_ISL_1397750, EPI_ISL_1397751 | ADIRONDACK MEDICAL CENTER | Wadsworth Center, New York State Department of Health | Alexis Russel; Daryl M. Lamson; Erasmus Schneider; Erica Lasek-Nesselquist; John Kelly; Jonathan Plitnick; Kirsten St. George; Matthew Shudt; Melissa A Leisner; Navjot Singh |
| EPI_ISL_1226485, EPI_ISL_1226511, EPI_ISL_1226563, EPI_ISL_1226706, EPI_ISL_1226819, EPI_ISL_1227029, EPI_ISL_1227031, EPI_ISL_1227032, EPI_ISL_1227208, EPI_ISL_1227209, EPI_ISL_1227232, EPI_ISL_1227233, EPI_ISL_1227235, EPI_ISL_1227263, EPI_ISL_1227264, EPI_ISL_1227265, EPI_ISL_1227266, EPI_ISL_1227267, EPI_ISL_1227268, EPI_ISL_1227269, EPI_ISL_1227270, EPI_ISL_1227271, EPI_ISL_1227461, EPI_ISL_1227462, EPI_ISL_1227558, EPI_ISL_1227572, EPI_ISL_1227573, EPI_ISL_1227790, EPI_ISL_1228219, EPI_ISL_1228220, EPI_ISL_1228221, EPI_ISL_1228275, EPI_ISL_1228276, EPI_ISL_1228277, EPI_ISL_1228279, EPI_ISL_1228282, EPI_ISL_1228283, EPI_ISL_1228430, EPI_ISL_1228477, EPI_ISL_1228478, EPI_ISL_1228479, EPI_ISL_1397542, EPI_ISL_1397543, EPI_ISL_1397544, EPI_ISL_1397545, EPI_ISL_1397551, EPI_ISL_1397834, EPI_ISL_1397835, EPI_ISL_1397837, EPI_ISL_1397843, EPI_ISL_1397853, EPI_ISL_1397854, EPI_ISL_1397855, EPI_ISL_1397856, EPI_ISL_1397858, EPI_ISL_1397861, EPI_ISL_1397866, EPI_ISL_1397867, EPI_ISL_1397868, EPI_ISL_1397869, EPI_ISL_1397961, EPI_ISL_1398021, EPI_ISL_1398022, EPI_ISL_1398023, EPI_ISL_1398024, EPI_ISL_1398025, EPI_ISL_1398026, EPI_ISL_1398027, EPI_ISL_1398147, EPI_ISL_1398148, EPI_ISL_1398149, EPI_ISL_1398150, EPI_ISL_1398151, EPI_ISL_1398152, EPI_ISL_1398153, EPI_ISL_1398274, EPI_ISL_1398275, EPI_ISL_1398276, EPI_ISL_1398277, EPI_ISL_1461295, EPI_ISL_1461296, EPI_ISL_1461297, EPI_ISL_1461298, EPI_ISL_1462506, EPI_ISL_1462508, EPI_ISL_1465615, EPI_ISL_1465616, EPI_ISL_1465634, EPI_ISL_1465635, EPI_ISL_1465636, EPI_ISL_1465639, EPI_ISL_1482455, EPI_ISL_1482456, EPI_ISL_1580700, EPI_ISL_1582154, EPI_ISL_1935630 | | | |
| see above | ALBANY MEDICAL CENTER | Wadsworth Center, New York State Department of Health | Alexis Russel; Alexis Russell; Catharine Prussing; Daryl M. Lamson; Erasmus Schneider; Erica Lasek-Nesselquist; John Kelly; Jonathan Plitnick; Kirsten St. George; Matthew Shudt; Melissa A Leisner; Navjot Singh |
| EPI_ISL_1445307, EPI_ISL_1445308, EPI_ISL_1445380, EPI_ISL_1445424, EPI_ISL_1445443, EPI_ISL_1445702, EPI_ISL_1445705, EPI_ISL_1445889, EPI_ISL_1445906, EPI_ISL_1445928, EPI_ISL_1445943, EPI_ISL_1446075, EPI_ISL_1446114, EPI_ISL_1446139, EPI_ISL_1446140, EPI_ISL_1446144, EPI_ISL_1446148, EPI_ISL_1446152, EPI_ISL_1479449, EPI_ISL_1479456, EPI_ISL_1479460, EPI_ISL_1479461, EPI_ISL_1479477, EPI_ISL_1493507, EPI_ISL_1493519, EPI_ISL_1493520, EPI_ISL_1493534, EPI_ISL_1493540, EPI_ISL_1493559, EPI_ISL_1550397, EPI_ISL_1550549, EPI_ISL_1561270 | | | |
| see above | Aegis Sciences Corporation | Centers for Disease Control and Prevention Division of Viral Diseases, Pathogen Discovery | Adrian Paskey; Alec Vest; Benjamin Rambo-Martin; Christopher Gulvick; Clinton R. Paden; Cyndi Clark; Dakota Howard; Darlene Wagner; Dhwani Batra; Dillon Nall; Duncan MacCannell; Ethan Sanders; Holly Houdeshell; Jason Caravas; Kara Moser; Matthew Hardison; Matthew Schmerer; Ola Kvalvaag; Patrick Campbell; Peter W. Cook; Rob Case; Scott Sammons; Shatavia Morrison; Shaun Westlund; Vikramsinha Ghorpade; Yvette Unoarumhi |
| EPI_ISL_1920105, EPI_ISL_1920109, EPI_ISL_1920112 | Albany Medical Center | Wadsworth Center, New York State Department of Health | Alexis Russell; Catharine Prussing; Daryl M. Lamson; Erasmus Schneider; Erica Lasek-Nesselquist; John Kelly; Jonathan Plitnick; Kirsten St. George; Matthew Shudt; Melissa A Leisner; Navjot Singh |
| EPI_ISL_1226551, EPI_ISL_1226556, EPI_ISL_1226571, EPI_ISL_1226572, EPI_ISL_1226574, EPI_ISL_1226577, EPI_ISL_1226583, EPI_ISL_1226622, EPI_ISL_1226645, EPI_ISL_1226646, EPI_ISL_1226658, EPI_ISL_1226669, EPI_ISL_1226932, EPI_ISL_1226985, EPI_ISL_1397694, EPI_ISL_1397695, EPI_ISL_1397696, EPI_ISL_1397697, EPI_ISL_1397698, EPI_ISL_1397699, EPI_ISL_1397912, EPI_ISL_1397913 | | | |
| see above | BIO-REFERENCE LABORATORIES | Wadsworth Center, New York State Department of Health | Alexis Russel; Daryl M. Lamson; Erasmus Schneider; Erica Lasek-Nesselquist; John Kelly; Jonathan Plitnick; Kirsten St. George; Matthew Shudt; Melissa A Leisner; Navjot Singh |
| EPI_ISL_1001460, EPI_ISL_1080751, EPI_ISL_1080756, EPI_ISL_1080759, EPI_ISL_1080761, EPI_ISL_1080767, EPI_ISL_1080768, EPI_ISL_1080769, EPI_ISL_1080772, EPI_ISL_1080773, EPI_ISL_1080775, EPI_ISL_1080777, EPI_ISL_1080778, EPI_ISL_1080779, EPI_ISL_1080782, EPI_ISL_1080794, EPI_ISL_1080795, EPI_ISL_1080802, EPI_ISL_1080804, EPI_ISL_1080805, EPI_ISL_1080806, EPI_ISL_1080807, EPI_ISL_1080808, EPI_ISL_1500570, EPI_ISL_1500571, EPI_ISL_1500572, EPI_ISL_1500573, EPI_ISL_1500574, EPI_ISL_1500575, EPI_ISL_1500576, EPI_ISL_1500577, EPI_ISL_1500578, EPI_ISL_1500579, EPI_ISL_1500580, EPI_ISL_1500581, EPI_ISL_1500582, EPI_ISL_1500583, EPI_ISL_1500584, EPI_ISL_1500585, EPI_ISL_1500586, EPI_ISL_1500587, EPI_ISL_1500588, EPI_ISL_1500589, EPI_ISL_1500590, EPI_ISL_1500591, EPI_ISL_1500592, EPI_ISL_1500593, EPI_ISL_1500594, EPI_ISL_1500595, EPI_ISL_1500596, EPI_ISL_1500597, EPI_ISL_1500598, EPI_ISL_1500599, EPI_ISL_1500600, EPI_ISL_1500601, EPI_ISL_1500602, EPI_ISL_1500603, EPI_ISL_1500604, EPI_ISL_1500605, EPI_ISL_1500606, EPI_ISL_1500607, EPI_ISL_1500608, EPI_ISL_1500609, EPI_ISL_1500610, EPI_ISL_1500611, EPI_ISL_1500612, EPI_ISL_1500613, EPI_ISL_1500614, EPI_ISL_1500615, EPI_ISL_1500616, EPI_ISL_1500617, EPI_ISL_1500618, EPI_ISL_1500619, EPI_ISL_1500620, EPI_ISL_1500621, EPI_ISL_1500622, EPI_ISL_1500623, EPI_ISL_1500625, EPI_ISL_1500626, EPI_ISL_1500627, EPI_ISL_1500628, EPI_ISL_1500629, EPI_ISL_1500630, EPI_ISL_1500631, EPI_ISL_1500632, EPI_ISL_1500633, EPI_ISL_1500634, EPI_ISL_1500635, EPI_ISL_1500636, EPI_ISL_1500637, EPI_ISL_1965849, EPI_ISL_1965851, EPI_ISL_1965852, EPI_ISL_1965853, EPI_ISL_1965854, EPI_ISL_1965855, EPI_ISL_1965856, EPI_ISL_1965857, EPI_ISL_1965858, EPI_ISL_1965859, EPI_ISL_1965860, EPI_ISL_1965861, EPI_ISL_1965862, EPI_ISL_1965863, EPI_ISL_1965864, EPI_ISL_1965865, EPI_ISL_1965866, EPI_ISL_1965867, EPI_ISL_1965868, EPI_ISL_1965869, EPI_ISL_1965870, EPI_ISL_1965871, EPI_ISL_1965872, EPI_ISL_1965873, EPI_ISL_1965874, EPI_ISL_1965875, EPI_ISL_1965876, EPI_ISL_1965877, EPI_ISL_1965878, EPI_ISL_1965879, EPI_ISL_1965880, EPI_ISL_1965881, EPI_ISL_1965882, EPI_ISL_1965883, EPI_ISL_1965885, EPI_ISL_1965886, EPI_ISL_1965887, EPI_ISL_1965888, EPI_ISL_1965889, EPI_ISL_1965890, EPI_ISL_1965891, EPI_ISL_1965892, EPI_ISL_1965893, EPI_ISL_1965894, EPI_ISL_1965895, EPI_ISL_1965896, EPI_ISL_1965897, EPI_ISL_1965898, EPI_ISL_1965899, EPI_ISL_1965900, EPI_ISL_1965901, EPI_ISL_1965902, EPI_ISL_1965903, EPI_ISL_1965904, EPI_ISL_1965905, EPI_ISL_1965906, EPI_ISL_1965907, EPI_ISL_1965908, EPI_ISL_1965909, EPI_ISL_1965910, EPI_ISL_1965911, EPI_ISL_1965912, EPI_ISL_1965913, EPI_ISL_1965914, EPI_ISL_1965915, EPI_ISL_1965916, EPI_ISL_1965917, EPI_ISL_1965924, EPI_ISL_1965925, EPI_ISL_1965926, EPI_ISL_1965927, EPI_ISL_1965928, EPI_ISL_1965931, EPI_ISL_1965932, EPI_ISL_1965933, EPI_ISL_1965934, EPI_ISL_1965935, EPI_ISL_1965936, EPI_ISL_1965939, EPI_ISL_1965940, EPI_ISL_1965941, EPI_ISL_1965942, EPI_ISL_1965944 | | | |
| see above | Clinical Microbiology Laboratory, NewYork Presbyterian Hospital/Columbia University Irving Medical Center | Uhlemann Laboratory, Columbia University Irving Medical Center | Anne Kelley; Anne-Catrin Uhlemann; David Ho; Hiroshi Mohri; Hiroshi Mori; Maya Tagliavia; Medini K. Annavajhala |
| EPI_ISL_1226461, EPI_ISL_1226518, EPI_ISL_1226637, EPI_ISL_1226638, EPI_ISL_1226728, EPI_ISL_1226759, EPI_ISL_1226765, EPI_ISL_1226805, EPI_ISL_1226806, EPI_ISL_1226807, EPI_ISL_1226930, EPI_ISL_1227002, EPI_ISL_1227003, EPI_ISL_1227004, EPI_ISL_1227068, EPI_ISL_1227069, EPI_ISL_1227088, EPI_ISL_1227089, EPI_ISL_1227096, EPI_ISL_1227166, EPI_ISL_1227222, EPI_ISL_1227401, EPI_ISL_1227402, EPI_ISL_1227406, EPI_ISL_1227407, EPI_ISL_1227408, EPI_ISL_1227409, EPI_ISL_1227416, EPI_ISL_1227435, EPI_ISL_1227536, EPI_ISL_1227934, EPI_ISL_1228576, EPI_ISL_1397636, EPI_ISL_1397637, EPI_ISL_1397638, EPI_ISL_1397639, EPI_ISL_1397640, EPI_ISL_1397641, EPI_ISL_1397642, EPI_ISL_1397643, EPI_ISL_1397644, EPI_ISL_1397645, EPI_ISL_1397646, EPI_ISL_1397647, EPI_ISL_1397648, EPI_ISL_1397649, EPI_ISL_1397824, EPI_ISL_1397825, EPI_ISL_1397826, EPI_ISL_1397827, EPI_ISL_1397828, EPI_ISL_1397886, EPI_ISL_1397887, EPI_ISL_1397888, EPI_ISL_1397889, EPI_ISL_1397890, EPI_ISL_1397891, EPI_ISL_1397892, EPI_ISL_1397893, EPI_ISL_1397894, EPI_ISL_1397895, EPI_ISL_1397896, EPI_ISL_1397897, EPI_ISL_1397898, EPI_ISL_1397899, EPI_ISL_1397900, EPI_ISL_1397901, EPI_ISL_1397902, EPI_ISL_1397903, EPI_ISL_1397904, EPI_ISL_1397905, EPI_ISL_1397906, EPI_ISL_1398144, EPI_ISL_1398145, EPI_ISL_1398146, EPI_ISL_1398163, EPI_ISL_1398164, EPI_ISL_1398165, EPI_ISL_1398166, EPI_ISL_1398167, EPI_ISL_1398168, EPI_ISL_1398169, EPI_ISL_1398170, EPI_ISL_1398171, EPI_ISL_1398172, EPI_ISL_1398173, EPI_ISL_1398174, EPI_ISL_1398175, EPI_ISL_1398176, EPI_ISL_1398177, EPI_ISL_1398178, EPI_ISL_1398179, EPI_ISL_1398180, EPI_ISL_1398181, EPI_ISL_1398182, EPI_ISL_1398183, EPI_ISL_1398184, EPI_ISL_1398185, EPI_ISL_1398186, EPI_ISL_1398187, EPI_ISL_1398188, EPI_ISL_1398189, EPI_ISL_1398190, EPI_ISL_1398191, EPI_ISL_1398192, EPI_ISL_1398193, EPI_ISL_1398194, EPI_ISL_1398195, EPI_ISL_1398196, EPI_ISL_1398197, EPI_ISL_1398198, EPI_ISL_1398199, EPI_ISL_1398200, EPI_ISL_1398201, EPI_ISL_1398202, EPI_ISL_1398203, EPI_ISL_1465633, EPI_ISL_1465693, EPI_ISL_1465694, EPI_ISL_1465695, EPI_ISL_1465803, EPI_ISL_1465804, EPI_ISL_1482277, EPI_ISL_1575472, EPI_ISL_1576820, EPI_ISL_1624010, EPI_ISL_1624011, EPI_ISL_1624404, EPI_ISL_1624405, EPI_ISL_1671160, EPI_ISL_1671176, EPI_ISL_1671210, EPI_ISL_1755943, EPI_ISL_1755944, EPI_ISL_1755945, EPI_ISL_1755946, EPI_ISL_1756326, EPI_ISL_1756328, EPI_ISL_1920078, EPI_ISL_1920094, EPI_ISL_1920137, EPI_ISL_1920139, EPI_ISL_1920140, EPI_ISL_1920141, EPI_ISL_1920143, EPI_ISL_1920144, EPI_ISL_1920146, EPI_ISL_1920148, EPI_ISL_1920149, EPI_ISL_1920156, EPI_ISL_1935625 | | | |
| see above | Columbia University Irving Medical Center | Wadsworth Center, New York State Department of Health | Alexis Russel; Alexis Russell; Catharine Prussing; Daryl M. Lamson; Erasmus Schneider; Erica Lasek-Nesselquist; John Kelly; Jonathan Plitnick; Kirsten St. George; Matthew Shudt; Melissa A Leisner; Navjot Singh |
| EPI_ISL_1228375, EPI_ISL_1228376, EPI_ISL_1228377 | DEPARTMENT OF VETERANS AFFAIR MEDICAL CENTER | Wadsworth Center, New York State Department of Health | Alexis Russel; Daryl M. Lamson; Erasmus Schneider; Erica Lasek-Nesselquist; John Kelly; Jonathan Plitnick; Kirsten St. George; Matthew Shudt; Melissa A Leisner; Navjot Singh |
| EPI_ISL_1017982, EPI_ISL_1017983, EPI_ISL_1017984, EPI_ISL_1017985, EPI_ISL_1018001, EPI_ISL_1064775, EPI_ISL_1064803, EPI_ISL_1064809, EPI_ISL_1064820, EPI_ISL_1064825, EPI_ISL_1064831, EPI_ISL_1064876, EPI_ISL_1064909, EPI_ISL_1200522, EPI_ISL_1200541, EPI_ISL_1200552, EPI_ISL_1200556, EPI_ISL_1200577, EPI_ISL_1200594, EPI_ISL_1200621, EPI_ISL_1200622, EPI_ISL_1200624, EPI_ISL_1200625, EPI_ISL_1261801, EPI_ISL_1261804, EPI_ISL_1261807, EPI_ISL_1261808, EPI_ISL_1261824, EPI_ISL_1261843, EPI_ISL_1261856, EPI_ISL_1261858, EPI_ISL_1261864, EPI_ISL_1337465, EPI_ISL_1337489, EPI_ISL_1337517 | | | |
| see above | DOHMH Central Harlem | New York City Public Health Laboratory | Jade Wang; et al. |
| EPI_ISL_1017978, EPI_ISL_1018011, EPI_ISL_1064766, EPI_ISL_1064779, EPI_ISL_1064782, EPI_ISL_1064797, EPI_ISL_1064872, EPI_ISL_1064895, EPI_ISL_1064896, EPI_ISL_1200518, EPI_ISL_1200528, EPI_ISL_1200529, EPI_ISL_1200530, EPI_ISL_1200534, EPI_ISL_1200560, EPI_ISL_1200564, EPI_ISL_1200574, EPI_ISL_1200584, EPI_ISL_1261817, EPI_ISL_1261852, EPI_ISL_1261855, EPI_ISL_1261862, EPI_ISL_1261863, EPI_ISL_1337464, EPI_ISL_1337500, EPI_ISL_1337542 | | | |
| see above | DOHMH Chelsea | New York City Public Health Laboratory | Jade Wang; et al. |
| EPI_ISL_1017975, EPI_ISL_1017986, EPI_ISL_1017987, EPI_ISL_1017988, EPI_ISL_1017995, EPI_ISL_1017996, EPI_ISL_1017997, EPI_ISL_1017998, EPI_ISL_1017999, EPI_ISL_1018005, EPI_ISL_1018027, EPI_ISL_1018028, EPI_ISL_1018029, EPI_ISL_1018030, EPI_ISL_1018031, EPI_ISL_1064757, EPI_ISL_1064760, EPI_ISL_1064761, EPI_ISL_1064767, EPI_ISL_1064784, EPI_ISL_1064786, EPI_ISL_1064789, EPI_ISL_1064808, EPI_ISL_1064811, EPI_ISL_1064847, EPI_ISL_1064870, EPI_ISL_1064871, EPI_ISL_1064873, EPI_ISL_1064874, EPI_ISL_1064888, EPI_ISL_1064893, EPI_ISL_1064897, EPI_ISL_1064905, EPI_ISL_1200533, EPI_ISL_1200540, EPI_ISL_1200545, EPI_ISL_1200546, EPI_ISL_1200565, EPI_ISL_1200568, EPI_ISL_1200569, EPI_ISL_1200570, EPI_ISL_1200591, EPI_ISL_1200592, EPI_ISL_1200593, EPI_ISL_1200597, EPI_ISL_1200598, EPI_ISL_1200603, EPI_ISL_1200613, EPI_ISL_1200614, EPI_ISL_1200626, EPI_ISL_1261806, EPI_ISL_1261810, EPI_ISL_1261812, EPI_ISL_1261813, EPI_ISL_1261832, EPI_ISL_1261833, EPI_ISL_1261835, EPI_ISL_1261836, EPI_ISL_1261837, EPI_ISL_1261844, EPI_ISL_1261859, EPI_ISL_1447142 | | | |
| see above | DOHMH Corona | New York City Public Health Laboratory | Jade Wang; et al. |
| EPI_ISL_1018021, EPI_ISL_1018024, EPI_ISL_1064777, EPI_ISL_1064791, EPI_ISL_1064798, EPI_ISL_1064824, EPI_ISL_1064835, EPI_ISL_1064883, EPI_ISL_1064890, EPI_ISL_1064904, EPI_ISL_1200524, EPI_ISL_1200543, EPI_ISL_1200554, EPI_ISL_1200571, EPI_ISL_1200605, EPI_ISL_1261803, EPI_ISL_1261821, EPI_ISL_1261825, EPI_ISL_1261834, EPI_ISL_1261861, EPI_ISL_1337488, EPI_ISL_1593729 | | | |
| see above | DOHMH Crown Heights | New York City Public Health Laboratory | Jade Wang; et al. |
| EPI_ISL_1064762, EPI_ISL_1064771, EPI_ISL_1064780, EPI_ISL_1064785, EPI_ISL_1064790, EPI_ISL_1064794, EPI_ISL_1064836, EPI_ISL_1064885, EPI_ISL_1064891, EPI_ISL_1200523, EPI_ISL_1200620, EPI_ISL_1261809, EPI_ISL_1261818, EPI_ISL_1261847, EPI_ISL_1261850, EPI_ISL_1337468, EPI_ISL_1337471, EPI_ISL_1337510 | | | |
| see above | DOHMH Fort Greene | New York City Public Health Laboratory | Jade Wang; et al. |
| EPI_ISL_1017976, EPI_ISL_1017977, EPI_ISL_1017992, EPI_ISL_1017993, EPI_ISL_1017994, EPI_ISL_1018012, EPI_ISL_1018013, EPI_ISL_1018014, EPI_ISL_1018015, EPI_ISL_1018016, EPI_ISL_1018017, EPI_ISL_1018018, EPI_ISL_1018019, EPI_ISL_1018020, EPI_ISL_1018022, EPI_ISL_1018023, EPI_ISL_1064755, EPI_ISL_1064759, EPI_ISL_1064781, EPI_ISL_1064788, EPI_ISL_1064807, EPI_ISL_1064819, EPI_ISL_1064830, EPI_ISL_1064837, EPI_ISL_1064838, EPI_ISL_1064839, EPI_ISL_1064840, EPI_ISL_1064848, EPI_ISL_1064858, EPI_ISL_1064865, EPI_ISL_1064866, EPI_ISL_1064867, EPI_ISL_1064868, EPI_ISL_1064869, EPI_ISL_1064884, EPI_ISL_1064886, EPI_ISL_1064892, EPI_ISL_1064906, EPI_ISL_1064907, EPI_ISL_1200520, EPI_ISL_1200521, EPI_ISL_1200535, EPI_ISL_1200536, EPI_ISL_1200542, EPI_ISL_1200544, EPI_ISL_1200547, EPI_ISL_1200551, EPI_ISL_1200553, EPI_ISL_1200563, EPI_ISL_1200572, EPI_ISL_1200573, EPI_ISL_1200581, EPI_ISL_1200583, EPI_ISL_1200585, EPI_ISL_1200590, EPI_ISL_1200595, EPI_ISL_1200596, EPI_ISL_1200615, EPI_ISL_1200616, EPI_ISL_1200617, EPI_ISL_1200618, EPI_ISL_1261790, EPI_ISL_1261791, EPI_ISL_1261792, EPI_ISL_1261793, EPI_ISL_1261794, EPI_ISL_1261802, EPI_ISL_1261815, EPI_ISL_1261820, EPI_ISL_1261822, EPI_ISL_1261831, EPI_ISL_1261857, EPI_ISL_1261860, EPI_ISL_1261866, EPI_ISL_1337501, EPI_ISL_1337525, EPI_ISL_1593730 | | | |
| see above | DOHMH Jamaica | New York City Public Health Laboratory | Jade Wang; et al. |
| EPI_ISL_1017974, EPI_ISL_1017980, EPI_ISL_1017981, EPI_ISL_1018000, EPI_ISL_1018006, EPI_ISL_1018007, EPI_ISL_1018008, EPI_ISL_1018009, EPI_ISL_1018010, EPI_ISL_1018025, EPI_ISL_1018026, EPI_ISL_1064787, EPI_ISL_1064801, EPI_ISL_1064802, EPI_ISL_1064812, EPI_ISL_1064834, EPI_ISL_1064841, EPI_ISL_1064842, EPI_ISL_1064849, EPI_ISL_1064850, EPI_ISL_1064861, EPI_ISL_1064862, EPI_ISL_1064875, EPI_ISL_1200550, EPI_ISL_1200557, EPI_ISL_1200558, EPI_ISL_1200559, EPI_ISL_1200567, EPI_ISL_1200575, EPI_ISL_1200578, EPI_ISL_1200579, EPI_ISL_1200580, EPI_ISL_1200582, EPI_ISL_1200588, EPI_ISL_1200589, EPI_ISL_1200600, EPI_ISL_1200604, EPI_ISL_1200612, EPI_ISL_1261814, EPI_ISL_1261826, EPI_ISL_1261838, EPI_ISL_1261839, EPI_ISL_1261848, EPI_ISL_1261851, EPI_ISL_1337463, EPI_ISL_1337473, EPI_ISL_1337481, EPI_ISL_1337485, EPI_ISL_1337523 | | | |

| | | | |
|---|---|---|---|
| see above | DOHMH Morrisania | New York City Public Health Laboratory | Jade Wang; et al. |
| EPI_ISL_1017969, EPI_ISL_1017970, EPI_ISL_1017979, EPI_ISL_1017991, EPI_ISL_1018002, EPI_ISL_1018003, EPI_ISL_1018004, EPI_ISL_1064756, EPI_ISL_1064769, EPI_ISL_1064799, EPI_ISL_1064805, EPI_ISL_1064806, EPI_ISL_1064821, EPI_ISL_1064822, EPI_ISL_1064826, EPI_ISL_1064859, EPI_ISL_1064860, EPI_ISL_1064887, EPI_ISL_1064894, EPI_ISL_1200538, EPI_ISL_1200548, EPI_ISL_1200566, EPI_ISL_1200586, EPI_ISL_1200587, EPI_ISL_1200601, EPI_ISL_1200602, EPI_ISL_1200619, EPI_ISL_1200623, EPI_ISL_1261798, EPI_ISL_1261805, EPI_ISL_1261823, EPI_ISL_1337509 | | | |
| see above | DOHMH PHL | New York City Public Health Laboratory | Jade Wang; et al. |
| EPI_ISL_1017971, EPI_ISL_1017972, EPI_ISL_1017973, EPI_ISL_1064823, EPI_ISL_1064908, EPI_ISL_1200525, EPI_ISL_1200537, EPI_ISL_1200606, EPI_ISL_1200611, EPI_ISL_1261795, EPI_ISL_1261799, EPI_ISL_1261816, EPI_ISL_1261840, EPI_ISL_1261841, EPI_ISL_1261842, EPI_ISL_1261845, EPI_ISL_1261849, EPI_ISL_1337526, EPI_ISL_1337543 | | | |
| see above | DOHMH Riverside | New York City Public Health Laboratory | Jade Wang; et al. |
| EPI_ISL_1121893, EPI_ISL_1121900, EPI_ISL_1121945, EPI_ISL_1121946, EPI_ISL_1121950 | Department of Health & Mental Hygiene Central Harlem | New York City Public Health Laboratory | Jade Wang; et al. |
| EPI_ISL_1121925, EPI_ISL_1121927, EPI_ISL_1121928, EPI_ISL_1121960 | Department of Health & Mental Hygiene Chelsea | New York City Public Health Laboratory | Jade Wang; et al. |
| EPI_ISL_1121887, EPI_ISL_1121899, EPI_ISL_1121905, EPI_ISL_1121906, EPI_ISL_1121907, EPI_ISL_1121908, EPI_ISL_1121932, EPI_ISL_1121933, EPI_ISL_1121934, EPI_ISL_1121935, EPI_ISL_1121936 | | | |
| see above | Department of Health & Mental Hygiene Corona | New York City Public Health Laboratory | Jade Wang; et al. |
| EPI_ISL_1121890, EPI_ISL_1121891, EPI_ISL_1121892, EPI_ISL_1121904, EPI_ISL_1121938, EPI_ISL_1121939 | Department of Health & Mental Hygiene Crown Heights | New York City Public Health Laboratory | Jade Wang; et al. |
| EPI_ISL_1121889, EPI_ISL_1121901, EPI_ISL_1121902, EPI_ISL_1121903, EPI_ISL_1121937 | Department of Health & Mental Hygiene Fort Greene | New York City Public Health Laboratory | Jade Wang; et al. |
| EPI_ISL_1121915, EPI_ISL_1121916, EPI_ISL_1121917, EPI_ISL_1121918, EPI_ISL_1121924, EPI_ISL_1121951, EPI_ISL_1121952, EPI_ISL_1121953, EPI_ISL_1121954, EPI_ISL_1121955, EPI_ISL_1121956, EPI_ISL_1121957, EPI_ISL_1121958, EPI_ISL_1121959 | | | |
| see above | Department of Health & Mental Hygiene Jamaica | New York City Public Health Laboratory | Jade Wang; et al. |
| EPI_ISL_1121888, EPI_ISL_1121913, EPI_ISL_1121919, EPI_ISL_1121920, EPI_ISL_1121921, EPI_ISL_1121922, EPI_ISL_1121923, EPI_ISL_1121926, EPI_ISL_1121940, EPI_ISL_1121941, EPI_ISL_1121942, EPI_ISL_1121944, EPI_ISL_1121947, EPI_ISL_1121948, EPI_ISL_1121949 | | | |
| see above | Department of Health & Mental Hygiene Morrisania | New York City Public Health Laboratory | Jade Wang; et al. |
| EPI_ISL_1121894, EPI_ISL_1121909, EPI_ISL_1121910, EPI_ISL_1121911, EPI_ISL_1121912, EPI_ISL_1121929, EPI_ISL_1121930, EPI_ISL_1121931 | | | |
| see above | Department of Health & Mental Hygiene PHL | New York City Public Health Laboratory | Jade Wang; et al. |
| EPI_ISL_1121914, EPI_ISL_1121943 | Department of Health & Mental Hygiene Riverside | New York City Public Health Laboratory | Jade Wang; et al. |
| EPI_ISL_1114470, EPI_ISL_1114471, EPI_ISL_1114492, EPI_ISL_1114493 | Department of Health and Mental Hygiene Central Harlem | New York City Public Health Laboratory | Jade Wang; et al. |
| EPI_ISL_1114500, EPI_ISL_1114501, EPI_ISL_1114509, EPI_ISL_1114524 | Department of Health and Mental Hygiene Chelsea | New York City Public Health Laboratory | Jade Wang; et al. |
| EPI_ISL_1114472, EPI_ISL_1114473, EPI_ISL_1114474, EPI_ISL_1114475, EPI_ISL_1114504, EPI_ISL_1114505, EPI_ISL_1114506, EPI_ISL_1114507, EPI_ISL_1114513, EPI_ISL_1114514, EPI_ISL_1114515 | | | |
| see above | Department of Health and Mental Hygiene Corona | New York City Public Health Laboratory | Jade Wang; et al. |
| EPI_ISL_1114485, EPI_ISL_1114499, EPI_ISL_1114502, EPI_ISL_1114516, EPI_ISL_1114517 | Department of Health and Mental Hygiene Crown Heights | New York City Public Health Laboratory | Jade Wang; et al. |
| EPI_ISL_1114494, EPI_ISL_1114495, EPI_ISL_1114496 | Department of Health and Mental Hygiene Fort Greene | New York City Public Health Laboratory | Jade Wang; et al. |
| EPI_ISL_1114477, EPI_ISL_1114478, EPI_ISL_1114479, EPI_ISL_1114480, EPI_ISL_1114481, EPI_ISL_1114508, EPI_ISL_1114518 | | | |
| see above | Department of Health and Mental Hygiene Jamaica | New York City Public Health Laboratory | Jade Wang; et al. |
| EPI_ISL_1114487, EPI_ISL_1114488, EPI_ISL_1114489, EPI_ISL_1114490, EPI_ISL_1114491 | Department of Health and Mental Hygiene Morrisania | New York City Public Health Laboratory | Jade Wang; et al. |
| EPI_ISL_1114469, EPI_ISL_1114476, EPI_ISL_1114482, EPI_ISL_1114483, EPI_ISL_1114484, EPI_ISL_1114486, EPI_ISL_1114510, EPI_ISL_1114519, EPI_ISL_1114520, EPI_ISL_1114521, EPI_ISL_1114522, EPI_ISL_1114523 | | | |
| see above | Department of Health and Mental Hygiene Public Health Laboratory | New York City Public Health Laboratory | Jade Wang; et al. |
| EPI_ISL_1114497, EPI_ISL_1114498, EPI_ISL_1114503 | Department of Health and Mental Hygiene Riverside | New York City Public Health Laboratory | Jade Wang; et al. |
| EPI_ISL_1017989, EPI_ISL_1017990, EPI_ISL_1064758, EPI_ISL_1064768, EPI_ISL_1064810, EPI_ISL_1064827, EPI_ISL_1064828, EPI_ISL_1064851, EPI_ISL_1064852, EPI_ISL_1064889, EPI_ISL_1064898, EPI_ISL_1114511, EPI_ISL_1114512, EPI_ISL_1200562, EPI_ISL_1200599, EPI_ISL_1261800, EPI_ISL_1261830, EPI_ISL_1261853, EPI_ISL_1261867, EPI_ISL_1337466, EPI_ISL_1337472 | | | |
| see above | Department of Homeless Services | New York City Public Health Laboratory | Jade Wang; et al. |
| EPI_ISL_1444909, EPI_ISL_1444910, EPI_ISL_1444911, EPI_ISL_1444913, EPI_ISL_1444914, EPI_ISL_1444915, EPI_ISL_1444916, EPI_ISL_1444917, EPI_ISL_1444918, EPI_ISL_1444923, EPI_ISL_1444924, EPI_ISL_1444925, EPI_ISL_1444926, EPI_ISL_1444927, EPI_ISL_1444928, EPI_ISL_1444929, EPI_ISL_1444930, EPI_ISL_1444931, EPI_ISL_1444932, EPI_ISL_1444933, EPI_ISL_1444934, EPI_ISL_1444935, EPI_ISL_1444936, EPI_ISL_1444937, EPI_ISL_1444938, EPI_ISL_1444939, EPI_ISL_1444940, EPI_ISL_1444941, EPI_ISL_1444942, EPI_ISL_1444943, EPI_ISL_1444944, EPI_ISL_1444945, EPI_ISL_1444946, EPI_ISL_1444947, EPI_ISL_1444948, EPI_ISL_1444949, EPI_ISL_1444950, EPI_ISL_1444951, EPI_ISL_1444952, EPI_ISL_1444953, EPI_ISL_1444954, EPI_ISL_1444955, EPI_ISL_1444956, EPI_ISL_1444957, EPI_ISL_1444958, EPI_ISL_1444959, EPI_ISL_1444960, EPI_ISL_1444961, EPI_ISL_1444962, EPI_ISL_1444963, EPI_ISL_1444964, EPI_ISL_1444965, EPI_ISL_1444966, EPI_ISL_1444967, EPI_ISL_1444969, EPI_ISL_1444970, EPI_ISL_1444971, EPI_ISL_1444972, EPI_ISL_1444973, EPI_ISL_1444974, EPI_ISL_1444975, EPI_ISL_1444976, EPI_ISL_1444977, EPI_ISL_1444978, EPI_ISL_1444979, EPI_ISL_1444980, EPI_ISL_1444981, EPI_ISL_1444982, EPI_ISL_1444983, EPI_ISL_1444984, EPI_ISL_1444985, EPI_ISL_1444986, EPI_ISL_1444987, EPI_ISL_1444988, EPI_ISL_1444989, EPI_ISL_1444990, EPI_ISL_1444991, EPI_ISL_1444992, EPI_ISL_1444993, EPI_ISL_1444994, EPI_ISL_1444995, EPI_ISL_1444996, EPI_ISL_1444997, EPI_ISL_1444998, EPI_ISL_1444999, EPI_ISL_1445000, EPI_ISL_1445001, EPI_ISL_1445002, EPI_ISL_1445003, EPI_ISL_1445004, EPI_ISL_1445005, EPI_ISL_1445006, EPI_ISL_1445007, EPI_ISL_1445008, EPI_ISL_1445009, EPI_ISL_1445010, EPI_ISL_1445011, EPI_ISL_1445012, EPI_ISL_1445013, EPI_ISL_1445014, EPI_ISL_1445015, EPI_ISL_1445016, EPI_ISL_1445017, EPI_ISL_1445018, EPI_ISL_1445019 | | | |
| see above | Erie County Public Health (ECPHL) | University at Buffalo Genomics and Bioinformatics Core | Alyssa Pohlman; Amanda Boccolucci; Brandon Marzullo; Donald Yergeau; Jennifer Surtees; Jonathan Bard; Natalie Lamb; Norma Nowak |
| EPI_ISL_1481835 | Fulgent Genetics | Centers for Disease Control and Prevention Division of Viral Diseases, Pathogen Discovery | Adrian Paskey; Becky Tsai; Benafsh Sapra; Benjamin Rambo-Martin; Christopher Gulvick; Clinton R. Paden; Dakota Howard; Darlene Wagner; Dhwani Batra; Doreen Ng; Duncan MacCannell; Harry Gao; James Xie; Jason Caravas; John Gao; Joseph Fierro; Kara Moser; Matthew Schmerer; Mickey Li; Peter W. Cook; Scott Sammons; Shatavia Morrison; Yan Meng; Yvette Unoarumhi |
| EPI_ISL_1228273, EPI_ISL_1228274, EPI_ISL_1228451, EPI_ISL_1228454, EPI_ISL_1228473, EPI_ISL_1228476, EPI_ISL_1397546, EPI_ISL_1397547, EPI_ISL_1397548, EPI_ISL_1397549, EPI_ISL_1397962, EPI_ISL_1397963, EPI_ISL_1397964, EPI_ISL_1397965, EPI_ISL_1398154, EPI_ISL_1398155, EPI_ISL_1398270, EPI_ISL_1461293, EPI_ISL_1465617, EPI_ISL_1755488 | | | |
| see above | GLENS FALLS HOSPITAL LABORATORY | Wadsworth Center, New York State Department of Health | Alexis Russel; Alexis Russell; Catharine Prussing; Daryl M. Lamson; Erasmus Schneider; Erica Lasek-Nesselquist; John Kelly; Jonathan Plitnick; Kirsten St. George; Matthew Shudt; Melissa A Leisner; Navjot Singh |

| | | | |
|---|---|---|---|
| EPI_ISL_1651225 | HOSPITAL FOR SPECIAL SURGERY | Wadsworth Center, New York State Department of Health | Alexis Russell; Catharine Prussing; Daryl M. Lamson; Erasmus Schneider; Erica Lasek-Nesselquist; John Kelly; Jonathan Plitnick; Kirsten St. George; Matthew Shudt; Melissa A Leisner; Navjot Singh |
| EPI_ISL_1200519 | HealthQuest Esoterics | New York City Public Health Laboratory | Jade Wang; et al. |
| EPI_ISL_1088840, EPI_ISL_1089387 | Helix / Illumina | Respiratory Viruses Branch, Division of Viral Diseases, Centers for Disease Control and Prevention | Alexandre Bolze; Ary Ascencio; Ben L. Rambo-Martin; Brad Sickler; Charlotte Rivera-Garcia; Christine Tran; Clinton R. Paden; Dakota Howard; David Becker; Dhwani Batra; Duncan MacCannell; Efren Sandoval; Eileen de Feo; Elizabeth Cirulli; Eric Allen; Geraint Levan; James Lu; Jan Antico; Jason Nguyen; Jimmy Ramirez; Jingtao Liu; Kelly Schiabor Barrett; Kim Gietzen; Magnus Isaksson; Marc Laurent; Matthew Tolentino; Nicole L. Washington; Peter W. Cook; Phil Febbo; Ryan Cho; Shannon Wickline; Sherry Wang; Simon White; Summer Galloway; Suxiang Tong; Tyler Cassens; William Lee |

EPI_ISL_1194777, EPI_ISL_1261929, EPI_ISL_1261938, EPI_ISL_1268329, EPI_ISL_1268980, EPI_ISL_1268992, EPI_ISL_1269067, EPI_ISL_1270016, EPI_ISL_1270051, EPI_ISL_1270165, EPI_ISL_1270213, EPI_ISL_1337041, EPI_ISL_1338149, EPI_ISL_1338229, EPI_ISL_1338263, EPI_ISL_1340378, EPI_ISL_1340381, EPI_ISL_1340392, EPI_ISL_1340441, EPI_ISL_1391780, EPI_ISL_1420931, EPI_ISL_1443911, EPI_ISL_1480025, EPI_ISL_1480043, EPI_ISL_1480059, EPI_ISL_1480155, EPI_ISL_1480290, EPI_ISL_1480429, EPI_ISL_1493799

| | | | |
|---|---|---|---|
| see above | Helix/Illumina | Centers for Disease Control and Prevention Division of Viral Diseases, Pathogen Discovery | Adrian Paskey; Alexandre Bolze; Ary Ascencio; Ben L. Rambo-Martin; Benjamin Rambo-Martin; Brad Sickler; Charlotte Rivera-Garcia; Christine Tran; Christopher Gulvick; Clinton R. Paden; Dakota Howard; Darlene Wagner; David Becker; Dhwani Batra; Duncan MacCannell; Efren Sandoval; Eileen de Feo; Elizabeth Cirulli; Eric Allen; Geraint Levan; James Lu; Jan Antico; Jason Caravas; Jason Nguyen; Jimmy Ramirez; Jingtao Liu; Kara Moser; Kelly Schiabor Barrett; Kim Gietzen; Magnus Isaksson; Marc Laurent; Matthew Schmerer; Matthew Tolentino; Nicole L. Washington; Peter W. Cook; Phil Febbo; Ryan Cho; Scott Sammons; Shannon Wickline; Shatavia Morrison; Sherry Wang; Simon White; Summer Galloway; Suxiang Tong; Tyler Cassens; William Lee; Yvette Unoarumhi |

EPI_ISL_1016565, EPI_ISL_1016602, EPI_ISL_1016643, EPI_ISL_1081047, EPI_ISL_1110651, EPI_ISL_1133941, EPI_ISL_1134090, EPI_ISL_1134455, EPI_ISL_1134696

| | | | |
|---|---|---|---|
| see above | Helix/Illumina | Respiratory Viruses Branch, Division of Viral Diseases, Centers for Disease Control and Prevention | Alexandre Bolze; Ary Ascencio; Ben L. Rambo-Martin; Brad Sickler; Charlotte Rivera-Garcia; Christine Tran; Clinton R. Paden; Dakota Howard; David Becker; Dhwani Batra; Duncan MacCannell; Efren Sandoval; Eileen de Feo; Elizabeth Cirulli; Eric Allen; Geraint Levan; James Lu; Jan Antico; Jason Nguyen; Jimmy Ramirez; Jingtao Liu; Kelly Schiabor Barrett; Kim Gietzen; Magnus Isaksson; Marc Laurent; Matthew Tolentino; Nicole L. Washington; Peter W. Cook; Phil Febbo; Ryan Cho; Shannon Wickline; Sherry Wang; Simon White; Summer Galloway; Suxiang Tong; Tyler Cassens; William Lee |
| EPI_ISL_1692677, EPI_ISL_1692705 | Infinity Biologix | Centers for Disease Control and Prevention Division of Viral Diseases, Pathogen Discovery | Adrian Paskey; Benjamin Rambo-Martin; Chirayu Goswami; Christian Bixby; Christopher Gulvick; Clinton R. Paden; Dakota Howard; Darlene Wagner; Dhwani Batra; Duncan MacCannell; Jason Caravas; Jonathan Schultz; Kara Moser; Matthew Schmerer; Peter W. Cook; Robin Grimwood; Russ Hager; Scott Sammons; Shatavia Morrison; Yihe Wang; Yvette Unoarumhi |

EPI_ISL_1226458, EPI_ISL_1226488, EPI_ISL_1226499, EPI_ISL_1226541, EPI_ISL_1226542, EPI_ISL_1226617, EPI_ISL_1226684, EPI_ISL_1226696, EPI_ISL_1226847, EPI_ISL_1226848, EPI_ISL_1227547, EPI_ISL_1227548, EPI_ISL_1227549, EPI_ISL_1227550, EPI_ISL_1227551, EPI_ISL_1227552, EPI_ISL_1227553, EPI_ISL_1227554, EPI_ISL_1227970, EPI_ISL_1227971, EPI_ISL_1227972, EPI_ISL_1228042

| | | | |
|---|---|---|---|
| see above | KALEIDA CENTER FOR LABORATORY MEDICINE | Wadsworth Center, New York State Department of Health | Alexis Russel; Daryl M. Lamson; Erasmus Schneider; Erica Lasek-Nesselquist; John Kelly; Jonathan Plitnick; Kirsten St. George; Matthew Shudt; Melissa A Leisner; Navjot Singh |

EPI_ISL_1193327, EPI_ISL_1193344, EPI_ISL_1193368, EPI_ISL_1220913, EPI_ISL_1220914, EPI_ISL_1220915, EPI_ISL_1220916, EPI_ISL_1220917, EPI_ISL_1220951, EPI_ISL_1220953, EPI_ISL_1221009, EPI_ISL_1221010, EPI_ISL_1221011, EPI_ISL_1221014, EPI_ISL_1221015, EPI_ISL_1221138, EPI_ISL_1221161, EPI_ISL_1221270, EPI_ISL_1221271, EPI_ISL_1221549, EPI_ISL_1221551, EPI_ISL_1221754, EPI_ISL_1221755, EPI_ISL_1221756, EPI_ISL_1221757, EPI_ISL_1221758, EPI_ISL_1221759, EPI_ISL_1221760, EPI_ISL_1221767, EPI_ISL_1221768, EPI_ISL_1221769, EPI_ISL_1221770, EPI_ISL_1221782, EPI_ISL_1222050, EPI_ISL_1222051, EPI_ISL_1222052, EPI_ISL_1222053, EPI_ISL_1222054, EPI_ISL_1222055, EPI_ISL_1222056, EPI_ISL_1222057, EPI_ISL_1222058, EPI_ISL_1222059, EPI_ISL_1222060, EPI_ISL_1222064, EPI_ISL_1222065, EPI_ISL_1222067, EPI_ISL_1222068, EPI_ISL_1222069, EPI_ISL_1222139, EPI_ISL_1222140, EPI_ISL_1222141, EPI_ISL_1222142, EPI_ISL_1222143, EPI_ISL_1291205, EPI_ISL_1291217, EPI_ISL_1291248, EPI_ISL_1291252, EPI_ISL_1291253, EPI_ISL_1298252, EPI_ISL_1298253, EPI_ISL_1298306, EPI_ISL_1298327, EPI_ISL_1298354, EPI_ISL_1298361, EPI_ISL_1319111, EPI_ISL_1319176, EPI_ISL_1319180, EPI_ISL_1319189, EPI_ISL_1319190, EPI_ISL_1319473, EPI_ISL_1319474, EPI_ISL_1319489, EPI_ISL_1319493, EPI_ISL_1319494, EPI_ISL_1319495, EPI_ISL_1319496, EPI_ISL_1319509, EPI_ISL_1319537, EPI_ISL_1319538, EPI_ISL_1319539, EPI_ISL_1319560, EPI_ISL_1319608, EPI_ISL_1319609, EPI_ISL_1319610, EPI_ISL_1319613, EPI_ISL_1319616, EPI_ISL_1319622, EPI_ISL_1319638, EPI_ISL_1319646, EPI_ISL_1319647, EPI_ISL_1319648, EPI_ISL_1319653, EPI_ISL_1319654, EPI_ISL_1319679, EPI_ISL_1319683, EPI_ISL_1319686, EPI_ISL_1319687, EPI_ISL_1319704, EPI_ISL_1319720, EPI_ISL_1319740, EPI_ISL_1319779, EPI_ISL_1319797, EPI_ISL_1319799, EPI_ISL_1319807, EPI_ISL_1319824, EPI_ISL_1319826, EPI_ISL_1319828, EPI_ISL_1319829, EPI_ISL_1319839, EPI_ISL_1319840, EPI_ISL_1319847, EPI_ISL_1319857, EPI_ISL_1319884, EPI_ISL_1319892, EPI_ISL_1319925, EPI_ISL_1319929, EPI_ISL_1319930, EPI_ISL_1319932, EPI_ISL_1319936, EPI_ISL_1319937, EPI_ISL_1319951, EPI_ISL_1319972, EPI_ISL_1320000, EPI_ISL_1320007, EPI_ISL_1320332, EPI_ISL_1320341, EPI_ISL_1320347, EPI_ISL_1320349, EPI_ISL_1320350, EPI_ISL_1320356, EPI_ISL_1320384, EPI_ISL_1320392, EPI_ISL_1320421, EPI_ISL_1320425, EPI_ISL_1320435, EPI_ISL_1320531, EPI_ISL_1320544, EPI_ISL_1320550, EPI_ISL_1320551, EPI_ISL_1320553, EPI_ISL_1320554, EPI_ISL_1320564, EPI_ISL_1320586, EPI_ISL_1320610, EPI_ISL_1320611, EPI_ISL_1320618, EPI_ISL_1320622, EPI_ISL_1320632, EPI_ISL_1320633, EPI_ISL_1320644, EPI_ISL_1320723, EPI_ISL_1320724, EPI_ISL_1320725, EPI_ISL_1320735, EPI_ISL_1320747, EPI_ISL_1320751, EPI_ISL_1320756, EPI_ISL_1320767, EPI_ISL_1320768, EPI_ISL_1320769, EPI_ISL_1320770, EPI_ISL_1320776, EPI_ISL_1320777, EPI_ISL_1320786, EPI_ISL_1320792, EPI_ISL_1320798, EPI_ISL_1320821, EPI_ISL_1320822, EPI_ISL_1320823, EPI_ISL_1320890, EPI_ISL_1320891, EPI_ISL_1320892, EPI_ISL_1320893, EPI_ISL_1320894, EPI_ISL_1320895, EPI_ISL_1320896, EPI_ISL_1320897, EPI_ISL_1320898, EPI_ISL_1320899, EPI_ISL_1320900, EPI_ISL_1320901, EPI_ISL_1320902, EPI_ISL_1320903, EPI_ISL_1320906, EPI_ISL_1320907, EPI_ISL_1320919, EPI_ISL_1320952, EPI_ISL_1320953, EPI_ISL_1320954, EPI_ISL_1320955, EPI_ISL_1320956, EPI_ISL_1320957, EPI_ISL_1320958, EPI_ISL_1320959, EPI_ISL_1320960, EPI_ISL_1320961, EPI_ISL_1320962, EPI_ISL_1320966, EPI_ISL_1320984, EPI_ISL_1321226, EPI_ISL_1338400, EPI_ISL_1338401, EPI_ISL_1338521, EPI_ISL_1338525, EPI_ISL_1338526, EPI_ISL_1338527, EPI_ISL_1338530, EPI_ISL_1338531, EPI_ISL_1338532, EPI_ISL_1338533, EPI_ISL_1338534, EPI_ISL_1338535, EPI_ISL_1338536, EPI_ISL_1338541, EPI_ISL_1338625, EPI_ISL_1338627, EPI_ISL_1338629, EPI_ISL_1338644, EPI_ISL_1338649, EPI_ISL_1338653, EPI_ISL_1338658, EPI_ISL_1338660, EPI_ISL_1338680, EPI_ISL_1338681, EPI_ISL_1338682, EPI_ISL_1338683, EPI_ISL_1338687, EPI_ISL_1338705, EPI_ISL_1338720, EPI_ISL_1338721, EPI_ISL_1338730, EPI_ISL_1338737, EPI_ISL_1338846, EPI_ISL_1338847, EPI_ISL_1339180, EPI_ISL_1339237, EPI_ISL_1339238, EPI_ISL_1339241, EPI_ISL_1339242, EPI_ISL_1339247, EPI_ISL_1339498, EPI_ISL_1339505, EPI_ISL_1339522, EPI_ISL_1339526, EPI_ISL_1339529, EPI_ISL_1339535, EPI_ISL_1339536, EPI_ISL_1339543, EPI_ISL_1339544, EPI_ISL_1339573, EPI_ISL_1339577, EPI_ISL_1339584, EPI_ISL_1339585, EPI_ISL_1339591, EPI_ISL_1339628, EPI_ISL_1339638, EPI_ISL_1339639, EPI_ISL_1339642, EPI_ISL_1421765, EPI_ISL_1421766, EPI_ISL_1421853, EPI_ISL_1421854, EPI_ISL_1421861, EPI_ISL_1421871, EPI_ISL_1421879, EPI_ISL_1421885, EPI_ISL_1421886, EPI_ISL_1421894, EPI_ISL_1421895, EPI_ISL_1421897, EPI_ISL_1421925, EPI_ISL_1421926, EPI_ISL_1421927, EPI_ISL_1421931, EPI_ISL_1421934, EPI_ISL_1421939, EPI_ISL_1421941, EPI_ISL_1421942, EPI_ISL_1421955, EPI_ISL_1421956, EPI_ISL_1421958, EPI_ISL_1421961, EPI_ISL_1421965, EPI_ISL_1421967, EPI_ISL_1421976, EPI_ISL_1421977, EPI_ISL_1421995, EPI_ISL_1421997, EPI_ISL_1422001, EPI_ISL_1422004, EPI_ISL_1422025, EPI_ISL_1422044, EPI_ISL_1422046, EPI_ISL_1422077, EPI_ISL_1422083, EPI_ISL_1422086, EPI_ISL_1422094, EPI_ISL_1422095, EPI_ISL_1422106, EPI_ISL_1422107, EPI_ISL_1422108, EPI_ISL_1422109, EPI_ISL_1422110, EPI_ISL_1422111, EPI_ISL_1422112, EPI_ISL_1422113, EPI_ISL_1422114, EPI_ISL_1422115, EPI_ISL_1422116, EPI_ISL_1422146, EPI_ISL_1422147, EPI_ISL_1422148, EPI_ISL_1422182, EPI_ISL_1422186, EPI_ISL_1422187, EPI_ISL_1461826, EPI_ISL_1461842, EPI_ISL_1462357, EPI_ISL_1462358, EPI_ISL_1462359, EPI_ISL_1462360, EPI_ISL_1462361, EPI_ISL_1462362, EPI_ISL_1462363, EPI_ISL_1462371, EPI_ISL_1462372, EPI_ISL_1462375, EPI_ISL_1462376, EPI_ISL_1462377, EPI_ISL_1462426, EPI_ISL_1462427, EPI_ISL_1462428, EPI_ISL_1462433, EPI_ISL_1462434, EPI_ISL_1463082, EPI_ISL_1480977, EPI_ISL_1481057, EPI_ISL_1481058, EPI_ISL_1481061, EPI_ISL_1481067, EPI_ISL_1481105, EPI_ISL_1481110, EPI_ISL_1481111, EPI_ISL_1481132, EPI_ISL_1481136, EPI_ISL_1481137, EPI_ISL_1481154, EPI_ISL_1481182, EPI_ISL_1481187, EPI_ISL_1481373, EPI_ISL_1481388, EPI_ISL_1481392, EPI_ISL_1481396, EPI_ISL_1481398, EPI_ISL_1481400, EPI_ISL_1481404, EPI_ISL_1481405, EPI_ISL_1481409, EPI_ISL_1481429, EPI_ISL_1481450, EPI_ISL_1481451, EPI_ISL_1481475, EPI_ISL_1481480, EPI_ISL_1481481, EPI_ISL_1548152, EPI_ISL_1548166, EPI_ISL_1548438, EPI_ISL_1548439, EPI_ISL_1548440, EPI_ISL_1548441, EPI_ISL_1548442, EPI_ISL_1548445, EPI_ISL_1548446, EPI_ISL_1548447, EPI_ISL_1548474, EPI_ISL_1548479

| | | | |
|---|---|---|---|
| see above | Laboratory Corporation of America | Centers for Disease Control and Prevention Division of Viral Diseases, Pathogen Discovery | Adrian Paskey; Amanda Douglas; Amanda Suchanek; Andrea Throop; Ayla Burns; Ben L. Rambo-Martin; Benjamin Rambo-Martin; Bobbi Croy; Brian Krueger; Brian Norvell; Christopher Gulvick; Christos Petropoulos; Clinton R. Paden; Craig Lukasik; Dakota Howard; Darlene Wagner; Debbie Boles; Dhwani Batra; Duncan MacCannell; Eyad Almasri; Eyad Almasri Debbie Boles; Goran Stevovic; Howard Engler; Hrushikesh Deshmukh; Jake Humphrey; Jana Schroth; Jason Caravas; Joe Voshell; John Pruitt; Jonathan Meltzer; Jonathan Williams; Kara Moser; Kimberly Wagner; Lax Iyer; Lyndon Tilson; Manoj Jain; Marcia Eisenberg; Mary Ann Cristobal; Mary Williamson; Matthew Schmerer; Michael Levandoski; Mike Sapeta; Mindy Nye; Minoo Agarwal; Mohan Kolli; Nuthawin Charoensri; Oren Cohen; Peter W. Cook; Prashant Gupta; Qian Zeng; Rama Ghatti; Scott Parker; Scott Ryan; Scott Sammons; Shatavia Morrison; Stanley Letovsky; Steven Ragan; Suresh Babu Selvaraju; Susan Countryman; Susan Hicks; Suxiang Tong; Suzanne Dale; Thomas Urban; Tim Kuphal; Tricia Zwiefelhofer; Vincent Drouillon; Yvette Unoarumhi |

EPI_ISL_1037306, EPI_ISL_1037640, EPI_ISL_1037650, EPI_ISL_1037790, EPI_ISL_1038074, EPI_ISL_1038179, EPI_ISL_1038223, EPI_ISL_1038235, EPI_ISL_1038406, EPI_ISL_1038784, EPI_ISL_1038785, EPI_ISL_1159904, EPI_ISL_1159919, EPI_ISL_1159978, EPI_ISL_1160029, EPI_ISL_1160030, EPI_ISL_1160031, EPI_ISL_1160038, EPI_ISL_1160045, EPI_ISL_1160097, EPI_ISL_1160115, EPI_ISL_1160181, EPI_ISL_1160251, EPI_ISL_1160298, EPI_ISL_1160318, EPI_ISL_1160325, EPI_ISL_1160377, EPI_ISL_1160400, EPI_ISL_1160444, EPI_ISL_1160465, EPI_ISL_1160491, EPI_ISL_1160494, EPI_ISL_1160513, EPI_ISL_1160545, EPI_ISL_1160546, EPI_ISL_1160556, EPI_ISL_1160565, EPI_ISL_1160590, EPI_ISL_1160593, EPI_ISL_1160600, EPI_ISL_1160628, EPI_ISL_1160656, EPI_ISL_1160697, EPI_ISL_1160700, EPI_ISL_1160705, EPI_ISL_1160708, EPI_ISL_1160711, EPI_ISL_1160732, EPI_ISL_1160733, EPI_ISL_1160735, EPI_ISL_1160751, EPI_ISL_1160802, EPI_ISL_1160815, EPI_ISL_1160844, EPI_ISL_1160846, EPI_ISL_1160847, EPI_ISL_1160905, EPI_ISL_1160914, EPI_ISL_1160919, EPI_ISL_1160925, EPI_ISL_1160943, EPI_ISL_1160944, EPI_ISL_1160953, EPI_ISL_1160966, EPI_ISL_1161138, EPI_ISL_1161139, EPI_ISL_1161140, EPI_ISL_1161141, EPI_ISL_1161144, EPI_ISL_1161146, EPI_ISL_1161157, EPI_ISL_1161168, EPI_ISL_1161190, EPI_ISL_1161363, EPI_ISL_1161367, EPI_ISL_1161380, EPI_ISL_1161381, EPI_ISL_1161382, EPI_ISL_1161440, EPI_ISL_1161448, EPI_ISL_1161451, EPI_ISL_1161468, EPI_ISL_1161473, EPI_ISL_1161530, EPI_ISL_1161558, EPI_ISL_1161621, EPI_ISL_1161627, EPI_ISL_1161631, EPI_ISL_1161673, EPI_ISL_1161680, EPI_ISL_1161690, EPI_ISL_1161764, EPI_ISL_1161777, EPI_ISL_1161778, EPI_ISL_1161780, EPI_ISL_1161782, EPI_ISL_1161800, EPI_ISL_1161808, EPI_ISL_1161818, EPI_ISL_1161824, EPI_ISL_1161828, EPI_ISL_1161890, EPI_ISL_1161909, EPI_ISL_1161921, EPI_ISL_1161941, EPI_ISL_1161956, EPI_ISL_1162030, EPI_ISL_1162073, EPI_ISL_1162080, EPI_ISL_1162091, EPI_ISL_1162111, EPI_ISL_1162132, EPI_ISL_1162147, EPI_ISL_1162153, EPI_ISL_1162165, EPI_ISL_1162230, EPI_ISL_1162268, EPI_ISL_1162277, EPI_ISL_1162294, EPI_ISL_1162297, EPI_ISL_1162306, EPI_ISL_1162348, EPI_ISL_1162387, EPI_ISL_1162403, EPI_ISL_1162478, EPI_ISL_1162484, EPI_ISL_1162519, EPI_ISL_1162531, EPI_ISL_1162532, EPI_ISL_1162603, EPI_ISL_1162626, EPI_ISL_1162663, EPI_ISL_1162664, EPI_ISL_1162682, EPI_ISL_1162685, EPI_ISL_1162699, EPI_ISL_1162773, EPI_ISL_1162784, EPI_ISL_1162806, EPI_ISL_1162816, EPI_ISL_1162827, EPI_ISL_1162836, EPI_ISL_1162845, EPI_ISL_1162860, EPI_ISL_1162881, EPI_ISL_1162883, EPI_ISL_1162912, EPI_ISL_1162922, EPI_ISL_1162928, EPI_ISL_1162942, EPI_ISL_1162953, EPI_ISL_1162996, EPI_ISL_1163009, EPI_ISL_1163010, EPI_ISL_1163012, EPI_ISL_1163024, EPI_ISL_1163043, EPI_ISL_1163082, EPI_ISL_1163132, EPI_ISL_1163158, EPI_ISL_1163161, EPI_ISL_1163198, EPI_ISL_1163227, EPI_ISL_1163228, EPI_ISL_1163229, EPI_ISL_1163234, EPI_ISL_1163235, EPI_ISL_1163268, EPI_ISL_1163272, EPI_ISL_1163292, EPI_ISL_1163293, EPI_ISL_1163294, EPI_ISL_1163295, EPI_ISL_1163349, EPI_ISL_1163373, EPI_ISL_1163387, EPI_ISL_1163393, EPI_ISL_1163440, EPI_ISL_1163441, EPI_ISL_1163450, EPI_ISL_1163463, EPI_ISL_1163483, EPI_ISL_1255301

| | | | |
|---|---|---|---|
| see above | Laboratory Corporation of America | Respiratory Viruses Branch, Division of Viral Diseases, Centers for Disease Control and Prevention | Amanda Douglas; Amanda Suchanek; Andrea Throop; Ayla Burns; Ben L. Rambo-Martin; Bobbi Croy; Brian Krueger; Brian Norvell; Christos Petropoulos; Clinton R. Paden; Craig Lukasik; Dakota Howard; Dhwani Batra; Duncan MacCannell; Eyad Almasri Debbie Boles; Goran Stevovic; Howard Engler; Hrushikesh Deshmukh; Jake Humphrey; Jana Schroth; Joe Voshell; John Pruitt; Jonathan Meltzer; Jonathan Williams; Kimberly Wagner; Lax Iyer; Lyndon Tilson; Manoj Jain; Marcia Eisenberg; Mary Ann Cristobal; Mary Williamson; Michael Levandoski; Mike Sapeta; Mindy Nye; Minoo Agarwal; Mohan Kolli; Nuthawin Charoensri; Oren Cohen; Peter W. Cook; Prashant Gupta; Qian Zeng; Rama Ghatti; Scott Parker; Scott Ryan; Stanley Letovsky; Steven Ragan; Suresh Babu Selvaraju; Susan Countryman; Susan Hicks; Suxiang Tong; Suzanne Dale; Thomas Urban; Tim Kuphal; Tricia Zwiefelhofer; Vincent Drouillon |

EPI_ISL_1016172, EPI_ISL_1016173, EPI_ISL_1016174, EPI_ISL_1016175, EPI_ISL_1016176, EPI_ISL_1016177, EPI_ISL_1016178, EPI_ISL_1016179, EPI_ISL_1016180, EPI_ISL_1016182, EPI_ISL_1016185, EPI_ISL_1016186, EPI_ISL_1016188, EPI_ISL_1016192, EPI_ISL_1016193, EPI_ISL_1016194, EPI_ISL_1016195, EPI_ISL_1016196, EPI_ISL_1016197, EPI_ISL_1016198, EPI_ISL_1016199, EPI_ISL_1016200, EPI_ISL_1016206, EPI_ISL_1016209, EPI_ISL_1016211, EPI_ISL_1016214, EPI_ISL_1016219, EPI_ISL_1226424, EPI_ISL_1226426, EPI_ISL_1226446, EPI_ISL_1226454, EPI_ISL_1226457, EPI_ISL_1226465, EPI_ISL_1226489, EPI_ISL_1226498, EPI_ISL_1226509, EPI_ISL_1226515, EPI_ISL_1226523, EPI_ISL_1226531, EPI_ISL_1226532, EPI_ISL_1226573, EPI_ISL_1226576, EPI_ISL_1226594, EPI_ISL_1226606, EPI_ISL_1226607, EPI_ISL_1226608, EPI_ISL_1226609, EPI_ISL_1226610, EPI_ISL_1226611, EPI_ISL_1226612, EPI_ISL_1226613, EPI_ISL_1226614, EPI_ISL_1226654, EPI_ISL_1226681, EPI_ISL_1226690, EPI_ISL_1226691, EPI_ISL_1226715, EPI_ISL_1226734, EPI_ISL_1226735, EPI_ISL_1226738, EPI_ISL_1226751, EPI_ISL_1226752, EPI_ISL_1226753, EPI_ISL_1226754, EPI_ISL_1226755, EPI_ISL_1226758, EPI_ISL_1226760, EPI_ISL_1226761, EPI_ISL_1226762, EPI_ISL_1226763, EPI_ISL_1226764, EPI_ISL_1226766, EPI_ISL_1226767, EPI_ISL_1226770, EPI_ISL_1226771, EPI_ISL_1226773, EPI_ISL_1226774, EPI_ISL_1226775, EPI_ISL_1226776, EPI_ISL_1226777, EPI_ISL_1226778, EPI_ISL_1226779, EPI_ISL_1226780, EPI_ISL_1226831, EPI_ISL_1226835, EPI_ISL_1226839, EPI_ISL_1226844, EPI_ISL_1226883, EPI_ISL_1226884, EPI_ISL_1226885, EPI_ISL_1226886, EPI_ISL_1226887, EPI_ISL_1226888, EPI_ISL_1226889, EPI_ISL_1226890, EPI_ISL_1226931, EPI_ISL_1226936, EPI_ISL_1226937, EPI_ISL_1226950, EPI_ISL_1226951, EPI_ISL_1226952, EPI_ISL_1226953, EPI_ISL_1226954, EPI_ISL_1226955, EPI_ISL_1226956, EPI_ISL_1226957, EPI_ISL_1226958, EPI_ISL_1226959, EPI_ISL_1226960, EPI_ISL_1226961, EPI_ISL_1226962, EPI_ISL_1226964, EPI_ISL_1226965, EPI_ISL_1226966, EPI_ISL_1226967, EPI_ISL_1226968, EPI_ISL_1226969, EPI_ISL_1226971, EPI_ISL_1226972, EPI_ISL_1226974, EPI_ISL_1226975, EPI_ISL_1226976, EPI_ISL_1226977, EPI_ISL_1226978, EPI_ISL_1226979, EPI_ISL_1227018, EPI_ISL_1227021, EPI_ISL_1227022, EPI_ISL_1227024, EPI_ISL_1227025, EPI_ISL_1227050, EPI_ISL_1227051, EPI_ISL_1227052, EPI_ISL_1227053, EPI_ISL_1227054, EPI_ISL_1227055, EPI_ISL_1227056, EPI_ISL_1227057, EPI_ISL_1227058, EPI_ISL_1227059, EPI_ISL_1227060, EPI_ISL_1227061, EPI_ISL_1227062, EPI_ISL_1227063, EPI_ISL_1227064, EPI_ISL_1227065, EPI_ISL_1227066, EPI_ISL_1227067, EPI_ISL_1227070, EPI_ISL_1227071, EPI_ISL_1227072, EPI_ISL_1227073, EPI_ISL_1227074, EPI_ISL_1227075, EPI_ISL_1227076, EPI_ISL_1227077, EPI_ISL_1227078, EPI_ISL_1227079, EPI_ISL_1227080, EPI_ISL_1227082, EPI_ISL_1227083, EPI_ISL_1227084, EPI_ISL_1227085, EPI_ISL_1227086, EPI_ISL_1227090, EPI_ISL_1227091, EPI_ISL_1227092, EPI_ISL_1227093, EPI_ISL_1227094, EPI_ISL_1227097, EPI_ISL_1227098, EPI_ISL_1227099, EPI_ISL_1227100, EPI_ISL_1227101, EPI_ISL_1227102, EPI_ISL_1227103, EPI_ISL_1227104, EPI_ISL_1227105, EPI_ISL_1227106, EPI_ISL_1227107, EPI_ISL_1227108, EPI_ISL_1227110, EPI_ISL_1227111, EPI_ISL_1227112, EPI_ISL_1227113, EPI_ISL_1227114, EPI_ISL_1227115, EPI_ISL_1227116, EPI_ISL_1227117, EPI_ISL_1227118, EPI_ISL_1227119, EPI_ISL_1227120, EPI_ISL_1227121, EPI_ISL_1227122, EPI_ISL_1227123, EPI_ISL_1227124, EPI_ISL_1227125, EPI_ISL_1227126, EPI_ISL_1227127, EPI_ISL_1227128, EPI_ISL_1227150, EPI_ISL_1227226, EPI_ISL_1227227, EPI_ISL_1227387, EPI_ISL_1227388, EPI_ISL_1227389, EPI_ISL_1227390, EPI_ISL_1227391, EPI_ISL_1227392, EPI_ISL_1227393, EPI_ISL_1227394, EPI_ISL_1227395, EPI_ISL_1227396, EPI_ISL_1227397, EPI_ISL_1227398, EPI_ISL_1227399, EPI_ISL_1227400, EPI_ISL_1227403, EPI_ISL_1227404, EPI_ISL_1227405, EPI_ISL_1227411, EPI_ISL_1227412, EPI_ISL_1227413, EPI_ISL_1227414, EPI_ISL_1227415, EPI_ISL_1227418, EPI_ISL_1227419, EPI_ISL_1227420, EPI_ISL_1227421, EPI_ISL_1227424, EPI_ISL_1227425, EPI_ISL_1227426, EPI_ISL_1227427, EPI_ISL_1227430, EPI_ISL_1227431, EPI_ISL_1227432, EPI_ISL_1227433, EPI_ISL_1227434, EPI_ISL_1227436, EPI_ISL_1227437, EPI_ISL_1227438, EPI_ISL_1227439, EPI_ISL_1227442, EPI_ISL_1227443, EPI_ISL_1227444, EPI_ISL_1227445, EPI_ISL_1227446, EPI_ISL_1227705, EPI_ISL_1227959, EPI_ISL_1228176, EPI_ISL_1228188, EPI_ISL_1228266, EPI_ISL_1228560, EPI_ISL_1228567, EPI_ISL_1228569, EPI_ISL_1228572, EPI_ISL_1228578, EPI_ISL_1228584, EPI_ISL_1397618, EPI_ISL_1397619, EPI_ISL_1397620, EPI_ISL_1397621, EPI_ISL_1397622, EPI_ISL_1397788, EPI_ISL_1397789, EPI_ISL_1397790, EPI_ISL_1397791, EPI_ISL_1397792, EPI_ISL_1397793, EPI_ISL_1397794, EPI_ISL_1397795, EPI_ISL_1397796, EPI_ISL_1397797, EPI_ISL_1397798, EPI_ISL_1397799, EPI_ISL_1397800, EPI_ISL_1397801, EPI_ISL_1397802, EPI_ISL_1397803, EPI_ISL_1397804, EPI_ISL_1397805, EPI_ISL_1397806, EPI_ISL_1397807, EPI_ISL_1397808, EPI_ISL_1397809, EPI_ISL_1397870, EPI_ISL_1397871, EPI_ISL_1397872, EPI_ISL_1397873, EPI_ISL_1397874, EPI_ISL_1397875, EPI_ISL_1397876, EPI_ISL_1397877, EPI_ISL_1397878, EPI_ISL_1397880, EPI_ISL_1397881, EPI_ISL_1397882, EPI_ISL_1397883, EPI_ISL_1397884, EPI_ISL_1397885, EPI_ISL_1398034, EPI_ISL_1398035, EPI_ISL_1398036, EPI_ISL_1398037, EPI_ISL_1398038, EPI_ISL_1398048, EPI_ISL_1398049, EPI_ISL_1398050, EPI_ISL_1398051, EPI_ISL_1398052, EPI_ISL_1398053, EPI_ISL_1398054, EPI_ISL_1398055, EPI_ISL_1398056, EPI_ISL_1398057, EPI_ISL_1398058, EPI_ISL_1398059, EPI_ISL_1398060, EPI_ISL_1398061, EPI_ISL_1398062, EPI_ISL_1398063, EPI_ISL_1398064, EPI_ISL_1398065, EPI_ISL_1398066, EPI_ISL_1398067, EPI_ISL_1398120, EPI_ISL_1398121, EPI_ISL_1398122, EPI_ISL_1398132, EPI_ISL_1398133, EPI_ISL_1398134, EPI_ISL_1398135, EPI_ISL_1398136, EPI_ISL_1398335, EPI_ISL_1398339, EPI_ISL_1462551, EPI_ISL_1465703, EPI_ISL_1465704, EPI_ISL_1465705, EPI_ISL_1465706, EPI_ISL_1465707, EPI_ISL_1465708, EPI_ISL_1465794, EPI_ISL_1465795, EPI_ISL_1465796, EPI_ISL_1465797, EPI_ISL_1465798, EPI_ISL_1465799, EPI_ISL_1465800, EPI_ISL_1465801, EPI_ISL_1575470, EPI_ISL_1576819, EPI_ISL_1623953, EPI_ISL_1623954, EPI_ISL_1623955, EPI_ISL_1623956, EPI_ISL_1623957, EPI_ISL_1623958, EPI_ISL_1623959, EPI_ISL_1623960, EPI_ISL_1623961, EPI_ISL_1623962, EPI_ISL_1623963, EPI_ISL_1623964, EPI_ISL_1623965, EPI_ISL_1623966, EPI_ISL_1623967, EPI_ISL_1623968, EPI_ISL_1623969, EPI_ISL_1623970, EPI_ISL_1623971, EPI_ISL_1623972, EPI_ISL_1623975, EPI_ISL_1623976, EPI_ISL_1623977, EPI_ISL_1623978, EPI_ISL_1623979, EPI_ISL_1623980, EPI_ISL_1623981, EPI_ISL_1623982, EPI_ISL_1623983, EPI_ISL_1623984, EPI_ISL_1623985, EPI_ISL_1623986, EPI_ISL_1623987, EPI_ISL_1623988, EPI_ISL_1623989, EPI_ISL_1651218, EPI_ISL_1671145, EPI_ISL_1671156, EPI_ISL_1671158, EPI_ISL_1671159, EPI_ISL_1671163, EPI_ISL_1671164, EPI_ISL_1671169, EPI_ISL_1671170, EPI_ISL_1671171, EPI_ISL_1671174, EPI_ISL_1671175, EPI_ISL_1671181, EPI_ISL_1671182, EPI_ISL_1671185, EPI_ISL_1671186, EPI_ISL_1671187, EPI_ISL_1671188, EPI_ISL_1671189, EPI_ISL_1671190, EPI_ISL_1671191, EPI_ISL_1671192, EPI_ISL_1671203, EPI_ISL_1671204, EPI_ISL_1671208, EPI_ISL_1740503, EPI_ISL_1740505, EPI_ISL_1755836, EPI_ISL_1755837, EPI_ISL_1755948, EPI_ISL_1755949, EPI_ISL_1755950, EPI_ISL_1755951, EPI_ISL_1755952, EPI_ISL_1755953, EPI_ISL_1755954, EPI_ISL_1755955, EPI_ISL_1755956, EPI_ISL_1755957, EPI_ISL_1755958, EPI_ISL_1755959, EPI_ISL_1755960, EPI_ISL_1755961, EPI_ISL_1755962, EPI_ISL_1755963, EPI_ISL_1755964, EPI_ISL_1789710, EPI_ISL_1789711, EPI_ISL_1789715, EPI_ISL_1855103, EPI_ISL_1855188, EPI_ISL_1855189, EPI_ISL_1855190, EPI_ISL_1855191, EPI_ISL_1855192, EPI_ISL_1855193, EPI_ISL_1855194, EPI_ISL_1855195, EPI_ISL_1855196, EPI_ISL_1855202, EPI_ISL_1855203, EPI_ISL_1855204, EPI_ISL_1855205, EPI_ISL_1855206, EPI_ISL_1855207, EPI_ISL_1855208, EPI_ISL_1855209, EPI_ISL_1855217, EPI_ISL_1855218, EPI_ISL_1855219, EPI_ISL_1935610, EPI_ISL_1935615, EPI_ISL_1935643, EPI_ISL_2098130, EPI_ISL_2134791

| | | | |
|---|---|---|---|
| see above | MONTEFIORE MEDICAL CENTER LABORATORIES | Wadsworth Center, New York State Department of Health | Alexis Russel; Alexis Russell; Catharine Prussing; Daryl M. Lamson; Erasmus Schneider; Erica Lasek-Nesselquist; John Kelly; Jonathan Plitnick; Kirsten St. George; Matthew Shudt; Melissa A Leisner; Navjot Singh |
| EPI_ISL_1465802 | MONTEFIORE NEW ROCHELLE HOSPITAL LABORATORY | Wadsworth Center, New York State Department of Health | Alexis Russell; Catharine Prussing; Daryl M. Lamson; Erasmus Schneider; Erica Lasek-Nesselquist; John Kelly; Jonathan Plitnick; Kirsten St. George; Matthew Shudt; Melissa A Leisner; Navjot Singh |

EPI_ISL_1049482, EPI_ISL_1300757, EPI_ISL_1300762, EPI_ISL_1300768, EPI_ISL_1300770, EPI_ISL_1300781, EPI_ISL_1300782, EPI_ISL_1300783, EPI_ISL_1300785, EPI_ISL_1300791, EPI_ISL_1300794, EPI_ISL_1300797, EPI_ISL_1300798, EPI_ISL_1300804, EPI_ISL_1300805, EPI_ISL_1300806, EPI_ISL_1300807, EPI_ISL_1300808, EPI_ISL_1300809, EPI_ISL_1300810, EPI_ISL_1300811, EPI_ISL_1300818, EPI_ISL_1300819, EPI_ISL_1300820, EPI_ISL_1300844, EPI_ISL_1300845, EPI_ISL_1300849, EPI_ISL_1300864, EPI_ISL_1300865, EPI_ISL_1300866, EPI_ISL_1300867, EPI_ISL_1300878, EPI_ISL_1300902, EPI_ISL_1300903, EPI_ISL_1300904, EPI_ISL_1300905, EPI_ISL_1300906, EPI_ISL_1300907, EPI_ISL_1300909, EPI_ISL_1300910, EPI_ISL_1300911, EPI_ISL_1300912, EPI_ISL_1300913, EPI_ISL_1300914, EPI_ISL_1300915, EPI_ISL_1300916, EPI_ISL_1300917, EPI_ISL_1300918, EPI_ISL_1300919, EPI_ISL_1300920, EPI_ISL_1300921, EPI_ISL_1300922, EPI_ISL_1300923, EPI_ISL_1300924, EPI_ISL_1300925, EPI_ISL_1300926, EPI_ISL_1300927, EPI_ISL_1300928, EPI_ISL_1300929, EPI_ISL_1300930, EPI_ISL_1300934, EPI_ISL_1300935, EPI_ISL_1301403, EPI_ISL_1301404, EPI_ISL_1301405, EPI_ISL_1301406, EPI_ISL_1301407, EPI_ISL_1301408, EPI_ISL_1301409, EPI_ISL_1301410, EPI_ISL_1301411, EPI_ISL_1301412, EPI_ISL_1301413, EPI_ISL_1301414, EPI_ISL_1301415, EPI_ISL_1301416, EPI_ISL_1301417, EPI_ISL_1301418, EPI_ISL_1301419, EPI_ISL_1301420, EPI_ISL_1301421, EPI_ISL_1301422, EPI_ISL_1301423, EPI_ISL_1301424, EPI_ISL_1301425, EPI_ISL_1301426, EPI_ISL_1301427, EPI_ISL_1301428, EPI_ISL_1301429, EPI_ISL_1301430, EPI_ISL_1301431, EPI_ISL_1301432, EPI_ISL_1301433, EPI_ISL_1301434, EPI_ISL_1301435, EPI_ISL_1301436, EPI_ISL_1301437, EPI_ISL_1301438, EPI_ISL_1301439, EPI_ISL_1708951, EPI_ISL_1708952, EPI_ISL_1708953, EPI_ISL_1708954, EPI_ISL_1708955, EPI_ISL_1708956, EPI_ISL_1708957, EPI_ISL_1708958, EPI_ISL_1708959, EPI_ISL_1708960, EPI_ISL_1708961, EPI_ISL_1708962, EPI_ISL_1708963, EPI_ISL_1708964, EPI_ISL_1708965, EPI_ISL_1708966, EPI_ISL_1708967, EPI_ISL_1708968, EPI_ISL_1708969, EPI_ISL_1708970, EPI_ISL_1708971, EPI_ISL_1708972, EPI_ISL_1708973, EPI_ISL_1708974, EPI_ISL_1708975, EPI_ISL_1708976, EPI_ISL_1708977, EPI_ISL_1708978, EPI_ISL_1708979, EPI_ISL_1708980, EPI_ISL_1708981, EPI_ISL_1708982, EPI_ISL_1708983, EPI_ISL_1708984, EPI_ISL_1708985, EPI_ISL_1708986, EPI_ISL_1708987, EPI_ISL_1708988, EPI_ISL_1708989, EPI_ISL_1708990, EPI_ISL_1708991, EPI_ISL_1708992, EPI_ISL_1708993, EPI_ISL_1708994, EPI_ISL_1708995, EPI_ISL_1708996, EPI_ISL_1708997, EPI_ISL_1708998, EPI_ISL_1708999, EPI_ISL_1709000, EPI_ISL_1709001, EPI_ISL_1709002, EPI_ISL_1709003, EPI_ISL_1709004, EPI_ISL_1709005, EPI_ISL_1709006, EPI_ISL_1709007, EPI_ISL_1709008, EPI_ISL_1709009, EPI_ISL_1709010, EPI_ISL_1709011, EPI_ISL_1709012, EPI_ISL_1709013, EPI_ISL_1709014, EPI_ISL_1709015, EPI_ISL_1709016, EPI_ISL_1709017, EPI_ISL_1709018, EPI_ISL_1709019, EPI_ISL_1709020, EPI_ISL_1709021, EPI_ISL_1709022, EPI_ISL_1709023, EPI_ISL_1709024, EPI_ISL_1709025, EPI_ISL_1709026, EPI_ISL_1709027, EPI_ISL_1709028, EPI_ISL_1709029, EPI_ISL_1709030, EPI_ISL_1709031, EPI_ISL_1709032, EPI_ISL_1709033, EPI_ISL_1709034, EPI_ISL_1709035, EPI_ISL_1709036, EPI_ISL_1709037, EPI_ISL_1709038, EPI_ISL_1709039, EPI_ISL_1709040, EPI_ISL_1709041, EPI_ISL_1709042, EPI_ISL_1709043, EPI_ISL_1709044, EPI_ISL_1709045, EPI_ISL_1709046, EPI_ISL_1709048, EPI_ISL_1709049, EPI_ISL_1709050, EPI_ISL_1709051, EPI_ISL_1709052, EPI_ISL_1709053, EPI_ISL_1709054, EPI_ISL_1709055, EPI_ISL_1709056, EPI_ISL_1709057, EPI_ISL_1709058, EPI_ISL_1709059, EPI_ISL_1709060, EPI_ISL_1709061, EPI_ISL_1709062, EPI_ISL_1709063, EPI_ISL_1709064, EPI_ISL_1709065, EPI_ISL_1709066, EPI_ISL_1709067, EPI_ISL_1709068, EPI_ISL_1709069, EPI_ISL_1709070, EPI_ISL_1709071, EPI_ISL_1709072, EPI_ISL_1709073, EPI_ISL_1709074, EPI_ISL_1709075, EPI_ISL_1709076, EPI_ISL_1709077, EPI_ISL_1709078, EPI_ISL_1709079, EPI_ISL_1709081, EPI_ISL_1709082, EPI_ISL_1709083, EPI_ISL_1709084, EPI_ISL_1709085, EPI_ISL_1709086, EPI_ISL_1709087, EPI_ISL_1709088, EPI_ISL_1709089, EPI_ISL_1709090, EPI_ISL_1709091, EPI_ISL_1709092, EPI_ISL_1709093, EPI_ISL_1709094, EPI_ISL_1709095, EPI_ISL_1709096, EPI_ISL_1709097, EPI_ISL_1709098, EPI_ISL_1709099, EPI_ISL_1709100, EPI_ISL_1709101, EPI_ISL_1709102, EPI_ISL_1709103, EPI_ISL_1709104, EPI_ISL_1709105, EPI_ISL_1709106, EPI_ISL_1709107, EPI_ISL_1709108, EPI_ISL_1709109, EPI_ISL_1709110, EPI_ISL_1709111, EPI_ISL_1709112, EPI_ISL_1709113, EPI_ISL_1709114, EPI_ISL_1709115, EPI_ISL_1709116, EPI_ISL_1709117, EPI_ISL_1709118, EPI_ISL_1709119, EPI_ISL_1709120, EPI_ISL_1709121, EPI_ISL_1709122, EPI_ISL_1709123, EPI_ISL_1709124, EPI_ISL_1709125, EPI_ISL_1709126, EPI_ISL_1709127, EPI_ISL_1709129, EPI_ISL_1709130, EPI_ISL_1709131, EPI_ISL_1709132, EPI_ISL_1709133, EPI_ISL_1709134, EPI_ISL_1709135, EPI_ISL_1709136, EPI_ISL_1709137, EPI_ISL_1709138, EPI_ISL_1709139, EPI_ISL_1709140, EPI_ISL_1709141, EPI_ISL_1709142, EPI_ISL_1709143, EPI_ISL_1709144, EPI_ISL_1709145, EPI_ISL_1709146, EPI_ISL_1709147, EPI_ISL_1709148, EPI_ISL_1709149, EPI_ISL_1709150, EPI_ISL_1709151, EPI_ISL_1709152, EPI_ISL_1709153, EPI_ISL_1709154, EPI_ISL_1709155, EPI_ISL_1709156, EPI_ISL_1709157, EPI_ISL_1709158, EPI_ISL_1709159, EPI_ISL_1709160, EPI_ISL_1709161, EPI_ISL_1709162, EPI_ISL_1709163, EPI_ISL_1709164, EPI_ISL_1709165, EPI_ISL_1709166, EPI_ISL_1709167, EPI_ISL_1709168, EPI_ISL_1709169, EPI_ISL_1709170, EPI_ISL_1709171, EPI_ISL_1709172, EPI_ISL_1709173, EPI_ISL_1709174, EPI_ISL_1709175, EPI_ISL_1709176, EPI_ISL_1709177, EPI_ISL_1709178, EPI_ISL_1709179, EPI_ISL_1709180, EPI_ISL_1709181, EPI_ISL_1709182, EPI_ISL_1709183, EPI_ISL_1709184, EPI_ISL_1709185, EPI_ISL_1709186, EPI_ISL_1709187, EPI_ISL_1709188, EPI_ISL_1709189, EPI_ISL_1709190, EPI_ISL_1709191, EPI_ISL_1709192, EPI_ISL_1709193, EPI_ISL_1709194, EPI_ISL_1709195, EPI_ISL_1709196, EPI_ISL_1709197, EPI_ISL_1709198, EPI_ISL_1709199, EPI_ISL_1709200, EPI_ISL_1709201, EPI_ISL_1709202, EPI_ISL_1709203, EPI_ISL_1709204, EPI_ISL_1709205,

| | | | |
|---|---|---|---|
| EPI_ISL_1709206, EPI_ISL_1709207, EPI_ISL_1709208, EPI_ISL_1709209, EPI_ISL_1709210, EPI_ISL_1709211, EPI_ISL_1709212, EPI_ISL_1709213, EPI_ISL_1709214, EPI_ISL_1709215, EPI_ISL_1709216, EPI_ISL_1709217, EPI_ISL_1709218, EPI_ISL_1709219, EPI_ISL_1709220, EPI_ISL_1709221, EPI_ISL_1709222, EPI_ISL_1709223, EPI_ISL_1709224, EPI_ISL_1709225, EPI_ISL_1709226, EPI_ISL_1709227, EPI_ISL_1709228, EPI_ISL_1709229, EPI_ISL_1709230, EPI_ISL_1709231, EPI_ISL_1709232, EPI_ISL_1709233, EPI_ISL_1709234, EPI_ISL_1709235, EPI_ISL_1709236, EPI_ISL_1709237, EPI_ISL_1709238, EPI_ISL_1709239, EPI_ISL_1709240, EPI_ISL_1709241, EPI_ISL_1709242, EPI_ISL_1709243, EPI_ISL_1709244, EPI_ISL_1709245, EPI_ISL_1709246, EPI_ISL_1709247, EPI_ISL_1709248, EPI_ISL_1709249, EPI_ISL_1709250, EPI_ISL_1709251, EPI_ISL_1709252, EPI_ISL_1709253, EPI_ISL_1709254, EPI_ISL_1709255, EPI_ISL_1709256, EPI_ISL_1709257, EPI_ISL_1709258, EPI_ISL_1709259, EPI_ISL_1709260, EPI_ISL_1709261, EPI_ISL_1709262, EPI_ISL_1709263, EPI_ISL_1709264, EPI_ISL_1709265, EPI_ISL_1709266, EPI_ISL_1709267, EPI_ISL_1709268, EPI_ISL_1709269, EPI_ISL_1709270, EPI_ISL_1709271, EPI_ISL_1709272, EPI_ISL_1709273, EPI_ISL_1709274, EPI_ISL_1709275, EPI_ISL_1709276, EPI_ISL_1709277, EPI_ISL_1709278, EPI_ISL_1709280, EPI_ISL_1709281, EPI_ISL_1709282, EPI_ISL_1709283, EPI_ISL_1709284, EPI_ISL_1709285, EPI_ISL_1709286, EPI_ISL_1709287, EPI_ISL_1709288, EPI_ISL_1709289, EPI_ISL_1709290, EPI_ISL_1709291, EPI_ISL_1709292, EPI_ISL_1709293, EPI_ISL_1709294, EPI_ISL_1709295, EPI_ISL_1709296, EPI_ISL_1709297, EPI_ISL_1709298, EPI_ISL_1709299 | | | |
| see above | MSHS Clinical Microbiology Laboratories | MSHS Pathogen Surveillance Program | Adolfo García-Sastre; Adriana van de Guchte; Ajay Obla; Alberto Paniz-Mondolfi; Ana S. Gonzalez-Reiche; Angela Amoako; Ashley Salimbangon; Betsaida Salom Melo; Bremy Alburquerque; Brianne Ciferri; Charles Gleason; Daniel Floda; Deena R. Altman; Denise Jurczyszak; Emilia Mia Sordillo; Gintaras Deikus; Giulio Kleiner; Gopi Patel; Hala Alshammary; Harm van Bakel; Irina Oussenko; Jayeeta Dutta; Juan Soto; Julia Matthews; Katherine Beach; Kathryn Twyman; Kayla Russo; Komal Srivastava; Levy Sominsky; Mahmoud Awawda; Marta Luksza; Matthew M. Hernandez; Melissa Gitman; Michael D. Nowak; Mitchell J. Sullivan; Nancy Francoeur; Robert Sebra; Sarah Schaefer; Shelcie Fabre; Shwetha Hara Sridhar; Viviana Simon; Ying-Chih Wang; Zenab Khan |
| EPI_ISL_1597696, EPI_ISL_1597697, EPI_ISL_1597698, EPI_ISL_1597699, EPI_ISL_1597700, EPI_ISL_1597701, EPI_ISL_1597702, EPI_ISL_1597703, EPI_ISL_1597706, EPI_ISL_1597711, EPI_ISL_1597712, EPI_ISL_1597713, EPI_ISL_1597714, EPI_ISL_1597715, EPI_ISL_1597716, EPI_ISL_1597717, EPI_ISL_1597725, EPI_ISL_1597726, EPI_ISL_1597727, EPI_ISL_1597730, EPI_ISL_1597731, EPI_ISL_1597732, EPI_ISL_1597733, EPI_ISL_1597745, EPI_ISL_1597746, EPI_ISL_1597747, EPI_ISL_1597748, EPI_ISL_1597749, EPI_ISL_1597750, EPI_ISL_1597751, EPI_ISL_1597752, EPI_ISL_1597753, EPI_ISL_1597754, EPI_ISL_1597755, EPI_ISL_1597756, EPI_ISL_1597757, EPI_ISL_1597758, EPI_ISL_1597759, EPI_ISL_1597760, EPI_ISL_1597761, EPI_ISL_1597762, EPI_ISL_1597763, EPI_ISL_1597764, EPI_ISL_1597765, EPI_ISL_1597766, EPI_ISL_1597767, EPI_ISL_1597768, EPI_ISL_1597769, EPI_ISL_1597770, EPI_ISL_1597771, EPI_ISL_1597772, EPI_ISL_1597773, EPI_ISL_1597774, EPI_ISL_1597775, EPI_ISL_1597776, EPI_ISL_1597777, EPI_ISL_1597778, EPI_ISL_1597779, EPI_ISL_1597780, EPI_ISL_1597781 | | | |
| see above | Montefiore Medical Center | Abbott | Amy Fox; Ana Olivo; Ana Vallari; Barbara Harris; Gavin Cloherty; Mary Rodgers; Todd Meyer; Yitz Goldstein |
| EPI_ISL_1919998, EPI_ISL_1920001, EPI_ISL_1920074, EPI_ISL_1920075, EPI_ISL_1920077, EPI_ISL_1920084, EPI_ISL_1920085, EPI_ISL_1920086, EPI_ISL_1920089, EPI_ISL_1920090, EPI_ISL_1920091, EPI_ISL_1920092, EPI_ISL_1920096, EPI_ISL_1920159, EPI_ISL_1920161 | | | |
| see above | Montefiore Medical Center Laboratories | Wadsworth Center, New York State Department of Health | Alexis Russell; Catharine Prussing; Daryl M. Lamson; Erasmus Schneider; Erica Lasek-Nesselquist; John Kelly; Jonathan Plitnick; Kirsten St. George; Matthew Shudt; Melissa A Leisner; Navjot Singh |
| EPI_ISL_1182091, EPI_ISL_1182092, EPI_ISL_1225664, EPI_ISL_1225678, EPI_ISL_1225699, EPI_ISL_1378825, EPI_ISL_1378826 | | | |
| see above | Murphy Medical Association | Grubaugh Lab - Yale School of Public Health | Anderson Brito; Annie Watkins; Caleb Neal; Chaney Kalinich; Chantal Vogels; Eva Laszlo; Isabell Ott; Jessica Rothman; Joseph Fauver; Mallery Breban; Mary Petrone; Nathan Grubaugh; Steven Murphy; Tara Alpert |
| EPI_ISL_1226420, EPI_ISL_1226798, EPI_ISL_1226815, EPI_ISL_1227014, EPI_ISL_1227026, EPI_ISL_1227870, EPI_ISL_1228174, EPI_ISL_1228175, EPI_ISL_1395750, EPI_ISL_1395753, EPI_ISL_1395754, EPI_ISL_1395756, EPI_ISL_1395757, EPI_ISL_1395758, EPI_ISL_1395760, EPI_ISL_1395763, EPI_ISL_1397650, EPI_ISL_1397651, EPI_ISL_1397652, EPI_ISL_1397653, EPI_ISL_1397654, EPI_ISL_1397655, EPI_ISL_1397656, EPI_ISL_1397657, EPI_ISL_1397658, EPI_ISL_1397659, EPI_ISL_1397660, EPI_ISL_1397661, EPI_ISL_1397662, EPI_ISL_1397663, EPI_ISL_1397664, EPI_ISL_1397665, EPI_ISL_1397666, EPI_ISL_1397667, EPI_ISL_1397668, EPI_ISL_1397669, EPI_ISL_1397670, EPI_ISL_1397671, EPI_ISL_1397672, EPI_ISL_1397673, EPI_ISL_1397674, EPI_ISL_1397675, EPI_ISL_1397676, EPI_ISL_1397677, EPI_ISL_1397678, EPI_ISL_1397679, EPI_ISL_1397680, EPI_ISL_1397681, EPI_ISL_1397682, EPI_ISL_1397683, EPI_ISL_1397684, EPI_ISL_1397685, EPI_ISL_1397686, EPI_ISL_1397687, EPI_ISL_1397688, EPI_ISL_1397689, EPI_ISL_1397690, EPI_ISL_1397691, EPI_ISL_1397692, EPI_ISL_1397693 | | | |
| see above | NORTH SHORE UNIVERSITY HOSPITAL | Wadsworth Center, New York State Department of Health | Alexis Russel; Daryl M. Lamson; Erasmus Schneider; Erica Lasek-Nesselquist; John Kelly; Jonathan Plitnick; Kirsten St. George; Matthew Shudt; Melissa A Leisner; Navjot Singh |
| EPI_ISL_1016136, EPI_ISL_1227860, EPI_ISL_1395707, EPI_ISL_1397829, EPI_ISL_1397830, EPI_ISL_1397907, EPI_ISL_1397908, EPI_ISL_1397909, EPI_ISL_1397910, EPI_ISL_1397911, EPI_ISL_1398068, EPI_ISL_1398069, EPI_ISL_1398070, EPI_ISL_1398071, EPI_ISL_1398072, EPI_ISL_1398073, EPI_ISL_1398074, EPI_ISL_1398075, EPI_ISL_1398076, EPI_ISL_1398077, EPI_ISL_1398078, EPI_ISL_1398079, EPI_ISL_1398080, EPI_ISL_1398081, EPI_ISL_1398082, EPI_ISL_1398083, EPI_ISL_1398084, EPI_ISL_1398085, EPI_ISL_1398086, EPI_ISL_1398087, EPI_ISL_1398088, EPI_ISL_1398089, EPI_ISL_1398090, EPI_ISL_1398091, EPI_ISL_1398092, EPI_ISL_1398093, EPI_ISL_1398094, EPI_ISL_1398095, EPI_ISL_1398096, EPI_ISL_1398097, EPI_ISL_1398098, EPI_ISL_1398099, EPI_ISL_1398100, EPI_ISL_1398101, EPI_ISL_1398102, EPI_ISL_1398103, EPI_ISL_1398104, EPI_ISL_1398105, EPI_ISL_1398106, EPI_ISL_1398107, EPI_ISL_1398108, EPI_ISL_1398109, EPI_ISL_1398110, EPI_ISL_1398111, EPI_ISL_1398112, EPI_ISL_1398113, EPI_ISL_1398114, EPI_ISL_1398115, EPI_ISL_1398116, EPI_ISL_1398117, EPI_ISL_1398118, EPI_ISL_1398119, EPI_ISL_1462561, EPI_ISL_1462566, EPI_ISL_1462569, EPI_ISL_1462570, EPI_ISL_1462576, EPI_ISL_1462584, EPI_ISL_1462585, EPI_ISL_1465757, EPI_ISL_1465758, EPI_ISL_1465759, EPI_ISL_1465761, EPI_ISL_1465835, EPI_ISL_1465836, EPI_ISL_1575469, EPI_ISL_1582156, EPI_ISL_1582405, EPI_ISL_1756327, EPI_ISL_1935609 | | | |
| see above | NORTHWELL HEALTH LABORATORIES | Wadsworth Center, New York State Department of Health | Alexis Russel; Alexis Russell; Catharine Prussing; Daryl M. Lamson; Erasmus Schneider; Erica Lasek-Nesselquist; John Kelly; Jonathan Plitnick; Kirsten St. George; Matthew Shudt; Melissa A Leisner; Navjot Singh |
| EPI_ISL_1273043, EPI_ISL_1273044, EPI_ISL_1273045, EPI_ISL_1273046, EPI_ISL_1273047, EPI_ISL_1318347, EPI_ISL_1318348, EPI_ISL_1318349, EPI_ISL_1446974, EPI_ISL_1446975, EPI_ISL_1447060, EPI_ISL_1447061, EPI_ISL_1447062, EPI_ISL_1678078 | | | |
| see above | NYC Department of Health and Mental Hygiene | Centers for Disease Control and Prevention Division of Viral Diseases, Pathogen Discovery | Alison Laufer Halpin; Anna Montmayeur; Anna Uehara; Ben L. Rambo-Martin; Clinton R. Paden; Dakota Howard; Darlene Wagner; Dave Wentworth; Dhwani Batra; Haibin Wang; Jasmine Padilla; Jing Zhang; Justin Lee; Katie Dillon; Krista Queen; Kristen Knipe; Kristine Lacek; Lori Rowe; Mark Burroughs; Matthew Schmerer; Mili Sheth; Peter Cook; Peter W. Cook; Rachel Marine; Sam Shepard; Sarah Nobles; Shoshona Le; Suxiang Tong; Vivien Dugan; Yan Li; Ying Tao; Yvette Unoarumhi |
| EPI_ISL_1700669, EPI_ISL_1700670, EPI_ISL_1700671, EPI_ISL_1700672 | NYC Department of Health and Mental Hygiene | Pathogen Discovery, Respiratory Viruses Branch, Division of Viral Diseases, Centers for Disease Control and Prevention | Adam Retchless; Anna Kelleher; Anna Montmayeur; Anna Uehara; Brian Lynch; Clinton R. Paden; Haibin Wang; Han Jia Justin Ng; Jing Zhang; Justin Lee; Krista Queen; Mark Burroughs; Peter Cook; Rachel Marine; Suxiang Tong; Yan Li; Ying Tao |
| EPI_ISL_1064800 | NYC HH Elmhurst Hospital Medical Center | New York City Public Health Laboratory | Jade Wang; et al. |
| EPI_ISL_1226419, EPI_ISL_1226430, EPI_ISL_1226440, EPI_ISL_1226441, EPI_ISL_1226443, EPI_ISL_1226459, EPI_ISL_1226500, EPI_ISL_1226507, EPI_ISL_1226510, EPI_ISL_1226521, EPI_ISL_1226545, EPI_ISL_1226546, EPI_ISL_1226547, EPI_ISL_1226548, EPI_ISL_1226549, EPI_ISL_1226554, EPI_ISL_1226560, EPI_ISL_1226561, EPI_ISL_1226562, EPI_ISL_1226564, EPI_ISL_1226565, EPI_ISL_1226566, EPI_ISL_1226580, EPI_ISL_1226581, EPI_ISL_1226582, EPI_ISL_1226595, EPI_ISL_1226600, EPI_ISL_1226620, EPI_ISL_1226621, EPI_ISL_1226625, EPI_ISL_1226636, EPI_ISL_1226639, EPI_ISL_1226640, EPI_ISL_1226641, EPI_ISL_1226642, EPI_ISL_1226643, EPI_ISL_1226644, EPI_ISL_1226647, EPI_ISL_1226648, EPI_ISL_1226651, EPI_ISL_1226652, EPI_ISL_1226660, EPI_ISL_1226661, EPI_ISL_1226673, EPI_ISL_1226685, EPI_ISL_1226692, EPI_ISL_1226711, EPI_ISL_1226718, EPI_ISL_1226719, EPI_ISL_1226729, EPI_ISL_1226739, EPI_ISL_1226768, EPI_ISL_1226769, EPI_ISL_1226790, EPI_ISL_1226791, EPI_ISL_1226792, EPI_ISL_1226793, EPI_ISL_1226794, EPI_ISL_1226795, EPI_ISL_1226796, EPI_ISL_1226797, EPI_ISL_1226808, EPI_ISL_1226809, EPI_ISL_1226810, EPI_ISL_1226813, EPI_ISL_1226814, EPI_ISL_1226816, EPI_ISL_1226817, EPI_ISL_1226832, EPI_ISL_1226833, EPI_ISL_1226854, EPI_ISL_1226855, EPI_ISL_1226862, EPI_ISL_1226867, EPI_ISL_1226869, EPI_ISL_1226893, EPI_ISL_1226894, EPI_ISL_1226895, EPI_ISL_1226896, EPI_ISL_1226900, EPI_ISL_1226916, EPI_ISL_1226917, EPI_ISL_1226918, EPI_ISL_1226919, EPI_ISL_1226920, EPI_ISL_1226921, EPI_ISL_1226922, EPI_ISL_1226923, EPI_ISL_1226935, EPI_ISL_1226963, EPI_ISL_1226982, EPI_ISL_1226983, EPI_ISL_1226984, EPI_ISL_1226987, EPI_ISL_1226998, EPI_ISL_1226999, EPI_ISL_1227001, EPI_ISL_1227005, EPI_ISL_1227006, EPI_ISL_1227009, EPI_ISL_1227010, EPI_ISL_1227011, EPI_ISL_1227012, EPI_ISL_1227013, EPI_ISL_1227015, EPI_ISL_1227027, EPI_ISL_1227028, EPI_ISL_1227081, EPI_ISL_1227087, EPI_ISL_1227095, EPI_ISL_1227109, EPI_ISL_1227137, EPI_ISL_1227138, EPI_ISL_1227139, EPI_ISL_1227140, EPI_ISL_1227141, EPI_ISL_1227142, EPI_ISL_1227143, EPI_ISL_1227144, EPI_ISL_1227145, EPI_ISL_1227146, EPI_ISL_1227147, EPI_ISL_1227148, EPI_ISL_1227149, EPI_ISL_1227155, EPI_ISL_1227156, EPI_ISL_1227157, EPI_ISL_1227158, EPI_ISL_1227159, EPI_ISL_1227160, EPI_ISL_1227161, EPI_ISL_1227167, EPI_ISL_1227168, EPI_ISL_1227169, EPI_ISL_1227170, EPI_ISL_1227171, EPI_ISL_1227172, EPI_ISL_1227184, EPI_ISL_1227186, EPI_ISL_1227187, EPI_ISL_1227188, EPI_ISL_1227190, EPI_ISL_1227191, EPI_ISL_1227192, EPI_ISL_1227193, EPI_ISL_1227194, EPI_ISL_1227195, EPI_ISL_1227196, EPI_ISL_1227197, EPI_ISL_1227198, EPI_ISL_1227199, EPI_ISL_1227200, EPI_ISL_1227201, EPI_ISL_1227202, EPI_ISL_1227203, EPI_ISL_1227204, EPI_ISL_1227212, EPI_ISL_1227213, EPI_ISL_1227221, EPI_ISL_1227238, EPI_ISL_1227244, EPI_ISL_1227422, EPI_ISL_1227423, EPI_ISL_1227428, EPI_ISL_1227429, EPI_ISL_1227440, EPI_ISL_1227680, EPI_ISL_1227681, EPI_ISL_1227682, EPI_ISL_1227683, EPI_ISL_1227686, EPI_ISL_1227687, EPI_ISL_1227688, EPI_ISL_1227689, EPI_ISL_1227690, EPI_ISL_1227691, EPI_ISL_1227692, EPI_ISL_1227693, EPI_ISL_1227694, EPI_ISL_1227695, EPI_ISL_1227696, EPI_ISL_1227697, EPI_ISL_1227698, EPI_ISL_1227699, EPI_ISL_1227700, EPI_ISL_1227701, EPI_ISL_1227702, EPI_ISL_1227703, EPI_ISL_1227704, EPI_ISL_1227706, EPI_ISL_1227861, EPI_ISL_1227862, EPI_ISL_1227863, EPI_ISL_1227864, EPI_ISL_1227865, EPI_ISL_1227866, EPI_ISL_1227868, EPI_ISL_1227869, EPI_ISL_1227871, EPI_ISL_1227872, EPI_ISL_1227873, EPI_ISL_1227874, EPI_ISL_1227875, EPI_ISL_1227876, EPI_ISL_1227877, EPI_ISL_1227878, EPI_ISL_1227879, EPI_ISL_1227938, EPI_ISL_1227939, EPI_ISL_1227940, EPI_ISL_1227941, EPI_ISL_1227942, EPI_ISL_1227946, EPI_ISL_1227947, EPI_ISL_1227948, EPI_ISL_1227949, EPI_ISL_1227950, EPI_ISL_1227951, EPI_ISL_1227952, EPI_ISL_1227953, EPI_ISL_1227954, EPI_ISL_1227955, EPI_ISL_1227956, EPI_ISL_1227957, EPI_ISL_1227958, EPI_ISL_1228177, EPI_ISL_1228178, EPI_ISL_1228179, EPI_ISL_1228180, EPI_ISL_1228182, EPI_ISL_1228185, EPI_ISL_1228186, EPI_ISL_1228187, EPI_ISL_1228189, EPI_ISL_1228190, EPI_ISL_1228191, EPI_ISL_1228192, EPI_ISL_1228193, EPI_ISL_1228194, EPI_ISL_1228195, EPI_ISL_1228196, EPI_ISL_1228197, EPI_ISL_1228198, EPI_ISL_1228199, EPI_ISL_1228200, EPI_ISL_1228201, EPI_ISL_1228202, EPI_ISL_1228203, EPI_ISL_1228204, EPI_ISL_1228205, EPI_ISL_1228206, EPI_ISL_1228207, EPI_ISL_1228208, EPI_ISL_1228209, EPI_ISL_1228210, EPI_ISL_1228211, EPI_ISL_1228212, EPI_ISL_1228213, EPI_ISL_1228214, EPI_ISL_1228215, EPI_ISL_1228216, EPI_ISL_1228217, EPI_ISL_1228237, EPI_ISL_1228238, EPI_ISL_1228239, EPI_ISL_1228265, EPI_ISL_1228369, EPI_ISL_1228370, EPI_ISL_1228371, EPI_ISL_1228372, EPI_ISL_1228373, EPI_ISL_1228563, EPI_ISL_1228570, EPI_ISL_1228574, EPI_ISL_1228575, EPI_ISL_1395706, EPI_ISL_1395708, EPI_ISL_1395709, EPI_ISL_1397623, EPI_ISL_1397624, EPI_ISL_1397625, EPI_ISL_1397626, EPI_ISL_1397627, EPI_ISL_1397628, EPI_ISL_1398284, EPI_ISL_1398285, EPI_ISL_1398286, EPI_ISL_1398287, EPI_ISL_1398288, EPI_ISL_1398289, EPI_ISL_1398290, EPI_ISL_1398291, EPI_ISL_1398292, EPI_ISL_1398293, EPI_ISL_1398294, EPI_ISL_1398295, EPI_ISL_1398296, EPI_ISL_1398297, EPI_ISL_1398298, EPI_ISL_1398299, EPI_ISL_1398300, EPI_ISL_1398301, EPI_ISL_1398302, EPI_ISL_1398303, EPI_ISL_1398304, EPI_ISL_1398305, EPI_ISL_1398306, EPI_ISL_1398307, EPI_ISL_1398308, EPI_ISL_1398309, EPI_ISL_1398310, EPI_ISL_1398311, EPI_ISL_1398318, EPI_ISL_1398319, EPI_ISL_1398320, EPI_ISL_1398321, EPI_ISL_1398322, EPI_ISL_1398323, EPI_ISL_1398324, EPI_ISL_1398325, EPI_ISL_1398326, EPI_ISL_1398327, EPI_ISL_1398328, EPI_ISL_1398329, EPI_ISL_1398330, EPI_ISL_1398340, EPI_ISL_1398341, EPI_ISL_1398342, EPI_ISL_1398343, EPI_ISL_1398344, EPI_ISL_1398345, EPI_ISL_1398346, EPI_ISL_1398347, EPI_ISL_1398348, EPI_ISL_1398349, EPI_ISL_1398350, EPI_ISL_1398351, EPI_ISL_1398352, EPI_ISL_1461319, EPI_ISL_1461320, EPI_ISL_1461321, EPI_ISL_1461323, EPI_ISL_1461324, EPI_ISL_1461325, EPI_ISL_1461326, EPI_ISL_1461327, EPI_ISL_1461328, EPI_ISL_1461329, EPI_ISL_1461330, EPI_ISL_1461331, EPI_ISL_1461332, EPI_ISL_1461333, EPI_ISL_1461334, EPI_ISL_1461335, EPI_ISL_1461336, EPI_ISL_1461337, EPI_ISL_1461338, EPI_ISL_1461339, EPI_ISL_1461340, EPI_ISL_1461341, EPI_ISL_1461342, EPI_ISL_1461343, EPI_ISL_1461344, EPI_ISL_1461345, EPI_ISL_1461346, EPI_ISL_1461347, EPI_ISL_1461348, EPI_ISL_1461349, EPI_ISL_1461350, EPI_ISL_1461351, EPI_ISL_1461352, EPI_ISL_1461353, EPI_ISL_1461354, EPI_ISL_1461355, EPI_ISL_1465714, EPI_ISL_1465715, EPI_ISL_1465716, EPI_ISL_1465717, EPI_ISL_1465718, EPI_ISL_1465719, EPI_ISL_1465720, EPI_ISL_1465721, EPI_ISL_1465722, EPI_ISL_1465805, EPI_ISL_1465806, EPI_ISL_1465807, EPI_ISL_1465808, EPI_ISL_1465809, EPI_ISL_1465810, EPI_ISL_1465811, EPI_ISL_1465812, EPI_ISL_1465813, EPI_ISL_1465814, EPI_ISL_1465815, EPI_ISL_1465816, EPI_ISL_1465817, EPI_ISL_1465818, EPI_ISL_1465819, EPI_ISL_1465820, EPI_ISL_1465821, EPI_ISL_1465822, EPI_ISL_1465823, EPI_ISL_1465824, EPI_ISL_1465825, EPI_ISL_1465826, EPI_ISL_1465827, EPI_ISL_1465828, EPI_ISL_1465829, EPI_ISL_1465830, EPI_ISL_1465831, EPI_ISL_1465832, EPI_ISL_1465833, EPI_ISL_1465834, EPI_ISL_1482287, EPI_ISL_1482288, EPI_ISL_1482289, EPI_ISL_1482290, EPI_ISL_1482291, EPI_ISL_1482292, EPI_ISL_1482293, EPI_ISL_1482294, EPI_ISL_1482295, EPI_ISL_1575459, EPI_ISL_1582400, EPI_ISL_1582401, EPI_ISL_1582402, EPI_ISL_1623973, EPI_ISL_1623974, EPI_ISL_1624083, EPI_ISL_1624084, EPI_ISL_1624085, EPI_ISL_1624086, EPI_ISL_1624087, EPI_ISL_1624088, EPI_ISL_1671141, EPI_ISL_1671142, EPI_ISL_1671148, EPI_ISL_1671149, EPI_ISL_1671150, EPI_ISL_1671151, EPI_ISL_1671152, EPI_ISL_1671153, EPI_ISL_1671154, EPI_ISL_1671155, EPI_ISL_1671157, EPI_ISL_1671167, EPI_ISL_1671168, EPI_ISL_1671172, EPI_ISL_1671173, EPI_ISL_1671177, EPI_ISL_1671178, EPI_ISL_1671179, EPI_ISL_1671180, EPI_ISL_1671183, EPI_ISL_1671184, EPI_ISL_1671195, EPI_ISL_1671196, EPI_ISL_1671197, EPI_ISL_1671198, EPI_ISL_1671199, EPI_ISL_1671200, EPI_ISL_1671205, EPI_ISL_1671206, EPI_ISL_1671207, EPI_ISL_1755445, EPI_ISL_1755446, EPI_ISL_1755447, EPI_ISL_1755448, EPI_ISL_1755449, EPI_ISL_1755450, EPI_ISL_1755451, EPI_ISL_1755452, EPI_ISL_1755453, EPI_ISL_1755454, EPI_ISL_1755455, EPI_ISL_1755456, EPI_ISL_1755457, EPI_ISL_1755458, EPI_ISL_1755459, EPI_ISL_1755460, EPI_ISL_1755461, EPI_ISL_1755462, EPI_ISL_1755463, EPI_ISL_1755464, EPI_ISL_1755465, EPI_ISL_1755466, EPI_ISL_1755467, EPI_ISL_1755468, EPI_ISL_1755469, EPI_ISL_1755470, EPI_ISL_1755471, EPI_ISL_1755472, EPI_ISL_1755473, EPI_ISL_1755474, EPI_ISL_1755475, EPI_ISL_1755476, EPI_ISL_1755477, EPI_ISL_1755478, EPI_ISL_1755479, EPI_ISL_1755480, EPI_ISL_1755481, EPI_ISL_1755482, EPI_ISL_1755483, EPI_ISL_1755484, EPI_ISL_1755485, EPI_ISL_1755486, EPI_ISL_1755491, EPI_ISL_1755492, EPI_ISL_1755493, EPI_ISL_1755494, EPI_ISL_1755495, EPI_ISL_1755496, EPI_ISL_1755497, EPI_ISL_1755498, EPI_ISL_1755499, EPI_ISL_1755500, EPI_ISL_1755501, EPI_ISL_1755941, EPI_ISL_1755942, EPI_ISL_1755947, EPI_ISL_1789700, EPI_ISL_1789701, EPI_ISL_1789702, EPI_ISL_1789703, EPI_ISL_1789704, EPI_ISL_1789705, EPI_ISL_1789708, EPI_ISL_1855186, EPI_ISL_1855187, EPI_ISL_1855197, EPI_ISL_1855198, EPI_ISL_1855199, EPI_ISL_1855200, EPI_ISL_1855201, EPI_ISL_1920070, EPI_ISL_1920071, EPI_ISL_1920072, EPI_ISL_1920073, EPI_ISL_1920079, EPI_ISL_1920080, EPI_ISL_1920083, EPI_ISL_1920087, EPI_ISL_1920088, EPI_ISL_1920095, EPI_ISL_1920163 | | | |
| see above | NYC Pandemic Response Lab | Wadsworth Center, New York State Department of Health | Alexis Russel; Alexis Russell; Catharine Prussing; Daryl M. Lamson; Erasmus Schneider; Erica Lasek-Nesselquist; John Kelly; Jonathan Plitnick; Kirsten St. George; Matthew Shudt; Melissa A Leisner; Navjot Singh |
| EPI_ISL_1379198, EPI_ISL_1379199, EPI_ISL_1379200, EPI_ISL_1379201, EPI_ISL_1379202, EPI_ISL_1379203, EPI_ISL_1379204, EPI_ISL_1379205, EPI_ISL_1379206, EPI_ISL_1379207, EPI_ISL_1379208, EPI_ISL_1379209, EPI_ISL_1379210, EPI_ISL_1379211, EPI_ISL_1379212, EPI_ISL_1379213, EPI_ISL_1379214, EPI_ISL_1379215, EPI_ISL_1379216, EPI_ISL_1379217, EPI_ISL_1379218, EPI_ISL_1379219, EPI_ISL_1379220, EPI_ISL_1379221, EPI_ISL_1379222, EPI_ISL_1379223, EPI_ISL_1379224, EPI_ISL_1379225, EPI_ISL_1379226, EPI_ISL_1379227, EPI_ISL_1379228, EPI_ISL_1379229, EPI_ISL_1379230, EPI_ISL_1379231, EPI_ISL_1379232, EPI_ISL_1379233, EPI_ISL_1379234, EPI_ISL_1379235, EPI_ISL_1379236, EPI_ISL_1379237, EPI_ISL_1379238, EPI_ISL_1379239, EPI_ISL_1379240, EPI_ISL_1379241, EPI_ISL_1379242, EPI_ISL_1379243, EPI_ISL_1379244, EPI_ISL_1379245, EPI_ISL_1379246, EPI_ISL_1379247, EPI_ISL_1379248, EPI_ISL_1379249, EPI_ISL_1379250, EPI_ISL_1379251, EPI_ISL_1379252, EPI_ISL_1379253, EPI_ISL_1379254, EPI_ISL_1379255, EPI_ISL_1379256, EPI_ISL_1379257, EPI_ISL_1379258, EPI_ISL_1379259, EPI_ISL_1379260, EPI_ISL_1379261, EPI_ISL_1379262, EPI_ISL_1379263, EPI_ISL_1379264, EPI_ISL_1379265, EPI_ISL_1379266, EPI_ISL_1379339, EPI_ISL_1379340, EPI_ISL_1379341, EPI_ISL_1379342, EPI_ISL_1379343, EPI_ISL_1379344, EPI_ISL_1379345, EPI_ISL_1379346, EPI_ISL_1379347, EPI_ISL_1379348, EPI_ISL_1379349, EPI_ISL_1379350, EPI_ISL_1379351, EPI_ISL_1379352, EPI_ISL_1379353, EPI_ISL_1379354, EPI_ISL_1379355, EPI_ISL_1379356, EPI_ISL_1379357, EPI_ISL_1379358, EPI_ISL_1379359, EPI_ISL_1379360, EPI_ISL_1379361, EPI_ISL_1379362, EPI_ISL_1379363, EPI_ISL_1379364, EPI_ISL_1379365, EPI_ISL_1379366, EPI_ISL_1379367, EPI_ISL_1379368, EPI_ISL_1379369, EPI_ISL_1379370, EPI_ISL_1379371, EPI_ISL_1379372, EPI_ISL_1379373, EPI_ISL_1379374, EPI_ISL_1379375, EPI_ISL_1379376, EPI_ISL_1379377, EPI_ISL_1379378, EPI_ISL_1379379, EPI_ISL_1379380, EPI_ISL_1379381, EPI_ISL_1379382, EPI_ISL_1379383, EPI_ISL_1379384, EPI_ISL_1379385, EPI_ISL_1379386, EPI_ISL_1379387, EPI_ISL_1379388, EPI_ISL_1379389, EPI_ISL_1379390, EPI_ISL_1968717, EPI_ISL_1968718, EPI_ISL_1968719, EPI_ISL_1968722, EPI_ISL_1968723, EPI_ISL_1968724, EPI_ISL_1968725 | | | |
| see above | NYP-WCM | New York Genome Center | Andre Corvelo; Arryn Craney; Chris Mason; Dayna M. Oschwald; Hanna Rennert; Lars F Westblade; Margaret Elizabeth Ross; Melissa Cushing; Michael Zody; Olivier Elemento; Priya Velu; Samantha Fennessey; Tom Maniatis |
| EPI_ISL_1272922, EPI_ISL_1272923, EPI_ISL_1272924, EPI_ISL_1272925, EPI_ISL_1272926, EPI_ISL_1272927, EPI_ISL_1695889, EPI_ISL_1695890, EPI_ISL_1695891, EPI_ISL_1695892 | | | |
| see above | NYSDOH Wadsworth Center, Virology Lab | Centers for Disease Control and Prevention Division of Viral Diseases, Pathogen Discovery | Alison Laufer Halpin; Anna Montmayeur; Anna Uehara; Ben L. Rambo-Martin; Clinton R. Paden; Dakota Howard; Darlene Wagner; Dave Wentworth; Dhwani Batra; Haibin Wang; Jasmine Padilla; Jing Zhang; Justin Lee; Katie Dillon; Krista Queen; Kristen Knipe; Kristine Lacek; Lori Rowe; Mark Burroughs; Matthew Schmerer; Mili Sheth; Peter Cook; Peter W. Cook; Rachel Marine; Sam Shepard; Sarah Nobles; Shoshona Le; Suxiang Tong; Vivien Dugan; Yan Li; Ying Tao; Yvette Unoarumhi |
| EPI_ISL_966245, EPI_ISL_1016013, EPI_ISL_1016014, EPI_ISL_1016015, EPI_ISL_1016016, EPI_ISL_1016017, EPI_ISL_1016018, EPI_ISL_1016075, EPI_ISL_1016100, EPI_ISL_1016170, EPI_ISL_1040853, EPI_ISL_1040854, EPI_ISL_1040855, EPI_ISL_1040856, EPI_ISL_1040857, EPI_ISL_1040858, EPI_ISL_1040859, EPI_ISL_1040861, EPI_ISL_1040862, EPI_ISL_1040863, EPI_ISL_1040864, EPI_ISL_1040865, EPI_ISL_1040866, EPI_ISL_1040867, EPI_ISL_1040870, EPI_ISL_1040871, EPI_ISL_1040872, EPI_ISL_1040873, EPI_ISL_1040874, EPI_ISL_1040875, EPI_ISL_1040876, EPI_ISL_1040877, EPI_ISL_1040878, EPI_ISL_1040879, EPI_ISL_1040882, EPI_ISL_1040883, EPI_ISL_1040884, EPI_ISL_1040885, EPI_ISL_1040886, EPI_ISL_1040887, EPI_ISL_1040888, EPI_ISL_1040889, EPI_ISL_1040890, EPI_ISL_1040891, EPI_ISL_1040892, EPI_ISL_1040893, EPI_ISL_1040894, EPI_ISL_1040895, EPI_ISL_1040896, EPI_ISL_1040897, EPI_ISL_1040898, EPI_ISL_1040899, EPI_ISL_1040900, EPI_ISL_1040901, EPI_ISL_1040904, EPI_ISL_1040905, EPI_ISL_1040906, EPI_ISL_1040908, EPI_ISL_1040909, EPI_ISL_1040910, EPI_ISL_1040911, EPI_ISL_1040913, EPI_ISL_1040914, EPI_ISL_1085093, EPI_ISL_1085161, EPI_ISL_1085166, EPI_ISL_1085168, EPI_ISL_1085171, EPI_ISL_1085175, EPI_ISL_1087272, EPI_ISL_1087273, EPI_ISL_1087274, EPI_ISL_1087275, EPI_ISL_1087277, EPI_ISL_1087278, EPI_ISL_1087279, EPI_ISL_1087280, EPI_ISL_1087281, EPI_ISL_1087282, EPI_ISL_1087283, EPI_ISL_1087284, EPI_ISL_1087285, EPI_ISL_1087287, EPI_ISL_1087288, EPI_ISL_1087289, EPI_ISL_1087290, EPI_ISL_1087291, EPI_ISL_1087293, EPI_ISL_1087294, EPI_ISL_1087295, EPI_ISL_1087296, EPI_ISL_1087297, EPI_ISL_1087298, EPI_ISL_1087299, EPI_ISL_1087300, EPI_ISL_1087301, EPI_ISL_1087313, EPI_ISL_1087314, EPI_ISL_1087315, EPI_ISL_1087316, EPI_ISL_1087317, EPI_ISL_1087318, EPI_ISL_1087319, EPI_ISL_1087320, EPI_ISL_1087321, EPI_ISL_1087322, EPI_ISL_1087323, EPI_ISL_1087324, EPI_ISL_1087325, EPI_ISL_1087326, EPI_ISL_1087327, EPI_ISL_1087328, EPI_ISL_1087329, EPI_ISL_1087330, EPI_ISL_1087331, EPI_ISL_1087332, EPI_ISL_1087333, EPI_ISL_1087334, EPI_ISL_1087335, EPI_ISL_1087336, EPI_ISL_1087337, EPI_ISL_1087338, EPI_ISL_1087339, EPI_ISL_1087340, EPI_ISL_1087341, EPI_ISL_1087342, EPI_ISL_1087343, EPI_ISL_1087344, EPI_ISL_1087345, EPI_ISL_1087346, EPI_ISL_1087347, EPI_ISL_1087348, EPI_ISL_1087349, EPI_ISL_1087350, EPI_ISL_1087351, EPI_ISL_1087352, EPI_ISL_1116476, EPI_ISL_1116477, EPI_ISL_1116478, EPI_ISL_1116479, EPI_ISL_1116480, EPI_ISL_1116481, EPI_ISL_1116482, EPI_ISL_1116483, EPI_ISL_1116484, EPI_ISL_1116485, EPI_ISL_1116486, EPI_ISL_1116487, EPI_ISL_1116488, EPI_ISL_1116489, EPI_ISL_1116490, EPI_ISL_1116491, EPI_ISL_1116492, EPI_ISL_1116493, EPI_ISL_1116494, EPI_ISL_1116495, EPI_ISL_1116496, EPI_ISL_1116497, EPI_ISL_1116498, EPI_ISL_1116499, EPI_ISL_1116500, EPI_ISL_1116501, EPI_ISL_1116502, EPI_ISL_1116504, EPI_ISL_1116505, EPI_ISL_1116506, EPI_ISL_1116507, EPI_ISL_1116508, EPI_ISL_1116509, EPI_ISL_1116510, EPI_ISL_1116511, EPI_ISL_1116512, EPI_ISL_1116513, EPI_ISL_1116514, EPI_ISL_1116515, EPI_ISL_1116516, EPI_ISL_1116517, EPI_ISL_1116518, EPI_ISL_1116519, EPI_ISL_1116520, EPI_ISL_1116521, EPI_ISL_1116522, EPI_ISL_1116523, EPI_ISL_1116524, EPI_ISL_1116525, EPI_ISL_1116526, EPI_ISL_1116527, EPI_ISL_1116528, EPI_ISL_1116529, EPI_ISL_1116530, EPI_ISL_1116531, EPI_ISL_1116532, EPI_ISL_1116533, EPI_ISL_1116534, EPI_ISL_1116535, EPI_ISL_1116536, EPI_ISL_1116537, EPI_ISL_1116538, EPI_ISL_1116539, EPI_ISL_1116540, EPI_ISL_1116541, EPI_ISL_1116542, EPI_ISL_1116543, EPI_ISL_1116544, EPI_ISL_1116545, EPI_ISL_1116546, EPI_ISL_1116547, EPI_ISL_1116548, EPI_ISL_1116549, EPI_ISL_1116550, EPI_ISL_1116551, EPI_ISL_1116552, EPI_ISL_1116553, EPI_ISL_1116554, EPI_ISL_1195744, EPI_ISL_1195745, EPI_ISL_1195746, EPI_ISL_1195747, EPI_ISL_1195748, EPI_ISL_1195749, EPI_ISL_1195750, EPI_ISL_1195751, EPI_ISL_1195752, EPI_ISL_1195753, EPI_ISL_1195754, EPI_ISL_1195755, EPI_ISL_1195756, EPI_ISL_1195757, EPI_ISL_1195758, EPI_ISL_1195759, EPI_ISL_1195760, EPI_ISL_1195761, EPI_ISL_1195762, EPI_ISL_1195763, EPI_ISL_1195764, EPI_ISL_1195765, EPI_ISL_1195766, EPI_ISL_1195767, EPI_ISL_1195768, EPI_ISL_1195770, EPI_ISL_1195771, EPI_ISL_1195772, EPI_ISL_1195773, EPI_ISL_1195774, EPI_ISL_1195775, EPI_ISL_1195776, EPI_ISL_1195777, EPI_ISL_1195778, EPI_ISL_1195779, EPI_ISL_1195780, EPI_ISL_1195781, EPI_ISL_1195782, EPI_ISL_1195783, EPI_ISL_1195784, EPI_ISL_1195785, EPI_ISL_1195786, EPI_ISL_1195787, EPI_ISL_1195788, EPI_ISL_1195789, EPI_ISL_1195790, EPI_ISL_1195791, EPI_ISL_1195792, EPI_ISL_1195793, EPI_ISL_1195794, EPI_ISL_1195795, EPI_ISL_1195796, EPI_ISL_1195798, EPI_ISL_1195799, EPI_ISL_1195801, EPI_ISL_1195802, EPI_ISL_1195803, EPI_ISL_1195804, EPI_ISL_1195805, EPI_ISL_1195806, EPI_ISL_1195807, EPI_ISL_1195808, EPI_ISL_1195809, EPI_ISL_1195810, EPI_ISL_1195811, EPI_ISL_1195812, EPI_ISL_1195813, EPI_ISL_1195814, EPI_ISL_1195815, EPI_ISL_1195816, EPI_ISL_1195817, EPI_ISL_1195818, EPI_ISL_1195819, EPI_ISL_1195820, EPI_ISL_1195821, EPI_ISL_1195822, EPI_ISL_1195823, EPI_ISL_1195824, EPI_ISL_1195825, EPI_ISL_1195826, EPI_ISL_1195827, EPI_ISL_1195828, EPI_ISL_1195829, EPI_ISL_1195830, EPI_ISL_1195831, EPI_ISL_1195832, EPI_ISL_1195833, EPI_ISL_1195834, EPI_ISL_1195835, EPI_ISL_1195836, EPI_ISL_1195837, EPI_ISL_1195838, EPI_ISL_1195839, EPI_ISL_1195840, EPI_ISL_1195841, EPI_ISL_1195842, EPI_ISL_1195843, EPI_ISL_1195845, EPI_ISL_1195846, EPI_ISL_1195848, EPI_ISL_1195849, EPI_ISL_1195850, EPI_ISL_1195851, EPI_ISL_1195852, EPI_ISL_1195853, EPI_ISL_1195854, EPI_ISL_1195855, EPI_ISL_1195856, EPI_ISL_1195857, EPI_ISL_1195858, EPI_ISL_1195859, EPI_ISL_1195860, EPI_ISL_1195861, EPI_ISL_1195862, EPI_ISL_1195864, EPI_ISL_1195865, EPI_ISL_1195866, EPI_ISL_1195867, EPI_ISL_1195868, EPI_ISL_1195869, EPI_ISL_1195870, EPI_ISL_1195871, EPI_ISL_1195872, EPI_ISL_1195873, EPI_ISL_1195874, EPI_ISL_1195875, EPI_ISL_1195876, EPI_ISL_1195877, EPI_ISL_1195878, EPI_ISL_1195879, EPI_ISL_1195880, EPI_ISL_1195881, EPI_ISL_1195882, EPI_ISL_1195883, EPI_ISL_1195884, EPI_ISL_1195885, EPI_ISL_1195886, EPI_ISL_1195887, EPI_ISL_1195888, EPI_ISL_1195889, EPI_ISL_1195890, EPI_ISL_1195892, EPI_ISL_1195893, EPI_ISL_1195894, EPI_ISL_1195895, EPI_ISL_1195896, EPI_ISL_1195897, EPI_ISL_1195898, EPI_ISL_1195899, EPI_ISL_1195900, EPI_ISL_1195901, EPI_ISL_1195902, EPI_ISL_1262501, EPI_ISL_1262502, EPI_ISL_1262503, EPI_ISL_1262504, EPI_ISL_1262505, EPI_ISL_1262506, EPI_ISL_1262507, EPI_ISL_1262508, EPI_ISL_1262509, EPI_ISL_1262510, EPI_ISL_1262511, EPI_ISL_1262512, EPI_ISL_1262513, EPI_ISL_1262514, EPI_ISL_1262515, EPI_ISL_1262516, EPI_ISL_1262517, EPI_ISL_1262518, EPI_ISL_1262519, EPI_ISL_1262520, EPI_ISL_1262521, EPI_ISL_1262522, EPI_ISL_1262523, EPI_ISL_1262524, EPI_ISL_1262525, EPI_ISL_1262526, EPI_ISL_1262527, EPI_ISL_1262528, EPI_ISL_1262529, EPI_ISL_1262530, EPI_ISL_1262531, EPI_ISL_1262532, EPI_ISL_1262533, EPI_ISL_1262534, EPI_ISL_1262535, EPI_ISL_1262536, EPI_ISL_1262537, EPI_ISL_1262538, EPI_ISL_1262539, EPI_ISL_1262540, EPI_ISL_1262541, EPI_ISL_1262542, EPI_ISL_1262543, EPI_ISL_1262544, EPI_ISL_1262545, EPI_ISL_1262546, EPI_ISL_1262547, EPI_ISL_1262548, EPI_ISL_1262549, EPI_ISL_1262550, EPI_ISL_1262551, EPI_ISL_1262552, EPI_ISL_1262553, EPI_ISL_1262554, EPI_ISL_1262555, EPI_ISL_1262556, EPI_ISL_1262557, EPI_ISL_1262558, EPI_ISL_1262559, EPI_ISL_1262560, EPI_ISL_1262561, EPI_ISL_1262562, EPI_ISL_1262563, EPI_ISL_1262564, EPI_ISL_1262565, EPI_ISL_1262566, EPI_ISL_1262567, EPI_ISL_1262568, EPI_ISL_1262569, EPI_ISL_1262570, EPI_ISL_1262571, EPI_ISL_1262572, EPI_ISL_1262573, EPI_ISL_1262574, EPI_ISL_1262575, EPI_ISL_1262576, EPI_ISL_1262577, EPI_ISL_1262578, EPI_ISL_1262579, EPI_ISL_1262580, EPI_ISL_1262581, EPI_ISL_1262582, EPI_ISL_1262583, EPI_ISL_1262584, EPI_ISL_1262585, EPI_ISL_1300175, EPI_ISL_1300176, EPI_ISL_1300177, EPI_ISL_1300178, EPI_ISL_1300179, EPI_ISL_1300180, EPI_ISL_1300181, EPI_ISL_1300183, EPI_ISL_1300184, EPI_ISL_1300185, EPI_ISL_1300186, EPI_ISL_1300187, EPI_ISL_1300188, EPI_ISL_1300189, EPI_ISL_1300190, EPI_ISL_1300191, EPI_ISL_1300192, EPI_ISL_1300193, EPI_ISL_1300194, EPI_ISL_1300195, EPI_ISL_1300196, EPI_ISL_1300197, EPI_ISL_1300198, EPI_ISL_1300199, EPI_ISL_1300200, EPI_ISL_1300201, EPI_ISL_1300202, EPI_ISL_1300203, EPI_ISL_1300204, EPI_ISL_1300205, EPI_ISL_1300206, EPI_ISL_1300207, EPI_ISL_1300208, EPI_ISL_1300209, EPI_ISL_1300210, EPI_ISL_1300211, EPI_ISL_1300212, EPI_ISL_1300213, EPI_ISL_1300214, EPI_ISL_1300215, EPI_ISL_1300216, EPI_ISL_1300217, EPI_ISL_1300218, EPI_ISL_1300219, EPI_ISL_1300220, EPI_ISL_1300221, EPI_ISL_1300222, EPI_ISL_1300223, EPI_ISL_1300224, EPI_ISL_1300225, EPI_ISL_1300226, EPI_ISL_1300227, EPI_ISL_1300228, EPI_ISL_1300229, EPI_ISL_1300230, EPI_ISL_1300231, EPI_ISL_1300232, EPI_ISL_1300233, EPI_ISL_1300234, EPI_ISL_1300235, EPI_ISL_1300236, EPI_ISL_1300237, EPI_ISL_1300238, EPI_ISL_1300239, EPI_ISL_1300240, EPI_ISL_1300241, EPI_ISL_1300242, EPI_ISL_1300243, EPI_ISL_1300244, EPI_ISL_1300245, EPI_ISL_1300246, EPI_ISL_1300247, EPI_ISL_1300248, EPI_ISL_1300249, EPI_ISL_1300250, EPI_ISL_1300251, EPI_ISL_1300252, EPI_ISL_1300253, | | | |

EPI_ISL_1300254, EPI_ISL_1300255, EPI_ISL_1300256, EPI_ISL_1300257, EPI_ISL_1300258, EPI_ISL_1300259, EPI_ISL_1300261, EPI_ISL_1300262, EPI_ISL_1300263, EPI_ISL_1300264, EPI_ISL_1300265, EPI_ISL_1300266, EPI_ISL_1300267, EPI_ISL_1300268, EPI_ISL_1300269, EPI_ISL_1300270, EPI_ISL_1300271, EPI_ISL_1300272, EPI_ISL_1300273, EPI_ISL_1300274, EPI_ISL_1300275, EPI_ISL_1300276,
EPI_ISL_1322746, EPI_ISL_1322747, EPI_ISL_1322748, EPI_ISL_1322749, EPI_ISL_1322750, EPI_ISL_1322752, EPI_ISL_1322753, EPI_ISL_1322754, EPI_ISL_1322757, EPI_ISL_1322758, EPI_ISL_1322759, EPI_ISL_1322760, EPI_ISL_1322761, EPI_ISL_1322790, EPI_ISL_1322791, EPI_ISL_1322792, EPI_ISL_1322795, EPI_ISL_1322797, EPI_ISL_1322800, EPI_ISL_1322801, EPI_ISL_1322802, EPI_ISL_1322803,
EPI_ISL_1322804, EPI_ISL_1322805, EPI_ISL_1322806, EPI_ISL_1322833, EPI_ISL_1423376, EPI_ISL_1423377, EPI_ISL_1423378, EPI_ISL_1423380, EPI_ISL_1423381, EPI_ISL_1423382, EPI_ISL_1423383, EPI_ISL_1423384, EPI_ISL_1423385, EPI_ISL_1423386, EPI_ISL_1423387, EPI_ISL_1423388, EPI_ISL_1423389, EPI_ISL_1423390, EPI_ISL_1423391, EPI_ISL_1423392, EPI_ISL_1423393, EPI_ISL_1423394,
EPI_ISL_1423395, EPI_ISL_1423396, EPI_ISL_1423397, EPI_ISL_1423398, EPI_ISL_1423399, EPI_ISL_1423400, EPI_ISL_1423401, EPI_ISL_1423402, EPI_ISL_1423403, EPI_ISL_1423404, EPI_ISL_1423405, EPI_ISL_1423406, EPI_ISL_1423407, EPI_ISL_1423408, EPI_ISL_1423409, EPI_ISL_1423410, EPI_ISL_1423411, EPI_ISL_1423413, EPI_ISL_1532785, EPI_ISL_1576945, EPI_ISL_1922862, EPI_ISL_1922872

| | | | |
|---|---|---|---|
| see above | NYU Langone Health | Departments of Pathology and Medicine, New York University School of Medicine | Adriana Heguy; Christian Marier; Dacia Dimartino; Emily Guzman; Gael Westby; Guiqing Wang; Paolo Cotzia; Paul Zappile; Peter Meyn; Sitharam Ramaswami; Yutong Zhang |

EPI_ISL_1016227, EPI_ISL_1016228, EPI_ISL_1016229, EPI_ISL_1016230, EPI_ISL_1016231, EPI_ISL_1016232, EPI_ISL_1016233, EPI_ISL_1016234, EPI_ISL_1016235, EPI_ISL_1016236, EPI_ISL_1016237, EPI_ISL_1016238, EPI_ISL_1016239, EPI_ISL_1016240, EPI_ISL_1016241, EPI_ISL_1016242, EPI_ISL_1016243, EPI_ISL_1016244, EPI_ISL_1016245, EPI_ISL_1016246, EPI_ISL_1016248, EPI_ISL_1226544,
EPI_ISL_1226559, EPI_ISL_1226624, EPI_ISL_1226757, EPI_ISL_1226811, EPI_ISL_1226812, EPI_ISL_1226843, EPI_ISL_1226850, EPI_ISL_1226915, EPI_ISL_1227000, EPI_ISL_1227164, EPI_ISL_1227165, EPI_ISL_1227182, EPI_ISL_1227183, EPI_ISL_1227185, EPI_ISL_1227410, EPI_ISL_1227417, EPI_ISL_1227679, EPI_ISL_1227684, EPI_ISL_1227685, EPI_ISL_1227867, EPI_ISL_1227935, EPI_ISL_1227936,
EPI_ISL_1227937, EPI_ISL_1227943, EPI_ISL_1227944, EPI_ISL_1227945, EPI_ISL_1228181, EPI_ISL_1228183, EPI_ISL_1228184, EPI_ISL_1228564, EPI_ISL_1228568, EPI_ISL_1855211, EPI_ISL_1855212, EPI_ISL_1855213, EPI_ISL_1855214

| | | | |
|---|---|---|---|
| see above | New York Presbyterian Hospital | Wadsworth Center, New York State Department of Health | Alexis Russel; Alexis Russell; Catharine Prussing; Daryl M. Lamson; Erasmus Schneider; Erica Lasek-Nesselquist; John Kelly; Jonathan Plitnick; Kirsten St. George; Matthew Shudt; Melissa A Leisner; Navjot Singh |
| EPI_ISL_1920132, EPI_ISL_1920151, EPI_ISL_1920169, EPI_ISL_1920170 | Northwell Health Laboratories | Wadsworth Center, New York State Department of Health | Alexis Russell; Catharine Prussing; Daryl M. Lamson; Erasmus Schneider; Erica Lasek-Nesselquist; John Kelly; Jonathan Plitnick; Kirsten St. George; Matthew Shudt; Melissa A Leisner; Navjot Singh |

EPI_ISL_966391, EPI_ISL_966400, EPI_ISL_966402, EPI_ISL_966411, EPI_ISL_966429, EPI_ISL_966430, EPI_ISL_966431, EPI_ISL_1018032, EPI_ISL_1018033, EPI_ISL_1018034, EPI_ISL_1018035, EPI_ISL_1018036, EPI_ISL_1018037, EPI_ISL_1018038, EPI_ISL_1018039, EPI_ISL_1018040, EPI_ISL_1018041, EPI_ISL_1018042, EPI_ISL_1018043, EPI_ISL_1018044, EPI_ISL_1018045, EPI_ISL_1018046,
EPI_ISL_1018047, EPI_ISL_1018048, EPI_ISL_1018049, EPI_ISL_1064751, EPI_ISL_1064752, EPI_ISL_1064753, EPI_ISL_1064754, EPI_ISL_1064763, EPI_ISL_1064764, EPI_ISL_1064765, EPI_ISL_1064770, EPI_ISL_1064772, EPI_ISL_1064773, EPI_ISL_1064774, EPI_ISL_1064776, EPI_ISL_1064778, EPI_ISL_1064783, EPI_ISL_1064792, EPI_ISL_1064793, EPI_ISL_1064795, EPI_ISL_1064796, EPI_ISL_1064804,
EPI_ISL_1064813, EPI_ISL_1064814, EPI_ISL_1064815, EPI_ISL_1064816, EPI_ISL_1064817, EPI_ISL_1064818, EPI_ISL_1064829, EPI_ISL_1064832, EPI_ISL_1064833, EPI_ISL_1064843, EPI_ISL_1064844, EPI_ISL_1064845, EPI_ISL_1064846, EPI_ISL_1064853, EPI_ISL_1064854, EPI_ISL_1064855, EPI_ISL_1064856, EPI_ISL_1064857, EPI_ISL_1064863, EPI_ISL_1064864, EPI_ISL_1064877, EPI_ISL_1064878,
EPI_ISL_1064879, EPI_ISL_1064880, EPI_ISL_1064881, EPI_ISL_1064882, EPI_ISL_1064899, EPI_ISL_1064900, EPI_ISL_1064901, EPI_ISL_1064902, EPI_ISL_1064903, EPI_ISL_1064910, EPI_ISL_1064911, EPI_ISL_1064912, EPI_ISL_1064913, EPI_ISL_1064914, EPI_ISL_1114525, EPI_ISL_1114526, EPI_ISL_1114527, EPI_ISL_1114528, EPI_ISL_1114529, EPI_ISL_1114530, EPI_ISL_1121895, EPI_ISL_1121896,
EPI_ISL_1121897, EPI_ISL_1121898, EPI_ISL_1200526, EPI_ISL_1200527, EPI_ISL_1200531, EPI_ISL_1200532, EPI_ISL_1200539, EPI_ISL_1200549, EPI_ISL_1200555, EPI_ISL_1200576, EPI_ISL_1200607, EPI_ISL_1200608, EPI_ISL_1200609, EPI_ISL_1200610, EPI_ISL_1200627, EPI_ISL_1261796, EPI_ISL_1261797, EPI_ISL_1261811, EPI_ISL_1261819, EPI_ISL_1261827, EPI_ISL_1261828, EPI_ISL_1261829,
EPI_ISL_1261846, EPI_ISL_1261854, EPI_ISL_1261865, EPI_ISL_1337518, EPI_ISL_1337519, EPI_ISL_1337524, EPI_ISL_1337527, EPI_ISL_1337541

| | | | |
|---|---|---|---|
| see above | OCME Office Of Chief Medical Examiner | New York City Public Health Laboratory | Jade Wang; et al. |

EPI_ISL_968080, EPI_ISL_984917, EPI_ISL_984918, EPI_ISL_984919, EPI_ISL_984920, EPI_ISL_984921, EPI_ISL_984922, EPI_ISL_984923, EPI_ISL_984924, EPI_ISL_984925, EPI_ISL_984926, EPI_ISL_984927, EPI_ISL_984928, EPI_ISL_984929, EPI_ISL_984930, EPI_ISL_984931, EPI_ISL_984932, EPI_ISL_984933, EPI_ISL_984934, EPI_ISL_984935, EPI_ISL_984936, EPI_ISL_984937, EPI_ISL_984938, EPI_ISL_984939,
EPI_ISL_984940, EPI_ISL_984941, EPI_ISL_984942, EPI_ISL_984943, EPI_ISL_984944, EPI_ISL_984945, EPI_ISL_984946, EPI_ISL_984947, EPI_ISL_994808, EPI_ISL_994809, EPI_ISL_994810, EPI_ISL_994811, EPI_ISL_994813, EPI_ISL_994814, EPI_ISL_994815, EPI_ISL_994821, EPI_ISL_994824, EPI_ISL_994825, EPI_ISL_994826, EPI_ISL_994829, EPI_ISL_994830, EPI_ISL_994831, EPI_ISL_994832, EPI_ISL_994833,
EPI_ISL_994834, EPI_ISL_994835, EPI_ISL_994837, EPI_ISL_994838, EPI_ISL_994839, EPI_ISL_994840, EPI_ISL_994841, EPI_ISL_994842, EPI_ISL_994843, EPI_ISL_994844, EPI_ISL_994845, EPI_ISL_994846, EPI_ISL_994847, EPI_ISL_994848, EPI_ISL_994849, EPI_ISL_994850, EPI_ISL_994851, EPI_ISL_994852, EPI_ISL_994853, EPI_ISL_994854, EPI_ISL_994855, EPI_ISL_994856, EPI_ISL_994857, EPI_ISL_994858,
EPI_ISL_994859, EPI_ISL_994860, EPI_ISL_994861, EPI_ISL_994862, EPI_ISL_994863, EPI_ISL_994864, EPI_ISL_994865, EPI_ISL_994866, EPI_ISL_994867, EPI_ISL_994868, EPI_ISL_994869, EPI_ISL_994870, EPI_ISL_994871, EPI_ISL_994872, EPI_ISL_994873, EPI_ISL_994874, EPI_ISL_994875, EPI_ISL_994876, EPI_ISL_994877, EPI_ISL_994878, EPI_ISL_994879, EPI_ISL_994880, EPI_ISL_994881, EPI_ISL_994882,
EPI_ISL_994883, EPI_ISL_994884, EPI_ISL_994885, EPI_ISL_994886, EPI_ISL_994887, EPI_ISL_994888, EPI_ISL_994889, EPI_ISL_994890, EPI_ISL_994891, EPI_ISL_994892, EPI_ISL_994893, EPI_ISL_994894, EPI_ISL_994895, EPI_ISL_994896, EPI_ISL_994897, EPI_ISL_994898, EPI_ISL_994899, EPI_ISL_994900, EPI_ISL_994901, EPI_ISL_994902, EPI_ISL_994903, EPI_ISL_994904, EPI_ISL_994905, EPI_ISL_994906,
EPI_ISL_994907, EPI_ISL_994908, EPI_ISL_994909, EPI_ISL_994910, EPI_ISL_994911, EPI_ISL_994912, EPI_ISL_994913, EPI_ISL_994914, EPI_ISL_994915, EPI_ISL_994916, EPI_ISL_994917, EPI_ISL_994918, EPI_ISL_994919, EPI_ISL_994920, EPI_ISL_994921, EPI_ISL_994922, EPI_ISL_994923, EPI_ISL_994924, EPI_ISL_994925, EPI_ISL_994926, EPI_ISL_994927, EPI_ISL_994928, EPI_ISL_994929, EPI_ISL_994930,
EPI_ISL_994931, EPI_ISL_994932, EPI_ISL_994933, EPI_ISL_994934, EPI_ISL_994935, EPI_ISL_994936, EPI_ISL_994937, EPI_ISL_994938, EPI_ISL_994939, EPI_ISL_994940, EPI_ISL_994941, EPI_ISL_994942, EPI_ISL_994943, EPI_ISL_994944, EPI_ISL_994945, EPI_ISL_994946, EPI_ISL_994947, EPI_ISL_994948, EPI_ISL_994949, EPI_ISL_994950, EPI_ISL_994951, EPI_ISL_994952, EPI_ISL_994953, EPI_ISL_994954,
EPI_ISL_994955, EPI_ISL_994956, EPI_ISL_994957, EPI_ISL_994958, EPI_ISL_994959, EPI_ISL_994960, EPI_ISL_994961, EPI_ISL_994962, EPI_ISL_994963, EPI_ISL_994964, EPI_ISL_994965, EPI_ISL_994966, EPI_ISL_994967, EPI_ISL_994968, EPI_ISL_994969, EPI_ISL_994970, EPI_ISL_994971, EPI_ISL_994972, EPI_ISL_994973, EPI_ISL_994974, EPI_ISL_994975, EPI_ISL_994976, EPI_ISL_994977, EPI_ISL_994978,
EPI_ISL_994979, EPI_ISL_994980, EPI_ISL_994981, EPI_ISL_994982, EPI_ISL_994983, EPI_ISL_994984, EPI_ISL_994985, EPI_ISL_994986, EPI_ISL_994987, EPI_ISL_994988, EPI_ISL_994989, EPI_ISL_994990, EPI_ISL_994991, EPI_ISL_994992, EPI_ISL_994993, EPI_ISL_994994, EPI_ISL_994995, EPI_ISL_994996, EPI_ISL_994997, EPI_ISL_994998, EPI_ISL_994999, EPI_ISL_995000, EPI_ISL_995001, EPI_ISL_995002,
EPI_ISL_995003, EPI_ISL_995004, EPI_ISL_995005, EPI_ISL_995006, EPI_ISL_995007, EPI_ISL_995008, EPI_ISL_995009, EPI_ISL_995010, EPI_ISL_995011, EPI_ISL_995012, EPI_ISL_995013, EPI_ISL_995014, EPI_ISL_995015, EPI_ISL_995016, EPI_ISL_995017, EPI_ISL_995018, EPI_ISL_995019, EPI_ISL_995020, EPI_ISL_995021, EPI_ISL_995022, EPI_ISL_995023, EPI_ISL_995024, EPI_ISL_995025, EPI_ISL_995026,
EPI_ISL_995027, EPI_ISL_995028, EPI_ISL_995029, EPI_ISL_995030, EPI_ISL_995031, EPI_ISL_995032, EPI_ISL_995033, EPI_ISL_995034, EPI_ISL_995035, EPI_ISL_995036, EPI_ISL_995037, EPI_ISL_995038, EPI_ISL_995039, EPI_ISL_995040, EPI_ISL_995041, EPI_ISL_995042, EPI_ISL_995043, EPI_ISL_995044, EPI_ISL_995045, EPI_ISL_995046, EPI_ISL_995047, EPI_ISL_995048, EPI_ISL_995049, EPI_ISL_995050,
EPI_ISL_995051, EPI_ISL_995052, EPI_ISL_995053, EPI_ISL_995054, EPI_ISL_995055, EPI_ISL_995056, EPI_ISL_995057, EPI_ISL_995058, EPI_ISL_995059, EPI_ISL_995060, EPI_ISL_995061, EPI_ISL_995062, EPI_ISL_995063, EPI_ISL_995064, EPI_ISL_995065, EPI_ISL_995066, EPI_ISL_995067, EPI_ISL_995068, EPI_ISL_995069, EPI_ISL_995070, EPI_ISL_995071, EPI_ISL_995072, EPI_ISL_995073, EPI_ISL_995074,
EPI_ISL_995075, EPI_ISL_995076, EPI_ISL_995077, EPI_ISL_995078, EPI_ISL_995079, EPI_ISL_995080, EPI_ISL_995081, EPI_ISL_995082, EPI_ISL_995083, EPI_ISL_995084, EPI_ISL_995085, EPI_ISL_995086, EPI_ISL_995087, EPI_ISL_995088, EPI_ISL_995089, EPI_ISL_995090, EPI_ISL_995091, EPI_ISL_995092, EPI_ISL_995093, EPI_ISL_995094, EPI_ISL_995095, EPI_ISL_995096, EPI_ISL_995097, EPI_ISL_995098,
EPI_ISL_995099, EPI_ISL_995100, EPI_ISL_995101, EPI_ISL_995102, EPI_ISL_995103, EPI_ISL_995104, EPI_ISL_995105, EPI_ISL_995106, EPI_ISL_995107, EPI_ISL_995108, EPI_ISL_995109, EPI_ISL_995110, EPI_ISL_995111, EPI_ISL_995112, EPI_ISL_995113, EPI_ISL_995114, EPI_ISL_995115, EPI_ISL_995116, EPI_ISL_995117, EPI_ISL_995118, EPI_ISL_995119, EPI_ISL_995120, EPI_ISL_995121, EPI_ISL_995122,
EPI_ISL_995123, EPI_ISL_995124, EPI_ISL_995125, EPI_ISL_995126, EPI_ISL_995127, EPI_ISL_995128, EPI_ISL_995129, EPI_ISL_995130, EPI_ISL_995131, EPI_ISL_995132, EPI_ISL_995133, EPI_ISL_995134, EPI_ISL_995135, EPI_ISL_995136, EPI_ISL_995137, EPI_ISL_995138, EPI_ISL_995139, EPI_ISL_995140, EPI_ISL_995141, EPI_ISL_995142, EPI_ISL_995143, EPI_ISL_995144, EPI_ISL_995145, EPI_ISL_995146,
EPI_ISL_995147, EPI_ISL_995148, EPI_ISL_995149, EPI_ISL_995150, EPI_ISL_995151, EPI_ISL_995152, EPI_ISL_995153, EPI_ISL_995154, EPI_ISL_995155, EPI_ISL_995156, EPI_ISL_995157, EPI_ISL_995158, EPI_ISL_995159, EPI_ISL_995160, EPI_ISL_995161, EPI_ISL_995162, EPI_ISL_995163, EPI_ISL_995164, EPI_ISL_995165, EPI_ISL_995166, EPI_ISL_995167, EPI_ISL_995168, EPI_ISL_995169, EPI_ISL_995170,
EPI_ISL_995171, EPI_ISL_1040480, EPI_ISL_1041213, EPI_ISL_1041214, EPI_ISL_1041215, EPI_ISL_1041216, EPI_ISL_1041217, EPI_ISL_1041218, EPI_ISL_1041219, EPI_ISL_1041220, EPI_ISL_1041221, EPI_ISL_1041222, EPI_ISL_1041223, EPI_ISL_1041224, EPI_ISL_1041225, EPI_ISL_1041226, EPI_ISL_1041227, EPI_ISL_1041228, EPI_ISL_1041229, EPI_ISL_1041230, EPI_ISL_1041231, EPI_ISL_1041232,
EPI_ISL_1041233, EPI_ISL_1041234, EPI_ISL_1041235, EPI_ISL_1041236, EPI_ISL_1041237, EPI_ISL_1041238, EPI_ISL_1041239, EPI_ISL_1041240, EPI_ISL_1041241, EPI_ISL_1041242, EPI_ISL_1041243, EPI_ISL_1041244, EPI_ISL_1041245, EPI_ISL_1041246, EPI_ISL_1041247, EPI_ISL_1041248, EPI_ISL_1041249, EPI_ISL_1041250, EPI_ISL_1041251, EPI_ISL_1041252, EPI_ISL_1041253, EPI_ISL_1041254,
EPI_ISL_1041255, EPI_ISL_1041256, EPI_ISL_1041257, EPI_ISL_1041258, EPI_ISL_1041259, EPI_ISL_1041260, EPI_ISL_1041261, EPI_ISL_1041262, EPI_ISL_1041263, EPI_ISL_1041264, EPI_ISL_1041265, EPI_ISL_1041266, EPI_ISL_1041267, EPI_ISL_1041268, EPI_ISL_1041269, EPI_ISL_1041270, EPI_ISL_1041271, EPI_ISL_1041272, EPI_ISL_1041273, EPI_ISL_1041274, EPI_ISL_1041275, EPI_ISL_1041276,
EPI_ISL_1041277, EPI_ISL_1041278, EPI_ISL_1041279, EPI_ISL_1041280, EPI_ISL_1041281, EPI_ISL_1041282, EPI_ISL_1041283, EPI_ISL_1041284, EPI_ISL_1041285, EPI_ISL_1041286, EPI_ISL_1041287, EPI_ISL_1041288, EPI_ISL_1041289, EPI_ISL_1041290, EPI_ISL_1041291, EPI_ISL_1041292, EPI_ISL_1041293, EPI_ISL_1041294, EPI_ISL_1041295, EPI_ISL_1041296, EPI_ISL_1041297, EPI_ISL_1041298,
EPI_ISL_1041299, EPI_ISL_1041300, EPI_ISL_1041301, EPI_ISL_1041302, EPI_ISL_1041303, EPI_ISL_1041304, EPI_ISL_1041305, EPI_ISL_1041306, EPI_ISL_1041307, EPI_ISL_1041308, EPI_ISL_1041309, EPI_ISL_1041310, EPI_ISL_1041311, EPI_ISL_1041312, EPI_ISL_1041313, EPI_ISL_1041314, EPI_ISL_1041315, EPI_ISL_1041316, EPI_ISL_1041317, EPI_ISL_1041318, EPI_ISL_1041319, EPI_ISL_1041320,
EPI_ISL_1041321, EPI_ISL_1041322, EPI_ISL_1041323, EPI_ISL_1041324, EPI_ISL_1041325, EPI_ISL_1041326, EPI_ISL_1041327, EPI_ISL_1041328, EPI_ISL_1041329, EPI_ISL_1041330, EPI_ISL_1041331, EPI_ISL_1041332, EPI_ISL_1041333, EPI_ISL_1041334, EPI_ISL_1041335, EPI_ISL_1041336, EPI_ISL_1041337, EPI_ISL_1041338, EPI_ISL_1041339, EPI_ISL_1041340, EPI_ISL_1041341, EPI_ISL_1041342,
EPI_ISL_1041343, EPI_ISL_1041344, EPI_ISL_1041345, EPI_ISL_1041346, EPI_ISL_1041347, EPI_ISL_1041348, EPI_ISL_1041349, EPI_ISL_1041350, EPI_ISL_1041351, EPI_ISL_1041352, EPI_ISL_1041353, EPI_ISL_1041354, EPI_ISL_1041355, EPI_ISL_1041356, EPI_ISL_1041357, EPI_ISL_1041358, EPI_ISL_1041359, EPI_ISL_1041360, EPI_ISL_1041361, EPI_ISL_1041362, EPI_ISL_1041363, EPI_ISL_1041364,
EPI_ISL_1041365, EPI_ISL_1041366, EPI_ISL_1041367, EPI_ISL_1041368, EPI_ISL_1041369, EPI_ISL_1041370, EPI_ISL_1041371, EPI_ISL_1041372, EPI_ISL_1041373, EPI_ISL_1041374, EPI_ISL_1041375, EPI_ISL_1041376, EPI_ISL_1041377, EPI_ISL_1041378, EPI_ISL_1041379, EPI_ISL_1041380, EPI_ISL_1041381, EPI_ISL_1041382, EPI_ISL_1041383, EPI_ISL_1041384, EPI_ISL_1041385, EPI_ISL_1041386,
EPI_ISL_1041387, EPI_ISL_1041388, EPI_ISL_1041389, EPI_ISL_1041390, EPI_ISL_1041391, EPI_ISL_1041392, EPI_ISL_1041393, EPI_ISL_1041394, EPI_ISL_1041395, EPI_ISL_1041396, EPI_ISL_1041397, EPI_ISL_1041398, EPI_ISL_1041399, EPI_ISL_1041400, EPI_ISL_1041401, EPI_ISL_1041402, EPI_ISL_1041403, EPI_ISL_1041404, EPI_ISL_1041405, EPI_ISL_1041406, EPI_ISL_1041407, EPI_ISL_1041408,
EPI_ISL_1041409, EPI_ISL_1041410, EPI_ISL_1041411, EPI_ISL_1041412, EPI_ISL_1041413, EPI_ISL_1041414, EPI_ISL_1041415, EPI_ISL_1041416, EPI_ISL_1041417, EPI_ISL_1041418, EPI_ISL_1041419, EPI_ISL_1041420, EPI_ISL_1041421, EPI_ISL_1041422, EPI_ISL_1041423, EPI_ISL_1041424, EPI_ISL_1041425, EPI_ISL_1041426, EPI_ISL_1041427, EPI_ISL_1041428, EPI_ISL_1041429, EPI_ISL_1041430,
EPI_ISL_1041431, EPI_ISL_1041432, EPI_ISL_1041433, EPI_ISL_1041434, EPI_ISL_1041435, EPI_ISL_1041436, EPI_ISL_1041437, EPI_ISL_1041438, EPI_ISL_1041439, EPI_ISL_1041440, EPI_ISL_1041441, EPI_ISL_1041442, EPI_ISL_1041443, EPI_ISL_1041444, EPI_ISL_1041445, EPI_ISL_1041446, EPI_ISL_1041447, EPI_ISL_1041448, EPI_ISL_1041449, EPI_ISL_1041450, EPI_ISL_1041451, EPI_ISL_1041452,
EPI_ISL_1041453, EPI_ISL_1041454, EPI_ISL_1041455, EPI_ISL_1041456, EPI_ISL_1041457, EPI_ISL_1041458, EPI_ISL_1041459, EPI_ISL_1041460, EPI_ISL_1041461, EPI_ISL_1041462, EPI_ISL_1041463, EPI_ISL_1041464, EPI_ISL_1041465, EPI_ISL_1041466, EPI_ISL_1041467, EPI_ISL_1041468, EPI_ISL_1041469, EPI_ISL_1041470, EPI_ISL_1041471, EPI_ISL_1041472, EPI_ISL_1041473, EPI_ISL_1041474,
EPI_ISL_1041475, EPI_ISL_1041476, EPI_ISL_1041477, EPI_ISL_1041478, EPI_ISL_1041479, EPI_ISL_1041480, EPI_ISL_1041481, EPI_ISL_1041482, EPI_ISL_1041483, EPI_ISL_1041484, EPI_ISL_1041485, EPI_ISL_1041486, EPI_ISL_1041487, EPI_ISL_1041488, EPI_ISL_1041489, EPI_ISL_1041490, EPI_ISL_1041491, EPI_ISL_1041492, EPI_ISL_1041493, EPI_ISL_1041494, EPI_ISL_1041495, EPI_ISL_1041496,
EPI_ISL_1041497, EPI_ISL_1041498, EPI_ISL_1041499, EPI_ISL_1041500, EPI_ISL_1041501, EPI_ISL_1041502, EPI_ISL_1041503, EPI_ISL_1041504, EPI_ISL_1041505, EPI_ISL_1041506, EPI_ISL_1041507, EPI_ISL_1041508, EPI_ISL_1041510, EPI_ISL_1041511, EPI_ISL_1041512, EPI_ISL_1041513, EPI_ISL_1041514, EPI_ISL_1041515, EPI_ISL_1041516, EPI_ISL_1041517, EPI_ISL_1041518, EPI_ISL_1041519,
EPI_ISL_1041520, EPI_ISL_1041521, EPI_ISL_1041522, EPI_ISL_1041523, EPI_ISL_1041524, EPI_ISL_1041525, EPI_ISL_1041526, EPI_ISL_1041527, EPI_ISL_1041528, EPI_ISL_1041529, EPI_ISL_1041530, EPI_ISL_1041531, EPI_ISL_1041532, EPI_ISL_1041533, EPI_ISL_1041534, EPI_ISL_1041535, EPI_ISL_1041536, EPI_ISL_1041537, EPI_ISL_1041538, EPI_ISL_1041539, EPI_ISL_1041540, EPI_ISL_1041541,
EPI_ISL_1041542, EPI_ISL_1041543, EPI_ISL_1041544, EPI_ISL_1041545, EPI_ISL_1041546, EPI_ISL_1041547, EPI_ISL_1041548, EPI_ISL_1041549, EPI_ISL_1041550, EPI_ISL_1041551, EPI_ISL_1041552, EPI_ISL_1041553, EPI_ISL_1041554, EPI_ISL_1041555, EPI_ISL_1041556, EPI_ISL_1041557, EPI_ISL_1041558, EPI_ISL_1041559, EPI_ISL_1041560, EPI_ISL_1041561, EPI_ISL_1041562, EPI_ISL_1041563,
EPI_ISL_1041564, EPI_ISL_1041565, EPI_ISL_1041566, EPI_ISL_1041567, EPI_ISL_1041568, EPI_ISL_1041569, EPI_ISL_1041570, EPI_ISL_1041571, EPI_ISL_1041572, EPI_ISL_1041573, EPI_ISL_1041574, EPI_ISL_1041575, EPI_ISL_1041576, EPI_ISL_1041577, EPI_ISL_1041578, EPI_ISL_1041579, EPI_ISL_1041580, EPI_ISL_1041581, EPI_ISL_1041582, EPI_ISL_1041583, EPI_ISL_1041584, EPI_ISL_1041585,
EPI_ISL_1041586, EPI_ISL_1041587, EPI_ISL_1041588, EPI_ISL_1041589, EPI_ISL_1041590, EPI_ISL_1041591, EPI_ISL_1041592, EPI_ISL_1041593, EPI_ISL_1041594, EPI_ISL_1041595, EPI_ISL_1041596, EPI_ISL_1041597, EPI_ISL_1041598, EPI_ISL_1041599, EPI_ISL_1041600, EPI_ISL_1041601, EPI_ISL_1041602, EPI_ISL_1041603, EPI_ISL_1041604, EPI_ISL_1041605, EPI_ISL_1041606, EPI_ISL_1041607,
EPI_ISL_1041608, EPI_ISL_1041609, EPI_ISL_1041610, EPI_ISL_1041611, EPI_ISL_1041612, EPI_ISL_1041613, EPI_ISL_1041614, EPI_ISL_1041615, EPI_ISL_1041616, EPI_ISL_1041617, EPI_ISL_1041618, EPI_ISL_1041619, EPI_ISL_1041620, EPI_ISL_1041621, EPI_ISL_1041622, EPI_ISL_1041623, EPI_ISL_1041624, EPI_ISL_1041625, EPI_ISL_1041626, EPI_ISL_1041627, EPI_ISL_1041628, EPI_ISL_1041629,
EPI_ISL_1041630, EPI_ISL_1041631, EPI_ISL_1041632, EPI_ISL_1041633, EPI_ISL_1041634, EPI_ISL_1041635, EPI_ISL_1041636, EPI_ISL_1041637, EPI_ISL_1041638, EPI_ISL_1041639, EPI_ISL_1041640, EPI_ISL_1041641, EPI_ISL_1041642, EPI_ISL_1041643, EPI_ISL_1041644, EPI_ISL_1041645, EPI_ISL_1041646, EPI_ISL_1041647, EPI_ISL_1041648, EPI_ISL_1041649, EPI_ISL_1041650, EPI_ISL_1041651,
EPI_ISL_1041652, EPI_ISL_1041653, EPI_ISL_1041654, EPI_ISL_1041655, EPI_ISL_1041656, EPI_ISL_1041657, EPI_ISL_1041658, EPI_ISL_1041659, EPI_ISL_1041660, EPI_ISL_1041661, EPI_ISL_1041662, EPI_ISL_1041663, EPI_ISL_1041664, EPI_ISL_1041665, EPI_ISL_1041666, EPI_ISL_1041667, EPI_ISL_1041668, EPI_ISL_1041669, EPI_ISL_1041670, EPI_ISL_1041671, EPI_ISL_1041672, EPI_ISL_1041673,
EPI_ISL_1041674, EPI_ISL_1041675, EPI_ISL_1041676, EPI_ISL_1041677, EPI_ISL_1041678, EPI_ISL_1041679, EPI_ISL_1041680, EPI_ISL_1041681, EPI_ISL_1041682, EPI_ISL_1041683, EPI_ISL_1041684, EPI_ISL_1041685, EPI_ISL_1041686, EPI_ISL_1041687, EPI_ISL_1041688, EPI_ISL_1041689, EPI_ISL_1041690, EPI_ISL_1041691, EPI_ISL_1041692, EPI_ISL_1041693, EPI_ISL_1041694, EPI_ISL_1041695,
EPI_ISL_1041696, EPI_ISL_1041697, EPI_ISL_1041698, EPI_ISL_1041699, EPI_ISL_1041700, EPI_ISL_1041701, EPI_ISL_1041702, EPI_ISL_1041703, EPI_ISL_1041704, EPI_ISL_1041705, EPI_ISL_1041706, EPI_ISL_1041707, EPI_ISL_1041708, EPI_ISL_1041709, EPI_ISL_1041710, EPI_ISL_1041711, EPI_ISL_1041712, EPI_ISL_1041713, EPI_ISL_1041714, EPI_ISL_1041715, EPI_ISL_1041716, EPI_ISL_1041717,
EPI_ISL_1041718, EPI_ISL_1041719, EPI_ISL_1041720, EPI_ISL_1041721, EPI_ISL_1041722, EPI_ISL_1041723, EPI_ISL_1041724, EPI_ISL_1041725, EPI_ISL_1041726, EPI_ISL_1041727, EPI_ISL_1041728, EPI_ISL_1041729, EPI_ISL_1041730, EPI_ISL_1041731, EPI_ISL_1041732, EPI_ISL_1041733, EPI_ISL_1041734, EPI_ISL_1041735, EPI_ISL_1041736, EPI_ISL_1041737, EPI_ISL_1041738, EPI_ISL_1041739,
EPI_ISL_1041740, EPI_ISL_1041741, EPI_ISL_1041742, EPI_ISL_1041743, EPI_ISL_1041744, EPI_ISL_1041745, EPI_ISL_1041746, EPI_ISL_1041747, EPI_ISL_1041748, EPI_ISL_1041749, EPI_ISL_1041750, EPI_ISL_1041751, EPI_ISL_1041752, EPI_ISL_1041753, EPI_ISL_1041754, EPI_ISL_1041755, EPI_ISL_1041756, EPI_ISL_1041757, EPI_ISL_1041758, EPI_ISL_1041759, EPI_ISL_1041760, EPI_ISL_1041761,
EPI_ISL_1041762, EPI_ISL_1041763, EPI_ISL_1041764, EPI_ISL_1041765, EPI_ISL_1041766, EPI_ISL_1041767, EPI_ISL_1041768, EPI_ISL_1041769, EPI_ISL_1041770, EPI_ISL_1041771, EPI_ISL_1041772, EPI_ISL_1041773, EPI_ISL_1041774, EPI_ISL_1041775, EPI_ISL_1041776, EPI_ISL_1041777, EPI_ISL_1041778, EPI_ISL_1041779, EPI_ISL_1041780, EPI_ISL_1041781, EPI_ISL_1041782, EPI_ISL_1041783,
EPI_ISL_1041784, EPI_ISL_1041785, EPI_ISL_1041786, EPI_ISL_1041787, EPI_ISL_1041788, EPI_ISL_1041789, EPI_ISL_1041790, EPI_ISL_1041791, EPI_ISL_1041792, EPI_ISL_1041793, EPI_ISL_1041794, EPI_ISL_1041795, EPI_ISL_1041796, EPI_ISL_1041797, EPI_ISL_1041798, EPI_ISL_1041799, EPI_ISL_1041800, EPI_ISL_1041801, EPI_ISL_1041802, EPI_ISL_1041803, EPI_ISL_1041804, EPI_ISL_1041805,
EPI_ISL_1041806, EPI_ISL_1041807, EPI_ISL_1041808, EPI_ISL_1041809, EPI_ISL_1041810, EPI_ISL_1041811, EPI_ISL_1041812, EPI_ISL_1041813, EPI_ISL_1041814, EPI_ISL_1041815, EPI_ISL_1041816, EPI_ISL_1041817, EPI_ISL_1041818, EPI_ISL_1041819, EPI_ISL_1041820, EPI_ISL_1041821, EPI_ISL_1041822, EPI_ISL_1041823, EPI_ISL_1041824, EPI_ISL_1041825, EPI_ISL_1041826, EPI_ISL_1041827,
EPI_ISL_1041828, EPI_ISL_1041829, EPI_ISL_1041830, EPI_ISL_1041831, EPI_ISL_1041832, EPI_ISL_1041833, EPI_ISL_1041834, EPI_ISL_1041835, EPI_ISL_1041836, EPI_ISL_1041837, EPI_ISL_1041838, EPI_ISL_1041839, EPI_ISL_1041840, EPI_ISL_1041841, EPI_ISL_1041842, EPI_ISL_1041843, EPI_ISL_1041844, EPI_ISL_1041845, EPI_ISL_1041846, EPI_ISL_1041847, EPI_ISL_1041848, EPI_ISL_1041849,
EPI_ISL_1041850, EPI_ISL_1041851, EPI_ISL_1041852, EPI_ISL_1041853, EPI_ISL_1041854, EPI_ISL_1041855, EPI_ISL_1041856, EPI_ISL_1041857, EPI_ISL_1041858, EPI_ISL_1041859, EPI_ISL_1041860, EPI_ISL_1041861, EPI_ISL_1041862, EPI_ISL_1041863, EPI_ISL_1041864, EPI_ISL_1041865, EPI_ISL_1041866, EPI_ISL_1041867, EPI_ISL_1041868, EPI_ISL_1041869, EPI_ISL_1041870, EPI_ISL_1041871,
EPI_ISL_1041872, EPI_ISL_1041873, EPI_ISL_1041874, EPI_ISL_1041875, EPI_ISL_1041876, EPI_ISL_1041877, EPI_ISL_1041878, EPI_ISL_1041879, EPI_ISL_1041880, EPI_ISL_1041881, EPI_ISL_1041882, EPI_ISL_1041883, EPI_ISL_1041884, EPI_ISL_1041885, EPI_ISL_1041886, EPI_ISL_1041887, EPI_ISL_1041888, EPI_ISL_1041889, EPI_ISL_1041890, EPI_ISL_1041891, EPI_ISL_1041892, EPI_ISL_1041893,
EPI_ISL_1041894, EPI_ISL_1041895, EPI_ISL_1041896, EPI_ISL_1041897, EPI_ISL_1041898, EPI_ISL_1041899, EPI_ISL_1041900, EPI_ISL_1041901, EPI_ISL_1041902, EPI_ISL_1041903, EPI_ISL_1041904, EPI_ISL_1041905, EPI_ISL_1041906, EPI_ISL_1041907, EPI_ISL_1041908, EPI_ISL_1041909, EPI_ISL_1041910, EPI_ISL_1041911, EPI_ISL_1041912, EPI_ISL_1041913, EPI_ISL_1041914, EPI_ISL_1041915,
EPI_ISL_1041916, EPI_ISL_1041917, EPI_ISL_1041918, EPI_ISL_1041919, EPI_ISL_1041920, EPI_ISL_1041921, EPI_ISL_1041922, EPI_ISL_1041923, EPI_ISL_1041924, EPI_ISL_1041925, EPI_ISL_1041926, EPI_ISL_1041927, EPI_ISL_1041928, EPI_ISL_1041929, EPI_ISL_1041930, EPI_ISL_1097781, EPI_ISL_1097782, EPI_ISL_1097783, EPI_ISL_1097784, EPI_ISL_1097785, EPI_ISL_1097786, EPI_ISL_1097787,
EPI_ISL_1097788, EPI_ISL_1097789, EPI_ISL_1097790, EPI_ISL_1097791, EPI_ISL_1097792, EPI_ISL_1097793, EPI_ISL_1097794, EPI_ISL_1097795, EPI_ISL_1097796, EPI_ISL_1097797, EPI_ISL_1097798, EPI_ISL_1097799, EPI_ISL_1097800, EPI_ISL_1097801, EPI_ISL_1097802, EPI_ISL_1097803, EPI_ISL_1097804, EPI_ISL_1097805, EPI_ISL_1097806, EPI_ISL_1097807, EPI_ISL_1097808, EPI_ISL_1097809,
EPI_ISL_1097810, EPI_ISL_1097811, EPI_ISL_1097812, EPI_ISL_1097813, EPI_ISL_1097814, EPI_ISL_1097815, EPI_ISL_1097816, EPI_ISL_1097817, EPI_ISL_1097818, EPI_ISL_1097819, EPI_ISL_1097820, EPI_ISL_1097821, EPI_ISL_1097822, EPI_ISL_1097823, EPI_ISL_1097824, EPI_ISL_1097825, EPI_ISL_1097826, EPI_ISL_1097827, EPI_ISL_1097828, EPI_ISL_1097829, EPI_ISL_1097830, EPI_ISL_1097831,
EPI_ISL_1097832, EPI_ISL_1097833, EPI_ISL_1097834, EPI_ISL_1097835, EPI_ISL_1097836, EPI_ISL_1097837, EPI_ISL_1097838, EPI_ISL_1097839, EPI_ISL_1097840, EPI_ISL_1097841, EPI_ISL_1097842, EPI_ISL_1097843, EPI_ISL_1097844, EPI_ISL_1097845, EPI_ISL_1097846, EPI_ISL_1097847, EPI_ISL_1097848, EPI_ISL_1097849, EPI_ISL_1097850, EPI_ISL_1097851, EPI_ISL_1097852, EPI_ISL_1097853,
EPI_ISL_1097854, EPI_ISL_1097855, EPI_ISL_1097856, EPI_ISL_1097857, EPI_ISL_1097858, EPI_ISL_1097859, EPI_ISL_1097860, EPI_ISL_1097861, EPI_ISL_1097862, EPI_ISL_1097863, EPI_ISL_1097864, EPI_ISL_1097865, EPI_ISL_1097866, EPI_ISL_1097867, EPI_ISL_1097868, EPI_ISL_1097869, EPI_ISL_1097870, EPI_ISL_1097871, EPI_ISL_1097872, EPI_ISL_1097873, EPI_ISL_1097874, EPI_ISL_1097875,
EPI_ISL_1097876, EPI_ISL_1097877, EPI_ISL_1097878, EPI_ISL_1097879, EPI_ISL_1097880, EPI_ISL_1097881, EPI_ISL_1097882, EPI_ISL_1097883, EPI_ISL_1097884, EPI_ISL_1097885, EPI_ISL_1097886, EPI_ISL_1097887, EPI_ISL_1097888, EPI_ISL_1097889, EPI_ISL_1097890, EPI_ISL_1097891, EPI_ISL_1097892, EPI_ISL_1097893, EPI_ISL_1097894, EPI_ISL_1097895, EPI_ISL_1097896, EPI_ISL_1097897,
EPI_ISL_1097898, EPI_ISL_1097899, EPI_ISL_1097900, EPI_ISL_1097901, EPI_ISL_1097902, EPI_ISL_1097903, EPI_ISL_1097904, EPI_ISL_1097905, EPI_ISL_1097906, EPI_ISL_1097907, EPI_ISL_1097908, EPI_ISL_1097909, EPI_ISL_1097910, EPI_ISL_1097911, EPI_ISL_1097912, EPI_ISL_1097913, EPI_ISL_1097914, EPI_ISL_1097915, EPI_ISL_1097916, EPI_ISL_1097917, EPI_ISL_1097918, EPI_ISL_1097919,
EPI_ISL_1097920, EPI_ISL_1097921, EPI_ISL_1097922, EPI_ISL_1097923, EPI_ISL_1097924, EPI_ISL_1097925, EPI_ISL_1097926, EPI_ISL_1097927, EPI_ISL_1097928, EPI_ISL_1097929, EPI_ISL_1097930, EPI_ISL_1097931, EPI_ISL_1097932, EPI_ISL_1097933, EPI_ISL_1097934, EPI_ISL_1097935, EPI_ISL_1097936, EPI_ISL_1097937, EPI_ISL_1097938, EPI_ISL_1097939, EPI_ISL_1097940, EPI_ISL_1097941,
EPI_ISL_1097942, EPI_ISL_1097943, EPI_ISL_1097944, EPI_ISL_1097945, EPI_ISL_1097946, EPI_ISL_1097947, EPI_ISL_1097948, EPI_ISL_1097949, EPI_ISL_1097950, EPI_ISL_1097951, EPI_ISL_1097952, EPI_ISL_1097953, EPI_ISL_1097954, EPI_ISL_1097955, EPI_ISL_1097956, EPI_ISL_1097957, EPI_ISL_1097958, EPI_ISL_1097959, EPI_ISL_1097960, EPI_ISL_1097961, EPI_ISL_1097962, EPI_ISL_1097963,
EPI_ISL_1097964, EPI_ISL_1097965, EPI_ISL_1097966, EPI_ISL_1097967, EPI_ISL_1097968, EPI_ISL_1097969, EPI_ISL_1097970, EPI_ISL_1097971, EPI_ISL_1097972, EPI_ISL_1097973, EPI_ISL_1097974, EPI_ISL_1097975, EPI_ISL_1097976, EPI_ISL_1097977, EPI_ISL_1097978, EPI_ISL_1097979, EPI_ISL_1097980, EPI_ISL_1097981, EPI_ISL_1097982, EPI_ISL_1097983, EPI_ISL_1097984, EPI_ISL_1097985,
EPI_ISL_1097986, EPI_ISL_1097987, EPI_ISL_1097988, EPI_ISL_1097989, EPI_ISL_1097990, EPI_ISL_1097991, EPI_ISL_1097992, EPI_ISL_1097993, EPI_ISL_1097994, EPI_ISL_1097995, EPI_ISL_1097996, EPI_ISL_1097997, EPI_ISL_1097998, EPI_ISL_1097999, EPI_ISL_1098000, EPI_ISL_1098001, EPI_ISL_1098002, EPI_ISL_1098003, EPI_ISL_1098004, EPI_ISL_1098005, EPI_ISL_1098006, EPI_ISL_1098007,
EPI_ISL_1098008, EPI_ISL_1098009, EPI_ISL_1098010, EPI_ISL_1098011, EPI_ISL_1098012, EPI_ISL_1098013, EPI_ISL_1098014, EPI_ISL_1098015, EPI_ISL_1098016, EPI_ISL_1098017, EPI_ISL_1098018, EPI_ISL_1098019, EPI_ISL_1098020, EPI_ISL_1098021, EPI_ISL_1098022, EPI_ISL_1098023, EPI_ISL_1098024, EPI_ISL_1098025, EPI_ISL_1098026, EPI_ISL_1098027, EPI_ISL_1098028, EPI_ISL_1098029,
EPI_ISL_1098030, EPI_ISL_1098031, EPI_ISL_1098032, EPI_ISL_1098033, EPI_ISL_1098034, EPI_ISL_1098035, EPI_ISL_1098036, EPI_ISL_1098037, EPI_ISL_1098038, EPI_ISL_1098039, EPI_ISL_1098040, EPI_ISL_1098041, EPI_ISL_1098042, EPI_ISL_1098043, EPI_ISL_1098044, EPI_ISL_1098045, EPI_ISL_1098046, EPI_ISL_1098047, EPI_ISL_1098048, EPI_ISL_1098049, EPI_ISL_1098050, EPI_ISL_1098051,
EPI_ISL_1098052, EPI_ISL_1098053, EPI_ISL_1098054, EPI_ISL_1098055, EPI_ISL_1098056, EPI_ISL_1098057, EPI_ISL_1098058, EPI_ISL_1098059, EPI_ISL_1098060, EPI_ISL_1098061, EPI_ISL_1098062, EPI_ISL_1098063, EPI_ISL_1098064, EPI_ISL_1098065, EPI_ISL_1098066, EPI_ISL_1098067, EPI_ISL_1098068, EPI_ISL_1098069, EPI_ISL_1098070, EPI_ISL_1098071, EPI_ISL_1098072, EPI_ISL_1098073,
EPI_ISL_1098074, EPI_ISL_1098075, EPI_ISL_1098076, EPI_ISL_1098077, EPI_ISL_1098078, EPI_ISL_1098079, EPI_ISL_1098080, EPI_ISL_1098081, EPI_ISL_1098082, EPI_ISL_1098083, EPI_ISL_1098084, EPI_ISL_1098085, EPI_ISL_1098086, EPI_ISL_1098087, EPI_ISL_1098088, EPI_ISL_1098089, EPI_ISL_1098090, EPI_ISL_1098091, EPI_ISL_1098092, EPI_ISL_1098093, EPI_ISL_1098094, EPI_ISL_1098095,
EPI_ISL_1098096, EPI_ISL_1098097, EPI_ISL_1098098, EPI_ISL_1098099, EPI_ISL_1098100, EPI_ISL_1098101, EPI_ISL_1098102, EPI_ISL_1098103, EPI_ISL_1098104, EPI_ISL_1098105, EPI_ISL_1098106, EPI_ISL_1098107, EPI_ISL_1098108, EPI_ISL_1098109, EPI_ISL_1098110, EPI_ISL_1098111, EPI_ISL_1098112, EPI_ISL_1098113, EPI_ISL_1098114, EPI_ISL_1098115, EPI_ISL_1098116, EPI_ISL_1098117,
EPI_ISL_1098118, EPI_ISL_1098119, EPI_ISL_1098120, EPI_ISL_1098121, EPI_ISL_1098122, EPI_ISL_1098123, EPI_ISL_1098124, EPI_ISL_1098125, EPI_ISL_1098126, EPI_ISL_1098127, EPI_ISL_1098128, EPI_ISL_1098129, EPI_ISL_1098130, EPI_ISL_1098131, EPI_ISL_1098132, EPI_ISL_1098133, EPI_ISL_1098134, EPI_ISL_1098135, EPI_ISL_1098136, EPI_ISL_1098137, EPI_ISL_1098138, EPI_ISL_1098139,
EPI_ISL_1098140, EPI_ISL_1098141, EPI_ISL_1098142, EPI_ISL_1098143, EPI_ISL_1098144, EPI_ISL_1098145, EPI_ISL_1098146, EPI_ISL_1098147, EPI_ISL_1098148, EPI_ISL_1098149, EPI_ISL_1098150, EPI_ISL_1098151, EPI_ISL_1098152, EPI_ISL_1098153, EPI_ISL_1098154, EPI_ISL_1098155, EPI_ISL_1098156, EPI_ISL_1098157, EPI_ISL_1098158, EPI_ISL_1098159, EPI_ISL_1098160, EPI_ISL_1098161,
EPI_ISL_1098162, EPI_ISL_1098163, EPI_ISL_1098164, EPI_ISL_1098165, EPI_ISL_1098166, EPI_ISL_1098167, EPI_ISL_1098168, EPI_ISL_1098169, EPI_ISL_1098170, EPI_ISL_1098171, EPI_ISL_1098172, EPI_ISL_1098173, EPI_ISL_1098174, EPI_ISL_1098175, EPI_ISL_1098176, EPI_ISL_1098177, EPI_ISL_1098178, EPI_ISL_1098179, EPI_ISL_1098180, EPI_ISL_1098181, EPI_ISL_1098182, EPI_ISL_1098183,
EPI_ISL_1098184, EPI_ISL_1098185, EPI_ISL_1098186, EPI_ISL_1098187, EPI_ISL_1098188, EPI_ISL_1098189, EPI_ISL_1098190, EPI_ISL_1098191, EPI_ISL_1098192, EPI_ISL_1098193, EPI_ISL_1098194, EPI_ISL_1098195, EPI_ISL_1098196, EPI_ISL_1098197, EPI_ISL_1098198, EPI_ISL_1098199, EPI_ISL_1098200, EPI_ISL_1098201, EPI_ISL_1098202, EPI_ISL_1098203, EPI_ISL_1098204, EPI_ISL_1098205,
EPI_ISL_1098206, EPI_ISL_1098207, EPI_ISL_1098208, EPI_ISL_1098209, EPI_ISL_1098210, EPI_ISL_1098211, EPI_ISL_1098212, EPI_ISL_1098213, EPI_ISL_1098214, EPI_ISL_1098215, EPI_ISL_1098216, EPI_ISL_1098217, EPI_ISL_1098218, EPI_ISL_1098219, EPI_ISL_1098220, EPI_ISL_1098221, EPI_ISL_1098222, EPI_ISL_1098223, EPI_ISL_1098224, EPI_ISL_1098225, EPI_ISL_1098226, EPI_ISL_1098227,
EPI_ISL_1098228, EPI_ISL_1098229, EPI_ISL_1098230, EPI_ISL_1098231, EPI_ISL_1098232, EPI_ISL_1098233, EPI_ISL_1098234, EPI_ISL_1098235, EPI_ISL_1098236, EPI_ISL_1098237, EPI_ISL_1098238, EPI_ISL_1098239, EPI_ISL_1098240, EPI_ISL_1098241, EPI_ISL_1098242, EPI_ISL_1098243, EPI_ISL_1098244, EPI_ISL_1098245, EPI_ISL_1098246, EPI_ISL_1098247, EPI_ISL_1098248, EPI_ISL_1098249,
EPI_ISL_1098250, EPI_ISL_1098251, EPI_ISL_1098252, EPI_ISL_1098253, EPI_ISL_1098254, EPI_ISL_1098255, EPI_ISL_1098256, EPI_ISL_1098257, EPI_ISL_1098258, EPI_ISL_1098259, EPI_ISL_1098260, EPI_ISL_1098261, EPI_ISL_1098262, EPI_ISL_1098263, EPI_ISL_1098264, EPI_ISL_1098265, EPI_ISL_1098266, EPI_ISL_1098267, EPI_ISL_1098268, EPI_ISL_1098269, EPI_ISL_1098270, EPI_ISL_1098271,
EPI_ISL_1098272, EPI_ISL_1098273, EPI_ISL_1098274, EPI_ISL_1098275, EPI_ISL_1098276, EPI_ISL_1098277, EPI_ISL_1098278, EPI_ISL_1098279, EPI_ISL_1098280, EPI_ISL_1098281, EPI_ISL_1098282, EPI_ISL_1098283, EPI_ISL_1098284, EPI_ISL_1098285, EPI_ISL_1098286, EPI_ISL_1098287, EPI_ISL_1098288, EPI_ISL_1098289, EPI_ISL_1098290, EPI_ISL_1098291, EPI_ISL_1098292, EPI_ISL_1098293,
EPI_ISL_1098294, EPI_ISL_1098295, EPI_ISL_1098296, EPI_ISL_1098297, EPI_ISL_1098298, EPI_ISL_1098299, EPI_ISL_1098300, EPI_ISL_1098301, EPI_ISL_1098302, EPI_ISL_1098303, EPI_ISL_1098304, EPI_ISL_1098305, EPI_ISL_1098306, EPI_ISL_1098307, EPI_ISL_1098308, EPI_ISL_1098309, EPI_ISL_1098310, EPI_ISL_1098311, EPI_ISL_1098312, EPI_ISL_1098313, EPI_ISL_1098314, EPI_ISL_1098315,
EPI_ISL_1098316, EPI_ISL_1098317, EPI_ISL_1098318, EPI_ISL_1098319, EPI_ISL_1098320, EPI_ISL_1098321, EPI_ISL_1098322, EPI_ISL_1098323, EPI_ISL_1098324, EPI_ISL_1098325, EPI_ISL_1098326, EPI_ISL_1098327, EPI_ISL_1098328, EPI_ISL_1098329, EPI_ISL_1098330, EPI_ISL_1098331, EPI_ISL_1098332, EPI_ISL_1098333, EPI_ISL_1098334, EPI_ISL_1098335, EPI_ISL_1098336, EPI_ISL_1098337,
EPI_ISL_1098338, EPI_ISL_1098339, EPI_ISL_1098340, EPI_ISL_1098341, EPI_ISL_1098342, EPI_ISL_1098343, EPI_ISL_1098344, EPI_ISL_1098345, EPI_ISL_1098346, EPI_ISL_1098347, EPI_ISL_1098348, EPI_ISL_1098349, EPI_ISL_1098350, EPI_ISL_1098351, EPI_ISL_1098352, EPI_ISL_1098353, EPI_ISL_1098354, EPI_ISL_1098355, EPI_ISL_1098356, EPI_ISL_1098357, EPI_ISL_1098358, EPI_ISL_1098359,
EPI_ISL_1098360, EPI_ISL_1098361, EPI_ISL_1098362, EPI_ISL_1098363, EPI_ISL_1098364, EPI_ISL_1098365, EPI_ISL_1098366, EPI_ISL_1098367, EPI_ISL_1098368, EPI_ISL_1098369, EPI_ISL_1098370, EPI_ISL_1098371, EPI_ISL_1098372, EPI_ISL_1098373, EPI_ISL_1098374, EPI_ISL_1098375, EPI_ISL_1098376, EPI_ISL_1098377, EPI_ISL_1098378, EPI_ISL_1098379, EPI_ISL_1098380, EPI_ISL_1098381,
EPI_ISL_1098382, EPI_ISL_1098383, EPI_ISL_1098384, EPI_ISL_1098385, EPI_ISL_1098386, EPI_ISL_1098387, EPI_ISL_1098388, EPI_ISL_1098389, EPI_ISL_1098390, EPI_ISL_1098391, EPI_ISL_1098392, EPI_ISL_1098393, EPI_ISL_1098394, EPI_ISL_1098395, EPI_ISL_1098396, EPI_ISL_1098397, EPI_ISL_1098398, EPI_ISL_1098399, EPI_ISL_1098400, EPI_ISL_1098401, EPI_ISL_1098402, EPI_ISL_1098403,
EPI_ISL_1098404, EPI_ISL_1098405, EPI_ISL_1098406, EPI_ISL_1098407, EPI_ISL_1098408, EPI_ISL_1098409, EPI_ISL_1098410, EPI_ISL_1098411, EPI_ISL_1098412, EPI_ISL_1098413, EPI_ISL_1098414, EPI_ISL_1098415, EPI_ISL_1098416, EPI_ISL_1098417, EPI_ISL_1098418, EPI_ISL_1098419, EPI_ISL_1098420, EPI_ISL_1098421, EPI_ISL_1098422, EPI_ISL_1098423, EPI_ISL_1098424, EPI_ISL_1098425,
EPI_ISL_1098426, EPI_ISL_1098427, EPI_ISL_1098428, EPI_ISL_1098429, EPI_ISL_1098430, EPI_ISL_1098431, EPI_ISL_1098432, EPI_ISL_1098433, EPI_ISL_1098434, EPI_ISL_1098435, EPI_ISL_1098436, EPI_ISL_1098437, EPI_ISL_1098438, EPI_ISL_1098439, EPI_ISL_1098440, EPI_ISL_1098441, EPI_ISL_1098442, EPI_ISL_1098443, EPI_ISL_1098444, EPI_ISL_1098445, EPI_ISL_1098446, EPI_ISL_1098447,
EPI_ISL_1098448, EPI_ISL_1098449, EPI_ISL_1098450, EPI_ISL_1098451, EPI_ISL_1098452, EPI_ISL_1098453, EPI_ISL_1098454, EPI_ISL_1098455, EPI_ISL_1098456, EPI_ISL_1098457, EPI_ISL_1098458, EPI_ISL_1098459, EPI_ISL_1098460, EPI_ISL_1098461, EPI_ISL_1098462, EPI_ISL_1098463, EPI_ISL_1098464, EPI_ISL_1098465, EPI_ISL_1098466, EPI_ISL_1098467, EPI_ISL_1098468, EPI_ISL_1098469,
EPI_ISL_1098470, EPI_ISL_1098471, EPI_ISL_1098472, EPI_ISL_1098473, EPI_ISL_1098474, EPI_ISL_1098475, EPI_ISL_1098476, EPI_ISL_1098477, EPI_ISL_1098478, EPI_ISL_1098479, EPI_ISL_1098480, EPI_ISL_1098481, EPI_ISL_1098482, EPI_ISL_1098483, EPI_ISL_1098484, EPI_ISL_1098485, EPI_ISL_1098486, EPI_ISL_1098487, EPI_ISL_1098488, EPI_ISL_1098489, EPI_ISL_1098490, EPI_ISL_1098491,
EPI_ISL_1098492, EPI_ISL_1098493, EPI_ISL_1098494, EPI_ISL_1098495, EPI_ISL_1098496, EPI_ISL_1098497, EPI_ISL_1098498, EPI_ISL_1098499, EPI_ISL_1098500, EPI_ISL_1098501, EPI_ISL_1098502, EPI_ISL_1098503, EPI_ISL_1098504, EPI_ISL_1098505, EPI_ISL_1098506, EPI_ISL_1098507, EPI_ISL_1098508, EPI_ISL_1098509, EPI_ISL_1098510, EPI_ISL_1098511, EPI_ISL_1098512, EPI_ISL_1098513,
EPI_ISL_1098514, EPI_ISL_1098515, EPI_ISL_1098516, EPI_ISL_1098517, EPI_ISL_1098518, EPI_ISL_1098519, EPI_ISL_1098520, EPI_ISL_1098521, EPI_ISL_1098522, EPI_ISL_1098523, EPI_ISL_1098524, EPI_ISL_1098525, EPI_ISL_1098526, EPI_ISL_1098527, EPI_ISL_1098528, EPI_ISL_1098529, EPI_ISL_1098530, EPI_ISL_1098531, EPI_ISL_1098532, EPI_ISL_1098533, EPI_ISL_1098534, EPI_ISL_1098535,

EPI_ISL_1098536, EPI_ISL_1098537, EPI_ISL_1098538, EPI_ISL_1098539, EPI_ISL_1098540, EPI_ISL_1098541, EPI_ISL_1098542, EPI_ISL_1098543, EPI_ISL_1098544, EPI_ISL_1098545, EPI_ISL_1098546, EPI_ISL_1098547, EPI_ISL_1098548, EPI_ISL_1098549, EPI_ISL_1098550, EPI_ISL_1098551, EPI_ISL_1098552, EPI_ISL_1098553, EPI_ISL_1098554, EPI_ISL_1098555, EPI_ISL_1098556, EPI_ISL_1098557, EPI_ISL_1098558, EPI_ISL_1098559, EPI_ISL_1098560, EPI_ISL_1098561, EPI_ISL_1098562, EPI_ISL_1098563, EPI_ISL_1098564, EPI_ISL_1098565, EPI_ISL_1098566, EPI_ISL_1098567, EPI_ISL_1098568, EPI_ISL_1098569, EPI_ISL_1098570, EPI_ISL_1098571, EPI_ISL_1098572, EPI_ISL_1098573, EPI_ISL_1098574, EPI_ISL_1098575, EPI_ISL_1098576, EPI_ISL_1098577, EPI_ISL_1098578, EPI_ISL_1098579, EPI_ISL_1098580, EPI_ISL_1098581, EPI_ISL_1098582, EPI_ISL_1098583, EPI_ISL_1098584, EPI_ISL_1098585, EPI_ISL_1098586, EPI_ISL_1098587, EPI_ISL_1098588, EPI_ISL_1098589, EPI_ISL_1098590, EPI_ISL_1098591, EPI_ISL_1098592, EPI_ISL_1098593, EPI_ISL_1098594, EPI_ISL_1098595, EPI_ISL_1098596, EPI_ISL_1098597, EPI_ISL_1098598, EPI_ISL_1098599, EPI_ISL_1098600, EPI_ISL_1098601, EPI_ISL_1172110, EPI_ISL_1172111, EPI_ISL_1172112, EPI_ISL_1172113, EPI_ISL_1172114, EPI_ISL_1172115, EPI_ISL_1172116, EPI_ISL_1172117, EPI_ISL_1172118, EPI_ISL_1172119, EPI_ISL_1172120, EPI_ISL_1172121, EPI_ISL_1172122, EPI_ISL_1172123, EPI_ISL_1172124, EPI_ISL_1172125, EPI_ISL_1172126, EPI_ISL_1172127, EPI_ISL_1172128, EPI_ISL_1172129, EPI_ISL_1172130, EPI_ISL_1172131, EPI_ISL_1172132, EPI_ISL_1172133, EPI_ISL_1172134, EPI_ISL_1172135, EPI_ISL_1172136, EPI_ISL_1172137, EPI_ISL_1172138, EPI_ISL_1172139, EPI_ISL_1172140, EPI_ISL_1172141, EPI_ISL_1172142, EPI_ISL_1172143, EPI_ISL_1172144, EPI_ISL_1172145, EPI_ISL_1172146, EPI_ISL_1172147, EPI_ISL_1172148, EPI_ISL_1172149, EPI_ISL_1172150, EPI_ISL_1172151, EPI_ISL_1172152, EPI_ISL_1172153, EPI_ISL_1172154, EPI_ISL_1172155, EPI_ISL_1172156, EPI_ISL_1172157, EPI_ISL_1172158, EPI_ISL_1172159, EPI_ISL_1172160, EPI_ISL_1172161, EPI_ISL_1172162, EPI_ISL_1172163, EPI_ISL_1172164, EPI_ISL_1172165, EPI_ISL_1172166, EPI_ISL_1172167, EPI_ISL_1172168, EPI_ISL_1172169, EPI_ISL_1172170, EPI_ISL_1172171, EPI_ISL_1172172, EPI_ISL_1172173, EPI_ISL_1172174, EPI_ISL_1172175, EPI_ISL_1172176, EPI_ISL_1172177, EPI_ISL_1172178, EPI_ISL_1172179, EPI_ISL_1172180, EPI_ISL_1172181, EPI_ISL_1172182, EPI_ISL_1172183, EPI_ISL_1172184, EPI_ISL_1172185, EPI_ISL_1172186, EPI_ISL_1172187, EPI_ISL_1172188, EPI_ISL_1172189, EPI_ISL_1172190, EPI_ISL_1172191, EPI_ISL_1172192, EPI_ISL_1172193, EPI_ISL_1172194, EPI_ISL_1172195, EPI_ISL_1172196, EPI_ISL_1172197, EPI_ISL_1172198, EPI_ISL_1172199, EPI_ISL_1172200, EPI_ISL_1172201, EPI_ISL_1172202, EPI_ISL_1172203, EPI_ISL_1172204, EPI_ISL_1172205, EPI_ISL_1172206, EPI_ISL_1172207, EPI_ISL_1172208, EPI_ISL_1172209, EPI_ISL_1172210, EPI_ISL_1172211, EPI_ISL_1172212, EPI_ISL_1172213, EPI_ISL_1172214, EPI_ISL_1172215, EPI_ISL_1172216, EPI_ISL_1172217, EPI_ISL_1172218, EPI_ISL_1172219, EPI_ISL_1172220, EPI_ISL_1172221, EPI_ISL_1172222, EPI_ISL_1172223, EPI_ISL_1172224, EPI_ISL_1172225, EPI_ISL_1172226, EPI_ISL_1172227, EPI_ISL_1172228, EPI_ISL_1172229, EPI_ISL_1172230, EPI_ISL_1172231, EPI_ISL_1172232, EPI_ISL_1172233, EPI_ISL_1172234, EPI_ISL_1172235, EPI_ISL_1172236, EPI_ISL_1172237, EPI_ISL_1172238, EPI_ISL_1172239, EPI_ISL_1172240, EPI_ISL_1172241, EPI_ISL_1172242, EPI_ISL_1172243, EPI_ISL_1172244, EPI_ISL_1172245, EPI_ISL_1172246, EPI_ISL_1172247, EPI_ISL_1172248, EPI_ISL_1172249, EPI_ISL_1172250, EPI_ISL_1172251, EPI_ISL_1172252, EPI_ISL_1172253, EPI_ISL_1172254, EPI_ISL_1172255, EPI_ISL_1172256, EPI_ISL_1172257, EPI_ISL_1172258, EPI_ISL_1172259, EPI_ISL_1172260, EPI_ISL_1172261, EPI_ISL_1172262, EPI_ISL_1172263, EPI_ISL_1172264, EPI_ISL_1172265, EPI_ISL_1172266, EPI_ISL_1172267, EPI_ISL_1172268, EPI_ISL_1172269, EPI_ISL_1172270, EPI_ISL_1172271, EPI_ISL_1172272, EPI_ISL_1172273, EPI_ISL_1172274, EPI_ISL_1172275, EPI_ISL_1172276, EPI_ISL_1172277, EPI_ISL_1172278, EPI_ISL_1172279, EPI_ISL_1172280, EPI_ISL_1172281, EPI_ISL_1172282, EPI_ISL_1172283, EPI_ISL_1172284, EPI_ISL_1172285, EPI_ISL_1172286, EPI_ISL_1172287, EPI_ISL_1172288, EPI_ISL_1172289, EPI_ISL_1172290, EPI_ISL_1172291, EPI_ISL_1172292, EPI_ISL_1172293, EPI_ISL_1172294, EPI_ISL_1172295, EPI_ISL_1172296, EPI_ISL_1172297, EPI_ISL_1172298, EPI_ISL_1172299, EPI_ISL_1172300, EPI_ISL_1172301, EPI_ISL_1172302, EPI_ISL_1172303, EPI_ISL_1172304, EPI_ISL_1172305, EPI_ISL_1172306, EPI_ISL_1172307, EPI_ISL_1172308, EPI_ISL_1172309, EPI_ISL_1172310, EPI_ISL_1172311, EPI_ISL_1172312, EPI_ISL_1172313, EPI_ISL_1172314, EPI_ISL_1172315, EPI_ISL_1172316, EPI_ISL_1172317, EPI_ISL_1172318, EPI_ISL_1172319, EPI_ISL_1172320, EPI_ISL_1172321, EPI_ISL_1172322, EPI_ISL_1172323, EPI_ISL_1172324, EPI_ISL_1172325, EPI_ISL_1172326, EPI_ISL_1172327, EPI_ISL_1172328, EPI_ISL_1172329, EPI_ISL_1172330, EPI_ISL_1172331, EPI_ISL_1172332, EPI_ISL_1172333, EPI_ISL_1172334, EPI_ISL_1172335, EPI_ISL_1172336, EPI_ISL_1172337, EPI_ISL_1172338, EPI_ISL_1172339, EPI_ISL_1172340, EPI_ISL_1172341, EPI_ISL_1172342, EPI_ISL_1172343, EPI_ISL_1172344, EPI_ISL_1172345, EPI_ISL_1172346, EPI_ISL_1172347, EPI_ISL_1172348, EPI_ISL_1172349, EPI_ISL_1172350, EPI_ISL_1172351, EPI_ISL_1172352, EPI_ISL_1172353, EPI_ISL_1172354, EPI_ISL_1172355, EPI_ISL_1172356, EPI_ISL_1172357, EPI_ISL_1172358, EPI_ISL_1172359, EPI_ISL_1172360, EPI_ISL_1172361, EPI_ISL_1172362, EPI_ISL_1172363, EPI_ISL_1172364, EPI_ISL_1172365, EPI_ISL_1172366, EPI_ISL_1172367, EPI_ISL_1172368, EPI_ISL_1172369, EPI_ISL_1172370, EPI_ISL_1172371, EPI_ISL_1172372, EPI_ISL_1172373, EPI_ISL_1172374, EPI_ISL_1172375, EPI_ISL_1172376, EPI_ISL_1172377, EPI_ISL_1172378, EPI_ISL_1172379, EPI_ISL_1172380, EPI_ISL_1172381, EPI_ISL_1172382, EPI_ISL_1172383, EPI_ISL_1172384, EPI_ISL_1172385, EPI_ISL_1172386, EPI_ISL_1172387, EPI_ISL_1172388, EPI_ISL_1172389, EPI_ISL_1172390, EPI_ISL_1172391, EPI_ISL_1172392, EPI_ISL_1172393, EPI_ISL_1172394, EPI_ISL_1172395, EPI_ISL_1172396, EPI_ISL_1172397, EPI_ISL_1172398, EPI_ISL_1172399, EPI_ISL_1172400, EPI_ISL_1172401, EPI_ISL_1172402, EPI_ISL_1172403, EPI_ISL_1172404, EPI_ISL_1172405, EPI_ISL_1172406, EPI_ISL_1172407, EPI_ISL_1172408, EPI_ISL_1172409, EPI_ISL_1172410, EPI_ISL_1172411, EPI_ISL_1172412, EPI_ISL_1172413, EPI_ISL_1172414, EPI_ISL_1172415, EPI_ISL_1172416, EPI_ISL_1172417, EPI_ISL_1172418, EPI_ISL_1172419, EPI_ISL_1172420, EPI_ISL_1172421, EPI_ISL_1172422, EPI_ISL_1172423, EPI_ISL_1172424, EPI_ISL_1172425, EPI_ISL_1172426, EPI_ISL_1172427, EPI_ISL_1172428, EPI_ISL_1172429, EPI_ISL_1172430, EPI_ISL_1172431, EPI_ISL_1172432, EPI_ISL_1172433, EPI_ISL_1172434, EPI_ISL_1172435, EPI_ISL_1172436, EPI_ISL_1172437, EPI_ISL_1172438, EPI_ISL_1172439, EPI_ISL_1172440, EPI_ISL_1172441, EPI_ISL_1172442, EPI_ISL_1172443, EPI_ISL_1172444, EPI_ISL_1172445, EPI_ISL_1172446, EPI_ISL_1172447, EPI_ISL_1172448, EPI_ISL_1172449, EPI_ISL_1172450, EPI_ISL_1172451, EPI_ISL_1172452, EPI_ISL_1172453, EPI_ISL_1172454, EPI_ISL_1172455, EPI_ISL_1172456, EPI_ISL_1172457, EPI_ISL_1172458, EPI_ISL_1172459, EPI_ISL_1172460, EPI_ISL_1172461, EPI_ISL_1172462, EPI_ISL_1172463, EPI_ISL_1172464, EPI_ISL_1172465, EPI_ISL_1172466, EPI_ISL_1172467, EPI_ISL_1172468, EPI_ISL_1172469, EPI_ISL_1172470, EPI_ISL_1172471, EPI_ISL_1172472, EPI_ISL_1172473, EPI_ISL_1172474, EPI_ISL_1172475, EPI_ISL_1172476, EPI_ISL_1172477, EPI_ISL_1172478, EPI_ISL_1172479, EPI_ISL_1172480, EPI_ISL_1172481, EPI_ISL_1172482, EPI_ISL_1172483, EPI_ISL_1172484, EPI_ISL_1172485, EPI_ISL_1172486, EPI_ISL_1172487, EPI_ISL_1172488, EPI_ISL_1172489, EPI_ISL_1172490, EPI_ISL_1172491, EPI_ISL_1172492, EPI_ISL_1172493, EPI_ISL_1172494, EPI_ISL_1172495, EPI_ISL_1172496, EPI_ISL_1172497, EPI_ISL_1172498, EPI_ISL_1172499, EPI_ISL_1172500, EPI_ISL_1172501, EPI_ISL_1172502, EPI_ISL_1172503, EPI_ISL_1172504, EPI_ISL_1172505, EPI_ISL_1172506, EPI_ISL_1172507, EPI_ISL_1172508, EPI_ISL_1172509, EPI_ISL_1172510, EPI_ISL_1172511, EPI_ISL_1172512, EPI_ISL_1172513, EPI_ISL_1172514, EPI_ISL_1172515, EPI_ISL_1172516, EPI_ISL_1172517, EPI_ISL_1172518, EPI_ISL_1172519, EPI_ISL_1172520, EPI_ISL_1172521, EPI_ISL_1172522, EPI_ISL_1172523, EPI_ISL_1172524, EPI_ISL_1172525, EPI_ISL_1172526, EPI_ISL_1172527, EPI_ISL_1172528, EPI_ISL_1172529, EPI_ISL_1172530, EPI_ISL_1172531, EPI_ISL_1172532, EPI_ISL_1172533, EPI_ISL_1172534, EPI_ISL_1172535, EPI_ISL_1172536, EPI_ISL_1172537, EPI_ISL_1172538, EPI_ISL_1172539, EPI_ISL_1172540, EPI_ISL_1172541, EPI_ISL_1172542, EPI_ISL_1172543, EPI_ISL_1172544, EPI_ISL_1172545, EPI_ISL_1172546, EPI_ISL_1172547, EPI_ISL_1172548, EPI_ISL_1172549, EPI_ISL_1172550, EPI_ISL_1172551, EPI_ISL_1172552, EPI_ISL_1172553, EPI_ISL_1172554, EPI_ISL_1172555, EPI_ISL_1172556, EPI_ISL_1172557, EPI_ISL_1172558, EPI_ISL_1172559, EPI_ISL_1172560, EPI_ISL_1172561, EPI_ISL_1172562, EPI_ISL_1172563, EPI_ISL_1172564, EPI_ISL_1172565, EPI_ISL_1172566, EPI_ISL_1172567, EPI_ISL_1172568, EPI_ISL_1172569, EPI_ISL_1172570, EPI_ISL_1172571, EPI_ISL_1172572, EPI_ISL_1172573, EPI_ISL_1172574, EPI_ISL_1172575, EPI_ISL_1172576, EPI_ISL_1172577, EPI_ISL_1172578, EPI_ISL_1172579, EPI_ISL_1172580, EPI_ISL_1172581, EPI_ISL_1172582, EPI_ISL_1172583, EPI_ISL_1172584, EPI_ISL_1172585, EPI_ISL_1172586, EPI_ISL_1172587, EPI_ISL_1172588, EPI_ISL_1172589, EPI_ISL_1172590, EPI_ISL_1172591, EPI_ISL_1172592, EPI_ISL_1172593, EPI_ISL_1172594, EPI_ISL_1172595, EPI_ISL_1172596, EPI_ISL_1172597, EPI_ISL_1172598, EPI_ISL_1172599, EPI_ISL_1172600, EPI_ISL_1172601, EPI_ISL_1172602, EPI_ISL_1172603, EPI_ISL_1172604, EPI_ISL_1172605, EPI_ISL_1172606, EPI_ISL_1172607, EPI_ISL_1172608, EPI_ISL_1172609, EPI_ISL_1172610, EPI_ISL_1172611, EPI_ISL_1172612, EPI_ISL_1172613, EPI_ISL_1172614, EPI_ISL_1172615, EPI_ISL_1172616, EPI_ISL_1172617, EPI_ISL_1172618, EPI_ISL_1172619, EPI_ISL_1172620, EPI_ISL_1172621, EPI_ISL_1172622, EPI_ISL_1172623, EPI_ISL_1172624, EPI_ISL_1172625, EPI_ISL_1172626, EPI_ISL_1172627, EPI_ISL_1172628, EPI_ISL_1172629, EPI_ISL_1172630, EPI_ISL_1172631, EPI_ISL_1172632, EPI_ISL_1172633, EPI_ISL_1172634, EPI_ISL_1172635, EPI_ISL_1172636, EPI_ISL_1172637, EPI_ISL_1172638, EPI_ISL_1172639, EPI_ISL_1172640, EPI_ISL_1172641, EPI_ISL_1172642, EPI_ISL_1172643, EPI_ISL_1172644, EPI_ISL_1172645, EPI_ISL_1172646, EPI_ISL_1172647, EPI_ISL_1172648, EPI_ISL_1172649, EPI_ISL_1172650, EPI_ISL_1172651, EPI_ISL_1172652, EPI_ISL_1172653, EPI_ISL_1172654, EPI_ISL_1172655, EPI_ISL_1172656, EPI_ISL_1172657, EPI_ISL_1172658, EPI_ISL_1172659, EPI_ISL_1172660, EPI_ISL_1172661, EPI_ISL_1172662, EPI_ISL_1172663, EPI_ISL_1172664, EPI_ISL_1172665, EPI_ISL_1172666, EPI_ISL_1172667, EPI_ISL_1172668, EPI_ISL_1172669, EPI_ISL_1172670, EPI_ISL_1172671, EPI_ISL_1172672, EPI_ISL_1172673, EPI_ISL_1172674, EPI_ISL_1172675, EPI_ISL_1172676, EPI_ISL_1172677, EPI_ISL_1172678, EPI_ISL_1172679, EPI_ISL_1172680, EPI_ISL_1172681, EPI_ISL_1172682, EPI_ISL_1172683, EPI_ISL_1172684, EPI_ISL_1172685, EPI_ISL_1172686, EPI_ISL_1172687, EPI_ISL_1172688, EPI_ISL_1172689, EPI_ISL_1172690, EPI_ISL_1172691, EPI_ISL_1172692, EPI_ISL_1172693, EPI_ISL_1172694, EPI_ISL_1172695, EPI_ISL_1172696, EPI_ISL_1172697, EPI_ISL_1172698, EPI_ISL_1172699, EPI_ISL_1172700, EPI_ISL_1172701, EPI_ISL_1172702, EPI_ISL_1172703, EPI_ISL_1172704, EPI_ISL_1172705, EPI_ISL_1172706, EPI_ISL_1172707, EPI_ISL_1172708, EPI_ISL_1172709, EPI_ISL_1172710, EPI_ISL_1172711, EPI_ISL_1172712, EPI_ISL_1172713, EPI_ISL_1172714, EPI_ISL_1172715, EPI_ISL_1172716, EPI_ISL_1172717, EPI_ISL_1172718, EPI_ISL_1172719, EPI_ISL_1172720, EPI_ISL_1172721, EPI_ISL_1172722, EPI_ISL_1172723, EPI_ISL_1172724, EPI_ISL_1172725, EPI_ISL_1172726, EPI_ISL_1172727, EPI_ISL_1172728, EPI_ISL_1172729, EPI_ISL_1172730, EPI_ISL_1172731, EPI_ISL_1172732, EPI_ISL_1172733, EPI_ISL_1172734, EPI_ISL_1172735, EPI_ISL_1172736, EPI_ISL_1172737, EPI_ISL_1172738, EPI_ISL_1172739, EPI_ISL_1172740, EPI_ISL_1172741, EPI_ISL_1172742, EPI_ISL_1172743, EPI_ISL_1172744, EPI_ISL_1172745, EPI_ISL_1172746, EPI_ISL_1172747, EPI_ISL_1172748, EPI_ISL_1172749, EPI_ISL_1172750, EPI_ISL_1172751, EPI_ISL_1172752, EPI_ISL_1172753, EPI_ISL_1172754, EPI_ISL_1172755, EPI_ISL_1172756, EPI_ISL_1172757, EPI_ISL_1172758, EPI_ISL_1172759, EPI_ISL_1172760, EPI_ISL_1172761, EPI_ISL_1172762, EPI_ISL_1172763, EPI_ISL_1172764, EPI_ISL_1172765, EPI_ISL_1172766, EPI_ISL_1172767, EPI_ISL_1172768, EPI_ISL_1172769, EPI_ISL_1172770, EPI_ISL_1172771, EPI_ISL_1172772, EPI_ISL_1172773, EPI_ISL_1172774, EPI_ISL_1172775, EPI_ISL_1172776, EPI_ISL_1172777, EPI_ISL_1172778, EPI_ISL_1172779, EPI_ISL_1172780, EPI_ISL_1172781, EPI_ISL_1172782, EPI_ISL_1172783, EPI_ISL_1172784, EPI_ISL_1172785, EPI_ISL_1172786, EPI_ISL_1172787, EPI_ISL_1172788, EPI_ISL_1172789, EPI_ISL_1172790, EPI_ISL_1172791, EPI_ISL_1172792, EPI_ISL_1172793, EPI_ISL_1172794, EPI_ISL_1172795, EPI_ISL_1172796, EPI_ISL_1172797, EPI_ISL_1172798, EPI_ISL_1172799, EPI_ISL_1172800, EPI_ISL_1172801, EPI_ISL_1172802, EPI_ISL_1172803, EPI_ISL_1172804, EPI_ISL_1172805, EPI_ISL_1172806, EPI_ISL_1172807, EPI_ISL_1172808, EPI_ISL_1172809, EPI_ISL_1172810, EPI_ISL_1172811, EPI_ISL_1172812, EPI_ISL_1172813, EPI_ISL_1172814, EPI_ISL_1172815, EPI_ISL_1172816, EPI_ISL_1172817, EPI_ISL_1172818, EPI_ISL_1172819, EPI_ISL_1172820, EPI_ISL_1172821, EPI_ISL_1172822, EPI_ISL_1172823, EPI_ISL_1172824, EPI_ISL_1172825, EPI_ISL_1172826, EPI_ISL_1172827, EPI_ISL_1172828, EPI_ISL_1172829, EPI_ISL_1172830, EPI_ISL_1172831, EPI_ISL_1172832, EPI_ISL_1172833, EPI_ISL_1172834, EPI_ISL_1172835, EPI_ISL_1172836, EPI_ISL_1172837, EPI_ISL_1172838, EPI_ISL_1172839, EPI_ISL_1172840, EPI_ISL_1172841, EPI_ISL_1172842, EPI_ISL_1172843, EPI_ISL_1172844, EPI_ISL_1172845, EPI_ISL_1172846, EPI_ISL_1172847, EPI_ISL_1172848, EPI_ISL_1172849, EPI_ISL_1172850, EPI_ISL_1172851, EPI_ISL_1172852, EPI_ISL_1172853, EPI_ISL_1172854, EPI_ISL_1172855, EPI_ISL_1172856, EPI_ISL_1172857, EPI_ISL_1172858, EPI_ISL_1172859, EPI_ISL_1172860, EPI_ISL_1172861, EPI_ISL_1172862, EPI_ISL_1172863, EPI_ISL_1172864, EPI_ISL_1172865, EPI_ISL_1172866, EPI_ISL_1172867, EPI_ISL_1172868, EPI_ISL_1172869, EPI_ISL_1172870, EPI_ISL_1172871, EPI_ISL_1172872, EPI_ISL_1172873, EPI_ISL_1172874, EPI_ISL_1172875, EPI_ISL_1172876, EPI_ISL_1172877, EPI_ISL_1172878, EPI_ISL_1172879, EPI_ISL_1172880, EPI_ISL_1172881, EPI_ISL_1172882, EPI_ISL_1172883, EPI_ISL_1172884, EPI_ISL_1172885, EPI_ISL_1172886, EPI_ISL_1172887, EPI_ISL_1172888, EPI_ISL_1172889, EPI_ISL_1172890, EPI_ISL_1172891, EPI_ISL_1172892, EPI_ISL_1172893, EPI_ISL_1172894, EPI_ISL_1172895, EPI_ISL_1172896, EPI_ISL_1172897, EPI_ISL_1172898, EPI_ISL_1172899, EPI_ISL_1172900, EPI_ISL_1172901, EPI_ISL_1172902, EPI_ISL_1172903, EPI_ISL_1172904, EPI_ISL_1172905, EPI_ISL_1172906, EPI_ISL_1172907, EPI_ISL_1172908, EPI_ISL_1172909, EPI_ISL_1172910, EPI_ISL_1172911, EPI_ISL_1172912, EPI_ISL_1172913, EPI_ISL_1172914, EPI_ISL_1172915, EPI_ISL_1172916, EPI_ISL_1172917, EPI_ISL_1172918, EPI_ISL_1172919, EPI_ISL_1172920, EPI_ISL_1172921, EPI_ISL_1172922, EPI_ISL_1172923, EPI_ISL_1172924, EPI_ISL_1172925, EPI_ISL_1172926, EPI_ISL_1172927, EPI_ISL_1172928, EPI_ISL_1172929, EPI_ISL_1172930, EPI_ISL_1172931, EPI_ISL_1172932, EPI_ISL_1172933, EPI_ISL_1172934, EPI_ISL_1172935, EPI_ISL_1172936, EPI_ISL_1172937, EPI_ISL_1172938, EPI_ISL_1172939, EPI_ISL_1172940, EPI_ISL_1172941, EPI_ISL_1172942, EPI_ISL_1172943, EPI_ISL_1172944, EPI_ISL_1172945, EPI_ISL_1172946, EPI_ISL_1172947, EPI_ISL_1172948, EPI_ISL_1172949, EPI_ISL_1172950, EPI_ISL_1172951, EPI_ISL_1172952, EPI_ISL_1172953, EPI_ISL_1172954, EPI_ISL_1172955, EPI_ISL_1172956, EPI_ISL_1172957, EPI_ISL_1172958, EPI_ISL_1172959, EPI_ISL_1172960, EPI_ISL_1172961, EPI_ISL_1172962, EPI_ISL_1172963, EPI_ISL_1172964, EPI_ISL_1172965, EPI_ISL_1172966, EPI_ISL_1172967, EPI_ISL_1172968, EPI_ISL_1172969, EPI_ISL_1172970, EPI_ISL_1172971, EPI_ISL_1172972, EPI_ISL_1172973, EPI_ISL_1172974, EPI_ISL_1172975, EPI_ISL_1172976, EPI_ISL_1172977, EPI_ISL_1172978, EPI_ISL_1172979, EPI_ISL_1172980, EPI_ISL_1172981, EPI_ISL_1172982, EPI_ISL_1172983, EPI_ISL_1172984, EPI_ISL_1172985, EPI_ISL_1172986, EPI_ISL_1172987, EPI_ISL_1172988, EPI_ISL_1172989, EPI_ISL_1172990, EPI_ISL_1172991, EPI_ISL_1172992, EPI_ISL_1172993, EPI_ISL_1172994, EPI_ISL_1172995, EPI_ISL_1172996, EPI_ISL_1172997, EPI_ISL_1172998, EPI_ISL_1172999, EPI_ISL_1173000, EPI_ISL_1173001, EPI_ISL_1173002, EPI_ISL_1173003, EPI_ISL_1173004, EPI_ISL_1173005, EPI_ISL_1173006, EPI_ISL_1173007, EPI_ISL_1173008, EPI_ISL_1173009, EPI_ISL_1173010, EPI_ISL_1173011, EPI_ISL_1173012, EPI_ISL_1173013, EPI_ISL_1173014, EPI_ISL_1173015, EPI_ISL_1173016, EPI_ISL_1173017, EPI_ISL_1173018, EPI_ISL_1173019, EPI_ISL_1173020, EPI_ISL_1173021, EPI_ISL_1173022, EPI_ISL_1173023, EPI_ISL_1173024, EPI_ISL_1173025, EPI_ISL_1173026, EPI_ISL_1173027, EPI_ISL_1173028, EPI_ISL_1173029, EPI_ISL_1173030, EPI_ISL_1173031, EPI_ISL_1173032, EPI_ISL_1173033, EPI_ISL_1173034, EPI_ISL_1173035, EPI_ISL_1173036, EPI_ISL_1173037, EPI_ISL_1173038, EPI_ISL_1173039, EPI_ISL_1173040, EPI_ISL_1173041, EPI_ISL_1173042, EPI_ISL_1173043, EPI_ISL_1173044, EPI_ISL_1173045, EPI_ISL_1173046, EPI_ISL_1173047, EPI_ISL_1173048, EPI_ISL_1173049, EPI_ISL_1173050, EPI_ISL_1173051, EPI_ISL_1173052, EPI_ISL_1173053, EPI_ISL_1173054, EPI_ISL_1173055, EPI_ISL_1173056, EPI_ISL_1173057, EPI_ISL_1173058, EPI_ISL_1173059, EPI_ISL_1173060, EPI_ISL_1173061, EPI_ISL_1173062, EPI_ISL_1173063, EPI_ISL_1173064, EPI_ISL_1173065, EPI_ISL_1173066, EPI_ISL_1173067, EPI_ISL_1173068, EPI_ISL_1173069, EPI_ISL_1173070, EPI_ISL_1173071, EPI_ISL_1173072, EPI_ISL_1173073, EPI_ISL_1173074, EPI_ISL_1173075, EPI_ISL_1173076, EPI_ISL_1173077, EPI_ISL_1173078, EPI_ISL_1173079, EPI_ISL_1173080, EPI_ISL_1173081, EPI_ISL_1173082, EPI_ISL_1173083, EPI_ISL_1173084, EPI_ISL_1173085, EPI_ISL_1173086, EPI_ISL_1173087, EPI_ISL_1173088, EPI_ISL_1173089, EPI_ISL_1173090, EPI_ISL_1173091, EPI_ISL_1173092, EPI_ISL_1173093, EPI_ISL_1173094, EPI_ISL_1173095, EPI_ISL_1173096, EPI_ISL_1173097, EPI_ISL_1173098, EPI_ISL_1173099, EPI_ISL_1173100, EPI_ISL_1173101, EPI_ISL_1173102, EPI_ISL_1173103, EPI_ISL_1173104, EPI_ISL_1173105, EPI_ISL_1173106, EPI_ISL_1173107, EPI_ISL_1173108, EPI_ISL_1173109, EPI_ISL_1173110, EPI_ISL_1173111, EPI_ISL_1173112, EPI_ISL_1173113, EPI_ISL_1173114, EPI_ISL_1173115, EPI_ISL_1173116, EPI_ISL_1173117, EPI_ISL_1173118, EPI_ISL_1173119, EPI_ISL_1173120, EPI_ISL_1173121, EPI_ISL_1173122, EPI_ISL_1173123, EPI_ISL_1173124, EPI_ISL_1173125, EPI_ISL_1173126, EPI_ISL_1173127, EPI_ISL_1173128, EPI_ISL_1173129, EPI_ISL_1173130, EPI_ISL_1173131, EPI_ISL_1173132, EPI_ISL_1173133, EPI_ISL_1173134, EPI_ISL_1173135, EPI_ISL_1173136, EPI_ISL_1173137, EPI_ISL_1173138, EPI_ISL_1173139, EPI_ISL_1173140, EPI_ISL_1173141, EPI_ISL_1173142, EPI_ISL_1173143, EPI_ISL_1173144, EPI_ISL_1173145, EPI_ISL_1173146, EPI_ISL_1173147, EPI_ISL_1173148, EPI_ISL_1173149, EPI_ISL_1173150, EPI_ISL_1173151, EPI_ISL_1173152, EPI_ISL_1173153, EPI_ISL_1173154, EPI_ISL_1173155, EPI_ISL_1173156, EPI_ISL_1173157, EPI_ISL_1173158, EPI_ISL_1258466, EPI_ISL_1258467, EPI_ISL_1258468, EPI_ISL_1258469, EPI_ISL_1258470, EPI_ISL_1258471, EPI_ISL_1258472, EPI_ISL_1258473, EPI_ISL_1258474, EPI_ISL_1258475, EPI_ISL_1258476, EPI_ISL_1258477, EPI_ISL_1258478, EPI_ISL_1258479, EPI_ISL_1258480, EPI_ISL_1258481, EPI_ISL_1258482, EPI_ISL_1258483, EPI_ISL_1258484, EPI_ISL_1258485, EPI_ISL_1258486, EPI_ISL_1258487, EPI_ISL_1258488, EPI_ISL_1258489, EPI_ISL_1258490, EPI_ISL_1258491, EPI_ISL_1258492, EPI_ISL_1258493, EPI_ISL_1258494, EPI_ISL_1258495, EPI_ISL_1258496, EPI_ISL_1258497, EPI_ISL_1258498, EPI_ISL_1258499, EPI_ISL_1258500, EPI_ISL_1258501, EPI_ISL_1258502, EPI_ISL_1258503, EPI_ISL_1258504, EPI_ISL_1258505, EPI_ISL_1258506, EPI_ISL_1258507, EPI_ISL_1258508, EPI_ISL_1258509, EPI_ISL_1258510, EPI_ISL_1258511, EPI_ISL_1258512, EPI_ISL_1258513, EPI_ISL_1258514, EPI_ISL_1258515, EPI_ISL_1258516, EPI_ISL_1258517, EPI_ISL_1258518, EPI_ISL_1258519, EPI_ISL_1258520, EPI_ISL_1258521, EPI_ISL_1258522, EPI_ISL_1258523, EPI_ISL_1258524, EPI_ISL_1258525, EPI_ISL_1258526, EPI_ISL_1258527, EPI_ISL_1258528, EPI_ISL_1258529, EPI_ISL_1258530, EPI_ISL_1258531, EPI_ISL_1258532, EPI_ISL_1258533, EPI_ISL_1258534, EPI_ISL_1258535, EPI_ISL_1258536, EPI_ISL_1258537, EPI_ISL_1258538, EPI_ISL_1258539, EPI_ISL_1258540, EPI_ISL_1258541, EPI_ISL_1258542, EPI_ISL_1258543, EPI_ISL_1258544, EPI_ISL_1258545, EPI_ISL_1258546, EPI_ISL_1258547, EPI_ISL_1258548, EPI_ISL_1258549, EPI_ISL_1258550, EPI_ISL_1258551, EPI_ISL_1258552, EPI_ISL_1258553, EPI_ISL_1258554, EPI_ISL_1258555, EPI_ISL_1258556, EPI_ISL_1258557, EPI_ISL_1258558, EPI_ISL_1258559, EPI_ISL_1258560, EPI_ISL_1258561, EPI_ISL_1258562, EPI_ISL_1258563, EPI_ISL_1258564, EPI_ISL_1258565, EPI_ISL_1258566, EPI_ISL_1258567, EPI_ISL_1258568, EPI_ISL_1258569, EPI_ISL_1258570, EPI_ISL_1258571, EPI_ISL_1258572, EPI_ISL_1258573, EPI_ISL_1258574, EPI_ISL_1258575, EPI_ISL_1258576, EPI_ISL_1258577, EPI_ISL_1258578, EPI_ISL_1258579, EPI_ISL_1258580, EPI_ISL_1258581, EPI_ISL_1258582, EPI_ISL_1258583, EPI_ISL_1258584, EPI_ISL_1258585, EPI_ISL_1258586, EPI_ISL_1258587, EPI_ISL_1258588, EPI_ISL_1258589, EPI_ISL_1258590, EPI_ISL_1258591, EPI_ISL_1258592, EPI_ISL_1258593, EPI_ISL_1258594, EPI_ISL_1258595, EPI_ISL_1258596, EPI_ISL_1258597, EPI_ISL_1258598, EPI_ISL_1258599, EPI_ISL_1258600, EPI_ISL_1258601, EPI_ISL_1258602, EPI_ISL_1258603, EPI_ISL_1258604, EPI_ISL_1258605, EPI_ISL_1258606, EPI_ISL_1258607, EPI_ISL_1258608, EPI_ISL_1258609, EPI_ISL_1258610, EPI_ISL_1258611, EPI_ISL_1258612, EPI_ISL_1258613, EPI_ISL_1258614, EPI_ISL_1258615, EPI_ISL_1258616, EPI_ISL_1258617, EPI_ISL_1258618, EPI_ISL_1258619, EPI_ISL_1258620, EPI_ISL_1258621, EPI_ISL_1258622, EPI_ISL_1258623, EPI_ISL_1258624, EPI_ISL_1258625, EPI_ISL_1258626, EPI_ISL_1258627, EPI_ISL_1258628, EPI_ISL_1258629, EPI_ISL_1258630, EPI_ISL_1258631, EPI_ISL_1258632, EPI_ISL_1258633, EPI_ISL_1258634, EPI_ISL_1258635, EPI_ISL_1258636, EPI_ISL_1258637, EPI_ISL_1258638, EPI_ISL_1258639, EPI_ISL_1258640, EPI_ISL_1258641, EPI_ISL_1258642, EPI_ISL_1258643, EPI_ISL_1258644, EPI_ISL_1258645, EPI_ISL_1258646, EPI_ISL_1258647, EPI_ISL_1258648, EPI_ISL_1258649, EPI_ISL_1258650, EPI_ISL_1258651, EPI_ISL_1258652, EPI_ISL_1258653, EPI_ISL_1258654, EPI_ISL_1258655, EPI_ISL_1258656, EPI_ISL_1258657, EPI_ISL_1258658, EPI_ISL_1258659, EPI_ISL_1258660, EPI_ISL_1258661, EPI_ISL_1258662, EPI_ISL_1258663, EPI_ISL_1258664, EPI_ISL_1258665, EPI_ISL_1258666, EPI_ISL_1258667, EPI_ISL_1258668, EPI_ISL_1258669, EPI_ISL_1258670, EPI_ISL_1258671, EPI_ISL_1258672, EPI_ISL_1258673, EPI_ISL_1258674, EPI_ISL_1258675, EPI_ISL_1258676, EPI_ISL_1258677, EPI_ISL_1258678, EPI_ISL_1258679, EPI_ISL_1258680, EPI_ISL_1258681, EPI_ISL_1258682, EPI_ISL_1258683, EPI_ISL_1258684, EPI_ISL_1258685, EPI_ISL_1258686, EPI_ISL_1258687, EPI_ISL_1258688, EPI_ISL_1258689, EPI_ISL_1258690, EPI_ISL_1258691, EPI_ISL_1258692, EPI_ISL_1258693, EPI_ISL_1258694, EPI_ISL_1258695, EPI_ISL_1258696, EPI_ISL_1258697, EPI_ISL_1258698, EPI_ISL_1258699, EPI_ISL_1258700, EPI_ISL_1258701, EPI_ISL_1258702, EPI_ISL_1258703, EPI_ISL_1258704, EPI_ISL_1258705, EPI_ISL_1258706, EPI_ISL_1258707, EPI_ISL_1258708, EPI_ISL_1258709, EPI_ISL_1258710, EPI_ISL_1258711, EPI_ISL_1258712, EPI_ISL_1258713, EPI_ISL_1258714, EPI_ISL_1258715, EPI_ISL_1258716, EPI_ISL_1258717, EPI_ISL_1258718, EPI_ISL_1258719, EPI_ISL_1258720, EPI_ISL_1258721, EPI_ISL_1258722, EPI_ISL_1258723, EPI_ISL_1258724, EPI_ISL_1258725, EPI_ISL_1258726, EPI_ISL_1258727, EPI_ISL_1258728, EPI_ISL_1258729, EPI_ISL_1258730, EPI_ISL_1258731, EPI_ISL_1258732, EPI_ISL_1258733, EPI_ISL_1258734, EPI_ISL_1258735, EPI_ISL_1258736, EPI_ISL_1258737, EPI_ISL_1258738, EPI_ISL_1258739, EPI_ISL_1258740, EPI_ISL_1258741, EPI_ISL_1258742, EPI_ISL_1258743, EPI_ISL_1258744, EPI_ISL_1258745, EPI_ISL_1258746, EPI_ISL_1258747, EPI_ISL_1258748, EPI_ISL_1258749, EPI_ISL_1258750, EPI_ISL_1258751, EPI_ISL_1258752, EPI_ISL_1258753, EPI_ISL_1258754, EPI_ISL_1258755, EPI_ISL_1258756, EPI_ISL_1258757, EPI_ISL_1258758, EPI_ISL_1258759, EPI_ISL_1258760, EPI_ISL_1258761, EPI_ISL_1258762, EPI_ISL_1258763, EPI_ISL_1258764, EPI_ISL_1258765, EPI_ISL_1258766, EPI_ISL_1258767, EPI_ISL_1258768, EPI_ISL_1258769, EPI_ISL_1258770, EPI_ISL_1258771, EPI_ISL_1258772, EPI_ISL_1258773, EPI_ISL_1258774, EPI_ISL_1258775, EPI_ISL_1258776, EPI_ISL_1258777, EPI_ISL_1258778, EPI_ISL_1258779, EPI_ISL_1258780, EPI_ISL_1258781, EPI_ISL_1258782, EPI_ISL_1258783, EPI_ISL_1258784, EPI_ISL_1258785, EPI_ISL_1258786, EPI_ISL_1258787, EPI_ISL_1258788, EPI_ISL_1258789, EPI_ISL_1258790, EPI_ISL_1258791, EPI_ISL_1258792, EPI_ISL_1258793, EPI_ISL_1258794, EPI_ISL_1258795, EPI_ISL_1258796, EPI_ISL_1258797, EPI_ISL_1258798, EPI_ISL_1258799, EPI_ISL_1258800, EPI_ISL_1258801, EPI_ISL_1258802, EPI_ISL_1258803, EPI_ISL_1258804, EPI_ISL_1258805, EPI_ISL_1258806, EPI_ISL_1258807, EPI_ISL_1258808, EPI_ISL_1258809, EPI_ISL_1258810, EPI_ISL_1258811, EPI_ISL_1258812, EPI_ISL_1258813, EPI_ISL_1258814, EPI_ISL_1258815, EPI_ISL_1258816, EPI_ISL_1258817, EPI_ISL_1258818, EPI_ISL_1258819, EPI_ISL_1258820, EPI_ISL_1258821, EPI_ISL_1258822, EPI_ISL_1258823, EPI_ISL_1258824, EPI_ISL_1258825, EPI_ISL_1258826, EPI_ISL_1258827, EPI_ISL_1258828, EPI_ISL_1258829, EPI_ISL_1258830, EPI_ISL_1258831, EPI_ISL_1258832, EPI_ISL_1258833, EPI_ISL_1258834, EPI_ISL_1258835, EPI_ISL_1258836, EPI_ISL_1258837, EPI_ISL_1258838, EPI_ISL_1258839, EPI_ISL_1258840, EPI_ISL_1258841, EPI_ISL_1258842, EPI_ISL_1258843, EPI_ISL_1258844, EPI_ISL_1258845, EPI_ISL_1258846, EPI_ISL_1258847, EPI_ISL_1258848, EPI_ISL_1258849, EPI_ISL_1258850, EPI_ISL_1258851, EPI_ISL_1258852, EPI_ISL_1258853, EPI_ISL_1258854, EPI_ISL_1258855, EPI_ISL_1258856, EPI_ISL_1258857, EPI_ISL_1258858, EPI_ISL_1258859, EPI_ISL_1258860, EPI_ISL_1258861, EPI_ISL_1258862, EPI_ISL_1258863, EPI_ISL_1258864, EPI_ISL_1258865, EPI_ISL_1258866, EPI_ISL_1258867, EPI_ISL_1258868, EPI_ISL_1258869, EPI_ISL_1258870, EPI_ISL_1258871, EPI_ISL_1258872, EPI_ISL_1258873, EPI_ISL_1258874, EPI_ISL_1258875, EPI_ISL_1258876, EPI_ISL_1258877, EPI_ISL_1258878, EPI_ISL_1258879, EPI_ISL_1258880, EPI_ISL_1258881, EPI_ISL_1258882, EPI_ISL_1258883, EPI_ISL_1258884, EPI_ISL_1258885, EPI_ISL_1258886, EPI_ISL_1258887, EPI_ISL_1258888, EPI_ISL_1258889, EPI_ISL_1258890, EPI_ISL_1258891, EPI_ISL_1258892, EPI_ISL_1258893, EPI_ISL_1258894, EPI_ISL_1258895, EPI_ISL_1258896, EPI_ISL_1258897, EPI_ISL_1258898, EPI_ISL_1258899, EPI_ISL_1258900, EPI_ISL_1258901, EPI_ISL_1258902, EPI_ISL_1258903, EPI_ISL_1258904, EPI_ISL_1258905, EPI_ISL_1258906, EPI_ISL_1258907, EPI_ISL_1258908, EPI_ISL_1258909, EPI_ISL_1258910, EPI_ISL_1258911, EPI_ISL_1258912, EPI_ISL_1258913, EPI_ISL_1258914, EPI_ISL_1258915, EPI_ISL_1258916, EPI_ISL_1258917, EPI_ISL_1258918, EPI_ISL_1258919, EPI_ISL_1258920, EPI_ISL_1258921, EPI_ISL_1258922, EPI_ISL_1258923, EPI_ISL_1258924, EPI_ISL_1258925, EPI_ISL_1258926, EPI_ISL_1258927, EPI_ISL_1258928, EPI_ISL_1258929, EPI_ISL_1258930, EPI_ISL_1258931, EPI_ISL_1258932, EPI_ISL_1258933, EPI_ISL_1258934, EPI_ISL_1258935, EPI_ISL_1258936, EPI_ISL_1258937, EPI_ISL_1258938, EPI_ISL_1258939, EPI_ISL_1258940, EPI_ISL_1258941, EPI_ISL_1258942, EPI_ISL_1258943, EPI_ISL_1258944, EPI_ISL_1258945, EPI_ISL_1258946, EPI_ISL_1258947, EPI_ISL_1258948, EPI_ISL_1258949, EPI_ISL_1258950, EPI_ISL_1258951, EPI_ISL_1258952, EPI_ISL_1258953, EPI_ISL_1258954, EPI_ISL_1258955, EPI_ISL_1258956, EPI_ISL_1258957, EPI_ISL_1258958, EPI_ISL_1258959, EPI_ISL_1258960, EPI_ISL_1258961, EPI_ISL_1258962, EPI_ISL_1258963, EPI_ISL_1258964, EPI_ISL_1258965, EPI_ISL_1258966, EPI_ISL_1258967, EPI_ISL_1258968, EPI_ISL_1258969, EPI_ISL_1258970, EPI_ISL_1258971, EPI_ISL_1258972, EPI_ISL_1258973, EPI_ISL_1258974, EPI_ISL_1258975, EPI_ISL_1258976, EPI_ISL_1258977, EPI_ISL_1258978, EPI_ISL_1258979, EPI_ISL_1258980, EPI_ISL_1258981, EPI_ISL_1258982, EPI_ISL_1258983, EPI_ISL_1258984, EPI_ISL_1258985, EPI_ISL_1258986, EPI_ISL_1258987, EPI_ISL_1307048, EPI_ISL_1307049, EPI_ISL_1307050, EPI_ISL_1307051, EPI_ISL_1307052, EPI_ISL_1307053, EPI_ISL_1307054, EPI_ISL_1307055, EPI_ISL_1307056, EPI_ISL_1307057, EPI_ISL_1307058, EPI_ISL_1307059, EPI_ISL_1307060, EPI_ISL_1307061, EPI_ISL_1307062, EPI_ISL_1307063, EPI_ISL_1307064, EPI_ISL_1307065, EPI_ISL_1307066, EPI_ISL_1307067, EPI_ISL_1307068, EPI_ISL_1307069, EPI_ISL_1307070, EPI_ISL_1307071, EPI_ISL_1307072, EPI_ISL_1307073, EPI_ISL_1307074, EPI_ISL_1307075, EPI_ISL_1307076, EPI_ISL_1307077, EPI_ISL_1307078, EPI_ISL_1307079, EPI_ISL_1307080, EPI_ISL_1307081, EPI_ISL_1307082, EPI_ISL_1307083, EPI_ISL_1307084, EPI_ISL_1307085, EPI_ISL_1307086, EPI_ISL_1307087, EPI_ISL_1307088, EPI_ISL_1307089, EPI_ISL_1307090, EPI_ISL_1307091, EPI_ISL_1307092, EPI_ISL_1307093, EPI_ISL_1307094, EPI_ISL_1307095, EPI_ISL_1307096, EPI_ISL_1307097, EPI_ISL_1307098, EPI_ISL_1307099, EPI_ISL_1307100, EPI_ISL_1307101, EPI_ISL_1307102, EPI_ISL_1307103, EPI_ISL_1307104, EPI_ISL_1307105, EPI_ISL_1307106, EPI_ISL_1307107, EPI_ISL_1307108, EPI_ISL_1307109, EPI_ISL_1307110, EPI_ISL_1307111, EPI_ISL_1307112, EPI_ISL_1307113, EPI_ISL_1307114, EPI_ISL_1307115, EPI_ISL_1307116, EPI_ISL_1307117, EPI_ISL_1307118, EPI_ISL_1307119, EPI_ISL_1307120, EPI_ISL_1307121, EPI_ISL_1307122, EPI_ISL_1307123, EPI_ISL_1307124, EPI_ISL_1307125, EPI_ISL_1307126, EPI_ISL_1307127, EPI_ISL_1307128, EPI_ISL_1307129, EPI_ISL_1307130, EPI_ISL_1307131, EPI_ISL_1307132, EPI_ISL_1307133, EPI_ISL_1307134, EPI_ISL_1307135, EPI_ISL_1307136, EPI_ISL_1307137, EPI_ISL_1307138, EPI_ISL_1307139, EPI_ISL_1307140, EPI_ISL_1307141, EPI_ISL_1307142, EPI_ISL_1307143, EPI_ISL_1307144, EPI_ISL_1307145, EPI_ISL_1307146, EPI_ISL_1307147, EPI_ISL_1307148, EPI_ISL_1307149, EPI_ISL_1307150, EPI_ISL_1307151, EPI_ISL_1307152, EPI_ISL_1307153, EPI_ISL_1307154, EPI_ISL_1307155, EPI_ISL_1307156, EPI_ISL_1307157, EPI_ISL_1307158, EPI_ISL_1307159, EPI_ISL_1307160, EPI_ISL_1307161, EPI_ISL_1307162, EPI_ISL_1307163, EPI_ISL_1307164, EPI_ISL_1307165, EPI_ISL_1307166, EPI_ISL_1307167, EPI_ISL_1307168, EPI_ISL_1307169, EPI_ISL_1307170, EPI_ISL_1307171, EPI_ISL_1307172, EPI_ISL_1307173, EPI_ISL_1307174, EPI_ISL_1307175, EPI_ISL_1307176, EPI_ISL_1307178, EPI_ISL_1307179, EPI_ISL_1307180, EPI_ISL_1307181, EPI_ISL_1307182, EPI_ISL_1307183, EPI_ISL_1307184, EPI_ISL_1307185, EPI_ISL_1307186, EPI_ISL_1307187, EPI_ISL_1307188, EPI_ISL_1307189, EPI_ISL_1307190, EPI_ISL_1307191, EPI_ISL_1307192, EPI_ISL_1307193, EPI_ISL_1307194, EPI_ISL_1307195, EPI_ISL_1307196, EPI_ISL_1307197, EPI_ISL_1307198, EPI_ISL_1307199, EPI_ISL_1307200, EPI_ISL_1307201, EPI_ISL_1307202, EPI_ISL_1307203, EPI_ISL_1307204, EPI_ISL_1307205, EPI_ISL_1307206, EPI_ISL_1307207, EPI_ISL_1307208, EPI_ISL_1307209, EPI_ISL_1307210, EPI_ISL_1307211, EPI_ISL_1307212, EPI_ISL_1307213, EPI_ISL_1307214, EPI_ISL_1307215, EPI_ISL_1307216, EPI_ISL_1307217, EPI_ISL_1307218, EPI_ISL_1307219, EPI_ISL_1307220, EPI_ISL_1307221, EPI_ISL_1307222, EPI_ISL_1307223, EPI_ISL_1307224, EPI_ISL_1307225, EPI_ISL_1307226, EPI_ISL_1307227, EPI_ISL_1307228, EPI_ISL_1307229, EPI_ISL_1307230, EPI_ISL_1307231, EPI_ISL_1307232, EPI_ISL_1307233, EPI_ISL_1307234, EPI_ISL_1307235, EPI_ISL_1307236, EPI_ISL_1307237, EPI_ISL_1307238, EPI_ISL_1307239, EPI_ISL_1307240, EPI_ISL_1307241, EPI_ISL_1307242, EPI_ISL_1307243, EPI_ISL_1307244, EPI_ISL_1307245, EPI_ISL_1307246, EPI_ISL_1307247, EPI_ISL_1307248, EPI_ISL_1307249, EPI_ISL_1307250, EPI_ISL_1307251, EPI_ISL_1307252, EPI_ISL_1307253, EPI_ISL_1307254, EPI_ISL_1307255, EPI_ISL_1307256, EPI_ISL_1307257, EPI_ISL_1307258, EPI_ISL_1307259, EPI_ISL_1307260, EPI_ISL_1307261, EPI_ISL_1307262, EPI_ISL_1307263, EPI_ISL_1307264, EPI_ISL_1307265, EPI_ISL_1307266, EPI_ISL_1307267, EPI_ISL_1307268, EPI_ISL_1307269, EPI_ISL_1307270, EPI_ISL_1307271, EPI_ISL_1307272, EPI_ISL_1307273, EPI_ISL_1307274, EPI_ISL_1307275, EPI_ISL_1307276, EPI_ISL_1307277, EPI_ISL_1307278, EPI_ISL_1307279, EPI_ISL_1307280, EPI_ISL_1307281, EPI_ISL_1307282, EPI_ISL_1307283, EPI_ISL_1307284, EPI_ISL_1307285, EPI_ISL_1307286, EPI_ISL_1307287, EPI_ISL_1307288, EPI_ISL_1307289, EPI_ISL_1307290, EPI_ISL_1307291, EPI_ISL_1307292, EPI_ISL_1307293, EPI_ISL_1307294, EPI_ISL_1307295, EPI_ISL_1307296, EPI_ISL_1307297, EPI_ISL_1307298, EPI_ISL_1307299, EPI_ISL_1307300, EPI_ISL_1307301, EPI_ISL_1307302, EPI_ISL_1307303, EPI_ISL_1307304, EPI_ISL_1307305, EPI_ISL_1307306, EPI_ISL_1307307, EPI_ISL_1307308, EPI_ISL_1307309, EPI_ISL_1307310, EPI_ISL_1307311, EPI_ISL_1307312, EPI_ISL_1307313, EPI_ISL_1307314, EPI_ISL_1307315, EPI_ISL_1307316, EPI_ISL_1307317, EPI_ISL_1307318, EPI_ISL_1307319, EPI_ISL_1307320, EPI_ISL_1307321, EPI_ISL_1307322, EPI_ISL_1307323, EPI_ISL_1307324, EPI_ISL_1307325, EPI_ISL_1307326, EPI_ISL_1307327, EPI_ISL_1307328, EPI_ISL_1307329, EPI_ISL_1307330, EPI_ISL_1307331, EPI_ISL_1307332, EPI_ISL_1307333, EPI_ISL_1307334, EPI_ISL_1307335, EPI_ISL_1307336, EPI_ISL_1307337, EPI_ISL_1307338, EPI_ISL_1307339, EPI_ISL_1307340, EPI_ISL_1307341, EPI_ISL_1307342, EPI_ISL_1307343, EPI_ISL_1307344, EPI_ISL_1307345, EPI_ISL_1307346, EPI_ISL_1307347, EPI_ISL_1307348, EPI_ISL_1307349, EPI_ISL_1307350, EPI_ISL_1307351, EPI_ISL_1307352, EPI_ISL_1307353, EPI_ISL_1307354, EPI_ISL_1307355, EPI_ISL_1307356, EPI_ISL_1307357, EPI_ISL_1307358, EPI_ISL_1307359, EPI_ISL_1307360, EPI_ISL_1307361, EPI_ISL_1307362, EPI_ISL_1307363, EPI_ISL_1307364, EPI_ISL_1307365, EPI_ISL_1307366, EPI_ISL_1307367, EPI_ISL_1307368, EPI_ISL_1307369, EPI_ISL_1307370, EPI_ISL_1307371, EPI_ISL_1307372, EPI_ISL_1307373, EPI_ISL_1307374, EPI_ISL_1307375, EPI_ISL_1307376, EPI_ISL_1307377, EPI_ISL_1307378, EPI_ISL_1307379, EPI_ISL_1307380, EPI_ISL_1307381, EPI_ISL_1307382, EPI_ISL_1307383, EPI_ISL_1307384, EPI_ISL_1307385, EPI_ISL_1307386, EPI_ISL_1307387, EPI_ISL_1307388, EPI_ISL_1307389, EPI_ISL_1307390, EPI_ISL_1307391, EPI_ISL_1307392, EPI_ISL_1307393, EPI_ISL_1307394, EPI_ISL_1307395, EPI_ISL_1307396, EPI_ISL_1307397, EPI_ISL_1307398, EPI_ISL_1307399, EPI_ISL_1307400, EPI_ISL_1307401, EPI_ISL_1307402, EPI_ISL_1307403, EPI_ISL_1307404, EPI_ISL_1307405, EPI_ISL_1307406, EPI_ISL_1307407, EPI_ISL_1307408, EPI_ISL_1307409, EPI_ISL_1307410, EPI_ISL_1307411, EPI_ISL_1307412, EPI_ISL_1307413, EPI_ISL_1307414, EPI_ISL_1307415, EPI_ISL_1307416, EPI_ISL_1307417, EPI_ISL_1307418, EPI_ISL_1307419, EPI_ISL_1307420, EPI_ISL_1307421, EPI_ISL_1307422, EPI_ISL_1307423, EPI_ISL_1307424, EPI_ISL_1307425, EPI_ISL_1307426, EPI_ISL_1307427, EPI_ISL_1307428, EPI_ISL_1307429, EPI_ISL_1307430, EPI_ISL_1307431, EPI_ISL_1307432, EPI_ISL_1307433, EPI_ISL_1307434, EPI_ISL_1307435, EPI_ISL_1307436, EPI_ISL_1307437, EPI_ISL_1307438, EPI_ISL_1307439, EPI_ISL_1307440, EPI_ISL_1307441, EPI_ISL_1307442, EPI_ISL_1307443, EPI_ISL_1307444, EPI_ISL_1307445, EPI_ISL_1307446, EPI_ISL_1307447, EPI_ISL_1307448, EPI_ISL_1307449, EPI_ISL_1307450, EPI_ISL_1307451, EPI_ISL_1307452, EPI_ISL_1307453, EPI_ISL_1307454, EPI_ISL_1307455, EPI_ISL_1307456, EPI_ISL_1307457, EPI_ISL_1307458, EPI_ISL_1307459, EPI_ISL_1307460, EPI_ISL_1307461, EPI_ISL_1307462, EPI_ISL_1307463, EPI_ISL_1307464, EPI_ISL_1307465, EPI_ISL_1307466, EPI_ISL_1307467, EPI_ISL_1307468, EPI_ISL_1307469, EPI_ISL_1307470, EPI_ISL_1307471, EPI_ISL_1307472, EPI_ISL_1307473, EPI_ISL_1307474, EPI_ISL_1307475, EPI_ISL_1307476, EPI_ISL_1307477, EPI_ISL_1307478, EPI_ISL_1307479, EPI_ISL_1307480, EPI_ISL_1307481, EPI_ISL_1307482, EPI_ISL_1307483, EPI_ISL_1307484, EPI_ISL_1307485, EPI_ISL_1307486, EPI_ISL_1307487, EPI_ISL_1307488, EPI_ISL_1307489, EPI_ISL_1307490, EPI_ISL_1307491, EPI_ISL_1307492, EPI_ISL_1307493, EPI_ISL_1307494, EPI_ISL_1307495, EPI_ISL_1307496, EPI_ISL_1307497, EPI_ISL_1307498, EPI_ISL_1307499, EPI_ISL_1307500, EPI_ISL_1307502, EPI_ISL_1307504, EPI_ISL_1307505, EPI_ISL_1307506, EPI_ISL_1307507, EPI_ISL_1307508, EPI_ISL_1307509, EPI_ISL_1307510, EPI_ISL_1307511, EPI_ISL_1307512, EPI_ISL_1307513, EPI_ISL_1307514, EPI_ISL_1307517, EPI_ISL_1307518, EPI_ISL_1307519, EPI_ISL_1307520, EPI_ISL_1307530, EPI_ISL_1307531, EPI_ISL_1307532, EPI_ISL_1307535, EPI_ISL_1307546, EPI_ISL_1307547, EPI_ISL_1307548, EPI_ISL_1307549, EPI_ISL_1307550, EPI_ISL_1307551, EPI_ISL_1307552, EPI_ISL_1307553, EPI_ISL_1307554, EPI_ISL_1307555, EPI_ISL_1307556, EPI_ISL_1307557, EPI_ISL_1307558, EPI_ISL_1307559, EPI_ISL_1307560, EPI_ISL_1307561, EPI_ISL_1307562, EPI_ISL_1307563, EPI_ISL_1307564, EPI_ISL_1307565, EPI_ISL_1307566, EPI_ISL_1307567, EPI_ISL_1307568, EPI_ISL_1307570, EPI_ISL_1307571, EPI_ISL_1307572, EPI_ISL_1307573, EPI_ISL_1307574, EPI_ISL_1307575, EPI_ISL_1307576, EPI_ISL_1307577, EPI_ISL_1307578, EPI_ISL_1307579, EPI_ISL_1307580, EPI_ISL_1307581, EPI_ISL_1307582, EPI_ISL_1307583, EPI_ISL_1307584, EPI_ISL_1307585, EPI_ISL_1307586, EPI_ISL_1307595, EPI_ISL_1307602, EPI_ISL_1307603, EPI_ISL_1307604, EPI_ISL_1307605, EPI_ISL_1307606, EPI_ISL_1307607, EPI_ISL_1307630, EPI_ISL_1307635

| | | | |
|---|---|---|---|
| see above | Pandemic Response Lab - NYC | Pandemic Response Lab, R&D | Cybill del Castillo; Dylan Law; Haiping Hao; Henry Lee; Jon Laurent; Melissa Hopkins; Michael Hammerling; Pradeep Bugga; Shinyoung Clair Kang; Sol Rey; William Ward |
| EPI_ISL_1633261 | Pathology and Laboratory Medicine Institute, Cleveland Clinic, Ohio, USA | Pathology and Laboratory Medicine Institute, Cleveland Clinic, Ohio, USA | Concetta Peck; Daniel H. Farkas; Daniel Rhoads; David Bosler; David Plunkett; Gary Procop; Jay Brock; Joy Nakitandwe; Kristen McDonnell; Yu-Wei Cheng; Zheng Jin Tu |

EPI_ISL_1193643, EPI_ISL_1193692, EPI_ISL_1193695, EPI_ISL_1193713, EPI_ISL_1193729, EPI_ISL_1193749, EPI_ISL_1193765, EPI_ISL_1193812, EPI_ISL_1193827, EPI_ISL_1193829, EPI_ISL_1193830, EPI_ISL_1193836, EPI_ISL_1193880, EPI_ISL_1193889, EPI_ISL_1193891, EPI_ISL_1193896, EPI_ISL_1193909, EPI_ISL_1194044, EPI_ISL_1194045, EPI_ISL_1194128, EPI_ISL_1194140, EPI_ISL_1194157, EPI_ISL_1194178, EPI_ISL_1194179, EPI_ISL_1194185, EPI_ISL_1194191, EPI_ISL_1194194, EPI_ISL_1194211, EPI_ISL_1194212, EPI_ISL_1194213, EPI_ISL_1194218, EPI_ISL_1194222, EPI_ISL_1194225, EPI_ISL_1194342, EPI_ISL_1194390, EPI_ISL_1194405, EPI_ISL_1194417, EPI_ISL_1194421, EPI_ISL_1194435, EPI_ISL_1194461, EPI_ISL_1220388, EPI_ISL_1220390, EPI_ISL_1220411, EPI_ISL_1220420, EPI_ISL_1220517, EPI_ISL_1220548, EPI_ISL_1220578, EPI_ISL_1220619, EPI_ISL_1220646, EPI_ISL_1220655, EPI_ISL_1220669, EPI_ISL_1220676, EPI_ISL_1220685, EPI_ISL_1220697, EPI_ISL_1220710, EPI_ISL_1220711, EPI_ISL_1252933, EPI_ISL_1252953, EPI_ISL_1252965, EPI_ISL_1252976, EPI_ISL_1253437, EPI_ISL_1254016, EPI_ISL_1254017, EPI_ISL_1254028, EPI_ISL_1254039, EPI_ISL_1254040, EPI_ISL_1254067, EPI_ISL_1254083, EPI_ISL_1254087, EPI_ISL_1254105, EPI_ISL_1254111, EPI_ISL_1254120, EPI_ISL_1254124, EPI_ISL_1254128, EPI_ISL_1254132, EPI_ISL_1254153, EPI_ISL_1254155, EPI_ISL_1254165, EPI_ISL_1254170, EPI_ISL_1254171, EPI_ISL_1254184, EPI_ISL_1254190, EPI_ISL_1254195, EPI_ISL_1254201, EPI_ISL_1254203, EPI_ISL_1254208, EPI_ISL_1254211, EPI_ISL_1254238, EPI_ISL_1254246, EPI_ISL_1254249, EPI_ISL_1254251, EPI_ISL_1254284, EPI_ISL_1254290, EPI_ISL_1254295, EPI_ISL_1254303, EPI_ISL_1254438, EPI_ISL_1254445, EPI_ISL_1254447, EPI_ISL_1265950, EPI_ISL_1265965, EPI_ISL_1265968, EPI_ISL_1265969, EPI_ISL_1265988, EPI_ISL_1265999, EPI_ISL_1266188, EPI_ISL_1266559, EPI_ISL_1266584, EPI_ISL_1266796, EPI_ISL_1266804, EPI_ISL_1267348,

| | | | |
|---|---|---|---|
| EPI_ISL_1267379, EPI_ISL_1267456, EPI_ISL_1291532, EPI_ISL_1291558, EPI_ISL_1291560, EPI_ISL_1291564, EPI_ISL_1297766, EPI_ISL_1297769, EPI_ISL_1298226, EPI_ISL_1298231, EPI_ISL_1298234, EPI_ISL_1298242, EPI_ISL_1314552, EPI_ISL_1314553, EPI_ISL_1314556, EPI_ISL_1314568, EPI_ISL_1314573, EPI_ISL_1314589, EPI_ISL_1314590, EPI_ISL_1314608, EPI_ISL_1314613, EPI_ISL_1314624, EPI_ISL_1314630, EPI_ISL_1314643, EPI_ISL_1314646, EPI_ISL_1314659, EPI_ISL_1314664, EPI_ISL_1314677, EPI_ISL_1314700, EPI_ISL_1314708, EPI_ISL_1314709, EPI_ISL_1314739, EPI_ISL_1314760, EPI_ISL_1314782, EPI_ISL_1314809, EPI_ISL_1314812, EPI_ISL_1314813, EPI_ISL_1314829, EPI_ISL_1314830, EPI_ISL_1314914, EPI_ISL_1314922, EPI_ISL_1335914, EPI_ISL_1335930, EPI_ISL_1336026, EPI_ISL_1336102, EPI_ISL_1339842, EPI_ISL_1340452, EPI_ISL_1340472, EPI_ISL_1340475, EPI_ISL_1340478, EPI_ISL_1340511, EPI_ISL_1340521, EPI_ISL_1340735, EPI_ISL_1367423, EPI_ISL_1367441, EPI_ISL_1367453, EPI_ISL_1367467, EPI_ISL_1367479, EPI_ISL_1422334, EPI_ISL_1422337, EPI_ISL_1422350, EPI_ISL_1422481, EPI_ISL_1422804, EPI_ISL_1422813, EPI_ISL_1422814, EPI_ISL_1422823, EPI_ISL_1422859, EPI_ISL_1422864, EPI_ISL_1422886, EPI_ISL_1422888, EPI_ISL_1422902, EPI_ISL_1422905, EPI_ISL_1422953, EPI_ISL_1422954, EPI_ISL_1422959, EPI_ISL_1422974, EPI_ISL_1480604, EPI_ISL_1480608, EPI_ISL_1480617, EPI_ISL_1480632, EPI_ISL_1480654, EPI_ISL_1480859, EPI_ISL_1480943, EPI_ISL_1480971, EPI_ISL_1480976, EPI_ISL_1551741 | | | |
| see above | Quest Diagnostics Incorporated | Centers for Disease Control and Prevention Division of Viral Diseases, Pathogen Discovery | A. Gerasimova; A. Perez; Adrian Paskey; B. Anderson; Ben L. Rambo-Martin; Benjamin Rambo-Martin; Christopher Gulvick; Clinton R. Paden; Dakota Howard; Darlene Wagner; Dhwani Batra; Duncan MacCannell; F. Lacbawan; I. A. Shlyakhter; Jason Caravas; K.E. Livingston; Kara Moser; L.E. Bernstein; M. Hua; Matthew Schmerer; P. Tanpaiboon; Peter W. Cook; R. M. Kagan; R. Owen; R. V. Rolando; S. H. Rosenthal; Scott Sammons; Shatavia Morrison; Suxiang Tong; Y. Liu; Yvette Unoarumhi |
| EPI_ISL_1086614, EPI_ISL_1086656, EPI_ISL_1087538, EPI_ISL_1087540, EPI_ISL_1087725, EPI_ISL_1087797, EPI_ISL_1087803, EPI_ISL_1087805, EPI_ISL_1087873, EPI_ISL_1087874, EPI_ISL_1087877, EPI_ISL_1087878, EPI_ISL_1087879, EPI_ISL_1087888, EPI_ISL_1087891, EPI_ISL_1087960, EPI_ISL_1087988, EPI_ISL_1087989, EPI_ISL_1087992, EPI_ISL_1088098, EPI_ISL_1088116, EPI_ISL_1088118, EPI_ISL_1088133, EPI_ISL_1088134, EPI_ISL_1088147, EPI_ISL_1088169, EPI_ISL_1088177, EPI_ISL_1088220, EPI_ISL_1088233, EPI_ISL_1090341, EPI_ISL_1090405, EPI_ISL_1090414, EPI_ISL_1090429, EPI_ISL_1090456, EPI_ISL_1090461, EPI_ISL_1090465, EPI_ISL_1090466, EPI_ISL_1090469, EPI_ISL_1090518, EPI_ISL_1090578, EPI_ISL_1090599, EPI_ISL_1090617, EPI_ISL_1090720, EPI_ISL_1090722, EPI_ISL_1090755, EPI_ISL_1090757, EPI_ISL_1090870, EPI_ISL_1090901, EPI_ISL_1090958, EPI_ISL_1113774, EPI_ISL_1113827, EPI_ISL_1113838, EPI_ISL_1113857, EPI_ISL_1114014, EPI_ISL_1114067, EPI_ISL_1114130, EPI_ISL_1114133, EPI_ISL_1114531, EPI_ISL_1114535, EPI_ISL_1114536, EPI_ISL_1114548, EPI_ISL_1114561, EPI_ISL_1114564, EPI_ISL_1114571, EPI_ISL_1114575, EPI_ISL_1114576, EPI_ISL_1114578, EPI_ISL_1114582, EPI_ISL_1114583, EPI_ISL_1114587, EPI_ISL_1114593, EPI_ISL_1114596, EPI_ISL_1114601, EPI_ISL_1114604, EPI_ISL_1114606, EPI_ISL_1114607, EPI_ISL_1114610, EPI_ISL_1114617, EPI_ISL_1114620, EPI_ISL_1114623, EPI_ISL_1114640, EPI_ISL_1114655, EPI_ISL_1114661, EPI_ISL_1114663, EPI_ISL_1114670, EPI_ISL_1135292, EPI_ISL_1135297, EPI_ISL_1135299, EPI_ISL_1139206, EPI_ISL_1139211, EPI_ISL_1139216, EPI_ISL_1139242, EPI_ISL_1139248, EPI_ISL_1139270, EPI_ISL_1139338, EPI_ISL_1139340, EPI_ISL_1139373, EPI_ISL_1139666, EPI_ISL_1139759, EPI_ISL_1139818, EPI_ISL_1158932, EPI_ISL_1158939, EPI_ISL_1158947, EPI_ISL_1158948, EPI_ISL_1158955, EPI_ISL_1158956, EPI_ISL_1158961, EPI_ISL_1158963, EPI_ISL_1158970, EPI_ISL_1158977, EPI_ISL_1158992, EPI_ISL_1158995, EPI_ISL_1158996, EPI_ISL_1158998, EPI_ISL_1159010, EPI_ISL_1159013, EPI_ISL_1159014, EPI_ISL_1159016, EPI_ISL_1159018, EPI_ISL_1159024, EPI_ISL_1159035, EPI_ISL_1159036, EPI_ISL_1159042, EPI_ISL_1159045, EPI_ISL_1159056, EPI_ISL_1159059, EPI_ISL_1159062, EPI_ISL_1159066, EPI_ISL_1290859 | | | |
| see above | Quest Diagnostics Incorporated | Respiratory Viruses Branch, Division of Viral Diseases, Centers for Disease Control and Prevention | A. Gerasimova; A. Perez; B. Anderson; Ben L. Rambo-Martin; Clinton R. Paden; Dakota Howard; Dhwani Batra; Duncan MacCannell; F. Lacbawan; I. A. Shlyakhter; K.E. Livingston; L.E. Bernstein; M. Hua; P. Tanpaiboon; Peter W. Cook; R. M. Kagan; R. Owen; R. V. Rolando; S. H. Rosenthal; Suxiang Tong; Y. Liu |
| EPI_ISL_1184135, EPI_ISL_1184136, EPI_ISL_1184137 | Rochester Regional Hospital | Multidrug-Resistant Organism Repository and Surveillance Network (MRSN) | Ana C. Ong; Brendan Corey; Brett E. Swierczewski; Edward E. Walsh; Emil Lesho; Francois Lebreton; Jason W. Bennett; and Patrick Mc Gann |
| EPI_ISL_1200561, EPI_ISL_1447143 | Rockefeller University Hospital | New York City Public Health Laboratory | Jade Wang; et al. |
| EPI_ISL_1227245, EPI_ISL_1228278, EPI_ISL_1398269, EPI_ISL_1582228, EPI_ISL_1624483, EPI_ISL_1624484, EPI_ISL_1651224, EPI_ISL_1671677, EPI_ISL_2098133 | | | |
| see above | SARATOGA HOSPITAL LABORATORY | Wadsworth Center, New York State Department of Health | Alexis Russel; Alexis Russell; Catharine Prussing; Daryl M. Lamson; Erasmus Schneider; Erica Lasek-Nesselquist; John Kelly; Jonathan Plitnick; Kirsten St. George; Matthew Shudt; Melissa A Leisner; Navjot Singh |
| EPI_ISL_1397831, EPI_ISL_1397832, EPI_ISL_1465844 | STONY BROOK UNIVERSITY HOSPITAL | Wadsworth Center, New York State Department of Health | Alexis Russel; Alexis Russell; Catharine Prussing; Daryl M. Lamson; Erasmus Schneider; Erica Lasek-Nesselquist; John Kelly; Jonathan Plitnick; Kirsten St. George; Matthew Shudt; Melissa A Leisner; Navjot Singh |
| EPI_ISL_1226593, EPI_ISL_1226683, EPI_ISL_1226732, EPI_ISL_1226872, EPI_ISL_1226873, EPI_ISL_1226981, EPI_ISL_1227129, EPI_ISL_1227173, EPI_ISL_1227239, EPI_ISL_1227240, EPI_ISL_1227241, EPI_ISL_1227447, EPI_ISL_1227463, EPI_ISL_1227464, EPI_ISL_1227465, EPI_ISL_1227641, EPI_ISL_1227642, EPI_ISL_1227708, EPI_ISL_1227726, EPI_ISL_1227727, EPI_ISL_1227991, EPI_ISL_1227992, EPI_ISL_1228005, EPI_ISL_1228006, EPI_ISL_1228007, EPI_ISL_1228008, EPI_ISL_1228009, EPI_ISL_1228010, EPI_ISL_1228011, EPI_ISL_1228012, EPI_ISL_1228013, EPI_ISL_1228014, EPI_ISL_1228015, EPI_ISL_1228016, EPI_ISL_1228017, EPI_ISL_1228018, EPI_ISL_1228019, EPI_ISL_1228020, EPI_ISL_1228021, EPI_ISL_1228022, EPI_ISL_1228293, EPI_ISL_1228380, EPI_ISL_1395751, EPI_ISL_1395752, EPI_ISL_1395755, EPI_ISL_1395759, EPI_ISL_1395761, EPI_ISL_1395762, EPI_ISL_1395764, EPI_ISL_1395765, EPI_ISL_1395767, EPI_ISL_1395770, EPI_ISL_1395772, EPI_ISL_1395774, EPI_ISL_1395775, EPI_ISL_1395776, EPI_ISL_1397592, EPI_ISL_1397593, EPI_ISL_1397594, EPI_ISL_1397595, EPI_ISL_1397596, EPI_ISL_1397597, EPI_ISL_1397598, EPI_ISL_1397599, EPI_ISL_1397600, EPI_ISL_1397601, EPI_ISL_1397602, EPI_ISL_1397603, EPI_ISL_1397604, EPI_ISL_1397605, EPI_ISL_1397606, EPI_ISL_1397607, EPI_ISL_1397608, EPI_ISL_1397609, EPI_ISL_1397610, EPI_ISL_1397611, EPI_ISL_1397629, EPI_ISL_1397706, EPI_ISL_1397784, EPI_ISL_1397785, EPI_ISL_1397786, EPI_ISL_1397787, EPI_ISL_1397968, EPI_ISL_1397969, EPI_ISL_1397970, EPI_ISL_1397983, EPI_ISL_1398002, EPI_ISL_1398003, EPI_ISL_1398004, EPI_ISL_1398156, EPI_ISL_1398157, EPI_ISL_1398158, EPI_ISL_1398159, EPI_ISL_1398160, EPI_ISL_1398161, EPI_ISL_1398162, EPI_ISL_1461294, EPI_ISL_1461310, EPI_ISL_1461311, EPI_ISL_1462500, EPI_ISL_1462502, EPI_ISL_1462503, EPI_ISL_1462505, EPI_ISL_1462536, EPI_ISL_1465638, EPI_ISL_1465689, EPI_ISL_1465690, EPI_ISL_1465691, EPI_ISL_1465692, EPI_ISL_1482274, EPI_ISL_1482275, EPI_ISL_1482276, EPI_ISL_1499447, EPI_ISL_1499451, EPI_ISL_1499452, EPI_ISL_1499453, EPI_ISL_1499454, EPI_ISL_1499455, EPI_ISL_1499456, EPI_ISL_1576812, EPI_ISL_1582149, EPI_ISL_1755487, EPI_ISL_1755489, EPI_ISL_1755490, EPI_ISL_2098631, EPI_ISL_2098632, EPI_ISL_2098633, EPI_ISL_2098634, EPI_ISL_2098635, EPI_ISL_2098636, EPI_ISL_2134798 | | | |
| see above | SUNY UPSTATE MEDICAL UNIVERSITY | Wadsworth Center, New York State Department of Health | Alexis Russel; Alexis Russell; Catharine Prussing; Daryl M. Lamson; Erasmus Schneider; Erica Lasek-Nesselquist; John Kelly; Jonathan Plitnick; Kirsten St. George; Matthew Shudt; Melissa A Leisner; Navjot Singh |
| EPI_ISL_1967686, EPI_ISL_1967688 | Stony Brook Medicine | New York Genome Center | Alaa B. Salaheldin; Andre Corvelo; Dayna M. Oschwald; Dimitri Gnatenko; Eric D. Spitzer; Jiang Chen; Josue M. Flores Acuna; Kenneth R. Shroyer; Margaret M. McGovern; Michael Zody; Richard R. Kew; Samantha Fennessey; Sarah A. Halbohn; Sarah A. Malik; Silvia G. Spitzer; Tom Maniatis |
| EPI_ISL_1016169, EPI_ISL_1016249, EPI_ISL_1016250, EPI_ISL_1016251, EPI_ISL_1016252, EPI_ISL_1016253, EPI_ISL_1016254, EPI_ISL_1016255, EPI_ISL_1226449, EPI_ISL_1226487, EPI_ISL_1226495, EPI_ISL_1226501, EPI_ISL_1226533, EPI_ISL_1226534, EPI_ISL_1226535, EPI_ISL_1226536, EPI_ISL_1226537, EPI_ISL_1226550, EPI_ISL_1226555, EPI_ISL_1226557, EPI_ISL_1226567, EPI_ISL_1226569, EPI_ISL_1226570, EPI_ISL_1226575, EPI_ISL_1226578, EPI_ISL_1226579, EPI_ISL_1226584, EPI_ISL_1226585, EPI_ISL_1226586, EPI_ISL_1226589, EPI_ISL_1226592, EPI_ISL_1226603, EPI_ISL_1226618, EPI_ISL_1226619, EPI_ISL_1226626, EPI_ISL_1226627, EPI_ISL_1226628, EPI_ISL_1226629, EPI_ISL_1226630, EPI_ISL_1226631, EPI_ISL_1226632, EPI_ISL_1226633, EPI_ISL_1226634, EPI_ISL_1226635, EPI_ISL_1226649, EPI_ISL_1226650, EPI_ISL_1226653, EPI_ISL_1226659, EPI_ISL_1226666, EPI_ISL_1226667, EPI_ISL_1226670, EPI_ISL_1226709, EPI_ISL_1226712, EPI_ISL_1227162, EPI_ISL_1397553, EPI_ISL_1397554, EPI_ISL_1397581, EPI_ISL_1397582, EPI_ISL_1397583, EPI_ISL_1397584, EPI_ISL_1397585, EPI_ISL_1397586, EPI_ISL_1397587, EPI_ISL_1397588, EPI_ISL_1397589, EPI_ISL_1397590, EPI_ISL_1397591, EPI_ISL_1397700, EPI_ISL_1397701, EPI_ISL_1397702, EPI_ISL_1397703, EPI_ISL_1397704, EPI_ISL_1397705, EPI_ISL_1397836, EPI_ISL_1397838, EPI_ISL_1397839, EPI_ISL_1397840, EPI_ISL_1397841, EPI_ISL_1397842, EPI_ISL_1397844, EPI_ISL_1397845, EPI_ISL_1397846, EPI_ISL_1397847, EPI_ISL_1397848, EPI_ISL_1397849, EPI_ISL_1397850, EPI_ISL_1397851, EPI_ISL_1397852, EPI_ISL_1397857, EPI_ISL_1397859, EPI_ISL_1397860, EPI_ISL_1397862, EPI_ISL_1397863, EPI_ISL_1397864, EPI_ISL_1397865, EPI_ISL_1461356, EPI_ISL_1461357, EPI_ISL_1461359, EPI_ISL_1461360, EPI_ISL_1461361, EPI_ISL_1461362, EPI_ISL_1461363, EPI_ISL_1461364, EPI_ISL_1461365, EPI_ISL_1461366, EPI_ISL_1461367, EPI_ISL_1461368, EPI_ISL_1461369, EPI_ISL_1461370, EPI_ISL_1461371, EPI_ISL_1461372, EPI_ISL_1461373, EPI_ISL_1461374, EPI_ISL_1461375, EPI_ISL_1461376, EPI_ISL_1461377, EPI_ISL_1462552, EPI_ISL_1576814, EPI_ISL_1582403 | | | |
| see above | TEMPUS LABS INC | Wadsworth Center, New York State Department of Health | Alexis Russel; Alexis Russell; Catharine Prussing; Daryl M. Lamson; Erasmus Schneider; Erica Lasek-Nesselquist; John Kelly; Jonathan Plitnick; Kirsten St. George; Matthew Shudt; Melissa A Leisner; Navjot Singh |
| EPI_ISL_1016130, EPI_ISL_1016131, EPI_ISL_1016132, EPI_ISL_1016133, EPI_ISL_1016137, EPI_ISL_1016139, EPI_ISL_1016154, EPI_ISL_1016155, EPI_ISL_1227210, EPI_ISL_1227640, EPI_ISL_1228051, EPI_ISL_1228291, EPI_ISL_1228453, EPI_ISL_1395766, EPI_ISL_1395768, EPI_ISL_1395769, EPI_ISL_1395771, EPI_ISL_1395773, EPI_ISL_1397612, EPI_ISL_1397613, EPI_ISL_1397614, EPI_ISL_1397615, EPI_ISL_1397616, EPI_ISL_1397617, EPI_ISL_1397783, EPI_ISL_1397966, EPI_ISL_1397967, EPI_ISL_1397988, EPI_ISL_1398259, EPI_ISL_1398260, EPI_ISL_1398261, EPI_ISL_1398262, EPI_ISL_1398263, EPI_ISL_1465674, EPI_ISL_1465675, EPI_ISL_1465760 | | | |
| see above | THE MARY IMOGENE BASSETT HOSPITAL | Wadsworth Center, New York State Department of Health | Alexis Russel; Alexis Russell; Catharine Prussing; Daryl M. Lamson; Erasmus Schneider; Erica Lasek-Nesselquist; John Kelly; Jonathan Plitnick; Kirsten St. George; Matthew Shudt; Melissa A Leisner; Navjot Singh |
| EPI_ISL_1378790, EPI_ISL_1378791, EPI_ISL_1378792, EPI_ISL_1378793, EPI_ISL_1378794, EPI_ISL_1378795, EPI_ISL_1378796 | | | |
| see above | Tempus | Grubaugh Lab - Yale School of Public Health | Anderson Brito; Annie Watkins; Chaney Kalinich; Chantal Vogels; Christopher E. Mason; Gaurav Khullar; Isabell Ott; Jessica Metti; Jessica Rothman; Joel T. Dudley; Joseph Fauver; Mallery Breban; Mary Petrone; Matthew J. MacKay; Megan Nash; Nathan Grubaugh; Nike Beaubier; Tara Alpert |
| EPI_ISL_983447, EPI_ISL_983448, EPI_ISL_983449, EPI_ISL_983450, EPI_ISL_983451, EPI_ISL_983452, EPI_ISL_1016101, EPI_ISL_1016102, EPI_ISL_1016103, EPI_ISL_1016104, EPI_ISL_1016105, EPI_ISL_1016106, EPI_ISL_1016107, EPI_ISL_1016108, EPI_ISL_1016109, EPI_ISL_1016110, EPI_ISL_1016118, EPI_ISL_1016119, EPI_ISL_1016120, EPI_ISL_1016121, EPI_ISL_1016122, EPI_ISL_1016123, EPI_ISL_1016124, EPI_ISL_1016125, EPI_ISL_1016126, EPI_ISL_1226421, EPI_ISL_1226432, EPI_ISL_1226478, EPI_ISL_1226480, EPI_ISL_1226486, EPI_ISL_1226514, EPI_ISL_1226516, EPI_ISL_1226517, EPI_ISL_1226520, EPI_ISL_1226552, EPI_ISL_1226598, EPI_ISL_1226602, EPI_ISL_1226623, EPI_ISL_1226897, EPI_ISL_1227454, EPI_ISL_1227455, EPI_ISL_1227563, EPI_ISL_1227716, EPI_ISL_1227717, EPI_ISL_1227718, EPI_ISL_1227719, EPI_ISL_1227759, EPI_ISL_1227760, EPI_ISL_1227761, EPI_ISL_1227762, EPI_ISL_1227763, EPI_ISL_1227764, EPI_ISL_1227765, EPI_ISL_1227766, EPI_ISL_1227767, EPI_ISL_1227768, EPI_ISL_1227769, EPI_ISL_1227770, EPI_ISL_1227771, EPI_ISL_1227772, EPI_ISL_1227773, EPI_ISL_1227774, EPI_ISL_1227775, EPI_ISL_1227776, EPI_ISL_1227777, EPI_ISL_1227778, EPI_ISL_1227779, EPI_ISL_1227780, EPI_ISL_1227781, EPI_ISL_1227782, EPI_ISL_1227783, EPI_ISL_1227784, EPI_ISL_1227785, EPI_ISL_1227786, EPI_ISL_1227787, EPI_ISL_1227788, EPI_ISL_1227789, EPI_ISL_1227973, EPI_ISL_1228028, EPI_ISL_1228029, EPI_ISL_1228030, EPI_ISL_1228031, EPI_ISL_1228041, EPI_ISL_1228043, EPI_ISL_1228044, EPI_ISL_1228045, EPI_ISL_1228116, EPI_ISL_1228306, EPI_ISL_1228481, EPI_ISL_1228482, EPI_ISL_1228483, EPI_ISL_1394629, EPI_ISL_1394630, EPI_ISL_1394631, EPI_ISL_1394632, EPI_ISL_1394633, EPI_ISL_1394634, EPI_ISL_1394635, EPI_ISL_1394636, EPI_ISL_1394637, EPI_ISL_1394638, EPI_ISL_1394639, EPI_ISL_1394640, EPI_ISL_1394641, EPI_ISL_1394643, EPI_ISL_1397552, EPI_ISL_1397555, EPI_ISL_1397556, EPI_ISL_1397557, EPI_ISL_1397558, EPI_ISL_1397559, EPI_ISL_1397560, EPI_ISL_1397561, EPI_ISL_1397562, EPI_ISL_1397563, EPI_ISL_1397564, EPI_ISL_1397565, EPI_ISL_1397566, EPI_ISL_1397567, EPI_ISL_1397568, EPI_ISL_1397569, EPI_ISL_1397570, EPI_ISL_1397571, EPI_ISL_1397572, EPI_ISL_1397573, EPI_ISL_1397574, EPI_ISL_1397575, EPI_ISL_1397576, EPI_ISL_1397577, EPI_ISL_1397578, EPI_ISL_1397579, EPI_ISL_1397580, EPI_ISL_1397630, EPI_ISL_1397631, EPI_ISL_1397632, EPI_ISL_1397633, EPI_ISL_1397634, EPI_ISL_1397635, EPI_ISL_1397810, EPI_ISL_1397811, EPI_ISL_1397812, EPI_ISL_1397813, EPI_ISL_1397814, EPI_ISL_1397815, EPI_ISL_1397816, EPI_ISL_1397817, EPI_ISL_1397818, EPI_ISL_1397819, EPI_ISL_1397820, EPI_ISL_1397821, EPI_ISL_1397822, EPI_ISL_1397823, EPI_ISL_1397971, EPI_ISL_1397972, EPI_ISL_1397973, EPI_ISL_1397974, EPI_ISL_1397975, EPI_ISL_1397976, EPI_ISL_1397977, EPI_ISL_1397978, EPI_ISL_1397979, EPI_ISL_1397980, EPI_ISL_1397981, EPI_ISL_1397982, EPI_ISL_1397984, EPI_ISL_1397985, EPI_ISL_1397986, EPI_ISL_1397987, EPI_ISL_1397989, EPI_ISL_1397990, EPI_ISL_1397991, EPI_ISL_1397992, EPI_ISL_1397993, EPI_ISL_1397994, EPI_ISL_1397995, EPI_ISL_1397996, EPI_ISL_1397997, EPI_ISL_1397998, EPI_ISL_1397999, EPI_ISL_1398000, EPI_ISL_1398001, EPI_ISL_1398005, EPI_ISL_1398006, EPI_ISL_1398007, EPI_ISL_1398008, EPI_ISL_1398009, EPI_ISL_1398010, EPI_ISL_1398011, EPI_ISL_1398012, EPI_ISL_1398013, EPI_ISL_1398014, EPI_ISL_1398015, EPI_ISL_1398016, EPI_ISL_1398017, EPI_ISL_1398018, EPI_ISL_1398019, EPI_ISL_1398020, EPI_ISL_1398123, EPI_ISL_1398124, EPI_ISL_1398125, EPI_ISL_1398126, EPI_ISL_1398127, EPI_ISL_1398128, EPI_ISL_1398129, EPI_ISL_1398130, EPI_ISL_1398131, EPI_ISL_1398137, EPI_ISL_1398138, EPI_ISL_1398139, EPI_ISL_1398140, EPI_ISL_1398141, EPI_ISL_1398142, EPI_ISL_1398143, EPI_ISL_1398271, EPI_ISL_1398272, EPI_ISL_1398273, EPI_ISL_1398312, EPI_ISL_1398313, EPI_ISL_1398314, EPI_ISL_1398315, EPI_ISL_1398316, EPI_ISL_1398317, EPI_ISL_1461290, EPI_ISL_1461291, EPI_ISL_1461292, EPI_ISL_1461300, EPI_ISL_1461301, EPI_ISL_1461302, EPI_ISL_1461303, EPI_ISL_1461304, EPI_ISL_1461305, EPI_ISL_1461306, EPI_ISL_1461307, EPI_ISL_1461308, EPI_ISL_1461309, EPI_ISL_1461318, EPI_ISL_1462509, EPI_ISL_1462510, EPI_ISL_1462512, EPI_ISL_1462513, EPI_ISL_1462515, EPI_ISL_1462516, EPI_ISL_1462518, EPI_ISL_1462519, EPI_ISL_1462521, EPI_ISL_1462523, EPI_ISL_1462524, EPI_ISL_1462525, EPI_ISL_1462527, EPI_ISL_1462528, EPI_ISL_1462530, EPI_ISL_1462531, EPI_ISL_1462533, EPI_ISL_1462534, EPI_ISL_1465618, EPI_ISL_1465619, EPI_ISL_1465620, EPI_ISL_1465621, EPI_ISL_1465623, EPI_ISL_1465624, EPI_ISL_1465625, EPI_ISL_1465626, EPI_ISL_1465627, EPI_ISL_1465628, EPI_ISL_1465629, EPI_ISL_1465630, EPI_ISL_1465631, EPI_ISL_1465632, EPI_ISL_1465640, EPI_ISL_1465641, EPI_ISL_1465642, EPI_ISL_1465643, EPI_ISL_1465644, EPI_ISL_1465645, EPI_ISL_1465646, EPI_ISL_1465647, EPI_ISL_1465648, EPI_ISL_1465649, EPI_ISL_1465650, EPI_ISL_1465651, EPI_ISL_1465652, EPI_ISL_1465653, EPI_ISL_1465654, EPI_ISL_1465655, EPI_ISL_1465656, EPI_ISL_1465657, EPI_ISL_1465658, EPI_ISL_1465659, EPI_ISL_1465660, EPI_ISL_1465661, EPI_ISL_1465662, EPI_ISL_1465663, EPI_ISL_1465664, EPI_ISL_1465667, EPI_ISL_1465696, EPI_ISL_1465726, EPI_ISL_1465727, EPI_ISL_1465728, EPI_ISL_1465729, EPI_ISL_1482278, EPI_ISL_1482279, EPI_ISL_1482280, EPI_ISL_1482281, EPI_ISL_1482282, EPI_ISL_1482283, EPI_ISL_1482284, EPI_ISL_1482285, EPI_ISL_1482286, EPI_ISL_1482299, EPI_ISL_1482302, EPI_ISL_1482303, EPI_ISL_1499396, EPI_ISL_1499397, EPI_ISL_1499398, EPI_ISL_1499399, EPI_ISL_1499400, EPI_ISL_1499401, EPI_ISL_1499402, EPI_ISL_1499403, EPI_ISL_1499408, EPI_ISL_1575446, EPI_ISL_1575447, EPI_ISL_1575448, EPI_ISL_1575450, EPI_ISL_1575451, EPI_ISL_1575452, EPI_ISL_1575455, EPI_ISL_1575456, EPI_ISL_1575457, EPI_ISL_1575458, EPI_ISL_1575461, EPI_ISL_1575473, EPI_ISL_1576823, EPI_ISL_1580671, EPI_ISL_1580672, EPI_ISL_1582151, EPI_ISL_1582398, EPI_ISL_1582404, EPI_ISL_1789706, EPI_ISL_1789707, EPI_ISL_1920127, EPI_ISL_1935648, EPI_ISL_2098501, EPI_ISL_2098660, EPI_ISL_2098661, EPI_ISL_2098662, EPI_ISL_2098663, EPI_ISL_2098664, EPI_ISL_2098665, EPI_ISL_2134792 | | | |
| see above | URMC LABS | Wadsworth Center, New York State Department of Health | Alexis Russel; Alexis Russell; Catharine Prussing; Daryl M. Lamson; Erasmus Schneider; Erica Lasek-Nesselquist; John Kelly; Jonathan Plitnick; Kirsten St. George; Matthew Shudt; Melissa A Leisner; Navjot Singh |
| EPI_ISL_1531475 | US Air Force School of Aerospace Medicine | US Air Force School of Aerospace Medicine | Amanda Javorina; Anthony Fries; Clarise Starr; Elizabeth Macias; Jennifer Meyer; Sarah Purves; William Buggele; William Gruner |
| EPI_ISL_1226438, EPI_ISL_1226496, EPI_ISL_1226497, EPI_ISL_1226538, EPI_ISL_1226539, EPI_ISL_1226558, EPI_ISL_1226568, EPI_ISL_1226616, EPI_ISL_1226655, EPI_ISL_1226656, EPI_ISL_1226657, EPI_ISL_1226668, EPI_ISL_1226678, EPI_ISL_1226679, EPI_ISL_1226695, EPI_ISL_1226723, EPI_ISL_1226756, EPI_ISL_1226772, EPI_ISL_1226783, EPI_ISL_1226784, EPI_ISL_1226785, EPI_ISL_1226786, EPI_ISL_1226787, EPI_ISL_1226788, EPI_ISL_1226818, EPI_ISL_1226821, EPI_ISL_1226829, EPI_ISL_1226830, EPI_ISL_1226834, EPI_ISL_1226836, EPI_ISL_1226837, EPI_ISL_1226838, EPI_ISL_1226840, EPI_ISL_1226841, EPI_ISL_1226863, EPI_ISL_1226870, EPI_ISL_1226925, EPI_ISL_1226970, EPI_ISL_1226973, EPI_ISL_1227019, EPI_ISL_1227020, EPI_ISL_1227023, EPI_ISL_1227176, EPI_ISL_1227177, EPI_ISL_1227189, EPI_ISL_1227207, EPI_ISL_1227217, EPI_ISL_1227218, EPI_ISL_1227219, EPI_ISL_1227220, EPI_ISL_1227223, EPI_ISL_1227224, EPI_ISL_1227225, EPI_ISL_1227228, EPI_ISL_1227229, EPI_ISL_1227247, EPI_ISL_1227248, EPI_ISL_1227441, EPI_ISL_1227530, EPI_ISL_1227531, EPI_ISL_1227532, EPI_ISL_1227533, EPI_ISL_1227534, EPI_ISL_1227535, EPI_ISL_1227537, EPI_ISL_1227538, EPI_ISL_1227539, EPI_ISL_1227540, EPI_ISL_1228120, EPI_ISL_1228121, EPI_ISL_1228263, EPI_ISL_1228264, EPI_ISL_1228267, EPI_ISL_1228374, EPI_ISL_1228422, EPI_ISL_1228423, EPI_ISL_1228424, EPI_ISL_1228425, EPI_ISL_1228426, EPI_ISL_1228427, EPI_ISL_1228428, EPI_ISL_1228429, EPI_ISL_1228431, EPI_ISL_1228432, EPI_ISL_1228433, EPI_ISL_1228561, EPI_ISL_1228562, EPI_ISL_1228565, EPI_ISL_1228566, EPI_ISL_1228571, EPI_ISL_1228573, EPI_ISL_1228577, EPI_ISL_1228579, EPI_ISL_1228580, EPI_ISL_1228581, EPI_ISL_1228582, EPI_ISL_1228583, EPI_ISL_1228585, EPI_ISL_1228586, EPI_ISL_1394599, EPI_ISL_1394603, EPI_ISL_1394604, EPI_ISL_1394605, EPI_ISL_1394606, EPI_ISL_1394607, EPI_ISL_1394608, EPI_ISL_1394609, EPI_ISL_1394610, EPI_ISL_1394611, EPI_ISL_1394612, EPI_ISL_1394613, EPI_ISL_1394614, EPI_ISL_1394615, EPI_ISL_1394616, EPI_ISL_1394617, EPI_ISL_1394618, EPI_ISL_1394619, EPI_ISL_1394620, EPI_ISL_1394621, EPI_ISL_1394622, EPI_ISL_1394623, EPI_ISL_1394624, EPI_ISL_1394625, EPI_ISL_1394626, EPI_ISL_1394627, EPI_ISL_1394628, EPI_ISL_1397550, EPI_ISL_1397914, EPI_ISL_1397915, EPI_ISL_1397916, EPI_ISL_1397917, EPI_ISL_1397918, EPI_ISL_1397919, EPI_ISL_1397920, EPI_ISL_1397921, EPI_ISL_1397922, EPI_ISL_1397923, EPI_ISL_1397924, EPI_ISL_1397925, EPI_ISL_1397926, EPI_ISL_1397927, EPI_ISL_1397928, EPI_ISL_1397929, EPI_ISL_1397930, EPI_ISL_1397931, EPI_ISL_1397932, EPI_ISL_1397933, EPI_ISL_1397934, EPI_ISL_1397935, EPI_ISL_1397936, EPI_ISL_1397937, EPI_ISL_1397938, EPI_ISL_1397939, EPI_ISL_1397940, EPI_ISL_1397941, EPI_ISL_1397942, EPI_ISL_1397943, EPI_ISL_1397944, EPI_ISL_1397945, EPI_ISL_1397946, EPI_ISL_1397947, EPI_ISL_1397948, EPI_ISL_1397949, EPI_ISL_1397950, EPI_ISL_1397951, EPI_ISL_1397952, EPI_ISL_1398028, EPI_ISL_1398029, EPI_ISL_1398030, EPI_ISL_1398031, EPI_ISL_1398032, EPI_ISL_1398033, EPI_ISL_1465622, EPI_ISL_1465637, EPI_ISL_1482394, EPI_ISL_1482395, EPI_ISL_1482539, EPI_ISL_1482540, EPI_ISL_1482541, EPI_ISL_1482542, EPI_ISL_1482543, EPI_ISL_1575453, EPI_ISL_1575454, EPI_ISL_1582308, EPI_ISL_1582309, EPI_ISL_1593872, EPI_ISL_1671144, EPI_ISL_1935612, EPI_ISL_1935614 | | | |
| see above | WESTCHESTER MEDICAL CENTER | Wadsworth Center, New York State Department of Health | Alexis Russel; Alexis Russell; Catharine Prussing; Daryl M. Lamson; Erasmus Schneider; Erica Lasek-Nesselquist; John Kelly; Jonathan Plitnick; Kirsten St. George; Matthew Shudt; Melissa A Leisner; Navjot Singh |
| EPI_ISL_1398268 | WHITE PLAINS HOSPITAL CENTER LABORATORY | Wadsworth Center, New York State Department of Health | Alexis Russel; Daryl M. Lamson; Erasmus Schneider; Erica Lasek-Nesselquist; John Kelly; Jonathan Plitnick; Kirsten St. George; Matthew Shudt; Melissa A Leisner; Navjot Singh |
| EPI_ISL_1226527, EPI_ISL_1226528, EPI_ISL_1226933, EPI_ISL_1227234, EPI_ISL_1227559, EPI_ISL_1227571, EPI_ISL_1227593, EPI_ISL_1227594, EPI_ISL_1227595, EPI_ISL_1227596, EPI_ISL_1227597, EPI_ISL_1227598, EPI_ISL_1227599, EPI_ISL_1227600, EPI_ISL_1227601, EPI_ISL_1227602, EPI_ISL_1227603, EPI_ISL_1227604, EPI_ISL_1227605, EPI_ISL_1227606, EPI_ISL_1227607, EPI_ISL_1227608, EPI_ISL_1227609, EPI_ISL_1227610, EPI_ISL_1227611, EPI_ISL_1227612, EPI_ISL_1227613, EPI_ISL_1227614, EPI_ISL_1227615, EPI_ISL_1227616, EPI_ISL_1228222, EPI_ISL_1228223, EPI_ISL_1228281, EPI_ISL_1228452, EPI_ISL_1228474, EPI_ISL_1228475, EPI_ISL_1397833 | | | |
| see above | Wadsworth Center, New York State Department of Health | Wadsworth Center, New York State Department of Health | Alexis Russel; Daryl M. Lamson; Erasmus Schneider; Erica Lasek-Nesselquist; John Kelly; Jonathan Plitnick; Kirsten St. George; Matthew Shudt; Melissa A Leisner; Navjot Singh |
| EPI_ISL_1968563, EPI_ISL_1968564, EPI_ISL_1968565, EPI_ISL_1968567, EPI_ISL_1968571, EPI_ISL_1968573, EPI_ISL_1968575, EPI_ISL_1968576, EPI_ISL_1968581, EPI_ISL_1968584, EPI_ISL_1968585, EPI_ISL_1968586, EPI_ISL_1968587, EPI_ISL_1968595, EPI_ISL_1968597, EPI_ISL_1968602, EPI_ISL_1968603, EPI_ISL_1968605, EPI_ISL_1968606, EPI_ISL_1968607, EPI_ISL_1968610, EPI_ISL_1968611, EPI_ISL_1968623, EPI_ISL_1968624, EPI_ISL_1968627, EPI_ISL_1971091, EPI_ISL_1971098, EPI_ISL_1971100, EPI_ISL_1971105, EPI_ISL_1971106, EPI_ISL_1971114, EPI_ISL_1971126, EPI_ISL_1971129, EPI_ISL_1971131, EPI_ISL_1971134, EPI_ISL_1971138, EPI_ISL_1971144, EPI_ISL_1971148, EPI_ISL_1971149, EPI_ISL_1971154, EPI_ISL_1971155, EPI_ISL_1971156, EPI_ISL_1971158, EPI_ISL_1971160, EPI_ISL_1971161, EPI_ISL_1971162, EPI_ISL_1971163, EPI_ISL_1971169, EPI_ISL_1971171, EPI_ISL_1971174, EPI_ISL_1971175, EPI_ISL_1971178, EPI_ISL_1971179, EPI_ISL_1971181, EPI_ISL_1971185, EPI_ISL_1971186, EPI_ISL_1971187, EPI_ISL_1971190, EPI_ISL_1971193, EPI_ISL_1971196, EPI_ISL_1971197, EPI_ISL_1971198, EPI_ISL_1971200, EPI_ISL_1971202, EPI_ISL_1971203, EPI_ISL_1971204, EPI_ISL_1971207, EPI_ISL_1971208, EPI_ISL_1971210, EPI_ISL_1971211, EPI_ISL_1971213, EPI_ISL_1971214, EPI_ISL_1971217, EPI_ISL_1971219, EPI_ISL_1971221, EPI_ISL_1971223, EPI_ISL_1971224, EPI_ISL_1971226, EPI_ISL_1971227, EPI_ISL_1971228, EPI_ISL_1971229, EPI_ISL_2031771 | | | |
| see above | Weill Cornell Medicine | New York Genome Center | Andre Corvelo; Arryn Craney; Chris Mason; Dayna M. Oschwald; Hanna Rennert; Lars F Westblade; Margaret Elizabeth Ross; Melissa Cushing; Michael Zody; Olivier Elemento; Priya Velu; Samantha Fennessey; Tom Maniatis |
| EPI_ISL_1920152, EPI_ISL_1920153, EPI_ISL_1920155, EPI_ISL_1920158, EPI_ISL_1920167, EPI_ISL_1920172, EPI_ISL_1920174 | | | |

| | | | |
|---|---|---|---|
| see above | Westchester Medical Center | Wadsworth Center, New York State Department of Health | Alexis Russell; Catharine Prussing; Daryl M. Lamson; Erasmus Schneider; Erica Lasek-Nesselquist; John Kelly; Jonathan Plitnick; Kirsten St. George; Matthew Shudt; Melissa A Leisner; Navjot Singh |
| EPI_ISL_1225669, EPI_ISL_1225706, EPI_ISL_1292992, EPI_ISL_1378687 | Yale Clinical Virology Lab | Grubaugh Lab - Yale School of Public Health | Anderson Brito; Annie Watkins; Chaney Kalinich; Chantal Vogels; Isabell Ott; Jessica Rothman; Joseph Fauver; Mallery Breban; Marie L. Landry; Mary Petrone; Nathan Grubaugh; Tara Alpert |
| EPI_ISL_2034813, EPI_ISL_2034818 | Yale Clinical Virology Lab | Yale Center for Genomic Analysis | Brooke Sullivan; Curt Scharfe; Irina Tikhonova; Kaya Bilguvar; Shrikant Mane |

We gratefully acknowledge the following Authors from the Originating laboratories responsible for obtaining the specimens, as well as the Submitting laboratories where the genome data were generated and shared via GISAID, on which this research is based.

All Submitters of data may be contacted directly via www.gisaid.org

Authors are sorted alphabetically.

| Accession ID | Originating Laboratory | Submitting Laboratory | Authors |
| --- | --- | --- | --- |
| EPI_ISL_1824033 | "NYSDOH Wadsworth Center, Virology Lab" | Centers for Disease Control and Prevention Division of Viral Diseases, Pathogen Discovery | Alison Laufer Halpin; Ben L. Rambo-Martin; Clinton R. Paden; Dakota Howard; Darlene Wagner; Dave Wentworth; Dhwani Batra; Jasmine Padilla; Justin Lee; Katie Dillon; Krista Queen; Kristen Knipe; Kristine Lacek; Mark Burroughs; Matthew Schmerer; Mili Sheth; Peter Cook; Sam Shepard; Sarah Nobles; Shoshona Le; Suxiang Tong; Vivien Dugan; Yvette Unoarumhi |
| EPI_ISL_1398278, EPI_ISL_1398279, EPI_ISL_1398280, EPI_ISL_1398281, EPI_ISL_1398282, EPI_ISL_1398283, EPI_ISL_1461299, EPI_ISL_1482457, EPI_ISL_1482458, EPI_ISL_1482459, EPI_ISL_1482460, EPI_ISL_1482461, EPI_ISL_1482462, EPI_ISL_1482463, EPI_ISL_1482464, EPI_ISL_1482465, EPI_ISL_1482466, EPI_ISL_1482467, EPI_ISL_1482468, EPI_ISL_1482469, EPI_ISL_1576824, EPI_ISL_1580741, EPI_ISL_1582218, EPI_ISL_1582260, EPI_ISL_1582407, EPI_ISL_1671313, EPI_ISL_1671315, EPI_ISL_1671317, EPI_ISL_1671318, EPI_ISL_1671319, EPI_ISL_1671320, EPI_ISL_1671323, EPI_ISL_1671324, EPI_ISL_1671325, EPI_ISL_1671326, EPI_ISL_1671327, EPI_ISL_1671337, EPI_ISL_1671354, EPI_ISL_1671357, EPI_ISL_1671358, EPI_ISL_1671462, EPI_ISL_1671623, EPI_ISL_1740509, EPI_ISL_1755838, EPI_ISL_1755839, EPI_ISL_1755840, EPI_ISL_1755841, EPI_ISL_1756329 | | | |
| see above | ALBANY MEDICAL CENTER | Wadsworth Center, New York State Department of Health | Alexis Russel; Alexis Russell; Catharine Prussing; Daryl M. Lamson; Erasmus Schneider; Erica Lasek-Nesselquist; John Kelly; Jonathan Plitnick; Kirsten St. George; Matthew Shudt; Melissa A Leisner; Navjot Singh |
| EPI_ISL_1445878, EPI_ISL_1445881, EPI_ISL_1446190, EPI_ISL_1479549, EPI_ISL_1479604, EPI_ISL_1479823, EPI_ISL_1491743, EPI_ISL_1491783, EPI_ISL_1491784, EPI_ISL_1491815, EPI_ISL_1491849, EPI_ISL_1491887, EPI_ISL_1491937, EPI_ISL_1513202, EPI_ISL_1513204, EPI_ISL_1513205, EPI_ISL_1513229, EPI_ISL_1513231, EPI_ISL_1513233, EPI_ISL_1513257, EPI_ISL_1513258, EPI_ISL_1513259, EPI_ISL_1513261, EPI_ISL_1513337, EPI_ISL_1513339, EPI_ISL_1513368, EPI_ISL_1513473, EPI_ISL_1513482, EPI_ISL_1513562, EPI_ISL_1513564, EPI_ISL_1513567, EPI_ISL_1513590, EPI_ISL_1513592, EPI_ISL_1525019, EPI_ISL_1525020, EPI_ISL_1525037, EPI_ISL_1525038, EPI_ISL_1525043, EPI_ISL_1525050, EPI_ISL_1525057, EPI_ISL_1525058, EPI_ISL_1525108, EPI_ISL_1525137, EPI_ISL_1525181, EPI_ISL_1525183, EPI_ISL_1525218, EPI_ISL_1525239, EPI_ISL_1525243, EPI_ISL_1550503, EPI_ISL_1550668, EPI_ISL_1550684, EPI_ISL_1550694, EPI_ISL_1550820, EPI_ISL_1550871, EPI_ISL_1550989, EPI_ISL_1551037, EPI_ISL_1551052, EPI_ISL_1551082, EPI_ISL_1551091, EPI_ISL_1551103, EPI_ISL_1551116, EPI_ISL_1551213, EPI_ISL_1551330, EPI_ISL_1551343, EPI_ISL_1551344, EPI_ISL_1551390, EPI_ISL_1551457, EPI_ISL_1551499, EPI_ISL_1551502, EPI_ISL_1551504, EPI_ISL_1551511, EPI_ISL_1551512, EPI_ISL_1551612, EPI_ISL_1551626, EPI_ISL_1559638, EPI_ISL_1559639, EPI_ISL_1559646, EPI_ISL_1559688, EPI_ISL_1559691, EPI_ISL_1559698, EPI_ISL_1559704, EPI_ISL_1559705, EPI_ISL_1559715, EPI_ISL_1559717, EPI_ISL_1559751, EPI_ISL_1559791, EPI_ISL_1559800, EPI_ISL_1559817, EPI_ISL_1559826, EPI_ISL_1559884, EPI_ISL_1559888, EPI_ISL_1559890, EPI_ISL_1559892, EPI_ISL_1559893, EPI_ISL_1559920, EPI_ISL_1559929, EPI_ISL_1559946, EPI_ISL_1559947, EPI_ISL_1559948, EPI_ISL_1559949, EPI_ISL_1560017, EPI_ISL_1560022, EPI_ISL_1560024, EPI_ISL_1560048, EPI_ISL_1560049, EPI_ISL_1560071, EPI_ISL_1560186, EPI_ISL_1560187, EPI_ISL_1560199, EPI_ISL_1560244, EPI_ISL_1560265, EPI_ISL_1560269, EPI_ISL_1560287, EPI_ISL_1560291, EPI_ISL_1560358, EPI_ISL_1560402, EPI_ISL_1560403, EPI_ISL_1560462, EPI_ISL_1560493, EPI_ISL_1560494, EPI_ISL_1560667, EPI_ISL_1560668, EPI_ISL_1560698, EPI_ISL_1560855, EPI_ISL_1560859, EPI_ISL_1560871, EPI_ISL_1560875, EPI_ISL_1560901, EPI_ISL_1560902, EPI_ISL_1560907, EPI_ISL_1560968, EPI_ISL_1560971, EPI_ISL_1561025, EPI_ISL_1561031, EPI_ISL_1561124, EPI_ISL_1561127, EPI_ISL_1561228, EPI_ISL_1561230, EPI_ISL_1561257, EPI_ISL_1561259, EPI_ISL_1561273, EPI_ISL_1561274, EPI_ISL_1561275, EPI_ISL_1561276, EPI_ISL_1561292, EPI_ISL_1561293, EPI_ISL_1561388, EPI_ISL_1561408, EPI_ISL_1561415, EPI_ISL_1561424, EPI_ISL_1561503, EPI_ISL_1561512, EPI_ISL_1561513, EPI_ISL_1561532, EPI_ISL_1561653, EPI_ISL_1561654, EPI_ISL_1561655, EPI_ISL_1561670, EPI_ISL_1561671, EPI_ISL_1561672, EPI_ISL_1561691, EPI_ISL_1561720, EPI_ISL_1561732, EPI_ISL_1561743, EPI_ISL_1561766, EPI_ISL_1561771, EPI_ISL_1561775, EPI_ISL_1561776, EPI_ISL_1561777, EPI_ISL_1561778, EPI_ISL_1561835, EPI_ISL_1561837, EPI_ISL_1561839, EPI_ISL_1561840, EPI_ISL_1561843, EPI_ISL_1561854, EPI_ISL_1561865, EPI_ISL_1561890, EPI_ISL_1561905, EPI_ISL_1561909, EPI_ISL_1561913, EPI_ISL_1562066, EPI_ISL_1562068, EPI_ISL_1562150, EPI_ISL_1562154, EPI_ISL_1562220, EPI_ISL_1562234, EPI_ISL_1562245, EPI_ISL_1562246, EPI_ISL_1562251, EPI_ISL_1562275, EPI_ISL_1562276, EPI_ISL_1562293, EPI_ISL_1562303, EPI_ISL_1562316, EPI_ISL_1562369, EPI_ISL_1562370, EPI_ISL_1562372, EPI_ISL_1562417, EPI_ISL_1562452, EPI_ISL_1562496, EPI_ISL_1562507, EPI_ISL_1562510, EPI_ISL_1562545, EPI_ISL_1562555, EPI_ISL_1562569, EPI_ISL_1562609, EPI_ISL_1562614, EPI_ISL_1562615, EPI_ISL_1562622, EPI_ISL_1562623, EPI_ISL_1562625, EPI_ISL_1562752, EPI_ISL_1562784, EPI_ISL_1562785, EPI_ISL_1562786, EPI_ISL_1562787, EPI_ISL_1562788, EPI_ISL_1562789, EPI_ISL_1562814, EPI_ISL_1562937, EPI_ISL_1562949, EPI_ISL_1562952, EPI_ISL_1562956, EPI_ISL_1562959, EPI_ISL_1562968, EPI_ISL_1562969, EPI_ISL_1562994, EPI_ISL_1563070, EPI_ISL_1563156, EPI_ISL_1563184, EPI_ISL_1563241, EPI_ISL_1563249, EPI_ISL_1563295, EPI_ISL_1563299, EPI_ISL_1563330, EPI_ISL_1563332, EPI_ISL_1563355, EPI_ISL_1563356, EPI_ISL_1563377, EPI_ISL_1563381, EPI_ISL_1563578, EPI_ISL_1563587, EPI_ISL_1563593, EPI_ISL_1563606, EPI_ISL_1563617, EPI_ISL_1563618, EPI_ISL_1649006, EPI_ISL_1649008, EPI_ISL_1649024, EPI_ISL_1649040, EPI_ISL_1649113, EPI_ISL_1649116, EPI_ISL_1649154, EPI_ISL_1649155, EPI_ISL_1649185, EPI_ISL_1649209, EPI_ISL_1649223, EPI_ISL_1649228, EPI_ISL_1649230, EPI_ISL_1649234, EPI_ISL_1649235, EPI_ISL_1649236, EPI_ISL_1649266, EPI_ISL_1649277, EPI_ISL_1649278, EPI_ISL_1649293, EPI_ISL_1649295, EPI_ISL_1649385, EPI_ISL_1649392, EPI_ISL_1649393, EPI_ISL_1649394, EPI_ISL_1649459, EPI_ISL_1649462, EPI_ISL_1649463, EPI_ISL_1649482, EPI_ISL_1649483, EPI_ISL_1649523, EPI_ISL_1649589, EPI_ISL_1649646, EPI_ISL_1649685, EPI_ISL_1649687, EPI_ISL_1649729, EPI_ISL_1649753, EPI_ISL_1649759, EPI_ISL_1649763, EPI_ISL_1649820, EPI_ISL_1649840, EPI_ISL_1649845, EPI_ISL_1650018, EPI_ISL_1650049, EPI_ISL_1650053, EPI_ISL_1650054, EPI_ISL_1650055, EPI_ISL_1650056, EPI_ISL_1650061, EPI_ISL_1650132, EPI_ISL_1650143, EPI_ISL_1650144, EPI_ISL_1650158, EPI_ISL_1650159, EPI_ISL_1650233, EPI_ISL_1650237, EPI_ISL_1650240, EPI_ISL_1650241, EPI_ISL_1650242, EPI_ISL_1650243, EPI_ISL_1650259, EPI_ISL_1650287, EPI_ISL_1650293, EPI_ISL_1650313, EPI_ISL_1650326, EPI_ISL_1650352, EPI_ISL_1650383, EPI_ISL_1650384, EPI_ISL_1650411, EPI_ISL_1650436, EPI_ISL_1650555, EPI_ISL_1650657, EPI_ISL_1650664, EPI_ISL_1650678, EPI_ISL_1650686, EPI_ISL_1650687, EPI_ISL_1650711, EPI_ISL_1650839, EPI_ISL_1650997, EPI_ISL_1651147, EPI_ISL_1651148, EPI_ISL_1667463, EPI_ISL_1667465, EPI_ISL_1667473, EPI_ISL_1667476, EPI_ISL_1667543, EPI_ISL_1667544, EPI_ISL_1667549, EPI_ISL_1667569, EPI_ISL_1667572, EPI_ISL_1667618, EPI_ISL_1667754, EPI_ISL_1667773, EPI_ISL_1667838, EPI_ISL_1667839, EPI_ISL_1667840, EPI_ISL_1667951, EPI_ISL_1668015, EPI_ISL_1668022, EPI_ISL_1668025, EPI_ISL_1668038, EPI_ISL_1668130, EPI_ISL_1668180, EPI_ISL_1668181, EPI_ISL_1668185, EPI_ISL_1668190, EPI_ISL_1668214, EPI_ISL_1668304, EPI_ISL_1668328, EPI_ISL_1668354, EPI_ISL_1668364, EPI_ISL_1668373, EPI_ISL_1668374, EPI_ISL_1668375, EPI_ISL_1685483, EPI_ISL_1685561, EPI_ISL_1685578, EPI_ISL_1685586, EPI_ISL_1685611, EPI_ISL_1685612, EPI_ISL_1685689, EPI_ISL_1685703, EPI_ISL_1685713, EPI_ISL_1685824, EPI_ISL_1685834, EPI_ISL_1685835, EPI_ISL_1685842, EPI_ISL_1685928, EPI_ISL_1685938, EPI_ISL_1685948, EPI_ISL_1685956, EPI_ISL_1685966, EPI_ISL_1685981, EPI_ISL_1685983, EPI_ISL_1685992, EPI_ISL_1686008, EPI_ISL_1686100, EPI_ISL_1686169, EPI_ISL_1686170, EPI_ISL_1686173, EPI_ISL_1686207, EPI_ISL_1686213, EPI_ISL_1686217, EPI_ISL_1686220, EPI_ISL_1686224, EPI_ISL_1686227, EPI_ISL_1686264, EPI_ISL_1686281, EPI_ISL_1686282, EPI_ISL_1686294, EPI_ISL_1686373, EPI_ISL_1686480, EPI_ISL_1686515, EPI_ISL_1686522, EPI_ISL_1686525, EPI_ISL_1686527, EPI_ISL_1686535, EPI_ISL_1686536, EPI_ISL_1686538, EPI_ISL_1686691, EPI_ISL_1686760, EPI_ISL_1686769, EPI_ISL_1686771, EPI_ISL_1686772, EPI_ISL_1686773, EPI_ISL_1686774, EPI_ISL_1686776, EPI_ISL_1686778, EPI_ISL_1686782, EPI_ISL_1686783, EPI_ISL_1686786, EPI_ISL_1686792, EPI_ISL_1686793, EPI_ISL_1686795, EPI_ISL_1686817, EPI_ISL_1686863, EPI_ISL_1686865, EPI_ISL_1686867, EPI_ISL_1686869, EPI_ISL_1686879, EPI_ISL_1686958, EPI_ISL_1686972, EPI_ISL_1686973, EPI_ISL_1686975, EPI_ISL_1687061, EPI_ISL_1687120, EPI_ISL_1687230, EPI_ISL_1687262, EPI_ISL_1687299, EPI_ISL_1687316, EPI_ISL_1687343, EPI_ISL_1687358, EPI_ISL_1687387, EPI_ISL_1687426, EPI_ISL_1687579, EPI_ISL_1687587, EPI_ISL_1687615, EPI_ISL_1687650, EPI_ISL_1687651, EPI_ISL_1687674, EPI_ISL_1687680, EPI_ISL_1687681, EPI_ISL_1687736, EPI_ISL_1687737, EPI_ISL_1687738, EPI_ISL_1687748, EPI_ISL_1687781, EPI_ISL_1687782, EPI_ISL_1687783, EPI_ISL_1687809, EPI_ISL_1687824, EPI_ISL_1687832, EPI_ISL_1687833, EPI_ISL_1687834, EPI_ISL_1687835, EPI_ISL_1687838, EPI_ISL_1687842, EPI_ISL_1687848, EPI_ISL_1687856, EPI_ISL_1687858, EPI_ISL_1687871, EPI_ISL_1688064, EPI_ISL_1688065, EPI_ISL_1688071, EPI_ISL_1688132, EPI_ISL_1688159, EPI_ISL_1688306, EPI_ISL_1688421, EPI_ISL_1688548, EPI_ISL_1688551, EPI_ISL_1688609, EPI_ISL_1688616, EPI_ISL_1688620, EPI_ISL_1688634, EPI_ISL_1688638, EPI_ISL_1688689, EPI_ISL_1688937, EPI_ISL_1688963, EPI_ISL_1689021, EPI_ISL_1689086, EPI_ISL_1689186, EPI_ISL_1689197, EPI_ISL_1689200, EPI_ISL_1689202, EPI_ISL_1689207, EPI_ISL_1689214, EPI_ISL_1689389, EPI_ISL_1689391, EPI_ISL_1689393, EPI_ISL_1689396, EPI_ISL_1689397, EPI_ISL_1689596, EPI_ISL_1689599, EPI_ISL_1689603, EPI_ISL_1689608, EPI_ISL_1689612, EPI_ISL_1689629, EPI_ISL_1689631, EPI_ISL_1689732, EPI_ISL_1689750, EPI_ISL_1689816, EPI_ISL_1689843, EPI_ISL_1689847, EPI_ISL_1689848, EPI_ISL_1689853, EPI_ISL_1689862, EPI_ISL_1689864, EPI_ISL_1689871, EPI_ISL_1690021, EPI_ISL_1690040, EPI_ISL_1690087, EPI_ISL_1690099, EPI_ISL_1690100, EPI_ISL_1690189, EPI_ISL_1690197, EPI_ISL_1690267, EPI_ISL_1690268, EPI_ISL_1695135, EPI_ISL_1695305, EPI_ISL_1695382, EPI_ISL_1695469, EPI_ISL_1695487, EPI_ISL_1695545, EPI_ISL_1695554, EPI_ISL_1695578, EPI_ISL_1701890, EPI_ISL_1701900, EPI_ISL_1701941, EPI_ISL_1701973, EPI_ISL_1701980, EPI_ISL_1701996, EPI_ISL_1701997, EPI_ISL_1702014, EPI_ISL_1702030, EPI_ISL_1702035, EPI_ISL_1702076, EPI_ISL_1702107, EPI_ISL_1702127, EPI_ISL_1702138, EPI_ISL_1702728, EPI_ISL_1702778, EPI_ISL_1702788, EPI_ISL_1702789, EPI_ISL_1702816, EPI_ISL_1702829, EPI_ISL_1702838, EPI_ISL_1702865, EPI_ISL_1702867, EPI_ISL_1702884, EPI_ISL_1702954, EPI_ISL_1702970, EPI_ISL_1735719, EPI_ISL_1735723, EPI_ISL_1735724, EPI_ISL_1735772, EPI_ISL_1735773, EPI_ISL_1735774, EPI_ISL_1735775, EPI_ISL_1735783, EPI_ISL_1735855, EPI_ISL_1735914, EPI_ISL_1735940, EPI_ISL_1736007, EPI_ISL_1736116, EPI_ISL_1736118, EPI_ISL_1736119, EPI_ISL_1736120, EPI_ISL_1736146, EPI_ISL_1736163, EPI_ISL_1736218, EPI_ISL_1736219, EPI_ISL_1736235, EPI_ISL_1736375, EPI_ISL_1736384, EPI_ISL_1736516, EPI_ISL_1736695, EPI_ISL_1736702, EPI_ISL_1737063, EPI_ISL_1737198, EPI_ISL_1737202, EPI_ISL_1737223, EPI_ISL_1737238, EPI_ISL_1737239, EPI_ISL_1737284, EPI_ISL_1737289, EPI_ISL_1737290, EPI_ISL_1737291, EPI_ISL_1737292, EPI_ISL_1737296, EPI_ISL_1737399, EPI_ISL_1737478, EPI_ISL_1737512, EPI_ISL_1737517, EPI_ISL_1737589, EPI_ISL_1737590, EPI_ISL_1738769, EPI_ISL_1753533, EPI_ISL_1753552, EPI_ISL_1753572, EPI_ISL_1753574, EPI_ISL_1753582, EPI_ISL_1834549, EPI_ISL_1834785, EPI_ISL_1834786, EPI_ISL_1834791, EPI_ISL_1834792, EPI_ISL_1834793, EPI_ISL_1834794, EPI_ISL_1834796, EPI_ISL_1834807, EPI_ISL_1834830, EPI_ISL_1834837, EPI_ISL_1834876, EPI_ISL_1834887, EPI_ISL_1834899, EPI_ISL_1834915, EPI_ISL_1834928, EPI_ISL_1834929, EPI_ISL_1834976, EPI_ISL_1835090, EPI_ISL_1835091, EPI_ISL_1835093, EPI_ISL_1835113, EPI_ISL_1835160, EPI_ISL_1835194, EPI_ISL_1835207, EPI_ISL_1835246, EPI_ISL_1835297, EPI_ISL_1835336, EPI_ISL_1835337, EPI_ISL_1835353, EPI_ISL_1835355, EPI_ISL_1835362, EPI_ISL_1836876, EPI_ISL_1900231, EPI_ISL_1900249, EPI_ISL_1900283, EPI_ISL_1925228, EPI_ISL_1925248, EPI_ISL_1925249, EPI_ISL_1925250, EPI_ISL_1925251, EPI_ISL_1925252, EPI_ISL_1925275, EPI_ISL_1925303, EPI_ISL_1925304, EPI_ISL_1925311, EPI_ISL_1925329, EPI_ISL_1925358, EPI_ISL_1925360, EPI_ISL_1925380, EPI_ISL_1925382, EPI_ISL_1925421, EPI_ISL_1925454, EPI_ISL_1925459, EPI_ISL_1925464, EPI_ISL_1925477, EPI_ISL_1925498, EPI_ISL_1925502, EPI_ISL_1925541, EPI_ISL_1925545, EPI_ISL_1925550, EPI_ISL_1925551, EPI_ISL_1925584, EPI_ISL_1925604, EPI_ISL_1925617 | | | |
| see above | Aegis Sciences Corporation | Centers for Disease Control and Prevention Division of Viral Diseases, Pathogen Discovery | Adrian Paskey; Alec Vest; Benjamin Rambo-Martin; Christopher Gulvick; Clinton R. Paden; Cyndi Clark; Dakota Howard; Darlene Wagner; Dhwani Batra; Dillon Nall; Duncan MacCannell; Ethan Sanders; Holly Houdeshell; Jason Caravas; Kara Moser; Matthew Hardison; Matthew Schmerer; Ola Kvalvaag; Patrick Campbell; Peter W. Cook; Rob Case; Scott Sammons; Shatavia Morrison; Shaun Westlund; Vikramsinha Ghorpade; Yvette Unoarumhi |
| EPI_ISL_1482408, EPI_ISL_1482409, EPI_ISL_1482446, EPI_ISL_1482447, EPI_ISL_1482448, EPI_ISL_1482449, EPI_ISL_1482450, EPI_ISL_1482451, EPI_ISL_1482452, EPI_ISL_1482453, EPI_ISL_1575477, EPI_ISL_1580667, EPI_ISL_1580668, EPI_ISL_1580669, EPI_ISL_1580670, EPI_ISL_1624409, EPI_ISL_1671444, EPI_ISL_1671445, EPI_ISL_1671446, EPI_ISL_1671447, EPI_ISL_1671454, EPI_ISL_1671455, EPI_ISL_1671456, EPI_ISL_1671457, EPI_ISL_1671476, EPI_ISL_1671495, EPI_ISL_1671496, EPI_ISL_1671596, EPI_ISL_1671618, EPI_ISL_1740510, EPI_ISL_1740511, EPI_ISL_1755849, EPI_ISL_1755850, EPI_ISL_1789712, EPI_ISL_2098126 | | | |
| see above | BOSTON HEART DIAGNOSTICS CORP | Wadsworth Center, New York State Department of Health | Alexis Russell; Catharine Prussing; Daryl M. Lamson; Erasmus Schneider; Erica Lasek-Nesselquist; John Kelly; Jonathan Plitnick; Kirsten St. George; Matthew Shudt; Melissa A Leisner; Navjot Singh |
| EPI_ISL_1413241, EPI_ISL_1413243, EPI_ISL_1413246, EPI_ISL_1413263, EPI_ISL_1413350, EPI_ISL_1516525, EPI_ISL_1516526, EPI_ISL_1516527, EPI_ISL_1516528, EPI_ISL_1516530, EPI_ISL_1516531, EPI_ISL_1516532, EPI_ISL_1516533, EPI_ISL_1516536, EPI_ISL_1516537, EPI_ISL_1516538, EPI_ISL_1516539, EPI_ISL_1516541, EPI_ISL_1516542, EPI_ISL_1516543, EPI_ISL_1516544, EPI_ISL_1516563, EPI_ISL_1516571, EPI_ISL_1578133, EPI_ISL_1578152, EPI_ISL_1578170, EPI_ISL_1578181, EPI_ISL_1578201, EPI_ISL_1578202, EPI_ISL_1578280, EPI_ISL_1578316, EPI_ISL_1578318, EPI_ISL_1578319, EPI_ISL_1578320, EPI_ISL_1578321, EPI_ISL_1578325, EPI_ISL_1578334, EPI_ISL_1578374, EPI_ISL_1578386, EPI_ISL_1578396, EPI_ISL_1578398, EPI_ISL_1578401, EPI_ISL_1578405, EPI_ISL_1578406, EPI_ISL_1578432, EPI_ISL_1710048, EPI_ISL_1710050, EPI_ISL_1710060, EPI_ISL_1710064, EPI_ISL_1710067, EPI_ISL_1710075, EPI_ISL_1710081, EPI_ISL_1710085, EPI_ISL_1710086, EPI_ISL_1710095, EPI_ISL_1710100, EPI_ISL_1710102, EPI_ISL_1710103, EPI_ISL_1710105, EPI_ISL_1710136, EPI_ISL_1710138 | | | |
| see above | Broad Institute Clinical Research Sequencing Platform | Infectious Disease Program, Broad Institute of Harvard and MIT | Adams, G.; B.L.; B.W.; Bauer, M.; Birren; Blumenstiel, B.; Brown, C.; Carter, A.; Chaluvadi, S.; D.J.; DeFelice, M.; DeRuff, K.; Dodge, S.; Gabriel, S.; Gallagher, G.; Gladden-Young, A.; Granger, B.; J.E.; K.J.; Lagerborg, K.; Larkin, K.; Lee, M.; Lemieux; Lennon, N.; Loreth, C.; Madoff, L.; McGovern, S.; Meldrim, J.; Normandin, E.; P.C.; Park; Pearlman, L.; Reilly, S.; Rudy, M.; Sabeti; Siddle; Smole, S.; Tomkins-Tinch, C.; Vicente, G.; and MacInnis |
| EPI_ISL_2101788, EPI_ISL_2101800 | Bronx Veterans Medical Research Foundation | Pathogen Discovery, Respiratory Viruses Branch, Division of Viral Diseases, Centers for Disease Control and Prevention | Adam Retchless; Anna Kelleher; Anna Uehara; Brian Lynch; Clinton R. Paden; Dhwani Batra; Haibin Wang; Han Jia Justin Ng; Jasmine Padilla; Jing Zhang; Justin Lee; Krista Queen; Mark Burroughs; Mili Sheth; Morgan Davis; Peter Cook; Rachel Marine; Sarah Nobles; Suxiang Tong; Tara Coalter; Yan Li; Ying Tao |
| EPI_ISL_1624487 | CHAMPLAIN VALLEY PHYSICIANS HOSPITAL MEDICAL CENTER | Wadsworth Center, New York State Department of Health | Alexis Russell; Catharine Prussing; Daryl M. Lamson; Erasmus Schneider; Erica Lasek-Nesselquist; John Kelly; Jonathan Plitnick; Kirsten St. George; Matthew Shudt; Melissa A Leisner; Navjot Singh |
| EPI_ISL_2036759 | CT Department of Public Health | CT Department of Public Health | Claire_Pearson; Tu_N_Nguyen |
| EPI_ISL_1965736, EPI_ISL_1965737, EPI_ISL_1965738, EPI_ISL_1965739, EPI_ISL_1965740, EPI_ISL_1965741, EPI_ISL_1965742, EPI_ISL_1965743, EPI_ISL_1965744, EPI_ISL_1965745, EPI_ISL_1965746, EPI_ISL_1965747, EPI_ISL_1965748, EPI_ISL_1965749, EPI_ISL_1965750, EPI_ISL_1965751, EPI_ISL_1965752, EPI_ISL_1965753, EPI_ISL_1965754, EPI_ISL_1965755, EPI_ISL_1965756, EPI_ISL_1965757, EPI_ISL_1965758, EPI_ISL_1965759, EPI_ISL_1965760, EPI_ISL_1965761, EPI_ISL_1965762, EPI_ISL_1965763, EPI_ISL_1965764, EPI_ISL_1965765, EPI_ISL_1965766, EPI_ISL_1965767, EPI_ISL_1965768, EPI_ISL_1965769, EPI_ISL_1965770, EPI_ISL_1965771, EPI_ISL_1965772, EPI_ISL_1965773, EPI_ISL_1965774, EPI_ISL_1965775, EPI_ISL_1965776, EPI_ISL_1965777, EPI_ISL_1965778, EPI_ISL_1965779, EPI_ISL_1965780, EPI_ISL_1965781, EPI_ISL_1965782, EPI_ISL_1965783, EPI_ISL_1965784, EPI_ISL_1965785, EPI_ISL_1965786, EPI_ISL_1965787, EPI_ISL_1965788, EPI_ISL_1965789, EPI_ISL_1965790, EPI_ISL_1965791, EPI_ISL_1965792, EPI_ISL_1965793, EPI_ISL_1965794, EPI_ISL_1965795, EPI_ISL_1965796, EPI_ISL_1965797, EPI_ISL_1965798, EPI_ISL_1965799, EPI_ISL_1965800, EPI_ISL_1965801, EPI_ISL_1965802, EPI_ISL_1965803, EPI_ISL_1965804, EPI_ISL_1965805, EPI_ISL_1965806, EPI_ISL_1965807, EPI_ISL_1965808, EPI_ISL_1965809, EPI_ISL_1965810, EPI_ISL_1965811, EPI_ISL_1965812, EPI_ISL_1965813, EPI_ISL_1965814, EPI_ISL_1965815, EPI_ISL_1965816, EPI_ISL_1965817, EPI_ISL_1965818, EPI_ISL_1965819, EPI_ISL_1965820, EPI_ISL_1965821, EPI_ISL_1965822, EPI_ISL_1965823, EPI_ISL_1965824, EPI_ISL_1965825, EPI_ISL_1965826, EPI_ISL_1965827, EPI_ISL_1965828, EPI_ISL_1965829, EPI_ISL_1965830, EPI_ISL_1965831, EPI_ISL_1965832, EPI_ISL_1965833, EPI_ISL_1965834, EPI_ISL_1965835, EPI_ISL_1965836, EPI_ISL_1965837, EPI_ISL_1965838, EPI_ISL_1965839, EPI_ISL_1965840, EPI_ISL_1965841, EPI_ISL_1965842, EPI_ISL_1965843, EPI_ISL_1965844, EPI_ISL_1965845, EPI_ISL_1965846, EPI_ISL_1965847, EPI_ISL_1965848, EPI_ISL_1965850 | | | |
| see above | Clinical Microbiology Laboratory, NewYork Presbyterian Hospital/Columbia University Irving Medical Center | Uhlemann Laboratory, Columbia University Irving Medical Center | Anne Kelley; Anne-Catrin Uhlemann; Maya Tagliavia; Medini K. Annavajhala |
| EPI_ISL_2031642, EPI_ISL_2031643, EPI_ISL_2031647 | Columbia University | Pathogen Discovery, Respiratory Viruses Branch, Division of Viral Diseases, Centers for Disease Control and Prevention | Adam Retchless; Anna Kelleher; Anna Uehara; Brian Lynch; Clinton R. Paden; Dhwani Batra; Haibin Wang; Han Jia Justin Ng; Jasmine Padilla; Jing Zhang; Justin Lee; Krista Queen; Mark Burroughs; Mili Sheth; Morgan Davis; Peter Cook; Rachel Marine; Sarah Nobles; Suxiang Tong; Tara Coalter; Yan Li; Ying Tao |
| EPI_ISL_1482410, EPI_ISL_1580687, EPI_ISL_1580688, EPI_ISL_1580689, EPI_ISL_1580690, EPI_ISL_1580691, EPI_ISL_1580692, EPI_ISL_1580693, EPI_ISL_1580694, EPI_ISL_1580695, EPI_ISL_1580696, EPI_ISL_1580697, EPI_ISL_1580698, EPI_ISL_1582214, EPI_ISL_1582215, EPI_ISL_1582216, EPI_ISL_1582217, EPI_ISL_1582261, EPI_ISL_1582262, EPI_ISL_1582263, EPI_ISL_1582264, EPI_ISL_1582265, EPI_ISL_1582266, EPI_ISL_1582267, EPI_ISL_1582268, EPI_ISL_1582269, EPI_ISL_1582270, EPI_ISL_1582271, EPI_ISL_1582337, EPI_ISL_1582368, EPI_ISL_1755854, EPI_ISL_1755855, EPI_ISL_1755856, EPI_ISL_1755857, EPI_ISL_1755858, EPI_ISL_1755859, EPI_ISL_1755860, EPI_ISL_1789713 | | | |
| see above | Columbia University Irving Medical Center | Wadsworth Center, New York State Department of Health | Alexis Russell; Catharine Prussing; Daryl M. Lamson; Erasmus Schneider; Erica Lasek-Nesselquist; John Kelly; Jonathan Plitnick; Kirsten St. George; Matthew Shudt; Melissa A Leisner; Navjot Singh |
| EPI_ISL_1337476, EPI_ISL_1337520, EPI_ISL_1337536, EPI_ISL_1337548, EPI_ISL_1337549, EPI_ISL_1447101, EPI_ISL_1447109, EPI_ISL_1447116, EPI_ISL_1447129, EPI_ISL_1447130, EPI_ISL_1447134, EPI_ISL_1447167, EPI_ISL_1447168, EPI_ISL_1447186, EPI_ISL_1593720, EPI_ISL_1593722, EPI_ISL_1593742, EPI_ISL_1593743, EPI_ISL_1593744, EPI_ISL_1593745, EPI_ISL_1593751, EPI_ISL_1593752, EPI_ISL_1593763, EPI_ISL_1593764, EPI_ISL_1593765, EPI_ISL_1593766, EPI_ISL_1593767, EPI_ISL_1694702, EPI_ISL_1694703, EPI_ISL_1694709 | | | |
| see above | DOHMH Central Harlem | New York City Public Health Laboratory | Jade Wang; et al. |
| EPI_ISL_1337479, EPI_ISL_1447093, EPI_ISL_1447096, EPI_ISL_1447118, EPI_ISL_1447123, EPI_ISL_1447133, EPI_ISL_1447156, EPI_ISL_1447157, EPI_ISL_1447169, EPI_ISL_1447193, EPI_ISL_1593733, EPI_ISL_1593754, EPI_ISL_1593755, EPI_ISL_1694697, EPI_ISL_1694698, EPI_ISL_1694706 | | | |
| see above | DOHMH Chelsea | New York City Public Health Laboratory | Jade Wang; et al. |
| EPI_ISL_1337467, EPI_ISL_1337475, EPI_ISL_1337477, EPI_ISL_1337484, EPI_ISL_1337491, EPI_ISL_1337507, EPI_ISL_1337511, EPI_ISL_1337512, EPI_ISL_1337514, EPI_ISL_1337515, EPI_ISL_1337531, EPI_ISL_1337551, EPI_ISL_1447088, EPI_ISL_1447095, EPI_ISL_1447126, EPI_ISL_1447136, EPI_ISL_1447137, EPI_ISL_1447139, EPI_ISL_1447140, EPI_ISL_1447141, EPI_ISL_1447144, EPI_ISL_1447147, EPI_ISL_1447155, EPI_ISL_1447160, EPI_ISL_1447165, EPI_ISL_1447171, EPI_ISL_1447172, EPI_ISL_1447185, EPI_ISL_1593721, EPI_ISL_1593737, EPI_ISL_1593746, EPI_ISL_1593759, EPI_ISL_1593760, EPI_ISL_1593761, EPI_ISL_1593768, EPI_ISL_1694707, EPI_ISL_1694710 | | | |

| | | | |
|---|---|---|---|
| see above | DOHMH Corona | New York City Public Health Laboratory | Jade Wang; et al. |
| EPI_ISL_1337474, EPI_ISL_1337490, EPI_ISL_1337492, EPI_ISL_1337493, EPI_ISL_1337494, EPI_ISL_1337499, EPI_ISL_1337528, EPI_ISL_1337534, EPI_ISL_1337535, EPI_ISL_1337544, EPI_ISL_1447089, EPI_ISL_1447113, EPI_ISL_1447128, EPI_ISL_1447148, EPI_ISL_1447161, EPI_ISL_1447162, EPI_ISL_1447166, EPI_ISL_1447170, EPI_ISL_1447191, EPI_ISL_1593741, EPI_ISL_1593749, EPI_ISL_1593769 | | | |
| see above | DOHMH Crown Heights | New York City Public Health Laboratory | Jade Wang; et al. |
| EPI_ISL_1447098, EPI_ISL_1447112, EPI_ISL_1447115, EPI_ISL_1447127, EPI_ISL_1447135, EPI_ISL_1447176, EPI_ISL_1447187, EPI_ISL_1593740, EPI_ISL_1593750, EPI_ISL_1593758, EPI_ISL_1593770 | | | |
| see above | DOHMH Fort Greene | New York City Public Health Laboratory | Jade Wang; et al. |
| EPI_ISL_1337469, EPI_ISL_1337486, EPI_ISL_1337487, EPI_ISL_1337495, EPI_ISL_1337496, EPI_ISL_1337497, EPI_ISL_1337498, EPI_ISL_1337504, EPI_ISL_1337505, EPI_ISL_1337513, EPI_ISL_1337521, EPI_ISL_1337522, EPI_ISL_1337532, EPI_ISL_1337533, EPI_ISL_1337537, EPI_ISL_1337538, EPI_ISL_1337550, EPI_ISL_1447092, EPI_ISL_1447094, EPI_ISL_1447103, EPI_ISL_1447106, EPI_ISL_1447117, EPI_ISL_1447120, EPI_ISL_1447121, EPI_ISL_1447122, EPI_ISL_1447124, EPI_ISL_1447125, EPI_ISL_1447131, EPI_ISL_1447132, EPI_ISL_1447146, EPI_ISL_1447151, EPI_ISL_1447152, EPI_ISL_1447153, EPI_ISL_1447158, EPI_ISL_1447159, EPI_ISL_1447163, EPI_ISL_1447164, EPI_ISL_1447175, EPI_ISL_1447179, EPI_ISL_1447180, EPI_ISL_1447188, EPI_ISL_1447189, EPI_ISL_1447190, EPI_ISL_1447192, EPI_ISL_1593732, EPI_ISL_1593735, EPI_ISL_1593738, EPI_ISL_1593739, EPI_ISL_1593747, EPI_ISL_1593748, EPI_ISL_1593762, EPI_ISL_1694699, EPI_ISL_1694708 | | | |
| see above | DOHMH Jamaica | New York City Public Health Laboratory | Jade Wang; et al. |
| EPI_ISL_1337470, EPI_ISL_1337478, EPI_ISL_1337502, EPI_ISL_1337506, EPI_ISL_1337540, EPI_ISL_1337545, EPI_ISL_1447087, EPI_ISL_1447090, EPI_ISL_1447097, EPI_ISL_1447108, EPI_ISL_1447110, EPI_ISL_1447181, EPI_ISL_1593753, EPI_ISL_1694704, EPI_ISL_1694705 | | | |
| see above | DOHMH Morrisania | New York City Public Health Laboratory | Jade Wang; et al. |
| EPI_ISL_1337508, EPI_ISL_1337516, EPI_ISL_1337530, EPI_ISL_1447099, EPI_ISL_1447107, EPI_ISL_1447145, EPI_ISL_1447149, EPI_ISL_1447150, EPI_ISL_1447184, EPI_ISL_1593731, EPI_ISL_1593734 | | | |
| see above | DOHMH PHL | New York City Public Health Laboratory | Jade Wang; et al. |
| EPI_ISL_1337480, EPI_ISL_1337483, EPI_ISL_1337503, EPI_ISL_1337547, EPI_ISL_1447105, EPI_ISL_1447154, EPI_ISL_1447173, EPI_ISL_1593736, EPI_ISL_1694696, EPI_ISL_1694700, EPI_ISL_1694701 | | | |
| see above | DOHMH Riverside | New York City Public Health Laboratory | Jade Wang; et al. |
| EPI_ISL_1447102, EPI_ISL_1447174, EPI_ISL_1447182, EPI_ISL_1593771, EPI_ISL_1804941, EPI_ISL_1804947 | Department of Homeless Services | New York City Public Health Laboratory | Jade Wang; et al. |
| EPI_ISL_1444912, EPI_ISL_1444919, EPI_ISL_1444920, EPI_ISL_1444921, EPI_ISL_1444922 | Erie County Public Health (ECPHL) | University at Buffalo Genomics and Bioinformatics Core | Alyssa Pohlman; Amanda Boccolucci; Brandon Marzullo; Donald Yergeau; Jennifer Surtees; Jonathan Bard; Natalie Lamb; Norma Nowak |
| EPI_ISL_1465853, EPI_ISL_1465854, EPI_ISL_1465855, EPI_ISL_1465856, EPI_ISL_1482333, EPI_ISL_1482334, EPI_ISL_1482335, EPI_ISL_1582406, EPI_ISL_1855241 | | | |
| see above | GLENS FALLS HOSPITAL LABORATORY | Wadsworth Center, New York State Department of Health | Alexis Russell; Catharine Prussing; Daryl M. Lamson; Erasmus Schneider; Erica Lasek-Nesselquist; John Kelly; Jonathan Plitnick; Kirsten St. George; Matthew Shudt; Melissa A Leisner; Navjot Singh |
| EPI_ISL_1465848, EPI_ISL_1465858, EPI_ISL_1582255, EPI_ISL_1624089, EPI_ISL_1624090 | GOOD SAMARITAN HOSPITAL LABORATORY | Wadsworth Center, New York State Department of Health | Alexis Russell; Catharine Prussing; Daryl M. Lamson; Erasmus Schneider; Erica Lasek-Nesselquist; John Kelly; Jonathan Plitnick; Kirsten St. George; Matthew Shudt; Melissa A Leisner; Navjot Singh |
| EPI_ISL_1920119, EPI_ISL_1920120 | Glens Falls Hospital Laboratory | Wadsworth Center, New York State Department of Health | Alexis Russell; Catharine Prussing; Daryl M. Lamson; Erasmus Schneider; Erica Lasek-Nesselquist; John Kelly; Jonathan Plitnick; Kirsten St. George; Matthew Shudt; Melissa A Leisner; Navjot Singh |
| EPI_ISL_1482470 | Good Samaritan Hospital Laboratory | Wadsworth Center, New York State Department of Health | Alexis Russell; Catharine Prussing; Daryl M. Lamson; Erasmus Schneider; Erica Lasek-Nesselquist; John Kelly; Jonathan Plitnick; Kirsten St. George; Matthew Shudt; Melissa A Leisner; Navjot Singh |
| EPI_ISL_1479386, EPI_ISL_1511926, EPI_ISL_1512010, EPI_ISL_1512135, EPI_ISL_1512243, EPI_ISL_1512415, EPI_ISL_1512449, EPI_ISL_1512483, EPI_ISL_1553421, EPI_ISL_1553555, EPI_ISL_1554114, EPI_ISL_1554306, EPI_ISL_1575510, EPI_ISL_1575846, EPI_ISL_1575913, EPI_ISL_1576513, EPI_ISL_1576617, EPI_ISL_1576668, EPI_ISL_1580918, EPI_ISL_1581703, EPI_ISL_1592417, EPI_ISL_1592426, EPI_ISL_1615131, EPI_ISL_1679008, EPI_ISL_1679009, EPI_ISL_1679029, EPI_ISL_1679426, EPI_ISL_1679621, EPI_ISL_1679640, EPI_ISL_1679734, EPI_ISL_1690309, EPI_ISL_1690352, EPI_ISL_1690546, EPI_ISL_1690890, EPI_ISL_1691214, EPI_ISL_1691266, EPI_ISL_1691275, EPI_ISL_1735142, EPI_ISL_1735643 | | | |
| see above | Helix/Illumina | Centers for Disease Control and Prevention Division of Viral Diseases, Pathogen Discovery | Adrian Paskey; Alexandre Bolze; Ary Ascencio; Benjamin Rambo-Martin; Brad Sickler; Charlotte Rivera-Garcia; Christine Tran; Christopher Gulvick; Clinton R. Paden; Dakota Howard; Darlene Wagner; David Becker; Dhwani Batra; Duncan MacCannell; Efren Sandoval; Eileen de Feo; Elizabeth Cirulli; Eric Allen; Geraint Levan; James Lu; Jan Antico; Jason Caravas; Jason Nguyen; Jimmy Ramirez; Jingtao Liu; Kara Moser; Kelly Schiabor Barrett; Kim Gietzen; Magnus Isaksson; Marc Laurent; Matthew Schmerer; Matthew Tolentino; Nicole L. Washington; Peter W. Cook; Phil Febbo; Ryan Cho; Scott Sammons; Shannon Wickline; Shatavia Morrison; Sherry Wang; Simon White; Tyler Cassens; William Lee; Yvette Unoarumhi |
| EPI_ISL_1465857 | ITHACA COLLEGE LABORATORY HAMMOND HEALTH CENTER | Wadsworth Center, New York State Department of Health | Alexis Russell; Catharine Prussing; Daryl M. Lamson; Erasmus Schneider; Erica Lasek-Nesselquist; John Kelly; Jonathan Plitnick; Kirsten St. George; Matthew Shudt; Melissa A Leisner; Navjot Singh |
| EPI_ISL_1512832, EPI_ISL_1512837, EPI_ISL_1512853, EPI_ISL_1512854, EPI_ISL_1512855, EPI_ISL_1512857, EPI_ISL_1512862, EPI_ISL_1512896, EPI_ISL_1512917, EPI_ISL_1512935, EPI_ISL_1512948, EPI_ISL_1512949, EPI_ISL_1512950, EPI_ISL_1512952, EPI_ISL_1513036, EPI_ISL_1513039, EPI_ISL_1513044, EPI_ISL_1513074, EPI_ISL_1513092, EPI_ISL_1513094, EPI_ISL_1513096, EPI_ISL_1513101, EPI_ISL_1513102, EPI_ISL_1513131, EPI_ISL_1587589, EPI_ISL_1587712, EPI_ISL_1587757, EPI_ISL_1587786, EPI_ISL_1587941, EPI_ISL_1587942, EPI_ISL_1587944, EPI_ISL_1587951, EPI_ISL_1587952, EPI_ISL_1587954, EPI_ISL_1587968, EPI_ISL_1587975, EPI_ISL_1588003, EPI_ISL_1588112, EPI_ISL_1588190, EPI_ISL_1588288, EPI_ISL_1588322, EPI_ISL_1588324, EPI_ISL_1588405, EPI_ISL_1588406, EPI_ISL_1588412, EPI_ISL_1588415, EPI_ISL_1588418, EPI_ISL_1588419, EPI_ISL_1588427, EPI_ISL_1588433, EPI_ISL_1691557, EPI_ISL_1691562, EPI_ISL_1691674, EPI_ISL_1691686, EPI_ISL_1691716, EPI_ISL_1691720, EPI_ISL_1691721, EPI_ISL_1691756, EPI_ISL_1691959, EPI_ISL_1691975, EPI_ISL_1691986, EPI_ISL_1691995, EPI_ISL_1692005, EPI_ISL_1692010, EPI_ISL_1692028, EPI_ISL_1692030, EPI_ISL_1692084, EPI_ISL_1692129, EPI_ISL_1692160, EPI_ISL_1692198, EPI_ISL_1692224, EPI_ISL_1692266, EPI_ISL_1692284, EPI_ISL_1692285, EPI_ISL_1692292, EPI_ISL_1692310, EPI_ISL_1692327, EPI_ISL_1692341, EPI_ISL_1692371, EPI_ISL_1692408, EPI_ISL_1692409, EPI_ISL_1692448, EPI_ISL_1692460, EPI_ISL_1692516, EPI_ISL_1692524, EPI_ISL_1692575, EPI_ISL_1692578, EPI_ISL_1692581, EPI_ISL_1692587, EPI_ISL_1692592, EPI_ISL_1692600, EPI_ISL_1692606, EPI_ISL_1692634, EPI_ISL_1692655, EPI_ISL_1692658, EPI_ISL_1692669, EPI_ISL_1692687, EPI_ISL_1692708, EPI_ISL_1693102, EPI_ISL_1693220, EPI_ISL_1693274, EPI_ISL_1693317, EPI_ISL_1693319, EPI_ISL_1693320, EPI_ISL_1693321, EPI_ISL_1693868, EPI_ISL_1694015, EPI_ISL_1694028, EPI_ISL_1694242, EPI_ISL_1694388, EPI_ISL_1694519, EPI_ISL_1702185, EPI_ISL_1702186, EPI_ISL_1702187, EPI_ISL_1702255, EPI_ISL_1702267, EPI_ISL_1702281, EPI_ISL_2013321 | | | |
| see above | Infinity Biologix | Centers for Disease Control and Prevention Division of Viral Diseases, Pathogen Discovery | Adrian Paskey; Benjamin Rambo-Martin; Chirayu Goswami; Christian Bixby; Christopher Gulvick; Clinton R. Paden; Dakota Howard; Darlene Wagner; Dhwani Batra; Duncan MacCannell; Jason Caravas; Jonathan Schultz; Kara Moser; Matthew Schmerer; Peter W. Cook; Robin Grimwood; Russ Hager; Scott Sammons; Shatavia Morrison; Yihe Wang; Yvette Unoarumhi |
| EPI_ISL_1461983, EPI_ISL_1461996, EPI_ISL_1461997, EPI_ISL_1461998, EPI_ISL_1462005, EPI_ISL_1462238, EPI_ISL_1462239, EPI_ISL_1462242, EPI_ISL_1462425, EPI_ISL_1462429, EPI_ISL_1462430, EPI_ISL_1462431, EPI_ISL_1462432, EPI_ISL_1462440, EPI_ISL_1462441, EPI_ISL_1462473, EPI_ISL_1462474, EPI_ISL_1462475, EPI_ISL_1462478, EPI_ISL_1462479, EPI_ISL_1462480, EPI_ISL_1462483, EPI_ISL_1462484, EPI_ISL_1462485, EPI_ISL_1462497, EPI_ISL_1462498, EPI_ISL_1462499, EPI_ISL_1462501, EPI_ISL_1462504, EPI_ISL_1462507, EPI_ISL_1462517, EPI_ISL_1462520, EPI_ISL_1462571, EPI_ISL_1462838, EPI_ISL_1463106, EPI_ISL_1463109, EPI_ISL_1463110, EPI_ISL_1463112, EPI_ISL_1463121, EPI_ISL_1463122, EPI_ISL_1463125, EPI_ISL_1463126, EPI_ISL_1463139, EPI_ISL_1463140, EPI_ISL_1463141, EPI_ISL_1463142, EPI_ISL_1463173, EPI_ISL_1463187, EPI_ISL_1463188, EPI_ISL_1463189, EPI_ISL_1463191, EPI_ISL_1463194, EPI_ISL_1463207, EPI_ISL_1463208, EPI_ISL_1463209, EPI_ISL_1463210, EPI_ISL_1463211, EPI_ISL_1463212, EPI_ISL_1463213, EPI_ISL_1463214, EPI_ISL_1463215, EPI_ISL_1463216, EPI_ISL_1463217, EPI_ISL_1463218, EPI_ISL_1463222, EPI_ISL_1463229, EPI_ISL_1463260, EPI_ISL_1463261, EPI_ISL_1463262, EPI_ISL_1463263, EPI_ISL_1463264, EPI_ISL_1463265, EPI_ISL_1463266, EPI_ISL_1463267, EPI_ISL_1463268, EPI_ISL_1463269, EPI_ISL_1463270, EPI_ISL_1463271, EPI_ISL_1463272, EPI_ISL_1463278, EPI_ISL_1463281, EPI_ISL_1463282, EPI_ISL_1463307, EPI_ISL_1463308, EPI_ISL_1463309, EPI_ISL_1463310, EPI_ISL_1463311, EPI_ISL_1463312, EPI_ISL_1463313, EPI_ISL_1463314, EPI_ISL_1463316, EPI_ISL_1463317, EPI_ISL_1463318, EPI_ISL_1463319, EPI_ISL_1463320, EPI_ISL_1463321, EPI_ISL_1463335, EPI_ISL_1463355, EPI_ISL_1463356, EPI_ISL_1463357, EPI_ISL_1463358, EPI_ISL_1463359, EPI_ISL_1463360, EPI_ISL_1463362, EPI_ISL_1463392, EPI_ISL_1463393, EPI_ISL_1463394, EPI_ISL_1463395, EPI_ISL_1463399, EPI_ISL_1463400, EPI_ISL_1513637, EPI_ISL_1513640, EPI_ISL_1513641, EPI_ISL_1513642, EPI_ISL_1513647, EPI_ISL_1513652, EPI_ISL_1513653, EPI_ISL_1513989, EPI_ISL_1513990, EPI_ISL_1513992, EPI_ISL_1513993, EPI_ISL_1513994, EPI_ISL_1513996, EPI_ISL_1513997, EPI_ISL_1514220, EPI_ISL_1514224, EPI_ISL_1514284, EPI_ISL_1514329, EPI_ISL_1514364, EPI_ISL_1514575, EPI_ISL_1514582, EPI_ISL_1514583, EPI_ISL_1514585, EPI_ISL_1514587, EPI_ISL_1514588, EPI_ISL_1514590, EPI_ISL_1514591, EPI_ISL_1514592, EPI_ISL_1514604, EPI_ISL_1514605, EPI_ISL_1514606, EPI_ISL_1514621, EPI_ISL_1514624, EPI_ISL_1514625, EPI_ISL_1514627, EPI_ISL_1514628, EPI_ISL_1514632, EPI_ISL_1514633, EPI_ISL_1514634, EPI_ISL_1514635, EPI_ISL_1514636, EPI_ISL_1514637, EPI_ISL_1514638, EPI_ISL_1514647, EPI_ISL_1514648, EPI_ISL_1514649, EPI_ISL_1514653, EPI_ISL_1514657, EPI_ISL_1514659, EPI_ISL_1514660, EPI_ISL_1514661, EPI_ISL_1514663, EPI_ISL_1514692, EPI_ISL_1514693, EPI_ISL_1514702, EPI_ISL_1514706, EPI_ISL_1514707, EPI_ISL_1514708, EPI_ISL_1514709, EPI_ISL_1514710, EPI_ISL_1514711, EPI_ISL_1514716, EPI_ISL_1514718, EPI_ISL_1514719, EPI_ISL_1514721, EPI_ISL_1515063, EPI_ISL_1515076, EPI_ISL_1515077, EPI_ISL_1515079, EPI_ISL_1515083, EPI_ISL_1515084, EPI_ISL_1515085, EPI_ISL_1515086, EPI_ISL_1515087, EPI_ISL_1515088, EPI_ISL_1515089, EPI_ISL_1515125, EPI_ISL_1515128, EPI_ISL_1515129, EPI_ISL_1515132, EPI_ISL_1515137, EPI_ISL_1515138, EPI_ISL_1515154, EPI_ISL_1515323, EPI_ISL_1515326, EPI_ISL_1515327, EPI_ISL_1515329, EPI_ISL_1515520, EPI_ISL_1548207, EPI_ISL_1548208, EPI_ISL_1548210, EPI_ISL_1548211, EPI_ISL_1548213, EPI_ISL_1548235, EPI_ISL_1548237, EPI_ISL_1548241, EPI_ISL_1548242, EPI_ISL_1548248, EPI_ISL_1548249, EPI_ISL_1548250, EPI_ISL_1548260, EPI_ISL_1548262, EPI_ISL_1548375, EPI_ISL_1548382, EPI_ISL_1548384, EPI_ISL_1548475, EPI_ISL_1548476, EPI_ISL_1548477, EPI_ISL_1548478, EPI_ISL_1548506, EPI_ISL_1548507, EPI_ISL_1548508, EPI_ISL_1548509, EPI_ISL_1548952, EPI_ISL_1548979, EPI_ISL_1548986, EPI_ISL_1548993, EPI_ISL_1549000, EPI_ISL_1549017, EPI_ISL_1549019, EPI_ISL_1549024, EPI_ISL_1549032, EPI_ISL_1549033, EPI_ISL_1549034, EPI_ISL_1549038, EPI_ISL_1549044, EPI_ISL_1549045, EPI_ISL_1549054, EPI_ISL_1549063, EPI_ISL_1549064, EPI_ISL_1549065, EPI_ISL_1549066, EPI_ISL_1549067, EPI_ISL_1549068, EPI_ISL_1549080, EPI_ISL_1549081, EPI_ISL_1549082, EPI_ISL_1549102, EPI_ISL_1549106, EPI_ISL_1549173, EPI_ISL_1549376, EPI_ISL_1549377, EPI_ISL_1549378, EPI_ISL_1549379, EPI_ISL_1549380, EPI_ISL_1549558, EPI_ISL_1549564, EPI_ISL_1549632, EPI_ISL_1549635, EPI_ISL_1549645, EPI_ISL_1549650, EPI_ISL_1549674, EPI_ISL_1549758, EPI_ISL_1549760, EPI_ISL_1549768, EPI_ISL_1549769, EPI_ISL_1549771, EPI_ISL_1549776, EPI_ISL_1549777, EPI_ISL_1549779, EPI_ISL_1549790, EPI_ISL_1549791, EPI_ISL_1549919, EPI_ISL_1549940, EPI_ISL_1549942, EPI_ISL_1549943, EPI_ISL_1550039, EPI_ISL_1550040, EPI_ISL_1609076, EPI_ISL_1609079, EPI_ISL_1609080, EPI_ISL_1609139, EPI_ISL_1609147, EPI_ISL_1609151, EPI_ISL_1609174, EPI_ISL_1609178, EPI_ISL_1609179, EPI_ISL_1609180, EPI_ISL_1609216, EPI_ISL_1609238, EPI_ISL_1609239, EPI_ISL_1609240, EPI_ISL_1609241, EPI_ISL_1609249, EPI_ISL_1609250, EPI_ISL_1609251, EPI_ISL_1609314, EPI_ISL_1609315, EPI_ISL_1609316, EPI_ISL_1609319, EPI_ISL_1609328, EPI_ISL_1609332, EPI_ISL_1609333, EPI_ISL_1609345, EPI_ISL_1609346, EPI_ISL_1609348, EPI_ISL_1609351, EPI_ISL_1609352, EPI_ISL_1609353, EPI_ISL_1609354, EPI_ISL_1609355, EPI_ISL_1609356, EPI_ISL_1609357, EPI_ISL_1609368, EPI_ISL_1609369, EPI_ISL_1609406, EPI_ISL_1609430, EPI_ISL_1609432, EPI_ISL_1609433, EPI_ISL_1609434, EPI_ISL_1609437, EPI_ISL_1609438, EPI_ISL_1609439, EPI_ISL_1609444, EPI_ISL_1609446, EPI_ISL_1609454, EPI_ISL_1609457, EPI_ISL_1609464, EPI_ISL_1609465, EPI_ISL_1609471, EPI_ISL_1609472, EPI_ISL_1609475, EPI_ISL_1609476, EPI_ISL_1609477, EPI_ISL_1609495, EPI_ISL_1609499, EPI_ISL_1609504, EPI_ISL_1609513, EPI_ISL_1609527, EPI_ISL_1609535, EPI_ISL_1609537, EPI_ISL_1609538, EPI_ISL_1609539, EPI_ISL_1609554, EPI_ISL_1609556, EPI_ISL_1609557, EPI_ISL_1609558, EPI_ISL_1609559, EPI_ISL_1609583, EPI_ISL_1609584, EPI_ISL_1609585, EPI_ISL_1609586, EPI_ISL_1609604, EPI_ISL_1609608, EPI_ISL_1609609, EPI_ISL_1609610, EPI_ISL_1609611, EPI_ISL_1609612, EPI_ISL_1609613, EPI_ISL_1609614, EPI_ISL_1609619, EPI_ISL_1609620, EPI_ISL_1609623, EPI_ISL_1609624, EPI_ISL_1609633, EPI_ISL_1609635, EPI_ISL_1609636, EPI_ISL_1609640, EPI_ISL_1609641, EPI_ISL_1609666, EPI_ISL_1609667, EPI_ISL_1609668, EPI_ISL_1609669, EPI_ISL_1609670, EPI_ISL_1609683, EPI_ISL_1609687, EPI_ISL_1609689, EPI_ISL_1609690, EPI_ISL_1609698, EPI_ISL_1609704, EPI_ISL_1609706, EPI_ISL_1609714, EPI_ISL_1609716, EPI_ISL_1609719, EPI_ISL_1609720, EPI_ISL_1609731, EPI_ISL_1609733, EPI_ISL_1609741, EPI_ISL_1609751, EPI_ISL_1609754, EPI_ISL_1609757, EPI_ISL_1609770, EPI_ISL_1609778, EPI_ISL_1609779, EPI_ISL_1609785, EPI_ISL_1609786, EPI_ISL_1609789, EPI_ISL_1609792, EPI_ISL_1609799, EPI_ISL_1609800, EPI_ISL_1609801, EPI_ISL_1609802, EPI_ISL_1609803, EPI_ISL_1609804, EPI_ISL_1609805, EPI_ISL_1609806, EPI_ISL_1609807, EPI_ISL_1609808, EPI_ISL_1609809, EPI_ISL_1609810, EPI_ISL_1609811, EPI_ISL_1609812, EPI_ISL_1609813, EPI_ISL_1609814, EPI_ISL_1609815, EPI_ISL_1609874, EPI_ISL_1609880, EPI_ISL_1609883, EPI_ISL_1609887, EPI_ISL_1609889, EPI_ISL_1609890, EPI_ISL_1609891, EPI_ISL_1609893, EPI_ISL_1609894, EPI_ISL_1609900, EPI_ISL_1609904, EPI_ISL_1609909, EPI_ISL_1609910, EPI_ISL_1609912, EPI_ISL_1609917, EPI_ISL_1609921, EPI_ISL_1609922, EPI_ISL_1609923, EPI_ISL_1609924, EPI_ISL_1609926, EPI_ISL_1609927, EPI_ISL_1609938, EPI_ISL_1609946, EPI_ISL_1609949, EPI_ISL_1609952, EPI_ISL_1609954, EPI_ISL_1609955, EPI_ISL_1609957, EPI_ISL_1609964, EPI_ISL_1609967, EPI_ISL_1610041, EPI_ISL_1610047, EPI_ISL_1610236, EPI_ISL_1610237, EPI_ISL_1610238, EPI_ISL_1610239, EPI_ISL_1610240, EPI_ISL_1610241, EPI_ISL_1610242, EPI_ISL_1610243, EPI_ISL_1610245, EPI_ISL_1610246, EPI_ISL_1610247, EPI_ISL_1610248, EPI_ISL_1610249, EPI_ISL_1610293, EPI_ISL_1610294, EPI_ISL_1610295, EPI_ISL_1610296, EPI_ISL_1610297, EPI_ISL_1610298, EPI_ISL_1610299, EPI_ISL_1610300, EPI_ISL_1610302, EPI_ISL_1610303, EPI_ISL_1610321, EPI_ISL_1610322, EPI_ISL_1610323, EPI_ISL_1610401, EPI_ISL_1610404, EPI_ISL_1610406, EPI_ISL_1610408, EPI_ISL_1610410, EPI_ISL_1610411, EPI_ISL_1610432, EPI_ISL_1610441, EPI_ISL_1610467, EPI_ISL_1610494, EPI_ISL_1610568, EPI_ISL_1610569, EPI_ISL_1610571, EPI_ISL_1610573, EPI_ISL_1610595, EPI_ISL_1610597, EPI_ISL_1610599, EPI_ISL_1610601, EPI_ISL_1610602, EPI_ISL_1610604, EPI_ISL_1610623, EPI_ISL_1610625, EPI_ISL_1610900, EPI_ISL_1610901, EPI_ISL_1610902, EPI_ISL_1610903, EPI_ISL_1610904, EPI_ISL_1610905, EPI_ISL_1610906, EPI_ISL_1610907, EPI_ISL_1610908, EPI_ISL_1610920, EPI_ISL_1610949, EPI_ISL_1610951, EPI_ISL_1610953, EPI_ISL_1611111, EPI_ISL_1611173, EPI_ISL_1611206, EPI_ISL_1611213, EPI_ISL_1611219, EPI_ISL_1611220, EPI_ISL_1611221, EPI_ISL_1611242, EPI_ISL_1611243, EPI_ISL_1611244, EPI_ISL_1611268, EPI_ISL_1611282, EPI_ISL_1611297, EPI_ISL_1611300, EPI_ISL_1611308, EPI_ISL_1611311, EPI_ISL_1611314, EPI_ISL_1611315, EPI_ISL_1611316, EPI_ISL_1611363, EPI_ISL_1611364, EPI_ISL_1611366, EPI_ISL_1611368, EPI_ISL_1611371, EPI_ISL_1611374, EPI_ISL_1611375, EPI_ISL_1611376, EPI_ISL_1611385, EPI_ISL_1611398, EPI_ISL_1611399, EPI_ISL_1611417, EPI_ISL_1611420, EPI_ISL_1611433, EPI_ISL_1611436, EPI_ISL_1611464, EPI_ISL_1611472, EPI_ISL_1611474, EPI_ISL_1611475, EPI_ISL_1611476, EPI_ISL_1611477, EPI_ISL_1611479, EPI_ISL_1611480, EPI_ISL_1611482, EPI_ISL_1611526, EPI_ISL_1611527, EPI_ISL_1611528, EPI_ISL_1611534, EPI_ISL_1611535, EPI_ISL_1611542, EPI_ISL_1611543, EPI_ISL_1611544, EPI_ISL_1611547, EPI_ISL_1611549, EPI_ISL_1611564, EPI_ISL_1611565, EPI_ISL_1611566, EPI_ISL_1611567, EPI_ISL_1611568, EPI_ISL_1611570, EPI_ISL_1611572, EPI_ISL_1611624, EPI_ISL_1611625, EPI_ISL_1611628, EPI_ISL_1611632, EPI_ISL_1611640, EPI_ISL_1611644, EPI_ISL_1611647, EPI_ISL_1611650, EPI_ISL_1611651, EPI_ISL_1611653, EPI_ISL_1611654, EPI_ISL_1611659, EPI_ISL_1611660, EPI_ISL_1611667, EPI_ISL_1611668, EPI_ISL_1611690, EPI_ISL_1611693, EPI_ISL_1611695, EPI_ISL_1611719, EPI_ISL_1611721, EPI_ISL_1611722, EPI_ISL_1611738, EPI_ISL_1611739, EPI_ISL_1611741, EPI_ISL_1611743, EPI_ISL_1611744, EPI_ISL_1611768, EPI_ISL_1611769, EPI_ISL_1611776, EPI_ISL_1611784, EPI_ISL_1611785, EPI_ISL_1611792, EPI_ISL_1611795, EPI_ISL_1611796, EPI_ISL_1611813, EPI_ISL_1611814, EPI_ISL_1611825, EPI_ISL_1611839, EPI_ISL_1611911, EPI_ISL_1611913, EPI_ISL_1611917, EPI_ISL_1611919, EPI_ISL_1611948, EPI_ISL_1611952, EPI_ISL_1611954, EPI_ISL_1611955, EPI_ISL_1611957, EPI_ISL_1611959, EPI_ISL_1680473, EPI_ISL_1680474, EPI_ISL_1680475, EPI_ISL_1680477, EPI_ISL_1680479, EPI_ISL_1680480, EPI_ISL_1680482, EPI_ISL_1680483, EPI_ISL_1680484, EPI_ISL_1680485, EPI_ISL_1680486, EPI_ISL_1680487, EPI_ISL_1680488, EPI_ISL_1680489, EPI_ISL_1680490, EPI_ISL_1680536, EPI_ISL_1680540, EPI_ISL_1680541, EPI_ISL_1680542, EPI_ISL_1680543, EPI_ISL_1680544, EPI_ISL_1680545, EPI_ISL_1680549, EPI_ISL_1680550, EPI_ISL_1680551, EPI_ISL_1680552, EPI_ISL_1680553, EPI_ISL_1680554, EPI_ISL_1680566, EPI_ISL_1680585, EPI_ISL_1680586, EPI_ISL_1680590, EPI_ISL_1680591, EPI_ISL_1680592, EPI_ISL_1680593, EPI_ISL_1680594, EPI_ISL_1680595, EPI_ISL_1680596, EPI_ISL_1680597, EPI_ISL_1680598, EPI_ISL_1680599, EPI_ISL_1680603, EPI_ISL_1680638, EPI_ISL_1680639, EPI_ISL_1680640, EPI_ISL_1680641, EPI_ISL_1680642, EPI_ISL_1680643, EPI_ISL_1680644, EPI_ISL_1680672, EPI_ISL_1680674, EPI_ISL_1680748, EPI_ISL_1680768, EPI_ISL_1682025, EPI_ISL_1682026, EPI_ISL_1682061, EPI_ISL_1682062, EPI_ISL_1682071, EPI_ISL_1682085, EPI_ISL_1682109, EPI_ISL_1682111, EPI_ISL_1682137, EPI_ISL_1682161, EPI_ISL_1682162, EPI_ISL_1682522, EPI_ISL_1682525, EPI_ISL_1682596, EPI_ISL_1682608, EPI_ISL_1682787, EPI_ISL_1682808, EPI_ISL_1682811, EPI_ISL_1682929, EPI_ISL_1682961, EPI_ISL_1683040, EPI_ISL_1683058, EPI_ISL_1683061, EPI_ISL_1683064, EPI_ISL_1683088, EPI_ISL_1683104, EPI_ISL_1683107, EPI_ISL_1683126, EPI_ISL_1683132, EPI_ISL_1683136, EPI_ISL_1683180, EPI_ISL_1683181, EPI_ISL_1683185, EPI_ISL_1683187, EPI_ISL_1683197, EPI_ISL_1683202, EPI_ISL_1683203, EPI_ISL_1683210, EPI_ISL_1683212, EPI_ISL_1683214, EPI_ISL_1683215, EPI_ISL_1683216, EPI_ISL_1683248, EPI_ISL_1683249, EPI_ISL_1683254, EPI_ISL_1683258, EPI_ISL_1683260, EPI_ISL_1683262, EPI_ISL_1683265, EPI_ISL_1683266, EPI_ISL_1683267, EPI_ISL_1683277, EPI_ISL_1683281, EPI_ISL_1683289, EPI_ISL_1683290, EPI_ISL_1683292, EPI_ISL_1683293, EPI_ISL_1683294, EPI_ISL_1683295, EPI_ISL_1683299, EPI_ISL_1683321, EPI_ISL_1683322, EPI_ISL_1683323, EPI_ISL_1683325, EPI_ISL_1683326, EPI_ISL_1683330, EPI_ISL_1683331, EPI_ISL_1683332, EPI_ISL_1683333, EPI_ISL_1683336, EPI_ISL_1683351, EPI_ISL_1683361, EPI_ISL_1683381, EPI_ISL_1683409, EPI_ISL_1683410, EPI_ISL_1683461, EPI_ISL_1683536, EPI_ISL_1695014, EPI_ISL_1695015 | | | |
| see above | Laboratory Corporation of America | Centers for Disease Control and Prevention Division of Viral Diseases, Pathogen Discovery | Adrian Paskey; Amanda Douglas; Amanda Suchanek; Andrea Throop; Ayla Burns; Benjamin Rambo-Martin; Bobbi Croy; Brian Krueger; Brian Norvell; Christopher Gulvick; Christos Petropoulos; Clinton R. Paden; Craig Lukasik; Dakota Howard; Darlene Wagner; Debbie Boles; Dhwani Batra; Duncan MacCannell; Eyad Almasri; Goran Stevovic; Howard Engler; Hrushikesh Deshmukh; Jake Humphrey; Jana Schroth; Jason Caravas; Joe Voshell; John Pruitt; Jonathan Meltzer; Jonathan Williams; Kara Moser; Kimberly Wagner; Lax Iyer; Lyndon Tilson; Manoj Jain; Marcia Eisenberg; Mary Ann Cristobal; Mary Williamson; Matthew Schmerer; Michael Levandoski; Mike Sapeta; Mindy Nye; Minoo Agarwal; Mohan Kolli; Nuthawin Charoensri; Oren Cohen; Peter W. Cook; Prashant Gupta; Qian Zeng; Rama Ghatti; Scott Parker; Scott Ryan; Scott Sammons; Shatavia Morrison; Stanley Letovsky; Steven Ragan; Suresh |

Babu Selvaraju; Susan Countryman; Susan Hicks; Suzanne Dale; Thomas Urban; Tim Kuphal; Tricia Zwiefelhofer; Vincent Drouillon; Yvette Unoarumhi

EPI_ISL_1398331, EPI_ISL_1398333, EPI_ISL_1398334, EPI_ISL_1398336, EPI_ISL_1398337, EPI_ISL_1398338, EPI_ISL_1465668, EPI_ISL_1465669, EPI_ISL_1465670, EPI_ISL_1465671, EPI_ISL_1465672, EPI_ISL_1465701, EPI_ISL_1465702, EPI_ISL_1465709, EPI_ISL_1465710, EPI_ISL_1465711, EPI_ISL_1465712, EPI_ISL_1465713, EPI_ISL_1576816, EPI_ISL_1580686, EPI_ISL_1582159, EPI_ISL_1582162, EPI_ISL_1582165, EPI_ISL_1582168, EPI_ISL_1582170, EPI_ISL_1582173, EPI_ISL_1582174, EPI_ISL_1582175, EPI_ISL_1582176, EPI_ISL_1582177, EPI_ISL_1582178, EPI_ISL_1582319, EPI_ISL_1582320, EPI_ISL_1582321, EPI_ISL_1593868, EPI_ISL_1593869, EPI_ISL_1593881, EPI_ISL_1671466, EPI_ISL_1671477, EPI_ISL_1671478, EPI_ISL_1671479, EPI_ISL_1671497, EPI_ISL_1671498, EPI_ISL_1671499, EPI_ISL_1671642, EPI_ISL_1671654, EPI_ISL_1671664, EPI_ISL_1740512, EPI_ISL_1740513, EPI_ISL_1755820, EPI_ISL_1755821, EPI_ISL_1755822, EPI_ISL_1755823, EPI_ISL_1755824, EPI_ISL_1755825, EPI_ISL_1755826, EPI_ISL_1755827, EPI_ISL_1755828, EPI_ISL_1755829, EPI_ISL_1755830, EPI_ISL_1755831, EPI_ISL_1755832, EPI_ISL_1755833, EPI_ISL_1755834, EPI_ISL_1855215, EPI_ISL_1855216

see above | MONTEFIORE MEDICAL CENTER LABORATORIES | Wadsworth Center, New York State Department of Health | Alexis Russel; Alexis Russell; Catharine Prussing; Daryl M. Lamson; Erasmus Schneider; Erica Lasek-Nesselquist; John Kelly; Jonathan Plitnick; Kirsten St. George; Matthew Shudt; Melissa A Leisner; Navjot Singh

EPI_ISL_1465849, EPI_ISL_1465850, EPI_ISL_1465851, EPI_ISL_1465852, EPI_ISL_1482336, EPI_ISL_1482337, EPI_ISL_1482338, EPI_ISL_1482339, EPI_ISL_1482340, EPI_ISL_1482341, EPI_ISL_1482342, EPI_ISL_1482343, EPI_ISL_1482344, EPI_ISL_1482345

see above | MONTEFIORE NEW ROCHELLE HOSPITAL LABORATORY | Wadsworth Center, New York State Department of Health | Alexis Russell; Catharine Prussing; Daryl M. Lamson; Erasmus Schneider; Erica Lasek-Nesselquist; John Kelly; Jonathan Plitnick; Kirsten St. George; Matthew Shudt; Melissa A Leisner; Navjot Singh

EPI_ISL_1709301, EPI_ISL_1709302, EPI_ISL_1709303, EPI_ISL_1709304, EPI_ISL_1709305, EPI_ISL_1709306, EPI_ISL_1709307, EPI_ISL_1709308, EPI_ISL_1709309, EPI_ISL_1709310, EPI_ISL_1709311, EPI_ISL_1709312, EPI_ISL_1709313, EPI_ISL_1709314, EPI_ISL_1709315, EPI_ISL_1709316, EPI_ISL_1709317, EPI_ISL_1709318, EPI_ISL_1709319, EPI_ISL_1709320, EPI_ISL_1709321, EPI_ISL_1709322, EPI_ISL_1709323, EPI_ISL_1709324, EPI_ISL_1709325, EPI_ISL_1709326, EPI_ISL_1709327, EPI_ISL_1709328, EPI_ISL_1709329, EPI_ISL_1709330, EPI_ISL_1709331, EPI_ISL_1709332, EPI_ISL_1709333, EPI_ISL_1709334, EPI_ISL_1709335, EPI_ISL_1709336, EPI_ISL_1709337, EPI_ISL_1709338, EPI_ISL_1709339, EPI_ISL_1709340, EPI_ISL_1709341, EPI_ISL_1709342, EPI_ISL_1709343, EPI_ISL_1709344, EPI_ISL_1709345, EPI_ISL_1709346, EPI_ISL_1709347, EPI_ISL_1709348, EPI_ISL_1709349, EPI_ISL_1709350, EPI_ISL_1709351, EPI_ISL_1709352, EPI_ISL_1709353, EPI_ISL_1709354, EPI_ISL_1709355, EPI_ISL_1709356, EPI_ISL_1709357, EPI_ISL_1709358, EPI_ISL_1709359, EPI_ISL_1709360, EPI_ISL_1709361, EPI_ISL_1709362, EPI_ISL_1709363, EPI_ISL_1709364, EPI_ISL_1709365, EPI_ISL_1709366, EPI_ISL_1709367, EPI_ISL_1709368, EPI_ISL_1709369, EPI_ISL_1709370, EPI_ISL_1709371, EPI_ISL_1709372, EPI_ISL_1709373, EPI_ISL_1709374, EPI_ISL_1709375, EPI_ISL_1709376, EPI_ISL_1709377, EPI_ISL_1709378, EPI_ISL_1709379, EPI_ISL_1709380, EPI_ISL_1709381, EPI_ISL_1709382, EPI_ISL_1709383, EPI_ISL_1709384, EPI_ISL_1709385, EPI_ISL_1709386, EPI_ISL_1709387, EPI_ISL_1709388, EPI_ISL_1709389, EPI_ISL_1709390, EPI_ISL_1709391, EPI_ISL_1709392, EPI_ISL_1709393, EPI_ISL_1709394, EPI_ISL_1709395, EPI_ISL_1709396, EPI_ISL_1709397, EPI_ISL_1709398, EPI_ISL_1709399, EPI_ISL_1709400, EPI_ISL_1709401, EPI_ISL_1709402, EPI_ISL_1709403, EPI_ISL_1709404, EPI_ISL_1709405, EPI_ISL_1709406, EPI_ISL_1709407, EPI_ISL_1709408, EPI_ISL_1709409, EPI_ISL_1709410, EPI_ISL_1709411, EPI_ISL_1709412, EPI_ISL_1709413, EPI_ISL_1709414, EPI_ISL_1709415, EPI_ISL_1709416, EPI_ISL_1709417, EPI_ISL_1709418, EPI_ISL_1709419, EPI_ISL_1709420, EPI_ISL_1709422, EPI_ISL_1709423, EPI_ISL_1709424, EPI_ISL_1709425, EPI_ISL_1709426, EPI_ISL_1709427, EPI_ISL_1709428, EPI_ISL_1709429, EPI_ISL_1709430, EPI_ISL_1709431, EPI_ISL_1709432, EPI_ISL_1709434, EPI_ISL_1709435, EPI_ISL_1709436, EPI_ISL_1709437, EPI_ISL_1709438, EPI_ISL_1709439, EPI_ISL_1709440, EPI_ISL_1709441, EPI_ISL_1709442, EPI_ISL_1709443, EPI_ISL_1709444, EPI_ISL_1709445, EPI_ISL_1709446, EPI_ISL_1709447, EPI_ISL_1709448, EPI_ISL_1709449, EPI_ISL_1709450, EPI_ISL_1709451, EPI_ISL_1709452, EPI_ISL_1709453, EPI_ISL_1709454, EPI_ISL_1709455, EPI_ISL_1709456, EPI_ISL_1709457, EPI_ISL_1709458, EPI_ISL_1709459, EPI_ISL_1709460, EPI_ISL_1709461, EPI_ISL_1709462, EPI_ISL_1709463, EPI_ISL_1709464, EPI_ISL_1709465, EPI_ISL_1709466, EPI_ISL_1709467, EPI_ISL_1709468, EPI_ISL_1709469, EPI_ISL_1709470, EPI_ISL_1709472, EPI_ISL_1709473, EPI_ISL_1709474, EPI_ISL_1709475, EPI_ISL_1709476, EPI_ISL_1709477, EPI_ISL_1709478, EPI_ISL_1709479, EPI_ISL_1709480, EPI_ISL_1709481, EPI_ISL_1709482, EPI_ISL_1709483, EPI_ISL_1709484, EPI_ISL_1709485, EPI_ISL_1709486, EPI_ISL_1709487, EPI_ISL_1709488, EPI_ISL_1709489, EPI_ISL_1709490, EPI_ISL_1709492, EPI_ISL_1709493, EPI_ISL_1709494, EPI_ISL_1709495, EPI_ISL_1709496, EPI_ISL_1709497, EPI_ISL_1709498, EPI_ISL_1709499, EPI_ISL_1709500, EPI_ISL_1709501, EPI_ISL_1709502, EPI_ISL_1709503, EPI_ISL_1709504, EPI_ISL_1709505, EPI_ISL_1709506, EPI_ISL_1709507, EPI_ISL_1709508, EPI_ISL_1709509, EPI_ISL_1709510, EPI_ISL_1709511, EPI_ISL_1709512, EPI_ISL_1709513, EPI_ISL_1709514, EPI_ISL_1709515, EPI_ISL_1709516, EPI_ISL_1709517, EPI_ISL_1709518, EPI_ISL_1709519, EPI_ISL_1709520, EPI_ISL_1709521, EPI_ISL_1709522, EPI_ISL_1709523, EPI_ISL_1709524, EPI_ISL_1709525, EPI_ISL_1709526, EPI_ISL_1709527, EPI_ISL_1709528, EPI_ISL_1709529, EPI_ISL_1709530, EPI_ISL_1709531, EPI_ISL_1709532, EPI_ISL_1709533, EPI_ISL_1709534, EPI_ISL_1709535, EPI_ISL_1709536, EPI_ISL_1709537, EPI_ISL_1709538, EPI_ISL_1709539, EPI_ISL_1709540, EPI_ISL_1709541, EPI_ISL_1709542, EPI_ISL_1709543, EPI_ISL_1709544, EPI_ISL_1709545, EPI_ISL_1709546, EPI_ISL_1709547, EPI_ISL_1709548, EPI_ISL_1709549, EPI_ISL_1709550, EPI_ISL_1709551, EPI_ISL_1709552, EPI_ISL_1709553, EPI_ISL_1709554, EPI_ISL_1709555, EPI_ISL_1709556, EPI_ISL_1709558, EPI_ISL_1709559, EPI_ISL_1709560, EPI_ISL_1709561, EPI_ISL_1709562, EPI_ISL_1709563, EPI_ISL_1709564, EPI_ISL_1709565, EPI_ISL_1709566, EPI_ISL_1709567, EPI_ISL_1709568, EPI_ISL_1709569, EPI_ISL_1709570, EPI_ISL_1709571, EPI_ISL_1709572, EPI_ISL_1709573, EPI_ISL_1709574, EPI_ISL_1709575, EPI_ISL_1709576, EPI_ISL_1709577, EPI_ISL_1709578, EPI_ISL_1709579, EPI_ISL_1709580, EPI_ISL_1709581, EPI_ISL_1709582, EPI_ISL_1709583, EPI_ISL_1709584, EPI_ISL_1709585, EPI_ISL_1709586, EPI_ISL_1709587, EPI_ISL_1709588, EPI_ISL_1709589, EPI_ISL_1709590, EPI_ISL_1709591, EPI_ISL_1709592, EPI_ISL_1709593, EPI_ISL_1709594, EPI_ISL_1709595, EPI_ISL_1709596, EPI_ISL_1709597, EPI_ISL_1709598, EPI_ISL_1709599, EPI_ISL_1709600, EPI_ISL_1709601, EPI_ISL_1709602, EPI_ISL_1709603, EPI_ISL_1709604, EPI_ISL_1709605, EPI_ISL_1709606, EPI_ISL_1709607, EPI_ISL_1709608, EPI_ISL_1709609, EPI_ISL_1709610, EPI_ISL_1709611, EPI_ISL_1709612, EPI_ISL_1709613, EPI_ISL_1709614, EPI_ISL_1709615, EPI_ISL_1709616, EPI_ISL_1709617, EPI_ISL_1709618, EPI_ISL_1709619, EPI_ISL_1709620, EPI_ISL_1709621, EPI_ISL_1709622, EPI_ISL_1709623, EPI_ISL_1709624, EPI_ISL_1709625, EPI_ISL_1709626, EPI_ISL_1709627, EPI_ISL_1709628, EPI_ISL_1709629, EPI_ISL_1709630, EPI_ISL_1709631, EPI_ISL_1709632, EPI_ISL_1709633, EPI_ISL_1709634, EPI_ISL_1709635, EPI_ISL_1709636, EPI_ISL_1709637, EPI_ISL_1709638, EPI_ISL_1709639, EPI_ISL_1709640, EPI_ISL_1709641, EPI_ISL_1709642, EPI_ISL_1709643, EPI_ISL_1709644, EPI_ISL_1709645, EPI_ISL_1709646, EPI_ISL_1709647, EPI_ISL_1709648, EPI_ISL_1709649, EPI_ISL_1709650, EPI_ISL_1709651, EPI_ISL_1709652, EPI_ISL_1709653, EPI_ISL_1709654, EPI_ISL_1709655, EPI_ISL_1709656, EPI_ISL_1709657, EPI_ISL_1709658, EPI_ISL_1709659, EPI_ISL_1709660, EPI_ISL_1709661, EPI_ISL_1709662, EPI_ISL_1709663, EPI_ISL_1709664, EPI_ISL_1709665, EPI_ISL_1709666, EPI_ISL_1709667, EPI_ISL_1709668, EPI_ISL_1709669, EPI_ISL_1709670, EPI_ISL_1709671, EPI_ISL_1709672, EPI_ISL_1709673, EPI_ISL_1709674, EPI_ISL_1709675, EPI_ISL_1709676, EPI_ISL_1709677, EPI_ISL_1709678, EPI_ISL_1709679, EPI_ISL_1709680, EPI_ISL_1709681, EPI_ISL_1709682, EPI_ISL_1709683, EPI_ISL_1709684, EPI_ISL_1709685, EPI_ISL_1709686, EPI_ISL_1709687, EPI_ISL_1709688, EPI_ISL_1709689, EPI_ISL_1709690, EPI_ISL_1709691, EPI_ISL_1709692, EPI_ISL_1709693, EPI_ISL_1709694, EPI_ISL_1709695, EPI_ISL_1709696, EPI_ISL_1709697, EPI_ISL_1709698, EPI_ISL_1709699, EPI_ISL_1709700, EPI_ISL_1709701, EPI_ISL_1709702, EPI_ISL_1709703, EPI_ISL_1709704, EPI_ISL_1709705, EPI_ISL_1709706, EPI_ISL_1709707, EPI_ISL_1709708, EPI_ISL_1709709, EPI_ISL_1709710, EPI_ISL_1709711, EPI_ISL_1709712, EPI_ISL_1709713, EPI_ISL_1709714, EPI_ISL_1709715, EPI_ISL_1709716, EPI_ISL_1709717, EPI_ISL_1709718, EPI_ISL_1709719, EPI_ISL_1709720, EPI_ISL_1709721, EPI_ISL_1709722, EPI_ISL_1709723, EPI_ISL_1709724, EPI_ISL_1709725, EPI_ISL_1709726, EPI_ISL_1709727, EPI_ISL_1709728, EPI_ISL_1709729, EPI_ISL_1709731, EPI_ISL_1709732, EPI_ISL_1709733, EPI_ISL_1709734, EPI_ISL_1709735, EPI_ISL_1709736, EPI_ISL_1709737, EPI_ISL_1709738, EPI_ISL_1709739, EPI_ISL_1709740, EPI_ISL_1709741, EPI_ISL_1709742, EPI_ISL_1709743, EPI_ISL_1709744, EPI_ISL_1709745, EPI_ISL_1709746, EPI_ISL_1709747, EPI_ISL_1709748, EPI_ISL_1709749, EPI_ISL_1709750, EPI_ISL_1709751, EPI_ISL_1709752, EPI_ISL_1709753, EPI_ISL_1709754, EPI_ISL_1709755, EPI_ISL_1709756, EPI_ISL_1709757, EPI_ISL_1709758, EPI_ISL_1709759, EPI_ISL_1709760, EPI_ISL_1709761, EPI_ISL_1709762, EPI_ISL_1709763, EPI_ISL_1709764, EPI_ISL_1709765, EPI_ISL_1709766, EPI_ISL_1709767, EPI_ISL_1709768, EPI_ISL_1709769, EPI_ISL_1709770, EPI_ISL_1709771, EPI_ISL_1709772, EPI_ISL_1709773, EPI_ISL_1709774, EPI_ISL_1709775, EPI_ISL_1709776, EPI_ISL_1709777, EPI_ISL_1709778, EPI_ISL_1709779, EPI_ISL_1709780, EPI_ISL_1709781, EPI_ISL_1709782, EPI_ISL_1709783, EPI_ISL_1709784, EPI_ISL_1709785, EPI_ISL_1709786, EPI_ISL_1709787, EPI_ISL_1709788, EPI_ISL_1709789, EPI_ISL_1709790, EPI_ISL_1709791, EPI_ISL_1709792, EPI_ISL_1709793, EPI_ISL_1709794, EPI_ISL_1709795, EPI_ISL_1709796, EPI_ISL_1709797, EPI_ISL_1709798, EPI_ISL_1709799, EPI_ISL_1709800, EPI_ISL_1709801, EPI_ISL_1709802, EPI_ISL_1709803, EPI_ISL_1709804, EPI_ISL_1709805, EPI_ISL_1709806, EPI_ISL_1709807, EPI_ISL_1709808, EPI_ISL_1709809, EPI_ISL_1709810, EPI_ISL_1709811, EPI_ISL_1709812, EPI_ISL_1709813, EPI_ISL_1709814, EPI_ISL_1709815, EPI_ISL_1709816, EPI_ISL_1709817, EPI_ISL_1709818, EPI_ISL_1709819, EPI_ISL_1709820, EPI_ISL_1709821, EPI_ISL_1709822, EPI_ISL_1709823, EPI_ISL_1709824, EPI_ISL_1709825, EPI_ISL_1709826, EPI_ISL_1709827, EPI_ISL_1709828, EPI_ISL_1709829, EPI_ISL_1709830, EPI_ISL_1709831, EPI_ISL_1709832, EPI_ISL_1709833, EPI_ISL_1709834, EPI_ISL_1709835, EPI_ISL_1709836, EPI_ISL_1709837, EPI_ISL_1709838, EPI_ISL_1709839, EPI_ISL_1709840, EPI_ISL_1709841, EPI_ISL_1709842, EPI_ISL_1709843, EPI_ISL_1709844, EPI_ISL_1709845, EPI_ISL_1709846, EPI_ISL_1709847, EPI_ISL_1709848, EPI_ISL_1709849, EPI_ISL_1709850, EPI_ISL_1709851, EPI_ISL_1709852, EPI_ISL_1709853, EPI_ISL_1709854, EPI_ISL_1709855, EPI_ISL_1709856, EPI_ISL_1709857, EPI_ISL_1709858, EPI_ISL_1709859, EPI_ISL_1709860, EPI_ISL_1709861, EPI_ISL_1709862, EPI_ISL_1709863, EPI_ISL_1709864, EPI_ISL_1709865, EPI_ISL_1709866, EPI_ISL_1709867, EPI_ISL_1709868, EPI_ISL_1709869, EPI_ISL_1709870, EPI_ISL_1709871, EPI_ISL_1709872, EPI_ISL_1709873, EPI_ISL_1709874, EPI_ISL_1709875, EPI_ISL_1709876, EPI_ISL_1709877, EPI_ISL_1709878, EPI_ISL_1709879, EPI_ISL_1709880, EPI_ISL_1709881, EPI_ISL_1709882, EPI_ISL_1709883, EPI_ISL_1709884, EPI_ISL_1709885, EPI_ISL_1709886, EPI_ISL_1709887, EPI_ISL_1709888, EPI_ISL_1709890, EPI_ISL_1709891, EPI_ISL_1709892, EPI_ISL_1709893, EPI_ISL_1709894, EPI_ISL_1709895, EPI_ISL_1709896, EPI_ISL_1709897, EPI_ISL_1709898, EPI_ISL_1709899, EPI_ISL_1709900, EPI_ISL_1709901, EPI_ISL_1709902, EPI_ISL_1709903, EPI_ISL_1709904, EPI_ISL_1709905, EPI_ISL_1709906, EPI_ISL_1709907, EPI_ISL_1709908, EPI_ISL_1709909, EPI_ISL_1709910, EPI_ISL_1709911, EPI_ISL_1709912

see above | MSHS Clinical Microbiology Laboratories | MSHS Pathogen Surveillance Program | Adolfo García-Sastre; Adriana van de Guchte; Ajay Obla; Alberto Paniz-Mondolfi; Ana S. Gonzalez-Reiche; Angela Amoako; Ashley Salimbangon; Betsaida Salom Melo; Bremy Alburquerque; Brianne Ciferri; Charles Gleason; Daniel Floda; Deena R. Altman; Denise Jurczyszak; Emilia Mia Sordillo; Gintaras Deikus; Giulio Kleiner; Gopi Patel; Hala Alshammary; Harm van Bakel; Irina Oussenko; Jayeeta Dutta; Juan Soto; Julia Matthews; Katherine Beach; Kathryn Twyman; Kayla Russo; Komal Srivastava; Levy Sominsky; Mahmoud Awawda; Marta Luksza; Matthew M. Hernandez; Melissa Gitman; Michael D. Nowak; Mitchell J. Sullivan; Nancy Francoeur; Robert Sebra; Sarah Schaefer; Shelcie Fabre; Shwetha Hara Sridhar; Viviana Simon; Ying-Chih Wang; Zenab Khan

EPI_ISL_2009117, EPI_ISL_2009125, EPI_ISL_2009127, EPI_ISL_2009131, EPI_ISL_2009132, EPI_ISL_2009137, EPI_ISL_2009138, EPI_ISL_2009140, EPI_ISL_2009144, EPI_ISL_2009150, EPI_ISL_2009152, EPI_ISL_2009158, EPI_ISL_2009166, EPI_ISL_2009167, EPI_ISL_2009168, EPI_ISL_2009169, EPI_ISL_2009176, EPI_ISL_2009177

see above | MSK Microbiology Lab | MSK Microbiology Lab | Esther Babady; Krupa Jani; Tracy McMillen

EPI_ISL_2011717 | Moderna Inc. | Moderna Inc. | Groves Dixon; Rolando Pajon; Yamuna Paila

EPI_ISL_1378824, EPI_ISL_1674703 | Murphy Medical Association | Grubaugh Lab - Yale School of Public Health | Anderson Brito; Annie Watkins; Caleb Neal; Chaney Kalinich; Chantal Vogels; Eva Laszlo; Isabel Ott; Isabell Ott; Jessica Rothman; Joseph Fauver; Mallery Breban; Mary Petrone; Nathan Grubaugh; Steven Murphy; Tara Alpert

EPI_ISL_1482346, EPI_ISL_1482347, EPI_ISL_1482348, EPI_ISL_1482349, EPI_ISL_1482350, EPI_ISL_1482351, EPI_ISL_1482352, EPI_ISL_1482353, EPI_ISL_1582256, EPI_ISL_1582257, EPI_ISL_1582258, EPI_ISL_1582259, EPI_ISL_1671448, EPI_ISL_1671452, EPI_ISL_1671453, EPI_ISL_1671493

see above | NORTH SHORE UNIVERSITY HOSPITAL | Wadsworth Center, New York State Department of Health | Alexis Russell; Catharine Prussing; Daryl M. Lamson; Erasmus Schneider; Erica Lasek-Nesselquist; John Kelly; Jonathan Plitnick; Kirsten St. George; Matthew Shudt; Melissa A Leisner; Navjot Singh

EPI_ISL_1462556, EPI_ISL_1462558, EPI_ISL_1462559, EPI_ISL_1462563, EPI_ISL_1462564, EPI_ISL_1462567, EPI_ISL_1462578, EPI_ISL_1462579, EPI_ISL_1462581, EPI_ISL_1462582, EPI_ISL_1462587, EPI_ISL_1462588, EPI_ISL_1462589, EPI_ISL_1462591, EPI_ISL_1462593, EPI_ISL_1462594, EPI_ISL_1462596, EPI_ISL_1462597, EPI_ISL_1462599, EPI_ISL_1462600, EPI_ISL_1462601, EPI_ISL_1462603, EPI_ISL_1462604, EPI_ISL_1462606, EPI_ISL_1462607, EPI_ISL_1462609, EPI_ISL_1462610, EPI_ISL_1462612, EPI_ISL_1462613, EPI_ISL_1462615, EPI_ISL_1462617, EPI_ISL_1462618, EPI_ISL_1462620, EPI_ISL_1462621, EPI_ISL_1462623, EPI_ISL_1462624, EPI_ISL_1465678, EPI_ISL_1465679, EPI_ISL_1465680, EPI_ISL_1465681, EPI_ISL_1465682, EPI_ISL_1465755, EPI_ISL_1465756, EPI_ISL_1465762, EPI_ISL_1465763, EPI_ISL_1465764, EPI_ISL_1465765, EPI_ISL_1465766, EPI_ISL_1465837, EPI_ISL_1482556, EPI_ISL_1482557, EPI_ISL_1482558, EPI_ISL_1482559, EPI_ISL_1482560, EPI_ISL_1482561, EPI_ISL_1482562, EPI_ISL_1482563, EPI_ISL_1482564, EPI_ISL_1482565, EPI_ISL_1482566, EPI_ISL_1482567, EPI_ISL_1482568, EPI_ISL_1482569, EPI_ISL_1482570, EPI_ISL_1482571, EPI_ISL_1482572, EPI_ISL_1482580, EPI_ISL_1482581, EPI_ISL_1482582, EPI_ISL_1482583, EPI_ISL_1482584, EPI_ISL_1482585, EPI_ISL_1482586, EPI_ISL_1482587, EPI_ISL_1482588, EPI_ISL_1482589, EPI_ISL_1482590, EPI_ISL_1482591, EPI_ISL_1482592, EPI_ISL_1482593, EPI_ISL_1482594, EPI_ISL_1482595, EPI_ISL_1482596, EPI_ISL_1482597, EPI_ISL_1482598, EPI_ISL_1482599, EPI_ISL_1482600, EPI_ISL_1482601, EPI_ISL_1482602, EPI_ISL_1482603, EPI_ISL_1482604, EPI_ISL_1482605, EPI_ISL_1482606, EPI_ISL_1482607, EPI_ISL_1482608, EPI_ISL_1482609, EPI_ISL_1482610, EPI_ISL_1482611, EPI_ISL_1482612, EPI_ISL_1482613, EPI_ISL_1482614, EPI_ISL_1482615, EPI_ISL_1482616, EPI_ISL_1575449, EPI_ISL_1575478, EPI_ISL_1576815, EPI_ISL_1576825, EPI_ISL_1576826, EPI_ISL_1580723, EPI_ISL_1580724, EPI_ISL_1580725, EPI_ISL_1580726, EPI_ISL_1580727, EPI_ISL_1580728, EPI_ISL_1580729, EPI_ISL_1580730, EPI_ISL_1580731, EPI_ISL_1580732, EPI_ISL_1580733, EPI_ISL_1580734, EPI_ISL_1580735, EPI_ISL_1580736, EPI_ISL_1580737, EPI_ISL_1580738, EPI_ISL_1582225, EPI_ISL_1582226, EPI_ISL_1582283, EPI_ISL_1582284, EPI_ISL_1582285, EPI_ISL_1582286, EPI_ISL_1582287, EPI_ISL_1582288, EPI_ISL_1582289, EPI_ISL_1582290, EPI_ISL_1582291, EPI_ISL_1582292, EPI_ISL_1582293, EPI_ISL_1582294, EPI_ISL_1582295, EPI_ISL_1582296, EPI_ISL_1582297, EPI_ISL_1582298, EPI_ISL_1582299, EPI_ISL_1582300, EPI_ISL_1582301, EPI_ISL_1582302, EPI_ISL_1582303, EPI_ISL_1582304, EPI_ISL_1582305, EPI_ISL_1582306, EPI_ISL_1582307, EPI_ISL_1582317, EPI_ISL_1582354, EPI_ISL_1582355, EPI_ISL_1582356, EPI_ISL_1582369, EPI_ISL_1582370, EPI_ISL_1582371, EPI_ISL_1582372, EPI_ISL_1582373, EPI_ISL_1582374, EPI_ISL_1582375, EPI_ISL_1582376, EPI_ISL_1582377, EPI_ISL_1582378, EPI_ISL_1582379, EPI_ISL_1582380, EPI_ISL_1582381, EPI_ISL_1582382, EPI_ISL_1582383, EPI_ISL_1582384, EPI_ISL_1582385, EPI_ISL_1582386, EPI_ISL_1582387, EPI_ISL_1582388, EPI_ISL_1582389, EPI_ISL_1593876, EPI_ISL_1593877, EPI_ISL_1593887, EPI_ISL_1671614, EPI_ISL_1671615, EPI_ISL_1671616, EPI_ISL_1671620, EPI_ISL_1671628, EPI_ISL_1671629, EPI_ISL_1671630, EPI_ISL_1671631, EPI_ISL_1671633, EPI_ISL_1671638, EPI_ISL_1671640, EPI_ISL_1671648, EPI_ISL_1671649, EPI_ISL_1671650, EPI_ISL_1671652, EPI_ISL_1671653, EPI_ISL_1671661, EPI_ISL_1671668, EPI_ISL_1671669, EPI_ISL_1671670, EPI_ISL_1671671, EPI_ISL_1740518, EPI_ISL_1740519, EPI_ISL_1756330

see above | NORTHWELL HEALTH LABORATORIES | Wadsworth Center, New York State Department of Health | Alexis Russell; Catharine Prussing; Daryl M. Lamson; Erasmus Schneider; Erica Lasek-Nesselquist; John Kelly; Jonathan Plitnick; Kirsten St. George; Matthew Shudt; Melissa A Leisner; Navjot Singh

EPI_ISL_1516205, EPI_ISL_1516206, EPI_ISL_1516207, EPI_ISL_1516208, EPI_ISL_1516209, EPI_ISL_1516210, EPI_ISL_1516211, EPI_ISL_1516212, EPI_ISL_1678056, EPI_ISL_1678057, EPI_ISL_1678058, EPI_ISL_1678059, EPI_ISL_1678060, EPI_ISL_1678061, EPI_ISL_1678062, EPI_ISL_1678063, EPI_ISL_1678064, EPI_ISL_1678065, EPI_ISL_1678066, EPI_ISL_1678067, EPI_ISL_1678068, EPI_ISL_1678069, EPI_ISL_1678070, EPI_ISL_1678071, EPI_ISL_1678072, EPI_ISL_1678073, EPI_ISL_1678074, EPI_ISL_1678075, EPI_ISL_1678076, EPI_ISL_1678077, EPI_ISL_1678079, EPI_ISL_1678080, EPI_ISL_1678257, EPI_ISL_1678258, EPI_ISL_1678259, EPI_ISL_1678260, EPI_ISL_1678261, EPI_ISL_1678262, EPI_ISL_1678263, EPI_ISL_1678264, EPI_ISL_1678265, EPI_ISL_1695970, EPI_ISL_1823946, EPI_ISL_1823948, EPI_ISL_1823950, EPI_ISL_1823953, EPI_ISL_1823955, EPI_ISL_1823956, EPI_ISL_1823957, EPI_ISL_1823958, EPI_ISL_1823961, EPI_ISL_1823963, EPI_ISL_1823964, EPI_ISL_1823965, EPI_ISL_1823967, EPI_ISL_1823968, EPI_ISL_1823969, EPI_ISL_1823970

see above | NYC Department of Health and Mental Hygiene | Centers for Disease Control and Prevention Division of Viral Diseases, Pathogen Discovery | Alison Laufer Halpin; Ben L. Rambo-Martin; Clinton R. Paden; Dakota Howard; Darlene Wagner; Dave Wentworth; Dhwani Batra; Jasmine Padilla; Justin Lee; Katie Dillon; Krista Queen; Kristen Knipe; Kristine Lacek; Mark Burroughs; Matthew Schmerer; Mili Sheth; Peter Cook; Sam Shepard; Sarah Nobles; Shoshona Le; Suxiang Tong; Vivien Dugan; Yvette Unoarumhi

EPI_ISL_1700673 | NYC Department of Health and Mental Hygiene | Pathogen Discovery, Respiratory Viruses Branch, Division of Viral Diseases, Centers for Disease Control and Prevention | Adam Retchless; Anna Kelleher; Anna Montmayeur; Anna Uehara; Brian Lynch; Clinton R. Paden; Haibin Wang; Han Jia Justin Ng; Jing Zhang; Justin Lee; Krista Queen; Mark Burroughs; Peter Cook; Rachel Marine; Suxiang Tong; Yan Li; Ying Tao

EPI_ISL_1582179, EPI_ISL_1582180, EPI_ISL_1582181, EPI_ISL_1582182, EPI_ISL_1582183, EPI_ISL_1582184, EPI_ISL_1582185, EPI_ISL_1582186, EPI_ISL_1582187, EPI_ISL_1582188, EPI_ISL_1582189, EPI_ISL_1582190, EPI_ISL_1582191, EPI_ISL_1582192, EPI_ISL_1582193, EPI_ISL_1582194, EPI_ISL_1582195, EPI_ISL_1582196, EPI_ISL_1582197, EPI_ISL_1582198, EPI_ISL_1582199, EPI_ISL_1582200, EPI_ISL_1582201, EPI_ISL_1582202, EPI_ISL_1582203, EPI_ISL_1582204, EPI_ISL_1582205, EPI_ISL_1582206, EPI_ISL_1582207, EPI_ISL_1582208, EPI_ISL_1582209, EPI_ISL_1582210, EPI_ISL_1582211, EPI_ISL_1582212, EPI_ISL_1582213, EPI_ISL_1582230, EPI_ISL_1582322, EPI_ISL_1582323, EPI_ISL_1582324, EPI_ISL_1582325, EPI_ISL_1582326, EPI_ISL_1582327, EPI_ISL_1582328, EPI_ISL_1582329, EPI_ISL_1582330, EPI_ISL_1582331, EPI_ISL_1582332, EPI_ISL_1582333, EPI_ISL_1582334, EPI_ISL_1582335, EPI_ISL_1582336, EPI_ISL_1593870, EPI_ISL_1593871, EPI_ISL_1593878, EPI_ISL_1593883, EPI_ISL_1605704, EPI_ISL_1854696, EPI_ISL_1854697, EPI_ISL_1854698, EPI_ISL_1854699, EPI_ISL_1854700, EPI_ISL_1854701, EPI_ISL_1854702, EPI_ISL_1854703, EPI_ISL_1854704, EPI_ISL_1854705, EPI_ISL_1854706, EPI_ISL_1854707, EPI_ISL_1854708, EPI_ISL_1854709, EPI_ISL_1854710, EPI_ISL_1854711, EPI_ISL_1854712, EPI_ISL_1854713, EPI_ISL_1854714, EPI_ISL_1854715, EPI_ISL_1854716, EPI_ISL_1854717, EPI_ISL_1854718, EPI_ISL_1854719, EPI_ISL_1854720, EPI_ISL_1854721, EPI_ISL_1854722, EPI_ISL_1854723, EPI_ISL_1854724, EPI_ISL_1854725, EPI_ISL_1854726, EPI_ISL_1854727, EPI_ISL_1854728, EPI_ISL_1854729, EPI_ISL_1854730, EPI_ISL_1854731, EPI_ISL_1854732, EPI_ISL_1854733, EPI_ISL_1854734, EPI_ISL_1854735, EPI_ISL_1935619, EPI_ISL_1935628

see above | NYC Pandemic Response Lab | Wadsworth Center, New York State Department of Health | Alexis Russell; Catharine Prussing; Daryl M. Lamson; Erasmus Schneider; Erica Lasek-Nesselquist; John Kelly; Jonathan Plitnick; Kirsten St. George; Matthew Shudt; Melissa A Leisner; Navjot Singh

EPI_ISL_1322745, EPI_ISL_1322751, EPI_ISL_1322755, EPI_ISL_1322756, EPI_ISL_1322762, EPI_ISL_1322763, EPI_ISL_1322764, EPI_ISL_1322765, EPI_ISL_1322766, EPI_ISL_1322767, EPI_ISL_1322768, EPI_ISL_1322769, EPI_ISL_1322770, EPI_ISL_1322771, EPI_ISL_1322772, EPI_ISL_1322773, EPI_ISL_1322774, EPI_ISL_1322775, EPI_ISL_1322776, EPI_ISL_1322777, EPI_ISL_1322778, EPI_ISL_1322779, EPI_ISL_1322780, EPI_ISL_1322781, EPI_ISL_1322782, EPI_ISL_1322783, EPI_ISL_1322784, EPI_ISL_1322785, EPI_ISL_1322786, EPI_ISL_1322787, EPI_ISL_1322788, EPI_ISL_1322789, EPI_ISL_1322793, EPI_ISL_1322794, EPI_ISL_1322796, EPI_ISL_1322798, EPI_ISL_1322799, EPI_ISL_1322807, EPI_ISL_1322808, EPI_ISL_1322809, EPI_ISL_1322810, EPI_ISL_1322811, EPI_ISL_1322812, EPI_ISL_1322813, EPI_ISL_1322814, EPI_ISL_1322815, EPI_ISL_1322816, EPI_ISL_1322817, EPI_ISL_1322818, EPI_ISL_1322819, EPI_ISL_1322821, EPI_ISL_1322822, EPI_ISL_1322823, EPI_ISL_1322824, EPI_ISL_1322825, EPI_ISL_1322826, EPI_ISL_1322827, EPI_ISL_1322828, EPI_ISL_1322829, EPI_ISL_1322830, EPI_ISL_1322831, EPI_ISL_1322832, EPI_ISL_1322834, EPI_ISL_1423412, EPI_ISL_1423414, EPI_ISL_1423415, EPI_ISL_1423416, EPI_ISL_1423417, EPI_ISL_1423418, EPI_ISL_1423419, EPI_ISL_1423420, EPI_ISL_1423421, EPI_ISL_1423422, EPI_ISL_1423423, EPI_ISL_1423424, EPI_ISL_1423425, EPI_ISL_1423426, EPI_ISL_1423427, EPI_ISL_1423428, EPI_ISL_1423429, EPI_ISL_1423430, EPI_ISL_1423431, EPI_ISL_1423432, EPI_ISL_1423433, EPI_ISL_1423434, EPI_ISL_1423435, EPI_ISL_1423436, EPI_ISL_1423437, EPI_ISL_1423438, EPI_ISL_1423439, EPI_ISL_1423440, EPI_ISL_1423441, EPI_ISL_1423442, EPI_ISL_1423443, EPI_ISL_1423444, EPI_ISL_1423447, EPI_ISL_1423448, EPI_ISL_1423449, EPI_ISL_1423450, EPI_ISL_1423451, EPI_ISL_1423452, EPI_ISL_1423453, EPI_ISL_1423454, EPI_ISL_1423455, EPI_ISL_1423456, EPI_ISL_1423457, EPI_ISL_1423458, EPI_ISL_1423459, EPI_ISL_1423460, EPI_ISL_1423461, EPI_ISL_1423462, EPI_ISL_1423463, EPI_ISL_1423464, EPI_ISL_1423465, EPI_ISL_1423466, EPI_ISL_1423467, EPI_ISL_1423468, EPI_ISL_1423469, EPI_ISL_1423470, EPI_ISL_1423471, EPI_ISL_1423472, EPI_ISL_1423473, EPI_ISL_1423474, EPI_ISL_1423475, EPI_ISL_1423476, EPI_ISL_1423477, EPI_ISL_1423478, EPI_ISL_1423479, EPI_ISL_1423480, EPI_ISL_1423481, EPI_ISL_1423483, EPI_ISL_1423484, EPI_ISL_1423485, EPI_ISL_1423486, EPI_ISL_1423487, EPI_ISL_1423488, EPI_ISL_1423489, EPI_ISL_1423490, EPI_ISL_1423492, EPI_ISL_1423493, EPI_ISL_1423494, EPI_ISL_1423495, EPI_ISL_1423496, EPI_ISL_1423497, EPI_ISL_1423498, EPI_ISL_1423499, EPI_ISL_1423500, EPI_ISL_1423501, EPI_ISL_1423502, EPI_ISL_1423503, EPI_ISL_1532731, EPI_ISL_1532732, EPI_ISL_1532733, EPI_ISL_1532734, EPI_ISL_1532735, EPI_ISL_1532736, EPI_ISL_1532737, EPI_ISL_1532738, EPI_ISL_1532739, EPI_ISL_1532740, EPI_ISL_1532741, EPI_ISL_1532742, EPI_ISL_1532743, EPI_ISL_1532744, EPI_ISL_1532746, EPI_ISL_1532747, EPI_ISL_1532748, EPI_ISL_1532749, EPI_ISL_1532750, EPI_ISL_1532751, EPI_ISL_1532752, EPI_ISL_1532753, EPI_ISL_1532754, EPI_ISL_1532755, EPI_ISL_1532756, EPI_ISL_1532757, EPI_ISL_1532758, EPI_ISL_1532759, EPI_ISL_1532760, EPI_ISL_1532761, EPI_ISL_1532762, EPI_ISL_1532763, EPI_ISL_1532765, EPI_ISL_1532766, EPI_ISL_1532767, EPI_ISL_1532769, EPI_ISL_1532770, EPI_ISL_1532771, EPI_ISL_1532772, EPI_ISL_1532773, EPI_ISL_1532774, EPI_ISL_1532775, EPI_ISL_1532776, EPI_ISL_1532777, EPI_ISL_1532778, EPI_ISL_1532779, EPI_ISL_1532780, EPI_ISL_1532781, EPI_ISL_1532783, EPI_ISL_1532784, EPI_ISL_1576862, EPI_ISL_1576863, EPI_ISL_1576864, EPI_ISL_1576865, EPI_ISL_1576866, EPI_ISL_1576867, EPI_ISL_1576868, EPI_ISL_1576869, EPI_ISL_1576870, EPI_ISL_1576871, EPI_ISL_1576872, EPI_ISL_1576873, EPI_ISL_1576874, EPI_ISL_1576875, EPI_ISL_1576876, EPI_ISL_1576877, EPI_ISL_1576878, EPI_ISL_1576879, EPI_ISL_1576880, EPI_ISL_1576881, EPI_ISL_1576882, EPI_ISL_1576883, EPI_ISL_1576884, EPI_ISL_1576885, EPI_ISL_1576886, EPI_ISL_1576887, EPI_ISL_1576888, EPI_ISL_1576889, EPI_ISL_1576890, EPI_ISL_1576891, EPI_ISL_1576892, EPI_ISL_1576893, EPI_ISL_1576894, EPI_ISL_1576895, EPI_ISL_1576896, EPI_ISL_1576897, EPI_ISL_1576898, EPI_ISL_1576899, EPI_ISL_1576900, EPI_ISL_1576902, EPI_ISL_1576904, EPI_ISL_1576906, EPI_ISL_1576907, EPI_ISL_1576908, EPI_ISL_1576909, EPI_ISL_1576910, EPI_ISL_1576911, EPI_ISL_1576912, EPI_ISL_1576913, EPI_ISL_1576914, EPI_ISL_1576915, EPI_ISL_1576916, EPI_ISL_1576917, EPI_ISL_1576918, EPI_ISL_1576919, EPI_ISL_1576920, EPI_ISL_1576921, EPI_ISL_1576922, EPI_ISL_1576923, EPI_ISL_1576924, EPI_ISL_1576925, EPI_ISL_1576926, EPI_ISL_1576927, EPI_ISL_1576928, EPI_ISL_1576929, EPI_ISL_1576931, EPI_ISL_1576932, EPI_ISL_1576936, EPI_ISL_1576946, EPI_ISL_1576948, EPI_ISL_1654574, EPI_ISL_1654575, EPI_ISL_1654576, EPI_ISL_1654577, EPI_ISL_1654578, EPI_ISL_1654579, EPI_ISL_1654580, EPI_ISL_1654581, EPI_ISL_1654582, EPI_ISL_1654583, EPI_ISL_1654584, EPI_ISL_1654585, EPI_ISL_1654587, EPI_ISL_1654588, EPI_ISL_1654589, EPI_ISL_1654590, EPI_ISL_1654591, EPI_ISL_1654592, EPI_ISL_1654593, EPI_ISL_1654594, EPI_ISL_1654595, EPI_ISL_1654596, EPI_ISL_1654597, EPI_ISL_1654599, EPI_ISL_1654600, EPI_ISL_1654601, EPI_ISL_1654602, EPI_ISL_1654603, EPI_ISL_1654604, EPI_ISL_1654605, EPI_ISL_1654606, EPI_ISL_1654607, EPI_ISL_1654608, EPI_ISL_1654609, EPI_ISL_1654610, EPI_ISL_1669821, EPI_ISL_1790045, EPI_ISL_1790046, EPI_ISL_1922854, EPI_ISL_1922855, EPI_ISL_1922856, EPI_ISL_1922860, EPI_ISL_1922864, EPI_ISL_1922865, EPI_ISL_1922868, EPI_ISL_1922869

see above | NYU Langone Health | Departments of Pathology and Medicine, New York University School of Medicine | Adriana Heguy; Christian Marier; Dacia Dimartino; Emily Guzman; Gael Westby; Guiqing Wang; Paolo Cotzia; Paul Zappile; Peter Meyn; Sitharam Ramaswami; Yutong Zhang

EPI_ISL_1920166, EPI_ISL_1920168 | Northwell Health Laboratories | Wadsworth Center, New York State Department of Health | Alexis Russell; Catharine Prussing; Daryl M. Lamson; Erasmus Schneider; Erica Lasek-Nesselquist; John Kelly; Jonathan Plitnick; Kirsten St. George; Matthew Shudt; Melissa A Leisner; Navjot Singh

EPI_ISL_1337482, EPI_ISL_1337529, EPI_ISL_1337539, EPI_ISL_1337546, EPI_ISL_1447091, EPI_ISL_1447100, EPI_ISL_1447104, EPI_ISL_1447111, EPI_ISL_1447114, EPI_ISL_1447119, EPI_ISL_1447138, EPI_ISL_1447177, EPI_ISL_1447178, EPI_ISL_1447183, EPI_ISL_1593756, EPI_ISL_1593757, EPI_ISL_1593772, EPI_ISL_1694721, EPI_ISL_1694722, EPI_ISL_1694723, EPI_ISL_1694724, EPI_ISL_1694725, EPI_ISL_1694726, EPI_ISL_1694727 see above | OCME Office Of Chief Medical Examiner | New York City Public Health Laboratory | Jade Wang; et al.

EPI_ISL_1306253, EPI_ISL_1306254, EPI_ISL_1306255, EPI_ISL_1306256, EPI_ISL_1306257, EPI_ISL_1306258, EPI_ISL_1306259, EPI_ISL_1306260, EPI_ISL_1306261, EPI_ISL_1306262, EPI_ISL_1306263, EPI_ISL_1306264, EPI_ISL_1306265, EPI_ISL_1306266, EPI_ISL_1306267, EPI_ISL_1306268, EPI_ISL_1306269, EPI_ISL_1306270, EPI_ISL_1306271, EPI_ISL_1306272, EPI_ISL_1306273, EPI_ISL_1306274,
EPI_ISL_1306275, EPI_ISL_1306276, EPI_ISL_1306277, EPI_ISL_1306278, EPI_ISL_1306279, EPI_ISL_1306280, EPI_ISL_1306281, EPI_ISL_1306282, EPI_ISL_1306283, EPI_ISL_1306284, EPI_ISL_1306285, EPI_ISL_1306286, EPI_ISL_1306287, EPI_ISL_1306288, EPI_ISL_1306289, EPI_ISL_1306290, EPI_ISL_1306291, EPI_ISL_1306292, EPI_ISL_1306293, EPI_ISL_1306294, EPI_ISL_1306295, EPI_ISL_1306296,
EPI_ISL_1306297, EPI_ISL_1306298, EPI_ISL_1306299, EPI_ISL_1306300, EPI_ISL_1306301, EPI_ISL_1306302, EPI_ISL_1306303, EPI_ISL_1306304, EPI_ISL_1306305, EPI_ISL_1306306, EPI_ISL_1306307, EPI_ISL_1306308, EPI_ISL_1306309, EPI_ISL_1306310, EPI_ISL_1306311, EPI_ISL_1306312, EPI_ISL_1306313, EPI_ISL_1306314, EPI_ISL_1306315, EPI_ISL_1306316, EPI_ISL_1306317, EPI_ISL_1306318,
EPI_ISL_1306319, EPI_ISL_1306320, EPI_ISL_1306321, EPI_ISL_1306322, EPI_ISL_1306323, EPI_ISL_1306324, EPI_ISL_1306325, EPI_ISL_1306326, EPI_ISL_1306327, EPI_ISL_1306328, EPI_ISL_1306329, EPI_ISL_1306330, EPI_ISL_1306331, EPI_ISL_1306332, EPI_ISL_1306333, EPI_ISL_1306334, EPI_ISL_1306335, EPI_ISL_1306336, EPI_ISL_1306337, EPI_ISL_1306338, EPI_ISL_1306339, EPI_ISL_1306340,
EPI_ISL_1306341, EPI_ISL_1306342, EPI_ISL_1306343, EPI_ISL_1306344, EPI_ISL_1306345, EPI_ISL_1306346, EPI_ISL_1306347, EPI_ISL_1306348, EPI_ISL_1306349, EPI_ISL_1306350, EPI_ISL_1306351, EPI_ISL_1306352, EPI_ISL_1306353, EPI_ISL_1306354, EPI_ISL_1306355, EPI_ISL_1306356, EPI_ISL_1306357, EPI_ISL_1306358, EPI_ISL_1306359, EPI_ISL_1306360, EPI_ISL_1306361, EPI_ISL_1306362,
EPI_ISL_1306363, EPI_ISL_1306364, EPI_ISL_1306365, EPI_ISL_1306366, EPI_ISL_1306367, EPI_ISL_1306368, EPI_ISL_1306369, EPI_ISL_1306370, EPI_ISL_1306371, EPI_ISL_1306372, EPI_ISL_1306373, EPI_ISL_1306374, EPI_ISL_1306375, EPI_ISL_1306376, EPI_ISL_1306377, EPI_ISL_1306378, EPI_ISL_1306379, EPI_ISL_1306380, EPI_ISL_1306381, EPI_ISL_1306382, EPI_ISL_1306383, EPI_ISL_1306384,
EPI_ISL_1306385, EPI_ISL_1306386, EPI_ISL_1306387, EPI_ISL_1306388, EPI_ISL_1306389, EPI_ISL_1306390, EPI_ISL_1306391, EPI_ISL_1306392, EPI_ISL_1306393, EPI_ISL_1306394, EPI_ISL_1306395, EPI_ISL_1306396, EPI_ISL_1306397, EPI_ISL_1306398, EPI_ISL_1306399, EPI_ISL_1306400, EPI_ISL_1306401, EPI_ISL_1306402, EPI_ISL_1306403, EPI_ISL_1306404, EPI_ISL_1306405, EPI_ISL_1306406,
EPI_ISL_1306407, EPI_ISL_1306408, EPI_ISL_1306409, EPI_ISL_1306410, EPI_ISL_1306411, EPI_ISL_1306412, EPI_ISL_1306413, EPI_ISL_1306414, EPI_ISL_1306415, EPI_ISL_1306416, EPI_ISL_1306417, EPI_ISL_1306418, EPI_ISL_1306419, EPI_ISL_1306420, EPI_ISL_1306421, EPI_ISL_1306422, EPI_ISL_1306423, EPI_ISL_1306424, EPI_ISL_1306425, EPI_ISL_1306426, EPI_ISL_1306427, EPI_ISL_1306428,
EPI_ISL_1306429, EPI_ISL_1306430, EPI_ISL_1306431, EPI_ISL_1306432, EPI_ISL_1306433, EPI_ISL_1306434, EPI_ISL_1306435, EPI_ISL_1306436, EPI_ISL_1306437, EPI_ISL_1306438, EPI_ISL_1306439, EPI_ISL_1306440, EPI_ISL_1306441, EPI_ISL_1306442, EPI_ISL_1306443, EPI_ISL_1306444, EPI_ISL_1306445, EPI_ISL_1306446, EPI_ISL_1306447, EPI_ISL_1306448, EPI_ISL_1306449, EPI_ISL_1306450,
EPI_ISL_1306451, EPI_ISL_1306452, EPI_ISL_1306453, EPI_ISL_1306454, EPI_ISL_1306455, EPI_ISL_1306456, EPI_ISL_1306457, EPI_ISL_1306458, EPI_ISL_1306459, EPI_ISL_1306460, EPI_ISL_1306461, EPI_ISL_1306462, EPI_ISL_1306463, EPI_ISL_1306464, EPI_ISL_1306465, EPI_ISL_1306466, EPI_ISL_1306467, EPI_ISL_1306468, EPI_ISL_1306469, EPI_ISL_1306470, EPI_ISL_1306471, EPI_ISL_1306472,
EPI_ISL_1306473, EPI_ISL_1306474, EPI_ISL_1306475, EPI_ISL_1306476, EPI_ISL_1306477, EPI_ISL_1306478, EPI_ISL_1306479, EPI_ISL_1306480, EPI_ISL_1306481, EPI_ISL_1306482, EPI_ISL_1306483, EPI_ISL_1306484, EPI_ISL_1306485, EPI_ISL_1306486, EPI_ISL_1306487, EPI_ISL_1306488, EPI_ISL_1306489, EPI_ISL_1306490, EPI_ISL_1306491, EPI_ISL_1306492, EPI_ISL_1306493, EPI_ISL_1306494,
EPI_ISL_1306495, EPI_ISL_1306496, EPI_ISL_1306497, EPI_ISL_1306498, EPI_ISL_1306499, EPI_ISL_1306500, EPI_ISL_1306501, EPI_ISL_1306502, EPI_ISL_1306503, EPI_ISL_1306504, EPI_ISL_1306505, EPI_ISL_1306506, EPI_ISL_1306507, EPI_ISL_1306508, EPI_ISL_1306509, EPI_ISL_1306510, EPI_ISL_1306511, EPI_ISL_1306512, EPI_ISL_1306513, EPI_ISL_1306514, EPI_ISL_1306515, EPI_ISL_1306516,
EPI_ISL_1306517, EPI_ISL_1306518, EPI_ISL_1306519, EPI_ISL_1306520, EPI_ISL_1306521, EPI_ISL_1306522, EPI_ISL_1306523, EPI_ISL_1306524, EPI_ISL_1306525, EPI_ISL_1306526, EPI_ISL_1306527, EPI_ISL_1306528, EPI_ISL_1306529, EPI_ISL_1306530, EPI_ISL_1306531, EPI_ISL_1306532, EPI_ISL_1306533, EPI_ISL_1306534, EPI_ISL_1306535, EPI_ISL_1306536, EPI_ISL_1306537, EPI_ISL_1306538,
EPI_ISL_1306539, EPI_ISL_1306540, EPI_ISL_1306541, EPI_ISL_1306542, EPI_ISL_1306543, EPI_ISL_1306544, EPI_ISL_1306545, EPI_ISL_1306546, EPI_ISL_1306547, EPI_ISL_1306548, EPI_ISL_1306549, EPI_ISL_1306550, EPI_ISL_1306551, EPI_ISL_1306552, EPI_ISL_1306553, EPI_ISL_1306554, EPI_ISL_1306555, EPI_ISL_1306556, EPI_ISL_1306557, EPI_ISL_1306558, EPI_ISL_1306559, EPI_ISL_1306560,
EPI_ISL_1306561, EPI_ISL_1306562, EPI_ISL_1306563, EPI_ISL_1306564, EPI_ISL_1306565, EPI_ISL_1306566, EPI_ISL_1306567, EPI_ISL_1306568, EPI_ISL_1306569, EPI_ISL_1306570, EPI_ISL_1306571, EPI_ISL_1306572, EPI_ISL_1306573, EPI_ISL_1306574, EPI_ISL_1306575, EPI_ISL_1306576, EPI_ISL_1306577, EPI_ISL_1306578, EPI_ISL_1306579, EPI_ISL_1306580, EPI_ISL_1306581, EPI_ISL_1306582,
EPI_ISL_1306583, EPI_ISL_1306584, EPI_ISL_1306585, EPI_ISL_1306586, EPI_ISL_1306587, EPI_ISL_1306588, EPI_ISL_1306589, EPI_ISL_1306590, EPI_ISL_1306591, EPI_ISL_1306592, EPI_ISL_1306593, EPI_ISL_1306594, EPI_ISL_1306595, EPI_ISL_1306596, EPI_ISL_1306597, EPI_ISL_1306598, EPI_ISL_1306599, EPI_ISL_1306600, EPI_ISL_1306601, EPI_ISL_1306602, EPI_ISL_1306603, EPI_ISL_1306604,
EPI_ISL_1306605, EPI_ISL_1306606, EPI_ISL_1306607, EPI_ISL_1306608, EPI_ISL_1306609, EPI_ISL_1306610, EPI_ISL_1306611, EPI_ISL_1306612, EPI_ISL_1306613, EPI_ISL_1306614, EPI_ISL_1306615, EPI_ISL_1306616, EPI_ISL_1306617, EPI_ISL_1306618, EPI_ISL_1306619, EPI_ISL_1306620, EPI_ISL_1306621, EPI_ISL_1306622, EPI_ISL_1306623, EPI_ISL_1306624, EPI_ISL_1306625, EPI_ISL_1306626,
EPI_ISL_1306627, EPI_ISL_1306628, EPI_ISL_1306629, EPI_ISL_1306630, EPI_ISL_1306631, EPI_ISL_1306632, EPI_ISL_1306633, EPI_ISL_1306634, EPI_ISL_1306635, EPI_ISL_1306636, EPI_ISL_1306637, EPI_ISL_1306638, EPI_ISL_1306639, EPI_ISL_1306640, EPI_ISL_1306641, EPI_ISL_1306642, EPI_ISL_1306643, EPI_ISL_1306644, EPI_ISL_1306645, EPI_ISL_1306646, EPI_ISL_1306647, EPI_ISL_1306648,
EPI_ISL_1306649, EPI_ISL_1306650, EPI_ISL_1306651, EPI_ISL_1306652, EPI_ISL_1306653, EPI_ISL_1306654, EPI_ISL_1306655, EPI_ISL_1306656, EPI_ISL_1306657, EPI_ISL_1306658, EPI_ISL_1306659, EPI_ISL_1306660, EPI_ISL_1306661, EPI_ISL_1306662, EPI_ISL_1306663, EPI_ISL_1306664, EPI_ISL_1306665, EPI_ISL_1306666, EPI_ISL_1306667, EPI_ISL_1306668, EPI_ISL_1306669, EPI_ISL_1306670,
EPI_ISL_1306671, EPI_ISL_1306672, EPI_ISL_1306673, EPI_ISL_1306674, EPI_ISL_1306675, EPI_ISL_1306676, EPI_ISL_1306677, EPI_ISL_1306678, EPI_ISL_1306679, EPI_ISL_1306680, EPI_ISL_1306681, EPI_ISL_1306682, EPI_ISL_1306683, EPI_ISL_1306684, EPI_ISL_1306685, EPI_ISL_1306686, EPI_ISL_1306687, EPI_ISL_1306688, EPI_ISL_1306689, EPI_ISL_1306690, EPI_ISL_1306691, EPI_ISL_1306692,
EPI_ISL_1306693, EPI_ISL_1306694, EPI_ISL_1306695, EPI_ISL_1306696, EPI_ISL_1306697, EPI_ISL_1306698, EPI_ISL_1306699, EPI_ISL_1306700, EPI_ISL_1306701, EPI_ISL_1306702, EPI_ISL_1306703, EPI_ISL_1306704, EPI_ISL_1306705, EPI_ISL_1306706, EPI_ISL_1306707, EPI_ISL_1306708, EPI_ISL_1306709, EPI_ISL_1306710, EPI_ISL_1306711, EPI_ISL_1306712, EPI_ISL_1306713, EPI_ISL_1306714,
EPI_ISL_1306715, EPI_ISL_1306716, EPI_ISL_1306717, EPI_ISL_1306718, EPI_ISL_1306719, EPI_ISL_1306720, EPI_ISL_1306721, EPI_ISL_1306722, EPI_ISL_1306723, EPI_ISL_1306724, EPI_ISL_1306725, EPI_ISL_1306726, EPI_ISL_1306727, EPI_ISL_1306728, EPI_ISL_1306729, EPI_ISL_1306730, EPI_ISL_1306731, EPI_ISL_1306732, EPI_ISL_1306733, EPI_ISL_1306734, EPI_ISL_1306735, EPI_ISL_1306736,
EPI_ISL_1306737, EPI_ISL_1306738, EPI_ISL_1306739, EPI_ISL_1306740, EPI_ISL_1306741, EPI_ISL_1306742, EPI_ISL_1306743, EPI_ISL_1306744, EPI_ISL_1306745, EPI_ISL_1306746, EPI_ISL_1306747, EPI_ISL_1306748, EPI_ISL_1306749, EPI_ISL_1306750, EPI_ISL_1306751, EPI_ISL_1306752, EPI_ISL_1306753, EPI_ISL_1306754, EPI_ISL_1306755, EPI_ISL_1306756, EPI_ISL_1306757, EPI_ISL_1306758,
EPI_ISL_1306759, EPI_ISL_1306760, EPI_ISL_1306761, EPI_ISL_1306762, EPI_ISL_1306763, EPI_ISL_1306764, EPI_ISL_1306765, EPI_ISL_1306766, EPI_ISL_1306767, EPI_ISL_1306768, EPI_ISL_1306769, EPI_ISL_1306770, EPI_ISL_1306771, EPI_ISL_1306772, EPI_ISL_1306773, EPI_ISL_1306774, EPI_ISL_1306775, EPI_ISL_1306776, EPI_ISL_1306777, EPI_ISL_1306778, EPI_ISL_1306779, EPI_ISL_1306780,
EPI_ISL_1306781, EPI_ISL_1306782, EPI_ISL_1306783, EPI_ISL_1306784, EPI_ISL_1306785, EPI_ISL_1306786, EPI_ISL_1306787, EPI_ISL_1306788, EPI_ISL_1306789, EPI_ISL_1306790, EPI_ISL_1306791, EPI_ISL_1306792, EPI_ISL_1306793, EPI_ISL_1306794, EPI_ISL_1306795, EPI_ISL_1306796, EPI_ISL_1306797, EPI_ISL_1306798, EPI_ISL_1306799, EPI_ISL_1306800, EPI_ISL_1306801, EPI_ISL_1306802,
EPI_ISL_1306803, EPI_ISL_1306804, EPI_ISL_1306805, EPI_ISL_1306806, EPI_ISL_1306807, EPI_ISL_1306808, EPI_ISL_1306809, EPI_ISL_1306810, EPI_ISL_1306811, EPI_ISL_1306812, EPI_ISL_1306813, EPI_ISL_1306814, EPI_ISL_1306815, EPI_ISL_1306816, EPI_ISL_1306817, EPI_ISL_1306818, EPI_ISL_1306819, EPI_ISL_1306820, EPI_ISL_1306821, EPI_ISL_1306822, EPI_ISL_1306823, EPI_ISL_1306824,
EPI_ISL_1306825, EPI_ISL_1306826, EPI_ISL_1306827, EPI_ISL_1306828, EPI_ISL_1306829, EPI_ISL_1306830, EPI_ISL_1306831, EPI_ISL_1306832, EPI_ISL_1306833, EPI_ISL_1306834, EPI_ISL_1306835, EPI_ISL_1306836, EPI_ISL_1306837, EPI_ISL_1306838, EPI_ISL_1306839, EPI_ISL_1306840, EPI_ISL_1306841, EPI_ISL_1306842, EPI_ISL_1306843, EPI_ISL_1306844, EPI_ISL_1306845, EPI_ISL_1306846,
EPI_ISL_1306847, EPI_ISL_1306848, EPI_ISL_1306849, EPI_ISL_1306850, EPI_ISL_1306851, EPI_ISL_1306852, EPI_ISL_1306853, EPI_ISL_1306854, EPI_ISL_1306855, EPI_ISL_1306856, EPI_ISL_1306857, EPI_ISL_1306858, EPI_ISL_1306859, EPI_ISL_1306860, EPI_ISL_1306861, EPI_ISL_1306862, EPI_ISL_1306863, EPI_ISL_1306864, EPI_ISL_1306865, EPI_ISL_1306866, EPI_ISL_1306867, EPI_ISL_1306868,
EPI_ISL_1306869, EPI_ISL_1306870, EPI_ISL_1306871, EPI_ISL_1306872, EPI_ISL_1306873, EPI_ISL_1306874, EPI_ISL_1306875, EPI_ISL_1306876, EPI_ISL_1306877, EPI_ISL_1306878, EPI_ISL_1306879, EPI_ISL_1306880, EPI_ISL_1306881, EPI_ISL_1306882, EPI_ISL_1306883, EPI_ISL_1306884, EPI_ISL_1306885, EPI_ISL_1306886, EPI_ISL_1306887, EPI_ISL_1306888, EPI_ISL_1306889, EPI_ISL_1306890,
EPI_ISL_1306891, EPI_ISL_1306892, EPI_ISL_1306893, EPI_ISL_1306894, EPI_ISL_1306895, EPI_ISL_1306896, EPI_ISL_1306897, EPI_ISL_1306898, EPI_ISL_1306899, EPI_ISL_1306900, EPI_ISL_1306901, EPI_ISL_1306902, EPI_ISL_1306903, EPI_ISL_1306904, EPI_ISL_1306905, EPI_ISL_1306906, EPI_ISL_1306907, EPI_ISL_1306908, EPI_ISL_1306909, EPI_ISL_1306910, EPI_ISL_1306911, EPI_ISL_1306912,
EPI_ISL_1306913, EPI_ISL_1306914, EPI_ISL_1306915, EPI_ISL_1306916, EPI_ISL_1306917, EPI_ISL_1306918, EPI_ISL_1306919, EPI_ISL_1306920, EPI_ISL_1306921, EPI_ISL_1306922, EPI_ISL_1306923, EPI_ISL_1306924, EPI_ISL_1306925, EPI_ISL_1306926, EPI_ISL_1306927, EPI_ISL_1306928, EPI_ISL_1306929, EPI_ISL_1306930, EPI_ISL_1306931, EPI_ISL_1306932, EPI_ISL_1306933, EPI_ISL_1306934,
EPI_ISL_1306935, EPI_ISL_1306936, EPI_ISL_1306937, EPI_ISL_1306938, EPI_ISL_1306939, EPI_ISL_1306940, EPI_ISL_1306941, EPI_ISL_1306942, EPI_ISL_1306943, EPI_ISL_1306944, EPI_ISL_1306945, EPI_ISL_1306946, EPI_ISL_1306947, EPI_ISL_1306948, EPI_ISL_1306949, EPI_ISL_1306950, EPI_ISL_1306951, EPI_ISL_1306952, EPI_ISL_1306953, EPI_ISL_1306954, EPI_ISL_1306955, EPI_ISL_1306956,
EPI_ISL_1306957, EPI_ISL_1306958, EPI_ISL_1306959, EPI_ISL_1306960, EPI_ISL_1306961, EPI_ISL_1306962, EPI_ISL_1306963, EPI_ISL_1306964, EPI_ISL_1306965, EPI_ISL_1306966, EPI_ISL_1306967, EPI_ISL_1306968, EPI_ISL_1306969, EPI_ISL_1306970, EPI_ISL_1306971, EPI_ISL_1306972, EPI_ISL_1306973, EPI_ISL_1306974, EPI_ISL_1306975, EPI_ISL_1306976, EPI_ISL_1306977, EPI_ISL_1306978,
EPI_ISL_1306979, EPI_ISL_1306980, EPI_ISL_1306981, EPI_ISL_1306982, EPI_ISL_1306983, EPI_ISL_1306984, EPI_ISL_1306985, EPI_ISL_1306986, EPI_ISL_1306987, EPI_ISL_1306988, EPI_ISL_1306989, EPI_ISL_1306990, EPI_ISL_1306991, EPI_ISL_1306992, EPI_ISL_1306993, EPI_ISL_1306994, EPI_ISL_1306995, EPI_ISL_1306996, EPI_ISL_1306997, EPI_ISL_1306998, EPI_ISL_1306999, EPI_ISL_1307000,
EPI_ISL_1307001, EPI_ISL_1307002, EPI_ISL_1307003, EPI_ISL_1307004, EPI_ISL_1307005, EPI_ISL_1307006, EPI_ISL_1307007, EPI_ISL_1307008, EPI_ISL_1307009, EPI_ISL_1307010, EPI_ISL_1307011, EPI_ISL_1307012, EPI_ISL_1307013, EPI_ISL_1307014, EPI_ISL_1307015, EPI_ISL_1307016, EPI_ISL_1307017, EPI_ISL_1307018, EPI_ISL_1307019, EPI_ISL_1307020, EPI_ISL_1307021, EPI_ISL_1307022,
EPI_ISL_1307023, EPI_ISL_1307024, EPI_ISL_1307025, EPI_ISL_1307026, EPI_ISL_1307027, EPI_ISL_1307028, EPI_ISL_1307029, EPI_ISL_1307030, EPI_ISL_1307031, EPI_ISL_1307032, EPI_ISL_1307033, EPI_ISL_1307034, EPI_ISL_1307035, EPI_ISL_1307036, EPI_ISL_1307037, EPI_ISL_1307038, EPI_ISL_1307039, EPI_ISL_1307040, EPI_ISL_1307041, EPI_ISL_1307042, EPI_ISL_1307043, EPI_ISL_1307044,
EPI_ISL_1307045, EPI_ISL_1307046, EPI_ISL_1307047, EPI_ISL_1307501, EPI_ISL_1307503, EPI_ISL_1307515, EPI_ISL_1307516, EPI_ISL_1307521, EPI_ISL_1307522, EPI_ISL_1307523, EPI_ISL_1307524, EPI_ISL_1307525, EPI_ISL_1307526, EPI_ISL_1307527, EPI_ISL_1307528, EPI_ISL_1307529, EPI_ISL_1307533, EPI_ISL_1307534, EPI_ISL_1307536, EPI_ISL_1307537, EPI_ISL_1307538, EPI_ISL_1307539,
EPI_ISL_1307540, EPI_ISL_1307541, EPI_ISL_1307542, EPI_ISL_1307543, EPI_ISL_1307544, EPI_ISL_1307545, EPI_ISL_1307569, EPI_ISL_1307587, EPI_ISL_1307588, EPI_ISL_1307589, EPI_ISL_1307590, EPI_ISL_1307591, EPI_ISL_1307592, EPI_ISL_1307593, EPI_ISL_1307594, EPI_ISL_1307596, EPI_ISL_1307597, EPI_ISL_1307598, EPI_ISL_1307599, EPI_ISL_1307600, EPI_ISL_1307601, EPI_ISL_1307608,
EPI_ISL_1307609, EPI_ISL_1307610, EPI_ISL_1307611, EPI_ISL_1307612, EPI_ISL_1307613, EPI_ISL_1307614, EPI_ISL_1307615, EPI_ISL_1307616, EPI_ISL_1307617, EPI_ISL_1307618, EPI_ISL_1307619, EPI_ISL_1307620, EPI_ISL_1307621, EPI_ISL_1307622, EPI_ISL_1307623, EPI_ISL_1307624, EPI_ISL_1307625, EPI_ISL_1307626, EPI_ISL_1307627, EPI_ISL_1307628, EPI_ISL_1307629, EPI_ISL_1307631,
EPI_ISL_1307632, EPI_ISL_1307633, EPI_ISL_1307634, EPI_ISL_1384885, EPI_ISL_1384886, EPI_ISL_1384887, EPI_ISL_1384888, EPI_ISL_1384889, EPI_ISL_1384890, EPI_ISL_1384891, EPI_ISL_1384892, EPI_ISL_1384893, EPI_ISL_1384894, EPI_ISL_1384895, EPI_ISL_1384896, EPI_ISL_1384897, EPI_ISL_1384898, EPI_ISL_1384899, EPI_ISL_1384900, EPI_ISL_1384901, EPI_ISL_1384902, EPI_ISL_1384903,
EPI_ISL_1384904, EPI_ISL_1384905, EPI_ISL_1384906, EPI_ISL_1384907, EPI_ISL_1384908, EPI_ISL_1384909, EPI_ISL_1384910, EPI_ISL_1384911, EPI_ISL_1384912, EPI_ISL_1384913, EPI_ISL_1384914, EPI_ISL_1384915, EPI_ISL_1384916, EPI_ISL_1384917, EPI_ISL_1384918, EPI_ISL_1384919, EPI_ISL_1384920, EPI_ISL_1384921, EPI_ISL_1384922, EPI_ISL_1384923, EPI_ISL_1384924, EPI_ISL_1384925,
EPI_ISL_1384926, EPI_ISL_1384927, EPI_ISL_1384928, EPI_ISL_1384929, EPI_ISL_1384930, EPI_ISL_1384931, EPI_ISL_1384932, EPI_ISL_1384933, EPI_ISL_1384934, EPI_ISL_1384935, EPI_ISL_1384936, EPI_ISL_1384937, EPI_ISL_1384938, EPI_ISL_1384939, EPI_ISL_1384940, EPI_ISL_1384941, EPI_ISL_1384942, EPI_ISL_1384943, EPI_ISL_1384944, EPI_ISL_1384945, EPI_ISL_1384946, EPI_ISL_1384947,
EPI_ISL_1384948, EPI_ISL_1384949, EPI_ISL_1384950, EPI_ISL_1384951, EPI_ISL_1384952, EPI_ISL_1384953, EPI_ISL_1384954, EPI_ISL_1384955, EPI_ISL_1384956, EPI_ISL_1384957, EPI_ISL_1384958, EPI_ISL_1384959, EPI_ISL_1384960, EPI_ISL_1384961, EPI_ISL_1384962, EPI_ISL_1384963, EPI_ISL_1384964, EPI_ISL_1384965, EPI_ISL_1384966, EPI_ISL_1384967, EPI_ISL_1384968, EPI_ISL_1384969,
EPI_ISL_1384970, EPI_ISL_1384971, EPI_ISL_1384972, EPI_ISL_1384973, EPI_ISL_1384974, EPI_ISL_1384975, EPI_ISL_1384976, EPI_ISL_1384977, EPI_ISL_1384978, EPI_ISL_1384979, EPI_ISL_1384980, EPI_ISL_1384981, EPI_ISL_1384982, EPI_ISL_1384983, EPI_ISL_1384984, EPI_ISL_1384985, EPI_ISL_1384986, EPI_ISL_1384987, EPI_ISL_1384988, EPI_ISL_1384989, EPI_ISL_1384990, EPI_ISL_1384991,
EPI_ISL_1384992, EPI_ISL_1384993, EPI_ISL_1384994, EPI_ISL_1384995, EPI_ISL_1384996, EPI_ISL_1384997, EPI_ISL_1384998, EPI_ISL_1384999, EPI_ISL_1385000, EPI_ISL_1385001, EPI_ISL_1385002, EPI_ISL_1385003, EPI_ISL_1385004, EPI_ISL_1385005, EPI_ISL_1385006, EPI_ISL_1385007, EPI_ISL_1385008, EPI_ISL_1385009, EPI_ISL_1385010, EPI_ISL_1385011, EPI_ISL_1385012, EPI_ISL_1385013,
EPI_ISL_1385014, EPI_ISL_1385015, EPI_ISL_1385016, EPI_ISL_1385017, EPI_ISL_1385018, EPI_ISL_1385019, EPI_ISL_1385020, EPI_ISL_1385021, EPI_ISL_1385022, EPI_ISL_1385023, EPI_ISL_1385024, EPI_ISL_1385025, EPI_ISL_1385026, EPI_ISL_1385027, EPI_ISL_1385028, EPI_ISL_1385029, EPI_ISL_1385030, EPI_ISL_1385031, EPI_ISL_1385032, EPI_ISL_1385033, EPI_ISL_1385034, EPI_ISL_1385035,
EPI_ISL_1385036, EPI_ISL_1385037, EPI_ISL_1385038, EPI_ISL_1385039, EPI_ISL_1385040, EPI_ISL_1385041, EPI_ISL_1385042, EPI_ISL_1385043, EPI_ISL_1385044, EPI_ISL_1385045, EPI_ISL_1385046, EPI_ISL_1385047, EPI_ISL_1385048, EPI_ISL_1385049, EPI_ISL_1385050, EPI_ISL_1385051, EPI_ISL_1385052, EPI_ISL_1385053, EPI_ISL_1385054, EPI_ISL_1385055, EPI_ISL_1385056, EPI_ISL_1385057,
EPI_ISL_1385058, EPI_ISL_1385059, EPI_ISL_1385060, EPI_ISL_1385061, EPI_ISL_1385062, EPI_ISL_1385063, EPI_ISL_1385064, EPI_ISL_1385065, EPI_ISL_1385066, EPI_ISL_1385067, EPI_ISL_1385068, EPI_ISL_1385069, EPI_ISL_1385070, EPI_ISL_1385071, EPI_ISL_1385072, EPI_ISL_1385073, EPI_ISL_1385074, EPI_ISL_1385075, EPI_ISL_1385076, EPI_ISL_1385077, EPI_ISL_1385078, EPI_ISL_1385079,
EPI_ISL_1385080, EPI_ISL_1385081, EPI_ISL_1385082, EPI_ISL_1385083, EPI_ISL_1385084, EPI_ISL_1385085, EPI_ISL_1385086, EPI_ISL_1385087, EPI_ISL_1385088, EPI_ISL_1385089, EPI_ISL_1385090, EPI_ISL_1385091, EPI_ISL_1385092, EPI_ISL_1385093, EPI_ISL_1385094, EPI_ISL_1385095, EPI_ISL_1385096, EPI_ISL_1385097, EPI_ISL_1385098, EPI_ISL_1385099, EPI_ISL_1385100, EPI_ISL_1385101,
EPI_ISL_1385102, EPI_ISL_1385103, EPI_ISL_1385104, EPI_ISL_1385105, EPI_ISL_1385106, EPI_ISL_1385107, EPI_ISL_1385108, EPI_ISL_1385109, EPI_ISL_1385110, EPI_ISL_1385111, EPI_ISL_1385112, EPI_ISL_1385113, EPI_ISL_1385114, EPI_ISL_1385115, EPI_ISL_1385116, EPI_ISL_1385117, EPI_ISL_1385118, EPI_ISL_1385119, EPI_ISL_1385120, EPI_ISL_1385121, EPI_ISL_1385122, EPI_ISL_1385123,
EPI_ISL_1385124, EPI_ISL_1385125, EPI_ISL_1385126, EPI_ISL_1385127, EPI_ISL_1385128, EPI_ISL_1385129, EPI_ISL_1385130, EPI_ISL_1385131, EPI_ISL_1385132, EPI_ISL_1385133, EPI_ISL_1385134, EPI_ISL_1385135, EPI_ISL_1385136, EPI_ISL_1385137, EPI_ISL_1385138, EPI_ISL_1385139, EPI_ISL_1385140, EPI_ISL_1385141, EPI_ISL_1385142, EPI_ISL_1385143, EPI_ISL_1385144, EPI_ISL_1385145,
EPI_ISL_1385146, EPI_ISL_1385147, EPI_ISL_1385148, EPI_ISL_1385149, EPI_ISL_1385150, EPI_ISL_1385151, EPI_ISL_1385152, EPI_ISL_1385153, EPI_ISL_1385154, EPI_ISL_1385155, EPI_ISL_1385156, EPI_ISL_1385157, EPI_ISL_1385158, EPI_ISL_1385159, EPI_ISL_1385160, EPI_ISL_1385161, EPI_ISL_1385162, EPI_ISL_1385163, EPI_ISL_1385164, EPI_ISL_1385165, EPI_ISL_1385166, EPI_ISL_1385167,
EPI_ISL_1385168, EPI_ISL_1385169, EPI_ISL_1385170, EPI_ISL_1385171, EPI_ISL_1385172, EPI_ISL_1385173, EPI_ISL_1385174, EPI_ISL_1385175, EPI_ISL_1385176, EPI_ISL_1385177, EPI_ISL_1385178, EPI_ISL_1385179, EPI_ISL_1385180, EPI_ISL_1385181, EPI_ISL_1385182, EPI_ISL_1385183, EPI_ISL_1385184, EPI_ISL_1385185, EPI_ISL_1385186, EPI_ISL_1385187, EPI_ISL_1385188, EPI_ISL_1385189,
EPI_ISL_1385190, EPI_ISL_1385191, EPI_ISL_1385192, EPI_ISL_1385193, EPI_ISL_1385194, EPI_ISL_1385195, EPI_ISL_1385196, EPI_ISL_1385197, EPI_ISL_1385198, EPI_ISL_1385199, EPI_ISL_1385200, EPI_ISL_1385201, EPI_ISL_1385202, EPI_ISL_1385203, EPI_ISL_1385204, EPI_ISL_1385205, EPI_ISL_1385206, EPI_ISL_1385207, EPI_ISL_1385208, EPI_ISL_1385209, EPI_ISL_1385210, EPI_ISL_1385211,
EPI_ISL_1385212, EPI_ISL_1385213, EPI_ISL_1385214, EPI_ISL_1385215, EPI_ISL_1385216, EPI_ISL_1385217, EPI_ISL_1385218, EPI_ISL_1385219, EPI_ISL_1385220, EPI_ISL_1385221, EPI_ISL_1385222, EPI_ISL_1385223, EPI_ISL_1385224, EPI_ISL_1385225, EPI_ISL_1385226, EPI_ISL_1385227, EPI_ISL_1385228, EPI_ISL_1385229, EPI_ISL_1385230, EPI_ISL_1385231, EPI_ISL_1385232, EPI_ISL_1385233,
EPI_ISL_1385234, EPI_ISL_1385235, EPI_ISL_1385236, EPI_ISL_1385237, EPI_ISL_1385238, EPI_ISL_1385239, EPI_ISL_1385240, EPI_ISL_1385241, EPI_ISL_1385242, EPI_ISL_1385243, EPI_ISL_1385244, EPI_ISL_1385245, EPI_ISL_1385246, EPI_ISL_1385247, EPI_ISL_1385248, EPI_ISL_1385249, EPI_ISL_1385250, EPI_ISL_1385251, EPI_ISL_1385252, EPI_ISL_1385253, EPI_ISL_1385254, EPI_ISL_1385255,
EPI_ISL_1385256, EPI_ISL_1385257, EPI_ISL_1385258, EPI_ISL_1385259, EPI_ISL_1385260, EPI_ISL_1385261, EPI_ISL_1385262, EPI_ISL_1385263, EPI_ISL_1385264, EPI_ISL_1385265, EPI_ISL_1385266, EPI_ISL_1385267, EPI_ISL_1385268, EPI_ISL_1385269, EPI_ISL_1385270, EPI_ISL_1385271, EPI_ISL_1385272, EPI_ISL_1385273, EPI_ISL_1385274, EPI_ISL_1385275, EPI_ISL_1385276, EPI_ISL_1385277,
EPI_ISL_1385278, EPI_ISL_1385279, EPI_ISL_1385280, EPI_ISL_1385281, EPI_ISL_1385282, EPI_ISL_1385283, EPI_ISL_1385284, EPI_ISL_1385285, EPI_ISL_1385286, EPI_ISL_1385287, EPI_ISL_1385288, EPI_ISL_1385289, EPI_ISL_1385290, EPI_ISL_1385291, EPI_ISL_1385292, EPI_ISL_1385293, EPI_ISL_1385294, EPI_ISL_1385295, EPI_ISL_1385296, EPI_ISL_1385297, EPI_ISL_1385298, EPI_ISL_1385299,
EPI_ISL_1385300, EPI_ISL_1385301, EPI_ISL_1385302, EPI_ISL_1385303, EPI_ISL_1385304, EPI_ISL_1385305, EPI_ISL_1385306, EPI_ISL_1385307, EPI_ISL_1385308, EPI_ISL_1385309, EPI_ISL_1385310, EPI_ISL_1385311, EPI_ISL_1385312, EPI_ISL_1385313, EPI_ISL_1385314, EPI_ISL_1385315, EPI_ISL_1385316, EPI_ISL_1385317, EPI_ISL_1385318, EPI_ISL_1385319, EPI_ISL_1385320, EPI_ISL_1385321,
EPI_ISL_1385322, EPI_ISL_1385323, EPI_ISL_1385324, EPI_ISL_1385325, EPI_ISL_1385326, EPI_ISL_1385327, EPI_ISL_1385328, EPI_ISL_1385329, EPI_ISL_1385330, EPI_ISL_1385331, EPI_ISL_1385332, EPI_ISL_1385333, EPI_ISL_1385334, EPI_ISL_1385335, EPI_ISL_1385336, EPI_ISL_1385337, EPI_ISL_1385338, EPI_ISL_1385339, EPI_ISL_1385340, EPI_ISL_1385341, EPI_ISL_1385342, EPI_ISL_1385343,
EPI_ISL_1385344, EPI_ISL_1385345, EPI_ISL_1385346, EPI_ISL_1385347, EPI_ISL_1385348, EPI_ISL_1385349, EPI_ISL_1385350, EPI_ISL_1385351, EPI_ISL_1385352, EPI_ISL_1385353, EPI_ISL_1385354, EPI_ISL_1385355, EPI_ISL_1385356, EPI_ISL_1385357, EPI_ISL_1385358, EPI_ISL_1385359, EPI_ISL_1385360, EPI_ISL_1385361, EPI_ISL_1385362, EPI_ISL_1385363, EPI_ISL_1385364, EPI_ISL_1385365,
EPI_ISL_1385366, EPI_ISL_1385367, EPI_ISL_1385368, EPI_ISL_1385369, EPI_ISL_1385370, EPI_ISL_1385371, EPI_ISL_1385372, EPI_ISL_1385373, EPI_ISL_1385374, EPI_ISL_1385375, EPI_ISL_1385376, EPI_ISL_1385377, EPI_ISL_1385378, EPI_ISL_1385379, EPI_ISL_1385380, EPI_ISL_1385381, EPI_ISL_1385382, EPI_ISL_1385383, EPI_ISL_1385384, EPI_ISL_1385385, EPI_ISL_1385386, EPI_ISL_1385387,
EPI_ISL_1385388, EPI_ISL_1385389, EPI_ISL_1385390, EPI_ISL_1385391, EPI_ISL_1385392, EPI_ISL_1385393, EPI_ISL_1385394, EPI_ISL_1385395, EPI_ISL_1385396, EPI_ISL_1385397, EPI_ISL_1385398, EPI_ISL_1385399, EPI_ISL_1385400, EPI_ISL_1385401, EPI_ISL_1385402, EPI_ISL_1385403, EPI_ISL_1385404, EPI_ISL_1385405, EPI_ISL_1385406, EPI_ISL_1385407, EPI_ISL_1385408, EPI_ISL_1385409,
EPI_ISL_1385410, EPI_ISL_1385411, EPI_ISL_1385412, EPI_ISL_1385413, EPI_ISL_1385414, EPI_ISL_1385415, EPI_ISL_1385416, EPI_ISL_1385417, EPI_ISL_1385418, EPI_ISL_1385419, EPI_ISL_1385420, EPI_ISL_1385421, EPI_ISL_1385422, EPI_ISL_1385423, EPI_ISL_1385424, EPI_ISL_1385425, EPI_ISL_1385426, EPI_ISL_1385427, EPI_ISL_1385428, EPI_ISL_1385429, EPI_ISL_1385430, EPI_ISL_1385431,
EPI_ISL_1385432, EPI_ISL_1385433, EPI_ISL_1385434, EPI_ISL_1385435, EPI_ISL_1385436, EPI_ISL_1385437, EPI_ISL_1385438, EPI_ISL_1385439, EPI_ISL_1385440, EPI_ISL_1385441, EPI_ISL_1385442, EPI_ISL_1385443, EPI_ISL_1385444, EPI_ISL_1385445, EPI_ISL_1385446, EPI_ISL_1385447, EPI_ISL_1385448, EPI_ISL_1385449, EPI_ISL_1385450, EPI_ISL_1385451, EPI_ISL_1385452, EPI_ISL_1385453,
EPI_ISL_1385454, EPI_ISL_1385455, EPI_ISL_1385456, EPI_ISL_1385457, EPI_ISL_1385458, EPI_ISL_1385459, EPI_ISL_1385460, EPI_ISL_1385461, EPI_ISL_1385462, EPI_ISL_1385463, EPI_ISL_1385464, EPI_ISL_1385465, EPI_ISL_1385466, EPI_ISL_1385467, EPI_ISL_1385468, EPI_ISL_1385469, EPI_ISL_1385470, EPI_ISL_1385471, EPI_ISL_1385472, EPI_ISL_1385473, EPI_ISL_1385474, EPI_ISL_1385475,
EPI_ISL_1385476, EPI_ISL_1385477, EPI_ISL_1385478, EPI_ISL_1385479, EPI_ISL_1385480, EPI_ISL_1385481, EPI_ISL_1385482, EPI_ISL_1385483, EPI_ISL_1385484, EPI_ISL_1385485, EPI_ISL_1385486, EPI_ISL_1385487, EPI_ISL_1385488, EPI_ISL_1385489, EPI_ISL_1385490, EPI_ISL_1385491, EPI_ISL_1385492, EPI_ISL_1385493, EPI_ISL_1385494, EPI_ISL_1385495, EPI_ISL_1385496, EPI_ISL_1385497,
EPI_ISL_1385498, EPI_ISL_1385499, EPI_ISL_1385500, EPI_ISL_1385501, EPI_ISL_1385502, EPI_ISL_1385503, EPI_ISL_1385504, EPI_ISL_1385505, EPI_ISL_1385506, EPI_ISL_1385507, EPI_ISL_1385508, EPI_ISL_1385509, EPI_ISL_1385510, EPI_ISL_1385511, EPI_ISL_1385512, EPI_ISL_1385513, EPI_ISL_1385514, EPI_ISL_1385515, EPI_ISL_1385516, EPI_ISL_1385517, EPI_ISL_1385518, EPI_ISL_1385519,
EPI_ISL_1385520, EPI_ISL_1385521, EPI_ISL_1385522, EPI_ISL_1385523, EPI_ISL_1385524, EPI_ISL_1385525, EPI_ISL_1385526, EPI_ISL_1385527, EPI_ISL_1385528, EPI_ISL_1385529, EPI_ISL_1385530, EPI_ISL_1385531, EPI_ISL_1385532, EPI_ISL_1385533, EPI_ISL_1385534, EPI_ISL_1385535, EPI_ISL_1385536, EPI_ISL_1385537, EPI_ISL_1385538, EPI_ISL_1385539, EPI_ISL_1385540, EPI_ISL_1385541,
EPI_ISL_1385542, EPI_ISL_1385543, EPI_ISL_1385544, EPI_ISL_1385545, EPI_ISL_1385546, EPI_ISL_1385547, EPI_ISL_1385548, EPI_ISL_1385549, EPI_ISL_1385550, EPI_ISL_1385551, EPI_ISL_1385552, EPI_ISL_1385553, EPI_ISL_1385554, EPI_ISL_1385555, EPI_ISL_1385556, EPI_ISL_1385557, EPI_ISL_1385558, EPI_ISL_1385559, EPI_ISL_1385560, EPI_ISL_1385561, EPI_ISL_1385562, EPI_ISL_1385563,
EPI_ISL_1385564, EPI_ISL_1385565, EPI_ISL_1385566, EPI_ISL_1385567, EPI_ISL_1385568, EPI_ISL_1385569, EPI_ISL_1385570, EPI_ISL_1385571, EPI_ISL_1385572, EPI_ISL_1385573, EPI_ISL_1385574, EPI_ISL_1385575, EPI_ISL_1385576, EPI_ISL_1385577, EPI_ISL_1385578, EPI_ISL_1385579, EPI_ISL_1385580, EPI_ISL_1385581, EPI_ISL_1385582, EPI_ISL_1385583, EPI_ISL_1385584, EPI_ISL_1385585,
EPI_ISL_1385586, EPI_ISL_1385587, EPI_ISL_1385588, EPI_ISL_1385589, EPI_ISL_1385590, EPI_ISL_1385591, EPI_ISL_1385592, EPI_ISL_1385593, EPI_ISL_1385594, EPI_ISL_1385595, EPI_ISL_1385596, EPI_ISL_1385597, EPI_ISL_1385598, EPI_ISL_1385599, EPI_ISL_1385600, EPI_ISL_1385601, EPI_ISL_1385602, EPI_ISL_1385603, EPI_ISL_1385604, EPI_ISL_1385605, EPI_ISL_1385606, EPI_ISL_1385607,
EPI_ISL_1385608, EPI_ISL_1385609, EPI_ISL_1385610, EPI_ISL_1385611, EPI_ISL_1385612, EPI_ISL_1385613, EPI_ISL_1385614, EPI_ISL_1385615, EPI_ISL_1385616, EPI_ISL_1385617, EPI_ISL_1385618, EPI_ISL_1385619, EPI_ISL_1385620, EPI_ISL_1385621, EPI_ISL_1385622, EPI_ISL_1385623, EPI_ISL_1385624, EPI_ISL_1385625, EPI_ISL_1385626, EPI_ISL_1385627, EPI_ISL_1385628, EPI_ISL_1385629,
EPI_ISL_1385630, EPI_ISL_1385631, EPI_ISL_1385632, EPI_ISL_1385633, EPI_ISL_1385634, EPI_ISL_1385635, EPI_ISL_1385636, EPI_ISL_1385637, EPI_ISL_1385638, EPI_ISL_1385639, EPI_ISL_1385640, EPI_ISL_1385641, EPI_ISL_1385642, EPI_ISL_1385643, EPI_ISL_1385644, EPI_ISL_1385645, EPI_ISL_1385646, EPI_ISL_1385647, EPI_ISL_1385648, EPI_ISL_1385649, EPI_ISL_1385650, EPI_ISL_1385651,
EPI_ISL_1385652, EPI_ISL_1385653, EPI_ISL_1385654, EPI_ISL_1385655, EPI_ISL_1385656, EPI_ISL_1385657, EPI_ISL_1385658, EPI_ISL_1385659, EPI_ISL_1385660, EPI_ISL_1385661, EPI_ISL_1385662, EPI_ISL_1385663, EPI_ISL_1385664, EPI_ISL_1385665, EPI_ISL_1385666, EPI_ISL_1385667, EPI_ISL_1385668, EPI_ISL_1385669, EPI_ISL_1385670, EPI_ISL_1385671, EPI_ISL_1385672, EPI_ISL_1385673,
EPI_ISL_1385674, EPI_ISL_1385675, EPI_ISL_1385676, EPI_ISL_1385677, EPI_ISL_1385678, EPI_ISL_1385679, EPI_ISL_1385680, EPI_ISL_1385681, EPI_ISL_1385682, EPI_ISL_1385683, EPI_ISL_1385684, EPI_ISL_1385685, EPI_ISL_1385686, EPI_ISL_1385687, EPI_ISL_1385688, EPI_ISL_1385689, EPI_ISL_1385690, EPI_ISL_1385691, EPI_ISL_1385692, EPI_ISL_1385693, EPI_ISL_1385694, EPI_ISL_1385695,
EPI_ISL_1385696, EPI_ISL_1385697, EPI_ISL_1385698, EPI_ISL_1385699, EPI_ISL_1385700, EPI_ISL_1385701, EPI_ISL_1385702, EPI_ISL_1385703, EPI_ISL_1385704, EPI_ISL_1385705, EPI_ISL_1385706, EPI_ISL_1385707, EPI_ISL_1385708, EPI_ISL_1385709, EPI_ISL_1385710, EPI_ISL_1385711, EPI_ISL_1385712, EPI_ISL_1385713, EPI_ISL_1385714, EPI_ISL_1385715, EPI_ISL_1385716, EPI_ISL_1385717,
EPI_ISL_1385718, EPI_ISL_1385719, EPI_ISL_1385720, EPI_ISL_1385721, EPI_ISL_1385722, EPI_ISL_1385723, EPI_ISL_1385724, EPI_ISL_1385725, EPI_ISL_1385726, EPI_ISL_1385727, EPI_ISL_1385728, EPI_ISL_1385729, EPI_ISL_1385730, EPI_ISL_1385731, EPI_ISL_1385732, EPI_ISL_1385733, EPI_ISL_1385734, EPI_ISL_1385735, EPI_ISL_1385736, EPI_ISL_1385737, EPI_ISL_1385738, EPI_ISL_1385739,
EPI_ISL_1385740, EPI_ISL_1385741, EPI_ISL_1385742, EPI_ISL_1385743, EPI_ISL_1385744, EPI_ISL_1385745, EPI_ISL_1385746, EPI_ISL_1385747, EPI_ISL_1385748, EPI_ISL_1385749, EPI_ISL_1385750, EPI_ISL_1385751, EPI_ISL_1385752, EPI_ISL_1385753, EPI_ISL_1385754, EPI_ISL_1385755, EPI_ISL_1385756, EPI_ISL_1385757, EPI_ISL_1385758, EPI_ISL_1385759, EPI_ISL_1385760, EPI_ISL_1385761,
EPI_ISL_1385762, EPI_ISL_1385763, EPI_ISL_1385764, EPI_ISL_1385765, EPI_ISL_1385766, EPI_ISL_1385767, EPI_ISL_1385768, EPI_ISL_1385769, EPI_ISL_1385770, EPI_ISL_1385771, EPI_ISL_1385772, EPI_ISL_1385773, EPI_ISL_1385774, EPI_ISL_1385775, EPI_ISL_1385776, EPI_ISL_1385777, EPI_ISL_1385778, EPI_ISL_1385779, EPI_ISL_1385780, EPI_ISL_1385781, EPI_ISL_1385782, EPI_ISL_1385783,
EPI_ISL_1470741, EPI_ISL_1470742, EPI_ISL_1470743, EPI_ISL_1470744, EPI_ISL_1470745, EPI_ISL_1470746, EPI_ISL_1470747, EPI_ISL_1470748, EPI_ISL_1470749, EPI_ISL_1470750, EPI_ISL_1470751, EPI_ISL_1470752, EPI_ISL_1470753, EPI_ISL_1470754, EPI_ISL_1470755, EPI_ISL_1470756, EPI_ISL_1470757, EPI_ISL_1470758, EPI_ISL_1470759, EPI_ISL_1470760, EPI_ISL_1470761, EPI_ISL_1470762,
EPI_ISL_1470763, EPI_ISL_1470764, EPI_ISL_1470765, EPI_ISL_1470766, EPI_ISL_1470767, EPI_ISL_1470768, EPI_ISL_1470769, EPI_ISL_1470770, EPI_ISL_1470771, EPI_ISL_1470772, EPI_ISL_1470773, EPI_ISL_1470774, EPI_ISL_1470775, EPI_ISL_1470776, EPI_ISL_1470777, EPI_ISL_1470778, EPI_ISL_1470779, EPI_ISL_1470780, EPI_ISL_1470781, EPI_ISL_1470782, EPI_ISL_1470783, EPI_ISL_1470784,
EPI_ISL_1470785, EPI_ISL_1470786, EPI_ISL_1470787, EPI_ISL_1470788, EPI_ISL_1470789, EPI_ISL_1470790, EPI_ISL_1470791, EPI_ISL_1470792, EPI_ISL_1470793, EPI_ISL_1470794, EPI_ISL_1470795, EPI_ISL_1470796, EPI_ISL_1470797, EPI_ISL_1470798, EPI_ISL_1470799, EPI_ISL_1470800, EPI_ISL_1470801, EPI_ISL_1470802, EPI_ISL_1470803, EPI_ISL_1470804, EPI_ISL_1470805, EPI_ISL_1470806,
EPI_ISL_1470807, EPI_ISL_1470808, EPI_ISL_1470809, EPI_ISL_1470810, EPI_ISL_1470811, EPI_ISL_1470812, EPI_ISL_1470813, EPI_ISL_1470814, EPI_ISL_1470815, EPI_ISL_1470816, EPI_ISL_1470817, EPI_ISL_1470818, EPI_ISL_1470819, EPI_ISL_1470820, EPI_ISL_1470821, EPI_ISL_1470822, EPI_ISL_1470823, EPI_ISL_1470824, EPI_ISL_1470825, EPI_ISL_1470826, EPI_ISL_1470827, EPI_ISL_1470828,
EPI_ISL_1470829, EPI_ISL_1470830, EPI_ISL_1470831, EPI_ISL_1470832, EPI_ISL_1470833, EPI_ISL_1470834, EPI_ISL_1470835, EPI_ISL_1470836, EPI_ISL_1470837, EPI_ISL_1470838, EPI_ISL_1470839, EPI_ISL_1470840, EPI_ISL_1470841, EPI_ISL_1470842, EPI_ISL_1470843, EPI_ISL_1470844, EPI_ISL_1470845, EPI_ISL_1470846, EPI_ISL_1470847, EPI_ISL_1470848, EPI_ISL_1470849, EPI_ISL_1470850,
EPI_ISL_1470851, EPI_ISL_1470852, EPI_ISL_1470853, EPI_ISL_1470854, EPI_ISL_1470855, EPI_ISL_1470856, EPI_ISL_1470857, EPI_ISL_1470858, EPI_ISL_1470859, EPI_ISL_1470860, EPI_ISL_1470861, EPI_ISL_1470862, EPI_ISL_1470863, EPI_ISL_1470864, EPI_ISL_1470865, EPI_ISL_1470866, EPI_ISL_1470867, EPI_ISL_1470868, EPI_ISL_1470869, EPI_ISL_1470870, EPI_ISL_1470871, EPI_ISL_1470872,
EPI_ISL_1470873, EPI_ISL_1470874, EPI_ISL_1470875, EPI_ISL_1470876, EPI_ISL_1470877, EPI_ISL_1470878, EPI_ISL_1470879, EPI_ISL_1470880, EPI_ISL_1470881, EPI_ISL_1470882, EPI_ISL_1470883, EPI_ISL_1470884, EPI_ISL_1470885, EPI_ISL_1470886, EPI_ISL_1470887, EPI_ISL_1470888, EPI_ISL_1470889, EPI_ISL_1470890, EPI_ISL_1470891, EPI_ISL_1470892, EPI_ISL_1470893, EPI_ISL_1470894,
EPI_ISL_1470895, EPI_ISL_1470896, EPI_ISL_1470897, EPI_ISL_1470898, EPI_ISL_1470899, EPI_ISL_1470900, EPI_ISL_1470901, EPI_ISL_1470902, EPI_ISL_1470903, EPI_ISL_1470904, EPI_ISL_1470905, EPI_ISL_1470906, EPI_ISL_1470907, EPI_ISL_1470908, EPI_ISL_1470909, EPI_ISL_1470910, EPI_ISL_1470911, EPI_ISL_1470912, EPI_ISL_1470913, EPI_ISL_1470914, EPI_ISL_1470915, EPI_ISL_1470916,
EPI_ISL_1470917, EPI_ISL_1470918, EPI_ISL_1470919, EPI_ISL_1470920, EPI_ISL_1470921, EPI_ISL_1470922, EPI_ISL_1470923, EPI_ISL_1470924, EPI_ISL_1470925, EPI_ISL_1470926, EPI_ISL_1470927, EPI_ISL_1470928, EPI_ISL_1470929, EPI_ISL_1470930, EPI_ISL_1470931, EPI_ISL_1470932, EPI_ISL_1470933, EPI_ISL_1470934, EPI_ISL_1470935, EPI_ISL_1470936, EPI_ISL_1470937, EPI_ISL_1470938,
EPI_ISL_1470939, EPI_ISL_1470940, EPI_ISL_1470941, EPI_ISL_1470942, EPI_ISL_1470943, EPI_ISL_1470944, EPI_ISL_1470945, EPI_ISL_1470946, EPI_ISL_1470947, EPI_ISL_1470948, EPI_ISL_1470949, EPI_ISL_1470950, EPI_ISL_1470951, EPI_ISL_1470952, EPI_ISL_1470953, EPI_ISL_1470954, EPI_ISL_1470955, EPI_ISL_1470956, EPI_ISL_1470957, EPI_ISL_1470958, EPI_ISL_1470959, EPI_ISL_1470960,
EPI_ISL_1470961, EPI_ISL_1470962, EPI_ISL_1470963, EPI_ISL_1470964, EPI_ISL_1470965, EPI_ISL_1470966, EPI_ISL_1470967, EPI_ISL_1470968, EPI_ISL_1470969, EPI_ISL_1470970, EPI_ISL_1470971, EPI_ISL_1470972, EPI_ISL_1470973, EPI_ISL_1470974, EPI_ISL_1470975, EPI_ISL_1470976, EPI_ISL_1470977, EPI_ISL_1470978, EPI_ISL_1470979, EPI_ISL_1470980, EPI_ISL_1470981, EPI_ISL_1470982,
EPI_ISL_1470983, EPI_ISL_1470984, EPI_ISL_1470985, EPI_ISL_1470986, EPI_ISL_1470987, EPI_ISL_1470988, EPI_ISL_1470989, EPI_ISL_1470990, EPI_ISL_1470991, EPI_ISL_1470992, EPI_ISL_1470993, EPI_ISL_1470994, EPI_ISL_1470995, EPI_ISL_1470996, EPI_ISL_1470997, EPI_ISL_1470998, EPI_ISL_1470999, EPI_ISL_1471000, EPI_ISL_1471001, EPI_ISL_1471002, EPI_ISL_1471003, EPI_ISL_1471004,
EPI_ISL_1471005, EPI_ISL_1471006, EPI_ISL_1471007, EPI_ISL_1471008, EPI_ISL_1471009, EPI_ISL_1471010, EPI_ISL_1471011, EPI_ISL_1471012, EPI_ISL_1471013, EPI_ISL_1471014, EPI_ISL_1471015, EPI_ISL_1471016, EPI_ISL_1471017, EPI_ISL_1471018, EPI_ISL_1471019, EPI_ISL_1471020, EPI_ISL_1471021, EPI_ISL_1471022, EPI_ISL_1471023, EPI_ISL_1471024, EPI_ISL_1471025, EPI_ISL_1471026,
EPI_ISL_1471027, EPI_ISL_1471028, EPI_ISL_1471029, EPI_ISL_1471030, EPI_ISL_1471031, EPI_ISL_1471032, EPI_ISL_1471033, EPI_ISL_1471034, EPI_ISL_1471035, EPI_ISL_1471036, EPI_ISL_1471037, EPI_ISL_1471038, EPI_ISL_1471039, EPI_ISL_1471040, EPI_ISL_1471041, EPI_ISL_1471042, EPI_ISL_1471043, EPI_ISL_1471044, EPI_ISL_1471045, EPI_ISL_1471046, EPI_ISL_1471047, EPI_ISL_1471048,
EPI_ISL_1471049, EPI_ISL_1471050, EPI_ISL_1471051, EPI_ISL_1471052, EPI_ISL_1471053, EPI_ISL_1471054, EPI_ISL_1471055, EPI_ISL_1471056, EPI_ISL_1471057, EPI_ISL_1471058, EPI_ISL_1471059, EPI_ISL_1471060, EPI_ISL_1471061, EPI_ISL_1471062, EPI_ISL_1471063, EPI_ISL_1471064, EPI_ISL_1471065, EPI_ISL_1471066, EPI_ISL_1471067, EPI_ISL_1471068, EPI_ISL_1471069, EPI_ISL_1471070,
EPI_ISL_1471071, EPI_ISL_1471072, EPI_ISL_1471073, EPI_ISL_1471074, EPI_ISL_1471075, EPI_ISL_1471076, EPI_ISL_1471077, EPI_ISL_1471078, EPI_ISL_1471079, EPI_ISL_1471080, EPI_ISL_1471081, EPI_ISL_1471082, EPI_ISL_1471083, EPI_ISL_1471084, EPI_ISL_1471085, EPI_ISL_1471086, EPI_ISL_1471087, EPI_ISL_1471088, EPI_ISL_1471089, EPI_ISL_1471090, EPI_ISL_1471091, EPI_ISL_1471092,
EPI_ISL_1471093, EPI_ISL_1471094, EPI_ISL_1471095, EPI_ISL_1471096, EPI_ISL_1471097, EPI_ISL_1471098, EPI_ISL_1471099, EPI_ISL_1471100, EPI_ISL_1471101, EPI_ISL_1471102, EPI_ISL_1471103, EPI_ISL_1471104, EPI_ISL_1471105, EPI_ISL_1471106, EPI_ISL_1471107, EPI_ISL_1471108, EPI_ISL_1471109, EPI_ISL_1471110, EPI_ISL_1471111, EPI_ISL_1471112, EPI_ISL_1471113, EPI_ISL_1471114,
EPI_ISL_1471115, EPI_ISL_1471116, EPI_ISL_1471117, EPI_ISL_1471118, EPI_ISL_1471119, EPI_ISL_1471120, EPI_ISL_1471121, EPI_ISL_1471122, EPI_ISL_1471123, EPI_ISL_1471124, EPI_ISL_1471125, EPI_ISL_1471126, EPI_ISL_1471127, EPI_ISL_1471128, EPI_ISL_1471129, EPI_ISL_1471130, EPI_ISL_1471131, EPI_ISL_1471132, EPI_ISL_1471133, EPI_ISL_1471134, EPI_ISL_1471135, EPI_ISL_1471136,
EPI_ISL_1471137, EPI_ISL_1471138, EPI_ISL_1471139, EPI_ISL_1471140, EPI_ISL_1471141, EPI_ISL_1471142, EPI_ISL_1471143, EPI_ISL_1471144, EPI_ISL_1471145, EPI_ISL_1471146, EPI_ISL_1471147, EPI_ISL_1471148, EPI_ISL_1471149, EPI_ISL_1471150, EPI_ISL_1471151, EPI_ISL_1471152, EPI_ISL_1471153, EPI_ISL_1471154, EPI_ISL_1471155, EPI_ISL_1471156, EPI_ISL_1471157, EPI_ISL_1471158,
EPI_ISL_1471159, EPI_ISL_1471160, EPI_ISL_1471161, EPI_ISL_1471162, EPI_ISL_1471163, EPI_ISL_1471164, EPI_ISL_1471165, EPI_ISL_1471166, EPI_ISL_1471167, EPI_ISL_1471168, EPI_ISL_1471169, EPI_ISL_1471170, EPI_ISL_1471171, EPI_ISL_1471172, EPI_ISL_1471173, EPI_ISL_1471174, EPI_ISL_1471175, EPI_ISL_1471176, EPI_ISL_1471177, EPI_ISL_1471178, EPI_ISL_1471179, EPI_ISL_1471180,
EPI_ISL_1471181, EPI_ISL_1471182, EPI_ISL_1471183, EPI_ISL_1471184, EPI_ISL_1471185, EPI_ISL_1471186, EPI_ISL_1471187, EPI_ISL_1471188, EPI_ISL_1471189, EPI_ISL_1471190, EPI_ISL_1471191, EPI_ISL_1471192, EPI_ISL_1471193, EPI_ISL_1471194, EPI_ISL_1471195, EPI_ISL_1471196, EPI_ISL_1471197, EPI_ISL_1471198, EPI_ISL_1471199, EPI_ISL_1471200, EPI_ISL_1471201, EPI_ISL_1471202,
EPI_ISL_1471203, EPI_ISL_1471204, EPI_ISL_1471205, EPI_ISL_1471206, EPI_ISL_1471207, EPI_ISL_1471208, EPI_ISL_1471209, EPI_ISL_1471210, EPI_ISL_1471211, EPI_ISL_1471212, EPI_ISL_1471213, EPI_ISL_1471214, EPI_ISL_1471215, EPI_ISL_1471216, EPI_ISL_1471217, EPI_ISL_1471218, EPI_ISL_1471219, EPI_ISL_1471220, EPI_ISL_1471221, EPI_ISL_1471222, EPI_ISL_1471223, EPI_ISL_1471224,
EPI_ISL_1471225, EPI_ISL_1471226, EPI_ISL_1471227, EPI_ISL_1471228, EPI_ISL_1471229, EPI_ISL_1471230, EPI_ISL_1471231, EPI_ISL_1471232, EPI_ISL_1471233, EPI_ISL_1471234, EPI_ISL_1471235, EPI_ISL_1471236, EPI_ISL_1471237, EPI_ISL_1471238, EPI_ISL_1471239, EPI_ISL_1471240, EPI_ISL_1471241, EPI_ISL_1471242, EPI_ISL_1471243, EPI_ISL_1471244, EPI_ISL_1471245, EPI_ISL_1471246,
EPI_ISL_1471247, EPI_ISL_1471248, EPI_ISL_1471249, EPI_ISL_1471250, EPI_ISL_1471251, EPI_ISL_1471252, EPI_ISL_1471253, EPI_ISL_1471254, EPI_ISL_1471255, EPI_ISL_1471256, EPI_ISL_1471257, EPI_ISL_1471258, EPI_ISL_1471259, EPI_ISL_1471260, EPI_ISL_1471261, EPI_ISL_1471262, EPI_ISL_1471263, EPI_ISL_1471264, EPI_ISL_1471265, EPI_ISL_1471266, EPI_ISL_1471267, EPI_ISL_1471268,
EPI_ISL_1471269, EPI_ISL_1471270, EPI_ISL_1471271, EPI_ISL_1471272, EPI_ISL_1471273, EPI_ISL_1471274, EPI_ISL_1471275, EPI_ISL_1471276, EPI_ISL_1471277, EPI_ISL_1471278, EPI_ISL_1471279, EPI_ISL_1471280, EPI_ISL_1471281, EPI_ISL_1471282, EPI_ISL_1471283, EPI_ISL_1471284, EPI_ISL_1471285, EPI_ISL_1471286, EPI_ISL_1471287, EPI_ISL_1471288, EPI_ISL_1471289, EPI_ISL_1471290,
EPI_ISL_1471291, EPI_ISL_1471292, EPI_ISL_1471293, EPI_ISL_1471294, EPI_ISL_1471295, EPI_ISL_1471296, EPI_ISL_1471297, EPI_ISL_1471298, EPI_ISL_1471299, EPI_ISL_1471300, EPI_ISL_1471301, EPI_ISL_1471302, EPI_ISL_1471303, EPI_ISL_1471304, EPI_ISL_1471305, EPI_ISL_1471306, EPI_ISL_1471307, EPI_ISL_1471308, EPI_ISL_1471309, EPI_ISL_1471310, EPI_ISL_1471311, EPI_ISL_1471312,
EPI_ISL_1471313, EPI_ISL_1471314, EPI_ISL_1471315, EPI_ISL_1471316, EPI_ISL_1471317, EPI_ISL_1471318, EPI_ISL_1471319, EPI_ISL_1471320, EPI_ISL_1471321, EPI_ISL_1471322, EPI_ISL_1471323, EPI_ISL_1471324, EPI_ISL_1471325, EPI_ISL_1471326, EPI_ISL_1471327, EPI_ISL_1471328, EPI_ISL_1471329, EPI_ISL_1471330, EPI_ISL_1471331, EPI_ISL_1471332, EPI_ISL_1471333, EPI_ISL_1471334,
EPI_ISL_1471335, EPI_ISL_1471336, EPI_ISL_1471337, EPI_ISL_1471338, EPI_ISL_1471339, EPI_ISL_1471340, EPI_ISL_1471341, EPI_ISL_1471342, EPI_ISL_1471343, EPI_ISL_1471344, EPI_ISL_1471345, EPI_ISL_1471346, EPI_ISL_1471347, EPI_ISL_1471348, EPI_ISL_1471349, EPI_ISL_1471350, EPI_ISL_1471351, EPI_ISL_1471352, EPI_ISL_1471353, EPI_ISL_1471354, EPI_ISL_1471355, EPI_ISL_1471356,

EPI_ISL_1471357, EPI_ISL_1471358, EPI_ISL_1471359, EPI_ISL_1471360, EPI_ISL_1471361, EPI_ISL_1471362, EPI_ISL_1471363, EPI_ISL_1471364, EPI_ISL_1471365, EPI_ISL_1471366, EPI_ISL_1471367, EPI_ISL_1471368, EPI_ISL_1471369, EPI_ISL_1471370, EPI_ISL_1471371, EPI_ISL_1471372, EPI_ISL_1471373, EPI_ISL_1471374, EPI_ISL_1471375, EPI_ISL_1471376, EPI_ISL_1471377, EPI_ISL_1471378,
EPI_ISL_1471379, EPI_ISL_1471380, EPI_ISL_1471381, EPI_ISL_1471382, EPI_ISL_1471383, EPI_ISL_1471384, EPI_ISL_1471385, EPI_ISL_1471386, EPI_ISL_1471387, EPI_ISL_1471388, EPI_ISL_1471389, EPI_ISL_1471390, EPI_ISL_1471391, EPI_ISL_1471392, EPI_ISL_1471393, EPI_ISL_1471394, EPI_ISL_1471395, EPI_ISL_1471396, EPI_ISL_1471397, EPI_ISL_1471398, EPI_ISL_1471399, EPI_ISL_1471400,
EPI_ISL_1471401, EPI_ISL_1471402, EPI_ISL_1471403, EPI_ISL_1471404, EPI_ISL_1471405, EPI_ISL_1471406, EPI_ISL_1471407, EPI_ISL_1471408, EPI_ISL_1471409, EPI_ISL_1471410, EPI_ISL_1471411, EPI_ISL_1471412, EPI_ISL_1471413, EPI_ISL_1471414, EPI_ISL_1471415, EPI_ISL_1471416, EPI_ISL_1471417, EPI_ISL_1471418, EPI_ISL_1471419, EPI_ISL_1471420, EPI_ISL_1471421, EPI_ISL_1471422,
EPI_ISL_1471423, EPI_ISL_1471424, EPI_ISL_1471425, EPI_ISL_1471426, EPI_ISL_1471427, EPI_ISL_1471428, EPI_ISL_1471429, EPI_ISL_1471430, EPI_ISL_1471431, EPI_ISL_1471432, EPI_ISL_1471433, EPI_ISL_1471434, EPI_ISL_1471435, EPI_ISL_1471436, EPI_ISL_1471437, EPI_ISL_1471438, EPI_ISL_1471439, EPI_ISL_1471440, EPI_ISL_1471441, EPI_ISL_1471442, EPI_ISL_1471443, EPI_ISL_1471444,
EPI_ISL_1471445, EPI_ISL_1471446, EPI_ISL_1471447, EPI_ISL_1471448, EPI_ISL_1471449, EPI_ISL_1471450, EPI_ISL_1471451, EPI_ISL_1471452, EPI_ISL_1471453, EPI_ISL_1471454, EPI_ISL_1471455, EPI_ISL_1471456, EPI_ISL_1471457, EPI_ISL_1471458, EPI_ISL_1471459, EPI_ISL_1471460, EPI_ISL_1471461, EPI_ISL_1471462, EPI_ISL_1471463, EPI_ISL_1471464, EPI_ISL_1471465, EPI_ISL_1471466,
EPI_ISL_1471467, EPI_ISL_1471468, EPI_ISL_1471469, EPI_ISL_1471470, EPI_ISL_1471471, EPI_ISL_1471472, EPI_ISL_1471473, EPI_ISL_1471474, EPI_ISL_1471475, EPI_ISL_1471476, EPI_ISL_1471477, EPI_ISL_1471478, EPI_ISL_1471479, EPI_ISL_1471480, EPI_ISL_1471481, EPI_ISL_1471482, EPI_ISL_1471483, EPI_ISL_1471484, EPI_ISL_1471485, EPI_ISL_1471486, EPI_ISL_1471487, EPI_ISL_1471488,
EPI_ISL_1471489, EPI_ISL_1471490, EPI_ISL_1471491, EPI_ISL_1471492, EPI_ISL_1471493, EPI_ISL_1471494, EPI_ISL_1471495, EPI_ISL_1471496, EPI_ISL_1471497, EPI_ISL_1471498, EPI_ISL_1471499, EPI_ISL_1471500, EPI_ISL_1471501, EPI_ISL_1471502, EPI_ISL_1471503, EPI_ISL_1471504, EPI_ISL_1471505, EPI_ISL_1471506, EPI_ISL_1471507, EPI_ISL_1471508, EPI_ISL_1471509, EPI_ISL_1471510,
EPI_ISL_1471511, EPI_ISL_1471512, EPI_ISL_1471513, EPI_ISL_1471514, EPI_ISL_1471515, EPI_ISL_1471516, EPI_ISL_1471517, EPI_ISL_1471518, EPI_ISL_1471519, EPI_ISL_1471520, EPI_ISL_1471521, EPI_ISL_1471522, EPI_ISL_1471523, EPI_ISL_1471524, EPI_ISL_1471525, EPI_ISL_1471526, EPI_ISL_1471527, EPI_ISL_1471528, EPI_ISL_1471529, EPI_ISL_1471530, EPI_ISL_1471531, EPI_ISL_1471532,
EPI_ISL_1471533, EPI_ISL_1471534, EPI_ISL_1471535, EPI_ISL_1471536, EPI_ISL_1471537, EPI_ISL_1471538, EPI_ISL_1471539, EPI_ISL_1471540, EPI_ISL_1471541, EPI_ISL_1471542, EPI_ISL_1471543, EPI_ISL_1471544, EPI_ISL_1471545, EPI_ISL_1471546, EPI_ISL_1471547, EPI_ISL_1471548, EPI_ISL_1471549, EPI_ISL_1471550, EPI_ISL_1471551, EPI_ISL_1471552, EPI_ISL_1471553, EPI_ISL_1471554,
EPI_ISL_1471555, EPI_ISL_1471556, EPI_ISL_1471557, EPI_ISL_1471558, EPI_ISL_1471559, EPI_ISL_1471560, EPI_ISL_1471561, EPI_ISL_1471562, EPI_ISL_1471563, EPI_ISL_1471564, EPI_ISL_1471565, EPI_ISL_1471566, EPI_ISL_1471567, EPI_ISL_1471568, EPI_ISL_1471569, EPI_ISL_1471570, EPI_ISL_1471571, EPI_ISL_1471572, EPI_ISL_1471573, EPI_ISL_1471574, EPI_ISL_1471575, EPI_ISL_1471576,
EPI_ISL_1471577, EPI_ISL_1471578, EPI_ISL_1471579, EPI_ISL_1471580, EPI_ISL_1471581, EPI_ISL_1471582, EPI_ISL_1471583, EPI_ISL_1471584, EPI_ISL_1471585, EPI_ISL_1471586, EPI_ISL_1471587, EPI_ISL_1471588, EPI_ISL_1471589, EPI_ISL_1471590, EPI_ISL_1471591, EPI_ISL_1471592, EPI_ISL_1471593, EPI_ISL_1471594, EPI_ISL_1471595, EPI_ISL_1471596, EPI_ISL_1471597, EPI_ISL_1471598,
EPI_ISL_1471599, EPI_ISL_1471600, EPI_ISL_1471601, EPI_ISL_1471602, EPI_ISL_1471603, EPI_ISL_1471604, EPI_ISL_1471605, EPI_ISL_1471606, EPI_ISL_1471607, EPI_ISL_1471608, EPI_ISL_1471609, EPI_ISL_1471610, EPI_ISL_1471611, EPI_ISL_1471612, EPI_ISL_1471613, EPI_ISL_1471614, EPI_ISL_1471615, EPI_ISL_1471616, EPI_ISL_1471617, EPI_ISL_1471618, EPI_ISL_1471619, EPI_ISL_1471620,
EPI_ISL_1471621, EPI_ISL_1471622, EPI_ISL_1471623, EPI_ISL_1471624, EPI_ISL_1471625, EPI_ISL_1471626, EPI_ISL_1471627, EPI_ISL_1471628, EPI_ISL_1471629, EPI_ISL_1471630, EPI_ISL_1471631, EPI_ISL_1471632, EPI_ISL_1471633, EPI_ISL_1471634, EPI_ISL_1471635, EPI_ISL_1471636, EPI_ISL_1471637, EPI_ISL_1471638, EPI_ISL_1471639, EPI_ISL_1471640, EPI_ISL_1471641, EPI_ISL_1471642,
EPI_ISL_1471643, EPI_ISL_1471644, EPI_ISL_1471645, EPI_ISL_1471646, EPI_ISL_1471647, EPI_ISL_1471648, EPI_ISL_1471649, EPI_ISL_1471650, EPI_ISL_1471651, EPI_ISL_1471652, EPI_ISL_1471653, EPI_ISL_1471654, EPI_ISL_1471655, EPI_ISL_1471656, EPI_ISL_1471657, EPI_ISL_1471658, EPI_ISL_1471659, EPI_ISL_1471660, EPI_ISL_1471661, EPI_ISL_1471662, EPI_ISL_1471663, EPI_ISL_1471664,
EPI_ISL_1471665, EPI_ISL_1471666, EPI_ISL_1471667, EPI_ISL_1471668, EPI_ISL_1471669, EPI_ISL_1471670, EPI_ISL_1471671, EPI_ISL_1471672, EPI_ISL_1471673, EPI_ISL_1471674, EPI_ISL_1471675, EPI_ISL_1471676, EPI_ISL_1471677, EPI_ISL_1471678, EPI_ISL_1471679, EPI_ISL_1471680, EPI_ISL_1471681, EPI_ISL_1471682, EPI_ISL_1471683, EPI_ISL_1471684, EPI_ISL_1471685, EPI_ISL_1471686,
EPI_ISL_1471687, EPI_ISL_1471688, EPI_ISL_1471689, EPI_ISL_1471690, EPI_ISL_1471691, EPI_ISL_1471692, EPI_ISL_1471693, EPI_ISL_1471694, EPI_ISL_1471695, EPI_ISL_1471696, EPI_ISL_1471697, EPI_ISL_1471698, EPI_ISL_1471699, EPI_ISL_1471700, EPI_ISL_1471701, EPI_ISL_1471702, EPI_ISL_1471703, EPI_ISL_1471704, EPI_ISL_1471705, EPI_ISL_1471706, EPI_ISL_1471707, EPI_ISL_1471708,
EPI_ISL_1471709, EPI_ISL_1471710, EPI_ISL_1471711, EPI_ISL_1471712, EPI_ISL_1471713, EPI_ISL_1471714, EPI_ISL_1471715, EPI_ISL_1471716, EPI_ISL_1471717, EPI_ISL_1471718, EPI_ISL_1471719, EPI_ISL_1471720, EPI_ISL_1471721, EPI_ISL_1471722, EPI_ISL_1471723, EPI_ISL_1471724, EPI_ISL_1471725, EPI_ISL_1471726, EPI_ISL_1471727, EPI_ISL_1471728, EPI_ISL_1471729, EPI_ISL_1471730,
EPI_ISL_1471731, EPI_ISL_1471732, EPI_ISL_1471733, EPI_ISL_1471734, EPI_ISL_1471735, EPI_ISL_1471736, EPI_ISL_1471737, EPI_ISL_1471738, EPI_ISL_1471739, EPI_ISL_1471740, EPI_ISL_1471741, EPI_ISL_1471742, EPI_ISL_1471743, EPI_ISL_1471744, EPI_ISL_1471745, EPI_ISL_1471746, EPI_ISL_1471747, EPI_ISL_1471748, EPI_ISL_1471749, EPI_ISL_1471750, EPI_ISL_1471751, EPI_ISL_1471752,
EPI_ISL_1471753, EPI_ISL_1471754, EPI_ISL_1471755, EPI_ISL_1471756, EPI_ISL_1471757, EPI_ISL_1471758, EPI_ISL_1471759, EPI_ISL_1471760, EPI_ISL_1471761, EPI_ISL_1471762, EPI_ISL_1471763, EPI_ISL_1471764, EPI_ISL_1471765, EPI_ISL_1471766, EPI_ISL_1471767, EPI_ISL_1471768, EPI_ISL_1471769, EPI_ISL_1471770, EPI_ISL_1471771, EPI_ISL_1471772, EPI_ISL_1471773, EPI_ISL_1471774,
EPI_ISL_1471775, EPI_ISL_1471776, EPI_ISL_1471777, EPI_ISL_1471778, EPI_ISL_1471779, EPI_ISL_1471780, EPI_ISL_1471781, EPI_ISL_1471782, EPI_ISL_1471783, EPI_ISL_1471784, EPI_ISL_1471785, EPI_ISL_1471786, EPI_ISL_1471787, EPI_ISL_1471788, EPI_ISL_1471789, EPI_ISL_1471790, EPI_ISL_1471791, EPI_ISL_1471792, EPI_ISL_1471793, EPI_ISL_1471794, EPI_ISL_1471795, EPI_ISL_1471796,
EPI_ISL_1471797, EPI_ISL_1471798, EPI_ISL_1471799, EPI_ISL_1471800, EPI_ISL_1471801, EPI_ISL_1471802, EPI_ISL_1471803, EPI_ISL_1471804, EPI_ISL_1471805, EPI_ISL_1471806, EPI_ISL_1471807, EPI_ISL_1471808, EPI_ISL_1471809, EPI_ISL_1471810, EPI_ISL_1471811, EPI_ISL_1471812, EPI_ISL_1471813, EPI_ISL_1471814, EPI_ISL_1471815, EPI_ISL_1471816, EPI_ISL_1471817, EPI_ISL_1471818,
EPI_ISL_1471819, EPI_ISL_1471820, EPI_ISL_1471821, EPI_ISL_1471822, EPI_ISL_1471823, EPI_ISL_1471824, EPI_ISL_1471825, EPI_ISL_1471826, EPI_ISL_1471827, EPI_ISL_1471828, EPI_ISL_1471829, EPI_ISL_1471830, EPI_ISL_1471831, EPI_ISL_1471832, EPI_ISL_1471833, EPI_ISL_1471834, EPI_ISL_1471835, EPI_ISL_1471836, EPI_ISL_1471837, EPI_ISL_1471838, EPI_ISL_1471839, EPI_ISL_1471840,
EPI_ISL_1471841, EPI_ISL_1471842, EPI_ISL_1471843, EPI_ISL_1471844, EPI_ISL_1471845, EPI_ISL_1471846, EPI_ISL_1471847, EPI_ISL_1471848, EPI_ISL_1471849, EPI_ISL_1471850, EPI_ISL_1471851, EPI_ISL_1471852, EPI_ISL_1471853, EPI_ISL_1471854, EPI_ISL_1471855, EPI_ISL_1471856, EPI_ISL_1471857, EPI_ISL_1471858, EPI_ISL_1471859, EPI_ISL_1471860, EPI_ISL_1471861, EPI_ISL_1471862,
EPI_ISL_1471863, EPI_ISL_1471864, EPI_ISL_1471865, EPI_ISL_1471866, EPI_ISL_1471867, EPI_ISL_1471868, EPI_ISL_1471869, EPI_ISL_1471870, EPI_ISL_1471871, EPI_ISL_1471872, EPI_ISL_1471873, EPI_ISL_1471874, EPI_ISL_1471875, EPI_ISL_1471876, EPI_ISL_1471877, EPI_ISL_1471878, EPI_ISL_1471879, EPI_ISL_1471880, EPI_ISL_1471881, EPI_ISL_1471882, EPI_ISL_1471883, EPI_ISL_1471884,
EPI_ISL_1471885, EPI_ISL_1471886, EPI_ISL_1471887, EPI_ISL_1471888, EPI_ISL_1471889, EPI_ISL_1471890, EPI_ISL_1471891, EPI_ISL_1471892, EPI_ISL_1471893, EPI_ISL_1471894, EPI_ISL_1471895, EPI_ISL_1471896, EPI_ISL_1471897, EPI_ISL_1471898, EPI_ISL_1471899, EPI_ISL_1471900, EPI_ISL_1471901, EPI_ISL_1471902, EPI_ISL_1471903, EPI_ISL_1471904, EPI_ISL_1471905, EPI_ISL_1471906,
EPI_ISL_1471907, EPI_ISL_1471908, EPI_ISL_1471909, EPI_ISL_1471910, EPI_ISL_1471911, EPI_ISL_1471912, EPI_ISL_1471913, EPI_ISL_1471914, EPI_ISL_1471915, EPI_ISL_1471916, EPI_ISL_1471917, EPI_ISL_1471918, EPI_ISL_1471919, EPI_ISL_1471920, EPI_ISL_1471921, EPI_ISL_1471922, EPI_ISL_1471923, EPI_ISL_1471924, EPI_ISL_1471925, EPI_ISL_1471926, EPI_ISL_1471927, EPI_ISL_1471928,
EPI_ISL_1471929, EPI_ISL_1471930, EPI_ISL_1471931, EPI_ISL_1471932, EPI_ISL_1471933, EPI_ISL_1471934, EPI_ISL_1471935, EPI_ISL_1471936, EPI_ISL_1471937, EPI_ISL_1471938, EPI_ISL_1471939, EPI_ISL_1471940, EPI_ISL_1471941, EPI_ISL_1471942, EPI_ISL_1471943, EPI_ISL_1471944, EPI_ISL_1471945, EPI_ISL_1471946, EPI_ISL_1471947, EPI_ISL_1471948, EPI_ISL_1471949, EPI_ISL_1471950,
EPI_ISL_1471951, EPI_ISL_1471952, EPI_ISL_1471953, EPI_ISL_1471954, EPI_ISL_1471955, EPI_ISL_1471956, EPI_ISL_1471957, EPI_ISL_1471958, EPI_ISL_1471959, EPI_ISL_1471960, EPI_ISL_1471961, EPI_ISL_1471962, EPI_ISL_1471963, EPI_ISL_1471964, EPI_ISL_1471965, EPI_ISL_1471966, EPI_ISL_1471967, EPI_ISL_1471968, EPI_ISL_1471969, EPI_ISL_1471970, EPI_ISL_1471971, EPI_ISL_1471972,
EPI_ISL_1471973, EPI_ISL_1471974, EPI_ISL_1471975, EPI_ISL_1471976, EPI_ISL_1471977, EPI_ISL_1471978, EPI_ISL_1471979, EPI_ISL_1471980, EPI_ISL_1471981, EPI_ISL_1471982, EPI_ISL_1471983, EPI_ISL_1471984, EPI_ISL_1471985, EPI_ISL_1471986, EPI_ISL_1471987, EPI_ISL_1471988, EPI_ISL_1471989, EPI_ISL_1471990, EPI_ISL_1471991, EPI_ISL_1471992, EPI_ISL_1471993, EPI_ISL_1471994,
EPI_ISL_1471995, EPI_ISL_1471996, EPI_ISL_1471997, EPI_ISL_1471998, EPI_ISL_1471999, EPI_ISL_1472000, EPI_ISL_1472001, EPI_ISL_1472002, EPI_ISL_1472003, EPI_ISL_1472004, EPI_ISL_1472005, EPI_ISL_1472006, EPI_ISL_1472007, EPI_ISL_1472008, EPI_ISL_1472009, EPI_ISL_1472010, EPI_ISL_1472011, EPI_ISL_1472012, EPI_ISL_1472013, EPI_ISL_1472014, EPI_ISL_1472015, EPI_ISL_1472016,
EPI_ISL_1472017, EPI_ISL_1472018, EPI_ISL_1472019, EPI_ISL_1472020, EPI_ISL_1472021, EPI_ISL_1472022, EPI_ISL_1472023, EPI_ISL_1472024, EPI_ISL_1472025, EPI_ISL_1472026, EPI_ISL_1472027, EPI_ISL_1472028, EPI_ISL_1472029, EPI_ISL_1472030, EPI_ISL_1472031, EPI_ISL_1472032, EPI_ISL_1472033, EPI_ISL_1472034, EPI_ISL_1472035, EPI_ISL_1472036, EPI_ISL_1472037, EPI_ISL_1472038,
EPI_ISL_1472039, EPI_ISL_1472040, EPI_ISL_1472041, EPI_ISL_1472042, EPI_ISL_1472043, EPI_ISL_1472044, EPI_ISL_1472045, EPI_ISL_1472046, EPI_ISL_1472047, EPI_ISL_1472048, EPI_ISL_1472049, EPI_ISL_1472050, EPI_ISL_1472051, EPI_ISL_1472052, EPI_ISL_1472053, EPI_ISL_1472054, EPI_ISL_1472055, EPI_ISL_1472056, EPI_ISL_1472057, EPI_ISL_1472058, EPI_ISL_1472059, EPI_ISL_1472060,
EPI_ISL_1472061, EPI_ISL_1472062, EPI_ISL_1472063, EPI_ISL_1472064, EPI_ISL_1472065, EPI_ISL_1472066, EPI_ISL_1472067, EPI_ISL_1472068, EPI_ISL_1472069, EPI_ISL_1472070, EPI_ISL_1472071, EPI_ISL_1472072, EPI_ISL_1472073, EPI_ISL_1472074, EPI_ISL_1472075, EPI_ISL_1472076, EPI_ISL_1542074, EPI_ISL_1542078, EPI_ISL_1542079, EPI_ISL_1542080, EPI_ISL_1542081, EPI_ISL_1542082,
EPI_ISL_1542083, EPI_ISL_1542084, EPI_ISL_1542085, EPI_ISL_1542086, EPI_ISL_1542088, EPI_ISL_1542090, EPI_ISL_1542091, EPI_ISL_1542092, EPI_ISL_1542093, EPI_ISL_1542097, EPI_ISL_1542098, EPI_ISL_1542099, EPI_ISL_1542100, EPI_ISL_1542101, EPI_ISL_1542103, EPI_ISL_1542104, EPI_ISL_1542106, EPI_ISL_1542107, EPI_ISL_1542109, EPI_ISL_1542113, EPI_ISL_1542114, EPI_ISL_1542115,
EPI_ISL_1542117, EPI_ISL_1542119, EPI_ISL_1542120, EPI_ISL_1542121, EPI_ISL_1542122, EPI_ISL_1542123, EPI_ISL_1542125, EPI_ISL_1542126, EPI_ISL_1542127, EPI_ISL_1542128, EPI_ISL_1542129, EPI_ISL_1542131, EPI_ISL_1542132, EPI_ISL_1542133, EPI_ISL_1542134, EPI_ISL_1542135, EPI_ISL_1542136, EPI_ISL_1542137, EPI_ISL_1542138, EPI_ISL_1542139, EPI_ISL_1542140, EPI_ISL_1542142,
EPI_ISL_1542144, EPI_ISL_1542145, EPI_ISL_1542146, EPI_ISL_1542147, EPI_ISL_1542148, EPI_ISL_1542149, EPI_ISL_1542150, EPI_ISL_1542151, EPI_ISL_1542152, EPI_ISL_1542153, EPI_ISL_1542154, EPI_ISL_1542155, EPI_ISL_1542158, EPI_ISL_1542159, EPI_ISL_1542160, EPI_ISL_1542161, EPI_ISL_1542162, EPI_ISL_1542163, EPI_ISL_1542164, EPI_ISL_1542167, EPI_ISL_1542168, EPI_ISL_1542169,
EPI_ISL_1542171, EPI_ISL_1542172, EPI_ISL_1542173, EPI_ISL_1542175, EPI_ISL_1542176, EPI_ISL_1542177, EPI_ISL_1542179, EPI_ISL_1542180, EPI_ISL_1542182, EPI_ISL_1542183, EPI_ISL_1542184, EPI_ISL_1542186, EPI_ISL_1542187, EPI_ISL_1542188, EPI_ISL_1542189, EPI_ISL_1542191, EPI_ISL_1542192, EPI_ISL_1542193, EPI_ISL_1542194, EPI_ISL_1542195, EPI_ISL_1542196, EPI_ISL_1542197,
EPI_ISL_1542198, EPI_ISL_1542199, EPI_ISL_1542201, EPI_ISL_1542202, EPI_ISL_1542204, EPI_ISL_1542205, EPI_ISL_1542207, EPI_ISL_1542209, EPI_ISL_1542210, EPI_ISL_1542211, EPI_ISL_1542212, EPI_ISL_1542213, EPI_ISL_1542214, EPI_ISL_1542215, EPI_ISL_1542216, EPI_ISL_1542218, EPI_ISL_1542219, EPI_ISL_1542220, EPI_ISL_1542222, EPI_ISL_1542223, EPI_ISL_1542224, EPI_ISL_1542225,
EPI_ISL_1542226, EPI_ISL_1542228, EPI_ISL_1542230, EPI_ISL_1542231, EPI_ISL_1542233, EPI_ISL_1542234, EPI_ISL_1542235, EPI_ISL_1542236, EPI_ISL_1542237, EPI_ISL_1542239, EPI_ISL_1542240, EPI_ISL_1542241, EPI_ISL_1542243, EPI_ISL_1542244, EPI_ISL_1542245, EPI_ISL_1542246, EPI_ISL_1542248, EPI_ISL_1542249, EPI_ISL_1542250, EPI_ISL_1542251, EPI_ISL_1542252, EPI_ISL_1542253,
EPI_ISL_1542254, EPI_ISL_1542256, EPI_ISL_1542257, EPI_ISL_1542258, EPI_ISL_1542259, EPI_ISL_1542261, EPI_ISL_1542262, EPI_ISL_1542263, EPI_ISL_1542264, EPI_ISL_1542265, EPI_ISL_1542266, EPI_ISL_1542267, EPI_ISL_1542268, EPI_ISL_1542269, EPI_ISL_1542270, EPI_ISL_1542271, EPI_ISL_1542272, EPI_ISL_1542274, EPI_ISL_1542275, EPI_ISL_1542276, EPI_ISL_1542277, EPI_ISL_1542278,
EPI_ISL_1542279, EPI_ISL_1542280, EPI_ISL_1542281, EPI_ISL_1542282, EPI_ISL_1542283, EPI_ISL_1542284, EPI_ISL_1542286, EPI_ISL_1542287, EPI_ISL_1542289, EPI_ISL_1542290, EPI_ISL_1542291, EPI_ISL_1542292, EPI_ISL_1542294, EPI_ISL_1542296, EPI_ISL_1542297, EPI_ISL_1542298, EPI_ISL_1542299, EPI_ISL_1542300, EPI_ISL_1542301, EPI_ISL_1542302, EPI_ISL_1542304, EPI_ISL_1542305,
EPI_ISL_1542306, EPI_ISL_1542308, EPI_ISL_1542309, EPI_ISL_1542310, EPI_ISL_1542311, EPI_ISL_1542313, EPI_ISL_1542315, EPI_ISL_1542316, EPI_ISL_1542317, EPI_ISL_1542319, EPI_ISL_1542320, EPI_ISL_1542321, EPI_ISL_1542323, EPI_ISL_1542324, EPI_ISL_1542325, EPI_ISL_1542326, EPI_ISL_1542327, EPI_ISL_1542328, EPI_ISL_1542330, EPI_ISL_1542331, EPI_ISL_1542334, EPI_ISL_1542335,
EPI_ISL_1542336, EPI_ISL_1542338, EPI_ISL_1542339, EPI_ISL_1542340, EPI_ISL_1542341, EPI_ISL_1542342, EPI_ISL_1542344, EPI_ISL_1542345, EPI_ISL_1542346, EPI_ISL_1542347, EPI_ISL_1542348, EPI_ISL_1542349, EPI_ISL_1542350, EPI_ISL_1542351, EPI_ISL_1542352, EPI_ISL_1542353, EPI_ISL_1542354, EPI_ISL_1542355, EPI_ISL_1542356, EPI_ISL_1542358, EPI_ISL_1542359, EPI_ISL_1542360,
EPI_ISL_1542361, EPI_ISL_1542362, EPI_ISL_1542363, EPI_ISL_1542364, EPI_ISL_1542365, EPI_ISL_1542367, EPI_ISL_1542368, EPI_ISL_1542369, EPI_ISL_1542370, EPI_ISL_1542373, EPI_ISL_1542374, EPI_ISL_1542375, EPI_ISL_1542376, EPI_ISL_1542377, EPI_ISL_1542378, EPI_ISL_1542379, EPI_ISL_1542381, EPI_ISL_1542382, EPI_ISL_1542383, EPI_ISL_1542384, EPI_ISL_1542385, EPI_ISL_1542386,
EPI_ISL_1542388, EPI_ISL_1542389, EPI_ISL_1542391, EPI_ISL_1542392, EPI_ISL_1542395, EPI_ISL_1542396, EPI_ISL_1542399, EPI_ISL_1542400, EPI_ISL_1542402, EPI_ISL_1542403, EPI_ISL_1542404, EPI_ISL_1542405, EPI_ISL_1542406, EPI_ISL_1542407, EPI_ISL_1542408, EPI_ISL_1542409, EPI_ISL_1542410, EPI_ISL_1542411, EPI_ISL_1542412, EPI_ISL_1542413, EPI_ISL_1542414, EPI_ISL_1542415,
EPI_ISL_1542416, EPI_ISL_1542417, EPI_ISL_1542418, EPI_ISL_1542419, EPI_ISL_1542420, EPI_ISL_1542421, EPI_ISL_1542423, EPI_ISL_1542424, EPI_ISL_1542425, EPI_ISL_1542426, EPI_ISL_1542428, EPI_ISL_1542429, EPI_ISL_1542431, EPI_ISL_1542432, EPI_ISL_1542433, EPI_ISL_1542434, EPI_ISL_1542435, EPI_ISL_1542436, EPI_ISL_1542437, EPI_ISL_1542438, EPI_ISL_1542439, EPI_ISL_1542440,
EPI_ISL_1542441, EPI_ISL_1542442, EPI_ISL_1542444, EPI_ISL_1542445, EPI_ISL_1542449, EPI_ISL_1542450, EPI_ISL_1542451, EPI_ISL_1542452, EPI_ISL_1542453, EPI_ISL_1542454, EPI_ISL_1542455, EPI_ISL_1542456, EPI_ISL_1542457, EPI_ISL_1542459, EPI_ISL_1542460, EPI_ISL_1542461, EPI_ISL_1542462, EPI_ISL_1542463, EPI_ISL_1542464, EPI_ISL_1542465, EPI_ISL_1542466, EPI_ISL_1542467,
EPI_ISL_1542468, EPI_ISL_1542469, EPI_ISL_1542470, EPI_ISL_1542471, EPI_ISL_1542472, EPI_ISL_1542473, EPI_ISL_1542474, EPI_ISL_1542475, EPI_ISL_1542476, EPI_ISL_1542477, EPI_ISL_1542479, EPI_ISL_1542480, EPI_ISL_1542482, EPI_ISL_1542483, EPI_ISL_1542484, EPI_ISL_1542485, EPI_ISL_1542487, EPI_ISL_1542488, EPI_ISL_1542489, EPI_ISL_1542490, EPI_ISL_1542491, EPI_ISL_1542492,
EPI_ISL_1542493, EPI_ISL_1542494, EPI_ISL_1542495, EPI_ISL_1542496, EPI_ISL_1542497, EPI_ISL_1542498, EPI_ISL_1542499, EPI_ISL_1542501, EPI_ISL_1542502, EPI_ISL_1542503, EPI_ISL_1542505, EPI_ISL_1542507, EPI_ISL_1542508, EPI_ISL_1542509, EPI_ISL_1542510, EPI_ISL_1542511, EPI_ISL_1542512, EPI_ISL_1542513, EPI_ISL_1542514, EPI_ISL_1542515, EPI_ISL_1542516, EPI_ISL_1542517,
EPI_ISL_1542518, EPI_ISL_1542519, EPI_ISL_1542521, EPI_ISL_1542522, EPI_ISL_1542523, EPI_ISL_1542524, EPI_ISL_1542526, EPI_ISL_1542527, EPI_ISL_1542528, EPI_ISL_1542529, EPI_ISL_1542530, EPI_ISL_1542531, EPI_ISL_1542532, EPI_ISL_1542533, EPI_ISL_1542534, EPI_ISL_1542535, EPI_ISL_1542537, EPI_ISL_1542539, EPI_ISL_1542540, EPI_ISL_1542542, EPI_ISL_1542546, EPI_ISL_1542547,
EPI_ISL_1542548, EPI_ISL_1542549, EPI_ISL_1542551, EPI_ISL_1542552, EPI_ISL_1542553, EPI_ISL_1542554, EPI_ISL_1542555, EPI_ISL_1542556, EPI_ISL_1542557, EPI_ISL_1542558, EPI_ISL_1542559, EPI_ISL_1542560, EPI_ISL_1542561, EPI_ISL_1542563, EPI_ISL_1542564, EPI_ISL_1542565, EPI_ISL_1542566, EPI_ISL_1542567, EPI_ISL_1542568, EPI_ISL_1542569, EPI_ISL_1542572, EPI_ISL_1542573,
EPI_ISL_1542575, EPI_ISL_1542576, EPI_ISL_1542577, EPI_ISL_1542578, EPI_ISL_1542579, EPI_ISL_1542580, EPI_ISL_1542581, EPI_ISL_1542582, EPI_ISL_1542583, EPI_ISL_1542584, EPI_ISL_1542585, EPI_ISL_1542586, EPI_ISL_1542587, EPI_ISL_1542588, EPI_ISL_1542589, EPI_ISL_1542590, EPI_ISL_1542592, EPI_ISL_1542593, EPI_ISL_1542594, EPI_ISL_1542595, EPI_ISL_1542597, EPI_ISL_1542598,
EPI_ISL_1542599, EPI_ISL_1542600, EPI_ISL_1542601, EPI_ISL_1542602, EPI_ISL_1542603, EPI_ISL_1542605, EPI_ISL_1542606, EPI_ISL_1542608, EPI_ISL_1542609, EPI_ISL_1542610, EPI_ISL_1542611, EPI_ISL_1542612, EPI_ISL_1542613, EPI_ISL_1542614, EPI_ISL_1542615, EPI_ISL_1542617, EPI_ISL_1542618, EPI_ISL_1542620, EPI_ISL_1542621, EPI_ISL_1542622, EPI_ISL_1542623, EPI_ISL_1542624,
EPI_ISL_1542625, EPI_ISL_1542627, EPI_ISL_1542628, EPI_ISL_1542629, EPI_ISL_1542630, EPI_ISL_1542631, EPI_ISL_1542632, EPI_ISL_1542633, EPI_ISL_1542634, EPI_ISL_1542635, EPI_ISL_1542636, EPI_ISL_1542637, EPI_ISL_1542638, EPI_ISL_1542639, EPI_ISL_1542640, EPI_ISL_1542641, EPI_ISL_1542643, EPI_ISL_1542644, EPI_ISL_1542646, EPI_ISL_1542648, EPI_ISL_1542649, EPI_ISL_1542650,
EPI_ISL_1542651, EPI_ISL_1542653, EPI_ISL_1542654, EPI_ISL_1542655, EPI_ISL_1542656, EPI_ISL_1542658, EPI_ISL_1542659, EPI_ISL_1542660, EPI_ISL_1542661, EPI_ISL_1542662, EPI_ISL_1542663, EPI_ISL_1542664, EPI_ISL_1542665, EPI_ISL_1542666, EPI_ISL_1542667, EPI_ISL_1542668, EPI_ISL_1542671, EPI_ISL_1542672, EPI_ISL_1542673, EPI_ISL_1542674, EPI_ISL_1542675, EPI_ISL_1542676,
EPI_ISL_1542677, EPI_ISL_1542678, EPI_ISL_1542679, EPI_ISL_1542681, EPI_ISL_1542682, EPI_ISL_1542683, EPI_ISL_1542685, EPI_ISL_1542686, EPI_ISL_1542687, EPI_ISL_1542688, EPI_ISL_1542689, EPI_ISL_1542690, EPI_ISL_1542691, EPI_ISL_1542693, EPI_ISL_1542694, EPI_ISL_1542696, EPI_ISL_1542698, EPI_ISL_1542699, EPI_ISL_1542700, EPI_ISL_1542701, EPI_ISL_1542702, EPI_ISL_1542703,
EPI_ISL_1542704, EPI_ISL_1542705, EPI_ISL_1542707, EPI_ISL_1542709, EPI_ISL_1542710, EPI_ISL_1542712, EPI_ISL_1542713, EPI_ISL_1542714, EPI_ISL_1542715, EPI_ISL_1542716, EPI_ISL_1542717, EPI_ISL_1542719, EPI_ISL_1542721, EPI_ISL_1542722, EPI_ISL_1542723, EPI_ISL_1542725, EPI_ISL_1542726, EPI_ISL_1542728, EPI_ISL_1542730, EPI_ISL_1542731, EPI_ISL_1542732, EPI_ISL_1542733,
EPI_ISL_1542734, EPI_ISL_1542735, EPI_ISL_1542736, EPI_ISL_1542737, EPI_ISL_1542738, EPI_ISL_1542739, EPI_ISL_1542740, EPI_ISL_1542741, EPI_ISL_1542742, EPI_ISL_1542743, EPI_ISL_1542744, EPI_ISL_1542747, EPI_ISL_1542748, EPI_ISL_1542749, EPI_ISL_1542750, EPI_ISL_1542751, EPI_ISL_1542753, EPI_ISL_1542754, EPI_ISL_1542756, EPI_ISL_1542757, EPI_ISL_1542759, EPI_ISL_1542760,
EPI_ISL_1542762, EPI_ISL_1542763, EPI_ISL_1542764, EPI_ISL_1542768, EPI_ISL_1542769, EPI_ISL_1542771, EPI_ISL_1542772, EPI_ISL_1542773, EPI_ISL_1542774, EPI_ISL_1542775, EPI_ISL_1542776, EPI_ISL_1542777, EPI_ISL_1542778, EPI_ISL_1542779, EPI_ISL_1542780, EPI_ISL_1542781, EPI_ISL_1542782, EPI_ISL_1542783, EPI_ISL_1542784, EPI_ISL_1542787, EPI_ISL_1542788, EPI_ISL_1542789,
EPI_ISL_1542790, EPI_ISL_1542791, EPI_ISL_1542792, EPI_ISL_1542793, EPI_ISL_1542794, EPI_ISL_1542795, EPI_ISL_1542796, EPI_ISL_1542797, EPI_ISL_1542798, EPI_ISL_1542799, EPI_ISL_1542800, EPI_ISL_1542801, EPI_ISL_1542802, EPI_ISL_1542803, EPI_ISL_1542804, EPI_ISL_1542805, EPI_ISL_1542806, EPI_ISL_1542807, EPI_ISL_1542809, EPI_ISL_1542810, EPI_ISL_1542811, EPI_ISL_1542812,
EPI_ISL_1542813, EPI_ISL_1542814, EPI_ISL_1542815, EPI_ISL_1542816, EPI_ISL_1542817, EPI_ISL_1542821, EPI_ISL_1542822, EPI_ISL_1542823, EPI_ISL_1542824, EPI_ISL_1542825, EPI_ISL_1542826, EPI_ISL_1542828, EPI_ISL_1542829, EPI_ISL_1542830, EPI_ISL_1542831, EPI_ISL_1542833, EPI_ISL_1542834, EPI_ISL_1542835, EPI_ISL_1542836, EPI_ISL_1542838, EPI_ISL_1542840, EPI_ISL_1542841,
EPI_ISL_1542842, EPI_ISL_1542843, EPI_ISL_1542844, EPI_ISL_1542845, EPI_ISL_1542846, EPI_ISL_1542847, EPI_ISL_1542849, EPI_ISL_1542850, EPI_ISL_1542851, EPI_ISL_1542852, EPI_ISL_1542854, EPI_ISL_1542855, EPI_ISL_1542856, EPI_ISL_1542857, EPI_ISL_1542859, EPI_ISL_1542860, EPI_ISL_1542861, EPI_ISL_1542862, EPI_ISL_1542863, EPI_ISL_1542864, EPI_ISL_1542865, EPI_ISL_1542867,
EPI_ISL_1542868, EPI_ISL_1542869, EPI_ISL_1542870, EPI_ISL_1542871, EPI_ISL_1542872, EPI_ISL_1542873, EPI_ISL_1542874, EPI_ISL_1542875, EPI_ISL_1542876, EPI_ISL_1542877, EPI_ISL_1542878, EPI_ISL_1542879, EPI_ISL_1542880, EPI_ISL_1542881, EPI_ISL_1542882, EPI_ISL_1542883, EPI_ISL_1542884, EPI_ISL_1542885, EPI_ISL_1542886, EPI_ISL_1542887, EPI_ISL_1542888, EPI_ISL_1542889,
EPI_ISL_1542890, EPI_ISL_1542891, EPI_ISL_1542892, EPI_ISL_1542893, EPI_ISL_1542895, EPI_ISL_1542897, EPI_ISL_1542900, EPI_ISL_1542901, EPI_ISL_1542902, EPI_ISL_1542903, EPI_ISL_1542904, EPI_ISL_1542905, EPI_ISL_1542906, EPI_ISL_1542907, EPI_ISL_1542908, EPI_ISL_1542909, EPI_ISL_1542910, EPI_ISL_1542911, EPI_ISL_1542912, EPI_ISL_1542913, EPI_ISL_1542914, EPI_ISL_1542915,
EPI_ISL_1542916, EPI_ISL_1542917, EPI_ISL_1542919, EPI_ISL_1542920, EPI_ISL_1542921, EPI_ISL_1542922, EPI_ISL_1542923, EPI_ISL_1542924, EPI_ISL_1542925, EPI_ISL_1542926, EPI_ISL_1542927, EPI_ISL_1542928, EPI_ISL_1542929, EPI_ISL_1542930, EPI_ISL_1542931, EPI_ISL_1542932, EPI_ISL_1542933, EPI_ISL_1542934, EPI_ISL_1542935, EPI_ISL_1542936, EPI_ISL_1542937, EPI_ISL_1542938,
EPI_ISL_1542939, EPI_ISL_1542940, EPI_ISL_1542941, EPI_ISL_1542942, EPI_ISL_1542943, EPI_ISL_1542944, EPI_ISL_1542945, EPI_ISL_1542946, EPI_ISL_1542947, EPI_ISL_1542948, EPI_ISL_1542949, EPI_ISL_1542950, EPI_ISL_1542951, EPI_ISL_1542952, EPI_ISL_1542953, EPI_ISL_1542954, EPI_ISL_1542955, EPI_ISL_1542956, EPI_ISL_1542957, EPI_ISL_1542958, EPI_ISL_1542959, EPI_ISL_1542960,
EPI_ISL_1542961, EPI_ISL_1542962, EPI_ISL_1542963, EPI_ISL_1542964, EPI_ISL_1542965, EPI_ISL_1542966, EPI_ISL_1542967, EPI_ISL_1542968, EPI_ISL_1542969, EPI_ISL_1542970, EPI_ISL_1542971, EPI_ISL_1542972, EPI_ISL_1542973, EPI_ISL_1542974, EPI_ISL_1542975, EPI_ISL_1542976, EPI_ISL_1542977, EPI_ISL_1542978, EPI_ISL_1542979, EPI_ISL_1542980, EPI_ISL_1542981, EPI_ISL_1542982,
EPI_ISL_1542983, EPI_ISL_1542984, EPI_ISL_1542985, EPI_ISL_1542986, EPI_ISL_1542987, EPI_ISL_1542988, EPI_ISL_1542989, EPI_ISL_1542990, EPI_ISL_1542991, EPI_ISL_1542992, EPI_ISL_1542993, EPI_ISL_1542994, EPI_ISL_1542995, EPI_ISL_1542996, EPI_ISL_1542997, EPI_ISL_1542998, EPI_ISL_1542999, EPI_ISL_1543000, EPI_ISL_1543001, EPI_ISL_1543002, EPI_ISL_1543003, EPI_ISL_1543004,
EPI_ISL_1543005, EPI_ISL_1543006, EPI_ISL_1543007, EPI_ISL_1543008, EPI_ISL_1543009, EPI_ISL_1543010, EPI_ISL_1543011, EPI_ISL_1543012, EPI_ISL_1543013, EPI_ISL_1543014, EPI_ISL_1543015, EPI_ISL_1543016, EPI_ISL_1543017, EPI_ISL_1543018, EPI_ISL_1543019, EPI_ISL_1543020, EPI_ISL_1543021, EPI_ISL_1543022, EPI_ISL_1543023, EPI_ISL_1543024, EPI_ISL_1543025, EPI_ISL_1543026,
EPI_ISL_1543027, EPI_ISL_1543028, EPI_ISL_1543029, EPI_ISL_1543030, EPI_ISL_1543031, EPI_ISL_1543032, EPI_ISL_1543033, EPI_ISL_1543034, EPI_ISL_1543035, EPI_ISL_1543036, EPI_ISL_1543037, EPI_ISL_1543038, EPI_ISL_1543039, EPI_ISL_1543040, EPI_ISL_1543041, EPI_ISL_1543042, EPI_ISL_1543043, EPI_ISL_1543044, EPI_ISL_1543045, EPI_ISL_1543046, EPI_ISL_1543047, EPI_ISL_1543048,
EPI_ISL_1543049, EPI_ISL_1543050, EPI_ISL_1543051, EPI_ISL_1543052, EPI_ISL_1543053, EPI_ISL_1543054, EPI_ISL_1543055, EPI_ISL_1543056, EPI_ISL_1543057, EPI_ISL_1543058, EPI_ISL_1543059, EPI_ISL_1543060, EPI_ISL_1543061, EPI_ISL_1543062, EPI_ISL_1543063, EPI_ISL_1543064, EPI_ISL_1543065, EPI_ISL_1543066, EPI_ISL_1543067, EPI_ISL_1543068, EPI_ISL_1543069, EPI_ISL_1543070,
EPI_ISL_1543071, EPI_ISL_1543072, EPI_ISL_1543073, EPI_ISL_1543074, EPI_ISL_1543075, EPI_ISL_1543076, EPI_ISL_1543077, EPI_ISL_1543078, EPI_ISL_1543079, EPI_ISL_1543080, EPI_ISL_1543081, EPI_ISL_1543082, EPI_ISL_1543083, EPI_ISL_1543084, EPI_ISL_1543085, EPI_ISL_1543086, EPI_ISL_1543087, EPI_ISL_1543088, EPI_ISL_1543089, EPI_ISL_1543090, EPI_ISL_1543091, EPI_ISL_1543092,
EPI_ISL_1543093, EPI_ISL_1543094, EPI_ISL_1543095, EPI_ISL_1543096, EPI_ISL_1543097, EPI_ISL_1543098, EPI_ISL_1543099, EPI_ISL_1543100, EPI_ISL_1543101, EPI_ISL_1543102, EPI_ISL_1543103, EPI_ISL_1543104, EPI_ISL_1543105, EPI_ISL_1543106, EPI_ISL_1543107, EPI_ISL_1543108, EPI_ISL_1543109, EPI_ISL_1543110, EPI_ISL_1543111, EPI_ISL_1543112, EPI_ISL_1543113, EPI_ISL_1543114,
EPI_ISL_1543115, EPI_ISL_1543116, EPI_ISL_1543117, EPI_ISL_1543118, EPI_ISL_1543119, EPI_ISL_1543120, EPI_ISL_1543121

| | | | |
|---|---|---|---|
| see above | Pandemic Response Lab - NYC | Pandemic Response Lab, R&D | Cybill del Castillo; Dylan Law; Haiping Hao; Henry Lee; Jon Laurent; Katharine Nelson; Melissa Hopkins; Michael Hammerling; Pradeep Bugga; Shinyoung Clair Kang; Sol Rey; William Ward |
| EPI_ISL_1625770 | Pathology and Laboratory Medicine Institute, Cleveland Clinic, Ohio, USA | Pathology and Laboratory Medicine Institute, Cleveland Clinic, Ohio, USA | Daniel H. Farkas; Daniel Rhoads; David Bosler; David Plunkett; Gary Procop; Jay Brock; Jennifer Starbuck; Joy Nakitandwe; Kristen McDonnell; Yu-Wei Cheng; Zheng Jin Tu |
| EPI_ISL_1694720 | Poplar Healthcare | New York City Public Health Laboratory | Jade Wang; et al. |

EPI_ISL_1479876, EPI_ISL_1479881, EPI_ISL_1479891, EPI_ISL_1479911, EPI_ISL_1491965, EPI_ISL_1491986, EPI_ISL_1491994, EPI_ISL_1492112, EPI_ISL_1493627, EPI_ISL_1511714, EPI_ISL_1511768, EPI_ISL_1511769, EPI_ISL_1511770, EPI_ISL_1511774, EPI_ISL_1511776, EPI_ISL_1511779, EPI_ISL_1511781, EPI_ISL_1511782, EPI_ISL_1511783, EPI_ISL_1511789, EPI_ISL_1511791, EPI_ISL_1511793,
EPI_ISL_1511794, EPI_ISL_1511797, EPI_ISL_1511802, EPI_ISL_1511805, EPI_ISL_1511809, EPI_ISL_1511813, EPI_ISL_1511814, EPI_ISL_1511815, EPI_ISL_1511816, EPI_ISL_1511817, EPI_ISL_1511820, EPI_ISL_1511821, EPI_ISL_1511822, EPI_ISL_1511824, EPI_ISL_1511829, EPI_ISL_1511830, EPI_ISL_1511831, EPI_ISL_1511832, EPI_ISL_1511834, EPI_ISL_1511836, EPI_ISL_1511838, EPI_ISL_1511842,
EPI_ISL_1511846, EPI_ISL_1511848, EPI_ISL_1511850, EPI_ISL_1511852, EPI_ISL_1511856, EPI_ISL_1511858, EPI_ISL_1551720, EPI_ISL_1551723, EPI_ISL_1551725, EPI_ISL_1551746, EPI_ISL_1551766, EPI_ISL_1551772, EPI_ISL_1552235, EPI_ISL_1552259, EPI_ISL_1552275, EPI_ISL_1552278, EPI_ISL_1552301, EPI_ISL_1552713, EPI_ISL_1552918, EPI_ISL_1553024, EPI_ISL_1553038, EPI_ISL_1553044,
EPI_ISL_1553049, EPI_ISL_1553053, EPI_ISL_1553056, EPI_ISL_1553061, EPI_ISL_1553064, EPI_ISL_1553069, EPI_ISL_1553070, EPI_ISL_1553071, EPI_ISL_1553075, EPI_ISL_1553076, EPI_ISL_1553080, EPI_ISL_1553082, EPI_ISL_1553085, EPI_ISL_1553086, EPI_ISL_1553087, EPI_ISL_1553092, EPI_ISL_1553093, EPI_ISL_1559420, EPI_ISL_1559430, EPI_ISL_1559442, EPI_ISL_1559557, EPI_ISL_1581871,
EPI_ISL_1581898, EPI_ISL_1581931, EPI_ISL_1581934, EPI_ISL_1581962, EPI_ISL_1582034, EPI_ISL_1582039, EPI_ISL_1582051, EPI_ISL_1582058, EPI_ISL_1582066, EPI_ISL_1582100, EPI_ISL_1582109, EPI_ISL_1582116, EPI_ISL_1582119, EPI_ISL_1582133, EPI_ISL_1582142, EPI_ISL_1582148, EPI_ISL_1582161, EPI_ISL_1614913, EPI_ISL_1614918, EPI_ISL_1614935, EPI_ISL_1614940, EPI_ISL_1614948,
EPI_ISL_1614957, EPI_ISL_1614967, EPI_ISL_1614977, EPI_ISL_1614981, EPI_ISL_1614988, EPI_ISL_1615021, EPI_ISL_1615032, EPI_ISL_1615037, EPI_ISL_1615039, EPI_ISL_1615047, EPI_ISL_1615061, EPI_ISL_1621729, EPI_ISL_1621744, EPI_ISL_1621747, EPI_ISL_1621749, EPI_ISL_1621786, EPI_ISL_1621789, EPI_ISL_1621807, EPI_ISL_1621812, EPI_ISL_1621815, EPI_ISL_1621818, EPI_ISL_1621850,
EPI_ISL_1621854, EPI_ISL_1621855, EPI_ISL_1621856, EPI_ISL_1621857, EPI_ISL_1621859, EPI_ISL_1621860, EPI_ISL_1621865, EPI_ISL_1621866, EPI_ISL_1621871, EPI_ISL_1621872, EPI_ISL_1621873, EPI_ISL_1621875, EPI_ISL_1621876, EPI_ISL_1621879, EPI_ISL_1621880, EPI_ISL_1621881, EPI_ISL_1621884, EPI_ISL_1621890, EPI_ISL_1621896, EPI_ISL_1621903, EPI_ISL_1621906, EPI_ISL_1621909,
EPI_ISL_1621913, EPI_ISL_1621915, EPI_ISL_1621921, EPI_ISL_1621924, EPI_ISL_1621925, EPI_ISL_1621928, EPI_ISL_1621941, EPI_ISL_1621942, EPI_ISL_1621945, EPI_ISL_1621946, EPI_ISL_1621947, EPI_ISL_1621956, EPI_ISL_1621958, EPI_ISL_1621962, EPI_ISL_1621988, EPI_ISL_1621991, EPI_ISL_1621992, EPI_ISL_1622001, EPI_ISL_1622003, EPI_ISL_1622005, EPI_ISL_1622007, EPI_ISL_1622008,
EPI_ISL_1622018, EPI_ISL_1622026, EPI_ISL_1622030, EPI_ISL_1622031, EPI_ISL_1622038, EPI_ISL_1622042, EPI_ISL_1622047, EPI_ISL_1622055, EPI_ISL_1622059, EPI_ISL_1622072, EPI_ISL_1622073, EPI_ISL_1622075, EPI_ISL_1622079, EPI_ISL_1622087, EPI_ISL_1622095, EPI_ISL_1622103, EPI_ISL_1622105, EPI_ISL_1622109, EPI_ISL_1648211, EPI_ISL_1648236, EPI_ISL_1648303, EPI_ISL_1648304,
EPI_ISL_1648314, EPI_ISL_1648317, EPI_ISL_1648320, EPI_ISL_1648327, EPI_ISL_1648341, EPI_ISL_1648358, EPI_ISL_1648362, EPI_ISL_1648365, EPI_ISL_1648384, EPI_ISL_1648386, EPI_ISL_1648392, EPI_ISL_1648397, EPI_ISL_1648401, EPI_ISL_1648402, EPI_ISL_1648417, EPI_ISL_1648437, EPI_ISL_1648461, EPI_ISL_1648499, EPI_ISL_1648538, EPI_ISL_1648558, EPI_ISL_1648561, EPI_ISL_1648586,
EPI_ISL_1648757, EPI_ISL_1648778, EPI_ISL_1648780, EPI_ISL_1648784, EPI_ISL_1648829, EPI_ISL_1648842, EPI_ISL_1648885, EPI_ISL_1648916, EPI_ISL_1668648, EPI_ISL_1668707, EPI_ISL_1668768, EPI_ISL_1669089, EPI_ISL_1691410, EPI_ISL_1691436, EPI_ISL_1691492, EPI_ISL_1694671, EPI_ISL_1694672, EPI_ISL_1694673, EPI_ISL_1695037, EPI_ISL_1695051, EPI_ISL_1695067, EPI_ISL_1695681,
EPI_ISL_1738235, EPI_ISL_1738591, EPI_ISL_1753625

| | | | |
|---|---|---|---|
| see above | Quest Diagnostics Incorporated | Centers for Disease Control and Prevention Division of Viral Diseases, Pathogen Discovery | A. Gerasimova; A. Perez; Adrian Paskey; B. Anderson; Ben L. Rambo-Martin; Benjamin Rambo-Martin; Christopher Gulvick; Clinton R. Paden; Dakota Howard; Darlene Wagner; Dhwani Batra; Duncan MacCannell; F. Lacbawan; I. A. Shlyakhter; Jason Caravas; K.E. Livingston; Kara Moser; L.E. Bernstein; M. Hua; Matthew Schmerer; P. Tanpaiboon; Peter W. Cook; R. M. Kagan; R. Owen; R. V. Rolando; S. H. Rosenthal; Scott Sammons; Shatavia Morrison; Suxiang Tong; Y. Liu; Yvette Unoarumhi |

EPI_ISL_1580739, EPI_ISL_1580740, EPI_ISL_1582227, EPI_ISL_1582229, EPI_ISL_1624485, EPI_ISL_1624486, EPI_ISL_1755992, EPI_ISL_1755993, EPI_ISL_1755994, EPI_ISL_1755995, EPI_ISL_1855220, EPI_ISL_2098625, EPI_ISL_2098627

| | | | |
|---|---|---|---|
| see above | SARATOGA HOSPITAL LABORATORY | Wadsworth Center, New York State Department of Health | Alexis Russell; Catharine Prussing; Daryl M. Lamson; Erasmus Schneider; Erica Lasek-Nesselquist; John Kelly; Jonathan Plitnick; Kirsten St. George; Matthew Shudt; Melissa A Leisner; Navjot Singh |
| EPI_ISL_1482454, EPI_ISL_1582272, EPI_ISL_1671468 | ST. FRANCIS HOSPITAL LABORATORY | Wadsworth Center, New York State Department of Health | Alexis Russell; Catharine Prussing; Daryl M. Lamson; Erasmus Schneider; Erica Lasek-Nesselquist; John Kelly; Jonathan Plitnick; Kirsten St. George; Matthew Shudt; Melissa A Leisner; Navjot Singh |

EPI_ISL_1465838, EPI_ISL_1465839, EPI_ISL_1465840, EPI_ISL_1465841, EPI_ISL_1465842, EPI_ISL_1465843, EPI_ISL_1465845, EPI_ISL_1465846, EPI_ISL_1465847, EPI_ISL_1582245, EPI_ISL_1582246, EPI_ISL_1582247, EPI_ISL_1582248, EPI_ISL_1582249, EPI_ISL_1582250, EPI_ISL_1582251, EPI_ISL_1582252, EPI_ISL_1582253, EPI_ISL_1582254, EPI_ISL_1671316, EPI_ISL_1671322, EPI_ISL_1671328,
EPI_ISL_1671329, EPI_ISL_1671330, EPI_ISL_1671336, EPI_ISL_1671349, EPI_ISL_1671356, EPI_ISL_1740508

| | | | |
|---|---|---|---|
| see above | STONY BROOK UNIVERSITY | Wadsworth Center, New York | Alexis Russell; Catharine Prussing; Daryl M. Lamson; Erasmus Schneider; Erica Lasek-Nesselquist; John Kelly; Jonathan Plitnick; Kirsten St. George; Matthew Shudt; Melissa A Leisner; Navjot Singh |

| | | | |
|---|---|---|---|
| | HOSPITAL | State Department of Health | |
| EPI_ISL_1461312, EPI_ISL_1461313, EPI_ISL_1461314, EPI_ISL_1461315, EPI_ISL_1461316, EPI_ISL_1461317, EPI_ISL_1462537, EPI_ISL_1462539, EPI_ISL_1462540, EPI_ISL_1462542, EPI_ISL_1462543, EPI_ISL_1462545, EPI_ISL_1462546, EPI_ISL_1462548, EPI_ISL_1462549, EPI_ISL_1465665, EPI_ISL_1465666, EPI_ISL_1465697, EPI_ISL_1465698, EPI_ISL_1465699, EPI_ISL_1465700, EPI_ISL_1482406, EPI_ISL_1482407, EPI_ISL_1575445, EPI_ISL_1576813, EPI_ISL_1580678, EPI_ISL_1580679, EPI_ISL_1580680, EPI_ISL_1580681, EPI_ISL_1580682, EPI_ISL_1580683, EPI_ISL_1580684, EPI_ISL_1580685, EPI_ISL_1582318, EPI_ISL_1582360, EPI_ISL_1582361, EPI_ISL_1582362, EPI_ISL_1582363, EPI_ISL_1582364, EPI_ISL_1582365, EPI_ISL_1582366, EPI_ISL_1582367, EPI_ISL_1582399, EPI_ISL_1624406, EPI_ISL_1624407, EPI_ISL_1624408, EPI_ISL_1624420, EPI_ISL_1671463, EPI_ISL_1671464, EPI_ISL_1671465, EPI_ISL_1671494, EPI_ISL_1671512, EPI_ISL_1671514, EPI_ISL_1671612, EPI_ISL_1755842, EPI_ISL_1755843, EPI_ISL_1755844, EPI_ISL_1755845, EPI_ISL_1755846, EPI_ISL_1755847, EPI_ISL_1755848, EPI_ISL_1789709 | | | |
| see above | SUNY UPSTATE MEDICAL UNIVERSITY | Wadsworth Center, New York State Department of Health | Alexis Russell; Catharine Prussing; Daryl M. Lamson; Erasmus Schneider; Erica Lasek-Nesselquist; John Kelly; Jonathan Plitnick; Kirsten St. George; Matthew Shudt; Melissa A Leisner; Navjot Singh |
| EPI_ISL_1967685, EPI_ISL_1967687 | Stony Brook Medicine | New York Genome Center | Alaa B. Salaheldin; Andre Corvelo; Dayna M. Oschwald; Dimitri Gnatenko; Eric D. Spitzer; Jiang Chen; Josue M. Flores Acuna; Kenneth R. Shroyer; Margaret M. McGovern; Michael Zody; Richard R. Kew; Samantha Fennessey; Sarah A. Halbohn; Sarah A. Malik; Silvia G. Spitzer; Tom Maniatis |
| EPI_ISL_1461358, EPI_ISL_1465859, EPI_ISL_1465860, EPI_ISL_1465861, EPI_ISL_1465862, EPI_ISL_1465863, EPI_ISL_1465864, EPI_ISL_1465865, EPI_ISL_1465866, EPI_ISL_1465867, EPI_ISL_1465868, EPI_ISL_1465869, EPI_ISL_1465870, EPI_ISL_1465871, EPI_ISL_1465872, EPI_ISL_1465873, EPI_ISL_1465874, EPI_ISL_1465875, EPI_ISL_1465876, EPI_ISL_1465877, EPI_ISL_1465878, EPI_ISL_1575471, EPI_ISL_1576821, EPI_ISL_1576822, EPI_ISL_1582232, EPI_ISL_1582233, EPI_ISL_1582234, EPI_ISL_1582235, EPI_ISL_1582236, EPI_ISL_1582237, EPI_ISL_1582238, EPI_ISL_1582239, EPI_ISL_1582240, EPI_ISL_1582241, EPI_ISL_1582242, EPI_ISL_1582243, EPI_ISL_1582244, EPI_ISL_1582357, EPI_ISL_1582358, EPI_ISL_1582359, EPI_ISL_1593873, EPI_ISL_1593874, EPI_ISL_1593875, EPI_ISL_1593884, EPI_ISL_1593885, EPI_ISL_1605699, EPI_ISL_1671312, EPI_ISL_1671314, EPI_ISL_1671331, EPI_ISL_1671332, EPI_ISL_1671333, EPI_ISL_1671334, EPI_ISL_1671335, EPI_ISL_1671338, EPI_ISL_1671339, EPI_ISL_1671340, EPI_ISL_1671342, EPI_ISL_1671343, EPI_ISL_1671344, EPI_ISL_1671345, EPI_ISL_1671346, EPI_ISL_1671347, EPI_ISL_1671348, EPI_ISL_1671352, EPI_ISL_1671353, EPI_ISL_1671355, EPI_ISL_1671643, EPI_ISL_1740506, EPI_ISL_1740507 | | | |
| see above | TEMPUS LABS INC | Wadsworth Center, New York State Department of Health | Alexis Russell; Catharine Prussing; Daryl M. Lamson; Erasmus Schneider; Erica Lasek-Nesselquist; John Kelly; Jonathan Plitnick; Kirsten St. George; Matthew Shudt; Melissa A Leisner; Navjot Singh |
| EPI_ISL_1462572, EPI_ISL_1462573, EPI_ISL_1462575, EPI_ISL_1465673, EPI_ISL_1465676, EPI_ISL_1465677 | THE MARY IMOGENE BASSETT HOSPITAL | Wadsworth Center, New York State Department of Health | Alexis Russell; Catharine Prussing; Daryl M. Lamson; Erasmus Schneider; Erica Lasek-Nesselquist; John Kelly; Jonathan Plitnick; Kirsten St. George; Matthew Shudt; Melissa A Leisner; Navjot Singh |
| EPI_ISL_1470626, EPI_ISL_1540718 | The Jackson Laboratory | The Jackson Laboratory | Adams M; Bergeron D; Kelly K; Li L; Long J; Omerza G; Renzette N |
| EPI_ISL_1465723, EPI_ISL_1465724, EPI_ISL_1465725, EPI_ISL_1465730, EPI_ISL_1465731, EPI_ISL_1465732, EPI_ISL_1465733, EPI_ISL_1465734, EPI_ISL_1465735, EPI_ISL_1465736, EPI_ISL_1465737, EPI_ISL_1465738, EPI_ISL_1465739, EPI_ISL_1465740, EPI_ISL_1465741, EPI_ISL_1465742, EPI_ISL_1465743, EPI_ISL_1465744, EPI_ISL_1465745, EPI_ISL_1465746, EPI_ISL_1465747, EPI_ISL_1465748, EPI_ISL_1465749, EPI_ISL_1465750, EPI_ISL_1465751, EPI_ISL_1465752, EPI_ISL_1465753, EPI_ISL_1465754, EPI_ISL_1482296, EPI_ISL_1482297, EPI_ISL_1482298, EPI_ISL_1482300, EPI_ISL_1482301, EPI_ISL_1482304, EPI_ISL_1482305, EPI_ISL_1482306, EPI_ISL_1482307, EPI_ISL_1482308, EPI_ISL_1482309, EPI_ISL_1482310, EPI_ISL_1482311, EPI_ISL_1482312, EPI_ISL_1482313, EPI_ISL_1482314, EPI_ISL_1482315, EPI_ISL_1482316, EPI_ISL_1482317, EPI_ISL_1482318, EPI_ISL_1482319, EPI_ISL_1482320, EPI_ISL_1482321, EPI_ISL_1482322, EPI_ISL_1482323, EPI_ISL_1482324, EPI_ISL_1482325, EPI_ISL_1482326, EPI_ISL_1482327, EPI_ISL_1482328, EPI_ISL_1482329, EPI_ISL_1482330, EPI_ISL_1482331, EPI_ISL_1482332, EPI_ISL_1482625, EPI_ISL_1482626, EPI_ISL_1575460, EPI_ISL_1575462, EPI_ISL_1575463, EPI_ISL_1575464, EPI_ISL_1575465, EPI_ISL_1575466, EPI_ISL_1575467, EPI_ISL_1575468, EPI_ISL_1575474, EPI_ISL_1575475, EPI_ISL_1575476, EPI_ISL_1576817, EPI_ISL_1576818, EPI_ISL_1580701, EPI_ISL_1580702, EPI_ISL_1580703, EPI_ISL_1580704, EPI_ISL_1580705, EPI_ISL_1580706, EPI_ISL_1580707, EPI_ISL_1580708, EPI_ISL_1580709, EPI_ISL_1580710, EPI_ISL_1580711, EPI_ISL_1580712, EPI_ISL_1580713, EPI_ISL_1580714, EPI_ISL_1580715, EPI_ISL_1580716, EPI_ISL_1580717, EPI_ISL_1580718, EPI_ISL_1580719, EPI_ISL_1580720, EPI_ISL_1580721, EPI_ISL_1580722, EPI_ISL_1582219, EPI_ISL_1582220, EPI_ISL_1582221, EPI_ISL_1582222, EPI_ISL_1582223, EPI_ISL_1582224, EPI_ISL_1582339, EPI_ISL_1582340, EPI_ISL_1582341, EPI_ISL_1582342, EPI_ISL_1582343, EPI_ISL_1582344, EPI_ISL_1582345, EPI_ISL_1582346, EPI_ISL_1582347, EPI_ISL_1582348, EPI_ISL_1582349, EPI_ISL_1582350, EPI_ISL_1582351, EPI_ISL_1582352, EPI_ISL_1582353, EPI_ISL_1593866, EPI_ISL_1593867, EPI_ISL_1593882, EPI_ISL_1624410, EPI_ISL_1624411, EPI_ISL_1624412, EPI_ISL_1624413, EPI_ISL_1624414, EPI_ISL_1624415, EPI_ISL_1624416, EPI_ISL_1624417, EPI_ISL_1624418, EPI_ISL_1624419, EPI_ISL_1624421, EPI_ISL_1624422, EPI_ISL_1624423, EPI_ISL_1624424, EPI_ISL_1624425, EPI_ISL_1624426, EPI_ISL_1624427, EPI_ISL_1624428, EPI_ISL_1624429, EPI_ISL_1624430, EPI_ISL_1624431, EPI_ISL_1624432, EPI_ISL_1624433, EPI_ISL_1624434, EPI_ISL_1624435, EPI_ISL_1624436, EPI_ISL_1624437, EPI_ISL_1624438, EPI_ISL_1624439, EPI_ISL_1624440, EPI_ISL_1624441, EPI_ISL_1624442, EPI_ISL_1624443, EPI_ISL_1624444, EPI_ISL_1624445, EPI_ISL_1624446, EPI_ISL_1624447, EPI_ISL_1624448, EPI_ISL_1624449, EPI_ISL_1624450, EPI_ISL_1624451, EPI_ISL_1624452, EPI_ISL_1624453, EPI_ISL_1624454, EPI_ISL_1624455, EPI_ISL_1624456, EPI_ISL_1624457, EPI_ISL_1624458, EPI_ISL_1624459, EPI_ISL_1624460, EPI_ISL_1624461, EPI_ISL_1624462, EPI_ISL_1624463, EPI_ISL_1624464, EPI_ISL_1624465, EPI_ISL_1624466, EPI_ISL_1624467, EPI_ISL_1624468, EPI_ISL_1624469, EPI_ISL_1624470, EPI_ISL_1624471, EPI_ISL_1624472, EPI_ISL_1624473, EPI_ISL_1624474, EPI_ISL_1624475, EPI_ISL_1624476, EPI_ISL_1624477, EPI_ISL_1624478, EPI_ISL_1624479, EPI_ISL_1624480, EPI_ISL_1624481, EPI_ISL_1624482, EPI_ISL_1651222, EPI_ISL_1651223, EPI_ISL_1651227, EPI_ISL_1651228, EPI_ISL_1671613, EPI_ISL_1671626, EPI_ISL_1671627, EPI_ISL_1671634, EPI_ISL_1671667, EPI_ISL_1740516, EPI_ISL_1740517, EPI_ISL_1755835, EPI_ISL_1755869, EPI_ISL_1755870, EPI_ISL_1755871, EPI_ISL_1755872, EPI_ISL_1755873, EPI_ISL_1755874, EPI_ISL_1755875, EPI_ISL_1755876, EPI_ISL_1755877, EPI_ISL_1755878, EPI_ISL_1755879, EPI_ISL_1755880, EPI_ISL_1755881, EPI_ISL_1755882, EPI_ISL_1755883, EPI_ISL_1755884, EPI_ISL_1755885, EPI_ISL_1755886, EPI_ISL_1755887, EPI_ISL_1755888, EPI_ISL_1755889, EPI_ISL_1755890, EPI_ISL_1755891, EPI_ISL_1755892, EPI_ISL_1755893, EPI_ISL_1755894, EPI_ISL_1755895, EPI_ISL_1755896, EPI_ISL_1755897, EPI_ISL_1755898, EPI_ISL_1755899, EPI_ISL_1755900, EPI_ISL_1755901, EPI_ISL_1755902, EPI_ISL_1755903, EPI_ISL_1755904, EPI_ISL_1755905, EPI_ISL_1755906, EPI_ISL_1755907, EPI_ISL_1920066, EPI_ISL_1920067, EPI_ISL_1920068, EPI_ISL_1920069, EPI_ISL_1920124, EPI_ISL_1920125, EPI_ISL_1920126, EPI_ISL_1920128, EPI_ISL_1920129, EPI_ISL_1920130, EPI_ISL_1920131, EPI_ISL_1920997, EPI_ISL_1920998, EPI_ISL_1920999, EPI_ISL_1921000, EPI_ISL_1921044, EPI_ISL_1921045, EPI_ISL_1921047, EPI_ISL_1921048, EPI_ISL_1921052, EPI_ISL_1921053, EPI_ISL_1921057, EPI_ISL_1921059, EPI_ISL_1921060, EPI_ISL_1921062, EPI_ISL_1921063, EPI_ISL_1921064, EPI_ISL_1921067, EPI_ISL_1921068, EPI_ISL_1921073, EPI_ISL_1935618, EPI_ISL_1935652, EPI_ISL_2098161, EPI_ISL_2098162, EPI_ISL_2098164, EPI_ISL_2098166, EPI_ISL_2098167, EPI_ISL_2098169, EPI_ISL_2098170, EPI_ISL_2098172, EPI_ISL_2098173, EPI_ISL_2098175, EPI_ISL_2098176, EPI_ISL_2098178, EPI_ISL_2098180, EPI_ISL_2098181, EPI_ISL_2098183, EPI_ISL_2098184, EPI_ISL_2098186, EPI_ISL_2098187, EPI_ISL_2098189, EPI_ISL_2098191, EPI_ISL_2098192, EPI_ISL_2098193, EPI_ISL_2098195, EPI_ISL_2098196, EPI_ISL_2098198, EPI_ISL_2098199, EPI_ISL_2098201, EPI_ISL_2098202, EPI_ISL_2098204, EPI_ISL_2098206, EPI_ISL_2098208, EPI_ISL_2098209, EPI_ISL_2098211, EPI_ISL_2098212, EPI_ISL_2098214, EPI_ISL_2098215, EPI_ISL_2098217, EPI_ISL_2098218, EPI_ISL_2098220, EPI_ISL_2098222, EPI_ISL_2098223, EPI_ISL_2098224, EPI_ISL_2098225, EPI_ISL_2098227, EPI_ISL_2098228, EPI_ISL_2134793, EPI_ISL_2134794 | | | |
| see above | URMC LABS | Wadsworth Center, New York State Department of Health | Alexis Russell; Catharine Prussing; Daryl M. Lamson; Erasmus Schneider; Erica Lasek-Nesselquist; John Kelly; Jonathan Plitnick; Kirsten St. George; Matthew Shudt; Melissa A Leisner; Navjot Singh |
| EPI_ISL_1462555, EPI_ISL_1482396, EPI_ISL_1482397, EPI_ISL_1482398, EPI_ISL_1482399, EPI_ISL_1482400, EPI_ISL_1482401, EPI_ISL_1482402, EPI_ISL_1482403, EPI_ISL_1482404, EPI_ISL_1482405, EPI_ISL_1482411, EPI_ISL_1482412, EPI_ISL_1482413, EPI_ISL_1482414, EPI_ISL_1482415, EPI_ISL_1482416, EPI_ISL_1482417, EPI_ISL_1482418, EPI_ISL_1482419, EPI_ISL_1482420, EPI_ISL_1482421, EPI_ISL_1482422, EPI_ISL_1482423, EPI_ISL_1482424, EPI_ISL_1482425, EPI_ISL_1482426, EPI_ISL_1482427, EPI_ISL_1482428, EPI_ISL_1482429, EPI_ISL_1482430, EPI_ISL_1482431, EPI_ISL_1482432, EPI_ISL_1482433, EPI_ISL_1482434, EPI_ISL_1482435, EPI_ISL_1482436, EPI_ISL_1482437, EPI_ISL_1482438, EPI_ISL_1482439, EPI_ISL_1482440, EPI_ISL_1482441, EPI_ISL_1482442, EPI_ISL_1482443, EPI_ISL_1482444, EPI_ISL_1482445, EPI_ISL_1482544, EPI_ISL_1482545, EPI_ISL_1482546, EPI_ISL_1482547, EPI_ISL_1482548, EPI_ISL_1482549, EPI_ISL_1482550, EPI_ISL_1482551, EPI_ISL_1482552, EPI_ISL_1482553, EPI_ISL_1482554, EPI_ISL_1482555, EPI_ISL_1482617, EPI_ISL_1482618, EPI_ISL_1482619, EPI_ISL_1482620, EPI_ISL_1482621, EPI_ISL_1482622, EPI_ISL_1482623, EPI_ISL_1482624, EPI_ISL_1580673, EPI_ISL_1580674, EPI_ISL_1580675, EPI_ISL_1580676, EPI_ISL_1580677, EPI_ISL_1580699, EPI_ISL_1582231, EPI_ISL_1582310, EPI_ISL_1582311, EPI_ISL_1582312, EPI_ISL_1582313, EPI_ISL_1582314, EPI_ISL_1582315, EPI_ISL_1582316, EPI_ISL_1582338, EPI_ISL_1593864, EPI_ISL_1593865, EPI_ISL_1671449, EPI_ISL_1671458, EPI_ISL_1671459, EPI_ISL_1671460, EPI_ISL_1671461, EPI_ISL_1671467, EPI_ISL_1671469, EPI_ISL_1671470, EPI_ISL_1671471, EPI_ISL_1671472, EPI_ISL_1671473, EPI_ISL_1671474, EPI_ISL_1671480, EPI_ISL_1671481, EPI_ISL_1671482, EPI_ISL_1671483, EPI_ISL_1671485, EPI_ISL_1671486, EPI_ISL_1671487, EPI_ISL_1671488, EPI_ISL_1671489, EPI_ISL_1671492, EPI_ISL_1671500, EPI_ISL_1671501, EPI_ISL_1671502, EPI_ISL_1671503, EPI_ISL_1671508, EPI_ISL_1671509, EPI_ISL_1671510, EPI_ISL_1671619, EPI_ISL_1671624, EPI_ISL_1671625, EPI_ISL_1671636, EPI_ISL_1671637, EPI_ISL_1671639, EPI_ISL_1671646, EPI_ISL_1671647, EPI_ISL_1671657, EPI_ISL_1671658, EPI_ISL_1671659, EPI_ISL_1671660, EPI_ISL_1671666, EPI_ISL_1740514, EPI_ISL_1755851, EPI_ISL_1755852, EPI_ISL_1755853, EPI_ISL_1755867, EPI_ISL_1755868, EPI_ISL_1755966, EPI_ISL_1755967 | | | |
| see above | WESTCHESTER MEDICAL CENTER | Wadsworth Center, New York State Department of Health | Alexis Russell; Catharine Prussing; Daryl M. Lamson; Erasmus Schneider; Erica Lasek-Nesselquist; John Kelly; Jonathan Plitnick; Kirsten St. George; Matthew Shudt; Melissa A Leisner; Navjot Singh |
| EPI_ISL_1482573, EPI_ISL_1482574, EPI_ISL_1482575, EPI_ISL_1482576, EPI_ISL_1482577, EPI_ISL_1482578, EPI_ISL_1482579, EPI_ISL_1582273, EPI_ISL_1582274, EPI_ISL_1582275, EPI_ISL_1582276, EPI_ISL_1582277, EPI_ISL_1582278, EPI_ISL_1582279, EPI_ISL_1582280, EPI_ISL_1582281, EPI_ISL_1582282, EPI_ISL_1593886, EPI_ISL_1671598, EPI_ISL_1671600, EPI_ISL_1671607, EPI_ISL_1671610, EPI_ISL_1671611, EPI_ISL_1671655, EPI_ISL_1671656 | | | |
| see above | WHITE PLAINS HOSPITAL CENTER LABORATORY | Wadsworth Center, New York State Department of Health | Alexis Russell; Catharine Prussing; Daryl M. Lamson; Erasmus Schneider; Erica Lasek-Nesselquist; John Kelly; Jonathan Plitnick; Kirsten St. George; Matthew Shudt; Melissa A Leisner; Navjot Singh |
| EPI_ISL_1967662, EPI_ISL_1967663, EPI_ISL_1967664, EPI_ISL_1967665, EPI_ISL_1967666, EPI_ISL_1967668, EPI_ISL_1967669, EPI_ISL_1967670, EPI_ISL_1967671, EPI_ISL_1967672, EPI_ISL_1967673, EPI_ISL_1967674, EPI_ISL_1967675, EPI_ISL_1967676, EPI_ISL_1967677, EPI_ISL_1967678, EPI_ISL_1967679, EPI_ISL_1967680, EPI_ISL_1967681, EPI_ISL_1967682, EPI_ISL_1967683, EPI_ISL_1967684, EPI_ISL_1971088, EPI_ISL_1971089, EPI_ISL_1971090, EPI_ISL_1971092, EPI_ISL_1971093, EPI_ISL_1971094, EPI_ISL_1971095, EPI_ISL_1971096, EPI_ISL_1971097, EPI_ISL_1971099, EPI_ISL_1971101, EPI_ISL_1971102, EPI_ISL_1971103, EPI_ISL_1971104, EPI_ISL_1971107, EPI_ISL_1971108, EPI_ISL_1971109, EPI_ISL_1971110, EPI_ISL_1971111, EPI_ISL_1971112, EPI_ISL_1971113, EPI_ISL_1971115, EPI_ISL_1971116, EPI_ISL_1971117, EPI_ISL_1971118, EPI_ISL_1971119, EPI_ISL_1971120, EPI_ISL_1971121, EPI_ISL_1971122, EPI_ISL_1971123, EPI_ISL_1971124, EPI_ISL_1971125, EPI_ISL_1971127, EPI_ISL_1971128, EPI_ISL_1971130, EPI_ISL_1971132, EPI_ISL_1971133, EPI_ISL_1971135, EPI_ISL_1971136, EPI_ISL_1971137, EPI_ISL_1971139, EPI_ISL_1971140, EPI_ISL_1971141, EPI_ISL_1971142, EPI_ISL_1971143, EPI_ISL_1971145, EPI_ISL_1971146, EPI_ISL_1971147, EPI_ISL_1971150, EPI_ISL_1971151, EPI_ISL_1971152, EPI_ISL_1971153, EPI_ISL_1971157, EPI_ISL_1971159, EPI_ISL_1971164, EPI_ISL_1971165, EPI_ISL_1971166, EPI_ISL_1971167, EPI_ISL_1971168, EPI_ISL_1971170, EPI_ISL_1971172, EPI_ISL_1971173, EPI_ISL_1971176, EPI_ISL_1971177, EPI_ISL_1971180, EPI_ISL_1971182, EPI_ISL_1971183, EPI_ISL_1971184, EPI_ISL_1971188, EPI_ISL_1971189, EPI_ISL_1971191, EPI_ISL_1971192, EPI_ISL_1971194, EPI_ISL_1971195, EPI_ISL_1971199, EPI_ISL_1971201, EPI_ISL_1971205, EPI_ISL_1971206, EPI_ISL_1971209, EPI_ISL_1971212, EPI_ISL_1971215, EPI_ISL_1971216, EPI_ISL_1971218, EPI_ISL_1971220, EPI_ISL_1971222, EPI_ISL_1971225, EPI_ISL_1971230, EPI_ISL_1971231, EPI_ISL_2031764, EPI_ISL_2031765, EPI_ISL_2031766, EPI_ISL_2031767, EPI_ISL_2086300, EPI_ISL_2086301, EPI_ISL_2086302, EPI_ISL_2086303 | | | |
| see above | Weill Cornell Medicine | New York Genome Center | Andre Corvelo; Arryn Craney; Chris Mason; Dayna M. Oschwald; Hanna Rennert; Lars F Westblade; Margaret Elizabeth Ross; Melissa Cushing; Michael Zody; Olivier Elemento; Priya Velu; Samantha Fennessey; Tom Maniatis |
| EPI_ISL_1920173 | Westchester Medical Center | Wadsworth Center, New York State Department of Health | Alexis Russell; Catharine Prussing; Daryl M. Lamson; Erasmus Schneider; Erica Lasek-Nesselquist; John Kelly; Jonathan Plitnick; Kirsten St. George; Matthew Shudt; Melissa A Leisner; Navjot Singh |
| EPI_ISL_1447572, EPI_ISL_1447577, EPI_ISL_1509180, EPI_ISL_1509238, EPI_ISL_1587343, EPI_ISL_1587390, EPI_ISL_1963661, EPI_ISL_1963667 | | | |
| see above | Yale Clinical Virology Lab | Grubaugh Lab - Yale School of Public Health | Anderson Brito; Annie Watkins; Chaney Kalinich; Chantal Vogels; Isabel Ott; Isabell Ott; Jessica Rothman; Joseph Fauver; Mallery Breban; Marie L. Landry; Mary Petrone; Nathan Grubaugh; Tara Alpert |
| EPI_ISL_1793502 | Yale Clinical Virology Lab | Yale Center for Genomic Analysis | Brooke Sullivan; Christopher Castaldi; Curt Scharfe; Irina Tikhonova; Kaya Bilguvar; Shrikant Mane |

We gratefully acknowledge the following Authors from the Originating laboratories responsible for obtaining the specimens, as well as the Submitting laboratories where the genome data were generated and shared via GISAID, on which this research is based.

All Submitters of data may be contacted directly via www.gisaid.org

Authors are sorted alphabetically.

| Accession ID | Originating Laboratory | Submitting Laboratory | Authors |
|---|---|---|---|
| EPI_ISL_1824034, EPI_ISL_1824035 | "NYSDOH Wadsworth Center, Virology Lab" | Centers for Disease Control and Prevention Division of Viral Diseases, Pathogen Discovery | Alison Laufer Halpin; Ben L. Rambo-Martin; Clinton R. Paden; Dakota Howard; Darlene Wagner; Dave Wentworth; Dhwani Batra; Jasmine Padilla; Justin Lee; Katie Dillon; Krista Queen; Kristen Knipe; Kristine Lacek; Mark Burroughs; Matthew Schmerer; Mili Sheth; Peter Cook; Sam Shepard; Sarah Nobles; Shoshona Le; Suxiang Tong; Vivien Dugan; Yvette Unoarumhi |
| EPI_ISL_1671321, EPI_ISL_1671341, EPI_ISL_1671350, EPI_ISL_1671351, EPI_ISL_1671359, EPI_ISL_1671450, EPI_ISL_1671451, EPI_ISL_1671599, EPI_ISL_1671645, EPI_ISL_1756001, EPI_ISL_1756002, EPI_ISL_1756003, EPI_ISL_1756004, EPI_ISL_1756005, EPI_ISL_1756006, EPI_ISL_1756007, EPI_ISL_1756008, EPI_ISL_1756009, EPI_ISL_1756010, EPI_ISL_1756011, EPI_ISL_1756012, EPI_ISL_1756013, EPI_ISL_1756014, EPI_ISL_1756015, EPI_ISL_1756016, EPI_ISL_1756017, EPI_ISL_1756018, EPI_ISL_1756019, EPI_ISL_1756020, EPI_ISL_1789716, EPI_ISL_1789717, EPI_ISL_1789718, EPI_ISL_1854692, EPI_ISL_1855116, EPI_ISL_1855117, EPI_ISL_1855118 | | | |
| see above | ALBANY MEDICAL CENTER | Wadsworth Center, New York State Department of Health | Alexis Russell; Catharine Prussing; Daryl M. Lamson; Erasmus Schneider; Erica Lasek-Nesselquist; John Kelly; Jonathan Plitnick; Kirsten St. George; Matthew Shudt; Melissa A Leisner; Navjot Singh |
| EPI_ISL_1667660, EPI_ISL_1667704, EPI_ISL_1667740, EPI_ISL_1667744, EPI_ISL_1667746, EPI_ISL_1667747, EPI_ISL_1685759, EPI_ISL_1687491, EPI_ISL_1687502, EPI_ISL_1687506, EPI_ISL_1687507, EPI_ISL_1687508, EPI_ISL_1689883, EPI_ISL_1689977, EPI_ISL_1690008, EPI_ISL_1690013, EPI_ISL_1690039, EPI_ISL_1690052, EPI_ISL_1690079, EPI_ISL_1690081, EPI_ISL_1690101, EPI_ISL_1690115, EPI_ISL_1690121, EPI_ISL_1690203, EPI_ISL_1690204, EPI_ISL_1690250, EPI_ISL_1701968, EPI_ISL_1701969, EPI_ISL_1736495, EPI_ISL_1736525, EPI_ISL_1736550, EPI_ISL_1736626, EPI_ISL_1736656, EPI_ISL_1736685, EPI_ISL_1736720, EPI_ISL_1736763, EPI_ISL_1736796, EPI_ISL_1736798, EPI_ISL_1736799, EPI_ISL_1736806, EPI_ISL_1736809, EPI_ISL_1736842, EPI_ISL_1736847, EPI_ISL_1736852, EPI_ISL_1736862, EPI_ISL_1736882, EPI_ISL_1736890, EPI_ISL_1736899, EPI_ISL_1736927, EPI_ISL_1736930, EPI_ISL_1737597, EPI_ISL_1737689, EPI_ISL_1737780, EPI_ISL_1737813, EPI_ISL_1737847, EPI_ISL_1753557, EPI_ISL_1834544, EPI_ISL_1834546, EPI_ISL_1834558, EPI_ISL_1834564, EPI_ISL_1834732, EPI_ISL_1835476, EPI_ISL_1835525, EPI_ISL_1835566, EPI_ISL_1835665, EPI_ISL_1835701, EPI_ISL_1835727, EPI_ISL_1835780, EPI_ISL_1835801, EPI_ISL_1835884, EPI_ISL_1835940, EPI_ISL_1835989, EPI_ISL_1836019, EPI_ISL_1836037, EPI_ISL_1836170, EPI_ISL_1836175, EPI_ISL_1836256, EPI_ISL_1836280, EPI_ISL_1836303, EPI_ISL_1836305, EPI_ISL_1836306, EPI_ISL_1836309, EPI_ISL_1836313, EPI_ISL_1836348, EPI_ISL_1836381, EPI_ISL_1836384, EPI_ISL_1836386, EPI_ISL_1836479, EPI_ISL_1836620, EPI_ISL_1836689, EPI_ISL_1836696, EPI_ISL_1836698, EPI_ISL_1836785, EPI_ISL_1836814, EPI_ISL_1836965, EPI_ISL_1837051, EPI_ISL_1837154, EPI_ISL_1837168, EPI_ISL_1837252, EPI_ISL_1837254, EPI_ISL_1837281, EPI_ISL_1837287, EPI_ISL_1837295, EPI_ISL_1837340, EPI_ISL_1837400, EPI_ISL_1837591, EPI_ISL_1837637, EPI_ISL_1837693, EPI_ISL_1837694, EPI_ISL_1837797, EPI_ISL_1837904, EPI_ISL_1837936, EPI_ISL_1837941, EPI_ISL_1837947, EPI_ISL_1838017, EPI_ISL_1838027, EPI_ISL_1838034, EPI_ISL_1838334, EPI_ISL_1838438, EPI_ISL_1900354, EPI_ISL_1900419, EPI_ISL_1922972, EPI_ISL_1922974, EPI_ISL_1922995, EPI_ISL_1923020, EPI_ISL_1923029, EPI_ISL_1923040, EPI_ISL_1923080, EPI_ISL_1923092, EPI_ISL_1923116, EPI_ISL_1923149, EPI_ISL_1942142, EPI_ISL_1994977, EPI_ISL_1994994, EPI_ISL_1995028, EPI_ISL_1995029, EPI_ISL_1995039, EPI_ISL_1995083, EPI_ISL_1995095, EPI_ISL_1995114, EPI_ISL_1995124, EPI_ISL_1995125, EPI_ISL_1995126, EPI_ISL_1995130, EPI_ISL_1995132, EPI_ISL_1995133, EPI_ISL_1995142, EPI_ISL_1995164, EPI_ISL_1995289, EPI_ISL_1995299, EPI_ISL_1995300, EPI_ISL_1995302, EPI_ISL_1995381, EPI_ISL_1995406, EPI_ISL_1995412, EPI_ISL_1995518, EPI_ISL_1995549, EPI_ISL_1995553, EPI_ISL_1995554, EPI_ISL_1995682, EPI_ISL_1995683, EPI_ISL_1995684, EPI_ISL_1995731, EPI_ISL_1995733, EPI_ISL_1995734, EPI_ISL_1995761, EPI_ISL_1995887, EPI_ISL_1995915, EPI_ISL_1995916, EPI_ISL_1995938, EPI_ISL_1995949, EPI_ISL_1995951, EPI_ISL_1995957, EPI_ISL_1996018, EPI_ISL_1996068, EPI_ISL_1996073, EPI_ISL_1996074, EPI_ISL_1996113, EPI_ISL_1996114, EPI_ISL_1996115, EPI_ISL_1996132, EPI_ISL_1996137, EPI_ISL_1996208, EPI_ISL_1996277, EPI_ISL_1996297, EPI_ISL_1996475, EPI_ISL_1996503, EPI_ISL_1996510, EPI_ISL_1996602, EPI_ISL_1996612, EPI_ISL_1996844, EPI_ISL_1996845, EPI_ISL_1996846, EPI_ISL_1996854, EPI_ISL_1996938, EPI_ISL_1996945, EPI_ISL_1996946, EPI_ISL_1996947, EPI_ISL_1996983, EPI_ISL_1996997, EPI_ISL_1997025, EPI_ISL_1997067, EPI_ISL_1997069, EPI_ISL_1997120, EPI_ISL_1997129, EPI_ISL_1997147, EPI_ISL_1997148, EPI_ISL_1997149, EPI_ISL_1997172, EPI_ISL_1997251, EPI_ISL_1997283, EPI_ISL_1997298, EPI_ISL_1997329, EPI_ISL_1997330, EPI_ISL_1997381, EPI_ISL_1997383, EPI_ISL_1997398, EPI_ISL_1997449, EPI_ISL_1997450, EPI_ISL_1997452, EPI_ISL_1997453, EPI_ISL_1997463, EPI_ISL_1997465, EPI_ISL_1997475, EPI_ISL_1997476, EPI_ISL_1997499, EPI_ISL_1997502, EPI_ISL_1997518, EPI_ISL_1997519, EPI_ISL_1997545, EPI_ISL_1997550, EPI_ISL_1997576, EPI_ISL_1997625, EPI_ISL_1997627, EPI_ISL_1997681, EPI_ISL_1997683, EPI_ISL_1997736, EPI_ISL_1997737, EPI_ISL_1997742, EPI_ISL_1997752, EPI_ISL_1997753, EPI_ISL_1997755, EPI_ISL_1997827, EPI_ISL_1997939, EPI_ISL_1997940, EPI_ISL_1997998, EPI_ISL_1998084, EPI_ISL_1998189, EPI_ISL_1998251, EPI_ISL_1998286, EPI_ISL_1998288, EPI_ISL_1998305, EPI_ISL_1998338, EPI_ISL_1998606, EPI_ISL_1998608, EPI_ISL_1998680, EPI_ISL_1998688, EPI_ISL_1998715, EPI_ISL_1998733, EPI_ISL_1998736, EPI_ISL_1998770, EPI_ISL_1999110, EPI_ISL_1999124, EPI_ISL_1999232, EPI_ISL_1999246, EPI_ISL_1999250, EPI_ISL_1999262, EPI_ISL_1999353, EPI_ISL_1999476, EPI_ISL_1999477, EPI_ISL_1999572, EPI_ISL_1999684, EPI_ISL_1999715, EPI_ISL_1999892, EPI_ISL_1999941, EPI_ISL_1999974, EPI_ISL_2000278, EPI_ISL_2000286, EPI_ISL_2000386, EPI_ISL_2000389, EPI_ISL_2000390, EPI_ISL_2000391, EPI_ISL_2000438, EPI_ISL_2000507, EPI_ISL_2000542, EPI_ISL_2035107, EPI_ISL_2035130, EPI_ISL_2035263, EPI_ISL_2040997 | | | |
| see above | Aegis Sciences Corporation | Centers for Disease Control and Prevention Division of Viral Diseases, Pathogen Discovery | Adrian Paskey; Alec Vest; Benjamin Rambo-Martin; Christopher Gulvick; Clinton R. Paden; Cyndi Clark; Dakota Howard; Darlene Wagner; Dhwani Batra; Dillon Nall; Duncan MacCannell; Ethan Sanders; Holly Houdeshell; Jason Caravas; Kara Moser; Matthew Hardison; Matthew Schmerer; Ola Kvalvaag; Patrick Campbell; Peter W. Cook; Rob Case; Scott Sammons; Shatavia Morrison; Shaun Westlund; Vikramsinha Ghorpade; Yvette Unoarumhi |
| EPI_ISL_1920060 | Albany Medical Center | Wadsworth Center, New York State Department of Health | Alexis Russell; Catharine Prussing; Daryl M. Lamson; Erasmus Schneider; Erica Lasek-Nesselquist; John Kelly; Jonathan Plitnick; Kirsten St. George; Matthew Shudt; Melissa A Leisner; Navjot Singh |
| EPI_ISL_1756356 | BIO-REFERENCE LABORATORIES | Wadsworth Center, New York State Department of Health | Alexis Russell; Catharine Prussing; Daryl M. Lamson; Erasmus Schneider; Erica Lasek-Nesselquist; John Kelly; Jonathan Plitnick; Kirsten St. George; Matthew Shudt; Melissa A Leisner; Navjot Singh |
| EPI_ISL_1855318 | BOSTON HEART DIAGNOSTICS CORP | Wadsworth Center, New York State Department of Health | Alexis Russell; Catharine Prussing; Daryl M. Lamson; Erasmus Schneider; Erica Lasek-Nesselquist; John Kelly; Jonathan Plitnick; Kirsten St. George; Matthew Shudt; Melissa A Leisner; Navjot Singh |
| EPI_ISL_1757072, EPI_ISL_1757073, EPI_ISL_1757074, EPI_ISL_1757076, EPI_ISL_1757077, EPI_ISL_1757089, EPI_ISL_1757091, EPI_ISL_1757092, EPI_ISL_1757279, EPI_ISL_1757284, EPI_ISL_1757286, EPI_ISL_1919090, EPI_ISL_1919091, EPI_ISL_1919092, EPI_ISL_1919093, EPI_ISL_1919094, EPI_ISL_1919095, EPI_ISL_1919096, EPI_ISL_1919097, EPI_ISL_1919098, EPI_ISL_1919099, EPI_ISL_1919100, EPI_ISL_1919101, EPI_ISL_1919102, EPI_ISL_1919103, EPI_ISL_1919104, EPI_ISL_1919105 | | | |
| see above | Broad Institute Clinical Research Sequencing Platform | Infectious Disease Program, Broad Institute of Harvard and MIT | Adams, G.; B.L.; B.W.; Bauer, M.; Birren; Blumenstiel, B.; Brown, C.; Carter, A.; Chaluvadi, S.; D.J.; DeFelice, M.; DeRuff, K.; Dodge, S.; Gabriel, S.; Gallagher, G.; Gladden-Young, A.; Granger, B.; J.E.; K.J.; Lagerborg, K.; Larkin, K.; Lee, M.; Lemieux; Lennon, N.; Loreth, C.; Madoff, L.; McGovern, S.; Meldrim, J.; Normandin, E.; P.C.; Park; Pearlman, L.; Reilly, S.; Rudy, M.; Sabeti; Siddle; Smole, S.; Tomkins-Tinch, C.; Vicente, G.; and MacInnis |
| EPI_ISL_1965723, EPI_ISL_1965724, EPI_ISL_1965725, EPI_ISL_1965726, EPI_ISL_1965727, EPI_ISL_1965728, EPI_ISL_1965729, EPI_ISL_1965730, EPI_ISL_1965731, EPI_ISL_1965732, EPI_ISL_1965733, EPI_ISL_1965734, EPI_ISL_1965735 | | | |
| see above | Clinical Microbiology Laboratory, NewYork Presbyterian Hospital/Columbia University Irving Medical Center | Uhlemann Laboratory, Columbia University Irving Medical Center | Anne Kelley; Anne-Catrin Uhlemann; Maya Tagliavia; Medini K. Annavajhala |
| EPI_ISL_1755861, EPI_ISL_1755862, EPI_ISL_1755863, EPI_ISL_1755864, EPI_ISL_1755865, EPI_ISL_1755866, EPI_ISL_1855286, EPI_ISL_1855287, EPI_ISL_1855288, EPI_ISL_1855289, EPI_ISL_1855290, EPI_ISL_1855291, EPI_ISL_1855292, EPI_ISL_1855293 | | | |
| see above | Columbia University Irving Medical Center | Wadsworth Center, New York State Department of Health | Alexis Russell; Catharine Prussing; Daryl M. Lamson; Erasmus Schneider; Erica Lasek-Nesselquist; John Kelly; Jonathan Plitnick; Kirsten St. George; Matthew Shudt; Melissa A Leisner; Navjot Singh |
| EPI_ISL_1694718, EPI_ISL_1694759, EPI_ISL_1694773, EPI_ISL_1804945 | DOHMH Central Harlem | New York City Public Health Laboratory | Jade Wang; et al. |
| EPI_ISL_1694730, EPI_ISL_1804946, EPI_ISL_1804950 | DOHMH Chelsea | New York City Public Health Laboratory | Jade Wang; et al. |
| EPI_ISL_1694713, EPI_ISL_1694714, EPI_ISL_1694734, EPI_ISL_1694735, EPI_ISL_1694750, EPI_ISL_1694751, EPI_ISL_1694752, EPI_ISL_1694753, EPI_ISL_1804943 | | | |
| see above | DOHMH Corona | New York City Public Health Laboratory | Jade Wang; et al. |
| EPI_ISL_1694711, EPI_ISL_1694728 | DOHMH Crown Heights | New York City Public Health Laboratory | Jade Wang; et al. |
| EPI_ISL_1694719, EPI_ISL_1694733, EPI_ISL_1694746, EPI_ISL_1694747 | DOHMH Fort Greene | New York City Public Health Laboratory | Jade Wang; et al. |
| EPI_ISL_1694712, EPI_ISL_1694736, EPI_ISL_1694737, EPI_ISL_1694738, EPI_ISL_1694739, EPI_ISL_1694740, EPI_ISL_1694754, EPI_ISL_1694755, EPI_ISL_1694756, EPI_ISL_1694757, EPI_ISL_1694758, EPI_ISL_1804942 | | | |
| see above | DOHMH Jamaica | New York City Public Health Laboratory | Jade Wang; et al. |
| EPI_ISL_1694717, EPI_ISL_1694729 | DOHMH Morrisania | New York City Public Health Laboratory | Jade Wang; et al. |
| EPI_ISL_1694715, EPI_ISL_1694716, EPI_ISL_1694741, EPI_ISL_1694742, EPI_ISL_1694743, EPI_ISL_1694744, EPI_ISL_1694745, EPI_ISL_1694748, EPI_ISL_1694749, EPI_ISL_1804944, EPI_ISL_1804951 | | | |
| see above | DOHMH PHL | New York City Public Health Laboratory | Jade Wang; et al. |
| EPI_ISL_1694760, EPI_ISL_1694761, EPI_ISL_1694762 | Department of Homeless Services | New York City Public Health Laboratory | Jade Wang; et al. |
| EPI_ISL_1937166, EPI_ISL_1937357 | Fulgent Genetics | Fulgent Genetics | Becky Tsai; Benafsh Sapra; Doreen Ng; Harry Gao; James Xie; John Gao; Joseph Fierro; Mickey Li; Yan Meng |
| EPI_ISL_1855144, EPI_ISL_1855146, EPI_ISL_1855147 | GLENS FALLS HOSPITAL LABORATORY | Wadsworth Center, New York State Department of Health | Alexis Russell; Catharine Prussing; Daryl M. Lamson; Erasmus Schneider; Erica Lasek-Nesselquist; John Kelly; Jonathan Plitnick; Kirsten St. George; Matthew Shudt; Melissa A Leisner; Navjot Singh |
| EPI_ISL_1734943, EPI_ISL_1796130, EPI_ISL_1796436, EPI_ISL_1796534, EPI_ISL_1796840, EPI_ISL_1797051, EPI_ISL_1797367, EPI_ISL_1803619, EPI_ISL_1803731, EPI_ISL_1803819, EPI_ISL_1906926, EPI_ISL_1907154, EPI_ISL_1907210, EPI_ISL_1907867 | | | |
| see above | Helix/Illumina | Centers for Disease | Adrian Paskey; Alexandre Bolze; Ary Ascencio; Benjamin Rambo-Martin; Brad Sickler; Charlotte Rivera-Garcia; Christine Tran; Christopher Gulvick; Clinton R. Paden; Dakota Howard; Darlene Wagner; David Becker; Dhwani Batra; Duncan MacCannell; Efren Sandoval; Eileen de Feo; Elizabeth Cirulli; Eric Allen; |

| | | | |
|---|---|---|---|
| | | Control and Prevention Division of Viral Diseases, Pathogen Discovery | Geraint Levan; James Lu; Jan Antico; Jason Caravas; Jason Nguyen; Jimmy Ramirez; Jingtao Liu; Kara Moser; Kelly Schiabor Barrett; Kim Gietzen; Magnus Isaksson; Marc Laurent; Matthew Schmerer; Matthew Tolentino; Nicole L. Washington; Peter W. Cook; Phil Febbo; Ryan Cho; Scott Sammons; Shannon Wickline; Shatavia Morrison; Sherry Wang; Simon White; Tyler Cassens; William Lee; Yvette Unoarumhi |
| EPI_ISL_1733367, EPI_ISL_1733640, EPI_ISL_1733721, EPI_ISL_1795506, EPI_ISL_1795589, EPI_ISL_1795597, EPI_ISL_1795605, EPI_ISL_1795615, EPI_ISL_1795620, EPI_ISL_1795621, EPI_ISL_1795622, EPI_ISL_1802835, EPI_ISL_1817992, EPI_ISL_2014189 | | | |
| see above | Infinity Biologix | Centers for Disease Control and Prevention Division of Viral Diseases, Pathogen Discovery | Adrian Paskey; Benjamin Rambo-Martin; Chirayu Goswami; Christian Bixby; Christopher Gulvick; Clinton R. Paden; Dakota Howard; Darlene Wagner; Dhwani Batra; Duncan MacCannell; Jason Caravas; Jonathan Schultz; Kara Moser; Matthew Schmerer; Peter W. Cook; Robin Grimwood; Russ Hager; Scott Sammons; Shatavia Morrison; Yihe Wang; Yvette Unoarumhi |
| EPI_ISL_1680747, EPI_ISL_1680765, EPI_ISL_1680767, EPI_ISL_1680769, EPI_ISL_1680802, EPI_ISL_1680847, EPI_ISL_1680848, EPI_ISL_1681168, EPI_ISL_1681169, EPI_ISL_1681170, EPI_ISL_1681171, EPI_ISL_1681172, EPI_ISL_1681173, EPI_ISL_1681187, EPI_ISL_1681188, EPI_ISL_1681190, EPI_ISL_1681193, EPI_ISL_1681195, EPI_ISL_1681249, EPI_ISL_1681250, EPI_ISL_1681251, EPI_ISL_1681309, EPI_ISL_1681310, EPI_ISL_1681311, EPI_ISL_1681314, EPI_ISL_1681315, EPI_ISL_1681316, EPI_ISL_1681318, EPI_ISL_1681321, EPI_ISL_1681370, EPI_ISL_1681371, EPI_ISL_1681372, EPI_ISL_1681409, EPI_ISL_1681415, EPI_ISL_1681416, EPI_ISL_1681417, EPI_ISL_1681418, EPI_ISL_1681419, EPI_ISL_1681420, EPI_ISL_1681444, EPI_ISL_1681445, EPI_ISL_1681446, EPI_ISL_1681447, EPI_ISL_1681448, EPI_ISL_1681451, EPI_ISL_1681452, EPI_ISL_1681453, EPI_ISL_1681461, EPI_ISL_1681495, EPI_ISL_1681496, EPI_ISL_1681497, EPI_ISL_1681498, EPI_ISL_1681499, EPI_ISL_1681500, EPI_ISL_1681542, EPI_ISL_1681543, EPI_ISL_1681544, EPI_ISL_1681545, EPI_ISL_1681546, EPI_ISL_1681547, EPI_ISL_1681548, EPI_ISL_1681549, EPI_ISL_1681550, EPI_ISL_1681551, EPI_ISL_1681552, EPI_ISL_1681644, EPI_ISL_1681645, EPI_ISL_1681646, EPI_ISL_1681647, EPI_ISL_1681648, EPI_ISL_1681649, EPI_ISL_1683358, EPI_ISL_1683413, EPI_ISL_1683458, EPI_ISL_1683464, EPI_ISL_1683485, EPI_ISL_1683494, EPI_ISL_1683497, EPI_ISL_1683499, EPI_ISL_1683501, EPI_ISL_1683503, EPI_ISL_1683504, EPI_ISL_1683531, EPI_ISL_1683532, EPI_ISL_1683537, EPI_ISL_1683542, EPI_ISL_1683543, EPI_ISL_1683558, EPI_ISL_1683566, EPI_ISL_1683599, EPI_ISL_1683617, EPI_ISL_1683634, EPI_ISL_1683635, EPI_ISL_1683661, EPI_ISL_1683662, EPI_ISL_1683663, EPI_ISL_1683664, EPI_ISL_1683666, EPI_ISL_1683667, EPI_ISL_1683668, EPI_ISL_1683669, EPI_ISL_1683670, EPI_ISL_1683672, EPI_ISL_1683675, EPI_ISL_1683701, EPI_ISL_1683749, EPI_ISL_1683756, EPI_ISL_1683760, EPI_ISL_1683768, EPI_ISL_1683769, EPI_ISL_1683772, EPI_ISL_1683799, EPI_ISL_1683800, EPI_ISL_1683808, EPI_ISL_1683811, EPI_ISL_1683833, EPI_ISL_1683835, EPI_ISL_1683836, EPI_ISL_1683837, EPI_ISL_1683839, EPI_ISL_1683909, EPI_ISL_1683910, EPI_ISL_1684063, EPI_ISL_1684065, EPI_ISL_1684071, EPI_ISL_1684084, EPI_ISL_1684085, EPI_ISL_1684153, EPI_ISL_1684162, EPI_ISL_1684164, EPI_ISL_1684165, EPI_ISL_1684167, EPI_ISL_1684168, EPI_ISL_1684173, EPI_ISL_1684174, EPI_ISL_1684177, EPI_ISL_1684211, EPI_ISL_1684212, EPI_ISL_1684213, EPI_ISL_1684214, EPI_ISL_1684253, EPI_ISL_1684254, EPI_ISL_1684255, EPI_ISL_1684256, EPI_ISL_1684261, EPI_ISL_1684263, EPI_ISL_1684265, EPI_ISL_1684266, EPI_ISL_1684267, EPI_ISL_1684271, EPI_ISL_1684276, EPI_ISL_1684277, EPI_ISL_1684279, EPI_ISL_1684287, EPI_ISL_1684289, EPI_ISL_1684303, EPI_ISL_1684305, EPI_ISL_1684307, EPI_ISL_1684541, EPI_ISL_1685066, EPI_ISL_1799429, EPI_ISL_1799495, EPI_ISL_1799693, EPI_ISL_1799740, EPI_ISL_1799741, EPI_ISL_1799744, EPI_ISL_1799746, EPI_ISL_1799747, EPI_ISL_1799769, EPI_ISL_1799781, EPI_ISL_1799784, EPI_ISL_1799790, EPI_ISL_1799840, EPI_ISL_1799841, EPI_ISL_1799842, EPI_ISL_1799877, EPI_ISL_1799938, EPI_ISL_1799950, EPI_ISL_1799970, EPI_ISL_1799974, EPI_ISL_1799975, EPI_ISL_1799980, EPI_ISL_1799983, EPI_ISL_1799984, EPI_ISL_1799985, EPI_ISL_1799986, EPI_ISL_1799992, EPI_ISL_1800344, EPI_ISL_1800349, EPI_ISL_1800355, EPI_ISL_1800357, EPI_ISL_1800359, EPI_ISL_1800402, EPI_ISL_1800404, EPI_ISL_1800405, EPI_ISL_1800406, EPI_ISL_1800407, EPI_ISL_1800409, EPI_ISL_1800413, EPI_ISL_1800455, EPI_ISL_1800457, EPI_ISL_1800460, EPI_ISL_1800461, EPI_ISL_1800470, EPI_ISL_1800471, EPI_ISL_1800472, EPI_ISL_1800473, EPI_ISL_1800475, EPI_ISL_1800479, EPI_ISL_1800488, EPI_ISL_1800489, EPI_ISL_1800490, EPI_ISL_1800491, EPI_ISL_1800492, EPI_ISL_1800513, EPI_ISL_1800515, EPI_ISL_1800516, EPI_ISL_1800517, EPI_ISL_1800527, EPI_ISL_1800533, EPI_ISL_1800541, EPI_ISL_1800542, EPI_ISL_1800666, EPI_ISL_1800695, EPI_ISL_1800701, EPI_ISL_1800709, EPI_ISL_1800747, EPI_ISL_1800753, EPI_ISL_1800765, EPI_ISL_1800766, EPI_ISL_1800773, EPI_ISL_1800774, EPI_ISL_1800778, EPI_ISL_1800780, EPI_ISL_1800781, EPI_ISL_1800859, EPI_ISL_1800860, EPI_ISL_1800866, EPI_ISL_1800889, EPI_ISL_1800891, EPI_ISL_1800892, EPI_ISL_1801293, EPI_ISL_1801320, EPI_ISL_1801321, EPI_ISL_1801323, EPI_ISL_1801324, EPI_ISL_1801325, EPI_ISL_1801327, EPI_ISL_1801403, EPI_ISL_1801437, EPI_ISL_1801487, EPI_ISL_1801528, EPI_ISL_1801663, EPI_ISL_1801664, EPI_ISL_1801666, EPI_ISL_1801667, EPI_ISL_1801668, EPI_ISL_1801669, EPI_ISL_1801670, EPI_ISL_1801671, EPI_ISL_1801672, EPI_ISL_1801673, EPI_ISL_1801762, EPI_ISL_1802513, EPI_ISL_1925986, EPI_ISL_2044080 | | | |
| see above | Laboratory Corporation of America | Centers for Disease Control and Prevention Division of Viral Diseases, Pathogen Discovery | Adrian Paskey; Amanda Douglas; Amanda Suchanek; Andrea Throop; Ayla Burns; Benjamin Rambo-Martin; Bobbi Croy; Brian Krueger; Brian Norvell; Christopher Gulvick; Christos Petropoulos; Clinton R. Paden; Craig Lukasik; Dakota Howard; Darlene Wagner; Debbie Boles; Dhwani Batra; Duncan MacCannell; Eyad Almasri; Goran Stevovic; Howard Engler; Hrushikesh Deshmukh; Jake Humphrey; Jana Schroth; Jason Caravas; Joe Voshell; John Pruitt; Jonathan Meltzer; Jonathan Williams; Kara Moser; Kimberly Wagner; Lax Iyer; Lyndon Tilson; Manoj Jain; Marcia Eisenberg; Mary Ann Cristobal; Mary Williamson; Matthew Schmerer; Michael Levandoski; Mike Sapeta; Mindy Nye; Minoo Agarwal; Mohan Kolli; Nuthawin Charoensri; Oren Cohen; Peter W. Cook; Prashant Gupta; Qian Zeng; Rama Ghatti; Scott Parker; Scott Ryan; Scott Sammons; Shatavia Morrison; Stanley Letovsky; Steven Ragan; Suresh Babu Selvaraju; Susan Countryman; Susan Hicks; Suzanne Dale; Thomas Urban; Tim Kuphal; Tricia Zwiefelhofer; Vincent Drouillon; Yvette Unoarumhi |
| EPI_ISL_1756342, EPI_ISL_1756343, EPI_ISL_1756344, EPI_ISL_1756345, EPI_ISL_1756346, EPI_ISL_1756347, EPI_ISL_1756348, EPI_ISL_1756349, EPI_ISL_1756350, EPI_ISL_1756351, EPI_ISL_1756352, EPI_ISL_1756353, EPI_ISL_1756354, EPI_ISL_1756355, EPI_ISL_1789720, EPI_ISL_1789721, EPI_ISL_1855324 | | | |
| see above | MONTEFIORE MEDICAL CENTER LABORATORIES | Wadsworth Center, New York State Department of Health | Alexis Russell; Catharine Prussing; Daryl M. Lamson; Erasmus Schneider; Erica Lasek-Nesselquist; John Kelly; Jonathan Plitnick; Kirsten St. George; Matthew Shudt; Melissa A Leisner; Navjot Singh |
| EPI_ISL_1709913, EPI_ISL_1709914, EPI_ISL_1709915, EPI_ISL_1709916, EPI_ISL_1709917, EPI_ISL_1709918, EPI_ISL_1709919, EPI_ISL_1709920, EPI_ISL_1709921, EPI_ISL_1709922, EPI_ISL_1709923, EPI_ISL_1709924, EPI_ISL_1709925, EPI_ISL_1709926, EPI_ISL_1709927, EPI_ISL_1709928, EPI_ISL_1709929, EPI_ISL_1709930, EPI_ISL_1709931, EPI_ISL_1709932, EPI_ISL_1709933, EPI_ISL_1709934, EPI_ISL_1709935, EPI_ISL_1709936, EPI_ISL_1709937, EPI_ISL_1709938, EPI_ISL_1709939, EPI_ISL_1709940, EPI_ISL_1709941, EPI_ISL_1709942, EPI_ISL_1709943, EPI_ISL_1709944, EPI_ISL_1709945, EPI_ISL_1709947, EPI_ISL_1709948, EPI_ISL_1709950, EPI_ISL_1709951, EPI_ISL_1709952, EPI_ISL_1709953, EPI_ISL_1709954, EPI_ISL_1709955, EPI_ISL_1709956, EPI_ISL_1709957, EPI_ISL_1709958, EPI_ISL_1709959, EPI_ISL_1709960, EPI_ISL_1709961, EPI_ISL_1709962, EPI_ISL_1709963, EPI_ISL_1709964, EPI_ISL_1709965, EPI_ISL_1709966, EPI_ISL_1709967, EPI_ISL_1709968, EPI_ISL_1709969, EPI_ISL_1709970, EPI_ISL_1709971, EPI_ISL_1709972, EPI_ISL_1709974, EPI_ISL_1709975, EPI_ISL_1709976, EPI_ISL_1709977, EPI_ISL_1709978, EPI_ISL_1709979, EPI_ISL_1709980, EPI_ISL_1709981, EPI_ISL_1709982, EPI_ISL_1709983, EPI_ISL_1709984, EPI_ISL_1709985, EPI_ISL_1709986, EPI_ISL_1709987, EPI_ISL_1709988, EPI_ISL_1709989, EPI_ISL_1709991, EPI_ISL_1709992, EPI_ISL_1709993, EPI_ISL_1709994, EPI_ISL_1709995, EPI_ISL_1709996, EPI_ISL_1709997, EPI_ISL_1709998, EPI_ISL_1709999, EPI_ISL_1710000, EPI_ISL_1710001, EPI_ISL_1710002, EPI_ISL_1710003, EPI_ISL_1710004, EPI_ISL_1710005, EPI_ISL_1710006, EPI_ISL_1710007, EPI_ISL_1710008, EPI_ISL_1710009, EPI_ISL_1710010, EPI_ISL_1710011, EPI_ISL_1710012, EPI_ISL_1710013, EPI_ISL_1710014 | | | |
| see above | MSHS Clinical Microbiology Laboratories | MSHS Pathogen Surveillance Program | Adolfo García-Sastre; Adriana van de Guchte; Ajay Obla; Alberto Paniz-Mondolfi; Ana S. Gonzalez-Reiche; Angela Amoako; Ashley Salimbangon; Betsaida Salom Melo; Bremy Alburquerque; Brianne Ciferri; Charles Gleason; Daniel Floda; Deena R. Altman; Denise Jurczyszak; Emilia Mia Sordillo; Gintaras Deikus; Giulio Kleiner; Gopi Patel; Hala Alshammary; Harm van Bakel; Irina Oussenko; Jayeeta Dutta; Juan Soto; Julia Matthews; Katherine Beach; Kathryn Twyman; Kayla Russo; Komal Srivastava; Levy Sominsky; Mahmoud Awawda; Marta Luksza; Matthew M. Hernandez; Melissa Gitman; Michael D. Nowak; Mitchell J. Sullivan; Nancy Francoeur; Robert Sebra; Sarah Schaefer; Shelcie Fabre; Shwetha Hara Sridhar; Viviana Simon; Ying-Chih Wang; Zenab Khan |
| EPI_ISL_2009122, EPI_ISL_2009123, EPI_ISL_2009133, EPI_ISL_2009134, EPI_ISL_2009139, EPI_ISL_2009141, EPI_ISL_2009142, EPI_ISL_2009143, EPI_ISL_2009160, EPI_ISL_2009165 | | | |
| see above | MSK Microbiology Lab | MSK Microbiology Lab | Esther Babady; Krupa Jani; Tracy McMillen |
| EPI_ISL_1919997, EPI_ISL_1919999, EPI_ISL_1920000, EPI_ISL_1920002, EPI_ISL_1920003, EPI_ISL_1920004, EPI_ISL_1920005, EPI_ISL_1920006 | | | |
| see above | Montefiore Medical Center Laboratories | Wadsworth Center, New York State Department of Health | Alexis Russell; Catharine Prussing; Daryl M. Lamson; Erasmus Schneider; Erica Lasek-Nesselquist; John Kelly; Jonathan Plitnick; Kirsten St. George; Matthew Shudt; Melissa A Leisner; Navjot Singh |
| EPI_ISL_1855349, EPI_ISL_1855353 | NORTH SHORE UNIVERSITY HOSPITAL | Wadsworth Center, New York State Department of Health | Alexis Russell; Catharine Prussing; Daryl M. Lamson; Erasmus Schneider; Erica Lasek-Nesselquist; John Kelly; Jonathan Plitnick; Kirsten St. George; Matthew Shudt; Melissa A Leisner; Navjot Singh |
| EPI_ISL_1671601, EPI_ISL_1671602, EPI_ISL_1671603, EPI_ISL_1671604, EPI_ISL_1671605, EPI_ISL_1671606, EPI_ISL_1671608, EPI_ISL_1671609, EPI_ISL_1671617, EPI_ISL_1671621, EPI_ISL_1671632, EPI_ISL_1671641, EPI_ISL_1671651, EPI_ISL_1671662, EPI_ISL_1671663, EPI_ISL_1671672, EPI_ISL_1671673, EPI_ISL_1671674, EPI_ISL_1671675, EPI_ISL_1671676, EPI_ISL_1740520, EPI_ISL_1740521, EPI_ISL_1740522, EPI_ISL_1935621, EPI_ISL_1935624, EPI_ISL_1935626, EPI_ISL_1935629 | | | |
| see above | NORTHWELL HEALTH LABORATORIES | Wadsworth Center, New York State Department of Health | Alexis Russell; Catharine Prussing; Daryl M. Lamson; Erasmus Schneider; Erica Lasek-Nesselquist; John Kelly; Jonathan Plitnick; Kirsten St. George; Matthew Shudt; Melissa A Leisner; Navjot Singh |
| EPI_ISL_1823944, EPI_ISL_1823945, EPI_ISL_1823947, EPI_ISL_1823949, EPI_ISL_1823951, EPI_ISL_1823952, EPI_ISL_1823954, EPI_ISL_1823959, EPI_ISL_1823960, EPI_ISL_1823962, EPI_ISL_1823966, EPI_ISL_1823971, EPI_ISL_1823972, EPI_ISL_1823973, EPI_ISL_1937596, EPI_ISL_1937598, EPI_ISL_1937600, EPI_ISL_1937601, EPI_ISL_1937603, EPI_ISL_1937605, EPI_ISL_1937606, EPI_ISL_1937608, EPI_ISL_1937610, EPI_ISL_1937611, EPI_ISL_1937613, EPI_ISL_1937615, EPI_ISL_1937616, EPI_ISL_1937618, EPI_ISL_1937620, EPI_ISL_1937622, EPI_ISL_1937623, EPI_ISL_2018422, EPI_ISL_2018423, EPI_ISL_2047506 | | | |
| see above | NYC Department of Health and Mental Hygiene | Centers for Disease Control and Prevention Division of Viral Diseases, Pathogen Discovery | Alison Laufer Halpin; Ben L. Rambo-Martin; Clinton R. Paden; Dakota Howard; Darlene Wagner; Dave Wentworth; Dhwani Batra; Jasmine Padilla; Justin Lee; Katie Dillon; Krista Queen; Kristen Knipe; Kristine Lacek; Mark Burroughs; Matthew Schmerer; Mili Sheth; Peter Cook; Sam Shepard; Sarah Nobles; Shoshona Le; Suxiang Tong; Vivien Dugan; Yvette Unoarumhi |
| EPI_ISL_1854736, EPI_ISL_1854737, EPI_ISL_1854738, EPI_ISL_1854739, EPI_ISL_1935631, EPI_ISL_1935636 | NYC Pandemic Response Lab | Wadsworth Center, New York State Department of Health | Alexis Russell; Catharine Prussing; Daryl M. Lamson; Erasmus Schneider; Erica Lasek-Nesselquist; John Kelly; Jonathan Plitnick; Kirsten St. George; Matthew Shudt; Melissa A Leisner; Navjot Singh |
| EPI_ISL_2018243, EPI_ISL_2018245, EPI_ISL_2018246 | NYSDOH Wadsworth Center, Virology Lab | Centers for Disease Control and Prevention Division of Viral Diseases, Pathogen Discovery | Alison Laufer Halpin; Ben L. Rambo-Martin; Clinton R. Paden; Dakota Howard; Darlene Wagner; Dave Wentworth; Dhwani Batra; Jasmine Padilla; Justin Lee; Katie Dillon; Krista Queen; Kristen Knipe; Kristine Lacek; Mark Burroughs; Matthew Schmerer; Mili Sheth; Peter Cook; Sam Shepard; Sarah Nobles; Shoshona Le; Suxiang Tong; Vivien Dugan; Yvette Unoarumhi |
| EPI_ISL_1576901, EPI_ISL_1576930, EPI_ISL_1576934, EPI_ISL_1576935, EPI_ISL_1576937, EPI_ISL_1576938, EPI_ISL_1576939, EPI_ISL_1576940, EPI_ISL_1576941, EPI_ISL_1576942, EPI_ISL_1576944, EPI_ISL_1654611, EPI_ISL_1654613, EPI_ISL_1654614, EPI_ISL_1654615, EPI_ISL_1654616, EPI_ISL_1654617, EPI_ISL_1654618, EPI_ISL_1654619, EPI_ISL_1654620, EPI_ISL_1654621, EPI_ISL_1654622, EPI_ISL_1654623, EPI_ISL_1654624, EPI_ISL_1654625, EPI_ISL_1654626, EPI_ISL_1654627, EPI_ISL_1654628, EPI_ISL_1654629, EPI_ISL_1654630, EPI_ISL_1654631, EPI_ISL_1654632, EPI_ISL_1654633, EPI_ISL_1654634, EPI_ISL_1654635, EPI_ISL_1654636, EPI_ISL_1654637, EPI_ISL_1654638, EPI_ISL_1654639, EPI_ISL_1654640, EPI_ISL_1654641, EPI_ISL_1654644, EPI_ISL_1654645, EPI_ISL_1654646, EPI_ISL_1654647, EPI_ISL_1669801, EPI_ISL_1669802, EPI_ISL_1669803, EPI_ISL_1789903, EPI_ISL_1789904, EPI_ISL_1789905, EPI_ISL_1789906, EPI_ISL_1789907, EPI_ISL_1789908, EPI_ISL_1789909, EPI_ISL_1789910, EPI_ISL_1789911, EPI_ISL_1789912, EPI_ISL_1789913, EPI_ISL_1789914, EPI_ISL_1789915, EPI_ISL_1789916, EPI_ISL_1789917, EPI_ISL_1789918, EPI_ISL_1789919, EPI_ISL_1789920, EPI_ISL_1789921, EPI_ISL_1789922, EPI_ISL_1789923, EPI_ISL_1789924, EPI_ISL_1789925, EPI_ISL_1789926, EPI_ISL_1789928, EPI_ISL_1789929, EPI_ISL_1789930, EPI_ISL_1789931, EPI_ISL_1789932, EPI_ISL_1789933, EPI_ISL_1789934, EPI_ISL_1789935, EPI_ISL_1789936, EPI_ISL_1789938, EPI_ISL_1789939, EPI_ISL_1789940, EPI_ISL_1789941, EPI_ISL_1789942, EPI_ISL_1789943, EPI_ISL_1789944, EPI_ISL_1789945, EPI_ISL_1789946, EPI_ISL_1789947, EPI_ISL_1789948, EPI_ISL_1789949, EPI_ISL_1789950, EPI_ISL_1789951, EPI_ISL_1789952, EPI_ISL_1789953, EPI_ISL_1789954, EPI_ISL_1789955, EPI_ISL_1789956, EPI_ISL_1789957, EPI_ISL_1789958, EPI_ISL_1789959, EPI_ISL_1789960, EPI_ISL_1789961, EPI_ISL_1789962, EPI_ISL_1789963, EPI_ISL_1789964, EPI_ISL_1789966, EPI_ISL_1789967, EPI_ISL_1789968, EPI_ISL_1789969, EPI_ISL_1789970, EPI_ISL_1789971, EPI_ISL_1789972, EPI_ISL_1789973, EPI_ISL_1789974, EPI_ISL_1789975, EPI_ISL_1789976, EPI_ISL_1789977, EPI_ISL_1789978, EPI_ISL_1789979, EPI_ISL_1790025, EPI_ISL_1790027, EPI_ISL_1790044, EPI_ISL_1790047, EPI_ISL_1922853, EPI_ISL_1922857, EPI_ISL_1922863, EPI_ISL_1922867, EPI_ISL_1922870 | | | |
| see above | NYU Langone Health | Departments of Pathology and Medicine, New York University School of Medicine | Adriana Heguy; Christian Marier; Dacia Dimartino; Emily Guzman; Gael Westby; Guiqing Wang; Paolo Cotzia; Paul Zappile; Peter Meyn; Sitharam Ramaswami; Yutong Zhang |
| EPI_ISL_1919988, EPI_ISL_1919989, EPI_ISL_1919993, EPI_ISL_1919994, EPI_ISL_1919995, EPI_ISL_1919996, EPI_ISL_1920011, EPI_ISL_1920012, EPI_ISL_1920013, EPI_ISL_1920015, EPI_ISL_1920016, EPI_ISL_1920018, EPI_ISL_1920019, EPI_ISL_1920020, EPI_ISL_1920021, EPI_ISL_1920022, EPI_ISL_1920023, EPI_ISL_1920024, EPI_ISL_1920025, EPI_ISL_1920027, EPI_ISL_1920028, EPI_ISL_1920031, EPI_ISL_1920032, EPI_ISL_1920033, EPI_ISL_1920034, EPI_ISL_1920036, EPI_ISL_1920037, EPI_ISL_1920038, EPI_ISL_1920039, EPI_ISL_1920040, EPI_ISL_1920042, EPI_ISL_1920045, EPI_ISL_1920101 | | | |
| see above | Northwell Health Laboratories | Wadsworth Center, New York State Department of Health | Alexis Russell; Catharine Prussing; Daryl M. Lamson; Erasmus Schneider; Erica Lasek-Nesselquist; John Kelly; Jonathan Plitnick; Kirsten St. George; Matthew Shudt; Melissa A Leisner; Navjot Singh |
| EPI_ISL_1704714 | Northwestern Memorial Hospital | Northwestern University - Ozer Lab | Chad J. Achenbach; Chao Qi; Egon A. Ozer; Judd F. Hultquist; Lacy M. Simons; Lawrence J. Jennings; Michael G. Ison; Ramon Lorenzo-Redondo; Taylor J. Dean |
| EPI_ISL_1694731, EPI_ISL_1694732, EPI_ISL_1694763, EPI_ISL_1694764, EPI_ISL_1694765, EPI_ISL_1694766, EPI_ISL_1694767, EPI_ISL_1694768, EPI_ISL_1694769, EPI_ISL_1694770, EPI_ISL_1694771, EPI_ISL_1694772, EPI_ISL_1804948, EPI_ISL_1804949 | | | |
| see above | OCME Office Of Chief Medical Examiner | New York City Public Health Laboratory | Jade Wang; et al. |
| EPI_ISL_1542075, EPI_ISL_1542076, EPI_ISL_1542077, EPI_ISL_1542087, EPI_ISL_1542089, EPI_ISL_1542094, EPI_ISL_1542095, EPI_ISL_1542096, EPI_ISL_1542102, EPI_ISL_1542105, EPI_ISL_1542108, EPI_ISL_1542110, EPI_ISL_1542111, EPI_ISL_1542112, EPI_ISL_1542116, EPI_ISL_1542118, EPI_ISL_1542124, EPI_ISL_1542130, EPI_ISL_1542141, EPI_ISL_1542143, EPI_ISL_1542156, EPI_ISL_1542157, EPI_ISL_1542165, EPI_ISL_1542166, EPI_ISL_1542170, EPI_ISL_1542174, EPI_ISL_1542178, EPI_ISL_1542181, EPI_ISL_1542185, EPI_ISL_1542190, EPI_ISL_1542200, EPI_ISL_1542203, EPI_ISL_1542206, EPI_ISL_1542208, EPI_ISL_1542217, EPI_ISL_1542221, EPI_ISL_1542227, EPI_ISL_1542229, EPI_ISL_1542232, EPI_ISL_1542238, EPI_ISL_1542242, EPI_ISL_1542247, EPI_ISL_1542255, EPI_ISL_1542260, EPI_ISL_1542273, EPI_ISL_1542285, EPI_ISL_1542288, EPI_ISL_1542293, EPI_ISL_1542295, EPI_ISL_1542303, EPI_ISL_1542307, EPI_ISL_1542312, EPI_ISL_1542314, EPI_ISL_1542318, EPI_ISL_1542322, EPI_ISL_1542329, EPI_ISL_1542332, EPI_ISL_1542333, EPI_ISL_1542337, EPI_ISL_1542343, EPI_ISL_1542357, EPI_ISL_1542366, EPI_ISL_1542371, EPI_ISL_1542372, EPI_ISL_1542380, EPI_ISL_1542387, EPI_ISL_1542390, EPI_ISL_1542393, EPI_ISL_1542394, EPI_ISL_1542397, EPI_ISL_1542398, EPI_ISL_1542401, EPI_ISL_1542422, EPI_ISL_1542427, EPI_ISL_1542430, EPI_ISL_1542443, EPI_ISL_1542446, EPI_ISL_1542447, EPI_ISL_1542448, EPI_ISL_1542458, EPI_ISL_1542478, EPI_ISL_1542481, EPI_ISL_1542486, EPI_ISL_1542500, EPI_ISL_1542504, EPI_ISL_1542506, EPI_ISL_1542520, EPI_ISL_1542525, EPI_ISL_1542536, EPI_ISL_1542538, EPI_ISL_1542541, EPI_ISL_1542543, EPI_ISL_1542544, EPI_ISL_1542545, EPI_ISL_1542550, EPI_ISL_1542562, EPI_ISL_1542570, EPI_ISL_1542571, EPI_ISL_1542574, EPI_ISL_1542591, EPI_ISL_1542596, EPI_ISL_1542604, EPI_ISL_1542607, EPI_ISL_1542616, EPI_ISL_1542619, EPI_ISL_1542626, EPI_ISL_1542642, EPI_ISL_1542645, EPI_ISL_1542647, EPI_ISL_1542652, EPI_ISL_1542657, EPI_ISL_1542669, EPI_ISL_1542670, EPI_ISL_1542680, EPI_ISL_1542684, EPI_ISL_1542692, EPI_ISL_1542695, EPI_ISL_1542697, EPI_ISL_1542706, EPI_ISL_1542708, EPI_ISL_1542711, EPI_ISL_1542718, EPI_ISL_1542720, EPI_ISL_1542724, EPI_ISL_1542727, EPI_ISL_1542729, EPI_ISL_1542745, EPI_ISL_1542746, EPI_ISL_1542752, EPI_ISL_1542755, EPI_ISL_1542758, EPI_ISL_1542761, EPI_ISL_1542765, EPI_ISL_1542766, EPI_ISL_1542767, EPI_ISL_1542770, EPI_ISL_1542785, EPI_ISL_1542786, EPI_ISL_1542808, EPI_ISL_1542818, EPI_ISL_1542819, EPI_ISL_1542820, EPI_ISL_1542827, EPI_ISL_1542832, EPI_ISL_1542837, EPI_ISL_1542839, EPI_ISL_1542848, EPI_ISL_1542853, EPI_ISL_1542858, EPI_ISL_1542866, EPI_ISL_1542894, EPI_ISL_1542896, EPI_ISL_1542898, EPI_ISL_1542899, EPI_ISL_1543122, EPI_ISL_1543123, EPI_ISL_1543124, EPI_ISL_1543125, EPI_ISL_1543126, EPI_ISL_1543127, EPI_ISL_1543128, EPI_ISL_1543129, EPI_ISL_1543130, EPI_ISL_1543131, EPI_ISL_1543132, EPI_ISL_1543133, EPI_ISL_1543134, EPI_ISL_1543135, EPI_ISL_1543136, EPI_ISL_1543137, EPI_ISL_1543138, EPI_ISL_1543139, EPI_ISL_1543140, EPI_ISL_1543141, EPI_ISL_1543142, EPI_ISL_1543143, EPI_ISL_1543144, EPI_ISL_1543145, EPI_ISL_1543146, EPI_ISL_1543147, EPI_ISL_1543148, EPI_ISL_1543149, EPI_ISL_1543150, EPI_ISL_1543151, EPI_ISL_1543152, EPI_ISL_1543153, EPI_ISL_1543154, EPI_ISL_1543155, EPI_ISL_1543156, EPI_ISL_1543157, EPI_ISL_1543158, EPI_ISL_1543159, EPI_ISL_1543160, EPI_ISL_1543161, EPI_ISL_1543162, EPI_ISL_1543163, EPI_ISL_1543164, EPI_ISL_1543165, EPI_ISL_1543166, EPI_ISL_1543167, EPI_ISL_1543168, EPI_ISL_1543169, EPI_ISL_1543170, EPI_ISL_1543171, EPI_ISL_1543172, EPI_ISL_1543173, EPI_ISL_1543174, EPI_ISL_1543175, EPI_ISL_1543176, EPI_ISL_1543177, EPI_ISL_1543178, EPI_ISL_1543179, EPI_ISL_1543180, EPI_ISL_1543181, EPI_ISL_1543182, EPI_ISL_1543183, EPI_ISL_1543184, EPI_ISL_1543185, EPI_ISL_1543186, EPI_ISL_1543187, EPI_ISL_1543188, EPI_ISL_1543189, EPI_ISL_1543190, EPI_ISL_1543191, EPI_ISL_1543192, EPI_ISL_1543193, EPI_ISL_1543194, EPI_ISL_1543195, EPI_ISL_1543196, EPI_ISL_1543197, EPI_ISL_1543198, EPI_ISL_1543199, EPI_ISL_1543200, EPI_ISL_1543201, EPI_ISL_1543202, EPI_ISL_1543203, EPI_ISL_1543204, EPI_ISL_1543205, EPI_ISL_1543206, EPI_ISL_1543207, EPI_ISL_1543208, EPI_ISL_1543209, | | | |

EPI_ISL_1543210, EPI_ISL_1543211, EPI_ISL_1543212, EPI_ISL_1543213, EPI_ISL_1543214, EPI_ISL_1543215, EPI_ISL_1543216, EPI_ISL_1543217, EPI_ISL_1543218, EPI_ISL_1543219, EPI_ISL_1543220, EPI_ISL_1543221, EPI_ISL_1543222, EPI_ISL_1543223, EPI_ISL_1543224, EPI_ISL_1543225, EPI_ISL_1543226, EPI_ISL_1543227, EPI_ISL_1543228, EPI_ISL_1543229, EPI_ISL_1543230, EPI_ISL_1543231,
EPI_ISL_1543232, EPI_ISL_1543233, EPI_ISL_1543234, EPI_ISL_1543235, EPI_ISL_1543236, EPI_ISL_1543237, EPI_ISL_1543238, EPI_ISL_1543239, EPI_ISL_1543240, EPI_ISL_1543241, EPI_ISL_1543242, EPI_ISL_1543243, EPI_ISL_1543244, EPI_ISL_1543245, EPI_ISL_1543246, EPI_ISL_1543247, EPI_ISL_1543248, EPI_ISL_1543249, EPI_ISL_1543250, EPI_ISL_1543251, EPI_ISL_1543252, EPI_ISL_1543253,
EPI_ISL_1543254, EPI_ISL_1543255, EPI_ISL_1543256, EPI_ISL_1543257, EPI_ISL_1543258, EPI_ISL_1543259, EPI_ISL_1543260, EPI_ISL_1543261, EPI_ISL_1543262, EPI_ISL_1543263, EPI_ISL_1543264, EPI_ISL_1543265, EPI_ISL_1543266, EPI_ISL_1543267, EPI_ISL_1543268, EPI_ISL_1543269, EPI_ISL_1543270, EPI_ISL_1543271, EPI_ISL_1543272, EPI_ISL_1543273, EPI_ISL_1543274, EPI_ISL_1543275,
EPI_ISL_1543276, EPI_ISL_1543277, EPI_ISL_1543278, EPI_ISL_1543279, EPI_ISL_1543280, EPI_ISL_1543281, EPI_ISL_1543282, EPI_ISL_1543283, EPI_ISL_1543284, EPI_ISL_1543285, EPI_ISL_1543286, EPI_ISL_1543287, EPI_ISL_1543288, EPI_ISL_1543289, EPI_ISL_1543290, EPI_ISL_1543291, EPI_ISL_1543292, EPI_ISL_1543293, EPI_ISL_1543294, EPI_ISL_1543295, EPI_ISL_1543296, EPI_ISL_1543297,
EPI_ISL_1543298, EPI_ISL_1543299, EPI_ISL_1543300, EPI_ISL_1543301, EPI_ISL_1543302, EPI_ISL_1543303, EPI_ISL_1543304, EPI_ISL_1543305, EPI_ISL_1543306, EPI_ISL_1543307, EPI_ISL_1543308, EPI_ISL_1543309, EPI_ISL_1543310, EPI_ISL_1543311, EPI_ISL_1543312, EPI_ISL_1543313, EPI_ISL_1543314, EPI_ISL_1543315, EPI_ISL_1543316, EPI_ISL_1543317, EPI_ISL_1543318, EPI_ISL_1543319,
EPI_ISL_1543320, EPI_ISL_1543321, EPI_ISL_1543322, EPI_ISL_1543323, EPI_ISL_1543324, EPI_ISL_1543325, EPI_ISL_1543326, EPI_ISL_1543327, EPI_ISL_1543328, EPI_ISL_1543329, EPI_ISL_1543330, EPI_ISL_1543331, EPI_ISL_1543332, EPI_ISL_1543333, EPI_ISL_1543334, EPI_ISL_1543335, EPI_ISL_1543336, EPI_ISL_1543337, EPI_ISL_1543338, EPI_ISL_1543339, EPI_ISL_1543340, EPI_ISL_1543341,
EPI_ISL_1543342, EPI_ISL_1543343, EPI_ISL_1543344, EPI_ISL_1543345, EPI_ISL_1543346, EPI_ISL_1543347, EPI_ISL_1543348, EPI_ISL_1543349, EPI_ISL_1543350, EPI_ISL_1543351, EPI_ISL_1543352, EPI_ISL_1543353, EPI_ISL_1543354, EPI_ISL_1543355, EPI_ISL_1543356, EPI_ISL_1543357, EPI_ISL_1543358, EPI_ISL_1543359, EPI_ISL_1543360, EPI_ISL_1543361, EPI_ISL_1543362, EPI_ISL_1543363,
EPI_ISL_1543364, EPI_ISL_1543365, EPI_ISL_1543366, EPI_ISL_1543367, EPI_ISL_1543368, EPI_ISL_1543369, EPI_ISL_1543370, EPI_ISL_1543371, EPI_ISL_1543372, EPI_ISL_1543373, EPI_ISL_1543374, EPI_ISL_1543375, EPI_ISL_1543376, EPI_ISL_1543377, EPI_ISL_1543378, EPI_ISL_1543379, EPI_ISL_1543380, EPI_ISL_1543381, EPI_ISL_1543382, EPI_ISL_1543383, EPI_ISL_1543384, EPI_ISL_1543385,
EPI_ISL_1543386, EPI_ISL_1543387, EPI_ISL_1543388, EPI_ISL_1543389, EPI_ISL_1543390, EPI_ISL_1543391, EPI_ISL_1543392, EPI_ISL_1543393, EPI_ISL_1543394, EPI_ISL_1543395, EPI_ISL_1543396, EPI_ISL_1543397, EPI_ISL_1543398, EPI_ISL_1543399, EPI_ISL_1543400, EPI_ISL_1543401, EPI_ISL_1543402, EPI_ISL_1543403, EPI_ISL_1543404, EPI_ISL_1543405, EPI_ISL_1543406, EPI_ISL_1543407,
EPI_ISL_1543408, EPI_ISL_1543409, EPI_ISL_1543410, EPI_ISL_1543411, EPI_ISL_1543412, EPI_ISL_1543413, EPI_ISL_1543414, EPI_ISL_1543415, EPI_ISL_1543416, EPI_ISL_1543417, EPI_ISL_1543418, EPI_ISL_1543419, EPI_ISL_1543420, EPI_ISL_1543421, EPI_ISL_1543422, EPI_ISL_1543423, EPI_ISL_1543424, EPI_ISL_1543425, EPI_ISL_1543426, EPI_ISL_1543427, EPI_ISL_1543428, EPI_ISL_1543429,
EPI_ISL_1543430, EPI_ISL_1543431, EPI_ISL_1543432, EPI_ISL_1543433, EPI_ISL_1543434, EPI_ISL_1543435, EPI_ISL_1543436, EPI_ISL_1543437, EPI_ISL_1543438, EPI_ISL_1543439, EPI_ISL_1543440, EPI_ISL_1543441, EPI_ISL_1543442, EPI_ISL_1543443, EPI_ISL_1543444, EPI_ISL_1543445, EPI_ISL_1543446, EPI_ISL_1543447, EPI_ISL_1543448, EPI_ISL_1543449, EPI_ISL_1543450, EPI_ISL_1543451,
EPI_ISL_1543452, EPI_ISL_1543453, EPI_ISL_1543454, EPI_ISL_1543455, EPI_ISL_1543456, EPI_ISL_1543457, EPI_ISL_1543458, EPI_ISL_1543459, EPI_ISL_1543460, EPI_ISL_1543461, EPI_ISL_1543462, EPI_ISL_1543463, EPI_ISL_1543464, EPI_ISL_1543465, EPI_ISL_1543466, EPI_ISL_1543467, EPI_ISL_1543468, EPI_ISL_1543469, EPI_ISL_1543470, EPI_ISL_1543471, EPI_ISL_1543472, EPI_ISL_1543473,
EPI_ISL_1543474, EPI_ISL_1543475, EPI_ISL_1543476, EPI_ISL_1543477, EPI_ISL_1543478, EPI_ISL_1543479, EPI_ISL_1543480, EPI_ISL_1543481, EPI_ISL_1543482, EPI_ISL_1543483, EPI_ISL_1543484, EPI_ISL_1543485, EPI_ISL_1543486, EPI_ISL_1543487, EPI_ISL_1543488, EPI_ISL_1543489, EPI_ISL_1543490, EPI_ISL_1543491, EPI_ISL_1543492, EPI_ISL_1543493, EPI_ISL_1543494, EPI_ISL_1543495,
EPI_ISL_1543496, EPI_ISL_1543497, EPI_ISL_1543498, EPI_ISL_1543499, EPI_ISL_1543500, EPI_ISL_1543501, EPI_ISL_1543502, EPI_ISL_1543503, EPI_ISL_1543504, EPI_ISL_1543505, EPI_ISL_1543506, EPI_ISL_1543507, EPI_ISL_1543508, EPI_ISL_1543509, EPI_ISL_1543510, EPI_ISL_1543511, EPI_ISL_1543512, EPI_ISL_1543513, EPI_ISL_1543514, EPI_ISL_1543515, EPI_ISL_1543516, EPI_ISL_1543517,
EPI_ISL_1543518, EPI_ISL_1543519, EPI_ISL_1543520, EPI_ISL_1543521, EPI_ISL_1543522, EPI_ISL_1543523, EPI_ISL_1543524, EPI_ISL_1543525, EPI_ISL_1543526, EPI_ISL_1543527, EPI_ISL_1543528, EPI_ISL_1543529, EPI_ISL_1543530, EPI_ISL_1543531, EPI_ISL_1543532, EPI_ISL_1543533, EPI_ISL_1543534, EPI_ISL_1543535, EPI_ISL_1543536, EPI_ISL_1543537, EPI_ISL_1543538, EPI_ISL_1543539,
EPI_ISL_1543540, EPI_ISL_1543541, EPI_ISL_1543542, EPI_ISL_1543543, EPI_ISL_1543544, EPI_ISL_1543545, EPI_ISL_1543546, EPI_ISL_1543547, EPI_ISL_1543548, EPI_ISL_1543549, EPI_ISL_1543550, EPI_ISL_1543551, EPI_ISL_1543552, EPI_ISL_1543553, EPI_ISL_1543554, EPI_ISL_1543555, EPI_ISL_1543556, EPI_ISL_1543557, EPI_ISL_1543558, EPI_ISL_1543559, EPI_ISL_1543560, EPI_ISL_1543561,
EPI_ISL_1543562, EPI_ISL_1543563, EPI_ISL_1543564, EPI_ISL_1543565, EPI_ISL_1543566, EPI_ISL_1543567, EPI_ISL_1543568, EPI_ISL_1543569, EPI_ISL_1543570, EPI_ISL_1543571, EPI_ISL_1543572, EPI_ISL_1543573, EPI_ISL_1543574, EPI_ISL_1543575, EPI_ISL_1543576, EPI_ISL_1543577, EPI_ISL_1543578, EPI_ISL_1543579, EPI_ISL_1543580, EPI_ISL_1543581, EPI_ISL_1543582, EPI_ISL_1543583,
EPI_ISL_1543584, EPI_ISL_1543585, EPI_ISL_1543586, EPI_ISL_1543587, EPI_ISL_1543588, EPI_ISL_1543589, EPI_ISL_1543590, EPI_ISL_1543591, EPI_ISL_1543592, EPI_ISL_1543593, EPI_ISL_1543594, EPI_ISL_1543595, EPI_ISL_1543596, EPI_ISL_1543597, EPI_ISL_1543598, EPI_ISL_1543599, EPI_ISL_1543600, EPI_ISL_1543601, EPI_ISL_1543602, EPI_ISL_1543603, EPI_ISL_1543604, EPI_ISL_1543605,
EPI_ISL_1543606, EPI_ISL_1543607, EPI_ISL_1543608, EPI_ISL_1543609, EPI_ISL_1543610, EPI_ISL_1543611, EPI_ISL_1543612, EPI_ISL_1543613, EPI_ISL_1543614, EPI_ISL_1543615, EPI_ISL_1543616, EPI_ISL_1543617, EPI_ISL_1543618, EPI_ISL_1543619, EPI_ISL_1543620, EPI_ISL_1543621, EPI_ISL_1543622, EPI_ISL_1543623, EPI_ISL_1543624, EPI_ISL_1543625, EPI_ISL_1543626, EPI_ISL_1543627,
EPI_ISL_1543628, EPI_ISL_1543629, EPI_ISL_1543630, EPI_ISL_1543631, EPI_ISL_1543632, EPI_ISL_1543633, EPI_ISL_1543634, EPI_ISL_1543635, EPI_ISL_1543636, EPI_ISL_1543637, EPI_ISL_1543638, EPI_ISL_1543639, EPI_ISL_1543640, EPI_ISL_1543641, EPI_ISL_1543642, EPI_ISL_1543643, EPI_ISL_1543644, EPI_ISL_1543645, EPI_ISL_1543646, EPI_ISL_1543647, EPI_ISL_1543648, EPI_ISL_1543649,
EPI_ISL_1543650, EPI_ISL_1543651, EPI_ISL_1543652, EPI_ISL_1543653, EPI_ISL_1543654, EPI_ISL_1543655, EPI_ISL_1543656, EPI_ISL_1543657, EPI_ISL_1543658, EPI_ISL_1543659, EPI_ISL_1543660, EPI_ISL_1543661, EPI_ISL_1543662, EPI_ISL_1543663, EPI_ISL_1543664, EPI_ISL_1543665, EPI_ISL_1543666, EPI_ISL_1543667, EPI_ISL_1543668, EPI_ISL_1543669, EPI_ISL_1543670, EPI_ISL_1543671,
EPI_ISL_1543672, EPI_ISL_1543673, EPI_ISL_1543674, EPI_ISL_1543675, EPI_ISL_1543676, EPI_ISL_1543677, EPI_ISL_1543678, EPI_ISL_1543679, EPI_ISL_1543680, EPI_ISL_1543681, EPI_ISL_1543682, EPI_ISL_1543683, EPI_ISL_1543684, EPI_ISL_1543685, EPI_ISL_1543686, EPI_ISL_1543687, EPI_ISL_1543688, EPI_ISL_1543689, EPI_ISL_1543690, EPI_ISL_1543691, EPI_ISL_1543692, EPI_ISL_1543693,
EPI_ISL_1543694, EPI_ISL_1543695, EPI_ISL_1543696, EPI_ISL_1543697, EPI_ISL_1543698, EPI_ISL_1543699, EPI_ISL_1543700, EPI_ISL_1543701, EPI_ISL_1543702, EPI_ISL_1543703, EPI_ISL_1543704, EPI_ISL_1543705, EPI_ISL_1543706, EPI_ISL_1543707, EPI_ISL_1543708, EPI_ISL_1543709, EPI_ISL_1543710, EPI_ISL_1543711, EPI_ISL_1543712, EPI_ISL_1543713, EPI_ISL_1543714, EPI_ISL_1543715,
EPI_ISL_1543716, EPI_ISL_1543717, EPI_ISL_1543718, EPI_ISL_1543719, EPI_ISL_1543720, EPI_ISL_1543721, EPI_ISL_1543722, EPI_ISL_1543723, EPI_ISL_1543724, EPI_ISL_1543725, EPI_ISL_1543726, EPI_ISL_1543727, EPI_ISL_1543728, EPI_ISL_1543729, EPI_ISL_1543730, EPI_ISL_1543731, EPI_ISL_1543732, EPI_ISL_1543733, EPI_ISL_1543734, EPI_ISL_1543735, EPI_ISL_1543736, EPI_ISL_1543737,
EPI_ISL_1543738, EPI_ISL_1543739, EPI_ISL_1543740, EPI_ISL_1543741, EPI_ISL_1543742, EPI_ISL_1543743, EPI_ISL_1543744, EPI_ISL_1543745, EPI_ISL_1543746, EPI_ISL_1543747, EPI_ISL_1543748, EPI_ISL_1543749, EPI_ISL_1543750, EPI_ISL_1543751, EPI_ISL_1543752, EPI_ISL_1543753, EPI_ISL_1543754, EPI_ISL_1543755, EPI_ISL_1543756, EPI_ISL_1543757, EPI_ISL_1543758, EPI_ISL_1543759,
EPI_ISL_1543760, EPI_ISL_1543761, EPI_ISL_1543762, EPI_ISL_1543763, EPI_ISL_1543764, EPI_ISL_1543765, EPI_ISL_1543766, EPI_ISL_1543767, EPI_ISL_1543768, EPI_ISL_1543769, EPI_ISL_1543770, EPI_ISL_1543771, EPI_ISL_1543772, EPI_ISL_1543773, EPI_ISL_1543774, EPI_ISL_1543775, EPI_ISL_1543776, EPI_ISL_1543777, EPI_ISL_1543778, EPI_ISL_1543779, EPI_ISL_1543780, EPI_ISL_1543781,
EPI_ISL_1543782, EPI_ISL_1543783, EPI_ISL_1543784, EPI_ISL_1543785, EPI_ISL_1543786, EPI_ISL_1543787, EPI_ISL_1543788, EPI_ISL_1543789, EPI_ISL_1543790, EPI_ISL_1543791, EPI_ISL_1543792, EPI_ISL_1543793, EPI_ISL_1543794, EPI_ISL_1543795, EPI_ISL_1543796, EPI_ISL_1543797, EPI_ISL_1543798, EPI_ISL_1543799, EPI_ISL_1543800, EPI_ISL_1543801, EPI_ISL_1543802, EPI_ISL_1543803,
EPI_ISL_1543804, EPI_ISL_1543805, EPI_ISL_1543806, EPI_ISL_1543807, EPI_ISL_1543808, EPI_ISL_1543809, EPI_ISL_1543810, EPI_ISL_1543811, EPI_ISL_1543812, EPI_ISL_1543813, EPI_ISL_1543814, EPI_ISL_1543815, EPI_ISL_1543816, EPI_ISL_1543817, EPI_ISL_1543818, EPI_ISL_1543819, EPI_ISL_1543820, EPI_ISL_1543821, EPI_ISL_1543822, EPI_ISL_1543823, EPI_ISL_1543824, EPI_ISL_1543825,
EPI_ISL_1543826, EPI_ISL_1543827, EPI_ISL_1543828, EPI_ISL_1543829, EPI_ISL_1543830, EPI_ISL_1543831, EPI_ISL_1543832, EPI_ISL_1543833, EPI_ISL_1543834, EPI_ISL_1543835, EPI_ISL_1543836, EPI_ISL_1543837, EPI_ISL_1543838, EPI_ISL_1543839, EPI_ISL_1543840, EPI_ISL_1543841, EPI_ISL_1543842, EPI_ISL_1543843, EPI_ISL_1543844, EPI_ISL_1543845, EPI_ISL_1543846, EPI_ISL_1543847,
EPI_ISL_1543848, EPI_ISL_1543849, EPI_ISL_1543850, EPI_ISL_1543851, EPI_ISL_1543852, EPI_ISL_1543853, EPI_ISL_1543854, EPI_ISL_1543855, EPI_ISL_1543856, EPI_ISL_1543857, EPI_ISL_1543858, EPI_ISL_1543859, EPI_ISL_1543860, EPI_ISL_1543861, EPI_ISL_1543862, EPI_ISL_1543863, EPI_ISL_1543864, EPI_ISL_1543865, EPI_ISL_1543866, EPI_ISL_1543867, EPI_ISL_1543868, EPI_ISL_1543869,
EPI_ISL_1543870, EPI_ISL_1543871, EPI_ISL_1543872, EPI_ISL_1543873, EPI_ISL_1543874, EPI_ISL_1543875, EPI_ISL_1543876, EPI_ISL_1543877, EPI_ISL_1543878, EPI_ISL_1543879, EPI_ISL_1543880, EPI_ISL_1543881, EPI_ISL_1543882, EPI_ISL_1543883, EPI_ISL_1543884, EPI_ISL_1543885, EPI_ISL_1543886, EPI_ISL_1543887, EPI_ISL_1543888, EPI_ISL_1543889, EPI_ISL_1543890, EPI_ISL_1543891,
EPI_ISL_1543892, EPI_ISL_1543893, EPI_ISL_1543894, EPI_ISL_1543895, EPI_ISL_1543896, EPI_ISL_1543897, EPI_ISL_1543898, EPI_ISL_1543899, EPI_ISL_1543900, EPI_ISL_1543901, EPI_ISL_1543902, EPI_ISL_1543903, EPI_ISL_1543904, EPI_ISL_1543905, EPI_ISL_1543906, EPI_ISL_1543907, EPI_ISL_1543908, EPI_ISL_1543909, EPI_ISL_1543910, EPI_ISL_1543911, EPI_ISL_1543912, EPI_ISL_1543913,
EPI_ISL_1543914, EPI_ISL_1543915, EPI_ISL_1543916, EPI_ISL_1543917, EPI_ISL_1543918, EPI_ISL_1543919, EPI_ISL_1543920, EPI_ISL_1633685, EPI_ISL_1633686, EPI_ISL_1633687, EPI_ISL_1633688, EPI_ISL_1633689, EPI_ISL_1633690, EPI_ISL_1633691, EPI_ISL_1633692, EPI_ISL_1633693, EPI_ISL_1633694, EPI_ISL_1633695, EPI_ISL_1633696, EPI_ISL_1633697, EPI_ISL_1633698, EPI_ISL_1633699,
EPI_ISL_1633700, EPI_ISL_1633701, EPI_ISL_1633702, EPI_ISL_1633703, EPI_ISL_1633704, EPI_ISL_1633705, EPI_ISL_1633706, EPI_ISL_1633707, EPI_ISL_1633708, EPI_ISL_1633709, EPI_ISL_1633710, EPI_ISL_1633711, EPI_ISL_1633712, EPI_ISL_1633713, EPI_ISL_1633714, EPI_ISL_1633715, EPI_ISL_1633716, EPI_ISL_1633717, EPI_ISL_1633718, EPI_ISL_1633719, EPI_ISL_1633720, EPI_ISL_1633721,
EPI_ISL_1633722, EPI_ISL_1633723, EPI_ISL_1633724, EPI_ISL_1633725, EPI_ISL_1633726, EPI_ISL_1633727, EPI_ISL_1633728, EPI_ISL_1633729, EPI_ISL_1633730, EPI_ISL_1633731, EPI_ISL_1633732, EPI_ISL_1633733, EPI_ISL_1633734, EPI_ISL_1633735, EPI_ISL_1633736, EPI_ISL_1633737, EPI_ISL_1633738, EPI_ISL_1633739, EPI_ISL_1633740, EPI_ISL_1633741, EPI_ISL_1633742, EPI_ISL_1633743,
EPI_ISL_1633744, EPI_ISL_1633745, EPI_ISL_1633746, EPI_ISL_1633747, EPI_ISL_1633748, EPI_ISL_1633749, EPI_ISL_1633750, EPI_ISL_1633751, EPI_ISL_1633752, EPI_ISL_1633753, EPI_ISL_1633754, EPI_ISL_1633755, EPI_ISL_1633756, EPI_ISL_1633757, EPI_ISL_1633758, EPI_ISL_1633759, EPI_ISL_1633760, EPI_ISL_1633761, EPI_ISL_1633762, EPI_ISL_1633763, EPI_ISL_1633764, EPI_ISL_1633765,
EPI_ISL_1633766, EPI_ISL_1633767, EPI_ISL_1633768, EPI_ISL_1633769, EPI_ISL_1633770, EPI_ISL_1633771, EPI_ISL_1633772, EPI_ISL_1633773, EPI_ISL_1633774, EPI_ISL_1633775, EPI_ISL_1633776, EPI_ISL_1633777, EPI_ISL_1633778, EPI_ISL_1633779, EPI_ISL_1633780, EPI_ISL_1633781, EPI_ISL_1633782, EPI_ISL_1633783, EPI_ISL_1633784, EPI_ISL_1633785, EPI_ISL_1633786, EPI_ISL_1633787,
EPI_ISL_1633788, EPI_ISL_1633789, EPI_ISL_1633790, EPI_ISL_1633791, EPI_ISL_1633792, EPI_ISL_1633793, EPI_ISL_1633794, EPI_ISL_1633795, EPI_ISL_1633796, EPI_ISL_1633797, EPI_ISL_1633798, EPI_ISL_1633799, EPI_ISL_1633800, EPI_ISL_1633801, EPI_ISL_1633802, EPI_ISL_1633803, EPI_ISL_1633804, EPI_ISL_1633805, EPI_ISL_1633806, EPI_ISL_1633807, EPI_ISL_1633808, EPI_ISL_1633809,
EPI_ISL_1633810, EPI_ISL_1633811, EPI_ISL_1633812, EPI_ISL_1633813, EPI_ISL_1633814, EPI_ISL_1633815, EPI_ISL_1633816, EPI_ISL_1633817, EPI_ISL_1633818, EPI_ISL_1633819, EPI_ISL_1633820, EPI_ISL_1633821, EPI_ISL_1633822, EPI_ISL_1633823, EPI_ISL_1633824, EPI_ISL_1633825, EPI_ISL_1633826, EPI_ISL_1633827, EPI_ISL_1633828, EPI_ISL_1633829, EPI_ISL_1633830, EPI_ISL_1633831,
EPI_ISL_1633832, EPI_ISL_1633833, EPI_ISL_1633834, EPI_ISL_1633835, EPI_ISL_1633836, EPI_ISL_1633837, EPI_ISL_1633838, EPI_ISL_1633839, EPI_ISL_1633840, EPI_ISL_1633841, EPI_ISL_1633842, EPI_ISL_1633843, EPI_ISL_1633844, EPI_ISL_1633845, EPI_ISL_1633846, EPI_ISL_1633847, EPI_ISL_1633848, EPI_ISL_1633849, EPI_ISL_1633850, EPI_ISL_1633851, EPI_ISL_1633852, EPI_ISL_1633853,
EPI_ISL_1633854, EPI_ISL_1633855, EPI_ISL_1633856, EPI_ISL_1633857, EPI_ISL_1633858, EPI_ISL_1633859, EPI_ISL_1633860, EPI_ISL_1633861, EPI_ISL_1633862, EPI_ISL_1633863, EPI_ISL_1633864, EPI_ISL_1633865, EPI_ISL_1633866, EPI_ISL_1633867, EPI_ISL_1633868, EPI_ISL_1633869, EPI_ISL_1633870, EPI_ISL_1633871, EPI_ISL_1633872, EPI_ISL_1633873, EPI_ISL_1633874, EPI_ISL_1633875,
EPI_ISL_1633876, EPI_ISL_1633877, EPI_ISL_1633878, EPI_ISL_1633879, EPI_ISL_1633880, EPI_ISL_1633881, EPI_ISL_1633882, EPI_ISL_1633883, EPI_ISL_1633884, EPI_ISL_1633885, EPI_ISL_1633886, EPI_ISL_1633887, EPI_ISL_1633888, EPI_ISL_1633889, EPI_ISL_1633890, EPI_ISL_1633891, EPI_ISL_1633892, EPI_ISL_1633893, EPI_ISL_1633894, EPI_ISL_1633895, EPI_ISL_1633896, EPI_ISL_1633897,
EPI_ISL_1633899, EPI_ISL_1633901, EPI_ISL_1633902, EPI_ISL_1633903, EPI_ISL_1633905, EPI_ISL_1633906, EPI_ISL_1633907, EPI_ISL_1633909, EPI_ISL_1633910, EPI_ISL_1633911, EPI_ISL_1633913, EPI_ISL_1633914, EPI_ISL_1633916, EPI_ISL_1633917, EPI_ISL_1633918, EPI_ISL_1633920, EPI_ISL_1633921, EPI_ISL_1633922, EPI_ISL_1633924, EPI_ISL_1633925, EPI_ISL_1633927, EPI_ISL_1633929,
EPI_ISL_1633930, EPI_ISL_1633931, EPI_ISL_1633932, EPI_ISL_1633934, EPI_ISL_1633935, EPI_ISL_1633937, EPI_ISL_1633938, EPI_ISL_1633939, EPI_ISL_1633940, EPI_ISL_1633942, EPI_ISL_1633943, EPI_ISL_1633944, EPI_ISL_1633946, EPI_ISL_1633947, EPI_ISL_1633949, EPI_ISL_1633951, EPI_ISL_1633953, EPI_ISL_1633954, EPI_ISL_1633956, EPI_ISL_1633958, EPI_ISL_1633959, EPI_ISL_1633960,
EPI_ISL_1633962, EPI_ISL_1633963, EPI_ISL_1633964, EPI_ISL_1633966, EPI_ISL_1633967, EPI_ISL_1633968, EPI_ISL_1633970, EPI_ISL_1633971, EPI_ISL_1633972, EPI_ISL_1633973, EPI_ISL_1633975, EPI_ISL_1633976, EPI_ISL_1633977, EPI_ISL_1633979, EPI_ISL_1633980, EPI_ISL_1633981, EPI_ISL_1633983, EPI_ISL_1633984, EPI_ISL_1633985, EPI_ISL_1633986, EPI_ISL_1633988, EPI_ISL_1633989,
EPI_ISL_1633990, EPI_ISL_1633992, EPI_ISL_1633993, EPI_ISL_1633994, EPI_ISL_1633996, EPI_ISL_1633997, EPI_ISL_1633998, EPI_ISL_1634000, EPI_ISL_1634001, EPI_ISL_1634002, EPI_ISL_1634004, EPI_ISL_1634005, EPI_ISL_1634007, EPI_ISL_1634008, EPI_ISL_1634009, EPI_ISL_1634011, EPI_ISL_1634012, EPI_ISL_1634014, EPI_ISL_1634016, EPI_ISL_1634018, EPI_ISL_1634019, EPI_ISL_1634021,
EPI_ISL_1634022, EPI_ISL_1634023, EPI_ISL_1634025, EPI_ISL_1634026, EPI_ISL_1634028, EPI_ISL_1634029, EPI_ISL_1634030, EPI_ISL_1634032, EPI_ISL_1634033, EPI_ISL_1634034, EPI_ISL_1634036, EPI_ISL_1634037, EPI_ISL_1634038, EPI_ISL_1634040, EPI_ISL_1634041, EPI_ISL_1634042, EPI_ISL_1634044, EPI_ISL_1634045, EPI_ISL_1634046, EPI_ISL_1634047, EPI_ISL_1634049, EPI_ISL_1634051,
EPI_ISL_1634052, EPI_ISL_1634053, EPI_ISL_1634054, EPI_ISL_1634056, EPI_ISL_1634058, EPI_ISL_1634060, EPI_ISL_1634061, EPI_ISL_1634062, EPI_ISL_1634064, EPI_ISL_1634065, EPI_ISL_1634066, EPI_ISL_1634068, EPI_ISL_1634069, EPI_ISL_1634071, EPI_ISL_1634072, EPI_ISL_1634074, EPI_ISL_1634076, EPI_ISL_1634077, EPI_ISL_1634079, EPI_ISL_1634080, EPI_ISL_1634082, EPI_ISL_1634083,
EPI_ISL_1634084, EPI_ISL_1634086, EPI_ISL_1634087, EPI_ISL_1634088, EPI_ISL_1634089, EPI_ISL_1634091, EPI_ISL_1634092, EPI_ISL_1634093, EPI_ISL_1634095, EPI_ISL_1634096, EPI_ISL_1634097, EPI_ISL_1634099, EPI_ISL_1634100, EPI_ISL_1634101, EPI_ISL_1634103, EPI_ISL_1634104, EPI_ISL_1634105, EPI_ISL_1634107, EPI_ISL_1634108, EPI_ISL_1634110, EPI_ISL_1634112, EPI_ISL_1634113,
EPI_ISL_1634114, EPI_ISL_1634116, EPI_ISL_1634118, EPI_ISL_1634120, EPI_ISL_1634121, EPI_ISL_1634123, EPI_ISL_1634124, EPI_ISL_1634125, EPI_ISL_1634126, EPI_ISL_1634128, EPI_ISL_1634130, EPI_ISL_1634132, EPI_ISL_1634133, EPI_ISL_1634135, EPI_ISL_1634136, EPI_ISL_1634137, EPI_ISL_1634139, EPI_ISL_1634140, EPI_ISL_1634141, EPI_ISL_1634150, EPI_ISL_1634153, EPI_ISL_1634155,
EPI_ISL_1634157, EPI_ISL_1634158, EPI_ISL_1634159, EPI_ISL_1634161, EPI_ISL_1634165, EPI_ISL_1634166, EPI_ISL_1634167, EPI_ISL_1634169, EPI_ISL_1634170, EPI_ISL_1634171, EPI_ISL_1634173, EPI_ISL_1634174, EPI_ISL_1634175, EPI_ISL_1634176, EPI_ISL_1634178, EPI_ISL_1634179, EPI_ISL_1634180, EPI_ISL_1634181, EPI_ISL_1634183, EPI_ISL_1634184, EPI_ISL_1634185, EPI_ISL_1634186,
EPI_ISL_1634187, EPI_ISL_1634188, EPI_ISL_1634189, EPI_ISL_1634191, EPI_ISL_1634192, EPI_ISL_1634194, EPI_ISL_1634195, EPI_ISL_1634196, EPI_ISL_1634197, EPI_ISL_1634198, EPI_ISL_1634199, EPI_ISL_1634200, EPI_ISL_1634201, EPI_ISL_1634202, EPI_ISL_1634204, EPI_ISL_1634205, EPI_ISL_1634206, EPI_ISL_1634207, EPI_ISL_1634208, EPI_ISL_1634210, EPI_ISL_1634211, EPI_ISL_1634212,
EPI_ISL_1634213, EPI_ISL_1634214, EPI_ISL_1634215, EPI_ISL_1634217, EPI_ISL_1634218, EPI_ISL_1634220, EPI_ISL_1634221, EPI_ISL_1634222, EPI_ISL_1634223, EPI_ISL_1634226, EPI_ISL_1634227, EPI_ISL_1634228, EPI_ISL_1634229, EPI_ISL_1634230, EPI_ISL_1634231, EPI_ISL_1634232, EPI_ISL_1634233, EPI_ISL_1634234, EPI_ISL_1634235, EPI_ISL_1634236, EPI_ISL_1634237, EPI_ISL_1634238,
EPI_ISL_1634239, EPI_ISL_1634240, EPI_ISL_1634241, EPI_ISL_1634242, EPI_ISL_1634243, EPI_ISL_1634244, EPI_ISL_1634245, EPI_ISL_1634246, EPI_ISL_1634247, EPI_ISL_1634248, EPI_ISL_1634250, EPI_ISL_1634251, EPI_ISL_1634252, EPI_ISL_1634253, EPI_ISL_1634255, EPI_ISL_1634256, EPI_ISL_1634258, EPI_ISL_1634259, EPI_ISL_1634260, EPI_ISL_1634261, EPI_ISL_1634262, EPI_ISL_1634263,
EPI_ISL_1634264, EPI_ISL_1634265, EPI_ISL_1634266, EPI_ISL_1634267, EPI_ISL_1634268, EPI_ISL_1634270, EPI_ISL_1634271, EPI_ISL_1634272, EPI_ISL_1634273, EPI_ISL_1634274, EPI_ISL_1634275, EPI_ISL_1634276, EPI_ISL_1634277, EPI_ISL_1634278, EPI_ISL_1634279, EPI_ISL_1634280, EPI_ISL_1634281, EPI_ISL_1634282, EPI_ISL_1634283, EPI_ISL_1634284, EPI_ISL_1634285, EPI_ISL_1634286,
EPI_ISL_1634287, EPI_ISL_1634288, EPI_ISL_1634289, EPI_ISL_1634290, EPI_ISL_1634291, EPI_ISL_1634292, EPI_ISL_1634293, EPI_ISL_1634294, EPI_ISL_1634295, EPI_ISL_1634296, EPI_ISL_1634297, EPI_ISL_1634298, EPI_ISL_1634300, EPI_ISL_1634301, EPI_ISL_1634302, EPI_ISL_1634303, EPI_ISL_1634305, EPI_ISL_1634306, EPI_ISL_1634307, EPI_ISL_1634308, EPI_ISL_1634309, EPI_ISL_1634310,
EPI_ISL_1634313, EPI_ISL_1634314, EPI_ISL_1634315, EPI_ISL_1634316, EPI_ISL_1634317, EPI_ISL_1634318, EPI_ISL_1634319, EPI_ISL_1634320, EPI_ISL_1634321, EPI_ISL_1634323, EPI_ISL_1634324, EPI_ISL_1634325, EPI_ISL_1634326, EPI_ISL_1634327, EPI_ISL_1634329, EPI_ISL_1634330, EPI_ISL_1634331, EPI_ISL_1634333, EPI_ISL_1634334, EPI_ISL_1634337, EPI_ISL_1634338, EPI_ISL_1635437,
EPI_ISL_1635438, EPI_ISL_1635439, EPI_ISL_1635440, EPI_ISL_1635441, EPI_ISL_1635442, EPI_ISL_1635443, EPI_ISL_1635444, EPI_ISL_1635445, EPI_ISL_1635446, EPI_ISL_1635447, EPI_ISL_1635448, EPI_ISL_1635449, EPI_ISL_1635450, EPI_ISL_1635451, EPI_ISL_1635452, EPI_ISL_1635453, EPI_ISL_1635454, EPI_ISL_1635455, EPI_ISL_1635456, EPI_ISL_1635457, EPI_ISL_1635458, EPI_ISL_1635459,
EPI_ISL_1635460, EPI_ISL_1635461, EPI_ISL_1635462, EPI_ISL_1635463, EPI_ISL_1635464, EPI_ISL_1635465, EPI_ISL_1635466, EPI_ISL_1635467, EPI_ISL_1635468, EPI_ISL_1635469, EPI_ISL_1635470, EPI_ISL_1635471, EPI_ISL_1635472, EPI_ISL_1635473, EPI_ISL_1635474, EPI_ISL_1635475, EPI_ISL_1635476, EPI_ISL_1635477, EPI_ISL_1635478, EPI_ISL_1635479, EPI_ISL_1635480, EPI_ISL_1635481,
EPI_ISL_1635482, EPI_ISL_1635483, EPI_ISL_1635484, EPI_ISL_1635485, EPI_ISL_1635486, EPI_ISL_1635487, EPI_ISL_1635488, EPI_ISL_1635489, EPI_ISL_1635490, EPI_ISL_1635491, EPI_ISL_1635492, EPI_ISL_1635493, EPI_ISL_1635494, EPI_ISL_1635495, EPI_ISL_1635496, EPI_ISL_1635497, EPI_ISL_1635498, EPI_ISL_1635499, EPI_ISL_1635500, EPI_ISL_1635501, EPI_ISL_1635502, EPI_ISL_1635503,
EPI_ISL_1635504, EPI_ISL_1635505, EPI_ISL_1635506, EPI_ISL_1635507, EPI_ISL_1635508, EPI_ISL_1635509, EPI_ISL_1635510, EPI_ISL_1635511, EPI_ISL_1635512, EPI_ISL_1635513, EPI_ISL_1635514, EPI_ISL_1635515, EPI_ISL_1635516, EPI_ISL_1635517, EPI_ISL_1635518, EPI_ISL_1635519, EPI_ISL_1635520, EPI_ISL_1635521, EPI_ISL_1635522, EPI_ISL_1635523, EPI_ISL_1635524, EPI_ISL_1635525,
EPI_ISL_1635526, EPI_ISL_1635527, EPI_ISL_1635528, EPI_ISL_1635529, EPI_ISL_1635530, EPI_ISL_1635531, EPI_ISL_1635532, EPI_ISL_1635533, EPI_ISL_1635534, EPI_ISL_1635535, EPI_ISL_1635536, EPI_ISL_1635537, EPI_ISL_1635538, EPI_ISL_1635539, EPI_ISL_1635540, EPI_ISL_1635541, EPI_ISL_1635542, EPI_ISL_1635543, EPI_ISL_1635544, EPI_ISL_1635545, EPI_ISL_1635546, EPI_ISL_1635547,
EPI_ISL_1635548, EPI_ISL_1635549, EPI_ISL_1635550, EPI_ISL_1635551, EPI_ISL_1635552, EPI_ISL_1635553, EPI_ISL_1635554, EPI_ISL_1635555, EPI_ISL_1635556, EPI_ISL_1635557, EPI_ISL_1635558, EPI_ISL_1635559, EPI_ISL_1635560, EPI_ISL_1635561, EPI_ISL_1635562, EPI_ISL_1635563, EPI_ISL_1635564, EPI_ISL_1635565, EPI_ISL_1635566, EPI_ISL_1635567, EPI_ISL_1635568, EPI_ISL_1635569,
EPI_ISL_1635570, EPI_ISL_1635571, EPI_ISL_1635572, EPI_ISL_1635573, EPI_ISL_1635574, EPI_ISL_1635575, EPI_ISL_1635576, EPI_ISL_1635577, EPI_ISL_1635578, EPI_ISL_1635579, EPI_ISL_1635580, EPI_ISL_1635581, EPI_ISL_1635582, EPI_ISL_1635583, EPI_ISL_1635584, EPI_ISL_1635585, EPI_ISL_1635586, EPI_ISL_1635587, EPI_ISL_1635588, EPI_ISL_1635589, EPI_ISL_1635590, EPI_ISL_1635591,
EPI_ISL_1635592, EPI_ISL_1635593, EPI_ISL_1635594, EPI_ISL_1635595, EPI_ISL_1635596, EPI_ISL_1635597, EPI_ISL_1635598, EPI_ISL_1635599, EPI_ISL_1635600, EPI_ISL_1635601, EPI_ISL_1635602, EPI_ISL_1635603, EPI_ISL_1635604, EPI_ISL_1635605, EPI_ISL_1635606, EPI_ISL_1635607, EPI_ISL_1635608, EPI_ISL_1635609, EPI_ISL_1635610, EPI_ISL_1635611, EPI_ISL_1635612, EPI_ISL_1635613,
EPI_ISL_1635614, EPI_ISL_1635615, EPI_ISL_1635616, EPI_ISL_1635617, EPI_ISL_1635618, EPI_ISL_1635619, EPI_ISL_1635620, EPI_ISL_1635621, EPI_ISL_1635622, EPI_ISL_1635623, EPI_ISL_1635624, EPI_ISL_1635625, EPI_ISL_1635626, EPI_ISL_1635627, EPI_ISL_1635628, EPI_ISL_1635629, EPI_ISL_1635630, EPI_ISL_1635631, EPI_ISL_1635632, EPI_ISL_1635633, EPI_ISL_1635634, EPI_ISL_1635635,
EPI_ISL_1635636, EPI_ISL_1635637, EPI_ISL_1635638, EPI_ISL_1635639, EPI_ISL_1635640, EPI_ISL_1635641, EPI_ISL_1635642, EPI_ISL_1635643, EPI_ISL_1635644, EPI_ISL_1635645, EPI_ISL_1635646, EPI_ISL_1635647, EPI_ISL_1635648, EPI_ISL_1635649, EPI_ISL_1635650, EPI_ISL_1635651, EPI_ISL_1635652, EPI_ISL_1635653, EPI_ISL_1635654, EPI_ISL_1635655, EPI_ISL_1635656, EPI_ISL_1635657,
EPI_ISL_1635658, EPI_ISL_1635659, EPI_ISL_1635660, EPI_ISL_1635661, EPI_ISL_1635662, EPI_ISL_1635663, EPI_ISL_1635664, EPI_ISL_1635665, EPI_ISL_1635667, EPI_ISL_1635668, EPI_ISL_1635669, EPI_ISL_1635670, EPI_ISL_1635671, EPI_ISL_1635672, EPI_ISL_1635673, EPI_ISL_1635674, EPI_ISL_1635675, EPI_ISL_1635676, EPI_ISL_1635677, EPI_ISL_1635678, EPI_ISL_1635679, EPI_ISL_1635680,
EPI_ISL_1635681, EPI_ISL_1635682, EPI_ISL_1635683, EPI_ISL_1635684, EPI_ISL_1635685, EPI_ISL_1635686, EPI_ISL_1635687, EPI_ISL_1635688, EPI_ISL_1635689, EPI_ISL_1635690, EPI_ISL_1635691, EPI_ISL_1635692, EPI_ISL_1635693, EPI_ISL_1635694, EPI_ISL_1635695, EPI_ISL_1635696, EPI_ISL_1635697, EPI_ISL_1635698, EPI_ISL_1635699, EPI_ISL_1635700, EPI_ISL_1635701, EPI_ISL_1635702,
EPI_ISL_1635703, EPI_ISL_1635704, EPI_ISL_1635705, EPI_ISL_1635706, EPI_ISL_1635707, EPI_ISL_1635708, EPI_ISL_1635709, EPI_ISL_1635710, EPI_ISL_1635711, EPI_ISL_1635712, EPI_ISL_1635713, EPI_ISL_1635714, EPI_ISL_1635715, EPI_ISL_1635716, EPI_ISL_1635717, EPI_ISL_1635718, EPI_ISL_1635719, EPI_ISL_1635720, EPI_ISL_1635721, EPI_ISL_1635722, EPI_ISL_1635723, EPI_ISL_1635724,
EPI_ISL_1635725, EPI_ISL_1635726, EPI_ISL_1635727, EPI_ISL_1635728, EPI_ISL_1635729, EPI_ISL_1635730, EPI_ISL_1635731, EPI_ISL_1635732, EPI_ISL_1635733, EPI_ISL_1635734, EPI_ISL_1635735, EPI_ISL_1635736, EPI_ISL_1635737, EPI_ISL_1635738, EPI_ISL_1635739, EPI_ISL_1635740, EPI_ISL_1635741, EPI_ISL_1635742, EPI_ISL_1635743, EPI_ISL_1635744, EPI_ISL_1635745, EPI_ISL_1635746,
EPI_ISL_1635747, EPI_ISL_1635748, EPI_ISL_1635749, EPI_ISL_1635750, EPI_ISL_1635751, EPI_ISL_1635752, EPI_ISL_1635753, EPI_ISL_1635755, EPI_ISL_1635756, EPI_ISL_1635757, EPI_ISL_1635758, EPI_ISL_1635759, EPI_ISL_1635760, EPI_ISL_1635761, EPI_ISL_1635762, EPI_ISL_1635763, EPI_ISL_1635764, EPI_ISL_1635765, EPI_ISL_1635766, EPI_ISL_1635767, EPI_ISL_1635768, EPI_ISL_1635769,
EPI_ISL_1635770, EPI_ISL_1635771, EPI_ISL_1635772, EPI_ISL_1635773, EPI_ISL_1635774, EPI_ISL_1635775, EPI_ISL_1635776, EPI_ISL_1635777, EPI_ISL_1635778, EPI_ISL_1635779, EPI_ISL_1635780, EPI_ISL_1635781, EPI_ISL_1635782, EPI_ISL_1635783, EPI_ISL_1635784, EPI_ISL_1635785, EPI_ISL_1635786, EPI_ISL_1635787, EPI_ISL_1635788, EPI_ISL_1635789, EPI_ISL_1635790, EPI_ISL_1635791,
EPI_ISL_1635792, EPI_ISL_1635793, EPI_ISL_1635794, EPI_ISL_1635795, EPI_ISL_1635796, EPI_ISL_1635797, EPI_ISL_1635798, EPI_ISL_1635799, EPI_ISL_1635800, EPI_ISL_1635801, EPI_ISL_1635802, EPI_ISL_1635803, EPI_ISL_1635804, EPI_ISL_1635805, EPI_ISL_1635806, EPI_ISL_1635807, EPI_ISL_1635808, EPI_ISL_1635809, EPI_ISL_1635810, EPI_ISL_1635811, EPI_ISL_1635812, EPI_ISL_1635813,
EPI_ISL_1635814, EPI_ISL_1635815, EPI_ISL_1635816, EPI_ISL_1635817, EPI_ISL_1635818, EPI_ISL_1635819, EPI_ISL_1635820, EPI_ISL_1635821, EPI_ISL_1635822, EPI_ISL_1635823, EPI_ISL_1635824, EPI_ISL_1635825, EPI_ISL_1635826, EPI_ISL_1635827, EPI_ISL_1635828, EPI_ISL_1635829, EPI_ISL_1635830, EPI_ISL_1635831, EPI_ISL_1635832, EPI_ISL_1635833, EPI_ISL_1635834, EPI_ISL_1635835,
EPI_ISL_1635836, EPI_ISL_1635837, EPI_ISL_1635838, EPI_ISL_1635839, EPI_ISL_1635840, EPI_ISL_1635841, EPI_ISL_1635842, EPI_ISL_1635843, EPI_ISL_1635844, EPI_ISL_1635845, EPI_ISL_1635846, EPI_ISL_1635847, EPI_ISL_1635848, EPI_ISL_1635849, EPI_ISL_1635850, EPI_ISL_1635851, EPI_ISL_1635852, EPI_ISL_1635853, EPI_ISL_1635854, EPI_ISL_1635855, EPI_ISL_1635856, EPI_ISL_1635857,
EPI_ISL_1635858, EPI_ISL_1635859, EPI_ISL_1635860, EPI_ISL_1635861, EPI_ISL_1635862, EPI_ISL_1635863, EPI_ISL_1635864, EPI_ISL_1635865, EPI_ISL_1635866, EPI_ISL_1635867, EPI_ISL_1635868, EPI_ISL_1635869, EPI_ISL_1635870, EPI_ISL_1635871, EPI_ISL_1635872, EPI_ISL_1635873, EPI_ISL_1635874, EPI_ISL_1635875, EPI_ISL_1635876, EPI_ISL_1635877, EPI_ISL_1635878, EPI_ISL_1635879,
EPI_ISL_1635880, EPI_ISL_1635881, EPI_ISL_1635882, EPI_ISL_1635883, EPI_ISL_1635884, EPI_ISL_1635885, EPI_ISL_1635886, EPI_ISL_1635887, EPI_ISL_1635888, EPI_ISL_1635889, EPI_ISL_1635890, EPI_ISL_1635891, EPI_ISL_1635892, EPI_ISL_1635893, EPI_ISL_1635894, EPI_ISL_1635895, EPI_ISL_1635896, EPI_ISL_1635897, EPI_ISL_1635898, EPI_ISL_1635899, EPI_ISL_1635900, EPI_ISL_1635901,
EPI_ISL_1635902, EPI_ISL_1635903, EPI_ISL_1635904, EPI_ISL_1635905, EPI_ISL_1635906, EPI_ISL_1635907, EPI_ISL_1635908, EPI_ISL_1635909, EPI_ISL_1635910, EPI_ISL_1635911, EPI_ISL_1635912, EPI_ISL_1635913, EPI_ISL_1635914, EPI_ISL_1635915, EPI_ISL_1635916, EPI_ISL_1635917, EPI_ISL_1635918, EPI_ISL_1635919, EPI_ISL_1635920, EPI_ISL_1635921, EPI_ISL_1635922, EPI_ISL_1635923,
EPI_ISL_1635924, EPI_ISL_1635925, EPI_ISL_1635926, EPI_ISL_1635927, EPI_ISL_1635928, EPI_ISL_1635929, EPI_ISL_1635930, EPI_ISL_1635931, EPI_ISL_1635932, EPI_ISL_1635933, EPI_ISL_1635934, EPI_ISL_1635935, EPI_ISL_1635936, EPI_ISL_1635937, EPI_ISL_1635938, EPI_ISL_1635939, EPI_ISL_1635940, EPI_ISL_1635941, EPI_ISL_1635942, EPI_ISL_1635943, EPI_ISL_1635944, EPI_ISL_1635945,
EPI_ISL_1635946, EPI_ISL_1635947, EPI_ISL_1635948, EPI_ISL_1635949, EPI_ISL_1635950, EPI_ISL_1635951, EPI_ISL_1635952, EPI_ISL_1635953, EPI_ISL_1635954, EPI_ISL_1635955, EPI_ISL_1635956, EPI_ISL_1635957, EPI_ISL_1635958, EPI_ISL_1635959, EPI_ISL_1635960, EPI_ISL_1635961, EPI_ISL_1635962, EPI_ISL_1635963, EPI_ISL_1635965, EPI_ISL_1635966, EPI_ISL_1635967, EPI_ISL_1635968,
EPI_ISL_1635969, EPI_ISL_1635970, EPI_ISL_1635971, EPI_ISL_1635972, EPI_ISL_1635973, EPI_ISL_1635974, EPI_ISL_1635975, EPI_ISL_1635976, EPI_ISL_1635977, EPI_ISL_1635978, EPI_ISL_1635979, EPI_ISL_1635980, EPI_ISL_1635981, EPI_ISL_1635982, EPI_ISL_1635983, EPI_ISL_1635984, EPI_ISL_1635985, EPI_ISL_1635986, EPI_ISL_1635987, EPI_ISL_1635988, EPI_ISL_1635989, EPI_ISL_1635990,
EPI_ISL_1635991, EPI_ISL_1635992, EPI_ISL_1635993, EPI_ISL_1635994, EPI_ISL_1635995, EPI_ISL_1635996, EPI_ISL_1635997, EPI_ISL_1635998, EPI_ISL_1635999, EPI_ISL_1636000, EPI_ISL_1636001, EPI_ISL_1636002, EPI_ISL_1636003, EPI_ISL_1636004, EPI_ISL_1636005, EPI_ISL_1636006, EPI_ISL_1636007, EPI_ISL_1636008, EPI_ISL_1636009, EPI_ISL_1636010, EPI_ISL_1636011, EPI_ISL_1636012,
EPI_ISL_1636013, EPI_ISL_1636014, EPI_ISL_1636015, EPI_ISL_1636016, EPI_ISL_1636017, EPI_ISL_1636018, EPI_ISL_1636019, EPI_ISL_1636020, EPI_ISL_1636021, EPI_ISL_1636022, EPI_ISL_1636023, EPI_ISL_1636024, EPI_ISL_1636025, EPI_ISL_1636026, EPI_ISL_1636027, EPI_ISL_1636028, EPI_ISL_1636029, EPI_ISL_1636030, EPI_ISL_1636031, EPI_ISL_1636032, EPI_ISL_1636033, EPI_ISL_1636034,
EPI_ISL_1636035, EPI_ISL_1636036, EPI_ISL_1636037, EPI_ISL_1636038, EPI_ISL_1636039, EPI_ISL_1636040, EPI_ISL_1636041, EPI_ISL_1636042, EPI_ISL_1636043, EPI_ISL_1636044, EPI_ISL_1636045, EPI_ISL_1636046, EPI_ISL_1636047, EPI_ISL_1636048, EPI_ISL_1636049, EPI_ISL_1636050, EPI_ISL_1636051, EPI_ISL_1636052, EPI_ISL_1636053, EPI_ISL_1636054, EPI_ISL_1636055, EPI_ISL_1636056,
EPI_ISL_1636057, EPI_ISL_1636058, EPI_ISL_1636059, EPI_ISL_1636060, EPI_ISL_1636061, EPI_ISL_1636062, EPI_ISL_1636063, EPI_ISL_1636064, EPI_ISL_1636065, EPI_ISL_1636066, EPI_ISL_1636067, EPI_ISL_1636068, EPI_ISL_1636069, EPI_ISL_1636070, EPI_ISL_1636071, EPI_ISL_1636072, EPI_ISL_1636073, EPI_ISL_1636074, EPI_ISL_1636075, EPI_ISL_1636076, EPI_ISL_1636077, EPI_ISL_1636078,
EPI_ISL_1636079, EPI_ISL_1636080, EPI_ISL_1636081, EPI_ISL_1636082, EPI_ISL_1636083, EPI_ISL_1636084, EPI_ISL_1636085, EPI_ISL_1636086, EPI_ISL_1636087, EPI_ISL_1636088, EPI_ISL_1636089, EPI_ISL_1636090, EPI_ISL_1636091, EPI_ISL_1636092, EPI_ISL_1636093, EPI_ISL_1636094, EPI_ISL_1636095, EPI_ISL_1636096, EPI_ISL_1636097, EPI_ISL_1636098, EPI_ISL_1636099, EPI_ISL_1636100,
EPI_ISL_1636101, EPI_ISL_1636102, EPI_ISL_1636103, EPI_ISL_1636104, EPI_ISL_1636105, EPI_ISL_1636106, EPI_ISL_1636107, EPI_ISL_1636108, EPI_ISL_1636109, EPI_ISL_1636111, EPI_ISL_1636112, EPI_ISL_1636113, EPI_ISL_1636114, EPI_ISL_1636115, EPI_ISL_1636116, EPI_ISL_1636117, EPI_ISL_1636118, EPI_ISL_1636119, EPI_ISL_1636120, EPI_ISL_1636121, EPI_ISL_1636122, EPI_ISL_1636123,
EPI_ISL_1636124, EPI_ISL_1636125, EPI_ISL_1636126, EPI_ISL_1636127, EPI_ISL_1636128, EPI_ISL_1636129, EPI_ISL_1636130, EPI_ISL_1636131, EPI_ISL_1636132, EPI_ISL_1636133, EPI_ISL_1636134, EPI_ISL_1636135, EPI_ISL_1636136, EPI_ISL_1636137, EPI_ISL_1636138, EPI_ISL_1636139, EPI_ISL_1636140, EPI_ISL_1636141, EPI_ISL_1636142, EPI_ISL_1636143, EPI_ISL_1636144, EPI_ISL_1636145,
EPI_ISL_1636146, EPI_ISL_1636147, EPI_ISL_1636148, EPI_ISL_1636149, EPI_ISL_1636150, EPI_ISL_1636151, EPI_ISL_1636152, EPI_ISL_1636153, EPI_ISL_1636154, EPI_ISL_1636155, EPI_ISL_1636156, EPI_ISL_1636157, EPI_ISL_1636158, EPI_ISL_1636159, EPI_ISL_1636160, EPI_ISL_1636161, EPI_ISL_1636162, EPI_ISL_1636163, EPI_ISL_1636164, EPI_ISL_1636165, EPI_ISL_1636166, EPI_ISL_1636167,
EPI_ISL_1636168, EPI_ISL_1636169, EPI_ISL_1636170, EPI_ISL_1636171, EPI_ISL_1636172, EPI_ISL_1636173, EPI_ISL_1636174, EPI_ISL_1636175, EPI_ISL_1636176, EPI_ISL_1636177, EPI_ISL_1636178, EPI_ISL_1636179, EPI_ISL_1636180, EPI_ISL_1636181, EPI_ISL_1636182, EPI_ISL_1636183, EPI_ISL_1636184, EPI_ISL_1636185, EPI_ISL_1636186, EPI_ISL_1636187, EPI_ISL_1636188, EPI_ISL_1636189,
EPI_ISL_1636190, EPI_ISL_1636191, EPI_ISL_1636192, EPI_ISL_1636193, EPI_ISL_1636194, EPI_ISL_1636195, EPI_ISL_1636196, EPI_ISL_1636197, EPI_ISL_1636198, EPI_ISL_1636199, EPI_ISL_1636200, EPI_ISL_1636201, EPI_ISL_1636202, EPI_ISL_1636203, EPI_ISL_1636204, EPI_ISL_1636205, EPI_ISL_1636206, EPI_ISL_1636207, EPI_ISL_1636208, EPI_ISL_1636209, EPI_ISL_1636210, EPI_ISL_1636211,
EPI_ISL_1636212, EPI_ISL_1636213, EPI_ISL_1636214, EPI_ISL_1636215, EPI_ISL_1636217, EPI_ISL_1636218, EPI_ISL_1636219, EPI_ISL_1636220, EPI_ISL_1636221, EPI_ISL_1636222, EPI_ISL_1636223, EPI_ISL_1636224, EPI_ISL_1636225, EPI_ISL_1636226, EPI_ISL_1636227, EPI_ISL_1636228, EPI_ISL_1636229, EPI_ISL_1636230, EPI_ISL_1636231, EPI_ISL_1636232, EPI_ISL_1636233, EPI_ISL_1636234,
EPI_ISL_1636235, EPI_ISL_1636236, EPI_ISL_1636237, EPI_ISL_1636238, EPI_ISL_1636239, EPI_ISL_1636240, EPI_ISL_1636241, EPI_ISL_1636242, EPI_ISL_1636243, EPI_ISL_1636244, EPI_ISL_1636245, EPI_ISL_1636246, EPI_ISL_1636247, EPI_ISL_1636248, EPI_ISL_1636249, EPI_ISL_1636250, EPI_ISL_1636251, EPI_ISL_1636252, EPI_ISL_1636253, EPI_ISL_1636254, EPI_ISL_1636255, EPI_ISL_1636256,
EPI_ISL_1636257, EPI_ISL_1636258, EPI_ISL_1636259, EPI_ISL_1636260, EPI_ISL_1636261, EPI_ISL_1636262, EPI_ISL_1636263, EPI_ISL_1636264, EPI_ISL_1636265, EPI_ISL_1636266, EPI_ISL_1636267, EPI_ISL_1636268, EPI_ISL_1636269, EPI_ISL_1636270, EPI_ISL_1636271, EPI_ISL_1636272, EPI_ISL_1636273, EPI_ISL_1636274, EPI_ISL_1636275, EPI_ISL_1636276, EPI_ISL_1636277, EPI_ISL_1636278,
EPI_ISL_1636279, EPI_ISL_1636280, EPI_ISL_1636281, EPI_ISL_1636282, EPI_ISL_1636283, EPI_ISL_1636284, EPI_ISL_1636285, EPI_ISL_1636286, EPI_ISL_1636287, EPI_ISL_1636288, EPI_ISL_1636289, EPI_ISL_1636290, EPI_ISL_1636291, EPI_ISL_1636292, EPI_ISL_1636293, EPI_ISL_1636294, EPI_ISL_1636295, EPI_ISL_1636296, EPI_ISL_1636297, EPI_ISL_1636298, EPI_ISL_1636299, EPI_ISL_1636300,
EPI_ISL_1636301, EPI_ISL_1636302, EPI_ISL_1636303, EPI_ISL_1636304, EPI_ISL_1636305, EPI_ISL_1636306, EPI_ISL_1636307, EPI_ISL_1636308, EPI_ISL_1636309, EPI_ISL_1636310, EPI_ISL_1636311, EPI_ISL_1636312, EPI_ISL_1636313, EPI_ISL_1636314, EPI_ISL_1636315, EPI_ISL_1636316, EPI_ISL_1636317, EPI_ISL_1636318, EPI_ISL_1636319, EPI_ISL_1636320, EPI_ISL_1636321, EPI_ISL_1636322,
EPI_ISL_1636323, EPI_ISL_1636324, EPI_ISL_1636325, EPI_ISL_1636326, EPI_ISL_1636327, EPI_ISL_1636328, EPI_ISL_1636329, EPI_ISL_1636330, EPI_ISL_1636331, EPI_ISL_1636332, EPI_ISL_1636333, EPI_ISL_1636334, EPI_ISL_1636335, EPI_ISL_1636336, EPI_ISL_1636337, EPI_ISL_1636338, EPI_ISL_1636339, EPI_ISL_1636340, EPI_ISL_1636341, EPI_ISL_1636342, EPI_ISL_1636343, EPI_ISL_1636344,
EPI_ISL_1636345, EPI_ISL_1636346, EPI_ISL_1636347, EPI_ISL_1636348, EPI_ISL_1636349, EPI_ISL_1636350, EPI_ISL_1636351, EPI_ISL_1636352, EPI_ISL_1636353, EPI_ISL_1636354, EPI_ISL_1636355, EPI_ISL_1636356, EPI_ISL_1636357, EPI_ISL_1636358, EPI_ISL_1636359, EPI_ISL_1636360, EPI_ISL_1636361, EPI_ISL_1636362, EPI_ISL_1636363, EPI_ISL_1636364, EPI_ISL_1636366, EPI_ISL_1636367,
EPI_ISL_1636368, EPI_ISL_1636369, EPI_ISL_1636370, EPI_ISL_1636371, EPI_ISL_1636372, EPI_ISL_1636373, EPI_ISL_1636374, EPI_ISL_1636375, EPI_ISL_1636376, EPI_ISL_1636377, EPI_ISL_1636378, EPI_ISL_1636379, EPI_ISL_1636380, EPI_ISL_1636381, EPI_ISL_1636382, EPI_ISL_1636383, EPI_ISL_1636384, EPI_ISL_1636385, EPI_ISL_1636386, EPI_ISL_1636387, EPI_ISL_1636388, EPI_ISL_1636389,
EPI_ISL_1636390, EPI_ISL_1636391, EPI_ISL_1636392, EPI_ISL_1636393, EPI_ISL_1636394, EPI_ISL_1636395, EPI_ISL_1636396, EPI_ISL_1636397, EPI_ISL_1717117, EPI_ISL_1717130, EPI_ISL_1717134, EPI_ISL_1717152, EPI_ISL_1717168, EPI_ISL_1717195, EPI_ISL_1717220, EPI_ISL_1717234, EPI_ISL_1717238, EPI_ISL_1717239, EPI_ISL_1717243, EPI_ISL_1717244, EPI_ISL_1717255, EPI_ISL_1717258,
EPI_ISL_1717259, EPI_ISL_1717266, EPI_ISL_1717267, EPI_ISL_1717272, EPI_ISL_1717276, EPI_ISL_1717282, EPI_ISL_1717301, EPI_ISL_1717306, EPI_ISL_1717314, EPI_ISL_1717326, EPI_ISL_1717329, EPI_ISL_1717352, EPI_ISL_1717358, EPI_ISL_1717365, EPI_ISL_1717386, EPI_ISL_1717396, EPI_ISL_1717428, EPI_ISL_1717434, EPI_ISL_1717466, EPI_ISL_1717495, EPI_ISL_1717509, EPI_ISL_1717510,
EPI_ISL_1717573, EPI_ISL_1717595, EPI_ISL_1717622, EPI_ISL_1717674

| | | | |
|---|---|---|---|
| see above | Pandemic Response Lab - NYC | Pandemic Response Lab, R&D | Cybill del Castillo; Dylan Law; Haiping Hao; Henry Lee; Jon Laurent; Katharine Nelson; Melissa Hopkins; Michael Hammerling; Pradeep Bugga; Shinyoung Clair Kang; Sol Rey; William Ward |

EPI_ISL_1702639, EPI_ISL_1738119, EPI_ISL_1738187, EPI_ISL_1738671, EPI_ISL_1738750, EPI_ISL_1753345, EPI_ISL_1753413, EPI_ISL_1753644, EPI_ISL_1761906, EPI_ISL_1761913, EPI_ISL_1761936, EPI_ISL_1761939, EPI_ISL_1761949, EPI_ISL_1761951, EPI_ISL_1761953, EPI_ISL_1761963, EPI_ISL_1761971, EPI_ISL_1761977, EPI_ISL_1761985, EPI_ISL_1762034, EPI_ISL_1762038, EPI_ISL_1798049,
EPI_ISL_1798098, EPI_ISL_1798112, EPI_ISL_1798113, EPI_ISL_1798146, EPI_ISL_1798164, EPI_ISL_1804820, EPI_ISL_1818070, EPI_ISL_1818295, EPI_ISL_1818327, EPI_ISL_1818331, EPI_ISL_1818346, EPI_ISL_1818349, EPI_ISL_1818401, EPI_ISL_1818422, EPI_ISL_1818519, EPI_ISL_1834218, EPI_ISL_1834241, EPI_ISL_1834304, EPI_ISL_1834326, EPI_ISL_1834346, EPI_ISL_1834359, EPI_ISL_1834371,
EPI_ISL_1834375, EPI_ISL_1834408, EPI_ISL_1834421, EPI_ISL_1834440, EPI_ISL_1838933, EPI_ISL_1838960, EPI_ISL_1838965, EPI_ISL_1838975, EPI_ISL_1838980, EPI_ISL_1838993, EPI_ISL_1838996, EPI_ISL_1839003, EPI_ISL_1839004, EPI_ISL_1839010, EPI_ISL_1839013, EPI_ISL_1839014, EPI_ISL_1839020, EPI_ISL_1839021, EPI_ISL_1839024, EPI_ISL_1839042, EPI_ISL_1839054, EPI_ISL_1839055,
EPI_ISL_1840412, EPI_ISL_1840423, EPI_ISL_1840431, EPI_ISL_1840435, EPI_ISL_1840458, EPI_ISL_1840526, EPI_ISL_1840528, EPI_ISL_1840531, EPI_ISL_1840546, EPI_ISL_1840559, EPI_ISL_1840561, EPI_ISL_1840567, EPI_ISL_1840570, EPI_ISL_1840602, EPI_ISL_1840604, EPI_ISL_1840623, EPI_ISL_1840625, EPI_ISL_1906183, EPI_ISL_1906197, EPI_ISL_1906206, EPI_ISL_1906210, EPI_ISL_1906215,

| | | | |
|---|---|---|---|
| EPI_ISL_1906217, EPI_ISL_1906223, EPI_ISL_1906244, EPI_ISL_1906253, EPI_ISL_1906258, EPI_ISL_1906268, EPI_ISL_1906270, EPI_ISL_1906272, EPI_ISL_1906274, EPI_ISL_1906276, EPI_ISL_1906286, EPI_ISL_1906290, EPI_ISL_1906308, EPI_ISL_1906318, EPI_ISL_1906329, EPI_ISL_1906345, EPI_ISL_1906347, EPI_ISL_1906349, EPI_ISL_1906353, EPI_ISL_1906361, EPI_ISL_1906372, EPI_ISL_1906400, EPI_ISL_1906402, EPI_ISL_1906407, EPI_ISL_1906420, EPI_ISL_1906422, EPI_ISL_1906437, EPI_ISL_1906440, EPI_ISL_1906452, EPI_ISL_1906455, EPI_ISL_1906461, EPI_ISL_1906474, EPI_ISL_1906480, EPI_ISL_1906482, EPI_ISL_1906486, EPI_ISL_1906491, EPI_ISL_1906504, EPI_ISL_1906511, EPI_ISL_1906516, EPI_ISL_1906523, EPI_ISL_1906526, EPI_ISL_1906530, EPI_ISL_1906532, EPI_ISL_1906538, EPI_ISL_1906542, EPI_ISL_1906546, EPI_ISL_1906549, EPI_ISL_1906555, EPI_ISL_1906557, EPI_ISL_1906562, EPI_ISL_1906564, EPI_ISL_1906567, EPI_ISL_1906573, EPI_ISL_1906593, EPI_ISL_1906595, EPI_ISL_1906600, EPI_ISL_1906619, EPI_ISL_1906623, EPI_ISL_1906634, EPI_ISL_1906639, EPI_ISL_1906657, EPI_ISL_1906659, EPI_ISL_1906665, EPI_ISL_1906714, EPI_ISL_1906721, EPI_ISL_1906747, EPI_ISL_1906761, EPI_ISL_1906774, EPI_ISL_1906781, EPI_ISL_1906788, EPI_ISL_1906797, EPI_ISL_1906806, EPI_ISL_1906814, EPI_ISL_1906823, EPI_ISL_1906828, EPI_ISL_1906832, EPI_ISL_1906844, EPI_ISL_1906850, EPI_ISL_1906851, EPI_ISL_1906852, EPI_ISL_1906855, EPI_ISL_1906857, EPI_ISL_1906858, EPI_ISL_1906861, EPI_ISL_1906866, EPI_ISL_1906868, EPI_ISL_1906872, EPI_ISL_1906879, EPI_ISL_1924286, EPI_ISL_1924290, EPI_ISL_1924295, EPI_ISL_1924297, EPI_ISL_1924298, EPI_ISL_1924300, EPI_ISL_1924301, EPI_ISL_1924308, EPI_ISL_1924311, EPI_ISL_1924316, EPI_ISL_1924318, EPI_ISL_1924321, EPI_ISL_1924322, EPI_ISL_1924323, EPI_ISL_1924328, EPI_ISL_1924330, EPI_ISL_1924332, EPI_ISL_1924340, EPI_ISL_1924350, EPI_ISL_1924353, EPI_ISL_1924358, EPI_ISL_1924361, EPI_ISL_1924362, EPI_ISL_1924364, EPI_ISL_1924365, EPI_ISL_1924371, EPI_ISL_1924374, EPI_ISL_1924376, EPI_ISL_1924377, EPI_ISL_1924378, EPI_ISL_1924379, EPI_ISL_1924381, EPI_ISL_1924382, EPI_ISL_1924385, EPI_ISL_1924400, EPI_ISL_1924407, EPI_ISL_1924409, EPI_ISL_1924410, EPI_ISL_1924417, EPI_ISL_1924420, EPI_ISL_1924422, EPI_ISL_1924423, EPI_ISL_1924424, EPI_ISL_1924427, EPI_ISL_1924429, EPI_ISL_1924430, EPI_ISL_1924514, EPI_ISL_1935569, EPI_ISL_1935574 | | | |
| see above | Quest Diagnostics Incorporated | Centers for Disease Control and Prevention Division of Viral Diseases, Pathogen Discovery | A. Gerasimova; A. Perez; Adrian Paskey; B. Anderson; Benjamin Rambo-Martin; Christopher Gulvick; Clinton R. Paden; Dakota Howard; Darlene Wagner; Dhwani Batra; Duncan MacCannell; F. Lacbawan; I. A. Shlyakhter; Jason Caravas; K.E. Livingston; Kara Moser; L.E. Bernstein; M. Hua; Matthew Schmerer; P. Tanpaiboon; Peter W. Cook; R. M. Kagan; R. Owen; R. V. Rolando; S. H. Rosenthal; Scott Sammons; Shatavia Morrison; Y. Liu; Yvette Unoarumhi |
| EPI_ISL_1755996, EPI_ISL_1755997, EPI_ISL_1755998, EPI_ISL_1755999, EPI_ISL_1855273 | SARATOGA HOSPITAL LABORATORY | Wadsworth Center, New York State Department of Health | Alexis Russell; Catharine Prussing; Daryl M. Lamson; Erasmus Schneider; Erica Lasek-Nesselquist; John Kelly; Jonathan Plitnick; Kirsten St. George; Matthew Shudt; Melissa A Leisner; Navjot Singh |
| EPI_ISL_1671484, EPI_ISL_1756368 | ST. FRANCIS HOSPITAL LABORATORY | Wadsworth Center, New York State Department of Health | Alexis Russell; Catharine Prussing; Daryl M. Lamson; Erasmus Schneider; Erica Lasek-Nesselquist; John Kelly; Jonathan Plitnick; Kirsten St. George; Matthew Shudt; Melissa A Leisner; Navjot Singh |
| EPI_ISL_1756000, EPI_ISL_1756331, EPI_ISL_1756332, EPI_ISL_1756333, EPI_ISL_1756334, EPI_ISL_1756335, EPI_ISL_1756336, EPI_ISL_1756337, EPI_ISL_1789719 | | | |
| see above | STONY BROOK UNIVERSITY HOSPITAL | Wadsworth Center, New York State Department of Health | Alexis Russell; Catharine Prussing; Daryl M. Lamson; Erasmus Schneider; Erica Lasek-Nesselquist; John Kelly; Jonathan Plitnick; Kirsten St. George; Matthew Shudt; Melissa A Leisner; Navjot Singh |
| EPI_ISL_1671513, EPI_ISL_1756338, EPI_ISL_1756339, EPI_ISL_1756340, EPI_ISL_1756341, EPI_ISL_1854691, EPI_ISL_1855105, EPI_ISL_1855106, EPI_ISL_1855107, EPI_ISL_1855108, EPI_ISL_1855109, EPI_ISL_1855110, EPI_ISL_1855111, EPI_ISL_1855112, EPI_ISL_1855113, EPI_ISL_1855114, EPI_ISL_1855115, EPI_ISL_1855221, EPI_ISL_1855222, EPI_ISL_1855223, EPI_ISL_1855224, EPI_ISL_1855225, EPI_ISL_1855354, EPI_ISL_1855355, EPI_ISL_1855356, EPI_ISL_1855357, EPI_ISL_1855358, EPI_ISL_1855359, EPI_ISL_1855360, EPI_ISL_1935639, EPI_ISL_1935640, EPI_ISL_1935641, EPI_ISL_1935642, EPI_ISL_1935646, EPI_ISL_1935647, EPI_ISL_1935649, EPI_ISL_1935651, EPI_ISL_2098135 | | | |
| see above | SUNY UPSTATE MEDICAL UNIVERSITY | Wadsworth Center, New York State Department of Health | Alexis Russell; Catharine Prussing; Daryl M. Lamson; Erasmus Schneider; Erica Lasek-Nesselquist; John Kelly; Jonathan Plitnick; Kirsten St. George; Matthew Shudt; Melissa A Leisner; Navjot Singh |
| EPI_ISL_1921011, EPI_ISL_1921012, EPI_ISL_1921013, EPI_ISL_1921038, EPI_ISL_1921039 | SUNY Upstate Medical University | Wadsworth Center, New York State Department of Health | Alexis Russell; Catharine Prussing; Daryl M. Lamson; Erasmus Schneider; Erica Lasek-Nesselquist; John Kelly; Jonathan Plitnick; Kirsten St. George; Matthew Shudt; Melissa A Leisner; Navjot Singh |
| EPI_ISL_1920029 | St. Francis Hospital Laboratory | Wadsworth Center, New York State Department of Health | Alexis Russell; Catharine Prussing; Daryl M. Lamson; Erasmus Schneider; Erica Lasek-Nesselquist; John Kelly; Jonathan Plitnick; Kirsten St. George; Matthew Shudt; Melissa A Leisner; Navjot Singh |
| EPI_ISL_1967689, EPI_ISL_1967690, EPI_ISL_1967691, EPI_ISL_1967692, EPI_ISL_1967693, EPI_ISL_1967694, EPI_ISL_1967695 | | | |
| see above | Stony Brook Medicine | New York Genome Center | Alaa B. Salaheldin; Andre Corvelo; Dayna M. Oschwald; Dimitri Gnatenko; Eric D. Spitzer; Jiang Chen; Josue M. Flores Acuna; Kenneth R. Shroyer; Margaret M. McGovern; Michael Zody; Richard R. Kew; Samantha Fennessey; Sarah A. Halbohn; Sarah A. Malik; Silvia G. Spitzer; Susan V. Donelan; Tom Maniatis |
| EPI_ISL_1920046, EPI_ISL_1920047 | Stony Brook University Hospital | Wadsworth Center, New York State Department of Health | Alexis Russell; Catharine Prussing; Daryl M. Lamson; Erasmus Schneider; Erica Lasek-Nesselquist; John Kelly; Jonathan Plitnick; Kirsten St. George; Matthew Shudt; Melissa A Leisner; Navjot Singh |
| EPI_ISL_1919990 | Suny Upstate Medical University | Wadsworth Center, New York State Department of Health | Alexis Russell; Catharine Prussing; Daryl M. Lamson; Erasmus Schneider; Erica Lasek-Nesselquist; John Kelly; Jonathan Plitnick; Kirsten St. George; Matthew Shudt; Melissa A Leisner; Navjot Singh |
| EPI_ISL_1755968, EPI_ISL_1755969, EPI_ISL_1755970, EPI_ISL_1755971, EPI_ISL_1755972, EPI_ISL_1755973, EPI_ISL_1755974, EPI_ISL_1755975, EPI_ISL_1755976, EPI_ISL_1755977, EPI_ISL_1755978, EPI_ISL_1755979, EPI_ISL_1755980, EPI_ISL_1755981, EPI_ISL_1755982, EPI_ISL_1755983, EPI_ISL_1755984, EPI_ISL_1755985, EPI_ISL_1755986, EPI_ISL_1755987, EPI_ISL_1755988, EPI_ISL_1755989, EPI_ISL_1755990, EPI_ISL_1755991, EPI_ISL_1855104, EPI_ISL_1855150, EPI_ISL_1855152, EPI_ISL_1855153, EPI_ISL_1855154, EPI_ISL_1855155, EPI_ISL_1855156, EPI_ISL_1855157, EPI_ISL_1855159, EPI_ISL_1855161, EPI_ISL_1855162, EPI_ISL_1855164, EPI_ISL_1855165, EPI_ISL_1855166, EPI_ISL_1855167, EPI_ISL_1855244, EPI_ISL_1855245, EPI_ISL_1855246, EPI_ISL_1855247, EPI_ISL_1855248, EPI_ISL_1935627, EPI_ISL_1935632 | | | |
| see above | TEMPUS LABS INC | Wadsworth Center, New York State Department of Health | Alexis Russell; Catharine Prussing; Daryl M. Lamson; Erasmus Schneider; Erica Lasek-Nesselquist; John Kelly; Jonathan Plitnick; Kirsten St. George; Matthew Shudt; Melissa A Leisner; Navjot Singh |
| EPI_ISL_1625677, EPI_ISL_1739440 | The Jackson Laboratory | The Jackson Laboratory | Adams M; Bergeron D; Kelly K; Li L; Long J; Omerza G; Renzette N |
| EPI_ISL_1756357, EPI_ISL_1756358, EPI_ISL_1756359, EPI_ISL_1756360, EPI_ISL_1756361, EPI_ISL_1756362, EPI_ISL_1756363, EPI_ISL_1756364, EPI_ISL_1756365, EPI_ISL_1756366, EPI_ISL_1756367, EPI_ISL_1854693, EPI_ISL_1854694, EPI_ISL_1854695, EPI_ISL_1855119, EPI_ISL_1855120, EPI_ISL_1855121, EPI_ISL_1855122, EPI_ISL_1855123, EPI_ISL_1855124, EPI_ISL_1855125, EPI_ISL_1855126, EPI_ISL_1855127, EPI_ISL_1855128, EPI_ISL_1855129, EPI_ISL_1855130, EPI_ISL_1855131, EPI_ISL_1855132, EPI_ISL_1855133, EPI_ISL_1855134, EPI_ISL_1855135, EPI_ISL_1855136, EPI_ISL_1855137, EPI_ISL_1855138, EPI_ISL_1855139, EPI_ISL_1855140, EPI_ISL_1855141, EPI_ISL_1855142, EPI_ISL_1855143, EPI_ISL_1855168, EPI_ISL_1855169, EPI_ISL_1855170, EPI_ISL_1855171, EPI_ISL_1855172, EPI_ISL_1855173, EPI_ISL_1855174, EPI_ISL_1855175, EPI_ISL_1855176, EPI_ISL_1855177, EPI_ISL_1855178, EPI_ISL_1855179, EPI_ISL_1855180, EPI_ISL_1855181, EPI_ISL_1855182, EPI_ISL_1855183, EPI_ISL_1855184, EPI_ISL_1855185, EPI_ISL_1855226, EPI_ISL_1855227, EPI_ISL_1855228, EPI_ISL_1855229, EPI_ISL_1855230, EPI_ISL_1855231, EPI_ISL_1855232, EPI_ISL_1855233, EPI_ISL_1855234, EPI_ISL_1855235, EPI_ISL_1855236, EPI_ISL_1855237, EPI_ISL_1855238, EPI_ISL_1855239, EPI_ISL_1855240, EPI_ISL_1855249, EPI_ISL_1855250, EPI_ISL_1855251, EPI_ISL_1855252, EPI_ISL_1855253, EPI_ISL_1855254, EPI_ISL_1855255, EPI_ISL_1855256, EPI_ISL_1855257, EPI_ISL_1855258, EPI_ISL_1855259, EPI_ISL_1855260, EPI_ISL_1855261, EPI_ISL_1855262, EPI_ISL_1855263, EPI_ISL_1855264, EPI_ISL_1855265, EPI_ISL_1855266, EPI_ISL_1855267, EPI_ISL_1855268, EPI_ISL_1855269, EPI_ISL_1855270, EPI_ISL_1855271, EPI_ISL_1855272, EPI_ISL_1855276, EPI_ISL_1855277, EPI_ISL_1855278, EPI_ISL_1855279, EPI_ISL_1855280, EPI_ISL_1855281, EPI_ISL_1855282, EPI_ISL_1855283, EPI_ISL_1855284, EPI_ISL_1855285, EPI_ISL_1855294, EPI_ISL_1855295, EPI_ISL_1855296, EPI_ISL_1855297, EPI_ISL_1855298, EPI_ISL_1855299, EPI_ISL_1855300, EPI_ISL_1855301, EPI_ISL_1855302, EPI_ISL_1855303, EPI_ISL_1855304, EPI_ISL_1855305, EPI_ISL_1855306, EPI_ISL_1855307, EPI_ISL_1855308, EPI_ISL_1855309, EPI_ISL_1921001, EPI_ISL_1921002, EPI_ISL_1921003, EPI_ISL_1921004, EPI_ISL_1921005, EPI_ISL_1921006, EPI_ISL_1921007, EPI_ISL_1921008, EPI_ISL_1921009, EPI_ISL_1921010, EPI_ISL_1921040, EPI_ISL_1921041, EPI_ISL_1921042, EPI_ISL_1921043, EPI_ISL_1921046, EPI_ISL_1921049, EPI_ISL_1921050, EPI_ISL_1921051, EPI_ISL_1921054, EPI_ISL_1921055, EPI_ISL_1921056, EPI_ISL_1921058, EPI_ISL_1921061, EPI_ISL_1921065, EPI_ISL_1921066, EPI_ISL_1921069, EPI_ISL_1921070, EPI_ISL_1921071, EPI_ISL_1921072, EPI_ISL_1921074, EPI_ISL_1921075, EPI_ISL_1921076, EPI_ISL_1921077, EPI_ISL_1921078, EPI_ISL_1921079, EPI_ISL_1935611, EPI_ISL_1935616, EPI_ISL_1935622, EPI_ISL_1935623, EPI_ISL_1935634, EPI_ISL_1935635, EPI_ISL_1935645, EPI_ISL_2098230, EPI_ISL_2098231, EPI_ISL_2134795 | | | |
| see above | URMC LABS | Wadsworth Center, New York State Department of Health | Alexis Russell; Catharine Prussing; Daryl M. Lamson; Erasmus Schneider; Erica Lasek-Nesselquist; John Kelly; Jonathan Plitnick; Kirsten St. George; Matthew Shudt; Melissa A Leisner; Navjot Singh |
| EPI_ISL_1671475, EPI_ISL_1671490, EPI_ISL_1671491, EPI_ISL_1671504, EPI_ISL_1671505, EPI_ISL_1671506, EPI_ISL_1671507, EPI_ISL_1671511, EPI_ISL_1671597, EPI_ISL_1671635, EPI_ISL_1740515, EPI_ISL_1755965, EPI_ISL_1756369, EPI_ISL_1756370, EPI_ISL_1756371, EPI_ISL_1756372, EPI_ISL_1756373, EPI_ISL_1756374, EPI_ISL_1756375, EPI_ISL_1756376, EPI_ISL_1756377, EPI_ISL_1756378, EPI_ISL_1756379, EPI_ISL_1756380, EPI_ISL_1756381, EPI_ISL_1756382, EPI_ISL_1756383, EPI_ISL_1756384, EPI_ISL_1756385, EPI_ISL_1756386, EPI_ISL_1756387, EPI_ISL_1756388, EPI_ISL_1756389, EPI_ISL_1756390, EPI_ISL_1756391, EPI_ISL_1756392, EPI_ISL_1756393, EPI_ISL_1756394, EPI_ISL_1756395, EPI_ISL_1756396, EPI_ISL_1756397, EPI_ISL_1756398, EPI_ISL_1756399, EPI_ISL_1756400, EPI_ISL_1756401, EPI_ISL_1756402, EPI_ISL_1756403, EPI_ISL_1756404, EPI_ISL_1756405, EPI_ISL_1756406, EPI_ISL_1756407, EPI_ISL_1756408, EPI_ISL_1789722, EPI_ISL_1789723, EPI_ISL_1855334 | | | |
| see above | WESTCHESTER MEDICAL CENTER | Wadsworth Center, New York State Department of Health | Alexis Russell; Catharine Prussing; Daryl M. Lamson; Erasmus Schneider; Erica Lasek-Nesselquist; John Kelly; Jonathan Plitnick; Kirsten St. George; Matthew Shudt; Melissa A Leisner; Navjot Singh |
| EPI_ISL_1671622, EPI_ISL_1671644, EPI_ISL_1671665 | WHITE PLAINS HOSPITAL CENTER LABORATORY | Wadsworth Center, New York State Department of Health | Alexis Russell; Catharine Prussing; Daryl M. Lamson; Erasmus Schneider; Erica Lasek-Nesselquist; John Kelly; Jonathan Plitnick; Kirsten St. George; Matthew Shudt; Melissa A Leisner; Navjot Singh |
| EPI_ISL_1920113, EPI_ISL_1920114, EPI_ISL_1920121 | Wadsworth Center, New York State Department of Health | Wadsworth Center, New York State Department of Health | Alexis Russell; Catharine Prussing; Daryl M. Lamson; Erasmus Schneider; Erica Lasek-Nesselquist; John Kelly; Jonathan Plitnick; Kirsten St. George; Matthew Shudt; Melissa A Leisner; Navjot Singh |
| EPI_ISL_1967667, EPI_ISL_1971369, EPI_ISL_1971370, EPI_ISL_1971371, EPI_ISL_1971372, EPI_ISL_1971373, EPI_ISL_1971374, EPI_ISL_1971375, EPI_ISL_1971376, EPI_ISL_1971377, EPI_ISL_1971378, EPI_ISL_1971379, EPI_ISL_1971380, EPI_ISL_1971381, EPI_ISL_1971382, EPI_ISL_1971383, EPI_ISL_1971384, EPI_ISL_1971385, EPI_ISL_1971386, EPI_ISL_1971387, EPI_ISL_1971388, EPI_ISL_1971389, EPI_ISL_1971390, EPI_ISL_1971391, EPI_ISL_1971392, EPI_ISL_1971393, EPI_ISL_1971394, EPI_ISL_1971395, EPI_ISL_1971396, EPI_ISL_1971397, EPI_ISL_1971398, EPI_ISL_1971399, EPI_ISL_1971400, EPI_ISL_1971401, EPI_ISL_1971402, EPI_ISL_1971403, EPI_ISL_1971404, EPI_ISL_1971405, EPI_ISL_1971406, EPI_ISL_2081128, EPI_ISL_2081130, EPI_ISL_2081131, EPI_ISL_2081132, EPI_ISL_2081133, EPI_ISL_2081134, EPI_ISL_2081135, EPI_ISL_2081136, EPI_ISL_2081137, EPI_ISL_2081138, EPI_ISL_2081139, EPI_ISL_2081140, EPI_ISL_2081141, EPI_ISL_2081142, EPI_ISL_2081143, EPI_ISL_2081144, EPI_ISL_2081146, EPI_ISL_2081147, EPI_ISL_2081148, EPI_ISL_2081149, EPI_ISL_2082729, EPI_ISL_2086266 | | | |
| see above | Weill Cornell Medicine | New York Genome Center | Andre Corvelo; Arryn Craney; Chris Mason; Dayna M. Oschwald; Hanna Rennert; Lars F Westblade; Margaret Elizabeth Ross; Melissa Cushing; Michael Zody; Olivier Elemento; Priya Velu; Samantha Fennessey; Tom Maniatis |
| EPI_ISL_2034886, EPI_ISL_2034934 | Yale Clinical Virology Lab | Yale Center for Genomic Analysis | Brooke Sullivan; Curt Scharfe; Irina Tikhonova; Kaya Bilguvar; Shrikant Mane |

We gratefully acknowledge the following Authors from the Originating laboratories responsible for obtaining the specimens, as well as the Submitting laboratories where the genome data were generated and shared via GISAID, on which this research is based.

All Submitters of data may be contacted directly via www.gisaid.org

Authors are sorted alphabetically.

| Accession ID | Originating Laboratory | Submitting Laboratory | Authors |
|---|---|---|---|
| EPI_ISL_1385784, EPI_ISL_1385785, EPI_ISL_1385786, EPI_ISL_1385789, EPI_ISL_1385790, EPI_ISL_1397958, EPI_ISL_1397959, EPI_ISL_1397960, EPI_ISL_1398039, EPI_ISL_1406391, EPI_ISL_1406393, EPI_ISL_1697282, EPI_ISL_1697283, EPI_ISL_1697284, EPI_ISL_1697288, EPI_ISL_1697367, EPI_ISL_1697368, EPI_ISL_1697369, EPI_ISL_1697370, EPI_ISL_1697371, EPI_ISL_1700780, EPI_ISL_1700781, EPI_ISL_1700782, EPI_ISL_1700786, EPI_ISL_1700787, EPI_ISL_1700882, EPI_ISL_1706278, EPI_ISL_1706361, EPI_ISL_1706363, EPI_ISL_1706369, EPI_ISL_1706371, EPI_ISL_1706372, EPI_ISL_1706581, EPI_ISL_1708744, EPI_ISL_1710244, EPI_ISL_1710353, EPI_ISL_1710506, EPI_ISL_1710507, EPI_ISL_1710508, EPI_ISL_1710509, EPI_ISL_1710510, EPI_ISL_1711669, EPI_ISL_1711670, EPI_ISL_1711671, EPI_ISL_1711672, EPI_ISL_1711673, EPI_ISL_1711674, EPI_ISL_1711675 | | | |
| see above | Department of Virology | Department of Virology | Massab Umair, Aamer Ikram, Muhammad Salman, Nazish Badar, Sana Tamim, Zaira Rehman, Abdul Ahad, Shannon Whitmer, Melissa Mobley, Austin Leach, Ketan Patel, Joel Montgomery, John Klena |
| EPI_ISL_2151345, EPI_ISL_2151346, EPI_ISL_2151347, EPI_ISL_2151348, EPI_ISL_2151349, EPI_ISL_2151350, EPI_ISL_2151351, EPI_ISL_2151352, EPI_ISL_2151353, EPI_ISL_2151354, EPI_ISL_2151355, EPI_ISL_2151356, EPI_ISL_2151357, EPI_ISL_2151358, EPI_ISL_2151359, EPI_ISL_2151360, EPI_ISL_2151361, EPI_ISL_2151362, EPI_ISL_2151363, EPI_ISL_2151364, EPI_ISL_2151365, EPI_ISL_2151366 | | | |
| see above | Jamil-ur-Rahman Center for Genome Research, Dr. Panjwani Center for Molecular Medicine and Drug Research | Jamil-ur-Rahman Center for Genome Research, Dr. Panjwani Center for Molecular Medicine and Drug Research | Irfan,M., Nisa,Z., Shakeel,M., Siddiqui,S., Ain,N., Hashmi,A.H., Khan,I.A. |
| EPI_ISL_709542 | National Institute of Blood Diseases (NIBD), Molecular Biology Lab | Genomics Lab NIBD | Samina Naz Mukry, Sayed Ali Raza, Shariq Ahmed, Aneeta Shahni, Gul Sufaida, Arshi Naz , Tahir Sultan Shamsi |
| EPI_ISL_757285, EPI_ISL_757286 | Department of Virology, Public Health Laboratories Division | Department of Virology, Public Health Laboratories Division | Massab Umair, Aamer Ikram, Muhammad Salman |
| EPI_ISL_766055 | Department of Virology | Department of Virology | Massab Umair, Muhammad Salman, Sana Tamim, Nazish Badar, Zaira Rehman, Adnan Khurshid, Samee Ullah, Abdul Ahad, Hamza Ahmed, Aamer Ikram |

We gratefully acknowledge the following Authors from the Originating laboratories responsible for obtaining the specimens, as well as the Submitting laboratories where the genome data were generated and shared via GISAID, on which this research is based.

All Submitters of data may be contacted directly via www.gisaid.org

Authors are sorted alphabetically.

| Accession ID | Originating Laboratory | Submitting Laboratory | Authors |
|---|---|---|---|
| EPI_ISL_1092325, EPI_ISL_1092326, EPI_ISL_1092327, EPI_ISL_1092328, EPI_ISL_1092329, EPI_ISL_1092330, EPI_ISL_1092331, EPI_ISL_1092332, EPI_ISL_1092333, EPI_ISL_1092334, EPI_ISL_1092335, EPI_ISL_1093148, EPI_ISL_1093150, EPI_ISL_1093152, EPI_ISL_1093153, EPI_ISL_1093154, EPI_ISL_1093155, EPI_ISL_1093156, EPI_ISL_1093157, EPI_ISL_1093158, EPI_ISL_1093159, EPI_ISL_1093160, EPI_ISL_1093161, EPI_ISL_1093162, EPI_ISL_1093163, EPI_ISL_1093164 | | | |
| see above | Laboratorio de Referencia Nacional de Virus Respiratorio. Instituto Nacional de Salud Perú | Laboratorio de Referencia Nacional de Biotecnología y Biología Molecular. Instituto Nacional de Salud Perú | Carlos Padilla Rojas, Karolyn Vega Chozo, Luis Barcena, Priscila Lope Pari, Omar Caceres Rey, Marco Galarza Perez, Maribel Huaringa Nuñez, Johanna Balbuena Torrez, Henri Bailon Calderon, Nancy Rojas Serrano |
| EPI_ISL_1111069, EPI_ISL_1111132, EPI_ISL_1111133, EPI_ISL_1111134, EPI_ISL_1111135, EPI_ISL_1111136, EPI_ISL_1111137, EPI_ISL_1111138, EPI_ISL_1111139, EPI_ISL_1111140, EPI_ISL_1111141, EPI_ISL_1111142, EPI_ISL_1111143, EPI_ISL_1111144, EPI_ISL_1111145, EPI_ISL_1111146, EPI_ISL_1111147, EPI_ISL_1111148, EPI_ISL_1111149, EPI_ISL_1111150, EPI_ISL_1111151, EPI_ISL_1111152, EPI_ISL_1111153, EPI_ISL_1111154, EPI_ISL_1111155, EPI_ISL_1111156, EPI_ISL_1111157, EPI_ISL_1111158, EPI_ISL_1111159, EPI_ISL_1111160, EPI_ISL_1111161, EPI_ISL_1111308, EPI_ISL_1111309, EPI_ISL_1111310, EPI_ISL_1111312, EPI_ISL_1111314, EPI_ISL_1111315, EPI_ISL_1111446, EPI_ISL_1111457, EPI_ISL_1111458, EPI_ISL_1111459, EPI_ISL_1111460, EPI_ISL_1111461, EPI_ISL_1111462, EPI_ISL_1111463, EPI_ISL_1111464, EPI_ISL_1111465, EPI_ISL_1111466, EPI_ISL_1111467, EPI_ISL_1111469, EPI_ISL_1111470, EPI_ISL_1111471, EPI_ISL_1111472, EPI_ISL_1111473, EPI_ISL_1111474, EPI_ISL_1111475, EPI_ISL_1111476, EPI_ISL_1111477, EPI_ISL_1111478, EPI_ISL_1111479, EPI_ISL_1111480, EPI_ISL_1111481, EPI_ISL_1111482, EPI_ISL_1111483, EPI_ISL_1111484, EPI_ISL_1111485, EPI_ISL_1111486, EPI_ISL_1111487, EPI_ISL_1111488, EPI_ISL_1111489, EPI_ISL_1111490, EPI_ISL_1111491, EPI_ISL_1111492, EPI_ISL_1111493, EPI_ISL_1111494, EPI_ISL_1111495, EPI_ISL_1111496, EPI_ISL_1111497, EPI_ISL_1111498, EPI_ISL_1111499, EPI_ISL_1111500, EPI_ISL_1111501, EPI_ISL_1111502, EPI_ISL_1111503, EPI_ISL_1111504, EPI_ISL_1111505 | | | |
| see above | Laboratorio de Referencia Nacional de Virus Respiratorio. Instituto Nacional de Salud Perú | Laboratorio de Referencia Nacional de Enteropatógenos. Instituto Nacional de Salud del Perú | Ronnie Gavilan Chavez, Junior Caro Castro, Willi Quino Sifuentes, Veronica Hurtado Vela, Iris Silva Molina, Fiorella Orellana Peralta |
| EPI_ISL_1137479, EPI_ISL_1137480, EPI_ISL_1137481, EPI_ISL_1137482, EPI_ISL_1137483, EPI_ISL_1137484, EPI_ISL_1137485, EPI_ISL_1137486, EPI_ISL_1137487, EPI_ISL_1137488, EPI_ISL_1137489, EPI_ISL_1137490, EPI_ISL_1137491, EPI_ISL_1138413, EPI_ISL_1138414, EPI_ISL_1138415, EPI_ISL_1138416, EPI_ISL_1138417, EPI_ISL_1138418, EPI_ISL_1138419, EPI_ISL_1138420, EPI_ISL_1138421, EPI_ISL_1138422 | | | |
| see above | Laboratorio de Referencia Nacional de Virus Respiratorio. Instituto Nacional de Salud Perú | Laboratorio de Referencia Nacional de Biotecnología y Biología Molecular. Instituto Nacional de Salud Perú | Carlos Padilla Rojas, Karolyn Vega Chozo, Luis Barcena, Priscila Lope Pari, Omar Caceres Rey, Marco Galarza Perez, Maribel Huaringa Nuñez, Johanna Balbuena Torrez, Henri Bailon Calderon, Nancy Rojas Serrano |
| EPI_ISL_1171630 | IAL Regional de Santos | Instituto Adolfo Lutz, Interdiciplinary Procedures Center, Strategic Laboratory | Claudio Tavares Sacchi, Claudia Regina Gonçalves, Erica Valessa Ramos Gomes, Karoline Rodrigues Campos, Caio Vinicius Dias Lopes |
| EPI_ISL_1532147, EPI_ISL_1532167, EPI_ISL_1532168, EPI_ISL_1532169, EPI_ISL_1532170, EPI_ISL_1532171, EPI_ISL_1532184, EPI_ISL_1532185, EPI_ISL_1532186, EPI_ISL_1532187, EPI_ISL_1532189, EPI_ISL_1532190, EPI_ISL_1532197, EPI_ISL_1532198, EPI_ISL_1532204, EPI_ISL_1532205, EPI_ISL_1532206, EPI_ISL_1532207, EPI_ISL_1532208, EPI_ISL_1532209, EPI_ISL_1532210, EPI_ISL_1532211, EPI_ISL_1532212, EPI_ISL_1532213, EPI_ISL_1532214, EPI_ISL_1532215, EPI_ISL_1532216, EPI_ISL_1532217, EPI_ISL_1532218, EPI_ISL_1532219, EPI_ISL_1532220, EPI_ISL_1532221, EPI_ISL_1532222, EPI_ISL_1532223, EPI_ISL_1532224, EPI_ISL_1532225, EPI_ISL_1532226, EPI_ISL_1532227, EPI_ISL_1532228, EPI_ISL_1532229, EPI_ISL_1532233, EPI_ISL_1534626, EPI_ISL_1534627, EPI_ISL_1534628, EPI_ISL_1534629, EPI_ISL_1534630, EPI_ISL_1534631, EPI_ISL_1534632, EPI_ISL_1534633, EPI_ISL_1534634, EPI_ISL_1534635, EPI_ISL_1534636, EPI_ISL_1534637, EPI_ISL_1534638, EPI_ISL_1534639, EPI_ISL_1534640, EPI_ISL_1534641, EPI_ISL_1534642, EPI_ISL_1534643, EPI_ISL_1534644, EPI_ISL_1534645, EPI_ISL_1534646, EPI_ISL_1534647, EPI_ISL_1534648, EPI_ISL_1534649, EPI_ISL_1534650, EPI_ISL_1534651, EPI_ISL_1534652, EPI_ISL_1534654, EPI_ISL_1534655, EPI_ISL_1534656, EPI_ISL_1534657, EPI_ISL_1534658, EPI_ISL_1534659, EPI_ISL_1534660, EPI_ISL_1534661, EPI_ISL_1534662, EPI_ISL_1534663, EPI_ISL_1534664, EPI_ISL_1534665, EPI_ISL_1534666, EPI_ISL_1534667, EPI_ISL_1534668, EPI_ISL_1534669, EPI_ISL_1534670, EPI_ISL_1534671, EPI_ISL_1534672, EPI_ISL_1534673, EPI_ISL_1534674, EPI_ISL_1534675, EPI_ISL_1534676, EPI_ISL_1534677, EPI_ISL_1534678, EPI_ISL_1534679, EPI_ISL_1534680, EPI_ISL_1534681, EPI_ISL_1534682, EPI_ISL_1534683, EPI_ISL_1534684, EPI_ISL_1534685, EPI_ISL_1534686, EPI_ISL_1534687, EPI_ISL_1534688, EPI_ISL_1534689 | | | |
| see above | Laboratorio de Referencia Nacional de Virus Respiratorio. Instituto Nacional de Salud Perú | Laboratorio de Referencia Nacional de Biotecnología y Biología Molecular. Instituto Nacional de Salud Perú | Carlos Padilla Rojas, Karolyn Vega Chozo, Luis Barcena, Priscila Lope Pari, Omar Caceres Rey, Marco Galarza Perez, Maribel Huaringa Nuñez, Johanna Balbuena Torrez, Henri Bailon Calderon, Nancy Rojas Serrano |
| EPI_ISL_1548074 | Laboratorio de Referencia Nacional de Virus Respiratorio. Instituto Nacional de Salud Peru | Laboratorio de Referencia Nacional de Biotecnología y Biología Molecular. Instituto Nacional de Salud Perú | Carlos Padilla Rojas, Karolyn Vega Chozo, Luis Barcena, Priscila Lope Pari, Omar Caceres Rey, Marco Galarza Perez, Maribel Huaringa Nuñez, Johanna Balbuena Torrez, Henri Bailon Calderon, Nancy Rojas Serrano |
| EPI_ISL_1563627 | Laboratorio de Referencia Nacional de Biotecnología y Biología Molecular. Instituto Nacional de Salud Peru | Laboratorio de Referencia Nacional de Biotecnología y Biología Molecular. Instituto Nacional de Salud Perú | Carlos Padilla Rojas, Karolyn Vega Chozo, Luis Barcena, Priscila Lope Pari, Omar Caceres Rey, Marco Galarza Perez, Maribel Huaringa Nuñez, Johanna Balbuena Torrez, Henri Bailon Calderon, Nancy Rojas Serrano |
| EPI_ISL_1575295, EPI_ISL_1575438, EPI_ISL_1575439, EPI_ISL_1575440 | Laboratorio de Referencia Nacional de Virus Respiratorio. Instituto Nacional de Salud Peru | Laboratorio de Referencia Nacional de Biotecnología y Biología Molecular. Instituto Nacional de Salud Peru | Carlos Padilla Rojas, Karolyn Vega Chozo, Luis Barcena, Priscila Lope Pari, Omar Caceres Rey, Marco Galarza Perez, Maribel Huaringa Nuñez, Johanna Balbuena Torrez, Henri Bailon Calderon, Nancy Rojas Serrano |
| EPI_ISL_1629762, EPI_ISL_1629763 | Hospital Carlos Alberto Seguín Escobedo - EsSalud | Laboratorio de Genómica Microbiana, Universidad Peruana Cayetano Heredia | Lenin Maturrano, Pablo Tsukayama, Alejandra Dávila-Barclay, Guillermo Salvatierra, Luis González, Pedro E. Romero, Diego Cuicapuza, Janet Huancachoque, Pool Marcos |
| EPI_ISL_1629764 | Instituto de Medicina Tropical Alexander Von Humboldt, Universidad Peruana Cayetano Heredia | Laboratorio de Genómica Microbiana, Universidad Peruana Cayetano Heredia | Lenin Maturrano, Pablo Tsukayama, Alejandra Dávila-Barclay, Guillermo Salvatierra, Luis González, Pedro E. Romero, Diego Cuicapuza, Janet Huancachoque, Pool Marcos |
| EPI_ISL_1629765, EPI_ISL_1629766, EPI_ISL_1629767, EPI_ISL_1629768, EPI_ISL_1629769, EPI_ISL_1629770, EPI_ISL_1629771, EPI_ISL_1629772, EPI_ISL_1629773, EPI_ISL_1629774, EPI_ISL_1629775, EPI_ISL_1629776, EPI_ISL_1629777, EPI_ISL_1629778, EPI_ISL_1629779, EPI_ISL_1629780, EPI_ISL_1629781, EPI_ISL_1629782 | | | |
| see above | Hospital Carlos Alberto Seguín Escobedo - EsSalud | Laboratorio de Genómica Microbiana, Universidad Peruana Cayetano Heredia | Lenin Maturrano, Pablo Tsukayama, Alejandra Dávila-Barclay, Guillermo Salvatierra, Luis González, Pedro E. Romero, Janet Huancachoque, Pool Marcos, Fernando Sánchez Fragoso, Lin Zevallos Cuarite |
| EPI_ISL_1664259, EPI_ISL_1701761, EPI_ISL_1701762, EPI_ISL_1701763, EPI_ISL_1701764, EPI_ISL_1701765, EPI_ISL_1701766, EPI_ISL_1701767, EPI_ISL_1701768, EPI_ISL_1701769, EPI_ISL_1701770, EPI_ISL_1701771, EPI_ISL_1701772, EPI_ISL_1701773, EPI_ISL_1701774, EPI_ISL_1701775 | | | |
| see above | Laboratorio de Referencia Nacional de Virus Respiratorio. Instituto Nacional de Salud Perú | Laboratorio de Referencia Nacional de Biotecnología y Biología Molecular. Instituto Nacional de Salud Perú | Carlos Padilla Rojas, Karolyn Vega Chozo, Luis Barcena, Priscila Lope Pari, Omar Caceres Rey, Marco Galarza Perez, Maribel Huaringa Nuñez, Johanna Balbuena Torrez, Henri Bailon Calderon, Nancy Rojas Serrano |
| EPI_ISL_729862, EPI_ISL_729863, EPI_ISL_729864, EPI_ISL_729865, EPI_ISL_729866, EPI_ISL_729867, EPI_ISL_729868 | Laboratorio de Referencia Nacional de Virus Respiratorios, Instituto Nacional de Salud Peru | Laboratorio de Genómica Microbiana, Universidad Peruana Cayetano Heredia | Pablo Tsukayama, Alejandra Dávila-Barclay, Luis González, Guillermo Salvatierra, Pedro E. Romero, Brenda Ayzanoa, Janet Huancachoque, Pool Marcos, Marco Galarza, Priscila Lope, Nancy Rojas |
| EPI_ISL_729869, EPI_ISL_729870, EPI_ISL_729871 | Instituto de Medicina Tropical, Universidad Nacional Toribio Rodríguez de Mendoza de Amazonas | Laboratorio de Genómica Microbiana, Universidad Peruana Cayetano Heredia | Pablo Tsukayama, Alejandra Dávila-Barclay, Luis González, Pedro E. Romero, Brenda Ayzanoa, Janet Huancachoque, Pool Marcos, Stella Chenet, Rafael Tapia, Cecilia Pajuelo, Carla Montenegro |
| EPI_ISL_729872, EPI_ISL_729873, EPI_ISL_729874, EPI_ISL_729875, EPI_ISL_729876, EPI_ISL_729877, EPI_ISL_729878, EPI_ISL_729879, EPI_ISL_729880, EPI_ISL_729881, EPI_ISL_729882, EPI_ISL_729883, EPI_ISL_729884, EPI_ISL_729885, EPI_ISL_729886, EPI_ISL_729887, EPI_ISL_729888, EPI_ISL_729889, EPI_ISL_729890, EPI_ISL_729891, EPI_ISL_729892, EPI_ISL_729893, EPI_ISL_729894, EPI_ISL_729895, EPI_ISL_729896, EPI_ISL_729897, EPI_ISL_729898, EPI_ISL_729899, EPI_ISL_729900, EPI_ISL_729901 | | | |
| see above | Laboratorio de Referencia Nacional de Virus Respiratorios, Instituto Nacional de Salud Peru | Laboratorio de Genómica Microbiana, Universidad Peruana Cayetano Heredia | Pablo Tsukayama, Alejandra Dávila-Barclay, Luis González, Guillermo Salvatierra, Pedro E. Romero, Brenda Ayzanoa, Janet Huancachoque, Pool Marcos, Marco Galarza, Priscila Lope, Nancy Rojas |
| EPI_ISL_729903, EPI_ISL_729904, EPI_ISL_729905, EPI_ISL_729906, EPI_ISL_729907, EPI_ISL_729908, EPI_ISL_729909, EPI_ISL_729911, EPI_ISL_729915, EPI_ISL_729916 | Instituto de Medicina Tropical, Universidad Nacional Toribio Rodríguez de Mendoza de Amazonas | Laboratorio de Genómica Microbiana, Universidad Peruana Cayetano Heredia | Pablo Tsukayama, Alejandra Dávila-Barclay, Luis González, Pedro E. Romero, Brenda Ayzanoa, Janet Huancachoque, Pool Marcos, Stella Chenet, Rafael Tapia, Cecilia Pajuelo, Carla Montenegro |
| EPI_ISL_812443, EPI_ISL_812444, EPI_ISL_812445, EPI_ISL_812446, EPI_ISL_812447, EPI_ISL_812448, EPI_ISL_812449, EPI_ISL_812450, EPI_ISL_812451, EPI_ISL_812452, EPI_ISL_812453, EPI_ISL_812454, EPI_ISL_812455, EPI_ISL_812456, EPI_ISL_812457, EPI_ISL_812458, EPI_ISL_812459, EPI_ISL_812460, EPI_ISL_812461, EPI_ISL_812462, EPI_ISL_812463, EPI_ISL_812464, EPI_ISL_812465, EPI_ISL_812466, EPI_ISL_812467, EPI_ISL_812468, EPI_ISL_812469, EPI_ISL_812470, EPI_ISL_812471, EPI_ISL_812472, EPI_ISL_812473, EPI_ISL_812474, EPI_ISL_812475, EPI_ISL_812476, EPI_ISL_812477, EPI_ISL_812478, EPI_ISL_812479, EPI_ISL_812480, EPI_ISL_812481, EPI_ISL_812482, EPI_ISL_812483, EPI_ISL_812484, EPI_ISL_812485, EPI_ISL_812486, EPI_ISL_812487, EPI_ISL_812488, EPI_ISL_812489, EPI_ISL_812490, EPI_ISL_812491, EPI_ISL_812492, EPI_ISL_812493, EPI_ISL_812494, EPI_ISL_812495, EPI_ISL_812496, EPI_ISL_812497, EPI_ISL_812498, EPI_ISL_812499, EPI_ISL_812500, EPI_ISL_812501, EPI_ISL_812502, EPI_ISL_812503, EPI_ISL_812504, EPI_ISL_812505, EPI_ISL_812506, EPI_ISL_812507, EPI_ISL_812508, EPI_ISL_812509, EPI_ISL_812510, EPI_ISL_812511, EPI_ISL_812512, EPI_ISL_812513, EPI_ISL_812514, EPI_ISL_812515, EPI_ISL_812516, EPI_ISL_812517, EPI_ISL_812712, EPI_ISL_812713, EPI_ISL_812714, EPI_ISL_812715, EPI_ISL_812716, EPI_ISL_812717, EPI_ISL_812718, EPI_ISL_812719, EPI_ISL_812720, EPI_ISL_812721 | | | |

| | | | |
|---|---|---|---|
| see above | Laboratorio de Referencia Nacional de Virus Respiratorios, Instituto Nacional de Salud Peru | Laboratorio de Genómica Microbiana, Universidad Peruana Cayetano Heredia | Pablo Tsukayama, Alejandra Dávila-Barclay, Guillermo Salvatierra, Luis González, Pedro E. Romero, Brenda Ayzanoa, Janet Huancachoque, Pool Marcos, Camila Castillo-Vilcahuamán, Oscar Escalante, Priscila Lope, Nancy Rojas |
| EPI_ISL_833038 | Laboratorio de Referencia Nacional de Virus Respiratorio. Instituto Nacional de Salud Perú | Laboratorio de Referencia Nacional de Biotecnología y Biología Molecular. Instituto Nacional de Salud Perú | Carlos Padilla Rojas, Luis Barcena, Karolyn Vega Chozo, Priscila Lope Pari, Omar Caceres Rey, Marco Galarza Perez, Maribel Huaringa Nuñez, Johanna Balbuena Torrez, Henri Bailon Calderon, Nancy Rojas Serrano |

We gratefully acknowledge the following Authors from the Originating laboratories responsible for obtaining the specimens, as well as the Submitting laboratories where the genome data were generated and shared via GISAID, on which this research is based.

All Submitters of data may be contacted directly via www.gisaid.org

Authors are sorted alphabetically.

| Accession ID | Originating Laboratory | Submitting Laboratory | Authors |
| --- | --- | --- | --- |
| EPI_ISL_1713378, EPI_ISL_1713379, EPI_ISL_1713380, EPI_ISL_1713381, EPI_ISL_1713383, EPI_ISL_1713384, EPI_ISL_1713385, EPI_ISL_1713387, EPI_ISL_1713388, EPI_ISL_1713389, EPI_ISL_1713390, EPI_ISL_1713391, EPI_ISL_1713392, EPI_ISL_1713393, EPI_ISL_1713394, EPI_ISL_1713395, EPI_ISL_1713396, EPI_ISL_1713397, EPI_ISL_1713398, EPI_ISL_1713399, EPI_ISL_1713400, EPI_ISL_1713401, EPI_ISL_1713402, EPI_ISL_1713403, EPI_ISL_1713404, EPI_ISL_1713405, EPI_ISL_1713406, EPI_ISL_1713407, EPI_ISL_1713408, EPI_ISL_1713409, EPI_ISL_1713410, EPI_ISL_1713411, EPI_ISL_1713412, EPI_ISL_1713413, EPI_ISL_1713414, EPI_ISL_1713415, EPI_ISL_1713416, EPI_ISL_1713417, EPI_ISL_1713418, EPI_ISL_1713419, EPI_ISL_1713421, EPI_ISL_1713422, EPI_ISL_1713423, EPI_ISL_1713424, EPI_ISL_1713425, EPI_ISL_1713426, EPI_ISL_1713427, EPI_ISL_1713428, EPI_ISL_1713429, EPI_ISL_1713430, EPI_ISL_1713440, EPI_ISL_1713441, EPI_ISL_1713442, EPI_ISL_1713448, EPI_ISL_1713449, EPI_ISL_1713450, EPI_ISL_1713451, EPI_ISL_1713452, EPI_ISL_1713453, EPI_ISL_1713454, EPI_ISL_1713455, EPI_ISL_1713456, EPI_ISL_1713457, EPI_ISL_1713458, EPI_ISL_1713459, EPI_ISL_1713460, EPI_ISL_1713461, EPI_ISL_1713462, EPI_ISL_1713463, EPI_ISL_1713464, EPI_ISL_1713465, EPI_ISL_1713466, EPI_ISL_1713467, EPI_ISL_1713484, EPI_ISL_1713485, EPI_ISL_1713486, EPI_ISL_1713487, EPI_ISL_1713488, EPI_ISL_1713489, EPI_ISL_1713490, EPI_ISL_1713491, EPI_ISL_1713492, EPI_ISL_1713493, EPI_ISL_1713494, EPI_ISL_1713495, EPI_ISL_1713496, EPI_ISL_1713497, EPI_ISL_1713498, EPI_ISL_1713499, EPI_ISL_1713500, EPI_ISL_1713501, EPI_ISL_1713503, EPI_ISL_1713504, EPI_ISL_1713505, EPI_ISL_1713506, EPI_ISL_1713507, EPI_ISL_1713508, EPI_ISL_1713509, EPI_ISL_1713510, EPI_ISL_1713511, EPI_ISL_1713512, EPI_ISL_1713513, EPI_ISL_1713514, EPI_ISL_1713515, EPI_ISL_1713516, EPI_ISL_1713517, EPI_ISL_1713518, EPI_ISL_1713519, EPI_ISL_1713520, EPI_ISL_1713521, EPI_ISL_1713524, EPI_ISL_1713525, EPI_ISL_1713526, EPI_ISL_1713527, EPI_ISL_1713528, EPI_ISL_1713529, EPI_ISL_1713530, EPI_ISL_1713531, EPI_ISL_1713532, EPI_ISL_1713533, EPI_ISL_1713534, EPI_ISL_1713535, EPI_ISL_1713536, EPI_ISL_1713537, EPI_ISL_1713538, EPI_ISL_1713539, EPI_ISL_1713541, EPI_ISL_1713542, EPI_ISL_1713543, EPI_ISL_1713544, EPI_ISL_1713545, EPI_ISL_1713546, EPI_ISL_1713547, EPI_ISL_1713548, EPI_ISL_1713549, EPI_ISL_1713551, EPI_ISL_1713561, EPI_ISL_1713565, EPI_ISL_1713567, EPI_ISL_1713568, EPI_ISL_1713569, EPI_ISL_1713570, EPI_ISL_1713571, EPI_ISL_1713572, EPI_ISL_1713573, EPI_ISL_1713574, EPI_ISL_1713575, EPI_ISL_1713576, EPI_ISL_1713577, EPI_ISL_1713578, EPI_ISL_1713579, EPI_ISL_1713580, EPI_ISL_1713581, EPI_ISL_1713582, EPI_ISL_1713583, EPI_ISL_1713584, EPI_ISL_1713585, EPI_ISL_1713586, EPI_ISL_1713587, EPI_ISL_1713588, EPI_ISL_1713589, EPI_ISL_1713590, EPI_ISL_1713591, EPI_ISL_1713592, EPI_ISL_1713593, EPI_ISL_1713594, EPI_ISL_1713595, EPI_ISL_1713596, EPI_ISL_1713597, EPI_ISL_1713598, EPI_ISL_1713599, EPI_ISL_1713600, EPI_ISL_1713601, EPI_ISL_1713602, EPI_ISL_1713603, EPI_ISL_1713604, EPI_ISL_1713605, EPI_ISL_1713606, EPI_ISL_1713607, EPI_ISL_1713608, EPI_ISL_1713609, EPI_ISL_1713610, EPI_ISL_1713611, EPI_ISL_1713612, EPI_ISL_1713613, EPI_ISL_1713614, EPI_ISL_1713615, EPI_ISL_1713616, EPI_ISL_1713617, EPI_ISL_1713618, EPI_ISL_1713619, EPI_ISL_1713620, EPI_ISL_1713621, EPI_ISL_1713622, EPI_ISL_1713623, EPI_ISL_1713624, EPI_ISL_1713625, EPI_ISL_1713626, EPI_ISL_1713627, EPI_ISL_1713628, EPI_ISL_1713629, EPI_ISL_1713630, EPI_ISL_1713631, EPI_ISL_1713632, EPI_ISL_1713633, EPI_ISL_1713634, EPI_ISL_1713635, EPI_ISL_1713636, EPI_ISL_1713637, EPI_ISL_1713638, EPI_ISL_1713639, EPI_ISL_1713640, EPI_ISL_1713641, EPI_ISL_1713642, EPI_ISL_1713643, EPI_ISL_1713644, EPI_ISL_1713645, EPI_ISL_1713646, EPI_ISL_1713647, EPI_ISL_1713648, EPI_ISL_1713649, EPI_ISL_1713650, EPI_ISL_1713651, EPI_ISL_1713652, EPI_ISL_1713653, EPI_ISL_1713654, EPI_ISL_1713655, EPI_ISL_1713656, EPI_ISL_1713657, EPI_ISL_1713658, EPI_ISL_1713659, EPI_ISL_1713660, EPI_ISL_1713661, EPI_ISL_1713662, EPI_ISL_1713663, EPI_ISL_1713664, EPI_ISL_1713665, EPI_ISL_1713666, EPI_ISL_1713667, EPI_ISL_1713668, EPI_ISL_1713669, EPI_ISL_1713670, EPI_ISL_1713671, EPI_ISL_1713672, EPI_ISL_1713673, EPI_ISL_1713674, EPI_ISL_1713675, EPI_ISL_1713676, EPI_ISL_1713677, EPI_ISL_1713678, EPI_ISL_1713679, EPI_ISL_1713680, EPI_ISL_1713681, EPI_ISL_1713682, EPI_ISL_1713683, EPI_ISL_1713684, EPI_ISL_1713685, EPI_ISL_1713686, EPI_ISL_1713687, EPI_ISL_1713688, EPI_ISL_1713689, EPI_ISL_1713690, EPI_ISL_1713691, EPI_ISL_1713692, EPI_ISL_1713693, EPI_ISL_1713694, EPI_ISL_1713695, EPI_ISL_1713696, EPI_ISL_1713697, EPI_ISL_1713698, EPI_ISL_1713699, EPI_ISL_1713700, EPI_ISL_1713701, EPI_ISL_1713702, EPI_ISL_1713703, EPI_ISL_1713704, EPI_ISL_1713705, EPI_ISL_1713706, EPI_ISL_1713707, EPI_ISL_1713708, EPI_ISL_1713709, EPI_ISL_1713710, EPI_ISL_1713711, EPI_ISL_1713712, EPI_ISL_1713713, EPI_ISL_1713714, EPI_ISL_1713715, EPI_ISL_1713716, EPI_ISL_1713717, EPI_ISL_1713718, EPI_ISL_1713719, EPI_ISL_1713720, EPI_ISL_1713721, EPI_ISL_1713722, EPI_ISL_1713723, EPI_ISL_1713724, EPI_ISL_1713725, EPI_ISL_1713726, EPI_ISL_1713727, EPI_ISL_1713728, EPI_ISL_1713729, EPI_ISL_1713730, EPI_ISL_1713731, EPI_ISL_1713732, EPI_ISL_1713733, EPI_ISL_1713734, EPI_ISL_1713735, EPI_ISL_1713736, EPI_ISL_1713737, EPI_ISL_1713738, EPI_ISL_1713739, EPI_ISL_1713740, EPI_ISL_1713741, EPI_ISL_1713742, EPI_ISL_1713743, EPI_ISL_1713744, EPI_ISL_1713745, EPI_ISL_1713746, EPI_ISL_1713747, EPI_ISL_1713748, EPI_ISL_1713749, EPI_ISL_1713750, EPI_ISL_1713751, EPI_ISL_1713752, EPI_ISL_1713753, EPI_ISL_1713754, EPI_ISL_1713755, EPI_ISL_1713756, EPI_ISL_1713757, EPI_ISL_1713758, EPI_ISL_1713759, EPI_ISL_1713760, EPI_ISL_1713761, EPI_ISL_1713762, EPI_ISL_1713763, EPI_ISL_1713764, EPI_ISL_1713765, EPI_ISL_1713766, EPI_ISL_1713767, EPI_ISL_1713768, EPI_ISL_1713769, EPI_ISL_1713770, EPI_ISL_1713771, EPI_ISL_1713772, EPI_ISL_1713773, EPI_ISL_1713774, EPI_ISL_1713775, EPI_ISL_1713776, EPI_ISL_1713777, EPI_ISL_1713778, EPI_ISL_1713779, EPI_ISL_1713780, EPI_ISL_1713781, EPI_ISL_1713782, EPI_ISL_1713783, EPI_ISL_1713784, EPI_ISL_1713785, EPI_ISL_1713786, EPI_ISL_1713787, EPI_ISL_1713788, EPI_ISL_1713789, EPI_ISL_1713790, EPI_ISL_1713791, EPI_ISL_1713792, EPI_ISL_1713793, EPI_ISL_1713794, EPI_ISL_1713795, EPI_ISL_1713796, EPI_ISL_1713797, EPI_ISL_1713798, EPI_ISL_1713799, EPI_ISL_1713800, EPI_ISL_1713801, EPI_ISL_1713802, EPI_ISL_1713803, EPI_ISL_1713804, EPI_ISL_1713805, EPI_ISL_1713806, EPI_ISL_1713807, EPI_ISL_1713808, EPI_ISL_1713809, EPI_ISL_1713810, EPI_ISL_1713811, EPI_ISL_1713812, EPI_ISL_1713813, EPI_ISL_1713814, EPI_ISL_1713815, EPI_ISL_1713816, EPI_ISL_1713817, EPI_ISL_1713818, EPI_ISL_1713819, EPI_ISL_1713820, EPI_ISL_1713821, EPI_ISL_1713822, EPI_ISL_1713823, EPI_ISL_1713824, EPI_ISL_1713825, EPI_ISL_1713826, EPI_ISL_1713827, EPI_ISL_1713828, EPI_ISL_1713829, EPI_ISL_1713830, EPI_ISL_1713831, EPI_ISL_1713832, EPI_ISL_1713833, EPI_ISL_1713834, EPI_ISL_1713835, EPI_ISL_1713836, EPI_ISL_1713837, EPI_ISL_1713838, EPI_ISL_1713839, EPI_ISL_1713840, EPI_ISL_1713841, EPI_ISL_1713842, EPI_ISL_1713843, EPI_ISL_1713844, EPI_ISL_1713845, EPI_ISL_1713846, EPI_ISL_1713847, EPI_ISL_1713848, EPI_ISL_1713849, EPI_ISL_1713850, EPI_ISL_1713851, EPI_ISL_1713852, EPI_ISL_1713853, EPI_ISL_1713854, EPI_ISL_1713855, EPI_ISL_1713856, EPI_ISL_1713857, EPI_ISL_1713858, EPI_ISL_1713859, EPI_ISL_1713860, EPI_ISL_1713861, EPI_ISL_1713862, EPI_ISL_1713863, EPI_ISL_1713864, EPI_ISL_1713865, EPI_ISL_1713866, EPI_ISL_1713867, EPI_ISL_1713868, EPI_ISL_1713869, EPI_ISL_1713870, EPI_ISL_1713871, EPI_ISL_1713872, EPI_ISL_1713873, EPI_ISL_1713874, EPI_ISL_1713875, EPI_ISL_1713876, EPI_ISL_1713877, EPI_ISL_1713878, EPI_ISL_1713879, EPI_ISL_1713880, EPI_ISL_1713881, EPI_ISL_1713882, EPI_ISL_1713883, EPI_ISL_1713884, EPI_ISL_1713885, EPI_ISL_1713886, EPI_ISL_1713887, EPI_ISL_1713888, EPI_ISL_1713889, EPI_ISL_1713890, EPI_ISL_1713891, EPI_ISL_1713892, EPI_ISL_1713893, EPI_ISL_1713894, EPI_ISL_1713895, EPI_ISL_1713896, EPI_ISL_1713897, EPI_ISL_1713898, EPI_ISL_1713899, EPI_ISL_1713900, EPI_ISL_1713901, EPI_ISL_1713902, EPI_ISL_1713903, EPI_ISL_1713904, EPI_ISL_1713905, EPI_ISL_1713906, EPI_ISL_1713907, EPI_ISL_1713908, EPI_ISL_1713909, EPI_ISL_1713910, EPI_ISL_1713911, EPI_ISL_1713912, EPI_ISL_1713913, EPI_ISL_1713914, EPI_ISL_1713915, EPI_ISL_1713916, EPI_ISL_1713917, EPI_ISL_1713918, EPI_ISL_1713919, EPI_ISL_1713920, EPI_ISL_1713921, EPI_ISL_1713922, EPI_ISL_1713923, EPI_ISL_1713924, EPI_ISL_1713925, EPI_ISL_1713926, EPI_ISL_1713927, EPI_ISL_1713928, EPI_ISL_1713929, EPI_ISL_1713931, EPI_ISL_1713932, EPI_ISL_1713933, EPI_ISL_1713934, EPI_ISL_1713935, EPI_ISL_1713936, EPI_ISL_1713937, EPI_ISL_1713938, EPI_ISL_1713940, EPI_ISL_1713941, EPI_ISL_1713942, EPI_ISL_1713944, EPI_ISL_1713945, EPI_ISL_1713946 | | | |
| see above | Ministry of Public Health / Hamad Medical Corporation | Biomedical Research Center (BRC), Qatar University / Qatar Genome Project (QGP) | BRC: Fatiha M. Benslimane, Heba A. Al-Khatib, Oal Al-Jamal, Dana Al-Batesh, Hadi M. Yassine, Asmaa A. Al-Thani. MOPH and HMC: Abdullatif Al-Khal, Muna A. S. Al-Maslamani, Mashael A. Al-Bader, Hamda Alromaihi, Roberto Bertollini, Peter V. Coyle, Einas A. E. Al-Kuwari, Hamad E. Al-Romaihi, Salih Al-Marri, Mohammed Al-Thani, Reham A. El-Kahlout. QBB: Tasneem Al-Hamad, Dina Elgakhlab QGP: Fatima H. Al-Kuwari, Chadi Saad |
| EPI_ISL_1714275, EPI_ISL_1714276, EPI_ISL_1714277, EPI_ISL_1714278, EPI_ISL_1714279, EPI_ISL_1714280, EPI_ISL_1714281, EPI_ISL_1714282, EPI_ISL_1714283, EPI_ISL_1714284, EPI_ISL_1714285, EPI_ISL_1714286, EPI_ISL_1714287, EPI_ISL_1714288, EPI_ISL_1714289, EPI_ISL_1714290, EPI_ISL_1714291, EPI_ISL_1714292, EPI_ISL_1714293, EPI_ISL_1714294, EPI_ISL_1714295, EPI_ISL_1714296, EPI_ISL_1714297, EPI_ISL_1714298, EPI_ISL_1714299, EPI_ISL_1714300, EPI_ISL_1714301, EPI_ISL_1714302, EPI_ISL_1714303, EPI_ISL_1714304, EPI_ISL_1714305, EPI_ISL_1714306, EPI_ISL_1714307, EPI_ISL_1714308, EPI_ISL_1714309, EPI_ISL_1714310, EPI_ISL_1714311, EPI_ISL_1714312, EPI_ISL_1714313, EPI_ISL_1714314, EPI_ISL_1714315, EPI_ISL_1714316, EPI_ISL_1714317, EPI_ISL_1714318, EPI_ISL_1714319, EPI_ISL_1714320, EPI_ISL_1714321, EPI_ISL_1714322, EPI_ISL_1714323, EPI_ISL_1714324, EPI_ISL_1714325, EPI_ISL_1714326, EPI_ISL_1714327, EPI_ISL_1714328, EPI_ISL_1714329, EPI_ISL_1714330, EPI_ISL_1714331, EPI_ISL_1714332, EPI_ISL_1714333, EPI_ISL_1714334, EPI_ISL_1714335, EPI_ISL_1714336, EPI_ISL_1714337, EPI_ISL_1714338, EPI_ISL_1714339, EPI_ISL_1714340, EPI_ISL_1714341, EPI_ISL_1714342, EPI_ISL_1714343, EPI_ISL_1714344, EPI_ISL_1714345, EPI_ISL_1714346, EPI_ISL_1714347, EPI_ISL_1714348, EPI_ISL_1714349, EPI_ISL_1714350, EPI_ISL_1714351, EPI_ISL_1714352, EPI_ISL_1714353 | | | |
| see above | Ministry of Public Health / Hamad Medical Corporation | Weill Cornell Medical College - Qatar (WCM-Q), Genomics Core Laboratory / Qatar Genome Project (QGP) | WCMQ: Ayeda A. Ahmed, Meryem Bensaad, Shameem Younuskunju, Yasmin Mohamoud, Laith Abu-Raddad, Joel A Malek. QGP: Fatima H. Al-Kuwari, Chadi Saad MOPH and HMC: Abdullatif Al-Khal, Muna A. S. Al-Maslamani, Mashael A. Al-Bader, Hamda Alromaihi, Roberto Bertollini, Peter V. Coyle, Einas A. E. Al-Kuwari, Hamad E. Al-Romaihi, Salih Al-Marri, Mohammed Al-Thani, Reham A. El-Kahlout. QBB: Tasneem Al-Hamad, Dina Elgakhlab |

We gratefully acknowledge the following Authors from the Originating laboratories responsible for obtaining the specimens, as well as the Submitting laboratories where the genome data were generated and shared via GISAID, on which this research is based.

All Submitters of data may be contacted directly via www.gisaid.org

Authors are sorted alphabetically.

| Accession ID | Originating Laboratory | Submitting Laboratory | Authors |
|---|---|---|---|
| EPI_ISL_1039057, EPI_ISL_1039058, EPI_ISL_1039059, EPI_ISL_1039060, EPI_ISL_1039061, EPI_ISL_1039062, EPI_ISL_1039063, EPI_ISL_1039064, EPI_ISL_1039065, EPI_ISL_1039066, EPI_ISL_1039067, EPI_ISL_1039068, EPI_ISL_1039069, EPI_ISL_1039070, EPI_ISL_1039071, EPI_ISL_1039072, EPI_ISL_1039073, EPI_ISL_1039074, EPI_ISL_1039075, EPI_ISL_1039076, EPI_ISL_1039077, EPI_ISL_1039078, EPI_ISL_1039079, EPI_ISL_1039080, EPI_ISL_1039081, EPI_ISL_1039082, EPI_ISL_1039083, EPI_ISL_1039084, EPI_ISL_1039085, EPI_ISL_1039086, EPI_ISL_1039087, EPI_ISL_1039088, EPI_ISL_1039089, EPI_ISL_1039090, EPI_ISL_1039091, EPI_ISL_1039092, EPI_ISL_1039093, EPI_ISL_1039094, EPI_ISL_1039095, EPI_ISL_1039096, EPI_ISL_1039097, EPI_ISL_1039098, EPI_ISL_1039099, EPI_ISL_1039100, EPI_ISL_1039101, EPI_ISL_1039102, EPI_ISL_1039103, EPI_ISL_1039104, EPI_ISL_1039105, EPI_ISL_1039106, EPI_ISL_1039107, EPI_ISL_1039108, EPI_ISL_1039109, EPI_ISL_1039110, EPI_ISL_1039111, EPI_ISL_1039112, EPI_ISL_1039113, EPI_ISL_1039114, EPI_ISL_1039115, EPI_ISL_1039116, EPI_ISL_1039117, EPI_ISL_1039118, EPI_ISL_1039119, EPI_ISL_1039120, EPI_ISL_1039121, EPI_ISL_1039122, EPI_ISL_1039123, EPI_ISL_1400492, EPI_ISL_1400525, EPI_ISL_1400552 | | | |
| see above | Center of Hygiene and Epidemiology in Belgorod Region | WHO National Influenza Centre Russian Federation | Andrey Komissarov; Anna Ivanova; Artem Fadeev; Daria Danilenko; Dmitry Bazhenov; Dmitry Lioznov; Elena Nabieva; Georgii Bazykin; Ksenia Safina; Kseniya Komissarova; Lyudmila Berdinskikh; Maria Pisareva; Maria Timofeeva; Tamila Musaeva; Veronika Eder |
| EPI_ISL_1859330, EPI_ISL_1859334, EPI_ISL_1859343, EPI_ISL_1859365, EPI_ISL_1859399, EPI_ISL_1859414, EPI_ISL_1859416, EPI_ISL_1859420, EPI_ISL_1859454, EPI_ISL_1859469, EPI_ISL_1859471, EPI_ISL_1859479, EPI_ISL_1859487, EPI_ISL_1859498, EPI_ISL_1859502, EPI_ISL_1859519, EPI_ISL_1859523, EPI_ISL_1859541, EPI_ISL_1859572, EPI_ISL_1859605, EPI_ISL_1859641, EPI_ISL_1859665, EPI_ISL_1859710, EPI_ISL_1859727, EPI_ISL_1859754, EPI_ISL_1859760 | | | |
| see above | City Clinical Hospital named after S.I. Spasokukotsky of Moscow Healthcare Department, Academic Consortium | FEDERAL RESEARCH AND CLINICAL CENTER OF PHYSICAL-CHEMICAL MEDICINE OF FEDERAL MEDICAL BIOLOGICAL AGENCY, Academic Consortium | Alexander Manolov; Alexander Pavlenko; Alexander Rumyantsev; Alexander Samoilov; Alexander Troitsky; Alexander Tulichev; Alexander Vasiliev; Alexey Volnukhin; Alla Petrova; Andrey Koritsky; Anna Blagonravova; Anna Varizhuk; Daria Boldyreva; Daria Kharlampieva; Daria Matyushkina; Dmitry Fedorov; Dmitry Pushkar; Ekaterina Moskaleva; Elena Galova; Elena Ilina; Elizaveta Orlova; Galina Trifonova; Gaukhar Yusubalieva; Ilshat Gafurov; Irina Shirokova; Konstantin Ivanov; Konstantin Kolontarev; Ksenia Klimina; Larisa Balykova; Lyubov Rychkova; Nadezhda Kolyshkina; Natalia Pozhenko; Oleg Ogarkov; Pavel Bashkirov; Polina Khromova; Polina Tikhonova; Ramiz Bakhtyev; Raushaniya Gaifullina; Sergey Rumyantsev; Svetlana Solovieva; Vadim Govorun; Valentin Manuvera; Varvara Musarova; Vasily Lazarev; Vladimir Baklaushev; Vladimir Rumyantsev; Vladimir Veselovsky; Vladislav Babenko; Vyacheslav Kuropatkin; Vyacheslav Sinkov; Yulia Galeeva; Yuri Udalov |
| EPI_ISL_1731043 | Collection of WHO National Influenza Center, N.F. Gamaleya National Research Center for Epidemiology and Microbiology of the Ministry of Health of the Russian Federation | N.F. Gamaleya Research Center for Epidemiology and Microbiology | Denis Logunov; Ekaterina Aksenova; Elena Burtseva; Kirill Krasnoslobodtsev; Marina Kunda; Natalia Ryzhova; Olga Voronina |
| EPI_ISL_1708511, EPI_ISL_1708512 | Collection of WHO National Influenza Center, N.F. Gamaleya National Research Center for Epidemiology and Microbiology of the Ministry of Health of the Russian Federation | Laboratory of genome analysis, N.F. Gamaleya National Research Center for Epidemiology and Microbiology of the Ministry of Health of the Russian Federation | Denis Logunov; Ekaterina Aksenova; Elena Burtseva; Kirill Krasnoslobodtsev; Marina Kunda; Natalia Ryzhova; Olga Voronina |
| EPI_ISL_1859417, EPI_ISL_1859570, EPI_ISL_1859654, EPI_ISL_1859685 | FEDERAL RESEARCH AND CLINICAL CENTER OF PHYSICAL-CHEMICAL MEDICINE OF FEDERAL MEDICAL BIOLOGICAL AGENCY, Academic Consortium | FEDERAL RESEARCH AND CLINICAL CENTER OF PHYSICAL-CHEMICAL MEDICINE OF FEDERAL MEDICAL BIOLOGICAL AGENCY, Academic Consortium | Alexander Manolov; Alexander Pavlenko; Alexander Rumyantsev; Alexander Samoilov; Alexander Troitsky; Alexander Tulichev; Alexander Vasiliev; Alexey Volnukhin; Alla Petrova; Andrey Koritsky; Anna Blagonravova; Anna Varizhuk; Daria Boldyreva; Daria Kharlampieva; Daria Matyushkina; Dmitry Fedorov; Dmitry Pushkar; Ekaterina Moskaleva; Elena Galova; Elena Ilina; Elizaveta Orlova; Galina Trifonova; Gaukhar Yusubalieva; Ilshat Gafurov; Irina Shirokova; Konstantin Ivanov; Konstantin Kolontarev; Ksenia Klimina; Larisa Balykova; Lyubov Rychkova; Nadezhda Kolyshkina; Natalia Pozhenko; Oleg Ogarkov; Pavel Bashkirov; Polina Khromova; Polina Tikhonova; Ramiz Bakhtyev; Raushaniya Gaifullina; Sergey Rumyantsev; Svetlana Solovieva; Vadim Govorun; Valentin Manuvera; Varvara Musarova; Vasily Lazarev; Vladimir Baklaushev; Vladimir Rumyantsev; Vladimir Veselovsky; Vladislav Babenko; Vyacheslav Kuropatkin; Vyacheslav Sinkov; Yulia Galeeva; Yuri Udalov |
| EPI_ISL_2086624, EPI_ISL_2086630, EPI_ISL_2086631, EPI_ISL_2086643, EPI_ISL_2086648, EPI_ISL_2086653, EPI_ISL_2086665, EPI_ISL_2086671, EPI_ISL_2086672, EPI_ISL_2086674, EPI_ISL_2086675, EPI_ISL_2096201 | | | |
| see above | FEDERAL STATE BUDGETARY INSTITUTION OF HEALTHCARE HOSPITAL OF THE RUSSIAN ACADEMY OF SCIENCES (TROITSK), Academic Consortium | FEDERAL RESEARCH AND CLINICAL CENTER OF PHYSICAL-CHEMICAL MEDICINE OF FEDERAL MEDICAL BIOLOGICAL AGENCY, Academic Consortium | Alexander Manolov; Alexander Pavlenko; Alexander Rumyantsev; Alexander Samoilov; Alexander Troitsky; Alexander Tulichev; Alexander Vasiliev; Alexey Volnukhin; Alla Petrova; Andrey Koritsky; Anna Blagonravova; Anna Varizhuk; Daria Boldyreva; Daria Kharlampieva; Daria Matyushkina; Dmitry Fedorov; Dmitry Pushkar; Ekaterina Moskaleva; Elena Galova; Elena Ilina; Elizaveta Orlova; Galina Trifonova; Gaukhar Yusubalieva; Ilshat Gafurov; Irina Shirokova; Konstantin Ivanov; Konstantin Kolontarev; Ksenia Klimina; Larisa Balykova; Lyubov Rychkova; Nadezhda Kolyshkina; Natalia Pozhenko; Oleg Ogarkov; Pavel Bashkirov; Polina Khromova; Polina Tikhonova; Ramiz Bakhtyev; Raushaniya Gaifullina; Sergey Rumyantsev; Svetlana Solovieva; Vadim Govorun; Valentin Manuvera; Varvara Musarova; Vasily Lazarev; Vladimir Baklaushev; Vladimir Rumyantsev; Vladimir Veselovsky; Vladislav Babenko; Vyacheslav Kuropatkin; Vyacheslav Sinkov; Yulia Galeeva; Yuri Udalov |
| EPI_ISL_1859410, EPI_ISL_1859758 | Federal State Budgetary Educational Institution of Higher Education B«Privolzhsky Research Medical UniversityB» of the Ministry of Health of the Russian Federation, Academic Consortium | FEDERAL RESEARCH AND CLINICAL CENTER OF PHYSICAL-CHEMICAL MEDICINE OF FEDERAL MEDICAL BIOLOGICAL AGENCY, Academic Consortium | Alexander Manolov; Alexander Pavlenko; Alexander Rumyantsev; Alexander Samoilov; Alexander Troitsky; Alexander Tulichev; Alexander Vasiliev; Alexey Volnukhin; Alla Petrova; Andrey Koritsky; Anna Blagonravova; Anna Varizhuk; Daria Boldyreva; Daria Kharlampieva; Daria Matyushkina; Dmitry Fedorov; Dmitry Pushkar; Ekaterina Moskaleva; Elena Galova; Elena Ilina; Elizaveta Orlova; Galina Trifonova; Gaukhar Yusubalieva; Ilshat Gafurov; Irina Shirokova; Konstantin Ivanov; Konstantin Kolontarev; Ksenia Klimina; Larisa Balykova; Lyubov Rychkova; Nadezhda Kolyshkina; Natalia Pozhenko; Oleg Ogarkov; Pavel Bashkirov; Polina Khromova; Polina Tikhonova; Ramiz Bakhtyev; Raushaniya Gaifullina; Sergey Rumyantsev; Svetlana Solovieva; Vadim Govorun; Valentin Manuvera; Varvara Musarova; Vasily Lazarev; Vladimir Baklaushev; Vladimir Rumyantsev; Vladimir Veselovsky; Vladislav Babenko; Vyacheslav Kuropatkin; Vyacheslav Sinkov; Yulia Galeeva; Yuri Udalov |
| EPI_ISL_2086623, EPI_ISL_2086636, EPI_ISL_2086638, EPI_ISL_2086639, EPI_ISL_2086646, EPI_ISL_2086649, EPI_ISL_2086650, EPI_ISL_2086651, EPI_ISL_2086657, EPI_ISL_2086659, EPI_ISL_2086666, EPI_ISL_2086673, EPI_ISL_2096200, EPI_ISL_2096202 | | | |
| see above | Federal State Public Scientific Institution «Scientific Centre for Family Health and Human Reproduction Problems», Academic Consortium | FEDERAL RESEARCH AND CLINICAL CENTER OF PHYSICAL-CHEMICAL MEDICINE OF FEDERAL MEDICAL BIOLOGICAL AGENCY, Academic Consortium | Alexander Manolov; Alexander Pavlenko; Alexander Rumyantsev; Alexander Samoilov; Alexander Troitsky; Alexander Tulichev; Alexander Vasiliev; Alexey Volnukhin; Alla Petrova; Andrey Koritsky; Anna Blagonravova; Anna Varizhuk; Daria Boldyreva; Daria Kharlampieva; Daria Matyushkina; Dmitry Fedorov; Dmitry Pushkar; Ekaterina Moskaleva; Elena Galova; Elena Ilina; Elizaveta Orlova; Galina Trifonova; Gaukhar Yusubalieva; Ilshat Gafurov; Irina Shirokova; Konstantin Ivanov; Konstantin Kolontarev; Ksenia Klimina; Larisa Balykova; Lyubov Rychkova; Nadezhda Kolyshkina; Natalia Pozhenko; Oleg Ogarkov; Pavel Bashkirov; Polina Khromova; Polina Tikhonova; Ramiz Bakhtyev; Raushaniya Gaifullina; Sergey Rumyantsev; Svetlana Solovieva; Vadim Govorun; Valentin Manuvera; Varvara Musarova; Vasily Lazarev; Vladimir Baklaushev; Vladimir Rumyantsev; Vladimir Veselovsky; Vladislav Babenko; Vyacheslav Kuropatkin; Vyacheslav Sinkov; Yulia Galeeva; Yuri Udalov |
| EPI_ISL_1259282, EPI_ISL_1259283 | Group for molecular genetics of pathogens | Group for molecular genetics of pathogens | Anna S. Gladkikh; Areg A.Totolian; Ekaterina A. Goncharova; Valerya A. Sbarzaglia; Vladimir G. Dedkov |
| EPI_ISL_699570, EPI_ISL_699571, EPI_ISL_699572, EPI_ISL_699573, EPI_ISL_735250, EPI_ISL_735251, EPI_ISL_735252, EPI_ISL_735253, EPI_ISL_735257, EPI_ISL_735258 | | | |
| see above | Group of Genetic Engineering and Biotechnology, Federal Budget Institution of Science 'Central Research Institute of Epidemiology' of The Federal Service on Customers' Rights Protection and Human Well-being Surveillance | Group of Genetic Engineering and Biotechnology, Federal Budget Institution of Science 'Central Research Institute of Epidemiology' of The Federal Service on Customers' Rights Protection and Human Well-being Surveillance | A.G.; A.S.; A.V.; Akimkin; Berlina; Bulanenko; Cherkashina; E.D.; E.V.; Golubeva; M.I.; O.Y.; Shipulina; Soloviova; Solovyova; Speranskaya; Tivanova; V.G.; V.P.; Valdokhina; Y.Y.; Zotova |
| EPI_ISL_596268, EPI_ISL_596269, EPI_ISL_596271, EPI_ISL_596272, EPI_ISL_596273, EPI_ISL_596274, EPI_ISL_596277, EPI_ISL_596278, EPI_ISL_596279, EPI_ISL_596280, EPI_ISL_596281, EPI_ISL_596282, EPI_ISL_596283, EPI_ISL_596284, EPI_ISL_596285, EPI_ISL_596286, EPI_ISL_596287, EPI_ISL_596288, EPI_ISL_596289, EPI_ISL_596290, EPI_ISL_596291, EPI_ISL_596292, EPI_ISL_596293, EPI_ISL_596294, EPI_ISL_596295, EPI_ISL_596296, EPI_ISL_596297, EPI_ISL_596298, EPI_ISL_596299, EPI_ISL_596300, EPI_ISL_596301, EPI_ISL_596302, EPI_ISL_596303, EPI_ISL_596304, EPI_ISL_596307, EPI_ISL_596308, EPI_ISL_596309, EPI_ISL_596315 | | | |
| see above | HELIX LCC | WHO National Influenza Centre Russian Federation | Andrey Komissarov; Anna Ivanova; Artem Fadeev; Daria Danilenko; Dmitry Bazhenov; Kseniya Komissarova |
| EPI_ISL_602332, EPI_ISL_602333, EPI_ISL_602334, EPI_ISL_602335, EPI_ISL_602336, EPI_ISL_602337, EPI_ISL_602338, EPI_ISL_602339, EPI_ISL_602340, EPI_ISL_602341, EPI_ISL_602342, EPI_ISL_602343, EPI_ISL_602344, EPI_ISL_602345, EPI_ISL_602346, EPI_ISL_602347, EPI_ISL_602348, EPI_ISL_602349, EPI_ISL_602350, EPI_ISL_602351, EPI_ISL_602352, EPI_ISL_602353, EPI_ISL_602354, EPI_ISL_602355, EPI_ISL_602356, EPI_ISL_602357, EPI_ISL_602358, EPI_ISL_602359, EPI_ISL_602360, EPI_ISL_602361, EPI_ISL_602362, EPI_ISL_602363, EPI_ISL_602364, EPI_ISL_602365, EPI_ISL_602366, EPI_ISL_602367, EPI_ISL_602368, EPI_ISL_602369, EPI_ISL_602370, EPI_ISL_602371, EPI_ISL_602372, EPI_ISL_602373, EPI_ISL_602374, EPI_ISL_602375, EPI_ISL_602376, EPI_ISL_602377, EPI_ISL_602378, EPI_ISL_602379, EPI_ISL_602380, EPI_ISL_602381, EPI_ISL_602382, EPI_ISL_602383, EPI_ISL_602384, EPI_ISL_602385, EPI_ISL_602386, EPI_ISL_602387, EPI_ISL_602388, EPI_ISL_602389, EPI_ISL_602390, EPI_ISL_602391, EPI_ISL_602392, EPI_ISL_602393, EPI_ISL_602394, EPI_ISL_602395, EPI_ISL_602396, EPI_ISL_602397, EPI_ISL_602398, EPI_ISL_602399, EPI_ISL_602400, EPI_ISL_602401, EPI_ISL_602402, EPI_ISL_602403, EPI_ISL_602404, EPI_ISL_602405, EPI_ISL_602406, EPI_ISL_602407, EPI_ISL_602408, EPI_ISL_602409, EPI_ISL_602410, EPI_ISL_602411, EPI_ISL_602417, EPI_ISL_602418, EPI_ISL_602419, EPI_ISL_602420, EPI_ISL_602421, EPI_ISL_602422, EPI_ISL_602423, EPI_ISL_602424, EPI_ISL_602425, EPI_ISL_602426, EPI_ISL_602427, EPI_ISL_602428, EPI_ISL_602429, EPI_ISL_602430, EPI_ISL_602431, EPI_ISL_602432, EPI_ISL_602433, EPI_ISL_602434, EPI_ISL_602435, EPI_ISL_602436, EPI_ISL_602437, EPI_ISL_602438, EPI_ISL_602439, EPI_ISL_602440, EPI_ISL_602441, EPI_ISL_602442, EPI_ISL_602443, EPI_ISL_602444, EPI_ISL_602445, EPI_ISL_602446, EPI_ISL_602447, EPI_ISL_602448, EPI_ISL_602449, EPI_ISL_602450, EPI_ISL_602451, EPI_ISL_602452, EPI_ISL_602453, EPI_ISL_602454, EPI_ISL_602455, EPI_ISL_602456, EPI_ISL_602457, EPI_ISL_602458, EPI_ISL_602459, EPI_ISL_602460, EPI_ISL_602461, EPI_ISL_639948, EPI_ISL_639949, EPI_ISL_639951, EPI_ISL_639952, EPI_ISL_639953, EPI_ISL_639954, EPI_ISL_639955, EPI_ISL_639956, EPI_ISL_639957, EPI_ISL_639958, EPI_ISL_639959, EPI_ISL_639960, EPI_ISL_639961, EPI_ISL_639962, EPI_ISL_639963, EPI_ISL_639964, EPI_ISL_639965, EPI_ISL_639966, EPI_ISL_639967, EPI_ISL_639968, EPI_ISL_639969, EPI_ISL_639970, EPI_ISL_639971, EPI_ISL_639972, EPI_ISL_639973, EPI_ISL_733023, EPI_ISL_733024, EPI_ISL_733025, EPI_ISL_733026, EPI_ISL_733027, EPI_ISL_733028, EPI_ISL_733029, EPI_ISL_733030, EPI_ISL_733031, EPI_ISL_733032, EPI_ISL_733033, EPI_ISL_733034, EPI_ISL_733035, EPI_ISL_733036, EPI_ISL_733037, EPI_ISL_733038, EPI_ISL_733039, EPI_ISL_733040, EPI_ISL_733041, EPI_ISL_733042, EPI_ISL_733043, EPI_ISL_733044, EPI_ISL_733045, EPI_ISL_733046, EPI_ISL_733047, EPI_ISL_733048, EPI_ISL_733049, EPI_ISL_733050, EPI_ISL_733051, EPI_ISL_733052, EPI_ISL_733053, EPI_ISL_733054, EPI_ISL_733055, EPI_ISL_733056, EPI_ISL_733057, EPI_ISL_733058, EPI_ISL_733059, EPI_ISL_733060, EPI_ISL_733061, EPI_ISL_733062, EPI_ISL_733063, EPI_ISL_733064, EPI_ISL_733065, EPI_ISL_733066, EPI_ISL_733067, EPI_ISL_733068, EPI_ISL_733069, EPI_ISL_733070, EPI_ISL_733071, EPI_ISL_733072, EPI_ISL_733073, EPI_ISL_733074, EPI_ISL_733075, EPI_ISL_733076, EPI_ISL_733079, EPI_ISL_733080, EPI_ISL_733081, EPI_ISL_733082, EPI_ISL_733083, EPI_ISL_733084, EPI_ISL_733085, EPI_ISL_733086, EPI_ISL_733087, EPI_ISL_733088, EPI_ISL_733089, EPI_ISL_733090, EPI_ISL_733091, EPI_ISL_733092, EPI_ISL_733093, EPI_ISL_733094, EPI_ISL_733095, EPI_ISL_733096, EPI_ISL_733097, EPI_ISL_733098, EPI_ISL_733099, EPI_ISL_733100, EPI_ISL_733101, EPI_ISL_733102, EPI_ISL_733103, EPI_ISL_733104, EPI_ISL_733105, EPI_ISL_733106, EPI_ISL_733107, EPI_ISL_733108, EPI_ISL_733109, EPI_ISL_733110, EPI_ISL_733111, EPI_ISL_733112, EPI_ISL_733113, EPI_ISL_733114, EPI_ISL_733115, EPI_ISL_733116, EPI_ISL_733117, EPI_ISL_733118, EPI_ISL_733119, EPI_ISL_733120, EPI_ISL_733121, EPI_ISL_733122, EPI_ISL_733123, EPI_ISL_733124, EPI_ISL_733125, EPI_ISL_733126, EPI_ISL_733127, EPI_ISL_733128, EPI_ISL_733129, EPI_ISL_733130, EPI_ISL_733131, EPI_ISL_733132, EPI_ISL_733133, EPI_ISL_733134, EPI_ISL_733135, EPI_ISL_733136, EPI_ISL_733137, EPI_ISL_733138, EPI_ISL_733139, EPI_ISL_733140, EPI_ISL_733141, EPI_ISL_733142, EPI_ISL_733143, EPI_ISL_733144, EPI_ISL_733145, EPI_ISL_733146, EPI_ISL_733147, EPI_ISL_733148, EPI_ISL_733149, EPI_ISL_733150, EPI_ISL_733152, EPI_ISL_733153, EPI_ISL_733301, EPI_ISL_733302, EPI_ISL_733303, EPI_ISL_733304, EPI_ISL_733305, EPI_ISL_733306, EPI_ISL_733307, EPI_ISL_733308, EPI_ISL_733309, EPI_ISL_733310, EPI_ISL_733311, EPI_ISL_733312, EPI_ISL_733313, EPI_ISL_733314, EPI_ISL_733315, EPI_ISL_733316, EPI_ISL_733317, EPI_ISL_733318, EPI_ISL_733319, EPI_ISL_733320, EPI_ISL_733321, EPI_ISL_733322, EPI_ISL_733323, EPI_ISL_733324, EPI_ISL_733325, EPI_ISL_733326, EPI_ISL_733327, EPI_ISL_733328, EPI_ISL_733329, EPI_ISL_733330, EPI_ISL_733331, EPI_ISL_733332, EPI_ISL_733333, EPI_ISL_733334, EPI_ISL_733335, EPI_ISL_733336, EPI_ISL_733337, EPI_ISL_733338, EPI_ISL_733339, EPI_ISL_733340, EPI_ISL_733341, EPI_ISL_733342, EPI_ISL_733343, EPI_ISL_733344, EPI_ISL_733345, EPI_ISL_733346, EPI_ISL_733347, EPI_ISL_733348, EPI_ISL_733349, EPI_ISL_733350, EPI_ISL_733351, EPI_ISL_733352, EPI_ISL_733353, EPI_ISL_733354, EPI_ISL_733355, EPI_ISL_733356, EPI_ISL_733357, EPI_ISL_733358, EPI_ISL_733359, EPI_ISL_733360, EPI_ISL_733361, EPI_ISL_733362, EPI_ISL_733363, EPI_ISL_733364, EPI_ISL_733365, EPI_ISL_733366, EPI_ISL_733367, EPI_ISL_733368, EPI_ISL_733369, EPI_ISL_733370, EPI_ISL_733371, EPI_ISL_733372, EPI_ISL_733373, EPI_ISL_733374, EPI_ISL_733375, EPI_ISL_733376, EPI_ISL_733377, EPI_ISL_733378, EPI_ISL_733379, EPI_ISL_733380, EPI_ISL_733381, EPI_ISL_733382, EPI_ISL_733383, EPI_ISL_733384, EPI_ISL_733385, EPI_ISL_733386, EPI_ISL_733387, EPI_ISL_733388, EPI_ISL_733389, EPI_ISL_733390, EPI_ISL_733391, EPI_ISL_733396, EPI_ISL_733397, EPI_ISL_733398, EPI_ISL_733399, EPI_ISL_733400, EPI_ISL_733401, EPI_ISL_733417, EPI_ISL_733418, EPI_ISL_733419, EPI_ISL_733420, EPI_ISL_733421, EPI_ISL_733422, EPI_ISL_733423, EPI_ISL_733424, EPI_ISL_733425, EPI_ISL_733426, EPI_ISL_733427, EPI_ISL_733428, EPI_ISL_733429, EPI_ISL_733430, EPI_ISL_733431, EPI_ISL_733432, EPI_ISL_733434, EPI_ISL_733435, EPI_ISL_733436, EPI_ISL_733437, EPI_ISL_733438, EPI_ISL_733439, EPI_ISL_733440, EPI_ISL_733441, EPI_ISL_733442, EPI_ISL_733443, EPI_ISL_733444, EPI_ISL_733445, EPI_ISL_733446, EPI_ISL_733447, EPI_ISL_733448, EPI_ISL_733449, EPI_ISL_733450, EPI_ISL_733451, EPI_ISL_733453, EPI_ISL_733454, EPI_ISL_733455, EPI_ISL_733456, EPI_ISL_733457, EPI_ISL_733458, EPI_ISL_872972, EPI_ISL_872973, EPI_ISL_872974, EPI_ISL_872975, EPI_ISL_872976, EPI_ISL_872977, EPI_ISL_872978, EPI_ISL_872979, EPI_ISL_872980, EPI_ISL_872981, EPI_ISL_872982, EPI_ISL_872983, EPI_ISL_872984, EPI_ISL_872985, EPI_ISL_872986, EPI_ISL_872987, EPI_ISL_872988, EPI_ISL_872989, EPI_ISL_872990, EPI_ISL_872991, EPI_ISL_872992, EPI_ISL_872993, EPI_ISL_872994, EPI_ISL_872995, EPI_ISL_872996, EPI_ISL_872997, EPI_ISL_872998, EPI_ISL_872999, EPI_ISL_873000, EPI_ISL_873001, EPI_ISL_873002, EPI_ISL_873003, EPI_ISL_873004, EPI_ISL_873005, EPI_ISL_873006, EPI_ISL_873007, EPI_ISL_873008, EPI_ISL_873009, EPI_ISL_873010, EPI_ISL_873011, EPI_ISL_873012, EPI_ISL_873013, EPI_ISL_873014, EPI_ISL_873015, EPI_ISL_873016, EPI_ISL_873017, EPI_ISL_873018, EPI_ISL_873019, EPI_ISL_873020, EPI_ISL_873021, EPI_ISL_873022, EPI_ISL_873023, EPI_ISL_873024, EPI_ISL_873025, EPI_ISL_873026, EPI_ISL_873027, EPI_ISL_873028, EPI_ISL_873029, EPI_ISL_873030, EPI_ISL_873031, EPI_ISL_873032, EPI_ISL_873033, EPI_ISL_873034, EPI_ISL_873035, EPI_ISL_873036, EPI_ISL_873037, EPI_ISL_873038, EPI_ISL_873039, EPI_ISL_1039124, EPI_ISL_1039125, EPI_ISL_1039126, EPI_ISL_1039127, EPI_ISL_1039128, EPI_ISL_1039129, EPI_ISL_1039130, EPI_ISL_1039131, EPI_ISL_1039132, EPI_ISL_1039133, EPI_ISL_1039134, EPI_ISL_1039135, EPI_ISL_1039136, EPI_ISL_1039137, EPI_ISL_1372318, EPI_ISL_1372319, EPI_ISL_1372320, EPI_ISL_1372321, EPI_ISL_1372322, EPI_ISL_1372323, EPI_ISL_1372324, EPI_ISL_1372325, EPI_ISL_1372326, EPI_ISL_1372327, EPI_ISL_1372328, EPI_ISL_1372329, EPI_ISL_1372330, EPI_ISL_1372331, EPI_ISL_1372332, EPI_ISL_1372333, EPI_ISL_1372334, EPI_ISL_1372335, EPI_ISL_1372336, EPI_ISL_1372337, EPI_ISL_1372338, EPI_ISL_1372339, EPI_ISL_1372340, EPI_ISL_1372341, EPI_ISL_1372342, EPI_ISL_1372343, EPI_ISL_1372344, EPI_ISL_1372345, EPI_ISL_1372346, EPI_ISL_1372347, EPI_ISL_1372348, EPI_ISL_1372349, EPI_ISL_1372350, EPI_ISL_1372351, EPI_ISL_1372352, EPI_ISL_1372353, EPI_ISL_1372354, EPI_ISL_1372355, EPI_ISL_1372356, EPI_ISL_1372357, EPI_ISL_1372358, EPI_ISL_1372359, EPI_ISL_1372360, EPI_ISL_1372361, EPI_ISL_1372362, EPI_ISL_1372363, EPI_ISL_1372364, EPI_ISL_1372365, EPI_ISL_1372366, EPI_ISL_1372367, EPI_ISL_1372368, EPI_ISL_1372369, EPI_ISL_1372370, EPI_ISL_1372371, EPI_ISL_1372372, EPI_ISL_1372373, EPI_ISL_1372374, EPI_ISL_1372375, EPI_ISL_1372376, EPI_ISL_1372377, EPI_ISL_1372378, EPI_ISL_1372379, EPI_ISL_1372380, EPI_ISL_1372381, EPI_ISL_1372382, EPI_ISL_1372383, EPI_ISL_1372384, EPI_ISL_1372385, EPI_ISL_1372386, EPI_ISL_1372387, EPI_ISL_1372388, EPI_ISL_1372389, EPI_ISL_1372390, EPI_ISL_1372391, EPI_ISL_1372392, EPI_ISL_1372393, EPI_ISL_1372394, EPI_ISL_1372395, EPI_ISL_1372396, EPI_ISL_1372397, EPI_ISL_1372398, EPI_ISL_1372399, EPI_ISL_1400490, EPI_ISL_1400493, EPI_ISL_1400495, EPI_ISL_1400498, EPI_ISL_1400501, EPI_ISL_1400502, EPI_ISL_1400503, EPI_ISL_1400505, EPI_ISL_1400509, EPI_ISL_1400510, EPI_ISL_1400511, EPI_ISL_1400512, EPI_ISL_1400513, EPI_ISL_1400515, EPI_ISL_1400516, EPI_ISL_1400518, EPI_ISL_1400519, EPI_ISL_1400520, EPI_ISL_1400521, EPI_ISL_1400526, EPI_ISL_1400528, EPI_ISL_1400530, EPI_ISL_1400531, EPI_ISL_1400541, EPI_ISL_1400542, EPI_ISL_1400543, EPI_ISL_1400544, EPI_ISL_1400545, EPI_ISL_1400553, EPI_ISL_1400554, EPI_ISL_1400555, EPI_ISL_1400556, EPI_ISL_1400562, EPI_ISL_1400563, EPI_ISL_1400564, EPI_ISL_1400565, EPI_ISL_1400566, EPI_ISL_1400567, EPI_ISL_1400568, EPI_ISL_1400571 | | | |
| see above | HELIX LLC | WHO National Influenza Centre Russian Federation | Andrey Komissarov; Anna Ivanova; Artem Fadeev; Daria Danilenko; Dmitry Bazhenov; Dmitry Lioznov; Elena Nabieva; Georgii Bazykin; Ksenia Safina; Kseniya Komissarova; Maria Pisareva; Maria Timofeeva; Mikhail Bakaev; Tamila Musaeva; Veronika Eder |
| EPI_ISL_872571, EPI_ISL_1039012, EPI_ISL_1039013, EPI_ISL_1039014, EPI_ISL_1039015, EPI_ISL_1039016, EPI_ISL_1039017, EPI_ISL_1039018, EPI_ISL_1039019, EPI_ISL_1039020, EPI_ISL_1039021, EPI_ISL_1039022, EPI_ISL_1039023, EPI_ISL_1039024, EPI_ISL_1039025, EPI_ISL_1039026, EPI_ISL_1039027, EPI_ISL_1039028, EPI_ISL_1039029, EPI_ISL_1039030, EPI_ISL_1039031, EPI_ISL_1039032, EPI_ISL_1039033, EPI_ISL_1039034, EPI_ISL_1039035, EPI_ISL_1039036, EPI_ISL_1039037, EPI_ISL_1039038, EPI_ISL_1039039, EPI_ISL_1039040, EPI_ISL_1039041, EPI_ISL_1039042, EPI_ISL_1039043, EPI_ISL_1039044, EPI_ISL_1039045, EPI_ISL_1039046, EPI_ISL_1039047, EPI_ISL_1039048, EPI_ISL_1039049, EPI_ISL_1039050, EPI_ISL_1039051, EPI_ISL_1039052, EPI_ISL_1039053, EPI_ISL_1039054, EPI_ISL_1039055, EPI_ISL_1039056, EPI_ISL_1400507, EPI_ISL_1400523, EPI_ISL_1400546, EPI_ISL_1400547, EPI_ISL_1400548, EPI_ISL_1400549, EPI_ISL_1400550, EPI_ISL_1400551, EPI_ISL_1400561 | | | |
| see above | Kamchatka Regional Children's Infectious Diseases Hospital | WHO National Influenza Centre Russian Federation | Andrey Komissarov; Anna Ivanova; Artem Fadeev; Daria Danilenko; Dmitry Bazhenov; Dmitry Lioznov; Elena Nabieva; Georgii Bazykin; Ksenia Safina; Kseniya Komissarova; Maria Pisareva; Maria Timofeeva; Mikhail Bakaev; Tamila Musaeva; Veronika Eder |
| EPI_ISL_1859323, EPI_ISL_1859337, EPI_ISL_1859341, EPI_ISL_1859345, EPI_ISL_1859356, EPI_ISL_1859367, EPI_ISL_1859368, EPI_ISL_1859372, EPI_ISL_1859376, EPI_ISL_1859378, EPI_ISL_1859382, EPI_ISL_1859384, EPI_ISL_1859385, EPI_ISL_1859387, EPI_ISL_1859401, EPI_ISL_1859406, EPI_ISL_1859424, EPI_ISL_1859427, EPI_ISL_1859433, EPI_ISL_1859443, EPI_ISL_1859450, EPI_ISL_1859475, EPI_ISL_1859477, EPI_ISL_1859481, EPI_ISL_1859513, EPI_ISL_1859515, EPI_ISL_1859521, EPI_ISL_1859530, EPI_ISL_1859547, EPI_ISL_1859562, EPI_ISL_1859600, EPI_ISL_1859602, EPI_ISL_1859621, EPI_ISL_1859626, EPI_ISL_1859628, EPI_ISL_1859658, EPI_ISL_1859673, EPI_ISL_1859675, EPI_ISL_1859677, EPI_ISL_1859681, EPI_ISL_1859683, EPI_ISL_1859687, EPI_ISL_1859689, EPI_ISL_1859692, EPI_ISL_1859696, EPI_ISL_1859700, EPI_ISL_1859721, EPI_ISL_1859729, EPI_ISL_1859733, EPI_ISL_1859736, EPI_ISL_1859738, EPI_ISL_1859746, EPI_ISL_1859764, EPI_ISL_2086625, EPI_ISL_2086626, EPI_ISL_2086627, EPI_ISL_2086628, EPI_ISL_2086629, EPI_ISL_2086632, EPI_ISL_2086633, EPI_ISL_2086634, EPI_ISL_2086635, EPI_ISL_2086637, EPI_ISL_2086640, EPI_ISL_2086642, EPI_ISL_2086644, EPI_ISL_2086645, EPI_ISL_2086652, EPI_ISL_2086654, EPI_ISL_2086655, EPI_ISL_2086656, EPI_ISL_2086658, EPI_ISL_2086660, EPI_ISL_2086661, EPI_ISL_2086662, EPI_ISL_2086663, EPI_ISL_2086664, EPI_ISL_2086668, EPI_ISL_2086669 | | | |
| see above | Kazan Federal University, Academic Consortium | FEDERAL RESEARCH AND CLINICAL CENTER OF PHYSICAL-CHEMICAL MEDICINE OF FEDERAL MEDICAL BIOLOGICAL AGENCY, Academic Consortium | Alexander Manolov; Alexander Pavlenko; Alexander Rumyantsev; Alexander Samoilov; Alexander Troitsky; Alexander Tulichev; Alexander Vasiliev; Alexey Volnukhin; Alla Petrova; Andrey Koritsky; Anna Blagonravova; Anna Varizhuk; Daria Boldyreva; Daria Kharlampieva; Daria Matyushkina; Dmitry Fedorov; Dmitry Pushkar; Ekaterina Moskaleva; Elena Galova; Elena Ilina; Elizaveta Orlova; Galina Trifonova; Gaukhar Yusubalieva; Ilshat Gafurov; Irina Shirokova; Konstantin Ivanov; Konstantin Kolontarev; Ksenia Klimina; Larisa Balykova; Lyubov Rychkova; Nadezhda Kolyshkina; Natalia Pozhenko; Oleg Ogarkov; Pavel Bashkirov; Polina Khromova; Polina Tikhonova; Ramiz Bakhtyev; Raushaniya Gaifullina; Sergey Rumyantsev; Svetlana Solovieva; Vadim Govorun; |

| | | | |
|---|---|---|---|
| | | | Valentin Manuvera; Varvara Musarova; Vasily Lazarev; Vladimir Baklaushev; Vladimir Rumyantsev; Vladimir Veselovsky; Vladislav Babenko; Vyacheslav Kuropatkin; Vyacheslav Sinkov; Yulia Galeeva; Yuri Udalov |
| EPI_ISL_869247, EPI_ISL_869250, EPI_ISL_869262, EPI_ISL_869263, EPI_ISL_869264, EPI_ISL_872570 | Krasnoyarsk Regional Clinical Hospital | WHO National Influenza Centre Russian Federation | Andrey Komissarov; Anna Ivanova; Artem Fadeev; Daria Danilenko; Dmitry Bazhenov; Dmitry Lioznov; Elena Nabieva; Georgii Bazykin; Ksenia Safina; Kseniya Komissarova; Mikhail Bakaev |
| EPI_ISL_1281568, EPI_ISL_1281569, EPI_ISL_1281571, EPI_ISL_1281573, EPI_ISL_1527390, EPI_ISL_1527391, EPI_ISL_1527392, EPI_ISL_1527393, EPI_ISL_1527394, EPI_ISL_1527395, EPI_ISL_1527396, EPI_ISL_1527397, EPI_ISL_1527398, EPI_ISL_1731069, EPI_ISL_1731070, EPI_ISL_1731071, EPI_ISL_1731072, EPI_ISL_1731073, EPI_ISL_1731074, EPI_ISL_1731075, EPI_ISL_1731076, EPI_ISL_1731077, EPI_ISL_1731078, EPI_ISL_1731079 | | | |
| see above | Laboratory of Extremely Dangerous Infections of Federal Budget Health Care Instute "Center of Hygiene and Epidemiology in Sakhalin region" | Group of Genomics and Postgenomic Technologies of Central Research Institute of Epidemiology | Akimkin VG; Arutiunian AE; Bulanenko VP; Byankina MA; Golubeva AG; Kaptelova VV; Kondrasheva LV; Kondrasheva LY; Korneenko EV; Saenko SS; Samoilov AE; Speranskaya AS; Tivanova EV; Valdokhina AV; Zagorodnov YL |
| EPI_ISL_1510606 | Laboratory of Extremely Dangerous Infections of Federal Budget Health Care Instute "Center of Hygiene and Epidemiology in Sverdlovsk region" | Group of Genomics and Postgenomic Technologies of Central Research Institute of Epidemiology | Akimkin VG; Berlina YY; Bulanenko VP; Cherkashina AS; Durasova AL; Egorova EA; Esaulkova AY; Golubeva AG; Kaptelova VV; Kondrasheva LY; Korneenko EV; Saenko SS; Samoilov AE; Shipulina OY; Snitkovskaya TE; Solovyeva ED; Speranskaya AS; Tivanova EV; Valdokhina AV; Zotova MI |
| EPI_ISL_1363115 | Laboratory of Polymerase Chain Reaction of Federal Budget Institution of Science "Central Research Institute of Epidemiology" of The Federal Service on Customers' Rights Protection and Human Well-being Surveillance | Group of Genomics and Postgenomic Technologies of Central Research Institute of Epidemiology | Akimkin VG; Berlina YY; Bulanenko VP; Cherkashina AS; Golubeva AG; Kaptelova VV; Kondrasheva LY; Korneenko EV; Saenko SS; Samoilov AE; Shipulina OY; Solovyeva ED; Speranskaya AS; Tivanova EV; Valdokhina AV; Zotova MI |
| EPI_ISL_984682, EPI_ISL_1082298, EPI_ISL_1082299, EPI_ISL_1082300, EPI_ISL_1082301, EPI_ISL_1082302, EPI_ISL_1082303, EPI_ISL_1082304, EPI_ISL_1082305, EPI_ISL_1082306, EPI_ISL_1082307, EPI_ISL_1082308, EPI_ISL_1082309, EPI_ISL_1082310, EPI_ISL_1082311, EPI_ISL_1082312, EPI_ISL_1082313, EPI_ISL_1082314, EPI_ISL_1082315, EPI_ISL_1082316, EPI_ISL_1082317, EPI_ISL_1082318, EPI_ISL_1082319, EPI_ISL_1365737 | | | |
| see above | Laboratory of Virology of Federal Budget Health Care Institution "Center of Hygiene and Epidemiology in Arkhangelsk region" | Group of Genomics and Postgenomic Technologies of Central Research Institute of Epidemiology | Akimkin VG; Bulanenko VP; Cherkashina AS; Golubeva AG; Kaptelova VV; Kondrasheva LV; Kondrasheva LY; Korneenko EV; Ponomareva YA; Saenko SS; Samoilov AE; Semyina LV; Shishko LA; Speranskaya AS; Tivanova EV; Valdokhina AV |
| EPI_ISL_984628, EPI_ISL_984629, EPI_ISL_984630, EPI_ISL_984631, EPI_ISL_984632, EPI_ISL_984633, EPI_ISL_984634, EPI_ISL_984635, EPI_ISL_984636, EPI_ISL_984637, EPI_ISL_984638, EPI_ISL_984639, EPI_ISL_984640, EPI_ISL_984641, EPI_ISL_984642, EPI_ISL_984643, EPI_ISL_984644, EPI_ISL_984645, EPI_ISL_984646, EPI_ISL_984647, EPI_ISL_984648, EPI_ISL_984649, EPI_ISL_984650, EPI_ISL_984651, EPI_ISL_984652, EPI_ISL_984653, EPI_ISL_984654, EPI_ISL_984655, EPI_ISL_984656, EPI_ISL_984657, EPI_ISL_984658 | | | |
| see above | Laboratory of Virology of Federal Budget Health Care Institution Center of Hygiene and Epidemiology in Arkhangelsk region | Group of Genomics and Postgenomic Technologies of Central Research Institute of Epidemiology | Akimkin VG; Bulanenko VP; Kaptelova VV; Kondrasheva LV; Korneenko EV; Ponomareva YA; Saenko SS; Samoilov AE; Semyina LV; Shishko LA; Speranskaya AS; Tivanova EV; Valdokhina AV |
| EPI_ISL_1527382, EPI_ISL_1527383, EPI_ISL_1527384, EPI_ISL_1527385, EPI_ISL_1527386, EPI_ISL_1527387, EPI_ISL_1527388, EPI_ISL_1527389 | | | |
| see above | Laboratory of Virology of Federal Budget Health Care Institution "Center of Hygiene and Epidemiology in Arkhangelsk region" | Group of Genomics and Postgenomic Technologies of Central Research Institute of Epidemiology | Akimkin VG; Bulanenko VP; Cherkashina AS; Golubeva AG; Kaptelova VV; Kondrasheva LY; Korneenko EV; Ponomareva YA; Saenko SS; Samoilov AE; Semyina LV; Shishko LA; Speranskaya AS; Tivanova EV; Valdokhina AV |
| EPI_ISL_717965, EPI_ISL_717966, EPI_ISL_717967, EPI_ISL_717968, EPI_ISL_717969, EPI_ISL_717970, EPI_ISL_717971, EPI_ISL_717972, EPI_ISL_717973, EPI_ISL_717974, EPI_ISL_732659, EPI_ISL_732660, EPI_ISL_732661, EPI_ISL_732662, EPI_ISL_732663, EPI_ISL_732664, EPI_ISL_732665, EPI_ISL_732666, EPI_ISL_732667, EPI_ISL_732668, EPI_ISL_732669, EPI_ISL_732670, EPI_ISL_732671, EPI_ISL_732672, EPI_ISL_732673, EPI_ISL_732674, EPI_ISL_732675, EPI_ISL_732676, EPI_ISL_745224, EPI_ISL_745225, EPI_ISL_745226, EPI_ISL_745227, EPI_ISL_745228, EPI_ISL_745229, EPI_ISL_745230, EPI_ISL_745231, EPI_ISL_745232, EPI_ISL_745233, EPI_ISL_745234, EPI_ISL_745235, EPI_ISL_745236, EPI_ISL_745237, EPI_ISL_745238, EPI_ISL_745239, EPI_ISL_745240, EPI_ISL_745241, EPI_ISL_745242, EPI_ISL_745243, EPI_ISL_745244, EPI_ISL_745245, EPI_ISL_745246, EPI_ISL_745247, EPI_ISL_745248, EPI_ISL_745249, EPI_ISL_745250, EPI_ISL_745251, EPI_ISL_745252, EPI_ISL_745253, EPI_ISL_745254, EPI_ISL_745255, EPI_ISL_745256, EPI_ISL_745257, EPI_ISL_745258, EPI_ISL_754196, EPI_ISL_754197, EPI_ISL_754198, EPI_ISL_754199, EPI_ISL_754200, EPI_ISL_754201, EPI_ISL_754202, EPI_ISL_754203, EPI_ISL_754204, EPI_ISL_754205, EPI_ISL_754206, EPI_ISL_754207, EPI_ISL_754208, EPI_ISL_754209, EPI_ISL_754210, EPI_ISL_754211, EPI_ISL_754212, EPI_ISL_754213, EPI_ISL_754214, EPI_ISL_1510607, EPI_ISL_1510616, EPI_ISL_1527364, EPI_ISL_1527365, EPI_ISL_1527366, EPI_ISL_1527367, EPI_ISL_1527368, EPI_ISL_1527369, EPI_ISL_1527370, EPI_ISL_1527371, EPI_ISL_1527372, EPI_ISL_1731067, EPI_ISL_1731068 | | | |
| see above | Molecular diagnostic laboratory of Federal Budget Institution of Science "Central Research Institute of Epidemiology" of The Federal Service on Customers' Rights Protection and Human Well-being Surveillance | Group of Genomics and Postgenomic Technologies of Central Research Institute of Epidemiology | Akimkin VG; Berlina YY; Bulanenko VP; Cherkashina AS; Dudorova AV; Golubeva AG; Kaptelova VV; Kondrasheva LY; Korneenko EV; Saenko SS; Samoilov AE; Shipulina OY; Solovyeva ED; Speranskaya AS; Tivanova EV; Valdokhina AV; Zotova MI |
| EPI_ISL_733483, EPI_ISL_733484, EPI_ISL_733485, EPI_ISL_733486, EPI_ISL_733487, EPI_ISL_733488, EPI_ISL_733489, EPI_ISL_733490, EPI_ISL_733491, EPI_ISL_733492, EPI_ISL_733493 | | | |
| see above | Omsk Research Institute of Natural Focal Infections | WHO National Influenza Centre Russian Federation | Aleksei Vasilenko; Andrey Komissarov; Anna Ivanova; Artem Fadeev; Daria Danilenko; Dmitry Bazhenov; Dmitry Lioznov; Ekaterina Gradoboeva; Elena Nabieva; Elena Poleshchuk; Georgii Bazykin; Ksenia Safina; Kseniya Komissarova; Valery Yakimenko |
| EPI_ISL_872643, EPI_ISL_1710849, EPI_ISL_1710850, EPI_ISL_1710851, EPI_ISL_1710852, EPI_ISL_1710853, EPI_ISL_1710854, EPI_ISL_1710855, EPI_ISL_1710856, EPI_ISL_1710857, EPI_ISL_1710858, EPI_ISL_1710859 | | | |
| see above | Pathogenic Microorganisms Variability Laboratory | Pathogenic Microorganisms Variability Laboratory | Alexander Gintsburg; Alexey Shchetinin; Andrei Siniavin; Andrey Pochtovyy; Anna Ignatieva; Denis Logunov; Elena Burtseva; Elena Shidlovskaya; Elizaveta Divisenko; Evgeniya Mukasheva; Evgeny Usachev; Kirill Krasnoslobotsev; Ludmila Kolobukhina; Maria Nikiforova; Nadezhda Kuznetsova; Olesya Venchakova; Olga Burgasova; Svetlana Smetanina; Svetlana Trushakova; Valeria Bacalin; Vladimir Gushchin |
| EPI_ISL_733185, EPI_ISL_733186, EPI_ISL_733190, EPI_ISL_733191, EPI_ISL_733192, EPI_ISL_733194, EPI_ISL_733203, EPI_ISL_733204, EPI_ISL_733205, EPI_ISL_733206, EPI_ISL_733207, EPI_ISL_733208, EPI_ISL_733214, EPI_ISL_733216, EPI_ISL_733217, EPI_ISL_733218, EPI_ISL_733219, EPI_ISL_733220, EPI_ISL_1039138, EPI_ISL_1039139, EPI_ISL_1039140, EPI_ISL_1039141, EPI_ISL_1039142, EPI_ISL_1039143, EPI_ISL_1039144, EPI_ISL_1039145, EPI_ISL_1039147, EPI_ISL_1039148, EPI_ISL_1039149, EPI_ISL_1039150, EPI_ISL_1039151, EPI_ISL_1039152, EPI_ISL_1039153, EPI_ISL_1039154, EPI_ISL_1039155, EPI_ISL_1372400, EPI_ISL_1372401, EPI_ISL_1372402, EPI_ISL_1372403, EPI_ISL_1372404, EPI_ISL_1372405, EPI_ISL_1372406, EPI_ISL_1372407, EPI_ISL_1372408, EPI_ISL_1372409, EPI_ISL_1400508, EPI_ISL_1400517, EPI_ISL_1400570, EPI_ISL_1491584, EPI_ISL_1491585, EPI_ISL_1491586, EPI_ISL_1491587, EPI_ISL_1491589, EPI_ISL_1491590, EPI_ISL_1491591, EPI_ISL_1491592, EPI_ISL_1491594, EPI_ISL_1491595, EPI_ISL_1491596, EPI_ISL_1491603, EPI_ISL_1491605, EPI_ISL_1491606, EPI_ISL_1491607, EPI_ISL_1491614, EPI_ISL_1491617, EPI_ISL_1491628, EPI_ISL_1491629, EPI_ISL_1491630, EPI_ISL_1491631, EPI_ISL_1491632, EPI_ISL_1491633, EPI_ISL_1491634, EPI_ISL_1491635, EPI_ISL_1491636, EPI_ISL_1491637, EPI_ISL_1491638, EPI_ISL_1491639, EPI_ISL_1491640, EPI_ISL_1491641, EPI_ISL_1491642, EPI_ISL_1491643, EPI_ISL_1491644, EPI_ISL_1491645, EPI_ISL_1491646, EPI_ISL_1491647, EPI_ISL_1491648, EPI_ISL_1491649, EPI_ISL_1491650, EPI_ISL_1491651, EPI_ISL_1491652, EPI_ISL_1491653, EPI_ISL_1491654, EPI_ISL_1491655, EPI_ISL_1491656, EPI_ISL_1491657, EPI_ISL_1491658, EPI_ISL_1491659, EPI_ISL_1491660, EPI_ISL_1491661, EPI_ISL_1491662, EPI_ISL_1491663, EPI_ISL_1491664, EPI_ISL_1491665, EPI_ISL_1491666, EPI_ISL_1491667, EPI_ISL_1491668, EPI_ISL_1491669, EPI_ISL_1491670 | | | |
| see above | Pathogenic Microorganisms Variability Laboratory | WHO National Influenza Centre Russian Federation | Alexander Gintsburg; Alexey Masharsky; Alexey Shchetinin; Andrey Komissarov; Andrey Pochtovyy; Anna Ivanova; Artem Fadeev; Artem Tkachuk; Daria Danilenko; Dmitry Bazhenov; Dmitry Lioznov; Elena Nabieva; Elena Shidlovskaya; Elena Vokalova; Elizaveta Divisenko; Evgeny Usachev; Georgii Bazykin; Ksenia Safina; Kseniya Komissarova; Maria Baturova; Maria Nikiforova; Maxim Rubalsky; Nadezhda Kuznetsova; Oleg Rubalsky; Sergey Alkhovsky; Tatyana Vishnevskaya; Vladimir Gushchin |
| EPI_ISL_661267 | Russian Academy of Sciences, Federal Research Center for Virology and Microbiology | Russian Academy of Sciences, Federal Research Center for Virology and Microbiology | Egorova, I.; Malogolovkin, A.; Nefedeva, M.; Titov, I. |
| EPI_ISL_733478, EPI_ISL_733479, EPI_ISL_733480, EPI_ISL_733481 | SOGAZ PROFMEDICINA LLC | WHO National Influenza Centre Russian Federation | Andrey Komissarov; Anna Ivanova; Artem Fadeev; Daria Danilenko; Dmitry Bazhenov; Dmitry Lioznov; Elena Nabieva; Georgii Bazykin; Ksenia Safina; Kseniya Komissarova |
| EPI_ISL_1731044, EPI_ISL_1731045 | State collection of viruses, N.F. Gamaleya National Research Center for Epidemiology and Microbiology of the Ministry of Health of the Russian Federation | N.F. Gamaleya Research Center for Epidemiology and Microbiology | Andrey Botikov; Anna Kovyrshina; Daria Grousova; Denis Logunov; Ekaterina Aksenova; Inna Dolzhikova; Marina Kunda; Natalia Ryzhova; Olga Voronina |
| EPI_ISL_1708513, EPI_ISL_1708514, EPI_ISL_1708515, EPI_ISL_1708516, EPI_ISL_1708519 | State collection of viruses, N.F. Gamaleya National Research Center for Epidemiology and Microbiology of the Ministry of Health of the Russian Federation | Laboratory of genome analysis, N.F. Gamaleya National Research Center for Epidemiology and Microbiology of the Ministry of Health of the Russian Federation | Andrey Botikov; Anna Kovyrshina; Daria Grousova; Denis Logunov; Ekaterina Aksenova; Inna Dolzhikova; Marina Kunda; Natalia Ryzhova; Olga Voronina |
| EPI_ISL_733228, EPI_ISL_733229, EPI_ISL_733230, EPI_ISL_733231, EPI_ISL_733232, EPI_ISL_733233, EPI_ISL_733234, EPI_ISL_733235, EPI_ISL_733236, EPI_ISL_733237, EPI_ISL_733238, EPI_ISL_733239, EPI_ISL_733240, EPI_ISL_733405, EPI_ISL_733406, EPI_ISL_733407, EPI_ISL_733408, EPI_ISL_733409, EPI_ISL_733410, EPI_ISL_733411, EPI_ISL_733412, EPI_ISL_733413, EPI_ISL_733414, EPI_ISL_733415, EPI_ISL_733416, EPI_ISL_770638, EPI_ISL_770642, EPI_ISL_770647, EPI_ISL_770648, EPI_ISL_770649, EPI_ISL_770650, EPI_ISL_770651, EPI_ISL_770652, EPI_ISL_770654, EPI_ISL_770655, EPI_ISL_770656, EPI_ISL_770657, EPI_ISL_770659, EPI_ISL_770662, EPI_ISL_770663, EPI_ISL_770664, EPI_ISL_770665, EPI_ISL_770667, EPI_ISL_770668, EPI_ISL_770671, EPI_ISL_770672, EPI_ISL_869242, EPI_ISL_869243, EPI_ISL_869244, EPI_ISL_869245, EPI_ISL_869246, EPI_ISL_869248, EPI_ISL_869249, EPI_ISL_869251, EPI_ISL_869252, EPI_ISL_869253, EPI_ISL_869254, EPI_ISL_869255, EPI_ISL_869256, EPI_ISL_869257, EPI_ISL_869258, EPI_ISL_869259, EPI_ISL_869260, EPI_ISL_869261, EPI_ISL_872569 | | | |
| see above | UMMC-Health | WHO National Influenza Centre Russian Federation | Andrey Komissarov; Anna Ivanova; Artem Fadeev; Daria Danilenko; Dmitry Bazhenov; Dmitry Lioznov; Elena Nabieva; Georgii Bazykin; Ksenia Safina; Kseniya Komissarova; Mikhail Bakaev; Tatiana Platonova |
| EPI_ISL_1363082, EPI_ISL_1363083, EPI_ISL_1363084, EPI_ISL_1363085, EPI_ISL_1363086, EPI_ISL_1363087, EPI_ISL_1363088, EPI_ISL_1363089, EPI_ISL_1363090, EPI_ISL_1363091, EPI_ISL_1363092, EPI_ISL_1363093, EPI_ISL_1363094, EPI_ISL_1363095, EPI_ISL_1363096, EPI_ISL_1363097, EPI_ISL_1363098, EPI_ISL_1363099, EPI_ISL_1363100, EPI_ISL_1363101, EPI_ISL_1363102, EPI_ISL_1363103, EPI_ISL_1363104, EPI_ISL_1363105, EPI_ISL_1363106, EPI_ISL_1363107, EPI_ISL_1363108, EPI_ISL_1363109, EPI_ISL_1363110, EPI_ISL_1363111, EPI_ISL_1363112, EPI_ISL_1363113 | | | |
| see above | Virological laboratory FBIH "The Center for Hygiene and Epidemiology in the Khabarovsk Region" | Group of Genomics and Postgenomic Technologies of Central Research Institute of Epidemiology | Akimkin VG; Bulanenko VP; Golubeva AG; Kaptelova VV; Kondrasheva LY; Korneenko EV; Lebedeva LA; Lukyanov AV; Saenko SS; Samoilov AE; Savosina LV; Shubina YA; Speranskaya AS; Tivanova EV; Valdokhina AV; Zhaleyko ZP |
| EPI_ISL_596232, EPI_ISL_596235, EPI_ISL_596236, EPI_ISL_596237, EPI_ISL_596238, EPI_ISL_596239, EPI_ISL_596240, EPI_ISL_596241, EPI_ISL_596242, EPI_ISL_596243, EPI_ISL_596244, EPI_ISL_596245, EPI_ISL_596246, EPI_ISL_596247, EPI_ISL_596248, EPI_ISL_596249, EPI_ISL_602413, EPI_ISL_602414, EPI_ISL_602415, EPI_ISL_602416, EPI_ISL_732974, EPI_ISL_732975, EPI_ISL_732976, EPI_ISL_732977, EPI_ISL_732978, EPI_ISL_732979, EPI_ISL_732980, EPI_ISL_732981, EPI_ISL_732982, EPI_ISL_732983, EPI_ISL_732984, EPI_ISL_732985, EPI_ISL_732986, EPI_ISL_732987, EPI_ISL_732988, EPI_ISL_732989, EPI_ISL_733154, EPI_ISL_733155, EPI_ISL_733183, EPI_ISL_733184, EPI_ISL_733241, EPI_ISL_733242, EPI_ISL_733243, EPI_ISL_733244, EPI_ISL_733245, EPI_ISL_733246, EPI_ISL_733247, EPI_ISL_733248, EPI_ISL_733249, EPI_ISL_733250, EPI_ISL_733251, EPI_ISL_733252, EPI_ISL_733253, EPI_ISL_733254, EPI_ISL_733255, EPI_ISL_733256, EPI_ISL_733257, EPI_ISL_733258, EPI_ISL_733259, EPI_ISL_733260, EPI_ISL_733261, EPI_ISL_733262, EPI_ISL_733263, EPI_ISL_733264, EPI_ISL_733265, EPI_ISL_733266, EPI_ISL_733267, EPI_ISL_733268, EPI_ISL_733269, EPI_ISL_733270, EPI_ISL_733271, EPI_ISL_733272, EPI_ISL_733273, EPI_ISL_733274, EPI_ISL_733275, EPI_ISL_733276, EPI_ISL_733277, EPI_ISL_733278, EPI_ISL_733279, EPI_ISL_733280, EPI_ISL_733281, EPI_ISL_733282, EPI_ISL_733283, EPI_ISL_733284, EPI_ISL_733285, EPI_ISL_733286, EPI_ISL_733287, EPI_ISL_733288, EPI_ISL_733289, EPI_ISL_733290, EPI_ISL_733291, EPI_ISL_733292, EPI_ISL_733293, EPI_ISL_733294, EPI_ISL_733295, EPI_ISL_733296, EPI_ISL_733297, EPI_ISL_733298, EPI_ISL_733299, EPI_ISL_733300, EPI_ISL_733394, EPI_ISL_733395, EPI_ISL_733403, EPI_ISL_733459, EPI_ISL_733460, EPI_ISL_733461, EPI_ISL_733462, EPI_ISL_733463, EPI_ISL_733464, EPI_ISL_733465, EPI_ISL_733466, EPI_ISL_733467, EPI_ISL_733468, EPI_ISL_733469, EPI_ISL_733470, EPI_ISL_733471, EPI_ISL_733472, EPI_ISL_733473, EPI_ISL_733474, EPI_ISL_733475, EPI_ISL_733476, EPI_ISL_733477, EPI_ISL_733494, EPI_ISL_733495, EPI_ISL_770633, EPI_ISL_770634, EPI_ISL_770635, EPI_ISL_770636, EPI_ISL_770637, EPI_ISL_770639, EPI_ISL_770640, EPI_ISL_770641, EPI_ISL_770643, EPI_ISL_770644, EPI_ISL_770645, EPI_ISL_770646, EPI_ISL_770653, EPI_ISL_770658, EPI_ISL_770660, EPI_ISL_770661, EPI_ISL_770666, EPI_ISL_770669, EPI_ISL_770673, EPI_ISL_770674, EPI_ISL_770675, EPI_ISL_770676, EPI_ISL_770677, EPI_ISL_770678, EPI_ISL_770679, EPI_ISL_770680, EPI_ISL_770681, EPI_ISL_770682, EPI_ISL_770683, EPI_ISL_770684, EPI_ISL_770685, EPI_ISL_770686, EPI_ISL_770687, EPI_ISL_811147, EPI_ISL_872898, EPI_ISL_872899, EPI_ISL_872900, EPI_ISL_872901, EPI_ISL_872902, EPI_ISL_872903, EPI_ISL_872904, EPI_ISL_872905, EPI_ISL_872906, EPI_ISL_872907, EPI_ISL_872908, EPI_ISL_872909, EPI_ISL_872910, EPI_ISL_872911, EPI_ISL_872912, EPI_ISL_872913, EPI_ISL_872914, EPI_ISL_872915, EPI_ISL_872916, EPI_ISL_872917, EPI_ISL_872918, EPI_ISL_872919, EPI_ISL_872920, EPI_ISL_872921, EPI_ISL_872922, EPI_ISL_872923, EPI_ISL_872924, EPI_ISL_872925, EPI_ISL_872926, EPI_ISL_872927, EPI_ISL_872928, EPI_ISL_872929, EPI_ISL_872930, EPI_ISL_872931, EPI_ISL_872932, EPI_ISL_872933, EPI_ISL_872934, EPI_ISL_872935, EPI_ISL_872936, EPI_ISL_872937, EPI_ISL_872938, EPI_ISL_872939, EPI_ISL_872940, EPI_ISL_872941, EPI_ISL_872942, EPI_ISL_872943, EPI_ISL_872944, EPI_ISL_872945, EPI_ISL_872946, EPI_ISL_872947, EPI_ISL_872948, EPI_ISL_872949, EPI_ISL_872950, EPI_ISL_872951, EPI_ISL_872952, EPI_ISL_872953, EPI_ISL_872954, EPI_ISL_872955, EPI_ISL_872956, EPI_ISL_872957, EPI_ISL_872958, EPI_ISL_872959, EPI_ISL_872960, EPI_ISL_872961, EPI_ISL_872962, EPI_ISL_872963, EPI_ISL_872964, EPI_ISL_872965, EPI_ISL_872966, EPI_ISL_872967, EPI_ISL_872968, EPI_ISL_872969, EPI_ISL_872970, EPI_ISL_872971, EPI_ISL_873040, EPI_ISL_1039008, EPI_ISL_1039009, EPI_ISL_1039010, EPI_ISL_1039011, EPI_ISL_1039156, EPI_ISL_1039157, EPI_ISL_1039158, EPI_ISL_1039159, EPI_ISL_1372283, EPI_ISL_1372284, EPI_ISL_1372285, EPI_ISL_1372286, EPI_ISL_1372287, EPI_ISL_1372288, EPI_ISL_1372300, EPI_ISL_1372301, EPI_ISL_1372302, EPI_ISL_1372303, EPI_ISL_1372304, EPI_ISL_1372305, EPI_ISL_1372306, EPI_ISL_1372307, EPI_ISL_1372308, EPI_ISL_1372309, EPI_ISL_1372310, EPI_ISL_1372311, EPI_ISL_1372312, EPI_ISL_1372313, EPI_ISL_1372314, EPI_ISL_1372315, EPI_ISL_1372316, EPI_ISL_1372317, EPI_ISL_1400489, EPI_ISL_1400491, EPI_ISL_1400494, EPI_ISL_1400497, EPI_ISL_1400500, EPI_ISL_1400504, EPI_ISL_1400506, EPI_ISL_1400514, EPI_ISL_1400522, EPI_ISL_1400524, EPI_ISL_1400529, EPI_ISL_1400532, EPI_ISL_1400533, EPI_ISL_1400534, EPI_ISL_1400535, EPI_ISL_1400536, EPI_ISL_1400537, EPI_ISL_1400538, EPI_ISL_1400539, EPI_ISL_1400540, EPI_ISL_1400559, EPI_ISL_1400560 | | | |
| see above | WHO National Influenza Centre Russian Federation | WHO National Influenza Centre Russian Federation | Andrey Komissarov; Anna Ivanova; Artem Fadeev; Daria Danilenko; Dmitry Bazhenov; Dmitry Lioznov; Elena Nabieva; Georgii Bazykin; Ksenia Safina; Kseniya Komissarova; Maria Pisareva; Maria Timofeeva; Mariia Sergeeva; Maxim Burlakov; Mikhail Bakaev; Tamila Musaeva; Veronika Eder |

We gratefully acknowledge the following Authors from the Originating laboratories responsible for obtaining the specimens, as well as the Submitting laboratories where the genome data were generated and shared via GISAID, on which this research is based.

All Submitters of data may be contacted directly via www.gisaid.org

Authors are sorted alphabetically.

| Accession ID | Originating Laboratory | Submitting Laboratory | Authors |
|---|---|---|---|
| EPI_ISL_640078, EPI_ISL_640089, EPI_ISL_640107, EPI_ISL_640115, EPI_ISL_640117, EPI_ISL_640127, EPI_ISL_700446, EPI_ISL_700555, EPI_ISL_1040736, EPI_ISL_1040737, EPI_ISL_1040741, EPI_ISL_1040743, EPI_ISL_1040744, EPI_ISL_1040769, EPI_ISL_1040770, EPI_ISL_1040771, EPI_ISL_1040772, EPI_ISL_1040773, EPI_ISL_1040790, EPI_ISL_1040791, EPI_ISL_1040792, EPI_ISL_1534327, EPI_ISL_1534328, EPI_ISL_1534329, EPI_ISL_1534330, EPI_ISL_1534374, EPI_ISL_1534401, EPI_ISL_1534418, EPI_ISL_2140653, EPI_ISL_2140666, EPI_ISL_2140674 | | | |
| see above | 2 Military Hospital wc MAA | NHLS/UCT | Arash Iranzadeh; Bruna Galvao; Carolyn Williamson; Deelan Doolabh; Diana Hardie; Emmanuel SJ; Innocent Mudau; Kruger Marais; Lynn Tyers; Marvin Hsiao; Stephen Korsman; Tegally H; de Oliveira T |
| EPI_ISL_960130, EPI_ISL_960151 | 2 Military Hospital wc MAA | National Health Laboratory Service/UCT | Arash Iranzadeh; Bruna Galvao; Carolyn Williamson; Deelan Doolabh; Diana Hardie; Innocent Mudau; Kruger Marais; Lynn Tyers; Marvin Hsiao; Stephen Korsman |
| EPI_ISL_602622, EPI_ISL_602624, EPI_ISL_602625, EPI_ISL_602626, EPI_ISL_602627, EPI_ISL_602628, EPI_ISL_602629, EPI_ISL_602630, EPI_ISL_602631, EPI_ISL_605784 | | | |
| see above | AHRI-Sigal | KRISP, KZN Research Innovation and Sequencing Platform | Cele S; Gazy I; Giandhari J; Karim F; Pillay S; Sigal A; Sigl A; Sigla; Tegally H; Wilkinson E; de Oliveira T |
| EPI_ISL_1229368 | AHRI-Sigal | KRISP, KZn Research Innovation and Sequencing Platform | Cele S; Gazy I; Giandhari J; Karim F; Pillay S; Sigal A; Tegally H; Wilkinson E; de Oliveira T |
| EPI_ISL_1534336 | Albertinia Clinic wc AAP | NHLS/UCT | Arash Iranzadeh; Bruna Galvao; Carolyn Williamson; Deelan Doolabh; Diana Hardie; Emmanuel SJ; Innocent Mudau; Kruger Marais; Lynn Tyers; Marvin Hsiao; Stephen Korsman; Tegally H; de Oliveira T |
| EPI_ISL_700475, EPI_ISL_700476, EPI_ISL_700521, EPI_ISL_700548, EPI_ISL_1040749, EPI_ISL_1534384 | Alma CDC wc AHC | NHLS/UCT | Arash Iranzadeh; Bruna Galvao; Carolyn Williamson; Deelan Doolabh; Diana Hardie; Emmanuel SJ; Houriiyah Tegally; Innocent Mudau; Kruger Marais; Lynn Tyers; Marvin Hsiao; Stephen Korsman; Tegally H; de Oliveira T |
| EPI_ISL_1534405 | Amalienstein Clinic wc AMS | NHLS/UCT | Arash Iranzadeh; Bruna Galvao; Carolyn Williamson; Deelan Doolabh; Diana Hardie; Emmanuel SJ; Innocent Mudau; Kruger Marais; Lynn Tyers; Marvin Hsiao; Stephen Korsman; Tegally H; de Oliveira T |
| EPI_ISL_660121, EPI_ISL_660122, EPI_ISL_660123, EPI_ISL_660124, EPI_ISL_660125, EPI_ISL_660126, EPI_ISL_660127, EPI_ISL_660128, EPI_ISL_660129 | | | |
| see above | Ampath | National Health Laboratory Service (NHLS), Tygerberg | Bronwyn Kleinhans; Davis M-A; Draper C; Eduan Wilkindon; Gert van Zyl; Houriiyah Tegally; Hsiao M; Kayla Delaney; Siegfried N; Susan Engelbrecht; Tulio de Oliveira; Williamson C; Wolfgang Preiser |
| EPI_ISL_860553 | Ampath-Netcare | KRISP, KZn Research Innovation and Sequencing Platform | Giandhari J; Khan S; Lessells R; Mdlalose K; Pillay S; Tegally H; Wilkinson E; York D; de Oliveira T |
| EPI_ISL_700553 | Beaufort West CDC wc BWC | NHLS/UCT | Arash Iranzadeh; Bruna Galvao; Carolyn Williamson; Deelan Doolabh; Diana Hardie; Innocent Mudau; Kruger Marais; Lynn Tyers; Marvin Hsiao; Stephen Korsman |
| EPI_ISL_640031, EPI_ISL_640047, EPI_ISL_700453 | Beaufort West Hospital wc BWH | NHLS/UCT | Arash Iranzadeh; Bruna Galvao; Carolyn Williamson; Deelan Doolabh; Diana Hardie; Innocent Mudau; Kruger Marais; Lynn Tyers; Marvin Hsiao; Stephen Korsman |
| EPI_ISL_960141 | Beaufort West Hospital wc BWH | National Health Laboratory Service/UCT | Arash Iranzadeh; Bruna Galvao; Carolyn Williamson; Deelan Doolabh; Diana Hardie; Innocent Mudau; Kruger Marais; Lynn Tyers; Marvin Hsiao; Stephen Korsman |
| EPI_ISL_745118, EPI_ISL_745134, EPI_ISL_745158, EPI_ISL_745159, EPI_ISL_745168, EPI_ISL_745169 | Bishop Lavis CDC wc BLP | National Health Laboratory Service (NHLS), Tygerberg | Bronwyn Kleinhans; Eduan Wilkindon; Gert van Zyl; Houriiyah Tegally; Kayla Delaney; Susan Engelbrecht; Tulio de Oliveira; Wolfgang Preiser |
| EPI_ISL_700589 | Bongolethu Clinic wc BLC | NHLS/UCT | Arash Iranzadeh; Bruna Galvao; Carolyn Williamson; Deelan Doolabh; Diana Hardie; Houriiyah Tegally; Innocent Mudau; Kruger Marais; Lynn Tyers; Marvin Hsiao; Stephen Korsman |
| EPI_ISL_640075 | Bothasig CDC wc BLD | NHLS/UCT | Arash Iranzadeh; Bruna Galvao; Carolyn Williamson; Deelan Doolabh; Diana Hardie; Innocent Mudau; Kruger Marais; Lynn Tyers; Marvin Hsiao; Stephen Korsman |
| EPI_ISL_745170, EPI_ISL_745190 | Brackengate Field Hospital COVID-19 wc BRG | National Health Laboratory Service (NHLS), Tygerberg | Bronwyn Kleinhans; Eduan Wilkindon; Gert van Zyl; Houriiyah Tegally; Kayla Delaney; Susan Engelbrecht; Tulio de Oliveira; Wolfgang Preiser |
| EPI_ISL_1534363 | Bredasdorp Clinic wc BDC | NHLS/UCT | Arash Iranzadeh; Bruna Galvao; Carolyn Williamson; Deelan Doolabh; Diana Hardie; Emmanuel SJ; Innocent Mudau; Kruger Marais; Lynn Tyers; Marvin Hsiao; Stephen Korsman; Tegally H; de Oliveira T |
| EPI_ISL_1040762 | Bredasdorp clinic | NHLS/UCT | Arash Iranzadeh; Bruna Galvao; Carolyn Williamson; Deelan Doolabh; Diana Hardie; Innocent Mudau; Kruger Marais; Lynn Tyers; Marvin Hsiao; Stephen Korsman |
| EPI_ISL_640036 | Bridgeton CDC wc BTC | NHLS/UCT | Arash Iranzadeh; Bruna Galvao; Carolyn Williamson; Deelan Doolabh; Diana Hardie; Innocent Mudau; Kruger Marais; Lynn Tyers; Marvin Hsiao; Stephen Korsman |
| EPI_ISL_1534396 | Brooklyn Chest Hospital wc BCH | NHLS/UCT | Arash Iranzadeh; Bruna Galvao; Carolyn Williamson; Deelan Doolabh; Diana Hardie; Emmanuel SJ; Innocent Mudau; Kruger Marais; Lynn Tyers; Marvin Hsiao; Stephen Korsman; Tegally H; de Oliveira T |
| EPI_ISL_2086877, EPI_ISL_2086878, EPI_ISL_2086879, EPI_ISL_2086880, EPI_ISL_2086881, EPI_ISL_2086882, EPI_ISL_2086883, EPI_ISL_2086884 | | | |
| see above | CHARLOTTE MAXEKE JOHANNESBURG ACADEMIC HOSPITAL | National Institute for Communicable Diseases of the National Health Laboratory Service | Amoako DG; Bhiman JN; Ismail A; Mahlangu B; Mohale T; Ntuli N; Scheepers C |
| EPI_ISL_2086904, EPI_ISL_2086905, EPI_ISL_2086906, EPI_ISL_2086907, EPI_ISL_2086908, EPI_ISL_2086910, EPI_ISL_2086911, EPI_ISL_2086912, EPI_ISL_2086913, EPI_ISL_2086915, EPI_ISL_2086921, EPI_ISL_2086922, EPI_ISL_2086923, EPI_ISL_2086924 | | | |
| see above | CHRIS HANI BARAGWANATH LABORATORY | National Institute for Communicable Diseases of the National Health Laboratory Service | Amoako DG; Bhiman JN; Ismail A; Mahlangu B; Mohale T; Ntuli N; Scheepers C |
| EPI_ISL_1534383, EPI_ISL_1534404 | Calitzdorp (Bergsig) Clinic wc CDC | NHLS/UCT | Arash Iranzadeh; Bruna Galvao; Carolyn Williamson; Deelan Doolabh; Diana Hardie; Emmanuel SJ; Innocent Mudau; Kruger Marais; Lynn Tyers; Marvin Hsiao; Stephen Korsman; Tegally H; de Oliveira T |
| EPI_ISL_745121 | Calvinia Hospital | National Health Laboratory Service (NHLS), Tygerberg | Bronwyn Kleinhans; Eduan Wilkindon; Gert van Zyl; Houriiyah Tegally; Kayla Delaney; Susan Engelbrecht; Tulio de Oliveira; Wolfgang Preiser |
| EPI_ISL_940075, EPI_ISL_940076, EPI_ISL_940077, EPI_ISL_940078, EPI_ISL_940079, EPI_ISL_940080, EPI_ISL_940081, EPI_ISL_940082, EPI_ISL_940083, EPI_ISL_940084, EPI_ISL_940085, EPI_ISL_940086, EPI_ISL_940087, EPI_ISL_940088, EPI_ISL_940089, EPI_ISL_940090, EPI_ISL_940091, EPI_ISL_940092, EPI_ISL_940093, EPI_ISL_940094, EPI_ISL_940095, EPI_ISL_940096, EPI_ISL_940097, EPI_ISL_940098, EPI_ISL_940099, EPI_ISL_940100, EPI_ISL_940101, EPI_ISL_940102, EPI_ISL_940103, EPI_ISL_940104, EPI_ISL_940106, EPI_ISL_940107, EPI_ISL_940109, EPI_ISL_940110, EPI_ISL_940111, EPI_ISL_940112, EPI_ISL_940113, EPI_ISL_940114, EPI_ISL_940115, EPI_ISL_940116, EPI_ISL_940117, EPI_ISL_940118, EPI_ISL_940119, EPI_ISL_940120, EPI_ISL_940121, EPI_ISL_940122, EPI_ISL_940123, EPI_ISL_940124, EPI_ISL_940126, EPI_ISL_940127, EPI_ISL_940128, EPI_ISL_940129, EPI_ISL_940130, EPI_ISL_940131, EPI_ISL_940132, EPI_ISL_940133, EPI_ISL_940134, EPI_ISL_940135, EPI_ISL_940136, EPI_ISL_940137, EPI_ISL_1239267, EPI_ISL_1239268, EPI_ISL_1239269, EPI_ISL_1239270, EPI_ISL_1239271, EPI_ISL_1239272, EPI_ISL_1239273, EPI_ISL_1239274, EPI_ISL_1239275, EPI_ISL_1239276, EPI_ISL_1239277, EPI_ISL_1239278, EPI_ISL_1239279, EPI_ISL_1239280, EPI_ISL_1239281, EPI_ISL_1239282, EPI_ISL_1239283, EPI_ISL_1239284, EPI_ISL_1239285, EPI_ISL_1239287, EPI_ISL_1239288, EPI_ISL_1239289, EPI_ISL_1239290, EPI_ISL_1239291, EPI_ISL_1239292, EPI_ISL_1239293, EPI_ISL_1239294, EPI_ISL_1239295, EPI_ISL_1239296, EPI_ISL_1239297, EPI_ISL_1239298, EPI_ISL_1239299, EPI_ISL_1239300, EPI_ISL_1239301, EPI_ISL_1239302, EPI_ISL_1239303, EPI_ISL_1239304, EPI_ISL_1239305, EPI_ISL_1239306, EPI_ISL_1239307, EPI_ISL_1239308, EPI_ISL_1239309, EPI_ISL_1239310, EPI_ISL_1239311, EPI_ISL_1239312, EPI_ISL_1239313, EPI_ISL_1239314, EPI_ISL_1239315, EPI_ISL_1239316, EPI_ISL_1239317, EPI_ISL_1239318, EPI_ISL_1239319, EPI_ISL_1239320, EPI_ISL_1239321, EPI_ISL_1239322, EPI_ISL_1239323, EPI_ISL_1239324, EPI_ISL_1239325, EPI_ISL_1239326, EPI_ISL_1239328, EPI_ISL_1239329 | | | |
| see above | Charlotte Maxeke Johannesburg Academic Hospital, National Health Laboratory Services, Gauteng, South Africa | National Institute for Communicable Diseases of the National Health Laboratory Service | Allam M; Amoako DG; Bhiman JN; Ismail A; Mahlangu B; Mohale T; Ntuli N; Scheepers C |
| EPI_ISL_1239330, EPI_ISL_1239331, EPI_ISL_1239332, EPI_ISL_1239333, EPI_ISL_1239334, EPI_ISL_1239335, EPI_ISL_1239336, EPI_ISL_1239337, EPI_ISL_1239338, EPI_ISL_1239339, EPI_ISL_1239340, EPI_ISL_1239341, EPI_ISL_1239342, EPI_ISL_1239343, EPI_ISL_1239344, EPI_ISL_1239345, EPI_ISL_1239346, EPI_ISL_1239347, EPI_ISL_1239348, EPI_ISL_1239349, EPI_ISL_1239350, EPI_ISL_1239351, EPI_ISL_1239352, EPI_ISL_1239353, EPI_ISL_1239354, EPI_ISL_1239355, EPI_ISL_1239356, EPI_ISL_1239357, EPI_ISL_1239358, EPI_ISL_1239360 | | | |
| see above | Chris Hani Baragwanath Hospital, National Health Laboratory Services, Gauteng, South Africa | National Institute for Communicable Diseases of the National Health Laboratory Service | Amoako DG; Bhiman JN; Ismail A; Mahlangu B; Mohale T; Ntuli N; Scheepers C |
| EPI_ISL_640068, EPI_ISL_700432, EPI_ISL_700519 | Clinic-in-Asla | NHLS/UCT | Arash Iranzadeh; Bruna Galvao; Carolyn Williamson; Deelan Doolabh; Diana Hardie; Houriiyah Tegally; Innocent Mudau; Kruger Marais; Lynn Tyers; Marvin Hsiao; Stephen Korsman |
| EPI_ISL_745128, EPI_ISL_745153 | CoVid EC Nelson Mandela Bay Metro | National Health Laboratory Service (NHLS), Tygerberg | Bronwyn Kleinhans; Eduan Wilkindon; Gert van Zyl; Houriiyah Tegally; Kayla Delaney; Susan Engelbrecht; Tulio de Oliveira; Wolfgang Preiser |
| EPI_ISL_1534340, EPI_ISL_1534355, EPI_ISL_1534395 | CoVid WC Garden Route | NHLS/UCT | Arash Iranzadeh; Bruna Galvao; Carolyn Williamson; Deelan Doolabh; Diana Hardie; Emmanuel SJ; Innocent Mudau; Kruger Marais; Lynn Tyers; Marvin Hsiao; Stephen Korsman; Tegally H; de Oliveira T |
| EPI_ISL_640015, EPI_ISL_640118, EPI_ISL_700414, EPI_ISL_700441, EPI_ISL_700467, EPI_ISL_700488, EPI_ISL_700492, EPI_ISL_700496, EPI_ISL_700504, EPI_ISL_700506, EPI_ISL_700533, EPI_ISL_700536, EPI_ISL_700538, EPI_ISL_700539, EPI_ISL_700554, EPI_ISL_700556, EPI_ISL_700557, EPI_ISL_700563, EPI_ISL_700581, EPI_ISL_700587, EPI_ISL_700590, EPI_ISL_700599, EPI_ISL_1534334, EPI_ISL_1534399, EPI_ISL_2140657, EPI_ISL_2140658, EPI_ISL_2140660, EPI_ISL_2140663 | | | |
| see above | Conville CDC wc CVC | NHLS/UCT | Arash Iranzadeh; Bruna Galvao; Carolyn Williamson; Deelan Doolabh; Diana Hardie; Emmanuel SJ; Houriiyah Tegally; Innocent Mudau; Kruger Marais; Lynn Tyers; Marvin Hsiao; Stephen Korsman; Tegally H; de Oliveira T |
| EPI_ISL_960119, EPI_ISL_960122, EPI_ISL_960127, EPI_ISL_960134 | Conville CDC wc CVC | National Health Laboratory Service/UCT | Arash Iranzadeh; Bruna Galvao; Carolyn Williamson; Deelan Doolabh; Diana Hardie; Innocent Mudau; Kruger Marais; Lynn Tyers; Marvin Hsiao; Stephen Korsman |
| EPI_ISL_696472, EPI_ISL_696492, EPI_ISL_696508, EPI_ISL_696509, EPI_ISL_696512 | Conville CDC wc CVC & NHLS/UCT | KRISP, KZN Research Innovation and Sequencing Platform | Arash Iranzadeh; Bruna Galvao; Carolyn Williamson; Deelan Doolabh; Diana Hardie; Emanuel James San; Houriiyah Tegally; Innocent Mudau; Jennifer Giandhari; Kruger Marais; Lynn Tyers; Marvin Hsiao; Stephen Korsman; Sureshnee Pillay; Tulio de Oliveira |
| EPI_ISL_700443, EPI_ISL_1534338, EPI_ISL_1534419 | Crags Clinic wc CRG | NHLS/UCT | Arash Iranzadeh; Bruna Galvao; Carolyn Williamson; Deelan Doolabh; Diana Hardie; Emmanuel SJ; Houriiyah Tegally; Innocent Mudau; Kruger Marais; Lynn Tyers; Marvin Hsiao; Stephen Korsman; Tegally H; de Oliveira T |
| EPI_ISL_696480 | Crags Clinic wc CRG & NHLS/UCT | KRISP, KZN Research Innovation and Sequencing Platform | Arash Iranzadeh; Bruna Galvao; Carolyn Williamson; Deelan Doolabh; Diana Hardie; Emanuel James San; Houriiyah Tegally; Innocent Mudau; Jennifer Giandhari; Kruger Marais; Lynn Tyers; Marvin Hsiao; Stephen Korsman; Sureshnee Pillay; Tulio de Oliveira |
| EPI_ISL_640029, EPI_ISL_640048, EPI_ISL_700420, EPI_ISL_700427, EPI_ISL_700448, EPI_ISL_700470, EPI_ISL_700564, EPI_ISL_700569, EPI_ISL_1534460 | | | |
| see above | D'Almeida Clinic wc DAL | NHLS/UCT | Arash Iranzadeh; Bruna Galvao; Carolyn Williamson; Deelan Doolabh; Diana Hardie; Emmanuel SJ; Houriiyah Tegally; Innocent Mudau; Kruger Marais; Lynn Tyers; Marvin Hsiao; Stephen Korsman; Tegally H; de Oliveira T |
| EPI_ISL_1534421 | DP Marais Hospital wc DPM | NHLS/UCT | Arash Iranzadeh; Bruna Galvao; Carolyn Williamson; Deelan Doolabh; Diana Hardie; Emmanuel SJ; Innocent Mudau; Kruger Marais; Lynn Tyers; Marvin Hsiao; Stephen Korsman; Tegally H; de Oliveira T |
| EPI_ISL_745171, EPI_ISL_745172 | Delft CHC wc DFP | National Health Laboratory Service (NHLS), Tygerberg | Bronwyn Kleinhans; Eduan Wilkindon; Gert van Zyl; Houriiyah Tegally; Kayla Delaney; Susan Engelbrecht; Tulio de Oliveira; Wolfgang Preiser |
| EPI_ISL_1502129, EPI_ISL_1502130, EPI_ISL_1502131, EPI_ISL_1502132, EPI_ISL_1502133, EPI_ISL_1502134, EPI_ISL_1502135, EPI_ISL_1502136, EPI_ISL_1502137, EPI_ISL_1502138, EPI_ISL_1502139, EPI_ISL_1502140, EPI_ISL_1502141, EPI_ISL_1502142, EPI_ISL_1502143, EPI_ISL_1502144, EPI_ISL_1502145, EPI_ISL_1591352, EPI_ISL_1591353, EPI_ISL_1591354, EPI_ISL_1591355, EPI_ISL_1591356, EPI_ISL_1591357, EPI_ISL_1591358, EPI_ISL_1591359, EPI_ISL_1591360, EPI_ISL_1591361, EPI_ISL_1591362, EPI_ISL_1591363, EPI_ISL_1591364, EPI_ISL_1591365, EPI_ISL_1591366, EPI_ISL_1591367, EPI_ISL_1591368, EPI_ISL_1591369, EPI_ISL_1591370, EPI_ISL_1591371, EPI_ISL_1591372, EPI_ISL_1591373, EPI_ISL_1591374, EPI_ISL_1591375, EPI_ISL_1591376, EPI_ISL_1591383, EPI_ISL_1591384, EPI_ISL_1591385, EPI_ISL_1591386, EPI_ISL_1591449, EPI_ISL_1608099, EPI_ISL_1608100, EPI_ISL_1608101 | | | |
| see above | Division of Medical Virology, National Health Laboratory Service (NHLS), Tygerberg | Division of Medical Virology, Stellenbosch University and | Bronwyn Kleinhans; Gert van Zyl; Susan Engelbrecht; Wolfgang Preiser |

| | | | |
|---|---|---|---|
| | Hospital / Stellenbosch University | NHLS Tygerberg Hospital | |
| EPI_ISL_960160 | Dr Abdurahman CDC wc DAC | National Health Laboratory Service/UCT | Arash Iranzadeh; Bruna Galvao; Carolyn Williamson; Deelan Doolabh; Diana Hardie; Innocent Mudau; Kruger Marais; Lynn Tyers; Marvin Hsiao; Stephen Korsman |
| EPI_ISL_745113, EPI_ISL_745132, EPI_ISL_745154 | Dr Ivan Toms Clinic wc IVT | National Health Laboratory Service (NHLS), Tygerberg | Bronwyn Kleinhans; Eduan Wilkindon; Gert van Zyl; Houriiyah Tegally; Kayla Delaney; Susan Engelbrecht; Tulio de Oliveira; Wolfgang Preiser |
| EPI_ISL_745112, EPI_ISL_745138, EPI_ISL_745139 | Dr Van Niekerk Hospital | National Health Laboratory Service (NHLS), Tygerberg | Bronwyn Kleinhans; Eduan Wilkindon; Gert van Zyl; Houriiyah Tegally; Kayla Delaney; Susan Engelbrecht; Tulio de Oliveira; Wolfgang Preiser |
| EPI_ISL_640091, EPI_ISL_640094, EPI_ISL_640095 | Du Noon CDC wc DNC | NHLS/UCT | Arash Iranzadeh; Bruna Galvao; Carolyn Williamson; Deelan Doolabh; Diana Hardie; Innocent Mudau; Kruger Marais; Lynn Tyers; Marvin Hsiao; Stephen Korsman |
| EPI_ISL_640024, EPI_ISL_700520, EPI_ISL_1534387 | Dysselsdorp Clinic wc DDC | NHLS/UCT | Arash Iranzadeh; Bruna Galvao; Carolyn Williamson; Deelan Doolabh; Diana Hardie; Emmanuel SJ; Houriiyah Tegally; Innocent Mudau; Kruger Marais; Lynn Tyers; Marvin Hsiao; Stephen Korsman; Tegally H; de Oliveira T |
| EPI_ISL_960096 | Dysselsdorp Clinic wc DDC | National Health Laboratory Service/UCT | Arash Iranzadeh; Bruna Galvao; Carolyn Williamson; Deelan Doolabh; Diana Hardie; Innocent Mudau; Kruger Marais; Lynn Tyers; Marvin Hsiao; Stephen Korsman |
| EPI_ISL_712077, EPI_ISL_712078, EPI_ISL_712092 | East London NHLS Laboratory, Eastern Cape, South Africa | National Institute for Communicable Diseases of the National Health Laboratory Service | Allam M; Bhiman JN; Ismail A; Mahlangu B; Mohale T; Ntuli N |
| EPI_ISL_745137, EPI_ISL_745151 | Edameni Clinic | National Health Laboratory Service (NHLS), Tygerberg | Bronwyn Kleinhans; Eduan Wilkindon; Gert van Zyl; Houriiyah Tegally; Kayla Delaney; Susan Engelbrecht; Tulio de Oliveira; Wolfgang Preiser |
| EPI_ISL_745119, EPI_ISL_745173, EPI_ISL_745174, EPI_ISL_745175, EPI_ISL_745176, EPI_ISL_745177, EPI_ISL_745178 | | | |
| see above | Eerste River Hospital wc ERH | National Health Laboratory Service (NHLS), Tygerberg | Bronwyn Kleinhans; Eduan Wilkindon; Gert van Zyl; Houriiyah Tegally; Kayla Delaney; Susan Engelbrecht; Tulio de Oliveira; Wolfgang Preiser |
| EPI_ISL_745110, EPI_ISL_745125, EPI_ISL_745162 | Elsies River CHC wc ERP | National Health Laboratory Service (NHLS), Tygerberg | Bronwyn Kleinhans; Eduan Wilkindon; Gert van Zyl; Houriiyah Tegally; Kayla Delaney; Susan Engelbrecht; Tulio de Oliveira; Wolfgang Preiser |
| EPI_ISL_2112529 | Epidemiology of Microbial Diseases, Yale School of Public Health | Epidemiology of Microbial Diseases, Yale School of Public Health | A.E.; A.F.; Alpert, T.; Breban; Brito; C.B.; C.C.; Fauver; Grubaugh; I.M.; Iwasaki, A.; J.E.; J.R.; Kalinich; Landry; Lucas, C.; M.E.; M.I.; M.L.; N.D.; Ott; Petrone; Rothman; Vogels; Watkins |
| EPI_ISL_640064 | Ethembeni Clinic [Prieska] | NHLS/UCT | Arash Iranzadeh; Bruna Galvao; Carolyn Williamson; Deelan Doolabh; Diana Hardie; Innocent Mudau; Kruger Marais; Lynn Tyers; Marvin Hsiao; Stephen Korsman |
| EPI_ISL_640033, EPI_ISL_640088, EPI_ISL_640090, EPI_ISL_640116, EPI_ISL_640121, EPI_ISL_640123, EPI_ISL_640124, EPI_ISL_640125, EPI_ISL_1040681, EPI_ISL_1040682, EPI_ISL_1040739, EPI_ISL_1040740, EPI_ISL_1040775, EPI_ISL_1040777, EPI_ISL_1040779, EPI_ISL_1040780, EPI_ISL_1534331, EPI_ISL_1534380, EPI_ISL_1534400, EPI_ISL_1817684, EPI_ISL_2140652, EPI_ISL_2140671, EPI_ISL_2140672 | | | |
| see above | False Bay Hospital wc FBH | NHLS/UCT | Arash Iranzadeh; Bruna Galvao; Carolyn Williamson; Deelan Doolabh; Diana Hardie; Emmanuel SJ; Innocent Mudau; Kruger Marais; Lynn Tyers; Marvin Hsiao; Stephen Korsman; Tegally H; de Oliveira T |
| EPI_ISL_960105, EPI_ISL_960110, EPI_ISL_960112 | False Bay Hospital wc FBH | National Health Laboratory Service/UCT | Arash Iranzadeh; Bruna Galvao; Carolyn Williamson; Deelan Doolabh; Diana Hardie; Innocent Mudau; Kruger Marais; Lynn Tyers; Marvin Hsiao; Stephen Korsman |
| EPI_ISL_640016, EPI_ISL_640027, EPI_ISL_700497, EPI_ISL_1534341, EPI_ISL_1534423, EPI_ISL_2140655 | George Hospital wc GRH | NHLS/UCT | Arash Iranzadeh; Bruna Galvao; Carolyn Williamson; Deelan Doolabh; Diana Hardie; Emmanuel SJ; Innocent Mudau; Kruger Marais; Lynn Tyers; Marvin Hsiao; Stephen Korsman; Tegally H; de Oliveira T |
| EPI_ISL_960118 | George Hospital wc GRH | National Health Laboratory Service/UCT | Arash Iranzadeh; Bruna Galvao; Carolyn Williamson; Deelan Doolabh; Diana Hardie; Innocent Mudau; Kruger Marais; Lynn Tyers; Marvin Hsiao; Stephen Korsman |
| EPI_ISL_696458, EPI_ISL_696471, EPI_ISL_696473, EPI_ISL_696481, EPI_ISL_696503 | George Hospital wc GRH & NHLS/UCT | KRISP, KZN Research Innovation and Sequencing Platform | Arash Iranzadeh; Bruna Galvao; Carolyn Williamson; Deelan Doolabh; Diana Hardie; Emanuel James San; Houriiyah Tegally; Innocent Mudau; Jennifer Giandhari; Kruger Marais; Lynn Tyers; Marvin Hsiao; Stephen Korsman; Sureshnee Pillay; Tulio de Oliveira |
| EPI_ISL_640049 | George Road Sat Clinic wc GWM | NHLS/UCT | Arash Iranzadeh; Bruna Galvao; Carolyn Williamson; Deelan Doolabh; Diana Hardie; Innocent Mudau; Kruger Marais; Lynn Tyers; Marvin Hsiao; Stephen Korsman |
| EPI_ISL_700543, EPI_ISL_700550 | Great Brak River Clinic wc GBC | NHLS/UCT | Arash Iranzadeh; Bruna Galvao; Carolyn Williamson; Deelan Doolabh; Diana Hardie; Houriiyah Tegally; Innocent Mudau; Kruger Marais; Lynn Tyers; Marvin Hsiao; Stephen Korsman |
| EPI_ISL_960149 | Great Brak River Clinic wc GBC | National Health Laboratory Service/UCT | Arash Iranzadeh; Bruna Galvao; Carolyn Williamson; Deelan Doolabh; Diana Hardie; Innocent Mudau; Kruger Marais; Lynn Tyers; Marvin Hsiao; Stephen Korsman |
| EPI_ISL_696466, EPI_ISL_696497, EPI_ISL_696501, EPI_ISL_696507 | Great Brak River Clinic wc GBC & NHLS/UCT | KRISP, KZN Research Innovation and Sequencing Platform | Arash Iranzadeh; Bruna Galvao; Carolyn Williamson; Deelan Doolabh; Diana Hardie; Emanuel James San; Houriiyah Tegally; Innocent Mudau; Jennifer Giandhari; Kruger Marais; Lynn Tyers; Marvin Hsiao; Stephen Korsman; Sureshnee Pillay; Tulio de Oliveira |
| EPI_ISL_640066, EPI_ISL_640067, EPI_ISL_640084, EPI_ISL_640111, EPI_ISL_640112, EPI_ISL_640119, EPI_ISL_640120, EPI_ISL_640122, EPI_ISL_640128, EPI_ISL_640129, EPI_ISL_640130, EPI_ISL_640131, EPI_ISL_640132, EPI_ISL_640133, EPI_ISL_640134, EPI_ISL_640135, EPI_ISL_640137, EPI_ISL_640138, EPI_ISL_640139, EPI_ISL_640140, EPI_ISL_700493, EPI_ISL_700507, EPI_ISL_700534, EPI_ISL_700577, EPI_ISL_1040644, EPI_ISL_1040645, EPI_ISL_1040646, EPI_ISL_1040648, EPI_ISL_1040649, EPI_ISL_1040650, EPI_ISL_1040651, EPI_ISL_1040652, EPI_ISL_1040653, EPI_ISL_1040654, EPI_ISL_1040656, EPI_ISL_1040657, EPI_ISL_1040658, EPI_ISL_1040659, EPI_ISL_1040660, EPI_ISL_1040661, EPI_ISL_1040663, EPI_ISL_1040668, EPI_ISL_1040672, EPI_ISL_1040673, EPI_ISL_1040676, EPI_ISL_1040683, EPI_ISL_1040684, EPI_ISL_1040685, EPI_ISL_1040686, EPI_ISL_1040687, EPI_ISL_1040689, EPI_ISL_1040690, EPI_ISL_1040691, EPI_ISL_1040692, EPI_ISL_1040693, EPI_ISL_1040696, EPI_ISL_1040697, EPI_ISL_1040698, EPI_ISL_1040699, EPI_ISL_1040700, EPI_ISL_1040704, EPI_ISL_1040705, EPI_ISL_1040706, EPI_ISL_1040707, EPI_ISL_1040708, EPI_ISL_1040711, EPI_ISL_1040714, EPI_ISL_1040716, EPI_ISL_1040720, EPI_ISL_1040722, EPI_ISL_1040723, EPI_ISL_1040725, EPI_ISL_1040726, EPI_ISL_1040727, EPI_ISL_1040728, EPI_ISL_1040729, EPI_ISL_1040731, EPI_ISL_1040733, EPI_ISL_1040735, EPI_ISL_1040738, EPI_ISL_1040745, EPI_ISL_1040747, EPI_ISL_1040748, EPI_ISL_1040750, EPI_ISL_1040751, EPI_ISL_1040752, EPI_ISL_1040753, EPI_ISL_1040754, EPI_ISL_1040755, EPI_ISL_1040756, EPI_ISL_1040758, EPI_ISL_1040759, EPI_ISL_1040763, EPI_ISL_1040765, EPI_ISL_1040767, EPI_ISL_1040768, EPI_ISL_1040781, EPI_ISL_1040782, EPI_ISL_1040783, EPI_ISL_1040785, EPI_ISL_1040786, EPI_ISL_1040787, EPI_ISL_1040789, EPI_ISL_1040795, EPI_ISL_1040796, EPI_ISL_1040800, EPI_ISL_1040801, EPI_ISL_1040802, EPI_ISL_1040804, EPI_ISL_1040807, EPI_ISL_1040808, EPI_ISL_1040811, EPI_ISL_1040812, EPI_ISL_1040813, EPI_ISL_1040815, EPI_ISL_1534313, EPI_ISL_1534315, EPI_ISL_1534316, EPI_ISL_1534317, EPI_ISL_1534318, EPI_ISL_1534319, EPI_ISL_1534320, EPI_ISL_1534322, EPI_ISL_1534323, EPI_ISL_1534324, EPI_ISL_1534325, EPI_ISL_1534326, EPI_ISL_1534345, EPI_ISL_1534346, EPI_ISL_1534347, EPI_ISL_1534348, EPI_ISL_1534349, EPI_ISL_1534350, EPI_ISL_1534351, EPI_ISL_1534352, EPI_ISL_1534362, EPI_ISL_1534376, EPI_ISL_1534389, EPI_ISL_1534390, EPI_ISL_1534391, EPI_ISL_1534392, EPI_ISL_1534393, EPI_ISL_1534394, EPI_ISL_1534411, EPI_ISL_1534412, EPI_ISL_1534413, EPI_ISL_1534414, EPI_ISL_1534415, EPI_ISL_1534416, EPI_ISL_1534455, EPI_ISL_1534457, EPI_ISL_1706553, EPI_ISL_1706554, EPI_ISL_1706555, EPI_ISL_1706556, EPI_ISL_1706557, EPI_ISL_1706558, EPI_ISL_1706559, EPI_ISL_1706560, EPI_ISL_1706561, EPI_ISL_1706570, EPI_ISL_1706571, EPI_ISL_1706576, EPI_ISL_1817557, EPI_ISL_1817668, EPI_ISL_1817683, EPI_ISL_1817686, EPI_ISL_1817750, EPI_ISL_1817751, EPI_ISL_1817753, EPI_ISL_1817754, EPI_ISL_2140675, EPI_ISL_2140676, EPI_ISL_2140677, EPI_ISL_2140678, EPI_ISL_2140679, EPI_ISL_2140680, EPI_ISL_2140681, EPI_ISL_2140683, EPI_ISL_2140684, EPI_ISL_2140685, EPI_ISL_2140686, EPI_ISL_2140687, EPI_ISL_2140688, EPI_ISL_2140689, EPI_ISL_2140690 | | | |
| see above | Groote Schuur Hospital wc GSH | NHLS/UCT | Arash Iranzadeh; Bruna Galvao; Carolyn Williamson; Deelan Doolabh; Diana Hardie; Emmanuel SJ; Innocent Mudau; Kruger Marais; Lynn Tyers; Marvin Hsiao; Stephen Korsman; Tegally H; de Oliveira T |
| EPI_ISL_960102, EPI_ISL_960103, EPI_ISL_960107, EPI_ISL_960109, EPI_ISL_960117, EPI_ISL_960126, EPI_ISL_960132, EPI_ISL_960133, EPI_ISL_960135, EPI_ISL_960136, EPI_ISL_960140, EPI_ISL_960142, EPI_ISL_960143, EPI_ISL_960145 | | | |
| see above | Groote Schuur Hospital wc GSH | National Health Laboratory Service/UCT | Arash Iranzadeh; Bruna Galvao; Carolyn Williamson; Deelan Doolabh; Diana Hardie; Innocent Mudau; Kruger Marais; Lynn Tyers; Marvin Hsiao; Stephen Korsman |
| EPI_ISL_696464 | Groote Schuur Hospital wc GSH & NHLS/UCT | KRISP, KZN Research Innovation and Sequencing Platform | Arash Iranzadeh; Bruna Galvao; Carolyn Williamson; Deelan Doolabh; Diana Hardie; Emanuel James San; Houriiyah Tegally; Innocent Mudau; Jennifer Giandhari; Kruger Marais; Lynn Tyers; Marvin Hsiao; Stephen Korsman; Sureshnee Pillay; Tulio de Oliveira |
| EPI_ISL_1040761 | Guguletu CHC | NHLS/UCT | Arash Iranzadeh; Bruna Galvao; Carolyn Williamson; Deelan Doolabh; Diana Hardie; Innocent Mudau; Kruger Marais; Lynn Tyers; Marvin Hsiao; Stephen Korsman |
| EPI_ISL_640069, EPI_ISL_640070, EPI_ISL_640071, EPI_ISL_1040666, EPI_ISL_1040667, EPI_ISL_1040669, EPI_ISL_1040671, EPI_ISL_1040675, EPI_ISL_1040678, EPI_ISL_1040710, EPI_ISL_1040713, EPI_ISL_1040715, EPI_ISL_1040718, EPI_ISL_1040721, EPI_ISL_1040724, EPI_ISL_1040732, EPI_ISL_1040734, EPI_ISL_1040793, EPI_ISL_1040805, EPI_ISL_1534397 | | | |
| see above | Guguletu CHC wc GDH | NHLS/UCT | Arash Iranzadeh; Bruna Galvao; Carolyn Williamson; Deelan Doolabh; Diana Hardie; Emmanuel SJ; Innocent Mudau; Kruger Marais; Lynn Tyers; Marvin Hsiao; Stephen Korsman; Tegally H; de Oliveira T |
| EPI_ISL_960154, EPI_ISL_960155, EPI_ISL_960156, EPI_ISL_960157, EPI_ISL_960158 | Guguletu CHC wc GDH | National Health Laboratory Service/UCT | Arash Iranzadeh; Bruna Galvao; Carolyn Williamson; Deelan Doolabh; Diana Hardie; Innocent Mudau; Kruger Marais; Lynn Tyers; Marvin Hsiao; Stephen Korsman |
| EPI_ISL_660137, EPI_ISL_660138, EPI_ISL_660139, EPI_ISL_660141, EPI_ISL_660142 | Hamadi | National Health Laboratory Service (NHLS), Tygerberg | Bronwyn Kleinhans; Davis M-A; Draper C; Eduan Wilkindon; Gert van Zyl; Houriiyah Tegally; Hsiao M; Kayla Delaney; Siegfried N; Susan Engelbrecht; Tulio de Oliveira; Williamson C; Wolfgang Preiser |
| EPI_ISL_1040694, EPI_ISL_1040695, EPI_ISL_1040717, EPI_ISL_1040730, EPI_ISL_1040764, EPI_ISL_1040797, EPI_ISL_1040809, EPI_ISL_1534314, EPI_ISL_1534437 | | | |
| see above | Hanover Park CHC wc HPH | NHLS/UCT | Arash Iranzadeh; Bruna Galvao; Carolyn Williamson; Deelan Doolabh; Diana Hardie; Emmanuel SJ; Innocent Mudau; Kruger Marais; Lynn Tyers; Marvin Hsiao; Stephen Korsman; Tegally H; de Oliveira T |
| EPI_ISL_640018, EPI_ISL_1534357, EPI_ISL_1534398 | Heidelberg Clinic wc HBC | NHLS/UCT | Arash Iranzadeh; Bruna Galvao; Carolyn Williamson; Deelan Doolabh; Diana Hardie; Emmanuel SJ; Innocent Mudau; Kruger Marais; Lynn Tyers; Marvin Hsiao; Stephen Korsman; Tegally H; de Oliveira T |
| EPI_ISL_1040719, EPI_ISL_1534402 | Heideveld CDC wc HVP | NHLS/UCT | Arash Iranzadeh; Bruna Galvao; Carolyn Williamson; Deelan Doolabh; Diana Hardie; Emmanuel SJ; Innocent Mudau; Kruger Marais; Lynn Tyers; Marvin Hsiao; Stephen Korsman; Tegally H; de Oliveira T |
| EPI_ISL_700445, EPI_ISL_700559, EPI_ISL_1040662, EPI_ISL_1040665, EPI_ISL_1040670, EPI_ISL_1040680, EPI_ISL_1040688, EPI_ISL_1040757, EPI_ISL_1040798, EPI_ISL_1040803, EPI_ISL_1040806, EPI_ISL_1040810, EPI_ISL_1534359 | | | |
| see above | Heideveld Emergency Centre | NHLS/UCT | Arash Iranzadeh; Bruna Galvao; Carolyn Williamson; Deelan Doolabh; Diana Hardie; Emmanuel SJ; Houriiyah Tegally; Innocent Mudau; Kruger Marais; Lynn Tyers; Marvin Hsiao; Stephen Korsman; Tegally H; de Oliveira T |
| EPI_ISL_960124, EPI_ISL_960153 | Heideveld Emergency Centre | National Health Laboratory Service/UCT | Arash Iranzadeh; Bruna Galvao; Carolyn Williamson; Deelan Doolabh; Diana Hardie; Innocent Mudau; Kruger Marais; Lynn Tyers; Marvin Hsiao; Stephen Korsman |
| EPI_ISL_700565, EPI_ISL_700571 | Herbertsdale Sat Clinic wc HBD | NHLS/UCT | Arash Iranzadeh; Bruna Galvao; Carolyn Williamson; Deelan Doolabh; Diana Hardie; Innocent Mudau; Kruger Marais; Lynn Tyers; Marvin Hsiao; Stephen Korsman |
| EPI_ISL_1534358 | Hillside Clinic wc BFW | NHLS/UCT | Arash Iranzadeh; Bruna Galvao; Carolyn Williamson; Deelan Doolabh; Diana Hardie; Emmanuel SJ; Innocent Mudau; Kruger Marais; Lynn Tyers; Marvin Hsiao; Stephen Korsman; Tegally H; de Oliveira T |
| EPI_ISL_700458, EPI_ISL_700524, EPI_ISL_700558 | Hornlee Clinic wc HLC | NHLS/UCT | Arash Iranzadeh; Bruna Galvao; Carolyn Williamson; Deelan Doolabh; Diana Hardie; Houriiyah Tegally; Innocent Mudau; Kruger Marais; Lynn Tyers; Marvin Hsiao; Stephen Korsman |
| EPI_ISL_696457, EPI_ISL_696479, EPI_ISL_696483, EPI_ISL_696486, EPI_ISL_696502 | Hornlee Clinic wc HLC & NHLS/UCT | KRISP, KZN Research Innovation and Sequencing Platform | Arash Iranzadeh; Bruna Galvao; Carolyn Williamson; Deelan Doolabh; Diana Hardie; Emanuel James San; Houriiyah Tegally; Innocent Mudau; Jennifer Giandhari; Kruger Marais; Lynn Tyers; Marvin Hsiao; Stephen Korsman; Sureshnee Pillay; Tulio de Oliveira |
| EPI_ISL_745135, EPI_ISL_745144 | Joe Slovo Hospital | National Health Laboratory Service (NHLS), Tygerberg | Bronwyn Kleinhans; Eduan Wilkindon; Gert van Zyl; Houriiyah Tegally; Kayla Delaney; Susan Engelbrecht; Tulio de Oliveira; Wolfgang Preiser |
| EPI_ISL_660221, EPI_ISL_660222, EPI_ISL_660228, EPI_ISL_660229, EPI_ISL_660230, EPI_ISL_660231, EPI_ISL_660232, EPI_ISL_660233, EPI_ISL_660256, EPI_ISL_660257, EPI_ISL_660258, EPI_ISL_660664 | | | |
| see above | KRISP, KZN Research Innovation and Sequencing Platform | KRISP, KZN Research Innovation and Sequencing Platform | Giandhari J; Khan S; Lessells R; Mdlalose K; Pillay S; Tegally H; Wilkinson E; York D; de Oliveira T |
| EPI_ISL_1040760 | Kensington CDC | NHLS/UCT | Arash Iranzadeh; Bruna Galvao; Carolyn Williamson; Deelan Doolabh; Diana Hardie; Innocent Mudau; Kruger Marais; Lynn Tyers; Marvin Hsiao; Stephen Korsman |
| EPI_ISL_640092, EPI_ISL_640093 | Kensington CDC wc KSC | NHLS/UCT | Arash Iranzadeh; Bruna Galvao; Carolyn Williamson; Deelan Doolabh; Diana Hardie; Innocent Mudau; Kruger Marais; Lynn Tyers; Marvin Hsiao; Stephen Korsman |
| EPI_ISL_696511 | Keurhoek Clinic wc KTT & NHLS/UCT | KRISP, KZN Research Innovation and Sequencing Platform | Arash Iranzadeh; Bruna Galvao; Carolyn Williamson; Deelan Doolabh; Diana Hardie; Emanuel James San; Houriiyah Tegally; Innocent Mudau; Jennifer Giandhari; Kruger Marais; Lynn Tyers; Marvin Hsiao; Stephen Korsman; Sureshnee Pillay; Tulio de Oliveira |
| EPI_ISL_700423, EPI_ISL_700464, EPI_ISL_700468, EPI_ISL_700485, EPI_ISL_700505, EPI_ISL_700540 | Khayelethu Clinic wc KLC | NHLS/UCT | Arash Iranzadeh; Bruna Galvao; Carolyn Williamson; Deelan Doolabh; Diana Hardie; Houriiyah Tegally; Innocent Mudau; Kruger Marais; Lynn Tyers; Marvin Hsiao; Stephen Korsman |
| EPI_ISL_696484, EPI_ISL_696505 | Khayelethu Clinic wc KLC & NHLS/UCT | KRISP, KZN Research Innovation and Sequencing Platform | Arash Iranzadeh; Bruna Galvao; Carolyn Williamson; Deelan Doolabh; Diana Hardie; Emanuel James San; Houriiyah Tegally; Innocent Mudau; Jennifer Giandhari; Kruger Marais; Lynn Tyers; Marvin Hsiao; Stephen Korsman; Sureshnee Pillay; Tulio de Oliveira |
| EPI_ISL_745111, EPI_ISL_745127, EPI_ISL_745163 | Khayelitsha (Site B) CHC wc KHC | National Health Laboratory Service (NHLS), Tygerberg | Bronwyn Kleinhans; Eduan Wilkindon; Gert van Zyl; Houriiyah Tegally; Kayla Delaney; Susan Engelbrecht; Tulio de Oliveira; Wolfgang Preiser |
| EPI_ISL_745114, EPI_ISL_745115, EPI_ISL_745116, EPI_ISL_745120, EPI_ISL_745122, EPI_ISL_745123, EPI_ISL_745145, EPI_ISL_745166 | | | |
| see above | Khayelitsha Hospital wc KHA | National Health Laboratory Service (NHLS), Tygerberg | Bronwyn Kleinhans; Eduan Wilkindon; Gert van Zyl; Houriiyah Tegally; Kayla Delaney; Susan Engelbrecht; Tulio de Oliveira; Wolfgang Preiser |

| | | | |
|---|---|---|---|
| EPI_ISL_1239225, EPI_ISL_1239226, EPI_ISL_1239227, EPI_ISL_1239228, EPI_ISL_1239229, EPI_ISL_1239230, EPI_ISL_1239231, EPI_ISL_1239232, EPI_ISL_1239233, EPI_ISL_1239234, EPI_ISL_1239235, EPI_ISL_1239236, EPI_ISL_1239237, EPI_ISL_1239238, EPI_ISL_1239239, EPI_ISL_1239240, EPI_ISL_1239241, EPI_ISL_1239242, EPI_ISL_1239243, EPI_ISL_1239244, EPI_ISL_1239245, EPI_ISL_1239246, EPI_ISL_1239247, EPI_ISL_1239248, EPI_ISL_1239249, EPI_ISL_1239250, EPI_ISL_1239251, EPI_ISL_1239252, EPI_ISL_1239253, EPI_ISL_1239254, EPI_ISL_1239255, EPI_ISL_1239256, EPI_ISL_1239257, EPI_ISL_1239258, EPI_ISL_1239259, EPI_ISL_1239260, EPI_ISL_1239261, EPI_ISL_1239262, EPI_ISL_1239263, EPI_ISL_1239264, EPI_ISL_1239265, EPI_ISL_1239266, EPI_ISL_1239458, EPI_ISL_1239460, EPI_ISL_1239461, EPI_ISL_1239463, EPI_ISL_1239466, EPI_ISL_1239467, EPI_ISL_1239468, EPI_ISL_1239471, EPI_ISL_1239474, EPI_ISL_1239475, EPI_ISL_1239476, EPI_ISL_1239477, EPI_ISL_1239478, EPI_ISL_1239480, EPI_ISL_1239481, EPI_ISL_1239483, EPI_ISL_1239485, EPI_ISL_1239486, EPI_ISL_1239488, EPI_ISL_1239489, EPI_ISL_1239508, EPI_ISL_1239512, EPI_ISL_1239518, EPI_ISL_1239519, EPI_ISL_1239520, EPI_ISL_1239522, EPI_ISL_1239527, EPI_ISL_1239529, EPI_ISL_1239531 | | | |
| see above | Kimberley Hospital, National Health Laboratory Services, Northern Cape, South Africa | National Institute for Communicable Diseases of the National Health Laboratory Service | Amoako DG; Bhiman JN; Ismail A; Mahlangu B; Mohale T; Ntuli N; Scheepers C |
| EPI_ISL_700433, EPI_ISL_700436, EPI_ISL_700440, EPI_ISL_700450, EPI_ISL_700469, EPI_ISL_700474, EPI_ISL_700526, EPI_ISL_700529, EPI_ISL_700531, EPI_ISL_700542, EPI_ISL_1534360 | | | |
| see above | Knysna CDC wc WLC | NHLS/UCT | Arash Iranzadeh; Bruna Galvao; Carolyn Williamson; Deelan Doolabh; Diana Hardie; Emmanuel SJ; Houriiyah Tegally; Innocent Mudau; Kruger Marais; Lynn Tyers; Marvin Hsiao; Stephen Korsman; Tegally H; de Oliveira T |
| EPI_ISL_696467, EPI_ISL_696495, EPI_ISL_696506 | Knysna CDC wc WLC & NHLS/UCT | KRISP, KZN Research Innovation and Sequencing Platform | Arash Iranzadeh; Bruna Galvao; Carolyn Williamson; Deelan Doolabh; Diana Hardie; Emanuel James San; Houriiyah Tegally; Innocent Mudau; Jennifer Giandhari; Kruger Marais; Lynn Tyers; Marvin Hsiao; Stephen Korsman; Sureshnee Pillay; Tulio de Oliveira |
| EPI_ISL_640022, EPI_ISL_640076, EPI_ISL_700419, EPI_ISL_700422, EPI_ISL_700426, EPI_ISL_700429, EPI_ISL_700456, EPI_ISL_700484, EPI_ISL_700501, EPI_ISL_700509, EPI_ISL_700510, EPI_ISL_700583, EPI_ISL_1534388, EPI_ISL_1534403 | | | |
| see above | Knysna Hospital wc KNY | NHLS/UCT | Arash Iranzadeh; Bruna Galvao; Carolyn Williamson; Deelan Doolabh; Diana Hardie; Emmanuel SJ; Houriiyah Tegally; Innocent Mudau; Kruger Marais; Lynn Tyers; Marvin Hsiao; Stephen Korsman; Tegally H; de Oliveira T |
| EPI_ISL_960120 | Knysna Hospital wc KNY | National Health Laboratory Service/UCT | Arash Iranzadeh; Bruna Galvao; Carolyn Williamson; Deelan Doolabh; Diana Hardie; Innocent Mudau; Kruger Marais; Lynn Tyers; Marvin Hsiao; Stephen Korsman |
| EPI_ISL_696463, EPI_ISL_696469, EPI_ISL_696477, EPI_ISL_696478, EPI_ISL_696482, EPI_ISL_696491 | Knysna Hospital wc KNY & NHLS/UCT | KRISP, KZN Research Innovation and Sequencing Platform | Arash Iranzadeh; Bruna Galvao; Carolyn Williamson; Deelan Doolabh; Diana Hardie; Emanuel James San; Houriiyah Tegally; Innocent Mudau; Jennifer Giandhari; Kruger Marais; Lynn Tyers; Marvin Hsiao; Stephen Korsman; Sureshnee Pillay; Tulio de Oliveira |
| EPI_ISL_745109, EPI_ISL_745131, EPI_ISL_745133, EPI_ISL_745136, EPI_ISL_745164, EPI_ISL_745179 | Kraaifontein CHC wc KFP | National Health Laboratory Service (NHLS), Tygerberg | Bronwyn Kleinhans; Eduan Wilkindon; Gert van Zyl; Houriiyah Tegally; Kayla Delaney; Susan Engelbrecht; Tulio de Oliveira; Wolfgang Preiser |
| EPI_ISL_2140670 | Kranshoek Clinic wc KSH | NHLS/UCT | Arash Iranzadeh; Bruna Galvao; Carolyn Williamson; Deelan Doolabh; Diana Hardie; Innocent Mudau; Kruger Marais; Lynn Tyers; Marvin Hsiao; Stephen Korsman |
| EPI_ISL_696460, EPI_ISL_696498 | Kranshoek Clinic wc KSH & NHLS/UCT | KRISP, KZN Research Innovation and Sequencing Platform | Arash Iranzadeh; Bruna Galvao; Carolyn Williamson; Deelan Doolabh; Diana Hardie; Emanuel James San; Houriiyah Tegally; Innocent Mudau; Jennifer Giandhari; Kruger Marais; Lynn Tyers; Marvin Hsiao; Stephen Korsman; Sureshnee Pillay; Tulio de Oliveira |
| EPI_ISL_640030, EPI_ISL_700532, EPI_ISL_1040674 | Kwamandlenkosi Clinic wc KMN | NHLS/UCT | Arash Iranzadeh; Bruna Galvao; Carolyn Williamson; Deelan Doolabh; Diana Hardie; Innocent Mudau; Kruger Marais; Lynn Tyers; Marvin Hsiao; Stephen Korsman |
| EPI_ISL_700437, EPI_ISL_700495, EPI_ISL_700535, EPI_ISL_700537, EPI_ISL_700541, EPI_ISL_700579, EPI_ISL_2140659, EPI_ISL_2140673 | | | |
| see above | Kwanokuthula CDC wc KWA | NHLS/UCT | Arash Iranzadeh; Bruna Galvao; Carolyn Williamson; Deelan Doolabh; Diana Hardie; Houriiyah Tegally; Innocent Mudau; Kruger Marais; Lynn Tyers; Marvin Hsiao; Stephen Korsman |
| EPI_ISL_960123 | Kwanokuthula CDC wc KWA | National Health Laboratory Service/UCT | Arash Iranzadeh; Bruna Galvao; Carolyn Williamson; Deelan Doolabh; Diana Hardie; Innocent Mudau; Kruger Marais; Lynn Tyers; Marvin Hsiao; Stephen Korsman |
| EPI_ISL_696465 | Kwanokuthula CDC wc KWA & NHLS/UCT | KRISP, KZN Research Innovation and Sequencing Platform | Arash Iranzadeh; Bruna Galvao; Carolyn Williamson; Deelan Doolabh; Diana Hardie; Emanuel James San; Houriiyah Tegally; Innocent Mudau; Jennifer Giandhari; Kruger Marais; Lynn Tyers; Marvin Hsiao; Stephen Korsman; Sureshnee Pillay; Tulio de Oliveira |
| EPI_ISL_700466 | Ladismith (Nissenville) Clinic wc LAF | NHLS/UCT | Arash Iranzadeh; Bruna Galvao; Carolyn Williamson; Deelan Doolabh; Diana Hardie; Innocent Mudau; Kruger Marais; Lynn Tyers; Marvin Hsiao; Stephen Korsman |
| EPI_ISL_745124, EPI_ISL_745152 | Laetitia Bam CHC | National Health Laboratory Service (NHLS), Tygerberg | Bronwyn Kleinhans; Eduan Wilkindon; Gert van Zyl; Houriiyah Tegally; Kayla Delaney; Susan Engelbrecht; Tulio de Oliveira; Wolfgang Preiser |
| EPI_ISL_660130, EPI_ISL_660131, EPI_ISL_660132, EPI_ISL_660133, EPI_ISL_660134, EPI_ISL_660135, EPI_ISL_660136 | | | |
| see above | Lancet | National Health Laboratory Service (NHLS), Tygerberg | Bronwyn Kleinhans; Davis M-A; Draper C; Eduan Wilkindon; Gert van Zyl; Houriiyah Tegally; Hsiao M; Kayla Delaney; Siegfried N; Susan Engelbrecht; Tulio de Oliveira; Williamson C; Wolfgang Preiser |
| EPI_ISL_622943, EPI_ISL_622946, EPI_ISL_622947, EPI_ISL_622948, EPI_ISL_622949, EPI_ISL_622950, EPI_ISL_622951, EPI_ISL_622952, EPI_ISL_622953, EPI_ISL_622955, EPI_ISL_622957, EPI_ISL_622959, EPI_ISL_622960, EPI_ISL_622961, EPI_ISL_622962, EPI_ISL_622963, EPI_ISL_622964, EPI_ISL_622965, EPI_ISL_622966, EPI_ISL_622967, EPI_ISL_622968, EPI_ISL_622969, EPI_ISL_622978, EPI_ISL_622983, EPI_ISL_622985, EPI_ISL_622986, EPI_ISL_622988, EPI_ISL_622990, EPI_ISL_622994, EPI_ISL_623064, EPI_ISL_623068, EPI_ISL_623073 | | | |
| see above | Lancet Laboratories | National Institute for Communicable Diseases of the National Health Laboratory Service | Allam M; Bhiman JN; Ismail A; Khumalo Z; Kwenda S; Mnyameni F; Mohale T; Mtshali P; Subramoney K |
| EPI_ISL_745130, EPI_ISL_745155, EPI_ISL_745160 | Langebaan Clinic wc LBC | National Health Laboratory Service (NHLS), Tygerberg | Bronwyn Kleinhans; Eduan Wilkindon; Gert van Zyl; Houriiyah Tegally; Kayla Delaney; Susan Engelbrecht; Tulio de Oliveira; Wolfgang Preiser |
| EPI_ISL_1040702, EPI_ISL_1040709 | Lentegeur Hospital wc LGH | NHLS/UCT | Arash Iranzadeh; Bruna Galvao; Carolyn Williamson; Deelan Doolabh; Diana Hardie; Innocent Mudau; Kruger Marais; Lynn Tyers; Marvin Hsiao; Stephen Korsman |
| EPI_ISL_736927, EPI_ISL_736930, EPI_ISL_736966, EPI_ISL_736967, EPI_ISL_736968, EPI_ISL_736995 | MDS | KRISP, KZN Research Innovation and Sequencing Platform | ChimukangaraB; Giandhari J; Khan S; Lessells R; Mdlalose K; Pillay S; Tegally H; Wilkinson E; York D; de Oliveira T |
| EPI_ISL_640074 | Mamre CDC wc MRC | NHLS/UCT | Arash Iranzadeh; Bruna Galvao; Carolyn Williamson; Deelan Doolabh; Diana Hardie; Innocent Mudau; Kruger Marais; Lynn Tyers; Marvin Hsiao; Stephen Korsman |
| EPI_ISL_495535, EPI_ISL_495536, EPI_ISL_495537, EPI_ISL_495538, EPI_ISL_495539, EPI_ISL_495540, EPI_ISL_495541, EPI_ISL_495542, EPI_ISL_515800, EPI_ISL_515801, EPI_ISL_515802, EPI_ISL_515803, EPI_ISL_515804, EPI_ISL_515805, EPI_ISL_515806, EPI_ISL_515807, EPI_ISL_515808, EPI_ISL_515809, EPI_ISL_515810, EPI_ISL_515811, EPI_ISL_515812, EPI_ISL_515813, EPI_ISL_515814, EPI_ISL_515815, EPI_ISL_515816, EPI_ISL_515817, EPI_ISL_515818, EPI_ISL_515819, EPI_ISL_515820, EPI_ISL_515821, EPI_ISL_515822, EPI_ISL_515823, EPI_ISL_515824, EPI_ISL_515825, EPI_ISL_515826, EPI_ISL_515827, EPI_ISL_515828, EPI_ISL_515829, EPI_ISL_515830, EPI_ISL_515831, EPI_ISL_515832, EPI_ISL_515833, EPI_ISL_515834, EPI_ISL_515835, EPI_ISL_515836, EPI_ISL_515837, EPI_ISL_515838, EPI_ISL_515839, EPI_ISL_515840, EPI_ISL_515841, EPI_ISL_515842, EPI_ISL_515843, EPI_ISL_515844, EPI_ISL_515845, EPI_ISL_515846, EPI_ISL_515847, EPI_ISL_515848, EPI_ISL_515849, EPI_ISL_515850, EPI_ISL_515851, EPI_ISL_515852, EPI_ISL_515853, EPI_ISL_515854, EPI_ISL_515855, EPI_ISL_515856, EPI_ISL_515857, EPI_ISL_515858, EPI_ISL_515859, EPI_ISL_515860, EPI_ISL_515861, EPI_ISL_515862, EPI_ISL_515863, EPI_ISL_515864, EPI_ISL_515865, EPI_ISL_515866, EPI_ISL_515867, EPI_ISL_515868, EPI_ISL_515869, EPI_ISL_515870, EPI_ISL_515871, EPI_ISL_515872, EPI_ISL_515873, EPI_ISL_515874, EPI_ISL_515875, EPI_ISL_515876, EPI_ISL_515877, EPI_ISL_515878, EPI_ISL_515879, EPI_ISL_515880, EPI_ISL_515881, EPI_ISL_515882, EPI_ISL_515883, EPI_ISL_515884, EPI_ISL_515885, EPI_ISL_515886, EPI_ISL_515887, EPI_ISL_515888, EPI_ISL_515889, EPI_ISL_515890, EPI_ISL_515891, EPI_ISL_515892, EPI_ISL_515893 | | | |
| see above | Medical Disagnostics Services (MDS) | KRISP, KZN Research Innovation and Sequencing Platform | Chimukangara B; ChimukangaraB; Giandhari J; Khan S; Lessells R; Mdlalose K; Pillay S; Tegally H; Wilkinson E; York D; de Oliveira T |
| EPI_ISL_640025 | Mitchells Plain CHC wc MHC | NHLS/UCT | Arash Iranzadeh; Bruna Galvao; Carolyn Williamson; Deelan Doolabh; Diana Hardie; Innocent Mudau; Kruger Marais; Lynn Tyers; Marvin Hsiao; Stephen Korsman |
| EPI_ISL_640020, EPI_ISL_640028, EPI_ISL_640065, EPI_ISL_700489, EPI_ISL_700572, EPI_ISL_700597, EPI_ISL_1040766, EPI_ISL_1040794, EPI_ISL_1534321, EPI_ISL_1534426, EPI_ISL_1534427, EPI_ISL_2140656, EPI_ISL_2140664, EPI_ISL_2140682 | | | |
| see above | Mitchells Plain Hospital wc MPH | NHLS/UCT | Arash Iranzadeh; Bruna Galvao; Carolyn Williamson; Deelan Doolabh; Diana Hardie; Emmanuel SJ; Innocent Mudau; Kruger Marais; Lynn Tyers; Marvin Hsiao; Stephen Korsman; Tegally H; de Oliveira T |
| EPI_ISL_960095, EPI_ISL_960097, EPI_ISL_960098, EPI_ISL_960100, EPI_ISL_960121, EPI_ISL_960125, EPI_ISL_960128, EPI_ISL_960129, EPI_ISL_960137, EPI_ISL_960148 | | | |
| see above | Mitchells Plain Hospital wc MPH | National Health Laboratory Service/UCT | Arash Iranzadeh; Bruna Galvao; Carolyn Williamson; Deelan Doolabh; Diana Hardie; Innocent Mudau; Kruger Marais; Lynn Tyers; Marvin Hsiao; Stephen Korsman |
| EPI_ISL_660259, EPI_ISL_660260, EPI_ISL_660261, EPI_ISL_660262, EPI_ISL_660263 | Molecualr Diagnostic Services (MDS) | KRISP, KZN Research Innovation and Sequencing Platform | Giandhari J; Khan S; Lessells R; Mdlalose K; Pillay S; Tegally H; Wilkinson E; York D; de Oliveira T |
| EPI_ISL_487329, EPI_ISL_487330, EPI_ISL_487331, EPI_ISL_487332, EPI_ISL_487333, EPI_ISL_487334, EPI_ISL_487335, EPI_ISL_487336, EPI_ISL_487337, EPI_ISL_487338, EPI_ISL_487339, EPI_ISL_487340, EPI_ISL_487341, EPI_ISL_1250507, EPI_ISL_1250509, EPI_ISL_1250512, EPI_ISL_1250514, EPI_ISL_1250515, EPI_ISL_1250517, EPI_ISL_1250518, EPI_ISL_1250520, EPI_ISL_1250522, EPI_ISL_1250523, EPI_ISL_1250525, EPI_ISL_1250526, EPI_ISL_1250528, EPI_ISL_1250529, EPI_ISL_1250531, EPI_ISL_1250533, EPI_ISL_1250535, EPI_ISL_1250537, EPI_ISL_1250538, EPI_ISL_1250540, EPI_ISL_1250542, EPI_ISL_1250543, EPI_ISL_1250545, EPI_ISL_1250547, EPI_ISL_1250548, EPI_ISL_1250550, EPI_ISL_1250551, EPI_ISL_1250552 | | | |
| see above | Molecular Diagnostics Services (MDS) | KRISP, KZN Research Innovation and Sequencing Platform | Chimukangara B; Emmanuel SJ; Giandhari J; Khan S; Lessells R; Maslo C; Mdlalose K; Pillay S; Sitharam L; Tegally H; Wilkinson E; York D; de Oliveira T |
| EPI_ISL_700479, EPI_ISL_1040701, EPI_ISL_1040712 | Mowbray Maternity Hospital wc MMH | NHLS/UCT | Arash Iranzadeh; Bruna Galvao; Carolyn Williamson; Deelan Doolabh; Diana Hardie; Innocent Mudau; Kruger Marais; Lynn Tyers; Marvin Hsiao; Stephen Korsman |
| EPI_ISL_960104 | Mowbray Maternity Hospital wc MMH | National Health Laboratory Service/UCT | Arash Iranzadeh; Bruna Galvao; Carolyn Williamson; Deelan Doolabh; Diana Hardie; Innocent Mudau; Kruger Marais; Lynn Tyers; Marvin Hsiao; Stephen Korsman |
| EPI_ISL_682323, EPI_ISL_682324, EPI_ISL_682325, EPI_ISL_682326, EPI_ISL_682327, EPI_ISL_682328, EPI_ISL_682329, EPI_ISL_682330, EPI_ISL_682331, EPI_ISL_682332, EPI_ISL_682333, EPI_ISL_682334, EPI_ISL_682335, EPI_ISL_682336, EPI_ISL_682337, EPI_ISL_682338, EPI_ISL_682339, EPI_ISL_682340, EPI_ISL_682341, EPI_ISL_682342, EPI_ISL_682343, EPI_ISL_682344, EPI_ISL_682345, EPI_ISL_682346, EPI_ISL_682347, EPI_ISL_682348, EPI_ISL_682349, EPI_ISL_682350, EPI_ISL_682351, EPI_ISL_682352, EPI_ISL_684047, EPI_ISL_912456, EPI_ISL_912457, EPI_ISL_912458, EPI_ISL_912459, EPI_ISL_912460, EPI_ISL_912461, EPI_ISL_912462, EPI_ISL_912463, EPI_ISL_912464, EPI_ISL_912465, EPI_ISL_912466, EPI_ISL_912467, EPI_ISL_912468, EPI_ISL_912469, EPI_ISL_912470, EPI_ISL_912471, EPI_ISL_912472, EPI_ISL_912473, EPI_ISL_912474, EPI_ISL_912475, EPI_ISL_912476, EPI_ISL_912477, EPI_ISL_912478, EPI_ISL_912479, EPI_ISL_912480, EPI_ISL_912481, EPI_ISL_912482, EPI_ISL_912483, EPI_ISL_912484, EPI_ISL_912485, EPI_ISL_912486, EPI_ISL_912487, EPI_ISL_912488, EPI_ISL_912489, EPI_ISL_912490, EPI_ISL_912491, EPI_ISL_912492, EPI_ISL_912493, EPI_ISL_912494, EPI_ISL_912495, EPI_ISL_912496, EPI_ISL_912497, EPI_ISL_912498, EPI_ISL_912499, EPI_ISL_912500, EPI_ISL_912501, EPI_ISL_912502, EPI_ISL_912503, EPI_ISL_912504, EPI_ISL_912505, EPI_ISL_912506, EPI_ISL_912507, EPI_ISL_912508, EPI_ISL_912509, EPI_ISL_912510, EPI_ISL_912511, EPI_ISL_912512, EPI_ISL_912513, EPI_ISL_912514, EPI_ISL_912515, EPI_ISL_912516, EPI_ISL_912517, EPI_ISL_912518, EPI_ISL_912519, EPI_ISL_912520, EPI_ISL_912521, EPI_ISL_912522, EPI_ISL_912523, EPI_ISL_912524, EPI_ISL_912525, EPI_ISL_912526, EPI_ISL_912527, EPI_ISL_912528, EPI_ISL_912529, EPI_ISL_912530, EPI_ISL_912531, EPI_ISL_912532, EPI_ISL_912533, EPI_ISL_912534, EPI_ISL_912535, EPI_ISL_912536, EPI_ISL_912537, EPI_ISL_912538, EPI_ISL_940147, EPI_ISL_940148, EPI_ISL_940149, EPI_ISL_940150, EPI_ISL_940151, EPI_ISL_940152, EPI_ISL_940153, EPI_ISL_940154, EPI_ISL_940155, EPI_ISL_940156, EPI_ISL_949618, EPI_ISL_949620, EPI_ISL_949622, EPI_ISL_949624, EPI_ISL_949626, EPI_ISL_949628, EPI_ISL_949630, EPI_ISL_949632, EPI_ISL_949634, EPI_ISL_1007984, EPI_ISL_1007985, EPI_ISL_1007986, EPI_ISL_1007987, EPI_ISL_1007988, EPI_ISL_1007989, EPI_ISL_1007990, EPI_ISL_1007991, EPI_ISL_1272300, EPI_ISL_1272301, EPI_ISL_1272302, EPI_ISL_1272303, EPI_ISL_1272304, EPI_ISL_1272305, EPI_ISL_1272306, EPI_ISL_1272307, EPI_ISL_1818892, EPI_ISL_1818893, EPI_ISL_1818894, EPI_ISL_1818895 | | | |
| see above | NHLS Universitas Academic | UFS Virology | D Goedhals; Emmanuel Ogunbayo; MM Nyaga; MT Mogotsi; P Nthiga; PA Bester; T de Oliveira |
| EPI_ISL_487277, EPI_ISL_487278, EPI_ISL_487279, EPI_ISL_487280, EPI_ISL_487281, EPI_ISL_487282, EPI_ISL_487283, EPI_ISL_487284, EPI_ISL_487285, EPI_ISL_487286, EPI_ISL_487287, EPI_ISL_487288, EPI_ISL_487289, EPI_ISL_487291, EPI_ISL_487292, EPI_ISL_487293, EPI_ISL_487294, EPI_ISL_487295, EPI_ISL_487298, EPI_ISL_487299, EPI_ISL_487300, EPI_ISL_487301, EPI_ISL_487302, EPI_ISL_487303, EPI_ISL_487313, EPI_ISL_487316, EPI_ISL_487318, EPI_ISL_487320, EPI_ISL_487321, EPI_ISL_487325, EPI_ISL_487328, EPI_ISL_487348, EPI_ISL_495516, EPI_ISL_495523, EPI_ISL_495524, EPI_ISL_495525, EPI_ISL_495526, EPI_ISL_495529, EPI_ISL_495532, EPI_ISL_495543, EPI_ISL_495544, EPI_ISL_495545, EPI_ISL_495546, EPI_ISL_495547, EPI_ISL_495548, EPI_ISL_495549, EPI_ISL_495550, EPI_ISL_495551, EPI_ISL_495552, EPI_ISL_495553, EPI_ISL_495554, EPI_ISL_495555, EPI_ISL_495556, EPI_ISL_495557, EPI_ISL_495558, EPI_ISL_495559, EPI_ISL_495560, EPI_ISL_495561, EPI_ISL_495562, EPI_ISL_498054, EPI_ISL_498055, EPI_ISL_498056, EPI_ISL_498057, EPI_ISL_498058, EPI_ISL_498059, EPI_ISL_498060, EPI_ISL_498061, EPI_ISL_498062, EPI_ISL_498063, EPI_ISL_498064, EPI_ISL_498065, EPI_ISL_498066, EPI_ISL_498067, EPI_ISL_498068, EPI_ISL_498069, EPI_ISL_498070, EPI_ISL_498071, EPI_ISL_498072, EPI_ISL_498073, EPI_ISL_498074, EPI_ISL_498075, EPI_ISL_498076, EPI_ISL_498077, EPI_ISL_498078, EPI_ISL_498079, EPI_ISL_498080, EPI_ISL_498081, EPI_ISL_498082, EPI_ISL_498083, EPI_ISL_498084, EPI_ISL_498085, EPI_ISL_498086, EPI_ISL_498087, EPI_ISL_498088, EPI_ISL_498089, EPI_ISL_498090, EPI_ISL_498091, EPI_ISL_498092, EPI_ISL_498093, EPI_ISL_498094, EPI_ISL_498095, EPI_ISL_498096, EPI_ISL_498097, EPI_ISL_498098, EPI_ISL_498099, EPI_ISL_498100, EPI_ISL_498101, EPI_ISL_498102, EPI_ISL_498103, EPI_ISL_498104, EPI_ISL_498105, EPI_ISL_498106, EPI_ISL_498107, EPI_ISL_498108, EPI_ISL_498109, EPI_ISL_498110, EPI_ISL_498111, EPI_ISL_498112, EPI_ISL_498113, EPI_ISL_498114, EPI_ISL_498115, EPI_ISL_498116, EPI_ISL_498117, EPI_ISL_498118, EPI_ISL_498119, EPI_ISL_498120, EPI_ISL_498121, EPI_ISL_498122, EPI_ISL_498123, EPI_ISL_498124, EPI_ISL_498125, EPI_ISL_498126, EPI_ISL_509223, EPI_ISL_509224, EPI_ISL_509225, EPI_ISL_509226, EPI_ISL_509227, EPI_ISL_509228, EPI_ISL_509229, EPI_ISL_509230, EPI_ISL_509231, EPI_ISL_509232, EPI_ISL_509233, EPI_ISL_509234, EPI_ISL_509235, EPI_ISL_509236, EPI_ISL_509237, EPI_ISL_509238, EPI_ISL_509239, EPI_ISL_509240, EPI_ISL_509241, EPI_ISL_509242, EPI_ISL_509243, EPI_ISL_509244, EPI_ISL_509245, EPI_ISL_509246, EPI_ISL_509247, EPI_ISL_509248, EPI_ISL_509249, EPI_ISL_509250, EPI_ISL_509251, EPI_ISL_509252, EPI_ISL_509253, EPI_ISL_509254, EPI_ISL_509255, EPI_ISL_509256, EPI_ISL_509257, EPI_ISL_509258, EPI_ISL_509259, EPI_ISL_509260, EPI_ISL_509261, EPI_ISL_509262, EPI_ISL_509263, EPI_ISL_509264, EPI_ISL_509265, EPI_ISL_509266, EPI_ISL_509267, EPI_ISL_509268, EPI_ISL_509269, EPI_ISL_509270, EPI_ISL_509271, EPI_ISL_509272, EPI_ISL_509273, EPI_ISL_509274, EPI_ISL_509275, EPI_ISL_509276, EPI_ISL_509277, EPI_ISL_509278, EPI_ISL_509279, EPI_ISL_509280, EPI_ISL_509281, EPI_ISL_509282, EPI_ISL_509283, EPI_ISL_509284, EPI_ISL_509285, EPI_ISL_509286, EPI_ISL_509287, EPI_ISL_509288, EPI_ISL_509289, EPI_ISL_509290, EPI_ISL_509291, EPI_ISL_509292, EPI_ISL_509293, EPI_ISL_509294, EPI_ISL_509295, EPI_ISL_509296, EPI_ISL_509297, EPI_ISL_509298, EPI_ISL_509299, EPI_ISL_509300, EPI_ISL_509301, EPI_ISL_509302, EPI_ISL_509303, EPI_ISL_509304, EPI_ISL_509305, EPI_ISL_509306, EPI_ISL_509307, EPI_ISL_509308, EPI_ISL_509309, EPI_ISL_509310, EPI_ISL_509311, EPI_ISL_509312, EPI_ISL_509313, EPI_ISL_509314, EPI_ISL_509315, EPI_ISL_509316, EPI_ISL_509317, EPI_ISL_509318, EPI_ISL_509319, EPI_ISL_509320, EPI_ISL_509321, EPI_ISL_509322, EPI_ISL_509323, EPI_ISL_509324, EPI_ISL_509325, EPI_ISL_509326, EPI_ISL_509327, EPI_ISL_509328, EPI_ISL_509329, EPI_ISL_509330, EPI_ISL_509331, EPI_ISL_509332, EPI_ISL_509333, EPI_ISL_509334, EPI_ISL_509335, EPI_ISL_509336, EPI_ISL_509337, EPI_ISL_509338, EPI_ISL_509339, EPI_ISL_509340, EPI_ISL_509341, EPI_ISL_509342, EPI_ISL_509343, EPI_ISL_509344, EPI_ISL_509345, EPI_ISL_509346, EPI_ISL_509347, EPI_ISL_509348, EPI_ISL_509349, EPI_ISL_509350, EPI_ISL_509351, EPI_ISL_509352, EPI_ISL_509353, EPI_ISL_509354, EPI_ISL_509355, EPI_ISL_509356, EPI_ISL_509357, EPI_ISL_509358, EPI_ISL_509359, EPI_ISL_509360, EPI_ISL_509361, EPI_ISL_509362, EPI_ISL_509363, EPI_ISL_509364, EPI_ISL_509365, EPI_ISL_509366, EPI_ISL_509367, EPI_ISL_509368, EPI_ISL_509369, EPI_ISL_509370, EPI_ISL_509371, EPI_ISL_515568, EPI_ISL_515569, EPI_ISL_515570, EPI_ISL_515571, EPI_ISL_515572, EPI_ISL_515573, EPI_ISL_515574, EPI_ISL_515575, EPI_ISL_515576, EPI_ISL_515577, EPI_ISL_515578, EPI_ISL_515579, EPI_ISL_515580, EPI_ISL_515581, EPI_ISL_515582, EPI_ISL_515583, EPI_ISL_515584, EPI_ISL_515585, EPI_ISL_515586, EPI_ISL_515587, EPI_ISL_515588, EPI_ISL_515589, EPI_ISL_515590, EPI_ISL_515591, EPI_ISL_515592, EPI_ISL_515593, EPI_ISL_515594, EPI_ISL_515595, EPI_ISL_515596, EPI_ISL_515597, EPI_ISL_515598, EPI_ISL_515599, EPI_ISL_515600, EPI_ISL_515601, EPI_ISL_515602, EPI_ISL_515603, EPI_ISL_515604, EPI_ISL_515605, EPI_ISL_515606, EPI_ISL_515607, EPI_ISL_515608, EPI_ISL_515609, EPI_ISL_515610, EPI_ISL_515611, EPI_ISL_515612, EPI_ISL_515613, EPI_ISL_515614, EPI_ISL_515615, EPI_ISL_515616, EPI_ISL_515617, EPI_ISL_515618, EPI_ISL_515619, EPI_ISL_515620, EPI_ISL_515621, EPI_ISL_515622, EPI_ISL_515623, EPI_ISL_515624, EPI_ISL_515625, EPI_ISL_515626, EPI_ISL_515627, EPI_ISL_515628, EPI_ISL_515629, EPI_ISL_515630, EPI_ISL_515631, EPI_ISL_515632, EPI_ISL_515633, EPI_ISL_515634, EPI_ISL_515635, EPI_ISL_515636, EPI_ISL_515637, EPI_ISL_515638, EPI_ISL_515639, EPI_ISL_515640, EPI_ISL_515641, EPI_ISL_515642, EPI_ISL_515643, EPI_ISL_515644, EPI_ISL_515645, EPI_ISL_515646, EPI_ISL_515647, EPI_ISL_515648, EPI_ISL_515649, EPI_ISL_515650, EPI_ISL_515651, EPI_ISL_515652, EPI_ISL_515653, EPI_ISL_515654, EPI_ISL_515655, EPI_ISL_515656, EPI_ISL_515657, EPI_ISL_515658, EPI_ISL_515659, EPI_ISL_515660, EPI_ISL_515661, EPI_ISL_515662, EPI_ISL_515663, EPI_ISL_515664, EPI_ISL_515665, EPI_ISL_515666, EPI_ISL_515667, EPI_ISL_515668, EPI_ISL_515669, EPI_ISL_515670, EPI_ISL_515671, EPI_ISL_515672, EPI_ISL_515673, EPI_ISL_515674, EPI_ISL_515675, EPI_ISL_515676, EPI_ISL_515677, EPI_ISL_515678, EPI_ISL_515679, EPI_ISL_515680, EPI_ISL_515681, EPI_ISL_515682, EPI_ISL_515683, EPI_ISL_515684, EPI_ISL_515685, EPI_ISL_515686, EPI_ISL_515687, EPI_ISL_515688, EPI_ISL_515689, EPI_ISL_515690, EPI_ISL_515691, EPI_ISL_515692, EPI_ISL_515693, EPI_ISL_515694, EPI_ISL_515695, EPI_ISL_515696, EPI_ISL_515697, EPI_ISL_515698, EPI_ISL_515699, EPI_ISL_515700, EPI_ISL_515701, EPI_ISL_515702, EPI_ISL_515703, EPI_ISL_515704, EPI_ISL_515705, EPI_ISL_515706, EPI_ISL_515707, EPI_ISL_515708, EPI_ISL_515709, EPI_ISL_515710, EPI_ISL_515711, EPI_ISL_515712, EPI_ISL_515713, EPI_ISL_515714, EPI_ISL_515715, EPI_ISL_515716, EPI_ISL_515717, EPI_ISL_515718, EPI_ISL_515719, EPI_ISL_515720, EPI_ISL_515721, EPI_ISL_515722, EPI_ISL_515723, EPI_ISL_515724, EPI_ISL_515725, EPI_ISL_515726, EPI_ISL_515727, EPI_ISL_515728, EPI_ISL_515729, EPI_ISL_515730, EPI_ISL_515731, EPI_ISL_515732, EPI_ISL_515733, EPI_ISL_515734, EPI_ISL_515735, EPI_ISL_515736, EPI_ISL_515737, EPI_ISL_515738, EPI_ISL_515739, EPI_ISL_515740, EPI_ISL_515741, EPI_ISL_515742, EPI_ISL_515743, EPI_ISL_515744, EPI_ISL_515745, EPI_ISL_515746, EPI_ISL_515747, EPI_ISL_515748, EPI_ISL_515749, EPI_ISL_515750, EPI_ISL_515751, EPI_ISL_515752, EPI_ISL_515753, EPI_ISL_515754, EPI_ISL_515755, EPI_ISL_515756, EPI_ISL_515757, EPI_ISL_515758, EPI_ISL_515759, EPI_ISL_515760, EPI_ISL_515761, EPI_ISL_515762, EPI_ISL_515763, EPI_ISL_515764, EPI_ISL_515765, EPI_ISL_515766, EPI_ISL_515767, EPI_ISL_515768, EPI_ISL_515769, EPI_ISL_515770, EPI_ISL_515771, EPI_ISL_515772, EPI_ISL_515773, EPI_ISL_515774, EPI_ISL_515775, EPI_ISL_515776, EPI_ISL_515777, EPI_ISL_515778, EPI_ISL_515779, EPI_ISL_515780, EPI_ISL_515781, EPI_ISL_515782, EPI_ISL_515783, EPI_ISL_515784, EPI_ISL_515785, EPI_ISL_515786, EPI_ISL_515787, EPI_ISL_515788, EPI_ISL_515789, EPI_ISL_515790, EPI_ISL_515791, EPI_ISL_515792, EPI_ISL_515793, EPI_ISL_515794, EPI_ISL_515795, EPI_ISL_515796, EPI_ISL_515797, EPI_ISL_515798, EPI_ISL_515799, EPI_ISL_518033, EPI_ISL_518034, EPI_ISL_518035, EPI_ISL_518036, EPI_ISL_518037, EPI_ISL_518038, EPI_ISL_518039, EPI_ISL_518040, EPI_ISL_518041, EPI_ISL_518042, EPI_ISL_518043, EPI_ISL_518044, EPI_ISL_518045, EPI_ISL_518046, EPI_ISL_518047, EPI_ISL_518048, EPI_ISL_518050, EPI_ISL_518051, EPI_ISL_518052, EPI_ISL_518053, EPI_ISL_529720, EPI_ISL_529721, EPI_ISL_529722, EPI_ISL_529723, EPI_ISL_529724, EPI_ISL_529725, EPI_ISL_529726, EPI_ISL_529727, EPI_ISL_529728, EPI_ISL_529729, EPI_ISL_529730, EPI_ISL_529731, EPI_ISL_529732, EPI_ISL_529733, EPI_ISL_529734, EPI_ISL_529735, EPI_ISL_529736, EPI_ISL_529737, EPI_ISL_529738, | | | |

| | | | |
|---|---|---|---|
| EPI_ISL_529739, EPI_ISL_529740, EPI_ISL_529741, EPI_ISL_529742, EPI_ISL_529743, EPI_ISL_529744, EPI_ISL_529745, EPI_ISL_529746, EPI_ISL_529747, EPI_ISL_529748, EPI_ISL_529749, EPI_ISL_529750, EPI_ISL_529751, EPI_ISL_529752, EPI_ISL_529753, EPI_ISL_529754, EPI_ISL_529755, EPI_ISL_529756, EPI_ISL_529757, EPI_ISL_529758, EPI_ISL_529759, EPI_ISL_529760, EPI_ISL_529761, EPI_ISL_529762, EPI_ISL_529763, EPI_ISL_529764, EPI_ISL_529765, EPI_ISL_529766, EPI_ISL_529767, EPI_ISL_529768, EPI_ISL_529769, EPI_ISL_529770, EPI_ISL_529771, EPI_ISL_529772, EPI_ISL_529773, EPI_ISL_529774, EPI_ISL_529775, EPI_ISL_529776, EPI_ISL_529777, EPI_ISL_529778, EPI_ISL_529779, EPI_ISL_529780, EPI_ISL_529781, EPI_ISL_529782, EPI_ISL_529783, EPI_ISL_529784, EPI_ISL_529785, EPI_ISL_529786, EPI_ISL_529787, EPI_ISL_529788, EPI_ISL_529789, EPI_ISL_529790, EPI_ISL_529791, EPI_ISL_529792, EPI_ISL_529793, EPI_ISL_529794, EPI_ISL_529795, EPI_ISL_529796, EPI_ISL_529797, EPI_ISL_529798, EPI_ISL_529799, EPI_ISL_529800, EPI_ISL_529801, EPI_ISL_529802, EPI_ISL_529803, EPI_ISL_535390, EPI_ISL_535391, EPI_ISL_535392, EPI_ISL_535393, EPI_ISL_535394, EPI_ISL_535395, EPI_ISL_535396, EPI_ISL_535397, EPI_ISL_535398, EPI_ISL_535399, EPI_ISL_535400, EPI_ISL_535401, EPI_ISL_535402, EPI_ISL_535403, EPI_ISL_535404, EPI_ISL_535405, EPI_ISL_535406, EPI_ISL_535407, EPI_ISL_535408, EPI_ISL_535409, EPI_ISL_535410, EPI_ISL_535411, EPI_ISL_535412, EPI_ISL_535413, EPI_ISL_535414, EPI_ISL_535415, EPI_ISL_535416, EPI_ISL_535417, EPI_ISL_535418, EPI_ISL_535419, EPI_ISL_535420, EPI_ISL_535421, EPI_ISL_535422, EPI_ISL_535423, EPI_ISL_535424, EPI_ISL_535425, EPI_ISL_535426, EPI_ISL_535427, EPI_ISL_535428, EPI_ISL_535429, EPI_ISL_535430, EPI_ISL_535431, EPI_ISL_535432, EPI_ISL_535433, EPI_ISL_535434, EPI_ISL_535435, EPI_ISL_535436, EPI_ISL_535437, EPI_ISL_535438, EPI_ISL_535439, EPI_ISL_535440, EPI_ISL_535441, EPI_ISL_535442, EPI_ISL_535443, EPI_ISL_535444, EPI_ISL_535445, EPI_ISL_535446, EPI_ISL_535447, EPI_ISL_535448, EPI_ISL_535449, EPI_ISL_535450, EPI_ISL_535451, EPI_ISL_535452, EPI_ISL_535453, EPI_ISL_535454, EPI_ISL_535455, EPI_ISL_535456, EPI_ISL_535457, EPI_ISL_535458, EPI_ISL_535459, EPI_ISL_535460, EPI_ISL_535461, EPI_ISL_535462, EPI_ISL_535463, EPI_ISL_535464, EPI_ISL_535465, EPI_ISL_535466, EPI_ISL_535467, EPI_ISL_535468, EPI_ISL_535469, EPI_ISL_535470, EPI_ISL_535471, EPI_ISL_535472, EPI_ISL_535473, EPI_ISL_535474, EPI_ISL_535475, EPI_ISL_535476, EPI_ISL_535477, EPI_ISL_535478, EPI_ISL_535479, EPI_ISL_535480, EPI_ISL_535481, EPI_ISL_535482, EPI_ISL_535483, EPI_ISL_535484, EPI_ISL_535485, EPI_ISL_535486, EPI_ISL_535487, EPI_ISL_535488, EPI_ISL_535496, EPI_ISL_535497, EPI_ISL_535498, EPI_ISL_535499, EPI_ISL_535500, EPI_ISL_535501, EPI_ISL_535502, EPI_ISL_535503, EPI_ISL_535504, EPI_ISL_535505, EPI_ISL_535506, EPI_ISL_535507, EPI_ISL_535508, EPI_ISL_535509, EPI_ISL_535510, EPI_ISL_535511, EPI_ISL_535512, EPI_ISL_535513, EPI_ISL_535514, EPI_ISL_535515, EPI_ISL_535516, EPI_ISL_535517, EPI_ISL_535518, EPI_ISL_535519, EPI_ISL_535520, EPI_ISL_535521, EPI_ISL_535522, EPI_ISL_535523, EPI_ISL_535524, EPI_ISL_535525, EPI_ISL_535526, EPI_ISL_535527, EPI_ISL_535528, EPI_ISL_535529, EPI_ISL_535530, EPI_ISL_535531, EPI_ISL_535532, EPI_ISL_535533, EPI_ISL_535534, EPI_ISL_535535, EPI_ISL_535536, EPI_ISL_535537, EPI_ISL_535538, EPI_ISL_535539, EPI_ISL_535540, EPI_ISL_535541, EPI_ISL_535542, EPI_ISL_535543, EPI_ISL_535544, EPI_ISL_535545, EPI_ISL_535546, EPI_ISL_535547, EPI_ISL_535548, EPI_ISL_535549, EPI_ISL_535550, EPI_ISL_535551, EPI_ISL_535552, EPI_ISL_535553, EPI_ISL_535554, EPI_ISL_535555, EPI_ISL_535556, EPI_ISL_535557, EPI_ISL_535558, EPI_ISL_535559, EPI_ISL_535560, EPI_ISL_535561, EPI_ISL_535562, EPI_ISL_535563, EPI_ISL_535564, EPI_ISL_535565, EPI_ISL_535566, EPI_ISL_535567, EPI_ISL_535568, EPI_ISL_535569, EPI_ISL_535570, EPI_ISL_535571, EPI_ISL_535572, EPI_ISL_602632, EPI_ISL_602633, EPI_ISL_602634, EPI_ISL_602635, EPI_ISL_602636, EPI_ISL_602637, EPI_ISL_602638, EPI_ISL_602639, EPI_ISL_602640, EPI_ISL_602641, EPI_ISL_602642, EPI_ISL_602643, EPI_ISL_602644, EPI_ISL_602645, EPI_ISL_602646, EPI_ISL_602647, EPI_ISL_602648, EPI_ISL_602649, EPI_ISL_602650, EPI_ISL_602651, EPI_ISL_602652, EPI_ISL_602653, EPI_ISL_602654, EPI_ISL_602655, EPI_ISL_602656, EPI_ISL_602657, EPI_ISL_602658, EPI_ISL_602659, EPI_ISL_602660, EPI_ISL_602661, EPI_ISL_602662, EPI_ISL_602663, EPI_ISL_602664, EPI_ISL_602665, EPI_ISL_602666, EPI_ISL_602667, EPI_ISL_602668, EPI_ISL_602669, EPI_ISL_602670, EPI_ISL_602671, EPI_ISL_602672, EPI_ISL_602673, EPI_ISL_602674, EPI_ISL_602675, EPI_ISL_602676, EPI_ISL_602677, EPI_ISL_602678, EPI_ISL_602679, EPI_ISL_602680, EPI_ISL_602681, EPI_ISL_602682, EPI_ISL_602683, EPI_ISL_602684, EPI_ISL_602685, EPI_ISL_602686, EPI_ISL_602687, EPI_ISL_602688, EPI_ISL_602689, EPI_ISL_602690, EPI_ISL_602691, EPI_ISL_602692, EPI_ISL_602693, EPI_ISL_602694, EPI_ISL_602695, EPI_ISL_602696, EPI_ISL_602697, EPI_ISL_602698, EPI_ISL_602699, EPI_ISL_602700, EPI_ISL_602701, EPI_ISL_602702, EPI_ISL_602703, EPI_ISL_602704, EPI_ISL_602705, EPI_ISL_602706, EPI_ISL_602707, EPI_ISL_602708, EPI_ISL_602709, EPI_ISL_602710, EPI_ISL_602711, EPI_ISL_602712, EPI_ISL_602713, EPI_ISL_602714, EPI_ISL_602715, EPI_ISL_602716, EPI_ISL_602717, EPI_ISL_602718, EPI_ISL_602719, EPI_ISL_602720, EPI_ISL_602721, EPI_ISL_602722, EPI_ISL_602723, EPI_ISL_602724, EPI_ISL_602725, EPI_ISL_602726, EPI_ISL_602727, EPI_ISL_602728, EPI_ISL_602729, EPI_ISL_602730, EPI_ISL_602731, EPI_ISL_602732, EPI_ISL_602733, EPI_ISL_602734, EPI_ISL_602735, EPI_ISL_602736, EPI_ISL_602737, EPI_ISL_602738, EPI_ISL_602739, EPI_ISL_602740, EPI_ISL_602741, EPI_ISL_602742, EPI_ISL_602743, EPI_ISL_602744, EPI_ISL_602745, EPI_ISL_602746, EPI_ISL_602747, EPI_ISL_602748, EPI_ISL_602749, EPI_ISL_602750, EPI_ISL_602751, EPI_ISL_602752, EPI_ISL_602753, EPI_ISL_602754, EPI_ISL_602755, EPI_ISL_602756, EPI_ISL_602757, EPI_ISL_602758, EPI_ISL_602759, EPI_ISL_602760, EPI_ISL_602761, EPI_ISL_602762, EPI_ISL_602763, EPI_ISL_602764, EPI_ISL_602765, EPI_ISL_602766, EPI_ISL_602767, EPI_ISL_602768, EPI_ISL_602769, EPI_ISL_602770, EPI_ISL_602771, EPI_ISL_602772, EPI_ISL_602773, EPI_ISL_602774, EPI_ISL_602775, EPI_ISL_602776, EPI_ISL_602777, EPI_ISL_602778, EPI_ISL_602779, EPI_ISL_602780, EPI_ISL_602781, EPI_ISL_602782, EPI_ISL_602783, EPI_ISL_602784, EPI_ISL_602785, EPI_ISL_602786, EPI_ISL_602787, EPI_ISL_602788, EPI_ISL_602789, EPI_ISL_602790, EPI_ISL_602791, EPI_ISL_602792, EPI_ISL_602793, EPI_ISL_602794, EPI_ISL_602795, EPI_ISL_602796, EPI_ISL_602797, EPI_ISL_602798, EPI_ISL_602799, EPI_ISL_602800, EPI_ISL_602801, EPI_ISL_602802, EPI_ISL_602803, EPI_ISL_602804, EPI_ISL_602805, EPI_ISL_602806, EPI_ISL_602807, EPI_ISL_602808, EPI_ISL_602809, EPI_ISL_602810, EPI_ISL_602811, EPI_ISL_602812, EPI_ISL_602813, EPI_ISL_602814, EPI_ISL_602815, EPI_ISL_602816, EPI_ISL_602817, EPI_ISL_602818, EPI_ISL_602819, EPI_ISL_602820, EPI_ISL_602821, EPI_ISL_602822, EPI_ISL_602823, EPI_ISL_602824, EPI_ISL_602825, EPI_ISL_602826, EPI_ISL_602827, EPI_ISL_602828, EPI_ISL_602829, EPI_ISL_602830, EPI_ISL_602831, EPI_ISL_602832, EPI_ISL_602833, EPI_ISL_602834, EPI_ISL_602835, EPI_ISL_602836, EPI_ISL_602837, EPI_ISL_602838, EPI_ISL_602839, EPI_ISL_602840, EPI_ISL_602841, EPI_ISL_602842, EPI_ISL_602843, EPI_ISL_602844, EPI_ISL_602845, EPI_ISL_602846, EPI_ISL_602847, EPI_ISL_602848, EPI_ISL_602849, EPI_ISL_602850, EPI_ISL_602851, EPI_ISL_602852, EPI_ISL_602853, EPI_ISL_602854, EPI_ISL_602855, EPI_ISL_602856, EPI_ISL_602857, EPI_ISL_602858, EPI_ISL_602859, EPI_ISL_602860, EPI_ISL_602861, EPI_ISL_602862, EPI_ISL_602863, EPI_ISL_602864, EPI_ISL_602865, EPI_ISL_602866, EPI_ISL_602867, EPI_ISL_602868, EPI_ISL_602869, EPI_ISL_602870, EPI_ISL_602871, EPI_ISL_602872, EPI_ISL_602873, EPI_ISL_602874, EPI_ISL_602875, EPI_ISL_602876, EPI_ISL_602877, EPI_ISL_602878, EPI_ISL_602879, EPI_ISL_602880, EPI_ISL_602881, EPI_ISL_602882, EPI_ISL_602883, EPI_ISL_602884, EPI_ISL_602885, EPI_ISL_602886, EPI_ISL_602887, EPI_ISL_602888, EPI_ISL_602889, EPI_ISL_602890, EPI_ISL_602891, EPI_ISL_602892, EPI_ISL_602893, EPI_ISL_602894, EPI_ISL_602895, EPI_ISL_602896, EPI_ISL_602897, EPI_ISL_602898, EPI_ISL_602899, EPI_ISL_602900, EPI_ISL_602901, EPI_ISL_602902, EPI_ISL_602903, EPI_ISL_602904, EPI_ISL_602905, EPI_ISL_602906, EPI_ISL_602907, EPI_ISL_602908, EPI_ISL_602909, EPI_ISL_602910, EPI_ISL_602911, EPI_ISL_602912, EPI_ISL_602913, EPI_ISL_602914, EPI_ISL_602915, EPI_ISL_602916, EPI_ISL_602917, EPI_ISL_602918, EPI_ISL_602919, EPI_ISL_602920, EPI_ISL_602921, EPI_ISL_602922, EPI_ISL_602923, EPI_ISL_602924, EPI_ISL_602925, EPI_ISL_602926, EPI_ISL_602927, EPI_ISL_602928, EPI_ISL_602929, EPI_ISL_602930, EPI_ISL_605785, EPI_ISL_605786, EPI_ISL_605787, EPI_ISL_605788, EPI_ISL_605789, EPI_ISL_605790, EPI_ISL_660159, EPI_ISL_660160, EPI_ISL_660161, EPI_ISL_660162, EPI_ISL_660163, EPI_ISL_660164, EPI_ISL_660165, EPI_ISL_660166, EPI_ISL_660167, EPI_ISL_660168, EPI_ISL_660169, EPI_ISL_660170, EPI_ISL_660171, EPI_ISL_660172, EPI_ISL_660173, EPI_ISL_660174, EPI_ISL_660175, EPI_ISL_660176, EPI_ISL_660177, EPI_ISL_660178, EPI_ISL_660179, EPI_ISL_660180, EPI_ISL_660181, EPI_ISL_660182, EPI_ISL_660183, EPI_ISL_660184, EPI_ISL_660185, EPI_ISL_660186, EPI_ISL_660187, EPI_ISL_660188, EPI_ISL_660189, EPI_ISL_660190, EPI_ISL_660191, EPI_ISL_660192, EPI_ISL_660193, EPI_ISL_660194, EPI_ISL_660195, EPI_ISL_660196, EPI_ISL_660197, EPI_ISL_660198, EPI_ISL_660199, EPI_ISL_660200, EPI_ISL_660201, EPI_ISL_660202, EPI_ISL_660203, EPI_ISL_660204, EPI_ISL_660205, EPI_ISL_660206, EPI_ISL_660207, EPI_ISL_660208, EPI_ISL_660209, EPI_ISL_660210, EPI_ISL_660211, EPI_ISL_660212, EPI_ISL_660213, EPI_ISL_660214, EPI_ISL_660215, EPI_ISL_660216, EPI_ISL_660217, EPI_ISL_660218, EPI_ISL_660219, EPI_ISL_660220, EPI_ISL_660223, EPI_ISL_660224, EPI_ISL_660225, EPI_ISL_660226, EPI_ISL_660227, EPI_ISL_660234, EPI_ISL_660235, EPI_ISL_660236, EPI_ISL_660237, EPI_ISL_660238, EPI_ISL_660239, EPI_ISL_660240, EPI_ISL_660241, EPI_ISL_660242, EPI_ISL_660243, EPI_ISL_660244, EPI_ISL_660245, EPI_ISL_660246, EPI_ISL_660247, EPI_ISL_660248, EPI_ISL_660249, EPI_ISL_660250, EPI_ISL_660251, EPI_ISL_660252, EPI_ISL_660253, EPI_ISL_660254, EPI_ISL_660255, EPI_ISL_660605, EPI_ISL_660606, EPI_ISL_660607, EPI_ISL_660608, EPI_ISL_660609, EPI_ISL_660610, EPI_ISL_660611, EPI_ISL_660612, EPI_ISL_660613, EPI_ISL_660614, EPI_ISL_660615, EPI_ISL_660616, EPI_ISL_660617, EPI_ISL_660618, EPI_ISL_660619, EPI_ISL_660620, EPI_ISL_660621, EPI_ISL_660622, EPI_ISL_660623, EPI_ISL_660624, EPI_ISL_660625, EPI_ISL_660626, EPI_ISL_660627, EPI_ISL_660628, EPI_ISL_660629, EPI_ISL_660630, EPI_ISL_660631, EPI_ISL_660632, EPI_ISL_660633, EPI_ISL_660634, EPI_ISL_660635, EPI_ISL_660636, EPI_ISL_660637, EPI_ISL_660638, EPI_ISL_660639, EPI_ISL_660640, EPI_ISL_660641, EPI_ISL_660642, EPI_ISL_660643, EPI_ISL_660644, EPI_ISL_660645, EPI_ISL_660646, EPI_ISL_660647, EPI_ISL_660648, EPI_ISL_660649, EPI_ISL_660650, EPI_ISL_660651, EPI_ISL_660652, EPI_ISL_660653, EPI_ISL_660654, EPI_ISL_660655, EPI_ISL_660656, EPI_ISL_660657, EPI_ISL_660658, EPI_ISL_660659, EPI_ISL_660660, EPI_ISL_660661, EPI_ISL_660662, EPI_ISL_660663, EPI_ISL_678566, EPI_ISL_678569, EPI_ISL_678570, EPI_ISL_678574, EPI_ISL_678575, EPI_ISL_678598, EPI_ISL_678599, EPI_ISL_678600, EPI_ISL_678601, EPI_ISL_678602, EPI_ISL_678603, EPI_ISL_678604, EPI_ISL_678605, EPI_ISL_678606, EPI_ISL_678607, EPI_ISL_678608, EPI_ISL_678609, EPI_ISL_678610, EPI_ISL_678611, EPI_ISL_678612, EPI_ISL_678613, EPI_ISL_678614, EPI_ISL_678615, EPI_ISL_678616, EPI_ISL_678617, EPI_ISL_678618, EPI_ISL_678619, EPI_ISL_678620, EPI_ISL_678621, EPI_ISL_678622, EPI_ISL_678623, EPI_ISL_678624, EPI_ISL_678625, EPI_ISL_678626, EPI_ISL_678627, EPI_ISL_678628, EPI_ISL_678629, EPI_ISL_678630, EPI_ISL_678631, EPI_ISL_678632, EPI_ISL_678633, EPI_ISL_678634, EPI_ISL_678635, EPI_ISL_678636, EPI_ISL_678637, EPI_ISL_678638, EPI_ISL_678639, EPI_ISL_678640, EPI_ISL_678641, EPI_ISL_678642, EPI_ISL_678643, EPI_ISL_736925, EPI_ISL_736926, EPI_ISL_736931, EPI_ISL_736932, EPI_ISL_736933, EPI_ISL_736934, EPI_ISL_736935, EPI_ISL_736938, EPI_ISL_736940, EPI_ISL_736941, EPI_ISL_736942, EPI_ISL_736943, EPI_ISL_736944, EPI_ISL_736945, EPI_ISL_736946, EPI_ISL_736947, EPI_ISL_736948, EPI_ISL_736949, EPI_ISL_736950, EPI_ISL_736951, EPI_ISL_736952, EPI_ISL_736953, EPI_ISL_736954, EPI_ISL_736955, EPI_ISL_736956, EPI_ISL_736957, EPI_ISL_736958, EPI_ISL_736959, EPI_ISL_736960, EPI_ISL_736969, EPI_ISL_736970, EPI_ISL_736971, EPI_ISL_736972, EPI_ISL_736973, EPI_ISL_736974, EPI_ISL_736975, EPI_ISL_736976, EPI_ISL_736977, EPI_ISL_736978, EPI_ISL_736979, EPI_ISL_736980, EPI_ISL_736984, EPI_ISL_736985, EPI_ISL_736986, EPI_ISL_736987, EPI_ISL_736988, EPI_ISL_736989, EPI_ISL_736990, EPI_ISL_736991, EPI_ISL_736992, EPI_ISL_736993, EPI_ISL_736994, EPI_ISL_825090, EPI_ISL_825091, EPI_ISL_825092, EPI_ISL_825093, EPI_ISL_825094, EPI_ISL_825095, EPI_ISL_825096, EPI_ISL_825097, EPI_ISL_825098, EPI_ISL_825099, EPI_ISL_825100, EPI_ISL_825101, EPI_ISL_825102, EPI_ISL_825103, EPI_ISL_825104, EPI_ISL_825105, EPI_ISL_825106, EPI_ISL_825107, EPI_ISL_825108, EPI_ISL_825109, EPI_ISL_825110, EPI_ISL_825111, EPI_ISL_825112, EPI_ISL_825113, EPI_ISL_825114, EPI_ISL_825115, EPI_ISL_825116, EPI_ISL_825117, EPI_ISL_825118, EPI_ISL_825119, EPI_ISL_825120, EPI_ISL_825121, EPI_ISL_825122, EPI_ISL_825123, EPI_ISL_825124, EPI_ISL_825125, EPI_ISL_825126, EPI_ISL_825127, EPI_ISL_825128, EPI_ISL_825129, EPI_ISL_825130, EPI_ISL_825131, EPI_ISL_825132, EPI_ISL_825133, EPI_ISL_825134, EPI_ISL_825135, EPI_ISL_825136, EPI_ISL_825137, EPI_ISL_825138, EPI_ISL_825139, EPI_ISL_825438, EPI_ISL_825439, EPI_ISL_825440, EPI_ISL_825441, EPI_ISL_825442, EPI_ISL_825443, EPI_ISL_825444, EPI_ISL_825445, EPI_ISL_825446, EPI_ISL_825447, EPI_ISL_825448, EPI_ISL_825449, EPI_ISL_825450, EPI_ISL_825451, EPI_ISL_825452, EPI_ISL_825453, EPI_ISL_825454, EPI_ISL_825455, EPI_ISL_825456, EPI_ISL_825457, EPI_ISL_825458, EPI_ISL_825459, EPI_ISL_825460, EPI_ISL_825461, EPI_ISL_825462, EPI_ISL_825463, EPI_ISL_825464, EPI_ISL_825465, EPI_ISL_825466, EPI_ISL_825467, EPI_ISL_825468, EPI_ISL_825469, EPI_ISL_825470, EPI_ISL_825471, EPI_ISL_825472, EPI_ISL_825473, EPI_ISL_825474, EPI_ISL_825475, EPI_ISL_825476, EPI_ISL_825477, EPI_ISL_825478, EPI_ISL_825479, EPI_ISL_825480, EPI_ISL_825481, EPI_ISL_825482, EPI_ISL_825483, EPI_ISL_825484, EPI_ISL_825485, EPI_ISL_825486, EPI_ISL_825487, EPI_ISL_825488, EPI_ISL_825489 | | | |
| see above | NHLS-IALCH | KRISP, KZN Research Innovation and Sequencing Platform | Cele S; Chimukangara B; ChimukangaraB; Gazy I; Giandhari J; Karim F; Khan S; Lessells R; Mdlalose K; Pillay S; Sigal A; Tegally H; Wilkinson E; York D; de Oliveira T |
| EPI_ISL_860554, EPI_ISL_860555, EPI_ISL_860556, EPI_ISL_860557, EPI_ISL_860558, EPI_ISL_860559, EPI_ISL_860560, EPI_ISL_860561, EPI_ISL_860562, EPI_ISL_860563, EPI_ISL_860564, EPI_ISL_860565, EPI_ISL_860566, EPI_ISL_860567, EPI_ISL_860568, EPI_ISL_860569, EPI_ISL_860570, EPI_ISL_860571, EPI_ISL_860572, EPI_ISL_860573, EPI_ISL_860574, EPI_ISL_860575, EPI_ISL_860576, EPI_ISL_860577, EPI_ISL_860578, EPI_ISL_860579, EPI_ISL_860580, EPI_ISL_860581, EPI_ISL_860582, EPI_ISL_860583, EPI_ISL_860584, EPI_ISL_860585, EPI_ISL_860586, EPI_ISL_860587, EPI_ISL_860588, EPI_ISL_860589, EPI_ISL_860590, EPI_ISL_860591, EPI_ISL_860592, EPI_ISL_860593, EPI_ISL_860594, EPI_ISL_860595, EPI_ISL_860596, EPI_ISL_860597, EPI_ISL_860598, EPI_ISL_860599, EPI_ISL_860600, EPI_ISL_860601, EPI_ISL_860602, EPI_ISL_860603, EPI_ISL_860604, EPI_ISL_860605, EPI_ISL_860606, EPI_ISL_860607, EPI_ISL_860608, EPI_ISL_860609, EPI_ISL_860610, EPI_ISL_860611, EPI_ISL_860612, EPI_ISL_860613, EPI_ISL_860614, EPI_ISL_860615, EPI_ISL_860616, EPI_ISL_860617, EPI_ISL_860618, EPI_ISL_860619, EPI_ISL_860620, EPI_ISL_860621, EPI_ISL_860622, EPI_ISL_860623, EPI_ISL_860624, EPI_ISL_860625, EPI_ISL_860626, EPI_ISL_860627, EPI_ISL_860628, EPI_ISL_860629, EPI_ISL_860630 | | | |
| see above | NHLS-IALCH | KRISP, KZn Research Innovation and Sequencing Platform | Giandhari J; Khan S; Lessells R; Mdlalose K; Pillay S; Tegally H; Wilkinson E; York D; de Oliveira T |
| EPI_ISL_736928, EPI_ISL_736929, EPI_ISL_736939, EPI_ISL_736981, EPI_ISL_736982, EPI_ISL_736983 | NHLS-UCT | KRISP, KZN Research Innovation and Sequencing Platform | Arash Iranzadeh; Bruna Galvao; Carolyn Williamson; Deelan Doolabh; Diana Hardie; Emanuel James San; Houriiyah Tegally; Innocent Mudau; Jennifer Giandhari; Kruger Marais; Lynn Tyers; Marvin Hsiao; Stephen Korsman; Sureshnee Pillay; Tulio de Oliveira |
| EPI_ISL_745188 | Nababeep Hospital | National Health Laboratory Service (NHLS), Tygerberg | Bronwyn Kleinhans; Eduan Wilkindon; Gert van Zyl; Houriiyah Tegally; Kayla Delaney; Susan Engelbrecht; Tulio de Oliveira; Wolfgang Preiser |
| EPI_ISL_622942, EPI_ISL_622944, EPI_ISL_622945, EPI_ISL_622958, EPI_ISL_622970, EPI_ISL_622972, EPI_ISL_622973, EPI_ISL_622974, EPI_ISL_622976, EPI_ISL_622977, EPI_ISL_622980, EPI_ISL_622981, EPI_ISL_622982, EPI_ISL_622987, EPI_ISL_622992, EPI_ISL_622995, EPI_ISL_622996, EPI_ISL_622998, EPI_ISL_622999, EPI_ISL_623000, EPI_ISL_623001, EPI_ISL_623002, EPI_ISL_623003, EPI_ISL_623004, EPI_ISL_623005, EPI_ISL_623006, EPI_ISL_623009, EPI_ISL_623010, EPI_ISL_623011, EPI_ISL_623012, EPI_ISL_623013, EPI_ISL_623015, EPI_ISL_623017, EPI_ISL_623018, EPI_ISL_623019, EPI_ISL_623021, EPI_ISL_623022, EPI_ISL_623024, EPI_ISL_623026, EPI_ISL_623027, EPI_ISL_623029, EPI_ISL_623030, EPI_ISL_623032, EPI_ISL_623033, EPI_ISL_623035, EPI_ISL_623036, EPI_ISL_623037, EPI_ISL_623038, EPI_ISL_623039, EPI_ISL_623040, EPI_ISL_623042, EPI_ISL_623043, EPI_ISL_623044, EPI_ISL_623045, EPI_ISL_623046, EPI_ISL_623047, EPI_ISL_623049, EPI_ISL_623051, EPI_ISL_623052, EPI_ISL_623053, EPI_ISL_623054, EPI_ISL_623055, EPI_ISL_623056, EPI_ISL_623057, EPI_ISL_623058, EPI_ISL_623059, EPI_ISL_623060, EPI_ISL_623061, EPI_ISL_623062, EPI_ISL_623066, EPI_ISL_623067, EPI_ISL_623069, EPI_ISL_623071, EPI_ISL_623072 | | | |
| see above | National Health Laboratory Service | National Institute for Communicable Diseases of the National Health Laboratory Service | Allam M; Bhiman JN; Ismail A; Khumalo Z; Kwenda S; Mnyameni F; Mohale T; Mtshali P; Subramoney K |
| EPI_ISL_634978, EPI_ISL_634979, EPI_ISL_634980, EPI_ISL_634981, EPI_ISL_634982, EPI_ISL_634983, EPI_ISL_634984, EPI_ISL_634985, EPI_ISL_634986, EPI_ISL_634987, EPI_ISL_634988, EPI_ISL_634989, EPI_ISL_634990, EPI_ISL_634991, EPI_ISL_634992, EPI_ISL_634993, EPI_ISL_634994, EPI_ISL_634995, EPI_ISL_634996, EPI_ISL_634997, EPI_ISL_634998, EPI_ISL_634999, EPI_ISL_635000, EPI_ISL_635001, EPI_ISL_635002, EPI_ISL_635003, EPI_ISL_635004, EPI_ISL_635005, EPI_ISL_635006, EPI_ISL_635007, EPI_ISL_635008, EPI_ISL_635009, EPI_ISL_635010, EPI_ISL_635011, EPI_ISL_635012, EPI_ISL_635013, EPI_ISL_635014, EPI_ISL_635015, EPI_ISL_635016, EPI_ISL_635017, EPI_ISL_635018, EPI_ISL_635019, EPI_ISL_635020, EPI_ISL_635021, EPI_ISL_635022, EPI_ISL_635023, EPI_ISL_635024, EPI_ISL_635025, EPI_ISL_635026, EPI_ISL_635027, EPI_ISL_635028, EPI_ISL_635029, EPI_ISL_635030, EPI_ISL_635031, EPI_ISL_635032, EPI_ISL_635033, EPI_ISL_635034, EPI_ISL_635035, EPI_ISL_635036, EPI_ISL_635037, EPI_ISL_635038, EPI_ISL_635039, EPI_ISL_635040, EPI_ISL_635041, EPI_ISL_635042, EPI_ISL_635043, EPI_ISL_635044, EPI_ISL_635045, EPI_ISL_635046, EPI_ISL_635047, EPI_ISL_635048, EPI_ISL_635049, EPI_ISL_635050, EPI_ISL_635051, EPI_ISL_635052, EPI_ISL_635053, EPI_ISL_635054, EPI_ISL_635055, EPI_ISL_635056, EPI_ISL_635057, EPI_ISL_635058 | | | |
| see above | National Health Laboratory Service - Inkosi Albert Luthuli Central Hospital (NHLS-IALCH) | KRISP, KZN Research Innovation and Sequencing Platform | Giandhari J; Khan S; Lessells R; Mdlalose K; Pillay S; Tegally H; Wilkinson E; York D; de Oliveira T |
| EPI_ISL_944124, EPI_ISL_944125, EPI_ISL_944126, EPI_ISL_944127, EPI_ISL_944128, EPI_ISL_944129, EPI_ISL_944130, EPI_ISL_944131, EPI_ISL_944132, EPI_ISL_944133, EPI_ISL_944134, EPI_ISL_944135, EPI_ISL_944136, EPI_ISL_944137, EPI_ISL_944138, EPI_ISL_944139, EPI_ISL_944140, EPI_ISL_944141, EPI_ISL_944142, EPI_ISL_944143, EPI_ISL_944144, EPI_ISL_944145, EPI_ISL_944146, EPI_ISL_944147, EPI_ISL_944148, EPI_ISL_944149, EPI_ISL_944150, EPI_ISL_944151, EPI_ISL_944152, EPI_ISL_944153, EPI_ISL_944154, EPI_ISL_944155, EPI_ISL_944156, EPI_ISL_944157, EPI_ISL_944158, EPI_ISL_944159, EPI_ISL_944160, EPI_ISL_944161, EPI_ISL_944162, EPI_ISL_944163, EPI_ISL_944164, EPI_ISL_944165, EPI_ISL_944166, EPI_ISL_944167, EPI_ISL_944168, EPI_ISL_944169, EPI_ISL_944170, EPI_ISL_944171, EPI_ISL_944172, EPI_ISL_944173, EPI_ISL_944174, EPI_ISL_944175, EPI_ISL_944176, EPI_ISL_944177, EPI_ISL_1250424, EPI_ISL_1250425, EPI_ISL_1250427, EPI_ISL_1250428, EPI_ISL_1250430, EPI_ISL_1250431, EPI_ISL_1250432, EPI_ISL_1250434, EPI_ISL_1250436, EPI_ISL_1250437, EPI_ISL_1250439, EPI_ISL_1250441, EPI_ISL_1250442, EPI_ISL_1250444, EPI_ISL_1250446, EPI_ISL_1250447, EPI_ISL_1250448, EPI_ISL_1250450, EPI_ISL_1250452, EPI_ISL_1250453, EPI_ISL_1250455, EPI_ISL_1250456, EPI_ISL_1250458, EPI_ISL_1250460, EPI_ISL_1250462, EPI_ISL_1250463, EPI_ISL_1250465, EPI_ISL_1250466, EPI_ISL_1250468, EPI_ISL_1250469, EPI_ISL_1250472, EPI_ISL_1250474, EPI_ISL_1250476, EPI_ISL_1250477, EPI_ISL_1250479, EPI_ISL_1250480, EPI_ISL_1250482, EPI_ISL_1250483, EPI_ISL_1250485, EPI_ISL_1250486, EPI_ISL_1250488, EPI_ISL_1250490, EPI_ISL_1250491, EPI_ISL_1250493, EPI_ISL_1250494, EPI_ISL_1250496, EPI_ISL_1250497, EPI_ISL_1250499, EPI_ISL_1250501, EPI_ISL_1250502, EPI_ISL_1250504, EPI_ISL_1250505, EPI_ISL_1366787, EPI_ISL_1366791, EPI_ISL_1366794, EPI_ISL_1366795, EPI_ISL_1366796, EPI_ISL_1366797, EPI_ISL_1366799, EPI_ISL_1366800, EPI_ISL_1366802, EPI_ISL_1366803, EPI_ISL_1366804, EPI_ISL_1366805, EPI_ISL_1366806, EPI_ISL_1366807, EPI_ISL_1366808, EPI_ISL_1366809, EPI_ISL_1366810, EPI_ISL_1366811, EPI_ISL_1366812, EPI_ISL_1366839, EPI_ISL_1817782, EPI_ISL_1817855, EPI_ISL_1817856 | | | |
| see above | National Health Laboratory Service, South Africa | KRISP, KZN Research Innovation and Sequencing Platform | Emmanuel SJ; Giandhari J; Khan S; Laguda-Akingba O; Lessells R; Maslo C; Mdlalose K; Naidoo Y; Pillay S; Ramphal U; Sitharam L; Tegally H; Wilkinson E; York D; de Oliveira T |
| EPI_ISL_1048431, EPI_ISL_1048432, EPI_ISL_1048433, EPI_ISL_1048434, EPI_ISL_1048435, EPI_ISL_1048436, EPI_ISL_1048437, EPI_ISL_1048438, EPI_ISL_1048439, EPI_ISL_1048440, EPI_ISL_1048441, EPI_ISL_1048442, EPI_ISL_1048443, EPI_ISL_1048444, EPI_ISL_1048445, EPI_ISL_1048446, EPI_ISL_1048447, EPI_ISL_1048448, EPI_ISL_1048449, EPI_ISL_1048450, EPI_ISL_1048451, EPI_ISL_1048452, EPI_ISL_1048453, EPI_ISL_1048454, EPI_ISL_1048455, EPI_ISL_1048456, EPI_ISL_1048457, EPI_ISL_1048458, EPI_ISL_1048459, EPI_ISL_1048460, EPI_ISL_1048461, EPI_ISL_1048462, EPI_ISL_1048463, EPI_ISL_1048464, EPI_ISL_1048465, EPI_ISL_1048466, EPI_ISL_1048467, EPI_ISL_1048468, EPI_ISL_1048469, EPI_ISL_1048470, EPI_ISL_1048471, EPI_ISL_1048472, EPI_ISL_1048473, EPI_ISL_1048474, EPI_ISL_1048475, EPI_ISL_1048476, EPI_ISL_1048477, EPI_ISL_1048478, EPI_ISL_1048479, EPI_ISL_1048480, EPI_ISL_1048481, EPI_ISL_1048482, EPI_ISL_1048483, EPI_ISL_1048484, EPI_ISL_1048485, EPI_ISL_1048486, EPI_ISL_1048487, EPI_ISL_1048488, EPI_ISL_1048489, EPI_ISL_1048490, EPI_ISL_1048491, EPI_ISL_1048492, EPI_ISL_1048493, EPI_ISL_1048494, EPI_ISL_1048495, EPI_ISL_1048496, EPI_ISL_1048497, EPI_ISL_1048498, EPI_ISL_1048499, EPI_ISL_1048500, EPI_ISL_1048507, EPI_ISL_1048508, EPI_ISL_1048518, EPI_ISL_1048520, EPI_ISL_1048535, EPI_ISL_1048537 | | | |
| see above | National Health Laboratory Service, South Africa | KRISP, KZn Research Innovation and Sequencing Platform | Emmanuel SJ; Giandhari J; Khan S; Laguda-Akingba O; Lessells R; Mdlalose K; Pillay S; Tegally H; Wilkinson E; York D; de Oliveira T |
| EPI_ISL_1363124, EPI_ISL_1363125, EPI_ISL_1363126, EPI_ISL_1363127, EPI_ISL_1363128, EPI_ISL_1363129, EPI_ISL_1363130, EPI_ISL_1363131, EPI_ISL_1363132, EPI_ISL_1363133, EPI_ISL_1363134, EPI_ISL_1363135, EPI_ISL_1363136, EPI_ISL_1363137, EPI_ISL_1363138, EPI_ISL_1363139, EPI_ISL_1363140, EPI_ISL_1363141, EPI_ISL_1363142, EPI_ISL_1363143, EPI_ISL_1363144, EPI_ISL_1363145, EPI_ISL_1363146, EPI_ISL_1363147, EPI_ISL_1363148, EPI_ISL_1363149, EPI_ISL_1363150, EPI_ISL_1372562, EPI_ISL_1372563 | | | |
| see above | National Health Laboratory Services | National Health Laboratory Services | Arshad Ismail; Florette Treurnicht; Kathleen Subramoney |
| EPI_ISL_514355, EPI_ISL_514356, EPI_ISL_514357, EPI_ISL_514358, EPI_ISL_514359, EPI_ISL_514360, EPI_ISL_514361, EPI_ISL_514362, EPI_ISL_514363, EPI_ISL_514364, EPI_ISL_514365, EPI_ISL_514366, EPI_ISL_514367, EPI_ISL_514368, EPI_ISL_514369, EPI_ISL_514370, EPI_ISL_514371, EPI_ISL_514372, EPI_ISL_514373, EPI_ISL_514374, EPI_ISL_514375, EPI_ISL_514376, EPI_ISL_514377, EPI_ISL_514378, EPI_ISL_514379, EPI_ISL_514380, EPI_ISL_514381, EPI_ISL_514382, EPI_ISL_514383, EPI_ISL_514384, EPI_ISL_514385, EPI_ISL_514386, EPI_ISL_514387, EPI_ISL_514388, EPI_ISL_514389, EPI_ISL_514390, EPI_ISL_514391, EPI_ISL_514392, EPI_ISL_514393, EPI_ISL_514394, EPI_ISL_514395, EPI_ISL_514396, EPI_ISL_514397, EPI_ISL_514398, EPI_ISL_514399, EPI_ISL_514400, EPI_ISL_514401, EPI_ISL_514402, EPI_ISL_514403, EPI_ISL_514404, EPI_ISL_514405, EPI_ISL_514406, EPI_ISL_514407, EPI_ISL_514408, EPI_ISL_514409, EPI_ISL_514410, EPI_ISL_514411, EPI_ISL_514412, EPI_ISL_514413, EPI_ISL_514414, EPI_ISL_514415, EPI_ISL_514416, EPI_ISL_514417, EPI_ISL_514418, EPI_ISL_514419, EPI_ISL_514420, EPI_ISL_514421, EPI_ISL_514422, EPI_ISL_514423, EPI_ISL_514424, EPI_ISL_515133, EPI_ISL_515134, EPI_ISL_515135, EPI_ISL_515136, EPI_ISL_515137, EPI_ISL_515138, EPI_ISL_515139, EPI_ISL_515140, EPI_ISL_515141, EPI_ISL_515142, EPI_ISL_515143, EPI_ISL_515144, EPI_ISL_515145, EPI_ISL_515146, EPI_ISL_515147, EPI_ISL_515148, EPI_ISL_515149, EPI_ISL_515150, EPI_ISL_515151, EPI_ISL_515152, EPI_ISL_515153, EPI_ISL_515154, EPI_ISL_515155, EPI_ISL_515156, EPI_ISL_515157, EPI_ISL_515158, EPI_ISL_515159, EPI_ISL_515160, EPI_ISL_515161, EPI_ISL_515162, EPI_ISL_515163, EPI_ISL_515164, EPI_ISL_515165, EPI_ISL_515166, EPI_ISL_515167, EPI_ISL_515168, EPI_ISL_515169, EPI_ISL_515170, EPI_ISL_515171, EPI_ISL_515172, EPI_ISL_515173, EPI_ISL_515174, EPI_ISL_515175, EPI_ISL_515176, EPI_ISL_515177, EPI_ISL_515178, EPI_ISL_515179, EPI_ISL_515180, EPI_ISL_622894, EPI_ISL_622895, EPI_ISL_622896, EPI_ISL_622897, EPI_ISL_622898, EPI_ISL_622899, EPI_ISL_622900, EPI_ISL_622901, EPI_ISL_622902, EPI_ISL_622903, EPI_ISL_622904, EPI_ISL_622905, EPI_ISL_622906, EPI_ISL_622907, EPI_ISL_622908, EPI_ISL_622909, EPI_ISL_622910, EPI_ISL_622911, EPI_ISL_622912, EPI_ISL_622914, EPI_ISL_622916, EPI_ISL_622917, EPI_ISL_622918, EPI_ISL_622919, EPI_ISL_622920, EPI_ISL_622921, EPI_ISL_622922, EPI_ISL_622923, EPI_ISL_622924, EPI_ISL_622925, EPI_ISL_622926, EPI_ISL_622927, EPI_ISL_622928, EPI_ISL_622929, EPI_ISL_622930, EPI_ISL_622931, EPI_ISL_622932, EPI_ISL_622933, EPI_ISL_622935, EPI_ISL_622936, EPI_ISL_622937, EPI_ISL_622938, EPI_ISL_622939, EPI_ISL_622940, EPI_ISL_622941 | | | |
| see above | National Institute for Communicable Diseases of the National Health Laboratory Service | National Institute for Communicable Diseases of the National Health Laboratory Service | Allam M; Bhiman JN; Ismail A; Khumalo Z; Kwenda S; Mnyameni F; Mohale T; Mtshali P; Subramoney K |
| EPI_ISL_939656, EPI_ISL_939657, EPI_ISL_939658, EPI_ISL_939659, EPI_ISL_940067, EPI_ISL_1239850, EPI_ISL_1239851, EPI_ISL_1239852, EPI_ISL_1239853, EPI_ISL_1239854, EPI_ISL_1239855, EPI_ISL_1239856, EPI_ISL_1239857, EPI_ISL_1239858, EPI_ISL_1239859, EPI_ISL_1239860, EPI_ISL_1239861, EPI_ISL_1239862, EPI_ISL_1239863, EPI_ISL_1239864, EPI_ISL_1239865, EPI_ISL_1239866, EPI_ISL_1239867, EPI_ISL_1239868, EPI_ISL_1239869, EPI_ISL_1239870, EPI_ISL_1239871, EPI_ISL_1239872, EPI_ISL_1239873, EPI_ISL_1239874, EPI_ISL_1239875, EPI_ISL_1239876, EPI_ISL_1239877, EPI_ISL_1239878, EPI_ISL_1239879, EPI_ISL_1239880, EPI_ISL_1239881, EPI_ISL_1239882, EPI_ISL_1827555, EPI_ISL_1827556, EPI_ISL_1827557, EPI_ISL_1827558, EPI_ISL_1827559, EPI_ISL_1827560, EPI_ISL_1827561, EPI_ISL_1827562, EPI_ISL_1827563, EPI_ISL_1827564, EPI_ISL_1827565, EPI_ISL_1827566, EPI_ISL_1827567, EPI_ISL_1827568, EPI_ISL_1827569, EPI_ISL_1827570, EPI_ISL_1827571, EPI_ISL_1827572, EPI_ISL_1827573, EPI_ISL_1827574, EPI_ISL_1827575, EPI_ISL_1827576, EPI_ISL_1827577, EPI_ISL_1827578, EPI_ISL_1827579, EPI_ISL_1827580, EPI_ISL_1827581, EPI_ISL_1827582, EPI_ISL_1827583, EPI_ISL_1827584, EPI_ISL_1827585, EPI_ISL_1827586, EPI_ISL_1827587, EPI_ISL_1827588, EPI_ISL_1827589, EPI_ISL_1827590, EPI_ISL_1827591, EPI_ISL_1827592, EPI_ISL_1827593, EPI_ISL_1827594, EPI_ISL_1827595, EPI_ISL_1827596, EPI_ISL_1827597, EPI_ISL_1827598, EPI_ISL_1827599, EPI_ISL_1827600, EPI_ISL_1827601, EPI_ISL_1827602, EPI_ISL_1827603, EPI_ISL_1827604, EPI_ISL_1827605, EPI_ISL_1827606, EPI_ISL_1827607, EPI_ISL_1827608, EPI_ISL_1827609, EPI_ISL_1827610, EPI_ISL_1827611, EPI_ISL_1827612, EPI_ISL_1827613, EPI_ISL_1827614, EPI_ISL_1827615, EPI_ISL_1827616, EPI_ISL_1827617, EPI_ISL_1827618, EPI_ISL_1827619, EPI_ISL_1827620, EPI_ISL_1827621, EPI_ISL_1827622, EPI_ISL_1827623, EPI_ISL_1827624, EPI_ISL_1827625, EPI_ISL_1827626, EPI_ISL_1827627, EPI_ISL_1827628, EPI_ISL_1827629, EPI_ISL_1827630, EPI_ISL_1827631, EPI_ISL_1827632, EPI_ISL_1827633, EPI_ISL_1914786, EPI_ISL_1914787, EPI_ISL_1914788, EPI_ISL_1914789, EPI_ISL_1914790, EPI_ISL_1914791, EPI_ISL_1914792, EPI_ISL_1914793, EPI_ISL_1914794, EPI_ISL_1914795, EPI_ISL_1914796, EPI_ISL_1914797, EPI_ISL_1914798, EPI_ISL_1914799, EPI_ISL_1914800, EPI_ISL_1914801, EPI_ISL_1914802, EPI_ISL_1914803, EPI_ISL_1914804, EPI_ISL_1914805, EPI_ISL_1914806, EPI_ISL_1914807, EPI_ISL_1914808, EPI_ISL_1914809, EPI_ISL_1914810, EPI_ISL_1914811, EPI_ISL_1914812, EPI_ISL_1914813, EPI_ISL_1914814, EPI_ISL_1914815, EPI_ISL_1914816, EPI_ISL_1914817, EPI_ISL_1914818, EPI_ISL_1914819, EPI_ISL_1914820, EPI_ISL_1914821, EPI_ISL_1914822, EPI_ISL_1914823, EPI_ISL_1914824, EPI_ISL_1914825, EPI_ISL_1914826, EPI_ISL_1914827, EPI_ISL_1914828, EPI_ISL_1914829, EPI_ISL_1914830, EPI_ISL_1914831, EPI_ISL_1914832, EPI_ISL_1914833, EPI_ISL_1914834, EPI_ISL_1914835, EPI_ISL_1914836, EPI_ISL_1914837, EPI_ISL_1914838, EPI_ISL_1914839, EPI_ISL_1914840, EPI_ISL_1914841, EPI_ISL_1914842, EPI_ISL_1914843, EPI_ISL_1914844, EPI_ISL_1914845, EPI_ISL_1914846, EPI_ISL_1914847, EPI_ISL_1914848, EPI_ISL_1914849, EPI_ISL_1914850, EPI_ISL_1914851, EPI_ISL_1914852, EPI_ISL_1914853, EPI_ISL_1914854, EPI_ISL_1914855, EPI_ISL_1914856, EPI_ISL_1914857, EPI_ISL_1914858, EPI_ISL_1914859, EPI_ISL_1914860, EPI_ISL_1914861, EPI_ISL_1914862, EPI_ISL_1914863, EPI_ISL_1914864, EPI_ISL_1914865, EPI_ISL_1914866, EPI_ISL_1914867, EPI_ISL_1914868, EPI_ISL_1914869, EPI_ISL_1914870, EPI_ISL_1914871, EPI_ISL_1914872, EPI_ISL_1914873, EPI_ISL_1914874, EPI_ISL_1914875, EPI_ISL_1914876, EPI_ISL_1914877, EPI_ISL_1914878, EPI_ISL_1914879, EPI_ISL_1914880, EPI_ISL_1914881, EPI_ISL_1914882, EPI_ISL_1914883, EPI_ISL_1914884, EPI_ISL_1914885, EPI_ISL_1914886, EPI_ISL_1914887, EPI_ISL_1914888, EPI_ISL_1914889, EPI_ISL_1914890, EPI_ISL_1914891, EPI_ISL_1914892, EPI_ISL_1914893, EPI_ISL_1914894, EPI_ISL_1914895, EPI_ISL_1914896, EPI_ISL_1914897, EPI_ISL_1914898, EPI_ISL_1914899, EPI_ISL_1914900, EPI_ISL_1914901, EPI_ISL_1914902, EPI_ISL_1914903, EPI_ISL_1914904, EPI_ISL_1914905, EPI_ISL_1914906, EPI_ISL_1914907, EPI_ISL_1914908, EPI_ISL_1914909, EPI_ISL_1914910, EPI_ISL_1914911, EPI_ISL_1914912, EPI_ISL_1914913, EPI_ISL_1914914, EPI_ISL_1914915, EPI_ISL_1914916, EPI_ISL_1914917, EPI_ISL_1914918, EPI_ISL_1914919, EPI_ISL_1914920, EPI_ISL_1914921, EPI_ISL_1914922, EPI_ISL_1914923, EPI_ISL_1914924, EPI_ISL_1914925, EPI_ISL_1914926, EPI_ISL_1914927 | | | |
| see above | National Institute for Communicable Diseases,National Health Laboratory Services, Gauteng, South Africa | National Institute for Communicable Diseases of the National Health Laboratory Service | Allam M; Amoako DG; Bhiman JN; Ismail A; Mahlangu B; Mohale T; Ntuli N; Scheepers C |
| EPI_ISL_678548, EPI_ISL_678549, EPI_ISL_678550, EPI_ISL_678551, EPI_ISL_678552, EPI_ISL_678553, EPI_ISL_678554, EPI_ISL_678555, EPI_ISL_678556, EPI_ISL_678557, EPI_ISL_678558, EPI_ISL_678559, EPI_ISL_678560, EPI_ISL_678561, EPI_ISL_678562, EPI_ISL_678563, EPI_ISL_678564, EPI_ISL_678565, EPI_ISL_678567, EPI_ISL_678568, EPI_ISL_678571, EPI_ISL_678572, EPI_ISL_678573, EPI_ISL_678576, EPI_ISL_678577, EPI_ISL_678578, EPI_ISL_678579, EPI_ISL_678580, EPI_ISL_678581, EPI_ISL_678582, EPI_ISL_678583, EPI_ISL_678584, EPI_ISL_678585, EPI_ISL_678586, EPI_ISL_678587, EPI_ISL_678588, EPI_ISL_678589, EPI_ISL_678590, EPI_ISL_678591, EPI_ISL_678592, EPI_ISL_678593, EPI_ISL_678594, EPI_ISL_678595, EPI_ISL_678596, EPI_ISL_678597, EPI_ISL_736936, EPI_ISL_736937, EPI_ISL_736961, EPI_ISL_736962, EPI_ISL_736963, EPI_ISL_736964, EPI_ISL_736965 | | | |
| see above | Netcare | KRISP, KZN Research Innovation and Sequencing Platform | ChimukangaraB; Giandhari J; Khan S; Lessells R; Mdlalose K; Pillay S; Tegally H; Wilkinson E; York D; de Oliveira T |

| | | | |
|---|---|---|---|
| EPI_ISL_1085951, EPI_ISL_1085952, EPI_ISL_1085953, EPI_ISL_1085954, EPI_ISL_1085955, EPI_ISL_1085956, EPI_ISL_1085957, EPI_ISL_1085958, EPI_ISL_1085959, EPI_ISL_1085960, EPI_ISL_1085961, EPI_ISL_1085962, EPI_ISL_1085963, EPI_ISL_1085964, EPI_ISL_1085965, EPI_ISL_1085966, EPI_ISL_1085967, EPI_ISL_1085968, EPI_ISL_1085969, EPI_ISL_1085970, EPI_ISL_1085971, EPI_ISL_1085972, EPI_ISL_1085973, EPI_ISL_1085974, EPI_ISL_1085975, EPI_ISL_1085976, EPI_ISL_1085977, EPI_ISL_1085978, EPI_ISL_1085979, EPI_ISL_1085980, EPI_ISL_1085981, EPI_ISL_1085982, EPI_ISL_1085983, EPI_ISL_1085984, EPI_ISL_1085985, EPI_ISL_1085986, EPI_ISL_1085987, EPI_ISL_1085988, EPI_ISL_1085989, EPI_ISL_1085990, EPI_ISL_1085991, EPI_ISL_1085992, EPI_ISL_1085993, EPI_ISL_1085994, EPI_ISL_1085995, EPI_ISL_1085996, EPI_ISL_1085997, EPI_ISL_1085998, EPI_ISL_1085999, EPI_ISL_1086000, EPI_ISL_1086001, EPI_ISL_1086002, EPI_ISL_1086003, EPI_ISL_1086004, EPI_ISL_1086005, EPI_ISL_1086006, EPI_ISL_1086007, EPI_ISL_1086008, EPI_ISL_1086009, EPI_ISL_1086010, EPI_ISL_1086011, EPI_ISL_1086012, EPI_ISL_1086013, EPI_ISL_1086014, EPI_ISL_1086015, EPI_ISL_1086016, EPI_ISL_1086017, EPI_ISL_1086018, EPI_ISL_1086019, EPI_ISL_1086020, EPI_ISL_1086021, EPI_ISL_1086022, EPI_ISL_1086023, EPI_ISL_1086024, EPI_ISL_1086025, EPI_ISL_1086026, EPI_ISL_1086027, EPI_ISL_1086028, EPI_ISL_1086029, EPI_ISL_1086030, EPI_ISL_1086031, EPI_ISL_1086032, EPI_ISL_1086033, EPI_ISL_1250356, EPI_ISL_1250358, EPI_ISL_1250359, EPI_ISL_1250361, EPI_ISL_1250362, EPI_ISL_1250364, EPI_ISL_1250366, EPI_ISL_1250367, EPI_ISL_1250369, EPI_ISL_1250370, EPI_ISL_1250371, EPI_ISL_1250373, EPI_ISL_1250375, EPI_ISL_1250376, EPI_ISL_1250378, EPI_ISL_1250379, EPI_ISL_1250381, EPI_ISL_1250383, EPI_ISL_1250385, EPI_ISL_1250386, EPI_ISL_1250388, EPI_ISL_1250390, EPI_ISL_1250392, EPI_ISL_1250393, EPI_ISL_1250394, EPI_ISL_1250396, EPI_ISL_1250397, EPI_ISL_1250399, EPI_ISL_1250401, EPI_ISL_1250402, EPI_ISL_1250404, EPI_ISL_1250405, EPI_ISL_1250407, EPI_ISL_1250409, EPI_ISL_1250410, EPI_ISL_1250412, EPI_ISL_1250413, EPI_ISL_1250414, EPI_ISL_1250416, EPI_ISL_1250417, EPI_ISL_1250419, EPI_ISL_1250420, EPI_ISL_1250422 | | | |
| see above | Netcare/AMPATH | KRISP, KZN Research Innovation and Sequencing Platform | Emmanuel SJ; Giandhari J; Khan S; Lessells R; Maslo C; Mdlalose K; Pillay S; Sitharam L; Tegally H; Wilkinson E; York D; de Oliveira T |
| EPI_ISL_700442, EPI_ISL_700444, EPI_ISL_2140669 | New Horizon Clinic wc NZC | NHLS/UCT | Arash Iranzadeh; Bruna Galvao; Carolyn Williamson; Deelan Doolabh; Diana Hardie; Houriiyah Tegally; Innocent Mudau; Kruger Marais; Lynn Tyers; Marvin Hsiao; Stephen Korsman |
| EPI_ISL_960116 | New Horizon Clinic wc NZC | National Health Laboratory Service/UCT | Arash Iranzadeh; Bruna Galvao; Carolyn Williamson; Deelan Doolabh; Diana Hardie; Innocent Mudau; Kruger Marais; Lynn Tyers; Marvin Hsiao; Stephen Korsman |
| EPI_ISL_696468, EPI_ISL_696489 | New Horizon Clinic wc NZC & NHLS/UCT | KRISP, KZN Research Innovation and Sequencing Platform | Arash Iranzadeh; Bruna Galvao; Carolyn Williamson; Deelan Doolabh; Diana Hardie; Emanuel James San; Houriiyah Tegally; Innocent Mudau; Jennifer Giandhari; Kruger Marais; Lynn Tyers; Marvin Hsiao; Stephen Korsman; Sureshnee Pillay; Tulio de Oliveira |
| EPI_ISL_1040703 | Nieuveldpark Clinic wc NVP | NHLS/UCT | Arash Iranzadeh; Bruna Galvao; Carolyn Williamson; Deelan Doolabh; Diana Hardie; Innocent Mudau; Kruger Marais; Lynn Tyers; Marvin Hsiao; Stephen Korsman |
| EPI_ISL_745142, EPI_ISL_745143 | Nomangesi Jayiya Clinic | National Health Laboratory Service (NHLS), Tygerberg | Bronwyn Kleinhans; Eduan Wilkindon; Gert van Zyl; Houriiyah Tegally; Kayla Delaney; Susan Engelbrecht; Tulio de Oliveira; Wolfgang Preiser |
| EPI_ISL_745126 | Okiep CHC | National Health Laboratory Service (NHLS), Tygerberg | Bronwyn Kleinhans; Eduan Wilkindon; Gert van Zyl; Houriiyah Tegally; Kayla Delaney; Susan Engelbrecht; Tulio de Oliveira; Wolfgang Preiser |
| EPI_ISL_640017, EPI_ISL_640023, EPI_ISL_640077, EPI_ISL_700417, EPI_ISL_700447, EPI_ISL_700575, EPI_ISL_700594, EPI_ISL_1534344, EPI_ISL_1534456, EPI_ISL_2140665 | | | |
| see above | Oudtshoorn Hospital wc OUD | NHLS/UCT | Arash Iranzadeh; Bruna Galvao; Carolyn Williamson; Deelan Doolabh; Diana Hardie; Emmanuel SJ; Innocent Mudau; Kruger Marais; Lynn Tyers; Marvin Hsiao; Stephen Korsman; Tegally H; de Oliveira T |
| EPI_ISL_960144, EPI_ISL_960146, EPI_ISL_960147 | Oudtshoorn Hospital wc OUD | National Health Laboratory Service/UCT | Arash Iranzadeh; Bruna Galvao; Carolyn Williamson; Deelan Doolabh; Diana Hardie; Innocent Mudau; Kruger Marais; Lynn Tyers; Marvin Hsiao; Stephen Korsman |
| EPI_ISL_2140667 | Oudtshoorn Sick Bay (SANDF) | NHLS/UCT | Arash Iranzadeh; Bruna Galvao; Carolyn Williamson; Deelan Doolabh; Diana Hardie; Innocent Mudau; Kruger Marais; Lynn Tyers; Marvin Hsiao; Stephen Korsman |
| EPI_ISL_700435, EPI_ISL_700439, EPI_ISL_700449, EPI_ISL_700451, EPI_ISL_700452, EPI_ISL_700478, EPI_ISL_700551, EPI_ISL_700586, EPI_ISL_1040655, EPI_ISL_1040679, EPI_ISL_1040799, EPI_ISL_1534425 | | | |
| see above | Pacaltsdorp Clinic wc PAC | NHLS/UCT | Arash Iranzadeh; Bruna Galvao; Carolyn Williamson; Deelan Doolabh; Diana Hardie; Emmanuel SJ; Houriiyah Tegally; Innocent Mudau; Kruger Marais; Lynn Tyers; Marvin Hsiao; Stephen Korsman; Tegally H; de Oliveira T |
| EPI_ISL_696453, EPI_ISL_696456, EPI_ISL_696459, EPI_ISL_696470, EPI_ISL_696474, EPI_ISL_696504, EPI_ISL_696513 | | | |
| see above | Pacaltsdorp Clinic wc PAC & NHLS/UCT | KRISP, KZN Research Innovation and Sequencing Platform | Arash Iranzadeh; Bruna Galvao; Carolyn Williamson; Deelan Doolabh; Diana Hardie; Emanuel James San; Houriiyah Tegally; Innocent Mudau; Jennifer Giandhari; Kruger Marais; Lynn Tyers; Marvin Hsiao; Stephen Korsman; Sureshnee Pillay; Tulio de Oliveira |
| EPI_ISL_660143, EPI_ISL_660144, EPI_ISL_660145, EPI_ISL_660146, EPI_ISL_660147, EPI_ISL_660148, EPI_ISL_660149, EPI_ISL_660150, EPI_ISL_660151, EPI_ISL_660152, EPI_ISL_660153, EPI_ISL_660154, EPI_ISL_660155, EPI_ISL_660156, EPI_ISL_660157, EPI_ISL_660158, EPI_ISL_912401 | | | |
| see above | PathCare | National Health Laboratory Service (NHLS), Tygerberg | Bronwyn Kleinhans; Davis M-A; Draper C; Eduan Wilkindon; Gert van Zyl; Houriiyah Tegally; Hsiao M; Jean Maritz; Kayla Delaney; Siegfried N; Susan Engelbrecht; Tulio de Oliveira; Williamson C; Wolfgang Preiser |
| EPI_ISL_960111 | Pelican Park CDC wc PAX | National Health Laboratory Service/UCT | Arash Iranzadeh; Bruna Galvao; Carolyn Williamson; Deelan Doolabh; Diana Hardie; Innocent Mudau; Kruger Marais; Lynn Tyers; Marvin Hsiao; Stephen Korsman |
| EPI_ISL_700430, EPI_ISL_700434, EPI_ISL_700500, EPI_ISL_700518, EPI_ISL_700528, EPI_ISL_700560, EPI_ISL_1534424 | | | |
| see above | Plettenberg Bay Clinic wc PLC | NHLS/UCT | Arash Iranzadeh; Bruna Galvao; Carolyn Williamson; Deelan Doolabh; Diana Hardie; Emmanuel SJ; Houriiyah Tegally; Innocent Mudau; Kruger Marais; Lynn Tyers; Marvin Hsiao; Stephen Korsman; Tegally H; de Oliveira T |
| EPI_ISL_960115 | Plettenberg Bay Clinic wc PLC | National Health Laboratory Service/UCT | Arash Iranzadeh; Bruna Galvao; Carolyn Williamson; Deelan Doolabh; Diana Hardie; Innocent Mudau; Kruger Marais; Lynn Tyers; Marvin Hsiao; Stephen Korsman |
| EPI_ISL_696454, EPI_ISL_696476 | Plettenberg Bay Clinic wc PLC & NHLS/UCT | KRISP, KZN Research Innovation and Sequencing Platform | Arash Iranzadeh; Bruna Galvao; Carolyn Williamson; Deelan Doolabh; Diana Hardie; Emanuel James San; Houriiyah Tegally; Innocent Mudau; Jennifer Giandhari; Kruger Marais; Lynn Tyers; Marvin Hsiao; Stephen Korsman; Sureshnee Pillay; Tulio de Oliveira |
| EPI_ISL_712070, EPI_ISL_712071, EPI_ISL_712072, EPI_ISL_712073, EPI_ISL_712074, EPI_ISL_712075, EPI_ISL_712076, EPI_ISL_712079, EPI_ISL_712080, EPI_ISL_712081, EPI_ISL_712082, EPI_ISL_712083, EPI_ISL_712084, EPI_ISL_712085, EPI_ISL_712086, EPI_ISL_712087, EPI_ISL_712088, EPI_ISL_712089, EPI_ISL_712090, EPI_ISL_712091, EPI_ISL_712093, EPI_ISL_712094, EPI_ISL_712095, EPI_ISL_712096, EPI_ISL_723468 | | | |
| see above | Port Elizabeth Provincial Hospital, National Health Laboratory Services, Eastern Cape, South Africa | National Institute for Communicable Diseases of the National Health Laboratory Service | Allam M; Bhiman JN; Ismail A; Mahlangu B; Mohale T; Ntuli N |
| EPI_ISL_745129, EPI_ISL_745187 | Port Nolloth Hospital | National Health Laboratory Service (NHLS), Tygerberg | Bronwyn Kleinhans; Eduan Wilkindon; Gert van Zyl; Houriiyah Tegally; Kayla Delaney; Susan Engelbrecht; Tulio de Oliveira; Wolfgang Preiser |
| EPI_ISL_2086931, EPI_ISL_2086932 | ROB FERREIRA LABORATORY | National Institute for Communicable Diseases of the National Health Laboratory Service | Amoako DG; Bhiman JN; Ismail A; Mahlangu B; Mohale T; Ntuli N; Scheepers C |
| EPI_ISL_640086, EPI_ISL_640136, EPI_ISL_1040742, EPI_ISL_1534342, EPI_ISL_1534343 | Red Cross Children's Hospital wc RXH | NHLS/UCT | Arash Iranzadeh; Bruna Galvao; Carolyn Williamson; Deelan Doolabh; Diana Hardie; Emmanuel SJ; Innocent Mudau; Kruger Marais; Lynn Tyers; Marvin Hsiao; Stephen Korsman; Tegally H; de Oliveira T |
| EPI_ISL_960106 | Red Cross Children's Hospital wc RXH | National Health Laboratory Service/UCT | Arash Iranzadeh; Bruna Galvao; Carolyn Williamson; Deelan Doolabh; Diana Hardie; Innocent Mudau; Kruger Marais; Lynn Tyers; Marvin Hsiao; Stephen Korsman |
| EPI_ISL_1040778, EPI_ISL_1534375 | Retreat CHC wc RHC | NHLS/UCT | Arash Iranzadeh; Bruna Galvao; Carolyn Williamson; Deelan Doolabh; Diana Hardie; Emmanuel SJ; Innocent Mudau; Kruger Marais; Lynn Tyers; Marvin Hsiao; Stephen Korsman; Tegally H; de Oliveira T |
| EPI_ISL_640035, EPI_ISL_700465, EPI_ISL_1534332, EPI_ISL_1534333, EPI_ISL_1534422 | Riversdale Clinic wc RAV | NHLS/UCT | Arash Iranzadeh; Bruna Galvao; Carolyn Williamson; Deelan Doolabh; Diana Hardie; Emmanuel SJ; Innocent Mudau; Kruger Marais; Lynn Tyers; Marvin Hsiao; Stephen Korsman; Tegally H; de Oliveira T |
| EPI_ISL_1239479, EPI_ISL_1239484, EPI_ISL_1239487, EPI_ISL_1239490, EPI_ISL_1239493, EPI_ISL_1239502, EPI_ISL_1239509, EPI_ISL_1239515, EPI_ISL_1239524, EPI_ISL_1239528, EPI_ISL_1239530 | | | |
| see above | Rob Ferreira Hospital, National Health Laboratory Services, Mpumalanga, South Africa | National Institute for Communicable Diseases of the National Health Laboratory Service | Amoako DG; Bhiman JN; Ismail A; Mahlangu B; Mohale T; Ntuli N; Scheepers C |
| EPI_ISL_745156 | SAS Saldanha | National Health Laboratory Service (NHLS), Tygerberg | Bronwyn Kleinhans; Eduan Wilkindon; Gert van Zyl; Houriiyah Tegally; Kayla Delaney; Susan Engelbrecht; Tulio de Oliveira; Wolfgang Preiser |
| EPI_ISL_745157, EPI_ISL_745161 | SAS Saldanha VPA | National Health Laboratory Service (NHLS), Tygerberg | Bronwyn Kleinhans; Eduan Wilkindon; Gert van Zyl; Houriiyah Tegally; Kayla Delaney; Susan Engelbrecht; Tulio de Oliveira; Wolfgang Preiser |
| EPI_ISL_2086909, EPI_ISL_2086914, EPI_ISL_2086916, EPI_ISL_2086917, EPI_ISL_2086918, EPI_ISL_2086919, EPI_ISL_2086920, EPI_ISL_2086925, EPI_ISL_2086926, EPI_ISL_2086927, EPI_ISL_2086928, EPI_ISL_2086929, EPI_ISL_2086930 | | | |
| see above | Sebokeng Lab | National Institute for Communicable Diseases of the National Health Laboratory Service | Amoako DG; Bhiman JN; Ismail A; Mahlangu B; Mohale T; Ntuli N; Scheepers C |
| EPI_ISL_640034, EPI_ISL_700588, EPI_ISL_1534339, EPI_ISL_1534356 | Sedgefield Clinic wc SGE | NHLS/UCT | Arash Iranzadeh; Bruna Galvao; Carolyn Williamson; Deelan Doolabh; Diana Hardie; Emmanuel SJ; Innocent Mudau; Kruger Marais; Lynn Tyers; Marvin Hsiao; Stephen Korsman; Tegally H; de Oliveira T |
| EPI_ISL_696462, EPI_ISL_696475, EPI_ISL_696487, EPI_ISL_696488, EPI_ISL_696510, EPI_ISL_696514, EPI_ISL_696515, EPI_ISL_696516, EPI_ISL_696517, EPI_ISL_696518 | | | |
| see above | Sedgefield Clinic wc SGE & NHLS/UCT | KRISP, KZN Research Innovation and Sequencing Platform | Arash Iranzadeh; Bruna Galvao; Carolyn Williamson; Deelan Doolabh; Diana Hardie; Emanuel James San; Houriiyah Tegally; Innocent Mudau; Jennifer Giandhari; Kruger Marais; Lynn Tyers; Marvin Hsiao; Stephen Korsman; Sureshnee Pillay; Tulio de Oliveira |
| EPI_ISL_1534335, EPI_ISL_1534417 | Still Bay Sat Clinic wc SFV | NHLS/UCT | Arash Iranzadeh; Bruna Galvao; Carolyn Williamson; Deelan Doolabh; Diana Hardie; Emmanuel SJ; Innocent Mudau; Kruger Marais; Lynn Tyers; Marvin Hsiao; Stephen Korsman; Tegally H; de Oliveira T |
| EPI_ISL_1040647 | Swellendam PHC | NHLS/UCT | Arash Iranzadeh; Bruna Galvao; Carolyn Williamson; Deelan Doolabh; Diana Hardie; Innocent Mudau; Kruger Marais; Lynn Tyers; Marvin Hsiao; Stephen Korsman |
| EPI_ISL_640019, EPI_ISL_640026, EPI_ISL_700424, EPI_ISL_700425, EPI_ISL_700428, EPI_ISL_700431, EPI_ISL_700438, EPI_ISL_700460, EPI_ISL_700461, EPI_ISL_700482, EPI_ISL_700483, EPI_ISL_700487, EPI_ISL_700490, EPI_ISL_700498, EPI_ISL_700502, EPI_ISL_2140654, EPI_ISL_2140661, EPI_ISL_2140662 | | | |
| see above | Thembalethu CDC wc THC | NHLS/UCT | Arash Iranzadeh; Bruna Galvao; Carolyn Williamson; Deelan Doolabh; Diana Hardie; Houriiyah Tegally; Innocent Mudau; Kruger Marais; Lynn Tyers; Marvin Hsiao; Stephen Korsman |
| EPI_ISL_960139 | Thembalethu CDC wc THC | National Health Laboratory Service/UCT | Arash Iranzadeh; Bruna Galvao; Carolyn Williamson; Deelan Doolabh; Diana Hardie; Innocent Mudau; Kruger Marais; Lynn Tyers; Marvin Hsiao; Stephen Korsman |
| EPI_ISL_696521 | Thembalethu CDC wc THC & NHLS/UC | KRISP, KZN Research Innovation and Sequencing Platform | Arash Iranzadeh; Bruna Galvao; Carolyn Williamson; Deelan Doolabh; Diana Hardie; Emanuel James San; Houriiyah Tegally; Innocent Mudau; Jennifer Giandhari; Kruger Marais; Lynn Tyers; Marvin Hsiao; Stephen Korsman; Sureshnee Pillay; Tulio de Oliveira |
| EPI_ISL_696452, EPI_ISL_696455, EPI_ISL_696461, EPI_ISL_696493, EPI_ISL_696494, EPI_ISL_696496, EPI_ISL_696499, EPI_ISL_696500, EPI_ISL_696519, EPI_ISL_696520 | | | |
| see above | Thembalethu CDC wc THC & NHLS/UCT | KRISP, KZN Research Innovation and Sequencing Platform | Arash Iranzadeh; Bruna Galvao; Carolyn Williamson; Deelan Doolabh; Diana Hardie; Emanuel James San; Houriiyah Tegally; Innocent Mudau; Jennifer Giandhari; Kruger Marais; Lynn Tyers; Marvin Hsiao; Stephen Korsman; Sureshnee Pillay; Tulio de Oliveira |
| EPI_ISL_696485, EPI_ISL_696490 | Touwsranten Clinic wc TST & NHLS/UCT | KRISP, KZN Research Innovation and Sequencing Platform | Arash Iranzadeh; Bruna Galvao; Carolyn Williamson; Deelan Doolabh; Diana Hardie; Emanuel James San; Houriiyah Tegally; Innocent Mudau; Jennifer Giandhari; Kruger Marais; Lynn Tyers; Marvin Hsiao; Stephen Korsman; Sureshnee Pillay; Tulio de Oliveira |
| EPI_ISL_1239459, EPI_ISL_1239462, EPI_ISL_1239464, EPI_ISL_1239465, EPI_ISL_1239469, EPI_ISL_1239470, EPI_ISL_1239472, EPI_ISL_1239473, EPI_ISL_1239482, EPI_ISL_1239491, EPI_ISL_1239492, EPI_ISL_1239494, EPI_ISL_1239495, EPI_ISL_1239496, EPI_ISL_1239497, EPI_ISL_1239498, EPI_ISL_1239499, EPI_ISL_1239500, EPI_ISL_1239501, EPI_ISL_1239503, EPI_ISL_1239504, EPI_ISL_1239505, EPI_ISL_1239506, EPI_ISL_1239507, EPI_ISL_1239510, EPI_ISL_1239511, EPI_ISL_1239513, EPI_ISL_1239514, EPI_ISL_1239516, EPI_ISL_1239517, EPI_ISL_1239521, EPI_ISL_1239523, EPI_ISL_1239525, EPI_ISL_1239526 | | | |
| see above | Tshepong Hospital, National Health Laboratory Services, North West, South Africa | National Institute for Communicable Diseases of the National Health Laboratory Service | Amoako DG; Bhiman JN; Ismail A; Mahlangu B; Mohale T; Ntuli N; Scheepers C |
| EPI_ISL_960152 | Tshwaragano Hospital | National Health Laboratory Service/UCT | Arash Iranzadeh; Bruna Galvao; Carolyn Williamson; Deelan Doolabh; Diana Hardie; Innocent Mudau; Kruger Marais; Lynn Tyers; Marvin Hsiao; Stephen Korsman |
| EPI_ISL_745140, EPI_ISL_745141, EPI_ISL_745147, EPI_ISL_745148, EPI_ISL_745149, EPI_ISL_745150, EPI_ISL_745180, EPI_ISL_745181, EPI_ISL_745182, EPI_ISL_745183, EPI_ISL_745184, EPI_ISL_745185 | | | |
| see above | Tygerberg Hospital wc TBH | National Health Laboratory Service (NHLS), Tygerberg | Bronwyn Kleinhans; Eduan Wilkindon; Gert van Zyl; Houriiyah Tegally; Kayla Delaney; Susan Engelbrecht; Tulio de Oliveira; Wolfgang Preiser |
| EPI_ISL_1324121, EPI_ISL_1324122, EPI_ISL_1324123, EPI_ISL_1324124, EPI_ISL_1324125, EPI_ISL_1324126, EPI_ISL_1324127, EPI_ISL_1324128, EPI_ISL_1324129, EPI_ISL_1324130, EPI_ISL_1324131 | | | |
| see above | UW Virology Lab | UW Virology Lab | Alexander Greninger; Hong Xie; Keith R Jerome; Lasata Shrestha; Margaret Mills; Meei-Li Huang; Michelle Lin; Noah Baker; Pavitra Roychoudhury; Saraswathi Sathees; Sean Ellis; Shah Mohamed Bakhash |

| | | | |
|---|---|---|---|
| EPI_ISL_940850, EPI_ISL_940851, EPI_ISL_940852, EPI_ISL_940853, EPI_ISL_940854, EPI_ISL_940855, EPI_ISL_940856, EPI_ISL_940857, EPI_ISL_940858, EPI_ISL_940859, EPI_ISL_940861, EPI_ISL_940862, EPI_ISL_940863, EPI_ISL_940864, EPI_ISL_940865, EPI_ISL_940866, EPI_ISL_940867, EPI_ISL_940868, EPI_ISL_940869, EPI_ISL_940870, EPI_ISL_940871, EPI_ISL_940872, EPI_ISL_940873, EPI_ISL_940874, EPI_ISL_940875, EPI_ISL_940876, EPI_ISL_940877, EPI_ISL_940878, EPI_ISL_940879, EPI_ISL_940880, EPI_ISL_940881, EPI_ISL_940882, EPI_ISL_940883, EPI_ISL_940884, EPI_ISL_940885, EPI_ISL_940886, EPI_ISL_940887, EPI_ISL_940888, EPI_ISL_940889 | | | |
| see above | Vaccines and Infectious Diseases Analytics Research Unit (VIDA) | KRISP, KZN Research Innovation and Sequencing Platform | Baillie Vicky; Giandhari Jennifer; Madhi Shabir; Naidoo Yeshnee; Pillay Sureshnee; Tegally Houriiyah; de Oliveira Tulio; du Plessis Jeanine |
| EPI_ISL_1132669, EPI_ISL_1132670, EPI_ISL_1132671, EPI_ISL_1132672, EPI_ISL_1132673, EPI_ISL_1132674, EPI_ISL_1132675, EPI_ISL_1132676, EPI_ISL_1132677, EPI_ISL_1132678, EPI_ISL_1132679, EPI_ISL_1132680, EPI_ISL_1132681, EPI_ISL_1132682, EPI_ISL_1132683, EPI_ISL_1132684, EPI_ISL_1132685, EPI_ISL_1132686, EPI_ISL_1132687, EPI_ISL_1132688, EPI_ISL_1132689, EPI_ISL_1132690, EPI_ISL_1132691, EPI_ISL_1132692, EPI_ISL_1132693, EPI_ISL_1132694, EPI_ISL_1132695, EPI_ISL_1132696, EPI_ISL_1132697, EPI_ISL_1132698, EPI_ISL_1132699, EPI_ISL_1132700, EPI_ISL_1132701, EPI_ISL_1132702, EPI_ISL_1132703, EPI_ISL_1132704, EPI_ISL_1132705, EPI_ISL_1132706, EPI_ISL_1132707, EPI_ISL_1132708, EPI_ISL_1132709, EPI_ISL_1132710, EPI_ISL_1132711, EPI_ISL_1132712, EPI_ISL_1132713, EPI_ISL_1132714, EPI_ISL_1132715, EPI_ISL_1132716, EPI_ISL_1132717, EPI_ISL_1132718, EPI_ISL_1132719, EPI_ISL_1132720, EPI_ISL_1132721, EPI_ISL_1132722, EPI_ISL_1132723, EPI_ISL_1132724, EPI_ISL_1132725, EPI_ISL_1132726, EPI_ISL_1132727, EPI_ISL_1132728 | | | |
| see above | Vaccines and Infectious Diseases Analytics Research Unit (VIDA) | KRISP, KZn Research Innovation and Sequencing Platform | Baillie Vicky; Giandhari Jennifer; Madhi Shabir; Naidoo Yeshnee; Pillay Sureshnee; Tegally Houriiyah; de Oliveira Tulio; du Plessis Jeanine |
| EPI_ISL_640037, EPI_ISL_640046, EPI_ISL_640114, EPI_ISL_1534381 | Valkenberg Hospital wc VBH | NHLS/UCT | Arash Iranzadeh; Bruna Galvao; Carolyn Williamson; Deelan Doolabh; Diana Hardie; Emmanuel SJ; Innocent Mudau; Kruger Marais; Lynn Tyers; Marvin Hsiao; Stephen Korsman; Tegally H; de Oliveira T |
| EPI_ISL_1534361 | Van Wyksdorp Sat Clinic wc VWS | NHLS/UCT | Arash Iranzadeh; Bruna Galvao; Carolyn Williamson; Deelan Doolabh; Diana Hardie; Emmanuel SJ; Innocent Mudau; Kruger Marais; Lynn Tyers; Marvin Hsiao; Stephen Korsman; Tegally H; de Oliveira T |
| EPI_ISL_640072, EPI_ISL_1040677, EPI_ISL_1534354 | Vanguard CHC wc VGC | NHLS/UCT | Arash Iranzadeh; Bruna Galvao; Carolyn Williamson; Deelan Doolabh; Diana Hardie; Emmanuel SJ; Innocent Mudau; Kruger Marais; Lynn Tyers; Marvin Hsiao; Stephen Korsman; Tegally H; de Oliveira T |
| EPI_ISL_960159 | Vanguard CHC wc VGC | National Health Laboratory Service/UCT | Arash Iranzadeh; Bruna Galvao; Carolyn Williamson; Deelan Doolabh; Diana Hardie; Innocent Mudau; Kruger Marais; Lynn Tyers; Marvin Hsiao; Stephen Korsman |
| EPI_ISL_640021, EPI_ISL_640032, EPI_ISL_640085, EPI_ISL_640087, EPI_ISL_640108, EPI_ISL_640109, EPI_ISL_640110, EPI_ISL_640113, EPI_ISL_640126, EPI_ISL_640141, EPI_ISL_700421, EPI_ISL_700546, EPI_ISL_700596, EPI_ISL_1040746, EPI_ISL_1040774, EPI_ISL_1040776, EPI_ISL_1040784, EPI_ISL_1040788, EPI_ISL_1040814, EPI_ISL_1534337, EPI_ISL_1534353, EPI_ISL_1534420, EPI_ISL_2140668 | | | |
| see above | Victoria Hospital wc VHW | NHLS/UCT | Arash Iranzadeh; Bruna Galvao; Carolyn Williamson; Deelan Doolabh; Diana Hardie; Emmanuel SJ; Innocent Mudau; Kruger Marais; Lynn Tyers; Marvin Hsiao; Stephen Korsman; Tegally H; de Oliveira T |
| EPI_ISL_960094, EPI_ISL_960099, EPI_ISL_960101, EPI_ISL_960108, EPI_ISL_960113, EPI_ISL_960114, EPI_ISL_960131, EPI_ISL_960138, EPI_ISL_960150 | | | |
| see above | Victoria Hospital wc VHW | National Health Laboratory Service/UCT | Arash Iranzadeh; Bruna Galvao; Carolyn Williamson; Deelan Doolabh; Diana Hardie; Innocent Mudau; Kruger Marais; Lynn Tyers; Marvin Hsiao; Stephen Korsman |
| EPI_ISL_745117, EPI_ISL_745165, EPI_ISL_745167, EPI_ISL_745189 | Vredenburg Hospital wc VBG | National Health Laboratory Service (NHLS), Tygerberg | Bronwyn Kleinhans; Eduan Wilkindon; Gert van Zyl; Houriiyah Tegally; Kayla Delaney; Susan Engelbrecht; Tulio de Oliveira; Wolfgang Preiser |
| EPI_ISL_745146 | Vredendal Hospital wc VRE | National Health Laboratory Service (NHLS), Tygerberg | Bronwyn Kleinhans; Eduan Wilkindon; Gert van Zyl; Houriiyah Tegally; Kayla Delaney; Susan Engelbrecht; Tulio de Oliveira; Wolfgang Preiser |
| EPI_ISL_745186 | Wallacedene Clinic wc WAL | National Health Laboratory Service (NHLS), Tygerberg | Bronwyn Kleinhans; Eduan Wilkindon; Gert van Zyl; Houriiyah Tegally; Kayla Delaney; Susan Engelbrecht; Tulio de Oliveira; Wolfgang Preiser |
| EPI_ISL_1040664 | Worcester Hospital | NHLS/UCT | Arash Iranzadeh; Bruna Galvao; Carolyn Williamson; Deelan Doolabh; Diana Hardie; Innocent Mudau; Kruger Marais; Lynn Tyers; Marvin Hsiao; Stephen Korsman |
| EPI_ISL_700416 | Zoar Clinic wc ZOA | NHLS/UCT | Arash Iranzadeh; Bruna Galvao; Carolyn Williamson; Deelan Doolabh; Diana Hardie; Innocent Mudau; Kruger Marais; Lynn Tyers; Marvin Hsiao; Stephen Korsman |

We gratefully acknowledge the following Authors from the Originating laboratories responsible for obtaining the specimens, as well as the Submitting laboratories where the genome data were generated and shared via GISAID, on which this research is based.

All Submitters of data may be contacted directly via www.gisaid.org

Authors are sorted alphabetically.

| Accession ID | Originating Laboratory | Submitting Laboratory | Authors |
|---|---|---|---|
| EPI_ISL_1064693, EPI_ISL_1168051 | A05 | The Public Health Agency of Sweden | Anna Risberg; Anna-Malin Linde; Carlo Berg; Karin Tegmark-Wisell; Maria Lind Karlberg; Mattias Haukland; Mia Brytting; Noura Walai; Oskar Karlsson Lindsjo; Petra Edquist; Petra Holmstrom; Reza Advani; Samuel Ohman; Sofia Stamouli |
| EPI_ISL_766699, EPI_ISL_766718, EPI_ISL_766719 | A05 Biomedicum | The Public Health Agency of Sweden | Department of Microbiology; The Public Health Agency of Sweden |
| EPI_ISL_1008401, EPI_ISL_1008402, EPI_ISL_1008403, EPI_ISL_1008404, EPI_ISL_1008405, EPI_ISL_1008406, EPI_ISL_1008407, EPI_ISL_1008408, EPI_ISL_1008409, EPI_ISL_1008410, EPI_ISL_1008411 | | | |
| see above | A05 Diagnostics | The Public Health Agency of Sweden | Anna Risberg; Anna-Malin Linde; Carlo Berg; Karin Tegmark-Wisell; Maria Lind Karlberg; Mattias Haukland; Mia Brytting; Noura Walai; Oskar Karlsson Lindsjo; Petra Edquist; Petra Holmstrom; Reza Advani; Sofia Stamouli |
| EPI_ISL_1008534, EPI_ISL_1008535, EPI_ISL_1008536, EPI_ISL_1008537, EPI_ISL_1008538, EPI_ISL_1008539, EPI_ISL_1231978, EPI_ISL_1231979 | | | |
| see above | A05 diagnostics | The Public Health Agency of Sweden | Anna Risberg; Anna-Malin Linde; Carlo Berg; Karin Tegmark-Wisell; Maria Lind Karlberg; Mattias Haukland; Mia Brytting; Noura Walai; Oskar Karlsson Lindsjo; Petra Edquist; Petra Holmstrom; Reza Advani; Sofia Stamouli |
| EPI_ISL_1008597 | A23 Lab AB | The Public Health Agency of Sweden | Anna Risberg; Anna-Malin Linde; Carlo Berg; Karin Tegmark-Wisell; Maria Lind Karlberg; Mattias Haukland; Mia Brytting; Noura Walai; Oskar Karlsson Lindsjo; Petra Edquist; Petra Holmstrom; Reza Advani; Sofia Stamouli |
| EPI_ISL_775550, EPI_ISL_775551, EPI_ISL_775552, EPI_ISL_775553, EPI_ISL_775554, EPI_ISL_775555, EPI_ISL_775556, EPI_ISL_775557, EPI_ISL_775558, EPI_ISL_775559, EPI_ISL_775560, EPI_ISL_775561, EPI_ISL_775562, EPI_ISL_775563, EPI_ISL_775564, EPI_ISL_775565, EPI_ISL_775566, EPI_ISL_775567, EPI_ISL_775568, EPI_ISL_775569, EPI_ISL_775570, EPI_ISL_775571, EPI_ISL_775575, EPI_ISL_788993, EPI_ISL_789000, EPI_ISL_789001, EPI_ISL_789002, EPI_ISL_789003, EPI_ISL_789004, EPI_ISL_789005, EPI_ISL_789006, EPI_ISL_789007, EPI_ISL_789008, EPI_ISL_789009, EPI_ISL_789010, EPI_ISL_789011, EPI_ISL_789018, EPI_ISL_789019, EPI_ISL_789020, EPI_ISL_789021, EPI_ISL_789022, EPI_ISL_789023, EPI_ISL_789024, EPI_ISL_831944, EPI_ISL_831947, EPI_ISL_831948, EPI_ISL_831949, EPI_ISL_831950, EPI_ISL_831951, EPI_ISL_831952 | | | |
| see above | ABC | The Public Health Agency of Sweden | Department of Microbiology; The Public Health Agency of Sweden |
| EPI_ISL_913291, EPI_ISL_913292, EPI_ISL_913293, EPI_ISL_913486 | ABC Labs | The Public Health Agency of Sweden | Anna Risberg; Anna-Malin Linde; Carlo Berg; Karin Tegmark-Wisell; Maria Lind Karlberg; Mattias Haukland; Mia Brytting; Noura Walai; Oskar Karlsson Lindsjo; Petra Edquist; Petra Holmstrom; Reza Advani; Sofia Stamouli |
| EPI_ISL_1008586, EPI_ISL_1008587, EPI_ISL_1008588 | ABC labs | The Public Health Agency of Sweden | Anna Risberg; Anna-Malin Linde; Carlo Berg; Karin Tegmark-Wisell; Maria Lind Karlberg; Mattias Haukland; Mia Brytting; Noura Walai; Oskar Karlsson Lindsjo; Petra Edquist; Petra Holmstrom; Reza Advani; Sofia Stamouli |
| EPI_ISL_1081843, EPI_ISL_1081844, EPI_ISL_1081845, EPI_ISL_1081846, EPI_ISL_1081847, EPI_ISL_1081848, EPI_ISL_1081849, EPI_ISL_1081850, EPI_ISL_1081851, EPI_ISL_1081852, EPI_ISL_1081853, EPI_ISL_1081854, EPI_ISL_1081855, EPI_ISL_1081856, EPI_ISL_1081857, EPI_ISL_1081858, EPI_ISL_1081859, EPI_ISL_1081860, EPI_ISL_1081861, EPI_ISL_1081862, EPI_ISL_1081863, EPI_ISL_1081864, EPI_ISL_1081865, EPI_ISL_1081866, EPI_ISL_1081867, EPI_ISL_1081868, EPI_ISL_1081869, EPI_ISL_1081870, EPI_ISL_1081871, EPI_ISL_1081872, EPI_ISL_1081873, EPI_ISL_1081874, EPI_ISL_1081875, EPI_ISL_1081876, EPI_ISL_1081877, EPI_ISL_1081878, EPI_ISL_1081879, EPI_ISL_1081880, EPI_ISL_1081881, EPI_ISL_1081882, EPI_ISL_1081883, EPI_ISL_1081884, EPI_ISL_1081885, EPI_ISL_1081886, EPI_ISL_1081887, EPI_ISL_1081888, EPI_ISL_1081889, EPI_ISL_1081890, EPI_ISL_1081891, EPI_ISL_1081892, EPI_ISL_1081893, EPI_ISL_1081894, EPI_ISL_1081895, EPI_ISL_1081896, EPI_ISL_1081897, EPI_ISL_1081898, EPI_ISL_1081899, EPI_ISL_1081900, EPI_ISL_1081901, EPI_ISL_1081902, EPI_ISL_1081903, EPI_ISL_1081904, EPI_ISL_1081905, EPI_ISL_1081906, EPI_ISL_1081907, EPI_ISL_1081908, EPI_ISL_1081909, EPI_ISL_1081910, EPI_ISL_1081911, EPI_ISL_1081912, EPI_ISL_1081913, EPI_ISL_1081914, EPI_ISL_1081915, EPI_ISL_1081916, EPI_ISL_1081917, EPI_ISL_1081918, EPI_ISL_1081919, EPI_ISL_1081920, EPI_ISL_1081921, EPI_ISL_1081923, EPI_ISL_1081925, EPI_ISL_1081927, EPI_ISL_1081929, EPI_ISL_1081931, EPI_ISL_1081933, EPI_ISL_1081936, EPI_ISL_1081938, EPI_ISL_1081944, EPI_ISL_1128779, EPI_ISL_1128782, EPI_ISL_1128784, EPI_ISL_1128786, EPI_ISL_1128789, EPI_ISL_1128791, EPI_ISL_1128794, EPI_ISL_1128796, EPI_ISL_1128798, EPI_ISL_1128801, EPI_ISL_1128803, EPI_ISL_1128805, EPI_ISL_1128808, EPI_ISL_1128810, EPI_ISL_1128812, EPI_ISL_1128815, EPI_ISL_1128817, EPI_ISL_1128819, EPI_ISL_1128822, EPI_ISL_1128824, EPI_ISL_1128826, EPI_ISL_1128829, EPI_ISL_1128831, EPI_ISL_1128833, EPI_ISL_1128835, EPI_ISL_1176391, EPI_ISL_1239168, EPI_ISL_1239169, EPI_ISL_1239170, EPI_ISL_1239171, EPI_ISL_1239172, EPI_ISL_1239173, EPI_ISL_1239174, EPI_ISL_1239175, EPI_ISL_1239176, EPI_ISL_1385876, EPI_ISL_1385889, EPI_ISL_1385890, EPI_ISL_1385891 | | | |
| see above | Clinical Microbiology, Infection Prevention and Control | Section for Molecular Diagnostics | Björn Hallström; Jonas Björkman |
| EPI_ISL_913275, EPI_ISL_913277, EPI_ISL_913281, EPI_ISL_913309, EPI_ISL_913378, EPI_ISL_913379, EPI_ISL_913380, EPI_ISL_913381, EPI_ISL_913382, EPI_ISL_913383, EPI_ISL_913384, EPI_ISL_913385, EPI_ISL_913386, EPI_ISL_913387, EPI_ISL_913388, EPI_ISL_913389, EPI_ISL_913390, EPI_ISL_913391, EPI_ISL_913392, EPI_ISL_913454, EPI_ISL_913459, EPI_ISL_934377, EPI_ISL_1008479, EPI_ISL_1008480, EPI_ISL_1008481, EPI_ISL_1008482, EPI_ISL_1008483, EPI_ISL_1008484, EPI_ISL_1008485, EPI_ISL_1008486, EPI_ISL_1008487, EPI_ISL_1008488, EPI_ISL_1008489, EPI_ISL_1064551, EPI_ISL_1064607, EPI_ISL_1064613, EPI_ISL_1064614, EPI_ISL_1064615, EPI_ISL_1064616, EPI_ISL_1064617, EPI_ISL_1064618, EPI_ISL_1064619, EPI_ISL_1064620, EPI_ISL_1168046 | | | |
| see above | Dynamic Code AB | The Public Health Agency of Sweden | Anna Risberg; Anna-Malin Linde; Carlo Berg; Karin Tegmark-Wisell; Maria Lind Karlberg; Mattias Haukland; Mia Brytting; Noura Walai; Oskar Karlsson Lindsjo; Petra Edquist; Petra Holmstrom; Reza Advani; Samuel Ohman; Sofia Stamouli |
| EPI_ISL_832005, EPI_ISL_913298, EPI_ISL_913299, EPI_ISL_913334, EPI_ISL_913335, EPI_ISL_913336, EPI_ISL_913370, EPI_ISL_934388, EPI_ISL_934390, EPI_ISL_934391, EPI_ISL_934392, EPI_ISL_934393, EPI_ISL_934394, EPI_ISL_934395, EPI_ISL_934396, EPI_ISL_934407, EPI_ISL_934408, EPI_ISL_934409, EPI_ISL_1008459, EPI_ISL_1008460, EPI_ISL_1008461, EPI_ISL_1008462, EPI_ISL_1008463, EPI_ISL_1008464, EPI_ISL_1008465, EPI_ISL_1008466, EPI_ISL_1008467, EPI_ISL_1008468, EPI_ISL_1008469, EPI_ISL_1008471, EPI_ISL_1008472, EPI_ISL_1008473, EPI_ISL_1008474, EPI_ISL_1008475, EPI_ISL_1008476, EPI_ISL_1008477, EPI_ISL_1008478, EPI_ISL_1008553, EPI_ISL_1008554, EPI_ISL_1008555, EPI_ISL_1008556, EPI_ISL_1008559, EPI_ISL_1008560, EPI_ISL_1008561, EPI_ISL_1008563, EPI_ISL_1008564, EPI_ISL_1008565, EPI_ISL_1008566, EPI_ISL_1008567, EPI_ISL_1008568, EPI_ISL_1008569, EPI_ISL_1008570, EPI_ISL_1008571, EPI_ISL_1008572, EPI_ISL_1008573, EPI_ISL_1008574, EPI_ISL_1008575, EPI_ISL_1008576, EPI_ISL_1008577, EPI_ISL_1008578, EPI_ISL_1008579, EPI_ISL_1008580, EPI_ISL_1008581, EPI_ISL_1008582, EPI_ISL_1008583, EPI_ISL_1008584, EPI_ISL_1008585, EPI_ISL_1008598, EPI_ISL_1008599, EPI_ISL_1008600, EPI_ISL_1064576, EPI_ISL_1064577, EPI_ISL_1064578, EPI_ISL_1064579, EPI_ISL_1064580, EPI_ISL_1064583, EPI_ISL_1064584, EPI_ISL_1064585, EPI_ISL_1064586, EPI_ISL_1064587, EPI_ISL_1064588, EPI_ISL_1064625, EPI_ISL_1064698, EPI_ISL_1064699, EPI_ISL_1168052, EPI_ISL_1168053, EPI_ISL_1168085, EPI_ISL_1190856, EPI_ISL_1190857, EPI_ISL_1190858, EPI_ISL_1190859, EPI_ISL_1190860, EPI_ISL_1190866, EPI_ISL_1190867, EPI_ISL_1190881, EPI_ISL_1190882, EPI_ISL_1190883, EPI_ISL_1190884, EPI_ISL_1231933, EPI_ISL_1231963, EPI_ISL_1231971, EPI_ISL_1231972, EPI_ISL_1231976, EPI_ISL_1231984, EPI_ISL_1231996, EPI_ISL_1232024, EPI_ISL_1232025, EPI_ISL_1232026, EPI_ISL_1232027 | | | |
| see above | Klinisk Mikrobiologi | The Public Health Agency of Sweden | Anna Risberg; Anna-Malin Linde; Carlo Berg; Department of Microbiology; Karin Tegmark-Wisell; Maria Lind Karlberg; Mattias Haukland; Mia Brytting; Noura Walai; Oskar Karlsson Lindsjo; Petra Edquist; Petra Holmstrom; Reza Advani; Samuel Ohman; Sofia Stamouli; The Public Health Agency of Sweden |
| EPI_ISL_766694, EPI_ISL_766714, EPI_ISL_775572, EPI_ISL_775573, EPI_ISL_775574, EPI_ISL_789044, EPI_ISL_831941, EPI_ISL_831942, EPI_ISL_831980, EPI_ISL_831981, EPI_ISL_913258, EPI_ISL_913259, EPI_ISL_913260, EPI_ISL_913261, EPI_ISL_913262, EPI_ISL_913264, EPI_ISL_913265, EPI_ISL_913268, EPI_ISL_913269, EPI_ISL_913270, EPI_ISL_913271, EPI_ISL_913272, EPI_ISL_913273, EPI_ISL_913274, EPI_ISL_913280, EPI_ISL_913283, EPI_ISL_913296, EPI_ISL_913297, EPI_ISL_913300, EPI_ISL_913307, EPI_ISL_913308, EPI_ISL_913311, EPI_ISL_913327, EPI_ISL_913328, EPI_ISL_913329, EPI_ISL_913359, EPI_ISL_913360, EPI_ISL_913361, EPI_ISL_913362, EPI_ISL_913367, EPI_ISL_913369, EPI_ISL_913371, EPI_ISL_913372, EPI_ISL_913373, EPI_ISL_913374, EPI_ISL_913453, EPI_ISL_913457, EPI_ISL_913458, EPI_ISL_913463, EPI_ISL_913465, EPI_ISL_913469, EPI_ISL_913470, EPI_ISL_913471, EPI_ISL_913485, EPI_ISL_913487, EPI_ISL_913488, EPI_ISL_913489, EPI_ISL_913490, EPI_ISL_913491, EPI_ISL_913492, EPI_ISL_913493, EPI_ISL_934324, EPI_ISL_934340, EPI_ISL_934343, EPI_ISL_934344, EPI_ISL_934345, EPI_ISL_934346, EPI_ISL_934347, EPI_ISL_934348, EPI_ISL_934349, EPI_ISL_934350, EPI_ISL_934351, EPI_ISL_934352, EPI_ISL_934353, EPI_ISL_934355, EPI_ISL_934356, EPI_ISL_934357, EPI_ISL_934358, EPI_ISL_934359, EPI_ISL_934360, EPI_ISL_934361, EPI_ISL_934362, EPI_ISL_934363, EPI_ISL_934375, EPI_ISL_934376, EPI_ISL_934378, EPI_ISL_934379, EPI_ISL_934380, EPI_ISL_934381, EPI_ISL_934382, EPI_ISL_934383, EPI_ISL_934384, EPI_ISL_934387, EPI_ISL_1008400, EPI_ISL_1008412, EPI_ISL_1008413, EPI_ISL_1008414, EPI_ISL_1008434, EPI_ISL_1008438, EPI_ISL_1008439, EPI_ISL_1008440, EPI_ISL_1008441, EPI_ISL_1008442, EPI_ISL_1008443, EPI_ISL_1008445, EPI_ISL_1008447, EPI_ISL_1008448, EPI_ISL_1008449, EPI_ISL_1008450, EPI_ISL_1008455, EPI_ISL_1008456, EPI_ISL_1008457, EPI_ISL_1008458, EPI_ISL_1008470, EPI_ISL_1008490, EPI_ISL_1008491, EPI_ISL_1008492, EPI_ISL_1008493, EPI_ISL_1008494, EPI_ISL_1008495, EPI_ISL_1008496, EPI_ISL_1008497, EPI_ISL_1008498, EPI_ISL_1008499, EPI_ISL_1008500, EPI_ISL_1008501, EPI_ISL_1008502, EPI_ISL_1008503, EPI_ISL_1008504, EPI_ISL_1008505, EPI_ISL_1008506, EPI_ISL_1008507, EPI_ISL_1008508, EPI_ISL_1008509, EPI_ISL_1008510, EPI_ISL_1008511, EPI_ISL_1008512, EPI_ISL_1008513, EPI_ISL_1008514, EPI_ISL_1008515, EPI_ISL_1008516, EPI_ISL_1008517, EPI_ISL_1008518, EPI_ISL_1008519, EPI_ISL_1008520, EPI_ISL_1008521, EPI_ISL_1008522, EPI_ISL_1008523, EPI_ISL_1008524, EPI_ISL_1008525, EPI_ISL_1008526, EPI_ISL_1008527, EPI_ISL_1008528, EPI_ISL_1008529, EPI_ISL_1008530, EPI_ISL_1008531, EPI_ISL_1008532, EPI_ISL_1008533, EPI_ISL_1008589, EPI_ISL_1008590, EPI_ISL_1008591, EPI_ISL_1008592, EPI_ISL_1008593, EPI_ISL_1008594, EPI_ISL_1008595, EPI_ISL_1008602, EPI_ISL_1064552, EPI_ISL_1064554, EPI_ISL_1064557, EPI_ISL_1064558, EPI_ISL_1064559, EPI_ISL_1064560, EPI_ISL_1064561, EPI_ISL_1064562, EPI_ISL_1064563, EPI_ISL_1064564, EPI_ISL_1064565, EPI_ISL_1064566, EPI_ISL_1064567, EPI_ISL_1064568, EPI_ISL_1064569, EPI_ISL_1064570, EPI_ISL_1064571, EPI_ISL_1064572, EPI_ISL_1064573, EPI_ISL_1064574, EPI_ISL_1064581, EPI_ISL_1064582, EPI_ISL_1064589, EPI_ISL_1064590, EPI_ISL_1064591, EPI_ISL_1064592, EPI_ISL_1064593, EPI_ISL_1064594, EPI_ISL_1064595, EPI_ISL_1064596, EPI_ISL_1064597, EPI_ISL_1064598, EPI_ISL_1064599, EPI_ISL_1064600, EPI_ISL_1064601, EPI_ISL_1064602, EPI_ISL_1064603, EPI_ISL_1064604, EPI_ISL_1064605, EPI_ISL_1064606, EPI_ISL_1064622, EPI_ISL_1064624, EPI_ISL_1064626, EPI_ISL_1064627, EPI_ISL_1064628, EPI_ISL_1064629, EPI_ISL_1064630, EPI_ISL_1064631, EPI_ISL_1064632, EPI_ISL_1064633, EPI_ISL_1064634, EPI_ISL_1064635, EPI_ISL_1064636, EPI_ISL_1064637, EPI_ISL_1064638, EPI_ISL_1064639, EPI_ISL_1064640, EPI_ISL_1064641, EPI_ISL_1064642, EPI_ISL_1064643, EPI_ISL_1064644, EPI_ISL_1064645, EPI_ISL_1064646, EPI_ISL_1064647, EPI_ISL_1064648, EPI_ISL_1064649, EPI_ISL_1064650, EPI_ISL_1064651, EPI_ISL_1064652, EPI_ISL_1064653, EPI_ISL_1064654, EPI_ISL_1064655, EPI_ISL_1064656, EPI_ISL_1064657, EPI_ISL_1064658, EPI_ISL_1064659, EPI_ISL_1064660, EPI_ISL_1064661, EPI_ISL_1064662, EPI_ISL_1064663, EPI_ISL_1064664, EPI_ISL_1064665, EPI_ISL_1064666, EPI_ISL_1064667, EPI_ISL_1064668, EPI_ISL_1064669, EPI_ISL_1064670, EPI_ISL_1064671, EPI_ISL_1064672, EPI_ISL_1064673, EPI_ISL_1064674, EPI_ISL_1064675, EPI_ISL_1064676, EPI_ISL_1064678, EPI_ISL_1064679, EPI_ISL_1064680, EPI_ISL_1064681, EPI_ISL_1064691, EPI_ISL_1064692, EPI_ISL_1064695, EPI_ISL_1064696, EPI_ISL_1064697, EPI_ISL_1064703, EPI_ISL_1064704, EPI_ISL_1064705, EPI_ISL_1064706, EPI_ISL_1064707, EPI_ISL_1168044, EPI_ISL_1168045, EPI_ISL_1168048, EPI_ISL_1168049, EPI_ISL_1168050, EPI_ISL_1168060, EPI_ISL_1168061, EPI_ISL_1168062, EPI_ISL_1168063, EPI_ISL_1168064, EPI_ISL_1168065, EPI_ISL_1168066, EPI_ISL_1168067, EPI_ISL_1168068, EPI_ISL_1168071, EPI_ISL_1168072, EPI_ISL_1168075, EPI_ISL_1168081, EPI_ISL_1168082, EPI_ISL_1168086, EPI_ISL_1168087, EPI_ISL_1168090, EPI_ISL_1168111, EPI_ISL_1168112, EPI_ISL_1168113, EPI_ISL_1168114, EPI_ISL_1168115, EPI_ISL_1168120, EPI_ISL_1168122, EPI_ISL_1168123, EPI_ISL_1168135, EPI_ISL_1168137, EPI_ISL_1168138, EPI_ISL_1168143, EPI_ISL_1168144, EPI_ISL_1168145, EPI_ISL_1168147, EPI_ISL_1168157, EPI_ISL_1190861, EPI_ISL_1190862, EPI_ISL_1190863, EPI_ISL_1190864, EPI_ISL_1190865, EPI_ISL_1190869, EPI_ISL_1190870, EPI_ISL_1190871, EPI_ISL_1190872, EPI_ISL_1190879, EPI_ISL_1231965, EPI_ISL_1231986, EPI_ISL_1231989, EPI_ISL_1231990, EPI_ISL_1232011, EPI_ISL_1300290, EPI_ISL_1300291, EPI_ISL_1300293 | | | |
| see above | Klinisk mikrobiologi | The Public Health Agency of Sweden | Anna Risberg; Anna-Malin Linde; Carlo Berg; Department of Microbiology; Karin Tegmark-Wisell; Maria Lind Karlberg; Mattias Haukland; Mia Brytting; Noura Walai; Oskar Karlsson Lindsjo; Petra Edquist; Petra Holmstrom; Reza Advani; Samuel Ohman; Sofia Stamouli; The Public Health Agency of Sweden |
| EPI_ISL_984717, EPI_ISL_984718, EPI_ISL_984719, EPI_ISL_984720, EPI_ISL_984721, EPI_ISL_984722, EPI_ISL_984723, EPI_ISL_984724, EPI_ISL_984725, EPI_ISL_984726, EPI_ISL_984727, EPI_ISL_984728, EPI_ISL_984729, EPI_ISL_984730, EPI_ISL_984731, EPI_ISL_984732, EPI_ISL_984733, EPI_ISL_984734, EPI_ISL_984735, EPI_ISL_984950, EPI_ISL_984951, EPI_ISL_984952, EPI_ISL_984953, EPI_ISL_984954, EPI_ISL_984955, EPI_ISL_984956, EPI_ISL_984957, EPI_ISL_984958, EPI_ISL_984959, EPI_ISL_984960, EPI_ISL_984961, EPI_ISL_984962, EPI_ISL_984963, EPI_ISL_984964, EPI_ISL_984965, EPI_ISL_984966, EPI_ISL_984967, EPI_ISL_984968, EPI_ISL_984969, EPI_ISL_984970, EPI_ISL_984971, EPI_ISL_984972, EPI_ISL_984973, EPI_ISL_984974, EPI_ISL_984975, EPI_ISL_984976, EPI_ISL_984977, EPI_ISL_984978, EPI_ISL_984979, EPI_ISL_985041, EPI_ISL_985042, EPI_ISL_985043, EPI_ISL_985044, EPI_ISL_985045, EPI_ISL_985046, EPI_ISL_985047, EPI_ISL_985048, EPI_ISL_985049, EPI_ISL_985050, EPI_ISL_985051, EPI_ISL_985052, EPI_ISL_985053, EPI_ISL_985054, EPI_ISL_985055, EPI_ISL_985056, EPI_ISL_985057, EPI_ISL_985320, EPI_ISL_985321, EPI_ISL_985322, EPI_ISL_985323, EPI_ISL_985324, EPI_ISL_985325, EPI_ISL_985326, EPI_ISL_985327, EPI_ISL_985328, EPI_ISL_985329, EPI_ISL_985330, EPI_ISL_985331, EPI_ISL_985332, EPI_ISL_985333, EPI_ISL_985334, EPI_ISL_985335, EPI_ISL_985336, EPI_ISL_985337, EPI_ISL_985338, EPI_ISL_985339, EPI_ISL_985340, EPI_ISL_985341, EPI_ISL_985342, EPI_ISL_985343, EPI_ISL_985344, EPI_ISL_985345, EPI_ISL_985346, EPI_ISL_985347, EPI_ISL_985348, EPI_ISL_985349, EPI_ISL_985350, EPI_ISL_985351, EPI_ISL_985352, EPI_ISL_985353, EPI_ISL_985354, EPI_ISL_985355, EPI_ISL_985356, EPI_ISL_985357, EPI_ISL_985358, EPI_ISL_985359, EPI_ISL_985360, EPI_ISL_985361, EPI_ISL_985362, EPI_ISL_985363, EPI_ISL_985364, EPI_ISL_985365, EPI_ISL_985366, EPI_ISL_985367, EPI_ISL_985368, EPI_ISL_985369, EPI_ISL_985370, EPI_ISL_985371, EPI_ISL_985372, EPI_ISL_985373, EPI_ISL_985374, EPI_ISL_985375, EPI_ISL_985376, EPI_ISL_985377, EPI_ISL_985378, EPI_ISL_985379, EPI_ISL_985380, EPI_ISL_985381, EPI_ISL_985382, EPI_ISL_985383, EPI_ISL_985384, EPI_ISL_985385, EPI_ISL_985386, EPI_ISL_985387, EPI_ISL_985388, EPI_ISL_985389, EPI_ISL_985390, EPI_ISL_985391, EPI_ISL_985392, EPI_ISL_1091497, EPI_ISL_1091501, EPI_ISL_1091502, EPI_ISL_1091511, EPI_ISL_1091512, EPI_ISL_1091514, EPI_ISL_1091516, EPI_ISL_1091517, EPI_ISL_1091518, EPI_ISL_1091519, EPI_ISL_1091520, EPI_ISL_1091521, EPI_ISL_1091533, EPI_ISL_1091534, EPI_ISL_1091535, EPI_ISL_1091536, EPI_ISL_1091537, EPI_ISL_1091538, EPI_ISL_1091539, EPI_ISL_1091569, EPI_ISL_1091570, EPI_ISL_1093492, EPI_ISL_1093493, EPI_ISL_1093494, EPI_ISL_1093495, EPI_ISL_1093496, EPI_ISL_1093497, EPI_ISL_1093498, EPI_ISL_1093499, EPI_ISL_1093500, EPI_ISL_1093501, EPI_ISL_1093502, EPI_ISL_1093503, EPI_ISL_1093504, EPI_ISL_1093505, EPI_ISL_1093506, EPI_ISL_1093507, EPI_ISL_1093508, EPI_ISL_1093509, EPI_ISL_1093510, EPI_ISL_1093511, EPI_ISL_1093512, EPI_ISL_1093513, EPI_ISL_1093514, EPI_ISL_1093515, EPI_ISL_1093516, EPI_ISL_1093517, EPI_ISL_1093518, EPI_ISL_1093519, EPI_ISL_1093520, EPI_ISL_1093521, EPI_ISL_1093522, EPI_ISL_1093523, EPI_ISL_1093524, EPI_ISL_1093525, EPI_ISL_1093526, EPI_ISL_1093527, EPI_ISL_1093528, EPI_ISL_1093529, EPI_ISL_1093530, EPI_ISL_1093531, EPI_ISL_1093532, EPI_ISL_1093533, EPI_ISL_1093534, EPI_ISL_1093535, EPI_ISL_1093536, EPI_ISL_1093537, EPI_ISL_1093538, EPI_ISL_1093539, EPI_ISL_1093540, EPI_ISL_1093541, EPI_ISL_1093542, EPI_ISL_1093543, EPI_ISL_1093544, EPI_ISL_1093545, EPI_ISL_1166539, EPI_ISL_1166540, EPI_ISL_1166541, EPI_ISL_1166542, EPI_ISL_1166543, EPI_ISL_1166544, EPI_ISL_1166545, EPI_ISL_1166546, EPI_ISL_1166547, EPI_ISL_1166548, EPI_ISL_1166549, EPI_ISL_1166550, EPI_ISL_1166551, EPI_ISL_1166552, EPI_ISL_1166553, EPI_ISL_1166554, EPI_ISL_1166555, EPI_ISL_1224950, EPI_ISL_1224951, EPI_ISL_1224952, EPI_ISL_1224953, EPI_ISL_1224954, EPI_ISL_1224955, EPI_ISL_1224956, EPI_ISL_1224957, EPI_ISL_1303381, EPI_ISL_1303382, EPI_ISL_1303383, EPI_ISL_1303384, EPI_ISL_1303385, EPI_ISL_1303386, EPI_ISL_1303387, EPI_ISL_1303388, EPI_ISL_1303389, EPI_ISL_1303390, EPI_ISL_1303391, EPI_ISL_1303392, EPI_ISL_1303393, EPI_ISL_1303394, EPI_ISL_1303395, EPI_ISL_1303396, EPI_ISL_1303397, EPI_ISL_1303399, EPI_ISL_1303400, EPI_ISL_1303401, EPI_ISL_1303402, EPI_ISL_1303403, EPI_ISL_1372903, EPI_ISL_1372909, EPI_ISL_1372910, EPI_ISL_1372920, EPI_ISL_1372931, EPI_ISL_1372943, EPI_ISL_1372944, EPI_ISL_1372961, EPI_ISL_1578117, EPI_ISL_1578118, EPI_ISL_2099890, EPI_ISL_2099891, EPI_ISL_2099892, EPI_ISL_2099893, EPI_ISL_2099894, EPI_ISL_2099895, EPI_ISL_2099896, EPI_ISL_2099897, EPI_ISL_2099898 | | | |
| see above | Klinisk mikrobiologi, Region Västerbotten | CBRN Defence and Security, Swedish Defence Research Agency | Andreas Sjödin; Anna-Lena Johansson; Annika Allard; Annika Osterman; Carolin Öhrman; Caroline Öhrman; David Sundell; Emelie Näslund Salomonsson; Ingrid Dacklin; Jonas Näslund; Kerstin Myrtennäs; Linda Karlsson; Malin Granberg; Mats Forsman; Stina Bäckman |
| EPI_ISL_766697, EPI_ISL_1064709 | Klinisk mikrobiologi, bakteriologi | The Public Health Agency of Sweden | Anna Risberg; Anna-Malin Linde; Carlo Berg; Department of Microbiology; Karin Tegmark-Wisell; Maria Lind Karlberg; Mattias Haukland; Mia Brytting; Noura Walai; Oskar Karlsson Lindsjo; Petra Edquist; Petra Holmstrom; Reza Advani; Samuel Ohman; Sofia Stamouli; The Public Health Agency of Sweden |
| EPI_ISL_1313180, EPI_ISL_1313280, EPI_ISL_1313281 | LaboratorieMedicinskt Centrum Gotland | Karolinska University Hospital | Annelie Bjerkner; Isak Sylvin; Jan Albert; Karolina Ininbergs; Lina Guerra Blomqvist; Lynda Eneh; Martin Ekman; Martina Wahlund; Robert Dyrdak; Sandra Broddesson; Tanja Normark; Tobias Allander; Valtteri Wirta; Zhibing Yun |
| EPI_ISL_1168094 | Laboratoriemedicin, Klinisk mikrobiologi | The Public Health Agency of Sweden | Anna Risberg; Anna-Malin Linde; Carlo Berg; Karin Tegmark-Wisell; Maria Lind Karlberg; Mattias Haukland; Mia Brytting; Noura Walai; Oskar Karlsson Lindsjo; Petra Edquist; Petra Holmstrom; Reza Advani; Sofia Stamouli |
| EPI_ISL_832003, EPI_ISL_832004, EPI_ISL_913312, EPI_ISL_913330, EPI_ISL_913331, EPI_ISL_913332, EPI_ISL_913333, EPI_ISL_1064700, EPI_ISL_1064701, EPI_ISL_1064702, EPI_ISL_1064710, EPI_ISL_1168054, EPI_ISL_1168055, EPI_ISL_1168056, EPI_ISL_1168057, EPI_ISL_1168058, EPI_ISL_1168059 | | | |
| see above | Mikrobiologen | The Public Health Agency of Sweden | Anna Risberg; Anna-Malin Linde; Carlo Berg; Department of Microbiology; Karin Tegmark-Wisell; Maria Lind Karlberg; Mattias Haukland; Mia Brytting; Noura Walai; Oskar Karlsson Lindsjo; Petra Edquist; Petra Holmstrom; Reza Advani; Samuel Ohman; Sofia Stamouli; The Public Health Agency of Sweden |
| EPI_ISL_1064608, EPI_ISL_1064609, EPI_ISL_1064610, EPI_ISL_1064611, EPI_ISL_1064612, EPI_ISL_1168096, EPI_ISL_1168097, EPI_ISL_1168098, EPI_ISL_1190873, EPI_ISL_1190874, EPI_ISL_1190875, EPI_ISL_1190876, EPI_ISL_1190877, EPI_ISL_1190878 | | | |
| see above | NPC | The Public Health Agency of Sweden | Anna Risberg; Anna-Malin Linde; Carlo Berg; Karin Tegmark-Wisell; Maria Lind Karlberg; Mattias Haukland; Mia Brytting; Noura Walai; Oskar Karlsson Lindsjo; Petra Edquist; Petra Holmstrom; Reza Advani; Samuel Ohman; Sofia Stamouli |
| EPI_ISL_934315, EPI_ISL_1008396, EPI_ISL_1008397, EPI_ISL_1008399 | Narhalsan Fjallbacka VC | The Public Health Agency of Sweden | Anna Risberg; Anna-Malin Linde; Carlo Berg; Karin Tegmark-Wisell; Maria Lind Karlberg; Mattias Haukland; Mia Brytting; Noura Walai; Oskar Karlsson Lindsjo; Petra Edquist; Petra Holmstrom; Reza Advani; Sofia Stamouli |
| EPI_ISL_1008398, EPI_ISL_1064555 | Omtanken Grimmered | The Public Health Agency of Sweden | Anna Risberg; Anna-Malin Linde; Carlo Berg; Karin Tegmark-Wisell; Maria Lind Karlberg; Mattias Haukland; Mia Brytting; Noura Walai; Oskar Karlsson Lindsjo; Petra Edquist; Petra Holmstrom; Reza Advani; Samuel Ohman; Sofia Stamouli |
| EPI_ISL_1492583, EPI_ISL_1492584, EPI_ISL_1492585, EPI_ISL_1492586, EPI_ISL_1492587, EPI_ISL_1492588, EPI_ISL_1492589, EPI_ISL_1492590, EPI_ISL_1492591, EPI_ISL_1492592, EPI_ISL_1492593, EPI_ISL_1492604, EPI_ISL_1492605, EPI_ISL_1492606, EPI_ISL_1492607, EPI_ISL_1492608, EPI_ISL_1492609, EPI_ISL_1492610, EPI_ISL_1492611, EPI_ISL_1492612, EPI_ISL_1492613, EPI_ISL_1492614, EPI_ISL_1622950 | | | |
| see above | Orebro University Hospital | Orebro University Hospital | Sundqvist M et al |
| EPI_ISL_1191588, EPI_ISL_1191589, EPI_ISL_1191590, EPI_ISL_1191591, EPI_ISL_1191592, EPI_ISL_1191593, EPI_ISL_1191594, EPI_ISL_1191595, EPI_ISL_1191596, EPI_ISL_1192468, EPI_ISL_1192469, EPI_ISL_1192470, EPI_ISL_1192471, EPI_ISL_1192472, EPI_ISL_1192473, EPI_ISL_1192474, EPI_ISL_1192475, EPI_ISL_1192476, EPI_ISL_1192477, EPI_ISL_1192478, EPI_ISL_1192479, EPI_ISL_1192480, EPI_ISL_1192481, EPI_ISL_1192482, EPI_ISL_1192483, EPI_ISL_1192484, EPI_ISL_1192485, EPI_ISL_1192486, EPI_ISL_1192487, EPI_ISL_1192488, EPI_ISL_1192489, EPI_ISL_1192490, EPI_ISL_1192491, EPI_ISL_1192492, EPI_ISL_1192493, EPI_ISL_1192494, EPI_ISL_1192495, EPI_ISL_1192496, EPI_ISL_1192497, EPI_ISL_1192498, EPI_ISL_1192499, EPI_ISL_1192500, EPI_ISL_1192501, EPI_ISL_1192502, EPI_ISL_1192503, EPI_ISL_1192504, EPI_ISL_1192505, EPI_ISL_1192506, EPI_ISL_1192507, EPI_ISL_1192508, EPI_ISL_1192509, EPI_ISL_1192510, EPI_ISL_1192511, EPI_ISL_1192512, EPI_ISL_1192513, EPI_ISL_1192514, EPI_ISL_1192515, EPI_ISL_1192516, EPI_ISL_1192517, EPI_ISL_1192518, EPI_ISL_1192519, EPI_ISL_1192520, EPI_ISL_1192521, EPI_ISL_1192522, EPI_ISL_1192523, EPI_ISL_1192524, EPI_ISL_1192525, EPI_ISL_1192526, EPI_ISL_1192527, EPI_ISL_1192528, EPI_ISL_1192529, EPI_ISL_1192530, EPI_ISL_1192531, EPI_ISL_1192532, EPI_ISL_1192533, EPI_ISL_1192534, EPI_ISL_1192535, EPI_ISL_1192536, EPI_ISL_1192537, EPI_ISL_1192538, EPI_ISL_1192539, EPI_ISL_1192540, EPI_ISL_1192541, EPI_ISL_1192542, EPI_ISL_1192543, EPI_ISL_1192544, EPI_ISL_1192545, EPI_ISL_1192546, | | | |

EPI_ISL_1192547, EPI_ISL_1192548, EPI_ISL_1192549, EPI_ISL_1192550, EPI_ISL_1192551, EPI_ISL_1192552, EPI_ISL_1192553, EPI_ISL_1192554, EPI_ISL_1192555, EPI_ISL_1192556, EPI_ISL_1192557, EPI_ISL_1192558, EPI_ISL_1192559, EPI_ISL_1192560, EPI_ISL_1192561, EPI_ISL_1192562, EPI_ISL_1192563, EPI_ISL_1192564, EPI_ISL_1192565, EPI_ISL_1192566, EPI_ISL_1192567, EPI_ISL_1192568,
EPI_ISL_1192569, EPI_ISL_1192570, EPI_ISL_1192571, EPI_ISL_1192572, EPI_ISL_1192573, EPI_ISL_1192574, EPI_ISL_1192575, EPI_ISL_1192576, EPI_ISL_1192577, EPI_ISL_1192578, EPI_ISL_1192579, EPI_ISL_1192580, EPI_ISL_1192581, EPI_ISL_1192582, EPI_ISL_1192583, EPI_ISL_1192584, EPI_ISL_1192585, EPI_ISL_1192586, EPI_ISL_1192587, EPI_ISL_1192588, EPI_ISL_1192589, EPI_ISL_1192590,
EPI_ISL_1192591, EPI_ISL_1192592, EPI_ISL_1192593, EPI_ISL_1192594, EPI_ISL_1192595, EPI_ISL_1192596, EPI_ISL_1192597, EPI_ISL_1192598, EPI_ISL_1192599, EPI_ISL_1192600, EPI_ISL_1192601, EPI_ISL_1192602, EPI_ISL_1192603, EPI_ISL_1192604, EPI_ISL_1192605, EPI_ISL_1192606, EPI_ISL_1192607, EPI_ISL_1192608, EPI_ISL_1192609, EPI_ISL_1192610, EPI_ISL_1192611, EPI_ISL_1192612,
EPI_ISL_1192613, EPI_ISL_1192614, EPI_ISL_1192615, EPI_ISL_1192616, EPI_ISL_1192617, EPI_ISL_1192618, EPI_ISL_1192619, EPI_ISL_1192620, EPI_ISL_1192621, EPI_ISL_1192622, EPI_ISL_1192623, EPI_ISL_1192624, EPI_ISL_1192625, EPI_ISL_1192626, EPI_ISL_1192627, EPI_ISL_1192628, EPI_ISL_1192629, EPI_ISL_1192630, EPI_ISL_1192631, EPI_ISL_1192632, EPI_ISL_1192633, EPI_ISL_1192634,
EPI_ISL_1192635, EPI_ISL_1192636, EPI_ISL_1192637, EPI_ISL_1192638, EPI_ISL_1192639, EPI_ISL_1192640, EPI_ISL_1192641, EPI_ISL_1192642, EPI_ISL_1192643, EPI_ISL_1192644, EPI_ISL_1192645, EPI_ISL_1192646, EPI_ISL_1192647, EPI_ISL_1192648, EPI_ISL_1192649, EPI_ISL_1192650, EPI_ISL_1192651, EPI_ISL_1192652, EPI_ISL_1192653, EPI_ISL_1192654, EPI_ISL_1192655, EPI_ISL_1192656,
EPI_ISL_1192657, EPI_ISL_1192658, EPI_ISL_1192659, EPI_ISL_1192660, EPI_ISL_1192661, EPI_ISL_1192662, EPI_ISL_1192663, EPI_ISL_1192664, EPI_ISL_1192665, EPI_ISL_1192666, EPI_ISL_1192667, EPI_ISL_1192668, EPI_ISL_1192669, EPI_ISL_1192670, EPI_ISL_1192671, EPI_ISL_1192672, EPI_ISL_1192673, EPI_ISL_1192674, EPI_ISL_1192675, EPI_ISL_1192676, EPI_ISL_1192677, EPI_ISL_1192678,
EPI_ISL_1192679, EPI_ISL_1192680, EPI_ISL_1192681, EPI_ISL_1192682, EPI_ISL_1192683, EPI_ISL_1192684, EPI_ISL_1192685, EPI_ISL_1192686, EPI_ISL_1192687, EPI_ISL_1192688, EPI_ISL_1192689, EPI_ISL_1192690, EPI_ISL_1192691, EPI_ISL_1192692, EPI_ISL_1192693, EPI_ISL_1192694, EPI_ISL_1192695, EPI_ISL_1192696, EPI_ISL_1192697, EPI_ISL_1192698, EPI_ISL_1192699, EPI_ISL_1192700,
EPI_ISL_1192701, EPI_ISL_1192702, EPI_ISL_1192703, EPI_ISL_1192704, EPI_ISL_1192705, EPI_ISL_1192706, EPI_ISL_1192707, EPI_ISL_1192708, EPI_ISL_1192709, EPI_ISL_1192710, EPI_ISL_1192711, EPI_ISL_1192712, EPI_ISL_1192713, EPI_ISL_1192714, EPI_ISL_1192715, EPI_ISL_1192716, EPI_ISL_1192717, EPI_ISL_1192718, EPI_ISL_1192719, EPI_ISL_1192720, EPI_ISL_1192721, EPI_ISL_1192722,
EPI_ISL_1192723, EPI_ISL_1192724, EPI_ISL_1192725, EPI_ISL_1192726, EPI_ISL_1192727, EPI_ISL_1192728, EPI_ISL_1192729, EPI_ISL_1192730, EPI_ISL_1192731, EPI_ISL_1192732, EPI_ISL_1192733, EPI_ISL_1192734, EPI_ISL_1192735, EPI_ISL_1192736, EPI_ISL_1192737, EPI_ISL_1192738, EPI_ISL_1192739, EPI_ISL_1192740, EPI_ISL_1192741, EPI_ISL_1192742, EPI_ISL_1192743, EPI_ISL_1192744,
EPI_ISL_1192745, EPI_ISL_1192746, EPI_ISL_1192747, EPI_ISL_1192748, EPI_ISL_1192749, EPI_ISL_1192750, EPI_ISL_1192751, EPI_ISL_1192752, EPI_ISL_1192753, EPI_ISL_1192754, EPI_ISL_1192755, EPI_ISL_1192756, EPI_ISL_1192757, EPI_ISL_1192758, EPI_ISL_1192759, EPI_ISL_1192760, EPI_ISL_1192761, EPI_ISL_1192762, EPI_ISL_1192763, EPI_ISL_1192764, EPI_ISL_1192765, EPI_ISL_1192766,
EPI_ISL_1192767, EPI_ISL_1192768, EPI_ISL_1192769, EPI_ISL_1192770, EPI_ISL_1192771, EPI_ISL_1192772, EPI_ISL_1192773, EPI_ISL_1192774, EPI_ISL_1192775, EPI_ISL_1192776, EPI_ISL_1192777, EPI_ISL_1192778, EPI_ISL_1192779, EPI_ISL_1192780, EPI_ISL_1192781, EPI_ISL_1192782, EPI_ISL_1192783, EPI_ISL_1192784, EPI_ISL_1192785, EPI_ISL_1192786, EPI_ISL_1192787, EPI_ISL_1192788,
EPI_ISL_1192789, EPI_ISL_1192790, EPI_ISL_1192791, EPI_ISL_1192792, EPI_ISL_1192793, EPI_ISL_1192794, EPI_ISL_1192795, EPI_ISL_1192796, EPI_ISL_1192797, EPI_ISL_1192798, EPI_ISL_1192799, EPI_ISL_1192800, EPI_ISL_1192801, EPI_ISL_1192802, EPI_ISL_1192803, EPI_ISL_1192804, EPI_ISL_1192805, EPI_ISL_1192806, EPI_ISL_1192807, EPI_ISL_1192808, EPI_ISL_1192809, EPI_ISL_1192810,
EPI_ISL_1192811, EPI_ISL_1192812, EPI_ISL_1192813, EPI_ISL_1192814, EPI_ISL_1192815, EPI_ISL_1192816, EPI_ISL_1192817, EPI_ISL_1192818, EPI_ISL_1192819, EPI_ISL_1192820, EPI_ISL_1192821, EPI_ISL_1192822, EPI_ISL_1192823, EPI_ISL_1192824, EPI_ISL_1192825, EPI_ISL_1192826, EPI_ISL_1192827, EPI_ISL_1192828, EPI_ISL_1192829, EPI_ISL_1192830, EPI_ISL_1192831, EPI_ISL_1192832,
EPI_ISL_1192833, EPI_ISL_1192834, EPI_ISL_1192835, EPI_ISL_1192836, EPI_ISL_1192837, EPI_ISL_1192838, EPI_ISL_1192839, EPI_ISL_1192840, EPI_ISL_1192841, EPI_ISL_1192842, EPI_ISL_1192843, EPI_ISL_1192844, EPI_ISL_1192845, EPI_ISL_1192846, EPI_ISL_1192847, EPI_ISL_1192848, EPI_ISL_1192849, EPI_ISL_1192850, EPI_ISL_1192851, EPI_ISL_1192852, EPI_ISL_1192853, EPI_ISL_1192854,
EPI_ISL_1192855, EPI_ISL_1192856, EPI_ISL_1192857, EPI_ISL_1192858, EPI_ISL_1192859, EPI_ISL_1192860, EPI_ISL_1192861, EPI_ISL_1192862, EPI_ISL_1192863, EPI_ISL_1192864, EPI_ISL_1192865, EPI_ISL_1192866, EPI_ISL_1192867, EPI_ISL_1192868, EPI_ISL_1192869, EPI_ISL_1192870, EPI_ISL_1192871, EPI_ISL_1192872, EPI_ISL_1192873, EPI_ISL_1192874, EPI_ISL_1192875, EPI_ISL_1192876,
EPI_ISL_1192877, EPI_ISL_1192878, EPI_ISL_1192879, EPI_ISL_1192880, EPI_ISL_1192881, EPI_ISL_1192882, EPI_ISL_1192883, EPI_ISL_1192884, EPI_ISL_1192885, EPI_ISL_1192886, EPI_ISL_1192887, EPI_ISL_1192888, EPI_ISL_1192889, EPI_ISL_1192890, EPI_ISL_1192891, EPI_ISL_1192892, EPI_ISL_1192893, EPI_ISL_1192894, EPI_ISL_1192895, EPI_ISL_1192896, EPI_ISL_1192897, EPI_ISL_1192898,
EPI_ISL_1192899, EPI_ISL_1192900, EPI_ISL_1192901, EPI_ISL_1192902, EPI_ISL_1192903, EPI_ISL_1192904, EPI_ISL_1192905, EPI_ISL_1192906, EPI_ISL_1192907, EPI_ISL_1192908, EPI_ISL_1192909, EPI_ISL_1192910, EPI_ISL_1192911, EPI_ISL_1192912, EPI_ISL_1192913, EPI_ISL_1192914, EPI_ISL_1192915, EPI_ISL_1192916, EPI_ISL_1192917, EPI_ISL_1192918, EPI_ISL_1192919, EPI_ISL_1192920,
EPI_ISL_1192921, EPI_ISL_1192922, EPI_ISL_1192923, EPI_ISL_1192924, EPI_ISL_1192925, EPI_ISL_1192926, EPI_ISL_1192927, EPI_ISL_1192928, EPI_ISL_1192929, EPI_ISL_1192930, EPI_ISL_1192931, EPI_ISL_1192932, EPI_ISL_1192933, EPI_ISL_1192934, EPI_ISL_1192935, EPI_ISL_1192936, EPI_ISL_1192937, EPI_ISL_1192938, EPI_ISL_1192939, EPI_ISL_1192940, EPI_ISL_1192941, EPI_ISL_1192942,
EPI_ISL_1192943, EPI_ISL_1192944, EPI_ISL_1192945, EPI_ISL_1192946, EPI_ISL_1192947, EPI_ISL_1192948, EPI_ISL_1192949, EPI_ISL_1192950, EPI_ISL_1192951, EPI_ISL_1192952, EPI_ISL_1192953, EPI_ISL_1192954, EPI_ISL_1192955, EPI_ISL_1192956, EPI_ISL_1192957, EPI_ISL_1192958, EPI_ISL_1192959, EPI_ISL_1192960, EPI_ISL_1192961, EPI_ISL_1192962, EPI_ISL_1192963, EPI_ISL_1192964,
EPI_ISL_1192965, EPI_ISL_1192966, EPI_ISL_1192967, EPI_ISL_1192968, EPI_ISL_1192969, EPI_ISL_1192970, EPI_ISL_1192971, EPI_ISL_1192972, EPI_ISL_1192973, EPI_ISL_1192974, EPI_ISL_1192975, EPI_ISL_1192976, EPI_ISL_1192977, EPI_ISL_1192978, EPI_ISL_1192979, EPI_ISL_1192980, EPI_ISL_1192981, EPI_ISL_1192982, EPI_ISL_1192983, EPI_ISL_1192984, EPI_ISL_1192985, EPI_ISL_1192986,
EPI_ISL_1192987, EPI_ISL_1192988, EPI_ISL_1192989, EPI_ISL_1192990, EPI_ISL_1192991, EPI_ISL_1192992, EPI_ISL_1192993, EPI_ISL_1192994, EPI_ISL_1192995, EPI_ISL_1192996, EPI_ISL_1192997, EPI_ISL_1192998, EPI_ISL_1192999, EPI_ISL_1193000, EPI_ISL_1193001, EPI_ISL_1193002, EPI_ISL_1193003, EPI_ISL_1193004, EPI_ISL_1193005, EPI_ISL_1193006, EPI_ISL_1193007, EPI_ISL_1193008,
EPI_ISL_1193009, EPI_ISL_1193010, EPI_ISL_1193011, EPI_ISL_1193012, EPI_ISL_1193013, EPI_ISL_1193014, EPI_ISL_1193015, EPI_ISL_1193016, EPI_ISL_1193017, EPI_ISL_1193018, EPI_ISL_1193019, EPI_ISL_1193020, EPI_ISL_1193021, EPI_ISL_1193022, EPI_ISL_1193023, EPI_ISL_1193024, EPI_ISL_1193025, EPI_ISL_1193026, EPI_ISL_1193027, EPI_ISL_1193028, EPI_ISL_1193029, EPI_ISL_1193030,
EPI_ISL_1193031, EPI_ISL_1193032, EPI_ISL_1193033, EPI_ISL_1193034, EPI_ISL_1193035, EPI_ISL_1193036, EPI_ISL_1193037, EPI_ISL_1193038, EPI_ISL_1193039, EPI_ISL_1193040, EPI_ISL_1193041, EPI_ISL_1193042, EPI_ISL_1193043, EPI_ISL_1193044, EPI_ISL_1193045, EPI_ISL_1193046, EPI_ISL_1193047, EPI_ISL_1193048, EPI_ISL_1193049, EPI_ISL_1193050, EPI_ISL_1193051, EPI_ISL_1193052,
EPI_ISL_1193053, EPI_ISL_1193054, EPI_ISL_1193055, EPI_ISL_1193056, EPI_ISL_1193057, EPI_ISL_1193058, EPI_ISL_1193059, EPI_ISL_1193060, EPI_ISL_1193061, EPI_ISL_1193062, EPI_ISL_1193063, EPI_ISL_1193064, EPI_ISL_1193065, EPI_ISL_1193066, EPI_ISL_1193067, EPI_ISL_1193068, EPI_ISL_1193069, EPI_ISL_1193070, EPI_ISL_1193071, EPI_ISL_1193072, EPI_ISL_1193073, EPI_ISL_1193074,
EPI_ISL_1193075, EPI_ISL_1193076, EPI_ISL_1193077, EPI_ISL_1193078, EPI_ISL_1193079, EPI_ISL_1193080, EPI_ISL_1193081, EPI_ISL_1193082, EPI_ISL_1193083, EPI_ISL_1193084, EPI_ISL_1193085, EPI_ISL_1193086, EPI_ISL_1193087, EPI_ISL_1193088, EPI_ISL_1193089, EPI_ISL_1193090, EPI_ISL_1193091, EPI_ISL_1193092, EPI_ISL_1193093, EPI_ISL_1193094, EPI_ISL_1193095, EPI_ISL_1193096,
EPI_ISL_1193097, EPI_ISL_1193098, EPI_ISL_1193099, EPI_ISL_1193100, EPI_ISL_1193101, EPI_ISL_1193102, EPI_ISL_1193103, EPI_ISL_1193104, EPI_ISL_1193105, EPI_ISL_1193106, EPI_ISL_1193107, EPI_ISL_1193108, EPI_ISL_1193109, EPI_ISL_1193110, EPI_ISL_1193111, EPI_ISL_1193112, EPI_ISL_1193113, EPI_ISL_1193114, EPI_ISL_1193115, EPI_ISL_1193116, EPI_ISL_1193117, EPI_ISL_1193118,
EPI_ISL_1193119, EPI_ISL_1193120, EPI_ISL_1193121, EPI_ISL_1193122, EPI_ISL_1193123, EPI_ISL_1193124, EPI_ISL_1193125, EPI_ISL_1193126, EPI_ISL_1193127, EPI_ISL_1193128, EPI_ISL_1193129, EPI_ISL_1193130, EPI_ISL_1193131, EPI_ISL_1193132, EPI_ISL_1193133, EPI_ISL_1193134, EPI_ISL_1193135, EPI_ISL_1193136, EPI_ISL_1193137, EPI_ISL_1193138, EPI_ISL_1193139, EPI_ISL_1193140,
EPI_ISL_1193141, EPI_ISL_1193142, EPI_ISL_1193143, EPI_ISL_1193144, EPI_ISL_1193145, EPI_ISL_1193146, EPI_ISL_1193147, EPI_ISL_1193148, EPI_ISL_1193149, EPI_ISL_1193150, EPI_ISL_1193151, EPI_ISL_1193152, EPI_ISL_1193153, EPI_ISL_1193154, EPI_ISL_1193155, EPI_ISL_1193156, EPI_ISL_1193157, EPI_ISL_1193158, EPI_ISL_1193159, EPI_ISL_1193160, EPI_ISL_1193161, EPI_ISL_1193162,
EPI_ISL_1193163, EPI_ISL_1193164, EPI_ISL_1193165, EPI_ISL_1193166, EPI_ISL_1193167, EPI_ISL_1193168, EPI_ISL_1193169, EPI_ISL_1193170, EPI_ISL_1193171, EPI_ISL_1193172, EPI_ISL_1193173, EPI_ISL_1193174, EPI_ISL_1193175, EPI_ISL_1193176, EPI_ISL_1193177, EPI_ISL_1193178, EPI_ISL_1193179, EPI_ISL_1193180, EPI_ISL_1193181, EPI_ISL_1193182, EPI_ISL_1193183, EPI_ISL_1193184,
EPI_ISL_1193185, EPI_ISL_1193186, EPI_ISL_1193187, EPI_ISL_1193188, EPI_ISL_1193189, EPI_ISL_1193190, EPI_ISL_1193191, EPI_ISL_1193192, EPI_ISL_1193193, EPI_ISL_1193194, EPI_ISL_1193195, EPI_ISL_1193196, EPI_ISL_1193197, EPI_ISL_1193198, EPI_ISL_1193199, EPI_ISL_1193200, EPI_ISL_1193201, EPI_ISL_1193202, EPI_ISL_1193203, EPI_ISL_1193204, EPI_ISL_1193205, EPI_ISL_1193206,
EPI_ISL_1193207, EPI_ISL_1193208, EPI_ISL_1193209, EPI_ISL_1193210, EPI_ISL_1193211, EPI_ISL_1193212, EPI_ISL_1193213, EPI_ISL_1193214, EPI_ISL_1193215, EPI_ISL_1193216, EPI_ISL_1193217, EPI_ISL_1193218, EPI_ISL_1193219, EPI_ISL_1193220, EPI_ISL_1193221, EPI_ISL_1193222, EPI_ISL_1193223, EPI_ISL_1193224, EPI_ISL_1193225, EPI_ISL_1193226, EPI_ISL_1193227, EPI_ISL_1193228,
EPI_ISL_1193229, EPI_ISL_1193230, EPI_ISL_1193231, EPI_ISL_1193232, EPI_ISL_1193233, EPI_ISL_1193234, EPI_ISL_1193235, EPI_ISL_1193236, EPI_ISL_1193237, EPI_ISL_1193238, EPI_ISL_1193239, EPI_ISL_1193240, EPI_ISL_1193241, EPI_ISL_1193242, EPI_ISL_1193243, EPI_ISL_1193244, EPI_ISL_1193245, EPI_ISL_1193246, EPI_ISL_1193247, EPI_ISL_1193248, EPI_ISL_1193249, EPI_ISL_1193250,
EPI_ISL_1193251, EPI_ISL_1193252, EPI_ISL_1193253, EPI_ISL_1193254, EPI_ISL_1193255, EPI_ISL_1193256, EPI_ISL_1193257, EPI_ISL_1193258, EPI_ISL_1193259, EPI_ISL_1197151, EPI_ISL_1197152, EPI_ISL_1197153, EPI_ISL_1197154, EPI_ISL_1197155, EPI_ISL_1197156, EPI_ISL_1197157, EPI_ISL_1197158, EPI_ISL_1197159, EPI_ISL_1197160, EPI_ISL_1197161, EPI_ISL_1197162, EPI_ISL_1197163,
EPI_ISL_1197164, EPI_ISL_1197165, EPI_ISL_1197166, EPI_ISL_1197167, EPI_ISL_1197168, EPI_ISL_1197169, EPI_ISL_1197170, EPI_ISL_1197171, EPI_ISL_1197172, EPI_ISL_1197173, EPI_ISL_1197174, EPI_ISL_1197175, EPI_ISL_1197176, EPI_ISL_1197177, EPI_ISL_1197178, EPI_ISL_1197179, EPI_ISL_1197180, EPI_ISL_1197181, EPI_ISL_1197182, EPI_ISL_1197183, EPI_ISL_1197184, EPI_ISL_1197185,
EPI_ISL_1197186, EPI_ISL_1197187, EPI_ISL_1197188, EPI_ISL_1197189, EPI_ISL_1197190, EPI_ISL_1197191, EPI_ISL_1197192, EPI_ISL_1197193, EPI_ISL_1197194, EPI_ISL_1197195, EPI_ISL_1197196, EPI_ISL_1197197, EPI_ISL_1197198, EPI_ISL_1197199, EPI_ISL_1197200, EPI_ISL_1197201, EPI_ISL_1197202, EPI_ISL_1197203, EPI_ISL_1197204, EPI_ISL_1197205, EPI_ISL_1197206, EPI_ISL_1197207,
EPI_ISL_1197208, EPI_ISL_1197209, EPI_ISL_1197210, EPI_ISL_1197211, EPI_ISL_1197212, EPI_ISL_1197213, EPI_ISL_1197214, EPI_ISL_1197215, EPI_ISL_1197216, EPI_ISL_1197217, EPI_ISL_1197218, EPI_ISL_1197219, EPI_ISL_1197220, EPI_ISL_1197221, EPI_ISL_1197222, EPI_ISL_1197223, EPI_ISL_1197224, EPI_ISL_1197225, EPI_ISL_1197226, EPI_ISL_1197227, EPI_ISL_1197228, EPI_ISL_1197229,
EPI_ISL_1197230, EPI_ISL_1197231, EPI_ISL_1197232, EPI_ISL_1197233, EPI_ISL_1197234, EPI_ISL_1197235, EPI_ISL_1197237, EPI_ISL_1197238, EPI_ISL_1197239, EPI_ISL_1197240, EPI_ISL_1197241, EPI_ISL_1197242, EPI_ISL_1197243, EPI_ISL_1197244, EPI_ISL_1197245, EPI_ISL_1197246, EPI_ISL_1197247, EPI_ISL_1197248, EPI_ISL_1197249, EPI_ISL_1197250, EPI_ISL_1197251, EPI_ISL_1197252,
EPI_ISL_1197253, EPI_ISL_1197254, EPI_ISL_1197255, EPI_ISL_1197256, EPI_ISL_1197257, EPI_ISL_1197258, EPI_ISL_1197259, EPI_ISL_1197260, EPI_ISL_1197261, EPI_ISL_1197262, EPI_ISL_1197263, EPI_ISL_1197264, EPI_ISL_1197265, EPI_ISL_1197266, EPI_ISL_1197267, EPI_ISL_1197268, EPI_ISL_1197269, EPI_ISL_1197270, EPI_ISL_1197271, EPI_ISL_1197272, EPI_ISL_1197273, EPI_ISL_1197274,
EPI_ISL_1197275, EPI_ISL_1197276, EPI_ISL_1197277, EPI_ISL_1197278, EPI_ISL_1197279, EPI_ISL_1197280, EPI_ISL_1197281, EPI_ISL_1197282, EPI_ISL_1197283, EPI_ISL_1197284, EPI_ISL_1197285, EPI_ISL_1197286, EPI_ISL_1197287, EPI_ISL_1197288, EPI_ISL_1197289, EPI_ISL_1197290, EPI_ISL_1197291, EPI_ISL_1197292, EPI_ISL_1197293, EPI_ISL_1197294, EPI_ISL_1197295, EPI_ISL_1197296,
EPI_ISL_1197297, EPI_ISL_1197298, EPI_ISL_1197299, EPI_ISL_1197300, EPI_ISL_1197301, EPI_ISL_1197302, EPI_ISL_1197303, EPI_ISL_1197304, EPI_ISL_1197305, EPI_ISL_1197306, EPI_ISL_1197307, EPI_ISL_1197308, EPI_ISL_1197309, EPI_ISL_1197310, EPI_ISL_1197311, EPI_ISL_1197312, EPI_ISL_1197313, EPI_ISL_1197314, EPI_ISL_1197315, EPI_ISL_1197316, EPI_ISL_1197317, EPI_ISL_1197318,
EPI_ISL_1197319, EPI_ISL_1197320, EPI_ISL_1197321, EPI_ISL_1197322, EPI_ISL_1197323, EPI_ISL_1197324, EPI_ISL_1197325, EPI_ISL_1197326, EPI_ISL_1197327, EPI_ISL_1197328, EPI_ISL_1197329, EPI_ISL_1197330, EPI_ISL_1197331, EPI_ISL_1197332, EPI_ISL_1197333, EPI_ISL_1197334, EPI_ISL_1197335, EPI_ISL_1197336, EPI_ISL_1197337, EPI_ISL_1197338, EPI_ISL_1197339, EPI_ISL_1197340,
EPI_ISL_1197341, EPI_ISL_1197342, EPI_ISL_1197343, EPI_ISL_1197344, EPI_ISL_1197345, EPI_ISL_1197346, EPI_ISL_1197347, EPI_ISL_1197348, EPI_ISL_1197349, EPI_ISL_1197350, EPI_ISL_1197351, EPI_ISL_1197352, EPI_ISL_1197353, EPI_ISL_1197354, EPI_ISL_1197355, EPI_ISL_1197356, EPI_ISL_1197357, EPI_ISL_1197358, EPI_ISL_1197359, EPI_ISL_1197360, EPI_ISL_1197361, EPI_ISL_1197362,
EPI_ISL_1197363, EPI_ISL_1197364, EPI_ISL_1197365, EPI_ISL_1197366, EPI_ISL_1197367, EPI_ISL_1197368, EPI_ISL_1197369, EPI_ISL_1197370, EPI_ISL_1197371, EPI_ISL_1197372, EPI_ISL_1197373, EPI_ISL_1197374, EPI_ISL_1197375, EPI_ISL_1197376, EPI_ISL_1197377, EPI_ISL_1197378, EPI_ISL_1197379, EPI_ISL_1197380, EPI_ISL_1197381, EPI_ISL_1197382, EPI_ISL_1197383, EPI_ISL_1197384,
EPI_ISL_1197385, EPI_ISL_1197386, EPI_ISL_1197387, EPI_ISL_1197388, EPI_ISL_1197389, EPI_ISL_1197390, EPI_ISL_1197391, EPI_ISL_1197392, EPI_ISL_1197393, EPI_ISL_1197394, EPI_ISL_1197395, EPI_ISL_1197396, EPI_ISL_1197397, EPI_ISL_1197398, EPI_ISL_1197399, EPI_ISL_1197400, EPI_ISL_1197401, EPI_ISL_1197402, EPI_ISL_1197403, EPI_ISL_1197404, EPI_ISL_1197405, EPI_ISL_1197406,
EPI_ISL_1197407, EPI_ISL_1197408, EPI_ISL_1197409, EPI_ISL_1197410, EPI_ISL_1197411, EPI_ISL_1197412, EPI_ISL_1197413, EPI_ISL_1197414, EPI_ISL_1197415, EPI_ISL_1197416, EPI_ISL_1197417, EPI_ISL_1197418, EPI_ISL_1197419, EPI_ISL_1197420, EPI_ISL_1197421, EPI_ISL_1197422, EPI_ISL_1197423, EPI_ISL_1197424, EPI_ISL_1197425, EPI_ISL_1197426, EPI_ISL_1197427, EPI_ISL_1197428,
EPI_ISL_1197429, EPI_ISL_1197430, EPI_ISL_1197431, EPI_ISL_1197432, EPI_ISL_1197433, EPI_ISL_1197434, EPI_ISL_1197435, EPI_ISL_1197436, EPI_ISL_1197437, EPI_ISL_1197438, EPI_ISL_1197439, EPI_ISL_1197440, EPI_ISL_1197441, EPI_ISL_1197442, EPI_ISL_1197443, EPI_ISL_1197444, EPI_ISL_1197445, EPI_ISL_1197446, EPI_ISL_1197447, EPI_ISL_1197448, EPI_ISL_1197449, EPI_ISL_1197450,
EPI_ISL_1197451, EPI_ISL_1197452, EPI_ISL_1197453, EPI_ISL_1197454, EPI_ISL_1197455, EPI_ISL_1197456, EPI_ISL_1197457, EPI_ISL_1197458, EPI_ISL_1197459, EPI_ISL_1197460, EPI_ISL_1197461, EPI_ISL_1197462, EPI_ISL_1197463, EPI_ISL_1197464, EPI_ISL_1197465, EPI_ISL_1197466, EPI_ISL_1197467, EPI_ISL_1197468, EPI_ISL_1197469, EPI_ISL_1197470, EPI_ISL_1197471, EPI_ISL_1197472,
EPI_ISL_1197473, EPI_ISL_1197474, EPI_ISL_1197475, EPI_ISL_1197476, EPI_ISL_1197477, EPI_ISL_1197478, EPI_ISL_1197479, EPI_ISL_1197480, EPI_ISL_1197481, EPI_ISL_1197482, EPI_ISL_1197483, EPI_ISL_1197484, EPI_ISL_1197485, EPI_ISL_1197486, EPI_ISL_1197487, EPI_ISL_1197488, EPI_ISL_1197489, EPI_ISL_1197490, EPI_ISL_1197491, EPI_ISL_1197492, EPI_ISL_1197493, EPI_ISL_1197494,
EPI_ISL_1197495, EPI_ISL_1197496, EPI_ISL_1197497, EPI_ISL_1197498, EPI_ISL_1197499, EPI_ISL_1197500, EPI_ISL_1197501, EPI_ISL_1197502, EPI_ISL_1197503, EPI_ISL_1197504, EPI_ISL_1197505, EPI_ISL_1197506, EPI_ISL_1197507, EPI_ISL_1197508, EPI_ISL_1197509, EPI_ISL_1197510, EPI_ISL_1197511, EPI_ISL_1197512, EPI_ISL_1197513, EPI_ISL_1197514, EPI_ISL_1197515, EPI_ISL_1197516,
EPI_ISL_1197517, EPI_ISL_1197518, EPI_ISL_1197519, EPI_ISL_1197520, EPI_ISL_1197521, EPI_ISL_1197522, EPI_ISL_1197523, EPI_ISL_1197524, EPI_ISL_1197525, EPI_ISL_1197526, EPI_ISL_1197527, EPI_ISL_1197528, EPI_ISL_1197529, EPI_ISL_1197530, EPI_ISL_1197531, EPI_ISL_1197532, EPI_ISL_1197533, EPI_ISL_1197534, EPI_ISL_1197535, EPI_ISL_1197536, EPI_ISL_1197537, EPI_ISL_1197538,
EPI_ISL_1197539, EPI_ISL_1197540, EPI_ISL_1197541, EPI_ISL_1197542, EPI_ISL_1197543, EPI_ISL_1197544, EPI_ISL_1197545, EPI_ISL_1197546, EPI_ISL_1197547, EPI_ISL_1197548, EPI_ISL_1197549, EPI_ISL_1197550, EPI_ISL_1197551, EPI_ISL_1197552, EPI_ISL_1197553, EPI_ISL_1197554, EPI_ISL_1197555, EPI_ISL_1197556, EPI_ISL_1197557, EPI_ISL_1197558, EPI_ISL_1197559, EPI_ISL_1197560,
EPI_ISL_1197561, EPI_ISL_1197562, EPI_ISL_1197563, EPI_ISL_1197564, EPI_ISL_1197565, EPI_ISL_1197566, EPI_ISL_1197567, EPI_ISL_1197568, EPI_ISL_1197569, EPI_ISL_1197570, EPI_ISL_1197571, EPI_ISL_1197572, EPI_ISL_1197573, EPI_ISL_1197574, EPI_ISL_1197575, EPI_ISL_1197576, EPI_ISL_1197577, EPI_ISL_1197578, EPI_ISL_1197579, EPI_ISL_1197580, EPI_ISL_1197581, EPI_ISL_1197582,
EPI_ISL_1197583, EPI_ISL_1197584, EPI_ISL_1197585, EPI_ISL_1197586, EPI_ISL_1197587, EPI_ISL_1197588, EPI_ISL_1197589, EPI_ISL_1197590, EPI_ISL_1197591, EPI_ISL_1197592, EPI_ISL_1197593, EPI_ISL_1197594, EPI_ISL_1197595, EPI_ISL_1197596, EPI_ISL_1197597, EPI_ISL_1197598, EPI_ISL_1197599, EPI_ISL_1197600, EPI_ISL_1197601, EPI_ISL_1197602, EPI_ISL_1197603, EPI_ISL_1197604,
EPI_ISL_1197605, EPI_ISL_1197606, EPI_ISL_1197607, EPI_ISL_1197608, EPI_ISL_1197609, EPI_ISL_1197610, EPI_ISL_1197611, EPI_ISL_1197612, EPI_ISL_1197613, EPI_ISL_1197614, EPI_ISL_1197615, EPI_ISL_1197616, EPI_ISL_1197617, EPI_ISL_1197618, EPI_ISL_1197619, EPI_ISL_1197620, EPI_ISL_1197621, EPI_ISL_1197622, EPI_ISL_1197623, EPI_ISL_1197624, EPI_ISL_1197625, EPI_ISL_1197626,
EPI_ISL_1197627, EPI_ISL_1197628, EPI_ISL_1197629, EPI_ISL_1197630, EPI_ISL_1197631, EPI_ISL_1197632, EPI_ISL_1197633, EPI_ISL_1197634, EPI_ISL_1197635, EPI_ISL_1197636, EPI_ISL_1197637, EPI_ISL_1197638, EPI_ISL_1197639, EPI_ISL_1197640, EPI_ISL_1197641, EPI_ISL_1197642, EPI_ISL_1197643, EPI_ISL_1197644, EPI_ISL_1197645, EPI_ISL_1197646, EPI_ISL_1197647, EPI_ISL_1197648,
EPI_ISL_1197649, EPI_ISL_1197650, EPI_ISL_1197651, EPI_ISL_1197652, EPI_ISL_1197653, EPI_ISL_1197654, EPI_ISL_1197655, EPI_ISL_1197656, EPI_ISL_1197657, EPI_ISL_1197658, EPI_ISL_1197659, EPI_ISL_1197660, EPI_ISL_1197661, EPI_ISL_1197662, EPI_ISL_1197663, EPI_ISL_1197664, EPI_ISL_1197665, EPI_ISL_1197666, EPI_ISL_1197667, EPI_ISL_1197668, EPI_ISL_1197669, EPI_ISL_1197670,
EPI_ISL_1197671, EPI_ISL_1197672, EPI_ISL_1197673, EPI_ISL_1197674, EPI_ISL_1197675, EPI_ISL_1197676, EPI_ISL_1197677, EPI_ISL_1197678, EPI_ISL_1197679, EPI_ISL_1197680, EPI_ISL_1197681, EPI_ISL_1197682, EPI_ISL_1197683, EPI_ISL_1197684, EPI_ISL_1197685, EPI_ISL_1197686, EPI_ISL_1197687, EPI_ISL_1197688, EPI_ISL_1197689, EPI_ISL_1197690, EPI_ISL_1197692, EPI_ISL_1197693,
EPI_ISL_1197694, EPI_ISL_1197695, EPI_ISL_1197696, EPI_ISL_1197697, EPI_ISL_1197698, EPI_ISL_1197699, EPI_ISL_1197700, EPI_ISL_1197701, EPI_ISL_1197702, EPI_ISL_1197703, EPI_ISL_1197704, EPI_ISL_1197705, EPI_ISL_1197706, EPI_ISL_1197707, EPI_ISL_1197708, EPI_ISL_1197709, EPI_ISL_1197710, EPI_ISL_1197711, EPI_ISL_1197712, EPI_ISL_1197713, EPI_ISL_1197714, EPI_ISL_1197715,
EPI_ISL_1197716, EPI_ISL_1197717, EPI_ISL_1197718, EPI_ISL_1197719, EPI_ISL_1197720, EPI_ISL_1197721, EPI_ISL_1197722, EPI_ISL_1197723, EPI_ISL_1197724, EPI_ISL_1197725, EPI_ISL_1197726, EPI_ISL_1197727, EPI_ISL_1197728, EPI_ISL_1197729, EPI_ISL_1197730, EPI_ISL_1197731, EPI_ISL_1197732, EPI_ISL_1197733, EPI_ISL_1197734, EPI_ISL_1197735, EPI_ISL_1197736, EPI_ISL_1197737,
EPI_ISL_1197738, EPI_ISL_1197739, EPI_ISL_1197740, EPI_ISL_1197741, EPI_ISL_1197742, EPI_ISL_1197743, EPI_ISL_1197744, EPI_ISL_1197745, EPI_ISL_1197746, EPI_ISL_1197747, EPI_ISL_1197748, EPI_ISL_1197749, EPI_ISL_1197750, EPI_ISL_1197751, EPI_ISL_1197752, EPI_ISL_1197753, EPI_ISL_1197754, EPI_ISL_1197755, EPI_ISL_1197756, EPI_ISL_1197757, EPI_ISL_1197758, EPI_ISL_1197759,
EPI_ISL_1197760, EPI_ISL_1197761, EPI_ISL_1197762, EPI_ISL_1197763, EPI_ISL_1197764, EPI_ISL_1197765, EPI_ISL_1197766, EPI_ISL_1197767, EPI_ISL_1197768, EPI_ISL_1197769, EPI_ISL_1197770, EPI_ISL_1197771, EPI_ISL_1197772, EPI_ISL_1197773, EPI_ISL_1197774, EPI_ISL_1197775, EPI_ISL_1197776, EPI_ISL_1197777, EPI_ISL_1197778, EPI_ISL_1197779, EPI_ISL_1197780, EPI_ISL_1197781,
EPI_ISL_1197782, EPI_ISL_1197783, EPI_ISL_1197784, EPI_ISL_1197785, EPI_ISL_1197786, EPI_ISL_1197787, EPI_ISL_1197788, EPI_ISL_1197789, EPI_ISL_1197790, EPI_ISL_1197791, EPI_ISL_1197792, EPI_ISL_1197793, EPI_ISL_1197794, EPI_ISL_1197795, EPI_ISL_1197796, EPI_ISL_1197797, EPI_ISL_1197798, EPI_ISL_1197799, EPI_ISL_1197800, EPI_ISL_1197801, EPI_ISL_1197802, EPI_ISL_1197803,
EPI_ISL_1197804, EPI_ISL_1197805, EPI_ISL_1197806, EPI_ISL_1197807, EPI_ISL_1197808, EPI_ISL_1197809, EPI_ISL_1197810, EPI_ISL_1197811, EPI_ISL_1197812, EPI_ISL_1197813, EPI_ISL_1197814, EPI_ISL_1197815, EPI_ISL_1197816, EPI_ISL_1197817, EPI_ISL_1197818, EPI_ISL_1197819, EPI_ISL_1197820, EPI_ISL_1197821, EPI_ISL_1197822, EPI_ISL_1197823, EPI_ISL_1197824, EPI_ISL_1197825,
EPI_ISL_1197826, EPI_ISL_1197827, EPI_ISL_1197828, EPI_ISL_1197829, EPI_ISL_1197830, EPI_ISL_1197831, EPI_ISL_1197832, EPI_ISL_1197833, EPI_ISL_1197834, EPI_ISL_1197835, EPI_ISL_1197836, EPI_ISL_1197837, EPI_ISL_1197838, EPI_ISL_1197839, EPI_ISL_1197840, EPI_ISL_1197841, EPI_ISL_1197842, EPI_ISL_1197843, EPI_ISL_1197844, EPI_ISL_1197845, EPI_ISL_1197846, EPI_ISL_1197847,
EPI_ISL_1197848, EPI_ISL_1197849, EPI_ISL_1197850, EPI_ISL_1197851, EPI_ISL_1197852, EPI_ISL_1197853, EPI_ISL_1197854, EPI_ISL_1197855, EPI_ISL_1197856, EPI_ISL_1197857, EPI_ISL_1197858, EPI_ISL_1197859, EPI_ISL_1197860, EPI_ISL_1197861, EPI_ISL_1197862, EPI_ISL_1197863, EPI_ISL_1197864, EPI_ISL_1197865, EPI_ISL_1197866, EPI_ISL_1197867, EPI_ISL_1197868, EPI_ISL_1197869,
EPI_ISL_1197870, EPI_ISL_1197871, EPI_ISL_1197872, EPI_ISL_1197873, EPI_ISL_1197874, EPI_ISL_1197875, EPI_ISL_1197876, EPI_ISL_1197877, EPI_ISL_1197878, EPI_ISL_1197879, EPI_ISL_1197880, EPI_ISL_1197881, EPI_ISL_1197882, EPI_ISL_1197883, EPI_ISL_1197884, EPI_ISL_1197885, EPI_ISL_1197886, EPI_ISL_1197887, EPI_ISL_1197888, EPI_ISL_1197889, EPI_ISL_1197890, EPI_ISL_1197891,
EPI_ISL_1197892, EPI_ISL_1197893, EPI_ISL_1197894, EPI_ISL_1197895, EPI_ISL_1197896, EPI_ISL_1197897, EPI_ISL_1197898, EPI_ISL_1197899, EPI_ISL_1197900, EPI_ISL_1197901, EPI_ISL_1197902, EPI_ISL_1197903, EPI_ISL_1197904, EPI_ISL_1197905, EPI_ISL_1197906, EPI_ISL_1197907, EPI_ISL_1197908, EPI_ISL_1197909, EPI_ISL_1197910, EPI_ISL_1197911, EPI_ISL_1197912, EPI_ISL_1197913,
EPI_ISL_1197914, EPI_ISL_1197915, EPI_ISL_1197916, EPI_ISL_1197917, EPI_ISL_1197918, EPI_ISL_1197919, EPI_ISL_1197920, EPI_ISL_1197921, EPI_ISL_1197922, EPI_ISL_1197923, EPI_ISL_1197924, EPI_ISL_1197925, EPI_ISL_1197926, EPI_ISL_1197927, EPI_ISL_1197928, EPI_ISL_1197929, EPI_ISL_1197930, EPI_ISL_1197931, EPI_ISL_1197932, EPI_ISL_1198009, EPI_ISL_1198010, EPI_ISL_1198011,
EPI_ISL_1198012, EPI_ISL_1198013, EPI_ISL_1198014, EPI_ISL_1198015, EPI_ISL_1198016, EPI_ISL_1198017, EPI_ISL_1198018, EPI_ISL_1198019, EPI_ISL_1198020, EPI_ISL_1198021, EPI_ISL_1198022, EPI_ISL_1198023, EPI_ISL_1198024, EPI_ISL_1198025, EPI_ISL_1198026, EPI_ISL_1198027, EPI_ISL_1198028, EPI_ISL_1198029, EPI_ISL_1198030, EPI_ISL_1198031, EPI_ISL_1198032, EPI_ISL_1198033,
EPI_ISL_1198034, EPI_ISL_1198035, EPI_ISL_1198036, EPI_ISL_1198037, EPI_ISL_1198038, EPI_ISL_1198039, EPI_ISL_1198040, EPI_ISL_1198041, EPI_ISL_1198042, EPI_ISL_1198043, EPI_ISL_1198044, EPI_ISL_1198045, EPI_ISL_1198046, EPI_ISL_1198047, EPI_ISL_1198048, EPI_ISL_1198049, EPI_ISL_1198050, EPI_ISL_1198051, EPI_ISL_1198052, EPI_ISL_1198053, EPI_ISL_1198054, EPI_ISL_1198055,
EPI_ISL_1198056, EPI_ISL_1198057, EPI_ISL_1198058, EPI_ISL_1198059, EPI_ISL_1198060, EPI_ISL_1198061, EPI_ISL_1198062, EPI_ISL_1198063, EPI_ISL_1198064, EPI_ISL_1198065, EPI_ISL_1198066, EPI_ISL_1198067, EPI_ISL_1198068, EPI_ISL_1198069, EPI_ISL_1198070, EPI_ISL_1198071, EPI_ISL_1198072, EPI_ISL_1198073, EPI_ISL_1198074, EPI_ISL_1198075, EPI_ISL_1198076, EPI_ISL_1198077,
EPI_ISL_1198078, EPI_ISL_1198079, EPI_ISL_1198080, EPI_ISL_1198081, EPI_ISL_1198082, EPI_ISL_1198083, EPI_ISL_1198084, EPI_ISL_1198085, EPI_ISL_1198086, EPI_ISL_1198087, EPI_ISL_1198088, EPI_ISL_1198089, EPI_ISL_1198090, EPI_ISL_1198091, EPI_ISL_1198092, EPI_ISL_1198093, EPI_ISL_1198094, EPI_ISL_1198095, EPI_ISL_1198096, EPI_ISL_1198097, EPI_ISL_1198098, EPI_ISL_1198099,
EPI_ISL_1198100, EPI_ISL_1198101, EPI_ISL_1198102, EPI_ISL_1198103, EPI_ISL_1198104, EPI_ISL_1198105, EPI_ISL_1198106, EPI_ISL_1198107, EPI_ISL_1198108, EPI_ISL_1198109, EPI_ISL_1198110, EPI_ISL_1198111, EPI_ISL_1198112, EPI_ISL_1198113, EPI_ISL_1198114, EPI_ISL_1198115, EPI_ISL_1198116, EPI_ISL_1198117, EPI_ISL_1198118, EPI_ISL_1198119, EPI_ISL_1198120, EPI_ISL_1198121,
EPI_ISL_1198122, EPI_ISL_1198123, EPI_ISL_1198124, EPI_ISL_1198125, EPI_ISL_1198126, EPI_ISL_1198127, EPI_ISL_1198128, EPI_ISL_1198129, EPI_ISL_1198130, EPI_ISL_1198131, EPI_ISL_1198132, EPI_ISL_1198133, EPI_ISL_1198134, EPI_ISL_1198135, EPI_ISL_1198136, EPI_ISL_1198137, EPI_ISL_1198138, EPI_ISL_1198139, EPI_ISL_1198140, EPI_ISL_1198141, EPI_ISL_1198142, EPI_ISL_1198143,
EPI_ISL_1198144, EPI_ISL_1198145, EPI_ISL_1198146, EPI_ISL_1198147, EPI_ISL_1198148, EPI_ISL_1198149, EPI_ISL_1198150, EPI_ISL_1198151, EPI_ISL_1198152, EPI_ISL_1198153, EPI_ISL_1198154, EPI_ISL_1198155, EPI_ISL_1198156, EPI_ISL_1198157, EPI_ISL_1198158, EPI_ISL_1198159, EPI_ISL_1198160, EPI_ISL_1198161, EPI_ISL_1198162, EPI_ISL_1198163, EPI_ISL_1198164, EPI_ISL_1198165,
EPI_ISL_1198166, EPI_ISL_1198167, EPI_ISL_1198168, EPI_ISL_1198169, EPI_ISL_1198170, EPI_ISL_1198171, EPI_ISL_1198172, EPI_ISL_1198173, EPI_ISL_1198174, EPI_ISL_1198175, EPI_ISL_1198176, EPI_ISL_1198177, EPI_ISL_1198178, EPI_ISL_1198179, EPI_ISL_1198180, EPI_ISL_1198181, EPI_ISL_1198182, EPI_ISL_1198183, EPI_ISL_1198184, EPI_ISL_1198185, EPI_ISL_1198186, EPI_ISL_1198187,
EPI_ISL_1198188, EPI_ISL_1198189, EPI_ISL_1198190, EPI_ISL_1198191, EPI_ISL_1198192, EPI_ISL_1198193, EPI_ISL_1198194, EPI_ISL_1198195, EPI_ISL_1198196, EPI_ISL_1198197, EPI_ISL_1198198, EPI_ISL_1198199, EPI_ISL_1198200, EPI_ISL_1198201, EPI_ISL_1198202, EPI_ISL_1198203, EPI_ISL_1198204, EPI_ISL_1198205, EPI_ISL_1198206, EPI_ISL_1198207, EPI_ISL_1198208, EPI_ISL_1198209,
EPI_ISL_1198210, EPI_ISL_1198211, EPI_ISL_1198212, EPI_ISL_1198213, EPI_ISL_1198214, EPI_ISL_1198215, EPI_ISL_1198216, EPI_ISL_1198217, EPI_ISL_1198218, EPI_ISL_1198219, EPI_ISL_1198220, EPI_ISL_1198221, EPI_ISL_1198222, EPI_ISL_1198223, EPI_ISL_1198224, EPI_ISL_1198225, EPI_ISL_1198226, EPI_ISL_1198227, EPI_ISL_1198228, EPI_ISL_1198229, EPI_ISL_1198230, EPI_ISL_1198231,
EPI_ISL_1198232, EPI_ISL_1198233, EPI_ISL_1198234, EPI_ISL_1198235, EPI_ISL_1198236, EPI_ISL_1198237, EPI_ISL_1198238, EPI_ISL_1198239, EPI_ISL_1198240, EPI_ISL_1198241, EPI_ISL_1198242, EPI_ISL_1198243, EPI_ISL_1198244, EPI_ISL_1198245, EPI_ISL_1198246, EPI_ISL_1198247, EPI_ISL_1198248, EPI_ISL_1198249, EPI_ISL_1198250, EPI_ISL_1198251, EPI_ISL_1198252, EPI_ISL_1198253,
EPI_ISL_1198254, EPI_ISL_1198255, EPI_ISL_1198256, EPI_ISL_1198257, EPI_ISL_1198258, EPI_ISL_1198259, EPI_ISL_1198260, EPI_ISL_1198261, EPI_ISL_1198262, EPI_ISL_1198263, EPI_ISL_1198264, EPI_ISL_1198265, EPI_ISL_1198266, EPI_ISL_1198267, EPI_ISL_1198268, EPI_ISL_1198269, EPI_ISL_1198270, EPI_ISL_1198271, EPI_ISL_1198272, EPI_ISL_1198273, EPI_ISL_1198274, EPI_ISL_1198275,
EPI_ISL_1198276, EPI_ISL_1198277, EPI_ISL_1198278, EPI_ISL_1198279, EPI_ISL_1198280, EPI_ISL_1198281, EPI_ISL_1198282, EPI_ISL_1198283, EPI_ISL_1198284, EPI_ISL_1198285, EPI_ISL_1198286, EPI_ISL_1198287, EPI_ISL_1198288, EPI_ISL_1198289, EPI_ISL_1198290, EPI_ISL_1198291, EPI_ISL_1198292, EPI_ISL_1198293, EPI_ISL_1198294, EPI_ISL_1198295, EPI_ISL_1198296, EPI_ISL_1198297,
EPI_ISL_1198298, EPI_ISL_1198299, EPI_ISL_1198300, EPI_ISL_1198301, EPI_ISL_1198302, EPI_ISL_1198303, EPI_ISL_1198304, EPI_ISL_1198305, EPI_ISL_1198306, EPI_ISL_1198307, EPI_ISL_1198308, EPI_ISL_1198309, EPI_ISL_1198310, EPI_ISL_1198311, EPI_ISL_1198312, EPI_ISL_1198313, EPI_ISL_1198314, EPI_ISL_1198315, EPI_ISL_1198316, EPI_ISL_1198317, EPI_ISL_1198318, EPI_ISL_1198319,
EPI_ISL_1198320, EPI_ISL_1198321, EPI_ISL_1198322, EPI_ISL_1198323, EPI_ISL_1198324, EPI_ISL_1198325, EPI_ISL_1198326, EPI_ISL_1198327, EPI_ISL_1198328, EPI_ISL_1198329, EPI_ISL_1198330, EPI_ISL_1198331, EPI_ISL_1198332, EPI_ISL_1198333, EPI_ISL_1198334, EPI_ISL_1198335, EPI_ISL_1198336, EPI_ISL_1198337, EPI_ISL_1198338, EPI_ISL_1198339, EPI_ISL_1198340, EPI_ISL_1198341,
EPI_ISL_1198342, EPI_ISL_1198343, EPI_ISL_1198344, EPI_ISL_1198345, EPI_ISL_1198346, EPI_ISL_1198347, EPI_ISL_1198348, EPI_ISL_1198349, EPI_ISL_1198350, EPI_ISL_1198351, EPI_ISL_1198352, EPI_ISL_1198353, EPI_ISL_1198354, EPI_ISL_1198355, EPI_ISL_1198356, EPI_ISL_1198357, EPI_ISL_1198358, EPI_ISL_1198359, EPI_ISL_1198360, EPI_ISL_1198361, EPI_ISL_1198362, EPI_ISL_1198363,
EPI_ISL_1198364, EPI_ISL_1198365, EPI_ISL_1198366, EPI_ISL_1198367, EPI_ISL_1198368, EPI_ISL_1198369, EPI_ISL_1198370, EPI_ISL_1198371, EPI_ISL_1198372, EPI_ISL_1198373, EPI_ISL_1198374, EPI_ISL_1198375, EPI_ISL_1198376, EPI_ISL_1198377, EPI_ISL_1198378, EPI_ISL_1198379, EPI_ISL_1198380, EPI_ISL_1198381, EPI_ISL_1198382, EPI_ISL_1198383, EPI_ISL_1198384, EPI_ISL_1198385,
EPI_ISL_1198386, EPI_ISL_1198387, EPI_ISL_1198388, EPI_ISL_1198389, EPI_ISL_1198390, EPI_ISL_1198391, EPI_ISL_1198392, EPI_ISL_1198393, EPI_ISL_1198394, EPI_ISL_1198395, EPI_ISL_1198396, EPI_ISL_1198397, EPI_ISL_1198398, EPI_ISL_1198399, EPI_ISL_1198400, EPI_ISL_1198401, EPI_ISL_1198402, EPI_ISL_1198403, EPI_ISL_1198404, EPI_ISL_1198405, EPI_ISL_1198406, EPI_ISL_1198407,
EPI_ISL_1198408, EPI_ISL_1198409, EPI_ISL_1198410, EPI_ISL_1198411, EPI_ISL_1198412, EPI_ISL_1198413, EPI_ISL_1198414, EPI_ISL_1198415, EPI_ISL_1198416, EPI_ISL_1198417, EPI_ISL_1198418, EPI_ISL_1198419, EPI_ISL_1198420, EPI_ISL_1198421, EPI_ISL_1198422, EPI_ISL_1198423, EPI_ISL_1198424, EPI_ISL_1198425, EPI_ISL_1198426, EPI_ISL_1198427, EPI_ISL_1198428, EPI_ISL_1198429,
EPI_ISL_1198430, EPI_ISL_1198431, EPI_ISL_1198432, EPI_ISL_1198433, EPI_ISL_1198434, EPI_ISL_1198435, EPI_ISL_1198436, EPI_ISL_1198437, EPI_ISL_1198438, EPI_ISL_1198439, EPI_ISL_1198440, EPI_ISL_1198441, EPI_ISL_1198442, EPI_ISL_1198443, EPI_ISL_1198444, EPI_ISL_1198445, EPI_ISL_1198446, EPI_ISL_1198447, EPI_ISL_1198448, EPI_ISL_1198449, EPI_ISL_1198450, EPI_ISL_1198451,
EPI_ISL_1198452, EPI_ISL_1198453, EPI_ISL_1198454, EPI_ISL_1198455, EPI_ISL_1198456, EPI_ISL_1198457, EPI_ISL_1198458, EPI_ISL_1198459, EPI_ISL_1198460, EPI_ISL_1198461, EPI_ISL_1198462, EPI_ISL_1198463, EPI_ISL_1198464, EPI_ISL_1198465, EPI_ISL_1198466, EPI_ISL_1198467, EPI_ISL_1198468, EPI_ISL_1198469, EPI_ISL_1198470, EPI_ISL_1198471, EPI_ISL_1198472, EPI_ISL_1198473,
EPI_ISL_1198474, EPI_ISL_1198475, EPI_ISL_1198476, EPI_ISL_1198477, EPI_ISL_1198478, EPI_ISL_1198479, EPI_ISL_1198480, EPI_ISL_1198481, EPI_ISL_1198482, EPI_ISL_1198483, EPI_ISL_1198484, EPI_ISL_1198485, EPI_ISL_1198486, EPI_ISL_1198487, EPI_ISL_1198488, EPI_ISL_1198489, EPI_ISL_1198490, EPI_ISL_1198491, EPI_ISL_1198492, EPI_ISL_1198493, EPI_ISL_1198494, EPI_ISL_1198495,
EPI_ISL_1198496, EPI_ISL_1198497, EPI_ISL_1198498, EPI_ISL_1198499, EPI_ISL_1198500, EPI_ISL_1198501, EPI_ISL_1198502, EPI_ISL_1198503, EPI_ISL_1198504, EPI_ISL_1198505, EPI_ISL_1198506, EPI_ISL_1198507, EPI_ISL_1198508, EPI_ISL_1198509, EPI_ISL_1198510, EPI_ISL_1198511, EPI_ISL_1198512, EPI_ISL_1198513, EPI_ISL_1198514, EPI_ISL_1198515, EPI_ISL_1198516, EPI_ISL_1198517,
EPI_ISL_1198518, EPI_ISL_1198519, EPI_ISL_1198520, EPI_ISL_1198521, EPI_ISL_1198522, EPI_ISL_1198523, EPI_ISL_1198524, EPI_ISL_1198525, EPI_ISL_1198526, EPI_ISL_1198527, EPI_ISL_1198528, EPI_ISL_1198529, EPI_ISL_1198530, EPI_ISL_1198531, EPI_ISL_1198532, EPI_ISL_1198533, EPI_ISL_1198534, EPI_ISL_1198535, EPI_ISL_1198536, EPI_ISL_1198537, EPI_ISL_1198538, EPI_ISL_1198539,
EPI_ISL_1198540, EPI_ISL_1198541, EPI_ISL_1198542, EPI_ISL_1198543, EPI_ISL_1198544, EPI_ISL_1198545, EPI_ISL_1198546, EPI_ISL_1198547, EPI_ISL_1198548, EPI_ISL_1198549, EPI_ISL_1198550, EPI_ISL_1198551, EPI_ISL_1198552, EPI_ISL_1198553, EPI_ISL_1198554, EPI_ISL_1198555, EPI_ISL_1198556, EPI_ISL_1198557, EPI_ISL_1198558, EPI_ISL_1198559, EPI_ISL_1198560, EPI_ISL_1198561,
EPI_ISL_1198562, EPI_ISL_1198563, EPI_ISL_1198564, EPI_ISL_1198565, EPI_ISL_1198566, EPI_ISL_1198567, EPI_ISL_1198568, EPI_ISL_1198569, EPI_ISL_1198570, EPI_ISL_1198571, EPI_ISL_1198572, EPI_ISL_1198573, EPI_ISL_1198574, EPI_ISL_1198575, EPI_ISL_1198576, EPI_ISL_1198577, EPI_ISL_1198578, EPI_ISL_1198579, EPI_ISL_1198580, EPI_ISL_1198581, EPI_ISL_1198582, EPI_ISL_1198583,
EPI_ISL_1198584, EPI_ISL_1198585, EPI_ISL_1198586, EPI_ISL_1198587, EPI_ISL_1198588, EPI_ISL_1198589, EPI_ISL_1198590, EPI_ISL_1198591, EPI_ISL_1198592, EPI_ISL_1198593, EPI_ISL_1198594, EPI_ISL_1198595, EPI_ISL_1198596, EPI_ISL_1198597, EPI_ISL_1198598, EPI_ISL_1198599, EPI_ISL_1198600, EPI_ISL_1198601, EPI_ISL_1198602, EPI_ISL_1198603, EPI_ISL_1198604, EPI_ISL_1198605,
EPI_ISL_1198606, EPI_ISL_1198607, EPI_ISL_1198608, EPI_ISL_1198609, EPI_ISL_1198610, EPI_ISL_1198611, EPI_ISL_1198612, EPI_ISL_1198613, EPI_ISL_1198614, EPI_ISL_1198615, EPI_ISL_1198616, EPI_ISL_1198617, EPI_ISL_1198618, EPI_ISL_1198619, EPI_ISL_1198620, EPI_ISL_1198621, EPI_ISL_1198622, EPI_ISL_1198623, EPI_ISL_1198624, EPI_ISL_1198625, EPI_ISL_1198626, EPI_ISL_1198627,
EPI_ISL_1198628, EPI_ISL_1198629, EPI_ISL_1198630, EPI_ISL_1198631, EPI_ISL_1198632, EPI_ISL_1198633, EPI_ISL_1198634, EPI_ISL_1198635, EPI_ISL_1198636, EPI_ISL_1198637, EPI_ISL_1198638, EPI_ISL_1198639, EPI_ISL_1198640, EPI_ISL_1198641, EPI_ISL_1198642, EPI_ISL_1198643, EPI_ISL_1198645, EPI_ISL_1198646, EPI_ISL_1198647, EPI_ISL_1198648, EPI_ISL_1198649, EPI_ISL_1198650,
EPI_ISL_1198651, EPI_ISL_1198652, EPI_ISL_1198653, EPI_ISL_1198654, EPI_ISL_1198655, EPI_ISL_1198656, EPI_ISL_1198657, EPI_ISL_1198658, EPI_ISL_1198659, EPI_ISL_1198660, EPI_ISL_1198661, EPI_ISL_1198662, EPI_ISL_1198663, EPI_ISL_1198664, EPI_ISL_1198665, EPI_ISL_1198666, EPI_ISL_1198667, EPI_ISL_1198668, EPI_ISL_1198669, EPI_ISL_1198670, EPI_ISL_1198671, EPI_ISL_1198672,
EPI_ISL_1198673, EPI_ISL_1198674, EPI_ISL_1198675, EPI_ISL_1198676, EPI_ISL_1198677, EPI_ISL_1198678, EPI_ISL_1198679, EPI_ISL_1198680, EPI_ISL_1198681, EPI_ISL_1198682, EPI_ISL_1198683, EPI_ISL_1198684, EPI_ISL_1198685, EPI_ISL_1198686, EPI_ISL_1198687, EPI_ISL_1198688, EPI_ISL_1198689, EPI_ISL_1198690, EPI_ISL_1198691, EPI_ISL_1198692, EPI_ISL_1198693, EPI_ISL_1198694,
EPI_ISL_1198695, EPI_ISL_1198696, EPI_ISL_1198697, EPI_ISL_1198699, EPI_ISL_1198700, EPI_ISL_1198701, EPI_ISL_1198702, EPI_ISL_1198703, EPI_ISL_1198704, EPI_ISL_1198705, EPI_ISL_1198706, EPI_ISL_1198707, EPI_ISL_1198708, EPI_ISL_1198709, EPI_ISL_1198710, EPI_ISL_1198711, EPI_ISL_1198712, EPI_ISL_1198713, EPI_ISL_1198714, EPI_ISL_1198715, EPI_ISL_1198716, EPI_ISL_1198717,
EPI_ISL_1198718, EPI_ISL_1198719, EPI_ISL_1198720, EPI_ISL_1198721, EPI_ISL_1198722, EPI_ISL_1198723, EPI_ISL_1198724, EPI_ISL_1198725, EPI_ISL_1198726, EPI_ISL_1198727, EPI_ISL_1198728, EPI_ISL_1198729, EPI_ISL_1198730, EPI_ISL_1198731, EPI_ISL_1198732, EPI_ISL_1198733, EPI_ISL_1198734, EPI_ISL_1198735, EPI_ISL_1198736, EPI_ISL_1198737, EPI_ISL_1198738, EPI_ISL_1198739,
EPI_ISL_1198740, EPI_ISL_1198741, EPI_ISL_1198742, EPI_ISL_1198743, EPI_ISL_1198744, EPI_ISL_1198745, EPI_ISL_1198746, EPI_ISL_1198747, EPI_ISL_1198748, EPI_ISL_1198749, EPI_ISL_1198750, EPI_ISL_1198751, EPI_ISL_1198752, EPI_ISL_1198753, EPI_ISL_1198754, EPI_ISL_1198755, EPI_ISL_1198756, EPI_ISL_1198757, EPI_ISL_1198758, EPI_ISL_1198759, EPI_ISL_1198760, EPI_ISL_1198761,
EPI_ISL_1198762, EPI_ISL_1198763, EPI_ISL_1198764, EPI_ISL_1198765, EPI_ISL_1198766, EPI_ISL_1198767, EPI_ISL_1198768, EPI_ISL_1198769, EPI_ISL_1198770, EPI_ISL_1198771, EPI_ISL_1198772, EPI_ISL_1198773, EPI_ISL_1198774, EPI_ISL_1198775, EPI_ISL_1198776, EPI_ISL_1198777, EPI_ISL_1198778, EPI_ISL_1198779, EPI_ISL_1198780, EPI_ISL_1198781, EPI_ISL_1198782, EPI_ISL_1198783,
EPI_ISL_1198784, EPI_ISL_1198785, EPI_ISL_1198786, EPI_ISL_1198787, EPI_ISL_1198788, EPI_ISL_1198789, EPI_ISL_1198790, EPI_ISL_1198791, EPI_ISL_1198792, EPI_ISL_1198852, EPI_ISL_1198853, EPI_ISL_1198854, EPI_ISL_1198855, EPI_ISL_1198856, EPI_ISL_1198857, EPI_ISL_1198858, EPI_ISL_1198859, EPI_ISL_1198860, EPI_ISL_1198861, EPI_ISL_1198862, EPI_ISL_1198863, EPI_ISL_1198864,
EPI_ISL_1198865, EPI_ISL_1198866, EPI_ISL_1198867, EPI_ISL_1198868, EPI_ISL_1198869, EPI_ISL_1198870, EPI_ISL_1198871, EPI_ISL_1198872, EPI_ISL_1198873, EPI_ISL_1198874, EPI_ISL_1198875, EPI_ISL_1198876, EPI_ISL_1198877, EPI_ISL_1198878, EPI_ISL_1198879, EPI_ISL_1198880, EPI_ISL_1198881, EPI_ISL_1198882, EPI_ISL_1198883, EPI_ISL_1198884, EPI_ISL_1198885, EPI_ISL_1198886,
EPI_ISL_1198887, EPI_ISL_1198888, EPI_ISL_1198889, EPI_ISL_1198890, EPI_ISL_1198891, EPI_ISL_1198892, EPI_ISL_1198893, EPI_ISL_1198894, EPI_ISL_1198895, EPI_ISL_1198896, EPI_ISL_1198897, EPI_ISL_1198898, EPI_ISL_1198899, EPI_ISL_1198900, EPI_ISL_1198901, EPI_ISL_1198902, EPI_ISL_1198903, EPI_ISL_1198904, EPI_ISL_1198905, EPI_ISL_1198906, EPI_ISL_1198907, EPI_ISL_1198908,
EPI_ISL_1198909, EPI_ISL_1198910, EPI_ISL_1198911, EPI_ISL_1198912, EPI_ISL_1198913, EPI_ISL_1198914, EPI_ISL_1198915, EPI_ISL_1198916, EPI_ISL_1198917, EPI_ISL_1198919, EPI_ISL_1198920, EPI_ISL_1198921, EPI_ISL_1198922, EPI_ISL_1198923, EPI_ISL_1198924, EPI_ISL_1198925, EPI_ISL_1198926, EPI_ISL_1198927, EPI_ISL_1198928, EPI_ISL_1198929, EPI_ISL_1198930, EPI_ISL_1198931,
EPI_ISL_1198932, EPI_ISL_1198933, EPI_ISL_1198934, EPI_ISL_1198935, EPI_ISL_1198936, EPI_ISL_1198937, EPI_ISL_1198938, EPI_ISL_1198939, EPI_ISL_1198940, EPI_ISL_1198941, EPI_ISL_1198942, EPI_ISL_1198943, EPI_ISL_1198944, EPI_ISL_1198945, EPI_ISL_1198946, EPI_ISL_1198947, EPI_ISL_1198948, EPI_ISL_1198949, EPI_ISL_1198950, EPI_ISL_1198951, EPI_ISL_1198952, EPI_ISL_1198953,
EPI_ISL_1198954, EPI_ISL_1198955, EPI_ISL_1198956, EPI_ISL_1198957, EPI_ISL_1198958, EPI_ISL_1198959, EPI_ISL_1198960, EPI_ISL_1198961, EPI_ISL_1198962, EPI_ISL_1198963, EPI_ISL_1198964, EPI_ISL_1198965, EPI_ISL_1198966, EPI_ISL_1198967, EPI_ISL_1198968, EPI_ISL_1198969, EPI_ISL_1198970, EPI_ISL_1198971, EPI_ISL_1198972, EPI_ISL_1198973, EPI_ISL_1198974, EPI_ISL_1198975,
EPI_ISL_1198976, EPI_ISL_1198977, EPI_ISL_1198978, EPI_ISL_1198979, EPI_ISL_1198980, EPI_ISL_1198981, EPI_ISL_1198982, EPI_ISL_1198983, EPI_ISL_1198984, EPI_ISL_1198985, EPI_ISL_1198986, EPI_ISL_1198987, EPI_ISL_1198988, EPI_ISL_1198989, EPI_ISL_1198990, EPI_ISL_1198991, EPI_ISL_1198992, EPI_ISL_1198993, EPI_ISL_1198994, EPI_ISL_1198995, EPI_ISL_1198996, EPI_ISL_1198997,

EPI_ISL_1198998, EPI_ISL_1198999, EPI_ISL_1199000, EPI_ISL_1199001, EPI_ISL_1199002, EPI_ISL_1199003, EPI_ISL_1199004, EPI_ISL_1199005, EPI_ISL_1199006, EPI_ISL_1199007, EPI_ISL_1199008, EPI_ISL_1199009, EPI_ISL_1199010, EPI_ISL_1199011, EPI_ISL_1199012, EPI_ISL_1199013, EPI_ISL_1199014, EPI_ISL_1199015, EPI_ISL_1199016, EPI_ISL_1199017, EPI_ISL_1199018, EPI_ISL_1199019,
EPI_ISL_1199020, EPI_ISL_1199021, EPI_ISL_1199022, EPI_ISL_1199023, EPI_ISL_1199024, EPI_ISL_1199025, EPI_ISL_1199026, EPI_ISL_1199027, EPI_ISL_1199028, EPI_ISL_1199029, EPI_ISL_1199030, EPI_ISL_1199031, EPI_ISL_1199032, EPI_ISL_1199033, EPI_ISL_1199034, EPI_ISL_1199035, EPI_ISL_1199036, EPI_ISL_1199037, EPI_ISL_1199038, EPI_ISL_1199039, EPI_ISL_1199040, EPI_ISL_1199041,
EPI_ISL_1199042, EPI_ISL_1199043, EPI_ISL_1199044, EPI_ISL_1199045, EPI_ISL_1199046, EPI_ISL_1199047, EPI_ISL_1199048, EPI_ISL_1199049, EPI_ISL_1199050, EPI_ISL_1199051, EPI_ISL_1199052, EPI_ISL_1199053, EPI_ISL_1199054, EPI_ISL_1199055, EPI_ISL_1199056, EPI_ISL_1199057, EPI_ISL_1199058, EPI_ISL_1199059, EPI_ISL_1199060, EPI_ISL_1199061, EPI_ISL_1199062, EPI_ISL_1199063,
EPI_ISL_1199064, EPI_ISL_1199065, EPI_ISL_1199066, EPI_ISL_1199067, EPI_ISL_1199068, EPI_ISL_1199069, EPI_ISL_1199070, EPI_ISL_1199071, EPI_ISL_1199072, EPI_ISL_1199073, EPI_ISL_1199074, EPI_ISL_1199075, EPI_ISL_1199076, EPI_ISL_1199077, EPI_ISL_1199078, EPI_ISL_1199079, EPI_ISL_1199080, EPI_ISL_1199081, EPI_ISL_1199082, EPI_ISL_1199083, EPI_ISL_1199084, EPI_ISL_1199085,
EPI_ISL_1199086, EPI_ISL_1199087, EPI_ISL_1199088, EPI_ISL_1199089, EPI_ISL_1199090, EPI_ISL_1199091, EPI_ISL_1199092, EPI_ISL_1199093, EPI_ISL_1199094, EPI_ISL_1199095, EPI_ISL_1199096, EPI_ISL_1199097, EPI_ISL_1199098, EPI_ISL_1199099, EPI_ISL_1199100, EPI_ISL_1199101, EPI_ISL_1199102, EPI_ISL_1199103, EPI_ISL_1199104, EPI_ISL_1199105, EPI_ISL_1199106, EPI_ISL_1199107,
EPI_ISL_1199108, EPI_ISL_1199109, EPI_ISL_1199110, EPI_ISL_1199111, EPI_ISL_1199112, EPI_ISL_1199113, EPI_ISL_1199114, EPI_ISL_1199115, EPI_ISL_1199116, EPI_ISL_1199117, EPI_ISL_1199118, EPI_ISL_1199119, EPI_ISL_1199120, EPI_ISL_1199121, EPI_ISL_1199122, EPI_ISL_1199123, EPI_ISL_1199124, EPI_ISL_1199125, EPI_ISL_1199126, EPI_ISL_1199127, EPI_ISL_1199128, EPI_ISL_1199129,
EPI_ISL_1199130, EPI_ISL_1199131, EPI_ISL_1199132, EPI_ISL_1199133, EPI_ISL_1199134, EPI_ISL_1199135, EPI_ISL_1199136, EPI_ISL_1199137, EPI_ISL_1199138, EPI_ISL_1199139, EPI_ISL_1199140, EPI_ISL_1199141, EPI_ISL_1199142, EPI_ISL_1199143, EPI_ISL_1199144, EPI_ISL_1199145, EPI_ISL_1199146, EPI_ISL_1199147, EPI_ISL_1199148, EPI_ISL_1199149, EPI_ISL_1199150, EPI_ISL_1199151,
EPI_ISL_1199152, EPI_ISL_1199153, EPI_ISL_1199154, EPI_ISL_1199155, EPI_ISL_1199156, EPI_ISL_1199157, EPI_ISL_1199158, EPI_ISL_1199159, EPI_ISL_1199160, EPI_ISL_1199161, EPI_ISL_1199162, EPI_ISL_1199163, EPI_ISL_1199164, EPI_ISL_1199165, EPI_ISL_1199166, EPI_ISL_1199167, EPI_ISL_1199168, EPI_ISL_1199169, EPI_ISL_1199170, EPI_ISL_1199171, EPI_ISL_1199172, EPI_ISL_1199173,
EPI_ISL_1199174, EPI_ISL_1199175, EPI_ISL_1199176, EPI_ISL_1199177, EPI_ISL_1199178, EPI_ISL_1199179, EPI_ISL_1199180, EPI_ISL_1199181, EPI_ISL_1199182, EPI_ISL_1199183, EPI_ISL_1199184, EPI_ISL_1199185, EPI_ISL_1199186, EPI_ISL_1199187, EPI_ISL_1199188, EPI_ISL_1199189, EPI_ISL_1199190, EPI_ISL_1199191, EPI_ISL_1199192, EPI_ISL_1199193, EPI_ISL_1199194, EPI_ISL_1199195,
EPI_ISL_1199196, EPI_ISL_1199197, EPI_ISL_1199198, EPI_ISL_1199199, EPI_ISL_1199200, EPI_ISL_1199201, EPI_ISL_1199202, EPI_ISL_1199203, EPI_ISL_1199204, EPI_ISL_1199205, EPI_ISL_1199206, EPI_ISL_1199207, EPI_ISL_1199208, EPI_ISL_1199209, EPI_ISL_1199210, EPI_ISL_1199211, EPI_ISL_1199212, EPI_ISL_1199213, EPI_ISL_1199214, EPI_ISL_1199215, EPI_ISL_1199216, EPI_ISL_1199217,
EPI_ISL_1199218, EPI_ISL_1199219, EPI_ISL_1199220, EPI_ISL_1199221, EPI_ISL_1199222, EPI_ISL_1199223, EPI_ISL_1199224, EPI_ISL_1199225, EPI_ISL_1199226, EPI_ISL_1199227, EPI_ISL_1199228, EPI_ISL_1199229, EPI_ISL_1199230, EPI_ISL_1199231, EPI_ISL_1199232, EPI_ISL_1199233, EPI_ISL_1199234, EPI_ISL_1199235, EPI_ISL_1199236, EPI_ISL_1199237, EPI_ISL_1199238, EPI_ISL_1199239,
EPI_ISL_1199240, EPI_ISL_1199241, EPI_ISL_1199242, EPI_ISL_1199243, EPI_ISL_1199244, EPI_ISL_1199245, EPI_ISL_1199246, EPI_ISL_1199247, EPI_ISL_1199248, EPI_ISL_1199249, EPI_ISL_1199250, EPI_ISL_1199251, EPI_ISL_1199252, EPI_ISL_1199253, EPI_ISL_1199254, EPI_ISL_1199255, EPI_ISL_1199256, EPI_ISL_1199257, EPI_ISL_1199258, EPI_ISL_1199259, EPI_ISL_1199260, EPI_ISL_1199261,
EPI_ISL_1199262, EPI_ISL_1199263, EPI_ISL_1199264, EPI_ISL_1199265, EPI_ISL_1199266, EPI_ISL_1199267, EPI_ISL_1199268, EPI_ISL_1199269, EPI_ISL_1199270, EPI_ISL_1199271, EPI_ISL_1199272, EPI_ISL_1199273, EPI_ISL_1199274, EPI_ISL_1199275, EPI_ISL_1199276, EPI_ISL_1199277, EPI_ISL_1199278, EPI_ISL_1199279, EPI_ISL_1199280, EPI_ISL_1199281, EPI_ISL_1199282, EPI_ISL_1199283,
EPI_ISL_1199284, EPI_ISL_1199285, EPI_ISL_1199286, EPI_ISL_1199287, EPI_ISL_1199288, EPI_ISL_1199289, EPI_ISL_1199290, EPI_ISL_1199291, EPI_ISL_1199292, EPI_ISL_1199293, EPI_ISL_1199294, EPI_ISL_1199295, EPI_ISL_1199296, EPI_ISL_1199297, EPI_ISL_1199298, EPI_ISL_1199299, EPI_ISL_1199300, EPI_ISL_1199301, EPI_ISL_1199302, EPI_ISL_1199303, EPI_ISL_1199304, EPI_ISL_1199305,
EPI_ISL_1199306, EPI_ISL_1199307, EPI_ISL_1199308, EPI_ISL_1199309, EPI_ISL_1199310, EPI_ISL_1199311, EPI_ISL_1199312, EPI_ISL_1199313, EPI_ISL_1199314, EPI_ISL_1199315, EPI_ISL_1199316, EPI_ISL_1199317, EPI_ISL_1199318, EPI_ISL_1199319, EPI_ISL_1199320, EPI_ISL_1199321, EPI_ISL_1199322, EPI_ISL_1199323, EPI_ISL_1199324, EPI_ISL_1199325, EPI_ISL_1199326, EPI_ISL_1199327,
EPI_ISL_1199328, EPI_ISL_1199329, EPI_ISL_1199330, EPI_ISL_1199331, EPI_ISL_1199332, EPI_ISL_1199333, EPI_ISL_1199334, EPI_ISL_1199335, EPI_ISL_1199336, EPI_ISL_1199337, EPI_ISL_1199338, EPI_ISL_1199339, EPI_ISL_1199340, EPI_ISL_1199341, EPI_ISL_1199342, EPI_ISL_1199343, EPI_ISL_1199344, EPI_ISL_1199345, EPI_ISL_1199346, EPI_ISL_1199347, EPI_ISL_1199348, EPI_ISL_1199349,
EPI_ISL_1199350, EPI_ISL_1199351, EPI_ISL_1199352, EPI_ISL_1199353, EPI_ISL_1199354, EPI_ISL_1199355, EPI_ISL_1199356, EPI_ISL_1199357, EPI_ISL_1199358, EPI_ISL_1199359, EPI_ISL_1199360, EPI_ISL_1199361, EPI_ISL_1199362, EPI_ISL_1199363, EPI_ISL_1199364, EPI_ISL_1199365, EPI_ISL_1199366, EPI_ISL_1199367, EPI_ISL_1199368, EPI_ISL_1199369, EPI_ISL_1199370, EPI_ISL_1199371,
EPI_ISL_1199372, EPI_ISL_1199373, EPI_ISL_1199374, EPI_ISL_1199375, EPI_ISL_1199376, EPI_ISL_1199377, EPI_ISL_1199378, EPI_ISL_1199379, EPI_ISL_1199380, EPI_ISL_1199381, EPI_ISL_1199382, EPI_ISL_1199383, EPI_ISL_1199384, EPI_ISL_1199385, EPI_ISL_1199387, EPI_ISL_1199388, EPI_ISL_1199389, EPI_ISL_1199390, EPI_ISL_1199391, EPI_ISL_1199392, EPI_ISL_1199393, EPI_ISL_1199394,
EPI_ISL_1199395, EPI_ISL_1199396, EPI_ISL_1199397, EPI_ISL_1199398, EPI_ISL_1199399, EPI_ISL_1199400, EPI_ISL_1199401, EPI_ISL_1199402, EPI_ISL_1199403, EPI_ISL_1199404, EPI_ISL_1199405, EPI_ISL_1199406, EPI_ISL_1199407, EPI_ISL_1199408, EPI_ISL_1199409, EPI_ISL_1199410, EPI_ISL_1199411, EPI_ISL_1199412, EPI_ISL_1199413, EPI_ISL_1199414, EPI_ISL_1199415, EPI_ISL_1199416,
EPI_ISL_1199417, EPI_ISL_1199418, EPI_ISL_1199419, EPI_ISL_1199420, EPI_ISL_1199421, EPI_ISL_1199422, EPI_ISL_1199423, EPI_ISL_1199424, EPI_ISL_1199425, EPI_ISL_1199426, EPI_ISL_1199427, EPI_ISL_1199428, EPI_ISL_1199429, EPI_ISL_1199430, EPI_ISL_1199431, EPI_ISL_1199432, EPI_ISL_1199433, EPI_ISL_1199434, EPI_ISL_1199435, EPI_ISL_1199436, EPI_ISL_1199437, EPI_ISL_1199438,
EPI_ISL_1199439, EPI_ISL_1199440, EPI_ISL_1199441, EPI_ISL_1199442, EPI_ISL_1199443, EPI_ISL_1199444, EPI_ISL_1199445, EPI_ISL_1199446, EPI_ISL_1199447, EPI_ISL_1199448, EPI_ISL_1199449, EPI_ISL_1199450, EPI_ISL_1199451, EPI_ISL_1199452, EPI_ISL_1199453, EPI_ISL_1199454, EPI_ISL_1199455, EPI_ISL_1199456, EPI_ISL_1199457, EPI_ISL_1199458, EPI_ISL_1199459, EPI_ISL_1199460,
EPI_ISL_1199461, EPI_ISL_1199462, EPI_ISL_1199463, EPI_ISL_1199464, EPI_ISL_1199465, EPI_ISL_1199466, EPI_ISL_1199467, EPI_ISL_1199468, EPI_ISL_1199469, EPI_ISL_1199470, EPI_ISL_1199471, EPI_ISL_1199472, EPI_ISL_1199473, EPI_ISL_1199474, EPI_ISL_1199475, EPI_ISL_1199476, EPI_ISL_1199477, EPI_ISL_1199478, EPI_ISL_1199479, EPI_ISL_1199480, EPI_ISL_1199481, EPI_ISL_1199482,
EPI_ISL_1199483, EPI_ISL_1199484, EPI_ISL_1199485, EPI_ISL_1199486, EPI_ISL_1199487, EPI_ISL_1199488, EPI_ISL_1199489, EPI_ISL_1199490, EPI_ISL_1199491, EPI_ISL_1199492, EPI_ISL_1199493, EPI_ISL_1199494, EPI_ISL_1199495, EPI_ISL_1199496, EPI_ISL_1199497, EPI_ISL_1199498, EPI_ISL_1199499, EPI_ISL_1199500, EPI_ISL_1199501, EPI_ISL_1199502, EPI_ISL_1199503, EPI_ISL_1199504,
EPI_ISL_1199505, EPI_ISL_1199506, EPI_ISL_1199507, EPI_ISL_1199508, EPI_ISL_1199509, EPI_ISL_1199510, EPI_ISL_1199511, EPI_ISL_1199512, EPI_ISL_1199513, EPI_ISL_1199514, EPI_ISL_1199515, EPI_ISL_1199516, EPI_ISL_1199517, EPI_ISL_1199518, EPI_ISL_1199519, EPI_ISL_1199520, EPI_ISL_1199521, EPI_ISL_1199522, EPI_ISL_1199523, EPI_ISL_1199524, EPI_ISL_1199525, EPI_ISL_1199526,
EPI_ISL_1199527, EPI_ISL_1199528, EPI_ISL_1199529, EPI_ISL_1199530, EPI_ISL_1199531, EPI_ISL_1199532, EPI_ISL_1199533, EPI_ISL_1199534, EPI_ISL_1199535, EPI_ISL_1199536, EPI_ISL_1199537, EPI_ISL_1199538, EPI_ISL_1199539, EPI_ISL_1199540, EPI_ISL_1199541, EPI_ISL_1199542, EPI_ISL_1199543, EPI_ISL_1199544, EPI_ISL_1199545, EPI_ISL_1199546, EPI_ISL_1199547, EPI_ISL_1199548,
EPI_ISL_1199549, EPI_ISL_1199550, EPI_ISL_1199551, EPI_ISL_1199552, EPI_ISL_1199553, EPI_ISL_1199554, EPI_ISL_1199555, EPI_ISL_1199556, EPI_ISL_1199557, EPI_ISL_1199558, EPI_ISL_1199559, EPI_ISL_1199560, EPI_ISL_1199561, EPI_ISL_1199562, EPI_ISL_1199563, EPI_ISL_1199564, EPI_ISL_1199565, EPI_ISL_1199566, EPI_ISL_1199567, EPI_ISL_1199568, EPI_ISL_1199569, EPI_ISL_1199570,
EPI_ISL_1199571, EPI_ISL_1199572, EPI_ISL_1199573, EPI_ISL_1199574, EPI_ISL_1199575, EPI_ISL_1199576, EPI_ISL_1199577, EPI_ISL_1199578, EPI_ISL_1199579, EPI_ISL_1199580, EPI_ISL_1199581, EPI_ISL_1199582, EPI_ISL_1199583, EPI_ISL_1199584, EPI_ISL_1199585, EPI_ISL_1199586, EPI_ISL_1199587, EPI_ISL_1199588, EPI_ISL_1199589, EPI_ISL_1199590, EPI_ISL_1199591, EPI_ISL_1199592,
EPI_ISL_1199593, EPI_ISL_1199594, EPI_ISL_1199595, EPI_ISL_1199596, EPI_ISL_1199597, EPI_ISL_1199598, EPI_ISL_1199599, EPI_ISL_1199600, EPI_ISL_1199601, EPI_ISL_1199602, EPI_ISL_1199603, EPI_ISL_1199604, EPI_ISL_1199605, EPI_ISL_1199606, EPI_ISL_1199607, EPI_ISL_1199608, EPI_ISL_1199609, EPI_ISL_1199610, EPI_ISL_1199611, EPI_ISL_1199612, EPI_ISL_1199613, EPI_ISL_1199614,
EPI_ISL_1199615, EPI_ISL_1199616, EPI_ISL_1199617, EPI_ISL_1199618, EPI_ISL_1199619, EPI_ISL_1199620, EPI_ISL_1199621, EPI_ISL_1199622, EPI_ISL_1199623, EPI_ISL_1199624, EPI_ISL_1199625, EPI_ISL_1199626, EPI_ISL_1199627, EPI_ISL_1199628, EPI_ISL_1199629, EPI_ISL_1199630, EPI_ISL_1199631, EPI_ISL_1199632, EPI_ISL_1199637, EPI_ISL_1199638, EPI_ISL_1199639, EPI_ISL_1199640,
EPI_ISL_1199641, EPI_ISL_1199642, EPI_ISL_1199643, EPI_ISL_1199644, EPI_ISL_1199645, EPI_ISL_1199646, EPI_ISL_1199647, EPI_ISL_1199648, EPI_ISL_1199649, EPI_ISL_1199650, EPI_ISL_1199651, EPI_ISL_1199652, EPI_ISL_1199653, EPI_ISL_1199654, EPI_ISL_1199655, EPI_ISL_1199656, EPI_ISL_1199657, EPI_ISL_1199658, EPI_ISL_1199659, EPI_ISL_1199660, EPI_ISL_1199661, EPI_ISL_1199662,
EPI_ISL_1199663, EPI_ISL_1199664, EPI_ISL_1199665, EPI_ISL_1199666, EPI_ISL_1199667, EPI_ISL_1199668, EPI_ISL_1199669, EPI_ISL_1199670, EPI_ISL_1199671, EPI_ISL_1199672, EPI_ISL_1199673, EPI_ISL_1199674, EPI_ISL_1199675, EPI_ISL_1199676, EPI_ISL_1199677, EPI_ISL_1199678, EPI_ISL_1199679, EPI_ISL_1199680, EPI_ISL_1199681, EPI_ISL_1199682, EPI_ISL_1199683, EPI_ISL_1199684,
EPI_ISL_1199685, EPI_ISL_1199686, EPI_ISL_1199687, EPI_ISL_1199688, EPI_ISL_1199689, EPI_ISL_1199690, EPI_ISL_1199691, EPI_ISL_1199692, EPI_ISL_1199693, EPI_ISL_1199694, EPI_ISL_1199695, EPI_ISL_1199696, EPI_ISL_1199697, EPI_ISL_1199698, EPI_ISL_1199699, EPI_ISL_1199700, EPI_ISL_1199701, EPI_ISL_1199702, EPI_ISL_1199703, EPI_ISL_1199704, EPI_ISL_1199705, EPI_ISL_1199706,
EPI_ISL_1199708, EPI_ISL_1199709, EPI_ISL_1199710, EPI_ISL_1199711, EPI_ISL_1199712, EPI_ISL_1199713, EPI_ISL_1199714, EPI_ISL_1199715, EPI_ISL_1199716, EPI_ISL_1199717, EPI_ISL_1199718, EPI_ISL_1199719, EPI_ISL_1199720, EPI_ISL_1199721, EPI_ISL_1199722, EPI_ISL_1199723, EPI_ISL_1199724, EPI_ISL_1199725, EPI_ISL_1199726, EPI_ISL_1199727, EPI_ISL_1199728, EPI_ISL_1199729,
EPI_ISL_1199730, EPI_ISL_1199731, EPI_ISL_1199732, EPI_ISL_1199733, EPI_ISL_1199734, EPI_ISL_1199735, EPI_ISL_1199736, EPI_ISL_1199737, EPI_ISL_1199738, EPI_ISL_1199739, EPI_ISL_1199740, EPI_ISL_1199741, EPI_ISL_1199742, EPI_ISL_1199743, EPI_ISL_1199744, EPI_ISL_1199745, EPI_ISL_1199746, EPI_ISL_1199747, EPI_ISL_1199748, EPI_ISL_1199749, EPI_ISL_1199750, EPI_ISL_1199751,
EPI_ISL_1199752, EPI_ISL_1199753, EPI_ISL_1199754, EPI_ISL_1199755, EPI_ISL_1199756, EPI_ISL_1199757, EPI_ISL_1199758, EPI_ISL_1199759, EPI_ISL_1199760, EPI_ISL_1199761, EPI_ISL_1199762, EPI_ISL_1199763, EPI_ISL_1199764, EPI_ISL_1199765, EPI_ISL_1199766, EPI_ISL_1199767, EPI_ISL_1199768, EPI_ISL_1199769, EPI_ISL_1199770, EPI_ISL_1199771, EPI_ISL_1199772, EPI_ISL_1199773,
EPI_ISL_1199774, EPI_ISL_1199775, EPI_ISL_1199776, EPI_ISL_1199777, EPI_ISL_1199778, EPI_ISL_1199779, EPI_ISL_1199780, EPI_ISL_1199781, EPI_ISL_1199782, EPI_ISL_1199783, EPI_ISL_1199784, EPI_ISL_1199785, EPI_ISL_1199786, EPI_ISL_1199787, EPI_ISL_1199788, EPI_ISL_1199789, EPI_ISL_1199790, EPI_ISL_1199791, EPI_ISL_1199792, EPI_ISL_1199793, EPI_ISL_1199794, EPI_ISL_1199795,
EPI_ISL_1199796, EPI_ISL_1199797, EPI_ISL_1199798, EPI_ISL_1199799, EPI_ISL_1199800, EPI_ISL_1199801, EPI_ISL_1199802, EPI_ISL_1199803, EPI_ISL_1199804, EPI_ISL_1199805, EPI_ISL_1199806, EPI_ISL_1199807, EPI_ISL_1199808, EPI_ISL_1199809, EPI_ISL_1199810, EPI_ISL_1199811, EPI_ISL_1199812, EPI_ISL_1199813, EPI_ISL_1199814, EPI_ISL_1199815, EPI_ISL_1199816, EPI_ISL_1199817,
EPI_ISL_1199818, EPI_ISL_1199819, EPI_ISL_1199820, EPI_ISL_1199821, EPI_ISL_1199822, EPI_ISL_1199823, EPI_ISL_1199824, EPI_ISL_1199825, EPI_ISL_1199826, EPI_ISL_1199827, EPI_ISL_1199828, EPI_ISL_1199829, EPI_ISL_1199830, EPI_ISL_1199831, EPI_ISL_1199832, EPI_ISL_1199833, EPI_ISL_1199834, EPI_ISL_1199835, EPI_ISL_1199836, EPI_ISL_1199837, EPI_ISL_1199838, EPI_ISL_1199839,
EPI_ISL_1199840, EPI_ISL_1199841, EPI_ISL_1199842, EPI_ISL_1199843, EPI_ISL_1199844, EPI_ISL_1199845, EPI_ISL_1199846, EPI_ISL_1199847, EPI_ISL_1199848, EPI_ISL_1199849, EPI_ISL_1199850, EPI_ISL_1199851, EPI_ISL_1199852, EPI_ISL_1199853, EPI_ISL_1199854, EPI_ISL_1199855, EPI_ISL_1199856, EPI_ISL_1199857, EPI_ISL_1199858, EPI_ISL_1199859, EPI_ISL_1199860, EPI_ISL_1199861,
EPI_ISL_1199862, EPI_ISL_1199863, EPI_ISL_1199864, EPI_ISL_1199865, EPI_ISL_1199866, EPI_ISL_1199867, EPI_ISL_1199868, EPI_ISL_1199869, EPI_ISL_1199870, EPI_ISL_1199871, EPI_ISL_1199872, EPI_ISL_1199873, EPI_ISL_1199874, EPI_ISL_1199875, EPI_ISL_1199876, EPI_ISL_1199877, EPI_ISL_1199878, EPI_ISL_1199879, EPI_ISL_1199880, EPI_ISL_1199881, EPI_ISL_1199882, EPI_ISL_1199883,
EPI_ISL_1199884, EPI_ISL_1199885, EPI_ISL_1199886, EPI_ISL_1199887, EPI_ISL_1199888, EPI_ISL_1199889, EPI_ISL_1199890, EPI_ISL_1199891, EPI_ISL_1199892, EPI_ISL_1199893, EPI_ISL_1199894, EPI_ISL_1199895, EPI_ISL_1199896, EPI_ISL_1199897, EPI_ISL_1199898, EPI_ISL_1199899, EPI_ISL_1199900, EPI_ISL_1199901, EPI_ISL_1199902, EPI_ISL_1199903, EPI_ISL_1199904, EPI_ISL_1199905,
EPI_ISL_1199906, EPI_ISL_1199907, EPI_ISL_1199908, EPI_ISL_1199909, EPI_ISL_1199910, EPI_ISL_1199911, EPI_ISL_1199912, EPI_ISL_1199913, EPI_ISL_1199914, EPI_ISL_1199915, EPI_ISL_1199916, EPI_ISL_1199917, EPI_ISL_1199918, EPI_ISL_1199919, EPI_ISL_1199920, EPI_ISL_1199921, EPI_ISL_1199922, EPI_ISL_1199923, EPI_ISL_1199924, EPI_ISL_1199925, EPI_ISL_1199926, EPI_ISL_1199927,
EPI_ISL_1199928, EPI_ISL_1199929, EPI_ISL_1199930, EPI_ISL_1199931, EPI_ISL_1199932, EPI_ISL_1199933, EPI_ISL_1199934, EPI_ISL_1199935, EPI_ISL_1199936, EPI_ISL_1199937, EPI_ISL_1199938, EPI_ISL_1199939, EPI_ISL_1199940, EPI_ISL_1199941, EPI_ISL_1199942, EPI_ISL_1199943, EPI_ISL_1199944, EPI_ISL_1199945, EPI_ISL_1199946, EPI_ISL_1199947, EPI_ISL_1199948, EPI_ISL_1199949,
EPI_ISL_1199950, EPI_ISL_1199951, EPI_ISL_1199952, EPI_ISL_1199953, EPI_ISL_1199954, EPI_ISL_1199955, EPI_ISL_1199956, EPI_ISL_1199957, EPI_ISL_1199958, EPI_ISL_1199959, EPI_ISL_1199960, EPI_ISL_1199961, EPI_ISL_1199962, EPI_ISL_1199963, EPI_ISL_1199964, EPI_ISL_1199965, EPI_ISL_1199966, EPI_ISL_1199967, EPI_ISL_1199968, EPI_ISL_1199969, EPI_ISL_1199970, EPI_ISL_1199971,
EPI_ISL_1199972, EPI_ISL_1199973, EPI_ISL_1199974, EPI_ISL_1199975, EPI_ISL_1199976, EPI_ISL_1199977, EPI_ISL_1199978, EPI_ISL_1199979, EPI_ISL_1199980, EPI_ISL_1199981, EPI_ISL_1199982, EPI_ISL_1199983, EPI_ISL_1199984, EPI_ISL_1199985, EPI_ISL_1199986, EPI_ISL_1199987, EPI_ISL_1199988, EPI_ISL_1199989, EPI_ISL_1199990, EPI_ISL_1199991, EPI_ISL_1199992, EPI_ISL_1199993,
EPI_ISL_1199994, EPI_ISL_1199995, EPI_ISL_1199996, EPI_ISL_1199997, EPI_ISL_1199998, EPI_ISL_1199999, EPI_ISL_1200000, EPI_ISL_1200001, EPI_ISL_1200002, EPI_ISL_1200003, EPI_ISL_1200005, EPI_ISL_1200006, EPI_ISL_1200007, EPI_ISL_1200008, EPI_ISL_1200009, EPI_ISL_1200010, EPI_ISL_1200011, EPI_ISL_1200012, EPI_ISL_1200013, EPI_ISL_1200014, EPI_ISL_1200015, EPI_ISL_1200016,
EPI_ISL_1200017, EPI_ISL_1200018, EPI_ISL_1200019, EPI_ISL_1200020, EPI_ISL_1200021, EPI_ISL_1200022, EPI_ISL_1200023, EPI_ISL_1200024, EPI_ISL_1200025, EPI_ISL_1200026, EPI_ISL_1200027, EPI_ISL_1200028, EPI_ISL_1200029, EPI_ISL_1200030, EPI_ISL_1200031, EPI_ISL_1200032, EPI_ISL_1200033, EPI_ISL_1200034, EPI_ISL_1200035, EPI_ISL_1200036, EPI_ISL_1200037, EPI_ISL_1200038,
EPI_ISL_1200039, EPI_ISL_1200040, EPI_ISL_1200041, EPI_ISL_1200042, EPI_ISL_1200043, EPI_ISL_1200044, EPI_ISL_1200045, EPI_ISL_1200046, EPI_ISL_1200047, EPI_ISL_1200048, EPI_ISL_1200049, EPI_ISL_1200050, EPI_ISL_1200051, EPI_ISL_1200052, EPI_ISL_1200053, EPI_ISL_1200054, EPI_ISL_1200055, EPI_ISL_1200056, EPI_ISL_1200057, EPI_ISL_1200058, EPI_ISL_1200059, EPI_ISL_1200060,
EPI_ISL_1200061, EPI_ISL_1200062, EPI_ISL_1200063, EPI_ISL_1200064, EPI_ISL_1200065, EPI_ISL_1200066, EPI_ISL_1200067, EPI_ISL_1200068, EPI_ISL_1200069, EPI_ISL_1200070, EPI_ISL_1200071, EPI_ISL_1200072, EPI_ISL_1200073, EPI_ISL_1200074, EPI_ISL_1200075, EPI_ISL_1200076, EPI_ISL_1200077, EPI_ISL_1200078, EPI_ISL_1200079, EPI_ISL_1200080, EPI_ISL_1200081, EPI_ISL_1200082,
EPI_ISL_1200083, EPI_ISL_1200084, EPI_ISL_1200085, EPI_ISL_1200087, EPI_ISL_1200088, EPI_ISL_1200089, EPI_ISL_1200090, EPI_ISL_1200091, EPI_ISL_1200092, EPI_ISL_1200093, EPI_ISL_1200094, EPI_ISL_1200095, EPI_ISL_1200096, EPI_ISL_1200097, EPI_ISL_1200098, EPI_ISL_1200099, EPI_ISL_1200100, EPI_ISL_1200101, EPI_ISL_1200102, EPI_ISL_1200103, EPI_ISL_1200104, EPI_ISL_1200105,
EPI_ISL_1200106, EPI_ISL_1200107, EPI_ISL_1200108, EPI_ISL_1200109, EPI_ISL_1200110, EPI_ISL_1200111, EPI_ISL_1200112, EPI_ISL_1200113, EPI_ISL_1200114, EPI_ISL_1200115, EPI_ISL_1200116, EPI_ISL_1200117, EPI_ISL_1200118, EPI_ISL_1200119, EPI_ISL_1200120, EPI_ISL_1200121, EPI_ISL_1200122, EPI_ISL_1200123, EPI_ISL_1200124, EPI_ISL_1200125, EPI_ISL_1200126, EPI_ISL_1200127,
EPI_ISL_1200128, EPI_ISL_1200129, EPI_ISL_1200130, EPI_ISL_1200131, EPI_ISL_1200132, EPI_ISL_1200133, EPI_ISL_1200134, EPI_ISL_1200135, EPI_ISL_1200136, EPI_ISL_1200137, EPI_ISL_1200138, EPI_ISL_1200139, EPI_ISL_1200140, EPI_ISL_1200141, EPI_ISL_1200142, EPI_ISL_1200143, EPI_ISL_1200144, EPI_ISL_1200145, EPI_ISL_1200146, EPI_ISL_1200147, EPI_ISL_1200148, EPI_ISL_1200149,
EPI_ISL_1200150, EPI_ISL_1200151, EPI_ISL_1200152, EPI_ISL_1200153, EPI_ISL_1200154, EPI_ISL_1200155, EPI_ISL_1200156, EPI_ISL_1200157, EPI_ISL_1200158, EPI_ISL_1200159, EPI_ISL_1200160, EPI_ISL_1200161, EPI_ISL_1200162, EPI_ISL_1200163, EPI_ISL_1200164, EPI_ISL_1200165, EPI_ISL_1200166, EPI_ISL_1200167, EPI_ISL_1200168, EPI_ISL_1200169, EPI_ISL_1200170, EPI_ISL_1200171,
EPI_ISL_1200172, EPI_ISL_1200173, EPI_ISL_1200174, EPI_ISL_1200175, EPI_ISL_1200176, EPI_ISL_1200177, EPI_ISL_1200178, EPI_ISL_1200179, EPI_ISL_1200180, EPI_ISL_1200181, EPI_ISL_1200182, EPI_ISL_1200183, EPI_ISL_1200184, EPI_ISL_1200185, EPI_ISL_1200186, EPI_ISL_1200187, EPI_ISL_1200188, EPI_ISL_1200189, EPI_ISL_1200190, EPI_ISL_1200191, EPI_ISL_1200192, EPI_ISL_1200193,
EPI_ISL_1200194, EPI_ISL_1200195, EPI_ISL_1200196, EPI_ISL_1200197, EPI_ISL_1200198, EPI_ISL_1200199, EPI_ISL_1200200, EPI_ISL_1200201, EPI_ISL_1200202, EPI_ISL_1200203, EPI_ISL_1200204, EPI_ISL_1200205, EPI_ISL_1200206, EPI_ISL_1200207, EPI_ISL_1200208, EPI_ISL_1200209, EPI_ISL_1200210, EPI_ISL_1200211, EPI_ISL_1200212, EPI_ISL_1200213, EPI_ISL_1200214, EPI_ISL_1200215,
EPI_ISL_1200216, EPI_ISL_1200217, EPI_ISL_1200218, EPI_ISL_1200219, EPI_ISL_1200220, EPI_ISL_1200221, EPI_ISL_1200222, EPI_ISL_1200223, EPI_ISL_1200224, EPI_ISL_1200225, EPI_ISL_1200226, EPI_ISL_1200227, EPI_ISL_1200228, EPI_ISL_1200229, EPI_ISL_1200230, EPI_ISL_1200231, EPI_ISL_1200232, EPI_ISL_1200233, EPI_ISL_1200234, EPI_ISL_1200235, EPI_ISL_1200236, EPI_ISL_1200237,
EPI_ISL_1200238, EPI_ISL_1200239, EPI_ISL_1200240, EPI_ISL_1200241, EPI_ISL_1200242, EPI_ISL_1200243, EPI_ISL_1200244, EPI_ISL_1200245, EPI_ISL_1200246, EPI_ISL_1200247, EPI_ISL_1200248, EPI_ISL_1200249, EPI_ISL_1200250, EPI_ISL_1200251, EPI_ISL_1200252, EPI_ISL_1200253, EPI_ISL_1200254, EPI_ISL_1200255, EPI_ISL_1200256, EPI_ISL_1200257, EPI_ISL_1200258, EPI_ISL_1200259,
EPI_ISL_1200260, EPI_ISL_1200261, EPI_ISL_1200262, EPI_ISL_1200263, EPI_ISL_1200264, EPI_ISL_1200265, EPI_ISL_1200266, EPI_ISL_1200267, EPI_ISL_1200268, EPI_ISL_1200269, EPI_ISL_1200270, EPI_ISL_1200271, EPI_ISL_1200272, EPI_ISL_1200273, EPI_ISL_1200274, EPI_ISL_1200275, EPI_ISL_1200276, EPI_ISL_1200277, EPI_ISL_1200278, EPI_ISL_1200279, EPI_ISL_1200280, EPI_ISL_1200281,
EPI_ISL_1200282, EPI_ISL_1200283, EPI_ISL_1200284, EPI_ISL_1200285, EPI_ISL_1200286, EPI_ISL_1200287, EPI_ISL_1200288, EPI_ISL_1200289, EPI_ISL_1200290, EPI_ISL_1200291, EPI_ISL_1200292, EPI_ISL_1200293, EPI_ISL_1200294, EPI_ISL_1200295, EPI_ISL_1200296, EPI_ISL_1200297, EPI_ISL_1200298, EPI_ISL_1200299, EPI_ISL_1200300, EPI_ISL_1200301, EPI_ISL_1200302, EPI_ISL_1200303,
EPI_ISL_1200304, EPI_ISL_1200305, EPI_ISL_1200306, EPI_ISL_1200307, EPI_ISL_1200308, EPI_ISL_1200309, EPI_ISL_1200310, EPI_ISL_1200311, EPI_ISL_1200312, EPI_ISL_1200313, EPI_ISL_1200314, EPI_ISL_1200315, EPI_ISL_1200316, EPI_ISL_1200317, EPI_ISL_1200318, EPI_ISL_1200319, EPI_ISL_1200320, EPI_ISL_1200321, EPI_ISL_1200322, EPI_ISL_1200323, EPI_ISL_1200324, EPI_ISL_1200325,
EPI_ISL_1200326, EPI_ISL_1200327, EPI_ISL_1200328, EPI_ISL_1200329, EPI_ISL_1200330, EPI_ISL_1200331, EPI_ISL_1200332, EPI_ISL_1200333, EPI_ISL_1200334, EPI_ISL_1200335, EPI_ISL_1200336, EPI_ISL_1200337, EPI_ISL_1200338, EPI_ISL_1200339, EPI_ISL_1200340, EPI_ISL_1200341, EPI_ISL_1200342, EPI_ISL_1200343, EPI_ISL_1200344, EPI_ISL_1200345, EPI_ISL_1200346, EPI_ISL_1200347,
EPI_ISL_1200348, EPI_ISL_1200349, EPI_ISL_1200350, EPI_ISL_1200351, EPI_ISL_1200352, EPI_ISL_1200353, EPI_ISL_1200354, EPI_ISL_1200355, EPI_ISL_1200356, EPI_ISL_1200357, EPI_ISL_1200358, EPI_ISL_1200359, EPI_ISL_1200360, EPI_ISL_1200361, EPI_ISL_1200362, EPI_ISL_1200363, EPI_ISL_1200364, EPI_ISL_1200365, EPI_ISL_1200366, EPI_ISL_1200367, EPI_ISL_1200368, EPI_ISL_1200369,
EPI_ISL_1200370, EPI_ISL_1200371, EPI_ISL_1200372, EPI_ISL_1200373, EPI_ISL_1200374, EPI_ISL_1200375, EPI_ISL_1200376, EPI_ISL_1200377, EPI_ISL_1200378, EPI_ISL_1200379, EPI_ISL_1200380, EPI_ISL_1200381, EPI_ISL_1200382, EPI_ISL_1200383, EPI_ISL_1200384, EPI_ISL_1200385, EPI_ISL_1200386, EPI_ISL_1200387, EPI_ISL_1200388, EPI_ISL_1200389, EPI_ISL_1200390, EPI_ISL_1200391,
EPI_ISL_1200392, EPI_ISL_1200393, EPI_ISL_1200394, EPI_ISL_1200395, EPI_ISL_1200396, EPI_ISL_1200397, EPI_ISL_1200398, EPI_ISL_1200399, EPI_ISL_1200400, EPI_ISL_1200401, EPI_ISL_1200402, EPI_ISL_1200403, EPI_ISL_1200404, EPI_ISL_1200405, EPI_ISL_1200406, EPI_ISL_1200407, EPI_ISL_1200408, EPI_ISL_1200409, EPI_ISL_1200410, EPI_ISL_1200411, EPI_ISL_1200412, EPI_ISL_1200413,
EPI_ISL_1200414, EPI_ISL_1200415, EPI_ISL_1200416, EPI_ISL_1200417, EPI_ISL_1200418, EPI_ISL_1200419, EPI_ISL_1200420, EPI_ISL_1200421, EPI_ISL_1200422, EPI_ISL_1200423, EPI_ISL_1200424, EPI_ISL_1200425, EPI_ISL_1200426, EPI_ISL_1200427, EPI_ISL_1200428, EPI_ISL_1200429, EPI_ISL_1200430, EPI_ISL_1200431, EPI_ISL_1200432, EPI_ISL_1200433, EPI_ISL_1200434, EPI_ISL_1200435,
EPI_ISL_1200436, EPI_ISL_1200437, EPI_ISL_1200438, EPI_ISL_1200439, EPI_ISL_1200440, EPI_ISL_1200441, EPI_ISL_1200442, EPI_ISL_1200443, EPI_ISL_1200444, EPI_ISL_1200445, EPI_ISL_1200446, EPI_ISL_1200447, EPI_ISL_1200448, EPI_ISL_1200449, EPI_ISL_1200450, EPI_ISL_1200451, EPI_ISL_1200452, EPI_ISL_1200453, EPI_ISL_1200454, EPI_ISL_1200455, EPI_ISL_1200456, EPI_ISL_1200457,
EPI_ISL_1200458, EPI_ISL_1200459, EPI_ISL_1200460, EPI_ISL_1200461, EPI_ISL_1200462, EPI_ISL_1200463, EPI_ISL_1200464, EPI_ISL_1200465, EPI_ISL_1200466, EPI_ISL_1200467, EPI_ISL_1200468, EPI_ISL_1200469, EPI_ISL_1200470, EPI_ISL_1200471, EPI_ISL_1200472, EPI_ISL_1200473, EPI_ISL_1200474, EPI_ISL_1200475, EPI_ISL_1200476, EPI_ISL_1200477, EPI_ISL_1200478, EPI_ISL_1200479,
EPI_ISL_1200480, EPI_ISL_1200481, EPI_ISL_1200482, EPI_ISL_1200483, EPI_ISL_1200484, EPI_ISL_1200485, EPI_ISL_1200486, EPI_ISL_1200487, EPI_ISL_1200488, EPI_ISL_1200489, EPI_ISL_1200490, EPI_ISL_1200491, EPI_ISL_1200492, EPI_ISL_1200493, EPI_ISL_1200494, EPI_ISL_1200495, EPI_ISL_1200496, EPI_ISL_1200497, EPI_ISL_1200498, EPI_ISL_1200499, EPI_ISL_1200500, EPI_ISL_1200501,
EPI_ISL_1200502, EPI_ISL_1200503, EPI_ISL_1200504, EPI_ISL_1200505, EPI_ISL_1200506, EPI_ISL_1200507, EPI_ISL_1200508, EPI_ISL_1200509, EPI_ISL_1200510, EPI_ISL_1200511, EPI_ISL_1200512, EPI_ISL_1200513, EPI_ISL_1200514, EPI_ISL_1289961, EPI_ISL_1289962, EPI_ISL_1289965, EPI_ISL_1289966, EPI_ISL_1289967, EPI_ISL_1289970, EPI_ISL_1289971, EPI_ISL_1289972, EPI_ISL_1289982,
EPI_ISL_1289984, EPI_ISL_1289987, EPI_ISL_1289989, EPI_ISL_1289993, EPI_ISL_1289997, EPI_ISL_1289999, EPI_ISL_1290002, EPI_ISL_1290004, EPI_ISL_1290007, EPI_ISL_1290009, EPI_ISL_1290012, EPI_ISL_1290015, EPI_ISL_1290018, EPI_ISL_1290019, EPI_ISL_1290021, EPI_ISL_1290022, EPI_ISL_1290024, EPI_ISL_1290025, EPI_ISL_1290028, EPI_ISL_1290030, EPI_ISL_1290031, EPI_ISL_1290033,
EPI_ISL_1290034, EPI_ISL_1290047, EPI_ISL_1290050, EPI_ISL_1290053, EPI_ISL_1290055, EPI_ISL_1290057, EPI_ISL_1290059, EPI_ISL_1290060, EPI_ISL_1290062, EPI_ISL_1290064, EPI_ISL_1290071, EPI_ISL_1290072, EPI_ISL_1290076, EPI_ISL_1290077, EPI_ISL_1290078, EPI_ISL_1290081, EPI_ISL_1290083, EPI_ISL_1290084, EPI_ISL_1290085, EPI_ISL_1290089, EPI_ISL_1290091, EPI_ISL_1290093,
EPI_ISL_1290095, EPI_ISL_1290097, EPI_ISL_1290100, EPI_ISL_1290101, EPI_ISL_1290102, EPI_ISL_1290103, EPI_ISL_1290111, EPI_ISL_1290112, EPI_ISL_1290113, EPI_ISL_1290118, EPI_ISL_1290121, EPI_ISL_1290122, EPI_ISL_1290124, EPI_ISL_1290125, EPI_ISL_1290127, EPI_ISL_1290132, EPI_ISL_1290134, EPI_ISL_1290135, EPI_ISL_1290136, EPI_ISL_1290137, EPI_ISL_1290139, EPI_ISL_1290140,
EPI_ISL_1290141, EPI_ISL_1290142, EPI_ISL_1290143, EPI_ISL_1290144, EPI_ISL_1290145, EPI_ISL_1290149, EPI_ISL_1290152, EPI_ISL_1290153, EPI_ISL_1290154, EPI_ISL_1290155, EPI_ISL_1290156, EPI_ISL_1290157, EPI_ISL_1290158, EPI_ISL_1290159, EPI_ISL_1290160, EPI_ISL_1290161, EPI_ISL_1290162, EPI_ISL_1290163, EPI_ISL_1290164, EPI_ISL_1290165, EPI_ISL_1290166, EPI_ISL_1290167,
EPI_ISL_1290168, EPI_ISL_1290169, EPI_ISL_1290170, EPI_ISL_1290171, EPI_ISL_1290172, EPI_ISL_1290173, EPI_ISL_1290174, EPI_ISL_1290175, EPI_ISL_1290176, EPI_ISL_1290177, EPI_ISL_1290178, EPI_ISL_1290179, EPI_ISL_1290180, EPI_ISL_1290181, EPI_ISL_1290182, EPI_ISL_1290183, EPI_ISL_1290184, EPI_ISL_1290185, EPI_ISL_1290186, EPI_ISL_1290187, EPI_ISL_1290188, EPI_ISL_1290189,
EPI_ISL_1290190, EPI_ISL_1290191, EPI_ISL_1290192, EPI_ISL_1290193, EPI_ISL_1290194, EPI_ISL_1290195, EPI_ISL_1290196, EPI_ISL_1290197, EPI_ISL_1290198, EPI_ISL_1290199, EPI_ISL_1290200, EPI_ISL_1290201, EPI_ISL_1290202, EPI_ISL_1290203, EPI_ISL_1290204, EPI_ISL_1290205, EPI_ISL_1290206, EPI_ISL_1290207, EPI_ISL_1290208, EPI_ISL_1290209, EPI_ISL_1290210, EPI_ISL_1290211,
EPI_ISL_1290212, EPI_ISL_1290213, EPI_ISL_1290214, EPI_ISL_1290215, EPI_ISL_1290216, EPI_ISL_1290217, EPI_ISL_1290218, EPI_ISL_1290219, EPI_ISL_1290220, EPI_ISL_1290221, EPI_ISL_1290222, EPI_ISL_1290223, EPI_ISL_1290224, EPI_ISL_1290225, EPI_ISL_1290226, EPI_ISL_1290227, EPI_ISL_1290228, EPI_ISL_1290229, EPI_ISL_1290230, EPI_ISL_1290231, EPI_ISL_1290232, EPI_ISL_1290233,
EPI_ISL_1290234, EPI_ISL_1290235, EPI_ISL_1290236, EPI_ISL_1290237, EPI_ISL_1290238, EPI_ISL_1290239, EPI_ISL_1290240, EPI_ISL_1290241, EPI_ISL_1290242, EPI_ISL_1290243, EPI_ISL_1290244, EPI_ISL_1290245, EPI_ISL_1290246, EPI_ISL_1290279, EPI_ISL_1290280, EPI_ISL_1290281, EPI_ISL_1290282, EPI_ISL_1290283, EPI_ISL_1290284, EPI_ISL_1290285, EPI_ISL_1290286, EPI_ISL_1290287,
EPI_ISL_1290294, EPI_ISL_1290295, EPI_ISL_1290296, EPI_ISL_1290297, EPI_ISL_1290298, EPI_ISL_1290299, EPI_ISL_1290300, EPI_ISL_1290301, EPI_ISL_1290302, EPI_ISL_1290303, EPI_ISL_1290304, EPI_ISL_1290305, EPI_ISL_1290306, EPI_ISL_1290307, EPI_ISL_1290308, EPI_ISL_1290309, EPI_ISL_1290310, EPI_ISL_1290311, EPI_ISL_1290312, EPI_ISL_1290313, EPI_ISL_1290314, EPI_ISL_1290315,
EPI_ISL_1290316, EPI_ISL_1290317, EPI_ISL_1290318, EPI_ISL_1290319, EPI_ISL_1290320, EPI_ISL_1290321, EPI_ISL_1290322, EPI_ISL_1290323, EPI_ISL_1290324, EPI_ISL_1290325, EPI_ISL_1290326, EPI_ISL_1290327, EPI_ISL_1290328, EPI_ISL_1290329, EPI_ISL_1290330, EPI_ISL_1290331, EPI_ISL_1290332, EPI_ISL_1290333, EPI_ISL_1290334, EPI_ISL_1290335, EPI_ISL_1290336, EPI_ISL_1290337,
EPI_ISL_1290338, EPI_ISL_1290339, EPI_ISL_1290340, EPI_ISL_1290341, EPI_ISL_1290342, EPI_ISL_1290343, EPI_ISL_1290344, EPI_ISL_1290359, EPI_ISL_1290360, EPI_ISL_1290361, EPI_ISL_1290362, EPI_ISL_1290363, EPI_ISL_1290364, EPI_ISL_1290365, EPI_ISL_1290366, EPI_ISL_1290367, EPI_ISL_1290368, EPI_ISL_1290369, EPI_ISL_1290370, EPI_ISL_1290371, EPI_ISL_1290372, EPI_ISL_1290373,
EPI_ISL_1290374, EPI_ISL_1290375, EPI_ISL_1290376, EPI_ISL_1290377, EPI_ISL_1290378, EPI_ISL_1290379, EPI_ISL_1290380, EPI_ISL_1290381, EPI_ISL_1290382, EPI_ISL_1290383, EPI_ISL_1290384, EPI_ISL_1290385, EPI_ISL_1290386, EPI_ISL_1290387, EPI_ISL_1290388, EPI_ISL_1290389, EPI_ISL_1290394, EPI_ISL_1290395, EPI_ISL_1290396, EPI_ISL_1290397, EPI_ISL_1290398, EPI_ISL_1290399,
EPI_ISL_1290405, EPI_ISL_1290406, EPI_ISL_1290408, EPI_ISL_1290409, EPI_ISL_1290415, EPI_ISL_1290416, EPI_ISL_1290417, EPI_ISL_1290418, EPI_ISL_1290419, EPI_ISL_1290420, EPI_ISL_1290421, EPI_ISL_1290422, EPI_ISL_1290423, EPI_ISL_1290424, EPI_ISL_1290425, EPI_ISL_1290426, EPI_ISL_1290427, EPI_ISL_1290461, EPI_ISL_1290462, EPI_ISL_1290463, EPI_ISL_1290464, EPI_ISL_1290465,
EPI_ISL_1290466, EPI_ISL_1290467, EPI_ISL_1290468, EPI_ISL_1290469, EPI_ISL_1290470, EPI_ISL_1290471, EPI_ISL_1290472, EPI_ISL_1290474, EPI_ISL_1290476, EPI_ISL_1290478, EPI_ISL_1290479, EPI_ISL_1290480, EPI_ISL_1290482, EPI_ISL_1290483, EPI_ISL_1290485, EPI_ISL_1290486, EPI_ISL_1290487, EPI_ISL_1290488, EPI_ISL_1290491, EPI_ISL_1290492, EPI_ISL_1290494, EPI_ISL_1290496,
EPI_ISL_1290497, EPI_ISL_1290501, EPI_ISL_1290503, EPI_ISL_1290504, EPI_ISL_1290508, EPI_ISL_1290513, EPI_ISL_1290517, EPI_ISL_1290522, EPI_ISL_1290524, EPI_ISL_1290528, EPI_ISL_1290529, EPI_ISL_1290530, EPI_ISL_1290531, EPI_ISL_1290532, EPI_ISL_1290535, EPI_ISL_1290536, EPI_ISL_1290537, EPI_ISL_1290538, EPI_ISL_1290539, EPI_ISL_1290540, EPI_ISL_1290541, EPI_ISL_1290542,
EPI_ISL_1290546, EPI_ISL_1290547, EPI_ISL_1290548, EPI_ISL_1290549, EPI_ISL_1290550, EPI_ISL_1290551, EPI_ISL_1290552, EPI_ISL_1290553, EPI_ISL_1290554, EPI_ISL_1290555, EPI_ISL_1290556, EPI_ISL_1290557, EPI_ISL_1290558, EPI_ISL_1290559, EPI_ISL_1290560, EPI_ISL_1290561, EPI_ISL_1290562, EPI_ISL_1290563, EPI_ISL_1290564, EPI_ISL_1290569, EPI_ISL_1290570, EPI_ISL_1290571,
EPI_ISL_1290573, EPI_ISL_1290574, EPI_ISL_1290575, EPI_ISL_1290576, EPI_ISL_1290577, EPI_ISL_1290578, EPI_ISL_1290579, EPI_ISL_1290580, EPI_ISL_1290581, EPI_ISL_1290582, EPI_ISL_1290587, EPI_ISL_1290588, EPI_ISL_1290589, EPI_ISL_1290590, EPI_ISL_1290591, EPI_ISL_1290592, EPI_ISL_1290593, EPI_ISL_1290594, EPI_ISL_1290595, EPI_ISL_1290596, EPI_ISL_1290597, EPI_ISL_1290598,
EPI_ISL_1290599, EPI_ISL_1290600, EPI_ISL_1290601, EPI_ISL_1290602, EPI_ISL_1290603, EPI_ISL_1290604, EPI_ISL_1290605, EPI_ISL_1290606, EPI_ISL_1290607, EPI_ISL_1290608, EPI_ISL_1290609, EPI_ISL_1290610, EPI_ISL_1290611, EPI_ISL_1290612, EPI_ISL_1290613, EPI_ISL_1290614, EPI_ISL_1290615, EPI_ISL_1290616, EPI_ISL_1290617, EPI_ISL_1290618, EPI_ISL_1290619, EPI_ISL_1290620,
EPI_ISL_1290621, EPI_ISL_1290622, EPI_ISL_1290623, EPI_ISL_1290624, EPI_ISL_1290625, EPI_ISL_1290626, EPI_ISL_1290627, EPI_ISL_1290628, EPI_ISL_1290629, EPI_ISL_1290630, EPI_ISL_1290631, EPI_ISL_1290632, EPI_ISL_1290633, EPI_ISL_1290634, EPI_ISL_1290635, EPI_ISL_1290636, EPI_ISL_1290637, EPI_ISL_1290638, EPI_ISL_1290639, EPI_ISL_1290640, EPI_ISL_1290641, EPI_ISL_1290642,
EPI_ISL_1290643, EPI_ISL_1290644, EPI_ISL_1290645, EPI_ISL_1290646, EPI_ISL_1290647, EPI_ISL_1290648, EPI_ISL_1290649, EPI_ISL_1290650, EPI_ISL_1290651, EPI_ISL_1290652, EPI_ISL_1290653, EPI_ISL_1290654, EPI_ISL_1290655, EPI_ISL_1290656, EPI_ISL_1290657, EPI_ISL_1290658, EPI_ISL_1290659, EPI_ISL_1290660, EPI_ISL_1290661, EPI_ISL_1290662, EPI_ISL_1290663, EPI_ISL_1290664,
EPI_ISL_1290665, EPI_ISL_1290666, EPI_ISL_1290667, EPI_ISL_1290668, EPI_ISL_1290669, EPI_ISL_1290670, EPI_ISL_1290671, EPI_ISL_1290672, EPI_ISL_1290673, EPI_ISL_1290674, EPI_ISL_1290675, EPI_ISL_1290676, EPI_ISL_1290677, EPI_ISL_1290678, EPI_ISL_1290679, EPI_ISL_1290680, EPI_ISL_1290681, EPI_ISL_1290682, EPI_ISL_1290683, EPI_ISL_1290684, EPI_ISL_1290685, EPI_ISL_1290686,
EPI_ISL_1363803, EPI_ISL_1363804, EPI_ISL_1363805, EPI_ISL_1363806, EPI_ISL_1363807, EPI_ISL_1363808, EPI_ISL_1363809, EPI_ISL_1363810, EPI_ISL_1363811, EPI_ISL_1363812, EPI_ISL_1363813, EPI_ISL_1363814, EPI_ISL_1363815, EPI_ISL_1363816, EPI_ISL_1363817, EPI_ISL_1363818, EPI_ISL_1363819, EPI_ISL_1363820, EPI_ISL_1363821, EPI_ISL_1363823, EPI_ISL_1363830, EPI_ISL_1363831,
EPI_ISL_1363832, EPI_ISL_1363833, EPI_ISL_1363834, EPI_ISL_1363835, EPI_ISL_1363836, EPI_ISL_1363837, EPI_ISL_1363838, EPI_ISL_1363839, EPI_ISL_1363840, EPI_ISL_1363841, EPI_ISL_1363842, EPI_ISL_1363843, EPI_ISL_1363844, EPI_ISL_1363845, EPI_ISL_1363846, EPI_ISL_1363847, EPI_ISL_1363848, EPI_ISL_1363849, EPI_ISL_1363850, EPI_ISL_1363851, EPI_ISL_1363852, EPI_ISL_1363853,
EPI_ISL_1363854, EPI_ISL_1363855, EPI_ISL_1363856, EPI_ISL_1363857, EPI_ISL_1363858, EPI_ISL_1363859, EPI_ISL_1363860, EPI_ISL_1363861, EPI_ISL_1363862, EPI_ISL_1363863, EPI_ISL_1363864, EPI_ISL_1363865, EPI_ISL_1363866, EPI_ISL_1363867, EPI_ISL_1363868, EPI_ISL_1363869, EPI_ISL_1363870, EPI_ISL_1363871, EPI_ISL_1363877, EPI_ISL_1363878, EPI_ISL_1363879, EPI_ISL_1363880,
EPI_ISL_1363881, EPI_ISL_1363896, EPI_ISL_1363897, EPI_ISL_1363898, EPI_ISL_1363899, EPI_ISL_1363900, EPI_ISL_1363901, EPI_ISL_1363902, EPI_ISL_1363903, EPI_ISL_1363904, EPI_ISL_1363905, EPI_ISL_1363906, EPI_ISL_1363907, EPI_ISL_1363908, EPI_ISL_1363909, EPI_ISL_1363910, EPI_ISL_1363911, EPI_ISL_1363912, EPI_ISL_1363913, EPI_ISL_1363914, EPI_ISL_1363915, EPI_ISL_1363916,
EPI_ISL_1363917, EPI_ISL_1363918, EPI_ISL_1363919, EPI_ISL_1363920, EPI_ISL_1363921, EPI_ISL_1363922, EPI_ISL_1363923, EPI_ISL_1363924, EPI_ISL_1363925, EPI_ISL_1363926, EPI_ISL_1363927, EPI_ISL_1363928, EPI_ISL_1363929, EPI_ISL_1363930, EPI_ISL_1363931, EPI_ISL_1363932, EPI_ISL_1363933, EPI_ISL_1363934, EPI_ISL_1363935, EPI_ISL_1363936, EPI_ISL_1363937, EPI_ISL_1363938,
EPI_ISL_1363939, EPI_ISL_1363940, EPI_ISL_1363941, EPI_ISL_1363942, EPI_ISL_1363943, EPI_ISL_1363944, EPI_ISL_1363945, EPI_ISL_1363946, EPI_ISL_1363947, EPI_ISL_1363948, EPI_ISL_1363958, EPI_ISL_1363959, EPI_ISL_1363960, EPI_ISL_1363961, EPI_ISL_1363962, EPI_ISL_1363963, EPI_ISL_1363964, EPI_ISL_1363965, EPI_ISL_1363966, EPI_ISL_1363967, EPI_ISL_1363968, EPI_ISL_1363969,
EPI_ISL_1363970, EPI_ISL_1363971, EPI_ISL_1363972, EPI_ISL_1363973, EPI_ISL_1363974, EPI_ISL_1363975, EPI_ISL_1363976, EPI_ISL_1363977, EPI_ISL_1363978, EPI_ISL_1363979, EPI_ISL_1363980, EPI_ISL_1363981, EPI_ISL_1363982, EPI_ISL_1363983, EPI_ISL_1363984, EPI_ISL_1363985, EPI_ISL_1363986, EPI_ISL_1363987, EPI_ISL_1363988, EPI_ISL_1363989, EPI_ISL_1363990, EPI_ISL_1363991,
EPI_ISL_1363992, EPI_ISL_1363996, EPI_ISL_1363997, EPI_ISL_1363998, EPI_ISL_1363999, EPI_ISL_1364000, EPI_ISL_1364001, EPI_ISL_1364002, EPI_ISL_1364003, EPI_ISL_1364004, EPI_ISL_1364005, EPI_ISL_1364006, EPI_ISL_1364007, EPI_ISL_1364008, EPI_ISL_1364009, EPI_ISL_1364010, EPI_ISL_1364011, EPI_ISL_1364012, EPI_ISL_1364013, EPI_ISL_1364014, EPI_ISL_1364015, EPI_ISL_1364016,
EPI_ISL_1364017, EPI_ISL_1364018, EPI_ISL_1364019, EPI_ISL_1364020, EPI_ISL_1364021, EPI_ISL_1364022, EPI_ISL_1364023, EPI_ISL_1364024, EPI_ISL_1364025, EPI_ISL_1364026, EPI_ISL_1364027, EPI_ISL_1364028, EPI_ISL_1364029, EPI_ISL_1364030, EPI_ISL_1364031, EPI_ISL_1364032, EPI_ISL_1364033, EPI_ISL_1364034, EPI_ISL_1364035, EPI_ISL_1364036, EPI_ISL_1364037, EPI_ISL_1364038,
EPI_ISL_1364039, EPI_ISL_1364040, EPI_ISL_1364041, EPI_ISL_1364042, EPI_ISL_1364046, EPI_ISL_1364047, EPI_ISL_1364048, EPI_ISL_1364049, EPI_ISL_1364050, EPI_ISL_1364051, EPI_ISL_1364052, EPI_ISL_1364053, EPI_ISL_1364054, EPI_ISL_1364055, EPI_ISL_1364056, EPI_ISL_1364057, EPI_ISL_1364058, EPI_ISL_1364059, EPI_ISL_1364060, EPI_ISL_1364061, EPI_ISL_1364062, EPI_ISL_1364071,
EPI_ISL_1364099, EPI_ISL_1364100, EPI_ISL_1364101, EPI_ISL_1364102, EPI_ISL_1364103, EPI_ISL_1364104, EPI_ISL_1364105, EPI_ISL_1364106, EPI_ISL_1364121, EPI_ISL_1364122, EPI_ISL_1364123, EPI_ISL_1364124, EPI_ISL_1364125, EPI_ISL_1364126, EPI_ISL_1364127, EPI_ISL_1364128, EPI_ISL_1364129, EPI_ISL_1364130, EPI_ISL_1364131, EPI_ISL_1364132, EPI_ISL_1364133, EPI_ISL_1364134,
EPI_ISL_1364135, EPI_ISL_1364136, EPI_ISL_1364137, EPI_ISL_1364138, EPI_ISL_1364139, EPI_ISL_1364140, EPI_ISL_1364141, EPI_ISL_1364142, EPI_ISL_1364143, EPI_ISL_1364144, EPI_ISL_1364145, EPI_ISL_1364146, EPI_ISL_1364147, EPI_ISL_1364148, EPI_ISL_1364159, EPI_ISL_1364160, EPI_ISL_1364161, EPI_ISL_1364162, EPI_ISL_1364163, EPI_ISL_1364164, EPI_ISL_1364165, EPI_ISL_1364166,
EPI_ISL_1364167, EPI_ISL_1364168, EPI_ISL_1364169, EPI_ISL_1364170, EPI_ISL_1364171, EPI_ISL_1364172, EPI_ISL_1364173, EPI_ISL_1364174, EPI_ISL_1364175, EPI_ISL_1364176, EPI_ISL_1364177, EPI_ISL_1364178, EPI_ISL_1364179, EPI_ISL_1364180, EPI_ISL_1364181, EPI_ISL_1364182, EPI_ISL_1364183, EPI_ISL_1364184, EPI_ISL_1364185, EPI_ISL_1364186, EPI_ISL_1364187, EPI_ISL_1364188,
EPI_ISL_1364190, EPI_ISL_1364191, EPI_ISL_1364192, EPI_ISL_1364193, EPI_ISL_1364194, EPI_ISL_1364195, EPI_ISL_1364196, EPI_ISL_1364197, EPI_ISL_1364198, EPI_ISL_1364199, EPI_ISL_1364200, EPI_ISL_1364201, EPI_ISL_1364202, EPI_ISL_1364203, EPI_ISL_1364204, EPI_ISL_1364205, EPI_ISL_1364206, EPI_ISL_1364207, EPI_ISL_1364208, EPI_ISL_1364209, EPI_ISL_1364210, EPI_ISL_1364211,
EPI_ISL_1364212, EPI_ISL_1364213, EPI_ISL_1364214, EPI_ISL_1364215, EPI_ISL_1364216, EPI_ISL_1364217, EPI_ISL_1364218, EPI_ISL_1364219, EPI_ISL_1364220, EPI_ISL_1364221, EPI_ISL_1364222, EPI_ISL_1364223, EPI_ISL_1364224, EPI_ISL_1364225, EPI_ISL_1364226, EPI_ISL_1364227, EPI_ISL_1364228, EPI_ISL_1364229, EPI_ISL_1364230, EPI_ISL_1364231, EPI_ISL_1364232, EPI_ISL_1364233,
EPI_ISL_1364234, EPI_ISL_1364235, EPI_ISL_1364236, EPI_ISL_1364237, EPI_ISL_1364238, EPI_ISL_1364239, EPI_ISL_1364240, EPI_ISL_1364241, EPI_ISL_1364242, EPI_ISL_1364243, EPI_ISL_1364244, EPI_ISL_1364245, EPI_ISL_1364246, EPI_ISL_1364262, EPI_ISL_1364263, EPI_ISL_1364265, EPI_ISL_1364266, EPI_ISL_1364268, EPI_ISL_1364277, EPI_ISL_1364278, EPI_ISL_1364279, EPI_ISL_1364280,
EPI_ISL_1364281, EPI_ISL_1364282, EPI_ISL_1364283, EPI_ISL_1364284, EPI_ISL_1364285, EPI_ISL_1364286, EPI_ISL_1364287, EPI_ISL_1364288, EPI_ISL_1364289, EPI_ISL_1364290, EPI_ISL_1364291, EPI_ISL_1364292, EPI_ISL_1364293, EPI_ISL_1364294, EPI_ISL_1364295, EPI_ISL_1364296, EPI_ISL_1364297, EPI_ISL_1364298, EPI_ISL_1364299, EPI_ISL_1364300, EPI_ISL_1364301, EPI_ISL_1364302,
EPI_ISL_1364303, EPI_ISL_1364304, EPI_ISL_1364305, EPI_ISL_1364314, EPI_ISL_1364315, EPI_ISL_1364316, EPI_ISL_1364322, EPI_ISL_1364341, EPI_ISL_1364342, EPI_ISL_1364343, EPI_ISL_1364344, EPI_ISL_1364345, EPI_ISL_1364346, EPI_ISL_1364347, EPI_ISL_1364348, EPI_ISL_1364349, EPI_ISL_1364350, EPI_ISL_1364351, EPI_ISL_1364352, EPI_ISL_1364353, EPI_ISL_1364354, EPI_ISL_1364355,
EPI_ISL_1364356, EPI_ISL_1364357, EPI_ISL_1364358, EPI_ISL_1364359, EPI_ISL_1364360, EPI_ISL_1364361, EPI_ISL_1364362, EPI_ISL_1364363, EPI_ISL_1364364, EPI_ISL_1364406, EPI_ISL_1364407, EPI_ISL_1364408, EPI_ISL_1364409, EPI_ISL_1364410, EPI_ISL_1364411, EPI_ISL_1364412, EPI_ISL_1364413, EPI_ISL_1364414, EPI_ISL_1364415, EPI_ISL_1364416, EPI_ISL_1364417, EPI_ISL_1364418,
EPI_ISL_1364419, EPI_ISL_1364420, EPI_ISL_1364421, EPI_ISL_1364422, EPI_ISL_1364423, EPI_ISL_1364424, EPI_ISL_1364425, EPI_ISL_1417446, EPI_ISL_1417448, EPI_ISL_1417462, EPI_ISL_1417472, EPI_ISL_1417494, EPI_ISL_1417495, EPI_ISL_1417497, EPI_ISL_1417500, EPI_ISL_1417502, EPI_ISL_1417506, EPI_ISL_1417536, EPI_ISL_1417568, EPI_ISL_1417572, EPI_ISL_1417577, EPI_ISL_1417586,

EPI_ISL_1417587, EPI_ISL_1417595, EPI_ISL_1417598, EPI_ISL_1417607, EPI_ISL_1417608, EPI_ISL_1417609, EPI_ISL_1417610, EPI_ISL_1417611, EPI_ISL_1417612, EPI_ISL_1417613, EPI_ISL_1417614, EPI_ISL_1417615, EPI_ISL_1417616, EPI_ISL_1417617, EPI_ISL_1417618, EPI_ISL_1417775, EPI_ISL_1417776, EPI_ISL_1417777, EPI_ISL_1417778, EPI_ISL_1417779, EPI_ISL_1417780, EPI_ISL_1417781,
EPI_ISL_1417782, EPI_ISL_1417783, EPI_ISL_1417784, EPI_ISL_1417785, EPI_ISL_1417786, EPI_ISL_1417787, EPI_ISL_1417788, EPI_ISL_1417789, EPI_ISL_1417790, EPI_ISL_1417791, EPI_ISL_1417792, EPI_ISL_1417793, EPI_ISL_1417794, EPI_ISL_1417795, EPI_ISL_1417796, EPI_ISL_1417797, EPI_ISL_1417798, EPI_ISL_1417799, EPI_ISL_1417800, EPI_ISL_1417801, EPI_ISL_1417802, EPI_ISL_1417803,
EPI_ISL_1417804, EPI_ISL_1417805, EPI_ISL_1417806, EPI_ISL_1417807, EPI_ISL_1417808, EPI_ISL_1417809, EPI_ISL_1417810, EPI_ISL_1417811, EPI_ISL_1417812, EPI_ISL_1417813, EPI_ISL_1417814, EPI_ISL_1417815, EPI_ISL_1417816, EPI_ISL_1417817, EPI_ISL_1417818, EPI_ISL_1417819, EPI_ISL_1417820, EPI_ISL_1417821, EPI_ISL_1417822, EPI_ISL_1417823, EPI_ISL_1417824, EPI_ISL_1417825,
EPI_ISL_1417826, EPI_ISL_1417827, EPI_ISL_1417828, EPI_ISL_1417829, EPI_ISL_1417830, EPI_ISL_1417831, EPI_ISL_1417832, EPI_ISL_1417833, EPI_ISL_1417834, EPI_ISL_1417835, EPI_ISL_1417836, EPI_ISL_1417837, EPI_ISL_1417838, EPI_ISL_1417839, EPI_ISL_1417840, EPI_ISL_1417841, EPI_ISL_1417842, EPI_ISL_1417843, EPI_ISL_1417844, EPI_ISL_1417845, EPI_ISL_1417846, EPI_ISL_1417847,
EPI_ISL_1417848, EPI_ISL_1417849, EPI_ISL_1417850, EPI_ISL_1417851, EPI_ISL_1417852, EPI_ISL_1417853, EPI_ISL_1417854, EPI_ISL_1417855, EPI_ISL_1417856, EPI_ISL_1417857, EPI_ISL_1417858, EPI_ISL_1417859, EPI_ISL_1417860, EPI_ISL_1417861, EPI_ISL_1417862, EPI_ISL_1417863, EPI_ISL_1417864, EPI_ISL_1417865, EPI_ISL_1417866, EPI_ISL_1417867, EPI_ISL_1417868, EPI_ISL_1417869,
EPI_ISL_1417870, EPI_ISL_1417871, EPI_ISL_1417872, EPI_ISL_1417873, EPI_ISL_1417874, EPI_ISL_1418000, EPI_ISL_1418001, EPI_ISL_1418002, EPI_ISL_1418003, EPI_ISL_1418004, EPI_ISL_1418005, EPI_ISL_1418006, EPI_ISL_1418007, EPI_ISL_1418008, EPI_ISL_1418009, EPI_ISL_1418010, EPI_ISL_1418011, EPI_ISL_1418012, EPI_ISL_1418013, EPI_ISL_1418014, EPI_ISL_1418015, EPI_ISL_1418016,
EPI_ISL_1418017, EPI_ISL_1418018, EPI_ISL_1418019, EPI_ISL_1418020, EPI_ISL_1418021, EPI_ISL_1418022, EPI_ISL_1418023, EPI_ISL_1418024, EPI_ISL_1418025, EPI_ISL_1418026, EPI_ISL_1418060, EPI_ISL_1418066, EPI_ISL_1418067, EPI_ISL_1418070, EPI_ISL_1418080, EPI_ISL_1418101, EPI_ISL_1418113, EPI_ISL_1418137, EPI_ISL_1418145, EPI_ISL_1418146, EPI_ISL_1418147, EPI_ISL_1418148,
EPI_ISL_1418149, EPI_ISL_1418150, EPI_ISL_1418151, EPI_ISL_1418185, EPI_ISL_1418186, EPI_ISL_1418332, EPI_ISL_1418337, EPI_ISL_1418339, EPI_ISL_1418341, EPI_ISL_1418348, EPI_ISL_1418349, EPI_ISL_1418362, EPI_ISL_1418366, EPI_ISL_1418370, EPI_ISL_1418372, EPI_ISL_1418373, EPI_ISL_1418374, EPI_ISL_1418376, EPI_ISL_1418516, EPI_ISL_1418517, EPI_ISL_1418518, EPI_ISL_1418519,
EPI_ISL_1418520, EPI_ISL_1418521, EPI_ISL_1418522, EPI_ISL_1418523, EPI_ISL_1418524, EPI_ISL_1418525, EPI_ISL_1418526, EPI_ISL_1418527, EPI_ISL_1418528, EPI_ISL_1418529, EPI_ISL_1418530, EPI_ISL_1418531, EPI_ISL_1418532, EPI_ISL_1418533, EPI_ISL_1418534, EPI_ISL_1418535, EPI_ISL_1418536, EPI_ISL_1418537, EPI_ISL_1418538, EPI_ISL_1418539, EPI_ISL_1418540, EPI_ISL_1418541,
EPI_ISL_1418602, EPI_ISL_1418603, EPI_ISL_1418604, EPI_ISL_1418605, EPI_ISL_1418606, EPI_ISL_1418607, EPI_ISL_1418608, EPI_ISL_1418609, EPI_ISL_1418610, EPI_ISL_1418611, EPI_ISL_1418612, EPI_ISL_1418613, EPI_ISL_1418614, EPI_ISL_1418615, EPI_ISL_1418616, EPI_ISL_1418617, EPI_ISL_1418618, EPI_ISL_1418622, EPI_ISL_1418623, EPI_ISL_1418631, EPI_ISL_1418632, EPI_ISL_1418633,
EPI_ISL_1418634, EPI_ISL_1418635, EPI_ISL_1418636, EPI_ISL_1418637, EPI_ISL_1418646, EPI_ISL_1418692, EPI_ISL_1418694, EPI_ISL_1418703, EPI_ISL_1418704, EPI_ISL_1418705, EPI_ISL_1418706, EPI_ISL_1418707, EPI_ISL_1418708, EPI_ISL_1418709, EPI_ISL_1418710, EPI_ISL_1418711, EPI_ISL_1418712, EPI_ISL_1418713, EPI_ISL_1418714, EPI_ISL_1418715, EPI_ISL_1418717, EPI_ISL_1418718,
EPI_ISL_1418719, EPI_ISL_1418723, EPI_ISL_1418724, EPI_ISL_1418725, EPI_ISL_1418726, EPI_ISL_1418727, EPI_ISL_1418728, EPI_ISL_1418729, EPI_ISL_1418730, EPI_ISL_1418731, EPI_ISL_1418732, EPI_ISL_1418733, EPI_ISL_1418734, EPI_ISL_1418735, EPI_ISL_1418736, EPI_ISL_1418737, EPI_ISL_1418738, EPI_ISL_1418739, EPI_ISL_1418740, EPI_ISL_1418741, EPI_ISL_1418742, EPI_ISL_1418743,
EPI_ISL_1418744, EPI_ISL_1418745, EPI_ISL_1418746, EPI_ISL_1418747, EPI_ISL_1418748, EPI_ISL_1418749, EPI_ISL_1418750, EPI_ISL_1418751, EPI_ISL_1418752, EPI_ISL_1418756, EPI_ISL_1418757, EPI_ISL_1418758, EPI_ISL_1418759, EPI_ISL_1418760, EPI_ISL_1418761, EPI_ISL_1418771, EPI_ISL_1418772, EPI_ISL_1418773, EPI_ISL_1418774, EPI_ISL_1418775, EPI_ISL_1418776, EPI_ISL_1418840,
EPI_ISL_1418841, EPI_ISL_1418842, EPI_ISL_1418843, EPI_ISL_1418844, EPI_ISL_1418845, EPI_ISL_1418846, EPI_ISL_1418847, EPI_ISL_1418848, EPI_ISL_1418849, EPI_ISL_1418850, EPI_ISL_1418851, EPI_ISL_1418852, EPI_ISL_1418853, EPI_ISL_1418854, EPI_ISL_1418855, EPI_ISL_1418856, EPI_ISL_1418857, EPI_ISL_1418858, EPI_ISL_1418859, EPI_ISL_1418860, EPI_ISL_1418861, EPI_ISL_1418862,
EPI_ISL_1418863, EPI_ISL_1418864, EPI_ISL_1418865, EPI_ISL_1418866, EPI_ISL_1418867, EPI_ISL_1418868, EPI_ISL_1418869, EPI_ISL_1418870, EPI_ISL_1418871, EPI_ISL_1418872, EPI_ISL_1418873, EPI_ISL_1418874, EPI_ISL_1418875, EPI_ISL_1418876, EPI_ISL_1418877, EPI_ISL_1418878, EPI_ISL_1418879, EPI_ISL_1418880, EPI_ISL_1418881, EPI_ISL_1418882, EPI_ISL_1418883, EPI_ISL_1418884,
EPI_ISL_1418885, EPI_ISL_1418886, EPI_ISL_1418887, EPI_ISL_1418888, EPI_ISL_1418889, EPI_ISL_1418890, EPI_ISL_1418891, EPI_ISL_1418892, EPI_ISL_1418893, EPI_ISL_1418894, EPI_ISL_1418895, EPI_ISL_1418896, EPI_ISL_1418897, EPI_ISL_1418898, EPI_ISL_1418899, EPI_ISL_1418900, EPI_ISL_1418901, EPI_ISL_1418902, EPI_ISL_1418903, EPI_ISL_1418904, EPI_ISL_1418905, EPI_ISL_1418906,
EPI_ISL_1418907, EPI_ISL_1418908, EPI_ISL_1418909, EPI_ISL_1418910, EPI_ISL_1418911, EPI_ISL_1418912, EPI_ISL_1418913, EPI_ISL_1418914, EPI_ISL_1418915, EPI_ISL_1418916, EPI_ISL_1418917, EPI_ISL_1418918, EPI_ISL_1418919, EPI_ISL_1418920, EPI_ISL_1418921, EPI_ISL_1418922, EPI_ISL_1418923, EPI_ISL_1418924, EPI_ISL_1418925, EPI_ISL_1418926, EPI_ISL_1418927, EPI_ISL_1418928,
EPI_ISL_1418929, EPI_ISL_1418930, EPI_ISL_1418931, EPI_ISL_1418932, EPI_ISL_1418933, EPI_ISL_1418934, EPI_ISL_1418935, EPI_ISL_1418936, EPI_ISL_1418937, EPI_ISL_1418938, EPI_ISL_1418939, EPI_ISL_1418940, EPI_ISL_1418941, EPI_ISL_1418942, EPI_ISL_1418943, EPI_ISL_1418944, EPI_ISL_1418945, EPI_ISL_1418946, EPI_ISL_1418947, EPI_ISL_1418948, EPI_ISL_1418949, EPI_ISL_1418950,
EPI_ISL_1418951, EPI_ISL_1418952, EPI_ISL_1418953, EPI_ISL_1418954, EPI_ISL_1418955, EPI_ISL_1418956, EPI_ISL_1418957, EPI_ISL_1418958, EPI_ISL_1418959, EPI_ISL_1418960, EPI_ISL_1418961, EPI_ISL_1418962, EPI_ISL_1418963, EPI_ISL_1418964, EPI_ISL_1418965, EPI_ISL_1418966, EPI_ISL_1418967, EPI_ISL_1418968, EPI_ISL_1418969, EPI_ISL_1418970, EPI_ISL_1418971, EPI_ISL_1418972,
EPI_ISL_1418973, EPI_ISL_1418974, EPI_ISL_1418975, EPI_ISL_1418976, EPI_ISL_1418977, EPI_ISL_1418978, EPI_ISL_1418979, EPI_ISL_1418980, EPI_ISL_1418981, EPI_ISL_1418982, EPI_ISL_1418983, EPI_ISL_1418984, EPI_ISL_1418985, EPI_ISL_1418986, EPI_ISL_1418987, EPI_ISL_1418988, EPI_ISL_1418989, EPI_ISL_1418990, EPI_ISL_1418991, EPI_ISL_1418992, EPI_ISL_1418993, EPI_ISL_1418994,
EPI_ISL_1418995, EPI_ISL_1418996, EPI_ISL_1418997, EPI_ISL_1418998, EPI_ISL_1418999, EPI_ISL_1419000, EPI_ISL_1419001, EPI_ISL_1419002, EPI_ISL_1419003, EPI_ISL_1419004, EPI_ISL_1419005, EPI_ISL_1419006, EPI_ISL_1419007, EPI_ISL_1419008, EPI_ISL_1419009, EPI_ISL_1419010, EPI_ISL_1419011, EPI_ISL_1419012, EPI_ISL_1419013, EPI_ISL_1419014, EPI_ISL_1419015, EPI_ISL_1419016,
EPI_ISL_1419017, EPI_ISL_1419018, EPI_ISL_1419019, EPI_ISL_1419020, EPI_ISL_1419021, EPI_ISL_1419022, EPI_ISL_1419023, EPI_ISL_1419024, EPI_ISL_1419025, EPI_ISL_1419026, EPI_ISL_1419027, EPI_ISL_1419028, EPI_ISL_1419029, EPI_ISL_1419030, EPI_ISL_1419031, EPI_ISL_1419032, EPI_ISL_1419033, EPI_ISL_1419034, EPI_ISL_1419035, EPI_ISL_1419036, EPI_ISL_1419037, EPI_ISL_1419038,
EPI_ISL_1419039, EPI_ISL_1419040, EPI_ISL_1419041, EPI_ISL_1419042, EPI_ISL_1419043, EPI_ISL_1419044, EPI_ISL_1419045, EPI_ISL_1419925, EPI_ISL_1419926, EPI_ISL_1419927, EPI_ISL_1419929, EPI_ISL_1419934, EPI_ISL_1419935, EPI_ISL_1419936, EPI_ISL_1419938, EPI_ISL_1419940, EPI_ISL_1419941, EPI_ISL_1419943, EPI_ISL_1419945, EPI_ISL_1419946, EPI_ISL_1419949, EPI_ISL_1419950,
EPI_ISL_1419953, EPI_ISL_1419956, EPI_ISL_1419961, EPI_ISL_1419962, EPI_ISL_1419966, EPI_ISL_1419969, EPI_ISL_1419971, EPI_ISL_1419973, EPI_ISL_1419975, EPI_ISL_1419977, EPI_ISL_1419980, EPI_ISL_1419983, EPI_ISL_1419984, EPI_ISL_1419985, EPI_ISL_1419986, EPI_ISL_1419989, EPI_ISL_1419990, EPI_ISL_1419992, EPI_ISL_1419993, EPI_ISL_1419995, EPI_ISL_1419996, EPI_ISL_1419997,
EPI_ISL_1419998, EPI_ISL_1420013, EPI_ISL_1420014, EPI_ISL_1420015, EPI_ISL_1420016, EPI_ISL_1420225, EPI_ISL_1420226, EPI_ISL_1420227, EPI_ISL_1420228, EPI_ISL_1420229, EPI_ISL_1420230, EPI_ISL_1420231, EPI_ISL_1420232, EPI_ISL_1420233, EPI_ISL_1420234, EPI_ISL_1420235, EPI_ISL_1420236, EPI_ISL_1420237, EPI_ISL_1420238, EPI_ISL_1420239, EPI_ISL_1420240, EPI_ISL_1420241,
EPI_ISL_1420242, EPI_ISL_1420243, EPI_ISL_1420244, EPI_ISL_1420245, EPI_ISL_1420246, EPI_ISL_1420247, EPI_ISL_1420248, EPI_ISL_1420249, EPI_ISL_1420250, EPI_ISL_1420251, EPI_ISL_1420252, EPI_ISL_1420253, EPI_ISL_1420254, EPI_ISL_1420255, EPI_ISL_1420256, EPI_ISL_1420257, EPI_ISL_1420258, EPI_ISL_1420259, EPI_ISL_1420260, EPI_ISL_1420261, EPI_ISL_1420262, EPI_ISL_1420263,
EPI_ISL_1420264, EPI_ISL_1420265, EPI_ISL_1420266, EPI_ISL_1420267, EPI_ISL_1420268, EPI_ISL_1420269, EPI_ISL_1420270, EPI_ISL_1420271, EPI_ISL_1420272, EPI_ISL_1420273, EPI_ISL_1420274, EPI_ISL_1420275, EPI_ISL_1420276, EPI_ISL_1420277, EPI_ISL_1420278, EPI_ISL_1420279, EPI_ISL_1420280, EPI_ISL_1420281, EPI_ISL_1420282, EPI_ISL_1420283, EPI_ISL_1420284, EPI_ISL_1420285,
EPI_ISL_1420286, EPI_ISL_1420287, EPI_ISL_1420288, EPI_ISL_1420289, EPI_ISL_1420290, EPI_ISL_1420291, EPI_ISL_1420292, EPI_ISL_1420293, EPI_ISL_1420294, EPI_ISL_1420295, EPI_ISL_1420296, EPI_ISL_1420297, EPI_ISL_1420298, EPI_ISL_1420299, EPI_ISL_1420300, EPI_ISL_1420301, EPI_ISL_1420302, EPI_ISL_1420303, EPI_ISL_1420304, EPI_ISL_1420305, EPI_ISL_1420306, EPI_ISL_1420307,
EPI_ISL_1420308, EPI_ISL_1420309, EPI_ISL_1420310, EPI_ISL_1420311, EPI_ISL_1420312, EPI_ISL_1420313, EPI_ISL_1420314, EPI_ISL_1420315, EPI_ISL_1420316, EPI_ISL_1420317, EPI_ISL_1420318, EPI_ISL_1420319, EPI_ISL_1420320, EPI_ISL_1420321, EPI_ISL_1420322, EPI_ISL_1420323, EPI_ISL_1420324, EPI_ISL_1420325, EPI_ISL_1420326, EPI_ISL_1420327, EPI_ISL_1420328, EPI_ISL_1420329,
EPI_ISL_1420330, EPI_ISL_1420331, EPI_ISL_1420332, EPI_ISL_1420333, EPI_ISL_1420334, EPI_ISL_1420335, EPI_ISL_1420336, EPI_ISL_1420337, EPI_ISL_1420338, EPI_ISL_1420339, EPI_ISL_1420340, EPI_ISL_1420341, EPI_ISL_1420342, EPI_ISL_1420343, EPI_ISL_1420344, EPI_ISL_1420345, EPI_ISL_1420346, EPI_ISL_1420347, EPI_ISL_1420348, EPI_ISL_1420349, EPI_ISL_1420350, EPI_ISL_1420351,
EPI_ISL_1420354, EPI_ISL_1420355, EPI_ISL_1420460, EPI_ISL_1420461, EPI_ISL_1420462, EPI_ISL_1420463, EPI_ISL_1420464, EPI_ISL_1420465, EPI_ISL_1420466, EPI_ISL_1420467, EPI_ISL_1420468, EPI_ISL_1420469, EPI_ISL_1420470, EPI_ISL_1420471, EPI_ISL_1420472, EPI_ISL_1420473, EPI_ISL_1420474, EPI_ISL_1420475, EPI_ISL_1420476, EPI_ISL_1420477, EPI_ISL_1420478, EPI_ISL_1420479,
EPI_ISL_1420480, EPI_ISL_1420481, EPI_ISL_1420482, EPI_ISL_1420483, EPI_ISL_1420484, EPI_ISL_1420485, EPI_ISL_1420486, EPI_ISL_1420487, EPI_ISL_1420488, EPI_ISL_1420489, EPI_ISL_1420490, EPI_ISL_1420491, EPI_ISL_1420492, EPI_ISL_1420493, EPI_ISL_1420494, EPI_ISL_1420495, EPI_ISL_1420496, EPI_ISL_1420497, EPI_ISL_1420498, EPI_ISL_1420499, EPI_ISL_1420500, EPI_ISL_1420501,
EPI_ISL_1420502, EPI_ISL_1420503, EPI_ISL_1420504, EPI_ISL_1420505, EPI_ISL_1420506, EPI_ISL_1420507, EPI_ISL_1420508, EPI_ISL_1420509, EPI_ISL_1420510, EPI_ISL_1420511, EPI_ISL_1420512, EPI_ISL_1420513, EPI_ISL_1420514, EPI_ISL_1420515, EPI_ISL_1420516, EPI_ISL_1420517, EPI_ISL_1420518, EPI_ISL_1420519, EPI_ISL_1420520, EPI_ISL_1420521, EPI_ISL_1420522, EPI_ISL_1420523,
EPI_ISL_1420524, EPI_ISL_1420525, EPI_ISL_1420526, EPI_ISL_1420551, EPI_ISL_1420552, EPI_ISL_1420553, EPI_ISL_1420554, EPI_ISL_1420555, EPI_ISL_1420556, EPI_ISL_1420557, EPI_ISL_1420558, EPI_ISL_1420560, EPI_ISL_1420561, EPI_ISL_1420563, EPI_ISL_1420565, EPI_ISL_1420566, EPI_ISL_1420567, EPI_ISL_1420568, EPI_ISL_1420596, EPI_ISL_1420597, EPI_ISL_1420598, EPI_ISL_1420599,
EPI_ISL_1420600, EPI_ISL_1420601, EPI_ISL_1420602, EPI_ISL_1420603, EPI_ISL_1420604, EPI_ISL_1420605, EPI_ISL_1420606, EPI_ISL_1420607, EPI_ISL_1420608, EPI_ISL_1420609, EPI_ISL_1420610, EPI_ISL_1420611, EPI_ISL_1420612, EPI_ISL_1420613, EPI_ISL_1420614, EPI_ISL_1420615, EPI_ISL_1420616, EPI_ISL_1420617, EPI_ISL_1420618, EPI_ISL_1420619, EPI_ISL_1420634, EPI_ISL_1599552,
EPI_ISL_1599553, EPI_ISL_1599554, EPI_ISL_1599555, EPI_ISL_1599556, EPI_ISL_1599557, EPI_ISL_1599558, EPI_ISL_1599559, EPI_ISL_1599560, EPI_ISL_1599561, EPI_ISL_1599562, EPI_ISL_1599563, EPI_ISL_1599564, EPI_ISL_1599565, EPI_ISL_1599566, EPI_ISL_1599567, EPI_ISL_1599568, EPI_ISL_1599569, EPI_ISL_1599570, EPI_ISL_1599571, EPI_ISL_1599572, EPI_ISL_1599573, EPI_ISL_1599574,
EPI_ISL_1599575, EPI_ISL_1599576, EPI_ISL_1599577, EPI_ISL_1599578, EPI_ISL_1599579, EPI_ISL_1599580, EPI_ISL_1599581, EPI_ISL_1599582, EPI_ISL_1599583, EPI_ISL_1599584, EPI_ISL_1599585, EPI_ISL_1599586, EPI_ISL_1599587, EPI_ISL_1599588, EPI_ISL_1599589, EPI_ISL_1599590, EPI_ISL_1599591, EPI_ISL_1599592, EPI_ISL_1599593, EPI_ISL_1599594, EPI_ISL_1599595, EPI_ISL_1599596,
EPI_ISL_1599597, EPI_ISL_1599598, EPI_ISL_1599599, EPI_ISL_1599600, EPI_ISL_1599601, EPI_ISL_1599602, EPI_ISL_1599603, EPI_ISL_1599604, EPI_ISL_1599605, EPI_ISL_1599606, EPI_ISL_1599607, EPI_ISL_1599608, EPI_ISL_1599609, EPI_ISL_1599610, EPI_ISL_1599611, EPI_ISL_1599612, EPI_ISL_1599613, EPI_ISL_1599614, EPI_ISL_1599615, EPI_ISL_1599616, EPI_ISL_1599617, EPI_ISL_1599618,
EPI_ISL_1599619, EPI_ISL_1599620, EPI_ISL_1599621, EPI_ISL_1599622, EPI_ISL_1599623, EPI_ISL_1599624, EPI_ISL_1599625, EPI_ISL_1599626, EPI_ISL_1599627, EPI_ISL_1599628, EPI_ISL_1599630, EPI_ISL_1599631, EPI_ISL_1599632, EPI_ISL_1599633, EPI_ISL_1599634, EPI_ISL_1599635, EPI_ISL_1599636, EPI_ISL_1599637, EPI_ISL_1599638, EPI_ISL_1599639, EPI_ISL_1599640, EPI_ISL_1599641,
EPI_ISL_1599642, EPI_ISL_1599643, EPI_ISL_1599644, EPI_ISL_1599645, EPI_ISL_1599646, EPI_ISL_1599647, EPI_ISL_1599648, EPI_ISL_1599649, EPI_ISL_1599650, EPI_ISL_1599651, EPI_ISL_1599652, EPI_ISL_1599653, EPI_ISL_1599654, EPI_ISL_1599655, EPI_ISL_1599656, EPI_ISL_1599657, EPI_ISL_1599658, EPI_ISL_1599659, EPI_ISL_1599660, EPI_ISL_1599661, EPI_ISL_1599662, EPI_ISL_1599663,
EPI_ISL_1599664, EPI_ISL_1599665, EPI_ISL_1599666, EPI_ISL_1599667, EPI_ISL_1599668, EPI_ISL_1599669, EPI_ISL_1599670, EPI_ISL_1599671, EPI_ISL_1599672, EPI_ISL_1599673, EPI_ISL_1599674, EPI_ISL_1599675, EPI_ISL_1599676, EPI_ISL_1599677, EPI_ISL_1599678, EPI_ISL_1599679, EPI_ISL_1599680, EPI_ISL_1599681, EPI_ISL_1599682, EPI_ISL_1599683, EPI_ISL_1599684, EPI_ISL_1599685,
EPI_ISL_1599686, EPI_ISL_1599687, EPI_ISL_1599688, EPI_ISL_1599689, EPI_ISL_1599690, EPI_ISL_1599691, EPI_ISL_1599692, EPI_ISL_1599693, EPI_ISL_1599694, EPI_ISL_1599695, EPI_ISL_1599696, EPI_ISL_1599697, EPI_ISL_1599698, EPI_ISL_1599699, EPI_ISL_1599700, EPI_ISL_1599701, EPI_ISL_1599702, EPI_ISL_1599703, EPI_ISL_1599704, EPI_ISL_1599705, EPI_ISL_1599706, EPI_ISL_1599707,
EPI_ISL_1599708, EPI_ISL_1599709, EPI_ISL_1599710, EPI_ISL_1599711, EPI_ISL_1599712, EPI_ISL_1599713, EPI_ISL_1599714, EPI_ISL_1599715, EPI_ISL_1599716, EPI_ISL_1599717, EPI_ISL_1599718, EPI_ISL_1599719, EPI_ISL_1599720, EPI_ISL_1599721, EPI_ISL_1599722, EPI_ISL_1599723, EPI_ISL_1599724, EPI_ISL_1599725, EPI_ISL_1599726, EPI_ISL_1599727, EPI_ISL_1599728, EPI_ISL_1599729,
EPI_ISL_1599730, EPI_ISL_1599731, EPI_ISL_1599732, EPI_ISL_1599733, EPI_ISL_1599734, EPI_ISL_1599735, EPI_ISL_1599736, EPI_ISL_1599737, EPI_ISL_1599738, EPI_ISL_1599739, EPI_ISL_1599740, EPI_ISL_1599742, EPI_ISL_1599743, EPI_ISL_1599744, EPI_ISL_1599745, EPI_ISL_1599746, EPI_ISL_1599747, EPI_ISL_1599748, EPI_ISL_1599749, EPI_ISL_1599750, EPI_ISL_1599751, EPI_ISL_1599752,
EPI_ISL_1599753, EPI_ISL_1599754, EPI_ISL_1599755, EPI_ISL_1599756, EPI_ISL_1599757, EPI_ISL_1599758, EPI_ISL_1599759, EPI_ISL_1599760, EPI_ISL_1599761, EPI_ISL_1599762, EPI_ISL_1599763, EPI_ISL_1599764, EPI_ISL_1599765, EPI_ISL_1599766, EPI_ISL_1599767, EPI_ISL_1599768, EPI_ISL_1599769, EPI_ISL_1599771, EPI_ISL_1599776, EPI_ISL_1599814, EPI_ISL_1599823, EPI_ISL_1599824,
EPI_ISL_1599825, EPI_ISL_1599826, EPI_ISL_1599827, EPI_ISL_1599828, EPI_ISL_1599829, EPI_ISL_1599836, EPI_ISL_1599837, EPI_ISL_1599838, EPI_ISL_1599839, EPI_ISL_1599868, EPI_ISL_1599869, EPI_ISL_1599870, EPI_ISL_1599871, EPI_ISL_1599872, EPI_ISL_1599873, EPI_ISL_1599874, EPI_ISL_1599875, EPI_ISL_1599876, EPI_ISL_1599877, EPI_ISL_1599878, EPI_ISL_1599879, EPI_ISL_1599880,
EPI_ISL_1599881, EPI_ISL_1599882, EPI_ISL_1599883, EPI_ISL_1599884, EPI_ISL_1599885, EPI_ISL_1599886, EPI_ISL_1599887, EPI_ISL_1599888, EPI_ISL_1599889, EPI_ISL_1599890, EPI_ISL_1599891, EPI_ISL_1599892, EPI_ISL_1599893, EPI_ISL_1599894, EPI_ISL_1599895, EPI_ISL_1599896, EPI_ISL_1599897, EPI_ISL_1599898, EPI_ISL_1599899, EPI_ISL_1599900, EPI_ISL_1599901, EPI_ISL_1599902,
EPI_ISL_1599907, EPI_ISL_1599908, EPI_ISL_1599909, EPI_ISL_1599910, EPI_ISL_1599911, EPI_ISL_1599912, EPI_ISL_1599913, EPI_ISL_1599914, EPI_ISL_1599915, EPI_ISL_1599916, EPI_ISL_1599917, EPI_ISL_1599918, EPI_ISL_1599919, EPI_ISL_1599920, EPI_ISL_1599921, EPI_ISL_1599922, EPI_ISL_1599923, EPI_ISL_1599924, EPI_ISL_1599925, EPI_ISL_1599926, EPI_ISL_1599927, EPI_ISL_1599928,
EPI_ISL_1599929, EPI_ISL_1599930, EPI_ISL_1599931, EPI_ISL_1599932, EPI_ISL_1599933, EPI_ISL_1599934, EPI_ISL_1599935, EPI_ISL_1599937, EPI_ISL_1599938, EPI_ISL_1599939, EPI_ISL_1599940, EPI_ISL_1599941, EPI_ISL_1599942, EPI_ISL_1599943, EPI_ISL_1599944, EPI_ISL_1599953, EPI_ISL_1599954, EPI_ISL_1599955, EPI_ISL_1599956, EPI_ISL_1599957, EPI_ISL_1599958, EPI_ISL_1599959,
EPI_ISL_1599960, EPI_ISL_1599961, EPI_ISL_1599962, EPI_ISL_1599963, EPI_ISL_1599964, EPI_ISL_1599965, EPI_ISL_1599966, EPI_ISL_1599967, EPI_ISL_1599968, EPI_ISL_1599969, EPI_ISL_1599970, EPI_ISL_1599971, EPI_ISL_1599972, EPI_ISL_1599973, EPI_ISL_1599974, EPI_ISL_1599975, EPI_ISL_1599976, EPI_ISL_1599977, EPI_ISL_1599978, EPI_ISL_1599979, EPI_ISL_1599980, EPI_ISL_1599981,
EPI_ISL_1599982, EPI_ISL_1599983, EPI_ISL_1599984, EPI_ISL_1599985, EPI_ISL_1599986, EPI_ISL_1599987, EPI_ISL_1599988, EPI_ISL_1599989, EPI_ISL_1599990, EPI_ISL_1599991, EPI_ISL_1599992, EPI_ISL_1599993, EPI_ISL_1599994, EPI_ISL_1599995, EPI_ISL_1599996, EPI_ISL_1599997, EPI_ISL_1599998, EPI_ISL_1599999, EPI_ISL_1600000, EPI_ISL_1600001, EPI_ISL_1600002, EPI_ISL_1600003,
EPI_ISL_1600004, EPI_ISL_1600005, EPI_ISL_1600006, EPI_ISL_1600007, EPI_ISL_1600008, EPI_ISL_1600009, EPI_ISL_1600010, EPI_ISL_1600011, EPI_ISL_1600012, EPI_ISL_1600013, EPI_ISL_1600014, EPI_ISL_1600015, EPI_ISL_1600016, EPI_ISL_1600017, EPI_ISL_1600018, EPI_ISL_1600019, EPI_ISL_1600020, EPI_ISL_1600021, EPI_ISL_1600022, EPI_ISL_1600023, EPI_ISL_1600024, EPI_ISL_1600025,
EPI_ISL_1600026, EPI_ISL_1600027, EPI_ISL_1600028, EPI_ISL_1600029, EPI_ISL_1600030, EPI_ISL_1600031, EPI_ISL_1600032, EPI_ISL_1600033, EPI_ISL_1600034, EPI_ISL_1600035, EPI_ISL_1600036, EPI_ISL_1600037, EPI_ISL_1600038, EPI_ISL_1600073, EPI_ISL_1600081, EPI_ISL_1600083, EPI_ISL_1600087, EPI_ISL_1600089, EPI_ISL_1600091, EPI_ISL_1600093, EPI_ISL_1600095, EPI_ISL_1600097,
EPI_ISL_1600099, EPI_ISL_1600102, EPI_ISL_1600104, EPI_ISL_1600106, EPI_ISL_1600108, EPI_ISL_1600110, EPI_ISL_1600112, EPI_ISL_1600114, EPI_ISL_1600116, EPI_ISL_1600118, EPI_ISL_1600120, EPI_ISL_1600122, EPI_ISL_1600124, EPI_ISL_1600126, EPI_ISL_1600128, EPI_ISL_1600130, EPI_ISL_1600132, EPI_ISL_1600207, EPI_ISL_1600208, EPI_ISL_1600209, EPI_ISL_1600210, EPI_ISL_1600211,
EPI_ISL_1600212, EPI_ISL_1600213, EPI_ISL_1600214, EPI_ISL_1600215, EPI_ISL_1600216, EPI_ISL_1600217, EPI_ISL_1600219, EPI_ISL_1600220, EPI_ISL_1600221, EPI_ISL_1600240, EPI_ISL_1600241, EPI_ISL_1600242, EPI_ISL_1600243, EPI_ISL_1600244, EPI_ISL_1600245, EPI_ISL_1600246, EPI_ISL_1600247, EPI_ISL_1600249, EPI_ISL_1600250, EPI_ISL_1600251, EPI_ISL_1600252, EPI_ISL_1600253,
EPI_ISL_1600254, EPI_ISL_1600255, EPI_ISL_1600256, EPI_ISL_1600257, EPI_ISL_1600258, EPI_ISL_1600260, EPI_ISL_1600261, EPI_ISL_1600262, EPI_ISL_1600263, EPI_ISL_1600264, EPI_ISL_1600265, EPI_ISL_1600266, EPI_ISL_1600267, EPI_ISL_1600268, EPI_ISL_1600272, EPI_ISL_1600275, EPI_ISL_1600276, EPI_ISL_1600277, EPI_ISL_1600278, EPI_ISL_1600279, EPI_ISL_1600280, EPI_ISL_1600281,
EPI_ISL_1600288, EPI_ISL_1600289, EPI_ISL_1600290, EPI_ISL_1600291, EPI_ISL_1600292, EPI_ISL_1600293, EPI_ISL_1600294, EPI_ISL_1600295, EPI_ISL_1600296, EPI_ISL_1600297, EPI_ISL_1600298, EPI_ISL_1600299, EPI_ISL_1600300, EPI_ISL_1600301, EPI_ISL_1600302, EPI_ISL_1600303, EPI_ISL_1600304, EPI_ISL_1600305, EPI_ISL_1600306, EPI_ISL_1600307, EPI_ISL_1600308, EPI_ISL_1600309,
EPI_ISL_1600310, EPI_ISL_1600311, EPI_ISL_1600312, EPI_ISL_1600313, EPI_ISL_1600314, EPI_ISL_1600315, EPI_ISL_1600316, EPI_ISL_1600317, EPI_ISL_1600318, EPI_ISL_1600319, EPI_ISL_1600320, EPI_ISL_1600321, EPI_ISL_1600322, EPI_ISL_1600323, EPI_ISL_1600324, EPI_ISL_1600325, EPI_ISL_1600326, EPI_ISL_1600327, EPI_ISL_1600328, EPI_ISL_1600330, EPI_ISL_1600331, EPI_ISL_1600332,
EPI_ISL_1600333, EPI_ISL_1600334, EPI_ISL_1600335, EPI_ISL_1600336, EPI_ISL_1600337, EPI_ISL_1600338, EPI_ISL_1600339, EPI_ISL_1600340, EPI_ISL_1600341, EPI_ISL_1600342, EPI_ISL_1600343, EPI_ISL_1600344, EPI_ISL_1600428, EPI_ISL_1600429, EPI_ISL_1600430, EPI_ISL_1600432, EPI_ISL_1600443, EPI_ISL_1600452, EPI_ISL_1600453, EPI_ISL_1600454, EPI_ISL_1600455, EPI_ISL_1600456,
EPI_ISL_1600457, EPI_ISL_1600458, EPI_ISL_1600459, EPI_ISL_1600499, EPI_ISL_1600500, EPI_ISL_1600501, EPI_ISL_1600502, EPI_ISL_1600503, EPI_ISL_1600504, EPI_ISL_1600505, EPI_ISL_1600506, EPI_ISL_1600507, EPI_ISL_1600541, EPI_ISL_1600542, EPI_ISL_1600578, EPI_ISL_1600579, EPI_ISL_1600580, EPI_ISL_1600581, EPI_ISL_1600587, EPI_ISL_1600588, EPI_ISL_1600589, EPI_ISL_1600590,
EPI_ISL_1600591, EPI_ISL_1600592, EPI_ISL_1600593, EPI_ISL_1600594, EPI_ISL_1600595, EPI_ISL_1600596, EPI_ISL_1600597, EPI_ISL_1600598, EPI_ISL_1600599, EPI_ISL_1600624, EPI_ISL_1600625, EPI_ISL_1600626, EPI_ISL_1600627, EPI_ISL_1600628, EPI_ISL_1600629, EPI_ISL_1617430, EPI_ISL_1617431, EPI_ISL_1617432, EPI_ISL_1617433, EPI_ISL_1617434, EPI_ISL_1617435, EPI_ISL_1617436,
EPI_ISL_1617437, EPI_ISL_1617438, EPI_ISL_1617439, EPI_ISL_1617440, EPI_ISL_1617441, EPI_ISL_1617442, EPI_ISL_1617443, EPI_ISL_1617444, EPI_ISL_1617445, EPI_ISL_1617446, EPI_ISL_1617447, EPI_ISL_1617448, EPI_ISL_1617449, EPI_ISL_1617450, EPI_ISL_1617451, EPI_ISL_1617452, EPI_ISL_1617453, EPI_ISL_1617454, EPI_ISL_1617455, EPI_ISL_1617456, EPI_ISL_1617457, EPI_ISL_1617458,
EPI_ISL_1617459, EPI_ISL_1617460, EPI_ISL_1617461, EPI_ISL_1617462, EPI_ISL_1617463, EPI_ISL_1617464, EPI_ISL_1617465, EPI_ISL_1617466, EPI_ISL_1617467, EPI_ISL_1617468, EPI_ISL_1617469, EPI_ISL_1617470, EPI_ISL_1617471, EPI_ISL_1617472, EPI_ISL_1617473, EPI_ISL_1617474, EPI_ISL_1617475, EPI_ISL_1617476, EPI_ISL_1617477, EPI_ISL_1617478, EPI_ISL_1617479, EPI_ISL_1617480,
EPI_ISL_1617481, EPI_ISL_1617482, EPI_ISL_1617483, EPI_ISL_1617484, EPI_ISL_1617485, EPI_ISL_1617486, EPI_ISL_1617487, EPI_ISL_1617488, EPI_ISL_1617489, EPI_ISL_1617490, EPI_ISL_1617491, EPI_ISL_1617492, EPI_ISL_1617493, EPI_ISL_1617494, EPI_ISL_1617495, EPI_ISL_1617496, EPI_ISL_1617497, EPI_ISL_1617498, EPI_ISL_1617499, EPI_ISL_1617500, EPI_ISL_1617501, EPI_ISL_1617502,
EPI_ISL_1617503, EPI_ISL_1617504, EPI_ISL_1617505, EPI_ISL_1617506, EPI_ISL_1617507, EPI_ISL_1617508, EPI_ISL_1617509, EPI_ISL_1617510, EPI_ISL_1617511, EPI_ISL_1617512, EPI_ISL_1617513, EPI_ISL_1617514, EPI_ISL_1617515, EPI_ISL_1617516, EPI_ISL_1617517, EPI_ISL_1617518, EPI_ISL_1617519, EPI_ISL_1617520, EPI_ISL_1617521, EPI_ISL_1617522, EPI_ISL_1617523, EPI_ISL_1617524,
EPI_ISL_1617525, EPI_ISL_1617526, EPI_ISL_1617527, EPI_ISL_1617528, EPI_ISL_1617529, EPI_ISL_1617530, EPI_ISL_1617531, EPI_ISL_1617532, EPI_ISL_1617533, EPI_ISL_1617534, EPI_ISL_1617535, EPI_ISL_1617536, EPI_ISL_1617537, EPI_ISL_1617538, EPI_ISL_1617539, EPI_ISL_1617540, EPI_ISL_1617541, EPI_ISL_1617542, EPI_ISL_1617543, EPI_ISL_1617544, EPI_ISL_1617545, EPI_ISL_1617546,
EPI_ISL_1617547, EPI_ISL_1617548, EPI_ISL_1617549, EPI_ISL_1617550, EPI_ISL_1617551, EPI_ISL_1617552, EPI_ISL_1617553, EPI_ISL_1617554, EPI_ISL_1617555, EPI_ISL_1617556, EPI_ISL_1617557, EPI_ISL_1617558, EPI_ISL_1617559, EPI_ISL_1617560, EPI_ISL_1617561, EPI_ISL_1617562, EPI_ISL_1617563, EPI_ISL_1617564, EPI_ISL_1617565, EPI_ISL_1617566, EPI_ISL_1617567, EPI_ISL_1617568,
EPI_ISL_1617569, EPI_ISL_1617570, EPI_ISL_1617571, EPI_ISL_1617572, EPI_ISL_1617573, EPI_ISL_1617574, EPI_ISL_1617575, EPI_ISL_1617576, EPI_ISL_1617577, EPI_ISL_1617578, EPI_ISL_1617579, EPI_ISL_1617580, EPI_ISL_1617581, EPI_ISL_1617626, EPI_ISL_1617627, EPI_ISL_1617628, EPI_ISL_1617629, EPI_ISL_1617630, EPI_ISL_1617631, EPI_ISL_1617632, EPI_ISL_1617633, EPI_ISL_1617634,
EPI_ISL_1617635, EPI_ISL_1617636, EPI_ISL_1617637, EPI_ISL_1617638, EPI_ISL_1617639, EPI_ISL_1617640, EPI_ISL_1617641, EPI_ISL_1617642, EPI_ISL_1617643, EPI_ISL_1617644, EPI_ISL_1617645, EPI_ISL_1617646, EPI_ISL_1617647, EPI_ISL_1617648, EPI_ISL_1617649, EPI_ISL_1617650, EPI_ISL_1617651, EPI_ISL_1617652, EPI_ISL_1617653, EPI_ISL_1617654, EPI_ISL_1617655, EPI_ISL_1617656,
EPI_ISL_1617657, EPI_ISL_1617658, EPI_ISL_1617659, EPI_ISL_1617660, EPI_ISL_1617661, EPI_ISL_1617662, EPI_ISL_1617663, EPI_ISL_1617664, EPI_ISL_1617665, EPI_ISL_1617666, EPI_ISL_1617667, EPI_ISL_1617668, EPI_ISL_1617669, EPI_ISL_1617670, EPI_ISL_1617671, EPI_ISL_1617672, EPI_ISL_1617673, EPI_ISL_1617674, EPI_ISL_1617675, EPI_ISL_1617676, EPI_ISL_1617677, EPI_ISL_1617678,
EPI_ISL_1617679, EPI_ISL_1617680, EPI_ISL_1617681, EPI_ISL_1617682, EPI_ISL_1617683, EPI_ISL_1617684, EPI_ISL_1617685, EPI_ISL_1617686, EPI_ISL_1617687, EPI_ISL_1617688, EPI_ISL_1617689, EPI_ISL_1617690, EPI_ISL_1617691, EPI_ISL_1617692, EPI_ISL_1617693, EPI_ISL_1617694, EPI_ISL_1617695, EPI_ISL_1617696, EPI_ISL_1617697, EPI_ISL_1617698, EPI_ISL_1617699, EPI_ISL_1617744,
EPI_ISL_1617783, EPI_ISL_1617908, EPI_ISL_1617909, EPI_ISL_1618082, EPI_ISL_1618083, EPI_ISL_1618084, EPI_ISL_1618085, EPI_ISL_1618086, EPI_ISL_1618087, EPI_ISL_1618088, EPI_ISL_1618089, EPI_ISL_1618090, EPI_ISL_1618091, EPI_ISL_1618092, EPI_ISL_1618093, EPI_ISL_1618094, EPI_ISL_1618095, EPI_ISL_1618096, EPI_ISL_1618097, EPI_ISL_1618098, EPI_ISL_1618099, EPI_ISL_1618100,
EPI_ISL_1618101, EPI_ISL_1618102, EPI_ISL_1618103, EPI_ISL_1618104, EPI_ISL_1618105, EPI_ISL_1618106, EPI_ISL_1618107, EPI_ISL_1618108, EPI_ISL_1618109, EPI_ISL_1618110, EPI_ISL_1618111, EPI_ISL_1618112, EPI_ISL_1618113, EPI_ISL_1618114, EPI_ISL_1618115, EPI_ISL_1618116, EPI_ISL_1618117, EPI_ISL_1618118, EPI_ISL_1618119, EPI_ISL_1618120, EPI_ISL_1618121, EPI_ISL_1618122,
EPI_ISL_1618123, EPI_ISL_1618124, EPI_ISL_1618125, EPI_ISL_1618126, EPI_ISL_1618127, EPI_ISL_1618128, EPI_ISL_1618129, EPI_ISL_1618130, EPI_ISL_1618131, EPI_ISL_1618132, EPI_ISL_1618133, EPI_ISL_1618134, EPI_ISL_1618135, EPI_ISL_1618136, EPI_ISL_1618137, EPI_ISL_1618138, EPI_ISL_1618139, EPI_ISL_1618140, EPI_ISL_1618141, EPI_ISL_1618142, EPI_ISL_1618143, EPI_ISL_1618144,
EPI_ISL_1618145, EPI_ISL_1618146, EPI_ISL_1618147, EPI_ISL_1618148, EPI_ISL_1618149, EPI_ISL_1618150, EPI_ISL_1618151, EPI_ISL_1618152, EPI_ISL_1618153, EPI_ISL_1618154, EPI_ISL_1618155, EPI_ISL_1618156, EPI_ISL_1618157, EPI_ISL_1618158, EPI_ISL_1618159, EPI_ISL_1618160, EPI_ISL_1618161, EPI_ISL_1618162, EPI_ISL_1618163, EPI_ISL_1618164, EPI_ISL_1618165, EPI_ISL_1618166,
EPI_ISL_1618167, EPI_ISL_1618168, EPI_ISL_1618169, EPI_ISL_1618170, EPI_ISL_1618171, EPI_ISL_1618172, EPI_ISL_1618173, EPI_ISL_1618174, EPI_ISL_1618175, EPI_ISL_1618176, EPI_ISL_1618177, EPI_ISL_1618178, EPI_ISL_1618179, EPI_ISL_1618180, EPI_ISL_1618181, EPI_ISL_1618182, EPI_ISL_1618183, EPI_ISL_1618184, EPI_ISL_1618185, EPI_ISL_1618186, EPI_ISL_1618187, EPI_ISL_1618188,
EPI_ISL_1618189, EPI_ISL_1618190, EPI_ISL_1618191, EPI_ISL_1618192, EPI_ISL_1618193, EPI_ISL_1618194, EPI_ISL_1618195, EPI_ISL_1618196, EPI_ISL_1618197, EPI_ISL_1618198, EPI_ISL_1618199, EPI_ISL_1618200, EPI_ISL_1618201, EPI_ISL_1618202, EPI_ISL_1618203, EPI_ISL_1618204, EPI_ISL_1618205, EPI_ISL_1618206, EPI_ISL_1618207, EPI_ISL_1618208, EPI_ISL_1618209, EPI_ISL_1618210,
EPI_ISL_1618211, EPI_ISL_1618212, EPI_ISL_1618213, EPI_ISL_1618214, EPI_ISL_1618215, EPI_ISL_1618216, EPI_ISL_1618217, EPI_ISL_1618218, EPI_ISL_1618219, EPI_ISL_1618220, EPI_ISL_1618221, EPI_ISL_1618222, EPI_ISL_1618223, EPI_ISL_1618273, EPI_ISL_1618274, EPI_ISL_1618275, EPI_ISL_1618276, EPI_ISL_1618277, EPI_ISL_1618278, EPI_ISL_1618279, EPI_ISL_1618280, EPI_ISL_1618281,
EPI_ISL_1618282, EPI_ISL_1618283, EPI_ISL_1618284, EPI_ISL_1618285, EPI_ISL_1618286, EPI_ISL_1618287, EPI_ISL_1618288, EPI_ISL_1618289, EPI_ISL_1618290, EPI_ISL_1618291, EPI_ISL_1618292, EPI_ISL_1618293, EPI_ISL_1618294, EPI_ISL_1618295, EPI_ISL_1618296, EPI_ISL_1618297, EPI_ISL_1618298, EPI_ISL_1618299, EPI_ISL_1618300, EPI_ISL_1618301, EPI_ISL_1618302, EPI_ISL_1618303,
EPI_ISL_1618304, EPI_ISL_1618305, EPI_ISL_1618306, EPI_ISL_1618307, EPI_ISL_1618308, EPI_ISL_1618309, EPI_ISL_1618310, EPI_ISL_1618311, EPI_ISL_1618312, EPI_ISL_1618313, EPI_ISL_1618314, EPI_ISL_1618315, EPI_ISL_1618316, EPI_ISL_1618317, EPI_ISL_1618318, EPI_ISL_1618319, EPI_ISL_1618320, EPI_ISL_1618321, EPI_ISL_1618322, EPI_ISL_1618323, EPI_ISL_1618324, EPI_ISL_1618325,
EPI_ISL_1618326, EPI_ISL_1618327, EPI_ISL_1618328, EPI_ISL_1618329, EPI_ISL_1618330, EPI_ISL_1618331, EPI_ISL_1618332, EPI_ISL_1618333, EPI_ISL_1618334, EPI_ISL_1618335, EPI_ISL_1618336, EPI_ISL_1618337, EPI_ISL_1618338, EPI_ISL_1618339, EPI_ISL_1618340, EPI_ISL_1618341, EPI_ISL_1618342, EPI_ISL_1618343, EPI_ISL_1618344, EPI_ISL_1618345, EPI_ISL_1618398, EPI_ISL_1618434,
EPI_ISL_1618435, EPI_ISL_1618436, EPI_ISL_1618437, EPI_ISL_1618438, EPI_ISL_1618439, EPI_ISL_1618440, EPI_ISL_1618441, EPI_ISL_1618442, EPI_ISL_1618443, EPI_ISL_1618444, EPI_ISL_1618445, EPI_ISL_1618446, EPI_ISL_1618447, EPI_ISL_1618448, EPI_ISL_1618449, EPI_ISL_1618450, EPI_ISL_1618451, EPI_ISL_1618452, EPI_ISL_1618453, EPI_ISL_1618454, EPI_ISL_1618455, EPI_ISL_1618456,
EPI_ISL_1618457, EPI_ISL_1618458, EPI_ISL_1618459, EPI_ISL_1618460, EPI_ISL_1618461, EPI_ISL_1618462, EPI_ISL_1618463, EPI_ISL_1618464, EPI_ISL_1618465, EPI_ISL_1618466, EPI_ISL_1618467, EPI_ISL_1618468, EPI_ISL_1618469, EPI_ISL_1618470, EPI_ISL_1618471, EPI_ISL_1618472, EPI_ISL_1618473, EPI_ISL_1618474, EPI_ISL_1618475, EPI_ISL_1618476, EPI_ISL_1618477, EPI_ISL_1618478,
EPI_ISL_1618479, EPI_ISL_1618480, EPI_ISL_1618481, EPI_ISL_1618482, EPI_ISL_1618483, EPI_ISL_1618484, EPI_ISL_1618485, EPI_ISL_1618486, EPI_ISL_1618487, EPI_ISL_1618488, EPI_ISL_1618489, EPI_ISL_1618490, EPI_ISL_1618491, EPI_ISL_1618492, EPI_ISL_1618493, EPI_ISL_1618494, EPI_ISL_1618495, EPI_ISL_1618496, EPI_ISL_1618497, EPI_ISL_1618498, EPI_ISL_1618499, EPI_ISL_1618500,
EPI_ISL_1618501, EPI_ISL_1618502, EPI_ISL_1618503, EPI_ISL_1618504, EPI_ISL_1618505, EPI_ISL_1618506, EPI_ISL_1618507, EPI_ISL_1618508, EPI_ISL_1618509, EPI_ISL_1618510, EPI_ISL_1618511, EPI_ISL_1618512, EPI_ISL_1618513, EPI_ISL_1618514, EPI_ISL_1618515, EPI_ISL_1618516, EPI_ISL_1618517, EPI_ISL_1618518, EPI_ISL_1618519, EPI_ISL_1618520, EPI_ISL_1618521, EPI_ISL_1618522,
EPI_ISL_1618523, EPI_ISL_1618524, EPI_ISL_1618525, EPI_ISL_1618550, EPI_ISL_1618554, EPI_ISL_1618641, EPI_ISL_1618642, EPI_ISL_1618643, EPI_ISL_1618644, EPI_ISL_1618645, EPI_ISL_1618646, EPI_ISL_1618647, EPI_ISL_1618648, EPI_ISL_1618652, EPI_ISL_1618653, EPI_ISL_1618657, EPI_ISL_1618658, EPI_ISL_1618659, EPI_ISL_1618660, EPI_ISL_1618661, EPI_ISL_1618662, EPI_ISL_1618663,
EPI_ISL_1618664, EPI_ISL_1618665, EPI_ISL_1618666, EPI_ISL_1618667, EPI_ISL_1618668, EPI_ISL_1618669, EPI_ISL_1618670, EPI_ISL_1618671, EPI_ISL_1618672, EPI_ISL_1618673, EPI_ISL_1618674, EPI_ISL_1618675, EPI_ISL_1618676, EPI_ISL_1618677, EPI_ISL_1618678, EPI_ISL_1618679, EPI_ISL_1618680, EPI_ISL_1618681, EPI_ISL_1618682, EPI_ISL_1618683, EPI_ISL_1618684, EPI_ISL_1618685,
EPI_ISL_1618686, EPI_ISL_1618687, EPI_ISL_1618688, EPI_ISL_1618689, EPI_ISL_1618690, EPI_ISL_1618691, EPI_ISL_1618692, EPI_ISL_1618693, EPI_ISL_1618694, EPI_ISL_1618695, EPI_ISL_1618696, EPI_ISL_1618697, EPI_ISL_1618698, EPI_ISL_1618699, EPI_ISL_1618700, EPI_ISL_1618701, EPI_ISL_1618702, EPI_ISL_1618703, EPI_ISL_1618704, EPI_ISL_1618705, EPI_ISL_1618706, EPI_ISL_1618707,
EPI_ISL_1618708, EPI_ISL_1618709, EPI_ISL_1618710, EPI_ISL_1618711, EPI_ISL_1618712, EPI_ISL_1618713, EPI_ISL_1618714, EPI_ISL_1618715, EPI_ISL_1618716, EPI_ISL_1618717, EPI_ISL_1618718, EPI_ISL_1618719, EPI_ISL_1618720, EPI_ISL_1618721, EPI_ISL_1618722, EPI_ISL_1618724, EPI_ISL_1618771, EPI_ISL_1618772, EPI_ISL_1618773, EPI_ISL_1618774, EPI_ISL_1618775, EPI_ISL_1618776,
EPI_ISL_1618777, EPI_ISL_1618778, EPI_ISL_1618779, EPI_ISL_1618780, EPI_ISL_1618781, EPI_ISL_1618782, EPI_ISL_1618783, EPI_ISL_1618784, EPI_ISL_1618785, EPI_ISL_1618786, EPI_ISL_1618787, EPI_ISL_1618788, EPI_ISL_1618789, EPI_ISL_1618790, EPI_ISL_1618791, EPI_ISL_1618792, EPI_ISL_1618793, EPI_ISL_1618794, EPI_ISL_1618795, EPI_ISL_1618796, EPI_ISL_1618797, EPI_ISL_1618798,
EPI_ISL_1618799, EPI_ISL_1618800, EPI_ISL_1618801, EPI_ISL_1618802, EPI_ISL_1618803, EPI_ISL_1618804, EPI_ISL_1618805, EPI_ISL_1618806, EPI_ISL_1618807, EPI_ISL_1618808, EPI_ISL_1618809, EPI_ISL_1618810, EPI_ISL_1618811, EPI_ISL_1618812, EPI_ISL_1618813, EPI_ISL_1618814, EPI_ISL_1618839, EPI_ISL_1618840, EPI_ISL_1618841, EPI_ISL_1618856, EPI_ISL_1618857, EPI_ISL_1618858,
EPI_ISL_1618859, EPI_ISL_1618860, EPI_ISL_1618861, EPI_ISL_1618862, EPI_ISL_1618863, EPI_ISL_1618865, EPI_ISL_1618866, EPI_ISL_1618867, EPI_ISL_1618868, EPI_ISL_1618869, EPI_ISL_1618870, EPI_ISL_1618871, EPI_ISL_1618872, EPI_ISL_1618873, EPI_ISL_1618874, EPI_ISL_1618875, EPI_ISL_1618876, EPI_ISL_1618877, EPI_ISL_1618878, EPI_ISL_1618879, EPI_ISL_1618880, EPI_ISL_1618881,
EPI_ISL_1618882, EPI_ISL_1618883, EPI_ISL_1618884, EPI_ISL_1618885, EPI_ISL_1618886, EPI_ISL_1618887, EPI_ISL_1618888, EPI_ISL_1618889, EPI_ISL_1618890, EPI_ISL_1618891, EPI_ISL_1618892, EPI_ISL_1618893, EPI_ISL_1618894, EPI_ISL_1618895, EPI_ISL_1618896, EPI_ISL_1618897, EPI_ISL_1618898, EPI_ISL_1618899, EPI_ISL_1618900, EPI_ISL_1618901, EPI_ISL_1618902, EPI_ISL_1618903,
EPI_ISL_1618904, EPI_ISL_1618905, EPI_ISL_1618906, EPI_ISL_1618907, EPI_ISL_1618908, EPI_ISL_1618909, EPI_ISL_1618910, EPI_ISL_1618911, EPI_ISL_1618912, EPI_ISL_1618913, EPI_ISL_1618914, EPI_ISL_1618915, EPI_ISL_1618916, EPI_ISL_1618917, EPI_ISL_1618918, EPI_ISL_1618919, EPI_ISL_1618920, EPI_ISL_1618921, EPI_ISL_1618922, EPI_ISL_1618923, EPI_ISL_1618924, EPI_ISL_1618925,
EPI_ISL_1618926, EPI_ISL_1618927, EPI_ISL_1618928, EPI_ISL_1618929, EPI_ISL_1618930, EPI_ISL_1618931, EPI_ISL_1618932, EPI_ISL_1618933, EPI_ISL_1618934, EPI_ISL_1618935, EPI_ISL_1618936, EPI_ISL_1618937, EPI_ISL_1618938, EPI_ISL_1618939, EPI_ISL_1618940, EPI_ISL_1618941, EPI_ISL_1618942, EPI_ISL_1618943, EPI_ISL_1618944, EPI_ISL_1618945, EPI_ISL_1618946, EPI_ISL_1618947,
EPI_ISL_1618948, EPI_ISL_1618949, EPI_ISL_1618950, EPI_ISL_1618951, EPI_ISL_1618952, EPI_ISL_1618953, EPI_ISL_1618954, EPI_ISL_1618955, EPI_ISL_1618956, EPI_ISL_1618957, EPI_ISL_1618958, EPI_ISL_1618959, EPI_ISL_1618960, EPI_ISL_1618961, EPI_ISL_1618962, EPI_ISL_1618963, EPI_ISL_1618964, EPI_ISL_1618965, EPI_ISL_1618966, EPI_ISL_1618967, EPI_ISL_1618968, EPI_ISL_1618969,
EPI_ISL_1618970, EPI_ISL_1618971, EPI_ISL_1618972, EPI_ISL_1618973, EPI_ISL_1618974, EPI_ISL_1618975, EPI_ISL_1618976, EPI_ISL_1618977, EPI_ISL_1618978, EPI_ISL_1618979, EPI_ISL_1618980, EPI_ISL_1618981, EPI_ISL_1618982, EPI_ISL_1618983, EPI_ISL_1618984, EPI_ISL_1618985, EPI_ISL_1618986, EPI_ISL_1618987, EPI_ISL_1618988, EPI_ISL_1618989, EPI_ISL_1618990, EPI_ISL_1618991,
EPI_ISL_1618992, EPI_ISL_1618993, EPI_ISL_1618994, EPI_ISL_1618995, EPI_ISL_1618996, EPI_ISL_1618997, EPI_ISL_1618998, EPI_ISL_1618999, EPI_ISL_1619000, EPI_ISL_1619001, EPI_ISL_1619002, EPI_ISL_1619003, EPI_ISL_1619004, EPI_ISL_1619005, EPI_ISL_1619006, EPI_ISL_1619007, EPI_ISL_1619008, EPI_ISL_1619009, EPI_ISL_1619010, EPI_ISL_1619011, EPI_ISL_1619012, EPI_ISL_1619013,
EPI_ISL_1619014, EPI_ISL_1619015, EPI_ISL_1619016, EPI_ISL_1619017, EPI_ISL_1619018, EPI_ISL_1619019, EPI_ISL_1619020, EPI_ISL_1619021, EPI_ISL_1619022, EPI_ISL_1619023, EPI_ISL_1619024, EPI_ISL_1619025, EPI_ISL_1619026, EPI_ISL_1619027, EPI_ISL_1619028, EPI_ISL_1619029, EPI_ISL_1619030, EPI_ISL_1619031, EPI_ISL_1619032, EPI_ISL_1619033, EPI_ISL_1619034, EPI_ISL_1619035,
EPI_ISL_1619036, EPI_ISL_1619037, EPI_ISL_1619038, EPI_ISL_1619039, EPI_ISL_1619040, EPI_ISL_1619041, EPI_ISL_1619042, EPI_ISL_1619043, EPI_ISL_1619044, EPI_ISL_1619045, EPI_ISL_1619046, EPI_ISL_1619047, EPI_ISL_1619048, EPI_ISL_1619049, EPI_ISL_1619050, EPI_ISL_1619051, EPI_ISL_1619052, EPI_ISL_1619053, EPI_ISL_1619054, EPI_ISL_1619055, EPI_ISL_1619056, EPI_ISL_1619057,
EPI_ISL_1619058, EPI_ISL_1619059, EPI_ISL_1619060, EPI_ISL_1619061, EPI_ISL_1619062, EPI_ISL_1619063, EPI_ISL_1619064, EPI_ISL_1619065, EPI_ISL_1619066, EPI_ISL_1619067, EPI_ISL_1619068, EPI_ISL_1619069, EPI_ISL_1619070, EPI_ISL_1619071, EPI_ISL_1619072, EPI_ISL_1619073, EPI_ISL_1619074, EPI_ISL_1619075, EPI_ISL_1619076, EPI_ISL_1619077, EPI_ISL_1619078, EPI_ISL_1619079,
EPI_ISL_1619080, EPI_ISL_1619081, EPI_ISL_1619082, EPI_ISL_1619083, EPI_ISL_1619084, EPI_ISL_1619085, EPI_ISL_1619086, EPI_ISL_1619087, EPI_ISL_1619088, EPI_ISL_1619089, EPI_ISL_1619090, EPI_ISL_1619091, EPI_ISL_1619092, EPI_ISL_1619093, EPI_ISL_1619094, EPI_ISL_1619095, EPI_ISL_1619096, EPI_ISL_1619097, EPI_ISL_1619098, EPI_ISL_1619099, EPI_ISL_1619100, EPI_ISL_1619101,
EPI_ISL_1619102, EPI_ISL_1619103, EPI_ISL_1619104, EPI_ISL_1619105, EPI_ISL_1619106, EPI_ISL_1619107, EPI_ISL_1619108, EPI_ISL_1619109, EPI_ISL_1619110, EPI_ISL_1619111, EPI_ISL_1619112, EPI_ISL_1619113, EPI_ISL_1619114, EPI_ISL_1619115, EPI_ISL_1619116, EPI_ISL_1619117, EPI_ISL_1619118, EPI_ISL_1619119, EPI_ISL_1619120, EPI_ISL_1619121, EPI_ISL_1619122, EPI_ISL_1619123,
EPI_ISL_1619124, EPI_ISL_1619125, EPI_ISL_1619126, EPI_ISL_1619127, EPI_ISL_1619128, EPI_ISL_1619129, EPI_ISL_1619130, EPI_ISL_1619131, EPI_ISL_1619132, EPI_ISL_1619133, EPI_ISL_1619134, EPI_ISL_1619135, EPI_ISL_1619136, EPI_ISL_1619137, EPI_ISL_1619138, EPI_ISL_1619139, EPI_ISL_1619140, EPI_ISL_1619141, EPI_ISL_1619142, EPI_ISL_1619143, EPI_ISL_1619144, EPI_ISL_1619145,
EPI_ISL_1619146, EPI_ISL_1619147, EPI_ISL_1619148, EPI_ISL_1619149, EPI_ISL_1619150, EPI_ISL_1619151, EPI_ISL_1619152, EPI_ISL_1619153, EPI_ISL_1619154, EPI_ISL_1619155, EPI_ISL_1619156, EPI_ISL_1619157, EPI_ISL_1619158, EPI_ISL_1619159, EPI_ISL_1619160, EPI_ISL_1619161, EPI_ISL_1619162, EPI_ISL_1619163, EPI_ISL_1619164, EPI_ISL_1619165, EPI_ISL_1619166, EPI_ISL_1619167,
EPI_ISL_1619168, EPI_ISL_1619169, EPI_ISL_1619170, EPI_ISL_1619171, EPI_ISL_1619172, EPI_ISL_1619173, EPI_ISL_1619174, EPI_ISL_1619175, EPI_ISL_1619176, EPI_ISL_1619177, EPI_ISL_1619178, EPI_ISL_1619179, EPI_ISL_1619180, EPI_ISL_1619181, EPI_ISL_1619182, EPI_ISL_1619183, EPI_ISL_1619184, EPI_ISL_1619185, EPI_ISL_1619186, EPI_ISL_1619187, EPI_ISL_1619188, EPI_ISL_1619189,
EPI_ISL_1619190, EPI_ISL_1619191, EPI_ISL_1619192, EPI_ISL_1619193, EPI_ISL_1619194, EPI_ISL_1619195, EPI_ISL_1619196, EPI_ISL_1619197, EPI_ISL_1619198, EPI_ISL_1619199, EPI_ISL_1619200, EPI_ISL_1619201, EPI_ISL_1619202, EPI_ISL_1619203, EPI_ISL_1619204, EPI_ISL_1619205, EPI_ISL_1619206, EPI_ISL_1619207, EPI_ISL_1619208, EPI_ISL_1619209, EPI_ISL_1619215, EPI_ISL_1619216,
EPI_ISL_1619229, EPI_ISL_1619491, EPI_ISL_1619492, EPI_ISL_1619493, EPI_ISL_1619494, EPI_ISL_1619495, EPI_ISL_1619496, EPI_ISL_1619497, EPI_ISL_1619498, EPI_ISL_1619499, EPI_ISL_1619500, EPI_ISL_1619501, EPI_ISL_1619502, EPI_ISL_1619503, EPI_ISL_1619504, EPI_ISL_1619505, EPI_ISL_1619506, EPI_ISL_1619507, EPI_ISL_1619508, EPI_ISL_1619509, EPI_ISL_1619510, EPI_ISL_1619511,
EPI_ISL_1619512, EPI_ISL_1619513, EPI_ISL_1619514, EPI_ISL_1619515, EPI_ISL_1619516, EPI_ISL_1619517, EPI_ISL_1619518, EPI_ISL_1619519, EPI_ISL_1619520, EPI_ISL_1619521, EPI_ISL_1619522, EPI_ISL_1619538, EPI_ISL_1619539, EPI_ISL_1619540, EPI_ISL_1619541, EPI_ISL_1619555, EPI_ISL_1619556, EPI_ISL_1619557, EPI_ISL_1619558, EPI_ISL_1619559, EPI_ISL_1619560, EPI_ISL_1619561,
EPI_ISL_1619562, EPI_ISL_1619563, EPI_ISL_1619564, EPI_ISL_1619565, EPI_ISL_1619566, EPI_ISL_1619567, EPI_ISL_1619568, EPI_ISL_1619569, EPI_ISL_1619570, EPI_ISL_1619571, EPI_ISL_1619572, EPI_ISL_1619573, EPI_ISL_1619574, EPI_ISL_1619575, EPI_ISL_1619576, EPI_ISL_1619577, EPI_ISL_1619578, EPI_ISL_1619579, EPI_ISL_1619580, EPI_ISL_1619581, EPI_ISL_1619582, EPI_ISL_1619583,

EPI_ISL_1619584, EPI_ISL_1619585, EPI_ISL_1619586, EPI_ISL_1619587, EPI_ISL_1619588, EPI_ISL_1619589, EPI_ISL_1619590, EPI_ISL_1619591, EPI_ISL_1619592, EPI_ISL_1619593, EPI_ISL_1619594, EPI_ISL_1619595, EPI_ISL_1619596, EPI_ISL_1619597, EPI_ISL_1619598, EPI_ISL_1619599, EPI_ISL_1619600, EPI_ISL_1619601, EPI_ISL_1619602, EPI_ISL_1619603, EPI_ISL_1619604, EPI_ISL_1619605, EPI_ISL_1619606, EPI_ISL_1619607, EPI_ISL_1619608, EPI_ISL_1619609, EPI_ISL_1619610, EPI_ISL_1619611, EPI_ISL_1619612, EPI_ISL_1619613, EPI_ISL_1619614, EPI_ISL_1619615, EPI_ISL_1619616, EPI_ISL_1619617, EPI_ISL_1619618, EPI_ISL_1619619, EPI_ISL_1619620, EPI_ISL_1619621, EPI_ISL_1619622, EPI_ISL_1619623, EPI_ISL_1619624, EPI_ISL_1619625, EPI_ISL_1619626, EPI_ISL_1619627, EPI_ISL_1619628, EPI_ISL_1619629, EPI_ISL_1619630, EPI_ISL_1619631, EPI_ISL_1619632, EPI_ISL_1619633, EPI_ISL_1619634, EPI_ISL_1619635, EPI_ISL_1619636, EPI_ISL_1619637, EPI_ISL_1619638, EPI_ISL_1619639, EPI_ISL_1619640, EPI_ISL_1619641, EPI_ISL_1619642, EPI_ISL_1619643, EPI_ISL_1619644, EPI_ISL_1619645, EPI_ISL_1619646, EPI_ISL_1619647, EPI_ISL_1619648, EPI_ISL_1619649, EPI_ISL_1619650, EPI_ISL_1619651, EPI_ISL_1619652, EPI_ISL_1619653, EPI_ISL_1619654, EPI_ISL_1619655, EPI_ISL_1619656, EPI_ISL_1619657, EPI_ISL_1619658, EPI_ISL_1619659, EPI_ISL_1619660, EPI_ISL_1619661, EPI_ISL_1619662, EPI_ISL_1619663, EPI_ISL_1619664, EPI_ISL_1619665, EPI_ISL_1619666, EPI_ISL_1619667, EPI_ISL_1619668, EPI_ISL_1619669, EPI_ISL_1619670, EPI_ISL_1619671, EPI_ISL_1619672, EPI_ISL_1619673, EPI_ISL_1619674, EPI_ISL_1619675, EPI_ISL_1619676, EPI_ISL_1619677, EPI_ISL_1619678, EPI_ISL_1619679, EPI_ISL_1619680, EPI_ISL_1619681, EPI_ISL_1619682, EPI_ISL_1619683, EPI_ISL_1619684, EPI_ISL_1619685, EPI_ISL_1619686, EPI_ISL_1619687, EPI_ISL_1619688, EPI_ISL_1619689, EPI_ISL_1619690, EPI_ISL_1619691, EPI_ISL_1619692, EPI_ISL_1619693, EPI_ISL_1619694, EPI_ISL_1619695, EPI_ISL_1619696, EPI_ISL_1619697, EPI_ISL_1619698, EPI_ISL_1619699, EPI_ISL_1619700, EPI_ISL_1619701, EPI_ISL_1619702, EPI_ISL_1619703, EPI_ISL_1619704, EPI_ISL_1619705, EPI_ISL_1619706, EPI_ISL_1619707, EPI_ISL_1619708, EPI_ISL_1619709, EPI_ISL_1619710, EPI_ISL_1619711, EPI_ISL_1619712, EPI_ISL_1619713, EPI_ISL_1619714, EPI_ISL_1619715, EPI_ISL_1619716, EPI_ISL_1619717, EPI_ISL_1619718, EPI_ISL_1619719, EPI_ISL_1619720, EPI_ISL_1619721, EPI_ISL_1619722, EPI_ISL_1619723, EPI_ISL_1619724, EPI_ISL_1619725, EPI_ISL_1619726, EPI_ISL_1619727, EPI_ISL_1619728, EPI_ISL_1619729, EPI_ISL_1619730, EPI_ISL_1619731, EPI_ISL_1619732, EPI_ISL_1619742, EPI_ISL_1619761, EPI_ISL_1619772, EPI_ISL_1619774, EPI_ISL_1619778, EPI_ISL_1659685, EPI_ISL_1659686, EPI_ISL_1659687, EPI_ISL_1659688, EPI_ISL_1659689, EPI_ISL_1659690, EPI_ISL_1659691, EPI_ISL_1659692, EPI_ISL_1659693, EPI_ISL_1659694, EPI_ISL_1659695, EPI_ISL_1659696, EPI_ISL_1659697, EPI_ISL_1659698, EPI_ISL_1659699, EPI_ISL_1659700, EPI_ISL_1659704, EPI_ISL_1659746, EPI_ISL_1659747, EPI_ISL_1659748, EPI_ISL_1659749, EPI_ISL_1659750, EPI_ISL_1659751, EPI_ISL_1659752, EPI_ISL_1659753, EPI_ISL_1659754, EPI_ISL_1659755, EPI_ISL_1659756, EPI_ISL_1659757, EPI_ISL_1659758, EPI_ISL_1659759, EPI_ISL_1659761, EPI_ISL_1659762, EPI_ISL_1659763, EPI_ISL_1659764, EPI_ISL_1659765, EPI_ISL_1659766, EPI_ISL_1659767, EPI_ISL_1659768, EPI_ISL_1659769, EPI_ISL_1659770, EPI_ISL_1659771, EPI_ISL_1659772, EPI_ISL_1659773, EPI_ISL_1659774, EPI_ISL_1659775, EPI_ISL_1659776, EPI_ISL_1659777, EPI_ISL_1659778, EPI_ISL_1659779, EPI_ISL_1659780, EPI_ISL_1659781, EPI_ISL_1659782, EPI_ISL_1659783, EPI_ISL_1659784, EPI_ISL_1659785, EPI_ISL_1659786, EPI_ISL_1659787, EPI_ISL_1659788, EPI_ISL_1659789, EPI_ISL_1659790, EPI_ISL_1659791, EPI_ISL_1659792, EPI_ISL_1659793, EPI_ISL_1659794, EPI_ISL_1659795, EPI_ISL_1659796, EPI_ISL_1659797, EPI_ISL_1659798, EPI_ISL_1659799, EPI_ISL_1659800, EPI_ISL_1659801, EPI_ISL_1659802, EPI_ISL_1659803, EPI_ISL_1659804, EPI_ISL_1659805, EPI_ISL_1659806, EPI_ISL_1659807, EPI_ISL_1659808, EPI_ISL_1659809, EPI_ISL_1659810, EPI_ISL_1659811, EPI_ISL_1659812, EPI_ISL_1659813, EPI_ISL_1659814, EPI_ISL_1659815, EPI_ISL_1659816, EPI_ISL_1659817, EPI_ISL_1659818, EPI_ISL_1659819, EPI_ISL_1659820, EPI_ISL_1659821, EPI_ISL_1659822, EPI_ISL_1659823, EPI_ISL_1659824, EPI_ISL_1659825, EPI_ISL_1659826, EPI_ISL_1659827, EPI_ISL_1659828, EPI_ISL_1659829, EPI_ISL_1659830, EPI_ISL_1659831, EPI_ISL_1659832, EPI_ISL_1659833, EPI_ISL_1659834, EPI_ISL_1659835, EPI_ISL_1659836, EPI_ISL_1659837, EPI_ISL_1659838, EPI_ISL_1659839, EPI_ISL_1659840, EPI_ISL_1659841, EPI_ISL_1659842, EPI_ISL_1659843, EPI_ISL_1659844, EPI_ISL_1659845, EPI_ISL_1659846, EPI_ISL_1659847, EPI_ISL_1659848, EPI_ISL_1659849, EPI_ISL_1659850, EPI_ISL_1659851, EPI_ISL_1659852, EPI_ISL_1659853, EPI_ISL_1659854, EPI_ISL_1659855, EPI_ISL_1659856, EPI_ISL_1659857, EPI_ISL_1659858, EPI_ISL_1659859, EPI_ISL_1659860, EPI_ISL_1659861, EPI_ISL_1659862, EPI_ISL_1659863, EPI_ISL_1659864, EPI_ISL_1659865, EPI_ISL_1659866, EPI_ISL_1659867, EPI_ISL_1659868, EPI_ISL_1659869, EPI_ISL_1659870, EPI_ISL_1659871, EPI_ISL_1659872, EPI_ISL_1659873, EPI_ISL_1659874, EPI_ISL_1659875, EPI_ISL_1659876, EPI_ISL_1659877, EPI_ISL_1659878, EPI_ISL_1659879, EPI_ISL_1659880, EPI_ISL_1659881, EPI_ISL_1659882, EPI_ISL_1659883, EPI_ISL_1659884, EPI_ISL_1659885, EPI_ISL_1659886, EPI_ISL_1659887, EPI_ISL_1659888, EPI_ISL_1659889, EPI_ISL_1659890, EPI_ISL_1659891, EPI_ISL_1659892, EPI_ISL_1659893, EPI_ISL_1659894, EPI_ISL_1659895, EPI_ISL_1659896, EPI_ISL_1659897, EPI_ISL_1659898, EPI_ISL_1659899, EPI_ISL_1659900, EPI_ISL_1659901, EPI_ISL_1659902, EPI_ISL_1659903, EPI_ISL_1659904, EPI_ISL_1659905, EPI_ISL_1659906, EPI_ISL_1659907, EPI_ISL_1659908, EPI_ISL_1659909, EPI_ISL_1659910, EPI_ISL_1659911, EPI_ISL_1659912, EPI_ISL_1659913, EPI_ISL_1659914, EPI_ISL_1659915, EPI_ISL_1659916, EPI_ISL_1659917, EPI_ISL_1659918, EPI_ISL_1659919, EPI_ISL_1659920, EPI_ISL_1659921, EPI_ISL_1659922, EPI_ISL_1659923, EPI_ISL_1659924, EPI_ISL_1659925, EPI_ISL_1659926, EPI_ISL_1659927, EPI_ISL_1659928, EPI_ISL_1659929, EPI_ISL_1659930, EPI_ISL_1659931, EPI_ISL_1659932, EPI_ISL_1659933, EPI_ISL_1659934, EPI_ISL_1659935, EPI_ISL_1659936, EPI_ISL_1659937, EPI_ISL_1659938, EPI_ISL_1659939, EPI_ISL_1659940, EPI_ISL_1659941, EPI_ISL_1659942, EPI_ISL_1659943, EPI_ISL_1659944, EPI_ISL_1659945, EPI_ISL_1659946, EPI_ISL_1659947, EPI_ISL_1659948, EPI_ISL_1659949, EPI_ISL_1659950, EPI_ISL_1659951, EPI_ISL_1659952, EPI_ISL_1659953, EPI_ISL_1659954, EPI_ISL_1659955, EPI_ISL_1659956, EPI_ISL_1659957, EPI_ISL_1659958, EPI_ISL_1659959, EPI_ISL_1659960, EPI_ISL_1659961, EPI_ISL_1659962, EPI_ISL_1659963, EPI_ISL_1659964, EPI_ISL_1659965, EPI_ISL_1659966, EPI_ISL_1659967, EPI_ISL_1659968, EPI_ISL_1659969, EPI_ISL_1659970, EPI_ISL_1659971, EPI_ISL_1659972, EPI_ISL_1659973, EPI_ISL_1659974, EPI_ISL_1659975, EPI_ISL_1659976, EPI_ISL_1659977, EPI_ISL_1659978, EPI_ISL_1659979, EPI_ISL_1659980, EPI_ISL_1659981, EPI_ISL_1659982, EPI_ISL_1659983, EPI_ISL_1659984, EPI_ISL_1659985, EPI_ISL_1659986, EPI_ISL_1659987, EPI_ISL_1659988, EPI_ISL_1659989, EPI_ISL_1659990, EPI_ISL_1659991, EPI_ISL_1676276, EPI_ISL_1747666, EPI_ISL_1747668, EPI_ISL_1747669, EPI_ISL_1747670, EPI_ISL_1747671, EPI_ISL_1747673, EPI_ISL_1747674, EPI_ISL_1747679, EPI_ISL_1747683, EPI_ISL_1747685, EPI_ISL_1747687, EPI_ISL_1747688, EPI_ISL_1747690, EPI_ISL_1747691, EPI_ISL_1747693, EPI_ISL_1747694, EPI_ISL_1747695, EPI_ISL_1747697, EPI_ISL_1747698, EPI_ISL_1747700, EPI_ISL_1747702, EPI_ISL_1747704, EPI_ISL_1747705, EPI_ISL_1747706, EPI_ISL_1747708, EPI_ISL_1747710, EPI_ISL_1747711, EPI_ISL_1747712, EPI_ISL_1747718, EPI_ISL_1747719, EPI_ISL_1747723, EPI_ISL_1747724, EPI_ISL_1747738, EPI_ISL_1747739, EPI_ISL_1747741, EPI_ISL_1747743, EPI_ISL_1747744, EPI_ISL_1747745, EPI_ISL_1747747, EPI_ISL_1747748, EPI_ISL_1747750, EPI_ISL_1747751, EPI_ISL_1747753, EPI_ISL_1747754, EPI_ISL_1747755, EPI_ISL_1747757, EPI_ISL_1747758, EPI_ISL_1747789, EPI_ISL_1747790, EPI_ISL_1747792, EPI_ISL_1747793, EPI_ISL_1747794, EPI_ISL_1747796, EPI_ISL_1747798, EPI_ISL_1747822, EPI_ISL_1747834, EPI_ISL_1747835, EPI_ISL_1747837, EPI_ISL_1747838, EPI_ISL_1747840, EPI_ISL_1747841, EPI_ISL_1747846, EPI_ISL_1747849, EPI_ISL_1747851, EPI_ISL_1747854, EPI_ISL_1747856, EPI_ISL_1747857, EPI_ISL_1747858, EPI_ISL_1747860, EPI_ISL_1747861, EPI_ISL_1747862, EPI_ISL_1747881, EPI_ISL_1747883, EPI_ISL_1747884, EPI_ISL_1747886, EPI_ISL_1747887, EPI_ISL_1747889, EPI_ISL_1747890, EPI_ISL_1747891, EPI_ISL_1747893, EPI_ISL_1747895, EPI_ISL_1747896, EPI_ISL_1747897, EPI_ISL_1747898, EPI_ISL_1747900, EPI_ISL_1747901, EPI_ISL_1747902, EPI_ISL_1747904, EPI_ISL_1747906, EPI_ISL_1747907, EPI_ISL_1747908, EPI_ISL_1747909, EPI_ISL_1747911, EPI_ISL_1747912, EPI_ISL_1747915, EPI_ISL_1747916, EPI_ISL_1747918, EPI_ISL_1747919, EPI_ISL_1747920, EPI_ISL_1747921, EPI_ISL_1747923, EPI_ISL_1747924, EPI_ISL_1747925, EPI_ISL_1747927, EPI_ISL_1747928, EPI_ISL_1747930, EPI_ISL_1747932, EPI_ISL_1747933, EPI_ISL_1747934, EPI_ISL_1747936, EPI_ISL_1747937, EPI_ISL_1747938, EPI_ISL_1747940, EPI_ISL_1747941, EPI_ISL_1747942, EPI_ISL_1747943, EPI_ISL_1747945, EPI_ISL_1747947, EPI_ISL_1747949, EPI_ISL_1747950, EPI_ISL_1747951, EPI_ISL_1747952, EPI_ISL_1747961, EPI_ISL_1747962, EPI_ISL_1747964, EPI_ISL_1747966, EPI_ISL_1747967, EPI_ISL_1747968, EPI_ISL_1747970, EPI_ISL_1747971, EPI_ISL_1747973, EPI_ISL_1747979, EPI_ISL_1747980, EPI_ISL_1747982, EPI_ISL_1747989, EPI_ISL_1747991, EPI_ISL_1748030, EPI_ISL_1748031, EPI_ISL_1748033, EPI_ISL_1748034, EPI_ISL_1748035, EPI_ISL_1748051, EPI_ISL_1748054, EPI_ISL_1748062, EPI_ISL_1748063, EPI_ISL_1748066, EPI_ISL_1748068, EPI_ISL_1748101, EPI_ISL_1748123, EPI_ISL_1748124, EPI_ISL_1748125, EPI_ISL_1748127, EPI_ISL_1748128, EPI_ISL_1748129, EPI_ISL_1748131, EPI_ISL_1748143, EPI_ISL_1748144, EPI_ISL_1748145, EPI_ISL_1748214, EPI_ISL_1748216, EPI_ISL_1748218, EPI_ISL_1748220, EPI_ISL_1748221, EPI_ISL_1807652, EPI_ISL_1807653, EPI_ISL_1807654, EPI_ISL_1807655, EPI_ISL_1807656, EPI_ISL_1807657, EPI_ISL_1807658, EPI_ISL_1807659, EPI_ISL_1807662, EPI_ISL_1807663, EPI_ISL_1807664, EPI_ISL_1807665, EPI_ISL_1807666, EPI_ISL_1807667, EPI_ISL_1807668, EPI_ISL_1807669, EPI_ISL_1807670, EPI_ISL_1807671, EPI_ISL_1807672, EPI_ISL_1807673, EPI_ISL_1807674, EPI_ISL_1807675, EPI_ISL_1807676, EPI_ISL_1807677, EPI_ISL_1807678, EPI_ISL_1807679, EPI_ISL_1807680, EPI_ISL_1807681, EPI_ISL_1807682, EPI_ISL_1807683, EPI_ISL_1807684, EPI_ISL_1807685, EPI_ISL_1807686, EPI_ISL_1807687, EPI_ISL_1807688, EPI_ISL_1807689, EPI_ISL_1807690, EPI_ISL_1807691, EPI_ISL_1807692, EPI_ISL_1807693, EPI_ISL_1807694, EPI_ISL_1807695, EPI_ISL_1807696, EPI_ISL_1807697, EPI_ISL_1807698, EPI_ISL_1807699, EPI_ISL_1807700, EPI_ISL_1807701, EPI_ISL_1807702, EPI_ISL_1807703, EPI_ISL_1807704, EPI_ISL_1807705, EPI_ISL_1807706, EPI_ISL_1807707, EPI_ISL_1807708, EPI_ISL_1807709, EPI_ISL_1807710, EPI_ISL_1807711, EPI_ISL_1807712, EPI_ISL_1807713, EPI_ISL_1807714, EPI_ISL_1807715, EPI_ISL_1807716, EPI_ISL_1807717, EPI_ISL_1807719, EPI_ISL_1807720, EPI_ISL_1807721, EPI_ISL_1807722, EPI_ISL_1807723, EPI_ISL_1807724, EPI_ISL_1807725, EPI_ISL_1807726, EPI_ISL_1807727, EPI_ISL_1807728, EPI_ISL_1807729, EPI_ISL_1807730, EPI_ISL_1807731, EPI_ISL_1807732, EPI_ISL_1807733, EPI_ISL_1807734, EPI_ISL_1807735, EPI_ISL_1807736, EPI_ISL_1807737, EPI_ISL_1807738, EPI_ISL_1807739, EPI_ISL_1807740, EPI_ISL_1807741, EPI_ISL_1807742, EPI_ISL_1807743, EPI_ISL_1807744, EPI_ISL_1807745, EPI_ISL_1807746, EPI_ISL_1807747, EPI_ISL_1807748, EPI_ISL_1807749, EPI_ISL_1807750, EPI_ISL_1807751, EPI_ISL_1809900, EPI_ISL_1809901, EPI_ISL_1809902, EPI_ISL_1809903, EPI_ISL_1809904, EPI_ISL_1809905, EPI_ISL_1809906, EPI_ISL_1809907, EPI_ISL_1809908, EPI_ISL_1809909, EPI_ISL_1809910, EPI_ISL_1809911, EPI_ISL_1809912, EPI_ISL_1809914, EPI_ISL_1809915, EPI_ISL_1809916, EPI_ISL_1809917, EPI_ISL_1809918, EPI_ISL_1809919, EPI_ISL_1809920, EPI_ISL_1809921, EPI_ISL_1809922, EPI_ISL_1809923, EPI_ISL_1809924, EPI_ISL_1809925, EPI_ISL_1809926, EPI_ISL_1809927, EPI_ISL_1809928, EPI_ISL_1809929, EPI_ISL_1809930, EPI_ISL_1809931, EPI_ISL_1809932, EPI_ISL_1809933, EPI_ISL_1809934, EPI_ISL_1809935, EPI_ISL_1809937, EPI_ISL_1809938, EPI_ISL_1809939, EPI_ISL_1809940, EPI_ISL_1809941, EPI_ISL_1809942, EPI_ISL_1809943, EPI_ISL_1809944, EPI_ISL_1809945, EPI_ISL_1809946, EPI_ISL_1809947, EPI_ISL_1809948, EPI_ISL_1809949, EPI_ISL_1809950, EPI_ISL_1809951, EPI_ISL_1809952, EPI_ISL_1809953, EPI_ISL_1809954, EPI_ISL_1809955, EPI_ISL_1809956, EPI_ISL_1809957, EPI_ISL_1809958, EPI_ISL_1809959, EPI_ISL_1809960, EPI_ISL_1809961, EPI_ISL_1809962, EPI_ISL_1809963, EPI_ISL_1809964, EPI_ISL_1809965, EPI_ISL_1809967, EPI_ISL_1809968, EPI_ISL_1809969, EPI_ISL_1809970, EPI_ISL_1809971, EPI_ISL_1809972, EPI_ISL_1809973, EPI_ISL_1809974, EPI_ISL_1809975, EPI_ISL_1809976, EPI_ISL_1809977, EPI_ISL_1809978, EPI_ISL_1809979, EPI_ISL_1809980, EPI_ISL_1809981, EPI_ISL_1809982, EPI_ISL_1809983, EPI_ISL_1809984, EPI_ISL_1809985, EPI_ISL_1809986, EPI_ISL_1809987, EPI_ISL_1809988, EPI_ISL_1809989, EPI_ISL_1809990, EPI_ISL_1831699, EPI_ISL_1831700, EPI_ISL_1831701, EPI_ISL_1831702, EPI_ISL_1831703, EPI_ISL_1831704, EPI_ISL_1831705, EPI_ISL_1831706, EPI_ISL_1831707, EPI_ISL_1831708, EPI_ISL_1831709, EPI_ISL_1831710, EPI_ISL_1831711, EPI_ISL_1831712, EPI_ISL_1831713, EPI_ISL_1831714, EPI_ISL_1831715, EPI_ISL_1831716, EPI_ISL_1831717, EPI_ISL_1831718, EPI_ISL_1831719, EPI_ISL_1831720, EPI_ISL_1831721, EPI_ISL_1831722, EPI_ISL_1831723, EPI_ISL_1831724, EPI_ISL_1831725, EPI_ISL_1831726, EPI_ISL_1831727, EPI_ISL_1831728, EPI_ISL_1831730, EPI_ISL_1831731, EPI_ISL_1831732, EPI_ISL_1831733, EPI_ISL_1831734, EPI_ISL_1831735, EPI_ISL_1831736, EPI_ISL_1831737, EPI_ISL_1831738, EPI_ISL_1831739, EPI_ISL_1831740, EPI_ISL_1831741, EPI_ISL_1831742, EPI_ISL_1831743, EPI_ISL_1831744, EPI_ISL_1831745, EPI_ISL_1831746, EPI_ISL_1831747, EPI_ISL_1831748, EPI_ISL_1831749, EPI_ISL_1831750, EPI_ISL_1831751, EPI_ISL_1831752, EPI_ISL_1831753, EPI_ISL_1831754, EPI_ISL_1831755, EPI_ISL_1831756, EPI_ISL_1831757, EPI_ISL_1831758, EPI_ISL_1831759, EPI_ISL_1831760, EPI_ISL_1831761, EPI_ISL_1831762, EPI_ISL_1831763, EPI_ISL_1831764, EPI_ISL_1831765, EPI_ISL_1831766, EPI_ISL_1831767, EPI_ISL_1831768, EPI_ISL_1831769, EPI_ISL_1831770, EPI_ISL_1831771, EPI_ISL_1831772, EPI_ISL_1831773, EPI_ISL_1831774, EPI_ISL_1831775, EPI_ISL_1831776, EPI_ISL_1831777, EPI_ISL_1831778, EPI_ISL_1831779, EPI_ISL_1831780, EPI_ISL_1831781, EPI_ISL_1831782, EPI_ISL_1831783, EPI_ISL_1831784, EPI_ISL_1831785, EPI_ISL_1831786, EPI_ISL_1831787, EPI_ISL_1831788, EPI_ISL_1831789, EPI_ISL_1831790, EPI_ISL_1831791, EPI_ISL_1831792, EPI_ISL_1831793, EPI_ISL_1831794, EPI_ISL_1831795, EPI_ISL_1831796, EPI_ISL_1831797, EPI_ISL_1831798, EPI_ISL_1831799, EPI_ISL_1831800, EPI_ISL_1831801, EPI_ISL_1831802, EPI_ISL_1831803, EPI_ISL_1831804, EPI_ISL_1831805, EPI_ISL_1831806, EPI_ISL_1831807, EPI_ISL_1831808, EPI_ISL_1831809, EPI_ISL_1831810, EPI_ISL_1831811, EPI_ISL_1831812, EPI_ISL_1831813, EPI_ISL_1831814, EPI_ISL_1831815, EPI_ISL_1831816, EPI_ISL_1831817, EPI_ISL_1831818, EPI_ISL_1831819, EPI_ISL_1831820, EPI_ISL_1831821, EPI_ISL_1831822, EPI_ISL_1831823, EPI_ISL_1831824, EPI_ISL_1831825, EPI_ISL_1831826, EPI_ISL_1831827, EPI_ISL_1831828, EPI_ISL_1831829, EPI_ISL_1831831, EPI_ISL_1831832, EPI_ISL_1831833, EPI_ISL_1831834, EPI_ISL_1831835, EPI_ISL_1831836, EPI_ISL_1831837, EPI_ISL_1831838, EPI_ISL_1831839, EPI_ISL_1831840, EPI_ISL_1831841, EPI_ISL_1831842, EPI_ISL_1831843, EPI_ISL_1831844, EPI_ISL_1831845, EPI_ISL_1831846, EPI_ISL_1831847, EPI_ISL_1831848, EPI_ISL_1831849, EPI_ISL_1831850, EPI_ISL_1831851, EPI_ISL_1831852, EPI_ISL_1831853, EPI_ISL_1831854, EPI_ISL_1831855, EPI_ISL_1831856, EPI_ISL_1831857, EPI_ISL_1831858, EPI_ISL_1831859, EPI_ISL_1831860, EPI_ISL_1831861, EPI_ISL_1831862, EPI_ISL_1831863, EPI_ISL_1831864, EPI_ISL_1831865, EPI_ISL_1831866, EPI_ISL_1831867, EPI_ISL_1831868, EPI_ISL_1831869, EPI_ISL_1831870, EPI_ISL_1831872, EPI_ISL_1831873, EPI_ISL_1831874, EPI_ISL_1831875, EPI_ISL_1831876, EPI_ISL_1831877, EPI_ISL_1831878, EPI_ISL_1831879, EPI_ISL_1831880, EPI_ISL_1831881, EPI_ISL_1831882, EPI_ISL_1831883, EPI_ISL_1831884, EPI_ISL_1831885, EPI_ISL_1831886, EPI_ISL_1831887, EPI_ISL_1831888, EPI_ISL_1831889, EPI_ISL_1831890, EPI_ISL_1831891, EPI_ISL_1831892, EPI_ISL_1831893, EPI_ISL_1831894, EPI_ISL_1831895, EPI_ISL_1831896, EPI_ISL_1831897, EPI_ISL_1831898, EPI_ISL_1831899, EPI_ISL_1831900, EPI_ISL_1831901, EPI_ISL_1831902, EPI_ISL_1831903, EPI_ISL_1831904, EPI_ISL_1831905, EPI_ISL_1831906, EPI_ISL_1831907, EPI_ISL_1831908, EPI_ISL_1831909, EPI_ISL_1831910, EPI_ISL_1831911, EPI_ISL_1831912, EPI_ISL_1831913, EPI_ISL_1831914, EPI_ISL_1831915, EPI_ISL_1831916, EPI_ISL_1831917, EPI_ISL_1831918, EPI_ISL_1831919, EPI_ISL_1831920, EPI_ISL_1831921, EPI_ISL_1831922, EPI_ISL_1831923, EPI_ISL_1831924, EPI_ISL_1831925, EPI_ISL_1831926, EPI_ISL_1831927, EPI_ISL_1831928, EPI_ISL_1831929, EPI_ISL_1831930, EPI_ISL_1831931, EPI_ISL_1831932, EPI_ISL_1831933, EPI_ISL_1831934, EPI_ISL_1831935, EPI_ISL_1831936, EPI_ISL_1831937, EPI_ISL_1831938, EPI_ISL_1831939, EPI_ISL_1831940, EPI_ISL_1831941, EPI_ISL_1831942, EPI_ISL_1831943, EPI_ISL_1831945, EPI_ISL_1831947, EPI_ISL_1831949, EPI_ISL_1831951, EPI_ISL_1831953, EPI_ISL_1831955, EPI_ISL_1831957, EPI_ISL_1831959, EPI_ISL_1831961, EPI_ISL_1831963, EPI_ISL_1831965, EPI_ISL_1831967, EPI_ISL_1831969, EPI_ISL_1831970, EPI_ISL_1831972, EPI_ISL_1831974, EPI_ISL_1831976, EPI_ISL_1831978, EPI_ISL_1831980, EPI_ISL_1831982, EPI_ISL_1831984, EPI_ISL_1831986, EPI_ISL_1831989, EPI_ISL_1831990, EPI_ISL_1831992, EPI_ISL_1831994, EPI_ISL_1831996, EPI_ISL_1831998, EPI_ISL_1832000, EPI_ISL_1832002, EPI_ISL_1832004, EPI_ISL_1832006, EPI_ISL_1832008, EPI_ISL_1832010, EPI_ISL_1832012, EPI_ISL_1832014, EPI_ISL_1832016, EPI_ISL_1832018, EPI_ISL_1832020, EPI_ISL_1832022, EPI_ISL_1832024, EPI_ISL_1832026, EPI_ISL_1832028, EPI_ISL_1832030, EPI_ISL_1832032, EPI_ISL_1832033, EPI_ISL_1832035, EPI_ISL_1832037, EPI_ISL_1832039, EPI_ISL_1832041, EPI_ISL_1832043, EPI_ISL_1832045, EPI_ISL_1832047, EPI_ISL_1832048, EPI_ISL_1832050, EPI_ISL_1832052, EPI_ISL_1832054, EPI_ISL_1832056, EPI_ISL_1832058, EPI_ISL_1832060, EPI_ISL_1832062, EPI_ISL_1832064, EPI_ISL_1832066, EPI_ISL_1832068, EPI_ISL_1832070, EPI_ISL_1832072, EPI_ISL_1832073, EPI_ISL_1832074, EPI_ISL_1832076, EPI_ISL_1832078, EPI_ISL_1832080, EPI_ISL_1832082, EPI_ISL_1832084, EPI_ISL_1832086, EPI_ISL_1832088, EPI_ISL_1832090, EPI_ISL_1832092, EPI_ISL_1832094, EPI_ISL_1832096, EPI_ISL_1832098, EPI_ISL_1832100, EPI_ISL_1832101, EPI_ISL_1832103, EPI_ISL_1832105, EPI_ISL_1832107, EPI_ISL_1832109, EPI_ISL_1832112, EPI_ISL_1832113, EPI_ISL_1832115, EPI_ISL_1832118, EPI_ISL_1832120, EPI_ISL_1832122, EPI_ISL_1832123, EPI_ISL_1832124, EPI_ISL_1832126, EPI_ISL_1832128, EPI_ISL_1832130, EPI_ISL_1832132, EPI_ISL_1832134, EPI_ISL_1832136, EPI_ISL_1832138, EPI_ISL_1832140, EPI_ISL_1832142, EPI_ISL_1832144, EPI_ISL_1832146, EPI_ISL_1832148, EPI_ISL_1832150, EPI_ISL_1832152, EPI_ISL_1832154, EPI_ISL_1832156, EPI_ISL_1832158, EPI_ISL_1832160, EPI_ISL_1832162, EPI_ISL_1832164, EPI_ISL_1832166, EPI_ISL_1832168, EPI_ISL_1832170, EPI_ISL_1832171, EPI_ISL_1832172, EPI_ISL_1832173, EPI_ISL_1832174, EPI_ISL_1832175, EPI_ISL_1832177, EPI_ISL_1832179, EPI_ISL_1832181, EPI_ISL_1832183, EPI_ISL_1832185, EPI_ISL_1832186, EPI_ISL_1832188, EPI_ISL_1832190, EPI_ISL_1832192, EPI_ISL_1832792, EPI_ISL_1899503, EPI_ISL_1900104, EPI_ISL_1900106, EPI_ISL_1900107, EPI_ISL_1900563, EPI_ISL_1900564, EPI_ISL_1900565, EPI_ISL_1900570, EPI_ISL_1900571, EPI_ISL_1900572, EPI_ISL_1900573, EPI_ISL_1900574, EPI_ISL_1900575, EPI_ISL_1900576, EPI_ISL_1900577, EPI_ISL_1900578, EPI_ISL_1900579, EPI_ISL_1900580, EPI_ISL_1900581, EPI_ISL_1900582, EPI_ISL_1900585, EPI_ISL_1900605, EPI_ISL_2098950, EPI_ISL_2098951, EPI_ISL_2098952, EPI_ISL_2098953, EPI_ISL_2098954, EPI_ISL_2098955, EPI_ISL_2099017, EPI_ISL_2099022, EPI_ISL_2099039, EPI_ISL_2099057, EPI_ISL_2099066, EPI_ISL_2099070, EPI_ISL_2099074, EPI_ISL_2099077, EPI_ISL_2099090, EPI_ISL_2099091, EPI_ISL_2099101, EPI_ISL_2099106, EPI_ISL_2099109, EPI_ISL_2099126, EPI_ISL_2099139, EPI_ISL_2099154, EPI_ISL_2099171

| | | | |
|---|---|---|---|
| see above | Swedish national genomic surveillance program of SARS-CoV-2 | The Public Health Agency of Sweden | Alma Brolund; Maria Lind Karlberg; Maximilian Riess; Swedish national genomic surveillance program of SARS-CoV-2 |

EPI_ISL_913256, EPI_ISL_913257, EPI_ISL_913276, EPI_ISL_913284, EPI_ISL_913301, EPI_ISL_913303, EPI_ISL_913304, EPI_ISL_913305, EPI_ISL_913456, EPI_ISL_913466, EPI_ISL_913467, EPI_ISL_934364, EPI_ISL_934365, EPI_ISL_934366, EPI_ISL_934367, EPI_ISL_934368, EPI_ISL_934369, EPI_ISL_934370, EPI_ISL_934371, EPI_ISL_934372, EPI_ISL_934373, EPI_ISL_934374, EPI_ISL_934397, EPI_ISL_934398, EPI_ISL_934399, EPI_ISL_934400, EPI_ISL_934401, EPI_ISL_934402, EPI_ISL_934403, EPI_ISL_934404, EPI_ISL_934410, EPI_ISL_934411, EPI_ISL_934412, EPI_ISL_1008451, EPI_ISL_1008452, EPI_ISL_1008453, EPI_ISL_1008454, EPI_ISL_1008596, EPI_ISL_1064623, EPI_ISL_1168037, EPI_ISL_1168047

| | | | |
|---|---|---|---|
| see above | Synlab Medilab, Mikrobiologi | The Public Health Agency of Sweden | Anna Risberg; Anna-Malin Linde; Carlo Berg; Karin Tegmark-Wisell; Maria Lind Karlberg; Mattias Haukland; Mia Brytting; Noura Walai; Oskar Karlsson Lindsjo; Petra Edquist; Petra Holmstrom; Reza Advani; Samuel Ohman; Sofia Stamouli |
| EPI_ISL_1008558 | TATAA Biocenter | The Public Health Agency of Sweden | Anna Risberg; Anna-Malin Linde; Carlo Berg; Karin Tegmark-Wisell; Maria Lind Karlberg; Mattias Haukland; Mia Brytting; Noura Walai; Oskar Karlsson Lindsjo; Petra Edquist; Petra Holmstrom; Reza Advani; Sofia Stamouli |

EPI_ISL_913263, EPI_ISL_913267, EPI_ISL_913286, EPI_ISL_913314, EPI_ISL_913375, EPI_ISL_913376, EPI_ISL_913377, EPI_ISL_913451, EPI_ISL_913452, EPI_ISL_913464, EPI_ISL_913468, EPI_ISL_913494, EPI_ISL_934406, EPI_ISL_1064682, EPI_ISL_1064683, EPI_ISL_1064684, EPI_ISL_1064708

| | | | |
|---|---|---|---|
| see above | The Public Health Agency of Sweden | The Public Health Agency of Sweden | Anna Risberg; Anna-Malin Linde; Carlo Berg; Karin Tegmark-Wisell; Maria Lind Karlberg; Mattias Haukland; Mia Brytting; Noura Walai; Oskar Karlsson Lindsjo; Petra Edquist; Petra Holmstrom; Reza Advani; Samuel Ohman; Sofia Stamouli |
| EPI_ISL_789054 | Unilabs AB | The Public Health Agency of Sweden | Department of Microbiology; The Public Health Agency of Sweden |

EPI_ISL_832006, EPI_ISL_832007, EPI_ISL_913266, EPI_ISL_913337, EPI_ISL_913338, EPI_ISL_913450, EPI_ISL_913455

| | | | |
|---|---|---|---|
| see above | Unilabs AB Skovde | The Public Health Agency of Sweden | Anna Risberg; Anna-Malin Linde; Carlo Berg; Department of Microbiology; Karin Tegmark-Wisell; Maria Lind Karlberg; Mattias Haukland; Mia Brytting; Noura Walai; Oskar Karlsson Lindsjo; Petra Edquist; Petra Holmstrom; Reza Advani; Sofia Stamouli; The Public Health Agency of Sweden |

EPI_ISL_1008446, EPI_ISL_1008540, EPI_ISL_1008541, EPI_ISL_1008542, EPI_ISL_1008543, EPI_ISL_1008544, EPI_ISL_1008545, EPI_ISL_1008546, EPI_ISL_1008547, EPI_ISL_1008549, EPI_ISL_1008550, EPI_ISL_1008551, EPI_ISL_1008557, EPI_ISL_1064575, EPI_ISL_1190868, EPI_ISL_1190880

| | | | |
|---|---|---|---|
| see above | Unilabs AB, Klinisk mikrobiologi | The Public Health Agency of Sweden | Anna Risberg; Anna-Malin Linde; Carlo Berg; Karin Tegmark-Wisell; Maria Lind Karlberg; Mattias Haukland; Mia Brytting; Noura Walai; Oskar Karlsson Lindsjo; Petra Edquist; Petra Holmstrom; Reza Advani; Samuel Ohman; Sofia Stamouli |
| EPI_ISL_1064550, EPI_ISL_1064677 | Unilabs AB, Klinisk mikrobiologi, | The Public Health Agency of Sweden | Anna Risberg; Anna-Malin Linde; Carlo Berg; Karin Tegmark-Wisell; Maria Lind Karlberg; Mattias Haukland; Mia Brytting; Noura Walai; Oskar Karlsson Lindsjo; Petra Edquist; Petra Holmstrom; Reza Advani; Samuel Ohman; Sofia Stamouli |
| EPI_ISL_1064685, EPI_ISL_1064686, EPI_ISL_1064687, EPI_ISL_1064688, EPI_ISL_1064689, EPI_ISL_1064690 | Unilabs Laboratoriemedicin | The Public Health Agency of Sweden | Anna Risberg; Anna-Malin Linde; Carlo Berg; Karin Tegmark-Wisell; Maria Lind Karlberg; Mattias Haukland; Mia Brytting; Noura Walai; Oskar Karlsson Lindsjo; Petra Edquist; Petra Holmstrom; Reza Advani; Samuel Ohman; Sofia Stamouli |
| EPI_ISL_831943, EPI_ISL_832008, EPI_ISL_913339, EPI_ISL_913340, EPI_ISL_913341, EPI_ISL_913342 | Unilabs, Mikrobiologiska laboratoriet | The Public Health Agency of Sweden | Anna Risberg; Anna-Malin Linde; Carlo Berg; Department of Microbiology; Karin Tegmark-Wisell; Maria Lind Karlberg; Mattias Haukland; Mia Brytting; Noura Walai; Oskar Karlsson Lindsjo; Petra Edquist; Petra Holmstrom; Reza Advani; Sofia Stamouli; The Public Health Agency of Sweden |

We gratefully acknowledge the following Authors from the Originating laboratories responsible for obtaining the specimens, as well as the Submitting laboratories where the genome data were generated and shared via GISAID, on which this research is based.

All Submitters of data may be contacted directly via www.gisaid.org

Authors are sorted alphabetically.

| Accession ID | Originating Laboratory | Submitting Laboratory | Authors |
|---|---|---|---|
| EPI_ISL_1168099, EPI_ISL_1168100, EPI_ISL_1168101, EPI_ISL_1168102, EPI_ISL_1168103, EPI_ISL_1168104, EPI_ISL_1168105, EPI_ISL_1168106, EPI_ISL_1231917, EPI_ISL_1231918, EPI_ISL_1231944, EPI_ISL_1231957, EPI_ISL_1231969, EPI_ISL_1231980, EPI_ISL_1231992, EPI_ISL_1231993 | | | |
| see above | A05 diagnostics | The Public Health Agency of Sweden | Anna Risberg; Anna-Malin Linde; Carlo Berg; Karin Tegmark-Wisell; Maria Lind Karlberg; Mattias Haukland; Mia Brytting; Noura Walai; Oskar Karlsson Lindsjo; Petra Edquist; Petra Holmstrom; Reza Advani; Sofia Stamouli |
| EPI_ISL_1168069, EPI_ISL_1168136 | ABC labs | The Public Health Agency of Sweden | Anna Risberg; Anna-Malin Linde; Carlo Berg; Karin Tegmark-Wisell; Maria Lind Karlberg; Mattias Haukland; Mia Brytting; Noura Walai; Oskar Karlsson Lindsjo; Petra Edquist; Petra Holmstrom; Reza Advani; Sofia Stamouli |
| EPI_ISL_1081940, EPI_ISL_1081942, EPI_ISL_1128837, EPI_ISL_1128839, EPI_ISL_1128842, EPI_ISL_1128844, EPI_ISL_1128846, EPI_ISL_1128849, EPI_ISL_1128851, EPI_ISL_1128853, EPI_ISL_1128855, EPI_ISL_1128858, EPI_ISL_1128860, EPI_ISL_1128862, EPI_ISL_1128866, EPI_ISL_1128868, EPI_ISL_1128870, EPI_ISL_1128873, EPI_ISL_1128875, EPI_ISL_1128878, EPI_ISL_1128880, EPI_ISL_1128882, EPI_ISL_1128884, EPI_ISL_1128887, EPI_ISL_1128889, EPI_ISL_1128892, EPI_ISL_1128894, EPI_ISL_1128896, EPI_ISL_1128899, EPI_ISL_1128901, EPI_ISL_1128903, EPI_ISL_1128906, EPI_ISL_1128908, EPI_ISL_1128909, EPI_ISL_1128911, EPI_ISL_1128913, EPI_ISL_1128914, EPI_ISL_1128916, EPI_ISL_1128918, EPI_ISL_1128919, EPI_ISL_1128921, EPI_ISL_1128923, EPI_ISL_1128925, EPI_ISL_1128928, EPI_ISL_1128930, EPI_ISL_1128932, EPI_ISL_1128935, EPI_ISL_1128937, EPI_ISL_1128939, EPI_ISL_1128941, EPI_ISL_1128944, EPI_ISL_1128946, EPI_ISL_1176356, EPI_ISL_1176357, EPI_ISL_1176358, EPI_ISL_1176359, EPI_ISL_1176360, EPI_ISL_1176361, EPI_ISL_1176362, EPI_ISL_1176363, EPI_ISL_1176364, EPI_ISL_1176365, EPI_ISL_1176366, EPI_ISL_1176367, EPI_ISL_1176368, EPI_ISL_1176369, EPI_ISL_1176370, EPI_ISL_1176371, EPI_ISL_1176372, EPI_ISL_1176373, EPI_ISL_1176374, EPI_ISL_1176375, EPI_ISL_1176376, EPI_ISL_1176377, EPI_ISL_1176378, EPI_ISL_1176379, EPI_ISL_1176380, EPI_ISL_1176381, EPI_ISL_1176382, EPI_ISL_1176383, EPI_ISL_1176384, EPI_ISL_1176387, EPI_ISL_1176388, EPI_ISL_1239146, EPI_ISL_1239156, EPI_ISL_1239177, EPI_ISL_1239178, EPI_ISL_1239179, EPI_ISL_1239180, EPI_ISL_1239181, EPI_ISL_1239182, EPI_ISL_1239183, EPI_ISL_1239184, EPI_ISL_1239194, EPI_ISL_1239199, EPI_ISL_1239200, EPI_ISL_1315430, EPI_ISL_1385885, EPI_ISL_1385886, EPI_ISL_1385887, EPI_ISL_1534099 | | | |
| see above | Clinical Microbiology, Infection Prevention and Control | Section for Molecular Diagnostics | Björn Hallström; Jonas Björkman |
| EPI_ISL_1313181, EPI_ISL_1313182, EPI_ISL_1313183, EPI_ISL_1313184, EPI_ISL_1313185, EPI_ISL_1313186, EPI_ISL_1313187, EPI_ISL_1313188, EPI_ISL_1313189, EPI_ISL_1313190, EPI_ISL_1313191, EPI_ISL_1313192, EPI_ISL_1313193, EPI_ISL_1313194, EPI_ISL_1313195, EPI_ISL_1313196, EPI_ISL_1313197, EPI_ISL_1313198, EPI_ISL_1313199, EPI_ISL_1313200, EPI_ISL_1313201, EPI_ISL_1313202, EPI_ISL_1313203, EPI_ISL_1313204, EPI_ISL_1313205, EPI_ISL_1313206, EPI_ISL_1313207, EPI_ISL_1313208, EPI_ISL_1313209, EPI_ISL_1313210, EPI_ISL_1313211, EPI_ISL_1313212, EPI_ISL_1313213, EPI_ISL_1313214, EPI_ISL_1313215, EPI_ISL_1313216, EPI_ISL_1313217, EPI_ISL_1313218, EPI_ISL_1313219, EPI_ISL_1313220, EPI_ISL_1313221, EPI_ISL_1313222, EPI_ISL_1313223, EPI_ISL_1313224, EPI_ISL_1313225, EPI_ISL_1313226, EPI_ISL_1313227, EPI_ISL_1313228, EPI_ISL_1313229, EPI_ISL_1313230, EPI_ISL_1313231, EPI_ISL_1313232, EPI_ISL_1313233, EPI_ISL_1313234, EPI_ISL_1313235, EPI_ISL_1313236, EPI_ISL_1313237, EPI_ISL_1313238, EPI_ISL_1313239, EPI_ISL_1313240, EPI_ISL_1313241, EPI_ISL_1313242, EPI_ISL_1313243, EPI_ISL_1313244, EPI_ISL_1313245, EPI_ISL_1313246, EPI_ISL_1313247, EPI_ISL_1313248, EPI_ISL_1313249, EPI_ISL_1313250, EPI_ISL_1313251, EPI_ISL_1313252, EPI_ISL_1313253, EPI_ISL_1313254, EPI_ISL_1313255, EPI_ISL_1313256, EPI_ISL_1313257, EPI_ISL_1313258, EPI_ISL_1313259, EPI_ISL_1313260, EPI_ISL_1313261, EPI_ISL_1313262, EPI_ISL_1313263, EPI_ISL_1313264, EPI_ISL_1313265, EPI_ISL_1313266, EPI_ISL_1313267, EPI_ISL_1313268, EPI_ISL_1313269, EPI_ISL_1313270, EPI_ISL_1313271, EPI_ISL_1313272, EPI_ISL_1313273, EPI_ISL_1313274, EPI_ISL_1313275, EPI_ISL_1313276, EPI_ISL_1313277, EPI_ISL_1313278, EPI_ISL_1313279, EPI_ISL_1313282, EPI_ISL_1313291, EPI_ISL_1313361, EPI_ISL_1313370, EPI_ISL_1313371, EPI_ISL_1313372, EPI_ISL_1313373, EPI_ISL_1313374, EPI_ISL_1313375, EPI_ISL_1313376, EPI_ISL_1313377, EPI_ISL_1313378, EPI_ISL_1313379, EPI_ISL_1313380, EPI_ISL_1313381, EPI_ISL_1313382, EPI_ISL_1313383, EPI_ISL_1313384, EPI_ISL_1313385, EPI_ISL_1313386, EPI_ISL_1313387, EPI_ISL_1313388, EPI_ISL_1313389, EPI_ISL_1313390, EPI_ISL_1313391, EPI_ISL_1313392, EPI_ISL_1313393, EPI_ISL_1313394, EPI_ISL_1313395, EPI_ISL_1313396, EPI_ISL_1313397, EPI_ISL_1313398, EPI_ISL_1313399, EPI_ISL_1313400, EPI_ISL_1313401, EPI_ISL_1313402, EPI_ISL_1313403, EPI_ISL_1313404, EPI_ISL_1313405, EPI_ISL_1313406, EPI_ISL_1313407, EPI_ISL_1313408, EPI_ISL_1313409, EPI_ISL_1313410, EPI_ISL_1313411, EPI_ISL_1313412, EPI_ISL_1313413, EPI_ISL_1313414, EPI_ISL_1313415, EPI_ISL_1313416, EPI_ISL_1313417, EPI_ISL_1313418, EPI_ISL_1313419, EPI_ISL_1313420, EPI_ISL_1313421, EPI_ISL_1313422, EPI_ISL_1313423, EPI_ISL_1313424, EPI_ISL_1313425, EPI_ISL_1313426, EPI_ISL_1313427, EPI_ISL_1313428, EPI_ISL_1313429, EPI_ISL_1313430, EPI_ISL_1313431, EPI_ISL_1313432, EPI_ISL_1313433, EPI_ISL_1313434, EPI_ISL_1313435, EPI_ISL_1313436, EPI_ISL_1313437, EPI_ISL_1313438, EPI_ISL_1313439, EPI_ISL_1313440, EPI_ISL_1313441, EPI_ISL_1313442, EPI_ISL_1313443, EPI_ISL_1313444, EPI_ISL_1313445, EPI_ISL_1313446, EPI_ISL_1313447, EPI_ISL_1313448, EPI_ISL_1313449, EPI_ISL_1313450, EPI_ISL_1313451, EPI_ISL_1313452, EPI_ISL_1313453, EPI_ISL_1313454, EPI_ISL_1313455, EPI_ISL_1313456, EPI_ISL_1313457, EPI_ISL_1313458, EPI_ISL_1313459, EPI_ISL_1313460, EPI_ISL_1313461, EPI_ISL_1313462, EPI_ISL_1313463, EPI_ISL_1313464, EPI_ISL_1313468, EPI_ISL_1313496, EPI_ISL_1313498, EPI_ISL_1313500, EPI_ISL_1313501, EPI_ISL_1313508, EPI_ISL_1313513, EPI_ISL_1360110 | | | |
| see above | Karolinska University Hospital | Karolinska University Hospital | Annelie Bjerkner; Isak Sylvin; Jan Albert; Karolina Ininbergs; Lina Guerra Blomqvist; Lynda Eneh; Martin Ekman; Martina Wahlund; Robert Dyrdak; Sandra Broddesson; Tanja Normark; Tobias Allander; Valtteri Wirta; Zhibing Yun |
| EPI_ISL_1168162, EPI_ISL_1168163, EPI_ISL_1168164, EPI_ISL_1168165, EPI_ISL_1168166, EPI_ISL_1168167, EPI_ISL_1231919, EPI_ISL_1231931, EPI_ISL_1231932, EPI_ISL_1231934, EPI_ISL_1231938, EPI_ISL_1231948, EPI_ISL_1231958, EPI_ISL_1231959, EPI_ISL_1231964, EPI_ISL_1231973, EPI_ISL_1231974, EPI_ISL_1231975, EPI_ISL_1231982, EPI_ISL_1231985, EPI_ISL_1231994, EPI_ISL_1231995, EPI_ISL_1232015, EPI_ISL_1232017 | | | |
| see above | Klinisk Mikrobiologi | The Public Health Agency of Sweden | Anna Risberg; Anna-Malin Linde; Carlo Berg; Karin Tegmark-Wisell; Maria Lind Karlberg; Mattias Haukland; Mia Brytting; Noura Walai; Oskar Karlsson Lindsjo; Petra Edquist; Petra Holmstrom; Reza Advani; Sofia Stamouli |
| EPI_ISL_1168070, EPI_ISL_1168073, EPI_ISL_1168074, EPI_ISL_1168076, EPI_ISL_1168077, EPI_ISL_1168078, EPI_ISL_1168079, EPI_ISL_1168080, EPI_ISL_1168083, EPI_ISL_1168084, EPI_ISL_1168088, EPI_ISL_1168089, EPI_ISL_1168091, EPI_ISL_1168092, EPI_ISL_1168093, EPI_ISL_1168107, EPI_ISL_1168108, EPI_ISL_1168109, EPI_ISL_1168110, EPI_ISL_1168116, EPI_ISL_1168117, EPI_ISL_1168118, EPI_ISL_1168119, EPI_ISL_1168121, EPI_ISL_1168124, EPI_ISL_1168125, EPI_ISL_1168126, EPI_ISL_1168127, EPI_ISL_1168128, EPI_ISL_1168129, EPI_ISL_1168130, EPI_ISL_1168131, EPI_ISL_1168132, EPI_ISL_1168133, EPI_ISL_1168134, EPI_ISL_1168139, EPI_ISL_1168140, EPI_ISL_1168141, EPI_ISL_1168142, EPI_ISL_1168146, EPI_ISL_1168148, EPI_ISL_1168149, EPI_ISL_1168150, EPI_ISL_1168151, EPI_ISL_1168152, EPI_ISL_1168153, EPI_ISL_1168154, EPI_ISL_1168155, EPI_ISL_1168156, EPI_ISL_1168158, EPI_ISL_1168159, EPI_ISL_1168160, EPI_ISL_1168161, EPI_ISL_1168168, EPI_ISL_1168169, EPI_ISL_1168170, EPI_ISL_1168171, EPI_ISL_1168172, EPI_ISL_1168173, EPI_ISL_1168174, EPI_ISL_1168175, EPI_ISL_1168176, EPI_ISL_1168177, EPI_ISL_1168178, EPI_ISL_1168179, EPI_ISL_1168180, EPI_ISL_1168181, EPI_ISL_1168182, EPI_ISL_1168183, EPI_ISL_1168184, EPI_ISL_1168185, EPI_ISL_1168186, EPI_ISL_1168187, EPI_ISL_1168188, EPI_ISL_1168189, EPI_ISL_1168190, EPI_ISL_1231920, EPI_ISL_1231921, EPI_ISL_1231922, EPI_ISL_1231923, EPI_ISL_1231940, EPI_ISL_1231941, EPI_ISL_1231942, EPI_ISL_1231943, EPI_ISL_1231947, EPI_ISL_1231951, EPI_ISL_1231966, EPI_ISL_1231967, EPI_ISL_1231983, EPI_ISL_1231997, EPI_ISL_1231998, EPI_ISL_1231999, EPI_ISL_1232000, EPI_ISL_1232001, EPI_ISL_1232002, EPI_ISL_1232003, EPI_ISL_1232008, EPI_ISL_1232009, EPI_ISL_1232010, EPI_ISL_1300277, EPI_ISL_1300278, EPI_ISL_1300279, EPI_ISL_1300280, EPI_ISL_1300281, EPI_ISL_1300282, EPI_ISL_1300283, EPI_ISL_1300284, EPI_ISL_1300292, EPI_ISL_1300297, EPI_ISL_1300298, EPI_ISL_1300299, EPI_ISL_1300308, EPI_ISL_1300325, EPI_ISL_1300326, EPI_ISL_1300328, EPI_ISL_1300331, EPI_ISL_1300333 | | | |
| see above | Klinisk mikrobiologi | The Public Health Agency of Sweden | Anna Risberg; Anna-Malin Linde; Carlo Berg; Karin Tegmark-Wisell; Maria Lind Karlberg; Mattias Haukland; Mia Brytting; Noura Walai; Oskar Karlsson Lindsjo; Petra Edquist; Petra Holmstrom; Reza Advani; Samuel Ohman; Sofia Stamouli |
| EPI_ISL_1091493, EPI_ISL_1091494, EPI_ISL_1091495, EPI_ISL_1091496, EPI_ISL_1091498, EPI_ISL_1091499, EPI_ISL_1091500, EPI_ISL_1091503, EPI_ISL_1091504, EPI_ISL_1091505, EPI_ISL_1091506, EPI_ISL_1091507, EPI_ISL_1091508, EPI_ISL_1091509, EPI_ISL_1091510, EPI_ISL_1091513, EPI_ISL_1091515, EPI_ISL_1091522, EPI_ISL_1091523, EPI_ISL_1091524, EPI_ISL_1091525, EPI_ISL_1091526, EPI_ISL_1091527, EPI_ISL_1091528, EPI_ISL_1091529, EPI_ISL_1091530, EPI_ISL_1091531, EPI_ISL_1091532, EPI_ISL_1091540, EPI_ISL_1091541, EPI_ISL_1091542, EPI_ISL_1091543, EPI_ISL_1091544, EPI_ISL_1091545, EPI_ISL_1091546, EPI_ISL_1091547, EPI_ISL_1091548, EPI_ISL_1091549, EPI_ISL_1091550, EPI_ISL_1091551, EPI_ISL_1091552, EPI_ISL_1091553, EPI_ISL_1091554, EPI_ISL_1091555, EPI_ISL_1091556, EPI_ISL_1091557, EPI_ISL_1091558, EPI_ISL_1091559, EPI_ISL_1091560, EPI_ISL_1091561, EPI_ISL_1091562, EPI_ISL_1091563, EPI_ISL_1091564, EPI_ISL_1091565, EPI_ISL_1091566, EPI_ISL_1091567, EPI_ISL_1091568, EPI_ISL_1091571, EPI_ISL_1091572, EPI_ISL_1091573, EPI_ISL_1091574, EPI_ISL_1091575, EPI_ISL_1166556, EPI_ISL_1166557, EPI_ISL_1166558, EPI_ISL_1166559, EPI_ISL_1166560, EPI_ISL_1166561, EPI_ISL_1166562, EPI_ISL_1166563, EPI_ISL_1166564, EPI_ISL_1166565, EPI_ISL_1166566, EPI_ISL_1166567, EPI_ISL_1166568, EPI_ISL_1166569, EPI_ISL_1166570, EPI_ISL_1166571, EPI_ISL_1166572, EPI_ISL_1166573, EPI_ISL_1166574, EPI_ISL_1166575, EPI_ISL_1166576, EPI_ISL_1166577, EPI_ISL_1166578, EPI_ISL_1166579, EPI_ISL_1166580, EPI_ISL_1166581, EPI_ISL_1166582, EPI_ISL_1166583, EPI_ISL_1166584, EPI_ISL_1166585, EPI_ISL_1166586, EPI_ISL_1166587, EPI_ISL_1166588, EPI_ISL_1166589, EPI_ISL_1166590, EPI_ISL_1166591, EPI_ISL_1166592, EPI_ISL_1166593, EPI_ISL_1166594, EPI_ISL_1166595, EPI_ISL_1166596, EPI_ISL_1166597, EPI_ISL_1166598, EPI_ISL_1166599, EPI_ISL_1166600, EPI_ISL_1166601, EPI_ISL_1166602, EPI_ISL_1166603, EPI_ISL_1166604, EPI_ISL_1166605, EPI_ISL_1166606, EPI_ISL_1166607, EPI_ISL_1166608, EPI_ISL_1166609, EPI_ISL_1166610, EPI_ISL_1166611, EPI_ISL_1166612, EPI_ISL_1166613, EPI_ISL_1166614, EPI_ISL_1224958, EPI_ISL_1224965, EPI_ISL_1224966, EPI_ISL_1224969, EPI_ISL_1303398, EPI_ISL_1372900, EPI_ISL_1372924, EPI_ISL_1372932, EPI_ISL_1372934, EPI_ISL_1578119, EPI_ISL_1578120 | | | |
| see above | Klinisk mikrobiologi, Region Västerbotten | CBRN Defence and Security, Swedish Defence Research Agency | Andreas Sjödin; Anna-Lena Johansson; Annika Allard; Annika Osterman; Caroline Öhrman; David Sundell; Emelie Näslund Salomonsson; Ingrid Dacklin; Jonas Näslund; Kerstin Myrtennäs; Linda Karlsson; Malin Granberg; Mats Forsman; Stina Bäckman |
| EPI_ISL_1231926 | Klinisk mikrobiologi, molekylar virologi | The Public Health Agency of Sweden | Anna Risberg; Anna-Malin Linde; Carlo Berg; Karin Tegmark-Wisell; Maria Lind Karlberg; Mattias Haukland; Mia Brytting; Noura Walai; Oskar Karlsson Lindsjo; Petra Edquist; Petra Holmstrom; Reza Advani; Sofia Stamouli |
| EPI_ISL_1313179 | LaboratorieMedicinskt Centrum Gotland | Karolinska University Hospital | Annelie Bjerkner; Isak Sylvin; Jan Albert; Karolina Ininbergs; Lina Guerra Blomqvist; Lynda Eneh; Martin Ekman; Martina Wahlund; Robert Dyrdak; Sandra Broddesson; Tanja Normark; Tobias Allander; Valtteri Wirta; Zhibing Yun |
| EPI_ISL_1232007 | Laboratoriemedicin, Klinisk mikrobiologi | The Public Health Agency of Sweden | Anna Risberg; Anna-Malin Linde; Carlo Berg; Karin Tegmark-Wisell; Maria Lind Karlberg; Mattias Haukland; Mia Brytting; Noura Walai; Oskar Karlsson Lindsjo; Petra Edquist; Petra Holmstrom; Reza Advani; Sofia Stamouli |
| EPI_ISL_1231915, EPI_ISL_1231927, EPI_ISL_1231928, EPI_ISL_1231929, EPI_ISL_1231930, EPI_ISL_1232012, EPI_ISL_1232013, EPI_ISL_1232014 | | | |
| see above | Mikrobiologen | The Public Health Agency of Sweden | Anna Risberg; Anna-Malin Linde; Carlo Berg; Karin Tegmark-Wisell; Maria Lind Karlberg; Mattias Haukland; Mia Brytting; Noura Walai; Oskar Karlsson Lindsjo; Petra Edquist; Petra Holmstrom; Reza Advani; Sofia Stamouli |
| EPI_ISL_1168095 | NPC | The Public Health Agency of Sweden | Anna Risberg; Anna-Malin Linde; Carlo Berg; Karin Tegmark-Wisell; Maria Lind Karlberg; Mattias Haukland; Mia Brytting; Noura Walai; Oskar Karlsson Lindsjo; Petra Edquist; Petra Holmstrom; Reza Advani; Sofia Stamouli |
| EPI_ISL_1492594, EPI_ISL_1492595, EPI_ISL_1492596, EPI_ISL_1492615, EPI_ISL_1492616, EPI_ISL_1492617 | Orebro University Hospital | Orebro University Hospital | Sundqvist M et al |
| EPI_ISL_1289963, EPI_ISL_1289964, EPI_ISL_1289968, EPI_ISL_1289969, EPI_ISL_1289973, EPI_ISL_1289974, EPI_ISL_1289975, EPI_ISL_1289976, EPI_ISL_1289977, EPI_ISL_1289978, EPI_ISL_1289979, EPI_ISL_1289980, EPI_ISL_1289981, EPI_ISL_1289983, EPI_ISL_1289985, EPI_ISL_1289986, EPI_ISL_1289988, EPI_ISL_1289990, EPI_ISL_1289991, EPI_ISL_1289992, EPI_ISL_1289994, EPI_ISL_1289995, EPI_ISL_1289996, EPI_ISL_1289998, EPI_ISL_1290000, EPI_ISL_1290001, EPI_ISL_1290003, EPI_ISL_1290005, EPI_ISL_1290006, EPI_ISL_1290008, EPI_ISL_1290010, EPI_ISL_1290011, EPI_ISL_1290013, EPI_ISL_1290014, EPI_ISL_1290016, EPI_ISL_1290017, EPI_ISL_1290020, EPI_ISL_1290023, EPI_ISL_1290026, EPI_ISL_1290027, EPI_ISL_1290029, EPI_ISL_1290032, EPI_ISL_1290035, EPI_ISL_1290036, EPI_ISL_1290037, EPI_ISL_1290038, EPI_ISL_1290039, EPI_ISL_1290040, EPI_ISL_1290041, EPI_ISL_1290042, EPI_ISL_1290043, EPI_ISL_1290044, EPI_ISL_1290045, EPI_ISL_1290046, EPI_ISL_1290048, EPI_ISL_1290049, EPI_ISL_1290051, EPI_ISL_1290052, EPI_ISL_1290054, EPI_ISL_1290056, EPI_ISL_1290058, EPI_ISL_1290061, EPI_ISL_1290063, EPI_ISL_1290065, EPI_ISL_1290066, EPI_ISL_1290067, EPI_ISL_1290068, EPI_ISL_1290069, EPI_ISL_1290070, EPI_ISL_1290073, EPI_ISL_1290074, EPI_ISL_1290075, EPI_ISL_1290079, EPI_ISL_1290080, EPI_ISL_1290082, EPI_ISL_1290086, EPI_ISL_1290088, EPI_ISL_1290090, EPI_ISL_1290092, EPI_ISL_1290094, EPI_ISL_1290096, EPI_ISL_1290098, EPI_ISL_1290099, EPI_ISL_1290104, EPI_ISL_1290105, EPI_ISL_1290106, EPI_ISL_1290107, EPI_ISL_1290108, EPI_ISL_1290109, EPI_ISL_1290110, EPI_ISL_1290114, EPI_ISL_1290115, EPI_ISL_1290116, EPI_ISL_1290117, EPI_ISL_1290119, EPI_ISL_1290120, EPI_ISL_1290123, EPI_ISL_1290126, EPI_ISL_1290128, EPI_ISL_1290129, EPI_ISL_1290130, EPI_ISL_1290131, EPI_ISL_1290133, EPI_ISL_1290138, EPI_ISL_1290146, EPI_ISL_1290147, EPI_ISL_1290148, EPI_ISL_1290247, EPI_ISL_1290248, EPI_ISL_1290249, EPI_ISL_1290250, EPI_ISL_1290251, EPI_ISL_1290252, EPI_ISL_1290253, EPI_ISL_1290254, EPI_ISL_1290255, EPI_ISL_1290256, EPI_ISL_1290257, EPI_ISL_1290258, EPI_ISL_1290259, EPI_ISL_1290260, EPI_ISL_1290261, EPI_ISL_1290262, EPI_ISL_1290263, EPI_ISL_1290264, EPI_ISL_1290265, EPI_ISL_1290266, EPI_ISL_1290267, EPI_ISL_1290268, EPI_ISL_1290269, EPI_ISL_1290270, EPI_ISL_1290271, EPI_ISL_1290272, EPI_ISL_1290273, EPI_ISL_1290274, EPI_ISL_1290275, EPI_ISL_1290276, EPI_ISL_1290277, EPI_ISL_1290278, EPI_ISL_1290288, EPI_ISL_1290289, EPI_ISL_1290290, EPI_ISL_1290291, EPI_ISL_1290292, EPI_ISL_1290293, EPI_ISL_1290345, EPI_ISL_1290346, EPI_ISL_1290347, EPI_ISL_1290348, EPI_ISL_1290349, EPI_ISL_1290350, EPI_ISL_1290351, EPI_ISL_1290352, EPI_ISL_1290353, EPI_ISL_1290354, EPI_ISL_1290355, EPI_ISL_1290356, EPI_ISL_1290357, EPI_ISL_1290358, EPI_ISL_1290390, EPI_ISL_1290391, EPI_ISL_1290392, EPI_ISL_1290393, EPI_ISL_1290400, EPI_ISL_1290401, EPI_ISL_1290402, EPI_ISL_1290403, EPI_ISL_1290404, EPI_ISL_1290407, EPI_ISL_1290410, EPI_ISL_1290411, EPI_ISL_1290412, EPI_ISL_1290413, EPI_ISL_1290414, EPI_ISL_1290428, EPI_ISL_1290429, EPI_ISL_1290430, EPI_ISL_1290431, EPI_ISL_1290432, EPI_ISL_1290433, EPI_ISL_1290434, EPI_ISL_1290435, EPI_ISL_1290436, EPI_ISL_1290437, EPI_ISL_1290438, EPI_ISL_1290439, EPI_ISL_1290440, EPI_ISL_1290441, EPI_ISL_1290442, EPI_ISL_1290443, EPI_ISL_1290444, EPI_ISL_1290445, EPI_ISL_1290446, EPI_ISL_1290447, EPI_ISL_1290448, EPI_ISL_1290449, EPI_ISL_1290450, EPI_ISL_1290451, EPI_ISL_1290452, EPI_ISL_1290453, EPI_ISL_1290454, EPI_ISL_1290455, EPI_ISL_1290456, EPI_ISL_1290457, EPI_ISL_1290458, EPI_ISL_1290459, EPI_ISL_1290460, EPI_ISL_1290473, EPI_ISL_1290475, EPI_ISL_1290477, EPI_ISL_1290481, EPI_ISL_1290484, EPI_ISL_1290489, EPI_ISL_1290490, EPI_ISL_1290493, EPI_ISL_1290495, EPI_ISL_1290498, EPI_ISL_1290499, EPI_ISL_1290500, EPI_ISL_1290502, EPI_ISL_1290505, EPI_ISL_1290506, EPI_ISL_1290507, EPI_ISL_1290509, EPI_ISL_1290510, EPI_ISL_1290511, EPI_ISL_1290512, EPI_ISL_1290514, EPI_ISL_1290515, EPI_ISL_1290516, EPI_ISL_1290518, EPI_ISL_1290519, EPI_ISL_1290520, EPI_ISL_1290521, EPI_ISL_1290523, EPI_ISL_1290525, EPI_ISL_1290526, EPI_ISL_1290527, EPI_ISL_1290533, EPI_ISL_1290565, EPI_ISL_1290566, EPI_ISL_1290567, EPI_ISL_1290572, EPI_ISL_1290583, EPI_ISL_1290687, EPI_ISL_1290688, EPI_ISL_1290689, EPI_ISL_1290690, EPI_ISL_1290691, EPI_ISL_1290692, EPI_ISL_1290693, EPI_ISL_1290694, EPI_ISL_1290695, EPI_ISL_1290696, EPI_ISL_1290697, EPI_ISL_1290698, EPI_ISL_1290699, EPI_ISL_1290700, EPI_ISL_1290701, EPI_ISL_1290702, EPI_ISL_1290703, EPI_ISL_1290704, EPI_ISL_1290705, EPI_ISL_1290706, EPI_ISL_1290707, EPI_ISL_1290708, EPI_ISL_1290709, EPI_ISL_1290710, EPI_ISL_1290711, EPI_ISL_1290712, EPI_ISL_1290713, EPI_ISL_1290714, EPI_ISL_1290715, EPI_ISL_1290716, EPI_ISL_1290717, EPI_ISL_1290718, EPI_ISL_1290719, EPI_ISL_1290720, EPI_ISL_1290721, EPI_ISL_1290722, EPI_ISL_1290723, EPI_ISL_1290724, EPI_ISL_1290725, EPI_ISL_1290726, EPI_ISL_1290727, EPI_ISL_1290728, EPI_ISL_1290729, EPI_ISL_1290730, EPI_ISL_1290731, EPI_ISL_1290732, EPI_ISL_1290733, EPI_ISL_1290734, EPI_ISL_1290735, EPI_ISL_1290736, EPI_ISL_1290737, EPI_ISL_1290738, EPI_ISL_1290739, EPI_ISL_1290740, EPI_ISL_1290741, EPI_ISL_1290742, EPI_ISL_1290743, EPI_ISL_1290744, EPI_ISL_1290745, EPI_ISL_1290746, EPI_ISL_1290747, EPI_ISL_1290748, EPI_ISL_1290749, EPI_ISL_1290750, EPI_ISL_1290751, EPI_ISL_1290752, EPI_ISL_1290753, EPI_ISL_1290754, EPI_ISL_1290755, EPI_ISL_1290756, EPI_ISL_1290757, EPI_ISL_1290758, EPI_ISL_1290759, EPI_ISL_1290760, EPI_ISL_1290761, EPI_ISL_1290762, EPI_ISL_1290763, EPI_ISL_1290764, EPI_ISL_1290765, EPI_ISL_1290766, EPI_ISL_1290767, EPI_ISL_1290768, EPI_ISL_1290769, EPI_ISL_1290770, EPI_ISL_1290771, EPI_ISL_1290772, EPI_ISL_1290773, EPI_ISL_1290774, EPI_ISL_1290775, EPI_ISL_1290776, EPI_ISL_1290777, EPI_ISL_1290778, EPI_ISL_1290779, EPI_ISL_1290780, EPI_ISL_1290781, EPI_ISL_1290782, EPI_ISL_1290783, EPI_ISL_1290784, EPI_ISL_1290785, EPI_ISL_1290786, EPI_ISL_1290787, EPI_ISL_1290788, EPI_ISL_1290789, EPI_ISL_1290790, EPI_ISL_1290791, EPI_ISL_1290792, EPI_ISL_1290793, EPI_ISL_1290794, EPI_ISL_1290795, EPI_ISL_1290796, EPI_ISL_1290797, EPI_ISL_1290798, EPI_ISL_1290799, EPI_ISL_1290800, EPI_ISL_1290801, EPI_ISL_1361935, EPI_ISL_1361936, EPI_ISL_1361937, EPI_ISL_1361938, EPI_ISL_1361939, EPI_ISL_1361940, EPI_ISL_1361941, EPI_ISL_1361942, EPI_ISL_1361943, EPI_ISL_1361944, EPI_ISL_1361945, EPI_ISL_1361946, EPI_ISL_1361947, EPI_ISL_1361948, EPI_ISL_1361949, EPI_ISL_1361950, EPI_ISL_1361951, EPI_ISL_1361952, EPI_ISL_1361953, EPI_ISL_1361954, EPI_ISL_1361955, EPI_ISL_1361956, EPI_ISL_1361957, EPI_ISL_1361958, EPI_ISL_1361959, EPI_ISL_1361960, EPI_ISL_1361961, EPI_ISL_1361962, EPI_ISL_1361963, EPI_ISL_1361964, EPI_ISL_1361965, EPI_ISL_1361966, EPI_ISL_1361967, EPI_ISL_1361968, EPI_ISL_1361969, EPI_ISL_1361970, EPI_ISL_1361971, EPI_ISL_1361972, EPI_ISL_1361973, EPI_ISL_1361974, EPI_ISL_1361975, EPI_ISL_1361976, EPI_ISL_1361977, EPI_ISL_1361978, EPI_ISL_1361979, EPI_ISL_1361980, EPI_ISL_1361981, EPI_ISL_1361982, EPI_ISL_1361983, EPI_ISL_1361984, EPI_ISL_1361985, EPI_ISL_1361986, EPI_ISL_1361987, EPI_ISL_1361988, EPI_ISL_1361989, EPI_ISL_1361990, EPI_ISL_1361991, EPI_ISL_1361992, EPI_ISL_1361993, EPI_ISL_1361994, EPI_ISL_1361995, EPI_ISL_1361996, EPI_ISL_1361997, EPI_ISL_1361998, EPI_ISL_1361999, EPI_ISL_1362000, EPI_ISL_1362001, EPI_ISL_1362002, EPI_ISL_1362003, EPI_ISL_1362004, EPI_ISL_1362005, EPI_ISL_1362006, EPI_ISL_1362007, EPI_ISL_1362008, EPI_ISL_1362009, EPI_ISL_1362010, EPI_ISL_1362011, EPI_ISL_1362012, EPI_ISL_1362013, EPI_ISL_1362014, EPI_ISL_1362015, EPI_ISL_1362016, EPI_ISL_1362017, EPI_ISL_1362018, EPI_ISL_1362019, EPI_ISL_1362020, EPI_ISL_1362021, EPI_ISL_1362022, EPI_ISL_1362023, EPI_ISL_1362024, EPI_ISL_1362025, EPI_ISL_1362026, EPI_ISL_1362027, EPI_ISL_1362028, EPI_ISL_1362029, EPI_ISL_1362030, EPI_ISL_1362031, EPI_ISL_1362032, EPI_ISL_1362033, EPI_ISL_1362034, EPI_ISL_1362035, EPI_ISL_1362036, EPI_ISL_1362037, EPI_ISL_1362038, EPI_ISL_1362039, EPI_ISL_1362040, EPI_ISL_1362041, EPI_ISL_1362042, EPI_ISL_1362043, EPI_ISL_1362044, EPI_ISL_1362045, EPI_ISL_1362046, EPI_ISL_1362047, EPI_ISL_1362048, EPI_ISL_1362049, EPI_ISL_1362050, EPI_ISL_1362051, EPI_ISL_1362052, EPI_ISL_1362053, EPI_ISL_1362054, EPI_ISL_1362055, EPI_ISL_1362056, EPI_ISL_1362057, EPI_ISL_1362058, EPI_ISL_1362059, EPI_ISL_1362060, EPI_ISL_1362061, EPI_ISL_1362062, EPI_ISL_1362063, EPI_ISL_1362064, EPI_ISL_1362065, EPI_ISL_1362066, EPI_ISL_1362067, EPI_ISL_1362068, EPI_ISL_1362069, EPI_ISL_1362070, EPI_ISL_1362071, EPI_ISL_1362072, EPI_ISL_1362073, EPI_ISL_1362074, EPI_ISL_1362075, EPI_ISL_1362076, EPI_ISL_1362077, EPI_ISL_1362078, EPI_ISL_1362079, EPI_ISL_1362080, EPI_ISL_1362081, EPI_ISL_1362082, EPI_ISL_1362083, EPI_ISL_1362084, EPI_ISL_1362085, EPI_ISL_1362086, EPI_ISL_1362087, EPI_ISL_1362088, EPI_ISL_1362089, EPI_ISL_1362090, EPI_ISL_1362091, EPI_ISL_1362092, EPI_ISL_1362093, EPI_ISL_1362094, EPI_ISL_1362095, EPI_ISL_1362096, EPI_ISL_1362097, EPI_ISL_1362098, EPI_ISL_1362099, EPI_ISL_1362100, EPI_ISL_1362101, EPI_ISL_1362102, EPI_ISL_1362103, EPI_ISL_1362104, EPI_ISL_1362105, EPI_ISL_1362106, EPI_ISL_1362107, EPI_ISL_1362108, EPI_ISL_1362109, EPI_ISL_1362110, EPI_ISL_1362111, EPI_ISL_1362112, EPI_ISL_1362113, EPI_ISL_1362114, EPI_ISL_1362115, EPI_ISL_1362116, EPI_ISL_1362117, EPI_ISL_1362118, EPI_ISL_1362119, EPI_ISL_1362120, EPI_ISL_1362121, EPI_ISL_1362122, EPI_ISL_1362123, EPI_ISL_1362124, EPI_ISL_1362125, EPI_ISL_1362126, EPI_ISL_1362127, EPI_ISL_1362128, EPI_ISL_1362129, EPI_ISL_1362130, EPI_ISL_1362131, EPI_ISL_1362132, EPI_ISL_1362133, EPI_ISL_1362134, EPI_ISL_1362135, EPI_ISL_1362136, EPI_ISL_1362137, EPI_ISL_1362138, EPI_ISL_1362139, EPI_ISL_1362140, EPI_ISL_1362141, EPI_ISL_1362142, EPI_ISL_1362143, EPI_ISL_1362144, EPI_ISL_1362145, EPI_ISL_1362146, EPI_ISL_1362147, EPI_ISL_1362148, EPI_ISL_1362149, EPI_ISL_1362150, EPI_ISL_1362151, EPI_ISL_1362152, EPI_ISL_1362153, EPI_ISL_1362154, EPI_ISL_1362155, EPI_ISL_1362156, EPI_ISL_1362157, EPI_ISL_1362158, EPI_ISL_1362159, EPI_ISL_1362160, EPI_ISL_1362161, EPI_ISL_1362162, EPI_ISL_1362163, EPI_ISL_1362164, EPI_ISL_1362165, EPI_ISL_1362166, EPI_ISL_1362167, EPI_ISL_1362168, EPI_ISL_1362169, EPI_ISL_1362170, EPI_ISL_1362171, EPI_ISL_1362172, EPI_ISL_1362173, EPI_ISL_1362174, EPI_ISL_1362175, EPI_ISL_1362176, EPI_ISL_1362177, EPI_ISL_1362178, EPI_ISL_1362179, EPI_ISL_1362180, EPI_ISL_1362181, EPI_ISL_1362182, EPI_ISL_1362183, EPI_ISL_1362184, EPI_ISL_1362185, EPI_ISL_1362186, EPI_ISL_1362187, EPI_ISL_1362188, EPI_ISL_1362189, EPI_ISL_1362190, EPI_ISL_1362191, EPI_ISL_1362192, EPI_ISL_1362193, EPI_ISL_1362194, EPI_ISL_1362195, EPI_ISL_1362196, EPI_ISL_1362197, EPI_ISL_1362198, EPI_ISL_1362199, EPI_ISL_1362200, EPI_ISL_1362201, EPI_ISL_1362202, EPI_ISL_1362203, EPI_ISL_1362204, EPI_ISL_1362205, EPI_ISL_1362206, EPI_ISL_1362207, EPI_ISL_1362208, EPI_ISL_1362209, EPI_ISL_1362210, EPI_ISL_1362211, EPI_ISL_1362212, EPI_ISL_1362213, EPI_ISL_1362214, EPI_ISL_1362215, EPI_ISL_1362216, EPI_ISL_1362217, EPI_ISL_1362218, EPI_ISL_1362219, EPI_ISL_1362220, EPI_ISL_1362221, EPI_ISL_1362222, EPI_ISL_1362223, EPI_ISL_1362224, EPI_ISL_1362225, EPI_ISL_1362226, EPI_ISL_1362227, EPI_ISL_1362228, EPI_ISL_1362229, EPI_ISL_1362230, EPI_ISL_1362231, EPI_ISL_1362232, EPI_ISL_1362233, EPI_ISL_1362234, EPI_ISL_1362235, EPI_ISL_1362236, EPI_ISL_1362237, EPI_ISL_1362238, EPI_ISL_1362239, EPI_ISL_1362240, EPI_ISL_1362241, EPI_ISL_1362242, EPI_ISL_1362243, EPI_ISL_1362244, EPI_ISL_1362245, EPI_ISL_1362246, EPI_ISL_1362247, EPI_ISL_1362248, EPI_ISL_1362249, EPI_ISL_1362250, EPI_ISL_1362251, EPI_ISL_1362252, EPI_ISL_1362253, EPI_ISL_1362254, EPI_ISL_1362255, EPI_ISL_1362256, EPI_ISL_1362257, EPI_ISL_1362258, EPI_ISL_1362259, EPI_ISL_1362260, EPI_ISL_1362261, EPI_ISL_1362262, EPI_ISL_1362263, EPI_ISL_1362264, EPI_ISL_1362265, EPI_ISL_1362266, EPI_ISL_1362267, EPI_ISL_1362268, EPI_ISL_1362269, EPI_ISL_1362270, EPI_ISL_1362271, EPI_ISL_1362272, EPI_ISL_1362273, EPI_ISL_1362274, EPI_ISL_1362275, EPI_ISL_1362276, EPI_ISL_1362277, EPI_ISL_1362278, EPI_ISL_1362279, EPI_ISL_1362280, EPI_ISL_1362281, EPI_ISL_1362282, EPI_ISL_1362283, EPI_ISL_1362284, EPI_ISL_1362285, EPI_ISL_1362286, EPI_ISL_1362287, EPI_ISL_1362288, EPI_ISL_1362289, EPI_ISL_1362290, EPI_ISL_1362291, EPI_ISL_1362292, EPI_ISL_1362293, EPI_ISL_1362294, EPI_ISL_1362295, EPI_ISL_1362296, EPI_ISL_1362297, EPI_ISL_1362298, EPI_ISL_1362299, EPI_ISL_1362300, EPI_ISL_1362301, | | | |

EPI_ISL_1362302, EPI_ISL_1362303, EPI_ISL_1362304, EPI_ISL_1362305, EPI_ISL_1362306, EPI_ISL_1362307, EPI_ISL_1362308, EPI_ISL_1362309, EPI_ISL_1362310, EPI_ISL_1362311, EPI_ISL_1362312, EPI_ISL_1362313, EPI_ISL_1362314, EPI_ISL_1362315, EPI_ISL_1362316, EPI_ISL_1362317, EPI_ISL_1362318, EPI_ISL_1362319, EPI_ISL_1362320, EPI_ISL_1362321, EPI_ISL_1362322, EPI_ISL_1362323, EPI_ISL_1362324, EPI_ISL_1362325, EPI_ISL_1362326, EPI_ISL_1362327, EPI_ISL_1362328, EPI_ISL_1362329, EPI_ISL_1362330, EPI_ISL_1362331, EPI_ISL_1362332, EPI_ISL_1362333, EPI_ISL_1362334, EPI_ISL_1362335, EPI_ISL_1362336, EPI_ISL_1362337, EPI_ISL_1362338, EPI_ISL_1362339, EPI_ISL_1362340, EPI_ISL_1362341, EPI_ISL_1362342, EPI_ISL_1362343, EPI_ISL_1362344, EPI_ISL_1362345, EPI_ISL_1362346, EPI_ISL_1362347, EPI_ISL_1362348, EPI_ISL_1362349, EPI_ISL_1362350, EPI_ISL_1362351, EPI_ISL_1362352, EPI_ISL_1362353, EPI_ISL_1362354, EPI_ISL_1362355, EPI_ISL_1362356, EPI_ISL_1362357, EPI_ISL_1362358, EPI_ISL_1362359, EPI_ISL_1362360, EPI_ISL_1362361, EPI_ISL_1362362, EPI_ISL_1362363, EPI_ISL_1362364, EPI_ISL_1362365, EPI_ISL_1362366, EPI_ISL_1362367, EPI_ISL_1362368, EPI_ISL_1362369, EPI_ISL_1362370, EPI_ISL_1362371, EPI_ISL_1362372, EPI_ISL_1362373, EPI_ISL_1362374, EPI_ISL_1362375, EPI_ISL_1362376, EPI_ISL_1362377, EPI_ISL_1362378, EPI_ISL_1362379, EPI_ISL_1362380, EPI_ISL_1362381, EPI_ISL_1362382, EPI_ISL_1362383, EPI_ISL_1362384, EPI_ISL_1362385, EPI_ISL_1362386, EPI_ISL_1362387, EPI_ISL_1362388, EPI_ISL_1362389, EPI_ISL_1362390, EPI_ISL_1362391, EPI_ISL_1362392, EPI_ISL_1362393, EPI_ISL_1362394, EPI_ISL_1362395, EPI_ISL_1362396, EPI_ISL_1362397, EPI_ISL_1362398, EPI_ISL_1362399, EPI_ISL_1362400, EPI_ISL_1362401, EPI_ISL_1362402, EPI_ISL_1362403, EPI_ISL_1362404, EPI_ISL_1362405, EPI_ISL_1362406, EPI_ISL_1362407, EPI_ISL_1362408, EPI_ISL_1362409, EPI_ISL_1362410, EPI_ISL_1362411, EPI_ISL_1362412, EPI_ISL_1362413, EPI_ISL_1362414, EPI_ISL_1362415, EPI_ISL_1362416, EPI_ISL_1362417, EPI_ISL_1362418, EPI_ISL_1362419, EPI_ISL_1362420, EPI_ISL_1362421, EPI_ISL_1362422, EPI_ISL_1362423, EPI_ISL_1362424, EPI_ISL_1362425, EPI_ISL_1362426, EPI_ISL_1362427, EPI_ISL_1362428, EPI_ISL_1362429, EPI_ISL_1362430, EPI_ISL_1362431, EPI_ISL_1362432, EPI_ISL_1362433, EPI_ISL_1362434, EPI_ISL_1362435, EPI_ISL_1362436, EPI_ISL_1362437, EPI_ISL_1362438, EPI_ISL_1362439, EPI_ISL_1362440, EPI_ISL_1362441, EPI_ISL_1362442, EPI_ISL_1362443, EPI_ISL_1362444, EPI_ISL_1362445, EPI_ISL_1362446, EPI_ISL_1362447, EPI_ISL_1362448, EPI_ISL_1362449, EPI_ISL_1362450, EPI_ISL_1362451, EPI_ISL_1362452, EPI_ISL_1362453, EPI_ISL_1362454, EPI_ISL_1362455, EPI_ISL_1362456, EPI_ISL_1362457, EPI_ISL_1362458, EPI_ISL_1362459, EPI_ISL_1362460, EPI_ISL_1362461, EPI_ISL_1362462, EPI_ISL_1362463, EPI_ISL_1362464, EPI_ISL_1362465, EPI_ISL_1362466, EPI_ISL_1362467, EPI_ISL_1362468, EPI_ISL_1362469, EPI_ISL_1362470, EPI_ISL_1362507, EPI_ISL_1362508, EPI_ISL_1362509, EPI_ISL_1362510, EPI_ISL_1362511, EPI_ISL_1362512, EPI_ISL_1362513, EPI_ISL_1362514, EPI_ISL_1362515, EPI_ISL_1362516, EPI_ISL_1362517, EPI_ISL_1362518, EPI_ISL_1362519, EPI_ISL_1362520, EPI_ISL_1362521, EPI_ISL_1362522, EPI_ISL_1362523, EPI_ISL_1362524, EPI_ISL_1362525, EPI_ISL_1362526, EPI_ISL_1362527, EPI_ISL_1362528, EPI_ISL_1362529, EPI_ISL_1362530, EPI_ISL_1362531, EPI_ISL_1362532, EPI_ISL_1362533, EPI_ISL_1362534, EPI_ISL_1362535, EPI_ISL_1362536, EPI_ISL_1362537, EPI_ISL_1362538, EPI_ISL_1362539, EPI_ISL_1362540, EPI_ISL_1362541, EPI_ISL_1362542, EPI_ISL_1362543, EPI_ISL_1362544, EPI_ISL_1362545, EPI_ISL_1362546, EPI_ISL_1362547, EPI_ISL_1362548, EPI_ISL_1362549, EPI_ISL_1362550, EPI_ISL_1362551, EPI_ISL_1363822, EPI_ISL_1363824, EPI_ISL_1363825, EPI_ISL_1363826, EPI_ISL_1363827, EPI_ISL_1363828, EPI_ISL_1363829, EPI_ISL_1363872, EPI_ISL_1363873, EPI_ISL_1363874, EPI_ISL_1363875, EPI_ISL_1363876, EPI_ISL_1363882, EPI_ISL_1363883, EPI_ISL_1363884, EPI_ISL_1363885, EPI_ISL_1363886, EPI_ISL_1363887, EPI_ISL_1363888, EPI_ISL_1363889, EPI_ISL_1363890, EPI_ISL_1363891, EPI_ISL_1363892, EPI_ISL_1363893, EPI_ISL_1363894, EPI_ISL_1363895, EPI_ISL_1363949, EPI_ISL_1363950, EPI_ISL_1363951, EPI_ISL_1363952, EPI_ISL_1363953, EPI_ISL_1363954, EPI_ISL_1363955, EPI_ISL_1363956, EPI_ISL_1363957, EPI_ISL_1363993, EPI_ISL_1363994, EPI_ISL_1363995, EPI_ISL_1364043, EPI_ISL_1364044, EPI_ISL_1364045, EPI_ISL_1364063, EPI_ISL_1364064, EPI_ISL_1364065, EPI_ISL_1364066, EPI_ISL_1364067, EPI_ISL_1364068, EPI_ISL_1364069, EPI_ISL_1364070, EPI_ISL_1364072, EPI_ISL_1364073, EPI_ISL_1364074, EPI_ISL_1364075, EPI_ISL_1364076, EPI_ISL_1364077, EPI_ISL_1364078, EPI_ISL_1364079, EPI_ISL_1364080, EPI_ISL_1364081, EPI_ISL_1364082, EPI_ISL_1364083, EPI_ISL_1364084, EPI_ISL_1364085, EPI_ISL_1364086, EPI_ISL_1364087, EPI_ISL_1364088, EPI_ISL_1364089, EPI_ISL_1364090, EPI_ISL_1364091, EPI_ISL_1364092, EPI_ISL_1364093, EPI_ISL_1364094, EPI_ISL_1364095, EPI_ISL_1364096, EPI_ISL_1364097, EPI_ISL_1364098, EPI_ISL_1364107, EPI_ISL_1364108, EPI_ISL_1364109, EPI_ISL_1364110, EPI_ISL_1364111, EPI_ISL_1364112, EPI_ISL_1364113, EPI_ISL_1364114, EPI_ISL_1364115, EPI_ISL_1364116, EPI_ISL_1364117, EPI_ISL_1364118, EPI_ISL_1364119, EPI_ISL_1364120, EPI_ISL_1364149, EPI_ISL_1364150, EPI_ISL_1364151, EPI_ISL_1364152, EPI_ISL_1364153, EPI_ISL_1364154, EPI_ISL_1364155, EPI_ISL_1364156, EPI_ISL_1364157, EPI_ISL_1364158, EPI_ISL_1364189, EPI_ISL_1364247, EPI_ISL_1364248, EPI_ISL_1364249, EPI_ISL_1364250, EPI_ISL_1364251, EPI_ISL_1364252, EPI_ISL_1364253, EPI_ISL_1364254, EPI_ISL_1364255, EPI_ISL_1364256, EPI_ISL_1364257, EPI_ISL_1364258, EPI_ISL_1364259, EPI_ISL_1364260, EPI_ISL_1364261, EPI_ISL_1364264, EPI_ISL_1364267, EPI_ISL_1364269, EPI_ISL_1364270, EPI_ISL_1364271, EPI_ISL_1364272, EPI_ISL_1364273, EPI_ISL_1364274, EPI_ISL_1364275, EPI_ISL_1364276, EPI_ISL_1364306, EPI_ISL_1364307, EPI_ISL_1364308, EPI_ISL_1364309, EPI_ISL_1364310, EPI_ISL_1364311, EPI_ISL_1364312, EPI_ISL_1364313, EPI_ISL_1364317, EPI_ISL_1364318, EPI_ISL_1364319, EPI_ISL_1364320, EPI_ISL_1364321, EPI_ISL_1364323, EPI_ISL_1364324, EPI_ISL_1364325, EPI_ISL_1364326, EPI_ISL_1364327, EPI_ISL_1364328, EPI_ISL_1364329, EPI_ISL_1364330, EPI_ISL_1364331, EPI_ISL_1364332, EPI_ISL_1364333, EPI_ISL_1364334, EPI_ISL_1364335, EPI_ISL_1364336, EPI_ISL_1364337, EPI_ISL_1364338, EPI_ISL_1364339, EPI_ISL_1364340, EPI_ISL_1364365, EPI_ISL_1364366, EPI_ISL_1364367, EPI_ISL_1364368, EPI_ISL_1364369, EPI_ISL_1364370, EPI_ISL_1364371, EPI_ISL_1364372, EPI_ISL_1364373, EPI_ISL_1364374, EPI_ISL_1364375, EPI_ISL_1364376, EPI_ISL_1364377, EPI_ISL_1364378, EPI_ISL_1364379, EPI_ISL_1364380, EPI_ISL_1364381, EPI_ISL_1364382, EPI_ISL_1364383, EPI_ISL_1364384, EPI_ISL_1364385, EPI_ISL_1364386, EPI_ISL_1364387, EPI_ISL_1364388, EPI_ISL_1364389, EPI_ISL_1364390, EPI_ISL_1364391, EPI_ISL_1364392, EPI_ISL_1364393, EPI_ISL_1364394, EPI_ISL_1364395, EPI_ISL_1364396, EPI_ISL_1364397, EPI_ISL_1364398, EPI_ISL_1364399, EPI_ISL_1364400, EPI_ISL_1364401, EPI_ISL_1364402, EPI_ISL_1364403, EPI_ISL_1364404, EPI_ISL_1364405, EPI_ISL_1364426, EPI_ISL_1364427, EPI_ISL_1364428, EPI_ISL_1417443, EPI_ISL_1417445, EPI_ISL_1417449, EPI_ISL_1417451, EPI_ISL_1417452, EPI_ISL_1417453, EPI_ISL_1417459, EPI_ISL_1417460, EPI_ISL_1417461, EPI_ISL_1417466, EPI_ISL_1417467, EPI_ISL_1417470, EPI_ISL_1417476, EPI_ISL_1417477, EPI_ISL_1417483, EPI_ISL_1417484, EPI_ISL_1417485, EPI_ISL_1417487, EPI_ISL_1417488, EPI_ISL_1417489, EPI_ISL_1417490, EPI_ISL_1417499, EPI_ISL_1417501, EPI_ISL_1417504, EPI_ISL_1417505, EPI_ISL_1417508, EPI_ISL_1417509, EPI_ISL_1417511, EPI_ISL_1417513, EPI_ISL_1417514, EPI_ISL_1417515, EPI_ISL_1417520, EPI_ISL_1417521, EPI_ISL_1417523, EPI_ISL_1417524, EPI_ISL_1417526, EPI_ISL_1417528, EPI_ISL_1417529, EPI_ISL_1417530, EPI_ISL_1417531, EPI_ISL_1417538, EPI_ISL_1417542, EPI_ISL_1417543, EPI_ISL_1417544, EPI_ISL_1417548, EPI_ISL_1417559, EPI_ISL_1417566, EPI_ISL_1417567, EPI_ISL_1417569, EPI_ISL_1417575, EPI_ISL_1417576, EPI_ISL_1417578, EPI_ISL_1417579, EPI_ISL_1417580, EPI_ISL_1417583, EPI_ISL_1417588, EPI_ISL_1417589, EPI_ISL_1417590, EPI_ISL_1417591, EPI_ISL_1417593, EPI_ISL_1417596, EPI_ISL_1417599, EPI_ISL_1417600, EPI_ISL_1417601, EPI_ISL_1417619, EPI_ISL_1417697, EPI_ISL_1417698, EPI_ISL_1417699, EPI_ISL_1417700, EPI_ISL_1417701, EPI_ISL_1417702, EPI_ISL_1417703, EPI_ISL_1417704, EPI_ISL_1417705, EPI_ISL_1417706, EPI_ISL_1417707, EPI_ISL_1417708, EPI_ISL_1417709, EPI_ISL_1417710, EPI_ISL_1417711, EPI_ISL_1417712, EPI_ISL_1417713, EPI_ISL_1417714, EPI_ISL_1417715, EPI_ISL_1417716, EPI_ISL_1417717, EPI_ISL_1417718, EPI_ISL_1417719, EPI_ISL_1417720, EPI_ISL_1417721, EPI_ISL_1417722, EPI_ISL_1417723, EPI_ISL_1417724, EPI_ISL_1417725, EPI_ISL_1417726, EPI_ISL_1417727, EPI_ISL_1417728, EPI_ISL_1417729, EPI_ISL_1417730, EPI_ISL_1417731, EPI_ISL_1417732, EPI_ISL_1417733, EPI_ISL_1417734, EPI_ISL_1417735, EPI_ISL_1417736, EPI_ISL_1417737, EPI_ISL_1417738, EPI_ISL_1417739, EPI_ISL_1417740, EPI_ISL_1417741, EPI_ISL_1417742, EPI_ISL_1417743, EPI_ISL_1417744, EPI_ISL_1417745, EPI_ISL_1417746, EPI_ISL_1417747, EPI_ISL_1417748, EPI_ISL_1417749, EPI_ISL_1417750, EPI_ISL_1417751, EPI_ISL_1417752, EPI_ISL_1417753, EPI_ISL_1417754, EPI_ISL_1417755, EPI_ISL_1417756, EPI_ISL_1417757, EPI_ISL_1417758, EPI_ISL_1417759, EPI_ISL_1417760, EPI_ISL_1417761, EPI_ISL_1417762, EPI_ISL_1417763, EPI_ISL_1417764, EPI_ISL_1417765, EPI_ISL_1417766, EPI_ISL_1417767, EPI_ISL_1417768, EPI_ISL_1417769, EPI_ISL_1417770, EPI_ISL_1417771, EPI_ISL_1417772, EPI_ISL_1417773, EPI_ISL_1417774, EPI_ISL_1417878, EPI_ISL_1417920, EPI_ISL_1417921, EPI_ISL_1417922, EPI_ISL_1417923, EPI_ISL_1417924, EPI_ISL_1417925, EPI_ISL_1417926, EPI_ISL_1417927, EPI_ISL_1417928, EPI_ISL_1417929, EPI_ISL_1417930, EPI_ISL_1417931, EPI_ISL_1417932, EPI_ISL_1417933, EPI_ISL_1417934, EPI_ISL_1417935, EPI_ISL_1417936, EPI_ISL_1417937, EPI_ISL_1417938, EPI_ISL_1417939, EPI_ISL_1417940, EPI_ISL_1417941, EPI_ISL_1417942, EPI_ISL_1417943, EPI_ISL_1417944, EPI_ISL_1417945, EPI_ISL_1417946, EPI_ISL_1417947, EPI_ISL_1417948, EPI_ISL_1417949, EPI_ISL_1417950, EPI_ISL_1417951, EPI_ISL_1417952, EPI_ISL_1417953, EPI_ISL_1417954, EPI_ISL_1417955, EPI_ISL_1417956, EPI_ISL_1417957, EPI_ISL_1417958, EPI_ISL_1417959, EPI_ISL_1417960, EPI_ISL_1417961, EPI_ISL_1417962, EPI_ISL_1417963, EPI_ISL_1417964, EPI_ISL_1417965, EPI_ISL_1417966, EPI_ISL_1417967, EPI_ISL_1417968, EPI_ISL_1417969, EPI_ISL_1417970, EPI_ISL_1417971, EPI_ISL_1417972, EPI_ISL_1417973, EPI_ISL_1417974, EPI_ISL_1417975, EPI_ISL_1417976, EPI_ISL_1417977, EPI_ISL_1417978, EPI_ISL_1417979, EPI_ISL_1417980, EPI_ISL_1417981, EPI_ISL_1417982, EPI_ISL_1417983, EPI_ISL_1417984, EPI_ISL_1417985, EPI_ISL_1417986, EPI_ISL_1417987, EPI_ISL_1417988, EPI_ISL_1417989, EPI_ISL_1417990, EPI_ISL_1417991, EPI_ISL_1417992, EPI_ISL_1417993, EPI_ISL_1417994, EPI_ISL_1417996, EPI_ISL_1417997, EPI_ISL_1417998, EPI_ISL_1417999, EPI_ISL_1418030, EPI_ISL_1418031, EPI_ISL_1418032, EPI_ISL_1418033, EPI_ISL_1418034, EPI_ISL_1418035, EPI_ISL_1418036, EPI_ISL_1418037, EPI_ISL_1418038, EPI_ISL_1418055, EPI_ISL_1418058, EPI_ISL_1418059, EPI_ISL_1418061, EPI_ISL_1418068, EPI_ISL_1418069, EPI_ISL_1418071, EPI_ISL_1418072, EPI_ISL_1418073, EPI_ISL_1418074, EPI_ISL_1418075, EPI_ISL_1418081, EPI_ISL_1418082, EPI_ISL_1418083, EPI_ISL_1418085, EPI_ISL_1418086, EPI_ISL_1418087, EPI_ISL_1418089, EPI_ISL_1418091, EPI_ISL_1418092, EPI_ISL_1418093, EPI_ISL_1418099, EPI_ISL_1418102, EPI_ISL_1418104, EPI_ISL_1418105, EPI_ISL_1418107, EPI_ISL_1418110, EPI_ISL_1418112, EPI_ISL_1418115, EPI_ISL_1418116, EPI_ISL_1418117, EPI_ISL_1418118, EPI_ISL_1418121, EPI_ISL_1418126, EPI_ISL_1418128, EPI_ISL_1418130, EPI_ISL_1418131, EPI_ISL_1418133, EPI_ISL_1418135, EPI_ISL_1418139, EPI_ISL_1418140, EPI_ISL_1418141, EPI_ISL_1418142, EPI_ISL_1418143, EPI_ISL_1418144, EPI_ISL_1418181, EPI_ISL_1418182, EPI_ISL_1418183, EPI_ISL_1418184, EPI_ISL_1418331, EPI_ISL_1418333, EPI_ISL_1418334, EPI_ISL_1418335, EPI_ISL_1418336, EPI_ISL_1418342, EPI_ISL_1418344, EPI_ISL_1418345, EPI_ISL_1418347, EPI_ISL_1418354, EPI_ISL_1418355, EPI_ISL_1418358, EPI_ISL_1418361, EPI_ISL_1418363, EPI_ISL_1418367, EPI_ISL_1418368, EPI_ISL_1418369, EPI_ISL_1418371, EPI_ISL_1418383, EPI_ISL_1418384, EPI_ISL_1418385, EPI_ISL_1418386, EPI_ISL_1418387, EPI_ISL_1418388, EPI_ISL_1418389, EPI_ISL_1418476, EPI_ISL_1418477, EPI_ISL_1418478, EPI_ISL_1418479, EPI_ISL_1418480, EPI_ISL_1418481, EPI_ISL_1418482, EPI_ISL_1418483, EPI_ISL_1418484, EPI_ISL_1418485, EPI_ISL_1418486, EPI_ISL_1418487, EPI_ISL_1418488, EPI_ISL_1418489, EPI_ISL_1418490, EPI_ISL_1418491, EPI_ISL_1418492, EPI_ISL_1418493, EPI_ISL_1418494, EPI_ISL_1418495, EPI_ISL_1418496, EPI_ISL_1418497, EPI_ISL_1418498, EPI_ISL_1418499, EPI_ISL_1418500, EPI_ISL_1418501, EPI_ISL_1418502, EPI_ISL_1418503, EPI_ISL_1418504, EPI_ISL_1418505, EPI_ISL_1418506, EPI_ISL_1418507, EPI_ISL_1418508, EPI_ISL_1418509, EPI_ISL_1418510, EPI_ISL_1418511, EPI_ISL_1418512, EPI_ISL_1418513, EPI_ISL_1418514, EPI_ISL_1418515, EPI_ISL_1418542, EPI_ISL_1418583, EPI_ISL_1418584, EPI_ISL_1418585, EPI_ISL_1418586, EPI_ISL_1418587, EPI_ISL_1418588, EPI_ISL_1418589, EPI_ISL_1418590, EPI_ISL_1418591, EPI_ISL_1418592, EPI_ISL_1418593, EPI_ISL_1418594, EPI_ISL_1418595, EPI_ISL_1418596, EPI_ISL_1418597, EPI_ISL_1418598, EPI_ISL_1418599, EPI_ISL_1418600, EPI_ISL_1418601, EPI_ISL_1418619, EPI_ISL_1418620, EPI_ISL_1418621, EPI_ISL_1418624, EPI_ISL_1418625, EPI_ISL_1418626, EPI_ISL_1418627, EPI_ISL_1418628, EPI_ISL_1418629, EPI_ISL_1418630, EPI_ISL_1418639, EPI_ISL_1418640, EPI_ISL_1418641, EPI_ISL_1418642, EPI_ISL_1418643, EPI_ISL_1418645, EPI_ISL_1418647, EPI_ISL_1418684, EPI_ISL_1418685, EPI_ISL_1418686, EPI_ISL_1418687, EPI_ISL_1418688, EPI_ISL_1418689, EPI_ISL_1418690, EPI_ISL_1418691, EPI_ISL_1418716, EPI_ISL_1418720, EPI_ISL_1418721, EPI_ISL_1418722, EPI_ISL_1418753, EPI_ISL_1418754, EPI_ISL_1418755, EPI_ISL_1418768, EPI_ISL_1418769, EPI_ISL_1418770, EPI_ISL_1418778, EPI_ISL_1418779, EPI_ISL_1418780, EPI_ISL_1418781, EPI_ISL_1418782, EPI_ISL_1418783, EPI_ISL_1418784, EPI_ISL_1418785, EPI_ISL_1418786, EPI_ISL_1418787, EPI_ISL_1418788, EPI_ISL_1418789, EPI_ISL_1418790, EPI_ISL_1418791, EPI_ISL_1418792, EPI_ISL_1418793, EPI_ISL_1418794, EPI_ISL_1418795, EPI_ISL_1418796, EPI_ISL_1418797, EPI_ISL_1418798, EPI_ISL_1418799, EPI_ISL_1418800, EPI_ISL_1418801, EPI_ISL_1418802, EPI_ISL_1418803, EPI_ISL_1418804, EPI_ISL_1418805, EPI_ISL_1418806, EPI_ISL_1418807, EPI_ISL_1418808, EPI_ISL_1418809, EPI_ISL_1418810, EPI_ISL_1418811, EPI_ISL_1418812, EPI_ISL_1418813, EPI_ISL_1418814, EPI_ISL_1418815, EPI_ISL_1418816, EPI_ISL_1418817, EPI_ISL_1418818, EPI_ISL_1418819, EPI_ISL_1418820, EPI_ISL_1418821, EPI_ISL_1418822, EPI_ISL_1418823, EPI_ISL_1418824, EPI_ISL_1418825, EPI_ISL_1418826, EPI_ISL_1418827, EPI_ISL_1418828, EPI_ISL_1418829, EPI_ISL_1418830, EPI_ISL_1418831, EPI_ISL_1418832, EPI_ISL_1418833, EPI_ISL_1418834, EPI_ISL_1418835, EPI_ISL_1418836, EPI_ISL_1418837, EPI_ISL_1418838, EPI_ISL_1418839, EPI_ISL_1419928, EPI_ISL_1419930, EPI_ISL_1419931, EPI_ISL_1419937, EPI_ISL_1419939, EPI_ISL_1419942, EPI_ISL_1419947, EPI_ISL_1419951, EPI_ISL_1419952, EPI_ISL_1419955, EPI_ISL_1419958, EPI_ISL_1419959, EPI_ISL_1419963, EPI_ISL_1419965, EPI_ISL_1419967, EPI_ISL_1419970, EPI_ISL_1419972, EPI_ISL_1419981, EPI_ISL_1419988, EPI_ISL_1419991, EPI_ISL_1420001, EPI_ISL_1420002, EPI_ISL_1420004, EPI_ISL_1420005, EPI_ISL_1420006, EPI_ISL_1420007, EPI_ISL_1420008, EPI_ISL_1420009, EPI_ISL_1420010, EPI_ISL_1420011, EPI_ISL_1420012, EPI_ISL_1420017, EPI_ISL_1420055, EPI_ISL_1420056, EPI_ISL_1420057, EPI_ISL_1420058, EPI_ISL_1420059, EPI_ISL_1420060, EPI_ISL_1420061, EPI_ISL_1420062, EPI_ISL_1420063, EPI_ISL_1420064, EPI_ISL_1420065, EPI_ISL_1420066, EPI_ISL_1420067, EPI_ISL_1420068, EPI_ISL_1420069, EPI_ISL_1420070, EPI_ISL_1420071, EPI_ISL_1420072, EPI_ISL_1420073, EPI_ISL_1420074, EPI_ISL_1420075, EPI_ISL_1420076, EPI_ISL_1420077, EPI_ISL_1420078, EPI_ISL_1420079, EPI_ISL_1420080, EPI_ISL_1420081, EPI_ISL_1420082, EPI_ISL_1420083, EPI_ISL_1420084, EPI_ISL_1420085, EPI_ISL_1420086, EPI_ISL_1420087, EPI_ISL_1420088, EPI_ISL_1420089, EPI_ISL_1420090, EPI_ISL_1420091, EPI_ISL_1420092, EPI_ISL_1420093, EPI_ISL_1420094, EPI_ISL_1420095, EPI_ISL_1420096, EPI_ISL_1420097, EPI_ISL_1420098, EPI_ISL_1420099, EPI_ISL_1420100, EPI_ISL_1420101, EPI_ISL_1420102, EPI_ISL_1420103, EPI_ISL_1420104, EPI_ISL_1420105, EPI_ISL_1420106, EPI_ISL_1420107, EPI_ISL_1420108, EPI_ISL_1420109, EPI_ISL_1420110, EPI_ISL_1420111, EPI_ISL_1420112, EPI_ISL_1420113, EPI_ISL_1420114, EPI_ISL_1420115, EPI_ISL_1420116, EPI_ISL_1420117, EPI_ISL_1420118, EPI_ISL_1420119, EPI_ISL_1420120, EPI_ISL_1420121, EPI_ISL_1420122, EPI_ISL_1420123, EPI_ISL_1420124, EPI_ISL_1420125, EPI_ISL_1420126, EPI_ISL_1420127, EPI_ISL_1420128, EPI_ISL_1420129, EPI_ISL_1420130, EPI_ISL_1420131, EPI_ISL_1420132, EPI_ISL_1420133, EPI_ISL_1420134, EPI_ISL_1420135, EPI_ISL_1420136, EPI_ISL_1420137, EPI_ISL_1420138, EPI_ISL_1420139, EPI_ISL_1420140, EPI_ISL_1420141, EPI_ISL_1420142, EPI_ISL_1420143, EPI_ISL_1420144, EPI_ISL_1420145, EPI_ISL_1420146, EPI_ISL_1420147, EPI_ISL_1420148, EPI_ISL_1420149, EPI_ISL_1420150, EPI_ISL_1420151, EPI_ISL_1420152, EPI_ISL_1420153, EPI_ISL_1420154, EPI_ISL_1420155, EPI_ISL_1420156, EPI_ISL_1420157, EPI_ISL_1420158, EPI_ISL_1420159, EPI_ISL_1420160, EPI_ISL_1420161, EPI_ISL_1420162, EPI_ISL_1420163, EPI_ISL_1420164, EPI_ISL_1420165, EPI_ISL_1420166, EPI_ISL_1420167, EPI_ISL_1420168, EPI_ISL_1420169, EPI_ISL_1420170, EPI_ISL_1420171, EPI_ISL_1420172, EPI_ISL_1420173, EPI_ISL_1420174, EPI_ISL_1420175, EPI_ISL_1420176, EPI_ISL_1420177, EPI_ISL_1420178, EPI_ISL_1420179, EPI_ISL_1420180, EPI_ISL_1420181, EPI_ISL_1420182, EPI_ISL_1420183, EPI_ISL_1420184, EPI_ISL_1420185, EPI_ISL_1420186, EPI_ISL_1420187, EPI_ISL_1420188, EPI_ISL_1420189, EPI_ISL_1420190, EPI_ISL_1420191, EPI_ISL_1420192, EPI_ISL_1420193, EPI_ISL_1420194, EPI_ISL_1420195, EPI_ISL_1420196, EPI_ISL_1420197, EPI_ISL_1420198, EPI_ISL_1420199, EPI_ISL_1420200, EPI_ISL_1420201, EPI_ISL_1420202, EPI_ISL_1420203, EPI_ISL_1420204, EPI_ISL_1420205, EPI_ISL_1420206, EPI_ISL_1420207, EPI_ISL_1420208, EPI_ISL_1420209, EPI_ISL_1420210, EPI_ISL_1420211, EPI_ISL_1420212, EPI_ISL_1420213, EPI_ISL_1420214, EPI_ISL_1420215, EPI_ISL_1420216, EPI_ISL_1420217, EPI_ISL_1420218, EPI_ISL_1420219, EPI_ISL_1420220, EPI_ISL_1420221, EPI_ISL_1420222, EPI_ISL_1420223, EPI_ISL_1420224, EPI_ISL_1420352, EPI_ISL_1420353, EPI_ISL_1420379, EPI_ISL_1420380, EPI_ISL_1420381, EPI_ISL_1420382, EPI_ISL_1420383, EPI_ISL_1420384, EPI_ISL_1420385, EPI_ISL_1420386, EPI_ISL_1420387, EPI_ISL_1420388, EPI_ISL_1420389, EPI_ISL_1420390, EPI_ISL_1420391, EPI_ISL_1420392, EPI_ISL_1420393, EPI_ISL_1420394, EPI_ISL_1420395, EPI_ISL_1420396, EPI_ISL_1420397, EPI_ISL_1420398, EPI_ISL_1420399, EPI_ISL_1420400, EPI_ISL_1420401, EPI_ISL_1420402, EPI_ISL_1420403, EPI_ISL_1420404, EPI_ISL_1420405, EPI_ISL_1420406, EPI_ISL_1420407, EPI_ISL_1420408, EPI_ISL_1420409, EPI_ISL_1420410, EPI_ISL_1420411, EPI_ISL_1420412, EPI_ISL_1420413, EPI_ISL_1420414, EPI_ISL_1420415, EPI_ISL_1420416, EPI_ISL_1420417, EPI_ISL_1420418, EPI_ISL_1420419, EPI_ISL_1420420, EPI_ISL_1420421, EPI_ISL_1420422, EPI_ISL_1420423, EPI_ISL_1420424, EPI_ISL_1420425, EPI_ISL_1420426, EPI_ISL_1420427, EPI_ISL_1420428, EPI_ISL_1420429, EPI_ISL_1420430, EPI_ISL_1420431, EPI_ISL_1420432, EPI_ISL_1420433, EPI_ISL_1420434, EPI_ISL_1420435, EPI_ISL_1420436, EPI_ISL_1420437, EPI_ISL_1420438, EPI_ISL_1420439, EPI_ISL_1420440, EPI_ISL_1420441, EPI_ISL_1420442, EPI_ISL_1420443, EPI_ISL_1420444, EPI_ISL_1420445, EPI_ISL_1420446, EPI_ISL_1420447, EPI_ISL_1420448, EPI_ISL_1420449, EPI_ISL_1420450, EPI_ISL_1420451, EPI_ISL_1420452, EPI_ISL_1420453, EPI_ISL_1420454, EPI_ISL_1420455, EPI_ISL_1420456, EPI_ISL_1420457, EPI_ISL_1420458, EPI_ISL_1420459, EPI_ISL_1420527, EPI_ISL_1420529, EPI_ISL_1420530, EPI_ISL_1420531, EPI_ISL_1420533, EPI_ISL_1420534, EPI_ISL_1420535, EPI_ISL_1420537, EPI_ISL_1420538, EPI_ISL_1420539, EPI_ISL_1420540, EPI_ISL_1420541, EPI_ISL_1420542, EPI_ISL_1420543, EPI_ISL_1420544, EPI_ISL_1420545, EPI_ISL_1420546, EPI_ISL_1420547, EPI_ISL_1420548, EPI_ISL_1420549, EPI_ISL_1420550, EPI_ISL_1420559, EPI_ISL_1420562, EPI_ISL_1420564, EPI_ISL_1420569, EPI_ISL_1420570, EPI_ISL_1420571, EPI_ISL_1420573, EPI_ISL_1420574, EPI_ISL_1420575, EPI_ISL_1420576, EPI_ISL_1420577, EPI_ISL_1420578, EPI_ISL_1420579, EPI_ISL_1420580, EPI_ISL_1420581, EPI_ISL_1420582, EPI_ISL_1420583, EPI_ISL_1420584, EPI_ISL_1420585, EPI_ISL_1420586, EPI_ISL_1420587, EPI_ISL_1420588, EPI_ISL_1420589, EPI_ISL_1420590, EPI_ISL_1420591, EPI_ISL_1420592, EPI_ISL_1420593, EPI_ISL_1420594, EPI_ISL_1420595, EPI_ISL_1420620, EPI_ISL_1420621, EPI_ISL_1420622, EPI_ISL_1420623, EPI_ISL_1420624, EPI_ISL_1420625, EPI_ISL_1420626, EPI_ISL_1420627, EPI_ISL_1420628, EPI_ISL_1420629, EPI_ISL_1420630, EPI_ISL_1420631, EPI_ISL_1420632, EPI_ISL_1420633, EPI_ISL_1599772, EPI_ISL_1599773, EPI_ISL_1599774, EPI_ISL_1599775, EPI_ISL_1599777, EPI_ISL_1599778, EPI_ISL_1599779, EPI_ISL_1599780, EPI_ISL_1599781, EPI_ISL_1599782, EPI_ISL_1599783, EPI_ISL_1599784, EPI_ISL_1599785, EPI_ISL_1599786, EPI_ISL_1599787, EPI_ISL_1599788, EPI_ISL_1599789, EPI_ISL_1599790, EPI_ISL_1599791, EPI_ISL_1599792, EPI_ISL_1599793, EPI_ISL_1599794, EPI_ISL_1599795, EPI_ISL_1599796, EPI_ISL_1599797, EPI_ISL_1599798, EPI_ISL_1599799, EPI_ISL_1599800, EPI_ISL_1599801, EPI_ISL_1599803, EPI_ISL_1599804, EPI_ISL_1599805, EPI_ISL_1599806, EPI_ISL_1599807, EPI_ISL_1599808, EPI_ISL_1599809, EPI_ISL_1599810, EPI_ISL_1599811, EPI_ISL_1599812, EPI_ISL_1599813, EPI_ISL_1599815, EPI_ISL_1599816, EPI_ISL_1599817, EPI_ISL_1599818, EPI_ISL_1599819, EPI_ISL_1599820, EPI_ISL_1599821, EPI_ISL_1599822, EPI_ISL_1599830, EPI_ISL_1599831, EPI_ISL_1599832, EPI_ISL_1599833, EPI_ISL_1599834, EPI_ISL_1599835, EPI_ISL_1599840, EPI_ISL_1599841, EPI_ISL_1599842, EPI_ISL_1599843, EPI_ISL_1599844, EPI_ISL_1599845, EPI_ISL_1599846, EPI_ISL_1599847, EPI_ISL_1599848, EPI_ISL_1599849, EPI_ISL_1599850, EPI_ISL_1599851, EPI_ISL_1599852, EPI_ISL_1599853, EPI_ISL_1599854, EPI_ISL_1599855, EPI_ISL_1599857, EPI_ISL_1599858, EPI_ISL_1599859, EPI_ISL_1599860, EPI_ISL_1599861, EPI_ISL_1599862, EPI_ISL_1599863, EPI_ISL_1599864, EPI_ISL_1599865, EPI_ISL_1599866, EPI_ISL_1599867, EPI_ISL_1599904, EPI_ISL_1599905, EPI_ISL_1599945, EPI_ISL_1599946, EPI_ISL_1599947, EPI_ISL_1599948, EPI_ISL_1599949, EPI_ISL_1599950, EPI_ISL_1599951, EPI_ISL_1599952, EPI_ISL_1600039, EPI_ISL_1600040, EPI_ISL_1600041, EPI_ISL_1600042, EPI_ISL_1600043, EPI_ISL_1600044, EPI_ISL_1600045, EPI_ISL_1600046, EPI_ISL_1600047, EPI_ISL_1600048, EPI_ISL_1600049, EPI_ISL_1600051, EPI_ISL_1600053, EPI_ISL_1600055, EPI_ISL_1600057, EPI_ISL_1600063, EPI_ISL_1600065, EPI_ISL_1600067, EPI_ISL_1600069, EPI_ISL_1600071, EPI_ISL_1600075, EPI_ISL_1600077, EPI_ISL_1600079, EPI_ISL_1600135, EPI_ISL_1600137, EPI_ISL_1600139, EPI_ISL_1600141, EPI_ISL_1600143, EPI_ISL_1600145, EPI_ISL_1600147, EPI_ISL_1600149, EPI_ISL_1600151, EPI_ISL_1600153, EPI_ISL_1600156, EPI_ISL_1600158, EPI_ISL_1600160, EPI_ISL_1600162, EPI_ISL_1600164, EPI_ISL_1600166, EPI_ISL_1600168, EPI_ISL_1600171, EPI_ISL_1600173, EPI_ISL_1600175, EPI_ISL_1600177, EPI_ISL_1600179, EPI_ISL_1600181, EPI_ISL_1600183, EPI_ISL_1600185, EPI_ISL_1600186, EPI_ISL_1600189, EPI_ISL_1600190, EPI_ISL_1600192, EPI_ISL_1600194, EPI_ISL_1600197, EPI_ISL_1600198, EPI_ISL_1600201, EPI_ISL_1600202, EPI_ISL_1600203, EPI_ISL_1600204, EPI_ISL_1600205, EPI_ISL_1600206, EPI_ISL_1600222, EPI_ISL_1600223, EPI_ISL_1600224, EPI_ISL_1600225, EPI_ISL_1600226, EPI_ISL_1600227, EPI_ISL_1600228, EPI_ISL_1600229, EPI_ISL_1600230, EPI_ISL_1600231, EPI_ISL_1600232, EPI_ISL_1600233, EPI_ISL_1600234, EPI_ISL_1600235, EPI_ISL_1600236, EPI_ISL_1600237, EPI_ISL_1600238, EPI_ISL_1600239, EPI_ISL_1600269, EPI_ISL_1600270, EPI_ISL_1600271, EPI_ISL_1600282, EPI_ISL_1600283, EPI_ISL_1600284, EPI_ISL_1600285, EPI_ISL_1600286, EPI_ISL_1600287, EPI_ISL_1600345, EPI_ISL_1600346, EPI_ISL_1600347, EPI_ISL_1600348, EPI_ISL_1600349, EPI_ISL_1600350, EPI_ISL_1600351, EPI_ISL_1600352, EPI_ISL_1600353, EPI_ISL_1600354, EPI_ISL_1600355, EPI_ISL_1600356, EPI_ISL_1600357, EPI_ISL_1600358, EPI_ISL_1600359, EPI_ISL_1600360, EPI_ISL_1600361, EPI_ISL_1600362, EPI_ISL_1600363, EPI_ISL_1600364, EPI_ISL_1600365, EPI_ISL_1600366, EPI_ISL_1600367, EPI_ISL_1600368, EPI_ISL_1600369, EPI_ISL_1600370, EPI_ISL_1600371, EPI_ISL_1600372, EPI_ISL_1600373, EPI_ISL_1600374, EPI_ISL_1600375, EPI_ISL_1600376, EPI_ISL_1600377, EPI_ISL_1600378, EPI_ISL_1600379, EPI_ISL_1600380, EPI_ISL_1600381, EPI_ISL_1600382, EPI_ISL_1600383, EPI_ISL_1600384, EPI_ISL_1600386, EPI_ISL_1600387, EPI_ISL_1600388, EPI_ISL_1600389, EPI_ISL_1600390, EPI_ISL_1600391, EPI_ISL_1600392, EPI_ISL_1600393, EPI_ISL_1600394, EPI_ISL_1600395, EPI_ISL_1600396, EPI_ISL_1600397, EPI_ISL_1600398, EPI_ISL_1600399, EPI_ISL_1600400, EPI_ISL_1600401, EPI_ISL_1600402, EPI_ISL_1600403, EPI_ISL_1600404, EPI_ISL_1600405, EPI_ISL_1600406, EPI_ISL_1600407, EPI_ISL_1600408, EPI_ISL_1600409, EPI_ISL_1600410, EPI_ISL_1600411, EPI_ISL_1600412, EPI_ISL_1600413, EPI_ISL_1600414, EPI_ISL_1600415, EPI_ISL_1600416, EPI_ISL_1600417, EPI_ISL_1600418, EPI_ISL_1600419, EPI_ISL_1600420, EPI_ISL_1600422, EPI_ISL_1600423, EPI_ISL_1600424, EPI_ISL_1600425, EPI_ISL_1600426, EPI_ISL_1600427, EPI_ISL_1600433, EPI_ISL_1600434, EPI_ISL_1600435, EPI_ISL_1600436, EPI_ISL_1600437, EPI_ISL_1600438, EPI_ISL_1600439, EPI_ISL_1600440, EPI_ISL_1600441, EPI_ISL_1600442, EPI_ISL_1600444, EPI_ISL_1600445, EPI_ISL_1600446, EPI_ISL_1600447, EPI_ISL_1600448, EPI_ISL_1600449, EPI_ISL_1600450, EPI_ISL_1600451, EPI_ISL_1600460, EPI_ISL_1600461, EPI_ISL_1600462, EPI_ISL_1600463, EPI_ISL_1600464, EPI_ISL_1600465, EPI_ISL_1600466, EPI_ISL_1600467, EPI_ISL_1600468, EPI_ISL_1600469, EPI_ISL_1600470, EPI_ISL_1600472, EPI_ISL_1600473, EPI_ISL_1600474, EPI_ISL_1600475, EPI_ISL_1600476, EPI_ISL_1600477, EPI_ISL_1600478, EPI_ISL_1600479, EPI_ISL_1600480, EPI_ISL_1600481, EPI_ISL_1600482, EPI_ISL_1600483, EPI_ISL_1600484, EPI_ISL_1600485, EPI_ISL_1600486, EPI_ISL_1600487, EPI_ISL_1600488, EPI_ISL_1600489, EPI_ISL_1600490, EPI_ISL_1600491, EPI_ISL_1600492, EPI_ISL_1600493, EPI_ISL_1600494, EPI_ISL_1600495, EPI_ISL_1600496, EPI_ISL_1600497, EPI_ISL_1600498, EPI_ISL_1600508, EPI_ISL_1600509, EPI_ISL_1600510, EPI_ISL_1600511, EPI_ISL_1600512, EPI_ISL_1600513, EPI_ISL_1600514, EPI_ISL_1600515, EPI_ISL_1600516, EPI_ISL_1600517, EPI_ISL_1600518, EPI_ISL_1600519, EPI_ISL_1600520, EPI_ISL_1600521, EPI_ISL_1600522, EPI_ISL_1600523, EPI_ISL_1600524, EPI_ISL_1600525, EPI_ISL_1600526, EPI_ISL_1600527, EPI_ISL_1600528, EPI_ISL_1600529, EPI_ISL_1600530, EPI_ISL_1600531, EPI_ISL_1600532, EPI_ISL_1600533, EPI_ISL_1600534, EPI_ISL_1600535, EPI_ISL_1600536, EPI_ISL_1600537, EPI_ISL_1600538, EPI_ISL_1600539, EPI_ISL_1600540, EPI_ISL_1600543, EPI_ISL_1600544, EPI_ISL_1600545, EPI_ISL_1600546, EPI_ISL_1600547, EPI_ISL_1600548, EPI_ISL_1600549, EPI_ISL_1600550, EPI_ISL_1600551, EPI_ISL_1600552, EPI_ISL_1600553, EPI_ISL_1600554, EPI_ISL_1600555, EPI_ISL_1600556, EPI_ISL_1600557, EPI_ISL_1600558, EPI_ISL_1600559, EPI_ISL_1600560, EPI_ISL_1600561, EPI_ISL_1600562, EPI_ISL_1600563, EPI_ISL_1600564, EPI_ISL_1600565, EPI_ISL_1600566, EPI_ISL_1600567, EPI_ISL_1600568, EPI_ISL_1600569, EPI_ISL_1600570, EPI_ISL_1600571, EPI_ISL_1600572, EPI_ISL_1600573, EPI_ISL_1600574, EPI_ISL_1600575, EPI_ISL_1600576, EPI_ISL_1600577, EPI_ISL_1600582, EPI_ISL_1600583, EPI_ISL_1600584, EPI_ISL_1600585, EPI_ISL_1600586, EPI_ISL_1600600, EPI_ISL_1600601, EPI_ISL_1600602, EPI_ISL_1600603, EPI_ISL_1600604, EPI_ISL_1600605, EPI_ISL_1600606, EPI_ISL_1600607, EPI_ISL_1600608, EPI_ISL_1600609, EPI_ISL_1600610, EPI_ISL_1600611, EPI_ISL_1600612, EPI_ISL_1600613, EPI_ISL_1600614, EPI_ISL_1600615, EPI_ISL_1600616, EPI_ISL_1600617, EPI_ISL_1600618, EPI_ISL_1600619, EPI_ISL_1600620, EPI_ISL_1600621, EPI_ISL_1600622, EPI_ISL_1600623, EPI_ISL_1600630, EPI_ISL_1600631, EPI_ISL_1600632, EPI_ISL_1600633, EPI_ISL_1600634, EPI_ISL_1600635, EPI_ISL_1600636, EPI_ISL_1600638, EPI_ISL_1600639, EPI_ISL_1600640, EPI_ISL_1600641, EPI_ISL_1600642, EPI_ISL_1600643, EPI_ISL_1600645, EPI_ISL_1600646, EPI_ISL_1600647, EPI_ISL_1600648, EPI_ISL_1600649, EPI_ISL_1600650, EPI_ISL_1600651, EPI_ISL_1600652, EPI_ISL_1600653, EPI_ISL_1600654, EPI_ISL_1600655, EPI_ISL_1600656, EPI_ISL_1600657, EPI_ISL_1600658, EPI_ISL_1600659, EPI_ISL_1600660, EPI_ISL_1600661, EPI_ISL_1600662, EPI_ISL_1600663, EPI_ISL_1600664, EPI_ISL_1600665, EPI_ISL_1600666, EPI_ISL_1600667, EPI_ISL_1600668, EPI_ISL_1600669, EPI_ISL_1600670, EPI_ISL_1600671, EPI_ISL_1600672, EPI_ISL_1600673, EPI_ISL_1600674, EPI_ISL_1600675, EPI_ISL_1600676, EPI_ISL_1600677, EPI_ISL_1600678, EPI_ISL_1600679, EPI_ISL_1600680, EPI_ISL_1600681, EPI_ISL_1600683, EPI_ISL_1600684, EPI_ISL_1600685, EPI_ISL_1600686, EPI_ISL_1600687, EPI_ISL_1600688, EPI_ISL_1600689, EPI_ISL_1600690, EPI_ISL_1600691, EPI_ISL_1600692, EPI_ISL_1600693, EPI_ISL_1600694, EPI_ISL_1600695, EPI_ISL_1600696, EPI_ISL_1600698, EPI_ISL_1600699, EPI_ISL_1600700, EPI_ISL_1600701, EPI_ISL_1600702, EPI_ISL_1600703, EPI_ISL_1600704, EPI_ISL_1600705, EPI_ISL_1600706, EPI_ISL_1600707, EPI_ISL_1600708, EPI_ISL_1600709, EPI_ISL_1600711, EPI_ISL_1600712, EPI_ISL_1600713, EPI_ISL_1600714, EPI_ISL_1600715, EPI_ISL_1600716, EPI_ISL_1600717, EPI_ISL_1600718, EPI_ISL_1600719, EPI_ISL_1600720, EPI_ISL_1600721, EPI_ISL_1600722, EPI_ISL_1600723, EPI_ISL_1600724, EPI_ISL_1600725, EPI_ISL_1600726, EPI_ISL_1600727, EPI_ISL_1600728, EPI_ISL_1600729, EPI_ISL_1600730, EPI_ISL_1600731, EPI_ISL_1600732, EPI_ISL_1600733, EPI_ISL_1600734, EPI_ISL_1600735, EPI_ISL_1600736, EPI_ISL_1600737, EPI_ISL_1600738, EPI_ISL_1600739, EPI_ISL_1600787, EPI_ISL_1600788, EPI_ISL_1600789, EPI_ISL_1600790, EPI_ISL_1600791, EPI_ISL_1600792, EPI_ISL_1600793, EPI_ISL_1600794, EPI_ISL_1600795, EPI_ISL_1600796, EPI_ISL_1600797, EPI_ISL_1600799, EPI_ISL_1600801, EPI_ISL_1600802, EPI_ISL_1600803, EPI_ISL_1600804, EPI_ISL_1600805, EPI_ISL_1600806, EPI_ISL_1600807, EPI_ISL_1600808, EPI_ISL_1600809, EPI_ISL_1600810, EPI_ISL_1600811, EPI_ISL_1600812, EPI_ISL_1600813, EPI_ISL_1600814, EPI_ISL_1600815, EPI_ISL_1600816, EPI_ISL_1600817, EPI_ISL_1600818, EPI_ISL_1600819, EPI_ISL_1600820, EPI_ISL_1600821, EPI_ISL_1600822, EPI_ISL_1600824, EPI_ISL_1600825, EPI_ISL_1600826, EPI_ISL_1600827, EPI_ISL_1600829, EPI_ISL_1600831, EPI_ISL_1600833, EPI_ISL_1600834, EPI_ISL_1600835, EPI_ISL_1600836, EPI_ISL_1600837, EPI_ISL_1600838, EPI_ISL_1600839, EPI_ISL_1600840, EPI_ISL_1600841, EPI_ISL_1600842, EPI_ISL_1600843, EPI_ISL_1600844, EPI_ISL_1600845, EPI_ISL_1600846, EPI_ISL_1600847, EPI_ISL_1600848, EPI_ISL_1600849, EPI_ISL_1600850, EPI_ISL_1600851, EPI_ISL_1600852, EPI_ISL_1600853, EPI_ISL_1600854, EPI_ISL_1600855, EPI_ISL_1600856, EPI_ISL_1600857, EPI_ISL_1600858, EPI_ISL_1600859, EPI_ISL_1600860, EPI_ISL_1600861, EPI_ISL_1600862, EPI_ISL_1600863, EPI_ISL_1600864, EPI_ISL_1600865, EPI_ISL_1600866, EPI_ISL_1600867, EPI_ISL_1600868, EPI_ISL_1600869, EPI_ISL_1600870, EPI_ISL_1600871, EPI_ISL_1600872, EPI_ISL_1600873, EPI_ISL_1600874, EPI_ISL_1600876, EPI_ISL_1600877, EPI_ISL_1600878, EPI_ISL_1600879, EPI_ISL_1600880, EPI_ISL_1600881, EPI_ISL_1600882, EPI_ISL_1600883, EPI_ISL_1600884, EPI_ISL_1600885, EPI_ISL_1600887, EPI_ISL_1600888, EPI_ISL_1600889, EPI_ISL_1600890, EPI_ISL_1600891, EPI_ISL_1600892, EPI_ISL_1600893, EPI_ISL_1600894, EPI_ISL_1600895, EPI_ISL_1600896, EPI_ISL_1600897, EPI_ISL_1600898, EPI_ISL_1600899, EPI_ISL_1600900, EPI_ISL_1600901, EPI_ISL_1600902, EPI_ISL_1600903, EPI_ISL_1600904, EPI_ISL_1600905, EPI_ISL_1600906, EPI_ISL_1600907, EPI_ISL_1600908, EPI_ISL_1600909, EPI_ISL_1600910, EPI_ISL_1600911, EPI_ISL_1600912, EPI_ISL_1600913, EPI_ISL_1600914, EPI_ISL_1600915, EPI_ISL_1600916, EPI_ISL_1600917, EPI_ISL_1600918, EPI_ISL_1600919, EPI_ISL_1600920, EPI_ISL_1600922, EPI_ISL_1600923, EPI_ISL_1600924, EPI_ISL_1600925, EPI_ISL_1600926, EPI_ISL_1600927, EPI_ISL_1600929, EPI_ISL_1600930, EPI_ISL_1600932, EPI_ISL_1600933, EPI_ISL_1600934, EPI_ISL_1600935, EPI_ISL_1600936, EPI_ISL_1600937, EPI_ISL_1600938, EPI_ISL_1600939, EPI_ISL_1600940, EPI_ISL_1600941, EPI_ISL_1600942, EPI_ISL_1600943, EPI_ISL_1600944, EPI_ISL_1600945, EPI_ISL_1600946, EPI_ISL_1600947, EPI_ISL_1600948, EPI_ISL_1600949, EPI_ISL_1600950, EPI_ISL_1600951, EPI_ISL_1600952, EPI_ISL_1600955, EPI_ISL_1600956, EPI_ISL_1600957, EPI_ISL_1600958, EPI_ISL_1600959, EPI_ISL_1600960, EPI_ISL_1600961, EPI_ISL_1600962, EPI_ISL_1600963, EPI_ISL_1600964, EPI_ISL_1600965, EPI_ISL_1600966, EPI_ISL_1600967, EPI_ISL_1600968, EPI_ISL_1600969, EPI_ISL_1600970, EPI_ISL_1600971, EPI_ISL_1600972, EPI_ISL_1600973, EPI_ISL_1600974, EPI_ISL_1600975, EPI_ISL_1600976, EPI_ISL_1600977, EPI_ISL_1600978, EPI_ISL_1600979, EPI_ISL_1600980, EPI_ISL_1600981, EPI_ISL_1600982, EPI_ISL_1600983, EPI_ISL_1600984, EPI_ISL_1600985, EPI_ISL_1600986, EPI_ISL_1600987, EPI_ISL_1600988, EPI_ISL_1600989, EPI_ISL_1600990, EPI_ISL_1600991, EPI_ISL_1600992, EPI_ISL_1600993, EPI_ISL_1600994, EPI_ISL_1600995, EPI_ISL_1600996, EPI_ISL_1600997, EPI_ISL_1600998, EPI_ISL_1600999, EPI_ISL_1601000, EPI_ISL_1601001, EPI_ISL_1601002, EPI_ISL_1601003, EPI_ISL_1601004, EPI_ISL_1601006, EPI_ISL_1601007, EPI_ISL_1601008, EPI_ISL_1601010, EPI_ISL_1601011, EPI_ISL_1601012, EPI_ISL_1601013, EPI_ISL_1601014, EPI_ISL_1601015, EPI_ISL_1601055, EPI_ISL_1601056, EPI_ISL_1601168, EPI_ISL_1601169, EPI_ISL_1601208, EPI_ISL_1601209, EPI_ISL_1601210, EPI_ISL_1601211, EPI_ISL_1601212, EPI_ISL_1601213, EPI_ISL_1601214, EPI_ISL_1601215, EPI_ISL_1601216, EPI_ISL_1601217, EPI_ISL_1601218, EPI_ISL_1601219, EPI_ISL_1601220, EPI_ISL_1601221, EPI_ISL_1601222, EPI_ISL_1601223, EPI_ISL_1601224, EPI_ISL_1601225, EPI_ISL_1601226, EPI_ISL_1601227, EPI_ISL_1601228, EPI_ISL_1601229, EPI_ISL_1601230, EPI_ISL_1601231, EPI_ISL_1601232, EPI_ISL_1601233, EPI_ISL_1601249, EPI_ISL_1601250, EPI_ISL_1601251, EPI_ISL_1601252, EPI_ISL_1601253, EPI_ISL_1601254, EPI_ISL_1601255, EPI_ISL_1601256, EPI_ISL_1601257, EPI_ISL_1601258, EPI_ISL_1601259, EPI_ISL_1601260, EPI_ISL_1601261, EPI_ISL_1601262, EPI_ISL_1601263, EPI_ISL_1601264, EPI_ISL_1601265, EPI_ISL_1601266, EPI_ISL_1601267, EPI_ISL_1601268, EPI_ISL_1601269, EPI_ISL_1601270, EPI_ISL_1601271, EPI_ISL_1601272, EPI_ISL_1601273, EPI_ISL_1601274, EPI_ISL_1601276, EPI_ISL_1601278, EPI_ISL_1601281, EPI_ISL_1601283, EPI_ISL_1601285, EPI_ISL_1601287, EPI_ISL_1601290, EPI_ISL_1601292, EPI_ISL_1601294, EPI_ISL_1601296, EPI_ISL_1601299, EPI_ISL_1601301, EPI_ISL_1601303, EPI_ISL_1601305, EPI_ISL_1601307, EPI_ISL_1601309, EPI_ISL_1601312, EPI_ISL_1601314, EPI_ISL_1601316, EPI_ISL_1601318, EPI_ISL_1601320, EPI_ISL_1601322, EPI_ISL_1601324, EPI_ISL_1601326, EPI_ISL_1601328, EPI_ISL_1601330, EPI_ISL_1601333, EPI_ISL_1601335, EPI_ISL_1601337, EPI_ISL_1601339, EPI_ISL_1601341, EPI_ISL_1601343, EPI_ISL_1601345, EPI_ISL_1601347, EPI_ISL_1601350, EPI_ISL_1601352, EPI_ISL_1601354, EPI_ISL_1601356, EPI_ISL_1601359, EPI_ISL_1601361, EPI_ISL_1601363, EPI_ISL_1601365, EPI_ISL_1601367, EPI_ISL_1601369, EPI_ISL_1601371, EPI_ISL_1601374, EPI_ISL_1601376, EPI_ISL_1601378, EPI_ISL_1601381, EPI_ISL_1601383, EPI_ISL_1601385, EPI_ISL_1601388, EPI_ISL_1601389, EPI_ISL_1601392, EPI_ISL_1601394, EPI_ISL_1601396, EPI_ISL_1601399, EPI_ISL_1601401, EPI_ISL_1601403, EPI_ISL_1601406, EPI_ISL_1601489, EPI_ISL_1601491, EPI_ISL_1601493, EPI_ISL_1601496, EPI_ISL_1601498, EPI_ISL_1601500, EPI_ISL_1601503, EPI_ISL_1601505, EPI_ISL_1601507, EPI_ISL_1601509, EPI_ISL_1601512, EPI_ISL_1601514, EPI_ISL_1601516, EPI_ISL_1601518, EPI_ISL_1601520, EPI_ISL_1601523, EPI_ISL_1601525, EPI_ISL_1601527, EPI_ISL_1601529, EPI_ISL_1601531, EPI_ISL_1601533, EPI_ISL_1601536, EPI_ISL_1601538, EPI_ISL_1601540, EPI_ISL_1601542, EPI_ISL_1601543, EPI_ISL_1601544, EPI_ISL_1601545, EPI_ISL_1601546, EPI_ISL_1601547, EPI_ISL_1601548, EPI_ISL_1601549, EPI_ISL_1601550, EPI_ISL_1601551, EPI_ISL_1601552, EPI_ISL_1601553, EPI_ISL_1601554, EPI_ISL_1601555, EPI_ISL_1601556, EPI_ISL_1601557, EPI_ISL_1601558, EPI_ISL_1601559, EPI_ISL_1601571, EPI_ISL_1601572, EPI_ISL_1601601, EPI_ISL_1601602, EPI_ISL_1601603, EPI_ISL_1601604, EPI_ISL_1601605, EPI_ISL_1601606, EPI_ISL_1601607, EPI_ISL_1601608, EPI_ISL_1601609, EPI_ISL_1601610, EPI_ISL_1601611, EPI_ISL_1601612, EPI_ISL_1601613, EPI_ISL_1601614, EPI_ISL_1601615, EPI_ISL_1601616, EPI_ISL_1601617, EPI_ISL_1601618, EPI_ISL_1601619, EPI_ISL_1601620, EPI_ISL_1601621, EPI_ISL_1601622, EPI_ISL_1601623, EPI_ISL_1601624, EPI_ISL_1601676, EPI_ISL_1601677, EPI_ISL_1601678, EPI_ISL_1601679, EPI_ISL_1601680, EPI_ISL_1601681, EPI_ISL_1601682, EPI_ISL_1601683, EPI_ISL_1601684, EPI_ISL_1601685, EPI_ISL_1601686, EPI_ISL_1601687, EPI_ISL_1601688, EPI_ISL_1601689, EPI_ISL_1601690, EPI_ISL_1601691, EPI_ISL_1601692, EPI_ISL_1601693, EPI_ISL_1601694, EPI_ISL_1601695, EPI_ISL_1601696, EPI_ISL_1601697, EPI_ISL_1601698, EPI_ISL_1601699, EPI_ISL_1601700, EPI_ISL_1601701, EPI_ISL_1601702, EPI_ISL_1601703, EPI_ISL_1601704, EPI_ISL_1601705, EPI_ISL_1601706, EPI_ISL_1601707, EPI_ISL_1601708, EPI_ISL_1601709, EPI_ISL_1601710, EPI_ISL_1601711, EPI_ISL_1601712, EPI_ISL_1601713, EPI_ISL_1601714, EPI_ISL_1601715, EPI_ISL_1601716, EPI_ISL_1601717, EPI_ISL_1601718, EPI_ISL_1601719, EPI_ISL_1601720, EPI_ISL_1601721, EPI_ISL_1601722, EPI_ISL_1601723, EPI_ISL_1601724, EPI_ISL_1601725, EPI_ISL_1601726, EPI_ISL_1601727, EPI_ISL_1601728, EPI_ISL_1601729, EPI_ISL_1601730, EPI_ISL_1601731, EPI_ISL_1601732, EPI_ISL_1601733, EPI_ISL_1601734, EPI_ISL_1601735, EPI_ISL_1601736, EPI_ISL_1601737, EPI_ISL_1601738, EPI_ISL_1601739, EPI_ISL_1601740, EPI_ISL_1601741, EPI_ISL_1601742, EPI_ISL_1601743, EPI_ISL_1601744, EPI_ISL_1601745, EPI_ISL_1601746, EPI_ISL_1601747, EPI_ISL_1601748, EPI_ISL_1601749, EPI_ISL_1601750, EPI_ISL_1601751, EPI_ISL_1601752, EPI_ISL_1601753, EPI_ISL_1601815, EPI_ISL_1601816, EPI_ISL_1601817, EPI_ISL_1601819, EPI_ISL_1601826, EPI_ISL_1601876, EPI_ISL_1601877, EPI_ISL_1601878, EPI_ISL_1601880, EPI_ISL_1601881, EPI_ISL_1601882, EPI_ISL_1601883, EPI_ISL_1601884, EPI_ISL_1601885, EPI_ISL_1601886, EPI_ISL_1601887, EPI_ISL_1601983, EPI_ISL_1601985, EPI_ISL_1601986, EPI_ISL_1601987, EPI_ISL_1601988, EPI_ISL_1601989, EPI_ISL_1601990, EPI_ISL_1602083, EPI_ISL_1602084, EPI_ISL_1602085, EPI_ISL_1602086, EPI_ISL_1602087, EPI_ISL_1602088, EPI_ISL_1602089, EPI_ISL_1602090, EPI_ISL_1602091, EPI_ISL_1602092, EPI_ISL_1602093, EPI_ISL_1602094, EPI_ISL_1602095, EPI_ISL_1602097, EPI_ISL_1602098, EPI_ISL_1602099, EPI_ISL_1602100, EPI_ISL_1602101, EPI_ISL_1602102, EPI_ISL_1602103, EPI_ISL_1602186, EPI_ISL_1602187, EPI_ISL_1602188, EPI_ISL_1602189, EPI_ISL_1602190, EPI_ISL_1602191, EPI_ISL_1602192, EPI_ISL_1602193, EPI_ISL_1602194, EPI_ISL_1602195, EPI_ISL_1602199, EPI_ISL_1602200, EPI_ISL_1602201, EPI_ISL_1602202, EPI_ISL_1602203, EPI_ISL_1602204, EPI_ISL_1602205, EPI_ISL_1602206, EPI_ISL_1602207, EPI_ISL_1602208, EPI_ISL_1602209, EPI_ISL_1602210, EPI_ISL_1602211, EPI_ISL_1602212, EPI_ISL_1602213, EPI_ISL_1602260, EPI_ISL_1602261, EPI_ISL_1602262, EPI_ISL_1602263, EPI_ISL_1602264, EPI_ISL_1602265, EPI_ISL_1602266, EPI_ISL_1602268, EPI_ISL_1602269, EPI_ISL_1602270, EPI_ISL_1602271, EPI_ISL_1602274, EPI_ISL_1602275, EPI_ISL_1602276, EPI_ISL_1602277, EPI_ISL_1602279, EPI_ISL_1602280, EPI_ISL_1602281, EPI_ISL_1602282, EPI_ISL_1602283, EPI_ISL_1602284, EPI_ISL_1602285, EPI_ISL_1602286, EPI_ISL_1602287, EPI_ISL_1602571, EPI_ISL_1602572, EPI_ISL_1602573, EPI_ISL_1602574, EPI_ISL_1602575, EPI_ISL_1602576, EPI_ISL_1602577, EPI_ISL_1602578, EPI_ISL_1602580, EPI_ISL_1602581, EPI_ISL_1602582, EPI_ISL_1602583, EPI_ISL_1602584, EPI_ISL_1602585, EPI_ISL_1602586, EPI_ISL_1602587, EPI_ISL_1602588, EPI_ISL_1602589, EPI_ISL_1602590, EPI_ISL_1602591, EPI_ISL_1602592, EPI_ISL_1602594, EPI_ISL_1602595, EPI_ISL_1602596, EPI_ISL_1602597, EPI_ISL_1602598, EPI_ISL_1602599, EPI_ISL_1602600, EPI_ISL_1602601, EPI_ISL_1602602, EPI_ISL_1602603, EPI_ISL_1602604, EPI_ISL_1602605, EPI_ISL_1602606, EPI_ISL_1602607, EPI_ISL_1602608, EPI_ISL_1602609, EPI_ISL_1602610, EPI_ISL_1602611, EPI_ISL_1602613, EPI_ISL_1602614, EPI_ISL_1602615, EPI_ISL_1602616, EPI_ISL_1602617, EPI_ISL_1602618, EPI_ISL_1602619, EPI_ISL_1602620, EPI_ISL_1602621, EPI_ISL_1602622, EPI_ISL_1602624, EPI_ISL_1603034, EPI_ISL_1603642, EPI_ISL_1603720, EPI_ISL_1603722, EPI_ISL_1603730, EPI_ISL_1603750, EPI_ISL_1606965, EPI_ISL_1617700, EPI_ISL_1617701, EPI_ISL_1617702, EPI_ISL_1617703, EPI_ISL_1617704, EPI_ISL_1617705, EPI_ISL_1617706, EPI_ISL_1617707, EPI_ISL_1617708, EPI_ISL_1617709, EPI_ISL_1617710, EPI_ISL_1617711, EPI_ISL_1617712, EPI_ISL_1617713, EPI_ISL_1617714, EPI_ISL_1617715, EPI_ISL_1617716, EPI_ISL_1617717, EPI_ISL_1617718, EPI_ISL_1617719, EPI_ISL_1617720, EPI_ISL_1617721, EPI_ISL_1617722, EPI_ISL_1617723, EPI_ISL_1617724, EPI_ISL_1617725, EPI_ISL_1617726, EPI_ISL_1617727, EPI_ISL_1617728, EPI_ISL_1617729, EPI_ISL_1617730, EPI_ISL_1617731, EPI_ISL_1617732, EPI_ISL_1617733, EPI_ISL_1617734,

EPI_ISL_1617735, EPI_ISL_1617736, EPI_ISL_1617737, EPI_ISL_1617738, EPI_ISL_1617739, EPI_ISL_1617740, EPI_ISL_1617741, EPI_ISL_1617742, EPI_ISL_1617743, EPI_ISL_1617745, EPI_ISL_1617746, EPI_ISL_1617747, EPI_ISL_1617748, EPI_ISL_1617749, EPI_ISL_1617750, EPI_ISL_1617751, EPI_ISL_1617752, EPI_ISL_1617753, EPI_ISL_1617754, EPI_ISL_1617755, EPI_ISL_1617756, EPI_ISL_1617757,
EPI_ISL_1617758, EPI_ISL_1617759, EPI_ISL_1617760, EPI_ISL_1617761, EPI_ISL_1617762, EPI_ISL_1617763, EPI_ISL_1617764, EPI_ISL_1617765, EPI_ISL_1617766, EPI_ISL_1617767, EPI_ISL_1617768, EPI_ISL_1617769, EPI_ISL_1617770, EPI_ISL_1617771, EPI_ISL_1617772, EPI_ISL_1617773, EPI_ISL_1617774, EPI_ISL_1617775, EPI_ISL_1617776, EPI_ISL_1617777, EPI_ISL_1617778, EPI_ISL_1617779,
EPI_ISL_1617780, EPI_ISL_1617781, EPI_ISL_1617782, EPI_ISL_1617784, EPI_ISL_1617785, EPI_ISL_1617786, EPI_ISL_1617787, EPI_ISL_1617788, EPI_ISL_1617789, EPI_ISL_1617790, EPI_ISL_1617791, EPI_ISL_1617792, EPI_ISL_1617793, EPI_ISL_1617794, EPI_ISL_1617795, EPI_ISL_1617796, EPI_ISL_1617797, EPI_ISL_1617798, EPI_ISL_1617799, EPI_ISL_1617800, EPI_ISL_1617801, EPI_ISL_1617802,
EPI_ISL_1617803, EPI_ISL_1617804, EPI_ISL_1617805, EPI_ISL_1617806, EPI_ISL_1617807, EPI_ISL_1617808, EPI_ISL_1617809, EPI_ISL_1617810, EPI_ISL_1617811, EPI_ISL_1617812, EPI_ISL_1617813, EPI_ISL_1617814, EPI_ISL_1617815, EPI_ISL_1617816, EPI_ISL_1617817, EPI_ISL_1617818, EPI_ISL_1617819, EPI_ISL_1617820, EPI_ISL_1617821, EPI_ISL_1617822, EPI_ISL_1617823, EPI_ISL_1617824,
EPI_ISL_1617825, EPI_ISL_1617826, EPI_ISL_1617827, EPI_ISL_1617828, EPI_ISL_1617829, EPI_ISL_1617830, EPI_ISL_1617831, EPI_ISL_1617832, EPI_ISL_1617833, EPI_ISL_1617834, EPI_ISL_1617835, EPI_ISL_1617836, EPI_ISL_1617837, EPI_ISL_1617838, EPI_ISL_1617839, EPI_ISL_1617840, EPI_ISL_1617841, EPI_ISL_1617842, EPI_ISL_1617843, EPI_ISL_1617844, EPI_ISL_1617845, EPI_ISL_1617846,
EPI_ISL_1617847, EPI_ISL_1617848, EPI_ISL_1617849, EPI_ISL_1617850, EPI_ISL_1617851, EPI_ISL_1617852, EPI_ISL_1617853, EPI_ISL_1617854, EPI_ISL_1617855, EPI_ISL_1617856, EPI_ISL_1617857, EPI_ISL_1617858, EPI_ISL_1617859, EPI_ISL_1617860, EPI_ISL_1617861, EPI_ISL_1617862, EPI_ISL_1617863, EPI_ISL_1617864, EPI_ISL_1617865, EPI_ISL_1617866, EPI_ISL_1617867, EPI_ISL_1617868,
EPI_ISL_1617869, EPI_ISL_1617870, EPI_ISL_1617871, EPI_ISL_1617872, EPI_ISL_1617873, EPI_ISL_1617874, EPI_ISL_1617875, EPI_ISL_1617876, EPI_ISL_1617877, EPI_ISL_1617878, EPI_ISL_1617879, EPI_ISL_1617880, EPI_ISL_1617881, EPI_ISL_1617882, EPI_ISL_1617883, EPI_ISL_1617884, EPI_ISL_1617885, EPI_ISL_1617886, EPI_ISL_1617887, EPI_ISL_1617888, EPI_ISL_1617889, EPI_ISL_1617890,
EPI_ISL_1617891, EPI_ISL_1617892, EPI_ISL_1617893, EPI_ISL_1617894, EPI_ISL_1617895, EPI_ISL_1617896, EPI_ISL_1617897, EPI_ISL_1617898, EPI_ISL_1617899, EPI_ISL_1617900, EPI_ISL_1617901, EPI_ISL_1617902, EPI_ISL_1617903, EPI_ISL_1617904, EPI_ISL_1617905, EPI_ISL_1617906, EPI_ISL_1617907, EPI_ISL_1617910, EPI_ISL_1617911, EPI_ISL_1617912, EPI_ISL_1617913, EPI_ISL_1617914,
EPI_ISL_1617915, EPI_ISL_1617916, EPI_ISL_1617917, EPI_ISL_1617918, EPI_ISL_1617919, EPI_ISL_1617920, EPI_ISL_1617921, EPI_ISL_1617922, EPI_ISL_1617923, EPI_ISL_1617924, EPI_ISL_1617925, EPI_ISL_1617926, EPI_ISL_1617927, EPI_ISL_1617928, EPI_ISL_1617929, EPI_ISL_1617930, EPI_ISL_1617931, EPI_ISL_1617932, EPI_ISL_1617933, EPI_ISL_1617934, EPI_ISL_1617935, EPI_ISL_1617936,
EPI_ISL_1617937, EPI_ISL_1617938, EPI_ISL_1617939, EPI_ISL_1617940, EPI_ISL_1617941, EPI_ISL_1617942, EPI_ISL_1617943, EPI_ISL_1617944, EPI_ISL_1617945, EPI_ISL_1617946, EPI_ISL_1617947, EPI_ISL_1617948, EPI_ISL_1617949, EPI_ISL_1617950, EPI_ISL_1617951, EPI_ISL_1617952, EPI_ISL_1617953, EPI_ISL_1617954, EPI_ISL_1617955, EPI_ISL_1617956, EPI_ISL_1617957, EPI_ISL_1617958,
EPI_ISL_1617959, EPI_ISL_1617960, EPI_ISL_1617961, EPI_ISL_1617962, EPI_ISL_1617963, EPI_ISL_1617964, EPI_ISL_1617965, EPI_ISL_1617966, EPI_ISL_1617967, EPI_ISL_1617968, EPI_ISL_1617969, EPI_ISL_1617970, EPI_ISL_1617971, EPI_ISL_1617972, EPI_ISL_1617973, EPI_ISL_1617974, EPI_ISL_1617975, EPI_ISL_1617976, EPI_ISL_1617977, EPI_ISL_1617978, EPI_ISL_1617979, EPI_ISL_1617980,
EPI_ISL_1617981, EPI_ISL_1617982, EPI_ISL_1617983, EPI_ISL_1617984, EPI_ISL_1617985, EPI_ISL_1617986, EPI_ISL_1617987, EPI_ISL_1617988, EPI_ISL_1617989, EPI_ISL_1617990, EPI_ISL_1617991, EPI_ISL_1617992, EPI_ISL_1617993, EPI_ISL_1617994, EPI_ISL_1617995, EPI_ISL_1617996, EPI_ISL_1617997, EPI_ISL_1617998, EPI_ISL_1617999, EPI_ISL_1618000, EPI_ISL_1618001, EPI_ISL_1618002,
EPI_ISL_1618003, EPI_ISL_1618004, EPI_ISL_1618005, EPI_ISL_1618006, EPI_ISL_1618007, EPI_ISL_1618008, EPI_ISL_1618009, EPI_ISL_1618010, EPI_ISL_1618011, EPI_ISL_1618012, EPI_ISL_1618013, EPI_ISL_1618014, EPI_ISL_1618015, EPI_ISL_1618016, EPI_ISL_1618017, EPI_ISL_1618018, EPI_ISL_1618019, EPI_ISL_1618020, EPI_ISL_1618021, EPI_ISL_1618024, EPI_ISL_1618025, EPI_ISL_1618026,
EPI_ISL_1618027, EPI_ISL_1618028, EPI_ISL_1618029, EPI_ISL_1618030, EPI_ISL_1618031, EPI_ISL_1618032, EPI_ISL_1618033, EPI_ISL_1618034, EPI_ISL_1618035, EPI_ISL_1618036, EPI_ISL_1618037, EPI_ISL_1618038, EPI_ISL_1618039, EPI_ISL_1618040, EPI_ISL_1618041, EPI_ISL_1618042, EPI_ISL_1618043, EPI_ISL_1618044, EPI_ISL_1618045, EPI_ISL_1618046, EPI_ISL_1618047, EPI_ISL_1618048,
EPI_ISL_1618049, EPI_ISL_1618050, EPI_ISL_1618051, EPI_ISL_1618052, EPI_ISL_1618053, EPI_ISL_1618054, EPI_ISL_1618055, EPI_ISL_1618056, EPI_ISL_1618057, EPI_ISL_1618058, EPI_ISL_1618059, EPI_ISL_1618060, EPI_ISL_1618061, EPI_ISL_1618062, EPI_ISL_1618063, EPI_ISL_1618064, EPI_ISL_1618065, EPI_ISL_1618066, EPI_ISL_1618067, EPI_ISL_1618068, EPI_ISL_1618069, EPI_ISL_1618070,
EPI_ISL_1618071, EPI_ISL_1618072, EPI_ISL_1618073, EPI_ISL_1618074, EPI_ISL_1618075, EPI_ISL_1618076, EPI_ISL_1618077, EPI_ISL_1618078, EPI_ISL_1618079, EPI_ISL_1618080, EPI_ISL_1618346, EPI_ISL_1618347, EPI_ISL_1618348, EPI_ISL_1618349, EPI_ISL_1618350, EPI_ISL_1618351, EPI_ISL_1618352, EPI_ISL_1618353, EPI_ISL_1618354, EPI_ISL_1618355, EPI_ISL_1618356, EPI_ISL_1618357,
EPI_ISL_1618358, EPI_ISL_1618359, EPI_ISL_1618360, EPI_ISL_1618361, EPI_ISL_1618362, EPI_ISL_1618363, EPI_ISL_1618364, EPI_ISL_1618365, EPI_ISL_1618366, EPI_ISL_1618367, EPI_ISL_1618368, EPI_ISL_1618369, EPI_ISL_1618370, EPI_ISL_1618371, EPI_ISL_1618372, EPI_ISL_1618373, EPI_ISL_1618374, EPI_ISL_1618375, EPI_ISL_1618376, EPI_ISL_1618377, EPI_ISL_1618378, EPI_ISL_1618379,
EPI_ISL_1618380, EPI_ISL_1618381, EPI_ISL_1618382, EPI_ISL_1618383, EPI_ISL_1618384, EPI_ISL_1618385, EPI_ISL_1618386, EPI_ISL_1618387, EPI_ISL_1618388, EPI_ISL_1618389, EPI_ISL_1618390, EPI_ISL_1618391, EPI_ISL_1618392, EPI_ISL_1618393, EPI_ISL_1618394, EPI_ISL_1618395, EPI_ISL_1618396, EPI_ISL_1618397, EPI_ISL_1618399, EPI_ISL_1618400, EPI_ISL_1618401, EPI_ISL_1618402,
EPI_ISL_1618403, EPI_ISL_1618404, EPI_ISL_1618405, EPI_ISL_1618406, EPI_ISL_1618407, EPI_ISL_1618408, EPI_ISL_1618409, EPI_ISL_1618410, EPI_ISL_1618411, EPI_ISL_1618412, EPI_ISL_1618413, EPI_ISL_1618414, EPI_ISL_1618415, EPI_ISL_1618416, EPI_ISL_1618417, EPI_ISL_1618418, EPI_ISL_1618419, EPI_ISL_1618420, EPI_ISL_1618421, EPI_ISL_1618422, EPI_ISL_1618423, EPI_ISL_1618424,
EPI_ISL_1618425, EPI_ISL_1618426, EPI_ISL_1618427, EPI_ISL_1618428, EPI_ISL_1618429, EPI_ISL_1618430, EPI_ISL_1618431, EPI_ISL_1618432, EPI_ISL_1618433, EPI_ISL_1618526, EPI_ISL_1618527, EPI_ISL_1618528, EPI_ISL_1618529, EPI_ISL_1618530, EPI_ISL_1618531, EPI_ISL_1618532, EPI_ISL_1618533, EPI_ISL_1618534, EPI_ISL_1618535, EPI_ISL_1618536, EPI_ISL_1618537, EPI_ISL_1618538,
EPI_ISL_1618539, EPI_ISL_1618540, EPI_ISL_1618541, EPI_ISL_1618542, EPI_ISL_1618543, EPI_ISL_1618544, EPI_ISL_1618545, EPI_ISL_1618546, EPI_ISL_1618547, EPI_ISL_1618548, EPI_ISL_1618549, EPI_ISL_1618551, EPI_ISL_1618552, EPI_ISL_1618553, EPI_ISL_1618555, EPI_ISL_1618556, EPI_ISL_1618557, EPI_ISL_1618558, EPI_ISL_1618559, EPI_ISL_1618560, EPI_ISL_1618561, EPI_ISL_1618562,
EPI_ISL_1618563, EPI_ISL_1618564, EPI_ISL_1618565, EPI_ISL_1618566, EPI_ISL_1618567, EPI_ISL_1618568, EPI_ISL_1618569, EPI_ISL_1618570, EPI_ISL_1618571, EPI_ISL_1618572, EPI_ISL_1618573, EPI_ISL_1618574, EPI_ISL_1618575, EPI_ISL_1618576, EPI_ISL_1618577, EPI_ISL_1618578, EPI_ISL_1618579, EPI_ISL_1618580, EPI_ISL_1618581, EPI_ISL_1618582, EPI_ISL_1618583, EPI_ISL_1618584,
EPI_ISL_1618585, EPI_ISL_1618586, EPI_ISL_1618587, EPI_ISL_1618588, EPI_ISL_1618589, EPI_ISL_1618590, EPI_ISL_1618591, EPI_ISL_1618592, EPI_ISL_1618593, EPI_ISL_1618594, EPI_ISL_1618595, EPI_ISL_1618596, EPI_ISL_1618597, EPI_ISL_1618598, EPI_ISL_1618599, EPI_ISL_1618600, EPI_ISL_1618601, EPI_ISL_1618602, EPI_ISL_1618603, EPI_ISL_1618604, EPI_ISL_1618605, EPI_ISL_1618606,
EPI_ISL_1618607, EPI_ISL_1618608, EPI_ISL_1618609, EPI_ISL_1618610, EPI_ISL_1618611, EPI_ISL_1618612, EPI_ISL_1618613, EPI_ISL_1618614, EPI_ISL_1618615, EPI_ISL_1618616, EPI_ISL_1618617, EPI_ISL_1618618, EPI_ISL_1618619, EPI_ISL_1618620, EPI_ISL_1618621, EPI_ISL_1618622, EPI_ISL_1618623, EPI_ISL_1618624, EPI_ISL_1618625, EPI_ISL_1618626, EPI_ISL_1618627, EPI_ISL_1618628,
EPI_ISL_1618629, EPI_ISL_1618630, EPI_ISL_1618631, EPI_ISL_1618632, EPI_ISL_1618633, EPI_ISL_1618634, EPI_ISL_1618635, EPI_ISL_1618636, EPI_ISL_1618637, EPI_ISL_1618638, EPI_ISL_1618639, EPI_ISL_1618640, EPI_ISL_1618723, EPI_ISL_1618725, EPI_ISL_1618726, EPI_ISL_1618727, EPI_ISL_1618728, EPI_ISL_1618729, EPI_ISL_1618730, EPI_ISL_1618731, EPI_ISL_1618732, EPI_ISL_1618733,
EPI_ISL_1618734, EPI_ISL_1618735, EPI_ISL_1618736, EPI_ISL_1618737, EPI_ISL_1618738, EPI_ISL_1618739, EPI_ISL_1618740, EPI_ISL_1618741, EPI_ISL_1618742, EPI_ISL_1618743, EPI_ISL_1618744, EPI_ISL_1618745, EPI_ISL_1618746, EPI_ISL_1618747, EPI_ISL_1618748, EPI_ISL_1618749, EPI_ISL_1618750, EPI_ISL_1618751, EPI_ISL_1618752, EPI_ISL_1618753, EPI_ISL_1618754, EPI_ISL_1618755,
EPI_ISL_1618756, EPI_ISL_1618757, EPI_ISL_1618758, EPI_ISL_1618759, EPI_ISL_1618760, EPI_ISL_1618761, EPI_ISL_1618762, EPI_ISL_1618763, EPI_ISL_1618764, EPI_ISL_1618765, EPI_ISL_1618766, EPI_ISL_1618767, EPI_ISL_1618768, EPI_ISL_1618769, EPI_ISL_1618770, EPI_ISL_1619210, EPI_ISL_1619211, EPI_ISL_1619212, EPI_ISL_1619213, EPI_ISL_1619214, EPI_ISL_1619217, EPI_ISL_1619218,
EPI_ISL_1619219, EPI_ISL_1619220, EPI_ISL_1619221, EPI_ISL_1619222, EPI_ISL_1619223, EPI_ISL_1619224, EPI_ISL_1619225, EPI_ISL_1619226, EPI_ISL_1619227, EPI_ISL_1619228, EPI_ISL_1619230, EPI_ISL_1619231, EPI_ISL_1619232, EPI_ISL_1619233, EPI_ISL_1619234, EPI_ISL_1619235, EPI_ISL_1619236, EPI_ISL_1619237, EPI_ISL_1619238, EPI_ISL_1619239, EPI_ISL_1619240, EPI_ISL_1619241,
EPI_ISL_1619242, EPI_ISL_1619243, EPI_ISL_1619244, EPI_ISL_1619245, EPI_ISL_1619246, EPI_ISL_1619247, EPI_ISL_1619248, EPI_ISL_1619249, EPI_ISL_1619250, EPI_ISL_1619251, EPI_ISL_1619252, EPI_ISL_1619253, EPI_ISL_1619254, EPI_ISL_1619255, EPI_ISL_1619256, EPI_ISL_1619257, EPI_ISL_1619258, EPI_ISL_1619259, EPI_ISL_1619260, EPI_ISL_1619261, EPI_ISL_1619262, EPI_ISL_1619263,
EPI_ISL_1619264, EPI_ISL_1619265, EPI_ISL_1619266, EPI_ISL_1619267, EPI_ISL_1619268, EPI_ISL_1619269, EPI_ISL_1619270, EPI_ISL_1619271, EPI_ISL_1619272, EPI_ISL_1619273, EPI_ISL_1619274, EPI_ISL_1619275, EPI_ISL_1619276, EPI_ISL_1619277, EPI_ISL_1619278, EPI_ISL_1619279, EPI_ISL_1619280, EPI_ISL_1619281, EPI_ISL_1619282, EPI_ISL_1619283, EPI_ISL_1619284, EPI_ISL_1619285,
EPI_ISL_1619286, EPI_ISL_1619287, EPI_ISL_1619288, EPI_ISL_1619289, EPI_ISL_1619290, EPI_ISL_1619291, EPI_ISL_1619292, EPI_ISL_1619293, EPI_ISL_1619294, EPI_ISL_1619295, EPI_ISL_1619296, EPI_ISL_1619297, EPI_ISL_1619298, EPI_ISL_1619299, EPI_ISL_1619300, EPI_ISL_1619301, EPI_ISL_1619302, EPI_ISL_1619303, EPI_ISL_1619304, EPI_ISL_1619305, EPI_ISL_1619306, EPI_ISL_1619307,
EPI_ISL_1619308, EPI_ISL_1619309, EPI_ISL_1619310, EPI_ISL_1619311, EPI_ISL_1619312, EPI_ISL_1619313, EPI_ISL_1619314, EPI_ISL_1619315, EPI_ISL_1619316, EPI_ISL_1619317, EPI_ISL_1619318, EPI_ISL_1619319, EPI_ISL_1619320, EPI_ISL_1619321, EPI_ISL_1619322, EPI_ISL_1619323, EPI_ISL_1619324, EPI_ISL_1619325, EPI_ISL_1619326, EPI_ISL_1619327, EPI_ISL_1619328, EPI_ISL_1619329,
EPI_ISL_1619330, EPI_ISL_1619331, EPI_ISL_1619332, EPI_ISL_1619333, EPI_ISL_1619334, EPI_ISL_1619335, EPI_ISL_1619336, EPI_ISL_1619337, EPI_ISL_1619338, EPI_ISL_1619339, EPI_ISL_1619340, EPI_ISL_1619341, EPI_ISL_1619342, EPI_ISL_1619343, EPI_ISL_1619344, EPI_ISL_1619345, EPI_ISL_1619346, EPI_ISL_1619347, EPI_ISL_1619348, EPI_ISL_1619349, EPI_ISL_1619350, EPI_ISL_1619351,
EPI_ISL_1619352, EPI_ISL_1619353, EPI_ISL_1619354, EPI_ISL_1619355, EPI_ISL_1619356, EPI_ISL_1619357, EPI_ISL_1619358, EPI_ISL_1619359, EPI_ISL_1619360, EPI_ISL_1619361, EPI_ISL_1619362, EPI_ISL_1619363, EPI_ISL_1619364, EPI_ISL_1619365, EPI_ISL_1619366, EPI_ISL_1619367, EPI_ISL_1619368, EPI_ISL_1619369, EPI_ISL_1619370, EPI_ISL_1619371, EPI_ISL_1619372, EPI_ISL_1619373,
EPI_ISL_1619374, EPI_ISL_1619375, EPI_ISL_1619376, EPI_ISL_1619377, EPI_ISL_1619378, EPI_ISL_1619379, EPI_ISL_1619380, EPI_ISL_1619381, EPI_ISL_1619382, EPI_ISL_1619383, EPI_ISL_1619384, EPI_ISL_1619385, EPI_ISL_1619386, EPI_ISL_1619387, EPI_ISL_1619388, EPI_ISL_1619389, EPI_ISL_1619390, EPI_ISL_1619391, EPI_ISL_1619392, EPI_ISL_1619393, EPI_ISL_1619394, EPI_ISL_1619395,
EPI_ISL_1619396, EPI_ISL_1619397, EPI_ISL_1619398, EPI_ISL_1619399, EPI_ISL_1619400, EPI_ISL_1619401, EPI_ISL_1619402, EPI_ISL_1619403, EPI_ISL_1619404, EPI_ISL_1619405, EPI_ISL_1619406, EPI_ISL_1619407, EPI_ISL_1619408, EPI_ISL_1619409, EPI_ISL_1619410, EPI_ISL_1619411, EPI_ISL_1619412, EPI_ISL_1619413, EPI_ISL_1619414, EPI_ISL_1619415, EPI_ISL_1619416, EPI_ISL_1619417,
EPI_ISL_1619418, EPI_ISL_1619419, EPI_ISL_1619420, EPI_ISL_1619421, EPI_ISL_1619422, EPI_ISL_1619423, EPI_ISL_1619424, EPI_ISL_1619425, EPI_ISL_1619426, EPI_ISL_1619427, EPI_ISL_1619428, EPI_ISL_1619429, EPI_ISL_1619430, EPI_ISL_1619431, EPI_ISL_1619432, EPI_ISL_1619433, EPI_ISL_1619434, EPI_ISL_1619435, EPI_ISL_1619436, EPI_ISL_1619437, EPI_ISL_1619438, EPI_ISL_1619439,
EPI_ISL_1619440, EPI_ISL_1619441, EPI_ISL_1619442, EPI_ISL_1619443, EPI_ISL_1619444, EPI_ISL_1619445, EPI_ISL_1619446, EPI_ISL_1619447, EPI_ISL_1619448, EPI_ISL_1619449, EPI_ISL_1619450, EPI_ISL_1619451, EPI_ISL_1619733, EPI_ISL_1619734, EPI_ISL_1619735, EPI_ISL_1619736, EPI_ISL_1619737, EPI_ISL_1619738, EPI_ISL_1619739, EPI_ISL_1619740, EPI_ISL_1619741, EPI_ISL_1619743,
EPI_ISL_1619744, EPI_ISL_1619745, EPI_ISL_1619746, EPI_ISL_1619747, EPI_ISL_1619748, EPI_ISL_1619749, EPI_ISL_1619750, EPI_ISL_1619751, EPI_ISL_1619752, EPI_ISL_1619753, EPI_ISL_1619754, EPI_ISL_1619755, EPI_ISL_1619756, EPI_ISL_1619757, EPI_ISL_1619758, EPI_ISL_1619759, EPI_ISL_1619760, EPI_ISL_1619762, EPI_ISL_1619763, EPI_ISL_1619764, EPI_ISL_1619765, EPI_ISL_1619766,
EPI_ISL_1619767, EPI_ISL_1619768, EPI_ISL_1619769, EPI_ISL_1619770, EPI_ISL_1619771, EPI_ISL_1619773, EPI_ISL_1619775, EPI_ISL_1619776, EPI_ISL_1619777, EPI_ISL_1619779, EPI_ISL_1619780, EPI_ISL_1619781, EPI_ISL_1619782, EPI_ISL_1619783, EPI_ISL_1619784, EPI_ISL_1619785, EPI_ISL_1619786, EPI_ISL_1619787, EPI_ISL_1619788, EPI_ISL_1619789, EPI_ISL_1619790, EPI_ISL_1619791,
EPI_ISL_1619792, EPI_ISL_1619793, EPI_ISL_1619794, EPI_ISL_1619795, EPI_ISL_1619796, EPI_ISL_1619797, EPI_ISL_1619798, EPI_ISL_1619799, EPI_ISL_1619800, EPI_ISL_1619801, EPI_ISL_1619802, EPI_ISL_1619803, EPI_ISL_1619804, EPI_ISL_1619805, EPI_ISL_1619806, EPI_ISL_1619807, EPI_ISL_1619808, EPI_ISL_1619809, EPI_ISL_1619810, EPI_ISL_1619811, EPI_ISL_1619812, EPI_ISL_1619813,
EPI_ISL_1619814, EPI_ISL_1619815, EPI_ISL_1619816, EPI_ISL_1619817, EPI_ISL_1619818, EPI_ISL_1619819, EPI_ISL_1619820, EPI_ISL_1619821, EPI_ISL_1619822, EPI_ISL_1619823, EPI_ISL_1619824, EPI_ISL_1619825, EPI_ISL_1619826, EPI_ISL_1619827, EPI_ISL_1619828, EPI_ISL_1619829, EPI_ISL_1619830, EPI_ISL_1619831, EPI_ISL_1619832, EPI_ISL_1619833, EPI_ISL_1619834, EPI_ISL_1619835,
EPI_ISL_1619836, EPI_ISL_1619837, EPI_ISL_1619838, EPI_ISL_1619839, EPI_ISL_1619840, EPI_ISL_1619841, EPI_ISL_1619842, EPI_ISL_1619843, EPI_ISL_1619844, EPI_ISL_1619845, EPI_ISL_1619846, EPI_ISL_1619847, EPI_ISL_1619848, EPI_ISL_1619849, EPI_ISL_1619850, EPI_ISL_1619851, EPI_ISL_1619852, EPI_ISL_1619853, EPI_ISL_1619854, EPI_ISL_1619855, EPI_ISL_1619856, EPI_ISL_1619857,
EPI_ISL_1619858, EPI_ISL_1619859, EPI_ISL_1619860, EPI_ISL_1619861, EPI_ISL_1619862, EPI_ISL_1619863, EPI_ISL_1619864, EPI_ISL_1619865, EPI_ISL_1619866, EPI_ISL_1619867, EPI_ISL_1619868, EPI_ISL_1619869, EPI_ISL_1619870, EPI_ISL_1619871, EPI_ISL_1619872, EPI_ISL_1619873, EPI_ISL_1619874, EPI_ISL_1619875, EPI_ISL_1619876, EPI_ISL_1619877, EPI_ISL_1619878, EPI_ISL_1619879,
EPI_ISL_1619880, EPI_ISL_1619881, EPI_ISL_1619882, EPI_ISL_1619883, EPI_ISL_1619884, EPI_ISL_1619885, EPI_ISL_1619886, EPI_ISL_1619887, EPI_ISL_1619888, EPI_ISL_1619889, EPI_ISL_1619890, EPI_ISL_1619891, EPI_ISL_1619892, EPI_ISL_1619893, EPI_ISL_1619894, EPI_ISL_1619895, EPI_ISL_1619896, EPI_ISL_1619897, EPI_ISL_1619898, EPI_ISL_1619899, EPI_ISL_1619900, EPI_ISL_1619901,
EPI_ISL_1619902, EPI_ISL_1619903, EPI_ISL_1619904, EPI_ISL_1619905, EPI_ISL_1619906, EPI_ISL_1619907, EPI_ISL_1619908, EPI_ISL_1619909, EPI_ISL_1619910, EPI_ISL_1619911, EPI_ISL_1619912, EPI_ISL_1619913, EPI_ISL_1619914, EPI_ISL_1619915, EPI_ISL_1619916, EPI_ISL_1619917, EPI_ISL_1619918, EPI_ISL_1619919, EPI_ISL_1619920, EPI_ISL_1619921, EPI_ISL_1619922, EPI_ISL_1619923,
EPI_ISL_1619924, EPI_ISL_1619925, EPI_ISL_1619926, EPI_ISL_1619927, EPI_ISL_1619928, EPI_ISL_1619929, EPI_ISL_1619930, EPI_ISL_1619931, EPI_ISL_1619932, EPI_ISL_1619933, EPI_ISL_1619934, EPI_ISL_1619935, EPI_ISL_1619936, EPI_ISL_1619937, EPI_ISL_1619938, EPI_ISL_1619939, EPI_ISL_1619940, EPI_ISL_1619941, EPI_ISL_1619942, EPI_ISL_1619943, EPI_ISL_1619944, EPI_ISL_1619945,
EPI_ISL_1619946, EPI_ISL_1619947, EPI_ISL_1619948, EPI_ISL_1619949, EPI_ISL_1619950, EPI_ISL_1619951, EPI_ISL_1619952, EPI_ISL_1619953, EPI_ISL_1619954, EPI_ISL_1619955, EPI_ISL_1619956, EPI_ISL_1619957, EPI_ISL_1619958, EPI_ISL_1619959, EPI_ISL_1619960, EPI_ISL_1619961, EPI_ISL_1619962, EPI_ISL_1619963, EPI_ISL_1619964, EPI_ISL_1619965, EPI_ISL_1619966, EPI_ISL_1619967,
EPI_ISL_1619968, EPI_ISL_1619969, EPI_ISL_1619970, EPI_ISL_1619971, EPI_ISL_1619972, EPI_ISL_1619973, EPI_ISL_1619974, EPI_ISL_1619975, EPI_ISL_1619976, EPI_ISL_1619977, EPI_ISL_1619978, EPI_ISL_1619979, EPI_ISL_1619980, EPI_ISL_1619981, EPI_ISL_1619982, EPI_ISL_1619983, EPI_ISL_1619984, EPI_ISL_1619985, EPI_ISL_1619986, EPI_ISL_1619987, EPI_ISL_1619988, EPI_ISL_1619989,
EPI_ISL_1619990, EPI_ISL_1619991, EPI_ISL_1619992, EPI_ISL_1619993, EPI_ISL_1619994, EPI_ISL_1619995, EPI_ISL_1619996, EPI_ISL_1619997, EPI_ISL_1619998, EPI_ISL_1619999, EPI_ISL_1620000, EPI_ISL_1620001, EPI_ISL_1620002, EPI_ISL_1620003, EPI_ISL_1620004, EPI_ISL_1620005, EPI_ISL_1620006, EPI_ISL_1620007, EPI_ISL_1620008, EPI_ISL_1620009, EPI_ISL_1620010, EPI_ISL_1620011,
EPI_ISL_1620012, EPI_ISL_1620013, EPI_ISL_1620014, EPI_ISL_1620015, EPI_ISL_1620016, EPI_ISL_1620017, EPI_ISL_1620018, EPI_ISL_1620019, EPI_ISL_1620020, EPI_ISL_1620021, EPI_ISL_1620022, EPI_ISL_1620023, EPI_ISL_1620024, EPI_ISL_1620025, EPI_ISL_1620026, EPI_ISL_1620027, EPI_ISL_1620028, EPI_ISL_1620029, EPI_ISL_1620030, EPI_ISL_1620031, EPI_ISL_1620032, EPI_ISL_1620033,
EPI_ISL_1620034, EPI_ISL_1620035, EPI_ISL_1620036, EPI_ISL_1620037, EPI_ISL_1620038, EPI_ISL_1620039, EPI_ISL_1620040, EPI_ISL_1620041, EPI_ISL_1620042, EPI_ISL_1620043, EPI_ISL_1620044, EPI_ISL_1620045, EPI_ISL_1620046, EPI_ISL_1620047, EPI_ISL_1620048, EPI_ISL_1620049, EPI_ISL_1620050, EPI_ISL_1620051, EPI_ISL_1620053, EPI_ISL_1620054, EPI_ISL_1620055, EPI_ISL_1620056,
EPI_ISL_1620057, EPI_ISL_1620058, EPI_ISL_1620059, EPI_ISL_1620060, EPI_ISL_1620061, EPI_ISL_1620062, EPI_ISL_1620063, EPI_ISL_1620064, EPI_ISL_1620065, EPI_ISL_1620066, EPI_ISL_1620067, EPI_ISL_1620068, EPI_ISL_1620069, EPI_ISL_1620070, EPI_ISL_1620071, EPI_ISL_1620072, EPI_ISL_1620073, EPI_ISL_1620074, EPI_ISL_1620075, EPI_ISL_1620076, EPI_ISL_1620077, EPI_ISL_1620078,
EPI_ISL_1620079, EPI_ISL_1620080, EPI_ISL_1620081, EPI_ISL_1620082, EPI_ISL_1620083, EPI_ISL_1620084, EPI_ISL_1620085, EPI_ISL_1620086, EPI_ISL_1620087, EPI_ISL_1620088, EPI_ISL_1620089, EPI_ISL_1620090, EPI_ISL_1620091, EPI_ISL_1620092, EPI_ISL_1620093, EPI_ISL_1620094, EPI_ISL_1620095, EPI_ISL_1620096, EPI_ISL_1620097, EPI_ISL_1620098, EPI_ISL_1620099, EPI_ISL_1620100,
EPI_ISL_1620101, EPI_ISL_1620102, EPI_ISL_1620103, EPI_ISL_1620104, EPI_ISL_1620105, EPI_ISL_1620106, EPI_ISL_1620107, EPI_ISL_1620108, EPI_ISL_1620109, EPI_ISL_1620110, EPI_ISL_1620111, EPI_ISL_1620112, EPI_ISL_1620113, EPI_ISL_1620114, EPI_ISL_1620115, EPI_ISL_1620116, EPI_ISL_1620117, EPI_ISL_1620118, EPI_ISL_1620119, EPI_ISL_1620120, EPI_ISL_1620121, EPI_ISL_1620122,
EPI_ISL_1620123, EPI_ISL_1620124, EPI_ISL_1620125, EPI_ISL_1620126, EPI_ISL_1620127, EPI_ISL_1620128, EPI_ISL_1620129, EPI_ISL_1659992, EPI_ISL_1659993, EPI_ISL_1659994, EPI_ISL_1659995, EPI_ISL_1659996, EPI_ISL_1747676, EPI_ISL_1747677, EPI_ISL_1747681, EPI_ISL_1747682, EPI_ISL_1747701, EPI_ISL_1747713, EPI_ISL_1747715, EPI_ISL_1747717, EPI_ISL_1747721, EPI_ISL_1747722,
EPI_ISL_1747726, EPI_ISL_1747727, EPI_ISL_1747729, EPI_ISL_1747730, EPI_ISL_1747731, EPI_ISL_1747733, EPI_ISL_1747734, EPI_ISL_1747735, EPI_ISL_1747737, EPI_ISL_1747760, EPI_ISL_1747761, EPI_ISL_1747763, EPI_ISL_1747764, EPI_ISL_1747765, EPI_ISL_1747767, EPI_ISL_1747769, EPI_ISL_1747770, EPI_ISL_1747771, EPI_ISL_1747773, EPI_ISL_1747774, EPI_ISL_1747775, EPI_ISL_1747777,
EPI_ISL_1747778, EPI_ISL_1747780, EPI_ISL_1747782, EPI_ISL_1747783, EPI_ISL_1747785, EPI_ISL_1747786, EPI_ISL_1747787, EPI_ISL_1747799, EPI_ISL_1747801, EPI_ISL_1747803, EPI_ISL_1747804, EPI_ISL_1747806, EPI_ISL_1747807, EPI_ISL_1747809, EPI_ISL_1747810, EPI_ISL_1747812, EPI_ISL_1747813, EPI_ISL_1747815, EPI_ISL_1747817, EPI_ISL_1747818, EPI_ISL_1747819, EPI_ISL_1747821,
EPI_ISL_1747824, EPI_ISL_1747825, EPI_ISL_1747827, EPI_ISL_1747828, EPI_ISL_1747830, EPI_ISL_1747831, EPI_ISL_1747832, EPI_ISL_1747843, EPI_ISL_1747844, EPI_ISL_1747848, EPI_ISL_1747852, EPI_ISL_1747863, EPI_ISL_1747865, EPI_ISL_1747866, EPI_ISL_1747867, EPI_ISL_1747869, EPI_ISL_1747871, EPI_ISL_1747872, EPI_ISL_1747873, EPI_ISL_1747875, EPI_ISL_1747876, EPI_ISL_1747878,
EPI_ISL_1747879, EPI_ISL_1747914, EPI_ISL_1747954, EPI_ISL_1747956, EPI_ISL_1747957, EPI_ISL_1747959, EPI_ISL_1747963, EPI_ISL_1747975, EPI_ISL_1747976, EPI_ISL_1747978, EPI_ISL_1747984, EPI_ISL_1747985, EPI_ISL_1747986, EPI_ISL_1747988, EPI_ISL_1747992, EPI_ISL_1747994, EPI_ISL_1747996, EPI_ISL_1747997, EPI_ISL_1747999, EPI_ISL_1748000, EPI_ISL_1748001, EPI_ISL_1748003,
EPI_ISL_1748004, EPI_ISL_1748006, EPI_ISL_1748008, EPI_ISL_1748009, EPI_ISL_1748011, EPI_ISL_1748012, EPI_ISL_1748013, EPI_ISL_1748015, EPI_ISL_1748025, EPI_ISL_1748027, EPI_ISL_1748029, EPI_ISL_1748036, EPI_ISL_1748038, EPI_ISL_1748040, EPI_ISL_1748041, EPI_ISL_1748043, EPI_ISL_1748044, EPI_ISL_1748046, EPI_ISL_1748047, EPI_ISL_1748049, EPI_ISL_1748050, EPI_ISL_1748053,
EPI_ISL_1748056, EPI_ISL_1748058, EPI_ISL_1748059, EPI_ISL_1748061, EPI_ISL_1748065, EPI_ISL_1748070, EPI_ISL_1748071, EPI_ISL_1748073, EPI_ISL_1748074, EPI_ISL_1748075, EPI_ISL_1748077, EPI_ISL_1748079, EPI_ISL_1748081, EPI_ISL_1748082, EPI_ISL_1748084, EPI_ISL_1748086, EPI_ISL_1748087, EPI_ISL_1748089, EPI_ISL_1748090, EPI_ISL_1748091, EPI_ISL_1748093, EPI_ISL_1748094,
EPI_ISL_1748096, EPI_ISL_1748097, EPI_ISL_1748099, EPI_ISL_1748103, EPI_ISL_1748104, EPI_ISL_1748105, EPI_ISL_1748106, EPI_ISL_1748108, EPI_ISL_1748109, EPI_ISL_1748110, EPI_ISL_1748111, EPI_ISL_1748113, EPI_ISL_1748115, EPI_ISL_1748116, EPI_ISL_1748118, EPI_ISL_1748119, EPI_ISL_1748120, EPI_ISL_1748121, EPI_ISL_1748132, EPI_ISL_1748133, EPI_ISL_1748135, EPI_ISL_1748136,
EPI_ISL_1748138, EPI_ISL_1748140, EPI_ISL_1748141, EPI_ISL_1748147, EPI_ISL_1748148, EPI_ISL_1748152, EPI_ISL_1748159, EPI_ISL_1748168, EPI_ISL_1748182, EPI_ISL_1748184, EPI_ISL_1748185, EPI_ISL_1748193, EPI_ISL_1748199, EPI_ISL_1748200, EPI_ISL_1748203, EPI_ISL_1748208, EPI_ISL_1748209, EPI_ISL_1748211, EPI_ISL_1748222, EPI_ISL_1748224, EPI_ISL_1748225, EPI_ISL_1748226,
EPI_ISL_1748227, EPI_ISL_1748229, EPI_ISL_1748230, EPI_ISL_1748237, EPI_ISL_1748239, EPI_ISL_1748252, EPI_ISL_1748253, EPI_ISL_1748255, EPI_ISL_1748335, EPI_ISL_1748447, EPI_ISL_1748467, EPI_ISL_1748508, EPI_ISL_1748510, EPI_ISL_1748635, EPI_ISL_1748636, EPI_ISL_1807375, EPI_ISL_1807376, EPI_ISL_1807377, EPI_ISL_1807378, EPI_ISL_1807379, EPI_ISL_1807380, EPI_ISL_1807381,
EPI_ISL_1807382, EPI_ISL_1807383, EPI_ISL_1807384, EPI_ISL_1807385, EPI_ISL_1807386, EPI_ISL_1807387, EPI_ISL_1807388, EPI_ISL_1807389, EPI_ISL_1807390, EPI_ISL_1807391, EPI_ISL_1807392, EPI_ISL_1807393, EPI_ISL_1807394, EPI_ISL_1807395, EPI_ISL_1807396, EPI_ISL_1807397, EPI_ISL_1807398, EPI_ISL_1807399, EPI_ISL_1807400, EPI_ISL_1807401, EPI_ISL_1807402, EPI_ISL_1807403,
EPI_ISL_1807404, EPI_ISL_1807405, EPI_ISL_1807406, EPI_ISL_1807407, EPI_ISL_1807408, EPI_ISL_1807409, EPI_ISL_1807410, EPI_ISL_1807411, EPI_ISL_1807412, EPI_ISL_1807413, EPI_ISL_1807414, EPI_ISL_1807415, EPI_ISL_1807416, EPI_ISL_1807417, EPI_ISL_1807418, EPI_ISL_1807419, EPI_ISL_1807420, EPI_ISL_1807421, EPI_ISL_1807422, EPI_ISL_1807423, EPI_ISL_1807424, EPI_ISL_1807425,
EPI_ISL_1807426, EPI_ISL_1807427, EPI_ISL_1807429, EPI_ISL_1807430, EPI_ISL_1807431, EPI_ISL_1807432, EPI_ISL_1807433, EPI_ISL_1807434, EPI_ISL_1807435, EPI_ISL_1807436, EPI_ISL_1807437, EPI_ISL_1807438, EPI_ISL_1807439, EPI_ISL_1807440, EPI_ISL_1807441, EPI_ISL_1807442, EPI_ISL_1807443, EPI_ISL_1807444, EPI_ISL_1807445, EPI_ISL_1807446, EPI_ISL_1807447, EPI_ISL_1807448,
EPI_ISL_1807449, EPI_ISL_1807450, EPI_ISL_1807451, EPI_ISL_1807452, EPI_ISL_1807453, EPI_ISL_1807454, EPI_ISL_1807455, EPI_ISL_1807456, EPI_ISL_1807457, EPI_ISL_1807458, EPI_ISL_1807459, EPI_ISL_1807460, EPI_ISL_1807461, EPI_ISL_1807462, EPI_ISL_1807463, EPI_ISL_1807464, EPI_ISL_1807465, EPI_ISL_1807466, EPI_ISL_1807467, EPI_ISL_1807468, EPI_ISL_1807469, EPI_ISL_1807470,
EPI_ISL_1807471, EPI_ISL_1807472, EPI_ISL_1807473, EPI_ISL_1807474, EPI_ISL_1807475, EPI_ISL_1807476, EPI_ISL_1807477, EPI_ISL_1807478, EPI_ISL_1807479, EPI_ISL_1807480, EPI_ISL_1807481, EPI_ISL_1807482, EPI_ISL_1807483, EPI_ISL_1807484, EPI_ISL_1807485, EPI_ISL_1807486, EPI_ISL_1807487, EPI_ISL_1807488, EPI_ISL_1807489, EPI_ISL_1807490, EPI_ISL_1807491, EPI_ISL_1807492,
EPI_ISL_1807493, EPI_ISL_1807494, EPI_ISL_1807495, EPI_ISL_1807496, EPI_ISL_1807497, EPI_ISL_1807498, EPI_ISL_1807499, EPI_ISL_1807500, EPI_ISL_1807501, EPI_ISL_1807503, EPI_ISL_1807504, EPI_ISL_1807505, EPI_ISL_1807506, EPI_ISL_1807507, EPI_ISL_1807508, EPI_ISL_1807509, EPI_ISL_1807510, EPI_ISL_1807511, EPI_ISL_1807512, EPI_ISL_1807513, EPI_ISL_1807514, EPI_ISL_1807515,
EPI_ISL_1807516, EPI_ISL_1807517, EPI_ISL_1807518, EPI_ISL_1807519, EPI_ISL_1807520, EPI_ISL_1807521, EPI_ISL_1807522, EPI_ISL_1807523, EPI_ISL_1807524, EPI_ISL_1807525, EPI_ISL_1807526, EPI_ISL_1807527, EPI_ISL_1807528, EPI_ISL_1807529, EPI_ISL_1807530, EPI_ISL_1807531, EPI_ISL_1807532, EPI_ISL_1807533, EPI_ISL_1807534, EPI_ISL_1807535, EPI_ISL_1807536, EPI_ISL_1807537,
EPI_ISL_1807538, EPI_ISL_1807539, EPI_ISL_1807540, EPI_ISL_1807541, EPI_ISL_1807542, EPI_ISL_1807543, EPI_ISL_1807544, EPI_ISL_1807545, EPI_ISL_1807546, EPI_ISL_1807547, EPI_ISL_1807548, EPI_ISL_1807549, EPI_ISL_1807550, EPI_ISL_1807551, EPI_ISL_1807552, EPI_ISL_1807553, EPI_ISL_1807554, EPI_ISL_1807555, EPI_ISL_1807556, EPI_ISL_1807557, EPI_ISL_1807558, EPI_ISL_1807559,
EPI_ISL_1807560, EPI_ISL_1807561, EPI_ISL_1807562, EPI_ISL_1807563, EPI_ISL_1807564, EPI_ISL_1807565, EPI_ISL_1807566, EPI_ISL_1807567, EPI_ISL_1807568, EPI_ISL_1807570, EPI_ISL_1807571, EPI_ISL_1807572, EPI_ISL_1807573, EPI_ISL_1807574, EPI_ISL_1807575, EPI_ISL_1807576, EPI_ISL_1807577, EPI_ISL_1807578, EPI_ISL_1807579, EPI_ISL_1807580, EPI_ISL_1807581, EPI_ISL_1807582,
EPI_ISL_1807583, EPI_ISL_1807584, EPI_ISL_1807585, EPI_ISL_1807586, EPI_ISL_1807587, EPI_ISL_1807588, EPI_ISL_1807589, EPI_ISL_1807590, EPI_ISL_1807591, EPI_ISL_1807592, EPI_ISL_1807593, EPI_ISL_1807594, EPI_ISL_1807595, EPI_ISL_1807596, EPI_ISL_1807597, EPI_ISL_1807598, EPI_ISL_1807599, EPI_ISL_1807600, EPI_ISL_1807601, EPI_ISL_1807602, EPI_ISL_1807603, EPI_ISL_1807604,
EPI_ISL_1807605, EPI_ISL_1807606, EPI_ISL_1807607, EPI_ISL_1807608, EPI_ISL_1807609, EPI_ISL_1807610, EPI_ISL_1807611, EPI_ISL_1807612, EPI_ISL_1807613, EPI_ISL_1807614, EPI_ISL_1807615, EPI_ISL_1807616, EPI_ISL_1807617, EPI_ISL_1807618, EPI_ISL_1807619, EPI_ISL_1807620, EPI_ISL_1807621, EPI_ISL_1807622, EPI_ISL_1807623, EPI_ISL_1807624, EPI_ISL_1807625, EPI_ISL_1807626,
EPI_ISL_1807627, EPI_ISL_1807628, EPI_ISL_1807629, EPI_ISL_1807630, EPI_ISL_1807631, EPI_ISL_1807632, EPI_ISL_1807633, EPI_ISL_1807634, EPI_ISL_1807635, EPI_ISL_1807636, EPI_ISL_1807637, EPI_ISL_1807638, EPI_ISL_1807639, EPI_ISL_1807640, EPI_ISL_1807641, EPI_ISL_1807642, EPI_ISL_1807643, EPI_ISL_1807644, EPI_ISL_1807645, EPI_ISL_1807646, EPI_ISL_1807647, EPI_ISL_1807648,
EPI_ISL_1807649, EPI_ISL_1807650, EPI_ISL_1807651, EPI_ISL_1807752, EPI_ISL_1807753, EPI_ISL_1807754, EPI_ISL_1807755, EPI_ISL_1807756, EPI_ISL_1807757, EPI_ISL_1807758, EPI_ISL_1807759, EPI_ISL_1807760, EPI_ISL_1807761, EPI_ISL_1807762, EPI_ISL_1807763, EPI_ISL_1807764, EPI_ISL_1807765, EPI_ISL_1807766, EPI_ISL_1807767, EPI_ISL_1807768, EPI_ISL_1807769, EPI_ISL_1807770,
EPI_ISL_1807771, EPI_ISL_1807772, EPI_ISL_1807773, EPI_ISL_1807775, EPI_ISL_1807776, EPI_ISL_1807777, EPI_ISL_1807778, EPI_ISL_1807779, EPI_ISL_1807780, EPI_ISL_1807781, EPI_ISL_1807782, EPI_ISL_1807783, EPI_ISL_1807784, EPI_ISL_1807785, EPI_ISL_1807786, EPI_ISL_1807787, EPI_ISL_1807788, EPI_ISL_1807789, EPI_ISL_1807790, EPI_ISL_1807791, EPI_ISL_1807793, EPI_ISL_1807794,
EPI_ISL_1807795, EPI_ISL_1807796, EPI_ISL_1807797, EPI_ISL_1807798, EPI_ISL_1807799, EPI_ISL_1807800, EPI_ISL_1807801, EPI_ISL_1807802, EPI_ISL_1807803, EPI_ISL_1807804, EPI_ISL_1807805, EPI_ISL_1807806, EPI_ISL_1807807, EPI_ISL_1807808, EPI_ISL_1807809, EPI_ISL_1807810, EPI_ISL_1807811, EPI_ISL_1807812, EPI_ISL_1807813, EPI_ISL_1807814, EPI_ISL_1807815, EPI_ISL_1807816,
EPI_ISL_1807817, EPI_ISL_1807818, EPI_ISL_1807819, EPI_ISL_1807820, EPI_ISL_1807822, EPI_ISL_1807823, EPI_ISL_1807824, EPI_ISL_1807825, EPI_ISL_1807826, EPI_ISL_1807827, EPI_ISL_1807828, EPI_ISL_1807829, EPI_ISL_1807830, EPI_ISL_1807831, EPI_ISL_1807832, EPI_ISL_1807833, EPI_ISL_1807834, EPI_ISL_1807835, EPI_ISL_1807836, EPI_ISL_1807837, EPI_ISL_1807840, EPI_ISL_1807841,
EPI_ISL_1807842, EPI_ISL_1807843, EPI_ISL_1807844, EPI_ISL_1807845, EPI_ISL_1807846, EPI_ISL_1807847, EPI_ISL_1807848, EPI_ISL_1807849, EPI_ISL_1807850, EPI_ISL_1807851, EPI_ISL_1807852, EPI_ISL_1807853, EPI_ISL_1807854, EPI_ISL_1807855, EPI_ISL_1807856, EPI_ISL_1807857, EPI_ISL_1807858, EPI_ISL_1807859, EPI_ISL_1807860, EPI_ISL_1807861, EPI_ISL_1807862, EPI_ISL_1807863,
EPI_ISL_1807864, EPI_ISL_1807865, EPI_ISL_1807866, EPI_ISL_1807867, EPI_ISL_1807868, EPI_ISL_1807869, EPI_ISL_1807870, EPI_ISL_1807871, EPI_ISL_1807872, EPI_ISL_1807873, EPI_ISL_1807874, EPI_ISL_1807875, EPI_ISL_1807876, EPI_ISL_1807877, EPI_ISL_1807878, EPI_ISL_1807879, EPI_ISL_1807880, EPI_ISL_1807881, EPI_ISL_1807882, EPI_ISL_1807883, EPI_ISL_1807884, EPI_ISL_1807885,
EPI_ISL_1807886, EPI_ISL_1807887, EPI_ISL_1807888, EPI_ISL_1807889, EPI_ISL_1807890, EPI_ISL_1807891, EPI_ISL_1807892, EPI_ISL_1807893, EPI_ISL_1807894, EPI_ISL_1807895, EPI_ISL_1807896, EPI_ISL_1807897, EPI_ISL_1807898, EPI_ISL_1807899, EPI_ISL_1807900, EPI_ISL_1807901, EPI_ISL_1807902, EPI_ISL_1807903, EPI_ISL_1807904, EPI_ISL_1807905, EPI_ISL_1807906, EPI_ISL_1807907,
EPI_ISL_1807908, EPI_ISL_1807909, EPI_ISL_1807910, EPI_ISL_1807911, EPI_ISL_1807912, EPI_ISL_1807913, EPI_ISL_1807914, EPI_ISL_1807915, EPI_ISL_1807916, EPI_ISL_1807917, EPI_ISL_1807918, EPI_ISL_1807919, EPI_ISL_1807920, EPI_ISL_1807921, EPI_ISL_1807922, EPI_ISL_1807923, EPI_ISL_1807924, EPI_ISL_1807925, EPI_ISL_1807926, EPI_ISL_1807927, EPI_ISL_1807928, EPI_ISL_1807929,
EPI_ISL_1807930, EPI_ISL_1807931, EPI_ISL_1807932, EPI_ISL_1807933, EPI_ISL_1807934, EPI_ISL_1807935, EPI_ISL_1807936, EPI_ISL_1807937, EPI_ISL_1807938, EPI_ISL_1807939, EPI_ISL_1807940, EPI_ISL_1807941, EPI_ISL_1807942, EPI_ISL_1807943, EPI_ISL_1807944, EPI_ISL_1807945, EPI_ISL_1807946, EPI_ISL_1807947, EPI_ISL_1807948, EPI_ISL_1807949, EPI_ISL_1807950, EPI_ISL_1807951,
EPI_ISL_1807952, EPI_ISL_1807953, EPI_ISL_1807955, EPI_ISL_1807956, EPI_ISL_1807957, EPI_ISL_1807958, EPI_ISL_1807959, EPI_ISL_1807960, EPI_ISL_1807961, EPI_ISL_1807962, EPI_ISL_1807963, EPI_ISL_1807964, EPI_ISL_1807965, EPI_ISL_1807966, EPI_ISL_1807967, EPI_ISL_1807968, EPI_ISL_1807969, EPI_ISL_1807970, EPI_ISL_1807971, EPI_ISL_1807972, EPI_ISL_1807973, EPI_ISL_1807974,
EPI_ISL_1807975, EPI_ISL_1807976, EPI_ISL_1807977, EPI_ISL_1807978, EPI_ISL_1807979, EPI_ISL_1807980, EPI_ISL_1807981, EPI_ISL_1807982, EPI_ISL_1807983, EPI_ISL_1807984, EPI_ISL_1807985, EPI_ISL_1807986, EPI_ISL_1807987, EPI_ISL_1807988, EPI_ISL_1807989, EPI_ISL_1807990, EPI_ISL_1807991, EPI_ISL_1807992, EPI_ISL_1807993, EPI_ISL_1807994, EPI_ISL_1807995, EPI_ISL_1807996,
EPI_ISL_1807997, EPI_ISL_1807998, EPI_ISL_1807999, EPI_ISL_1808000, EPI_ISL_1808002, EPI_ISL_1808003, EPI_ISL_1808004, EPI_ISL_1808005, EPI_ISL_1808006, EPI_ISL_1808007, EPI_ISL_1808008, EPI_ISL_1808009, EPI_ISL_1808010, EPI_ISL_1808011, EPI_ISL_1808012, EPI_ISL_1808013, EPI_ISL_1808014, EPI_ISL_1808015, EPI_ISL_1808016, EPI_ISL_1808017, EPI_ISL_1808018, EPI_ISL_1808019,
EPI_ISL_1808020, EPI_ISL_1808021, EPI_ISL_1808022, EPI_ISL_1808023, EPI_ISL_1808024, EPI_ISL_1808025, EPI_ISL_1808026, EPI_ISL_1808027, EPI_ISL_1808028, EPI_ISL_1808029, EPI_ISL_1808030, EPI_ISL_1808031, EPI_ISL_1808032, EPI_ISL_1808033, EPI_ISL_1808034, EPI_ISL_1808035, EPI_ISL_1808036, EPI_ISL_1808037, EPI_ISL_1808038, EPI_ISL_1808039, EPI_ISL_1808040, EPI_ISL_1808041,
EPI_ISL_1808042, EPI_ISL_1808043, EPI_ISL_1808044, EPI_ISL_1808045, EPI_ISL_1808046, EPI_ISL_1808047, EPI_ISL_1808048, EPI_ISL_1808049, EPI_ISL_1808050, EPI_ISL_1808051, EPI_ISL_1808052, EPI_ISL_1808053, EPI_ISL_1808054, EPI_ISL_1808055, EPI_ISL_1808056, EPI_ISL_1808057, EPI_ISL_1808058, EPI_ISL_1808059, EPI_ISL_1808060, EPI_ISL_1808061, EPI_ISL_1808063, EPI_ISL_1808064,
EPI_ISL_1808066, EPI_ISL_1808067, EPI_ISL_1808068, EPI_ISL_1808069, EPI_ISL_1808070, EPI_ISL_1808071, EPI_ISL_1808072, EPI_ISL_1808073, EPI_ISL_1808074, EPI_ISL_1808075, EPI_ISL_1808076, EPI_ISL_1808077, EPI_ISL_1808078, EPI_ISL_1808079, EPI_ISL_1808080, EPI_ISL_1808081, EPI_ISL_1808082, EPI_ISL_1808083, EPI_ISL_1808084, EPI_ISL_1808085, EPI_ISL_1808086, EPI_ISL_1808087,
EPI_ISL_1808088, EPI_ISL_1808089, EPI_ISL_1808090, EPI_ISL_1808091, EPI_ISL_1808092, EPI_ISL_1808093, EPI_ISL_1808094, EPI_ISL_1808095, EPI_ISL_1808096, EPI_ISL_1808097, EPI_ISL_1808098, EPI_ISL_1808099, EPI_ISL_1808100, EPI_ISL_1808101, EPI_ISL_1808102, EPI_ISL_1808103, EPI_ISL_1808104, EPI_ISL_1808105, EPI_ISL_1808106, EPI_ISL_1808107, EPI_ISL_1808108, EPI_ISL_1808109,
EPI_ISL_1808110, EPI_ISL_1808111, EPI_ISL_1808112, EPI_ISL_1808113, EPI_ISL_1808114, EPI_ISL_1808115, EPI_ISL_1808116, EPI_ISL_1808117, EPI_ISL_1808119, EPI_ISL_1808120, EPI_ISL_1808121, EPI_ISL_1808122, EPI_ISL_1808123, EPI_ISL_1808124, EPI_ISL_1808125, EPI_ISL_1808126, EPI_ISL_1808796, EPI_ISL_1808798, EPI_ISL_1808799, EPI_ISL_1808800, EPI_ISL_1808801, EPI_ISL_1808803,
EPI_ISL_1808804, EPI_ISL_1808806, EPI_ISL_1808807, EPI_ISL_1808808, EPI_ISL_1808809, EPI_ISL_1808812, EPI_ISL_1808813, EPI_ISL_1808815, EPI_ISL_1809812, EPI_ISL_1809813, EPI_ISL_1809814, EPI_ISL_1809815, EPI_ISL_1809816, EPI_ISL_1809817, EPI_ISL_1809818, EPI_ISL_1809819, EPI_ISL_1809820, EPI_ISL_1809821, EPI_ISL_1809822, EPI_ISL_1809823, EPI_ISL_1809824, EPI_ISL_1809825,
EPI_ISL_1809826, EPI_ISL_1809827, EPI_ISL_1809828, EPI_ISL_1809829, EPI_ISL_1809830, EPI_ISL_1809831, EPI_ISL_1809832, EPI_ISL_1809833, EPI_ISL_1809834, EPI_ISL_1809835, EPI_ISL_1809836, EPI_ISL_1809837, EPI_ISL_1809838, EPI_ISL_1809839, EPI_ISL_1809840, EPI_ISL_1809841, EPI_ISL_1809842, EPI_ISL_1809843, EPI_ISL_1809844, EPI_ISL_1809845, EPI_ISL_1809846, EPI_ISL_1809847,
EPI_ISL_1809848, EPI_ISL_1809849, EPI_ISL_1809850, EPI_ISL_1809851, EPI_ISL_1809852, EPI_ISL_1809853, EPI_ISL_1809854, EPI_ISL_1809855, EPI_ISL_1809856, EPI_ISL_1809857, EPI_ISL_1809858, EPI_ISL_1809859, EPI_ISL_1809860, EPI_ISL_1809861, EPI_ISL_1809862, EPI_ISL_1809863, EPI_ISL_1809864, EPI_ISL_1809865, EPI_ISL_1809866, EPI_ISL_1809867, EPI_ISL_1809868, EPI_ISL_1809869,
EPI_ISL_1809870, EPI_ISL_1809871, EPI_ISL_1809872, EPI_ISL_1809873, EPI_ISL_1809874, EPI_ISL_1809875, EPI_ISL_1809876, EPI_ISL_1809877, EPI_ISL_1809878, EPI_ISL_1809880, EPI_ISL_1809881, EPI_ISL_1809882, EPI_ISL_1809883, EPI_ISL_1809884, EPI_ISL_1809885, EPI_ISL_1809886, EPI_ISL_1809887, EPI_ISL_1809888, EPI_ISL_1809889, EPI_ISL_1809890, EPI_ISL_1809891, EPI_ISL_1809892,
EPI_ISL_1809893, EPI_ISL_1809894, EPI_ISL_1809895, EPI_ISL_1809896, EPI_ISL_1809897, EPI_ISL_1809898, EPI_ISL_1809899, EPI_ISL_1809991, EPI_ISL_1809992, EPI_ISL_1809993, EPI_ISL_1809994, EPI_ISL_1809995, EPI_ISL_1809996, EPI_ISL_1809997, EPI_ISL_1809998, EPI_ISL_1809999, EPI_ISL_1810000, EPI_ISL_1810001, EPI_ISL_1810002, EPI_ISL_1810003, EPI_ISL_1810004, EPI_ISL_1810005,
EPI_ISL_1810006, EPI_ISL_1810007, EPI_ISL_1810008, EPI_ISL_1810009, EPI_ISL_1810010, EPI_ISL_1810011, EPI_ISL_1810012, EPI_ISL_1810013, EPI_ISL_1810014, EPI_ISL_1810015, EPI_ISL_1810016, EPI_ISL_1810017, EPI_ISL_1810018, EPI_ISL_1810019, EPI_ISL_1810020, EPI_ISL_1810021, EPI_ISL_1810022, EPI_ISL_1810023, EPI_ISL_1810024, EPI_ISL_1810025, EPI_ISL_1810026, EPI_ISL_1810027,
EPI_ISL_1810028, EPI_ISL_1810029, EPI_ISL_1810030, EPI_ISL_1810031, EPI_ISL_1810032, EPI_ISL_1810033, EPI_ISL_1810034, EPI_ISL_1810035, EPI_ISL_1810036, EPI_ISL_1810037, EPI_ISL_1810038, EPI_ISL_1810039, EPI_ISL_1810040, EPI_ISL_1810041, EPI_ISL_1810042, EPI_ISL_1810043, EPI_ISL_1810044, EPI_ISL_1810045, EPI_ISL_1810046, EPI_ISL_1810047, EPI_ISL_1810048, EPI_ISL_1810049,
EPI_ISL_1810050, EPI_ISL_1810051, EPI_ISL_1810052, EPI_ISL_1810053, EPI_ISL_1810054, EPI_ISL_1810055, EPI_ISL_1810056, EPI_ISL_1810057, EPI_ISL_1810058, EPI_ISL_1810059, EPI_ISL_1810060, EPI_ISL_1810061, EPI_ISL_1810062, EPI_ISL_1810063, EPI_ISL_1810064, EPI_ISL_1810065, EPI_ISL_1810066, EPI_ISL_1810067, EPI_ISL_1810068, EPI_ISL_1810069, EPI_ISL_1810070, EPI_ISL_1810071,
EPI_ISL_1810072, EPI_ISL_1810073, EPI_ISL_1810074, EPI_ISL_1810075, EPI_ISL_1810076, EPI_ISL_1810077, EPI_ISL_1810078, EPI_ISL_1810079, EPI_ISL_1810080, EPI_ISL_1810081, EPI_ISL_1810082, EPI_ISL_1810083, EPI_ISL_1810084, EPI_ISL_1810085, EPI_ISL_1810086, EPI_ISL_1810087, EPI_ISL_1810088, EPI_ISL_1810089, EPI_ISL_1810090, EPI_ISL_1810091, EPI_ISL_1810092, EPI_ISL_1810093,
EPI_ISL_1810094, EPI_ISL_1810095, EPI_ISL_1810096, EPI_ISL_1810097, EPI_ISL_1810098, EPI_ISL_1810099, EPI_ISL_1810100, EPI_ISL_1810101, EPI_ISL_1810102, EPI_ISL_1810103, EPI_ISL_1810104, EPI_ISL_1810105, EPI_ISL_1810106, EPI_ISL_1810107, EPI_ISL_1810108, EPI_ISL_1810109, EPI_ISL_1810110, EPI_ISL_1810111, EPI_ISL_1810112, EPI_ISL_1810113, EPI_ISL_1810114, EPI_ISL_1810115,
EPI_ISL_1810116, EPI_ISL_1810117, EPI_ISL_1810118, EPI_ISL_1810119, EPI_ISL_1810120, EPI_ISL_1810121, EPI_ISL_1810122, EPI_ISL_1810123, EPI_ISL_1810124, EPI_ISL_1810125, EPI_ISL_1810126, EPI_ISL_1810127, EPI_ISL_1810128, EPI_ISL_1810129, EPI_ISL_1810130, EPI_ISL_1810131, EPI_ISL_1810132, EPI_ISL_1810133, EPI_ISL_1810134, EPI_ISL_1810135, EPI_ISL_1810136, EPI_ISL_1810137,
EPI_ISL_1810138, EPI_ISL_1810139, EPI_ISL_1810140, EPI_ISL_1810141, EPI_ISL_1810142, EPI_ISL_1810143, EPI_ISL_1810144, EPI_ISL_1810145, EPI_ISL_1810146, EPI_ISL_1810147, EPI_ISL_1810148, EPI_ISL_1810149, EPI_ISL_1810150, EPI_ISL_1810151, EPI_ISL_1810152, EPI_ISL_1810153, EPI_ISL_1810154, EPI_ISL_1810155, EPI_ISL_1810156, EPI_ISL_1810157, EPI_ISL_1810158, EPI_ISL_1810159,
EPI_ISL_1810160, EPI_ISL_1810161, EPI_ISL_1810162, EPI_ISL_1810163, EPI_ISL_1810164, EPI_ISL_1810165, EPI_ISL_1810166, EPI_ISL_1810167, EPI_ISL_1810168, EPI_ISL_1810169, EPI_ISL_1810170, EPI_ISL_1810171, EPI_ISL_1810172, EPI_ISL_1810173, EPI_ISL_1810174, EPI_ISL_1810175, EPI_ISL_1810176, EPI_ISL_1810177, EPI_ISL_1810178, EPI_ISL_1810179, EPI_ISL_1810180, EPI_ISL_1810181,
EPI_ISL_1810182, EPI_ISL_1810183, EPI_ISL_1810184, EPI_ISL_1810185, EPI_ISL_1810186, EPI_ISL_1810187, EPI_ISL_1810188, EPI_ISL_1810189, EPI_ISL_1810190, EPI_ISL_1810191, EPI_ISL_1810192, EPI_ISL_1810193, EPI_ISL_1810194, EPI_ISL_1810195, EPI_ISL_1810196, EPI_ISL_1810197, EPI_ISL_1810198, EPI_ISL_1810199, EPI_ISL_1810200, EPI_ISL_1810201, EPI_ISL_1810202, EPI_ISL_1810203,
EPI_ISL_1810204, EPI_ISL_1810205, EPI_ISL_1810206, EPI_ISL_1810207, EPI_ISL_1810208, EPI_ISL_1810209, EPI_ISL_1810210, EPI_ISL_1810211, EPI_ISL_1810212, EPI_ISL_1810213, EPI_ISL_1810214, EPI_ISL_1810215, EPI_ISL_1810216, EPI_ISL_1810217, EPI_ISL_1810218, EPI_ISL_1810219, EPI_ISL_1810220, EPI_ISL_1810221, EPI_ISL_1810222, EPI_ISL_1810223, EPI_ISL_1810224, EPI_ISL_1810225,
EPI_ISL_1810226, EPI_ISL_1810227, EPI_ISL_1810228, EPI_ISL_1810229, EPI_ISL_1810230, EPI_ISL_1810231, EPI_ISL_1810232, EPI_ISL_1810233, EPI_ISL_1810234, EPI_ISL_1810235, EPI_ISL_1810236, EPI_ISL_1810237, EPI_ISL_1810238, EPI_ISL_1810239, EPI_ISL_1810240, EPI_ISL_1810241, EPI_ISL_1810242, EPI_ISL_1810243, EPI_ISL_1810244, EPI_ISL_1810245, EPI_ISL_1810246, EPI_ISL_1810247,

| | | | |
|---|---|---|---|
| EPI_ISL_1810248, EPI_ISL_1810249, EPI_ISL_1810250, EPI_ISL_1810251, EPI_ISL_1810252, EPI_ISL_1810253, EPI_ISL_1810254, EPI_ISL_1810255, EPI_ISL_1810256, EPI_ISL_1810257, EPI_ISL_1810258, EPI_ISL_1810665, EPI_ISL_1810666, EPI_ISL_1810667, EPI_ISL_1810668, EPI_ISL_1810669, EPI_ISL_1810670, EPI_ISL_1810671, EPI_ISL_1810672, EPI_ISL_1810673, EPI_ISL_1810674, EPI_ISL_1810675, EPI_ISL_1810676, EPI_ISL_1810677, EPI_ISL_1810678, EPI_ISL_1810679, EPI_ISL_1810680, EPI_ISL_1810681, EPI_ISL_1810682, EPI_ISL_1810683, EPI_ISL_1810684, EPI_ISL_1810685, EPI_ISL_1810686, EPI_ISL_1810687, EPI_ISL_1810688, EPI_ISL_1810689, EPI_ISL_1810690, EPI_ISL_1810691, EPI_ISL_1810692, EPI_ISL_1810693, EPI_ISL_1810694, EPI_ISL_1810695, EPI_ISL_1810696, EPI_ISL_1810697, EPI_ISL_1810698, EPI_ISL_1810699, EPI_ISL_1810700, EPI_ISL_1810701, EPI_ISL_1810702, EPI_ISL_1810703, EPI_ISL_1810704, EPI_ISL_1810705, EPI_ISL_1810706, EPI_ISL_1810707, EPI_ISL_1810708, EPI_ISL_1810709, EPI_ISL_1810710, EPI_ISL_1810712, EPI_ISL_1810713, EPI_ISL_1810714, EPI_ISL_1810715, EPI_ISL_1810716, EPI_ISL_1810717, EPI_ISL_1810718, EPI_ISL_1810720, EPI_ISL_1810721, EPI_ISL_1831698, EPI_ISL_1832347, EPI_ISL_1832348, EPI_ISL_1832349, EPI_ISL_1832350, EPI_ISL_1832351, EPI_ISL_1832352, EPI_ISL_1832354, EPI_ISL_1832356, EPI_ISL_1832358, EPI_ISL_1832360, EPI_ISL_1832362, EPI_ISL_1832364, EPI_ISL_1832365, EPI_ISL_1832367, EPI_ISL_1832369, EPI_ISL_1832371, EPI_ISL_1832373, EPI_ISL_1832375, EPI_ISL_1832377, EPI_ISL_1832379, EPI_ISL_1832381, EPI_ISL_1832382, EPI_ISL_1832384, EPI_ISL_1832386, EPI_ISL_1832388, EPI_ISL_1832390, EPI_ISL_1832392, EPI_ISL_1832394, EPI_ISL_1832396, EPI_ISL_1832398, EPI_ISL_1832400, EPI_ISL_1832402, EPI_ISL_1832404, EPI_ISL_1832406, EPI_ISL_1832407, EPI_ISL_1832409, EPI_ISL_1832411, EPI_ISL_1832413, EPI_ISL_1832417, EPI_ISL_1832419, EPI_ISL_1832421, EPI_ISL_1832423, EPI_ISL_1832425, EPI_ISL_1832427, EPI_ISL_1832428, EPI_ISL_1832430, EPI_ISL_1832432, EPI_ISL_1832434, EPI_ISL_1832437, EPI_ISL_1832439, EPI_ISL_1832440, EPI_ISL_1832442, EPI_ISL_1832444, EPI_ISL_1832446, EPI_ISL_1832448, EPI_ISL_1832450, EPI_ISL_1832452, EPI_ISL_1832454, EPI_ISL_1832456, EPI_ISL_1832458, EPI_ISL_1832460, EPI_ISL_1832462, EPI_ISL_1832464, EPI_ISL_1832465, EPI_ISL_1832467, EPI_ISL_1832469, EPI_ISL_1832472, EPI_ISL_1832473, EPI_ISL_1832475, EPI_ISL_1832477, EPI_ISL_1832479, EPI_ISL_1832481, EPI_ISL_1832483, EPI_ISL_1832487, EPI_ISL_1832489, EPI_ISL_1832491, EPI_ISL_1832493, EPI_ISL_1832495, EPI_ISL_1832497, EPI_ISL_1832499, EPI_ISL_1832501, EPI_ISL_1832503, EPI_ISL_1832504, EPI_ISL_1832505, EPI_ISL_1832506, EPI_ISL_1832508, EPI_ISL_1832509, EPI_ISL_1832511, EPI_ISL_1832513, EPI_ISL_1832515, EPI_ISL_1832795, EPI_ISL_1832806, EPI_ISL_1832808, EPI_ISL_1832819, EPI_ISL_1832831, EPI_ISL_1832853, EPI_ISL_1897916, EPI_ISL_1897973, EPI_ISL_1897974, EPI_ISL_1900095, EPI_ISL_1900108, EPI_ISL_1900109, EPI_ISL_1900110, EPI_ISL_1900111, EPI_ISL_1900112, EPI_ISL_1900113, EPI_ISL_1903537, EPI_ISL_1903846, EPI_ISL_1903847, EPI_ISL_1903848, EPI_ISL_1903849, EPI_ISL_1903850, EPI_ISL_1903851, EPI_ISL_1903852, EPI_ISL_1903853, EPI_ISL_1903854, EPI_ISL_1903855, EPI_ISL_1903856, EPI_ISL_1903857, EPI_ISL_1903858, EPI_ISL_1903859, EPI_ISL_1903860, EPI_ISL_1903861, EPI_ISL_1903862, EPI_ISL_1903863, EPI_ISL_1904175, EPI_ISL_1904176, EPI_ISL_1904177, EPI_ISL_1904178, EPI_ISL_1904179, EPI_ISL_1904180, EPI_ISL_1904181, EPI_ISL_1904182, EPI_ISL_1904183, EPI_ISL_1904184, EPI_ISL_1904185, EPI_ISL_1904186, EPI_ISL_1904187, EPI_ISL_1904188, EPI_ISL_1904189, EPI_ISL_1904190, EPI_ISL_1904191, EPI_ISL_1904192, EPI_ISL_1904193, EPI_ISL_1904194, EPI_ISL_1904195, EPI_ISL_1904257, EPI_ISL_1904258, EPI_ISL_1904259, EPI_ISL_1904260, EPI_ISL_1904261, EPI_ISL_1904262, EPI_ISL_1904263, EPI_ISL_1904264, EPI_ISL_1904265, EPI_ISL_1904266, EPI_ISL_1904267, EPI_ISL_1904268, EPI_ISL_1904269, EPI_ISL_1904270, EPI_ISL_1904271, EPI_ISL_1904272, EPI_ISL_1904273, EPI_ISL_1904274, EPI_ISL_1904275, EPI_ISL_1904276, EPI_ISL_1904277, EPI_ISL_1904278, EPI_ISL_1904279, EPI_ISL_1904280, EPI_ISL_1904281, EPI_ISL_1904282, EPI_ISL_1904283, EPI_ISL_1904284, EPI_ISL_1904285, EPI_ISL_1904286, EPI_ISL_1904287, EPI_ISL_1904288, EPI_ISL_1904289, EPI_ISL_1904290, EPI_ISL_1904291, EPI_ISL_1904292, EPI_ISL_1904294 | | | |
| see above | Swedish national genomic surveillance program of SARS-CoV-2 | The Public Health Agency of Sweden | Alma Brolund; Maria Lind Karlberg; Maximilian Riess; Swedish national genomic surveillance program of SARS-CoV-2 |
| EPI_ISL_1300286 | The Public Health Agency of Sweden | The Public Health Agency of Sweden | Anna Risberg; Anna-Malin Linde; Carlo Berg; Karin Tegmark-Wisell; Maria Lind Karlberg; Mattias Haukland; Mia Brytting; Noura Walai; Oskar Karlsson Lindsjo; Petra Edquist; Petra Holmstrom; Reza Advani; Samuel Ohman; Sofia Stamouli |
| EPI_ISL_1231924, EPI_ISL_1231925, EPI_ISL_1231945, EPI_ISL_1231946, EPI_ISL_1231952, EPI_ISL_1231981, EPI_ISL_1232004, EPI_ISL_1232005, EPI_ISL_1232006 | | | |
| see above | Unilabs Klinisk kemi, akutlaboratoriet | The Public Health Agency of Sweden | Anna Risberg; Anna-Malin Linde; Carlo Berg; Karin Tegmark-Wisell; Maria Lind Karlberg; Mattias Haukland; Mia Brytting; Noura Walai; Oskar Karlsson Lindsjo; Petra Edquist; Petra Holmstrom; Reza Advani; Sofia Stamouli |

We gratefully acknowledge the following Authors from the Originating laboratories responsible for obtaining the specimens, as well as the Submitting laboratories where the genome data were generated and shared via GISAID, on which this research is based.

All Submitters of data may be contacted directly via www.gisaid.org

Authors are sorted alphabetically.

| Accession ID | Originating Laboratory | Submitting Laboratory | Authors |
| --- | --- | --- | --- |
| EPI_ISL_730567, EPI_ISL_730569, EPI_ISL_730570, EPI_ISL_730575, EPI_ISL_730577, EPI_ISL_730620, EPI_ISL_984736, EPI_ISL_984737, EPI_ISL_984738, EPI_ISL_984739, EPI_ISL_984740, EPI_ISL_984741, EPI_ISL_984742, EPI_ISL_984743, EPI_ISL_984744, EPI_ISL_984745, EPI_ISL_984746, EPI_ISL_984747, EPI_ISL_984748, EPI_ISL_984749, EPI_ISL_984750, EPI_ISL_984751, EPI_ISL_984752, EPI_ISL_984980, EPI_ISL_984981, EPI_ISL_984982, EPI_ISL_984983, EPI_ISL_984984, EPI_ISL_984985, EPI_ISL_984986, EPI_ISL_984987, EPI_ISL_984988, EPI_ISL_984989, EPI_ISL_984990, EPI_ISL_984991, EPI_ISL_984992, EPI_ISL_984993, EPI_ISL_984994 | | | |
| see above | Gazi University Faculty of Medicine, Medical Virology Laboratory | Gazi University Faculty of Medicine, Medical Virology Laboratory | Erdem Şahin; Gülendam Bozdayı; Hager Muftah; Işıl Fidan; Kayhan Çağlar; Murat Dizbay; Selin Yiğit; Shaknoza Sarzhanova; Özlem Güzel Tunçcan |
| EPI_ISL_833141, EPI_ISL_833142, EPI_ISL_833143, EPI_ISL_833144, EPI_ISL_833145, EPI_ISL_833146, EPI_ISL_833147, EPI_ISL_833148, EPI_ISL_885146, EPI_ISL_955153, EPI_ISL_955154 | | | |
| see above | Genomic Laboratory (GLAB) (Conjoint lab of Health Directorate of Istanbul and Istanbul Technical University) | Genomic Laboratory (GLAB), Istanbul Technical University | Gizem Dinler Doganay; Ilker Karacan; Levent Doganay; Nisan Denizce Can; Pari Sharifli; Payam Zolfagharian; Tugba Kizilboga Akgun |
| EPI_ISL_1663583, EPI_ISL_2004078, EPI_ISL_2004089, EPI_ISL_2004090, EPI_ISL_2004097, EPI_ISL_2004098, EPI_ISL_2004099, EPI_ISL_2015096, EPI_ISL_2015099, EPI_ISL_2015100, EPI_ISL_2015121, EPI_ISL_2015139, EPI_ISL_2015140, EPI_ISL_2015142, EPI_ISL_2015143 | | | |
| see above | IU-Cerrahpasa, Cerrahpasa School of Medicine, COVID-19 Lab | IU-Cerrahpasa, Cerrahpasa School of Medicine, COVID-19 Lab | Kenan Midilli; Mert Kuskucu; Yesim Tuyji Tok |
| EPI_ISL_2003980 | Istanbul University-Cerrahpasa, Cerrahpasa Medical School, COVID Lab | Istanbul University-Cerrahpasa, Cerrahpasa COVID Laboratory | Kenan Midilli; Mert Ahmet Kuskucu; Yesim Tuyji Tok |
| EPI_ISL_1061413 | Istanbul University-Cerrahpasa, Cerrahpasa School of Medicine, COVID-19 Laboratory | Istanbul University-Cerrahpasa, Cerrahpasa School of Medicine, COVID-19 Laboratory | Ebru Yucebag; Fusun Can; Haluk Eraksoy; Kenan Midilli; Mert Ahmet Kuskucu; Okan Kadir Nohut; Serap Şimşek Yavuz; Yesim Tuyji Tok; Zarifa Abullayeva |
| EPI_ISL_811136, EPI_ISL_811137, EPI_ISL_811138, EPI_ISL_811139, EPI_ISL_811140, EPI_ISL_811141, EPI_ISL_811142, EPI_ISL_811143, EPI_ISL_812761, EPI_ISL_812762, EPI_ISL_812763, EPI_ISL_812764, EPI_ISL_812765, EPI_ISL_812766, EPI_ISL_812767, EPI_ISL_812768, EPI_ISL_812769, EPI_ISL_812770, EPI_ISL_812771, EPI_ISL_812772, EPI_ISL_812773, EPI_ISL_812774, EPI_ISL_812775, EPI_ISL_812776, EPI_ISL_812777, EPI_ISL_812778, EPI_ISL_812779, EPI_ISL_812780, EPI_ISL_812781, EPI_ISL_812873, EPI_ISL_812874, EPI_ISL_812875, EPI_ISL_812876, EPI_ISL_812877, EPI_ISL_812878, EPI_ISL_812879, EPI_ISL_812880, EPI_ISL_812881, EPI_ISL_812882, EPI_ISL_812883, EPI_ISL_812884, EPI_ISL_812885, EPI_ISL_812886, EPI_ISL_812887, EPI_ISL_812888, EPI_ISL_812889, EPI_ISL_812890, EPI_ISL_812891, EPI_ISL_812892, EPI_ISL_812893, EPI_ISL_812894, EPI_ISL_812895, EPI_ISL_812896, EPI_ISL_812897, EPI_ISL_812898, EPI_ISL_812899, EPI_ISL_812900, EPI_ISL_812901, EPI_ISL_812902, EPI_ISL_812903, EPI_ISL_812904, EPI_ISL_812905, EPI_ISL_812906, EPI_ISL_812907, EPI_ISL_812908, EPI_ISL_812909, EPI_ISL_812910, EPI_ISL_812911, EPI_ISL_812912, EPI_ISL_812913, EPI_ISL_812914, EPI_ISL_812915, EPI_ISL_812916, EPI_ISL_812917, EPI_ISL_812918, EPI_ISL_812919, EPI_ISL_812920, EPI_ISL_812921, EPI_ISL_814068, EPI_ISL_894246, EPI_ISL_894247, EPI_ISL_894248, EPI_ISL_894249, EPI_ISL_894250, EPI_ISL_894251, EPI_ISL_894252, EPI_ISL_894253, EPI_ISL_894254, EPI_ISL_894255, EPI_ISL_894256, EPI_ISL_894257, EPI_ISL_894258, EPI_ISL_894259, EPI_ISL_894260, EPI_ISL_894261, EPI_ISL_894262, EPI_ISL_894263, EPI_ISL_894264, EPI_ISL_894265, EPI_ISL_894266, EPI_ISL_894267, EPI_ISL_894268, EPI_ISL_894269, EPI_ISL_894270, EPI_ISL_894271, EPI_ISL_894272, EPI_ISL_894273, EPI_ISL_894274, EPI_ISL_894275, EPI_ISL_894276, EPI_ISL_903347, EPI_ISL_906049, EPI_ISL_935048, EPI_ISL_935049, EPI_ISL_935050, EPI_ISL_935051, EPI_ISL_935052, EPI_ISL_935053, EPI_ISL_935054, EPI_ISL_935055, EPI_ISL_935056, EPI_ISL_935057, EPI_ISL_935058, EPI_ISL_935059, EPI_ISL_935060, EPI_ISL_935061, EPI_ISL_935062, EPI_ISL_935063, EPI_ISL_935064, EPI_ISL_935065, EPI_ISL_935066, EPI_ISL_935067, EPI_ISL_935068, EPI_ISL_935069, EPI_ISL_935070, EPI_ISL_935071, EPI_ISL_935072, EPI_ISL_935073, EPI_ISL_935074, EPI_ISL_935075, EPI_ISL_935076, EPI_ISL_935077, EPI_ISL_935078, EPI_ISL_935079, EPI_ISL_935080, EPI_ISL_935081, EPI_ISL_935082, EPI_ISL_935083, EPI_ISL_935084, EPI_ISL_935085, EPI_ISL_935086, EPI_ISL_935087, EPI_ISL_935088, EPI_ISL_935089, EPI_ISL_935090, EPI_ISL_935091, EPI_ISL_935092, EPI_ISL_935093, EPI_ISL_935094, EPI_ISL_935095, EPI_ISL_935096, EPI_ISL_935097, EPI_ISL_935098, EPI_ISL_935099, EPI_ISL_935100, EPI_ISL_935101, EPI_ISL_935102, EPI_ISL_939627, EPI_ISL_940633, EPI_ISL_940634, EPI_ISL_940635, EPI_ISL_940636, EPI_ISL_940637, EPI_ISL_940638, EPI_ISL_940639, EPI_ISL_940640, EPI_ISL_940641, EPI_ISL_940642, EPI_ISL_940643, EPI_ISL_940644, EPI_ISL_940645, EPI_ISL_940646, EPI_ISL_940647, EPI_ISL_940648, EPI_ISL_940649, EPI_ISL_940650, EPI_ISL_940651, EPI_ISL_940652, EPI_ISL_940653, EPI_ISL_940654, EPI_ISL_940655, EPI_ISL_940656, EPI_ISL_940657, EPI_ISL_940658, EPI_ISL_940659, EPI_ISL_940660, EPI_ISL_940661, EPI_ISL_940662, EPI_ISL_940663, EPI_ISL_940664, EPI_ISL_940665, EPI_ISL_940666, EPI_ISL_940667, EPI_ISL_940668, EPI_ISL_940669, EPI_ISL_940670, EPI_ISL_940671, EPI_ISL_940672, EPI_ISL_940673, EPI_ISL_940674, EPI_ISL_940675, EPI_ISL_940676, EPI_ISL_940677, EPI_ISL_940678, EPI_ISL_940679, EPI_ISL_940680, EPI_ISL_940681, EPI_ISL_940682, EPI_ISL_940709, EPI_ISL_940710, EPI_ISL_940711, EPI_ISL_940712, EPI_ISL_940713, EPI_ISL_983409, EPI_ISL_983410, EPI_ISL_983411, EPI_ISL_983412, EPI_ISL_983413, EPI_ISL_983414, EPI_ISL_983415, EPI_ISL_983416, EPI_ISL_983417, EPI_ISL_983418, EPI_ISL_983419, EPI_ISL_983420, EPI_ISL_983421, EPI_ISL_983422, EPI_ISL_983423, EPI_ISL_983424, EPI_ISL_983425, EPI_ISL_983426, EPI_ISL_983427, EPI_ISL_983428, EPI_ISL_983429, EPI_ISL_983430, EPI_ISL_983431, EPI_ISL_983432, EPI_ISL_983433, EPI_ISL_983434, EPI_ISL_983435, EPI_ISL_983436, EPI_ISL_983437, EPI_ISL_984301, EPI_ISL_1008616, EPI_ISL_1008617, EPI_ISL_1008618, EPI_ISL_1008619, EPI_ISL_1008620, EPI_ISL_1008621, EPI_ISL_1008622, EPI_ISL_1008623, EPI_ISL_1008624, EPI_ISL_1008625, EPI_ISL_1008626, EPI_ISL_1008627, EPI_ISL_1008628, EPI_ISL_1008629, EPI_ISL_1008630, EPI_ISL_1008631, EPI_ISL_1008632, EPI_ISL_1008633, EPI_ISL_1008634, EPI_ISL_1008635, EPI_ISL_1008636, EPI_ISL_1008637, EPI_ISL_1008638, EPI_ISL_1008639, EPI_ISL_1008640, EPI_ISL_1008641, EPI_ISL_1008642, EPI_ISL_1008643, EPI_ISL_1008644, EPI_ISL_1008645, EPI_ISL_1008646, EPI_ISL_1008647, EPI_ISL_1008648, EPI_ISL_1008649, EPI_ISL_1008650, EPI_ISL_1008651, EPI_ISL_1008652, EPI_ISL_1008653, EPI_ISL_1008654, EPI_ISL_1008655, EPI_ISL_1008656, EPI_ISL_1020411, EPI_ISL_1020412, EPI_ISL_1020413, EPI_ISL_1020414, EPI_ISL_1020415, EPI_ISL_1020416, EPI_ISL_1020417, EPI_ISL_1020418, EPI_ISL_1020419, EPI_ISL_1020420, EPI_ISL_1020421, EPI_ISL_1020422, EPI_ISL_1020423, EPI_ISL_1020424, EPI_ISL_1020425, EPI_ISL_1020426, EPI_ISL_1020427, EPI_ISL_1020428, EPI_ISL_1020429, EPI_ISL_1020430, EPI_ISL_1020431, EPI_ISL_1020432, EPI_ISL_1020433, EPI_ISL_1020434, EPI_ISL_1020435, EPI_ISL_1020436, EPI_ISL_1020437, EPI_ISL_1020438, EPI_ISL_1020439, EPI_ISL_1020440, EPI_ISL_1020441, EPI_ISL_1020442, EPI_ISL_1020443, EPI_ISL_1020444, EPI_ISL_1020445, EPI_ISL_1020446, EPI_ISL_1020447, EPI_ISL_1020448, EPI_ISL_1020449, EPI_ISL_1020450, EPI_ISL_1020451, EPI_ISL_1020452, EPI_ISL_1020453, EPI_ISL_1020454, EPI_ISL_1020455, EPI_ISL_1020456, EPI_ISL_1020457, EPI_ISL_1020458, EPI_ISL_1020459, EPI_ISL_1020460, EPI_ISL_1020461, EPI_ISL_1020462, EPI_ISL_1020463, EPI_ISL_1020464, EPI_ISL_1020465, EPI_ISL_1020466, EPI_ISL_1020467, EPI_ISL_1020468, EPI_ISL_1020469, EPI_ISL_1020470, EPI_ISL_1020471, EPI_ISL_1020472, EPI_ISL_1020473, EPI_ISL_1020474, EPI_ISL_1020475, EPI_ISL_1020476, EPI_ISL_1020477, EPI_ISL_1020478, EPI_ISL_1020479, EPI_ISL_1020480, EPI_ISL_1020481, EPI_ISL_1020482, EPI_ISL_1020483, EPI_ISL_1020484, EPI_ISL_1020485, EPI_ISL_1034231, EPI_ISL_1034232, EPI_ISL_1034233, EPI_ISL_1034234, EPI_ISL_1034235, EPI_ISL_1034236, EPI_ISL_1034237, EPI_ISL_1034238, EPI_ISL_1034239, EPI_ISL_1034240, EPI_ISL_1034241, EPI_ISL_1034242, EPI_ISL_1034243, EPI_ISL_1034244, EPI_ISL_1034245, EPI_ISL_1034246, EPI_ISL_1034247, EPI_ISL_1034248, EPI_ISL_1034249, EPI_ISL_1034250, EPI_ISL_1034251, EPI_ISL_1034252, EPI_ISL_1034253, EPI_ISL_1034254, EPI_ISL_1034255, EPI_ISL_1034256, EPI_ISL_1034257, EPI_ISL_1034258, EPI_ISL_1034259, EPI_ISL_1063916, EPI_ISL_1063917, EPI_ISL_1063918, EPI_ISL_1063919, EPI_ISL_1063920, EPI_ISL_1063921, EPI_ISL_1063922, EPI_ISL_1063923, EPI_ISL_1063924, EPI_ISL_1063925, EPI_ISL_1063926, EPI_ISL_1063927, EPI_ISL_1063928, EPI_ISL_1063929, EPI_ISL_1063930, EPI_ISL_1063931, EPI_ISL_1063932, EPI_ISL_1063933, EPI_ISL_1063934, EPI_ISL_1063935, EPI_ISL_1063936, EPI_ISL_1063937, EPI_ISL_1063938, EPI_ISL_1063939, EPI_ISL_1063940, EPI_ISL_1063941, EPI_ISL_1063942, EPI_ISL_1063943, EPI_ISL_1063944, EPI_ISL_1063945, EPI_ISL_1063946, EPI_ISL_1063947, EPI_ISL_1063948, EPI_ISL_1063949, EPI_ISL_1063950, EPI_ISL_1063951, EPI_ISL_1063952, EPI_ISL_1063953, EPI_ISL_1063954, EPI_ISL_1063955, EPI_ISL_1063956, EPI_ISL_1063957, EPI_ISL_1063958, EPI_ISL_1063959, EPI_ISL_1063960, EPI_ISL_1063961, EPI_ISL_1063962, EPI_ISL_1063963, EPI_ISL_1063964, EPI_ISL_1063965, EPI_ISL_1063966, EPI_ISL_1063967, EPI_ISL_1063968, EPI_ISL_1063969, EPI_ISL_1063970, EPI_ISL_1063971, EPI_ISL_1063972, EPI_ISL_1063973, EPI_ISL_1063974, EPI_ISL_1063975, EPI_ISL_1063976, EPI_ISL_1063977, EPI_ISL_1063978, EPI_ISL_1063979, EPI_ISL_1063980, EPI_ISL_1063981, EPI_ISL_1063982, EPI_ISL_1063983, EPI_ISL_1063984, EPI_ISL_1063985, EPI_ISL_1063986, EPI_ISL_1063988, EPI_ISL_1063989, EPI_ISL_1063990, EPI_ISL_1063991, EPI_ISL_1063992, EPI_ISL_1073641, EPI_ISL_1073642, EPI_ISL_1073643, EPI_ISL_1073644, EPI_ISL_1073645, EPI_ISL_1073646, EPI_ISL_1073647, EPI_ISL_1073648, EPI_ISL_1073649, EPI_ISL_1073650, EPI_ISL_1073651, EPI_ISL_1073652, EPI_ISL_1073653, EPI_ISL_1073654, EPI_ISL_1073655, EPI_ISL_1073656, EPI_ISL_1073657, EPI_ISL_1073658, EPI_ISL_1073659, EPI_ISL_1073660, EPI_ISL_1073661, EPI_ISL_1073662, EPI_ISL_1073663, EPI_ISL_1073664, EPI_ISL_1073665, EPI_ISL_1073666, EPI_ISL_1073667, EPI_ISL_1073668, EPI_ISL_1073669, EPI_ISL_1073670, EPI_ISL_1073671, EPI_ISL_1073672, EPI_ISL_1073673, EPI_ISL_1073674, EPI_ISL_1073675, EPI_ISL_1073676, EPI_ISL_1073677, EPI_ISL_1073678, EPI_ISL_1073679, EPI_ISL_1073680, EPI_ISL_1073681, EPI_ISL_1073682, EPI_ISL_1073683, EPI_ISL_1073684, EPI_ISL_1073685, EPI_ISL_1073686, EPI_ISL_1073687, EPI_ISL_1073688, EPI_ISL_1073689, EPI_ISL_1073690, EPI_ISL_1073691, EPI_ISL_1073692, EPI_ISL_1073693, EPI_ISL_1073694, EPI_ISL_1073695, EPI_ISL_1073696, EPI_ISL_1073697, EPI_ISL_1073698, EPI_ISL_1073699, EPI_ISL_1073700, EPI_ISL_1073701, EPI_ISL_1073702, EPI_ISL_1073703, EPI_ISL_1073704, EPI_ISL_1073705, EPI_ISL_1073706, EPI_ISL_1073707, EPI_ISL_1073708, EPI_ISL_1073709, EPI_ISL_1073710, EPI_ISL_1073711, EPI_ISL_1073712, EPI_ISL_1073713, EPI_ISL_1073714, EPI_ISL_1073715, EPI_ISL_1073716, EPI_ISL_1073717, EPI_ISL_1073718, EPI_ISL_1073719, EPI_ISL_1073720, EPI_ISL_1073721, EPI_ISL_1073722, EPI_ISL_1073723, EPI_ISL_1073724, EPI_ISL_1073725, EPI_ISL_1073726, EPI_ISL_1073727, EPI_ISL_1073728, EPI_ISL_1073729, EPI_ISL_1073730, EPI_ISL_1073731, EPI_ISL_1073732, EPI_ISL_1073733, EPI_ISL_1073734, EPI_ISL_1073735, EPI_ISL_1073736, EPI_ISL_1073737, EPI_ISL_1073738, EPI_ISL_1073739, EPI_ISL_1073740, EPI_ISL_1073741, EPI_ISL_1073742, EPI_ISL_1073743, EPI_ISL_1073744, EPI_ISL_1073745, EPI_ISL_1073746, EPI_ISL_1073747, EPI_ISL_1073748, EPI_ISL_1073749, EPI_ISL_1073750, EPI_ISL_1073751, EPI_ISL_1073752, EPI_ISL_1073753, EPI_ISL_1073754, EPI_ISL_1073755, EPI_ISL_1073756, EPI_ISL_1073757, EPI_ISL_1073758, EPI_ISL_1073759, EPI_ISL_1073760, EPI_ISL_1073761, EPI_ISL_1073762, EPI_ISL_1073763, EPI_ISL_1073764, EPI_ISL_1073765, EPI_ISL_1073766, EPI_ISL_1073767, EPI_ISL_1073768, EPI_ISL_1073769, EPI_ISL_1073770, EPI_ISL_1073771, EPI_ISL_1073772, EPI_ISL_1073773, EPI_ISL_1073774, EPI_ISL_1073775, EPI_ISL_1073776, EPI_ISL_1073777, EPI_ISL_1073778, EPI_ISL_1073779, EPI_ISL_1073780, EPI_ISL_1073781, EPI_ISL_1073782, EPI_ISL_1073783, EPI_ISL_1073784, EPI_ISL_1073785, EPI_ISL_1073786, EPI_ISL_1073787, EPI_ISL_1073788, EPI_ISL_1073789, EPI_ISL_1073790, EPI_ISL_1073791, EPI_ISL_1073792, EPI_ISL_1073793, EPI_ISL_1073794, EPI_ISL_1073795, EPI_ISL_1073796, EPI_ISL_1073797, EPI_ISL_1073798, EPI_ISL_1073799, EPI_ISL_1073800, EPI_ISL_1073801, EPI_ISL_1073802, EPI_ISL_1073803, EPI_ISL_1073804, EPI_ISL_1073805, EPI_ISL_1073806, EPI_ISL_1073807, EPI_ISL_1073808, EPI_ISL_1073809, EPI_ISL_1073810, EPI_ISL_1073811, EPI_ISL_1073812, EPI_ISL_1082388, EPI_ISL_1082389, EPI_ISL_1082390, EPI_ISL_1082391, EPI_ISL_1082392, EPI_ISL_1082393, EPI_ISL_1082394, EPI_ISL_1082395, EPI_ISL_1082396, EPI_ISL_1082397, EPI_ISL_1082398, EPI_ISL_1082399, EPI_ISL_1082400, EPI_ISL_1082401, EPI_ISL_1082402, EPI_ISL_1082403, EPI_ISL_1082404, EPI_ISL_1082405, EPI_ISL_1082406, EPI_ISL_1082407, EPI_ISL_1082408, EPI_ISL_1082409, EPI_ISL_1082410, EPI_ISL_1082411, EPI_ISL_1082412, EPI_ISL_1082413, EPI_ISL_1082414, EPI_ISL_1082415, EPI_ISL_1082416, EPI_ISL_1082417, EPI_ISL_1082418, EPI_ISL_1082419, EPI_ISL_1082420, EPI_ISL_1082421, EPI_ISL_1082422, EPI_ISL_1082423, EPI_ISL_1082424, EPI_ISL_1082425, EPI_ISL_1082426, EPI_ISL_1082427, EPI_ISL_1082428, EPI_ISL_1082429, EPI_ISL_1082430, EPI_ISL_1082431, EPI_ISL_1082432, EPI_ISL_1082433, EPI_ISL_1082434, EPI_ISL_1082435, EPI_ISL_1082436, EPI_ISL_1082437, EPI_ISL_1082438, EPI_ISL_1082439, EPI_ISL_1082440, EPI_ISL_1082441, EPI_ISL_1082442, EPI_ISL_1082443, EPI_ISL_1082444, EPI_ISL_1082445, EPI_ISL_1082446, EPI_ISL_1082447, EPI_ISL_1082448, EPI_ISL_1095517, EPI_ISL_1095518, EPI_ISL_1095519, EPI_ISL_1095520, EPI_ISL_1095521, EPI_ISL_1095522, EPI_ISL_1095523, EPI_ISL_1095524, EPI_ISL_1095525, EPI_ISL_1095526, EPI_ISL_1095527, EPI_ISL_1095528, EPI_ISL_1095529, EPI_ISL_1095530, EPI_ISL_1095531, EPI_ISL_1095532, EPI_ISL_1095533, EPI_ISL_1095534, EPI_ISL_1095535, EPI_ISL_1095536, EPI_ISL_1095537, EPI_ISL_1095538, EPI_ISL_1095539, EPI_ISL_1095540, EPI_ISL_1095541, EPI_ISL_1095542, EPI_ISL_1095543, EPI_ISL_1095544, EPI_ISL_1095545, EPI_ISL_1095546, EPI_ISL_1095547, EPI_ISL_1095548, EPI_ISL_1095549, EPI_ISL_1095550, EPI_ISL_1095551, EPI_ISL_1095552, EPI_ISL_1095553, EPI_ISL_1095554, EPI_ISL_1095555, EPI_ISL_1095556, EPI_ISL_1095557, EPI_ISL_1095558, EPI_ISL_1095559, EPI_ISL_1095560, EPI_ISL_1095561, EPI_ISL_1095562, EPI_ISL_1095563, EPI_ISL_1095564, EPI_ISL_1095565, EPI_ISL_1095566, EPI_ISL_1095567, EPI_ISL_1095568, EPI_ISL_1095569, EPI_ISL_1095570, EPI_ISL_1095571, EPI_ISL_1095572, EPI_ISL_1095573, EPI_ISL_1095574, EPI_ISL_1095575, EPI_ISL_1095576, EPI_ISL_1095577, EPI_ISL_1095578, EPI_ISL_1095579, EPI_ISL_1095580, EPI_ISL_1095581, EPI_ISL_1095582, EPI_ISL_1095583, EPI_ISL_1095584, EPI_ISL_1095585, EPI_ISL_1095586, EPI_ISL_1095587, EPI_ISL_1095588, EPI_ISL_1095589, EPI_ISL_1095590, EPI_ISL_1095591, EPI_ISL_1095592, EPI_ISL_1095593, EPI_ISL_1095594, EPI_ISL_1095595, EPI_ISL_1095596, EPI_ISL_1095597, EPI_ISL_1095598, EPI_ISL_1095599, EPI_ISL_1095600, EPI_ISL_1095601, EPI_ISL_1095602, EPI_ISL_1095603, EPI_ISL_1095604, EPI_ISL_1095605, EPI_ISL_1095606, EPI_ISL_1095607, EPI_ISL_1095608, EPI_ISL_1095609, EPI_ISL_1096030, EPI_ISL_1096031, EPI_ISL_1096032, EPI_ISL_1096033, EPI_ISL_1096034, EPI_ISL_1096035, EPI_ISL_1096036, EPI_ISL_1096037, EPI_ISL_1096038, EPI_ISL_1096039, EPI_ISL_1096040, EPI_ISL_1096041, EPI_ISL_1096042, EPI_ISL_1096043, EPI_ISL_1096044, EPI_ISL_1096045, EPI_ISL_1096046, EPI_ISL_1096047, EPI_ISL_1096048, EPI_ISL_1096049, EPI_ISL_1096050, EPI_ISL_1096051, EPI_ISL_1096052, EPI_ISL_1096053, EPI_ISL_1096054, EPI_ISL_1096055, EPI_ISL_1096056, EPI_ISL_1096057, EPI_ISL_1096058, EPI_ISL_1096059, EPI_ISL_1096060, EPI_ISL_1096061, EPI_ISL_1096062, EPI_ISL_1096063, EPI_ISL_1096064, EPI_ISL_1096065, EPI_ISL_1096066, EPI_ISL_1096067, EPI_ISL_1096068, EPI_ISL_1096069, EPI_ISL_1096070, EPI_ISL_1096071, EPI_ISL_1096072, EPI_ISL_1096073, EPI_ISL_1096074, EPI_ISL_1096075, EPI_ISL_1096076, EPI_ISL_1096077, EPI_ISL_1096078, EPI_ISL_1096079, EPI_ISL_1096080, EPI_ISL_1096081, EPI_ISL_1096082, EPI_ISL_1096083, EPI_ISL_1096084, EPI_ISL_1096085, EPI_ISL_1096086, EPI_ISL_1096087, EPI_ISL_1096088, EPI_ISL_1096089, EPI_ISL_1096090, EPI_ISL_1096091, EPI_ISL_1096092, EPI_ISL_1096093, EPI_ISL_1096094, EPI_ISL_1096095, EPI_ISL_1096096, EPI_ISL_1096097, EPI_ISL_1096098, EPI_ISL_1096099, EPI_ISL_1096100, EPI_ISL_1096101, EPI_ISL_1096102, EPI_ISL_1096103, EPI_ISL_1096104, EPI_ISL_1096105, EPI_ISL_1096106, EPI_ISL_1096107, EPI_ISL_1096108, EPI_ISL_1096109, EPI_ISL_1096110, EPI_ISL_1096111, EPI_ISL_1096112, EPI_ISL_1096113, EPI_ISL_1096114, EPI_ISL_1096115, EPI_ISL_1096116, EPI_ISL_1096117, EPI_ISL_1096118, EPI_ISL_1096119, EPI_ISL_1097030, EPI_ISL_1097031, EPI_ISL_1097032, EPI_ISL_1097033, EPI_ISL_1097034, EPI_ISL_1097035, EPI_ISL_1097036, EPI_ISL_1097037, EPI_ISL_1097038, EPI_ISL_1097039, EPI_ISL_1097040, EPI_ISL_1097041, EPI_ISL_1097042, EPI_ISL_1097043, EPI_ISL_1097044, EPI_ISL_1097045, EPI_ISL_1097046, EPI_ISL_1097047, EPI_ISL_1097048, EPI_ISL_1097049, EPI_ISL_1097050, EPI_ISL_1097051, EPI_ISL_1097052, EPI_ISL_1097053, EPI_ISL_1097054, EPI_ISL_1097055, EPI_ISL_1097056, EPI_ISL_1097057, EPI_ISL_1097058, EPI_ISL_1097059, EPI_ISL_1097060, EPI_ISL_1097061, EPI_ISL_1097062, EPI_ISL_1097063, EPI_ISL_1097064, EPI_ISL_1097065, EPI_ISL_1097066, EPI_ISL_1097067, EPI_ISL_1097068, EPI_ISL_1097069, EPI_ISL_1097070, EPI_ISL_1097071, EPI_ISL_1097072, EPI_ISL_1097073, EPI_ISL_1097074, EPI_ISL_1097075, EPI_ISL_1097076, EPI_ISL_1097077, EPI_ISL_1097078, EPI_ISL_1097079, EPI_ISL_1097080, EPI_ISL_1097081, EPI_ISL_1097082, EPI_ISL_1097083, EPI_ISL_1097084, EPI_ISL_1097085, EPI_ISL_1097086, EPI_ISL_1097087, EPI_ISL_1097088, EPI_ISL_1097089, EPI_ISL_1097090, EPI_ISL_1097091, EPI_ISL_1097092, EPI_ISL_1097093, EPI_ISL_1097094, EPI_ISL_1097095, EPI_ISL_1097096, EPI_ISL_1097097, EPI_ISL_1097098, EPI_ISL_1097099, EPI_ISL_1097100, EPI_ISL_1097101, EPI_ISL_1097102, EPI_ISL_1097103, EPI_ISL_1097104, EPI_ISL_1097105, EPI_ISL_1097106, EPI_ISL_1097107, EPI_ISL_1097108, EPI_ISL_1097109, EPI_ISL_1097110, EPI_ISL_1097111, EPI_ISL_1097112, EPI_ISL_1097113, EPI_ISL_1097114, EPI_ISL_1097115, EPI_ISL_1097116, EPI_ISL_1097117, EPI_ISL_1097118, EPI_ISL_1097119, EPI_ISL_1097120, EPI_ISL_1097121, EPI_ISL_1097122, EPI_ISL_1097123, EPI_ISL_1097124, EPI_ISL_1097125, EPI_ISL_1097126, EPI_ISL_1097127, EPI_ISL_1097128, EPI_ISL_1097129, EPI_ISL_1097130, EPI_ISL_1097131, EPI_ISL_1097132, EPI_ISL_1097133, EPI_ISL_1097134, EPI_ISL_1097135, EPI_ISL_1097136, EPI_ISL_1097137, EPI_ISL_1097138, EPI_ISL_1097139, EPI_ISL_1097140, EPI_ISL_1097141, EPI_ISL_1097142, EPI_ISL_1097143, EPI_ISL_1097144, EPI_ISL_1097145, EPI_ISL_1097146, EPI_ISL_1097147, EPI_ISL_1097148, EPI_ISL_1097149, EPI_ISL_1097150, EPI_ISL_1097151, EPI_ISL_1097152, EPI_ISL_1097153, EPI_ISL_1097154, EPI_ISL_1097155, EPI_ISL_1097156, EPI_ISL_1097157, EPI_ISL_1097158, EPI_ISL_1097159, EPI_ISL_1097160, EPI_ISL_1097161, EPI_ISL_1097162, EPI_ISL_1097163, EPI_ISL_1097164, EPI_ISL_1097165, EPI_ISL_1097166, EPI_ISL_1097167, EPI_ISL_1097168, EPI_ISL_1097169, EPI_ISL_1097170, EPI_ISL_1097171, EPI_ISL_1097172, EPI_ISL_1097173, EPI_ISL_1097174, EPI_ISL_1097175, EPI_ISL_1097176, EPI_ISL_1097177, EPI_ISL_1097178, EPI_ISL_1097179, EPI_ISL_1097180, EPI_ISL_1097181, EPI_ISL_1097182, EPI_ISL_1097183, EPI_ISL_1097184, EPI_ISL_1097185, EPI_ISL_1097186, EPI_ISL_1097187, EPI_ISL_1097188, EPI_ISL_1097189, EPI_ISL_1097190, EPI_ISL_1097191, EPI_ISL_1097192, EPI_ISL_1097193, EPI_ISL_1097194, EPI_ISL_1097195, EPI_ISL_1097196, EPI_ISL_1097197, EPI_ISL_1097198, EPI_ISL_1097199, EPI_ISL_1097200, EPI_ISL_1097201, EPI_ISL_1097202, EPI_ISL_1097203, EPI_ISL_1097204, EPI_ISL_1097205, EPI_ISL_1097206, EPI_ISL_1097207, EPI_ISL_1097208, EPI_ISL_1097209, EPI_ISL_1097210, EPI_ISL_1097211, EPI_ISL_1097212, EPI_ISL_1097213, EPI_ISL_1097214, EPI_ISL_1097215, EPI_ISL_1097216, EPI_ISL_1097217, EPI_ISL_1097218, EPI_ISL_1097219, EPI_ISL_1097220, EPI_ISL_1097221, EPI_ISL_1097222, EPI_ISL_1097223, EPI_ISL_1097224, EPI_ISL_1097225, EPI_ISL_1097226, EPI_ISL_1097227, EPI_ISL_1097228, EPI_ISL_1097229, EPI_ISL_1097230, EPI_ISL_1097231, EPI_ISL_1097232, EPI_ISL_1097233, EPI_ISL_1097234, EPI_ISL_1097235, EPI_ISL_1097236, EPI_ISL_1097237, EPI_ISL_1097238, EPI_ISL_1097239, EPI_ISL_1097240, EPI_ISL_1097241, EPI_ISL_1097242, EPI_ISL_1097243, EPI_ISL_1097244, EPI_ISL_1097245, EPI_ISL_1097246, EPI_ISL_1097247, EPI_ISL_1097248, EPI_ISL_1097249, EPI_ISL_1097250, EPI_ISL_1097251, EPI_ISL_1097252, EPI_ISL_1097253, EPI_ISL_1097254, EPI_ISL_1097255, EPI_ISL_1097256, EPI_ISL_1097257, EPI_ISL_1097258, EPI_ISL_1097259, EPI_ISL_1097260, EPI_ISL_1097261, EPI_ISL_1097262, EPI_ISL_1097263, EPI_ISL_1097264, EPI_ISL_1097265, EPI_ISL_1097266, EPI_ISL_1097267, EPI_ISL_1097268, EPI_ISL_1097269, EPI_ISL_1097270, EPI_ISL_1097271, EPI_ISL_1097272, EPI_ISL_1097273, EPI_ISL_1097274, EPI_ISL_1097275, EPI_ISL_1097276, EPI_ISL_1097277, EPI_ISL_1097278, EPI_ISL_1097279, EPI_ISL_1097280, EPI_ISL_1097281, EPI_ISL_1097282, EPI_ISL_1097283, EPI_ISL_1097284, EPI_ISL_1097285, EPI_ISL_1097286, EPI_ISL_1097287, EPI_ISL_1097288, EPI_ISL_1097289, EPI_ISL_1097290, EPI_ISL_1097291, EPI_ISL_1097292, EPI_ISL_1097293, EPI_ISL_1097294, EPI_ISL_1097295, EPI_ISL_1097296, EPI_ISL_1097297, EPI_ISL_1097298, EPI_ISL_1097299, EPI_ISL_1097300, EPI_ISL_1097301, EPI_ISL_1097302, EPI_ISL_1097303, EPI_ISL_1097304, EPI_ISL_1097305, EPI_ISL_1097306, EPI_ISL_1097307, EPI_ISL_1117597, EPI_ISL_1117598, EPI_ISL_1117599, EPI_ISL_1117600, EPI_ISL_1117601, EPI_ISL_1117602, EPI_ISL_1117603, EPI_ISL_1117604, EPI_ISL_1117605, EPI_ISL_1117606, EPI_ISL_1117607, EPI_ISL_1117608, EPI_ISL_1117609, EPI_ISL_1117610, EPI_ISL_1117611, EPI_ISL_1117612, EPI_ISL_1117613, EPI_ISL_1117614, EPI_ISL_1117615, EPI_ISL_1117616, EPI_ISL_1117617, EPI_ISL_1117618, EPI_ISL_1117619, EPI_ISL_1117620, EPI_ISL_1117621, EPI_ISL_1117622, EPI_ISL_1117623, EPI_ISL_1117624, EPI_ISL_1117625, EPI_ISL_1117626, EPI_ISL_1117627, EPI_ISL_1117628, EPI_ISL_1117629, EPI_ISL_1117630, EPI_ISL_1117631, EPI_ISL_1117632, EPI_ISL_1117633, EPI_ISL_1117634, EPI_ISL_1117635, EPI_ISL_1117636, EPI_ISL_1117637, EPI_ISL_1117638, EPI_ISL_1117639, EPI_ISL_1117640, EPI_ISL_1117641, EPI_ISL_1117642, EPI_ISL_1117643, EPI_ISL_1117644, EPI_ISL_1117645, EPI_ISL_1117646, EPI_ISL_1117647, EPI_ISL_1117648, EPI_ISL_1117649, EPI_ISL_1117650, EPI_ISL_1117651, EPI_ISL_1117652, EPI_ISL_1117653, EPI_ISL_1117654, EPI_ISL_1117655, EPI_ISL_1117656, EPI_ISL_1117657, EPI_ISL_1117658, EPI_ISL_1117659, EPI_ISL_1117660, EPI_ISL_1164466, EPI_ISL_1164467, EPI_ISL_1164468, EPI_ISL_1164469, EPI_ISL_1164470, EPI_ISL_1164471, EPI_ISL_1164472, EPI_ISL_1164473, EPI_ISL_1164474, EPI_ISL_1164475, EPI_ISL_1164476, EPI_ISL_1164477, EPI_ISL_1164478, EPI_ISL_1164479, EPI_ISL_1164480, EPI_ISL_1164481, EPI_ISL_1164482, EPI_ISL_1164483, EPI_ISL_1167568, EPI_ISL_1167569, EPI_ISL_1167570, EPI_ISL_1167571, EPI_ISL_1167572, EPI_ISL_1167573, EPI_ISL_1167574, EPI_ISL_1167575, EPI_ISL_1167576, EPI_ISL_1167577, EPI_ISL_1167578, EPI_ISL_1167579, EPI_ISL_1167580, EPI_ISL_1167581, EPI_ISL_1167582, EPI_ISL_1167583, EPI_ISL_1167584, EPI_ISL_1167585, EPI_ISL_1167586, EPI_ISL_1167587, EPI_ISL_1167588, EPI_ISL_1167589, EPI_ISL_1167590, EPI_ISL_1167591, EPI_ISL_1167592, EPI_ISL_1167593, EPI_ISL_1167594, EPI_ISL_1167595, EPI_ISL_1167596, EPI_ISL_1167597, EPI_ISL_1167598, EPI_ISL_1167599, EPI_ISL_1167600, EPI_ISL_1167601, EPI_ISL_1167602, EPI_ISL_1167603, EPI_ISL_1167604, EPI_ISL_1167605, EPI_ISL_1167606, EPI_ISL_1167607, EPI_ISL_1167608, EPI_ISL_1167609, EPI_ISL_1167610, EPI_ISL_1167611, EPI_ISL_1167612, EPI_ISL_1167613, EPI_ISL_1167614, EPI_ISL_1167615, EPI_ISL_1167616, EPI_ISL_1167617, EPI_ISL_1167618, EPI_ISL_1167619, EPI_ISL_1167620, EPI_ISL_1167621, EPI_ISL_1167622, EPI_ISL_1167623, EPI_ISL_1167624, EPI_ISL_1167625, EPI_ISL_1167626, EPI_ISL_1167627, EPI_ISL_1167628, EPI_ISL_1167629, EPI_ISL_1167630, EPI_ISL_1167631, EPI_ISL_1167632, EPI_ISL_1167633, EPI_ISL_1167634, EPI_ISL_1167635, EPI_ISL_1167636, EPI_ISL_1167637, EPI_ISL_1167638, EPI_ISL_1167639, EPI_ISL_1167640, EPI_ISL_1167641, EPI_ISL_1167642, EPI_ISL_1167643, EPI_ISL_1167644, EPI_ISL_1167645, EPI_ISL_1167646, EPI_ISL_1167647, EPI_ISL_1167648, EPI_ISL_1167649, EPI_ISL_1167650, EPI_ISL_1167651, EPI_ISL_1167652, EPI_ISL_1167653, EPI_ISL_1167654, EPI_ISL_1167655, EPI_ISL_1167656, EPI_ISL_1167657, EPI_ISL_1167658, EPI_ISL_1167659, EPI_ISL_1167660, EPI_ISL_1167661, EPI_ISL_1167662, EPI_ISL_1167663, EPI_ISL_1169975, EPI_ISL_1169976, EPI_ISL_1169977, EPI_ISL_1169978, EPI_ISL_1169979, EPI_ISL_1169980, EPI_ISL_1169981, EPI_ISL_1169982, EPI_ISL_1169983, EPI_ISL_1169984, EPI_ISL_1169985, EPI_ISL_1169986, EPI_ISL_1169987, EPI_ISL_1169988, EPI_ISL_1169989, EPI_ISL_1169990, EPI_ISL_1169991, EPI_ISL_1169992, EPI_ISL_1169993, EPI_ISL_1169994, EPI_ISL_1169995, EPI_ISL_1169996, EPI_ISL_1169997, EPI_ISL_1169998, EPI_ISL_1169999, EPI_ISL_1170000, EPI_ISL_1170001, EPI_ISL_1170002, EPI_ISL_1170003, EPI_ISL_1170004, EPI_ISL_1170005, EPI_ISL_1170006, EPI_ISL_1170007, EPI_ISL_1170008, EPI_ISL_1170009, EPI_ISL_1170010, EPI_ISL_1170011, EPI_ISL_1170012, EPI_ISL_1170013, EPI_ISL_1170014, EPI_ISL_1170015, EPI_ISL_1170016, EPI_ISL_1170017, EPI_ISL_1170018, EPI_ISL_1170019, EPI_ISL_1170020, EPI_ISL_1170021, EPI_ISL_1170022, EPI_ISL_1170023, EPI_ISL_1170024, EPI_ISL_1170025, EPI_ISL_1170026, EPI_ISL_1170027, EPI_ISL_1170028, EPI_ISL_1170029, EPI_ISL_1170030, EPI_ISL_1170031, EPI_ISL_1170032, EPI_ISL_1170033, EPI_ISL_1170034, EPI_ISL_1170035, EPI_ISL_1170036, EPI_ISL_1170037, EPI_ISL_1379944, EPI_ISL_1379945, EPI_ISL_1379946, EPI_ISL_1379947, EPI_ISL_1379948, EPI_ISL_1379949, EPI_ISL_1379950, EPI_ISL_1379951, EPI_ISL_1379952, EPI_ISL_1379953, EPI_ISL_1379954, EPI_ISL_1379955, EPI_ISL_1386007, EPI_ISL_1386009, EPI_ISL_1386011, EPI_ISL_1386013, EPI_ISL_1386014, EPI_ISL_1386016, EPI_ISL_1386018, EPI_ISL_1386020, EPI_ISL_1386021, EPI_ISL_1386023, EPI_ISL_1386025, EPI_ISL_1386027, EPI_ISL_1386028, EPI_ISL_1386030, EPI_ISL_1386032, EPI_ISL_1386033, EPI_ISL_1760125 | | | |
| see above | Ministry of Health Turkey | Ministry of Health Turkey | Ayse Basak Altas; Ayşe Başak Altaş; Fatma Bayrakdar; Gulay Korukluoglu; Gülay Korukluoğlu; Suleyman Yalcin; Süleyman Yalcin; Yasemin Cosgun; Yasemin Coşgun |
| EPI_ISL_1224941, EPI_ISL_1225156, EPI_ISL_1225157, EPI_ISL_1225158, EPI_ISL_1225159, EPI_ISL_1225160, EPI_ISL_1225161 | | | |
| see above | Turkish Society of Internal Medicine | Gen Era Diagnostics Corporation Life Science Research and Molecular Diagnostics | Serhat Ünal |

We gratefully acknowledge the following Authors from the Originating laboratories responsible for obtaining the specimens, as well as the Submitting laboratories where the genome data were generated and shared via GISAID, on which this research is based.

All Submitters of data may be contacted directly via www.gisaid.org

Authors are sorted alphabetically.

| Accession ID | Originating Laboratory | Submitting Laboratory | Authors |
|---|---|---|---|
| EPI_ISL_859561, EPI_ISL_859562, EPI_ISL_859563, EPI_ISL_859564, EPI_ISL_859565, EPI_ISL_859566, EPI_ISL_859567, EPI_ISL_859568, EPI_ISL_859569, EPI_ISL_859570, EPI_ISL_859571, EPI_ISL_859572, EPI_ISL_859573, EPI_ISL_859574, EPI_ISL_859575, EPI_ISL_859576, EPI_ISL_859577, EPI_ISL_859601, EPI_ISL_859602, EPI_ISL_859603, EPI_ISL_859604, EPI_ISL_859605, EPI_ISL_859607, EPI_ISL_859609, EPI_ISL_859610, EPI_ISL_859619, EPI_ISL_859622, EPI_ISL_859637, EPI_ISL_859656, EPI_ISL_859657, EPI_ISL_859658, EPI_ISL_859659, EPI_ISL_859660, EPI_ISL_859661, EPI_ISL_859662, EPI_ISL_859663, EPI_ISL_859664, EPI_ISL_859665, EPI_ISL_859666, EPI_ISL_859667, EPI_ISL_859668, EPI_ISL_859669, EPI_ISL_859670, EPI_ISL_859671, EPI_ISL_859672, EPI_ISL_859673, EPI_ISL_859674, EPI_ISL_859675, EPI_ISL_859676, EPI_ISL_859677, EPI_ISL_859678, EPI_ISL_859679, EPI_ISL_859680, EPI_ISL_859681, EPI_ISL_859682, EPI_ISL_859683, EPI_ISL_859684, EPI_ISL_859685, EPI_ISL_859686, EPI_ISL_859687, EPI_ISL_859688, EPI_ISL_859689, EPI_ISL_859690, EPI_ISL_859691, EPI_ISL_859692, EPI_ISL_859693, EPI_ISL_859694, EPI_ISL_859695, EPI_ISL_859696, EPI_ISL_859697, EPI_ISL_859698, EPI_ISL_859699, EPI_ISL_859700, EPI_ISL_859701, EPI_ISL_859702, EPI_ISL_859703, EPI_ISL_859704, EPI_ISL_859705, EPI_ISL_859706, EPI_ISL_859707, EPI_ISL_859708, EPI_ISL_859709, EPI_ISL_859710, EPI_ISL_859711, EPI_ISL_859712, EPI_ISL_859713, EPI_ISL_859714, EPI_ISL_859715, EPI_ISL_859716, EPI_ISL_859717, EPI_ISL_859718, EPI_ISL_859719, EPI_ISL_859720, EPI_ISL_859721, EPI_ISL_859722, EPI_ISL_859723, EPI_ISL_859724, EPI_ISL_859725, EPI_ISL_859726, EPI_ISL_859727, EPI_ISL_859728, EPI_ISL_859729, EPI_ISL_859730, EPI_ISL_859731, EPI_ISL_859732, EPI_ISL_859733, EPI_ISL_859734, EPI_ISL_859735, EPI_ISL_859736, EPI_ISL_859737, EPI_ISL_859738, EPI_ISL_859739, EPI_ISL_859740, EPI_ISL_859741, EPI_ISL_859742, EPI_ISL_859743, EPI_ISL_859744, EPI_ISL_859745, EPI_ISL_859746, EPI_ISL_859747, EPI_ISL_859748, EPI_ISL_859749, EPI_ISL_859750, EPI_ISL_859751, EPI_ISL_859752, EPI_ISL_859753, EPI_ISL_859754, EPI_ISL_859755, EPI_ISL_859756, EPI_ISL_859757, EPI_ISL_859758, EPI_ISL_859759, EPI_ISL_859760, EPI_ISL_859761, EPI_ISL_859762, EPI_ISL_859763, EPI_ISL_859764, EPI_ISL_859765, EPI_ISL_859766, EPI_ISL_859767, EPI_ISL_859768, EPI_ISL_859769, EPI_ISL_859770, EPI_ISL_859771, EPI_ISL_859772, EPI_ISL_859773, EPI_ISL_859774, EPI_ISL_859775, EPI_ISL_859776, EPI_ISL_859777, EPI_ISL_859778, EPI_ISL_859779, EPI_ISL_859780, EPI_ISL_859781, EPI_ISL_859782, EPI_ISL_859783, EPI_ISL_859784, EPI_ISL_859785, EPI_ISL_859786, EPI_ISL_859787, EPI_ISL_859788, EPI_ISL_859789, EPI_ISL_859790, EPI_ISL_859791, EPI_ISL_859792, EPI_ISL_859793, EPI_ISL_859794, EPI_ISL_859795, EPI_ISL_859796, EPI_ISL_859797, EPI_ISL_859798, EPI_ISL_859799, EPI_ISL_859800, EPI_ISL_859801, EPI_ISL_859802, EPI_ISL_859803, EPI_ISL_859804, EPI_ISL_859805, EPI_ISL_859806, EPI_ISL_859807, EPI_ISL_859808, EPI_ISL_859809, EPI_ISL_859810, EPI_ISL_859811, EPI_ISL_859812, EPI_ISL_859813, EPI_ISL_859814, EPI_ISL_859815, EPI_ISL_859816, EPI_ISL_859817, EPI_ISL_859818, EPI_ISL_859819, EPI_ISL_859820, EPI_ISL_859821, EPI_ISL_859822, EPI_ISL_859823, EPI_ISL_859824, EPI_ISL_859825, EPI_ISL_859826, EPI_ISL_859827, EPI_ISL_859828, EPI_ISL_859829, EPI_ISL_859830, EPI_ISL_859831, EPI_ISL_859832, EPI_ISL_859833, EPI_ISL_859834, EPI_ISL_859835, EPI_ISL_859836, EPI_ISL_859837, EPI_ISL_859838, EPI_ISL_859839, EPI_ISL_859840, EPI_ISL_859841, EPI_ISL_859842, EPI_ISL_859843, EPI_ISL_859844, EPI_ISL_859845, EPI_ISL_859846, EPI_ISL_859847, EPI_ISL_859848, EPI_ISL_859849, EPI_ISL_859850, EPI_ISL_859851, EPI_ISL_859852, EPI_ISL_859853, EPI_ISL_859854, EPI_ISL_859855, EPI_ISL_859856, EPI_ISL_859857, EPI_ISL_859858, EPI_ISL_859859, EPI_ISL_859860, EPI_ISL_859861, EPI_ISL_859862, EPI_ISL_859863, EPI_ISL_859864, EPI_ISL_859865, EPI_ISL_859866, EPI_ISL_859867, EPI_ISL_859868, EPI_ISL_859869, EPI_ISL_859870, EPI_ISL_859871, EPI_ISL_859872, EPI_ISL_859873, EPI_ISL_859874, EPI_ISL_859875, EPI_ISL_859876, EPI_ISL_859877, EPI_ISL_859878, EPI_ISL_859879, EPI_ISL_859880, EPI_ISL_859881, EPI_ISL_859882, EPI_ISL_859883, EPI_ISL_859884, EPI_ISL_859885, EPI_ISL_859886, EPI_ISL_859887, EPI_ISL_859888, EPI_ISL_859889, EPI_ISL_859890, EPI_ISL_859891, EPI_ISL_859892, EPI_ISL_859893, EPI_ISL_859894, EPI_ISL_859895, EPI_ISL_859896, EPI_ISL_859897, EPI_ISL_859898, EPI_ISL_859899, EPI_ISL_859900, EPI_ISL_859901, EPI_ISL_859902, EPI_ISL_859903, EPI_ISL_859904, EPI_ISL_859905, EPI_ISL_859906, EPI_ISL_859907, EPI_ISL_859908, EPI_ISL_859909, EPI_ISL_859910, EPI_ISL_859911, EPI_ISL_859912, EPI_ISL_859913, EPI_ISL_859914, EPI_ISL_859915, EPI_ISL_859916, EPI_ISL_859917, EPI_ISL_859918, EPI_ISL_859919, EPI_ISL_859920, EPI_ISL_859921, EPI_ISL_859922, EPI_ISL_859923, EPI_ISL_859924, EPI_ISL_859925, EPI_ISL_859926, EPI_ISL_859927, EPI_ISL_859928, EPI_ISL_859929, EPI_ISL_859930, EPI_ISL_859931, EPI_ISL_859932, EPI_ISL_859933, EPI_ISL_859934, EPI_ISL_859935, EPI_ISL_860090, EPI_ISL_860091 | | | |
| see above | BTC, Khalifa University | BTC, Khalifa University | Al Safar et al |
| EPI_ISL_582642, EPI_ISL_582643, EPI_ISL_582644, EPI_ISL_582645, EPI_ISL_582646, EPI_ISL_582647, EPI_ISL_582648, EPI_ISL_582649, EPI_ISL_582650, EPI_ISL_582651, EPI_ISL_582652, EPI_ISL_582653, EPI_ISL_582654, EPI_ISL_582655, EPI_ISL_582656, EPI_ISL_582657, EPI_ISL_582658, EPI_ISL_582659, EPI_ISL_582660, EPI_ISL_582661, EPI_ISL_582662, EPI_ISL_582663, EPI_ISL_582664, EPI_ISL_582665, EPI_ISL_582666, EPI_ISL_582667, EPI_ISL_582668, EPI_ISL_582669, EPI_ISL_582670, EPI_ISL_582671, EPI_ISL_582672, EPI_ISL_582673, EPI_ISL_582674, EPI_ISL_582675, EPI_ISL_582676, EPI_ISL_582677, EPI_ISL_582678, EPI_ISL_582679, EPI_ISL_582680, EPI_ISL_582681, EPI_ISL_582682, EPI_ISL_582683, EPI_ISL_582684, EPI_ISL_582685, EPI_ISL_582686, EPI_ISL_582687, EPI_ISL_582688, EPI_ISL_582689, EPI_ISL_582690, EPI_ISL_582691 | | | |
| see above | Sheikh Khalifa Medical City | Molecular/Surveillance lab Sheikh Khalifa Medical City | Amirtharaj Francis; Hala Imambaccus; Hiba Saud; Sahar Almarzooqi; Sajeed Abdul; Stefan Weber |

We gratefully acknowledge the following Authors from the Originating laboratories responsible for obtaining the specimens, as well as the Submitting laboratories where the genome data were generated and shared via GISAID, on which this research is based.

All Submitters of data may be contacted directly via www.gisaid.org

Authors are sorted alphabetically.

| Accession ID | Originating Laboratory | Submitting Laboratory | Authors |
|---|---|---|---|
| EPI_ISL_842849, EPI_ISL_842850, EPI_ISL_842851, EPI_ISL_842853, EPI_ISL_842854, EPI_ISL_842855, EPI_ISL_842856, EPI_ISL_842857 | | | |
| see above | Barts Health NHS Trust | COVID-19 Genomics UK (COG-UK) Consortium | BROAD; Beatrix; CUTINO-MOGUEL; Claire; David; Dola; HARRINGTON; KELE; Maria-Teresa; OWOYEMI; Raghavendran; SHYLINI |
| EPI_ISL_868821, EPI_ISL_1054835, EPI_ISL_1054836, EPI_ISL_1054837, EPI_ISL_1054838, EPI_ISL_1054839, EPI_ISL_1054840, EPI_ISL_1054841, EPI_ISL_1054842, EPI_ISL_1054843, EPI_ISL_1054844, EPI_ISL_1054845, EPI_ISL_1054846, EPI_ISL_1054847, EPI_ISL_1054848, EPI_ISL_1054850, EPI_ISL_1054851, EPI_ISL_1054855, EPI_ISL_1054856, EPI_ISL_1054857, EPI_ISL_1054858, EPI_ISL_1054859, EPI_ISL_1833917, EPI_ISL_1833918, EPI_ISL_1833919, EPI_ISL_1833920, EPI_ISL_1833922, EPI_ISL_1833925, EPI_ISL_1833926, EPI_ISL_1833930, EPI_ISL_1833932, EPI_ISL_1833934, EPI_ISL_1833935, EPI_ISL_1833937, EPI_ISL_1833938, EPI_ISL_1833939, EPI_ISL_1833940, EPI_ISL_1833941, EPI_ISL_1833943, EPI_ISL_1833944, EPI_ISL_1833948, EPI_ISL_2128544, EPI_ISL_2128566, EPI_ISL_2128572, EPI_ISL_2128579, EPI_ISL_2128586, EPI_ISL_2128598 | | | |
| see above | Bioinformatics and Biostatistics Lab, Advanced Sequencing Facility | COVID-19 Genomics UK (COG-UK) Consortium | Aengus Stewart; Chelsea Sawyer; Harshil Patel; Jerome Nicod; Laura Cubitt; Margaret Crawford |
| EPI_ISL_595100, EPI_ISL_595101, EPI_ISL_595102, EPI_ISL_595103, EPI_ISL_595104, EPI_ISL_595105, EPI_ISL_595106, EPI_ISL_595107, EPI_ISL_595108, EPI_ISL_595109, EPI_ISL_595110, EPI_ISL_595111, EPI_ISL_595112, EPI_ISL_595113, EPI_ISL_595114, EPI_ISL_611589, EPI_ISL_611689, EPI_ISL_611841, EPI_ISL_611927, EPI_ISL_611957, EPI_ISL_611958, EPI_ISL_611959, EPI_ISL_611962, EPI_ISL_611971, EPI_ISL_613288, EPI_ISL_613289, EPI_ISL_613290, EPI_ISL_613291, EPI_ISL_613292, EPI_ISL_613293, EPI_ISL_613302, EPI_ISL_613303, EPI_ISL_613304, EPI_ISL_613305, EPI_ISL_613306, EPI_ISL_637305, EPI_ISL_637369, EPI_ISL_637370, EPI_ISL_637371, EPI_ISL_637372, EPI_ISL_637453, EPI_ISL_637496, EPI_ISL_637497, EPI_ISL_637610, EPI_ISL_637659, EPI_ISL_637782, EPI_ISL_637783, EPI_ISL_637932, EPI_ISL_638002, EPI_ISL_638455, EPI_ISL_638456, EPI_ISL_638457, EPI_ISL_638458, EPI_ISL_639007, EPI_ISL_650168, EPI_ISL_650221, EPI_ISL_650401, EPI_ISL_650566, EPI_ISL_650567, EPI_ISL_650568, EPI_ISL_650569, EPI_ISL_650570, EPI_ISL_650571, EPI_ISL_650584, EPI_ISL_650585, EPI_ISL_650711, EPI_ISL_650947, EPI_ISL_650949, EPI_ISL_650964, EPI_ISL_650965, EPI_ISL_650966, EPI_ISL_651067, EPI_ISL_651099, EPI_ISL_652103, EPI_ISL_652104, EPI_ISL_652105, EPI_ISL_652106, EPI_ISL_652107, EPI_ISL_652108, EPI_ISL_652109, EPI_ISL_652110, EPI_ISL_665391, EPI_ISL_665652, EPI_ISL_665849, EPI_ISL_727705, EPI_ISL_727707, EPI_ISL_727712, EPI_ISL_741348, EPI_ISL_742260, EPI_ISL_867936, EPI_ISL_867937, EPI_ISL_867938, EPI_ISL_867939, EPI_ISL_867940, EPI_ISL_867954, EPI_ISL_867958, EPI_ISL_867959, EPI_ISL_867960, EPI_ISL_867961, EPI_ISL_867971, EPI_ISL_867972, EPI_ISL_867973, EPI_ISL_867974, EPI_ISL_867976, EPI_ISL_867977, EPI_ISL_867978, EPI_ISL_867979, EPI_ISL_867980, EPI_ISL_867981, EPI_ISL_1105656 | | | |
| see above | Centre for Enzyme Innovation, University of Portsmouth / Translational Research Laboratory, Portsmouth Hospitals NHS Trust | COVID-19 Genomics UK (COG-UK) Consortium | Allyson Lloyd; Angela Beckett; Anoop Chauhan; Christopher Fearn; Ethan Butcher; Garry Scarlett; Kate Cook; Katie Loveson; Kelly Bicknell; Robert Impey; Salman Goudarzi; Samuel Robson; Sarah Wyllie; Scott Elliott; Sharon Glaysher; Yann Bourgeois |
| EPI_ISL_584286, EPI_ISL_584287, EPI_ISL_584288, EPI_ISL_584289, EPI_ISL_584290, EPI_ISL_584291, EPI_ISL_584294, EPI_ISL_584295, EPI_ISL_584296, EPI_ISL_584297, EPI_ISL_584298, EPI_ISL_584299, EPI_ISL_584300, EPI_ISL_584301, EPI_ISL_584302, EPI_ISL_584304, EPI_ISL_584305, EPI_ISL_584306, EPI_ISL_584307, EPI_ISL_584309, EPI_ISL_584310, EPI_ISL_584311, EPI_ISL_584312, EPI_ISL_584313, EPI_ISL_584314, EPI_ISL_584316, EPI_ISL_584317, EPI_ISL_584318, EPI_ISL_584321, EPI_ISL_584322, EPI_ISL_584323, EPI_ISL_612007, EPI_ISL_612034, EPI_ISL_612262, EPI_ISL_612263, EPI_ISL_612264, EPI_ISL_612265, EPI_ISL_612266, EPI_ISL_612267, EPI_ISL_612268, EPI_ISL_612269, EPI_ISL_612270, EPI_ISL_612271, EPI_ISL_612272, EPI_ISL_612273, EPI_ISL_637287, EPI_ISL_637318, EPI_ISL_637331, EPI_ISL_637362, EPI_ISL_637393, EPI_ISL_637398, EPI_ISL_637500, EPI_ISL_637519, EPI_ISL_637583, EPI_ISL_637668, EPI_ISL_637672, EPI_ISL_637673, EPI_ISL_637681, EPI_ISL_637682, EPI_ISL_637683, EPI_ISL_637684, EPI_ISL_637810, EPI_ISL_637811, EPI_ISL_637812, EPI_ISL_637813, EPI_ISL_637814, EPI_ISL_637815, EPI_ISL_637816, EPI_ISL_637817, EPI_ISL_637818, EPI_ISL_637819, EPI_ISL_637820, EPI_ISL_637821, EPI_ISL_637822, EPI_ISL_637823, EPI_ISL_637824, EPI_ISL_637825, EPI_ISL_637826, EPI_ISL_637827, EPI_ISL_637828, EPI_ISL_637829, EPI_ISL_637830, EPI_ISL_637831, EPI_ISL_637832, EPI_ISL_637833, EPI_ISL_637834, EPI_ISL_637835, EPI_ISL_637836, EPI_ISL_637837, EPI_ISL_637838, EPI_ISL_637839, EPI_ISL_637840, EPI_ISL_637841, EPI_ISL_637842, EPI_ISL_637843, EPI_ISL_637844, EPI_ISL_637845, EPI_ISL_637846, EPI_ISL_637847, EPI_ISL_637848, EPI_ISL_637849, EPI_ISL_637850, EPI_ISL_637851, EPI_ISL_637852, EPI_ISL_637906, EPI_ISL_637916, EPI_ISL_638146, EPI_ISL_638169, EPI_ISL_638731, EPI_ISL_638732, EPI_ISL_638733, EPI_ISL_638734, EPI_ISL_638735, EPI_ISL_638936, EPI_ISL_638937, EPI_ISL_638938, EPI_ISL_638939, EPI_ISL_638940, EPI_ISL_638941, EPI_ISL_638942, EPI_ISL_638943, EPI_ISL_842110, EPI_ISL_952943, EPI_ISL_952944, EPI_ISL_952945, EPI_ISL_952946, EPI_ISL_952947, EPI_ISL_952948, EPI_ISL_952949 | | | |
| see above | Department of Pathology, University of Cambridge | COVID-19 Genomics UK (COG-UK) Consortium | Aminu S. Jahun; Grant Hall; Ian Goodfellow; Iliana Georgana; Malte Pinckert; Martin D. Curran; Myra Hosmillo; Rhys Izu; Surendra Parmar; Yasmin Chaudhry |
| EPI_ISL_586612, EPI_ISL_586617, EPI_ISL_586621, EPI_ISL_586709, EPI_ISL_586907, EPI_ISL_587085, EPI_ISL_587265, EPI_ISL_587977, EPI_ISL_588007, EPI_ISL_588026, EPI_ISL_597633, EPI_ISL_597984, EPI_ISL_598163, EPI_ISL_609209, EPI_ISL_645240, EPI_ISL_645473, EPI_ISL_649219, EPI_ISL_649234, EPI_ISL_649261, EPI_ISL_649309, EPI_ISL_649312, EPI_ISL_649321, EPI_ISL_649368, EPI_ISL_649389, EPI_ISL_822317, EPI_ISL_892226, EPI_ISL_918125 | | | |
| see above | Lighthouse Lab in Alderley Park | Wellcome Sanger Institute for the COVID-19 Genomics UK (COG-UK) Consortium | Cordelia Langford; David K. Jackson; Dominic Kwiatkowski; Ewan Harrison; Ian Johnston; Jacquelyn Wynn; John Sillitoe on behalf of the Wellcome Sanger Institute COVID-19 Surveillance Team; John Sillitoe on behalf of the Wellcome Sanger Institute COVID-19 Surveillance Team (http://www.sanger.ac.uk/covid-team); Mairead Hyland; Roberto Amato; Sonia Goncalves; The Lighthouse Lab in Alderley Park and Alex Alderton |
| EPI_ISL_586572, EPI_ISL_586573, EPI_ISL_586574, EPI_ISL_586575, EPI_ISL_586576, EPI_ISL_586577, EPI_ISL_586578, EPI_ISL_586579, EPI_ISL_586580, EPI_ISL_586581, EPI_ISL_586582, EPI_ISL_586583, EPI_ISL_586584, EPI_ISL_586585, EPI_ISL_586586, EPI_ISL_586587, EPI_ISL_586588, EPI_ISL_586589, EPI_ISL_586590, EPI_ISL_586591, EPI_ISL_586592, EPI_ISL_586593, EPI_ISL_586594, EPI_ISL_586595, EPI_ISL_586596, EPI_ISL_586597, EPI_ISL_586598, EPI_ISL_586599, EPI_ISL_586600, EPI_ISL_586601, EPI_ISL_586602, EPI_ISL_586603, EPI_ISL_586604, EPI_ISL_586605, EPI_ISL_586606, EPI_ISL_586607, EPI_ISL_586608, EPI_ISL_586609, EPI_ISL_586610, EPI_ISL_586611, EPI_ISL_586613, EPI_ISL_586614, EPI_ISL_586615, EPI_ISL_586616, EPI_ISL_586618, EPI_ISL_586619, EPI_ISL_586620, EPI_ISL_586622, EPI_ISL_586623, EPI_ISL_586624, EPI_ISL_586625, EPI_ISL_586626, EPI_ISL_586627, EPI_ISL_586628, EPI_ISL_586629, EPI_ISL_586630, EPI_ISL_586631, EPI_ISL_586632, EPI_ISL_586633, EPI_ISL_586634, EPI_ISL_586635, EPI_ISL_586636, EPI_ISL_586637, EPI_ISL_586638, EPI_ISL_586639, EPI_ISL_586640, EPI_ISL_586641, EPI_ISL_586642, EPI_ISL_586643, EPI_ISL_586644, EPI_ISL_586645, EPI_ISL_586646, EPI_ISL_586647, EPI_ISL_586648, EPI_ISL_586649, EPI_ISL_586650, EPI_ISL_586651, EPI_ISL_586652, EPI_ISL_586653, EPI_ISL_586654, EPI_ISL_586655, EPI_ISL_586656, EPI_ISL_586657, EPI_ISL_586658, EPI_ISL_586659, EPI_ISL_586660, EPI_ISL_586661, EPI_ISL_586662, EPI_ISL_586663, EPI_ISL_586664, EPI_ISL_586665, EPI_ISL_586666, EPI_ISL_586667, EPI_ISL_586668, EPI_ISL_586669, EPI_ISL_586670, EPI_ISL_586671, EPI_ISL_586672, EPI_ISL_586673, EPI_ISL_586674, EPI_ISL_586675, EPI_ISL_586676, EPI_ISL_586677, EPI_ISL_586678, EPI_ISL_586679, EPI_ISL_586680, EPI_ISL_586681, EPI_ISL_586682, EPI_ISL_586683, EPI_ISL_586684, EPI_ISL_586685, EPI_ISL_586686, EPI_ISL_586687, EPI_ISL_586688, EPI_ISL_586689, EPI_ISL_586690, EPI_ISL_586691, EPI_ISL_586692, EPI_ISL_586693, EPI_ISL_586694, EPI_ISL_586695, EPI_ISL_586696, EPI_ISL_586697, EPI_ISL_586698, EPI_ISL_586699, EPI_ISL_586700, EPI_ISL_586701, EPI_ISL_586702, EPI_ISL_586703, EPI_ISL_586704, EPI_ISL_586705, EPI_ISL_586706, EPI_ISL_586707, EPI_ISL_586708, EPI_ISL_586710, EPI_ISL_586711, EPI_ISL_586712, EPI_ISL_586713, EPI_ISL_586714, EPI_ISL_586715, EPI_ISL_586716, EPI_ISL_586717, EPI_ISL_586718, EPI_ISL_586719, EPI_ISL_586720, EPI_ISL_586721, EPI_ISL_586722, EPI_ISL_586723, EPI_ISL_586724, EPI_ISL_586725, EPI_ISL_586726, EPI_ISL_586727, EPI_ISL_586728, EPI_ISL_586729, EPI_ISL_586730, EPI_ISL_586731, EPI_ISL_586732, EPI_ISL_586733, EPI_ISL_586734, EPI_ISL_586735, EPI_ISL_586736, EPI_ISL_586737, EPI_ISL_586738, EPI_ISL_586739, EPI_ISL_586740, EPI_ISL_586741, EPI_ISL_586742, EPI_ISL_586743, EPI_ISL_586744, EPI_ISL_586745, EPI_ISL_586746, EPI_ISL_586747, EPI_ISL_586748, EPI_ISL_586749, EPI_ISL_586750, EPI_ISL_586751, EPI_ISL_586752, EPI_ISL_586753, EPI_ISL_586754, EPI_ISL_586755, EPI_ISL_586756, EPI_ISL_586757, EPI_ISL_586758, EPI_ISL_586759, EPI_ISL_586760, EPI_ISL_586761, EPI_ISL_586762, EPI_ISL_586763, EPI_ISL_586764, EPI_ISL_586765, EPI_ISL_586766, EPI_ISL_586767, EPI_ISL_586768, EPI_ISL_586769, EPI_ISL_586770, EPI_ISL_586771, EPI_ISL_586772, EPI_ISL_586773, EPI_ISL_586774, EPI_ISL_586775, EPI_ISL_586776, EPI_ISL_586777, EPI_ISL_586778, EPI_ISL_586779, EPI_ISL_586780, EPI_ISL_586781, EPI_ISL_586782, EPI_ISL_586783, EPI_ISL_586784, EPI_ISL_586785, EPI_ISL_586786, EPI_ISL_586787, EPI_ISL_586788, EPI_ISL_586789, EPI_ISL_586790, EPI_ISL_586791, EPI_ISL_586792, EPI_ISL_586793, EPI_ISL_586794, EPI_ISL_586795, EPI_ISL_586796, EPI_ISL_586797, EPI_ISL_586798, EPI_ISL_586799, EPI_ISL_586800, EPI_ISL_586801, EPI_ISL_586802, EPI_ISL_586803, EPI_ISL_586804, EPI_ISL_586805, EPI_ISL_586806, EPI_ISL_586807, EPI_ISL_586808, EPI_ISL_586809, EPI_ISL_586810, EPI_ISL_586811, EPI_ISL_586812, EPI_ISL_586813, EPI_ISL_586814, EPI_ISL_586815, EPI_ISL_586816, EPI_ISL_586817, EPI_ISL_586818, EPI_ISL_586819, EPI_ISL_586820, EPI_ISL_586821, EPI_ISL_586822, EPI_ISL_586823, EPI_ISL_586824, EPI_ISL_586825, EPI_ISL_586826, EPI_ISL_586827, EPI_ISL_586828, EPI_ISL_586829, EPI_ISL_586830, EPI_ISL_586831, EPI_ISL_586832, EPI_ISL_586833, EPI_ISL_586834, EPI_ISL_586835, EPI_ISL_586836, EPI_ISL_586837, EPI_ISL_586838, EPI_ISL_586839, EPI_ISL_586840, EPI_ISL_586841, EPI_ISL_586842, EPI_ISL_586843, EPI_ISL_586844, EPI_ISL_586845, EPI_ISL_586846, EPI_ISL_586847, EPI_ISL_586848, EPI_ISL_586849, EPI_ISL_586850, EPI_ISL_586851, EPI_ISL_586852, EPI_ISL_586853, EPI_ISL_586854, EPI_ISL_586855, EPI_ISL_586856, EPI_ISL_586857, EPI_ISL_586858, EPI_ISL_586859, EPI_ISL_586860, EPI_ISL_586861, EPI_ISL_586862, EPI_ISL_586863, EPI_ISL_586864, EPI_ISL_586865, EPI_ISL_586866, EPI_ISL_586867, EPI_ISL_586868, EPI_ISL_586869, EPI_ISL_586870, EPI_ISL_586871, EPI_ISL_586872, EPI_ISL_586873, EPI_ISL_586874, EPI_ISL_586875, EPI_ISL_586876, EPI_ISL_586877, EPI_ISL_586878, EPI_ISL_586879, EPI_ISL_586880, EPI_ISL_586881, EPI_ISL_586882, EPI_ISL_586883, EPI_ISL_586884, EPI_ISL_586885, EPI_ISL_586886, EPI_ISL_586887, EPI_ISL_586888, EPI_ISL_586889, EPI_ISL_586890, EPI_ISL_586891, EPI_ISL_586892, EPI_ISL_586893, EPI_ISL_586894, EPI_ISL_586895, EPI_ISL_586896, EPI_ISL_586897, EPI_ISL_586898, EPI_ISL_586899, EPI_ISL_586900, EPI_ISL_586901, EPI_ISL_586902, EPI_ISL_586903, EPI_ISL_586904, EPI_ISL_586905, EPI_ISL_586906, EPI_ISL_586908, EPI_ISL_586909, EPI_ISL_586910, EPI_ISL_586911, EPI_ISL_586912, EPI_ISL_586913, EPI_ISL_586914, EPI_ISL_586915, EPI_ISL_586916, EPI_ISL_586917, EPI_ISL_586918, EPI_ISL_586919, EPI_ISL_586920, EPI_ISL_586921, EPI_ISL_586922, EPI_ISL_586923, EPI_ISL_586924, EPI_ISL_586925, EPI_ISL_586926, EPI_ISL_586927, EPI_ISL_586928, EPI_ISL_586929, EPI_ISL_586930, EPI_ISL_586931, EPI_ISL_586932, EPI_ISL_586933, EPI_ISL_586934, EPI_ISL_586935, EPI_ISL_586936, EPI_ISL_586937, EPI_ISL_586938, EPI_ISL_586939, EPI_ISL_586940, EPI_ISL_586941, EPI_ISL_586942, EPI_ISL_586943, EPI_ISL_586944, EPI_ISL_586945, EPI_ISL_586946, EPI_ISL_586947, EPI_ISL_586948, EPI_ISL_586949, EPI_ISL_586950, EPI_ISL_586951, EPI_ISL_586952, EPI_ISL_586953, EPI_ISL_586954, EPI_ISL_586955, EPI_ISL_586956, EPI_ISL_586957, EPI_ISL_586958, EPI_ISL_586959, EPI_ISL_586960, EPI_ISL_586961, EPI_ISL_586962, EPI_ISL_586963, EPI_ISL_586964, EPI_ISL_586965, EPI_ISL_586966, EPI_ISL_586967, EPI_ISL_586968, EPI_ISL_586969, EPI_ISL_586970, EPI_ISL_586971, EPI_ISL_586972, EPI_ISL_586973, EPI_ISL_586974, EPI_ISL_586975, EPI_ISL_586976, EPI_ISL_586977, EPI_ISL_586978, EPI_ISL_586979, EPI_ISL_586980, EPI_ISL_586981, EPI_ISL_586982, EPI_ISL_586983, EPI_ISL_586984, EPI_ISL_586985, EPI_ISL_586986, EPI_ISL_586987, EPI_ISL_586988, EPI_ISL_586989, EPI_ISL_586990, EPI_ISL_586991, EPI_ISL_586992, EPI_ISL_586993, EPI_ISL_586994, EPI_ISL_586995, EPI_ISL_586996, EPI_ISL_586997, EPI_ISL_586998, EPI_ISL_586999, EPI_ISL_587000, EPI_ISL_587001, EPI_ISL_587002, EPI_ISL_587003, EPI_ISL_587004, EPI_ISL_587005, EPI_ISL_587006, EPI_ISL_587007, EPI_ISL_587008, EPI_ISL_587009, EPI_ISL_587010, EPI_ISL_587011, EPI_ISL_587012, EPI_ISL_587013, EPI_ISL_587014, EPI_ISL_587015, EPI_ISL_587016, EPI_ISL_587017, EPI_ISL_587018, EPI_ISL_587019, EPI_ISL_587020, EPI_ISL_587021, EPI_ISL_587022, EPI_ISL_587023, EPI_ISL_587024, EPI_ISL_587025, EPI_ISL_587026, EPI_ISL_587027, EPI_ISL_587028, EPI_ISL_587029, EPI_ISL_587030, EPI_ISL_587031, EPI_ISL_587032, EPI_ISL_587033, EPI_ISL_587034, EPI_ISL_587035, EPI_ISL_587036, EPI_ISL_587037, EPI_ISL_587038, EPI_ISL_587039, EPI_ISL_587040, EPI_ISL_587041, EPI_ISL_587042, EPI_ISL_587043, EPI_ISL_587044, EPI_ISL_587045, EPI_ISL_587046, EPI_ISL_587047, EPI_ISL_587048, EPI_ISL_587049, EPI_ISL_587050, EPI_ISL_587051, EPI_ISL_587052, EPI_ISL_587053, EPI_ISL_587054, EPI_ISL_587055, EPI_ISL_587056, EPI_ISL_587057, EPI_ISL_587058, EPI_ISL_587059, EPI_ISL_587060, EPI_ISL_587061, EPI_ISL_587062, EPI_ISL_587063, EPI_ISL_587064, EPI_ISL_587065, EPI_ISL_587066, EPI_ISL_587067, EPI_ISL_587068, EPI_ISL_587069, EPI_ISL_587070, EPI_ISL_587071, EPI_ISL_587072, EPI_ISL_587073, EPI_ISL_587074, EPI_ISL_587075, EPI_ISL_587076, EPI_ISL_587077, EPI_ISL_587078, EPI_ISL_587079, EPI_ISL_587080, EPI_ISL_587081, EPI_ISL_587082, EPI_ISL_587083, EPI_ISL_587084, EPI_ISL_587086, EPI_ISL_587087, EPI_ISL_587088, EPI_ISL_587089, EPI_ISL_587090, EPI_ISL_587091, EPI_ISL_587092, EPI_ISL_587093, EPI_ISL_587094, EPI_ISL_587095, EPI_ISL_587096, EPI_ISL_587097, EPI_ISL_587098, EPI_ISL_587099, EPI_ISL_587100, EPI_ISL_587101, EPI_ISL_587102, EPI_ISL_587103, EPI_ISL_587104, EPI_ISL_587105, EPI_ISL_587106, EPI_ISL_587107, EPI_ISL_587108, EPI_ISL_587109, EPI_ISL_587110, EPI_ISL_587111, EPI_ISL_587112, EPI_ISL_587113, EPI_ISL_587114, EPI_ISL_587115, EPI_ISL_587116, EPI_ISL_587117, EPI_ISL_587118, EPI_ISL_587119, EPI_ISL_587120, EPI_ISL_587121, EPI_ISL_587122, EPI_ISL_587123, EPI_ISL_587124, EPI_ISL_587125, EPI_ISL_587126, EPI_ISL_587127, EPI_ISL_587128, EPI_ISL_587129, EPI_ISL_587130, EPI_ISL_587131, EPI_ISL_587132, EPI_ISL_587133, EPI_ISL_587134, EPI_ISL_587135, EPI_ISL_587136, EPI_ISL_587137, EPI_ISL_587138, EPI_ISL_587139, EPI_ISL_587140, EPI_ISL_587141, EPI_ISL_587142, EPI_ISL_587143, EPI_ISL_587144, EPI_ISL_587145, EPI_ISL_587146, EPI_ISL_587147, EPI_ISL_587148, EPI_ISL_587149, EPI_ISL_587150, EPI_ISL_587151, EPI_ISL_587152, EPI_ISL_587153, EPI_ISL_587154, EPI_ISL_587155, EPI_ISL_587156, EPI_ISL_587157, EPI_ISL_587158, EPI_ISL_587159, EPI_ISL_587160, EPI_ISL_587161, EPI_ISL_587162, EPI_ISL_587163, EPI_ISL_587164, EPI_ISL_587165, EPI_ISL_587166, EPI_ISL_587167, EPI_ISL_587168, EPI_ISL_587169, EPI_ISL_587170, EPI_ISL_587171, EPI_ISL_587172, EPI_ISL_587173, EPI_ISL_587174, EPI_ISL_587175, EPI_ISL_587176, EPI_ISL_587177, EPI_ISL_587178, EPI_ISL_587179, EPI_ISL_587180, EPI_ISL_587181, EPI_ISL_587182, EPI_ISL_587183, EPI_ISL_587184, EPI_ISL_587185, EPI_ISL_587186, EPI_ISL_587187, EPI_ISL_587188, EPI_ISL_587189, EPI_ISL_587190, EPI_ISL_587191, EPI_ISL_587192, EPI_ISL_587193, EPI_ISL_587194, EPI_ISL_587195, EPI_ISL_587196, EPI_ISL_587197, EPI_ISL_587198, EPI_ISL_587199, EPI_ISL_587200, EPI_ISL_587201, EPI_ISL_587202, EPI_ISL_587203, EPI_ISL_587204, EPI_ISL_587205, EPI_ISL_587206, EPI_ISL_587207, EPI_ISL_587208, EPI_ISL_587209, EPI_ISL_587210, EPI_ISL_587211, EPI_ISL_587212, EPI_ISL_587213, EPI_ISL_587214, EPI_ISL_587215, EPI_ISL_587216, EPI_ISL_587217, EPI_ISL_587218, EPI_ISL_587219, EPI_ISL_587220, EPI_ISL_587221, EPI_ISL_587222, EPI_ISL_587223, EPI_ISL_587224, EPI_ISL_587225, EPI_ISL_587226, EPI_ISL_587227, EPI_ISL_587228, EPI_ISL_587229, EPI_ISL_587230, EPI_ISL_587231, EPI_ISL_587232, EPI_ISL_587233, EPI_ISL_587234, EPI_ISL_587235, EPI_ISL_587236, EPI_ISL_587237, EPI_ISL_587238, EPI_ISL_587239, EPI_ISL_587240, EPI_ISL_587241, EPI_ISL_587242, EPI_ISL_587243, EPI_ISL_587244, EPI_ISL_587245, EPI_ISL_587246, EPI_ISL_587247, EPI_ISL_587248, EPI_ISL_587249, EPI_ISL_587250, EPI_ISL_587251, EPI_ISL_587252, EPI_ISL_587253, EPI_ISL_587254, EPI_ISL_587255, EPI_ISL_587256, EPI_ISL_587257, EPI_ISL_587258, EPI_ISL_587259, EPI_ISL_587260, EPI_ISL_587261, EPI_ISL_587262, EPI_ISL_587263, EPI_ISL_587264, EPI_ISL_587266, EPI_ISL_587267, EPI_ISL_587268, EPI_ISL_587269, EPI_ISL_587270, EPI_ISL_587271, EPI_ISL_587272, EPI_ISL_587273, EPI_ISL_587274, EPI_ISL_587275, EPI_ISL_587276, EPI_ISL_587277, EPI_ISL_587278, EPI_ISL_587279, EPI_ISL_587280, EPI_ISL_587281, EPI_ISL_587282, EPI_ISL_587283, EPI_ISL_587284, EPI_ISL_587285, EPI_ISL_587286, EPI_ISL_587287, EPI_ISL_587288, EPI_ISL_587289, EPI_ISL_587290, EPI_ISL_587291, EPI_ISL_587292, EPI_ISL_587293, EPI_ISL_587294, EPI_ISL_587295, EPI_ISL_587296, EPI_ISL_587297, EPI_ISL_587298, EPI_ISL_587299, EPI_ISL_587300, EPI_ISL_587301, EPI_ISL_587302, EPI_ISL_587711, EPI_ISL_587712, EPI_ISL_587713, EPI_ISL_587714, EPI_ISL_587715, EPI_ISL_587716, EPI_ISL_587717, EPI_ISL_587718, EPI_ISL_587719, EPI_ISL_587720, EPI_ISL_587721, EPI_ISL_587722, EPI_ISL_587723, EPI_ISL_587724, EPI_ISL_587725, EPI_ISL_587726, EPI_ISL_587727, EPI_ISL_587728, EPI_ISL_587729, EPI_ISL_587730, EPI_ISL_587731, EPI_ISL_587732, EPI_ISL_587733, EPI_ISL_587734, EPI_ISL_587735, EPI_ISL_587736, EPI_ISL_587737, EPI_ISL_587738, EPI_ISL_587739, EPI_ISL_587740, EPI_ISL_587741, EPI_ISL_587742, EPI_ISL_587743, EPI_ISL_587744, EPI_ISL_587745, EPI_ISL_587746, EPI_ISL_587747, EPI_ISL_587748, EPI_ISL_587749, EPI_ISL_587750, EPI_ISL_587751, EPI_ISL_587752, EPI_ISL_587753, EPI_ISL_587754, EPI_ISL_587755, EPI_ISL_587756, EPI_ISL_587757, EPI_ISL_587758, EPI_ISL_587759, EPI_ISL_587760, EPI_ISL_587761, EPI_ISL_587762, EPI_ISL_587763, EPI_ISL_587764, EPI_ISL_587765, EPI_ISL_587766, EPI_ISL_587767, EPI_ISL_587768, EPI_ISL_587769, EPI_ISL_587770, EPI_ISL_587771, EPI_ISL_587772, EPI_ISL_587773, EPI_ISL_587774, EPI_ISL_587775, EPI_ISL_587776, EPI_ISL_587777, EPI_ISL_587778, EPI_ISL_587779, EPI_ISL_587780, EPI_ISL_587781, EPI_ISL_587782, EPI_ISL_587783, EPI_ISL_587784, EPI_ISL_587785, EPI_ISL_587786, EPI_ISL_587787, EPI_ISL_587788, EPI_ISL_587789, EPI_ISL_587790, EPI_ISL_587791, EPI_ISL_587792, EPI_ISL_587793, EPI_ISL_587794, EPI_ISL_587795, EPI_ISL_587796, EPI_ISL_587797, EPI_ISL_587798, EPI_ISL_587799, EPI_ISL_587800, EPI_ISL_587801, EPI_ISL_587802, EPI_ISL_587803, EPI_ISL_587804, EPI_ISL_587805, EPI_ISL_587806, EPI_ISL_587807, EPI_ISL_587808, EPI_ISL_587809, EPI_ISL_587810, EPI_ISL_587811, EPI_ISL_587812, EPI_ISL_587813, EPI_ISL_587814, EPI_ISL_587815, EPI_ISL_587816, EPI_ISL_587817, EPI_ISL_587818, EPI_ISL_587819, EPI_ISL_587820, EPI_ISL_587821, EPI_ISL_587822, EPI_ISL_587823, EPI_ISL_587824, EPI_ISL_587825, EPI_ISL_587826, EPI_ISL_587827, EPI_ISL_587828, EPI_ISL_587829, EPI_ISL_587830, EPI_ISL_587831, EPI_ISL_587832, EPI_ISL_587833, EPI_ISL_587834, EPI_ISL_587835, EPI_ISL_587836, EPI_ISL_587837, EPI_ISL_587838, EPI_ISL_587839, EPI_ISL_587840, EPI_ISL_587841, EPI_ISL_587842, EPI_ISL_587843, EPI_ISL_587844, EPI_ISL_587845, EPI_ISL_587846, EPI_ISL_587847, EPI_ISL_587848, EPI_ISL_587849, EPI_ISL_587850, EPI_ISL_587851, EPI_ISL_587852, EPI_ISL_587853, EPI_ISL_587854, EPI_ISL_587855, EPI_ISL_587856, EPI_ISL_587857, EPI_ISL_587858, EPI_ISL_587859, EPI_ISL_587860, EPI_ISL_587861, EPI_ISL_587862, EPI_ISL_587863, EPI_ISL_587864, EPI_ISL_587865, EPI_ISL_587866, EPI_ISL_587867, EPI_ISL_587868, EPI_ISL_587869, EPI_ISL_587870, EPI_ISL_587871, EPI_ISL_587872, EPI_ISL_587873, EPI_ISL_587874, EPI_ISL_587875, EPI_ISL_587876, EPI_ISL_587877, EPI_ISL_587878, EPI_ISL_587879, EPI_ISL_587880, EPI_ISL_587881, EPI_ISL_587882, EPI_ISL_587883, EPI_ISL_587884, EPI_ISL_587885, EPI_ISL_587886, EPI_ISL_587887, EPI_ISL_587888, EPI_ISL_587889, EPI_ISL_587890, EPI_ISL_587891, EPI_ISL_587892, EPI_ISL_587893, EPI_ISL_587894, EPI_ISL_587895, EPI_ISL_587896, EPI_ISL_587897, EPI_ISL_587898, EPI_ISL_587899, EPI_ISL_587900, EPI_ISL_587901, EPI_ISL_587902, EPI_ISL_587903, EPI_ISL_587904, EPI_ISL_587905, EPI_ISL_587906, EPI_ISL_587907, EPI_ISL_587908, EPI_ISL_587909, EPI_ISL_587910, EPI_ISL_587911, EPI_ISL_587912, EPI_ISL_587913, EPI_ISL_587914, EPI_ISL_587915, EPI_ISL_587916, EPI_ISL_587917, EPI_ISL_587918, EPI_ISL_587919, EPI_ISL_587920, EPI_ISL_587921, EPI_ISL_587922, EPI_ISL_587923, EPI_ISL_587924, EPI_ISL_587925, EPI_ISL_587926, EPI_ISL_587927, EPI_ISL_587928, EPI_ISL_587929, EPI_ISL_587930, EPI_ISL_587931, EPI_ISL_587932, EPI_ISL_587933, EPI_ISL_587934, EPI_ISL_587935, EPI_ISL_587936, EPI_ISL_587937, EPI_ISL_587938, EPI_ISL_587939, EPI_ISL_587940, EPI_ISL_587941, EPI_ISL_587942, EPI_ISL_587943, EPI_ISL_587944, EPI_ISL_587945, EPI_ISL_587946, EPI_ISL_587947, EPI_ISL_587948, EPI_ISL_587949, EPI_ISL_587950, EPI_ISL_587951, EPI_ISL_587952, EPI_ISL_587953, EPI_ISL_587954, EPI_ISL_587955, EPI_ISL_587956, EPI_ISL_587957, EPI_ISL_587958, EPI_ISL_587959, EPI_ISL_587960, EPI_ISL_587961, EPI_ISL_587962, EPI_ISL_587963, EPI_ISL_587964, EPI_ISL_587965, EPI_ISL_587966, EPI_ISL_587967, EPI_ISL_587968, EPI_ISL_587969, EPI_ISL_587970, EPI_ISL_587971, EPI_ISL_587972, EPI_ISL_587973, EPI_ISL_587974, EPI_ISL_587975, EPI_ISL_587976, EPI_ISL_587978, EPI_ISL_587979, EPI_ISL_587980, EPI_ISL_587981, EPI_ISL_587982, EPI_ISL_587983, EPI_ISL_587984, EPI_ISL_587985, EPI_ISL_587986, EPI_ISL_587987, EPI_ISL_587988, EPI_ISL_587989, EPI_ISL_587990, EPI_ISL_587991, EPI_ISL_587992, EPI_ISL_587993, EPI_ISL_587994, EPI_ISL_587995, EPI_ISL_587996, EPI_ISL_587997, EPI_ISL_587998, EPI_ISL_587999, EPI_ISL_588000, EPI_ISL_588001, EPI_ISL_588002, EPI_ISL_588003, EPI_ISL_588004, EPI_ISL_588005, EPI_ISL_588006, EPI_ISL_588008, EPI_ISL_588009, EPI_ISL_588010, EPI_ISL_588011, EPI_ISL_588012, EPI_ISL_588013, EPI_ISL_588014, EPI_ISL_588015, EPI_ISL_588016, EPI_ISL_588017, EPI_ISL_588018, EPI_ISL_588019, EPI_ISL_588020, EPI_ISL_588021, EPI_ISL_588022, EPI_ISL_588023, EPI_ISL_588024, EPI_ISL_588025, EPI_ISL_588027, EPI_ISL_588028, EPI_ISL_588029, EPI_ISL_588030, EPI_ISL_588031, EPI_ISL_588032, EPI_ISL_588033, EPI_ISL_588034, EPI_ISL_588035, EPI_ISL_588036, EPI_ISL_588037, EPI_ISL_588038, EPI_ISL_588039, EPI_ISL_588040, EPI_ISL_588041, EPI_ISL_588042, EPI_ISL_588043, EPI_ISL_588044, EPI_ISL_588045, EPI_ISL_588046, EPI_ISL_588047, EPI_ISL_588048, EPI_ISL_588049, EPI_ISL_588050, EPI_ISL_588051, EPI_ISL_588052, EPI_ISL_588053, EPI_ISL_588054, EPI_ISL_588055, EPI_ISL_588056, EPI_ISL_588057, EPI_ISL_588058, EPI_ISL_588059, EPI_ISL_597523, EPI_ISL_597524, EPI_ISL_597525, EPI_ISL_597526, EPI_ISL_597527, EPI_ISL_597528, EPI_ISL_597529, EPI_ISL_597530, EPI_ISL_597531, EPI_ISL_597532, EPI_ISL_597533, EPI_ISL_597534, EPI_ISL_597535, EPI_ISL_597536, EPI_ISL_597537, EPI_ISL_597538, EPI_ISL_597539, EPI_ISL_597540, EPI_ISL_597541, EPI_ISL_597542, EPI_ISL_597543, EPI_ISL_597544, EPI_ISL_597545, EPI_ISL_597546, EPI_ISL_597547, EPI_ISL_597548, EPI_ISL_597549, EPI_ISL_597550, EPI_ISL_597551, EPI_ISL_597552, EPI_ISL_597553, EPI_ISL_597554, EPI_ISL_597555, EPI_ISL_597556, EPI_ISL_597557, EPI_ISL_597558, EPI_ISL_597559, EPI_ISL_597560, EPI_ISL_597561, EPI_ISL_597562, EPI_ISL_597563, EPI_ISL_597564, EPI_ISL_597565, EPI_ISL_597566, EPI_ISL_597567, EPI_ISL_597568, EPI_ISL_597569, EPI_ISL_597570, EPI_ISL_597571, EPI_ISL_597572, EPI_ISL_597573, EPI_ISL_597574, EPI_ISL_597575, EPI_ISL_597576, EPI_ISL_597577, EPI_ISL_597578, EPI_ISL_597579, EPI_ISL_597580, EPI_ISL_597581, EPI_ISL_597582, EPI_ISL_597583, EPI_ISL_597584, EPI_ISL_597585, EPI_ISL_597586, EPI_ISL_597587, EPI_ISL_597588, EPI_ISL_597589, EPI_ISL_597590, EPI_ISL_597591, EPI_ISL_597592, EPI_ISL_597593, EPI_ISL_597594, EPI_ISL_597595, EPI_ISL_597596, EPI_ISL_597597, EPI_ISL_597598, EPI_ISL_597599, EPI_ISL_597600, EPI_ISL_597601, EPI_ISL_597602, EPI_ISL_597603, EPI_ISL_597604, EPI_ISL_597605, EPI_ISL_597606, EPI_ISL_597607, EPI_ISL_597608, EPI_ISL_597609, EPI_ISL_597610, EPI_ISL_597611, EPI_ISL_597612, EPI_ISL_597613, EPI_ISL_597614, EPI_ISL_597615, EPI_ISL_597616, EPI_ISL_597617, EPI_ISL_597618, EPI_ISL_597619, EPI_ISL_597620, EPI_ISL_597621, EPI_ISL_597622, EPI_ISL_597623, EPI_ISL_597624, EPI_ISL_597625, EPI_ISL_597626, EPI_ISL_597627, EPI_ISL_597628, EPI_ISL_597629, EPI_ISL_597630, EPI_ISL_597631, EPI_ISL_597632, EPI_ISL_597634, EPI_ISL_597635, EPI_ISL_597636, EPI_ISL_597637, EPI_ISL_597638, EPI_ISL_597639, EPI_ISL_597640, EPI_ISL_597641, EPI_ISL_597642, EPI_ISL_597643, EPI_ISL_597644, EPI_ISL_597645, EPI_ISL_597646, EPI_ISL_597647, EPI_ISL_597648, EPI_ISL_597649, EPI_ISL_597650, EPI_ISL_597651, EPI_ISL_597652, EPI_ISL_597653, EPI_ISL_597654, EPI_ISL_597655, EPI_ISL_597656, EPI_ISL_597657, EPI_ISL_597658, EPI_ISL_597659, EPI_ISL_597660, EPI_ISL_597661, EPI_ISL_597662, EPI_ISL_597663, EPI_ISL_597664, EPI_ISL_597665, EPI_ISL_597666, EPI_ISL_597667, EPI_ISL_597668, EPI_ISL_597669, EPI_ISL_597670, EPI_ISL_597671, EPI_ISL_597672, EPI_ISL_597673, EPI_ISL_597674, EPI_ISL_597675, EPI_ISL_597676, EPI_ISL_597677, EPI_ISL_597678, EPI_ISL_597679, EPI_ISL_597680, EPI_ISL_597681, EPI_ISL_597682, EPI_ISL_597683, EPI_ISL_597684, EPI_ISL_597685, EPI_ISL_597686, EPI_ISL_597687, EPI_ISL_597688, EPI_ISL_597689, EPI_ISL_597690, EPI_ISL_597691, EPI_ISL_597692, EPI_ISL_597693, EPI_ISL_597694, EPI_ISL_597695, EPI_ISL_597696, EPI_ISL_597697, EPI_ISL_597698, EPI_ISL_597699, EPI_ISL_597700, EPI_ISL_597701, EPI_ISL_597702, EPI_ISL_597703, EPI_ISL_597704, EPI_ISL_597705, EPI_ISL_597706, EPI_ISL_597707, EPI_ISL_597708, EPI_ISL_597709, EPI_ISL_597710, EPI_ISL_597711, EPI_ISL_597712, EPI_ISL_597713, EPI_ISL_597714, EPI_ISL_597715, EPI_ISL_597716, EPI_ISL_597717, EPI_ISL_597718, EPI_ISL_597719, EPI_ISL_597720, EPI_ISL_597721, EPI_ISL_597722, EPI_ISL_597723, EPI_ISL_597724, EPI_ISL_597725, EPI_ISL_597726, EPI_ISL_597727, EPI_ISL_597728, EPI_ISL_597729, EPI_ISL_597730, EPI_ISL_597731, EPI_ISL_597732, EPI_ISL_597733, EPI_ISL_597734, EPI_ISL_597735, EPI_ISL_597736, EPI_ISL_597737, EPI_ISL_597738, EPI_ISL_597739, EPI_ISL_597740, EPI_ISL_597741, EPI_ISL_597742, EPI_ISL_597743, EPI_ISL_597744, EPI_ISL_597745, EPI_ISL_597746, EPI_ISL_597747, EPI_ISL_597748, EPI_ISL_597749, EPI_ISL_597750, EPI_ISL_597751, EPI_ISL_597752, EPI_ISL_597753, EPI_ISL_597754, EPI_ISL_597755, EPI_ISL_597756, EPI_ISL_597757, EPI_ISL_597758, EPI_ISL_597759, EPI_ISL_597760, EPI_ISL_597761, EPI_ISL_597762, EPI_ISL_597763, EPI_ISL_597764, EPI_ISL_597765, EPI_ISL_597766, EPI_ISL_597767, EPI_ISL_597768, EPI_ISL_597769, EPI_ISL_597770, EPI_ISL_597771, EPI_ISL_597772, EPI_ISL_597773, EPI_ISL_597774, EPI_ISL_597775, EPI_ISL_597776, EPI_ISL_597777, EPI_ISL_597778, EPI_ISL_597779, EPI_ISL_597780, EPI_ISL_597781, EPI_ISL_597782, EPI_ISL_597783, EPI_ISL_597784, EPI_ISL_597785, EPI_ISL_597786, EPI_ISL_597787, EPI_ISL_597788, EPI_ISL_597789, EPI_ISL_597790, EPI_ISL_597791, EPI_ISL_597792, EPI_ISL_597793, EPI_ISL_597794, EPI_ISL_597795, EPI_ISL_597796, EPI_ISL_597797, EPI_ISL_597798, EPI_ISL_597799, EPI_ISL_597800, EPI_ISL_597801, EPI_ISL_597802, EPI_ISL_597803, EPI_ISL_597804, EPI_ISL_597805, EPI_ISL_597806, EPI_ISL_597807, EPI_ISL_597808, EPI_ISL_597809, EPI_ISL_597810, EPI_ISL_597811, EPI_ISL_597812, EPI_ISL_597813, EPI_ISL_597814, EPI_ISL_597815, EPI_ISL_597816, EPI_ISL_597817, EPI_ISL_597818, EPI_ISL_597819, EPI_ISL_597820, EPI_ISL_597821, EPI_ISL_597822, EPI_ISL_597823, EPI_ISL_597824, EPI_ISL_597825, EPI_ISL_597826, EPI_ISL_597827, EPI_ISL_597828, EPI_ISL_597829, EPI_ISL_597830, EPI_ISL_597831, EPI_ISL_597832, EPI_ISL_597833, EPI_ISL_597834, EPI_ISL_597835, EPI_ISL_597836, EPI_ISL_597837, EPI_ISL_597838, EPI_ISL_597839, EPI_ISL_597840, EPI_ISL_597841, EPI_ISL_597842, EPI_ISL_597843, EPI_ISL_597844, EPI_ISL_597845, EPI_ISL_597846, EPI_ISL_597847, EPI_ISL_597848, EPI_ISL_597849, EPI_ISL_597850, EPI_ISL_597851, EPI_ISL_597852, EPI_ISL_597853, EPI_ISL_597854, EPI_ISL_597855, EPI_ISL_597856, EPI_ISL_597857, EPI_ISL_597858, EPI_ISL_597859, EPI_ISL_597860, EPI_ISL_597861, EPI_ISL_597862, EPI_ISL_597863, EPI_ISL_597864, EPI_ISL_597865, EPI_ISL_597866, EPI_ISL_597867, EPI_ISL_597868, EPI_ISL_597869, EPI_ISL_597870, EPI_ISL_597871, EPI_ISL_597872, EPI_ISL_597873, EPI_ISL_597874, EPI_ISL_597875, EPI_ISL_597876, EPI_ISL_597877, EPI_ISL_597878, EPI_ISL_597879, EPI_ISL_597880, EPI_ISL_597881, EPI_ISL_597882, EPI_ISL_597883, EPI_ISL_597884, EPI_ISL_597885, EPI_ISL_597886, EPI_ISL_597887, EPI_ISL_597888, EPI_ISL_597889, EPI_ISL_597890, EPI_ISL_597891, EPI_ISL_597892, EPI_ISL_597893, EPI_ISL_597894, EPI_ISL_597895, EPI_ISL_597896, EPI_ISL_597897, EPI_ISL_597898, EPI_ISL_597899, EPI_ISL_597900, EPI_ISL_597901, EPI_ISL_597902, EPI_ISL_597903, EPI_ISL_597904, EPI_ISL_597905, EPI_ISL_597906, EPI_ISL_597907, EPI_ISL_597908, EPI_ISL_597909, EPI_ISL_597910, EPI_ISL_597911, EPI_ISL_597912, EPI_ISL_597913, EPI_ISL_597914, EPI_ISL_597915, EPI_ISL_597916, EPI_ISL_597917, EPI_ISL_597918, EPI_ISL_597919, EPI_ISL_597920, EPI_ISL_597921, EPI_ISL_597922, EPI_ISL_597923, EPI_ISL_597924, EPI_ISL_597925, EPI_ISL_597926, EPI_ISL_597927, EPI_ISL_597928, EPI_ISL_597929, EPI_ISL_597930, EPI_ISL_597931, EPI_ISL_597932, EPI_ISL_597933, EPI_ISL_597934, EPI_ISL_597935, EPI_ISL_597936, EPI_ISL_597937, EPI_ISL_597938, EPI_ISL_597939, EPI_ISL_597940, EPI_ISL_597941, EPI_ISL_597942, EPI_ISL_597943, EPI_ISL_597944, EPI_ISL_597945, EPI_ISL_597946, EPI_ISL_597947, EPI_ISL_597948, EPI_ISL_597949, EPI_ISL_597950, EPI_ISL_597951, EPI_ISL_597952, EPI_ISL_597953, EPI_ISL_597954, EPI_ISL_597955, EPI_ISL_597956, EPI_ISL_597957, EPI_ISL_597958, EPI_ISL_597959, EPI_ISL_597960, EPI_ISL_597961, EPI_ISL_597962, EPI_ISL_597963, EPI_ISL_597964, EPI_ISL_597965, EPI_ISL_597966, EPI_ISL_597967, EPI_ISL_597968, EPI_ISL_597969, EPI_ISL_597970, EPI_ISL_597971, EPI_ISL_597972, EPI_ISL_597973, EPI_ISL_597974, EPI_ISL_597975, EPI_ISL_597976, EPI_ISL_597977, EPI_ISL_597978, EPI_ISL_597979, EPI_ISL_597980, EPI_ISL_597981, EPI_ISL_597982, EPI_ISL_597983, EPI_ISL_597985, EPI_ISL_597986, EPI_ISL_597987, EPI_ISL_597988, EPI_ISL_597989, EPI_ISL_597990, EPI_ISL_597991, EPI_ISL_597992, EPI_ISL_597993, EPI_ISL_597994, EPI_ISL_597995, EPI_ISL_597996, EPI_ISL_597997, EPI_ISL_597998, EPI_ISL_597999, EPI_ISL_598000, EPI_ISL_598001, EPI_ISL_598002, EPI_ISL_598003, EPI_ISL_598004, EPI_ISL_598005, EPI_ISL_598006, EPI_ISL_598007, EPI_ISL_598008, EPI_ISL_598009, EPI_ISL_598010, EPI_ISL_598011, EPI_ISL_598012, EPI_ISL_598013, EPI_ISL_598014, EPI_ISL_598015, EPI_ISL_598016, EPI_ISL_598017, EPI_ISL_598018, EPI_ISL_598019, EPI_ISL_598020, EPI_ISL_598021, EPI_ISL_598022, EPI_ISL_598023, EPI_ISL_598024, EPI_ISL_598025, EPI_ISL_598026, EPI_ISL_598027, EPI_ISL_598028, EPI_ISL_598029, EPI_ISL_598030, EPI_ISL_598031, EPI_ISL_598032, EPI_ISL_598033, EPI_ISL_598034, EPI_ISL_598035, EPI_ISL_598036, EPI_ISL_598037, EPI_ISL_598038, | | | |

EPI_ISL_598039, EPI_ISL_598040, EPI_ISL_598041, EPI_ISL_598042, EPI_ISL_598043, EPI_ISL_598044, EPI_ISL_598045, EPI_ISL_598046, EPI_ISL_598047, EPI_ISL_598048, EPI_ISL_598049, EPI_ISL_598050, EPI_ISL_598051, EPI_ISL_598052, EPI_ISL_598053, EPI_ISL_598054, EPI_ISL_598055, EPI_ISL_598056, EPI_ISL_598057, EPI_ISL_598058, EPI_ISL_598059, EPI_ISL_598060, EPI_ISL_598061, EPI_ISL_598062, EPI_ISL_598063, EPI_ISL_598064, EPI_ISL_598065, EPI_ISL_598066, EPI_ISL_598067, EPI_ISL_598068, EPI_ISL_598069, EPI_ISL_598070, EPI_ISL_598071, EPI_ISL_598072, EPI_ISL_598073, EPI_ISL_598074, EPI_ISL_598075, EPI_ISL_598076, EPI_ISL_598077, EPI_ISL_598078, EPI_ISL_598079, EPI_ISL_598080, EPI_ISL_598081, EPI_ISL_598082, EPI_ISL_598083, EPI_ISL_598084, EPI_ISL_598085, EPI_ISL_598086, EPI_ISL_598087, EPI_ISL_598088, EPI_ISL_598089, EPI_ISL_598090, EPI_ISL_598091, EPI_ISL_598092, EPI_ISL_598093, EPI_ISL_598094, EPI_ISL_598095, EPI_ISL_598096, EPI_ISL_598097, EPI_ISL_598098, EPI_ISL_598099, EPI_ISL_598100, EPI_ISL_598101, EPI_ISL_598102, EPI_ISL_598103, EPI_ISL_598104, EPI_ISL_598105, EPI_ISL_598106, EPI_ISL_598107, EPI_ISL_598108, EPI_ISL_598109, EPI_ISL_598110, EPI_ISL_598111, EPI_ISL_598112, EPI_ISL_598113, EPI_ISL_598114, EPI_ISL_598115, EPI_ISL_598116, EPI_ISL_598117, EPI_ISL_598118, EPI_ISL_598119, EPI_ISL_598120, EPI_ISL_598121, EPI_ISL_598122, EPI_ISL_598123, EPI_ISL_598124, EPI_ISL_598125, EPI_ISL_598126, EPI_ISL_598127, EPI_ISL_598128, EPI_ISL_598129, EPI_ISL_598130, EPI_ISL_598131, EPI_ISL_598132, EPI_ISL_598133, EPI_ISL_598134, EPI_ISL_598135, EPI_ISL_598136, EPI_ISL_598137, EPI_ISL_598138, EPI_ISL_598139, EPI_ISL_598140, EPI_ISL_598141, EPI_ISL_598142, EPI_ISL_598143, EPI_ISL_598144, EPI_ISL_598145, EPI_ISL_598146, EPI_ISL_598147, EPI_ISL_598148, EPI_ISL_598149, EPI_ISL_598150, EPI_ISL_598151, EPI_ISL_598152, EPI_ISL_598153, EPI_ISL_598154, EPI_ISL_598155, EPI_ISL_598156, EPI_ISL_598157, EPI_ISL_598158, EPI_ISL_598159, EPI_ISL_598160, EPI_ISL_598161, EPI_ISL_598162, EPI_ISL_598164, EPI_ISL_598165, EPI_ISL_598166, EPI_ISL_598167, EPI_ISL_598168, EPI_ISL_598169, EPI_ISL_598170, EPI_ISL_598171, EPI_ISL_598172, EPI_ISL_598173, EPI_ISL_598174, EPI_ISL_598175, EPI_ISL_598176, EPI_ISL_598177, EPI_ISL_598178, EPI_ISL_598179, EPI_ISL_598180, EPI_ISL_598181, EPI_ISL_598182, EPI_ISL_598183, EPI_ISL_598184, EPI_ISL_598185, EPI_ISL_598186, EPI_ISL_598187, EPI_ISL_598188, EPI_ISL_598189, EPI_ISL_598190, EPI_ISL_598191, EPI_ISL_598192, EPI_ISL_598193, EPI_ISL_598194, EPI_ISL_598195, EPI_ISL_598196, EPI_ISL_598197, EPI_ISL_598198, EPI_ISL_598199, EPI_ISL_598200, EPI_ISL_598201, EPI_ISL_598202, EPI_ISL_598203, EPI_ISL_598204, EPI_ISL_598205, EPI_ISL_598206, EPI_ISL_598207, EPI_ISL_598208, EPI_ISL_598209, EPI_ISL_598210, EPI_ISL_598211, EPI_ISL_598212, EPI_ISL_598213, EPI_ISL_598214, EPI_ISL_598215, EPI_ISL_598216, EPI_ISL_598217, EPI_ISL_598218, EPI_ISL_598219, EPI_ISL_598220, EPI_ISL_598221, EPI_ISL_598222, EPI_ISL_598223, EPI_ISL_598224, EPI_ISL_598225, EPI_ISL_598226, EPI_ISL_598227, EPI_ISL_598228, EPI_ISL_598229, EPI_ISL_598230, EPI_ISL_598231, EPI_ISL_598232, EPI_ISL_598233, EPI_ISL_598234, EPI_ISL_598235, EPI_ISL_598236, EPI_ISL_598237, EPI_ISL_598238, EPI_ISL_598239, EPI_ISL_598240, EPI_ISL_598241, EPI_ISL_598242, EPI_ISL_598243, EPI_ISL_598244, EPI_ISL_598245, EPI_ISL_598246, EPI_ISL_598247, EPI_ISL_598248, EPI_ISL_598824, EPI_ISL_598825, EPI_ISL_598826, EPI_ISL_598827, EPI_ISL_598828, EPI_ISL_598829, EPI_ISL_598830, EPI_ISL_600521, EPI_ISL_600523, EPI_ISL_600525, EPI_ISL_600526, EPI_ISL_600528, EPI_ISL_600529, EPI_ISL_600531, EPI_ISL_600532, EPI_ISL_600534, EPI_ISL_600536, EPI_ISL_600537, EPI_ISL_608425, EPI_ISL_608440, EPI_ISL_608518, EPI_ISL_608622, EPI_ISL_608690, EPI_ISL_608864, EPI_ISL_608896, EPI_ISL_608909, EPI_ISL_608914, EPI_ISL_608916, EPI_ISL_608920, EPI_ISL_608934, EPI_ISL_608963, EPI_ISL_608973, EPI_ISL_608976, EPI_ISL_609004, EPI_ISL_609047, EPI_ISL_609060, EPI_ISL_609092, EPI_ISL_609124, EPI_ISL_609153, EPI_ISL_609207, EPI_ISL_609208, EPI_ISL_609210, EPI_ISL_609211, EPI_ISL_609212, EPI_ISL_609213, EPI_ISL_609214, EPI_ISL_609215, EPI_ISL_609216, EPI_ISL_609217, EPI_ISL_609218, EPI_ISL_625422, EPI_ISL_625423, EPI_ISL_625425, EPI_ISL_625427, EPI_ISL_625429, EPI_ISL_625433, EPI_ISL_625435, EPI_ISL_625436, EPI_ISL_630359, EPI_ISL_630369, EPI_ISL_630399, EPI_ISL_630414, EPI_ISL_630417, EPI_ISL_630441, EPI_ISL_630444, EPI_ISL_630506, EPI_ISL_630512, EPI_ISL_630550, EPI_ISL_630644, EPI_ISL_630682, EPI_ISL_630693, EPI_ISL_630701, EPI_ISL_630971, EPI_ISL_630976, EPI_ISL_630994, EPI_ISL_631023, EPI_ISL_631057, EPI_ISL_631089, EPI_ISL_631111, EPI_ISL_631116, EPI_ISL_631127, EPI_ISL_631128, EPI_ISL_631163, EPI_ISL_631173, EPI_ISL_631175, EPI_ISL_631202, EPI_ISL_631212, EPI_ISL_631222, EPI_ISL_631228, EPI_ISL_631241, EPI_ISL_634754

| | | | |
|---|---|---|---|
| see above | Lighthouse Lab in Alderley Park | Wellcome Sanger Institute for the COVID-19 Genomics UK (COG-UK) consortium | Cordelia Langford; David K. Jackson; Dominic Kwiatkowski; Ewan Harrison; Ian Johnston; Jacquelyn Wynn; John Sillitoe on behalf of the Wellcome Sanger Institute COVID-19 Surveillance Team; John Sillitoe on behalf of the Wellcome Sanger Institute COVID-19 Surveillance Team (http://www.sanger.ac.uk/covid-team); Mairead Hyland; Roberto Amato; Sonia Goncalves; The Lighthouse Lab in Alderley Park and Alex Alderton |

EPI_ISL_596969, EPI_ISL_597074, EPI_ISL_597099, EPI_ISL_597189, EPI_ISL_609378, EPI_ISL_609393, EPI_ISL_609450, EPI_ISL_609472, EPI_ISL_609527, EPI_ISL_609534, EPI_ISL_609600, EPI_ISL_609746, EPI_ISL_647213, EPI_ISL_822310, EPI_ISL_822311, EPI_ISL_822312, EPI_ISL_822313, EPI_ISL_822314, EPI_ISL_822315, EPI_ISL_822316, EPI_ISL_994653

| | | | |
|---|---|---|---|
| see above | Lighthouse Lab in Cambridge | Wellcome Sanger Institute for the COVID-19 Genomics UK (COG-UK) Consortium | Cordelia Langford; David K. Jackson; Dominic Kwiatkowski; Ewan Harrison; Ian Johnston; John Sillitoe on behalf of the Wellcome Sanger Institute COVID-19 Surveillance Team; Rob Howes; Roberto Amato; Sonia Goncalves; The Lighthouse Lab in Cambridge and Alex Alderton |

EPI_ISL_596928, EPI_ISL_596929, EPI_ISL_596930, EPI_ISL_596931, EPI_ISL_596932, EPI_ISL_596933, EPI_ISL_596934, EPI_ISL_596935, EPI_ISL_596936, EPI_ISL_596937, EPI_ISL_596938, EPI_ISL_596939, EPI_ISL_596940, EPI_ISL_596941, EPI_ISL_596942, EPI_ISL_596943, EPI_ISL_596944, EPI_ISL_596945, EPI_ISL_596946, EPI_ISL_596947, EPI_ISL_596948, EPI_ISL_596949, EPI_ISL_596950, EPI_ISL_596951, EPI_ISL_596952, EPI_ISL_596953, EPI_ISL_596954, EPI_ISL_596955, EPI_ISL_596956, EPI_ISL_596957, EPI_ISL_596958, EPI_ISL_596959, EPI_ISL_596960, EPI_ISL_596961, EPI_ISL_596962, EPI_ISL_596963, EPI_ISL_596965, EPI_ISL_596966, EPI_ISL_596967, EPI_ISL_596968, EPI_ISL_596970, EPI_ISL_596971, EPI_ISL_596972, EPI_ISL_596973, EPI_ISL_596974, EPI_ISL_596975, EPI_ISL_596976, EPI_ISL_596977, EPI_ISL_596978, EPI_ISL_596979, EPI_ISL_596980, EPI_ISL_596981, EPI_ISL_596982, EPI_ISL_596983, EPI_ISL_596984, EPI_ISL_596985, EPI_ISL_596986, EPI_ISL_596987, EPI_ISL_596988, EPI_ISL_596989, EPI_ISL_596990, EPI_ISL_596991, EPI_ISL_596992, EPI_ISL_596993, EPI_ISL_596994, EPI_ISL_596995, EPI_ISL_596996, EPI_ISL_596997, EPI_ISL_596998, EPI_ISL_596999, EPI_ISL_597000, EPI_ISL_597001, EPI_ISL_597002, EPI_ISL_597003, EPI_ISL_597004, EPI_ISL_597005, EPI_ISL_597006, EPI_ISL_597007, EPI_ISL_597008, EPI_ISL_597009, EPI_ISL_597010, EPI_ISL_597011, EPI_ISL_597012, EPI_ISL_597013, EPI_ISL_597014, EPI_ISL_597015, EPI_ISL_597016, EPI_ISL_597017, EPI_ISL_597018, EPI_ISL_597019, EPI_ISL_597020, EPI_ISL_597021, EPI_ISL_597022, EPI_ISL_597023, EPI_ISL_597024, EPI_ISL_597025, EPI_ISL_597026, EPI_ISL_597027, EPI_ISL_597028, EPI_ISL_597029, EPI_ISL_597030, EPI_ISL_597031, EPI_ISL_597032, EPI_ISL_597033, EPI_ISL_597034, EPI_ISL_597035, EPI_ISL_597036, EPI_ISL_597037, EPI_ISL_597038, EPI_ISL_597039, EPI_ISL_597040, EPI_ISL_597041, EPI_ISL_597042, EPI_ISL_597043, EPI_ISL_597044, EPI_ISL_597045, EPI_ISL_597046, EPI_ISL_597047, EPI_ISL_597048, EPI_ISL_597049, EPI_ISL_597050, EPI_ISL_597051, EPI_ISL_597052, EPI_ISL_597053, EPI_ISL_597054, EPI_ISL_597055, EPI_ISL_597056, EPI_ISL_597057, EPI_ISL_597058, EPI_ISL_597059, EPI_ISL_597060, EPI_ISL_597061, EPI_ISL_597062, EPI_ISL_597063, EPI_ISL_597064, EPI_ISL_597065, EPI_ISL_597066, EPI_ISL_597067, EPI_ISL_597068, EPI_ISL_597069, EPI_ISL_597070, EPI_ISL_597071, EPI_ISL_597072, EPI_ISL_597073, EPI_ISL_597075, EPI_ISL_597076, EPI_ISL_597077, EPI_ISL_597078, EPI_ISL_597079, EPI_ISL_597080, EPI_ISL_597081, EPI_ISL_597082, EPI_ISL_597083, EPI_ISL_597084, EPI_ISL_597085, EPI_ISL_597086, EPI_ISL_597087, EPI_ISL_597088, EPI_ISL_597089, EPI_ISL_597090, EPI_ISL_597091, EPI_ISL_597092, EPI_ISL_597093, EPI_ISL_597094, EPI_ISL_597095, EPI_ISL_597096, EPI_ISL_597097, EPI_ISL_597098, EPI_ISL_597100, EPI_ISL_597101, EPI_ISL_597102, EPI_ISL_597103, EPI_ISL_597104, EPI_ISL_597105, EPI_ISL_597106, EPI_ISL_597107, EPI_ISL_597108, EPI_ISL_597109, EPI_ISL_597110, EPI_ISL_597111, EPI_ISL_597112, EPI_ISL_597113, EPI_ISL_597114, EPI_ISL_597115, EPI_ISL_597116, EPI_ISL_597117, EPI_ISL_597118, EPI_ISL_597119, EPI_ISL_597120, EPI_ISL_597121, EPI_ISL_597122, EPI_ISL_597123, EPI_ISL_597124, EPI_ISL_597125, EPI_ISL_597126, EPI_ISL_597127, EPI_ISL_597128, EPI_ISL_597129, EPI_ISL_597130, EPI_ISL_597131, EPI_ISL_597132, EPI_ISL_597133, EPI_ISL_597134, EPI_ISL_597135, EPI_ISL_597136, EPI_ISL_597137, EPI_ISL_597138, EPI_ISL_597139, EPI_ISL_597140, EPI_ISL_597141, EPI_ISL_597142, EPI_ISL_597143, EPI_ISL_597144, EPI_ISL_597145, EPI_ISL_597146, EPI_ISL_597147, EPI_ISL_597148, EPI_ISL_597149, EPI_ISL_597150, EPI_ISL_597151, EPI_ISL_597152, EPI_ISL_597153, EPI_ISL_597154, EPI_ISL_597155, EPI_ISL_597156, EPI_ISL_597157, EPI_ISL_597158, EPI_ISL_597159, EPI_ISL_597160, EPI_ISL_597161, EPI_ISL_597162, EPI_ISL_597163, EPI_ISL_597164, EPI_ISL_597165, EPI_ISL_597166, EPI_ISL_597167, EPI_ISL_597168, EPI_ISL_597169, EPI_ISL_597170, EPI_ISL_597171, EPI_ISL_597172, EPI_ISL_597173, EPI_ISL_597174, EPI_ISL_597175, EPI_ISL_597176, EPI_ISL_597177, EPI_ISL_597178, EPI_ISL_597179, EPI_ISL_597180, EPI_ISL_597181, EPI_ISL_597182, EPI_ISL_597183, EPI_ISL_597184, EPI_ISL_597185, EPI_ISL_597186, EPI_ISL_597187, EPI_ISL_597188, EPI_ISL_597190, EPI_ISL_597191, EPI_ISL_597192, EPI_ISL_597193, EPI_ISL_597194, EPI_ISL_597196, EPI_ISL_597197, EPI_ISL_597198, EPI_ISL_597199, EPI_ISL_597200, EPI_ISL_597201, EPI_ISL_597202, EPI_ISL_597203, EPI_ISL_597204, EPI_ISL_597205, EPI_ISL_597206, EPI_ISL_597207, EPI_ISL_597208, EPI_ISL_597209, EPI_ISL_597210, EPI_ISL_597211, EPI_ISL_597212, EPI_ISL_597213, EPI_ISL_597214, EPI_ISL_597215, EPI_ISL_597217, EPI_ISL_597218, EPI_ISL_597219, EPI_ISL_597220, EPI_ISL_597221, EPI_ISL_597222, EPI_ISL_597223, EPI_ISL_597224, EPI_ISL_597225, EPI_ISL_597226, EPI_ISL_597227, EPI_ISL_597228, EPI_ISL_597230, EPI_ISL_597231, EPI_ISL_597232, EPI_ISL_597233, EPI_ISL_597237, EPI_ISL_597238, EPI_ISL_597239, EPI_ISL_597241, EPI_ISL_597243, EPI_ISL_597246, EPI_ISL_597247, EPI_ISL_597251, EPI_ISL_597254, EPI_ISL_597255, EPI_ISL_597256, EPI_ISL_597257, EPI_ISL_597258, EPI_ISL_597259, EPI_ISL_597262, EPI_ISL_597265, EPI_ISL_597266, EPI_ISL_597269, EPI_ISL_597270, EPI_ISL_597271, EPI_ISL_597273, EPI_ISL_597274, EPI_ISL_597279, EPI_ISL_597282, EPI_ISL_597283, EPI_ISL_597284, EPI_ISL_597285, EPI_ISL_597286, EPI_ISL_597287, EPI_ISL_597289, EPI_ISL_597293, EPI_ISL_597294, EPI_ISL_597295, EPI_ISL_597297, EPI_ISL_597300, EPI_ISL_597301, EPI_ISL_597304, EPI_ISL_597305, EPI_ISL_597306, EPI_ISL_597307, EPI_ISL_597308, EPI_ISL_597309, EPI_ISL_597310, EPI_ISL_597316, EPI_ISL_597317, EPI_ISL_597319, EPI_ISL_597320, EPI_ISL_597321, EPI_ISL_597323, EPI_ISL_597324, EPI_ISL_597325, EPI_ISL_597327, EPI_ISL_597331, EPI_ISL_597335, EPI_ISL_597337, EPI_ISL_597340, EPI_ISL_597341, EPI_ISL_597342, EPI_ISL_597343, EPI_ISL_597344, EPI_ISL_597345, EPI_ISL_597346, EPI_ISL_597349, EPI_ISL_597358, EPI_ISL_597361, EPI_ISL_597365, EPI_ISL_597366, EPI_ISL_597367, EPI_ISL_597368, EPI_ISL_597371, EPI_ISL_597372, EPI_ISL_597373, EPI_ISL_597374, EPI_ISL_597375, EPI_ISL_597377, EPI_ISL_597378, EPI_ISL_597380, EPI_ISL_597381, EPI_ISL_597388, EPI_ISL_597390, EPI_ISL_597392, EPI_ISL_597396, EPI_ISL_597397, EPI_ISL_597398, EPI_ISL_597399, EPI_ISL_597401, EPI_ISL_597402, EPI_ISL_597403, EPI_ISL_597405, EPI_ISL_597412, EPI_ISL_597414, EPI_ISL_597418, EPI_ISL_597424, EPI_ISL_597425, EPI_ISL_597426, EPI_ISL_597427, EPI_ISL_597428, EPI_ISL_597432, EPI_ISL_597435, EPI_ISL_597437, EPI_ISL_597438, EPI_ISL_597441, EPI_ISL_597445, EPI_ISL_597446, EPI_ISL_597449, EPI_ISL_597450, EPI_ISL_597451, EPI_ISL_597453, EPI_ISL_597458, EPI_ISL_597459, EPI_ISL_597460, EPI_ISL_597461, EPI_ISL_597462, EPI_ISL_597464, EPI_ISL_597466, EPI_ISL_597467, EPI_ISL_597468, EPI_ISL_597469, EPI_ISL_597470, EPI_ISL_597472, EPI_ISL_597473, EPI_ISL_597476, EPI_ISL_597477, EPI_ISL_597478, EPI_ISL_597479, EPI_ISL_597480, EPI_ISL_597483, EPI_ISL_597487, EPI_ISL_597488, EPI_ISL_597489, EPI_ISL_597490, EPI_ISL_597494, EPI_ISL_597495, EPI_ISL_597497, EPI_ISL_597502, EPI_ISL_597504, EPI_ISL_597505, EPI_ISL_597507, EPI_ISL_597508, EPI_ISL_597510, EPI_ISL_597512, EPI_ISL_597517, EPI_ISL_597519, EPI_ISL_597520, EPI_ISL_608400, EPI_ISL_608429, EPI_ISL_608544, EPI_ISL_608552, EPI_ISL_608573, EPI_ISL_608611, EPI_ISL_608867, EPI_ISL_608873, EPI_ISL_608875, EPI_ISL_608949, EPI_ISL_608974, EPI_ISL_609014, EPI_ISL_609050, EPI_ISL_609067, EPI_ISL_609097, EPI_ISL_609101, EPI_ISL_609142, EPI_ISL_609145, EPI_ISL_609159, EPI_ISL_609160, EPI_ISL_609161, EPI_ISL_609162, EPI_ISL_609163, EPI_ISL_609164, EPI_ISL_609165, EPI_ISL_609166, EPI_ISL_609167, EPI_ISL_609168, EPI_ISL_609169, EPI_ISL_609170, EPI_ISL_609171, EPI_ISL_609172, EPI_ISL_609173, EPI_ISL_609174, EPI_ISL_609175, EPI_ISL_609176, EPI_ISL_609177, EPI_ISL_609180, EPI_ISL_609181, EPI_ISL_609183, EPI_ISL_609188, EPI_ISL_609193, EPI_ISL_609195, EPI_ISL_609196, EPI_ISL_609200, EPI_ISL_609201, EPI_ISL_609203, EPI_ISL_609204, EPI_ISL_609205, EPI_ISL_609308, EPI_ISL_609309, EPI_ISL_609310, EPI_ISL_609311, EPI_ISL_609312, EPI_ISL_609313, EPI_ISL_609314, EPI_ISL_609315, EPI_ISL_609316, EPI_ISL_609317, EPI_ISL_609318, EPI_ISL_609319, EPI_ISL_609320, EPI_ISL_609321, EPI_ISL_609322, EPI_ISL_609323, EPI_ISL_609324, EPI_ISL_609325, EPI_ISL_609326, EPI_ISL_609327, EPI_ISL_609328, EPI_ISL_609329, EPI_ISL_609330, EPI_ISL_609331, EPI_ISL_609332, EPI_ISL_609333, EPI_ISL_609334, EPI_ISL_609335, EPI_ISL_609336, EPI_ISL_609337, EPI_ISL_609338, EPI_ISL_609339, EPI_ISL_609340, EPI_ISL_609341, EPI_ISL_609342, EPI_ISL_609343, EPI_ISL_609344, EPI_ISL_609345, EPI_ISL_609346, EPI_ISL_609347, EPI_ISL_609348, EPI_ISL_609349, EPI_ISL_609350, EPI_ISL_609351, EPI_ISL_609352, EPI_ISL_609353, EPI_ISL_609354, EPI_ISL_609355, EPI_ISL_609356, EPI_ISL_609357, EPI_ISL_609358, EPI_ISL_609359, EPI_ISL_609360, EPI_ISL_609361, EPI_ISL_609362, EPI_ISL_609363, EPI_ISL_609364, EPI_ISL_609365, EPI_ISL_609366, EPI_ISL_609367, EPI_ISL_609368, EPI_ISL_609369, EPI_ISL_609370, EPI_ISL_609371, EPI_ISL_609372, EPI_ISL_609373, EPI_ISL_609374, EPI_ISL_609375, EPI_ISL_609376, EPI_ISL_609377, EPI_ISL_609379, EPI_ISL_609380, EPI_ISL_609381, EPI_ISL_609382, EPI_ISL_609383, EPI_ISL_609384, EPI_ISL_609385, EPI_ISL_609386, EPI_ISL_609387, EPI_ISL_609388, EPI_ISL_609389, EPI_ISL_609390, EPI_ISL_609391, EPI_ISL_609392, EPI_ISL_609394, EPI_ISL_609395, EPI_ISL_609396, EPI_ISL_609397, EPI_ISL_609398, EPI_ISL_609399, EPI_ISL_609400, EPI_ISL_609401, EPI_ISL_609402, EPI_ISL_609403, EPI_ISL_609404, EPI_ISL_609405, EPI_ISL_609406, EPI_ISL_609407, EPI_ISL_609408, EPI_ISL_609409, EPI_ISL_609410, EPI_ISL_609411, EPI_ISL_609412, EPI_ISL_609413, EPI_ISL_609414, EPI_ISL_609415, EPI_ISL_609416, EPI_ISL_609417, EPI_ISL_609418, EPI_ISL_609419, EPI_ISL_609420, EPI_ISL_609421, EPI_ISL_609422, EPI_ISL_609423, EPI_ISL_609424, EPI_ISL_609425, EPI_ISL_609426, EPI_ISL_609427, EPI_ISL_609428, EPI_ISL_609429, EPI_ISL_609430, EPI_ISL_609431, EPI_ISL_609432, EPI_ISL_609433, EPI_ISL_609434, EPI_ISL_609435, EPI_ISL_609436, EPI_ISL_609437, EPI_ISL_609438, EPI_ISL_609439, EPI_ISL_609440, EPI_ISL_609441, EPI_ISL_609442, EPI_ISL_609443, EPI_ISL_609444, EPI_ISL_609445, EPI_ISL_609446, EPI_ISL_609447, EPI_ISL_609448, EPI_ISL_609449, EPI_ISL_609451, EPI_ISL_609452, EPI_ISL_609453, EPI_ISL_609454, EPI_ISL_609455, EPI_ISL_609456, EPI_ISL_609457, EPI_ISL_609458, EPI_ISL_609459, EPI_ISL_609460, EPI_ISL_609461, EPI_ISL_609462, EPI_ISL_609463, EPI_ISL_609464, EPI_ISL_609465, EPI_ISL_609466, EPI_ISL_609467, EPI_ISL_609468, EPI_ISL_609469, EPI_ISL_609470, EPI_ISL_609471, EPI_ISL_609473, EPI_ISL_609474, EPI_ISL_609475, EPI_ISL_609476, EPI_ISL_609477, EPI_ISL_609478, EPI_ISL_609479, EPI_ISL_609480, EPI_ISL_609481, EPI_ISL_609482, EPI_ISL_609483, EPI_ISL_609484, EPI_ISL_609485, EPI_ISL_609486, EPI_ISL_609487, EPI_ISL_609488, EPI_ISL_609489, EPI_ISL_609490, EPI_ISL_609491, EPI_ISL_609492, EPI_ISL_609493, EPI_ISL_609494, EPI_ISL_609495, EPI_ISL_609496, EPI_ISL_609497, EPI_ISL_609498, EPI_ISL_609499, EPI_ISL_609500, EPI_ISL_609501, EPI_ISL_609502, EPI_ISL_609503, EPI_ISL_609504, EPI_ISL_609505, EPI_ISL_609506, EPI_ISL_609507, EPI_ISL_609508, EPI_ISL_609509, EPI_ISL_609510, EPI_ISL_609511, EPI_ISL_609512, EPI_ISL_609513, EPI_ISL_609514, EPI_ISL_609515, EPI_ISL_609516, EPI_ISL_609517, EPI_ISL_609518, EPI_ISL_609519, EPI_ISL_609520, EPI_ISL_609521, EPI_ISL_609522, EPI_ISL_609523, EPI_ISL_609524, EPI_ISL_609525, EPI_ISL_609526, EPI_ISL_609528, EPI_ISL_609529, EPI_ISL_609530, EPI_ISL_609531, EPI_ISL_609532, EPI_ISL_609533, EPI_ISL_609535, EPI_ISL_609536, EPI_ISL_609537, EPI_ISL_609538, EPI_ISL_609539, EPI_ISL_609540, EPI_ISL_609541, EPI_ISL_609542, EPI_ISL_609543, EPI_ISL_609544, EPI_ISL_609545, EPI_ISL_609546, EPI_ISL_609547, EPI_ISL_609548, EPI_ISL_609549, EPI_ISL_609550, EPI_ISL_609551, EPI_ISL_609552, EPI_ISL_609553, EPI_ISL_609554, EPI_ISL_609555, EPI_ISL_609556, EPI_ISL_609557, EPI_ISL_609558, EPI_ISL_609559, EPI_ISL_609560, EPI_ISL_609561, EPI_ISL_609562, EPI_ISL_609563, EPI_ISL_609564, EPI_ISL_609565, EPI_ISL_609566, EPI_ISL_609567, EPI_ISL_609568, EPI_ISL_609569, EPI_ISL_609570, EPI_ISL_609571, EPI_ISL_609572, EPI_ISL_609573, EPI_ISL_609574, EPI_ISL_609575, EPI_ISL_609576, EPI_ISL_609577, EPI_ISL_609578, EPI_ISL_609579, EPI_ISL_609580, EPI_ISL_609581, EPI_ISL_609582, EPI_ISL_609583, EPI_ISL_609584, EPI_ISL_609585, EPI_ISL_609586, EPI_ISL_609587, EPI_ISL_609588, EPI_ISL_609589, EPI_ISL_609590, EPI_ISL_609591, EPI_ISL_609592, EPI_ISL_609593, EPI_ISL_609594, EPI_ISL_609595, EPI_ISL_609596, EPI_ISL_609597, EPI_ISL_609598, EPI_ISL_609599, EPI_ISL_609601, EPI_ISL_609602, EPI_ISL_609603, EPI_ISL_609604, EPI_ISL_609605, EPI_ISL_609606, EPI_ISL_609607, EPI_ISL_609608, EPI_ISL_609609, EPI_ISL_609610, EPI_ISL_609611, EPI_ISL_609612, EPI_ISL_609613, EPI_ISL_609614, EPI_ISL_609615, EPI_ISL_609616, EPI_ISL_609617, EPI_ISL_609618, EPI_ISL_609619, EPI_ISL_609620, EPI_ISL_609621, EPI_ISL_609622, EPI_ISL_609623, EPI_ISL_609624, EPI_ISL_609625, EPI_ISL_609626, EPI_ISL_609627, EPI_ISL_609628, EPI_ISL_609629, EPI_ISL_609630, EPI_ISL_609631, EPI_ISL_609632, EPI_ISL_609633, EPI_ISL_609634, EPI_ISL_609635, EPI_ISL_609636, EPI_ISL_609637, EPI_ISL_609638, EPI_ISL_609639, EPI_ISL_609640, EPI_ISL_609641, EPI_ISL_609642, EPI_ISL_609643, EPI_ISL_609644, EPI_ISL_609645, EPI_ISL_609646, EPI_ISL_609647, EPI_ISL_609648, EPI_ISL_609649, EPI_ISL_609650, EPI_ISL_609651, EPI_ISL_609652, EPI_ISL_609653, EPI_ISL_609654, EPI_ISL_609655, EPI_ISL_609656, EPI_ISL_609657, EPI_ISL_609658, EPI_ISL_609659, EPI_ISL_609660, EPI_ISL_609661, EPI_ISL_609662, EPI_ISL_609663, EPI_ISL_609664, EPI_ISL_609665, EPI_ISL_609666, EPI_ISL_609667, EPI_ISL_609668, EPI_ISL_609669, EPI_ISL_609670, EPI_ISL_609671, EPI_ISL_609672, EPI_ISL_609673, EPI_ISL_609674, EPI_ISL_609675, EPI_ISL_609676, EPI_ISL_609677, EPI_ISL_609678, EPI_ISL_609679, EPI_ISL_609680, EPI_ISL_609681, EPI_ISL_609682, EPI_ISL_609683, EPI_ISL_609684, EPI_ISL_609685, EPI_ISL_609686, EPI_ISL_609687, EPI_ISL_609688, EPI_ISL_609689, EPI_ISL_609690, EPI_ISL_609691, EPI_ISL_609692, EPI_ISL_609693, EPI_ISL_609694, EPI_ISL_609695, EPI_ISL_609696, EPI_ISL_609697, EPI_ISL_609698, EPI_ISL_609699, EPI_ISL_609700, EPI_ISL_609701, EPI_ISL_609702, EPI_ISL_609703, EPI_ISL_609704, EPI_ISL_609705, EPI_ISL_609706, EPI_ISL_609707, EPI_ISL_609708, EPI_ISL_609709, EPI_ISL_609710, EPI_ISL_609711, EPI_ISL_609712, EPI_ISL_609713, EPI_ISL_609714, EPI_ISL_609715, EPI_ISL_609716, EPI_ISL_609717, EPI_ISL_609718, EPI_ISL_609719, EPI_ISL_609720, EPI_ISL_609721, EPI_ISL_609722, EPI_ISL_609723, EPI_ISL_609724, EPI_ISL_609725, EPI_ISL_609726, EPI_ISL_609727, EPI_ISL_609728, EPI_ISL_609729, EPI_ISL_609730, EPI_ISL_609731, EPI_ISL_609732, EPI_ISL_609733, EPI_ISL_609734, EPI_ISL_609735, EPI_ISL_609736, EPI_ISL_609737, EPI_ISL_609738, EPI_ISL_609739, EPI_ISL_609740, EPI_ISL_609741, EPI_ISL_609742, EPI_ISL_609743, EPI_ISL_609744, EPI_ISL_609745, EPI_ISL_609747, EPI_ISL_609748, EPI_ISL_609749, EPI_ISL_609750, EPI_ISL_609751, EPI_ISL_609752, EPI_ISL_609753, EPI_ISL_609754, EPI_ISL_609755, EPI_ISL_609756, EPI_ISL_609757, EPI_ISL_609758, EPI_ISL_609759, EPI_ISL_609760, EPI_ISL_609761, EPI_ISL_609762, EPI_ISL_609763, EPI_ISL_609764, EPI_ISL_609765, EPI_ISL_609766, EPI_ISL_609767, EPI_ISL_609768, EPI_ISL_609769, EPI_ISL_609770, EPI_ISL_609771, EPI_ISL_609772, EPI_ISL_609773, EPI_ISL_609774, EPI_ISL_609775, EPI_ISL_609776, EPI_ISL_609777, EPI_ISL_609778, EPI_ISL_609779, EPI_ISL_609780, EPI_ISL_609781, EPI_ISL_609782, EPI_ISL_609783, EPI_ISL_609784, EPI_ISL_609785, EPI_ISL_609786, EPI_ISL_609787, EPI_ISL_609788, EPI_ISL_609789, EPI_ISL_609790, EPI_ISL_609791, EPI_ISL_609792, EPI_ISL_609793, EPI_ISL_609794, EPI_ISL_609795, EPI_ISL_609796, EPI_ISL_609797, EPI_ISL_609798, EPI_ISL_609799, EPI_ISL_609800, EPI_ISL_610657, EPI_ISL_610726, EPI_ISL_610785, EPI_ISL_610914, EPI_ISL_625300, EPI_ISL_625432, EPI_ISL_625437, EPI_ISL_625438, EPI_ISL_625439, EPI_ISL_625440, EPI_ISL_625441, EPI_ISL_625442, EPI_ISL_625443, EPI_ISL_625444, EPI_ISL_625445, EPI_ISL_625446, EPI_ISL_625447, EPI_ISL_625448, EPI_ISL_625449, EPI_ISL_625450, EPI_ISL_625451, EPI_ISL_625452, EPI_ISL_625453

| | | | |
|---|---|---|---|
| see above | Lighthouse Lab in Cambridge | Wellcome Sanger Institute for the COVID-19 Genomics UK (COG-UK) consortium | Cordelia Langford; David K. Jackson; Dominic Kwiatkowski; Ewan Harrison; Ian Johnston; John Sillitoe on behalf of the Wellcome Sanger Institute COVID-19 Surveillance Team; John Sillitoe on behalf of the Wellcome Sanger Institute COVID-19 Surveillance Team (http://www.sanger.ac.uk/covid-team); Rob Howes; Roberto Amato; Sonia Goncalves; The Lighthouse Lab in Cambridge and Alex Alderton |

EPI_ISL_587303, EPI_ISL_587386, EPI_ISL_587477, EPI_ISL_588075, EPI_ISL_588125, EPI_ISL_588128, EPI_ISL_588175, EPI_ISL_598970, EPI_ISL_599010, EPI_ISL_599077, EPI_ISL_599166, EPI_ISL_599183, EPI_ISL_599194, EPI_ISL_599201, EPI_ISL_599270, EPI_ISL_599314, EPI_ISL_600139, EPI_ISL_600221, EPI_ISL_822318, EPI_ISL_822319, EPI_ISL_822320

| | | | |
|---|---|---|---|
| see above | Lighthouse Lab in Glasgow | Wellcome Sanger Institute for the COVID-19 Genomics UK (COG-UK) Consortium | Anna Dominiczak and Alex Alderton; Carol Clugston; Cordelia Langford; David Gray; David K. Jackson; Dominic Kwiatkowski; Ewan Harrison; Harper VanSteenhouse; Ian Johnston; John Sillitoe on behalf of the Wellcome Sanger Institute COVID-19 Surveillance Team; Roberto Amato; Sonia Goncalves; Yumi Kasai |

EPI_ISL_587304, EPI_ISL_587305, EPI_ISL_587306, EPI_ISL_587307, EPI_ISL_587308, EPI_ISL_587309, EPI_ISL_587310, EPI_ISL_587312, EPI_ISL_587313, EPI_ISL_587314, EPI_ISL_587316, EPI_ISL_587317, EPI_ISL_587318, EPI_ISL_587319, EPI_ISL_587320, EPI_ISL_587321, EPI_ISL_587322, EPI_ISL_587323, EPI_ISL_587324, EPI_ISL_587325, EPI_ISL_587326, EPI_ISL_587327, EPI_ISL_587328, EPI_ISL_587329, EPI_ISL_587330, EPI_ISL_587331, EPI_ISL_587332, EPI_ISL_587333, EPI_ISL_587334, EPI_ISL_587335, EPI_ISL_587336, EPI_ISL_587337, EPI_ISL_587338, EPI_ISL_587339, EPI_ISL_587340, EPI_ISL_587341, EPI_ISL_587342, EPI_ISL_587343, EPI_ISL_587344, EPI_ISL_587345, EPI_ISL_587346, EPI_ISL_587347, EPI_ISL_587348, EPI_ISL_587349, EPI_ISL_587350, EPI_ISL_587351, EPI_ISL_587352, EPI_ISL_587353, EPI_ISL_587354, EPI_ISL_587355, EPI_ISL_587356, EPI_ISL_587357, EPI_ISL_587358, EPI_ISL_587359, EPI_ISL_587360, EPI_ISL_587361, EPI_ISL_587362, EPI_ISL_587363, EPI_ISL_587364, EPI_ISL_587365, EPI_ISL_587366, EPI_ISL_587367, EPI_ISL_587368, EPI_ISL_587369, EPI_ISL_587370, EPI_ISL_587371, EPI_ISL_587372, EPI_ISL_587373, EPI_ISL_587374, EPI_ISL_587375, EPI_ISL_587376, EPI_ISL_587377, EPI_ISL_587378, EPI_ISL_587379, EPI_ISL_587380, EPI_ISL_587381, EPI_ISL_587382, EPI_ISL_587383, EPI_ISL_587384, EPI_ISL_587385, EPI_ISL_587387, EPI_ISL_587388, EPI_ISL_587389, EPI_ISL_587390, EPI_ISL_587391, EPI_ISL_587392, EPI_ISL_587393, EPI_ISL_587394, EPI_ISL_587395, EPI_ISL_587396, EPI_ISL_587397, EPI_ISL_587398, EPI_ISL_587399, EPI_ISL_587400, EPI_ISL_587401, EPI_ISL_587402, EPI_ISL_587403, EPI_ISL_587404, EPI_ISL_587405, EPI_ISL_587406, EPI_ISL_587407, EPI_ISL_587408, EPI_ISL_587409, EPI_ISL_587410, EPI_ISL_587411, EPI_ISL_587412, EPI_ISL_587413, EPI_ISL_587414, EPI_ISL_587415, EPI_ISL_587416, EPI_ISL_587417, EPI_ISL_587418, EPI_ISL_587419, EPI_ISL_587420, EPI_ISL_587421, EPI_ISL_587422, EPI_ISL_587423, EPI_ISL_587424, EPI_ISL_587425, EPI_ISL_587426, EPI_ISL_587427, EPI_ISL_587428, EPI_ISL_587429, EPI_ISL_587430, EPI_ISL_587431, EPI_ISL_587432, EPI_ISL_587433, EPI_ISL_587434, EPI_ISL_587435, EPI_ISL_587436, EPI_ISL_587437, EPI_ISL_587438, EPI_ISL_587439, EPI_ISL_587440, EPI_ISL_587441, EPI_ISL_587442, EPI_ISL_587443, EPI_ISL_587444, EPI_ISL_587445, EPI_ISL_587446, EPI_ISL_587447, EPI_ISL_587448, EPI_ISL_587449, EPI_ISL_587450, EPI_ISL_587451, EPI_ISL_587453, EPI_ISL_587454, EPI_ISL_587455, EPI_ISL_587456, EPI_ISL_587457, EPI_ISL_587458, EPI_ISL_587459, EPI_ISL_587460, EPI_ISL_587461, EPI_ISL_587462, EPI_ISL_587463, EPI_ISL_587464, EPI_ISL_587465, EPI_ISL_587466, EPI_ISL_587467, EPI_ISL_587468, EPI_ISL_587469, EPI_ISL_587470, EPI_ISL_587471, EPI_ISL_587472, EPI_ISL_587473, EPI_ISL_587474, EPI_ISL_587475, EPI_ISL_587476, EPI_ISL_587478, EPI_ISL_587479, EPI_ISL_587480, EPI_ISL_587481, EPI_ISL_587482, EPI_ISL_587483, EPI_ISL_587484, EPI_ISL_587485, EPI_ISL_587486, EPI_ISL_587487, EPI_ISL_587488, EPI_ISL_587489, EPI_ISL_587490, EPI_ISL_587491, EPI_ISL_587492, EPI_ISL_587493, EPI_ISL_587494, EPI_ISL_587495, EPI_ISL_587496, EPI_ISL_587497, EPI_ISL_587498, EPI_ISL_587499, EPI_ISL_587500, EPI_ISL_587501, EPI_ISL_587502, EPI_ISL_587503, EPI_ISL_587504, EPI_ISL_587505, EPI_ISL_587506, EPI_ISL_587507, EPI_ISL_587508, EPI_ISL_587509, EPI_ISL_587510, EPI_ISL_587511, EPI_ISL_587512, EPI_ISL_587513, EPI_ISL_587514, EPI_ISL_587515, EPI_ISL_587516, EPI_ISL_587517, EPI_ISL_587518, EPI_ISL_587519, EPI_ISL_587520, EPI_ISL_587521, EPI_ISL_587522, EPI_ISL_587523, EPI_ISL_587524, EPI_ISL_587525, EPI_ISL_587526, EPI_ISL_587527, EPI_ISL_587528, EPI_ISL_587529, EPI_ISL_587530, EPI_ISL_587531, EPI_ISL_587532, EPI_ISL_587533, EPI_ISL_587534, EPI_ISL_587535, EPI_ISL_587536, EPI_ISL_587537, EPI_ISL_587538, EPI_ISL_587539, EPI_ISL_587540, EPI_ISL_587541, EPI_ISL_587542, EPI_ISL_587543, EPI_ISL_587544, EPI_ISL_587545, EPI_ISL_587546, EPI_ISL_587547, EPI_ISL_587548, EPI_ISL_587549, EPI_ISL_587550, EPI_ISL_587551, EPI_ISL_587552, EPI_ISL_587553, EPI_ISL_587554, EPI_ISL_587555, EPI_ISL_587556, EPI_ISL_587557, EPI_ISL_587558, EPI_ISL_587559, EPI_ISL_587560, EPI_ISL_587561, EPI_ISL_587562, EPI_ISL_587563, EPI_ISL_587564, EPI_ISL_587565, EPI_ISL_587566, EPI_ISL_587567, EPI_ISL_587568, EPI_ISL_587569, EPI_ISL_587570, EPI_ISL_587571, EPI_ISL_587572, EPI_ISL_587573, EPI_ISL_587574, EPI_ISL_587575, EPI_ISL_587576, EPI_ISL_587577, EPI_ISL_587578, EPI_ISL_587579, EPI_ISL_587580, EPI_ISL_587581, EPI_ISL_587582, EPI_ISL_587583, EPI_ISL_587584, EPI_ISL_587585, EPI_ISL_587586, EPI_ISL_587587, EPI_ISL_587588, EPI_ISL_587589, EPI_ISL_587590, EPI_ISL_587591, EPI_ISL_587592, EPI_ISL_587593, EPI_ISL_587594, EPI_ISL_587595, EPI_ISL_587596, EPI_ISL_587597, EPI_ISL_587598, EPI_ISL_587599, EPI_ISL_587600, EPI_ISL_587601, EPI_ISL_587602, EPI_ISL_587603, EPI_ISL_587604, EPI_ISL_587605, EPI_ISL_587606, EPI_ISL_587607, EPI_ISL_587608, EPI_ISL_587609, EPI_ISL_587610, EPI_ISL_587611, EPI_ISL_587612, EPI_ISL_587613, EPI_ISL_587614, EPI_ISL_587615, EPI_ISL_587616, EPI_ISL_587617, EPI_ISL_587618, EPI_ISL_587619, EPI_ISL_587620, EPI_ISL_587621, EPI_ISL_587622, EPI_ISL_587623, EPI_ISL_587624, EPI_ISL_587625, EPI_ISL_587626, EPI_ISL_587627, EPI_ISL_587628, EPI_ISL_587629, EPI_ISL_587630, EPI_ISL_587631, EPI_ISL_587632, EPI_ISL_587633, EPI_ISL_587634, EPI_ISL_587635, EPI_ISL_587636, EPI_ISL_587637, EPI_ISL_587638, EPI_ISL_587639, EPI_ISL_587640, EPI_ISL_587641, EPI_ISL_587642, EPI_ISL_587643, EPI_ISL_587644, EPI_ISL_587645, EPI_ISL_587646, EPI_ISL_587647, EPI_ISL_587648, EPI_ISL_587649, EPI_ISL_587650, EPI_ISL_587651, EPI_ISL_587652, EPI_ISL_587653, EPI_ISL_587654, EPI_ISL_587655, EPI_ISL_587656, EPI_ISL_587657, EPI_ISL_587658, EPI_ISL_587659, EPI_ISL_587660, EPI_ISL_587661, EPI_ISL_587662, EPI_ISL_587663, EPI_ISL_587664, EPI_ISL_587665, EPI_ISL_587666, EPI_ISL_587667, EPI_ISL_587668, EPI_ISL_587669, EPI_ISL_587670, EPI_ISL_587671, EPI_ISL_587672, EPI_ISL_587673, EPI_ISL_587674, EPI_ISL_587675, EPI_ISL_587676, EPI_ISL_587677, EPI_ISL_587678, EPI_ISL_587679, EPI_ISL_587680, EPI_ISL_587681, EPI_ISL_587682, EPI_ISL_587683, EPI_ISL_587684, EPI_ISL_587685, EPI_ISL_587686, EPI_ISL_587687, EPI_ISL_587688, EPI_ISL_587689, EPI_ISL_587690, EPI_ISL_587691, EPI_ISL_587692, EPI_ISL_587693, EPI_ISL_587694, EPI_ISL_587695, EPI_ISL_587696, EPI_ISL_587697, EPI_ISL_587698, EPI_ISL_587699, EPI_ISL_587700, EPI_ISL_587701, EPI_ISL_587702, EPI_ISL_587703, EPI_ISL_587704, EPI_ISL_587705, EPI_ISL_587706, EPI_ISL_587707, EPI_ISL_587708, EPI_ISL_587709, EPI_ISL_587710, EPI_ISL_588060, EPI_ISL_588061, EPI_ISL_588062, EPI_ISL_588063, EPI_ISL_588065, EPI_ISL_588066, EPI_ISL_588067, EPI_ISL_588068, EPI_ISL_588069, EPI_ISL_588070, EPI_ISL_588072, EPI_ISL_588073, EPI_ISL_588074, EPI_ISL_588076, EPI_ISL_588077, EPI_ISL_588078, EPI_ISL_588079, EPI_ISL_588080, EPI_ISL_588081, EPI_ISL_588082, EPI_ISL_588083, EPI_ISL_588084, EPI_ISL_588086, EPI_ISL_588087, EPI_ISL_588088, EPI_ISL_588089, EPI_ISL_588090, EPI_ISL_588091, EPI_ISL_588092, EPI_ISL_588093, EPI_ISL_588094, EPI_ISL_588095, EPI_ISL_588096, EPI_ISL_588097, EPI_ISL_588098, EPI_ISL_588099, EPI_ISL_588101, EPI_ISL_588102, EPI_ISL_588103, EPI_ISL_588104, EPI_ISL_588105, EPI_ISL_588106, EPI_ISL_588107, EPI_ISL_588108, EPI_ISL_588109, EPI_ISL_588110, EPI_ISL_588111, EPI_ISL_588112, EPI_ISL_588113, EPI_ISL_588114, EPI_ISL_588115, EPI_ISL_588116, EPI_ISL_588117, EPI_ISL_588118, EPI_ISL_588119, EPI_ISL_588120, EPI_ISL_588121, EPI_ISL_588122, EPI_ISL_588123, EPI_ISL_588124, EPI_ISL_588126, EPI_ISL_588127, EPI_ISL_588129, EPI_ISL_588130, EPI_ISL_588131, EPI_ISL_588132, EPI_ISL_588133, EPI_ISL_588134, EPI_ISL_588135, EPI_ISL_588136, EPI_ISL_588137, EPI_ISL_588138, EPI_ISL_588139, EPI_ISL_588140, EPI_ISL_588141, EPI_ISL_588142, EPI_ISL_588143, EPI_ISL_588144, EPI_ISL_588145, EPI_ISL_588146, EPI_ISL_588147, EPI_ISL_588148, EPI_ISL_588149, EPI_ISL_588150, EPI_ISL_588151, EPI_ISL_588152, EPI_ISL_588153, EPI_ISL_588154, EPI_ISL_588155, EPI_ISL_588156, EPI_ISL_588157, EPI_ISL_588158, EPI_ISL_588159, EPI_ISL_588160, EPI_ISL_588161, EPI_ISL_588162, EPI_ISL_588163, EPI_ISL_588164, EPI_ISL_588165, EPI_ISL_588168, EPI_ISL_588169, EPI_ISL_588171, EPI_ISL_588173, EPI_ISL_588174, EPI_ISL_588176, EPI_ISL_588177, EPI_ISL_588178, EPI_ISL_588179, EPI_ISL_588180, EPI_ISL_588182, EPI_ISL_588183, EPI_ISL_588184, EPI_ISL_588186, EPI_ISL_588187, EPI_ISL_588188, EPI_ISL_588191, EPI_ISL_588192, EPI_ISL_588193, EPI_ISL_588194, EPI_ISL_588195, EPI_ISL_588196, EPI_ISL_588197, EPI_ISL_588198, EPI_ISL_588199, EPI_ISL_588201, EPI_ISL_588202, EPI_ISL_588203, EPI_ISL_588204, EPI_ISL_588207, EPI_ISL_588209, EPI_ISL_588210, EPI_ISL_588211, EPI_ISL_588212, EPI_ISL_588213, EPI_ISL_588214, EPI_ISL_588215, EPI_ISL_588216, EPI_ISL_588217, EPI_ISL_588218, EPI_ISL_588219, EPI_ISL_588220, EPI_ISL_588221, EPI_ISL_588222, EPI_ISL_588223, EPI_ISL_588224, EPI_ISL_588225, EPI_ISL_588226, EPI_ISL_588227, EPI_ISL_588228, EPI_ISL_588229, EPI_ISL_588230, EPI_ISL_588232, EPI_ISL_588234, EPI_ISL_588235, EPI_ISL_588236, EPI_ISL_588237, EPI_ISL_588238, EPI_ISL_588239, EPI_ISL_588240, EPI_ISL_588241, EPI_ISL_588242, EPI_ISL_588243, EPI_ISL_588244, EPI_ISL_588245, EPI_ISL_588246, EPI_ISL_588247, EPI_ISL_588248, EPI_ISL_588251, EPI_ISL_588252, EPI_ISL_588253, EPI_ISL_588254, EPI_ISL_588256, EPI_ISL_588257, EPI_ISL_588258, EPI_ISL_588259, EPI_ISL_588261, EPI_ISL_588262, EPI_ISL_588263, EPI_ISL_588264, EPI_ISL_588265, EPI_ISL_588266, EPI_ISL_588267, EPI_ISL_588269, EPI_ISL_588270, EPI_ISL_588272, EPI_ISL_588273, EPI_ISL_588274, EPI_ISL_597333, EPI_ISL_597359, EPI_ISL_597501, EPI_ISL_598831, EPI_ISL_598832, EPI_ISL_598833, EPI_ISL_598834, EPI_ISL_598835, EPI_ISL_598836, EPI_ISL_598837, EPI_ISL_598838, EPI_ISL_598839, EPI_ISL_598840, EPI_ISL_598841, EPI_ISL_598842, EPI_ISL_598843, EPI_ISL_598844, EPI_ISL_598845, EPI_ISL_598846, EPI_ISL_598847, EPI_ISL_598848, EPI_ISL_598849, EPI_ISL_598850, EPI_ISL_598851, EPI_ISL_598852, EPI_ISL_598853, EPI_ISL_598854, EPI_ISL_598855, EPI_ISL_598856, EPI_ISL_598857, EPI_ISL_598858, EPI_ISL_598859, EPI_ISL_598860, EPI_ISL_598861, EPI_ISL_598862, EPI_ISL_598863, EPI_ISL_598864, EPI_ISL_598865, EPI_ISL_598866, EPI_ISL_598867, EPI_ISL_598868, EPI_ISL_598869, EPI_ISL_598870, EPI_ISL_598871, EPI_ISL_598872, EPI_ISL_598873, EPI_ISL_598874, EPI_ISL_598875, EPI_ISL_598876, EPI_ISL_598877, EPI_ISL_598878, EPI_ISL_598879, EPI_ISL_598880, EPI_ISL_598881, EPI_ISL_598882, EPI_ISL_598883, EPI_ISL_598884, EPI_ISL_598885, EPI_ISL_598886, EPI_ISL_598887, EPI_ISL_598888, EPI_ISL_598889, EPI_ISL_598890, EPI_ISL_598891, EPI_ISL_598892, EPI_ISL_598893, EPI_ISL_598894, EPI_ISL_598895, EPI_ISL_598896, EPI_ISL_598897, EPI_ISL_598898, EPI_ISL_598899, EPI_ISL_598900, EPI_ISL_598901, EPI_ISL_598902, EPI_ISL_598903, EPI_ISL_598904, EPI_ISL_598905, EPI_ISL_598906, EPI_ISL_598907, EPI_ISL_598908, EPI_ISL_598909, EPI_ISL_598910, EPI_ISL_598911, EPI_ISL_598912, EPI_ISL_598913, EPI_ISL_598914, EPI_ISL_598915, EPI_ISL_598916, EPI_ISL_598917, EPI_ISL_598918, EPI_ISL_598919, EPI_ISL_598920, EPI_ISL_598921, EPI_ISL_598922, EPI_ISL_598923, EPI_ISL_598924, EPI_ISL_598925, EPI_ISL_598926, EPI_ISL_598927, EPI_ISL_598928, EPI_ISL_598929, EPI_ISL_598930, EPI_ISL_598931, EPI_ISL_598932, EPI_ISL_598933, EPI_ISL_598934, EPI_ISL_598935, EPI_ISL_598936, EPI_ISL_598937, EPI_ISL_598938, EPI_ISL_598939, EPI_ISL_598940, EPI_ISL_598941, EPI_ISL_598942, EPI_ISL_598943, EPI_ISL_598944, EPI_ISL_598945, EPI_ISL_598946, EPI_ISL_598947, EPI_ISL_598948, EPI_ISL_598949, EPI_ISL_598950, EPI_ISL_598951, EPI_ISL_598952, EPI_ISL_598953, EPI_ISL_598954, EPI_ISL_598955, EPI_ISL_598956, EPI_ISL_598957, EPI_ISL_598958, EPI_ISL_598959, EPI_ISL_598960, EPI_ISL_598961, EPI_ISL_598962, EPI_ISL_598963, EPI_ISL_598964, EPI_ISL_598965, EPI_ISL_598966, EPI_ISL_598967, EPI_ISL_598968, EPI_ISL_598969, EPI_ISL_598971, EPI_ISL_598972, EPI_ISL_598973, EPI_ISL_598974, EPI_ISL_598975, EPI_ISL_598976, EPI_ISL_598977, EPI_ISL_598978, EPI_ISL_598979, EPI_ISL_598980, EPI_ISL_598981, EPI_ISL_598982, EPI_ISL_598983, EPI_ISL_598984, EPI_ISL_598985, EPI_ISL_598986, EPI_ISL_598987, EPI_ISL_598988, EPI_ISL_598989, EPI_ISL_598990, EPI_ISL_598991, EPI_ISL_598992, EPI_ISL_598993, EPI_ISL_598994, EPI_ISL_598995, EPI_ISL_598996, EPI_ISL_598997, EPI_ISL_598998, EPI_ISL_598999, EPI_ISL_599000, EPI_ISL_599001, EPI_ISL_599002, EPI_ISL_599003, EPI_ISL_599004, EPI_ISL_599005, EPI_ISL_599006, EPI_ISL_599007, EPI_ISL_599008, EPI_ISL_599009, EPI_ISL_599011, EPI_ISL_599012, EPI_ISL_599013, EPI_ISL_599014, EPI_ISL_599015, EPI_ISL_599016, EPI_ISL_599017, EPI_ISL_599018, EPI_ISL_599019, EPI_ISL_599020, EPI_ISL_599021, EPI_ISL_599022, EPI_ISL_599023, EPI_ISL_599024, EPI_ISL_599025, EPI_ISL_599026, EPI_ISL_599027, EPI_ISL_599028, EPI_ISL_599029, EPI_ISL_599030, EPI_ISL_599031, EPI_ISL_599032, EPI_ISL_599033, EPI_ISL_599034, EPI_ISL_599035, EPI_ISL_599036, EPI_ISL_599037, EPI_ISL_599038, EPI_ISL_599039, EPI_ISL_599040, EPI_ISL_599041, EPI_ISL_599042, EPI_ISL_599043, EPI_ISL_599044, EPI_ISL_599045, EPI_ISL_599046, EPI_ISL_599047, EPI_ISL_599048, EPI_ISL_599049, EPI_ISL_599050, EPI_ISL_599051, EPI_ISL_599052, EPI_ISL_599053, EPI_ISL_599054, EPI_ISL_599055, EPI_ISL_599056, EPI_ISL_599057, EPI_ISL_599058, EPI_ISL_599059, EPI_ISL_599060, EPI_ISL_599061, EPI_ISL_599062, EPI_ISL_599063, EPI_ISL_599064, EPI_ISL_599065, EPI_ISL_599066, EPI_ISL_599067, EPI_ISL_599068, EPI_ISL_599069, EPI_ISL_599070, EPI_ISL_599071, EPI_ISL_599072, EPI_ISL_599073, EPI_ISL_599074, EPI_ISL_599075, EPI_ISL_599076, EPI_ISL_599078, EPI_ISL_599079, EPI_ISL_599080, EPI_ISL_599081, EPI_ISL_599082, EPI_ISL_599083, EPI_ISL_599084, EPI_ISL_599085, EPI_ISL_599086, EPI_ISL_599087, EPI_ISL_599088, EPI_ISL_599089, EPI_ISL_599090, EPI_ISL_599091, EPI_ISL_599092, EPI_ISL_599093, EPI_ISL_599094, EPI_ISL_599095, EPI_ISL_599096, EPI_ISL_599097, EPI_ISL_599098, EPI_ISL_599099, EPI_ISL_599100, EPI_ISL_599101, EPI_ISL_599102, EPI_ISL_599103, EPI_ISL_599104, EPI_ISL_599105, EPI_ISL_599106, EPI_ISL_599107, EPI_ISL_599108, EPI_ISL_599109, EPI_ISL_599110, EPI_ISL_599111, EPI_ISL_599112, EPI_ISL_599113, EPI_ISL_599114, EPI_ISL_599115, EPI_ISL_599116, EPI_ISL_599117, EPI_ISL_599118, EPI_ISL_599119, EPI_ISL_599120, EPI_ISL_599121, EPI_ISL_599122, EPI_ISL_599123, EPI_ISL_599124, EPI_ISL_599125, EPI_ISL_599126, EPI_ISL_599127, EPI_ISL_599128, EPI_ISL_599129, EPI_ISL_599130, EPI_ISL_599131, EPI_ISL_599132, EPI_ISL_599133, EPI_ISL_599134, EPI_ISL_599135, EPI_ISL_599136, EPI_ISL_599137, EPI_ISL_599138, EPI_ISL_599139, EPI_ISL_599140, EPI_ISL_599141, EPI_ISL_599142, EPI_ISL_599143, EPI_ISL_599144, EPI_ISL_599145, EPI_ISL_599146, EPI_ISL_599147, EPI_ISL_599148, EPI_ISL_599149, EPI_ISL_599150, EPI_ISL_599151, EPI_ISL_599152, EPI_ISL_599153,

EPI_ISL_599154, EPI_ISL_599155, EPI_ISL_599156, EPI_ISL_599157, EPI_ISL_599158, EPI_ISL_599159, EPI_ISL_599160, EPI_ISL_599161, EPI_ISL_599162, EPI_ISL_599163, EPI_ISL_599164, EPI_ISL_599165, EPI_ISL_599167, EPI_ISL_599168, EPI_ISL_599169, EPI_ISL_599170, EPI_ISL_599171, EPI_ISL_599172, EPI_ISL_599173, EPI_ISL_599174, EPI_ISL_599175, EPI_ISL_599176, EPI_ISL_599177, EPI_ISL_599178, EPI_ISL_599179, EPI_ISL_599180, EPI_ISL_599181, EPI_ISL_599182, EPI_ISL_599184, EPI_ISL_599185, EPI_ISL_599186, EPI_ISL_599187, EPI_ISL_599188, EPI_ISL_599189, EPI_ISL_599190, EPI_ISL_599191, EPI_ISL_599192, EPI_ISL_599193, EPI_ISL_599195, EPI_ISL_599196, EPI_ISL_599197, EPI_ISL_599198, EPI_ISL_599199, EPI_ISL_599200, EPI_ISL_599202, EPI_ISL_599203, EPI_ISL_599204, EPI_ISL_599205, EPI_ISL_599206, EPI_ISL_599207, EPI_ISL_599208, EPI_ISL_599209, EPI_ISL_599210, EPI_ISL_599211, EPI_ISL_599212, EPI_ISL_599213, EPI_ISL_599214, EPI_ISL_599215, EPI_ISL_599216, EPI_ISL_599217, EPI_ISL_599218, EPI_ISL_599219, EPI_ISL_599220, EPI_ISL_599221, EPI_ISL_599222, EPI_ISL_599223, EPI_ISL_599224, EPI_ISL_599225, EPI_ISL_599226, EPI_ISL_599227, EPI_ISL_599228, EPI_ISL_599229, EPI_ISL_599230, EPI_ISL_599231, EPI_ISL_599232, EPI_ISL_599233, EPI_ISL_599234, EPI_ISL_599235, EPI_ISL_599236, EPI_ISL_599237, EPI_ISL_599238, EPI_ISL_599239, EPI_ISL_599240, EPI_ISL_599241, EPI_ISL_599242, EPI_ISL_599243, EPI_ISL_599244, EPI_ISL_599245, EPI_ISL_599246, EPI_ISL_599247, EPI_ISL_599248, EPI_ISL_599249, EPI_ISL_599250, EPI_ISL_599251, EPI_ISL_599252, EPI_ISL_599253, EPI_ISL_599254, EPI_ISL_599255, EPI_ISL_599256, EPI_ISL_599257, EPI_ISL_599258, EPI_ISL_599259, EPI_ISL_599260, EPI_ISL_599261, EPI_ISL_599262, EPI_ISL_599263, EPI_ISL_599264, EPI_ISL_599265, EPI_ISL_599266, EPI_ISL_599267, EPI_ISL_599268, EPI_ISL_599269, EPI_ISL_599271, EPI_ISL_599272, EPI_ISL_599273, EPI_ISL_599274, EPI_ISL_599275, EPI_ISL_599276, EPI_ISL_599277, EPI_ISL_599278, EPI_ISL_599279, EPI_ISL_599280, EPI_ISL_599281, EPI_ISL_599282, EPI_ISL_599283, EPI_ISL_599284, EPI_ISL_599285, EPI_ISL_599286, EPI_ISL_599287, EPI_ISL_599288, EPI_ISL_599289, EPI_ISL_599290, EPI_ISL_599291, EPI_ISL_599292, EPI_ISL_599293, EPI_ISL_599294, EPI_ISL_599295, EPI_ISL_599296, EPI_ISL_599297, EPI_ISL_599298, EPI_ISL_599299, EPI_ISL_599300, EPI_ISL_599301, EPI_ISL_599302, EPI_ISL_599303, EPI_ISL_599304, EPI_ISL_599305, EPI_ISL_599306, EPI_ISL_599307, EPI_ISL_599308, EPI_ISL_599309, EPI_ISL_599310, EPI_ISL_599311, EPI_ISL_599312, EPI_ISL_599313, EPI_ISL_599315, EPI_ISL_599316, EPI_ISL_599317, EPI_ISL_599318, EPI_ISL_599319, EPI_ISL_599320, EPI_ISL_599321, EPI_ISL_599322, EPI_ISL_599323, EPI_ISL_599324, EPI_ISL_599325, EPI_ISL_599326, EPI_ISL_599327, EPI_ISL_599328, EPI_ISL_599329, EPI_ISL_599330, EPI_ISL_599331, EPI_ISL_599332, EPI_ISL_599333, EPI_ISL_599334, EPI_ISL_599335, EPI_ISL_599336, EPI_ISL_599337, EPI_ISL_599338, EPI_ISL_599339, EPI_ISL_599340, EPI_ISL_599341, EPI_ISL_599342, EPI_ISL_599343, EPI_ISL_599344, EPI_ISL_599345, EPI_ISL_599346, EPI_ISL_599347, EPI_ISL_599348, EPI_ISL_599349, EPI_ISL_599350, EPI_ISL_599351, EPI_ISL_599352, EPI_ISL_599353, EPI_ISL_599354, EPI_ISL_599355, EPI_ISL_599356, EPI_ISL_599357, EPI_ISL_599358, EPI_ISL_599359, EPI_ISL_599360, EPI_ISL_599361, EPI_ISL_599362, EPI_ISL_599363, EPI_ISL_599364, EPI_ISL_599365, EPI_ISL_599366, EPI_ISL_599367, EPI_ISL_599368, EPI_ISL_599369, EPI_ISL_599370, EPI_ISL_599371, EPI_ISL_599372, EPI_ISL_599373, EPI_ISL_599374, EPI_ISL_599375, EPI_ISL_599376, EPI_ISL_599377, EPI_ISL_599378, EPI_ISL_599379, EPI_ISL_599380, EPI_ISL_599381, EPI_ISL_599382, EPI_ISL_599383, EPI_ISL_599384, EPI_ISL_599385, EPI_ISL_599386, EPI_ISL_599387, EPI_ISL_599388, EPI_ISL_599389, EPI_ISL_599390, EPI_ISL_599391, EPI_ISL_599392, EPI_ISL_599393, EPI_ISL_599394, EPI_ISL_599395, EPI_ISL_599396, EPI_ISL_599397, EPI_ISL_599398, EPI_ISL_599399, EPI_ISL_599400, EPI_ISL_599401, EPI_ISL_599402, EPI_ISL_599403, EPI_ISL_599404, EPI_ISL_599405, EPI_ISL_599406, EPI_ISL_599407, EPI_ISL_599408, EPI_ISL_599409, EPI_ISL_599410, EPI_ISL_599411, EPI_ISL_599412, EPI_ISL_599413, EPI_ISL_599414, EPI_ISL_599415, EPI_ISL_599416, EPI_ISL_599417, EPI_ISL_599418, EPI_ISL_599419, EPI_ISL_599420, EPI_ISL_599421, EPI_ISL_599422, EPI_ISL_599423, EPI_ISL_599424, EPI_ISL_599425, EPI_ISL_599426, EPI_ISL_599427, EPI_ISL_599428, EPI_ISL_599429, EPI_ISL_599430, EPI_ISL_599431, EPI_ISL_599432, EPI_ISL_599433, EPI_ISL_599434, EPI_ISL_599435, EPI_ISL_599436, EPI_ISL_599437, EPI_ISL_599438, EPI_ISL_599439, EPI_ISL_599440, EPI_ISL_599441, EPI_ISL_599442, EPI_ISL_599443, EPI_ISL_599444, EPI_ISL_599445, EPI_ISL_599446, EPI_ISL_599447, EPI_ISL_599448, EPI_ISL_599449, EPI_ISL_599450, EPI_ISL_599451, EPI_ISL_599452, EPI_ISL_599453, EPI_ISL_599454, EPI_ISL_599455, EPI_ISL_599456, EPI_ISL_599457, EPI_ISL_599458, EPI_ISL_599459, EPI_ISL_599460, EPI_ISL_599461, EPI_ISL_599462, EPI_ISL_599463, EPI_ISL_599464, EPI_ISL_599465, EPI_ISL_599466, EPI_ISL_599467, EPI_ISL_599468, EPI_ISL_599469, EPI_ISL_599470, EPI_ISL_599471, EPI_ISL_599472, EPI_ISL_599473, EPI_ISL_599474, EPI_ISL_599475, EPI_ISL_599476, EPI_ISL_599477, EPI_ISL_599478, EPI_ISL_599479, EPI_ISL_599480, EPI_ISL_599481, EPI_ISL_599482, EPI_ISL_599483, EPI_ISL_600128, EPI_ISL_600129, EPI_ISL_600130, EPI_ISL_600131, EPI_ISL_600132, EPI_ISL_600133, EPI_ISL_600134, EPI_ISL_600135, EPI_ISL_600136, EPI_ISL_600137, EPI_ISL_600138, EPI_ISL_600140, EPI_ISL_600141, EPI_ISL_600142, EPI_ISL_600143, EPI_ISL_600144, EPI_ISL_600145, EPI_ISL_600146, EPI_ISL_600147, EPI_ISL_600148, EPI_ISL_600149, EPI_ISL_600150, EPI_ISL_600151, EPI_ISL_600152, EPI_ISL_600153, EPI_ISL_600154, EPI_ISL_600155, EPI_ISL_600156, EPI_ISL_600157, EPI_ISL_600158, EPI_ISL_600159, EPI_ISL_600160, EPI_ISL_600161, EPI_ISL_600162, EPI_ISL_600163, EPI_ISL_600164, EPI_ISL_600165, EPI_ISL_600166, EPI_ISL_600167, EPI_ISL_600168, EPI_ISL_600169, EPI_ISL_600170, EPI_ISL_600171, EPI_ISL_600172, EPI_ISL_600173, EPI_ISL_600174, EPI_ISL_600175, EPI_ISL_600176, EPI_ISL_600177, EPI_ISL_600178, EPI_ISL_600179, EPI_ISL_600180, EPI_ISL_600181, EPI_ISL_600182, EPI_ISL_600183, EPI_ISL_600184, EPI_ISL_600185, EPI_ISL_600186, EPI_ISL_600187, EPI_ISL_600188, EPI_ISL_600189, EPI_ISL_600190, EPI_ISL_600191, EPI_ISL_600192, EPI_ISL_600193, EPI_ISL_600194, EPI_ISL_600195, EPI_ISL_600196, EPI_ISL_600197, EPI_ISL_600198, EPI_ISL_600199, EPI_ISL_600200, EPI_ISL_600201, EPI_ISL_600202, EPI_ISL_600203, EPI_ISL_600204, EPI_ISL_600205, EPI_ISL_600206, EPI_ISL_600207, EPI_ISL_600208, EPI_ISL_600209, EPI_ISL_600210, EPI_ISL_600211, EPI_ISL_600212, EPI_ISL_600213, EPI_ISL_600214, EPI_ISL_600215, EPI_ISL_600216, EPI_ISL_600217, EPI_ISL_600218, EPI_ISL_600219, EPI_ISL_600220, EPI_ISL_600222, EPI_ISL_600223, EPI_ISL_600224, EPI_ISL_600225, EPI_ISL_600226, EPI_ISL_600227, EPI_ISL_600228, EPI_ISL_600229, EPI_ISL_600230, EPI_ISL_600231, EPI_ISL_600232, EPI_ISL_600233, EPI_ISL_600234, EPI_ISL_600235, EPI_ISL_600236, EPI_ISL_600237, EPI_ISL_600238, EPI_ISL_600239, EPI_ISL_600240, EPI_ISL_600241, EPI_ISL_600242, EPI_ISL_600243, EPI_ISL_600244, EPI_ISL_600245, EPI_ISL_600246, EPI_ISL_600247, EPI_ISL_600248, EPI_ISL_600249, EPI_ISL_600250, EPI_ISL_600251, EPI_ISL_600252, EPI_ISL_600253, EPI_ISL_600254, EPI_ISL_600255, EPI_ISL_600256, EPI_ISL_600257, EPI_ISL_600258, EPI_ISL_600259, EPI_ISL_600260, EPI_ISL_600261, EPI_ISL_600262, EPI_ISL_600263, EPI_ISL_600264, EPI_ISL_600265, EPI_ISL_600266, EPI_ISL_600267, EPI_ISL_600268, EPI_ISL_600269, EPI_ISL_600270, EPI_ISL_600271, EPI_ISL_600272, EPI_ISL_600273, EPI_ISL_600274, EPI_ISL_600275, EPI_ISL_600276, EPI_ISL_600277, EPI_ISL_600278, EPI_ISL_600279, EPI_ISL_600280, EPI_ISL_600281, EPI_ISL_600282, EPI_ISL_600283, EPI_ISL_600284, EPI_ISL_600285, EPI_ISL_600286, EPI_ISL_600287, EPI_ISL_600288, EPI_ISL_600289, EPI_ISL_600290, EPI_ISL_600291, EPI_ISL_600292, EPI_ISL_600293, EPI_ISL_600294, EPI_ISL_600295, EPI_ISL_600296, EPI_ISL_600297, EPI_ISL_600298, EPI_ISL_600299, EPI_ISL_600300, EPI_ISL_600301, EPI_ISL_600302, EPI_ISL_600303, EPI_ISL_600304, EPI_ISL_600305, EPI_ISL_600306, EPI_ISL_600307, EPI_ISL_600308, EPI_ISL_600309, EPI_ISL_600310, EPI_ISL_600311, EPI_ISL_600312, EPI_ISL_600313, EPI_ISL_600314, EPI_ISL_600315, EPI_ISL_600316, EPI_ISL_600317, EPI_ISL_600318, EPI_ISL_600319, EPI_ISL_600320, EPI_ISL_600321, EPI_ISL_600322, EPI_ISL_600323, EPI_ISL_600324, EPI_ISL_600325, EPI_ISL_600326, EPI_ISL_600327, EPI_ISL_600328, EPI_ISL_600329, EPI_ISL_600330, EPI_ISL_600331, EPI_ISL_600332, EPI_ISL_600333, EPI_ISL_600334, EPI_ISL_600335, EPI_ISL_600336, EPI_ISL_600337, EPI_ISL_600338, EPI_ISL_600339, EPI_ISL_600340, EPI_ISL_600341, EPI_ISL_600342, EPI_ISL_600343, EPI_ISL_600344, EPI_ISL_600345, EPI_ISL_600346, EPI_ISL_600347, EPI_ISL_600348, EPI_ISL_600349, EPI_ISL_600350, EPI_ISL_600351, EPI_ISL_600352, EPI_ISL_600353, EPI_ISL_600354, EPI_ISL_600355, EPI_ISL_600356, EPI_ISL_600357, EPI_ISL_600358, EPI_ISL_600359, EPI_ISL_600360, EPI_ISL_600361, EPI_ISL_600362, EPI_ISL_600363, EPI_ISL_600364, EPI_ISL_600365, EPI_ISL_600366, EPI_ISL_600367, EPI_ISL_600368, EPI_ISL_600369, EPI_ISL_600370, EPI_ISL_600371, EPI_ISL_600372, EPI_ISL_600373, EPI_ISL_600374, EPI_ISL_600375, EPI_ISL_600376, EPI_ISL_600377, EPI_ISL_600378, EPI_ISL_600379, EPI_ISL_600380, EPI_ISL_600381, EPI_ISL_600382, EPI_ISL_600383, EPI_ISL_600384, EPI_ISL_600385, EPI_ISL_600386, EPI_ISL_600387, EPI_ISL_600388, EPI_ISL_600389, EPI_ISL_600390, EPI_ISL_600391, EPI_ISL_600392, EPI_ISL_600393, EPI_ISL_600394, EPI_ISL_600395, EPI_ISL_600396, EPI_ISL_600397, EPI_ISL_600398, EPI_ISL_600399, EPI_ISL_600400, EPI_ISL_600401, EPI_ISL_600402, EPI_ISL_600403, EPI_ISL_600404, EPI_ISL_600405, EPI_ISL_600406, EPI_ISL_600407, EPI_ISL_600408, EPI_ISL_600409, EPI_ISL_600410, EPI_ISL_600411, EPI_ISL_600412, EPI_ISL_600413, EPI_ISL_600414, EPI_ISL_600415, EPI_ISL_600416, EPI_ISL_600417, EPI_ISL_600418, EPI_ISL_600419, EPI_ISL_600420, EPI_ISL_600421, EPI_ISL_600422, EPI_ISL_600423, EPI_ISL_600424, EPI_ISL_600425, EPI_ISL_600426, EPI_ISL_600427, EPI_ISL_600447, EPI_ISL_600448, EPI_ISL_600450, EPI_ISL_600452, EPI_ISL_600453, EPI_ISL_600455, EPI_ISL_600457, EPI_ISL_600458, EPI_ISL_600460, EPI_ISL_600462, EPI_ISL_600464, EPI_ISL_600465, EPI_ISL_600467, EPI_ISL_600469, EPI_ISL_600470, EPI_ISL_600472, EPI_ISL_600474, EPI_ISL_600475, EPI_ISL_600477, EPI_ISL_600479, EPI_ISL_600481, EPI_ISL_600482, EPI_ISL_600483, EPI_ISL_600485, EPI_ISL_600486, EPI_ISL_600488, EPI_ISL_600490, EPI_ISL_600491, EPI_ISL_600493, EPI_ISL_600494, EPI_ISL_600496, EPI_ISL_600497, EPI_ISL_600499, EPI_ISL_600501, EPI_ISL_600503, EPI_ISL_600504, EPI_ISL_600506, EPI_ISL_600508, EPI_ISL_600509, EPI_ISL_600511, EPI_ISL_600513, EPI_ISL_600514, EPI_ISL_600516, EPI_ISL_600518, EPI_ISL_600520, EPI_ISL_600539, EPI_ISL_600540, EPI_ISL_600542, EPI_ISL_600544, EPI_ISL_600545, EPI_ISL_600547, EPI_ISL_600548, EPI_ISL_600550, EPI_ISL_600551, EPI_ISL_600553, EPI_ISL_600555, EPI_ISL_600556, EPI_ISL_600558, EPI_ISL_600560, EPI_ISL_600561, EPI_ISL_600563, EPI_ISL_600565, EPI_ISL_600566, EPI_ISL_600568, EPI_ISL_600569, EPI_ISL_600571, EPI_ISL_600572, EPI_ISL_600573, EPI_ISL_600574, EPI_ISL_600575, EPI_ISL_600576, EPI_ISL_600577, EPI_ISL_600578, EPI_ISL_600579, EPI_ISL_600580, EPI_ISL_600581, EPI_ISL_600582, EPI_ISL_600583, EPI_ISL_600584, EPI_ISL_600585, EPI_ISL_600586, EPI_ISL_600587, EPI_ISL_600588, EPI_ISL_600589, EPI_ISL_600590, EPI_ISL_600591, EPI_ISL_600592, EPI_ISL_600593, EPI_ISL_600594, EPI_ISL_600595, EPI_ISL_600596, EPI_ISL_600597, EPI_ISL_600598, EPI_ISL_600599, EPI_ISL_600600, EPI_ISL_600601, EPI_ISL_600602, EPI_ISL_600603, EPI_ISL_600604, EPI_ISL_600605, EPI_ISL_600606, EPI_ISL_600607, EPI_ISL_600608, EPI_ISL_600609, EPI_ISL_600610, EPI_ISL_600611, EPI_ISL_600612, EPI_ISL_600613, EPI_ISL_600614, EPI_ISL_600616, EPI_ISL_600617, EPI_ISL_600618

| | | | |
|---|---|---|---|
| see above | Lighthouse Lab in Glasgow | Wellcome Sanger Institute for the COVID-19 Genomics UK (COG-UK) consortium | Anna Dominiczak and Alex Alderton; Carol Clugston; Cordelia Langford; David Gray; David K. Jackson; Dominic Kwiatkowski; Ewan Harrison; Harper VanSteenhouse; Ian Johnston; John Sillitoe on behalf of the Wellcome Sanger Institute COVID-19 Surveillance Team; John Sillitoe on behalf of the Wellcome Sanger Institute COVID-19 Surveillance Team (http://www.sanger.ac.uk/covid-team); Roberto Amato; Sonia Goncalves; Yumi Kasai |

EPI_ISL_598249, EPI_ISL_598305, EPI_ISL_598330, EPI_ISL_598331, EPI_ISL_598361, EPI_ISL_598408, EPI_ISL_598433, EPI_ISL_598436, EPI_ISL_598606, EPI_ISL_598638, EPI_ISL_598667, EPI_ISL_598821, EPI_ISL_599845, EPI_ISL_599888, EPI_ISL_599948, EPI_ISL_599960, EPI_ISL_600056, EPI_ISL_608748, EPI_ISL_608834, EPI_ISL_609230, EPI_ISL_609267, EPI_ISL_609302, EPI_ISL_709942, EPI_ISL_709944, EPI_ISL_709947, EPI_ISL_822309

| | | | |
|---|---|---|---|
| see above | Lighthouse Lab in Milton Keynes | Wellcome Sanger Institute for the COVID-19 Genomics UK (COG-UK) Consortium | Cordelia Langford; David K. Jackson; Dominic Kwiatkowski; Ewan Harrison; Ian Johnston; John Sillitoe on behalf of the Wellcome Sanger Institute COVID-19 Surveillance Team; John Sillitoe on behalf of the Wellcome Sanger Institute COVID-19 Surveillance Team (http://www.sanger.ac.uk/covid-team); Roberto Amato; Sonia Goncalves; The Lighthouse Lab in Milton Keynes and Alex Alderton |

EPI_ISL_580826, EPI_ISL_598250, EPI_ISL_598251, EPI_ISL_598252, EPI_ISL_598253, EPI_ISL_598254, EPI_ISL_598255, EPI_ISL_598256, EPI_ISL_598257, EPI_ISL_598258, EPI_ISL_598259, EPI_ISL_598260, EPI_ISL_598261, EPI_ISL_598262, EPI_ISL_598263, EPI_ISL_598264, EPI_ISL_598265, EPI_ISL_598266, EPI_ISL_598267, EPI_ISL_598268, EPI_ISL_598269, EPI_ISL_598270, EPI_ISL_598271, EPI_ISL_598272, EPI_ISL_598273, EPI_ISL_598274, EPI_ISL_598275, EPI_ISL_598276, EPI_ISL_598277, EPI_ISL_598278, EPI_ISL_598279, EPI_ISL_598280, EPI_ISL_598281, EPI_ISL_598282, EPI_ISL_598283, EPI_ISL_598284, EPI_ISL_598285, EPI_ISL_598286, EPI_ISL_598287, EPI_ISL_598288, EPI_ISL_598289, EPI_ISL_598290, EPI_ISL_598291, EPI_ISL_598292, EPI_ISL_598293, EPI_ISL_598294, EPI_ISL_598295, EPI_ISL_598296, EPI_ISL_598297, EPI_ISL_598298, EPI_ISL_598299, EPI_ISL_598300, EPI_ISL_598301, EPI_ISL_598302, EPI_ISL_598303, EPI_ISL_598304, EPI_ISL_598306, EPI_ISL_598307, EPI_ISL_598308, EPI_ISL_598309, EPI_ISL_598310, EPI_ISL_598311, EPI_ISL_598312, EPI_ISL_598313, EPI_ISL_598314, EPI_ISL_598315, EPI_ISL_598316, EPI_ISL_598317, EPI_ISL_598318, EPI_ISL_598319, EPI_ISL_598320, EPI_ISL_598321, EPI_ISL_598322, EPI_ISL_598323, EPI_ISL_598324, EPI_ISL_598325, EPI_ISL_598326, EPI_ISL_598327, EPI_ISL_598328, EPI_ISL_598329, EPI_ISL_598332, EPI_ISL_598333, EPI_ISL_598334, EPI_ISL_598335, EPI_ISL_598336, EPI_ISL_598337, EPI_ISL_598338, EPI_ISL_598339, EPI_ISL_598340, EPI_ISL_598341, EPI_ISL_598342, EPI_ISL_598343, EPI_ISL_598344, EPI_ISL_598345, EPI_ISL_598346, EPI_ISL_598347, EPI_ISL_598348, EPI_ISL_598349, EPI_ISL_598350, EPI_ISL_598351, EPI_ISL_598352, EPI_ISL_598353, EPI_ISL_598354, EPI_ISL_598355, EPI_ISL_598356, EPI_ISL_598357, EPI_ISL_598358, EPI_ISL_598359, EPI_ISL_598360, EPI_ISL_598362, EPI_ISL_598363, EPI_ISL_598364, EPI_ISL_598365, EPI_ISL_598366, EPI_ISL_598367, EPI_ISL_598368, EPI_ISL_598369, EPI_ISL_598370, EPI_ISL_598371, EPI_ISL_598372, EPI_ISL_598373, EPI_ISL_598374, EPI_ISL_598375, EPI_ISL_598376, EPI_ISL_598377, EPI_ISL_598378, EPI_ISL_598379, EPI_ISL_598380, EPI_ISL_598381, EPI_ISL_598382, EPI_ISL_598383, EPI_ISL_598384, EPI_ISL_598385, EPI_ISL_598386, EPI_ISL_598387, EPI_ISL_598388, EPI_ISL_598389, EPI_ISL_598390, EPI_ISL_598391, EPI_ISL_598392, EPI_ISL_598393, EPI_ISL_598394, EPI_ISL_598395, EPI_ISL_598396, EPI_ISL_598397, EPI_ISL_598398, EPI_ISL_598399, EPI_ISL_598400, EPI_ISL_598401, EPI_ISL_598402, EPI_ISL_598403, EPI_ISL_598404, EPI_ISL_598405, EPI_ISL_598406, EPI_ISL_598407, EPI_ISL_598409, EPI_ISL_598410, EPI_ISL_598411, EPI_ISL_598412, EPI_ISL_598413, EPI_ISL_598414, EPI_ISL_598415, EPI_ISL_598416, EPI_ISL_598417, EPI_ISL_598418, EPI_ISL_598419, EPI_ISL_598420, EPI_ISL_598421, EPI_ISL_598422, EPI_ISL_598423, EPI_ISL_598424, EPI_ISL_598425, EPI_ISL_598426, EPI_ISL_598427, EPI_ISL_598428, EPI_ISL_598429, EPI_ISL_598430, EPI_ISL_598431, EPI_ISL_598432, EPI_ISL_598434, EPI_ISL_598435, EPI_ISL_598437, EPI_ISL_598438, EPI_ISL_598439, EPI_ISL_598440, EPI_ISL_598441, EPI_ISL_598442, EPI_ISL_598443, EPI_ISL_598444, EPI_ISL_598445, EPI_ISL_598446, EPI_ISL_598447, EPI_ISL_598448, EPI_ISL_598449, EPI_ISL_598450, EPI_ISL_598451, EPI_ISL_598452, EPI_ISL_598453, EPI_ISL_598454, EPI_ISL_598455, EPI_ISL_598456, EPI_ISL_598457, EPI_ISL_598458, EPI_ISL_598459, EPI_ISL_598460, EPI_ISL_598461, EPI_ISL_598462, EPI_ISL_598463, EPI_ISL_598464, EPI_ISL_598465, EPI_ISL_598466, EPI_ISL_598467, EPI_ISL_598468, EPI_ISL_598469, EPI_ISL_598470, EPI_ISL_598471, EPI_ISL_598472, EPI_ISL_598473, EPI_ISL_598474, EPI_ISL_598475, EPI_ISL_598476, EPI_ISL_598477, EPI_ISL_598478, EPI_ISL_598479, EPI_ISL_598480, EPI_ISL_598481, EPI_ISL_598482, EPI_ISL_598483, EPI_ISL_598484, EPI_ISL_598485, EPI_ISL_598486, EPI_ISL_598487, EPI_ISL_598488, EPI_ISL_598489, EPI_ISL_598490, EPI_ISL_598491, EPI_ISL_598492, EPI_ISL_598493, EPI_ISL_598494, EPI_ISL_598495, EPI_ISL_598496, EPI_ISL_598497, EPI_ISL_598498, EPI_ISL_598499, EPI_ISL_598500, EPI_ISL_598501, EPI_ISL_598502, EPI_ISL_598503, EPI_ISL_598504, EPI_ISL_598505, EPI_ISL_598506, EPI_ISL_598507, EPI_ISL_598508, EPI_ISL_598509, EPI_ISL_598510, EPI_ISL_598511, EPI_ISL_598512, EPI_ISL_598513, EPI_ISL_598514, EPI_ISL_598515, EPI_ISL_598516, EPI_ISL_598517, EPI_ISL_598518, EPI_ISL_598519, EPI_ISL_598520, EPI_ISL_598521, EPI_ISL_598522, EPI_ISL_598523, EPI_ISL_598524, EPI_ISL_598525, EPI_ISL_598526, EPI_ISL_598527, EPI_ISL_598528, EPI_ISL_598529, EPI_ISL_598530, EPI_ISL_598531, EPI_ISL_598532, EPI_ISL_598533, EPI_ISL_598534, EPI_ISL_598535, EPI_ISL_598536, EPI_ISL_598537, EPI_ISL_598538, EPI_ISL_598539, EPI_ISL_598540, EPI_ISL_598541, EPI_ISL_598542, EPI_ISL_598543, EPI_ISL_598544, EPI_ISL_598545, EPI_ISL_598546, EPI_ISL_598547, EPI_ISL_598548, EPI_ISL_598549, EPI_ISL_598550, EPI_ISL_598551, EPI_ISL_598552, EPI_ISL_598553, EPI_ISL_598554, EPI_ISL_598555, EPI_ISL_598556, EPI_ISL_598557, EPI_ISL_598558, EPI_ISL_598559, EPI_ISL_598560, EPI_ISL_598561, EPI_ISL_598562, EPI_ISL_598563, EPI_ISL_598564, EPI_ISL_598565, EPI_ISL_598566, EPI_ISL_598567, EPI_ISL_598568, EPI_ISL_598569, EPI_ISL_598570, EPI_ISL_598571, EPI_ISL_598572, EPI_ISL_598573, EPI_ISL_598574, EPI_ISL_598575, EPI_ISL_598576, EPI_ISL_598577, EPI_ISL_598578, EPI_ISL_598579, EPI_ISL_598580, EPI_ISL_598581, EPI_ISL_598582, EPI_ISL_598583, EPI_ISL_598584, EPI_ISL_598585, EPI_ISL_598586, EPI_ISL_598587, EPI_ISL_598588, EPI_ISL_598589, EPI_ISL_598590, EPI_ISL_598591, EPI_ISL_598592, EPI_ISL_598593, EPI_ISL_598594, EPI_ISL_598595, EPI_ISL_598596, EPI_ISL_598597, EPI_ISL_598598, EPI_ISL_598599, EPI_ISL_598600, EPI_ISL_598601, EPI_ISL_598602, EPI_ISL_598603, EPI_ISL_598604, EPI_ISL_598605, EPI_ISL_598607, EPI_ISL_598608, EPI_ISL_598609, EPI_ISL_598610, EPI_ISL_598611, EPI_ISL_598612, EPI_ISL_598613, EPI_ISL_598614, EPI_ISL_598615, EPI_ISL_598616, EPI_ISL_598617, EPI_ISL_598618, EPI_ISL_598619, EPI_ISL_598620, EPI_ISL_598621, EPI_ISL_598622, EPI_ISL_598623, EPI_ISL_598624, EPI_ISL_598625, EPI_ISL_598626, EPI_ISL_598627, EPI_ISL_598628, EPI_ISL_598629, EPI_ISL_598630, EPI_ISL_598631, EPI_ISL_598632, EPI_ISL_598633, EPI_ISL_598634, EPI_ISL_598635, EPI_ISL_598636, EPI_ISL_598637, EPI_ISL_598639, EPI_ISL_598640, EPI_ISL_598641, EPI_ISL_598642, EPI_ISL_598643, EPI_ISL_598644, EPI_ISL_598645, EPI_ISL_598646, EPI_ISL_598647, EPI_ISL_598648, EPI_ISL_598649, EPI_ISL_598650, EPI_ISL_598651, EPI_ISL_598652, EPI_ISL_598653, EPI_ISL_598654, EPI_ISL_598655, EPI_ISL_598656, EPI_ISL_598657, EPI_ISL_598658, EPI_ISL_598659, EPI_ISL_598660, EPI_ISL_598661, EPI_ISL_598662, EPI_ISL_598663, EPI_ISL_598664, EPI_ISL_598665, EPI_ISL_598666, EPI_ISL_598668, EPI_ISL_598669, EPI_ISL_598670, EPI_ISL_598671, EPI_ISL_598672, EPI_ISL_598673, EPI_ISL_598674, EPI_ISL_598675, EPI_ISL_598676, EPI_ISL_598677, EPI_ISL_598678, EPI_ISL_598679, EPI_ISL_598680, EPI_ISL_598681, EPI_ISL_598682, EPI_ISL_598683, EPI_ISL_598684, EPI_ISL_598685, EPI_ISL_598686, EPI_ISL_598687, EPI_ISL_598688, EPI_ISL_598689, EPI_ISL_598690, EPI_ISL_598691, EPI_ISL_598692, EPI_ISL_598693, EPI_ISL_598694, EPI_ISL_598695, EPI_ISL_598696, EPI_ISL_598697, EPI_ISL_598698, EPI_ISL_598699, EPI_ISL_598700, EPI_ISL_598701, EPI_ISL_598702, EPI_ISL_598703, EPI_ISL_598704, EPI_ISL_598705, EPI_ISL_598706, EPI_ISL_598707, EPI_ISL_598708, EPI_ISL_598709, EPI_ISL_598710, EPI_ISL_598711, EPI_ISL_598712, EPI_ISL_598713, EPI_ISL_598714, EPI_ISL_598715, EPI_ISL_598716, EPI_ISL_598717, EPI_ISL_598718, EPI_ISL_598719, EPI_ISL_598720, EPI_ISL_598721, EPI_ISL_598722, EPI_ISL_598723, EPI_ISL_598724, EPI_ISL_598725, EPI_ISL_598726, EPI_ISL_598727, EPI_ISL_598728, EPI_ISL_598729, EPI_ISL_598730, EPI_ISL_598731, EPI_ISL_598732, EPI_ISL_598733, EPI_ISL_598734, EPI_ISL_598735, EPI_ISL_598736, EPI_ISL_598737, EPI_ISL_598738, EPI_ISL_598739, EPI_ISL_598740, EPI_ISL_598741, EPI_ISL_598742, EPI_ISL_598743, EPI_ISL_598744, EPI_ISL_598745, EPI_ISL_598746, EPI_ISL_598747, EPI_ISL_598748, EPI_ISL_598749, EPI_ISL_598750, EPI_ISL_598751, EPI_ISL_598752, EPI_ISL_598753, EPI_ISL_598754, EPI_ISL_598755, EPI_ISL_598756, EPI_ISL_598757, EPI_ISL_598758, EPI_ISL_598759, EPI_ISL_598760, EPI_ISL_598761, EPI_ISL_598762, EPI_ISL_598763, EPI_ISL_598764, EPI_ISL_598765, EPI_ISL_598766, EPI_ISL_598767, EPI_ISL_598768, EPI_ISL_598769, EPI_ISL_598770, EPI_ISL_598771, EPI_ISL_598772, EPI_ISL_598773, EPI_ISL_598774, EPI_ISL_598775, EPI_ISL_598776, EPI_ISL_598777, EPI_ISL_598778, EPI_ISL_598779, EPI_ISL_598780, EPI_ISL_598781, EPI_ISL_598782, EPI_ISL_598783, EPI_ISL_598784, EPI_ISL_598785, EPI_ISL_598786, EPI_ISL_598787, EPI_ISL_598788, EPI_ISL_598789, EPI_ISL_598790, EPI_ISL_598791, EPI_ISL_598792, EPI_ISL_598793, EPI_ISL_598794, EPI_ISL_598795, EPI_ISL_598796, EPI_ISL_598797, EPI_ISL_598798, EPI_ISL_598799, EPI_ISL_598800, EPI_ISL_598801, EPI_ISL_598802, EPI_ISL_598803, EPI_ISL_598804, EPI_ISL_598805, EPI_ISL_598806, EPI_ISL_598807, EPI_ISL_598808, EPI_ISL_598809, EPI_ISL_598810, EPI_ISL_598811, EPI_ISL_598812, EPI_ISL_598813, EPI_ISL_598814, EPI_ISL_598815, EPI_ISL_598816, EPI_ISL_598817, EPI_ISL_598818, EPI_ISL_598819, EPI_ISL_598820, EPI_ISL_598822, EPI_ISL_598823, EPI_ISL_599818, EPI_ISL_599819, EPI_ISL_599820, EPI_ISL_599821, EPI_ISL_599822, EPI_ISL_599823, EPI_ISL_599824, EPI_ISL_599825, EPI_ISL_599826, EPI_ISL_599827, EPI_ISL_599828, EPI_ISL_599829, EPI_ISL_599830, EPI_ISL_599831, EPI_ISL_599832, EPI_ISL_599833, EPI_ISL_599834, EPI_ISL_599835, EPI_ISL_599836, EPI_ISL_599837, EPI_ISL_599838, EPI_ISL_599839, EPI_ISL_599840, EPI_ISL_599841, EPI_ISL_599842, EPI_ISL_599843, EPI_ISL_599844, EPI_ISL_599846, EPI_ISL_599847, EPI_ISL_599848, EPI_ISL_599849, EPI_ISL_599850, EPI_ISL_599851, EPI_ISL_599852, EPI_ISL_599853, EPI_ISL_599854, EPI_ISL_599855, EPI_ISL_599856, EPI_ISL_599857, EPI_ISL_599858, EPI_ISL_599859, EPI_ISL_599860, EPI_ISL_599861, EPI_ISL_599862, EPI_ISL_599863, EPI_ISL_599864, EPI_ISL_599865, EPI_ISL_599866, EPI_ISL_599867, EPI_ISL_599868, EPI_ISL_599869, EPI_ISL_599870, EPI_ISL_599871, EPI_ISL_599872, EPI_ISL_599873, EPI_ISL_599874, EPI_ISL_599875, EPI_ISL_599876, EPI_ISL_599877, EPI_ISL_599878, EPI_ISL_599879, EPI_ISL_599880, EPI_ISL_599881, EPI_ISL_599882, EPI_ISL_599883, EPI_ISL_599884, EPI_ISL_599885, EPI_ISL_599886, EPI_ISL_599887, EPI_ISL_599889, EPI_ISL_599890, EPI_ISL_599891, EPI_ISL_599892, EPI_ISL_599893, EPI_ISL_599894, EPI_ISL_599895, EPI_ISL_599896, EPI_ISL_599897, EPI_ISL_599898, EPI_ISL_599899, EPI_ISL_599900, EPI_ISL_599901, EPI_ISL_599902, EPI_ISL_599903, EPI_ISL_599904, EPI_ISL_599905, EPI_ISL_599906, EPI_ISL_599907, EPI_ISL_599908, EPI_ISL_599909, EPI_ISL_599910, EPI_ISL_599911, EPI_ISL_599912, EPI_ISL_599913, EPI_ISL_599914, EPI_ISL_599915, EPI_ISL_599916, EPI_ISL_599917, EPI_ISL_599918, EPI_ISL_599919, EPI_ISL_599920, EPI_ISL_599921, EPI_ISL_599922, EPI_ISL_599923, EPI_ISL_599924, EPI_ISL_599925, EPI_ISL_599926, EPI_ISL_599927, EPI_ISL_599928, EPI_ISL_599929, EPI_ISL_599930, EPI_ISL_599931, EPI_ISL_599932, EPI_ISL_599933, EPI_ISL_599934, EPI_ISL_599935, EPI_ISL_599936, EPI_ISL_599937, EPI_ISL_599938, EPI_ISL_599939, EPI_ISL_599940, EPI_ISL_599941, EPI_ISL_599942, EPI_ISL_599943, EPI_ISL_599944, EPI_ISL_599945, EPI_ISL_599946, EPI_ISL_599947, EPI_ISL_599949, EPI_ISL_599950, EPI_ISL_599951, EPI_ISL_599952, EPI_ISL_599953, EPI_ISL_599954, EPI_ISL_599955, EPI_ISL_599956, EPI_ISL_599957, EPI_ISL_599958, EPI_ISL_599959, EPI_ISL_599961, EPI_ISL_599962, EPI_ISL_599963, EPI_ISL_599964, EPI_ISL_599965, EPI_ISL_599966, EPI_ISL_599967, EPI_ISL_599968, EPI_ISL_599969, EPI_ISL_599970, EPI_ISL_599971, EPI_ISL_599972, EPI_ISL_599973, EPI_ISL_599974, EPI_ISL_599975, EPI_ISL_599976, EPI_ISL_599977, EPI_ISL_599978, EPI_ISL_599979, EPI_ISL_599980, EPI_ISL_599981, EPI_ISL_599982, EPI_ISL_599983, EPI_ISL_599984, EPI_ISL_599985, EPI_ISL_599986, EPI_ISL_599987, EPI_ISL_599988, EPI_ISL_599989, EPI_ISL_599990, EPI_ISL_599991, EPI_ISL_599992, EPI_ISL_599993, EPI_ISL_599994, EPI_ISL_599995, EPI_ISL_599996, EPI_ISL_599997, EPI_ISL_599998, EPI_ISL_599999, EPI_ISL_600000, EPI_ISL_600001, EPI_ISL_600002, EPI_ISL_600003, EPI_ISL_600004, EPI_ISL_600005, EPI_ISL_600006, EPI_ISL_600007, EPI_ISL_600008, EPI_ISL_600009, EPI_ISL_600010, EPI_ISL_600011, EPI_ISL_600012, EPI_ISL_600013, EPI_ISL_600014, EPI_ISL_600015, EPI_ISL_600016, EPI_ISL_600017, EPI_ISL_600018, EPI_ISL_600019, EPI_ISL_600020, EPI_ISL_600021, EPI_ISL_600022, EPI_ISL_600023, EPI_ISL_600024, EPI_ISL_600025, EPI_ISL_600026, EPI_ISL_600027, EPI_ISL_600028, EPI_ISL_600029, EPI_ISL_600030, EPI_ISL_600031, EPI_ISL_600032, EPI_ISL_600033, EPI_ISL_600034, EPI_ISL_600035, EPI_ISL_600036, EPI_ISL_600037, EPI_ISL_600038, EPI_ISL_600039, EPI_ISL_600040, EPI_ISL_600041, EPI_ISL_600042, EPI_ISL_600043, EPI_ISL_600044, EPI_ISL_600045, EPI_ISL_600046, EPI_ISL_600047, EPI_ISL_600048, EPI_ISL_600049, EPI_ISL_600050, EPI_ISL_600051, EPI_ISL_600052, EPI_ISL_600053, EPI_ISL_600054, EPI_ISL_600055, EPI_ISL_600057, EPI_ISL_600058, EPI_ISL_600059, EPI_ISL_600060, EPI_ISL_600061, EPI_ISL_600062, EPI_ISL_600063, EPI_ISL_600064, EPI_ISL_600065, EPI_ISL_600066, EPI_ISL_600067, EPI_ISL_600068, EPI_ISL_600069, EPI_ISL_600070, EPI_ISL_600071, EPI_ISL_600072, EPI_ISL_600073, EPI_ISL_600074, EPI_ISL_600075, EPI_ISL_600076, EPI_ISL_600077, EPI_ISL_600078, EPI_ISL_600079, EPI_ISL_600080, EPI_ISL_600081, EPI_ISL_600082, EPI_ISL_600083, EPI_ISL_600084, EPI_ISL_600085, EPI_ISL_600086, EPI_ISL_600087, EPI_ISL_600088, EPI_ISL_600089, EPI_ISL_600090, EPI_ISL_600091, EPI_ISL_600092, EPI_ISL_600093, EPI_ISL_600094, EPI_ISL_600095, EPI_ISL_600096, EPI_ISL_600097, EPI_ISL_600098, EPI_ISL_600099, EPI_ISL_600100, EPI_ISL_600101, EPI_ISL_600102, EPI_ISL_600103, EPI_ISL_600104, EPI_ISL_600105, EPI_ISL_600106, EPI_ISL_600107, EPI_ISL_600108, EPI_ISL_600109, EPI_ISL_600110, EPI_ISL_600111, EPI_ISL_600112, EPI_ISL_600113, EPI_ISL_600114, EPI_ISL_600115, EPI_ISL_600116, EPI_ISL_600117, EPI_ISL_600118, EPI_ISL_600119, EPI_ISL_600120, EPI_ISL_600121, EPI_ISL_600122, EPI_ISL_600123, EPI_ISL_600124, EPI_ISL_600125, EPI_ISL_600126, EPI_ISL_600127, EPI_ISL_608697, EPI_ISL_608698, EPI_ISL_608699, EPI_ISL_608700, EPI_ISL_608701, EPI_ISL_608702, EPI_ISL_608703, EPI_ISL_608704, EPI_ISL_608705, EPI_ISL_608706, EPI_ISL_608708, EPI_ISL_608709, EPI_ISL_608710, EPI_ISL_608711, EPI_ISL_608712, EPI_ISL_608713, EPI_ISL_608714, EPI_ISL_608715, EPI_ISL_608716, EPI_ISL_608717, EPI_ISL_608718, EPI_ISL_608719, EPI_ISL_608720, EPI_ISL_608721, EPI_ISL_608722, EPI_ISL_608723, EPI_ISL_608724, EPI_ISL_608725, EPI_ISL_608726, EPI_ISL_608727, EPI_ISL_608728, EPI_ISL_608729, EPI_ISL_608730, EPI_ISL_608731, EPI_ISL_608732, EPI_ISL_608733, EPI_ISL_608734, EPI_ISL_608735, EPI_ISL_608736, EPI_ISL_608737, EPI_ISL_608738, EPI_ISL_608739, EPI_ISL_608740, EPI_ISL_608741, EPI_ISL_608742, EPI_ISL_608743, EPI_ISL_608744, EPI_ISL_608745, EPI_ISL_608746, EPI_ISL_608747, EPI_ISL_608749, EPI_ISL_608750, EPI_ISL_608752, EPI_ISL_608753, EPI_ISL_608754, EPI_ISL_608755, EPI_ISL_608756, EPI_ISL_608757, EPI_ISL_608758, EPI_ISL_608759, EPI_ISL_608760, EPI_ISL_608761, EPI_ISL_608762, EPI_ISL_608763, EPI_ISL_608764, EPI_ISL_608765, EPI_ISL_608766, EPI_ISL_608767, EPI_ISL_608768, EPI_ISL_608769, EPI_ISL_608770, EPI_ISL_608771, EPI_ISL_608772, EPI_ISL_608773, EPI_ISL_608774, EPI_ISL_608775, EPI_ISL_608776, EPI_ISL_608777, EPI_ISL_608778, EPI_ISL_608779, EPI_ISL_608780, EPI_ISL_608781, EPI_ISL_608782, EPI_ISL_608783, EPI_ISL_608784, EPI_ISL_608786, EPI_ISL_608787, EPI_ISL_608788, EPI_ISL_608789, EPI_ISL_608790, EPI_ISL_608791, EPI_ISL_608792, EPI_ISL_608793, EPI_ISL_608794, EPI_ISL_608795, EPI_ISL_608796, EPI_ISL_608797, EPI_ISL_608798, EPI_ISL_608799, EPI_ISL_608800, EPI_ISL_608801, EPI_ISL_608802, EPI_ISL_608803, EPI_ISL_608804, EPI_ISL_608805, EPI_ISL_608806, EPI_ISL_608807, EPI_ISL_608808, EPI_ISL_608809, EPI_ISL_608810, EPI_ISL_608811, EPI_ISL_608812, EPI_ISL_608813, EPI_ISL_608814, EPI_ISL_608815, EPI_ISL_608816, EPI_ISL_608817, EPI_ISL_608818, EPI_ISL_608819, EPI_ISL_608820, EPI_ISL_608821, EPI_ISL_608822, EPI_ISL_608823, EPI_ISL_608824, EPI_ISL_608825, EPI_ISL_608826, EPI_ISL_608827, EPI_ISL_608828, EPI_ISL_608829, EPI_ISL_608830, EPI_ISL_608831, EPI_ISL_608832, EPI_ISL_608833, EPI_ISL_608835, EPI_ISL_608836, EPI_ISL_608837, EPI_ISL_608838, EPI_ISL_608839, EPI_ISL_608840, EPI_ISL_608841, EPI_ISL_608842, EPI_ISL_608843, EPI_ISL_608844, EPI_ISL_608846, EPI_ISL_608848, EPI_ISL_608850, EPI_ISL_608854, EPI_ISL_608857, EPI_ISL_608859, EPI_ISL_608861, EPI_ISL_608870, EPI_ISL_608871, EPI_ISL_608893, EPI_ISL_608901, EPI_ISL_608905, EPI_ISL_608936, EPI_ISL_608940, EPI_ISL_608951, EPI_ISL_608954, EPI_ISL_608975, EPI_ISL_608977, EPI_ISL_608978, EPI_ISL_608986, EPI_ISL_609001, EPI_ISL_609006, EPI_ISL_609008, EPI_ISL_609015, EPI_ISL_609021, EPI_ISL_609024, EPI_ISL_609026, EPI_ISL_609027, EPI_ISL_609029, EPI_ISL_609030, EPI_ISL_609036, EPI_ISL_609038, EPI_ISL_609042, EPI_ISL_609044, EPI_ISL_609046, EPI_ISL_609049, EPI_ISL_609058, EPI_ISL_609059, EPI_ISL_609062, EPI_ISL_609063, EPI_ISL_609066, EPI_ISL_609071, EPI_ISL_609076, EPI_ISL_609084, EPI_ISL_609108, EPI_ISL_609110, EPI_ISL_609115, EPI_ISL_609116, EPI_ISL_609117, EPI_ISL_609126, EPI_ISL_609128, EPI_ISL_609136, EPI_ISL_609137, EPI_ISL_609138, EPI_ISL_609139, EPI_ISL_609140, EPI_ISL_609149, EPI_ISL_609151, EPI_ISL_609155, EPI_ISL_609219, EPI_ISL_609220, EPI_ISL_609221, EPI_ISL_609222, EPI_ISL_609223, EPI_ISL_609224, EPI_ISL_609225, EPI_ISL_609226, EPI_ISL_609227, EPI_ISL_609228, EPI_ISL_609229, EPI_ISL_609231, EPI_ISL_609232, EPI_ISL_609233, EPI_ISL_609234, EPI_ISL_609235, EPI_ISL_609236, EPI_ISL_609237, EPI_ISL_609238, EPI_ISL_609239, EPI_ISL_609240, EPI_ISL_609241, EPI_ISL_609242, EPI_ISL_609243, EPI_ISL_609244, EPI_ISL_609245, EPI_ISL_609246, EPI_ISL_609247, EPI_ISL_609248, EPI_ISL_609249, EPI_ISL_609250, EPI_ISL_609251, EPI_ISL_609252, EPI_ISL_609253, EPI_ISL_609254, EPI_ISL_609255, EPI_ISL_609256, EPI_ISL_609257, EPI_ISL_609258, EPI_ISL_609259, EPI_ISL_609260, EPI_ISL_609261, EPI_ISL_609262, EPI_ISL_609263, EPI_ISL_609264, EPI_ISL_609265, EPI_ISL_609266, EPI_ISL_609268, EPI_ISL_609269, EPI_ISL_609270, EPI_ISL_609271, EPI_ISL_609272, EPI_ISL_609273, EPI_ISL_609274, EPI_ISL_609275, EPI_ISL_609276, EPI_ISL_609277, EPI_ISL_609278, EPI_ISL_609279, EPI_ISL_609280, EPI_ISL_609281, EPI_ISL_609282, EPI_ISL_609283, EPI_ISL_609284, EPI_ISL_609285, EPI_ISL_609286, EPI_ISL_609287, EPI_ISL_609288, EPI_ISL_609289, EPI_ISL_609290, EPI_ISL_609291, EPI_ISL_609292, EPI_ISL_609293, EPI_ISL_609294, EPI_ISL_609295, EPI_ISL_609296, EPI_ISL_609297, EPI_ISL_609298, EPI_ISL_609299, EPI_ISL_609300, EPI_ISL_609301, EPI_ISL_609303, EPI_ISL_609304, EPI_ISL_609305, EPI_ISL_609306, EPI_ISL_609307, EPI_ISL_625405, EPI_ISL_625406, EPI_ISL_625407, EPI_ISL_625408, EPI_ISL_625409, EPI_ISL_625410, EPI_ISL_625411, EPI_ISL_625412, EPI_ISL_625413, EPI_ISL_625414, EPI_ISL_625415, EPI_ISL_625416, EPI_ISL_625417, EPI_ISL_625418, EPI_ISL_625419, EPI_ISL_625420, EPI_ISL_625421, EPI_ISL_625424, EPI_ISL_625426, EPI_ISL_625428, EPI_ISL_625430

| | | | |
|---|---|---|---|
| see above | Lighthouse Lab in Milton Keynes | Wellcome Sanger Institute for the COVID-19 Genomics UK (COG-UK) consortium | Cordelia Langford; David K. Jackson; Dominic Kwiatkowski; Ewan Harrison; Ian Johnston; John Sillitoe on behalf of the Wellcome Sanger Institute COVID-19 Surveillance Team; John Sillitoe on behalf of the Wellcome Sanger Institute COVID-19 Surveillance Team (http://www.sanger.ac.uk/covid-team); Roberto Amato; Sonia Goncalves; The Lighthouse Lab in Milton Keynes and Alex Alderton |

EPI_ISL_584599, EPI_ISL_584601, EPI_ISL_584605, EPI_ISL_594850, EPI_ISL_594852, EPI_ISL_594853, EPI_ISL_594854, EPI_ISL_594855, EPI_ISL_594856, EPI_ISL_594857, EPI_ISL_594858, EPI_ISL_594859, EPI_ISL_594860, EPI_ISL_595799, EPI_ISL_595800, EPI_ISL_595801, EPI_ISL_595822, EPI_ISL_611515, EPI_ISL_611517, EPI_ISL_611522, EPI_ISL_611542, EPI_ISL_611543, EPI_ISL_611568, EPI_ISL_611580, EPI_ISL_611600, EPI_ISL_611603, EPI_ISL_611630, EPI_ISL_611635, EPI_ISL_611642, EPI_ISL_611658, EPI_ISL_611664, EPI_ISL_611667, EPI_ISL_611675, EPI_ISL_611695, EPI_ISL_611721, EPI_ISL_611724, EPI_ISL_611728, EPI_ISL_611729, EPI_ISL_611730, EPI_ISL_611731, EPI_ISL_611736, EPI_ISL_611737, EPI_ISL_611764, EPI_ISL_611765, EPI_ISL_611784, EPI_ISL_611785, EPI_ISL_611790, EPI_ISL_611815, EPI_ISL_611827, EPI_ISL_611851, EPI_ISL_611857, EPI_ISL_611885, EPI_ISL_611908, EPI_ISL_611910, EPI_ISL_611911, EPI_ISL_611914, EPI_ISL_611921, EPI_ISL_611978, EPI_ISL_611981, EPI_ISL_611985, EPI_ISL_611995, EPI_ISL_611999, EPI_ISL_612001, EPI_ISL_612009, EPI_ISL_612010, EPI_ISL_612021, EPI_ISL_612024, EPI_ISL_612035, EPI_ISL_612036, EPI_ISL_612039, EPI_ISL_612059, EPI_ISL_612060, EPI_ISL_612064, EPI_ISL_612066, EPI_ISL_612071, EPI_ISL_612080, EPI_ISL_612088, EPI_ISL_612101, EPI_ISL_612111, EPI_ISL_612112, EPI_ISL_612433, EPI_ISL_612434, EPI_ISL_612435, EPI_ISL_626665, EPI_ISL_626697, EPI_ISL_626709, EPI_ISL_626766, EPI_ISL_626789, EPI_ISL_626994, EPI_ISL_627004, EPI_ISL_627017, EPI_ISL_627127, EPI_ISL_627131, EPI_ISL_627135, EPI_ISL_627136, EPI_ISL_627137, EPI_ISL_627152, EPI_ISL_627174, EPI_ISL_627177, EPI_ISL_627178, EPI_ISL_627179, EPI_ISL_627188, EPI_ISL_813657, EPI_ISL_813658, EPI_ISL_813659, EPI_ISL_813660, EPI_ISL_813662, EPI_ISL_813663, EPI_ISL_813664, EPI_ISL_813665, EPI_ISL_813666, EPI_ISL_813667, EPI_ISL_813668, EPI_ISL_813669, EPI_ISL_813670, EPI_ISL_813671, EPI_ISL_813672, EPI_ISL_813673, EPI_ISL_813674, EPI_ISL_813675, EPI_ISL_813676, EPI_ISL_1104308, EPI_ISL_1636735, EPI_ISL_1636736, EPI_ISL_1636737, EPI_ISL_1636738

| | | | |
|---|---|---|---|
| see above | Liverpool Clinical Laboratories | COVID-19 Genomics UK (COG-UK) Consortium | A Alrezaihi; Alessandro Gerada; Alistair Darby; Angela Cowell; Anita Lucaci; Anu Chawla; Cassie Olateju; Catherine Hartley; Charlotte Nelson; Ecaterina Vamos; Elaine O'Toole; Eleanor G Bentley; Ghada T Shawli; Hermione Webster; Isabel García-Dorival; James Johnson; James P Stewart; Jenifer Manson; Joanne Watts; Jones Benjamin; Jordan J Clark; Julian Hiscox; L Luu; Lucille Rainbow; M Almsaud; Margaret Hughes; Mark Whitehead; Matthew Gemmell; Miren Iturriza-Gomara; Muhannad Alruwaili; N.P Randle; Neil Swainston; PKF Gilmore; Parul Sharma; Rebekah Penrice-Randal; Richard Eccles; Richard Gregory; Sam Haldenby; Steve Paterson; Stuart D Armstrong; Trevor Ian Robinson; Ximeng Han |

EPI_ISL_584695, EPI_ISL_584696, EPI_ISL_584697, EPI_ISL_584698, EPI_ISL_584699, EPI_ISL_584700, EPI_ISL_584701, EPI_ISL_584702, EPI_ISL_584703, EPI_ISL_584704, EPI_ISL_584705, EPI_ISL_584706, EPI_ISL_584707, EPI_ISL_584708, EPI_ISL_584709, EPI_ISL_584710, EPI_ISL_584711, EPI_ISL_584712, EPI_ISL_584713, EPI_ISL_584714, EPI_ISL_584715, EPI_ISL_594987, EPI_ISL_594988, EPI_ISL_594989, EPI_ISL_594990, EPI_ISL_594991, EPI_ISL_594992, EPI_ISL_594993, EPI_ISL_594994, EPI_ISL_594995, EPI_ISL_594996, EPI_ISL_594997, EPI_ISL_594998, EPI_ISL_594999, EPI_ISL_595000, EPI_ISL_595001, EPI_ISL_595002, EPI_ISL_595003, EPI_ISL_595004, EPI_ISL_595005, EPI_ISL_595006, EPI_ISL_595007, EPI_ISL_595008, EPI_ISL_595009, EPI_ISL_595010, EPI_ISL_595011, EPI_ISL_595012, EPI_ISL_595013, EPI_ISL_595014, EPI_ISL_595015, EPI_ISL_595016, EPI_ISL_595017, EPI_ISL_595018, EPI_ISL_595019, EPI_ISL_595020, EPI_ISL_595021, EPI_ISL_595022, EPI_ISL_595023, EPI_ISL_595024, EPI_ISL_595025, EPI_ISL_595026, EPI_ISL_595027, EPI_ISL_595028, EPI_ISL_595029, EPI_ISL_595030, EPI_ISL_595031, EPI_ISL_595032, EPI_ISL_595033, EPI_ISL_595034, EPI_ISL_595035, EPI_ISL_595036, EPI_ISL_595037, EPI_ISL_595038, EPI_ISL_595039, EPI_ISL_595040, EPI_ISL_595041, EPI_ISL_595042, EPI_ISL_595043, EPI_ISL_595044, EPI_ISL_595045, EPI_ISL_611524, EPI_ISL_611572, EPI_ISL_611574, EPI_ISL_611576, EPI_ISL_611577, EPI_ISL_611591, EPI_ISL_611643, EPI_ISL_611644, EPI_ISL_611645, EPI_ISL_611646, EPI_ISL_611647, EPI_ISL_611648, EPI_ISL_611657, EPI_ISL_611668, EPI_ISL_611746, EPI_ISL_611756, EPI_ISL_611757, EPI_ISL_611758, EPI_ISL_611759, EPI_ISL_611760, EPI_ISL_611767, EPI_ISL_611768, EPI_ISL_611769, EPI_ISL_611770, EPI_ISL_611839, EPI_ISL_611846, EPI_ISL_611864, EPI_ISL_611868, EPI_ISL_611917, EPI_ISL_611930, EPI_ISL_611941, EPI_ISL_611942, EPI_ISL_611943, EPI_ISL_611944, EPI_ISL_611945, EPI_ISL_611946, EPI_ISL_611947, EPI_ISL_611948, EPI_ISL_611949, EPI_ISL_611950,

EPI_ISL_611951, EPI_ISL_611952, EPI_ISL_611953, EPI_ISL_611980, EPI_ISL_612008, EPI_ISL_612027, EPI_ISL_612030, EPI_ISL_612054, EPI_ISL_612085, EPI_ISL_612097, EPI_ISL_612100, EPI_ISL_612104, EPI_ISL_612110, EPI_ISL_612134, EPI_ISL_612523, EPI_ISL_612524, EPI_ISL_612525, EPI_ISL_612526, EPI_ISL_612527, EPI_ISL_612528, EPI_ISL_612529, EPI_ISL_612530, EPI_ISL_612531, EPI_ISL_612532, EPI_ISL_612533, EPI_ISL_612534, EPI_ISL_612535, EPI_ISL_612536, EPI_ISL_612537, EPI_ISL_612538, EPI_ISL_612539, EPI_ISL_612540, EPI_ISL_612541, EPI_ISL_612542, EPI_ISL_612543, EPI_ISL_612544, EPI_ISL_612545, EPI_ISL_612549, EPI_ISL_612556, EPI_ISL_612557, EPI_ISL_612558, EPI_ISL_612559, EPI_ISL_612560, EPI_ISL_612561, EPI_ISL_612562, EPI_ISL_612563, EPI_ISL_612564, EPI_ISL_612565, EPI_ISL_612566, EPI_ISL_637793, EPI_ISL_637794, EPI_ISL_637902, EPI_ISL_637994, EPI_ISL_638560, EPI_ISL_638561, EPI_ISL_638562, EPI_ISL_638563, EPI_ISL_638565, EPI_ISL_638566, EPI_ISL_638567, EPI_ISL_638568, EPI_ISL_638569, EPI_ISL_650241, EPI_ISL_650348, EPI_ISL_650394, EPI_ISL_650640, EPI_ISL_650684, EPI_ISL_651062, EPI_ISL_651063, EPI_ISL_651117, EPI_ISL_651126, EPI_ISL_651345, EPI_ISL_652343, EPI_ISL_652344, EPI_ISL_665278, EPI_ISL_665349, EPI_ISL_665409, EPI_ISL_665540, EPI_ISL_665621, EPI_ISL_665676, EPI_ISL_665680, EPI_ISL_665718, EPI_ISL_665744, EPI_ISL_665787, EPI_ISL_665868, EPI_ISL_665869, EPI_ISL_665870, EPI_ISL_665871, EPI_ISL_665872, EPI_ISL_665873, EPI_ISL_665874, EPI_ISL_665875, EPI_ISL_724967, EPI_ISL_724968, EPI_ISL_724969, EPI_ISL_724978, EPI_ISL_724979, EPI_ISL_839732

| | | | |
|---|---|---|---|
| see above | Northumbria University / South Tees Hospitals NHS Foundation Trust / North Cumbria Integrated Care NHS Foundation Trust / North Tees and Hartlepool NHS Foundation Trust / Newcastle Hospitals NHS Foundation Trust | COVID-19 Genomics UK (COG-UK) Consortium | Andrew Nelson; Brendan Payne; Clive Graham; Darren L Smith; Debra Padgett; Edward Barton; Emma Swindells; Garren Scott; Gary Black; Gary Eltringham; Giles S Holt; Greg R Young; Jane Greenaway; Jennifer Collins; John Allan; Joshua Loh; Lynn Dover; Matthew Bashton; Mohammad A Tariq; Paul Baker; Sarah Essex; Steve Liggett; Wen C Yew; Yusri Taha |

EPI_ISL_595327, EPI_ISL_627079, EPI_ISL_627085, EPI_ISL_627250, EPI_ISL_627251, EPI_ISL_627255, EPI_ISL_627256, EPI_ISL_627257, EPI_ISL_627258, EPI_ISL_627259, EPI_ISL_627263, EPI_ISL_627281, EPI_ISL_627282, EPI_ISL_627283, EPI_ISL_627284, EPI_ISL_627285, EPI_ISL_627286, EPI_ISL_627290, EPI_ISL_627294, EPI_ISL_627297, EPI_ISL_627299, EPI_ISL_627303, EPI_ISL_627305, EPI_ISL_628334, EPI_ISL_628335, EPI_ISL_628336, EPI_ISL_628337, EPI_ISL_628338, EPI_ISL_628339, EPI_ISL_628340, EPI_ISL_628341, EPI_ISL_628342, EPI_ISL_628343, EPI_ISL_628344, EPI_ISL_628345, EPI_ISL_628346, EPI_ISL_628347, EPI_ISL_628348, EPI_ISL_628349, EPI_ISL_628350, EPI_ISL_628351, EPI_ISL_628352, EPI_ISL_628353, EPI_ISL_628354, EPI_ISL_628355, EPI_ISL_628356, EPI_ISL_628357, EPI_ISL_628358, EPI_ISL_628359, EPI_ISL_628360, EPI_ISL_628361, EPI_ISL_628362, EPI_ISL_628363, EPI_ISL_628364, EPI_ISL_628365, EPI_ISL_628366, EPI_ISL_628367, EPI_ISL_628368, EPI_ISL_628414, EPI_ISL_628415, EPI_ISL_628416, EPI_ISL_628417, EPI_ISL_628418, EPI_ISL_628419, EPI_ISL_628420, EPI_ISL_628422, EPI_ISL_628423, EPI_ISL_628424, EPI_ISL_628425, EPI_ISL_628426, EPI_ISL_628427, EPI_ISL_628428, EPI_ISL_628429, EPI_ISL_628430, EPI_ISL_628431, EPI_ISL_628432, EPI_ISL_628433, EPI_ISL_628434, EPI_ISL_628435, EPI_ISL_628436, EPI_ISL_628437, EPI_ISL_628438, EPI_ISL_628439, EPI_ISL_628440, EPI_ISL_628441, EPI_ISL_628442, EPI_ISL_628443, EPI_ISL_628444, EPI_ISL_628445, EPI_ISL_628446, EPI_ISL_628447, EPI_ISL_628448, EPI_ISL_628449, EPI_ISL_628450, EPI_ISL_628451, EPI_ISL_628452, EPI_ISL_628453, EPI_ISL_628454, EPI_ISL_628455, EPI_ISL_628456, EPI_ISL_628457, EPI_ISL_628458, EPI_ISL_628459, EPI_ISL_628460, EPI_ISL_628461, EPI_ISL_628462, EPI_ISL_628463, EPI_ISL_628464, EPI_ISL_628465, EPI_ISL_628466, EPI_ISL_628467, EPI_ISL_628468, EPI_ISL_628469, EPI_ISL_628470, EPI_ISL_628471, EPI_ISL_628472, EPI_ISL_628473, EPI_ISL_628474, EPI_ISL_628475, EPI_ISL_628476, EPI_ISL_628477, EPI_ISL_628478, EPI_ISL_628479, EPI_ISL_628480, EPI_ISL_628481, EPI_ISL_628482, EPI_ISL_628483, EPI_ISL_628484, EPI_ISL_628485, EPI_ISL_628486, EPI_ISL_628487, EPI_ISL_628488, EPI_ISL_628489, EPI_ISL_628490, EPI_ISL_628491, EPI_ISL_628492, EPI_ISL_628493, EPI_ISL_628494, EPI_ISL_628495, EPI_ISL_628496, EPI_ISL_628497, EPI_ISL_628498, EPI_ISL_628499, EPI_ISL_628500, EPI_ISL_628501, EPI_ISL_628502, EPI_ISL_628503, EPI_ISL_628504, EPI_ISL_628505, EPI_ISL_628506, EPI_ISL_628507, EPI_ISL_628508, EPI_ISL_628509, EPI_ISL_628510, EPI_ISL_628511, EPI_ISL_628512, EPI_ISL_628513, EPI_ISL_628514, EPI_ISL_628515, EPI_ISL_628516, EPI_ISL_628517, EPI_ISL_628518, EPI_ISL_628519, EPI_ISL_628520, EPI_ISL_628521, EPI_ISL_628522, EPI_ISL_628523, EPI_ISL_628524, EPI_ISL_628525, EPI_ISL_628526, EPI_ISL_628527, EPI_ISL_628528, EPI_ISL_628529, EPI_ISL_628530, EPI_ISL_628531, EPI_ISL_628532, EPI_ISL_628533, EPI_ISL_628534, EPI_ISL_628535, EPI_ISL_628536, EPI_ISL_628537, EPI_ISL_628538, EPI_ISL_628539, EPI_ISL_628540, EPI_ISL_628541, EPI_ISL_628542, EPI_ISL_628544, EPI_ISL_628545, EPI_ISL_628546, EPI_ISL_628547, EPI_ISL_628548, EPI_ISL_628549, EPI_ISL_628550, EPI_ISL_628551, EPI_ISL_628552, EPI_ISL_628553, EPI_ISL_628554, EPI_ISL_628555, EPI_ISL_628556, EPI_ISL_628557, EPI_ISL_628558, EPI_ISL_628559, EPI_ISL_628560, EPI_ISL_628561, EPI_ISL_628562, EPI_ISL_628563, EPI_ISL_628564, EPI_ISL_628565, EPI_ISL_628566, EPI_ISL_628567, EPI_ISL_628568, EPI_ISL_628569, EPI_ISL_628570, EPI_ISL_628571, EPI_ISL_628572, EPI_ISL_628573, EPI_ISL_628574, EPI_ISL_628575, EPI_ISL_628576, EPI_ISL_628577, EPI_ISL_628578, EPI_ISL_628579, EPI_ISL_628580, EPI_ISL_628581, EPI_ISL_628582, EPI_ISL_628583, EPI_ISL_628584, EPI_ISL_628585, EPI_ISL_628586, EPI_ISL_628587, EPI_ISL_628588, EPI_ISL_628589, EPI_ISL_628590, EPI_ISL_628591, EPI_ISL_628592, EPI_ISL_628593, EPI_ISL_628594, EPI_ISL_628595, EPI_ISL_628596, EPI_ISL_628597, EPI_ISL_628598, EPI_ISL_628599, EPI_ISL_628600, EPI_ISL_628601, EPI_ISL_628602, EPI_ISL_628603, EPI_ISL_628604, EPI_ISL_628605, EPI_ISL_628606, EPI_ISL_628607, EPI_ISL_628608, EPI_ISL_628609, EPI_ISL_628610, EPI_ISL_628611, EPI_ISL_628612, EPI_ISL_628613, EPI_ISL_628614, EPI_ISL_628615, EPI_ISL_628616, EPI_ISL_628617, EPI_ISL_628618, EPI_ISL_638736, EPI_ISL_638744, EPI_ISL_705443, EPI_ISL_705464, EPI_ISL_705465, EPI_ISL_705466, EPI_ISL_705476, EPI_ISL_705499, EPI_ISL_705500, EPI_ISL_706891, EPI_ISL_706892, EPI_ISL_706893, EPI_ISL_706894, EPI_ISL_706895, EPI_ISL_706896, EPI_ISL_706897, EPI_ISL_706898, EPI_ISL_706899, EPI_ISL_706900, EPI_ISL_706901, EPI_ISL_706902, EPI_ISL_706903, EPI_ISL_706904, EPI_ISL_706905, EPI_ISL_706906, EPI_ISL_706907, EPI_ISL_706908, EPI_ISL_706909, EPI_ISL_706910, EPI_ISL_706911, EPI_ISL_706912, EPI_ISL_706913, EPI_ISL_706914, EPI_ISL_706915, EPI_ISL_706916, EPI_ISL_706917, EPI_ISL_706918, EPI_ISL_706919, EPI_ISL_706920, EPI_ISL_706921, EPI_ISL_706922, EPI_ISL_706923, EPI_ISL_706924, EPI_ISL_706925, EPI_ISL_706926, EPI_ISL_706927, EPI_ISL_706928, EPI_ISL_706929, EPI_ISL_741760, EPI_ISL_741761, EPI_ISL_741762, EPI_ISL_741763, EPI_ISL_741764, EPI_ISL_741765, EPI_ISL_842107, EPI_ISL_842111, EPI_ISL_842112, EPI_ISL_842113, EPI_ISL_842116, EPI_ISL_842117, EPI_ISL_842118, EPI_ISL_952942

| | | | |
|---|---|---|---|
| see above | Oxford Viromics, NDM, University of Oxford; Oxford University Hospitals; Basingstoke and North Hampshire Hospital | COVID-19 Genomics UK (COG-UK) Consortium | Alex Mobbs; Amy Trebes; Anita Justice; Catrin Moore; Christophe Fraser; David Bonsall; David Buck; Emma Wise; George Macintyre; Jessica Lynch; John Todd; Mariateresa de Cesare; Matilde Mori; Monique Andersson; Nathan Moore; Nick Cortes; Robert Shaw; Stephen Kidd; Tanya Golubchik; Timothy Peto |

EPI_ISL_577097, EPI_ISL_577098, EPI_ISL_577099, EPI_ISL_577100, EPI_ISL_577101, EPI_ISL_577103, EPI_ISL_577104, EPI_ISL_577105, EPI_ISL_577106, EPI_ISL_577107, EPI_ISL_577108, EPI_ISL_577109, EPI_ISL_584716, EPI_ISL_584717, EPI_ISL_584718, EPI_ISL_584721, EPI_ISL_584722, EPI_ISL_584723, EPI_ISL_584724, EPI_ISL_584725, EPI_ISL_584726, EPI_ISL_584727, EPI_ISL_584729, EPI_ISL_584730, EPI_ISL_584731, EPI_ISL_584732, EPI_ISL_584733, EPI_ISL_584735, EPI_ISL_584736, EPI_ISL_584737, EPI_ISL_584738, EPI_ISL_584739, EPI_ISL_584740, EPI_ISL_584741, EPI_ISL_584742, EPI_ISL_584743, EPI_ISL_584744, EPI_ISL_584745, EPI_ISL_584746, EPI_ISL_584747, EPI_ISL_584748, EPI_ISL_584749, EPI_ISL_584906, EPI_ISL_584907, EPI_ISL_584908, EPI_ISL_584909, EPI_ISL_584910, EPI_ISL_584911, EPI_ISL_584912, EPI_ISL_584913, EPI_ISL_584914, EPI_ISL_584915, EPI_ISL_584916, EPI_ISL_584917, EPI_ISL_584918, EPI_ISL_584919, EPI_ISL_584920, EPI_ISL_584921, EPI_ISL_584922, EPI_ISL_584923, EPI_ISL_584924, EPI_ISL_584925, EPI_ISL_584926, EPI_ISL_584927, EPI_ISL_584928, EPI_ISL_584929, EPI_ISL_584930, EPI_ISL_584931, EPI_ISL_584932, EPI_ISL_584933, EPI_ISL_584934, EPI_ISL_584935, EPI_ISL_584936, EPI_ISL_584937, EPI_ISL_584938, EPI_ISL_584939, EPI_ISL_584940, EPI_ISL_584941, EPI_ISL_584942, EPI_ISL_584943, EPI_ISL_584944, EPI_ISL_584945, EPI_ISL_584946, EPI_ISL_584947, EPI_ISL_584948, EPI_ISL_584949, EPI_ISL_584950, EPI_ISL_584951, EPI_ISL_584952, EPI_ISL_584953, EPI_ISL_584954, EPI_ISL_584955, EPI_ISL_584956, EPI_ISL_584957, EPI_ISL_584958, EPI_ISL_584959, EPI_ISL_584960, EPI_ISL_584961, EPI_ISL_584962, EPI_ISL_584963, EPI_ISL_584964, EPI_ISL_584965, EPI_ISL_584966, EPI_ISL_584967, EPI_ISL_584968, EPI_ISL_584969, EPI_ISL_584970, EPI_ISL_584971, EPI_ISL_584972, EPI_ISL_584973, EPI_ISL_584974, EPI_ISL_584975, EPI_ISL_584976, EPI_ISL_584977, EPI_ISL_584978, EPI_ISL_584979, EPI_ISL_584980, EPI_ISL_584981, EPI_ISL_584982, EPI_ISL_584983, EPI_ISL_584984, EPI_ISL_584985, EPI_ISL_584986, EPI_ISL_584987, EPI_ISL_584988, EPI_ISL_584989, EPI_ISL_584990, EPI_ISL_584991, EPI_ISL_584992, EPI_ISL_584993, EPI_ISL_584994, EPI_ISL_584995, EPI_ISL_584996, EPI_ISL_584997, EPI_ISL_584998, EPI_ISL_584999, EPI_ISL_585000, EPI_ISL_585001, EPI_ISL_585002, EPI_ISL_585003, EPI_ISL_585004, EPI_ISL_585005, EPI_ISL_585006, EPI_ISL_585007, EPI_ISL_585008, EPI_ISL_585260, EPI_ISL_585583, EPI_ISL_585584, EPI_ISL_585585, EPI_ISL_585586, EPI_ISL_585587, EPI_ISL_585588, EPI_ISL_585589, EPI_ISL_585590, EPI_ISL_585591, EPI_ISL_585592, EPI_ISL_585593, EPI_ISL_585594, EPI_ISL_585596, EPI_ISL_585598, EPI_ISL_585599, EPI_ISL_585600, EPI_ISL_585601, EPI_ISL_585602, EPI_ISL_585603, EPI_ISL_585604, EPI_ISL_585605, EPI_ISL_585606, EPI_ISL_585607, EPI_ISL_585608, EPI_ISL_585609, EPI_ISL_585610, EPI_ISL_585611, EPI_ISL_585612, EPI_ISL_585613, EPI_ISL_585614, EPI_ISL_585615, EPI_ISL_585616, EPI_ISL_585617, EPI_ISL_585618, EPI_ISL_585619, EPI_ISL_586240, EPI_ISL_586241, EPI_ISL_586242, EPI_ISL_595115, EPI_ISL_595116, EPI_ISL_595117, EPI_ISL_595118, EPI_ISL_595119, EPI_ISL_595120, EPI_ISL_595121, EPI_ISL_595122, EPI_ISL_595123, EPI_ISL_595124, EPI_ISL_595125, EPI_ISL_595126, EPI_ISL_595127, EPI_ISL_595128, EPI_ISL_595129, EPI_ISL_595130, EPI_ISL_595131, EPI_ISL_595132, EPI_ISL_595133, EPI_ISL_595134, EPI_ISL_595135, EPI_ISL_595136, EPI_ISL_595137, EPI_ISL_595138, EPI_ISL_595139, EPI_ISL_595140, EPI_ISL_595141, EPI_ISL_595142, EPI_ISL_595143, EPI_ISL_595144, EPI_ISL_595145, EPI_ISL_595146, EPI_ISL_595147, EPI_ISL_595148, EPI_ISL_595149, EPI_ISL_595150, EPI_ISL_595151, EPI_ISL_595152, EPI_ISL_595153, EPI_ISL_595154, EPI_ISL_595155, EPI_ISL_595156, EPI_ISL_595157, EPI_ISL_595158, EPI_ISL_595159, EPI_ISL_595160, EPI_ISL_595161, EPI_ISL_595162, EPI_ISL_595163, EPI_ISL_595164, EPI_ISL_595165, EPI_ISL_595166, EPI_ISL_595167, EPI_ISL_595168, EPI_ISL_595169, EPI_ISL_595170, EPI_ISL_595171, EPI_ISL_595172, EPI_ISL_595174, EPI_ISL_595175, EPI_ISL_595176, EPI_ISL_595200, EPI_ISL_595837, EPI_ISL_595838, EPI_ISL_595839, EPI_ISL_595840, EPI_ISL_595841, EPI_ISL_595842, EPI_ISL_595843, EPI_ISL_595844, EPI_ISL_595845, EPI_ISL_595846, EPI_ISL_595847, EPI_ISL_595848, EPI_ISL_595849, EPI_ISL_595850, EPI_ISL_595851, EPI_ISL_595852, EPI_ISL_595853, EPI_ISL_595854, EPI_ISL_595855, EPI_ISL_595856, EPI_ISL_595857, EPI_ISL_595858, EPI_ISL_595859, EPI_ISL_595860, EPI_ISL_595861, EPI_ISL_595862, EPI_ISL_595863, EPI_ISL_595864, EPI_ISL_595865, EPI_ISL_595866, EPI_ISL_595867, EPI_ISL_595868, EPI_ISL_595869, EPI_ISL_595870, EPI_ISL_595871, EPI_ISL_595872, EPI_ISL_595873, EPI_ISL_595874, EPI_ISL_595875, EPI_ISL_595876, EPI_ISL_595877, EPI_ISL_595878, EPI_ISL_595879, EPI_ISL_595880, EPI_ISL_595881, EPI_ISL_595882, EPI_ISL_595883, EPI_ISL_595884, EPI_ISL_595885, EPI_ISL_595886, EPI_ISL_595887, EPI_ISL_595888, EPI_ISL_595889, EPI_ISL_595890, EPI_ISL_595891, EPI_ISL_595892, EPI_ISL_595893, EPI_ISL_595894, EPI_ISL_595895, EPI_ISL_595896, EPI_ISL_595897, EPI_ISL_595898, EPI_ISL_595899, EPI_ISL_595900, EPI_ISL_595901, EPI_ISL_595902, EPI_ISL_595903, EPI_ISL_595904, EPI_ISL_595905, EPI_ISL_595906, EPI_ISL_595907, EPI_ISL_595908, EPI_ISL_595909, EPI_ISL_595910, EPI_ISL_595911, EPI_ISL_595912, EPI_ISL_595913, EPI_ISL_595914, EPI_ISL_595915, EPI_ISL_595916, EPI_ISL_595917, EPI_ISL_595918, EPI_ISL_595919, EPI_ISL_595920, EPI_ISL_595921, EPI_ISL_595922, EPI_ISL_595923, EPI_ISL_595924, EPI_ISL_595925, EPI_ISL_595926, EPI_ISL_595927, EPI_ISL_595928, EPI_ISL_595929, EPI_ISL_595930, EPI_ISL_595931, EPI_ISL_595932, EPI_ISL_595933, EPI_ISL_595934, EPI_ISL_595935, EPI_ISL_595936, EPI_ISL_595937, EPI_ISL_595938, EPI_ISL_595939, EPI_ISL_595940, EPI_ISL_595941, EPI_ISL_595942, EPI_ISL_595943, EPI_ISL_595944, EPI_ISL_595945, EPI_ISL_595946, EPI_ISL_595947, EPI_ISL_595948, EPI_ISL_595949, EPI_ISL_595950, EPI_ISL_595951, EPI_ISL_595952, EPI_ISL_595953, EPI_ISL_595954, EPI_ISL_595955, EPI_ISL_595956, EPI_ISL_595957, EPI_ISL_595958, EPI_ISL_595959, EPI_ISL_595960, EPI_ISL_595961, EPI_ISL_595962, EPI_ISL_595963, EPI_ISL_595964, EPI_ISL_595965, EPI_ISL_595966, EPI_ISL_595967, EPI_ISL_595968, EPI_ISL_595969, EPI_ISL_595970, EPI_ISL_595971, EPI_ISL_595972, EPI_ISL_595973, EPI_ISL_595974, EPI_ISL_595975, EPI_ISL_595976, EPI_ISL_595977, EPI_ISL_595978, EPI_ISL_595979, EPI_ISL_595980, EPI_ISL_595981, EPI_ISL_595982, EPI_ISL_595983, EPI_ISL_595984, EPI_ISL_595985, EPI_ISL_595986, EPI_ISL_595987, EPI_ISL_595988, EPI_ISL_595989, EPI_ISL_595990, EPI_ISL_595991, EPI_ISL_595992, EPI_ISL_595993, EPI_ISL_595994, EPI_ISL_595995, EPI_ISL_595996, EPI_ISL_595997, EPI_ISL_595998, EPI_ISL_595999, EPI_ISL_596000, EPI_ISL_596001, EPI_ISL_596002, EPI_ISL_596003, EPI_ISL_596004, EPI_ISL_596005, EPI_ISL_596006, EPI_ISL_596007, EPI_ISL_596008, EPI_ISL_596009, EPI_ISL_596010, EPI_ISL_596011, EPI_ISL_596012, EPI_ISL_596013, EPI_ISL_596014, EPI_ISL_596015, EPI_ISL_596016, EPI_ISL_596017, EPI_ISL_596018, EPI_ISL_596019, EPI_ISL_596020, EPI_ISL_596021, EPI_ISL_596022, EPI_ISL_596023, EPI_ISL_596024, EPI_ISL_596025, EPI_ISL_596026, EPI_ISL_596027, EPI_ISL_596028, EPI_ISL_596029, EPI_ISL_596030, EPI_ISL_596031, EPI_ISL_596032, EPI_ISL_596033, EPI_ISL_596034, EPI_ISL_596035, EPI_ISL_596036, EPI_ISL_596037, EPI_ISL_596038, EPI_ISL_596039, EPI_ISL_596040, EPI_ISL_596041, EPI_ISL_596042, EPI_ISL_596043, EPI_ISL_596044, EPI_ISL_596045, EPI_ISL_596046, EPI_ISL_596047, EPI_ISL_596048, EPI_ISL_596049, EPI_ISL_596050, EPI_ISL_596051, EPI_ISL_596052, EPI_ISL_596053, EPI_ISL_596054, EPI_ISL_596055, EPI_ISL_596056, EPI_ISL_596057, EPI_ISL_596058, EPI_ISL_596059, EPI_ISL_596060, EPI_ISL_596061, EPI_ISL_596062, EPI_ISL_596063, EPI_ISL_596064, EPI_ISL_596065, EPI_ISL_596066, EPI_ISL_596067, EPI_ISL_596068, EPI_ISL_596069, EPI_ISL_596070, EPI_ISL_596071, EPI_ISL_596072, EPI_ISL_596073, EPI_ISL_596074, EPI_ISL_596075, EPI_ISL_596076, EPI_ISL_596077, EPI_ISL_596078, EPI_ISL_596079, EPI_ISL_596080, EPI_ISL_596081, EPI_ISL_596082, EPI_ISL_596083, EPI_ISL_596084, EPI_ISL_596085, EPI_ISL_596086, EPI_ISL_596087, EPI_ISL_596088, EPI_ISL_596089, EPI_ISL_596090, EPI_ISL_596091, EPI_ISL_596092, EPI_ISL_596093, EPI_ISL_596094, EPI_ISL_596095, EPI_ISL_596096, EPI_ISL_596097, EPI_ISL_596098, EPI_ISL_596099, EPI_ISL_596100, EPI_ISL_596101, EPI_ISL_596102, EPI_ISL_596103, EPI_ISL_596104, EPI_ISL_596105, EPI_ISL_596106, EPI_ISL_596107, EPI_ISL_596108, EPI_ISL_596109, EPI_ISL_596110, EPI_ISL_596111, EPI_ISL_596112, EPI_ISL_596113, EPI_ISL_596114, EPI_ISL_596115, EPI_ISL_596145, EPI_ISL_596146, EPI_ISL_596147, EPI_ISL_596148, EPI_ISL_596149, EPI_ISL_596150, EPI_ISL_596151, EPI_ISL_596152, EPI_ISL_596153, EPI_ISL_596154, EPI_ISL_596155, EPI_ISL_596156, EPI_ISL_596157, EPI_ISL_596158, EPI_ISL_596159, EPI_ISL_596160, EPI_ISL_596161, EPI_ISL_596162, EPI_ISL_596163, EPI_ISL_596164, EPI_ISL_596165, EPI_ISL_596166, EPI_ISL_596167, EPI_ISL_596168, EPI_ISL_596169, EPI_ISL_596170, EPI_ISL_596171, EPI_ISL_596172, EPI_ISL_596173, EPI_ISL_596174, EPI_ISL_596175, EPI_ISL_596176, EPI_ISL_596177, EPI_ISL_596178, EPI_ISL_596179, EPI_ISL_596180, EPI_ISL_596181, EPI_ISL_596182, EPI_ISL_596184, EPI_ISL_596185, EPI_ISL_596186, EPI_ISL_596187, EPI_ISL_596188, EPI_ISL_665296, EPI_ISL_665505, EPI_ISL_665656, EPI_ISL_665671, EPI_ISL_665702, EPI_ISL_665850, EPI_ISL_665957, EPI_ISL_665958, EPI_ISL_665959, EPI_ISL_665960, EPI_ISL_665961, EPI_ISL_665962, EPI_ISL_665963, EPI_ISL_725159, EPI_ISL_725161, EPI_ISL_725165, EPI_ISL_725169, EPI_ISL_725170, EPI_ISL_725172, EPI_ISL_725173, EPI_ISL_725187, EPI_ISL_842100, EPI_ISL_842101, EPI_ISL_842102, EPI_ISL_842103, EPI_ISL_842104, EPI_ISL_842105, EPI_ISL_842106, EPI_ISL_842109, EPI_ISL_842114, EPI_ISL_842115, EPI_ISL_924077, EPI_ISL_1046890, EPI_ISL_1046891, EPI_ISL_1046893, EPI_ISL_1046894, EPI_ISL_1046895, EPI_ISL_1046896, EPI_ISL_1046897, EPI_ISL_1046898, EPI_ISL_1046899, EPI_ISL_1046900, EPI_ISL_1046901, EPI_ISL_1046902, EPI_ISL_1046903, EPI_ISL_1046904, EPI_ISL_1046905, EPI_ISL_1046906, EPI_ISL_1046907, EPI_ISL_1046908, EPI_ISL_1046910, EPI_ISL_1046912, EPI_ISL_1046913, EPI_ISL_1046914, EPI_ISL_1046915, EPI_ISL_1046916, EPI_ISL_1046917, EPI_ISL_1046918, EPI_ISL_1046919, EPI_ISL_1046920, EPI_ISL_1046921, EPI_ISL_1046924, EPI_ISL_1046925, EPI_ISL_1046927, EPI_ISL_1046928, EPI_ISL_1046930, EPI_ISL_1046932, EPI_ISL_1046933, EPI_ISL_1046934, EPI_ISL_1046935, EPI_ISL_1046936, EPI_ISL_1046937, EPI_ISL_1046938, EPI_ISL_1046939, EPI_ISL_1046940, EPI_ISL_1046941, EPI_ISL_1046942, EPI_ISL_1046943, EPI_ISL_1046944, EPI_ISL_1046945, EPI_ISL_1046947, EPI_ISL_1046948, EPI_ISL_1046950

| | | | |
|---|---|---|---|
| see above | Quadram Institute Bioscience | COVID-19 Genomics UK (COG-UK) Consortium | Alexander J Trotter; Alison E. Mather; Alp Aydin; Ana P. Tedim; Anastasia Kolyva; Andrew Bell; Andrew J. Page; Claire Stuart; Dave J. Baker; Gemma L. Kay; John Wain; Justin O'Grady; Leonardo de Oliveira Martins; Lizzie Meadows; Maria Diaz; Mark Webber; Muhammed Yasir; Nabil-Fareed Alikhan; Ngozi Elumogo; Nicholas M. Thomson; Rachael Stanley; Rachel Gilroy; Reenesh Prakash; Samir Dervisevic; Samuel Bloomfield; Steven Rudder; Thanh Le-Viet |

EPI_ISL_572444, EPI_ISL_572460, EPI_ISL_572476, EPI_ISL_572582, EPI_ISL_572598, EPI_ISL_572603, EPI_ISL_572706, EPI_ISL_572833, EPI_ISL_573466, EPI_ISL_573467, EPI_ISL_573468, EPI_ISL_573469, EPI_ISL_573470, EPI_ISL_573471, EPI_ISL_573472, EPI_ISL_573473, EPI_ISL_573474, EPI_ISL_573475, EPI_ISL_573476, EPI_ISL_573477, EPI_ISL_577110, EPI_ISL_577111, EPI_ISL_577112, EPI_ISL_577113, EPI_ISL_577114, EPI_ISL_577115, EPI_ISL_577116, EPI_ISL_577117, EPI_ISL_577118, EPI_ISL_577119, EPI_ISL_577120, EPI_ISL_577121, EPI_ISL_577122, EPI_ISL_577123, EPI_ISL_577124, EPI_ISL_577125, EPI_ISL_577126, EPI_ISL_577127, EPI_ISL_577128, EPI_ISL_577129, EPI_ISL_577130, EPI_ISL_577131, EPI_ISL_577132, EPI_ISL_577133, EPI_ISL_577134, EPI_ISL_577135, EPI_ISL_577136, EPI_ISL_577137, EPI_ISL_577138, EPI_ISL_577139, EPI_ISL_577140, EPI_ISL_577141, EPI_ISL_577142, EPI_ISL_577143, EPI_ISL_577144, EPI_ISL_577145, EPI_ISL_577146, EPI_ISL_577147, EPI_ISL_577148, EPI_ISL_577149, EPI_ISL_577150, EPI_ISL_577151, EPI_ISL_577152, EPI_ISL_577153, EPI_ISL_577154, EPI_ISL_577155, EPI_ISL_577156, EPI_ISL_577157, EPI_ISL_577158, EPI_ISL_577159, EPI_ISL_577160, EPI_ISL_577161, EPI_ISL_577162, EPI_ISL_577163, EPI_ISL_577164, EPI_ISL_577165, EPI_ISL_577166, EPI_ISL_577167, EPI_ISL_577168, EPI_ISL_577169, EPI_ISL_577170, EPI_ISL_577171, EPI_ISL_577172, EPI_ISL_577173, EPI_ISL_577174, EPI_ISL_577175, EPI_ISL_577176, EPI_ISL_577177, EPI_ISL_577178, EPI_ISL_577179, EPI_ISL_577180, EPI_ISL_577181, EPI_ISL_577182, EPI_ISL_577183, EPI_ISL_577184, EPI_ISL_577185, EPI_ISL_577186, EPI_ISL_577187, EPI_ISL_577188, EPI_ISL_577189, EPI_ISL_577190, EPI_ISL_577191, EPI_ISL_577192, EPI_ISL_584750, EPI_ISL_584751, EPI_ISL_584752, EPI_ISL_584753, EPI_ISL_584754, EPI_ISL_584755, EPI_ISL_584756, EPI_ISL_584757, EPI_ISL_584758, EPI_ISL_584759, EPI_ISL_584760, EPI_ISL_595046, EPI_ISL_595047, EPI_ISL_595048, EPI_ISL_595049, EPI_ISL_595050, EPI_ISL_595051, EPI_ISL_595052, EPI_ISL_595053, EPI_ISL_595054, EPI_ISL_595055, EPI_ISL_595056, EPI_ISL_595057, EPI_ISL_595058, EPI_ISL_595059, EPI_ISL_595060, EPI_ISL_595061, EPI_ISL_595062, EPI_ISL_595063, EPI_ISL_595064, EPI_ISL_595065, EPI_ISL_595066, EPI_ISL_595067, EPI_ISL_595068, EPI_ISL_595069, EPI_ISL_595070, EPI_ISL_595071, EPI_ISL_595072, EPI_ISL_595073, EPI_ISL_595074, EPI_ISL_595075, EPI_ISL_595076, EPI_ISL_595077, EPI_ISL_595078, EPI_ISL_595079, EPI_ISL_595080, EPI_ISL_595081, EPI_ISL_595082, EPI_ISL_595083, EPI_ISL_595084, EPI_ISL_595085, EPI_ISL_595086, EPI_ISL_595087, EPI_ISL_595088, EPI_ISL_595089, EPI_ISL_595090, EPI_ISL_595091, EPI_ISL_595092, EPI_ISL_595093, EPI_ISL_595094, EPI_ISL_595095, EPI_ISL_595096, EPI_ISL_595097, EPI_ISL_595098, EPI_ISL_595099, EPI_ISL_665940

| | | | |
|---|---|---|---|
| see above | Queens Medical Centre, Clinical Microbiology Department / DeepSeq Nottingham | COVID-19 Genomics UK (COG-UK) Consortium | Christopher Moore; Fei Sang; Gemma Clark; Hannah Howson-Wells; Johnny Debebe; Jonathan Ball; Joseph Chappell; Manjinder Khakh; Matthew Carlisle; Matthew Loose; Michelle M Lister; Nadine Holmes; Patrick McClure; Theocharis Tsoleridis; Vicki M Fleming; Victoria Wright; Wendy Smith |

EPI_ISL_626828, EPI_ISL_626859, EPI_ISL_626953, EPI_ISL_627000, EPI_ISL_627008, EPI_ISL_627038, EPI_ISL_627070, EPI_ISL_627158, EPI_ISL_627199, EPI_ISL_627531, EPI_ISL_627533, EPI_ISL_627535, EPI_ISL_627536, EPI_ISL_627537, EPI_ISL_627538, EPI_ISL_627539, EPI_ISL_627540, EPI_ISL_627541, EPI_ISL_627542, EPI_ISL_627543, EPI_ISL_627544, EPI_ISL_627545, EPI_ISL_627546, EPI_ISL_627547, EPI_ISL_627548, EPI_ISL_627549, EPI_ISL_627550, EPI_ISL_627551, EPI_ISL_627552, EPI_ISL_627553, EPI_ISL_627554, EPI_ISL_627555, EPI_ISL_627556, EPI_ISL_627557, EPI_ISL_627558, EPI_ISL_680245, EPI_ISL_680246, EPI_ISL_680247, EPI_ISL_680249, EPI_ISL_680250, EPI_ISL_680254, EPI_ISL_680255, EPI_ISL_680256, EPI_ISL_680257, EPI_ISL_680282, EPI_ISL_680290, EPI_ISL_680291, EPI_ISL_680292, EPI_ISL_680293, EPI_ISL_680294, EPI_ISL_680295, EPI_ISL_680296, EPI_ISL_680297, EPI_ISL_680298, EPI_ISL_680299, EPI_ISL_680306, EPI_ISL_680307, EPI_ISL_680308, EPI_ISL_680309, EPI_ISL_680310, EPI_ISL_680311, EPI_ISL_680312, EPI_ISL_680313, EPI_ISL_680314, EPI_ISL_680315, EPI_ISL_680316, EPI_ISL_680317, EPI_ISL_680318, EPI_ISL_680319, EPI_ISL_680320, EPI_ISL_680321, EPI_ISL_680322, EPI_ISL_680323, EPI_ISL_680324, EPI_ISL_680325, EPI_ISL_680326, EPI_ISL_680327, EPI_ISL_680328, EPI_ISL_680436, EPI_ISL_680437, EPI_ISL_680438, EPI_ISL_680439, EPI_ISL_680441

| | | | |
|---|---|---|---|
| see above | Regional Virus Laboratory, Belfast Health and Social Care Trust | COVID-19 Genomics UK (COG-UK) Consortium | Alison Watt; Ciara Cox; Conall McCaughey; David Simpson; Derek Fairley; James McKenna; Mairead Connor; Susan Feeney; Tanya Curran; Zoltan Molnar |

EPI_ISL_637117, EPI_ISL_649817, EPI_ISL_686535, EPI_ISL_686536, EPI_ISL_686537, EPI_ISL_686539, EPI_ISL_686540, EPI_ISL_686597

| | | | |
|---|---|---|---|
| see above | Respiratory Virus Unit, Microbiology Services Colindale, Public Health England | COVID-19 Genomics UK (COG-UK) Consortium | PHE Covid Sequencing Team |

EPI_ISL_583973, EPI_ISL_583974, EPI_ISL_583975, EPI_ISL_583976, EPI_ISL_583977, EPI_ISL_583978, EPI_ISL_583979, EPI_ISL_583980, EPI_ISL_583981, EPI_ISL_583985, EPI_ISL_583986, EPI_ISL_583987, EPI_ISL_583988, EPI_ISL_583989, EPI_ISL_583990, EPI_ISL_583992, EPI_ISL_584066, EPI_ISL_593804, EPI_ISL_593810, EPI_ISL_603189, EPI_ISL_603190, EPI_ISL_603192, EPI_ISL_603201, EPI_ISL_603212, EPI_ISL_603215, EPI_ISL_609827, EPI_ISL_609828, EPI_ISL_609829, EPI_ISL_609830, EPI_ISL_609831, EPI_ISL_609832, EPI_ISL_609833, EPI_ISL_609834, EPI_ISL_609835, EPI_ISL_609836, EPI_ISL_609837, EPI_ISL_609838, EPI_ISL_609839, EPI_ISL_609840, EPI_ISL_609841, EPI_ISL_609842, EPI_ISL_609843, EPI_ISL_609844, EPI_ISL_609845, EPI_ISL_609846, EPI_ISL_609847, EPI_ISL_609848, EPI_ISL_609849, EPI_ISL_609850, EPI_ISL_609851, EPI_ISL_609852, EPI_ISL_609853, EPI_ISL_609854, EPI_ISL_609855, EPI_ISL_609856, EPI_ISL_609857, EPI_ISL_609858, EPI_ISL_609859, EPI_ISL_609860, EPI_ISL_609869, EPI_ISL_609871, EPI_ISL_609872, EPI_ISL_609874, EPI_ISL_609881, EPI_ISL_609882, EPI_ISL_609884, EPI_ISL_613587, EPI_ISL_613588, EPI_ISL_613589, EPI_ISL_613633, EPI_ISL_622833, EPI_ISL_622834, EPI_ISL_622835, EPI_ISL_622836, EPI_ISL_622837, EPI_ISL_622838, EPI_ISL_622839, EPI_ISL_622840, EPI_ISL_622841, EPI_ISL_622842, EPI_ISL_622843, EPI_ISL_622846, EPI_ISL_622872, EPI_ISL_622892, EPI_ISL_622893, EPI_ISL_632796, EPI_ISL_636688, EPI_ISL_636689, EPI_ISL_636690, EPI_ISL_636691, EPI_ISL_636692, EPI_ISL_636693, EPI_ISL_636694, EPI_ISL_636695, EPI_ISL_636696, EPI_ISL_636697, EPI_ISL_636698, EPI_ISL_636699, EPI_ISL_636700, EPI_ISL_636701, EPI_ISL_636702, EPI_ISL_636703

| | | | |
|---|---|---|---|
| see above | Respiratory Virus Unit, Microbiology Services Colindale, Public Health England | Respiratory Virus Unit, Microbiology Services Colindale, Public Health England | PHE Covid Sequencing Team |

EPI_ISL_733580, EPI_ISL_733581, EPI_ISL_733583, EPI_ISL_733584, EPI_ISL_733586, EPI_ISL_733587, EPI_ISL_733588, EPI_ISL_804261, EPI_ISL_804262, EPI_ISL_804263, EPI_ISL_804264, EPI_ISL_825493, EPI_ISL_825497, EPI_ISL_825575, EPI_ISL_825576, EPI_ISL_825577, EPI_ISL_825578, EPI_ISL_825579, EPI_ISL_860685, EPI_ISL_860689

| | | | |
|---|---|---|---|
| see above | Respiratory Virus Unit, National Infection Service, Public Health England | COVID-19 Genomics UK (COG-UK) Consortium | PHE Covid Sequencing Team |

EPI_ISL_664352, EPI_ISL_664361, EPI_ISL_664479, EPI_ISL_664505, EPI_ISL_665160, EPI_ISL_665183, EPI_ISL_665184, EPI_ISL_665185, EPI_ISL_665186, EPI_ISL_665187, EPI_ISL_665193, EPI_ISL_665194, EPI_ISL_665195, EPI_ISL_665196, EPI_ISL_665223, EPI_ISL_665241, EPI_ISL_679412, EPI_ISL_679413, EPI_ISL_1105305, EPI_ISL_1105312, EPI_ISL_1105318, EPI_ISL_1105326

| | | | |
|---|---|---|---|
| see above | University College London Hospital | COVID-19 Genomics UK (COG-UK) Consortium | Catherine Houlihan; Dan Frampton; Judith Heaney; Matthew Byott; Moira Spyer and Eleni Nastouli; Stuart Kirk |

EPI_ISL_594862, EPI_ISL_594863, EPI_ISL_594864, EPI_ISL_594865, EPI_ISL_594866, EPI_ISL_594867, EPI_ISL_594868, EPI_ISL_594869, EPI_ISL_594870, EPI_ISL_594871, EPI_ISL_594872, EPI_ISL_594873, EPI_ISL_594874, EPI_ISL_594875, EPI_ISL_594876, EPI_ISL_594877, EPI_ISL_594879, EPI_ISL_594880, EPI_ISL_594881, EPI_ISL_594883, EPI_ISL_594884, EPI_ISL_594888, EPI_ISL_612103, EPI_ISL_612198, EPI_ISL_612457, EPI_ISL_627104, EPI_ISL_627108, EPI_ISL_627110, EPI_ISL_627472, EPI_ISL_627473, EPI_ISL_627474, EPI_ISL_627475, EPI_ISL_627476, EPI_ISL_627477, EPI_ISL_627478, EPI_ISL_627479, EPI_ISL_627480, EPI_ISL_627481, EPI_ISL_627491, EPI_ISL_637995, EPI_ISL_679550, EPI_ISL_679551, EPI_ISL_679552, EPI_ISL_679553, EPI_ISL_679554, EPI_ISL_679555, EPI_ISL_679556, EPI_ISL_679557, EPI_ISL_679558, EPI_ISL_679559, EPI_ISL_679560, EPI_ISL_679561, EPI_ISL_679562, EPI_ISL_679563, EPI_ISL_679564, EPI_ISL_679565, EPI_ISL_679566, EPI_ISL_679567, EPI_ISL_679568, EPI_ISL_679569, EPI_ISL_679570, EPI_ISL_679571, EPI_ISL_679572, EPI_ISL_679573, EPI_ISL_679574, EPI_ISL_679627, EPI_ISL_679628, EPI_ISL_702690, EPI_ISL_703179, EPI_ISL_703332, EPI_ISL_704046, EPI_ISL_704084, EPI_ISL_704163, EPI_ISL_704193, EPI_ISL_705425, EPI_ISL_1637132

| | | | |
|---|---|---|---|
| see above | University College London, Great Ormond Street Hospital for Children NHS Foundation Trust, Imperial College Healthcare NHS Trust | COVID-19 Genomics UK (COG-UK) Consortium | Alison Holmes; Charlotte Williams; Helena Tutill; Jacqueline Findlay; James Price; Judith Breuer; Julianne Brown; Kathryn Harris; Leysa Forrest; Mark Kristiansen; Paola Niola; Paola Resende Silva; Patricia Dyal; Paul Randell; Rachel Williams; Samuel Weeks; Sergi Castellano; Sunando Roy; Tony Brooks; Yasmin Panchbhaya |

EPI_ISL_594484, EPI_ISL_594536, EPI_ISL_594537, EPI_ISL_594538, EPI_ISL_594539, EPI_ISL_594540, EPI_ISL_594541, EPI_ISL_594542, EPI_ISL_594543, EPI_ISL_594544, EPI_ISL_594545, EPI_ISL_594546, EPI_ISL_594547, EPI_ISL_594548, EPI_ISL_594549, EPI_ISL_594550, EPI_ISL_594551, EPI_ISL_594552, EPI_ISL_594553, EPI_ISL_594554, EPI_ISL_594555, EPI_ISL_594556, EPI_ISL_594557, EPI_ISL_594558, EPI_ISL_594559, EPI_ISL_594560, EPI_ISL_594561, EPI_ISL_594562, EPI_ISL_594563, EPI_ISL_594564, EPI_ISL_594565, EPI_ISL_594566, EPI_ISL_594567, EPI_ISL_594568, EPI_ISL_594569, EPI_ISL_594570, EPI_ISL_594571, EPI_ISL_594572, EPI_ISL_594573, EPI_ISL_594574, EPI_ISL_594575, EPI_ISL_594576, EPI_ISL_594577, EPI_ISL_594578, EPI_ISL_594579, EPI_ISL_594580, EPI_ISL_594581, EPI_ISL_594582, EPI_ISL_594583, EPI_ISL_594584, EPI_ISL_594585, EPI_ISL_594586, EPI_ISL_594587, EPI_ISL_594588, EPI_ISL_594589, EPI_ISL_594590, EPI_ISL_594591, EPI_ISL_594592, EPI_ISL_594593, EPI_ISL_594594, EPI_ISL_594595, EPI_ISL_594596, EPI_ISL_594597, EPI_ISL_594598, EPI_ISL_594599, EPI_ISL_594600, EPI_ISL_594601, EPI_ISL_594602, EPI_ISL_594603, EPI_ISL_594604, EPI_ISL_594605, EPI_ISL_594606, EPI_ISL_594607, EPI_ISL_594608, EPI_ISL_594609, EPI_ISL_594610, EPI_ISL_594611, EPI_ISL_594612, EPI_ISL_594613, EPI_ISL_594614, EPI_ISL_594615, EPI_ISL_594616, EPI_ISL_611556, EPI_ISL_611671, EPI_ISL_611696, EPI_ISL_611700, EPI_ISL_611776, EPI_ISL_611814, EPI_ISL_611847, EPI_ISL_611859, EPI_ISL_612015, EPI_ISL_612047, EPI_ISL_612073, EPI_ISL_612219, EPI_ISL_612220, EPI_ISL_612221, EPI_ISL_612222, EPI_ISL_612223, EPI_ISL_612224, EPI_ISL_612225, EPI_ISL_612226, EPI_ISL_612228, EPI_ISL_612229, EPI_ISL_612230, EPI_ISL_612231, EPI_ISL_612232, EPI_ISL_612233, EPI_ISL_612234, EPI_ISL_612235, EPI_ISL_612236, EPI_ISL_612237, EPI_ISL_612238, EPI_ISL_612239, EPI_ISL_612240, EPI_ISL_612241, EPI_ISL_612243, EPI_ISL_612244, EPI_ISL_612245, EPI_ISL_612246, EPI_ISL_612247, EPI_ISL_612248, EPI_ISL_612249, EPI_ISL_612250, EPI_ISL_612251, EPI_ISL_612252, EPI_ISL_612253, EPI_ISL_612254, EPI_ISL_612255, EPI_ISL_612256, EPI_ISL_612257, EPI_ISL_612258

| | | | |
|---|---|---|---|
| see above | University of Birmingham | COVID-19 Genomics UK | Alex Richter; Andrew Bosworth; Andrew Bosworth. Queen Elizabeth Hospital: Anna Casey; Andrew D Beggs PHE Heartlands Lab: Husam Osman; Charlotte Poxon; Institute of Microbiology; Joanne Stockton; Josh Quick; Kasun Wanigasooriya; Mike Kidd; |

(COG-UK) Consortium | Nicholas Loman. University of Birmingham Testing Laboratory: Celina M Whalley; Oliver Pickles; Radoslaw Poplawski; Samuel Nicholls; University of Birmingham: Claire McMurray; Will Rowe

EPI_ISL_584883, EPI_ISL_584884, EPI_ISL_584885, EPI_ISL_584886, EPI_ISL_584887, EPI_ISL_584888, EPI_ISL_584889, EPI_ISL_584890, EPI_ISL_584891, EPI_ISL_584892, EPI_ISL_584893, EPI_ISL_584894, EPI_ISL_584895, EPI_ISL_584896, EPI_ISL_584897, EPI_ISL_584898, EPI_ISL_584899, EPI_ISL_584900, EPI_ISL_584901, EPI_ISL_584902, EPI_ISL_584903, EPI_ISL_584904, EPI_ISL_584905, EPI_ISL_585533, EPI_ISL_585534, EPI_ISL_585535, EPI_ISL_585536, EPI_ISL_585537, EPI_ISL_585538, EPI_ISL_585539, EPI_ISL_585540, EPI_ISL_585541, EPI_ISL_585542, EPI_ISL_585543, EPI_ISL_585544, EPI_ISL_585545, EPI_ISL_585546, EPI_ISL_585547, EPI_ISL_585548, EPI_ISL_585549, EPI_ISL_585550, EPI_ISL_585551, EPI_ISL_585552, EPI_ISL_585553, EPI_ISL_585554, EPI_ISL_585555, EPI_ISL_585556, EPI_ISL_585557, EPI_ISL_585558, EPI_ISL_585559, EPI_ISL_585560, EPI_ISL_585561, EPI_ISL_585562, EPI_ISL_585563, EPI_ISL_585564, EPI_ISL_585565, EPI_ISL_585566, EPI_ISL_585567, EPI_ISL_585568, EPI_ISL_585569, EPI_ISL_585570, EPI_ISL_585571, EPI_ISL_585572, EPI_ISL_585573, EPI_ISL_585574, EPI_ISL_585575, EPI_ISL_585576, EPI_ISL_585577, EPI_ISL_585578, EPI_ISL_585579, EPI_ISL_585580, EPI_ISL_585581, EPI_ISL_585582, EPI_ISL_611599, EPI_ISL_611836, EPI_ISL_611837, EPI_ISL_611939, EPI_ISL_611940, EPI_ISL_612118, EPI_ISL_612130, EPI_ISL_612379, EPI_ISL_612380, EPI_ISL_612381, EPI_ISL_612382, EPI_ISL_612383, EPI_ISL_612384, EPI_ISL_612385, EPI_ISL_612386, EPI_ISL_612387, EPI_ISL_612388, EPI_ISL_612389, EPI_ISL_612390, EPI_ISL_612391, EPI_ISL_612392, EPI_ISL_612393, EPI_ISL_626668, EPI_ISL_626669, EPI_ISL_626671, EPI_ISL_626682, EPI_ISL_626729, EPI_ISL_626731, EPI_ISL_626793, EPI_ISL_626794, EPI_ISL_626800, EPI_ISL_626804, EPI_ISL_626810, EPI_ISL_626890, EPI_ISL_626891, EPI_ISL_626939, EPI_ISL_626944, EPI_ISL_627065, EPI_ISL_627067, EPI_ISL_627102, EPI_ISL_627112, EPI_ISL_627143, EPI_ISL_627325, EPI_ISL_627326, EPI_ISL_627327, EPI_ISL_627328, EPI_ISL_627429, EPI_ISL_627435, EPI_ISL_627436, EPI_ISL_627438, EPI_ISL_627439, EPI_ISL_665273, EPI_ISL_665276, EPI_ISL_665277, EPI_ISL_665279, EPI_ISL_665282, EPI_ISL_665324, EPI_ISL_665325, EPI_ISL_665326, EPI_ISL_665332, EPI_ISL_665333, EPI_ISL_665417, EPI_ISL_665419, EPI_ISL_665435, EPI_ISL_665514, EPI_ISL_665515, EPI_ISL_665547, EPI_ISL_665548, EPI_ISL_665549, EPI_ISL_665570, EPI_ISL_665571, EPI_ISL_665663, EPI_ISL_665666, EPI_ISL_665675, EPI_ISL_665686, EPI_ISL_665694, EPI_ISL_665700, EPI_ISL_665721, EPI_ISL_665734, EPI_ISL_665738, EPI_ISL_665760, EPI_ISL_665851, EPI_ISL_665852, EPI_ISL_665853, EPI_ISL_665854, EPI_ISL_665855, EPI_ISL_665856, EPI_ISL_665857, EPI_ISL_665858, EPI_ISL_665859, EPI_ISL_665860, EPI_ISL_665861, EPI_ISL_665862, EPI_ISL_665863

see above | University of Exeter | COVID-19 Genomics UK (COG-UK) Consortium | Aaron Jeffries; Audrey Farbos; Ben Temperton; Jane Masoli; Joanna Warwick-Dugdale; Michelle Michelsen; Robyn Manley; Stephen Michell

EPI_ISL_585460, EPI_ISL_585461, EPI_ISL_585462, EPI_ISL_585463, EPI_ISL_585464, EPI_ISL_585465, EPI_ISL_585466, EPI_ISL_585467, EPI_ISL_585468, EPI_ISL_585469, EPI_ISL_585470, EPI_ISL_585471, EPI_ISL_585472, EPI_ISL_585473, EPI_ISL_585474, EPI_ISL_585475, EPI_ISL_585476, EPI_ISL_585477, EPI_ISL_585478, EPI_ISL_585479, EPI_ISL_585480, EPI_ISL_585481, EPI_ISL_585482, EPI_ISL_585483, EPI_ISL_585484, EPI_ISL_585485, EPI_ISL_585486, EPI_ISL_585488, EPI_ISL_585489, EPI_ISL_585490, EPI_ISL_585491, EPI_ISL_585492, EPI_ISL_585493, EPI_ISL_585494, EPI_ISL_585495, EPI_ISL_585496, EPI_ISL_585497, EPI_ISL_585498, EPI_ISL_585499, EPI_ISL_594816, EPI_ISL_594817, EPI_ISL_594818, EPI_ISL_594819, EPI_ISL_594820, EPI_ISL_594821, EPI_ISL_594822, EPI_ISL_594823, EPI_ISL_594824, EPI_ISL_594825, EPI_ISL_594826, EPI_ISL_594827, EPI_ISL_594828, EPI_ISL_594829, EPI_ISL_594830, EPI_ISL_594831, EPI_ISL_594832, EPI_ISL_594833, EPI_ISL_594834, EPI_ISL_611685, EPI_ISL_611707, EPI_ISL_611989, EPI_ISL_612126, EPI_ISL_612320, EPI_ISL_612321, EPI_ISL_612322, EPI_ISL_612323, EPI_ISL_612324, EPI_ISL_612326, EPI_ISL_612327, EPI_ISL_612328, EPI_ISL_612329, EPI_ISL_612330, EPI_ISL_612331, EPI_ISL_612332, EPI_ISL_612333, EPI_ISL_612334, EPI_ISL_612335, EPI_ISL_612336, EPI_ISL_612337, EPI_ISL_612338, EPI_ISL_612339, EPI_ISL_612340, EPI_ISL_612347, EPI_ISL_650832, EPI_ISL_651520, EPI_ISL_651957, EPI_ISL_680469, EPI_ISL_680470, EPI_ISL_680471, EPI_ISL_680472, EPI_ISL_680473, EPI_ISL_680474, EPI_ISL_680475, EPI_ISL_680476, EPI_ISL_680495, EPI_ISL_680496, EPI_ISL_703622, EPI_ISL_703782, EPI_ISL_703784, EPI_ISL_705761, EPI_ISL_705763, EPI_ISL_705778

see above | Virology Department, Royal Infirmary of Edinburgh, NHS Lothian / School of Biological Sciences, University of Edinburgh / Institute of Genetics and Molecular Medicine, University of Edinburgh | COVID-19 Genomics UK (COG-UK) Consortium | Balcaza C; Colquhoun R; Dewar R; Gallagher M; Hill V; Jackson B; McCrone JT; McHugh M; O'Toole Á; Rambaut A; Rooke S; Scher E; Templeton K; Williams TC; Yu X

EPI_ISL_585009, EPI_ISL_585010, EPI_ISL_585011, EPI_ISL_585012, EPI_ISL_585014, EPI_ISL_585016, EPI_ISL_585018, EPI_ISL_585019, EPI_ISL_585020, EPI_ISL_585021, EPI_ISL_585023, EPI_ISL_585027, EPI_ISL_585028, EPI_ISL_585029, EPI_ISL_585030, EPI_ISL_585033, EPI_ISL_585034, EPI_ISL_585039, EPI_ISL_585042, EPI_ISL_585046, EPI_ISL_585047, EPI_ISL_585049, EPI_ISL_585050, EPI_ISL_585053, EPI_ISL_585055, EPI_ISL_585058, EPI_ISL_585059, EPI_ISL_585061, EPI_ISL_585062, EPI_ISL_585065, EPI_ISL_585066, EPI_ISL_585067, EPI_ISL_585068, EPI_ISL_585069, EPI_ISL_585070, EPI_ISL_585074, EPI_ISL_585075, EPI_ISL_585078, EPI_ISL_585079, EPI_ISL_585080, EPI_ISL_585081, EPI_ISL_585082, EPI_ISL_585083, EPI_ISL_585084, EPI_ISL_585087, EPI_ISL_585091, EPI_ISL_595802, EPI_ISL_595803, EPI_ISL_595804, EPI_ISL_595805, EPI_ISL_595806, EPI_ISL_595807, EPI_ISL_595808, EPI_ISL_595809, EPI_ISL_595810, EPI_ISL_595811, EPI_ISL_595812, EPI_ISL_595813, EPI_ISL_595814, EPI_ISL_595815, EPI_ISL_595816, EPI_ISL_595817, EPI_ISL_595818, EPI_ISL_595819, EPI_ISL_595820, EPI_ISL_595821, EPI_ISL_595823, EPI_ISL_595824, EPI_ISL_595825, EPI_ISL_595826, EPI_ISL_595827, EPI_ISL_595828, EPI_ISL_595829, EPI_ISL_595830, EPI_ISL_595831, EPI_ISL_595832, EPI_ISL_595833, EPI_ISL_595834, EPI_ISL_595835, EPI_ISL_595836, EPI_ISL_596191, EPI_ISL_596192, EPI_ISL_596194, EPI_ISL_596195, EPI_ISL_596196, EPI_ISL_596197, EPI_ISL_596198, EPI_ISL_596199, EPI_ISL_596200, EPI_ISL_596201, EPI_ISL_596202, EPI_ISL_596203, EPI_ISL_596204, EPI_ISL_596205, EPI_ISL_596206, EPI_ISL_596207, EPI_ISL_596208, EPI_ISL_596209, EPI_ISL_596210, EPI_ISL_596211, EPI_ISL_596212, EPI_ISL_596213, EPI_ISL_596214, EPI_ISL_596215, EPI_ISL_596218, EPI_ISL_596219, EPI_ISL_596220, EPI_ISL_596221, EPI_ISL_596222, EPI_ISL_596223, EPI_ISL_596224, EPI_ISL_596225, EPI_ISL_596226, EPI_ISL_596227, EPI_ISL_611536, EPI_ISL_611679, EPI_ISL_611733, EPI_ISL_611789, EPI_ISL_611800, EPI_ISL_611829, EPI_ISL_611988, EPI_ISL_612020, EPI_ISL_613358, EPI_ISL_613360, EPI_ISL_613362, EPI_ISL_613364, EPI_ISL_613371, EPI_ISL_613372, EPI_ISL_613379, EPI_ISL_613380, EPI_ISL_613386, EPI_ISL_613387, EPI_ISL_613389, EPI_ISL_613392, EPI_ISL_613399, EPI_ISL_613403, EPI_ISL_613405, EPI_ISL_626664, EPI_ISL_626679, EPI_ISL_626680, EPI_ISL_626710, EPI_ISL_626734, EPI_ISL_626739, EPI_ISL_626743, EPI_ISL_626747, EPI_ISL_626752, EPI_ISL_626754, EPI_ISL_626757, EPI_ISL_626771, EPI_ISL_626781, EPI_ISL_626786, EPI_ISL_626791, EPI_ISL_626809, EPI_ISL_626817, EPI_ISL_626821, EPI_ISL_626823, EPI_ISL_626827, EPI_ISL_626830, EPI_ISL_626852, EPI_ISL_626896, EPI_ISL_626898, EPI_ISL_626899, EPI_ISL_626900, EPI_ISL_626901, EPI_ISL_626902, EPI_ISL_626903, EPI_ISL_626906, EPI_ISL_626907, EPI_ISL_626908, EPI_ISL_626909, EPI_ISL_626911, EPI_ISL_626912, EPI_ISL_626913, EPI_ISL_626915, EPI_ISL_626916, EPI_ISL_626918, EPI_ISL_626922, EPI_ISL_626923, EPI_ISL_626924, EPI_ISL_626929, EPI_ISL_626932, EPI_ISL_626938, EPI_ISL_626947, EPI_ISL_626948, EPI_ISL_626949, EPI_ISL_626950, EPI_ISL_626951, EPI_ISL_627019, EPI_ISL_627047, EPI_ISL_627103, EPI_ISL_627130, EPI_ISL_627164, EPI_ISL_627184, EPI_ISL_627191, EPI_ISL_627252, EPI_ISL_627253, EPI_ISL_627254, EPI_ISL_627264, EPI_ISL_627280, EPI_ISL_627298, EPI_ISL_627313, EPI_ISL_627492, EPI_ISL_627493, EPI_ISL_627494, EPI_ISL_627495, EPI_ISL_627496, EPI_ISL_627497, EPI_ISL_627498, EPI_ISL_627499, EPI_ISL_627500, EPI_ISL_627501, EPI_ISL_627502, EPI_ISL_627503, EPI_ISL_627504, EPI_ISL_627505, EPI_ISL_627506, EPI_ISL_627507, EPI_ISL_627508, EPI_ISL_627509, EPI_ISL_627510, EPI_ISL_627511, EPI_ISL_627512, EPI_ISL_627513, EPI_ISL_627514, EPI_ISL_627515, EPI_ISL_627516, EPI_ISL_627517, EPI_ISL_628369, EPI_ISL_628370, EPI_ISL_628371, EPI_ISL_628372, EPI_ISL_628373, EPI_ISL_628374, EPI_ISL_628375, EPI_ISL_628376, EPI_ISL_628377, EPI_ISL_628378, EPI_ISL_628379, EPI_ISL_628380, EPI_ISL_628381, EPI_ISL_628382, EPI_ISL_628383, EPI_ISL_628384, EPI_ISL_628385, EPI_ISL_628386, EPI_ISL_628387, EPI_ISL_628388, EPI_ISL_628389, EPI_ISL_628390, EPI_ISL_628391, EPI_ISL_628392, EPI_ISL_628393, EPI_ISL_628394, EPI_ISL_628395, EPI_ISL_628396, EPI_ISL_628397, EPI_ISL_628398, EPI_ISL_628399, EPI_ISL_628400, EPI_ISL_628401, EPI_ISL_628402, EPI_ISL_628403, EPI_ISL_628404, EPI_ISL_628405, EPI_ISL_628406, EPI_ISL_628407, EPI_ISL_628408, EPI_ISL_628409, EPI_ISL_628410, EPI_ISL_628411, EPI_ISL_628412, EPI_ISL_628413, EPI_ISL_628421, EPI_ISL_637559, EPI_ISL_638017, EPI_ISL_638882, EPI_ISL_650244, EPI_ISL_650283, EPI_ISL_650372, EPI_ISL_650663, EPI_ISL_651243, EPI_ISL_653071, EPI_ISL_653074, EPI_ISL_653075, EPI_ISL_653086, EPI_ISL_653087, EPI_ISL_653093, EPI_ISL_653097, EPI_ISL_679986, EPI_ISL_679987, EPI_ISL_679988, EPI_ISL_679989, EPI_ISL_679990, EPI_ISL_679991, EPI_ISL_679992, EPI_ISL_679993, EPI_ISL_679994, EPI_ISL_679995, EPI_ISL_679996, EPI_ISL_679997, EPI_ISL_679998, EPI_ISL_679999, EPI_ISL_680000, EPI_ISL_680001, EPI_ISL_680002, EPI_ISL_680003, EPI_ISL_680004, EPI_ISL_680005, EPI_ISL_680006, EPI_ISL_680007, EPI_ISL_680008, EPI_ISL_680009, EPI_ISL_680010, EPI_ISL_680011, EPI_ISL_680012, EPI_ISL_680013, EPI_ISL_680014, EPI_ISL_680015, EPI_ISL_680556, EPI_ISL_680557, EPI_ISL_680558, EPI_ISL_680559, EPI_ISL_680560, EPI_ISL_680561, EPI_ISL_816362, EPI_ISL_924418, EPI_ISL_924426, EPI_ISL_1000650, EPI_ISL_1000793

see above | Virology Department, Sheffield Teaching Hospitals NHS Foundation Trust/Department of Infection, Immunity and Cardiovascular Disease, The Medical School, University of Sheffield | COVID-19 Genomics UK (COG-UK) Consortium | Adri Angyal; Alex Keeley; Benjamin Lindsey; Cariad Evans; Danielle Groves; Dave Partridge; Katie Johnson; Laura Carrilero; Luke Green; Matthew Parker; Matthew Wyles; Mehmet Yavuz; Mohammad Raza; Nikki Smith; Paul Parsons; Rachel Tucker; Rebecca Brown; Thushan de Silva

EPI_ISL_577504, EPI_ISL_577507, EPI_ISL_577513, EPI_ISL_577526, EPI_ISL_577530, EPI_ISL_577531, EPI_ISL_585630, EPI_ISL_585631, EPI_ISL_585632, EPI_ISL_585633, EPI_ISL_585634, EPI_ISL_585635, EPI_ISL_585636, EPI_ISL_585637, EPI_ISL_585638, EPI_ISL_585639, EPI_ISL_585640, EPI_ISL_585641, EPI_ISL_585642, EPI_ISL_585643, EPI_ISL_585644, EPI_ISL_585645, EPI_ISL_585646, EPI_ISL_585647, EPI_ISL_585648, EPI_ISL_585649, EPI_ISL_585650, EPI_ISL_585651, EPI_ISL_585652, EPI_ISL_585654, EPI_ISL_585655, EPI_ISL_585656, EPI_ISL_585657, EPI_ISL_585658, EPI_ISL_585659, EPI_ISL_585660, EPI_ISL_585661, EPI_ISL_585662, EPI_ISL_585663, EPI_ISL_585664, EPI_ISL_585665, EPI_ISL_585666, EPI_ISL_585667, EPI_ISL_585668, EPI_ISL_585669, EPI_ISL_585670, EPI_ISL_585671, EPI_ISL_585672, EPI_ISL_585673, EPI_ISL_585674, EPI_ISL_585675, EPI_ISL_585676, EPI_ISL_585677, EPI_ISL_585678, EPI_ISL_585679, EPI_ISL_585680, EPI_ISL_585681, EPI_ISL_585682, EPI_ISL_585683, EPI_ISL_585684, EPI_ISL_585685, EPI_ISL_585686, EPI_ISL_585687, EPI_ISL_585688, EPI_ISL_585689, EPI_ISL_585690, EPI_ISL_585691, EPI_ISL_585692, EPI_ISL_585693, EPI_ISL_585694, EPI_ISL_585695, EPI_ISL_585696, EPI_ISL_585697, EPI_ISL_585698, EPI_ISL_585699, EPI_ISL_585700, EPI_ISL_585701, EPI_ISL_585702, EPI_ISL_585703, EPI_ISL_585704, EPI_ISL_585705, EPI_ISL_585706, EPI_ISL_585707, EPI_ISL_585708, EPI_ISL_585709, EPI_ISL_585710, EPI_ISL_585711, EPI_ISL_585712, EPI_ISL_585713, EPI_ISL_585714, EPI_ISL_585715, EPI_ISL_585716, EPI_ISL_585717, EPI_ISL_585718, EPI_ISL_585719, EPI_ISL_585721, EPI_ISL_585722, EPI_ISL_585723, EPI_ISL_585724, EPI_ISL_585725, EPI_ISL_585726, EPI_ISL_585727, EPI_ISL_585728, EPI_ISL_585729, EPI_ISL_585730, EPI_ISL_585731, EPI_ISL_585732, EPI_ISL_585733, EPI_ISL_585734, EPI_ISL_585735, EPI_ISL_585736, EPI_ISL_585737, EPI_ISL_585738, EPI_ISL_585739, EPI_ISL_585740, EPI_ISL_585741, EPI_ISL_585742, EPI_ISL_585743, EPI_ISL_585744, EPI_ISL_585745, EPI_ISL_585746, EPI_ISL_585747, EPI_ISL_585748, EPI_ISL_585749, EPI_ISL_585750, EPI_ISL_585751, EPI_ISL_585752, EPI_ISL_585753, EPI_ISL_585754, EPI_ISL_585755, EPI_ISL_585756, EPI_ISL_585757, EPI_ISL_585758, EPI_ISL_585759, EPI_ISL_585760, EPI_ISL_585762, EPI_ISL_585763, EPI_ISL_585764, EPI_ISL_585765, EPI_ISL_585766, EPI_ISL_585767, EPI_ISL_585768, EPI_ISL_585769, EPI_ISL_585770, EPI_ISL_585771, EPI_ISL_585772, EPI_ISL_585773, EPI_ISL_585774, EPI_ISL_585775, EPI_ISL_585776, EPI_ISL_585777, EPI_ISL_585778, EPI_ISL_585779, EPI_ISL_585780, EPI_ISL_585781, EPI_ISL_585782, EPI_ISL_585783, EPI_ISL_585784, EPI_ISL_585785, EPI_ISL_585786, EPI_ISL_585787, EPI_ISL_585788, EPI_ISL_585789, EPI_ISL_585790, EPI_ISL_585791, EPI_ISL_585792, EPI_ISL_585793, EPI_ISL_585794, EPI_ISL_585795, EPI_ISL_585796, EPI_ISL_585798, EPI_ISL_585799, EPI_ISL_585800, EPI_ISL_585801, EPI_ISL_585802, EPI_ISL_585803, EPI_ISL_585804, EPI_ISL_585805, EPI_ISL_585806, EPI_ISL_585807, EPI_ISL_585808, EPI_ISL_585809, EPI_ISL_585810, EPI_ISL_585811, EPI_ISL_585812, EPI_ISL_585813, EPI_ISL_585814, EPI_ISL_585815, EPI_ISL_585816, EPI_ISL_585817, EPI_ISL_585818, EPI_ISL_585819, EPI_ISL_585820, EPI_ISL_585821, EPI_ISL_585822, EPI_ISL_585823, EPI_ISL_585824, EPI_ISL_585825, EPI_ISL_585826, EPI_ISL_585827, EPI_ISL_585828, EPI_ISL_585829, EPI_ISL_585830, EPI_ISL_585831, EPI_ISL_585832, EPI_ISL_585833, EPI_ISL_585834, EPI_ISL_585835, EPI_ISL_585836, EPI_ISL_585837, EPI_ISL_585838, EPI_ISL_585839, EPI_ISL_585840, EPI_ISL_585841, EPI_ISL_585842, EPI_ISL_585843, EPI_ISL_585844, EPI_ISL_585845, EPI_ISL_585846, EPI_ISL_585847, EPI_ISL_585848, EPI_ISL_585849, EPI_ISL_585850, EPI_ISL_585851, EPI_ISL_585852, EPI_ISL_585853, EPI_ISL_585854, EPI_ISL_585855, EPI_ISL_585856, EPI_ISL_585857, EPI_ISL_585858, EPI_ISL_585859, EPI_ISL_585860, EPI_ISL_585861, EPI_ISL_585862, EPI_ISL_585863, EPI_ISL_585864, EPI_ISL_585865, EPI_ISL_585866, EPI_ISL_585867, EPI_ISL_585868, EPI_ISL_585869, EPI_ISL_585870, EPI_ISL_585871, EPI_ISL_585872, EPI_ISL_585873, EPI_ISL_585874, EPI_ISL_585875, EPI_ISL_585876, EPI_ISL_585877, EPI_ISL_585878, EPI_ISL_585879, EPI_ISL_585880, EPI_ISL_585881, EPI_ISL_585882, EPI_ISL_585883, EPI_ISL_585884, EPI_ISL_585885, EPI_ISL_585887, EPI_ISL_585888, EPI_ISL_585889, EPI_ISL_585890, EPI_ISL_585891, EPI_ISL_585892, EPI_ISL_585893, EPI_ISL_585894, EPI_ISL_585895, EPI_ISL_585896, EPI_ISL_585897, EPI_ISL_585898, EPI_ISL_585899, EPI_ISL_585901, EPI_ISL_585902, EPI_ISL_585903, EPI_ISL_585904, EPI_ISL_585905, EPI_ISL_585906, EPI_ISL_585907, EPI_ISL_585908, EPI_ISL_585909, EPI_ISL_585910, EPI_ISL_585911, EPI_ISL_585912, EPI_ISL_585913, EPI_ISL_585914, EPI_ISL_585915, EPI_ISL_585916, EPI_ISL_585917, EPI_ISL_585918, EPI_ISL_585919, EPI_ISL_585920, EPI_ISL_585921, EPI_ISL_585922, EPI_ISL_585923, EPI_ISL_585924, EPI_ISL_585925, EPI_ISL_585926, EPI_ISL_585927, EPI_ISL_585928, EPI_ISL_585929, EPI_ISL_585930, EPI_ISL_585931, EPI_ISL_585932, EPI_ISL_585933, EPI_ISL_585934, EPI_ISL_585935, EPI_ISL_585936, EPI_ISL_585937, EPI_ISL_585938, EPI_ISL_585939, EPI_ISL_585940, EPI_ISL_585941, EPI_ISL_585942, EPI_ISL_585943, EPI_ISL_585944, EPI_ISL_585945, EPI_ISL_585946, EPI_ISL_585947, EPI_ISL_585948, EPI_ISL_585949, EPI_ISL_585950, EPI_ISL_585951, EPI_ISL_585952, EPI_ISL_585953, EPI_ISL_585954, EPI_ISL_585955, EPI_ISL_585956, EPI_ISL_585957, EPI_ISL_585958, EPI_ISL_585959, EPI_ISL_585960, EPI_ISL_585961, EPI_ISL_585962, EPI_ISL_585963, EPI_ISL_585964, EPI_ISL_585965, EPI_ISL_585966, EPI_ISL_585967, EPI_ISL_585968, EPI_ISL_585969, EPI_ISL_585970, EPI_ISL_585971, EPI_ISL_585972, EPI_ISL_585973, EPI_ISL_585974, EPI_ISL_585975, EPI_ISL_585976, EPI_ISL_585978, EPI_ISL_585979, EPI_ISL_585980, EPI_ISL_585981, EPI_ISL_585982, EPI_ISL_585983, EPI_ISL_585984, EPI_ISL_585985, EPI_ISL_585986, EPI_ISL_585987, EPI_ISL_585988, EPI_ISL_585989, EPI_ISL_585990, EPI_ISL_585991, EPI_ISL_585992, EPI_ISL_585993, EPI_ISL_585994, EPI_ISL_585995, EPI_ISL_585996, EPI_ISL_585997, EPI_ISL_585998, EPI_ISL_585999, EPI_ISL_586000, EPI_ISL_586001, EPI_ISL_586002, EPI_ISL_586003, EPI_ISL_586004, EPI_ISL_586005, EPI_ISL_586006, EPI_ISL_586007, EPI_ISL_586008, EPI_ISL_586009, EPI_ISL_586010, EPI_ISL_586011, EPI_ISL_586012, EPI_ISL_586013, EPI_ISL_586014, EPI_ISL_586015, EPI_ISL_586016, EPI_ISL_586017, EPI_ISL_586018, EPI_ISL_586019, EPI_ISL_586020, EPI_ISL_586021, EPI_ISL_586022, EPI_ISL_586023, EPI_ISL_586024, EPI_ISL_586026, EPI_ISL_586027, EPI_ISL_586028, EPI_ISL_586030, EPI_ISL_586031, EPI_ISL_586032, EPI_ISL_586033, EPI_ISL_586034, EPI_ISL_586036, EPI_ISL_586037, EPI_ISL_586038, EPI_ISL_586039, EPI_ISL_586041, EPI_ISL_586043, EPI_ISL_586044, EPI_ISL_586045, EPI_ISL_586047, EPI_ISL_586048, EPI_ISL_586050, EPI_ISL_586051, EPI_ISL_586052, EPI_ISL_586053, EPI_ISL_586054, EPI_ISL_586055, EPI_ISL_586056, EPI_ISL_586057, EPI_ISL_586058, EPI_ISL_586060, EPI_ISL_586061, EPI_ISL_586062, EPI_ISL_586063, EPI_ISL_586064, EPI_ISL_586065, EPI_ISL_586066, EPI_ISL_586067, EPI_ISL_586068, EPI_ISL_586069, EPI_ISL_586070, EPI_ISL_586071, EPI_ISL_586072, EPI_ISL_586073, EPI_ISL_586074, EPI_ISL_586076, EPI_ISL_586077, EPI_ISL_586078, EPI_ISL_586079, EPI_ISL_586081, EPI_ISL_586082, EPI_ISL_586083, EPI_ISL_586084, EPI_ISL_586087, EPI_ISL_586088, EPI_ISL_586089, EPI_ISL_586091, EPI_ISL_586092, EPI_ISL_586093, EPI_ISL_586094, EPI_ISL_586095, EPI_ISL_586096, EPI_ISL_586097, EPI_ISL_586098, EPI_ISL_586099, EPI_ISL_586100, EPI_ISL_586101, EPI_ISL_586102, EPI_ISL_586103, EPI_ISL_586105, EPI_ISL_586106, EPI_ISL_586108, EPI_ISL_586110, EPI_ISL_586114, EPI_ISL_586115, EPI_ISL_586116, EPI_ISL_586117, EPI_ISL_586118, EPI_ISL_586119, EPI_ISL_586120, EPI_ISL_586121, EPI_ISL_586122, EPI_ISL_586124, EPI_ISL_586127, EPI_ISL_586128, EPI_ISL_586129, EPI_ISL_586132, EPI_ISL_586133, EPI_ISL_586134, EPI_ISL_586135, EPI_ISL_586136, EPI_ISL_586137, EPI_ISL_586138, EPI_ISL_586139, EPI_ISL_586142, EPI_ISL_586143, EPI_ISL_586144, EPI_ISL_586145, EPI_ISL_586146, EPI_ISL_586147, EPI_ISL_586148, EPI_ISL_586150, EPI_ISL_586151, EPI_ISL_586152, EPI_ISL_586154, EPI_ISL_586155, EPI_ISL_586156, EPI_ISL_586157, EPI_ISL_586158, EPI_ISL_586159, EPI_ISL_586160, EPI_ISL_586161, EPI_ISL_586162, EPI_ISL_586163, EPI_ISL_586164, EPI_ISL_586166, EPI_ISL_586167, EPI_ISL_586168, EPI_ISL_586169, EPI_ISL_586171, EPI_ISL_586173, EPI_ISL_586174, EPI_ISL_586175, EPI_ISL_586176, EPI_ISL_586177, EPI_ISL_586178, EPI_ISL_586179, EPI_ISL_586180, EPI_ISL_586182, EPI_ISL_586183, EPI_ISL_586184, EPI_ISL_586185, EPI_ISL_586186, EPI_ISL_586187, EPI_ISL_586189, EPI_ISL_586191, EPI_ISL_586192, EPI_ISL_586193, EPI_ISL_586195, EPI_ISL_586196, EPI_ISL_586197, EPI_ISL_586198, EPI_ISL_586199, EPI_ISL_586200, EPI_ISL_586201, EPI_ISL_586202, EPI_ISL_586203, EPI_ISL_586204, EPI_ISL_586205, EPI_ISL_586206, EPI_ISL_586207, EPI_ISL_586208, EPI_ISL_586210, EPI_ISL_586211, EPI_ISL_586213, EPI_ISL_586214, EPI_ISL_586215, EPI_ISL_586216, EPI_ISL_586217, EPI_ISL_586218, EPI_ISL_586219, EPI_ISL_586220, EPI_ISL_586221, EPI_ISL_586222, EPI_ISL_586223, EPI_ISL_586224, EPI_ISL_586225, EPI_ISL_586226, EPI_ISL_586227, EPI_ISL_586228, EPI_ISL_586229, EPI_ISL_586230, EPI_ISL_586232, EPI_ISL_586233, EPI_ISL_586234, EPI_ISL_586235, EPI_ISL_586237, EPI_ISL_586239, EPI_ISL_595329, EPI_ISL_595330, EPI_ISL_595334, EPI_ISL_595336, EPI_ISL_595338, EPI_ISL_595340, EPI_ISL_595351, EPI_ISL_595352, EPI_ISL_595353, EPI_ISL_595354, EPI_ISL_595370, EPI_ISL_595373, EPI_ISL_595374, EPI_ISL_595375, EPI_ISL_595377, EPI_ISL_595380, EPI_ISL_595383, EPI_ISL_595385, EPI_ISL_595386, EPI_ISL_595387, EPI_ISL_595391, EPI_ISL_595393, EPI_ISL_595398, EPI_ISL_595403, EPI_ISL_595405, EPI_ISL_595406, EPI_ISL_595407, EPI_ISL_595411, EPI_ISL_595412, EPI_ISL_595413, EPI_ISL_595414, EPI_ISL_595418, EPI_ISL_595420, EPI_ISL_595421, EPI_ISL_595426, EPI_ISL_595433, EPI_ISL_595434, EPI_ISL_595435, EPI_ISL_595436, EPI_ISL_595437, EPI_ISL_595439, EPI_ISL_595441, EPI_ISL_595442, EPI_ISL_595446, EPI_ISL_595447, EPI_ISL_595448, EPI_ISL_595451, EPI_ISL_595452, EPI_ISL_595454, EPI_ISL_595455, EPI_ISL_595457, EPI_ISL_595459, EPI_ISL_595461, EPI_ISL_595465, EPI_ISL_595468, EPI_ISL_595469, EPI_ISL_595470, EPI_ISL_595472, EPI_ISL_595476, EPI_ISL_595478, EPI_ISL_595479, EPI_ISL_595480, EPI_ISL_595481, EPI_ISL_595482, EPI_ISL_595483, EPI_ISL_595488, EPI_ISL_595489, EPI_ISL_595491, EPI_ISL_595492, EPI_ISL_595494, EPI_ISL_595496, EPI_ISL_595502, EPI_ISL_595503, EPI_ISL_595505, EPI_ISL_595507, EPI_ISL_595512, EPI_ISL_595513, EPI_ISL_595514, EPI_ISL_595523, EPI_ISL_595524, EPI_ISL_595525, EPI_ISL_595528, EPI_ISL_595530, EPI_ISL_595534, EPI_ISL_595538, EPI_ISL_595544, EPI_ISL_595545, EPI_ISL_595546, EPI_ISL_595549, EPI_ISL_595550, EPI_ISL_595554, EPI_ISL_595555, EPI_ISL_595557, EPI_ISL_595558, EPI_ISL_595561, EPI_ISL_595566, EPI_ISL_595568, EPI_ISL_595573, EPI_ISL_595574, EPI_ISL_595575, EPI_ISL_595579, EPI_ISL_595588, EPI_ISL_595593, EPI_ISL_595595, EPI_ISL_595596, EPI_ISL_595598, EPI_ISL_595599, EPI_ISL_595601, EPI_ISL_595602, EPI_ISL_595605, EPI_ISL_595606, EPI_ISL_595608, EPI_ISL_595609, EPI_ISL_595611, EPI_ISL_595615, EPI_ISL_611636, EPI_ISL_611683, EPI_ISL_611832, EPI_ISL_611834, EPI_ISL_611849, EPI_ISL_611912, EPI_ISL_611968, EPI_ISL_611969, EPI_ISL_611970, EPI_ISL_612048, EPI_ISL_612053, EPI_ISL_612117, EPI_ISL_612121, EPI_ISL_612122, EPI_ISL_612132, EPI_ISL_612133, EPI_ISL_612138, EPI_ISL_612139, EPI_ISL_612141, EPI_ISL_612142, EPI_ISL_612144, EPI_ISL_612145, EPI_ISL_612162, EPI_ISL_612163, EPI_ISL_612165, EPI_ISL_612169, EPI_ISL_612171, EPI_ISL_612184, EPI_ISL_612200, EPI_ISL_612201, EPI_ISL_612458, EPI_ISL_612459, EPI_ISL_612460, EPI_ISL_612461, EPI_ISL_612462, EPI_ISL_612463, EPI_ISL_612464, EPI_ISL_612465, EPI_ISL_612466, EPI_ISL_612467, EPI_ISL_612468, EPI_ISL_612469, EPI_ISL_612470, EPI_ISL_612471, EPI_ISL_612472, EPI_ISL_612473, EPI_ISL_612474, EPI_ISL_612475, EPI_ISL_612476, EPI_ISL_612477, EPI_ISL_612478, EPI_ISL_612479, EPI_ISL_612480, EPI_ISL_612481, EPI_ISL_612482, EPI_ISL_612483, EPI_ISL_612484, EPI_ISL_612485, EPI_ISL_612486, EPI_ISL_612487, EPI_ISL_612488, EPI_ISL_612489, EPI_ISL_612490, EPI_ISL_612491, EPI_ISL_612492, EPI_ISL_612493, EPI_ISL_612494, EPI_ISL_612495, EPI_ISL_612496, EPI_ISL_612497, EPI_ISL_612498, EPI_ISL_612499, EPI_ISL_612500, EPI_ISL_612501, EPI_ISL_612502, EPI_ISL_612503, EPI_ISL_612504, EPI_ISL_612505, EPI_ISL_612506, EPI_ISL_612507, EPI_ISL_612508, EPI_ISL_612509, EPI_ISL_612510, EPI_ISL_612511, EPI_ISL_612512, EPI_ISL_612513, EPI_ISL_612514, EPI_ISL_612515, EPI_ISL_612516, EPI_ISL_612517, EPI_ISL_612518, EPI_ISL_612519, EPI_ISL_612520, EPI_ISL_612521, EPI_ISL_612522, EPI_ISL_612607, EPI_ISL_612608, EPI_ISL_612609, EPI_ISL_612616, EPI_ISL_612627, EPI_ISL_612628, EPI_ISL_612636, EPI_ISL_612639, EPI_ISL_612642, EPI_ISL_612643, EPI_ISL_612644, EPI_ISL_612646, EPI_ISL_612647, EPI_ISL_612650, EPI_ISL_612651, EPI_ISL_612657, EPI_ISL_612658, EPI_ISL_612662, EPI_ISL_612663, EPI_ISL_612666, EPI_ISL_612668, EPI_ISL_612669, EPI_ISL_612670, EPI_ISL_612671, EPI_ISL_612678, EPI_ISL_612680, EPI_ISL_612684, EPI_ISL_612687, EPI_ISL_612688, EPI_ISL_612690, EPI_ISL_612691, EPI_ISL_612693, EPI_ISL_612695, EPI_ISL_612696, EPI_ISL_612697, EPI_ISL_612700, EPI_ISL_612704, EPI_ISL_612706, EPI_ISL_612707, EPI_ISL_612708, EPI_ISL_612710, EPI_ISL_612711, EPI_ISL_612714, EPI_ISL_612716, EPI_ISL_612720, EPI_ISL_612721, EPI_ISL_612725, EPI_ISL_612726, EPI_ISL_612727, EPI_ISL_612728, EPI_ISL_612731, EPI_ISL_612732, EPI_ISL_612733, EPI_ISL_612737, EPI_ISL_612738, EPI_ISL_612739, EPI_ISL_612745, EPI_ISL_612746, EPI_ISL_612747, EPI_ISL_612751, EPI_ISL_612753, EPI_ISL_612754, EPI_ISL_612755, EPI_ISL_612756, EPI_ISL_612760, EPI_ISL_612762, EPI_ISL_612765, EPI_ISL_612767, EPI_ISL_612768, EPI_ISL_612769, EPI_ISL_612770, EPI_ISL_612773, EPI_ISL_612774, EPI_ISL_612782, EPI_ISL_612786, EPI_ISL_612787, EPI_ISL_612788, EPI_ISL_612789, EPI_ISL_612790, EPI_ISL_612791, EPI_ISL_612794, EPI_ISL_612795, EPI_ISL_612796, EPI_ISL_612797, EPI_ISL_612799, EPI_ISL_612803, EPI_ISL_612804, EPI_ISL_612808, EPI_ISL_612809, EPI_ISL_612811, EPI_ISL_612812, EPI_ISL_612813, EPI_ISL_612814, EPI_ISL_612816, EPI_ISL_612817, EPI_ISL_612818, EPI_ISL_612819, EPI_ISL_612821, EPI_ISL_612824, EPI_ISL_612826, EPI_ISL_612828, EPI_ISL_612829, EPI_ISL_612830, EPI_ISL_612833, EPI_ISL_612836, EPI_ISL_612838, EPI_ISL_612839, EPI_ISL_612850, EPI_ISL_612851, EPI_ISL_612854, EPI_ISL_612862, EPI_ISL_612867, EPI_ISL_612870, EPI_ISL_612880, EPI_ISL_612881, EPI_ISL_612887, EPI_ISL_612897, EPI_ISL_612901, EPI_ISL_612905, EPI_ISL_612911, EPI_ISL_612913, EPI_ISL_612920, EPI_ISL_612923, EPI_ISL_612925, EPI_ISL_612928, EPI_ISL_612938, EPI_ISL_612944, EPI_ISL_612960, EPI_ISL_612964, EPI_ISL_612967, EPI_ISL_612978, EPI_ISL_612985, EPI_ISL_613010, EPI_ISL_613014, EPI_ISL_613020, EPI_ISL_613045, EPI_ISL_613078, EPI_ISL_613098, EPI_ISL_613100, EPI_ISL_613105, EPI_ISL_613114, EPI_ISL_613136, EPI_ISL_613151, EPI_ISL_613168, EPI_ISL_626733, EPI_ISL_626833, EPI_ISL_626850, EPI_ISL_626869, EPI_ISL_626875, EPI_ISL_627012, EPI_ISL_627023, EPI_ISL_627026, EPI_ISL_627097, EPI_ISL_627115, EPI_ISL_627155, EPI_ISL_627203, EPI_ISL_627212, EPI_ISL_627219, EPI_ISL_627306, EPI_ISL_627307, EPI_ISL_627308, EPI_ISL_627482, EPI_ISL_627483, EPI_ISL_627484, EPI_ISL_627485, EPI_ISL_627486, EPI_ISL_627487, EPI_ISL_627488, EPI_ISL_627489, EPI_ISL_627490, EPI_ISL_627735, EPI_ISL_627743, EPI_ISL_627759, EPI_ISL_627780, EPI_ISL_627783, EPI_ISL_627799, EPI_ISL_627806, EPI_ISL_627828, EPI_ISL_627832, EPI_ISL_627849, EPI_ISL_627871, EPI_ISL_627872, EPI_ISL_627875, EPI_ISL_627880, EPI_ISL_627905, EPI_ISL_637346, EPI_ISL_637347, EPI_ISL_637400, EPI_ISL_637437, EPI_ISL_637458, EPI_ISL_637524, EPI_ISL_637537, EPI_ISL_637596, EPI_ISL_637667, EPI_ISL_637899, EPI_ISL_637997, EPI_ISL_638041, EPI_ISL_638084, EPI_ISL_638085, EPI_ISL_638682, EPI_ISL_638683, EPI_ISL_638684, EPI_ISL_638685, EPI_ISL_638686, EPI_ISL_638687, EPI_ISL_638688, EPI_ISL_638689, EPI_ISL_638690, EPI_ISL_638691, EPI_ISL_638692, EPI_ISL_638693, EPI_ISL_638694, EPI_ISL_638695, EPI_ISL_638696, EPI_ISL_638697, EPI_ISL_638698, EPI_ISL_638699, EPI_ISL_638700, EPI_ISL_638701, EPI_ISL_638702, EPI_ISL_638703, EPI_ISL_638704, EPI_ISL_638705, EPI_ISL_638706, EPI_ISL_638707, EPI_ISL_638708, EPI_ISL_638709, EPI_ISL_638710, EPI_ISL_638711, EPI_ISL_638712, EPI_ISL_638713, EPI_ISL_638714, EPI_ISL_638715, EPI_ISL_638716, EPI_ISL_638717, EPI_ISL_638718, EPI_ISL_638719, EPI_ISL_638720, EPI_ISL_638721, EPI_ISL_638722, EPI_ISL_638723, EPI_ISL_638724, EPI_ISL_638725, EPI_ISL_638726, EPI_ISL_638727, EPI_ISL_638728, EPI_ISL_638729, EPI_ISL_638730, EPI_ISL_638928, EPI_ISL_638929, EPI_ISL_638930, EPI_ISL_638931, EPI_ISL_638932, EPI_ISL_638933, EPI_ISL_638934, EPI_ISL_638935, EPI_ISL_639626, EPI_ISL_680612, EPI_ISL_680614, EPI_ISL_741347

see above | Wales Specialist Virology Centre Sequencing lab: Pathogen Genomics Unit | COVID-19 Genomics UK (COG-UK) Consortium | Alec Birchley; Alexander Adams; Amy Gaskin; Angela Marchbank; Bree Gatica-Wilcox; Catherine Moore; Jason Coombes; Joanne Watkins; Joel Southgate; Johnathan Evans; Laura Gifford; Lauren Gilbert; Lee Graham; Malorie Perry; Matthew Bull; Nicole Pacchiarini; Sally Corden; Sara Kumziene-Summerhayes; Sara Rey; Sarah Taylor; Simon Cottrell; Sophie Jones; Tom Connor

EPI_ISL_585425, EPI_ISL_594645, EPI_ISL_594646, EPI_ISL_594647, EPI_ISL_594648, EPI_ISL_594649, EPI_ISL_594650, EPI_ISL_594651, EPI_ISL_594652, EPI_ISL_594653, EPI_ISL_594654, EPI_ISL_594655, EPI_ISL_594656, EPI_ISL_594657, EPI_ISL_594658, EPI_ISL_594659, EPI_ISL_594660, EPI_ISL_594661, EPI_ISL_594662, EPI_ISL_594663, EPI_ISL_594664, EPI_ISL_594665, EPI_ISL_594666, EPI_ISL_594667, EPI_ISL_594668, EPI_ISL_594669, EPI_ISL_594670, EPI_ISL_594671, EPI_ISL_594672, EPI_ISL_594673, EPI_ISL_594674, EPI_ISL_594675, EPI_ISL_594676, EPI_ISL_594677, EPI_ISL_594678, EPI_ISL_594679, EPI_ISL_594680, EPI_ISL_594681, EPI_ISL_594682, EPI_ISL_594683, EPI_ISL_594684, EPI_ISL_594685, EPI_ISL_594686, EPI_ISL_594687, EPI_ISL_594688, EPI_ISL_594689, EPI_ISL_594690, EPI_ISL_594691, EPI_ISL_594692, EPI_ISL_594693, EPI_ISL_594694, EPI_ISL_594695, EPI_ISL_594696, EPI_ISL_594697, EPI_ISL_594698, EPI_ISL_594699, EPI_ISL_594700, EPI_ISL_594701, EPI_ISL_594702, EPI_ISL_594703, EPI_ISL_594704, EPI_ISL_594705, EPI_ISL_594706, EPI_ISL_594707, EPI_ISL_594708, EPI_ISL_594709, EPI_ISL_594710, EPI_ISL_594711, EPI_ISL_594712, EPI_ISL_594713, EPI_ISL_594714, EPI_ISL_594715, EPI_ISL_594716, EPI_ISL_594717, EPI_ISL_594718, EPI_ISL_594719, EPI_ISL_594720, EPI_ISL_594721, EPI_ISL_594722, EPI_ISL_594723, EPI_ISL_594724, EPI_ISL_594725, EPI_ISL_594726, EPI_ISL_594727, EPI_ISL_594728, EPI_ISL_594729, EPI_ISL_594730, EPI_ISL_594731, EPI_ISL_594732, EPI_ISL_594733, EPI_ISL_594734, EPI_ISL_594735, EPI_ISL_594736, EPI_ISL_594737, EPI_ISL_594738, EPI_ISL_594739, EPI_ISL_594740, EPI_ISL_594741, EPI_ISL_594745, EPI_ISL_594746, EPI_ISL_594747, EPI_ISL_594748, EPI_ISL_594749, EPI_ISL_594750, EPI_ISL_594752, EPI_ISL_594753, EPI_ISL_594754, EPI_ISL_594755, EPI_ISL_594760, EPI_ISL_594761, EPI_ISL_594762, EPI_ISL_594763, EPI_ISL_594764, EPI_ISL_594765, EPI_ISL_594766, EPI_ISL_594767, EPI_ISL_594768, EPI_ISL_594769, EPI_ISL_594770, EPI_ISL_594771, EPI_ISL_594772, EPI_ISL_594773, EPI_ISL_594774, EPI_ISL_594775, EPI_ISL_594776, EPI_ISL_594777, EPI_ISL_594778, EPI_ISL_594779, EPI_ISL_594780, EPI_ISL_594782, EPI_ISL_594783, EPI_ISL_594784, EPI_ISL_594785, EPI_ISL_594792, EPI_ISL_611533, EPI_ISL_611547, EPI_ISL_611548, EPI_ISL_611610, EPI_ISL_611611, EPI_ISL_611660, EPI_ISL_611709, EPI_ISL_611712, EPI_ISL_611713, EPI_ISL_611818, EPI_ISL_611820, EPI_ISL_611852, EPI_ISL_611867, EPI_ISL_611877, EPI_ISL_611878, EPI_ISL_611879, EPI_ISL_611880, EPI_ISL_611881, EPI_ISL_611890, EPI_ISL_612290, EPI_ISL_612299, EPI_ISL_612300, EPI_ISL_612301, EPI_ISL_612302, EPI_ISL_612303, EPI_ISL_612304, EPI_ISL_612305, EPI_ISL_612306, EPI_ISL_612307, EPI_ISL_612308, EPI_ISL_612309, EPI_ISL_612310, EPI_ISL_612311, EPI_ISL_612312, EPI_ISL_612313, EPI_ISL_626933, EPI_ISL_626970, EPI_ISL_627014, EPI_ISL_627117, EPI_ISL_650292, EPI_ISL_650490, EPI_ISL_650497, EPI_ISL_650503, EPI_ISL_650517, EPI_ISL_650518, EPI_ISL_650519, EPI_ISL_650523, EPI_ISL_650712, EPI_ISL_650762, EPI_ISL_650853, EPI_ISL_650927, EPI_ISL_651330, EPI_ISL_651493, EPI_ISL_651498, EPI_ISL_651583, EPI_ISL_651733, EPI_ISL_651734, EPI_ISL_651735, EPI_ISL_651736, EPI_ISL_651737, EPI_ISL_651738, EPI_ISL_651739, EPI_ISL_651785, EPI_ISL_651786, EPI_ISL_651787, EPI_ISL_651788, EPI_ISL_651789, EPI_ISL_651955, EPI_ISL_651956

see above | West of Scotland Specialist Virology Centre, NHSGGC / MRC-University of Glasgow Centre for Virus Research | COVID-19 Genomics UK (COG-UK) Consortium | Alasdair MacLean; Alice Broos; Ana da Silva Filipe; Antonia Ho; Daniel Mair; David L Robertson; Elihu Aranday-Cortes; Emma Thomson; Guy Mollett; Igor Starinskij; James Shepherd; Jenna Nichols; Joseph Hughes; Kathy Li; Kathy Smollett; Kyriaki Nomikou; Lily Tong; Marc Niebel; Matthew Holden; Natasha Jesudason; Natasha Johnson; Patawee Asamaphan; Rachel Blacow; Rajiv Shah; Richard Orton; Rory Gunson; Sarah McDonald; Sharif Shaaban; Sreenu Vattipally; Stephen Carmichael

We gratefully acknowledge the following Authors from the Originating laboratories responsible for obtaining the specimens, as well as the Submitting laboratories where the genome data were generated and shared via GISAID, on which this research is based.

All Submitters of data may be contacted directly via www.gisaid.org

Authors are sorted alphabetically.

| Accession ID | Originating Laboratory | Submitting Laboratory | Authors |
|---|---|---|---|
| EPI_ISL_842862 | Barts Health NHS Trust | COVID-19 Genomics UK (COG-UK) Consortium | BROAD; Beatrix; CUTINO-MOGUEL; Claire; David; Dola; HARRINGTON; KELE; Maria-Teresa; OWOYEMI; Raghavendran; SHYLINI |
| EPI_ISL_868789, EPI_ISL_868790, EPI_ISL_868791, EPI_ISL_868793, EPI_ISL_868795, EPI_ISL_868797, EPI_ISL_868798, EPI_ISL_868799, EPI_ISL_868800, EPI_ISL_868801, EPI_ISL_868802, EPI_ISL_868804, EPI_ISL_868805, EPI_ISL_868808, EPI_ISL_868810, EPI_ISL_868813, EPI_ISL_868815, EPI_ISL_868816, EPI_ISL_868817, EPI_ISL_868820, EPI_ISL_868822, EPI_ISL_868823, EPI_ISL_868824, EPI_ISL_868825, EPI_ISL_868826, EPI_ISL_868827, EPI_ISL_868828, EPI_ISL_868829, EPI_ISL_868830, EPI_ISL_868832, EPI_ISL_868833, EPI_ISL_868834, EPI_ISL_868835, EPI_ISL_868836, EPI_ISL_868837, EPI_ISL_868838, EPI_ISL_868839, EPI_ISL_868840, EPI_ISL_868841, EPI_ISL_1054853, EPI_ISL_1054854, EPI_ISL_1054860, EPI_ISL_1476758, EPI_ISL_2128542, EPI_ISL_2128546, EPI_ISL_2128548, EPI_ISL_2128550, EPI_ISL_2128556, EPI_ISL_2128560, EPI_ISL_2128562, EPI_ISL_2128570, EPI_ISL_2128574, EPI_ISL_2128576, EPI_ISL_2128578, EPI_ISL_2128584, EPI_ISL_2128587, EPI_ISL_2128588, EPI_ISL_2128592, EPI_ISL_2128594 | | | |
| see above | Bioinformatics and Biostatistics Lab, Advanced Sequencing Facility | COVID-19 Genomics UK (COG-UK) Consortium | Aengus Stewart; Chelsea Sawyer; Harshil Patel; Jerome Nicod; Laura Cubitt; Margaret Crawford |
| EPI_ISL_611526, EPI_ISL_611532, EPI_ISL_611535, EPI_ISL_611537, EPI_ISL_611538, EPI_ISL_611585, EPI_ISL_611590, EPI_ISL_611592, EPI_ISL_611638, EPI_ISL_611754, EPI_ISL_611755, EPI_ISL_611761, EPI_ISL_611763, EPI_ISL_611772, EPI_ISL_611773, EPI_ISL_611916, EPI_ISL_611928, EPI_ISL_611934, EPI_ISL_611960, EPI_ISL_611983, EPI_ISL_611998, EPI_ISL_612068, EPI_ISL_612069, EPI_ISL_612082, EPI_ISL_612089, EPI_ISL_612092, EPI_ISL_612094, EPI_ISL_613294, EPI_ISL_613295, EPI_ISL_613296, EPI_ISL_613297, EPI_ISL_613298, EPI_ISL_626684, EPI_ISL_650957, EPI_ISL_652622, EPI_ISL_652623, EPI_ISL_727706, EPI_ISL_727709, EPI_ISL_867933, EPI_ISL_867935, EPI_ISL_867955, EPI_ISL_867956, EPI_ISL_867957, EPI_ISL_867969, EPI_ISL_867970, EPI_ISL_867975, EPI_ISL_923228, EPI_ISL_923259, EPI_ISL_923264, EPI_ISL_923269, EPI_ISL_923270, EPI_ISL_923271, EPI_ISL_923272, EPI_ISL_923274, EPI_ISL_923282, EPI_ISL_923283, EPI_ISL_923285, EPI_ISL_923286, EPI_ISL_923288, EPI_ISL_923299, EPI_ISL_1000236 | | | |
| see above | Centre for Enzyme Innovation, University of Portsmouth / Translational Research Laboratory, Portsmouth Hospitals NHS Trust | COVID-19 Genomics UK (COG-UK) Consortium | Allyson Lloyd; Angela Beckett; Anoop Chauhan; Christopher Fearn; Ethan Butcher; Garry Scarlett; Kate Cook; Katie Loveson; Kelly Bicknell; Robert Impey; Salman Goudarzi; Samuel Robson; Sarah Wyllie; Scott Elliott; Sharon Glaysher; Yann Bourgeois |
| EPI_ISL_611593, EPI_ISL_611938, EPI_ISL_611964, EPI_ISL_611965, EPI_ISL_611966, EPI_ISL_611967, EPI_ISL_612099, EPI_ISL_612106, EPI_ISL_612123, EPI_ISL_612186, EPI_ISL_612274, EPI_ISL_612275, EPI_ISL_612276, EPI_ISL_612277, EPI_ISL_612278, EPI_ISL_612279, EPI_ISL_612280, EPI_ISL_612281, EPI_ISL_612282, EPI_ISL_612283, EPI_ISL_637260, EPI_ISL_637264, EPI_ISL_637267, EPI_ISL_637268, EPI_ISL_637269, EPI_ISL_637270, EPI_ISL_637271, EPI_ISL_637272, EPI_ISL_637276, EPI_ISL_637304, EPI_ISL_637308, EPI_ISL_637309, EPI_ISL_637310, EPI_ISL_637314, EPI_ISL_637315, EPI_ISL_637316, EPI_ISL_637317, EPI_ISL_637348, EPI_ISL_637353, EPI_ISL_637355, EPI_ISL_637356, EPI_ISL_637357, EPI_ISL_637360, EPI_ISL_637363, EPI_ISL_637364, EPI_ISL_637365, EPI_ISL_637366, EPI_ISL_637367, EPI_ISL_637368, EPI_ISL_637384, EPI_ISL_637386, EPI_ISL_637388, EPI_ISL_637389, EPI_ISL_637391, EPI_ISL_637394, EPI_ISL_637396, EPI_ISL_637397, EPI_ISL_637403, EPI_ISL_637404, EPI_ISL_637405, EPI_ISL_637406, EPI_ISL_637407, EPI_ISL_637408, EPI_ISL_637424, EPI_ISL_637426, EPI_ISL_637435, EPI_ISL_637451, EPI_ISL_637457, EPI_ISL_637460, EPI_ISL_637461, EPI_ISL_637462, EPI_ISL_637465, EPI_ISL_637472, EPI_ISL_637473, EPI_ISL_637474, EPI_ISL_637475, EPI_ISL_637476, EPI_ISL_637477, EPI_ISL_637478, EPI_ISL_637479, EPI_ISL_637480, EPI_ISL_637481, EPI_ISL_637482, EPI_ISL_637483, EPI_ISL_637484, EPI_ISL_637485, EPI_ISL_637486, EPI_ISL_637487, EPI_ISL_637488, EPI_ISL_637489, EPI_ISL_637490, EPI_ISL_637501, EPI_ISL_637503, EPI_ISL_637504, EPI_ISL_637507, EPI_ISL_637508, EPI_ISL_637510, EPI_ISL_637512, EPI_ISL_637513, EPI_ISL_637514, EPI_ISL_637515, EPI_ISL_637516, EPI_ISL_637517, EPI_ISL_637518, EPI_ISL_637527, EPI_ISL_637532, EPI_ISL_637534, EPI_ISL_637548, EPI_ISL_637552, EPI_ISL_637560, EPI_ISL_637613, EPI_ISL_637628, EPI_ISL_637629, EPI_ISL_637630, EPI_ISL_637631, EPI_ISL_637638, EPI_ISL_637641, EPI_ISL_637642, EPI_ISL_637643, EPI_ISL_637648, EPI_ISL_637649, EPI_ISL_637650, EPI_ISL_637651, EPI_ISL_637652, EPI_ISL_637653, EPI_ISL_637654, EPI_ISL_637655, EPI_ISL_637656, EPI_ISL_637657, EPI_ISL_637658, EPI_ISL_637676, EPI_ISL_637677, EPI_ISL_637678, EPI_ISL_637680, EPI_ISL_637685, EPI_ISL_637686, EPI_ISL_637687, EPI_ISL_637688, EPI_ISL_637689, EPI_ISL_637690, EPI_ISL_637691, EPI_ISL_637692, EPI_ISL_637693, EPI_ISL_637695, EPI_ISL_637697, EPI_ISL_637700, EPI_ISL_637701, EPI_ISL_637702, EPI_ISL_637703, EPI_ISL_637704, EPI_ISL_637705, EPI_ISL_637706, EPI_ISL_637707, EPI_ISL_637721, EPI_ISL_637722, EPI_ISL_637723, EPI_ISL_637724, EPI_ISL_637725, EPI_ISL_637728, EPI_ISL_637729, EPI_ISL_637730, EPI_ISL_637731, EPI_ISL_637732, EPI_ISL_637733, EPI_ISL_637734, EPI_ISL_637735, EPI_ISL_637736, EPI_ISL_637737, EPI_ISL_637739, EPI_ISL_637740, EPI_ISL_637741, EPI_ISL_637742, EPI_ISL_637743, EPI_ISL_637744, EPI_ISL_637745, EPI_ISL_637746, EPI_ISL_637747, EPI_ISL_637748, EPI_ISL_637756, EPI_ISL_637757, EPI_ISL_637758, EPI_ISL_637759, EPI_ISL_637760, EPI_ISL_637761, EPI_ISL_637762, EPI_ISL_637763, EPI_ISL_637764, EPI_ISL_637765, EPI_ISL_637766, EPI_ISL_637767, EPI_ISL_637768, EPI_ISL_637769, EPI_ISL_637770, EPI_ISL_637771, EPI_ISL_637772, EPI_ISL_637773, EPI_ISL_637774, EPI_ISL_637775, EPI_ISL_637776, EPI_ISL_637777, EPI_ISL_637778, EPI_ISL_637779, EPI_ISL_637859, EPI_ISL_637873, EPI_ISL_637877, EPI_ISL_637890, EPI_ISL_637936, EPI_ISL_637938, EPI_ISL_637939, EPI_ISL_637940, EPI_ISL_637942, EPI_ISL_637963, EPI_ISL_637967, EPI_ISL_637974, EPI_ISL_637977, EPI_ISL_637980, EPI_ISL_637982, EPI_ISL_637983, EPI_ISL_638000, EPI_ISL_638001, EPI_ISL_638008, EPI_ISL_638013, EPI_ISL_638014, EPI_ISL_638018, EPI_ISL_638019, EPI_ISL_638036, EPI_ISL_638038, EPI_ISL_638039, EPI_ISL_638067, EPI_ISL_638068, EPI_ISL_638072, EPI_ISL_638138, EPI_ISL_638177, EPI_ISL_638178, EPI_ISL_638179, EPI_ISL_638180, EPI_ISL_638181, EPI_ISL_638182, EPI_ISL_638183, EPI_ISL_638184, EPI_ISL_638185, EPI_ISL_638186, EPI_ISL_638190, EPI_ISL_638191, EPI_ISL_638192, EPI_ISL_638195, EPI_ISL_638197, EPI_ISL_638198, EPI_ISL_638199, EPI_ISL_638200, EPI_ISL_638201, EPI_ISL_638202, EPI_ISL_638203, EPI_ISL_638204, EPI_ISL_638205, EPI_ISL_638293, EPI_ISL_638294, EPI_ISL_638295, EPI_ISL_638296, EPI_ISL_638297, EPI_ISL_638298, EPI_ISL_638299, EPI_ISL_638302, EPI_ISL_638303, EPI_ISL_638304, EPI_ISL_638305, EPI_ISL_638306, EPI_ISL_638307, EPI_ISL_638308, EPI_ISL_638309, EPI_ISL_638310, EPI_ISL_638311, EPI_ISL_638312, EPI_ISL_638313, EPI_ISL_638314, EPI_ISL_638316, EPI_ISL_638317, EPI_ISL_638318, EPI_ISL_638319, EPI_ISL_638320, EPI_ISL_638321, EPI_ISL_638322, EPI_ISL_638323, EPI_ISL_638363, EPI_ISL_638372, EPI_ISL_638373, EPI_ISL_638374, EPI_ISL_638375, EPI_ISL_638376, EPI_ISL_638377, EPI_ISL_638378, EPI_ISL_638379, EPI_ISL_638380, EPI_ISL_638381, EPI_ISL_638382, EPI_ISL_638383, EPI_ISL_638384, EPI_ISL_638386, EPI_ISL_638387, EPI_ISL_638388, EPI_ISL_638389, EPI_ISL_638390, EPI_ISL_638391, EPI_ISL_638392, EPI_ISL_638393, EPI_ISL_638394, EPI_ISL_638395, EPI_ISL_638396, EPI_ISL_638397, EPI_ISL_638398, EPI_ISL_638399, EPI_ISL_638400, EPI_ISL_638444, EPI_ISL_638445, EPI_ISL_638446, EPI_ISL_638447, EPI_ISL_638448, EPI_ISL_705537, EPI_ISL_705541 | | | |
| see above | Department of Pathology, University of Cambridge | COVID-19 Genomics UK (COG-UK) Consortium | Aminu S. Jahun; Grant Hall; Ian Goodfellow; Iliana Georgana; Malte Pinckert; Martin D. Curran; Myra Hosmillo; Surendra Parmar; Yasmin Chaudhry |
| EPI_ISL_606948, EPI_ISL_606960, EPI_ISL_607189, EPI_ISL_607200, EPI_ISL_607894, EPI_ISL_608147, EPI_ISL_608216, EPI_ISL_608310, EPI_ISL_610332, EPI_ISL_610502, EPI_ISL_610503, EPI_ISL_630474, EPI_ISL_630533, EPI_ISL_630547, EPI_ISL_644118, EPI_ISL_649426, EPI_ISL_649457, EPI_ISL_826480, EPI_ISL_826481, EPI_ISL_826482, EPI_ISL_826483, EPI_ISL_826484, EPI_ISL_826485, EPI_ISL_826515, EPI_ISL_826517, EPI_ISL_826519 | | | |
| see above | Lighthouse Lab in Alderley Park | Wellcome Sanger Institute for the COVID-19 Genomics UK (COG-UK) Consortium | Cordelia Langford; David K. Jackson; Dominic Kwiatkowski; Ewan Harrison; Ian Johnston; Jacquelyn Wynn; John Sillitoe on behalf of the Wellcome Sanger Institute COVID-19 Surveillance Team; John Sillitoe on behalf of the Wellcome Sanger Institute COVID-19 Surveillance Team (http://www.sanger.ac.uk/covid-team); Mairead Hyland; Roberto Amato; Sonia Goncalves; The Lighthouse Lab in Alderley Park and Alex Alderton |
| EPI_ISL_605932, EPI_ISL_605933, EPI_ISL_605934, EPI_ISL_605935, EPI_ISL_605936, EPI_ISL_605937, EPI_ISL_605938, EPI_ISL_605939, EPI_ISL_605940, EPI_ISL_605941, EPI_ISL_605942, EPI_ISL_605943, EPI_ISL_605944, EPI_ISL_605945, EPI_ISL_605946, EPI_ISL_605947, EPI_ISL_605948, EPI_ISL_605949, EPI_ISL_605950, EPI_ISL_605951, EPI_ISL_605952, EPI_ISL_605953, EPI_ISL_605954, EPI_ISL_605955, EPI_ISL_605956, EPI_ISL_605957, EPI_ISL_605958, EPI_ISL_605959, EPI_ISL_605960, EPI_ISL_605961, EPI_ISL_605962, EPI_ISL_605963, EPI_ISL_605964, EPI_ISL_605965, EPI_ISL_605966, EPI_ISL_605967, EPI_ISL_605968, EPI_ISL_605969, EPI_ISL_605970, EPI_ISL_605971, EPI_ISL_605972, EPI_ISL_605973, EPI_ISL_605974, EPI_ISL_605975, EPI_ISL_605976, EPI_ISL_605977, EPI_ISL_605978, EPI_ISL_605979, EPI_ISL_605980, EPI_ISL_605981, EPI_ISL_605982, EPI_ISL_605983, EPI_ISL_605984, EPI_ISL_605985, EPI_ISL_605986, EPI_ISL_605987, EPI_ISL_605988, EPI_ISL_605989, EPI_ISL_605990, EPI_ISL_605991, EPI_ISL_605992, EPI_ISL_605993, EPI_ISL_605994, EPI_ISL_605995, EPI_ISL_605996, EPI_ISL_605997, EPI_ISL_605998, EPI_ISL_605999, EPI_ISL_606000, EPI_ISL_606001, EPI_ISL_606002, EPI_ISL_606003, EPI_ISL_606004, EPI_ISL_606005, EPI_ISL_606006, EPI_ISL_606007, EPI_ISL_606008, EPI_ISL_606009, EPI_ISL_606010, EPI_ISL_606011, EPI_ISL_606012, EPI_ISL_606013, EPI_ISL_606014, EPI_ISL_606015, EPI_ISL_606016, EPI_ISL_606017, EPI_ISL_606018, EPI_ISL_606019, EPI_ISL_606020, EPI_ISL_606021, EPI_ISL_606022, EPI_ISL_606023, EPI_ISL_606024, EPI_ISL_606025, EPI_ISL_606026, EPI_ISL_606027, EPI_ISL_606028, EPI_ISL_606029, EPI_ISL_606030, EPI_ISL_606031, EPI_ISL_606032, EPI_ISL_606033, EPI_ISL_606034, EPI_ISL_606035, EPI_ISL_606036, EPI_ISL_606037, EPI_ISL_606038, EPI_ISL_606039, EPI_ISL_606040, EPI_ISL_606041, EPI_ISL_606042, EPI_ISL_606043, EPI_ISL_606044, EPI_ISL_606045, EPI_ISL_606046, EPI_ISL_606047, EPI_ISL_606048, EPI_ISL_606049, EPI_ISL_606050, EPI_ISL_606051, EPI_ISL_606052, EPI_ISL_606053, EPI_ISL_606054, EPI_ISL_606055, EPI_ISL_606056, EPI_ISL_606057, EPI_ISL_606058, EPI_ISL_606059, EPI_ISL_606060, EPI_ISL_606061, EPI_ISL_606062, EPI_ISL_606063, EPI_ISL_606064, EPI_ISL_606065, EPI_ISL_606066, EPI_ISL_606067, EPI_ISL_606068, EPI_ISL_606069, EPI_ISL_606070, EPI_ISL_606071, EPI_ISL_606072, EPI_ISL_606073, EPI_ISL_606074, EPI_ISL_606075, EPI_ISL_606076, EPI_ISL_606077, EPI_ISL_606078, EPI_ISL_606079, EPI_ISL_606080, EPI_ISL_606081, EPI_ISL_606082, EPI_ISL_606083, EPI_ISL_606084, EPI_ISL_606085, EPI_ISL_606086, EPI_ISL_606087, EPI_ISL_606088, EPI_ISL_606089, EPI_ISL_606090, EPI_ISL_606091, EPI_ISL_606092, EPI_ISL_606093, EPI_ISL_606094, EPI_ISL_606095, EPI_ISL_606096, EPI_ISL_606097, EPI_ISL_606098, EPI_ISL_606099, EPI_ISL_606100, EPI_ISL_606101, EPI_ISL_606102, EPI_ISL_606103, EPI_ISL_606104, EPI_ISL_606105, EPI_ISL_606106, EPI_ISL_606107, EPI_ISL_606108, EPI_ISL_606109, EPI_ISL_606110, EPI_ISL_606111, EPI_ISL_606112, EPI_ISL_606113, EPI_ISL_606114, EPI_ISL_606115, EPI_ISL_606116, EPI_ISL_606117, EPI_ISL_606118, EPI_ISL_606119, EPI_ISL_606120, EPI_ISL_606121, EPI_ISL_606122, EPI_ISL_606123, EPI_ISL_606124, EPI_ISL_606125, EPI_ISL_606126, EPI_ISL_606127, EPI_ISL_606128, EPI_ISL_606129, EPI_ISL_606130, EPI_ISL_606131, EPI_ISL_606132, EPI_ISL_606133, EPI_ISL_606134, EPI_ISL_606135, EPI_ISL_606136, EPI_ISL_606137, EPI_ISL_606138, EPI_ISL_606139, EPI_ISL_606140, EPI_ISL_606141, EPI_ISL_606142, EPI_ISL_606143, EPI_ISL_606144, EPI_ISL_606145, EPI_ISL_606146, EPI_ISL_606147, EPI_ISL_606148, EPI_ISL_606149, EPI_ISL_606150, EPI_ISL_606151, EPI_ISL_606152, EPI_ISL_606153, EPI_ISL_606154, EPI_ISL_606155, EPI_ISL_606156, EPI_ISL_606157, EPI_ISL_606158, EPI_ISL_606159, EPI_ISL_606160, EPI_ISL_606161, EPI_ISL_606162, EPI_ISL_606163, EPI_ISL_606164, EPI_ISL_606165, EPI_ISL_606166, EPI_ISL_606167, EPI_ISL_606168, EPI_ISL_606169, EPI_ISL_606170, EPI_ISL_606171, EPI_ISL_606172, EPI_ISL_606173, EPI_ISL_606174, EPI_ISL_606175, EPI_ISL_606176, EPI_ISL_606177, EPI_ISL_606178, EPI_ISL_606179, EPI_ISL_606180, EPI_ISL_606181, EPI_ISL_606182, EPI_ISL_606183, EPI_ISL_606184, EPI_ISL_606185, EPI_ISL_606186, EPI_ISL_606187, EPI_ISL_606188, EPI_ISL_606189, EPI_ISL_606190, EPI_ISL_606191, EPI_ISL_606192, EPI_ISL_606193, EPI_ISL_606194, EPI_ISL_606195, EPI_ISL_606196, EPI_ISL_606197, EPI_ISL_606198, EPI_ISL_606199, EPI_ISL_606200, EPI_ISL_606201, EPI_ISL_606202, EPI_ISL_606203, EPI_ISL_606204, EPI_ISL_606205, EPI_ISL_606206, EPI_ISL_606207, EPI_ISL_606208, EPI_ISL_606209, EPI_ISL_606210, EPI_ISL_606211, EPI_ISL_606212, EPI_ISL_606213, EPI_ISL_606214, EPI_ISL_606215, EPI_ISL_606216, EPI_ISL_606217, EPI_ISL_606218, EPI_ISL_606219, EPI_ISL_606220, EPI_ISL_606221, EPI_ISL_606222, EPI_ISL_606223, EPI_ISL_606224, EPI_ISL_606225, EPI_ISL_606226, EPI_ISL_606227, EPI_ISL_606228, EPI_ISL_606229, EPI_ISL_606230, EPI_ISL_606231, EPI_ISL_606232, EPI_ISL_606233, EPI_ISL_606234, EPI_ISL_606235, EPI_ISL_606236, EPI_ISL_606237, EPI_ISL_606238, EPI_ISL_606239, EPI_ISL_606240, EPI_ISL_606241, EPI_ISL_606242, EPI_ISL_606243, EPI_ISL_606244, EPI_ISL_606245, EPI_ISL_606246, EPI_ISL_606247, EPI_ISL_606248, EPI_ISL_606249, EPI_ISL_606250, EPI_ISL_606251, EPI_ISL_606252, EPI_ISL_606253, EPI_ISL_606254, EPI_ISL_606255, EPI_ISL_606256, EPI_ISL_606257, EPI_ISL_606258, EPI_ISL_606259, EPI_ISL_606260, EPI_ISL_606261, EPI_ISL_606262, EPI_ISL_606263, EPI_ISL_606264, EPI_ISL_606265, EPI_ISL_606266, EPI_ISL_606267, EPI_ISL_606268, EPI_ISL_606269, EPI_ISL_606270, EPI_ISL_606271, EPI_ISL_606272, EPI_ISL_606273, EPI_ISL_606274, EPI_ISL_606275, EPI_ISL_606276, EPI_ISL_606277, EPI_ISL_606278, EPI_ISL_606279, EPI_ISL_606280, EPI_ISL_606281, EPI_ISL_606282, EPI_ISL_606283, EPI_ISL_606284, EPI_ISL_606285, EPI_ISL_606286, EPI_ISL_606287, EPI_ISL_606288, EPI_ISL_606289, EPI_ISL_606290, EPI_ISL_606291, EPI_ISL_606292, EPI_ISL_606293, EPI_ISL_606294, EPI_ISL_606295, EPI_ISL_606924, EPI_ISL_606925, EPI_ISL_606926, EPI_ISL_606927, EPI_ISL_606928, EPI_ISL_606929, EPI_ISL_606930, EPI_ISL_606931, EPI_ISL_606932, EPI_ISL_606933, EPI_ISL_606934, EPI_ISL_606935, EPI_ISL_606936, EPI_ISL_606937, EPI_ISL_606938, EPI_ISL_606939, EPI_ISL_606940, EPI_ISL_606941, EPI_ISL_606942, EPI_ISL_606943, EPI_ISL_606944, EPI_ISL_606945, EPI_ISL_606946, EPI_ISL_606947, EPI_ISL_606949, EPI_ISL_606950, EPI_ISL_606951, EPI_ISL_606952, EPI_ISL_606953, EPI_ISL_606954, EPI_ISL_606955, EPI_ISL_606956, EPI_ISL_606957, EPI_ISL_606958, EPI_ISL_606959, EPI_ISL_606961, EPI_ISL_606962, EPI_ISL_606963, EPI_ISL_606964, EPI_ISL_606965, EPI_ISL_606966, EPI_ISL_606967, EPI_ISL_606968, EPI_ISL_606969, EPI_ISL_606970, EPI_ISL_606971, EPI_ISL_606972, EPI_ISL_606973, EPI_ISL_606974, EPI_ISL_606975, EPI_ISL_606976, EPI_ISL_606977, EPI_ISL_606978, EPI_ISL_606979, EPI_ISL_606980, EPI_ISL_606981, EPI_ISL_606982, EPI_ISL_606983, EPI_ISL_606984, EPI_ISL_606985, EPI_ISL_606986, EPI_ISL_606987, EPI_ISL_606988, EPI_ISL_606989, EPI_ISL_606990, EPI_ISL_606991, EPI_ISL_606992, EPI_ISL_606993, EPI_ISL_606994, EPI_ISL_606995, EPI_ISL_606996, EPI_ISL_606997, EPI_ISL_606998, EPI_ISL_606999, EPI_ISL_607000, EPI_ISL_607001, EPI_ISL_607002, EPI_ISL_607003, EPI_ISL_607004, EPI_ISL_607005, EPI_ISL_607006, EPI_ISL_607007, EPI_ISL_607008, EPI_ISL_607009, EPI_ISL_607010, EPI_ISL_607011, EPI_ISL_607012, EPI_ISL_607013, EPI_ISL_607014, EPI_ISL_607015, EPI_ISL_607016, EPI_ISL_607017, EPI_ISL_607018, EPI_ISL_607019, EPI_ISL_607020, EPI_ISL_607021, EPI_ISL_607022, EPI_ISL_607023, EPI_ISL_607024, EPI_ISL_607025, EPI_ISL_607026, EPI_ISL_607027, EPI_ISL_607028, EPI_ISL_607029, EPI_ISL_607030, EPI_ISL_607031, EPI_ISL_607032, EPI_ISL_607033, EPI_ISL_607034, EPI_ISL_607035, EPI_ISL_607036, EPI_ISL_607037, EPI_ISL_607038, EPI_ISL_607039, EPI_ISL_607040, EPI_ISL_607041, EPI_ISL_607042, EPI_ISL_607043, EPI_ISL_607044, EPI_ISL_607045, EPI_ISL_607046, EPI_ISL_607047, EPI_ISL_607048, EPI_ISL_607049, EPI_ISL_607050, EPI_ISL_607051, EPI_ISL_607052, EPI_ISL_607053, EPI_ISL_607054, EPI_ISL_607055, EPI_ISL_607056, EPI_ISL_607057, EPI_ISL_607058, EPI_ISL_607059, EPI_ISL_607060, EPI_ISL_607061, EPI_ISL_607062, EPI_ISL_607063, EPI_ISL_607064, EPI_ISL_607065, EPI_ISL_607066, EPI_ISL_607067, EPI_ISL_607068, EPI_ISL_607069, EPI_ISL_607070, EPI_ISL_607071, EPI_ISL_607072, EPI_ISL_607073, EPI_ISL_607074, EPI_ISL_607075, EPI_ISL_607076, EPI_ISL_607077, EPI_ISL_607078, EPI_ISL_607079, EPI_ISL_607080, EPI_ISL_607081, EPI_ISL_607082, EPI_ISL_607083, EPI_ISL_607084, EPI_ISL_607085, EPI_ISL_607086, EPI_ISL_607087, EPI_ISL_607088, EPI_ISL_607089, EPI_ISL_607090, EPI_ISL_607091, EPI_ISL_607092, EPI_ISL_607093, EPI_ISL_607094, EPI_ISL_607095, EPI_ISL_607096, EPI_ISL_607097, EPI_ISL_607098, EPI_ISL_607099, EPI_ISL_607100, EPI_ISL_607101, EPI_ISL_607102, EPI_ISL_607103, EPI_ISL_607104, EPI_ISL_607105, EPI_ISL_607106, EPI_ISL_607107, EPI_ISL_607108, EPI_ISL_607109, EPI_ISL_607110, EPI_ISL_607111, EPI_ISL_607112, EPI_ISL_607113, EPI_ISL_607114, EPI_ISL_607115, EPI_ISL_607116, EPI_ISL_607117, EPI_ISL_607118, EPI_ISL_607119, EPI_ISL_607120, EPI_ISL_607121, EPI_ISL_607122, EPI_ISL_607123, EPI_ISL_607124, EPI_ISL_607125, EPI_ISL_607126, EPI_ISL_607127, EPI_ISL_607128, EPI_ISL_607129, EPI_ISL_607130, EPI_ISL_607131, EPI_ISL_607132, EPI_ISL_607133, EPI_ISL_607134, EPI_ISL_607135, EPI_ISL_607136, EPI_ISL_607137, EPI_ISL_607138, EPI_ISL_607139, EPI_ISL_607140, EPI_ISL_607141, EPI_ISL_607142, EPI_ISL_607143, EPI_ISL_607144, EPI_ISL_607145, EPI_ISL_607146, EPI_ISL_607147, EPI_ISL_607148, EPI_ISL_607149, EPI_ISL_607150, EPI_ISL_607151, EPI_ISL_607152, EPI_ISL_607153, EPI_ISL_607154, EPI_ISL_607155, EPI_ISL_607156, EPI_ISL_607157, EPI_ISL_607158, EPI_ISL_607159, EPI_ISL_607160, EPI_ISL_607161, EPI_ISL_607162, EPI_ISL_607163, EPI_ISL_607164, EPI_ISL_607165, EPI_ISL_607166, EPI_ISL_607167, EPI_ISL_607168, EPI_ISL_607169, EPI_ISL_607170, EPI_ISL_607171, EPI_ISL_607172, EPI_ISL_607173, EPI_ISL_607174, EPI_ISL_607175, EPI_ISL_607176, EPI_ISL_607177, EPI_ISL_607178, EPI_ISL_607179, EPI_ISL_607180, EPI_ISL_607181, EPI_ISL_607182, EPI_ISL_607183, EPI_ISL_607184, EPI_ISL_607185, EPI_ISL_607186, EPI_ISL_607187, EPI_ISL_607188, EPI_ISL_607190, EPI_ISL_607191, EPI_ISL_607192, EPI_ISL_607193, EPI_ISL_607194, EPI_ISL_607195, EPI_ISL_607196, EPI_ISL_607197, EPI_ISL_607198, EPI_ISL_607199, EPI_ISL_607201, EPI_ISL_607202, EPI_ISL_607203, EPI_ISL_607204, EPI_ISL_607205, EPI_ISL_607206, EPI_ISL_607207, EPI_ISL_607208, EPI_ISL_607209, EPI_ISL_607210, EPI_ISL_607211, EPI_ISL_607212, EPI_ISL_607213, EPI_ISL_607214, EPI_ISL_607215, EPI_ISL_607216, EPI_ISL_607217, EPI_ISL_607218, EPI_ISL_607219, EPI_ISL_607220, EPI_ISL_607221, EPI_ISL_607222, EPI_ISL_607223, EPI_ISL_607224, EPI_ISL_607225, EPI_ISL_607226, EPI_ISL_607227, EPI_ISL_607228, EPI_ISL_607229, EPI_ISL_607230, EPI_ISL_607231, EPI_ISL_607232, EPI_ISL_607233, EPI_ISL_607234, EPI_ISL_607235, EPI_ISL_607236, EPI_ISL_607237, EPI_ISL_607238, EPI_ISL_607239, EPI_ISL_607240, EPI_ISL_607241, EPI_ISL_607242, EPI_ISL_607243, EPI_ISL_607244, EPI_ISL_607245, EPI_ISL_607246, EPI_ISL_607247, EPI_ISL_607248, EPI_ISL_607249, EPI_ISL_607250, EPI_ISL_607251, EPI_ISL_607252, EPI_ISL_607253, EPI_ISL_607254, EPI_ISL_607255, EPI_ISL_607256, EPI_ISL_607257, EPI_ISL_607258, EPI_ISL_607259, EPI_ISL_607260, EPI_ISL_607261, EPI_ISL_607262, EPI_ISL_607263, EPI_ISL_607264, EPI_ISL_607265, EPI_ISL_607266, EPI_ISL_607267, EPI_ISL_607268, EPI_ISL_607269, EPI_ISL_607270, EPI_ISL_607271, EPI_ISL_607272, EPI_ISL_607273, EPI_ISL_607274, EPI_ISL_607275, EPI_ISL_607276, EPI_ISL_607277, EPI_ISL_607278, EPI_ISL_607279, EPI_ISL_607280, EPI_ISL_607281, EPI_ISL_607282, EPI_ISL_607283, EPI_ISL_607284, EPI_ISL_607285, EPI_ISL_607286, EPI_ISL_607287, EPI_ISL_607288, EPI_ISL_607289, EPI_ISL_607290, EPI_ISL_607623, EPI_ISL_607624, EPI_ISL_607625, EPI_ISL_607626, EPI_ISL_607627, EPI_ISL_607628, EPI_ISL_607629, EPI_ISL_607630, EPI_ISL_607631, EPI_ISL_607632, EPI_ISL_607633, EPI_ISL_607634, EPI_ISL_607635, EPI_ISL_607636, EPI_ISL_607637, EPI_ISL_607638, EPI_ISL_607639, EPI_ISL_607640, EPI_ISL_607641, EPI_ISL_607642, EPI_ISL_607643, EPI_ISL_607644, EPI_ISL_607645, EPI_ISL_607646, EPI_ISL_607647, EPI_ISL_607648, EPI_ISL_607649, EPI_ISL_607650, EPI_ISL_607651, EPI_ISL_607652, EPI_ISL_607653, EPI_ISL_607654, EPI_ISL_607655, EPI_ISL_607656, EPI_ISL_607657, EPI_ISL_607658, EPI_ISL_607659, EPI_ISL_607660, EPI_ISL_607661, EPI_ISL_607662, EPI_ISL_607663, EPI_ISL_607664, EPI_ISL_607665, EPI_ISL_607666, EPI_ISL_607667, EPI_ISL_607668, EPI_ISL_607669, EPI_ISL_607670, EPI_ISL_607671, EPI_ISL_607672, EPI_ISL_607673, EPI_ISL_607674, EPI_ISL_607675, EPI_ISL_607676, EPI_ISL_607677, EPI_ISL_607678, EPI_ISL_607679, EPI_ISL_607680, EPI_ISL_607681, EPI_ISL_607682, EPI_ISL_607683, EPI_ISL_607684, EPI_ISL_607685, EPI_ISL_607686, EPI_ISL_607687, EPI_ISL_607688, EPI_ISL_607689, EPI_ISL_607690, EPI_ISL_607691, EPI_ISL_607692, EPI_ISL_607693, EPI_ISL_607694, EPI_ISL_607695, EPI_ISL_607696, EPI_ISL_607697, EPI_ISL_607698, EPI_ISL_607699, EPI_ISL_607700, EPI_ISL_607701, EPI_ISL_607702, EPI_ISL_607703, EPI_ISL_607704, EPI_ISL_607705, EPI_ISL_607706, EPI_ISL_607707, EPI_ISL_607708, EPI_ISL_607709, EPI_ISL_607710, EPI_ISL_607711, EPI_ISL_607712, EPI_ISL_607713, EPI_ISL_607714, EPI_ISL_607715, EPI_ISL_607716, EPI_ISL_607717, EPI_ISL_607718, EPI_ISL_607719, EPI_ISL_607720, EPI_ISL_607721, EPI_ISL_607722, EPI_ISL_607723, EPI_ISL_607724, EPI_ISL_607725, EPI_ISL_607726, EPI_ISL_607727, EPI_ISL_607728, EPI_ISL_607729, EPI_ISL_607730, EPI_ISL_607731, EPI_ISL_607732, EPI_ISL_607733, EPI_ISL_607734, EPI_ISL_607735, EPI_ISL_607736, EPI_ISL_607737, EPI_ISL_607738, EPI_ISL_607739, EPI_ISL_607740, EPI_ISL_607741, EPI_ISL_607742, EPI_ISL_607743, EPI_ISL_607744, EPI_ISL_607745, EPI_ISL_607746, EPI_ISL_607747, EPI_ISL_607748, EPI_ISL_607749, EPI_ISL_607750, EPI_ISL_607751, EPI_ISL_607752, EPI_ISL_607753, EPI_ISL_607754, EPI_ISL_607755, EPI_ISL_607756, EPI_ISL_607757, EPI_ISL_607758, EPI_ISL_607759, EPI_ISL_607760, EPI_ISL_607761, EPI_ISL_607762, EPI_ISL_607763, EPI_ISL_607764, EPI_ISL_607765, EPI_ISL_607766, EPI_ISL_607767, EPI_ISL_607768, EPI_ISL_607769, EPI_ISL_607770, EPI_ISL_607771, EPI_ISL_607772, EPI_ISL_607773, EPI_ISL_607774, EPI_ISL_607775, EPI_ISL_607776, EPI_ISL_607777, EPI_ISL_607778, EPI_ISL_607779, EPI_ISL_607780, EPI_ISL_607781, EPI_ISL_607782, EPI_ISL_607783, EPI_ISL_607784, EPI_ISL_607785, EPI_ISL_607786, EPI_ISL_607787, EPI_ISL_607788, EPI_ISL_607789, EPI_ISL_607790, EPI_ISL_607791, EPI_ISL_607792, EPI_ISL_607793, EPI_ISL_607794, EPI_ISL_607795, EPI_ISL_607796, EPI_ISL_607797, EPI_ISL_607798, EPI_ISL_607799, EPI_ISL_607800, EPI_ISL_607801, EPI_ISL_607802, EPI_ISL_607803, EPI_ISL_607804, EPI_ISL_607805, EPI_ISL_607806, EPI_ISL_607807, EPI_ISL_607808, EPI_ISL_607809, EPI_ISL_607810, EPI_ISL_607811, EPI_ISL_607812, EPI_ISL_607813, EPI_ISL_607814, EPI_ISL_607815, EPI_ISL_607816, EPI_ISL_607817, EPI_ISL_607818, EPI_ISL_607819, EPI_ISL_607820, EPI_ISL_607821, EPI_ISL_607822, EPI_ISL_607823, EPI_ISL_607824, EPI_ISL_607825, EPI_ISL_607826, EPI_ISL_607827, EPI_ISL_607828, EPI_ISL_607829, EPI_ISL_607830, EPI_ISL_607831, EPI_ISL_607832, EPI_ISL_607833, EPI_ISL_607834, EPI_ISL_607835, EPI_ISL_607836, EPI_ISL_607837, EPI_ISL_607838, EPI_ISL_607839, EPI_ISL_607840, EPI_ISL_607841, EPI_ISL_607842, EPI_ISL_607843, EPI_ISL_607844, EPI_ISL_607845, EPI_ISL_607846, EPI_ISL_607847, EPI_ISL_607848, EPI_ISL_607849, EPI_ISL_607850, EPI_ISL_607851, EPI_ISL_607852, EPI_ISL_607853, EPI_ISL_607854, EPI_ISL_607855, EPI_ISL_607856, EPI_ISL_607857, EPI_ISL_607858, EPI_ISL_607859, EPI_ISL_607860, EPI_ISL_607861, EPI_ISL_607862, EPI_ISL_607863, EPI_ISL_607864, EPI_ISL_607865, EPI_ISL_607866, EPI_ISL_607867, EPI_ISL_607868, EPI_ISL_607869, EPI_ISL_607870, EPI_ISL_607871, EPI_ISL_607872, EPI_ISL_607873, EPI_ISL_607874, EPI_ISL_607875, EPI_ISL_607876, EPI_ISL_607877, EPI_ISL_607878, EPI_ISL_607879, EPI_ISL_607880, EPI_ISL_607881, EPI_ISL_607882, EPI_ISL_607883, EPI_ISL_607884, EPI_ISL_607885, EPI_ISL_607886, EPI_ISL_607887, EPI_ISL_607888, EPI_ISL_607889, EPI_ISL_607890, EPI_ISL_607891, EPI_ISL_607892, EPI_ISL_607893, EPI_ISL_607895, EPI_ISL_607896, EPI_ISL_607897, EPI_ISL_607898, EPI_ISL_607899, EPI_ISL_607900, EPI_ISL_607901, EPI_ISL_607902, EPI_ISL_607903, EPI_ISL_607904, EPI_ISL_607905, EPI_ISL_607906, EPI_ISL_607907, EPI_ISL_607908, EPI_ISL_607909, EPI_ISL_607910, EPI_ISL_607911, EPI_ISL_607912, EPI_ISL_607913, EPI_ISL_607914, EPI_ISL_607915, EPI_ISL_607916, EPI_ISL_607917, EPI_ISL_607918, EPI_ISL_607919, EPI_ISL_607920, EPI_ISL_607921, EPI_ISL_607922, EPI_ISL_607923, EPI_ISL_607924, EPI_ISL_607925, EPI_ISL_607926, EPI_ISL_607927, EPI_ISL_607928, EPI_ISL_607929, EPI_ISL_607930, EPI_ISL_607931, EPI_ISL_607932, EPI_ISL_607933, EPI_ISL_607934, EPI_ISL_607935, EPI_ISL_607936, EPI_ISL_607937, EPI_ISL_607938, EPI_ISL_607939, EPI_ISL_607940, EPI_ISL_607941, EPI_ISL_607942, EPI_ISL_607943, EPI_ISL_607944, EPI_ISL_607945, EPI_ISL_607946, EPI_ISL_607947, EPI_ISL_607948, EPI_ISL_607949, EPI_ISL_607950, EPI_ISL_607951, EPI_ISL_607952, EPI_ISL_607953, EPI_ISL_607954, EPI_ISL_607955, EPI_ISL_607956, EPI_ISL_607957, EPI_ISL_607958, EPI_ISL_607959, EPI_ISL_607960, EPI_ISL_607961, EPI_ISL_607962, EPI_ISL_607963, EPI_ISL_607964, EPI_ISL_607965, EPI_ISL_607966, EPI_ISL_607967, EPI_ISL_607968, EPI_ISL_607969, EPI_ISL_607970, EPI_ISL_607971, EPI_ISL_607972, EPI_ISL_607973, EPI_ISL_607974, EPI_ISL_607975, EPI_ISL_607976, EPI_ISL_607977, EPI_ISL_607978, EPI_ISL_607979, EPI_ISL_607980, EPI_ISL_607981, EPI_ISL_607982, EPI_ISL_607983, EPI_ISL_607984, EPI_ISL_607985, EPI_ISL_607986, EPI_ISL_607987, EPI_ISL_607988, EPI_ISL_607989, EPI_ISL_607990, EPI_ISL_607991, EPI_ISL_607992, EPI_ISL_607993, EPI_ISL_607994, EPI_ISL_607995, EPI_ISL_607996, EPI_ISL_607997, EPI_ISL_607998, EPI_ISL_607999, EPI_ISL_608000, EPI_ISL_608001, EPI_ISL_608002, EPI_ISL_608003, EPI_ISL_608004, EPI_ISL_608005, EPI_ISL_608006, EPI_ISL_608007, EPI_ISL_608008, EPI_ISL_608009, EPI_ISL_608010, EPI_ISL_608011, EPI_ISL_608012, EPI_ISL_608013, EPI_ISL_608014, EPI_ISL_608015, EPI_ISL_608016, EPI_ISL_608017, EPI_ISL_608018, EPI_ISL_608019, EPI_ISL_608020, EPI_ISL_608021, EPI_ISL_608022, EPI_ISL_608023, EPI_ISL_608024, EPI_ISL_608025, EPI_ISL_608026, EPI_ISL_608027, EPI_ISL_608028, EPI_ISL_608029, EPI_ISL_608030, EPI_ISL_608031, EPI_ISL_608032, EPI_ISL_608033, EPI_ISL_608034, EPI_ISL_608035, EPI_ISL_608036, EPI_ISL_608037, EPI_ISL_608038, EPI_ISL_608039, EPI_ISL_608040, EPI_ISL_608041, EPI_ISL_608042, EPI_ISL_608043, EPI_ISL_608044, EPI_ISL_608045, EPI_ISL_608046, EPI_ISL_608047, EPI_ISL_608048, EPI_ISL_608049, EPI_ISL_608050, EPI_ISL_608051, EPI_ISL_608052, EPI_ISL_608053, EPI_ISL_608054, EPI_ISL_608055, EPI_ISL_608056, EPI_ISL_608057, EPI_ISL_608058, EPI_ISL_608059, EPI_ISL_608060, EPI_ISL_608061, EPI_ISL_608062, EPI_ISL_608063, EPI_ISL_608064, EPI_ISL_608065, EPI_ISL_608066, EPI_ISL_608067, EPI_ISL_608068, EPI_ISL_608069, EPI_ISL_608070, EPI_ISL_608071, EPI_ISL_608072, EPI_ISL_608073, EPI_ISL_608074, EPI_ISL_608075, EPI_ISL_608076, EPI_ISL_608077, EPI_ISL_608078, EPI_ISL_608079, EPI_ISL_608080, EPI_ISL_608081, EPI_ISL_608082, EPI_ISL_608083, EPI_ISL_608084, EPI_ISL_608085, EPI_ISL_608086, EPI_ISL_608087, EPI_ISL_608088, EPI_ISL_608089, EPI_ISL_608090, EPI_ISL_608091, EPI_ISL_608092, EPI_ISL_608093, EPI_ISL_608094, EPI_ISL_608095, EPI_ISL_608096, EPI_ISL_608097, EPI_ISL_608098, EPI_ISL_608099, EPI_ISL_608100, EPI_ISL_608101, EPI_ISL_608102, EPI_ISL_608103, EPI_ISL_608104, EPI_ISL_608105, EPI_ISL_608106, EPI_ISL_608107, EPI_ISL_608108, EPI_ISL_608109, EPI_ISL_608110, EPI_ISL_608111, EPI_ISL_608112, EPI_ISL_608113, EPI_ISL_608114, EPI_ISL_608115, EPI_ISL_608116, EPI_ISL_608117, EPI_ISL_608118, EPI_ISL_608119, EPI_ISL_608120, EPI_ISL_608121, EPI_ISL_608122, EPI_ISL_608123, EPI_ISL_608124, EPI_ISL_608125, EPI_ISL_608126, EPI_ISL_608127, EPI_ISL_608128, EPI_ISL_608129, EPI_ISL_608130, EPI_ISL_608131, EPI_ISL_608132, EPI_ISL_608133, EPI_ISL_608134, EPI_ISL_608135, EPI_ISL_608136, EPI_ISL_608137, EPI_ISL_608138, EPI_ISL_608139, EPI_ISL_608140, EPI_ISL_608141, EPI_ISL_608142, EPI_ISL_608143, EPI_ISL_608144, EPI_ISL_608145, EPI_ISL_608146, EPI_ISL_608148, EPI_ISL_608149, EPI_ISL_608150, EPI_ISL_608151, EPI_ISL_608152, EPI_ISL_608153, EPI_ISL_608154, EPI_ISL_608155, EPI_ISL_608156, EPI_ISL_608157, EPI_ISL_608158, EPI_ISL_608159, EPI_ISL_608160, EPI_ISL_608161, EPI_ISL_608162, EPI_ISL_608163, EPI_ISL_608164, EPI_ISL_608165, EPI_ISL_608166, EPI_ISL_608167, EPI_ISL_608168, EPI_ISL_608169, EPI_ISL_608170, EPI_ISL_608171, EPI_ISL_608172, EPI_ISL_608173, EPI_ISL_608174, EPI_ISL_608175, EPI_ISL_608176, EPI_ISL_608177, EPI_ISL_608178, EPI_ISL_608179, EPI_ISL_608180, EPI_ISL_608181, EPI_ISL_608182, EPI_ISL_608183, EPI_ISL_608184, EPI_ISL_608185, EPI_ISL_608186, EPI_ISL_608187, EPI_ISL_608188, EPI_ISL_608189, EPI_ISL_608190, EPI_ISL_608191, EPI_ISL_608192, EPI_ISL_608193, EPI_ISL_608194, EPI_ISL_608195, EPI_ISL_608196, EPI_ISL_608197, EPI_ISL_608198, EPI_ISL_608199, EPI_ISL_608200, EPI_ISL_608201, EPI_ISL_608202, EPI_ISL_608203, EPI_ISL_608204, EPI_ISL_608205, EPI_ISL_608206, EPI_ISL_608207, EPI_ISL_608208, EPI_ISL_608209, EPI_ISL_608210, EPI_ISL_608211, EPI_ISL_608212, EPI_ISL_608213, EPI_ISL_608214, EPI_ISL_608215, EPI_ISL_608217, EPI_ISL_608218, EPI_ISL_608219, EPI_ISL_608220, EPI_ISL_608221, EPI_ISL_608222, EPI_ISL_608223, EPI_ISL_608224, EPI_ISL_608225, EPI_ISL_608226, EPI_ISL_608227, EPI_ISL_608228, EPI_ISL_608229, EPI_ISL_608230, EPI_ISL_608231, EPI_ISL_608232, EPI_ISL_608233, EPI_ISL_608234, EPI_ISL_608235, EPI_ISL_608236, EPI_ISL_608237, EPI_ISL_608238, EPI_ISL_608239, EPI_ISL_608240, EPI_ISL_608241, EPI_ISL_608242, EPI_ISL_608243, EPI_ISL_608244, EPI_ISL_608245, EPI_ISL_608246, EPI_ISL_608247, EPI_ISL_608248, EPI_ISL_608249, EPI_ISL_608250, EPI_ISL_608251, EPI_ISL_608252, EPI_ISL_608253, EPI_ISL_608254, EPI_ISL_608255, EPI_ISL_608256, EPI_ISL_608257, EPI_ISL_608258, EPI_ISL_608259, EPI_ISL_608260, EPI_ISL_608261, EPI_ISL_608262, EPI_ISL_608263, EPI_ISL_608264, EPI_ISL_608265, EPI_ISL_608266, EPI_ISL_608267, EPI_ISL_608268, EPI_ISL_608269, EPI_ISL_608270, EPI_ISL_608271, EPI_ISL_608272, EPI_ISL_608273, EPI_ISL_608274, EPI_ISL_608275, EPI_ISL_608276, EPI_ISL_608277, EPI_ISL_608278, EPI_ISL_608279, EPI_ISL_608280, EPI_ISL_608281, EPI_ISL_608282, EPI_ISL_608283, EPI_ISL_608284, EPI_ISL_608285, EPI_ISL_608286, EPI_ISL_608287, EPI_ISL_608288, EPI_ISL_608289, EPI_ISL_608290, EPI_ISL_608291, EPI_ISL_608292, EPI_ISL_608293, EPI_ISL_608294, EPI_ISL_608295, EPI_ISL_608296, EPI_ISL_608297, EPI_ISL_608298, EPI_ISL_608299, EPI_ISL_608300, EPI_ISL_608301, EPI_ISL_608302, EPI_ISL_608303, EPI_ISL_608304, EPI_ISL_608305, EPI_ISL_608306, EPI_ISL_608307, EPI_ISL_608308, EPI_ISL_608309, EPI_ISL_608311, EPI_ISL_608312, EPI_ISL_608313, EPI_ISL_608314, EPI_ISL_608315, EPI_ISL_608316, EPI_ISL_608317, EPI_ISL_608318, EPI_ISL_608319, EPI_ISL_608320, EPI_ISL_608321, EPI_ISL_608322, EPI_ISL_608323, EPI_ISL_608324, EPI_ISL_608325, EPI_ISL_608326, EPI_ISL_608327, EPI_ISL_608328, EPI_ISL_608329, EPI_ISL_608330, EPI_ISL_608331, EPI_ISL_608332, EPI_ISL_608333, EPI_ISL_608334, EPI_ISL_608335, EPI_ISL_608336, EPI_ISL_608337, EPI_ISL_608338, EPI_ISL_608339, EPI_ISL_608340, EPI_ISL_608341, EPI_ISL_608342, EPI_ISL_608343, EPI_ISL_608344, EPI_ISL_608345, EPI_ISL_608346, EPI_ISL_608350, EPI_ISL_608351, EPI_ISL_608352, EPI_ISL_608355, EPI_ISL_608356, EPI_ISL_608358, EPI_ISL_608361, EPI_ISL_608362, EPI_ISL_608364, EPI_ISL_608365, EPI_ISL_608366, EPI_ISL_608367, EPI_ISL_608369, EPI_ISL_608371, EPI_ISL_608372, EPI_ISL_608374, EPI_ISL_608375, EPI_ISL_608376, EPI_ISL_608377, EPI_ISL_608378, EPI_ISL_608379, EPI_ISL_608380, EPI_ISL_608383, EPI_ISL_608384, EPI_ISL_608385, EPI_ISL_608386, EPI_ISL_608387, EPI_ISL_608390, EPI_ISL_608399, EPI_ISL_608401, EPI_ISL_608403, EPI_ISL_608405, EPI_ISL_608406, EPI_ISL_608409, EPI_ISL_608413, EPI_ISL_608414, EPI_ISL_608415, EPI_ISL_608418, EPI_ISL_608419, EPI_ISL_608420, EPI_ISL_608421, | | | |

EPI_ISL_608422, EPI_ISL_608431, EPI_ISL_608434, EPI_ISL_608435, EPI_ISL_608438, EPI_ISL_608441, EPI_ISL_608443, EPI_ISL_608444, EPI_ISL_608445, EPI_ISL_608448, EPI_ISL_608452, EPI_ISL_608453, EPI_ISL_608454, EPI_ISL_608457, EPI_ISL_608460, EPI_ISL_608461, EPI_ISL_608462, EPI_ISL_608465, EPI_ISL_608468, EPI_ISL_608469, EPI_ISL_608470, EPI_ISL_608473, EPI_ISL_608474, EPI_ISL_608479, EPI_ISL_608481, EPI_ISL_608483, EPI_ISL_608487, EPI_ISL_608488, EPI_ISL_608489, EPI_ISL_608491, EPI_ISL_608496, EPI_ISL_608499, EPI_ISL_608500, EPI_ISL_608502, EPI_ISL_608503, EPI_ISL_608505, EPI_ISL_608511, EPI_ISL_608512, EPI_ISL_608513, EPI_ISL_608516, EPI_ISL_608517, EPI_ISL_608519, EPI_ISL_608520, EPI_ISL_608526, EPI_ISL_608528, EPI_ISL_608533, EPI_ISL_608534, EPI_ISL_608539, EPI_ISL_608541, EPI_ISL_608542, EPI_ISL_608543, EPI_ISL_608548, EPI_ISL_608549, EPI_ISL_608551, EPI_ISL_608553, EPI_ISL_608554, EPI_ISL_608555, EPI_ISL_608556, EPI_ISL_608562, EPI_ISL_608563, EPI_ISL_608564, EPI_ISL_608567, EPI_ISL_608569, EPI_ISL_608574, EPI_ISL_608575, EPI_ISL_608579, EPI_ISL_608580, EPI_ISL_608581, EPI_ISL_608582, EPI_ISL_608583, EPI_ISL_608584, EPI_ISL_608586, EPI_ISL_608587, EPI_ISL_608589, EPI_ISL_608590, EPI_ISL_608591, EPI_ISL_608592, EPI_ISL_608593, EPI_ISL_608594, EPI_ISL_608595, EPI_ISL_608599, EPI_ISL_608600, EPI_ISL_608601, EPI_ISL_608603, EPI_ISL_608605, EPI_ISL_608606, EPI_ISL_608609, EPI_ISL_608610, EPI_ISL_608614, EPI_ISL_608617, EPI_ISL_608621, EPI_ISL_608623, EPI_ISL_608625, EPI_ISL_608630, EPI_ISL_608634, EPI_ISL_608635, EPI_ISL_608639, EPI_ISL_608640, EPI_ISL_608646, EPI_ISL_608647, EPI_ISL_608648, EPI_ISL_608649, EPI_ISL_608652, EPI_ISL_608655, EPI_ISL_608657, EPI_ISL_608658, EPI_ISL_608659, EPI_ISL_608661, EPI_ISL_608662, EPI_ISL_608663, EPI_ISL_608665, EPI_ISL_608667, EPI_ISL_608668, EPI_ISL_608669, EPI_ISL_608670, EPI_ISL_608672, EPI_ISL_608674, EPI_ISL_608675, EPI_ISL_608676, EPI_ISL_608677, EPI_ISL_608680, EPI_ISL_608684, EPI_ISL_608685, EPI_ISL_608686, EPI_ISL_608687, EPI_ISL_608691, EPI_ISL_608692, EPI_ISL_608694, EPI_ISL_608847, EPI_ISL_608849, EPI_ISL_608851, EPI_ISL_608853, EPI_ISL_608855, EPI_ISL_608856, EPI_ISL_608858, EPI_ISL_608860, EPI_ISL_608862, EPI_ISL_608863, EPI_ISL_608865, EPI_ISL_608866, EPI_ISL_608869, EPI_ISL_608872, EPI_ISL_608874, EPI_ISL_608877, EPI_ISL_608878, EPI_ISL_608880, EPI_ISL_608883, EPI_ISL_608884, EPI_ISL_608885, EPI_ISL_608886, EPI_ISL_608887, EPI_ISL_608888, EPI_ISL_608889, EPI_ISL_608890, EPI_ISL_608891, EPI_ISL_608895, EPI_ISL_608897, EPI_ISL_608898, EPI_ISL_608899, EPI_ISL_608900, EPI_ISL_608903, EPI_ISL_608904, EPI_ISL_608911, EPI_ISL_608913, EPI_ISL_608915, EPI_ISL_608917, EPI_ISL_608919, EPI_ISL_608921, EPI_ISL_608922, EPI_ISL_608925, EPI_ISL_608926, EPI_ISL_608928, EPI_ISL_608929, EPI_ISL_608930, EPI_ISL_608931, EPI_ISL_608932, EPI_ISL_608933, EPI_ISL_608938, EPI_ISL_608941, EPI_ISL_608942, EPI_ISL_608944, EPI_ISL_608945, EPI_ISL_608946, EPI_ISL_608948, EPI_ISL_608950, EPI_ISL_608952, EPI_ISL_608955, EPI_ISL_608956, EPI_ISL_608957, EPI_ISL_608961, EPI_ISL_608962, EPI_ISL_608964, EPI_ISL_608966, EPI_ISL_608970, EPI_ISL_608971, EPI_ISL_608972, EPI_ISL_608979, EPI_ISL_608980, EPI_ISL_608981, EPI_ISL_608983, EPI_ISL_608984, EPI_ISL_608985, EPI_ISL_608988, EPI_ISL_608989, EPI_ISL_608991, EPI_ISL_608992, EPI_ISL_608993, EPI_ISL_608994, EPI_ISL_608995, EPI_ISL_608998, EPI_ISL_608999, EPI_ISL_609000, EPI_ISL_609002, EPI_ISL_609007, EPI_ISL_609009, EPI_ISL_609010, EPI_ISL_609011, EPI_ISL_609012, EPI_ISL_609013, EPI_ISL_609016, EPI_ISL_609019, EPI_ISL_609020, EPI_ISL_609022, EPI_ISL_609028, EPI_ISL_609031, EPI_ISL_609032, EPI_ISL_609034, EPI_ISL_609035, EPI_ISL_609039, EPI_ISL_609040, EPI_ISL_609041, EPI_ISL_609043, EPI_ISL_609051, EPI_ISL_609052, EPI_ISL_609053, EPI_ISL_609055, EPI_ISL_609056, EPI_ISL_609057, EPI_ISL_609061, EPI_ISL_609064, EPI_ISL_609065, EPI_ISL_609068, EPI_ISL_609069, EPI_ISL_609070, EPI_ISL_609073, EPI_ISL_609074, EPI_ISL_609075, EPI_ISL_609078, EPI_ISL_609079, EPI_ISL_609080, EPI_ISL_609083, EPI_ISL_609085, EPI_ISL_609087, EPI_ISL_609089, EPI_ISL_609090, EPI_ISL_609093, EPI_ISL_609094, EPI_ISL_609095, EPI_ISL_609096, EPI_ISL_609098, EPI_ISL_609102, EPI_ISL_609103, EPI_ISL_609104, EPI_ISL_609105, EPI_ISL_609106, EPI_ISL_609107, EPI_ISL_609109, EPI_ISL_609112, EPI_ISL_609120, EPI_ISL_609125, EPI_ISL_609127, EPI_ISL_609130, EPI_ISL_609132, EPI_ISL_609133, EPI_ISL_609134, EPI_ISL_609135, EPI_ISL_609141, EPI_ISL_609144, EPI_ISL_609146, EPI_ISL_609147, EPI_ISL_609148, EPI_ISL_609150, EPI_ISL_609152, EPI_ISL_609154, EPI_ISL_609157, EPI_ISL_610286, EPI_ISL_610287, EPI_ISL_610288, EPI_ISL_610289, EPI_ISL_610290, EPI_ISL_610291, EPI_ISL_610292, EPI_ISL_610293, EPI_ISL_610294, EPI_ISL_610295, EPI_ISL_610296, EPI_ISL_610297, EPI_ISL_610298, EPI_ISL_610299, EPI_ISL_610300, EPI_ISL_610301, EPI_ISL_610302, EPI_ISL_610303, EPI_ISL_610304, EPI_ISL_610305, EPI_ISL_610306, EPI_ISL_610307, EPI_ISL_610308, EPI_ISL_610309, EPI_ISL_610310, EPI_ISL_610311, EPI_ISL_610312, EPI_ISL_610313, EPI_ISL_610314, EPI_ISL_610315, EPI_ISL_610316, EPI_ISL_610317, EPI_ISL_610318, EPI_ISL_610319, EPI_ISL_610320, EPI_ISL_610321, EPI_ISL_610322, EPI_ISL_610323, EPI_ISL_610324, EPI_ISL_610325, EPI_ISL_610326, EPI_ISL_610327, EPI_ISL_610328, EPI_ISL_610329, EPI_ISL_610330, EPI_ISL_610331, EPI_ISL_610333, EPI_ISL_610334, EPI_ISL_610335, EPI_ISL_610336, EPI_ISL_610337, EPI_ISL_610338, EPI_ISL_610339, EPI_ISL_610340, EPI_ISL_610341, EPI_ISL_610343, EPI_ISL_610344, EPI_ISL_610345, EPI_ISL_610346, EPI_ISL_610347, EPI_ISL_610348, EPI_ISL_610349, EPI_ISL_610350, EPI_ISL_610351, EPI_ISL_610352, EPI_ISL_610353, EPI_ISL_610354, EPI_ISL_610355, EPI_ISL_610356, EPI_ISL_610357, EPI_ISL_610358, EPI_ISL_610359, EPI_ISL_610360, EPI_ISL_610361, EPI_ISL_610362, EPI_ISL_610363, EPI_ISL_610364, EPI_ISL_610365, EPI_ISL_610366, EPI_ISL_610367, EPI_ISL_610368, EPI_ISL_610369, EPI_ISL_610370, EPI_ISL_610371, EPI_ISL_610372, EPI_ISL_610373, EPI_ISL_610374, EPI_ISL_610375, EPI_ISL_610376, EPI_ISL_610377, EPI_ISL_610378, EPI_ISL_610379, EPI_ISL_610380, EPI_ISL_610381, EPI_ISL_610382, EPI_ISL_610383, EPI_ISL_610384, EPI_ISL_610385, EPI_ISL_610386, EPI_ISL_610387, EPI_ISL_610388, EPI_ISL_610389, EPI_ISL_610390, EPI_ISL_610391, EPI_ISL_610392, EPI_ISL_610393, EPI_ISL_610394, EPI_ISL_610395, EPI_ISL_610396, EPI_ISL_610397, EPI_ISL_610398, EPI_ISL_610399, EPI_ISL_610400, EPI_ISL_610401, EPI_ISL_610402, EPI_ISL_610403, EPI_ISL_610404, EPI_ISL_610405, EPI_ISL_610406, EPI_ISL_610407, EPI_ISL_610408, EPI_ISL_610409, EPI_ISL_610410, EPI_ISL_610411, EPI_ISL_610412, EPI_ISL_610413, EPI_ISL_610414, EPI_ISL_610415, EPI_ISL_610416, EPI_ISL_610417, EPI_ISL_610418, EPI_ISL_610419, EPI_ISL_610420, EPI_ISL_610421, EPI_ISL_610422, EPI_ISL_610423, EPI_ISL_610424, EPI_ISL_610425, EPI_ISL_610426, EPI_ISL_610427, EPI_ISL_610428, EPI_ISL_610429, EPI_ISL_610430, EPI_ISL_610431, EPI_ISL_610432, EPI_ISL_610433, EPI_ISL_610434, EPI_ISL_610436, EPI_ISL_610437, EPI_ISL_610438, EPI_ISL_610439, EPI_ISL_610440, EPI_ISL_610441, EPI_ISL_610442, EPI_ISL_610443, EPI_ISL_610444, EPI_ISL_610445, EPI_ISL_610446, EPI_ISL_610447, EPI_ISL_610448, EPI_ISL_610449, EPI_ISL_610450, EPI_ISL_610451, EPI_ISL_610452, EPI_ISL_610453, EPI_ISL_610454, EPI_ISL_610455, EPI_ISL_610456, EPI_ISL_610457, EPI_ISL_610458, EPI_ISL_610459, EPI_ISL_610460, EPI_ISL_610461, EPI_ISL_610462, EPI_ISL_610463, EPI_ISL_610464, EPI_ISL_610465, EPI_ISL_610466, EPI_ISL_610467, EPI_ISL_610468, EPI_ISL_610469, EPI_ISL_610470, EPI_ISL_610471, EPI_ISL_610472, EPI_ISL_610473, EPI_ISL_610474, EPI_ISL_610475, EPI_ISL_610476, EPI_ISL_610477, EPI_ISL_610478, EPI_ISL_610479, EPI_ISL_610480, EPI_ISL_610481, EPI_ISL_610482, EPI_ISL_610483, EPI_ISL_610484, EPI_ISL_610485, EPI_ISL_610486, EPI_ISL_610487, EPI_ISL_610488, EPI_ISL_610489, EPI_ISL_610490, EPI_ISL_610491, EPI_ISL_610492, EPI_ISL_610493, EPI_ISL_610494, EPI_ISL_610495, EPI_ISL_610496, EPI_ISL_610497, EPI_ISL_610498, EPI_ISL_610499, EPI_ISL_610500, EPI_ISL_610501, EPI_ISL_610504, EPI_ISL_610505, EPI_ISL_610506, EPI_ISL_610507, EPI_ISL_610508, EPI_ISL_610509, EPI_ISL_610510, EPI_ISL_610511, EPI_ISL_610512, EPI_ISL_610513, EPI_ISL_610514, EPI_ISL_610515, EPI_ISL_610516, EPI_ISL_610517, EPI_ISL_610518, EPI_ISL_610519, EPI_ISL_610520, EPI_ISL_610521, EPI_ISL_610522, EPI_ISL_610523, EPI_ISL_610524, EPI_ISL_610525, EPI_ISL_610526, EPI_ISL_610527, EPI_ISL_610528, EPI_ISL_610529, EPI_ISL_610530, EPI_ISL_610531, EPI_ISL_610532, EPI_ISL_610533, EPI_ISL_610534, EPI_ISL_610535, EPI_ISL_610536, EPI_ISL_610537, EPI_ISL_610538, EPI_ISL_610539, EPI_ISL_610540, EPI_ISL_610541, EPI_ISL_610542, EPI_ISL_610543, EPI_ISL_610544, EPI_ISL_610545, EPI_ISL_610546, EPI_ISL_610547, EPI_ISL_610548, EPI_ISL_610549, EPI_ISL_610550, EPI_ISL_610551, EPI_ISL_610552, EPI_ISL_610553, EPI_ISL_610554, EPI_ISL_610555, EPI_ISL_610556, EPI_ISL_610557, EPI_ISL_610558, EPI_ISL_610559, EPI_ISL_610560, EPI_ISL_610561, EPI_ISL_610562, EPI_ISL_610563, EPI_ISL_610564, EPI_ISL_610565, EPI_ISL_610566, EPI_ISL_610567, EPI_ISL_610568, EPI_ISL_610569, EPI_ISL_610570, EPI_ISL_610571, EPI_ISL_610572, EPI_ISL_610573, EPI_ISL_610574, EPI_ISL_610575, EPI_ISL_610576, EPI_ISL_610577, EPI_ISL_610578, EPI_ISL_610579, EPI_ISL_610580, EPI_ISL_610581, EPI_ISL_610582, EPI_ISL_610583, EPI_ISL_610584, EPI_ISL_610585, EPI_ISL_610586, EPI_ISL_610587, EPI_ISL_610588, EPI_ISL_610589, EPI_ISL_610590, EPI_ISL_610591, EPI_ISL_610592, EPI_ISL_610593, EPI_ISL_610594, EPI_ISL_610595, EPI_ISL_610596, EPI_ISL_610597, EPI_ISL_610598, EPI_ISL_610599, EPI_ISL_610600, EPI_ISL_610601, EPI_ISL_610602, EPI_ISL_610603, EPI_ISL_610604, EPI_ISL_610605, EPI_ISL_610606, EPI_ISL_610607, EPI_ISL_610608, EPI_ISL_610609, EPI_ISL_610610, EPI_ISL_610611, EPI_ISL_610612, EPI_ISL_610613, EPI_ISL_610614, EPI_ISL_610615, EPI_ISL_610616, EPI_ISL_610617, EPI_ISL_610618, EPI_ISL_610619, EPI_ISL_610620, EPI_ISL_610621, EPI_ISL_610622, EPI_ISL_610623, EPI_ISL_610624, EPI_ISL_610625, EPI_ISL_610626, EPI_ISL_610627, EPI_ISL_610628, EPI_ISL_610923, EPI_ISL_610924, EPI_ISL_610925, EPI_ISL_610929, EPI_ISL_610930, EPI_ISL_610931, EPI_ISL_610934, EPI_ISL_610935, EPI_ISL_610936, EPI_ISL_610937, EPI_ISL_610940, EPI_ISL_610941, EPI_ISL_610942, EPI_ISL_610943, EPI_ISL_610945, EPI_ISL_610949, EPI_ISL_610950, EPI_ISL_610951, EPI_ISL_610952, EPI_ISL_610953, EPI_ISL_610955, EPI_ISL_610956, EPI_ISL_610957, EPI_ISL_610958, EPI_ISL_610959, EPI_ISL_610961, EPI_ISL_610962, EPI_ISL_610964, EPI_ISL_610965, EPI_ISL_610966, EPI_ISL_610968, EPI_ISL_610969, EPI_ISL_610970, EPI_ISL_610971, EPI_ISL_610972, EPI_ISL_610973, EPI_ISL_610974, EPI_ISL_610975, EPI_ISL_610976, EPI_ISL_610977, EPI_ISL_610978, EPI_ISL_610979, EPI_ISL_610981, EPI_ISL_610983, EPI_ISL_610985, EPI_ISL_610986, EPI_ISL_610987, EPI_ISL_610988, EPI_ISL_610991, EPI_ISL_610992, EPI_ISL_610993, EPI_ISL_610994, EPI_ISL_610995, EPI_ISL_610996, EPI_ISL_610997, EPI_ISL_610998, EPI_ISL_610999, EPI_ISL_611001, EPI_ISL_611002, EPI_ISL_611003, EPI_ISL_611006, EPI_ISL_611007, EPI_ISL_611008, EPI_ISL_611009, EPI_ISL_611010, EPI_ISL_611011, EPI_ISL_611012, EPI_ISL_611013, EPI_ISL_611017, EPI_ISL_611018, EPI_ISL_611019, EPI_ISL_611020, EPI_ISL_611021, EPI_ISL_611022, EPI_ISL_611024, EPI_ISL_611025, EPI_ISL_611026, EPI_ISL_611027, EPI_ISL_611028, EPI_ISL_611030, EPI_ISL_611031, EPI_ISL_611032, EPI_ISL_611033, EPI_ISL_611035, EPI_ISL_611036, EPI_ISL_611037, EPI_ISL_611038, EPI_ISL_611039, EPI_ISL_611040, EPI_ISL_611042, EPI_ISL_611043, EPI_ISL_611044, EPI_ISL_611047, EPI_ISL_611048, EPI_ISL_611049, EPI_ISL_611050, EPI_ISL_611051, EPI_ISL_611052, EPI_ISL_611053, EPI_ISL_611058, EPI_ISL_611060, EPI_ISL_611061, EPI_ISL_611062, EPI_ISL_611063, EPI_ISL_611064, EPI_ISL_611065, EPI_ISL_611066, EPI_ISL_611067, EPI_ISL_611068, EPI_ISL_611069, EPI_ISL_611070, EPI_ISL_611071, EPI_ISL_611072, EPI_ISL_611073, EPI_ISL_611074, EPI_ISL_611075, EPI_ISL_611076, EPI_ISL_611077, EPI_ISL_611078, EPI_ISL_611079, EPI_ISL_611080, EPI_ISL_611081, EPI_ISL_611082, EPI_ISL_611083, EPI_ISL_611084, EPI_ISL_611085, EPI_ISL_611086, EPI_ISL_611087, EPI_ISL_611088, EPI_ISL_611089, EPI_ISL_611091, EPI_ISL_611092, EPI_ISL_611094, EPI_ISL_611095, EPI_ISL_611096, EPI_ISL_611097, EPI_ISL_611098, EPI_ISL_611099, EPI_ISL_611100, EPI_ISL_611101, EPI_ISL_611103, EPI_ISL_611105, EPI_ISL_611107, EPI_ISL_611108, EPI_ISL_611109, EPI_ISL_611110, EPI_ISL_611111, EPI_ISL_611114, EPI_ISL_611115, EPI_ISL_611116, EPI_ISL_611117, EPI_ISL_611118, EPI_ISL_611119, EPI_ISL_611120, EPI_ISL_611122, EPI_ISL_611123, EPI_ISL_611124, EPI_ISL_611125, EPI_ISL_611126, EPI_ISL_611127, EPI_ISL_611128, EPI_ISL_611129, EPI_ISL_611130, EPI_ISL_611133, EPI_ISL_611136, EPI_ISL_611137, EPI_ISL_611138, EPI_ISL_611139, EPI_ISL_611141, EPI_ISL_611142, EPI_ISL_611144, EPI_ISL_611145, EPI_ISL_611146, EPI_ISL_611147, EPI_ISL_611148, EPI_ISL_611149, EPI_ISL_611150, EPI_ISL_611151, EPI_ISL_611152, EPI_ISL_611153, EPI_ISL_611154, EPI_ISL_611155, EPI_ISL_611156, EPI_ISL_611157, EPI_ISL_611158, EPI_ISL_611159, EPI_ISL_611160, EPI_ISL_611161, EPI_ISL_611162, EPI_ISL_611164, EPI_ISL_611165, EPI_ISL_611166, EPI_ISL_611167, EPI_ISL_611168, EPI_ISL_611169, EPI_ISL_611170, EPI_ISL_611171, EPI_ISL_611172, EPI_ISL_611173, EPI_ISL_611174, EPI_ISL_611175, EPI_ISL_611176, EPI_ISL_611177, EPI_ISL_611178, EPI_ISL_611179, EPI_ISL_611180, EPI_ISL_611181, EPI_ISL_611182, EPI_ISL_611184, EPI_ISL_611185, EPI_ISL_611186, EPI_ISL_611187, EPI_ISL_611189, EPI_ISL_611190, EPI_ISL_611191, EPI_ISL_611192, EPI_ISL_611193, EPI_ISL_611195, EPI_ISL_611196, EPI_ISL_611198, EPI_ISL_611199, EPI_ISL_611200, EPI_ISL_611202, EPI_ISL_611204, EPI_ISL_611205, EPI_ISL_611206, EPI_ISL_611208, EPI_ISL_611210, EPI_ISL_611211, EPI_ISL_611214, EPI_ISL_611215, EPI_ISL_611216, EPI_ISL_611218, EPI_ISL_611219, EPI_ISL_611220, EPI_ISL_611221, EPI_ISL_611222, EPI_ISL_611223, EPI_ISL_611224, EPI_ISL_611225, EPI_ISL_611226, EPI_ISL_611227, EPI_ISL_611228, EPI_ISL_611229, EPI_ISL_611231, EPI_ISL_611233, EPI_ISL_611234, EPI_ISL_611236, EPI_ISL_611237, EPI_ISL_611238, EPI_ISL_611239, EPI_ISL_611240, EPI_ISL_611241, EPI_ISL_611242, EPI_ISL_611244, EPI_ISL_611245, EPI_ISL_611246, EPI_ISL_611247, EPI_ISL_611248, EPI_ISL_611249, EPI_ISL_611250, EPI_ISL_611251, EPI_ISL_611252, EPI_ISL_611253, EPI_ISL_611254, EPI_ISL_611256, EPI_ISL_611257, EPI_ISL_611258, EPI_ISL_611259, EPI_ISL_611260, EPI_ISL_611261, EPI_ISL_611262, EPI_ISL_611263, EPI_ISL_611264, EPI_ISL_611265, EPI_ISL_611266, EPI_ISL_611268, EPI_ISL_611269, EPI_ISL_611270, EPI_ISL_611271, EPI_ISL_611272, EPI_ISL_611274, EPI_ISL_611275, EPI_ISL_611276, EPI_ISL_611278, EPI_ISL_611279, EPI_ISL_611280, EPI_ISL_611281, EPI_ISL_611282, EPI_ISL_611283, EPI_ISL_611284, EPI_ISL_611285, EPI_ISL_624560, EPI_ISL_624562, EPI_ISL_624563, EPI_ISL_624565, EPI_ISL_624567, EPI_ISL_624568, EPI_ISL_624576, EPI_ISL_624577, EPI_ISL_624581, EPI_ISL_624584, EPI_ISL_624587, EPI_ISL_624589, EPI_ISL_624590, EPI_ISL_624591, EPI_ISL_624593, EPI_ISL_624595, EPI_ISL_624598, EPI_ISL_624599, EPI_ISL_624603, EPI_ISL_624604, EPI_ISL_624605, EPI_ISL_624606, EPI_ISL_624608, EPI_ISL_624610, EPI_ISL_624611, EPI_ISL_624613, EPI_ISL_624617, EPI_ISL_624624, EPI_ISL_624625, EPI_ISL_624626, EPI_ISL_624629, EPI_ISL_624630, EPI_ISL_624634, EPI_ISL_624637, EPI_ISL_624641, EPI_ISL_624643, EPI_ISL_624645, EPI_ISL_624647, EPI_ISL_624649, EPI_ISL_624650, EPI_ISL_624651, EPI_ISL_624652, EPI_ISL_624654, EPI_ISL_624656, EPI_ISL_624657, EPI_ISL_624658, EPI_ISL_624659, EPI_ISL_624663, EPI_ISL_624667, EPI_ISL_624672, EPI_ISL_624674, EPI_ISL_624675, EPI_ISL_624677, EPI_ISL_624678, EPI_ISL_624680, EPI_ISL_624681, EPI_ISL_624682, EPI_ISL_624683, EPI_ISL_624687, EPI_ISL_624693, EPI_ISL_624694, EPI_ISL_624696, EPI_ISL_624697, EPI_ISL_624703, EPI_ISL_624704, EPI_ISL_624707, EPI_ISL_624708, EPI_ISL_624709, EPI_ISL_624710, EPI_ISL_624711, EPI_ISL_624713, EPI_ISL_624715, EPI_ISL_624716, EPI_ISL_624717, EPI_ISL_624718, EPI_ISL_624721, EPI_ISL_624722, EPI_ISL_624723, EPI_ISL_624725, EPI_ISL_624726, EPI_ISL_624727, EPI_ISL_624728, EPI_ISL_624731, EPI_ISL_624732, EPI_ISL_624742, EPI_ISL_624745, EPI_ISL_624746, EPI_ISL_624747, EPI_ISL_624749, EPI_ISL_624753, EPI_ISL_624754, EPI_ISL_624756, EPI_ISL_624759, EPI_ISL_624762, EPI_ISL_624764, EPI_ISL_624765, EPI_ISL_624769, EPI_ISL_624770, EPI_ISL_624773, EPI_ISL_624774, EPI_ISL_624778, EPI_ISL_624779, EPI_ISL_624781, EPI_ISL_624783, EPI_ISL_624785, EPI_ISL_624786, EPI_ISL_624789, EPI_ISL_624790, EPI_ISL_624791, EPI_ISL_624792, EPI_ISL_624793, EPI_ISL_624796, EPI_ISL_624800, EPI_ISL_624805, EPI_ISL_624810, EPI_ISL_624814, EPI_ISL_624816, EPI_ISL_624820, EPI_ISL_624821, EPI_ISL_624824, EPI_ISL_624826, EPI_ISL_624831, EPI_ISL_624836, EPI_ISL_624838, EPI_ISL_624840, EPI_ISL_624841, EPI_ISL_624843, EPI_ISL_624844, EPI_ISL_624846, EPI_ISL_624847, EPI_ISL_624849, EPI_ISL_624850, EPI_ISL_624851, EPI_ISL_624852, EPI_ISL_624856, EPI_ISL_624858, EPI_ISL_624859, EPI_ISL_624863, EPI_ISL_624869, EPI_ISL_624871, EPI_ISL_624872, EPI_ISL_624874, EPI_ISL_624880, EPI_ISL_624881, EPI_ISL_624883, EPI_ISL_624885, EPI_ISL_624889, EPI_ISL_624895, EPI_ISL_624896, EPI_ISL_624897, EPI_ISL_624898, EPI_ISL_624901, EPI_ISL_624903, EPI_ISL_624905, EPI_ISL_624906, EPI_ISL_625267, EPI_ISL_625268, EPI_ISL_625269, EPI_ISL_625270, EPI_ISL_625271, EPI_ISL_625272, EPI_ISL_625273, EPI_ISL_625274, EPI_ISL_625275, EPI_ISL_625276, EPI_ISL_625277, EPI_ISL_625278, EPI_ISL_625279, EPI_ISL_625280, EPI_ISL_625396, EPI_ISL_625397, EPI_ISL_625398, EPI_ISL_625399, EPI_ISL_625400, EPI_ISL_625401, EPI_ISL_625402, EPI_ISL_625403, EPI_ISL_625404, EPI_ISL_625431, EPI_ISL_625434, EPI_ISL_630348, EPI_ISL_630349, EPI_ISL_630350, EPI_ISL_630351, EPI_ISL_630352, EPI_ISL_630353, EPI_ISL_630354, EPI_ISL_630355, EPI_ISL_630356, EPI_ISL_630357, EPI_ISL_630358, EPI_ISL_630360, EPI_ISL_630361, EPI_ISL_630362, EPI_ISL_630363, EPI_ISL_630364, EPI_ISL_630365, EPI_ISL_630366, EPI_ISL_630367, EPI_ISL_630368, EPI_ISL_630370, EPI_ISL_630371, EPI_ISL_630372, EPI_ISL_630373, EPI_ISL_630374, EPI_ISL_630375, EPI_ISL_630376, EPI_ISL_630377, EPI_ISL_630378, EPI_ISL_630379, EPI_ISL_630380, EPI_ISL_630381, EPI_ISL_630382, EPI_ISL_630383, EPI_ISL_630384, EPI_ISL_630385, EPI_ISL_630386, EPI_ISL_630387, EPI_ISL_630388, EPI_ISL_630389, EPI_ISL_630390, EPI_ISL_630391, EPI_ISL_630392, EPI_ISL_630393, EPI_ISL_630394, EPI_ISL_630395, EPI_ISL_630396, EPI_ISL_630397, EPI_ISL_630398, EPI_ISL_630400, EPI_ISL_630401, EPI_ISL_630402, EPI_ISL_630403, EPI_ISL_630404, EPI_ISL_630405, EPI_ISL_630406, EPI_ISL_630407, EPI_ISL_630408, EPI_ISL_630409, EPI_ISL_630410, EPI_ISL_630411, EPI_ISL_630412, EPI_ISL_630413, EPI_ISL_630415, EPI_ISL_630416, EPI_ISL_630418, EPI_ISL_630419, EPI_ISL_630420, EPI_ISL_630421, EPI_ISL_630422, EPI_ISL_630423, EPI_ISL_630424, EPI_ISL_630425, EPI_ISL_630426, EPI_ISL_630427, EPI_ISL_630428, EPI_ISL_630429, EPI_ISL_630430, EPI_ISL_630431, EPI_ISL_630432, EPI_ISL_630433, EPI_ISL_630434, EPI_ISL_630435, EPI_ISL_630436, EPI_ISL_630437, EPI_ISL_630438, EPI_ISL_630439, EPI_ISL_630440, EPI_ISL_630442, EPI_ISL_630443, EPI_ISL_630445, EPI_ISL_630446, EPI_ISL_630447, EPI_ISL_630448, EPI_ISL_630449, EPI_ISL_630450, EPI_ISL_630451, EPI_ISL_630452, EPI_ISL_630453, EPI_ISL_630454, EPI_ISL_630455, EPI_ISL_630456, EPI_ISL_630457, EPI_ISL_630458, EPI_ISL_630459, EPI_ISL_630460, EPI_ISL_630461, EPI_ISL_630462, EPI_ISL_630463, EPI_ISL_630464, EPI_ISL_630465, EPI_ISL_630466, EPI_ISL_630467, EPI_ISL_630468, EPI_ISL_630469, EPI_ISL_630470, EPI_ISL_630471, EPI_ISL_630472, EPI_ISL_630473, EPI_ISL_630475, EPI_ISL_630476, EPI_ISL_630477, EPI_ISL_630478, EPI_ISL_630479, EPI_ISL_630480, EPI_ISL_630481, EPI_ISL_630482, EPI_ISL_630483, EPI_ISL_630484, EPI_ISL_630485, EPI_ISL_630486, EPI_ISL_630487, EPI_ISL_630488, EPI_ISL_630489, EPI_ISL_630490, EPI_ISL_630491, EPI_ISL_630492, EPI_ISL_630493, EPI_ISL_630494, EPI_ISL_630495, EPI_ISL_630496, EPI_ISL_630497, EPI_ISL_630498, EPI_ISL_630499, EPI_ISL_630500, EPI_ISL_630501, EPI_ISL_630502, EPI_ISL_630503, EPI_ISL_630504, EPI_ISL_630505, EPI_ISL_630507, EPI_ISL_630508, EPI_ISL_630509, EPI_ISL_630510, EPI_ISL_630511, EPI_ISL_630513, EPI_ISL_630514, EPI_ISL_630515, EPI_ISL_630516, EPI_ISL_630517, EPI_ISL_630518, EPI_ISL_630519, EPI_ISL_630520, EPI_ISL_630521, EPI_ISL_630522, EPI_ISL_630523, EPI_ISL_630524, EPI_ISL_630525, EPI_ISL_630526, EPI_ISL_630527, EPI_ISL_630528, EPI_ISL_630529, EPI_ISL_630530, EPI_ISL_630531, EPI_ISL_630532, EPI_ISL_630534, EPI_ISL_630535, EPI_ISL_630536, EPI_ISL_630537, EPI_ISL_630538, EPI_ISL_630539, EPI_ISL_630540, EPI_ISL_630541, EPI_ISL_630542, EPI_ISL_630543, EPI_ISL_630544, EPI_ISL_630545, EPI_ISL_630546, EPI_ISL_630548, EPI_ISL_630549, EPI_ISL_630551, EPI_ISL_630552, EPI_ISL_630553, EPI_ISL_630554, EPI_ISL_630555, EPI_ISL_630556, EPI_ISL_630557, EPI_ISL_630558, EPI_ISL_630559, EPI_ISL_630560, EPI_ISL_630561, EPI_ISL_630562, EPI_ISL_630563, EPI_ISL_630564, EPI_ISL_630565, EPI_ISL_630566, EPI_ISL_630567, EPI_ISL_630568, EPI_ISL_630569, EPI_ISL_630570, EPI_ISL_630571, EPI_ISL_630572, EPI_ISL_630573, EPI_ISL_630574, EPI_ISL_630575, EPI_ISL_630576, EPI_ISL_630577, EPI_ISL_630578, EPI_ISL_630579, EPI_ISL_630580, EPI_ISL_630581, EPI_ISL_630582, EPI_ISL_630583, EPI_ISL_630584, EPI_ISL_630585, EPI_ISL_630586, EPI_ISL_630587, EPI_ISL_630588, EPI_ISL_630589, EPI_ISL_630590, EPI_ISL_630591, EPI_ISL_630592, EPI_ISL_630593, EPI_ISL_630594, EPI_ISL_630595, EPI_ISL_630596, EPI_ISL_630597, EPI_ISL_630598, EPI_ISL_630599, EPI_ISL_630600, EPI_ISL_630601, EPI_ISL_630602, EPI_ISL_630603, EPI_ISL_630604, EPI_ISL_630605, EPI_ISL_630606, EPI_ISL_630607, EPI_ISL_630608, EPI_ISL_630609, EPI_ISL_630610, EPI_ISL_630611, EPI_ISL_630612, EPI_ISL_630613, EPI_ISL_630614, EPI_ISL_630615, EPI_ISL_630616, EPI_ISL_630617, EPI_ISL_630618, EPI_ISL_630619, EPI_ISL_630620, EPI_ISL_630621, EPI_ISL_630622, EPI_ISL_630623, EPI_ISL_630624, EPI_ISL_630625, EPI_ISL_630626, EPI_ISL_630627, EPI_ISL_630628, EPI_ISL_630629, EPI_ISL_630630, EPI_ISL_630631, EPI_ISL_630632, EPI_ISL_630633, EPI_ISL_630634, EPI_ISL_630635, EPI_ISL_630636, EPI_ISL_630637, EPI_ISL_630638, EPI_ISL_630639, EPI_ISL_630640, EPI_ISL_630641, EPI_ISL_630642, EPI_ISL_630643, EPI_ISL_630645, EPI_ISL_630646, EPI_ISL_630647, EPI_ISL_630648, EPI_ISL_630649, EPI_ISL_630650, EPI_ISL_630651, EPI_ISL_630652, EPI_ISL_630653, EPI_ISL_630654, EPI_ISL_630655, EPI_ISL_630656, EPI_ISL_630657, EPI_ISL_630658, EPI_ISL_630659, EPI_ISL_630660, EPI_ISL_630661, EPI_ISL_630662, EPI_ISL_630663, EPI_ISL_630664, EPI_ISL_630665, EPI_ISL_630666, EPI_ISL_630667, EPI_ISL_630668, EPI_ISL_630669, EPI_ISL_630670, EPI_ISL_630671, EPI_ISL_630672, EPI_ISL_630673, EPI_ISL_630674, EPI_ISL_630675, EPI_ISL_630676, EPI_ISL_630677, EPI_ISL_630678, EPI_ISL_630679, EPI_ISL_630680, EPI_ISL_630681, EPI_ISL_630683, EPI_ISL_630684, EPI_ISL_630685, EPI_ISL_630686, EPI_ISL_630687, EPI_ISL_630688, EPI_ISL_630689, EPI_ISL_630690, EPI_ISL_630691, EPI_ISL_630692, EPI_ISL_630694, EPI_ISL_630695, EPI_ISL_630696, EPI_ISL_630697, EPI_ISL_630698, EPI_ISL_630699, EPI_ISL_630700, EPI_ISL_630975, EPI_ISL_630982, EPI_ISL_630983, EPI_ISL_630985, EPI_ISL_630992, EPI_ISL_630999, EPI_ISL_631011, EPI_ISL_631012, EPI_ISL_631014, EPI_ISL_631018, EPI_ISL_631027, EPI_ISL_631031, EPI_ISL_631033, EPI_ISL_631035, EPI_ISL_631041, EPI_ISL_631044, EPI_ISL_631046, EPI_ISL_631050, EPI_ISL_631056, EPI_ISL_631059, EPI_ISL_631060, EPI_ISL_631074, EPI_ISL_631081, EPI_ISL_631084, EPI_ISL_631093, EPI_ISL_631096, EPI_ISL_631099, EPI_ISL_631107, EPI_ISL_631113, EPI_ISL_631131, EPI_ISL_631133, EPI_ISL_631150, EPI_ISL_631165, EPI_ISL_631184, EPI_ISL_631189, EPI_ISL_631190, EPI_ISL_631215, EPI_ISL_631217, EPI_ISL_631231, EPI_ISL_631250, EPI_ISL_631265, EPI_ISL_631269, EPI_ISL_634755, EPI_ISL_634756

| | | | |
|---|---|---|---|
| see above | Lighthouse Lab in Alderley Park | Wellcome Sanger Institute for the COVID-19 Genomics UK (COG-UK) consortium | Cordelia Langford; David K. Jackson; Dominic Kwiatkowski; Ewan Harrison; Ian Johnston; Jacquelyn Wynn; John Sillitoe on behalf of the Wellcome Sanger Institute COVID-19 Surveillance Team; John Sillitoe on behalf of the Wellcome Sanger Institute COVID-19 Surveillance Team (http://www.sanger.ac.uk/covid-team); Mairead Hyland; Roberto Amato; Sonia Goncalves; The Lighthouse Lab in Alderley Park and Alex Alderton |

EPI_ISL_608347, EPI_ISL_608455, EPI_ISL_608681, EPI_ISL_608907, EPI_ISL_609121, EPI_ISL_610702, EPI_ISL_610779, EPI_ISL_610843, EPI_ISL_625289, EPI_ISL_625312, EPI_ISL_647697, EPI_ISL_826518, EPI_ISL_892225

| | | | |
|---|---|---|---|
| see above | Lighthouse Lab in Cambridge | Wellcome Sanger Institute for the COVID-19 Genomics UK (COG-UK) Consortium | Cordelia Langford; David K. Jackson; Dominic Kwiatkowski; Ewan Harrison; Ian Johnston; John Sillitoe on behalf of the Wellcome Sanger Institute COVID-19 Surveillance Team; John Sillitoe on behalf of the Wellcome Sanger Institute COVID-19 Surveillance Team (http://www.sanger.ac.uk/covid-team); Rob Howes; Roberto Amato; Sonia Goncalves; The Lighthouse Lab in Cambridge and Alex Alderton |

EPI_ISL_596964, EPI_ISL_597195, EPI_ISL_597216, EPI_ISL_597229, EPI_ISL_597248, EPI_ISL_597250, EPI_ISL_597264, EPI_ISL_597350, EPI_ISL_597354, EPI_ISL_608348, EPI_ISL_608349, EPI_ISL_608353, EPI_ISL_608354, EPI_ISL_608357, EPI_ISL_608359, EPI_ISL_608360, EPI_ISL_608363, EPI_ISL_608368, EPI_ISL_608370, EPI_ISL_608373, EPI_ISL_608381, EPI_ISL_608382, EPI_ISL_608388, EPI_ISL_608389, EPI_ISL_608391, EPI_ISL_608392, EPI_ISL_608393, EPI_ISL_608394, EPI_ISL_608395, EPI_ISL_608396, EPI_ISL_608397, EPI_ISL_608398, EPI_ISL_608402, EPI_ISL_608404, EPI_ISL_608407, EPI_ISL_608408, EPI_ISL_608410, EPI_ISL_608411, EPI_ISL_608416, EPI_ISL_608417, EPI_ISL_608423, EPI_ISL_608424, EPI_ISL_608426, EPI_ISL_608428, EPI_ISL_608430, EPI_ISL_608432, EPI_ISL_608433, EPI_ISL_608436, EPI_ISL_608437, EPI_ISL_608439, EPI_ISL_608442, EPI_ISL_608446, EPI_ISL_608447, EPI_ISL_608449, EPI_ISL_608450, EPI_ISL_608451, EPI_ISL_608456, EPI_ISL_608458, EPI_ISL_608459, EPI_ISL_608463, EPI_ISL_608464, EPI_ISL_608466, EPI_ISL_608467, EPI_ISL_608471, EPI_ISL_608472, EPI_ISL_608475, EPI_ISL_608476, EPI_ISL_608477, EPI_ISL_608478, EPI_ISL_608480, EPI_ISL_608482, EPI_ISL_608484, EPI_ISL_608485, EPI_ISL_608486, EPI_ISL_608490, EPI_ISL_608492, EPI_ISL_608493, EPI_ISL_608494, EPI_ISL_608495, EPI_ISL_608497, EPI_ISL_608498, EPI_ISL_608501, EPI_ISL_608504, EPI_ISL_608506, EPI_ISL_608507, EPI_ISL_608508, EPI_ISL_608509, EPI_ISL_608510, EPI_ISL_608514, EPI_ISL_608515, EPI_ISL_608521, EPI_ISL_608522, EPI_ISL_608523, EPI_ISL_608524, EPI_ISL_608525, EPI_ISL_608527, EPI_ISL_608529, EPI_ISL_608530, EPI_ISL_608531, EPI_ISL_608532, EPI_ISL_608535, EPI_ISL_608536, EPI_ISL_608537, EPI_ISL_608538, EPI_ISL_608540, EPI_ISL_608545, EPI_ISL_608546, EPI_ISL_608547, EPI_ISL_608550, EPI_ISL_608557, EPI_ISL_608558, EPI_ISL_608559, EPI_ISL_608560, EPI_ISL_608561, EPI_ISL_608565, EPI_ISL_608566, EPI_ISL_608568, EPI_ISL_608570, EPI_ISL_608571, EPI_ISL_608572, EPI_ISL_608576, EPI_ISL_608577, EPI_ISL_608578, EPI_ISL_608585, EPI_ISL_608588, EPI_ISL_608596, EPI_ISL_608597, EPI_ISL_608598, EPI_ISL_608602, EPI_ISL_608604, EPI_ISL_608607, EPI_ISL_608608, EPI_ISL_608612, EPI_ISL_608613, EPI_ISL_608615, EPI_ISL_608616, EPI_ISL_608618, EPI_ISL_608619, EPI_ISL_608620, EPI_ISL_608624, EPI_ISL_608626, EPI_ISL_608627, EPI_ISL_608628, EPI_ISL_608629, EPI_ISL_608631, EPI_ISL_608632, EPI_ISL_608633, EPI_ISL_608636, EPI_ISL_608637, EPI_ISL_608638, EPI_ISL_608641, EPI_ISL_608642, EPI_ISL_608643, EPI_ISL_608644, EPI_ISL_608645, EPI_ISL_608650, EPI_ISL_608651, EPI_ISL_608653, EPI_ISL_608654, EPI_ISL_608656, EPI_ISL_608660, EPI_ISL_608664, EPI_ISL_608666, EPI_ISL_608671, EPI_ISL_608673, EPI_ISL_608678, EPI_ISL_608679, EPI_ISL_608682, EPI_ISL_608683, EPI_ISL_608688, EPI_ISL_608689, EPI_ISL_608690, EPI_ISL_608693, EPI_ISL_608695, EPI_ISL_608696, EPI_ISL_608845, EPI_ISL_608852, EPI_ISL_608868, EPI_ISL_608876, EPI_ISL_608879, EPI_ISL_608881, EPI_ISL_608892, EPI_ISL_608902, EPI_ISL_608906, EPI_ISL_608908, EPI_ISL_608910, EPI_ISL_608912, EPI_ISL_608918, EPI_ISL_608923, EPI_ISL_608924, EPI_ISL_608927, EPI_ISL_608935, EPI_ISL_608937, EPI_ISL_608939, EPI_ISL_608943, EPI_ISL_608947, EPI_ISL_608953, EPI_ISL_608958, EPI_ISL_608959, EPI_ISL_608960, EPI_ISL_608965, EPI_ISL_608967, EPI_ISL_608968, EPI_ISL_608969, EPI_ISL_608982, EPI_ISL_608987, EPI_ISL_608990, EPI_ISL_608996, EPI_ISL_608997, EPI_ISL_609003, EPI_ISL_609005, EPI_ISL_609018, EPI_ISL_609023, EPI_ISL_609025, EPI_ISL_609033, EPI_ISL_609037, EPI_ISL_609045, EPI_ISL_609048, EPI_ISL_609054, EPI_ISL_609072, EPI_ISL_609077, EPI_ISL_609081, EPI_ISL_609082, EPI_ISL_609086, EPI_ISL_609088, EPI_ISL_609091, EPI_ISL_609099, EPI_ISL_609100, EPI_ISL_609111, EPI_ISL_609113, EPI_ISL_609114, EPI_ISL_609118, EPI_ISL_609119, EPI_ISL_609122, EPI_ISL_609123, EPI_ISL_609129, EPI_ISL_609131, EPI_ISL_609143, EPI_ISL_609156, EPI_ISL_609158, EPI_ISL_610629, EPI_ISL_610630, EPI_ISL_610631, EPI_ISL_610632, EPI_ISL_610634, EPI_ISL_610635, EPI_ISL_610636, EPI_ISL_610637, EPI_ISL_610638, EPI_ISL_610639, EPI_ISL_610640, EPI_ISL_610641, EPI_ISL_610642, EPI_ISL_610643, EPI_ISL_610644, EPI_ISL_610645, EPI_ISL_610646, EPI_ISL_610647, EPI_ISL_610648, EPI_ISL_610649, EPI_ISL_610650, EPI_ISL_610651, EPI_ISL_610652, EPI_ISL_610653, EPI_ISL_610654, EPI_ISL_610655, EPI_ISL_610656, EPI_ISL_610658, EPI_ISL_610659, EPI_ISL_610660, EPI_ISL_610661, EPI_ISL_610662, EPI_ISL_610663, EPI_ISL_610664, EPI_ISL_610665, EPI_ISL_610666, EPI_ISL_610667, EPI_ISL_610668, EPI_ISL_610669, EPI_ISL_610670, EPI_ISL_610671, EPI_ISL_610672, EPI_ISL_610673, EPI_ISL_610674, EPI_ISL_610675, EPI_ISL_610676, EPI_ISL_610677, EPI_ISL_610678, EPI_ISL_610679, EPI_ISL_610680, EPI_ISL_610681, EPI_ISL_610682, EPI_ISL_610683, EPI_ISL_610684, EPI_ISL_610685, EPI_ISL_610686, EPI_ISL_610687, EPI_ISL_610688, EPI_ISL_610689, EPI_ISL_610690, EPI_ISL_610691, EPI_ISL_610692, EPI_ISL_610693, EPI_ISL_610694, EPI_ISL_610695, EPI_ISL_610696, EPI_ISL_610697, EPI_ISL_610698, EPI_ISL_610699, EPI_ISL_610700, EPI_ISL_610701, EPI_ISL_610703, EPI_ISL_610704, EPI_ISL_610705, EPI_ISL_610706, EPI_ISL_610707, EPI_ISL_610708, EPI_ISL_610709, EPI_ISL_610710, EPI_ISL_610711, EPI_ISL_610712, EPI_ISL_610713, EPI_ISL_610714, EPI_ISL_610715, EPI_ISL_610716, EPI_ISL_610717, EPI_ISL_610718, EPI_ISL_610719, EPI_ISL_610720, EPI_ISL_610721, EPI_ISL_610722, EPI_ISL_610723, EPI_ISL_610724, EPI_ISL_610725, EPI_ISL_610727, EPI_ISL_610728, EPI_ISL_610729, EPI_ISL_610730, EPI_ISL_610731, EPI_ISL_610732, EPI_ISL_610733, EPI_ISL_610734, EPI_ISL_610735, EPI_ISL_610736, EPI_ISL_610737, EPI_ISL_610738, EPI_ISL_610739, EPI_ISL_610740, EPI_ISL_610741, EPI_ISL_610742, EPI_ISL_610743, EPI_ISL_610744, EPI_ISL_610745, EPI_ISL_610746, EPI_ISL_610747, EPI_ISL_610748, EPI_ISL_610749, EPI_ISL_610750, EPI_ISL_610751, EPI_ISL_610752, EPI_ISL_610753, EPI_ISL_610754, EPI_ISL_610755, EPI_ISL_610756, EPI_ISL_610757, EPI_ISL_610758, EPI_ISL_610759, EPI_ISL_610760, EPI_ISL_610761, EPI_ISL_610762, EPI_ISL_610763, EPI_ISL_610764, EPI_ISL_610765, EPI_ISL_610766, EPI_ISL_610767, EPI_ISL_610768, EPI_ISL_610769, EPI_ISL_610770, EPI_ISL_610771, EPI_ISL_610772, EPI_ISL_610773, EPI_ISL_610774, EPI_ISL_610775, EPI_ISL_610776, EPI_ISL_610777, EPI_ISL_610778, EPI_ISL_610780, EPI_ISL_610781, EPI_ISL_610782, EPI_ISL_610783, EPI_ISL_610784, EPI_ISL_610786, EPI_ISL_610787, EPI_ISL_610788, EPI_ISL_610789, EPI_ISL_610790, EPI_ISL_610791, EPI_ISL_610792, EPI_ISL_610793, EPI_ISL_610794, EPI_ISL_610795, EPI_ISL_610796, EPI_ISL_610797, EPI_ISL_610798, EPI_ISL_610799, EPI_ISL_610800, EPI_ISL_610801, EPI_ISL_610802, EPI_ISL_610803, EPI_ISL_610804, EPI_ISL_610805, EPI_ISL_610806, EPI_ISL_610807, EPI_ISL_610808, EPI_ISL_610809, EPI_ISL_610810, EPI_ISL_610811, EPI_ISL_610812, EPI_ISL_610813, EPI_ISL_610814, EPI_ISL_610815, EPI_ISL_610816, EPI_ISL_610817, EPI_ISL_610818, EPI_ISL_610819, EPI_ISL_610820, EPI_ISL_610821, EPI_ISL_610822, EPI_ISL_610823, EPI_ISL_610824, EPI_ISL_610825, EPI_ISL_610826, EPI_ISL_610827, EPI_ISL_610828, EPI_ISL_610829, EPI_ISL_610830, EPI_ISL_610831, EPI_ISL_610832, EPI_ISL_610833, EPI_ISL_610834, EPI_ISL_610835, EPI_ISL_610836, EPI_ISL_610837, EPI_ISL_610838, EPI_ISL_610839, EPI_ISL_610840, EPI_ISL_610841, EPI_ISL_610842, EPI_ISL_610844, EPI_ISL_610845, EPI_ISL_610846, EPI_ISL_610847, EPI_ISL_610848, EPI_ISL_610849, EPI_ISL_610850, EPI_ISL_610851, EPI_ISL_610852, EPI_ISL_610853, EPI_ISL_610854, EPI_ISL_610855, EPI_ISL_610856, EPI_ISL_610857, EPI_ISL_610858, EPI_ISL_610859, EPI_ISL_610860, EPI_ISL_610861, EPI_ISL_610862, EPI_ISL_610863, EPI_ISL_610864, EPI_ISL_610865, EPI_ISL_610866, EPI_ISL_610867, EPI_ISL_610868, EPI_ISL_610869, EPI_ISL_610870, EPI_ISL_610871, EPI_ISL_610872, EPI_ISL_610873, EPI_ISL_610874, EPI_ISL_610875, EPI_ISL_610876, EPI_ISL_610877, EPI_ISL_610878, EPI_ISL_610879, EPI_ISL_610880, EPI_ISL_610881, EPI_ISL_610882, EPI_ISL_610883, EPI_ISL_610884, EPI_ISL_610885, EPI_ISL_610886, EPI_ISL_610887, EPI_ISL_610888, EPI_ISL_610889, EPI_ISL_610890, EPI_ISL_610891, EPI_ISL_610892, EPI_ISL_610893, EPI_ISL_610894, EPI_ISL_610895, EPI_ISL_610896, EPI_ISL_610897, EPI_ISL_610898, EPI_ISL_610899, EPI_ISL_610900, EPI_ISL_610901, EPI_ISL_610902, EPI_ISL_610903, EPI_ISL_610904, EPI_ISL_610905, EPI_ISL_610906, EPI_ISL_610907, EPI_ISL_610908, EPI_ISL_610909, EPI_ISL_610910, EPI_ISL_610911, EPI_ISL_610912, EPI_ISL_610913, EPI_ISL_610915, EPI_ISL_610916, EPI_ISL_610917, EPI_ISL_610918, EPI_ISL_610919, EPI_ISL_610920, EPI_ISL_610921, EPI_ISL_625281, EPI_ISL_625282, EPI_ISL_625283, EPI_ISL_625284, EPI_ISL_625285, EPI_ISL_625286, EPI_ISL_625287, EPI_ISL_625288, EPI_ISL_625290, EPI_ISL_625291, EPI_ISL_625292, EPI_ISL_625293, EPI_ISL_625294, EPI_ISL_625295, EPI_ISL_625296, EPI_ISL_625297, EPI_ISL_625298, EPI_ISL_625299, EPI_ISL_625301, EPI_ISL_625302, EPI_ISL_625303, EPI_ISL_625304, EPI_ISL_625305, EPI_ISL_625306, EPI_ISL_625307, EPI_ISL_625308, EPI_ISL_625309, EPI_ISL_625310, EPI_ISL_625311, EPI_ISL_625313, EPI_ISL_625314

| | | | |
|---|---|---|---|
| see above | Lighthouse Lab in Cambridge | Wellcome Sanger Institute for the COVID-19 Genomics UK (COG-UK) consortium | Cordelia Langford; David K. Jackson; Dominic Kwiatkowski; Ewan Harrison; Ian Johnston; John Sillitoe on behalf of the Wellcome Sanger Institute COVID-19 Surveillance Team; John Sillitoe on behalf of the Wellcome Sanger Institute COVID-19 Surveillance Team (http://www.sanger.ac.uk/covid-team); Rob Howes; Roberto Amato; Sonia Goncalves; The Lighthouse Lab in Cambridge and Alex Alderton |

EPI_ISL_609187, EPI_ISL_609192, EPI_ISL_643044, EPI_ISL_643045, EPI_ISL_643046, EPI_ISL_643047, EPI_ISL_643048, EPI_ISL_643049, EPI_ISL_643050, EPI_ISL_643051, EPI_ISL_643052, EPI_ISL_643053, EPI_ISL_643054, EPI_ISL_643055, EPI_ISL_643056, EPI_ISL_643057, EPI_ISL_643058, EPI_ISL_643059, EPI_ISL_643060, EPI_ISL_643061, EPI_ISL_643062, EPI_ISL_643063, EPI_ISL_643064, EPI_ISL_643065, EPI_ISL_643066, EPI_ISL_643067, EPI_ISL_643068, EPI_ISL_643069, EPI_ISL_643070, EPI_ISL_643071, EPI_ISL_643072, EPI_ISL_643073, EPI_ISL_643074, EPI_ISL_643075, EPI_ISL_643076, EPI_ISL_643077, EPI_ISL_643078, EPI_ISL_643079, EPI_ISL_643080, EPI_ISL_643081, EPI_ISL_643082, EPI_ISL_643083, EPI_ISL_643084, EPI_ISL_643085, EPI_ISL_643086, EPI_ISL_643087, EPI_ISL_643088, EPI_ISL_643089, EPI_ISL_643090, EPI_ISL_643091, EPI_ISL_643092, EPI_ISL_643093, EPI_ISL_643094, EPI_ISL_643095, EPI_ISL_643096, EPI_ISL_643097, EPI_ISL_643098, EPI_ISL_643099, EPI_ISL_643100, EPI_ISL_643101, EPI_ISL_643102, EPI_ISL_643103, EPI_ISL_643104, EPI_ISL_643105, EPI_ISL_643106, EPI_ISL_643107, EPI_ISL_643108, EPI_ISL_643109, EPI_ISL_643110, EPI_ISL_643111, EPI_ISL_643112, EPI_ISL_643113, EPI_ISL_643114, EPI_ISL_643115, EPI_ISL_643116, EPI_ISL_643117, EPI_ISL_643118, EPI_ISL_643119, EPI_ISL_643120, EPI_ISL_643121, EPI_ISL_643122, EPI_ISL_643123, EPI_ISL_643124, EPI_ISL_643125, EPI_ISL_643126, EPI_ISL_643127, EPI_ISL_643128, EPI_ISL_643129, EPI_ISL_643130, EPI_ISL_643131, EPI_ISL_643132, EPI_ISL_643133, EPI_ISL_643134, EPI_ISL_643135, EPI_ISL_643136, EPI_ISL_643137, EPI_ISL_643138, EPI_ISL_643139, EPI_ISL_643140, EPI_ISL_643141, EPI_ISL_643142, EPI_ISL_643143, EPI_ISL_643144, EPI_ISL_643145, EPI_ISL_643146, EPI_ISL_643147, EPI_ISL_643148, EPI_ISL_643149, EPI_ISL_643150, EPI_ISL_643151, EPI_ISL_643152, EPI_ISL_643153, EPI_ISL_643154, EPI_ISL_643155, EPI_ISL_643156, EPI_ISL_643157, EPI_ISL_643158, EPI_ISL_643159, EPI_ISL_643160, EPI_ISL_643161, EPI_ISL_643162, EPI_ISL_643163, EPI_ISL_643164, EPI_ISL_643165, EPI_ISL_643166, EPI_ISL_643167, EPI_ISL_643168, EPI_ISL_643169, EPI_ISL_643170, EPI_ISL_643171, EPI_ISL_643172, EPI_ISL_643173, EPI_ISL_643174, EPI_ISL_643175, EPI_ISL_643176, EPI_ISL_643177, EPI_ISL_643178, EPI_ISL_643179, EPI_ISL_643180, EPI_ISL_643181, EPI_ISL_643182, EPI_ISL_643183, EPI_ISL_643184, EPI_ISL_643185, EPI_ISL_643186, EPI_ISL_643187, EPI_ISL_643188, EPI_ISL_643189, EPI_ISL_643190, EPI_ISL_643191, EPI_ISL_643192, EPI_ISL_643193, EPI_ISL_643194, EPI_ISL_643195, EPI_ISL_643196, EPI_ISL_643197, EPI_ISL_643198, EPI_ISL_643199, EPI_ISL_643200, EPI_ISL_643201, EPI_ISL_643202, EPI_ISL_643203, EPI_ISL_643204, EPI_ISL_643205, EPI_ISL_643206, EPI_ISL_643207, EPI_ISL_643208, EPI_ISL_643209, EPI_ISL_643210, EPI_ISL_643211, EPI_ISL_643212, EPI_ISL_643213, EPI_ISL_643214, EPI_ISL_643215, EPI_ISL_643216, EPI_ISL_643217, EPI_ISL_643218, EPI_ISL_643219, EPI_ISL_643220, EPI_ISL_643221, EPI_ISL_643222, EPI_ISL_643223, EPI_ISL_643224, EPI_ISL_643225, EPI_ISL_643226, EPI_ISL_643227, EPI_ISL_643228, EPI_ISL_643229, EPI_ISL_643230, EPI_ISL_643231, EPI_ISL_643232, EPI_ISL_643233, EPI_ISL_643234, EPI_ISL_643235, EPI_ISL_643236, EPI_ISL_643237, EPI_ISL_643238, EPI_ISL_643239, EPI_ISL_643240, EPI_ISL_643241, EPI_ISL_643242, EPI_ISL_643243, EPI_ISL_643244, EPI_ISL_643245, EPI_ISL_643246, EPI_ISL_643247, EPI_ISL_643248, EPI_ISL_643249, EPI_ISL_643250, EPI_ISL_643251, EPI_ISL_643252, EPI_ISL_643253, EPI_ISL_643254, EPI_ISL_643255, EPI_ISL_643256, EPI_ISL_643257, EPI_ISL_643258, EPI_ISL_643259, EPI_ISL_643260, EPI_ISL_643261, EPI_ISL_643262, EPI_ISL_643263, EPI_ISL_643264, EPI_ISL_643265, EPI_ISL_643266, EPI_ISL_643267, EPI_ISL_643268, EPI_ISL_643269, EPI_ISL_643270, EPI_ISL_643271, EPI_ISL_643272, EPI_ISL_643273, EPI_ISL_643274, EPI_ISL_643275, EPI_ISL_643276, EPI_ISL_643277, EPI_ISL_643278, EPI_ISL_643279, EPI_ISL_643280, EPI_ISL_643281, EPI_ISL_643282, EPI_ISL_643283, EPI_ISL_643284, EPI_ISL_643285, EPI_ISL_643286, EPI_ISL_643287, EPI_ISL_643288, EPI_ISL_643290, EPI_ISL_643291, EPI_ISL_643292, EPI_ISL_643293, EPI_ISL_643294, EPI_ISL_643295, EPI_ISL_643296, EPI_ISL_643297, EPI_ISL_643298, EPI_ISL_643299, EPI_ISL_643300, EPI_ISL_643301, EPI_ISL_643302, EPI_ISL_643303, EPI_ISL_643304, EPI_ISL_643305, EPI_ISL_643306, EPI_ISL_643307, EPI_ISL_643308, EPI_ISL_643309, EPI_ISL_643310, EPI_ISL_643311, EPI_ISL_643312, EPI_ISL_643313, EPI_ISL_643314, EPI_ISL_647854, EPI_ISL_647855, EPI_ISL_647856, EPI_ISL_647857, EPI_ISL_647858, EPI_ISL_647859, EPI_ISL_647860, EPI_ISL_649556, EPI_ISL_649557, EPI_ISL_649558, EPI_ISL_649559, EPI_ISL_649560, EPI_ISL_649561, EPI_ISL_649562, EPI_ISL_649563,

| | | | |
|---|---|---|---|
| EPI_ISL_649564, EPI_ISL_649565, EPI_ISL_649566, EPI_ISL_649567, EPI_ISL_649568, EPI_ISL_721536, EPI_ISL_721537, EPI_ISL_721538, EPI_ISL_721539, EPI_ISL_721540, EPI_ISL_721541, EPI_ISL_721544, EPI_ISL_721556, EPI_ISL_721558, EPI_ISL_721559, EPI_ISL_721561, EPI_ISL_721562, EPI_ISL_721563, EPI_ISL_721564, EPI_ISL_721565, EPI_ISL_721567 | | | |
| see above | Lighthouse Lab in Glasgow | Wellcome Sanger Institute for the COVID-19 Genomics UK (COG-UK) Consortium | Anna Dominiczak and Alex Alderton; Carol Clugston; Cordelia Langford; David Gray; David K. Jackson; Dominic Kwiatkowski; Ewan Harrison; Harper VanSteenhouse; Ian Johnston; John Sillitoe on behalf of the Wellcome Sanger Institute COVID-19 Surveillance Team; John Sillitoe on behalf of the Wellcome Sanger Institute COVID-19 Surveillance Team (http://www.sanger.ac.uk/covid-team); Roberto Amato; Sonia Goncalves; Yumi Kasai |
| EPI_ISL_597234, EPI_ISL_597235, EPI_ISL_597236, EPI_ISL_597240, EPI_ISL_597242, EPI_ISL_597244, EPI_ISL_597245, EPI_ISL_597249, EPI_ISL_597252, EPI_ISL_597260, EPI_ISL_597261, EPI_ISL_597263, EPI_ISL_597267, EPI_ISL_597268, EPI_ISL_597272, EPI_ISL_597275, EPI_ISL_597276, EPI_ISL_597277, EPI_ISL_597278, EPI_ISL_597280, EPI_ISL_597281, EPI_ISL_597288, EPI_ISL_597290, EPI_ISL_597291, EPI_ISL_597292, EPI_ISL_597296, EPI_ISL_597298, EPI_ISL_597299, EPI_ISL_597302, EPI_ISL_597303, EPI_ISL_597311, EPI_ISL_597312, EPI_ISL_597313, EPI_ISL_597314, EPI_ISL_597315, EPI_ISL_597318, EPI_ISL_597322, EPI_ISL_597326, EPI_ISL_597328, EPI_ISL_597329, EPI_ISL_597330, EPI_ISL_597332, EPI_ISL_597334, EPI_ISL_597336, EPI_ISL_597338, EPI_ISL_597339, EPI_ISL_597347, EPI_ISL_597348, EPI_ISL_597351, EPI_ISL_597352, EPI_ISL_597353, EPI_ISL_597355, EPI_ISL_597356, EPI_ISL_597357, EPI_ISL_597360, EPI_ISL_597362, EPI_ISL_597363, EPI_ISL_597364, EPI_ISL_597369, EPI_ISL_597370, EPI_ISL_597376, EPI_ISL_597379, EPI_ISL_597382, EPI_ISL_597383, EPI_ISL_597384, EPI_ISL_597385, EPI_ISL_597386, EPI_ISL_597387, EPI_ISL_597389, EPI_ISL_597391, EPI_ISL_597393, EPI_ISL_597394, EPI_ISL_597395, EPI_ISL_597400, EPI_ISL_597404, EPI_ISL_597406, EPI_ISL_597407, EPI_ISL_597408, EPI_ISL_597409, EPI_ISL_597410, EPI_ISL_597411, EPI_ISL_597413, EPI_ISL_597415, EPI_ISL_597416, EPI_ISL_597417, EPI_ISL_597419, EPI_ISL_597420, EPI_ISL_597421, EPI_ISL_597422, EPI_ISL_597423, EPI_ISL_597429, EPI_ISL_597430, EPI_ISL_597431, EPI_ISL_597433, EPI_ISL_597434, EPI_ISL_597436, EPI_ISL_597439, EPI_ISL_597440, EPI_ISL_597442, EPI_ISL_597443, EPI_ISL_597444, EPI_ISL_597447, EPI_ISL_597448, EPI_ISL_597452, EPI_ISL_597454, EPI_ISL_597455, EPI_ISL_597456, EPI_ISL_597457, EPI_ISL_597463, EPI_ISL_597465, EPI_ISL_597471, EPI_ISL_597474, EPI_ISL_597475, EPI_ISL_597481, EPI_ISL_597482, EPI_ISL_597484, EPI_ISL_597485, EPI_ISL_597486, EPI_ISL_597491, EPI_ISL_597492, EPI_ISL_597493, EPI_ISL_597496, EPI_ISL_597498, EPI_ISL_597499, EPI_ISL_597500, EPI_ISL_597503, EPI_ISL_597506, EPI_ISL_597509, EPI_ISL_597511, EPI_ISL_597513, EPI_ISL_597514, EPI_ISL_597515, EPI_ISL_597516, EPI_ISL_597518, EPI_ISL_597521, EPI_ISL_597522, EPI_ISL_609178, EPI_ISL_609179, EPI_ISL_609182, EPI_ISL_609184, EPI_ISL_609185, EPI_ISL_609186, EPI_ISL_609189, EPI_ISL_609190, EPI_ISL_609191, EPI_ISL_609194, EPI_ISL_609197, EPI_ISL_609198, EPI_ISL_609199, EPI_ISL_609202, EPI_ISL_609206, EPI_ISL_611298, EPI_ISL_611308, EPI_ISL_611318, EPI_ISL_611321, EPI_ISL_611346, EPI_ISL_611350, EPI_ISL_611372, EPI_ISL_611374, EPI_ISL_611389, EPI_ISL_611403, EPI_ISL_611406, EPI_ISL_611410, EPI_ISL_611422, EPI_ISL_611426, EPI_ISL_611435, EPI_ISL_611438, EPI_ISL_611439, EPI_ISL_611443, EPI_ISL_611454, EPI_ISL_611484, EPI_ISL_623895, EPI_ISL_623924, EPI_ISL_623954, EPI_ISL_624188, EPI_ISL_625324, EPI_ISL_625382, EPI_ISL_630702, EPI_ISL_630704, EPI_ISL_630965 | | | |
| see above | Lighthouse Lab in Glasgow | Wellcome Sanger Institute for the COVID-19 Genomics UK (COG-UK) consortium | Anna Dominiczak and Alex Alderton; Carol Clugston; Cordelia Langford; David Gray; David K. Jackson; Dominic Kwiatkowski; Ewan Harrison; Harper VanSteenhouse; Ian Johnston; John Sillitoe on behalf of the Wellcome Sanger Institute COVID-19 Surveillance Team; John Sillitoe on behalf of the Wellcome Sanger Institute COVID-19 Surveillance Team (http://www.sanger.ac.uk/covid-team); Roberto Amato; Sonia Goncalves; Yumi Kasai |
| EPI_ISL_606311, EPI_ISL_606314, EPI_ISL_606353, EPI_ISL_606402, EPI_ISL_606415, EPI_ISL_606462, EPI_ISL_606516, EPI_ISL_606524, EPI_ISL_606588, EPI_ISL_606608, EPI_ISL_606654, EPI_ISL_606797, EPI_ISL_607448, EPI_ISL_607461, EPI_ISL_607516, EPI_ISL_607550, EPI_ISL_607572, EPI_ISL_607579, EPI_ISL_607587, EPI_ISL_644087, EPI_ISL_649386, EPI_ISL_649424, EPI_ISL_709911, EPI_ISL_709914, EPI_ISL_709932, EPI_ISL_709933, EPI_ISL_709934, EPI_ISL_709935, EPI_ISL_709936, EPI_ISL_709937, EPI_ISL_709938, EPI_ISL_709939, EPI_ISL_709940, EPI_ISL_709941, EPI_ISL_709943, EPI_ISL_709945, EPI_ISL_709946, EPI_ISL_756377, EPI_ISL_892224 | | | |
| see above | Lighthouse Lab in Milton Keynes | Wellcome Sanger Institute for the COVID-19 Genomics UK (COG-UK) Consortium | Cordelia Langford; David K. Jackson; Dominic Kwiatkowski; Ewan Harrison; Ian Johnston; John Sillitoe on behalf of the Wellcome Sanger Institute COVID-19 Surveillance Team; John Sillitoe on behalf of the Wellcome Sanger Institute COVID-19 Surveillance Team (http://www.sanger.ac.uk/covid-team); Roberto Amato; Sonia Goncalves; The Lighthouse Lab in Milton Keynes and Alex Alderton |
| EPI_ISL_606296, EPI_ISL_606297, EPI_ISL_606298, EPI_ISL_606299, EPI_ISL_606300, EPI_ISL_606301, EPI_ISL_606302, EPI_ISL_606303, EPI_ISL_606304, EPI_ISL_606305, EPI_ISL_606306, EPI_ISL_606307, EPI_ISL_606308, EPI_ISL_606309, EPI_ISL_606310, EPI_ISL_606312, EPI_ISL_606313, EPI_ISL_606315, EPI_ISL_606316, EPI_ISL_606317, EPI_ISL_606318, EPI_ISL_606319, EPI_ISL_606320, EPI_ISL_606321, EPI_ISL_606322, EPI_ISL_606323, EPI_ISL_606324, EPI_ISL_606325, EPI_ISL_606326, EPI_ISL_606327, EPI_ISL_606328, EPI_ISL_606329, EPI_ISL_606330, EPI_ISL_606331, EPI_ISL_606332, EPI_ISL_606333, EPI_ISL_606334, EPI_ISL_606335, EPI_ISL_606336, EPI_ISL_606337, EPI_ISL_606338, EPI_ISL_606339, EPI_ISL_606340, EPI_ISL_606341, EPI_ISL_606342, EPI_ISL_606343, EPI_ISL_606344, EPI_ISL_606345, EPI_ISL_606346, EPI_ISL_606347, EPI_ISL_606348, EPI_ISL_606349, EPI_ISL_606350, EPI_ISL_606351, EPI_ISL_606352, EPI_ISL_606354, EPI_ISL_606355, EPI_ISL_606356, EPI_ISL_606357, EPI_ISL_606358, EPI_ISL_606359, EPI_ISL_606360, EPI_ISL_606361, EPI_ISL_606362, EPI_ISL_606363, EPI_ISL_606364, EPI_ISL_606365, EPI_ISL_606366, EPI_ISL_606367, EPI_ISL_606368, EPI_ISL_606369, EPI_ISL_606370, EPI_ISL_606371, EPI_ISL_606372, EPI_ISL_606373, EPI_ISL_606374, EPI_ISL_606375, EPI_ISL_606376, EPI_ISL_606377, EPI_ISL_606378, EPI_ISL_606379, EPI_ISL_606380, EPI_ISL_606381, EPI_ISL_606382, EPI_ISL_606383, EPI_ISL_606384, EPI_ISL_606385, EPI_ISL_606386, EPI_ISL_606387, EPI_ISL_606388, EPI_ISL_606389, EPI_ISL_606390, EPI_ISL_606391, EPI_ISL_606392, EPI_ISL_606393, EPI_ISL_606394, EPI_ISL_606395, EPI_ISL_606396, EPI_ISL_606397, EPI_ISL_606398, EPI_ISL_606399, EPI_ISL_606400, EPI_ISL_606401, EPI_ISL_606403, EPI_ISL_606404, EPI_ISL_606405, EPI_ISL_606406, EPI_ISL_606407, EPI_ISL_606408, EPI_ISL_606409, EPI_ISL_606410, EPI_ISL_606411, EPI_ISL_606412, EPI_ISL_606413, EPI_ISL_606414, EPI_ISL_606416, EPI_ISL_606417, EPI_ISL_606418, EPI_ISL_606419, EPI_ISL_606420, EPI_ISL_606421, EPI_ISL_606422, EPI_ISL_606423, EPI_ISL_606424, EPI_ISL_606425, EPI_ISL_606426, EPI_ISL_606427, EPI_ISL_606428, EPI_ISL_606429, EPI_ISL_606430, EPI_ISL_606431, EPI_ISL_606432, EPI_ISL_606433, EPI_ISL_606434, EPI_ISL_606435, EPI_ISL_606436, EPI_ISL_606437, EPI_ISL_606438, EPI_ISL_606439, EPI_ISL_606440, EPI_ISL_606441, EPI_ISL_606442, EPI_ISL_606443, EPI_ISL_606444, EPI_ISL_606445, EPI_ISL_606446, EPI_ISL_606447, EPI_ISL_606448, EPI_ISL_606449, EPI_ISL_606450, EPI_ISL_606451, EPI_ISL_606452, EPI_ISL_606453, EPI_ISL_606454, EPI_ISL_606455, EPI_ISL_606456, EPI_ISL_606457, EPI_ISL_606458, EPI_ISL_606459, EPI_ISL_606460, EPI_ISL_606461, EPI_ISL_606463, EPI_ISL_606464, EPI_ISL_606465, EPI_ISL_606466, EPI_ISL_606467, EPI_ISL_606468, EPI_ISL_606469, EPI_ISL_606470, EPI_ISL_606471, EPI_ISL_606472, EPI_ISL_606473, EPI_ISL_606474, EPI_ISL_606475, EPI_ISL_606476, EPI_ISL_606477, EPI_ISL_606478, EPI_ISL_606479, EPI_ISL_606480, EPI_ISL_606481, EPI_ISL_606482, EPI_ISL_606483, EPI_ISL_606484, EPI_ISL_606485, EPI_ISL_606486, EPI_ISL_606487, EPI_ISL_606488, EPI_ISL_606489, EPI_ISL_606490, EPI_ISL_606491, EPI_ISL_606492, EPI_ISL_606493, EPI_ISL_606494, EPI_ISL_606495, EPI_ISL_606496, EPI_ISL_606497, EPI_ISL_606498, EPI_ISL_606499, EPI_ISL_606500, EPI_ISL_606501, EPI_ISL_606502, EPI_ISL_606503, EPI_ISL_606504, EPI_ISL_606505, EPI_ISL_606506, EPI_ISL_606507, EPI_ISL_606508, EPI_ISL_606509, EPI_ISL_606510, EPI_ISL_606511, EPI_ISL_606512, EPI_ISL_606513, EPI_ISL_606514, EPI_ISL_606515, EPI_ISL_606517, EPI_ISL_606518, EPI_ISL_606519, EPI_ISL_606520, EPI_ISL_606521, EPI_ISL_606522, EPI_ISL_606523, EPI_ISL_606525, EPI_ISL_606526, EPI_ISL_606527, EPI_ISL_606528, EPI_ISL_606529, EPI_ISL_606530, EPI_ISL_606531, EPI_ISL_606532, EPI_ISL_606533, EPI_ISL_606534, EPI_ISL_606535, EPI_ISL_606536, EPI_ISL_606537, EPI_ISL_606538, EPI_ISL_606539, EPI_ISL_606540, EPI_ISL_606541, EPI_ISL_606542, EPI_ISL_606543, EPI_ISL_606544, EPI_ISL_606545, EPI_ISL_606546, EPI_ISL_606547, EPI_ISL_606548, EPI_ISL_606549, EPI_ISL_606550, EPI_ISL_606551, EPI_ISL_606552, EPI_ISL_606553, EPI_ISL_606554, EPI_ISL_606555, EPI_ISL_606556, EPI_ISL_606557, EPI_ISL_606558, EPI_ISL_606559, EPI_ISL_606560, EPI_ISL_606561, EPI_ISL_606562, EPI_ISL_606563, EPI_ISL_606564, EPI_ISL_606565, EPI_ISL_606566, EPI_ISL_606567, EPI_ISL_606568, EPI_ISL_606569, EPI_ISL_606570, EPI_ISL_606571, EPI_ISL_606572, EPI_ISL_606573, EPI_ISL_606574, EPI_ISL_606575, EPI_ISL_606576, EPI_ISL_606577, EPI_ISL_606578, EPI_ISL_606579, EPI_ISL_606580, EPI_ISL_606581, EPI_ISL_606582, EPI_ISL_606583, EPI_ISL_606584, EPI_ISL_606585, EPI_ISL_606586, EPI_ISL_606587, EPI_ISL_606589, EPI_ISL_606590, EPI_ISL_606591, EPI_ISL_606592, EPI_ISL_606593, EPI_ISL_606594, EPI_ISL_606595, EPI_ISL_606596, EPI_ISL_606597, EPI_ISL_606598, EPI_ISL_606599, EPI_ISL_606600, EPI_ISL_606601, EPI_ISL_606602, EPI_ISL_606603, EPI_ISL_606604, EPI_ISL_606605, EPI_ISL_606606, EPI_ISL_606607, EPI_ISL_606609, EPI_ISL_606610, EPI_ISL_606611, EPI_ISL_606612, EPI_ISL_606613, EPI_ISL_606614, EPI_ISL_606615, EPI_ISL_606616, EPI_ISL_606617, EPI_ISL_606618, EPI_ISL_606619, EPI_ISL_606620, EPI_ISL_606621, EPI_ISL_606622, EPI_ISL_606623, EPI_ISL_606624, EPI_ISL_606625, EPI_ISL_606626, EPI_ISL_606627, EPI_ISL_606628, EPI_ISL_606629, EPI_ISL_606630, EPI_ISL_606631, EPI_ISL_606632, EPI_ISL_606633, EPI_ISL_606634, EPI_ISL_606635, EPI_ISL_606636, EPI_ISL_606637, EPI_ISL_606638, EPI_ISL_606639, EPI_ISL_606640, EPI_ISL_606641, EPI_ISL_606642, EPI_ISL_606643, EPI_ISL_606644, EPI_ISL_606645, EPI_ISL_606646, EPI_ISL_606647, EPI_ISL_606648, EPI_ISL_606649, EPI_ISL_606650, EPI_ISL_606651, EPI_ISL_606652, EPI_ISL_606653, EPI_ISL_606655, EPI_ISL_606656, EPI_ISL_606657, EPI_ISL_606658, EPI_ISL_606659, EPI_ISL_606660, EPI_ISL_606661, EPI_ISL_606662, EPI_ISL_606663, EPI_ISL_606664, EPI_ISL_606665, EPI_ISL_606666, EPI_ISL_606667, EPI_ISL_606668, EPI_ISL_606669, EPI_ISL_606670, EPI_ISL_606671, EPI_ISL_606672, EPI_ISL_606673, EPI_ISL_606674, EPI_ISL_606675, EPI_ISL_606676, EPI_ISL_606677, EPI_ISL_606678, EPI_ISL_606679, EPI_ISL_606680, EPI_ISL_606681, EPI_ISL_606682, EPI_ISL_606683, EPI_ISL_606684, EPI_ISL_606685, EPI_ISL_606686, EPI_ISL_606687, EPI_ISL_606688, EPI_ISL_606689, EPI_ISL_606690, EPI_ISL_606691, EPI_ISL_606692, EPI_ISL_606693, EPI_ISL_606694, EPI_ISL_606695, EPI_ISL_606696, EPI_ISL_606697, EPI_ISL_606698, EPI_ISL_606699, EPI_ISL_606700, EPI_ISL_606701, EPI_ISL_606702, EPI_ISL_606703, EPI_ISL_606704, EPI_ISL_606705, EPI_ISL_606706, EPI_ISL_606707, EPI_ISL_606708, EPI_ISL_606709, EPI_ISL_606710, EPI_ISL_606711, EPI_ISL_606712, EPI_ISL_606713, EPI_ISL_606714, EPI_ISL_606715, EPI_ISL_606716, EPI_ISL_606717, EPI_ISL_606718, EPI_ISL_606719, EPI_ISL_606720, EPI_ISL_606721, EPI_ISL_606722, EPI_ISL_606723, EPI_ISL_606724, EPI_ISL_606725, EPI_ISL_606726, EPI_ISL_606727, EPI_ISL_606728, EPI_ISL_606729, EPI_ISL_606730, EPI_ISL_606731, EPI_ISL_606732, EPI_ISL_606733, EPI_ISL_606734, EPI_ISL_606735, EPI_ISL_606736, EPI_ISL_606737, EPI_ISL_606738, EPI_ISL_606739, EPI_ISL_606740, EPI_ISL_606741, EPI_ISL_606742, EPI_ISL_606743, EPI_ISL_606744, EPI_ISL_606745, EPI_ISL_606746, EPI_ISL_606747, EPI_ISL_606748, EPI_ISL_606749, EPI_ISL_606750, EPI_ISL_606751, EPI_ISL_606752, EPI_ISL_606753, EPI_ISL_606754, EPI_ISL_606755, EPI_ISL_606756, EPI_ISL_606757, EPI_ISL_606758, EPI_ISL_606759, EPI_ISL_606760, EPI_ISL_606761, EPI_ISL_606762, EPI_ISL_606763, EPI_ISL_606764, EPI_ISL_606765, EPI_ISL_606766, EPI_ISL_606767, EPI_ISL_606768, EPI_ISL_606769, EPI_ISL_606770, EPI_ISL_606771, EPI_ISL_606772, EPI_ISL_606773, EPI_ISL_606774, EPI_ISL_606775, EPI_ISL_606776, EPI_ISL_606777, EPI_ISL_606778, EPI_ISL_606779, EPI_ISL_606780, EPI_ISL_606781, EPI_ISL_606782, EPI_ISL_606783, EPI_ISL_606784, EPI_ISL_606785, EPI_ISL_606786, EPI_ISL_606787, EPI_ISL_606788, EPI_ISL_606789, EPI_ISL_606790, EPI_ISL_606791, EPI_ISL_606792, EPI_ISL_606793, EPI_ISL_606794, EPI_ISL_606795, EPI_ISL_606796, EPI_ISL_606798, EPI_ISL_606799, EPI_ISL_606800, EPI_ISL_606801, EPI_ISL_606802, EPI_ISL_606803, EPI_ISL_606804, EPI_ISL_606805, EPI_ISL_606806, EPI_ISL_606807, EPI_ISL_606808, EPI_ISL_606809, EPI_ISL_606810, EPI_ISL_606811, EPI_ISL_606812, EPI_ISL_606813, EPI_ISL_606814, EPI_ISL_606815, EPI_ISL_606816, EPI_ISL_606817, EPI_ISL_606818, EPI_ISL_606819, EPI_ISL_606820, EPI_ISL_606821, EPI_ISL_606822, EPI_ISL_606823, EPI_ISL_606824, EPI_ISL_606825, EPI_ISL_606826, EPI_ISL_606827, EPI_ISL_606828, EPI_ISL_606829, EPI_ISL_606830, EPI_ISL_606831, EPI_ISL_606832, EPI_ISL_606833, EPI_ISL_606834, EPI_ISL_606835, EPI_ISL_606836, EPI_ISL_606837, EPI_ISL_606838, EPI_ISL_606839, EPI_ISL_606840, EPI_ISL_606841, EPI_ISL_606842, EPI_ISL_606843, EPI_ISL_606844, EPI_ISL_606845, EPI_ISL_606846, EPI_ISL_606847, EPI_ISL_606848, EPI_ISL_606849, EPI_ISL_606850, EPI_ISL_606851, EPI_ISL_606852, EPI_ISL_606853, EPI_ISL_606854, EPI_ISL_606855, EPI_ISL_606856, EPI_ISL_606857, EPI_ISL_606858, EPI_ISL_606859, EPI_ISL_606860, EPI_ISL_606861, EPI_ISL_606862, EPI_ISL_606863, EPI_ISL_606864, EPI_ISL_606865, EPI_ISL_606866, EPI_ISL_606867, EPI_ISL_606868, EPI_ISL_606869, EPI_ISL_606870, EPI_ISL_606871, EPI_ISL_606872, EPI_ISL_606873, EPI_ISL_606874, EPI_ISL_606875, EPI_ISL_606876, EPI_ISL_606877, EPI_ISL_606878, EPI_ISL_606879, EPI_ISL_606880, EPI_ISL_606881, EPI_ISL_606882, EPI_ISL_606883, EPI_ISL_606884, EPI_ISL_606885, EPI_ISL_606886, EPI_ISL_606887, EPI_ISL_606888, EPI_ISL_606889, EPI_ISL_606890, EPI_ISL_606891, EPI_ISL_606892, EPI_ISL_606893, EPI_ISL_606894, EPI_ISL_606895, EPI_ISL_606896, EPI_ISL_606897, EPI_ISL_606898, EPI_ISL_606899, EPI_ISL_606900, EPI_ISL_606901, EPI_ISL_606902, EPI_ISL_606903, EPI_ISL_606904, EPI_ISL_606905, EPI_ISL_606906, EPI_ISL_606907, EPI_ISL_606908, EPI_ISL_606909, EPI_ISL_606910, EPI_ISL_606911, EPI_ISL_606912, EPI_ISL_606913, EPI_ISL_606914, EPI_ISL_606915, EPI_ISL_606916, EPI_ISL_606917, EPI_ISL_606918, EPI_ISL_606919, EPI_ISL_606920, EPI_ISL_606921, EPI_ISL_606922, EPI_ISL_606923, EPI_ISL_607291, EPI_ISL_607292, EPI_ISL_607293, EPI_ISL_607294, EPI_ISL_607295, EPI_ISL_607296, EPI_ISL_607297, EPI_ISL_607298, EPI_ISL_607299, EPI_ISL_607300, EPI_ISL_607301, EPI_ISL_607302, EPI_ISL_607303, EPI_ISL_607304, EPI_ISL_607305, EPI_ISL_607306, EPI_ISL_607307, EPI_ISL_607308, EPI_ISL_607309, EPI_ISL_607310, EPI_ISL_607311, EPI_ISL_607312, EPI_ISL_607313, EPI_ISL_607314, EPI_ISL_607315, EPI_ISL_607316, EPI_ISL_607317, EPI_ISL_607318, EPI_ISL_607319, EPI_ISL_607320, EPI_ISL_607321, EPI_ISL_607322, EPI_ISL_607323, EPI_ISL_607324, EPI_ISL_607325, EPI_ISL_607326, EPI_ISL_607327, EPI_ISL_607328, EPI_ISL_607329, EPI_ISL_607330, EPI_ISL_607331, EPI_ISL_607332, EPI_ISL_607333, EPI_ISL_607334, EPI_ISL_607335, EPI_ISL_607336, EPI_ISL_607337, EPI_ISL_607338, EPI_ISL_607339, EPI_ISL_607340, EPI_ISL_607341, EPI_ISL_607342, EPI_ISL_607343, EPI_ISL_607344, EPI_ISL_607345, EPI_ISL_607346, EPI_ISL_607347, EPI_ISL_607348, EPI_ISL_607349, EPI_ISL_607350, EPI_ISL_607351, EPI_ISL_607352, EPI_ISL_607353, EPI_ISL_607354, EPI_ISL_607355, EPI_ISL_607356, EPI_ISL_607357, EPI_ISL_607358, EPI_ISL_607359, EPI_ISL_607360, EPI_ISL_607361, EPI_ISL_607362, EPI_ISL_607363, EPI_ISL_607364, EPI_ISL_607365, EPI_ISL_607366, EPI_ISL_607367, EPI_ISL_607368, EPI_ISL_607369, EPI_ISL_607370, EPI_ISL_607371, EPI_ISL_607372, EPI_ISL_607373, EPI_ISL_607374, EPI_ISL_607375, EPI_ISL_607376, EPI_ISL_607377, EPI_ISL_607378, EPI_ISL_607379, EPI_ISL_607380, EPI_ISL_607381, EPI_ISL_607382, EPI_ISL_607383, EPI_ISL_607384, EPI_ISL_607385, EPI_ISL_607386, EPI_ISL_607387, EPI_ISL_607388, EPI_ISL_607389, EPI_ISL_607390, EPI_ISL_607391, EPI_ISL_607392, EPI_ISL_607393, EPI_ISL_607394, EPI_ISL_607395, EPI_ISL_607396, EPI_ISL_607397, EPI_ISL_607398, EPI_ISL_607399, EPI_ISL_607400, EPI_ISL_607401, EPI_ISL_607402, EPI_ISL_607403, EPI_ISL_607404, EPI_ISL_607405, EPI_ISL_607406, EPI_ISL_607407, EPI_ISL_607408, EPI_ISL_607409, EPI_ISL_607410, EPI_ISL_607411, EPI_ISL_607412, EPI_ISL_607413, EPI_ISL_607414, EPI_ISL_607415, EPI_ISL_607416, EPI_ISL_607417, EPI_ISL_607418, EPI_ISL_607419, EPI_ISL_607420, EPI_ISL_607421, EPI_ISL_607422, EPI_ISL_607423, EPI_ISL_607424, EPI_ISL_607425, EPI_ISL_607426, EPI_ISL_607427, EPI_ISL_607428, EPI_ISL_607429, EPI_ISL_607430, EPI_ISL_607431, EPI_ISL_607432, EPI_ISL_607433, EPI_ISL_607434, EPI_ISL_607435, EPI_ISL_607436, EPI_ISL_607437, EPI_ISL_607438, EPI_ISL_607439, EPI_ISL_607440, EPI_ISL_607441, EPI_ISL_607442, EPI_ISL_607443, EPI_ISL_607444, EPI_ISL_607445, EPI_ISL_607446, EPI_ISL_607447, EPI_ISL_607449, EPI_ISL_607450, EPI_ISL_607451, EPI_ISL_607452, EPI_ISL_607453, EPI_ISL_607454, EPI_ISL_607455, EPI_ISL_607456, EPI_ISL_607457, EPI_ISL_607458, EPI_ISL_607459, EPI_ISL_607460, EPI_ISL_607462, EPI_ISL_607463, EPI_ISL_607464, EPI_ISL_607465, EPI_ISL_607466, EPI_ISL_607467, EPI_ISL_607468, EPI_ISL_607469, EPI_ISL_607470, EPI_ISL_607471, EPI_ISL_607472, EPI_ISL_607473, EPI_ISL_607474, EPI_ISL_607475, EPI_ISL_607476, EPI_ISL_607477, EPI_ISL_607478, EPI_ISL_607479, EPI_ISL_607480, EPI_ISL_607481, EPI_ISL_607482, EPI_ISL_607483, EPI_ISL_607484, EPI_ISL_607485, EPI_ISL_607486, EPI_ISL_607487, EPI_ISL_607488, EPI_ISL_607489, EPI_ISL_607490, EPI_ISL_607491, EPI_ISL_607492, EPI_ISL_607493, EPI_ISL_607494, EPI_ISL_607495, EPI_ISL_607496, EPI_ISL_607497, EPI_ISL_607498, EPI_ISL_607499, EPI_ISL_607500, EPI_ISL_607501, EPI_ISL_607502, EPI_ISL_607503, EPI_ISL_607504, EPI_ISL_607505, EPI_ISL_607506, EPI_ISL_607507, EPI_ISL_607508, EPI_ISL_607509, EPI_ISL_607510, EPI_ISL_607511, EPI_ISL_607512, EPI_ISL_607513, EPI_ISL_607514, EPI_ISL_607515, EPI_ISL_607517, EPI_ISL_607518, EPI_ISL_607519, EPI_ISL_607520, EPI_ISL_607521, EPI_ISL_607522, EPI_ISL_607523, EPI_ISL_607524, EPI_ISL_607525, EPI_ISL_607526, EPI_ISL_607527, EPI_ISL_607528, EPI_ISL_607529, EPI_ISL_607530, EPI_ISL_607531, EPI_ISL_607532, EPI_ISL_607533, EPI_ISL_607534, EPI_ISL_607535, EPI_ISL_607536, EPI_ISL_607537, EPI_ISL_607538, EPI_ISL_607539, EPI_ISL_607540, EPI_ISL_607541, EPI_ISL_607542, EPI_ISL_607543, EPI_ISL_607544, EPI_ISL_607545, EPI_ISL_607546, EPI_ISL_607547, EPI_ISL_607548, EPI_ISL_607549, EPI_ISL_607551, EPI_ISL_607552, EPI_ISL_607553, EPI_ISL_607554, EPI_ISL_607555, EPI_ISL_607556, EPI_ISL_607557, EPI_ISL_607558, EPI_ISL_607559, EPI_ISL_607560, EPI_ISL_607561, EPI_ISL_607562, EPI_ISL_607563, EPI_ISL_607564, EPI_ISL_607565, EPI_ISL_607566, EPI_ISL_607567, EPI_ISL_607568, EPI_ISL_607569, EPI_ISL_607570, EPI_ISL_607571, EPI_ISL_607573, EPI_ISL_607574, EPI_ISL_607575, EPI_ISL_607576, EPI_ISL_607577, EPI_ISL_607578, EPI_ISL_607580, EPI_ISL_607581, EPI_ISL_607582, EPI_ISL_607583, EPI_ISL_607584, EPI_ISL_607585, EPI_ISL_607586, EPI_ISL_607588, EPI_ISL_607589, EPI_ISL_607590, EPI_ISL_607591, EPI_ISL_607592, EPI_ISL_607593, EPI_ISL_607594, EPI_ISL_607595, EPI_ISL_607596, EPI_ISL_607597, EPI_ISL_607598, EPI_ISL_607599, EPI_ISL_607600, EPI_ISL_607601, EPI_ISL_607602, EPI_ISL_607603, EPI_ISL_607604, EPI_ISL_607605, EPI_ISL_607606, EPI_ISL_607607, EPI_ISL_607608, EPI_ISL_607609, EPI_ISL_607610, EPI_ISL_607611, EPI_ISL_607612, EPI_ISL_607613, EPI_ISL_607614, EPI_ISL_607615, EPI_ISL_607616, EPI_ISL_607617, EPI_ISL_607618, EPI_ISL_607619, EPI_ISL_607620, EPI_ISL_607621, EPI_ISL_607622, EPI_ISL_608707, EPI_ISL_608751, EPI_ISL_608785, EPI_ISL_608882, EPI_ISL_608894, EPI_ISL_609017, EPI_ISL_623212, EPI_ISL_623218, EPI_ISL_623237, EPI_ISL_623238, EPI_ISL_623262, EPI_ISL_623293, EPI_ISL_623320, EPI_ISL_623331, EPI_ISL_623338, EPI_ISL_623340, EPI_ISL_623367, EPI_ISL_623377, EPI_ISL_623421, EPI_ISL_623434, EPI_ISL_623478, EPI_ISL_623486, EPI_ISL_623496, EPI_ISL_623499, EPI_ISL_629181, EPI_ISL_629216, EPI_ISL_629280, EPI_ISL_629382, EPI_ISL_629395, EPI_ISL_629483, EPI_ISL_629563, EPI_ISL_630048, EPI_ISL_630088, EPI_ISL_630104, EPI_ISL_630120, EPI_ISL_630139, EPI_ISL_630160, EPI_ISL_630174, EPI_ISL_630250, EPI_ISL_630272, EPI_ISL_630292, EPI_ISL_630293, EPI_ISL_630302, EPI_ISL_631108, EPI_ISL_631251, EPI_ISL_634310, EPI_ISL_634388, EPI_ISL_634392, EPI_ISL_634424 | | | |
| see above | Lighthouse Lab in Milton Keynes | Wellcome Sanger Institute for the COVID-19 Genomics UK (COG-UK) consortium | Cordelia Langford; David K. Jackson; Dominic Kwiatkowski; Ewan Harrison; Ian Johnston; John Sillitoe on behalf of the Wellcome Sanger Institute COVID-19 Surveillance Team; John Sillitoe on behalf of the Wellcome Sanger Institute COVID-19 Surveillance Team (http://www.sanger.ac.uk/covid-team); Roberto Amato; Sonia Goncalves; The Lighthouse Lab in Milton Keynes and Alex Alderton |
| EPI_ISL_611516, EPI_ISL_611570, EPI_ISL_611571, EPI_ISL_611613, EPI_ISL_611634, EPI_ISL_611741, EPI_ISL_611750, EPI_ISL_611786, EPI_ISL_611889, EPI_ISL_612003, EPI_ISL_612019, EPI_ISL_612041, EPI_ISL_612043, EPI_ISL_612044, EPI_ISL_612061, EPI_ISL_612072, EPI_ISL_612114, EPI_ISL_626661, EPI_ISL_626694, EPI_ISL_626712, EPI_ISL_626714, EPI_ISL_626736, EPI_ISL_626742, EPI_ISL_626760, EPI_ISL_626807, EPI_ISL_626815, EPI_ISL_626832, EPI_ISL_626866, EPI_ISL_626905, EPI_ISL_627106, EPI_ISL_627107, EPI_ISL_627114, EPI_ISL_627123, EPI_ISL_627125, EPI_ISL_627132, EPI_ISL_627139, EPI_ISL_627141, EPI_ISL_627153, EPI_ISL_627156, EPI_ISL_627162, EPI_ISL_627171, EPI_ISL_627176, EPI_ISL_627181, EPI_ISL_627183, EPI_ISL_627185, EPI_ISL_627445, EPI_ISL_627446, EPI_ISL_627447, EPI_ISL_627448, EPI_ISL_627449, EPI_ISL_637914, EPI_ISL_650114, EPI_ISL_650183, EPI_ISL_650284, EPI_ISL_650331, EPI_ISL_650336, EPI_ISL_650547, EPI_ISL_650561, EPI_ISL_650619, EPI_ISL_650654, EPI_ISL_650658, EPI_ISL_650671, EPI_ISL_650672, EPI_ISL_650755, EPI_ISL_650892, EPI_ISL_650904, EPI_ISL_650928, EPI_ISL_650935, EPI_ISL_650936, EPI_ISL_650946, EPI_ISL_651162, EPI_ISL_651192, EPI_ISL_651270, EPI_ISL_651295, EPI_ISL_651311, EPI_ISL_651344, EPI_ISL_651358, EPI_ISL_651403, EPI_ISL_652078, EPI_ISL_652079, EPI_ISL_652080, EPI_ISL_652081, EPI_ISL_652082, EPI_ISL_652083, EPI_ISL_652084, EPI_ISL_652086, EPI_ISL_1104309, EPI_ISL_1104310 | | | |
| see above | Liverpool Clinical Laboratories | COVID-19 Genomics UK (COG-UK) Consortium | A Alrezaihi; Alessandro Gerada; Alistair Darby; Angela Cowell; Anita Lucaci; Anu Chawla; Cassie Olateju; Catherine Hartley; Charlotte Nelson; Ecaterina Vamos; Elaine O'Toole; Eleanor G Bentley; Ghada T Shawli; Isabel García-Dorival; James Johnson; James P Stewart; Jenifer Manson; Joanne Watts; Jones Benjamin; Jordan J Clark; Julian Hiscox; L Luu; Lucille Rainbow; M Almsaud; Margaret Hughes; Mark Whitehead; Matthew Gemmell; Miren Iturriza-Gomara; Muhannad Alruwaili; N.P Randle; Neil Swainston; PKF Gilmore; Parul Sharma; Rebekah Penrice-Randal; Richard Eccles; Richard Gregory; Sam Haldenby; Steve Paterson; Stuart D Armstrong; Trevor Ian Robinson; Ximeng Han |
| EPI_ISL_611649, EPI_ISL_611752, EPI_ISL_612026, EPI_ISL_612567, EPI_ISL_612568, EPI_ISL_612569, EPI_ISL_612570, EPI_ISL_637283, EPI_ISL_637312, EPI_ISL_637322, EPI_ISL_637352, EPI_ISL_637376, EPI_ISL_637463, EPI_ISL_637520, EPI_ISL_637521, EPI_ISL_637581, EPI_ISL_637784, EPI_ISL_637785, EPI_ISL_637786, EPI_ISL_637787, EPI_ISL_637795, EPI_ISL_637796, EPI_ISL_637797, EPI_ISL_637979, EPI_ISL_637996, EPI_ISL_638009, EPI_ISL_638010, EPI_ISL_638555, EPI_ISL_638556, EPI_ISL_638564, EPI_ISL_638570, EPI_ISL_638571, EPI_ISL_638572, EPI_ISL_638573, EPI_ISL_638574, EPI_ISL_638575, EPI_ISL_638576, EPI_ISL_638577, EPI_ISL_638578, EPI_ISL_638579, EPI_ISL_638580, EPI_ISL_650125, EPI_ISL_650147, EPI_ISL_650148, EPI_ISL_650156, EPI_ISL_650246, EPI_ISL_650247, EPI_ISL_650279, EPI_ISL_650282, EPI_ISL_650351, EPI_ISL_650355, EPI_ISL_650428, EPI_ISL_650572, EPI_ISL_650586, EPI_ISL_650587, EPI_ISL_650588, EPI_ISL_650624, EPI_ISL_650625, EPI_ISL_650785, EPI_ISL_650950, EPI_ISL_650967, EPI_ISL_650968, EPI_ISL_651013, EPI_ISL_651314, EPI_ISL_651354, EPI_ISL_651369, EPI_ISL_651388, EPI_ISL_652111, EPI_ISL_652112, EPI_ISL_652113, EPI_ISL_652114, EPI_ISL_652115, EPI_ISL_652116, EPI_ISL_665262, EPI_ISL_665264, EPI_ISL_665272, EPI_ISL_665275, EPI_ISL_665303, EPI_ISL_665305, EPI_ISL_665308, EPI_ISL_665310, EPI_ISL_665311, EPI_ISL_665312, EPI_ISL_665313, EPI_ISL_665314, EPI_ISL_665319, EPI_ISL_665323, EPI_ISL_665350, EPI_ISL_665368, EPI_ISL_665379, EPI_ISL_665387, EPI_ISL_665392, EPI_ISL_665393, EPI_ISL_665394, EPI_ISL_665395, EPI_ISL_665396, EPI_ISL_665402, EPI_ISL_665403, EPI_ISL_665404, EPI_ISL_665405, EPI_ISL_665408, EPI_ISL_665412, EPI_ISL_665432, EPI_ISL_665490, EPI_ISL_665491, EPI_ISL_665492, EPI_ISL_665493, EPI_ISL_665494, EPI_ISL_665495, EPI_ISL_665499, EPI_ISL_665500, EPI_ISL_665501, EPI_ISL_665502, EPI_ISL_665506, EPI_ISL_665600, EPI_ISL_665617, EPI_ISL_665618, EPI_ISL_665626, EPI_ISL_665630, EPI_ISL_665667, EPI_ISL_665701, EPI_ISL_665706, EPI_ISL_665735, EPI_ISL_665964, EPI_ISL_665965, EPI_ISL_665966, EPI_ISL_665967, EPI_ISL_665968, EPI_ISL_665969, EPI_ISL_665970, EPI_ISL_665983, EPI_ISL_665986, EPI_ISL_665999, EPI_ISL_666084, EPI_ISL_666085, EPI_ISL_666086, EPI_ISL_666087, EPI_ISL_666591, EPI_ISL_705225, EPI_ISL_724966, EPI_ISL_724975, EPI_ISL_724977, EPI_ISL_819448, EPI_ISL_819449, EPI_ISL_1637222, EPI_ISL_1637223 | | | |
| see above | Northumbria University / South Tees Hospitals NHS Foundation Trust / North Cumbria Integrated Care NHS Foundation Trust / North Tees and Hartlepool NHS Foundation Trust / Newcastle Hospitals NHS Foundation Trust | COVID-19 Genomics UK (COG-UK) Consortium | Andrew Nelson; Brendan Payne; Clive Graham; Darren L Smith; Debra Padgett; Edward Barton; Emma Swindells; Garren Scott; Gary Black; Gary Eltringham; Giles S Holt; Greg R Young; Jane Greenaway; Jennifer Collins; John Allan; Joshua Loh; Lynn Dover; Matthew Bashton; Mohammad A Tariq; Paul Baker; Sarah Essex; Steve Liggett; Wen C Yew; Yusri Taha |
| EPI_ISL_1546791 | Originating lab: Wales Specialist Virology Centre Sequencing lab: Pathogen Genomics Unit | Public Health Wales Microbiology Cardiff Wales Specialist Virology Centre | Alec Birchley; Alexander Adams; Amy Gaskin; Angela Marchbank; Bree Gatica-Wilcox; Catherine Moore; Jason Coombes; Joanne Watkins; Joel Southgate; Johnathan Evans; Laura Gifford; Lauren Gilbert; Lee Graham; Malorie Perry; Matthew Bull; Nicole Pacchiarini; Sally Corden; Sara Kumziene-Summerhayes; Sara Rey; Sarah Taylor; Simon Cottrell; Sophie Jones; Tom Connor |
| EPI_ISL_627260, EPI_ISL_627261, EPI_ISL_627262, EPI_ISL_627287, EPI_ISL_627288, EPI_ISL_627289, EPI_ISL_627292, EPI_ISL_627302, EPI_ISL_628619, EPI_ISL_628620, EPI_ISL_628621, EPI_ISL_628622, EPI_ISL_628623, EPI_ISL_628624, EPI_ISL_628625, EPI_ISL_628626, EPI_ISL_628627, EPI_ISL_628628, EPI_ISL_628629, EPI_ISL_628630, EPI_ISL_628631, EPI_ISL_628632, EPI_ISL_628633, EPI_ISL_628634, EPI_ISL_628635, EPI_ISL_628636, EPI_ISL_628637, EPI_ISL_628638, EPI_ISL_628639, EPI_ISL_628640, EPI_ISL_628641, EPI_ISL_628642, EPI_ISL_628643, EPI_ISL_628644, EPI_ISL_628645, EPI_ISL_628646, EPI_ISL_628647, EPI_ISL_628648, EPI_ISL_628649, EPI_ISL_628650, EPI_ISL_628651, EPI_ISL_628652, EPI_ISL_628653, EPI_ISL_628654, EPI_ISL_628655, EPI_ISL_628656, EPI_ISL_628657, EPI_ISL_628658, EPI_ISL_628659, EPI_ISL_628660, EPI_ISL_628661, EPI_ISL_628662, EPI_ISL_628663, EPI_ISL_628664, EPI_ISL_628665, EPI_ISL_628666, EPI_ISL_628667, EPI_ISL_628668, EPI_ISL_628669, EPI_ISL_628670, EPI_ISL_628671, EPI_ISL_628672, EPI_ISL_628673, EPI_ISL_628674, EPI_ISL_628675, EPI_ISL_628676, EPI_ISL_628677, EPI_ISL_628678, EPI_ISL_628679, EPI_ISL_628680, EPI_ISL_628681, EPI_ISL_628682, EPI_ISL_628683, EPI_ISL_628684, EPI_ISL_628685, EPI_ISL_628686, EPI_ISL_628687, EPI_ISL_628688, EPI_ISL_628689, EPI_ISL_628690, EPI_ISL_628691, EPI_ISL_628692, EPI_ISL_628693, EPI_ISL_628694, EPI_ISL_628695, EPI_ISL_628696, EPI_ISL_628697, EPI_ISL_628698, EPI_ISL_628699, EPI_ISL_628700, EPI_ISL_637855, EPI_ISL_637858, EPI_ISL_637861, EPI_ISL_637867, EPI_ISL_637871, EPI_ISL_638037, EPI_ISL_638073, EPI_ISL_638074, EPI_ISL_638075, EPI_ISL_638076, EPI_ISL_638077, EPI_ISL_638078, EPI_ISL_638086, EPI_ISL_638087, EPI_ISL_638088, EPI_ISL_638089, EPI_ISL_638144, EPI_ISL_638145, EPI_ISL_638164, EPI_ISL_638170, EPI_ISL_638171, EPI_ISL_638173, EPI_ISL_638175, EPI_ISL_638176, EPI_ISL_638207, EPI_ISL_638459, EPI_ISL_638460, EPI_ISL_638461, EPI_ISL_638462, EPI_ISL_638463, EPI_ISL_638464, EPI_ISL_638465, EPI_ISL_638466, EPI_ISL_638467, EPI_ISL_638468, EPI_ISL_638469, EPI_ISL_638470, EPI_ISL_638471, EPI_ISL_638472, EPI_ISL_638473, EPI_ISL_638474, EPI_ISL_638475, EPI_ISL_638476, EPI_ISL_638477, EPI_ISL_638478, EPI_ISL_638479, EPI_ISL_638480, EPI_ISL_638481, EPI_ISL_638482, EPI_ISL_638483, EPI_ISL_638484, EPI_ISL_638485, EPI_ISL_638486, EPI_ISL_638487, EPI_ISL_638488, EPI_ISL_638489, EPI_ISL_638490, EPI_ISL_638491, EPI_ISL_638492, EPI_ISL_638493, EPI_ISL_638494, EPI_ISL_638495, EPI_ISL_638496, EPI_ISL_638497, EPI_ISL_638498, EPI_ISL_638499, EPI_ISL_638500, EPI_ISL_638501, EPI_ISL_638502, EPI_ISL_638503, EPI_ISL_638504, EPI_ISL_638505, EPI_ISL_638506, EPI_ISL_638507, EPI_ISL_638508, EPI_ISL_638509, EPI_ISL_638510, EPI_ISL_638511, EPI_ISL_638512, EPI_ISL_638513, EPI_ISL_638514, EPI_ISL_638515, EPI_ISL_638516, EPI_ISL_638517, EPI_ISL_638518, EPI_ISL_638519, EPI_ISL_638520, EPI_ISL_638521, EPI_ISL_638522, EPI_ISL_638523, EPI_ISL_638524, EPI_ISL_638525, EPI_ISL_638526, EPI_ISL_638527, EPI_ISL_638528, EPI_ISL_638529, EPI_ISL_638530, EPI_ISL_638531, EPI_ISL_638737, EPI_ISL_638738, EPI_ISL_638739, EPI_ISL_638740, EPI_ISL_638741, EPI_ISL_638742, EPI_ISL_638743, EPI_ISL_638745, EPI_ISL_638746, EPI_ISL_638747, EPI_ISL_638748, EPI_ISL_638749, EPI_ISL_638750, EPI_ISL_638751, EPI_ISL_638752, EPI_ISL_638753, EPI_ISL_638754, EPI_ISL_638755, EPI_ISL_638756, EPI_ISL_638757, EPI_ISL_638758, EPI_ISL_638759, EPI_ISL_638760, EPI_ISL_638761, EPI_ISL_638762, EPI_ISL_638763, EPI_ISL_638764, EPI_ISL_638765, EPI_ISL_638766, EPI_ISL_638767, EPI_ISL_638768, EPI_ISL_638769, EPI_ISL_638770, EPI_ISL_638771, EPI_ISL_638772, EPI_ISL_638773, EPI_ISL_638774, EPI_ISL_638775, EPI_ISL_638776, EPI_ISL_638777, EPI_ISL_638778, EPI_ISL_638779, EPI_ISL_638780, EPI_ISL_638781, EPI_ISL_638782, EPI_ISL_638783, EPI_ISL_638784, EPI_ISL_638785, EPI_ISL_638786, EPI_ISL_638787, EPI_ISL_638788, EPI_ISL_638789, EPI_ISL_638790, EPI_ISL_638791, EPI_ISL_638792, EPI_ISL_638793, EPI_ISL_638794, EPI_ISL_638795, EPI_ISL_638796, EPI_ISL_638797, EPI_ISL_638798, EPI_ISL_638799, EPI_ISL_638800, EPI_ISL_638801, EPI_ISL_638802, EPI_ISL_638803, EPI_ISL_638804, EPI_ISL_638805, EPI_ISL_638806, EPI_ISL_638807, EPI_ISL_638808, EPI_ISL_638809, EPI_ISL_638810, EPI_ISL_638811, EPI_ISL_638812, EPI_ISL_638813, EPI_ISL_638814, EPI_ISL_638815, EPI_ISL_638816, EPI_ISL_638817, EPI_ISL_638818, EPI_ISL_638819, EPI_ISL_638820, EPI_ISL_638821, EPI_ISL_638822, EPI_ISL_638823, EPI_ISL_638824, EPI_ISL_638825, EPI_ISL_638826, EPI_ISL_638827, EPI_ISL_638828, EPI_ISL_638829, EPI_ISL_638830, EPI_ISL_638831, EPI_ISL_638832, EPI_ISL_638833, EPI_ISL_638834, EPI_ISL_638835, EPI_ISL_638836, EPI_ISL_638837, EPI_ISL_638838, EPI_ISL_638839, EPI_ISL_638840, EPI_ISL_638841, EPI_ISL_638842, EPI_ISL_638843, EPI_ISL_638844, EPI_ISL_638845, EPI_ISL_638846, EPI_ISL_638847, EPI_ISL_638848, EPI_ISL_638849, EPI_ISL_638850, EPI_ISL_638851, EPI_ISL_638852, EPI_ISL_638853, EPI_ISL_638854, EPI_ISL_638855, EPI_ISL_638856, EPI_ISL_638857, EPI_ISL_638858, EPI_ISL_638859, EPI_ISL_638860, EPI_ISL_638861, EPI_ISL_638862, EPI_ISL_638863, EPI_ISL_638864, EPI_ISL_638865, EPI_ISL_638866, EPI_ISL_638867, EPI_ISL_638868, EPI_ISL_638869, EPI_ISL_638870, EPI_ISL_638871, EPI_ISL_638872, EPI_ISL_638873, EPI_ISL_638874, EPI_ISL_638875, EPI_ISL_638876, EPI_ISL_638894, EPI_ISL_638944, EPI_ISL_638945, EPI_ISL_638946, EPI_ISL_638947, EPI_ISL_638948, EPI_ISL_638949, EPI_ISL_638950, EPI_ISL_638951, EPI_ISL_638952, EPI_ISL_638953, EPI_ISL_638954, EPI_ISL_638955, EPI_ISL_638956, EPI_ISL_638957, EPI_ISL_638958, EPI_ISL_638959, EPI_ISL_638960, EPI_ISL_638961, EPI_ISL_638962, EPI_ISL_638963, EPI_ISL_638964, EPI_ISL_638965, EPI_ISL_638966, EPI_ISL_638967, EPI_ISL_638968, EPI_ISL_638969, EPI_ISL_638970, EPI_ISL_638971, EPI_ISL_638972, EPI_ISL_638973, EPI_ISL_638974, EPI_ISL_638975, EPI_ISL_638976, EPI_ISL_638977, EPI_ISL_638978, EPI_ISL_638979, EPI_ISL_638980, EPI_ISL_638981, EPI_ISL_638982, EPI_ISL_638983, EPI_ISL_638984, EPI_ISL_638985, EPI_ISL_638986, EPI_ISL_638987, EPI_ISL_638988, EPI_ISL_638989, EPI_ISL_638990, EPI_ISL_638991, EPI_ISL_638992, EPI_ISL_638993, EPI_ISL_638994, EPI_ISL_638995, EPI_ISL_638996, EPI_ISL_638997, EPI_ISL_638998, EPI_ISL_638999, EPI_ISL_639000, EPI_ISL_639001, EPI_ISL_639627, EPI_ISL_639628, EPI_ISL_639629, EPI_ISL_639630, EPI_ISL_652121, EPI_ISL_652122, EPI_ISL_652761, EPI_ISL_652917, EPI_ISL_652919, EPI_ISL_652947, EPI_ISL_652983, EPI_ISL_653010, EPI_ISL_653026, EPI_ISL_704594, EPI_ISL_704812, EPI_ISL_704845, EPI_ISL_704866, EPI_ISL_704869, EPI_ISL_705414, EPI_ISL_705417, EPI_ISL_705444, EPI_ISL_705473, EPI_ISL_705489, EPI_ISL_705498, EPI_ISL_705972, EPI_ISL_705973, EPI_ISL_705974, EPI_ISL_705975, EPI_ISL_705976, EPI_ISL_705977, EPI_ISL_705978, EPI_ISL_705979, EPI_ISL_705980, EPI_ISL_705981, EPI_ISL_705982, EPI_ISL_705983, EPI_ISL_705984, EPI_ISL_705985, EPI_ISL_705986, EPI_ISL_705987, EPI_ISL_705988, EPI_ISL_705989, EPI_ISL_705990, EPI_ISL_705991, EPI_ISL_705992, EPI_ISL_705993, EPI_ISL_705994, EPI_ISL_705995, EPI_ISL_705996, EPI_ISL_705997, EPI_ISL_705998, EPI_ISL_705999, EPI_ISL_706000, EPI_ISL_706001, EPI_ISL_706002, EPI_ISL_706003, EPI_ISL_706004, EPI_ISL_706005, EPI_ISL_706006, EPI_ISL_706007, EPI_ISL_706008, EPI_ISL_706009, EPI_ISL_706010, EPI_ISL_706011, EPI_ISL_706012, EPI_ISL_706930, EPI_ISL_706931, EPI_ISL_706932, EPI_ISL_706933, EPI_ISL_706934, | | | |

| | | | |
|---|---|---|---|
| EPI_ISL_706935, EPI_ISL_706936, EPI_ISL_706937, EPI_ISL_706938, EPI_ISL_706939, EPI_ISL_706940, EPI_ISL_706941, EPI_ISL_706942, EPI_ISL_706943, EPI_ISL_706944, EPI_ISL_706945, EPI_ISL_706946, EPI_ISL_706947, EPI_ISL_706948, EPI_ISL_706949, EPI_ISL_706950, EPI_ISL_706951, EPI_ISL_706952, EPI_ISL_706953, EPI_ISL_706954, EPI_ISL_706955, EPI_ISL_706956, EPI_ISL_706957, EPI_ISL_706958, EPI_ISL_706959, EPI_ISL_706960, EPI_ISL_706961, EPI_ISL_706962, EPI_ISL_706963, EPI_ISL_706964, EPI_ISL_706965, EPI_ISL_706966, EPI_ISL_706967, EPI_ISL_706968, EPI_ISL_706969, EPI_ISL_706970, EPI_ISL_706971, EPI_ISL_706972, EPI_ISL_706973, EPI_ISL_706974, EPI_ISL_741349, EPI_ISL_741350, EPI_ISL_741351, EPI_ISL_741352, EPI_ISL_741353, EPI_ISL_741354, EPI_ISL_741355, EPI_ISL_741766, EPI_ISL_741767, EPI_ISL_741768, EPI_ISL_741769, EPI_ISL_741770, EPI_ISL_741771, EPI_ISL_741773, EPI_ISL_842119, EPI_ISL_842120, EPI_ISL_842122, EPI_ISL_842123, EPI_ISL_842124, EPI_ISL_842125, EPI_ISL_842126, EPI_ISL_842127, EPI_ISL_842128, EPI_ISL_842129, EPI_ISL_842130, EPI_ISL_842131, EPI_ISL_842132, EPI_ISL_842133, EPI_ISL_842134, EPI_ISL_842135, EPI_ISL_842136, EPI_ISL_842138, EPI_ISL_842140, EPI_ISL_842143, EPI_ISL_842144, EPI_ISL_842145, EPI_ISL_842147, EPI_ISL_842148, EPI_ISL_842149, EPI_ISL_842150, EPI_ISL_842151, EPI_ISL_842152, EPI_ISL_842154, EPI_ISL_842155, EPI_ISL_842156, EPI_ISL_842157, EPI_ISL_842158, EPI_ISL_842159, EPI_ISL_842160, EPI_ISL_842162, EPI_ISL_842163, EPI_ISL_842165, EPI_ISL_842166, EPI_ISL_842167, EPI_ISL_842168, EPI_ISL_842169, EPI_ISL_842170, EPI_ISL_842171, EPI_ISL_842175, EPI_ISL_842178, EPI_ISL_842179, EPI_ISL_842180, EPI_ISL_842181, EPI_ISL_842184, EPI_ISL_842186, EPI_ISL_842187, EPI_ISL_842188, EPI_ISL_842190, EPI_ISL_842191, EPI_ISL_842192, EPI_ISL_842194, EPI_ISL_842195, EPI_ISL_842196, EPI_ISL_842197, EPI_ISL_842198, EPI_ISL_842199, EPI_ISL_842200, EPI_ISL_924078 | | | |
| see above | Oxford Viromics, NDM, University of Oxford; Oxford University Hospitals; Basingstoke and North Hampshire Hospital | COVID-19 Genomics UK (COG-UK) Consortium | Alex Mobbs; Amy Trebes; Anita Justice; Catrin Moore; Christophe Fraser; David Bonsall; David Buck; Emma Wise; George Macintyre; Jessica Lynch; John Todd; Mariateresa de Cesare; Matilde Mori; Monique Andersson; Nathan Moore; Nick Cortes; Robert Shaw; Stephen Kidd; Tanya Golubchik; Timothy Peto |
| EPI_ISL_584734, EPI_ISL_595173, EPI_ISL_595177, EPI_ISL_595178, EPI_ISL_595179, EPI_ISL_595180, EPI_ISL_595181, EPI_ISL_595182, EPI_ISL_595183, EPI_ISL_595184, EPI_ISL_595185, EPI_ISL_595186, EPI_ISL_595187, EPI_ISL_595188, EPI_ISL_595189, EPI_ISL_595190, EPI_ISL_595191, EPI_ISL_595192, EPI_ISL_595193, EPI_ISL_595194, EPI_ISL_595195, EPI_ISL_595196, EPI_ISL_595197, EPI_ISL_595198, EPI_ISL_595199, EPI_ISL_595201, EPI_ISL_595202, EPI_ISL_595203, EPI_ISL_595204, EPI_ISL_595205, EPI_ISL_595206, EPI_ISL_595207, EPI_ISL_595208, EPI_ISL_595210, EPI_ISL_595211, EPI_ISL_595212, EPI_ISL_595213, EPI_ISL_595214, EPI_ISL_595215, EPI_ISL_595216, EPI_ISL_595217, EPI_ISL_595218, EPI_ISL_595219, EPI_ISL_595220, EPI_ISL_595221, EPI_ISL_595222, EPI_ISL_595223, EPI_ISL_595224, EPI_ISL_595225, EPI_ISL_595226, EPI_ISL_595227, EPI_ISL_595228, EPI_ISL_595229, EPI_ISL_595230, EPI_ISL_595231, EPI_ISL_596116, EPI_ISL_596117, EPI_ISL_596118, EPI_ISL_596119, EPI_ISL_596120, EPI_ISL_596121, EPI_ISL_596122, EPI_ISL_596123, EPI_ISL_596124, EPI_ISL_596125, EPI_ISL_596126, EPI_ISL_596127, EPI_ISL_596128, EPI_ISL_596129, EPI_ISL_596130, EPI_ISL_596131, EPI_ISL_596132, EPI_ISL_596133, EPI_ISL_596134, EPI_ISL_596135, EPI_ISL_596136, EPI_ISL_596137, EPI_ISL_596138, EPI_ISL_596139, EPI_ISL_596140, EPI_ISL_596141, EPI_ISL_596142, EPI_ISL_596143, EPI_ISL_596144, EPI_ISL_596183, EPI_ISL_596189, EPI_ISL_596190, EPI_ISL_626656, EPI_ISL_626657, EPI_ISL_626660, EPI_ISL_626663, EPI_ISL_626681, EPI_ISL_626693, EPI_ISL_626698, EPI_ISL_626700, EPI_ISL_626701, EPI_ISL_626704, EPI_ISL_626705, EPI_ISL_626737, EPI_ISL_626741, EPI_ISL_626744, EPI_ISL_626745, EPI_ISL_626751, EPI_ISL_626753, EPI_ISL_626756, EPI_ISL_626759, EPI_ISL_626765, EPI_ISL_626779, EPI_ISL_626790, EPI_ISL_626795, EPI_ISL_626808, EPI_ISL_626819, EPI_ISL_626860, EPI_ISL_626868, EPI_ISL_626893, EPI_ISL_626894, EPI_ISL_626914, EPI_ISL_626954, EPI_ISL_626956, EPI_ISL_626964, EPI_ISL_627009, EPI_ISL_627016, EPI_ISL_627024, EPI_ISL_627041, EPI_ISL_627077, EPI_ISL_627080, EPI_ISL_627098, EPI_ISL_627099, EPI_ISL_627148, EPI_ISL_627170, EPI_ISL_627180, EPI_ISL_627187, EPI_ISL_627198, EPI_ISL_627213, EPI_ISL_627300, EPI_ISL_627570, EPI_ISL_627571, EPI_ISL_627572, EPI_ISL_627573, EPI_ISL_627574, EPI_ISL_627575, EPI_ISL_627576, EPI_ISL_627577, EPI_ISL_627578, EPI_ISL_627579, EPI_ISL_627580, EPI_ISL_627581, EPI_ISL_627582, EPI_ISL_627583, EPI_ISL_627586, EPI_ISL_627587, EPI_ISL_627588, EPI_ISL_627589, EPI_ISL_627590, EPI_ISL_627591, EPI_ISL_627592, EPI_ISL_627593, EPI_ISL_627594, EPI_ISL_627595, EPI_ISL_627596, EPI_ISL_627602, EPI_ISL_627603, EPI_ISL_627607, EPI_ISL_627612, EPI_ISL_627613, EPI_ISL_627615, EPI_ISL_627616, EPI_ISL_627627, EPI_ISL_627630, EPI_ISL_627631, EPI_ISL_627632, EPI_ISL_627633, EPI_ISL_627634, EPI_ISL_627635, EPI_ISL_627637, EPI_ISL_627638, EPI_ISL_627639, EPI_ISL_627640, EPI_ISL_627643, EPI_ISL_627644, EPI_ISL_627652, EPI_ISL_627656, EPI_ISL_627664, EPI_ISL_627665, EPI_ISL_665411, EPI_ISL_665445, EPI_ISL_725162, EPI_ISL_725163, EPI_ISL_725168, EPI_ISL_725191, EPI_ISL_866820, EPI_ISL_866821 | | | |
| see above | Quadram Institute Bioscience | COVID-19 Genomics UK (COG-UK) Consortium | Alexander J Trotter; Alison E. Mather; Alp Aydin; Ana P. Tedim; Anastasia Kolyva; Andrew Bell; Andrew J. Page; Claire Stuart; Dave J. Baker; Gemma L. Kay; John Wain; Justin O'Grady; Leonardo de Oliveira Martins; Lizzie Meadows; Maria Diaz; Mark Webber; Muhammed Yasir; Nabil-Fareed Alikhan; Ngozi Elumogo; Nicholas M. Thomson; Rachael Stanley; Rachel Gilroy; Reenesh Prakash; Samir Dervisevic; Samuel Bloomfield; Steven Rudder; Thanh Le-Viet |
| EPI_ISL_584761, EPI_ISL_584762, EPI_ISL_584763, EPI_ISL_584764, EPI_ISL_584765, EPI_ISL_584766, EPI_ISL_584767, EPI_ISL_584768, EPI_ISL_584769, EPI_ISL_584770, EPI_ISL_584771, EPI_ISL_584772, EPI_ISL_584773, EPI_ISL_584774, EPI_ISL_584775, EPI_ISL_584776, EPI_ISL_584777, EPI_ISL_584778, EPI_ISL_584779, EPI_ISL_584780, EPI_ISL_584781, EPI_ISL_584782, EPI_ISL_584783, EPI_ISL_584784, EPI_ISL_584785, EPI_ISL_584786, EPI_ISL_584787, EPI_ISL_584788, EPI_ISL_584789, EPI_ISL_584790, EPI_ISL_584791, EPI_ISL_584792, EPI_ISL_584793, EPI_ISL_584794, EPI_ISL_584795, EPI_ISL_584796, EPI_ISL_584797, EPI_ISL_584798, EPI_ISL_584799, EPI_ISL_584800, EPI_ISL_584801, EPI_ISL_584802, EPI_ISL_584803, EPI_ISL_584804, EPI_ISL_584805, EPI_ISL_584806, EPI_ISL_584807, EPI_ISL_584808, EPI_ISL_595232, EPI_ISL_595233, EPI_ISL_595234, EPI_ISL_595235, EPI_ISL_595236, EPI_ISL_595237, EPI_ISL_595238, EPI_ISL_595239, EPI_ISL_595240, EPI_ISL_595241, EPI_ISL_595242, EPI_ISL_595243, EPI_ISL_595244, EPI_ISL_595245, EPI_ISL_595246, EPI_ISL_595247, EPI_ISL_595248, EPI_ISL_595249, EPI_ISL_595250, EPI_ISL_595251, EPI_ISL_595252, EPI_ISL_595253, EPI_ISL_595254, EPI_ISL_595255, EPI_ISL_595256, EPI_ISL_595257, EPI_ISL_595258, EPI_ISL_595259, EPI_ISL_595260, EPI_ISL_595261, EPI_ISL_595262, EPI_ISL_595263, EPI_ISL_595264, EPI_ISL_595265, EPI_ISL_595266, EPI_ISL_595267, EPI_ISL_595268, EPI_ISL_595269, EPI_ISL_595270, EPI_ISL_595271, EPI_ISL_595272, EPI_ISL_595273, EPI_ISL_595274, EPI_ISL_595275, EPI_ISL_595276, EPI_ISL_595277, EPI_ISL_595278, EPI_ISL_595279, EPI_ISL_595280, EPI_ISL_595281, EPI_ISL_595282, EPI_ISL_595283, EPI_ISL_595284, EPI_ISL_595285, EPI_ISL_595286, EPI_ISL_595287, EPI_ISL_595288, EPI_ISL_595289, EPI_ISL_595290, EPI_ISL_595291, EPI_ISL_595292, EPI_ISL_595293, EPI_ISL_595294, EPI_ISL_595295, EPI_ISL_595296, EPI_ISL_595297, EPI_ISL_595298, EPI_ISL_595299, EPI_ISL_595300, EPI_ISL_595301, EPI_ISL_595302, EPI_ISL_595303, EPI_ISL_595304, EPI_ISL_595305, EPI_ISL_595306, EPI_ISL_595307, EPI_ISL_595308, EPI_ISL_595309, EPI_ISL_595310, EPI_ISL_595311, EPI_ISL_595312, EPI_ISL_595313, EPI_ISL_595314, EPI_ISL_595315, EPI_ISL_595316, EPI_ISL_595317, EPI_ISL_595318, EPI_ISL_595319, EPI_ISL_595320, EPI_ISL_595321, EPI_ISL_595322, EPI_ISL_595323, EPI_ISL_611546, EPI_ISL_611604, EPI_ISL_611680, EPI_ISL_611753, EPI_ISL_611810, EPI_ISL_611812, EPI_ISL_611813, EPI_ISL_611919, EPI_ISL_611954, EPI_ISL_611963, EPI_ISL_612032, EPI_ISL_612077, EPI_ISL_612107, EPI_ISL_612158, EPI_ISL_612159, EPI_ISL_612181, EPI_ISL_612182, EPI_ISL_612183, EPI_ISL_612197, EPI_ISL_612571, EPI_ISL_612572, EPI_ISL_612573, EPI_ISL_613307, EPI_ISL_613308, EPI_ISL_613309, EPI_ISL_613310, EPI_ISL_613311, EPI_ISL_613312, EPI_ISL_613313, EPI_ISL_613314, EPI_ISL_613315, EPI_ISL_613316, EPI_ISL_613317, EPI_ISL_613318, EPI_ISL_613319, EPI_ISL_613320, EPI_ISL_613321, EPI_ISL_613322, EPI_ISL_613323, EPI_ISL_613324, EPI_ISL_613325, EPI_ISL_613326, EPI_ISL_613327, EPI_ISL_613328, EPI_ISL_613329, EPI_ISL_613330, EPI_ISL_613331, EPI_ISL_613332, EPI_ISL_613333, EPI_ISL_613334, EPI_ISL_613335, EPI_ISL_613336, EPI_ISL_613337, EPI_ISL_613338, EPI_ISL_613339, EPI_ISL_613340, EPI_ISL_613341, EPI_ISL_613342, EPI_ISL_613343, EPI_ISL_613344, EPI_ISL_613345, EPI_ISL_613346, EPI_ISL_613347, EPI_ISL_613348, EPI_ISL_613349, EPI_ISL_613350, EPI_ISL_613351, EPI_ISL_613352, EPI_ISL_613353, EPI_ISL_613354, EPI_ISL_613355, EPI_ISL_627666, EPI_ISL_638584, EPI_ISL_638585, EPI_ISL_651071, EPI_ISL_652627, EPI_ISL_665971, EPI_ISL_665972, EPI_ISL_665973, EPI_ISL_665974, EPI_ISL_665975, EPI_ISL_665976, EPI_ISL_665978, EPI_ISL_665979, EPI_ISL_665980, EPI_ISL_665981, EPI_ISL_839981 | | | |
| see above | Queens Medical Centre, Clinical Microbiology Department / DeepSeq Nottingham | COVID-19 Genomics UK (COG-UK) Consortium | Christopher Moore; Fei Sang; Gemma Clark; Hannah Howson-Wells; Johnny Debebe; Jonathan Ball; Joseph Chappell; Manjinder Khakh; Matthew Carlisle; Matthew Loose; Michelle M Lister; Nadine Holmes; Patrick McClure; Theocharis Tsoleridis; Vicki M Fleming; Victoria Wright; Wendy Smith |
| EPI_ISL_627020, EPI_ISL_627046, EPI_ISL_627518, EPI_ISL_627519, EPI_ISL_627520, EPI_ISL_627521, EPI_ISL_627522, EPI_ISL_627523, EPI_ISL_627524, EPI_ISL_627525, EPI_ISL_627526, EPI_ISL_627527, EPI_ISL_627528, EPI_ISL_627529, EPI_ISL_627530, EPI_ISL_627532, EPI_ISL_627534, EPI_ISL_627559, EPI_ISL_627560, EPI_ISL_627561, EPI_ISL_627562, EPI_ISL_627563, EPI_ISL_627564, EPI_ISL_627565, EPI_ISL_627566, EPI_ISL_627567, EPI_ISL_627568, EPI_ISL_627569, EPI_ISL_650415, EPI_ISL_650524, EPI_ISL_650535, EPI_ISL_650543, EPI_ISL_650598, EPI_ISL_650810, EPI_ISL_650888, EPI_ISL_650901, EPI_ISL_651128, EPI_ISL_651215, EPI_ISL_651329, EPI_ISL_651387, EPI_ISL_651404, EPI_ISL_651501, EPI_ISL_652305, EPI_ISL_652307, EPI_ISL_652309, EPI_ISL_652310, EPI_ISL_652315, EPI_ISL_652330, EPI_ISL_652331, EPI_ISL_652332, EPI_ISL_652333, EPI_ISL_652334, EPI_ISL_652335, EPI_ISL_652336, EPI_ISL_680206, EPI_ISL_680208, EPI_ISL_680210, EPI_ISL_680212, EPI_ISL_680214, EPI_ISL_680217, EPI_ISL_680220, EPI_ISL_680221, EPI_ISL_680222, EPI_ISL_680225, EPI_ISL_680329, EPI_ISL_680331, EPI_ISL_680332, EPI_ISL_680333, EPI_ISL_680334, EPI_ISL_680335, EPI_ISL_680336, EPI_ISL_680337, EPI_ISL_680338, EPI_ISL_680339, EPI_ISL_680340, EPI_ISL_680341, EPI_ISL_680342, EPI_ISL_680343, EPI_ISL_680344, EPI_ISL_680345, EPI_ISL_680346, EPI_ISL_680347, EPI_ISL_680348, EPI_ISL_680349, EPI_ISL_680350, EPI_ISL_680351, EPI_ISL_680352, EPI_ISL_680353, EPI_ISL_680354, EPI_ISL_680355, EPI_ISL_680356, EPI_ISL_680357, EPI_ISL_680358, EPI_ISL_680359, EPI_ISL_680360, EPI_ISL_680361, EPI_ISL_680362, EPI_ISL_680363, EPI_ISL_680364, EPI_ISL_680365, EPI_ISL_680366, EPI_ISL_680367, EPI_ISL_680368, EPI_ISL_680369, EPI_ISL_680370, EPI_ISL_680371, EPI_ISL_680372, EPI_ISL_680373, EPI_ISL_680374, EPI_ISL_680375, EPI_ISL_680376, EPI_ISL_680377, EPI_ISL_680378, EPI_ISL_680379, EPI_ISL_680380, EPI_ISL_680381, EPI_ISL_680382, EPI_ISL_680383, EPI_ISL_680384, EPI_ISL_680385, EPI_ISL_680386, EPI_ISL_680387, EPI_ISL_680388, EPI_ISL_680389, EPI_ISL_680390, EPI_ISL_680391, EPI_ISL_680392, EPI_ISL_680393, EPI_ISL_680394, EPI_ISL_680395, EPI_ISL_680396, EPI_ISL_680397, EPI_ISL_680398, EPI_ISL_680399, EPI_ISL_680400, EPI_ISL_680401, EPI_ISL_680402, EPI_ISL_680403, EPI_ISL_680404, EPI_ISL_680405, EPI_ISL_680406, EPI_ISL_680407, EPI_ISL_680408, EPI_ISL_680409, EPI_ISL_680410, EPI_ISL_680411, EPI_ISL_680412, EPI_ISL_680413, EPI_ISL_680414, EPI_ISL_680415, EPI_ISL_680416, EPI_ISL_680417, EPI_ISL_680418, EPI_ISL_680419, EPI_ISL_680420, EPI_ISL_680421, EPI_ISL_680422, EPI_ISL_680423, EPI_ISL_680424, EPI_ISL_680425, EPI_ISL_680426, EPI_ISL_680427, EPI_ISL_680428, EPI_ISL_680429, EPI_ISL_680430, EPI_ISL_680431, EPI_ISL_680432, EPI_ISL_680433, EPI_ISL_680434, EPI_ISL_680435, EPI_ISL_763388, EPI_ISL_763395, EPI_ISL_763405, EPI_ISL_763408, EPI_ISL_763417, EPI_ISL_763446, EPI_ISL_763450, EPI_ISL_763524, EPI_ISL_763573, EPI_ISL_763583, EPI_ISL_763667, EPI_ISL_763719, EPI_ISL_763729, EPI_ISL_764127, EPI_ISL_764154, EPI_ISL_764173, EPI_ISL_764206, EPI_ISL_764262, EPI_ISL_764263, EPI_ISL_764387, EPI_ISL_764388, EPI_ISL_764389, EPI_ISL_764390, EPI_ISL_764391, EPI_ISL_764392, EPI_ISL_764393, EPI_ISL_764394, EPI_ISL_764395, EPI_ISL_764403, EPI_ISL_764404, EPI_ISL_764405, EPI_ISL_764406, EPI_ISL_764407, EPI_ISL_764408, EPI_ISL_764409, EPI_ISL_764410, EPI_ISL_764411, EPI_ISL_764412, EPI_ISL_764413, EPI_ISL_764414, EPI_ISL_764415, EPI_ISL_764416, EPI_ISL_764417, EPI_ISL_764418, EPI_ISL_764419, EPI_ISL_764420, EPI_ISL_764421, EPI_ISL_764422, EPI_ISL_764423, EPI_ISL_764424, EPI_ISL_764425, EPI_ISL_764426, EPI_ISL_764427, EPI_ISL_764428, EPI_ISL_764429, EPI_ISL_764430 | | | |
| see above | Regional Virus Laboratory, Belfast Health and Social Care Trust | COVID-19 Genomics UK (COG-UK) Consortium | Alison Watt; Ciara Cox; Conall McCaughey; David Simpson; Derek Fairley; James McKenna; Mairead Connor; Susan Feeney; Tanya Curran; Zoltan Molnar |
| EPI_ISL_637114, EPI_ISL_637115, EPI_ISL_637116, EPI_ISL_637155, EPI_ISL_637156, EPI_ISL_637157, EPI_ISL_649808, EPI_ISL_649809, EPI_ISL_649810, EPI_ISL_649811, EPI_ISL_649812, EPI_ISL_649813, EPI_ISL_649814, EPI_ISL_649815, EPI_ISL_649816, EPI_ISL_649819, EPI_ISL_649820, EPI_ISL_649821, EPI_ISL_649822, EPI_ISL_649824, EPI_ISL_649828, EPI_ISL_649829, EPI_ISL_649830, EPI_ISL_660546, EPI_ISL_660742, EPI_ISL_660743, EPI_ISL_660744, EPI_ISL_660745, EPI_ISL_666769, EPI_ISL_668444, EPI_ISL_678751 | | | |
| see above | Respiratory Virus Unit, Microbiology Services Colindale, Public Health England | COVID-19 Genomics UK (COG-UK) Consortium | PHE Covid Sequencing Team |
| EPI_ISL_593998, EPI_ISL_603188, EPI_ISL_603191, EPI_ISL_603193, EPI_ISL_603194, EPI_ISL_603195, EPI_ISL_603196, EPI_ISL_603197, EPI_ISL_603198, EPI_ISL_603199, EPI_ISL_603200, EPI_ISL_603202, EPI_ISL_603203, EPI_ISL_603204, EPI_ISL_603205, EPI_ISL_603206, EPI_ISL_603207, EPI_ISL_603208, EPI_ISL_603209, EPI_ISL_603210, EPI_ISL_603211, EPI_ISL_603213, EPI_ISL_603214, EPI_ISL_609861, EPI_ISL_609862, EPI_ISL_609863, EPI_ISL_609864, EPI_ISL_609865, EPI_ISL_609866, EPI_ISL_609867, EPI_ISL_609868, EPI_ISL_609870, EPI_ISL_609873, EPI_ISL_609875, EPI_ISL_609876, EPI_ISL_609877, EPI_ISL_609878, EPI_ISL_609879, EPI_ISL_609880, EPI_ISL_609883, EPI_ISL_609885, EPI_ISL_609886, EPI_ISL_609887, EPI_ISL_609888, EPI_ISL_609889, EPI_ISL_609890, EPI_ISL_609891, EPI_ISL_609892, EPI_ISL_609893, EPI_ISL_609894, EPI_ISL_609895, EPI_ISL_609896, EPI_ISL_609897, EPI_ISL_609898, EPI_ISL_609899, EPI_ISL_609900, EPI_ISL_609901, EPI_ISL_609902, EPI_ISL_609903, EPI_ISL_609904, EPI_ISL_609905, EPI_ISL_609906, EPI_ISL_609907, EPI_ISL_609908, EPI_ISL_609909, EPI_ISL_613565, EPI_ISL_613568, EPI_ISL_613569, EPI_ISL_613571, EPI_ISL_613572, EPI_ISL_613573, EPI_ISL_613574, EPI_ISL_613575, EPI_ISL_613576, EPI_ISL_613577, EPI_ISL_613578, EPI_ISL_613579, EPI_ISL_613580, EPI_ISL_613581, EPI_ISL_613582, EPI_ISL_613583, EPI_ISL_613584, EPI_ISL_613585, EPI_ISL_613586, EPI_ISL_613590, EPI_ISL_613591, EPI_ISL_613592, EPI_ISL_613593, EPI_ISL_613594, EPI_ISL_613595, EPI_ISL_613596, EPI_ISL_613597, EPI_ISL_613598, EPI_ISL_613599, EPI_ISL_613600, EPI_ISL_613601, EPI_ISL_613602, EPI_ISL_613603, EPI_ISL_613604, EPI_ISL_613605, EPI_ISL_613606, EPI_ISL_613607, EPI_ISL_613608, EPI_ISL_613609, EPI_ISL_613610, EPI_ISL_613611, EPI_ISL_613612, EPI_ISL_613613, EPI_ISL_613614, EPI_ISL_613615, EPI_ISL_613616, EPI_ISL_613617, EPI_ISL_613618, EPI_ISL_613619, EPI_ISL_613620, EPI_ISL_613621, EPI_ISL_613622, EPI_ISL_613623, EPI_ISL_613624, EPI_ISL_613625, EPI_ISL_613626, EPI_ISL_613627, EPI_ISL_613628, EPI_ISL_613629, EPI_ISL_613630, EPI_ISL_613631, EPI_ISL_613632, EPI_ISL_613957, EPI_ISL_622844, EPI_ISL_622845, EPI_ISL_622847, EPI_ISL_622848, EPI_ISL_622849, EPI_ISL_622850, EPI_ISL_622851, EPI_ISL_622852, EPI_ISL_622853, EPI_ISL_622854, EPI_ISL_622855, EPI_ISL_622856, EPI_ISL_622857, EPI_ISL_622858, EPI_ISL_622859, EPI_ISL_622860, EPI_ISL_622861, EPI_ISL_622862, EPI_ISL_622863, EPI_ISL_622864, EPI_ISL_622865, EPI_ISL_622866, EPI_ISL_622867, EPI_ISL_622868, EPI_ISL_622869, EPI_ISL_622870, EPI_ISL_622871, EPI_ISL_622873, EPI_ISL_622874, EPI_ISL_622875, EPI_ISL_622876, EPI_ISL_622877, EPI_ISL_622880, EPI_ISL_622882, EPI_ISL_622884, EPI_ISL_622885, EPI_ISL_622887, EPI_ISL_622888, EPI_ISL_622889, EPI_ISL_622890, EPI_ISL_636705, EPI_ISL_636706, EPI_ISL_636707, EPI_ISL_636708, EPI_ISL_636709, EPI_ISL_636710, EPI_ISL_636711, EPI_ISL_636712, EPI_ISL_636713, EPI_ISL_636714, EPI_ISL_636715, EPI_ISL_636736 | | | |
| see above | Respiratory Virus Unit, Microbiology Services Colindale, Public Health England | Respiratory Virus Unit, Microbiology Services Colindale, Public Health England | PHE Covid Sequencing Team |
| EPI_ISL_825492, EPI_ISL_825495, EPI_ISL_825496, EPI_ISL_825580, EPI_ISL_825581, EPI_ISL_825582, EPI_ISL_837078 | | | |
| see above | Respiratory Virus Unit, National Infection Service, Public Health England | COVID-19 Genomics UK (COG-UK) Consortium | PHE Covid Sequencing Team |
| EPI_ISL_664340, EPI_ISL_664367, EPI_ISL_664498, EPI_ISL_664500, EPI_ISL_664507, EPI_ISL_664672, EPI_ISL_665197, EPI_ISL_665198, EPI_ISL_665199, EPI_ISL_665200, EPI_ISL_665201, EPI_ISL_665202, EPI_ISL_665209, EPI_ISL_665210, EPI_ISL_665211, EPI_ISL_665242, EPI_ISL_665822, EPI_ISL_679414, EPI_ISL_679415, EPI_ISL_1104846, EPI_ISL_1105319, EPI_ISL_1105335 | | | |
| see above | University College London Hospital | COVID-19 Genomics UK (COG-UK) Consortium | Catherine Houlihan; Dan Frampton; Judith Heaney; Matthew Byott; Moira Spyer and Eleni Nastouli; Stuart Kirk |
| EPI_ISL_594878, EPI_ISL_594882, EPI_ISL_594885, EPI_ISL_594886, EPI_ISL_594887, EPI_ISL_611523, EPI_ISL_611558, EPI_ISL_611559, EPI_ISL_611595, EPI_ISL_611596, EPI_ISL_611597, EPI_ISL_611622, EPI_ISL_611623, EPI_ISL_611665, EPI_ISL_611822, EPI_ISL_611996, EPI_ISL_612014, EPI_ISL_612078, EPI_ISL_612079, EPI_ISL_612450, EPI_ISL_612452, EPI_ISL_612453, EPI_ISL_612454, EPI_ISL_627082, EPI_ISL_627175, EPI_ISL_627450, EPI_ISL_627451, EPI_ISL_627452, EPI_ISL_627453, EPI_ISL_627454, EPI_ISL_627455, EPI_ISL_627456, EPI_ISL_627457, EPI_ISL_627458, EPI_ISL_627459, EPI_ISL_627460, EPI_ISL_627461, EPI_ISL_627462, EPI_ISL_627463, EPI_ISL_627464, EPI_ISL_651132, EPI_ISL_1637145, EPI_ISL_1637147, EPI_ISL_1637149, EPI_ISL_1637155, EPI_ISL_1637156 | | | |
| see above | University College London, Great Ormond Street Hospital for Children NHS Foundation Trust, Imperial College Healthcare NHS Trust | COVID-19 Genomics UK (COG-UK) Consortium | Alison Holmes; Charlotte Williams; Helena Tutill; Jacqueline Findlay; James Price; Judith Breuer; Julianne Brown; Kathryn Harris; Leysa Forrest; Mark Kristiansen; Paola Niola; Paola Resende Silva; Patricia Dyal; Paul Randell; Rachel Williams; Samuel Weeks; Sergi Castellano; Sunando Roy; Tony Brooks; Yasmin Panchbhaya |
| EPI_ISL_611550, EPI_ISL_611554, EPI_ISL_611608, EPI_ISL_611612, EPI_ISL_611692, EPI_ISL_611705, EPI_ISL_611825, EPI_ISL_611855, EPI_ISL_611856, EPI_ISL_611858, EPI_ISL_611860, EPI_ISL_612011, EPI_ISL_612207, EPI_ISL_612208, EPI_ISL_612209, EPI_ISL_612210, EPI_ISL_612211, EPI_ISL_612212, EPI_ISL_612213, EPI_ISL_612214, EPI_ISL_612215, EPI_ISL_612216, EPI_ISL_612217, EPI_ISL_612218, EPI_ISL_612227, EPI_ISL_612242, EPI_ISL_612259, EPI_ISL_612260, EPI_ISL_651591, EPI_ISL_679132 | | | |
| see above | University of Birmingham | COVID-19 Genomics UK (COG-UK) Consortium | Alex Richter; Andrew Bosworth; Andrew Bosworth. Queen Elizabeth Hospital: Anna Casey; Andrew D Beggs PHE Heartlands Lab: Husam Osman; Charlotte Poxon; Institute of Microbiology; Joanne Stockton; Josh Quick; Kasun Wanigasooriya; Mike Kidd; Nicholas Loman. University of Birmingham Testing Laboratory: Celina M Whalley; Oliver Pickles; Radoslaw Poplawski; Samuel Nicholls; University of Birmingham: Claire McMurray; Will Rowe |
| EPI_ISL_611838, EPI_ISL_611987, EPI_ISL_612400, EPI_ISL_626662, EPI_ISL_626666, EPI_ISL_626672, EPI_ISL_626673, EPI_ISL_626676, EPI_ISL_626683, EPI_ISL_626685, EPI_ISL_626686, EPI_ISL_626715, EPI_ISL_626716, EPI_ISL_626717, EPI_ISL_626718, EPI_ISL_626719, EPI_ISL_626720, EPI_ISL_626721, EPI_ISL_626722, EPI_ISL_626723, EPI_ISL_626724, EPI_ISL_626725, EPI_ISL_626726, EPI_ISL_626727, EPI_ISL_626728, EPI_ISL_626772, EPI_ISL_626773, EPI_ISL_626774, EPI_ISL_626775, EPI_ISL_626776, EPI_ISL_626785, EPI_ISL_626797, EPI_ISL_626798, EPI_ISL_626799, EPI_ISL_626801, EPI_ISL_626803, EPI_ISL_626805, EPI_ISL_626813, EPI_ISL_626882, EPI_ISL_626883, EPI_ISL_626884, EPI_ISL_626885, EPI_ISL_626886, EPI_ISL_626887, EPI_ISL_626888, EPI_ISL_626910, EPI_ISL_626925, EPI_ISL_626940, EPI_ISL_626941, EPI_ISL_626942, EPI_ISL_626943, EPI_ISL_626945, EPI_ISL_627031, EPI_ISL_627058, EPI_ISL_627059, EPI_ISL_627060, EPI_ISL_627061, EPI_ISL_627062, EPI_ISL_627111, EPI_ISL_627118, EPI_ISL_627145, EPI_ISL_627149, EPI_ISL_627159, EPI_ISL_627163, EPI_ISL_627166, EPI_ISL_627182, EPI_ISL_627186, EPI_ISL_627189, EPI_ISL_627190, EPI_ISL_627196, EPI_ISL_627210, EPI_ISL_627291, EPI_ISL_627314, EPI_ISL_627315, EPI_ISL_627316, EPI_ISL_627317, EPI_ISL_627318, EPI_ISL_627319, EPI_ISL_627320, EPI_ISL_627321, EPI_ISL_627322, EPI_ISL_627323, EPI_ISL_627324, EPI_ISL_627424, EPI_ISL_627425, EPI_ISL_627426, EPI_ISL_627427, EPI_ISL_627428, EPI_ISL_627431, EPI_ISL_627432, EPI_ISL_627433, EPI_ISL_650259, EPI_ISL_650444, EPI_ISL_650642, EPI_ISL_651074, EPI_ISL_651547, EPI_ISL_652676, EPI_ISL_652679, EPI_ISL_652689, EPI_ISL_652701, EPI_ISL_652709 | | | |
| see above | University of Exeter | COVID-19 Genomics UK (COG-UK) Consortium | Aaron Jeffries; Audrey Farbos; Ben Temperton; Jane Masoli; Joanna Warwick-Dugdale; Michelle Michelsen; Robyn Manley; Stephen Michell |
| EPI_ISL_585487, EPI_ISL_585500, EPI_ISL_585501, EPI_ISL_594835, EPI_ISL_594836, EPI_ISL_594837, EPI_ISL_594838, EPI_ISL_594839, EPI_ISL_594840, EPI_ISL_594841, EPI_ISL_594842, EPI_ISL_594843, EPI_ISL_594844, EPI_ISL_594845, EPI_ISL_594846, EPI_ISL_594847, EPI_ISL_594848, EPI_ISL_611544, EPI_ISL_611549, EPI_ISL_611842, EPI_ISL_611850, EPI_ISL_611894, EPI_ISL_611924, EPI_ISL_612037, EPI_ISL_612074, EPI_ISL_612127, EPI_ISL_612128, EPI_ISL_612129, EPI_ISL_612187, EPI_ISL_612188, EPI_ISL_612189, EPI_ISL_612314, EPI_ISL_612315, EPI_ISL_612316, EPI_ISL_612317, EPI_ISL_612318, EPI_ISL_612319, EPI_ISL_612341, EPI_ISL_612342, EPI_ISL_612343, EPI_ISL_612344, EPI_ISL_612345, EPI_ISL_612346, EPI_ISL_612348, EPI_ISL_612349, EPI_ISL_612350, EPI_ISL_612351, EPI_ISL_612352, EPI_ISL_612353, EPI_ISL_612354, EPI_ISL_612355, EPI_ISL_612356, EPI_ISL_612357, EPI_ISL_612358, EPI_ISL_612359, EPI_ISL_612360, EPI_ISL_612361, EPI_ISL_612362, EPI_ISL_612363, EPI_ISL_612364, EPI_ISL_612365, EPI_ISL_612366, EPI_ISL_612367, EPI_ISL_612368, EPI_ISL_612369, EPI_ISL_612370, EPI_ISL_612371, EPI_ISL_612372, EPI_ISL_612373, EPI_ISL_612374, EPI_ISL_650531, EPI_ISL_650833, EPI_ISL_651958, EPI_ISL_651959, EPI_ISL_651971, EPI_ISL_652062, EPI_ISL_680477, EPI_ISL_680478, EPI_ISL_680479, EPI_ISL_680480, EPI_ISL_680481, EPI_ISL_680482, EPI_ISL_680483, EPI_ISL_680484, EPI_ISL_680485, EPI_ISL_680511, EPI_ISL_680512, EPI_ISL_703144, EPI_ISL_703198, EPI_ISL_705391, EPI_ISL_705762, EPI_ISL_705779 | | | |
| see above | Virology Department, Royal Infirmary of Edinburgh, NHS Lothian / School of Biological Sciences, University of Edinburgh / Institute of Genetics and Molecular Medicine, University of Edinburgh | COVID-19 Genomics UK (COG-UK) Consortium | Balcaza C; Colquhoun R; Dewar R; Gallagher M; Hill V; Jackson B; McCrone JT; McHugh M; O'Toole Á; Rambaut A; Rooke S; Scher E; Templeton K; Williams TC; Yu X |
| EPI_ISL_596193, EPI_ISL_596216, EPI_ISL_596217, EPI_ISL_611520, EPI_ISL_611525, EPI_ISL_611528, EPI_ISL_611541, EPI_ISL_611545, EPI_ISL_611553, EPI_ISL_611583, EPI_ISL_611584, EPI_ISL_611594, EPI_ISL_611602, EPI_ISL_611606, EPI_ISL_611650, EPI_ISL_611651, EPI_ISL_611655, EPI_ISL_611656, EPI_ISL_611661, EPI_ISL_611669, EPI_ISL_611678, EPI_ISL_611681, EPI_ISL_611682, EPI_ISL_611690, EPI_ISL_611698, EPI_ISL_611699, EPI_ISL_611744, EPI_ISL_611745, EPI_ISL_611774, EPI_ISL_611775, EPI_ISL_611777, EPI_ISL_611778, EPI_ISL_611779, EPI_ISL_611780, EPI_ISL_611781, EPI_ISL_611782, EPI_ISL_611783, EPI_ISL_611787, EPI_ISL_611788, EPI_ISL_611793, EPI_ISL_611794, EPI_ISL_611795, EPI_ISL_611797, EPI_ISL_611798, EPI_ISL_611801, EPI_ISL_611802, EPI_ISL_611804, EPI_ISL_611805, EPI_ISL_611806, EPI_ISL_611816, EPI_ISL_611817, EPI_ISL_611861, EPI_ISL_611862, EPI_ISL_611920, EPI_ISL_611936, EPI_ISL_611979, EPI_ISL_611997, EPI_ISL_612005, EPI_ISL_612022, EPI_ISL_612023, EPI_ISL_612033, EPI_ISL_612050, EPI_ISL_612055, EPI_ISL_612058, EPI_ISL_612062, EPI_ISL_612067, EPI_ISL_612084, EPI_ISL_612102, EPI_ISL_612196, EPI_ISL_613356, EPI_ISL_613357, EPI_ISL_613359, EPI_ISL_613361, EPI_ISL_613363, EPI_ISL_613365, EPI_ISL_613367, EPI_ISL_613368, EPI_ISL_613369, EPI_ISL_613370, EPI_ISL_613373, EPI_ISL_613374, EPI_ISL_613375, EPI_ISL_613376, EPI_ISL_613377, EPI_ISL_613378, EPI_ISL_613381, EPI_ISL_613382, EPI_ISL_613383, EPI_ISL_613384, EPI_ISL_613385, EPI_ISL_613388, EPI_ISL_613390, EPI_ISL_613391, EPI_ISL_613393, EPI_ISL_613394, EPI_ISL_613395, EPI_ISL_613396, EPI_ISL_613397, EPI_ISL_613398, EPI_ISL_613400, EPI_ISL_613401, EPI_ISL_613402, EPI_ISL_613404, EPI_ISL_613406, EPI_ISL_613407, EPI_ISL_613408, EPI_ISL_613409, EPI_ISL_626735, EPI_ISL_626824, EPI_ISL_637285, EPI_ISL_637290, EPI_ISL_637538, EPI_ISL_637561, EPI_ISL_637621, EPI_ISL_637876, EPI_ISL_637879, EPI_ISL_637881, EPI_ISL_637887, EPI_ISL_637897, EPI_ISL_637901, EPI_ISL_637944, EPI_ISL_637946, EPI_ISL_637973, EPI_ISL_637992, EPI_ISL_638878, EPI_ISL_638890, EPI_ISL_638892, EPI_ISL_650680, EPI_ISL_651076, EPI_ISL_651530, EPI_ISL_653063, EPI_ISL_653069, EPI_ISL_653077, EPI_ISL_653084, EPI_ISL_653092, EPI_ISL_924341, EPI_ISL_924419, EPI_ISL_924420, EPI_ISL_924424 | | | |
| see above | Virology Department, Sheffield Teaching Hospitals NHS Foundation Trust/Department of Infection, Immunity and Cardiovascular Disease, The Medical School, University of Sheffield | COVID-19 Genomics UK (COG-UK) Consortium | Adri Angyal; Alex Keeley; Benjamin Lindsey; Cariad Evans; Danielle Groves; Dave Partridge; Katie Johnson; Laura Carrilero; Luke Green; Matthew Parker; Matthew Wyles; Mehmet Yavuz; Mohammad Raza; Nikki Smith; Paul Parsons; Rachel Tucker; Rebecca Brown; Thushan de Silva |
| EPI_ISL_586025, EPI_ISL_586029, EPI_ISL_586035, EPI_ISL_586040, EPI_ISL_586042, EPI_ISL_586046, EPI_ISL_586049, EPI_ISL_586059, EPI_ISL_586075, EPI_ISL_586085, EPI_ISL_586086, EPI_ISL_586090, EPI_ISL_586104, EPI_ISL_586107, EPI_ISL_586109, EPI_ISL_586111, EPI_ISL_586112, EPI_ISL_586113, EPI_ISL_586123, EPI_ISL_586125, EPI_ISL_586126, EPI_ISL_586130, EPI_ISL_586141, EPI_ISL_586149, EPI_ISL_586153, EPI_ISL_586165, EPI_ISL_586172, EPI_ISL_586181, EPI_ISL_586190, EPI_ISL_586194, EPI_ISL_586209, EPI_ISL_586231, EPI_ISL_586236, EPI_ISL_586238, EPI_ISL_595331, EPI_ISL_595332, EPI_ISL_595333, EPI_ISL_595335, EPI_ISL_595337, EPI_ISL_595339, EPI_ISL_595341, EPI_ISL_595342, EPI_ISL_595343, EPI_ISL_595344, EPI_ISL_595345, EPI_ISL_595346, EPI_ISL_595347, EPI_ISL_595348, EPI_ISL_595349, EPI_ISL_595350, EPI_ISL_595355, EPI_ISL_595356, EPI_ISL_595357, EPI_ISL_595358, EPI_ISL_595359, EPI_ISL_595360, EPI_ISL_595361, EPI_ISL_595362, EPI_ISL_595363, EPI_ISL_595364, EPI_ISL_595365, EPI_ISL_595366, EPI_ISL_595367, EPI_ISL_595368, EPI_ISL_595372, EPI_ISL_595378, EPI_ISL_595379, EPI_ISL_595384, EPI_ISL_595389, EPI_ISL_595390, EPI_ISL_595392, EPI_ISL_595394, EPI_ISL_595395, EPI_ISL_595396, EPI_ISL_595399, EPI_ISL_595400, EPI_ISL_595401, EPI_ISL_595409, EPI_ISL_595410, EPI_ISL_595415, EPI_ISL_595416, EPI_ISL_595419, EPI_ISL_595422, EPI_ISL_595423, EPI_ISL_595425, EPI_ISL_595428, EPI_ISL_595429, EPI_ISL_595431, EPI_ISL_595432, EPI_ISL_595438, EPI_ISL_595440, EPI_ISL_595443, EPI_ISL_595445, EPI_ISL_595449, EPI_ISL_595450, EPI_ISL_595453, | | | |

EPI_ISL_595456, EPI_ISL_595458, EPI_ISL_595460, EPI_ISL_595462, EPI_ISL_595463, EPI_ISL_595464, EPI_ISL_595466, EPI_ISL_595467, EPI_ISL_595474, EPI_ISL_595477, EPI_ISL_595484, EPI_ISL_595485, EPI_ISL_595486, EPI_ISL_595487, EPI_ISL_595490, EPI_ISL_595499, EPI_ISL_595506, EPI_ISL_595508, EPI_ISL_595509, EPI_ISL_595510, EPI_ISL_595511, EPI_ISL_595515, EPI_ISL_595516, EPI_ISL_595517, EPI_ISL_595518, EPI_ISL_595519, EPI_ISL_595521, EPI_ISL_595522, EPI_ISL_595526, EPI_ISL_595529, EPI_ISL_595531, EPI_ISL_595533, EPI_ISL_595535, EPI_ISL_595536, EPI_ISL_595537, EPI_ISL_595539, EPI_ISL_595540, EPI_ISL_595541, EPI_ISL_595542, EPI_ISL_595543, EPI_ISL_595547, EPI_ISL_595548, EPI_ISL_595552, EPI_ISL_595553, EPI_ISL_595560, EPI_ISL_595563, EPI_ISL_595567, EPI_ISL_595569, EPI_ISL_595570, EPI_ISL_595577, EPI_ISL_595578, EPI_ISL_595581, EPI_ISL_595582, EPI_ISL_595584, EPI_ISL_595585, EPI_ISL_595587, EPI_ISL_595589, EPI_ISL_595590, EPI_ISL_595594, EPI_ISL_595597, EPI_ISL_595600, EPI_ISL_595603, EPI_ISL_595604, EPI_ISL_595613, EPI_ISL_595614, EPI_ISL_595616, EPI_ISL_611518, EPI_ISL_611534, EPI_ISL_611560, EPI_ISL_611561, EPI_ISL_611562, EPI_ISL_611564, EPI_ISL_611573, EPI_ISL_611578, EPI_ISL_611581, EPI_ISL_611588, EPI_ISL_611601, EPI_ISL_611609, EPI_ISL_611626, EPI_ISL_611637, EPI_ISL_611652, EPI_ISL_611663, EPI_ISL_611673, EPI_ISL_611674, EPI_ISL_611686, EPI_ISL_611687, EPI_ISL_611688, EPI_ISL_611691, EPI_ISL_611704, EPI_ISL_611711, EPI_ISL_611725, EPI_ISL_611732, EPI_ISL_611738, EPI_ISL_611740, EPI_ISL_611742, EPI_ISL_611748, EPI_ISL_611749, EPI_ISL_611792, EPI_ISL_611809, EPI_ISL_611824, EPI_ISL_611828, EPI_ISL_611835, EPI_ISL_611854, EPI_ISL_611863, EPI_ISL_611865, EPI_ISL_611887, EPI_ISL_611888, EPI_ISL_611896, EPI_ISL_611900, EPI_ISL_611901, EPI_ISL_611902, EPI_ISL_611906, EPI_ISL_611907, EPI_ISL_611909, EPI_ISL_611913, EPI_ISL_611922, EPI_ISL_611923, EPI_ISL_611932, EPI_ISL_611972, EPI_ISL_611973, EPI_ISL_611975, EPI_ISL_611982, EPI_ISL_611986, EPI_ISL_611992, EPI_ISL_611993, EPI_ISL_612002, EPI_ISL_612006, EPI_ISL_612012, EPI_ISL_612025, EPI_ISL_612029, EPI_ISL_612031, EPI_ISL_612038, EPI_ISL_612045, EPI_ISL_612049, EPI_ISL_612052, EPI_ISL_612076, EPI_ISL_612087, EPI_ISL_612091, EPI_ISL_612093, EPI_ISL_612098, EPI_ISL_612109, EPI_ISL_612119, EPI_ISL_612135, EPI_ISL_612136, EPI_ISL_612137, EPI_ISL_612140, EPI_ISL_612143, EPI_ISL_612148, EPI_ISL_612150, EPI_ISL_612151, EPI_ISL_612153, EPI_ISL_612154, EPI_ISL_612160, EPI_ISL_612161, EPI_ISL_612164, EPI_ISL_612166, EPI_ISL_612167, EPI_ISL_612168, EPI_ISL_612170, EPI_ISL_612172, EPI_ISL_612173, EPI_ISL_612174, EPI_ISL_612175, EPI_ISL_612176, EPI_ISL_612177, EPI_ISL_612178, EPI_ISL_612179, EPI_ISL_612180, EPI_ISL_612194, EPI_ISL_612195, EPI_ISL_612202, EPI_ISL_612610, EPI_ISL_612611, EPI_ISL_612612, EPI_ISL_612613, EPI_ISL_612614, EPI_ISL_612615, EPI_ISL_612617, EPI_ISL_612618, EPI_ISL_612619, EPI_ISL_612620, EPI_ISL_612621, EPI_ISL_612622, EPI_ISL_612623, EPI_ISL_612624, EPI_ISL_612625, EPI_ISL_612626, EPI_ISL_612629, EPI_ISL_612630, EPI_ISL_612631, EPI_ISL_612632, EPI_ISL_612633, EPI_ISL_612634, EPI_ISL_612635, EPI_ISL_612637, EPI_ISL_612638, EPI_ISL_612640, EPI_ISL_612641, EPI_ISL_612645, EPI_ISL_612648, EPI_ISL_612649, EPI_ISL_612652, EPI_ISL_612653, EPI_ISL_612654, EPI_ISL_612655, EPI_ISL_612656, EPI_ISL_612659, EPI_ISL_612660, EPI_ISL_612661, EPI_ISL_612664, EPI_ISL_612665, EPI_ISL_612667, EPI_ISL_612672, EPI_ISL_612673, EPI_ISL_612674, EPI_ISL_612675, EPI_ISL_612676, EPI_ISL_612677, EPI_ISL_612679, EPI_ISL_612681, EPI_ISL_612682, EPI_ISL_612683, EPI_ISL_612685, EPI_ISL_612686, EPI_ISL_612689, EPI_ISL_612692, EPI_ISL_612694, EPI_ISL_612698, EPI_ISL_612699, EPI_ISL_612701, EPI_ISL_612702, EPI_ISL_612703, EPI_ISL_612705, EPI_ISL_612709, EPI_ISL_612712, EPI_ISL_612713, EPI_ISL_612715, EPI_ISL_612717, EPI_ISL_612718, EPI_ISL_612719, EPI_ISL_612723, EPI_ISL_612724, EPI_ISL_612729, EPI_ISL_612730, EPI_ISL_612734, EPI_ISL_612735, EPI_ISL_612736, EPI_ISL_612740, EPI_ISL_612741, EPI_ISL_612742, EPI_ISL_612743, EPI_ISL_612744, EPI_ISL_612748, EPI_ISL_612749, EPI_ISL_612750, EPI_ISL_612752, EPI_ISL_612757, EPI_ISL_612758, EPI_ISL_612759, EPI_ISL_612761, EPI_ISL_612763, EPI_ISL_612764, EPI_ISL_612766, EPI_ISL_612771, EPI_ISL_612772, EPI_ISL_612775, EPI_ISL_612776, EPI_ISL_612777, EPI_ISL_612778, EPI_ISL_612779, EPI_ISL_612780, EPI_ISL_612781, EPI_ISL_612783, EPI_ISL_612784, EPI_ISL_612785, EPI_ISL_612792, EPI_ISL_612793, EPI_ISL_612798, EPI_ISL_612800, EPI_ISL_612801, EPI_ISL_612802, EPI_ISL_612805, EPI_ISL_612806, EPI_ISL_612807, EPI_ISL_612810, EPI_ISL_612815, EPI_ISL_612820, EPI_ISL_612822, EPI_ISL_612823, EPI_ISL_612825, EPI_ISL_612827, EPI_ISL_612831, EPI_ISL_612832, EPI_ISL_612834, EPI_ISL_612835, EPI_ISL_612837, EPI_ISL_612840, EPI_ISL_612841, EPI_ISL_612842, EPI_ISL_612846, EPI_ISL_612847, EPI_ISL_612853, EPI_ISL_612856, EPI_ISL_612858, EPI_ISL_612860, EPI_ISL_612863, EPI_ISL_612864, EPI_ISL_612866, EPI_ISL_612869, EPI_ISL_612871, EPI_ISL_612872, EPI_ISL_612873, EPI_ISL_612874, EPI_ISL_612876, EPI_ISL_612879, EPI_ISL_612883, EPI_ISL_612884, EPI_ISL_612885, EPI_ISL_612886, EPI_ISL_612888, EPI_ISL_612889, EPI_ISL_612890, EPI_ISL_612893, EPI_ISL_612894, EPI_ISL_612896, EPI_ISL_612898, EPI_ISL_612900, EPI_ISL_612904, EPI_ISL_612906, EPI_ISL_612907, EPI_ISL_612909, EPI_ISL_612910, EPI_ISL_612912, EPI_ISL_612914, EPI_ISL_612916, EPI_ISL_612917, EPI_ISL_612919, EPI_ISL_612921, EPI_ISL_612926, EPI_ISL_612929, EPI_ISL_612930, EPI_ISL_612931, EPI_ISL_612935, EPI_ISL_612937, EPI_ISL_612939, EPI_ISL_612941, EPI_ISL_612945, EPI_ISL_612946, EPI_ISL_612947, EPI_ISL_612948, EPI_ISL_612949, EPI_ISL_612950, EPI_ISL_612951, EPI_ISL_612955, EPI_ISL_612956, EPI_ISL_612959, EPI_ISL_612961, EPI_ISL_612965, EPI_ISL_612966, EPI_ISL_612970, EPI_ISL_612971, EPI_ISL_612973, EPI_ISL_612974, EPI_ISL_612977, EPI_ISL_612979, EPI_ISL_612980, EPI_ISL_612981, EPI_ISL_612982, EPI_ISL_612983, EPI_ISL_612984, EPI_ISL_612987, EPI_ISL_612989, EPI_ISL_612990, EPI_ISL_612993, EPI_ISL_612996, EPI_ISL_612999, EPI_ISL_613000, EPI_ISL_613001, EPI_ISL_613002, EPI_ISL_613004, EPI_ISL_613005, EPI_ISL_613006, EPI_ISL_613007, EPI_ISL_613008, EPI_ISL_613009, EPI_ISL_613011, EPI_ISL_613012, EPI_ISL_613013, EPI_ISL_613015, EPI_ISL_613016, EPI_ISL_613017, EPI_ISL_613018, EPI_ISL_613019, EPI_ISL_613022, EPI_ISL_613023, EPI_ISL_613024, EPI_ISL_613027, EPI_ISL_613028, EPI_ISL_613029, EPI_ISL_613030, EPI_ISL_613031, EPI_ISL_613032, EPI_ISL_613033, EPI_ISL_613034, EPI_ISL_613035, EPI_ISL_613036, EPI_ISL_613037, EPI_ISL_613038, EPI_ISL_613039, EPI_ISL_613040, EPI_ISL_613041, EPI_ISL_613042, EPI_ISL_613046, EPI_ISL_613047, EPI_ISL_613048, EPI_ISL_613049, EPI_ISL_613050, EPI_ISL_613051, EPI_ISL_613052, EPI_ISL_613053, EPI_ISL_613054, EPI_ISL_613055, EPI_ISL_613056, EPI_ISL_613058, EPI_ISL_613059, EPI_ISL_613060, EPI_ISL_613062, EPI_ISL_613063, EPI_ISL_613064, EPI_ISL_613065, EPI_ISL_613066, EPI_ISL_613067, EPI_ISL_613069, EPI_ISL_613070, EPI_ISL_613071, EPI_ISL_613072, EPI_ISL_613073, EPI_ISL_613074, EPI_ISL_613075, EPI_ISL_613076, EPI_ISL_613077, EPI_ISL_613079, EPI_ISL_613080, EPI_ISL_613081, EPI_ISL_613082, EPI_ISL_613083, EPI_ISL_613084, EPI_ISL_613085, EPI_ISL_613086, EPI_ISL_613087, EPI_ISL_613088, EPI_ISL_613089, EPI_ISL_613091, EPI_ISL_613092, EPI_ISL_613093, EPI_ISL_613094, EPI_ISL_613095, EPI_ISL_613096, EPI_ISL_613097, EPI_ISL_613099, EPI_ISL_613102, EPI_ISL_613103, EPI_ISL_613104, EPI_ISL_613106, EPI_ISL_613107, EPI_ISL_613108, EPI_ISL_613109, EPI_ISL_613110, EPI_ISL_613111, EPI_ISL_613112, EPI_ISL_613113, EPI_ISL_613115, EPI_ISL_613116, EPI_ISL_613117, EPI_ISL_613118, EPI_ISL_613119, EPI_ISL_613120, EPI_ISL_613121, EPI_ISL_613122, EPI_ISL_613123, EPI_ISL_613124, EPI_ISL_613126, EPI_ISL_613127, EPI_ISL_613128, EPI_ISL_613129, EPI_ISL_613130, EPI_ISL_613131, EPI_ISL_613132, EPI_ISL_613133, EPI_ISL_613134, EPI_ISL_613135, EPI_ISL_613139, EPI_ISL_613140, EPI_ISL_613142, EPI_ISL_613143, EPI_ISL_613144, EPI_ISL_613145, EPI_ISL_613146, EPI_ISL_613147, EPI_ISL_613150, EPI_ISL_613152, EPI_ISL_613153, EPI_ISL_613154, EPI_ISL_613156, EPI_ISL_613157, EPI_ISL_613158, EPI_ISL_613159, EPI_ISL_613160, EPI_ISL_613161, EPI_ISL_613162, EPI_ISL_613163, EPI_ISL_613164, EPI_ISL_613165, EPI_ISL_613166, EPI_ISL_613167, EPI_ISL_613169, EPI_ISL_613170, EPI_ISL_613171, EPI_ISL_613172, EPI_ISL_613173, EPI_ISL_613174, EPI_ISL_613175, EPI_ISL_613176, EPI_ISL_613179, EPI_ISL_613180, EPI_ISL_613181, EPI_ISL_613182, EPI_ISL_613183, EPI_ISL_613184, EPI_ISL_613187, EPI_ISL_613188, EPI_ISL_613190, EPI_ISL_613197, EPI_ISL_613203, EPI_ISL_613204, EPI_ISL_613216, EPI_ISL_613220, EPI_ISL_613225, EPI_ISL_613226, EPI_ISL_613229, EPI_ISL_613231, EPI_ISL_613241, EPI_ISL_613250, EPI_ISL_613254, EPI_ISL_613256, EPI_ISL_613258, EPI_ISL_613265, EPI_ISL_613269, EPI_ISL_613271, EPI_ISL_613275, EPI_ISL_613278, EPI_ISL_613279, EPI_ISL_613280, EPI_ISL_626780, EPI_ISL_626787, EPI_ISL_626820, EPI_ISL_626862, EPI_ISL_626921, EPI_ISL_627002, EPI_ISL_627010, EPI_ISL_627027, EPI_ISL_627032, EPI_ISL_627043, EPI_ISL_627048, EPI_ISL_627050, EPI_ISL_627078, EPI_ISL_627088, EPI_ISL_627089, EPI_ISL_627090, EPI_ISL_627091, EPI_ISL_627092, EPI_ISL_627093, EPI_ISL_627094, EPI_ISL_627095, EPI_ISL_627113, EPI_ISL_627120, EPI_ISL_627154, EPI_ISL_627161, EPI_ISL_627168, EPI_ISL_627197, EPI_ISL_627222, EPI_ISL_627266, EPI_ISL_627267, EPI_ISL_627269, EPI_ISL_627270, EPI_ISL_627271, EPI_ISL_627274, EPI_ISL_627275, EPI_ISL_627276, EPI_ISL_627293, EPI_ISL_627733, EPI_ISL_627738, EPI_ISL_627739, EPI_ISL_627740, EPI_ISL_627741, EPI_ISL_627746, EPI_ISL_627747, EPI_ISL_627749, EPI_ISL_627750, EPI_ISL_627752, EPI_ISL_627753, EPI_ISL_627754, EPI_ISL_627755, EPI_ISL_627760, EPI_ISL_627767, EPI_ISL_627768, EPI_ISL_627769, EPI_ISL_627770, EPI_ISL_627778, EPI_ISL_627784, EPI_ISL_627786, EPI_ISL_627788, EPI_ISL_627792, EPI_ISL_627793, EPI_ISL_627794, EPI_ISL_627802, EPI_ISL_627803, EPI_ISL_627807, EPI_ISL_627809, EPI_ISL_627810, EPI_ISL_627817, EPI_ISL_627818, EPI_ISL_627820, EPI_ISL_627821, EPI_ISL_627822, EPI_ISL_627825, EPI_ISL_627826, EPI_ISL_627829, EPI_ISL_627831, EPI_ISL_627833, EPI_ISL_627834, EPI_ISL_627836, EPI_ISL_627839, EPI_ISL_627840, EPI_ISL_627842, EPI_ISL_627843, EPI_ISL_627844, EPI_ISL_627845, EPI_ISL_627846, EPI_ISL_627847, EPI_ISL_627848, EPI_ISL_627854, EPI_ISL_627855, EPI_ISL_627858, EPI_ISL_627860, EPI_ISL_627862, EPI_ISL_627868, EPI_ISL_627869, EPI_ISL_627877, EPI_ISL_627878, EPI_ISL_627882, EPI_ISL_627887, EPI_ISL_627888, EPI_ISL_627890, EPI_ISL_627891, EPI_ISL_627892, EPI_ISL_627897, EPI_ISL_627902, EPI_ISL_627904, EPI_ISL_627906, EPI_ISL_627907, EPI_ISL_627909, EPI_ISL_627910, EPI_ISL_627912, EPI_ISL_627918, EPI_ISL_627920, EPI_ISL_627932, EPI_ISL_627933, EPI_ISL_627934, EPI_ISL_627936, EPI_ISL_627942, EPI_ISL_627955, EPI_ISL_627976, EPI_ISL_627977, EPI_ISL_627978, EPI_ISL_627979, EPI_ISL_627980, EPI_ISL_627983, EPI_ISL_627994, EPI_ISL_627997, EPI_ISL_628001, EPI_ISL_628011, EPI_ISL_628012, EPI_ISL_628020, EPI_ISL_628022, EPI_ISL_628026, EPI_ISL_628030, EPI_ISL_628034, EPI_ISL_628037, EPI_ISL_628044, EPI_ISL_628045, EPI_ISL_628059, EPI_ISL_628064, EPI_ISL_628073, EPI_ISL_628083, EPI_ISL_628102, EPI_ISL_628111, EPI_ISL_628112, EPI_ISL_628119, EPI_ISL_628121, EPI_ISL_628128, EPI_ISL_628133, EPI_ISL_628143, EPI_ISL_628176, EPI_ISL_628223, EPI_ISL_628284, EPI_ISL_637256, EPI_ISL_637258, EPI_ISL_637327, EPI_ISL_637445, EPI_ISL_637448, EPI_ISL_637449, EPI_ISL_637544, EPI_ISL_637546, EPI_ISL_637601, EPI_ISL_637606, EPI_ISL_637616, EPI_ISL_637618, EPI_ISL_637619, EPI_ISL_637882, EPI_ISL_637962, EPI_ISL_638051, EPI_ISL_638096, EPI_ISL_638099, EPI_ISL_638153, EPI_ISL_639012, EPI_ISL_639013, EPI_ISL_639014, EPI_ISL_639023, EPI_ISL_639030, EPI_ISL_639046, EPI_ISL_639050, EPI_ISL_639052, EPI_ISL_639063, EPI_ISL_639064, EPI_ISL_639065, EPI_ISL_639071, EPI_ISL_639072, EPI_ISL_639073, EPI_ISL_639081, EPI_ISL_639083, EPI_ISL_639086, EPI_ISL_639087, EPI_ISL_639094, EPI_ISL_639095, EPI_ISL_639097, EPI_ISL_639100, EPI_ISL_639104, EPI_ISL_639107, EPI_ISL_639130, EPI_ISL_639134, EPI_ISL_639135, EPI_ISL_639136, EPI_ISL_639141, EPI_ISL_639143, EPI_ISL_639147, EPI_ISL_639149, EPI_ISL_639150, EPI_ISL_639151, EPI_ISL_639153, EPI_ISL_639163, EPI_ISL_639165, EPI_ISL_639168, EPI_ISL_639169, EPI_ISL_639170, EPI_ISL_639172, EPI_ISL_639177, EPI_ISL_639184, EPI_ISL_639187, EPI_ISL_639455, EPI_ISL_639522, EPI_ISL_639554, EPI_ISL_666195, EPI_ISL_666205, EPI_ISL_666288, EPI_ISL_666323, EPI_ISL_680580, EPI_ISL_680581, EPI_ISL_680589, EPI_ISL_680611, EPI_ISL_680613, EPI_ISL_680615, EPI_ISL_704918, EPI_ISL_705229, EPI_ISL_705433, EPI_ISL_705463, EPI_ISL_705488, EPI_ISL_706470, EPI_ISL_706603, EPI_ISL_706660, EPI_ISL_706661, EPI_ISL_727180

| | | | |
|---|---|---|---|
| see above | Wales Specialist Virology Centre Sequencing lab: Pathogen Genomics Unit | COVID-19 Genomics UK (COG-UK) Consortium | Alec Birchley; Alexander Adams; Amy Gaskin; Angela Marchbank; Bree Gatica-Wilcox; Catherine Moore; Jason Coombes; Joanne Watkins; Joel Southgate; Johnathan Evans; Laura Gifford; Lauren Gilbert; Lee Graham; Malorie Perry; Matthew Bull; Nicole Pacchiarini; Sally Corden; Sara Kumziene-Summerhayes; Sara Rey; Sarah Taylor; Simon Cottrell; Sophie Jones; Tom Connor |

EPI_ISL_594742, EPI_ISL_594743, EPI_ISL_594744, EPI_ISL_594751, EPI_ISL_594756, EPI_ISL_594757, EPI_ISL_594758, EPI_ISL_594759, EPI_ISL_594781, EPI_ISL_594786, EPI_ISL_594787, EPI_ISL_594788, EPI_ISL_594789, EPI_ISL_594790, EPI_ISL_594791, EPI_ISL_594793, EPI_ISL_594794, EPI_ISL_594795, EPI_ISL_594796, EPI_ISL_594797, EPI_ISL_594798, EPI_ISL_594799, EPI_ISL_594800, EPI_ISL_594801, EPI_ISL_594802, EPI_ISL_594803, EPI_ISL_594804, EPI_ISL_594805, EPI_ISL_594806, EPI_ISL_594807, EPI_ISL_594808, EPI_ISL_594809, EPI_ISL_594810, EPI_ISL_594811, EPI_ISL_594812, EPI_ISL_594813, EPI_ISL_594814, EPI_ISL_594815, EPI_ISL_626687, EPI_ISL_626758, EPI_ISL_626812, EPI_ISL_626818, EPI_ISL_626822, EPI_ISL_626835, EPI_ISL_626837, EPI_ISL_626838, EPI_ISL_626839, EPI_ISL_626840, EPI_ISL_626841, EPI_ISL_626842, EPI_ISL_626844, EPI_ISL_626845, EPI_ISL_626847, EPI_ISL_626848, EPI_ISL_626855, EPI_ISL_626873, EPI_ISL_626897, EPI_ISL_626904, EPI_ISL_626917, EPI_ISL_626927, EPI_ISL_626934, EPI_ISL_626935, EPI_ISL_626958, EPI_ISL_626962, EPI_ISL_626963, EPI_ISL_626967, EPI_ISL_626968, EPI_ISL_626971, EPI_ISL_626972, EPI_ISL_626973, EPI_ISL_626974, EPI_ISL_626975, EPI_ISL_626976, EPI_ISL_626977, EPI_ISL_626978, EPI_ISL_626979, EPI_ISL_626980, EPI_ISL_626981, EPI_ISL_626982, EPI_ISL_626983, EPI_ISL_626984, EPI_ISL_626985, EPI_ISL_626986, EPI_ISL_626987, EPI_ISL_626988, EPI_ISL_626989, EPI_ISL_626990, EPI_ISL_626991, EPI_ISL_626992, EPI_ISL_627045, EPI_ISL_627049, EPI_ISL_627063, EPI_ISL_627173, EPI_ISL_627195, EPI_ISL_627200, EPI_ISL_627201, EPI_ISL_627202, EPI_ISL_627368, EPI_ISL_627369, EPI_ISL_627370, EPI_ISL_627371, EPI_ISL_627372, EPI_ISL_627377, EPI_ISL_627378, EPI_ISL_627380, EPI_ISL_627381, EPI_ISL_627382, EPI_ISL_627383, EPI_ISL_627384, EPI_ISL_627385, EPI_ISL_627386, EPI_ISL_627387, EPI_ISL_627389, EPI_ISL_627390, EPI_ISL_627392, EPI_ISL_627393, EPI_ISL_627394, EPI_ISL_627395, EPI_ISL_627396, EPI_ISL_627397, EPI_ISL_627399, EPI_ISL_627400, EPI_ISL_627401, EPI_ISL_627402, EPI_ISL_627403, EPI_ISL_627404, EPI_ISL_627405, EPI_ISL_627406, EPI_ISL_627407, EPI_ISL_627408, EPI_ISL_627409, EPI_ISL_627410, EPI_ISL_627411, EPI_ISL_627412, EPI_ISL_627413, EPI_ISL_627414, EPI_ISL_627415, EPI_ISL_627416, EPI_ISL_627417, EPI_ISL_627418, EPI_ISL_627419, EPI_ISL_627420, EPI_ISL_627421, EPI_ISL_627422, EPI_ISL_627423, EPI_ISL_650730, EPI_ISL_651814, EPI_ISL_651815, EPI_ISL_651816, EPI_ISL_651817, EPI_ISL_651818

| | | | |
|---|---|---|---|
| see above | West of Scotland Specialist Virology Centre, NHSGGC / MRC-University of Glasgow Centre for Virus Research | COVID-19 Genomics UK (COG-UK) Consortium | Alasdair MacLean; Alice Broos; Ana da Silva Filipe; Antonia Ho; Daniel Mair; David L Robertson; Elihu Aranday-Cortes; Emma Thomson; Guy Mollett; Igor Starinskij; James Shepherd; Jenna Nichols; Joseph Hughes; Kathy Li; Kathy Smollett; Kyriaki Nomikou; Lily Tong; Marc Niebel; Matthew Holden; Natasha Jesudason; Natasha Johnson; Patawee Asamaphan; Rachel Blacow; Rajiv Shah; Richard Orton; Rory Gunson; Sarah McDonald; Sharif Shaaban; Sreenu Vattipally; Stephen Carmichael |

We gratefully acknowledge the following Authors from the Originating laboratories responsible for obtaining the specimens, as well as the Submitting laboratories where the genome data were generated and shared via GISAID, on which this research is based.

All Submitters of data may be contacted directly via www.gisaid.org

Authors are sorted alphabetically.

| Accession ID | Originating Laboratory | Submitting Laboratory | Authors |
|---|---|---|---|
| EPI_ISL_842863, EPI_ISL_842866 | Barts Health NHS Trust | COVID-19 Genomics UK (COG-UK) Consortium | BROAD; Beatrix; CUTINO-MOGUEL; Claire; David; Dola; HARRINGTON; KELE; Maria-Teresa; OWOYEMI; Raghavendran; SHYLINI |
| EPI_ISL_868938, EPI_ISL_868939, EPI_ISL_868940, EPI_ISL_868941, EPI_ISL_868942, EPI_ISL_868943, EPI_ISL_868944, EPI_ISL_868945, EPI_ISL_868946, EPI_ISL_868947, EPI_ISL_868948, EPI_ISL_924605, EPI_ISL_924606, EPI_ISL_924607, EPI_ISL_924608, EPI_ISL_924609, EPI_ISL_924610, EPI_ISL_924611, EPI_ISL_924612, EPI_ISL_924613, EPI_ISL_924614, EPI_ISL_924615, EPI_ISL_924616, EPI_ISL_924617, EPI_ISL_924618, EPI_ISL_924619, EPI_ISL_924620, EPI_ISL_924621, EPI_ISL_925014, EPI_ISL_925015, EPI_ISL_925016, EPI_ISL_925017, EPI_ISL_925018, EPI_ISL_925019, EPI_ISL_925020, EPI_ISL_925021, EPI_ISL_925022, EPI_ISL_925023, EPI_ISL_925024, EPI_ISL_925025, EPI_ISL_925026, EPI_ISL_925027, EPI_ISL_925028, EPI_ISL_925029, EPI_ISL_925030, EPI_ISL_1476754, EPI_ISL_1476850, EPI_ISL_2128590 | | | |
| see above | Bioinformatics and Biostatistics Lab, Advanced Sequencing Facility | COVID-19 Genomics UK (COG-UK) Consortium | Aengus Stewart; Chelsea Sawyer; Harshil Patel; Jerome Nicod; Laura Cubitt; Margaret Crawford |
| EPI_ISL_626677, EPI_ISL_626678, EPI_ISL_626689, EPI_ISL_626738, EPI_ISL_626778, EPI_ISL_626792, EPI_ISL_626878, EPI_ISL_626879, EPI_ISL_626880, EPI_ISL_626895, EPI_ISL_626936, EPI_ISL_627056, EPI_ISL_627057, EPI_ISL_627116, EPI_ISL_627128, EPI_ISL_627144, EPI_ISL_628320, EPI_ISL_628321, EPI_ISL_628322, EPI_ISL_628325, EPI_ISL_628326, EPI_ISL_628327, EPI_ISL_628328, EPI_ISL_628329, EPI_ISL_628330, EPI_ISL_628331, EPI_ISL_628332, EPI_ISL_628333, EPI_ISL_650248, EPI_ISL_650249, EPI_ISL_650578, EPI_ISL_650600, EPI_ISL_650601, EPI_ISL_650602, EPI_ISL_650603, EPI_ISL_650982, EPI_ISL_651068, EPI_ISL_651277, EPI_ISL_652624, EPI_ISL_652625, EPI_ISL_652626, EPI_ISL_665487, EPI_ISL_665488, EPI_ISL_680016, EPI_ISL_680017, EPI_ISL_680018, EPI_ISL_680019, EPI_ISL_680020, EPI_ISL_680021, EPI_ISL_680022, EPI_ISL_680023, EPI_ISL_680024, EPI_ISL_680025, EPI_ISL_680026, EPI_ISL_680027, EPI_ISL_680028, EPI_ISL_680029, EPI_ISL_680030, EPI_ISL_680031, EPI_ISL_680032, EPI_ISL_680033, EPI_ISL_680034, EPI_ISL_680035, EPI_ISL_680036, EPI_ISL_680562, EPI_ISL_680563, EPI_ISL_680564, EPI_ISL_680565, EPI_ISL_680566, EPI_ISL_680567, EPI_ISL_680568, EPI_ISL_680569, EPI_ISL_680570, EPI_ISL_727703, EPI_ISL_727704, EPI_ISL_727710, EPI_ISL_727711, EPI_ISL_727774, EPI_ISL_727775, EPI_ISL_727776, EPI_ISL_727777, EPI_ISL_727778, EPI_ISL_727779, EPI_ISL_727780, EPI_ISL_727781, EPI_ISL_727782, EPI_ISL_727783, EPI_ISL_727784, EPI_ISL_727785, EPI_ISL_727786, EPI_ISL_727787, EPI_ISL_727788, EPI_ISL_727789, EPI_ISL_727790, EPI_ISL_727791, EPI_ISL_727792, EPI_ISL_727793, EPI_ISL_727794, EPI_ISL_727795, EPI_ISL_727796, EPI_ISL_727797, EPI_ISL_727798, EPI_ISL_727799, EPI_ISL_727800, EPI_ISL_727801, EPI_ISL_727802, EPI_ISL_727803, EPI_ISL_727804, EPI_ISL_727805, EPI_ISL_727806, EPI_ISL_741357, EPI_ISL_741358, EPI_ISL_741772, EPI_ISL_867930, EPI_ISL_867931, EPI_ISL_867932, EPI_ISL_867934, EPI_ISL_867941, EPI_ISL_867942, EPI_ISL_923273, EPI_ISL_923277, EPI_ISL_923278, EPI_ISL_923289, EPI_ISL_923297, EPI_ISL_952436, EPI_ISL_1108835, EPI_ISL_1730865 | | | |
| see above | Centre for Enzyme Innovation, University of Portsmouth / Translational Research Laboratory, Portsmouth Hospitals NHS Trust | COVID-19 Genomics UK (COG-UK) Consortium | Allyson Lloyd; Angela Beckett; Anoop Chauhan; Christopher Fearn; Ethan Butcher; Garry Scarlett; Kate Cook; Katie Loveson; Kelly Bicknell; Robert Impey; Salman Goudarzi; Samuel Robson; Sarah Wyllie; Scott Elliott; Sharon Glaysher; Yann Bourgeois |
| EPI_ISL_637294, EPI_ISL_637399, EPI_ISL_637410, EPI_ISL_637418, EPI_ISL_637420, EPI_ISL_637433, EPI_ISL_637464, EPI_ISL_637468, EPI_ISL_637469, EPI_ISL_637470, EPI_ISL_637471, EPI_ISL_637505, EPI_ISL_637506, EPI_ISL_637509, EPI_ISL_637511, EPI_ISL_637529, EPI_ISL_637530, EPI_ISL_637566, EPI_ISL_637567, EPI_ISL_637624, EPI_ISL_637636, EPI_ISL_637637, EPI_ISL_637640, EPI_ISL_637694, EPI_ISL_637696, EPI_ISL_637698, EPI_ISL_637699, EPI_ISL_637727, EPI_ISL_637738, EPI_ISL_638027, EPI_ISL_638040, EPI_ISL_638066, EPI_ISL_638165, EPI_ISL_638166, EPI_ISL_638167, EPI_ISL_638187, EPI_ISL_638188, EPI_ISL_638189, EPI_ISL_638193, EPI_ISL_638194, EPI_ISL_638196, EPI_ISL_638324, EPI_ISL_638325, EPI_ISL_638327, EPI_ISL_638328, EPI_ISL_638329, EPI_ISL_638330, EPI_ISL_638332, EPI_ISL_638333, EPI_ISL_638334, EPI_ISL_638335, EPI_ISL_638337, EPI_ISL_638338, EPI_ISL_638339, EPI_ISL_638340, EPI_ISL_638341, EPI_ISL_638342, EPI_ISL_638343, EPI_ISL_638344, EPI_ISL_638345, EPI_ISL_638346, EPI_ISL_638347, EPI_ISL_638348, EPI_ISL_638349, EPI_ISL_638350, EPI_ISL_638351, EPI_ISL_638352, EPI_ISL_638353, EPI_ISL_638354, EPI_ISL_638355, EPI_ISL_638356, EPI_ISL_638357, EPI_ISL_638358, EPI_ISL_638359, EPI_ISL_638360, EPI_ISL_638361, EPI_ISL_638362, EPI_ISL_638364, EPI_ISL_638365, EPI_ISL_638366, EPI_ISL_638367, EPI_ISL_638368, EPI_ISL_638369, EPI_ISL_638370, EPI_ISL_638371, EPI_ISL_638385, EPI_ISL_650122, EPI_ISL_650127, EPI_ISL_650129, EPI_ISL_650130, EPI_ISL_650133, EPI_ISL_650149, EPI_ISL_650200, EPI_ISL_650217, EPI_ISL_650228, EPI_ISL_650236, EPI_ISL_650239, EPI_ISL_650243, EPI_ISL_650262, EPI_ISL_650265, EPI_ISL_650356, EPI_ISL_650376, EPI_ISL_650383, EPI_ISL_650391, EPI_ISL_650392, EPI_ISL_650398, EPI_ISL_650400, EPI_ISL_650553, EPI_ISL_650564, EPI_ISL_650573, EPI_ISL_650574, EPI_ISL_650589, EPI_ISL_650590, EPI_ISL_650591, EPI_ISL_650592, EPI_ISL_650593, EPI_ISL_650594, EPI_ISL_650595, EPI_ISL_650609, EPI_ISL_650626, EPI_ISL_650627, EPI_ISL_650628, EPI_ISL_650629, EPI_ISL_650630, EPI_ISL_650631, EPI_ISL_650632, EPI_ISL_650661, EPI_ISL_650666, EPI_ISL_650697, EPI_ISL_650698, EPI_ISL_650699, EPI_ISL_650700, EPI_ISL_650910, EPI_ISL_650922, EPI_ISL_650951, EPI_ISL_650952, EPI_ISL_650959, EPI_ISL_650969, EPI_ISL_650970, EPI_ISL_650971, EPI_ISL_650972, EPI_ISL_650973, EPI_ISL_650983, EPI_ISL_650985, EPI_ISL_650986, EPI_ISL_650994, EPI_ISL_651014, EPI_ISL_651015, EPI_ISL_651016, EPI_ISL_651017, EPI_ISL_651018, EPI_ISL_651019, EPI_ISL_651020, EPI_ISL_651021, EPI_ISL_651022, EPI_ISL_651023, EPI_ISL_651024, EPI_ISL_651025, EPI_ISL_651026, EPI_ISL_651027, EPI_ISL_651028, EPI_ISL_651029, EPI_ISL_651030, EPI_ISL_651031, EPI_ISL_651032, EPI_ISL_651033, EPI_ISL_651034, EPI_ISL_651035, EPI_ISL_651036, EPI_ISL_651037, EPI_ISL_651038, EPI_ISL_651039, EPI_ISL_651040, EPI_ISL_651041, EPI_ISL_651042, EPI_ISL_651043, EPI_ISL_651044, EPI_ISL_651045, EPI_ISL_651046, EPI_ISL_651047, EPI_ISL_651080, EPI_ISL_651081, EPI_ISL_651172, EPI_ISL_651176, EPI_ISL_651186, EPI_ISL_651202, EPI_ISL_651235, EPI_ISL_651278, EPI_ISL_651280, EPI_ISL_651281, EPI_ISL_651283, EPI_ISL_651284, EPI_ISL_651302, EPI_ISL_651331, EPI_ISL_651339, EPI_ISL_651346, EPI_ISL_651348, EPI_ISL_651360, EPI_ISL_651361, EPI_ISL_651362, EPI_ISL_651365, EPI_ISL_651370, EPI_ISL_651435, EPI_ISL_651436, EPI_ISL_651488, EPI_ISL_651653, EPI_ISL_651654, EPI_ISL_651655, EPI_ISL_651656, EPI_ISL_651657, EPI_ISL_651660, EPI_ISL_651662, EPI_ISL_651675, EPI_ISL_652157, EPI_ISL_652158, EPI_ISL_652159, EPI_ISL_652160, EPI_ISL_652161, EPI_ISL_652162, EPI_ISL_652163, EPI_ISL_652164, EPI_ISL_652165, EPI_ISL_652166, EPI_ISL_652167, EPI_ISL_652168, EPI_ISL_652169, EPI_ISL_652170, EPI_ISL_652171, EPI_ISL_652172, EPI_ISL_652173, EPI_ISL_652174, EPI_ISL_652175, EPI_ISL_652176, EPI_ISL_652177, EPI_ISL_652178, EPI_ISL_652179, EPI_ISL_704470, EPI_ISL_704473, EPI_ISL_705538, EPI_ISL_705540, EPI_ISL_705544, EPI_ISL_705546, EPI_ISL_705550 | | | |
| see above | Department of Pathology, University of Cambridge | COVID-19 Genomics UK (COG-UK) Consortium | Aminu S. Jahun; Grant Hall; Ian Goodfellow; Iliana Georgana; Malte Pinckert; Martin D. Curran; Myra Hosmillo; Surendra Parmar; Yasmin Chaudhry |
| EPI_ISL_611093, EPI_ISL_623864, EPI_ISL_623904, EPI_ISL_623974, EPI_ISL_623983, EPI_ISL_624094, EPI_ISL_624244, EPI_ISL_624291, EPI_ISL_624350, EPI_ISL_624354, EPI_ISL_624909, EPI_ISL_625213, EPI_ISL_625240, EPI_ISL_630780, EPI_ISL_630891, EPI_ISL_644095, EPI_ISL_644096, EPI_ISL_644097, EPI_ISL_644098, EPI_ISL_644100, EPI_ISL_644103, EPI_ISL_644104, EPI_ISL_644105, EPI_ISL_644106, EPI_ISL_644108, EPI_ISL_644109, EPI_ISL_644110, EPI_ISL_644111, EPI_ISL_644113, EPI_ISL_645297, EPI_ISL_645418, EPI_ISL_645452, EPI_ISL_645459, EPI_ISL_645464, EPI_ISL_645502, EPI_ISL_645503, EPI_ISL_649220, EPI_ISL_649257, EPI_ISL_649305, EPI_ISL_649369, EPI_ISL_649370, EPI_ISL_826486, EPI_ISL_826487, EPI_ISL_826488, EPI_ISL_826489, EPI_ISL_826490, EPI_ISL_826491, EPI_ISL_826492, EPI_ISL_826493, EPI_ISL_826494, EPI_ISL_826495, EPI_ISL_826496, EPI_ISL_826497, EPI_ISL_826498, EPI_ISL_826499, EPI_ISL_826500, EPI_ISL_826501, EPI_ISL_826502, EPI_ISL_826503, EPI_ISL_826504, EPI_ISL_826505, EPI_ISL_826506, EPI_ISL_826507, EPI_ISL_826508, EPI_ISL_826509, EPI_ISL_826510, EPI_ISL_826511, EPI_ISL_826512, EPI_ISL_826513, EPI_ISL_826514, EPI_ISL_892221, EPI_ISL_892222, EPI_ISL_892223, EPI_ISL_2004453 | | | |
| see above | Lighthouse Lab in Alderley Park | Wellcome Sanger Institute for the COVID-19 Genomics UK (COG-UK) Consortium | Cordelia Langford; David K. Jackson; Dominic Kwiatkowski; Ewan Harrison; Ian Johnston; Jacquelyn Wynn; Jeffrey Barrett; John Sillitoe on behalf of the Wellcome Sanger Institute COVID-19 Surveillance Team; John Sillitoe on behalf of the Wellcome Sanger Institute COVID-19 Surveillance Team (http://www.sanger.ac.uk/covid-team); Mairead Hyland; Roberto Amato; Sonia Goncalves; The Lighthouse Lab in Alderley Park and Alex Alderton |
| EPI_ISL_610922, EPI_ISL_610926, EPI_ISL_610927, EPI_ISL_610928, EPI_ISL_610932, EPI_ISL_610933, EPI_ISL_610938, EPI_ISL_610939, EPI_ISL_610944, EPI_ISL_610946, EPI_ISL_610947, EPI_ISL_610948, EPI_ISL_610954, EPI_ISL_610960, EPI_ISL_610963, EPI_ISL_610967, EPI_ISL_610980, EPI_ISL_610982, EPI_ISL_610984, EPI_ISL_610989, EPI_ISL_610990, EPI_ISL_611000, EPI_ISL_611004, EPI_ISL_611005, EPI_ISL_611014, EPI_ISL_611015, EPI_ISL_611016, EPI_ISL_611023, EPI_ISL_611029, EPI_ISL_611034, EPI_ISL_611041, EPI_ISL_611045, EPI_ISL_611046, EPI_ISL_611054, EPI_ISL_611055, EPI_ISL_611056, EPI_ISL_611057, EPI_ISL_611059, EPI_ISL_611090, EPI_ISL_611102, EPI_ISL_611104, EPI_ISL_611106, EPI_ISL_611112, EPI_ISL_611113, EPI_ISL_611121, EPI_ISL_611131, EPI_ISL_611132, EPI_ISL_611134, EPI_ISL_611135, EPI_ISL_611140, EPI_ISL_611143, EPI_ISL_611163, EPI_ISL_611183, EPI_ISL_611188, EPI_ISL_611194, EPI_ISL_611197, EPI_ISL_611201, EPI_ISL_611203, EPI_ISL_611207, EPI_ISL_611209, EPI_ISL_611212, EPI_ISL_611213, EPI_ISL_611217, EPI_ISL_611230, EPI_ISL_611232, EPI_ISL_611235, EPI_ISL_611243, EPI_ISL_611255, EPI_ISL_611267, EPI_ISL_611273, EPI_ISL_611277, EPI_ISL_611286, EPI_ISL_623858, EPI_ISL_623859, EPI_ISL_623860, EPI_ISL_623861, EPI_ISL_623862, EPI_ISL_623866, EPI_ISL_623867, EPI_ISL_623869, EPI_ISL_623870, EPI_ISL_623871, EPI_ISL_623875, EPI_ISL_623877, EPI_ISL_623878, EPI_ISL_623879, EPI_ISL_623880, EPI_ISL_623881, EPI_ISL_623882, EPI_ISL_623885, EPI_ISL_623887, EPI_ISL_623888, EPI_ISL_623889, EPI_ISL_623890, EPI_ISL_623891, EPI_ISL_623892, EPI_ISL_623894, EPI_ISL_623896, EPI_ISL_623897, EPI_ISL_623898, EPI_ISL_623899, EPI_ISL_623900, EPI_ISL_623901, EPI_ISL_623902, EPI_ISL_623905, EPI_ISL_623906, EPI_ISL_623909, EPI_ISL_623910, EPI_ISL_623912, EPI_ISL_623913, EPI_ISL_623914, EPI_ISL_623915, EPI_ISL_623918, EPI_ISL_623919, EPI_ISL_623920, EPI_ISL_623922, EPI_ISL_623923, EPI_ISL_623925, EPI_ISL_623926, EPI_ISL_623927, EPI_ISL_623928, EPI_ISL_623929, EPI_ISL_623931, EPI_ISL_623932, EPI_ISL_623935, EPI_ISL_623936, EPI_ISL_623937, EPI_ISL_623938, EPI_ISL_623939, EPI_ISL_623940, EPI_ISL_623941, EPI_ISL_623943, EPI_ISL_623945, EPI_ISL_623946, EPI_ISL_623947, EPI_ISL_623948, EPI_ISL_623950, EPI_ISL_623951, EPI_ISL_623952, EPI_ISL_623953, EPI_ISL_623955, EPI_ISL_623957, EPI_ISL_623958, EPI_ISL_623960, EPI_ISL_623961, EPI_ISL_623962, EPI_ISL_623964, EPI_ISL_623965, EPI_ISL_623966, EPI_ISL_623967, EPI_ISL_623968, EPI_ISL_623969, EPI_ISL_623970, EPI_ISL_623971, EPI_ISL_623972, EPI_ISL_623973, EPI_ISL_623978, EPI_ISL_623979, EPI_ISL_623980, EPI_ISL_623981, EPI_ISL_623982, EPI_ISL_623984, EPI_ISL_623985, EPI_ISL_623987, EPI_ISL_623988, EPI_ISL_623989, EPI_ISL_623992, EPI_ISL_623993, EPI_ISL_623996, EPI_ISL_623997, EPI_ISL_623998, EPI_ISL_623999, EPI_ISL_624000, EPI_ISL_624001, EPI_ISL_624002, EPI_ISL_624003, EPI_ISL_624004, EPI_ISL_624005, EPI_ISL_624006, EPI_ISL_624007, EPI_ISL_624008, EPI_ISL_624009, EPI_ISL_624011, EPI_ISL_624013, EPI_ISL_624014, EPI_ISL_624015, EPI_ISL_624016, EPI_ISL_624017, EPI_ISL_624020, EPI_ISL_624021, EPI_ISL_624023, EPI_ISL_624024, EPI_ISL_624025, EPI_ISL_624026, EPI_ISL_624030, EPI_ISL_624031, EPI_ISL_624033, EPI_ISL_624035, EPI_ISL_624036, EPI_ISL_624037, EPI_ISL_624038, EPI_ISL_624039, EPI_ISL_624040, EPI_ISL_624041, EPI_ISL_624042, EPI_ISL_624045, EPI_ISL_624046, EPI_ISL_624047, EPI_ISL_624048, EPI_ISL_624050, EPI_ISL_624051, EPI_ISL_624052, EPI_ISL_624053, EPI_ISL_624054, EPI_ISL_624055, EPI_ISL_624056, EPI_ISL_624057, EPI_ISL_624058, EPI_ISL_624059, EPI_ISL_624060, EPI_ISL_624061, EPI_ISL_624062, EPI_ISL_624063, EPI_ISL_624064, EPI_ISL_624065, EPI_ISL_624066, EPI_ISL_624068, EPI_ISL_624069, EPI_ISL_624070, EPI_ISL_624071, EPI_ISL_624072, EPI_ISL_624074, EPI_ISL_624075, EPI_ISL_624076, EPI_ISL_624078, EPI_ISL_624080, EPI_ISL_624081, EPI_ISL_624082, EPI_ISL_624083, EPI_ISL_624084, EPI_ISL_624085, EPI_ISL_624086, EPI_ISL_624087, EPI_ISL_624088, EPI_ISL_624089, EPI_ISL_624090, EPI_ISL_624091, EPI_ISL_624093, EPI_ISL_624095, EPI_ISL_624096, EPI_ISL_624097, EPI_ISL_624098, EPI_ISL_624099, EPI_ISL_624100, EPI_ISL_624101, EPI_ISL_624102, EPI_ISL_624103, EPI_ISL_624104, EPI_ISL_624106, EPI_ISL_624107, EPI_ISL_624108, EPI_ISL_624109, EPI_ISL_624110, EPI_ISL_624111, EPI_ISL_624113, EPI_ISL_624115, EPI_ISL_624116, EPI_ISL_624117, EPI_ISL_624118, EPI_ISL_624119, EPI_ISL_624121, EPI_ISL_624123, EPI_ISL_624124, EPI_ISL_624125, EPI_ISL_624126, EPI_ISL_624127, EPI_ISL_624129, EPI_ISL_624130, EPI_ISL_624131, EPI_ISL_624133, EPI_ISL_624134, EPI_ISL_624135, EPI_ISL_624136, EPI_ISL_624137, EPI_ISL_624138, EPI_ISL_624139, EPI_ISL_624141, EPI_ISL_624142, EPI_ISL_624143, EPI_ISL_624144, EPI_ISL_624146, EPI_ISL_624149, EPI_ISL_624150, EPI_ISL_624151, EPI_ISL_624152, EPI_ISL_624153, EPI_ISL_624154, EPI_ISL_624155, EPI_ISL_624156, EPI_ISL_624157, EPI_ISL_624159, EPI_ISL_624160, EPI_ISL_624161, EPI_ISL_624162, EPI_ISL_624166, EPI_ISL_624167, EPI_ISL_624168, EPI_ISL_624169, EPI_ISL_624170, EPI_ISL_624171, EPI_ISL_624172, EPI_ISL_624173, EPI_ISL_624174, EPI_ISL_624175, EPI_ISL_624177, EPI_ISL_624178, EPI_ISL_624179, EPI_ISL_624180, EPI_ISL_624182, EPI_ISL_624183, EPI_ISL_624184, EPI_ISL_624185, EPI_ISL_624186, EPI_ISL_624187, EPI_ISL_624190, EPI_ISL_624191, EPI_ISL_624192, EPI_ISL_624193, EPI_ISL_624194, EPI_ISL_624195, EPI_ISL_624196, EPI_ISL_624198, EPI_ISL_624199, EPI_ISL_624200, EPI_ISL_624201, EPI_ISL_624202, EPI_ISL_624203, EPI_ISL_624204, EPI_ISL_624205, EPI_ISL_624206, EPI_ISL_624207, EPI_ISL_624208, EPI_ISL_624209, EPI_ISL_624210, EPI_ISL_624211, EPI_ISL_624212, EPI_ISL_624213, EPI_ISL_624214, EPI_ISL_624215, EPI_ISL_624216, EPI_ISL_624217, EPI_ISL_624218, EPI_ISL_624219, EPI_ISL_624220, EPI_ISL_624221, EPI_ISL_624222, EPI_ISL_624223, EPI_ISL_624224, EPI_ISL_624225, EPI_ISL_624226, EPI_ISL_624227, EPI_ISL_624228, EPI_ISL_624229, EPI_ISL_624230, EPI_ISL_624231, EPI_ISL_624232, EPI_ISL_624233, EPI_ISL_624234, EPI_ISL_624235, EPI_ISL_624236, EPI_ISL_624237, EPI_ISL_624238, EPI_ISL_624239, EPI_ISL_624240, EPI_ISL_624241, EPI_ISL_624242, EPI_ISL_624243, EPI_ISL_624245, EPI_ISL_624246, EPI_ISL_624247, EPI_ISL_624248, EPI_ISL_624249, EPI_ISL_624250, EPI_ISL_624251, EPI_ISL_624252, EPI_ISL_624253, EPI_ISL_624254, EPI_ISL_624255, EPI_ISL_624256, EPI_ISL_624257, EPI_ISL_624258, EPI_ISL_624259, EPI_ISL_624260, EPI_ISL_624261, EPI_ISL_624262, EPI_ISL_624263, EPI_ISL_624264, EPI_ISL_624265, EPI_ISL_624266, EPI_ISL_624267, EPI_ISL_624268, EPI_ISL_624269, EPI_ISL_624270, EPI_ISL_624271, EPI_ISL_624272, EPI_ISL_624273, EPI_ISL_624274, EPI_ISL_624275, EPI_ISL_624276, EPI_ISL_624277, EPI_ISL_624278, EPI_ISL_624279, EPI_ISL_624280, EPI_ISL_624281, EPI_ISL_624282, EPI_ISL_624283, EPI_ISL_624284, EPI_ISL_624285, EPI_ISL_624286, EPI_ISL_624287, EPI_ISL_624288, EPI_ISL_624289, EPI_ISL_624290, EPI_ISL_624292, EPI_ISL_624293, EPI_ISL_624294, EPI_ISL_624295, EPI_ISL_624296, EPI_ISL_624297, EPI_ISL_624298, EPI_ISL_624299, EPI_ISL_624300, EPI_ISL_624301, EPI_ISL_624302, EPI_ISL_624303, EPI_ISL_624304, EPI_ISL_624305, EPI_ISL_624306, EPI_ISL_624307, EPI_ISL_624308, EPI_ISL_624309, EPI_ISL_624310, EPI_ISL_624311, EPI_ISL_624312, EPI_ISL_624313, EPI_ISL_624314, EPI_ISL_624315, EPI_ISL_624316, EPI_ISL_624317, EPI_ISL_624318, EPI_ISL_624319, EPI_ISL_624320, EPI_ISL_624321, EPI_ISL_624322, EPI_ISL_624323, EPI_ISL_624324, EPI_ISL_624325, EPI_ISL_624326, EPI_ISL_624327, EPI_ISL_624328, EPI_ISL_624329, EPI_ISL_624330, EPI_ISL_624331, EPI_ISL_624332, EPI_ISL_624333, EPI_ISL_624334, EPI_ISL_624335, EPI_ISL_624336, EPI_ISL_624337, EPI_ISL_624338, EPI_ISL_624339, EPI_ISL_624340, EPI_ISL_624341, EPI_ISL_624342, EPI_ISL_624343, EPI_ISL_624344, EPI_ISL_624345, EPI_ISL_624346, EPI_ISL_624347, EPI_ISL_624348, EPI_ISL_624349, EPI_ISL_624351, EPI_ISL_624352, EPI_ISL_624353, EPI_ISL_624355, EPI_ISL_624356, EPI_ISL_624357, EPI_ISL_624358, EPI_ISL_624359, EPI_ISL_624360, EPI_ISL_624361, EPI_ISL_624362, EPI_ISL_624363, EPI_ISL_624364, EPI_ISL_624365, EPI_ISL_624366, EPI_ISL_624367, EPI_ISL_624368, EPI_ISL_624369, EPI_ISL_624370, EPI_ISL_624371, EPI_ISL_624372, EPI_ISL_624373, EPI_ISL_624374, EPI_ISL_624375, EPI_ISL_624376, EPI_ISL_624377, EPI_ISL_624378, EPI_ISL_624379, EPI_ISL_624380, EPI_ISL_624381, EPI_ISL_624382, EPI_ISL_624383, EPI_ISL_624384, EPI_ISL_624385, EPI_ISL_624386, EPI_ISL_624387, EPI_ISL_624388, EPI_ISL_624389, EPI_ISL_624390, EPI_ISL_624391, EPI_ISL_624392, EPI_ISL_624393, EPI_ISL_624394, EPI_ISL_624395, EPI_ISL_624396, EPI_ISL_624397, EPI_ISL_624398, EPI_ISL_624399, EPI_ISL_624400, EPI_ISL_624401, EPI_ISL_624402, EPI_ISL_624403, EPI_ISL_624404, EPI_ISL_624405, EPI_ISL_624406, EPI_ISL_624407, EPI_ISL_624408, EPI_ISL_624409, EPI_ISL_624410, EPI_ISL_624411, EPI_ISL_624412, EPI_ISL_624413, EPI_ISL_624414, EPI_ISL_624415, EPI_ISL_624416, EPI_ISL_624417, EPI_ISL_624418, EPI_ISL_624419, EPI_ISL_624420, EPI_ISL_624421, EPI_ISL_624422, EPI_ISL_624423, EPI_ISL_624424, EPI_ISL_624425, EPI_ISL_624426, EPI_ISL_624427, EPI_ISL_624428, EPI_ISL_624429, EPI_ISL_624430, EPI_ISL_624431, EPI_ISL_624432, EPI_ISL_624433, EPI_ISL_624434, EPI_ISL_624435, EPI_ISL_624436, EPI_ISL_624437, EPI_ISL_624438, EPI_ISL_624439, EPI_ISL_624440, EPI_ISL_624441, EPI_ISL_624442, EPI_ISL_624443, EPI_ISL_624444, EPI_ISL_624445, EPI_ISL_624446, EPI_ISL_624447, EPI_ISL_624448, EPI_ISL_624449, EPI_ISL_624450, EPI_ISL_624451, EPI_ISL_624452, EPI_ISL_624453, EPI_ISL_624454, EPI_ISL_624455, EPI_ISL_624456, EPI_ISL_624457, EPI_ISL_624458, EPI_ISL_624459, EPI_ISL_624460, EPI_ISL_624461, EPI_ISL_624462, EPI_ISL_624463, EPI_ISL_624464, EPI_ISL_624465, EPI_ISL_624466, EPI_ISL_624467, EPI_ISL_624468, EPI_ISL_624469, EPI_ISL_624470, EPI_ISL_624471, EPI_ISL_624472, EPI_ISL_624473, EPI_ISL_624474, EPI_ISL_624475, EPI_ISL_624476, EPI_ISL_624477, EPI_ISL_624478, EPI_ISL_624479, EPI_ISL_624480, EPI_ISL_624481, EPI_ISL_624482, EPI_ISL_624483, EPI_ISL_624484, EPI_ISL_624485, EPI_ISL_624486, EPI_ISL_624487, EPI_ISL_624488, EPI_ISL_624489, EPI_ISL_624490, EPI_ISL_624491, EPI_ISL_624492, EPI_ISL_624493, EPI_ISL_624494, EPI_ISL_624495, EPI_ISL_624496, EPI_ISL_624497, EPI_ISL_624498, EPI_ISL_624499, EPI_ISL_624500, EPI_ISL_624501, EPI_ISL_624502, EPI_ISL_624503, EPI_ISL_624504, EPI_ISL_624505, EPI_ISL_624506, EPI_ISL_624507, EPI_ISL_624508, EPI_ISL_624509, EPI_ISL_624510, EPI_ISL_624511, EPI_ISL_624512, EPI_ISL_624513, EPI_ISL_624514, EPI_ISL_624515, EPI_ISL_624516, EPI_ISL_624517, EPI_ISL_624518, EPI_ISL_624519, EPI_ISL_624520, EPI_ISL_624521, EPI_ISL_624522, EPI_ISL_624523, EPI_ISL_624524, EPI_ISL_624525, EPI_ISL_624526, EPI_ISL_624527, EPI_ISL_624528, EPI_ISL_624529, EPI_ISL_624530, EPI_ISL_624531, EPI_ISL_624532, EPI_ISL_624533, EPI_ISL_624534, EPI_ISL_624535, EPI_ISL_624536, EPI_ISL_624537, EPI_ISL_624538, EPI_ISL_624539, EPI_ISL_624540, EPI_ISL_624541, EPI_ISL_624542, EPI_ISL_624543, EPI_ISL_624544, EPI_ISL_624545, EPI_ISL_624546, EPI_ISL_624547, EPI_ISL_624548, EPI_ISL_624549, EPI_ISL_624550, EPI_ISL_624551, EPI_ISL_624552, EPI_ISL_624553, EPI_ISL_624554, EPI_ISL_624555, EPI_ISL_624556, EPI_ISL_624557, EPI_ISL_624558, EPI_ISL_624559, EPI_ISL_624561, EPI_ISL_624564, EPI_ISL_624566, EPI_ISL_624569, EPI_ISL_624570, EPI_ISL_624571, EPI_ISL_624572, EPI_ISL_624573, EPI_ISL_624574, EPI_ISL_624575, EPI_ISL_624578, EPI_ISL_624579, EPI_ISL_624580, EPI_ISL_624582, EPI_ISL_624583, EPI_ISL_624585, EPI_ISL_624586, EPI_ISL_624588, EPI_ISL_624592, EPI_ISL_624594, EPI_ISL_624596, EPI_ISL_624597, EPI_ISL_624600, EPI_ISL_624601, EPI_ISL_624602, EPI_ISL_624607, EPI_ISL_624609, EPI_ISL_624612, EPI_ISL_624614, EPI_ISL_624615, EPI_ISL_624616, EPI_ISL_624618, EPI_ISL_624619, EPI_ISL_624620, EPI_ISL_624621, EPI_ISL_624622, EPI_ISL_624623, EPI_ISL_624627, EPI_ISL_624628, EPI_ISL_624631, EPI_ISL_624632, EPI_ISL_624633, EPI_ISL_624635, EPI_ISL_624636, EPI_ISL_624638, EPI_ISL_624639, EPI_ISL_624640, EPI_ISL_624642, EPI_ISL_624644, EPI_ISL_624646, EPI_ISL_624648, EPI_ISL_624653, EPI_ISL_624655, EPI_ISL_624660, EPI_ISL_624661, EPI_ISL_624662, EPI_ISL_624664, EPI_ISL_624665, EPI_ISL_624666, EPI_ISL_624668, EPI_ISL_624669, EPI_ISL_624670, EPI_ISL_624671, EPI_ISL_624673, EPI_ISL_624676, EPI_ISL_624679, EPI_ISL_624684, EPI_ISL_624685, EPI_ISL_624686, EPI_ISL_624688, EPI_ISL_624689, EPI_ISL_624690, EPI_ISL_624691, EPI_ISL_624692, EPI_ISL_624695, EPI_ISL_624698, EPI_ISL_624699, EPI_ISL_624700, EPI_ISL_624701, EPI_ISL_624702, EPI_ISL_624705, EPI_ISL_624706, EPI_ISL_624712, EPI_ISL_624714, EPI_ISL_624719, EPI_ISL_624720, EPI_ISL_624724, EPI_ISL_624729, EPI_ISL_624730, EPI_ISL_624733, EPI_ISL_624734, EPI_ISL_624735, EPI_ISL_624736, EPI_ISL_624737, EPI_ISL_624738, EPI_ISL_624739, EPI_ISL_624740, EPI_ISL_624741, EPI_ISL_624743, EPI_ISL_624744, EPI_ISL_624748, EPI_ISL_624750, EPI_ISL_624751, EPI_ISL_624752, EPI_ISL_624755, EPI_ISL_624757, EPI_ISL_624758, EPI_ISL_624760, EPI_ISL_624761, EPI_ISL_624763, EPI_ISL_624766, EPI_ISL_624767, EPI_ISL_624768, EPI_ISL_624771, EPI_ISL_624772, EPI_ISL_624775, EPI_ISL_624776, EPI_ISL_624777, EPI_ISL_624782, EPI_ISL_624784, EPI_ISL_624787, EPI_ISL_624788, EPI_ISL_624794, EPI_ISL_624795, EPI_ISL_624797, EPI_ISL_624798, EPI_ISL_624799, EPI_ISL_624801, EPI_ISL_624802, EPI_ISL_624803, EPI_ISL_624804, EPI_ISL_624806, EPI_ISL_624807, EPI_ISL_624808, EPI_ISL_624809, EPI_ISL_624811, EPI_ISL_624812, EPI_ISL_624813, EPI_ISL_624815, EPI_ISL_624817, EPI_ISL_624818, EPI_ISL_624819, EPI_ISL_624822, EPI_ISL_624823, EPI_ISL_624825, EPI_ISL_624827, EPI_ISL_624828, EPI_ISL_624829, EPI_ISL_624830, EPI_ISL_624832, EPI_ISL_624833, EPI_ISL_624834, EPI_ISL_624835, EPI_ISL_624837, EPI_ISL_624839, EPI_ISL_624842, EPI_ISL_624845, EPI_ISL_624848, EPI_ISL_624853, EPI_ISL_624854, EPI_ISL_624855, EPI_ISL_624857, EPI_ISL_624860, EPI_ISL_624861, EPI_ISL_624862, EPI_ISL_624864, EPI_ISL_624865, EPI_ISL_624866, EPI_ISL_624867, EPI_ISL_624868, EPI_ISL_624870, EPI_ISL_624873, EPI_ISL_624875, EPI_ISL_624876, EPI_ISL_624877, EPI_ISL_624878, EPI_ISL_624879, EPI_ISL_624882, EPI_ISL_624886, EPI_ISL_624887, EPI_ISL_624888, EPI_ISL_624890, EPI_ISL_624891, EPI_ISL_624892, EPI_ISL_624893, EPI_ISL_624894, EPI_ISL_624900, EPI_ISL_624902, EPI_ISL_624904, EPI_ISL_624907, EPI_ISL_624908, EPI_ISL_624910, EPI_ISL_624911, EPI_ISL_624912, EPI_ISL_624913, EPI_ISL_624914, EPI_ISL_624915, EPI_ISL_624916, EPI_ISL_624917, EPI_ISL_624918, EPI_ISL_624919, EPI_ISL_624920, EPI_ISL_624921, EPI_ISL_624922, EPI_ISL_624923, EPI_ISL_624924, EPI_ISL_624925, EPI_ISL_624926, EPI_ISL_624927, EPI_ISL_624928, EPI_ISL_624929, EPI_ISL_624930, EPI_ISL_624931, EPI_ISL_624932, EPI_ISL_624933, EPI_ISL_624934, EPI_ISL_624935, EPI_ISL_624936, EPI_ISL_624937, EPI_ISL_624938, EPI_ISL_624939, EPI_ISL_624940, EPI_ISL_624941, EPI_ISL_624942, EPI_ISL_624943, EPI_ISL_624944, EPI_ISL_624945, EPI_ISL_624946, EPI_ISL_624947, EPI_ISL_624948, EPI_ISL_624949, EPI_ISL_624950, EPI_ISL_624951, EPI_ISL_624952, EPI_ISL_624953, EPI_ISL_624954, EPI_ISL_624955, EPI_ISL_624956, EPI_ISL_624957, EPI_ISL_624958, EPI_ISL_624959, EPI_ISL_624960, EPI_ISL_624961, EPI_ISL_624962, EPI_ISL_624963, EPI_ISL_624964, EPI_ISL_624965, EPI_ISL_624966, EPI_ISL_624967, EPI_ISL_624968, EPI_ISL_624969, EPI_ISL_624970, EPI_ISL_624971, EPI_ISL_624972, EPI_ISL_624973, EPI_ISL_624974, EPI_ISL_624975, EPI_ISL_624976, EPI_ISL_624977, EPI_ISL_624978, EPI_ISL_624979, EPI_ISL_624980, EPI_ISL_624981, EPI_ISL_624982, EPI_ISL_624983, EPI_ISL_624984, EPI_ISL_624985, EPI_ISL_624986, EPI_ISL_624987, EPI_ISL_624988, EPI_ISL_624989, EPI_ISL_624990, EPI_ISL_624991, EPI_ISL_624992, EPI_ISL_624993, EPI_ISL_624994, EPI_ISL_624995, EPI_ISL_624996, EPI_ISL_624997, EPI_ISL_624998, EPI_ISL_624999, EPI_ISL_625000, EPI_ISL_625001, EPI_ISL_625002, EPI_ISL_625003, EPI_ISL_625004, EPI_ISL_625005, EPI_ISL_625006, EPI_ISL_625007, EPI_ISL_625008, EPI_ISL_625009, EPI_ISL_625010, EPI_ISL_625011, EPI_ISL_625012, EPI_ISL_625013, EPI_ISL_625014, EPI_ISL_625015, EPI_ISL_625016, EPI_ISL_625017, EPI_ISL_625018, EPI_ISL_625019, EPI_ISL_625020, EPI_ISL_625021, EPI_ISL_625022, EPI_ISL_625023, EPI_ISL_625024, EPI_ISL_625025, EPI_ISL_625026, EPI_ISL_625027, EPI_ISL_625028, EPI_ISL_625029, EPI_ISL_625030, EPI_ISL_625031, EPI_ISL_625032, EPI_ISL_625033, EPI_ISL_625034, EPI_ISL_625035, EPI_ISL_625036, EPI_ISL_625037, EPI_ISL_625038, EPI_ISL_625039, EPI_ISL_625040, EPI_ISL_625041, EPI_ISL_625042, EPI_ISL_625043, EPI_ISL_625044, EPI_ISL_625045, EPI_ISL_625046, EPI_ISL_625047, EPI_ISL_625048, EPI_ISL_625049, EPI_ISL_625050, EPI_ISL_625051, EPI_ISL_625052, EPI_ISL_625053, EPI_ISL_625054, EPI_ISL_625055, EPI_ISL_625056, EPI_ISL_625057, EPI_ISL_625058, EPI_ISL_625059, EPI_ISL_625060, EPI_ISL_625061, EPI_ISL_625062, EPI_ISL_625063, EPI_ISL_625064, EPI_ISL_625065, EPI_ISL_625066, EPI_ISL_625067, EPI_ISL_625068, EPI_ISL_625069, EPI_ISL_625070, EPI_ISL_625071, EPI_ISL_625072, EPI_ISL_625073, EPI_ISL_625074, EPI_ISL_625075, EPI_ISL_625076, EPI_ISL_625077, EPI_ISL_625078, EPI_ISL_625079, EPI_ISL_625080, EPI_ISL_625081, EPI_ISL_625082, EPI_ISL_625083, EPI_ISL_625084, EPI_ISL_625085, EPI_ISL_625086, EPI_ISL_625087, EPI_ISL_625088, EPI_ISL_625089, EPI_ISL_625090, EPI_ISL_625091, EPI_ISL_625092, EPI_ISL_625093, EPI_ISL_625094, EPI_ISL_625095, EPI_ISL_625096, EPI_ISL_625097, EPI_ISL_625098, EPI_ISL_625099, EPI_ISL_625100, EPI_ISL_625101, EPI_ISL_625102, EPI_ISL_625103, EPI_ISL_625104, EPI_ISL_625105, EPI_ISL_625106, EPI_ISL_625107, EPI_ISL_625108, EPI_ISL_625109, EPI_ISL_625110, EPI_ISL_625111, EPI_ISL_625112, EPI_ISL_625113, EPI_ISL_625114, EPI_ISL_625115, EPI_ISL_625116, EPI_ISL_625117, EPI_ISL_625118, EPI_ISL_625119, EPI_ISL_625120, EPI_ISL_625121, EPI_ISL_625122, EPI_ISL_625123, EPI_ISL_625124, EPI_ISL_625125, EPI_ISL_625126, EPI_ISL_625127, EPI_ISL_625128, EPI_ISL_625129, EPI_ISL_625130, EPI_ISL_625131, EPI_ISL_625132, EPI_ISL_625133, EPI_ISL_625134, EPI_ISL_625135, EPI_ISL_625136, EPI_ISL_625137, EPI_ISL_625138, EPI_ISL_625139, EPI_ISL_625140, EPI_ISL_625141, EPI_ISL_625142, EPI_ISL_625143, EPI_ISL_625144, EPI_ISL_625145, EPI_ISL_625146, EPI_ISL_625147, EPI_ISL_625148, EPI_ISL_625149, EPI_ISL_625150, EPI_ISL_625151, EPI_ISL_625152, EPI_ISL_625153, EPI_ISL_625154, EPI_ISL_625155, EPI_ISL_625156, EPI_ISL_625157, EPI_ISL_625158, EPI_ISL_625159, EPI_ISL_625160, EPI_ISL_625161, EPI_ISL_625162, EPI_ISL_625163, EPI_ISL_625164, EPI_ISL_625165, EPI_ISL_625166, EPI_ISL_625167, EPI_ISL_625168, EPI_ISL_625169, EPI_ISL_625170, EPI_ISL_625171, EPI_ISL_625172, EPI_ISL_625173, EPI_ISL_625174, EPI_ISL_625175, EPI_ISL_625176, EPI_ISL_625177, EPI_ISL_625178, EPI_ISL_625179, EPI_ISL_625180, EPI_ISL_625181, EPI_ISL_625182, EPI_ISL_625183, EPI_ISL_625184, EPI_ISL_625185, EPI_ISL_625186, EPI_ISL_625187, EPI_ISL_625188, EPI_ISL_625189, EPI_ISL_625190, EPI_ISL_625191, EPI_ISL_625192, EPI_ISL_625193, EPI_ISL_625194, EPI_ISL_625195, EPI_ISL_625196, EPI_ISL_625197, EPI_ISL_625198, EPI_ISL_625199, EPI_ISL_625200, EPI_ISL_625201, EPI_ISL_625202, EPI_ISL_625203, EPI_ISL_625204, EPI_ISL_625205, EPI_ISL_625206, EPI_ISL_625207, EPI_ISL_625208, EPI_ISL_625209, EPI_ISL_625210, EPI_ISL_625211, EPI_ISL_625212, EPI_ISL_625214, EPI_ISL_625215, EPI_ISL_625216, EPI_ISL_625217, EPI_ISL_625218, EPI_ISL_625219, EPI_ISL_625220, EPI_ISL_625221, EPI_ISL_625222, EPI_ISL_625223, EPI_ISL_625224, EPI_ISL_625225, EPI_ISL_625226, EPI_ISL_625227, EPI_ISL_625228, EPI_ISL_625229, EPI_ISL_625230, EPI_ISL_625231, EPI_ISL_625232, EPI_ISL_625233, EPI_ISL_625234, EPI_ISL_625235, EPI_ISL_625236, EPI_ISL_625237, EPI_ISL_625238, EPI_ISL_625239, EPI_ISL_625241, EPI_ISL_625242, EPI_ISL_625243, EPI_ISL_625244, EPI_ISL_625245, EPI_ISL_625246, EPI_ISL_625247, EPI_ISL_625248, EPI_ISL_625249, EPI_ISL_625250, EPI_ISL_625251, EPI_ISL_625252, EPI_ISL_625253, EPI_ISL_625254, EPI_ISL_625255, EPI_ISL_625256, EPI_ISL_625257, EPI_ISL_625258, EPI_ISL_625259, EPI_ISL_625260, EPI_ISL_625261, EPI_ISL_625262, EPI_ISL_625263, EPI_ISL_625264, EPI_ISL_625265, EPI_ISL_625266, EPI_ISL_630706, EPI_ISL_630707, EPI_ISL_630708, EPI_ISL_630709, EPI_ISL_630710, EPI_ISL_630711, EPI_ISL_630713, EPI_ISL_630714, EPI_ISL_630715, EPI_ISL_630716, EPI_ISL_630717, EPI_ISL_630720, EPI_ISL_630721, EPI_ISL_630722, EPI_ISL_630723, EPI_ISL_630724, EPI_ISL_630726, EPI_ISL_630727, EPI_ISL_630728, EPI_ISL_630729, EPI_ISL_630730, EPI_ISL_630732, EPI_ISL_630734, EPI_ISL_630735, EPI_ISL_630736, EPI_ISL_630737, EPI_ISL_630738, EPI_ISL_630739, EPI_ISL_630740, EPI_ISL_630742, EPI_ISL_630743, EPI_ISL_630744, EPI_ISL_630746, EPI_ISL_630747, EPI_ISL_630748, EPI_ISL_630750, EPI_ISL_630751, EPI_ISL_630752, EPI_ISL_630753, EPI_ISL_630754, EPI_ISL_630755, EPI_ISL_630756, EPI_ISL_630757, EPI_ISL_630758, EPI_ISL_630759, EPI_ISL_630760, EPI_ISL_630762, EPI_ISL_630763, EPI_ISL_630765, EPI_ISL_630766, EPI_ISL_630767, EPI_ISL_630768, EPI_ISL_630769, EPI_ISL_630770, EPI_ISL_630771, EPI_ISL_630772, EPI_ISL_630773, EPI_ISL_630774, EPI_ISL_630776, EPI_ISL_630777, EPI_ISL_630779, EPI_ISL_630781, EPI_ISL_630783, EPI_ISL_630784, EPI_ISL_630785, EPI_ISL_630786, EPI_ISL_630788, EPI_ISL_630789, EPI_ISL_630790, EPI_ISL_630791, EPI_ISL_630792, EPI_ISL_630793, EPI_ISL_630794, EPI_ISL_630795, EPI_ISL_630796, EPI_ISL_630797, EPI_ISL_630798, EPI_ISL_630799, EPI_ISL_630800, EPI_ISL_630801, EPI_ISL_630802, EPI_ISL_630803, EPI_ISL_630805, EPI_ISL_630806, EPI_ISL_630807, EPI_ISL_630808, EPI_ISL_630809, EPI_ISL_630810, EPI_ISL_630811, EPI_ISL_630812, EPI_ISL_630814, EPI_ISL_630816, EPI_ISL_630817, EPI_ISL_630818, EPI_ISL_630819, EPI_ISL_630820, EPI_ISL_630821, EPI_ISL_630822, EPI_ISL_630823, EPI_ISL_630825, EPI_ISL_630826, EPI_ISL_630828, EPI_ISL_630829, EPI_ISL_630830, EPI_ISL_630831, EPI_ISL_630832, EPI_ISL_630834, EPI_ISL_630835, EPI_ISL_630836, EPI_ISL_630837, EPI_ISL_630839, EPI_ISL_630840, EPI_ISL_630841, EPI_ISL_630842, EPI_ISL_630843, EPI_ISL_630844, EPI_ISL_630845, EPI_ISL_630846, EPI_ISL_630847, EPI_ISL_630848, EPI_ISL_630849, EPI_ISL_630852, EPI_ISL_630854, EPI_ISL_630855, EPI_ISL_630856, EPI_ISL_630857, EPI_ISL_630858, EPI_ISL_630860, EPI_ISL_630861, EPI_ISL_630863, EPI_ISL_630865, EPI_ISL_630866, EPI_ISL_630867, EPI_ISL_630868, EPI_ISL_630869, EPI_ISL_630874, EPI_ISL_630877, EPI_ISL_630878, EPI_ISL_630879, EPI_ISL_630880, EPI_ISL_630881, EPI_ISL_630882, EPI_ISL_630884, EPI_ISL_630885, EPI_ISL_630886, EPI_ISL_630887, EPI_ISL_630889, EPI_ISL_630890, EPI_ISL_630892, EPI_ISL_630893, EPI_ISL_630894, EPI_ISL_630895, EPI_ISL_630896, EPI_ISL_630897, EPI_ISL_630898, EPI_ISL_630899, EPI_ISL_630901, EPI_ISL_630902, EPI_ISL_630903, EPI_ISL_630904, EPI_ISL_630905, EPI_ISL_630906, EPI_ISL_630907, EPI_ISL_630908, EPI_ISL_630909, EPI_ISL_630910, EPI_ISL_630911, EPI_ISL_630912, EPI_ISL_630913, EPI_ISL_630914, EPI_ISL_630916, EPI_ISL_630917, EPI_ISL_630918, EPI_ISL_630920, EPI_ISL_630921, EPI_ISL_630922, EPI_ISL_630924, EPI_ISL_630925, EPI_ISL_630927, EPI_ISL_630928, EPI_ISL_630929, EPI_ISL_630930, EPI_ISL_630932, EPI_ISL_630934, EPI_ISL_630935, EPI_ISL_630936, EPI_ISL_630937, EPI_ISL_630938, EPI_ISL_630939, EPI_ISL_630940, EPI_ISL_630942, EPI_ISL_630943, EPI_ISL_630944, EPI_ISL_630945, EPI_ISL_630946, EPI_ISL_630947, EPI_ISL_630950, EPI_ISL_630952, EPI_ISL_630953, EPI_ISL_630954, EPI_ISL_630956, EPI_ISL_630958, EPI_ISL_630960, EPI_ISL_630961, EPI_ISL_630962, | | | |

EPI_ISL_630963, EPI_ISL_630964, EPI_ISL_630966, EPI_ISL_630967, EPI_ISL_630968, EPI_ISL_630969, EPI_ISL_630970, EPI_ISL_634757, EPI_ISL_634758, EPI_ISL_634759, EPI_ISL_634760, EPI_ISL_634761, EPI_ISL_634762, EPI_ISL_634763, EPI_ISL_634764, EPI_ISL_634765, EPI_ISL_634766, EPI_ISL_634767, EPI_ISL_634768, EPI_ISL_634769, EPI_ISL_634770, EPI_ISL_634771, EPI_ISL_634772, EPI_ISL_634773, EPI_ISL_634774, EPI_ISL_634775, EPI_ISL_634776, EPI_ISL_634777, EPI_ISL_634778, EPI_ISL_634779, EPI_ISL_634780, EPI_ISL_634781, EPI_ISL_634782, EPI_ISL_634783, EPI_ISL_634784, EPI_ISL_634785, EPI_ISL_634786, EPI_ISL_634787, EPI_ISL_634788

| | | | |
|---|---|---|---|
| see above | Lighthouse Lab in Alderley Park | Wellcome Sanger Institute for the COVID-19 Genomics UK (COG-UK) consortium | Cordelia Langford; David K. Jackson; Dominic Kwiatkowski; Ewan Harrison; Ian Johnston; Jacquelyn Wynn; John Sillitoe on behalf of the Wellcome Sanger Institute COVID-19 Surveillance Team; John Sillitoe on behalf of the Wellcome Sanger Institute COVID-19 Surveillance Team (http://www.sanger.ac.uk/covid-team); Mairead Hyland; Roberto Amato; Sonia Goncalves; The Lighthouse Lab in Alderley Park and Alex Alderton |

EPI_ISL_623477, EPI_ISL_623781, EPI_ISL_631159, EPI_ISL_633165, EPI_ISL_633221, EPI_ISL_643330, EPI_ISL_643413, EPI_ISL_643768, EPI_ISL_644115, EPI_ISL_644117, EPI_ISL_647900, EPI_ISL_647901, EPI_ISL_647906, EPI_ISL_647908, EPI_ISL_647909, EPI_ISL_647913, EPI_ISL_647916

| | | | |
|---|---|---|---|
| see above | Lighthouse Lab in Cambridge | Wellcome Sanger Institute for the COVID-19 Genomics UK (COG-UK) Consortium | Cordelia Langford; David K. Jackson; Dominic Kwiatkowski; Ewan Harrison; Ian Johnston; John Sillitoe on behalf of the Wellcome Sanger Institute COVID-19 Surveillance Team; John Sillitoe on behalf of the Wellcome Sanger Institute COVID-19 Surveillance Team (http://www.sanger.ac.uk/covid-team); Rob Howes; Roberto Amato; Sonia Goncalves; The Lighthouse Lab in Cambridge and Alex Alderton |

EPI_ISL_623220, EPI_ISL_623222, EPI_ISL_623234, EPI_ISL_623245, EPI_ISL_623247, EPI_ISL_623248, EPI_ISL_623256, EPI_ISL_623268, EPI_ISL_623273, EPI_ISL_623276, EPI_ISL_623281, EPI_ISL_623286, EPI_ISL_623288, EPI_ISL_623301, EPI_ISL_623308, EPI_ISL_623312, EPI_ISL_623321, EPI_ISL_623323, EPI_ISL_623330, EPI_ISL_623333, EPI_ISL_623334, EPI_ISL_623337, EPI_ISL_623343, EPI_ISL_623344, EPI_ISL_623345, EPI_ISL_623346, EPI_ISL_623347, EPI_ISL_623359, EPI_ISL_623360, EPI_ISL_623363, EPI_ISL_623364, EPI_ISL_623366, EPI_ISL_623370, EPI_ISL_623385, EPI_ISL_623386, EPI_ISL_623390, EPI_ISL_623392, EPI_ISL_623393, EPI_ISL_623397, EPI_ISL_623398, EPI_ISL_623404, EPI_ISL_623409, EPI_ISL_623411, EPI_ISL_623424, EPI_ISL_623425, EPI_ISL_623426, EPI_ISL_623427, EPI_ISL_623433, EPI_ISL_623437, EPI_ISL_623440, EPI_ISL_623444, EPI_ISL_623445, EPI_ISL_623448, EPI_ISL_623449, EPI_ISL_623451, EPI_ISL_623474, EPI_ISL_623475, EPI_ISL_623482, EPI_ISL_623485, EPI_ISL_623488, EPI_ISL_623489, EPI_ISL_623490, EPI_ISL_623494, EPI_ISL_623495, EPI_ISL_623520, EPI_ISL_623527, EPI_ISL_623528, EPI_ISL_623531, EPI_ISL_623532, EPI_ISL_623537, EPI_ISL_623539, EPI_ISL_623544, EPI_ISL_623545, EPI_ISL_623547, EPI_ISL_623548, EPI_ISL_623550, EPI_ISL_623551, EPI_ISL_623555, EPI_ISL_623556, EPI_ISL_623557, EPI_ISL_623558, EPI_ISL_623559, EPI_ISL_623560, EPI_ISL_623562, EPI_ISL_623567, EPI_ISL_623568, EPI_ISL_623569, EPI_ISL_623570, EPI_ISL_623572, EPI_ISL_623574, EPI_ISL_623575, EPI_ISL_623576, EPI_ISL_623578, EPI_ISL_623581, EPI_ISL_623584, EPI_ISL_623585, EPI_ISL_623587, EPI_ISL_623592, EPI_ISL_623593, EPI_ISL_623595, EPI_ISL_623600, EPI_ISL_623605, EPI_ISL_623608, EPI_ISL_623609, EPI_ISL_623611, EPI_ISL_623612, EPI_ISL_623616, EPI_ISL_623617, EPI_ISL_623619, EPI_ISL_623626, EPI_ISL_623630, EPI_ISL_623633, EPI_ISL_623639, EPI_ISL_623640, EPI_ISL_623641, EPI_ISL_623643, EPI_ISL_623644, EPI_ISL_623645, EPI_ISL_623647, EPI_ISL_623650, EPI_ISL_623651, EPI_ISL_623652, EPI_ISL_623653, EPI_ISL_623654, EPI_ISL_623655, EPI_ISL_623657, EPI_ISL_623658, EPI_ISL_623661, EPI_ISL_623662, EPI_ISL_623663, EPI_ISL_623664, EPI_ISL_623665, EPI_ISL_623666, EPI_ISL_623667, EPI_ISL_623674, EPI_ISL_623675, EPI_ISL_623679, EPI_ISL_623680, EPI_ISL_623683, EPI_ISL_623688, EPI_ISL_623690, EPI_ISL_623694, EPI_ISL_623700, EPI_ISL_623702, EPI_ISL_623703, EPI_ISL_623710, EPI_ISL_623712, EPI_ISL_623716, EPI_ISL_623717, EPI_ISL_623721, EPI_ISL_623724, EPI_ISL_623725, EPI_ISL_623726, EPI_ISL_623731, EPI_ISL_623733, EPI_ISL_623735, EPI_ISL_623736, EPI_ISL_623740, EPI_ISL_623741, EPI_ISL_623742, EPI_ISL_623746, EPI_ISL_623749, EPI_ISL_623750, EPI_ISL_623751, EPI_ISL_623754, EPI_ISL_623758, EPI_ISL_623759, EPI_ISL_623762, EPI_ISL_623763, EPI_ISL_623764, EPI_ISL_623772, EPI_ISL_623774, EPI_ISL_623775, EPI_ISL_623776, EPI_ISL_623785, EPI_ISL_623788, EPI_ISL_623790, EPI_ISL_623791, EPI_ISL_623793, EPI_ISL_623794, EPI_ISL_623796, EPI_ISL_623797, EPI_ISL_623798, EPI_ISL_623799, EPI_ISL_623800, EPI_ISL_623802, EPI_ISL_623806, EPI_ISL_623808, EPI_ISL_623809, EPI_ISL_623810, EPI_ISL_623811, EPI_ISL_623812, EPI_ISL_623815, EPI_ISL_623816, EPI_ISL_623822, EPI_ISL_623823, EPI_ISL_623828, EPI_ISL_623829, EPI_ISL_623831, EPI_ISL_623832, EPI_ISL_623833, EPI_ISL_623834, EPI_ISL_623838, EPI_ISL_623840, EPI_ISL_623841, EPI_ISL_623843, EPI_ISL_623844, EPI_ISL_623846, EPI_ISL_623847, EPI_ISL_623848, EPI_ISL_623849, EPI_ISL_623854, EPI_ISL_623855, EPI_ISL_623856, EPI_ISL_623857, EPI_ISL_630972, EPI_ISL_630977, EPI_ISL_630979, EPI_ISL_630984, EPI_ISL_630988, EPI_ISL_630996, EPI_ISL_630997, EPI_ISL_631001, EPI_ISL_631003, EPI_ISL_631005, EPI_ISL_631008, EPI_ISL_631016, EPI_ISL_631017, EPI_ISL_631025, EPI_ISL_631028, EPI_ISL_631038, EPI_ISL_631039, EPI_ISL_631045, EPI_ISL_631053, EPI_ISL_631055, EPI_ISL_631062, EPI_ISL_631063, EPI_ISL_631069, EPI_ISL_631078, EPI_ISL_631080, EPI_ISL_631094, EPI_ISL_631101, EPI_ISL_631105, EPI_ISL_631109, EPI_ISL_631117, EPI_ISL_631118, EPI_ISL_631123, EPI_ISL_631129, EPI_ISL_631132, EPI_ISL_631135, EPI_ISL_631137, EPI_ISL_631142, EPI_ISL_631144, EPI_ISL_631148, EPI_ISL_631149, EPI_ISL_631154, EPI_ISL_631155, EPI_ISL_631160, EPI_ISL_631164, EPI_ISL_631171, EPI_ISL_631174, EPI_ISL_631177, EPI_ISL_631180, EPI_ISL_631181, EPI_ISL_631185, EPI_ISL_631191, EPI_ISL_631192, EPI_ISL_631194, EPI_ISL_631199, EPI_ISL_631204, EPI_ISL_631208, EPI_ISL_631219, EPI_ISL_631220, EPI_ISL_631221, EPI_ISL_631224, EPI_ISL_631225, EPI_ISL_631235, EPI_ISL_631237, EPI_ISL_631245, EPI_ISL_631254, EPI_ISL_631257, EPI_ISL_631262, EPI_ISL_631266, EPI_ISL_631268, EPI_ISL_631270, EPI_ISL_631271, EPI_ISL_631272, EPI_ISL_631273, EPI_ISL_631274, EPI_ISL_633100, EPI_ISL_633101, EPI_ISL_633102, EPI_ISL_633104, EPI_ISL_633105, EPI_ISL_633106, EPI_ISL_633107, EPI_ISL_633108, EPI_ISL_633109, EPI_ISL_633110, EPI_ISL_633114, EPI_ISL_633116, EPI_ISL_633117, EPI_ISL_633120, EPI_ISL_633121, EPI_ISL_633122, EPI_ISL_633123, EPI_ISL_633124, EPI_ISL_633126, EPI_ISL_633127, EPI_ISL_633128, EPI_ISL_633129, EPI_ISL_633130, EPI_ISL_633131, EPI_ISL_633135, EPI_ISL_633136, EPI_ISL_633140, EPI_ISL_633141, EPI_ISL_633142, EPI_ISL_633147, EPI_ISL_633148, EPI_ISL_633149, EPI_ISL_633150, EPI_ISL_633153, EPI_ISL_633155, EPI_ISL_633156, EPI_ISL_633157, EPI_ISL_633159, EPI_ISL_633161, EPI_ISL_633163, EPI_ISL_633164, EPI_ISL_633170, EPI_ISL_633172, EPI_ISL_633173, EPI_ISL_633176, EPI_ISL_633177, EPI_ISL_633183, EPI_ISL_633186, EPI_ISL_633187, EPI_ISL_633188, EPI_ISL_633189, EPI_ISL_633192, EPI_ISL_633194, EPI_ISL_633197, EPI_ISL_633198, EPI_ISL_633199, EPI_ISL_633200, EPI_ISL_633205, EPI_ISL_633208, EPI_ISL_633211, EPI_ISL_633212, EPI_ISL_633214, EPI_ISL_633215, EPI_ISL_633216, EPI_ISL_633217, EPI_ISL_633218, EPI_ISL_633219, EPI_ISL_633220, EPI_ISL_633223, EPI_ISL_633224, EPI_ISL_633225, EPI_ISL_633227, EPI_ISL_633229, EPI_ISL_633231, EPI_ISL_633234, EPI_ISL_633235, EPI_ISL_633236, EPI_ISL_633238, EPI_ISL_633239, EPI_ISL_633242, EPI_ISL_633245, EPI_ISL_633246, EPI_ISL_633247, EPI_ISL_633248, EPI_ISL_633249, EPI_ISL_633250, EPI_ISL_633252, EPI_ISL_633253, EPI_ISL_633254, EPI_ISL_633255, EPI_ISL_633257, EPI_ISL_633258, EPI_ISL_633260, EPI_ISL_633263, EPI_ISL_633264, EPI_ISL_633265, EPI_ISL_633266, EPI_ISL_633267, EPI_ISL_633270, EPI_ISL_633273, EPI_ISL_633277, EPI_ISL_633278, EPI_ISL_633281, EPI_ISL_633284, EPI_ISL_633286, EPI_ISL_633288, EPI_ISL_633291, EPI_ISL_633292, EPI_ISL_633294, EPI_ISL_633295, EPI_ISL_633297, EPI_ISL_633298, EPI_ISL_633299, EPI_ISL_633300, EPI_ISL_633302, EPI_ISL_633308, EPI_ISL_633311, EPI_ISL_633313, EPI_ISL_633314, EPI_ISL_633315, EPI_ISL_633319, EPI_ISL_633321, EPI_ISL_633322, EPI_ISL_633324, EPI_ISL_633325, EPI_ISL_633329, EPI_ISL_633330, EPI_ISL_633333, EPI_ISL_633334, EPI_ISL_633336, EPI_ISL_633337, EPI_ISL_633340, EPI_ISL_633341, EPI_ISL_633344, EPI_ISL_633345, EPI_ISL_633346, EPI_ISL_633347, EPI_ISL_633349, EPI_ISL_633350, EPI_ISL_633356, EPI_ISL_633357, EPI_ISL_633359, EPI_ISL_633360, EPI_ISL_633363, EPI_ISL_633365, EPI_ISL_633366, EPI_ISL_633369, EPI_ISL_633370, EPI_ISL_633372, EPI_ISL_633373, EPI_ISL_633374, EPI_ISL_633375, EPI_ISL_633378, EPI_ISL_633380, EPI_ISL_633383, EPI_ISL_633385, EPI_ISL_633388, EPI_ISL_633389, EPI_ISL_633390, EPI_ISL_633393, EPI_ISL_633395, EPI_ISL_633397, EPI_ISL_633398, EPI_ISL_633399, EPI_ISL_633403, EPI_ISL_633405, EPI_ISL_633407, EPI_ISL_633408, EPI_ISL_634789, EPI_ISL_634790, EPI_ISL_634791, EPI_ISL_634792, EPI_ISL_634793, EPI_ISL_634794, EPI_ISL_634795, EPI_ISL_634796, EPI_ISL_634797, EPI_ISL_634798, EPI_ISL_634799, EPI_ISL_634800, EPI_ISL_634801, EPI_ISL_634802, EPI_ISL_634803, EPI_ISL_634804, EPI_ISL_634805, EPI_ISL_634806, EPI_ISL_634807, EPI_ISL_634808, EPI_ISL_634809, EPI_ISL_634810, EPI_ISL_634811, EPI_ISL_634812, EPI_ISL_634813, EPI_ISL_634814, EPI_ISL_634815, EPI_ISL_634816

| | | | |
|---|---|---|---|
| see above | Lighthouse Lab in Cambridge | Wellcome Sanger Institute for the COVID-19 Genomics UK (COG-UK) consortium | Cordelia Langford; David K. Jackson; Dominic Kwiatkowski; Ewan Harrison; Ian Johnston; John Sillitoe on behalf of the Wellcome Sanger Institute COVID-19 Surveillance Team; John Sillitoe on behalf of the Wellcome Sanger Institute COVID-19 Surveillance Team (http://www.sanger.ac.uk/covid-team); Rob Howes; Roberto Amato; Sonia Goncalves; The Lighthouse Lab in Cambridge and Alex Alderton |

EPI_ISL_611304, EPI_ISL_611378, EPI_ISL_624197, EPI_ISL_629940, EPI_ISL_633118, EPI_ISL_633282, EPI_ISL_633361, EPI_ISL_633492, EPI_ISL_633577, EPI_ISL_633653, EPI_ISL_633700, EPI_ISL_633760, EPI_ISL_633782, EPI_ISL_633799, EPI_ISL_634156, EPI_ISL_634678, EPI_ISL_643992, EPI_ISL_643993, EPI_ISL_643994, EPI_ISL_643995, EPI_ISL_643996, EPI_ISL_643997, EPI_ISL_643998, EPI_ISL_643999, EPI_ISL_644000, EPI_ISL_644001, EPI_ISL_644002, EPI_ISL_644003, EPI_ISL_644004, EPI_ISL_644005, EPI_ISL_644006, EPI_ISL_644007, EPI_ISL_644008, EPI_ISL_644009, EPI_ISL_644010, EPI_ISL_644011, EPI_ISL_644012, EPI_ISL_644013, EPI_ISL_644014, EPI_ISL_644015, EPI_ISL_644016, EPI_ISL_644017, EPI_ISL_644018, EPI_ISL_644019, EPI_ISL_644020, EPI_ISL_644021, EPI_ISL_644022, EPI_ISL_644023, EPI_ISL_644024, EPI_ISL_644025, EPI_ISL_644026, EPI_ISL_644027, EPI_ISL_644028, EPI_ISL_644029, EPI_ISL_644030, EPI_ISL_644031, EPI_ISL_644032, EPI_ISL_644033, EPI_ISL_644034, EPI_ISL_644035, EPI_ISL_644036, EPI_ISL_644037, EPI_ISL_644038, EPI_ISL_644039, EPI_ISL_644040, EPI_ISL_644041, EPI_ISL_644042, EPI_ISL_644043, EPI_ISL_644044, EPI_ISL_644045, EPI_ISL_644046, EPI_ISL_644047, EPI_ISL_644048, EPI_ISL_644049, EPI_ISL_644050, EPI_ISL_644051, EPI_ISL_644093, EPI_ISL_644094, EPI_ISL_644099, EPI_ISL_644101, EPI_ISL_644102, EPI_ISL_644107, EPI_ISL_644112, EPI_ISL_647873, EPI_ISL_647874, EPI_ISL_647875, EPI_ISL_647877, EPI_ISL_647878, EPI_ISL_647879, EPI_ISL_647880, EPI_ISL_647881, EPI_ISL_647882, EPI_ISL_647883, EPI_ISL_647884, EPI_ISL_647885, EPI_ISL_647886, EPI_ISL_647887, EPI_ISL_647888, EPI_ISL_647889, EPI_ISL_647890, EPI_ISL_647891, EPI_ISL_647892, EPI_ISL_647893, EPI_ISL_647894, EPI_ISL_647895, EPI_ISL_647896, EPI_ISL_647897, EPI_ISL_647898, EPI_ISL_647899, EPI_ISL_647902, EPI_ISL_647903, EPI_ISL_647904, EPI_ISL_647905, EPI_ISL_647907, EPI_ISL_647910, EPI_ISL_647911, EPI_ISL_647912, EPI_ISL_647914, EPI_ISL_647915, EPI_ISL_647917, EPI_ISL_647918, EPI_ISL_647919, EPI_ISL_647920, EPI_ISL_647921, EPI_ISL_647922, EPI_ISL_647923, EPI_ISL_647924, EPI_ISL_647925, EPI_ISL_647926, EPI_ISL_647927, EPI_ISL_647928, EPI_ISL_647929, EPI_ISL_647930, EPI_ISL_647931, EPI_ISL_647932, EPI_ISL_647933, EPI_ISL_647934, EPI_ISL_647935, EPI_ISL_647936, EPI_ISL_647937, EPI_ISL_647938, EPI_ISL_647939, EPI_ISL_647940, EPI_ISL_647941, EPI_ISL_647942, EPI_ISL_647943, EPI_ISL_647944, EPI_ISL_647945, EPI_ISL_647946, EPI_ISL_647947, EPI_ISL_647948, EPI_ISL_647949, EPI_ISL_647950, EPI_ISL_647951, EPI_ISL_647952, EPI_ISL_647953, EPI_ISL_647954, EPI_ISL_647955, EPI_ISL_647956, EPI_ISL_647957, EPI_ISL_647958, EPI_ISL_647959, EPI_ISL_647960, EPI_ISL_647961, EPI_ISL_647962, EPI_ISL_647963, EPI_ISL_647964, EPI_ISL_647965, EPI_ISL_647966, EPI_ISL_647967, EPI_ISL_647968, EPI_ISL_649634, EPI_ISL_649635, EPI_ISL_649636, EPI_ISL_649637, EPI_ISL_649638, EPI_ISL_649639, EPI_ISL_649640, EPI_ISL_649641, EPI_ISL_649642, EPI_ISL_649643, EPI_ISL_649644, EPI_ISL_649645, EPI_ISL_649646, EPI_ISL_649647, EPI_ISL_649648, EPI_ISL_649649, EPI_ISL_649650, EPI_ISL_649651, EPI_ISL_649652, EPI_ISL_649653, EPI_ISL_649654, EPI_ISL_649655, EPI_ISL_649656, EPI_ISL_649657, EPI_ISL_649658, EPI_ISL_649659, EPI_ISL_649660, EPI_ISL_649662, EPI_ISL_649663, EPI_ISL_649665, EPI_ISL_649666, EPI_ISL_649667, EPI_ISL_649668, EPI_ISL_649670, EPI_ISL_649671, EPI_ISL_649672, EPI_ISL_649673, EPI_ISL_649674, EPI_ISL_649675, EPI_ISL_649676, EPI_ISL_649677, EPI_ISL_649678, EPI_ISL_649679, EPI_ISL_649680, EPI_ISL_649681, EPI_ISL_649682, EPI_ISL_649683, EPI_ISL_649684, EPI_ISL_649685, EPI_ISL_649686, EPI_ISL_649687, EPI_ISL_649688, EPI_ISL_649689, EPI_ISL_649690, EPI_ISL_649691, EPI_ISL_649692, EPI_ISL_649693, EPI_ISL_649694, EPI_ISL_649695, EPI_ISL_649696, EPI_ISL_649697, EPI_ISL_649698, EPI_ISL_649699, EPI_ISL_649700, EPI_ISL_649701, EPI_ISL_649702, EPI_ISL_649703, EPI_ISL_649704, EPI_ISL_649705, EPI_ISL_649706, EPI_ISL_649707, EPI_ISL_649708, EPI_ISL_649709, EPI_ISL_649711, EPI_ISL_649712, EPI_ISL_649713, EPI_ISL_649714, EPI_ISL_649715, EPI_ISL_649716, EPI_ISL_649717, EPI_ISL_649718, EPI_ISL_649719, EPI_ISL_649720, EPI_ISL_649721, EPI_ISL_649722, EPI_ISL_649723, EPI_ISL_649724, EPI_ISL_649725, EPI_ISL_649726, EPI_ISL_649727, EPI_ISL_649728, EPI_ISL_649729, EPI_ISL_649730, EPI_ISL_649731, EPI_ISL_649732, EPI_ISL_649733, EPI_ISL_649734, EPI_ISL_649735, EPI_ISL_649736, EPI_ISL_649737, EPI_ISL_649738, EPI_ISL_649739, EPI_ISL_649740, EPI_ISL_649741, EPI_ISL_649742, EPI_ISL_649743, EPI_ISL_649744, EPI_ISL_649745, EPI_ISL_649746, EPI_ISL_649747, EPI_ISL_649748, EPI_ISL_649749, EPI_ISL_649750, EPI_ISL_649751, EPI_ISL_649752, EPI_ISL_649753, EPI_ISL_649754, EPI_ISL_649755, EPI_ISL_649756, EPI_ISL_649757, EPI_ISL_649758, EPI_ISL_649759, EPI_ISL_649760, EPI_ISL_649761, EPI_ISL_649762, EPI_ISL_649763, EPI_ISL_649764, EPI_ISL_649765, EPI_ISL_649766, EPI_ISL_649767, EPI_ISL_649768, EPI_ISL_649769, EPI_ISL_649770, EPI_ISL_649771, EPI_ISL_649772, EPI_ISL_649773, EPI_ISL_649774, EPI_ISL_649775, EPI_ISL_649776, EPI_ISL_649777, EPI_ISL_649778, EPI_ISL_649779, EPI_ISL_649780, EPI_ISL_649781, EPI_ISL_649782, EPI_ISL_649783, EPI_ISL_662746, EPI_ISL_662856, EPI_ISL_721542, EPI_ISL_721543, EPI_ISL_721545, EPI_ISL_721546, EPI_ISL_721547, EPI_ISL_721548, EPI_ISL_721549, EPI_ISL_721550, EPI_ISL_721551, EPI_ISL_721552, EPI_ISL_721553, EPI_ISL_721554, EPI_ISL_721555, EPI_ISL_721557, EPI_ISL_721560, EPI_ISL_721566, EPI_ISL_826516

| | | | |
|---|---|---|---|
| see above | Lighthouse Lab in Glasgow | Wellcome Sanger Institute for the COVID-19 Genomics UK (COG-UK) Consortium | Anna Dominiczak and Alex Alderton; Carol Clugston; Cordelia Langford; David Gray; David K. Jackson; Dominic Kwiatkowski; Ewan Harrison; Harper VanSteenhouse; Ian Johnston; John Sillitoe on behalf of the Wellcome Sanger Institute COVID-19 Surveillance Team; John Sillitoe on behalf of the Wellcome Sanger Institute COVID-19 Surveillance Team (http://www.sanger.ac.uk/covid-team); Roberto Amato; Sonia Goncalves; Yumi Kasai |

EPI_ISL_611287, EPI_ISL_611288, EPI_ISL_611289, EPI_ISL_611290, EPI_ISL_611291, EPI_ISL_611292, EPI_ISL_611293, EPI_ISL_611294, EPI_ISL_611295, EPI_ISL_611296, EPI_ISL_611297, EPI_ISL_611299, EPI_ISL_611300, EPI_ISL_611301, EPI_ISL_611302, EPI_ISL_611303, EPI_ISL_611305, EPI_ISL_611306, EPI_ISL_611307, EPI_ISL_611309, EPI_ISL_611310, EPI_ISL_611311, EPI_ISL_611312, EPI_ISL_611313, EPI_ISL_611314, EPI_ISL_611315, EPI_ISL_611316, EPI_ISL_611317, EPI_ISL_611319, EPI_ISL_611320, EPI_ISL_611322, EPI_ISL_611323, EPI_ISL_611324, EPI_ISL_611325, EPI_ISL_611326, EPI_ISL_611327, EPI_ISL_611328, EPI_ISL_611329, EPI_ISL_611330, EPI_ISL_611331, EPI_ISL_611332, EPI_ISL_611333, EPI_ISL_611334, EPI_ISL_611335, EPI_ISL_611336, EPI_ISL_611337, EPI_ISL_611338, EPI_ISL_611339, EPI_ISL_611340, EPI_ISL_611341, EPI_ISL_611342, EPI_ISL_611343, EPI_ISL_611344, EPI_ISL_611345, EPI_ISL_611347, EPI_ISL_611348, EPI_ISL_611349, EPI_ISL_611351, EPI_ISL_611352, EPI_ISL_611353, EPI_ISL_611354, EPI_ISL_611355, EPI_ISL_611356, EPI_ISL_611357, EPI_ISL_611358, EPI_ISL_611359, EPI_ISL_611360, EPI_ISL_611361, EPI_ISL_611362, EPI_ISL_611363, EPI_ISL_611364, EPI_ISL_611365, EPI_ISL_611366, EPI_ISL_611367, EPI_ISL_611368, EPI_ISL_611369, EPI_ISL_611370, EPI_ISL_611371, EPI_ISL_611373, EPI_ISL_611375, EPI_ISL_611376, EPI_ISL_611377, EPI_ISL_611379, EPI_ISL_611380, EPI_ISL_611381, EPI_ISL_611382, EPI_ISL_611383, EPI_ISL_611384, EPI_ISL_611385, EPI_ISL_611386, EPI_ISL_611387, EPI_ISL_611388, EPI_ISL_611390, EPI_ISL_611391, EPI_ISL_611392, EPI_ISL_611393, EPI_ISL_611394, EPI_ISL_611395, EPI_ISL_611396, EPI_ISL_611397, EPI_ISL_611398, EPI_ISL_611399, EPI_ISL_611400, EPI_ISL_611401, EPI_ISL_611402, EPI_ISL_611404, EPI_ISL_611405, EPI_ISL_611407, EPI_ISL_611408, EPI_ISL_611409, EPI_ISL_611411, EPI_ISL_611412, EPI_ISL_611413, EPI_ISL_611414, EPI_ISL_611415, EPI_ISL_611416, EPI_ISL_611417, EPI_ISL_611418, EPI_ISL_611419, EPI_ISL_611420, EPI_ISL_611421, EPI_ISL_611423, EPI_ISL_611424, EPI_ISL_611425, EPI_ISL_611427, EPI_ISL_611428, EPI_ISL_611429, EPI_ISL_611430, EPI_ISL_611431, EPI_ISL_611432, EPI_ISL_611433, EPI_ISL_611434, EPI_ISL_611436, EPI_ISL_611437, EPI_ISL_611440, EPI_ISL_611441, EPI_ISL_611442, EPI_ISL_611444, EPI_ISL_611445, EPI_ISL_611446, EPI_ISL_611447, EPI_ISL_611448, EPI_ISL_611449, EPI_ISL_611450, EPI_ISL_611451, EPI_ISL_611452, EPI_ISL_611453, EPI_ISL_611455, EPI_ISL_611456, EPI_ISL_611457, EPI_ISL_611458, EPI_ISL_611459, EPI_ISL_611460, EPI_ISL_611461, EPI_ISL_611462, EPI_ISL_611463, EPI_ISL_611464, EPI_ISL_611465, EPI_ISL_611466, EPI_ISL_611467, EPI_ISL_611468, EPI_ISL_611469, EPI_ISL_611470, EPI_ISL_611471, EPI_ISL_611472, EPI_ISL_611473, EPI_ISL_611474, EPI_ISL_611475, EPI_ISL_611476, EPI_ISL_611477, EPI_ISL_611478, EPI_ISL_611479, EPI_ISL_611480, EPI_ISL_611481, EPI_ISL_611482, EPI_ISL_611483, EPI_ISL_623863, EPI_ISL_623865, EPI_ISL_623868, EPI_ISL_623872, EPI_ISL_623873, EPI_ISL_623874, EPI_ISL_623876, EPI_ISL_623883, EPI_ISL_623884, EPI_ISL_623886, EPI_ISL_623893, EPI_ISL_623903, EPI_ISL_623907, EPI_ISL_623908, EPI_ISL_623911, EPI_ISL_623916, EPI_ISL_623917, EPI_ISL_623921, EPI_ISL_623930, EPI_ISL_623933, EPI_ISL_623934, EPI_ISL_623942, EPI_ISL_623944, EPI_ISL_623949, EPI_ISL_623956, EPI_ISL_623959, EPI_ISL_623963, EPI_ISL_623975, EPI_ISL_623976, EPI_ISL_623977, EPI_ISL_623986, EPI_ISL_623990, EPI_ISL_623991, EPI_ISL_623994, EPI_ISL_623995, EPI_ISL_624010, EPI_ISL_624012, EPI_ISL_624018, EPI_ISL_624019, EPI_ISL_624022, EPI_ISL_624027, EPI_ISL_624028, EPI_ISL_624029, EPI_ISL_624032, EPI_ISL_624034, EPI_ISL_624043, EPI_ISL_624044, EPI_ISL_624049, EPI_ISL_624067, EPI_ISL_624073, EPI_ISL_624077, EPI_ISL_624079, EPI_ISL_624092, EPI_ISL_624105, EPI_ISL_624112, EPI_ISL_624114, EPI_ISL_624120, EPI_ISL_624122, EPI_ISL_624128, EPI_ISL_624132, EPI_ISL_624140, EPI_ISL_624145, EPI_ISL_624147, EPI_ISL_624148, EPI_ISL_624158, EPI_ISL_624163, EPI_ISL_624164, EPI_ISL_624165, EPI_ISL_624176, EPI_ISL_624181, EPI_ISL_624189, EPI_ISL_625315, EPI_ISL_625316, EPI_ISL_625317, EPI_ISL_625318, EPI_ISL_625319, EPI_ISL_625320, EPI_ISL_625321, EPI_ISL_625322, EPI_ISL_625323, EPI_ISL_625325, EPI_ISL_625326, EPI_ISL_625327, EPI_ISL_625328, EPI_ISL_625329, EPI_ISL_625330, EPI_ISL_625331, EPI_ISL_625332, EPI_ISL_625333, EPI_ISL_625334, EPI_ISL_625335, EPI_ISL_625336, EPI_ISL_625337, EPI_ISL_625338, EPI_ISL_625339, EPI_ISL_625340, EPI_ISL_625341, EPI_ISL_625342, EPI_ISL_625343, EPI_ISL_625344, EPI_ISL_625345, EPI_ISL_625346, EPI_ISL_625347, EPI_ISL_625348, EPI_ISL_625349, EPI_ISL_625350, EPI_ISL_625351, EPI_ISL_625352, EPI_ISL_625353, EPI_ISL_625354, EPI_ISL_625355, EPI_ISL_625356, EPI_ISL_625357, EPI_ISL_625358, EPI_ISL_625359, EPI_ISL_625360, EPI_ISL_625361, EPI_ISL_625362, EPI_ISL_625363, EPI_ISL_625364, EPI_ISL_625365, EPI_ISL_625366, EPI_ISL_625367, EPI_ISL_625368, EPI_ISL_625369, EPI_ISL_625370, EPI_ISL_625371, EPI_ISL_625372, EPI_ISL_625373, EPI_ISL_625374, EPI_ISL_625375, EPI_ISL_625376, EPI_ISL_625377, EPI_ISL_625378, EPI_ISL_625379, EPI_ISL_625380, EPI_ISL_625381, EPI_ISL_625383, EPI_ISL_625384, EPI_ISL_625385, EPI_ISL_625386, EPI_ISL_625387, EPI_ISL_625388, EPI_ISL_625389, EPI_ISL_625390, EPI_ISL_625391, EPI_ISL_625392, EPI_ISL_625393, EPI_ISL_625394, EPI_ISL_625395, EPI_ISL_629754, EPI_ISL_629755, EPI_ISL_629756, EPI_ISL_629757, EPI_ISL_629758, EPI_ISL_629759, EPI_ISL_629760, EPI_ISL_629761, EPI_ISL_629762, EPI_ISL_629763, EPI_ISL_629764, EPI_ISL_629765, EPI_ISL_629766, EPI_ISL_629767, EPI_ISL_629768, EPI_ISL_629769, EPI_ISL_629770, EPI_ISL_629771, EPI_ISL_629772, EPI_ISL_629773, EPI_ISL_629774, EPI_ISL_629775, EPI_ISL_629776, EPI_ISL_629777, EPI_ISL_629778, EPI_ISL_629779, EPI_ISL_629780, EPI_ISL_629781, EPI_ISL_629782, EPI_ISL_629783, EPI_ISL_629784, EPI_ISL_629785, EPI_ISL_629786, EPI_ISL_629787, EPI_ISL_629788, EPI_ISL_629789, EPI_ISL_629790, EPI_ISL_629791, EPI_ISL_629792, EPI_ISL_629793, EPI_ISL_629794, EPI_ISL_629795, EPI_ISL_629796, EPI_ISL_629797, EPI_ISL_629798, EPI_ISL_629799, EPI_ISL_629800, EPI_ISL_629801, EPI_ISL_629802, EPI_ISL_629803, EPI_ISL_629804, EPI_ISL_629805, EPI_ISL_629806, EPI_ISL_629807, EPI_ISL_629808, EPI_ISL_629809, EPI_ISL_629810, EPI_ISL_629811, EPI_ISL_629812, EPI_ISL_629813, EPI_ISL_629814, EPI_ISL_629815, EPI_ISL_629816, EPI_ISL_629817, EPI_ISL_629818, EPI_ISL_629819, EPI_ISL_629820, EPI_ISL_629821, EPI_ISL_629822, EPI_ISL_629823, EPI_ISL_629824, EPI_ISL_629825, EPI_ISL_629826, EPI_ISL_629827, EPI_ISL_629828, EPI_ISL_629829, EPI_ISL_629830, EPI_ISL_629831, EPI_ISL_629832, EPI_ISL_629833, EPI_ISL_629834, EPI_ISL_629835, EPI_ISL_629836, EPI_ISL_629837, EPI_ISL_629838, EPI_ISL_629839, EPI_ISL_629840, EPI_ISL_629841, EPI_ISL_629842, EPI_ISL_629843, EPI_ISL_629844, EPI_ISL_629845, EPI_ISL_629846, EPI_ISL_629847, EPI_ISL_629848, EPI_ISL_629849, EPI_ISL_629850, EPI_ISL_629851, EPI_ISL_629852, EPI_ISL_629853, EPI_ISL_629854, EPI_ISL_629855, EPI_ISL_629856, EPI_ISL_629857, EPI_ISL_629858, EPI_ISL_629859, EPI_ISL_629860, EPI_ISL_629861, EPI_ISL_629862, EPI_ISL_629863, EPI_ISL_629864, EPI_ISL_629865, EPI_ISL_629866, EPI_ISL_629867, EPI_ISL_629868, EPI_ISL_629869, EPI_ISL_629870, EPI_ISL_629871, EPI_ISL_629872, EPI_ISL_629873, EPI_ISL_629874, EPI_ISL_629875, EPI_ISL_629876, EPI_ISL_629877, EPI_ISL_629878, EPI_ISL_629879, EPI_ISL_629880, EPI_ISL_629881, EPI_ISL_629882, EPI_ISL_629883, EPI_ISL_629884, EPI_ISL_629885, EPI_ISL_629886, EPI_ISL_629887, EPI_ISL_629888, EPI_ISL_629889, EPI_ISL_629890, EPI_ISL_629891, EPI_ISL_629892, EPI_ISL_629893, EPI_ISL_629894, EPI_ISL_629895, EPI_ISL_629896, EPI_ISL_629897, EPI_ISL_629898, EPI_ISL_629899, EPI_ISL_629900, EPI_ISL_629901, EPI_ISL_629902, EPI_ISL_629903, EPI_ISL_629904, EPI_ISL_629905, EPI_ISL_629906, EPI_ISL_629907, EPI_ISL_629908, EPI_ISL_629909, EPI_ISL_629910, EPI_ISL_629911, EPI_ISL_629912, EPI_ISL_629913, EPI_ISL_629914, EPI_ISL_629915, EPI_ISL_629916, EPI_ISL_629917, EPI_ISL_629918, EPI_ISL_629919, EPI_ISL_629920, EPI_ISL_629921, EPI_ISL_629922, EPI_ISL_629923, EPI_ISL_629924, EPI_ISL_629925, EPI_ISL_629926, EPI_ISL_629927, EPI_ISL_629928, EPI_ISL_629929, EPI_ISL_629930, EPI_ISL_629931, EPI_ISL_629932, EPI_ISL_629933, EPI_ISL_629934, EPI_ISL_629935, EPI_ISL_629936, EPI_ISL_629937, EPI_ISL_629938, EPI_ISL_629939, EPI_ISL_629941, EPI_ISL_629942, EPI_ISL_629943, EPI_ISL_629944, EPI_ISL_629945, EPI_ISL_629946, EPI_ISL_629947, EPI_ISL_629948, EPI_ISL_629949, EPI_ISL_629950, EPI_ISL_629951, EPI_ISL_629952, EPI_ISL_629953, EPI_ISL_629954, EPI_ISL_629955, EPI_ISL_629956, EPI_ISL_629957, EPI_ISL_629958, EPI_ISL_629959, EPI_ISL_629960, EPI_ISL_629961, EPI_ISL_629962, EPI_ISL_629963, EPI_ISL_629964, EPI_ISL_629965, EPI_ISL_629966, EPI_ISL_629967, EPI_ISL_629968, EPI_ISL_629969, EPI_ISL_629970, EPI_ISL_629971, EPI_ISL_629972, EPI_ISL_629973, EPI_ISL_629974, EPI_ISL_629975, EPI_ISL_629976, EPI_ISL_629977, EPI_ISL_629978, EPI_ISL_629979, EPI_ISL_629980, EPI_ISL_629981, EPI_ISL_629982, EPI_ISL_629983, EPI_ISL_629984, EPI_ISL_629985, EPI_ISL_629986, EPI_ISL_629987, EPI_ISL_629988, EPI_ISL_629989, EPI_ISL_629990, EPI_ISL_629991, EPI_ISL_629992, EPI_ISL_629993, EPI_ISL_629994, EPI_ISL_629995, EPI_ISL_629996, EPI_ISL_629997, EPI_ISL_629998, EPI_ISL_629999, EPI_ISL_630000, EPI_ISL_630001, EPI_ISL_630002, EPI_ISL_630003, EPI_ISL_630004, EPI_ISL_630005, EPI_ISL_630006, EPI_ISL_630703, EPI_ISL_630705, EPI_ISL_630712, EPI_ISL_630718, EPI_ISL_630719, EPI_ISL_630725, EPI_ISL_630731, EPI_ISL_630733, EPI_ISL_630741, EPI_ISL_630745, EPI_ISL_630749, EPI_ISL_630761, EPI_ISL_630764, EPI_ISL_630775, EPI_ISL_630778, EPI_ISL_630782, EPI_ISL_630787, EPI_ISL_630804, EPI_ISL_630813, EPI_ISL_630815, EPI_ISL_630824, EPI_ISL_630827, EPI_ISL_630833, EPI_ISL_630838, EPI_ISL_630850, EPI_ISL_630851, EPI_ISL_630853, EPI_ISL_630859, EPI_ISL_630862, EPI_ISL_630864, EPI_ISL_630870, EPI_ISL_630871, EPI_ISL_630872, EPI_ISL_630873, EPI_ISL_630875, EPI_ISL_630876, EPI_ISL_630883, EPI_ISL_630888, EPI_ISL_630900, EPI_ISL_630915, EPI_ISL_630919, EPI_ISL_630923, EPI_ISL_630926, EPI_ISL_630931, EPI_ISL_630933, EPI_ISL_630941, EPI_ISL_630948, EPI_ISL_630949, EPI_ISL_630951, EPI_ISL_630955, EPI_ISL_630957, EPI_ISL_630959, EPI_ISL_633103, EPI_ISL_633111, EPI_ISL_633112, EPI_ISL_633113, EPI_ISL_633115, EPI_ISL_633119, EPI_ISL_633125, EPI_ISL_633132, EPI_ISL_633133, EPI_ISL_633134, EPI_ISL_633137, EPI_ISL_633138, EPI_ISL_633139, EPI_ISL_633143, EPI_ISL_633144, EPI_ISL_633145, EPI_ISL_633146, EPI_ISL_633151, EPI_ISL_633152, EPI_ISL_633154, EPI_ISL_633158, EPI_ISL_633160, EPI_ISL_633162, EPI_ISL_633166, EPI_ISL_633167, EPI_ISL_633168, EPI_ISL_633169, EPI_ISL_633171, EPI_ISL_633174, EPI_ISL_633175, EPI_ISL_633178, EPI_ISL_633179, EPI_ISL_633180, EPI_ISL_633181, EPI_ISL_633182, EPI_ISL_633184, EPI_ISL_633185, EPI_ISL_633190, EPI_ISL_633191, EPI_ISL_633193, EPI_ISL_633195, EPI_ISL_633196, EPI_ISL_633201, EPI_ISL_633202, EPI_ISL_633203, EPI_ISL_633204, EPI_ISL_633206, EPI_ISL_633207, EPI_ISL_633209, EPI_ISL_633210, EPI_ISL_633213, EPI_ISL_633222, EPI_ISL_633226, EPI_ISL_633228, EPI_ISL_633230, EPI_ISL_633232, EPI_ISL_633233, EPI_ISL_633237, EPI_ISL_633240, EPI_ISL_633241, EPI_ISL_633243, EPI_ISL_633244, EPI_ISL_633251, EPI_ISL_633256, EPI_ISL_633259, EPI_ISL_633261, EPI_ISL_633262, EPI_ISL_633268, EPI_ISL_633269, EPI_ISL_633271, EPI_ISL_633272, EPI_ISL_633274, EPI_ISL_633276, EPI_ISL_633279, EPI_ISL_633280, EPI_ISL_633283, EPI_ISL_633285, EPI_ISL_633287, EPI_ISL_633289, EPI_ISL_633290, EPI_ISL_633293, EPI_ISL_633296, EPI_ISL_633301, EPI_ISL_633303, EPI_ISL_633304, EPI_ISL_633305, EPI_ISL_633306, EPI_ISL_633307, EPI_ISL_633309, EPI_ISL_633310, EPI_ISL_633312, EPI_ISL_633316, EPI_ISL_633317, EPI_ISL_633320, EPI_ISL_633323, EPI_ISL_633326, EPI_ISL_633327, EPI_ISL_633328, EPI_ISL_633331, EPI_ISL_633332, EPI_ISL_633335, EPI_ISL_633338, EPI_ISL_633339, EPI_ISL_633342, EPI_ISL_633343, EPI_ISL_633348, EPI_ISL_633351, EPI_ISL_633352, EPI_ISL_633353, EPI_ISL_633354, EPI_ISL_633355, EPI_ISL_633358, EPI_ISL_633362, EPI_ISL_633364, EPI_ISL_633367, EPI_ISL_633368, EPI_ISL_633371, EPI_ISL_633376, EPI_ISL_633377, EPI_ISL_633379, EPI_ISL_633381, EPI_ISL_633382, EPI_ISL_633384, EPI_ISL_633386, EPI_ISL_633387, EPI_ISL_633391, EPI_ISL_633394, EPI_ISL_633396, EPI_ISL_633400, EPI_ISL_633401, EPI_ISL_633402, EPI_ISL_633404, EPI_ISL_633406, EPI_ISL_633409, EPI_ISL_633410, EPI_ISL_633411, EPI_ISL_633412, EPI_ISL_633413, EPI_ISL_633414, EPI_ISL_633415, EPI_ISL_633416, EPI_ISL_633417, EPI_ISL_633418, EPI_ISL_633419, EPI_ISL_633420, EPI_ISL_633421, EPI_ISL_633422, EPI_ISL_633423, EPI_ISL_633424, EPI_ISL_633425, EPI_ISL_633426, EPI_ISL_633427, EPI_ISL_633428, EPI_ISL_633429, EPI_ISL_633430, EPI_ISL_633431, EPI_ISL_633432, EPI_ISL_633433, EPI_ISL_633434, EPI_ISL_633435, EPI_ISL_633436, EPI_ISL_633437, EPI_ISL_633438, EPI_ISL_633439, EPI_ISL_633440, EPI_ISL_633441, EPI_ISL_633442, EPI_ISL_633443, EPI_ISL_633444, EPI_ISL_633445, EPI_ISL_633446, EPI_ISL_633447, EPI_ISL_633448, EPI_ISL_633449, EPI_ISL_633450, EPI_ISL_633451, EPI_ISL_633452, EPI_ISL_633453, EPI_ISL_633454, EPI_ISL_633455, EPI_ISL_633456, EPI_ISL_633457, EPI_ISL_633458, EPI_ISL_633459, EPI_ISL_633460, EPI_ISL_633461, EPI_ISL_633462, EPI_ISL_633463, EPI_ISL_633464, EPI_ISL_633465, EPI_ISL_633466, EPI_ISL_633467, EPI_ISL_633468, EPI_ISL_633469, EPI_ISL_633470, EPI_ISL_633471, EPI_ISL_633472, EPI_ISL_633473, EPI_ISL_633474, EPI_ISL_633475, EPI_ISL_633476, EPI_ISL_633477, EPI_ISL_633478, EPI_ISL_633479, EPI_ISL_633480, EPI_ISL_633481, EPI_ISL_633482, EPI_ISL_633483, EPI_ISL_633484, EPI_ISL_633485, EPI_ISL_633486, EPI_ISL_633487, EPI_ISL_633488, EPI_ISL_633489, EPI_ISL_633490, EPI_ISL_633491, EPI_ISL_633493, EPI_ISL_633494, EPI_ISL_633495, EPI_ISL_633496, EPI_ISL_633497, EPI_ISL_633498, EPI_ISL_633499, EPI_ISL_633500, EPI_ISL_633501, EPI_ISL_633502, EPI_ISL_633503, EPI_ISL_633504, EPI_ISL_633505, EPI_ISL_633506, EPI_ISL_633507, EPI_ISL_633508, EPI_ISL_633509, EPI_ISL_633510, EPI_ISL_633511, EPI_ISL_633512, EPI_ISL_633513, EPI_ISL_633514, EPI_ISL_633515, EPI_ISL_633516, EPI_ISL_633517, EPI_ISL_633518, EPI_ISL_633519, EPI_ISL_633520, EPI_ISL_633521, EPI_ISL_633522, EPI_ISL_633523, EPI_ISL_633524, EPI_ISL_633525, EPI_ISL_633526, EPI_ISL_633527, EPI_ISL_633528, EPI_ISL_633529, EPI_ISL_633530, EPI_ISL_633531, EPI_ISL_633532, EPI_ISL_633533, EPI_ISL_633534, EPI_ISL_633535, EPI_ISL_633536, EPI_ISL_633537, EPI_ISL_633538, EPI_ISL_633539, EPI_ISL_633540, EPI_ISL_633541, EPI_ISL_633542, EPI_ISL_633543, EPI_ISL_633544, EPI_ISL_633545, EPI_ISL_633546, EPI_ISL_633547, EPI_ISL_633548, EPI_ISL_633549, EPI_ISL_633550, EPI_ISL_633551, EPI_ISL_633552, EPI_ISL_633553, EPI_ISL_633554, EPI_ISL_633555, EPI_ISL_633556, EPI_ISL_633557, EPI_ISL_633558, EPI_ISL_633559, EPI_ISL_633560, EPI_ISL_633561, EPI_ISL_633562, EPI_ISL_633563, EPI_ISL_633564, EPI_ISL_633565, EPI_ISL_633566, EPI_ISL_633567, EPI_ISL_633568, EPI_ISL_633569, EPI_ISL_633570, EPI_ISL_633571, EPI_ISL_633572, EPI_ISL_633573, EPI_ISL_633574, EPI_ISL_633575, EPI_ISL_633576, EPI_ISL_633578, EPI_ISL_633579, EPI_ISL_633580, EPI_ISL_633581, EPI_ISL_633582, EPI_ISL_633583, EPI_ISL_633584, EPI_ISL_633585, EPI_ISL_633586, EPI_ISL_633587, EPI_ISL_633588, EPI_ISL_633589, EPI_ISL_633590, EPI_ISL_633592, EPI_ISL_633593, EPI_ISL_633594, EPI_ISL_633595, EPI_ISL_633596, EPI_ISL_633597, EPI_ISL_633598, EPI_ISL_633599, EPI_ISL_633600, EPI_ISL_633601, EPI_ISL_633602, EPI_ISL_633603, EPI_ISL_633604, EPI_ISL_633605, EPI_ISL_633606, EPI_ISL_633607, EPI_ISL_633608, EPI_ISL_633609, EPI_ISL_633610, EPI_ISL_633611, EPI_ISL_633612, EPI_ISL_633613, EPI_ISL_633614, EPI_ISL_633615, EPI_ISL_633616, EPI_ISL_633617, EPI_ISL_633618, EPI_ISL_633619, EPI_ISL_633620, EPI_ISL_633621, EPI_ISL_633622, EPI_ISL_633623, EPI_ISL_633624, EPI_ISL_633625, EPI_ISL_633626, EPI_ISL_633627, EPI_ISL_633628, EPI_ISL_633629, EPI_ISL_633630, EPI_ISL_633631, EPI_ISL_633632, EPI_ISL_633633, EPI_ISL_633634, EPI_ISL_633635, EPI_ISL_633636, EPI_ISL_633637, EPI_ISL_633638, EPI_ISL_633639, EPI_ISL_633640, EPI_ISL_633641, EPI_ISL_633642, EPI_ISL_633643, EPI_ISL_633644, EPI_ISL_633645, EPI_ISL_633646, EPI_ISL_633647, EPI_ISL_633648, EPI_ISL_633649, EPI_ISL_633650, EPI_ISL_633651, EPI_ISL_633652, EPI_ISL_633654, EPI_ISL_633655, EPI_ISL_633656, EPI_ISL_633657, EPI_ISL_633658, EPI_ISL_633659, EPI_ISL_633660, EPI_ISL_633661, EPI_ISL_633662, EPI_ISL_633663, EPI_ISL_633664, EPI_ISL_633665, EPI_ISL_633666, EPI_ISL_633667, EPI_ISL_633668, EPI_ISL_633669, EPI_ISL_633670, EPI_ISL_633671, EPI_ISL_633672, EPI_ISL_633673, EPI_ISL_633674, EPI_ISL_633675, EPI_ISL_633676, EPI_ISL_633677, EPI_ISL_633678, EPI_ISL_633679, EPI_ISL_633680, EPI_ISL_633682, EPI_ISL_633683, EPI_ISL_633684, EPI_ISL_633685, EPI_ISL_633686, EPI_ISL_633687, EPI_ISL_633688, EPI_ISL_633689, EPI_ISL_633690, EPI_ISL_633691, EPI_ISL_633692, EPI_ISL_633693, EPI_ISL_633694, EPI_ISL_633695, EPI_ISL_633696, EPI_ISL_633697, EPI_ISL_633698, EPI_ISL_633699, EPI_ISL_633701, EPI_ISL_633702, EPI_ISL_633703, EPI_ISL_633704, EPI_ISL_633705, EPI_ISL_633706, EPI_ISL_633707, EPI_ISL_633708, EPI_ISL_633709, EPI_ISL_633710, EPI_ISL_633711, EPI_ISL_633712, EPI_ISL_633713, EPI_ISL_633714, EPI_ISL_633715, EPI_ISL_633716, EPI_ISL_633717, EPI_ISL_633718, EPI_ISL_633719, EPI_ISL_633720, EPI_ISL_633721, EPI_ISL_633722, EPI_ISL_633723, EPI_ISL_633724, EPI_ISL_633725, EPI_ISL_633726, EPI_ISL_633727, EPI_ISL_633728, EPI_ISL_633729, EPI_ISL_633730, EPI_ISL_633731, EPI_ISL_633732, EPI_ISL_633733, EPI_ISL_633734, EPI_ISL_633735, EPI_ISL_633736, EPI_ISL_633737, EPI_ISL_633738, EPI_ISL_633739, EPI_ISL_633740, EPI_ISL_633741, EPI_ISL_633742, EPI_ISL_633743, EPI_ISL_633744, EPI_ISL_633745, EPI_ISL_633746, EPI_ISL_633747, EPI_ISL_633748, EPI_ISL_633749, EPI_ISL_633750, EPI_ISL_633751, EPI_ISL_633752, EPI_ISL_633753, EPI_ISL_633754, EPI_ISL_633755, EPI_ISL_633756, EPI_ISL_633757, EPI_ISL_633758, EPI_ISL_633759, EPI_ISL_633761, EPI_ISL_633762, EPI_ISL_633763, EPI_ISL_633764, EPI_ISL_633765, EPI_ISL_633766, EPI_ISL_633767, EPI_ISL_633768, EPI_ISL_633769, EPI_ISL_633770, EPI_ISL_633771, EPI_ISL_633772, EPI_ISL_633773, EPI_ISL_633774, EPI_ISL_633775, EPI_ISL_633776, EPI_ISL_633777, EPI_ISL_633778, EPI_ISL_633779, EPI_ISL_633780, EPI_ISL_633781, EPI_ISL_633783, EPI_ISL_633784, EPI_ISL_633785, EPI_ISL_633786, EPI_ISL_633787, EPI_ISL_633788, EPI_ISL_633789, EPI_ISL_633790, EPI_ISL_633791, EPI_ISL_633792, EPI_ISL_633793, EPI_ISL_633794, EPI_ISL_633795, EPI_ISL_633796, EPI_ISL_633797, EPI_ISL_633798, EPI_ISL_633800, EPI_ISL_633801, EPI_ISL_633802, EPI_ISL_633803, EPI_ISL_633804, EPI_ISL_633805, EPI_ISL_633806, EPI_ISL_633807, EPI_ISL_633808, EPI_ISL_633809, EPI_ISL_633810, EPI_ISL_633811, EPI_ISL_633812, EPI_ISL_633813, EPI_ISL_633814, EPI_ISL_633815, EPI_ISL_633816, EPI_ISL_633817, EPI_ISL_633818, EPI_ISL_633819, EPI_ISL_633820, EPI_ISL_633821, EPI_ISL_633822, EPI_ISL_633823, EPI_ISL_633824, EPI_ISL_633825, EPI_ISL_633826, EPI_ISL_633827, EPI_ISL_633828, EPI_ISL_633829, EPI_ISL_633830, EPI_ISL_633831, EPI_ISL_633832, EPI_ISL_633833, EPI_ISL_633834, EPI_ISL_633835, EPI_ISL_633836, EPI_ISL_633837, EPI_ISL_633838, EPI_ISL_633839, EPI_ISL_633840, EPI_ISL_633841, EPI_ISL_633842, EPI_ISL_633843, EPI_ISL_633844, EPI_ISL_633845, EPI_ISL_633846, EPI_ISL_633847, EPI_ISL_633848, EPI_ISL_633849, EPI_ISL_633850, EPI_ISL_633851, EPI_ISL_633852, EPI_ISL_633853, EPI_ISL_633854, EPI_ISL_633855, EPI_ISL_633856, EPI_ISL_633857, EPI_ISL_633858, EPI_ISL_633859, EPI_ISL_633860, EPI_ISL_633861, EPI_ISL_633862, EPI_ISL_633863, EPI_ISL_633864, EPI_ISL_633865, EPI_ISL_633866, EPI_ISL_633867, EPI_ISL_633868, EPI_ISL_633869, EPI_ISL_633870, EPI_ISL_633871, EPI_ISL_633872, EPI_ISL_633873, EPI_ISL_633874, EPI_ISL_633875, EPI_ISL_633876, EPI_ISL_633877, EPI_ISL_633878, EPI_ISL_633879, EPI_ISL_633880, EPI_ISL_633881, EPI_ISL_633882, EPI_ISL_633883, EPI_ISL_633884, EPI_ISL_633885, EPI_ISL_633886, EPI_ISL_633887, EPI_ISL_633888, EPI_ISL_633889, EPI_ISL_633890, EPI_ISL_633891, EPI_ISL_633892, EPI_ISL_633893, EPI_ISL_633894, EPI_ISL_633895, EPI_ISL_633896, EPI_ISL_633897, EPI_ISL_633898, EPI_ISL_633899, EPI_ISL_633900, EPI_ISL_633901, EPI_ISL_633902, EPI_ISL_633903, EPI_ISL_633904, EPI_ISL_633905, EPI_ISL_633906, EPI_ISL_633907, EPI_ISL_633908, EPI_ISL_633909, EPI_ISL_633910, EPI_ISL_633911, EPI_ISL_633912, EPI_ISL_633913, EPI_ISL_633914, EPI_ISL_633915, EPI_ISL_633916, EPI_ISL_633917, EPI_ISL_633918, EPI_ISL_633919, EPI_ISL_633920, EPI_ISL_633921, EPI_ISL_633922, EPI_ISL_633923, EPI_ISL_633924, EPI_ISL_633925, EPI_ISL_633926, EPI_ISL_633927, EPI_ISL_633928, EPI_ISL_633929, EPI_ISL_633930, EPI_ISL_633931, EPI_ISL_633932, EPI_ISL_633933, EPI_ISL_633934, EPI_ISL_633935, EPI_ISL_633936, EPI_ISL_633937, EPI_ISL_633938, EPI_ISL_633939, EPI_ISL_633940, EPI_ISL_633941, EPI_ISL_633942, EPI_ISL_633943, EPI_ISL_633944, EPI_ISL_633945, EPI_ISL_633946, EPI_ISL_633947, EPI_ISL_633948, EPI_ISL_633949, EPI_ISL_633950, EPI_ISL_633951, EPI_ISL_633952, EPI_ISL_633953, EPI_ISL_633954, EPI_ISL_633955, EPI_ISL_633956, EPI_ISL_633957, EPI_ISL_633958, EPI_ISL_633959, EPI_ISL_633960, EPI_ISL_633961, EPI_ISL_633962, EPI_ISL_633963, EPI_ISL_633964, EPI_ISL_633965, EPI_ISL_633966, EPI_ISL_633967, EPI_ISL_633968, EPI_ISL_633969, EPI_ISL_633970, EPI_ISL_633971, EPI_ISL_633972, EPI_ISL_633973, EPI_ISL_633974, EPI_ISL_633975, EPI_ISL_633976, EPI_ISL_633977, EPI_ISL_633978, EPI_ISL_633979, EPI_ISL_633980, EPI_ISL_633981, EPI_ISL_633982, EPI_ISL_633983, EPI_ISL_633984, EPI_ISL_633985, EPI_ISL_633986, EPI_ISL_633987, EPI_ISL_633988, EPI_ISL_633989, EPI_ISL_633990, EPI_ISL_633991, EPI_ISL_633992, EPI_ISL_633993, EPI_ISL_633994, EPI_ISL_633995, EPI_ISL_633996, EPI_ISL_633997, EPI_ISL_633998, EPI_ISL_633999, EPI_ISL_634000, EPI_ISL_634001, EPI_ISL_634002, EPI_ISL_634003, EPI_ISL_634004, EPI_ISL_634005, EPI_ISL_634006, EPI_ISL_634007, EPI_ISL_634008, EPI_ISL_634009, EPI_ISL_634010, EPI_ISL_634011, EPI_ISL_634012, EPI_ISL_634013, EPI_ISL_634014, EPI_ISL_634015, EPI_ISL_634016, EPI_ISL_634017, EPI_ISL_634018, EPI_ISL_634019, EPI_ISL_634020, EPI_ISL_634021, EPI_ISL_634022,

| | | | |
|---|---|---|---|
| EPI_ISL_634023, EPI_ISL_634024, EPI_ISL_634025, EPI_ISL_634026, EPI_ISL_634027, EPI_ISL_634028, EPI_ISL_634029, EPI_ISL_634030, EPI_ISL_634031, EPI_ISL_634032, EPI_ISL_634033, EPI_ISL_634034, EPI_ISL_634035, EPI_ISL_634036, EPI_ISL_634037, EPI_ISL_634038, EPI_ISL_634039, EPI_ISL_634040, EPI_ISL_634041, EPI_ISL_634042, EPI_ISL_634043, EPI_ISL_634044, EPI_ISL_634045, EPI_ISL_634046, EPI_ISL_634047, EPI_ISL_634048, EPI_ISL_634049, EPI_ISL_634050, EPI_ISL_634051, EPI_ISL_634052, EPI_ISL_634053, EPI_ISL_634054, EPI_ISL_634055, EPI_ISL_634056, EPI_ISL_634057, EPI_ISL_634058, EPI_ISL_634059, EPI_ISL_634060, EPI_ISL_634061, EPI_ISL_634062, EPI_ISL_634063, EPI_ISL_634064, EPI_ISL_634065, EPI_ISL_634066, EPI_ISL_634067, EPI_ISL_634068, EPI_ISL_634069, EPI_ISL_634070, EPI_ISL_634071, EPI_ISL_634072, EPI_ISL_634073, EPI_ISL_634074, EPI_ISL_634075, EPI_ISL_634076, EPI_ISL_634077, EPI_ISL_634078, EPI_ISL_634079, EPI_ISL_634080, EPI_ISL_634081, EPI_ISL_634082, EPI_ISL_634083, EPI_ISL_634084, EPI_ISL_634085, EPI_ISL_634086, EPI_ISL_634087, EPI_ISL_634088, EPI_ISL_634089, EPI_ISL_634090, EPI_ISL_634091, EPI_ISL_634092, EPI_ISL_634093, EPI_ISL_634094, EPI_ISL_634095, EPI_ISL_634096, EPI_ISL_634097, EPI_ISL_634098, EPI_ISL_634099, EPI_ISL_634100, EPI_ISL_634101, EPI_ISL_634102, EPI_ISL_634103, EPI_ISL_634104, EPI_ISL_634105, EPI_ISL_634106, EPI_ISL_634107, EPI_ISL_634108, EPI_ISL_634109, EPI_ISL_634110, EPI_ISL_634111, EPI_ISL_634112, EPI_ISL_634113, EPI_ISL_634114, EPI_ISL_634115, EPI_ISL_634116, EPI_ISL_634117, EPI_ISL_634118, EPI_ISL_634119, EPI_ISL_634120, EPI_ISL_634121, EPI_ISL_634122, EPI_ISL_634123, EPI_ISL_634124, EPI_ISL_634125, EPI_ISL_634126, EPI_ISL_634127, EPI_ISL_634128, EPI_ISL_634129, EPI_ISL_634130, EPI_ISL_634131, EPI_ISL_634132, EPI_ISL_634133, EPI_ISL_634134, EPI_ISL_634135, EPI_ISL_634136, EPI_ISL_634137, EPI_ISL_634138, EPI_ISL_634139, EPI_ISL_634140, EPI_ISL_634141, EPI_ISL_634142, EPI_ISL_634143, EPI_ISL_634144, EPI_ISL_634145, EPI_ISL_634146, EPI_ISL_634147, EPI_ISL_634148, EPI_ISL_634149, EPI_ISL_634150, EPI_ISL_634151, EPI_ISL_634152, EPI_ISL_634153, EPI_ISL_634154, EPI_ISL_634155, EPI_ISL_634157, EPI_ISL_634158, EPI_ISL_634159, EPI_ISL_634160, EPI_ISL_634161, EPI_ISL_634162, EPI_ISL_634163, EPI_ISL_634164, EPI_ISL_634165, EPI_ISL_634166, EPI_ISL_634167, EPI_ISL_634168, EPI_ISL_634169, EPI_ISL_634170, EPI_ISL_634171, EPI_ISL_634172, EPI_ISL_634173, EPI_ISL_634174, EPI_ISL_634175, EPI_ISL_634176, EPI_ISL_634177, EPI_ISL_634178, EPI_ISL_634179, EPI_ISL_634180, EPI_ISL_634181, EPI_ISL_634182, EPI_ISL_634183, EPI_ISL_634184, EPI_ISL_634185, EPI_ISL_634186, EPI_ISL_634187, EPI_ISL_634188, EPI_ISL_634189, EPI_ISL_634190, EPI_ISL_634191, EPI_ISL_634192, EPI_ISL_634193, EPI_ISL_634194, EPI_ISL_634195, EPI_ISL_634196, EPI_ISL_634197, EPI_ISL_634198, EPI_ISL_634199, EPI_ISL_634200, EPI_ISL_634201, EPI_ISL_634202, EPI_ISL_634203, EPI_ISL_634204, EPI_ISL_634205, EPI_ISL_634206, EPI_ISL_634207, EPI_ISL_634208, EPI_ISL_634209, EPI_ISL_634210, EPI_ISL_634211, EPI_ISL_634212, EPI_ISL_634213, EPI_ISL_634214, EPI_ISL_634215, EPI_ISL_634216, EPI_ISL_634217, EPI_ISL_634218, EPI_ISL_634219, EPI_ISL_634220, EPI_ISL_634221, EPI_ISL_634222, EPI_ISL_634223, EPI_ISL_634224, EPI_ISL_634225, EPI_ISL_634226, EPI_ISL_634227, EPI_ISL_634228, EPI_ISL_634229, EPI_ISL_634230, EPI_ISL_634231, EPI_ISL_634232, EPI_ISL_634233, EPI_ISL_634234, EPI_ISL_634235, EPI_ISL_634236, EPI_ISL_634237, EPI_ISL_634238, EPI_ISL_634239, EPI_ISL_634240, EPI_ISL_634241, EPI_ISL_634242, EPI_ISL_634243, EPI_ISL_634244, EPI_ISL_634245, EPI_ISL_634246, EPI_ISL_634247, EPI_ISL_634248, EPI_ISL_634249, EPI_ISL_634250, EPI_ISL_634251, EPI_ISL_634252, EPI_ISL_634253, EPI_ISL_634254, EPI_ISL_634255, EPI_ISL_634520, EPI_ISL_634521, EPI_ISL_634522, EPI_ISL_634523, EPI_ISL_634524, EPI_ISL_634525, EPI_ISL_634526, EPI_ISL_634527, EPI_ISL_634528, EPI_ISL_634529, EPI_ISL_634530, EPI_ISL_634531, EPI_ISL_634532, EPI_ISL_634533, EPI_ISL_634534, EPI_ISL_634535, EPI_ISL_634536, EPI_ISL_634537, EPI_ISL_634538, EPI_ISL_634539, EPI_ISL_634540, EPI_ISL_634541, EPI_ISL_634542, EPI_ISL_634543, EPI_ISL_634544, EPI_ISL_634545, EPI_ISL_634546, EPI_ISL_634547, EPI_ISL_634548, EPI_ISL_634549, EPI_ISL_634550, EPI_ISL_634551, EPI_ISL_634552, EPI_ISL_634553, EPI_ISL_634554, EPI_ISL_634555, EPI_ISL_634556, EPI_ISL_634557, EPI_ISL_634558, EPI_ISL_634559, EPI_ISL_634560, EPI_ISL_634561, EPI_ISL_634562, EPI_ISL_634563, EPI_ISL_634564, EPI_ISL_634565, EPI_ISL_634566, EPI_ISL_634567, EPI_ISL_634568, EPI_ISL_634569, EPI_ISL_634570, EPI_ISL_634571, EPI_ISL_634572, EPI_ISL_634573, EPI_ISL_634574, EPI_ISL_634575, EPI_ISL_634576, EPI_ISL_634577, EPI_ISL_634578, EPI_ISL_634579, EPI_ISL_634580, EPI_ISL_634581, EPI_ISL_634582, EPI_ISL_634583, EPI_ISL_634584, EPI_ISL_634585, EPI_ISL_634586, EPI_ISL_634587, EPI_ISL_634588, EPI_ISL_634589, EPI_ISL_634590, EPI_ISL_634591, EPI_ISL_634592, EPI_ISL_634593, EPI_ISL_634594, EPI_ISL_634595, EPI_ISL_634596, EPI_ISL_634597, EPI_ISL_634598, EPI_ISL_634599, EPI_ISL_634600, EPI_ISL_634601, EPI_ISL_634602, EPI_ISL_634603, EPI_ISL_634604, EPI_ISL_634605, EPI_ISL_634606, EPI_ISL_634607, EPI_ISL_634608, EPI_ISL_634609, EPI_ISL_634610, EPI_ISL_634611, EPI_ISL_634612, EPI_ISL_634613, EPI_ISL_634614, EPI_ISL_634615, EPI_ISL_634616, EPI_ISL_634617, EPI_ISL_634618, EPI_ISL_634619, EPI_ISL_634620, EPI_ISL_634621, EPI_ISL_634622, EPI_ISL_634623, EPI_ISL_634624, EPI_ISL_634625, EPI_ISL_634626, EPI_ISL_634627, EPI_ISL_634628, EPI_ISL_634629, EPI_ISL_634630, EPI_ISL_634631, EPI_ISL_634632, EPI_ISL_634633, EPI_ISL_634634, EPI_ISL_634635, EPI_ISL_634636, EPI_ISL_634637, EPI_ISL_634638, EPI_ISL_634639, EPI_ISL_634640, EPI_ISL_634641, EPI_ISL_634642, EPI_ISL_634643, EPI_ISL_634644, EPI_ISL_634645, EPI_ISL_634646, EPI_ISL_634647, EPI_ISL_634648, EPI_ISL_634649, EPI_ISL_634650, EPI_ISL_634651, EPI_ISL_634652, EPI_ISL_634653, EPI_ISL_634654, EPI_ISL_634655, EPI_ISL_634656, EPI_ISL_634657, EPI_ISL_634658, EPI_ISL_634659, EPI_ISL_634660, EPI_ISL_634661, EPI_ISL_634662, EPI_ISL_634663, EPI_ISL_634664, EPI_ISL_634665, EPI_ISL_634666, EPI_ISL_634667, EPI_ISL_634668, EPI_ISL_634669, EPI_ISL_634670, EPI_ISL_634671, EPI_ISL_634672, EPI_ISL_634673, EPI_ISL_634674, EPI_ISL_634675, EPI_ISL_634676, EPI_ISL_634677, EPI_ISL_634679, EPI_ISL_634680, EPI_ISL_634681, EPI_ISL_634682, EPI_ISL_634683, EPI_ISL_634684, EPI_ISL_634685, EPI_ISL_634686, EPI_ISL_634687, EPI_ISL_634688, EPI_ISL_634689, EPI_ISL_634690, EPI_ISL_634691, EPI_ISL_634692, EPI_ISL_634693, EPI_ISL_634694, EPI_ISL_634695, EPI_ISL_634696, EPI_ISL_634697, EPI_ISL_634698, EPI_ISL_634699, EPI_ISL_634700, EPI_ISL_634701, EPI_ISL_634702, EPI_ISL_634703, EPI_ISL_634704, EPI_ISL_634705, EPI_ISL_634706, EPI_ISL_634707, EPI_ISL_634708, EPI_ISL_634709, EPI_ISL_634710, EPI_ISL_634711, EPI_ISL_634712, EPI_ISL_634713, EPI_ISL_634714, EPI_ISL_634715, EPI_ISL_634716, EPI_ISL_634717, EPI_ISL_634718, EPI_ISL_634719, EPI_ISL_634720, EPI_ISL_634721, EPI_ISL_634722, EPI_ISL_634723, EPI_ISL_634724, EPI_ISL_634725, EPI_ISL_634726, EPI_ISL_634727, EPI_ISL_634728, EPI_ISL_634729, EPI_ISL_634730, EPI_ISL_634731, EPI_ISL_634732, EPI_ISL_634733, EPI_ISL_634734, EPI_ISL_634735, EPI_ISL_634736, EPI_ISL_634737, EPI_ISL_634738, EPI_ISL_634739, EPI_ISL_634740, EPI_ISL_634741, EPI_ISL_634742, EPI_ISL_634743, EPI_ISL_634744, EPI_ISL_634745, EPI_ISL_634746, EPI_ISL_634747, EPI_ISL_634748, EPI_ISL_634749, EPI_ISL_634750, EPI_ISL_634751, EPI_ISL_634752, EPI_ISL_634753 | | | |
| see above | Lighthouse Lab in Glasgow | Wellcome Sanger Institute for the COVID-19 Genomics UK (COG-UK) consortium | Anna Dominiczak and Alex Alderton; Carol Clugston; Cordelia Langford; David Gray; David K. Jackson; Dominic Kwiatkowski; Ewan Harrison; Harper VanSteenhouse; Ian Johnston; John Sillitoe on behalf of the Wellcome Sanger Institute COVID-19 Surveillance Team; John Sillitoe on behalf of the Wellcome Sanger Institute COVID-19 Surveillance Team (http://www.sanger.ac.uk/covid-team); Roberto Amato; Sonia Goncalves; Yumi Kasai |
| EPI_ISL_623214, EPI_ISL_623259, EPI_ISL_623298, EPI_ISL_623311, EPI_ISL_623313, EPI_ISL_623428, EPI_ISL_623456, EPI_ISL_623484, EPI_ISL_623515, EPI_ISL_623522, EPI_ISL_623602, EPI_ISL_629138, EPI_ISL_629199, EPI_ISL_629219, EPI_ISL_629245, EPI_ISL_629409, EPI_ISL_629490, EPI_ISL_629512, EPI_ISL_629526, EPI_ISL_629580, EPI_ISL_629590, EPI_ISL_629592, EPI_ISL_629727, EPI_ISL_630191, EPI_ISL_630219, EPI_ISL_630249, EPI_ISL_631090, EPI_ISL_631102, EPI_ISL_634266, EPI_ISL_634341, EPI_ISL_634376, EPI_ISL_634384, EPI_ISL_634385, EPI_ISL_634406, EPI_ISL_634428, EPI_ISL_634465, EPI_ISL_634483, EPI_ISL_634499, EPI_ISL_642761, EPI_ISL_642762, EPI_ISL_642763, EPI_ISL_642764, EPI_ISL_642765, EPI_ISL_642766, EPI_ISL_642767, EPI_ISL_642768, EPI_ISL_642769, EPI_ISL_642770, EPI_ISL_642771, EPI_ISL_642772, EPI_ISL_642773, EPI_ISL_642774, EPI_ISL_642775, EPI_ISL_642776, EPI_ISL_642777, EPI_ISL_642778, EPI_ISL_642779, EPI_ISL_642780, EPI_ISL_642781, EPI_ISL_642782, EPI_ISL_642783, EPI_ISL_642784, EPI_ISL_642785, EPI_ISL_642786, EPI_ISL_642787, EPI_ISL_642788, EPI_ISL_642789, EPI_ISL_642790, EPI_ISL_642791, EPI_ISL_642792, EPI_ISL_642793, EPI_ISL_642794, EPI_ISL_642795, EPI_ISL_642796, EPI_ISL_642797, EPI_ISL_642798, EPI_ISL_642799, EPI_ISL_642800, EPI_ISL_642801, EPI_ISL_642802, EPI_ISL_642803, EPI_ISL_642804, EPI_ISL_642805, EPI_ISL_642806, EPI_ISL_642807, EPI_ISL_642808, EPI_ISL_642809, EPI_ISL_642810, EPI_ISL_642811, EPI_ISL_642812, EPI_ISL_642813, EPI_ISL_642814, EPI_ISL_642815, EPI_ISL_642816, EPI_ISL_642817, EPI_ISL_642818, EPI_ISL_642819, EPI_ISL_642820, EPI_ISL_642821, EPI_ISL_642822, EPI_ISL_642823, EPI_ISL_642824, EPI_ISL_642825, EPI_ISL_642826, EPI_ISL_642827, EPI_ISL_642828, EPI_ISL_642829, EPI_ISL_642830, EPI_ISL_642831, EPI_ISL_642832, EPI_ISL_642833, EPI_ISL_642834, EPI_ISL_642835, EPI_ISL_642836, EPI_ISL_642837, EPI_ISL_642838, EPI_ISL_642839, EPI_ISL_642840, EPI_ISL_642841, EPI_ISL_642842, EPI_ISL_642843, EPI_ISL_642844, EPI_ISL_642845, EPI_ISL_642846, EPI_ISL_642847, EPI_ISL_642848, EPI_ISL_642849, EPI_ISL_642850, EPI_ISL_642851, EPI_ISL_642852, EPI_ISL_642853, EPI_ISL_642854, EPI_ISL_642855, EPI_ISL_642856, EPI_ISL_642857, EPI_ISL_642858, EPI_ISL_642859, EPI_ISL_642860, EPI_ISL_642861, EPI_ISL_642862, EPI_ISL_642863, EPI_ISL_642864, EPI_ISL_642865, EPI_ISL_642866, EPI_ISL_642867, EPI_ISL_642868, EPI_ISL_642869, EPI_ISL_642870, EPI_ISL_642871, EPI_ISL_642872, EPI_ISL_642873, EPI_ISL_642874, EPI_ISL_642875, EPI_ISL_642876, EPI_ISL_642877, EPI_ISL_642878, EPI_ISL_642879, EPI_ISL_642880, EPI_ISL_642881, EPI_ISL_642882, EPI_ISL_642883, EPI_ISL_642884, EPI_ISL_642885, EPI_ISL_642886, EPI_ISL_642887, EPI_ISL_642888, EPI_ISL_642889, EPI_ISL_642890, EPI_ISL_642891, EPI_ISL_642892, EPI_ISL_642893, EPI_ISL_642894, EPI_ISL_642895, EPI_ISL_642896, EPI_ISL_642897, EPI_ISL_642898, EPI_ISL_642899, EPI_ISL_642900, EPI_ISL_642901, EPI_ISL_642902, EPI_ISL_642903, EPI_ISL_642904, EPI_ISL_642905, EPI_ISL_642906, EPI_ISL_642907, EPI_ISL_642908, EPI_ISL_642909, EPI_ISL_642910, EPI_ISL_642911, EPI_ISL_642912, EPI_ISL_642913, EPI_ISL_642914, EPI_ISL_642915, EPI_ISL_642916, EPI_ISL_642917, EPI_ISL_642918, EPI_ISL_642919, EPI_ISL_642920, EPI_ISL_642921, EPI_ISL_642922, EPI_ISL_642923, EPI_ISL_642924, EPI_ISL_642925, EPI_ISL_642926, EPI_ISL_642927, EPI_ISL_642928, EPI_ISL_642929, EPI_ISL_642930, EPI_ISL_642931, EPI_ISL_642932, EPI_ISL_642933, EPI_ISL_642934, EPI_ISL_642935, EPI_ISL_642936, EPI_ISL_642937, EPI_ISL_642938, EPI_ISL_642939, EPI_ISL_642940, EPI_ISL_642941, EPI_ISL_642942, EPI_ISL_642943, EPI_ISL_642944, EPI_ISL_642945, EPI_ISL_642946, EPI_ISL_642947, EPI_ISL_642948, EPI_ISL_642949, EPI_ISL_642950, EPI_ISL_642951, EPI_ISL_642952, EPI_ISL_642953, EPI_ISL_642954, EPI_ISL_642955, EPI_ISL_642956, EPI_ISL_642957, EPI_ISL_642958, EPI_ISL_642959, EPI_ISL_642960, EPI_ISL_642961, EPI_ISL_642962, EPI_ISL_642963, EPI_ISL_642964, EPI_ISL_642965, EPI_ISL_642966, EPI_ISL_642967, EPI_ISL_642968, EPI_ISL_642969, EPI_ISL_642970, EPI_ISL_642971, EPI_ISL_642972, EPI_ISL_642973, EPI_ISL_642974, EPI_ISL_642975, EPI_ISL_642976, EPI_ISL_642977, EPI_ISL_642978, EPI_ISL_642979, EPI_ISL_642980, EPI_ISL_642981, EPI_ISL_642982, EPI_ISL_642983, EPI_ISL_642984, EPI_ISL_642985, EPI_ISL_642986, EPI_ISL_642987, EPI_ISL_642988, EPI_ISL_642989, EPI_ISL_642990, EPI_ISL_642991, EPI_ISL_642992, EPI_ISL_642993, EPI_ISL_642994, EPI_ISL_642995, EPI_ISL_642996, EPI_ISL_642997, EPI_ISL_642998, EPI_ISL_642999, EPI_ISL_643000, EPI_ISL_643001, EPI_ISL_643002, EPI_ISL_643003, EPI_ISL_643004, EPI_ISL_643005, EPI_ISL_643006, EPI_ISL_643007, EPI_ISL_643008, EPI_ISL_643009, EPI_ISL_643010, EPI_ISL_643011, EPI_ISL_643012, EPI_ISL_643013, EPI_ISL_643014, EPI_ISL_643015, EPI_ISL_643016, EPI_ISL_643017, EPI_ISL_643018, EPI_ISL_643019, EPI_ISL_643020, EPI_ISL_643021, EPI_ISL_643022, EPI_ISL_643023, EPI_ISL_643024, EPI_ISL_643025, EPI_ISL_643026, EPI_ISL_643027, EPI_ISL_643028, EPI_ISL_643029, EPI_ISL_643030, EPI_ISL_643031, EPI_ISL_643032, EPI_ISL_643033, EPI_ISL_643034, EPI_ISL_643035, EPI_ISL_643036, EPI_ISL_643037, EPI_ISL_643038, EPI_ISL_643039, EPI_ISL_643040, EPI_ISL_643041, EPI_ISL_643042, EPI_ISL_643043, EPI_ISL_643983, EPI_ISL_643984, EPI_ISL_643985, EPI_ISL_643986, EPI_ISL_643987, EPI_ISL_643988, EPI_ISL_643989, EPI_ISL_643990, EPI_ISL_643991, EPI_ISL_644052, EPI_ISL_644053, EPI_ISL_644054, EPI_ISL_644055, EPI_ISL_644056, EPI_ISL_644057, EPI_ISL_644058, EPI_ISL_644059, EPI_ISL_644060, EPI_ISL_644061, EPI_ISL_644062, EPI_ISL_644063, EPI_ISL_644064, EPI_ISL_644065, EPI_ISL_644066, EPI_ISL_644067, EPI_ISL_644068, EPI_ISL_644069, EPI_ISL_644070, EPI_ISL_644071, EPI_ISL_644072, EPI_ISL_644073, EPI_ISL_644074, EPI_ISL_644075, EPI_ISL_644076, EPI_ISL_644077, EPI_ISL_644078, EPI_ISL_644079, EPI_ISL_644080, EPI_ISL_644081, EPI_ISL_644082, EPI_ISL_644083, EPI_ISL_644084, EPI_ISL_644085, EPI_ISL_644086, EPI_ISL_644088, EPI_ISL_644089, EPI_ISL_644090, EPI_ISL_644091, EPI_ISL_644092, EPI_ISL_644114, EPI_ISL_644116, EPI_ISL_644119, EPI_ISL_644120, EPI_ISL_647807, EPI_ISL_647808, EPI_ISL_647809, EPI_ISL_647810, EPI_ISL_647811, EPI_ISL_647812, EPI_ISL_647813, EPI_ISL_647814, EPI_ISL_647815, EPI_ISL_647816, EPI_ISL_647817, EPI_ISL_647818, EPI_ISL_647819, EPI_ISL_647820, EPI_ISL_647821, EPI_ISL_647822, EPI_ISL_647823, EPI_ISL_647824, EPI_ISL_647825, EPI_ISL_647826, EPI_ISL_647827, EPI_ISL_647828, EPI_ISL_647829, EPI_ISL_647830, EPI_ISL_647831, EPI_ISL_647832, EPI_ISL_647833, EPI_ISL_647834, EPI_ISL_647835, EPI_ISL_647836, EPI_ISL_647837, EPI_ISL_647838, EPI_ISL_647839, EPI_ISL_647840, EPI_ISL_647841, EPI_ISL_647842, EPI_ISL_647843, EPI_ISL_647844, EPI_ISL_647845, EPI_ISL_647846, EPI_ISL_647847, EPI_ISL_647848, EPI_ISL_647849, EPI_ISL_647850, EPI_ISL_647851, EPI_ISL_647852, EPI_ISL_647853, EPI_ISL_649192, EPI_ISL_649205, EPI_ISL_649255, EPI_ISL_649283, EPI_ISL_649320, EPI_ISL_649362, EPI_ISL_649405, EPI_ISL_649479, EPI_ISL_649548, EPI_ISL_649549, EPI_ISL_649550, EPI_ISL_649551, EPI_ISL_649552, EPI_ISL_649553, EPI_ISL_649554, EPI_ISL_649555, EPI_ISL_709906, EPI_ISL_709907, EPI_ISL_709908, EPI_ISL_709909, EPI_ISL_709910, EPI_ISL_709912, EPI_ISL_709913, EPI_ISL_709915, EPI_ISL_709916, EPI_ISL_709917, EPI_ISL_709918, EPI_ISL_709919, EPI_ISL_709920, EPI_ISL_709921, EPI_ISL_709922, EPI_ISL_709923, EPI_ISL_709924, EPI_ISL_709925, EPI_ISL_709926, EPI_ISL_709927, EPI_ISL_709928, EPI_ISL_709929, EPI_ISL_709930, EPI_ISL_709931 | | | |
| see above | Lighthouse Lab in Milton Keynes | Wellcome Sanger Institute for the COVID-19 Genomics UK (COG-UK) Consortium | Cordelia Langford; David K. Jackson; Dominic Kwiatkowski; Ewan Harrison; Ian Johnston; John Sillitoe on behalf of the Wellcome Sanger Institute COVID-19 Surveillance Team; John Sillitoe on behalf of the Wellcome Sanger Institute COVID-19 Surveillance Team (http://www.sanger.ac.uk/covid-team); Roberto Amato; Sonia Goncalves; The Lighthouse Lab in Milton Keynes and Alex Alderton |
| EPI_ISL_623211, EPI_ISL_623213, EPI_ISL_623215, EPI_ISL_623216, EPI_ISL_623217, EPI_ISL_623219, EPI_ISL_623221, EPI_ISL_623223, EPI_ISL_623224, EPI_ISL_623225, EPI_ISL_623226, EPI_ISL_623227, EPI_ISL_623228, EPI_ISL_623229, EPI_ISL_623230, EPI_ISL_623231, EPI_ISL_623232, EPI_ISL_623233, EPI_ISL_623235, EPI_ISL_623236, EPI_ISL_623239, EPI_ISL_623240, EPI_ISL_623241, EPI_ISL_623242, EPI_ISL_623243, EPI_ISL_623244, EPI_ISL_623246, EPI_ISL_623249, EPI_ISL_623250, EPI_ISL_623251, EPI_ISL_623252, EPI_ISL_623253, EPI_ISL_623254, EPI_ISL_623255, EPI_ISL_623257, EPI_ISL_623258, EPI_ISL_623260, EPI_ISL_623261, EPI_ISL_623263, EPI_ISL_623264, EPI_ISL_623265, EPI_ISL_623266, EPI_ISL_623267, EPI_ISL_623269, EPI_ISL_623270, EPI_ISL_623271, EPI_ISL_623272, EPI_ISL_623274, EPI_ISL_623275, EPI_ISL_623277, EPI_ISL_623278, EPI_ISL_623279, EPI_ISL_623280, EPI_ISL_623282, EPI_ISL_623283, EPI_ISL_623284, EPI_ISL_623285, EPI_ISL_623287, EPI_ISL_623289, EPI_ISL_623290, EPI_ISL_623291, EPI_ISL_623292, EPI_ISL_623294, EPI_ISL_623295, EPI_ISL_623296, EPI_ISL_623297, EPI_ISL_623299, EPI_ISL_623300, EPI_ISL_623302, EPI_ISL_623303, EPI_ISL_623304, EPI_ISL_623305, EPI_ISL_623306, EPI_ISL_623307, EPI_ISL_623309, EPI_ISL_623310, EPI_ISL_623314, EPI_ISL_623315, EPI_ISL_623316, EPI_ISL_623317, EPI_ISL_623318, EPI_ISL_623319, EPI_ISL_623322, EPI_ISL_623324, EPI_ISL_623325, EPI_ISL_623326, EPI_ISL_623327, EPI_ISL_623328, EPI_ISL_623329, EPI_ISL_623332, EPI_ISL_623335, EPI_ISL_623336, EPI_ISL_623339, EPI_ISL_623341, EPI_ISL_623342, EPI_ISL_623348, EPI_ISL_623349, EPI_ISL_623350, EPI_ISL_623351, EPI_ISL_623352, EPI_ISL_623353, EPI_ISL_623354, EPI_ISL_623355, EPI_ISL_623356, EPI_ISL_623357, EPI_ISL_623358, EPI_ISL_623361, EPI_ISL_623362, EPI_ISL_623365, EPI_ISL_623368, EPI_ISL_623369, EPI_ISL_623371, EPI_ISL_623372, EPI_ISL_623373, EPI_ISL_623374, EPI_ISL_623375, EPI_ISL_623376, EPI_ISL_623378, EPI_ISL_623379, EPI_ISL_623380, EPI_ISL_623381, EPI_ISL_623382, EPI_ISL_623383, EPI_ISL_623384, EPI_ISL_623387, EPI_ISL_623388, EPI_ISL_623389, EPI_ISL_623391, EPI_ISL_623394, EPI_ISL_623395, EPI_ISL_623396, EPI_ISL_623399, EPI_ISL_623400, EPI_ISL_623401, EPI_ISL_623402, EPI_ISL_623403, EPI_ISL_623405, EPI_ISL_623406, EPI_ISL_623407, EPI_ISL_623408, EPI_ISL_623410, EPI_ISL_623412, EPI_ISL_623413, EPI_ISL_623414, EPI_ISL_623415, EPI_ISL_623416, EPI_ISL_623417, EPI_ISL_623418, EPI_ISL_623419, EPI_ISL_623420, EPI_ISL_623422, EPI_ISL_623423, EPI_ISL_623429, EPI_ISL_623430, EPI_ISL_623431, EPI_ISL_623432, EPI_ISL_623435, EPI_ISL_623436, EPI_ISL_623438, EPI_ISL_623439, EPI_ISL_623441, EPI_ISL_623442, EPI_ISL_623443, EPI_ISL_623446, EPI_ISL_623447, EPI_ISL_623450, EPI_ISL_623452, EPI_ISL_623453, EPI_ISL_623454, EPI_ISL_623455, EPI_ISL_623457, EPI_ISL_623458, EPI_ISL_623459, EPI_ISL_623460, EPI_ISL_623461, EPI_ISL_623462, EPI_ISL_623463, EPI_ISL_623464, EPI_ISL_623465, EPI_ISL_623466, EPI_ISL_623467, EPI_ISL_623468, EPI_ISL_623469, EPI_ISL_623470, EPI_ISL_623471, EPI_ISL_623472, EPI_ISL_623473, EPI_ISL_623476, EPI_ISL_623479, EPI_ISL_623480, EPI_ISL_623481, EPI_ISL_623483, EPI_ISL_623487, EPI_ISL_623491, EPI_ISL_623492, EPI_ISL_623493, EPI_ISL_623497, EPI_ISL_623498, EPI_ISL_623500, EPI_ISL_623501, EPI_ISL_623502, EPI_ISL_623503, EPI_ISL_623504, EPI_ISL_623505, EPI_ISL_623506, EPI_ISL_623507, EPI_ISL_623508, EPI_ISL_623509, EPI_ISL_623510, EPI_ISL_623511, EPI_ISL_623512, EPI_ISL_623513, EPI_ISL_623514, EPI_ISL_623516, EPI_ISL_623517, EPI_ISL_623518, EPI_ISL_623519, EPI_ISL_623521, EPI_ISL_623523, EPI_ISL_623524, EPI_ISL_623525, EPI_ISL_623526, EPI_ISL_623529, EPI_ISL_623530, EPI_ISL_623533, EPI_ISL_623534, EPI_ISL_623535, EPI_ISL_623536, EPI_ISL_623538, EPI_ISL_623540, EPI_ISL_623541, EPI_ISL_623542, EPI_ISL_623543, EPI_ISL_623546, EPI_ISL_623549, EPI_ISL_623552, EPI_ISL_623553, EPI_ISL_623554, EPI_ISL_623561, EPI_ISL_623563, EPI_ISL_623564, EPI_ISL_623565, EPI_ISL_623566, EPI_ISL_623571, EPI_ISL_623573, EPI_ISL_623577, EPI_ISL_623579, EPI_ISL_623580, EPI_ISL_623582, EPI_ISL_623583, EPI_ISL_623586, EPI_ISL_623588, EPI_ISL_623589, EPI_ISL_623590, EPI_ISL_623591, EPI_ISL_623594, EPI_ISL_623596, EPI_ISL_623597, EPI_ISL_623598, EPI_ISL_623599, EPI_ISL_623601, EPI_ISL_623603, EPI_ISL_623604, EPI_ISL_623606, EPI_ISL_623607, EPI_ISL_623610, EPI_ISL_623613, EPI_ISL_623614, EPI_ISL_623615, EPI_ISL_623618, EPI_ISL_623620, EPI_ISL_623621, EPI_ISL_623622, EPI_ISL_623623, EPI_ISL_623624, EPI_ISL_623625, EPI_ISL_623627, EPI_ISL_623628, EPI_ISL_623629, EPI_ISL_623631, EPI_ISL_623632, EPI_ISL_623634, EPI_ISL_623635, EPI_ISL_623636, EPI_ISL_623637, EPI_ISL_623638, EPI_ISL_623642, EPI_ISL_623646, EPI_ISL_623648, EPI_ISL_623649, EPI_ISL_623656, EPI_ISL_623659, EPI_ISL_623660, EPI_ISL_623668, EPI_ISL_623669, EPI_ISL_623670, EPI_ISL_623671, EPI_ISL_623672, EPI_ISL_623673, EPI_ISL_623676, EPI_ISL_623677, EPI_ISL_623678, EPI_ISL_623681, EPI_ISL_623682, EPI_ISL_623684, EPI_ISL_623685, EPI_ISL_623686, EPI_ISL_623687, EPI_ISL_623689, EPI_ISL_623691, EPI_ISL_623692, EPI_ISL_623693, EPI_ISL_623695, EPI_ISL_623696, EPI_ISL_623697, EPI_ISL_623698, EPI_ISL_623699, EPI_ISL_623701, EPI_ISL_623704, EPI_ISL_623705, EPI_ISL_623706, EPI_ISL_623707, EPI_ISL_623708, EPI_ISL_623709, EPI_ISL_623711, EPI_ISL_623713, EPI_ISL_623714, EPI_ISL_623715, EPI_ISL_623718, EPI_ISL_623719, EPI_ISL_623720, EPI_ISL_623722, EPI_ISL_623723, EPI_ISL_623727, EPI_ISL_623728, EPI_ISL_623729, EPI_ISL_623730, EPI_ISL_623732, EPI_ISL_623734, EPI_ISL_623737, EPI_ISL_623738, EPI_ISL_623739, EPI_ISL_623743, EPI_ISL_623744, EPI_ISL_623745, EPI_ISL_623747, EPI_ISL_623748, EPI_ISL_623752, EPI_ISL_623753, EPI_ISL_623755, EPI_ISL_623756, EPI_ISL_623757, EPI_ISL_623760, EPI_ISL_623761, EPI_ISL_623765, EPI_ISL_623766, EPI_ISL_623767, EPI_ISL_623768, EPI_ISL_623769, EPI_ISL_623770, EPI_ISL_623771, EPI_ISL_623773, EPI_ISL_623777, EPI_ISL_623778, EPI_ISL_623779, EPI_ISL_623780, EPI_ISL_623782, EPI_ISL_623783, EPI_ISL_623784, EPI_ISL_623786, EPI_ISL_623787, EPI_ISL_623789, EPI_ISL_623792, EPI_ISL_623795, EPI_ISL_623801, EPI_ISL_623803, EPI_ISL_623804, EPI_ISL_623805, EPI_ISL_623807, EPI_ISL_623813, EPI_ISL_623814, EPI_ISL_623817, EPI_ISL_623818, EPI_ISL_623819, EPI_ISL_623820, EPI_ISL_623821, EPI_ISL_623824, EPI_ISL_623825, EPI_ISL_623826, EPI_ISL_623827, EPI_ISL_623830, EPI_ISL_623835, EPI_ISL_623836, EPI_ISL_623837, EPI_ISL_623839, EPI_ISL_623842, EPI_ISL_623845, EPI_ISL_623850, EPI_ISL_623851, EPI_ISL_623852, EPI_ISL_623853, EPI_ISL_629109, EPI_ISL_629110, EPI_ISL_629111, EPI_ISL_629112, EPI_ISL_629113, EPI_ISL_629114, EPI_ISL_629115, EPI_ISL_629116, EPI_ISL_629117, EPI_ISL_629118, EPI_ISL_629119, EPI_ISL_629120, EPI_ISL_629121, EPI_ISL_629122, EPI_ISL_629123, EPI_ISL_629124, EPI_ISL_629125, EPI_ISL_629126, EPI_ISL_629127, EPI_ISL_629128, EPI_ISL_629129, EPI_ISL_629130, EPI_ISL_629131, EPI_ISL_629132, EPI_ISL_629133, EPI_ISL_629134, EPI_ISL_629135, EPI_ISL_629136, EPI_ISL_629137, EPI_ISL_629139, EPI_ISL_629140, EPI_ISL_629141, EPI_ISL_629142, EPI_ISL_629143, EPI_ISL_629144, EPI_ISL_629145, EPI_ISL_629146, EPI_ISL_629147, EPI_ISL_629148, EPI_ISL_629149, EPI_ISL_629150, EPI_ISL_629151, EPI_ISL_629152, EPI_ISL_629153, EPI_ISL_629154, EPI_ISL_629155, EPI_ISL_629156, EPI_ISL_629157, EPI_ISL_629158, EPI_ISL_629159, EPI_ISL_629160, EPI_ISL_629161, EPI_ISL_629162, EPI_ISL_629163, EPI_ISL_629164, EPI_ISL_629165, EPI_ISL_629166, EPI_ISL_629167, EPI_ISL_629168, EPI_ISL_629169, EPI_ISL_629170, EPI_ISL_629171, EPI_ISL_629172, EPI_ISL_629173, EPI_ISL_629174, EPI_ISL_629175, EPI_ISL_629176, EPI_ISL_629177, EPI_ISL_629178, EPI_ISL_629179, EPI_ISL_629180, EPI_ISL_629182, EPI_ISL_629183, EPI_ISL_629184, EPI_ISL_629185, EPI_ISL_629186, EPI_ISL_629187, EPI_ISL_629188, EPI_ISL_629189, EPI_ISL_629190, EPI_ISL_629191, EPI_ISL_629192, EPI_ISL_629193, EPI_ISL_629194, EPI_ISL_629195, EPI_ISL_629196, EPI_ISL_629197, EPI_ISL_629198, EPI_ISL_629200, EPI_ISL_629201, EPI_ISL_629202, EPI_ISL_629203, EPI_ISL_629204, EPI_ISL_629205, EPI_ISL_629206, EPI_ISL_629207, EPI_ISL_629208, EPI_ISL_629209, EPI_ISL_629210, EPI_ISL_629211, EPI_ISL_629212, EPI_ISL_629213, EPI_ISL_629214, EPI_ISL_629215, EPI_ISL_629217, EPI_ISL_629218, EPI_ISL_629220, EPI_ISL_629221, EPI_ISL_629222, EPI_ISL_629223, EPI_ISL_629224, EPI_ISL_629225, EPI_ISL_629226, EPI_ISL_629227, EPI_ISL_629228, EPI_ISL_629229, EPI_ISL_629230, EPI_ISL_629231, EPI_ISL_629232, EPI_ISL_629233, EPI_ISL_629234, EPI_ISL_629235, EPI_ISL_629236, EPI_ISL_629237, EPI_ISL_629238, EPI_ISL_629239, EPI_ISL_629240, EPI_ISL_629241, EPI_ISL_629242, EPI_ISL_629243, EPI_ISL_629244, EPI_ISL_629246, EPI_ISL_629247, EPI_ISL_629248, EPI_ISL_629249, EPI_ISL_629250, EPI_ISL_629251, EPI_ISL_629252, EPI_ISL_629253, EPI_ISL_629254, EPI_ISL_629255, EPI_ISL_629256, EPI_ISL_629257, EPI_ISL_629258, EPI_ISL_629259, EPI_ISL_629260, EPI_ISL_629261, EPI_ISL_629262, EPI_ISL_629263, EPI_ISL_629264, EPI_ISL_629265, EPI_ISL_629266, EPI_ISL_629267, EPI_ISL_629268, EPI_ISL_629269, EPI_ISL_629270, EPI_ISL_629271, EPI_ISL_629272, EPI_ISL_629273, EPI_ISL_629274, EPI_ISL_629275, EPI_ISL_629276, EPI_ISL_629277, EPI_ISL_629278, EPI_ISL_629279, EPI_ISL_629281, EPI_ISL_629282, EPI_ISL_629283, EPI_ISL_629284, EPI_ISL_629285, EPI_ISL_629286, EPI_ISL_629287, EPI_ISL_629288, EPI_ISL_629289, EPI_ISL_629290, EPI_ISL_629291, EPI_ISL_629292, EPI_ISL_629293, EPI_ISL_629294, EPI_ISL_629295, EPI_ISL_629296, EPI_ISL_629297, EPI_ISL_629298, EPI_ISL_629299, EPI_ISL_629300, EPI_ISL_629301, EPI_ISL_629302, EPI_ISL_629303, EPI_ISL_629304, EPI_ISL_629305, EPI_ISL_629306, EPI_ISL_629307, EPI_ISL_629308, EPI_ISL_629309, EPI_ISL_629310, EPI_ISL_629311, EPI_ISL_629312, EPI_ISL_629313, EPI_ISL_629314, EPI_ISL_629315, EPI_ISL_629316, EPI_ISL_629317, EPI_ISL_629318, EPI_ISL_629319, EPI_ISL_629320, EPI_ISL_629321, EPI_ISL_629322, EPI_ISL_629323, EPI_ISL_629324, EPI_ISL_629325, EPI_ISL_629326, EPI_ISL_629327, EPI_ISL_629328, EPI_ISL_629329, EPI_ISL_629330, EPI_ISL_629331, EPI_ISL_629332, EPI_ISL_629333, EPI_ISL_629334, EPI_ISL_629335, EPI_ISL_629336, EPI_ISL_629337, EPI_ISL_629338, EPI_ISL_629339, EPI_ISL_629340, EPI_ISL_629341, EPI_ISL_629342, EPI_ISL_629343, EPI_ISL_629344, EPI_ISL_629345, EPI_ISL_629346, EPI_ISL_629347, EPI_ISL_629348, EPI_ISL_629349, EPI_ISL_629350, EPI_ISL_629351, EPI_ISL_629352, EPI_ISL_629353, EPI_ISL_629354, EPI_ISL_629355, EPI_ISL_629356, EPI_ISL_629357, EPI_ISL_629358, EPI_ISL_629359, EPI_ISL_629360, EPI_ISL_629361, EPI_ISL_629362, EPI_ISL_629363, EPI_ISL_629364, EPI_ISL_629365, EPI_ISL_629366, EPI_ISL_629367, EPI_ISL_629368, EPI_ISL_629369, EPI_ISL_629370, EPI_ISL_629371, EPI_ISL_629372, EPI_ISL_629373, EPI_ISL_629374, EPI_ISL_629375, EPI_ISL_629376, EPI_ISL_629377, EPI_ISL_629378, EPI_ISL_629379, EPI_ISL_629380, EPI_ISL_629381, EPI_ISL_629383, EPI_ISL_629384, EPI_ISL_629385, EPI_ISL_629386, EPI_ISL_629387, EPI_ISL_629388, EPI_ISL_629389, EPI_ISL_629390, EPI_ISL_629391, EPI_ISL_629392, EPI_ISL_629393, EPI_ISL_629394, EPI_ISL_629396, EPI_ISL_629397, EPI_ISL_629398, EPI_ISL_629399, EPI_ISL_629400, EPI_ISL_629401, EPI_ISL_629402, EPI_ISL_629403, EPI_ISL_629404, EPI_ISL_629405, EPI_ISL_629406, EPI_ISL_629407, EPI_ISL_629408, EPI_ISL_629410, EPI_ISL_629411, EPI_ISL_629412, EPI_ISL_629413, EPI_ISL_629414, EPI_ISL_629415, EPI_ISL_629416, EPI_ISL_629417, EPI_ISL_629418, EPI_ISL_629419, EPI_ISL_629420, EPI_ISL_629421, EPI_ISL_629422, EPI_ISL_629423, EPI_ISL_629424, EPI_ISL_629425, EPI_ISL_629426, EPI_ISL_629427, EPI_ISL_629428, EPI_ISL_629429, EPI_ISL_629430, EPI_ISL_629431, EPI_ISL_629432, EPI_ISL_629433, EPI_ISL_629434, EPI_ISL_629435, EPI_ISL_629436, EPI_ISL_629437, EPI_ISL_629438, EPI_ISL_629439, EPI_ISL_629440, EPI_ISL_629441, EPI_ISL_629442, EPI_ISL_629443, EPI_ISL_629444, EPI_ISL_629445, EPI_ISL_629446, EPI_ISL_629447, EPI_ISL_629448, EPI_ISL_629449, EPI_ISL_629450, EPI_ISL_629451, EPI_ISL_629452, EPI_ISL_629453, EPI_ISL_629454, EPI_ISL_629455, EPI_ISL_629456, EPI_ISL_629457, EPI_ISL_629458, EPI_ISL_629459, EPI_ISL_629460, EPI_ISL_629461, EPI_ISL_629462, EPI_ISL_629463, EPI_ISL_629464, EPI_ISL_629465, EPI_ISL_629466, EPI_ISL_629467, EPI_ISL_629468, EPI_ISL_629469, EPI_ISL_629470, EPI_ISL_629471, EPI_ISL_629472, EPI_ISL_629473, EPI_ISL_629474, EPI_ISL_629475, EPI_ISL_629476, EPI_ISL_629477, EPI_ISL_629478, EPI_ISL_629479, EPI_ISL_629480, EPI_ISL_629481, EPI_ISL_629482, EPI_ISL_629484, EPI_ISL_629485, EPI_ISL_629486, EPI_ISL_629487, EPI_ISL_629488, EPI_ISL_629489, EPI_ISL_629491, EPI_ISL_629492, EPI_ISL_629493, EPI_ISL_629494, EPI_ISL_629495, EPI_ISL_629496, EPI_ISL_629497, EPI_ISL_629498, EPI_ISL_629499, EPI_ISL_629500, EPI_ISL_629501, EPI_ISL_629502, EPI_ISL_629503, EPI_ISL_629504, EPI_ISL_629505, EPI_ISL_629506, EPI_ISL_629507, EPI_ISL_629508, EPI_ISL_629509, EPI_ISL_629510, EPI_ISL_629511, EPI_ISL_629513, EPI_ISL_629514, EPI_ISL_629515, EPI_ISL_629516, EPI_ISL_629517, EPI_ISL_629518, EPI_ISL_629519, EPI_ISL_629520, EPI_ISL_629521, EPI_ISL_629522, EPI_ISL_629523, EPI_ISL_629524, EPI_ISL_629525, EPI_ISL_629527, EPI_ISL_629528, EPI_ISL_629529, EPI_ISL_629530, EPI_ISL_629531, EPI_ISL_629532, EPI_ISL_629533, EPI_ISL_629534, EPI_ISL_629535, EPI_ISL_629536, EPI_ISL_629537, EPI_ISL_629538, EPI_ISL_629539, EPI_ISL_629540, EPI_ISL_629541, EPI_ISL_629542, EPI_ISL_629543, EPI_ISL_629544, EPI_ISL_629545, EPI_ISL_629546, EPI_ISL_629547, EPI_ISL_629548, EPI_ISL_629549, EPI_ISL_629550, EPI_ISL_629551, EPI_ISL_629552, EPI_ISL_629553, EPI_ISL_629554, EPI_ISL_629555, EPI_ISL_629556, EPI_ISL_629557, EPI_ISL_629558, EPI_ISL_629559, EPI_ISL_629560, EPI_ISL_629561, EPI_ISL_629562, EPI_ISL_629564, EPI_ISL_629565, EPI_ISL_629566, EPI_ISL_629567, EPI_ISL_629568, EPI_ISL_629569, EPI_ISL_629570, EPI_ISL_629571, EPI_ISL_629572, EPI_ISL_629573, EPI_ISL_629574, EPI_ISL_629575, EPI_ISL_629576, EPI_ISL_629577, EPI_ISL_629578, EPI_ISL_629579, EPI_ISL_629581, EPI_ISL_629582, EPI_ISL_629583, EPI_ISL_629584, EPI_ISL_629585, EPI_ISL_629586, EPI_ISL_629587, EPI_ISL_629588, EPI_ISL_629589, EPI_ISL_629591, EPI_ISL_629593, EPI_ISL_629594, EPI_ISL_629595, EPI_ISL_629596, EPI_ISL_629597, EPI_ISL_629598, EPI_ISL_629599, EPI_ISL_629600, EPI_ISL_629601, EPI_ISL_629602, EPI_ISL_629603, EPI_ISL_629604, EPI_ISL_629605, EPI_ISL_629606, EPI_ISL_629607, EPI_ISL_629608, EPI_ISL_629609, EPI_ISL_629610, EPI_ISL_629611, EPI_ISL_629612, EPI_ISL_629613, EPI_ISL_629614, EPI_ISL_629615, EPI_ISL_629616, EPI_ISL_629617, EPI_ISL_629618, EPI_ISL_629619, EPI_ISL_629620, EPI_ISL_629621, EPI_ISL_629622, EPI_ISL_629623, EPI_ISL_629624, EPI_ISL_629625, EPI_ISL_629626, EPI_ISL_629627, EPI_ISL_629628, EPI_ISL_629629, EPI_ISL_629630, EPI_ISL_629631, EPI_ISL_629632, EPI_ISL_629633, EPI_ISL_629634, EPI_ISL_629635, EPI_ISL_629636, EPI_ISL_629637, EPI_ISL_629638, EPI_ISL_629639, EPI_ISL_629640, EPI_ISL_629641, EPI_ISL_629642, EPI_ISL_629643, EPI_ISL_629644, EPI_ISL_629645, EPI_ISL_629646, EPI_ISL_629647, EPI_ISL_629648, EPI_ISL_629649, EPI_ISL_629650, EPI_ISL_629651, EPI_ISL_629652, EPI_ISL_629653, EPI_ISL_629654, EPI_ISL_629655, EPI_ISL_629656, EPI_ISL_629657, EPI_ISL_629658, EPI_ISL_629659, EPI_ISL_629660, EPI_ISL_629661, EPI_ISL_629662, EPI_ISL_629663, EPI_ISL_629664, EPI_ISL_629665, EPI_ISL_629666, EPI_ISL_629667, EPI_ISL_629668, EPI_ISL_629669, EPI_ISL_629670, EPI_ISL_629671, EPI_ISL_629672, EPI_ISL_629673, EPI_ISL_629674, EPI_ISL_629675, EPI_ISL_629676, EPI_ISL_629677, EPI_ISL_629678, EPI_ISL_629679, EPI_ISL_629680, EPI_ISL_629681, EPI_ISL_629682, EPI_ISL_629683, EPI_ISL_629684, EPI_ISL_629685, EPI_ISL_629686, EPI_ISL_629687, EPI_ISL_629688, EPI_ISL_629689, EPI_ISL_629690, EPI_ISL_629691, EPI_ISL_629692, EPI_ISL_629693, EPI_ISL_629694, EPI_ISL_629695, EPI_ISL_629696, EPI_ISL_629697, EPI_ISL_629698, EPI_ISL_629699, EPI_ISL_629700, EPI_ISL_629701, EPI_ISL_629702, EPI_ISL_629703, EPI_ISL_629704, EPI_ISL_629705, EPI_ISL_629706, EPI_ISL_629707, EPI_ISL_629708, EPI_ISL_629709, EPI_ISL_629710, EPI_ISL_629711, EPI_ISL_629712, EPI_ISL_629713, EPI_ISL_629714, EPI_ISL_629715, EPI_ISL_629716, EPI_ISL_629717, EPI_ISL_629718, EPI_ISL_629719, EPI_ISL_629720, EPI_ISL_629721, EPI_ISL_629722, EPI_ISL_629723, EPI_ISL_629724, EPI_ISL_629725, EPI_ISL_629726, EPI_ISL_629728, EPI_ISL_629729, EPI_ISL_629730, EPI_ISL_629731, EPI_ISL_629732, EPI_ISL_629733, EPI_ISL_629734, EPI_ISL_629735, EPI_ISL_629736, EPI_ISL_629737, EPI_ISL_629738, EPI_ISL_629739, EPI_ISL_629740, EPI_ISL_629741, EPI_ISL_629742, EPI_ISL_629743, EPI_ISL_629744, EPI_ISL_629745, EPI_ISL_629746, EPI_ISL_629747, EPI_ISL_629748, EPI_ISL_629749, EPI_ISL_629750, EPI_ISL_629751, EPI_ISL_629752, EPI_ISL_629753, EPI_ISL_630007, EPI_ISL_630008, EPI_ISL_630009, EPI_ISL_630010, EPI_ISL_630011, EPI_ISL_630012, EPI_ISL_630013, EPI_ISL_630014, EPI_ISL_630015, EPI_ISL_630016, EPI_ISL_630017, EPI_ISL_630018, EPI_ISL_630019, EPI_ISL_630020, EPI_ISL_630021, EPI_ISL_630022, EPI_ISL_630023, EPI_ISL_630024, EPI_ISL_630025, EPI_ISL_630026, EPI_ISL_630027, EPI_ISL_630028, EPI_ISL_630029, EPI_ISL_630030, EPI_ISL_630031, EPI_ISL_630032, EPI_ISL_630033, EPI_ISL_630034, EPI_ISL_630035, EPI_ISL_630036, EPI_ISL_630037, EPI_ISL_630038, EPI_ISL_630039, EPI_ISL_630040, EPI_ISL_630041, EPI_ISL_630042, EPI_ISL_630043, EPI_ISL_630044, EPI_ISL_630045, EPI_ISL_630046, EPI_ISL_630047, EPI_ISL_630049, EPI_ISL_630050, EPI_ISL_630051, EPI_ISL_630052, EPI_ISL_630053, EPI_ISL_630054, EPI_ISL_630055, EPI_ISL_630056, EPI_ISL_630057, EPI_ISL_630058, EPI_ISL_630059, EPI_ISL_630060, EPI_ISL_630061, EPI_ISL_630062, EPI_ISL_630063, EPI_ISL_630064, EPI_ISL_630065, EPI_ISL_630066, EPI_ISL_630067, EPI_ISL_630068, EPI_ISL_630069, EPI_ISL_630070, EPI_ISL_630071, EPI_ISL_630072, EPI_ISL_630073, EPI_ISL_630074, EPI_ISL_630075, EPI_ISL_630076, EPI_ISL_630077, EPI_ISL_630078, EPI_ISL_630079, EPI_ISL_630080, EPI_ISL_630081, EPI_ISL_630082, EPI_ISL_630083, EPI_ISL_630084, EPI_ISL_630085, EPI_ISL_630086, EPI_ISL_630087, EPI_ISL_630089, EPI_ISL_630090, EPI_ISL_630091, EPI_ISL_630092, EPI_ISL_630093, EPI_ISL_630094, EPI_ISL_630095, EPI_ISL_630096, EPI_ISL_630097, EPI_ISL_630098, EPI_ISL_630099, EPI_ISL_630100, EPI_ISL_630101, EPI_ISL_630102, EPI_ISL_630103, EPI_ISL_630105, EPI_ISL_630106, EPI_ISL_630107, EPI_ISL_630108, EPI_ISL_630109, EPI_ISL_630110, EPI_ISL_630111, EPI_ISL_630112, EPI_ISL_630113, EPI_ISL_630114, EPI_ISL_630115, EPI_ISL_630116, EPI_ISL_630117, EPI_ISL_630118, EPI_ISL_630119, EPI_ISL_630121, EPI_ISL_630122, EPI_ISL_630123, EPI_ISL_630124, EPI_ISL_630125, EPI_ISL_630126, EPI_ISL_630127, EPI_ISL_630128, EPI_ISL_630129, EPI_ISL_630130, EPI_ISL_630131, EPI_ISL_630132, EPI_ISL_630133, EPI_ISL_630134, EPI_ISL_630135, EPI_ISL_630136, EPI_ISL_630137, EPI_ISL_630138, EPI_ISL_630140, EPI_ISL_630141, EPI_ISL_630142, EPI_ISL_630143, EPI_ISL_630144, EPI_ISL_630145, EPI_ISL_630146, EPI_ISL_630147, EPI_ISL_630148, EPI_ISL_630149, EPI_ISL_630150, EPI_ISL_630151, EPI_ISL_630152, EPI_ISL_630153, EPI_ISL_630154, EPI_ISL_630155, EPI_ISL_630156, EPI_ISL_630157, EPI_ISL_630158, EPI_ISL_630159, EPI_ISL_630161, EPI_ISL_630162, EPI_ISL_630163, EPI_ISL_630164, EPI_ISL_630165, EPI_ISL_630166, EPI_ISL_630167, EPI_ISL_630168, EPI_ISL_630169, EPI_ISL_630170, EPI_ISL_630171, EPI_ISL_630172, EPI_ISL_630173, EPI_ISL_630175, EPI_ISL_630176, EPI_ISL_630177, EPI_ISL_630178, EPI_ISL_630179, EPI_ISL_630180, EPI_ISL_630181, EPI_ISL_630182, EPI_ISL_630183, EPI_ISL_630184, EPI_ISL_630185, EPI_ISL_630186, EPI_ISL_630187, EPI_ISL_630188, EPI_ISL_630189, EPI_ISL_630190, EPI_ISL_630192, EPI_ISL_630193, EPI_ISL_630194, EPI_ISL_630195, EPI_ISL_630196, EPI_ISL_630197, EPI_ISL_630198, EPI_ISL_630199, EPI_ISL_630200, EPI_ISL_630201, EPI_ISL_630202, EPI_ISL_630203, EPI_ISL_630204, EPI_ISL_630205, EPI_ISL_630206, EPI_ISL_630207, EPI_ISL_630208, EPI_ISL_630209, EPI_ISL_630210, EPI_ISL_630211, EPI_ISL_630212, EPI_ISL_630213, EPI_ISL_630214, EPI_ISL_630215, EPI_ISL_630216, EPI_ISL_630217, EPI_ISL_630218, EPI_ISL_630220, EPI_ISL_630221, EPI_ISL_630222, EPI_ISL_630223, EPI_ISL_630224, EPI_ISL_630225, EPI_ISL_630226, EPI_ISL_630227, EPI_ISL_630228, EPI_ISL_630229, EPI_ISL_630230, EPI_ISL_630231, EPI_ISL_630232, EPI_ISL_630233, EPI_ISL_630234, EPI_ISL_630235, EPI_ISL_630236, EPI_ISL_630237, EPI_ISL_630238, EPI_ISL_630239, EPI_ISL_630240, EPI_ISL_630241, EPI_ISL_630242, EPI_ISL_630243, EPI_ISL_630244, EPI_ISL_630245, EPI_ISL_630246, EPI_ISL_630247, EPI_ISL_630248, EPI_ISL_630251, EPI_ISL_630252, EPI_ISL_630253, EPI_ISL_630254, EPI_ISL_630255, EPI_ISL_630256, EPI_ISL_630257, EPI_ISL_630258, EPI_ISL_630259, EPI_ISL_630260, EPI_ISL_630261, EPI_ISL_630262, EPI_ISL_630263, EPI_ISL_630264, EPI_ISL_630265, EPI_ISL_630266, EPI_ISL_630267, EPI_ISL_630268, EPI_ISL_630269, EPI_ISL_630270, EPI_ISL_630271, EPI_ISL_630273, EPI_ISL_630274, EPI_ISL_630275, EPI_ISL_630276, EPI_ISL_630277, EPI_ISL_630278, EPI_ISL_630279, EPI_ISL_630280, EPI_ISL_630281, EPI_ISL_630282, EPI_ISL_630283, EPI_ISL_630284, EPI_ISL_630285, EPI_ISL_630286, EPI_ISL_630287, EPI_ISL_630288, EPI_ISL_630289, EPI_ISL_630290, EPI_ISL_630291, EPI_ISL_630294, EPI_ISL_630295, EPI_ISL_630296, EPI_ISL_630297, EPI_ISL_630298, EPI_ISL_630299, EPI_ISL_630300, EPI_ISL_630301, EPI_ISL_630303, EPI_ISL_630304, EPI_ISL_630305, EPI_ISL_630306, EPI_ISL_630307, EPI_ISL_630308, EPI_ISL_630309, EPI_ISL_630310, EPI_ISL_630311, EPI_ISL_630312, EPI_ISL_630313, EPI_ISL_630314, EPI_ISL_630315, EPI_ISL_630316, EPI_ISL_630317, EPI_ISL_630318, EPI_ISL_630319, EPI_ISL_630320, EPI_ISL_630321, EPI_ISL_630322, EPI_ISL_630323, EPI_ISL_630324, EPI_ISL_630325, EPI_ISL_630326, EPI_ISL_630327, EPI_ISL_630328, EPI_ISL_630329, EPI_ISL_630330, EPI_ISL_630331, EPI_ISL_630332, EPI_ISL_630333, EPI_ISL_630334, EPI_ISL_630335, EPI_ISL_630336, EPI_ISL_630337, EPI_ISL_630338, EPI_ISL_630339, EPI_ISL_630340, EPI_ISL_630341, EPI_ISL_630342, EPI_ISL_630343, EPI_ISL_630344, EPI_ISL_630345, EPI_ISL_630346, EPI_ISL_630347, EPI_ISL_630973, EPI_ISL_630974, EPI_ISL_630978, EPI_ISL_630980, EPI_ISL_630981, EPI_ISL_630986, EPI_ISL_630987, EPI_ISL_630989, EPI_ISL_630990, EPI_ISL_630991, EPI_ISL_630993, EPI_ISL_630995, EPI_ISL_630998, EPI_ISL_631000, EPI_ISL_631002, EPI_ISL_631004, EPI_ISL_631006, EPI_ISL_631007, EPI_ISL_631009, EPI_ISL_631010, EPI_ISL_631013, EPI_ISL_631015, EPI_ISL_631019, EPI_ISL_631020, EPI_ISL_631021, EPI_ISL_631022, EPI_ISL_631024, EPI_ISL_631026, EPI_ISL_631029, EPI_ISL_631030, EPI_ISL_631032, EPI_ISL_631034, EPI_ISL_631036, EPI_ISL_631037, EPI_ISL_631040, EPI_ISL_631042, EPI_ISL_631043, EPI_ISL_631047, EPI_ISL_631048, EPI_ISL_631049, EPI_ISL_631051, EPI_ISL_631052, EPI_ISL_631054, EPI_ISL_631058, EPI_ISL_631061, EPI_ISL_631064, EPI_ISL_631065, EPI_ISL_631066, EPI_ISL_631067, EPI_ISL_631068, EPI_ISL_631070, EPI_ISL_631071, EPI_ISL_631072, EPI_ISL_631073, EPI_ISL_631075, EPI_ISL_631076, EPI_ISL_631077, EPI_ISL_631079, EPI_ISL_631082, EPI_ISL_631083, EPI_ISL_631085, EPI_ISL_631086, EPI_ISL_631087, EPI_ISL_631088, EPI_ISL_631091, EPI_ISL_631092, EPI_ISL_631095, EPI_ISL_631097, EPI_ISL_631098, EPI_ISL_631100, EPI_ISL_631103, EPI_ISL_631104, EPI_ISL_631106, EPI_ISL_631110, EPI_ISL_631112, EPI_ISL_631114, EPI_ISL_631115, EPI_ISL_631119, EPI_ISL_631120, EPI_ISL_631121, EPI_ISL_631122, EPI_ISL_631124, EPI_ISL_631125, EPI_ISL_631126, EPI_ISL_631130, EPI_ISL_631134, EPI_ISL_631136, EPI_ISL_631138, EPI_ISL_631139, EPI_ISL_631140, EPI_ISL_631141, EPI_ISL_631143, EPI_ISL_631145, EPI_ISL_631146, EPI_ISL_631147, EPI_ISL_631151, EPI_ISL_631152, EPI_ISL_631153, EPI_ISL_631156, EPI_ISL_631157, EPI_ISL_631158, EPI_ISL_631161, EPI_ISL_631162, EPI_ISL_631166, EPI_ISL_631167, EPI_ISL_631168, EPI_ISL_631169, EPI_ISL_631170, EPI_ISL_631172, EPI_ISL_631176, EPI_ISL_631178, EPI_ISL_631179, EPI_ISL_631182, EPI_ISL_631183, EPI_ISL_631186, EPI_ISL_631187, EPI_ISL_631188, EPI_ISL_631193, EPI_ISL_631195, EPI_ISL_631196, EPI_ISL_631197, EPI_ISL_631198, EPI_ISL_631200, EPI_ISL_631201, EPI_ISL_631203, EPI_ISL_631205, EPI_ISL_631206, EPI_ISL_631207, EPI_ISL_631209, EPI_ISL_631210, EPI_ISL_631211, EPI_ISL_631213, EPI_ISL_631214, EPI_ISL_631216, EPI_ISL_631218, EPI_ISL_631223, EPI_ISL_631226, EPI_ISL_631227, EPI_ISL_631229, EPI_ISL_631230, EPI_ISL_631232, EPI_ISL_631233, EPI_ISL_631234, EPI_ISL_631236, EPI_ISL_631238, EPI_ISL_631239, EPI_ISL_631240, EPI_ISL_631242, EPI_ISL_631243, EPI_ISL_631244, EPI_ISL_631246, EPI_ISL_631247, EPI_ISL_631248, EPI_ISL_631249, EPI_ISL_631252, EPI_ISL_631253, EPI_ISL_631255, EPI_ISL_631256, EPI_ISL_631258, | | | |

EPI_ISL_631259, EPI_ISL_631260, EPI_ISL_631261, EPI_ISL_631263, EPI_ISL_631264, EPI_ISL_631267, EPI_ISL_631275, EPI_ISL_634256, EPI_ISL_634257, EPI_ISL_634258, EPI_ISL_634259, EPI_ISL_634260, EPI_ISL_634261, EPI_ISL_634262, EPI_ISL_634263, EPI_ISL_634264, EPI_ISL_634265, EPI_ISL_634267, EPI_ISL_634268, EPI_ISL_634269, EPI_ISL_634270, EPI_ISL_634271, EPI_ISL_634272, EPI_ISL_634273, EPI_ISL_634274, EPI_ISL_634275, EPI_ISL_634276, EPI_ISL_634277, EPI_ISL_634278, EPI_ISL_634279, EPI_ISL_634280, EPI_ISL_634281, EPI_ISL_634282, EPI_ISL_634283, EPI_ISL_634284, EPI_ISL_634285, EPI_ISL_634286, EPI_ISL_634287, EPI_ISL_634288, EPI_ISL_634289, EPI_ISL_634290, EPI_ISL_634291, EPI_ISL_634292, EPI_ISL_634293, EPI_ISL_634294, EPI_ISL_634295, EPI_ISL_634296, EPI_ISL_634297, EPI_ISL_634298, EPI_ISL_634299, EPI_ISL_634300, EPI_ISL_634301, EPI_ISL_634302, EPI_ISL_634303, EPI_ISL_634304, EPI_ISL_634305, EPI_ISL_634306, EPI_ISL_634307, EPI_ISL_634308, EPI_ISL_634309, EPI_ISL_634311, EPI_ISL_634312, EPI_ISL_634313, EPI_ISL_634314, EPI_ISL_634315, EPI_ISL_634316, EPI_ISL_634317, EPI_ISL_634318, EPI_ISL_634319, EPI_ISL_634320, EPI_ISL_634321, EPI_ISL_634322, EPI_ISL_634323, EPI_ISL_634324, EPI_ISL_634325, EPI_ISL_634326, EPI_ISL_634327, EPI_ISL_634328, EPI_ISL_634329, EPI_ISL_634330, EPI_ISL_634331, EPI_ISL_634332, EPI_ISL_634333, EPI_ISL_634334, EPI_ISL_634335, EPI_ISL_634336, EPI_ISL_634337, EPI_ISL_634338, EPI_ISL_634339, EPI_ISL_634340, EPI_ISL_634342, EPI_ISL_634343, EPI_ISL_634344, EPI_ISL_634345, EPI_ISL_634346, EPI_ISL_634347, EPI_ISL_634348, EPI_ISL_634349, EPI_ISL_634350, EPI_ISL_634351, EPI_ISL_634352, EPI_ISL_634353, EPI_ISL_634354, EPI_ISL_634355, EPI_ISL_634356, EPI_ISL_634357, EPI_ISL_634358, EPI_ISL_634359, EPI_ISL_634360, EPI_ISL_634361, EPI_ISL_634362, EPI_ISL_634363, EPI_ISL_634364, EPI_ISL_634365, EPI_ISL_634366, EPI_ISL_634367, EPI_ISL_634368, EPI_ISL_634369, EPI_ISL_634370, EPI_ISL_634371, EPI_ISL_634372, EPI_ISL_634373, EPI_ISL_634374, EPI_ISL_634375, EPI_ISL_634377, EPI_ISL_634378, EPI_ISL_634379, EPI_ISL_634380, EPI_ISL_634381, EPI_ISL_634382, EPI_ISL_634383, EPI_ISL_634386, EPI_ISL_634387, EPI_ISL_634389, EPI_ISL_634390, EPI_ISL_634391, EPI_ISL_634393, EPI_ISL_634394, EPI_ISL_634395, EPI_ISL_634396, EPI_ISL_634397, EPI_ISL_634398, EPI_ISL_634399, EPI_ISL_634400, EPI_ISL_634401, EPI_ISL_634402, EPI_ISL_634403, EPI_ISL_634404, EPI_ISL_634405, EPI_ISL_634407, EPI_ISL_634408, EPI_ISL_634409, EPI_ISL_634410, EPI_ISL_634411, EPI_ISL_634412, EPI_ISL_634413, EPI_ISL_634414, EPI_ISL_634415, EPI_ISL_634416, EPI_ISL_634417, EPI_ISL_634418, EPI_ISL_634419, EPI_ISL_634420, EPI_ISL_634421, EPI_ISL_634422, EPI_ISL_634423, EPI_ISL_634425, EPI_ISL_634426, EPI_ISL_634427, EPI_ISL_634429, EPI_ISL_634430, EPI_ISL_634431, EPI_ISL_634432, EPI_ISL_634433, EPI_ISL_634434, EPI_ISL_634435, EPI_ISL_634436, EPI_ISL_634437, EPI_ISL_634438, EPI_ISL_634439, EPI_ISL_634440, EPI_ISL_634441, EPI_ISL_634442, EPI_ISL_634443, EPI_ISL_634444, EPI_ISL_634445, EPI_ISL_634446, EPI_ISL_634447, EPI_ISL_634448, EPI_ISL_634449, EPI_ISL_634450, EPI_ISL_634451, EPI_ISL_634452, EPI_ISL_634453, EPI_ISL_634454, EPI_ISL_634455, EPI_ISL_634456, EPI_ISL_634457, EPI_ISL_634458, EPI_ISL_634459, EPI_ISL_634460, EPI_ISL_634461, EPI_ISL_634462, EPI_ISL_634463, EPI_ISL_634464, EPI_ISL_634466, EPI_ISL_634467, EPI_ISL_634468, EPI_ISL_634469, EPI_ISL_634470, EPI_ISL_634471, EPI_ISL_634472, EPI_ISL_634473, EPI_ISL_634474, EPI_ISL_634475, EPI_ISL_634476, EPI_ISL_634477, EPI_ISL_634478, EPI_ISL_634479, EPI_ISL_634480, EPI_ISL_634481, EPI_ISL_634482, EPI_ISL_634484, EPI_ISL_634485, EPI_ISL_634486, EPI_ISL_634487, EPI_ISL_634488, EPI_ISL_634489, EPI_ISL_634490, EPI_ISL_634491, EPI_ISL_634492, EPI_ISL_634493, EPI_ISL_634494, EPI_ISL_634495, EPI_ISL_634496, EPI_ISL_634497, EPI_ISL_634498, EPI_ISL_634500, EPI_ISL_634501, EPI_ISL_634502, EPI_ISL_634503, EPI_ISL_634504, EPI_ISL_634505, EPI_ISL_634506, EPI_ISL_634507, EPI_ISL_634508, EPI_ISL_634509, EPI_ISL_634510, EPI_ISL_634511, EPI_ISL_634512, EPI_ISL_634513, EPI_ISL_634514, EPI_ISL_634515, EPI_ISL_634516, EPI_ISL_634517, EPI_ISL_634518, EPI_ISL_634519

| | | | |
|---|---|---|---|
| see above | Lighthouse Lab in Milton Keynes | Wellcome Sanger Institute for the COVID-19 Genomics UK (COG-UK) consortium | Cordelia Langford; David K. Jackson; Dominic Kwiatkowski; Ewan Harrison; Ian Johnston; John Sillitoe on behalf of the Wellcome Sanger Institute COVID-19 Surveillance Team; John Sillitoe on behalf of the Wellcome Sanger Institute COVID-19 Surveillance Team (http://www.sanger.ac.uk/covid-team); Roberto Amato; Sonia Goncalves; The Lighthouse Lab in Milton Keynes and Alex Alderton |

EPI_ISL_627109, EPI_ISL_627133, EPI_ISL_627138, EPI_ISL_637262, EPI_ISL_637265, EPI_ISL_637293, EPI_ISL_637295, EPI_ISL_637296, EPI_ISL_637303, EPI_ISL_637306, EPI_ISL_637313, EPI_ISL_637323, EPI_ISL_637325, EPI_ISL_637329, EPI_ISL_637337, EPI_ISL_637338, EPI_ISL_637339, EPI_ISL_637350, EPI_ISL_637390, EPI_ISL_637401, EPI_ISL_637438, EPI_ISL_637439, EPI_ISL_637443, EPI_ISL_637444, EPI_ISL_637874, EPI_ISL_637891, EPI_ISL_637893, EPI_ISL_637908, EPI_ISL_637910, EPI_ISL_637911, EPI_ISL_637917, EPI_ISL_637923, EPI_ISL_637925, EPI_ISL_637926, EPI_ISL_637929, EPI_ISL_637930, EPI_ISL_637933, EPI_ISL_637934, EPI_ISL_637941, EPI_ISL_637948, EPI_ISL_637950, EPI_ISL_637954, EPI_ISL_637959, EPI_ISL_637960, EPI_ISL_637961, EPI_ISL_637965, EPI_ISL_637968, EPI_ISL_637969, EPI_ISL_637970, EPI_ISL_637972, EPI_ISL_637981, EPI_ISL_637984, EPI_ISL_637987, EPI_ISL_637989, EPI_ISL_637990, EPI_ISL_637993, EPI_ISL_638004, EPI_ISL_638005, EPI_ISL_638015, EPI_ISL_638021, EPI_ISL_638023, EPI_ISL_639002, EPI_ISL_650166, EPI_ISL_650192, EPI_ISL_650194, EPI_ISL_650196, EPI_ISL_650198, EPI_ISL_650199, EPI_ISL_650202, EPI_ISL_650205, EPI_ISL_650242, EPI_ISL_650275, EPI_ISL_650315, EPI_ISL_650318, EPI_ISL_650324, EPI_ISL_650326, EPI_ISL_650341, EPI_ISL_650366, EPI_ISL_650381, EPI_ISL_650419, EPI_ISL_650440, EPI_ISL_650448, EPI_ISL_650479, EPI_ISL_650515, EPI_ISL_650529, EPI_ISL_650532, EPI_ISL_650537, EPI_ISL_650544, EPI_ISL_650655, EPI_ISL_650656, EPI_ISL_650657, EPI_ISL_650688, EPI_ISL_650694, EPI_ISL_650710, EPI_ISL_650714, EPI_ISL_650782, EPI_ISL_650847, EPI_ISL_650877, EPI_ISL_650881, EPI_ISL_650882, EPI_ISL_650886, EPI_ISL_650887, EPI_ISL_650889, EPI_ISL_650891, EPI_ISL_650893, EPI_ISL_650897, EPI_ISL_650908, EPI_ISL_650920, EPI_ISL_650924, EPI_ISL_650926, EPI_ISL_650932, EPI_ISL_650933, EPI_ISL_650934, EPI_ISL_650939, EPI_ISL_650945, EPI_ISL_651095, EPI_ISL_651114, EPI_ISL_651115, EPI_ISL_651129, EPI_ISL_651131, EPI_ISL_651138, EPI_ISL_651145, EPI_ISL_651149, EPI_ISL_651153, EPI_ISL_651161, EPI_ISL_651165, EPI_ISL_651170, EPI_ISL_651178, EPI_ISL_651196, EPI_ISL_651199, EPI_ISL_651205, EPI_ISL_651208, EPI_ISL_651225, EPI_ISL_651236, EPI_ISL_651288, EPI_ISL_651296, EPI_ISL_651312, EPI_ISL_651328, EPI_ISL_651368, EPI_ISL_651375, EPI_ISL_651376, EPI_ISL_651391, EPI_ISL_651396, EPI_ISL_651400, EPI_ISL_651407, EPI_ISL_651429, EPI_ISL_651499, EPI_ISL_652073, EPI_ISL_652074, EPI_ISL_652075, EPI_ISL_652076, EPI_ISL_652077, EPI_ISL_652085, EPI_ISL_652087, EPI_ISL_652088, EPI_ISL_664164, EPI_ISL_664232, EPI_ISL_664278, EPI_ISL_664301, EPI_ISL_664333, EPI_ISL_664338, EPI_ISL_664345, EPI_ISL_664349, EPI_ISL_664355, EPI_ISL_664364, EPI_ISL_664424, EPI_ISL_664443, EPI_ISL_664444, EPI_ISL_664445, EPI_ISL_664468, EPI_ISL_664469, EPI_ISL_664501, EPI_ISL_664523, EPI_ISL_664653, EPI_ISL_664705, EPI_ISL_664732, EPI_ISL_665128, EPI_ISL_665129, EPI_ISL_665130, EPI_ISL_665133, EPI_ISL_665139, EPI_ISL_679390, EPI_ISL_679391, EPI_ISL_702849, EPI_ISL_952951, EPI_ISL_952952, EPI_ISL_1104311, EPI_ISL_1636789, EPI_ISL_1636790, EPI_ISL_1636791, EPI_ISL_1636792, EPI_ISL_1636793, EPI_ISL_1636794, EPI_ISL_1636795, EPI_ISL_1636796, EPI_ISL_1636797, EPI_ISL_1636798, EPI_ISL_1636799, EPI_ISL_1636800, EPI_ISL_1636801, EPI_ISL_1636802, EPI_ISL_1636803, EPI_ISL_1636804, EPI_ISL_1636805, EPI_ISL_1636806, EPI_ISL_1636807, EPI_ISL_1636808, EPI_ISL_1636809, EPI_ISL_1636810, EPI_ISL_1636811, EPI_ISL_1636812, EPI_ISL_1636813

| | | | |
|---|---|---|---|
| see above | Liverpool Clinical Laboratories | COVID-19 Genomics UK (COG-UK) Consortium | A Alrezaihi; Alessandro Gerada; Alistair Darby; Angela Cowell; Anita Lucaci; Anu Chawla; Cassie Olateju; Catherine Hartley; Charlotte Nelson; Ecaterina Vamos; Elaine O'Toole; Eleanor G Bentley; Ghada T Shawli; Hermione Webster; Isabel García-Dorival; James Johnson; James P Stewart; Jenifer Manson; Joanne Watts; Jones Benjamin; Jordan J Clark; Julian Hiscox; L Luu; Lucille Rainbow; M Almsaud; Margaret Hughes; Mark Whitehead; Matthew Gemmell; Miren Iturriza-Gomara; Muhannad Alruwaili; N.P Randle; Neil Swainston; PKF Gilmore; Parul Sharma; Rebekah Penrice-Randal; Richard Eccles; Richard Gregory; Sam Haldenby; Steve Paterson; Stuart D Armstrong; Trevor Ian Robinson; Ximeng Han |

EPI_ISL_637311, EPI_ISL_637373, EPI_ISL_637374, EPI_ISL_637375, EPI_ISL_637377, EPI_ISL_637455, EPI_ISL_637522, EPI_ISL_637788, EPI_ISL_637789, EPI_ISL_637790, EPI_ISL_637791, EPI_ISL_637792, EPI_ISL_637883, EPI_ISL_637943, EPI_ISL_638557, EPI_ISL_638558, EPI_ISL_638559, EPI_ISL_650142, EPI_ISL_650359, EPI_ISL_650393, EPI_ISL_650703, EPI_ISL_651060, EPI_ISL_651061, EPI_ISL_652337, EPI_ISL_652338, EPI_ISL_652339, EPI_ISL_652340, EPI_ISL_652341, EPI_ISL_652342, EPI_ISL_665280, EPI_ISL_665300, EPI_ISL_665317, EPI_ISL_665322, EPI_ISL_665329, EPI_ISL_665388, EPI_ISL_665389, EPI_ISL_665397, EPI_ISL_665398, EPI_ISL_665407, EPI_ISL_665489, EPI_ISL_665620, EPI_ISL_665627, EPI_ISL_665628, EPI_ISL_665631, EPI_ISL_665681, EPI_ISL_665739, EPI_ISL_665784, EPI_ISL_665984, EPI_ISL_665985, EPI_ISL_665987, EPI_ISL_665988, EPI_ISL_665989, EPI_ISL_665990, EPI_ISL_665991, EPI_ISL_665992, EPI_ISL_665993, EPI_ISL_665994, EPI_ISL_665995, EPI_ISL_665996, EPI_ISL_665997, EPI_ISL_665998, EPI_ISL_666000, EPI_ISL_666001, EPI_ISL_666002, EPI_ISL_666088, EPI_ISL_666089, EPI_ISL_666090, EPI_ISL_666091, EPI_ISL_666092, EPI_ISL_666093, EPI_ISL_666094, EPI_ISL_666095, EPI_ISL_666096, EPI_ISL_666097, EPI_ISL_666098, EPI_ISL_666099, EPI_ISL_666100, EPI_ISL_702525, EPI_ISL_705226, EPI_ISL_705302, EPI_ISL_724964, EPI_ISL_819450, EPI_ISL_819451, EPI_ISL_819452, EPI_ISL_819453, EPI_ISL_1637212, EPI_ISL_1637217, EPI_ISL_1637218, EPI_ISL_1637219

| | | | |
|---|---|---|---|
| see above | Northumbria University / South Tees Hospitals NHS Foundation Trust / North Cumbria Integrated Care NHS Foundation Trust / North Tees and Hartlepool NHS Foundation Trust / Newcastle Hospitals NHS Foundation Trust | COVID-19 Genomics UK (COG-UK) Consortium | Andrew Nelson; Brendan Payne; Clive Graham; Darren L Smith; Debra Padgett; Edward Barton; Emma Swindells; Garren Scott; Gary Black; Gary Eltringham; Giles S Holt; Greg R Young; Jane Greenaway; Jennifer Collins; John Allan; Joshua Loh; Lynn Dover; Matthew Bashton; Mohammad A Tariq; Paul Baker; Sarah Essex; Steve Liggett; Wen C Yew; Yusri Taha |
| EPI_ISL_1546783, EPI_ISL_1546794 | Originating lab: Wales Specialist Virology Centre Sequencing lab: Pathogen Genomics Unit | Public Health Wales Microbiology Cardiff Wales Specialist Virology Centre | Alec Birchley; Alexander Adams; Amy Gaskin; Angela Marchbank; Bree Gatica-Wilcox; Catherine Moore; Jason Coombes; Joanne Watkins; Joel Southgate; Johnathan Evans; Laura Gifford; Lauren Gilbert; Lee Graham; Malorie Perry; Matthew Bull; Nicole Pacchiarini; Sally Corden; Sara Kumziene-Summerhayes; Sara Rey; Sarah Taylor; Simon Cottrell; Sophie Jones; Tom Connor |

EPI_ISL_651086, EPI_ISL_651087, EPI_ISL_651088, EPI_ISL_651103, EPI_ISL_651104, EPI_ISL_651105, EPI_ISL_651106, EPI_ISL_651417, EPI_ISL_651421, EPI_ISL_651431, EPI_ISL_651432, EPI_ISL_651433, EPI_ISL_651469, EPI_ISL_651470, EPI_ISL_651471, EPI_ISL_651472, EPI_ISL_651473, EPI_ISL_651474, EPI_ISL_651475, EPI_ISL_651476, EPI_ISL_651477, EPI_ISL_651478, EPI_ISL_651479, EPI_ISL_651480, EPI_ISL_651481, EPI_ISL_651482, EPI_ISL_651483, EPI_ISL_651485, EPI_ISL_651505, EPI_ISL_651506, EPI_ISL_651507, EPI_ISL_651508, EPI_ISL_651509, EPI_ISL_651510, EPI_ISL_651511, EPI_ISL_651513, EPI_ISL_651517, EPI_ISL_651525, EPI_ISL_651529, EPI_ISL_651541, EPI_ISL_651544, EPI_ISL_651545, EPI_ISL_651548, EPI_ISL_651549, EPI_ISL_651586, EPI_ISL_651587, EPI_ISL_652117, EPI_ISL_652118, EPI_ISL_652119, EPI_ISL_652120, EPI_ISL_652123, EPI_ISL_652124, EPI_ISL_652125, EPI_ISL_652126, EPI_ISL_652127, EPI_ISL_652128, EPI_ISL_652129, EPI_ISL_652130, EPI_ISL_652131, EPI_ISL_652132, EPI_ISL_652133, EPI_ISL_652134, EPI_ISL_652135, EPI_ISL_652136, EPI_ISL_652137, EPI_ISL_652138, EPI_ISL_652139, EPI_ISL_652140, EPI_ISL_652141, EPI_ISL_652142, EPI_ISL_652143, EPI_ISL_652144, EPI_ISL_652145, EPI_ISL_652146, EPI_ISL_652147, EPI_ISL_652148, EPI_ISL_652149, EPI_ISL_652150, EPI_ISL_652711, EPI_ISL_652712, EPI_ISL_652713, EPI_ISL_652714, EPI_ISL_652715, EPI_ISL_652716, EPI_ISL_652717, EPI_ISL_652718, EPI_ISL_652719, EPI_ISL_652720, EPI_ISL_652721, EPI_ISL_652722, EPI_ISL_652723, EPI_ISL_652724, EPI_ISL_652725, EPI_ISL_652726, EPI_ISL_652727, EPI_ISL_652728, EPI_ISL_652729, EPI_ISL_652730, EPI_ISL_652731, EPI_ISL_652732, EPI_ISL_652733, EPI_ISL_652734, EPI_ISL_652735, EPI_ISL_652736, EPI_ISL_652737, EPI_ISL_652738, EPI_ISL_652739, EPI_ISL_652740, EPI_ISL_652741, EPI_ISL_652742, EPI_ISL_652743, EPI_ISL_652744, EPI_ISL_652745, EPI_ISL_652746, EPI_ISL_652747, EPI_ISL_652748, EPI_ISL_652749, EPI_ISL_652750, EPI_ISL_652751, EPI_ISL_652752, EPI_ISL_652753, EPI_ISL_652754, EPI_ISL_652755, EPI_ISL_652756, EPI_ISL_652757, EPI_ISL_652758, EPI_ISL_652759, EPI_ISL_652760, EPI_ISL_652762, EPI_ISL_652763, EPI_ISL_652764, EPI_ISL_652765, EPI_ISL_652766, EPI_ISL_652767, EPI_ISL_652768, EPI_ISL_652769, EPI_ISL_652770, EPI_ISL_652771, EPI_ISL_652772, EPI_ISL_652773, EPI_ISL_652774, EPI_ISL_652775, EPI_ISL_652776, EPI_ISL_652777, EPI_ISL_652778, EPI_ISL_652779, EPI_ISL_652780, EPI_ISL_652781, EPI_ISL_652782, EPI_ISL_652783, EPI_ISL_652784, EPI_ISL_652785, EPI_ISL_652786, EPI_ISL_652787, EPI_ISL_652788, EPI_ISL_652789, EPI_ISL_652790, EPI_ISL_652791, EPI_ISL_652792, EPI_ISL_652793, EPI_ISL_652794, EPI_ISL_652795, EPI_ISL_652796, EPI_ISL_652797, EPI_ISL_652798, EPI_ISL_652799, EPI_ISL_652800, EPI_ISL_652801, EPI_ISL_652802, EPI_ISL_652803, EPI_ISL_652804, EPI_ISL_652805, EPI_ISL_652806, EPI_ISL_652807, EPI_ISL_652808, EPI_ISL_652809, EPI_ISL_652810, EPI_ISL_652811, EPI_ISL_652812, EPI_ISL_652813, EPI_ISL_652814, EPI_ISL_652815, EPI_ISL_652816, EPI_ISL_652817, EPI_ISL_652818, EPI_ISL_652819, EPI_ISL_652820, EPI_ISL_652821, EPI_ISL_652822, EPI_ISL_652823, EPI_ISL_652824, EPI_ISL_652825, EPI_ISL_652826, EPI_ISL_652827, EPI_ISL_652828, EPI_ISL_652829, EPI_ISL_652830, EPI_ISL_652831, EPI_ISL_652832, EPI_ISL_652833, EPI_ISL_652834, EPI_ISL_652835, EPI_ISL_652836, EPI_ISL_652837, EPI_ISL_652838, EPI_ISL_652839, EPI_ISL_652840, EPI_ISL_652841, EPI_ISL_652842, EPI_ISL_652843, EPI_ISL_652844, EPI_ISL_652845, EPI_ISL_652846, EPI_ISL_652847, EPI_ISL_652848, EPI_ISL_652849, EPI_ISL_652850, EPI_ISL_652851, EPI_ISL_652852, EPI_ISL_652853, EPI_ISL_652854, EPI_ISL_652855, EPI_ISL_652856, EPI_ISL_652857, EPI_ISL_652858, EPI_ISL_652859, EPI_ISL_652860, EPI_ISL_652861, EPI_ISL_652862, EPI_ISL_652863, EPI_ISL_652864, EPI_ISL_652865, EPI_ISL_652866, EPI_ISL_652867, EPI_ISL_652868, EPI_ISL_652869, EPI_ISL_652870, EPI_ISL_652871, EPI_ISL_652872, EPI_ISL_652873, EPI_ISL_652874, EPI_ISL_652875, EPI_ISL_652876, EPI_ISL_652877, EPI_ISL_652878, EPI_ISL_652879, EPI_ISL_652880, EPI_ISL_652881, EPI_ISL_652882, EPI_ISL_652883, EPI_ISL_652884, EPI_ISL_652885, EPI_ISL_652886, EPI_ISL_652887, EPI_ISL_652888, EPI_ISL_652889, EPI_ISL_652890, EPI_ISL_652891, EPI_ISL_652892, EPI_ISL_652893, EPI_ISL_652894, EPI_ISL_652895, EPI_ISL_652896, EPI_ISL_652897, EPI_ISL_652898, EPI_ISL_652899, EPI_ISL_652900, EPI_ISL_652901, EPI_ISL_652902, EPI_ISL_652903, EPI_ISL_652904, EPI_ISL_652905, EPI_ISL_652906, EPI_ISL_652907, EPI_ISL_652908, EPI_ISL_652909, EPI_ISL_652910, EPI_ISL_652911, EPI_ISL_652912, EPI_ISL_652913, EPI_ISL_652914, EPI_ISL_652915, EPI_ISL_652916, EPI_ISL_652918, EPI_ISL_652920, EPI_ISL_652921, EPI_ISL_652922, EPI_ISL_652923, EPI_ISL_652924, EPI_ISL_652925, EPI_ISL_652926, EPI_ISL_652927, EPI_ISL_652928, EPI_ISL_652929, EPI_ISL_652930, EPI_ISL_652931, EPI_ISL_652932, EPI_ISL_652933, EPI_ISL_652934, EPI_ISL_652935, EPI_ISL_652936, EPI_ISL_652937, EPI_ISL_652938, EPI_ISL_652939, EPI_ISL_652940, EPI_ISL_652941, EPI_ISL_652942, EPI_ISL_652943, EPI_ISL_652944, EPI_ISL_652945, EPI_ISL_652946, EPI_ISL_652948, EPI_ISL_652949, EPI_ISL_652950, EPI_ISL_652951, EPI_ISL_652952, EPI_ISL_652953, EPI_ISL_652954, EPI_ISL_652955, EPI_ISL_652956, EPI_ISL_652957, EPI_ISL_652958, EPI_ISL_652959, EPI_ISL_652960, EPI_ISL_652961, EPI_ISL_652962, EPI_ISL_652963, EPI_ISL_652964, EPI_ISL_652965, EPI_ISL_652966, EPI_ISL_652967, EPI_ISL_652968, EPI_ISL_652969, EPI_ISL_652970, EPI_ISL_652971, EPI_ISL_652972, EPI_ISL_652973, EPI_ISL_652974, EPI_ISL_652975, EPI_ISL_652976, EPI_ISL_652977, EPI_ISL_652978, EPI_ISL_652979, EPI_ISL_652980, EPI_ISL_652981, EPI_ISL_652982, EPI_ISL_652984, EPI_ISL_652985, EPI_ISL_652986, EPI_ISL_652987, EPI_ISL_652988, EPI_ISL_652989, EPI_ISL_652990, EPI_ISL_652991, EPI_ISL_652992, EPI_ISL_652993, EPI_ISL_652994, EPI_ISL_652995, EPI_ISL_652996, EPI_ISL_652997, EPI_ISL_652998, EPI_ISL_652999, EPI_ISL_653000, EPI_ISL_653001, EPI_ISL_653002, EPI_ISL_653003, EPI_ISL_653004, EPI_ISL_653005, EPI_ISL_653006, EPI_ISL_653007, EPI_ISL_653008, EPI_ISL_653009, EPI_ISL_653011, EPI_ISL_653012, EPI_ISL_653013, EPI_ISL_653014, EPI_ISL_653015, EPI_ISL_653016, EPI_ISL_653017, EPI_ISL_653018, EPI_ISL_653019, EPI_ISL_653020, EPI_ISL_653021, EPI_ISL_653022, EPI_ISL_653023, EPI_ISL_653024, EPI_ISL_653025, EPI_ISL_741356, EPI_ISL_741774, EPI_ISL_741775, EPI_ISL_741776, EPI_ISL_839360

| | | | |
|---|---|---|---|
| see above | Oxford Viromics, NDM, University of Oxford; Oxford University Hospitals; Basingstoke and North Hampshire Hospital | COVID-19 Genomics UK (COG-UK) Consortium | Alex Mobbs; Amy Trebes; Anita Justice; Catrin Moore; Christophe Fraser; David Bonsall; David Buck; Emma Wise; George Macintyre; Jessica Lynch; John Todd; Mariateresa de Cesare; Matilde Mori; Monique Andersson; Nathan Moore; Nick Cortes; Robert Shaw; Stephen Kidd; Tanya Golubchik; Timothy Peto |

EPI_ISL_626667, EPI_ISL_626699, EPI_ISL_626702, EPI_ISL_626755, EPI_ISL_626764, EPI_ISL_626782, EPI_ISL_626814, EPI_ISL_626854, EPI_ISL_626864, EPI_ISL_626955, EPI_ISL_626997, EPI_ISL_626999, EPI_ISL_627005, EPI_ISL_627036, EPI_ISL_627071, EPI_ISL_627072, EPI_ISL_627073, EPI_ISL_627074, EPI_ISL_627075, EPI_ISL_627076, EPI_ISL_627081, EPI_ISL_627083, EPI_ISL_627084, EPI_ISL_627121, EPI_ISL_627151, EPI_ISL_627160, EPI_ISL_627193, EPI_ISL_627214, EPI_ISL_627584, EPI_ISL_627597, EPI_ISL_627598, EPI_ISL_627599, EPI_ISL_627600, EPI_ISL_627601, EPI_ISL_627604, EPI_ISL_627605, EPI_ISL_627606, EPI_ISL_627608, EPI_ISL_627609, EPI_ISL_627610, EPI_ISL_627611, EPI_ISL_627614, EPI_ISL_627617, EPI_ISL_627618, EPI_ISL_627619, EPI_ISL_627620, EPI_ISL_627621, EPI_ISL_627622, EPI_ISL_627623, EPI_ISL_627624, EPI_ISL_627625, EPI_ISL_627626, EPI_ISL_627628, EPI_ISL_627629, EPI_ISL_627636, EPI_ISL_627641, EPI_ISL_627642, EPI_ISL_627645, EPI_ISL_627646, EPI_ISL_627647, EPI_ISL_627648, EPI_ISL_627649, EPI_ISL_627650, EPI_ISL_627651, EPI_ISL_627653, EPI_ISL_627654, EPI_ISL_627655, EPI_ISL_627657, EPI_ISL_627658, EPI_ISL_627659, EPI_ISL_627660, EPI_ISL_627661, EPI_ISL_627662, EPI_ISL_627663, EPI_ISL_637286, EPI_ISL_637864, EPI_ISL_650116, EPI_ISL_650117, EPI_ISL_650181, EPI_ISL_650190, EPI_ISL_650224, EPI_ISL_650309, EPI_ISL_650319, EPI_ISL_650322, EPI_ISL_650404, EPI_ISL_650409, EPI_ISL_650446, EPI_ISL_650464, EPI_ISL_650480, EPI_ISL_650516, EPI_ISL_650528, EPI_ISL_650541, EPI_ISL_650552, EPI_ISL_650559, EPI_ISL_650673, EPI_ISL_650716, EPI_ISL_650765, EPI_ISL_650829, EPI_ISL_650878, EPI_ISL_650879, EPI_ISL_650883, EPI_ISL_650894, EPI_ISL_650902, EPI_ISL_650912, EPI_ISL_650913, EPI_ISL_650917, EPI_ISL_650918, EPI_ISL_650931, EPI_ISL_650954, EPI_ISL_650976, EPI_ISL_651079, EPI_ISL_651082, EPI_ISL_651089, EPI_ISL_651113, EPI_ISL_651116, EPI_ISL_651124, EPI_ISL_651147, EPI_ISL_651151, EPI_ISL_651167, EPI_ISL_651168, EPI_ISL_651185, EPI_ISL_651194, EPI_ISL_651211, EPI_ISL_651214, EPI_ISL_651244, EPI_ISL_651250, EPI_ISL_651257, EPI_ISL_651258, EPI_ISL_651261, EPI_ISL_651262, EPI_ISL_651324, EPI_ISL_651338, EPI_ISL_651351, EPI_ISL_651397, EPI_ISL_651434, EPI_ISL_651515, EPI_ISL_651526, EPI_ISL_651588, EPI_ISL_652151, EPI_ISL_652152, EPI_ISL_652153, EPI_ISL_652154, EPI_ISL_652155, EPI_ISL_652156, EPI_ISL_652216, EPI_ISL_652217, EPI_ISL_652218, EPI_ISL_652219, EPI_ISL_652220, EPI_ISL_652221, EPI_ISL_652222, EPI_ISL_652223, EPI_ISL_652224, EPI_ISL_652225, EPI_ISL_652226, EPI_ISL_652227, EPI_ISL_652228, EPI_ISL_652229, EPI_ISL_652230, EPI_ISL_652231, EPI_ISL_652232, EPI_ISL_652233, EPI_ISL_652235, EPI_ISL_652236, EPI_ISL_652237, EPI_ISL_652238, EPI_ISL_652239, EPI_ISL_652240, EPI_ISL_652259, EPI_ISL_652261, EPI_ISL_652262, EPI_ISL_652263, EPI_ISL_652348, EPI_ISL_665298, EPI_ISL_665507, EPI_ISL_665729, EPI_ISL_665865, EPI_ISL_725264, EPI_ISL_725270, EPI_ISL_725274, EPI_ISL_725278, EPI_ISL_725287, EPI_ISL_725289, EPI_ISL_866803, EPI_ISL_866819, EPI_ISL_866825

| | | | |
|---|---|---|---|
| see above | Quadram Institute Bioscience | COVID-19 Genomics UK (COG-UK) Consortium | Alexander J Trotter; Alison E. Mather; Alp Aydin; Ana P. Tedim; Anastasia Kolyva; Andrew Bell; Andrew J. Page; Claire Stuart; Dave J. Baker; Gemma L. Kay; John Wain; Justin O'Grady; Leonardo de Oliveira Martins; Lizzie Meadows; Maria Diaz; Mark Webber; Muhammed Yasir; Nabil-Fareed Alikhan; Ngozi Elumogo; Nicholas M. Thomson; Rachael Stanley; Rachel Gilroy; Reenesh Prakash; Samir Dervisevic; Samuel Bloomfield; Steven Rudder; Thanh Le-Viet |

EPI_ISL_595324, EPI_ISL_595325, EPI_ISL_611582, EPI_ISL_611659, EPI_ISL_611662, EPI_ISL_611670, EPI_ISL_611676, EPI_ISL_611677, EPI_ISL_611791, EPI_ISL_611831, EPI_ISL_611833, EPI_ISL_611840, EPI_ISL_611955, EPI_ISL_611956, EPI_ISL_612192, EPI_ISL_612574, EPI_ISL_612575, EPI_ISL_612576, EPI_ISL_612577, EPI_ISL_612578, EPI_ISL_612579, EPI_ISL_612580, EPI_ISL_612581, EPI_ISL_612582, EPI_ISL_612583, EPI_ISL_612584, EPI_ISL_612585, EPI_ISL_612586, EPI_ISL_612587, EPI_ISL_612588, EPI_ISL_612589, EPI_ISL_612590, EPI_ISL_612591, EPI_ISL_612592, EPI_ISL_612593, EPI_ISL_612594, EPI_ISL_612595, EPI_ISL_612596, EPI_ISL_612597, EPI_ISL_612598, EPI_ISL_612599, EPI_ISL_612600, EPI_ISL_612601, EPI_ISL_612602, EPI_ISL_612603, EPI_ISL_612604, EPI_ISL_612605, EPI_ISL_612606, EPI_ISL_626919, EPI_ISL_627667, EPI_ISL_627668, EPI_ISL_627669, EPI_ISL_627670, EPI_ISL_627671, EPI_ISL_627672, EPI_ISL_627673, EPI_ISL_627674, EPI_ISL_627675, EPI_ISL_627676, EPI_ISL_627679, EPI_ISL_650136, EPI_ISL_650150, EPI_ISL_650154, EPI_ISL_650157, EPI_ISL_650347, EPI_ISL_650386, EPI_ISL_650449, EPI_ISL_650556, EPI_ISL_650558, EPI_ISL_650611, EPI_ISL_650641, EPI_ISL_650701, EPI_ISL_650846, EPI_ISL_650937, EPI_ISL_651072, EPI_ISL_651073, EPI_ISL_651096, EPI_ISL_651232, EPI_ISL_651242, EPI_ISL_651249, EPI_ISL_651274, EPI_ISL_651285, EPI_ISL_651293, EPI_ISL_651309, EPI_ISL_651386, EPI_ISL_651466, EPI_ISL_651504, EPI_ISL_651528, EPI_ISL_651584, EPI_ISL_651585, EPI_ISL_652184, EPI_ISL_652185, EPI_ISL_652186, EPI_ISL_652187, EPI_ISL_652188, EPI_ISL_652189, EPI_ISL_652190, EPI_ISL_652191, EPI_ISL_652192, EPI_ISL_652193, EPI_ISL_652194, EPI_ISL_652195, EPI_ISL_652196, EPI_ISL_652197, EPI_ISL_652198, EPI_ISL_652199, EPI_ISL_652200, EPI_ISL_652201, EPI_ISL_652202, EPI_ISL_652203, EPI_ISL_652204, EPI_ISL_652205, EPI_ISL_652206, EPI_ISL_652207, EPI_ISL_652208, EPI_ISL_652209, EPI_ISL_652210, EPI_ISL_652211, EPI_ISL_652212, EPI_ISL_652213, EPI_ISL_652214, EPI_ISL_652215, EPI_ISL_652628, EPI_ISL_652629, EPI_ISL_652630, EPI_ISL_652631, EPI_ISL_652632, EPI_ISL_652633, EPI_ISL_652634, EPI_ISL_652635, EPI_ISL_652636, EPI_ISL_652637, EPI_ISL_652638, EPI_ISL_652639, EPI_ISL_652640, EPI_ISL_652641, EPI_ISL_652642, EPI_ISL_652643, EPI_ISL_652644, EPI_ISL_652645, EPI_ISL_652646, EPI_ISL_652647, EPI_ISL_652648, EPI_ISL_652649, EPI_ISL_652650, EPI_ISL_652651, EPI_ISL_652652, EPI_ISL_652653, EPI_ISL_652654, EPI_ISL_652655, EPI_ISL_652656, EPI_ISL_652657, EPI_ISL_652658, EPI_ISL_652659, EPI_ISL_652660, EPI_ISL_652661, EPI_ISL_652662, EPI_ISL_652663, EPI_ISL_652664, EPI_ISL_652665, EPI_ISL_652666, EPI_ISL_652667, EPI_ISL_652668, EPI_ISL_652669, EPI_ISL_652670, EPI_ISL_652671, EPI_ISL_652672, EPI_ISL_652673, EPI_ISL_652674, EPI_ISL_665699, EPI_ISL_665761, EPI_ISL_665977, EPI_ISL_665982, EPI_ISL_679924, EPI_ISL_679925, EPI_ISL_679926, EPI_ISL_839982, EPI_ISL_839983, EPI_ISL_839984, EPI_ISL_866889

| | | | |
|---|---|---|---|
| see above | Queens Medical Centre, Clinical Microbiology Department / DeepSeq Nottingham | COVID-19 Genomics UK (COG-UK) Consortium | Christopher Moore; Fei Sang; Gemma Clark; Hannah Howson-Wells; Johnny Debebe; Jonathan Ball; Joseph Chappell; Manjinder Khakh; Matthew Carlisle; Matthew Loose; Michelle M Lister; Nadine Holmes; Patrick McClure; Theocharis Tsoleridis; Vicki M Fleming; Victoria Wright; Wendy Smith |

EPI_ISL_650270, EPI_ISL_650329, EPI_ISL_650334, EPI_ISL_650378, EPI_ISL_650452, EPI_ISL_650513, EPI_ISL_650898, EPI_ISL_650923, EPI_ISL_650930, EPI_ISL_651097, EPI_ISL_651318, EPI_ISL_651410, EPI_ISL_652304, EPI_ISL_652306, EPI_ISL_652308, EPI_ISL_652311, EPI_ISL_652312, EPI_ISL_652313, EPI_ISL_652314, EPI_ISL_652316, EPI_ISL_652317, EPI_ISL_652318, EPI_ISL_652319, EPI_ISL_652320, EPI_ISL_652321, EPI_ISL_652322, EPI_ISL_652323, EPI_ISL_652324, EPI_ISL_652325, EPI_ISL_652326, EPI_ISL_652327, EPI_ISL_652328, EPI_ISL_652329, EPI_ISL_680198, EPI_ISL_680199, EPI_ISL_680200, EPI_ISL_680201, EPI_ISL_680202, EPI_ISL_680203, EPI_ISL_680204, EPI_ISL_680205, EPI_ISL_680207, EPI_ISL_680209, EPI_ISL_680211, EPI_ISL_680213, EPI_ISL_680215, EPI_ISL_680216, EPI_ISL_680218, EPI_ISL_680219, EPI_ISL_680223, EPI_ISL_680224, EPI_ISL_680330, EPI_ISL_742123, EPI_ISL_742124, EPI_ISL_742125, EPI_ISL_763367, EPI_ISL_763400, EPI_ISL_763401, EPI_ISL_763440, EPI_ISL_763475, EPI_ISL_763482, EPI_ISL_763596, EPI_ISL_763646, EPI_ISL_764143, EPI_ISL_764148, EPI_ISL_764187, EPI_ISL_764261, EPI_ISL_764396, EPI_ISL_764397, EPI_ISL_764398, EPI_ISL_764399, EPI_ISL_764400, EPI_ISL_764401, EPI_ISL_764402, EPI_ISL_764441, EPI_ISL_764442, EPI_ISL_764443, EPI_ISL_764444, EPI_ISL_921025, EPI_ISL_921121

| | | | |
|---|---|---|---|
| see above | Regional Virus Laboratory, Belfast Health and Social Care Trust | COVID-19 Genomics UK (COG-UK) Consortium | Alison Watt; Ciara Cox; Conall McCaughey; David Simpson; Derek Fairley; James McKenna; Mairead Connor; Susan Feeney; Tanya Curran; Zoltan Molnar |

EPI_ISL_637118, EPI_ISL_637119, EPI_ISL_637120, EPI_ISL_637121, EPI_ISL_637122, EPI_ISL_637123, EPI_ISL_637124, EPI_ISL_637125, EPI_ISL_637126, EPI_ISL_637127, EPI_ISL_637128, EPI_ISL_637129, EPI_ISL_637130, EPI_ISL_637131, EPI_ISL_637132, EPI_ISL_637133, EPI_ISL_637134, EPI_ISL_637135, EPI_ISL_637136, EPI_ISL_637137, EPI_ISL_637138, EPI_ISL_637139, EPI_ISL_637140, EPI_ISL_637141, EPI_ISL_637142, EPI_ISL_637143, EPI_ISL_637144, EPI_ISL_637145, EPI_ISL_637146, EPI_ISL_637147, EPI_ISL_637148, EPI_ISL_637149, EPI_ISL_637150, EPI_ISL_637151, EPI_ISL_637152, EPI_ISL_637154, EPI_ISL_637167, EPI_ISL_639821, EPI_ISL_649818, EPI_ISL_649823, EPI_ISL_649825, EPI_ISL_649826, EPI_ISL_649827, EPI_ISL_649831, EPI_ISL_649832, EPI_ISL_649833, EPI_ISL_649834, EPI_ISL_649835, EPI_ISL_649836, EPI_ISL_649837, EPI_ISL_649838, EPI_ISL_649839, EPI_ISL_649840, EPI_ISL_649841, EPI_ISL_649842, EPI_ISL_649843, EPI_ISL_649844, EPI_ISL_649845, EPI_ISL_649846, EPI_ISL_649847, EPI_ISL_649848, EPI_ISL_649849, EPI_ISL_649850, EPI_ISL_649851, EPI_ISL_649852, EPI_ISL_649853, EPI_ISL_649854, EPI_ISL_649855, EPI_ISL_649856, EPI_ISL_649857, EPI_ISL_649858, EPI_ISL_649859, EPI_ISL_649860, EPI_ISL_649861, EPI_ISL_649862, EPI_ISL_649863, EPI_ISL_649864, EPI_ISL_649865, EPI_ISL_649866, EPI_ISL_649867, EPI_ISL_649868, EPI_ISL_649869, EPI_ISL_649870, EPI_ISL_649871, EPI_ISL_649872, EPI_ISL_649873, EPI_ISL_649874, EPI_ISL_649875, EPI_ISL_649876, EPI_ISL_649877, EPI_ISL_649878, EPI_ISL_649879, EPI_ISL_649880, EPI_ISL_649881, EPI_ISL_649882, EPI_ISL_649883, EPI_ISL_649884, EPI_ISL_649885, EPI_ISL_649886, EPI_ISL_649887, EPI_ISL_649888, EPI_ISL_653933, EPI_ISL_653935, EPI_ISL_653936, EPI_ISL_653939, EPI_ISL_653940, EPI_ISL_653941, EPI_ISL_653943, EPI_ISL_653944, EPI_ISL_653945, EPI_ISL_653946, EPI_ISL_653948, EPI_ISL_653949, EPI_ISL_653955, EPI_ISL_653956, EPI_ISL_653957, EPI_ISL_653958, EPI_ISL_653959, EPI_ISL_653960, EPI_ISL_653961, EPI_ISL_653962, EPI_ISL_653963, EPI_ISL_653964, EPI_ISL_653965, EPI_ISL_653966, EPI_ISL_653967, EPI_ISL_653968, EPI_ISL_653969, EPI_ISL_653973, EPI_ISL_653975, EPI_ISL_653977, EPI_ISL_653978, EPI_ISL_660547, EPI_ISL_660548, EPI_ISL_660549, EPI_ISL_660550, EPI_ISL_660601, EPI_ISL_660602, EPI_ISL_660603, EPI_ISL_660746, EPI_ISL_660747, EPI_ISL_660748, EPI_ISL_660749, EPI_ISL_660750, EPI_ISL_660751, EPI_ISL_660752, EPI_ISL_668445, EPI_ISL_668446, EPI_ISL_678662

| | | | |
|---|---|---|---|
| see above | Respiratory Virus Unit, Microbiology Services Colindale, Public Health England | COVID-19 Genomics UK (COG-UK) Consortium | PHE Covid Sequencing Team |

EPI_ISL_613634, EPI_ISL_613635, EPI_ISL_622878, EPI_ISL_622879, EPI_ISL_622881, EPI_ISL_622883, EPI_ISL_622886, EPI_ISL_622891, EPI_ISL_636716, EPI_ISL_636717, EPI_ISL_636719, EPI_ISL_636720, EPI_ISL_636721, EPI_ISL_636722, EPI_ISL_636723, EPI_ISL_636725, EPI_ISL_636726, EPI_ISL_636727, EPI_ISL_636729, EPI_ISL_636730

| | | | |
|---|---|---|---|
| see above | Respiratory Virus Unit, Microbiology Services Colindale, Public Health England | Respiratory Virus Unit, Microbiology Services Colindale, Public Health England | PHE Covid Sequencing Team |

EPI_ISL_766083, EPI_ISL_825548, EPI_ISL_825583, EPI_ISL_825584, EPI_ISL_825585, EPI_ISL_825587, EPI_ISL_825588

| | | | |
|---|---|---|---|
| see above | Respiratory Virus Unit, National Infection Service, Public Health England | COVID-19 Genomics UK (COG-UK) Consortium | PHE Covid Sequencing Team |

EPI_ISL_664147, EPI_ISL_664335, EPI_ISL_664342, EPI_ISL_664356, EPI_ISL_664472, EPI_ISL_664517, EPI_ISL_664711, EPI_ISL_664724, EPI_ISL_664753, EPI_ISL_664754, EPI_ISL_664839, EPI_ISL_665203, EPI_ISL_665204, EPI_ISL_665205, EPI_ISL_665212, EPI_ISL_665213, EPI_ISL_665214, EPI_ISL_665215, EPI_ISL_665216, EPI_ISL_665217, EPI_ISL_665218, EPI_ISL_665219, EPI_ISL_665220, EPI_ISL_665221, EPI_ISL_665222, EPI_ISL_665224, EPI_ISL_665225, EPI_ISL_665226, EPI_ISL_665227, EPI_ISL_665228, EPI_ISL_665229, EPI_ISL_665230, EPI_ISL_665231, EPI_ISL_665232, EPI_ISL_665233, EPI_ISL_665234, EPI_ISL_665235, EPI_ISL_665236, EPI_ISL_665597

| | | | |
|---|---|---|---|
| see above | University College London Hospital | COVID-19 Genomics UK (COG-UK) Consortium | Catherine Houlihan; Dan Frampton; Judith Heaney; Matthew Byott; Moira Spyer and Eleni Nastouli; Stuart Kirk |

EPI_ISL_611527, EPI_ISL_611823, EPI_ISL_612000, EPI_ISL_612105, EPI_ISL_612451, EPI_ISL_637895, EPI_ISL_637912, EPI_ISL_638003, EPI_ISL_638031, EPI_ISL_638032, EPI_ISL_651240, EPI_ISL_651253, EPI_ISL_665674, EPI_ISL_665695, EPI_ISL_665711, EPI_ISL_679457, EPI_ISL_1637134, EPI_ISL_1730334

| | | | |
|---|---|---|---|
| see above | University College London, Great Ormond Street Hospital for Children NHS Foundation Trust, Imperial College Healthcare NHS Trust | COVID-19 Genomics UK (COG-UK) Consortium | Alison Holmes; Charlotte Williams; Helena Tutill; Jacqueline Findlay; James Price; Judith Breuer; Julianne Brown; Kathryn Harris; Leysa Forrest; Mark Kristiansen; Paola Niola; Paola Resende Silva; Patricia Dyal; Paul Randell; Rachel Williams; Samuel Weeks; Sergi Castellano; Sunando Roy; Tony Brooks; Yasmin Panchbhaya |

EPI_ISL_650172, EPI_ISL_650271, EPI_ISL_650418, EPI_ISL_650463, EPI_ISL_650790, EPI_ISL_650824, EPI_ISL_651287, EPI_ISL_651413, EPI_ISL_651597, EPI_ISL_651598, EPI_ISL_651599, EPI_ISL_651602, EPI_ISL_651604, EPI_ISL_651610, EPI_ISL_651611, EPI_ISL_651612, EPI_ISL_651613, EPI_ISL_651614, EPI_ISL_651615, EPI_ISL_651616, EPI_ISL_651617, EPI_ISL_651618, EPI_ISL_651620, EPI_ISL_651621, EPI_ISL_812971, EPI_ISL_812972, EPI_ISL_812973, EPI_ISL_812974, EPI_ISL_812975, EPI_ISL_812976, EPI_ISL_812977, EPI_ISL_812978, EPI_ISL_812979, EPI_ISL_812980, EPI_ISL_812981, EPI_ISL_812982, EPI_ISL_812983

| | | | |
|---|---|---|---|
| see above | University of Birmingham | COVID-19 Genomics UK (COG-UK) Consortium | Alex Richter; Andrew Bosworth; Andrew Bosworth. Queen Elizabeth Hospital: Anna Casey; Andrew D Beggs PHE Heartlands Lab: Husam Osman; Charlotte Poxon; Institute of Microbiology; Joanne Stockton; Josh Quick; Kasun Wanigasooriya; Mike Kidd; Nicholas Loman. University of Birmingham Testing Laboratory: Celina M Whalley; Oliver Pickles; Radoslaw Poplawski; Samuel Nicholls; University of Birmingham: Claire McMurray; Will Rowe |

EPI_ISL_626713, EPI_ISL_626730, EPI_ISL_626777, EPI_ISL_626802, EPI_ISL_626889, EPI_ISL_626946, EPI_ISL_627194, EPI_ISL_627430, EPI_ISL_627434, EPI_ISL_650225, EPI_ISL_650260, EPI_ISL_650261, EPI_ISL_650296, EPI_ISL_650360, EPI_ISL_650361, EPI_ISL_650445, EPI_ISL_650643, EPI_ISL_650644, EPI_ISL_650645, EPI_ISL_650788, EPI_ISL_651075, EPI_ISL_651143, EPI_ISL_651231, EPI_ISL_651300, EPI_ISL_651372, EPI_ISL_651384, EPI_ISL_651467, EPI_ISL_651468, EPI_ISL_652675, EPI_ISL_652677, EPI_ISL_652678, EPI_ISL_652680, EPI_ISL_652681, EPI_ISL_652682, EPI_ISL_652683, EPI_ISL_652684, EPI_ISL_652685, EPI_ISL_652686, EPI_ISL_652687, EPI_ISL_652688, EPI_ISL_652690, EPI_ISL_652691, EPI_ISL_652692, EPI_ISL_652693, EPI_ISL_652694, EPI_ISL_652695, EPI_ISL_652696, EPI_ISL_652697, EPI_ISL_652698, EPI_ISL_652699, EPI_ISL_652700, EPI_ISL_652702, EPI_ISL_652703, EPI_ISL_652704, EPI_ISL_652705, EPI_ISL_652706, EPI_ISL_652707, EPI_ISL_652708, EPI_ISL_652710, EPI_ISL_664123, EPI_ISL_664124, EPI_ISL_664127, EPI_ISL_664128, EPI_ISL_664129, EPI_ISL_664208, EPI_ISL_664209, EPI_ISL_664215, EPI_ISL_664218, EPI_ISL_664288, EPI_ISL_664318, EPI_ISL_664331, EPI_ISL_664371, EPI_ISL_664423, EPI_ISL_664458, EPI_ISL_664474, EPI_ISL_664491, EPI_ISL_664603, EPI_ISL_664612, EPI_ISL_664683, EPI_ISL_664686, EPI_ISL_665054, EPI_ISL_665055, EPI_ISL_665056, EPI_ISL_665057, EPI_ISL_665058, EPI_ISL_665059, EPI_ISL_665060, EPI_ISL_665061, EPI_ISL_665062, EPI_ISL_665064, EPI_ISL_665065, EPI_ISL_665422, EPI_ISL_665719, EPI_ISL_997307, EPI_ISL_997308, EPI_ISL_1247536

| | | | |
|---|---|---|---|
| see above | University of Exeter | COVID-19 Genomics UK (COG-UK) Consortium | Aaron Jeffries; Audrey Farbos; Ben Temperton; Jane Masoli; Joanna Warwick-Dugdale; Michelle Michelsen; Robyn Manley; Stephen Michell |
| EPI_ISL_1104230, EPI_ISL_1104233, EPI_ISL_1104260, EPI_ISL_1104261, EPI_ISL_1177690 | Virology Department, Royal Infirmary of Edinburgh, NHS Lothian / School of Biological Sciences, University of Edinburgh | COVID-19 Genomics UK (COG-UK) Consortium | Colquhoun R; Cotton S; Dewar R; Hill V; Jackson B; McCrone JT; McHugh M; O'Toole Á; Rambaut A; Rooke S; Scher E; Templeton K; Yu X |

EPI_ISL_650138, EPI_ISL_650152, EPI_ISL_650159, EPI_ISL_650165, EPI_ISL_650167, EPI_ISL_650173, EPI_ISL_650177, EPI_ISL_650179, EPI_ISL_650180, EPI_ISL_650182, EPI_ISL_650191, EPI_ISL_650220, EPI_ISL_650255, EPI_ISL_650263, EPI_ISL_650264, EPI_ISL_650277, EPI_ISL_650280, EPI_ISL_650300, EPI_ISL_650301, EPI_ISL_650302, EPI_ISL_650303, EPI_ISL_650304, EPI_ISL_650305, EPI_ISL_650306, EPI_ISL_650307, EPI_ISL_650308, EPI_ISL_650426, EPI_ISL_650499, EPI_ISL_650500, EPI_ISL_650618, EPI_ISL_650690, EPI_ISL_650775, EPI_ISL_650776, EPI_ISL_650777, EPI_ISL_650778, EPI_ISL_650793, EPI_ISL_650794, EPI_ISL_650795, EPI_ISL_650796, EPI_ISL_650797, EPI_ISL_650798, EPI_ISL_650814, EPI_ISL_650815, EPI_ISL_650819, EPI_ISL_650834, EPI_ISL_651012, EPI_ISL_651120, EPI_ISL_651171, EPI_ISL_651221, EPI_ISL_651228, EPI_ISL_651299, EPI_ISL_651301, EPI_ISL_651316, EPI_ISL_651320, EPI_ISL_651322, EPI_ISL_651343, EPI_ISL_651350, EPI_ISL_651366, EPI_ISL_651395, EPI_ISL_651425, EPI_ISL_651426, EPI_ISL_651521, EPI_ISL_651522, EPI_ISL_651960, EPI_ISL_651961, EPI_ISL_651962, EPI_ISL_651963, EPI_ISL_651964, EPI_ISL_651965, EPI_ISL_651966, EPI_ISL_651967, EPI_ISL_651968, EPI_ISL_651969, EPI_ISL_651970, EPI_ISL_651972, EPI_ISL_651973, EPI_ISL_651974, EPI_ISL_651975, EPI_ISL_651976, EPI_ISL_651977, EPI_ISL_651978, EPI_ISL_651979, EPI_ISL_651980, EPI_ISL_651981, EPI_ISL_651982, EPI_ISL_651983, EPI_ISL_651984, EPI_ISL_651985, EPI_ISL_651986, EPI_ISL_651987, EPI_ISL_651988, EPI_ISL_651989, EPI_ISL_651990, EPI_ISL_651991, EPI_ISL_651992, EPI_ISL_651993, EPI_ISL_651994, EPI_ISL_651995, EPI_ISL_651996, EPI_ISL_651997, EPI_ISL_651998, EPI_ISL_651999, EPI_ISL_652000, EPI_ISL_652001, EPI_ISL_652002, EPI_ISL_652003, EPI_ISL_652004, EPI_ISL_652005, EPI_ISL_652006, EPI_ISL_652007, EPI_ISL_652008, EPI_ISL_652009, EPI_ISL_652010, EPI_ISL_652011, EPI_ISL_652012, EPI_ISL_652015, EPI_ISL_652041, EPI_ISL_652042, EPI_ISL_652043, EPI_ISL_652044, EPI_ISL_652045, EPI_ISL_652046, EPI_ISL_652047, EPI_ISL_652048, EPI_ISL_652063, EPI_ISL_680498, EPI_ISL_680499, EPI_ISL_680500, EPI_ISL_680501, EPI_ISL_680502, EPI_ISL_680503, EPI_ISL_680504, EPI_ISL_680505, EPI_ISL_680506, EPI_ISL_680514, EPI_ISL_703787

| | | | |
|---|---|---|---|
| see above | Virology Department, Royal Infirmary of Edinburgh, NHS Lothian / School of Biological Sciences, University of Edinburgh / Institute of Genetics and Molecular Medicine, University of Edinburgh | COVID-19 Genomics UK (COG-UK) Consortium | Balcaza C; Colquhoun R; Dewar R; Gallagher M; Hill V; Jackson B; McCrone JT; McHugh M; O'Toole Á; Rambaut A; Rooke S; Scher E; Templeton K; Williams TC; Yu X |

EPI_ISL_611587, EPI_ISL_611598, EPI_ISL_611799, EPI_ISL_611803, EPI_ISL_611830, EPI_ISL_611869, EPI_ISL_613366, EPI_ISL_637266, EPI_ISL_637320, EPI_ISL_637326, EPI_ISL_637361, EPI_ISL_637381, EPI_ISL_637409, EPI_ISL_637531, EPI_ISL_637535, EPI_ISL_637539, EPI_ISL_637540, EPI_ISL_637541, EPI_ISL_637553, EPI_ISL_637580, EPI_ISL_637594, EPI_ISL_637862, EPI_ISL_637863, EPI_ISL_637905, EPI_ISL_638090, EPI_ISL_638168, EPI_ISL_638532, EPI_ISL_638533, EPI_ISL_638534, EPI_ISL_638535, EPI_ISL_638536, EPI_ISL_638537, EPI_ISL_638538, EPI_ISL_638539, EPI_ISL_638540, EPI_ISL_638541, EPI_ISL_638542, EPI_ISL_638543, EPI_ISL_638544, EPI_ISL_638545, EPI_ISL_638546, EPI_ISL_638547, EPI_ISL_638548, EPI_ISL_638549, EPI_ISL_638550, EPI_ISL_638551, EPI_ISL_638552, EPI_ISL_638553, EPI_ISL_638554, EPI_ISL_639008, EPI_ISL_639009, EPI_ISL_639010, EPI_ISL_650124, EPI_ISL_650209, EPI_ISL_650237, EPI_ISL_650362, EPI_ISL_650363, EPI_ISL_650367, EPI_ISL_650368, EPI_ISL_650369, EPI_ISL_650374, EPI_ISL_650384, EPI_ISL_650407, EPI_ISL_650423, EPI_ISL_650425, EPI_ISL_650488, EPI_ISL_650554, EPI_ISL_650650, EPI_ISL_650651, EPI_ISL_650652, EPI_ISL_650653, EPI_ISL_650659, EPI_ISL_650674, EPI_ISL_650675, EPI_ISL_650676, EPI_ISL_650678, EPI_ISL_650718, EPI_ISL_651184, EPI_ISL_651341, EPI_ISL_651437, EPI_ISL_652180, EPI_ISL_652181, EPI_ISL_652182, EPI_ISL_652183, EPI_ISL_653083, EPI_ISL_653085, EPI_ISL_653095, EPI_ISL_665864, EPI_ISL_703674, EPI_ISL_816317, EPI_ISL_842348, EPI_ISL_842349, EPI_ISL_842350, EPI_ISL_842351, EPI_ISL_842354, EPI_ISL_924423

| | | | |
|---|---|---|---|
| see above | Virology Department, Sheffield Teaching Hospitals NHS Foundation Trust/Department of Infection, Immunity and Cardiovascular Disease, The Medical School, University of Sheffield | COVID-19 Genomics UK (COG-UK) Consortium | Adri Angyal; Alex Keeley; Benjamin Lindsey; Cariad Evans; Danielle Groves; Dave Partridge; Katie Johnson; Laura Carrilero; Luke Green; Matthew Parker; Matthew Wyles; Mehmet Yavuz; Mohammad Raza; Nikki Smith; Paul Parsons; Rachel Tucker; Rebecca Brown; Thushan de Silva |

EPI_ISL_611519, EPI_ISL_611529, EPI_ISL_611530, EPI_ISL_611563, EPI_ISL_611566, EPI_ISL_611586, EPI_ISL_611605, EPI_ISL_611624, EPI_ISL_611625, EPI_ISL_611633, EPI_ISL_611640, EPI_ISL_611666, EPI_ISL_611684, EPI_ISL_611697, EPI_ISL_611706, EPI_ISL_611722, EPI_ISL_611726, EPI_ISL_611735, EPI_ISL_611819, EPI_ISL_611903, EPI_ISL_611904, EPI_ISL_611905, EPI_ISL_611926, EPI_ISL_611931, EPI_ISL_611994, EPI_ISL_612013, EPI_ISL_612018, EPI_ISL_612040, EPI_ISL_612081, EPI_ISL_612083, EPI_ISL_612116, EPI_ISL_612120, EPI_ISL_612146, EPI_ISL_612147, EPI_ISL_612149, EPI_ISL_612152, EPI_ISL_612155, EPI_ISL_612156, EPI_ISL_612157, EPI_ISL_612185, EPI_ISL_612193, EPI_ISL_612199, EPI_ISL_612203, EPI_ISL_612204, EPI_ISL_612205, EPI_ISL_612206, EPI_ISL_612843, EPI_ISL_612844, EPI_ISL_612845, EPI_ISL_612848, EPI_ISL_612849, EPI_ISL_612852, EPI_ISL_612855, EPI_ISL_612857, EPI_ISL_612859, EPI_ISL_612861, EPI_ISL_612865, EPI_ISL_612868, EPI_ISL_612875, EPI_ISL_612877, EPI_ISL_612878, EPI_ISL_612882, EPI_ISL_612891, EPI_ISL_612892, EPI_ISL_612895, EPI_ISL_612899, EPI_ISL_612902, EPI_ISL_612903, EPI_ISL_612908, EPI_ISL_612915, EPI_ISL_612918, EPI_ISL_612922, EPI_ISL_612924, EPI_ISL_612927, EPI_ISL_612932, EPI_ISL_612933, EPI_ISL_612934, EPI_ISL_612936, EPI_ISL_612940, EPI_ISL_612942, EPI_ISL_612943, EPI_ISL_612952, EPI_ISL_612953, EPI_ISL_612954, EPI_ISL_612957, EPI_ISL_612958, EPI_ISL_612962, EPI_ISL_612963, EPI_ISL_612969, EPI_ISL_612972, EPI_ISL_612975, EPI_ISL_612976, EPI_ISL_612986, EPI_ISL_612988, EPI_ISL_612991, EPI_ISL_612992, EPI_ISL_612994, EPI_ISL_612995, EPI_ISL_612997, EPI_ISL_612998, EPI_ISL_613021, EPI_ISL_613025, EPI_ISL_613026, EPI_ISL_613043, EPI_ISL_613044, EPI_ISL_613057, EPI_ISL_613068, EPI_ISL_613090, EPI_ISL_613101, EPI_ISL_613125, EPI_ISL_613137, EPI_ISL_613138, EPI_ISL_613141, EPI_ISL_613148, EPI_ISL_613155, EPI_ISL_613178, EPI_ISL_613185, EPI_ISL_613186, EPI_ISL_613189, EPI_ISL_613191, EPI_ISL_613192, EPI_ISL_613193, EPI_ISL_613194, EPI_ISL_613195, EPI_ISL_613196, EPI_ISL_613198, EPI_ISL_613199, EPI_ISL_613200, EPI_ISL_613201, EPI_ISL_613202, EPI_ISL_613205, EPI_ISL_613206, EPI_ISL_613207, EPI_ISL_613208, EPI_ISL_613209, EPI_ISL_613210, EPI_ISL_613211, EPI_ISL_613212, EPI_ISL_613213, EPI_ISL_613214, EPI_ISL_613215, EPI_ISL_613217, EPI_ISL_613218, EPI_ISL_613219, EPI_ISL_613221, EPI_ISL_613222, EPI_ISL_613223, EPI_ISL_613224, EPI_ISL_613227, EPI_ISL_613228, EPI_ISL_613230, EPI_ISL_613232, EPI_ISL_613233, EPI_ISL_613234, EPI_ISL_613235, EPI_ISL_613236, EPI_ISL_613237, EPI_ISL_613238, EPI_ISL_613239, EPI_ISL_613240, EPI_ISL_613242, EPI_ISL_613243, EPI_ISL_613244, EPI_ISL_613245, EPI_ISL_613246, EPI_ISL_613247, EPI_ISL_613248, EPI_ISL_613249, EPI_ISL_613251, EPI_ISL_613252, EPI_ISL_613253, EPI_ISL_613255, EPI_ISL_613257, EPI_ISL_613259, EPI_ISL_613260, EPI_ISL_613261, EPI_ISL_613262, EPI_ISL_613263, EPI_ISL_613264, EPI_ISL_613266, EPI_ISL_613267, EPI_ISL_613268, EPI_ISL_613270, EPI_ISL_613272, EPI_ISL_613273, EPI_ISL_613274, EPI_ISL_613276, EPI_ISL_613277, EPI_ISL_613281, EPI_ISL_626658, EPI_ISL_626674, EPI_ISL_626675, EPI_ISL_626696, EPI_ISL_626703, EPI_ISL_626706, EPI_ISL_626707, EPI_ISL_626708, EPI_ISL_626711, EPI_ISL_626746, EPI_ISL_626748, EPI_ISL_626761, EPI_ISL_626762, EPI_ISL_626763, EPI_ISL_626767, EPI_ISL_626768, EPI_ISL_626769, EPI_ISL_626783, EPI_ISL_626784, EPI_ISL_626788, EPI_ISL_626796, EPI_ISL_626829, EPI_ISL_626836, EPI_ISL_626853, EPI_ISL_626856, EPI_ISL_626857, EPI_ISL_626861, EPI_ISL_626863, EPI_ISL_626865, EPI_ISL_626867, EPI_ISL_626870, EPI_ISL_626871, EPI_ISL_626872, EPI_ISL_626874, EPI_ISL_626876, EPI_ISL_626877, EPI_ISL_626920, EPI_ISL_626928, EPI_ISL_626931, EPI_ISL_626959, EPI_ISL_626960, EPI_ISL_626961, EPI_ISL_626965, EPI_ISL_626993, EPI_ISL_626996, EPI_ISL_626998, EPI_ISL_627001, EPI_ISL_627003, EPI_ISL_627006, EPI_ISL_627007, EPI_ISL_627011, EPI_ISL_627013, EPI_ISL_627015, EPI_ISL_627018, EPI_ISL_627021, EPI_ISL_627022, EPI_ISL_627025, EPI_ISL_627028, EPI_ISL_627029, EPI_ISL_627033, EPI_ISL_627034, EPI_ISL_627035, EPI_ISL_627039, EPI_ISL_627042, EPI_ISL_627044, EPI_ISL_627051, EPI_ISL_627054, EPI_ISL_627055, EPI_ISL_627069, EPI_ISL_627086, EPI_ISL_627087, EPI_ISL_627096, EPI_ISL_627100, EPI_ISL_627101, EPI_ISL_627105, EPI_ISL_627119, EPI_ISL_627122, EPI_ISL_627124, EPI_ISL_627126, EPI_ISL_627129, EPI_ISL_627134, EPI_ISL_627140, EPI_ISL_627157, EPI_ISL_627167, EPI_ISL_627169, EPI_ISL_627172, EPI_ISL_627192, EPI_ISL_627204, EPI_ISL_627205, EPI_ISL_627206, EPI_ISL_627207, EPI_ISL_627217, EPI_ISL_627218, EPI_ISL_627221, EPI_ISL_627223, EPI_ISL_627224, EPI_ISL_627225, EPI_ISL_627226, EPI_ISL_627227, EPI_ISL_627228, EPI_ISL_627229, EPI_ISL_627230, EPI_ISL_627231, EPI_ISL_627232, EPI_ISL_627233, EPI_ISL_627234, EPI_ISL_627235, EPI_ISL_627236, EPI_ISL_627237, EPI_ISL_627238, EPI_ISL_627239, EPI_ISL_627240, EPI_ISL_627241, EPI_ISL_627242, EPI_ISL_627243, EPI_ISL_627244, EPI_ISL_627245, EPI_ISL_627246, EPI_ISL_627247, EPI_ISL_627248, EPI_ISL_627249, EPI_ISL_627265, EPI_ISL_627268, EPI_ISL_627272, EPI_ISL_627273, EPI_ISL_627277, EPI_ISL_627278, EPI_ISL_627279, EPI_ISL_627295, EPI_ISL_627296, EPI_ISL_627301, EPI_ISL_627732, EPI_ISL_627734, EPI_ISL_627736, EPI_ISL_627737, EPI_ISL_627742, EPI_ISL_627744, EPI_ISL_627745, EPI_ISL_627751, EPI_ISL_627756, EPI_ISL_627757, EPI_ISL_627758, EPI_ISL_627761, EPI_ISL_627762, EPI_ISL_627763, EPI_ISL_627764, EPI_ISL_627765, EPI_ISL_627766, EPI_ISL_627771, EPI_ISL_627772, EPI_ISL_627773, EPI_ISL_627774, EPI_ISL_627775, EPI_ISL_627776, EPI_ISL_627777, EPI_ISL_627779, EPI_ISL_627781, EPI_ISL_627782, EPI_ISL_627787, EPI_ISL_627789, EPI_ISL_627790, EPI_ISL_627791, EPI_ISL_627795, EPI_ISL_627796, EPI_ISL_627797, EPI_ISL_627798, EPI_ISL_627800, EPI_ISL_627801, EPI_ISL_627804, EPI_ISL_627808, EPI_ISL_627812, EPI_ISL_627813, EPI_ISL_627814, EPI_ISL_627815, EPI_ISL_627816, EPI_ISL_627819, EPI_ISL_627823, EPI_ISL_627827, EPI_ISL_627830, EPI_ISL_627835, EPI_ISL_627837, EPI_ISL_627838, EPI_ISL_627841, EPI_ISL_627850, EPI_ISL_627851, EPI_ISL_627852, EPI_ISL_627853, EPI_ISL_627856, EPI_ISL_627857, EPI_ISL_627859, EPI_ISL_627861, EPI_ISL_627863, EPI_ISL_627864, EPI_ISL_627866, EPI_ISL_627867, EPI_ISL_627870, EPI_ISL_627874, EPI_ISL_627876, EPI_ISL_627879, EPI_ISL_627881, EPI_ISL_627883, EPI_ISL_627884, EPI_ISL_627885, EPI_ISL_627886, EPI_ISL_627889, EPI_ISL_627893, EPI_ISL_627894, EPI_ISL_627895, EPI_ISL_627896, EPI_ISL_627898, EPI_ISL_627900, EPI_ISL_627901, EPI_ISL_627903, EPI_ISL_627908, EPI_ISL_627911, EPI_ISL_627913, EPI_ISL_627914, EPI_ISL_627915, EPI_ISL_627916, EPI_ISL_627917, EPI_ISL_627919, EPI_ISL_627921, EPI_ISL_627922, EPI_ISL_627923, EPI_ISL_627924, EPI_ISL_627925, EPI_ISL_627926, EPI_ISL_627927, EPI_ISL_627928, EPI_ISL_627929, EPI_ISL_627930, EPI_ISL_627931, EPI_ISL_627935, EPI_ISL_627937, EPI_ISL_627938, EPI_ISL_627939, EPI_ISL_627940, EPI_ISL_627941, EPI_ISL_627943, EPI_ISL_627944, EPI_ISL_627945, EPI_ISL_627946, EPI_ISL_627947, EPI_ISL_627948, EPI_ISL_627949, EPI_ISL_627950, EPI_ISL_627951, EPI_ISL_627952, EPI_ISL_627953, EPI_ISL_627954, EPI_ISL_627956, EPI_ISL_627957, EPI_ISL_627958, EPI_ISL_627959, EPI_ISL_627960, EPI_ISL_627961, EPI_ISL_627962, EPI_ISL_627963, EPI_ISL_627964, EPI_ISL_627965, EPI_ISL_627966, EPI_ISL_627967, EPI_ISL_627968, EPI_ISL_627969, EPI_ISL_627970, EPI_ISL_627971, EPI_ISL_627972, EPI_ISL_627973, EPI_ISL_627974, EPI_ISL_627975, EPI_ISL_627981, EPI_ISL_627982, EPI_ISL_627984, EPI_ISL_627985, EPI_ISL_627986, EPI_ISL_627987, EPI_ISL_627988, EPI_ISL_627989, EPI_ISL_627990, EPI_ISL_627991, EPI_ISL_627992, EPI_ISL_627993, EPI_ISL_627995, EPI_ISL_627996, EPI_ISL_627998, EPI_ISL_627999, EPI_ISL_628000, EPI_ISL_628002, EPI_ISL_628003, EPI_ISL_628004, EPI_ISL_628005, EPI_ISL_628006, EPI_ISL_628007, EPI_ISL_628008, EPI_ISL_628009, EPI_ISL_628010, EPI_ISL_628013, EPI_ISL_628014, EPI_ISL_628015, EPI_ISL_628017, EPI_ISL_628018, EPI_ISL_628019, EPI_ISL_628021, EPI_ISL_628023, EPI_ISL_628024, EPI_ISL_628025, EPI_ISL_628027, EPI_ISL_628028, EPI_ISL_628029, EPI_ISL_628031, EPI_ISL_628032, EPI_ISL_628033, EPI_ISL_628035, EPI_ISL_628036, EPI_ISL_628038, EPI_ISL_628039, EPI_ISL_628040, EPI_ISL_628041, EPI_ISL_628042, EPI_ISL_628043, EPI_ISL_628046, EPI_ISL_628047, EPI_ISL_628048, EPI_ISL_628049, EPI_ISL_628050, EPI_ISL_628051, EPI_ISL_628052, EPI_ISL_628053, EPI_ISL_628054, EPI_ISL_628055, EPI_ISL_628056, EPI_ISL_628057, EPI_ISL_628058, EPI_ISL_628060, EPI_ISL_628061, EPI_ISL_628062, EPI_ISL_628063, EPI_ISL_628065, EPI_ISL_628066, EPI_ISL_628067, EPI_ISL_628068, EPI_ISL_628069, EPI_ISL_628070, EPI_ISL_628071, EPI_ISL_628072, EPI_ISL_628074, EPI_ISL_628075, EPI_ISL_628076, EPI_ISL_628077, EPI_ISL_628078, EPI_ISL_628079, EPI_ISL_628080, EPI_ISL_628081, EPI_ISL_628082, EPI_ISL_628084, EPI_ISL_628085, EPI_ISL_628086, EPI_ISL_628087, EPI_ISL_628088, EPI_ISL_628089, EPI_ISL_628090, EPI_ISL_628091, EPI_ISL_628092, EPI_ISL_628093, EPI_ISL_628094, EPI_ISL_628095, EPI_ISL_628096, EPI_ISL_628097, EPI_ISL_628098, EPI_ISL_628099, EPI_ISL_628100, EPI_ISL_628101, EPI_ISL_628103, EPI_ISL_628104, EPI_ISL_628105, EPI_ISL_628106, EPI_ISL_628107, EPI_ISL_628108, EPI_ISL_628109, EPI_ISL_628110, EPI_ISL_628113, EPI_ISL_628114, EPI_ISL_628115, EPI_ISL_628116, EPI_ISL_628117, EPI_ISL_628118, EPI_ISL_628120, EPI_ISL_628122, EPI_ISL_628123, EPI_ISL_628124, EPI_ISL_628125, EPI_ISL_628126, EPI_ISL_628127, EPI_ISL_628129, EPI_ISL_628130, EPI_ISL_628131, EPI_ISL_628132, EPI_ISL_628134, EPI_ISL_628135, EPI_ISL_628136, EPI_ISL_628137, EPI_ISL_628138, EPI_ISL_628139, EPI_ISL_628140, EPI_ISL_628141, EPI_ISL_628142, EPI_ISL_628144, EPI_ISL_628145, EPI_ISL_628146, EPI_ISL_628147, EPI_ISL_628148, EPI_ISL_628149, EPI_ISL_628150, EPI_ISL_628151, EPI_ISL_628152, EPI_ISL_628153, EPI_ISL_628154, EPI_ISL_628155, EPI_ISL_628156, EPI_ISL_628157, EPI_ISL_628158, EPI_ISL_628159, EPI_ISL_628160, EPI_ISL_628161, EPI_ISL_628162, EPI_ISL_628163, EPI_ISL_628164, EPI_ISL_628165, EPI_ISL_628166, EPI_ISL_628167, EPI_ISL_628168, EPI_ISL_628169, EPI_ISL_628170, EPI_ISL_628171, EPI_ISL_628172, EPI_ISL_628173, EPI_ISL_628174, EPI_ISL_628175, EPI_ISL_628177, EPI_ISL_628178, EPI_ISL_628179, EPI_ISL_628180, EPI_ISL_628181, EPI_ISL_628182, EPI_ISL_628183, EPI_ISL_628184, EPI_ISL_628185, EPI_ISL_628186, EPI_ISL_628187, EPI_ISL_628188, EPI_ISL_628189, EPI_ISL_628190, EPI_ISL_628191, EPI_ISL_628192, EPI_ISL_628193, EPI_ISL_628194, EPI_ISL_628195, EPI_ISL_628196, EPI_ISL_628197, EPI_ISL_628198, EPI_ISL_628199, EPI_ISL_628200, EPI_ISL_628201, EPI_ISL_628202, EPI_ISL_628203, EPI_ISL_628204, EPI_ISL_628205, EPI_ISL_628206, EPI_ISL_628207, EPI_ISL_628208, EPI_ISL_628209, EPI_ISL_628210, EPI_ISL_628211, EPI_ISL_628212, EPI_ISL_628213, EPI_ISL_628214, EPI_ISL_628215, EPI_ISL_628216, EPI_ISL_628217, EPI_ISL_628218, EPI_ISL_628219, EPI_ISL_628220, EPI_ISL_628221, EPI_ISL_628222, EPI_ISL_628224, EPI_ISL_628225, EPI_ISL_628226, EPI_ISL_628227, EPI_ISL_628228, EPI_ISL_628229, EPI_ISL_628230, EPI_ISL_628231, EPI_ISL_628232, EPI_ISL_628233, EPI_ISL_628234, EPI_ISL_628235, EPI_ISL_628236, EPI_ISL_628237, EPI_ISL_628238, EPI_ISL_628239, EPI_ISL_628240, EPI_ISL_628241, EPI_ISL_628242, EPI_ISL_628243, EPI_ISL_628244, EPI_ISL_628245, EPI_ISL_628246, EPI_ISL_628247, EPI_ISL_628248, EPI_ISL_628249, EPI_ISL_628250, EPI_ISL_628251, EPI_ISL_628252, EPI_ISL_628253, EPI_ISL_628254, EPI_ISL_628255, EPI_ISL_628256, EPI_ISL_628257, EPI_ISL_628258, EPI_ISL_628259, EPI_ISL_628260, EPI_ISL_628261, EPI_ISL_628262, EPI_ISL_628263, EPI_ISL_628264, EPI_ISL_628265, EPI_ISL_628266, EPI_ISL_628267, EPI_ISL_628268, EPI_ISL_628269, EPI_ISL_628270, EPI_ISL_628271, EPI_ISL_628272, EPI_ISL_628273, EPI_ISL_628274, EPI_ISL_628275, EPI_ISL_628276, EPI_ISL_628277, EPI_ISL_628278, EPI_ISL_628279, EPI_ISL_628280, EPI_ISL_628281, EPI_ISL_628282, EPI_ISL_628283, EPI_ISL_628285, EPI_ISL_628286, EPI_ISL_628287, EPI_ISL_628288, EPI_ISL_628289, EPI_ISL_628290, EPI_ISL_628291, EPI_ISL_628292, EPI_ISL_628293, EPI_ISL_628294, EPI_ISL_628295, EPI_ISL_628296, EPI_ISL_628297, EPI_ISL_628298, EPI_ISL_628299, EPI_ISL_628300, EPI_ISL_628301, EPI_ISL_628302, EPI_ISL_628303, EPI_ISL_628304, EPI_ISL_628305, EPI_ISL_628306, EPI_ISL_628307, EPI_ISL_628308, EPI_ISL_628309, EPI_ISL_628310, EPI_ISL_628311, EPI_ISL_637385, EPI_ISL_637543, EPI_ISL_637599, EPI_ISL_637604, EPI_ISL_637607, EPI_ISL_637611, EPI_ISL_637612, EPI_ISL_637614, EPI_ISL_637622, EPI_ISL_637627, EPI_ISL_637854, EPI_ISL_637870, EPI_ISL_637884, EPI_ISL_637885, EPI_ISL_637886, EPI_ISL_637894, EPI_ISL_637900, EPI_ISL_637904, EPI_ISL_637919, EPI_ISL_637924, EPI_ISL_637952, EPI_ISL_637956, EPI_ISL_637957, EPI_ISL_637964, EPI_ISL_638028, EPI_ISL_638042, EPI_ISL_638043, EPI_ISL_638045, EPI_ISL_638048, EPI_ISL_638049, EPI_ISL_638053, EPI_ISL_638054, EPI_ISL_638093, EPI_ISL_638102, EPI_ISL_638110, EPI_ISL_638117, EPI_ISL_638118, EPI_ISL_638119, EPI_ISL_638122, EPI_ISL_638126, EPI_ISL_638127, EPI_ISL_638130, EPI_ISL_638147, EPI_ISL_638148, EPI_ISL_638149, EPI_ISL_638150, EPI_ISL_638151, EPI_ISL_638152, EPI_ISL_638154, EPI_ISL_638156, EPI_ISL_638157, EPI_ISL_638160, EPI_ISL_639015, EPI_ISL_639019, EPI_ISL_639020, EPI_ISL_639022, EPI_ISL_639024, EPI_ISL_639025, EPI_ISL_639028, EPI_ISL_639033, EPI_ISL_639036, EPI_ISL_639038, EPI_ISL_639039, EPI_ISL_639042, EPI_ISL_639043, EPI_ISL_639044, EPI_ISL_639048, EPI_ISL_639049, EPI_ISL_639051, EPI_ISL_639054, EPI_ISL_639058, EPI_ISL_639059, EPI_ISL_639061, EPI_ISL_639066, EPI_ISL_639068, EPI_ISL_639069, EPI_ISL_639070, EPI_ISL_639074, EPI_ISL_639075, EPI_ISL_639076, EPI_ISL_639079, EPI_ISL_639084, EPI_ISL_639089, EPI_ISL_639093, EPI_ISL_639098, EPI_ISL_639099, EPI_ISL_639101, EPI_ISL_639102, EPI_ISL_639103, EPI_ISL_639110, EPI_ISL_639112, EPI_ISL_639113, EPI_ISL_639114, EPI_ISL_639116, EPI_ISL_639117, EPI_ISL_639118, EPI_ISL_639120, EPI_ISL_639123, EPI_ISL_639124, EPI_ISL_639126, EPI_ISL_639127, EPI_ISL_639128, EPI_ISL_639129, EPI_ISL_639132, EPI_ISL_639137, EPI_ISL_639138, EPI_ISL_639140, EPI_ISL_639142, EPI_ISL_639144, EPI_ISL_639145, EPI_ISL_639152, EPI_ISL_639154, EPI_ISL_639159, EPI_ISL_639162, EPI_ISL_639171, EPI_ISL_639178, EPI_ISL_639180, EPI_ISL_639185, EPI_ISL_639186, EPI_ISL_639191, EPI_ISL_639192, EPI_ISL_639194, EPI_ISL_639207, EPI_ISL_639209, EPI_ISL_639210, EPI_ISL_639214, EPI_ISL_639223, EPI_ISL_639224, EPI_ISL_639226, EPI_ISL_639229, EPI_ISL_639230, EPI_ISL_639231, EPI_ISL_639232, EPI_ISL_639236, EPI_ISL_639242, EPI_ISL_639245, EPI_ISL_639251, EPI_ISL_639257, EPI_ISL_639260, EPI_ISL_639264, EPI_ISL_639265, EPI_ISL_639266, EPI_ISL_639268, EPI_ISL_639278, EPI_ISL_639279, EPI_ISL_639281, EPI_ISL_639283, EPI_ISL_639285, EPI_ISL_639286, EPI_ISL_639287, EPI_ISL_639291, EPI_ISL_639295, EPI_ISL_639297, EPI_ISL_639298, EPI_ISL_639304, EPI_ISL_639307, EPI_ISL_639312, EPI_ISL_639320, EPI_ISL_639326, EPI_ISL_639336, EPI_ISL_639337, EPI_ISL_639340, EPI_ISL_639343, EPI_ISL_639348, EPI_ISL_639349, EPI_ISL_639350, EPI_ISL_639352, EPI_ISL_639354, EPI_ISL_639357, EPI_ISL_639364, EPI_ISL_639370, EPI_ISL_639373, EPI_ISL_639380, EPI_ISL_639382, EPI_ISL_639389, EPI_ISL_639391, EPI_ISL_639406, EPI_ISL_639411, EPI_ISL_639415, EPI_ISL_639419, EPI_ISL_639429, EPI_ISL_639430, EPI_ISL_639435, EPI_ISL_639442, EPI_ISL_639446, EPI_ISL_639451, EPI_ISL_639454, EPI_ISL_639458, EPI_ISL_639470, EPI_ISL_639471, EPI_ISL_639478, EPI_ISL_639487, EPI_ISL_639502, EPI_ISL_639505, EPI_ISL_639511, EPI_ISL_639513, EPI_ISL_639519, EPI_ISL_639533, EPI_ISL_639538, EPI_ISL_639543, EPI_ISL_639546, EPI_ISL_639550, EPI_ISL_639552, EPI_ISL_639564, EPI_ISL_639570, EPI_ISL_639573, EPI_ISL_639575, EPI_ISL_639579, EPI_ISL_639582, EPI_ISL_639583, EPI_ISL_639589, EPI_ISL_639590, EPI_ISL_639595, EPI_ISL_639598, EPI_ISL_639613, EPI_ISL_639619, EPI_ISL_639622, EPI_ISL_650660, EPI_ISL_650695, EPI_ISL_650914, EPI_ISL_651207, EPI_ISL_651415, EPI_ISL_651420, EPI_ISL_651441, EPI_ISL_651443, EPI_ISL_651444, EPI_ISL_651445, EPI_ISL_651446, EPI_ISL_651447, EPI_ISL_651448, EPI_ISL_651449, EPI_ISL_651451, EPI_ISL_651455, EPI_ISL_651457, EPI_ISL_651465, EPI_ISL_651503, EPI_ISL_652414, EPI_ISL_652415, EPI_ISL_652416, EPI_ISL_652417, EPI_ISL_652418, EPI_ISL_652419, EPI_ISL_652420, EPI_ISL_652421, EPI_ISL_652422, EPI_ISL_652423, EPI_ISL_652424, EPI_ISL_652425, EPI_ISL_652426, EPI_ISL_652427, EPI_ISL_652428, EPI_ISL_652429, EPI_ISL_652430, EPI_ISL_652431, EPI_ISL_652433, EPI_ISL_652434, EPI_ISL_652435, EPI_ISL_652436, EPI_ISL_652437, EPI_ISL_652438, EPI_ISL_652439, EPI_ISL_652440, EPI_ISL_652441, EPI_ISL_652442, EPI_ISL_652443, EPI_ISL_652444, EPI_ISL_652445, EPI_ISL_652446, EPI_ISL_652447, EPI_ISL_652448, EPI_ISL_652449, EPI_ISL_652450, EPI_ISL_652451, EPI_ISL_652452, EPI_ISL_652453, EPI_ISL_652454, EPI_ISL_652456, EPI_ISL_652457, EPI_ISL_652458, EPI_ISL_652459, EPI_ISL_652460, EPI_ISL_652461, EPI_ISL_652463, EPI_ISL_652464, EPI_ISL_652465, EPI_ISL_652467, EPI_ISL_652468, EPI_ISL_652469, EPI_ISL_652471, EPI_ISL_652472, EPI_ISL_652473, EPI_ISL_652475, EPI_ISL_652476, EPI_ISL_652477, EPI_ISL_652478, EPI_ISL_652479, EPI_ISL_652480, EPI_ISL_652483, EPI_ISL_652484, EPI_ISL_652485, EPI_ISL_652486, EPI_ISL_652488, EPI_ISL_652489, EPI_ISL_652490, EPI_ISL_652491, EPI_ISL_652492, EPI_ISL_652493, EPI_ISL_652494, EPI_ISL_652495, EPI_ISL_652503, EPI_ISL_652536, EPI_ISL_652543, EPI_ISL_652546, EPI_ISL_652555, EPI_ISL_652568, EPI_ISL_652598, EPI_ISL_652603, EPI_ISL_652606, EPI_ISL_652609, EPI_ISL_666140, EPI_ISL_666181, EPI_ISL_666244, EPI_ISL_666350, EPI_ISL_666352, EPI_ISL_666355, EPI_ISL_666364, EPI_ISL_680616, EPI_ISL_680617, EPI_ISL_680618, EPI_ISL_680619, EPI_ISL_705459, EPI_ISL_725999, EPI_ISL_742283, EPI_ISL_742284, EPI_ISL_742285, EPI_ISL_742302, EPI_ISL_742303, EPI_ISL_742304, EPI_ISL_742305, EPI_ISL_742306, EPI_ISL_742307

| | | | |
|---|---|---|---|
| see above | Wales Specialist Virology Centre Sequencing lab: Pathogen Genomics Unit | COVID-19 Genomics UK (COG-UK) Consortium | Alec Birchley; Alexander Adams; Amy Gaskin; Angela Marchbank; Bree Gatica-Wilcox; Catherine Moore; Jason Coombes; Joanne Watkins; Joel Southgate; Johnathan Evans; Laura Gifford; Lauren Gilbert; Lee Graham; Malorie Perry; Matthew Bull; Nicole Pacchiarini; Sally Corden; Sara Kumziene-Summerhayes; Sara Rey; Sarah Taylor; Simon Cottrell; Sophie Jones; Tom Connor |

EPI_ISL_626691, EPI_ISL_626692, EPI_ISL_626749, EPI_ISL_626831, EPI_ISL_626846, EPI_ISL_626849, EPI_ISL_626851, EPI_ISL_626957, EPI_ISL_626995, EPI_ISL_627030, EPI_ISL_627165, EPI_ISL_627209, EPI_ISL_627379, EPI_ISL_627388, EPI_ISL_627391, EPI_ISL_627398, EPI_ISL_650175, EPI_ISL_650312, EPI_ISL_650313, EPI_ISL_650328, EPI_ISL_650412, EPI_ISL_650438, EPI_ISL_650477, EPI_ISL_650483, EPI_ISL_650484, EPI_ISL_650491, EPI_ISL_650492, EPI_ISL_650494, EPI_ISL_650498, EPI_ISL_650504, EPI_ISL_650505, EPI_ISL_650509, EPI_ISL_650510, EPI_ISL_650520, EPI_ISL_650521, EPI_ISL_650525, EPI_ISL_650526, EPI_ISL_650725, EPI_ISL_650731, EPI_ISL_650732, EPI_ISL_650734, EPI_ISL_650739, EPI_ISL_650745, EPI_ISL_650746, EPI_ISL_650747, EPI_ISL_650763, EPI_ISL_650802, EPI_ISL_650816, EPI_ISL_650838, EPI_ISL_650839, EPI_ISL_650840, EPI_ISL_650841, EPI_ISL_650842, EPI_ISL_650843, EPI_ISL_650845, EPI_ISL_650848, EPI_ISL_650854, EPI_ISL_650855, EPI_ISL_650856, EPI_ISL_650857, EPI_ISL_650858, EPI_ISL_650859, EPI_ISL_650860, EPI_ISL_650861, EPI_ISL_650862, EPI_ISL_650863, EPI_ISL_650872, EPI_ISL_650873, EPI_ISL_651160, EPI_ISL_651310, EPI_ISL_651402, EPI_ISL_651494, EPI_ISL_651495, EPI_ISL_651531, EPI_ISL_651532, EPI_ISL_651533, EPI_ISL_651740, EPI_ISL_651741, EPI_ISL_651742, EPI_ISL_651743, EPI_ISL_651744, EPI_ISL_651745, EPI_ISL_651746, EPI_ISL_651747, EPI_ISL_651748, EPI_ISL_651749, EPI_ISL_651750, EPI_ISL_651751, EPI_ISL_651752, EPI_ISL_651753, EPI_ISL_651754, EPI_ISL_651755, EPI_ISL_651756, EPI_ISL_651757, EPI_ISL_651758, EPI_ISL_651759, EPI_ISL_651760, EPI_ISL_651761, EPI_ISL_651762, EPI_ISL_651763, EPI_ISL_651764, EPI_ISL_651765, EPI_ISL_651766, EPI_ISL_651767, EPI_ISL_651768, EPI_ISL_651769, EPI_ISL_651770, EPI_ISL_651771, EPI_ISL_651772, EPI_ISL_651773, EPI_ISL_651774, EPI_ISL_651775, EPI_ISL_651776, EPI_ISL_651777, EPI_ISL_651778, EPI_ISL_651779, EPI_ISL_651780, EPI_ISL_651781, EPI_ISL_651782, EPI_ISL_651783, EPI_ISL_651784, EPI_ISL_651819, EPI_ISL_651820, EPI_ISL_651832, EPI_ISL_651833, EPI_ISL_651834, EPI_ISL_651835, EPI_ISL_651836, EPI_ISL_651837, EPI_ISL_651838, EPI_ISL_651839, EPI_ISL_651840, EPI_ISL_651845, EPI_ISL_651846, EPI_ISL_651847, EPI_ISL_651874, EPI_ISL_651875, EPI_ISL_651876, EPI_ISL_651877, EPI_ISL_651878, EPI_ISL_651879, EPI_ISL_651880, EPI_ISL_651881, EPI_ISL_651884, EPI_ISL_703771, EPI_ISL_997067

| | | | |
|---|---|---|---|
| see above | West of Scotland Specialist Virology Centre, NHSGGC / MRC-University of Glasgow Centre for Virus Research | COVID-19 Genomics UK (COG-UK) Consortium | Alasdair MacLean; Alice Broos; Ana da Silva Filipe; Antonia Ho; Daniel Mair; David L Robertson; Elihu Aranday-Cortes; Emma Thomson; Guy Mollett; Igor Starinskij; James Shepherd; Jenna Nichols; Joseph Hughes; Kathy Li; Kathy Smollett; Kyriaki Nomikou; Lily Tong; Marc Niebel; Matthew Holden; Natasha Jesudason; Natasha Johnson; Patawee Asamaphan; Rachel Blacow; Rajiv Shah; Richard Orton; Rory Gunson; Sarah McDonald; Sharif Shaaban; Sreenu Vattipally; Stephen Carmichael |

We gratefully acknowledge the following Authors from the Originating laboratories responsible for obtaining the specimens, as well as the Submitting laboratories where the genome data were generated and shared via GISAID, on which this research is based.

All Submitters of data may be contacted directly via www.gisaid.org

Authors are sorted alphabetically.

| Accession ID | Originating Laboratory | Submitting Laboratory | Authors |
|---|---|---|---|
| EPI_ISL_842865, EPI_ISL_842867, EPI_ISL_842868, EPI_ISL_842903, EPI_ISL_842939, EPI_ISL_1636884, EPI_ISL_1636894, EPI_ISL_1636895, EPI_ISL_1636896, EPI_ISL_1636897, EPI_ISL_1636898, EPI_ISL_1636899, EPI_ISL_1636900, EPI_ISL_1636901, EPI_ISL_1636902, EPI_ISL_1636903, EPI_ISL_1636904, EPI_ISL_1636905, EPI_ISL_1636906, EPI_ISL_1636907, EPI_ISL_1636908, EPI_ISL_1636909, EPI_ISL_1636910, EPI_ISL_1636911, EPI_ISL_1636912, EPI_ISL_1636985 | | | |
| see above | Barts Health NHS Trust | COVID-19 Genomics UK (COG-UK) Consortium | BROAD; Beatrix; CUTINO-MOGUEL; Claire; David; Dola; HARRINGTON; KELE; KULASEGARAN-SHYLINI; Maria-Teresa; OWOYEMI; Raghavendran; SHYLINI |
| EPI_ISL_868792, EPI_ISL_868794, EPI_ISL_868796, EPI_ISL_868803, EPI_ISL_868806, EPI_ISL_868807, EPI_ISL_868811, EPI_ISL_868812, EPI_ISL_868814, EPI_ISL_868819, EPI_ISL_1476853 | | | |
| see above | Bioinformatics and Biostatistics Lab, Advanced Sequencing Facility | COVID-19 Genomics UK (COG-UK) Consortium | Aengus Stewart; Chelsea Sawyer; Harshil Patel; Jerome Nicod; Laura Cubitt; Margaret Crawford |
| EPI_ISL_628323, EPI_ISL_628324, EPI_ISL_650379, EPI_ISL_650388, EPI_ISL_650575, EPI_ISL_650576, EPI_ISL_650604, EPI_ISL_650605, EPI_ISL_650608, EPI_ISL_650941, EPI_ISL_650980, EPI_ISL_651069, EPI_ISL_651187, EPI_ISL_652620, EPI_ISL_652621, EPI_ISL_664137, EPI_ISL_664191, EPI_ISL_664302, EPI_ISL_664374, EPI_ISL_664375, EPI_ISL_664376, EPI_ISL_664377, EPI_ISL_664492, EPI_ISL_664526, EPI_ISL_664556, EPI_ISL_664557, EPI_ISL_664558, EPI_ISL_664559, EPI_ISL_664679, EPI_ISL_664690, EPI_ISL_664746, EPI_ISL_664763, EPI_ISL_664767, EPI_ISL_664840, EPI_ISL_664841, EPI_ISL_664842, EPI_ISL_664843, EPI_ISL_664844, EPI_ISL_664845, EPI_ISL_664846, EPI_ISL_664847, EPI_ISL_664848, EPI_ISL_664849, EPI_ISL_664850, EPI_ISL_664851, EPI_ISL_664852, EPI_ISL_664853, EPI_ISL_664854, EPI_ISL_664855, EPI_ISL_665338, EPI_ISL_665347, EPI_ISL_665723, EPI_ISL_665726, EPI_ISL_666107, EPI_ISL_666108, EPI_ISL_666109, EPI_ISL_678831, EPI_ISL_680578, EPI_ISL_723521, EPI_ISL_723522, EPI_ISL_723523, EPI_ISL_723524, EPI_ISL_723525, EPI_ISL_723526, EPI_ISL_723527, EPI_ISL_723528, EPI_ISL_723529, EPI_ISL_723530, EPI_ISL_723531, EPI_ISL_723532, EPI_ISL_723533, EPI_ISL_723534, EPI_ISL_723535, EPI_ISL_723536, EPI_ISL_723537, EPI_ISL_723538, EPI_ISL_723539, EPI_ISL_723540, EPI_ISL_723541, EPI_ISL_723542, EPI_ISL_723543, EPI_ISL_723544, EPI_ISL_723545, EPI_ISL_723546, EPI_ISL_723547, EPI_ISL_723548, EPI_ISL_723549, EPI_ISL_723550, EPI_ISL_723551, EPI_ISL_723552, EPI_ISL_723553, EPI_ISL_723554, EPI_ISL_723555, EPI_ISL_723556, EPI_ISL_723557, EPI_ISL_727702, EPI_ISL_727708, EPI_ISL_727750, EPI_ISL_867944, EPI_ISL_867963, EPI_ISL_867964, EPI_ISL_867965, EPI_ISL_867967, EPI_ISL_923224, EPI_ISL_1730868, EPI_ISL_1979968 | | | |
| see above | Centre for Enzyme Innovation, University of Portsmouth / Translational Research Laboratory, Portsmouth Hospitals NHS Trust | COVID-19 Genomics UK (COG-UK) Consortium | Allyson Lloyd; Angela Beckett; Anoop Chauhan; Christopher Fearn; Ethan Butcher; Garry Scarlett; Kate Cook; Katie Loveson; Kelly Bicknell; Robert Impey; Salman Goudarzi; Samuel Robson; Sarah Wyllie; Scott Elliott; Sharon Glaysher; Yann Bourgeois |
| EPI_ISL_650143, EPI_ISL_650232, EPI_ISL_650240, EPI_ISL_650273, EPI_ISL_650353, EPI_ISL_650364, EPI_ISL_650365, EPI_ISL_650390, EPI_ISL_650397, EPI_ISL_650447, EPI_ISL_650481, EPI_ISL_650562, EPI_ISL_650580, EPI_ISL_650583, EPI_ISL_650596, EPI_ISL_650614, EPI_ISL_650615, EPI_ISL_650616, EPI_ISL_650617, EPI_ISL_650633, EPI_ISL_650664, EPI_ISL_650682, EPI_ISL_650768, EPI_ISL_650774, EPI_ISL_650836, EPI_ISL_650899, EPI_ISL_650916, EPI_ISL_650978, EPI_ISL_650990, EPI_ISL_650991, EPI_ISL_650993, EPI_ISL_651001, EPI_ISL_651002, EPI_ISL_651003, EPI_ISL_651004, EPI_ISL_651005, EPI_ISL_651006, EPI_ISL_651007, EPI_ISL_651008, EPI_ISL_651049, EPI_ISL_651050, EPI_ISL_651051, EPI_ISL_651052, EPI_ISL_651122, EPI_ISL_651136, EPI_ISL_651269, EPI_ISL_651276, EPI_ISL_651292, EPI_ISL_651363, EPI_ISL_651383, EPI_ISL_651423, EPI_ISL_651489, EPI_ISL_651490, EPI_ISL_651492, EPI_ISL_651542, EPI_ISL_651556, EPI_ISL_651557, EPI_ISL_651560, EPI_ISL_651562, EPI_ISL_651564, EPI_ISL_651568, EPI_ISL_651570, EPI_ISL_651571, EPI_ISL_651572, EPI_ISL_651574, EPI_ISL_651576, EPI_ISL_651578, EPI_ISL_651580, EPI_ISL_651581, EPI_ISL_651659, EPI_ISL_651663, EPI_ISL_651664, EPI_ISL_651665, EPI_ISL_651666, EPI_ISL_651667, EPI_ISL_651668, EPI_ISL_651669, EPI_ISL_651670, EPI_ISL_651671, EPI_ISL_651672, EPI_ISL_651673, EPI_ISL_651674, EPI_ISL_651676, EPI_ISL_651677, EPI_ISL_651679, EPI_ISL_651680, EPI_ISL_651681, EPI_ISL_651682, EPI_ISL_651683, EPI_ISL_651684, EPI_ISL_651685, EPI_ISL_651686, EPI_ISL_651687, EPI_ISL_651688, EPI_ISL_651689, EPI_ISL_651690, EPI_ISL_651691, EPI_ISL_651692, EPI_ISL_651693, EPI_ISL_651694, EPI_ISL_651695, EPI_ISL_651696, EPI_ISL_651697, EPI_ISL_651698, EPI_ISL_651699, EPI_ISL_651700, EPI_ISL_651702, EPI_ISL_651703, EPI_ISL_651704, EPI_ISL_651705, EPI_ISL_651706, EPI_ISL_651707, EPI_ISL_651711, EPI_ISL_651712, EPI_ISL_651713, EPI_ISL_652286, EPI_ISL_652287, EPI_ISL_652288, EPI_ISL_664167, EPI_ISL_664217, EPI_ISL_664422, EPI_ISL_664532, EPI_ISL_664542, EPI_ISL_664568, EPI_ISL_664569, EPI_ISL_664573, EPI_ISL_664574, EPI_ISL_664576, EPI_ISL_664577, EPI_ISL_664582, EPI_ISL_664599, EPI_ISL_664721, EPI_ISL_664941, EPI_ISL_664943, EPI_ISL_664944, EPI_ISL_664945, EPI_ISL_664969, EPI_ISL_679140, EPI_ISL_680037, EPI_ISL_680038, EPI_ISL_680039, EPI_ISL_680040, EPI_ISL_680041, EPI_ISL_680042, EPI_ISL_680043, EPI_ISL_680044, EPI_ISL_680045, EPI_ISL_680046, EPI_ISL_680047, EPI_ISL_680048, EPI_ISL_680049, EPI_ISL_680050, EPI_ISL_680051, EPI_ISL_680052, EPI_ISL_680053, EPI_ISL_680054, EPI_ISL_680055, EPI_ISL_680056, EPI_ISL_680057, EPI_ISL_680058, EPI_ISL_680059, EPI_ISL_680060, EPI_ISL_680061, EPI_ISL_680062, EPI_ISL_680063, EPI_ISL_680064, EPI_ISL_680065, EPI_ISL_680066, EPI_ISL_680067, EPI_ISL_680068, EPI_ISL_680069, EPI_ISL_680070, EPI_ISL_680071, EPI_ISL_680072, EPI_ISL_680073, EPI_ISL_680074, EPI_ISL_680075, EPI_ISL_680076, EPI_ISL_680077, EPI_ISL_680078, EPI_ISL_680079, EPI_ISL_680080, EPI_ISL_680081, EPI_ISL_680082, EPI_ISL_680083, EPI_ISL_680084, EPI_ISL_680085, EPI_ISL_680086, EPI_ISL_680087, EPI_ISL_680088, EPI_ISL_680089, EPI_ISL_680090, EPI_ISL_680091, EPI_ISL_680092, EPI_ISL_680093, EPI_ISL_680094, EPI_ISL_680095, EPI_ISL_680096, EPI_ISL_680097, EPI_ISL_680098, EPI_ISL_680099, EPI_ISL_680100, EPI_ISL_680571, EPI_ISL_680572, EPI_ISL_680573, EPI_ISL_680574, EPI_ISL_680575, EPI_ISL_680576, EPI_ISL_680577, EPI_ISL_681260, EPI_ISL_705502, EPI_ISL_705547, EPI_ISL_705548 | | | |
| see above | Department of Pathology, University of Cambridge | COVID-19 Genomics UK (COG-UK) Consortium | Aminu S. Jahun; Grant Hall; Ian Goodfellow; Iliana Georgana; Malte Pinckert; Martin D. Curran; Myra Hosmillo; Surendra Parmar; Yasmin Chaudhry |
| EPI_ISL_645266, EPI_ISL_645362, EPI_ISL_645368, EPI_ISL_645530, EPI_ISL_645531, EPI_ISL_645532, EPI_ISL_645533, EPI_ISL_645534, EPI_ISL_645535, EPI_ISL_645536, EPI_ISL_645537, EPI_ISL_645538, EPI_ISL_645539, EPI_ISL_645540, EPI_ISL_645541, EPI_ISL_645542, EPI_ISL_645543, EPI_ISL_645544, EPI_ISL_645545, EPI_ISL_645546, EPI_ISL_645547, EPI_ISL_645548, EPI_ISL_645549, EPI_ISL_645550, EPI_ISL_645551, EPI_ISL_645552, EPI_ISL_645553, EPI_ISL_645554, EPI_ISL_645555, EPI_ISL_645556, EPI_ISL_645557, EPI_ISL_645558, EPI_ISL_645559, EPI_ISL_645560, EPI_ISL_645561, EPI_ISL_645562, EPI_ISL_645563, EPI_ISL_645564, EPI_ISL_645565, EPI_ISL_645566, EPI_ISL_645567, EPI_ISL_645568, EPI_ISL_645569, EPI_ISL_645570, EPI_ISL_645571, EPI_ISL_645572, EPI_ISL_645573, EPI_ISL_645574, EPI_ISL_645575, EPI_ISL_645576, EPI_ISL_645577, EPI_ISL_645578, EPI_ISL_645579, EPI_ISL_645580, EPI_ISL_645581, EPI_ISL_645582, EPI_ISL_645583, EPI_ISL_645584, EPI_ISL_645585, EPI_ISL_645586, EPI_ISL_645587, EPI_ISL_645588, EPI_ISL_645589, EPI_ISL_645590, EPI_ISL_645591, EPI_ISL_645592, EPI_ISL_645593, EPI_ISL_645594, EPI_ISL_645595, EPI_ISL_645596, EPI_ISL_645597, EPI_ISL_645598, EPI_ISL_645599, EPI_ISL_645600, EPI_ISL_645601, EPI_ISL_645602, EPI_ISL_645603, EPI_ISL_645604, EPI_ISL_645605, EPI_ISL_645606, EPI_ISL_645607, EPI_ISL_645608, EPI_ISL_645609, EPI_ISL_645610, EPI_ISL_645611, EPI_ISL_645612, EPI_ISL_645613, EPI_ISL_645614, EPI_ISL_645615, EPI_ISL_645616, EPI_ISL_645617, EPI_ISL_645618, EPI_ISL_645619, EPI_ISL_645620, EPI_ISL_645621, EPI_ISL_645622, EPI_ISL_645623, EPI_ISL_645624, EPI_ISL_645625, EPI_ISL_645626, EPI_ISL_645627, EPI_ISL_645628, EPI_ISL_645629, EPI_ISL_645630, EPI_ISL_645631, EPI_ISL_645632, EPI_ISL_645633, EPI_ISL_645634, EPI_ISL_645635, EPI_ISL_645636, EPI_ISL_645637, EPI_ISL_645638, EPI_ISL_645639, EPI_ISL_645640, EPI_ISL_645641, EPI_ISL_645642, EPI_ISL_645643, EPI_ISL_645644, EPI_ISL_645645, EPI_ISL_645646, EPI_ISL_645647, EPI_ISL_645648, EPI_ISL_645649, EPI_ISL_645650, EPI_ISL_645651, EPI_ISL_645652, EPI_ISL_645653, EPI_ISL_645654, EPI_ISL_645655, EPI_ISL_645656, EPI_ISL_645657, EPI_ISL_645658, EPI_ISL_645659, EPI_ISL_645660, EPI_ISL_645661, EPI_ISL_645662, EPI_ISL_645663, EPI_ISL_645664, EPI_ISL_645665, EPI_ISL_645666, EPI_ISL_645667, EPI_ISL_645668, EPI_ISL_645669, EPI_ISL_645670, EPI_ISL_645671, EPI_ISL_645672, EPI_ISL_645673, EPI_ISL_645674, EPI_ISL_645675, EPI_ISL_645676, EPI_ISL_645677, EPI_ISL_645678, EPI_ISL_645679, EPI_ISL_645680, EPI_ISL_645681, EPI_ISL_645682, EPI_ISL_645683, EPI_ISL_645684, EPI_ISL_645685, EPI_ISL_645686, EPI_ISL_645687, EPI_ISL_645688, EPI_ISL_645689, EPI_ISL_645690, EPI_ISL_645691, EPI_ISL_645692, EPI_ISL_645693, EPI_ISL_645694, EPI_ISL_645695, EPI_ISL_645696, EPI_ISL_645697, EPI_ISL_645698, EPI_ISL_645699, EPI_ISL_645700, EPI_ISL_645701, EPI_ISL_645702, EPI_ISL_645703, EPI_ISL_645704, EPI_ISL_645705, EPI_ISL_645706, EPI_ISL_645707, EPI_ISL_645708, EPI_ISL_645709, EPI_ISL_645710, EPI_ISL_645711, EPI_ISL_645712, EPI_ISL_645713, EPI_ISL_645714, EPI_ISL_645715, EPI_ISL_645716, EPI_ISL_645717, EPI_ISL_645718, EPI_ISL_645719, EPI_ISL_645720, EPI_ISL_645721, EPI_ISL_645722, EPI_ISL_645723, EPI_ISL_645724, EPI_ISL_645725, EPI_ISL_645726, EPI_ISL_645727, EPI_ISL_645728, EPI_ISL_645729, EPI_ISL_645730, EPI_ISL_645731, EPI_ISL_645732, EPI_ISL_645733, EPI_ISL_645734, EPI_ISL_645735, EPI_ISL_645736, EPI_ISL_645737, EPI_ISL_645738, EPI_ISL_645739, EPI_ISL_645740, EPI_ISL_645741, EPI_ISL_645742, EPI_ISL_645743, EPI_ISL_645744, EPI_ISL_645745, EPI_ISL_645746, EPI_ISL_645747, EPI_ISL_645748, EPI_ISL_645749, EPI_ISL_645750, EPI_ISL_645751, EPI_ISL_645752, EPI_ISL_645753, EPI_ISL_645754, EPI_ISL_645755, EPI_ISL_645756, EPI_ISL_645757, EPI_ISL_645758, EPI_ISL_645759, EPI_ISL_645760, EPI_ISL_645761, EPI_ISL_645762, EPI_ISL_645763, EPI_ISL_645764, EPI_ISL_645765, EPI_ISL_645766, EPI_ISL_645767, EPI_ISL_645768, EPI_ISL_645769, EPI_ISL_645770, EPI_ISL_645771, EPI_ISL_645772, EPI_ISL_645773, EPI_ISL_645774, EPI_ISL_645775, EPI_ISL_645776, EPI_ISL_645777, EPI_ISL_645778, EPI_ISL_645779, EPI_ISL_645780, EPI_ISL_645781, EPI_ISL_645782, EPI_ISL_645783, EPI_ISL_645784, EPI_ISL_645785, EPI_ISL_645786, EPI_ISL_645787, EPI_ISL_645788, EPI_ISL_645789, EPI_ISL_645790, EPI_ISL_645791, EPI_ISL_645792, EPI_ISL_645793, EPI_ISL_645794, EPI_ISL_645795, EPI_ISL_645796, EPI_ISL_645797, EPI_ISL_645798, EPI_ISL_645799, EPI_ISL_645800, EPI_ISL_645801, EPI_ISL_645802, EPI_ISL_645803, EPI_ISL_645804, EPI_ISL_645805, EPI_ISL_645806, EPI_ISL_645807, EPI_ISL_645808, EPI_ISL_645809, EPI_ISL_645810, EPI_ISL_645811, EPI_ISL_645812, EPI_ISL_645813, EPI_ISL_645814, EPI_ISL_645815, EPI_ISL_645816, EPI_ISL_645817, EPI_ISL_645818, EPI_ISL_645819, EPI_ISL_645820, EPI_ISL_645821, EPI_ISL_645822, EPI_ISL_645823, EPI_ISL_645824, EPI_ISL_645825, EPI_ISL_645827, EPI_ISL_645830, EPI_ISL_645831, EPI_ISL_645833, EPI_ISL_645834, EPI_ISL_645835, EPI_ISL_645837, EPI_ISL_645839, EPI_ISL_645841, EPI_ISL_645842, EPI_ISL_645843, EPI_ISL_645844, EPI_ISL_645845, EPI_ISL_645846, EPI_ISL_645847, EPI_ISL_645848, EPI_ISL_645849, EPI_ISL_645853, EPI_ISL_645854, EPI_ISL_645855, EPI_ISL_645856, EPI_ISL_645857, EPI_ISL_645858, EPI_ISL_645859, EPI_ISL_645860, EPI_ISL_645861, EPI_ISL_645862, EPI_ISL_645863, EPI_ISL_645864, EPI_ISL_645865, EPI_ISL_645867, EPI_ISL_645870, EPI_ISL_645871, EPI_ISL_645872, EPI_ISL_645873, EPI_ISL_645874, EPI_ISL_645875, EPI_ISL_645876, EPI_ISL_645878, EPI_ISL_645879, EPI_ISL_645881, EPI_ISL_645883, EPI_ISL_645884, EPI_ISL_645887, EPI_ISL_645888, EPI_ISL_645889, EPI_ISL_645890, EPI_ISL_645891, EPI_ISL_645892, EPI_ISL_645893, EPI_ISL_645894, EPI_ISL_645895, EPI_ISL_645898, EPI_ISL_645900, EPI_ISL_645901, EPI_ISL_645903, EPI_ISL_645904, EPI_ISL_645906, EPI_ISL_645907, EPI_ISL_645909, EPI_ISL_645910, EPI_ISL_645911, EPI_ISL_645912, EPI_ISL_645913, EPI_ISL_645914, EPI_ISL_645915, EPI_ISL_645919, EPI_ISL_645921, EPI_ISL_645922, EPI_ISL_645924, EPI_ISL_645928, EPI_ISL_645929, EPI_ISL_645931, EPI_ISL_645932, EPI_ISL_645933, EPI_ISL_645934, EPI_ISL_645935, EPI_ISL_645936, EPI_ISL_645937, EPI_ISL_645938, EPI_ISL_645939, EPI_ISL_645940, EPI_ISL_645941, EPI_ISL_645942, EPI_ISL_645943, EPI_ISL_645944, EPI_ISL_645946, EPI_ISL_645947, EPI_ISL_645948, EPI_ISL_645950, EPI_ISL_645951, EPI_ISL_645952, EPI_ISL_645953, EPI_ISL_645954, EPI_ISL_645955, EPI_ISL_645956, EPI_ISL_645957, EPI_ISL_645960, EPI_ISL_645961, EPI_ISL_645963, EPI_ISL_645964, EPI_ISL_645965, EPI_ISL_645966, EPI_ISL_645967, EPI_ISL_645969, EPI_ISL_645971, EPI_ISL_645972, EPI_ISL_645973, EPI_ISL_645974, EPI_ISL_645975, EPI_ISL_645976, EPI_ISL_645977, EPI_ISL_645978, EPI_ISL_645982, EPI_ISL_645984, EPI_ISL_645985, EPI_ISL_645986, EPI_ISL_645988, EPI_ISL_645990, EPI_ISL_645991, EPI_ISL_645996, EPI_ISL_645997, EPI_ISL_645999, EPI_ISL_646000, EPI_ISL_646001, EPI_ISL_646002, EPI_ISL_646003, EPI_ISL_646004, EPI_ISL_646005, EPI_ISL_646006, EPI_ISL_646007, EPI_ISL_646008, EPI_ISL_646010, EPI_ISL_646011, EPI_ISL_646012, EPI_ISL_646013, EPI_ISL_646015, EPI_ISL_646016, EPI_ISL_646019, EPI_ISL_646020, EPI_ISL_646021, EPI_ISL_646023, EPI_ISL_646024, EPI_ISL_646025, EPI_ISL_646026, EPI_ISL_646029, EPI_ISL_646033, EPI_ISL_646036, EPI_ISL_646037, EPI_ISL_646038, EPI_ISL_646040, EPI_ISL_646042, EPI_ISL_646043, EPI_ISL_646044, EPI_ISL_646045, EPI_ISL_646048, EPI_ISL_646049, EPI_ISL_646050, EPI_ISL_646051, EPI_ISL_646053, EPI_ISL_646054, EPI_ISL_646055, EPI_ISL_646057, EPI_ISL_646059, EPI_ISL_646060, EPI_ISL_646061, EPI_ISL_646062, EPI_ISL_646063, EPI_ISL_646064, EPI_ISL_646065, EPI_ISL_646066, EPI_ISL_646068, EPI_ISL_646069, EPI_ISL_646071, EPI_ISL_646072, EPI_ISL_646073, EPI_ISL_646074, EPI_ISL_646075, EPI_ISL_646077, EPI_ISL_646078, EPI_ISL_646079, EPI_ISL_646080, EPI_ISL_646081, EPI_ISL_646082, EPI_ISL_646087, EPI_ISL_646088, EPI_ISL_646090, EPI_ISL_646092, EPI_ISL_646093, EPI_ISL_646094, EPI_ISL_646095, EPI_ISL_646096, EPI_ISL_646097, EPI_ISL_646098, EPI_ISL_646099, EPI_ISL_646100, EPI_ISL_646103, EPI_ISL_646104, EPI_ISL_646106, EPI_ISL_646107, EPI_ISL_646108, EPI_ISL_646109, EPI_ISL_646110, EPI_ISL_646111, EPI_ISL_646112, EPI_ISL_646113, EPI_ISL_646114, EPI_ISL_646116, EPI_ISL_646117, EPI_ISL_646118, EPI_ISL_646119, EPI_ISL_646120, EPI_ISL_646121, EPI_ISL_646123, EPI_ISL_646124, EPI_ISL_646125, EPI_ISL_646126, EPI_ISL_646127, EPI_ISL_646130, EPI_ISL_646132, EPI_ISL_646133, EPI_ISL_646134, EPI_ISL_646137, EPI_ISL_646138, EPI_ISL_646148, EPI_ISL_646150, EPI_ISL_646155, EPI_ISL_646156, EPI_ISL_646157, EPI_ISL_646162, EPI_ISL_646166, EPI_ISL_646169, EPI_ISL_646170, EPI_ISL_646173, EPI_ISL_646180, EPI_ISL_646182, EPI_ISL_646185, EPI_ISL_646189, EPI_ISL_646191, EPI_ISL_646193, EPI_ISL_646198, EPI_ISL_646199, EPI_ISL_646204, EPI_ISL_646207, EPI_ISL_646208, EPI_ISL_646212, EPI_ISL_646215, EPI_ISL_646223, EPI_ISL_646224, EPI_ISL_646231, EPI_ISL_646235, EPI_ISL_646242, EPI_ISL_646245, EPI_ISL_646248, EPI_ISL_646249, EPI_ISL_646252, EPI_ISL_646261, EPI_ISL_646268, EPI_ISL_646269, EPI_ISL_646271, EPI_ISL_646276, EPI_ISL_646277, EPI_ISL_646278, EPI_ISL_646279, EPI_ISL_646283, EPI_ISL_646291, EPI_ISL_646297, EPI_ISL_646299, EPI_ISL_646303, EPI_ISL_646305, EPI_ISL_646307, EPI_ISL_646309, EPI_ISL_646313, EPI_ISL_646314, EPI_ISL_646318, EPI_ISL_646320, EPI_ISL_646323, EPI_ISL_646327, EPI_ISL_646334, EPI_ISL_646342, EPI_ISL_646343, EPI_ISL_646345, EPI_ISL_646346, EPI_ISL_646347, EPI_ISL_646349, EPI_ISL_646350, EPI_ISL_646354, EPI_ISL_646356, EPI_ISL_646362, EPI_ISL_646363, EPI_ISL_646366, EPI_ISL_646369, EPI_ISL_646370, EPI_ISL_646373, EPI_ISL_646375, EPI_ISL_646376, EPI_ISL_646377, EPI_ISL_646382, EPI_ISL_646385, EPI_ISL_646388, EPI_ISL_646397, EPI_ISL_646402, EPI_ISL_646412, EPI_ISL_646418, EPI_ISL_646419, EPI_ISL_646420, EPI_ISL_646426, EPI_ISL_646428, EPI_ISL_646429, EPI_ISL_646430, EPI_ISL_646431, EPI_ISL_646432, EPI_ISL_646434, EPI_ISL_646441, EPI_ISL_646442, EPI_ISL_646443, EPI_ISL_646448, EPI_ISL_646454, EPI_ISL_646463, EPI_ISL_646466, EPI_ISL_646467, EPI_ISL_646468, EPI_ISL_646476, EPI_ISL_646477, EPI_ISL_646478, EPI_ISL_646479, EPI_ISL_646485, EPI_ISL_646486, EPI_ISL_646487, EPI_ISL_646488, EPI_ISL_646489, EPI_ISL_646490, EPI_ISL_646491, EPI_ISL_646492, EPI_ISL_646493, EPI_ISL_646494, EPI_ISL_646497, EPI_ISL_646500, EPI_ISL_646502, EPI_ISL_646505, EPI_ISL_646506, EPI_ISL_646508, EPI_ISL_646509, EPI_ISL_646510, EPI_ISL_646511, EPI_ISL_646512, EPI_ISL_646513, EPI_ISL_646514, EPI_ISL_646515, EPI_ISL_646516, EPI_ISL_646518, EPI_ISL_646520, EPI_ISL_646521, EPI_ISL_646522, EPI_ISL_646525, EPI_ISL_646526, EPI_ISL_646528, EPI_ISL_646529, EPI_ISL_646532, EPI_ISL_646533, EPI_ISL_646535, EPI_ISL_646537, EPI_ISL_646538, EPI_ISL_646540, EPI_ISL_646541, EPI_ISL_646542, EPI_ISL_646543, EPI_ISL_646544, EPI_ISL_646545, EPI_ISL_646546, EPI_ISL_646548, EPI_ISL_646549, EPI_ISL_646551, EPI_ISL_646555, EPI_ISL_646557, EPI_ISL_646558, EPI_ISL_646559, EPI_ISL_646560, EPI_ISL_646561, EPI_ISL_646563, EPI_ISL_646564, EPI_ISL_646566, EPI_ISL_646567, EPI_ISL_646568, EPI_ISL_646569, EPI_ISL_646571, EPI_ISL_646572, EPI_ISL_646573, EPI_ISL_646576, EPI_ISL_646577, EPI_ISL_646580, EPI_ISL_646581, EPI_ISL_646583, EPI_ISL_646585, EPI_ISL_646586, EPI_ISL_646591, EPI_ISL_646592, EPI_ISL_646594, EPI_ISL_646595, EPI_ISL_646597, EPI_ISL_646599, EPI_ISL_646600, EPI_ISL_646601, EPI_ISL_646603, EPI_ISL_646605, EPI_ISL_646606, EPI_ISL_646607, EPI_ISL_646610, EPI_ISL_646612, EPI_ISL_646613, EPI_ISL_646614, EPI_ISL_646616, EPI_ISL_646618, EPI_ISL_646621, EPI_ISL_646623, EPI_ISL_646625, EPI_ISL_646626, EPI_ISL_646628, EPI_ISL_646631, EPI_ISL_646635, EPI_ISL_646638, EPI_ISL_646639, EPI_ISL_646640, EPI_ISL_646642, EPI_ISL_646648, EPI_ISL_646651, EPI_ISL_646652, EPI_ISL_646653, EPI_ISL_646655, EPI_ISL_646658, EPI_ISL_646660, EPI_ISL_646662, EPI_ISL_646664, EPI_ISL_646665, EPI_ISL_646666, EPI_ISL_646668, EPI_ISL_646671, EPI_ISL_646673, EPI_ISL_646675, EPI_ISL_646676, EPI_ISL_646678, EPI_ISL_646683, EPI_ISL_646684, EPI_ISL_646686, EPI_ISL_646687, EPI_ISL_646689, EPI_ISL_646691, EPI_ISL_646692, EPI_ISL_646693, EPI_ISL_646695, EPI_ISL_646696, EPI_ISL_646697, EPI_ISL_646698, EPI_ISL_646702, EPI_ISL_646706, EPI_ISL_646707, EPI_ISL_646708, EPI_ISL_646710, EPI_ISL_646711, EPI_ISL_646712, EPI_ISL_646713, EPI_ISL_646715, EPI_ISL_646716, EPI_ISL_646718, EPI_ISL_646719, EPI_ISL_646721, EPI_ISL_646722, EPI_ISL_646724, EPI_ISL_646727, EPI_ISL_646731, EPI_ISL_646734, EPI_ISL_646736, EPI_ISL_646737, EPI_ISL_646738, EPI_ISL_646739, EPI_ISL_646740, EPI_ISL_646741, EPI_ISL_646743, EPI_ISL_646744, EPI_ISL_646746, EPI_ISL_646747, EPI_ISL_646749, EPI_ISL_646751, EPI_ISL_646752, EPI_ISL_646753, EPI_ISL_646755, EPI_ISL_646756, EPI_ISL_646757, EPI_ISL_646758, EPI_ISL_646760, EPI_ISL_646761, EPI_ISL_646762, EPI_ISL_646763, EPI_ISL_646767, EPI_ISL_646768, EPI_ISL_646769, EPI_ISL_646770, EPI_ISL_646771, EPI_ISL_646773, EPI_ISL_646776, EPI_ISL_646782, EPI_ISL_646784, EPI_ISL_646787, EPI_ISL_646789, EPI_ISL_646790, EPI_ISL_646791, EPI_ISL_646793, EPI_ISL_646795, EPI_ISL_646796, EPI_ISL_646797, EPI_ISL_646799, EPI_ISL_646800, EPI_ISL_646801, EPI_ISL_646802, EPI_ISL_646803, EPI_ISL_646804, EPI_ISL_646805, EPI_ISL_646806, EPI_ISL_646807, EPI_ISL_646808, EPI_ISL_646809, EPI_ISL_646811, EPI_ISL_646812, EPI_ISL_646813, EPI_ISL_646814, EPI_ISL_646815, EPI_ISL_646816, EPI_ISL_646817, EPI_ISL_646819, EPI_ISL_646820, EPI_ISL_646821, EPI_ISL_646822, EPI_ISL_646823, EPI_ISL_646824, EPI_ISL_646827, EPI_ISL_646828, EPI_ISL_646829, EPI_ISL_646830, EPI_ISL_646831, EPI_ISL_646832, EPI_ISL_646833, EPI_ISL_646834, EPI_ISL_646835, EPI_ISL_646836, EPI_ISL_646838, EPI_ISL_646839, EPI_ISL_646841, EPI_ISL_646842, EPI_ISL_646843, EPI_ISL_646844, EPI_ISL_646845, EPI_ISL_646846, EPI_ISL_646847, EPI_ISL_646848, EPI_ISL_646849, EPI_ISL_646851, EPI_ISL_646852, EPI_ISL_646855, EPI_ISL_646856, EPI_ISL_646857, EPI_ISL_646858, EPI_ISL_646859, EPI_ISL_646860, EPI_ISL_646861, EPI_ISL_646862, EPI_ISL_646863, EPI_ISL_646864, EPI_ISL_646865, EPI_ISL_646866, EPI_ISL_646867, EPI_ISL_646871, EPI_ISL_646873, EPI_ISL_646875, EPI_ISL_646876, EPI_ISL_646878, EPI_ISL_646879, EPI_ISL_646881, EPI_ISL_646882, EPI_ISL_646883, EPI_ISL_646884, EPI_ISL_646886, EPI_ISL_646887, EPI_ISL_646888, EPI_ISL_646889, EPI_ISL_646890, EPI_ISL_646893, EPI_ISL_646894, EPI_ISL_646895, EPI_ISL_646896, EPI_ISL_646897, EPI_ISL_646898, EPI_ISL_646899, EPI_ISL_646900, EPI_ISL_646901, EPI_ISL_646903, EPI_ISL_646904, EPI_ISL_646906, EPI_ISL_646907, EPI_ISL_646908, EPI_ISL_646909, EPI_ISL_646910, EPI_ISL_646911, EPI_ISL_646913, EPI_ISL_646914, EPI_ISL_646916, EPI_ISL_646917, EPI_ISL_646918, EPI_ISL_646919, EPI_ISL_646920, EPI_ISL_646922, EPI_ISL_646923, EPI_ISL_646924, EPI_ISL_646925, EPI_ISL_646926, EPI_ISL_646927, EPI_ISL_646928, EPI_ISL_646930, EPI_ISL_646931, EPI_ISL_646932, EPI_ISL_646933, EPI_ISL_646934, EPI_ISL_646935, EPI_ISL_646938, EPI_ISL_646940, EPI_ISL_646941, EPI_ISL_646942, EPI_ISL_646943, EPI_ISL_646944, EPI_ISL_646945, EPI_ISL_646946, EPI_ISL_646947, EPI_ISL_646949, EPI_ISL_646950, EPI_ISL_646951, EPI_ISL_646952, EPI_ISL_646953, EPI_ISL_646954, EPI_ISL_646956, EPI_ISL_646957, EPI_ISL_646958, EPI_ISL_646959, EPI_ISL_646961, EPI_ISL_646962, EPI_ISL_646963, EPI_ISL_646965, EPI_ISL_646966, EPI_ISL_646968, EPI_ISL_646969, EPI_ISL_646970, EPI_ISL_646971, EPI_ISL_646972, EPI_ISL_646973, EPI_ISL_646978, EPI_ISL_646979, EPI_ISL_646980, EPI_ISL_646981, EPI_ISL_646982, EPI_ISL_646984, EPI_ISL_646985, EPI_ISL_646986, EPI_ISL_646987, EPI_ISL_646989, EPI_ISL_646990, EPI_ISL_646991, EPI_ISL_646993, EPI_ISL_646994, EPI_ISL_646995, EPI_ISL_646997, EPI_ISL_646998, EPI_ISL_646999, EPI_ISL_647000, EPI_ISL_647002, EPI_ISL_647004, EPI_ISL_647005, EPI_ISL_647007, EPI_ISL_647008, EPI_ISL_647009, EPI_ISL_647010, EPI_ISL_647011, EPI_ISL_647012, EPI_ISL_647013, EPI_ISL_647015, EPI_ISL_647016, EPI_ISL_647017, EPI_ISL_647018, EPI_ISL_647019, EPI_ISL_647020, EPI_ISL_647021, EPI_ISL_647022, EPI_ISL_647024, EPI_ISL_647025, EPI_ISL_647026, EPI_ISL_647027, EPI_ISL_647028, EPI_ISL_647029, EPI_ISL_647030, EPI_ISL_647033, EPI_ISL_647034, EPI_ISL_647036, EPI_ISL_647037, EPI_ISL_647039, EPI_ISL_647040, EPI_ISL_647041, EPI_ISL_647042, EPI_ISL_647043, EPI_ISL_647044, EPI_ISL_647046, EPI_ISL_647047, EPI_ISL_647048, EPI_ISL_647049, EPI_ISL_647051, EPI_ISL_647052, EPI_ISL_647054, EPI_ISL_647055, EPI_ISL_647056, EPI_ISL_647057, EPI_ISL_647058, EPI_ISL_647060, EPI_ISL_647062, EPI_ISL_647063, EPI_ISL_647064, EPI_ISL_647065, EPI_ISL_647066, EPI_ISL_647068, EPI_ISL_647069, EPI_ISL_647070, EPI_ISL_647071, EPI_ISL_647072, EPI_ISL_647074, EPI_ISL_647075, EPI_ISL_647077, EPI_ISL_647078, EPI_ISL_647079, EPI_ISL_647080, EPI_ISL_647082, EPI_ISL_647083, EPI_ISL_647084, EPI_ISL_647085, EPI_ISL_647086, EPI_ISL_647088, EPI_ISL_647089, EPI_ISL_647091, EPI_ISL_647092, EPI_ISL_647093, EPI_ISL_647094, EPI_ISL_647095, EPI_ISL_647096, EPI_ISL_647098, EPI_ISL_647099, EPI_ISL_647100, EPI_ISL_647101, EPI_ISL_647103, EPI_ISL_647104, EPI_ISL_647106, EPI_ISL_647107, EPI_ISL_647108, EPI_ISL_647111, EPI_ISL_647112, EPI_ISL_647113, EPI_ISL_647116, EPI_ISL_647118, EPI_ISL_647119, EPI_ISL_647120, EPI_ISL_647122, EPI_ISL_647468, EPI_ISL_647469, EPI_ISL_647473, EPI_ISL_647474, EPI_ISL_647477, EPI_ISL_647497, EPI_ISL_647499, EPI_ISL_647503, EPI_ISL_647506, EPI_ISL_647511, EPI_ISL_647513, EPI_ISL_647526, EPI_ISL_647528, EPI_ISL_647535, EPI_ISL_647536, EPI_ISL_647541, EPI_ISL_647546, EPI_ISL_647550, EPI_ISL_647553, EPI_ISL_647558, EPI_ISL_647560, EPI_ISL_647572, EPI_ISL_647573, EPI_ISL_647576, EPI_ISL_647579, EPI_ISL_647580, EPI_ISL_647584, EPI_ISL_647591, EPI_ISL_647594, EPI_ISL_647603, EPI_ISL_647606, EPI_ISL_647608, EPI_ISL_647612, EPI_ISL_647615, EPI_ISL_647618, EPI_ISL_647624, EPI_ISL_647630, EPI_ISL_647632, EPI_ISL_647634, EPI_ISL_647635, EPI_ISL_647638, EPI_ISL_647643, EPI_ISL_647650, EPI_ISL_647655, EPI_ISL_647659, EPI_ISL_647667, EPI_ISL_647668, EPI_ISL_647672, EPI_ISL_647677, EPI_ISL_647682, EPI_ISL_647683, EPI_ISL_647689, EPI_ISL_647693, EPI_ISL_647695, EPI_ISL_647698, EPI_ISL_647700, EPI_ISL_647704, EPI_ISL_647705, EPI_ISL_647708, EPI_ISL_647711, EPI_ISL_647726, EPI_ISL_647733, EPI_ISL_647741, EPI_ISL_647750, EPI_ISL_647752, EPI_ISL_647762, EPI_ISL_647771, EPI_ISL_647776, EPI_ISL_647780, EPI_ISL_647786, EPI_ISL_647787, EPI_ISL_647792, EPI_ISL_649228, EPI_ISL_649252, EPI_ISL_649354, EPI_ISL_649432, EPI_ISL_649483, EPI_ISL_649484, EPI_ISL_649485, EPI_ISL_649486, EPI_ISL_649487, EPI_ISL_649488, EPI_ISL_649489, EPI_ISL_649490, EPI_ISL_649491, EPI_ISL_710908, EPI_ISL_710909, EPI_ISL_710910, EPI_ISL_710911, EPI_ISL_710912, EPI_ISL_710913, EPI_ISL_710914, EPI_ISL_710915, EPI_ISL_710916, EPI_ISL_710917, EPI_ISL_710918, EPI_ISL_710919, EPI_ISL_710920, EPI_ISL_710921, EPI_ISL_710922, EPI_ISL_710923, EPI_ISL_710924, EPI_ISL_710925, EPI_ISL_710926, EPI_ISL_710927, EPI_ISL_710928, EPI_ISL_710929, EPI_ISL_710930, EPI_ISL_710931, EPI_ISL_710932, EPI_ISL_710933, EPI_ISL_710934, EPI_ISL_710935, EPI_ISL_710936, EPI_ISL_710937, EPI_ISL_710938, EPI_ISL_710939, EPI_ISL_710940, EPI_ISL_710941, EPI_ISL_710942, EPI_ISL_710943, EPI_ISL_710944, EPI_ISL_710945, EPI_ISL_710946, EPI_ISL_710947, EPI_ISL_710948, EPI_ISL_710949, EPI_ISL_710950, EPI_ISL_710951, EPI_ISL_710952, EPI_ISL_710953, EPI_ISL_710954, EPI_ISL_710955, EPI_ISL_710956, EPI_ISL_710957, EPI_ISL_710958, EPI_ISL_710959, EPI_ISL_710960, EPI_ISL_710961, EPI_ISL_710962, EPI_ISL_710963, EPI_ISL_710964, EPI_ISL_710965, EPI_ISL_710966, EPI_ISL_710967, EPI_ISL_710968, EPI_ISL_710969, EPI_ISL_710970, EPI_ISL_710971, EPI_ISL_710972, EPI_ISL_710973, EPI_ISL_710974, EPI_ISL_710975, EPI_ISL_710976, EPI_ISL_710977, EPI_ISL_710978, EPI_ISL_710979, EPI_ISL_710980, EPI_ISL_710981, EPI_ISL_710982, EPI_ISL_710983, EPI_ISL_710984, EPI_ISL_710985, EPI_ISL_710986, EPI_ISL_710987, EPI_ISL_710988, EPI_ISL_710989, EPI_ISL_710990, EPI_ISL_710991, EPI_ISL_710992, EPI_ISL_710993, EPI_ISL_710994, EPI_ISL_711008, EPI_ISL_711009, EPI_ISL_711010, EPI_ISL_711011, EPI_ISL_711012, EPI_ISL_711013, EPI_ISL_711015, EPI_ISL_711016, EPI_ISL_711017, EPI_ISL_711018, EPI_ISL_711019, EPI_ISL_711020, EPI_ISL_711021, EPI_ISL_711022, EPI_ISL_711027, EPI_ISL_711028, EPI_ISL_711029, EPI_ISL_711030, EPI_ISL_711031, EPI_ISL_711032, EPI_ISL_711033, EPI_ISL_711034, EPI_ISL_711035, EPI_ISL_711036, EPI_ISL_711037, EPI_ISL_711039, EPI_ISL_711040, EPI_ISL_711041, EPI_ISL_711042, EPI_ISL_711043, EPI_ISL_711044, EPI_ISL_711045, EPI_ISL_711046, EPI_ISL_711047, EPI_ISL_892220 | | | |
| see above | Lighthouse Lab in Alderley Park | Wellcome Sanger Institute for the COVID-19 Genomics UK (COG-UK) Consortium | Cordelia Langford; David K. Jackson; Dominic Kwiatkowski; Ewan Harrison; Ian Johnston; Jacquelyn Wynn; John Sillitoe on behalf of the Wellcome Sanger Institute COVID-19 Surveillance Team; John Sillitoe on behalf of the Wellcome Sanger Institute COVID-19 Surveillance Team (http://www.sanger.ac.uk/covid-team); Mairead Hyland; Roberto Amato; Sonia Goncalves; The Lighthouse Lab in Alderley Park and Alex Alderton |
| EPI_ISL_643315, EPI_ISL_643316, EPI_ISL_643317, EPI_ISL_643318, EPI_ISL_643319, EPI_ISL_643320, EPI_ISL_643321, EPI_ISL_643322, EPI_ISL_643323, EPI_ISL_643324, EPI_ISL_643325, EPI_ISL_643326, EPI_ISL_643327, EPI_ISL_643328, EPI_ISL_643329, EPI_ISL_643331, EPI_ISL_643332, EPI_ISL_643333, EPI_ISL_643334, EPI_ISL_643335, EPI_ISL_643336, EPI_ISL_643337, EPI_ISL_643338, EPI_ISL_643339, EPI_ISL_643340, EPI_ISL_643341, EPI_ISL_643342, EPI_ISL_643343, EPI_ISL_643344, EPI_ISL_643345, EPI_ISL_643346, EPI_ISL_643347, EPI_ISL_643348, EPI_ISL_643349, EPI_ISL_643350, EPI_ISL_643351, EPI_ISL_643352, EPI_ISL_643353, EPI_ISL_643354, EPI_ISL_643355, EPI_ISL_643356, EPI_ISL_643357, EPI_ISL_643358, EPI_ISL_643359, EPI_ISL_643360, EPI_ISL_643361, EPI_ISL_643362, EPI_ISL_643363, EPI_ISL_643364, EPI_ISL_643365, EPI_ISL_643366, EPI_ISL_643367, EPI_ISL_643368, EPI_ISL_643369, EPI_ISL_643370, EPI_ISL_643371, EPI_ISL_643372, EPI_ISL_643373, EPI_ISL_643374, EPI_ISL_643375, EPI_ISL_643376, EPI_ISL_643377, EPI_ISL_643378, EPI_ISL_643379, EPI_ISL_643380, EPI_ISL_643381, EPI_ISL_643382, EPI_ISL_643383, EPI_ISL_643384, EPI_ISL_643385, EPI_ISL_643386, EPI_ISL_643387, EPI_ISL_643388, EPI_ISL_643389, EPI_ISL_643390, EPI_ISL_643391, EPI_ISL_643392, EPI_ISL_643393, EPI_ISL_643394, EPI_ISL_643395, EPI_ISL_643396, EPI_ISL_643397, EPI_ISL_643398, EPI_ISL_643399, EPI_ISL_643400, EPI_ISL_643401, EPI_ISL_643402, EPI_ISL_643403, EPI_ISL_643404, EPI_ISL_643405, EPI_ISL_643406, EPI_ISL_643407, EPI_ISL_643408, EPI_ISL_643409, EPI_ISL_643410, EPI_ISL_643411, EPI_ISL_643412, EPI_ISL_643414, EPI_ISL_643415, EPI_ISL_643416, EPI_ISL_643417, EPI_ISL_643418, EPI_ISL_643419, EPI_ISL_643420, EPI_ISL_643421, EPI_ISL_643422, EPI_ISL_643423, EPI_ISL_643424, EPI_ISL_643425, EPI_ISL_643426, EPI_ISL_643427, EPI_ISL_643428, EPI_ISL_643429, EPI_ISL_643430, EPI_ISL_643431, EPI_ISL_643432, EPI_ISL_643433, EPI_ISL_643434, EPI_ISL_643435, EPI_ISL_643436, EPI_ISL_643437, EPI_ISL_643438, EPI_ISL_643439, EPI_ISL_643440, EPI_ISL_643441, EPI_ISL_643442, EPI_ISL_643443, EPI_ISL_643444, EPI_ISL_643445, EPI_ISL_643446, EPI_ISL_643447, EPI_ISL_643448, EPI_ISL_643449, EPI_ISL_643450, EPI_ISL_643451, EPI_ISL_643452, EPI_ISL_643453, EPI_ISL_643454, EPI_ISL_643455, EPI_ISL_643456, EPI_ISL_643457, EPI_ISL_643458, EPI_ISL_643459, EPI_ISL_643460, EPI_ISL_643461, EPI_ISL_643462, EPI_ISL_643463, EPI_ISL_643464, EPI_ISL_643465, EPI_ISL_643466, EPI_ISL_643467, EPI_ISL_643468, EPI_ISL_643469, EPI_ISL_643470, EPI_ISL_643471, EPI_ISL_643472, EPI_ISL_643473, EPI_ISL_643474, EPI_ISL_643475, EPI_ISL_643476, EPI_ISL_643477, EPI_ISL_643478, EPI_ISL_643479, EPI_ISL_643480, EPI_ISL_643481, EPI_ISL_643482, EPI_ISL_643483, EPI_ISL_643484, EPI_ISL_643485, EPI_ISL_643486, EPI_ISL_643487, EPI_ISL_643488, EPI_ISL_643489, EPI_ISL_643490, EPI_ISL_643491, EPI_ISL_643492, EPI_ISL_643493, EPI_ISL_643494, EPI_ISL_643495, EPI_ISL_643496, EPI_ISL_643497, EPI_ISL_643498, EPI_ISL_643499, EPI_ISL_643500, EPI_ISL_643501, EPI_ISL_643502, EPI_ISL_643503, EPI_ISL_643504, EPI_ISL_643505, EPI_ISL_643506, EPI_ISL_643507, EPI_ISL_643508, EPI_ISL_643509, EPI_ISL_643510, EPI_ISL_643511, EPI_ISL_643512, EPI_ISL_643513, EPI_ISL_643514, EPI_ISL_643515, EPI_ISL_643516, EPI_ISL_643517, EPI_ISL_643518, EPI_ISL_643519, EPI_ISL_643520, EPI_ISL_643521, EPI_ISL_643522, EPI_ISL_643523, EPI_ISL_643524, EPI_ISL_643525, EPI_ISL_643526, EPI_ISL_643527, EPI_ISL_643528, EPI_ISL_643529, EPI_ISL_643530, EPI_ISL_643531, EPI_ISL_643532, EPI_ISL_643533, EPI_ISL_643534, EPI_ISL_643535, EPI_ISL_643536, EPI_ISL_643537, EPI_ISL_643538, EPI_ISL_643539, EPI_ISL_643540, EPI_ISL_643541, EPI_ISL_643542, EPI_ISL_643543, EPI_ISL_643544, EPI_ISL_643545, EPI_ISL_643546, EPI_ISL_643547, EPI_ISL_643548, EPI_ISL_643549, EPI_ISL_643550, EPI_ISL_643551, EPI_ISL_643552, EPI_ISL_643553, EPI_ISL_643554, EPI_ISL_643555, EPI_ISL_643556, EPI_ISL_643557, EPI_ISL_643558, EPI_ISL_643559, EPI_ISL_643560, EPI_ISL_643561, EPI_ISL_643562, EPI_ISL_643563, EPI_ISL_643564, EPI_ISL_643565, EPI_ISL_643566, EPI_ISL_643567, EPI_ISL_643568, EPI_ISL_643569, EPI_ISL_643570, EPI_ISL_643571, EPI_ISL_643572, EPI_ISL_643573, EPI_ISL_643574, EPI_ISL_643575, EPI_ISL_643576, EPI_ISL_643577, EPI_ISL_643578, EPI_ISL_643579, EPI_ISL_643580, EPI_ISL_643581, EPI_ISL_643582, EPI_ISL_643583, EPI_ISL_643584, EPI_ISL_643585, EPI_ISL_643586, EPI_ISL_643587, EPI_ISL_643588, EPI_ISL_643589, EPI_ISL_643590, EPI_ISL_643591, EPI_ISL_643592, EPI_ISL_643593, EPI_ISL_643594, EPI_ISL_643595, EPI_ISL_643596, EPI_ISL_643597, EPI_ISL_643598, EPI_ISL_643599, EPI_ISL_643600, EPI_ISL_643601, EPI_ISL_643602, EPI_ISL_643603, EPI_ISL_643604, EPI_ISL_643605, EPI_ISL_643606, EPI_ISL_643607, EPI_ISL_643608, EPI_ISL_643609, EPI_ISL_643610, EPI_ISL_643611, EPI_ISL_643612, EPI_ISL_643613, EPI_ISL_643614, EPI_ISL_643615, EPI_ISL_643616, EPI_ISL_643617, EPI_ISL_643618, EPI_ISL_643619, EPI_ISL_643620, EPI_ISL_643621, EPI_ISL_643622, EPI_ISL_643623, EPI_ISL_643624, EPI_ISL_643625, EPI_ISL_643626, EPI_ISL_643627, EPI_ISL_643628, | | | |

| | | | |
|---|---|---|---|
| EPI_ISL_643629, EPI_ISL_643630, EPI_ISL_643631, EPI_ISL_643632, EPI_ISL_643633, EPI_ISL_643634, EPI_ISL_643635, EPI_ISL_643636, EPI_ISL_643637, EPI_ISL_643638, EPI_ISL_643639, EPI_ISL_643640, EPI_ISL_643641, EPI_ISL_643642, EPI_ISL_643643, EPI_ISL_643644, EPI_ISL_643645, EPI_ISL_643646, EPI_ISL_643647, EPI_ISL_643648, EPI_ISL_643649, EPI_ISL_643650, EPI_ISL_643651, EPI_ISL_643652, EPI_ISL_643653, EPI_ISL_643654, EPI_ISL_643655, EPI_ISL_643656, EPI_ISL_643657, EPI_ISL_643658, EPI_ISL_643659, EPI_ISL_643660, EPI_ISL_643661, EPI_ISL_643662, EPI_ISL_643663, EPI_ISL_643664, EPI_ISL_643665, EPI_ISL_643666, EPI_ISL_643667, EPI_ISL_643668, EPI_ISL_643669, EPI_ISL_643670, EPI_ISL_643671, EPI_ISL_643672, EPI_ISL_643673, EPI_ISL_643674, EPI_ISL_643675, EPI_ISL_643676, EPI_ISL_643677, EPI_ISL_643678, EPI_ISL_643679, EPI_ISL_643680, EPI_ISL_643681, EPI_ISL_643682, EPI_ISL_643683, EPI_ISL_643684, EPI_ISL_643685, EPI_ISL_643686, EPI_ISL_643687, EPI_ISL_643688, EPI_ISL_643689, EPI_ISL_643690, EPI_ISL_643691, EPI_ISL_643692, EPI_ISL_643693, EPI_ISL_643694, EPI_ISL_643695, EPI_ISL_643696, EPI_ISL_643697, EPI_ISL_643698, EPI_ISL_643699, EPI_ISL_643700, EPI_ISL_643701, EPI_ISL_643702, EPI_ISL_643703, EPI_ISL_643704, EPI_ISL_643705, EPI_ISL_643706, EPI_ISL_643707, EPI_ISL_643708, EPI_ISL_643709, EPI_ISL_643710, EPI_ISL_643711, EPI_ISL_643712, EPI_ISL_643713, EPI_ISL_643714, EPI_ISL_643715, EPI_ISL_643716, EPI_ISL_643717, EPI_ISL_643718, EPI_ISL_643719, EPI_ISL_643720, EPI_ISL_643721, EPI_ISL_643722, EPI_ISL_643723, EPI_ISL_643724, EPI_ISL_643725, EPI_ISL_643726, EPI_ISL_643727, EPI_ISL_643728, EPI_ISL_643729, EPI_ISL_643730, EPI_ISL_643731, EPI_ISL_643732, EPI_ISL_643733, EPI_ISL_643734, EPI_ISL_643735, EPI_ISL_643736, EPI_ISL_643737, EPI_ISL_643738, EPI_ISL_643739, EPI_ISL_643740, EPI_ISL_643741, EPI_ISL_643742, EPI_ISL_643743, EPI_ISL_643744, EPI_ISL_643745, EPI_ISL_643746, EPI_ISL_643747, EPI_ISL_643748, EPI_ISL_643749, EPI_ISL_643750, EPI_ISL_643751, EPI_ISL_643752, EPI_ISL_643753, EPI_ISL_643754, EPI_ISL_643755, EPI_ISL_643756, EPI_ISL_643757, EPI_ISL_643758, EPI_ISL_643759, EPI_ISL_643760, EPI_ISL_643761, EPI_ISL_643762, EPI_ISL_643763, EPI_ISL_643764, EPI_ISL_643765, EPI_ISL_643766, EPI_ISL_643767, EPI_ISL_643769, EPI_ISL_643770, EPI_ISL_643771, EPI_ISL_643772, EPI_ISL_643773, EPI_ISL_643774, EPI_ISL_643775, EPI_ISL_643776, EPI_ISL_643777, EPI_ISL_643778, EPI_ISL_643779, EPI_ISL_643780, EPI_ISL_643781, EPI_ISL_643782, EPI_ISL_643783, EPI_ISL_643784, EPI_ISL_643785, EPI_ISL_643786, EPI_ISL_643787, EPI_ISL_643788, EPI_ISL_643789, EPI_ISL_643790, EPI_ISL_643791, EPI_ISL_643792, EPI_ISL_643793, EPI_ISL_643794, EPI_ISL_643795, EPI_ISL_643796, EPI_ISL_643797, EPI_ISL_643798, EPI_ISL_643799, EPI_ISL_643800, EPI_ISL_643801, EPI_ISL_643802, EPI_ISL_643803, EPI_ISL_643804, EPI_ISL_643805, EPI_ISL_643806, EPI_ISL_643807, EPI_ISL_643808, EPI_ISL_643809, EPI_ISL_643810, EPI_ISL_643811, EPI_ISL_643812, EPI_ISL_643813, EPI_ISL_643814, EPI_ISL_643815, EPI_ISL_643816, EPI_ISL_643817, EPI_ISL_643818, EPI_ISL_643819, EPI_ISL_643820, EPI_ISL_643821, EPI_ISL_643822, EPI_ISL_643823, EPI_ISL_643824, EPI_ISL_643825, EPI_ISL_643826, EPI_ISL_643827, EPI_ISL_643828, EPI_ISL_643829, EPI_ISL_643830, EPI_ISL_643831, EPI_ISL_643832, EPI_ISL_643833, EPI_ISL_643834, EPI_ISL_643835, EPI_ISL_643836, EPI_ISL_643837, EPI_ISL_643838, EPI_ISL_643839, EPI_ISL_643840, EPI_ISL_643841, EPI_ISL_643842, EPI_ISL_643843, EPI_ISL_643844, EPI_ISL_643845, EPI_ISL_643846, EPI_ISL_643847, EPI_ISL_643848, EPI_ISL_643849, EPI_ISL_643850, EPI_ISL_643851, EPI_ISL_643852, EPI_ISL_643853, EPI_ISL_643854, EPI_ISL_643855, EPI_ISL_643856, EPI_ISL_643857, EPI_ISL_643858, EPI_ISL_643859, EPI_ISL_643860, EPI_ISL_643861, EPI_ISL_643862, EPI_ISL_643863, EPI_ISL_643864, EPI_ISL_643865, EPI_ISL_643866, EPI_ISL_643867, EPI_ISL_643868, EPI_ISL_643869, EPI_ISL_643870, EPI_ISL_643871, EPI_ISL_643872, EPI_ISL_643873, EPI_ISL_643874, EPI_ISL_643875, EPI_ISL_643876, EPI_ISL_643877, EPI_ISL_643878, EPI_ISL_643879, EPI_ISL_643880, EPI_ISL_643881, EPI_ISL_643882, EPI_ISL_643883, EPI_ISL_643884, EPI_ISL_643885, EPI_ISL_643886, EPI_ISL_643887, EPI_ISL_643888, EPI_ISL_643889, EPI_ISL_643890, EPI_ISL_643891, EPI_ISL_643892, EPI_ISL_643893, EPI_ISL_643894, EPI_ISL_643895, EPI_ISL_643896, EPI_ISL_643897, EPI_ISL_643898, EPI_ISL_643899, EPI_ISL_643900, EPI_ISL_643901, EPI_ISL_643902, EPI_ISL_643903, EPI_ISL_643904, EPI_ISL_643905, EPI_ISL_643906, EPI_ISL_643907, EPI_ISL_643908, EPI_ISL_643909, EPI_ISL_643910, EPI_ISL_643911, EPI_ISL_643912, EPI_ISL_643913, EPI_ISL_643914, EPI_ISL_643915, EPI_ISL_643916, EPI_ISL_643917, EPI_ISL_643918, EPI_ISL_643919, EPI_ISL_643920, EPI_ISL_643921, EPI_ISL_643922, EPI_ISL_643923, EPI_ISL_643924, EPI_ISL_643925, EPI_ISL_643926, EPI_ISL_643927, EPI_ISL_643928, EPI_ISL_643929, EPI_ISL_643930, EPI_ISL_643931, EPI_ISL_643932, EPI_ISL_643933, EPI_ISL_643934, EPI_ISL_643935, EPI_ISL_643936, EPI_ISL_643937, EPI_ISL_643938, EPI_ISL_643939, EPI_ISL_643940, EPI_ISL_643941, EPI_ISL_643942, EPI_ISL_643943, EPI_ISL_643944, EPI_ISL_643945, EPI_ISL_643946, EPI_ISL_643947, EPI_ISL_643948, EPI_ISL_643949, EPI_ISL_643950, EPI_ISL_643951, EPI_ISL_643952, EPI_ISL_643953, EPI_ISL_643954, EPI_ISL_643955, EPI_ISL_643956, EPI_ISL_643957, EPI_ISL_643958, EPI_ISL_643959, EPI_ISL_643960, EPI_ISL_643961, EPI_ISL_643962, EPI_ISL_643963, EPI_ISL_643964, EPI_ISL_643965, EPI_ISL_643966, EPI_ISL_643967, EPI_ISL_643968, EPI_ISL_643969, EPI_ISL_643970, EPI_ISL_643971, EPI_ISL_643972, EPI_ISL_643973, EPI_ISL_643974, EPI_ISL_643975, EPI_ISL_643976, EPI_ISL_643977, EPI_ISL_643978, EPI_ISL_643979, EPI_ISL_643980, EPI_ISL_643981, EPI_ISL_643982, EPI_ISL_645322, EPI_ISL_645414, EPI_ISL_645826, EPI_ISL_645828, EPI_ISL_645829, EPI_ISL_645832, EPI_ISL_645836, EPI_ISL_645838, EPI_ISL_645840, EPI_ISL_645850, EPI_ISL_645851, EPI_ISL_645852, EPI_ISL_645866, EPI_ISL_645868, EPI_ISL_645869, EPI_ISL_645877, EPI_ISL_645880, EPI_ISL_645882, EPI_ISL_645885, EPI_ISL_645886, EPI_ISL_645896, EPI_ISL_645897, EPI_ISL_645899, EPI_ISL_645902, EPI_ISL_645905, EPI_ISL_645908, EPI_ISL_645916, EPI_ISL_645917, EPI_ISL_645918, EPI_ISL_645920, EPI_ISL_645923, EPI_ISL_645925, EPI_ISL_645926, EPI_ISL_645927, EPI_ISL_645930, EPI_ISL_645945, EPI_ISL_645949, EPI_ISL_645958, EPI_ISL_645959, EPI_ISL_645962, EPI_ISL_645968, EPI_ISL_645970, EPI_ISL_645979, EPI_ISL_645980, EPI_ISL_645981, EPI_ISL_645983, EPI_ISL_645987, EPI_ISL_645989, EPI_ISL_645992, EPI_ISL_645993, EPI_ISL_645994, EPI_ISL_645995, EPI_ISL_645998, EPI_ISL_646009, EPI_ISL_646014, EPI_ISL_646017, EPI_ISL_646018, EPI_ISL_646022, EPI_ISL_646027, EPI_ISL_646028, EPI_ISL_646030, EPI_ISL_646031, EPI_ISL_646032, EPI_ISL_646034, EPI_ISL_646035, EPI_ISL_646039, EPI_ISL_646041, EPI_ISL_646046, EPI_ISL_646047, EPI_ISL_646052, EPI_ISL_646056, EPI_ISL_646058, EPI_ISL_646067, EPI_ISL_646070, EPI_ISL_646076, EPI_ISL_646083, EPI_ISL_646084, EPI_ISL_646085, EPI_ISL_646086, EPI_ISL_646089, EPI_ISL_646091, EPI_ISL_646101, EPI_ISL_646102, EPI_ISL_646105, EPI_ISL_646115, EPI_ISL_646122, EPI_ISL_646128, EPI_ISL_646129, EPI_ISL_646131, EPI_ISL_646135, EPI_ISL_646136, EPI_ISL_646139, EPI_ISL_647123, EPI_ISL_647124, EPI_ISL_647125, EPI_ISL_647126, EPI_ISL_647127, EPI_ISL_647128, EPI_ISL_647129, EPI_ISL_647130, EPI_ISL_647131, EPI_ISL_647132, EPI_ISL_647133, EPI_ISL_647134, EPI_ISL_647135, EPI_ISL_647136, EPI_ISL_647137, EPI_ISL_647138, EPI_ISL_647139, EPI_ISL_647140, EPI_ISL_647141, EPI_ISL_647142, EPI_ISL_647143, EPI_ISL_647144, EPI_ISL_647145, EPI_ISL_647146, EPI_ISL_647147, EPI_ISL_647148, EPI_ISL_647149, EPI_ISL_647150, EPI_ISL_647151, EPI_ISL_647152, EPI_ISL_647153, EPI_ISL_647154, EPI_ISL_647155, EPI_ISL_647156, EPI_ISL_647157, EPI_ISL_647158, EPI_ISL_647159, EPI_ISL_647160, EPI_ISL_647161, EPI_ISL_647162, EPI_ISL_647163, EPI_ISL_647164, EPI_ISL_647165, EPI_ISL_647166, EPI_ISL_647167, EPI_ISL_647168, EPI_ISL_647169, EPI_ISL_647170, EPI_ISL_647171, EPI_ISL_647172, EPI_ISL_647173, EPI_ISL_647174, EPI_ISL_647175, EPI_ISL_647176, EPI_ISL_647177, EPI_ISL_647178, EPI_ISL_647179, EPI_ISL_647180, EPI_ISL_647181, EPI_ISL_647182, EPI_ISL_647183, EPI_ISL_647184, EPI_ISL_647185, EPI_ISL_647186, EPI_ISL_647187, EPI_ISL_647188, EPI_ISL_647189, EPI_ISL_647190, EPI_ISL_647191, EPI_ISL_647192, EPI_ISL_647193, EPI_ISL_647194, EPI_ISL_647195, EPI_ISL_647196, EPI_ISL_647197, EPI_ISL_647198, EPI_ISL_647199, EPI_ISL_647200, EPI_ISL_647201, EPI_ISL_647202, EPI_ISL_647203, EPI_ISL_647204, EPI_ISL_647205, EPI_ISL_647206, EPI_ISL_647207, EPI_ISL_647208, EPI_ISL_647209, EPI_ISL_647210, EPI_ISL_647211, EPI_ISL_647212, EPI_ISL_647214, EPI_ISL_647215, EPI_ISL_647216, EPI_ISL_647217, EPI_ISL_647218, EPI_ISL_647219, EPI_ISL_647220, EPI_ISL_647221, EPI_ISL_647222, EPI_ISL_647223, EPI_ISL_647224, EPI_ISL_647225, EPI_ISL_647226, EPI_ISL_647227, EPI_ISL_647228, EPI_ISL_647229, EPI_ISL_647230, EPI_ISL_647231, EPI_ISL_647232, EPI_ISL_647233, EPI_ISL_647234, EPI_ISL_647235, EPI_ISL_647236, EPI_ISL_647237, EPI_ISL_647238, EPI_ISL_647239, EPI_ISL_647240, EPI_ISL_647241, EPI_ISL_647242, EPI_ISL_647243, EPI_ISL_647244, EPI_ISL_647245, EPI_ISL_647246, EPI_ISL_647247, EPI_ISL_647248, EPI_ISL_647249, EPI_ISL_647250, EPI_ISL_647251, EPI_ISL_647252, EPI_ISL_647253, EPI_ISL_647254, EPI_ISL_647255, EPI_ISL_647256, EPI_ISL_647257, EPI_ISL_647258, EPI_ISL_647259, EPI_ISL_647260, EPI_ISL_647261, EPI_ISL_647262, EPI_ISL_647263, EPI_ISL_647264, EPI_ISL_647265, EPI_ISL_647266, EPI_ISL_647267, EPI_ISL_647268, EPI_ISL_647269, EPI_ISL_647270, EPI_ISL_647271, EPI_ISL_647272, EPI_ISL_647273, EPI_ISL_647274, EPI_ISL_647275, EPI_ISL_647276, EPI_ISL_647277, EPI_ISL_647278, EPI_ISL_647279, EPI_ISL_647280, EPI_ISL_647281, EPI_ISL_647282, EPI_ISL_647283, EPI_ISL_647284, EPI_ISL_647285, EPI_ISL_647286, EPI_ISL_647287, EPI_ISL_647288, EPI_ISL_647289, EPI_ISL_647290, EPI_ISL_647291, EPI_ISL_647292, EPI_ISL_647293, EPI_ISL_647294, EPI_ISL_647295, EPI_ISL_647296, EPI_ISL_647297, EPI_ISL_647298, EPI_ISL_647299, EPI_ISL_647300, EPI_ISL_647301, EPI_ISL_647302, EPI_ISL_647303, EPI_ISL_647304, EPI_ISL_647305, EPI_ISL_647306, EPI_ISL_647307, EPI_ISL_647308, EPI_ISL_647309, EPI_ISL_647310, EPI_ISL_647311, EPI_ISL_647312, EPI_ISL_647313, EPI_ISL_647314, EPI_ISL_647315, EPI_ISL_647316, EPI_ISL_647317, EPI_ISL_647318, EPI_ISL_647319, EPI_ISL_647320, EPI_ISL_647321, EPI_ISL_647322, EPI_ISL_647323, EPI_ISL_647324, EPI_ISL_647325, EPI_ISL_647326, EPI_ISL_647327, EPI_ISL_647328, EPI_ISL_647329, EPI_ISL_647330, EPI_ISL_647331, EPI_ISL_647332, EPI_ISL_647333, EPI_ISL_647334, EPI_ISL_647335, EPI_ISL_647336, EPI_ISL_647337, EPI_ISL_647338, EPI_ISL_647339, EPI_ISL_647340, EPI_ISL_647341, EPI_ISL_647342, EPI_ISL_647343, EPI_ISL_647344, EPI_ISL_647345, EPI_ISL_647346, EPI_ISL_647347, EPI_ISL_647348, EPI_ISL_647349, EPI_ISL_647350, EPI_ISL_647351, EPI_ISL_647352, EPI_ISL_647353, EPI_ISL_647354, EPI_ISL_647355, EPI_ISL_647356, EPI_ISL_647357, EPI_ISL_647358, EPI_ISL_647359, EPI_ISL_647360, EPI_ISL_647361, EPI_ISL_647362, EPI_ISL_647363, EPI_ISL_647364, EPI_ISL_647365, EPI_ISL_647366, EPI_ISL_647367, EPI_ISL_647368, EPI_ISL_647369, EPI_ISL_647370, EPI_ISL_647371, EPI_ISL_647372, EPI_ISL_647373, EPI_ISL_647374, EPI_ISL_647375, EPI_ISL_647376, EPI_ISL_647377, EPI_ISL_647378, EPI_ISL_647379, EPI_ISL_647380, EPI_ISL_647381, EPI_ISL_647382, EPI_ISL_647383, EPI_ISL_647384, EPI_ISL_647385, EPI_ISL_647386, EPI_ISL_647387, EPI_ISL_647388, EPI_ISL_647389, EPI_ISL_647390, EPI_ISL_647391, EPI_ISL_647392, EPI_ISL_647393, EPI_ISL_647394, EPI_ISL_647395, EPI_ISL_647396, EPI_ISL_647397, EPI_ISL_647398, EPI_ISL_647399, EPI_ISL_647400, EPI_ISL_647401, EPI_ISL_647402, EPI_ISL_647403, EPI_ISL_647404, EPI_ISL_647405, EPI_ISL_647406, EPI_ISL_647407, EPI_ISL_647408, EPI_ISL_647409, EPI_ISL_647410, EPI_ISL_647411, EPI_ISL_647412, EPI_ISL_647413, EPI_ISL_647414, EPI_ISL_647415, EPI_ISL_647416, EPI_ISL_647417, EPI_ISL_647418, EPI_ISL_647419, EPI_ISL_647420, EPI_ISL_647421, EPI_ISL_647422, EPI_ISL_647423, EPI_ISL_647424, EPI_ISL_647425, EPI_ISL_647426, EPI_ISL_647427, EPI_ISL_647428, EPI_ISL_647429, EPI_ISL_647430, EPI_ISL_647431, EPI_ISL_647432, EPI_ISL_647433, EPI_ISL_647434, EPI_ISL_647435, EPI_ISL_647436, EPI_ISL_647437, EPI_ISL_647438, EPI_ISL_647439, EPI_ISL_647440, EPI_ISL_647441, EPI_ISL_647442, EPI_ISL_647443, EPI_ISL_647444, EPI_ISL_647445, EPI_ISL_647446, EPI_ISL_647447, EPI_ISL_647448, EPI_ISL_647449, EPI_ISL_647450, EPI_ISL_647451, EPI_ISL_647452, EPI_ISL_647453, EPI_ISL_647454, EPI_ISL_647455, EPI_ISL_647456, EPI_ISL_647457, EPI_ISL_647458, EPI_ISL_647459, EPI_ISL_647460, EPI_ISL_647461, EPI_ISL_647462, EPI_ISL_647463, EPI_ISL_647464, EPI_ISL_647465, EPI_ISL_647466, EPI_ISL_647467, EPI_ISL_647470, EPI_ISL_647471, EPI_ISL_647472, EPI_ISL_647475, EPI_ISL_647476, EPI_ISL_647478, EPI_ISL_647479, EPI_ISL_647480, EPI_ISL_647481, EPI_ISL_647482, EPI_ISL_647483, EPI_ISL_647484, EPI_ISL_647485, EPI_ISL_647486, EPI_ISL_647487, EPI_ISL_647488, EPI_ISL_647489, EPI_ISL_647490, EPI_ISL_647491, EPI_ISL_647492, EPI_ISL_647493, EPI_ISL_647494, EPI_ISL_647495, EPI_ISL_647496, EPI_ISL_647498, EPI_ISL_647500, EPI_ISL_647501, EPI_ISL_647502, EPI_ISL_647504, EPI_ISL_647505, EPI_ISL_647507, EPI_ISL_647508, EPI_ISL_647509, EPI_ISL_647510, EPI_ISL_647512, EPI_ISL_647514, EPI_ISL_647515, EPI_ISL_647516, EPI_ISL_647517, EPI_ISL_647518, EPI_ISL_647519, EPI_ISL_647520, EPI_ISL_647521, EPI_ISL_647522, EPI_ISL_647523, EPI_ISL_647524, EPI_ISL_647525, EPI_ISL_647527, EPI_ISL_647529, EPI_ISL_647530, EPI_ISL_647531, EPI_ISL_647532, EPI_ISL_647533, EPI_ISL_647534, EPI_ISL_647537, EPI_ISL_647538, EPI_ISL_647539, EPI_ISL_647540, EPI_ISL_647542, EPI_ISL_647543, EPI_ISL_647544, EPI_ISL_647545, EPI_ISL_647547, EPI_ISL_647548, EPI_ISL_647549, EPI_ISL_647551, EPI_ISL_647552, EPI_ISL_647554, EPI_ISL_647555, EPI_ISL_647556, EPI_ISL_647557, EPI_ISL_647559, EPI_ISL_647561, EPI_ISL_647562, EPI_ISL_647563, EPI_ISL_647564, EPI_ISL_647565, EPI_ISL_647566, EPI_ISL_647567, EPI_ISL_647568, EPI_ISL_647569, EPI_ISL_647570, EPI_ISL_647571, EPI_ISL_647574, EPI_ISL_647575, EPI_ISL_647577, EPI_ISL_647578, EPI_ISL_647581, EPI_ISL_647582, EPI_ISL_647583, EPI_ISL_647585, EPI_ISL_647586, EPI_ISL_647587, EPI_ISL_647588, EPI_ISL_647589, EPI_ISL_647590, EPI_ISL_647592, EPI_ISL_647593, EPI_ISL_647595, EPI_ISL_647596, EPI_ISL_647597, EPI_ISL_647598, EPI_ISL_647599, EPI_ISL_647600, EPI_ISL_647601, EPI_ISL_647602, EPI_ISL_647604, EPI_ISL_647605, EPI_ISL_647607, EPI_ISL_647609, EPI_ISL_647610, EPI_ISL_647611, EPI_ISL_647613, EPI_ISL_647614, EPI_ISL_647616, EPI_ISL_647617, EPI_ISL_647619, EPI_ISL_647620, EPI_ISL_647621, EPI_ISL_647622, EPI_ISL_647623, EPI_ISL_647625, EPI_ISL_647626, EPI_ISL_647627, EPI_ISL_647628, EPI_ISL_647629, EPI_ISL_647631, EPI_ISL_647633, EPI_ISL_647636, EPI_ISL_647637, EPI_ISL_647639, EPI_ISL_647640, EPI_ISL_647641, EPI_ISL_647642, EPI_ISL_647644, EPI_ISL_647645, EPI_ISL_647646, EPI_ISL_647647, EPI_ISL_647648, EPI_ISL_647649, EPI_ISL_647651, EPI_ISL_647652, EPI_ISL_647653, EPI_ISL_647654, EPI_ISL_647656, EPI_ISL_647657, EPI_ISL_647658, EPI_ISL_647660, EPI_ISL_647661, EPI_ISL_647662, EPI_ISL_647663, EPI_ISL_647664, EPI_ISL_647665, EPI_ISL_647666, EPI_ISL_647669, EPI_ISL_647670, EPI_ISL_647671, EPI_ISL_647673, EPI_ISL_647674, EPI_ISL_647675, EPI_ISL_647676, EPI_ISL_647678, EPI_ISL_647679, EPI_ISL_647680, EPI_ISL_647681, EPI_ISL_647684, EPI_ISL_647685, EPI_ISL_647686, EPI_ISL_647687, EPI_ISL_647688, EPI_ISL_647690, EPI_ISL_647691, EPI_ISL_647692, EPI_ISL_647694, EPI_ISL_647696, EPI_ISL_647699, EPI_ISL_647701, EPI_ISL_647702, EPI_ISL_647703, EPI_ISL_647706, EPI_ISL_647707, EPI_ISL_647709, EPI_ISL_647710, EPI_ISL_647712, EPI_ISL_647713, EPI_ISL_647714, EPI_ISL_647715, EPI_ISL_647716, EPI_ISL_647717, EPI_ISL_647718, EPI_ISL_647719, EPI_ISL_647720, EPI_ISL_647721, EPI_ISL_647722, EPI_ISL_647723, EPI_ISL_647724, EPI_ISL_647725, EPI_ISL_647727, EPI_ISL_647728, EPI_ISL_647729, EPI_ISL_647730, EPI_ISL_647731, EPI_ISL_647732, EPI_ISL_647734, EPI_ISL_647735, EPI_ISL_647736, EPI_ISL_647737, EPI_ISL_647738, EPI_ISL_647739, EPI_ISL_647740, EPI_ISL_647742, EPI_ISL_647743, EPI_ISL_647744, EPI_ISL_647745, EPI_ISL_647746, EPI_ISL_647747, EPI_ISL_647748, EPI_ISL_647749, EPI_ISL_647751, EPI_ISL_647753, EPI_ISL_647754, EPI_ISL_647755, EPI_ISL_647756, EPI_ISL_647757, EPI_ISL_647758, EPI_ISL_647759, EPI_ISL_647760, EPI_ISL_647761, EPI_ISL_647763, EPI_ISL_647764, EPI_ISL_647765, EPI_ISL_647766, EPI_ISL_647767, EPI_ISL_647768, EPI_ISL_647769, EPI_ISL_647770, EPI_ISL_647772, EPI_ISL_647773, EPI_ISL_647774, EPI_ISL_647775, EPI_ISL_647777, EPI_ISL_647778, EPI_ISL_647779, EPI_ISL_647781, EPI_ISL_647782, EPI_ISL_647783, EPI_ISL_647784, EPI_ISL_647785, EPI_ISL_647788, EPI_ISL_647789, EPI_ISL_647790, EPI_ISL_647791, EPI_ISL_647793, EPI_ISL_647794, EPI_ISL_647795, EPI_ISL_647796, EPI_ISL_649492, EPI_ISL_649493, EPI_ISL_649625, EPI_ISL_649626, EPI_ISL_649627, EPI_ISL_649628, EPI_ISL_649629, EPI_ISL_649630, EPI_ISL_649631, EPI_ISL_649632, EPI_ISL_649633, EPI_ISL_659135, EPI_ISL_659163, EPI_ISL_659167, EPI_ISL_659345, EPI_ISL_659387, EPI_ISL_659395, EPI_ISL_659542, EPI_ISL_672912, EPI_ISL_672913, EPI_ISL_672914, EPI_ISL_672915, EPI_ISL_672916, EPI_ISL_672917, EPI_ISL_672918, EPI_ISL_672919, EPI_ISL_672920, EPI_ISL_672921, EPI_ISL_672922, EPI_ISL_672923, EPI_ISL_672924, EPI_ISL_672925, EPI_ISL_672926, EPI_ISL_672927, EPI_ISL_672928, EPI_ISL_672929, EPI_ISL_672930, EPI_ISL_672931, EPI_ISL_672932, EPI_ISL_672933, EPI_ISL_672934, EPI_ISL_672935, EPI_ISL_672936, EPI_ISL_672937, EPI_ISL_672938, EPI_ISL_672939, EPI_ISL_672940, EPI_ISL_672941, EPI_ISL_672942, EPI_ISL_672943, EPI_ISL_672944, EPI_ISL_672945, EPI_ISL_672946, EPI_ISL_672947, EPI_ISL_672948, EPI_ISL_672949, EPI_ISL_672950, EPI_ISL_672951, EPI_ISL_672952, EPI_ISL_672953, EPI_ISL_672954, EPI_ISL_672955, EPI_ISL_672956, EPI_ISL_672957, EPI_ISL_672958, EPI_ISL_672959, EPI_ISL_672960, EPI_ISL_672961, EPI_ISL_672962, EPI_ISL_672963, EPI_ISL_672964, EPI_ISL_672965, EPI_ISL_672966, EPI_ISL_672967, EPI_ISL_672968, EPI_ISL_672969, EPI_ISL_672970, EPI_ISL_672971, EPI_ISL_672972, EPI_ISL_672973, EPI_ISL_672974, EPI_ISL_672975, EPI_ISL_672976, EPI_ISL_672977, EPI_ISL_672978, EPI_ISL_672979, EPI_ISL_672980, EPI_ISL_672981, EPI_ISL_672982, EPI_ISL_672983, EPI_ISL_672984, EPI_ISL_672985, EPI_ISL_672986, EPI_ISL_672987, EPI_ISL_672988, EPI_ISL_672989, EPI_ISL_672990, EPI_ISL_672991, EPI_ISL_672992, EPI_ISL_672993, EPI_ISL_672994, EPI_ISL_672995, EPI_ISL_672996, EPI_ISL_672997, EPI_ISL_672998, EPI_ISL_672999, EPI_ISL_673000, EPI_ISL_673001, EPI_ISL_673002, EPI_ISL_673003, EPI_ISL_673004, EPI_ISL_673005, EPI_ISL_673006, EPI_ISL_673007, EPI_ISL_673008, EPI_ISL_673009, EPI_ISL_673010, EPI_ISL_673011, EPI_ISL_673012, EPI_ISL_673013, EPI_ISL_673014, EPI_ISL_673015, EPI_ISL_673016, EPI_ISL_673017, EPI_ISL_673018, EPI_ISL_673019, EPI_ISL_673020, EPI_ISL_673021, EPI_ISL_673022, EPI_ISL_673023, EPI_ISL_673024, EPI_ISL_673025, EPI_ISL_673026, EPI_ISL_673027, EPI_ISL_673028, EPI_ISL_673029, EPI_ISL_673030, EPI_ISL_673031, EPI_ISL_673032, EPI_ISL_673033, EPI_ISL_673034, EPI_ISL_673035, EPI_ISL_673036, EPI_ISL_673037, EPI_ISL_673038, EPI_ISL_673039, EPI_ISL_673040, EPI_ISL_673041, EPI_ISL_673042, EPI_ISL_673043, EPI_ISL_673044, EPI_ISL_673045, EPI_ISL_673046, EPI_ISL_673047, EPI_ISL_673048, EPI_ISL_673049, EPI_ISL_673050, EPI_ISL_673051, EPI_ISL_673052, EPI_ISL_673053, EPI_ISL_673054, EPI_ISL_673055, EPI_ISL_673056, EPI_ISL_673057, EPI_ISL_673058, EPI_ISL_673059, EPI_ISL_673060, EPI_ISL_673061, EPI_ISL_673062, EPI_ISL_673063, EPI_ISL_673064, EPI_ISL_673065, EPI_ISL_673066, EPI_ISL_673067, EPI_ISL_673068, EPI_ISL_673069, EPI_ISL_673070, EPI_ISL_673071, EPI_ISL_673072, EPI_ISL_673073, EPI_ISL_673074, EPI_ISL_673075, EPI_ISL_673076, EPI_ISL_673077, EPI_ISL_673078, EPI_ISL_673079, EPI_ISL_673080, EPI_ISL_673081, EPI_ISL_673082, EPI_ISL_673083, EPI_ISL_673084, EPI_ISL_673085, EPI_ISL_673086, EPI_ISL_673087, EPI_ISL_673088, EPI_ISL_673089, EPI_ISL_673090, EPI_ISL_673091, EPI_ISL_673092, EPI_ISL_673093, EPI_ISL_673094, EPI_ISL_673095, EPI_ISL_673096, EPI_ISL_673097, EPI_ISL_673098, EPI_ISL_673099, EPI_ISL_673100, EPI_ISL_673101, EPI_ISL_673102, EPI_ISL_673103, EPI_ISL_673104, EPI_ISL_673105, EPI_ISL_673106, EPI_ISL_673107, EPI_ISL_673108, EPI_ISL_673109, EPI_ISL_673110, EPI_ISL_673111, EPI_ISL_673112, EPI_ISL_673113, EPI_ISL_673114, EPI_ISL_673115, EPI_ISL_673116, EPI_ISL_673117, EPI_ISL_673118, EPI_ISL_673119, EPI_ISL_673120, EPI_ISL_673121, EPI_ISL_673122, EPI_ISL_673123, EPI_ISL_673124, EPI_ISL_673125, EPI_ISL_673126, EPI_ISL_673127, EPI_ISL_673128, EPI_ISL_673129, EPI_ISL_673130, EPI_ISL_673131, EPI_ISL_673132, EPI_ISL_673133, EPI_ISL_673134, EPI_ISL_673135, EPI_ISL_673136, EPI_ISL_673137, EPI_ISL_673138, EPI_ISL_673139, EPI_ISL_673140, EPI_ISL_673141, EPI_ISL_673142, EPI_ISL_673143, EPI_ISL_673144, EPI_ISL_673145, EPI_ISL_673146, EPI_ISL_673147, EPI_ISL_673148, EPI_ISL_673149, EPI_ISL_673150, EPI_ISL_673151, EPI_ISL_673152, EPI_ISL_673153, EPI_ISL_673154, EPI_ISL_673155, EPI_ISL_673156, EPI_ISL_673157, EPI_ISL_673158, EPI_ISL_673159, EPI_ISL_673160, EPI_ISL_673161, EPI_ISL_673162, EPI_ISL_673163, EPI_ISL_673164, EPI_ISL_673165, EPI_ISL_673166, EPI_ISL_673167, EPI_ISL_673168, EPI_ISL_673169, EPI_ISL_673170, EPI_ISL_673171, EPI_ISL_673172, EPI_ISL_673173, EPI_ISL_673174, EPI_ISL_673175, EPI_ISL_673176, EPI_ISL_673177, EPI_ISL_673178, EPI_ISL_673179, EPI_ISL_673180, EPI_ISL_673181, EPI_ISL_673182, EPI_ISL_673183, EPI_ISL_673184, EPI_ISL_673185, EPI_ISL_673186, EPI_ISL_673187, EPI_ISL_673188, EPI_ISL_673189, EPI_ISL_673190, EPI_ISL_673191, EPI_ISL_673192, EPI_ISL_673193, EPI_ISL_673194, EPI_ISL_673195, EPI_ISL_673196, EPI_ISL_673197, EPI_ISL_673198, EPI_ISL_673199, EPI_ISL_673200, EPI_ISL_673201, EPI_ISL_673202, EPI_ISL_673203, EPI_ISL_673204, EPI_ISL_673205, EPI_ISL_673206, EPI_ISL_673207, EPI_ISL_673208, EPI_ISL_673209, EPI_ISL_673210, EPI_ISL_673211, EPI_ISL_673212, EPI_ISL_673213, EPI_ISL_673214, EPI_ISL_673215, EPI_ISL_673216, EPI_ISL_673217, EPI_ISL_673218, EPI_ISL_673219, EPI_ISL_673220, EPI_ISL_673221, EPI_ISL_673222, EPI_ISL_673223, EPI_ISL_673224, EPI_ISL_673225, EPI_ISL_673226, EPI_ISL_673227, EPI_ISL_673228, EPI_ISL_673229, EPI_ISL_673230, EPI_ISL_673231, EPI_ISL_673232, EPI_ISL_673233, EPI_ISL_673234, EPI_ISL_673235, EPI_ISL_681656, EPI_ISL_681657, EPI_ISL_710836, EPI_ISL_711023, EPI_ISL_711024, EPI_ISL_711025, EPI_ISL_711026, EPI_ISL_711038 | | | |
| see above | Lighthouse Lab in Cambridge | Wellcome Sanger Institute for the COVID-19 Genomics UK (COG-UK) Consortium | Cordelia Langford; David K. Jackson; Dominic Kwiatkowski; Ewan Harrison; Ian Johnston; John Sillitoe on behalf of the Wellcome Sanger Institute COVID-19 Surveillance Team; John Sillitoe on behalf of the Wellcome Sanger Institute COVID-19 Surveillance Team (http://www.sanger.ac.uk/covid-team); Rob Howes; Roberto Amato; Sonia Goncalves; The Lighthouse Lab in Cambridge and Alex Alderton |
| EPI_ISL_633275 | Lighthouse Lab in Cambridge | Wellcome Sanger Institute for the COVID-19 Genomics UK (COG-UK) consortium | Cordelia Langford; David K. Jackson; Dominic Kwiatkowski; Ewan Harrison; Ian Johnston; John Sillitoe on behalf of the Wellcome Sanger Institute COVID-19 Surveillance Team; Rob Howes; Roberto Amato; Sonia Goncalves; The Lighthouse Lab in Cambridge and Alex Alderton |
| EPI_ISL_641607, EPI_ISL_641608, EPI_ISL_641609, EPI_ISL_641610, EPI_ISL_641611, EPI_ISL_641612, EPI_ISL_641613, EPI_ISL_641614, EPI_ISL_641615, EPI_ISL_641616, EPI_ISL_641617, EPI_ISL_641618, EPI_ISL_641619, EPI_ISL_641620, EPI_ISL_641621, EPI_ISL_641622, EPI_ISL_641623, EPI_ISL_641624, EPI_ISL_641625, EPI_ISL_641626, EPI_ISL_641627, EPI_ISL_641628, EPI_ISL_641629, EPI_ISL_641630, EPI_ISL_641631, EPI_ISL_641632, EPI_ISL_641633, EPI_ISL_641634, EPI_ISL_641635, EPI_ISL_641636, EPI_ISL_641637, EPI_ISL_641638, EPI_ISL_641639, EPI_ISL_641640, EPI_ISL_641641, EPI_ISL_641642, EPI_ISL_641643, EPI_ISL_641644, EPI_ISL_641645, EPI_ISL_641646, EPI_ISL_641647, EPI_ISL_641648, EPI_ISL_641649, EPI_ISL_641650, EPI_ISL_641651, EPI_ISL_641652, EPI_ISL_641653, EPI_ISL_641654, EPI_ISL_641655, EPI_ISL_641656, EPI_ISL_641657, EPI_ISL_641658, EPI_ISL_641659, EPI_ISL_641660, EPI_ISL_641661, EPI_ISL_641662, EPI_ISL_641663, EPI_ISL_641664, EPI_ISL_641665, EPI_ISL_641666, EPI_ISL_641667, EPI_ISL_641668, EPI_ISL_641669, EPI_ISL_641670, EPI_ISL_641671, EPI_ISL_641672, EPI_ISL_641673, EPI_ISL_641674, EPI_ISL_641675, EPI_ISL_641676, EPI_ISL_641677, EPI_ISL_641678, EPI_ISL_641679, EPI_ISL_641680, EPI_ISL_641681, EPI_ISL_641682, EPI_ISL_641683, EPI_ISL_641684, EPI_ISL_641685, EPI_ISL_641686, EPI_ISL_641687, EPI_ISL_641688, EPI_ISL_641689, EPI_ISL_641690, EPI_ISL_641691, EPI_ISL_641692, EPI_ISL_641693, EPI_ISL_641694, EPI_ISL_641695, EPI_ISL_641696, EPI_ISL_641697, EPI_ISL_641698, EPI_ISL_641699, EPI_ISL_641700, EPI_ISL_641701, EPI_ISL_641702, EPI_ISL_641703, EPI_ISL_641704, EPI_ISL_641705, EPI_ISL_641706, EPI_ISL_641707, EPI_ISL_641708, EPI_ISL_641709, EPI_ISL_641710, EPI_ISL_641711, EPI_ISL_641712, EPI_ISL_641713, EPI_ISL_641714, EPI_ISL_641715, EPI_ISL_641716, EPI_ISL_641717, EPI_ISL_641718, EPI_ISL_641719, EPI_ISL_641720, EPI_ISL_641721, EPI_ISL_641722, EPI_ISL_641723, EPI_ISL_641724, EPI_ISL_641725, EPI_ISL_641726, EPI_ISL_641727, EPI_ISL_641728, EPI_ISL_641729, EPI_ISL_641730, EPI_ISL_641731, EPI_ISL_641732, EPI_ISL_641733, EPI_ISL_641734, EPI_ISL_641735, EPI_ISL_641736, EPI_ISL_641737, EPI_ISL_641738, EPI_ISL_641739, EPI_ISL_641740, EPI_ISL_641741, EPI_ISL_641742, EPI_ISL_641743, EPI_ISL_641744, EPI_ISL_641745, EPI_ISL_641746, EPI_ISL_641747, EPI_ISL_641748, EPI_ISL_641749, EPI_ISL_641750, EPI_ISL_641751, EPI_ISL_641752, EPI_ISL_641753, EPI_ISL_641754, EPI_ISL_641755, EPI_ISL_641756, EPI_ISL_641757, EPI_ISL_641758, EPI_ISL_641759, EPI_ISL_641760, EPI_ISL_641761, EPI_ISL_641762, EPI_ISL_641763, EPI_ISL_641764, EPI_ISL_641765, EPI_ISL_641766, EPI_ISL_641767, EPI_ISL_641768, EPI_ISL_641769, EPI_ISL_641770, EPI_ISL_641771, EPI_ISL_641772, EPI_ISL_641773, EPI_ISL_641774, EPI_ISL_641775, EPI_ISL_641776, EPI_ISL_641777, EPI_ISL_641778, EPI_ISL_641779, EPI_ISL_641780, EPI_ISL_641781, EPI_ISL_641782, EPI_ISL_641783, EPI_ISL_641784, EPI_ISL_641785, EPI_ISL_641786, EPI_ISL_641787, EPI_ISL_641788, EPI_ISL_641789, EPI_ISL_641790, EPI_ISL_641791, EPI_ISL_641792, EPI_ISL_641793, EPI_ISL_641794, EPI_ISL_641795, EPI_ISL_641796, EPI_ISL_641797, EPI_ISL_641798, EPI_ISL_641799, EPI_ISL_641800, EPI_ISL_641801, EPI_ISL_641802, EPI_ISL_641803, EPI_ISL_641804, EPI_ISL_641805, EPI_ISL_641806, EPI_ISL_641807, EPI_ISL_641808, EPI_ISL_641809, EPI_ISL_641810, EPI_ISL_641811, EPI_ISL_641812, EPI_ISL_641813, EPI_ISL_641814, EPI_ISL_641815, EPI_ISL_641816, EPI_ISL_641817, EPI_ISL_641818, EPI_ISL_641819, EPI_ISL_641820, EPI_ISL_641821, EPI_ISL_641822, EPI_ISL_641823, EPI_ISL_641824, EPI_ISL_641825, EPI_ISL_641826, EPI_ISL_641827, EPI_ISL_641828, EPI_ISL_641829, EPI_ISL_641830, EPI_ISL_641831, EPI_ISL_641832, EPI_ISL_641833, EPI_ISL_641834, EPI_ISL_641835, EPI_ISL_641836, EPI_ISL_641837, EPI_ISL_641838, EPI_ISL_641839, EPI_ISL_641840, EPI_ISL_641841, EPI_ISL_641842, EPI_ISL_641843, EPI_ISL_641844, EPI_ISL_641845, EPI_ISL_641846, EPI_ISL_641847, EPI_ISL_641848, EPI_ISL_641849, EPI_ISL_641850, EPI_ISL_641851, EPI_ISL_641852, EPI_ISL_641853, EPI_ISL_641854, EPI_ISL_641855, EPI_ISL_641856, EPI_ISL_641857, EPI_ISL_641858, EPI_ISL_641859, EPI_ISL_641860, EPI_ISL_641861, EPI_ISL_641862, EPI_ISL_641863, EPI_ISL_641864, EPI_ISL_641865, EPI_ISL_641866, EPI_ISL_641867, EPI_ISL_641868, EPI_ISL_641869, EPI_ISL_641870, EPI_ISL_641871, EPI_ISL_641872, EPI_ISL_641873, EPI_ISL_641874, EPI_ISL_641875, EPI_ISL_641876, EPI_ISL_641878, EPI_ISL_641879, EPI_ISL_641880, EPI_ISL_641881, EPI_ISL_641882, EPI_ISL_641883, EPI_ISL_641884, EPI_ISL_641885, EPI_ISL_641886, EPI_ISL_641887, EPI_ISL_641888, EPI_ISL_641889, EPI_ISL_641890, EPI_ISL_641891, EPI_ISL_641892, EPI_ISL_641893, EPI_ISL_641894, EPI_ISL_641895, EPI_ISL_641896, EPI_ISL_641897, EPI_ISL_641898, EPI_ISL_641899, EPI_ISL_641900, EPI_ISL_641901, EPI_ISL_641902, EPI_ISL_641903, EPI_ISL_641904, EPI_ISL_641905, EPI_ISL_641906, EPI_ISL_641907, EPI_ISL_641908, EPI_ISL_641909, EPI_ISL_641910, EPI_ISL_641911, EPI_ISL_641912, EPI_ISL_641913, EPI_ISL_641914, EPI_ISL_641915, EPI_ISL_641916, EPI_ISL_641917, EPI_ISL_641918, EPI_ISL_641919, EPI_ISL_641920, EPI_ISL_641921, EPI_ISL_641922, EPI_ISL_641923, EPI_ISL_641924, EPI_ISL_641925, EPI_ISL_641926, EPI_ISL_641927, EPI_ISL_641928, EPI_ISL_641929, EPI_ISL_641930, EPI_ISL_641931, EPI_ISL_641932, EPI_ISL_641933, EPI_ISL_641934, EPI_ISL_641935, EPI_ISL_641936, EPI_ISL_641937, EPI_ISL_641938, EPI_ISL_641939, EPI_ISL_641940, EPI_ISL_641941, EPI_ISL_641942, EPI_ISL_641943, EPI_ISL_641944, EPI_ISL_641945, EPI_ISL_641946, EPI_ISL_641947, EPI_ISL_641948, EPI_ISL_641949, EPI_ISL_641950, EPI_ISL_641951, EPI_ISL_641952, EPI_ISL_641953, EPI_ISL_641954, EPI_ISL_641955, EPI_ISL_641956, EPI_ISL_641957, EPI_ISL_641958, EPI_ISL_641959, EPI_ISL_641960, EPI_ISL_641961, EPI_ISL_641962, EPI_ISL_641963, EPI_ISL_641964, EPI_ISL_641965, EPI_ISL_641966, EPI_ISL_641967, EPI_ISL_641968, EPI_ISL_641969, EPI_ISL_641970, EPI_ISL_641971, EPI_ISL_641972, EPI_ISL_641973, EPI_ISL_641974, EPI_ISL_641975, EPI_ISL_641976, EPI_ISL_641977, EPI_ISL_641978, EPI_ISL_641979, EPI_ISL_641980, EPI_ISL_641981, EPI_ISL_641982, EPI_ISL_641983, EPI_ISL_641984, EPI_ISL_641985, EPI_ISL_641986, EPI_ISL_641987, EPI_ISL_641988, EPI_ISL_641989, EPI_ISL_641990, EPI_ISL_641991, EPI_ISL_641992, EPI_ISL_641993, EPI_ISL_641994, EPI_ISL_641995, EPI_ISL_641996, EPI_ISL_641997, EPI_ISL_641998, EPI_ISL_641999, EPI_ISL_642000, EPI_ISL_642001, EPI_ISL_642002, EPI_ISL_642003, EPI_ISL_642004, EPI_ISL_642005, EPI_ISL_642006, EPI_ISL_642007, EPI_ISL_642008, EPI_ISL_642009, EPI_ISL_642010, EPI_ISL_642011, EPI_ISL_642012, EPI_ISL_642013, EPI_ISL_642014, EPI_ISL_642015, EPI_ISL_642016, EPI_ISL_642017, EPI_ISL_642018, EPI_ISL_642019, EPI_ISL_642020, EPI_ISL_642021, EPI_ISL_642022, EPI_ISL_642023, EPI_ISL_642024, EPI_ISL_642025, EPI_ISL_642026, EPI_ISL_642027, EPI_ISL_642028, EPI_ISL_642029, EPI_ISL_642030, EPI_ISL_642031, EPI_ISL_642032, EPI_ISL_642033, EPI_ISL_642034, EPI_ISL_642035, EPI_ISL_642036, EPI_ISL_642037, EPI_ISL_642038, EPI_ISL_642039, EPI_ISL_642040, EPI_ISL_642041, EPI_ISL_642042, EPI_ISL_642043, EPI_ISL_642044, EPI_ISL_642045, EPI_ISL_642046, EPI_ISL_642047, EPI_ISL_642048, EPI_ISL_642049, EPI_ISL_642050, EPI_ISL_642051, EPI_ISL_642052, EPI_ISL_642053, EPI_ISL_642054, EPI_ISL_642055, EPI_ISL_642056, EPI_ISL_642057, EPI_ISL_642058, EPI_ISL_642059, EPI_ISL_642060, EPI_ISL_642061, EPI_ISL_642062, EPI_ISL_642063, EPI_ISL_642064, EPI_ISL_642065, EPI_ISL_642066, EPI_ISL_642067, EPI_ISL_642068, EPI_ISL_642069, EPI_ISL_642070, EPI_ISL_642071, EPI_ISL_642072, EPI_ISL_642073, EPI_ISL_642074, EPI_ISL_642075, EPI_ISL_642076, EPI_ISL_642077, EPI_ISL_642078, EPI_ISL_642079, EPI_ISL_642080, EPI_ISL_642081, EPI_ISL_642082, EPI_ISL_642083, EPI_ISL_642084, EPI_ISL_642085, EPI_ISL_642086, EPI_ISL_642087, EPI_ISL_642088, EPI_ISL_642089, EPI_ISL_642090, EPI_ISL_642091, EPI_ISL_642092, EPI_ISL_642093, EPI_ISL_642094, EPI_ISL_642095, EPI_ISL_642096, EPI_ISL_642097, EPI_ISL_642098, EPI_ISL_642099, EPI_ISL_642100, EPI_ISL_642101, EPI_ISL_642102, EPI_ISL_642103, EPI_ISL_642104, EPI_ISL_642105, EPI_ISL_642106, EPI_ISL_642107, EPI_ISL_642108, EPI_ISL_642109, EPI_ISL_642110, EPI_ISL_642111, EPI_ISL_642112, EPI_ISL_642113, EPI_ISL_642114, EPI_ISL_642115, EPI_ISL_642116, EPI_ISL_642117, EPI_ISL_642118, EPI_ISL_642119, EPI_ISL_642120, EPI_ISL_642121, EPI_ISL_642122, EPI_ISL_642123, EPI_ISL_642124, EPI_ISL_642125, EPI_ISL_642126, EPI_ISL_642127, EPI_ISL_642128, EPI_ISL_642129, EPI_ISL_642130, EPI_ISL_642131, EPI_ISL_642132, EPI_ISL_642133, EPI_ISL_642134, EPI_ISL_642135, EPI_ISL_642136, EPI_ISL_642137, EPI_ISL_642138, EPI_ISL_642139, EPI_ISL_642140, EPI_ISL_642141, EPI_ISL_642142, EPI_ISL_642143, EPI_ISL_642144, EPI_ISL_642145, EPI_ISL_642146, EPI_ISL_642147, EPI_ISL_642148, EPI_ISL_642149, EPI_ISL_642150, EPI_ISL_642151, EPI_ISL_642152, EPI_ISL_642153, EPI_ISL_642154, EPI_ISL_642155, EPI_ISL_642156, EPI_ISL_642157, EPI_ISL_642158, EPI_ISL_642159, EPI_ISL_642160, EPI_ISL_642161, EPI_ISL_642162, EPI_ISL_642163, EPI_ISL_642164, EPI_ISL_642165, EPI_ISL_642166, EPI_ISL_642167, EPI_ISL_642168, EPI_ISL_642169, EPI_ISL_642170, EPI_ISL_642171, EPI_ISL_642172, EPI_ISL_642173, EPI_ISL_642174, EPI_ISL_642175, EPI_ISL_642176, EPI_ISL_642177, EPI_ISL_642178, EPI_ISL_642179, EPI_ISL_642180, EPI_ISL_642181, EPI_ISL_642182, EPI_ISL_642183, EPI_ISL_642184, EPI_ISL_642185, EPI_ISL_642186, EPI_ISL_642187, EPI_ISL_642188, EPI_ISL_642189, EPI_ISL_642190, EPI_ISL_642191, EPI_ISL_642192, EPI_ISL_642193, EPI_ISL_642194, EPI_ISL_642195, EPI_ISL_642196, EPI_ISL_642197, EPI_ISL_642198, EPI_ISL_642199, EPI_ISL_642200, EPI_ISL_642201, EPI_ISL_642202, EPI_ISL_642203, EPI_ISL_642204, EPI_ISL_642205, EPI_ISL_642206, EPI_ISL_642207, EPI_ISL_642208, EPI_ISL_642209, EPI_ISL_642210, EPI_ISL_642211, EPI_ISL_642212, EPI_ISL_642213, EPI_ISL_642214, EPI_ISL_642215, EPI_ISL_642216, EPI_ISL_642217, EPI_ISL_642218, EPI_ISL_642219, EPI_ISL_642220, EPI_ISL_642221, EPI_ISL_642222, EPI_ISL_642223, EPI_ISL_642224, EPI_ISL_642225, EPI_ISL_642226, EPI_ISL_642227, EPI_ISL_642228, EPI_ISL_642229, EPI_ISL_642230, EPI_ISL_642231, EPI_ISL_642232, EPI_ISL_642233, EPI_ISL_642234, EPI_ISL_642235, EPI_ISL_642236, EPI_ISL_642237, EPI_ISL_642238, EPI_ISL_642239, EPI_ISL_642240, EPI_ISL_642241, EPI_ISL_642242, EPI_ISL_642243, EPI_ISL_642244, EPI_ISL_642245, EPI_ISL_642246, EPI_ISL_642247, EPI_ISL_642248, EPI_ISL_642249, EPI_ISL_642250, EPI_ISL_642251, EPI_ISL_642252, EPI_ISL_642253, EPI_ISL_642254, EPI_ISL_642255, EPI_ISL_642256, EPI_ISL_642257, EPI_ISL_642258, EPI_ISL_642259, EPI_ISL_642260, EPI_ISL_642261, EPI_ISL_642262, EPI_ISL_642263, EPI_ISL_642264, EPI_ISL_642265, EPI_ISL_642266, EPI_ISL_642267, EPI_ISL_642268, EPI_ISL_642269, EPI_ISL_642270, EPI_ISL_642271, EPI_ISL_642272, EPI_ISL_642273, EPI_ISL_642274, EPI_ISL_642275, EPI_ISL_642276, EPI_ISL_642277, EPI_ISL_642278, EPI_ISL_642279, EPI_ISL_642280, EPI_ISL_642281, EPI_ISL_642282, EPI_ISL_642283, EPI_ISL_642284, EPI_ISL_642285, EPI_ISL_642286, EPI_ISL_642287, EPI_ISL_642288, EPI_ISL_642289, EPI_ISL_642290, EPI_ISL_642291, EPI_ISL_642292, EPI_ISL_642293, EPI_ISL_642294, EPI_ISL_642295, EPI_ISL_642296, EPI_ISL_642297, EPI_ISL_642298, EPI_ISL_642299, EPI_ISL_642300, EPI_ISL_642301, EPI_ISL_642302, EPI_ISL_642303, EPI_ISL_642304, EPI_ISL_642305, EPI_ISL_642306, EPI_ISL_642307, EPI_ISL_642308, EPI_ISL_642309, EPI_ISL_642310, EPI_ISL_642311, EPI_ISL_642312, EPI_ISL_642313, EPI_ISL_642314, EPI_ISL_642315, EPI_ISL_642316, EPI_ISL_642317, EPI_ISL_642318, EPI_ISL_642319, EPI_ISL_642320, EPI_ISL_642321, EPI_ISL_642322, EPI_ISL_642323, EPI_ISL_642324, EPI_ISL_642325, EPI_ISL_642326, EPI_ISL_642327, EPI_ISL_642328, EPI_ISL_642329, EPI_ISL_642330, EPI_ISL_642331, EPI_ISL_642332, EPI_ISL_642333, EPI_ISL_642334, EPI_ISL_642335, EPI_ISL_642336, EPI_ISL_642337, EPI_ISL_642338, EPI_ISL_642339, EPI_ISL_642340, EPI_ISL_642341, EPI_ISL_642342, EPI_ISL_642343, EPI_ISL_642344, EPI_ISL_642345, EPI_ISL_642346, EPI_ISL_642347, EPI_ISL_642348, EPI_ISL_642349, EPI_ISL_642350, EPI_ISL_642351, EPI_ISL_642352, EPI_ISL_642353, EPI_ISL_642354, EPI_ISL_642355, EPI_ISL_642356, EPI_ISL_642357, EPI_ISL_642358, EPI_ISL_642359, EPI_ISL_642360, EPI_ISL_642361, EPI_ISL_642362, EPI_ISL_642363, EPI_ISL_642364, EPI_ISL_642365, EPI_ISL_642366, EPI_ISL_642367, EPI_ISL_642368, EPI_ISL_642369, EPI_ISL_642370, EPI_ISL_642371, EPI_ISL_642372, EPI_ISL_642373, EPI_ISL_642374, EPI_ISL_642375, EPI_ISL_642376, EPI_ISL_642377, EPI_ISL_642378, EPI_ISL_642379, EPI_ISL_642380, EPI_ISL_642381, EPI_ISL_642382, EPI_ISL_642383, EPI_ISL_642384, EPI_ISL_642385, EPI_ISL_642386, EPI_ISL_642387, EPI_ISL_642388, EPI_ISL_642389, EPI_ISL_642390, EPI_ISL_642391, EPI_ISL_642392, EPI_ISL_642393, EPI_ISL_642394, EPI_ISL_642395, EPI_ISL_642396, EPI_ISL_642397, EPI_ISL_642398, EPI_ISL_642399, EPI_ISL_642400, EPI_ISL_642401, EPI_ISL_642402, EPI_ISL_642403, EPI_ISL_642404, EPI_ISL_642405, EPI_ISL_642406, EPI_ISL_642407, EPI_ISL_642408, EPI_ISL_642409, EPI_ISL_642410, EPI_ISL_642411, EPI_ISL_642412, EPI_ISL_642413, EPI_ISL_642414, EPI_ISL_642415, EPI_ISL_642416, EPI_ISL_642417, EPI_ISL_642418, EPI_ISL_642419, EPI_ISL_642420, EPI_ISL_642421, EPI_ISL_642422, EPI_ISL_642423, EPI_ISL_642424, EPI_ISL_642425, EPI_ISL_642426, EPI_ISL_642427, EPI_ISL_642428, EPI_ISL_642429, EPI_ISL_642430, EPI_ISL_642431, EPI_ISL_642432, EPI_ISL_642433, EPI_ISL_642434, EPI_ISL_642435, EPI_ISL_642436, EPI_ISL_642437, EPI_ISL_642438, EPI_ISL_642439, EPI_ISL_642440, EPI_ISL_642441, EPI_ISL_642442, EPI_ISL_642443, EPI_ISL_642444, EPI_ISL_642445, EPI_ISL_642446, EPI_ISL_642447, EPI_ISL_642448, EPI_ISL_642449, EPI_ISL_642450, EPI_ISL_642451, EPI_ISL_642452, EPI_ISL_642453, EPI_ISL_642454, EPI_ISL_642455, EPI_ISL_642456, EPI_ISL_642457, EPI_ISL_642458, EPI_ISL_642459, EPI_ISL_642460, EPI_ISL_642461, EPI_ISL_642462, EPI_ISL_642463, EPI_ISL_642464, EPI_ISL_642465, EPI_ISL_642466, EPI_ISL_642467, EPI_ISL_642468, EPI_ISL_642469, EPI_ISL_642470, EPI_ISL_642471, EPI_ISL_642472, EPI_ISL_642473, EPI_ISL_642474, EPI_ISL_642475, EPI_ISL_642476, EPI_ISL_642477, EPI_ISL_642478, EPI_ISL_642479, EPI_ISL_642480, EPI_ISL_642481, EPI_ISL_642482, EPI_ISL_642483, EPI_ISL_642484, EPI_ISL_642485, EPI_ISL_642486, EPI_ISL_642487, EPI_ISL_642488, EPI_ISL_642489, EPI_ISL_642490, EPI_ISL_642491, EPI_ISL_642492, EPI_ISL_642493, EPI_ISL_642494, EPI_ISL_642495, EPI_ISL_642496, EPI_ISL_642497, EPI_ISL_642498, EPI_ISL_642499, EPI_ISL_642500, EPI_ISL_642501, EPI_ISL_642502, EPI_ISL_642503, EPI_ISL_642504, EPI_ISL_642505, EPI_ISL_642506, EPI_ISL_642507, EPI_ISL_642508, EPI_ISL_642509, EPI_ISL_642510, EPI_ISL_642511, EPI_ISL_642512, EPI_ISL_642513, EPI_ISL_642514, EPI_ISL_642515, EPI_ISL_642516, EPI_ISL_642517, EPI_ISL_642518, EPI_ISL_642519, EPI_ISL_642520, EPI_ISL_642521, EPI_ISL_642522, EPI_ISL_642523, EPI_ISL_642524, EPI_ISL_642525, EPI_ISL_642526, EPI_ISL_642527, EPI_ISL_642528, EPI_ISL_642529, EPI_ISL_642530, EPI_ISL_642531, EPI_ISL_642532, EPI_ISL_642533, EPI_ISL_642534, EPI_ISL_642535, EPI_ISL_642536, EPI_ISL_642537, EPI_ISL_642538, EPI_ISL_642539, EPI_ISL_642540, EPI_ISL_642541, EPI_ISL_642542, EPI_ISL_642543, EPI_ISL_642544, EPI_ISL_642545, EPI_ISL_642546, EPI_ISL_642547, EPI_ISL_642548, EPI_ISL_642549, EPI_ISL_642550, EPI_ISL_642551, EPI_ISL_642552, EPI_ISL_642553, EPI_ISL_642554, EPI_ISL_642555, EPI_ISL_642556, EPI_ISL_642557, EPI_ISL_642558, EPI_ISL_642559, EPI_ISL_642560, EPI_ISL_642561, EPI_ISL_642562, EPI_ISL_642563, EPI_ISL_642564, EPI_ISL_642565, EPI_ISL_642566, EPI_ISL_642567, EPI_ISL_642568, EPI_ISL_642569, EPI_ISL_642570, EPI_ISL_642571, EPI_ISL_642572, EPI_ISL_642573, EPI_ISL_642574, EPI_ISL_642575, EPI_ISL_642576, EPI_ISL_642577, EPI_ISL_642578, EPI_ISL_642579, EPI_ISL_642580, EPI_ISL_642581, EPI_ISL_642582, EPI_ISL_642583, EPI_ISL_642584, EPI_ISL_642585, EPI_ISL_642586, EPI_ISL_642587, EPI_ISL_642588, EPI_ISL_642589, EPI_ISL_642590, EPI_ISL_642591, EPI_ISL_642592, EPI_ISL_642593, EPI_ISL_642594, EPI_ISL_642595, EPI_ISL_642596, EPI_ISL_642597, EPI_ISL_642598, EPI_ISL_642599, EPI_ISL_642600, EPI_ISL_642601, EPI_ISL_642602, EPI_ISL_642603, EPI_ISL_642604, EPI_ISL_642605, EPI_ISL_642606, EPI_ISL_642607, EPI_ISL_642608, EPI_ISL_642609, EPI_ISL_642610, EPI_ISL_642611, EPI_ISL_642612, EPI_ISL_642613, EPI_ISL_642614, EPI_ISL_642615, EPI_ISL_642616, EPI_ISL_642617, EPI_ISL_642618, EPI_ISL_642619, EPI_ISL_642620, EPI_ISL_642621, EPI_ISL_642622, EPI_ISL_642623, EPI_ISL_642624, EPI_ISL_642625, EPI_ISL_642626, EPI_ISL_642627, EPI_ISL_642628, EPI_ISL_642629, EPI_ISL_642630, EPI_ISL_642631, EPI_ISL_642632, EPI_ISL_642633, EPI_ISL_642634, EPI_ISL_642635, EPI_ISL_642636, EPI_ISL_642637, EPI_ISL_642638, EPI_ISL_642639, EPI_ISL_642640, EPI_ISL_642641, EPI_ISL_642642, EPI_ISL_642643, EPI_ISL_642644, EPI_ISL_642645, EPI_ISL_642646, EPI_ISL_642647, EPI_ISL_642648, EPI_ISL_642649, EPI_ISL_642650, EPI_ISL_642651, EPI_ISL_642652, EPI_ISL_642653, EPI_ISL_642654, EPI_ISL_642655, EPI_ISL_642656, EPI_ISL_642657, EPI_ISL_642658, EPI_ISL_642659, EPI_ISL_642660, EPI_ISL_642661, EPI_ISL_642662, EPI_ISL_642663, | | | |

| | | | |
|---|---|---|---|
| EPI_ISL_642664, EPI_ISL_642665, EPI_ISL_642666, EPI_ISL_642667, EPI_ISL_642668, EPI_ISL_642669, EPI_ISL_642670, EPI_ISL_642671, EPI_ISL_642672, EPI_ISL_642673, EPI_ISL_642674, EPI_ISL_642675, EPI_ISL_642676, EPI_ISL_642677, EPI_ISL_642678, EPI_ISL_642679, EPI_ISL_642680, EPI_ISL_642681, EPI_ISL_642682, EPI_ISL_642683, EPI_ISL_642684, EPI_ISL_642685, EPI_ISL_642686, EPI_ISL_642687, EPI_ISL_642688, EPI_ISL_642689, EPI_ISL_642690, EPI_ISL_642691, EPI_ISL_642692, EPI_ISL_642693, EPI_ISL_642694, EPI_ISL_642695, EPI_ISL_642696, EPI_ISL_642697, EPI_ISL_642698, EPI_ISL_642699, EPI_ISL_642700, EPI_ISL_642701, EPI_ISL_642702, EPI_ISL_642703, EPI_ISL_642704, EPI_ISL_642705, EPI_ISL_642706, EPI_ISL_642707, EPI_ISL_642708, EPI_ISL_642709, EPI_ISL_642710, EPI_ISL_642711, EPI_ISL_642712, EPI_ISL_642713, EPI_ISL_642714, EPI_ISL_642715, EPI_ISL_642716, EPI_ISL_642717, EPI_ISL_642718, EPI_ISL_642719, EPI_ISL_642720, EPI_ISL_642721, EPI_ISL_642722, EPI_ISL_642723, EPI_ISL_642724, EPI_ISL_642725, EPI_ISL_642726, EPI_ISL_642727, EPI_ISL_642728, EPI_ISL_642729, EPI_ISL_642730, EPI_ISL_642731, EPI_ISL_642732, EPI_ISL_642733, EPI_ISL_642734, EPI_ISL_642735, EPI_ISL_642736, EPI_ISL_642737, EPI_ISL_642738, EPI_ISL_642739, EPI_ISL_642740, EPI_ISL_642741, EPI_ISL_642742, EPI_ISL_642743, EPI_ISL_642744, EPI_ISL_642745, EPI_ISL_642746, EPI_ISL_642747, EPI_ISL_642748, EPI_ISL_642749, EPI_ISL_642750, EPI_ISL_642751, EPI_ISL_642752, EPI_ISL_642753, EPI_ISL_642754, EPI_ISL_642755, EPI_ISL_642756, EPI_ISL_642757, EPI_ISL_642758, EPI_ISL_642759, EPI_ISL_642760, EPI_ISL_646140, EPI_ISL_646141, EPI_ISL_646142, EPI_ISL_646143, EPI_ISL_646144, EPI_ISL_646145, EPI_ISL_646146, EPI_ISL_646147, EPI_ISL_646149, EPI_ISL_646151, EPI_ISL_646152, EPI_ISL_646153, EPI_ISL_646154, EPI_ISL_646158, EPI_ISL_646159, EPI_ISL_646160, EPI_ISL_646161, EPI_ISL_646163, EPI_ISL_646164, EPI_ISL_646165, EPI_ISL_646167, EPI_ISL_646168, EPI_ISL_646171, EPI_ISL_646172, EPI_ISL_646174, EPI_ISL_646175, EPI_ISL_646176, EPI_ISL_646177, EPI_ISL_646178, EPI_ISL_646179, EPI_ISL_646181, EPI_ISL_646183, EPI_ISL_646184, EPI_ISL_646186, EPI_ISL_646187, EPI_ISL_646188, EPI_ISL_646190, EPI_ISL_646192, EPI_ISL_646194, EPI_ISL_646195, EPI_ISL_646196, EPI_ISL_646197, EPI_ISL_646200, EPI_ISL_646201, EPI_ISL_646202, EPI_ISL_646203, EPI_ISL_646205, EPI_ISL_646206, EPI_ISL_646209, EPI_ISL_646210, EPI_ISL_646211, EPI_ISL_646213, EPI_ISL_646214, EPI_ISL_646216, EPI_ISL_646217, EPI_ISL_646218, EPI_ISL_646219, EPI_ISL_646220, EPI_ISL_646221, EPI_ISL_646222, EPI_ISL_646225, EPI_ISL_646226, EPI_ISL_646227, EPI_ISL_646228, EPI_ISL_646229, EPI_ISL_646230, EPI_ISL_646232, EPI_ISL_646233, EPI_ISL_646234, EPI_ISL_646236, EPI_ISL_646237, EPI_ISL_646238, EPI_ISL_646239, EPI_ISL_646240, EPI_ISL_646241, EPI_ISL_646243, EPI_ISL_646244, EPI_ISL_646246, EPI_ISL_646247, EPI_ISL_646250, EPI_ISL_646251, EPI_ISL_646253, EPI_ISL_646254, EPI_ISL_646255, EPI_ISL_646256, EPI_ISL_646257, EPI_ISL_646258, EPI_ISL_646259, EPI_ISL_646260, EPI_ISL_646262, EPI_ISL_646263, EPI_ISL_646264, EPI_ISL_646265, EPI_ISL_646266, EPI_ISL_646267, EPI_ISL_646270, EPI_ISL_646272, EPI_ISL_646273, EPI_ISL_646274, EPI_ISL_646275, EPI_ISL_646280, EPI_ISL_646281, EPI_ISL_646282, EPI_ISL_646284, EPI_ISL_646285, EPI_ISL_646286, EPI_ISL_646287, EPI_ISL_646288, EPI_ISL_646289, EPI_ISL_646290, EPI_ISL_646292, EPI_ISL_646293, EPI_ISL_646294, EPI_ISL_646295, EPI_ISL_646296, EPI_ISL_646298, EPI_ISL_646300, EPI_ISL_646301, EPI_ISL_646302, EPI_ISL_646304, EPI_ISL_646306, EPI_ISL_646308, EPI_ISL_646310, EPI_ISL_646311, EPI_ISL_646312, EPI_ISL_646315, EPI_ISL_646316, EPI_ISL_646317, EPI_ISL_646319, EPI_ISL_646321, EPI_ISL_646322, EPI_ISL_646324, EPI_ISL_646325, EPI_ISL_646326, EPI_ISL_646328, EPI_ISL_646329, EPI_ISL_646330, EPI_ISL_646331, EPI_ISL_646332, EPI_ISL_646333, EPI_ISL_646335, EPI_ISL_646336, EPI_ISL_646337, EPI_ISL_646338, EPI_ISL_646339, EPI_ISL_646340, EPI_ISL_646341, EPI_ISL_646344, EPI_ISL_646348, EPI_ISL_646351, EPI_ISL_646352, EPI_ISL_646353, EPI_ISL_646355, EPI_ISL_646357, EPI_ISL_646358, EPI_ISL_646359, EPI_ISL_646360, EPI_ISL_646361, EPI_ISL_646364, EPI_ISL_646365, EPI_ISL_646367, EPI_ISL_646368, EPI_ISL_646371, EPI_ISL_646372, EPI_ISL_646374, EPI_ISL_646378, EPI_ISL_646379, EPI_ISL_646380, EPI_ISL_646381, EPI_ISL_646383, EPI_ISL_646384, EPI_ISL_646386, EPI_ISL_646387, EPI_ISL_646389, EPI_ISL_646390, EPI_ISL_646391, EPI_ISL_646392, EPI_ISL_646393, EPI_ISL_646394, EPI_ISL_646395, EPI_ISL_646396, EPI_ISL_646398, EPI_ISL_646399, EPI_ISL_646400, EPI_ISL_646401, EPI_ISL_646403, EPI_ISL_646404, EPI_ISL_646405, EPI_ISL_646406, EPI_ISL_646407, EPI_ISL_646408, EPI_ISL_646409, EPI_ISL_646410, EPI_ISL_646411, EPI_ISL_646413, EPI_ISL_646414, EPI_ISL_646415, EPI_ISL_646416, EPI_ISL_646417, EPI_ISL_646421, EPI_ISL_646422, EPI_ISL_646423, EPI_ISL_646424, EPI_ISL_646425, EPI_ISL_646427, EPI_ISL_646433, EPI_ISL_646435, EPI_ISL_646436, EPI_ISL_646437, EPI_ISL_646438, EPI_ISL_646439, EPI_ISL_646440, EPI_ISL_646444, EPI_ISL_646445, EPI_ISL_646446, EPI_ISL_646447, EPI_ISL_646449, EPI_ISL_646450, EPI_ISL_646451, EPI_ISL_646452, EPI_ISL_646453, EPI_ISL_646455, EPI_ISL_646456, EPI_ISL_646457, EPI_ISL_646458, EPI_ISL_646459, EPI_ISL_646460, EPI_ISL_646461, EPI_ISL_646462, EPI_ISL_646464, EPI_ISL_646465, EPI_ISL_646469, EPI_ISL_646470, EPI_ISL_646471, EPI_ISL_646472, EPI_ISL_646473, EPI_ISL_646474, EPI_ISL_646475, EPI_ISL_646480, EPI_ISL_646481, EPI_ISL_646482, EPI_ISL_646483, EPI_ISL_646484, EPI_ISL_646495, EPI_ISL_646496, EPI_ISL_646498, EPI_ISL_646499, EPI_ISL_646501, EPI_ISL_646503, EPI_ISL_646504, EPI_ISL_646507, EPI_ISL_646517, EPI_ISL_646519, EPI_ISL_646523, EPI_ISL_646524, EPI_ISL_646527, EPI_ISL_646530, EPI_ISL_646531, EPI_ISL_646534, EPI_ISL_646536, EPI_ISL_646539, EPI_ISL_646547, EPI_ISL_646550, EPI_ISL_646552, EPI_ISL_646553, EPI_ISL_646554, EPI_ISL_646556, EPI_ISL_646562, EPI_ISL_646565, EPI_ISL_646570, EPI_ISL_646574, EPI_ISL_646575, EPI_ISL_646578, EPI_ISL_646579, EPI_ISL_646582, EPI_ISL_646584, EPI_ISL_646587, EPI_ISL_646588, EPI_ISL_646589, EPI_ISL_646590, EPI_ISL_646593, EPI_ISL_646596, EPI_ISL_646598, EPI_ISL_646602, EPI_ISL_646604, EPI_ISL_646608, EPI_ISL_646609, EPI_ISL_646611, EPI_ISL_646615, EPI_ISL_646617, EPI_ISL_646619, EPI_ISL_646620, EPI_ISL_646622, EPI_ISL_646624, EPI_ISL_646627, EPI_ISL_646629, EPI_ISL_646630, EPI_ISL_646632, EPI_ISL_646633, EPI_ISL_646634, EPI_ISL_646636, EPI_ISL_646637, EPI_ISL_646641, EPI_ISL_646643, EPI_ISL_646644, EPI_ISL_646645, EPI_ISL_646646, EPI_ISL_646647, EPI_ISL_646649, EPI_ISL_646650, EPI_ISL_646654, EPI_ISL_646656, EPI_ISL_646657, EPI_ISL_646659, EPI_ISL_646661, EPI_ISL_646663, EPI_ISL_646667, EPI_ISL_646669, EPI_ISL_646670, EPI_ISL_646672, EPI_ISL_646674, EPI_ISL_646677, EPI_ISL_646679, EPI_ISL_646680, EPI_ISL_646681, EPI_ISL_646682, EPI_ISL_646685, EPI_ISL_646688, EPI_ISL_646690, EPI_ISL_646694, EPI_ISL_646699, EPI_ISL_646700, EPI_ISL_646701, EPI_ISL_646703, EPI_ISL_646704, EPI_ISL_646705, EPI_ISL_646709, EPI_ISL_646714, EPI_ISL_646717, EPI_ISL_646720, EPI_ISL_646723, EPI_ISL_646725, EPI_ISL_646726, EPI_ISL_646728, EPI_ISL_646729, EPI_ISL_646730, EPI_ISL_646732, EPI_ISL_646733, EPI_ISL_646735, EPI_ISL_646742, EPI_ISL_646745, EPI_ISL_646748, EPI_ISL_646750, EPI_ISL_646754, EPI_ISL_646759, EPI_ISL_646764, EPI_ISL_646765, EPI_ISL_646766, EPI_ISL_646772, EPI_ISL_646774, EPI_ISL_646775, EPI_ISL_646777, EPI_ISL_646778, EPI_ISL_646779, EPI_ISL_646780, EPI_ISL_646781, EPI_ISL_646783, EPI_ISL_646785, EPI_ISL_646786, EPI_ISL_646788, EPI_ISL_646792, EPI_ISL_646794, EPI_ISL_646798, EPI_ISL_646810, EPI_ISL_646818, EPI_ISL_646825, EPI_ISL_646826, EPI_ISL_646837, EPI_ISL_646840, EPI_ISL_646850, EPI_ISL_646853, EPI_ISL_646854, EPI_ISL_646868, EPI_ISL_646869, EPI_ISL_646870, EPI_ISL_646872, EPI_ISL_646877, EPI_ISL_646880, EPI_ISL_646885, EPI_ISL_646891, EPI_ISL_646892, EPI_ISL_646902, EPI_ISL_646905, EPI_ISL_646912, EPI_ISL_646915, EPI_ISL_646921, EPI_ISL_646929, EPI_ISL_646936, EPI_ISL_646937, EPI_ISL_646939, EPI_ISL_646948, EPI_ISL_646955, EPI_ISL_646960, EPI_ISL_646964, EPI_ISL_646967, EPI_ISL_646974, EPI_ISL_646975, EPI_ISL_646976, EPI_ISL_646977, EPI_ISL_646983, EPI_ISL_646988, EPI_ISL_646992, EPI_ISL_646996, EPI_ISL_647001, EPI_ISL_647003, EPI_ISL_647006, EPI_ISL_647014, EPI_ISL_647023, EPI_ISL_647031, EPI_ISL_647032, EPI_ISL_647035, EPI_ISL_647038, EPI_ISL_647045, EPI_ISL_647050, EPI_ISL_647053, EPI_ISL_647059, EPI_ISL_647061, EPI_ISL_647067, EPI_ISL_647073, EPI_ISL_647076, EPI_ISL_647081, EPI_ISL_647087, EPI_ISL_647090, EPI_ISL_647097, EPI_ISL_647102, EPI_ISL_647105, EPI_ISL_647109, EPI_ISL_647114, EPI_ISL_647115, EPI_ISL_647117, EPI_ISL_647121, EPI_ISL_647797, EPI_ISL_647798, EPI_ISL_647799, EPI_ISL_647800, EPI_ISL_647801, EPI_ISL_647802, EPI_ISL_647803, EPI_ISL_647804, EPI_ISL_647805, EPI_ISL_647806, EPI_ISL_647861, EPI_ISL_647862, EPI_ISL_647863, EPI_ISL_647864, EPI_ISL_647865, EPI_ISL_647866, EPI_ISL_647867, EPI_ISL_647868, EPI_ISL_647869, EPI_ISL_647870, EPI_ISL_647871, EPI_ISL_647872, EPI_ISL_647876, EPI_ISL_649494, EPI_ISL_649495, EPI_ISL_649496, EPI_ISL_649497, EPI_ISL_649498, EPI_ISL_649499, EPI_ISL_649500, EPI_ISL_649501, EPI_ISL_649502, EPI_ISL_649503, EPI_ISL_649504, EPI_ISL_649505, EPI_ISL_649506, EPI_ISL_649507, EPI_ISL_649508, EPI_ISL_649509, EPI_ISL_649510, EPI_ISL_649511, EPI_ISL_649512, EPI_ISL_649513, EPI_ISL_649514, EPI_ISL_649515, EPI_ISL_649516, EPI_ISL_649517, EPI_ISL_649518, EPI_ISL_649519, EPI_ISL_649520, EPI_ISL_649521, EPI_ISL_649522, EPI_ISL_649523, EPI_ISL_649524, EPI_ISL_649525, EPI_ISL_649526, EPI_ISL_649527, EPI_ISL_649528, EPI_ISL_649529, EPI_ISL_649530, EPI_ISL_649531, EPI_ISL_649532, EPI_ISL_649533, EPI_ISL_649534, EPI_ISL_649535, EPI_ISL_649536, EPI_ISL_649537, EPI_ISL_649538, EPI_ISL_649539, EPI_ISL_649540, EPI_ISL_649541, EPI_ISL_649542, EPI_ISL_649543, EPI_ISL_649544, EPI_ISL_649545, EPI_ISL_649546, EPI_ISL_649547, EPI_ISL_649569, EPI_ISL_649570, EPI_ISL_649571, EPI_ISL_649572, EPI_ISL_649573, EPI_ISL_649574, EPI_ISL_649575, EPI_ISL_649576, EPI_ISL_649577, EPI_ISL_649578, EPI_ISL_649579, EPI_ISL_649580, EPI_ISL_649581, EPI_ISL_649582, EPI_ISL_649583, EPI_ISL_649584, EPI_ISL_649585, EPI_ISL_649586, EPI_ISL_649587, EPI_ISL_649588, EPI_ISL_649589, EPI_ISL_649590, EPI_ISL_649591, EPI_ISL_649592, EPI_ISL_649593, EPI_ISL_649594, EPI_ISL_649595, EPI_ISL_649596, EPI_ISL_649597, EPI_ISL_649598, EPI_ISL_649599, EPI_ISL_649600, EPI_ISL_649601, EPI_ISL_649602, EPI_ISL_649603, EPI_ISL_649604, EPI_ISL_649605, EPI_ISL_649606, EPI_ISL_649607, EPI_ISL_649608, EPI_ISL_649609, EPI_ISL_649610, EPI_ISL_649611, EPI_ISL_649612, EPI_ISL_649613, EPI_ISL_649614, EPI_ISL_649615, EPI_ISL_649616, EPI_ISL_649617, EPI_ISL_649618, EPI_ISL_649619, EPI_ISL_649620, EPI_ISL_649621, EPI_ISL_649622, EPI_ISL_649623, EPI_ISL_649624, EPI_ISL_649661, EPI_ISL_649664, EPI_ISL_649669, EPI_ISL_649710, EPI_ISL_659744, EPI_ISL_659745, EPI_ISL_659746, EPI_ISL_659747, EPI_ISL_659748, EPI_ISL_659749, EPI_ISL_659750, EPI_ISL_659751, EPI_ISL_659752, EPI_ISL_659753, EPI_ISL_659754, EPI_ISL_659755, EPI_ISL_659756, EPI_ISL_659757, EPI_ISL_659758, EPI_ISL_659759, EPI_ISL_659760, EPI_ISL_659761, EPI_ISL_659762, EPI_ISL_659763, EPI_ISL_659764, EPI_ISL_659765, EPI_ISL_659766, EPI_ISL_659767, EPI_ISL_659768, EPI_ISL_659769, EPI_ISL_659770, EPI_ISL_659771, EPI_ISL_659772, EPI_ISL_659773, EPI_ISL_659774, EPI_ISL_659775, EPI_ISL_659776, EPI_ISL_659777, EPI_ISL_659778, EPI_ISL_659779, EPI_ISL_659780, EPI_ISL_659781, EPI_ISL_659782, EPI_ISL_659783, EPI_ISL_659784, EPI_ISL_659785, EPI_ISL_659786, EPI_ISL_659787, EPI_ISL_659788, EPI_ISL_659789, EPI_ISL_659790, EPI_ISL_659791, EPI_ISL_659792, EPI_ISL_659793, EPI_ISL_659794, EPI_ISL_659795, EPI_ISL_659796, EPI_ISL_659797, EPI_ISL_659798, EPI_ISL_659799, EPI_ISL_659800, EPI_ISL_659801, EPI_ISL_659802, EPI_ISL_659803, EPI_ISL_659804, EPI_ISL_659805, EPI_ISL_659806, EPI_ISL_659807, EPI_ISL_659808, EPI_ISL_659809, EPI_ISL_659810, EPI_ISL_659811, EPI_ISL_659812, EPI_ISL_659813, EPI_ISL_659814, EPI_ISL_659815, EPI_ISL_659816, EPI_ISL_659817, EPI_ISL_659818, EPI_ISL_659819, EPI_ISL_659820, EPI_ISL_659821, EPI_ISL_659822, EPI_ISL_659823, EPI_ISL_659824, EPI_ISL_659825, EPI_ISL_659826, EPI_ISL_659827, EPI_ISL_659828, EPI_ISL_659829, EPI_ISL_659830, EPI_ISL_659831, EPI_ISL_659832, EPI_ISL_659833, EPI_ISL_659834, EPI_ISL_659835, EPI_ISL_659836, EPI_ISL_659837, EPI_ISL_659838, EPI_ISL_659839, EPI_ISL_659840, EPI_ISL_659841, EPI_ISL_659842, EPI_ISL_659843, EPI_ISL_659844, EPI_ISL_659845, EPI_ISL_659846, EPI_ISL_659847, EPI_ISL_659848, EPI_ISL_659849, EPI_ISL_659850, EPI_ISL_659851, EPI_ISL_659852, EPI_ISL_659853, EPI_ISL_659854, EPI_ISL_659855, EPI_ISL_659856, EPI_ISL_659857, EPI_ISL_659858, EPI_ISL_659859, EPI_ISL_659860, EPI_ISL_659861, EPI_ISL_659862, EPI_ISL_659863, EPI_ISL_659864, EPI_ISL_659865, EPI_ISL_659866, EPI_ISL_659867, EPI_ISL_659868, EPI_ISL_659869, EPI_ISL_659870, EPI_ISL_659871, EPI_ISL_659872, EPI_ISL_659873, EPI_ISL_659874, EPI_ISL_659875, EPI_ISL_659876, EPI_ISL_659877, EPI_ISL_659878, EPI_ISL_659879, EPI_ISL_659880, EPI_ISL_659881, EPI_ISL_659882, EPI_ISL_659883, EPI_ISL_659884, EPI_ISL_659885, EPI_ISL_659886, EPI_ISL_659887, EPI_ISL_659888, EPI_ISL_659889, EPI_ISL_659890, EPI_ISL_659891, EPI_ISL_659892, EPI_ISL_659893, EPI_ISL_659894, EPI_ISL_659895, EPI_ISL_659896, EPI_ISL_659897, EPI_ISL_659898, EPI_ISL_659899, EPI_ISL_659900, EPI_ISL_659901, EPI_ISL_659902, EPI_ISL_659903, EPI_ISL_659904, EPI_ISL_659905, EPI_ISL_659906, EPI_ISL_659907, EPI_ISL_659908, EPI_ISL_659909, EPI_ISL_659910, EPI_ISL_659911, EPI_ISL_659912, EPI_ISL_659913, EPI_ISL_659914, EPI_ISL_659915, EPI_ISL_659916, EPI_ISL_659917, EPI_ISL_659918, EPI_ISL_659919, EPI_ISL_659920, EPI_ISL_659921, EPI_ISL_659922, EPI_ISL_659923, EPI_ISL_659924, EPI_ISL_659925, EPI_ISL_659926, EPI_ISL_659927, EPI_ISL_659928, EPI_ISL_659929, EPI_ISL_659930, EPI_ISL_659931, EPI_ISL_659932, EPI_ISL_659933, EPI_ISL_659934, EPI_ISL_659935, EPI_ISL_659936, EPI_ISL_659937, EPI_ISL_659938, EPI_ISL_659939, EPI_ISL_659940, EPI_ISL_659941, EPI_ISL_659942, EPI_ISL_659943, EPI_ISL_659944, EPI_ISL_659945, EPI_ISL_659946, EPI_ISL_659947, EPI_ISL_659948, EPI_ISL_659949, EPI_ISL_659950, EPI_ISL_659951, EPI_ISL_659952, EPI_ISL_659953, EPI_ISL_659954, EPI_ISL_659955, EPI_ISL_659956, EPI_ISL_659957, EPI_ISL_659958, EPI_ISL_659959, EPI_ISL_659960, EPI_ISL_659961, EPI_ISL_659962, EPI_ISL_659963, EPI_ISL_659964, EPI_ISL_659965, EPI_ISL_659966, EPI_ISL_659967, EPI_ISL_659968, EPI_ISL_659969, EPI_ISL_659970, EPI_ISL_659971, EPI_ISL_659972, EPI_ISL_659973, EPI_ISL_659974, EPI_ISL_659975, EPI_ISL_659976, EPI_ISL_659977, EPI_ISL_659978, EPI_ISL_659979, EPI_ISL_659980, EPI_ISL_659981, EPI_ISL_659982, EPI_ISL_659983, EPI_ISL_659984, EPI_ISL_659985, EPI_ISL_659986, EPI_ISL_659987, EPI_ISL_659988, EPI_ISL_659989, EPI_ISL_659990, EPI_ISL_659991, EPI_ISL_659992, EPI_ISL_659993, EPI_ISL_659994, EPI_ISL_659995, EPI_ISL_659996, EPI_ISL_659997, EPI_ISL_659998, EPI_ISL_659999, EPI_ISL_660000, EPI_ISL_660001, EPI_ISL_660002, EPI_ISL_660003, EPI_ISL_660004, EPI_ISL_660005, EPI_ISL_660006, EPI_ISL_660007, EPI_ISL_660008, EPI_ISL_660009, EPI_ISL_660010, EPI_ISL_660011, EPI_ISL_660012, EPI_ISL_660013, EPI_ISL_660014, EPI_ISL_660015, EPI_ISL_660016, EPI_ISL_660017, EPI_ISL_660018, EPI_ISL_660019, EPI_ISL_660020, EPI_ISL_660021, EPI_ISL_660022, EPI_ISL_660023, EPI_ISL_660024, EPI_ISL_660025, EPI_ISL_660026, EPI_ISL_660027, EPI_ISL_660028, EPI_ISL_660029, EPI_ISL_660030, EPI_ISL_660031, EPI_ISL_660032, EPI_ISL_660033, EPI_ISL_660034, EPI_ISL_660035, EPI_ISL_660036, EPI_ISL_660037, EPI_ISL_660038, EPI_ISL_660039, EPI_ISL_660040, EPI_ISL_660041, EPI_ISL_660042, EPI_ISL_660043, EPI_ISL_660044, EPI_ISL_660045, EPI_ISL_660046, EPI_ISL_660047, EPI_ISL_660048, EPI_ISL_660049, EPI_ISL_660050, EPI_ISL_660051, EPI_ISL_660052, EPI_ISL_660053, EPI_ISL_660054, EPI_ISL_660055, EPI_ISL_660056, EPI_ISL_660057, EPI_ISL_660058, EPI_ISL_660059, EPI_ISL_660060, EPI_ISL_660061, EPI_ISL_660062, EPI_ISL_660063, EPI_ISL_660064, EPI_ISL_660065, EPI_ISL_660066, EPI_ISL_660067, EPI_ISL_662654, EPI_ISL_662655, EPI_ISL_662656, EPI_ISL_662657, EPI_ISL_662658, EPI_ISL_662659, EPI_ISL_662660, EPI_ISL_662661, EPI_ISL_662662, EPI_ISL_662663, EPI_ISL_662664, EPI_ISL_662665, EPI_ISL_662666, EPI_ISL_662667, EPI_ISL_662668, EPI_ISL_662669, EPI_ISL_662670, EPI_ISL_662671, EPI_ISL_662672, EPI_ISL_662673, EPI_ISL_662674, EPI_ISL_662675, EPI_ISL_662676, EPI_ISL_662677, EPI_ISL_662678, EPI_ISL_662679, EPI_ISL_662680, EPI_ISL_662681, EPI_ISL_662682, EPI_ISL_662683, EPI_ISL_662684, EPI_ISL_662685, EPI_ISL_662686, EPI_ISL_662687, EPI_ISL_662688, EPI_ISL_662689, EPI_ISL_662690, EPI_ISL_662691, EPI_ISL_662692, EPI_ISL_662693, EPI_ISL_662694, EPI_ISL_662695, EPI_ISL_662696, EPI_ISL_662697, EPI_ISL_662698, EPI_ISL_662699, EPI_ISL_662700, EPI_ISL_662701, EPI_ISL_662702, EPI_ISL_662703, EPI_ISL_662704, EPI_ISL_662705, EPI_ISL_662706, EPI_ISL_662707, EPI_ISL_662708, EPI_ISL_662709, EPI_ISL_662710, EPI_ISL_662711, EPI_ISL_662712, EPI_ISL_662713, EPI_ISL_662714, EPI_ISL_662715, EPI_ISL_662716, EPI_ISL_662717, EPI_ISL_662718, EPI_ISL_662719, EPI_ISL_662720, EPI_ISL_662721, EPI_ISL_662722, EPI_ISL_662723, EPI_ISL_662724, EPI_ISL_662725, EPI_ISL_662726, EPI_ISL_662727, EPI_ISL_662728, EPI_ISL_662729, EPI_ISL_662730, EPI_ISL_662731, EPI_ISL_662732, EPI_ISL_662733, EPI_ISL_662734, EPI_ISL_662735, EPI_ISL_662736, EPI_ISL_662737, EPI_ISL_662738, EPI_ISL_662739, EPI_ISL_662740, EPI_ISL_662741, EPI_ISL_662742, EPI_ISL_662743, EPI_ISL_662744, EPI_ISL_662745, EPI_ISL_662747, EPI_ISL_662748, EPI_ISL_662749, EPI_ISL_662750, EPI_ISL_662751, EPI_ISL_662752, EPI_ISL_662753, EPI_ISL_662754, EPI_ISL_662755, EPI_ISL_662756, EPI_ISL_662757, EPI_ISL_662758, EPI_ISL_662759, EPI_ISL_662760, EPI_ISL_662761, EPI_ISL_662762, EPI_ISL_662763, EPI_ISL_662764, EPI_ISL_662765, EPI_ISL_662766, EPI_ISL_662767, EPI_ISL_662768, EPI_ISL_662769, EPI_ISL_662770, EPI_ISL_662771, EPI_ISL_662772, EPI_ISL_662773, EPI_ISL_662774, EPI_ISL_662775, EPI_ISL_662776, EPI_ISL_662777, EPI_ISL_662778, EPI_ISL_662779, EPI_ISL_662780, EPI_ISL_662781, EPI_ISL_662782, EPI_ISL_662783, EPI_ISL_662784, EPI_ISL_662785, EPI_ISL_662786, EPI_ISL_662787, EPI_ISL_662788, EPI_ISL_662789, EPI_ISL_662790, EPI_ISL_662791, EPI_ISL_662792, EPI_ISL_662793, EPI_ISL_662794, EPI_ISL_662795, EPI_ISL_662796, EPI_ISL_662797, EPI_ISL_662798, EPI_ISL_662799, EPI_ISL_662800, EPI_ISL_662801, EPI_ISL_662802, EPI_ISL_662803, EPI_ISL_662804, EPI_ISL_662805, EPI_ISL_662806, EPI_ISL_662807, EPI_ISL_662808, EPI_ISL_662809, EPI_ISL_662810, EPI_ISL_662811, EPI_ISL_662812, EPI_ISL_662813, EPI_ISL_662814, EPI_ISL_662815, EPI_ISL_662816, EPI_ISL_662817, EPI_ISL_662818, EPI_ISL_662819, EPI_ISL_662820, EPI_ISL_662821, EPI_ISL_662822, EPI_ISL_662823, EPI_ISL_662824, EPI_ISL_662825, EPI_ISL_662826, EPI_ISL_662827, EPI_ISL_662828, EPI_ISL_662829, EPI_ISL_662830, EPI_ISL_662831, EPI_ISL_662832, EPI_ISL_662833, EPI_ISL_662834, EPI_ISL_662835, EPI_ISL_662836, EPI_ISL_662837, EPI_ISL_662838, EPI_ISL_662839, EPI_ISL_662840, EPI_ISL_662841, EPI_ISL_662842, EPI_ISL_662843, EPI_ISL_662844, EPI_ISL_662845, EPI_ISL_662846, EPI_ISL_662847, EPI_ISL_662848, EPI_ISL_662849, EPI_ISL_662850, EPI_ISL_662851, EPI_ISL_662852, EPI_ISL_662853, EPI_ISL_662854, EPI_ISL_662855, EPI_ISL_662857, EPI_ISL_662858, EPI_ISL_662859, EPI_ISL_662860, EPI_ISL_662861, EPI_ISL_662862, EPI_ISL_662863, EPI_ISL_662864, EPI_ISL_662865, EPI_ISL_662866, EPI_ISL_662867, EPI_ISL_662868, EPI_ISL_662869, EPI_ISL_662870, EPI_ISL_662871, EPI_ISL_662872, EPI_ISL_662873, EPI_ISL_662874, EPI_ISL_662875, EPI_ISL_662876, EPI_ISL_662877, EPI_ISL_662878, EPI_ISL_662879, EPI_ISL_662880, EPI_ISL_662881, EPI_ISL_662882, EPI_ISL_662883, EPI_ISL_662884, EPI_ISL_662885, EPI_ISL_662886, EPI_ISL_662887, EPI_ISL_662888, EPI_ISL_662889, EPI_ISL_662890, EPI_ISL_662891, EPI_ISL_662892, EPI_ISL_662893, EPI_ISL_662894, EPI_ISL_662895, EPI_ISL_662896, EPI_ISL_662897, EPI_ISL_662898, EPI_ISL_662899, EPI_ISL_662900, EPI_ISL_662901, EPI_ISL_662902, EPI_ISL_662903, EPI_ISL_662904, EPI_ISL_662905, EPI_ISL_662906, EPI_ISL_662907, EPI_ISL_662908, EPI_ISL_662909, EPI_ISL_662910, EPI_ISL_662911, EPI_ISL_662912, EPI_ISL_662913, EPI_ISL_662914, EPI_ISL_662915, EPI_ISL_662916, EPI_ISL_662917, EPI_ISL_662918, EPI_ISL_662919, EPI_ISL_662920, EPI_ISL_662921, EPI_ISL_662922, EPI_ISL_662923, EPI_ISL_662924, EPI_ISL_662925, EPI_ISL_662926, EPI_ISL_662927, EPI_ISL_662928, EPI_ISL_662929, EPI_ISL_662930, EPI_ISL_662931, EPI_ISL_662932, EPI_ISL_662933, EPI_ISL_662934, EPI_ISL_662935, EPI_ISL_662936, EPI_ISL_662937, EPI_ISL_662938, EPI_ISL_662939, EPI_ISL_662940, EPI_ISL_662941, EPI_ISL_662942, EPI_ISL_662943, EPI_ISL_662944, EPI_ISL_662945, EPI_ISL_662946, EPI_ISL_662947, EPI_ISL_662948, EPI_ISL_662949, EPI_ISL_662950, EPI_ISL_662951, EPI_ISL_662952, EPI_ISL_662953, EPI_ISL_662954, EPI_ISL_662955, EPI_ISL_662956, EPI_ISL_662957, EPI_ISL_662958, EPI_ISL_662959, EPI_ISL_662960, EPI_ISL_662961, EPI_ISL_662962, EPI_ISL_662963, EPI_ISL_662964, EPI_ISL_662965, EPI_ISL_662966, EPI_ISL_662967, EPI_ISL_662968, EPI_ISL_662969, EPI_ISL_662970, EPI_ISL_662971, EPI_ISL_662972, EPI_ISL_662973, EPI_ISL_662974, EPI_ISL_662975, EPI_ISL_662976, EPI_ISL_662977, EPI_ISL_662978, EPI_ISL_662979, EPI_ISL_662980, EPI_ISL_662981, EPI_ISL_662982, EPI_ISL_662983, EPI_ISL_662984, EPI_ISL_662985, EPI_ISL_662986, EPI_ISL_662987, EPI_ISL_662988, EPI_ISL_662989, EPI_ISL_662990, EPI_ISL_662991, EPI_ISL_662992, EPI_ISL_662993, EPI_ISL_662994, EPI_ISL_662995, EPI_ISL_662996, EPI_ISL_662997, EPI_ISL_662998, EPI_ISL_662999, EPI_ISL_663000, EPI_ISL_663001, EPI_ISL_663002, EPI_ISL_663003, EPI_ISL_663004, EPI_ISL_663005, EPI_ISL_663006, EPI_ISL_663007, EPI_ISL_663008, EPI_ISL_663009, EPI_ISL_663010, EPI_ISL_663011, EPI_ISL_663012, EPI_ISL_663013, EPI_ISL_663014, EPI_ISL_663015, EPI_ISL_663016, EPI_ISL_663017, EPI_ISL_663018, EPI_ISL_663019, EPI_ISL_663020, EPI_ISL_663021, EPI_ISL_663022, EPI_ISL_663023, EPI_ISL_663024, EPI_ISL_663025, EPI_ISL_663026, EPI_ISL_663027, EPI_ISL_663028, EPI_ISL_663029, EPI_ISL_663030, EPI_ISL_663031, EPI_ISL_663032, EPI_ISL_663033, EPI_ISL_663034, EPI_ISL_663035, EPI_ISL_663036, EPI_ISL_663037, EPI_ISL_663038, EPI_ISL_663039, EPI_ISL_663040, EPI_ISL_663041, EPI_ISL_663042, EPI_ISL_663043, EPI_ISL_663044, EPI_ISL_663045, EPI_ISL_663046, EPI_ISL_663047, EPI_ISL_663048, EPI_ISL_663049, EPI_ISL_663050, EPI_ISL_663051, EPI_ISL_663052, EPI_ISL_663053, EPI_ISL_663054, EPI_ISL_663055, EPI_ISL_663056, EPI_ISL_663057, EPI_ISL_663058, EPI_ISL_663059, EPI_ISL_663060, EPI_ISL_663061, EPI_ISL_663062, EPI_ISL_663063, EPI_ISL_663064, EPI_ISL_663065, EPI_ISL_663066, EPI_ISL_663067, EPI_ISL_663068, EPI_ISL_663069, EPI_ISL_663070, EPI_ISL_663071, EPI_ISL_663072, EPI_ISL_663073, EPI_ISL_663074, EPI_ISL_663075, EPI_ISL_663076, EPI_ISL_663077, EPI_ISL_663078, EPI_ISL_663079, EPI_ISL_663080, EPI_ISL_663081, EPI_ISL_663082, EPI_ISL_663083, EPI_ISL_663084, EPI_ISL_663085, EPI_ISL_663086, EPI_ISL_663087, EPI_ISL_663088, EPI_ISL_663089, EPI_ISL_663090, EPI_ISL_663091, EPI_ISL_663092, EPI_ISL_663093, EPI_ISL_663094, EPI_ISL_663095, EPI_ISL_663096, EPI_ISL_663097, EPI_ISL_663098, EPI_ISL_663099, EPI_ISL_663100, EPI_ISL_663101, EPI_ISL_663102, EPI_ISL_663103, EPI_ISL_663104, EPI_ISL_663105, EPI_ISL_663106, EPI_ISL_663107, EPI_ISL_663108, EPI_ISL_663109, EPI_ISL_663110, EPI_ISL_663111, EPI_ISL_663112, EPI_ISL_663113, EPI_ISL_663114, EPI_ISL_663115, EPI_ISL_663116, EPI_ISL_663117, EPI_ISL_663118, EPI_ISL_663119, EPI_ISL_663120, EPI_ISL_663121, EPI_ISL_663122, EPI_ISL_663123, EPI_ISL_663124, EPI_ISL_663125, EPI_ISL_663126, EPI_ISL_663127, EPI_ISL_663128, EPI_ISL_663129, EPI_ISL_663130, EPI_ISL_663131, EPI_ISL_663132, EPI_ISL_663133, EPI_ISL_663134, EPI_ISL_663135, EPI_ISL_663136, EPI_ISL_663137, EPI_ISL_663138, EPI_ISL_663139, EPI_ISL_663140, EPI_ISL_663141, EPI_ISL_663142, EPI_ISL_663143, EPI_ISL_663144, EPI_ISL_663145, EPI_ISL_663146, EPI_ISL_663147, EPI_ISL_663148, EPI_ISL_663149, EPI_ISL_663150, EPI_ISL_663151, EPI_ISL_663152, EPI_ISL_663153, EPI_ISL_663154, EPI_ISL_663155, EPI_ISL_663156, EPI_ISL_663157, EPI_ISL_663158, EPI_ISL_663159, EPI_ISL_663160, EPI_ISL_663161, EPI_ISL_663162, EPI_ISL_663163, EPI_ISL_663164, EPI_ISL_663165, EPI_ISL_663166, EPI_ISL_663167, EPI_ISL_663168, EPI_ISL_663169, EPI_ISL_663170, EPI_ISL_663171, EPI_ISL_663172, EPI_ISL_663173, EPI_ISL_663174, EPI_ISL_663175, EPI_ISL_663176, EPI_ISL_663177, EPI_ISL_663178, EPI_ISL_663179, EPI_ISL_663180, EPI_ISL_663181, EPI_ISL_663182, EPI_ISL_663183, EPI_ISL_663184, EPI_ISL_663185, EPI_ISL_663186, EPI_ISL_663187, EPI_ISL_663188, EPI_ISL_663189, EPI_ISL_663190, EPI_ISL_663191, EPI_ISL_663192, EPI_ISL_663193, EPI_ISL_663194, EPI_ISL_663195, EPI_ISL_663196, EPI_ISL_663197, EPI_ISL_663198, EPI_ISL_663199, EPI_ISL_663200, EPI_ISL_663201, EPI_ISL_663202, EPI_ISL_663203, EPI_ISL_663204, EPI_ISL_663205, EPI_ISL_676395, EPI_ISL_676396, EPI_ISL_676397, EPI_ISL_676398, EPI_ISL_676399, EPI_ISL_676400, EPI_ISL_676401, EPI_ISL_676402, EPI_ISL_676403, EPI_ISL_676404, EPI_ISL_676405, EPI_ISL_676406, EPI_ISL_676407, EPI_ISL_676408, EPI_ISL_676409, EPI_ISL_676410, EPI_ISL_676411, EPI_ISL_676412, EPI_ISL_676413, EPI_ISL_676414, EPI_ISL_676415, EPI_ISL_676416, EPI_ISL_676417, EPI_ISL_676418, EPI_ISL_676419, EPI_ISL_676420, EPI_ISL_676421, EPI_ISL_676422, EPI_ISL_676423, EPI_ISL_676424, EPI_ISL_676425, EPI_ISL_676426, EPI_ISL_676427, EPI_ISL_676428, EPI_ISL_676429, EPI_ISL_676430, EPI_ISL_676431, EPI_ISL_676432, EPI_ISL_676433, EPI_ISL_676434, EPI_ISL_676435, EPI_ISL_676436, EPI_ISL_676437, EPI_ISL_676438, EPI_ISL_676439, EPI_ISL_676440, EPI_ISL_676441, EPI_ISL_676442, EPI_ISL_676443, EPI_ISL_676444, EPI_ISL_676445, EPI_ISL_676446, EPI_ISL_676447, EPI_ISL_676448, EPI_ISL_676449, EPI_ISL_676450, EPI_ISL_676451, EPI_ISL_676452, EPI_ISL_676453, EPI_ISL_676454, EPI_ISL_676455, EPI_ISL_676456, EPI_ISL_676457, EPI_ISL_676458, EPI_ISL_676459, EPI_ISL_676460, EPI_ISL_676461, EPI_ISL_676462, EPI_ISL_676463, EPI_ISL_676464, EPI_ISL_676465, EPI_ISL_676466, EPI_ISL_676467, EPI_ISL_676468, EPI_ISL_676469, EPI_ISL_676470, EPI_ISL_676471, EPI_ISL_676472, EPI_ISL_676473, EPI_ISL_676474, EPI_ISL_676475, EPI_ISL_676476, EPI_ISL_676477, EPI_ISL_676478, EPI_ISL_676479, EPI_ISL_676480, EPI_ISL_676481, EPI_ISL_676482, EPI_ISL_676483, EPI_ISL_676484, EPI_ISL_676485, EPI_ISL_676486, EPI_ISL_676487, EPI_ISL_676488, EPI_ISL_710903, EPI_ISL_710904, EPI_ISL_710905, EPI_ISL_710995, EPI_ISL_710996, EPI_ISL_710997, EPI_ISL_710998, EPI_ISL_710999, EPI_ISL_711000, EPI_ISL_711001, EPI_ISL_711002, EPI_ISL_711003, EPI_ISL_711004, EPI_ISL_711005, EPI_ISL_711006, EPI_ISL_711007, EPI_ISL_711014, EPI_ISL_711048, EPI_ISL_711049, EPI_ISL_711051, EPI_ISL_711052, EPI_ISL_711053, EPI_ISL_711054, EPI_ISL_721534, EPI_ISL_721535 | | | |
| see above | Lighthouse Lab in Glasgow | Wellcome Sanger Institute for the COVID-19 Genomics UK (COG-UK) Consortium | Anna Dominiczak and Alex Alderton; Carol Clugston; Cordelia Langford; David Gray; David K. Jackson; Dominic Kwiatkowski; Ewan Harrison; Harper VanSteenhouse; Ian Johnston; John Sillitoe on behalf of the Wellcome Sanger Institute COVID-19 Surveillance Team; John Sillitoe on behalf of the Wellcome Sanger Institute COVID-19 Surveillance Team (http://www.sanger.ac.uk/covid-team); Roberto Amato; Sonia Goncalves; Yumi Kasai |
| EPI_ISL_633318, EPI_ISL_633392, EPI_ISL_633591, EPI_ISL_633681 | Lighthouse Lab in Glasgow | Wellcome Sanger Institute for the COVID-19 Genomics UK (COG-UK) consortium | Anna Dominiczak and Alex Alderton; Carol Clugston; Cordelia Langford; David Gray; David K. Jackson; Dominic Kwiatkowski; Ewan Harrison; Harper VanSteenhouse; Ian Johnston; John Sillitoe on behalf of the Wellcome Sanger Institute COVID-19 Surveillance Team; Roberto Amato; Sonia Goncalves; Yumi Kasai |
| EPI_ISL_645221, EPI_ISL_645226, EPI_ISL_645241, EPI_ISL_645243, EPI_ISL_645244, EPI_ISL_645263, EPI_ISL_645264, EPI_ISL_645285, EPI_ISL_645296, EPI_ISL_645300, EPI_ISL_645302, EPI_ISL_645308, EPI_ISL_645319, EPI_ISL_645324, EPI_ISL_645327, EPI_ISL_645329, EPI_ISL_645345, EPI_ISL_645354, EPI_ISL_645360, EPI_ISL_645367, EPI_ISL_645369, EPI_ISL_645374, EPI_ISL_645375, EPI_ISL_645383, EPI_ISL_645391, EPI_ISL_645393, EPI_ISL_645395, EPI_ISL_645399, EPI_ISL_645405, EPI_ISL_645406, EPI_ISL_645421, EPI_ISL_645424, EPI_ISL_645438, EPI_ISL_645441, EPI_ISL_645453, EPI_ISL_645477, EPI_ISL_645489, EPI_ISL_645513, EPI_ISL_645518, EPI_ISL_645520, EPI_ISL_656731, EPI_ISL_656744, EPI_ISL_656811, EPI_ISL_656827, EPI_ISL_656860, EPI_ISL_656865, EPI_ISL_656879, EPI_ISL_656950, EPI_ISL_657023, EPI_ISL_657361, EPI_ISL_657472, EPI_ISL_657488, EPI_ISL_657532, EPI_ISL_657640, EPI_ISL_657665, EPI_ISL_657712, EPI_ISL_657751, EPI_ISL_657770, EPI_ISL_657779, EPI_ISL_657784, EPI_ISL_657785, EPI_ISL_657788, EPI_ISL_657789, EPI_ISL_657821, EPI_ISL_657832, EPI_ISL_657856, EPI_ISL_657861, EPI_ISL_657869, EPI_ISL_657894, EPI_ISL_657898, EPI_ISL_657962, EPI_ISL_657973, EPI_ISL_657991, EPI_ISL_658000, EPI_ISL_658015, EPI_ISL_658035, EPI_ISL_658047, EPI_ISL_658079, EPI_ISL_658105, EPI_ISL_658111, EPI_ISL_658115, EPI_ISL_658143, EPI_ISL_658158, EPI_ISL_658261, EPI_ISL_658268, EPI_ISL_658306, EPI_ISL_658391, EPI_ISL_658395, EPI_ISL_658741, EPI_ISL_658742, EPI_ISL_658743, EPI_ISL_658746, EPI_ISL_658747, EPI_ISL_658748, EPI_ISL_658749, EPI_ISL_658751, EPI_ISL_658754, EPI_ISL_658755, EPI_ISL_658757, EPI_ISL_658758, EPI_ISL_658761, EPI_ISL_658762, EPI_ISL_658765, EPI_ISL_658766, EPI_ISL_658770, EPI_ISL_658771, EPI_ISL_658772, EPI_ISL_658773, EPI_ISL_658774, EPI_ISL_658776, EPI_ISL_658777, EPI_ISL_658778, EPI_ISL_658779, EPI_ISL_658780, EPI_ISL_658782, EPI_ISL_658783, EPI_ISL_658784, EPI_ISL_658785, EPI_ISL_658786, EPI_ISL_658789, EPI_ISL_658790, EPI_ISL_658791, EPI_ISL_658792, EPI_ISL_658793, EPI_ISL_658794, EPI_ISL_658796, EPI_ISL_658797, EPI_ISL_658798, EPI_ISL_658799, EPI_ISL_658801, EPI_ISL_658802, EPI_ISL_658803, EPI_ISL_658804, EPI_ISL_658805, EPI_ISL_658806, EPI_ISL_658808, EPI_ISL_658810, EPI_ISL_658811, EPI_ISL_658812, EPI_ISL_658813, EPI_ISL_658814, EPI_ISL_658815, EPI_ISL_658817, EPI_ISL_658818, EPI_ISL_658819, EPI_ISL_658820, EPI_ISL_658821, EPI_ISL_658822, EPI_ISL_658823, EPI_ISL_658824, EPI_ISL_658825, EPI_ISL_658826, EPI_ISL_658827, EPI_ISL_658828, EPI_ISL_658830, EPI_ISL_658831, EPI_ISL_658832, EPI_ISL_658834, EPI_ISL_658835, EPI_ISL_658839, EPI_ISL_658840, EPI_ISL_658841, EPI_ISL_658842, EPI_ISL_658843, EPI_ISL_658844, EPI_ISL_658845, EPI_ISL_658847, EPI_ISL_658848, EPI_ISL_658849, EPI_ISL_658850, EPI_ISL_658851, EPI_ISL_658853, EPI_ISL_658854, EPI_ISL_658855, EPI_ISL_658856, EPI_ISL_658857, EPI_ISL_658858, EPI_ISL_658859, EPI_ISL_658860, EPI_ISL_658861, EPI_ISL_658862, EPI_ISL_658867, EPI_ISL_658869, EPI_ISL_658871, EPI_ISL_658872, EPI_ISL_658874, EPI_ISL_658876, EPI_ISL_658879, EPI_ISL_658882, EPI_ISL_658884, EPI_ISL_658887, EPI_ISL_658889, EPI_ISL_658890, EPI_ISL_658892, EPI_ISL_658894, EPI_ISL_658895, EPI_ISL_658897, EPI_ISL_658898, EPI_ISL_658902, EPI_ISL_658903, EPI_ISL_658906, EPI_ISL_658907, EPI_ISL_658908, EPI_ISL_658910, EPI_ISL_658911, EPI_ISL_658915, EPI_ISL_658917, EPI_ISL_658918, EPI_ISL_658920, EPI_ISL_658922, EPI_ISL_658924, EPI_ISL_658926, EPI_ISL_658927, EPI_ISL_658928, EPI_ISL_658929, EPI_ISL_658932, EPI_ISL_658934, EPI_ISL_658935, EPI_ISL_658937, EPI_ISL_658938, EPI_ISL_658939, EPI_ISL_658940, EPI_ISL_658941, EPI_ISL_658942, EPI_ISL_658943, EPI_ISL_658944, EPI_ISL_658945, EPI_ISL_658946, EPI_ISL_658947, EPI_ISL_658949, EPI_ISL_658950, EPI_ISL_658951, EPI_ISL_658952, EPI_ISL_658953, EPI_ISL_658955, EPI_ISL_658956, EPI_ISL_658957, EPI_ISL_658958, EPI_ISL_658960, EPI_ISL_658961, EPI_ISL_658962, EPI_ISL_658964, EPI_ISL_658965, EPI_ISL_658966, EPI_ISL_658967, EPI_ISL_658968, EPI_ISL_658969, EPI_ISL_658972, EPI_ISL_658973, EPI_ISL_658974, EPI_ISL_658975, EPI_ISL_658977, EPI_ISL_658978, EPI_ISL_658980, EPI_ISL_658981, EPI_ISL_658982, EPI_ISL_658983, EPI_ISL_658984, EPI_ISL_658985, EPI_ISL_658986, EPI_ISL_658987, EPI_ISL_658988, EPI_ISL_658989, EPI_ISL_658990, EPI_ISL_658991, EPI_ISL_658992, EPI_ISL_658993, EPI_ISL_658994, EPI_ISL_658997, EPI_ISL_658998, EPI_ISL_658999, EPI_ISL_659001, EPI_ISL_659002, EPI_ISL_659003, EPI_ISL_659004, EPI_ISL_659006, EPI_ISL_659008, EPI_ISL_659009, EPI_ISL_659010, EPI_ISL_659012, EPI_ISL_659013, EPI_ISL_659014, EPI_ISL_659015, EPI_ISL_659016, EPI_ISL_659020, EPI_ISL_659021, EPI_ISL_659022, EPI_ISL_659023, EPI_ISL_659024, EPI_ISL_659026, EPI_ISL_659027, EPI_ISL_659029, EPI_ISL_659030, EPI_ISL_659031, EPI_ISL_659032, EPI_ISL_659033, EPI_ISL_659034, EPI_ISL_659035, EPI_ISL_659036, EPI_ISL_659037, EPI_ISL_659038, EPI_ISL_659039, EPI_ISL_659041, EPI_ISL_659042, EPI_ISL_659043, EPI_ISL_659044, EPI_ISL_659046, EPI_ISL_659047, EPI_ISL_659048, EPI_ISL_659049, EPI_ISL_659051, EPI_ISL_659052, EPI_ISL_659055, EPI_ISL_659056, EPI_ISL_659057, EPI_ISL_659058, EPI_ISL_659059, EPI_ISL_659060, EPI_ISL_659061, EPI_ISL_659062, EPI_ISL_659063, EPI_ISL_659064, EPI_ISL_659065, EPI_ISL_659066, EPI_ISL_659067, EPI_ISL_659068, EPI_ISL_659069, EPI_ISL_659074, EPI_ISL_659075, EPI_ISL_659078, EPI_ISL_659081, EPI_ISL_659085, EPI_ISL_659086, EPI_ISL_659087, EPI_ISL_659088, EPI_ISL_659089, EPI_ISL_659092, EPI_ISL_659095, EPI_ISL_659096, EPI_ISL_662637, EPI_ISL_662638, EPI_ISL_662639, EPI_ISL_662640, EPI_ISL_662641, EPI_ISL_662642, EPI_ISL_662643, EPI_ISL_662644, EPI_ISL_662645, EPI_ISL_662646, EPI_ISL_709901, EPI_ISL_709903, EPI_ISL_709904, EPI_ISL_709905, EPI_ISL_710907 | | | |
| see above | Lighthouse Lab in Milton Keynes | Wellcome Sanger Institute for the COVID-19 Genomics UK (COG-UK) Consortium | Cordelia Langford; David K. Jackson; Dominic Kwiatkowski; Ewan Harrison; Ian Johnston; John Sillitoe on behalf of the Wellcome Sanger Institute COVID-19 Surveillance Team; Roberto Amato; Sonia Goncalves; The Lighthouse Lab in Milton Keynes and Alex Alderton |
| EPI_ISL_637257, EPI_ISL_637259, EPI_ISL_637263, EPI_ISL_637274, EPI_ISL_637336, EPI_ISL_637434, EPI_ISL_637617, EPI_ISL_637898, EPI_ISL_637945, EPI_ISL_637976, EPI_ISL_638011, EPI_ISL_638025, EPI_ISL_638449, EPI_ISL_638450, EPI_ISL_638451, EPI_ISL_638452, EPI_ISL_638453, EPI_ISL_650115, EPI_ISL_650139, EPI_ISL_650160, EPI_ISL_650161, EPI_ISL_650162, EPI_ISL_650184, EPI_ISL_650189, EPI_ISL_650256, EPI_ISL_650257, EPI_ISL_650268, EPI_ISL_650269, EPI_ISL_650278, EPI_ISL_650299, EPI_ISL_650453, EPI_ISL_650496, EPI_ISL_650563, EPI_ISL_651123, EPI_ISL_651146, EPI_ISL_651148, EPI_ISL_651155, EPI_ISL_651175, EPI_ISL_651181, EPI_ISL_651197, EPI_ISL_651213, EPI_ISL_651223, EPI_ISL_651234, EPI_ISL_651239, EPI_ISL_651248, EPI_ISL_651251, EPI_ISL_651265, EPI_ISL_651271, EPI_ISL_651272, EPI_ISL_651273, EPI_ISL_651306, EPI_ISL_651307, EPI_ISL_651347, EPI_ISL_651355, EPI_ISL_664111, EPI_ISL_664134, EPI_ISL_664138, EPI_ISL_664142, EPI_ISL_664203, EPI_ISL_664219, EPI_ISL_664229, EPI_ISL_664230, EPI_ISL_664231, EPI_ISL_664336, EPI_ISL_664339, EPI_ISL_664343, EPI_ISL_664348, EPI_ISL_664354, EPI_ISL_664362, EPI_ISL_664448, EPI_ISL_664475, EPI_ISL_664486, EPI_ISL_664489, EPI_ISL_664513, EPI_ISL_664519, EPI_ISL_664524, EPI_ISL_664606, EPI_ISL_664614, EPI_ISL_664623, EPI_ISL_664632, EPI_ISL_664637, EPI_ISL_664649, EPI_ISL_664652, EPI_ISL_664664, EPI_ISL_664665, EPI_ISL_664667, EPI_ISL_664668, EPI_ISL_664670, EPI_ISL_664693, EPI_ISL_664695, EPI_ISL_664726, EPI_ISL_664733, EPI_ISL_665131, EPI_ISL_665132, EPI_ISL_665134, EPI_ISL_665135, EPI_ISL_665136, EPI_ISL_665137, EPI_ISL_665138, EPI_ISL_665140, EPI_ISL_665142, EPI_ISL_665294, EPI_ISL_665327, EPI_ISL_665328, EPI_ISL_665384, EPI_ISL_665414, EPI_ISL_665662, EPI_ISL_665683, EPI_ISL_665704, EPI_ISL_665707, EPI_ISL_666022, EPI_ISL_702512, EPI_ISL_702541, EPI_ISL_702556, EPI_ISL_702566, EPI_ISL_702569, EPI_ISL_702667, EPI_ISL_702697, EPI_ISL_702758, EPI_ISL_702779, EPI_ISL_702792, EPI_ISL_702877, EPI_ISL_702897, EPI_ISL_702953, EPI_ISL_703085, EPI_ISL_703412, EPI_ISL_703711, EPI_ISL_703881, EPI_ISL_703958, EPI_ISL_704057, EPI_ISL_704129, EPI_ISL_704201, EPI_ISL_704934, EPI_ISL_704937, EPI_ISL_705064, EPI_ISL_705198, EPI_ISL_705252, EPI_ISL_705259, EPI_ISL_705263, EPI_ISL_705284, EPI_ISL_705301, EPI_ISL_705314, EPI_ISL_705318, EPI_ISL_705322, EPI_ISL_705323, EPI_ISL_705340, EPI_ISL_705348, EPI_ISL_705360, EPI_ISL_705362, EPI_ISL_705393, EPI_ISL_705402, EPI_ISL_705407, EPI_ISL_705411, EPI_ISL_865164, EPI_ISL_865165, EPI_ISL_865166, EPI_ISL_865167, EPI_ISL_865168, EPI_ISL_865170, EPI_ISL_1104312, EPI_ISL_1104313, EPI_ISL_1104314, EPI_ISL_1104315, EPI_ISL_1104316, EPI_ISL_1104317 | | | |
| see above | Liverpool Clinical Laboratories | COVID-19 Genomics UK (COG-UK) Consortium | A Alrezaihi; Alessandro Gerada; Alistair Darby; Angela Cowell; Anita Lucaci; Anu Chawla; Cassie Olateju; Catherine Hartley; Charlotte Nelson; Ecaterina Vamos; Elaine O'Toole; Eleanor G Bentley; Ghada T Shawli; Isabel García-Dorival; James Johnson; James P Stewart; Jenifer Manson; Joanne Watts; Jones Benjamin; Jordan J Clark; Julian Hiscox; L Luu; Lucille Rainbow; M Almsaud; Margaret Hughes; Mark Whitehead; Matthew Gemmell; Miren Iturriza-Gomara; Muhannad Alruwaili; N.P Randle; Neil Swainston; PKF Gilmore; Parul Sharma; Rebekah Penrice-Randal; Richard Eccles; Richard Gregory; Sam Haldenby; Steve Paterson; Stuart D Armstrong; Trevor Ian Robinson; Ximeng Han |
| EPI_ISL_637280, EPI_ISL_637523, EPI_ISL_637605, EPI_ISL_664107, EPI_ISL_664121, EPI_ISL_664190, EPI_ISL_664192, EPI_ISL_664238, EPI_ISL_664240, EPI_ISL_664241, EPI_ISL_664242, EPI_ISL_664243, EPI_ISL_664244, EPI_ISL_664245, EPI_ISL_664246, EPI_ISL_664247, EPI_ISL_664360, EPI_ISL_664373, EPI_ISL_664385, EPI_ISL_664386, EPI_ISL_664387, EPI_ISL_664388, EPI_ISL_664389, EPI_ISL_664390, EPI_ISL_664391, EPI_ISL_664392, EPI_ISL_664393, EPI_ISL_664394, EPI_ISL_664395, EPI_ISL_664396, EPI_ISL_664503, EPI_ISL_664541, EPI_ISL_664544, EPI_ISL_664560, EPI_ISL_664561, EPI_ISL_664562, EPI_ISL_664563, EPI_ISL_664564, EPI_ISL_664639, EPI_ISL_664696, EPI_ISL_664736, EPI_ISL_664856, EPI_ISL_665261, EPI_ISL_665271, EPI_ISL_665307, EPI_ISL_665318, EPI_ISL_665390, EPI_ISL_665399, | | | |

| | | | |
|---|---|---|---|
| EPI_ISL_665400, EPI_ISL_665401, EPI_ISL_665410, EPI_ISL_665496, EPI_ISL_665497, EPI_ISL_665498, EPI_ISL_665503, EPI_ISL_665509, EPI_ISL_665629, EPI_ISL_665651, EPI_ISL_665673, EPI_ISL_665679, EPI_ISL_665687, EPI_ISL_665717, EPI_ISL_665736, EPI_ISL_665742, EPI_ISL_666003, EPI_ISL_666004, EPI_ISL_666005, EPI_ISL_666006, EPI_ISL_666007, EPI_ISL_666008, EPI_ISL_666009, EPI_ISL_666010, EPI_ISL_666011, EPI_ISL_666012, EPI_ISL_666013, EPI_ISL_666014, EPI_ISL_666015, EPI_ISL_666016, EPI_ISL_666017, EPI_ISL_666018, EPI_ISL_666019, EPI_ISL_666020, EPI_ISL_666021, EPI_ISL_666101, EPI_ISL_666102, EPI_ISL_666103, EPI_ISL_666104, EPI_ISL_666105, EPI_ISL_666106, EPI_ISL_666592, EPI_ISL_702505, EPI_ISL_702784, EPI_ISL_702820, EPI_ISL_702990, EPI_ISL_703002, EPI_ISL_704120, EPI_ISL_704166, EPI_ISL_704579, EPI_ISL_705105, EPI_ISL_705219, EPI_ISL_705224, EPI_ISL_705279, EPI_ISL_705280, EPI_ISL_705281, EPI_ISL_705293, EPI_ISL_705355, EPI_ISL_706276, EPI_ISL_706353, EPI_ISL_706354, EPI_ISL_724963, EPI_ISL_724965, EPI_ISL_724974, EPI_ISL_724980, EPI_ISL_724981, EPI_ISL_724982, EPI_ISL_819454, EPI_ISL_819455, EPI_ISL_819456, EPI_ISL_819457, EPI_ISL_1637213, EPI_ISL_1637214, EPI_ISL_1637216, EPI_ISL_1637221 | | | |
| see above | Northumbria University / South Tees Hospitals NHS Foundation Trust / North Cumbria Integrated Care NHS Foundation Trust / North Tees and Hartlepool NHS Foundation Trust / Newcastle Hospitals NHS Foundation Trust | COVID-19 Genomics UK (COG-UK) Consortium | Andrew Nelson; Brendan Payne; Clive Graham; Darren L Smith; Debra Padgett; Edward Barton; Emma Swindells; Garren Scott; Gary Black; Gary Eltringham; Giles S Holt; Greg R Young; Jane Greenaway; Jennifer Collins; John Allan; Joshua Loh; Lynn Dover; Matthew Bashton; Mohammad A Tariq; Paul Baker; Sarah Essex; Steve Liggett; Wen C Yew; Yusri Taha |
| EPI_ISL_1249308 | Originating lab: Wales Specialist Virology Centre Sequencing lab: Pathogen Genomics Unit | Public Health Wales Microbiology Cardiff Wales Specialist Virology Centre | Alec Birchley; Alexander Adams; Amy Gaskin; Angela Marchbank; Bree Gatica-Wilcox; Catherine Moore; Jason Coombes; Joanne Watkins; Joel Southgate; Johnathan Evans; Laura Gifford; Lauren Gilbert; Lee Graham; Malorie Perry; Matthew Bull; Nicole Pacchiarini; Sally Corden; Sara Kumziene-Summerhayes; Sara Rey; Sarah Taylor; Simon Cottrell; Sophie Jones; Tom Connor |
| EPI_ISL_679629, EPI_ISL_679630, EPI_ISL_679631, EPI_ISL_679632, EPI_ISL_679633, EPI_ISL_679634, EPI_ISL_679635, EPI_ISL_679636, EPI_ISL_679637, EPI_ISL_679638, EPI_ISL_679639, EPI_ISL_679640, EPI_ISL_679641, EPI_ISL_679642, EPI_ISL_679643, EPI_ISL_679644, EPI_ISL_679645, EPI_ISL_679646, EPI_ISL_679647, EPI_ISL_679648, EPI_ISL_679649, EPI_ISL_679650, EPI_ISL_679651, EPI_ISL_679652, EPI_ISL_679653, EPI_ISL_679654, EPI_ISL_679655, EPI_ISL_679656, EPI_ISL_679657, EPI_ISL_679658, EPI_ISL_679659, EPI_ISL_679660, EPI_ISL_679661, EPI_ISL_679662, EPI_ISL_679663, EPI_ISL_679664, EPI_ISL_679665, EPI_ISL_679666, EPI_ISL_679667, EPI_ISL_679668, EPI_ISL_679669, EPI_ISL_679670, EPI_ISL_679671, EPI_ISL_679672, EPI_ISL_679673, EPI_ISL_679674, EPI_ISL_679675, EPI_ISL_679676, EPI_ISL_679677, EPI_ISL_679678, EPI_ISL_679679, EPI_ISL_679680, EPI_ISL_679681, EPI_ISL_679682, EPI_ISL_679683, EPI_ISL_679684, EPI_ISL_679685, EPI_ISL_679686, EPI_ISL_679687, EPI_ISL_679688, EPI_ISL_679689, EPI_ISL_679690, EPI_ISL_679691, EPI_ISL_679692, EPI_ISL_679693, EPI_ISL_679694, EPI_ISL_679695, EPI_ISL_679696, EPI_ISL_679697, EPI_ISL_679698, EPI_ISL_679699, EPI_ISL_679700, EPI_ISL_679701, EPI_ISL_679702, EPI_ISL_679703, EPI_ISL_679704, EPI_ISL_679705, EPI_ISL_679706, EPI_ISL_679707, EPI_ISL_679708, EPI_ISL_679709, EPI_ISL_679710, EPI_ISL_679711, EPI_ISL_679712, EPI_ISL_679713, EPI_ISL_679714, EPI_ISL_679715, EPI_ISL_679716, EPI_ISL_679717, EPI_ISL_679718, EPI_ISL_679719, EPI_ISL_679720, EPI_ISL_679721, EPI_ISL_679722, EPI_ISL_679723, EPI_ISL_679724, EPI_ISL_679725, EPI_ISL_679726, EPI_ISL_679727, EPI_ISL_679728, EPI_ISL_679729, EPI_ISL_679730, EPI_ISL_679731, EPI_ISL_679732, EPI_ISL_679733, EPI_ISL_679734, EPI_ISL_679735, EPI_ISL_679736, EPI_ISL_679737, EPI_ISL_679738, EPI_ISL_679739, EPI_ISL_679740, EPI_ISL_679741, EPI_ISL_679742, EPI_ISL_679743, EPI_ISL_679744, EPI_ISL_679745, EPI_ISL_679746, EPI_ISL_679747, EPI_ISL_679748, EPI_ISL_679749, EPI_ISL_679750, EPI_ISL_679751, EPI_ISL_679752, EPI_ISL_679753, EPI_ISL_679754, EPI_ISL_679755, EPI_ISL_679756, EPI_ISL_679757, EPI_ISL_679758, EPI_ISL_679759, EPI_ISL_679760, EPI_ISL_679761, EPI_ISL_679762, EPI_ISL_679763, EPI_ISL_679764, EPI_ISL_679765, EPI_ISL_679766, EPI_ISL_679767, EPI_ISL_679768, EPI_ISL_679769, EPI_ISL_679770, EPI_ISL_679771, EPI_ISL_679772, EPI_ISL_679773, EPI_ISL_679774, EPI_ISL_679775, EPI_ISL_679776, EPI_ISL_679777, EPI_ISL_679778, EPI_ISL_679779, EPI_ISL_679780, EPI_ISL_679781, EPI_ISL_679782, EPI_ISL_679783, EPI_ISL_679784, EPI_ISL_679785, EPI_ISL_679786, EPI_ISL_679787, EPI_ISL_679788, EPI_ISL_679789, EPI_ISL_679790, EPI_ISL_679791, EPI_ISL_679792, EPI_ISL_679793, EPI_ISL_679794, EPI_ISL_679795, EPI_ISL_679796, EPI_ISL_679797, EPI_ISL_679798, EPI_ISL_679799, EPI_ISL_679800, EPI_ISL_679801, EPI_ISL_679802, EPI_ISL_679803, EPI_ISL_679804, EPI_ISL_679805, EPI_ISL_679806, EPI_ISL_679807, EPI_ISL_679808, EPI_ISL_679809, EPI_ISL_679810, EPI_ISL_679811, EPI_ISL_679812, EPI_ISL_679813, EPI_ISL_679814, EPI_ISL_679815, EPI_ISL_679816, EPI_ISL_679817, EPI_ISL_679818, EPI_ISL_679819, EPI_ISL_679820, EPI_ISL_679821, EPI_ISL_679822, EPI_ISL_679823, EPI_ISL_679824, EPI_ISL_679825, EPI_ISL_679826, EPI_ISL_679827, EPI_ISL_679828, EPI_ISL_679829, EPI_ISL_679830, EPI_ISL_679831, EPI_ISL_679832, EPI_ISL_679833, EPI_ISL_679834, EPI_ISL_679835, EPI_ISL_679836, EPI_ISL_679837, EPI_ISL_679838, EPI_ISL_679839, EPI_ISL_679840, EPI_ISL_679841, EPI_ISL_679842, EPI_ISL_679843, EPI_ISL_679844, EPI_ISL_679845, EPI_ISL_679846, EPI_ISL_679847, EPI_ISL_679848, EPI_ISL_679849, EPI_ISL_679850, EPI_ISL_679851, EPI_ISL_680579, EPI_ISL_704676, EPI_ISL_704679, EPI_ISL_704682, EPI_ISL_704685, EPI_ISL_704687, EPI_ISL_704690, EPI_ISL_704692, EPI_ISL_704695, EPI_ISL_704698, EPI_ISL_704701, EPI_ISL_704703, EPI_ISL_704706, EPI_ISL_704815, EPI_ISL_704817, EPI_ISL_704836, EPI_ISL_704839, EPI_ISL_704861, EPI_ISL_704864, EPI_ISL_704872, EPI_ISL_704874, EPI_ISL_704877, EPI_ISL_704879, EPI_ISL_704882, EPI_ISL_704885, EPI_ISL_705426, EPI_ISL_705427, EPI_ISL_705428, EPI_ISL_705429, EPI_ISL_705430, EPI_ISL_705450, EPI_ISL_705451, EPI_ISL_705452, EPI_ISL_705453, EPI_ISL_705454, EPI_ISL_705455, EPI_ISL_705456, EPI_ISL_705467, EPI_ISL_705468, EPI_ISL_705469, EPI_ISL_705470, EPI_ISL_705474, EPI_ISL_705475, EPI_ISL_705477, EPI_ISL_705478, EPI_ISL_705479, EPI_ISL_705480, EPI_ISL_705481, EPI_ISL_705482, EPI_ISL_705483, EPI_ISL_705484, EPI_ISL_705485, EPI_ISL_705493, EPI_ISL_705497, EPI_ISL_705526, EPI_ISL_706013, EPI_ISL_706014, EPI_ISL_706015, EPI_ISL_706016, EPI_ISL_706017, EPI_ISL_706018, EPI_ISL_706019, EPI_ISL_706020, EPI_ISL_706021, EPI_ISL_706022, EPI_ISL_706023, EPI_ISL_706024, EPI_ISL_706025, EPI_ISL_706026, EPI_ISL_706027, EPI_ISL_706028, EPI_ISL_706029, EPI_ISL_706030, EPI_ISL_706031, EPI_ISL_706032, EPI_ISL_706033, EPI_ISL_706034, EPI_ISL_706035, EPI_ISL_706036, EPI_ISL_706037, EPI_ISL_706038, EPI_ISL_706039, EPI_ISL_706040, EPI_ISL_706041, EPI_ISL_706042, EPI_ISL_706043, EPI_ISL_706044, EPI_ISL_706045, EPI_ISL_706046, EPI_ISL_706047, EPI_ISL_706048, EPI_ISL_706049, EPI_ISL_706050, EPI_ISL_706051, EPI_ISL_706052, EPI_ISL_706053, EPI_ISL_706054, EPI_ISL_706055, EPI_ISL_706056, EPI_ISL_706057, EPI_ISL_706058, EPI_ISL_706059, EPI_ISL_706060, EPI_ISL_706061, EPI_ISL_706062, EPI_ISL_706063, EPI_ISL_706064, EPI_ISL_706065, EPI_ISL_706066, EPI_ISL_706067, EPI_ISL_706068, EPI_ISL_706069, EPI_ISL_706070, EPI_ISL_706071, EPI_ISL_706072, EPI_ISL_706073, EPI_ISL_706074, EPI_ISL_706075, EPI_ISL_706076, EPI_ISL_706077, EPI_ISL_706078, EPI_ISL_706079, EPI_ISL_706080, EPI_ISL_706081, EPI_ISL_706082, EPI_ISL_706083, EPI_ISL_706084, EPI_ISL_706085, EPI_ISL_706086, EPI_ISL_706087, EPI_ISL_706088, EPI_ISL_706089, EPI_ISL_706090, EPI_ISL_706091, EPI_ISL_706092, EPI_ISL_706093, EPI_ISL_706094, EPI_ISL_706095, EPI_ISL_706096, EPI_ISL_706097, EPI_ISL_706098, EPI_ISL_706099, EPI_ISL_706100, EPI_ISL_706101, EPI_ISL_706102, EPI_ISL_706103, EPI_ISL_706104, EPI_ISL_706105, EPI_ISL_706106, EPI_ISL_706107, EPI_ISL_706108, EPI_ISL_706109, EPI_ISL_706110, EPI_ISL_706111, EPI_ISL_706112, EPI_ISL_706113, EPI_ISL_706114, EPI_ISL_706115, EPI_ISL_706116, EPI_ISL_706117, EPI_ISL_706118, EPI_ISL_706119, EPI_ISL_706120, EPI_ISL_706121, EPI_ISL_706122, EPI_ISL_706123, EPI_ISL_706124, EPI_ISL_706125, EPI_ISL_706126, EPI_ISL_706127, EPI_ISL_706129, EPI_ISL_706130, EPI_ISL_706131, EPI_ISL_706132, EPI_ISL_706133, EPI_ISL_706134, EPI_ISL_706135, EPI_ISL_706136, EPI_ISL_706137, EPI_ISL_706138, EPI_ISL_706139, EPI_ISL_706140, EPI_ISL_706141, EPI_ISL_706142, EPI_ISL_706143, EPI_ISL_706144, EPI_ISL_706145, EPI_ISL_706146, EPI_ISL_706147, EPI_ISL_706148, EPI_ISL_706149, EPI_ISL_706150, EPI_ISL_706151, EPI_ISL_706152, EPI_ISL_706153, EPI_ISL_706154, EPI_ISL_706155, EPI_ISL_706156, EPI_ISL_706157, EPI_ISL_706158, EPI_ISL_706159, EPI_ISL_706160, EPI_ISL_706161, EPI_ISL_706162, EPI_ISL_706163, EPI_ISL_706164, EPI_ISL_706165, EPI_ISL_706166, EPI_ISL_706167, EPI_ISL_706168, EPI_ISL_706169, EPI_ISL_706170, EPI_ISL_706171, EPI_ISL_706172, EPI_ISL_706173, EPI_ISL_706174, EPI_ISL_706175, EPI_ISL_706176, EPI_ISL_706177, EPI_ISL_706178, EPI_ISL_706179, EPI_ISL_706180, EPI_ISL_706181, EPI_ISL_706182, EPI_ISL_706183, EPI_ISL_706184, EPI_ISL_706185, EPI_ISL_706186, EPI_ISL_706187, EPI_ISL_706188, EPI_ISL_706189, EPI_ISL_706190, EPI_ISL_706191, EPI_ISL_706192, EPI_ISL_706193, EPI_ISL_706195, EPI_ISL_706196, EPI_ISL_706197, EPI_ISL_706198, EPI_ISL_706199, EPI_ISL_706200, EPI_ISL_706201, EPI_ISL_706202, EPI_ISL_706203, EPI_ISL_706204, EPI_ISL_706205, EPI_ISL_706206, EPI_ISL_706207, EPI_ISL_706208, EPI_ISL_706209, EPI_ISL_706210, EPI_ISL_706211, EPI_ISL_706212, EPI_ISL_706213, EPI_ISL_706214, EPI_ISL_706215, EPI_ISL_706216, EPI_ISL_706217, EPI_ISL_706218, EPI_ISL_706219, EPI_ISL_706220, EPI_ISL_706221, EPI_ISL_706222, EPI_ISL_706223, EPI_ISL_706224, EPI_ISL_706225, EPI_ISL_706226, EPI_ISL_706227, EPI_ISL_706228, EPI_ISL_706229, EPI_ISL_706230, EPI_ISL_706231, EPI_ISL_706232, EPI_ISL_706233, EPI_ISL_706234, EPI_ISL_706235, EPI_ISL_706236, EPI_ISL_706237, EPI_ISL_706238, EPI_ISL_706239, EPI_ISL_706240, EPI_ISL_706241, EPI_ISL_706242, EPI_ISL_706243, EPI_ISL_706244, EPI_ISL_706245, EPI_ISL_706246, EPI_ISL_706247, EPI_ISL_706248, EPI_ISL_706249, EPI_ISL_706250, EPI_ISL_706251, EPI_ISL_706252, EPI_ISL_706253, EPI_ISL_706254, EPI_ISL_706255, EPI_ISL_706256, EPI_ISL_706257, EPI_ISL_706258, EPI_ISL_706259, EPI_ISL_706260, EPI_ISL_706261, EPI_ISL_706262, EPI_ISL_706263, EPI_ISL_706264, EPI_ISL_706265, EPI_ISL_706266, EPI_ISL_706267, EPI_ISL_706268, EPI_ISL_706269, EPI_ISL_706270, EPI_ISL_706271, EPI_ISL_706272, EPI_ISL_706273, EPI_ISL_706274, EPI_ISL_706275, EPI_ISL_741362, EPI_ISL_741363, EPI_ISL_741364, EPI_ISL_741365, EPI_ISL_741366, EPI_ISL_741367, EPI_ISL_741368, EPI_ISL_741369, EPI_ISL_741370, EPI_ISL_741371, EPI_ISL_741372, EPI_ISL_741373, EPI_ISL_741374, EPI_ISL_741375, EPI_ISL_741376, EPI_ISL_741377, EPI_ISL_741378, EPI_ISL_741379, EPI_ISL_741380, EPI_ISL_741381, EPI_ISL_741382, EPI_ISL_741383, EPI_ISL_741384, EPI_ISL_741385, EPI_ISL_741386, EPI_ISL_741387, EPI_ISL_741388, EPI_ISL_741389, EPI_ISL_741390, EPI_ISL_741391, EPI_ISL_741392, EPI_ISL_741393, EPI_ISL_741394, EPI_ISL_741395, EPI_ISL_741396, EPI_ISL_741397, EPI_ISL_741398, EPI_ISL_741399, EPI_ISL_741400, EPI_ISL_741401, EPI_ISL_741402, EPI_ISL_741403, EPI_ISL_741404, EPI_ISL_741405, EPI_ISL_741406, EPI_ISL_741407, EPI_ISL_741408, EPI_ISL_741409, EPI_ISL_741410, EPI_ISL_741411, EPI_ISL_741412, EPI_ISL_741413, EPI_ISL_741414, EPI_ISL_741415, EPI_ISL_741416, EPI_ISL_741417, EPI_ISL_741418, EPI_ISL_741419, EPI_ISL_741420, EPI_ISL_742261, EPI_ISL_840202, EPI_ISL_950232, EPI_ISL_1309733, EPI_ISL_1833045 | | | |
| see above | Oxford Viromics, NDM, University of Oxford; Oxford University Hospitals; Basingstoke and North Hampshire Hospital | COVID-19 Genomics UK (COG-UK) Consortium | Alex Mobbs; Amy Trebes; Anita Justice; Catrin Moore; Christophe Fraser; David Bonsall; David Buck; Emma Wise; George Macintyre; Jessica Lynch; John Todd; Mariateresa de Cesare; Matilde Mori; Monique Andersson; Nathan Moore; Nick Cortes; Robert Shaw; Stephen Kidd; Tanya Golubchik; Timothy Peto |
| EPI_ISL_637284, EPI_ISL_637301, EPI_ISL_637359, EPI_ISL_637442, EPI_ISL_637536, EPI_ISL_637550, EPI_ISL_637557, EPI_ISL_637588, EPI_ISL_637589, EPI_ISL_637602, EPI_ISL_637857, EPI_ISL_637865, EPI_ISL_637866, EPI_ISL_637878, EPI_ISL_637922, EPI_ISL_638033, EPI_ISL_638581, EPI_ISL_638582, EPI_ISL_638583, EPI_ISL_650120, EPI_ISL_650128, EPI_ISL_650197, EPI_ISL_650214, EPI_ISL_650323, EPI_ISL_650325, EPI_ISL_650330, EPI_ISL_650335, EPI_ISL_650402, EPI_ISL_650413, EPI_ISL_650417, EPI_ISL_650476, EPI_ISL_650522, EPI_ISL_650545, EPI_ISL_650681, EPI_ISL_650683, EPI_ISL_650737, EPI_ISL_650756, EPI_ISL_650758, EPI_ISL_650761, EPI_ISL_650766, EPI_ISL_650817, EPI_ISL_650828, EPI_ISL_650885, EPI_ISL_650895, EPI_ISL_650911, EPI_ISL_650938, EPI_ISL_650940, EPI_ISL_650944, EPI_ISL_650953, EPI_ISL_650955, EPI_ISL_650958, EPI_ISL_650974, EPI_ISL_650975, EPI_ISL_650977, EPI_ISL_651048, EPI_ISL_651093, EPI_ISL_651100, EPI_ISL_651101, EPI_ISL_651110, EPI_ISL_651119, EPI_ISL_651133, EPI_ISL_651134, EPI_ISL_651142, EPI_ISL_651156, EPI_ISL_651173, EPI_ISL_651209, EPI_ISL_651216, EPI_ISL_651226, EPI_ISL_651247, EPI_ISL_651252, EPI_ISL_651255, EPI_ISL_651267, EPI_ISL_651268, EPI_ISL_651279, EPI_ISL_651298, EPI_ISL_651317, EPI_ISL_651337, EPI_ISL_651349, EPI_ISL_651357, EPI_ISL_651411, EPI_ISL_651412, EPI_ISL_651418, EPI_ISL_651419, EPI_ISL_651438, EPI_ISL_651484, EPI_ISL_651512, EPI_ISL_651516, EPI_ISL_652241, EPI_ISL_652242, EPI_ISL_652243, EPI_ISL_652244, EPI_ISL_652245, EPI_ISL_652246, EPI_ISL_652247, EPI_ISL_652248, EPI_ISL_652249, EPI_ISL_652250, EPI_ISL_652251, EPI_ISL_652252, EPI_ISL_652253, EPI_ISL_652254, EPI_ISL_652255, EPI_ISL_652256, EPI_ISL_652257, EPI_ISL_652258, EPI_ISL_652260, EPI_ISL_652264, EPI_ISL_652265, EPI_ISL_652266, EPI_ISL_652267, EPI_ISL_652268, EPI_ISL_652269, EPI_ISL_652270, EPI_ISL_652271, EPI_ISL_652272, EPI_ISL_652273, EPI_ISL_652274, EPI_ISL_652275, EPI_ISL_652276, EPI_ISL_652277, EPI_ISL_652278, EPI_ISL_652279, EPI_ISL_652280, EPI_ISL_652281, EPI_ISL_652282, EPI_ISL_652283, EPI_ISL_652284, EPI_ISL_652285, EPI_ISL_652349, EPI_ISL_652356, EPI_ISL_652358, EPI_ISL_652362, EPI_ISL_652363, EPI_ISL_653027, EPI_ISL_653028, EPI_ISL_653029, EPI_ISL_653030, EPI_ISL_653031, EPI_ISL_653032, EPI_ISL_653033, EPI_ISL_653034, EPI_ISL_653035, EPI_ISL_653036, EPI_ISL_653037, EPI_ISL_653038, EPI_ISL_653039, EPI_ISL_653040, EPI_ISL_653041, EPI_ISL_653042, EPI_ISL_653043, EPI_ISL_653044, EPI_ISL_653045, EPI_ISL_653046, EPI_ISL_653047, EPI_ISL_653048, EPI_ISL_653049, EPI_ISL_653050, EPI_ISL_653051, EPI_ISL_653052, EPI_ISL_653053, EPI_ISL_653054, EPI_ISL_653055, EPI_ISL_653056, EPI_ISL_653057, EPI_ISL_653058, EPI_ISL_665263, EPI_ISL_665269, EPI_ISL_665292, EPI_ISL_665337, EPI_ISL_665351, EPI_ISL_665354, EPI_ISL_665355, EPI_ISL_665360, EPI_ISL_665371, EPI_ISL_665406, EPI_ISL_665444, EPI_ISL_665454, EPI_ISL_665456, EPI_ISL_665457, EPI_ISL_665458, EPI_ISL_665461, EPI_ISL_665467, EPI_ISL_665481, EPI_ISL_665520, EPI_ISL_665533, EPI_ISL_665568, EPI_ISL_665581, EPI_ISL_665583, EPI_ISL_665587, EPI_ISL_665591, EPI_ISL_665657, EPI_ISL_665672, EPI_ISL_665684, EPI_ISL_665731, EPI_ISL_665747, EPI_ISL_665788, EPI_ISL_665866, EPI_ISL_665867, EPI_ISL_665877, EPI_ISL_665878, EPI_ISL_665879, EPI_ISL_665884, EPI_ISL_665885, EPI_ISL_665887, EPI_ISL_665888, EPI_ISL_665889, EPI_ISL_665890, EPI_ISL_665891, EPI_ISL_665892, EPI_ISL_665893, EPI_ISL_665895, EPI_ISL_665896, EPI_ISL_725246, EPI_ISL_725266, EPI_ISL_725271, EPI_ISL_725277, EPI_ISL_725279, EPI_ISL_741567 | | | |
| see above | Quadram Institute Bioscience | COVID-19 Genomics UK (COG-UK) Consortium | Alexander J Trotter; Alison E. Mather; Alp Aydin; Ana P. Tedim; Anastasia Kolyva; Andrew Bell; Andrew J. Page; Claire Stuart; Dave J. Baker; Gemma L. Kay; John Wain; Justin O'Grady; Leonardo de Oliveira Martins; Lizzie Meadows; Maria Diaz; Mark Webber; Muhammed Yasir; Nabil-Fareed Alikhan; Ngozi Elumogo; Nicholas M. Thomson; Rachael Stanley; Rachel Gilroy; Reenesh Prakash; Samir Dervisevic; Samuel Bloomfield; Steven Rudder; Thanh Le-Viet |
| EPI_ISL_626659, EPI_ISL_626670, EPI_ISL_626688, EPI_ISL_626695, EPI_ISL_626740, EPI_ISL_626750, EPI_ISL_626770, EPI_ISL_626811, EPI_ISL_626816, EPI_ISL_626834, EPI_ISL_626881, EPI_ISL_627053, EPI_ISL_627068, EPI_ISL_627147, EPI_ISL_627215, EPI_ISL_627216, EPI_ISL_627304, EPI_ISL_627309, EPI_ISL_627310, EPI_ISL_627677, EPI_ISL_627678, EPI_ISL_627680, EPI_ISL_627681, EPI_ISL_627682, EPI_ISL_627683, EPI_ISL_627684, EPI_ISL_627685, EPI_ISL_627686, EPI_ISL_627687, EPI_ISL_627688, EPI_ISL_627689, EPI_ISL_627690, EPI_ISL_627691, EPI_ISL_627692, EPI_ISL_627693, EPI_ISL_627694, EPI_ISL_627695, EPI_ISL_627696, EPI_ISL_627697, EPI_ISL_627698, EPI_ISL_627699, EPI_ISL_627700, EPI_ISL_627701, EPI_ISL_627702, EPI_ISL_627703, EPI_ISL_627704, EPI_ISL_627705, EPI_ISL_627706, EPI_ISL_627707, EPI_ISL_627708, EPI_ISL_627709, EPI_ISL_627710, EPI_ISL_627711, EPI_ISL_627712, EPI_ISL_627713, EPI_ISL_627714, EPI_ISL_627715, EPI_ISL_627716, EPI_ISL_627717, EPI_ISL_627718, EPI_ISL_627719, EPI_ISL_627720, EPI_ISL_627721, EPI_ISL_627722, EPI_ISL_627723, EPI_ISL_627724, EPI_ISL_627725, EPI_ISL_627726, EPI_ISL_627727, EPI_ISL_627728, EPI_ISL_627729, EPI_ISL_637300, EPI_ISL_637343, EPI_ISL_637419, EPI_ISL_637660, EPI_ISL_638586, EPI_ISL_638589, EPI_ISL_638590, EPI_ISL_638591, EPI_ISL_651398, EPI_ISL_679927, EPI_ISL_679928, EPI_ISL_679929, EPI_ISL_679930, EPI_ISL_679931, EPI_ISL_704581, EPI_ISL_741609, EPI_ISL_839985, EPI_ISL_1978965 | | | |
| see above | Queens Medical Centre, Clinical Microbiology Department / DeepSeq Nottingham | COVID-19 Genomics UK (COG-UK) Consortium | Christopher Moore; Emily Park; Fei Sang; Gemma Clark; Hannah Howson-Wells; Jack Hill; Jayasree Dey; Johnny Debebe; Jonathan Ball; Joseph Chappell; Manjinder Khakh; Matthew Carlisle; Matthew Loose; Michelle M Lister; Nadine Holmes; Patrick McClure; Theocharis Tsoleridis; Timothy Byaruhanga; Vicki M Fleming; Victoria Wright; Wendy Smith |
| EPI_ISL_763723, EPI_ISL_764432, EPI_ISL_764435 | Regional Virus Laboratory, Belfast Health and Social Care Trust | COVID-19 Genomics UK (COG-UK) Consortium | Alison Watt; Ciara Cox; Conall McCaughey; David Simpson; Derek Fairley; James McKenna; Mairead Connor; Susan Feeney; Tanya Curran; Zoltan Molnar |
| EPI_ISL_637158, EPI_ISL_637159, EPI_ISL_637160, EPI_ISL_637161, EPI_ISL_637162, EPI_ISL_637163, EPI_ISL_637164, EPI_ISL_637165, EPI_ISL_637166, EPI_ISL_637168, EPI_ISL_637169, EPI_ISL_637170, EPI_ISL_637171, EPI_ISL_637172, EPI_ISL_637173, EPI_ISL_637174, EPI_ISL_637175, EPI_ISL_637176, EPI_ISL_637177, EPI_ISL_637178, EPI_ISL_637179, EPI_ISL_637180, EPI_ISL_637181, EPI_ISL_637182, EPI_ISL_637183, EPI_ISL_637184, EPI_ISL_639822, EPI_ISL_649786, EPI_ISL_649787, EPI_ISL_649788, EPI_ISL_649789, EPI_ISL_649790, EPI_ISL_649792, EPI_ISL_649793, EPI_ISL_649794, EPI_ISL_649795, EPI_ISL_649796, EPI_ISL_649797, EPI_ISL_649799, EPI_ISL_649800, EPI_ISL_649801, EPI_ISL_649802, EPI_ISL_649803, EPI_ISL_649806, EPI_ISL_649807, EPI_ISL_649890, EPI_ISL_653937, EPI_ISL_653938, EPI_ISL_653942, EPI_ISL_653947, EPI_ISL_653951, EPI_ISL_653952, EPI_ISL_653953, EPI_ISL_653954, EPI_ISL_653970, EPI_ISL_653971, EPI_ISL_653972, EPI_ISL_653974, EPI_ISL_653976, EPI_ISL_653979, EPI_ISL_653980, EPI_ISL_653981, EPI_ISL_653982, EPI_ISL_653983, EPI_ISL_653984, EPI_ISL_653985, EPI_ISL_653986, EPI_ISL_653987, EPI_ISL_653988, EPI_ISL_653989, EPI_ISL_653990, EPI_ISL_653991, EPI_ISL_653992, EPI_ISL_653993, EPI_ISL_653994, EPI_ISL_653995, EPI_ISL_653996, EPI_ISL_653997, EPI_ISL_653998, EPI_ISL_653999, EPI_ISL_654000, EPI_ISL_654001, EPI_ISL_654002, EPI_ISL_654003, EPI_ISL_654004, EPI_ISL_654005, EPI_ISL_654006, EPI_ISL_654007, EPI_ISL_654008, EPI_ISL_654009, EPI_ISL_654010, EPI_ISL_654011, EPI_ISL_654012, EPI_ISL_654013, EPI_ISL_654014, EPI_ISL_654015, EPI_ISL_660604, EPI_ISL_660753, EPI_ISL_660754, EPI_ISL_660755, EPI_ISL_660756, EPI_ISL_660757, EPI_ISL_660758, EPI_ISL_660759, EPI_ISL_660760, EPI_ISL_660761, EPI_ISL_660762, EPI_ISL_660763, EPI_ISL_660764, EPI_ISL_660765, EPI_ISL_660766, EPI_ISL_660767, EPI_ISL_660768, EPI_ISL_660769, EPI_ISL_660770, EPI_ISL_660771, EPI_ISL_660772, EPI_ISL_660773, EPI_ISL_660774, EPI_ISL_660775, EPI_ISL_660776, EPI_ISL_660777, EPI_ISL_660778, EPI_ISL_660779, EPI_ISL_660780, EPI_ISL_660781, EPI_ISL_660782, EPI_ISL_660783, EPI_ISL_660784, EPI_ISL_660785, EPI_ISL_660786, EPI_ISL_660787, EPI_ISL_660788, EPI_ISL_660789, EPI_ISL_660790, EPI_ISL_660791, EPI_ISL_660792, EPI_ISL_660793, EPI_ISL_666726, EPI_ISL_666729, EPI_ISL_666732, EPI_ISL_666733, EPI_ISL_666734, EPI_ISL_666735, EPI_ISL_666736, EPI_ISL_666740, EPI_ISL_666741, EPI_ISL_666770, EPI_ISL_678644, EPI_ISL_678645, EPI_ISL_678646, EPI_ISL_678647, EPI_ISL_678648, EPI_ISL_678649, EPI_ISL_678650, EPI_ISL_678651, EPI_ISL_678652, EPI_ISL_678653, EPI_ISL_678654, EPI_ISL_678655, EPI_ISL_678656, EPI_ISL_678657, EPI_ISL_678658, EPI_ISL_678659, EPI_ISL_678660, EPI_ISL_678661, EPI_ISL_678663, EPI_ISL_678664, EPI_ISL_678665, EPI_ISL_678666, EPI_ISL_678667, EPI_ISL_678668, EPI_ISL_678669, EPI_ISL_678670, EPI_ISL_678671, EPI_ISL_678672, EPI_ISL_678673, EPI_ISL_678674, EPI_ISL_678675, EPI_ISL_678676, EPI_ISL_678677, EPI_ISL_678678, EPI_ISL_678679, EPI_ISL_678680, EPI_ISL_678683, EPI_ISL_678684, EPI_ISL_678688, EPI_ISL_678694, EPI_ISL_678700, EPI_ISL_678704, EPI_ISL_678787, EPI_ISL_678788, EPI_ISL_678789, EPI_ISL_678790, EPI_ISL_678791, EPI_ISL_678792, EPI_ISL_678793, EPI_ISL_678794, EPI_ISL_678795, EPI_ISL_678796, EPI_ISL_678797, EPI_ISL_678798, EPI_ISL_678799, EPI_ISL_678800, EPI_ISL_678801, EPI_ISL_678802, EPI_ISL_678804, EPI_ISL_678805, EPI_ISL_678806, EPI_ISL_678807, EPI_ISL_678808, EPI_ISL_678809, EPI_ISL_678811, EPI_ISL_678812, EPI_ISL_678813, EPI_ISL_678814, EPI_ISL_678815, EPI_ISL_678816, EPI_ISL_678817, EPI_ISL_678818, EPI_ISL_678819, EPI_ISL_678820, EPI_ISL_678821, EPI_ISL_678822, EPI_ISL_678823, EPI_ISL_678824, EPI_ISL_678827, EPI_ISL_681323, EPI_ISL_681324, EPI_ISL_683491, EPI_ISL_683492, EPI_ISL_686541, EPI_ISL_686542, EPI_ISL_686543, EPI_ISL_686544, EPI_ISL_686545 | | | |
| see above | Respiratory Virus Unit, Microbiology Services Colindale, Public Health England | COVID-19 Genomics UK (COG-UK) Consortium | PHE Covid Sequencing Team |
| EPI_ISL_636732, EPI_ISL_636733 | Respiratory Virus Unit, Microbiology Services Colindale, Public Health England | Respiratory Virus Unit, Microbiology Services Colindale, Public Health England | PHE Covid Sequencing Team |
| EPI_ISL_825549, EPI_ISL_825589, EPI_ISL_825590, EPI_ISL_825591, EPI_ISL_825592, EPI_ISL_825593, EPI_ISL_825594 | | | |
| see above | Respiratory Virus Unit, National Infection Service, Public Health England | COVID-19 Genomics UK (COG-UK) Consortium | PHE Covid Sequencing Team |
| EPI_ISL_664228, EPI_ISL_664359, EPI_ISL_664372, EPI_ISL_664734, EPI_ISL_664755, EPI_ISL_665237, EPI_ISL_665238, EPI_ISL_665239, EPI_ISL_665243, EPI_ISL_665244, EPI_ISL_665245, EPI_ISL_665246, EPI_ISL_665247, EPI_ISL_665248, EPI_ISL_665249, EPI_ISL_665250, EPI_ISL_665251, EPI_ISL_665252, EPI_ISL_665253, EPI_ISL_665382, EPI_ISL_665530, EPI_ISL_665805, EPI_ISL_665806 | | | |
| see above | University College London Hospital | COVID-19 Genomics UK (COG-UK) Consortium | Catherine Houlihan; Dan Frampton; Judith Heaney; Matthew Byott; Moira Spyer and Eleni Nastouli; Stuart Kirk |
| EPI_ISL_637446, EPI_ISL_637937, EPI_ISL_637999, EPI_ISL_638022, EPI_ISL_650123, EPI_ISL_651179, EPI_ISL_651201, EPI_ISL_665335, EPI_ISL_665428, EPI_ISL_665703, EPI_ISL_665720, EPI_ISL_679447, EPI_ISL_679448, EPI_ISL_679449, EPI_ISL_679450, EPI_ISL_679451, EPI_ISL_679452, EPI_ISL_679453, EPI_ISL_679454, EPI_ISL_679455, EPI_ISL_679456, EPI_ISL_679458, EPI_ISL_679459, EPI_ISL_679460, EPI_ISL_679461, EPI_ISL_679462, EPI_ISL_679463, EPI_ISL_679464, EPI_ISL_679465, EPI_ISL_679466, EPI_ISL_679467, EPI_ISL_679468, EPI_ISL_679469, EPI_ISL_679470, EPI_ISL_679471, EPI_ISL_679472, EPI_ISL_679473, EPI_ISL_679474, EPI_ISL_679475, EPI_ISL_679476, EPI_ISL_679477, EPI_ISL_679479, EPI_ISL_679492, EPI_ISL_702831, EPI_ISL_705236, EPI_ISL_705268, EPI_ISL_705338, EPI_ISL_705395, EPI_ISL_724617, EPI_ISL_724620, EPI_ISL_724625, EPI_ISL_724629, EPI_ISL_724635, EPI_ISL_724638, EPI_ISL_724655, EPI_ISL_724658, EPI_ISL_724659, EPI_ISL_724660, EPI_ISL_724662, EPI_ISL_724664, EPI_ISL_724673, EPI_ISL_724675, EPI_ISL_724678, EPI_ISL_724679, EPI_ISL_724682, EPI_ISL_724683, EPI_ISL_724684, EPI_ISL_724694, EPI_ISL_724697, EPI_ISL_724705, EPI_ISL_724708, EPI_ISL_724716, EPI_ISL_724719, EPI_ISL_724725, EPI_ISL_724740, EPI_ISL_724753, EPI_ISL_724759, EPI_ISL_1637138, EPI_ISL_1637139, EPI_ISL_1637140, EPI_ISL_1637142, EPI_ISL_1637143, EPI_ISL_1637144, EPI_ISL_1637148 | | | |
| see above | University College London, Great Ormond Street Hospital for Children NHS Foundation Trust, Imperial College Healthcare NHS Trust | COVID-19 Genomics UK (COG-UK) Consortium | Alison Holmes; Charlotte Williams; Helena Tutill; Jacqueline Findlay; James Price; Judith Breuer; Julianne Brown; Kathryn Harris; Leysa Forrest; Mark Kristiansen; Paola Niola; Paola Resende Silva; Patricia Dyal; Paul Randell; Rachel Williams; Samuel Weeks; Sergi Castellano; Sunando Roy; Tony Brooks; Yasmin Panchbhaya |
| EPI_ISL_650153, EPI_ISL_650155, EPI_ISL_650158, EPI_ISL_650171, EPI_ISL_650176, EPI_ISL_650233, EPI_ISL_650234, EPI_ISL_650235, EPI_ISL_650250, EPI_ISL_650251, EPI_ISL_650252, EPI_ISL_650253, EPI_ISL_650274, EPI_ISL_650281, EPI_ISL_650354, EPI_ISL_650405, EPI_ISL_650410, EPI_ISL_650416, EPI_ISL_650436, EPI_ISL_650437, EPI_ISL_650613, EPI_ISL_650647, EPI_ISL_650665, EPI_ISL_650668, EPI_ISL_650692, EPI_ISL_650693, EPI_ISL_650724, EPI_ISL_650729, EPI_ISL_650740, EPI_ISL_650741, EPI_ISL_650742, EPI_ISL_650744, EPI_ISL_650769, EPI_ISL_650770, EPI_ISL_650771, EPI_ISL_650772, EPI_ISL_650773, EPI_ISL_650791, EPI_ISL_650792, EPI_ISL_650822, EPI_ISL_650823, EPI_ISL_650849, EPI_ISL_650948, EPI_ISL_650995, EPI_ISL_650996, EPI_ISL_650997, EPI_ISL_650998, EPI_ISL_651127, EPI_ISL_651137, EPI_ISL_651140, EPI_ISL_651190, EPI_ISL_651193, EPI_ISL_651227, EPI_ISL_651256, EPI_ISL_651282, EPI_ISL_651304, EPI_ISL_651305, EPI_ISL_651321, EPI_ISL_651378, EPI_ISL_651393, EPI_ISL_651405, EPI_ISL_651486, EPI_ISL_651519, EPI_ISL_651592, EPI_ISL_651600, EPI_ISL_651601, EPI_ISL_651603, EPI_ISL_651605, EPI_ISL_651606, EPI_ISL_651607, EPI_ISL_651608, EPI_ISL_651609, EPI_ISL_651619, EPI_ISL_651622, EPI_ISL_651623, EPI_ISL_651624, EPI_ISL_651625, EPI_ISL_651626, EPI_ISL_651627, EPI_ISL_651628, EPI_ISL_651629, EPI_ISL_651632, EPI_ISL_679130, EPI_ISL_679131, EPI_ISL_679133, EPI_ISL_812984, EPI_ISL_812985, EPI_ISL_812986, EPI_ISL_812987, EPI_ISL_812988, EPI_ISL_812989, EPI_ISL_812990, EPI_ISL_812991, EPI_ISL_812992, EPI_ISL_812993, EPI_ISL_812994, EPI_ISL_812995, EPI_ISL_812996, EPI_ISL_812997, EPI_ISL_812998, EPI_ISL_812999, EPI_ISL_813000, EPI_ISL_813001, EPI_ISL_813002, EPI_ISL_813003, EPI_ISL_813004, EPI_ISL_813005, EPI_ISL_813006, EPI_ISL_813007, EPI_ISL_813008, EPI_ISL_813009, EPI_ISL_813010, EPI_ISL_813011 | | | |
| see above | University of Birmingham | COVID-19 Genomics UK (COG-UK) Consortium | Alex Richter; Andrew Bosworth; Andrew Bosworth. Queen Elizabeth Hospital: Anna Casey; Andrew D Beggs PHE Heartlands Lab: Husam Osman; Charlotte Poxon; Institute of Microbiology; Joanne Stockton; Josh Quick; Kasun Wanigasooriya; Mike Kidd; Nicholas Loman. University of Birmingham Testing Laboratory: Celina M Whalley; Oliver Pickles; Radoslaw Poplawski; Samuel Nicholls; University of Birmingham: Claire McMurray; Will Rowe |
| EPI_ISL_650141, EPI_ISL_650151, EPI_ISL_650163, EPI_ISL_650215, EPI_ISL_650219, EPI_ISL_650222, EPI_ISL_650226, EPI_ISL_650258, EPI_ISL_650352, EPI_ISL_650357, EPI_ISL_650358, EPI_ISL_650377, EPI_ISL_650399, EPI_ISL_650414, EPI_ISL_650565, EPI_ISL_650597, EPI_ISL_650606, EPI_ISL_650607, EPI_ISL_650610, EPI_ISL_650634, EPI_ISL_650635, EPI_ISL_650636, EPI_ISL_650637, EPI_ISL_650638, EPI_ISL_650639, EPI_ISL_650667, EPI_ISL_650702, EPI_ISL_650783, EPI_ISL_650784, EPI_ISL_651053, EPI_ISL_651054, EPI_ISL_651055, EPI_ISL_651056, EPI_ISL_651057, EPI_ISL_651058, EPI_ISL_651059, EPI_ISL_651083, EPI_ISL_651084, EPI_ISL_651085, EPI_ISL_651130, EPI_ISL_651177, EPI_ISL_651183, EPI_ISL_651238, EPI_ISL_651241, EPI_ISL_651289, EPI_ISL_651290, EPI_ISL_651373, EPI_ISL_651439, EPI_ISL_651527, EPI_ISL_652289, EPI_ISL_652290, EPI_ISL_652291, EPI_ISL_652292, EPI_ISL_652293, EPI_ISL_652294, EPI_ISL_652295, EPI_ISL_652296, EPI_ISL_652297, EPI_ISL_652298, EPI_ISL_652299, EPI_ISL_652300, EPI_ISL_652301, EPI_ISL_652302, EPI_ISL_652303, EPI_ISL_664117, EPI_ISL_664162, EPI_ISL_664170, EPI_ISL_664205, EPI_ISL_664206, EPI_ISL_664207, EPI_ISL_664289, EPI_ISL_664295, EPI_ISL_664319, EPI_ISL_664327, EPI_ISL_664328, EPI_ISL_664330, EPI_ISL_664398, EPI_ISL_664399, EPI_ISL_664439, EPI_ISL_664465, EPI_ISL_664473, EPI_ISL_664592, EPI_ISL_664650, EPI_ISL_664691, EPI_ISL_664699, EPI_ISL_664716, EPI_ISL_664720, EPI_ISL_664737, EPI_ISL_664740, EPI_ISL_665046, EPI_ISL_665048, EPI_ISL_665049, EPI_ISL_665050, EPI_ISL_665052, EPI_ISL_665053, EPI_ISL_665067, EPI_ISL_741359, EPI_ISL_741360, EPI_ISL_741361, EPI_ISL_742117, EPI_ISL_997309, EPI_ISL_997310, EPI_ISL_997311, EPI_ISL_997312, EPI_ISL_1247548 | | | |
| see above | University of Exeter | COVID-19 Genomics UK (COG- | Aaron Jeffries; Audrey Farbos; Ben Temperton; Jane Masoli; Joanna Warwick-Dugdale; Michelle Michelsen; Robyn Manley; Stephen Michell |

| | | | |
|---|---|---|---|
| | | UK) Consortium | |
| EPI_ISL_1104259 | Virology Department, Royal Infirmary of Edinburgh, NHS Lothian / School of Biological Sciences, University of Edinburgh | COVID-19 Genomics UK (COG-UK) Consortium | Colquhoun R; Cotton S; Dewar R; Hill V; Jackson B; McCrone JT; McHugh M; O'Toole Á; Rambaut A; Rooke S; Scher E; Templeton K; Yu X |
| EPI_ISL_650135, EPI_ISL_650267, EPI_ISL_650286, EPI_ISL_650311, EPI_ISL_650427, EPI_ISL_650433, EPI_ISL_650435, EPI_ISL_650439, EPI_ISL_650456, EPI_ISL_650478, EPI_ISL_650495, EPI_ISL_650501, EPI_ISL_650757, EPI_ISL_650779, EPI_ISL_650780, EPI_ISL_650781, EPI_ISL_650799, EPI_ISL_650800, EPI_ISL_650812, EPI_ISL_650813, EPI_ISL_650826, EPI_ISL_651141, EPI_ISL_651158, EPI_ISL_651169, EPI_ISL_651188, EPI_ISL_651229, EPI_ISL_651230, EPI_ISL_651294, EPI_ISL_651367, EPI_ISL_651385, EPI_ISL_651394, EPI_ISL_651427, EPI_ISL_651428, EPI_ISL_651523, EPI_ISL_651524, EPI_ISL_652013, EPI_ISL_652014, EPI_ISL_652016, EPI_ISL_652017, EPI_ISL_652018, EPI_ISL_652019, EPI_ISL_652020, EPI_ISL_652021, EPI_ISL_652022, EPI_ISL_652023, EPI_ISL_652024, EPI_ISL_652025, EPI_ISL_652026, EPI_ISL_652027, EPI_ISL_652028, EPI_ISL_652029, EPI_ISL_652030, EPI_ISL_652031, EPI_ISL_652032, EPI_ISL_652033, EPI_ISL_652034, EPI_ISL_652035, EPI_ISL_652036, EPI_ISL_652037, EPI_ISL_652038, EPI_ISL_652039, EPI_ISL_652040, EPI_ISL_652049, EPI_ISL_652050, EPI_ISL_652051, EPI_ISL_652052, EPI_ISL_652053, EPI_ISL_652054, EPI_ISL_652057, EPI_ISL_652058, EPI_ISL_652059, EPI_ISL_652060, EPI_ISL_652061, EPI_ISL_652064, EPI_ISL_652065, EPI_ISL_652066, EPI_ISL_665552, EPI_ISL_665553, EPI_ISL_665554, EPI_ISL_666060, EPI_ISL_666061, EPI_ISL_666062, EPI_ISL_666063, EPI_ISL_666064, EPI_ISL_666065, EPI_ISL_666066, EPI_ISL_666067, EPI_ISL_666068, EPI_ISL_666069, EPI_ISL_666071, EPI_ISL_666073, EPI_ISL_680507, EPI_ISL_680508, EPI_ISL_680509, EPI_ISL_680513, EPI_ISL_680515, EPI_ISL_680516, EPI_ISL_680517, EPI_ISL_680518, EPI_ISL_680519, EPI_ISL_680520, EPI_ISL_680521, EPI_ISL_680522, EPI_ISL_680523, EPI_ISL_680524, EPI_ISL_680525, EPI_ISL_680526, EPI_ISL_680527, EPI_ISL_680528, EPI_ISL_680529, EPI_ISL_680530, EPI_ISL_680531, EPI_ISL_680532, EPI_ISL_680533, EPI_ISL_680534, EPI_ISL_680535, EPI_ISL_680536, EPI_ISL_680537, EPI_ISL_680544, EPI_ISL_680545, EPI_ISL_702937, EPI_ISL_703863, EPI_ISL_705771, EPI_ISL_705772, EPI_ISL_705798, EPI_ISL_705799, EPI_ISL_764356, EPI_ISL_814681, EPI_ISL_814682, EPI_ISL_814683, EPI_ISL_814684, EPI_ISL_814685, EPI_ISL_814686, EPI_ISL_814687, EPI_ISL_838313, EPI_ISL_949647, EPI_ISL_949648, EPI_ISL_949649 | | | |
| see above | Virology Department, Royal Infirmary of Edinburgh, NHS Lothian / School of Biological Sciences, University of Edinburgh / Institute of Genetics and Molecular Medicine, University of Edinburgh | COVID-19 Genomics UK (COG-UK) Consortium | Balcaza C; Colquhoun R; Dewar R; Gallagher M; Hill V; Jackson B; McCrone JT; McHugh M; O'Toole Á; Rambaut A; Rooke S; Scher E; Templeton K; Williams TC; Yu X |
| EPI_ISL_637273, EPI_ISL_637291, EPI_ISL_637297, EPI_ISL_637358, EPI_ISL_637436, EPI_ISL_637533, EPI_ISL_637584, EPI_ISL_637585, EPI_ISL_637872, EPI_ISL_637985, EPI_ISL_638024, EPI_ISL_638877, EPI_ISL_638879, EPI_ISL_638880, EPI_ISL_638881, EPI_ISL_638883, EPI_ISL_638884, EPI_ISL_638885, EPI_ISL_638886, EPI_ISL_638887, EPI_ISL_638888, EPI_ISL_638889, EPI_ISL_638891, EPI_ISL_638893, EPI_ISL_650121, EPI_ISL_650131, EPI_ISL_650208, EPI_ISL_650210, EPI_ISL_650349, EPI_ISL_650370, EPI_ISL_650395, EPI_ISL_650408, EPI_ISL_650422, EPI_ISL_650646, EPI_ISL_650662, EPI_ISL_650670, EPI_ISL_650679, EPI_ISL_650689, EPI_ISL_650691, EPI_ISL_650705, EPI_ISL_650706, EPI_ISL_650713, EPI_ISL_650715, EPI_ISL_650807, EPI_ISL_650808, EPI_ISL_651078, EPI_ISL_651109, EPI_ISL_651121, EPI_ISL_651152, EPI_ISL_651224, EPI_ISL_651233, EPI_ISL_651291, EPI_ISL_651374, EPI_ISL_651414, EPI_ISL_651514, EPI_ISL_651518, EPI_ISL_651715, EPI_ISL_651716, EPI_ISL_651717, EPI_ISL_651718, EPI_ISL_651719, EPI_ISL_651720, EPI_ISL_651721, EPI_ISL_651722, EPI_ISL_651723, EPI_ISL_651724, EPI_ISL_651725, EPI_ISL_651726, EPI_ISL_651727, EPI_ISL_651728, EPI_ISL_651729, EPI_ISL_651730, EPI_ISL_651731, EPI_ISL_651732, EPI_ISL_653059, EPI_ISL_653060, EPI_ISL_653061, EPI_ISL_653062, EPI_ISL_653064, EPI_ISL_653065, EPI_ISL_653066, EPI_ISL_653067, EPI_ISL_653068, EPI_ISL_653072, EPI_ISL_653073, EPI_ISL_653081, EPI_ISL_653082, EPI_ISL_653088, EPI_ISL_653090, EPI_ISL_653091, EPI_ISL_653094, EPI_ISL_653096, EPI_ISL_664283, EPI_ISL_664298, EPI_ISL_664370, EPI_ISL_664425, EPI_ISL_664426, EPI_ISL_664431, EPI_ISL_664437, EPI_ISL_664600, EPI_ISL_664602, EPI_ISL_664604, EPI_ISL_664630, EPI_ISL_664689, EPI_ISL_664713, EPI_ISL_664752, EPI_ISL_665023, EPI_ISL_665024, EPI_ISL_665025, EPI_ISL_665026, EPI_ISL_665027, EPI_ISL_665028, EPI_ISL_665029, EPI_ISL_665030, EPI_ISL_665031, EPI_ISL_665032, EPI_ISL_665033, EPI_ISL_665034, EPI_ISL_665035, EPI_ISL_665036, EPI_ISL_665037, EPI_ISL_665038, EPI_ISL_665039, EPI_ISL_665040, EPI_ISL_665041, EPI_ISL_665042, EPI_ISL_665043, EPI_ISL_665044, EPI_ISL_665045, EPI_ISL_679903, EPI_ISL_679904, EPI_ISL_679905, EPI_ISL_679906, EPI_ISL_679907, EPI_ISL_679908, EPI_ISL_679909, EPI_ISL_679910, EPI_ISL_679911, EPI_ISL_679912, EPI_ISL_679913, EPI_ISL_679914, EPI_ISL_680131, EPI_ISL_703048, EPI_ISL_727990, EPI_ISL_727992, EPI_ISL_727993, EPI_ISL_727994, EPI_ISL_727995, EPI_ISL_842347, EPI_ISL_842352, EPI_ISL_842353, EPI_ISL_924421, EPI_ISL_924425, EPI_ISL_924429 | | | |
| see above | Virology Department, Sheffield Teaching Hospitals NHS Foundation Trust/Department of Infection, Immunity and Cardiovascular Disease, The Medical School, University of Sheffield | COVID-19 Genomics UK (COG-UK) Consortium | Adri Angyal; Alex Keeley; Benjamin Lindsey; Cariad Evans; Danielle Groves; Dave Partridge; Katie Johnson; Laura Carrilero; Luke Green; Matthew Parker; Matthew Wyles; Mehmet Yavuz; Mohammad Raza; Nikki Smith; Paul Parsons; Rachel Tucker; Rebecca Brown; Thushan de Silva |
| EPI_ISL_637288, EPI_ISL_637289, EPI_ISL_637302, EPI_ISL_637319, EPI_ISL_637330, EPI_ISL_637332, EPI_ISL_637349, EPI_ISL_637378, EPI_ISL_637379, EPI_ISL_637380, EPI_ISL_637395, EPI_ISL_637412, EPI_ISL_637422, EPI_ISL_637447, EPI_ISL_637450, EPI_ISL_637452, EPI_ISL_637454, EPI_ISL_637525, EPI_ISL_637542, EPI_ISL_637551, EPI_ISL_637556, EPI_ISL_637586, EPI_ISL_637587, EPI_ISL_637593, EPI_ISL_637597, EPI_ISL_637600, EPI_ISL_637603, EPI_ISL_637608, EPI_ISL_637609, EPI_ISL_637615, EPI_ISL_637626, EPI_ISL_637675, EPI_ISL_637868, EPI_ISL_637869, EPI_ISL_637896, EPI_ISL_637907, EPI_ISL_637909, EPI_ISL_637915, EPI_ISL_637918, EPI_ISL_637927, EPI_ISL_637928, EPI_ISL_637958, EPI_ISL_637971, EPI_ISL_637998, EPI_ISL_638007, EPI_ISL_638012, EPI_ISL_638034, EPI_ISL_638035, EPI_ISL_638044, EPI_ISL_638046, EPI_ISL_638047, EPI_ISL_638052, EPI_ISL_638055, EPI_ISL_638056, EPI_ISL_638057, EPI_ISL_638058, EPI_ISL_638059, EPI_ISL_638060, EPI_ISL_638061, EPI_ISL_638062, EPI_ISL_638063, EPI_ISL_638064, EPI_ISL_638091, EPI_ISL_638092, EPI_ISL_638094, EPI_ISL_638095, EPI_ISL_638097, EPI_ISL_638098, EPI_ISL_638100, EPI_ISL_638101, EPI_ISL_638103, EPI_ISL_638104, EPI_ISL_638105, EPI_ISL_638106, EPI_ISL_638107, EPI_ISL_638108, EPI_ISL_638109, EPI_ISL_638111, EPI_ISL_638112, EPI_ISL_638113, EPI_ISL_638114, EPI_ISL_638115, EPI_ISL_638116, EPI_ISL_638120, EPI_ISL_638121, EPI_ISL_638123, EPI_ISL_638124, EPI_ISL_638125, EPI_ISL_638128, EPI_ISL_638129, EPI_ISL_638155, EPI_ISL_638158, EPI_ISL_638159, EPI_ISL_638161, EPI_ISL_638162, EPI_ISL_638163, EPI_ISL_638172, EPI_ISL_638206, EPI_ISL_639011, EPI_ISL_639016, EPI_ISL_639017, EPI_ISL_639018, EPI_ISL_639021, EPI_ISL_639026, EPI_ISL_639027, EPI_ISL_639029, EPI_ISL_639031, EPI_ISL_639032, EPI_ISL_639034, EPI_ISL_639035, EPI_ISL_639037, EPI_ISL_639040, EPI_ISL_639041, EPI_ISL_639045, EPI_ISL_639047, EPI_ISL_639053, EPI_ISL_639055, EPI_ISL_639056, EPI_ISL_639057, EPI_ISL_639060, EPI_ISL_639062, EPI_ISL_639067, EPI_ISL_639077, EPI_ISL_639078, EPI_ISL_639080, EPI_ISL_639082, EPI_ISL_639085, EPI_ISL_639088, EPI_ISL_639090, EPI_ISL_639091, EPI_ISL_639092, EPI_ISL_639096, EPI_ISL_639105, EPI_ISL_639106, EPI_ISL_639108, EPI_ISL_639109, EPI_ISL_639111, EPI_ISL_639115, EPI_ISL_639119, EPI_ISL_639121, EPI_ISL_639122, EPI_ISL_639125, EPI_ISL_639131, EPI_ISL_639133, EPI_ISL_639139, EPI_ISL_639146, EPI_ISL_639148, EPI_ISL_639155, EPI_ISL_639156, EPI_ISL_639157, EPI_ISL_639158, EPI_ISL_639160, EPI_ISL_639161, EPI_ISL_639164, EPI_ISL_639166, EPI_ISL_639167, EPI_ISL_639173, EPI_ISL_639174, EPI_ISL_639175, EPI_ISL_639176, EPI_ISL_639179, EPI_ISL_639182, EPI_ISL_639183, EPI_ISL_639188, EPI_ISL_639189, EPI_ISL_639190, EPI_ISL_639193, EPI_ISL_639195, EPI_ISL_639196, EPI_ISL_639197, EPI_ISL_639198, EPI_ISL_639199, EPI_ISL_639200, EPI_ISL_639201, EPI_ISL_639202, EPI_ISL_639203, EPI_ISL_639204, EPI_ISL_639205, EPI_ISL_639206, EPI_ISL_639208, EPI_ISL_639211, EPI_ISL_639212, EPI_ISL_639213, EPI_ISL_639215, EPI_ISL_639216, EPI_ISL_639217, EPI_ISL_639218, EPI_ISL_639219, EPI_ISL_639220, EPI_ISL_639221, EPI_ISL_639222, EPI_ISL_639225, EPI_ISL_639227, EPI_ISL_639228, EPI_ISL_639233, EPI_ISL_639234, EPI_ISL_639235, EPI_ISL_639237, EPI_ISL_639238, EPI_ISL_639239, EPI_ISL_639240, EPI_ISL_639241, EPI_ISL_639243, EPI_ISL_639244, EPI_ISL_639246, EPI_ISL_639247, EPI_ISL_639248, EPI_ISL_639249, EPI_ISL_639250, EPI_ISL_639252, EPI_ISL_639253, EPI_ISL_639254, EPI_ISL_639255, EPI_ISL_639256, EPI_ISL_639258, EPI_ISL_639259, EPI_ISL_639261, EPI_ISL_639262, EPI_ISL_639263, EPI_ISL_639267, EPI_ISL_639269, EPI_ISL_639270, EPI_ISL_639271, EPI_ISL_639272, EPI_ISL_639273, EPI_ISL_639274, EPI_ISL_639275, EPI_ISL_639276, EPI_ISL_639277, EPI_ISL_639280, EPI_ISL_639282, EPI_ISL_639284, EPI_ISL_639288, EPI_ISL_639289, EPI_ISL_639290, EPI_ISL_639292, EPI_ISL_639293, EPI_ISL_639294, EPI_ISL_639296, EPI_ISL_639299, EPI_ISL_639300, EPI_ISL_639301, EPI_ISL_639302, EPI_ISL_639303, EPI_ISL_639305, EPI_ISL_639306, EPI_ISL_639308, EPI_ISL_639309, EPI_ISL_639310, EPI_ISL_639311, EPI_ISL_639313, EPI_ISL_639314, EPI_ISL_639315, EPI_ISL_639316, EPI_ISL_639317, EPI_ISL_639318, EPI_ISL_639319, EPI_ISL_639321, EPI_ISL_639322, EPI_ISL_639323, EPI_ISL_639324, EPI_ISL_639325, EPI_ISL_639327, EPI_ISL_639328, EPI_ISL_639329, EPI_ISL_639330, EPI_ISL_639331, EPI_ISL_639332, EPI_ISL_639333, EPI_ISL_639334, EPI_ISL_639335, EPI_ISL_639338, EPI_ISL_639339, EPI_ISL_639341, EPI_ISL_639342, EPI_ISL_639344, EPI_ISL_639345, EPI_ISL_639346, EPI_ISL_639347, EPI_ISL_639351, EPI_ISL_639353, EPI_ISL_639355, EPI_ISL_639356, EPI_ISL_639358, EPI_ISL_639359, EPI_ISL_639360, EPI_ISL_639361, EPI_ISL_639362, EPI_ISL_639363, EPI_ISL_639365, EPI_ISL_639366, EPI_ISL_639367, EPI_ISL_639368, EPI_ISL_639369, EPI_ISL_639371, EPI_ISL_639372, EPI_ISL_639374, EPI_ISL_639375, EPI_ISL_639376, EPI_ISL_639377, EPI_ISL_639378, EPI_ISL_639379, EPI_ISL_639381, EPI_ISL_639383, EPI_ISL_639384, EPI_ISL_639385, EPI_ISL_639386, EPI_ISL_639387, EPI_ISL_639388, EPI_ISL_639390, EPI_ISL_639392, EPI_ISL_639393, EPI_ISL_639394, EPI_ISL_639395, EPI_ISL_639396, EPI_ISL_639397, EPI_ISL_639398, EPI_ISL_639399, EPI_ISL_639400, EPI_ISL_639401, EPI_ISL_639402, EPI_ISL_639403, EPI_ISL_639404, EPI_ISL_639405, EPI_ISL_639407, EPI_ISL_639408, EPI_ISL_639409, EPI_ISL_639410, EPI_ISL_639412, EPI_ISL_639413, EPI_ISL_639414, EPI_ISL_639416, EPI_ISL_639417, EPI_ISL_639418, EPI_ISL_639420, EPI_ISL_639421, EPI_ISL_639422, EPI_ISL_639423, EPI_ISL_639424, EPI_ISL_639425, EPI_ISL_639426, EPI_ISL_639427, EPI_ISL_639428, EPI_ISL_639431, EPI_ISL_639432, EPI_ISL_639433, EPI_ISL_639434, EPI_ISL_639436, EPI_ISL_639437, EPI_ISL_639438, EPI_ISL_639439, EPI_ISL_639440, EPI_ISL_639441, EPI_ISL_639443, EPI_ISL_639444, EPI_ISL_639445, EPI_ISL_639447, EPI_ISL_639448, EPI_ISL_639449, EPI_ISL_639450, EPI_ISL_639452, EPI_ISL_639453, EPI_ISL_639456, EPI_ISL_639457, EPI_ISL_639459, EPI_ISL_639460, EPI_ISL_639461, EPI_ISL_639462, EPI_ISL_639463, EPI_ISL_639464, EPI_ISL_639465, EPI_ISL_639466, EPI_ISL_639467, EPI_ISL_639468, EPI_ISL_639469, EPI_ISL_639472, EPI_ISL_639473, EPI_ISL_639474, EPI_ISL_639475, EPI_ISL_639476, EPI_ISL_639477, EPI_ISL_639479, EPI_ISL_639480, EPI_ISL_639481, EPI_ISL_639482, EPI_ISL_639483, EPI_ISL_639484, EPI_ISL_639485, EPI_ISL_639486, EPI_ISL_639488, EPI_ISL_639489, EPI_ISL_639490, EPI_ISL_639491, EPI_ISL_639492, EPI_ISL_639493, EPI_ISL_639494, EPI_ISL_639495, EPI_ISL_639496, EPI_ISL_639497, EPI_ISL_639498, EPI_ISL_639499, EPI_ISL_639500, EPI_ISL_639501, EPI_ISL_639503, EPI_ISL_639504, EPI_ISL_639506, EPI_ISL_639507, EPI_ISL_639508, EPI_ISL_639509, EPI_ISL_639510, EPI_ISL_639512, EPI_ISL_639514, EPI_ISL_639515, EPI_ISL_639516, EPI_ISL_639517, EPI_ISL_639518, EPI_ISL_639520, EPI_ISL_639521, EPI_ISL_639523, EPI_ISL_639524, EPI_ISL_639525, EPI_ISL_639526, EPI_ISL_639527, EPI_ISL_639528, EPI_ISL_639529, EPI_ISL_639530, EPI_ISL_639531, EPI_ISL_639532, EPI_ISL_639534, EPI_ISL_639535, EPI_ISL_639536, EPI_ISL_639537, EPI_ISL_639539, EPI_ISL_639540, EPI_ISL_639541, EPI_ISL_639542, EPI_ISL_639544, EPI_ISL_639545, EPI_ISL_639547, EPI_ISL_639548, EPI_ISL_639549, EPI_ISL_639551, EPI_ISL_639553, EPI_ISL_639555, EPI_ISL_639556, EPI_ISL_639557, EPI_ISL_639558, EPI_ISL_639559, EPI_ISL_639560, EPI_ISL_639561, EPI_ISL_639562, EPI_ISL_639563, EPI_ISL_639565, EPI_ISL_639566, EPI_ISL_639567, EPI_ISL_639568, EPI_ISL_639569, EPI_ISL_639571, EPI_ISL_639572, EPI_ISL_639574, EPI_ISL_639576, EPI_ISL_639577, EPI_ISL_639578, EPI_ISL_639580, EPI_ISL_639581, EPI_ISL_639584, EPI_ISL_639585, EPI_ISL_639586, EPI_ISL_639587, EPI_ISL_639588, EPI_ISL_639591, EPI_ISL_639592, EPI_ISL_639593, EPI_ISL_639594, EPI_ISL_639596, EPI_ISL_639597, EPI_ISL_639599, EPI_ISL_639600, EPI_ISL_639601, EPI_ISL_639602, EPI_ISL_639603, EPI_ISL_639604, EPI_ISL_639605, EPI_ISL_639606, EPI_ISL_639607, EPI_ISL_639608, EPI_ISL_639609, EPI_ISL_639610, EPI_ISL_639611, EPI_ISL_639612, EPI_ISL_639614, EPI_ISL_639615, EPI_ISL_639616, EPI_ISL_639617, EPI_ISL_639618, EPI_ISL_639620, EPI_ISL_639621, EPI_ISL_639623, EPI_ISL_650204, EPI_ISL_650206, EPI_ISL_650213, EPI_ISL_650320, EPI_ISL_650338, EPI_ISL_650339, EPI_ISL_650340, EPI_ISL_650431, EPI_ISL_650549, EPI_ISL_650551, EPI_ISL_650787, EPI_ISL_650884, EPI_ISL_650896, EPI_ISL_650905, EPI_ISL_650906, EPI_ISL_650919, EPI_ISL_650929, EPI_ISL_651108, EPI_ISL_651112, EPI_ISL_651154, EPI_ISL_651191, EPI_ISL_651264, EPI_ISL_651416, EPI_ISL_651440, EPI_ISL_651442, EPI_ISL_651450, EPI_ISL_651452, EPI_ISL_651453, EPI_ISL_651454, EPI_ISL_651456, EPI_ISL_651458, EPI_ISL_651459, EPI_ISL_651460, EPI_ISL_651461, EPI_ISL_651462, EPI_ISL_651463, EPI_ISL_651464, EPI_ISL_651502, EPI_ISL_651543, EPI_ISL_651546, EPI_ISL_652432, EPI_ISL_652455, EPI_ISL_652462, EPI_ISL_652466, EPI_ISL_652470, EPI_ISL_652474, EPI_ISL_652481, EPI_ISL_652482, EPI_ISL_652487, EPI_ISL_652496, EPI_ISL_652497, EPI_ISL_652498, EPI_ISL_652499, EPI_ISL_652500, EPI_ISL_652501, EPI_ISL_652502, EPI_ISL_652504, EPI_ISL_652505, EPI_ISL_652506, EPI_ISL_652507, EPI_ISL_652508, EPI_ISL_652509, EPI_ISL_652510, EPI_ISL_652511, EPI_ISL_652512, EPI_ISL_652513, EPI_ISL_652514, EPI_ISL_652515, EPI_ISL_652516, EPI_ISL_652517, EPI_ISL_652518, EPI_ISL_652519, EPI_ISL_652520, EPI_ISL_652521, EPI_ISL_652522, EPI_ISL_652523, EPI_ISL_652524, EPI_ISL_652525, EPI_ISL_652526, EPI_ISL_652527, EPI_ISL_652528, EPI_ISL_652529, EPI_ISL_652530, EPI_ISL_652531, EPI_ISL_652532, EPI_ISL_652533, EPI_ISL_652534, EPI_ISL_652535, EPI_ISL_652537, EPI_ISL_652538, EPI_ISL_652539, EPI_ISL_652540, EPI_ISL_652541, EPI_ISL_652542, EPI_ISL_652544, EPI_ISL_652545, EPI_ISL_652547, EPI_ISL_652548, EPI_ISL_652549, EPI_ISL_652550, EPI_ISL_652551, EPI_ISL_652552, EPI_ISL_652553, EPI_ISL_652554, EPI_ISL_652556, EPI_ISL_652557, EPI_ISL_652558, EPI_ISL_652559, EPI_ISL_652560, EPI_ISL_652561, EPI_ISL_652562, EPI_ISL_652563, EPI_ISL_652564, EPI_ISL_652565, EPI_ISL_652566, EPI_ISL_652567, EPI_ISL_652569, EPI_ISL_652570, EPI_ISL_652571, EPI_ISL_652572, EPI_ISL_652573, EPI_ISL_652574, EPI_ISL_652575, EPI_ISL_652576, EPI_ISL_652577, EPI_ISL_652578, EPI_ISL_652579, EPI_ISL_652580, EPI_ISL_652581, EPI_ISL_652582, EPI_ISL_652583, EPI_ISL_652584, EPI_ISL_652585, EPI_ISL_652586, EPI_ISL_652587, EPI_ISL_652588, EPI_ISL_652589, EPI_ISL_652590, EPI_ISL_652591, EPI_ISL_652592, EPI_ISL_652593, EPI_ISL_652594, EPI_ISL_652595, EPI_ISL_652596, EPI_ISL_652597, EPI_ISL_652599, EPI_ISL_652600, EPI_ISL_652601, EPI_ISL_652602, EPI_ISL_652604, EPI_ISL_652605, EPI_ISL_652607, EPI_ISL_652608, EPI_ISL_652610, EPI_ISL_652611, EPI_ISL_652612, EPI_ISL_652613, EPI_ISL_652614, EPI_ISL_652615, EPI_ISL_652616, EPI_ISL_652617, EPI_ISL_652618, EPI_ISL_665281, EPI_ISL_665288, EPI_ISL_665302, EPI_ISL_665315, EPI_ISL_665331, EPI_ISL_665373, EPI_ISL_665451, EPI_ISL_665459, EPI_ISL_665468, EPI_ISL_665469, EPI_ISL_665473, EPI_ISL_665474, EPI_ISL_665504, EPI_ISL_665511, EPI_ISL_665516, EPI_ISL_665526, EPI_ISL_665527, EPI_ISL_665573, EPI_ISL_665576, EPI_ISL_665582, EPI_ISL_665584, EPI_ISL_665585, EPI_ISL_665588, EPI_ISL_665589, EPI_ISL_665602, EPI_ISL_665604, EPI_ISL_665607, EPI_ISL_665608, EPI_ISL_665653, EPI_ISL_665655, EPI_ISL_665659, EPI_ISL_665660, EPI_ISL_665677, EPI_ISL_665691, EPI_ISL_665713, EPI_ISL_665743, EPI_ISL_665750, EPI_ISL_665751, EPI_ISL_665752, EPI_ISL_665753, EPI_ISL_665754, EPI_ISL_665755, EPI_ISL_665756, EPI_ISL_665764, EPI_ISL_665768, EPI_ISL_665771, EPI_ISL_665783, EPI_ISL_665794, EPI_ISL_665795, EPI_ISL_665799, EPI_ISL_665800, EPI_ISL_665802, EPI_ISL_665803, EPI_ISL_666111, EPI_ISL_666112, EPI_ISL_666113, EPI_ISL_666115, EPI_ISL_666117, EPI_ISL_666118, EPI_ISL_666119, EPI_ISL_666120, EPI_ISL_666123, EPI_ISL_666124, EPI_ISL_666125, EPI_ISL_666127, EPI_ISL_666128, EPI_ISL_666131, EPI_ISL_666132, EPI_ISL_666134, EPI_ISL_666135, EPI_ISL_666136, EPI_ISL_666137, EPI_ISL_666139, EPI_ISL_666141, EPI_ISL_666142, EPI_ISL_666144, EPI_ISL_666147, EPI_ISL_666148, EPI_ISL_666149, EPI_ISL_666150, EPI_ISL_666151, EPI_ISL_666153, EPI_ISL_666157, EPI_ISL_666158, EPI_ISL_666159, EPI_ISL_666160, EPI_ISL_666161, EPI_ISL_666162, EPI_ISL_666163, EPI_ISL_666164, EPI_ISL_666165, EPI_ISL_666166, EPI_ISL_666168, EPI_ISL_666169, EPI_ISL_666171, EPI_ISL_666172, EPI_ISL_666174, EPI_ISL_666175, EPI_ISL_666177, EPI_ISL_666179, EPI_ISL_666180, EPI_ISL_666182, EPI_ISL_666184, EPI_ISL_666185, EPI_ISL_666192, EPI_ISL_666193, EPI_ISL_666194, EPI_ISL_666197, EPI_ISL_666199, EPI_ISL_666200, EPI_ISL_666204, EPI_ISL_666207, EPI_ISL_666208, EPI_ISL_666209, EPI_ISL_666210, EPI_ISL_666211, EPI_ISL_666212, EPI_ISL_666213, EPI_ISL_666214, EPI_ISL_666218, EPI_ISL_666220, EPI_ISL_666221, EPI_ISL_666222, EPI_ISL_666223, EPI_ISL_666224, EPI_ISL_666226, EPI_ISL_666227, EPI_ISL_666229, EPI_ISL_666230, EPI_ISL_666231, EPI_ISL_666232, EPI_ISL_666234, EPI_ISL_666235, EPI_ISL_666237, EPI_ISL_666238, EPI_ISL_666240, EPI_ISL_666241, EPI_ISL_666242, EPI_ISL_666243, EPI_ISL_666245, EPI_ISL_666247, EPI_ISL_666248, EPI_ISL_666250, EPI_ISL_666252, EPI_ISL_666253, EPI_ISL_666254, EPI_ISL_666258, EPI_ISL_666261, EPI_ISL_666263, EPI_ISL_666264, EPI_ISL_666265, EPI_ISL_666266, EPI_ISL_666267, EPI_ISL_666268, EPI_ISL_666270, EPI_ISL_666273, EPI_ISL_666274, EPI_ISL_666276, EPI_ISL_666277, EPI_ISL_666280, EPI_ISL_666283, EPI_ISL_666285, EPI_ISL_666286, EPI_ISL_666287, EPI_ISL_666291, EPI_ISL_666292, EPI_ISL_666298, EPI_ISL_666301, EPI_ISL_666305, EPI_ISL_666307, EPI_ISL_666310, EPI_ISL_666312, EPI_ISL_666313, EPI_ISL_666315, EPI_ISL_666318, EPI_ISL_666319, EPI_ISL_666320, EPI_ISL_666321, EPI_ISL_666322, EPI_ISL_666325, EPI_ISL_666327, EPI_ISL_666328, EPI_ISL_666329, EPI_ISL_666330, EPI_ISL_666331, EPI_ISL_666332, EPI_ISL_666334, EPI_ISL_666335, EPI_ISL_666337, EPI_ISL_666338, EPI_ISL_666340, EPI_ISL_666342, EPI_ISL_666343, EPI_ISL_666344, EPI_ISL_666345, EPI_ISL_666346, EPI_ISL_666347, EPI_ISL_666349, EPI_ISL_666351, EPI_ISL_666354, EPI_ISL_666356, EPI_ISL_666358, EPI_ISL_666360, EPI_ISL_666361, EPI_ISL_666362, EPI_ISL_666365, EPI_ISL_666367, EPI_ISL_666371, EPI_ISL_666376, EPI_ISL_666377, EPI_ISL_666379, EPI_ISL_666383, EPI_ISL_666396, EPI_ISL_666397, EPI_ISL_666399, EPI_ISL_666401, EPI_ISL_666405, EPI_ISL_666406, EPI_ISL_666407, EPI_ISL_666409, EPI_ISL_666411, EPI_ISL_666412, EPI_ISL_666417, EPI_ISL_666418, EPI_ISL_666424, EPI_ISL_666425, EPI_ISL_666426, EPI_ISL_666430, EPI_ISL_666437, EPI_ISL_666443, EPI_ISL_666451, EPI_ISL_666452, EPI_ISL_666454, EPI_ISL_666457, EPI_ISL_666458, EPI_ISL_666459, EPI_ISL_666460, EPI_ISL_666461, EPI_ISL_666464, EPI_ISL_666465, EPI_ISL_666467, EPI_ISL_666470, EPI_ISL_666474, EPI_ISL_666476, EPI_ISL_666480, EPI_ISL_666481, EPI_ISL_666482, EPI_ISL_666483, EPI_ISL_666491, EPI_ISL_666493, EPI_ISL_666497, EPI_ISL_666498, EPI_ISL_666499, EPI_ISL_666502, EPI_ISL_666504, EPI_ISL_666505, EPI_ISL_666509, EPI_ISL_666513, EPI_ISL_666514, EPI_ISL_666516, EPI_ISL_666518, EPI_ISL_666520, EPI_ISL_666525, EPI_ISL_666532, EPI_ISL_666536, EPI_ISL_666537, EPI_ISL_666539, EPI_ISL_666547, EPI_ISL_666548, EPI_ISL_666550, EPI_ISL_666551, EPI_ISL_666554, EPI_ISL_666555, EPI_ISL_666565, EPI_ISL_666567, EPI_ISL_666580, EPI_ISL_666581, EPI_ISL_666583, EPI_ISL_666585, EPI_ISL_666586, EPI_ISL_666588, EPI_ISL_666589, EPI_ISL_680582, EPI_ISL_680585, EPI_ISL_680587, EPI_ISL_680590, EPI_ISL_680620, EPI_ISL_680621, EPI_ISL_680622, EPI_ISL_680623, EPI_ISL_680624, EPI_ISL_680625, EPI_ISL_680626, EPI_ISL_680627, EPI_ISL_680628, EPI_ISL_680629, EPI_ISL_680630, EPI_ISL_680631, EPI_ISL_680632, EPI_ISL_680633, EPI_ISL_680684, EPI_ISL_680685, EPI_ISL_680714, EPI_ISL_680715, EPI_ISL_680716, EPI_ISL_680717, EPI_ISL_680718, EPI_ISL_680719, EPI_ISL_680720, EPI_ISL_680721, EPI_ISL_680722, EPI_ISL_680723, EPI_ISL_680724, EPI_ISL_680725, EPI_ISL_680726, EPI_ISL_680727, EPI_ISL_680728, EPI_ISL_680729, EPI_ISL_680730, EPI_ISL_680731, EPI_ISL_680732, EPI_ISL_680733, EPI_ISL_680734, EPI_ISL_680735, EPI_ISL_680736, EPI_ISL_680737, EPI_ISL_680738, EPI_ISL_680740, EPI_ISL_680959, EPI_ISL_680960, EPI_ISL_680961, EPI_ISL_680962, EPI_ISL_680963, EPI_ISL_680964, EPI_ISL_680965, EPI_ISL_680966, EPI_ISL_680967, EPI_ISL_680968, EPI_ISL_681028, EPI_ISL_681029, EPI_ISL_681030, EPI_ISL_681031, EPI_ISL_681032, EPI_ISL_681033, EPI_ISL_681034, EPI_ISL_681035, EPI_ISL_681036, EPI_ISL_681037, EPI_ISL_681038, EPI_ISL_681039, EPI_ISL_702890, EPI_ISL_702896, EPI_ISL_703227, EPI_ISL_703299, EPI_ISL_703346, EPI_ISL_703627, EPI_ISL_703642, EPI_ISL_703855, EPI_ISL_703902, EPI_ISL_704017, EPI_ISL_704171, EPI_ISL_704320, EPI_ISL_705265, EPI_ISL_705390, EPI_ISL_705461, EPI_ISL_705462, EPI_ISL_706442, EPI_ISL_706471, EPI_ISL_706472, EPI_ISL_706473, EPI_ISL_706474, EPI_ISL_706475, EPI_ISL_706476, EPI_ISL_706477, EPI_ISL_706478, EPI_ISL_706479, EPI_ISL_706480, EPI_ISL_706481, EPI_ISL_706482, EPI_ISL_706483, EPI_ISL_706659, EPI_ISL_706666, EPI_ISL_706667, EPI_ISL_706668, EPI_ISL_706669, EPI_ISL_706670, EPI_ISL_706671, EPI_ISL_706672, EPI_ISL_706673, EPI_ISL_706674, EPI_ISL_706679, EPI_ISL_706680, EPI_ISL_706681, EPI_ISL_706682, EPI_ISL_726017, EPI_ISL_742300, EPI_ISL_742301, EPI_ISL_742308, EPI_ISL_742309, EPI_ISL_742310, EPI_ISL_742311, EPI_ISL_742312, EPI_ISL_742313, EPI_ISL_742314, EPI_ISL_742315, EPI_ISL_742316, EPI_ISL_814836 | | | |
| see above | Wales Specialist Virology Centre Sequencing lab: Pathogen Genomics Unit | COVID-19 Genomics UK (COG-UK) Consortium | Alec Birchley; Alexander Adams; Amy Gaskin; Angela Marchbank; Bree Gatica-Wilcox; Catherine Moore; Jason Coombes; Joanne Watkins; Joel Southgate; Johnathan Evans; Laura Gifford; Lauren Gilbert; Lee Graham; Malorie Perry; Matthew Bull; Nicole Pacchiarini; Sally Corden; Sara Kumziene-Summerhayes; Sara Rey; Sarah Taylor; Simon Cottrell; Sophie Jones; Tom Connor |
| EPI_ISL_650146, EPI_ISL_650164, EPI_ISL_650293, EPI_ISL_650297, EPI_ISL_650298, EPI_ISL_650310, EPI_ISL_650316, EPI_ISL_650375, EPI_ISL_650424, EPI_ISL_650482, EPI_ISL_650506, EPI_ISL_650507, EPI_ISL_650508, EPI_ISL_650511, EPI_ISL_650514, EPI_ISL_650538, EPI_ISL_650550, EPI_ISL_650709, EPI_ISL_650727, EPI_ISL_650733, EPI_ISL_650735, EPI_ISL_650736, EPI_ISL_650748, EPI_ISL_650749, EPI_ISL_650750, EPI_ISL_650751, EPI_ISL_650752, EPI_ISL_650753, EPI_ISL_650754, EPI_ISL_650759, EPI_ISL_650764, EPI_ISL_650789, EPI_ISL_650804, EPI_ISL_650805, EPI_ISL_650837, EPI_ISL_650864, EPI_ISL_650865, EPI_ISL_650866, EPI_ISL_650867, EPI_ISL_650868, EPI_ISL_650869, EPI_ISL_650870, EPI_ISL_650871, EPI_ISL_650874, EPI_ISL_650875, EPI_ISL_650925, EPI_ISL_650963, EPI_ISL_651125, EPI_ISL_651303, EPI_ISL_651315, EPI_ISL_651359, EPI_ISL_651380, EPI_ISL_651406, EPI_ISL_651534, EPI_ISL_651535, EPI_ISL_651536, EPI_ISL_651537, EPI_ISL_651811, EPI_ISL_651812, EPI_ISL_651813, EPI_ISL_651821, EPI_ISL_651822, EPI_ISL_651823, EPI_ISL_651824, EPI_ISL_651825, EPI_ISL_651826, EPI_ISL_651827, EPI_ISL_651828, EPI_ISL_651829, EPI_ISL_651830, EPI_ISL_651831, EPI_ISL_651841, EPI_ISL_651842, EPI_ISL_651843, EPI_ISL_651844, EPI_ISL_651848, EPI_ISL_651849, EPI_ISL_651850, EPI_ISL_651851, EPI_ISL_651852, EPI_ISL_651853, EPI_ISL_651854, EPI_ISL_651855, EPI_ISL_651856, EPI_ISL_651857, EPI_ISL_651858, EPI_ISL_651859, EPI_ISL_651860, EPI_ISL_651861, EPI_ISL_651862, EPI_ISL_651863, EPI_ISL_651864, EPI_ISL_651865, EPI_ISL_651866, EPI_ISL_651867, EPI_ISL_651868, EPI_ISL_651869, EPI_ISL_651870, EPI_ISL_651871, EPI_ISL_651872, EPI_ISL_651873, EPI_ISL_651882, EPI_ISL_651883, EPI_ISL_651887, EPI_ISL_651890, EPI_ISL_651891, EPI_ISL_651892, EPI_ISL_651900, EPI_ISL_651928, EPI_ISL_651930, EPI_ISL_651946, EPI_ISL_680442, EPI_ISL_680443, EPI_ISL_680444, EPI_ISL_703755, EPI_ISL_704089, EPI_ISL_704952, EPI_ISL_705316 | | | |
| see above | West of Scotland Specialist Virology Centre, NHSGGC / MRC-University of Glasgow Centre for Virus Research | COVID-19 Genomics UK (COG-UK) Consortium | Alasdair MacLean; Alice Broos; Ana da Silva Filipe; Antonia Ho; Daniel Mair; David L Robertson; Emma Thomson; Guy Mollett; James Shepherd; Jenna Nichols; Joseph Hughes; Kathy Li; Kathy Smollett; Kyriaki Nomikou; Lily Tong; Matthew Holden; Natasha Johnson; Rachel Blacow; Richard Orton; Rory Gunson; Sarah McDonald; Sharif Shaaban; Sreenu Vattipally; Stephen Carmichael |

We gratefully acknowledge the following Authors from the Originating laboratories responsible for obtaining the specimens, as well as the Submitting laboratories where the genome data were generated and shared via GISAID, on which this research is based.

All Submitters of data may be contacted directly via www.gisaid.org

Authors are sorted alphabetically.

| Accession ID | Originating Laboratory | Submitting Laboratory | Authors |
|---|---|---|---|
| EPI_ISL_842869, EPI_ISL_842870, EPI_ISL_842871, EPI_ISL_842872, EPI_ISL_842873, EPI_ISL_842874, EPI_ISL_842875, EPI_ISL_842876, EPI_ISL_842877, EPI_ISL_842878, EPI_ISL_842879, EPI_ISL_842880, EPI_ISL_842881, EPI_ISL_842882, EPI_ISL_842883, EPI_ISL_842884, EPI_ISL_842885, EPI_ISL_842886, EPI_ISL_842887, EPI_ISL_842888, EPI_ISL_842889, EPI_ISL_842890, EPI_ISL_842891, EPI_ISL_842892, EPI_ISL_842893, EPI_ISL_842894, EPI_ISL_842895, EPI_ISL_842896, EPI_ISL_842897, EPI_ISL_842898, EPI_ISL_842902, EPI_ISL_842904, EPI_ISL_842905, EPI_ISL_842906, EPI_ISL_842907, EPI_ISL_842908, EPI_ISL_842909, EPI_ISL_842919, EPI_ISL_842920, EPI_ISL_842921, EPI_ISL_842922, EPI_ISL_842932, EPI_ISL_949770, EPI_ISL_1048105, EPI_ISL_1636885, EPI_ISL_1636886, EPI_ISL_1636887, EPI_ISL_1636913, EPI_ISL_1636915, EPI_ISL_1636916, EPI_ISL_1636917, EPI_ISL_1636918, EPI_ISL_1636987 | | | |
| see above | Barts Health NHS Trust | COVID-19 Genomics UK (COG-UK) Consortium | BROAD; Beatrix; CUTINO-MOGUEL; Claire; David; Dola; HARRINGTON; KELE; KULASEGARAN-SHYLINI; Maria-Teresa; OWOYEMI; Raghavendran; SHYLINI |
| EPI_ISL_1476854 | Bioinformatics and Biostatistics Lab, Advanced Sequencing Facility | COVID-19 Genomics UK (COG-UK) Consortium | Aengus Stewart; Chelsea Sawyer; Harshil Patel; Jerome Nicod; Laura Cubitt; Margaret Crawford |
| EPI_ISL_650577, EPI_ISL_650581, EPI_ISL_650956, EPI_ISL_650981, EPI_ISL_651066, EPI_ISL_665348, EPI_ISL_727697, EPI_ISL_727698, EPI_ISL_727699, EPI_ISL_727700, EPI_ISL_727701, EPI_ISL_727735, EPI_ISL_727736, EPI_ISL_727737, EPI_ISL_727739, EPI_ISL_727740, EPI_ISL_727741, EPI_ISL_727742, EPI_ISL_727743, EPI_ISL_727744, EPI_ISL_727745, EPI_ISL_727746, EPI_ISL_727747, EPI_ISL_727748, EPI_ISL_727749, EPI_ISL_741724, EPI_ISL_741725, EPI_ISL_741726, EPI_ISL_741727, EPI_ISL_741728, EPI_ISL_741729, EPI_ISL_741735, EPI_ISL_741742, EPI_ISL_741745, EPI_ISL_741746, EPI_ISL_741753, EPI_ISL_741755, EPI_ISL_867943, EPI_ISL_867945, EPI_ISL_867962, EPI_ISL_923213, EPI_ISL_923216, EPI_ISL_923226, EPI_ISL_952403, EPI_ISL_1000280, EPI_ISL_1730876 | | | |
| see above | Centre for Enzyme Innovation, University of Portsmouth / Translational Research Laboratory, Portsmouth Hospitals NHS Trust | COVID-19 Genomics UK (COG-UK) Consortium | Allyson Lloyd; Angela Beckett; Anoop Chauhan; Christopher Fearn; Ethan Butcher; Garry Scarlett; Kate Cook; Katie Loveson; Kelly Bicknell; Robert Impey; Salman Goudarzi; Samuel Robson; Sarah Wyllie; Scott Elliott; Sharon Glaysher; Yann Bourgeois |
| EPI_ISL_650396, EPI_ISL_650612, EPI_ISL_650707, EPI_ISL_650708, EPI_ISL_650962, EPI_ISL_650988, EPI_ISL_650989, EPI_ISL_650992, EPI_ISL_651009, EPI_ISL_651010, EPI_ISL_651550, EPI_ISL_651551, EPI_ISL_651552, EPI_ISL_651553, EPI_ISL_651554, EPI_ISL_651555, EPI_ISL_651558, EPI_ISL_651559, EPI_ISL_651561, EPI_ISL_651563, EPI_ISL_651565, EPI_ISL_651566, EPI_ISL_651567, EPI_ISL_651569, EPI_ISL_651573, EPI_ISL_651575, EPI_ISL_651577, EPI_ISL_651579, EPI_ISL_651582, EPI_ISL_651701, EPI_ISL_651708, EPI_ISL_651709, EPI_ISL_651710, EPI_ISL_651714, EPI_ISL_664109, EPI_ISL_664126, EPI_ISL_664141, EPI_ISL_664143, EPI_ISL_664152, EPI_ISL_664157, EPI_ISL_664160, EPI_ISL_664161, EPI_ISL_664174, EPI_ISL_664177, EPI_ISL_664178, EPI_ISL_664180, EPI_ISL_664181, EPI_ISL_664182, EPI_ISL_664185, EPI_ISL_664198, EPI_ISL_664213, EPI_ISL_664221, EPI_ISL_664237, EPI_ISL_664249, EPI_ISL_664282, EPI_ISL_664286, EPI_ISL_664306, EPI_ISL_664307, EPI_ISL_664308, EPI_ISL_664312, EPI_ISL_664313, EPI_ISL_664314, EPI_ISL_664322, EPI_ISL_664323, EPI_ISL_664324, EPI_ISL_664378, EPI_ISL_664379, EPI_ISL_664397, EPI_ISL_664421, EPI_ISL_664428, EPI_ISL_664432, EPI_ISL_664433, EPI_ISL_664434, EPI_ISL_664453, EPI_ISL_664454, EPI_ISL_664514, EPI_ISL_664518, EPI_ISL_664528, EPI_ISL_664533, EPI_ISL_664534, EPI_ISL_664550, EPI_ISL_664551, EPI_ISL_664567, EPI_ISL_664570, EPI_ISL_664571, EPI_ISL_664572, EPI_ISL_664575, EPI_ISL_664578, EPI_ISL_664579, EPI_ISL_664580, EPI_ISL_664581, EPI_ISL_664584, EPI_ISL_664585, EPI_ISL_664586, EPI_ISL_664587, EPI_ISL_664598, EPI_ISL_664622, EPI_ISL_664625, EPI_ISL_664627, EPI_ISL_664631, EPI_ISL_664647, EPI_ISL_664655, EPI_ISL_664659, EPI_ISL_664660, EPI_ISL_664662, EPI_ISL_664666, EPI_ISL_664680, EPI_ISL_664684, EPI_ISL_664697, EPI_ISL_664719, EPI_ISL_664801, EPI_ISL_664802, EPI_ISL_664803, EPI_ISL_664804, EPI_ISL_664805, EPI_ISL_664806, EPI_ISL_664807, EPI_ISL_664808, EPI_ISL_664809, EPI_ISL_664810, EPI_ISL_664811, EPI_ISL_664812, EPI_ISL_664813, EPI_ISL_664814, EPI_ISL_664815, EPI_ISL_664816, EPI_ISL_664817, EPI_ISL_664818, EPI_ISL_664819, EPI_ISL_664820, EPI_ISL_664821, EPI_ISL_664822, EPI_ISL_664823, EPI_ISL_664824, EPI_ISL_664825, EPI_ISL_664826, EPI_ISL_664827, EPI_ISL_664828, EPI_ISL_664829, EPI_ISL_664830, EPI_ISL_664831, EPI_ISL_664832, EPI_ISL_664834, EPI_ISL_664942, EPI_ISL_664946, EPI_ISL_664947, EPI_ISL_664948, EPI_ISL_664949, EPI_ISL_664950, EPI_ISL_664951, EPI_ISL_664952, EPI_ISL_664953, EPI_ISL_664954, EPI_ISL_664955, EPI_ISL_664956, EPI_ISL_664957, EPI_ISL_664958, EPI_ISL_664959, EPI_ISL_664960, EPI_ISL_664961, EPI_ISL_664962, EPI_ISL_664963, EPI_ISL_664964, EPI_ISL_664965, EPI_ISL_664966, EPI_ISL_664967, EPI_ISL_664968, EPI_ISL_664970, EPI_ISL_664971, EPI_ISL_664972, EPI_ISL_664973, EPI_ISL_664974, EPI_ISL_664975, EPI_ISL_664976, EPI_ISL_664977, EPI_ISL_664978, EPI_ISL_664979, EPI_ISL_664989, EPI_ISL_664990, EPI_ISL_664992, EPI_ISL_664993, EPI_ISL_664999, EPI_ISL_665000, EPI_ISL_665001, EPI_ISL_665002, EPI_ISL_665003, EPI_ISL_678832, EPI_ISL_678833, EPI_ISL_678834, EPI_ISL_678835, EPI_ISL_678836, EPI_ISL_678837, EPI_ISL_678838, EPI_ISL_678839, EPI_ISL_678840, EPI_ISL_678841, EPI_ISL_678842, EPI_ISL_678843, EPI_ISL_678844, EPI_ISL_678845, EPI_ISL_678846, EPI_ISL_678847, EPI_ISL_678848, EPI_ISL_678849, EPI_ISL_678850, EPI_ISL_678851, EPI_ISL_678852, EPI_ISL_678853, EPI_ISL_678854, EPI_ISL_678855, EPI_ISL_678856, EPI_ISL_678857, EPI_ISL_678858, EPI_ISL_678859, EPI_ISL_678860, EPI_ISL_678861, EPI_ISL_678862, EPI_ISL_678863, EPI_ISL_678864, EPI_ISL_678865, EPI_ISL_678866, EPI_ISL_678867, EPI_ISL_678868, EPI_ISL_678869, EPI_ISL_678870, EPI_ISL_678871, EPI_ISL_678872, EPI_ISL_678873, EPI_ISL_678874, EPI_ISL_678875, EPI_ISL_678876, EPI_ISL_678877, EPI_ISL_678878, EPI_ISL_678879, EPI_ISL_678880, EPI_ISL_678881, EPI_ISL_678882, EPI_ISL_678883, EPI_ISL_678884, EPI_ISL_678885, EPI_ISL_678886, EPI_ISL_678887, EPI_ISL_678888, EPI_ISL_678889, EPI_ISL_678890, EPI_ISL_678891, EPI_ISL_678892, EPI_ISL_678893, EPI_ISL_678894, EPI_ISL_678895, EPI_ISL_678896, EPI_ISL_678897, EPI_ISL_678898, EPI_ISL_678899, EPI_ISL_678900, EPI_ISL_678901, EPI_ISL_678902, EPI_ISL_678903, EPI_ISL_678904, EPI_ISL_678905, EPI_ISL_678906, EPI_ISL_678907, EPI_ISL_678908, EPI_ISL_678909, EPI_ISL_678910, EPI_ISL_678911, EPI_ISL_678912, EPI_ISL_678913, EPI_ISL_678914, EPI_ISL_678915, EPI_ISL_679134, EPI_ISL_679135, EPI_ISL_679136, EPI_ISL_679137, EPI_ISL_679138, EPI_ISL_679139, EPI_ISL_679141, EPI_ISL_679142, EPI_ISL_679143, EPI_ISL_679144, EPI_ISL_679145, EPI_ISL_679146, EPI_ISL_679147, EPI_ISL_679148, EPI_ISL_679149, EPI_ISL_679150, EPI_ISL_679151, EPI_ISL_679152, EPI_ISL_679153, EPI_ISL_679154, EPI_ISL_679155, EPI_ISL_679158, EPI_ISL_702474, EPI_ISL_702723, EPI_ISL_702763, EPI_ISL_702782, EPI_ISL_702787, EPI_ISL_702834, EPI_ISL_702847, EPI_ISL_702860, EPI_ISL_702971, EPI_ISL_702977, EPI_ISL_703093, EPI_ISL_703115, EPI_ISL_703406, EPI_ISL_703432, EPI_ISL_703435, EPI_ISL_703438, EPI_ISL_703440, EPI_ISL_703459, EPI_ISL_703462, EPI_ISL_703465, EPI_ISL_703468, EPI_ISL_703471, EPI_ISL_703473, EPI_ISL_703513, EPI_ISL_703516, EPI_ISL_703519, EPI_ISL_703521, EPI_ISL_703715, EPI_ISL_704076, EPI_ISL_704219, EPI_ISL_704222, EPI_ISL_704225, EPI_ISL_704273, EPI_ISL_704276, EPI_ISL_704280, EPI_ISL_704283, EPI_ISL_704286, EPI_ISL_704389, EPI_ISL_704392, EPI_ISL_704395, EPI_ISL_704427, EPI_ISL_704430, EPI_ISL_704433, EPI_ISL_704435, EPI_ISL_704438, EPI_ISL_704441, EPI_ISL_704443, EPI_ISL_704446, EPI_ISL_704449, EPI_ISL_704452, EPI_ISL_704454, EPI_ISL_704457, EPI_ISL_704460, EPI_ISL_704462, EPI_ISL_704465, EPI_ISL_704467, EPI_ISL_704957, EPI_ISL_705140, EPI_ISL_705257, EPI_ISL_705290, EPI_ISL_705333, EPI_ISL_705346, EPI_ISL_705365, EPI_ISL_705382, EPI_ISL_705529, EPI_ISL_705530, EPI_ISL_705531, EPI_ISL_705532, EPI_ISL_705533, EPI_ISL_705534, EPI_ISL_705535 | | | |
| see above | Department of Pathology, University of Cambridge | COVID-19 Genomics UK (COG-UK) Consortium | Aminu S. Jahun; Grant Hall; Ian Goodfellow; Iliana Georgana; Malte Pinckert; Martin D. Curran; Myra Hosmillo; Surendra Parmar; Yasmin Chaudhry |
| EPI_ISL_649193, EPI_ISL_649194, EPI_ISL_649195, EPI_ISL_649196, EPI_ISL_649197, EPI_ISL_649198, EPI_ISL_649199, EPI_ISL_649200, EPI_ISL_649201, EPI_ISL_649202, EPI_ISL_649203, EPI_ISL_649204, EPI_ISL_649206, EPI_ISL_649207, EPI_ISL_649208, EPI_ISL_649209, EPI_ISL_649210, EPI_ISL_649211, EPI_ISL_649212, EPI_ISL_649213, EPI_ISL_649214, EPI_ISL_649215, EPI_ISL_649216, EPI_ISL_649217, EPI_ISL_649218, EPI_ISL_649221, EPI_ISL_649222, EPI_ISL_649223, EPI_ISL_649224, EPI_ISL_649225, EPI_ISL_649226, EPI_ISL_649227, EPI_ISL_649229, EPI_ISL_649230, EPI_ISL_649231, EPI_ISL_649232, EPI_ISL_649233, EPI_ISL_649235, EPI_ISL_649236, EPI_ISL_649237, EPI_ISL_649238, EPI_ISL_649239, EPI_ISL_649240, EPI_ISL_649241, EPI_ISL_649242, EPI_ISL_649243, EPI_ISL_649244, EPI_ISL_649245, EPI_ISL_649246, EPI_ISL_649247, EPI_ISL_649248, EPI_ISL_649249, EPI_ISL_649250, EPI_ISL_649251, EPI_ISL_649253, EPI_ISL_649254, EPI_ISL_649256, EPI_ISL_649258, EPI_ISL_649259, EPI_ISL_649260, EPI_ISL_649262, EPI_ISL_649263, EPI_ISL_649264, EPI_ISL_649265, EPI_ISL_649266, EPI_ISL_649267, EPI_ISL_649268, EPI_ISL_649269, EPI_ISL_649270, EPI_ISL_649271, EPI_ISL_649272, EPI_ISL_649273, EPI_ISL_649274, EPI_ISL_649275, EPI_ISL_649276, EPI_ISL_649277, EPI_ISL_649278, EPI_ISL_649279, EPI_ISL_649280, EPI_ISL_649281, EPI_ISL_649282, EPI_ISL_649284, EPI_ISL_649285, EPI_ISL_649286, EPI_ISL_649287, EPI_ISL_649288, EPI_ISL_649289, EPI_ISL_649290, EPI_ISL_649291, EPI_ISL_649292, EPI_ISL_649293, EPI_ISL_649294, EPI_ISL_649295, EPI_ISL_649296, EPI_ISL_649297, EPI_ISL_649298, EPI_ISL_649299, EPI_ISL_649300, EPI_ISL_649301, EPI_ISL_649302, EPI_ISL_649303, EPI_ISL_649304, EPI_ISL_649306, EPI_ISL_649307, EPI_ISL_649308, EPI_ISL_649310, EPI_ISL_649311, EPI_ISL_649313, EPI_ISL_649314, EPI_ISL_649315, EPI_ISL_649316, EPI_ISL_649317, EPI_ISL_649318, EPI_ISL_649319, EPI_ISL_649322, EPI_ISL_649323, EPI_ISL_649324, EPI_ISL_649325, EPI_ISL_649326, EPI_ISL_649327, EPI_ISL_649328, EPI_ISL_649329, EPI_ISL_649330, EPI_ISL_649331, EPI_ISL_649332, EPI_ISL_649333, EPI_ISL_649334, EPI_ISL_649335, EPI_ISL_649336, EPI_ISL_649337, EPI_ISL_649338, EPI_ISL_649339, EPI_ISL_649340, EPI_ISL_649341, EPI_ISL_649342, EPI_ISL_649343, EPI_ISL_649344, EPI_ISL_649345, EPI_ISL_649346, EPI_ISL_649347, EPI_ISL_649348, EPI_ISL_649349, EPI_ISL_649350, EPI_ISL_649351, EPI_ISL_649352, EPI_ISL_649353, EPI_ISL_649355, EPI_ISL_649357, EPI_ISL_649358, EPI_ISL_649359, EPI_ISL_649360, EPI_ISL_649361, EPI_ISL_649363, EPI_ISL_649364, EPI_ISL_649365, EPI_ISL_649366, EPI_ISL_649367, EPI_ISL_649371, EPI_ISL_649372, EPI_ISL_649373, EPI_ISL_649374, EPI_ISL_649375, EPI_ISL_649376, EPI_ISL_649377, EPI_ISL_649378, EPI_ISL_649379, EPI_ISL_649380, EPI_ISL_649381, EPI_ISL_649382, EPI_ISL_649383, EPI_ISL_649384, EPI_ISL_649385, EPI_ISL_649387, EPI_ISL_649388, EPI_ISL_649390, EPI_ISL_649391, EPI_ISL_649392, EPI_ISL_649393, EPI_ISL_649394, EPI_ISL_649395, EPI_ISL_649396, EPI_ISL_649397, EPI_ISL_649398, EPI_ISL_649399, EPI_ISL_649400, EPI_ISL_649401, EPI_ISL_649402, EPI_ISL_649403, EPI_ISL_649404, EPI_ISL_649406, EPI_ISL_649407, EPI_ISL_649408, EPI_ISL_649409, EPI_ISL_649410, EPI_ISL_649411, EPI_ISL_649412, EPI_ISL_649413, EPI_ISL_649414, EPI_ISL_649415, EPI_ISL_649416, EPI_ISL_649417, EPI_ISL_649418, EPI_ISL_649419, EPI_ISL_649420, EPI_ISL_649421, EPI_ISL_649422, EPI_ISL_649423, EPI_ISL_649425, EPI_ISL_649427, EPI_ISL_649428, EPI_ISL_649429, EPI_ISL_649430, EPI_ISL_649431, EPI_ISL_649433, EPI_ISL_649434, EPI_ISL_649435, EPI_ISL_649436, EPI_ISL_649437, EPI_ISL_649438, EPI_ISL_649439, EPI_ISL_649440, EPI_ISL_649441, EPI_ISL_649442, EPI_ISL_649443, EPI_ISL_649444, EPI_ISL_649445, EPI_ISL_649446, EPI_ISL_649447, EPI_ISL_649448, EPI_ISL_649449, EPI_ISL_649450, EPI_ISL_649451, EPI_ISL_649452, EPI_ISL_649453, EPI_ISL_649454, EPI_ISL_649455, EPI_ISL_649456, EPI_ISL_649458, EPI_ISL_649459, EPI_ISL_649460, EPI_ISL_649461, EPI_ISL_649462, EPI_ISL_649463, EPI_ISL_649464, EPI_ISL_649465, EPI_ISL_649466, EPI_ISL_649467, EPI_ISL_649468, EPI_ISL_649469, EPI_ISL_649470, EPI_ISL_649471, EPI_ISL_649472, EPI_ISL_649473, EPI_ISL_649474, EPI_ISL_649475, EPI_ISL_649476, EPI_ISL_649477, EPI_ISL_649478, EPI_ISL_649480, EPI_ISL_649481, EPI_ISL_655273, EPI_ISL_655274, EPI_ISL_655275, EPI_ISL_655276, EPI_ISL_655277, EPI_ISL_655278, EPI_ISL_655279, EPI_ISL_655280, EPI_ISL_655281, EPI_ISL_655282, EPI_ISL_655283, EPI_ISL_655284, EPI_ISL_655285, EPI_ISL_655286, EPI_ISL_655287, EPI_ISL_655288, EPI_ISL_655289, EPI_ISL_655290, EPI_ISL_655291, EPI_ISL_655292, EPI_ISL_655293, EPI_ISL_655294, EPI_ISL_655295, EPI_ISL_655296, EPI_ISL_655297, EPI_ISL_655298, EPI_ISL_655299, EPI_ISL_655300, EPI_ISL_655301, EPI_ISL_655302, EPI_ISL_655303, EPI_ISL_655304, EPI_ISL_655305, EPI_ISL_655306, EPI_ISL_655307, EPI_ISL_655308, EPI_ISL_655309, EPI_ISL_655310, EPI_ISL_655311, EPI_ISL_655312, EPI_ISL_655313, EPI_ISL_655314, EPI_ISL_655315, EPI_ISL_655316, EPI_ISL_655317, EPI_ISL_655318, EPI_ISL_655319, EPI_ISL_655320, EPI_ISL_655321, EPI_ISL_655322, EPI_ISL_655323, EPI_ISL_655324, EPI_ISL_655325, EPI_ISL_655326, EPI_ISL_655327, EPI_ISL_655328, EPI_ISL_655329, EPI_ISL_655330, EPI_ISL_655331, EPI_ISL_655332, EPI_ISL_655333, EPI_ISL_655334, EPI_ISL_655335, EPI_ISL_655336, EPI_ISL_655337, EPI_ISL_655338, EPI_ISL_655339, EPI_ISL_655340, EPI_ISL_655341, EPI_ISL_655342, EPI_ISL_655343, EPI_ISL_655344, EPI_ISL_655345, EPI_ISL_655346, EPI_ISL_655347, EPI_ISL_655348, EPI_ISL_655349, EPI_ISL_655350, EPI_ISL_655351, EPI_ISL_655352, EPI_ISL_655353, EPI_ISL_655354, EPI_ISL_655355, EPI_ISL_655356, EPI_ISL_655357, EPI_ISL_655358, EPI_ISL_655359, EPI_ISL_655360, EPI_ISL_655361, EPI_ISL_655362, EPI_ISL_655363, EPI_ISL_655364, EPI_ISL_655365, EPI_ISL_655366, EPI_ISL_655367, EPI_ISL_655368, EPI_ISL_655369, EPI_ISL_655370, EPI_ISL_655371, EPI_ISL_655372, EPI_ISL_655373, EPI_ISL_655374, EPI_ISL_655375, EPI_ISL_655376, EPI_ISL_655377, EPI_ISL_655378, EPI_ISL_655379, EPI_ISL_655380, EPI_ISL_655381, EPI_ISL_655382, EPI_ISL_655383, EPI_ISL_655384, EPI_ISL_655385, EPI_ISL_655386, EPI_ISL_655387, EPI_ISL_655388, EPI_ISL_655389, EPI_ISL_655390, EPI_ISL_655391, EPI_ISL_655392, EPI_ISL_655393, EPI_ISL_655394, EPI_ISL_655395, EPI_ISL_655396, EPI_ISL_655397, EPI_ISL_655398, EPI_ISL_655399, EPI_ISL_655400, EPI_ISL_655401, EPI_ISL_655402, EPI_ISL_655403, EPI_ISL_655404, EPI_ISL_655405, EPI_ISL_655406, EPI_ISL_655407, EPI_ISL_655408, EPI_ISL_655409, EPI_ISL_655410, EPI_ISL_655411, EPI_ISL_655412, EPI_ISL_655413, EPI_ISL_655414, EPI_ISL_655415, EPI_ISL_655416, EPI_ISL_655417, EPI_ISL_655418, EPI_ISL_655419, EPI_ISL_655420, EPI_ISL_655421, EPI_ISL_655422, EPI_ISL_655423, EPI_ISL_655424, EPI_ISL_655425, EPI_ISL_655426, EPI_ISL_655427, EPI_ISL_655428, EPI_ISL_655429, EPI_ISL_655430, EPI_ISL_655431, EPI_ISL_655432, EPI_ISL_655433, EPI_ISL_655434, EPI_ISL_655435, EPI_ISL_655436, EPI_ISL_655437, EPI_ISL_655438, EPI_ISL_655439, EPI_ISL_655440, EPI_ISL_655441, EPI_ISL_655442, EPI_ISL_655443, EPI_ISL_655444, EPI_ISL_655445, EPI_ISL_655446, EPI_ISL_655447, EPI_ISL_655448, EPI_ISL_655449, EPI_ISL_655450, EPI_ISL_655451, EPI_ISL_655452, EPI_ISL_655453, EPI_ISL_655454, EPI_ISL_655455, EPI_ISL_655456, EPI_ISL_655457, EPI_ISL_655458, EPI_ISL_655459, EPI_ISL_655460, EPI_ISL_655461, EPI_ISL_655462, EPI_ISL_655463, EPI_ISL_655464, EPI_ISL_655465, EPI_ISL_655466, EPI_ISL_655467, EPI_ISL_655468, EPI_ISL_655469, EPI_ISL_655470, EPI_ISL_655471, EPI_ISL_655472, EPI_ISL_655473, EPI_ISL_655474, EPI_ISL_655475, EPI_ISL_655476, EPI_ISL_655477, EPI_ISL_655478, EPI_ISL_655479, EPI_ISL_655480, EPI_ISL_655481, EPI_ISL_655482, EPI_ISL_655483, EPI_ISL_655484, EPI_ISL_655485, EPI_ISL_655486, EPI_ISL_655487, EPI_ISL_655488, EPI_ISL_655489, EPI_ISL_655490, EPI_ISL_655491, EPI_ISL_655492, EPI_ISL_655493, EPI_ISL_655494, EPI_ISL_655495, EPI_ISL_655496, EPI_ISL_655497, EPI_ISL_655498, EPI_ISL_655499, EPI_ISL_655500, EPI_ISL_655501, EPI_ISL_655502, EPI_ISL_655503, EPI_ISL_655504, EPI_ISL_655505, EPI_ISL_655506, EPI_ISL_655507, EPI_ISL_655508, EPI_ISL_655509, EPI_ISL_655510, EPI_ISL_655511, EPI_ISL_655512, EPI_ISL_655513, EPI_ISL_655514, EPI_ISL_655515, EPI_ISL_655516, EPI_ISL_655517, EPI_ISL_655518, EPI_ISL_655519, EPI_ISL_655520, EPI_ISL_655521, EPI_ISL_655522, EPI_ISL_655523, EPI_ISL_655524, EPI_ISL_655525, EPI_ISL_655526, EPI_ISL_655527, EPI_ISL_655528, EPI_ISL_655529, EPI_ISL_655530, EPI_ISL_655531, EPI_ISL_655532, EPI_ISL_655533, EPI_ISL_655534, EPI_ISL_655535, EPI_ISL_655536, EPI_ISL_655537, EPI_ISL_655538, EPI_ISL_655539, EPI_ISL_655540, EPI_ISL_655541, EPI_ISL_655542, EPI_ISL_655543, EPI_ISL_655544, EPI_ISL_655545, EPI_ISL_655546, EPI_ISL_655547, EPI_ISL_655548, EPI_ISL_655549, EPI_ISL_655550, EPI_ISL_655551, EPI_ISL_655552, EPI_ISL_655553, EPI_ISL_655554, EPI_ISL_655555, EPI_ISL_655556, EPI_ISL_655557, EPI_ISL_655558, EPI_ISL_655559, EPI_ISL_655560, EPI_ISL_655561, EPI_ISL_655562, EPI_ISL_655563, EPI_ISL_655564, EPI_ISL_655565, EPI_ISL_655566, EPI_ISL_655567, EPI_ISL_655568, EPI_ISL_655569, EPI_ISL_655570, EPI_ISL_655571, EPI_ISL_655572, EPI_ISL_655573, EPI_ISL_655574, EPI_ISL_655575, EPI_ISL_655576, EPI_ISL_655577, EPI_ISL_655578, EPI_ISL_655579, EPI_ISL_655580, EPI_ISL_655581, EPI_ISL_655582, EPI_ISL_655583, EPI_ISL_655584, EPI_ISL_655585, EPI_ISL_655586, EPI_ISL_655587, EPI_ISL_655588, EPI_ISL_655589, EPI_ISL_655590, EPI_ISL_655591, EPI_ISL_655592, EPI_ISL_655593, EPI_ISL_655594, EPI_ISL_655595, EPI_ISL_655596, EPI_ISL_655597, EPI_ISL_655598, EPI_ISL_655599, EPI_ISL_655600, EPI_ISL_655601, EPI_ISL_655602, EPI_ISL_655603, EPI_ISL_655604, EPI_ISL_655605, EPI_ISL_655606, EPI_ISL_655607, EPI_ISL_655608, EPI_ISL_655609, EPI_ISL_655610, EPI_ISL_655611, EPI_ISL_655612, EPI_ISL_655613, EPI_ISL_655614, EPI_ISL_655615, EPI_ISL_655616, EPI_ISL_655617, EPI_ISL_655618, EPI_ISL_655619, EPI_ISL_655620, EPI_ISL_655621, EPI_ISL_655622, EPI_ISL_655623, EPI_ISL_655624, EPI_ISL_655625, EPI_ISL_655626, EPI_ISL_655627, EPI_ISL_655628, EPI_ISL_655629, EPI_ISL_655630, EPI_ISL_655631, EPI_ISL_655632, EPI_ISL_655633, EPI_ISL_655634, EPI_ISL_655635, EPI_ISL_655636, EPI_ISL_655637, EPI_ISL_655638, EPI_ISL_655639, EPI_ISL_655640, EPI_ISL_655641, EPI_ISL_655642, EPI_ISL_655643, EPI_ISL_655644, EPI_ISL_655645, EPI_ISL_655646, EPI_ISL_655647, EPI_ISL_655648, EPI_ISL_655649, EPI_ISL_655650, EPI_ISL_655652, EPI_ISL_655653, EPI_ISL_655654, EPI_ISL_655655, EPI_ISL_655656, EPI_ISL_655657, EPI_ISL_655658, EPI_ISL_655659, EPI_ISL_655661, EPI_ISL_655663, EPI_ISL_655664, EPI_ISL_655665, EPI_ISL_655666, EPI_ISL_655667, EPI_ISL_655668, EPI_ISL_655669, EPI_ISL_655670, EPI_ISL_655671, EPI_ISL_655673, EPI_ISL_655674, EPI_ISL_655675, EPI_ISL_655676, EPI_ISL_655678, EPI_ISL_655679, EPI_ISL_655680, EPI_ISL_655681, EPI_ISL_655682, EPI_ISL_655683, EPI_ISL_655684, EPI_ISL_655685, EPI_ISL_655686, EPI_ISL_655687, EPI_ISL_655688, EPI_ISL_655689, EPI_ISL_655690, EPI_ISL_655691, EPI_ISL_655692, EPI_ISL_655693, EPI_ISL_655696, EPI_ISL_655698, EPI_ISL_655700, EPI_ISL_655701, EPI_ISL_655702, EPI_ISL_655703, EPI_ISL_655704, EPI_ISL_655705, EPI_ISL_655706, EPI_ISL_655707, EPI_ISL_655708, EPI_ISL_655709, EPI_ISL_655710, EPI_ISL_655712, EPI_ISL_655713, EPI_ISL_655714, EPI_ISL_655715, EPI_ISL_655716, EPI_ISL_655717, EPI_ISL_655718, EPI_ISL_655720, EPI_ISL_655721, EPI_ISL_655722, EPI_ISL_655724, EPI_ISL_655725, EPI_ISL_655726, EPI_ISL_655728, EPI_ISL_655729, EPI_ISL_655730, EPI_ISL_655731, EPI_ISL_655732, EPI_ISL_655733, EPI_ISL_655734, EPI_ISL_655736, EPI_ISL_655738, EPI_ISL_655740, EPI_ISL_655741, EPI_ISL_655742, EPI_ISL_655743, EPI_ISL_655746, EPI_ISL_655747, EPI_ISL_655748, EPI_ISL_655749, EPI_ISL_655750, EPI_ISL_655751, EPI_ISL_655752, EPI_ISL_655753, EPI_ISL_655754, EPI_ISL_655755, EPI_ISL_655757, EPI_ISL_655758, EPI_ISL_655759, EPI_ISL_655760, EPI_ISL_655761, EPI_ISL_655763, EPI_ISL_655764, EPI_ISL_655765, EPI_ISL_655766, EPI_ISL_655767, EPI_ISL_655771, EPI_ISL_655772, EPI_ISL_655774, EPI_ISL_655775, EPI_ISL_655776, EPI_ISL_655777, EPI_ISL_655778, EPI_ISL_655781, EPI_ISL_655782, EPI_ISL_655784, EPI_ISL_655785, EPI_ISL_655786, EPI_ISL_655787, EPI_ISL_655789, EPI_ISL_655790, EPI_ISL_655791, EPI_ISL_655792, EPI_ISL_655793, EPI_ISL_655794, EPI_ISL_655795, EPI_ISL_655796, EPI_ISL_655797, EPI_ISL_655799, EPI_ISL_655800, EPI_ISL_655801, EPI_ISL_655802, EPI_ISL_655803, EPI_ISL_655805, EPI_ISL_655808, EPI_ISL_655810, EPI_ISL_655811, EPI_ISL_655812, EPI_ISL_655813, EPI_ISL_655814, EPI_ISL_655815, EPI_ISL_655816, EPI_ISL_655818, EPI_ISL_655820, EPI_ISL_655821, EPI_ISL_655823, EPI_ISL_655824, EPI_ISL_655825, EPI_ISL_655826, EPI_ISL_655827, EPI_ISL_655829, EPI_ISL_655830, EPI_ISL_655831, EPI_ISL_655833, EPI_ISL_655834, EPI_ISL_655835, EPI_ISL_655836, EPI_ISL_655837, EPI_ISL_655838, EPI_ISL_655839, EPI_ISL_655840, EPI_ISL_655842, EPI_ISL_655843, EPI_ISL_655844, EPI_ISL_655845, EPI_ISL_655846, EPI_ISL_655848, EPI_ISL_655849, EPI_ISL_655851, EPI_ISL_655852, EPI_ISL_655853, EPI_ISL_655855, EPI_ISL_655856, EPI_ISL_655857, EPI_ISL_655858, EPI_ISL_655859, EPI_ISL_655860, EPI_ISL_655862, EPI_ISL_655863, EPI_ISL_655866, EPI_ISL_655867, EPI_ISL_655869, EPI_ISL_655870, EPI_ISL_655871, EPI_ISL_655872, EPI_ISL_655875, EPI_ISL_655876, EPI_ISL_655877, EPI_ISL_655878, EPI_ISL_655880, EPI_ISL_655881, EPI_ISL_655882, EPI_ISL_655884, EPI_ISL_655885, EPI_ISL_655886, EPI_ISL_655887, EPI_ISL_655888, EPI_ISL_655889, EPI_ISL_655890, EPI_ISL_655892, EPI_ISL_655893, EPI_ISL_655894, EPI_ISL_655895, EPI_ISL_655896, EPI_ISL_655897, EPI_ISL_655898, EPI_ISL_655899, EPI_ISL_655900, EPI_ISL_655901, EPI_ISL_655902, EPI_ISL_655903, EPI_ISL_655904, EPI_ISL_655905, EPI_ISL_655906, EPI_ISL_655908, EPI_ISL_655909, EPI_ISL_655910, EPI_ISL_655913, EPI_ISL_655914, EPI_ISL_655915, EPI_ISL_655916, EPI_ISL_655917, EPI_ISL_655919, EPI_ISL_655921, EPI_ISL_655922, EPI_ISL_655923, EPI_ISL_655925, EPI_ISL_655926, EPI_ISL_655927, EPI_ISL_655928, EPI_ISL_655930, EPI_ISL_655931, EPI_ISL_655932, EPI_ISL_655934, EPI_ISL_655935, EPI_ISL_655936, EPI_ISL_655937, EPI_ISL_655938, EPI_ISL_655939, EPI_ISL_655940, EPI_ISL_655941, EPI_ISL_655942, EPI_ISL_655943, EPI_ISL_655944, EPI_ISL_655945, EPI_ISL_655946, EPI_ISL_655949, EPI_ISL_655950, EPI_ISL_655951, EPI_ISL_655952, EPI_ISL_655953, EPI_ISL_655954, EPI_ISL_655955, EPI_ISL_655956, EPI_ISL_655958, EPI_ISL_655960, EPI_ISL_655961, EPI_ISL_655962, EPI_ISL_655963, EPI_ISL_655964, EPI_ISL_655965, EPI_ISL_655966, EPI_ISL_655968, EPI_ISL_655969, EPI_ISL_655970, EPI_ISL_655972, EPI_ISL_655974, EPI_ISL_655976, EPI_ISL_655977, EPI_ISL_655978, EPI_ISL_655979, EPI_ISL_655980, EPI_ISL_655981, EPI_ISL_655982, EPI_ISL_655983, EPI_ISL_655984, EPI_ISL_655985, EPI_ISL_655986, EPI_ISL_655987, EPI_ISL_655988, EPI_ISL_655989, EPI_ISL_655990, EPI_ISL_655991, EPI_ISL_655992, EPI_ISL_655993, EPI_ISL_655994, EPI_ISL_655995, EPI_ISL_655996, EPI_ISL_655997, EPI_ISL_655998, EPI_ISL_655999, EPI_ISL_656000, EPI_ISL_656001, EPI_ISL_656002, EPI_ISL_656003, EPI_ISL_656004, EPI_ISL_656005, EPI_ISL_656006, EPI_ISL_656007, EPI_ISL_656008, EPI_ISL_656009, EPI_ISL_656010, EPI_ISL_656011, EPI_ISL_656012, EPI_ISL_656013, EPI_ISL_656014, EPI_ISL_656015, EPI_ISL_656016, EPI_ISL_656017, EPI_ISL_656018, EPI_ISL_656019, EPI_ISL_656020, EPI_ISL_656021, EPI_ISL_656022, EPI_ISL_656023, EPI_ISL_656024, EPI_ISL_656025, EPI_ISL_656026, EPI_ISL_656027, EPI_ISL_656028, EPI_ISL_656029, EPI_ISL_656030, EPI_ISL_656031, EPI_ISL_656032, EPI_ISL_656033, EPI_ISL_656034, EPI_ISL_656035, EPI_ISL_656036, EPI_ISL_656037, EPI_ISL_656038, EPI_ISL_656039, EPI_ISL_656040, EPI_ISL_656041, EPI_ISL_656042, EPI_ISL_656043, EPI_ISL_656044, EPI_ISL_656045, EPI_ISL_656046, EPI_ISL_656047, EPI_ISL_656048, EPI_ISL_656049, EPI_ISL_656050, EPI_ISL_656051, EPI_ISL_656052, EPI_ISL_656053, EPI_ISL_656054, EPI_ISL_656055, EPI_ISL_656056, EPI_ISL_656057, EPI_ISL_656058, EPI_ISL_656059, EPI_ISL_656060, EPI_ISL_656061, EPI_ISL_656062, EPI_ISL_656063, EPI_ISL_656064, EPI_ISL_656065, EPI_ISL_656066, EPI_ISL_656067, EPI_ISL_656068, EPI_ISL_656069, EPI_ISL_656070, EPI_ISL_656071, EPI_ISL_656072, EPI_ISL_656073, EPI_ISL_656074, EPI_ISL_656075, EPI_ISL_656076, EPI_ISL_656077, EPI_ISL_656078, EPI_ISL_656079, EPI_ISL_656080, EPI_ISL_656081, EPI_ISL_656082, EPI_ISL_656083, EPI_ISL_656084, EPI_ISL_656085, EPI_ISL_656086, EPI_ISL_656087, EPI_ISL_656088, EPI_ISL_656089, EPI_ISL_656090, EPI_ISL_656091, EPI_ISL_656092, EPI_ISL_656093, EPI_ISL_656094, EPI_ISL_656095, EPI_ISL_656096, EPI_ISL_656097, EPI_ISL_656098, EPI_ISL_656099, EPI_ISL_656100, EPI_ISL_656101, EPI_ISL_656102, EPI_ISL_656103, EPI_ISL_656104, EPI_ISL_656105, EPI_ISL_656106, EPI_ISL_656107, EPI_ISL_656108, EPI_ISL_656109, EPI_ISL_656110, EPI_ISL_656111, EPI_ISL_656112, EPI_ISL_656113, EPI_ISL_656114, EPI_ISL_656115, EPI_ISL_656116, EPI_ISL_656117, EPI_ISL_656118, EPI_ISL_656119, EPI_ISL_656120, EPI_ISL_656121, EPI_ISL_656122, EPI_ISL_656123, EPI_ISL_656124, EPI_ISL_656125, EPI_ISL_656126, EPI_ISL_656127, EPI_ISL_656128, EPI_ISL_656129, EPI_ISL_656130, EPI_ISL_656131, EPI_ISL_656132, EPI_ISL_656133, EPI_ISL_656134, EPI_ISL_656135, EPI_ISL_656136, EPI_ISL_656137, EPI_ISL_656138, EPI_ISL_656139, EPI_ISL_656140, EPI_ISL_656141, EPI_ISL_656142, EPI_ISL_656143, EPI_ISL_656144, EPI_ISL_656145, EPI_ISL_656146, EPI_ISL_656147, EPI_ISL_656148, EPI_ISL_656149, EPI_ISL_656150, EPI_ISL_656151, EPI_ISL_656152, EPI_ISL_656153, EPI_ISL_656154, EPI_ISL_656155, EPI_ISL_656156, EPI_ISL_656157, EPI_ISL_656158, EPI_ISL_656159, EPI_ISL_656160, EPI_ISL_656161, EPI_ISL_656162, EPI_ISL_656163, EPI_ISL_656164, EPI_ISL_656165, EPI_ISL_656166, EPI_ISL_656167, EPI_ISL_656168, EPI_ISL_656169, EPI_ISL_656170, EPI_ISL_656171, EPI_ISL_656172, EPI_ISL_656173, EPI_ISL_656174, EPI_ISL_656175, EPI_ISL_656176, EPI_ISL_656177, EPI_ISL_656178, EPI_ISL_656179, EPI_ISL_656180, EPI_ISL_656181, EPI_ISL_656182, EPI_ISL_656183, EPI_ISL_656184, EPI_ISL_656185, EPI_ISL_656186, EPI_ISL_656187, EPI_ISL_656188, EPI_ISL_656189, EPI_ISL_656190, EPI_ISL_656191, EPI_ISL_656192, EPI_ISL_656193, EPI_ISL_656194, EPI_ISL_656195, EPI_ISL_656196, EPI_ISL_656197, EPI_ISL_656198, EPI_ISL_656199, EPI_ISL_656200, EPI_ISL_656201, EPI_ISL_656202, EPI_ISL_656203, EPI_ISL_656204, EPI_ISL_656205, EPI_ISL_656206, EPI_ISL_656207, EPI_ISL_656208, EPI_ISL_656209, EPI_ISL_656210, EPI_ISL_656211, EPI_ISL_656212, EPI_ISL_656213, EPI_ISL_656214, EPI_ISL_656215, EPI_ISL_656216, EPI_ISL_656217, EPI_ISL_656218, EPI_ISL_656219, EPI_ISL_656220, EPI_ISL_656221, EPI_ISL_656222, EPI_ISL_656223, EPI_ISL_656224, EPI_ISL_656225, EPI_ISL_656226, EPI_ISL_656227, EPI_ISL_656228, EPI_ISL_656229, EPI_ISL_656230, EPI_ISL_656231, EPI_ISL_656232, EPI_ISL_656233, EPI_ISL_656234, EPI_ISL_656235, EPI_ISL_656236, EPI_ISL_656237, EPI_ISL_656238, EPI_ISL_656239, EPI_ISL_656240, EPI_ISL_656241, EPI_ISL_656242, EPI_ISL_656243, EPI_ISL_656244, EPI_ISL_656245, EPI_ISL_656246, EPI_ISL_656247, EPI_ISL_656248, EPI_ISL_656249, EPI_ISL_656250, EPI_ISL_656251, EPI_ISL_656252, EPI_ISL_656253, EPI_ISL_656254, EPI_ISL_656255, EPI_ISL_656256, EPI_ISL_656257, EPI_ISL_656258, EPI_ISL_656259, EPI_ISL_656260, EPI_ISL_656261, EPI_ISL_656262, EPI_ISL_656263, EPI_ISL_656264, EPI_ISL_656265, EPI_ISL_656266, EPI_ISL_656267, EPI_ISL_656268, EPI_ISL_656269, EPI_ISL_656270, EPI_ISL_656271, EPI_ISL_656272, EPI_ISL_656273, EPI_ISL_656274, EPI_ISL_656275, EPI_ISL_656276, EPI_ISL_656277, EPI_ISL_656278, EPI_ISL_656279, EPI_ISL_656280, EPI_ISL_656281, EPI_ISL_656282, EPI_ISL_656283, EPI_ISL_656284, EPI_ISL_656285, EPI_ISL_656286, EPI_ISL_656287, EPI_ISL_656288, EPI_ISL_656289, EPI_ISL_656290, EPI_ISL_656291, EPI_ISL_656292, EPI_ISL_656293, EPI_ISL_656294, EPI_ISL_656295, EPI_ISL_656296, EPI_ISL_656297, EPI_ISL_656298, EPI_ISL_656299, EPI_ISL_656300, EPI_ISL_656301, EPI_ISL_656302, EPI_ISL_656303, EPI_ISL_656304, EPI_ISL_656305, EPI_ISL_656306, EPI_ISL_656307, EPI_ISL_656308, EPI_ISL_656309, EPI_ISL_656310, EPI_ISL_656311, EPI_ISL_656312, EPI_ISL_656313, EPI_ISL_656314, EPI_ISL_656315, EPI_ISL_656316, EPI_ISL_656317, EPI_ISL_656318, EPI_ISL_656319, EPI_ISL_656320, EPI_ISL_656321, EPI_ISL_656322, EPI_ISL_656323, EPI_ISL_656324, EPI_ISL_656325, EPI_ISL_656326, EPI_ISL_656327, EPI_ISL_656328, EPI_ISL_656329, EPI_ISL_656330, EPI_ISL_656331, EPI_ISL_656332, EPI_ISL_656333, EPI_ISL_656334, EPI_ISL_656335, EPI_ISL_656336, EPI_ISL_656337, EPI_ISL_656338, EPI_ISL_656339, EPI_ISL_656340, EPI_ISL_656341, EPI_ISL_656342, EPI_ISL_656343, EPI_ISL_656344, EPI_ISL_656345, EPI_ISL_656346, EPI_ISL_656347, EPI_ISL_656348, EPI_ISL_656349, EPI_ISL_656350, EPI_ISL_656351, EPI_ISL_656352, EPI_ISL_656353, EPI_ISL_656354, EPI_ISL_656355, EPI_ISL_656356, EPI_ISL_656357, EPI_ISL_656358, EPI_ISL_656359, EPI_ISL_656361, EPI_ISL_656362, EPI_ISL_656363, EPI_ISL_656364, EPI_ISL_656365, EPI_ISL_656366, EPI_ISL_656367, EPI_ISL_656368, EPI_ISL_656369, EPI_ISL_656370, EPI_ISL_656371, EPI_ISL_656372, EPI_ISL_656373, EPI_ISL_656374, EPI_ISL_656375, EPI_ISL_656376, EPI_ISL_656377, EPI_ISL_656378, EPI_ISL_656379, EPI_ISL_656380, EPI_ISL_656382, EPI_ISL_656383, EPI_ISL_656384, EPI_ISL_656385, EPI_ISL_656386, EPI_ISL_656387, EPI_ISL_656388, EPI_ISL_656389, EPI_ISL_656390, EPI_ISL_656391, EPI_ISL_656392, EPI_ISL_656393, EPI_ISL_656394, EPI_ISL_656395, EPI_ISL_656396, EPI_ISL_656397, EPI_ISL_656398, EPI_ISL_656399, EPI_ISL_656400, EPI_ISL_656401, EPI_ISL_656402, EPI_ISL_656403, EPI_ISL_656404, EPI_ISL_656405, EPI_ISL_656406, EPI_ISL_656407, EPI_ISL_656408, EPI_ISL_656409, EPI_ISL_656410, EPI_ISL_656411, EPI_ISL_656412, EPI_ISL_656413, EPI_ISL_656414, EPI_ISL_656415, EPI_ISL_656416, EPI_ISL_656417, EPI_ISL_656418, EPI_ISL_656419, EPI_ISL_656420, EPI_ISL_656421, EPI_ISL_656422, EPI_ISL_656423, EPI_ISL_656424, EPI_ISL_656425, EPI_ISL_656426, EPI_ISL_656427, EPI_ISL_656428, EPI_ISL_656429, EPI_ISL_656430, EPI_ISL_656431, EPI_ISL_656432, EPI_ISL_656433, EPI_ISL_656434, EPI_ISL_656435, EPI_ISL_656436, EPI_ISL_656437, EPI_ISL_656438, EPI_ISL_656439, EPI_ISL_656440, EPI_ISL_656441, EPI_ISL_656442, EPI_ISL_656443, EPI_ISL_656444, EPI_ISL_656445, EPI_ISL_656446, EPI_ISL_656447, EPI_ISL_656448, EPI_ISL_656449, EPI_ISL_656450, EPI_ISL_656451, EPI_ISL_656452, EPI_ISL_656453, EPI_ISL_656454, EPI_ISL_656455, EPI_ISL_656456, EPI_ISL_656457, EPI_ISL_656458, EPI_ISL_656459, EPI_ISL_656460, EPI_ISL_656461, EPI_ISL_656462, EPI_ISL_656463, EPI_ISL_656464, EPI_ISL_656465, EPI_ISL_656466, EPI_ISL_656467, EPI_ISL_656468, EPI_ISL_656469, EPI_ISL_656470, EPI_ISL_656472, EPI_ISL_656473, EPI_ISL_656474, EPI_ISL_656475, EPI_ISL_656476, EPI_ISL_656477, EPI_ISL_656478, EPI_ISL_656479, EPI_ISL_656480, EPI_ISL_656481, EPI_ISL_656482, EPI_ISL_656483, EPI_ISL_656484, EPI_ISL_656485, EPI_ISL_656486, EPI_ISL_656487, EPI_ISL_656488, EPI_ISL_656489, EPI_ISL_656490, EPI_ISL_656491, EPI_ISL_656492, EPI_ISL_656493, EPI_ISL_656494, EPI_ISL_656495, EPI_ISL_656496, EPI_ISL_656497, EPI_ISL_656498, EPI_ISL_656499, EPI_ISL_656500, EPI_ISL_656501, EPI_ISL_656502, EPI_ISL_656503, EPI_ISL_656505, EPI_ISL_656506, EPI_ISL_656507, EPI_ISL_656508, EPI_ISL_656509, EPI_ISL_656510, EPI_ISL_656511, EPI_ISL_656512, EPI_ISL_656513, EPI_ISL_656514, EPI_ISL_656515, EPI_ISL_656516, EPI_ISL_656517, EPI_ISL_656518, EPI_ISL_656519, EPI_ISL_656520, EPI_ISL_656521, EPI_ISL_656522, EPI_ISL_656523, EPI_ISL_656524, EPI_ISL_656525, EPI_ISL_656526, EPI_ISL_656527, EPI_ISL_656528, EPI_ISL_656529, EPI_ISL_656530, EPI_ISL_656531, EPI_ISL_656532, EPI_ISL_656533, EPI_ISL_656534, EPI_ISL_656535, EPI_ISL_656536, EPI_ISL_656537, EPI_ISL_656538, EPI_ISL_656539, EPI_ISL_656540, EPI_ISL_656541, EPI_ISL_656542, EPI_ISL_656543, EPI_ISL_656544, EPI_ISL_656545, EPI_ISL_656546, EPI_ISL_656547, EPI_ISL_656548, EPI_ISL_656549, EPI_ISL_656550, EPI_ISL_656551, EPI_ISL_656552, EPI_ISL_656553, EPI_ISL_656554, EPI_ISL_656555, EPI_ISL_656556, EPI_ISL_656557, EPI_ISL_656558, EPI_ISL_656559, EPI_ISL_656560, EPI_ISL_656561, EPI_ISL_656562, EPI_ISL_656563, EPI_ISL_656564, EPI_ISL_656565, EPI_ISL_656566, EPI_ISL_656567, EPI_ISL_656568, EPI_ISL_656569, EPI_ISL_656570, EPI_ISL_656571, EPI_ISL_656572, EPI_ISL_656573, EPI_ISL_656574, EPI_ISL_656575, EPI_ISL_656576, EPI_ISL_656577, EPI_ISL_656578, EPI_ISL_656579, EPI_ISL_656580, EPI_ISL_656581, EPI_ISL_656582, EPI_ISL_656583, EPI_ISL_656584, EPI_ISL_656585, EPI_ISL_656586, EPI_ISL_656587, EPI_ISL_656588, EPI_ISL_656589, EPI_ISL_656590, EPI_ISL_656591, EPI_ISL_656592, EPI_ISL_656593, EPI_ISL_656594, EPI_ISL_656595, EPI_ISL_656596, EPI_ISL_656597, EPI_ISL_656598, EPI_ISL_656599, EPI_ISL_656600, EPI_ISL_656601, EPI_ISL_656602, EPI_ISL_656603, EPI_ISL_656604, EPI_ISL_656605, EPI_ISL_656606, EPI_ISL_656607, EPI_ISL_656608, EPI_ISL_656609, EPI_ISL_656610, EPI_ISL_656611, EPI_ISL_656612, EPI_ISL_656613, EPI_ISL_656614, EPI_ISL_656615, EPI_ISL_656616, EPI_ISL_656617, EPI_ISL_656618, EPI_ISL_656619, EPI_ISL_656620, EPI_ISL_656621, EPI_ISL_656622, EPI_ISL_656623, EPI_ISL_656624, EPI_ISL_656625, EPI_ISL_656626, EPI_ISL_656627, EPI_ISL_656628, EPI_ISL_656629, EPI_ISL_656630, EPI_ISL_656631, EPI_ISL_656632, EPI_ISL_656633, EPI_ISL_656634, EPI_ISL_656635, EPI_ISL_656636, EPI_ISL_656637, EPI_ISL_656638, EPI_ISL_656639, EPI_ISL_656640, EPI_ISL_656641, EPI_ISL_656642, EPI_ISL_656643, EPI_ISL_656644, EPI_ISL_656645, EPI_ISL_656646, EPI_ISL_656647, EPI_ISL_656648, EPI_ISL_656649, EPI_ISL_656650, EPI_ISL_656651, EPI_ISL_656652, EPI_ISL_656653, EPI_ISL_656654, EPI_ISL_656655, EPI_ISL_656656, EPI_ISL_656657, EPI_ISL_656658, EPI_ISL_656659, EPI_ISL_656660, EPI_ISL_656661, EPI_ISL_656662, EPI_ISL_656663, EPI_ISL_656664, EPI_ISL_656665, EPI_ISL_656666, EPI_ISL_656667, EPI_ISL_656668, EPI_ISL_656669, EPI_ISL_656670, EPI_ISL_656671, EPI_ISL_656672, EPI_ISL_656673, EPI_ISL_656674, EPI_ISL_656675, EPI_ISL_656676, EPI_ISL_656677, EPI_ISL_656678, EPI_ISL_656679, EPI_ISL_656680, EPI_ISL_656681, EPI_ISL_656682, EPI_ISL_656683, EPI_ISL_656684, EPI_ISL_662623, EPI_ISL_662624, EPI_ISL_662625, EPI_ISL_662626, EPI_ISL_662628, EPI_ISL_662629, EPI_ISL_662630, | | | |

EPI_ISL_662647, EPI_ISL_662648, EPI_ISL_662649, EPI_ISL_662650, EPI_ISL_674223, EPI_ISL_674418, EPI_ISL_675483, EPI_ISL_675491, EPI_ISL_675494, EPI_ISL_675514, EPI_ISL_675537, EPI_ISL_675546, EPI_ISL_675559, EPI_ISL_675589, EPI_ISL_675605, EPI_ISL_675633, EPI_ISL_675643, EPI_ISL_675656, EPI_ISL_675681, EPI_ISL_675685, EPI_ISL_675720, EPI_ISL_675721, EPI_ISL_675746, EPI_ISL_675855, EPI_ISL_675925, EPI_ISL_710865, EPI_ISL_710866, EPI_ISL_710869, EPI_ISL_710870, EPI_ISL_710871, EPI_ISL_710873, EPI_ISL_710874, EPI_ISL_710875, EPI_ISL_710876, EPI_ISL_710887, EPI_ISL_710888, EPI_ISL_710889, EPI_ISL_710890, EPI_ISL_710891, EPI_ISL_710892, EPI_ISL_710893, EPI_ISL_710894, EPI_ISL_710895, EPI_ISL_710896, EPI_ISL_710897, EPI_ISL_710898, EPI_ISL_710899, EPI_ISL_710900, EPI_ISL_710901, EPI_ISL_710902, EPI_ISL_782843, EPI_ISL_892218, EPI_ISL_892219

| | | | |
|---|---|---|---|
| see above | Lighthouse Lab in Alderley Park | Wellcome Sanger Institute for the COVID-19 Genomics UK (COG-UK) Consortium | Cordelia Langford; David K. Jackson; Dominic Kwiatkowski; Ewan Harrison; Ian Johnston; Jacquelyn Wynn; John Sillitoe on behalf of the Wellcome Sanger Institute COVID-19 Surveillance Team; John Sillitoe on behalf of the Wellcome Sanger Institute COVID-19 Surveillance Team (http://www.sanger.ac.uk/covid-team); Mairead Hyland; Roberto Amato; Sonia Goncalves; The Lighthouse Lab in Alderley Park and Alex Alderton |

EPI_ISL_645219, EPI_ISL_645220, EPI_ISL_645222, EPI_ISL_645223, EPI_ISL_645224, EPI_ISL_645225, EPI_ISL_645227, EPI_ISL_645228, EPI_ISL_645229, EPI_ISL_645230, EPI_ISL_645231, EPI_ISL_645232, EPI_ISL_645233, EPI_ISL_645234, EPI_ISL_645235, EPI_ISL_645236, EPI_ISL_645237, EPI_ISL_645238, EPI_ISL_645239, EPI_ISL_645242, EPI_ISL_645245, EPI_ISL_645246, EPI_ISL_645247, EPI_ISL_645248, EPI_ISL_645249, EPI_ISL_645250, EPI_ISL_645251, EPI_ISL_645252, EPI_ISL_645253, EPI_ISL_645254, EPI_ISL_645255, EPI_ISL_645256, EPI_ISL_645257, EPI_ISL_645258, EPI_ISL_645259, EPI_ISL_645260, EPI_ISL_645261, EPI_ISL_645262, EPI_ISL_645265, EPI_ISL_645267, EPI_ISL_645268, EPI_ISL_645269, EPI_ISL_645270, EPI_ISL_645271, EPI_ISL_645272, EPI_ISL_645273, EPI_ISL_645274, EPI_ISL_645275, EPI_ISL_645276, EPI_ISL_645277, EPI_ISL_645278, EPI_ISL_645279, EPI_ISL_645280, EPI_ISL_645281, EPI_ISL_645282, EPI_ISL_645283, EPI_ISL_645284, EPI_ISL_645286, EPI_ISL_645287, EPI_ISL_645288, EPI_ISL_645289, EPI_ISL_645290, EPI_ISL_645291, EPI_ISL_645292, EPI_ISL_645293, EPI_ISL_645294, EPI_ISL_645295, EPI_ISL_645298, EPI_ISL_645299, EPI_ISL_645301, EPI_ISL_645304, EPI_ISL_645305, EPI_ISL_645306, EPI_ISL_645307, EPI_ISL_645309, EPI_ISL_645310, EPI_ISL_645311, EPI_ISL_645312, EPI_ISL_645313, EPI_ISL_645314, EPI_ISL_645315, EPI_ISL_645316, EPI_ISL_645317, EPI_ISL_645318, EPI_ISL_645320, EPI_ISL_645321, EPI_ISL_645323, EPI_ISL_645325, EPI_ISL_645326, EPI_ISL_645328, EPI_ISL_645330, EPI_ISL_645331, EPI_ISL_645332, EPI_ISL_645333, EPI_ISL_645334, EPI_ISL_645335, EPI_ISL_645336, EPI_ISL_645337, EPI_ISL_645338, EPI_ISL_645339, EPI_ISL_645340, EPI_ISL_645341, EPI_ISL_645342, EPI_ISL_645343, EPI_ISL_645344, EPI_ISL_645346, EPI_ISL_645347, EPI_ISL_645348, EPI_ISL_645349, EPI_ISL_645350, EPI_ISL_645351, EPI_ISL_645352, EPI_ISL_645353, EPI_ISL_645355, EPI_ISL_645356, EPI_ISL_645357, EPI_ISL_645358, EPI_ISL_645359, EPI_ISL_645361, EPI_ISL_645363, EPI_ISL_645364, EPI_ISL_645365, EPI_ISL_645366, EPI_ISL_645370, EPI_ISL_645371, EPI_ISL_645372, EPI_ISL_645373, EPI_ISL_645376, EPI_ISL_645377, EPI_ISL_645378, EPI_ISL_645379, EPI_ISL_645380, EPI_ISL_645381, EPI_ISL_645382, EPI_ISL_645384, EPI_ISL_645385, EPI_ISL_645386, EPI_ISL_645387, EPI_ISL_645388, EPI_ISL_645389, EPI_ISL_645390, EPI_ISL_645392, EPI_ISL_645394, EPI_ISL_645396, EPI_ISL_645397, EPI_ISL_645398, EPI_ISL_645400, EPI_ISL_645401, EPI_ISL_645402, EPI_ISL_645403, EPI_ISL_645404, EPI_ISL_645407, EPI_ISL_645408, EPI_ISL_645409, EPI_ISL_645410, EPI_ISL_645411, EPI_ISL_645412, EPI_ISL_645413, EPI_ISL_645415, EPI_ISL_645416, EPI_ISL_645417, EPI_ISL_645419, EPI_ISL_645420, EPI_ISL_645422, EPI_ISL_645423, EPI_ISL_645425, EPI_ISL_645426, EPI_ISL_645427, EPI_ISL_645428, EPI_ISL_645429, EPI_ISL_645430, EPI_ISL_645431, EPI_ISL_645432, EPI_ISL_645433, EPI_ISL_645434, EPI_ISL_645435, EPI_ISL_645436, EPI_ISL_645437, EPI_ISL_645439, EPI_ISL_645440, EPI_ISL_645442, EPI_ISL_645443, EPI_ISL_645444, EPI_ISL_645445, EPI_ISL_645446, EPI_ISL_645447, EPI_ISL_645448, EPI_ISL_645449, EPI_ISL_645450, EPI_ISL_645451, EPI_ISL_645454, EPI_ISL_645455, EPI_ISL_645456, EPI_ISL_645457, EPI_ISL_645458, EPI_ISL_645460, EPI_ISL_645461, EPI_ISL_645462, EPI_ISL_645463, EPI_ISL_645465, EPI_ISL_645466, EPI_ISL_645467, EPI_ISL_645468, EPI_ISL_645469, EPI_ISL_645470, EPI_ISL_645471, EPI_ISL_645472, EPI_ISL_645474, EPI_ISL_645475, EPI_ISL_645476, EPI_ISL_645478, EPI_ISL_645479, EPI_ISL_645480, EPI_ISL_645481, EPI_ISL_645482, EPI_ISL_645483, EPI_ISL_645484, EPI_ISL_645485, EPI_ISL_645486, EPI_ISL_645487, EPI_ISL_645488, EPI_ISL_645490, EPI_ISL_645491, EPI_ISL_645492, EPI_ISL_645493, EPI_ISL_645494, EPI_ISL_645495, EPI_ISL_645496, EPI_ISL_645497, EPI_ISL_645498, EPI_ISL_645500, EPI_ISL_645501, EPI_ISL_645504, EPI_ISL_645505, EPI_ISL_645506, EPI_ISL_645507, EPI_ISL_645509, EPI_ISL_645510, EPI_ISL_645511, EPI_ISL_645512, EPI_ISL_645514, EPI_ISL_645515, EPI_ISL_645516, EPI_ISL_645519, EPI_ISL_645521, EPI_ISL_645522, EPI_ISL_645523, EPI_ISL_645524, EPI_ISL_645525, EPI_ISL_645526, EPI_ISL_645527, EPI_ISL_645528, EPI_ISL_645529, EPI_ISL_649482, EPI_ISL_656686, EPI_ISL_656687, EPI_ISL_656688, EPI_ISL_656689, EPI_ISL_656690, EPI_ISL_656691, EPI_ISL_656692, EPI_ISL_656693, EPI_ISL_656694, EPI_ISL_656695, EPI_ISL_656696, EPI_ISL_656697, EPI_ISL_656698, EPI_ISL_656699, EPI_ISL_656700, EPI_ISL_656701, EPI_ISL_656702, EPI_ISL_656703, EPI_ISL_656704, EPI_ISL_656706, EPI_ISL_656707, EPI_ISL_656708, EPI_ISL_656709, EPI_ISL_656710, EPI_ISL_656711, EPI_ISL_656712, EPI_ISL_656713, EPI_ISL_656714, EPI_ISL_656715, EPI_ISL_656716, EPI_ISL_656717, EPI_ISL_656718, EPI_ISL_656719, EPI_ISL_656720, EPI_ISL_656721, EPI_ISL_656722, EPI_ISL_656723, EPI_ISL_656724, EPI_ISL_656725, EPI_ISL_656726, EPI_ISL_656727, EPI_ISL_656729, EPI_ISL_656730, EPI_ISL_656734, EPI_ISL_656735, EPI_ISL_656736, EPI_ISL_656737, EPI_ISL_656738, EPI_ISL_656739, EPI_ISL_656740, EPI_ISL_656741, EPI_ISL_656742, EPI_ISL_656743, EPI_ISL_656747, EPI_ISL_656748, EPI_ISL_656749, EPI_ISL_656750, EPI_ISL_656751, EPI_ISL_656752, EPI_ISL_656753, EPI_ISL_656754, EPI_ISL_656755, EPI_ISL_656756, EPI_ISL_656758, EPI_ISL_656759, EPI_ISL_656760, EPI_ISL_656761, EPI_ISL_656762, EPI_ISL_656763, EPI_ISL_656764, EPI_ISL_656765, EPI_ISL_656766, EPI_ISL_656767, EPI_ISL_656768, EPI_ISL_656769, EPI_ISL_656770, EPI_ISL_656771, EPI_ISL_656772, EPI_ISL_656773, EPI_ISL_656775, EPI_ISL_656776, EPI_ISL_656780, EPI_ISL_656781, EPI_ISL_656782, EPI_ISL_656783, EPI_ISL_656784, EPI_ISL_656785, EPI_ISL_656786, EPI_ISL_656787, EPI_ISL_656790, EPI_ISL_656791, EPI_ISL_656792, EPI_ISL_656793, EPI_ISL_656794, EPI_ISL_656795, EPI_ISL_656796, EPI_ISL_656797, EPI_ISL_656800, EPI_ISL_656801, EPI_ISL_656802, EPI_ISL_656803, EPI_ISL_656804, EPI_ISL_656805, EPI_ISL_656806, EPI_ISL_656809, EPI_ISL_656812, EPI_ISL_656813, EPI_ISL_656814, EPI_ISL_656815, EPI_ISL_656816, EPI_ISL_656817, EPI_ISL_656818, EPI_ISL_656819, EPI_ISL_656820, EPI_ISL_656821, EPI_ISL_656822, EPI_ISL_656823, EPI_ISL_656825, EPI_ISL_656826, EPI_ISL_656828, EPI_ISL_656830, EPI_ISL_656831, EPI_ISL_656832, EPI_ISL_656833, EPI_ISL_656836, EPI_ISL_656837, EPI_ISL_656839, EPI_ISL_656842, EPI_ISL_656843, EPI_ISL_656844, EPI_ISL_656845, EPI_ISL_656847, EPI_ISL_656849, EPI_ISL_656850, EPI_ISL_656851, EPI_ISL_656852, EPI_ISL_656853, EPI_ISL_656854, EPI_ISL_656855, EPI_ISL_656856, EPI_ISL_656857, EPI_ISL_656858, EPI_ISL_656859, EPI_ISL_656863, EPI_ISL_656864, EPI_ISL_656866, EPI_ISL_656868, EPI_ISL_656870, EPI_ISL_656872, EPI_ISL_656873, EPI_ISL_656874, EPI_ISL_656875, EPI_ISL_656876, EPI_ISL_656878, EPI_ISL_656880, EPI_ISL_656881, EPI_ISL_656882, EPI_ISL_656883, EPI_ISL_656884, EPI_ISL_656885, EPI_ISL_656886, EPI_ISL_656888, EPI_ISL_656889, EPI_ISL_656890, EPI_ISL_656891, EPI_ISL_656894, EPI_ISL_656895, EPI_ISL_656896, EPI_ISL_656897, EPI_ISL_656898, EPI_ISL_656899, EPI_ISL_656900, EPI_ISL_656901, EPI_ISL_656902, EPI_ISL_656903, EPI_ISL_656904, EPI_ISL_656905, EPI_ISL_656907, EPI_ISL_656908, EPI_ISL_656909, EPI_ISL_656911, EPI_ISL_656912, EPI_ISL_656914, EPI_ISL_656915, EPI_ISL_656916, EPI_ISL_656917, EPI_ISL_656918, EPI_ISL_656919, EPI_ISL_656921, EPI_ISL_656923, EPI_ISL_656925, EPI_ISL_656926, EPI_ISL_656927, EPI_ISL_656928, EPI_ISL_656929, EPI_ISL_656931, EPI_ISL_656933, EPI_ISL_656934, EPI_ISL_656937, EPI_ISL_656939, EPI_ISL_656940, EPI_ISL_656941, EPI_ISL_656943, EPI_ISL_656946, EPI_ISL_656948, EPI_ISL_656949, EPI_ISL_656951, EPI_ISL_656952, EPI_ISL_656953, EPI_ISL_656954, EPI_ISL_656956, EPI_ISL_656959, EPI_ISL_656960, EPI_ISL_656963, EPI_ISL_656964, EPI_ISL_656967, EPI_ISL_656968, EPI_ISL_656971, EPI_ISL_656972, EPI_ISL_656975, EPI_ISL_656977, EPI_ISL_656978, EPI_ISL_656981, EPI_ISL_656983, EPI_ISL_656984, EPI_ISL_656985, EPI_ISL_656986, EPI_ISL_656988, EPI_ISL_656989, EPI_ISL_656990, EPI_ISL_656992, EPI_ISL_656993, EPI_ISL_656994, EPI_ISL_656995, EPI_ISL_656996, EPI_ISL_656998, EPI_ISL_656999, EPI_ISL_657000, EPI_ISL_657001, EPI_ISL_657002, EPI_ISL_657005, EPI_ISL_657006, EPI_ISL_657007, EPI_ISL_657009, EPI_ISL_657010, EPI_ISL_657012, EPI_ISL_657013, EPI_ISL_657015, EPI_ISL_657018, EPI_ISL_657019, EPI_ISL_657020, EPI_ISL_657022, EPI_ISL_657024, EPI_ISL_657025, EPI_ISL_657356, EPI_ISL_657357, EPI_ISL_657358, EPI_ISL_657362, EPI_ISL_657363, EPI_ISL_657366, EPI_ISL_657368, EPI_ISL_657374, EPI_ISL_657376, EPI_ISL_657378, EPI_ISL_657380, EPI_ISL_657383, EPI_ISL_657387, EPI_ISL_657388, EPI_ISL_657389, EPI_ISL_657394, EPI_ISL_657395, EPI_ISL_657397, EPI_ISL_657399, EPI_ISL_657402, EPI_ISL_657403, EPI_ISL_657404, EPI_ISL_657407, EPI_ISL_657409, EPI_ISL_657412, EPI_ISL_657415, EPI_ISL_657421, EPI_ISL_657422, EPI_ISL_657423, EPI_ISL_657426, EPI_ISL_657427, EPI_ISL_657431, EPI_ISL_657432, EPI_ISL_657433, EPI_ISL_657434, EPI_ISL_657435, EPI_ISL_657436, EPI_ISL_657437, EPI_ISL_657442, EPI_ISL_657444, EPI_ISL_657445, EPI_ISL_657447, EPI_ISL_657448, EPI_ISL_657449, EPI_ISL_657454, EPI_ISL_657456, EPI_ISL_657461, EPI_ISL_657468, EPI_ISL_657469, EPI_ISL_657470, EPI_ISL_657471, EPI_ISL_657474, EPI_ISL_657475, EPI_ISL_657476, EPI_ISL_657477, EPI_ISL_657478, EPI_ISL_657481, EPI_ISL_657482, EPI_ISL_657489, EPI_ISL_657492, EPI_ISL_657496, EPI_ISL_657499, EPI_ISL_657501, EPI_ISL_657502, EPI_ISL_657504, EPI_ISL_657506, EPI_ISL_657507, EPI_ISL_657508, EPI_ISL_657511, EPI_ISL_657514, EPI_ISL_657516, EPI_ISL_657517, EPI_ISL_657520, EPI_ISL_657521, EPI_ISL_657522, EPI_ISL_657523, EPI_ISL_657524, EPI_ISL_657525, EPI_ISL_657528, EPI_ISL_657529, EPI_ISL_657530, EPI_ISL_657534, EPI_ISL_657537, EPI_ISL_657538, EPI_ISL_657539, EPI_ISL_657540, EPI_ISL_657541, EPI_ISL_657545, EPI_ISL_657546, EPI_ISL_657548, EPI_ISL_657549, EPI_ISL_657551, EPI_ISL_657552, EPI_ISL_657559, EPI_ISL_657561, EPI_ISL_657562, EPI_ISL_657565, EPI_ISL_657566, EPI_ISL_657569, EPI_ISL_657572, EPI_ISL_657573, EPI_ISL_657576, EPI_ISL_657578, EPI_ISL_657580, EPI_ISL_657582, EPI_ISL_657586, EPI_ISL_657588, EPI_ISL_657590, EPI_ISL_657592, EPI_ISL_657595, EPI_ISL_657597, EPI_ISL_657598, EPI_ISL_657600, EPI_ISL_657601, EPI_ISL_657603, EPI_ISL_657604, EPI_ISL_657612, EPI_ISL_657613, EPI_ISL_657617, EPI_ISL_657618, EPI_ISL_657619, EPI_ISL_657620, EPI_ISL_657621, EPI_ISL_657622, EPI_ISL_657623, EPI_ISL_657624, EPI_ISL_657626, EPI_ISL_657628, EPI_ISL_657630, EPI_ISL_657633, EPI_ISL_657634, EPI_ISL_657637, EPI_ISL_657638, EPI_ISL_657641, EPI_ISL_657643, EPI_ISL_657645, EPI_ISL_657646, EPI_ISL_657649, EPI_ISL_657652, EPI_ISL_657653, EPI_ISL_657654, EPI_ISL_657655, EPI_ISL_657656, EPI_ISL_657658, EPI_ISL_657659, EPI_ISL_657660, EPI_ISL_657664, EPI_ISL_657668, EPI_ISL_657669, EPI_ISL_657670, EPI_ISL_657671, EPI_ISL_657672, EPI_ISL_657673, EPI_ISL_657674, EPI_ISL_657675, EPI_ISL_657677, EPI_ISL_657678, EPI_ISL_657679, EPI_ISL_657684, EPI_ISL_657685, EPI_ISL_657686, EPI_ISL_657687, EPI_ISL_657689, EPI_ISL_657690, EPI_ISL_657691, EPI_ISL_657693, EPI_ISL_657697, EPI_ISL_657698, EPI_ISL_657699, EPI_ISL_657701, EPI_ISL_657704, EPI_ISL_657707, EPI_ISL_657708, EPI_ISL_657713, EPI_ISL_657720, EPI_ISL_657721, EPI_ISL_657723, EPI_ISL_657724, EPI_ISL_657725, EPI_ISL_657726, EPI_ISL_657728, EPI_ISL_657731, EPI_ISL_657736, EPI_ISL_657737, EPI_ISL_657739, EPI_ISL_657744, EPI_ISL_657746, EPI_ISL_657747, EPI_ISL_657749, EPI_ISL_657750, EPI_ISL_657754, EPI_ISL_657756, EPI_ISL_657759, EPI_ISL_657760, EPI_ISL_657762, EPI_ISL_657763, EPI_ISL_657765, EPI_ISL_657766, EPI_ISL_657767, EPI_ISL_657769, EPI_ISL_657771, EPI_ISL_657780, EPI_ISL_657782, EPI_ISL_657786, EPI_ISL_657787, EPI_ISL_657790, EPI_ISL_657793, EPI_ISL_657795, EPI_ISL_657798, EPI_ISL_657799, EPI_ISL_657801, EPI_ISL_657802, EPI_ISL_657803, EPI_ISL_657804, EPI_ISL_657805, EPI_ISL_657806, EPI_ISL_657807, EPI_ISL_657809, EPI_ISL_657810, EPI_ISL_657813, EPI_ISL_657815, EPI_ISL_657816, EPI_ISL_657817, EPI_ISL_657818, EPI_ISL_657824, EPI_ISL_657825, EPI_ISL_657826, EPI_ISL_657828, EPI_ISL_657831, EPI_ISL_657833, EPI_ISL_657834, EPI_ISL_657837, EPI_ISL_657838, EPI_ISL_657839, EPI_ISL_657841, EPI_ISL_657845, EPI_ISL_657846, EPI_ISL_657847, EPI_ISL_657849, EPI_ISL_657851, EPI_ISL_657852, EPI_ISL_657853, EPI_ISL_657855, EPI_ISL_657857, EPI_ISL_657860, EPI_ISL_657863, EPI_ISL_657864, EPI_ISL_657866, EPI_ISL_657867, EPI_ISL_657868, EPI_ISL_657871, EPI_ISL_657873, EPI_ISL_657875, EPI_ISL_657876, EPI_ISL_657877, EPI_ISL_657880, EPI_ISL_657881, EPI_ISL_657882, EPI_ISL_657883, EPI_ISL_657884, EPI_ISL_657885, EPI_ISL_657888, EPI_ISL_657890, EPI_ISL_657891, EPI_ISL_657893, EPI_ISL_657895, EPI_ISL_657896, EPI_ISL_657897, EPI_ISL_657899, EPI_ISL_657900, EPI_ISL_657902, EPI_ISL_657903, EPI_ISL_657904, EPI_ISL_657907, EPI_ISL_657908, EPI_ISL_657909, EPI_ISL_657911, EPI_ISL_657912, EPI_ISL_657914, EPI_ISL_657917, EPI_ISL_657920, EPI_ISL_657924, EPI_ISL_657926, EPI_ISL_657931, EPI_ISL_657934, EPI_ISL_657935, EPI_ISL_657936, EPI_ISL_657937, EPI_ISL_657939, EPI_ISL_657942, EPI_ISL_657943, EPI_ISL_657946, EPI_ISL_657947, EPI_ISL_657949, EPI_ISL_657951, EPI_ISL_657952, EPI_ISL_657953, EPI_ISL_657957, EPI_ISL_657960, EPI_ISL_657964, EPI_ISL_657966, EPI_ISL_657967, EPI_ISL_657968, EPI_ISL_657974, EPI_ISL_657976, EPI_ISL_657978, EPI_ISL_657983, EPI_ISL_657986, EPI_ISL_657987, EPI_ISL_657989, EPI_ISL_657990, EPI_ISL_657992, EPI_ISL_657994, EPI_ISL_657998, EPI_ISL_658001, EPI_ISL_658007, EPI_ISL_658008, EPI_ISL_658010, EPI_ISL_658011, EPI_ISL_658012, EPI_ISL_658013, EPI_ISL_658014, EPI_ISL_658019, EPI_ISL_658020, EPI_ISL_658024, EPI_ISL_658028, EPI_ISL_658029, EPI_ISL_658030, EPI_ISL_658031, EPI_ISL_658032, EPI_ISL_658033, EPI_ISL_658034, EPI_ISL_658036, EPI_ISL_658037, EPI_ISL_658041, EPI_ISL_658043, EPI_ISL_658045, EPI_ISL_658046, EPI_ISL_658048, EPI_ISL_658052, EPI_ISL_658055, EPI_ISL_658056, EPI_ISL_658063, EPI_ISL_658064, EPI_ISL_658069, EPI_ISL_658071, EPI_ISL_658072, EPI_ISL_658073, EPI_ISL_658074, EPI_ISL_658075, EPI_ISL_658076, EPI_ISL_658077, EPI_ISL_658078, EPI_ISL_658080, EPI_ISL_658083, EPI_ISL_658084, EPI_ISL_658085, EPI_ISL_658086, EPI_ISL_658089, EPI_ISL_658092, EPI_ISL_658093, EPI_ISL_658095, EPI_ISL_658096, EPI_ISL_658097, EPI_ISL_658098, EPI_ISL_658099, EPI_ISL_658100, EPI_ISL_658101, EPI_ISL_658104, EPI_ISL_658110, EPI_ISL_658116, EPI_ISL_658119, EPI_ISL_658120, EPI_ISL_658122, EPI_ISL_658124, EPI_ISL_658125, EPI_ISL_658128, EPI_ISL_658131, EPI_ISL_658133, EPI_ISL_658134, EPI_ISL_658138, EPI_ISL_658142, EPI_ISL_658144, EPI_ISL_658148, EPI_ISL_658153, EPI_ISL_658156, EPI_ISL_658157, EPI_ISL_658159, EPI_ISL_658160, EPI_ISL_658161, EPI_ISL_658162, EPI_ISL_658163, EPI_ISL_658164, EPI_ISL_658166, EPI_ISL_658167, EPI_ISL_658171, EPI_ISL_658177, EPI_ISL_658178, EPI_ISL_658179, EPI_ISL_658181, EPI_ISL_658186, EPI_ISL_658187, EPI_ISL_658188, EPI_ISL_658189, EPI_ISL_658190, EPI_ISL_658191, EPI_ISL_658192, EPI_ISL_658194, EPI_ISL_658195, EPI_ISL_658196, EPI_ISL_658197, EPI_ISL_658198, EPI_ISL_658199, EPI_ISL_658200, EPI_ISL_658201, EPI_ISL_658202, EPI_ISL_658203, EPI_ISL_658206, EPI_ISL_658207, EPI_ISL_658208, EPI_ISL_658209, EPI_ISL_658212, EPI_ISL_658214, EPI_ISL_658215, EPI_ISL_658218, EPI_ISL_658220, EPI_ISL_658221, EPI_ISL_658224, EPI_ISL_658226, EPI_ISL_658227, EPI_ISL_658228, EPI_ISL_658231, EPI_ISL_658234, EPI_ISL_658236, EPI_ISL_658239, EPI_ISL_658242, EPI_ISL_658246, EPI_ISL_658252, EPI_ISL_658256, EPI_ISL_658258, EPI_ISL_658260, EPI_ISL_658263, EPI_ISL_658266, EPI_ISL_658270, EPI_ISL_658271, EPI_ISL_658272, EPI_ISL_658274, EPI_ISL_658275, EPI_ISL_658276, EPI_ISL_658278, EPI_ISL_658279, EPI_ISL_658285, EPI_ISL_658286, EPI_ISL_658287, EPI_ISL_658292, EPI_ISL_658299, EPI_ISL_658304, EPI_ISL_658305, EPI_ISL_658307, EPI_ISL_658310, EPI_ISL_658311, EPI_ISL_658312, EPI_ISL_658313, EPI_ISL_658314, EPI_ISL_658316, EPI_ISL_658317, EPI_ISL_658320, EPI_ISL_658321, EPI_ISL_658323, EPI_ISL_658326, EPI_ISL_658327, EPI_ISL_658330, EPI_ISL_658331, EPI_ISL_658332, EPI_ISL_658334, EPI_ISL_658335, EPI_ISL_658337, EPI_ISL_658340, EPI_ISL_658341, EPI_ISL_658343, EPI_ISL_658345, EPI_ISL_658348, EPI_ISL_658351, EPI_ISL_658352, EPI_ISL_658355, EPI_ISL_658356, EPI_ISL_658360, EPI_ISL_658361, EPI_ISL_658362, EPI_ISL_658363, EPI_ISL_658364, EPI_ISL_658366, EPI_ISL_658367, EPI_ISL_658370, EPI_ISL_658371, EPI_ISL_658376, EPI_ISL_658381, EPI_ISL_658382, EPI_ISL_658383, EPI_ISL_658387, EPI_ISL_658388, EPI_ISL_658390, EPI_ISL_658394, EPI_ISL_658396, EPI_ISL_658397, EPI_ISL_658398, EPI_ISL_658402, EPI_ISL_658744, EPI_ISL_658745, EPI_ISL_658750, EPI_ISL_658752, EPI_ISL_658753, EPI_ISL_658756, EPI_ISL_658759, EPI_ISL_658760, EPI_ISL_658763, EPI_ISL_658764, EPI_ISL_658767, EPI_ISL_658768, EPI_ISL_658769, EPI_ISL_658775, EPI_ISL_658781, EPI_ISL_658787, EPI_ISL_658788, EPI_ISL_658795, EPI_ISL_658800, EPI_ISL_658807, EPI_ISL_658809, EPI_ISL_658816, EPI_ISL_658829, EPI_ISL_658833, EPI_ISL_658836, EPI_ISL_658837, EPI_ISL_658838, EPI_ISL_658846, EPI_ISL_658852, EPI_ISL_658864, EPI_ISL_658866, EPI_ISL_658877, EPI_ISL_658881, EPI_ISL_658885, EPI_ISL_658900, EPI_ISL_658905, EPI_ISL_658909, EPI_ISL_658912, EPI_ISL_658913, EPI_ISL_658914, EPI_ISL_658916, EPI_ISL_658919, EPI_ISL_658921, EPI_ISL_658923, EPI_ISL_658925, EPI_ISL_658930, EPI_ISL_658931, EPI_ISL_658933, EPI_ISL_658936, EPI_ISL_658948, EPI_ISL_658954, EPI_ISL_658959, EPI_ISL_658963, EPI_ISL_658970, EPI_ISL_658971, EPI_ISL_658976, EPI_ISL_658979, EPI_ISL_658995, EPI_ISL_658996, EPI_ISL_659000, EPI_ISL_659005, EPI_ISL_659007, EPI_ISL_659011, EPI_ISL_659017, EPI_ISL_659018, EPI_ISL_659019, EPI_ISL_659025, EPI_ISL_659028, EPI_ISL_659040, EPI_ISL_659045, EPI_ISL_659050, EPI_ISL_659053, EPI_ISL_659054, EPI_ISL_659070, EPI_ISL_659071, EPI_ISL_659072, EPI_ISL_659073, EPI_ISL_659076, EPI_ISL_659079, EPI_ISL_659080, EPI_ISL_659082, EPI_ISL_659083, EPI_ISL_659084, EPI_ISL_659090, EPI_ISL_659091, EPI_ISL_659093, EPI_ISL_659094, EPI_ISL_659097, EPI_ISL_659098, EPI_ISL_659099, EPI_ISL_659100, EPI_ISL_659101, EPI_ISL_659102, EPI_ISL_659103, EPI_ISL_659104, EPI_ISL_659106, EPI_ISL_659107, EPI_ISL_659108, EPI_ISL_659109, EPI_ISL_659110, EPI_ISL_659111, EPI_ISL_659112, EPI_ISL_659113, EPI_ISL_659114, EPI_ISL_659115, EPI_ISL_659116, EPI_ISL_659117, EPI_ISL_659118, EPI_ISL_659119, EPI_ISL_659120, EPI_ISL_659121, EPI_ISL_659122, EPI_ISL_659123, EPI_ISL_659124, EPI_ISL_659125, EPI_ISL_659126, EPI_ISL_659127, EPI_ISL_659128, EPI_ISL_659130, EPI_ISL_659131, EPI_ISL_659132, EPI_ISL_659133, EPI_ISL_659134, EPI_ISL_659136, EPI_ISL_659137, EPI_ISL_659138, EPI_ISL_659139, EPI_ISL_659140, EPI_ISL_659141, EPI_ISL_659142, EPI_ISL_659143, EPI_ISL_659144, EPI_ISL_659145, EPI_ISL_659146, EPI_ISL_659147, EPI_ISL_659148, EPI_ISL_659149, EPI_ISL_659150, EPI_ISL_659151, EPI_ISL_659152, EPI_ISL_659153, EPI_ISL_659154, EPI_ISL_659155, EPI_ISL_659156, EPI_ISL_659157, EPI_ISL_659158, EPI_ISL_659159, EPI_ISL_659160, EPI_ISL_659161, EPI_ISL_659162, EPI_ISL_659164, EPI_ISL_659165, EPI_ISL_659166, EPI_ISL_659168, EPI_ISL_659169, EPI_ISL_659170, EPI_ISL_659171, EPI_ISL_659172, EPI_ISL_659173, EPI_ISL_659174, EPI_ISL_659175, EPI_ISL_659176, EPI_ISL_659177, EPI_ISL_659178, EPI_ISL_659179, EPI_ISL_659180, EPI_ISL_659181, EPI_ISL_659182, EPI_ISL_659183, EPI_ISL_659184, EPI_ISL_659185, EPI_ISL_659187, EPI_ISL_659188, EPI_ISL_659189, EPI_ISL_659190, EPI_ISL_659191, EPI_ISL_659192, EPI_ISL_659193, EPI_ISL_659194, EPI_ISL_659195, EPI_ISL_659196, EPI_ISL_659197, EPI_ISL_659198, EPI_ISL_659199, EPI_ISL_659200, EPI_ISL_659201, EPI_ISL_659202, EPI_ISL_659203, EPI_ISL_659204, EPI_ISL_659205, EPI_ISL_659206, EPI_ISL_659207, EPI_ISL_659208, EPI_ISL_659209, EPI_ISL_659210, EPI_ISL_659211, EPI_ISL_659212, EPI_ISL_659213, EPI_ISL_659214, EPI_ISL_659215, EPI_ISL_659216, EPI_ISL_659217, EPI_ISL_659218, EPI_ISL_659219, EPI_ISL_659220, EPI_ISL_659221, EPI_ISL_659222, EPI_ISL_659223, EPI_ISL_659224, EPI_ISL_659225, EPI_ISL_659226, EPI_ISL_659227, EPI_ISL_659228, EPI_ISL_659229, EPI_ISL_659230, EPI_ISL_659231, EPI_ISL_659232, EPI_ISL_659233, EPI_ISL_659234, EPI_ISL_659235, EPI_ISL_659236, EPI_ISL_659237, EPI_ISL_659238, EPI_ISL_659239, EPI_ISL_659240, EPI_ISL_659241, EPI_ISL_659242, EPI_ISL_659243, EPI_ISL_659244, EPI_ISL_659245, EPI_ISL_659246, EPI_ISL_659247, EPI_ISL_659248, EPI_ISL_659249, EPI_ISL_659250, EPI_ISL_659251, EPI_ISL_659252, EPI_ISL_659254, EPI_ISL_659255, EPI_ISL_659256, EPI_ISL_659257, EPI_ISL_659258, EPI_ISL_659259, EPI_ISL_659260, EPI_ISL_659261, EPI_ISL_659262, EPI_ISL_659263, EPI_ISL_659264, EPI_ISL_659265, EPI_ISL_659266, EPI_ISL_659267, EPI_ISL_659268, EPI_ISL_659269, EPI_ISL_659270, EPI_ISL_659271, EPI_ISL_659272, EPI_ISL_659273, EPI_ISL_659274, EPI_ISL_659275, EPI_ISL_659276, EPI_ISL_659277, EPI_ISL_659278, EPI_ISL_659279, EPI_ISL_659280, EPI_ISL_659281, EPI_ISL_659282, EPI_ISL_659283, EPI_ISL_659284, EPI_ISL_659285, EPI_ISL_659286, EPI_ISL_659287, EPI_ISL_659288, EPI_ISL_659289, EPI_ISL_659290, EPI_ISL_659291, EPI_ISL_659292, EPI_ISL_659293, EPI_ISL_659294, EPI_ISL_659295, EPI_ISL_659296, EPI_ISL_659297, EPI_ISL_659298, EPI_ISL_659299, EPI_ISL_659300, EPI_ISL_659301, EPI_ISL_659302, EPI_ISL_659303, EPI_ISL_659304, EPI_ISL_659305, EPI_ISL_659306, EPI_ISL_659307, EPI_ISL_659308, EPI_ISL_659309, EPI_ISL_659310, EPI_ISL_659311, EPI_ISL_659312, EPI_ISL_659313, EPI_ISL_659314, EPI_ISL_659315, EPI_ISL_659316, EPI_ISL_659317, EPI_ISL_659318, EPI_ISL_659319, EPI_ISL_659320, EPI_ISL_659321, EPI_ISL_659322, EPI_ISL_659323, EPI_ISL_659324, EPI_ISL_659325, EPI_ISL_659326, EPI_ISL_659327, EPI_ISL_659328, EPI_ISL_659329, EPI_ISL_659330, EPI_ISL_659331, EPI_ISL_659332, EPI_ISL_659333, EPI_ISL_659334, EPI_ISL_659335, EPI_ISL_659336, EPI_ISL_659337, EPI_ISL_659338, EPI_ISL_659339, EPI_ISL_659340, EPI_ISL_659341, EPI_ISL_659342, EPI_ISL_659343, EPI_ISL_659344, EPI_ISL_659346, EPI_ISL_659347, EPI_ISL_659348, EPI_ISL_659349, EPI_ISL_659350, EPI_ISL_659351, EPI_ISL_659352, EPI_ISL_659353, EPI_ISL_659354, EPI_ISL_659355, EPI_ISL_659356, EPI_ISL_659357, EPI_ISL_659358, EPI_ISL_659359, EPI_ISL_659360, EPI_ISL_659361, EPI_ISL_659362, EPI_ISL_659363, EPI_ISL_659364, EPI_ISL_659365, EPI_ISL_659366, EPI_ISL_659367, EPI_ISL_659368, EPI_ISL_659369, EPI_ISL_659930, EPI_ISL_659371, EPI_ISL_659372, EPI_ISL_659373, EPI_ISL_659374, EPI_ISL_659375, EPI_ISL_659376, EPI_ISL_659377, EPI_ISL_659378, EPI_ISL_659379, EPI_ISL_659380, EPI_ISL_659381, EPI_ISL_659382, EPI_ISL_659383, EPI_ISL_659384, EPI_ISL_659385, EPI_ISL_659386, EPI_ISL_659388, EPI_ISL_659389, EPI_ISL_659390, EPI_ISL_659391, EPI_ISL_659392, EPI_ISL_659393, EPI_ISL_659394, EPI_ISL_659396, EPI_ISL_659397, EPI_ISL_659398, EPI_ISL_659399, EPI_ISL_659400, EPI_ISL_659401, EPI_ISL_659402, EPI_ISL_659403, EPI_ISL_659404, EPI_ISL_659405, EPI_ISL_659406, EPI_ISL_659407, EPI_ISL_659408, EPI_ISL_659409, EPI_ISL_659410, EPI_ISL_659411, EPI_ISL_659412, EPI_ISL_659413, EPI_ISL_659414, EPI_ISL_659415, EPI_ISL_659416, EPI_ISL_659417, EPI_ISL_659418, EPI_ISL_659419, EPI_ISL_659420, EPI_ISL_659421, EPI_ISL_659422, EPI_ISL_659423, EPI_ISL_659424, EPI_ISL_659425, EPI_ISL_659426, EPI_ISL_659427, EPI_ISL_659428, EPI_ISL_659429, EPI_ISL_659430, EPI_ISL_659431, EPI_ISL_659432, EPI_ISL_659433, EPI_ISL_659434, EPI_ISL_659435, EPI_ISL_659436, EPI_ISL_659437, EPI_ISL_659438, EPI_ISL_659439, EPI_ISL_659440, EPI_ISL_659441, EPI_ISL_659442, EPI_ISL_659443, EPI_ISL_659444, EPI_ISL_659445, EPI_ISL_659446, EPI_ISL_659447, EPI_ISL_659448, EPI_ISL_659449, EPI_ISL_659450, EPI_ISL_659451, EPI_ISL_659452, EPI_ISL_659453, EPI_ISL_659454, EPI_ISL_659455, EPI_ISL_659456, EPI_ISL_659457, EPI_ISL_659458, EPI_ISL_659459, EPI_ISL_659460, EPI_ISL_659461, EPI_ISL_659462, EPI_ISL_659463, EPI_ISL_659464, EPI_ISL_659465, EPI_ISL_659466, EPI_ISL_659467, EPI_ISL_659468, EPI_ISL_659469, EPI_ISL_659470, EPI_ISL_659471, EPI_ISL_659472, EPI_ISL_659473, EPI_ISL_659474, EPI_ISL_659475, EPI_ISL_659476, EPI_ISL_659477, EPI_ISL_659478, EPI_ISL_659479, EPI_ISL_659480, EPI_ISL_659481, EPI_ISL_659482, EPI_ISL_659483, EPI_ISL_659484, EPI_ISL_659485, EPI_ISL_659486, EPI_ISL_659487, EPI_ISL_659488, EPI_ISL_659489, EPI_ISL_659490, EPI_ISL_659491, EPI_ISL_659492, EPI_ISL_659493, EPI_ISL_659494, EPI_ISL_659495, EPI_ISL_659496, EPI_ISL_659497, EPI_ISL_659498, EPI_ISL_659499, EPI_ISL_659500, EPI_ISL_659501, EPI_ISL_659502, EPI_ISL_659503, EPI_ISL_659504, EPI_ISL_659505, EPI_ISL_659506, EPI_ISL_659507, EPI_ISL_659508, EPI_ISL_659509, EPI_ISL_659510, EPI_ISL_659511, EPI_ISL_659512, EPI_ISL_659513, EPI_ISL_659514, EPI_ISL_659515, EPI_ISL_659516, EPI_ISL_659517, EPI_ISL_659518, EPI_ISL_659519, EPI_ISL_659520, EPI_ISL_659521, EPI_ISL_659522, EPI_ISL_659523, EPI_ISL_659524, EPI_ISL_659525, EPI_ISL_659526, EPI_ISL_659527, EPI_ISL_659528, EPI_ISL_659529, EPI_ISL_659530, EPI_ISL_659531, EPI_ISL_659532, EPI_ISL_659533, EPI_ISL_659534, EPI_ISL_659535, EPI_ISL_659536, EPI_ISL_659537, EPI_ISL_659538, EPI_ISL_659539, EPI_ISL_659540, EPI_ISL_659541, EPI_ISL_659543, EPI_ISL_659544, EPI_ISL_659545, EPI_ISL_659546, EPI_ISL_659547, EPI_ISL_659548, EPI_ISL_659549, EPI_ISL_659550, EPI_ISL_659551, EPI_ISL_659552, EPI_ISL_659553, EPI_ISL_659554, EPI_ISL_659555, EPI_ISL_659556, EPI_ISL_659557, EPI_ISL_659558, EPI_ISL_659559, EPI_ISL_659560, EPI_ISL_659561, EPI_ISL_659562, EPI_ISL_659563, EPI_ISL_659564, EPI_ISL_659565, EPI_ISL_659566, EPI_ISL_659567, EPI_ISL_659568, EPI_ISL_659569, EPI_ISL_659570, EPI_ISL_659571, EPI_ISL_659572, EPI_ISL_659573, EPI_ISL_659574, EPI_ISL_659575, EPI_ISL_659576, EPI_ISL_659577, EPI_ISL_659578, EPI_ISL_659579, EPI_ISL_659580, EPI_ISL_659581, EPI_ISL_659582, EPI_ISL_659583, EPI_ISL_659584, EPI_ISL_659585, EPI_ISL_659586, EPI_ISL_659587, EPI_ISL_659588, EPI_ISL_659589, EPI_ISL_659590, EPI_ISL_659591, EPI_ISL_659593, EPI_ISL_659594, EPI_ISL_659595, EPI_ISL_659596, EPI_ISL_659597, EPI_ISL_659598, EPI_ISL_659599, EPI_ISL_659600, EPI_ISL_659601, EPI_ISL_659602, EPI_ISL_659603, EPI_ISL_659604, EPI_ISL_659605, EPI_ISL_659606, EPI_ISL_659607, EPI_ISL_659608, EPI_ISL_659609, EPI_ISL_659610, EPI_ISL_659611, EPI_ISL_659612, EPI_ISL_659613, EPI_ISL_659614, EPI_ISL_659615, EPI_ISL_659616, EPI_ISL_659617, EPI_ISL_659618, EPI_ISL_659619, EPI_ISL_659620, EPI_ISL_659621, EPI_ISL_659622, EPI_ISL_659623, EPI_ISL_659624, EPI_ISL_659625, EPI_ISL_659626, EPI_ISL_659627, EPI_ISL_659628, EPI_ISL_659629, EPI_ISL_659630, EPI_ISL_659631, EPI_ISL_659632, EPI_ISL_659634, EPI_ISL_659635, EPI_ISL_659636, EPI_ISL_659637, EPI_ISL_659638, EPI_ISL_659639, EPI_ISL_659640, EPI_ISL_659641, EPI_ISL_659642, EPI_ISL_659643, EPI_ISL_659644, EPI_ISL_659645, EPI_ISL_659646, EPI_ISL_659647, EPI_ISL_659648, EPI_ISL_659649, EPI_ISL_659650, EPI_ISL_659651, EPI_ISL_659652, EPI_ISL_659653, EPI_ISL_659654, EPI_ISL_659655, EPI_ISL_659656, EPI_ISL_659657, EPI_ISL_659658, EPI_ISL_659659, EPI_ISL_659660, EPI_ISL_659661, EPI_ISL_659662, EPI_ISL_659663, EPI_ISL_659664, EPI_ISL_659665, EPI_ISL_659666, EPI_ISL_659667, EPI_ISL_659668, EPI_ISL_659669, EPI_ISL_659670, EPI_ISL_659671, EPI_ISL_659672, EPI_ISL_659673, EPI_ISL_659674, EPI_ISL_659675, EPI_ISL_659676, EPI_ISL_659677, EPI_ISL_659678, EPI_ISL_659679, EPI_ISL_659680, EPI_ISL_659681, EPI_ISL_659682, EPI_ISL_659683, EPI_ISL_659684, EPI_ISL_659685, EPI_ISL_659686, EPI_ISL_659687, EPI_ISL_659688, EPI_ISL_659689, EPI_ISL_659690, EPI_ISL_659691, EPI_ISL_659692, EPI_ISL_659693, EPI_ISL_659694, EPI_ISL_659695, EPI_ISL_659696, EPI_ISL_659697, EPI_ISL_659698, EPI_ISL_659699, EPI_ISL_659700, EPI_ISL_659701, EPI_ISL_659702, EPI_ISL_659703, EPI_ISL_659704, EPI_ISL_659705, EPI_ISL_659706, EPI_ISL_659707, EPI_ISL_659708, EPI_ISL_659709, EPI_ISL_659710, EPI_ISL_659711, EPI_ISL_659712, EPI_ISL_659713, EPI_ISL_659714, EPI_ISL_659715, EPI_ISL_659716, EPI_ISL_659717, EPI_ISL_659718, EPI_ISL_659719, EPI_ISL_659720, EPI_ISL_659721, EPI_ISL_659722, EPI_ISL_659723, EPI_ISL_659724, EPI_ISL_659725, EPI_ISL_659726, EPI_ISL_659727, EPI_ISL_659728, EPI_ISL_659729, EPI_ISL_659730, EPI_ISL_659731, EPI_ISL_659732, EPI_ISL_659733, EPI_ISL_659734, EPI_ISL_659735, EPI_ISL_659736, EPI_ISL_659737, EPI_ISL_659738, EPI_ISL_659739, EPI_ISL_659740, EPI_ISL_659741, EPI_ISL_659742, EPI_ISL_659743, EPI_ISL_661640, EPI_ISL_661647, EPI_ISL_661651, EPI_ISL_661652, EPI_ISL_661654, EPI_ISL_661664, EPI_ISL_661675, EPI_ISL_661684, EPI_ISL_661687, EPI_ISL_661691, EPI_ISL_661695, EPI_ISL_661696, EPI_ISL_661710, EPI_ISL_661711, EPI_ISL_661712, EPI_ISL_661713, EPI_ISL_661717, EPI_ISL_661721, EPI_ISL_661729, EPI_ISL_661732, EPI_ISL_661734, EPI_ISL_661741, EPI_ISL_661742, EPI_ISL_661743, EPI_ISL_661747, EPI_ISL_661749, EPI_ISL_661751, EPI_ISL_661752, EPI_ISL_661761, EPI_ISL_661778, EPI_ISL_661781, EPI_ISL_661789, EPI_ISL_661791, EPI_ISL_661797, EPI_ISL_661799, EPI_ISL_661801, EPI_ISL_661805, EPI_ISL_661811, EPI_ISL_661815, EPI_ISL_661817, EPI_ISL_661830, EPI_ISL_661833, EPI_ISL_661834, EPI_ISL_661844, EPI_ISL_661845, EPI_ISL_661863, EPI_ISL_661869, EPI_ISL_661874, EPI_ISL_661876, EPI_ISL_661881, EPI_ISL_661883, EPI_ISL_661886, EPI_ISL_661887, EPI_ISL_661895, EPI_ISL_661897, EPI_ISL_661900, EPI_ISL_661902, EPI_ISL_661903, EPI_ISL_661917, EPI_ISL_661918, EPI_ISL_661926, EPI_ISL_661930, EPI_ISL_661941, EPI_ISL_661947, EPI_ISL_661953, EPI_ISL_661954, EPI_ISL_662631, EPI_ISL_662632, EPI_ISL_662635, EPI_ISL_662651, EPI_ISL_662652, EPI_ISL_662653, EPI_ISL_667829, EPI_ISL_667861, EPI_ISL_667865, EPI_ISL_667870, EPI_ISL_667902, EPI_ISL_667906, EPI_ISL_667933, EPI_ISL_667955, EPI_ISL_667961, EPI_ISL_667988, EPI_ISL_667991, EPI_ISL_668027, EPI_ISL_668071, EPI_ISL_668072, EPI_ISL_668080, EPI_ISL_673310, EPI_ISL_673385, EPI_ISL_673441, EPI_ISL_673447, EPI_ISL_673851, EPI_ISL_676383, EPI_ISL_710878, EPI_ISL_783367, EPI_ISL_783368, EPI_ISL_783370, EPI_ISL_783374, EPI_ISL_783376, EPI_ISL_783377, EPI_ISL_783379, EPI_ISL_783383, EPI_ISL_783394, EPI_ISL_783398, EPI_ISL_783399, EPI_ISL_783400, EPI_ISL_783408, EPI_ISL_783410, EPI_ISL_783412, EPI_ISL_783413, EPI_ISL_783423, EPI_ISL_783425, EPI_ISL_783426, EPI_ISL_783427, EPI_ISL_783428, EPI_ISL_783429, EPI_ISL_783433, EPI_ISL_783445, EPI_ISL_783449, EPI_ISL_783454, EPI_ISL_783457, EPI_ISL_783462, EPI_ISL_783464, EPI_ISL_783465, EPI_ISL_783475, EPI_ISL_783478, EPI_ISL_783490, EPI_ISL_783501, EPI_ISL_783515, EPI_ISL_783516, EPI_ISL_783517, EPI_ISL_783518, EPI_ISL_783522, EPI_ISL_783525, EPI_ISL_783526, EPI_ISL_783534

| | | | |
|---|---|---|---|
| see above | Lighthouse Lab in Cambridge | Wellcome Sanger Institute for the COVID-19 Genomics UK (COG-UK) Consortium | Cordelia Langford; David K. Jackson; Dominic Kwiatkowski; Ewan Harrison; Ian Johnston; John Sillitoe on behalf of the Wellcome Sanger Institute COVID-19 Surveillance Team; John Sillitoe on behalf of the Wellcome Sanger Institute COVID-19 Surveillance Team (http://www.sanger.ac.uk/covid-team); Rob Howes; Roberto Amato; Sonia Goncalves; The Lighthouse Lab in Cambridge and Alex Alderton |

EPI_ISL_641877, EPI_ISL_646874, EPI_ISL_654959, EPI_ISL_654960, EPI_ISL_654961, EPI_ISL_654962, EPI_ISL_654963, EPI_ISL_654964, EPI_ISL_654965, EPI_ISL_654966, EPI_ISL_654967, EPI_ISL_654968, EPI_ISL_654969, EPI_ISL_654970, EPI_ISL_654971, EPI_ISL_654972, EPI_ISL_654973, EPI_ISL_654974, EPI_ISL_654975, EPI_ISL_654976, EPI_ISL_654977, EPI_ISL_654978, EPI_ISL_654979, EPI_ISL_654980, EPI_ISL_654981, EPI_ISL_654982, EPI_ISL_654983, EPI_ISL_654984, EPI_ISL_654985, EPI_ISL_654986, EPI_ISL_654987, EPI_ISL_654988, EPI_ISL_654989, EPI_ISL_654990, EPI_ISL_654991, EPI_ISL_654992, EPI_ISL_654993, EPI_ISL_654994, EPI_ISL_654996, EPI_ISL_654997, EPI_ISL_654998, EPI_ISL_654999, EPI_ISL_655000, EPI_ISL_655001, EPI_ISL_655002, EPI_ISL_655003, EPI_ISL_655004, EPI_ISL_655005, EPI_ISL_655006, EPI_ISL_655007, EPI_ISL_655008, EPI_ISL_655009, EPI_ISL_655010, EPI_ISL_655011, EPI_ISL_655012, EPI_ISL_655013, EPI_ISL_655014, EPI_ISL_655015, EPI_ISL_655016, EPI_ISL_655017, EPI_ISL_655018, EPI_ISL_655019, EPI_ISL_655020, EPI_ISL_655021, EPI_ISL_655022, EPI_ISL_655023, EPI_ISL_655024, EPI_ISL_655025, EPI_ISL_655026, EPI_ISL_655027, EPI_ISL_655028, EPI_ISL_655029, EPI_ISL_655030, EPI_ISL_655031, EPI_ISL_655032, EPI_ISL_655033, EPI_ISL_655034, EPI_ISL_655035, EPI_ISL_655036, EPI_ISL_655037, EPI_ISL_655038, EPI_ISL_655039, EPI_ISL_655040, EPI_ISL_655041, EPI_ISL_655042, EPI_ISL_655044, EPI_ISL_655045, EPI_ISL_655046, EPI_ISL_655047, EPI_ISL_655048, EPI_ISL_655049, EPI_ISL_655050, EPI_ISL_655051, EPI_ISL_655052, EPI_ISL_655053, EPI_ISL_655054, EPI_ISL_655055, EPI_ISL_655056, EPI_ISL_655057, EPI_ISL_655058, EPI_ISL_655059, EPI_ISL_655060, EPI_ISL_655061, EPI_ISL_655062, EPI_ISL_655063, EPI_ISL_655064, EPI_ISL_655065, EPI_ISL_655066, EPI_ISL_655067, EPI_ISL_655068, EPI_ISL_655069, EPI_ISL_655070, EPI_ISL_655071, EPI_ISL_655072, EPI_ISL_655073, EPI_ISL_655074, EPI_ISL_655075, EPI_ISL_655076, EPI_ISL_655077, EPI_ISL_655078, EPI_ISL_655079, EPI_ISL_655080, EPI_ISL_655081, EPI_ISL_655082, EPI_ISL_655083, EPI_ISL_655084, EPI_ISL_655085, EPI_ISL_655086, EPI_ISL_655087, EPI_ISL_655088, EPI_ISL_655089, EPI_ISL_655090, EPI_ISL_655091, EPI_ISL_655092, EPI_ISL_655093, EPI_ISL_655094, EPI_ISL_655095, EPI_ISL_655096, EPI_ISL_655097, EPI_ISL_655098, EPI_ISL_655099, EPI_ISL_655100, EPI_ISL_655101, EPI_ISL_655102, EPI_ISL_655103, EPI_ISL_655104, EPI_ISL_655105, EPI_ISL_655106, EPI_ISL_655107, EPI_ISL_655108, EPI_ISL_655109, EPI_ISL_655110, EPI_ISL_655111, EPI_ISL_655112, EPI_ISL_655113, EPI_ISL_655114, EPI_ISL_655115, EPI_ISL_655116, EPI_ISL_655117, EPI_ISL_655118, EPI_ISL_655119, EPI_ISL_655120, EPI_ISL_655121, EPI_ISL_655122, EPI_ISL_655123, EPI_ISL_655124, EPI_ISL_655125, EPI_ISL_655126, EPI_ISL_655127, EPI_ISL_655128, EPI_ISL_655129, EPI_ISL_655130, EPI_ISL_655131, EPI_ISL_655132, EPI_ISL_655133, EPI_ISL_655134, EPI_ISL_655135, EPI_ISL_655136, EPI_ISL_655137, EPI_ISL_655138, EPI_ISL_655139, EPI_ISL_655140, EPI_ISL_655141, EPI_ISL_655142, EPI_ISL_655143, EPI_ISL_655144, EPI_ISL_655145, EPI_ISL_655146, EPI_ISL_655147, EPI_ISL_655148, EPI_ISL_655149, EPI_ISL_655150, EPI_ISL_655151, EPI_ISL_655152, EPI_ISL_655153, EPI_ISL_655154, EPI_ISL_655155, EPI_ISL_655156, EPI_ISL_655157, EPI_ISL_655158, EPI_ISL_655159, EPI_ISL_655160, EPI_ISL_655161, EPI_ISL_655162, EPI_ISL_655163, EPI_ISL_655164, EPI_ISL_655165, EPI_ISL_655166, EPI_ISL_655167, EPI_ISL_655168, EPI_ISL_655169, EPI_ISL_655170, EPI_ISL_655171, EPI_ISL_655172, EPI_ISL_655173, EPI_ISL_655174, EPI_ISL_655175, EPI_ISL_655176, EPI_ISL_655177, EPI_ISL_655178, EPI_ISL_655179, EPI_ISL_655180, EPI_ISL_655181, EPI_ISL_655182, EPI_ISL_655183, EPI_ISL_655184, EPI_ISL_655185, EPI_ISL_655186, EPI_ISL_655187, EPI_ISL_655188, EPI_ISL_655189, EPI_ISL_655190, EPI_ISL_655191, EPI_ISL_655192, EPI_ISL_655193, EPI_ISL_655194, EPI_ISL_655195, EPI_ISL_655196, EPI_ISL_655197, EPI_ISL_655198, EPI_ISL_655199, EPI_ISL_655200, EPI_ISL_655201, EPI_ISL_655202, EPI_ISL_655203, EPI_ISL_655204, EPI_ISL_655205, EPI_ISL_655206, EPI_ISL_655207, EPI_ISL_655208, EPI_ISL_655209, EPI_ISL_655210, EPI_ISL_655211, EPI_ISL_655212, EPI_ISL_655213, EPI_ISL_655214, EPI_ISL_655215, EPI_ISL_655216, EPI_ISL_655217, EPI_ISL_655218, EPI_ISL_655219, EPI_ISL_655220, EPI_ISL_655221, EPI_ISL_655222, EPI_ISL_655223, EPI_ISL_655224, EPI_ISL_655225, EPI_ISL_655226, EPI_ISL_655227, EPI_ISL_655228, EPI_ISL_655229, EPI_ISL_655230, EPI_ISL_655231, EPI_ISL_655232, EPI_ISL_655233, EPI_ISL_655234, EPI_ISL_655235, EPI_ISL_655236, EPI_ISL_655237, EPI_ISL_655238, EPI_ISL_655239, EPI_ISL_655240, EPI_ISL_655241, EPI_ISL_655242, EPI_ISL_655243, EPI_ISL_655244, EPI_ISL_655245, EPI_ISL_655246, EPI_ISL_655247, EPI_ISL_655248, EPI_ISL_655249, EPI_ISL_655250, EPI_ISL_655251, EPI_ISL_655252, EPI_ISL_655253, EPI_ISL_655254, EPI_ISL_655255, EPI_ISL_655256, EPI_ISL_655257, EPI_ISL_655258, EPI_ISL_655259, EPI_ISL_655260, EPI_ISL_655261, EPI_ISL_655262, EPI_ISL_655263, EPI_ISL_655264, EPI_ISL_655265, EPI_ISL_655266, EPI_ISL_655267, EPI_ISL_655268, EPI_ISL_655269, EPI_ISL_655270, EPI_ISL_655271, EPI_ISL_655272, EPI_ISL_655651, EPI_ISL_655660, EPI_ISL_655662, EPI_ISL_655672, EPI_ISL_655677, EPI_ISL_655694, EPI_ISL_655695, EPI_ISL_655697, EPI_ISL_655699, EPI_ISL_655711, EPI_ISL_655719, EPI_ISL_655723, EPI_ISL_655727, EPI_ISL_655735, EPI_ISL_655737, EPI_ISL_655739, EPI_ISL_655744, EPI_ISL_655745, EPI_ISL_655756, EPI_ISL_655762, EPI_ISL_655768, EPI_ISL_655769, EPI_ISL_655770, EPI_ISL_655773, EPI_ISL_655779, EPI_ISL_655780, EPI_ISL_655783, EPI_ISL_655788, EPI_ISL_655798, EPI_ISL_655804, EPI_ISL_655806, EPI_ISL_655807, EPI_ISL_655809, EPI_ISL_655817, EPI_ISL_655819, EPI_ISL_655822, EPI_ISL_655828, EPI_ISL_655832, EPI_ISL_655841, EPI_ISL_655847, EPI_ISL_655850, EPI_ISL_655854, EPI_ISL_655861, EPI_ISL_655864, EPI_ISL_655865, EPI_ISL_655868, EPI_ISL_655873, EPI_ISL_655874, EPI_ISL_655879, EPI_ISL_655883, EPI_ISL_655891, EPI_ISL_655907, EPI_ISL_655911, EPI_ISL_655912, EPI_ISL_655918, EPI_ISL_655920, EPI_ISL_655924, EPI_ISL_655929, EPI_ISL_655933, EPI_ISL_655947, EPI_ISL_655948, EPI_ISL_655957, EPI_ISL_655959, EPI_ISL_655967, EPI_ISL_655971, EPI_ISL_655973, EPI_ISL_655975, EPI_ISL_657027, EPI_ISL_657028, EPI_ISL_657029, EPI_ISL_657030, EPI_ISL_657031, EPI_ISL_657032, EPI_ISL_657033, EPI_ISL_657034, EPI_ISL_657035, EPI_ISL_657036, EPI_ISL_657037, EPI_ISL_657038, EPI_ISL_657039, EPI_ISL_657040, EPI_ISL_657041, EPI_ISL_657042, EPI_ISL_657043, EPI_ISL_657044, EPI_ISL_657045, EPI_ISL_657046, EPI_ISL_657047, EPI_ISL_657048, EPI_ISL_657049, EPI_ISL_657050, EPI_ISL_657051, EPI_ISL_657052, EPI_ISL_657053, EPI_ISL_657054, EPI_ISL_657055, EPI_ISL_657056, EPI_ISL_657057, EPI_ISL_657058, EPI_ISL_657059, EPI_ISL_657060, EPI_ISL_657061, EPI_ISL_657062, EPI_ISL_657063, EPI_ISL_657064, EPI_ISL_657065, EPI_ISL_657066, EPI_ISL_657067, EPI_ISL_657068, EPI_ISL_657069, EPI_ISL_657070, EPI_ISL_657071, EPI_ISL_657072, EPI_ISL_657073, EPI_ISL_657074, EPI_ISL_657075, EPI_ISL_657076, EPI_ISL_657077, EPI_ISL_657078, EPI_ISL_657079, EPI_ISL_657080, EPI_ISL_657081, EPI_ISL_657082, EPI_ISL_657083, EPI_ISL_657084, EPI_ISL_657085, EPI_ISL_657086, EPI_ISL_657087, EPI_ISL_657088, EPI_ISL_657089, EPI_ISL_657090, EPI_ISL_657091, EPI_ISL_657092, EPI_ISL_657093, EPI_ISL_657094, EPI_ISL_657095, EPI_ISL_657096, EPI_ISL_657097, EPI_ISL_657098, EPI_ISL_657099, EPI_ISL_657100, EPI_ISL_657101, EPI_ISL_657102, EPI_ISL_657103, EPI_ISL_657104, EPI_ISL_657105, EPI_ISL_657106, EPI_ISL_657107, EPI_ISL_657108, EPI_ISL_657109, EPI_ISL_657110, EPI_ISL_657111, EPI_ISL_657112, EPI_ISL_657113, EPI_ISL_657114, EPI_ISL_657115, EPI_ISL_657116, EPI_ISL_657117, EPI_ISL_657118, EPI_ISL_657119, EPI_ISL_657120, EPI_ISL_657121, EPI_ISL_657122, EPI_ISL_657123,

| | | | |
|---|---|---|---|
| EPI_ISL_657124, EPI_ISL_657125, EPI_ISL_657126, EPI_ISL_657127, EPI_ISL_657128, EPI_ISL_657129, EPI_ISL_657130, EPI_ISL_657131, EPI_ISL_657132, EPI_ISL_657133, EPI_ISL_657134, EPI_ISL_657135, EPI_ISL_657136, EPI_ISL_657137, EPI_ISL_657138, EPI_ISL_657139, EPI_ISL_657140, EPI_ISL_657141, EPI_ISL_657142, EPI_ISL_657143, EPI_ISL_657144, EPI_ISL_657145, EPI_ISL_657146, EPI_ISL_657147, EPI_ISL_657148, EPI_ISL_657149, EPI_ISL_657150, EPI_ISL_657151, EPI_ISL_657152, EPI_ISL_657153, EPI_ISL_657154, EPI_ISL_657155, EPI_ISL_657156, EPI_ISL_657157, EPI_ISL_657158, EPI_ISL_657159, EPI_ISL_657160, EPI_ISL_657161, EPI_ISL_657162, EPI_ISL_657163, EPI_ISL_657164, EPI_ISL_657165, EPI_ISL_657166, EPI_ISL_657167, EPI_ISL_657168, EPI_ISL_657169, EPI_ISL_657170, EPI_ISL_657171, EPI_ISL_657172, EPI_ISL_657173, EPI_ISL_657174, EPI_ISL_657175, EPI_ISL_657176, EPI_ISL_657177, EPI_ISL_657178, EPI_ISL_657179, EPI_ISL_657180, EPI_ISL_657181, EPI_ISL_657182, EPI_ISL_657183, EPI_ISL_657184, EPI_ISL_657185, EPI_ISL_657186, EPI_ISL_657187, EPI_ISL_657188, EPI_ISL_657189, EPI_ISL_657190, EPI_ISL_657191, EPI_ISL_657192, EPI_ISL_657193, EPI_ISL_657194, EPI_ISL_657195, EPI_ISL_657196, EPI_ISL_657197, EPI_ISL_657198, EPI_ISL_657199, EPI_ISL_657200, EPI_ISL_657201, EPI_ISL_657202, EPI_ISL_657203, EPI_ISL_657204, EPI_ISL_657205, EPI_ISL_657206, EPI_ISL_657207, EPI_ISL_657208, EPI_ISL_657209, EPI_ISL_657210, EPI_ISL_657211, EPI_ISL_657212, EPI_ISL_657213, EPI_ISL_657214, EPI_ISL_657215, EPI_ISL_657216, EPI_ISL_657217, EPI_ISL_657218, EPI_ISL_657219, EPI_ISL_657220, EPI_ISL_657221, EPI_ISL_657222, EPI_ISL_657223, EPI_ISL_657224, EPI_ISL_657225, EPI_ISL_657226, EPI_ISL_657227, EPI_ISL_657228, EPI_ISL_657229, EPI_ISL_657230, EPI_ISL_657231, EPI_ISL_657232, EPI_ISL_657233, EPI_ISL_657234, EPI_ISL_657235, EPI_ISL_657236, EPI_ISL_657237, EPI_ISL_657238, EPI_ISL_657239, EPI_ISL_657240, EPI_ISL_657241, EPI_ISL_657242, EPI_ISL_657243, EPI_ISL_657244, EPI_ISL_657245, EPI_ISL_657246, EPI_ISL_657247, EPI_ISL_657248, EPI_ISL_657249, EPI_ISL_657250, EPI_ISL_657251, EPI_ISL_657252, EPI_ISL_657253, EPI_ISL_657254, EPI_ISL_657255, EPI_ISL_657256, EPI_ISL_657257, EPI_ISL_657258, EPI_ISL_657259, EPI_ISL_657260, EPI_ISL_657261, EPI_ISL_657262, EPI_ISL_657263, EPI_ISL_657264, EPI_ISL_657265, EPI_ISL_657266, EPI_ISL_657267, EPI_ISL_657268, EPI_ISL_657269, EPI_ISL_657270, EPI_ISL_657271, EPI_ISL_657272, EPI_ISL_657273, EPI_ISL_657274, EPI_ISL_657275, EPI_ISL_657276, EPI_ISL_657277, EPI_ISL_657278, EPI_ISL_657279, EPI_ISL_657280, EPI_ISL_657281, EPI_ISL_657282, EPI_ISL_657283, EPI_ISL_657284, EPI_ISL_657285, EPI_ISL_657286, EPI_ISL_657287, EPI_ISL_657288, EPI_ISL_657289, EPI_ISL_657290, EPI_ISL_657291, EPI_ISL_657292, EPI_ISL_657293, EPI_ISL_657294, EPI_ISL_657295, EPI_ISL_657296, EPI_ISL_657297, EPI_ISL_657298, EPI_ISL_657299, EPI_ISL_657300, EPI_ISL_657301, EPI_ISL_657302, EPI_ISL_657303, EPI_ISL_657304, EPI_ISL_657305, EPI_ISL_657306, EPI_ISL_657307, EPI_ISL_657308, EPI_ISL_657309, EPI_ISL_657310, EPI_ISL_657311, EPI_ISL_657312, EPI_ISL_657313, EPI_ISL_657314, EPI_ISL_657315, EPI_ISL_657316, EPI_ISL_657317, EPI_ISL_657318, EPI_ISL_657319, EPI_ISL_657320, EPI_ISL_657321, EPI_ISL_657322, EPI_ISL_657323, EPI_ISL_657324, EPI_ISL_657325, EPI_ISL_657326, EPI_ISL_657327, EPI_ISL_657328, EPI_ISL_657329, EPI_ISL_657330, EPI_ISL_657331, EPI_ISL_657332, EPI_ISL_657333, EPI_ISL_657334, EPI_ISL_657335, EPI_ISL_657336, EPI_ISL_657337, EPI_ISL_657338, EPI_ISL_657339, EPI_ISL_657340, EPI_ISL_657341, EPI_ISL_657342, EPI_ISL_657343, EPI_ISL_657344, EPI_ISL_657345, EPI_ISL_657346, EPI_ISL_657347, EPI_ISL_657348, EPI_ISL_657349, EPI_ISL_657350, EPI_ISL_657351, EPI_ISL_657352, EPI_ISL_657353, EPI_ISL_657354, EPI_ISL_658404, EPI_ISL_658405, EPI_ISL_658406, EPI_ISL_658407, EPI_ISL_658408, EPI_ISL_658409, EPI_ISL_658410, EPI_ISL_658411, EPI_ISL_658412, EPI_ISL_658413, EPI_ISL_658414, EPI_ISL_658415, EPI_ISL_658416, EPI_ISL_658417, EPI_ISL_658418, EPI_ISL_658419, EPI_ISL_658420, EPI_ISL_658421, EPI_ISL_658422, EPI_ISL_658423, EPI_ISL_658424, EPI_ISL_658425, EPI_ISL_658426, EPI_ISL_658427, EPI_ISL_658428, EPI_ISL_658429, EPI_ISL_658430, EPI_ISL_658431, EPI_ISL_658432, EPI_ISL_658433, EPI_ISL_658434, EPI_ISL_658435, EPI_ISL_658436, EPI_ISL_658437, EPI_ISL_658438, EPI_ISL_658439, EPI_ISL_658440, EPI_ISL_658441, EPI_ISL_658442, EPI_ISL_658443, EPI_ISL_658444, EPI_ISL_658445, EPI_ISL_658446, EPI_ISL_658447, EPI_ISL_658448, EPI_ISL_658449, EPI_ISL_658450, EPI_ISL_658451, EPI_ISL_658452, EPI_ISL_658453, EPI_ISL_658454, EPI_ISL_658455, EPI_ISL_658456, EPI_ISL_658457, EPI_ISL_658458, EPI_ISL_658459, EPI_ISL_658460, EPI_ISL_658461, EPI_ISL_658462, EPI_ISL_658463, EPI_ISL_658464, EPI_ISL_658465, EPI_ISL_658466, EPI_ISL_658467, EPI_ISL_658468, EPI_ISL_658469, EPI_ISL_658470, EPI_ISL_658471, EPI_ISL_658472, EPI_ISL_658473, EPI_ISL_658474, EPI_ISL_658475, EPI_ISL_658476, EPI_ISL_658477, EPI_ISL_658478, EPI_ISL_658479, EPI_ISL_658480, EPI_ISL_658481, EPI_ISL_658482, EPI_ISL_658483, EPI_ISL_658484, EPI_ISL_658485, EPI_ISL_658486, EPI_ISL_658487, EPI_ISL_658488, EPI_ISL_658489, EPI_ISL_658490, EPI_ISL_658491, EPI_ISL_658492, EPI_ISL_658493, EPI_ISL_658494, EPI_ISL_658495, EPI_ISL_658496, EPI_ISL_658497, EPI_ISL_658498, EPI_ISL_658499, EPI_ISL_658500, EPI_ISL_658501, EPI_ISL_658502, EPI_ISL_658503, EPI_ISL_658504, EPI_ISL_658505, EPI_ISL_658506, EPI_ISL_658507, EPI_ISL_658508, EPI_ISL_658509, EPI_ISL_658510, EPI_ISL_658511, EPI_ISL_658512, EPI_ISL_658513, EPI_ISL_658514, EPI_ISL_658515, EPI_ISL_658516, EPI_ISL_658517, EPI_ISL_658518, EPI_ISL_658519, EPI_ISL_658520, EPI_ISL_658521, EPI_ISL_658522, EPI_ISL_658523, EPI_ISL_658524, EPI_ISL_658525, EPI_ISL_658526, EPI_ISL_658527, EPI_ISL_658528, EPI_ISL_658529, EPI_ISL_658530, EPI_ISL_658531, EPI_ISL_658532, EPI_ISL_658533, EPI_ISL_658534, EPI_ISL_658535, EPI_ISL_658536, EPI_ISL_658537, EPI_ISL_658538, EPI_ISL_658539, EPI_ISL_658540, EPI_ISL_658541, EPI_ISL_658542, EPI_ISL_658543, EPI_ISL_658544, EPI_ISL_658545, EPI_ISL_658546, EPI_ISL_658547, EPI_ISL_658548, EPI_ISL_658549, EPI_ISL_658550, EPI_ISL_658551, EPI_ISL_658552, EPI_ISL_658553, EPI_ISL_658554, EPI_ISL_658555, EPI_ISL_658556, EPI_ISL_658557, EPI_ISL_658558, EPI_ISL_658559, EPI_ISL_658560, EPI_ISL_658561, EPI_ISL_658562, EPI_ISL_658563, EPI_ISL_658564, EPI_ISL_658565, EPI_ISL_658566, EPI_ISL_658567, EPI_ISL_658568, EPI_ISL_658569, EPI_ISL_658570, EPI_ISL_658571, EPI_ISL_658572, EPI_ISL_658573, EPI_ISL_658574, EPI_ISL_658575, EPI_ISL_658576, EPI_ISL_658577, EPI_ISL_658578, EPI_ISL_658579, EPI_ISL_658580, EPI_ISL_658581, EPI_ISL_658582, EPI_ISL_658583, EPI_ISL_658584, EPI_ISL_658585, EPI_ISL_658586, EPI_ISL_658587, EPI_ISL_658588, EPI_ISL_658589, EPI_ISL_658590, EPI_ISL_658591, EPI_ISL_658592, EPI_ISL_658593, EPI_ISL_658594, EPI_ISL_658595, EPI_ISL_658596, EPI_ISL_658597, EPI_ISL_658598, EPI_ISL_658599, EPI_ISL_658600, EPI_ISL_658601, EPI_ISL_658602, EPI_ISL_658603, EPI_ISL_658604, EPI_ISL_658605, EPI_ISL_658606, EPI_ISL_658607, EPI_ISL_658608, EPI_ISL_658609, EPI_ISL_658610, EPI_ISL_658611, EPI_ISL_658612, EPI_ISL_658613, EPI_ISL_658614, EPI_ISL_658615, EPI_ISL_658616, EPI_ISL_658617, EPI_ISL_658618, EPI_ISL_658619, EPI_ISL_658620, EPI_ISL_658621, EPI_ISL_658622, EPI_ISL_658623, EPI_ISL_658624, EPI_ISL_658625, EPI_ISL_658626, EPI_ISL_658627, EPI_ISL_658628, EPI_ISL_658629, EPI_ISL_658630, EPI_ISL_658631, EPI_ISL_658632, EPI_ISL_658633, EPI_ISL_658634, EPI_ISL_658635, EPI_ISL_658636, EPI_ISL_658637, EPI_ISL_658638, EPI_ISL_658639, EPI_ISL_658640, EPI_ISL_658641, EPI_ISL_658642, EPI_ISL_658643, EPI_ISL_658644, EPI_ISL_658645, EPI_ISL_658646, EPI_ISL_658647, EPI_ISL_658648, EPI_ISL_658649, EPI_ISL_658650, EPI_ISL_658651, EPI_ISL_658652, EPI_ISL_658653, EPI_ISL_658654, EPI_ISL_658655, EPI_ISL_658656, EPI_ISL_658657, EPI_ISL_658658, EPI_ISL_658659, EPI_ISL_658660, EPI_ISL_658661, EPI_ISL_658662, EPI_ISL_658663, EPI_ISL_658664, EPI_ISL_658665, EPI_ISL_658666, EPI_ISL_658667, EPI_ISL_658668, EPI_ISL_658669, EPI_ISL_658670, EPI_ISL_658671, EPI_ISL_658672, EPI_ISL_658673, EPI_ISL_658674, EPI_ISL_658675, EPI_ISL_658676, EPI_ISL_658677, EPI_ISL_658678, EPI_ISL_658679, EPI_ISL_658680, EPI_ISL_658681, EPI_ISL_658682, EPI_ISL_658683, EPI_ISL_658684, EPI_ISL_658685, EPI_ISL_658686, EPI_ISL_658687, EPI_ISL_658688, EPI_ISL_658689, EPI_ISL_658690, EPI_ISL_658691, EPI_ISL_658692, EPI_ISL_658693, EPI_ISL_658694, EPI_ISL_658695, EPI_ISL_658696, EPI_ISL_658697, EPI_ISL_658698, EPI_ISL_658699, EPI_ISL_658700, EPI_ISL_658701, EPI_ISL_658702, EPI_ISL_658703, EPI_ISL_658704, EPI_ISL_658705, EPI_ISL_658706, EPI_ISL_658707, EPI_ISL_658708, EPI_ISL_658709, EPI_ISL_658710, EPI_ISL_658711, EPI_ISL_658712, EPI_ISL_658713, EPI_ISL_658714, EPI_ISL_658715, EPI_ISL_658716, EPI_ISL_658717, EPI_ISL_658718, EPI_ISL_658719, EPI_ISL_658720, EPI_ISL_658721, EPI_ISL_658722, EPI_ISL_658723, EPI_ISL_658724, EPI_ISL_658725, EPI_ISL_658726, EPI_ISL_658727, EPI_ISL_658728, EPI_ISL_658729, EPI_ISL_658730, EPI_ISL_658731, EPI_ISL_658732, EPI_ISL_658733, EPI_ISL_658734, EPI_ISL_658735, EPI_ISL_658736, EPI_ISL_658737, EPI_ISL_658738, EPI_ISL_658739, EPI_ISL_658740, EPI_ISL_661314, EPI_ISL_661315, EPI_ISL_661316, EPI_ISL_661317, EPI_ISL_661318, EPI_ISL_661319, EPI_ISL_661320, EPI_ISL_661321, EPI_ISL_661322, EPI_ISL_661323, EPI_ISL_661324, EPI_ISL_661325, EPI_ISL_661326, EPI_ISL_661327, EPI_ISL_661328, EPI_ISL_661329, EPI_ISL_661330, EPI_ISL_661331, EPI_ISL_661332, EPI_ISL_661333, EPI_ISL_661334, EPI_ISL_661335, EPI_ISL_661336, EPI_ISL_661337, EPI_ISL_661338, EPI_ISL_661339, EPI_ISL_661340, EPI_ISL_661341, EPI_ISL_661342, EPI_ISL_661343, EPI_ISL_661344, EPI_ISL_661345, EPI_ISL_661346, EPI_ISL_661347, EPI_ISL_661348, EPI_ISL_661349, EPI_ISL_661350, EPI_ISL_661351, EPI_ISL_661352, EPI_ISL_661353, EPI_ISL_661354, EPI_ISL_661355, EPI_ISL_661356, EPI_ISL_661357, EPI_ISL_661358, EPI_ISL_661359, EPI_ISL_661360, EPI_ISL_661361, EPI_ISL_661362, EPI_ISL_661363, EPI_ISL_661364, EPI_ISL_661365, EPI_ISL_661366, EPI_ISL_661367, EPI_ISL_661368, EPI_ISL_661369, EPI_ISL_661370, EPI_ISL_661371, EPI_ISL_661372, EPI_ISL_661373, EPI_ISL_661374, EPI_ISL_661375, EPI_ISL_661376, EPI_ISL_661377, EPI_ISL_661378, EPI_ISL_661379, EPI_ISL_661380, EPI_ISL_661381, EPI_ISL_661382, EPI_ISL_661383, EPI_ISL_661384, EPI_ISL_661385, EPI_ISL_661386, EPI_ISL_661387, EPI_ISL_661388, EPI_ISL_661389, EPI_ISL_661390, EPI_ISL_661391, EPI_ISL_661392, EPI_ISL_661393, EPI_ISL_661394, EPI_ISL_661395, EPI_ISL_661396, EPI_ISL_661397, EPI_ISL_661398, EPI_ISL_661399, EPI_ISL_661400, EPI_ISL_661401, EPI_ISL_661402, EPI_ISL_661403, EPI_ISL_661404, EPI_ISL_661405, EPI_ISL_661406, EPI_ISL_661407, EPI_ISL_661408, EPI_ISL_661409, EPI_ISL_661410, EPI_ISL_661411, EPI_ISL_661412, EPI_ISL_661413, EPI_ISL_661414, EPI_ISL_661415, EPI_ISL_661416, EPI_ISL_661417, EPI_ISL_661418, EPI_ISL_661419, EPI_ISL_661420, EPI_ISL_661421, EPI_ISL_661422, EPI_ISL_661423, EPI_ISL_661424, EPI_ISL_661425, EPI_ISL_661426, EPI_ISL_661427, EPI_ISL_661428, EPI_ISL_661429, EPI_ISL_661430, EPI_ISL_661431, EPI_ISL_661432, EPI_ISL_661433, EPI_ISL_661434, EPI_ISL_661435, EPI_ISL_661436, EPI_ISL_661437, EPI_ISL_661438, EPI_ISL_661439, EPI_ISL_661440, EPI_ISL_661441, EPI_ISL_661442, EPI_ISL_661443, EPI_ISL_661444, EPI_ISL_661445, EPI_ISL_661446, EPI_ISL_661447, EPI_ISL_661448, EPI_ISL_661449, EPI_ISL_661450, EPI_ISL_661451, EPI_ISL_661452, EPI_ISL_661453, EPI_ISL_661454, EPI_ISL_661455, EPI_ISL_661456, EPI_ISL_661457, EPI_ISL_661458, EPI_ISL_661459, EPI_ISL_661460, EPI_ISL_661461, EPI_ISL_661462, EPI_ISL_661463, EPI_ISL_661464, EPI_ISL_661465, EPI_ISL_661466, EPI_ISL_661467, EPI_ISL_661468, EPI_ISL_661469, EPI_ISL_661470, EPI_ISL_661471, EPI_ISL_661472, EPI_ISL_661473, EPI_ISL_661474, EPI_ISL_661475, EPI_ISL_661476, EPI_ISL_661477, EPI_ISL_661478, EPI_ISL_661479, EPI_ISL_661480, EPI_ISL_661481, EPI_ISL_661482, EPI_ISL_661483, EPI_ISL_661484, EPI_ISL_661485, EPI_ISL_661486, EPI_ISL_661487, EPI_ISL_661488, EPI_ISL_661489, EPI_ISL_661490, EPI_ISL_661491, EPI_ISL_661492, EPI_ISL_661493, EPI_ISL_661494, EPI_ISL_661495, EPI_ISL_661496, EPI_ISL_661497, EPI_ISL_661498, EPI_ISL_661499, EPI_ISL_661500, EPI_ISL_661501, EPI_ISL_661502, EPI_ISL_661503, EPI_ISL_661504, EPI_ISL_661505, EPI_ISL_661506, EPI_ISL_661507, EPI_ISL_661508, EPI_ISL_661509, EPI_ISL_661510, EPI_ISL_661511, EPI_ISL_661512, EPI_ISL_661513, EPI_ISL_661514, EPI_ISL_661515, EPI_ISL_661516, EPI_ISL_661517, EPI_ISL_661518, EPI_ISL_661519, EPI_ISL_661520, EPI_ISL_661521, EPI_ISL_661522, EPI_ISL_661523, EPI_ISL_661524, EPI_ISL_661525, EPI_ISL_661526, EPI_ISL_661527, EPI_ISL_661528, EPI_ISL_661529, EPI_ISL_661530, EPI_ISL_661531, EPI_ISL_661532, EPI_ISL_661533, EPI_ISL_661534, EPI_ISL_661535, EPI_ISL_661536, EPI_ISL_661537, EPI_ISL_661538, EPI_ISL_661539, EPI_ISL_661540, EPI_ISL_661541, EPI_ISL_661542, EPI_ISL_661543, EPI_ISL_661544, EPI_ISL_661545, EPI_ISL_661546, EPI_ISL_661547, EPI_ISL_661548, EPI_ISL_661549, EPI_ISL_661550, EPI_ISL_661551, EPI_ISL_661552, EPI_ISL_661553, EPI_ISL_661554, EPI_ISL_661555, EPI_ISL_661556, EPI_ISL_661557, EPI_ISL_661558, EPI_ISL_661559, EPI_ISL_661560, EPI_ISL_661561, EPI_ISL_661562, EPI_ISL_661563, EPI_ISL_661564, EPI_ISL_661565, EPI_ISL_661566, EPI_ISL_661567, EPI_ISL_661568, EPI_ISL_661569, EPI_ISL_661570, EPI_ISL_661571, EPI_ISL_661572, EPI_ISL_661573, EPI_ISL_661574, EPI_ISL_661575, EPI_ISL_661576, EPI_ISL_661577, EPI_ISL_661578, EPI_ISL_661579, EPI_ISL_661580, EPI_ISL_661581, EPI_ISL_661582, EPI_ISL_661583, EPI_ISL_661584, EPI_ISL_661585, EPI_ISL_661586, EPI_ISL_661587, EPI_ISL_661588, EPI_ISL_661589, EPI_ISL_661590, EPI_ISL_661591, EPI_ISL_661592, EPI_ISL_661593, EPI_ISL_661594, EPI_ISL_661595, EPI_ISL_661596, EPI_ISL_661597, EPI_ISL_661598, EPI_ISL_661599, EPI_ISL_661600, EPI_ISL_661601, EPI_ISL_661602, EPI_ISL_661603, EPI_ISL_661604, EPI_ISL_661605, EPI_ISL_661606, EPI_ISL_661607, EPI_ISL_661608, EPI_ISL_661609, EPI_ISL_661610, EPI_ISL_661611, EPI_ISL_661612, EPI_ISL_661613, EPI_ISL_661614, EPI_ISL_661615, EPI_ISL_661616, EPI_ISL_661617, EPI_ISL_661618, EPI_ISL_661619, EPI_ISL_661620, EPI_ISL_661621, EPI_ISL_661622, EPI_ISL_661623, EPI_ISL_661624, EPI_ISL_661625, EPI_ISL_661626, EPI_ISL_661627, EPI_ISL_661628, EPI_ISL_661629, EPI_ISL_661630, EPI_ISL_661631, EPI_ISL_661957, EPI_ISL_661959, EPI_ISL_661960, EPI_ISL_661963, EPI_ISL_661967, EPI_ISL_661969, EPI_ISL_661971, EPI_ISL_661972, EPI_ISL_661973, EPI_ISL_661975, EPI_ISL_661976, EPI_ISL_661979, EPI_ISL_661981, EPI_ISL_661984, EPI_ISL_661987, EPI_ISL_661993, EPI_ISL_661994, EPI_ISL_661996, EPI_ISL_661997, EPI_ISL_662003, EPI_ISL_662005, EPI_ISL_662007, EPI_ISL_662009, EPI_ISL_662011, EPI_ISL_662012, EPI_ISL_662016, EPI_ISL_662021, EPI_ISL_662023, EPI_ISL_662024, EPI_ISL_662026, EPI_ISL_662027, EPI_ISL_662028, EPI_ISL_662030, EPI_ISL_662033, EPI_ISL_662035, EPI_ISL_662037, EPI_ISL_662039, EPI_ISL_662040, EPI_ISL_662041, EPI_ISL_662042, EPI_ISL_662044, EPI_ISL_662052, EPI_ISL_662054, EPI_ISL_662059, EPI_ISL_662064, EPI_ISL_662066, EPI_ISL_662069, EPI_ISL_662072, EPI_ISL_662073, EPI_ISL_662075, EPI_ISL_662076, EPI_ISL_662077, EPI_ISL_662082, EPI_ISL_662083, EPI_ISL_662084, EPI_ISL_662085, EPI_ISL_662087, EPI_ISL_662088, EPI_ISL_662089, EPI_ISL_662090, EPI_ISL_662091, EPI_ISL_662092, EPI_ISL_662094, EPI_ISL_662095, EPI_ISL_662096, EPI_ISL_662102, EPI_ISL_662103, EPI_ISL_662104, EPI_ISL_662109, EPI_ISL_662110, EPI_ISL_662111, EPI_ISL_662112, EPI_ISL_662113, EPI_ISL_662117, EPI_ISL_662119, EPI_ISL_662121, EPI_ISL_662125, EPI_ISL_662126, EPI_ISL_662127, EPI_ISL_662128, EPI_ISL_662130, EPI_ISL_662134, EPI_ISL_662136, EPI_ISL_662139, EPI_ISL_662142, EPI_ISL_662144, EPI_ISL_662147, EPI_ISL_662149, EPI_ISL_662152, EPI_ISL_662153, EPI_ISL_662154, EPI_ISL_662155, EPI_ISL_662157, EPI_ISL_662158, EPI_ISL_662161, EPI_ISL_662168, EPI_ISL_662171, EPI_ISL_662172, EPI_ISL_662180, EPI_ISL_662185, EPI_ISL_662188, EPI_ISL_662189, EPI_ISL_662190, EPI_ISL_662193, EPI_ISL_662195, EPI_ISL_662197, EPI_ISL_662199, EPI_ISL_662201, EPI_ISL_662204, EPI_ISL_662205, EPI_ISL_662210, EPI_ISL_662211, EPI_ISL_662213, EPI_ISL_662216, EPI_ISL_662220, EPI_ISL_662221, EPI_ISL_662222, EPI_ISL_662224, EPI_ISL_662225, EPI_ISL_662226, EPI_ISL_662230, EPI_ISL_662232, EPI_ISL_662241, EPI_ISL_662242, EPI_ISL_662243, EPI_ISL_662246, EPI_ISL_662250, EPI_ISL_662251, EPI_ISL_662253, EPI_ISL_662256, EPI_ISL_662257, EPI_ISL_662259, EPI_ISL_662261, EPI_ISL_662262, EPI_ISL_662264, EPI_ISL_662268, EPI_ISL_662271, EPI_ISL_662277, EPI_ISL_662278, EPI_ISL_662280, EPI_ISL_662282, EPI_ISL_662283, EPI_ISL_662284, EPI_ISL_662287, EPI_ISL_662288, EPI_ISL_662289, EPI_ISL_662291, EPI_ISL_662293, EPI_ISL_662294, EPI_ISL_662295, EPI_ISL_662297, EPI_ISL_662298, EPI_ISL_662299, EPI_ISL_662301, EPI_ISL_662302, EPI_ISL_662303, EPI_ISL_662304, EPI_ISL_662306, EPI_ISL_662307, EPI_ISL_662310, EPI_ISL_662311, EPI_ISL_662313, EPI_ISL_662314, EPI_ISL_662315, EPI_ISL_662316, EPI_ISL_662317, EPI_ISL_662318, EPI_ISL_662319, EPI_ISL_662321, EPI_ISL_662322, EPI_ISL_662323, EPI_ISL_662324, EPI_ISL_662325, EPI_ISL_662326, EPI_ISL_662328, EPI_ISL_662329, EPI_ISL_662330, EPI_ISL_662331, EPI_ISL_662332, EPI_ISL_662333, EPI_ISL_662335, EPI_ISL_662336, EPI_ISL_662337, EPI_ISL_662338, EPI_ISL_662339, EPI_ISL_662340, EPI_ISL_662341, EPI_ISL_662345, EPI_ISL_662346, EPI_ISL_662347, EPI_ISL_662348, EPI_ISL_662349, EPI_ISL_662350, EPI_ISL_662351, EPI_ISL_662353, EPI_ISL_662355, EPI_ISL_662357, EPI_ISL_662360, EPI_ISL_662361, EPI_ISL_662362, EPI_ISL_662363, EPI_ISL_662364, EPI_ISL_662366, EPI_ISL_662367, EPI_ISL_662368, EPI_ISL_662370, EPI_ISL_662371, EPI_ISL_662372, EPI_ISL_662376, EPI_ISL_662377, EPI_ISL_662378, EPI_ISL_662379, EPI_ISL_662380, EPI_ISL_662383, EPI_ISL_662384, EPI_ISL_662386, EPI_ISL_662387, EPI_ISL_662388, EPI_ISL_662392, EPI_ISL_662395, EPI_ISL_662396, EPI_ISL_662397, EPI_ISL_662398, EPI_ISL_662399, EPI_ISL_662400, EPI_ISL_662401, EPI_ISL_662402, EPI_ISL_662404, EPI_ISL_662406, EPI_ISL_662407, EPI_ISL_662408, EPI_ISL_662409, EPI_ISL_662413, EPI_ISL_662414, EPI_ISL_662416, EPI_ISL_662417, EPI_ISL_662420, EPI_ISL_662421, EPI_ISL_662422, EPI_ISL_662423, EPI_ISL_662424, EPI_ISL_662425, EPI_ISL_662426, EPI_ISL_662428, EPI_ISL_662430, EPI_ISL_662432, EPI_ISL_662434, EPI_ISL_662435, EPI_ISL_662436, EPI_ISL_662437, EPI_ISL_662438, EPI_ISL_662439, EPI_ISL_662440, EPI_ISL_662441, EPI_ISL_662442, EPI_ISL_662443, EPI_ISL_662445, EPI_ISL_662447, EPI_ISL_662448, EPI_ISL_662449, EPI_ISL_662450, EPI_ISL_662451, EPI_ISL_662453, EPI_ISL_662454, EPI_ISL_662455, EPI_ISL_662456, EPI_ISL_662458, EPI_ISL_662459, EPI_ISL_662460, EPI_ISL_662461, EPI_ISL_662464, EPI_ISL_662465, EPI_ISL_662466, EPI_ISL_662467, EPI_ISL_662469, EPI_ISL_662470, EPI_ISL_662471, EPI_ISL_662473, EPI_ISL_662475, EPI_ISL_662476, EPI_ISL_662479, EPI_ISL_662481, EPI_ISL_662482, EPI_ISL_662483, EPI_ISL_662484, EPI_ISL_662485, EPI_ISL_662490, EPI_ISL_662491, EPI_ISL_662493, EPI_ISL_662494, EPI_ISL_662495, EPI_ISL_662497, EPI_ISL_662498, EPI_ISL_662500, EPI_ISL_662501, EPI_ISL_662503, EPI_ISL_662504, EPI_ISL_662505, EPI_ISL_662506, EPI_ISL_662509, EPI_ISL_662510, EPI_ISL_662511, EPI_ISL_662512, EPI_ISL_662513, EPI_ISL_662515, EPI_ISL_662516, EPI_ISL_662517, EPI_ISL_662519, EPI_ISL_662521, EPI_ISL_662522, EPI_ISL_662523, EPI_ISL_662524, EPI_ISL_662525, EPI_ISL_662527, EPI_ISL_662528, EPI_ISL_662529, EPI_ISL_662530, EPI_ISL_662531, EPI_ISL_662532, EPI_ISL_662533, EPI_ISL_662534, EPI_ISL_662536, EPI_ISL_662537, EPI_ISL_662538, EPI_ISL_662539, EPI_ISL_662540, EPI_ISL_662541, EPI_ISL_662542, EPI_ISL_662543, EPI_ISL_662544, EPI_ISL_662545, EPI_ISL_662546, EPI_ISL_662547, EPI_ISL_662548, EPI_ISL_662550, EPI_ISL_662551, EPI_ISL_662552, EPI_ISL_662554, EPI_ISL_662555, EPI_ISL_662556, EPI_ISL_662557, EPI_ISL_662559, EPI_ISL_662560, EPI_ISL_662561, EPI_ISL_662562, EPI_ISL_662563, EPI_ISL_662565, EPI_ISL_662566, EPI_ISL_662567, EPI_ISL_662568, EPI_ISL_662569, EPI_ISL_662570, EPI_ISL_662571, EPI_ISL_662572, EPI_ISL_662573, EPI_ISL_662574, EPI_ISL_662577, EPI_ISL_662582, EPI_ISL_662584, EPI_ISL_662585, EPI_ISL_662586, EPI_ISL_662587, EPI_ISL_662589, EPI_ISL_662590, EPI_ISL_662591, EPI_ISL_662592, EPI_ISL_662594, EPI_ISL_662595, EPI_ISL_662596, EPI_ISL_662598, EPI_ISL_662599, EPI_ISL_662600, EPI_ISL_662601, EPI_ISL_662602, EPI_ISL_662603, EPI_ISL_662606, EPI_ISL_662607, EPI_ISL_662608, EPI_ISL_662611, EPI_ISL_662612, EPI_ISL_662613, EPI_ISL_662615, EPI_ISL_662618, EPI_ISL_662619, EPI_ISL_662620, EPI_ISL_662627, EPI_ISL_668348, EPI_ISL_676313, EPI_ISL_676314, EPI_ISL_676315, EPI_ISL_676316, EPI_ISL_676317, EPI_ISL_676318, EPI_ISL_676319, EPI_ISL_676320, EPI_ISL_676321, EPI_ISL_676322, EPI_ISL_676323, EPI_ISL_676324, EPI_ISL_676325, EPI_ISL_676326, EPI_ISL_676327, EPI_ISL_676328, EPI_ISL_676329, EPI_ISL_676330, EPI_ISL_676331, EPI_ISL_676332, EPI_ISL_676333, EPI_ISL_676334, EPI_ISL_676335, EPI_ISL_676336, EPI_ISL_676337, EPI_ISL_676338, EPI_ISL_676339, EPI_ISL_676340, EPI_ISL_676341, EPI_ISL_676342, EPI_ISL_676343, EPI_ISL_676344, EPI_ISL_676345, EPI_ISL_676346, EPI_ISL_676347, EPI_ISL_676348, EPI_ISL_676349, EPI_ISL_676350, EPI_ISL_676351, EPI_ISL_676352, EPI_ISL_676359, EPI_ISL_676360, EPI_ISL_676361, EPI_ISL_676362, EPI_ISL_676363, EPI_ISL_676364, EPI_ISL_676365, EPI_ISL_676366, EPI_ISL_676367, EPI_ISL_676368, EPI_ISL_676369, EPI_ISL_676370, EPI_ISL_676371, EPI_ISL_676372, EPI_ISL_676373, EPI_ISL_676374, EPI_ISL_676375, EPI_ISL_676376, EPI_ISL_676377, EPI_ISL_676378, EPI_ISL_676379, EPI_ISL_676380, EPI_ISL_676381, EPI_ISL_676382, EPI_ISL_676384, EPI_ISL_676385, EPI_ISL_676386, EPI_ISL_676387, EPI_ISL_676388, EPI_ISL_676389, EPI_ISL_676390, EPI_ISL_676391, EPI_ISL_676392, EPI_ISL_676393, EPI_ISL_676394, EPI_ISL_710862, EPI_ISL_710863, EPI_ISL_710864, EPI_ISL_710867, EPI_ISL_710868, EPI_ISL_710872, EPI_ISL_710877, EPI_ISL_710879, EPI_ISL_710880, EPI_ISL_710881, EPI_ISL_710882, EPI_ISL_710883, EPI_ISL_710884, EPI_ISL_710885, EPI_ISL_710886 | | | |
| see above | Lighthouse Lab in Glasgow | Wellcome Sanger Institute for the COVID-19 Genomics UK (COG-UK) Consortium | Anna Dominiczak and Alex Alderton; Carol Clugston; Cordelia Langford; David Gray; David K. Jackson; Dominic Kwiatkowski; Ewan Harrison; Harper VanSteenhouse; Ian Johnston; John Sillitoe on behalf of the Wellcome Sanger Institute COVID-19 Surveillance Team; John Sillitoe on behalf of the Wellcome Sanger Institute COVID-19 Surveillance Team (http://www.sanger.ac.uk/covid-team); Roberto Amato; Sonia Goncalves; Yumi Kasai |
| EPI_ISL_651886 | Lighthouse Lab in Glasgow / MRC-University of Glasgow Centre for Virus Research | COVID-19 Genomics UK (COG-UK) Consortium | Alasdair MacLean; Alice Broos; Ana da Silva Filipe; Anna Dominiczak; Antonia Ho; Carol Clugston; Daniel Mair; David Gray; David L Robertson; Emma Thomson; Harper VanSteenhouse; James Shepherd; Jenna Nichols; Joseph Hughes; Kathy Li; Kathy Smollett; Kyriaki Nomikou; Lily Tong; Matthew Holden; Natasha Johnson; Richard Orton; Rory Gunson; Sarah McDonald; Sharif Shaaban; Sreenu Vattipally; Stephen Carmichael; Yumi Kasai |
| EPI_ISL_656685, EPI_ISL_656705, EPI_ISL_656728, EPI_ISL_656732, EPI_ISL_656733, EPI_ISL_656745, EPI_ISL_656746, EPI_ISL_656757, EPI_ISL_656774, EPI_ISL_656777, EPI_ISL_656778, EPI_ISL_656779, EPI_ISL_656788, EPI_ISL_656789, EPI_ISL_656798, EPI_ISL_656799, EPI_ISL_656807, EPI_ISL_656808, EPI_ISL_656810, EPI_ISL_656824, EPI_ISL_656829, EPI_ISL_656834, EPI_ISL_656835, EPI_ISL_656838, EPI_ISL_656840, EPI_ISL_656841, EPI_ISL_656846, EPI_ISL_656848, EPI_ISL_656861, EPI_ISL_656862, EPI_ISL_656867, EPI_ISL_656869, EPI_ISL_656871, EPI_ISL_656877, EPI_ISL_656887, EPI_ISL_656892, EPI_ISL_656893, EPI_ISL_656906, EPI_ISL_656910, EPI_ISL_656913, EPI_ISL_656920, EPI_ISL_656922, EPI_ISL_656924, EPI_ISL_656930, EPI_ISL_656932, EPI_ISL_656935, EPI_ISL_656936, EPI_ISL_656938, EPI_ISL_656942, EPI_ISL_656945, EPI_ISL_656947, EPI_ISL_656955, EPI_ISL_656957, EPI_ISL_656958, EPI_ISL_656961, EPI_ISL_656962, EPI_ISL_656965, EPI_ISL_656966, EPI_ISL_656969, EPI_ISL_656973, EPI_ISL_656974, EPI_ISL_656976, EPI_ISL_656979, EPI_ISL_656980, EPI_ISL_656982, EPI_ISL_656987, EPI_ISL_656991, EPI_ISL_656997, EPI_ISL_657003, EPI_ISL_657004, EPI_ISL_657008, EPI_ISL_657011, EPI_ISL_657014, EPI_ISL_657016, EPI_ISL_657017, EPI_ISL_657021, EPI_ISL_657026, EPI_ISL_657355, EPI_ISL_657359, EPI_ISL_657360, EPI_ISL_657364, EPI_ISL_657365, EPI_ISL_657367, EPI_ISL_657369, EPI_ISL_657370, EPI_ISL_657371, EPI_ISL_657372, EPI_ISL_657373, EPI_ISL_657375, EPI_ISL_657377, EPI_ISL_657379, EPI_ISL_657381, EPI_ISL_657382, EPI_ISL_657384, EPI_ISL_657385, EPI_ISL_657386, EPI_ISL_657390, EPI_ISL_657391, EPI_ISL_657392, EPI_ISL_657393, EPI_ISL_657396, EPI_ISL_657398, EPI_ISL_657400, EPI_ISL_657401, EPI_ISL_657405, EPI_ISL_657406, EPI_ISL_657408, EPI_ISL_657410, EPI_ISL_657411, EPI_ISL_657413, EPI_ISL_657414, EPI_ISL_657416, EPI_ISL_657417, EPI_ISL_657418, EPI_ISL_657419, EPI_ISL_657420, EPI_ISL_657424, EPI_ISL_657425, EPI_ISL_657428, EPI_ISL_657429, EPI_ISL_657430, EPI_ISL_657438, EPI_ISL_657439, EPI_ISL_657440, EPI_ISL_657441, EPI_ISL_657443, EPI_ISL_657446, EPI_ISL_657451, EPI_ISL_657453, EPI_ISL_657457, EPI_ISL_657459, EPI_ISL_657462, EPI_ISL_657464, EPI_ISL_657473, EPI_ISL_657479, EPI_ISL_657480, EPI_ISL_657483, EPI_ISL_657484, EPI_ISL_657485, EPI_ISL_657486, EPI_ISL_657487, EPI_ISL_657490, EPI_ISL_657491, EPI_ISL_657493, EPI_ISL_657494, EPI_ISL_657495, EPI_ISL_657497, EPI_ISL_657500, EPI_ISL_657503, EPI_ISL_657505, EPI_ISL_657509, EPI_ISL_657510, EPI_ISL_657512, EPI_ISL_657513, EPI_ISL_657515, EPI_ISL_657518, EPI_ISL_657519, EPI_ISL_657526, EPI_ISL_657527, EPI_ISL_657531, EPI_ISL_657533, EPI_ISL_657535, EPI_ISL_657536, EPI_ISL_657542, EPI_ISL_657543, EPI_ISL_657544, EPI_ISL_657547, EPI_ISL_657550, EPI_ISL_657553, EPI_ISL_657554, EPI_ISL_657555, EPI_ISL_657556, EPI_ISL_657557, EPI_ISL_657558, EPI_ISL_657560, EPI_ISL_657563, EPI_ISL_657564, EPI_ISL_657567, EPI_ISL_657568, EPI_ISL_657570, EPI_ISL_657571, EPI_ISL_657574, EPI_ISL_657575, EPI_ISL_657577, EPI_ISL_657579, EPI_ISL_657581, EPI_ISL_657583, EPI_ISL_657584, EPI_ISL_657585, EPI_ISL_657587, EPI_ISL_657589, EPI_ISL_657591, EPI_ISL_657593, EPI_ISL_657594, EPI_ISL_657596, EPI_ISL_657599, EPI_ISL_657602, EPI_ISL_657605, EPI_ISL_657606, EPI_ISL_657607, EPI_ISL_657608, EPI_ISL_657609, EPI_ISL_657610, EPI_ISL_657611, EPI_ISL_657614, EPI_ISL_657615, EPI_ISL_657616, EPI_ISL_657625, EPI_ISL_657627, EPI_ISL_657629, EPI_ISL_657631, EPI_ISL_657632, EPI_ISL_657635, EPI_ISL_657636, EPI_ISL_657639, EPI_ISL_657642, EPI_ISL_657644, EPI_ISL_657647, EPI_ISL_657648, EPI_ISL_657650, EPI_ISL_657651, EPI_ISL_657657, EPI_ISL_657661, EPI_ISL_657662, EPI_ISL_657663, EPI_ISL_657666, EPI_ISL_657667, EPI_ISL_657676, EPI_ISL_657680, EPI_ISL_657681, EPI_ISL_657682, EPI_ISL_657683, EPI_ISL_657688, EPI_ISL_657692, EPI_ISL_657694, EPI_ISL_657695, EPI_ISL_657696, EPI_ISL_657700, EPI_ISL_657702, EPI_ISL_657703, EPI_ISL_657705, EPI_ISL_657706, EPI_ISL_657709, EPI_ISL_657710, EPI_ISL_657711, EPI_ISL_657714, EPI_ISL_657715, EPI_ISL_657716, EPI_ISL_657717, EPI_ISL_657718, EPI_ISL_657719, EPI_ISL_657722, EPI_ISL_657727, EPI_ISL_657729, EPI_ISL_657730, EPI_ISL_657732, EPI_ISL_657733, EPI_ISL_657734, EPI_ISL_657735, EPI_ISL_657738, EPI_ISL_657740, EPI_ISL_657741, EPI_ISL_657742, EPI_ISL_657743, EPI_ISL_657745, EPI_ISL_657748, EPI_ISL_657752, EPI_ISL_657753, EPI_ISL_657755, EPI_ISL_657757, EPI_ISL_657758, EPI_ISL_657761, EPI_ISL_657768, EPI_ISL_657772, EPI_ISL_657773, EPI_ISL_657774, EPI_ISL_657775, EPI_ISL_657776, EPI_ISL_657777, EPI_ISL_657778, EPI_ISL_657781, EPI_ISL_657783, EPI_ISL_657791, EPI_ISL_657792, EPI_ISL_657794, EPI_ISL_657796, EPI_ISL_657797, EPI_ISL_657800, EPI_ISL_657808, EPI_ISL_657811, EPI_ISL_657812, EPI_ISL_657814, EPI_ISL_657819, EPI_ISL_657820, EPI_ISL_657822, EPI_ISL_657823, EPI_ISL_657827, EPI_ISL_657829, EPI_ISL_657830, EPI_ISL_657835, EPI_ISL_657836, EPI_ISL_657840, EPI_ISL_657842, EPI_ISL_657843, EPI_ISL_657844, EPI_ISL_657848, EPI_ISL_657850, EPI_ISL_657854, EPI_ISL_657858, EPI_ISL_657859, EPI_ISL_657862, EPI_ISL_657865, EPI_ISL_657870, EPI_ISL_657872, EPI_ISL_657874, EPI_ISL_657878, EPI_ISL_657879, EPI_ISL_657886, EPI_ISL_657887, EPI_ISL_657889, EPI_ISL_657892, EPI_ISL_657901, EPI_ISL_657905, EPI_ISL_657906, EPI_ISL_657910, EPI_ISL_657913, EPI_ISL_657915, EPI_ISL_657916, EPI_ISL_657918, EPI_ISL_657919, EPI_ISL_657921, EPI_ISL_657922, EPI_ISL_657923, EPI_ISL_657925, EPI_ISL_657927, EPI_ISL_657928, EPI_ISL_657929, EPI_ISL_657930, EPI_ISL_657932, EPI_ISL_657938, EPI_ISL_657940, EPI_ISL_657941, EPI_ISL_657944, EPI_ISL_657948, EPI_ISL_657950, EPI_ISL_657954, EPI_ISL_657955, EPI_ISL_657956, EPI_ISL_657958, EPI_ISL_657959, EPI_ISL_657961, EPI_ISL_657963, EPI_ISL_657965, EPI_ISL_657969, EPI_ISL_657970, EPI_ISL_657971, EPI_ISL_657972, EPI_ISL_657975, EPI_ISL_657977, EPI_ISL_657979, EPI_ISL_657980, EPI_ISL_657981, EPI_ISL_657982, EPI_ISL_657984, EPI_ISL_657985, EPI_ISL_657988, EPI_ISL_657993, EPI_ISL_657995, EPI_ISL_657996, EPI_ISL_657997, EPI_ISL_657999, EPI_ISL_658002, EPI_ISL_658003, EPI_ISL_658004, EPI_ISL_658005, EPI_ISL_658006, EPI_ISL_658009, EPI_ISL_658016, EPI_ISL_658017, EPI_ISL_658018, EPI_ISL_658021, EPI_ISL_658022, EPI_ISL_658023, EPI_ISL_658025, EPI_ISL_658026, EPI_ISL_658027, EPI_ISL_658038, EPI_ISL_658039, EPI_ISL_658040, EPI_ISL_658042, EPI_ISL_658044, EPI_ISL_658049, EPI_ISL_658050, EPI_ISL_658051, EPI_ISL_658053, EPI_ISL_658057, EPI_ISL_658058, EPI_ISL_658059, EPI_ISL_658060, EPI_ISL_658061, EPI_ISL_658062, EPI_ISL_658065, EPI_ISL_658066, EPI_ISL_658067, EPI_ISL_658068, EPI_ISL_658070, EPI_ISL_658081, EPI_ISL_658082, EPI_ISL_658087, EPI_ISL_658088, EPI_ISL_658090, EPI_ISL_658091, EPI_ISL_658094, EPI_ISL_658102, EPI_ISL_658103, EPI_ISL_658106, EPI_ISL_658107, EPI_ISL_658108, EPI_ISL_658109, EPI_ISL_658112, EPI_ISL_658113, EPI_ISL_658114, EPI_ISL_658117, EPI_ISL_658118, EPI_ISL_658121, EPI_ISL_658123, EPI_ISL_658126, EPI_ISL_658127, EPI_ISL_658129, EPI_ISL_658130, EPI_ISL_658132, EPI_ISL_658135, EPI_ISL_658136, EPI_ISL_658137, EPI_ISL_658139, EPI_ISL_658140, EPI_ISL_658141, EPI_ISL_658145, EPI_ISL_658146, EPI_ISL_658147, EPI_ISL_658149, EPI_ISL_658150, EPI_ISL_658151, EPI_ISL_658152, EPI_ISL_658154, EPI_ISL_658155, EPI_ISL_658165, EPI_ISL_658168, EPI_ISL_658169, EPI_ISL_658170, EPI_ISL_658172, EPI_ISL_658173, EPI_ISL_658174, EPI_ISL_658175, EPI_ISL_658176, EPI_ISL_658180, EPI_ISL_658182, EPI_ISL_658183, EPI_ISL_658184, EPI_ISL_658185, EPI_ISL_658193, EPI_ISL_658204, EPI_ISL_658205, EPI_ISL_658210, EPI_ISL_658211, EPI_ISL_658213, EPI_ISL_658216, EPI_ISL_658217, EPI_ISL_658219, EPI_ISL_658222, EPI_ISL_658223, EPI_ISL_658225, EPI_ISL_658229, EPI_ISL_658230, EPI_ISL_658232, EPI_ISL_658233, EPI_ISL_658235, EPI_ISL_658237, EPI_ISL_658238, EPI_ISL_658240, EPI_ISL_658241, EPI_ISL_658243, EPI_ISL_658244, EPI_ISL_658245, EPI_ISL_658247, EPI_ISL_658248, EPI_ISL_658249, EPI_ISL_658250, EPI_ISL_658251, EPI_ISL_658253, EPI_ISL_658254, EPI_ISL_658255, EPI_ISL_658257, EPI_ISL_658259, EPI_ISL_658262, EPI_ISL_658264, EPI_ISL_658265, EPI_ISL_658267, EPI_ISL_658269, EPI_ISL_658273, EPI_ISL_658277, EPI_ISL_658280, EPI_ISL_658281, EPI_ISL_658282, EPI_ISL_658283, EPI_ISL_658284, EPI_ISL_658288, EPI_ISL_658289, EPI_ISL_658290, EPI_ISL_658291, EPI_ISL_658293, EPI_ISL_658294, EPI_ISL_658295, EPI_ISL_658296, EPI_ISL_658297, EPI_ISL_658298, EPI_ISL_658300, EPI_ISL_658301, EPI_ISL_658302, EPI_ISL_658303, EPI_ISL_658308, EPI_ISL_658309, EPI_ISL_658315, EPI_ISL_658318, EPI_ISL_658319, EPI_ISL_658322, EPI_ISL_658324, EPI_ISL_658325, EPI_ISL_658328, EPI_ISL_658329, EPI_ISL_658333, EPI_ISL_658336, EPI_ISL_658338, EPI_ISL_658339, EPI_ISL_658342, EPI_ISL_658344, EPI_ISL_658346, EPI_ISL_658347, EPI_ISL_658349, EPI_ISL_658350, EPI_ISL_658353, EPI_ISL_658354, EPI_ISL_658357, EPI_ISL_658358, EPI_ISL_658359, EPI_ISL_658365, EPI_ISL_658368, EPI_ISL_658369, EPI_ISL_658372, EPI_ISL_658373, EPI_ISL_658374, EPI_ISL_658375, EPI_ISL_658377, EPI_ISL_658378, EPI_ISL_658379, EPI_ISL_658380, EPI_ISL_658384, EPI_ISL_658385, EPI_ISL_658386, EPI_ISL_658389, EPI_ISL_658392, EPI_ISL_658393, EPI_ISL_658399, EPI_ISL_658400, EPI_ISL_658401, EPI_ISL_658403, EPI_ISL_662633, EPI_ISL_662634, EPI_ISL_662636, EPI_ISL_673995, EPI_ISL_674832, EPI_ISL_709886, EPI_ISL_709893, EPI_ISL_709899, EPI_ISL_709900, EPI_ISL_709902 | | | |
| see above | Lighthouse Lab in Milton Keynes | Wellcome Sanger Institute for the COVID-19 Genomics UK (COG-UK) Consortium | Cordelia Langford; David K. Jackson; Dominic Kwiatkowski; Ewan Harrison; Ian Johnston; John Sillitoe on behalf of the Wellcome Sanger Institute COVID-19 Surveillance Team; Roberto Amato; Sonia Goncalves; The Lighthouse Lab in Milton Keynes and Alex Alderton |
| EPI_ISL_650112, EPI_ISL_650132, EPI_ISL_650140, EPI_ISL_650203, EPI_ISL_650207, EPI_ISL_650272, EPI_ISL_650290, EPI_ISL_650321, EPI_ISL_650333, EPI_ISL_650337, EPI_ISL_650342, EPI_ISL_650344, EPI_ISL_650346, EPI_ISL_650382, EPI_ISL_650420, EPI_ISL_650434, EPI_ISL_650441, EPI_ISL_650442, EPI_ISL_650473, EPI_ISL_650486, EPI_ISL_650527, EPI_ISL_650533, EPI_ISL_650539, EPI_ISL_650540, EPI_ISL_650542, EPI_ISL_650560, EPI_ISL_650620, EPI_ISL_650621, EPI_ISL_650622, EPI_ISL_650623, EPI_ISL_650801, EPI_ISL_650809, EPI_ISL_650831, EPI_ISL_650907, EPI_ISL_650909, EPI_ISL_650921, EPI_ISL_651135, EPI_ISL_651163, EPI_ISL_651212, EPI_ISL_651219, EPI_ISL_651245, EPI_ISL_651259, EPI_ISL_651263, EPI_ISL_651297, EPI_ISL_651319, EPI_ISL_651325, EPI_ISL_651327, EPI_ISL_651334, EPI_ISL_651336, EPI_ISL_651364, EPI_ISL_651430, EPI_ISL_651500, EPI_ISL_652089, EPI_ISL_652090, EPI_ISL_652091, EPI_ISL_652092, EPI_ISL_652093, EPI_ISL_652094, EPI_ISL_652095, EPI_ISL_652096, EPI_ISL_652097, EPI_ISL_652098, EPI_ISL_652099, EPI_ISL_652100, EPI_ISL_652101, EPI_ISL_652102, EPI_ISL_664341, EPI_ISL_664436, EPI_ISL_664440, EPI_ISL_664481, EPI_ISL_664539, EPI_ISL_664615, EPI_ISL_664620, EPI_ISL_664621, EPI_ISL_664624, EPI_ISL_664634, EPI_ISL_664640, EPI_ISL_664648, EPI_ISL_664654, EPI_ISL_664663, EPI_ISL_664685, EPI_ISL_664700, EPI_ISL_664727, EPI_ISL_664729, EPI_ISL_664730, EPI_ISL_665113, EPI_ISL_665141, EPI_ISL_679381, EPI_ISL_679382, EPI_ISL_679383, EPI_ISL_679384, EPI_ISL_679385, EPI_ISL_679386, EPI_ISL_679387, EPI_ISL_679388, EPI_ISL_679392, EPI_ISL_702498, EPI_ISL_702536, EPI_ISL_702582, EPI_ISL_702587, EPI_ISL_702597, EPI_ISL_702647, EPI_ISL_702656, EPI_ISL_702664, EPI_ISL_702672, EPI_ISL_702687, EPI_ISL_702708, EPI_ISL_702710, EPI_ISL_702721, EPI_ISL_702726, EPI_ISL_702739, EPI_ISL_702753, EPI_ISL_702756, EPI_ISL_702766, EPI_ISL_702774, EPI_ISL_702828, EPI_ISL_702854, EPI_ISL_702865, EPI_ISL_702869, EPI_ISL_702888, EPI_ISL_702900, EPI_ISL_702913, EPI_ISL_702920, EPI_ISL_702922, EPI_ISL_702928, EPI_ISL_702939, EPI_ISL_702966, EPI_ISL_703046, EPI_ISL_703059, EPI_ISL_703121, EPI_ISL_703150, EPI_ISL_703158, EPI_ISL_703195, EPI_ISL_703225, EPI_ISL_703230, EPI_ISL_703254, EPI_ISL_703256, EPI_ISL_703281, EPI_ISL_703284, EPI_ISL_703308, EPI_ISL_703316, EPI_ISL_703417, EPI_ISL_703530, EPI_ISL_703596, EPI_ISL_703652, EPI_ISL_703663, EPI_ISL_703712, EPI_ISL_703725, EPI_ISL_703731, EPI_ISL_703774, EPI_ISL_703888, EPI_ISL_703935, EPI_ISL_703990, EPI_ISL_704012, EPI_ISL_704020, EPI_ISL_704028, EPI_ISL_704062, EPI_ISL_704065, EPI_ISL_704068, EPI_ISL_704079, EPI_ISL_704097, EPI_ISL_704231, EPI_ISL_704970, EPI_ISL_704986, EPI_ISL_705013, EPI_ISL_705040, EPI_ISL_705182, EPI_ISL_705190, EPI_ISL_705218, EPI_ISL_705238, EPI_ISL_705260, EPI_ISL_705264, EPI_ISL_705275, EPI_ISL_705277, EPI_ISL_705298, EPI_ISL_705312, EPI_ISL_705320, EPI_ISL_705330, EPI_ISL_705344, EPI_ISL_705354, EPI_ISL_705359, EPI_ISL_705363, EPI_ISL_705406, EPI_ISL_705409, EPI_ISL_705423, EPI_ISL_705424, EPI_ISL_705449, EPI_ISL_705525, EPI_ISL_705806, EPI_ISL_705807, EPI_ISL_705808, EPI_ISL_705865, EPI_ISL_705866, EPI_ISL_705867, EPI_ISL_705868, EPI_ISL_705869, EPI_ISL_705870, EPI_ISL_705871, EPI_ISL_705872, EPI_ISL_705873, EPI_ISL_705874, EPI_ISL_705876, EPI_ISL_705878, EPI_ISL_705879, EPI_ISL_705880, EPI_ISL_705881, EPI_ISL_705882, EPI_ISL_705883, EPI_ISL_705884, EPI_ISL_705885, EPI_ISL_705895, EPI_ISL_705896, EPI_ISL_705897, EPI_ISL_705898, EPI_ISL_705899, EPI_ISL_705900, EPI_ISL_705901, EPI_ISL_705902, EPI_ISL_705903, EPI_ISL_705904, EPI_ISL_705905, EPI_ISL_705906, EPI_ISL_705907, EPI_ISL_705908, EPI_ISL_705909, EPI_ISL_705910, EPI_ISL_705911, EPI_ISL_705912, EPI_ISL_705913, EPI_ISL_705914, EPI_ISL_705915, EPI_ISL_705916, EPI_ISL_705917, EPI_ISL_705918, EPI_ISL_705920, EPI_ISL_865169, EPI_ISL_865171, EPI_ISL_1104318, EPI_ISL_1104319, EPI_ISL_1177806, EPI_ISL_1177807, EPI_ISL_1177808, EPI_ISL_1177809, EPI_ISL_1177810, EPI_ISL_1177811, EPI_ISL_1177812, EPI_ISL_1177813, EPI_ISL_1177814, EPI_ISL_1177815, EPI_ISL_1177816, EPI_ISL_1177817, EPI_ISL_1177818, EPI_ISL_1177819, EPI_ISL_1177820, EPI_ISL_1177823, EPI_ISL_1636814, EPI_ISL_1636815 | | | |
| see above | Liverpool Clinical Laboratories | COVID-19 Genomics UK (COG-UK) Consortium | A Alrezaihi; Alessandro Gerada; Alistair Darby; Angela Cowell; Anita Lucaci; Anu Chawla; Cassie Olateju; Catherine Hartley; Charlotte Nelson; Ecaterina Vamos; Elaine O'Toole; Eleanor G Bentley; Ghada T Shawli; Hermione Webster; Isabel García-Dorival; James Johnson; James P Stewart; Jenifer Manson; Joanne Watts; Jones Benjamin; Jordan J Clark; Julian Hiscox; L Luu; Lucille Rainbow; M Almsaud; Margaret Hughes; Mark Whitehead; Matthew Gemmell; Miren Iturriza-Gomara; Muhannad Alruwaili; N.P Randle; Neil Swainston; PKF Gilmore; Parul Sharma; Rebekah Penrice-Randal; Richard Eccles; Richard Gregory; Sam Haldenby; Steve Paterson; Stuart D Armstrong; Trevor Ian Robinson; Ximeng Han |
| EPI_ISL_637382, EPI_ISL_637383, EPI_ISL_637988, EPI_ISL_702809, EPI_ISL_702835, EPI_ISL_702982, EPI_ISL_703005, EPI_ISL_703533, EPI_ISL_703535, EPI_ISL_703701, EPI_ISL_704567, EPI_ISL_705212, EPI_ISL_705341, EPI_ISL_706277, EPI_ISL_706278, EPI_ISL_706286, EPI_ISL_706288, EPI_ISL_706289, EPI_ISL_706290, EPI_ISL_706291, EPI_ISL_706292, EPI_ISL_706293, EPI_ISL_706294, EPI_ISL_706295, EPI_ISL_706296, EPI_ISL_723558, EPI_ISL_723559, EPI_ISL_723560, EPI_ISL_723561, EPI_ISL_723562, EPI_ISL_723563, EPI_ISL_723564, EPI_ISL_723565, EPI_ISL_723566, EPI_ISL_723567, EPI_ISL_723568, EPI_ISL_723569, EPI_ISL_723570, EPI_ISL_723571, EPI_ISL_723572, EPI_ISL_723573, EPI_ISL_723574, EPI_ISL_723575, EPI_ISL_723576, EPI_ISL_723577, EPI_ISL_723578, EPI_ISL_723579, EPI_ISL_723580, EPI_ISL_723581, EPI_ISL_723582, EPI_ISL_723583, EPI_ISL_723584, EPI_ISL_723585, EPI_ISL_723586, EPI_ISL_723587, EPI_ISL_723588, EPI_ISL_723589, EPI_ISL_723590, EPI_ISL_723591, EPI_ISL_723592, EPI_ISL_723593, EPI_ISL_723594, EPI_ISL_723595, EPI_ISL_723596, EPI_ISL_723597, EPI_ISL_723598, EPI_ISL_723599, EPI_ISL_723600, EPI_ISL_723601, EPI_ISL_723602, EPI_ISL_723603, EPI_ISL_723604, EPI_ISL_723605, EPI_ISL_723606, EPI_ISL_723607, EPI_ISL_723608, EPI_ISL_723609, EPI_ISL_723610, EPI_ISL_723611, EPI_ISL_723612, EPI_ISL_723613, EPI_ISL_723614, EPI_ISL_723615, EPI_ISL_723616, EPI_ISL_723617, EPI_ISL_723618, EPI_ISL_723619, EPI_ISL_723620, EPI_ISL_723621, EPI_ISL_723622, EPI_ISL_723623, EPI_ISL_723624, EPI_ISL_723625, EPI_ISL_723626, EPI_ISL_723627, EPI_ISL_723628, | | | |

| | | | |
|---|---|---|---|
| EPI_ISL_723629, EPI_ISL_723630, EPI_ISL_724971, EPI_ISL_724972, EPI_ISL_724983, EPI_ISL_724984, EPI_ISL_724985, EPI_ISL_724986, EPI_ISL_724987, EPI_ISL_724988, EPI_ISL_724989, EPI_ISL_724990, EPI_ISL_724991, EPI_ISL_724992, EPI_ISL_724993, EPI_ISL_724994, EPI_ISL_724995, EPI_ISL_724996, EPI_ISL_724997, EPI_ISL_724998, EPI_ISL_724999, EPI_ISL_725000, EPI_ISL_725001, EPI_ISL_819460, EPI_ISL_819461, EPI_ISL_1637215, EPI_ISL_1637220 | | | |
| see above | Northumbria University / South Tees Hospitals NHS Foundation Trust / North Cumbria Integrated Care NHS Foundation Trust / North Tees and Hartlepool NHS Foundation Trust / Newcastle Hospitals NHS Foundation Trust | COVID-19 Genomics UK (COG-UK) Consortium | Andrew Nelson; Brendan Payne; Clive Graham; Darren L Smith; Debra Padgett; Edward Barton; Emma Swindells; Garren Scott; Gary Black; Gary Eltringham; Giles S Holt; Greg R Young; Jane Greenaway; Jennifer Collins; John Allan; Joshua Loh; Lynn Dover; Matthew Bashton; Mohammad A Tariq; Paul Baker; Sarah Essex; Steve Liggett; Wen C Yew; Yusri Taha |
| EPI_ISL_706128, EPI_ISL_723631, EPI_ISL_723632, EPI_ISL_723633, EPI_ISL_723634, EPI_ISL_723635, EPI_ISL_723636, EPI_ISL_723637, EPI_ISL_723638, EPI_ISL_723639, EPI_ISL_723640, EPI_ISL_723641, EPI_ISL_723642, EPI_ISL_723643, EPI_ISL_723644, EPI_ISL_723645, EPI_ISL_723646, EPI_ISL_723647, EPI_ISL_723648, EPI_ISL_723649, EPI_ISL_723650, EPI_ISL_723651, EPI_ISL_723652, EPI_ISL_723653, EPI_ISL_723654, EPI_ISL_723655, EPI_ISL_723656, EPI_ISL_723657, EPI_ISL_723658, EPI_ISL_723659, EPI_ISL_723660, EPI_ISL_723661, EPI_ISL_723662, EPI_ISL_723663, EPI_ISL_723664, EPI_ISL_723665, EPI_ISL_723666, EPI_ISL_723667, EPI_ISL_723668, EPI_ISL_723669, EPI_ISL_723670, EPI_ISL_723671, EPI_ISL_723672, EPI_ISL_723729, EPI_ISL_723964, EPI_ISL_723967, EPI_ISL_723982, EPI_ISL_724016, EPI_ISL_724021, EPI_ISL_724022, EPI_ISL_724023, EPI_ISL_724024, EPI_ISL_724025, EPI_ISL_724043, EPI_ISL_741032, EPI_ISL_741033, EPI_ISL_741054, EPI_ISL_741055, EPI_ISL_741085, EPI_ISL_741105, EPI_ISL_741107, EPI_ISL_741108, EPI_ISL_741109, EPI_ISL_741110, EPI_ISL_741150, EPI_ISL_839362, EPI_ISL_839363, EPI_ISL_839364, EPI_ISL_839365, EPI_ISL_839367, EPI_ISL_839368, EPI_ISL_839369, EPI_ISL_839370, EPI_ISL_839371, EPI_ISL_839372, EPI_ISL_839373, EPI_ISL_839374, EPI_ISL_839375, EPI_ISL_839376, EPI_ISL_839377, EPI_ISL_839378, EPI_ISL_839379, EPI_ISL_839380, EPI_ISL_839381, EPI_ISL_839382, EPI_ISL_839383, EPI_ISL_839384, EPI_ISL_839386, EPI_ISL_839387, EPI_ISL_839388, EPI_ISL_839389, EPI_ISL_839390, EPI_ISL_839391, EPI_ISL_839392, EPI_ISL_839393, EPI_ISL_839394, EPI_ISL_839395, EPI_ISL_839396, EPI_ISL_839397, EPI_ISL_839398, EPI_ISL_839399, EPI_ISL_839400, EPI_ISL_839401, EPI_ISL_839402, EPI_ISL_839403, EPI_ISL_839404, EPI_ISL_839405, EPI_ISL_839406, EPI_ISL_839407, EPI_ISL_839408, EPI_ISL_839409, EPI_ISL_839410, EPI_ISL_839411, EPI_ISL_839413, EPI_ISL_839414, EPI_ISL_839415, EPI_ISL_839416, EPI_ISL_839417, EPI_ISL_839418, EPI_ISL_839419, EPI_ISL_839420, EPI_ISL_839421, EPI_ISL_839422, EPI_ISL_839423, EPI_ISL_839424, EPI_ISL_839425, EPI_ISL_839426, EPI_ISL_839427, EPI_ISL_839428, EPI_ISL_839429, EPI_ISL_839430, EPI_ISL_839431, EPI_ISL_839432, EPI_ISL_839433, EPI_ISL_839434, EPI_ISL_839435, EPI_ISL_839436, EPI_ISL_839438, EPI_ISL_839439, EPI_ISL_839440, EPI_ISL_839441, EPI_ISL_839442, EPI_ISL_839443, EPI_ISL_839444, EPI_ISL_839445, EPI_ISL_839446, EPI_ISL_839447, EPI_ISL_839448, EPI_ISL_839449, EPI_ISL_839450, EPI_ISL_839451, EPI_ISL_839452, EPI_ISL_839453, EPI_ISL_839454, EPI_ISL_839455, EPI_ISL_839456, EPI_ISL_839457, EPI_ISL_839458, EPI_ISL_839459, EPI_ISL_839460, EPI_ISL_839461, EPI_ISL_839462, EPI_ISL_839463, EPI_ISL_839464, EPI_ISL_839465, EPI_ISL_839466, EPI_ISL_839467, EPI_ISL_839468, EPI_ISL_839469, EPI_ISL_839470, EPI_ISL_839471, EPI_ISL_839472, EPI_ISL_839473, EPI_ISL_839474, EPI_ISL_839475, EPI_ISL_839476, EPI_ISL_839477, EPI_ISL_839478, EPI_ISL_839479, EPI_ISL_839480, EPI_ISL_839481, EPI_ISL_839482, EPI_ISL_839483, EPI_ISL_840234, EPI_ISL_840235, EPI_ISL_840236, EPI_ISL_840237, EPI_ISL_840238, EPI_ISL_840239, EPI_ISL_840240, EPI_ISL_840241, EPI_ISL_840242, EPI_ISL_840243, EPI_ISL_840244, EPI_ISL_840245, EPI_ISL_840246, EPI_ISL_1309734 | | | |
| see above | Oxford Viromics, NDM, University of Oxford; Oxford University Hospitals; Basingstoke and North Hampshire Hospital | COVID-19 Genomics UK (COG-UK) Consortium | Alex Mobbs; Amy Trebes; Anita Justice; Catrin Moore; Christophe Fraser; David Bonsall; David Buck; Emma Wise; George Macintyre; Jessica Lynch; John Todd; Mariateresa de Cesare; Matilde Mori; Monique Andersson; Nathan Moore; Nick Cortes; Robert Shaw; Stephen Kidd; Tanya Golubchik; Timothy Peto |
| EPI_ISL_650113, EPI_ISL_650118, EPI_ISL_650145, EPI_ISL_650185, EPI_ISL_650188, EPI_ISL_650195, EPI_ISL_650211, EPI_ISL_650227, EPI_ISL_650229, EPI_ISL_650230, EPI_ISL_650285, EPI_ISL_650287, EPI_ISL_650294, EPI_ISL_650327, EPI_ISL_650332, EPI_ISL_650343, EPI_ISL_650345, EPI_ISL_650371, EPI_ISL_650421, EPI_ISL_650429, EPI_ISL_650457, EPI_ISL_650458, EPI_ISL_650461, EPI_ISL_650465, EPI_ISL_650466, EPI_ISL_650467, EPI_ISL_650474, EPI_ISL_650475, EPI_ISL_650485, EPI_ISL_650487, EPI_ISL_650489, EPI_ISL_650534, EPI_ISL_650555, EPI_ISL_650685, EPI_ISL_650686, EPI_ISL_650719, EPI_ISL_650720, EPI_ISL_650722, EPI_ISL_650818, EPI_ISL_650880, EPI_ISL_650890, EPI_ISL_650900, EPI_ISL_650903, EPI_ISL_650915, EPI_ISL_650943, EPI_ISL_651090, EPI_ISL_651091, EPI_ISL_651092, EPI_ISL_651094, EPI_ISL_651102, EPI_ISL_651118, EPI_ISL_651139, EPI_ISL_651150, EPI_ISL_651157, EPI_ISL_651159, EPI_ISL_651182, EPI_ISL_651198, EPI_ISL_651203, EPI_ISL_651204, EPI_ISL_651206, EPI_ISL_651210, EPI_ISL_651217, EPI_ISL_651220, EPI_ISL_651254, EPI_ISL_651326, EPI_ISL_651332, EPI_ISL_651333, EPI_ISL_651335, EPI_ISL_651340, EPI_ISL_651352, EPI_ISL_651371, EPI_ISL_651392, EPI_ISL_652350, EPI_ISL_652351, EPI_ISL_652352, EPI_ISL_652353, EPI_ISL_652354, EPI_ISL_652355, EPI_ISL_652357, EPI_ISL_652359, EPI_ISL_652360, EPI_ISL_652361, EPI_ISL_665265, EPI_ISL_665293, EPI_ISL_665306, EPI_ISL_665342, EPI_ISL_665343, EPI_ISL_665346, EPI_ISL_665353, EPI_ISL_665356, EPI_ISL_665357, EPI_ISL_665358, EPI_ISL_665359, EPI_ISL_665364, EPI_ISL_665372, EPI_ISL_665375, EPI_ISL_665431, EPI_ISL_665437, EPI_ISL_665446, EPI_ISL_665447, EPI_ISL_665448, EPI_ISL_665449, EPI_ISL_665542, EPI_ISL_665567, EPI_ISL_665577, EPI_ISL_665590, EPI_ISL_665593, EPI_ISL_665682, EPI_ISL_665880, EPI_ISL_665881, EPI_ISL_665882, EPI_ISL_665883, EPI_ISL_665886, EPI_ISL_665894, EPI_ISL_725113, EPI_ISL_725116, EPI_ISL_725117, EPI_ISL_725121, EPI_ISL_725128, EPI_ISL_725130, EPI_ISL_725136, EPI_ISL_725166, EPI_ISL_725167, EPI_ISL_725185, EPI_ISL_725190, EPI_ISL_725193, EPI_ISL_725267, EPI_ISL_725282, EPI_ISL_725294, EPI_ISL_725345, EPI_ISL_741552 | | | |
| see above | Quadram Institute Bioscience | COVID-19 Genomics UK (COG-UK) Consortium | Alexander J Trotter; Alison E. Mather; Alp Aydin; Ana P. Tedim; Anastasia Kolyva; Andrew Bell; Andrew J. Page; Claire Stuart; Dave J. Baker; Gemma L. Kay; John Wain; Justin O'Grady; Leonardo de Oliveira Martins; Lizzie Meadows; Maria Diaz; Mark Webber; Muhammed Yasir; Nabil-Fareed Alikhan; Ngozi Elumogo; Nicholas M. Thomson; Rachael Stanley; Rachel Gilroy; Reenesh Prakash; Samir Dervisevic; Samuel Bloomfield; Steven Rudder; Thanh Le-Viet |
| EPI_ISL_637298, EPI_ISL_637342, EPI_ISL_637344, EPI_ISL_637427, EPI_ISL_637555, EPI_ISL_637558, EPI_ISL_637582, EPI_ISL_637661, EPI_ISL_637798, EPI_ISL_637799, EPI_ISL_637800, EPI_ISL_637935, EPI_ISL_638587, EPI_ISL_638588, EPI_ISL_638592, EPI_ISL_638593, EPI_ISL_638594, EPI_ISL_638595, EPI_ISL_638596, EPI_ISL_638597, EPI_ISL_638598, EPI_ISL_638599, EPI_ISL_638600, EPI_ISL_638601, EPI_ISL_638602, EPI_ISL_638603, EPI_ISL_638604, EPI_ISL_638605, EPI_ISL_638606, EPI_ISL_638607, EPI_ISL_638608, EPI_ISL_638609, EPI_ISL_638610, EPI_ISL_638611, EPI_ISL_638612, EPI_ISL_638613, EPI_ISL_650295, EPI_ISL_650738, EPI_ISL_652364, EPI_ISL_652365, EPI_ISL_652366, EPI_ISL_652367, EPI_ISL_652368, EPI_ISL_652369, EPI_ISL_652370, EPI_ISL_652371, EPI_ISL_652372, EPI_ISL_665441, EPI_ISL_665650, EPI_ISL_665898, EPI_ISL_665899, EPI_ISL_665941, EPI_ISL_679932, EPI_ISL_704315, EPI_ISL_819458, EPI_ISL_819459, EPI_ISL_866901, EPI_ISL_866902, EPI_ISL_866903, EPI_ISL_950636, EPI_ISL_950637 | | | |
| see above | Queens Medical Centre, Clinical Microbiology Department / DeepSeq Nottingham | COVID-19 Genomics UK (COG-UK) Consortium | Christopher Moore; Fei Sang; Gemma Clark; Hannah Howson-Wells; Johnny Debebe; Jonathan Ball; Joseph Chappell; Manjinder Khakh; Matthew Carlisle; Matthew Loose; Michelle M Lister; Nadine Holmes; Patrick McClure; Theocharis Tsoleridis; Vicki M Fleming; Victoria Wright; Wendy Smith |
| EPI_ISL_742118, EPI_ISL_742119, EPI_ISL_742120, EPI_ISL_742121, EPI_ISL_742122, EPI_ISL_742126, EPI_ISL_742127, EPI_ISL_742128, EPI_ISL_742140, EPI_ISL_742141, EPI_ISL_742142, EPI_ISL_742143, EPI_ISL_742144, EPI_ISL_742145, EPI_ISL_742146, EPI_ISL_742147, EPI_ISL_742148, EPI_ISL_742149, EPI_ISL_742150, EPI_ISL_742151, EPI_ISL_742152, EPI_ISL_742153, EPI_ISL_742154, EPI_ISL_742155, EPI_ISL_742156, EPI_ISL_742157, EPI_ISL_742158, EPI_ISL_742159, EPI_ISL_742160, EPI_ISL_742161, EPI_ISL_742162, EPI_ISL_742163, EPI_ISL_742164, EPI_ISL_742165, EPI_ISL_742166, EPI_ISL_742167, EPI_ISL_742168, EPI_ISL_742170, EPI_ISL_742171, EPI_ISL_742173, EPI_ISL_763373, EPI_ISL_763642, EPI_ISL_764431, EPI_ISL_764433, EPI_ISL_764440, EPI_ISL_921122 | | | |
| see above | Regional Virus Laboratory, Belfast Health and Social Care Trust | COVID-19 Genomics UK (COG-UK) Consortium | Alison Watt; Ciara Cox; Conall McCaughey; David Simpson; Derek Fairley; James McKenna; Mairead Connor; Susan Feeney; Tanya Curran; Zoltan Molnar |
| EPI_ISL_664020, EPI_ISL_664021, EPI_ISL_664022, EPI_ISL_664023, EPI_ISL_664024, EPI_ISL_664025, EPI_ISL_664026, EPI_ISL_664027, EPI_ISL_664028, EPI_ISL_664029, EPI_ISL_664030, EPI_ISL_664031, EPI_ISL_664032, EPI_ISL_664033, EPI_ISL_664034, EPI_ISL_664035, EPI_ISL_664036, EPI_ISL_664037, EPI_ISL_664038, EPI_ISL_664039, EPI_ISL_664040, EPI_ISL_664041, EPI_ISL_664042, EPI_ISL_664043, EPI_ISL_664044, EPI_ISL_664045, EPI_ISL_664049, EPI_ISL_664050, EPI_ISL_664053, EPI_ISL_664054, EPI_ISL_664055, EPI_ISL_664056, EPI_ISL_664057, EPI_ISL_664058, EPI_ISL_664060, EPI_ISL_664061, EPI_ISL_664062, EPI_ISL_664063, EPI_ISL_664064, EPI_ISL_664065, EPI_ISL_664066, EPI_ISL_664067, EPI_ISL_664068, EPI_ISL_664069, EPI_ISL_664070, EPI_ISL_664071, EPI_ISL_664072, EPI_ISL_664073, EPI_ISL_664074, EPI_ISL_664075, EPI_ISL_664076, EPI_ISL_664077, EPI_ISL_664078, EPI_ISL_664079, EPI_ISL_664080, EPI_ISL_664081, EPI_ISL_664082, EPI_ISL_664083, EPI_ISL_664084, EPI_ISL_664085, EPI_ISL_664086, EPI_ISL_664087, EPI_ISL_664088, EPI_ISL_664093, EPI_ISL_664094, EPI_ISL_664095, EPI_ISL_664096, EPI_ISL_664098, EPI_ISL_664099, EPI_ISL_664100, EPI_ISL_666727, EPI_ISL_666728, EPI_ISL_666730, EPI_ISL_666738, EPI_ISL_666739, EPI_ISL_666742, EPI_ISL_666743, EPI_ISL_666744, EPI_ISL_666745, EPI_ISL_666746, EPI_ISL_666747, EPI_ISL_666748, EPI_ISL_666749, EPI_ISL_666750, EPI_ISL_666751, EPI_ISL_666752, EPI_ISL_666753, EPI_ISL_666754, EPI_ISL_666755, EPI_ISL_666756, EPI_ISL_666757, EPI_ISL_666758, EPI_ISL_666759, EPI_ISL_666760, EPI_ISL_666761, EPI_ISL_666762, EPI_ISL_666763, EPI_ISL_666764, EPI_ISL_666765, EPI_ISL_666766, EPI_ISL_666767, EPI_ISL_678681, EPI_ISL_678682, EPI_ISL_678685, EPI_ISL_678686, EPI_ISL_678687, EPI_ISL_678689, EPI_ISL_678690, EPI_ISL_678691, EPI_ISL_678692, EPI_ISL_678693, EPI_ISL_678695, EPI_ISL_678696, EPI_ISL_678697, EPI_ISL_678698, EPI_ISL_678699, EPI_ISL_678701, EPI_ISL_678702, EPI_ISL_678703, EPI_ISL_678705, EPI_ISL_683452, EPI_ISL_683454, EPI_ISL_683455, EPI_ISL_683458, EPI_ISL_683477, EPI_ISL_683481, EPI_ISL_683493, EPI_ISL_683494, EPI_ISL_683495, EPI_ISL_683496, EPI_ISL_683497, EPI_ISL_683498, EPI_ISL_683499, EPI_ISL_683500, EPI_ISL_683501, EPI_ISL_683502, EPI_ISL_683503, EPI_ISL_683504, EPI_ISL_683505, EPI_ISL_683506, EPI_ISL_683507, EPI_ISL_683508, EPI_ISL_683509, EPI_ISL_683510, EPI_ISL_683511, EPI_ISL_683512, EPI_ISL_683513, EPI_ISL_683514, EPI_ISL_683515, EPI_ISL_683516, EPI_ISL_683517, EPI_ISL_683518, EPI_ISL_683523, EPI_ISL_686108, EPI_ISL_686546, EPI_ISL_686547, EPI_ISL_686548, EPI_ISL_686549, EPI_ISL_686550, EPI_ISL_686551, EPI_ISL_686552, EPI_ISL_686553, EPI_ISL_686554, EPI_ISL_686555, EPI_ISL_686556, EPI_ISL_686557, EPI_ISL_686558, EPI_ISL_686559, EPI_ISL_686560, EPI_ISL_686561, EPI_ISL_686562, EPI_ISL_686563, EPI_ISL_686564, EPI_ISL_686565, EPI_ISL_686566, EPI_ISL_686567, EPI_ISL_686568, EPI_ISL_686569, EPI_ISL_686570, EPI_ISL_686571, EPI_ISL_686572, EPI_ISL_686573, EPI_ISL_686574, EPI_ISL_686575, EPI_ISL_686576, EPI_ISL_686577, EPI_ISL_686578, EPI_ISL_686579, EPI_ISL_693393, EPI_ISL_693400, EPI_ISL_693408, EPI_ISL_693409, EPI_ISL_693410, EPI_ISL_693411, EPI_ISL_693414, EPI_ISL_693415, EPI_ISL_693419, EPI_ISL_693420, EPI_ISL_693422, EPI_ISL_693433, EPI_ISL_693485 | | | |
| see above | Respiratory Virus Unit, Microbiology Services Colindale, Public Health England | COVID-19 Genomics UK (COG-UK) Consortium | PHE Covid Sequencing Team |
| EPI_ISL_636734, EPI_ISL_636735 | Respiratory Virus Unit, Microbiology Services Colindale, Public Health England | Respiratory Virus Unit, Microbiology Services Colindale, Public Health England | PHE Covid Sequencing Team |
| EPI_ISL_762997, EPI_ISL_766101, EPI_ISL_766263, EPI_ISL_766264, EPI_ISL_766265, EPI_ISL_766266, EPI_ISL_804258, EPI_ISL_825595, EPI_ISL_825596 | | | |
| see above | Respiratory Virus Unit, National Infection Service, Public Health England | COVID-19 Genomics UK (COG-UK) Consortium | PHE Covid Sequencing Team |
| EPI_ISL_665297, EPI_ISL_665299, EPI_ISL_665352, EPI_ISL_665374, EPI_ISL_665376, EPI_ISL_665466, EPI_ISL_665517, EPI_ISL_665572, EPI_ISL_665634, EPI_ISL_665635, EPI_ISL_665644, EPI_ISL_665759, EPI_ISL_665785, EPI_ISL_665801, EPI_ISL_665807, EPI_ISL_665808, EPI_ISL_665809, EPI_ISL_665810, EPI_ISL_665811, EPI_ISL_665812, EPI_ISL_665813, EPI_ISL_665814, EPI_ISL_665815, EPI_ISL_665823, EPI_ISL_665826, EPI_ISL_665827, EPI_ISL_665828, EPI_ISL_665829, EPI_ISL_665830, EPI_ISL_866048, EPI_ISL_920209, EPI_ISL_920213, EPI_ISL_920529, EPI_ISL_920530, EPI_ISL_1104883, EPI_ISL_1104889, EPI_ISL_1104892, EPI_ISL_1104895, EPI_ISL_1104923, EPI_ISL_1104924 | | | |
| see above | University College London Hospital | COVID-19 Genomics UK (COG-UK) Consortium | Catherine Houlihan; Dan Frampton; Judith Heaney; Matthew Byott; Moira Spyer and Eleni Nastouli; Stuart Kirk |
| EPI_ISL_665266, EPI_ISL_665270, EPI_ISL_665339, EPI_ISL_665378, EPI_ISL_665413, EPI_ISL_665661, EPI_ISL_665664, EPI_ISL_665669, EPI_ISL_665688, EPI_ISL_665705, EPI_ISL_665709, EPI_ISL_665710, EPI_ISL_665715, EPI_ISL_665716, EPI_ISL_665733, EPI_ISL_665741, EPI_ISL_679478, EPI_ISL_679480, EPI_ISL_679481, EPI_ISL_679482, EPI_ISL_679483, EPI_ISL_679484, EPI_ISL_679485, EPI_ISL_679486, EPI_ISL_679487, EPI_ISL_679488, EPI_ISL_679489, EPI_ISL_679490, EPI_ISL_679491, EPI_ISL_679493, EPI_ISL_679494, EPI_ISL_679495, EPI_ISL_679496, EPI_ISL_679497, EPI_ISL_679498, EPI_ISL_679499, EPI_ISL_679500, EPI_ISL_679502, EPI_ISL_679505, EPI_ISL_679621, EPI_ISL_679622, EPI_ISL_679623, EPI_ISL_702839, EPI_ISL_703745, EPI_ISL_704994, EPI_ISL_705035, EPI_ISL_705171, EPI_ISL_705205, EPI_ISL_705243, EPI_ISL_705307, EPI_ISL_705308, EPI_ISL_705309, EPI_ISL_724540, EPI_ISL_724594, EPI_ISL_724611, EPI_ISL_724613, EPI_ISL_724614, EPI_ISL_724626, EPI_ISL_724628, EPI_ISL_724630, EPI_ISL_724631, EPI_ISL_724632, EPI_ISL_724633, EPI_ISL_724636, EPI_ISL_724637, EPI_ISL_724639, EPI_ISL_724642, EPI_ISL_724648, EPI_ISL_724649, EPI_ISL_724650, EPI_ISL_724651, EPI_ISL_724652, EPI_ISL_724656, EPI_ISL_724661, EPI_ISL_724665, EPI_ISL_724666, EPI_ISL_724674, EPI_ISL_724677, EPI_ISL_724680, EPI_ISL_724681, EPI_ISL_724685, EPI_ISL_724686, EPI_ISL_724687, EPI_ISL_724689, EPI_ISL_724690, EPI_ISL_724692, EPI_ISL_724693, EPI_ISL_724695, EPI_ISL_724699, EPI_ISL_724707, EPI_ISL_724709, EPI_ISL_724710, EPI_ISL_724713, EPI_ISL_724715, EPI_ISL_724717, EPI_ISL_724718, EPI_ISL_724720, EPI_ISL_724722, EPI_ISL_724723, EPI_ISL_724724, EPI_ISL_724734, EPI_ISL_724735, EPI_ISL_724736, EPI_ISL_724737, EPI_ISL_724738, EPI_ISL_724739, EPI_ISL_724743, EPI_ISL_724744, EPI_ISL_724745, EPI_ISL_724746, EPI_ISL_724747, EPI_ISL_724748, EPI_ISL_724749, EPI_ISL_724750, EPI_ISL_724751, EPI_ISL_724752, EPI_ISL_724754, EPI_ISL_724756, EPI_ISL_724758, EPI_ISL_724776, EPI_ISL_724777, EPI_ISL_865684, EPI_ISL_865686, EPI_ISL_1386874, EPI_ISL_1637152 | | | |
| see above | University College London, Great Ormond Street Hospital for Children NHS Foundation Trust, Imperial College Healthcare NHS Trust | COVID-19 Genomics UK (COG-UK) Consortium | Alison Holmes; Charlotte Williams; Helena Tutill; Jacqueline Findlay; James Price; Judith Breuer; Julianne Brown; Kathryn Harris; Leysa Forrest; Mark Kristiansen; Paola Niola; Paola Resende Silva; Patricia Dyal; Paul Randell; Rachel Williams; Samuel Weeks; Sergi Castellano; Sunando Roy; Tony Brooks; Yasmin Panchbhaya |
| EPI_ISL_650119, EPI_ISL_650126, EPI_ISL_650137, EPI_ISL_650144, EPI_ISL_650212, EPI_ISL_650216, EPI_ISL_650218, EPI_ISL_650245, EPI_ISL_650254, EPI_ISL_650350, EPI_ISL_650373, EPI_ISL_650432, EPI_ISL_650582, EPI_ISL_650648, EPI_ISL_650717, EPI_ISL_650743, EPI_ISL_650760, EPI_ISL_650825, EPI_ISL_650999, EPI_ISL_651000, EPI_ISL_651098, EPI_ISL_651144, EPI_ISL_651174, EPI_ISL_651195, EPI_ISL_651266, EPI_ISL_651275, EPI_ISL_651342, EPI_ISL_651353, EPI_ISL_651379, EPI_ISL_651381, EPI_ISL_651390, EPI_ISL_651422, EPI_ISL_651630, EPI_ISL_651631, EPI_ISL_651633, EPI_ISL_651635, EPI_ISL_651636, EPI_ISL_651637, EPI_ISL_651638, EPI_ISL_651639, EPI_ISL_651640, EPI_ISL_651641, EPI_ISL_651642, EPI_ISL_651643, EPI_ISL_651644, EPI_ISL_651645, EPI_ISL_651646, EPI_ISL_651647, EPI_ISL_651648, EPI_ISL_651649, EPI_ISL_651650, EPI_ISL_651651, EPI_ISL_651652, EPI_ISL_679008, EPI_ISL_679009, EPI_ISL_679010, EPI_ISL_679011, EPI_ISL_679012, EPI_ISL_679013, EPI_ISL_679014, EPI_ISL_679015, EPI_ISL_679016, EPI_ISL_679017, EPI_ISL_679018, EPI_ISL_679019, EPI_ISL_679020, EPI_ISL_679021, EPI_ISL_679022, EPI_ISL_679023, EPI_ISL_679024, EPI_ISL_679025, EPI_ISL_679026, EPI_ISL_679027, EPI_ISL_679028, EPI_ISL_679029, EPI_ISL_679030, EPI_ISL_679031, EPI_ISL_679032, EPI_ISL_679033, EPI_ISL_679034, EPI_ISL_679035, EPI_ISL_679036, EPI_ISL_679037, EPI_ISL_679038, EPI_ISL_679039, EPI_ISL_679040, EPI_ISL_679041, EPI_ISL_679042, EPI_ISL_679043, EPI_ISL_679044, EPI_ISL_679045, EPI_ISL_679046, EPI_ISL_679047, EPI_ISL_679048, EPI_ISL_679049, EPI_ISL_679050, EPI_ISL_679053, EPI_ISL_679054, EPI_ISL_679055, EPI_ISL_679056, EPI_ISL_679057, EPI_ISL_679058, EPI_ISL_679059, EPI_ISL_679060, EPI_ISL_679061, EPI_ISL_679062, EPI_ISL_679063, EPI_ISL_679064, EPI_ISL_679065, EPI_ISL_679066, EPI_ISL_679067, EPI_ISL_679068, EPI_ISL_679069, EPI_ISL_679070, EPI_ISL_679071, EPI_ISL_679072, EPI_ISL_679073, EPI_ISL_679074, EPI_ISL_679075, EPI_ISL_679076, EPI_ISL_679077, EPI_ISL_679078, EPI_ISL_679079, EPI_ISL_679080, EPI_ISL_679081, EPI_ISL_679082, EPI_ISL_679083, EPI_ISL_679084, EPI_ISL_679085, EPI_ISL_679086, EPI_ISL_679087, EPI_ISL_679088, EPI_ISL_679089, EPI_ISL_679090, EPI_ISL_679091, EPI_ISL_679092, EPI_ISL_679093, EPI_ISL_679094, EPI_ISL_679095, EPI_ISL_679096, EPI_ISL_679097, EPI_ISL_679098, EPI_ISL_679099, EPI_ISL_679100, EPI_ISL_679101, EPI_ISL_679102, EPI_ISL_679103, EPI_ISL_679129, EPI_ISL_813012, EPI_ISL_813013, EPI_ISL_813014, EPI_ISL_813015, EPI_ISL_813016, EPI_ISL_813017, EPI_ISL_813018, EPI_ISL_813019, EPI_ISL_813020, EPI_ISL_813021, EPI_ISL_813022, EPI_ISL_813024, EPI_ISL_813029, EPI_ISL_949327, EPI_ISL_949328, EPI_ISL_949329, EPI_ISL_949330, EPI_ISL_949331, EPI_ISL_949332, EPI_ISL_949333, EPI_ISL_949334, EPI_ISL_949337, EPI_ISL_949338, EPI_ISL_949339, EPI_ISL_949340, EPI_ISL_949341, EPI_ISL_949342 | | | |
| see above | University of Birmingham | COVID-19 Genomics UK (COG-UK) Consortium | Alex Richter; Andrew Bosworth; Andrew Bosworth. Queen Elizabeth Hospital: Anna Casey; Andrew D Beggs PHE Heartlands Lab: Husam Osman; Charlotte Poxon; Institute of Microbiology; Joanne Stockton; Josh Quick; Kasun Wanigasooriya; Mike Kidd; Nicholas Loman. University of Birmingham Testing Laboratory: Celina M Whalley; Oliver Pickles; Radoslaw Poplawski; Samuel Nicholls; University of Birmingham: Claire McMurray; Will Rowe |
| EPI_ISL_664114, EPI_ISL_664115, EPI_ISL_664116, EPI_ISL_664136, EPI_ISL_664144, EPI_ISL_664145, EPI_ISL_664153, EPI_ISL_664173, EPI_ISL_664183, EPI_ISL_664184, EPI_ISL_664193, EPI_ISL_664194, EPI_ISL_664195, EPI_ISL_664202, EPI_ISL_664210, EPI_ISL_664211, EPI_ISL_664212, EPI_ISL_664214, EPI_ISL_664216, EPI_ISL_664239, EPI_ISL_664250, EPI_ISL_664251, EPI_ISL_664252, EPI_ISL_664253, EPI_ISL_664254, EPI_ISL_664255, EPI_ISL_664256, EPI_ISL_664257, EPI_ISL_664258, EPI_ISL_664259, EPI_ISL_664260, EPI_ISL_664261, EPI_ISL_664262, EPI_ISL_664263, EPI_ISL_664264, EPI_ISL_664265, EPI_ISL_664266, EPI_ISL_664267, EPI_ISL_664268, EPI_ISL_664269, EPI_ISL_664270, EPI_ISL_664271, EPI_ISL_664272, EPI_ISL_664273, EPI_ISL_664274, EPI_ISL_664275, EPI_ISL_664279, EPI_ISL_664284, EPI_ISL_664285, EPI_ISL_664287, EPI_ISL_664292, EPI_ISL_664293, EPI_ISL_664294, EPI_ISL_664296, EPI_ISL_664297, EPI_ISL_664365, EPI_ISL_664400, EPI_ISL_664401, EPI_ISL_664402, EPI_ISL_664403, EPI_ISL_664404, EPI_ISL_664405, EPI_ISL_664406, EPI_ISL_664407, EPI_ISL_664408, EPI_ISL_664409, EPI_ISL_664410, EPI_ISL_664411, EPI_ISL_664412, EPI_ISL_664413, EPI_ISL_664414, EPI_ISL_664415, EPI_ISL_664416, EPI_ISL_664417, EPI_ISL_664418, EPI_ISL_664419, EPI_ISL_664420, EPI_ISL_664429, EPI_ISL_664459, EPI_ISL_664509, EPI_ISL_664545, EPI_ISL_664594, EPI_ISL_664595, EPI_ISL_664596, EPI_ISL_664597, EPI_ISL_664608, EPI_ISL_664611, EPI_ISL_664626, EPI_ISL_664629, EPI_ISL_664644, EPI_ISL_664674, EPI_ISL_664682, EPI_ISL_664687, EPI_ISL_664709, EPI_ISL_664715, EPI_ISL_664718, EPI_ISL_664722, EPI_ISL_665066, EPI_ISL_665068, EPI_ISL_665069, EPI_ISL_665070, EPI_ISL_665071, EPI_ISL_665072, EPI_ISL_665073, EPI_ISL_665074, EPI_ISL_665075, EPI_ISL_665076, EPI_ISL_665077, EPI_ISL_665078, EPI_ISL_665079, EPI_ISL_665080, EPI_ISL_665081, EPI_ISL_665082, EPI_ISL_665083, EPI_ISL_665084, EPI_ISL_665085, EPI_ISL_665086, EPI_ISL_665087, EPI_ISL_665088, EPI_ISL_665089, EPI_ISL_665090, EPI_ISL_665091, EPI_ISL_665092, EPI_ISL_665093, EPI_ISL_665094, EPI_ISL_665095, EPI_ISL_665096, EPI_ISL_665097, EPI_ISL_665098, EPI_ISL_665099, EPI_ISL_665100, EPI_ISL_665101, EPI_ISL_665102, EPI_ISL_665103, EPI_ISL_665104, EPI_ISL_665105, EPI_ISL_665106, EPI_ISL_665107, EPI_ISL_665108, EPI_ISL_665109, EPI_ISL_665110, EPI_ISL_665111, EPI_ISL_1247533, EPI_ISL_1247543, EPI_ISL_1247549, EPI_ISL_1545810, EPI_ISL_1545812 | | | |
| see above | University of Exeter | COVID-19 Genomics UK (COG-UK) Consortium | Aaron Jeffries; Audrey Farbos; Ben Temperton; Jane Masoli; Joanna Warwick-Dugdale; Michelle Michelsen; Robyn Manley; Stephen Michell |
| EPI_ISL_997121 | Virology Department, Royal Infirmary of Edinburgh, NHS Lothian / School of Biological Sciences, University of Edinburgh | COVID-19 Genomics UK (COG-UK) Consortium | Colquhoun R; Cotton S; Dewar R; Hill V; Jackson B; McCrone JT; McHugh M; O'Toole Á; Rambaut A; Rooke S; Scher E; Templeton K; Yu X |
| EPI_ISL_652055, EPI_ISL_652056, EPI_ISL_665340, EPI_ISL_665555, EPI_ISL_665563, EPI_ISL_665646, EPI_ISL_665725, EPI_ISL_665762, EPI_ISL_665790, EPI_ISL_665791, EPI_ISL_666070, EPI_ISL_666072, EPI_ISL_666074, EPI_ISL_666075, EPI_ISL_666076, EPI_ISL_666077, EPI_ISL_666078, EPI_ISL_666079, EPI_ISL_666080, EPI_ISL_666081, EPI_ISL_666082, EPI_ISL_666083, EPI_ISL_680510, EPI_ISL_680538, EPI_ISL_680546, EPI_ISL_680547, EPI_ISL_702528, EPI_ISL_702594, EPI_ISL_703779, EPI_ISL_703950, EPI_ISL_704187, EPI_ISL_705751, EPI_ISL_705752, EPI_ISL_705753, EPI_ISL_705754, EPI_ISL_705755, EPI_ISL_705756, EPI_ISL_705757, EPI_ISL_705758, EPI_ISL_705759, EPI_ISL_705760, EPI_ISL_705764, EPI_ISL_705765, EPI_ISL_705766, EPI_ISL_705767, EPI_ISL_705770, EPI_ISL_705773, EPI_ISL_705800, EPI_ISL_705801, EPI_ISL_742202, EPI_ISL_742243, EPI_ISL_742245, EPI_ISL_742246, EPI_ISL_814333, EPI_ISL_814546, EPI_ISL_814569, EPI_ISL_814570, EPI_ISL_838314, EPI_ISL_865162 | | | |
| see above | Virology Department, Royal Infirmary of Edinburgh, NHS Lothian / School of Biological Sciences, University of Edinburgh / Institute of Genetics and Molecular Medicine, University of Edinburgh | COVID-19 Genomics UK (COG-UK) Consortium | Balcaza C; Colquhoun R; Dewar R; Gallagher M; Hill V; Jackson B; McCrone JT; McHugh M; O'Toole Á; Rambaut A; Rooke S; Scher E; Templeton K; Williams TC; Yu X |
| EPI_ISL_650134, EPI_ISL_650406, EPI_ISL_651077, EPI_ISL_653070, EPI_ISL_653076, EPI_ISL_653078, EPI_ISL_653079, EPI_ISL_653080, EPI_ISL_653089, EPI_ISL_679852, EPI_ISL_679853, EPI_ISL_679854, EPI_ISL_679855, EPI_ISL_679856, EPI_ISL_679857, EPI_ISL_679858, EPI_ISL_679859, EPI_ISL_679860, EPI_ISL_679861, EPI_ISL_679862, EPI_ISL_679863, EPI_ISL_679864, EPI_ISL_679865, EPI_ISL_679866, EPI_ISL_679867, EPI_ISL_679868, EPI_ISL_679869, EPI_ISL_679870, EPI_ISL_679871, EPI_ISL_679872, EPI_ISL_679873, EPI_ISL_679874, EPI_ISL_679875, EPI_ISL_679876, EPI_ISL_679877, EPI_ISL_679878, EPI_ISL_679879, EPI_ISL_679880, EPI_ISL_679881, EPI_ISL_679882, EPI_ISL_679883, EPI_ISL_679884, EPI_ISL_679885, EPI_ISL_679886, EPI_ISL_679887, EPI_ISL_679888, EPI_ISL_679889, EPI_ISL_679890, EPI_ISL_679891, EPI_ISL_679892, EPI_ISL_679893, EPI_ISL_679894, EPI_ISL_679895, EPI_ISL_679896, EPI_ISL_679897, EPI_ISL_679898, EPI_ISL_679899, EPI_ISL_679900, EPI_ISL_679901, EPI_ISL_679902, EPI_ISL_679915, EPI_ISL_679916, EPI_ISL_679917, EPI_ISL_679918, EPI_ISL_679919, EPI_ISL_679920, EPI_ISL_679921, EPI_ISL_679922, EPI_ISL_679923, EPI_ISL_680101, EPI_ISL_680117, EPI_ISL_680120, EPI_ISL_680122, EPI_ISL_680123, EPI_ISL_680126, EPI_ISL_680127, EPI_ISL_680129, EPI_ISL_680130, EPI_ISL_680133, EPI_ISL_680139, EPI_ISL_680145, EPI_ISL_680149, EPI_ISL_680152, EPI_ISL_680156, EPI_ISL_680159, EPI_ISL_680160, EPI_ISL_680162, EPI_ISL_680163, EPI_ISL_680165, EPI_ISL_680166, EPI_ISL_680169, EPI_ISL_680172, EPI_ISL_680176, EPI_ISL_680177, EPI_ISL_680179, EPI_ISL_680182, EPI_ISL_680184, EPI_ISL_680188, EPI_ISL_680189, EPI_ISL_680191, EPI_ISL_680195, EPI_ISL_680197, EPI_ISL_727997, EPI_ISL_816510, EPI_ISL_816574, EPI_ISL_816645, EPI_ISL_924315, EPI_ISL_924385, EPI_ISL_924422, EPI_ISL_924427 | | | |
| see above | Virology Department, Sheffield Teaching Hospitals NHS Foundation Trust/Department of Infection, Immunity and Cardiovascular Disease, The Medical School, University of Sheffield | COVID-19 Genomics UK (COG-UK) Consortium | Adri Angyal; Alex Keeley; Benjamin Lindsey; Cariad Evans; Danielle Groves; Dave Partridge; Katie Johnson; Laura Carrilero; Luke Green; Matthew Parker; Matthew Wyles; Mehmet Yavuz; Mohammad Raza; Nikki Smith; Paul Parsons; Rachel Tucker; Rebecca Brown; Thushan de Silva |

| | | | |
|---|---|---|---|
| EPI_ISL_637625, EPI_ISL_650536, EPI_ISL_652619, EPI_ISL_665284, EPI_ISL_665289, EPI_ISL_665309, EPI_ISL_665361, EPI_ISL_665386, EPI_ISL_665433, EPI_ISL_665442, EPI_ISL_665455, EPI_ISL_665460, EPI_ISL_665462, EPI_ISL_665464, EPI_ISL_665539, EPI_ISL_665546, EPI_ISL_665556, EPI_ISL_665557, EPI_ISL_665558, EPI_ISL_665569, EPI_ISL_665575, EPI_ISL_665586, EPI_ISL_665592, EPI_ISL_665595, EPI_ISL_665598, EPI_ISL_665605, EPI_ISL_665606, EPI_ISL_665613, EPI_ISL_665619, EPI_ISL_665642, EPI_ISL_665647, EPI_ISL_665654, EPI_ISL_665658, EPI_ISL_665670, EPI_ISL_665690, EPI_ISL_665708, EPI_ISL_665722, EPI_ISL_665728, EPI_ISL_665763, EPI_ISL_665765, EPI_ISL_665766, EPI_ISL_665767, EPI_ISL_665769, EPI_ISL_665770, EPI_ISL_665772, EPI_ISL_665773, EPI_ISL_665774, EPI_ISL_665775, EPI_ISL_665776, EPI_ISL_665777, EPI_ISL_665778, EPI_ISL_665779, EPI_ISL_665780, EPI_ISL_665781, EPI_ISL_665782, EPI_ISL_665793, EPI_ISL_665796, EPI_ISL_665797, EPI_ISL_665798, EPI_ISL_666110, EPI_ISL_666114, EPI_ISL_666116, EPI_ISL_666121, EPI_ISL_666122, EPI_ISL_666126, EPI_ISL_666129, EPI_ISL_666130, EPI_ISL_666133, EPI_ISL_666138, EPI_ISL_666143, EPI_ISL_666145, EPI_ISL_666146, EPI_ISL_666152, EPI_ISL_666154, EPI_ISL_666155, EPI_ISL_666156, EPI_ISL_666167, EPI_ISL_666170, EPI_ISL_666173, EPI_ISL_666176, EPI_ISL_666178, EPI_ISL_666183, EPI_ISL_666186, EPI_ISL_666187, EPI_ISL_666188, EPI_ISL_666189, EPI_ISL_666190, EPI_ISL_666191, EPI_ISL_666196, EPI_ISL_666198, EPI_ISL_666201, EPI_ISL_666202, EPI_ISL_666203, EPI_ISL_666206, EPI_ISL_666215, EPI_ISL_666216, EPI_ISL_666217, EPI_ISL_666219, EPI_ISL_666225, EPI_ISL_666228, EPI_ISL_666233, EPI_ISL_666236, EPI_ISL_666239, EPI_ISL_666246, EPI_ISL_666249, EPI_ISL_666251, EPI_ISL_666255, EPI_ISL_666256, EPI_ISL_666257, EPI_ISL_666259, EPI_ISL_666260, EPI_ISL_666262, EPI_ISL_666269, EPI_ISL_666271, EPI_ISL_666272, EPI_ISL_666275, EPI_ISL_666278, EPI_ISL_666279, EPI_ISL_666281, EPI_ISL_666282, EPI_ISL_666284, EPI_ISL_666289, EPI_ISL_666290, EPI_ISL_666293, EPI_ISL_666294, EPI_ISL_666295, EPI_ISL_666296, EPI_ISL_666297, EPI_ISL_666299, EPI_ISL_666300, EPI_ISL_666302, EPI_ISL_666303, EPI_ISL_666304, EPI_ISL_666306, EPI_ISL_666308, EPI_ISL_666309, EPI_ISL_666311, EPI_ISL_666314, EPI_ISL_666316, EPI_ISL_666317, EPI_ISL_666324, EPI_ISL_666326, EPI_ISL_666333, EPI_ISL_666336, EPI_ISL_666339, EPI_ISL_666341, EPI_ISL_666348, EPI_ISL_666353, EPI_ISL_666357, EPI_ISL_666359, EPI_ISL_666363, EPI_ISL_666366, EPI_ISL_666368, EPI_ISL_666369, EPI_ISL_666370, EPI_ISL_666372, EPI_ISL_666373, EPI_ISL_666374, EPI_ISL_666375, EPI_ISL_666378, EPI_ISL_666380, EPI_ISL_666381, EPI_ISL_666382, EPI_ISL_666384, EPI_ISL_666385, EPI_ISL_666386, EPI_ISL_666387, EPI_ISL_666388, EPI_ISL_666389, EPI_ISL_666390, EPI_ISL_666391, EPI_ISL_666392, EPI_ISL_666393, EPI_ISL_666394, EPI_ISL_666395, EPI_ISL_666398, EPI_ISL_666400, EPI_ISL_666402, EPI_ISL_666403, EPI_ISL_666404, EPI_ISL_666408, EPI_ISL_666410, EPI_ISL_666413, EPI_ISL_666414, EPI_ISL_666415, EPI_ISL_666416, EPI_ISL_666419, EPI_ISL_666420, EPI_ISL_666421, EPI_ISL_666422, EPI_ISL_666423, EPI_ISL_666427, EPI_ISL_666428, EPI_ISL_666429, EPI_ISL_666431, EPI_ISL_666432, EPI_ISL_666433, EPI_ISL_666434, EPI_ISL_666435, EPI_ISL_666436, EPI_ISL_666438, EPI_ISL_666439, EPI_ISL_666440, EPI_ISL_666441, EPI_ISL_666442, EPI_ISL_666444, EPI_ISL_666445, EPI_ISL_666446, EPI_ISL_666447, EPI_ISL_666448, EPI_ISL_666449, EPI_ISL_666450, EPI_ISL_666453, EPI_ISL_666455, EPI_ISL_666456, EPI_ISL_666462, EPI_ISL_666463, EPI_ISL_666466, EPI_ISL_666468, EPI_ISL_666469, EPI_ISL_666471, EPI_ISL_666472, EPI_ISL_666473, EPI_ISL_666475, EPI_ISL_666477, EPI_ISL_666478, EPI_ISL_666479, EPI_ISL_666484, EPI_ISL_666485, EPI_ISL_666486, EPI_ISL_666487, EPI_ISL_666488, EPI_ISL_666489, EPI_ISL_666490, EPI_ISL_666492, EPI_ISL_666494, EPI_ISL_666495, EPI_ISL_666496, EPI_ISL_666500, EPI_ISL_666501, EPI_ISL_666503, EPI_ISL_666507, EPI_ISL_666508, EPI_ISL_666510, EPI_ISL_666511, EPI_ISL_666512, EPI_ISL_666515, EPI_ISL_666517, EPI_ISL_666519, EPI_ISL_666521, EPI_ISL_666522, EPI_ISL_666523, EPI_ISL_666524, EPI_ISL_666526, EPI_ISL_666527, EPI_ISL_666528, EPI_ISL_666529, EPI_ISL_666530, EPI_ISL_666531, EPI_ISL_666533, EPI_ISL_666534, EPI_ISL_666535, EPI_ISL_666538, EPI_ISL_666540, EPI_ISL_666541, EPI_ISL_666542, EPI_ISL_666543, EPI_ISL_666544, EPI_ISL_666545, EPI_ISL_666546, EPI_ISL_666549, EPI_ISL_666552, EPI_ISL_666553, EPI_ISL_666556, EPI_ISL_666557, EPI_ISL_666558, EPI_ISL_666559, EPI_ISL_666560, EPI_ISL_666561, EPI_ISL_666562, EPI_ISL_666563, EPI_ISL_666564, EPI_ISL_666566, EPI_ISL_666569, EPI_ISL_666570, EPI_ISL_666571, EPI_ISL_666572, EPI_ISL_666573, EPI_ISL_666574, EPI_ISL_666575, EPI_ISL_666576, EPI_ISL_666577, EPI_ISL_666578, EPI_ISL_666579, EPI_ISL_666582, EPI_ISL_666584, EPI_ISL_666587, EPI_ISL_666590, EPI_ISL_680583, EPI_ISL_680584, EPI_ISL_680586, EPI_ISL_680588, EPI_ISL_680606, EPI_ISL_680634, EPI_ISL_680635, EPI_ISL_680636, EPI_ISL_680637, EPI_ISL_680638, EPI_ISL_680639, EPI_ISL_680671, EPI_ISL_680672, EPI_ISL_680673, EPI_ISL_680674, EPI_ISL_680675, EPI_ISL_680676, EPI_ISL_680677, EPI_ISL_680678, EPI_ISL_680679, EPI_ISL_680680, EPI_ISL_680683, EPI_ISL_680689, EPI_ISL_680690, EPI_ISL_680691, EPI_ISL_680692, EPI_ISL_680693, EPI_ISL_680694, EPI_ISL_680695, EPI_ISL_680696, EPI_ISL_680697, EPI_ISL_680698, EPI_ISL_680701, EPI_ISL_680713, EPI_ISL_680739, EPI_ISL_680741, EPI_ISL_680742, EPI_ISL_680743, EPI_ISL_680744, EPI_ISL_680745, EPI_ISL_680746, EPI_ISL_680748, EPI_ISL_680749, EPI_ISL_680750, EPI_ISL_680751, EPI_ISL_680752, EPI_ISL_680753, EPI_ISL_680754, EPI_ISL_680755, EPI_ISL_680756, EPI_ISL_680757, EPI_ISL_680758, EPI_ISL_680759, EPI_ISL_680760, EPI_ISL_680761, EPI_ISL_680762, EPI_ISL_680763, EPI_ISL_680764, EPI_ISL_680765, EPI_ISL_680766, EPI_ISL_680767, EPI_ISL_680768, EPI_ISL_680769, EPI_ISL_680770, EPI_ISL_680771, EPI_ISL_680772, EPI_ISL_680773, EPI_ISL_680774, EPI_ISL_680775, EPI_ISL_680776, EPI_ISL_680777, EPI_ISL_680778, EPI_ISL_680779, EPI_ISL_680780, EPI_ISL_680781, EPI_ISL_680782, EPI_ISL_680783, EPI_ISL_680784, EPI_ISL_680785, EPI_ISL_680786, EPI_ISL_680787, EPI_ISL_680788, EPI_ISL_680789, EPI_ISL_680790, EPI_ISL_680791, EPI_ISL_680792, EPI_ISL_680793, EPI_ISL_680794, EPI_ISL_680795, EPI_ISL_680796, EPI_ISL_680797, EPI_ISL_680798, EPI_ISL_680799, EPI_ISL_680800, EPI_ISL_680801, EPI_ISL_680802, EPI_ISL_680803, EPI_ISL_680804, EPI_ISL_680805, EPI_ISL_680806, EPI_ISL_680807, EPI_ISL_680808, EPI_ISL_680809, EPI_ISL_680810, EPI_ISL_680811, EPI_ISL_680812, EPI_ISL_680813, EPI_ISL_680814, EPI_ISL_680815, EPI_ISL_680816, EPI_ISL_680817, EPI_ISL_680818, EPI_ISL_680819, EPI_ISL_680822, EPI_ISL_680823, EPI_ISL_680824, EPI_ISL_680825, EPI_ISL_680826, EPI_ISL_680827, EPI_ISL_680828, EPI_ISL_680829, EPI_ISL_680830, EPI_ISL_680831, EPI_ISL_680832, EPI_ISL_680833, EPI_ISL_680834, EPI_ISL_680835, EPI_ISL_680836, EPI_ISL_680837, EPI_ISL_680838, EPI_ISL_680840, EPI_ISL_680841, EPI_ISL_680842, EPI_ISL_680843, EPI_ISL_680844, EPI_ISL_680845, EPI_ISL_680846, EPI_ISL_680847, EPI_ISL_680852, EPI_ISL_680853, EPI_ISL_680854, EPI_ISL_680855, EPI_ISL_680857, EPI_ISL_680953, EPI_ISL_680954, EPI_ISL_680955, EPI_ISL_680956, EPI_ISL_680957, EPI_ISL_680958, EPI_ISL_680969, EPI_ISL_680970, EPI_ISL_680971, EPI_ISL_680972, EPI_ISL_680973, EPI_ISL_680974, EPI_ISL_680975, EPI_ISL_680976, EPI_ISL_680977, EPI_ISL_680978, EPI_ISL_680979, EPI_ISL_680980, EPI_ISL_680981, EPI_ISL_680982, EPI_ISL_680983, EPI_ISL_680984, EPI_ISL_680985, EPI_ISL_680986, EPI_ISL_680987, EPI_ISL_680988, EPI_ISL_680989, EPI_ISL_680990, EPI_ISL_680991, EPI_ISL_680992, EPI_ISL_680993, EPI_ISL_680994, EPI_ISL_680995, EPI_ISL_680996, EPI_ISL_680997, EPI_ISL_680998, EPI_ISL_680999, EPI_ISL_681000, EPI_ISL_681001, EPI_ISL_681002, EPI_ISL_681003, EPI_ISL_681004, EPI_ISL_681005, EPI_ISL_681006, EPI_ISL_681007, EPI_ISL_681008, EPI_ISL_681009, EPI_ISL_681010, EPI_ISL_681011, EPI_ISL_681012, EPI_ISL_681018, EPI_ISL_681021, EPI_ISL_681022, EPI_ISL_681023, EPI_ISL_681024, EPI_ISL_681025, EPI_ISL_681026, EPI_ISL_681027, EPI_ISL_681040, EPI_ISL_681041, EPI_ISL_681042, EPI_ISL_681043, EPI_ISL_681044, EPI_ISL_681045, EPI_ISL_681046, EPI_ISL_681047, EPI_ISL_681048, EPI_ISL_681049, EPI_ISL_681050, EPI_ISL_681051, EPI_ISL_681052, EPI_ISL_681053, EPI_ISL_681054, EPI_ISL_681055, EPI_ISL_681056, EPI_ISL_681057, EPI_ISL_681058, EPI_ISL_681059, EPI_ISL_681060, EPI_ISL_681061, EPI_ISL_681062, EPI_ISL_681063, EPI_ISL_681064, EPI_ISL_681065, EPI_ISL_681066, EPI_ISL_681067, EPI_ISL_681068, EPI_ISL_681069, EPI_ISL_681070, EPI_ISL_681071, EPI_ISL_681072, EPI_ISL_681073, EPI_ISL_681074, EPI_ISL_681075, EPI_ISL_681076, EPI_ISL_681077, EPI_ISL_681078, EPI_ISL_681079, EPI_ISL_681080, EPI_ISL_681081, EPI_ISL_681082, EPI_ISL_681083, EPI_ISL_681084, EPI_ISL_681085, EPI_ISL_681086, EPI_ISL_681087, EPI_ISL_681088, EPI_ISL_681089, EPI_ISL_681090, EPI_ISL_681091, EPI_ISL_681092, EPI_ISL_681093, EPI_ISL_681094, EPI_ISL_681095, EPI_ISL_681096, EPI_ISL_681097, EPI_ISL_681098, EPI_ISL_681099, EPI_ISL_681100, EPI_ISL_681101, EPI_ISL_681102, EPI_ISL_681103, EPI_ISL_681104, EPI_ISL_681105, EPI_ISL_681106, EPI_ISL_681107, EPI_ISL_681108, EPI_ISL_681109, EPI_ISL_681110, EPI_ISL_681111, EPI_ISL_681112, EPI_ISL_681113, EPI_ISL_681114, EPI_ISL_681115, EPI_ISL_681116, EPI_ISL_681117, EPI_ISL_681118, EPI_ISL_681119, EPI_ISL_681120, EPI_ISL_681121, EPI_ISL_681122, EPI_ISL_681123, EPI_ISL_681124, EPI_ISL_681125, EPI_ISL_681126, EPI_ISL_681127, EPI_ISL_681128, EPI_ISL_681129, EPI_ISL_681130, EPI_ISL_681131, EPI_ISL_681132, EPI_ISL_681133, EPI_ISL_681134, EPI_ISL_681135, EPI_ISL_681136, EPI_ISL_681137, EPI_ISL_681138, EPI_ISL_681139, EPI_ISL_681140, EPI_ISL_681141, EPI_ISL_681142, EPI_ISL_681143, EPI_ISL_681144, EPI_ISL_681145, EPI_ISL_681146, EPI_ISL_681147, EPI_ISL_681148, EPI_ISL_681149, EPI_ISL_681150, EPI_ISL_681151, EPI_ISL_681152, EPI_ISL_681153, EPI_ISL_681154, EPI_ISL_681155, EPI_ISL_681156, EPI_ISL_681157, EPI_ISL_681158, EPI_ISL_681159, EPI_ISL_681160, EPI_ISL_681161, EPI_ISL_681162, EPI_ISL_681163, EPI_ISL_681164, EPI_ISL_681165, EPI_ISL_681166, EPI_ISL_681167, EPI_ISL_681168, EPI_ISL_681169, EPI_ISL_681170, EPI_ISL_681171, EPI_ISL_681172, EPI_ISL_681173, EPI_ISL_681174, EPI_ISL_681175, EPI_ISL_681176, EPI_ISL_681177, EPI_ISL_681178, EPI_ISL_681179, EPI_ISL_681180, EPI_ISL_681181, EPI_ISL_681182, EPI_ISL_681183, EPI_ISL_681184, EPI_ISL_681185, EPI_ISL_681186, EPI_ISL_681187, EPI_ISL_681188, EPI_ISL_681189, EPI_ISL_681190, EPI_ISL_681191, EPI_ISL_681192, EPI_ISL_681193, EPI_ISL_681194, EPI_ISL_681195, EPI_ISL_681196, EPI_ISL_681197, EPI_ISL_681198, EPI_ISL_681199, EPI_ISL_681200, EPI_ISL_681201, EPI_ISL_681202, EPI_ISL_681203, EPI_ISL_681204, EPI_ISL_681205, EPI_ISL_681206, EPI_ISL_681207, EPI_ISL_681208, EPI_ISL_681209, EPI_ISL_681210, EPI_ISL_681211, EPI_ISL_681212, EPI_ISL_681213, EPI_ISL_681214, EPI_ISL_681215, EPI_ISL_681216, EPI_ISL_681217, EPI_ISL_681218, EPI_ISL_681219, EPI_ISL_681220, EPI_ISL_681221, EPI_ISL_681222, EPI_ISL_681223, EPI_ISL_681224, EPI_ISL_681225, EPI_ISL_681226, EPI_ISL_681227, EPI_ISL_681228, EPI_ISL_681229, EPI_ISL_681230, EPI_ISL_681231, EPI_ISL_681232, EPI_ISL_681233, EPI_ISL_681234, EPI_ISL_681235, EPI_ISL_681236, EPI_ISL_681237, EPI_ISL_681238, EPI_ISL_681239, EPI_ISL_681240, EPI_ISL_681241, EPI_ISL_681242, EPI_ISL_681243, EPI_ISL_681244, EPI_ISL_681245, EPI_ISL_681247, EPI_ISL_681248, EPI_ISL_681249, EPI_ISL_681250, EPI_ISL_681251, EPI_ISL_681252, EPI_ISL_681253, EPI_ISL_681254, EPI_ISL_681255, EPI_ISL_703129, EPI_ISL_703443, EPI_ISL_703868, EPI_ISL_703908, EPI_ISL_705015, EPI_ISL_705070, EPI_ISL_705166, EPI_ISL_705328, EPI_ISL_705418, EPI_ISL_705419, EPI_ISL_705436, EPI_ISL_705437, EPI_ISL_705438, EPI_ISL_705460, EPI_ISL_705527, EPI_ISL_706438, EPI_ISL_706443, EPI_ISL_706450, EPI_ISL_706484, EPI_ISL_706485, EPI_ISL_706486, EPI_ISL_706487, EPI_ISL_706488, EPI_ISL_706489, EPI_ISL_706490, EPI_ISL_706491, EPI_ISL_706493, EPI_ISL_706495, EPI_ISL_706496, EPI_ISL_706589, EPI_ISL_706590, EPI_ISL_706591, EPI_ISL_706592, EPI_ISL_706593, EPI_ISL_706594, EPI_ISL_706595, EPI_ISL_706604, EPI_ISL_706605, EPI_ISL_706606, EPI_ISL_706614, EPI_ISL_706615, EPI_ISL_706616, EPI_ISL_706617, EPI_ISL_706618, EPI_ISL_706636, EPI_ISL_706637, EPI_ISL_706663, EPI_ISL_706664, EPI_ISL_706665, EPI_ISL_706675, EPI_ISL_706676, EPI_ISL_706677, EPI_ISL_706678, EPI_ISL_706683, EPI_ISL_706684, EPI_ISL_706685, EPI_ISL_706686, EPI_ISL_706687, EPI_ISL_706688, EPI_ISL_706689, EPI_ISL_706690, EPI_ISL_706691, EPI_ISL_706692, EPI_ISL_706693, EPI_ISL_706694, EPI_ISL_706695, EPI_ISL_706696, EPI_ISL_706697, EPI_ISL_706698, EPI_ISL_706699, EPI_ISL_706700, EPI_ISL_706701, EPI_ISL_706702, EPI_ISL_706703, EPI_ISL_706704, EPI_ISL_706705, EPI_ISL_706706, EPI_ISL_706707, EPI_ISL_706708, EPI_ISL_706709, EPI_ISL_706710, EPI_ISL_706711, EPI_ISL_706712, EPI_ISL_706713, EPI_ISL_706714, EPI_ISL_706715, EPI_ISL_706716, EPI_ISL_706717, EPI_ISL_706718, EPI_ISL_706719, EPI_ISL_706720, EPI_ISL_706721, EPI_ISL_706722, EPI_ISL_706723, EPI_ISL_706724, EPI_ISL_706725, EPI_ISL_706726, EPI_ISL_706727, EPI_ISL_706728, EPI_ISL_706729, EPI_ISL_706730, EPI_ISL_706731, EPI_ISL_706732, EPI_ISL_706733, EPI_ISL_706734, EPI_ISL_706735, EPI_ISL_706736, EPI_ISL_706737, EPI_ISL_706738, EPI_ISL_706739, EPI_ISL_706740, EPI_ISL_706741, EPI_ISL_706742, EPI_ISL_706743, EPI_ISL_706744, EPI_ISL_706745, EPI_ISL_706746, EPI_ISL_706747, EPI_ISL_706748, EPI_ISL_706749, EPI_ISL_706750, EPI_ISL_706751, EPI_ISL_706752, EPI_ISL_706753, EPI_ISL_706754, EPI_ISL_706755, EPI_ISL_706756, EPI_ISL_706757, EPI_ISL_706758, EPI_ISL_706759, EPI_ISL_706760, EPI_ISL_706761, EPI_ISL_706762, EPI_ISL_706763, EPI_ISL_725670, EPI_ISL_725985, EPI_ISL_725986, EPI_ISL_725987, EPI_ISL_725988, EPI_ISL_725989, EPI_ISL_726018, EPI_ISL_726033, EPI_ISL_726034, EPI_ISL_726035, EPI_ISL_726036, EPI_ISL_726037, EPI_ISL_726038, EPI_ISL_726039, EPI_ISL_726040, EPI_ISL_726041, EPI_ISL_726042, EPI_ISL_726044, EPI_ISL_742281, EPI_ISL_742282, EPI_ISL_742317, EPI_ISL_742318, EPI_ISL_742319, EPI_ISL_742320, EPI_ISL_742321, EPI_ISL_742323, EPI_ISL_742324, EPI_ISL_822345 | | | |
| see above | Wales Specialist Virology Centre Sequencing lab: Pathogen Genomics Unit | COVID-19 Genomics UK (COG-UK) Consortium | Alec Birchley; Alexander Adams; Amy Gaskin; Angela Marchbank; Bree Gatica-Wilcox; Catherine Moore; Jason Coombes; Joanne Watkins; Joel Southgate; Johnathan Evans; Laura Gifford; Lauren Gilbert; Lee Graham; Malorie Perry; Matthew Bull; Nicole Pacchiarini; Sally Corden; Sara Kumziene-Summerhayes; Sara Rey; Sarah Taylor; Simon Cottrell; Sophie Jones; Tom Connor |
| EPI_ISL_650451, EPI_ISL_650649, EPI_ISL_650726, EPI_ISL_650728, EPI_ISL_650806, EPI_ISL_650876, EPI_ISL_651011, EPI_ISL_651377, EPI_ISL_651496, EPI_ISL_651497, EPI_ISL_651538, EPI_ISL_651539, EPI_ISL_651540, EPI_ISL_651885, EPI_ISL_651888, EPI_ISL_651889, EPI_ISL_651893, EPI_ISL_651894, EPI_ISL_651895, EPI_ISL_651896, EPI_ISL_651897, EPI_ISL_651898, EPI_ISL_651899, EPI_ISL_651901, EPI_ISL_651902, EPI_ISL_651903, EPI_ISL_651904, EPI_ISL_651905, EPI_ISL_651906, EPI_ISL_651907, EPI_ISL_651908, EPI_ISL_651909, EPI_ISL_651910, EPI_ISL_651911, EPI_ISL_651912, EPI_ISL_651913, EPI_ISL_651914, EPI_ISL_651915, EPI_ISL_651916, EPI_ISL_651917, EPI_ISL_651918, EPI_ISL_651919, EPI_ISL_651920, EPI_ISL_651921, EPI_ISL_651922, EPI_ISL_651923, EPI_ISL_651924, EPI_ISL_651925, EPI_ISL_651926, EPI_ISL_651927, EPI_ISL_651929, EPI_ISL_651931, EPI_ISL_651932, EPI_ISL_651933, EPI_ISL_651934, EPI_ISL_651935, EPI_ISL_651936, EPI_ISL_651937, EPI_ISL_651938, EPI_ISL_651939, EPI_ISL_651940, EPI_ISL_651941, EPI_ISL_651942, EPI_ISL_651943, EPI_ISL_651944, EPI_ISL_651945, EPI_ISL_651947, EPI_ISL_651948, EPI_ISL_651949, EPI_ISL_651950, EPI_ISL_651951, EPI_ISL_651952, EPI_ISL_651953, EPI_ISL_651954, EPI_ISL_665283, EPI_ISL_665522, EPI_ISL_665523, EPI_ISL_665529, EPI_ISL_665535, EPI_ISL_665541, EPI_ISL_665564, EPI_ISL_665565, EPI_ISL_665566, EPI_ISL_665578, EPI_ISL_665640, EPI_ISL_665792, EPI_ISL_665804, EPI_ISL_666023, EPI_ISL_666024, EPI_ISL_666025, EPI_ISL_666026, EPI_ISL_666027, EPI_ISL_666028, EPI_ISL_680445, EPI_ISL_680446, EPI_ISL_680447, EPI_ISL_680448, EPI_ISL_703190, EPI_ISL_703192, EPI_ISL_703287, EPI_ISL_703718, EPI_ISL_703829, EPI_ISL_703831, EPI_ISL_703834, EPI_ISL_703837, EPI_ISL_704206, EPI_ISL_705636, EPI_ISL_705637, EPI_ISL_919253, EPI_ISL_919254 | | | |
| see above | West of Scotland Specialist Virology Centre, NHSGGC / MRC-University of Glasgow Centre for Virus Research | COVID-19 Genomics UK (COG-UK) Consortium | Alasdair MacLean; Alice Broos; Ana da Silva Filipe; Antonia Ho; Daniel Mair; David L Robertson; Emma Thomson; Guy Mollett; James Shepherd; Jenna Nichols; Joseph Hughes; Kathy Li; Kathy Smollett; Kyriaki Nomikou; Lily Tong; Matthew Holden; Natasha Johnson; Rachel Blacow; Richard Orton; Rory Gunson; Sarah McDonald; Sharif Shaaban; Sreenu Vattipally; Stephen Carmichael |

We gratefully acknowledge the following Authors from the Originating laboratories responsible for obtaining the specimens, as well as the Submitting laboratories where the genome data were generated and shared via GISAID, on which this research is based.

All Submitters of data may be contacted directly via www.gisaid.org

Authors are sorted alphabetically.

| Accession ID | Originating Laboratory | Submitting Laboratory | Authors |
|---|---|---|---|
| EPI_ISL_842899, EPI_ISL_842900, EPI_ISL_842901, EPI_ISL_842914, EPI_ISL_842915, EPI_ISL_842923, EPI_ISL_842924, EPI_ISL_1048102, EPI_ISL_1636919, EPI_ISL_1636920, EPI_ISL_1636921, EPI_ISL_1636922, EPI_ISL_1636923, EPI_ISL_1636924, EPI_ISL_1636925, EPI_ISL_1636926 | | | |
| see above | Barts Health NHS Trust | COVID-19 Genomics UK (COG-UK) Consortium | BROAD; Beatrix; CUTINO-MOGUEL; Claire; David; Dola; HARRINGTON; KELE; KULASEGARAN-SHYLINI; Maria-Teresa; OWOYEMI; Raghavendran; SHYLINI |
| EPI_ISL_868758, EPI_ISL_868759, EPI_ISL_868761, EPI_ISL_868762, EPI_ISL_868764, EPI_ISL_868765, EPI_ISL_868767, EPI_ISL_868768, EPI_ISL_868769, EPI_ISL_868771, EPI_ISL_868774, EPI_ISL_868775, EPI_ISL_868777, EPI_ISL_868778, EPI_ISL_868779, EPI_ISL_868785, EPI_ISL_868787, EPI_ISL_1476756, EPI_ISL_1476757 | | | |
| see above | Bioinformatics and Biostatistics Lab, Advanced Sequencing Facility | COVID-19 Genomics UK (COG-UK) Consortium | Aengus Stewart; Chelsea Sawyer; Harshil Patel; Jerome Nicod; Laura Cubitt; Margaret Crawford |
| EPI_ISL_665616, EPI_ISL_665625, EPI_ISL_727738, EPI_ISL_741719, EPI_ISL_741720, EPI_ISL_741721, EPI_ISL_741722, EPI_ISL_741723, EPI_ISL_741743, EPI_ISL_741744, EPI_ISL_867966, EPI_ISL_923214, EPI_ISL_923215, EPI_ISL_952428, EPI_ISL_952431, EPI_ISL_1107755, EPI_ISL_1730878, EPI_ISL_1730880, EPI_ISL_1730881 | | | |
| see above | Centre for Enzyme Innovation, University of Portsmouth / Translational Research Laboratory, Portsmouth Hospitals NHS Trust | COVID-19 Genomics UK (COG-UK) Consortium | Allyson Lloyd; Angela Beckett; Anoop Chauhan; Christopher Fearn; Ethan Butcher; Garry Scarlett; Kate Cook; Katie Loveson; Kelly Bicknell; Robert Impey; Salman Goudarzi; Samuel Robson; Sarah Wyllie; Scott Elliott; Sharon Glaysher; Yann Bourgeois |
| EPI_ISL_664108, EPI_ISL_664112, EPI_ISL_664118, EPI_ISL_664119, EPI_ISL_664130, EPI_ISL_664131, EPI_ISL_664132, EPI_ISL_664133, EPI_ISL_664139, EPI_ISL_664140, EPI_ISL_664155, EPI_ISL_664156, EPI_ISL_664165, EPI_ISL_664166, EPI_ISL_664168, EPI_ISL_664172, EPI_ISL_664175, EPI_ISL_664176, EPI_ISL_664179, EPI_ISL_664201, EPI_ISL_664277, EPI_ISL_664280, EPI_ISL_664291, EPI_ISL_664303, EPI_ISL_664304, EPI_ISL_664305, EPI_ISL_664309, EPI_ISL_664310, EPI_ISL_664311, EPI_ISL_664315, EPI_ISL_664317, EPI_ISL_664320, EPI_ISL_664321, EPI_ISL_664325, EPI_ISL_664346, EPI_ISL_664380, EPI_ISL_664451, EPI_ISL_664455, EPI_ISL_664470, EPI_ISL_664516, EPI_ISL_664527, EPI_ISL_664529, EPI_ISL_664530, EPI_ISL_664531, EPI_ISL_664535, EPI_ISL_664536, EPI_ISL_664537, EPI_ISL_664543, EPI_ISL_664547, EPI_ISL_664548, EPI_ISL_664549, EPI_ISL_664552, EPI_ISL_664553, EPI_ISL_664554, EPI_ISL_664555, EPI_ISL_664588, EPI_ISL_664589, EPI_ISL_664590, EPI_ISL_664601, EPI_ISL_664616, EPI_ISL_664656, EPI_ISL_664676, EPI_ISL_664677, EPI_ISL_664698, EPI_ISL_664708, EPI_ISL_664717, EPI_ISL_664731, EPI_ISL_664739, EPI_ISL_664748, EPI_ISL_664749, EPI_ISL_664765, EPI_ISL_664766, EPI_ISL_664768, EPI_ISL_664769, EPI_ISL_664770, EPI_ISL_664771, EPI_ISL_664772, EPI_ISL_664773, EPI_ISL_664774, EPI_ISL_664775, EPI_ISL_664776, EPI_ISL_664777, EPI_ISL_664778, EPI_ISL_664779, EPI_ISL_664780, EPI_ISL_664781, EPI_ISL_664782, EPI_ISL_664783, EPI_ISL_664784, EPI_ISL_664785, EPI_ISL_664786, EPI_ISL_664787, EPI_ISL_664788, EPI_ISL_664789, EPI_ISL_664790, EPI_ISL_664791, EPI_ISL_664792, EPI_ISL_664793, EPI_ISL_664794, EPI_ISL_664795, EPI_ISL_664796, EPI_ISL_664797, EPI_ISL_664798, EPI_ISL_664799, EPI_ISL_664800, EPI_ISL_664833, EPI_ISL_664835, EPI_ISL_664836, EPI_ISL_664980, EPI_ISL_664981, EPI_ISL_664982, EPI_ISL_664983, EPI_ISL_664984, EPI_ISL_664985, EPI_ISL_664986, EPI_ISL_664988, EPI_ISL_664994, EPI_ISL_664995, EPI_ISL_664996, EPI_ISL_664997, EPI_ISL_664998, EPI_ISL_665004, EPI_ISL_665005, EPI_ISL_665006, EPI_ISL_665007, EPI_ISL_665008, EPI_ISL_665009, EPI_ISL_665010, EPI_ISL_665011, EPI_ISL_665012, EPI_ISL_665022, EPI_ISL_679156, EPI_ISL_679157, EPI_ISL_679159, EPI_ISL_679167, EPI_ISL_679169, EPI_ISL_679173, EPI_ISL_679177, EPI_ISL_679178, EPI_ISL_679179, EPI_ISL_679180, EPI_ISL_679181, EPI_ISL_679182, EPI_ISL_679183, EPI_ISL_679184, EPI_ISL_679185, EPI_ISL_679186, EPI_ISL_679188, EPI_ISL_679190, EPI_ISL_679192, EPI_ISL_679194, EPI_ISL_679196, EPI_ISL_679197, EPI_ISL_679198, EPI_ISL_679204, EPI_ISL_679205, EPI_ISL_679206, EPI_ISL_679207, EPI_ISL_679208, EPI_ISL_679209, EPI_ISL_679210, EPI_ISL_679211, EPI_ISL_679212, EPI_ISL_679213, EPI_ISL_679214, EPI_ISL_679215, EPI_ISL_679216, EPI_ISL_679217, EPI_ISL_679219, EPI_ISL_679220, EPI_ISL_679221, EPI_ISL_679223, EPI_ISL_679224, EPI_ISL_679226, EPI_ISL_679228, EPI_ISL_679230, EPI_ISL_679232, EPI_ISL_679234, EPI_ISL_679236, EPI_ISL_679238, EPI_ISL_679240, EPI_ISL_679242, EPI_ISL_679244, EPI_ISL_679246, EPI_ISL_679248 | | | |
| see above | Department of Pathology, University of Cambridge | COVID-19 Genomics UK (COG-UK) Consortium | Aminu S. Jahun; Grant Hall; Ian Goodfellow; Iliana Georgana; Malte Pinckert; Martin D. Curran; Myra Hosmillo; Surendra Parmar; Yasmin Chaudhry |
| EPI_ISL_661956, EPI_ISL_661958, EPI_ISL_661961, EPI_ISL_661962, EPI_ISL_661964, EPI_ISL_661965, EPI_ISL_661966, EPI_ISL_661968, EPI_ISL_661970, EPI_ISL_661974, EPI_ISL_661977, EPI_ISL_661978, EPI_ISL_661980, EPI_ISL_661982, EPI_ISL_661983, EPI_ISL_661985, EPI_ISL_661986, EPI_ISL_661988, EPI_ISL_661989, EPI_ISL_661990, EPI_ISL_661991, EPI_ISL_661992, EPI_ISL_661995, EPI_ISL_661998, EPI_ISL_661999, EPI_ISL_662000, EPI_ISL_662001, EPI_ISL_662002, EPI_ISL_662004, EPI_ISL_662006, EPI_ISL_662008, EPI_ISL_662010, EPI_ISL_662013, EPI_ISL_662014, EPI_ISL_662015, EPI_ISL_662017, EPI_ISL_662018, EPI_ISL_662019, EPI_ISL_662020, EPI_ISL_662022, EPI_ISL_662025, EPI_ISL_662029, EPI_ISL_662031, EPI_ISL_662032, EPI_ISL_662034, EPI_ISL_662036, EPI_ISL_662038, EPI_ISL_662043, EPI_ISL_662045, EPI_ISL_662046, EPI_ISL_662047, EPI_ISL_662048, EPI_ISL_662049, EPI_ISL_662050, EPI_ISL_662051, EPI_ISL_662053, EPI_ISL_662055, EPI_ISL_662056, EPI_ISL_662057, EPI_ISL_662058, EPI_ISL_662060, EPI_ISL_662061, EPI_ISL_662062, EPI_ISL_662063, EPI_ISL_662065, EPI_ISL_662067, EPI_ISL_662068, EPI_ISL_662070, EPI_ISL_662071, EPI_ISL_662074, EPI_ISL_662078, EPI_ISL_662079, EPI_ISL_662080, EPI_ISL_662081, EPI_ISL_662086, EPI_ISL_662093, EPI_ISL_662097, EPI_ISL_662098, EPI_ISL_662099, EPI_ISL_662100, EPI_ISL_662101, EPI_ISL_662105, EPI_ISL_662106, EPI_ISL_662107, EPI_ISL_662108, EPI_ISL_662114, EPI_ISL_662115, EPI_ISL_662116, EPI_ISL_662118, EPI_ISL_662120, EPI_ISL_662122, EPI_ISL_662123, EPI_ISL_662124, EPI_ISL_662129, EPI_ISL_662131, EPI_ISL_662132, EPI_ISL_662133, EPI_ISL_662135, EPI_ISL_662137, EPI_ISL_662138, EPI_ISL_662140, EPI_ISL_662141, EPI_ISL_662143, EPI_ISL_662145, EPI_ISL_662146, EPI_ISL_662148, EPI_ISL_662150, EPI_ISL_662151, EPI_ISL_662156, EPI_ISL_662159, EPI_ISL_662160, EPI_ISL_662162, EPI_ISL_662163, EPI_ISL_662164, EPI_ISL_662165, EPI_ISL_662166, EPI_ISL_662167, EPI_ISL_662169, EPI_ISL_662170, EPI_ISL_662173, EPI_ISL_662174, EPI_ISL_662175, EPI_ISL_662176, EPI_ISL_662177, EPI_ISL_662178, EPI_ISL_662179, EPI_ISL_662181, EPI_ISL_662182, EPI_ISL_662183, EPI_ISL_662184, EPI_ISL_662186, EPI_ISL_662187, EPI_ISL_662191, EPI_ISL_662192, EPI_ISL_662194, EPI_ISL_662196, EPI_ISL_662198, EPI_ISL_662200, EPI_ISL_662202, EPI_ISL_662203, EPI_ISL_662206, EPI_ISL_662207, EPI_ISL_662208, EPI_ISL_662209, EPI_ISL_662212, EPI_ISL_662214, EPI_ISL_662215, EPI_ISL_662217, EPI_ISL_662218, EPI_ISL_662219, EPI_ISL_662223, EPI_ISL_662227, EPI_ISL_662228, EPI_ISL_662229, EPI_ISL_662231, EPI_ISL_662233, EPI_ISL_662234, EPI_ISL_662235, EPI_ISL_662236, EPI_ISL_662237, EPI_ISL_662238, EPI_ISL_662239, EPI_ISL_662240, EPI_ISL_662244, EPI_ISL_662245, EPI_ISL_662247, EPI_ISL_662248, EPI_ISL_662249, EPI_ISL_662252, EPI_ISL_662254, EPI_ISL_662255, EPI_ISL_662258, EPI_ISL_662260, EPI_ISL_662263, EPI_ISL_662265, EPI_ISL_662266, EPI_ISL_662267, EPI_ISL_662269, EPI_ISL_662270, EPI_ISL_662272, EPI_ISL_662273, EPI_ISL_662274, EPI_ISL_662275, EPI_ISL_662276, EPI_ISL_662279, EPI_ISL_662281, EPI_ISL_662285, EPI_ISL_662286, EPI_ISL_662290, EPI_ISL_662292, EPI_ISL_662296, EPI_ISL_662300, EPI_ISL_662305, EPI_ISL_662308, EPI_ISL_662309, EPI_ISL_662312, EPI_ISL_662320, EPI_ISL_662327, EPI_ISL_662334, EPI_ISL_662342, EPI_ISL_662343, EPI_ISL_662344, EPI_ISL_662352, EPI_ISL_662354, EPI_ISL_662356, EPI_ISL_662358, EPI_ISL_662359, EPI_ISL_662365, EPI_ISL_662369, EPI_ISL_662373, EPI_ISL_662374, EPI_ISL_662375, EPI_ISL_662381, EPI_ISL_662382, EPI_ISL_662385, EPI_ISL_662389, EPI_ISL_662390, EPI_ISL_662391, EPI_ISL_662393, EPI_ISL_662394, EPI_ISL_662403, EPI_ISL_662405, EPI_ISL_662410, EPI_ISL_662411, EPI_ISL_662412, EPI_ISL_662415, EPI_ISL_662418, EPI_ISL_662419, EPI_ISL_662427, EPI_ISL_662429, EPI_ISL_662431, EPI_ISL_662433, EPI_ISL_662444, EPI_ISL_662446, EPI_ISL_662452, EPI_ISL_662457, EPI_ISL_662462, EPI_ISL_662463, EPI_ISL_662468, EPI_ISL_662472, EPI_ISL_662474, EPI_ISL_662477, EPI_ISL_662478, EPI_ISL_662480, EPI_ISL_662486, EPI_ISL_662487, EPI_ISL_662488, EPI_ISL_662489, EPI_ISL_662492, EPI_ISL_662496, EPI_ISL_662499, EPI_ISL_662502, EPI_ISL_662507, EPI_ISL_662508, EPI_ISL_662514, EPI_ISL_662518, EPI_ISL_662520, EPI_ISL_662526, EPI_ISL_662535, EPI_ISL_662549, EPI_ISL_662553, EPI_ISL_662558, EPI_ISL_662564, EPI_ISL_662575, EPI_ISL_662576, EPI_ISL_662578, EPI_ISL_662579, EPI_ISL_662580, EPI_ISL_662581, EPI_ISL_662583, EPI_ISL_662588, EPI_ISL_662593, EPI_ISL_662597, EPI_ISL_662604, EPI_ISL_662605, EPI_ISL_662609, EPI_ISL_662610, EPI_ISL_662614, EPI_ISL_662616, EPI_ISL_662617, EPI_ISL_662621, EPI_ISL_662622, EPI_ISL_674159, EPI_ISL_674160, EPI_ISL_674161, EPI_ISL_674162, EPI_ISL_674164, EPI_ISL_674165, EPI_ISL_674168, EPI_ISL_674169, EPI_ISL_674170, EPI_ISL_674171, EPI_ISL_674172, EPI_ISL_674174, EPI_ISL_674175, EPI_ISL_674176, EPI_ISL_674177, EPI_ISL_674179, EPI_ISL_674181, EPI_ISL_674182, EPI_ISL_674186, EPI_ISL_674188, EPI_ISL_674190, EPI_ISL_674192, EPI_ISL_674193, EPI_ISL_674194, EPI_ISL_674195, EPI_ISL_674197, EPI_ISL_674199, EPI_ISL_674201, EPI_ISL_674203, EPI_ISL_674204, EPI_ISL_674206, EPI_ISL_674207, EPI_ISL_674208, EPI_ISL_674211, EPI_ISL_674217, EPI_ISL_674222, EPI_ISL_674224, EPI_ISL_674226, EPI_ISL_674227, EPI_ISL_674228, EPI_ISL_674229, EPI_ISL_674231, EPI_ISL_674233, EPI_ISL_674234, EPI_ISL_674240, EPI_ISL_674241, EPI_ISL_674242, EPI_ISL_674243, EPI_ISL_674245, EPI_ISL_674249, EPI_ISL_674253, EPI_ISL_674254, EPI_ISL_674255, EPI_ISL_674260, EPI_ISL_674261, EPI_ISL_674264, EPI_ISL_674267, EPI_ISL_674268, EPI_ISL_674272, EPI_ISL_674276, EPI_ISL_674277, EPI_ISL_674281, EPI_ISL_674283, EPI_ISL_674284, EPI_ISL_674285, EPI_ISL_674286, EPI_ISL_674287, EPI_ISL_674292, EPI_ISL_674294, EPI_ISL_674295, EPI_ISL_674296, EPI_ISL_674297, EPI_ISL_674299, EPI_ISL_674300, EPI_ISL_674301, EPI_ISL_674305, EPI_ISL_674308, EPI_ISL_674310, EPI_ISL_674311, EPI_ISL_674313, EPI_ISL_674317, EPI_ISL_674318, EPI_ISL_674319, EPI_ISL_674322, EPI_ISL_674323, EPI_ISL_674324, EPI_ISL_674328, EPI_ISL_674329, EPI_ISL_674332, EPI_ISL_674334, EPI_ISL_674337, EPI_ISL_674339, EPI_ISL_674340, EPI_ISL_674341, EPI_ISL_674343, EPI_ISL_674345, EPI_ISL_674346, EPI_ISL_674347, EPI_ISL_674351, EPI_ISL_674352, EPI_ISL_674354, EPI_ISL_674357, EPI_ISL_674360, EPI_ISL_674361, EPI_ISL_674362, EPI_ISL_674364, EPI_ISL_674366, EPI_ISL_674367, EPI_ISL_674368, EPI_ISL_674369, EPI_ISL_674370, EPI_ISL_674372, EPI_ISL_674374, EPI_ISL_674375, EPI_ISL_674376, EPI_ISL_674380, EPI_ISL_674383, EPI_ISL_674385, EPI_ISL_674389, EPI_ISL_674390, EPI_ISL_674391, EPI_ISL_674392, EPI_ISL_674394, EPI_ISL_674396, EPI_ISL_674397, EPI_ISL_674399, EPI_ISL_674400, EPI_ISL_674402, EPI_ISL_674404, EPI_ISL_674406, EPI_ISL_674408, EPI_ISL_674409, EPI_ISL_674411, EPI_ISL_674412, EPI_ISL_674413, EPI_ISL_674416, EPI_ISL_674419, EPI_ISL_674420, EPI_ISL_674421, EPI_ISL_674424, EPI_ISL_674428, EPI_ISL_674429, EPI_ISL_674431, EPI_ISL_674432, EPI_ISL_674434, EPI_ISL_674435, EPI_ISL_674436, EPI_ISL_674438, EPI_ISL_674439, EPI_ISL_674442, EPI_ISL_674445, EPI_ISL_674446, EPI_ISL_674447, EPI_ISL_674451, EPI_ISL_674452, EPI_ISL_674454, EPI_ISL_674455, EPI_ISL_674456, EPI_ISL_674459, EPI_ISL_674462, EPI_ISL_674465, EPI_ISL_674466, EPI_ISL_674467, EPI_ISL_674469, EPI_ISL_674470, EPI_ISL_674471, EPI_ISL_674472, EPI_ISL_674474, EPI_ISL_674476, EPI_ISL_674477, EPI_ISL_674478, EPI_ISL_674479, EPI_ISL_674484, EPI_ISL_674485, EPI_ISL_674486, EPI_ISL_674487, EPI_ISL_674488, EPI_ISL_674490, EPI_ISL_674493, EPI_ISL_674495, EPI_ISL_675130, EPI_ISL_675133, EPI_ISL_675135, EPI_ISL_675136, EPI_ISL_675137, EPI_ISL_675138, EPI_ISL_675139, EPI_ISL_675140, EPI_ISL_675141, EPI_ISL_675142, EPI_ISL_675143, EPI_ISL_675144, EPI_ISL_675146, EPI_ISL_675147, EPI_ISL_675148, EPI_ISL_675149, EPI_ISL_675150, EPI_ISL_675151, EPI_ISL_675152, EPI_ISL_675154, EPI_ISL_675155, EPI_ISL_675156, EPI_ISL_675157, EPI_ISL_675158, EPI_ISL_675159, EPI_ISL_675161, EPI_ISL_675162, EPI_ISL_675163, EPI_ISL_675164, EPI_ISL_675166, EPI_ISL_675167, EPI_ISL_675168, EPI_ISL_675169, EPI_ISL_675171, EPI_ISL_675172, EPI_ISL_675173, EPI_ISL_675174, EPI_ISL_675175, EPI_ISL_675176, EPI_ISL_675177, EPI_ISL_675180, EPI_ISL_675181, EPI_ISL_675182, EPI_ISL_675184, EPI_ISL_675186, EPI_ISL_675187, EPI_ISL_675189, EPI_ISL_675190, EPI_ISL_675191, EPI_ISL_675192, EPI_ISL_675193, EPI_ISL_675194, EPI_ISL_675195, EPI_ISL_675196, EPI_ISL_675197, EPI_ISL_675198, EPI_ISL_675199, EPI_ISL_675200, EPI_ISL_675201, EPI_ISL_675202, EPI_ISL_675203, EPI_ISL_675204, EPI_ISL_675205, EPI_ISL_675206, EPI_ISL_675207, EPI_ISL_675208, EPI_ISL_675209, EPI_ISL_675211, EPI_ISL_675213, EPI_ISL_675214, EPI_ISL_675215, EPI_ISL_675216, EPI_ISL_675218, EPI_ISL_675220, EPI_ISL_675221, EPI_ISL_675222, EPI_ISL_675223, EPI_ISL_675225, EPI_ISL_675226, EPI_ISL_675229, EPI_ISL_675230, EPI_ISL_675231, EPI_ISL_675232, EPI_ISL_675233, EPI_ISL_675234, EPI_ISL_675235, EPI_ISL_675237, EPI_ISL_675239, EPI_ISL_675240, EPI_ISL_675242, EPI_ISL_675245, EPI_ISL_675246, EPI_ISL_675247, EPI_ISL_675248, EPI_ISL_675249, EPI_ISL_675250, EPI_ISL_675251, EPI_ISL_675252, EPI_ISL_675253, EPI_ISL_675254, EPI_ISL_675255, EPI_ISL_675256, EPI_ISL_675257, EPI_ISL_675258, EPI_ISL_675259, EPI_ISL_675261, EPI_ISL_675262, EPI_ISL_675263, EPI_ISL_675265, EPI_ISL_675266, EPI_ISL_675267, EPI_ISL_675268, EPI_ISL_675269, EPI_ISL_675270, EPI_ISL_675271, EPI_ISL_675272, EPI_ISL_675273, EPI_ISL_675274, EPI_ISL_675278, EPI_ISL_675279, EPI_ISL_675280, EPI_ISL_675281, EPI_ISL_675283, EPI_ISL_675287, EPI_ISL_675288, EPI_ISL_675289, EPI_ISL_675290, EPI_ISL_675291, EPI_ISL_675293, EPI_ISL_675294, EPI_ISL_675295, EPI_ISL_675296, EPI_ISL_675297, EPI_ISL_675298, EPI_ISL_675299, EPI_ISL_675300, EPI_ISL_675302, EPI_ISL_675303, EPI_ISL_675304, EPI_ISL_675310, EPI_ISL_675312, EPI_ISL_675314, EPI_ISL_675315, EPI_ISL_675316, EPI_ISL_675321, EPI_ISL_675322, EPI_ISL_675323, EPI_ISL_675324, EPI_ISL_675325, EPI_ISL_675327, EPI_ISL_675328, EPI_ISL_675329, EPI_ISL_675330, EPI_ISL_675333, EPI_ISL_675334, EPI_ISL_675335, EPI_ISL_675336, EPI_ISL_675337, EPI_ISL_675338, EPI_ISL_675339, EPI_ISL_675341, EPI_ISL_675342, EPI_ISL_675343, EPI_ISL_675344, EPI_ISL_675345, EPI_ISL_675346, EPI_ISL_675347, EPI_ISL_675348, EPI_ISL_675349, EPI_ISL_675350, EPI_ISL_675351, EPI_ISL_675352, EPI_ISL_675354, EPI_ISL_675355, EPI_ISL_675356, EPI_ISL_675358, EPI_ISL_675359, EPI_ISL_675360, EPI_ISL_675361, EPI_ISL_675362, EPI_ISL_675363, EPI_ISL_675365, EPI_ISL_675366, EPI_ISL_675367, EPI_ISL_675369, EPI_ISL_675370, EPI_ISL_675372, EPI_ISL_675373, EPI_ISL_675374, EPI_ISL_675375, EPI_ISL_675376, EPI_ISL_675377, EPI_ISL_675378, EPI_ISL_675379, EPI_ISL_675380, EPI_ISL_675381, EPI_ISL_675382, EPI_ISL_675383, EPI_ISL_675384, EPI_ISL_675385, EPI_ISL_675387, EPI_ISL_675388, EPI_ISL_675390, EPI_ISL_675391, EPI_ISL_675392, EPI_ISL_675394, EPI_ISL_675395, EPI_ISL_675396, EPI_ISL_675398, EPI_ISL_675400, EPI_ISL_675401, EPI_ISL_675402, EPI_ISL_675403, EPI_ISL_675405, EPI_ISL_675406, EPI_ISL_675407, EPI_ISL_675408, EPI_ISL_675409, EPI_ISL_675410, EPI_ISL_675411, EPI_ISL_675412, EPI_ISL_675413, EPI_ISL_675414, EPI_ISL_675415, EPI_ISL_675416, EPI_ISL_675418, EPI_ISL_675419, EPI_ISL_675421, EPI_ISL_675422, EPI_ISL_675425, EPI_ISL_675426, EPI_ISL_675427, EPI_ISL_675428, EPI_ISL_675429, EPI_ISL_675430, EPI_ISL_675433, EPI_ISL_675434, EPI_ISL_675435, EPI_ISL_675437, EPI_ISL_675438, EPI_ISL_675440, EPI_ISL_675441, EPI_ISL_675442, EPI_ISL_675445, EPI_ISL_675447, EPI_ISL_675448, EPI_ISL_675449, EPI_ISL_675450, EPI_ISL_675451, EPI_ISL_675452, EPI_ISL_675453, EPI_ISL_675454, EPI_ISL_675455, EPI_ISL_675456, EPI_ISL_675457, EPI_ISL_675458, EPI_ISL_675459, EPI_ISL_675460, EPI_ISL_675461, EPI_ISL_675463, EPI_ISL_675464, EPI_ISL_675465, EPI_ISL_675466, EPI_ISL_675468, EPI_ISL_675469, EPI_ISL_675470, EPI_ISL_675471, EPI_ISL_675473, EPI_ISL_675474, EPI_ISL_675476, EPI_ISL_675477, EPI_ISL_675478, EPI_ISL_675479, EPI_ISL_675481, EPI_ISL_675482, EPI_ISL_675484, EPI_ISL_675487, EPI_ISL_675488, EPI_ISL_675489, EPI_ISL_675492, EPI_ISL_675493, EPI_ISL_675497, EPI_ISL_675498, EPI_ISL_675501, EPI_ISL_675502, EPI_ISL_675508, EPI_ISL_675509, EPI_ISL_675510, EPI_ISL_675511, EPI_ISL_675512, EPI_ISL_675515, EPI_ISL_675516, EPI_ISL_675517, EPI_ISL_675519, EPI_ISL_675520, EPI_ISL_675522, EPI_ISL_675523, EPI_ISL_675524, EPI_ISL_675525, EPI_ISL_675526, EPI_ISL_675527, EPI_ISL_675529, EPI_ISL_675530, EPI_ISL_675531, EPI_ISL_675532, EPI_ISL_675534, EPI_ISL_675536, EPI_ISL_675539, EPI_ISL_675541, EPI_ISL_675543, EPI_ISL_675544, EPI_ISL_675545, EPI_ISL_675548, EPI_ISL_675549, EPI_ISL_675551, EPI_ISL_675552, EPI_ISL_675553, EPI_ISL_675554, EPI_ISL_675557, EPI_ISL_675561, EPI_ISL_675562, EPI_ISL_675563, EPI_ISL_675567, EPI_ISL_675568, EPI_ISL_675570, EPI_ISL_675571, EPI_ISL_675573, EPI_ISL_675574, EPI_ISL_675575, EPI_ISL_675577, EPI_ISL_675579, EPI_ISL_675580, EPI_ISL_675581, EPI_ISL_675582, EPI_ISL_675583, EPI_ISL_675584, EPI_ISL_675585, EPI_ISL_675586, EPI_ISL_675587, EPI_ISL_675588, EPI_ISL_675590, EPI_ISL_675591, EPI_ISL_675592, EPI_ISL_675593, EPI_ISL_675594, EPI_ISL_675595, EPI_ISL_675597, EPI_ISL_675598, EPI_ISL_675599, EPI_ISL_675600, EPI_ISL_675603, EPI_ISL_675604, EPI_ISL_675607, EPI_ISL_675608, EPI_ISL_675609, EPI_ISL_675610, EPI_ISL_675611, EPI_ISL_675612, EPI_ISL_675613, EPI_ISL_675614, EPI_ISL_675615, EPI_ISL_675616, EPI_ISL_675617, EPI_ISL_675618, EPI_ISL_675619, EPI_ISL_675620, EPI_ISL_675622, EPI_ISL_675623, EPI_ISL_675624, EPI_ISL_675625, EPI_ISL_675626, EPI_ISL_675627, EPI_ISL_675628, EPI_ISL_675629, EPI_ISL_675630, EPI_ISL_675632, EPI_ISL_675635, EPI_ISL_675636, EPI_ISL_675637, EPI_ISL_675638, EPI_ISL_675639, EPI_ISL_675640, EPI_ISL_675641, EPI_ISL_675642, EPI_ISL_675645, EPI_ISL_675646, EPI_ISL_675647, EPI_ISL_675648, EPI_ISL_675649, EPI_ISL_675650, EPI_ISL_675651, EPI_ISL_675653, EPI_ISL_675654, EPI_ISL_675655, EPI_ISL_675657, EPI_ISL_675658, EPI_ISL_675659, EPI_ISL_675662, EPI_ISL_675664, EPI_ISL_675665, EPI_ISL_675666, EPI_ISL_675668, EPI_ISL_675669, EPI_ISL_675670, EPI_ISL_675671, EPI_ISL_675672, EPI_ISL_675673, EPI_ISL_675674, EPI_ISL_675675, EPI_ISL_675677, EPI_ISL_675678, EPI_ISL_675679, EPI_ISL_675680, EPI_ISL_675683, EPI_ISL_675684, EPI_ISL_675686, EPI_ISL_675687, EPI_ISL_675688, EPI_ISL_675689, EPI_ISL_675691, EPI_ISL_675692, EPI_ISL_675696, EPI_ISL_675697, EPI_ISL_675699, EPI_ISL_675700, EPI_ISL_675701, EPI_ISL_675702, EPI_ISL_675703, EPI_ISL_675705, EPI_ISL_675706, EPI_ISL_675707, EPI_ISL_675708, EPI_ISL_675710, EPI_ISL_675715, EPI_ISL_675716, EPI_ISL_675719, EPI_ISL_675735, EPI_ISL_675739, EPI_ISL_675740, EPI_ISL_675750, EPI_ISL_675751, EPI_ISL_675756, EPI_ISL_675757, EPI_ISL_675759, EPI_ISL_675760, EPI_ISL_675766, EPI_ISL_675770, EPI_ISL_675771, EPI_ISL_675774, EPI_ISL_675775, EPI_ISL_675776, EPI_ISL_675777, EPI_ISL_675779, EPI_ISL_675785, EPI_ISL_675788, EPI_ISL_675794, EPI_ISL_675798, EPI_ISL_675799, EPI_ISL_675800, EPI_ISL_675802, EPI_ISL_675803, EPI_ISL_675806, EPI_ISL_675810, EPI_ISL_675812, EPI_ISL_675813, EPI_ISL_675817, EPI_ISL_675818, EPI_ISL_675820, EPI_ISL_675822, EPI_ISL_675823, EPI_ISL_675825, EPI_ISL_675826, EPI_ISL_675829, EPI_ISL_675833, EPI_ISL_675839, EPI_ISL_675847, EPI_ISL_675848, EPI_ISL_675852, EPI_ISL_675868, EPI_ISL_675878, EPI_ISL_675880, EPI_ISL_675882, EPI_ISL_675890, EPI_ISL_675894, EPI_ISL_675897, EPI_ISL_675901, EPI_ISL_675902, EPI_ISL_675903, EPI_ISL_675908, EPI_ISL_675913, EPI_ISL_675916, EPI_ISL_675920, EPI_ISL_675924, EPI_ISL_675929, EPI_ISL_675930, EPI_ISL_675934, EPI_ISL_675937, EPI_ISL_675939, EPI_ISL_675942, EPI_ISL_675944, EPI_ISL_675952, EPI_ISL_675954, EPI_ISL_675962, EPI_ISL_675965, EPI_ISL_675968, EPI_ISL_675973, EPI_ISL_675974, EPI_ISL_675976, EPI_ISL_675978, EPI_ISL_675980, EPI_ISL_675982, EPI_ISL_675986, EPI_ISL_675987, EPI_ISL_675988, EPI_ISL_675993, EPI_ISL_676000, EPI_ISL_676002, EPI_ISL_676006, EPI_ISL_676010, EPI_ISL_681661, EPI_ISL_681662, EPI_ISL_681665, EPI_ISL_710837, EPI_ISL_915911 | | | |
| see above | Lighthouse Lab in Alderley Park | Wellcome Sanger Institute for the COVID-19 Genomics UK (COG-UK) Consortium | Cordelia Langford; David K. Jackson; Dominic Kwiatkowski; Ewan Harrison; Ian Johnston; Jacquelyn Wynn; John Sillitoe on behalf of the Wellcome Sanger Institute COVID-19 Surveillance Team; Mairead Hyland; Roberto Amato; Sonia Goncalves; The Lighthouse Lab in Alderley Park and Alex Alderton |
| EPI_ISL_661638, EPI_ISL_661641, EPI_ISL_661653, EPI_ISL_661656, EPI_ISL_661679, EPI_ISL_661680, EPI_ISL_661693, EPI_ISL_661697, EPI_ISL_661702, EPI_ISL_661726, EPI_ISL_661759, EPI_ISL_661775, EPI_ISL_661779, EPI_ISL_661783, EPI_ISL_661785, EPI_ISL_661836, EPI_ISL_661853, EPI_ISL_661854, EPI_ISL_661856, EPI_ISL_661873, EPI_ISL_661925, EPI_ISL_661928, EPI_ISL_661944, EPI_ISL_661951, EPI_ISL_667811, EPI_ISL_667812, EPI_ISL_667813, EPI_ISL_667815, EPI_ISL_667817, EPI_ISL_667818, EPI_ISL_667819, EPI_ISL_667820, EPI_ISL_667821, EPI_ISL_667823, EPI_ISL_667826, EPI_ISL_667827, EPI_ISL_667830, EPI_ISL_667833, EPI_ISL_667834, EPI_ISL_667835, EPI_ISL_667837, EPI_ISL_667839, EPI_ISL_667840, EPI_ISL_667841, EPI_ISL_667842, EPI_ISL_667845, EPI_ISL_667846, EPI_ISL_667848, EPI_ISL_667852, EPI_ISL_667853, EPI_ISL_667854, EPI_ISL_667856, EPI_ISL_667858, EPI_ISL_667862, EPI_ISL_667869, EPI_ISL_667871, EPI_ISL_667874, EPI_ISL_667875, EPI_ISL_667877, EPI_ISL_667878, EPI_ISL_667880, EPI_ISL_667882, EPI_ISL_667884, EPI_ISL_667886, EPI_ISL_667890, EPI_ISL_667892, EPI_ISL_667894, EPI_ISL_667896, EPI_ISL_667897, EPI_ISL_667899, EPI_ISL_667901, EPI_ISL_667903, EPI_ISL_667905, EPI_ISL_667908, EPI_ISL_667909, EPI_ISL_667910, EPI_ISL_667911, EPI_ISL_667913, EPI_ISL_667914, EPI_ISL_667916, EPI_ISL_667921, EPI_ISL_667923, EPI_ISL_667926, EPI_ISL_667932, EPI_ISL_667934, EPI_ISL_667935, EPI_ISL_667936, EPI_ISL_667937, EPI_ISL_667942, EPI_ISL_667944, EPI_ISL_667946, EPI_ISL_667949, EPI_ISL_667951, EPI_ISL_667953, EPI_ISL_667954, EPI_ISL_667956, EPI_ISL_667964, EPI_ISL_667965, EPI_ISL_667968, EPI_ISL_667971, EPI_ISL_667974, EPI_ISL_667979, EPI_ISL_667981, EPI_ISL_667984, EPI_ISL_667985, EPI_ISL_667993, EPI_ISL_667994, EPI_ISL_667995, EPI_ISL_667999, EPI_ISL_668002, EPI_ISL_668003, EPI_ISL_668007, EPI_ISL_668008, EPI_ISL_668009, EPI_ISL_668011, EPI_ISL_668012, EPI_ISL_668014, EPI_ISL_668015, EPI_ISL_668016, EPI_ISL_668017, EPI_ISL_668018, EPI_ISL_668019, EPI_ISL_668023, EPI_ISL_668026, EPI_ISL_668037, EPI_ISL_668039, EPI_ISL_668040, EPI_ISL_668041, EPI_ISL_668042, EPI_ISL_668043, EPI_ISL_668046, EPI_ISL_668047, EPI_ISL_668051, EPI_ISL_668058, EPI_ISL_668059, EPI_ISL_668061, EPI_ISL_668065, EPI_ISL_668068, EPI_ISL_668069, EPI_ISL_668070, EPI_ISL_668076, EPI_ISL_668077, EPI_ISL_668079, EPI_ISL_668081, EPI_ISL_668085, EPI_ISL_668086, EPI_ISL_668087, EPI_ISL_668088, EPI_ISL_668089, EPI_ISL_668090, EPI_ISL_668091, EPI_ISL_668092, EPI_ISL_668097, EPI_ISL_668099, EPI_ISL_668100, EPI_ISL_668101, EPI_ISL_668103, EPI_ISL_668104, EPI_ISL_668108, EPI_ISL_668109, EPI_ISL_668110, EPI_ISL_668112, EPI_ISL_668115, EPI_ISL_668122, EPI_ISL_668123, EPI_ISL_668127, EPI_ISL_668128, EPI_ISL_668133, EPI_ISL_668136, EPI_ISL_668137, EPI_ISL_668138, EPI_ISL_668140, EPI_ISL_668141, EPI_ISL_668148, EPI_ISL_668149, EPI_ISL_668153, EPI_ISL_668154, EPI_ISL_668155, EPI_ISL_668163, EPI_ISL_668166, EPI_ISL_668169, EPI_ISL_668171, EPI_ISL_668172, EPI_ISL_668180, EPI_ISL_668188, EPI_ISL_668190, EPI_ISL_668192, EPI_ISL_668194, EPI_ISL_668200, EPI_ISL_668201, EPI_ISL_668212, EPI_ISL_668219, EPI_ISL_668222, EPI_ISL_668227, EPI_ISL_668237, EPI_ISL_668239, EPI_ISL_668251, EPI_ISL_668252, EPI_ISL_668254, EPI_ISL_668255, EPI_ISL_668261, EPI_ISL_668262, EPI_ISL_668265, EPI_ISL_668267, EPI_ISL_668275, EPI_ISL_668280, EPI_ISL_668281, EPI_ISL_668283, EPI_ISL_668289, EPI_ISL_668294, EPI_ISL_668296, EPI_ISL_668297, EPI_ISL_668298, EPI_ISL_668299, EPI_ISL_668313, EPI_ISL_668315, EPI_ISL_668316, EPI_ISL_668317, EPI_ISL_668319, EPI_ISL_668322, EPI_ISL_668329, EPI_ISL_668336, EPI_ISL_668341, EPI_ISL_668342, EPI_ISL_668345, EPI_ISL_668353, EPI_ISL_668355, EPI_ISL_668365, EPI_ISL_668368, EPI_ISL_668369, EPI_ISL_668373, EPI_ISL_668374, EPI_ISL_668376, EPI_ISL_668379, EPI_ISL_668382, EPI_ISL_668385, EPI_ISL_668388, EPI_ISL_668389, EPI_ISL_673236, EPI_ISL_673237, EPI_ISL_673238, EPI_ISL_673239, EPI_ISL_673240, EPI_ISL_673241, EPI_ISL_673242, EPI_ISL_673243, EPI_ISL_673244, EPI_ISL_673245, EPI_ISL_673246, EPI_ISL_673247, EPI_ISL_673248, EPI_ISL_673249, EPI_ISL_673250, EPI_ISL_673251, EPI_ISL_673252, EPI_ISL_673253, EPI_ISL_673254, EPI_ISL_673255, EPI_ISL_673256, EPI_ISL_673257, EPI_ISL_673258, EPI_ISL_673259, EPI_ISL_673260, EPI_ISL_673261, EPI_ISL_673262, EPI_ISL_673263, EPI_ISL_673264, EPI_ISL_673265, EPI_ISL_673266, EPI_ISL_673267, EPI_ISL_673268, EPI_ISL_673269, EPI_ISL_673270, EPI_ISL_673271, EPI_ISL_673272, EPI_ISL_673273, EPI_ISL_673274, EPI_ISL_673275, EPI_ISL_673276, EPI_ISL_673277, EPI_ISL_673278, EPI_ISL_673279, EPI_ISL_673280, EPI_ISL_673281, EPI_ISL_673282, EPI_ISL_673283, EPI_ISL_673284, EPI_ISL_673285, EPI_ISL_673286, EPI_ISL_673287, EPI_ISL_673288, EPI_ISL_673289, EPI_ISL_673290, EPI_ISL_673291, EPI_ISL_673292, EPI_ISL_673293, EPI_ISL_673294, EPI_ISL_673295, EPI_ISL_673296, EPI_ISL_673297, EPI_ISL_673298, EPI_ISL_673299, EPI_ISL_673300, EPI_ISL_673301, EPI_ISL_673302, EPI_ISL_673303, EPI_ISL_673304, EPI_ISL_673305, EPI_ISL_673306, EPI_ISL_673307, EPI_ISL_673308, EPI_ISL_673309, EPI_ISL_673311, EPI_ISL_673312, EPI_ISL_673313, EPI_ISL_673314, EPI_ISL_673315, EPI_ISL_673316, EPI_ISL_673317, EPI_ISL_673318, EPI_ISL_673319, EPI_ISL_673320, EPI_ISL_673321, EPI_ISL_673322, EPI_ISL_673323, EPI_ISL_673324, EPI_ISL_673325, EPI_ISL_673326, EPI_ISL_673327, EPI_ISL_673328, EPI_ISL_673329, EPI_ISL_673330, EPI_ISL_673331, EPI_ISL_673332, EPI_ISL_673333, EPI_ISL_673334, EPI_ISL_673335, EPI_ISL_673336, EPI_ISL_673337, EPI_ISL_673338, EPI_ISL_673339, EPI_ISL_673340, EPI_ISL_673341, EPI_ISL_673342, EPI_ISL_673343, EPI_ISL_673344, EPI_ISL_673345, EPI_ISL_673346, EPI_ISL_673347, EPI_ISL_673348, EPI_ISL_673349, EPI_ISL_673350, EPI_ISL_673351, EPI_ISL_673352, EPI_ISL_673353, EPI_ISL_673354, EPI_ISL_673355, EPI_ISL_673356, EPI_ISL_673357, EPI_ISL_673358, EPI_ISL_673359, EPI_ISL_673360, EPI_ISL_673361, EPI_ISL_673362, EPI_ISL_673363, EPI_ISL_673364, EPI_ISL_673365, EPI_ISL_673366, EPI_ISL_673367, EPI_ISL_673368, EPI_ISL_673369, EPI_ISL_673370, EPI_ISL_673371, EPI_ISL_673372, EPI_ISL_673373, EPI_ISL_673374, EPI_ISL_673375, EPI_ISL_673376, EPI_ISL_673377, EPI_ISL_673378, EPI_ISL_673379, EPI_ISL_673380, EPI_ISL_673381, EPI_ISL_673382, EPI_ISL_673383, EPI_ISL_673384, EPI_ISL_673386, EPI_ISL_673387, EPI_ISL_673388, EPI_ISL_673389, EPI_ISL_673390, EPI_ISL_673391, EPI_ISL_673392, EPI_ISL_673393, EPI_ISL_673394, EPI_ISL_673395, EPI_ISL_673396, EPI_ISL_673397, EPI_ISL_673398, EPI_ISL_673399, EPI_ISL_673400, EPI_ISL_673401, EPI_ISL_673402, EPI_ISL_673403, EPI_ISL_673404, EPI_ISL_673405, EPI_ISL_673406, EPI_ISL_673407, EPI_ISL_673408, EPI_ISL_673409, EPI_ISL_673410, EPI_ISL_673411, EPI_ISL_673412, EPI_ISL_673413, EPI_ISL_673414, EPI_ISL_673415, EPI_ISL_673416, EPI_ISL_673417, EPI_ISL_673418, EPI_ISL_673419, EPI_ISL_673420, EPI_ISL_673421, EPI_ISL_673422, EPI_ISL_673423, EPI_ISL_673424, EPI_ISL_673425, EPI_ISL_673426, EPI_ISL_673427, EPI_ISL_673428, EPI_ISL_673429, EPI_ISL_673430, EPI_ISL_673431, EPI_ISL_673432, EPI_ISL_673433, EPI_ISL_673434, EPI_ISL_673435, EPI_ISL_673436, EPI_ISL_673437, EPI_ISL_673438, EPI_ISL_673439, EPI_ISL_673440, EPI_ISL_673442, EPI_ISL_673443, EPI_ISL_673444, EPI_ISL_673445, EPI_ISL_673446, EPI_ISL_673448, EPI_ISL_673449, EPI_ISL_673450, EPI_ISL_673451, EPI_ISL_673452, EPI_ISL_673453, EPI_ISL_673454, EPI_ISL_673455, EPI_ISL_673456, EPI_ISL_673457, EPI_ISL_673458, EPI_ISL_673459, EPI_ISL_673460, EPI_ISL_673461, EPI_ISL_673462, EPI_ISL_673463, EPI_ISL_673464, EPI_ISL_673465, EPI_ISL_673466, EPI_ISL_673467, EPI_ISL_673468, EPI_ISL_673469, EPI_ISL_673470, EPI_ISL_673471, EPI_ISL_673472, EPI_ISL_673473, EPI_ISL_673474, EPI_ISL_673475, EPI_ISL_673476, EPI_ISL_673477, EPI_ISL_673478, EPI_ISL_673479, EPI_ISL_673480, EPI_ISL_673481, EPI_ISL_673482, EPI_ISL_673483, EPI_ISL_673484, EPI_ISL_673485, EPI_ISL_673486, EPI_ISL_673487, EPI_ISL_673488, EPI_ISL_673489, EPI_ISL_673490, EPI_ISL_673491, EPI_ISL_673492, EPI_ISL_673493, EPI_ISL_673494, EPI_ISL_673495, EPI_ISL_673496, EPI_ISL_673497, EPI_ISL_673498, EPI_ISL_673499, EPI_ISL_673500, EPI_ISL_673501, EPI_ISL_673502, EPI_ISL_673503, EPI_ISL_673504, EPI_ISL_673505, EPI_ISL_673506, EPI_ISL_673507, EPI_ISL_673508, EPI_ISL_673509, EPI_ISL_673510, EPI_ISL_673511, EPI_ISL_673512, EPI_ISL_673513, EPI_ISL_673514, EPI_ISL_673515, EPI_ISL_673516, EPI_ISL_673517, EPI_ISL_673518, EPI_ISL_673519, EPI_ISL_673520, EPI_ISL_673521, EPI_ISL_673522, EPI_ISL_673523, EPI_ISL_673524, EPI_ISL_673525, EPI_ISL_673526, EPI_ISL_673527, EPI_ISL_673528, EPI_ISL_673529, EPI_ISL_673530, EPI_ISL_673531, EPI_ISL_673532, EPI_ISL_673533, EPI_ISL_673534, EPI_ISL_673535, EPI_ISL_673537, EPI_ISL_673538, EPI_ISL_673539, EPI_ISL_673540, EPI_ISL_673541, EPI_ISL_673542, EPI_ISL_673544, EPI_ISL_673545, EPI_ISL_673546, EPI_ISL_673547, EPI_ISL_673548, EPI_ISL_673549, EPI_ISL_673550, EPI_ISL_673551, EPI_ISL_673552, EPI_ISL_673553, EPI_ISL_673554, EPI_ISL_673555, EPI_ISL_673556, EPI_ISL_673557, EPI_ISL_673558, EPI_ISL_673559, EPI_ISL_673560, EPI_ISL_673561, EPI_ISL_673562, EPI_ISL_673563, EPI_ISL_673565, EPI_ISL_673567, EPI_ISL_673568, EPI_ISL_673569, EPI_ISL_673570, EPI_ISL_673571, EPI_ISL_673572, EPI_ISL_673573, EPI_ISL_673574, EPI_ISL_673575, EPI_ISL_673576, EPI_ISL_673578, EPI_ISL_673579, EPI_ISL_673580, EPI_ISL_673581, EPI_ISL_673582, EPI_ISL_673583, EPI_ISL_673584, EPI_ISL_673585, EPI_ISL_673586, EPI_ISL_673587, EPI_ISL_673588, EPI_ISL_673589, EPI_ISL_673590, EPI_ISL_673591, EPI_ISL_673593, EPI_ISL_673595, EPI_ISL_673596, EPI_ISL_673597, EPI_ISL_673600, EPI_ISL_673602, EPI_ISL_673604, EPI_ISL_673605, EPI_ISL_673606, EPI_ISL_673607, EPI_ISL_673608, EPI_ISL_673609, EPI_ISL_673610, EPI_ISL_673611, EPI_ISL_673612, EPI_ISL_673613, EPI_ISL_673614, EPI_ISL_673615, EPI_ISL_673617, EPI_ISL_673618, EPI_ISL_673619, EPI_ISL_673620, EPI_ISL_673622, EPI_ISL_673623, EPI_ISL_673624, EPI_ISL_673625, EPI_ISL_673626, EPI_ISL_673627, EPI_ISL_673628, EPI_ISL_673629, EPI_ISL_673630, EPI_ISL_673631, EPI_ISL_673632, EPI_ISL_673635, EPI_ISL_673636, EPI_ISL_673637, EPI_ISL_673638, EPI_ISL_673639, EPI_ISL_673640, EPI_ISL_673641, EPI_ISL_673643, EPI_ISL_673644, EPI_ISL_673647, EPI_ISL_673648, EPI_ISL_673649, EPI_ISL_673650, EPI_ISL_673651, EPI_ISL_673653, EPI_ISL_673654, EPI_ISL_673656, EPI_ISL_673658, EPI_ISL_673660, EPI_ISL_673661, EPI_ISL_673662, EPI_ISL_673664, EPI_ISL_673665, EPI_ISL_673666, EPI_ISL_673667, EPI_ISL_673668, EPI_ISL_673669, EPI_ISL_673670, EPI_ISL_673671, EPI_ISL_673672, EPI_ISL_673674, EPI_ISL_673675, EPI_ISL_673677, EPI_ISL_673678, EPI_ISL_673679, EPI_ISL_673680, EPI_ISL_673681, EPI_ISL_673682, EPI_ISL_673683, EPI_ISL_673684, EPI_ISL_673685, EPI_ISL_673687, EPI_ISL_673689, EPI_ISL_673690, EPI_ISL_673692, EPI_ISL_673693, EPI_ISL_673695, EPI_ISL_673696, EPI_ISL_673697, EPI_ISL_673698, EPI_ISL_673699, EPI_ISL_673700, EPI_ISL_673701, EPI_ISL_673703, EPI_ISL_673704, EPI_ISL_673705, EPI_ISL_673706, EPI_ISL_673707, EPI_ISL_673710, EPI_ISL_673711, EPI_ISL_673712, EPI_ISL_673713, EPI_ISL_673716, EPI_ISL_673717, EPI_ISL_673719, EPI_ISL_673720, EPI_ISL_673721, EPI_ISL_673723, EPI_ISL_673724, EPI_ISL_673725, EPI_ISL_673727, EPI_ISL_673728, EPI_ISL_673731, EPI_ISL_673732, EPI_ISL_673734, EPI_ISL_673735, EPI_ISL_673736, EPI_ISL_673737, EPI_ISL_673739, EPI_ISL_673740, EPI_ISL_673741, EPI_ISL_673742, EPI_ISL_673743, EPI_ISL_673744, EPI_ISL_673747, EPI_ISL_673748, EPI_ISL_673749, EPI_ISL_673751, EPI_ISL_673752, EPI_ISL_673753, EPI_ISL_673754, EPI_ISL_673755, | | | |

EPI_ISL_673756, EPI_ISL_673757, EPI_ISL_673758, EPI_ISL_673759, EPI_ISL_673761, EPI_ISL_673762, EPI_ISL_673763, EPI_ISL_673766, EPI_ISL_673768, EPI_ISL_673769, EPI_ISL_673771, EPI_ISL_673772, EPI_ISL_673773, EPI_ISL_673774, EPI_ISL_673775, EPI_ISL_673776, EPI_ISL_673777, EPI_ISL_673779, EPI_ISL_673780, EPI_ISL_673781, EPI_ISL_673782, EPI_ISL_673783, EPI_ISL_673784, EPI_ISL_673786, EPI_ISL_673788, EPI_ISL_673789, EPI_ISL_673791, EPI_ISL_673792, EPI_ISL_673793, EPI_ISL_673794, EPI_ISL_673795, EPI_ISL_673797, EPI_ISL_673799, EPI_ISL_673800, EPI_ISL_673802, EPI_ISL_673804, EPI_ISL_673805, EPI_ISL_673806, EPI_ISL_673807, EPI_ISL_673808, EPI_ISL_673809, EPI_ISL_673810, EPI_ISL_673811, EPI_ISL_673815, EPI_ISL_673816, EPI_ISL_673817, EPI_ISL_673818, EPI_ISL_673819, EPI_ISL_673820, EPI_ISL_673823, EPI_ISL_673824, EPI_ISL_673825, EPI_ISL_673826, EPI_ISL_673830, EPI_ISL_673831, EPI_ISL_673836, EPI_ISL_673837, EPI_ISL_673838, EPI_ISL_673839, EPI_ISL_673841, EPI_ISL_673842, EPI_ISL_673843, EPI_ISL_673844, EPI_ISL_673845, EPI_ISL_673846, EPI_ISL_673847, EPI_ISL_673849, EPI_ISL_673850, EPI_ISL_673852, EPI_ISL_673853, EPI_ISL_673854, EPI_ISL_673855, EPI_ISL_673858, EPI_ISL_673860, EPI_ISL_673861, EPI_ISL_673862, EPI_ISL_673863, EPI_ISL_673864, EPI_ISL_673866, EPI_ISL_673868, EPI_ISL_673869, EPI_ISL_673870, EPI_ISL_674826, EPI_ISL_674828, EPI_ISL_674834, EPI_ISL_674837, EPI_ISL_674838, EPI_ISL_674840, EPI_ISL_674845, EPI_ISL_674846, EPI_ISL_674847, EPI_ISL_674849, EPI_ISL_674850, EPI_ISL_674851, EPI_ISL_674859, EPI_ISL_674862, EPI_ISL_674866, EPI_ISL_674867, EPI_ISL_674868, EPI_ISL_674871, EPI_ISL_674873, EPI_ISL_674876, EPI_ISL_674883, EPI_ISL_674893, EPI_ISL_674896, EPI_ISL_674899, EPI_ISL_674906, EPI_ISL_674908, EPI_ISL_674909, EPI_ISL_674910, EPI_ISL_674913, EPI_ISL_674917, EPI_ISL_674919, EPI_ISL_674920, EPI_ISL_674922, EPI_ISL_674923, EPI_ISL_674924, EPI_ISL_674926, EPI_ISL_674933, EPI_ISL_674936, EPI_ISL_674939, EPI_ISL_674940, EPI_ISL_674944, EPI_ISL_674946, EPI_ISL_674949, EPI_ISL_674950, EPI_ISL_674951, EPI_ISL_674952, EPI_ISL_674953, EPI_ISL_674957, EPI_ISL_674959, EPI_ISL_674962, EPI_ISL_674972, EPI_ISL_674973, EPI_ISL_674977, EPI_ISL_674983, EPI_ISL_674988, EPI_ISL_674992, EPI_ISL_674995, EPI_ISL_674998, EPI_ISL_675006, EPI_ISL_675008, EPI_ISL_675009, EPI_ISL_675012, EPI_ISL_675018, EPI_ISL_675024, EPI_ISL_675026, EPI_ISL_675027, EPI_ISL_675033, EPI_ISL_675038, EPI_ISL_675039, EPI_ISL_675040, EPI_ISL_675042, EPI_ISL_675046, EPI_ISL_675053, EPI_ISL_675056, EPI_ISL_675064, EPI_ISL_675071, EPI_ISL_675075, EPI_ISL_675080, EPI_ISL_675082, EPI_ISL_675087, EPI_ISL_675094, EPI_ISL_675104, EPI_ISL_675107, EPI_ISL_675110, EPI_ISL_675117, EPI_ISL_675120, EPI_ISL_675126, EPI_ISL_681658, EPI_ISL_681659, EPI_ISL_681671, EPI_ISL_681672, EPI_ISL_704750, EPI_ISL_710835, EPI_ISL_783369, EPI_ISL_783371, EPI_ISL_783372, EPI_ISL_783373, EPI_ISL_783375, EPI_ISL_783378, EPI_ISL_783380, EPI_ISL_783381, EPI_ISL_783382, EPI_ISL_783384, EPI_ISL_783385, EPI_ISL_783386, EPI_ISL_783387, EPI_ISL_783388, EPI_ISL_783389, EPI_ISL_783390, EPI_ISL_783391, EPI_ISL_783392, EPI_ISL_783395, EPI_ISL_783396, EPI_ISL_783397, EPI_ISL_783401, EPI_ISL_783402, EPI_ISL_783403, EPI_ISL_783404, EPI_ISL_783405, EPI_ISL_783406, EPI_ISL_783407, EPI_ISL_783409, EPI_ISL_783411, EPI_ISL_783414, EPI_ISL_783415, EPI_ISL_783416, EPI_ISL_783417, EPI_ISL_783418, EPI_ISL_783419, EPI_ISL_783420, EPI_ISL_783421, EPI_ISL_783422, EPI_ISL_783424, EPI_ISL_783430, EPI_ISL_783431, EPI_ISL_783432, EPI_ISL_783434, EPI_ISL_783435, EPI_ISL_783436, EPI_ISL_783437, EPI_ISL_783438, EPI_ISL_783439, EPI_ISL_783440, EPI_ISL_783441, EPI_ISL_783442, EPI_ISL_783443, EPI_ISL_783444, EPI_ISL_783446, EPI_ISL_783447, EPI_ISL_783448, EPI_ISL_783450, EPI_ISL_783451, EPI_ISL_783452, EPI_ISL_783453, EPI_ISL_783455, EPI_ISL_783456, EPI_ISL_783458, EPI_ISL_783459, EPI_ISL_783460, EPI_ISL_783461, EPI_ISL_783463, EPI_ISL_783466, EPI_ISL_783467, EPI_ISL_783468, EPI_ISL_783469, EPI_ISL_783470, EPI_ISL_783471, EPI_ISL_783472, EPI_ISL_783473, EPI_ISL_783474, EPI_ISL_783476, EPI_ISL_783477, EPI_ISL_783479, EPI_ISL_783480, EPI_ISL_783481, EPI_ISL_783482, EPI_ISL_783483, EPI_ISL_783484, EPI_ISL_783485, EPI_ISL_783486, EPI_ISL_783487, EPI_ISL_783488, EPI_ISL_783489, EPI_ISL_783491, EPI_ISL_783492, EPI_ISL_783493, EPI_ISL_783494, EPI_ISL_783495, EPI_ISL_783496, EPI_ISL_783497, EPI_ISL_783498, EPI_ISL_783499, EPI_ISL_783500, EPI_ISL_783502, EPI_ISL_783503, EPI_ISL_783504, EPI_ISL_783505, EPI_ISL_783506, EPI_ISL_783507, EPI_ISL_783508, EPI_ISL_783509, EPI_ISL_783510, EPI_ISL_783511, EPI_ISL_783512, EPI_ISL_783513, EPI_ISL_783514, EPI_ISL_783519, EPI_ISL_783520, EPI_ISL_783521, EPI_ISL_783523, EPI_ISL_783524, EPI_ISL_783527, EPI_ISL_783528, EPI_ISL_783530, EPI_ISL_783531, EPI_ISL_783532, EPI_ISL_783533, EPI_ISL_783535, EPI_ISL_801055, EPI_ISL_801148, EPI_ISL_859555, EPI_ISL_859560

| | | | |
|---|---|---|---|
| see above | Lighthouse Lab in Cambridge | Wellcome Sanger Institute for the COVID-19 Genomics UK (COG-UK) Consortium | Cordelia Langford; David K. Jackson; Dominic Kwiatkowski; Ewan Harrison; Ian Johnston; John Sillitoe on behalf of the Wellcome Sanger Institute COVID-19 Surveillance Team; John Sillitoe on behalf of the Wellcome Sanger Institute COVID-19 Surveillance Team (http://www.sanger.ac.uk/covid-team); Rob Howes; Roberto Amato; Sonia Goncalves; The Lighthouse Lab in Cambridge and Alex Alderton |

EPI_ISL_647110, EPI_ISL_661632, EPI_ISL_661633, EPI_ISL_661634, EPI_ISL_661635, EPI_ISL_661636, EPI_ISL_661637, EPI_ISL_661639, EPI_ISL_661642, EPI_ISL_661643, EPI_ISL_661644, EPI_ISL_661645, EPI_ISL_661646, EPI_ISL_661648, EPI_ISL_661649, EPI_ISL_661650, EPI_ISL_661655, EPI_ISL_661657, EPI_ISL_661658, EPI_ISL_661659, EPI_ISL_661660, EPI_ISL_661661, EPI_ISL_661662, EPI_ISL_661663, EPI_ISL_661665, EPI_ISL_661666, EPI_ISL_661667, EPI_ISL_661668, EPI_ISL_661669, EPI_ISL_661670, EPI_ISL_661671, EPI_ISL_661672, EPI_ISL_661673, EPI_ISL_661674, EPI_ISL_661676, EPI_ISL_661677, EPI_ISL_661678, EPI_ISL_661681, EPI_ISL_661682, EPI_ISL_661683, EPI_ISL_661685, EPI_ISL_661686, EPI_ISL_661688, EPI_ISL_661689, EPI_ISL_661690, EPI_ISL_661692, EPI_ISL_661694, EPI_ISL_661698, EPI_ISL_661699, EPI_ISL_661700, EPI_ISL_661701, EPI_ISL_661703, EPI_ISL_661704, EPI_ISL_661705, EPI_ISL_661706, EPI_ISL_661707, EPI_ISL_661708, EPI_ISL_661709, EPI_ISL_661714, EPI_ISL_661715, EPI_ISL_661716, EPI_ISL_661718, EPI_ISL_661719, EPI_ISL_661720, EPI_ISL_661722, EPI_ISL_661723, EPI_ISL_661724, EPI_ISL_661725, EPI_ISL_661727, EPI_ISL_661728, EPI_ISL_661730, EPI_ISL_661731, EPI_ISL_661733, EPI_ISL_661736, EPI_ISL_661737, EPI_ISL_661738, EPI_ISL_661739, EPI_ISL_661740, EPI_ISL_661744, EPI_ISL_661745, EPI_ISL_661746, EPI_ISL_661748, EPI_ISL_661750, EPI_ISL_661753, EPI_ISL_661755, EPI_ISL_661756, EPI_ISL_661757, EPI_ISL_661758, EPI_ISL_661760, EPI_ISL_661762, EPI_ISL_661763, EPI_ISL_661764, EPI_ISL_661765, EPI_ISL_661766, EPI_ISL_661767, EPI_ISL_661768, EPI_ISL_661769, EPI_ISL_661770, EPI_ISL_661771, EPI_ISL_661772, EPI_ISL_661773, EPI_ISL_661774, EPI_ISL_661776, EPI_ISL_661777, EPI_ISL_661780, EPI_ISL_661782, EPI_ISL_661784, EPI_ISL_661786, EPI_ISL_661787, EPI_ISL_661788, EPI_ISL_661790, EPI_ISL_661792, EPI_ISL_661793, EPI_ISL_661794, EPI_ISL_661795, EPI_ISL_661796, EPI_ISL_661798, EPI_ISL_661800, EPI_ISL_661802, EPI_ISL_661803, EPI_ISL_661804, EPI_ISL_661806, EPI_ISL_661807, EPI_ISL_661808, EPI_ISL_661809, EPI_ISL_661810, EPI_ISL_661812, EPI_ISL_661813, EPI_ISL_661814, EPI_ISL_661816, EPI_ISL_661818, EPI_ISL_661819, EPI_ISL_661820, EPI_ISL_661821, EPI_ISL_661822, EPI_ISL_661823, EPI_ISL_661824, EPI_ISL_661825, EPI_ISL_661826, EPI_ISL_661827, EPI_ISL_661828, EPI_ISL_661829, EPI_ISL_661831, EPI_ISL_661832, EPI_ISL_661835, EPI_ISL_661837, EPI_ISL_661838, EPI_ISL_661839, EPI_ISL_661840, EPI_ISL_661841, EPI_ISL_661842, EPI_ISL_661843, EPI_ISL_661846, EPI_ISL_661847, EPI_ISL_661848, EPI_ISL_661849, EPI_ISL_661850, EPI_ISL_661851, EPI_ISL_661852, EPI_ISL_661855, EPI_ISL_661857, EPI_ISL_661858, EPI_ISL_661859, EPI_ISL_661860, EPI_ISL_661861, EPI_ISL_661862, EPI_ISL_661864, EPI_ISL_661865, EPI_ISL_661866, EPI_ISL_661867, EPI_ISL_661868, EPI_ISL_661870, EPI_ISL_661871, EPI_ISL_661872, EPI_ISL_661875, EPI_ISL_661877, EPI_ISL_661878, EPI_ISL_661879, EPI_ISL_661880, EPI_ISL_661882, EPI_ISL_661884, EPI_ISL_661885, EPI_ISL_661888, EPI_ISL_661889, EPI_ISL_661890, EPI_ISL_661891, EPI_ISL_661892, EPI_ISL_661893, EPI_ISL_661894, EPI_ISL_661896, EPI_ISL_661898, EPI_ISL_661899, EPI_ISL_661901, EPI_ISL_661904, EPI_ISL_661905, EPI_ISL_661906, EPI_ISL_661907, EPI_ISL_661908, EPI_ISL_661909, EPI_ISL_661910, EPI_ISL_661911, EPI_ISL_661912, EPI_ISL_661913, EPI_ISL_661914, EPI_ISL_661915, EPI_ISL_661916, EPI_ISL_661919, EPI_ISL_661920, EPI_ISL_661921, EPI_ISL_661922, EPI_ISL_661923, EPI_ISL_661924, EPI_ISL_661927, EPI_ISL_661929, EPI_ISL_661931, EPI_ISL_661932, EPI_ISL_661933, EPI_ISL_661934, EPI_ISL_661935, EPI_ISL_661936, EPI_ISL_661937, EPI_ISL_661938, EPI_ISL_661939, EPI_ISL_661940, EPI_ISL_661942, EPI_ISL_661943, EPI_ISL_661945, EPI_ISL_661946, EPI_ISL_661948, EPI_ISL_661949, EPI_ISL_661950, EPI_ISL_661952, EPI_ISL_661955, EPI_ISL_667814, EPI_ISL_667822, EPI_ISL_667824, EPI_ISL_667825, EPI_ISL_667828, EPI_ISL_667831, EPI_ISL_667832, EPI_ISL_667836, EPI_ISL_667838, EPI_ISL_667843, EPI_ISL_667844, EPI_ISL_667847, EPI_ISL_667849, EPI_ISL_667850, EPI_ISL_667851, EPI_ISL_667855, EPI_ISL_667857, EPI_ISL_667859, EPI_ISL_667860, EPI_ISL_667863, EPI_ISL_667864, EPI_ISL_667866, EPI_ISL_667867, EPI_ISL_667868, EPI_ISL_667872, EPI_ISL_667873, EPI_ISL_667876, EPI_ISL_667879, EPI_ISL_667881, EPI_ISL_667883, EPI_ISL_667885, EPI_ISL_667887, EPI_ISL_667888, EPI_ISL_667889, EPI_ISL_667891, EPI_ISL_667893, EPI_ISL_667895, EPI_ISL_667898, EPI_ISL_667900, EPI_ISL_667904, EPI_ISL_667907, EPI_ISL_667912, EPI_ISL_667915, EPI_ISL_667917, EPI_ISL_667918, EPI_ISL_667919, EPI_ISL_667920, EPI_ISL_667922, EPI_ISL_667924, EPI_ISL_667925, EPI_ISL_667927, EPI_ISL_667928, EPI_ISL_667929, EPI_ISL_667930, EPI_ISL_667931, EPI_ISL_667938, EPI_ISL_667939, EPI_ISL_667940, EPI_ISL_667941, EPI_ISL_667943, EPI_ISL_667947, EPI_ISL_667948, EPI_ISL_667950, EPI_ISL_667952, EPI_ISL_667957, EPI_ISL_667958, EPI_ISL_667959, EPI_ISL_667960, EPI_ISL_667962, EPI_ISL_667963, EPI_ISL_667966, EPI_ISL_667967, EPI_ISL_667969, EPI_ISL_667970, EPI_ISL_667972, EPI_ISL_667973, EPI_ISL_667975, EPI_ISL_667976, EPI_ISL_667977, EPI_ISL_667978, EPI_ISL_667980, EPI_ISL_667982, EPI_ISL_667983, EPI_ISL_667986, EPI_ISL_667987, EPI_ISL_667989, EPI_ISL_667990, EPI_ISL_667992, EPI_ISL_667996, EPI_ISL_667997, EPI_ISL_667998, EPI_ISL_668000, EPI_ISL_668001, EPI_ISL_668004, EPI_ISL_668005, EPI_ISL_668006, EPI_ISL_668010, EPI_ISL_668013, EPI_ISL_668020, EPI_ISL_668021, EPI_ISL_668022, EPI_ISL_668024, EPI_ISL_668028, EPI_ISL_668029, EPI_ISL_668030, EPI_ISL_668031, EPI_ISL_668032, EPI_ISL_668033, EPI_ISL_668034, EPI_ISL_668035, EPI_ISL_668036, EPI_ISL_668038, EPI_ISL_668044, EPI_ISL_668045, EPI_ISL_668049, EPI_ISL_668050, EPI_ISL_668052, EPI_ISL_668053, EPI_ISL_668054, EPI_ISL_668055, EPI_ISL_668056, EPI_ISL_668057, EPI_ISL_668060, EPI_ISL_668062, EPI_ISL_668063, EPI_ISL_668064, EPI_ISL_668066, EPI_ISL_668067, EPI_ISL_668073, EPI_ISL_668074, EPI_ISL_668075, EPI_ISL_668078, EPI_ISL_668082, EPI_ISL_668083, EPI_ISL_668084, EPI_ISL_668093, EPI_ISL_668094, EPI_ISL_668096, EPI_ISL_668098, EPI_ISL_668102, EPI_ISL_668105, EPI_ISL_668106, EPI_ISL_668107, EPI_ISL_668111, EPI_ISL_668113, EPI_ISL_668114, EPI_ISL_668116, EPI_ISL_668117, EPI_ISL_668118, EPI_ISL_668119, EPI_ISL_668120, EPI_ISL_668121, EPI_ISL_668124, EPI_ISL_668125, EPI_ISL_668126, EPI_ISL_668129, EPI_ISL_668130, EPI_ISL_668131, EPI_ISL_668132, EPI_ISL_668134, EPI_ISL_668135, EPI_ISL_668139, EPI_ISL_668142, EPI_ISL_668143, EPI_ISL_668144, EPI_ISL_668145, EPI_ISL_668146, EPI_ISL_668147, EPI_ISL_668150, EPI_ISL_668151, EPI_ISL_668152, EPI_ISL_668156, EPI_ISL_668157, EPI_ISL_668158, EPI_ISL_668159, EPI_ISL_668160, EPI_ISL_668161, EPI_ISL_668162, EPI_ISL_668164, EPI_ISL_668165, EPI_ISL_668167, EPI_ISL_668168, EPI_ISL_668170, EPI_ISL_668173, EPI_ISL_668174, EPI_ISL_668175, EPI_ISL_668176, EPI_ISL_668177, EPI_ISL_668178, EPI_ISL_668179, EPI_ISL_668181, EPI_ISL_668182, EPI_ISL_668183, EPI_ISL_668184, EPI_ISL_668185, EPI_ISL_668186, EPI_ISL_668187, EPI_ISL_668189, EPI_ISL_668191, EPI_ISL_668193, EPI_ISL_668195, EPI_ISL_668196, EPI_ISL_668197, EPI_ISL_668198, EPI_ISL_668199, EPI_ISL_668202, EPI_ISL_668203, EPI_ISL_668204, EPI_ISL_668205, EPI_ISL_668206, EPI_ISL_668207, EPI_ISL_668208, EPI_ISL_668209, EPI_ISL_668210, EPI_ISL_668213, EPI_ISL_668214, EPI_ISL_668215, EPI_ISL_668216, EPI_ISL_668217, EPI_ISL_668218, EPI_ISL_668220, EPI_ISL_668221, EPI_ISL_668223, EPI_ISL_668224, EPI_ISL_668225, EPI_ISL_668226, EPI_ISL_668228, EPI_ISL_668229, EPI_ISL_668230, EPI_ISL_668231, EPI_ISL_668232, EPI_ISL_668233, EPI_ISL_668234, EPI_ISL_668235, EPI_ISL_668236, EPI_ISL_668238, EPI_ISL_668240, EPI_ISL_668241, EPI_ISL_668242, EPI_ISL_668243, EPI_ISL_668244, EPI_ISL_668245, EPI_ISL_668246, EPI_ISL_668247, EPI_ISL_668248, EPI_ISL_668249, EPI_ISL_668250, EPI_ISL_668253, EPI_ISL_668256, EPI_ISL_668257, EPI_ISL_668258, EPI_ISL_668259, EPI_ISL_668260, EPI_ISL_668263, EPI_ISL_668264, EPI_ISL_668266, EPI_ISL_668268, EPI_ISL_668269, EPI_ISL_668270, EPI_ISL_668271, EPI_ISL_668272, EPI_ISL_668273, EPI_ISL_668274, EPI_ISL_668276, EPI_ISL_668277, EPI_ISL_668278, EPI_ISL_668279, EPI_ISL_668282, EPI_ISL_668284, EPI_ISL_668285, EPI_ISL_668286, EPI_ISL_668287, EPI_ISL_668288, EPI_ISL_668290, EPI_ISL_668291, EPI_ISL_668292, EPI_ISL_668293, EPI_ISL_668295, EPI_ISL_668300, EPI_ISL_668301, EPI_ISL_668302, EPI_ISL_668303, EPI_ISL_668304, EPI_ISL_668305, EPI_ISL_668306, EPI_ISL_668307, EPI_ISL_668308, EPI_ISL_668309, EPI_ISL_668310, EPI_ISL_668311, EPI_ISL_668312, EPI_ISL_668314, EPI_ISL_668318, EPI_ISL_668320, EPI_ISL_668321, EPI_ISL_668323, EPI_ISL_668324, EPI_ISL_668325, EPI_ISL_668326, EPI_ISL_668327, EPI_ISL_668328, EPI_ISL_668330, EPI_ISL_668331, EPI_ISL_668332, EPI_ISL_668333, EPI_ISL_668334, EPI_ISL_668335, EPI_ISL_668337, EPI_ISL_668338, EPI_ISL_668339, EPI_ISL_668340, EPI_ISL_668343, EPI_ISL_668344, EPI_ISL_668346, EPI_ISL_668347, EPI_ISL_668349, EPI_ISL_668350, EPI_ISL_668351, EPI_ISL_668352, EPI_ISL_668356, EPI_ISL_668357, EPI_ISL_668358, EPI_ISL_668359, EPI_ISL_668360, EPI_ISL_668361, EPI_ISL_668362, EPI_ISL_668363, EPI_ISL_668364, EPI_ISL_668366, EPI_ISL_668367, EPI_ISL_668370, EPI_ISL_668371, EPI_ISL_668372, EPI_ISL_668375, EPI_ISL_668377, EPI_ISL_668378, EPI_ISL_668380, EPI_ISL_668381, EPI_ISL_668383, EPI_ISL_668384, EPI_ISL_668386, EPI_ISL_668387, EPI_ISL_674498, EPI_ISL_674499, EPI_ISL_674501, EPI_ISL_674504, EPI_ISL_674514, EPI_ISL_674516, EPI_ISL_674522, EPI_ISL_674527, EPI_ISL_674528, EPI_ISL_674530, EPI_ISL_674543, EPI_ISL_674544, EPI_ISL_674545, EPI_ISL_674550, EPI_ISL_674558, EPI_ISL_674563, EPI_ISL_674565, EPI_ISL_674569, EPI_ISL_674585, EPI_ISL_674589, EPI_ISL_674590, EPI_ISL_674592, EPI_ISL_674595, EPI_ISL_674603, EPI_ISL_674605, EPI_ISL_674609, EPI_ISL_674613, EPI_ISL_674625, EPI_ISL_674629, EPI_ISL_674635, EPI_ISL_674652, EPI_ISL_674653, EPI_ISL_674655, EPI_ISL_674657, EPI_ISL_674658, EPI_ISL_674661, EPI_ISL_674662, EPI_ISL_674667, EPI_ISL_674670, EPI_ISL_674674, EPI_ISL_674678, EPI_ISL_674685, EPI_ISL_674690, EPI_ISL_674691, EPI_ISL_674692, EPI_ISL_674698, EPI_ISL_674707, EPI_ISL_674714, EPI_ISL_674716, EPI_ISL_674717, EPI_ISL_674722, EPI_ISL_674723, EPI_ISL_674724, EPI_ISL_674725, EPI_ISL_674736, EPI_ISL_674740, EPI_ISL_674743, EPI_ISL_674744, EPI_ISL_674745, EPI_ISL_674746, EPI_ISL_674747, EPI_ISL_674749, EPI_ISL_674751, EPI_ISL_674752, EPI_ISL_674756, EPI_ISL_674758, EPI_ISL_674762, EPI_ISL_674763, EPI_ISL_674772, EPI_ISL_674773, EPI_ISL_674779, EPI_ISL_674781, EPI_ISL_674783, EPI_ISL_674786, EPI_ISL_674791, EPI_ISL_674792, EPI_ISL_674794, EPI_ISL_674795, EPI_ISL_674804, EPI_ISL_674808, EPI_ISL_674809, EPI_ISL_676353, EPI_ISL_676354, EPI_ISL_676355, EPI_ISL_676356, EPI_ISL_676357, EPI_ISL_676358, EPI_ISL_703486, EPI_ISL_708843, EPI_ISL_710838, EPI_ISL_710839, EPI_ISL_710840, EPI_ISL_710841, EPI_ISL_710842, EPI_ISL_710843, EPI_ISL_710844, EPI_ISL_710845, EPI_ISL_710846, EPI_ISL_710847, EPI_ISL_710848, EPI_ISL_710849, EPI_ISL_710850, EPI_ISL_710851, EPI_ISL_710852, EPI_ISL_710853, EPI_ISL_710854, EPI_ISL_710855, EPI_ISL_710856, EPI_ISL_710857, EPI_ISL_710858, EPI_ISL_710859, EPI_ISL_710860, EPI_ISL_710861, EPI_ISL_719776, EPI_ISL_719801, EPI_ISL_719820, EPI_ISL_719837, EPI_ISL_719841, EPI_ISL_719849, EPI_ISL_719852, EPI_ISL_719858, EPI_ISL_719886, EPI_ISL_719888, EPI_ISL_719918, EPI_ISL_719921, EPI_ISL_719926, EPI_ISL_719934, EPI_ISL_719935, EPI_ISL_719939, EPI_ISL_719968, EPI_ISL_720916, EPI_ISL_720927, EPI_ISL_720928, EPI_ISL_720940, EPI_ISL_720946, EPI_ISL_720952, EPI_ISL_720954, EPI_ISL_720958, EPI_ISL_720962, EPI_ISL_720969, EPI_ISL_720980, EPI_ISL_720981, EPI_ISL_720983, EPI_ISL_720987, EPI_ISL_721000, EPI_ISL_721009, EPI_ISL_721021, EPI_ISL_721027, EPI_ISL_721061, EPI_ISL_721062, EPI_ISL_721070, EPI_ISL_721072, EPI_ISL_721074, EPI_ISL_721077, EPI_ISL_721092, EPI_ISL_721095, EPI_ISL_721109, EPI_ISL_721119, EPI_ISL_721131, EPI_ISL_721133, EPI_ISL_721163, EPI_ISL_721167, EPI_ISL_721170, EPI_ISL_757202, EPI_ISL_801231, EPI_ISL_801232, EPI_ISL_801233, EPI_ISL_801234, EPI_ISL_801235, EPI_ISL_801236, EPI_ISL_801237, EPI_ISL_801238, EPI_ISL_801239, EPI_ISL_801240, EPI_ISL_801241, EPI_ISL_801242, EPI_ISL_801243, EPI_ISL_801244, EPI_ISL_801245, EPI_ISL_801246, EPI_ISL_801247, EPI_ISL_801248, EPI_ISL_801249, EPI_ISL_801250, EPI_ISL_801251, EPI_ISL_801252, EPI_ISL_801253, EPI_ISL_801254, EPI_ISL_801255, EPI_ISL_801256, EPI_ISL_801257, EPI_ISL_801258, EPI_ISL_801259, EPI_ISL_801260, EPI_ISL_801261, EPI_ISL_801262, EPI_ISL_801263, EPI_ISL_801264, EPI_ISL_801265, EPI_ISL_801266, EPI_ISL_801267, EPI_ISL_801268, EPI_ISL_801269, EPI_ISL_801270, EPI_ISL_801271, EPI_ISL_801272, EPI_ISL_801273, EPI_ISL_801274, EPI_ISL_801275, EPI_ISL_801276, EPI_ISL_801277, EPI_ISL_801278, EPI_ISL_801279, EPI_ISL_801280, EPI_ISL_801281, EPI_ISL_801282, EPI_ISL_801283, EPI_ISL_801284, EPI_ISL_801285, EPI_ISL_801286, EPI_ISL_801287, EPI_ISL_801288, EPI_ISL_801289, EPI_ISL_801290, EPI_ISL_801291, EPI_ISL_801292, EPI_ISL_801293, EPI_ISL_801294, EPI_ISL_801295, EPI_ISL_801296, EPI_ISL_801297, EPI_ISL_801298, EPI_ISL_801299, EPI_ISL_801300, EPI_ISL_801301, EPI_ISL_801302, EPI_ISL_801303, EPI_ISL_801304, EPI_ISL_801305, EPI_ISL_801306, EPI_ISL_801307, EPI_ISL_801308, EPI_ISL_801309, EPI_ISL_801310, EPI_ISL_801311, EPI_ISL_801312, EPI_ISL_801313, EPI_ISL_801314, EPI_ISL_801315, EPI_ISL_801316, EPI_ISL_801317, EPI_ISL_801318, EPI_ISL_801319, EPI_ISL_801320, EPI_ISL_801321, EPI_ISL_801322, EPI_ISL_801323, EPI_ISL_801324, EPI_ISL_801325, EPI_ISL_801326, EPI_ISL_801327, EPI_ISL_801328, EPI_ISL_801329, EPI_ISL_801330, EPI_ISL_801331, EPI_ISL_801332, EPI_ISL_801333, EPI_ISL_801334, EPI_ISL_801335, EPI_ISL_801336, EPI_ISL_801337, EPI_ISL_801338, EPI_ISL_801339, EPI_ISL_801340, EPI_ISL_801341, EPI_ISL_801342, EPI_ISL_801343, EPI_ISL_801344, EPI_ISL_801345, EPI_ISL_801346, EPI_ISL_801347, EPI_ISL_801348, EPI_ISL_801349, EPI_ISL_801350, EPI_ISL_801351, EPI_ISL_801352, EPI_ISL_801353, EPI_ISL_801354, EPI_ISL_801355, EPI_ISL_801356, EPI_ISL_801357, EPI_ISL_801358, EPI_ISL_801359, EPI_ISL_801360, EPI_ISL_801361, EPI_ISL_801362, EPI_ISL_801363, EPI_ISL_801364, EPI_ISL_801365, EPI_ISL_801366, EPI_ISL_801367, EPI_ISL_801368, EPI_ISL_801369, EPI_ISL_801370, EPI_ISL_801371, EPI_ISL_801372, EPI_ISL_801373, EPI_ISL_801374, EPI_ISL_801375, EPI_ISL_801376, EPI_ISL_801377, EPI_ISL_801378, EPI_ISL_801379, EPI_ISL_801380, EPI_ISL_801381, EPI_ISL_801382, EPI_ISL_801383, EPI_ISL_801384, EPI_ISL_801385

| | | | |
|---|---|---|---|
| see above | Lighthouse Lab in Glasgow | Wellcome Sanger Institute for the COVID-19 Genomics UK (COG-UK) Consortium | Anna Dominiczak and Alex Alderton; Carol Clugston; Cordelia Langford; David Gray; David K. Jackson; Dominic Kwiatkowski; Ewan Harrison; Harper VanSteenhouse; Ian Johnston; John Sillitoe on behalf of the Wellcome Sanger Institute COVID-19 Surveillance Team; John Sillitoe on behalf of the Wellcome Sanger Institute COVID-19 Surveillance Team (http://www.sanger.ac.uk/covid-team); Roberto Amato; Sonia Goncalves; Yumi Kasai |

EPI_ISL_673566, EPI_ISL_673577, EPI_ISL_673592, EPI_ISL_673594, EPI_ISL_673599, EPI_ISL_673601, EPI_ISL_673603, EPI_ISL_673616, EPI_ISL_673621, EPI_ISL_673633, EPI_ISL_673634, EPI_ISL_673642, EPI_ISL_673645, EPI_ISL_673646, EPI_ISL_673652, EPI_ISL_673655, EPI_ISL_673657, EPI_ISL_673659, EPI_ISL_673663, EPI_ISL_673673, EPI_ISL_673676, EPI_ISL_673686, EPI_ISL_673688, EPI_ISL_673691, EPI_ISL_673696, EPI_ISL_673702, EPI_ISL_673708, EPI_ISL_673709, EPI_ISL_673715, EPI_ISL_673722, EPI_ISL_673726, EPI_ISL_673729, EPI_ISL_673730, EPI_ISL_673733, EPI_ISL_673738, EPI_ISL_673745, EPI_ISL_673746, EPI_ISL_673750, EPI_ISL_673760, EPI_ISL_673764, EPI_ISL_673765, EPI_ISL_673767, EPI_ISL_673770, EPI_ISL_673778, EPI_ISL_673785, EPI_ISL_673787, EPI_ISL_673790, EPI_ISL_673796, EPI_ISL_673798, EPI_ISL_673801, EPI_ISL_673803, EPI_ISL_673812, EPI_ISL_673813, EPI_ISL_673814, EPI_ISL_673821, EPI_ISL_673822, EPI_ISL_673827, EPI_ISL_673828, EPI_ISL_673829, EPI_ISL_673832, EPI_ISL_673833, EPI_ISL_673834, EPI_ISL_673835, EPI_ISL_673840, EPI_ISL_673848, EPI_ISL_673851, EPI_ISL_673856, EPI_ISL_673857, EPI_ISL_673859, EPI_ISL_673865, EPI_ISL_673867, EPI_ISL_673871, EPI_ISL_673872, EPI_ISL_673873, EPI_ISL_673874, EPI_ISL_673875, EPI_ISL_673876, EPI_ISL_673877, EPI_ISL_673878, EPI_ISL_673879, EPI_ISL_673880, EPI_ISL_673881, EPI_ISL_673882, EPI_ISL_673883, EPI_ISL_673884, EPI_ISL_673885, EPI_ISL_673886, EPI_ISL_673887, EPI_ISL_673888, EPI_ISL_673889, EPI_ISL_673890, EPI_ISL_673891, EPI_ISL_673892, EPI_ISL_673893, EPI_ISL_673894, EPI_ISL_673895, EPI_ISL_673896, EPI_ISL_673897, EPI_ISL_673898, EPI_ISL_673899, EPI_ISL_673900, EPI_ISL_673901, EPI_ISL_673902, EPI_ISL_673903, EPI_ISL_673904, EPI_ISL_673905, EPI_ISL_673906, EPI_ISL_673907, EPI_ISL_673908, EPI_ISL_673909, EPI_ISL_673910, EPI_ISL_673911, EPI_ISL_673912, EPI_ISL_673913, EPI_ISL_673914, EPI_ISL_673915, EPI_ISL_673916, EPI_ISL_673917, EPI_ISL_673918, EPI_ISL_673919, EPI_ISL_673920, EPI_ISL_673921, EPI_ISL_673922, EPI_ISL_673923, EPI_ISL_673924, EPI_ISL_673925, EPI_ISL_673926, EPI_ISL_673927, EPI_ISL_673928, EPI_ISL_673929, EPI_ISL_673930, EPI_ISL_673931, EPI_ISL_673932, EPI_ISL_673933, EPI_ISL_673934, EPI_ISL_673935, EPI_ISL_673936, EPI_ISL_673937, EPI_ISL_673938, EPI_ISL_673939, EPI_ISL_673940, EPI_ISL_673941, EPI_ISL_673942, EPI_ISL_673943, EPI_ISL_673944, EPI_ISL_673945, EPI_ISL_673946, EPI_ISL_673947, EPI_ISL_673948, EPI_ISL_673949, EPI_ISL_673950, EPI_ISL_673951, EPI_ISL_673952, EPI_ISL_673953, EPI_ISL_673954, EPI_ISL_673955, EPI_ISL_673956, EPI_ISL_673957, EPI_ISL_673959, EPI_ISL_673960, EPI_ISL_673961, EPI_ISL_673962, EPI_ISL_673963, EPI_ISL_673964, EPI_ISL_673966, EPI_ISL_673967, EPI_ISL_673968, EPI_ISL_673969, EPI_ISL_673970, EPI_ISL_673971, EPI_ISL_673972, EPI_ISL_673973, EPI_ISL_673974, EPI_ISL_673975, EPI_ISL_673976, EPI_ISL_673977, EPI_ISL_673978, EPI_ISL_673979, EPI_ISL_673980, EPI_ISL_673981, EPI_ISL_673982, EPI_ISL_673983, EPI_ISL_673984, EPI_ISL_673985, EPI_ISL_673986, EPI_ISL_673987, EPI_ISL_673988, EPI_ISL_673989, EPI_ISL_673990, EPI_ISL_673991, EPI_ISL_673992, EPI_ISL_673993, EPI_ISL_673994, EPI_ISL_673996, EPI_ISL_673997, EPI_ISL_673998, EPI_ISL_673999, EPI_ISL_674000, EPI_ISL_674001, EPI_ISL_674002, EPI_ISL_674003, EPI_ISL_674004, EPI_ISL_674005, EPI_ISL_674006, EPI_ISL_674007, EPI_ISL_674008, EPI_ISL_674009, EPI_ISL_674010, EPI_ISL_674011, EPI_ISL_674012, EPI_ISL_674013, EPI_ISL_674014, EPI_ISL_674015, EPI_ISL_674016, EPI_ISL_674017, EPI_ISL_674018, EPI_ISL_674019, EPI_ISL_674020, EPI_ISL_674021, EPI_ISL_674022, EPI_ISL_674023, EPI_ISL_674024, EPI_ISL_674025, EPI_ISL_674026, EPI_ISL_674027, EPI_ISL_674028, EPI_ISL_674029, EPI_ISL_674030, EPI_ISL_674031, EPI_ISL_674032, EPI_ISL_674033, EPI_ISL_674034, EPI_ISL_674035, EPI_ISL_674036, EPI_ISL_674037, EPI_ISL_674038, EPI_ISL_674039, EPI_ISL_674040, EPI_ISL_674041, EPI_ISL_674042, EPI_ISL_674043, EPI_ISL_674044, EPI_ISL_674045, EPI_ISL_674046, EPI_ISL_674047, EPI_ISL_674048, EPI_ISL_674049, EPI_ISL_674050, EPI_ISL_674051, EPI_ISL_674052, EPI_ISL_674053, EPI_ISL_674054, EPI_ISL_674055, EPI_ISL_674056, EPI_ISL_674057, EPI_ISL_674058, EPI_ISL_674059, EPI_ISL_674060, EPI_ISL_674061, EPI_ISL_674062, EPI_ISL_674063, EPI_ISL_674064, EPI_ISL_674065, EPI_ISL_674066, EPI_ISL_674067, EPI_ISL_674068, EPI_ISL_674069, EPI_ISL_674070, EPI_ISL_674071, EPI_ISL_674072, EPI_ISL_674073, EPI_ISL_674074, EPI_ISL_674075, EPI_ISL_674076, EPI_ISL_674077, EPI_ISL_674078, EPI_ISL_674079, EPI_ISL_674080, EPI_ISL_674081, EPI_ISL_674082, EPI_ISL_674083, EPI_ISL_674084, EPI_ISL_674085, EPI_ISL_674086, EPI_ISL_674087, EPI_ISL_674088, EPI_ISL_674089, EPI_ISL_674090, EPI_ISL_674091, EPI_ISL_674092, EPI_ISL_674093, EPI_ISL_674094, EPI_ISL_674095, EPI_ISL_674096, EPI_ISL_674097, EPI_ISL_674098, EPI_ISL_674099, EPI_ISL_674100, EPI_ISL_674101, EPI_ISL_674102, EPI_ISL_674103, EPI_ISL_674104, EPI_ISL_674105, EPI_ISL_674106, EPI_ISL_674107, EPI_ISL_674108, EPI_ISL_674109, EPI_ISL_674110, EPI_ISL_674111, EPI_ISL_674112, EPI_ISL_674113, EPI_ISL_674114, EPI_ISL_674115, EPI_ISL_674116, EPI_ISL_674117, EPI_ISL_674118, EPI_ISL_674119, EPI_ISL_674120, EPI_ISL_674121, EPI_ISL_674122, EPI_ISL_674123, EPI_ISL_674124, EPI_ISL_674125, EPI_ISL_674126, EPI_ISL_674127, EPI_ISL_674128, EPI_ISL_674129, EPI_ISL_674130, EPI_ISL_674131, EPI_ISL_674132, EPI_ISL_674133, EPI_ISL_674134, EPI_ISL_674135, EPI_ISL_674136, EPI_ISL_674137, EPI_ISL_674138, EPI_ISL_674139, EPI_ISL_674140, EPI_ISL_674141, EPI_ISL_674143, EPI_ISL_674144, EPI_ISL_674145, EPI_ISL_674146, EPI_ISL_674147, EPI_ISL_674148, EPI_ISL_674149, EPI_ISL_674150, EPI_ISL_674151, EPI_ISL_674152, EPI_ISL_674153, EPI_ISL_674154, EPI_ISL_674155, EPI_ISL_674156, EPI_ISL_674157, EPI_ISL_674163, EPI_ISL_674166, EPI_ISL_674167, EPI_ISL_674173, EPI_ISL_674178, EPI_ISL_674180, EPI_ISL_674183, EPI_ISL_674184, EPI_ISL_674185, EPI_ISL_674187, EPI_ISL_674189, EPI_ISL_674191, EPI_ISL_674196, EPI_ISL_674198, EPI_ISL_674200, EPI_ISL_674202, EPI_ISL_674205, EPI_ISL_674209, EPI_ISL_674210, EPI_ISL_674212, EPI_ISL_674213, EPI_ISL_674214, EPI_ISL_674215, EPI_ISL_674216, EPI_ISL_674218, EPI_ISL_674219, EPI_ISL_674220, EPI_ISL_674221, EPI_ISL_674230, EPI_ISL_674232, EPI_ISL_674235, EPI_ISL_674236, EPI_ISL_674237, EPI_ISL_674238, EPI_ISL_674239, EPI_ISL_674244, EPI_ISL_674246, EPI_ISL_674247, EPI_ISL_674248, EPI_ISL_674251, EPI_ISL_674252, EPI_ISL_674256, EPI_ISL_674257, EPI_ISL_674258, EPI_ISL_674259, EPI_ISL_674262, EPI_ISL_674263, EPI_ISL_674265, EPI_ISL_674266, EPI_ISL_674269, EPI_ISL_674270, EPI_ISL_674271, EPI_ISL_674273, EPI_ISL_674274, EPI_ISL_674275, EPI_ISL_674278, EPI_ISL_674279, EPI_ISL_674280, EPI_ISL_674282, EPI_ISL_674288, EPI_ISL_674289, EPI_ISL_674290, EPI_ISL_674291, EPI_ISL_674293, EPI_ISL_674298, EPI_ISL_674302, EPI_ISL_674303, EPI_ISL_674304, EPI_ISL_674306, EPI_ISL_674307, EPI_ISL_674309, EPI_ISL_674312, EPI_ISL_674314, EPI_ISL_674315, EPI_ISL_674316, EPI_ISL_674320, EPI_ISL_674321, EPI_ISL_674325, EPI_ISL_674326, EPI_ISL_674327, EPI_ISL_674330, EPI_ISL_674331, EPI_ISL_674333, EPI_ISL_674335, EPI_ISL_674336, EPI_ISL_674338, EPI_ISL_674342, EPI_ISL_674344, EPI_ISL_674348, EPI_ISL_674349, EPI_ISL_674350, EPI_ISL_674353, EPI_ISL_674355, EPI_ISL_674356, EPI_ISL_674358, EPI_ISL_674359, EPI_ISL_674363, EPI_ISL_674365, EPI_ISL_674371, EPI_ISL_674373, EPI_ISL_674377, EPI_ISL_674378, EPI_ISL_674379, EPI_ISL_674381, EPI_ISL_674382, EPI_ISL_674384, EPI_ISL_674386, EPI_ISL_674387, EPI_ISL_674388, EPI_ISL_674393, EPI_ISL_674395, EPI_ISL_674398, EPI_ISL_674401, EPI_ISL_674403, EPI_ISL_674405, EPI_ISL_674407, EPI_ISL_674410, EPI_ISL_674414, EPI_ISL_674415, EPI_ISL_674417, EPI_ISL_674422, EPI_ISL_674423, EPI_ISL_674425, EPI_ISL_674426, EPI_ISL_674427, EPI_ISL_674430, EPI_ISL_674433, EPI_ISL_674437, EPI_ISL_674440, EPI_ISL_674441, EPI_ISL_674443, EPI_ISL_674444, EPI_ISL_674448, EPI_ISL_674449, EPI_ISL_674453, EPI_ISL_674457, EPI_ISL_674458, EPI_ISL_674460, EPI_ISL_674461, EPI_ISL_674463, EPI_ISL_674464, EPI_ISL_674468, EPI_ISL_674473, EPI_ISL_674475, EPI_ISL_674480, EPI_ISL_674481, EPI_ISL_674482, EPI_ISL_674483, EPI_ISL_674489, EPI_ISL_674491, EPI_ISL_674492, EPI_ISL_674494, EPI_ISL_674496, EPI_ISL_674497, EPI_ISL_674500, EPI_ISL_674502, EPI_ISL_674503, EPI_ISL_674505, EPI_ISL_674506, EPI_ISL_674507, EPI_ISL_674508, EPI_ISL_674509, EPI_ISL_674510, EPI_ISL_674511, EPI_ISL_674512, EPI_ISL_674513, EPI_ISL_674515, EPI_ISL_674517, EPI_ISL_674518, EPI_ISL_674519, EPI_ISL_674520, EPI_ISL_674521, EPI_ISL_674523, EPI_ISL_674524, EPI_ISL_674525, EPI_ISL_674526, EPI_ISL_674529, EPI_ISL_674531, EPI_ISL_674532, EPI_ISL_674533, EPI_ISL_674534, EPI_ISL_674535, EPI_ISL_674536, EPI_ISL_674537, EPI_ISL_674538, EPI_ISL_674539, EPI_ISL_674540, EPI_ISL_674541, EPI_ISL_674542, EPI_ISL_674546, EPI_ISL_674547, EPI_ISL_674548, EPI_ISL_674549, EPI_ISL_674551, EPI_ISL_674552, EPI_ISL_674553, EPI_ISL_674554, EPI_ISL_674555, EPI_ISL_674556, EPI_ISL_674557, EPI_ISL_674559, EPI_ISL_674560, EPI_ISL_674561, EPI_ISL_674562, EPI_ISL_674564, EPI_ISL_674566, EPI_ISL_674567, EPI_ISL_674568, EPI_ISL_674570, EPI_ISL_674571, EPI_ISL_674572, EPI_ISL_674573, EPI_ISL_674574, EPI_ISL_674575, EPI_ISL_674576, EPI_ISL_674577, EPI_ISL_674578, EPI_ISL_674579, EPI_ISL_674580, EPI_ISL_674581, EPI_ISL_674582, EPI_ISL_674583, EPI_ISL_674584, EPI_ISL_674586, EPI_ISL_674587, EPI_ISL_674588, EPI_ISL_674591, EPI_ISL_674593, EPI_ISL_674594, EPI_ISL_674596, EPI_ISL_674597, EPI_ISL_674598, EPI_ISL_674599, EPI_ISL_674600, EPI_ISL_674601, EPI_ISL_674602, EPI_ISL_674604, EPI_ISL_674606, EPI_ISL_674607, EPI_ISL_674608, EPI_ISL_674610, EPI_ISL_674611, EPI_ISL_674612, EPI_ISL_674614, EPI_ISL_674615, EPI_ISL_674616, EPI_ISL_674617, EPI_ISL_674618, EPI_ISL_674619, EPI_ISL_674620, EPI_ISL_674621, EPI_ISL_674622, EPI_ISL_674623, EPI_ISL_674624, EPI_ISL_674626, EPI_ISL_674627, EPI_ISL_674628, EPI_ISL_674630, EPI_ISL_674631, EPI_ISL_674632, EPI_ISL_674633, EPI_ISL_674634, EPI_ISL_674636, EPI_ISL_674637, EPI_ISL_674638, EPI_ISL_674639, EPI_ISL_674640, EPI_ISL_674641, EPI_ISL_674642, EPI_ISL_674643, EPI_ISL_674644, EPI_ISL_674645, EPI_ISL_674646, EPI_ISL_674647, EPI_ISL_674648, EPI_ISL_674649, EPI_ISL_674650, EPI_ISL_674651, EPI_ISL_674654, EPI_ISL_674656, EPI_ISL_674659, EPI_ISL_674660, EPI_ISL_674663, EPI_ISL_674664, EPI_ISL_674665, EPI_ISL_674666, EPI_ISL_674668, EPI_ISL_674669, EPI_ISL_674671, EPI_ISL_674672, EPI_ISL_674673, EPI_ISL_674675, EPI_ISL_674676, EPI_ISL_674677, EPI_ISL_674679, EPI_ISL_674680, EPI_ISL_674681, EPI_ISL_674682, EPI_ISL_674683, EPI_ISL_674684, EPI_ISL_674686, EPI_ISL_674687, EPI_ISL_674688, EPI_ISL_674689, EPI_ISL_674693, EPI_ISL_674694, EPI_ISL_674695, EPI_ISL_674696, EPI_ISL_674697, EPI_ISL_674699, EPI_ISL_674700, EPI_ISL_674701, EPI_ISL_674702, EPI_ISL_674703, EPI_ISL_674704, EPI_ISL_674705, EPI_ISL_674706, EPI_ISL_674708, EPI_ISL_674709, EPI_ISL_674710, EPI_ISL_674711, EPI_ISL_674712, EPI_ISL_674713, EPI_ISL_674715, EPI_ISL_674718, EPI_ISL_674719, EPI_ISL_674720, EPI_ISL_674721, EPI_ISL_674726, EPI_ISL_674727, EPI_ISL_674728, EPI_ISL_674729, EPI_ISL_674730, EPI_ISL_674731, EPI_ISL_674732, EPI_ISL_674733, EPI_ISL_674734, EPI_ISL_674735, EPI_ISL_674737, EPI_ISL_674738, EPI_ISL_674739, EPI_ISL_674741, EPI_ISL_674742, EPI_ISL_674748, EPI_ISL_674750, EPI_ISL_674753, EPI_ISL_674754, EPI_ISL_674755, EPI_ISL_674757, EPI_ISL_674759, EPI_ISL_674760, EPI_ISL_674761, EPI_ISL_674764, EPI_ISL_674765, EPI_ISL_674766, EPI_ISL_674767, EPI_ISL_674768, EPI_ISL_674769, EPI_ISL_674770, EPI_ISL_674771, EPI_ISL_674774, EPI_ISL_674775, EPI_ISL_674776, EPI_ISL_674777, EPI_ISL_674778, EPI_ISL_674780, EPI_ISL_674782, EPI_ISL_674784, EPI_ISL_674785, EPI_ISL_674787, EPI_ISL_674788, EPI_ISL_674789, EPI_ISL_674790, EPI_ISL_674793, EPI_ISL_674796, EPI_ISL_674797, EPI_ISL_674798, EPI_ISL_674799, EPI_ISL_674800, EPI_ISL_674801, EPI_ISL_674802, EPI_ISL_674803, EPI_ISL_674805, EPI_ISL_674806, EPI_ISL_674807, EPI_ISL_674810, EPI_ISL_674811, EPI_ISL_674812, EPI_ISL_674813, EPI_ISL_674814, EPI_ISL_674815, EPI_ISL_674816, EPI_ISL_674817, EPI_ISL_674818, EPI_ISL_674819, EPI_ISL_674820, EPI_ISL_674821, EPI_ISL_674822, EPI_ISL_674823, EPI_ISL_674824, EPI_ISL_674825, EPI_ISL_674827, EPI_ISL_674829, EPI_ISL_674830, EPI_ISL_674831, EPI_ISL_674833, EPI_ISL_674835, EPI_ISL_674836, EPI_ISL_674839, EPI_ISL_674841, EPI_ISL_674842, EPI_ISL_674843, EPI_ISL_674844, EPI_ISL_674848, EPI_ISL_674852, EPI_ISL_674853, EPI_ISL_674854, EPI_ISL_674855, EPI_ISL_674856, EPI_ISL_674857, EPI_ISL_674858, EPI_ISL_674860, EPI_ISL_674861, EPI_ISL_674864, EPI_ISL_674865, EPI_ISL_674869, EPI_ISL_674870, EPI_ISL_674872, EPI_ISL_674874, EPI_ISL_674875, EPI_ISL_674877, EPI_ISL_674878, EPI_ISL_674879, EPI_ISL_674880, EPI_ISL_674882, EPI_ISL_674884, EPI_ISL_674885, EPI_ISL_674886, EPI_ISL_674887, EPI_ISL_674888, EPI_ISL_674889, EPI_ISL_674890, EPI_ISL_674891, EPI_ISL_674892, EPI_ISL_674894, EPI_ISL_674895, EPI_ISL_674897, EPI_ISL_674898, EPI_ISL_674900, EPI_ISL_674901, EPI_ISL_674902, EPI_ISL_674903, EPI_ISL_674904, EPI_ISL_674905, EPI_ISL_674907, EPI_ISL_674911, EPI_ISL_674912, EPI_ISL_674914, EPI_ISL_674915, EPI_ISL_674916, EPI_ISL_674918, EPI_ISL_674921, EPI_ISL_674925, EPI_ISL_674927, EPI_ISL_674928, EPI_ISL_674929, EPI_ISL_674930, EPI_ISL_674931, EPI_ISL_674932, EPI_ISL_674934, EPI_ISL_674935, EPI_ISL_674937, EPI_ISL_674938, EPI_ISL_674941, EPI_ISL_674942, EPI_ISL_674943, EPI_ISL_674945, EPI_ISL_674947, EPI_ISL_674948, EPI_ISL_674954, EPI_ISL_674955, EPI_ISL_674956, EPI_ISL_674958, EPI_ISL_674960, EPI_ISL_674961, EPI_ISL_674963, EPI_ISL_674964, EPI_ISL_674965, EPI_ISL_674966, EPI_ISL_674967, EPI_ISL_674968, EPI_ISL_674969, EPI_ISL_674970, EPI_ISL_674971, EPI_ISL_674974, EPI_ISL_674975, EPI_ISL_674976, EPI_ISL_674978, EPI_ISL_674979, EPI_ISL_674980, EPI_ISL_674981, EPI_ISL_674982, EPI_ISL_674984, EPI_ISL_674985, EPI_ISL_674986, EPI_ISL_674987, EPI_ISL_674989, EPI_ISL_674990, EPI_ISL_674991, EPI_ISL_674993, EPI_ISL_674994, EPI_ISL_674996, EPI_ISL_674997, EPI_ISL_674999, EPI_ISL_675000, EPI_ISL_675001, EPI_ISL_675002, EPI_ISL_675003, EPI_ISL_675004, EPI_ISL_675005, EPI_ISL_675007, EPI_ISL_675010, EPI_ISL_675011, EPI_ISL_675013, EPI_ISL_675014, EPI_ISL_675015, EPI_ISL_675016, EPI_ISL_675017, EPI_ISL_675019, EPI_ISL_675020, EPI_ISL_675021, EPI_ISL_675023, EPI_ISL_675025, EPI_ISL_675028, EPI_ISL_675029, EPI_ISL_675030, EPI_ISL_675031, EPI_ISL_675032, EPI_ISL_675034, EPI_ISL_675035, EPI_ISL_675036, EPI_ISL_675037, EPI_ISL_675041, EPI_ISL_675043, EPI_ISL_675044, EPI_ISL_675045, EPI_ISL_675047, EPI_ISL_675048, EPI_ISL_675049, EPI_ISL_675050, EPI_ISL_675051, EPI_ISL_675052, EPI_ISL_675054, EPI_ISL_675055, EPI_ISL_675057, EPI_ISL_675058, EPI_ISL_675059, EPI_ISL_675060, EPI_ISL_675061, EPI_ISL_675062, EPI_ISL_675063, EPI_ISL_675065, EPI_ISL_675066, EPI_ISL_675067, EPI_ISL_675068, EPI_ISL_675069, EPI_ISL_675070, EPI_ISL_675072, EPI_ISL_675073, EPI_ISL_675074, EPI_ISL_675076, EPI_ISL_675077, EPI_ISL_675078, EPI_ISL_675079, EPI_ISL_675081, EPI_ISL_675083, EPI_ISL_675084, EPI_ISL_675085, EPI_ISL_675086, EPI_ISL_675088, EPI_ISL_675089, EPI_ISL_675090, EPI_ISL_675091, EPI_ISL_675092, EPI_ISL_675093, EPI_ISL_675095, EPI_ISL_675096, EPI_ISL_675097, EPI_ISL_675098, EPI_ISL_675099, EPI_ISL_675100, EPI_ISL_675101, EPI_ISL_675102, EPI_ISL_675103, EPI_ISL_675105, EPI_ISL_675106, EPI_ISL_675108, EPI_ISL_675109, EPI_ISL_675111, EPI_ISL_675112, EPI_ISL_675113, EPI_ISL_675114, EPI_ISL_675115, EPI_ISL_675116, EPI_ISL_675118, EPI_ISL_675119, EPI_ISL_675121, EPI_ISL_675122, EPI_ISL_675123, EPI_ISL_675124, EPI_ISL_675125, EPI_ISL_675127, EPI_ISL_675128, EPI_ISL_675129, EPI_ISL_675131, EPI_ISL_675132, EPI_ISL_675134, EPI_ISL_675145, EPI_ISL_675153, EPI_ISL_675160, EPI_ISL_675165, EPI_ISL_675170, EPI_ISL_675178, EPI_ISL_675179, EPI_ISL_675183, EPI_ISL_675185, EPI_ISL_675188, EPI_ISL_675210, EPI_ISL_675212, EPI_ISL_675217, EPI_ISL_675219, EPI_ISL_675224, EPI_ISL_675227, EPI_ISL_675228, EPI_ISL_675236, EPI_ISL_675238, EPI_ISL_675241, EPI_ISL_675243, EPI_ISL_675244, EPI_ISL_675260, EPI_ISL_675264, EPI_ISL_675275, EPI_ISL_675276, EPI_ISL_675277, EPI_ISL_675282, EPI_ISL_675284, EPI_ISL_675285, EPI_ISL_675286, EPI_ISL_675292, EPI_ISL_675301, EPI_ISL_675305, EPI_ISL_675306, EPI_ISL_675307, EPI_ISL_675308, EPI_ISL_675309, EPI_ISL_675311, EPI_ISL_675313, EPI_ISL_675317, EPI_ISL_675318, EPI_ISL_675319, EPI_ISL_675320, EPI_ISL_675326, EPI_ISL_675331, EPI_ISL_675332, EPI_ISL_675340, EPI_ISL_675353, EPI_ISL_675357, EPI_ISL_675364, EPI_ISL_675368, EPI_ISL_675371, EPI_ISL_675386, EPI_ISL_675389, EPI_ISL_675393, EPI_ISL_675397, EPI_ISL_675399, EPI_ISL_675404, EPI_ISL_675417, EPI_ISL_675420, EPI_ISL_675423, EPI_ISL_675424, EPI_ISL_675431, EPI_ISL_675432, EPI_ISL_675436, EPI_ISL_675439, EPI_ISL_675443, EPI_ISL_675444, EPI_ISL_675446, EPI_ISL_675462, EPI_ISL_675467, EPI_ISL_675472, EPI_ISL_675475, EPI_ISL_675480, EPI_ISL_675485, EPI_ISL_675486, EPI_ISL_675490, EPI_ISL_675495, EPI_ISL_675496, EPI_ISL_675499, EPI_ISL_675500, EPI_ISL_675503, EPI_ISL_675504, EPI_ISL_675505, EPI_ISL_675506, EPI_ISL_675507, EPI_ISL_675513, EPI_ISL_675518, EPI_ISL_675528, EPI_ISL_675533, EPI_ISL_675535, EPI_ISL_675538, EPI_ISL_675540, EPI_ISL_675542, EPI_ISL_675547, EPI_ISL_675550, EPI_ISL_675555, EPI_ISL_675556, EPI_ISL_675558, EPI_ISL_675560, EPI_ISL_675564, EPI_ISL_675565, EPI_ISL_675566, EPI_ISL_675569, EPI_ISL_675572, EPI_ISL_675576, EPI_ISL_675578, EPI_ISL_675591, EPI_ISL_675601, EPI_ISL_675602, EPI_ISL_675606, EPI_ISL_675621, EPI_ISL_675631, EPI_ISL_675634, EPI_ISL_675644, EPI_ISL_675652, EPI_ISL_675660, EPI_ISL_675661, EPI_ISL_675663, EPI_ISL_675667, EPI_ISL_675676, EPI_ISL_675682, EPI_ISL_675690, EPI_ISL_675694, EPI_ISL_675695, EPI_ISL_675698, EPI_ISL_675704, EPI_ISL_675709, EPI_ISL_675711, EPI_ISL_675712, EPI_ISL_675713, EPI_ISL_675714, EPI_ISL_675717, EPI_ISL_675718, EPI_ISL_675722, EPI_ISL_675723, EPI_ISL_675724, EPI_ISL_675725, EPI_ISL_675726, EPI_ISL_675727, EPI_ISL_675728, EPI_ISL_675729, EPI_ISL_675730, EPI_ISL_675731, EPI_ISL_675732, EPI_ISL_675733, EPI_ISL_675734, EPI_ISL_675736, EPI_ISL_675737, EPI_ISL_675738, EPI_ISL_675741, EPI_ISL_675743, EPI_ISL_675744, EPI_ISL_675745, EPI_ISL_675747, EPI_ISL_675748, EPI_ISL_675749, EPI_ISL_675752, EPI_ISL_675753, EPI_ISL_675754, EPI_ISL_675755, EPI_ISL_675758, EPI_ISL_675761, EPI_ISL_675762, EPI_ISL_675763, EPI_ISL_675764, EPI_ISL_675765, EPI_ISL_675767, EPI_ISL_675768, EPI_ISL_675769, EPI_ISL_675772, EPI_ISL_675773, EPI_ISL_675778, EPI_ISL_675780, EPI_ISL_675781, EPI_ISL_675782, EPI_ISL_675783, EPI_ISL_675784, EPI_ISL_675786, EPI_ISL_675787, EPI_ISL_675789, EPI_ISL_675790, EPI_ISL_675791, EPI_ISL_675792, EPI_ISL_675793, EPI_ISL_675795, EPI_ISL_675796, EPI_ISL_675797, EPI_ISL_675801, EPI_ISL_675804, EPI_ISL_675805, EPI_ISL_675807, EPI_ISL_675808, EPI_ISL_675809, EPI_ISL_675811, EPI_ISL_675815, EPI_ISL_675816, EPI_ISL_675821, EPI_ISL_675824, EPI_ISL_675827, EPI_ISL_675828, EPI_ISL_675830, EPI_ISL_675831, EPI_ISL_675832, EPI_ISL_675834, EPI_ISL_675835, EPI_ISL_675837, EPI_ISL_675838, EPI_ISL_675840, EPI_ISL_675841, EPI_ISL_675842, EPI_ISL_675844, EPI_ISL_675845, EPI_ISL_675849, EPI_ISL_675850, EPI_ISL_675851, EPI_ISL_675853, EPI_ISL_675854, EPI_ISL_675856, EPI_ISL_675857, EPI_ISL_675858, EPI_ISL_675859, EPI_ISL_675860, EPI_ISL_675861, EPI_ISL_675862, EPI_ISL_675863, EPI_ISL_675864, EPI_ISL_675865, EPI_ISL_675866, EPI_ISL_675867, EPI_ISL_675869, EPI_ISL_675870, EPI_ISL_675871, EPI_ISL_675872, EPI_ISL_675873, EPI_ISL_675874, EPI_ISL_675875,

EPI_ISL_675876, EPI_ISL_675877, EPI_ISL_675879, EPI_ISL_675881, EPI_ISL_675883, EPI_ISL_675884, EPI_ISL_675885, EPI_ISL_675886, EPI_ISL_675887, EPI_ISL_675888, EPI_ISL_675889, EPI_ISL_675891, EPI_ISL_675893, EPI_ISL_675895, EPI_ISL_675896, EPI_ISL_675898, EPI_ISL_675899, EPI_ISL_675900, EPI_ISL_675904, EPI_ISL_675905, EPI_ISL_675906, EPI_ISL_675907, EPI_ISL_675909, EPI_ISL_675910, EPI_ISL_675911, EPI_ISL_675912, EPI_ISL_675914, EPI_ISL_675915, EPI_ISL_675917, EPI_ISL_675918, EPI_ISL_675919, EPI_ISL_675921, EPI_ISL_675922, EPI_ISL_675926, EPI_ISL_675927, EPI_ISL_675928, EPI_ISL_675931, EPI_ISL_675932, EPI_ISL_675933, EPI_ISL_675935, EPI_ISL_675936, EPI_ISL_675938, EPI_ISL_675940, EPI_ISL_675941, EPI_ISL_675943, EPI_ISL_675945, EPI_ISL_675946, EPI_ISL_675947, EPI_ISL_675948, EPI_ISL_675949, EPI_ISL_675950, EPI_ISL_675951, EPI_ISL_675953, EPI_ISL_675955, EPI_ISL_675956, EPI_ISL_675957, EPI_ISL_675958, EPI_ISL_675959, EPI_ISL_675960, EPI_ISL_675961, EPI_ISL_675963, EPI_ISL_675964, EPI_ISL_675966, EPI_ISL_675967, EPI_ISL_675969, EPI_ISL_675970, EPI_ISL_675971, EPI_ISL_675972, EPI_ISL_675975, EPI_ISL_675977, EPI_ISL_675979, EPI_ISL_675981, EPI_ISL_675983, EPI_ISL_675984, EPI_ISL_675985, EPI_ISL_675989, EPI_ISL_675990, EPI_ISL_675991, EPI_ISL_675992, EPI_ISL_675994, EPI_ISL_675995, EPI_ISL_675996, EPI_ISL_675997, EPI_ISL_675998, EPI_ISL_675999, EPI_ISL_676001, EPI_ISL_676003, EPI_ISL_676004, EPI_ISL_676005, EPI_ISL_676007, EPI_ISL_676008, EPI_ISL_676009, EPI_ISL_676011, EPI_ISL_676012, EPI_ISL_676013, EPI_ISL_676014, EPI_ISL_676015, EPI_ISL_676016, EPI_ISL_676017, EPI_ISL_676018, EPI_ISL_676019, EPI_ISL_676020, EPI_ISL_676021, EPI_ISL_676022, EPI_ISL_676023, EPI_ISL_676024, EPI_ISL_676025, EPI_ISL_676026, EPI_ISL_676027, EPI_ISL_676028, EPI_ISL_676029, EPI_ISL_676030, EPI_ISL_676031, EPI_ISL_676032, EPI_ISL_676033, EPI_ISL_676034, EPI_ISL_676036, EPI_ISL_676037, EPI_ISL_676038, EPI_ISL_676039, EPI_ISL_676040, EPI_ISL_676041, EPI_ISL_676042, EPI_ISL_676043, EPI_ISL_676044, EPI_ISL_676045, EPI_ISL_676046, EPI_ISL_676047, EPI_ISL_676048, EPI_ISL_676049, EPI_ISL_676050, EPI_ISL_676051, EPI_ISL_676052, EPI_ISL_676053, EPI_ISL_676054, EPI_ISL_676055, EPI_ISL_676056, EPI_ISL_676057, EPI_ISL_676058, EPI_ISL_676059, EPI_ISL_676060, EPI_ISL_676061, EPI_ISL_676062, EPI_ISL_676063, EPI_ISL_676064, EPI_ISL_676065, EPI_ISL_676066, EPI_ISL_676067, EPI_ISL_676068, EPI_ISL_676069, EPI_ISL_676070, EPI_ISL_676071, EPI_ISL_676072, EPI_ISL_676073, EPI_ISL_676074, EPI_ISL_676075, EPI_ISL_676076, EPI_ISL_676077, EPI_ISL_676078, EPI_ISL_676079, EPI_ISL_676080, EPI_ISL_676081, EPI_ISL_676082, EPI_ISL_676083, EPI_ISL_676084, EPI_ISL_676085, EPI_ISL_676086, EPI_ISL_676087, EPI_ISL_676088, EPI_ISL_676089, EPI_ISL_676090, EPI_ISL_676091, EPI_ISL_676092, EPI_ISL_676093, EPI_ISL_676094, EPI_ISL_676095, EPI_ISL_676096, EPI_ISL_676097, EPI_ISL_676098, EPI_ISL_676099, EPI_ISL_676100, EPI_ISL_676101, EPI_ISL_676102, EPI_ISL_676103, EPI_ISL_676104, EPI_ISL_676105, EPI_ISL_676106, EPI_ISL_676107, EPI_ISL_676108, EPI_ISL_676109, EPI_ISL_676110, EPI_ISL_676111, EPI_ISL_676112, EPI_ISL_676113, EPI_ISL_676114, EPI_ISL_676115, EPI_ISL_676116, EPI_ISL_676117, EPI_ISL_676118, EPI_ISL_676119, EPI_ISL_676120, EPI_ISL_676121, EPI_ISL_676122, EPI_ISL_676123, EPI_ISL_676124, EPI_ISL_676125, EPI_ISL_676126, EPI_ISL_676127, EPI_ISL_676128, EPI_ISL_676129, EPI_ISL_676130, EPI_ISL_676131, EPI_ISL_676132, EPI_ISL_676133, EPI_ISL_676134, EPI_ISL_676135, EPI_ISL_676136, EPI_ISL_676137, EPI_ISL_676138, EPI_ISL_676139, EPI_ISL_676140, EPI_ISL_676141, EPI_ISL_676142, EPI_ISL_676143, EPI_ISL_676144, EPI_ISL_676145, EPI_ISL_676146, EPI_ISL_676147, EPI_ISL_676148, EPI_ISL_676149, EPI_ISL_676150, EPI_ISL_676151, EPI_ISL_676152, EPI_ISL_676153, EPI_ISL_676154, EPI_ISL_676155, EPI_ISL_676156, EPI_ISL_676157, EPI_ISL_676158, EPI_ISL_676159, EPI_ISL_676160, EPI_ISL_676161, EPI_ISL_676162, EPI_ISL_676163, EPI_ISL_676164, EPI_ISL_676165, EPI_ISL_676166, EPI_ISL_676167, EPI_ISL_676168, EPI_ISL_676169, EPI_ISL_676170, EPI_ISL_676171, EPI_ISL_676172, EPI_ISL_676173, EPI_ISL_676174, EPI_ISL_676175, EPI_ISL_676176, EPI_ISL_676177, EPI_ISL_676178, EPI_ISL_676179, EPI_ISL_676180, EPI_ISL_676181, EPI_ISL_676182, EPI_ISL_676183, EPI_ISL_676184, EPI_ISL_676185, EPI_ISL_676186, EPI_ISL_676187, EPI_ISL_676188, EPI_ISL_676189, EPI_ISL_676190, EPI_ISL_676191, EPI_ISL_676192, EPI_ISL_676193, EPI_ISL_676194, EPI_ISL_676195, EPI_ISL_676196, EPI_ISL_676197, EPI_ISL_676198, EPI_ISL_676199, EPI_ISL_676200, EPI_ISL_676201, EPI_ISL_676202, EPI_ISL_676203, EPI_ISL_676204, EPI_ISL_676205, EPI_ISL_676206, EPI_ISL_676207, EPI_ISL_676208, EPI_ISL_676209, EPI_ISL_676210, EPI_ISL_676211, EPI_ISL_676212, EPI_ISL_676213, EPI_ISL_676214, EPI_ISL_676215, EPI_ISL_676216, EPI_ISL_676217, EPI_ISL_676218, EPI_ISL_676219, EPI_ISL_676220, EPI_ISL_676221, EPI_ISL_676222, EPI_ISL_676223, EPI_ISL_676224, EPI_ISL_676225, EPI_ISL_676226, EPI_ISL_676227, EPI_ISL_676228, EPI_ISL_676229, EPI_ISL_676230, EPI_ISL_676231, EPI_ISL_676232, EPI_ISL_676233, EPI_ISL_676234, EPI_ISL_676235, EPI_ISL_676236, EPI_ISL_676237, EPI_ISL_676238, EPI_ISL_676239, EPI_ISL_676240, EPI_ISL_676241, EPI_ISL_676242, EPI_ISL_676243, EPI_ISL_676244, EPI_ISL_676245, EPI_ISL_676246, EPI_ISL_676247, EPI_ISL_676248, EPI_ISL_676249, EPI_ISL_676250, EPI_ISL_676251, EPI_ISL_676252, EPI_ISL_676253, EPI_ISL_676254, EPI_ISL_676255, EPI_ISL_676256, EPI_ISL_676257, EPI_ISL_676258, EPI_ISL_676259, EPI_ISL_676260, EPI_ISL_676261, EPI_ISL_676262, EPI_ISL_676263, EPI_ISL_676264, EPI_ISL_676265, EPI_ISL_676266, EPI_ISL_676267, EPI_ISL_676268, EPI_ISL_676269, EPI_ISL_676270, EPI_ISL_676271, EPI_ISL_676272, EPI_ISL_676273, EPI_ISL_676274, EPI_ISL_676275, EPI_ISL_676276, EPI_ISL_676277, EPI_ISL_676278, EPI_ISL_676279, EPI_ISL_676280, EPI_ISL_676281, EPI_ISL_676282, EPI_ISL_676283, EPI_ISL_676284, EPI_ISL_676285, EPI_ISL_676286, EPI_ISL_676287, EPI_ISL_676288, EPI_ISL_676289, EPI_ISL_676290, EPI_ISL_676291, EPI_ISL_676292, EPI_ISL_676293, EPI_ISL_676294, EPI_ISL_676295, EPI_ISL_676296, EPI_ISL_676297, EPI_ISL_676298, EPI_ISL_676299, EPI_ISL_676300, EPI_ISL_676301, EPI_ISL_676302, EPI_ISL_676303, EPI_ISL_676304, EPI_ISL_676305, EPI_ISL_676306, EPI_ISL_676307, EPI_ISL_676308, EPI_ISL_676309, EPI_ISL_676310, EPI_ISL_676311, EPI_ISL_676312, EPI_ISL_681327, EPI_ISL_681329, EPI_ISL_681333, EPI_ISL_681334, EPI_ISL_681340, EPI_ISL_681341, EPI_ISL_681343, EPI_ISL_681345, EPI_ISL_681347, EPI_ISL_681349, EPI_ISL_681351, EPI_ISL_681352, EPI_ISL_681355, EPI_ISL_681359, EPI_ISL_681363, EPI_ISL_681367, EPI_ISL_681370, EPI_ISL_681373, EPI_ISL_681374, EPI_ISL_681377, EPI_ISL_681381, EPI_ISL_681382, EPI_ISL_681384, EPI_ISL_681387, EPI_ISL_681390, EPI_ISL_681391, EPI_ISL_681394, EPI_ISL_681400, EPI_ISL_681402, EPI_ISL_681403, EPI_ISL_681408, EPI_ISL_681409, EPI_ISL_681410, EPI_ISL_681411, EPI_ISL_681412, EPI_ISL_681413, EPI_ISL_681414, EPI_ISL_681420, EPI_ISL_681423, EPI_ISL_681429, EPI_ISL_681430, EPI_ISL_681433, EPI_ISL_681435, EPI_ISL_681440, EPI_ISL_681441, EPI_ISL_681443, EPI_ISL_681449, EPI_ISL_681452, EPI_ISL_681453, EPI_ISL_681454, EPI_ISL_681455, EPI_ISL_681458, EPI_ISL_681464, EPI_ISL_681465, EPI_ISL_681470, EPI_ISL_681472, EPI_ISL_681474, EPI_ISL_681478, EPI_ISL_681483, EPI_ISL_681486, EPI_ISL_681487, EPI_ISL_681489, EPI_ISL_681490, EPI_ISL_681491, EPI_ISL_681493, EPI_ISL_681497, EPI_ISL_681500, EPI_ISL_681507, EPI_ISL_681509, EPI_ISL_681512, EPI_ISL_681513, EPI_ISL_681516, EPI_ISL_681519, EPI_ISL_681520, EPI_ISL_681523, EPI_ISL_681524, EPI_ISL_681526, EPI_ISL_681528, EPI_ISL_681529, EPI_ISL_681531, EPI_ISL_681532, EPI_ISL_681533, EPI_ISL_681534, EPI_ISL_681539, EPI_ISL_681542, EPI_ISL_681545, EPI_ISL_681547, EPI_ISL_681549, EPI_ISL_681550, EPI_ISL_681551, EPI_ISL_681556, EPI_ISL_681557, EPI_ISL_681558, EPI_ISL_681559, EPI_ISL_681560, EPI_ISL_681561, EPI_ISL_681562, EPI_ISL_681563, EPI_ISL_681564, EPI_ISL_681565, EPI_ISL_681566, EPI_ISL_681568, EPI_ISL_681569, EPI_ISL_681574, EPI_ISL_681575, EPI_ISL_681577, EPI_ISL_681578, EPI_ISL_681579, EPI_ISL_681583, EPI_ISL_681587, EPI_ISL_681590, EPI_ISL_681592, EPI_ISL_681595, EPI_ISL_681597, EPI_ISL_681599, EPI_ISL_681606, EPI_ISL_681607, EPI_ISL_681610, EPI_ISL_681614, EPI_ISL_681617, EPI_ISL_681618, EPI_ISL_681620, EPI_ISL_681621, EPI_ISL_681623, EPI_ISL_681624, EPI_ISL_681625, EPI_ISL_681629, EPI_ISL_681630, EPI_ISL_681633, EPI_ISL_681634, EPI_ISL_681635, EPI_ISL_681636, EPI_ISL_681638, EPI_ISL_681639, EPI_ISL_681641, EPI_ISL_681642, EPI_ISL_681643, EPI_ISL_681646, EPI_ISL_681647, EPI_ISL_681651, EPI_ISL_681655, EPI_ISL_681660, EPI_ISL_681663, EPI_ISL_681664, EPI_ISL_681666, EPI_ISL_681667, EPI_ISL_681668, EPI_ISL_681669, EPI_ISL_681670, EPI_ISL_705191, EPI_ISL_705192, EPI_ISL_705194, EPI_ISL_705196, EPI_ISL_705197, EPI_ISL_705199, EPI_ISL_705202, EPI_ISL_709882, EPI_ISL_709883, EPI_ISL_709884, EPI_ISL_709885, EPI_ISL_709887, EPI_ISL_709888, EPI_ISL_709889, EPI_ISL_709890, EPI_ISL_709891, EPI_ISL_709892, EPI_ISL_709894, EPI_ISL_709895, EPI_ISL_709896, EPI_ISL_709897, EPI_ISL_709898, EPI_ISL_719987, EPI_ISL_719990, EPI_ISL_719991, EPI_ISL_719992, EPI_ISL_719994, EPI_ISL_719995, EPI_ISL_719997, EPI_ISL_720001, EPI_ISL_720003, EPI_ISL_720004, EPI_ISL_720005, EPI_ISL_720006, EPI_ISL_720007, EPI_ISL_720008, EPI_ISL_720010, EPI_ISL_720011, EPI_ISL_720012, EPI_ISL_720014, EPI_ISL_720016, EPI_ISL_720017, EPI_ISL_720021, EPI_ISL_720026, EPI_ISL_720029, EPI_ISL_720030, EPI_ISL_720032, EPI_ISL_720035, EPI_ISL_720039, EPI_ISL_720040, EPI_ISL_720041, EPI_ISL_720047, EPI_ISL_720049, EPI_ISL_720050, EPI_ISL_720054, EPI_ISL_720055, EPI_ISL_720056, EPI_ISL_720057, EPI_ISL_720061, EPI_ISL_720062, EPI_ISL_720063, EPI_ISL_720065, EPI_ISL_720066, EPI_ISL_720067, EPI_ISL_720068, EPI_ISL_720069, EPI_ISL_720072, EPI_ISL_720074, EPI_ISL_720076, EPI_ISL_720078, EPI_ISL_720081, EPI_ISL_720089, EPI_ISL_720092, EPI_ISL_720093, EPI_ISL_720095, EPI_ISL_720098, EPI_ISL_720108, EPI_ISL_720109, EPI_ISL_720112, EPI_ISL_720113, EPI_ISL_720114, EPI_ISL_720115, EPI_ISL_720117, EPI_ISL_720119, EPI_ISL_720122, EPI_ISL_720124, EPI_ISL_720125, EPI_ISL_720127, EPI_ISL_720133, EPI_ISL_720136, EPI_ISL_720137, EPI_ISL_720143, EPI_ISL_720144, EPI_ISL_720145, EPI_ISL_720147, EPI_ISL_720148, EPI_ISL_720149, EPI_ISL_720150, EPI_ISL_720151, EPI_ISL_720153, EPI_ISL_720158, EPI_ISL_720159, EPI_ISL_720161, EPI_ISL_720163, EPI_ISL_720165, EPI_ISL_720167, EPI_ISL_720168, EPI_ISL_720169, EPI_ISL_720170, EPI_ISL_720172, EPI_ISL_720173, EPI_ISL_720174, EPI_ISL_720175, EPI_ISL_720176, EPI_ISL_720178, EPI_ISL_720179, EPI_ISL_720180, EPI_ISL_720184, EPI_ISL_720185, EPI_ISL_720186, EPI_ISL_720188, EPI_ISL_720193, EPI_ISL_720195, EPI_ISL_720197, EPI_ISL_720198, EPI_ISL_720204, EPI_ISL_720209, EPI_ISL_720210, EPI_ISL_720218, EPI_ISL_720222, EPI_ISL_720224, EPI_ISL_720229, EPI_ISL_720231, EPI_ISL_720235, EPI_ISL_720236, EPI_ISL_720237, EPI_ISL_720238, EPI_ISL_720240, EPI_ISL_720245, EPI_ISL_720246, EPI_ISL_720251, EPI_ISL_720255, EPI_ISL_720262, EPI_ISL_720263, EPI_ISL_720267, EPI_ISL_720268, EPI_ISL_720273, EPI_ISL_720275, EPI_ISL_720276, EPI_ISL_720277, EPI_ISL_720278, EPI_ISL_720281, EPI_ISL_720282, EPI_ISL_720286, EPI_ISL_720288, EPI_ISL_720289, EPI_ISL_720292, EPI_ISL_720293, EPI_ISL_720294, EPI_ISL_720295, EPI_ISL_720296, EPI_ISL_720300, EPI_ISL_720305, EPI_ISL_720306, EPI_ISL_720313, EPI_ISL_720315, EPI_ISL_720319, EPI_ISL_720322, EPI_ISL_720323, EPI_ISL_720326, EPI_ISL_720327, EPI_ISL_720331, EPI_ISL_720332, EPI_ISL_720333, EPI_ISL_720335, EPI_ISL_720337, EPI_ISL_720339, EPI_ISL_720340, EPI_ISL_720345, EPI_ISL_720346, EPI_ISL_720347, EPI_ISL_720349, EPI_ISL_720350, EPI_ISL_720351, EPI_ISL_720353, EPI_ISL_720354, EPI_ISL_720356, EPI_ISL_720358, EPI_ISL_720359, EPI_ISL_720361, EPI_ISL_720362, EPI_ISL_720363, EPI_ISL_720367, EPI_ISL_720368, EPI_ISL_720369, EPI_ISL_720371, EPI_ISL_720372, EPI_ISL_720373, EPI_ISL_720374, EPI_ISL_720375, EPI_ISL_720381, EPI_ISL_720385, EPI_ISL_720387, EPI_ISL_720390, EPI_ISL_720391, EPI_ISL_720394, EPI_ISL_720396, EPI_ISL_720401, EPI_ISL_720406, EPI_ISL_720408, EPI_ISL_720409, EPI_ISL_720412, EPI_ISL_720413, EPI_ISL_720414, EPI_ISL_720415, EPI_ISL_720416, EPI_ISL_720418, EPI_ISL_720420, EPI_ISL_720421, EPI_ISL_720422, EPI_ISL_720425, EPI_ISL_720431, EPI_ISL_720434, EPI_ISL_720436, EPI_ISL_720437, EPI_ISL_720442, EPI_ISL_720448, EPI_ISL_720451, EPI_ISL_720453, EPI_ISL_720454, EPI_ISL_720460, EPI_ISL_720461, EPI_ISL_720463, EPI_ISL_720466, EPI_ISL_720467, EPI_ISL_720469, EPI_ISL_720472, EPI_ISL_720475, EPI_ISL_720478, EPI_ISL_720482, EPI_ISL_720483, EPI_ISL_720486, EPI_ISL_720489, EPI_ISL_720490, EPI_ISL_720493, EPI_ISL_720494, EPI_ISL_720498, EPI_ISL_720499, EPI_ISL_720502, EPI_ISL_720504, EPI_ISL_720512, EPI_ISL_720514, EPI_ISL_720519, EPI_ISL_720521, EPI_ISL_720522, EPI_ISL_720524, EPI_ISL_720525, EPI_ISL_720527, EPI_ISL_720529, EPI_ISL_720532, EPI_ISL_720533, EPI_ISL_720537, EPI_ISL_720539, EPI_ISL_720541, EPI_ISL_720543, EPI_ISL_720545, EPI_ISL_720548, EPI_ISL_720550, EPI_ISL_720551, EPI_ISL_720558, EPI_ISL_720559, EPI_ISL_720561, EPI_ISL_720563, EPI_ISL_720565, EPI_ISL_720571, EPI_ISL_720573, EPI_ISL_720574, EPI_ISL_720576, EPI_ISL_720577, EPI_ISL_720578, EPI_ISL_720579, EPI_ISL_720580, EPI_ISL_720581, EPI_ISL_720584, EPI_ISL_720585, EPI_ISL_720588, EPI_ISL_720594, EPI_ISL_720597, EPI_ISL_720602, EPI_ISL_720605, EPI_ISL_720607, EPI_ISL_720609, EPI_ISL_720614, EPI_ISL_720619, EPI_ISL_720621, EPI_ISL_720622, EPI_ISL_720623, EPI_ISL_720627, EPI_ISL_720629, EPI_ISL_720630, EPI_ISL_720631, EPI_ISL_720634, EPI_ISL_720637, EPI_ISL_720638, EPI_ISL_720641, EPI_ISL_720646, EPI_ISL_720647, EPI_ISL_720649, EPI_ISL_720651, EPI_ISL_720652, EPI_ISL_720654, EPI_ISL_720659, EPI_ISL_720661, EPI_ISL_720664, EPI_ISL_731251, EPI_ISL_731252, EPI_ISL_783071, EPI_ISL_783072, EPI_ISL_783075, EPI_ISL_783077, EPI_ISL_783078, EPI_ISL_783083, EPI_ISL_783085, EPI_ISL_783087, EPI_ISL_783090, EPI_ISL_783099, EPI_ISL_783105, EPI_ISL_783106, EPI_ISL_783110, EPI_ISL_783112, EPI_ISL_783113, EPI_ISL_783114, EPI_ISL_783124, EPI_ISL_783129, EPI_ISL_783132, EPI_ISL_783136, EPI_ISL_783142, EPI_ISL_783148, EPI_ISL_783149, EPI_ISL_783150, EPI_ISL_783153, EPI_ISL_783154, EPI_ISL_783155, EPI_ISL_783157, EPI_ISL_783158, EPI_ISL_783159, EPI_ISL_783161, EPI_ISL_783162, EPI_ISL_783165, EPI_ISL_783168, EPI_ISL_783170, EPI_ISL_783171, EPI_ISL_783174, EPI_ISL_783177, EPI_ISL_783178, EPI_ISL_783179, EPI_ISL_783184, EPI_ISL_783189, EPI_ISL_783191, EPI_ISL_783195, EPI_ISL_783198, EPI_ISL_783200, EPI_ISL_783201, EPI_ISL_783206, EPI_ISL_783207, EPI_ISL_783215, EPI_ISL_783221, EPI_ISL_783224, EPI_ISL_783225, EPI_ISL_783226, EPI_ISL_783230, EPI_ISL_783231, EPI_ISL_783235, EPI_ISL_783237, EPI_ISL_783238, EPI_ISL_783240, EPI_ISL_783241, EPI_ISL_783243, EPI_ISL_783245, EPI_ISL_783246, EPI_ISL_783247, EPI_ISL_783250, EPI_ISL_783253, EPI_ISL_783255, EPI_ISL_783257, EPI_ISL_783261, EPI_ISL_783263, EPI_ISL_783267, EPI_ISL_783268, EPI_ISL_783269, EPI_ISL_783270, EPI_ISL_783279, EPI_ISL_783281, EPI_ISL_783283, EPI_ISL_783284, EPI_ISL_783291, EPI_ISL_783293, EPI_ISL_783296, EPI_ISL_783298, EPI_ISL_783303, EPI_ISL_783306, EPI_ISL_783308, EPI_ISL_783311, EPI_ISL_783312, EPI_ISL_783314, EPI_ISL_783316, EPI_ISL_783317, EPI_ISL_783321, EPI_ISL_783322, EPI_ISL_783324, EPI_ISL_783329, EPI_ISL_783330, EPI_ISL_783332, EPI_ISL_783334, EPI_ISL_783336, EPI_ISL_783337, EPI_ISL_783339, EPI_ISL_783340, EPI_ISL_783342, EPI_ISL_783344, EPI_ISL_783345, EPI_ISL_783347, EPI_ISL_783348, EPI_ISL_783351, EPI_ISL_783354, EPI_ISL_783355, EPI_ISL_783359, EPI_ISL_783360, EPI_ISL_783365, EPI_ISL_801183, EPI_ISL_801184, EPI_ISL_801185, EPI_ISL_801186, EPI_ISL_801187, EPI_ISL_801188, EPI_ISL_801189, EPI_ISL_801190, EPI_ISL_801191, EPI_ISL_801192, EPI_ISL_801195, EPI_ISL_801196, EPI_ISL_801198, EPI_ISL_801201, EPI_ISL_801202, EPI_ISL_801203, EPI_ISL_801205, EPI_ISL_801206, EPI_ISL_801207, EPI_ISL_801209, EPI_ISL_801211, EPI_ISL_801213, EPI_ISL_801214, EPI_ISL_801215, EPI_ISL_801216, EPI_ISL_801217, EPI_ISL_801218, EPI_ISL_801220, EPI_ISL_801221, EPI_ISL_801222, EPI_ISL_801224, EPI_ISL_801226, EPI_ISL_801227, EPI_ISL_801228, EPI_ISL_801229, EPI_ISL_801230, EPI_ISL_892217, EPI_ISL_933495

| | | | |
|---|---|---|---|
| see above | Lighthouse Lab in Milton Keynes | Wellcome Sanger Institute for the COVID-19 Genomics UK (COG-UK) Consortium | Cordelia Langford; David K. Jackson; Dominic Kwiatkowski; Ewan Harrison; Ian Johnston; John Sillitoe on behalf of the Wellcome Sanger Institute COVID-19 Surveillance Team; Roberto Amato; Sonia Goncalves; The Lighthouse Lab in Milton Keynes and Alex Alderton |

EPI_ISL_664113, EPI_ISL_664146, EPI_ISL_664151, EPI_ISL_664163, EPI_ISL_664204, EPI_ISL_664220, EPI_ISL_664223, EPI_ISL_664299, EPI_ISL_664332, EPI_ISL_664337, EPI_ISL_664350, EPI_ISL_664363, EPI_ISL_664368, EPI_ISL_664430, EPI_ISL_664435, EPI_ISL_664478, EPI_ISL_664480, EPI_ISL_664487, EPI_ISL_664496, EPI_ISL_664511, EPI_ISL_664607, EPI_ISL_664613, EPI_ISL_664619, EPI_ISL_664633, EPI_ISL_664643, EPI_ISL_664646, EPI_ISL_664671, EPI_ISL_664675, EPI_ISL_664678, EPI_ISL_664681, EPI_ISL_664692, EPI_ISL_664710, EPI_ISL_664738, EPI_ISL_665114, EPI_ISL_665115, EPI_ISL_665116, EPI_ISL_665117, EPI_ISL_665118, EPI_ISL_665119, EPI_ISL_665120, EPI_ISL_665121, EPI_ISL_665122, EPI_ISL_665123, EPI_ISL_665124, EPI_ISL_665125, EPI_ISL_665126, EPI_ISL_665127, EPI_ISL_665696, EPI_ISL_679389, EPI_ISL_702571, EPI_ISL_702591, EPI_ISL_702600, EPI_ISL_702612, EPI_ISL_702628, EPI_ISL_702636, EPI_ISL_702642, EPI_ISL_702684, EPI_ISL_702729, EPI_ISL_702769, EPI_ISL_702838, EPI_ISL_702887, EPI_ISL_702917, EPI_ISL_702947, EPI_ISL_702980, EPI_ISL_703051, EPI_ISL_703061, EPI_ISL_703080, EPI_ISL_703174, EPI_ISL_703219, EPI_ISL_703235, EPI_ISL_703272, EPI_ISL_703364, EPI_ISL_703367, EPI_ISL_703372, EPI_ISL_703375, EPI_ISL_703387, EPI_ISL_703403, EPI_ISL_703577, EPI_ISL_703624, EPI_ISL_703698, EPI_ISL_703726, EPI_ISL_703729, EPI_ISL_703730, EPI_ISL_703732, EPI_ISL_703826, EPI_ISL_703884, EPI_ISL_703891, EPI_ISL_703937, EPI_ISL_703952, EPI_ISL_703961, EPI_ISL_703992, EPI_ISL_703995, EPI_ISL_704022, EPI_ISL_704033, EPI_ISL_704049, EPI_ISL_704108, EPI_ISL_704168, EPI_ISL_704209, EPI_ISL_704947, EPI_ISL_704950, EPI_ISL_705086, EPI_ISL_705168, EPI_ISL_705179, EPI_ISL_705227, EPI_ISL_705231, EPI_ISL_705240, EPI_ISL_705271, EPI_ISL_705305, EPI_ISL_705326, EPI_ISL_705404, EPI_ISL_705408, EPI_ISL_705446, EPI_ISL_705447, EPI_ISL_705448, EPI_ISL_705875, EPI_ISL_705877, EPI_ISL_705886, EPI_ISL_705887, EPI_ISL_705888, EPI_ISL_705889, EPI_ISL_705890, EPI_ISL_705891, EPI_ISL_705892, EPI_ISL_705894, EPI_ISL_705919, EPI_ISL_705929, EPI_ISL_705930, EPI_ISL_705931, EPI_ISL_705932, EPI_ISL_705933, EPI_ISL_705934, EPI_ISL_705935, EPI_ISL_705936, EPI_ISL_705937, EPI_ISL_705938, EPI_ISL_705939, EPI_ISL_865172, EPI_ISL_865173, EPI_ISL_1177821, EPI_ISL_1177822, EPI_ISL_1636816

| | | | |
|---|---|---|---|
| see above | Liverpool Clinical Laboratories | COVID-19 Genomics UK (COG-UK) Consortium | A Alrezaihi; Alessandro Gerada; Alistair Darby; Angela Cowell; Anita Lucaci; Anu Chawla; Cassie Olateju; Catherine Hartley; Charlotte Nelson; Ecaterina Vamos; Elaine O'Toole; Eleanor G Bentley; Ghada T Shawli; Hermione Webster; Isabel Garcia-Dorival; James Johnson; James P Stewart; Jenifer Manson; Joanne Watts; Jones Benjamin; Jordan J Clark; Julian Hiscox; L Luu; Lucille Rainbow; M Almsaud; Margaret Hughes; Mark Whitehead; Matthew Gemmell; Miren Iturriza-Gomara; Muhannad Alruwaili; N.P Randle; Neil Swainston; PKF Gilmore; Parul Sharma; Rebekah Penrice-Randal; Richard Eccles; Richard Gregory; Sam Haldenby; Steve Paterson; Stuart D Armstrong; Trevor Ian Robinson; Ximeng Han |

EPI_ISL_702533, EPI_ISL_702547, EPI_ISL_702812, EPI_ISL_702823, EPI_ISL_702836, EPI_ISL_702855, EPI_ISL_702959, EPI_ISL_703007, EPI_ISL_703034, EPI_ISL_703064, EPI_ISL_703390, EPI_ISL_703537, EPI_ISL_703540, EPI_ISL_703542, EPI_ISL_703599, EPI_ISL_703728, EPI_ISL_704387, EPI_ISL_704570, EPI_ISL_704577, EPI_ISL_704578, EPI_ISL_705102, EPI_ISL_705137, EPI_ISL_705148, EPI_ISL_705266, EPI_ISL_705295, EPI_ISL_705353, EPI_ISL_705361, EPI_ISL_705367, EPI_ISL_705381, EPI_ISL_705458, EPI_ISL_706279, EPI_ISL_706297, EPI_ISL_706298, EPI_ISL_706299, EPI_ISL_706300, EPI_ISL_706301, EPI_ISL_706302, EPI_ISL_706303, EPI_ISL_706304, EPI_ISL_706305, EPI_ISL_706306, EPI_ISL_706307, EPI_ISL_706308, EPI_ISL_706309, EPI_ISL_706310, EPI_ISL_706311, EPI_ISL_706313, EPI_ISL_706314, EPI_ISL_706316, EPI_ISL_706332, EPI_ISL_706333, EPI_ISL_706334, EPI_ISL_706335, EPI_ISL_706336, EPI_ISL_706337, EPI_ISL_706338, EPI_ISL_706339, EPI_ISL_706341, EPI_ISL_706342, EPI_ISL_706343, EPI_ISL_706344, EPI_ISL_706345, EPI_ISL_706346, EPI_ISL_706347, EPI_ISL_706348, EPI_ISL_724973, EPI_ISL_725002, EPI_ISL_725003, EPI_ISL_725004, EPI_ISL_725005, EPI_ISL_725006, EPI_ISL_725007, EPI_ISL_725008, EPI_ISL_725009, EPI_ISL_725010, EPI_ISL_725011, EPI_ISL_921676

| | | | |
|---|---|---|---|
| see above | Northumbria University / South Tees Hospitals NHS Foundation Trust / North Cumbria Integrated Care NHS Foundation Trust / North Tees and Hartlepool NHS Foundation Trust / Newcastle Hospitals NHS Foundation Trust | COVID-19 Genomics UK (COG-UK) Consortium | Andrew Nelson; Brendan Payne; Clive Graham; Darren L Smith; Debra Padgett; Edward Barton; Emma Swindells; Garren Scott; Gary Black; Gary Eltringham; Giles S Holt; Greg R Young; Jane Greenaway; Jennifer Collins; John Allan; Joshua Loh; Lynn Dover; Matthew Bashton; Mohammad A Tariq; Paul Baker; Sarah Essex; Steve Liggett; Wen C Yew; Yusri Taha |

EPI_ISL_723674, EPI_ISL_723675, EPI_ISL_723676, EPI_ISL_723677, EPI_ISL_723678, EPI_ISL_723679, EPI_ISL_723680, EPI_ISL_723681, EPI_ISL_723682, EPI_ISL_723683, EPI_ISL_723684, EPI_ISL_723685, EPI_ISL_723686, EPI_ISL_723687, EPI_ISL_723688, EPI_ISL_723689, EPI_ISL_723690, EPI_ISL_723691, EPI_ISL_723692, EPI_ISL_723693, EPI_ISL_723694, EPI_ISL_723695, EPI_ISL_723696, EPI_ISL_723697, EPI_ISL_723698, EPI_ISL_723699, EPI_ISL_723700, EPI_ISL_723701, EPI_ISL_723702, EPI_ISL_723703, EPI_ISL_723704, EPI_ISL_723705, EPI_ISL_723706, EPI_ISL_723707, EPI_ISL_723708, EPI_ISL_723709, EPI_ISL_723710, EPI_ISL_723711, EPI_ISL_723712, EPI_ISL_723713, EPI_ISL_723714, EPI_ISL_723715, EPI_ISL_723716, EPI_ISL_723717, EPI_ISL_723718, EPI_ISL_723719, EPI_ISL_723720, EPI_ISL_723721, EPI_ISL_723722, EPI_ISL_723723, EPI_ISL_723724, EPI_ISL_723725, EPI_ISL_723726, EPI_ISL_723727, EPI_ISL_723728, EPI_ISL_723730, EPI_ISL_723731, EPI_ISL_723732, EPI_ISL_723733, EPI_ISL_723734, EPI_ISL_723735, EPI_ISL_723736, EPI_ISL_723737, EPI_ISL_723738, EPI_ISL_723739, EPI_ISL_723740, EPI_ISL_723741, EPI_ISL_723742, EPI_ISL_723743, EPI_ISL_723744, EPI_ISL_723745, EPI_ISL_723746, EPI_ISL_723747, EPI_ISL_723748, EPI_ISL_723749, EPI_ISL_723750, EPI_ISL_723751, EPI_ISL_723752, EPI_ISL_723753, EPI_ISL_723754, EPI_ISL_723755, EPI_ISL_723756, EPI_ISL_723757, EPI_ISL_723758, EPI_ISL_723759, EPI_ISL_723760, EPI_ISL_723761, EPI_ISL_723762, EPI_ISL_723763, EPI_ISL_723764, EPI_ISL_723765, EPI_ISL_723766, EPI_ISL_723767, EPI_ISL_723768, EPI_ISL_723769, EPI_ISL_723770, EPI_ISL_723771, EPI_ISL_723772, EPI_ISL_723773, EPI_ISL_723774, EPI_ISL_723775, EPI_ISL_723776, EPI_ISL_723777, EPI_ISL_723778, EPI_ISL_723779, EPI_ISL_723780, EPI_ISL_723781, EPI_ISL_723782, EPI_ISL_723783, EPI_ISL_723784, EPI_ISL_723785, EPI_ISL_723786, EPI_ISL_723787, EPI_ISL_723788, EPI_ISL_723789, EPI_ISL_723790, EPI_ISL_723791, EPI_ISL_723792, EPI_ISL_723793, EPI_ISL_723794, EPI_ISL_723795, EPI_ISL_723796, EPI_ISL_723797, EPI_ISL_723798, EPI_ISL_723799, EPI_ISL_723800, EPI_ISL_723801, EPI_ISL_723802, EPI_ISL_723803, EPI_ISL_723804, EPI_ISL_723805, EPI_ISL_723806, EPI_ISL_723807, EPI_ISL_723808, EPI_ISL_723809, EPI_ISL_723810, EPI_ISL_723811, EPI_ISL_723812, EPI_ISL_723813, EPI_ISL_723814, EPI_ISL_723815, EPI_ISL_723816, EPI_ISL_723817, EPI_ISL_723818, EPI_ISL_723819, EPI_ISL_723820, EPI_ISL_723821, EPI_ISL_723822, EPI_ISL_723823, EPI_ISL_723824, EPI_ISL_723825, EPI_ISL_723826, EPI_ISL_723827, EPI_ISL_723828, EPI_ISL_723829, EPI_ISL_723830, EPI_ISL_723831, EPI_ISL_723832, EPI_ISL_723833, EPI_ISL_723834, EPI_ISL_723835, EPI_ISL_723836, EPI_ISL_723837, EPI_ISL_723838, EPI_ISL_723839, EPI_ISL_723840, EPI_ISL_723841, EPI_ISL_723842, EPI_ISL_723843, EPI_ISL_723844, EPI_ISL_723845, EPI_ISL_723846, EPI_ISL_723847, EPI_ISL_723848, EPI_ISL_723849, EPI_ISL_723850, EPI_ISL_723949, EPI_ISL_723950, EPI_ISL_723951, EPI_ISL_723952, EPI_ISL_723953, EPI_ISL_723954, EPI_ISL_723955, EPI_ISL_723956, EPI_ISL_723957, EPI_ISL_723958, EPI_ISL_723959, EPI_ISL_723960, EPI_ISL_723961, EPI_ISL_723962, EPI_ISL_723963, EPI_ISL_723965, EPI_ISL_723966, EPI_ISL_723968, EPI_ISL_723969, EPI_ISL_723970, EPI_ISL_723971, EPI_ISL_723972, EPI_ISL_723973, EPI_ISL_723974, EPI_ISL_723975, EPI_ISL_723976, EPI_ISL_723977, EPI_ISL_723978, EPI_ISL_723979, EPI_ISL_723980, EPI_ISL_723983, EPI_ISL_723984, EPI_ISL_723986, EPI_ISL_723987, EPI_ISL_723989, EPI_ISL_723990, EPI_ISL_723991, EPI_ISL_723992, EPI_ISL_723993, EPI_ISL_723994, EPI_ISL_723995, EPI_ISL_723996, EPI_ISL_723997, EPI_ISL_723998, EPI_ISL_723999, EPI_ISL_724000, EPI_ISL_724001, EPI_ISL_724002, EPI_ISL_724003, EPI_ISL_724004, EPI_ISL_724005, EPI_ISL_724006, EPI_ISL_724007, EPI_ISL_724008, EPI_ISL_724009, EPI_ISL_724010, EPI_ISL_724011, EPI_ISL_724012, EPI_ISL_724013, EPI_ISL_724014, EPI_ISL_724015, EPI_ISL_724017, EPI_ISL_724018, EPI_ISL_724019, EPI_ISL_724020, EPI_ISL_724026, EPI_ISL_724027, EPI_ISL_724028, EPI_ISL_724029, EPI_ISL_724030, EPI_ISL_724031, EPI_ISL_724032, EPI_ISL_724034, EPI_ISL_724035, EPI_ISL_724036, EPI_ISL_724037, EPI_ISL_724038, EPI_ISL_724039, EPI_ISL_724040, EPI_ISL_724041, EPI_ISL_724042, EPI_ISL_724044, EPI_ISL_724045, EPI_ISL_724046, EPI_ISL_724047, EPI_ISL_724048, EPI_ISL_724049, EPI_ISL_724050, EPI_ISL_724051, EPI_ISL_724052, EPI_ISL_724053, EPI_ISL_724054, EPI_ISL_724055, EPI_ISL_724056, EPI_ISL_724057, EPI_ISL_724058, EPI_ISL_724059, EPI_ISL_724060, EPI_ISL_724061, EPI_ISL_724062, EPI_ISL_724778, EPI_ISL_724779, EPI_ISL_724780, EPI_ISL_724781, EPI_ISL_724782, EPI_ISL_724783, EPI_ISL_724784, EPI_ISL_724785, EPI_ISL_724786, EPI_ISL_724787, EPI_ISL_724788, EPI_ISL_724789, EPI_ISL_724790, EPI_ISL_724791, EPI_ISL_724792, EPI_ISL_724793, EPI_ISL_724794, EPI_ISL_724795, EPI_ISL_724796, EPI_ISL_724797, EPI_ISL_724798, EPI_ISL_724799, EPI_ISL_724800, EPI_ISL_724801, EPI_ISL_724802, EPI_ISL_724803, EPI_ISL_724804, EPI_ISL_724805, EPI_ISL_724806, EPI_ISL_724807, EPI_ISL_724808, EPI_ISL_724809, EPI_ISL_724810, EPI_ISL_724811, EPI_ISL_724812, EPI_ISL_724813, EPI_ISL_724814, EPI_ISL_724815, EPI_ISL_724816, EPI_ISL_724817, EPI_ISL_724818, EPI_ISL_724819, EPI_ISL_724820, EPI_ISL_724821, EPI_ISL_724822, EPI_ISL_724823, EPI_ISL_724824, EPI_ISL_724825, EPI_ISL_724826, EPI_ISL_724827, EPI_ISL_724828, EPI_ISL_724829, EPI_ISL_724830, EPI_ISL_724831, EPI_ISL_724832, EPI_ISL_724833, EPI_ISL_724834, EPI_ISL_724835, EPI_ISL_727807, EPI_ISL_727808, EPI_ISL_727809, EPI_ISL_727810, EPI_ISL_727811, EPI_ISL_727813, EPI_ISL_727814, EPI_ISL_727815, EPI_ISL_727816, EPI_ISL_727817, EPI_ISL_727818, EPI_ISL_727819, EPI_ISL_727820, EPI_ISL_727821, EPI_ISL_727822, EPI_ISL_727823, EPI_ISL_727824, EPI_ISL_727825, EPI_ISL_727826, EPI_ISL_727827, EPI_ISL_727828, EPI_ISL_727829, EPI_ISL_727830, EPI_ISL_727831, EPI_ISL_727832, EPI_ISL_727833, EPI_ISL_727834, EPI_ISL_727835, EPI_ISL_727836, EPI_ISL_727837, EPI_ISL_727838, EPI_ISL_727839, EPI_ISL_727840, EPI_ISL_727841, EPI_ISL_727842, EPI_ISL_727843, EPI_ISL_727844, EPI_ISL_727845, EPI_ISL_727846, EPI_ISL_727847, EPI_ISL_727848, EPI_ISL_727849, EPI_ISL_727850, EPI_ISL_727851, EPI_ISL_727852, EPI_ISL_727853, EPI_ISL_727854, EPI_ISL_727855, EPI_ISL_727856, EPI_ISL_727857, EPI_ISL_727858, EPI_ISL_727859, EPI_ISL_727860, EPI_ISL_740897, EPI_ISL_740898, EPI_ISL_740899, EPI_ISL_740900, EPI_ISL_740901, EPI_ISL_740902, EPI_ISL_740903, EPI_ISL_740904, EPI_ISL_740905, EPI_ISL_740906, EPI_ISL_740907, EPI_ISL_740908, EPI_ISL_740909, EPI_ISL_740910, EPI_ISL_740911, EPI_ISL_740912, EPI_ISL_740913, EPI_ISL_740914, EPI_ISL_740915, EPI_ISL_740916, EPI_ISL_740917, EPI_ISL_740918, EPI_ISL_740919, EPI_ISL_740920, EPI_ISL_740921, EPI_ISL_740922, EPI_ISL_740923, EPI_ISL_740924, EPI_ISL_740925, EPI_ISL_740926, EPI_ISL_740927, EPI_ISL_740928, EPI_ISL_740929, EPI_ISL_740930, EPI_ISL_740931, EPI_ISL_740932, EPI_ISL_740933, EPI_ISL_740934, EPI_ISL_740935, EPI_ISL_740936, EPI_ISL_740937, EPI_ISL_740938, EPI_ISL_740939, EPI_ISL_740940, EPI_ISL_740941, EPI_ISL_740942, EPI_ISL_740943, EPI_ISL_740944, EPI_ISL_740945, EPI_ISL_740946, EPI_ISL_740947, EPI_ISL_740948, EPI_ISL_740949, EPI_ISL_740950, EPI_ISL_740951, EPI_ISL_740952, EPI_ISL_741018, EPI_ISL_741019, EPI_ISL_741020, EPI_ISL_741021, EPI_ISL_741022, EPI_ISL_741023, EPI_ISL_741024, EPI_ISL_741025, EPI_ISL_741026, EPI_ISL_741027, EPI_ISL_741028, EPI_ISL_741029, EPI_ISL_741030, EPI_ISL_741031, EPI_ISL_741034, EPI_ISL_741035, EPI_ISL_741036, EPI_ISL_741037, EPI_ISL_741038, EPI_ISL_741039, EPI_ISL_741040, EPI_ISL_741041, EPI_ISL_741042, EPI_ISL_741043, EPI_ISL_741044, EPI_ISL_741045, EPI_ISL_741046, EPI_ISL_741047, EPI_ISL_741048, EPI_ISL_741049, EPI_ISL_741050, EPI_ISL_741051, EPI_ISL_741052, EPI_ISL_741053, EPI_ISL_741056, EPI_ISL_741057, EPI_ISL_741058, EPI_ISL_741059, EPI_ISL_741060, EPI_ISL_741061, EPI_ISL_741062, EPI_ISL_741063, EPI_ISL_741064, EPI_ISL_741065, EPI_ISL_741066, EPI_ISL_741067, EPI_ISL_741068, EPI_ISL_741069, EPI_ISL_741070, EPI_ISL_741071, EPI_ISL_741072, EPI_ISL_741073, EPI_ISL_741074, EPI_ISL_741075, EPI_ISL_741076, EPI_ISL_741077, EPI_ISL_741078, EPI_ISL_741079, EPI_ISL_741080, EPI_ISL_741081, EPI_ISL_741082, EPI_ISL_741083, EPI_ISL_741084, EPI_ISL_741086, EPI_ISL_741087, EPI_ISL_741088, EPI_ISL_741089, EPI_ISL_741090, EPI_ISL_741091, EPI_ISL_741092, EPI_ISL_741093, EPI_ISL_741094, EPI_ISL_741095, EPI_ISL_741096, EPI_ISL_741097, EPI_ISL_741098, EPI_ISL_741099, EPI_ISL_741100, EPI_ISL_741101, EPI_ISL_741102, EPI_ISL_741103, EPI_ISL_741104, EPI_ISL_741106, EPI_ISL_741111, EPI_ISL_741112, EPI_ISL_741113, EPI_ISL_741114, EPI_ISL_741115, EPI_ISL_741116, EPI_ISL_741117, EPI_ISL_741118, EPI_ISL_741119, EPI_ISL_741120, EPI_ISL_741121, EPI_ISL_741122, EPI_ISL_741123, EPI_ISL_741124, EPI_ISL_741125, EPI_ISL_741126, EPI_ISL_741127, EPI_ISL_741128, EPI_ISL_741129, EPI_ISL_741130, EPI_ISL_741131, EPI_ISL_741132, EPI_ISL_741133, EPI_ISL_741134, EPI_ISL_741135, EPI_ISL_741136, EPI_ISL_741137, EPI_ISL_741138, EPI_ISL_741139, EPI_ISL_741140, EPI_ISL_741141, EPI_ISL_741142, EPI_ISL_741143, EPI_ISL_741144, EPI_ISL_741145, EPI_ISL_741146, EPI_ISL_741147, EPI_ISL_741148, EPI_ISL_741149, EPI_ISL_741151, EPI_ISL_741152, EPI_ISL_741153, EPI_ISL_741154, EPI_ISL_741155, EPI_ISL_741156, EPI_ISL_741157, EPI_ISL_741158, EPI_ISL_741159, EPI_ISL_741160, EPI_ISL_741161, EPI_ISL_741162, EPI_ISL_741163, EPI_ISL_741164, EPI_ISL_741165, EPI_ISL_741166, EPI_ISL_741167, EPI_ISL_741168, EPI_ISL_741169, EPI_ISL_741170, EPI_ISL_741171, EPI_ISL_741172, EPI_ISL_741173, EPI_ISL_741174, EPI_ISL_741175, EPI_ISL_741176, EPI_ISL_741177, EPI_ISL_741178, EPI_ISL_741179, EPI_ISL_741180, EPI_ISL_741181, EPI_ISL_741182, EPI_ISL_741183, EPI_ISL_741184, EPI_ISL_741185, EPI_ISL_741777, EPI_ISL_741778, EPI_ISL_741779, EPI_ISL_741780, EPI_ISL_741781, EPI_ISL_741782, EPI_ISL_741783, EPI_ISL_741784, EPI_ISL_741785, EPI_ISL_741786, EPI_ISL_741787, EPI_ISL_741788, EPI_ISL_741789, EPI_ISL_741790, EPI_ISL_741791, EPI_ISL_741792, EPI_ISL_741793, EPI_ISL_741794, EPI_ISL_741795, EPI_ISL_741796, EPI_ISL_741797, EPI_ISL_741798, EPI_ISL_741799, EPI_ISL_741800, EPI_ISL_741801, EPI_ISL_741802, EPI_ISL_741803, EPI_ISL_741804, EPI_ISL_741805, EPI_ISL_741806, EPI_ISL_741807, EPI_ISL_741808, EPI_ISL_741809, EPI_ISL_741810, EPI_ISL_741811, EPI_ISL_741812, EPI_ISL_741813, EPI_ISL_741814, EPI_ISL_741815, EPI_ISL_741816, EPI_ISL_741817, EPI_ISL_741818, EPI_ISL_741819, EPI_ISL_741820, EPI_ISL_741821, EPI_ISL_741822, EPI_ISL_741823, EPI_ISL_741824, EPI_ISL_741825, EPI_ISL_741826, EPI_ISL_741827, EPI_ISL_741828, EPI_ISL_741829, EPI_ISL_741830, EPI_ISL_741831, EPI_ISL_741832, EPI_ISL_741833, EPI_ISL_741834, EPI_ISL_741835, EPI_ISL_741836, EPI_ISL_741837, EPI_ISL_741838, EPI_ISL_741839, EPI_ISL_741840, EPI_ISL_741841, EPI_ISL_741842, EPI_ISL_741843, EPI_ISL_741844, EPI_ISL_741846, EPI_ISL_741847, EPI_ISL_741848, EPI_ISL_741849, EPI_ISL_741850, EPI_ISL_741851, EPI_ISL_741852, EPI_ISL_741853, EPI_ISL_741854, EPI_ISL_741855, EPI_ISL_741856, EPI_ISL_741857, EPI_ISL_741858, EPI_ISL_741859, EPI_ISL_741860, EPI_ISL_741861, EPI_ISL_741862, EPI_ISL_741863, EPI_ISL_741864, EPI_ISL_741865, EPI_ISL_741866, EPI_ISL_741867, EPI_ISL_741868, EPI_ISL_741869, EPI_ISL_741870, EPI_ISL_741871, EPI_ISL_741872, EPI_ISL_741873, EPI_ISL_741874, EPI_ISL_741875, EPI_ISL_741876, EPI_ISL_741877, EPI_ISL_741878, EPI_ISL_741879, EPI_ISL_741880, EPI_ISL_741881, EPI_ISL_741882, EPI_ISL_741883, EPI_ISL_741884, EPI_ISL_741885, EPI_ISL_741886, EPI_ISL_741887, EPI_ISL_741888, EPI_ISL_741889, EPI_ISL_741890, EPI_ISL_741891, EPI_ISL_741892, EPI_ISL_741893, EPI_ISL_741894, EPI_ISL_741895, EPI_ISL_741896, EPI_ISL_741897, EPI_ISL_741898, EPI_ISL_741899, EPI_ISL_741900, EPI_ISL_741901, EPI_ISL_741902, EPI_ISL_741903, EPI_ISL_741904, EPI_ISL_741905, EPI_ISL_741906, EPI_ISL_741907, EPI_ISL_741908, EPI_ISL_741909, EPI_ISL_741910, EPI_ISL_741911, EPI_ISL_741912, EPI_ISL_741913, EPI_ISL_741914, EPI_ISL_741915, EPI_ISL_741916, EPI_ISL_741917, EPI_ISL_741918, EPI_ISL_741919, EPI_ISL_741920, EPI_ISL_741921, EPI_ISL_741922, EPI_ISL_741923, EPI_ISL_741924, EPI_ISL_741925, EPI_ISL_741926, EPI_ISL_741927, EPI_ISL_741928, EPI_ISL_741929, EPI_ISL_741930, EPI_ISL_741931, EPI_ISL_741932, EPI_ISL_741933, EPI_ISL_741934, EPI_ISL_741935, EPI_ISL_742247, EPI_ISL_742248, EPI_ISL_742249, EPI_ISL_742250, EPI_ISL_742251, EPI_ISL_742252, EPI_ISL_742253, EPI_ISL_742254, EPI_ISL_742255, EPI_ISL_742256, EPI_ISL_742257, EPI_ISL_742258, EPI_ISL_742259, EPI_ISL_840186, EPI_ISL_840187, EPI_ISL_840194, EPI_ISL_840195, EPI_ISL_840196, EPI_ISL_840197, EPI_ISL_840198, EPI_ISL_840199, EPI_ISL_840201, EPI_ISL_840228, EPI_ISL_840229, EPI_ISL_840230, EPI_ISL_840231, EPI_ISL_840232, EPI_ISL_840233, EPI_ISL_1833044

| | | | |
|---|---|---|---|
| see above | Oxford Viromics, NDM, University of Oxford; Oxford University Hospitals; Basingstoke and North Hampshire Hospital | COVID-19 Genomics UK (COG-UK) Consortium | Alex Mobbs; Amy Trebes; Anita Justice; Catrin Moore; Christophe Fraser; David Bonsall; David Buck; Emma Wise; George Macintyre; Jessica Lynch; John Todd; Mariateresa de Cesare; Matilde Mori; Monique Andersson; Nathan Moore; Nick Cortes; Robert Shaw; Stephen Kidd; Tanya Golubchik; Timothy Peto |

EPI_ISL_650170, EPI_ISL_650174, EPI_ISL_650193, EPI_ISL_650201, EPI_ISL_650231, EPI_ISL_650288, EPI_ISL_650289, EPI_ISL_650317, EPI_ISL_650385, EPI_ISL_650387, EPI_ISL_650454, EPI_ISL_650455, EPI_ISL_650459, EPI_ISL_650460, EPI_ISL_650462, EPI_ISL_650468, EPI_ISL_650469, EPI_ISL_650470, EPI_ISL_650472, EPI_ISL_650493, EPI_ISL_650502, EPI_ISL_650512, EPI_ISL_650530, EPI_ISL_650546, EPI_ISL_650548, EPI_ISL_650557, EPI_ISL_650677, EPI_ISL_650687, EPI_ISL_650696, EPI_ISL_650721, EPI_ISL_651111, EPI_ISL_651180, EPI_ISL_651218, EPI_ISL_651237, EPI_ISL_651246, EPI_ISL_651323, EPI_ISL_651399, EPI_ISL_651409, EPI_ISL_664110, EPI_ISL_664122, EPI_ISL_664135, EPI_ISL_664154, EPI_ISL_664158, EPI_ISL_664159, EPI_ISL_664169, EPI_ISL_664186, EPI_ISL_664222, EPI_ISL_664224, EPI_ISL_664226, EPI_ISL_664227, EPI_ISL_664235, EPI_ISL_664248, EPI_ISL_664290, EPI_ISL_664300, EPI_ISL_664334, EPI_ISL_664344, EPI_ISL_664347, EPI_ISL_664351, EPI_ISL_664357, EPI_ISL_664358, EPI_ISL_664366, EPI_ISL_664369, EPI_ISL_664441, EPI_ISL_664442, EPI_ISL_664450, EPI_ISL_664452, EPI_ISL_664460, EPI_ISL_664461, EPI_ISL_664462, EPI_ISL_664463, EPI_ISL_664464, EPI_ISL_664467, EPI_ISL_664476, EPI_ISL_664477, EPI_ISL_664482, EPI_ISL_664483, EPI_ISL_664484, EPI_ISL_664485, EPI_ISL_664488, EPI_ISL_664495, EPI_ISL_664502, EPI_ISL_664504, EPI_ISL_664506, EPI_ISL_664508, EPI_ISL_664515, EPI_ISL_664520, EPI_ISL_664522, EPI_ISL_664565, EPI_ISL_664566, EPI_ISL_664617, EPI_ISL_664628, EPI_ISL_664636, EPI_ISL_664638, EPI_ISL_664641, EPI_ISL_664642, EPI_ISL_664645, EPI_ISL_664657, EPI_ISL_664688, EPI_ISL_664704, EPI_ISL_664706, EPI_ISL_664714, EPI_ISL_664725, EPI_ISL_664735, EPI_ISL_664741, EPI_ISL_664743, EPI_ISL_664747, EPI_ISL_664756, EPI_ISL_664757, EPI_ISL_664758, EPI_ISL_664759, EPI_ISL_664760, EPI_ISL_664761, EPI_ISL_664764, EPI_ISL_664838, EPI_ISL_664857, EPI_ISL_664858, EPI_ISL_664859, EPI_ISL_664860, EPI_ISL_664861, EPI_ISL_664862, EPI_ISL_664863, EPI_ISL_664864, EPI_ISL_664865, EPI_ISL_664866, EPI_ISL_664867, EPI_ISL_664868, EPI_ISL_664869, EPI_ISL_664870, EPI_ISL_664871, EPI_ISL_664872, EPI_ISL_664873, EPI_ISL_664874, EPI_ISL_664875, EPI_ISL_664876, EPI_ISL_664877, EPI_ISL_664878, EPI_ISL_664879, EPI_ISL_664880, EPI_ISL_664881, EPI_ISL_664882, EPI_ISL_664883, EPI_ISL_664884, EPI_ISL_664885, EPI_ISL_664886, EPI_ISL_664887, EPI_ISL_664888, EPI_ISL_664889, EPI_ISL_664890, EPI_ISL_664891, EPI_ISL_664892, EPI_ISL_664893, EPI_ISL_664894, EPI_ISL_664895, EPI_ISL_664896, EPI_ISL_664897, EPI_ISL_664898, EPI_ISL_664899, EPI_ISL_664900, EPI_ISL_664901, EPI_ISL_664902, EPI_ISL_664905, EPI_ISL_664906, EPI_ISL_664907, EPI_ISL_664908, EPI_ISL_664909, EPI_ISL_664910, EPI_ISL_664911, EPI_ISL_664912, EPI_ISL_664913, EPI_ISL_664914, EPI_ISL_664915, EPI_ISL_664916, EPI_ISL_664917, EPI_ISL_664918, EPI_ISL_664919, EPI_ISL_664920, EPI_ISL_664921, EPI_ISL_664922, EPI_ISL_664923, EPI_ISL_664924, EPI_ISL_664925, EPI_ISL_664926, EPI_ISL_664927, EPI_ISL_664928, EPI_ISL_664929, EPI_ISL_664930, EPI_ISL_664931, EPI_ISL_664932, EPI_ISL_664933, EPI_ISL_664934,

| | | | |
|---|---|---|---|
| EPI_ISL_664935, EPI_ISL_664936, EPI_ISL_664937, EPI_ISL_664938, EPI_ISL_664939, EPI_ISL_664940, EPI_ISL_665370, EPI_ISL_665427, EPI_ISL_665438, EPI_ISL_665560, EPI_ISL_665638, EPI_ISL_665693, EPI_ISL_665746, EPI_ISL_678916, EPI_ISL_678917, EPI_ISL_678918, EPI_ISL_678919, EPI_ISL_723673, EPI_ISL_724125, EPI_ISL_724126, EPI_ISL_724188, EPI_ISL_725112, EPI_ISL_725114, EPI_ISL_725115, EPI_ISL_725118, EPI_ISL_725119, EPI_ISL_725120, EPI_ISL_725122, EPI_ISL_725123, EPI_ISL_725124, EPI_ISL_725125, EPI_ISL_725126, EPI_ISL_725127, EPI_ISL_725129, EPI_ISL_725131, EPI_ISL_725132, EPI_ISL_725133, EPI_ISL_725134, EPI_ISL_725135, EPI_ISL_725137, EPI_ISL_725164, EPI_ISL_725171, EPI_ISL_725175, EPI_ISL_725176, EPI_ISL_725177, EPI_ISL_725179, EPI_ISL_725180, EPI_ISL_725181, EPI_ISL_725182, EPI_ISL_725184, EPI_ISL_725195, EPI_ISL_725196, EPI_ISL_725197, EPI_ISL_725198, EPI_ISL_725202, EPI_ISL_725204, EPI_ISL_725211, EPI_ISL_725212, EPI_ISL_725214, EPI_ISL_725216, EPI_ISL_725218, EPI_ISL_725222, EPI_ISL_725227, EPI_ISL_725228, EPI_ISL_725230, EPI_ISL_725231, EPI_ISL_725233, EPI_ISL_725237, EPI_ISL_725238, EPI_ISL_725239, EPI_ISL_725242, EPI_ISL_725245, EPI_ISL_725247, EPI_ISL_725248, EPI_ISL_725249, EPI_ISL_725250, EPI_ISL_725254, EPI_ISL_725255, EPI_ISL_725256, EPI_ISL_725257, EPI_ISL_725258, EPI_ISL_725261, EPI_ISL_725263, EPI_ISL_725265, EPI_ISL_725268, EPI_ISL_725269, EPI_ISL_725272, EPI_ISL_725273, EPI_ISL_725276, EPI_ISL_725280, EPI_ISL_725283, EPI_ISL_725285, EPI_ISL_725286, EPI_ISL_725290, EPI_ISL_725291, EPI_ISL_725292, EPI_ISL_725293, EPI_ISL_725295, EPI_ISL_741529, EPI_ISL_741571, EPI_ISL_866805, EPI_ISL_866818, EPI_ISL_866826 | | | |
| see above | Quadram Institute Bioscience | COVID-19 Genomics UK (COG-UK) Consortium | Alexander J Trotter; Alison E. Mather; Alp Aydin; Ana P. Tedim; Anastasia Kolyva; Andrew Bell; Andrew J. Page; Claire Stuart; Dave J. Baker; Gemma L. Kay; John Wain; Justin O'Grady; Leonardo de Oliveira Martins; Lizzie Meadows; Maria Diaz; Mark Webber; Muhammed Yasir; Nabil-Fareed Alikhan; Ngozi Elumogo; Nicholas M. Thomson; Rachael Stanley; Rachel Gilroy; Reenesh Prakash; Samir Dervisevic; Samuel Bloomfield; Steven Rudder; Thanh Le-Viet |
| EPI_ISL_650178, EPI_ISL_650186, EPI_ISL_650187, EPI_ISL_650238, EPI_ISL_650266, EPI_ISL_650276, EPI_ISL_650291, EPI_ISL_650314, EPI_ISL_650430, EPI_ISL_650443, EPI_ISL_650450, EPI_ISL_650471, EPI_ISL_650599, EPI_ISL_650723, EPI_ISL_650767, EPI_ISL_650786, EPI_ISL_650803, EPI_ISL_650811, EPI_ISL_650820, EPI_ISL_650821, EPI_ISL_650830, EPI_ISL_650835, EPI_ISL_650979, EPI_ISL_650987, EPI_ISL_651065, EPI_ISL_651107, EPI_ISL_651166, EPI_ISL_651189, EPI_ISL_651260, EPI_ISL_651286, EPI_ISL_651308, EPI_ISL_651313, EPI_ISL_651389, EPI_ISL_651401, EPI_ISL_651408, EPI_ISL_652373, EPI_ISL_652374, EPI_ISL_652375, EPI_ISL_652376, EPI_ISL_652377, EPI_ISL_652378, EPI_ISL_652379, EPI_ISL_652380, EPI_ISL_652381, EPI_ISL_652382, EPI_ISL_652383, EPI_ISL_652384, EPI_ISL_652385, EPI_ISL_652386, EPI_ISL_652387, EPI_ISL_652388, EPI_ISL_652389, EPI_ISL_652390, EPI_ISL_652391, EPI_ISL_652392, EPI_ISL_652393, EPI_ISL_652394, EPI_ISL_652395, EPI_ISL_652396, EPI_ISL_652397, EPI_ISL_652398, EPI_ISL_652399, EPI_ISL_652400, EPI_ISL_652401, EPI_ISL_652402, EPI_ISL_652403, EPI_ISL_652404, EPI_ISL_652405, EPI_ISL_652406, EPI_ISL_652407, EPI_ISL_652408, EPI_ISL_652409, EPI_ISL_652410, EPI_ISL_652411, EPI_ISL_652412, EPI_ISL_652413, EPI_ISL_665420, EPI_ISL_665426, EPI_ISL_665443, EPI_ISL_665543, EPI_ISL_665550, EPI_ISL_665551, EPI_ISL_665561, EPI_ISL_665622, EPI_ISL_665900, EPI_ISL_665901, EPI_ISL_665902, EPI_ISL_665903, EPI_ISL_665904, EPI_ISL_665905, EPI_ISL_665942, EPI_ISL_678920, EPI_ISL_678921, EPI_ISL_678922, EPI_ISL_678923, EPI_ISL_678924, EPI_ISL_678925, EPI_ISL_678926, EPI_ISL_678927, EPI_ISL_678928, EPI_ISL_678929, EPI_ISL_678930, EPI_ISL_678931, EPI_ISL_678932, EPI_ISL_678933, EPI_ISL_678934, EPI_ISL_678935, EPI_ISL_678936, EPI_ISL_678937, EPI_ISL_678938, EPI_ISL_678939, EPI_ISL_678940, EPI_ISL_678941, EPI_ISL_678942, EPI_ISL_678943, EPI_ISL_678944, EPI_ISL_678945, EPI_ISL_678946, EPI_ISL_678947, EPI_ISL_678948, EPI_ISL_678949, EPI_ISL_678950, EPI_ISL_678951, EPI_ISL_678952, EPI_ISL_678953, EPI_ISL_678954, EPI_ISL_678955, EPI_ISL_678956, EPI_ISL_678957, EPI_ISL_678958, EPI_ISL_678959, EPI_ISL_678960, EPI_ISL_678961, EPI_ISL_678962, EPI_ISL_678963, EPI_ISL_678964, EPI_ISL_678965, EPI_ISL_678966, EPI_ISL_678967, EPI_ISL_678968, EPI_ISL_678969, EPI_ISL_678970, EPI_ISL_678971, EPI_ISL_678972, EPI_ISL_678973, EPI_ISL_678974, EPI_ISL_678975, EPI_ISL_678976, EPI_ISL_678977, EPI_ISL_678978, EPI_ISL_678979, EPI_ISL_678980, EPI_ISL_678981, EPI_ISL_678982, EPI_ISL_678983, EPI_ISL_678984, EPI_ISL_678985, EPI_ISL_678986, EPI_ISL_678987, EPI_ISL_678988, EPI_ISL_678989, EPI_ISL_678990, EPI_ISL_678991, EPI_ISL_678992, EPI_ISL_678993, EPI_ISL_678994, EPI_ISL_678995, EPI_ISL_678996, EPI_ISL_678997, EPI_ISL_678998, EPI_ISL_678999, EPI_ISL_679000, EPI_ISL_679001, EPI_ISL_679002, EPI_ISL_679003, EPI_ISL_679004, EPI_ISL_679005, EPI_ISL_679006, EPI_ISL_679007, EPI_ISL_679308, EPI_ISL_838343, EPI_ISL_838349, EPI_ISL_839986 | | | |
| see above | Queens Medical Centre, Clinical Microbiology Department / DeepSeq Nottingham | COVID-19 Genomics UK (COG-UK) Consortium | Christopher Moore; Fei Sang; Gemma Clark; Hannah Howson-Wells; Johnny Debebe; Jonathan Ball; Joseph Chappell; Manjinder Khakh; Matthew Carlisle; Matthew Loose; Michelle M Lister; Nadine Holmes; Patrick McClure; Theocharis Tsoleridis; Vicki M Fleming; Victoria Wright; Wendy Smith |
| EPI_ISL_742129, EPI_ISL_742130, EPI_ISL_742131, EPI_ISL_742132, EPI_ISL_742133, EPI_ISL_742134, EPI_ISL_742135, EPI_ISL_742136, EPI_ISL_742137, EPI_ISL_742138, EPI_ISL_742139, EPI_ISL_763463, EPI_ISL_764434, EPI_ISL_764436, EPI_ISL_764437, EPI_ISL_764438, EPI_ISL_764439 | | | |
| see above | Regional Virus Laboratory, Belfast Health and Social Care Trust | COVID-19 Genomics UK (COG-UK) Consortium | Alison Watt; Ciara Cox; Conall McCaughey; David Simpson; Derek Fairley; James McKenna; Mairead Connor; Susan Feeney; Tanya Curran; Zoltan Molnar |
| EPI_ISL_644162, EPI_ISL_645133, EPI_ISL_645134, EPI_ISL_645135, EPI_ISL_645136, EPI_ISL_649108, EPI_ISL_649110, EPI_ISL_649889, EPI_ISL_649891, EPI_ISL_649892, EPI_ISL_649936, EPI_ISL_649937, EPI_ISL_666768, EPI_ISL_668442, EPI_ISL_668443, EPI_ISL_678706, EPI_ISL_678707, EPI_ISL_678708, EPI_ISL_678709, EPI_ISL_678710, EPI_ISL_678711, EPI_ISL_678712, EPI_ISL_678713, EPI_ISL_678714, EPI_ISL_678715, EPI_ISL_678716, EPI_ISL_678717, EPI_ISL_678718, EPI_ISL_678719, EPI_ISL_678720, EPI_ISL_678721, EPI_ISL_678722, EPI_ISL_678723, EPI_ISL_678724, EPI_ISL_678725, EPI_ISL_678726, EPI_ISL_678728, EPI_ISL_678741, EPI_ISL_678760, EPI_ISL_683457, EPI_ISL_683461, EPI_ISL_683479, EPI_ISL_683480, EPI_ISL_683485, EPI_ISL_683486, EPI_ISL_683487, EPI_ISL_683488, EPI_ISL_683519, EPI_ISL_683520, EPI_ISL_683521, EPI_ISL_683522, EPI_ISL_683524, EPI_ISL_683525, EPI_ISL_683526, EPI_ISL_683527, EPI_ISL_683528, EPI_ISL_683529, EPI_ISL_683530, EPI_ISL_683531, EPI_ISL_683532, EPI_ISL_683533, EPI_ISL_683534, EPI_ISL_683535, EPI_ISL_683536, EPI_ISL_683537, EPI_ISL_683538, EPI_ISL_683539, EPI_ISL_683540, EPI_ISL_683541, EPI_ISL_683542, EPI_ISL_683543, EPI_ISL_683544, EPI_ISL_683545, EPI_ISL_686580, EPI_ISL_686581, EPI_ISL_686582, EPI_ISL_686583, EPI_ISL_686584, EPI_ISL_686585, EPI_ISL_686586, EPI_ISL_686587, EPI_ISL_686588, EPI_ISL_686589, EPI_ISL_686590, EPI_ISL_686591, EPI_ISL_686592, EPI_ISL_686593, EPI_ISL_686594, EPI_ISL_686595, EPI_ISL_686596, EPI_ISL_686598, EPI_ISL_686599, EPI_ISL_686600, EPI_ISL_686601, EPI_ISL_686602, EPI_ISL_686603, EPI_ISL_686604, EPI_ISL_686605, EPI_ISL_686606, EPI_ISL_686607, EPI_ISL_686608, EPI_ISL_686609, EPI_ISL_686610, EPI_ISL_693392, EPI_ISL_693394, EPI_ISL_693395, EPI_ISL_693396, EPI_ISL_693397, EPI_ISL_693398, EPI_ISL_693399, EPI_ISL_693401, EPI_ISL_693402, EPI_ISL_693403, EPI_ISL_693404, EPI_ISL_693405, EPI_ISL_693407, EPI_ISL_693412, EPI_ISL_693413, EPI_ISL_693416, EPI_ISL_693417, EPI_ISL_693418, EPI_ISL_693421, EPI_ISL_693423, EPI_ISL_693424, EPI_ISL_693425, EPI_ISL_693426, EPI_ISL_693427, EPI_ISL_693428, EPI_ISL_693429, EPI_ISL_693430, EPI_ISL_693431, EPI_ISL_693432, EPI_ISL_693434, EPI_ISL_693435, EPI_ISL_693436, EPI_ISL_693437, EPI_ISL_693438, EPI_ISL_693439, EPI_ISL_693440, EPI_ISL_693441, EPI_ISL_693442, EPI_ISL_693443, EPI_ISL_693444, EPI_ISL_693445, EPI_ISL_693446, EPI_ISL_693447, EPI_ISL_693448, EPI_ISL_693449, EPI_ISL_693450, EPI_ISL_693451, EPI_ISL_693452, EPI_ISL_693453, EPI_ISL_693454, EPI_ISL_693455, EPI_ISL_693456, EPI_ISL_693457, EPI_ISL_693458, EPI_ISL_693459, EPI_ISL_693460, EPI_ISL_693461, EPI_ISL_693462, EPI_ISL_693463, EPI_ISL_693464, EPI_ISL_693465, EPI_ISL_693466, EPI_ISL_693467, EPI_ISL_693469, EPI_ISL_693470, EPI_ISL_693484, EPI_ISL_693486, EPI_ISL_693487 | | | |
| see above | Respiratory Virus Unit, Microbiology Services Colindale, Public Health England | COVID-19 Genomics UK (COG-UK) Consortium | PHE Covid Sequencing Team |
| EPI_ISL_636974, EPI_ISL_637097 | Respiratory Virus Unit, Microbiology Services Colindale, Public Health England | Respiratory Virus Unit, Microbiology Services Colindale, Public Health England | PHE Covid Sequencing Team |
| EPI_ISL_766076, EPI_ISL_766077, EPI_ISL_766094, EPI_ISL_766098, EPI_ISL_766267, EPI_ISL_766268, EPI_ISL_766269, EPI_ISL_766270, EPI_ISL_766271, EPI_ISL_766272, EPI_ISL_766273, EPI_ISL_769864, EPI_ISL_769865, EPI_ISL_825597, EPI_ISL_825598, EPI_ISL_825599, EPI_ISL_825600, EPI_ISL_825601, EPI_ISL_825602, EPI_ISL_825603, EPI_ISL_825604 | | | |
| see above | Respiratory Virus Unit, National Infection Service, Public Health England | COVID-19 Genomics UK (COG-UK) Consortium | PHE Covid Sequencing Team |
| EPI_ISL_665287, EPI_ISL_665320, EPI_ISL_665334, EPI_ISL_665525, EPI_ISL_665559, EPI_ISL_665580, EPI_ISL_665611, EPI_ISL_665632, EPI_ISL_665636, EPI_ISL_665637, EPI_ISL_665643, EPI_ISL_665645, EPI_ISL_665648, EPI_ISL_665649, EPI_ISL_665757, EPI_ISL_665758, EPI_ISL_665786, EPI_ISL_665816, EPI_ISL_665817, EPI_ISL_665818, EPI_ISL_665819, EPI_ISL_665820, EPI_ISL_665821, EPI_ISL_665824, EPI_ISL_665825, EPI_ISL_665831, EPI_ISL_665832, EPI_ISL_665833, EPI_ISL_665834, EPI_ISL_665835, EPI_ISL_665836, EPI_ISL_665837, EPI_ISL_665838, EPI_ISL_665839, EPI_ISL_665840, EPI_ISL_665841, EPI_ISL_665842, EPI_ISL_665843, EPI_ISL_665844, EPI_ISL_665845, EPI_ISL_665846, EPI_ISL_665847, EPI_ISL_665848, EPI_ISL_679430, EPI_ISL_679431, EPI_ISL_679434, EPI_ISL_705949, EPI_ISL_866053, EPI_ISL_866054, EPI_ISL_920268, EPI_ISL_920278, EPI_ISL_920376, EPI_ISL_920577, EPI_ISL_920586, EPI_ISL_1104841, EPI_ISL_1104842, EPI_ISL_1104854, EPI_ISL_1104869, EPI_ISL_1104877, EPI_ISL_1104885, EPI_ISL_1104896, EPI_ISL_1104897, EPI_ISL_1104905, EPI_ISL_1104906, EPI_ISL_1104907, EPI_ISL_1104915, EPI_ISL_1104929, EPI_ISL_1104948, EPI_ISL_1104968, EPI_ISL_1105083 | | | |
| see above | University College London Hospital | COVID-19 Genomics UK (COG-UK) Consortium | Catherine Houlihan; Dan Frampton; Judith Heaney; Matthew Byott; Moira Spyer and Eleni Nastouli; Stuart Kirk |
| EPI_ISL_679501, EPI_ISL_679503, EPI_ISL_679504, EPI_ISL_679506, EPI_ISL_679507, EPI_ISL_679508, EPI_ISL_679509, EPI_ISL_679510, EPI_ISL_679511, EPI_ISL_679512, EPI_ISL_679513, EPI_ISL_679514, EPI_ISL_679515, EPI_ISL_679516, EPI_ISL_679517, EPI_ISL_679518, EPI_ISL_679519, EPI_ISL_679520, EPI_ISL_679521, EPI_ISL_679522, EPI_ISL_679523, EPI_ISL_679524, EPI_ISL_679525, EPI_ISL_679526, EPI_ISL_679527, EPI_ISL_679528, EPI_ISL_679529, EPI_ISL_679530, EPI_ISL_679531, EPI_ISL_679532, EPI_ISL_679533, EPI_ISL_679534, EPI_ISL_679535, EPI_ISL_679536, EPI_ISL_679537, EPI_ISL_679538, EPI_ISL_679539, EPI_ISL_679540, EPI_ISL_679541, EPI_ISL_679542, EPI_ISL_679543, EPI_ISL_679544, EPI_ISL_679545, EPI_ISL_679546, EPI_ISL_679547, EPI_ISL_679548, EPI_ISL_679575, EPI_ISL_679576, EPI_ISL_679577, EPI_ISL_679578, EPI_ISL_679579, EPI_ISL_679580, EPI_ISL_679584, EPI_ISL_679587, EPI_ISL_679624, EPI_ISL_679625, EPI_ISL_679626, EPI_ISL_702930, EPI_ISL_703319, EPI_ISL_703335, EPI_ISL_703383, EPI_ISL_703706, EPI_ISL_703739, EPI_ISL_703767, EPI_ISL_703879, EPI_ISL_704086, EPI_ISL_704092, EPI_ISL_704312, EPI_ISL_704372, EPI_ISL_705018, EPI_ISL_705143, EPI_ISL_705368, EPI_ISL_705397, EPI_ISL_705961, EPI_ISL_705962, EPI_ISL_705964, EPI_ISL_705965, EPI_ISL_705966, EPI_ISL_705969, EPI_ISL_705971, EPI_ISL_724610, EPI_ISL_724612, EPI_ISL_724615, EPI_ISL_724616, EPI_ISL_724618, EPI_ISL_724619, EPI_ISL_724621, EPI_ISL_724622, EPI_ISL_724623, EPI_ISL_724624, EPI_ISL_724627, EPI_ISL_724634, EPI_ISL_724640, EPI_ISL_724641, EPI_ISL_724643, EPI_ISL_724644, EPI_ISL_724645, EPI_ISL_724646, EPI_ISL_724647, EPI_ISL_724653, EPI_ISL_724654, EPI_ISL_724657, EPI_ISL_724663, EPI_ISL_724667, EPI_ISL_724668, EPI_ISL_724669, EPI_ISL_724670, EPI_ISL_724671, EPI_ISL_724672, EPI_ISL_724676, EPI_ISL_724688, EPI_ISL_724696, EPI_ISL_724700, EPI_ISL_724701, EPI_ISL_724702, EPI_ISL_724703, EPI_ISL_724704, EPI_ISL_724706, EPI_ISL_724711, EPI_ISL_724712, EPI_ISL_724714, EPI_ISL_724721, EPI_ISL_724726, EPI_ISL_724727, EPI_ISL_724728, EPI_ISL_724729, EPI_ISL_724730, EPI_ISL_724731, EPI_ISL_724732, EPI_ISL_724733, EPI_ISL_724741, EPI_ISL_724742, EPI_ISL_724755, EPI_ISL_724757, EPI_ISL_724760, EPI_ISL_724761, EPI_ISL_724762, EPI_ISL_724763, EPI_ISL_724764, EPI_ISL_724765, EPI_ISL_724766, EPI_ISL_724767, EPI_ISL_724768, EPI_ISL_724769, EPI_ISL_724770, EPI_ISL_724771, EPI_ISL_724772, EPI_ISL_724773, EPI_ISL_724774, EPI_ISL_724775, EPI_ISL_865683, EPI_ISL_865685, EPI_ISL_865695, EPI_ISL_865696, EPI_ISL_865697, EPI_ISL_865698, EPI_ISL_1637150, EPI_ISL_1637151, EPI_ISL_1637154 | | | |
| see above | University College London, Great Ormond Street Hospital for Children NHS Foundation Trust, Imperial College Healthcare NHS Trust | COVID-19 Genomics UK (COG-UK) Consortium | Alison Holmes; Charlotte Williams; Helena Tutill; Jacqueline Findlay; James Price; Judith Breuer; Julianne Brown; Kathryn Harris; Leysa Forrest; Mark Kristiansen; Paola Niola; Paola Resende Silva; Patricia Dyal; Paul Randell; Rachel Williams; Samuel Weeks; Sergi Castellano; Sunando Roy; Tony Brooks; Yasmin Panchbhaya |
| EPI_ISL_679104, EPI_ISL_679105, EPI_ISL_679106, EPI_ISL_679107, EPI_ISL_679108, EPI_ISL_679109, EPI_ISL_679110, EPI_ISL_679111, EPI_ISL_679112, EPI_ISL_679113, EPI_ISL_679114, EPI_ISL_679115, EPI_ISL_679116, EPI_ISL_679117, EPI_ISL_679118, EPI_ISL_679119, EPI_ISL_679120, EPI_ISL_679121, EPI_ISL_679122, EPI_ISL_679123, EPI_ISL_679124, EPI_ISL_679125, EPI_ISL_679126, EPI_ISL_679127, EPI_ISL_679128, EPI_ISL_813023, EPI_ISL_813025, EPI_ISL_813026, EPI_ISL_813027, EPI_ISL_813028, EPI_ISL_918564, EPI_ISL_918665, EPI_ISL_949335, EPI_ISL_949336, EPI_ISL_949343, EPI_ISL_949344, EPI_ISL_949347, EPI_ISL_949348, EPI_ISL_949349, EPI_ISL_949350, EPI_ISL_949351, EPI_ISL_949352, EPI_ISL_949353, EPI_ISL_949354, EPI_ISL_949355, EPI_ISL_949356, EPI_ISL_949357, EPI_ISL_949358, EPI_ISL_949359, EPI_ISL_949360, EPI_ISL_949361, EPI_ISL_949362, EPI_ISL_949363, EPI_ISL_949364, EPI_ISL_949365, EPI_ISL_949366, EPI_ISL_949367, EPI_ISL_949368, EPI_ISL_949369, EPI_ISL_949370, EPI_ISL_949371, EPI_ISL_949372, EPI_ISL_949373, EPI_ISL_949374, EPI_ISL_949375, EPI_ISL_949376, EPI_ISL_949377, EPI_ISL_949378, EPI_ISL_949379, EPI_ISL_949380, EPI_ISL_949381, EPI_ISL_949382, EPI_ISL_949383, EPI_ISL_949384, EPI_ISL_949385, EPI_ISL_949386, EPI_ISL_949387, EPI_ISL_949388, EPI_ISL_949389, EPI_ISL_949390, EPI_ISL_1474495 | | | |
| see above | University of Birmingham | COVID-19 Genomics UK (COG-UK) Consortium | Alex Richter; Andrew Bosworth; Andrew Bosworth. Queen Elizabeth Hospital: Anna Casey; Andrew D Beggs PHE Heartlands Lab: Husam Osman; Charlotte Poxon; Institute of Microbiology; Joanne Stockton; Josh Quick; Kasun Wanigasooriya; Mike Kidd; Nicholas Loman. University of Birmingham Testing Laboratory: Celina M Whalley; Oliver Pickles; Radoslaw Poplawski; Samuel Nicholls; University of Birmingham: Claire McMurray; Will Rowe |
| EPI_ISL_664276, EPI_ISL_664609, EPI_ISL_665112, EPI_ISL_997313, EPI_ISL_997314, EPI_ISL_997320, EPI_ISL_997321, EPI_ISL_1247537, EPI_ISL_1247540, EPI_ISL_1247542, EPI_ISL_1247545, EPI_ISL_1247546 | | | |
| see above | University of Exeter | COVID-19 Genomics UK (COG-UK) Consortium | Aaron Jeffries; Audrey Farbos; Ben Temperton; Jane Masoli; Joanna Warwick-Dugdale; Michelle Michelsen; Robyn Manley; Stephen Michell |
| EPI_ISL_680539, EPI_ISL_680540, EPI_ISL_680541, EPI_ISL_680542, EPI_ISL_680543, EPI_ISL_680548, EPI_ISL_680549, EPI_ISL_680550, EPI_ISL_680551, EPI_ISL_680552, EPI_ISL_680553, EPI_ISL_680554, EPI_ISL_703810, EPI_ISL_705768, EPI_ISL_705769, EPI_ISL_705803, EPI_ISL_742244, EPI_ISL_814534, EPI_ISL_814571, EPI_ISL_814678 | | | |
| see above | Virology Department, Royal Infirmary of Edinburgh, NHS Lothian / School of Biological Sciences, University of Edinburgh / Institute of Genetics and Molecular Medicine, University of Edinburgh | COVID-19 Genomics UK (COG-UK) Consortium | Balcaza C; Colquhoun R; Dewar R; Gallagher M; Hill V; Jackson B; McCrone JT; McHugh M; O'Toole Á; Rambaut A; Rooke S; Scher E; Templeton K; Williams TC; Yu X |
| EPI_ISL_680102, EPI_ISL_680103, EPI_ISL_680104, EPI_ISL_680105, EPI_ISL_680106, EPI_ISL_680107, EPI_ISL_680108, EPI_ISL_680116, EPI_ISL_680119, EPI_ISL_680124, EPI_ISL_680125, EPI_ISL_680132, EPI_ISL_680138, EPI_ISL_680140, EPI_ISL_680143, EPI_ISL_680151, EPI_ISL_680153, EPI_ISL_680157, EPI_ISL_680161, EPI_ISL_680164, EPI_ISL_680167, EPI_ISL_680173, EPI_ISL_680174, EPI_ISL_680175, EPI_ISL_680180, EPI_ISL_680185, EPI_ISL_680193, EPI_ISL_680196, EPI_ISL_705276, EPI_ISL_705374, EPI_ISL_706988, EPI_ISL_706991, EPI_ISL_706994, EPI_ISL_727991, EPI_ISL_816453, EPI_ISL_842204 | | | |
| see above | Virology Department, Sheffield Teaching Hospitals NHS Foundation Trust/Department of Infection, Immunity and Cardiovascular Disease, The Medical School, University of Sheffield | COVID-19 Genomics UK (COG-UK) Consortium | Adri Angyal; Alex Keeley; Benjamin Lindsey; Cariad Evans; Danielle Groves; Dave Partridge; Katie Johnson; Laura Carrilero; Luke Green; Matthew Parker; Matthew Wyles; Mehmet Yavuz; Mohammad Raza; Nikki Smith; Paul Parsons; Rachel Tucker; Rebecca Brown; Thushan de Silva |
| EPI_ISL_680591, EPI_ISL_680592, EPI_ISL_680593, EPI_ISL_680594, EPI_ISL_680595, EPI_ISL_680596, EPI_ISL_680597, EPI_ISL_680598, EPI_ISL_680599, EPI_ISL_680600, EPI_ISL_680601, EPI_ISL_680602, EPI_ISL_680603, EPI_ISL_680604, EPI_ISL_680605, EPI_ISL_680607, EPI_ISL_680608, EPI_ISL_680609, EPI_ISL_680610, EPI_ISL_680640, EPI_ISL_680641, EPI_ISL_680642, EPI_ISL_680643, EPI_ISL_680644, EPI_ISL_680645, EPI_ISL_680646, EPI_ISL_680647, EPI_ISL_680648, EPI_ISL_680649, EPI_ISL_680650, EPI_ISL_680651, EPI_ISL_680652, EPI_ISL_680653, EPI_ISL_680654, EPI_ISL_680655, EPI_ISL_680656, EPI_ISL_680657, EPI_ISL_680658, EPI_ISL_680659, EPI_ISL_680660, EPI_ISL_680661, EPI_ISL_680662, EPI_ISL_680663, EPI_ISL_680664, EPI_ISL_680665, EPI_ISL_680666, EPI_ISL_680667, EPI_ISL_680668, EPI_ISL_680669, EPI_ISL_680670, EPI_ISL_680681, EPI_ISL_680682, EPI_ISL_680686, EPI_ISL_680687, EPI_ISL_680688, EPI_ISL_680699, EPI_ISL_680700, EPI_ISL_680702, EPI_ISL_680703, EPI_ISL_680704, EPI_ISL_680705, EPI_ISL_680706, EPI_ISL_680707, EPI_ISL_680708, EPI_ISL_680709, EPI_ISL_680710, EPI_ISL_680711, EPI_ISL_680712, EPI_ISL_680747, EPI_ISL_680820, EPI_ISL_680821, EPI_ISL_680839, EPI_ISL_680848, EPI_ISL_680849, EPI_ISL_680850, EPI_ISL_680851, EPI_ISL_680856, EPI_ISL_680858, EPI_ISL_680859, EPI_ISL_680860, EPI_ISL_680861, EPI_ISL_680862, EPI_ISL_680863, EPI_ISL_680864, EPI_ISL_680865, EPI_ISL_680866, EPI_ISL_680867, EPI_ISL_680868, EPI_ISL_680869, EPI_ISL_680870, EPI_ISL_680871, EPI_ISL_680872, EPI_ISL_680873, EPI_ISL_680874, EPI_ISL_680875, EPI_ISL_680876, EPI_ISL_680877, EPI_ISL_680878, EPI_ISL_680879, EPI_ISL_680880, EPI_ISL_680881, EPI_ISL_680882, EPI_ISL_680883, EPI_ISL_680884, EPI_ISL_680885, EPI_ISL_680886, EPI_ISL_680887, EPI_ISL_680888, EPI_ISL_680889, EPI_ISL_680890, EPI_ISL_680891, EPI_ISL_680892, EPI_ISL_680893, EPI_ISL_680894, EPI_ISL_680895, EPI_ISL_680896, EPI_ISL_680897, EPI_ISL_680898, EPI_ISL_680899, EPI_ISL_680900, EPI_ISL_680901, EPI_ISL_680902, EPI_ISL_680903, EPI_ISL_680904, EPI_ISL_680905, EPI_ISL_680906, EPI_ISL_680907, EPI_ISL_680908, EPI_ISL_680909, EPI_ISL_680910, EPI_ISL_680911, EPI_ISL_680912, EPI_ISL_680913, EPI_ISL_680914, EPI_ISL_680915, EPI_ISL_680916, EPI_ISL_680917, EPI_ISL_680918, EPI_ISL_680919, EPI_ISL_680920, EPI_ISL_680921, EPI_ISL_680922, EPI_ISL_680923, EPI_ISL_680924, EPI_ISL_680925, EPI_ISL_680926, EPI_ISL_680927, EPI_ISL_680928, EPI_ISL_680929, EPI_ISL_680930, EPI_ISL_680931, EPI_ISL_680932, EPI_ISL_680933, EPI_ISL_680934, EPI_ISL_680935, EPI_ISL_680936, EPI_ISL_680937, EPI_ISL_680938, EPI_ISL_680939, EPI_ISL_680940, EPI_ISL_680941, EPI_ISL_680942, EPI_ISL_680943, EPI_ISL_680944, EPI_ISL_680945, EPI_ISL_680946, EPI_ISL_680947, EPI_ISL_680948, EPI_ISL_680949, EPI_ISL_680950, EPI_ISL_680951, EPI_ISL_680952, EPI_ISL_681013, EPI_ISL_681014, EPI_ISL_681015, EPI_ISL_681016, EPI_ISL_681017, EPI_ISL_681019, EPI_ISL_681020, EPI_ISL_681246, EPI_ISL_681256, EPI_ISL_681257, EPI_ISL_681258, EPI_ISL_681259, EPI_ISL_702553, EPI_ISL_702692, EPI_ISL_702742, EPI_ISL_702817, EPI_ISL_702857, EPI_ISL_702859, EPI_ISL_702863, EPI_ISL_702864, EPI_ISL_702895, EPI_ISL_702905, EPI_ISL_703066, EPI_ISL_703139, EPI_ISL_703203, EPI_ISL_703251, EPI_ISL_703259, EPI_ISL_703278, EPI_ISL_703302, EPI_ISL_703321, EPI_ISL_703330, EPI_ISL_703343, EPI_ISL_703351, EPI_ISL_703359, EPI_ISL_703392, EPI_ISL_703395, EPI_ISL_703398, EPI_ISL_703614, EPI_ISL_703721, EPI_ISL_703722, EPI_ISL_703760, EPI_ISL_703768, EPI_ISL_703858, EPI_ISL_703860, EPI_ISL_703870, EPI_ISL_703873, EPI_ISL_703899, EPI_ISL_703918, EPI_ISL_703940, EPI_ISL_703942, EPI_ISL_703987, EPI_ISL_704014, EPI_ISL_704030, EPI_ISL_704038, EPI_ISL_704044, EPI_ISL_704081, EPI_ISL_704095, EPI_ISL_704112, EPI_ISL_704126, EPI_ISL_704161, EPI_ISL_704317, EPI_ISL_704931, EPI_ISL_704981, EPI_ISL_705239, EPI_ISL_705306, EPI_ISL_705327, EPI_ISL_705366, EPI_ISL_705384, EPI_ISL_705412, EPI_ISL_705413, EPI_ISL_705420, EPI_ISL_705434, EPI_ISL_705439, EPI_ISL_705440, EPI_ISL_705442, EPI_ISL_705445, EPI_ISL_706439, EPI_ISL_706440, EPI_ISL_706441, EPI_ISL_706444, EPI_ISL_706446, EPI_ISL_706447, EPI_ISL_706448, EPI_ISL_706449, EPI_ISL_706451, EPI_ISL_706452, EPI_ISL_706453, EPI_ISL_706454, EPI_ISL_706455, EPI_ISL_706456, EPI_ISL_706457, EPI_ISL_706458, EPI_ISL_706492, EPI_ISL_706494, EPI_ISL_706497, EPI_ISL_706498, EPI_ISL_706499, EPI_ISL_706500, EPI_ISL_706501, EPI_ISL_706502, EPI_ISL_706503, EPI_ISL_706504, EPI_ISL_706505, EPI_ISL_706506, EPI_ISL_706507, EPI_ISL_706508, EPI_ISL_706509, EPI_ISL_706510, EPI_ISL_706511, EPI_ISL_706512, EPI_ISL_706513, EPI_ISL_706514, EPI_ISL_706515, EPI_ISL_706516, EPI_ISL_706517, EPI_ISL_706518, EPI_ISL_706519, EPI_ISL_706520, EPI_ISL_706521, EPI_ISL_706522, EPI_ISL_706523, EPI_ISL_706524, EPI_ISL_706525, EPI_ISL_706526, EPI_ISL_706527, EPI_ISL_706528, EPI_ISL_706529, EPI_ISL_706530, EPI_ISL_706531, EPI_ISL_706532, EPI_ISL_706533, EPI_ISL_706534, EPI_ISL_706535, EPI_ISL_706536, EPI_ISL_706537, EPI_ISL_706538, EPI_ISL_706539, EPI_ISL_706540, EPI_ISL_706541, EPI_ISL_706542, EPI_ISL_706543, EPI_ISL_706544, EPI_ISL_706545, EPI_ISL_706546, EPI_ISL_706547, EPI_ISL_706548, EPI_ISL_706549, EPI_ISL_706550, EPI_ISL_706551, EPI_ISL_706552, EPI_ISL_706553, EPI_ISL_706554, EPI_ISL_706555, EPI_ISL_706556, EPI_ISL_706557, EPI_ISL_706558, EPI_ISL_706559, EPI_ISL_706560, EPI_ISL_706561, EPI_ISL_706562, EPI_ISL_706563, EPI_ISL_706564, EPI_ISL_706565, EPI_ISL_706566, EPI_ISL_706567, EPI_ISL_706568, EPI_ISL_706569, EPI_ISL_706570, EPI_ISL_706571, EPI_ISL_706572, EPI_ISL_706573, EPI_ISL_706574, EPI_ISL_706575, EPI_ISL_706596, EPI_ISL_706597, EPI_ISL_706598, EPI_ISL_706599, EPI_ISL_706600, EPI_ISL_706601, EPI_ISL_706607, EPI_ISL_706608, EPI_ISL_706609, EPI_ISL_706613, EPI_ISL_706619, EPI_ISL_706620, EPI_ISL_706621, EPI_ISL_706622, EPI_ISL_706623, EPI_ISL_706624, EPI_ISL_706625, EPI_ISL_706626, EPI_ISL_706627, EPI_ISL_706628, EPI_ISL_706629, EPI_ISL_706630, EPI_ISL_706631, EPI_ISL_706632, EPI_ISL_706633, EPI_ISL_706634, EPI_ISL_706635, EPI_ISL_706638, EPI_ISL_706639, EPI_ISL_706640, EPI_ISL_706641, EPI_ISL_706642, EPI_ISL_706643, EPI_ISL_706644, EPI_ISL_706645, EPI_ISL_706648, EPI_ISL_706649, EPI_ISL_706650, EPI_ISL_706651, EPI_ISL_706658, EPI_ISL_706662, EPI_ISL_706764, EPI_ISL_706765, EPI_ISL_706766, EPI_ISL_706767, EPI_ISL_706768, EPI_ISL_706769, EPI_ISL_706770, EPI_ISL_706771, EPI_ISL_706772, EPI_ISL_706773, EPI_ISL_706774, EPI_ISL_706775, EPI_ISL_706776, EPI_ISL_706777, EPI_ISL_706778, EPI_ISL_706779, EPI_ISL_706780, EPI_ISL_706781, EPI_ISL_706782, EPI_ISL_706783, EPI_ISL_706784, EPI_ISL_706785, EPI_ISL_706786, EPI_ISL_706787, EPI_ISL_706788, EPI_ISL_706789, EPI_ISL_706790, EPI_ISL_706791, EPI_ISL_706792, EPI_ISL_706793, EPI_ISL_706794, EPI_ISL_706795, EPI_ISL_706796, EPI_ISL_706797, EPI_ISL_706798, EPI_ISL_706799, EPI_ISL_706800, EPI_ISL_706801, EPI_ISL_706802, EPI_ISL_706803, EPI_ISL_706804, EPI_ISL_706805, EPI_ISL_706806, EPI_ISL_706807, EPI_ISL_706808, EPI_ISL_706809, EPI_ISL_706810, EPI_ISL_706811, EPI_ISL_706812, EPI_ISL_706813, EPI_ISL_706814, EPI_ISL_706815, EPI_ISL_706816, EPI_ISL_706817, EPI_ISL_706818, EPI_ISL_706819, EPI_ISL_706820, EPI_ISL_706821, EPI_ISL_706822, EPI_ISL_706823, EPI_ISL_706824, EPI_ISL_706825, EPI_ISL_706826, EPI_ISL_706827, EPI_ISL_706828, EPI_ISL_706829, EPI_ISL_706830, EPI_ISL_706831, EPI_ISL_706832, EPI_ISL_706833, EPI_ISL_706834, EPI_ISL_706835, EPI_ISL_706836, EPI_ISL_706837, EPI_ISL_706838, EPI_ISL_706840, EPI_ISL_706841, EPI_ISL_706842, EPI_ISL_706843, EPI_ISL_706844, EPI_ISL_706845, EPI_ISL_706846, EPI_ISL_706847, EPI_ISL_706848, EPI_ISL_706849, EPI_ISL_706850, EPI_ISL_706851, EPI_ISL_706852, EPI_ISL_725990, EPI_ISL_725991, EPI_ISL_725992, EPI_ISL_725993, EPI_ISL_725994, EPI_ISL_725995, EPI_ISL_725996, EPI_ISL_725998, EPI_ISL_726005, EPI_ISL_726010, EPI_ISL_726011, EPI_ISL_726019, EPI_ISL_726020, EPI_ISL_726021, EPI_ISL_726043, EPI_ISL_726045, EPI_ISL_726046, EPI_ISL_726047, EPI_ISL_726048, EPI_ISL_726049, EPI_ISL_726050, EPI_ISL_726051, EPI_ISL_726052, EPI_ISL_726054, EPI_ISL_726055, EPI_ISL_726056, EPI_ISL_726057, EPI_ISL_726058, EPI_ISL_726059, EPI_ISL_726060, EPI_ISL_726061, EPI_ISL_726062, EPI_ISL_726063, EPI_ISL_726064, EPI_ISL_726065, EPI_ISL_726066, EPI_ISL_726067, EPI_ISL_726068, EPI_ISL_726069, EPI_ISL_726070, EPI_ISL_726071, EPI_ISL_726072, EPI_ISL_726073, EPI_ISL_726074, EPI_ISL_726075, EPI_ISL_726076, EPI_ISL_726077, EPI_ISL_726078, EPI_ISL_726079, EPI_ISL_726080, EPI_ISL_726081, EPI_ISL_726082, EPI_ISL_726083, EPI_ISL_726086, EPI_ISL_726087, EPI_ISL_726088, EPI_ISL_726089, EPI_ISL_726090, EPI_ISL_726091, EPI_ISL_726092, EPI_ISL_726093, EPI_ISL_726094, EPI_ISL_726095, EPI_ISL_726096, EPI_ISL_726099, EPI_ISL_726213, EPI_ISL_726253, EPI_ISL_726254, EPI_ISL_742276, EPI_ISL_742278, EPI_ISL_742279, EPI_ISL_742280, EPI_ISL_742322, EPI_ISL_742325, EPI_ISL_742326, EPI_ISL_742327, EPI_ISL_742328, EPI_ISL_742329, EPI_ISL_742330, EPI_ISL_742331, EPI_ISL_742332, EPI_ISL_742333, EPI_ISL_742334, EPI_ISL_742335, EPI_ISL_742336, EPI_ISL_742337, EPI_ISL_742338, EPI_ISL_742339, EPI_ISL_742340, EPI_ISL_742341, EPI_ISL_742342, EPI_ISL_742343, EPI_ISL_742344, EPI_ISL_742345, EPI_ISL_742346, EPI_ISL_742347, EPI_ISL_742348, EPI_ISL_742349, EPI_ISL_742350, EPI_ISL_742559, EPI_ISL_742586, EPI_ISL_742587 | | | |
| see above | Wales Specialist Virology Centre Sequencing lab: Pathogen Genomics Unit | COVID-19 Genomics UK (COG-UK) Consortium | Alec Birchley; Alexander Adams; Amy Gaskin; Angela Marchbank; Bree Gatica-Wilcox; Catherine Moore; Jason Coombes; Joanne Watkins; Joel Southgate; Johnathan Evans; Laura Gifford; Lauren Gilbert; Lee Graham; Malorie Perry; Matthew Bull; Nicole Pacchiarini; Sally Corden; Sara Kumziene-Summerhayes; Sara Rey; Sarah Taylor; Simon Cottrell; Sophie Jones; Tom Connor |
| EPI_ISL_665286, EPI_ISL_665321, EPI_ISL_665363, EPI_ISL_665385, EPI_ISL_665416, EPI_ISL_665425, EPI_ISL_665436, EPI_ISL_665439, EPI_ISL_665521, EPI_ISL_665524, EPI_ISL_665531, EPI_ISL_665532, EPI_ISL_665536, EPI_ISL_665537, EPI_ISL_665538, EPI_ISL_665579, EPI_ISL_665609, EPI_ISL_665633, EPI_ISL_665641, EPI_ISL_665685, EPI_ISL_665727, EPI_ISL_665748, EPI_ISL_666029, EPI_ISL_666030, EPI_ISL_666031, EPI_ISL_666032, EPI_ISL_666033, EPI_ISL_666034, EPI_ISL_666035, EPI_ISL_666036, EPI_ISL_666037, EPI_ISL_666038, EPI_ISL_666039, EPI_ISL_666040, EPI_ISL_666041, EPI_ISL_666042, EPI_ISL_666043, EPI_ISL_666044, EPI_ISL_666045, EPI_ISL_666046, EPI_ISL_666047, EPI_ISL_666048, EPI_ISL_666049, EPI_ISL_666050, EPI_ISL_666051, EPI_ISL_666052, EPI_ISL_666053, EPI_ISL_666054, EPI_ISL_666055, EPI_ISL_666056, EPI_ISL_666057, EPI_ISL_666058, EPI_ISL_666059, EPI_ISL_680449, EPI_ISL_680450, EPI_ISL_680451, EPI_ISL_702618, EPI_ISL_702867, EPI_ISL_702892, EPI_ISL_703249, EPI_ISL_703269, EPI_ISL_703748, EPI_ISL_703749, EPI_ISL_703839, EPI_ISL_703911, EPI_ISL_704595, EPI_ISL_705124, EPI_ISL_705528, EPI_ISL_705638, EPI_ISL_705639, EPI_ISL_705640, EPI_ISL_705641, EPI_ISL_705642, EPI_ISL_705643, EPI_ISL_705644, EPI_ISL_705645, EPI_ISL_705646, EPI_ISL_705647, EPI_ISL_705648, EPI_ISL_705650, EPI_ISL_705651, EPI_ISL_705652, EPI_ISL_705653, EPI_ISL_705654, EPI_ISL_705655, EPI_ISL_705656, EPI_ISL_705657, EPI_ISL_705658, EPI_ISL_705659, EPI_ISL_705660, EPI_ISL_705661, EPI_ISL_705662, EPI_ISL_705663, EPI_ISL_705664, EPI_ISL_705711, EPI_ISL_705730, | | | |

| | | | | |
|---|---|---|---|---|
| EPI_ISL_724212 | | | | |
| | see above | West of Scotland Specialist Virology Centre, NHSGGC / MRC-University of Glasgow Centre for Virus Research | COVID-19 Genomics UK (COG-UK) Consortium | Alasdair MacLean; Alice Broos; Ana da Silva Filipe; Antonia Ho; Daniel Mair; David L Robertson; Emma Thomson; Guy Mollett; James Shepherd; Jenna Nichols; Joseph Hughes; Kathy Li; Kathy Smollett; Kyriaki Nomikou; Lily Tong; Matthew Holden; Natasha Johnson; Rachel Blacow; Richard Orton; Rory Gunson; Sarah McDonald; Sharif Shaaban; Sreenu Vattipally; Stephen Carmichael |

We gratefully acknowledge the following Authors from the Originating laboratories responsible for obtaining the specimens, as well as the Submitting laboratories where the genome data were generated and shared via GISAID, on which this research is based.

All Submitters of data may be contacted directly via www.gisaid.org

Authors are sorted alphabetically.

| Accession ID | Originating Laboratory | Submitting Laboratory | Authors |
|---|---|---|---|
| EPI_ISL_842910, EPI_ISL_842911, EPI_ISL_842912, EPI_ISL_842913, EPI_ISL_842916, EPI_ISL_842917, EPI_ISL_842918, EPI_ISL_842925, EPI_ISL_842926, EPI_ISL_842927, EPI_ISL_842928, EPI_ISL_842929, EPI_ISL_842930, EPI_ISL_842931, EPI_ISL_842933, EPI_ISL_842934, EPI_ISL_842935, EPI_ISL_842936, EPI_ISL_842937, EPI_ISL_842938, EPI_ISL_842940, EPI_ISL_842941, EPI_ISL_842942, EPI_ISL_842943, EPI_ISL_842944, EPI_ISL_842945, EPI_ISL_842946, EPI_ISL_842947, EPI_ISL_842948, EPI_ISL_842949, EPI_ISL_842950, EPI_ISL_842951, EPI_ISL_842952, EPI_ISL_842953, EPI_ISL_842954, EPI_ISL_842955, EPI_ISL_842956, EPI_ISL_842957, EPI_ISL_842958, EPI_ISL_842959, EPI_ISL_842960, EPI_ISL_842961, EPI_ISL_842962, EPI_ISL_842963, EPI_ISL_842964, EPI_ISL_842965, EPI_ISL_842967, EPI_ISL_842968, EPI_ISL_842969, EPI_ISL_842970, EPI_ISL_842971, EPI_ISL_842972, EPI_ISL_842973, EPI_ISL_842974, EPI_ISL_842975, EPI_ISL_842976, EPI_ISL_842977, EPI_ISL_842978, EPI_ISL_842979, EPI_ISL_842980, EPI_ISL_842981, EPI_ISL_842984, EPI_ISL_842985, EPI_ISL_842986, EPI_ISL_842987, EPI_ISL_842988, EPI_ISL_842989, EPI_ISL_842990, EPI_ISL_842991, EPI_ISL_842992, EPI_ISL_842993, EPI_ISL_842994, EPI_ISL_842995, EPI_ISL_842996, EPI_ISL_842997, EPI_ISL_842998, EPI_ISL_842999, EPI_ISL_843000, EPI_ISL_843001, EPI_ISL_843002, EPI_ISL_843003, EPI_ISL_843004, EPI_ISL_843005, EPI_ISL_843007, EPI_ISL_843013, EPI_ISL_843014, EPI_ISL_843015, EPI_ISL_843016, EPI_ISL_843017, EPI_ISL_843018, EPI_ISL_843019, EPI_ISL_843020, EPI_ISL_843041, EPI_ISL_1636888, EPI_ISL_1636889, EPI_ISL_1636890, EPI_ISL_1636914, EPI_ISL_1636927, EPI_ISL_1636928, EPI_ISL_1636929, EPI_ISL_1636930, EPI_ISL_1636986, EPI_ISL_1975099 | | | |
| see above | Barts Health NHS Trust | COVID-19 Genomics UK (COG-UK) Consortium | BROAD; Beatrix; CUTINO-MOGUEL; Claire; David; Dola; HARRINGTON; KELE; KULASEGARAN-SHYLINI; Maria-Teresa; OWOYEMI; Raghavendran; SHYLINI |
| EPI_ISL_868760, EPI_ISL_868763, EPI_ISL_868766, EPI_ISL_868772, EPI_ISL_868773, EPI_ISL_868776, EPI_ISL_868780, EPI_ISL_868781, EPI_ISL_868782, EPI_ISL_868783, EPI_ISL_868784, EPI_ISL_868788, EPI_ISL_868878, EPI_ISL_868879, EPI_ISL_868880, EPI_ISL_868881, EPI_ISL_868882, EPI_ISL_868883, EPI_ISL_868884, EPI_ISL_868885, EPI_ISL_868886, EPI_ISL_868887, EPI_ISL_868888, EPI_ISL_868890, EPI_ISL_868891, EPI_ISL_868892, EPI_ISL_868893, EPI_ISL_868894, EPI_ISL_868895, EPI_ISL_868896, EPI_ISL_868897, EPI_ISL_868898, EPI_ISL_868899, EPI_ISL_868900, EPI_ISL_868985, EPI_ISL_868986, EPI_ISL_868988, EPI_ISL_868989, EPI_ISL_868990, EPI_ISL_868991, EPI_ISL_868992, EPI_ISL_868994, EPI_ISL_868997, EPI_ISL_868998, EPI_ISL_869002, EPI_ISL_869003, EPI_ISL_869004, EPI_ISL_869007, EPI_ISL_869009, EPI_ISL_869010, EPI_ISL_869011, EPI_ISL_869012, EPI_ISL_869013, EPI_ISL_924783, EPI_ISL_924784, EPI_ISL_924785, EPI_ISL_924786, EPI_ISL_924788, EPI_ISL_924789, EPI_ISL_924790, EPI_ISL_924791, EPI_ISL_924792, EPI_ISL_924793, EPI_ISL_1476752 | | | |
| see above | Bioinformatics and Biostatistics Lab, Advanced Sequencing Facility | COVID-19 Genomics UK (COG-UK) Consortium | Aengus Stewart; Chelsea Sawyer; Harshil Patel; Jerome Nicod; Laura Cubitt; Margaret Crawford |
| EPI_ISL_679971, EPI_ISL_727734, EPI_ISL_741716, EPI_ISL_741717, EPI_ISL_741718, EPI_ISL_741738, EPI_ISL_741739, EPI_ISL_741740, EPI_ISL_741741, EPI_ISL_741757, EPI_ISL_741758, EPI_ISL_741759, EPI_ISL_867950, EPI_ISL_867951, EPI_ISL_952390, EPI_ISL_952399, EPI_ISL_952404, EPI_ISL_1000265, EPI_ISL_1000277 | | | |
| see above | Centre for Enzyme Innovation, University of Portsmouth / Translational Research Laboratory, Portsmouth Hospitals NHS Trust | COVID-19 Genomics UK (COG-UK) Consortium | Allyson Lloyd; Angela Beckett; Anoop Chauhan; Christopher Fearn; Ethan Butcher; Garry Scarlett; Kate Cook; Katie Loveson; Kelly Bicknell; Robert Impey; Salman Goudarzi; Samuel Robson; Sarah Wyllie; Scott Elliott; Sharon Glaysher; Yann Bourgeois |
| EPI_ISL_664120, EPI_ISL_664149, EPI_ISL_664150, EPI_ISL_664199, EPI_ISL_664200, EPI_ISL_664234, EPI_ISL_664316, EPI_ISL_664326, EPI_ISL_664381, EPI_ISL_664382, EPI_ISL_664383, EPI_ISL_664384, EPI_ISL_664446, EPI_ISL_664538, EPI_ISL_664546, EPI_ISL_664591, EPI_ISL_664610, EPI_ISL_664635, EPI_ISL_665013, EPI_ISL_665014, EPI_ISL_665015, EPI_ISL_665016, EPI_ISL_665017, EPI_ISL_665018, EPI_ISL_665019, EPI_ISL_665020, EPI_ISL_665021, EPI_ISL_679160, EPI_ISL_679161, EPI_ISL_679162, EPI_ISL_679163, EPI_ISL_679164, EPI_ISL_679165, EPI_ISL_679166, EPI_ISL_679168, EPI_ISL_679170, EPI_ISL_679171, EPI_ISL_679172, EPI_ISL_679174, EPI_ISL_679175, EPI_ISL_679176, EPI_ISL_679187, EPI_ISL_679189, EPI_ISL_679191, EPI_ISL_679193, EPI_ISL_679195, EPI_ISL_679199, EPI_ISL_679200, EPI_ISL_679201, EPI_ISL_679202, EPI_ISL_679203, EPI_ISL_679218, EPI_ISL_702489, EPI_ISL_702500, EPI_ISL_702561, EPI_ISL_702703, EPI_ISL_702734, EPI_ISL_702848, EPI_ISL_702876, EPI_ISL_702883, EPI_ISL_702886, EPI_ISL_702891, EPI_ISL_703032, EPI_ISL_703076, EPI_ISL_703118, EPI_ISL_703127, EPI_ISL_703213, EPI_ISL_703238, EPI_ISL_703451, EPI_ISL_703457, EPI_ISL_703476, EPI_ISL_703479, EPI_ISL_703508, EPI_ISL_703524, EPI_ISL_703619, EPI_ISL_703629, EPI_ISL_703632, EPI_ISL_703635, EPI_ISL_703679, EPI_ISL_703693, EPI_ISL_703708, EPI_ISL_703709, EPI_ISL_703710, EPI_ISL_703713, EPI_ISL_703716, EPI_ISL_703724, EPI_ISL_703741, EPI_ISL_703742, EPI_ISL_703747, EPI_ISL_704060, EPI_ISL_704115, EPI_ISL_704203, EPI_ISL_704247, EPI_ISL_704250, EPI_ISL_704252, EPI_ISL_704255, EPI_ISL_704257, EPI_ISL_704269, EPI_ISL_704289, EPI_ISL_704291, EPI_ISL_704294, EPI_ISL_704297, EPI_ISL_704299, EPI_ISL_704302, EPI_ISL_704305, EPI_ISL_704307, EPI_ISL_704309, EPI_ISL_704328, EPI_ISL_704331, EPI_ISL_704333, EPI_ISL_704336, EPI_ISL_704339, EPI_ISL_704341, EPI_ISL_704344, EPI_ISL_704347, EPI_ISL_704350, EPI_ISL_704353, EPI_ISL_704356, EPI_ISL_704359, EPI_ISL_704361, EPI_ISL_704364, EPI_ISL_704366, EPI_ISL_704369, EPI_ISL_704374, EPI_ISL_704377, EPI_ISL_704379, EPI_ISL_704382, EPI_ISL_704384, EPI_ISL_704478, EPI_ISL_704481, EPI_ISL_704484, EPI_ISL_704486, EPI_ISL_704489, EPI_ISL_704492, EPI_ISL_704494, EPI_ISL_704497, EPI_ISL_704499, EPI_ISL_704502, EPI_ISL_704505, EPI_ISL_704507, EPI_ISL_704510, EPI_ISL_704513, EPI_ISL_704516, EPI_ISL_704518, EPI_ISL_704521, EPI_ISL_704524, EPI_ISL_704526, EPI_ISL_704529, EPI_ISL_704532, EPI_ISL_704535, EPI_ISL_704537, EPI_ISL_704540, EPI_ISL_704543, EPI_ISL_704545, EPI_ISL_704548, EPI_ISL_704551, EPI_ISL_704554, EPI_ISL_704557, EPI_ISL_704606, EPI_ISL_704610, EPI_ISL_704611, EPI_ISL_704612, EPI_ISL_704613, EPI_ISL_704616, EPI_ISL_704617, EPI_ISL_704618, EPI_ISL_704640, EPI_ISL_704641, EPI_ISL_704643, EPI_ISL_704646, EPI_ISL_704648, EPI_ISL_704651, EPI_ISL_704654, EPI_ISL_704656, EPI_ISL_704659, EPI_ISL_704662, EPI_ISL_704664, EPI_ISL_704834, EPI_ISL_705081, EPI_ISL_705110, EPI_ISL_705217, EPI_ISL_705289, EPI_ISL_705339, EPI_ISL_705376, EPI_ISL_705410, EPI_ISL_705536, EPI_ISL_705551, EPI_ISL_705555, EPI_ISL_705556, EPI_ISL_705557, EPI_ISL_705559, EPI_ISL_705560, EPI_ISL_705561, EPI_ISL_705562, EPI_ISL_705563, EPI_ISL_705564, EPI_ISL_705565, EPI_ISL_705566, EPI_ISL_705629, EPI_ISL_705630, EPI_ISL_705631, EPI_ISL_705632, EPI_ISL_705633, EPI_ISL_705634, EPI_ISL_705635, EPI_ISL_723852, EPI_ISL_723853, EPI_ISL_723854, EPI_ISL_723855, EPI_ISL_723856, EPI_ISL_723857, EPI_ISL_723858, EPI_ISL_723859, EPI_ISL_723860, EPI_ISL_723861, EPI_ISL_723862, EPI_ISL_723866, EPI_ISL_723867, EPI_ISL_723869, EPI_ISL_723870, EPI_ISL_723872, EPI_ISL_723873, EPI_ISL_723874, EPI_ISL_723875, EPI_ISL_723876, EPI_ISL_723877, EPI_ISL_723878, EPI_ISL_723879, EPI_ISL_723880, EPI_ISL_723881, EPI_ISL_723882, EPI_ISL_723883, EPI_ISL_723896, EPI_ISL_740953, EPI_ISL_740954, EPI_ISL_763790, EPI_ISL_763873, EPI_ISL_764302, EPI_ISL_764327 | | | |
| see above | Department of Pathology, University of Cambridge | COVID-19 Genomics UK (COG-UK) Consortium | Aminu S. Jahun; Grant Hall; Ian Goodfellow; Iliana Georgana; Malte Pinckert; Martin D. Curran; Myra Hosmillo; Surendra Parmar; Yasmin Chaudhry |
| EPI_ISL_693332, EPI_ISL_693333, EPI_ISL_701673, EPI_ISL_701674, EPI_ISL_701675, EPI_ISL_701676, EPI_ISL_701677, EPI_ISL_701678, EPI_ISL_701679, EPI_ISL_701680, EPI_ISL_701681, EPI_ISL_701682, EPI_ISL_701683, EPI_ISL_701684, EPI_ISL_701685, EPI_ISL_701686, EPI_ISL_701687, EPI_ISL_701688, EPI_ISL_701689, EPI_ISL_701690, EPI_ISL_701691, EPI_ISL_701692, EPI_ISL_701693, EPI_ISL_701694, EPI_ISL_701695, EPI_ISL_701696, EPI_ISL_701697, EPI_ISL_701698, EPI_ISL_701699, EPI_ISL_701700, EPI_ISL_701701, EPI_ISL_701702, EPI_ISL_701703, EPI_ISL_701704, EPI_ISL_701705, EPI_ISL_701706, EPI_ISL_701707, EPI_ISL_701708, EPI_ISL_701709, EPI_ISL_701710, EPI_ISL_701711, EPI_ISL_701712, EPI_ISL_701713, EPI_ISL_701714, EPI_ISL_701715, EPI_ISL_701716, EPI_ISL_701717, EPI_ISL_701718, EPI_ISL_701719, EPI_ISL_701720, EPI_ISL_701721, EPI_ISL_701722, EPI_ISL_701723, EPI_ISL_701725, EPI_ISL_701726, EPI_ISL_701727, EPI_ISL_701728, EPI_ISL_701729, EPI_ISL_701730, EPI_ISL_701731, EPI_ISL_701732, EPI_ISL_701733, EPI_ISL_701734, EPI_ISL_701735, EPI_ISL_701736, EPI_ISL_701737, EPI_ISL_701738, EPI_ISL_701739, EPI_ISL_701740, EPI_ISL_701741, EPI_ISL_701742, EPI_ISL_701743, EPI_ISL_701744, EPI_ISL_701745, EPI_ISL_701746, EPI_ISL_701747, EPI_ISL_701748, EPI_ISL_701749, EPI_ISL_701750, EPI_ISL_701751, EPI_ISL_701752, EPI_ISL_701753, EPI_ISL_701754, EPI_ISL_701755, EPI_ISL_701756, EPI_ISL_701757, EPI_ISL_701758, EPI_ISL_701759, EPI_ISL_701760, EPI_ISL_701761, EPI_ISL_701762, EPI_ISL_701763, EPI_ISL_701764, EPI_ISL_701765, EPI_ISL_701766, EPI_ISL_701767, EPI_ISL_701768, EPI_ISL_701769, EPI_ISL_701770, EPI_ISL_701771, EPI_ISL_701772, EPI_ISL_701773, EPI_ISL_701774, EPI_ISL_701775, EPI_ISL_701776, EPI_ISL_701777, EPI_ISL_701778, EPI_ISL_701779, EPI_ISL_701780, EPI_ISL_701781, EPI_ISL_701782, EPI_ISL_701783, EPI_ISL_701784, EPI_ISL_701785, EPI_ISL_701786, EPI_ISL_701787, EPI_ISL_701788, EPI_ISL_701789, EPI_ISL_701790, EPI_ISL_701791, EPI_ISL_701792, EPI_ISL_701793, EPI_ISL_701794, EPI_ISL_701795, EPI_ISL_701796, EPI_ISL_701797, EPI_ISL_701798, EPI_ISL_701799, EPI_ISL_701800, EPI_ISL_701801, EPI_ISL_701802, EPI_ISL_701803, EPI_ISL_701804, EPI_ISL_701805, EPI_ISL_701806, EPI_ISL_701807, EPI_ISL_701808, EPI_ISL_701809, EPI_ISL_701810, EPI_ISL_701811, EPI_ISL_701812, EPI_ISL_701813, EPI_ISL_701814, EPI_ISL_701815, EPI_ISL_701816, EPI_ISL_701817, EPI_ISL_701818, EPI_ISL_701819, EPI_ISL_701820, EPI_ISL_701821, EPI_ISL_701822, EPI_ISL_701823, EPI_ISL_701824, EPI_ISL_701825, EPI_ISL_701826, EPI_ISL_701827, EPI_ISL_701828, EPI_ISL_701829, EPI_ISL_701830, EPI_ISL_701831, EPI_ISL_701832, EPI_ISL_701833, EPI_ISL_701834, EPI_ISL_701835, EPI_ISL_701836, EPI_ISL_701837, EPI_ISL_701838, EPI_ISL_701839, EPI_ISL_701840, EPI_ISL_701841, EPI_ISL_701842, EPI_ISL_701843, EPI_ISL_701844, EPI_ISL_701845, EPI_ISL_701846, EPI_ISL_701847, EPI_ISL_701848, EPI_ISL_701849, EPI_ISL_701850, EPI_ISL_701851, EPI_ISL_701852, EPI_ISL_701853, EPI_ISL_701854, EPI_ISL_701855, EPI_ISL_701856, EPI_ISL_701857, EPI_ISL_701858, EPI_ISL_701859, EPI_ISL_701860, EPI_ISL_701861, EPI_ISL_701862, EPI_ISL_701863, EPI_ISL_701864, EPI_ISL_701865, EPI_ISL_701866, EPI_ISL_701867, EPI_ISL_701868, EPI_ISL_701869, EPI_ISL_701870, EPI_ISL_701871, EPI_ISL_701872, EPI_ISL_701873, EPI_ISL_701874, EPI_ISL_701875, EPI_ISL_701876, EPI_ISL_701877, EPI_ISL_701878, EPI_ISL_701879, EPI_ISL_701880, EPI_ISL_701881, EPI_ISL_701882, EPI_ISL_701883, EPI_ISL_701884, EPI_ISL_701885, EPI_ISL_701886, EPI_ISL_701887, EPI_ISL_701888, EPI_ISL_701889, EPI_ISL_701890, EPI_ISL_701891, EPI_ISL_701892, EPI_ISL_701893, EPI_ISL_701894, EPI_ISL_701895, EPI_ISL_701896, EPI_ISL_701897, EPI_ISL_701898, EPI_ISL_701899, EPI_ISL_701900, EPI_ISL_701901, EPI_ISL_701902, EPI_ISL_701903, EPI_ISL_701904, EPI_ISL_701905, EPI_ISL_701906, EPI_ISL_701907, EPI_ISL_701908, EPI_ISL_701909, EPI_ISL_701910, EPI_ISL_701911, EPI_ISL_701912, EPI_ISL_701913, EPI_ISL_701914, EPI_ISL_701915, EPI_ISL_701916, EPI_ISL_701917, EPI_ISL_701918, EPI_ISL_701919, EPI_ISL_701920, EPI_ISL_701921, EPI_ISL_701922, EPI_ISL_701923, EPI_ISL_701924, EPI_ISL_701925, EPI_ISL_701926, EPI_ISL_701927, EPI_ISL_701928, EPI_ISL_701929, EPI_ISL_701930, EPI_ISL_701931, EPI_ISL_701932, EPI_ISL_701933, EPI_ISL_701934, EPI_ISL_701935, EPI_ISL_701936, EPI_ISL_701937, EPI_ISL_701938, EPI_ISL_701939, EPI_ISL_701940, EPI_ISL_701941, EPI_ISL_701942, EPI_ISL_701943, EPI_ISL_701944, EPI_ISL_701945, EPI_ISL_701946, EPI_ISL_701947, EPI_ISL_701948, EPI_ISL_701949, EPI_ISL_701950, EPI_ISL_701951, EPI_ISL_701952, EPI_ISL_701953, EPI_ISL_701954, EPI_ISL_701955, EPI_ISL_701956, EPI_ISL_701957, EPI_ISL_701958, EPI_ISL_701959, EPI_ISL_701960, EPI_ISL_701962, EPI_ISL_701963, EPI_ISL_701964, EPI_ISL_701965, EPI_ISL_701966, EPI_ISL_701967, EPI_ISL_701968, EPI_ISL_701969, EPI_ISL_701970, EPI_ISL_701971, EPI_ISL_701972, EPI_ISL_701973, EPI_ISL_701974, EPI_ISL_701975, EPI_ISL_701976, EPI_ISL_701977, EPI_ISL_701978, EPI_ISL_701979, EPI_ISL_701980, EPI_ISL_701981, EPI_ISL_701982, EPI_ISL_701983, EPI_ISL_701984, EPI_ISL_701985, EPI_ISL_701986, EPI_ISL_701987, EPI_ISL_701988, EPI_ISL_701989, EPI_ISL_701990, EPI_ISL_701991, EPI_ISL_701992, EPI_ISL_701993, EPI_ISL_701994, EPI_ISL_701995, EPI_ISL_701996, EPI_ISL_701997, EPI_ISL_701998, EPI_ISL_701999, EPI_ISL_702000, EPI_ISL_702001, EPI_ISL_702002, EPI_ISL_702003, EPI_ISL_702004, EPI_ISL_702005, EPI_ISL_702006, EPI_ISL_702007, EPI_ISL_702008, EPI_ISL_702009, EPI_ISL_702010, EPI_ISL_702011, EPI_ISL_702012, EPI_ISL_702013, EPI_ISL_702014, EPI_ISL_702015, EPI_ISL_702016, EPI_ISL_702017, EPI_ISL_702018, EPI_ISL_702019, EPI_ISL_702020, EPI_ISL_702021, EPI_ISL_702022, EPI_ISL_702370, EPI_ISL_702371, EPI_ISL_702372, EPI_ISL_702373, EPI_ISL_702374, EPI_ISL_702375, EPI_ISL_702376, EPI_ISL_702377, EPI_ISL_702378, EPI_ISL_702379, EPI_ISL_702380, EPI_ISL_702381, EPI_ISL_702382, EPI_ISL_702383, EPI_ISL_702384, EPI_ISL_702386, EPI_ISL_702387, EPI_ISL_702388, EPI_ISL_702389, EPI_ISL_702390, EPI_ISL_702391, EPI_ISL_702392, EPI_ISL_702393, EPI_ISL_702394, EPI_ISL_702395, EPI_ISL_702396, EPI_ISL_702397, EPI_ISL_702398, EPI_ISL_702399, EPI_ISL_702400, EPI_ISL_702401, EPI_ISL_702402, EPI_ISL_702403, EPI_ISL_702404, EPI_ISL_702405, EPI_ISL_702406, EPI_ISL_702407, EPI_ISL_702408, EPI_ISL_702409, EPI_ISL_702410, EPI_ISL_702412, EPI_ISL_702413, EPI_ISL_702414, EPI_ISL_702415, EPI_ISL_702416, EPI_ISL_702417, EPI_ISL_702418, EPI_ISL_702419, EPI_ISL_702420, EPI_ISL_702421, EPI_ISL_702422, EPI_ISL_702423, EPI_ISL_702424, EPI_ISL_702425, EPI_ISL_702426, EPI_ISL_702427, EPI_ISL_702428, EPI_ISL_702429, EPI_ISL_702430, EPI_ISL_702431, EPI_ISL_702432, EPI_ISL_702433, EPI_ISL_702434, EPI_ISL_702435, EPI_ISL_702436, EPI_ISL_702437, EPI_ISL_702439, EPI_ISL_702440, EPI_ISL_702441, EPI_ISL_702442, EPI_ISL_702443, EPI_ISL_702444, EPI_ISL_702445, EPI_ISL_702446, EPI_ISL_702447, EPI_ISL_702448, EPI_ISL_702449, EPI_ISL_702450, EPI_ISL_702451, EPI_ISL_702452, EPI_ISL_702453, EPI_ISL_702454, EPI_ISL_702455, EPI_ISL_702456, EPI_ISL_702457, EPI_ISL_702458, EPI_ISL_702459, EPI_ISL_702460, EPI_ISL_702461, EPI_ISL_702462, EPI_ISL_702463, EPI_ISL_702465, EPI_ISL_702466, EPI_ISL_702467, EPI_ISL_702468, EPI_ISL_702470, EPI_ISL_702472, EPI_ISL_702473, EPI_ISL_702475, EPI_ISL_702477, EPI_ISL_702479, EPI_ISL_702480, EPI_ISL_702482, EPI_ISL_702483, EPI_ISL_702485, EPI_ISL_702486, EPI_ISL_702488, EPI_ISL_702490, EPI_ISL_702491, EPI_ISL_702493, EPI_ISL_702494, EPI_ISL_702496, EPI_ISL_702497, EPI_ISL_702499, EPI_ISL_702501, EPI_ISL_702502, EPI_ISL_702503, EPI_ISL_702506, EPI_ISL_702508, EPI_ISL_702509, EPI_ISL_702511, EPI_ISL_702513, EPI_ISL_702514, EPI_ISL_702516, EPI_ISL_702517, EPI_ISL_702519, EPI_ISL_702521, EPI_ISL_702522, EPI_ISL_702524, EPI_ISL_702526, EPI_ISL_702527, EPI_ISL_702529, EPI_ISL_702530, EPI_ISL_702532, EPI_ISL_702534, EPI_ISL_702535, EPI_ISL_702537, EPI_ISL_702538, EPI_ISL_702540, EPI_ISL_702542, EPI_ISL_702543, EPI_ISL_702545, EPI_ISL_702546, EPI_ISL_702548, EPI_ISL_702551, EPI_ISL_702552, EPI_ISL_702554, EPI_ISL_702555, EPI_ISL_702557, EPI_ISL_702559, EPI_ISL_702563, EPI_ISL_702565, EPI_ISL_702567, EPI_ISL_702568, EPI_ISL_702570, EPI_ISL_702572, EPI_ISL_702573, EPI_ISL_702575, EPI_ISL_702576, EPI_ISL_702578, EPI_ISL_702580, EPI_ISL_702581, EPI_ISL_702583, EPI_ISL_702584, EPI_ISL_702586, EPI_ISL_702588, EPI_ISL_702589, EPI_ISL_702592, EPI_ISL_702593, EPI_ISL_702595, EPI_ISL_702596, EPI_ISL_702598, EPI_ISL_702599, EPI_ISL_702601, EPI_ISL_702603, EPI_ISL_702604, EPI_ISL_702606, EPI_ISL_702608, EPI_ISL_702609, EPI_ISL_702611, EPI_ISL_702613, EPI_ISL_702614, EPI_ISL_702616, EPI_ISL_702617, EPI_ISL_702619, EPI_ISL_702621, EPI_ISL_702622, EPI_ISL_702624, EPI_ISL_702625, EPI_ISL_702627, EPI_ISL_702629, EPI_ISL_702630, EPI_ISL_702632, EPI_ISL_702634, EPI_ISL_702635, EPI_ISL_702637, EPI_ISL_702638, EPI_ISL_702640, EPI_ISL_702641, EPI_ISL_702643, EPI_ISL_702645, EPI_ISL_702646, EPI_ISL_702648, EPI_ISL_702650, EPI_ISL_702651, EPI_ISL_702653, EPI_ISL_702654, EPI_ISL_702657, EPI_ISL_702658, EPI_ISL_702660, EPI_ISL_702662, EPI_ISL_702663, EPI_ISL_702665, EPI_ISL_702666, EPI_ISL_702668, EPI_ISL_702670, EPI_ISL_702671, EPI_ISL_702673, EPI_ISL_702675, EPI_ISL_702676, EPI_ISL_702678, EPI_ISL_702680, EPI_ISL_702681, EPI_ISL_702683, EPI_ISL_702685, EPI_ISL_702686, EPI_ISL_702688, EPI_ISL_702689, EPI_ISL_702691, EPI_ISL_702693, EPI_ISL_702695, EPI_ISL_702696, EPI_ISL_702699, EPI_ISL_702701, EPI_ISL_702702, EPI_ISL_702704, EPI_ISL_702706, EPI_ISL_702707, EPI_ISL_702709, EPI_ISL_702711, EPI_ISL_702712, EPI_ISL_702714, EPI_ISL_702715, EPI_ISL_702717, EPI_ISL_702719, EPI_ISL_702720, EPI_ISL_702722, EPI_ISL_702724, EPI_ISL_702725, EPI_ISL_702727, EPI_ISL_702728, EPI_ISL_702730, EPI_ISL_702732, EPI_ISL_702733, EPI_ISL_702735, EPI_ISL_702737, EPI_ISL_702738, EPI_ISL_702740, EPI_ISL_702741, EPI_ISL_702743, EPI_ISL_702745, EPI_ISL_702746, EPI_ISL_702748, EPI_ISL_702750, EPI_ISL_702752, EPI_ISL_702754, EPI_ISL_702755, EPI_ISL_702757, EPI_ISL_702759, EPI_ISL_702760, EPI_ISL_702762, EPI_ISL_702764, EPI_ISL_702765, EPI_ISL_702767, EPI_ISL_702768, EPI_ISL_702770, EPI_ISL_702772, EPI_ISL_702773, EPI_ISL_702775, EPI_ISL_702777, EPI_ISL_702778, EPI_ISL_702780, EPI_ISL_702781, EPI_ISL_702783, EPI_ISL_702785, EPI_ISL_702786, EPI_ISL_702788, EPI_ISL_702789, EPI_ISL_702791, EPI_ISL_702793, EPI_ISL_702796, EPI_ISL_702797, EPI_ISL_702799, EPI_ISL_702800, EPI_ISL_702802, EPI_ISL_702804, EPI_ISL_702805, EPI_ISL_702807, EPI_ISL_702808, EPI_ISL_702810, EPI_ISL_702811, EPI_ISL_702813, EPI_ISL_702814, EPI_ISL_702816, EPI_ISL_702818, EPI_ISL_702819, EPI_ISL_702821, EPI_ISL_702822, EPI_ISL_702898, EPI_ISL_702899, EPI_ISL_702901, EPI_ISL_702903, EPI_ISL_702904, EPI_ISL_702906, EPI_ISL_702908, EPI_ISL_702909, EPI_ISL_702912, EPI_ISL_702914, EPI_ISL_702916, EPI_ISL_702918, EPI_ISL_702919, EPI_ISL_702921, EPI_ISL_702923, EPI_ISL_702924, EPI_ISL_702926, EPI_ISL_702927, EPI_ISL_702929, EPI_ISL_702931, EPI_ISL_702933, EPI_ISL_702934, EPI_ISL_702936, EPI_ISL_702938, EPI_ISL_702940, EPI_ISL_702942, EPI_ISL_702943, EPI_ISL_702945, EPI_ISL_702946, EPI_ISL_702948, EPI_ISL_702951, EPI_ISL_702952, EPI_ISL_702954, EPI_ISL_702955, EPI_ISL_702957, EPI_ISL_702958, EPI_ISL_702960, EPI_ISL_702962, EPI_ISL_702963, EPI_ISL_702965, EPI_ISL_702967, EPI_ISL_702968, EPI_ISL_702970, EPI_ISL_702972, EPI_ISL_702973, EPI_ISL_702975, EPI_ISL_702976, EPI_ISL_702978, EPI_ISL_702979, EPI_ISL_702981, EPI_ISL_702983, EPI_ISL_702984, EPI_ISL_702986, EPI_ISL_702988, EPI_ISL_702989, EPI_ISL_702991, EPI_ISL_702993, EPI_ISL_702995, EPI_ISL_702997, EPI_ISL_702998, EPI_ISL_703000, EPI_ISL_703001, EPI_ISL_703003, EPI_ISL_703004, EPI_ISL_703006, EPI_ISL_703008, EPI_ISL_703009, EPI_ISL_703011, EPI_ISL_703012, EPI_ISL_703014, EPI_ISL_703016, EPI_ISL_703017, EPI_ISL_703019, EPI_ISL_703021, EPI_ISL_703022, EPI_ISL_703024, EPI_ISL_703025, EPI_ISL_703027, EPI_ISL_703028, EPI_ISL_703030, EPI_ISL_703031, EPI_ISL_703033, EPI_ISL_703035, EPI_ISL_703037, EPI_ISL_703039, EPI_ISL_703041, EPI_ISL_703042, EPI_ISL_703044, EPI_ISL_703045, EPI_ISL_703047, EPI_ISL_703049, EPI_ISL_703050, EPI_ISL_703052, EPI_ISL_703054, EPI_ISL_703055, EPI_ISL_703057, EPI_ISL_703058, EPI_ISL_703060, EPI_ISL_703062, EPI_ISL_703063, EPI_ISL_703065, EPI_ISL_703067, EPI_ISL_703068, EPI_ISL_703070, EPI_ISL_703072, EPI_ISL_703073, EPI_ISL_703075, EPI_ISL_703078, EPI_ISL_703079, EPI_ISL_703081, EPI_ISL_703083, EPI_ISL_703084, EPI_ISL_703086, EPI_ISL_703087, EPI_ISL_703089, EPI_ISL_703091, EPI_ISL_703092, EPI_ISL_703094, EPI_ISL_703095, EPI_ISL_703097, EPI_ISL_703098, EPI_ISL_703100, EPI_ISL_703101, EPI_ISL_703103, EPI_ISL_703104, EPI_ISL_703106, EPI_ISL_703108, EPI_ISL_703109, EPI_ISL_703111, EPI_ISL_703112, EPI_ISL_703114, EPI_ISL_703116, EPI_ISL_703117, EPI_ISL_703119, EPI_ISL_703120, EPI_ISL_703122, EPI_ISL_703123, EPI_ISL_703125, EPI_ISL_703126, EPI_ISL_703128, EPI_ISL_703130, EPI_ISL_703131, EPI_ISL_703134, EPI_ISL_703135, EPI_ISL_703137, EPI_ISL_703138, EPI_ISL_703140, EPI_ISL_703142, EPI_ISL_703143, EPI_ISL_703145, EPI_ISL_703146, EPI_ISL_703148, EPI_ISL_703149, EPI_ISL_703151, EPI_ISL_703153, EPI_ISL_703154, EPI_ISL_703156, EPI_ISL_703157, EPI_ISL_703159, EPI_ISL_703160, EPI_ISL_703161, EPI_ISL_703163, EPI_ISL_703164, EPI_ISL_703166, EPI_ISL_703168, EPI_ISL_703169, EPI_ISL_703171, EPI_ISL_703173, EPI_ISL_703175, EPI_ISL_703177, EPI_ISL_703178, EPI_ISL_703180, EPI_ISL_703181, EPI_ISL_703183, EPI_ISL_703185, EPI_ISL_703186, EPI_ISL_703188, EPI_ISL_703189, EPI_ISL_703191, EPI_ISL_703193, EPI_ISL_703194, EPI_ISL_703196, EPI_ISL_703197, EPI_ISL_703199, EPI_ISL_703201, EPI_ISL_703202, EPI_ISL_703204, EPI_ISL_703205, EPI_ISL_703207, EPI_ISL_703209, EPI_ISL_703210, EPI_ISL_703212, EPI_ISL_703214, EPI_ISL_703215, EPI_ISL_703217, EPI_ISL_703218, EPI_ISL_703220, EPI_ISL_703221, EPI_ISL_703223, EPI_ISL_703224, EPI_ISL_703226, EPI_ISL_703228, EPI_ISL_703229, EPI_ISL_703231, EPI_ISL_703232, EPI_ISL_703234, EPI_ISL_703236, EPI_ISL_703237, EPI_ISL_703239, EPI_ISL_703241, EPI_ISL_703242, EPI_ISL_703244, EPI_ISL_703245, EPI_ISL_703247, EPI_ISL_703248, EPI_ISL_703250, EPI_ISL_703252, EPI_ISL_703253, EPI_ISL_703255, EPI_ISL_703257, EPI_ISL_703258, EPI_ISL_703260, EPI_ISL_703265, EPI_ISL_703266, EPI_ISL_703268, EPI_ISL_703271, EPI_ISL_703274, EPI_ISL_703276, EPI_ISL_703277, EPI_ISL_703282, EPI_ISL_703293, EPI_ISL_703294, EPI_ISL_703298, EPI_ISL_703301, EPI_ISL_703306, EPI_ISL_703307, EPI_ISL_703311, EPI_ISL_703312, EPI_ISL_703314, EPI_ISL_703317, EPI_ISL_703318, EPI_ISL_703322, EPI_ISL_703326, EPI_ISL_703328, EPI_ISL_703329, EPI_ISL_703333, EPI_ISL_703336, EPI_ISL_703339, EPI_ISL_703341, EPI_ISL_703342, EPI_ISL_703344, EPI_ISL_703347, EPI_ISL_703357, EPI_ISL_703363, EPI_ISL_703365, EPI_ISL_703368, EPI_ISL_703373, EPI_ISL_703374, EPI_ISL_703376, EPI_ISL_703386, EPI_ISL_703388, EPI_ISL_703394, EPI_ISL_703396, EPI_ISL_703397, EPI_ISL_703402, EPI_ISL_703404, EPI_ISL_703405, EPI_ISL_703413, EPI_ISL_703416, EPI_ISL_703418, EPI_ISL_703420, EPI_ISL_703425, EPI_ISL_703433, EPI_ISL_703434, EPI_ISL_703436, EPI_ISL_703437, EPI_ISL_703441, EPI_ISL_703448, EPI_ISL_703450, EPI_ISL_703452, EPI_ISL_703455, EPI_ISL_703460, EPI_ISL_703461, EPI_ISL_703464, EPI_ISL_703467, EPI_ISL_703470, EPI_ISL_703475, EPI_ISL_703477, EPI_ISL_703478, EPI_ISL_703480, EPI_ISL_703483, EPI_ISL_703485, EPI_ISL_703488, EPI_ISL_703490, EPI_ISL_703493, EPI_ISL_703494, EPI_ISL_703498, EPI_ISL_703503, EPI_ISL_703504, EPI_ISL_703507, EPI_ISL_703509, EPI_ISL_703518, EPI_ISL_703520, EPI_ISL_703523, EPI_ISL_703526, EPI_ISL_703528, EPI_ISL_703531, EPI_ISL_703532, EPI_ISL_703536, EPI_ISL_703539, EPI_ISL_703543, EPI_ISL_703544, EPI_ISL_703547, EPI_ISL_703549, EPI_ISL_703552, EPI_ISL_703555, EPI_ISL_703558, EPI_ISL_703563, EPI_ISL_703565, EPI_ISL_703566, EPI_ISL_703568, EPI_ISL_703570, EPI_ISL_703571, EPI_ISL_703573, EPI_ISL_703574, EPI_ISL_703576, EPI_ISL_703580, EPI_ISL_703581, EPI_ISL_703586, EPI_ISL_703588, EPI_ISL_703589, EPI_ISL_703591, EPI_ISL_703592, EPI_ISL_703594, EPI_ISL_703595, EPI_ISL_703597, EPI_ISL_703602, EPI_ISL_703603, EPI_ISL_703605, EPI_ISL_703613, EPI_ISL_703615, EPI_ISL_703618, EPI_ISL_703620, EPI_ISL_703623, EPI_ISL_703625, EPI_ISL_703628, EPI_ISL_703630, EPI_ISL_703637, EPI_ISL_703638, EPI_ISL_703646, EPI_ISL_703648, EPI_ISL_703662, EPI_ISL_703664, EPI_ISL_703665, EPI_ISL_703667, EPI_ISL_703670, EPI_ISL_703675, EPI_ISL_703678, EPI_ISL_703680, EPI_ISL_703681, EPI_ISL_703683, EPI_ISL_703686, EPI_ISL_703688, EPI_ISL_703689, EPI_ISL_703691, EPI_ISL_703692, EPI_ISL_703696, EPI_ISL_703699, EPI_ISL_703702, EPI_ISL_703705, EPI_ISL_703707, EPI_ISL_703777, EPI_ISL_703778, EPI_ISL_703780, EPI_ISL_703781, EPI_ISL_703783, EPI_ISL_703785, EPI_ISL_703788, EPI_ISL_703791, EPI_ISL_703793, EPI_ISL_703795, EPI_ISL_703796, EPI_ISL_703798, EPI_ISL_703803, EPI_ISL_703804, EPI_ISL_703809, EPI_ISL_703812, EPI_ISL_703814, EPI_ISL_703817, EPI_ISL_710699, EPI_ISL_710700, EPI_ISL_710701, EPI_ISL_710702, EPI_ISL_710703, EPI_ISL_710704, EPI_ISL_710705, EPI_ISL_710706, EPI_ISL_710707, EPI_ISL_710708, EPI_ISL_710709, EPI_ISL_710710, EPI_ISL_710711, EPI_ISL_710712, EPI_ISL_710713, EPI_ISL_710714, EPI_ISL_710715, EPI_ISL_710716, EPI_ISL_710717, EPI_ISL_710718, EPI_ISL_710719, EPI_ISL_710720, EPI_ISL_710721, EPI_ISL_710722, EPI_ISL_710723, EPI_ISL_710724, EPI_ISL_710725, EPI_ISL_710726, EPI_ISL_710727, EPI_ISL_710728, EPI_ISL_710729, EPI_ISL_710730, EPI_ISL_710731, EPI_ISL_710732, EPI_ISL_710733, EPI_ISL_710734, EPI_ISL_710735, EPI_ISL_710736, EPI_ISL_710737, EPI_ISL_710738, EPI_ISL_710739, EPI_ISL_710740, EPI_ISL_710741, EPI_ISL_710742, EPI_ISL_710743, EPI_ISL_710744, EPI_ISL_710745, EPI_ISL_710746, EPI_ISL_710747, EPI_ISL_710749, EPI_ISL_710750, EPI_ISL_710751, EPI_ISL_710752, EPI_ISL_721185, EPI_ISL_721186, EPI_ISL_721187, EPI_ISL_721188, EPI_ISL_721189, EPI_ISL_721190, EPI_ISL_721191, EPI_ISL_721192, EPI_ISL_721193, EPI_ISL_721194, EPI_ISL_721195, EPI_ISL_721196, EPI_ISL_721197, EPI_ISL_721198, EPI_ISL_721199, EPI_ISL_721200, EPI_ISL_721201, EPI_ISL_721202, EPI_ISL_721203, EPI_ISL_721204, EPI_ISL_721205, EPI_ISL_721206, EPI_ISL_721207, EPI_ISL_721208, EPI_ISL_721209, EPI_ISL_721210, EPI_ISL_721211, EPI_ISL_721212, EPI_ISL_721213, EPI_ISL_721214, EPI_ISL_721215, EPI_ISL_721216, EPI_ISL_721217, EPI_ISL_721218, EPI_ISL_721219, EPI_ISL_721220, EPI_ISL_721221, EPI_ISL_721222, EPI_ISL_721223, EPI_ISL_721224, EPI_ISL_721225, EPI_ISL_721226, EPI_ISL_721227, EPI_ISL_721228, EPI_ISL_721229, EPI_ISL_721230, EPI_ISL_721231, EPI_ISL_721232, EPI_ISL_721233, EPI_ISL_721234, EPI_ISL_721235, EPI_ISL_721236, EPI_ISL_721237, EPI_ISL_721238, EPI_ISL_721239, EPI_ISL_721240, EPI_ISL_721241, EPI_ISL_721242, EPI_ISL_721243, EPI_ISL_721244, EPI_ISL_721245, EPI_ISL_721246, EPI_ISL_721247, EPI_ISL_721248, EPI_ISL_721249, EPI_ISL_721250, EPI_ISL_721251, EPI_ISL_721252, EPI_ISL_721253, EPI_ISL_721254, EPI_ISL_721255, EPI_ISL_721256, EPI_ISL_721257, EPI_ISL_721258, EPI_ISL_721259, EPI_ISL_721260, EPI_ISL_721261, EPI_ISL_721262, EPI_ISL_721263, EPI_ISL_721264, EPI_ISL_721265, EPI_ISL_721266, EPI_ISL_721267, EPI_ISL_721268, EPI_ISL_721269, EPI_ISL_721270, EPI_ISL_721271, EPI_ISL_721272, EPI_ISL_721273, EPI_ISL_721274, EPI_ISL_721275, EPI_ISL_721276, EPI_ISL_721277, EPI_ISL_721278, EPI_ISL_721279, EPI_ISL_721280, EPI_ISL_721281, EPI_ISL_721282, EPI_ISL_721283, EPI_ISL_721284, EPI_ISL_721285, EPI_ISL_721286, EPI_ISL_721287, EPI_ISL_721288, EPI_ISL_721289, EPI_ISL_721290, EPI_ISL_721291, EPI_ISL_721292, EPI_ISL_721293, EPI_ISL_721294, EPI_ISL_721295, EPI_ISL_721296, EPI_ISL_721297, EPI_ISL_721298, EPI_ISL_721299, EPI_ISL_721300, EPI_ISL_721301, EPI_ISL_721302, EPI_ISL_721303, EPI_ISL_721304, EPI_ISL_721305, EPI_ISL_721306, EPI_ISL_721307, EPI_ISL_721308, EPI_ISL_721309, EPI_ISL_721310, EPI_ISL_721311, EPI_ISL_721312, EPI_ISL_721313, EPI_ISL_721314, EPI_ISL_721315, EPI_ISL_721316, EPI_ISL_721317, EPI_ISL_721318, EPI_ISL_721319, EPI_ISL_721320, EPI_ISL_721321, EPI_ISL_721322, EPI_ISL_721323, EPI_ISL_721324, EPI_ISL_721325, EPI_ISL_721326, EPI_ISL_721327, EPI_ISL_721328, EPI_ISL_721329, EPI_ISL_721330, EPI_ISL_721331, EPI_ISL_721332, EPI_ISL_721333, EPI_ISL_721334, EPI_ISL_721335, EPI_ISL_721336, EPI_ISL_721337, EPI_ISL_721338, EPI_ISL_721339, EPI_ISL_721340, EPI_ISL_721341, EPI_ISL_721342, EPI_ISL_721343, EPI_ISL_721344, EPI_ISL_721345, EPI_ISL_721346, EPI_ISL_721347, EPI_ISL_721348, EPI_ISL_721349, EPI_ISL_721350, EPI_ISL_721351, EPI_ISL_721352, EPI_ISL_721353, EPI_ISL_721354, EPI_ISL_721355, EPI_ISL_721356, EPI_ISL_721357, EPI_ISL_721358, EPI_ISL_721359, EPI_ISL_721360, EPI_ISL_721361, EPI_ISL_721362, EPI_ISL_721363, EPI_ISL_721364, EPI_ISL_721365, EPI_ISL_721366, EPI_ISL_721367, EPI_ISL_721368, EPI_ISL_721369, EPI_ISL_721370, EPI_ISL_721371, EPI_ISL_721372, EPI_ISL_721373, EPI_ISL_721374, EPI_ISL_721375, EPI_ISL_721376, EPI_ISL_721377, EPI_ISL_721378, EPI_ISL_721379, EPI_ISL_721380, EPI_ISL_721381, EPI_ISL_721382, EPI_ISL_721383, EPI_ISL_721384, EPI_ISL_721385, EPI_ISL_721386, EPI_ISL_721387, EPI_ISL_721388, EPI_ISL_721389, EPI_ISL_721390, EPI_ISL_721391, EPI_ISL_721392, EPI_ISL_721393, EPI_ISL_721394, EPI_ISL_721395, EPI_ISL_721396, EPI_ISL_721397, EPI_ISL_721398, EPI_ISL_721399, EPI_ISL_721400, EPI_ISL_721401, EPI_ISL_721402, EPI_ISL_721403, EPI_ISL_721404, EPI_ISL_721405, EPI_ISL_721406, EPI_ISL_721407, EPI_ISL_721408, EPI_ISL_721409, EPI_ISL_721410, EPI_ISL_721411, EPI_ISL_721412, EPI_ISL_721413, EPI_ISL_721414, EPI_ISL_721415, EPI_ISL_721416, EPI_ISL_721417, EPI_ISL_721418, EPI_ISL_721419, EPI_ISL_721420, EPI_ISL_721421, EPI_ISL_721422, EPI_ISL_721423, EPI_ISL_721424, EPI_ISL_721425, EPI_ISL_721426, EPI_ISL_721427, EPI_ISL_721428, EPI_ISL_721429, EPI_ISL_721430, EPI_ISL_721431, EPI_ISL_721432, EPI_ISL_721433, EPI_ISL_721434, EPI_ISL_721435, EPI_ISL_721436, EPI_ISL_721437, EPI_ISL_721438, EPI_ISL_721439, EPI_ISL_721440, EPI_ISL_721441, EPI_ISL_721442, EPI_ISL_721443, EPI_ISL_721444, EPI_ISL_721445, EPI_ISL_721446, EPI_ISL_721447, EPI_ISL_721448, EPI_ISL_721449, EPI_ISL_721450, EPI_ISL_721451, EPI_ISL_721452, EPI_ISL_721453, EPI_ISL_721454, EPI_ISL_721455, EPI_ISL_721456, EPI_ISL_721457, EPI_ISL_721458, EPI_ISL_721459, EPI_ISL_721460, EPI_ISL_721461, EPI_ISL_721462, EPI_ISL_721463, EPI_ISL_721464, EPI_ISL_721465, EPI_ISL_721466, EPI_ISL_721467, EPI_ISL_721468, EPI_ISL_721469, EPI_ISL_721470, EPI_ISL_721471, EPI_ISL_721472, EPI_ISL_721473, EPI_ISL_721474, EPI_ISL_721475, EPI_ISL_721476, EPI_ISL_721477, EPI_ISL_721478, EPI_ISL_721479, EPI_ISL_721480, EPI_ISL_721481, EPI_ISL_721482, EPI_ISL_721483, EPI_ISL_721484, EPI_ISL_721485, EPI_ISL_721486, EPI_ISL_721487, EPI_ISL_721488, EPI_ISL_721489, EPI_ISL_721490, EPI_ISL_721491, EPI_ISL_721492, EPI_ISL_721493, EPI_ISL_721494, EPI_ISL_721495, EPI_ISL_721496, EPI_ISL_721497, EPI_ISL_721498, EPI_ISL_721499, EPI_ISL_721500, EPI_ISL_721501, EPI_ISL_721502, EPI_ISL_721503, EPI_ISL_721504, EPI_ISL_721505, EPI_ISL_721506, EPI_ISL_721507, EPI_ISL_721508, EPI_ISL_721509, EPI_ISL_721510, EPI_ISL_721511, EPI_ISL_721512, EPI_ISL_721513, EPI_ISL_721514, EPI_ISL_721515, EPI_ISL_721516, EPI_ISL_767319, EPI_ISL_767320, EPI_ISL_767321, EPI_ISL_767322, EPI_ISL_767323, EPI_ISL_767324, EPI_ISL_767325, EPI_ISL_767326, EPI_ISL_767327, EPI_ISL_767328, EPI_ISL_767329, EPI_ISL_767330, EPI_ISL_767331, EPI_ISL_767332, EPI_ISL_767333, EPI_ISL_915440, EPI_ISL_915441, EPI_ISL_915442, EPI_ISL_915443, EPI_ISL_915444, EPI_ISL_915445, EPI_ISL_915446, EPI_ISL_915447, EPI_ISL_915448, EPI_ISL_915449, EPI_ISL_915450, EPI_ISL_915451, EPI_ISL_915452, EPI_ISL_915453, EPI_ISL_915454, EPI_ISL_915455, EPI_ISL_915456, EPI_ISL_915457, EPI_ISL_915458, EPI_ISL_915459, EPI_ISL_915460, EPI_ISL_915461, EPI_ISL_915462, EPI_ISL_915463, EPI_ISL_915464, EPI_ISL_915465, EPI_ISL_915466, EPI_ISL_915467, EPI_ISL_915468, EPI_ISL_915469, EPI_ISL_915470, EPI_ISL_915471, EPI_ISL_915472, EPI_ISL_915473, EPI_ISL_915474, EPI_ISL_915475, EPI_ISL_915476, EPI_ISL_915477, EPI_ISL_915478, EPI_ISL_915479, EPI_ISL_915480, EPI_ISL_915481, EPI_ISL_915482, EPI_ISL_915483, EPI_ISL_915484, EPI_ISL_915485, EPI_ISL_915486, EPI_ISL_915487, EPI_ISL_915488, EPI_ISL_915489, EPI_ISL_915490, EPI_ISL_915491, EPI_ISL_915492, EPI_ISL_915493, EPI_ISL_915494, EPI_ISL_915495, EPI_ISL_915496, EPI_ISL_915497, EPI_ISL_915498, EPI_ISL_915499, EPI_ISL_915500, EPI_ISL_915501, EPI_ISL_915502, EPI_ISL_915503, EPI_ISL_915504, EPI_ISL_915505, EPI_ISL_915506, EPI_ISL_915507, EPI_ISL_915508, EPI_ISL_915509, EPI_ISL_915510, EPI_ISL_915511, EPI_ISL_915512, EPI_ISL_915513, EPI_ISL_915514, EPI_ISL_915515, EPI_ISL_915516, EPI_ISL_915517, EPI_ISL_915518, EPI_ISL_915519, EPI_ISL_915520, EPI_ISL_915521, EPI_ISL_915522, EPI_ISL_915523, EPI_ISL_915524, EPI_ISL_915525, EPI_ISL_915526, EPI_ISL_915527, EPI_ISL_915528, EPI_ISL_915529, EPI_ISL_915530, EPI_ISL_915531, EPI_ISL_915532, EPI_ISL_915533, EPI_ISL_915534, EPI_ISL_915535, EPI_ISL_915536, EPI_ISL_915537, EPI_ISL_915538, EPI_ISL_915539, EPI_ISL_915540, EPI_ISL_915541, EPI_ISL_915542, EPI_ISL_915543, EPI_ISL_915544, EPI_ISL_915545, EPI_ISL_915546, EPI_ISL_915547, EPI_ISL_915548, EPI_ISL_915549, EPI_ISL_915550, EPI_ISL_915551, EPI_ISL_915552, EPI_ISL_915553, EPI_ISL_915554, EPI_ISL_915555, EPI_ISL_915556, EPI_ISL_915557, EPI_ISL_915558, EPI_ISL_915559, EPI_ISL_915560, EPI_ISL_915561, EPI_ISL_915562, EPI_ISL_915563, EPI_ISL_915564, EPI_ISL_915565, EPI_ISL_915566, EPI_ISL_915567, EPI_ISL_915568, EPI_ISL_915569, EPI_ISL_915570, EPI_ISL_915571, EPI_ISL_915572, EPI_ISL_915573, EPI_ISL_915574, EPI_ISL_915575, EPI_ISL_915576, EPI_ISL_915577, EPI_ISL_915578, EPI_ISL_915579, EPI_ISL_915580, EPI_ISL_915581, EPI_ISL_915582, EPI_ISL_915583, EPI_ISL_915584, EPI_ISL_915585, EPI_ISL_915586, EPI_ISL_915587, EPI_ISL_915588, EPI_ISL_915589, EPI_ISL_915590, EPI_ISL_915591, EPI_ISL_915592, EPI_ISL_915593, EPI_ISL_915594, EPI_ISL_915595, EPI_ISL_915596, EPI_ISL_915597, EPI_ISL_915598, EPI_ISL_915599, EPI_ISL_915600, EPI_ISL_915601, EPI_ISL_915602, EPI_ISL_915603, EPI_ISL_915604, EPI_ISL_915605, EPI_ISL_915606, EPI_ISL_915607, EPI_ISL_915608, EPI_ISL_915609, EPI_ISL_915610, EPI_ISL_915611, EPI_ISL_915612, EPI_ISL_915613, EPI_ISL_915614, EPI_ISL_915615, EPI_ISL_915616, EPI_ISL_915617, EPI_ISL_915618, EPI_ISL_915619, EPI_ISL_915620, EPI_ISL_915621, EPI_ISL_915622, EPI_ISL_915623, EPI_ISL_915624, | | | |

EPI_ISL_915625, EPI_ISL_915626, EPI_ISL_915627, EPI_ISL_915628, EPI_ISL_915629, EPI_ISL_915630, EPI_ISL_915631, EPI_ISL_915632, EPI_ISL_915633, EPI_ISL_915634, EPI_ISL_915635, EPI_ISL_915636, EPI_ISL_915637, EPI_ISL_915638, EPI_ISL_915639, EPI_ISL_915640, EPI_ISL_915641, EPI_ISL_915642, EPI_ISL_915643, EPI_ISL_915644, EPI_ISL_915645, EPI_ISL_915646, EPI_ISL_915647, EPI_ISL_915648,
EPI_ISL_915649, EPI_ISL_915650, EPI_ISL_915651, EPI_ISL_915652, EPI_ISL_915653, EPI_ISL_915654, EPI_ISL_915655, EPI_ISL_915656, EPI_ISL_915657, EPI_ISL_915658, EPI_ISL_915659, EPI_ISL_915660, EPI_ISL_915661, EPI_ISL_915662, EPI_ISL_915663, EPI_ISL_915664, EPI_ISL_915665, EPI_ISL_915666, EPI_ISL_915667, EPI_ISL_915668, EPI_ISL_915669, EPI_ISL_915670, EPI_ISL_915671, EPI_ISL_915672,
EPI_ISL_915673, EPI_ISL_915674, EPI_ISL_915675, EPI_ISL_915676, EPI_ISL_915677, EPI_ISL_915678, EPI_ISL_915679, EPI_ISL_915680, EPI_ISL_915681, EPI_ISL_915682, EPI_ISL_915683, EPI_ISL_915684, EPI_ISL_915685, EPI_ISL_915686, EPI_ISL_915687, EPI_ISL_915688, EPI_ISL_915689, EPI_ISL_915690, EPI_ISL_915691, EPI_ISL_915692, EPI_ISL_915693, EPI_ISL_915694, EPI_ISL_915695, EPI_ISL_915696,
EPI_ISL_915697, EPI_ISL_915698, EPI_ISL_915699, EPI_ISL_915700, EPI_ISL_915701, EPI_ISL_915702, EPI_ISL_915703, EPI_ISL_915704, EPI_ISL_915705, EPI_ISL_915706, EPI_ISL_915707, EPI_ISL_915708, EPI_ISL_915709, EPI_ISL_915710, EPI_ISL_915711, EPI_ISL_915712, EPI_ISL_915713, EPI_ISL_915714, EPI_ISL_915715, EPI_ISL_915716, EPI_ISL_915717, EPI_ISL_915718, EPI_ISL_915719, EPI_ISL_915720,
EPI_ISL_915721, EPI_ISL_915722, EPI_ISL_915723, EPI_ISL_915724, EPI_ISL_915725, EPI_ISL_915726, EPI_ISL_915727, EPI_ISL_915728, EPI_ISL_915729, EPI_ISL_915730, EPI_ISL_915731, EPI_ISL_915732, EPI_ISL_915733, EPI_ISL_915734, EPI_ISL_915735, EPI_ISL_915736, EPI_ISL_915737, EPI_ISL_915738, EPI_ISL_915739, EPI_ISL_915740, EPI_ISL_915741, EPI_ISL_915742, EPI_ISL_915743, EPI_ISL_915744,
EPI_ISL_915745, EPI_ISL_915746, EPI_ISL_915747, EPI_ISL_915748, EPI_ISL_915749, EPI_ISL_915750, EPI_ISL_915751, EPI_ISL_915752, EPI_ISL_915753, EPI_ISL_915754, EPI_ISL_915755, EPI_ISL_915756, EPI_ISL_915757, EPI_ISL_915758, EPI_ISL_915759, EPI_ISL_915760, EPI_ISL_915761, EPI_ISL_915762, EPI_ISL_915763, EPI_ISL_915764, EPI_ISL_915765, EPI_ISL_915766, EPI_ISL_915767, EPI_ISL_915768,
EPI_ISL_915769, EPI_ISL_915770, EPI_ISL_915771, EPI_ISL_915772, EPI_ISL_915773, EPI_ISL_915774, EPI_ISL_915775, EPI_ISL_915776, EPI_ISL_915777, EPI_ISL_915778, EPI_ISL_915779, EPI_ISL_915780, EPI_ISL_915781, EPI_ISL_915782, EPI_ISL_915783, EPI_ISL_915784, EPI_ISL_915785, EPI_ISL_915786, EPI_ISL_915787, EPI_ISL_915788, EPI_ISL_915789, EPI_ISL_915790, EPI_ISL_915791, EPI_ISL_915792,
EPI_ISL_915793, EPI_ISL_915794, EPI_ISL_915795, EPI_ISL_915796, EPI_ISL_915797, EPI_ISL_915798, EPI_ISL_915799, EPI_ISL_915800, EPI_ISL_915801, EPI_ISL_915802, EPI_ISL_915803, EPI_ISL_915804, EPI_ISL_915805, EPI_ISL_915806, EPI_ISL_915807, EPI_ISL_915808, EPI_ISL_915809, EPI_ISL_915810, EPI_ISL_915811, EPI_ISL_915812, EPI_ISL_915813, EPI_ISL_915814, EPI_ISL_915815, EPI_ISL_915816,
EPI_ISL_915817, EPI_ISL_915818, EPI_ISL_915819, EPI_ISL_915820, EPI_ISL_915821, EPI_ISL_915822, EPI_ISL_915823, EPI_ISL_915824, EPI_ISL_915825, EPI_ISL_915826, EPI_ISL_915827, EPI_ISL_915828, EPI_ISL_915829, EPI_ISL_915830, EPI_ISL_915831, EPI_ISL_915832, EPI_ISL_915833, EPI_ISL_915834, EPI_ISL_915835, EPI_ISL_915836, EPI_ISL_915837, EPI_ISL_915838, EPI_ISL_915839, EPI_ISL_915840,
EPI_ISL_915841, EPI_ISL_915842, EPI_ISL_915843, EPI_ISL_915844, EPI_ISL_915845, EPI_ISL_915846, EPI_ISL_915847, EPI_ISL_915848, EPI_ISL_915849, EPI_ISL_915850, EPI_ISL_915851, EPI_ISL_915852, EPI_ISL_915853, EPI_ISL_915854, EPI_ISL_915855, EPI_ISL_915856, EPI_ISL_915857, EPI_ISL_915858, EPI_ISL_915859, EPI_ISL_915860, EPI_ISL_915861, EPI_ISL_915862, EPI_ISL_915863, EPI_ISL_915864,
EPI_ISL_915865, EPI_ISL_915866, EPI_ISL_915867, EPI_ISL_915868, EPI_ISL_915869, EPI_ISL_915870, EPI_ISL_915871, EPI_ISL_915872, EPI_ISL_915873, EPI_ISL_915874, EPI_ISL_915875, EPI_ISL_915876, EPI_ISL_915877, EPI_ISL_915878, EPI_ISL_915879, EPI_ISL_915880, EPI_ISL_915881, EPI_ISL_915882, EPI_ISL_915883, EPI_ISL_915884, EPI_ISL_915885, EPI_ISL_915886, EPI_ISL_915887, EPI_ISL_915888,
EPI_ISL_915889, EPI_ISL_915890, EPI_ISL_915891, EPI_ISL_915892, EPI_ISL_915893, EPI_ISL_915894, EPI_ISL_915895, EPI_ISL_915896, EPI_ISL_915897, EPI_ISL_915898, EPI_ISL_915899, EPI_ISL_915900, EPI_ISL_915901, EPI_ISL_915902, EPI_ISL_915903, EPI_ISL_915904, EPI_ISL_915905, EPI_ISL_915906, EPI_ISL_915907, EPI_ISL_915908, EPI_ISL_915909, EPI_ISL_915910, EPI_ISL_915912, EPI_ISL_915913,
EPI_ISL_915914, EPI_ISL_915915, EPI_ISL_915916, EPI_ISL_915917, EPI_ISL_915918, EPI_ISL_915919, EPI_ISL_915920, EPI_ISL_915921, EPI_ISL_915922, EPI_ISL_915923, EPI_ISL_915924, EPI_ISL_915925, EPI_ISL_915926, EPI_ISL_915927, EPI_ISL_915928, EPI_ISL_915929, EPI_ISL_915930, EPI_ISL_915931, EPI_ISL_915932, EPI_ISL_915933, EPI_ISL_915934, EPI_ISL_915935, EPI_ISL_915936, EPI_ISL_915937,
EPI_ISL_915938, EPI_ISL_915939, EPI_ISL_915940, EPI_ISL_915941, EPI_ISL_915942, EPI_ISL_915943, EPI_ISL_915944, EPI_ISL_915945, EPI_ISL_915946, EPI_ISL_915947, EPI_ISL_915948, EPI_ISL_915949, EPI_ISL_915950, EPI_ISL_915951, EPI_ISL_915952, EPI_ISL_915953, EPI_ISL_915954, EPI_ISL_915955, EPI_ISL_915956, EPI_ISL_915957, EPI_ISL_915958, EPI_ISL_915959, EPI_ISL_915960, EPI_ISL_915961,
EPI_ISL_915962, EPI_ISL_915963, EPI_ISL_915964, EPI_ISL_915965, EPI_ISL_915966, EPI_ISL_915967, EPI_ISL_915968, EPI_ISL_915969, EPI_ISL_915970, EPI_ISL_915971, EPI_ISL_915972, EPI_ISL_915973, EPI_ISL_915974, EPI_ISL_915975, EPI_ISL_915976, EPI_ISL_915977, EPI_ISL_915978, EPI_ISL_915979, EPI_ISL_915980, EPI_ISL_915981, EPI_ISL_915982, EPI_ISL_915983, EPI_ISL_915984, EPI_ISL_915985,
EPI_ISL_915986, EPI_ISL_915987, EPI_ISL_915988, EPI_ISL_915989, EPI_ISL_915990, EPI_ISL_915991, EPI_ISL_915992, EPI_ISL_915993, EPI_ISL_915994, EPI_ISL_915995, EPI_ISL_915996, EPI_ISL_915997, EPI_ISL_915998, EPI_ISL_915999, EPI_ISL_916000, EPI_ISL_916001, EPI_ISL_916002, EPI_ISL_916003, EPI_ISL_916004, EPI_ISL_916005, EPI_ISL_916006, EPI_ISL_916007, EPI_ISL_916008, EPI_ISL_916009,
EPI_ISL_916010, EPI_ISL_916011, EPI_ISL_916012, EPI_ISL_916013, EPI_ISL_916014, EPI_ISL_916015, EPI_ISL_916016, EPI_ISL_916017, EPI_ISL_916018, EPI_ISL_916019, EPI_ISL_916020, EPI_ISL_916021, EPI_ISL_916022, EPI_ISL_916023, EPI_ISL_916024, EPI_ISL_916025, EPI_ISL_916026, EPI_ISL_916027, EPI_ISL_916028, EPI_ISL_916029, EPI_ISL_916030, EPI_ISL_916031, EPI_ISL_916032, EPI_ISL_916033,
EPI_ISL_916034, EPI_ISL_916035, EPI_ISL_916036, EPI_ISL_916037, EPI_ISL_916038, EPI_ISL_916039, EPI_ISL_916040, EPI_ISL_916041, EPI_ISL_916042, EPI_ISL_916043, EPI_ISL_916044, EPI_ISL_916045, EPI_ISL_916046, EPI_ISL_916047, EPI_ISL_916048, EPI_ISL_916049, EPI_ISL_916050, EPI_ISL_916051, EPI_ISL_916052, EPI_ISL_916053, EPI_ISL_916054, EPI_ISL_916055, EPI_ISL_916056, EPI_ISL_916057,
EPI_ISL_916058, EPI_ISL_916059, EPI_ISL_916060, EPI_ISL_916061, EPI_ISL_916062, EPI_ISL_916063, EPI_ISL_916064, EPI_ISL_916065, EPI_ISL_916066, EPI_ISL_916067, EPI_ISL_916068, EPI_ISL_916069, EPI_ISL_916070, EPI_ISL_916071, EPI_ISL_916072, EPI_ISL_916073, EPI_ISL_916074, EPI_ISL_916075, EPI_ISL_916076, EPI_ISL_916077, EPI_ISL_916078, EPI_ISL_916079, EPI_ISL_916080, EPI_ISL_916081,
EPI_ISL_916082, EPI_ISL_916083, EPI_ISL_916084, EPI_ISL_916085, EPI_ISL_916086, EPI_ISL_916087, EPI_ISL_916088, EPI_ISL_916089, EPI_ISL_916090, EPI_ISL_916091, EPI_ISL_916092, EPI_ISL_916093, EPI_ISL_916094, EPI_ISL_916095, EPI_ISL_916096, EPI_ISL_916097, EPI_ISL_916098, EPI_ISL_916099, EPI_ISL_916100, EPI_ISL_916101, EPI_ISL_916102, EPI_ISL_916103, EPI_ISL_916104, EPI_ISL_916105,
EPI_ISL_916106, EPI_ISL_916107, EPI_ISL_916108, EPI_ISL_916109, EPI_ISL_916110, EPI_ISL_916111, EPI_ISL_916112, EPI_ISL_916113, EPI_ISL_916114, EPI_ISL_916115, EPI_ISL_916116, EPI_ISL_916117, EPI_ISL_916118, EPI_ISL_916120, EPI_ISL_916121, EPI_ISL_916122, EPI_ISL_916123, EPI_ISL_916124, EPI_ISL_916125, EPI_ISL_916126, EPI_ISL_916127, EPI_ISL_916128, EPI_ISL_916129, EPI_ISL_916130,
EPI_ISL_916131, EPI_ISL_916132, EPI_ISL_916133, EPI_ISL_916134, EPI_ISL_916135, EPI_ISL_916136, EPI_ISL_916137, EPI_ISL_916138, EPI_ISL_916139, EPI_ISL_916140, EPI_ISL_916141, EPI_ISL_916142, EPI_ISL_916143, EPI_ISL_916144, EPI_ISL_916145, EPI_ISL_916146, EPI_ISL_916147, EPI_ISL_916148, EPI_ISL_916149, EPI_ISL_916150, EPI_ISL_916152, EPI_ISL_916153, EPI_ISL_916154, EPI_ISL_916155,
EPI_ISL_916156, EPI_ISL_916157, EPI_ISL_916158, EPI_ISL_916159, EPI_ISL_916160, EPI_ISL_916161, EPI_ISL_916162, EPI_ISL_916163, EPI_ISL_916165, EPI_ISL_916166, EPI_ISL_916167, EPI_ISL_916168, EPI_ISL_916169, EPI_ISL_916170, EPI_ISL_916171, EPI_ISL_916172, EPI_ISL_916173, EPI_ISL_916174, EPI_ISL_916175, EPI_ISL_916176, EPI_ISL_916177, EPI_ISL_916178, EPI_ISL_916179, EPI_ISL_991309,
EPI_ISL_991310, EPI_ISL_991311, EPI_ISL_991312, EPI_ISL_991313, EPI_ISL_991314, EPI_ISL_991315, EPI_ISL_991316, EPI_ISL_991317, EPI_ISL_991318, EPI_ISL_991319, EPI_ISL_991320, EPI_ISL_991321, EPI_ISL_991322, EPI_ISL_991323, EPI_ISL_991324, EPI_ISL_991325, EPI_ISL_991326, EPI_ISL_991327, EPI_ISL_991328, EPI_ISL_991329, EPI_ISL_991330, EPI_ISL_991331, EPI_ISL_991332, EPI_ISL_991333,
EPI_ISL_991334, EPI_ISL_991335, EPI_ISL_991336, EPI_ISL_991337, EPI_ISL_991338, EPI_ISL_991339, EPI_ISL_991340, EPI_ISL_991341, EPI_ISL_991342, EPI_ISL_991343, EPI_ISL_991344, EPI_ISL_991345, EPI_ISL_991346, EPI_ISL_991347, EPI_ISL_991348, EPI_ISL_991349, EPI_ISL_991350

| | | | |
|---|---|---|---|
| see above | Lighthouse Lab in Alderley Park | Wellcome Sanger Institute for the COVID-19 Genomics UK (COG-UK) Consortium | Cordelia Langford; David K. Jackson; Dominic Kwiatkowski; Ewan Harrison; Ian Johnston; Jacquelyn Wynn; John Sillitoe on behalf of the Wellcome Sanger Institute COVID-19 Surveillance Team; Mairead Hyland; Roberto Amato; Sonia Goncalves; The Lighthouse Lab in Alderley Park and Alex Alderton |

EPI_ISL_702023, EPI_ISL_702024, EPI_ISL_702026, EPI_ISL_702029, EPI_ISL_702030, EPI_ISL_702038, EPI_ISL_702039, EPI_ISL_702040, EPI_ISL_702047, EPI_ISL_702051, EPI_ISL_702052, EPI_ISL_702053, EPI_ISL_702058, EPI_ISL_702059, EPI_ISL_702060, EPI_ISL_702064, EPI_ISL_702065, EPI_ISL_702066, EPI_ISL_702067, EPI_ISL_702070, EPI_ISL_702073, EPI_ISL_702074, EPI_ISL_702079, EPI_ISL_702083,
EPI_ISL_702084, EPI_ISL_702086, EPI_ISL_702088, EPI_ISL_702089, EPI_ISL_702092, EPI_ISL_702095, EPI_ISL_702101, EPI_ISL_702102, EPI_ISL_702103, EPI_ISL_702104, EPI_ISL_702107, EPI_ISL_702108, EPI_ISL_702110, EPI_ISL_702111, EPI_ISL_702113, EPI_ISL_702118, EPI_ISL_702125, EPI_ISL_702127, EPI_ISL_702128, EPI_ISL_702129, EPI_ISL_702131, EPI_ISL_702133, EPI_ISL_702134, EPI_ISL_702135,
EPI_ISL_702136, EPI_ISL_702137, EPI_ISL_702142, EPI_ISL_702143, EPI_ISL_702146, EPI_ISL_702149, EPI_ISL_702150, EPI_ISL_702152, EPI_ISL_702153, EPI_ISL_702154, EPI_ISL_702155, EPI_ISL_702157, EPI_ISL_702160, EPI_ISL_702161, EPI_ISL_702167, EPI_ISL_702168, EPI_ISL_702171, EPI_ISL_702172, EPI_ISL_702177, EPI_ISL_702178, EPI_ISL_702179, EPI_ISL_702181, EPI_ISL_702184, EPI_ISL_702185,
EPI_ISL_702186, EPI_ISL_702187, EPI_ISL_702189, EPI_ISL_702190, EPI_ISL_702191, EPI_ISL_702194, EPI_ISL_702198, EPI_ISL_702201, EPI_ISL_702203, EPI_ISL_702205, EPI_ISL_702206, EPI_ISL_702208, EPI_ISL_702209, EPI_ISL_702211, EPI_ISL_702213, EPI_ISL_702216, EPI_ISL_702222, EPI_ISL_702223, EPI_ISL_702225, EPI_ISL_702230, EPI_ISL_702232, EPI_ISL_702236, EPI_ISL_702237, EPI_ISL_702240,
EPI_ISL_702242, EPI_ISL_702243, EPI_ISL_702245, EPI_ISL_702250, EPI_ISL_702251, EPI_ISL_702252, EPI_ISL_702256, EPI_ISL_702259, EPI_ISL_702260, EPI_ISL_702261, EPI_ISL_702263, EPI_ISL_702264, EPI_ISL_702265, EPI_ISL_702266, EPI_ISL_702267, EPI_ISL_702268, EPI_ISL_702269, EPI_ISL_702270, EPI_ISL_702271, EPI_ISL_702277, EPI_ISL_702282, EPI_ISL_702283, EPI_ISL_702285, EPI_ISL_702286,
EPI_ISL_702287, EPI_ISL_702289, EPI_ISL_702291, EPI_ISL_702294, EPI_ISL_702295, EPI_ISL_702297, EPI_ISL_702302, EPI_ISL_702304, EPI_ISL_702306, EPI_ISL_702310, EPI_ISL_702312, EPI_ISL_702317, EPI_ISL_702318, EPI_ISL_702319, EPI_ISL_702321, EPI_ISL_702322, EPI_ISL_702324, EPI_ISL_702325, EPI_ISL_702326, EPI_ISL_702329, EPI_ISL_702330, EPI_ISL_702331, EPI_ISL_702332, EPI_ISL_702334,
EPI_ISL_702338, EPI_ISL_702339, EPI_ISL_702340, EPI_ISL_702342, EPI_ISL_702343, EPI_ISL_702345, EPI_ISL_702346, EPI_ISL_702347, EPI_ISL_702348, EPI_ISL_702349, EPI_ISL_702350, EPI_ISL_702351, EPI_ISL_702354, EPI_ISL_702355, EPI_ISL_702356, EPI_ISL_702357, EPI_ISL_702362, EPI_ISL_702363, EPI_ISL_702364, EPI_ISL_702365, EPI_ISL_702366, EPI_ISL_702367, EPI_ISL_704688, EPI_ISL_704689,
EPI_ISL_704691, EPI_ISL_704693, EPI_ISL_704694, EPI_ISL_704696, EPI_ISL_704697, EPI_ISL_704699, EPI_ISL_704700, EPI_ISL_704702, EPI_ISL_704704, EPI_ISL_704705, EPI_ISL_704707, EPI_ISL_704708, EPI_ISL_704710, EPI_ISL_704712, EPI_ISL_704713, EPI_ISL_704715, EPI_ISL_704717, EPI_ISL_704718, EPI_ISL_704720, EPI_ISL_704721, EPI_ISL_704723, EPI_ISL_704724, EPI_ISL_704726, EPI_ISL_704727,
EPI_ISL_704729, EPI_ISL_704730, EPI_ISL_704732, EPI_ISL_704733, EPI_ISL_704735, EPI_ISL_704737, EPI_ISL_704738, EPI_ISL_704740, EPI_ISL_704742, EPI_ISL_704743, EPI_ISL_704745, EPI_ISL_704747, EPI_ISL_704748, EPI_ISL_704751, EPI_ISL_704753, EPI_ISL_704755, EPI_ISL_704756, EPI_ISL_704758, EPI_ISL_704760, EPI_ISL_704761, EPI_ISL_704763, EPI_ISL_704764, EPI_ISL_704766, EPI_ISL_704767,
EPI_ISL_704769, EPI_ISL_704771, EPI_ISL_704773, EPI_ISL_704774, EPI_ISL_704776, EPI_ISL_704778, EPI_ISL_704779, EPI_ISL_704781, EPI_ISL_704782, EPI_ISL_704784, EPI_ISL_704785, EPI_ISL_704787, EPI_ISL_704789, EPI_ISL_704790, EPI_ISL_704792, EPI_ISL_704793, EPI_ISL_704795, EPI_ISL_704796, EPI_ISL_704798, EPI_ISL_704800, EPI_ISL_704801, EPI_ISL_704803, EPI_ISL_704805, EPI_ISL_704806,
EPI_ISL_704808, EPI_ISL_704810, EPI_ISL_704811, EPI_ISL_704813, EPI_ISL_704814, EPI_ISL_704816, EPI_ISL_704818, EPI_ISL_704819, EPI_ISL_704821, EPI_ISL_704822, EPI_ISL_704824, EPI_ISL_704825, EPI_ISL_704827, EPI_ISL_704829, EPI_ISL_704830, EPI_ISL_704832, EPI_ISL_704833, EPI_ISL_704835, EPI_ISL_704837, EPI_ISL_704838, EPI_ISL_704840, EPI_ISL_704841, EPI_ISL_704843, EPI_ISL_704844,
EPI_ISL_704846, EPI_ISL_704848, EPI_ISL_704849, EPI_ISL_704851, EPI_ISL_704852, EPI_ISL_704854, EPI_ISL_704856, EPI_ISL_704857, EPI_ISL_704859, EPI_ISL_704860, EPI_ISL_704862, EPI_ISL_704863, EPI_ISL_704865, EPI_ISL_704867, EPI_ISL_704868, EPI_ISL_704870, EPI_ISL_704871, EPI_ISL_704873, EPI_ISL_704875, EPI_ISL_704876, EPI_ISL_704878, EPI_ISL_704880, EPI_ISL_704881, EPI_ISL_704883,
EPI_ISL_704884, EPI_ISL_704886, EPI_ISL_704888, EPI_ISL_704889, EPI_ISL_704891, EPI_ISL_704893, EPI_ISL_704894, EPI_ISL_704896, EPI_ISL_704898, EPI_ISL_704899, EPI_ISL_704901, EPI_ISL_704902, EPI_ISL_704904, EPI_ISL_704906, EPI_ISL_704907, EPI_ISL_704909, EPI_ISL_704910, EPI_ISL_704912, EPI_ISL_704913, EPI_ISL_704915, EPI_ISL_704917, EPI_ISL_704919, EPI_ISL_704920, EPI_ISL_704922,
EPI_ISL_704924, EPI_ISL_704925, EPI_ISL_704927, EPI_ISL_704928, EPI_ISL_704930, EPI_ISL_704932, EPI_ISL_704933, EPI_ISL_704935, EPI_ISL_704936, EPI_ISL_704938, EPI_ISL_704940, EPI_ISL_704941, EPI_ISL_704943, EPI_ISL_704944, EPI_ISL_704946, EPI_ISL_704948, EPI_ISL_704949, EPI_ISL_704951, EPI_ISL_704953, EPI_ISL_704954, EPI_ISL_704956, EPI_ISL_704958, EPI_ISL_704959, EPI_ISL_704961,
EPI_ISL_704962, EPI_ISL_704964, EPI_ISL_704966, EPI_ISL_704967, EPI_ISL_704969, EPI_ISL_704971, EPI_ISL_704972, EPI_ISL_704974, EPI_ISL_704975, EPI_ISL_704977, EPI_ISL_704979, EPI_ISL_704980, EPI_ISL_704982, EPI_ISL_704984, EPI_ISL_704985, EPI_ISL_704987, EPI_ISL_704988, EPI_ISL_704990, EPI_ISL_704992, EPI_ISL_704993, EPI_ISL_704995, EPI_ISL_704996, EPI_ISL_704998, EPI_ISL_705000,
EPI_ISL_705001, EPI_ISL_705003, EPI_ISL_705004, EPI_ISL_705006, EPI_ISL_705008, EPI_ISL_705009, EPI_ISL_705011, EPI_ISL_705012, EPI_ISL_705014, EPI_ISL_705016, EPI_ISL_705017, EPI_ISL_705019, EPI_ISL_705020, EPI_ISL_705022, EPI_ISL_705023, EPI_ISL_705025, EPI_ISL_705027, EPI_ISL_705028, EPI_ISL_705029, EPI_ISL_705031, EPI_ISL_705033, EPI_ISL_705034, EPI_ISL_705036, EPI_ISL_705037,
EPI_ISL_705039, EPI_ISL_705041, EPI_ISL_705042, EPI_ISL_705044, EPI_ISL_705045, EPI_ISL_705047, EPI_ISL_705049, EPI_ISL_705050, EPI_ISL_705052, EPI_ISL_705053, EPI_ISL_705055, EPI_ISL_705057, EPI_ISL_705058, EPI_ISL_705060, EPI_ISL_705061, EPI_ISL_705063, EPI_ISL_705065, EPI_ISL_705066, EPI_ISL_705068, EPI_ISL_705069, EPI_ISL_705071, EPI_ISL_705073, EPI_ISL_705074, EPI_ISL_705076,
EPI_ISL_705077, EPI_ISL_705079, EPI_ISL_705080, EPI_ISL_705082, EPI_ISL_705084, EPI_ISL_705085, EPI_ISL_705087, EPI_ISL_705089, EPI_ISL_705090, EPI_ISL_705092, EPI_ISL_705093, EPI_ISL_705095, EPI_ISL_705097, EPI_ISL_705098, EPI_ISL_705100, EPI_ISL_705101, EPI_ISL_705103, EPI_ISL_705104, EPI_ISL_705106, EPI_ISL_705107, EPI_ISL_705109, EPI_ISL_705111, EPI_ISL_705112, EPI_ISL_705114,
EPI_ISL_705116, EPI_ISL_705117, EPI_ISL_705119, EPI_ISL_705120, EPI_ISL_705122, EPI_ISL_705123, EPI_ISL_705125, EPI_ISL_705127, EPI_ISL_705128, EPI_ISL_705130, EPI_ISL_705131, EPI_ISL_705133, EPI_ISL_705135, EPI_ISL_705136, EPI_ISL_705138, EPI_ISL_705139, EPI_ISL_705141, EPI_ISL_705142, EPI_ISL_705144, EPI_ISL_705146, EPI_ISL_705147, EPI_ISL_705149, EPI_ISL_705151, EPI_ISL_705152,
EPI_ISL_705154, EPI_ISL_705155, EPI_ISL_705157, EPI_ISL_705159, EPI_ISL_705160, EPI_ISL_705162, EPI_ISL_705164, EPI_ISL_705165, EPI_ISL_705167, EPI_ISL_705169, EPI_ISL_705170, EPI_ISL_705172, EPI_ISL_705173, EPI_ISL_705175, EPI_ISL_705177, EPI_ISL_705178, EPI_ISL_705180, EPI_ISL_705181, EPI_ISL_705183, EPI_ISL_705185, EPI_ISL_705186, EPI_ISL_705188, EPI_ISL_705189, EPI_ISL_710821,
EPI_ISL_710822, EPI_ISL_710823, EPI_ISL_710824, EPI_ISL_710825, EPI_ISL_710826, EPI_ISL_710827, EPI_ISL_710828, EPI_ISL_710829, EPI_ISL_800220, EPI_ISL_800885, EPI_ISL_800886, EPI_ISL_800887, EPI_ISL_800888, EPI_ISL_800889, EPI_ISL_800890, EPI_ISL_800891, EPI_ISL_800892, EPI_ISL_800893, EPI_ISL_800894, EPI_ISL_800895, EPI_ISL_800896, EPI_ISL_800897, EPI_ISL_800898, EPI_ISL_800899,
EPI_ISL_800900, EPI_ISL_800901, EPI_ISL_800902, EPI_ISL_800903, EPI_ISL_800904, EPI_ISL_800905, EPI_ISL_800906, EPI_ISL_800907, EPI_ISL_800908, EPI_ISL_800909, EPI_ISL_800910, EPI_ISL_800911, EPI_ISL_800912, EPI_ISL_800913, EPI_ISL_800914, EPI_ISL_800915, EPI_ISL_800916, EPI_ISL_800917, EPI_ISL_800918, EPI_ISL_800919, EPI_ISL_800920, EPI_ISL_800921, EPI_ISL_800922, EPI_ISL_800923,
EPI_ISL_800924, EPI_ISL_800925, EPI_ISL_800926, EPI_ISL_800927, EPI_ISL_800928, EPI_ISL_800929, EPI_ISL_800930, EPI_ISL_800931, EPI_ISL_800932, EPI_ISL_800933, EPI_ISL_800934, EPI_ISL_800935, EPI_ISL_800936, EPI_ISL_800937, EPI_ISL_800938, EPI_ISL_800939, EPI_ISL_800940, EPI_ISL_800941, EPI_ISL_800942, EPI_ISL_800943, EPI_ISL_800944, EPI_ISL_800945, EPI_ISL_800946, EPI_ISL_800947,
EPI_ISL_800948, EPI_ISL_800949, EPI_ISL_800950, EPI_ISL_800951, EPI_ISL_800952, EPI_ISL_800953, EPI_ISL_800954, EPI_ISL_800955, EPI_ISL_800956, EPI_ISL_800957, EPI_ISL_800958, EPI_ISL_800959, EPI_ISL_800960, EPI_ISL_800961, EPI_ISL_800962, EPI_ISL_800963, EPI_ISL_800964, EPI_ISL_800965, EPI_ISL_800966, EPI_ISL_800967, EPI_ISL_800968, EPI_ISL_800969, EPI_ISL_800970, EPI_ISL_800971,
EPI_ISL_800972, EPI_ISL_800973, EPI_ISL_800974, EPI_ISL_800975, EPI_ISL_800976, EPI_ISL_800977, EPI_ISL_800978, EPI_ISL_800979, EPI_ISL_800980, EPI_ISL_800981, EPI_ISL_800982, EPI_ISL_800983, EPI_ISL_800984, EPI_ISL_800985, EPI_ISL_800986, EPI_ISL_800987, EPI_ISL_800988, EPI_ISL_800989, EPI_ISL_800990, EPI_ISL_800991, EPI_ISL_800992, EPI_ISL_800993, EPI_ISL_800994, EPI_ISL_800995,
EPI_ISL_800996, EPI_ISL_800997, EPI_ISL_800998, EPI_ISL_800999, EPI_ISL_801000, EPI_ISL_801001, EPI_ISL_801002, EPI_ISL_801003, EPI_ISL_801004, EPI_ISL_801005, EPI_ISL_801006, EPI_ISL_801007, EPI_ISL_801008, EPI_ISL_801009, EPI_ISL_801010, EPI_ISL_801011, EPI_ISL_801012, EPI_ISL_801013, EPI_ISL_801014, EPI_ISL_801015, EPI_ISL_801016, EPI_ISL_801017, EPI_ISL_801018, EPI_ISL_801019,
EPI_ISL_801020, EPI_ISL_801021, EPI_ISL_801022, EPI_ISL_801023, EPI_ISL_801024, EPI_ISL_801025, EPI_ISL_801026, EPI_ISL_801027, EPI_ISL_801028, EPI_ISL_801029, EPI_ISL_801030, EPI_ISL_801031, EPI_ISL_801032, EPI_ISL_801033, EPI_ISL_801034, EPI_ISL_801035, EPI_ISL_801036, EPI_ISL_801037, EPI_ISL_801038, EPI_ISL_801039, EPI_ISL_801040, EPI_ISL_801041, EPI_ISL_801042, EPI_ISL_801043,
EPI_ISL_801044, EPI_ISL_801045, EPI_ISL_801046, EPI_ISL_801047, EPI_ISL_801048, EPI_ISL_801049, EPI_ISL_801050, EPI_ISL_801051, EPI_ISL_801052, EPI_ISL_801053, EPI_ISL_801054, EPI_ISL_801056, EPI_ISL_801057, EPI_ISL_801058, EPI_ISL_801059, EPI_ISL_801060, EPI_ISL_801061, EPI_ISL_801062, EPI_ISL_801063, EPI_ISL_801064, EPI_ISL_801065, EPI_ISL_801066, EPI_ISL_801067, EPI_ISL_801068,
EPI_ISL_801069, EPI_ISL_801070, EPI_ISL_801071, EPI_ISL_801072, EPI_ISL_801073, EPI_ISL_801074, EPI_ISL_801075, EPI_ISL_801076, EPI_ISL_801077, EPI_ISL_801078, EPI_ISL_801079, EPI_ISL_801080, EPI_ISL_801081, EPI_ISL_801082, EPI_ISL_801083, EPI_ISL_801084, EPI_ISL_801085, EPI_ISL_801086, EPI_ISL_801087, EPI_ISL_801088, EPI_ISL_801089, EPI_ISL_801090, EPI_ISL_801091, EPI_ISL_801092,
EPI_ISL_801093, EPI_ISL_801094, EPI_ISL_801095, EPI_ISL_801097, EPI_ISL_801098, EPI_ISL_801099, EPI_ISL_801100, EPI_ISL_801101, EPI_ISL_801102, EPI_ISL_801103, EPI_ISL_801104, EPI_ISL_801105, EPI_ISL_801106, EPI_ISL_801107, EPI_ISL_801108, EPI_ISL_801109, EPI_ISL_801110, EPI_ISL_801111, EPI_ISL_801112, EPI_ISL_801113, EPI_ISL_801114, EPI_ISL_801115, EPI_ISL_801116, EPI_ISL_801117,
EPI_ISL_801118, EPI_ISL_801119, EPI_ISL_801120, EPI_ISL_801121, EPI_ISL_801122, EPI_ISL_801123, EPI_ISL_801124, EPI_ISL_801125, EPI_ISL_801126, EPI_ISL_801127, EPI_ISL_801128, EPI_ISL_801129, EPI_ISL_801130, EPI_ISL_801131, EPI_ISL_801132, EPI_ISL_801133, EPI_ISL_801134, EPI_ISL_801135, EPI_ISL_801136, EPI_ISL_801137, EPI_ISL_801138, EPI_ISL_801139, EPI_ISL_801140, EPI_ISL_801141,
EPI_ISL_801142, EPI_ISL_801143, EPI_ISL_801144, EPI_ISL_801145, EPI_ISL_801146, EPI_ISL_801147, EPI_ISL_801149, EPI_ISL_801150, EPI_ISL_801151, EPI_ISL_801152, EPI_ISL_801153, EPI_ISL_801154, EPI_ISL_801155, EPI_ISL_801156, EPI_ISL_801157, EPI_ISL_801158, EPI_ISL_801159, EPI_ISL_801160, EPI_ISL_801161, EPI_ISL_801162, EPI_ISL_801163, EPI_ISL_801164, EPI_ISL_801165, EPI_ISL_801166,
EPI_ISL_801167, EPI_ISL_801168, EPI_ISL_801169, EPI_ISL_801170, EPI_ISL_801171, EPI_ISL_801172, EPI_ISL_801173, EPI_ISL_801174, EPI_ISL_801175, EPI_ISL_801176, EPI_ISL_801177, EPI_ISL_801178, EPI_ISL_859554, EPI_ISL_859556, EPI_ISL_859557, EPI_ISL_859558, EPI_ISL_859559

| | | | |
|---|---|---|---|
| see above | Lighthouse Lab in Cambridge | Wellcome Sanger Institute for the COVID-19 Genomics UK (COG-UK) Consortium | Cordelia Langford; David K. Jackson; Dominic Kwiatkowski; Ewan Harrison; Ian Johnston; John Sillitoe on behalf of the Wellcome Sanger Institute COVID-19 Surveillance Team; Rob Howes; Roberto Amato; Sonia Goncalves; The Lighthouse Lab in Cambridge and Alex Alderton |

EPI_ISL_672752, EPI_ISL_672753, EPI_ISL_672754, EPI_ISL_672755, EPI_ISL_672756, EPI_ISL_672757, EPI_ISL_672758, EPI_ISL_672759, EPI_ISL_672760, EPI_ISL_672761, EPI_ISL_672762, EPI_ISL_672763, EPI_ISL_672764, EPI_ISL_672765, EPI_ISL_672766, EPI_ISL_672767, EPI_ISL_672768, EPI_ISL_672769, EPI_ISL_672770, EPI_ISL_672771, EPI_ISL_672772, EPI_ISL_672773, EPI_ISL_672774, EPI_ISL_672775,
EPI_ISL_672776, EPI_ISL_672777, EPI_ISL_672778, EPI_ISL_672779, EPI_ISL_672780, EPI_ISL_672781, EPI_ISL_672782, EPI_ISL_672783, EPI_ISL_672784, EPI_ISL_672785, EPI_ISL_672786, EPI_ISL_672787, EPI_ISL_672788, EPI_ISL_672789, EPI_ISL_672790, EPI_ISL_672791, EPI_ISL_672792, EPI_ISL_672793, EPI_ISL_672794, EPI_ISL_672795, EPI_ISL_672796, EPI_ISL_672797, EPI_ISL_672798, EPI_ISL_672799,
EPI_ISL_672800, EPI_ISL_672801, EPI_ISL_672802, EPI_ISL_672803, EPI_ISL_672804, EPI_ISL_672805, EPI_ISL_672806, EPI_ISL_672807, EPI_ISL_672808, EPI_ISL_672809, EPI_ISL_672810, EPI_ISL_672811, EPI_ISL_672812, EPI_ISL_672813, EPI_ISL_672814, EPI_ISL_672815, EPI_ISL_672816, EPI_ISL_672817, EPI_ISL_672818, EPI_ISL_672819, EPI_ISL_672820, EPI_ISL_672821, EPI_ISL_672822, EPI_ISL_672823,
EPI_ISL_672824, EPI_ISL_672825, EPI_ISL_672826, EPI_ISL_672827, EPI_ISL_672828, EPI_ISL_672829, EPI_ISL_672830, EPI_ISL_672831, EPI_ISL_672832, EPI_ISL_672833, EPI_ISL_672834, EPI_ISL_672835, EPI_ISL_672836, EPI_ISL_672837, EPI_ISL_672838, EPI_ISL_672839, EPI_ISL_672840, EPI_ISL_672841, EPI_ISL_672842, EPI_ISL_672843, EPI_ISL_672844, EPI_ISL_672845, EPI_ISL_672846, EPI_ISL_672847,
EPI_ISL_672848, EPI_ISL_672849, EPI_ISL_672850, EPI_ISL_672851, EPI_ISL_672852, EPI_ISL_672853, EPI_ISL_672854, EPI_ISL_672855, EPI_ISL_672856, EPI_ISL_672857, EPI_ISL_672858, EPI_ISL_672859, EPI_ISL_672860, EPI_ISL_672861, EPI_ISL_672862, EPI_ISL_672863, EPI_ISL_672864, EPI_ISL_672865, EPI_ISL_672866, EPI_ISL_672867, EPI_ISL_672868, EPI_ISL_672869, EPI_ISL_672870, EPI_ISL_672871,
EPI_ISL_672872, EPI_ISL_672873, EPI_ISL_672874, EPI_ISL_672875, EPI_ISL_672876, EPI_ISL_672877, EPI_ISL_672878, EPI_ISL_672879, EPI_ISL_672880, EPI_ISL_672881, EPI_ISL_672882, EPI_ISL_672883, EPI_ISL_672884, EPI_ISL_672885, EPI_ISL_672886, EPI_ISL_672887, EPI_ISL_672888, EPI_ISL_672889, EPI_ISL_672890, EPI_ISL_672891, EPI_ISL_672892, EPI_ISL_672893, EPI_ISL_672894, EPI_ISL_672895,
EPI_ISL_672896, EPI_ISL_672897, EPI_ISL_672898, EPI_ISL_672899, EPI_ISL_672900, EPI_ISL_672901, EPI_ISL_672902, EPI_ISL_672903, EPI_ISL_672904, EPI_ISL_672905, EPI_ISL_672906, EPI_ISL_672907, EPI_ISL_672908, EPI_ISL_672909, EPI_ISL_672910, EPI_ISL_672911, EPI_ISL_703262, EPI_ISL_703263, EPI_ISL_703270, EPI_ISL_703273, EPI_ISL_703280, EPI_ISL_703283, EPI_ISL_703285, EPI_ISL_703286,
EPI_ISL_703288, EPI_ISL_703290, EPI_ISL_703291, EPI_ISL_703296, EPI_ISL_703300, EPI_ISL_703303, EPI_ISL_703304, EPI_ISL_703309, EPI_ISL_703315, EPI_ISL_703320, EPI_ISL_703323, EPI_ISL_703325, EPI_ISL_703331, EPI_ISL_703334, EPI_ISL_703337, EPI_ISL_703345, EPI_ISL_703349, EPI_ISL_703350, EPI_ISL_703352, EPI_ISL_703354, EPI_ISL_703355, EPI_ISL_703358, EPI_ISL_703360, EPI_ISL_703362,
EPI_ISL_703366, EPI_ISL_703370, EPI_ISL_703371, EPI_ISL_703378, EPI_ISL_703379, EPI_ISL_703381, EPI_ISL_703382, EPI_ISL_703384, EPI_ISL_703389, EPI_ISL_703391, EPI_ISL_703393, EPI_ISL_703399, EPI_ISL_703401, EPI_ISL_703407, EPI_ISL_703408, EPI_ISL_703410, EPI_ISL_703411, EPI_ISL_703415, EPI_ISL_703422, EPI_ISL_703423, EPI_ISL_703426, EPI_ISL_703428, EPI_ISL_703429, EPI_ISL_703431,
EPI_ISL_703439, EPI_ISL_703442, EPI_ISL_703444, EPI_ISL_703445, EPI_ISL_703447, EPI_ISL_703453, EPI_ISL_703456, EPI_ISL_703458, EPI_ISL_703463, EPI_ISL_703466, EPI_ISL_703469, EPI_ISL_703472, EPI_ISL_703474, EPI_ISL_703482, EPI_ISL_703491, EPI_ISL_703496, EPI_ISL_703499, EPI_ISL_703501, EPI_ISL_703506, EPI_ISL_703510, EPI_ISL_703512, EPI_ISL_703514, EPI_ISL_703515, EPI_ISL_703517,
EPI_ISL_703522, EPI_ISL_703525, EPI_ISL_703529, EPI_ISL_703534, EPI_ISL_703538, EPI_ISL_703541, EPI_ISL_703546, EPI_ISL_703550, EPI_ISL_703553, EPI_ISL_703557, EPI_ISL_703560, EPI_ISL_703561, EPI_ISL_703578, EPI_ISL_703583, EPI_ISL_703584, EPI_ISL_703598, EPI_ISL_703600, EPI_ISL_703607, EPI_ISL_703608, EPI_ISL_703610, EPI_ISL_703612, EPI_ISL_703617, EPI_ISL_703621, EPI_ISL_703626,
EPI_ISL_703631, EPI_ISL_703633, EPI_ISL_703634, EPI_ISL_703640, EPI_ISL_703641, EPI_ISL_703643, EPI_ISL_703645, EPI_ISL_703650, EPI_ISL_703651, EPI_ISL_703653, EPI_ISL_703654, EPI_ISL_703656, EPI_ISL_703657, EPI_ISL_703659, EPI_ISL_703661, EPI_ISL_703669, EPI_ISL_703672, EPI_ISL_703673, EPI_ISL_703677, EPI_ISL_703685, EPI_ISL_703694, EPI_ISL_703697, EPI_ISL_703700, EPI_ISL_703703,
EPI_ISL_703786, EPI_ISL_703790, EPI_ISL_703799, EPI_ISL_703801, EPI_ISL_703806, EPI_ISL_703808, EPI_ISL_703811, EPI_ISL_703816, EPI_ISL_703819, EPI_ISL_703820, EPI_ISL_703822, EPI_ISL_703824, EPI_ISL_703825, EPI_ISL_703827, EPI_ISL_703828, EPI_ISL_703830, EPI_ISL_703832, EPI_ISL_703833, EPI_ISL_703835, EPI_ISL_703836, EPI_ISL_703838, EPI_ISL_703840, EPI_ISL_703841, EPI_ISL_703843,
EPI_ISL_703844, EPI_ISL_703846, EPI_ISL_703848, EPI_ISL_703849, EPI_ISL_703851, EPI_ISL_703852, EPI_ISL_703854, EPI_ISL_703856, EPI_ISL_703857, EPI_ISL_703859, EPI_ISL_703861, EPI_ISL_703862, EPI_ISL_703864, EPI_ISL_703866, EPI_ISL_703867, EPI_ISL_703869, EPI_ISL_703871, EPI_ISL_703872, EPI_ISL_703874, EPI_ISL_703875, EPI_ISL_703877, EPI_ISL_703878, EPI_ISL_703880, EPI_ISL_703882,
EPI_ISL_703883, EPI_ISL_703885, EPI_ISL_703887, EPI_ISL_703889, EPI_ISL_703890, EPI_ISL_703892, EPI_ISL_703893, EPI_ISL_703895, EPI_ISL_703896, EPI_ISL_703898, EPI_ISL_703900, EPI_ISL_703901, EPI_ISL_703903, EPI_ISL_703904, EPI_ISL_703906, EPI_ISL_703907, EPI_ISL_703909, EPI_ISL_703910, EPI_ISL_703912, EPI_ISL_703914, EPI_ISL_703915, EPI_ISL_703917, EPI_ISL_703919, EPI_ISL_703920,
EPI_ISL_703922, EPI_ISL_703923, EPI_ISL_703925, EPI_ISL_703926, EPI_ISL_703928, EPI_ISL_703930, EPI_ISL_703931, EPI_ISL_703933, EPI_ISL_703934, EPI_ISL_703936, EPI_ISL_703938, EPI_ISL_703939, EPI_ISL_703941, EPI_ISL_703943, EPI_ISL_703944, EPI_ISL_703946, EPI_ISL_703948, EPI_ISL_703949, EPI_ISL_703951, EPI_ISL_703953, EPI_ISL_703954, EPI_ISL_703956, EPI_ISL_703957, EPI_ISL_703959,
EPI_ISL_703960, EPI_ISL_703962, EPI_ISL_703964, EPI_ISL_703965, EPI_ISL_703967, EPI_ISL_703969, EPI_ISL_703970, EPI_ISL_703972, EPI_ISL_703973, EPI_ISL_703975, EPI_ISL_703976, EPI_ISL_703978, EPI_ISL_703980, EPI_ISL_703981, EPI_ISL_703983, EPI_ISL_703984, EPI_ISL_703986, EPI_ISL_703988, EPI_ISL_703989, EPI_ISL_703991, EPI_ISL_703993, EPI_ISL_703994, EPI_ISL_703996, EPI_ISL_703997,
EPI_ISL_703999, EPI_ISL_704000, EPI_ISL_704002, EPI_ISL_704004, EPI_ISL_704005, EPI_ISL_704007, EPI_ISL_704008, EPI_ISL_704010, EPI_ISL_704011, EPI_ISL_704013, EPI_ISL_704015, EPI_ISL_704016, EPI_ISL_704018, EPI_ISL_704019, EPI_ISL_704021, EPI_ISL_704023, EPI_ISL_704024, EPI_ISL_704026, EPI_ISL_704027, EPI_ISL_704029, EPI_ISL_704031, EPI_ISL_704032, EPI_ISL_704034, EPI_ISL_704035,
EPI_ISL_704037, EPI_ISL_704039, EPI_ISL_704040, EPI_ISL_704042, EPI_ISL_704043, EPI_ISL_704045, EPI_ISL_704047, EPI_ISL_704048, EPI_ISL_704050, EPI_ISL_704051, EPI_ISL_704053, EPI_ISL_704055, EPI_ISL_704056, EPI_ISL_704058, EPI_ISL_704059, EPI_ISL_704061, EPI_ISL_704063, EPI_ISL_704064, EPI_ISL_704066, EPI_ISL_704067, EPI_ISL_704069, EPI_ISL_704071, EPI_ISL_704072, EPI_ISL_704074,
EPI_ISL_704075, EPI_ISL_704077, EPI_ISL_704078, EPI_ISL_704080, EPI_ISL_704082, EPI_ISL_704083, EPI_ISL_704085, EPI_ISL_704087, EPI_ISL_704088, EPI_ISL_704090, EPI_ISL_704091, EPI_ISL_704093, EPI_ISL_704094, EPI_ISL_704096, EPI_ISL_704098, EPI_ISL_704099, EPI_ISL_704101, EPI_ISL_704103, EPI_ISL_704104, EPI_ISL_704106, EPI_ISL_704107, EPI_ISL_704109, EPI_ISL_704110, EPI_ISL_704113,
EPI_ISL_704114, EPI_ISL_704116, EPI_ISL_704117, EPI_ISL_704119, EPI_ISL_704121, EPI_ISL_704122, EPI_ISL_704124, EPI_ISL_704125, EPI_ISL_704127, EPI_ISL_704128, EPI_ISL_704130, EPI_ISL_704132, EPI_ISL_704133, EPI_ISL_704135, EPI_ISL_704136, EPI_ISL_704138, EPI_ISL_704140, EPI_ISL_704141, EPI_ISL_704143, EPI_ISL_704144, EPI_ISL_704146, EPI_ISL_704147, EPI_ISL_704149, EPI_ISL_704151,
EPI_ISL_704152, EPI_ISL_704154, EPI_ISL_704155, EPI_ISL_704157, EPI_ISL_704159, EPI_ISL_704160, EPI_ISL_704162, EPI_ISL_704164, EPI_ISL_704165, EPI_ISL_704167, EPI_ISL_704169, EPI_ISL_704170, EPI_ISL_704172, EPI_ISL_704173, EPI_ISL_704175, EPI_ISL_704177, EPI_ISL_704178, EPI_ISL_704180, EPI_ISL_704182, EPI_ISL_704183, EPI_ISL_704185, EPI_ISL_704186, EPI_ISL_704188, EPI_ISL_704189,
EPI_ISL_704191, EPI_ISL_704192, EPI_ISL_704194, EPI_ISL_704196, EPI_ISL_704197, EPI_ISL_704199, EPI_ISL_704200, EPI_ISL_704202, EPI_ISL_704204, EPI_ISL_704205, EPI_ISL_704207, EPI_ISL_704208, EPI_ISL_704210, EPI_ISL_704211, EPI_ISL_704212, EPI_ISL_704213, EPI_ISL_704214, EPI_ISL_704215, EPI_ISL_704216, EPI_ISL_704218, EPI_ISL_704220, EPI_ISL_704221, EPI_ISL_704223, EPI_ISL_704224,
EPI_ISL_704226, EPI_ISL_704227, EPI_ISL_704229, EPI_ISL_704230, EPI_ISL_704232, EPI_ISL_704234, EPI_ISL_704235, EPI_ISL_704237, EPI_ISL_704239, EPI_ISL_704240, EPI_ISL_704242, EPI_ISL_704244, EPI_ISL_704245, EPI_ISL_704246, EPI_ISL_704248, EPI_ISL_704249, EPI_ISL_704251, EPI_ISL_704253, EPI_ISL_704254, EPI_ISL_704256, EPI_ISL_704258, EPI_ISL_704259, EPI_ISL_704261, EPI_ISL_704263,
EPI_ISL_704264, EPI_ISL_704265, EPI_ISL_704267, EPI_ISL_704268, EPI_ISL_704270, EPI_ISL_704272, EPI_ISL_704274, EPI_ISL_704275, EPI_ISL_704277, EPI_ISL_704278, EPI_ISL_704279, EPI_ISL_704281, EPI_ISL_704282, EPI_ISL_704284, EPI_ISL_704285, EPI_ISL_704287, EPI_ISL_704288, EPI_ISL_704290, EPI_ISL_704292, EPI_ISL_704293, EPI_ISL_704295, EPI_ISL_704296, EPI_ISL_704298, EPI_ISL_704300,
EPI_ISL_704301, EPI_ISL_704303, EPI_ISL_704304, EPI_ISL_704306, EPI_ISL_704308, EPI_ISL_704310, EPI_ISL_704311, EPI_ISL_704313, EPI_ISL_704314, EPI_ISL_704316, EPI_ISL_704318, EPI_ISL_704319, EPI_ISL_704321, EPI_ISL_704323, EPI_ISL_704324, EPI_ISL_704326, EPI_ISL_704327, EPI_ISL_704329, EPI_ISL_704330, EPI_ISL_704332, EPI_ISL_704334, EPI_ISL_704335, EPI_ISL_704337, EPI_ISL_704338,
EPI_ISL_704340, EPI_ISL_704342, EPI_ISL_704343, EPI_ISL_704345, EPI_ISL_704346, EPI_ISL_704348, EPI_ISL_704349, EPI_ISL_704351, EPI_ISL_704352, EPI_ISL_704354, EPI_ISL_704355, EPI_ISL_704357, EPI_ISL_704358, EPI_ISL_704360, EPI_ISL_704362, EPI_ISL_704363, EPI_ISL_704365, EPI_ISL_704367, EPI_ISL_704368, EPI_ISL_704370, EPI_ISL_704371, EPI_ISL_704373, EPI_ISL_704375, EPI_ISL_704376,
EPI_ISL_704378, EPI_ISL_704380, EPI_ISL_704381, EPI_ISL_704383, EPI_ISL_704385, EPI_ISL_704386, EPI_ISL_704388, EPI_ISL_704390, EPI_ISL_704391, EPI_ISL_704393, EPI_ISL_704394, EPI_ISL_704396, EPI_ISL_704397, EPI_ISL_704399, EPI_ISL_704400, EPI_ISL_704402, EPI_ISL_704404, EPI_ISL_704405, EPI_ISL_704407, EPI_ISL_704409, EPI_ISL_704410, EPI_ISL_704412, EPI_ISL_704414, EPI_ISL_704415,
EPI_ISL_704417, EPI_ISL_704418, EPI_ISL_704420, EPI_ISL_704421, EPI_ISL_704423, EPI_ISL_704424, EPI_ISL_704426, EPI_ISL_704428, EPI_ISL_704429, EPI_ISL_704431, EPI_ISL_704432, EPI_ISL_704434, EPI_ISL_704436, EPI_ISL_704437, EPI_ISL_704439, EPI_ISL_704440, EPI_ISL_704442, EPI_ISL_704444, EPI_ISL_704445, EPI_ISL_704447, EPI_ISL_704448, EPI_ISL_704450, EPI_ISL_704451, EPI_ISL_704453,
EPI_ISL_704455, EPI_ISL_704456, EPI_ISL_704458, EPI_ISL_704459, EPI_ISL_704461, EPI_ISL_704463, EPI_ISL_704464, EPI_ISL_704466, EPI_ISL_704468, EPI_ISL_704469, EPI_ISL_704471, EPI_ISL_704472, EPI_ISL_704474, EPI_ISL_704476, EPI_ISL_704477, EPI_ISL_704479, EPI_ISL_704480, EPI_ISL_704482, EPI_ISL_704483, EPI_ISL_704485, EPI_ISL_704487, EPI_ISL_704488, EPI_ISL_704490, EPI_ISL_704491,
EPI_ISL_704493, EPI_ISL_704495, EPI_ISL_704496, EPI_ISL_704498, EPI_ISL_704500, EPI_ISL_704501, EPI_ISL_704503, EPI_ISL_704504, EPI_ISL_704506, EPI_ISL_704508, EPI_ISL_704509, EPI_ISL_704511, EPI_ISL_704512, EPI_ISL_704514, EPI_ISL_704515, EPI_ISL_704517, EPI_ISL_704519, EPI_ISL_704520, EPI_ISL_704522, EPI_ISL_704523, EPI_ISL_704525, EPI_ISL_704527, EPI_ISL_704528, EPI_ISL_704530,
EPI_ISL_704531, EPI_ISL_704533, EPI_ISL_704534, EPI_ISL_704536, EPI_ISL_704538, EPI_ISL_704539, EPI_ISL_704541, EPI_ISL_704542, EPI_ISL_704544, EPI_ISL_704546, EPI_ISL_704547, EPI_ISL_704549, EPI_ISL_704550, EPI_ISL_704552, EPI_ISL_704553, EPI_ISL_704555, EPI_ISL_704556, EPI_ISL_704558, EPI_ISL_704560, EPI_ISL_704561, EPI_ISL_704563, EPI_ISL_704565, EPI_ISL_704566, EPI_ISL_704568,
EPI_ISL_704569, EPI_ISL_704571, EPI_ISL_704642, EPI_ISL_704644, EPI_ISL_704645, EPI_ISL_704647, EPI_ISL_704649, EPI_ISL_704650, EPI_ISL_704652, EPI_ISL_704653, EPI_ISL_704655, EPI_ISL_704657, EPI_ISL_704658, EPI_ISL_704660, EPI_ISL_704661, EPI_ISL_704663, EPI_ISL_704665, EPI_ISL_704667, EPI_ISL_704668, EPI_ISL_704670, EPI_ISL_704672, EPI_ISL_704673, EPI_ISL_704675, EPI_ISL_704677,
EPI_ISL_704678, EPI_ISL_704680, EPI_ISL_704681, EPI_ISL_704683, EPI_ISL_704684, EPI_ISL_704686, EPI_ISL_710748, EPI_ISL_710753, EPI_ISL_710754, EPI_ISL_710755, EPI_ISL_710756, EPI_ISL_710757, EPI_ISL_710758, EPI_ISL_710759, EPI_ISL_710760, EPI_ISL_710761, EPI_ISL_710762, EPI_ISL_710763, EPI_ISL_710764, EPI_ISL_710765, EPI_ISL_710766, EPI_ISL_710767, EPI_ISL_710768, EPI_ISL_710769,
EPI_ISL_710770, EPI_ISL_710771, EPI_ISL_710772, EPI_ISL_710773, EPI_ISL_710774, EPI_ISL_710775, EPI_ISL_710776, EPI_ISL_710777, EPI_ISL_710778, EPI_ISL_710779, EPI_ISL_710780, EPI_ISL_710781, EPI_ISL_710782, EPI_ISL_710783, EPI_ISL_710784, EPI_ISL_710785, EPI_ISL_710786, EPI_ISL_710787, EPI_ISL_710788, EPI_ISL_710789, EPI_ISL_710790, EPI_ISL_710791, EPI_ISL_710792, EPI_ISL_710793,
EPI_ISL_710794, EPI_ISL_710795, EPI_ISL_710796, EPI_ISL_710797, EPI_ISL_710798, EPI_ISL_710799, EPI_ISL_710800, EPI_ISL_710801, EPI_ISL_710802, EPI_ISL_710803, EPI_ISL_710804, EPI_ISL_710805, EPI_ISL_710806, EPI_ISL_710807, EPI_ISL_710808, EPI_ISL_710809, EPI_ISL_710810, EPI_ISL_710811, EPI_ISL_710812, EPI_ISL_710813, EPI_ISL_710814, EPI_ISL_710815, EPI_ISL_710816, EPI_ISL_710817,
EPI_ISL_710818, EPI_ISL_710819, EPI_ISL_710820, EPI_ISL_710830, EPI_ISL_710831, EPI_ISL_710832, EPI_ISL_710833, EPI_ISL_710834, EPI_ISL_719605, EPI_ISL_719606, EPI_ISL_719607, EPI_ISL_719608, EPI_ISL_719609, EPI_ISL_719610, EPI_ISL_719611, EPI_ISL_719612, EPI_ISL_719613, EPI_ISL_719614, EPI_ISL_719615, EPI_ISL_719616, EPI_ISL_719617, EPI_ISL_719618, EPI_ISL_719619, EPI_ISL_719620,
EPI_ISL_719621, EPI_ISL_719622, EPI_ISL_719623, EPI_ISL_719624, EPI_ISL_719625, EPI_ISL_719626, EPI_ISL_719627, EPI_ISL_719628, EPI_ISL_719629, EPI_ISL_719630, EPI_ISL_719631, EPI_ISL_719632, EPI_ISL_719633, EPI_ISL_719634, EPI_ISL_719635, EPI_ISL_719636, EPI_ISL_719637, EPI_ISL_719638, EPI_ISL_719639, EPI_ISL_719640, EPI_ISL_719641, EPI_ISL_719642, EPI_ISL_719643, EPI_ISL_719644,
EPI_ISL_719645, EPI_ISL_719646, EPI_ISL_719647, EPI_ISL_719648, EPI_ISL_719649, EPI_ISL_719650, EPI_ISL_719651, EPI_ISL_719652, EPI_ISL_719653, EPI_ISL_719654, EPI_ISL_719655, EPI_ISL_719656, EPI_ISL_719657, EPI_ISL_719658, EPI_ISL_719659, EPI_ISL_719660, EPI_ISL_719661, EPI_ISL_719662, EPI_ISL_719663, EPI_ISL_719664, EPI_ISL_719665, EPI_ISL_719666, EPI_ISL_719667, EPI_ISL_719668,
EPI_ISL_719669, EPI_ISL_719670, EPI_ISL_719671, EPI_ISL_719672, EPI_ISL_719673, EPI_ISL_719674, EPI_ISL_719675, EPI_ISL_719676, EPI_ISL_719677, EPI_ISL_719678, EPI_ISL_719679, EPI_ISL_719680, EPI_ISL_719681, EPI_ISL_719682, EPI_ISL_719683, EPI_ISL_719684, EPI_ISL_719685, EPI_ISL_719686, EPI_ISL_719687, EPI_ISL_719688, EPI_ISL_719689, EPI_ISL_719690, EPI_ISL_719691, EPI_ISL_719692,
EPI_ISL_719693, EPI_ISL_719694, EPI_ISL_719695, EPI_ISL_719696, EPI_ISL_719697, EPI_ISL_719698, EPI_ISL_719699, EPI_ISL_719700, EPI_ISL_719701, EPI_ISL_719702, EPI_ISL_719703, EPI_ISL_719704, EPI_ISL_719705, EPI_ISL_719706, EPI_ISL_719707, EPI_ISL_719708, EPI_ISL_719709, EPI_ISL_719710, EPI_ISL_719711, EPI_ISL_719712, EPI_ISL_719713, EPI_ISL_719714, EPI_ISL_719715, EPI_ISL_719716,
EPI_ISL_719717, EPI_ISL_719719, EPI_ISL_719720, EPI_ISL_719721, EPI_ISL_719722, EPI_ISL_719723, EPI_ISL_719724, EPI_ISL_719725, EPI_ISL_719726, EPI_ISL_719727, EPI_ISL_719728, EPI_ISL_719729, EPI_ISL_719730, EPI_ISL_719731, EPI_ISL_719732, EPI_ISL_719733, EPI_ISL_719734, EPI_ISL_719735, EPI_ISL_719736, EPI_ISL_719737, EPI_ISL_719738, EPI_ISL_719739, EPI_ISL_719740, EPI_ISL_719741,
EPI_ISL_719742, EPI_ISL_719743, EPI_ISL_719744, EPI_ISL_719745, EPI_ISL_719746, EPI_ISL_719747, EPI_ISL_719748, EPI_ISL_719749, EPI_ISL_719750, EPI_ISL_719751, EPI_ISL_719752, EPI_ISL_719753, EPI_ISL_719754, EPI_ISL_719755, EPI_ISL_719756, EPI_ISL_719757, EPI_ISL_719758, EPI_ISL_719759, EPI_ISL_719760, EPI_ISL_719761, EPI_ISL_719762, EPI_ISL_719763, EPI_ISL_719764, EPI_ISL_719765,
EPI_ISL_719766, EPI_ISL_719767, EPI_ISL_719768, EPI_ISL_719769, EPI_ISL_719770, EPI_ISL_719771, EPI_ISL_719772, EPI_ISL_719773, EPI_ISL_719774, EPI_ISL_719775, EPI_ISL_719777, EPI_ISL_719778, EPI_ISL_719779, EPI_ISL_719780, EPI_ISL_719781, EPI_ISL_719782, EPI_ISL_719783, EPI_ISL_719784, EPI_ISL_719785, EPI_ISL_719786, EPI_ISL_719787, EPI_ISL_719788, EPI_ISL_719789, EPI_ISL_719790,

EPI_ISL_719791, EPI_ISL_719792, EPI_ISL_719793, EPI_ISL_719794, EPI_ISL_719795, EPI_ISL_719796, EPI_ISL_719797, EPI_ISL_719798, EPI_ISL_719799, EPI_ISL_719800, EPI_ISL_719802, EPI_ISL_719803, EPI_ISL_719804, EPI_ISL_719805, EPI_ISL_719806, EPI_ISL_719807, EPI_ISL_719808, EPI_ISL_719809, EPI_ISL_719810, EPI_ISL_719811, EPI_ISL_719812, EPI_ISL_719813, EPI_ISL_719814, EPI_ISL_719815, EPI_ISL_719816, EPI_ISL_719817, EPI_ISL_719818, EPI_ISL_719819, EPI_ISL_719821, EPI_ISL_719822, EPI_ISL_719823, EPI_ISL_719824, EPI_ISL_719825, EPI_ISL_719826, EPI_ISL_719827, EPI_ISL_719828, EPI_ISL_719829, EPI_ISL_719830, EPI_ISL_719831, EPI_ISL_719832, EPI_ISL_719833, EPI_ISL_719834, EPI_ISL_719835, EPI_ISL_719836, EPI_ISL_719838, EPI_ISL_719839, EPI_ISL_719840, EPI_ISL_719842, EPI_ISL_719843, EPI_ISL_719844, EPI_ISL_719845, EPI_ISL_719846, EPI_ISL_719847, EPI_ISL_719848, EPI_ISL_719850, EPI_ISL_719851, EPI_ISL_719853, EPI_ISL_719854, EPI_ISL_719855, EPI_ISL_719856, EPI_ISL_719857, EPI_ISL_719859, EPI_ISL_719860, EPI_ISL_719861, EPI_ISL_719862, EPI_ISL_719863, EPI_ISL_719864, EPI_ISL_719865, EPI_ISL_719866, EPI_ISL_719867, EPI_ISL_719868, EPI_ISL_719869, EPI_ISL_719870, EPI_ISL_719871, EPI_ISL_719872, EPI_ISL_719873, EPI_ISL_719874, EPI_ISL_719875, EPI_ISL_719876, EPI_ISL_719877, EPI_ISL_719878, EPI_ISL_719879, EPI_ISL_719880, EPI_ISL_719881, EPI_ISL_719882, EPI_ISL_719883, EPI_ISL_719884, EPI_ISL_719885, EPI_ISL_719887, EPI_ISL_719889, EPI_ISL_719890, EPI_ISL_719891, EPI_ISL_719892, EPI_ISL_719893, EPI_ISL_719894, EPI_ISL_719895, EPI_ISL_719896, EPI_ISL_719897, EPI_ISL_719898, EPI_ISL_719899, EPI_ISL_719900, EPI_ISL_719901, EPI_ISL_719902, EPI_ISL_719903, EPI_ISL_719904, EPI_ISL_719905, EPI_ISL_719906, EPI_ISL_719907, EPI_ISL_719908, EPI_ISL_719909, EPI_ISL_719910, EPI_ISL_719911, EPI_ISL_719912, EPI_ISL_719913, EPI_ISL_719914, EPI_ISL_719915, EPI_ISL_719916, EPI_ISL_719917, EPI_ISL_719919, EPI_ISL_719920, EPI_ISL_719922, EPI_ISL_719923, EPI_ISL_719924, EPI_ISL_719925, EPI_ISL_719927, EPI_ISL_719928, EPI_ISL_719929, EPI_ISL_719930, EPI_ISL_719931, EPI_ISL_719932, EPI_ISL_719933, EPI_ISL_719936, EPI_ISL_719937, EPI_ISL_719938, EPI_ISL_719940, EPI_ISL_719941, EPI_ISL_719942, EPI_ISL_719943, EPI_ISL_719944, EPI_ISL_719945, EPI_ISL_719946, EPI_ISL_719947, EPI_ISL_719948, EPI_ISL_719949, EPI_ISL_719950, EPI_ISL_719951, EPI_ISL_719952, EPI_ISL_719953, EPI_ISL_719954, EPI_ISL_719955, EPI_ISL_719956, EPI_ISL_719957, EPI_ISL_719958, EPI_ISL_719959, EPI_ISL_719960, EPI_ISL_719961, EPI_ISL_719962, EPI_ISL_719963, EPI_ISL_719964, EPI_ISL_719965, EPI_ISL_719966, EPI_ISL_719967, EPI_ISL_719969, EPI_ISL_719970, EPI_ISL_719971, EPI_ISL_719972, EPI_ISL_719973, EPI_ISL_719974, EPI_ISL_719975, EPI_ISL_719976, EPI_ISL_719977, EPI_ISL_719978, EPI_ISL_719979, EPI_ISL_719980, EPI_ISL_719981, EPI_ISL_719982, EPI_ISL_719983, EPI_ISL_719984, EPI_ISL_719985, EPI_ISL_719986, EPI_ISL_720909, EPI_ISL_720910, EPI_ISL_720911, EPI_ISL_720912, EPI_ISL_720913, EPI_ISL_720914, EPI_ISL_720915, EPI_ISL_720917, EPI_ISL_720918, EPI_ISL_720919, EPI_ISL_720920, EPI_ISL_720921, EPI_ISL_720922, EPI_ISL_720923, EPI_ISL_720924, EPI_ISL_720925, EPI_ISL_720926, EPI_ISL_720929, EPI_ISL_720930, EPI_ISL_720931, EPI_ISL_720932, EPI_ISL_720933, EPI_ISL_720934, EPI_ISL_720935, EPI_ISL_720936, EPI_ISL_720937, EPI_ISL_720938, EPI_ISL_720939, EPI_ISL_720941, EPI_ISL_720942, EPI_ISL_720943, EPI_ISL_720944, EPI_ISL_720947, EPI_ISL_720948, EPI_ISL_720949, EPI_ISL_720950, EPI_ISL_720951, EPI_ISL_720953, EPI_ISL_720955, EPI_ISL_720956, EPI_ISL_720957, EPI_ISL_720959, EPI_ISL_720960, EPI_ISL_720961, EPI_ISL_720963, EPI_ISL_720964, EPI_ISL_720965, EPI_ISL_720966, EPI_ISL_720967, EPI_ISL_720968, EPI_ISL_720970, EPI_ISL_720971, EPI_ISL_720972, EPI_ISL_720973, EPI_ISL_720974, EPI_ISL_720975, EPI_ISL_720976, EPI_ISL_720977, EPI_ISL_720978, EPI_ISL_720979, EPI_ISL_720982, EPI_ISL_720984, EPI_ISL_720985, EPI_ISL_720986, EPI_ISL_720988, EPI_ISL_720989, EPI_ISL_720990, EPI_ISL_720991, EPI_ISL_720992, EPI_ISL_720993, EPI_ISL_720994, EPI_ISL_720995, EPI_ISL_720996, EPI_ISL_720997, EPI_ISL_720998, EPI_ISL_720999, EPI_ISL_721001, EPI_ISL_721002, EPI_ISL_721003, EPI_ISL_721004, EPI_ISL_721005, EPI_ISL_721006, EPI_ISL_721007, EPI_ISL_721008, EPI_ISL_721010, EPI_ISL_721011, EPI_ISL_721012, EPI_ISL_721013, EPI_ISL_721014, EPI_ISL_721015, EPI_ISL_721016, EPI_ISL_721017, EPI_ISL_721018, EPI_ISL_721019, EPI_ISL_721020, EPI_ISL_721022, EPI_ISL_721023, EPI_ISL_721024, EPI_ISL_721025, EPI_ISL_721026, EPI_ISL_721028, EPI_ISL_721029, EPI_ISL_721030, EPI_ISL_721031, EPI_ISL_721032, EPI_ISL_721033, EPI_ISL_721034, EPI_ISL_721035, EPI_ISL_721036, EPI_ISL_721037, EPI_ISL_721038, EPI_ISL_721039, EPI_ISL_721040, EPI_ISL_721041, EPI_ISL_721043, EPI_ISL_721044, EPI_ISL_721045, EPI_ISL_721046, EPI_ISL_721047, EPI_ISL_721048, EPI_ISL_721049, EPI_ISL_721050, EPI_ISL_721051, EPI_ISL_721052, EPI_ISL_721053, EPI_ISL_721054, EPI_ISL_721055, EPI_ISL_721056, EPI_ISL_721057, EPI_ISL_721058, EPI_ISL_721059, EPI_ISL_721060, EPI_ISL_721063, EPI_ISL_721064, EPI_ISL_721065, EPI_ISL_721066, EPI_ISL_721067, EPI_ISL_721068, EPI_ISL_721069, EPI_ISL_721071, EPI_ISL_721073, EPI_ISL_721075, EPI_ISL_721076, EPI_ISL_721078, EPI_ISL_721079, EPI_ISL_721080, EPI_ISL_721081, EPI_ISL_721082, EPI_ISL_721083, EPI_ISL_721084, EPI_ISL_721085, EPI_ISL_721086, EPI_ISL_721087, EPI_ISL_721088, EPI_ISL_721089, EPI_ISL_721090, EPI_ISL_721091, EPI_ISL_721093, EPI_ISL_721094, EPI_ISL_721096, EPI_ISL_721097, EPI_ISL_721098, EPI_ISL_721099, EPI_ISL_721100, EPI_ISL_721101, EPI_ISL_721102, EPI_ISL_721103, EPI_ISL_721104, EPI_ISL_721105, EPI_ISL_721106, EPI_ISL_721107, EPI_ISL_721108, EPI_ISL_721110, EPI_ISL_721111, EPI_ISL_721112, EPI_ISL_721113, EPI_ISL_721114, EPI_ISL_721115, EPI_ISL_721116, EPI_ISL_721117, EPI_ISL_721118, EPI_ISL_721120, EPI_ISL_721121, EPI_ISL_721122, EPI_ISL_721123, EPI_ISL_721124, EPI_ISL_721125, EPI_ISL_721126, EPI_ISL_721127, EPI_ISL_721128, EPI_ISL_721129, EPI_ISL_721130, EPI_ISL_721132, EPI_ISL_721134, EPI_ISL_721135, EPI_ISL_721136, EPI_ISL_721137, EPI_ISL_721138, EPI_ISL_721139, EPI_ISL_721140, EPI_ISL_721141, EPI_ISL_721142, EPI_ISL_721143, EPI_ISL_721144, EPI_ISL_721145, EPI_ISL_721146, EPI_ISL_721147, EPI_ISL_721148, EPI_ISL_721149, EPI_ISL_721150, EPI_ISL_721151, EPI_ISL_721152, EPI_ISL_721153, EPI_ISL_721154, EPI_ISL_721155, EPI_ISL_721156, EPI_ISL_721157, EPI_ISL_721158, EPI_ISL_721159, EPI_ISL_721160, EPI_ISL_721161, EPI_ISL_721162, EPI_ISL_721164, EPI_ISL_721165, EPI_ISL_721166, EPI_ISL_721168, EPI_ISL_721169, EPI_ISL_721171, EPI_ISL_721172, EPI_ISL_721173, EPI_ISL_721174, EPI_ISL_721175, EPI_ISL_721176, EPI_ISL_721177, EPI_ISL_721178, EPI_ISL_721179, EPI_ISL_721180, EPI_ISL_721181, EPI_ISL_721182, EPI_ISL_721183, EPI_ISL_721184, EPI_ISL_721533, EPI_ISL_728300, EPI_ISL_731894, EPI_ISL_757187, EPI_ISL_757188, EPI_ISL_757189, EPI_ISL_757190, EPI_ISL_757191, EPI_ISL_757192, EPI_ISL_757193, EPI_ISL_757194, EPI_ISL_757195, EPI_ISL_757196, EPI_ISL_757197, EPI_ISL_757198, EPI_ISL_757199, EPI_ISL_757200, EPI_ISL_757201, EPI_ISL_757203, EPI_ISL_757204, EPI_ISL_757205, EPI_ISL_757206, EPI_ISL_757207, EPI_ISL_757208, EPI_ISL_757209, EPI_ISL_757210, EPI_ISL_757211, EPI_ISL_757212, EPI_ISL_757213, EPI_ISL_757214, EPI_ISL_757215, EPI_ISL_757216, EPI_ISL_757217, EPI_ISL_757218, EPI_ISL_757219, EPI_ISL_757220, EPI_ISL_757221, EPI_ISL_757222, EPI_ISL_757223, EPI_ISL_757224, EPI_ISL_757273, EPI_ISL_757274, EPI_ISL_757275, EPI_ISL_757276, EPI_ISL_757277, EPI_ISL_757278, EPI_ISL_757279, EPI_ISL_757280, EPI_ISL_757281, EPI_ISL_757282, EPI_ISL_757283, EPI_ISL_757284, EPI_ISL_760490

| | | | |
|---|---|---|---|
| see above | Lighthouse Lab in Glasgow | Wellcome Sanger Institute for the COVID-19 Genomics UK (COG-UK) Consortium | Anna Dominiczak and Alex Alderton; Carol Clugston; Cordelia Langford; David Gray; David K. Jackson; Dominic Kwiatkowski; Ewan Harrison; Harper VanSteenhouse; Ian Johnston; John Sillitoe on behalf of the Wellcome Sanger Institute COVID-19 Surveillance Team; Roberto Amato; Sonia Goncalves; Yumi Kasai |

EPI_ISL_674863, EPI_ISL_681325, EPI_ISL_681326, EPI_ISL_681328, EPI_ISL_681330, EPI_ISL_681331, EPI_ISL_681332, EPI_ISL_681335, EPI_ISL_681336, EPI_ISL_681337, EPI_ISL_681338, EPI_ISL_681339, EPI_ISL_681344, EPI_ISL_681346, EPI_ISL_681348, EPI_ISL_681350, EPI_ISL_681353, EPI_ISL_681354, EPI_ISL_681356, EPI_ISL_681357, EPI_ISL_681358, EPI_ISL_681360, EPI_ISL_681361, EPI_ISL_681362, EPI_ISL_681364, EPI_ISL_681365, EPI_ISL_681366, EPI_ISL_681368, EPI_ISL_681369, EPI_ISL_681371, EPI_ISL_681372, EPI_ISL_681375, EPI_ISL_681376, EPI_ISL_681378, EPI_ISL_681379, EPI_ISL_681380, EPI_ISL_681383, EPI_ISL_681385, EPI_ISL_681386, EPI_ISL_681388, EPI_ISL_681389, EPI_ISL_681392, EPI_ISL_681393, EPI_ISL_681395, EPI_ISL_681396, EPI_ISL_681397, EPI_ISL_681398, EPI_ISL_681399, EPI_ISL_681401, EPI_ISL_681404, EPI_ISL_681405, EPI_ISL_681406, EPI_ISL_681407, EPI_ISL_681415, EPI_ISL_681416, EPI_ISL_681417, EPI_ISL_681418, EPI_ISL_681419, EPI_ISL_681421, EPI_ISL_681422, EPI_ISL_681424, EPI_ISL_681425, EPI_ISL_681426, EPI_ISL_681427, EPI_ISL_681428, EPI_ISL_681431, EPI_ISL_681432, EPI_ISL_681434, EPI_ISL_681436, EPI_ISL_681437, EPI_ISL_681438, EPI_ISL_681439, EPI_ISL_681442, EPI_ISL_681444, EPI_ISL_681445, EPI_ISL_681446, EPI_ISL_681447, EPI_ISL_681448, EPI_ISL_681450, EPI_ISL_681451, EPI_ISL_681456, EPI_ISL_681457, EPI_ISL_681459, EPI_ISL_681460, EPI_ISL_681461, EPI_ISL_681462, EPI_ISL_681463, EPI_ISL_681466, EPI_ISL_681467, EPI_ISL_681468, EPI_ISL_681469, EPI_ISL_681471, EPI_ISL_681473, EPI_ISL_681475, EPI_ISL_681476, EPI_ISL_681477, EPI_ISL_681479, EPI_ISL_681480, EPI_ISL_681481, EPI_ISL_681482, EPI_ISL_681484, EPI_ISL_681485, EPI_ISL_681488, EPI_ISL_681492, EPI_ISL_681494, EPI_ISL_681495, EPI_ISL_681496, EPI_ISL_681498, EPI_ISL_681499, EPI_ISL_681501, EPI_ISL_681502, EPI_ISL_681503, EPI_ISL_681504, EPI_ISL_681505, EPI_ISL_681506, EPI_ISL_681508, EPI_ISL_681510, EPI_ISL_681511, EPI_ISL_681514, EPI_ISL_681515, EPI_ISL_681517, EPI_ISL_681518, EPI_ISL_681521, EPI_ISL_681522, EPI_ISL_681525, EPI_ISL_681527, EPI_ISL_681530, EPI_ISL_681535, EPI_ISL_681536, EPI_ISL_681537, EPI_ISL_681538, EPI_ISL_681540, EPI_ISL_681541, EPI_ISL_681543, EPI_ISL_681544, EPI_ISL_681548, EPI_ISL_681552, EPI_ISL_681553, EPI_ISL_681554, EPI_ISL_681555, EPI_ISL_681567, EPI_ISL_681570, EPI_ISL_681571, EPI_ISL_681572, EPI_ISL_681573, EPI_ISL_681576, EPI_ISL_681580, EPI_ISL_681581, EPI_ISL_681582, EPI_ISL_681584, EPI_ISL_681585, EPI_ISL_681586, EPI_ISL_681588, EPI_ISL_681589, EPI_ISL_681591, EPI_ISL_681593, EPI_ISL_681594, EPI_ISL_681596, EPI_ISL_681598, EPI_ISL_681600, EPI_ISL_681601, EPI_ISL_681602, EPI_ISL_681603, EPI_ISL_681604, EPI_ISL_681605, EPI_ISL_681608, EPI_ISL_681609, EPI_ISL_681611, EPI_ISL_681612, EPI_ISL_681613, EPI_ISL_681615, EPI_ISL_681616, EPI_ISL_681619, EPI_ISL_681622, EPI_ISL_681626, EPI_ISL_681627, EPI_ISL_681628, EPI_ISL_681631, EPI_ISL_681632, EPI_ISL_681637, EPI_ISL_681640, EPI_ISL_681644, EPI_ISL_681645, EPI_ISL_681648, EPI_ISL_681649, EPI_ISL_681650, EPI_ISL_681652, EPI_ISL_681653, EPI_ISL_681654, EPI_ISL_705200, EPI_ISL_709567, EPI_ISL_709568, EPI_ISL_709569, EPI_ISL_709570, EPI_ISL_709571, EPI_ISL_709572, EPI_ISL_709573, EPI_ISL_709574, EPI_ISL_709575, EPI_ISL_709576, EPI_ISL_709577, EPI_ISL_709578, EPI_ISL_709579, EPI_ISL_709580, EPI_ISL_709581, EPI_ISL_709582, EPI_ISL_709583, EPI_ISL_709584, EPI_ISL_709585, EPI_ISL_709586, EPI_ISL_709587, EPI_ISL_709588, EPI_ISL_709589, EPI_ISL_709590, EPI_ISL_709591, EPI_ISL_709592, EPI_ISL_709593, EPI_ISL_709594, EPI_ISL_709595, EPI_ISL_709596, EPI_ISL_709597, EPI_ISL_709598, EPI_ISL_709599, EPI_ISL_709600, EPI_ISL_709601, EPI_ISL_709602, EPI_ISL_709603, EPI_ISL_709604, EPI_ISL_709605, EPI_ISL_709606, EPI_ISL_709607, EPI_ISL_709608, EPI_ISL_709609, EPI_ISL_709610, EPI_ISL_709611, EPI_ISL_709612, EPI_ISL_709613, EPI_ISL_709614, EPI_ISL_709615, EPI_ISL_709616, EPI_ISL_709617, EPI_ISL_709618, EPI_ISL_709619, EPI_ISL_709620, EPI_ISL_709621, EPI_ISL_709622, EPI_ISL_709623, EPI_ISL_709624, EPI_ISL_709625, EPI_ISL_709626, EPI_ISL_709627, EPI_ISL_709628, EPI_ISL_709629, EPI_ISL_709630, EPI_ISL_709631, EPI_ISL_709632, EPI_ISL_709633, EPI_ISL_709634, EPI_ISL_709635, EPI_ISL_709636, EPI_ISL_709637, EPI_ISL_709638, EPI_ISL_709639, EPI_ISL_709640, EPI_ISL_709641, EPI_ISL_709642, EPI_ISL_709643, EPI_ISL_709644, EPI_ISL_709645, EPI_ISL_709646, EPI_ISL_709647, EPI_ISL_709648, EPI_ISL_709649, EPI_ISL_709650, EPI_ISL_709651, EPI_ISL_709652, EPI_ISL_709653, EPI_ISL_709654, EPI_ISL_709655, EPI_ISL_709656, EPI_ISL_709657, EPI_ISL_709658, EPI_ISL_709659, EPI_ISL_709660, EPI_ISL_709661, EPI_ISL_709662, EPI_ISL_709663, EPI_ISL_709664, EPI_ISL_709665, EPI_ISL_709666, EPI_ISL_709667, EPI_ISL_709668, EPI_ISL_709669, EPI_ISL_709670, EPI_ISL_709671, EPI_ISL_709672, EPI_ISL_709673, EPI_ISL_709674, EPI_ISL_709675, EPI_ISL_709676, EPI_ISL_709677, EPI_ISL_709678, EPI_ISL_709679, EPI_ISL_709680, EPI_ISL_709681, EPI_ISL_709682, EPI_ISL_709683, EPI_ISL_709684, EPI_ISL_709685, EPI_ISL_709686, EPI_ISL_709687, EPI_ISL_709688, EPI_ISL_709689, EPI_ISL_709690, EPI_ISL_709691, EPI_ISL_709692, EPI_ISL_709693, EPI_ISL_709694, EPI_ISL_709695, EPI_ISL_709696, EPI_ISL_709697, EPI_ISL_709698, EPI_ISL_709699, EPI_ISL_709700, EPI_ISL_709701, EPI_ISL_709702, EPI_ISL_709703, EPI_ISL_709704, EPI_ISL_709705, EPI_ISL_709706, EPI_ISL_709707, EPI_ISL_709708, EPI_ISL_709709, EPI_ISL_709710, EPI_ISL_709711, EPI_ISL_709712, EPI_ISL_709713, EPI_ISL_709714, EPI_ISL_709715, EPI_ISL_709716, EPI_ISL_709717, EPI_ISL_709718, EPI_ISL_709719, EPI_ISL_709720, EPI_ISL_709721, EPI_ISL_709722, EPI_ISL_709723, EPI_ISL_709724, EPI_ISL_709725, EPI_ISL_709726, EPI_ISL_709727, EPI_ISL_709728, EPI_ISL_709729, EPI_ISL_709730, EPI_ISL_709731, EPI_ISL_709732, EPI_ISL_709733, EPI_ISL_709734, EPI_ISL_709735, EPI_ISL_709736, EPI_ISL_709737, EPI_ISL_709738, EPI_ISL_709739, EPI_ISL_709740, EPI_ISL_709741, EPI_ISL_709742, EPI_ISL_709743, EPI_ISL_709744, EPI_ISL_709745, EPI_ISL_709746, EPI_ISL_709747, EPI_ISL_709748, EPI_ISL_709749, EPI_ISL_709750, EPI_ISL_709751, EPI_ISL_709752, EPI_ISL_709753, EPI_ISL_709754, EPI_ISL_709755, EPI_ISL_709756, EPI_ISL_709757, EPI_ISL_709758, EPI_ISL_709759, EPI_ISL_709760, EPI_ISL_709761, EPI_ISL_709762, EPI_ISL_709763, EPI_ISL_709764, EPI_ISL_709765, EPI_ISL_709766, EPI_ISL_709767, EPI_ISL_709768, EPI_ISL_709769, EPI_ISL_709770, EPI_ISL_709771, EPI_ISL_709772, EPI_ISL_709773, EPI_ISL_709774, EPI_ISL_709775, EPI_ISL_709776, EPI_ISL_709777, EPI_ISL_709778, EPI_ISL_709779, EPI_ISL_709780, EPI_ISL_709781, EPI_ISL_709782, EPI_ISL_709783, EPI_ISL_709784, EPI_ISL_709785, EPI_ISL_709786, EPI_ISL_709787, EPI_ISL_709788, EPI_ISL_709789, EPI_ISL_709790, EPI_ISL_709791, EPI_ISL_709792, EPI_ISL_709793, EPI_ISL_709794, EPI_ISL_709795, EPI_ISL_709796, EPI_ISL_709797, EPI_ISL_709798, EPI_ISL_709799, EPI_ISL_719988, EPI_ISL_719989, EPI_ISL_719993, EPI_ISL_719996, EPI_ISL_719998, EPI_ISL_719999, EPI_ISL_720000, EPI_ISL_720002, EPI_ISL_720009, EPI_ISL_720013, EPI_ISL_720015, EPI_ISL_720018, EPI_ISL_720019, EPI_ISL_720020, EPI_ISL_720022, EPI_ISL_720023, EPI_ISL_720024, EPI_ISL_720025, EPI_ISL_720027, EPI_ISL_720028, EPI_ISL_720031, EPI_ISL_720033, EPI_ISL_720034, EPI_ISL_720036, EPI_ISL_720037, EPI_ISL_720038, EPI_ISL_720042, EPI_ISL_720043, EPI_ISL_720044, EPI_ISL_720046, EPI_ISL_720048, EPI_ISL_720051, EPI_ISL_720052, EPI_ISL_720053, EPI_ISL_720058, EPI_ISL_720059, EPI_ISL_720060, EPI_ISL_720064, EPI_ISL_720070, EPI_ISL_720071, EPI_ISL_720073, EPI_ISL_720075, EPI_ISL_720077, EPI_ISL_720079, EPI_ISL_720080, EPI_ISL_720082, EPI_ISL_720083, EPI_ISL_720084, EPI_ISL_720085, EPI_ISL_720086, EPI_ISL_720087, EPI_ISL_720088, EPI_ISL_720090, EPI_ISL_720091, EPI_ISL_720094, EPI_ISL_720096, EPI_ISL_720097, EPI_ISL_720099, EPI_ISL_720100, EPI_ISL_720101, EPI_ISL_720102, EPI_ISL_720103, EPI_ISL_720104, EPI_ISL_720105, EPI_ISL_720106, EPI_ISL_720107, EPI_ISL_720110, EPI_ISL_720111, EPI_ISL_720116, EPI_ISL_720118, EPI_ISL_720120, EPI_ISL_720121, EPI_ISL_720123, EPI_ISL_720126, EPI_ISL_720129, EPI_ISL_720130, EPI_ISL_720131, EPI_ISL_720132, EPI_ISL_720134, EPI_ISL_720135, EPI_ISL_720138, EPI_ISL_720139, EPI_ISL_720140, EPI_ISL_720141, EPI_ISL_720152, EPI_ISL_720154, EPI_ISL_720155, EPI_ISL_720156, EPI_ISL_720157, EPI_ISL_720162, EPI_ISL_720164, EPI_ISL_720166, EPI_ISL_720171, EPI_ISL_720177, EPI_ISL_720181, EPI_ISL_720182, EPI_ISL_720183, EPI_ISL_720187, EPI_ISL_720189, EPI_ISL_720190, EPI_ISL_720191, EPI_ISL_720192, EPI_ISL_720194, EPI_ISL_720196, EPI_ISL_720199, EPI_ISL_720200, EPI_ISL_720201, EPI_ISL_720202, EPI_ISL_720203, EPI_ISL_720205, EPI_ISL_720206, EPI_ISL_720207, EPI_ISL_720208, EPI_ISL_720211, EPI_ISL_720212, EPI_ISL_720213, EPI_ISL_720214, EPI_ISL_720215, EPI_ISL_720216, EPI_ISL_720217, EPI_ISL_720218, EPI_ISL_720219, EPI_ISL_720221, EPI_ISL_720223, EPI_ISL_720225, EPI_ISL_720226, EPI_ISL_720227, EPI_ISL_720228, EPI_ISL_720230, EPI_ISL_720232, EPI_ISL_720233, EPI_ISL_720234, EPI_ISL_720239, EPI_ISL_720241, EPI_ISL_720242, EPI_ISL_720243, EPI_ISL_720244, EPI_ISL_720247, EPI_ISL_720248, EPI_ISL_720249, EPI_ISL_720250, EPI_ISL_720252, EPI_ISL_720253, EPI_ISL_720254, EPI_ISL_720256, EPI_ISL_720258, EPI_ISL_720259, EPI_ISL_720260, EPI_ISL_720261, EPI_ISL_720264, EPI_ISL_720265, EPI_ISL_720266, EPI_ISL_720269, EPI_ISL_720270, EPI_ISL_720271, EPI_ISL_720272, EPI_ISL_720274, EPI_ISL_720279, EPI_ISL_720280, EPI_ISL_720283, EPI_ISL_720284, EPI_ISL_720285, EPI_ISL_720287, EPI_ISL_720290, EPI_ISL_720291, EPI_ISL_720297, EPI_ISL_720298, EPI_ISL_720299, EPI_ISL_720301, EPI_ISL_720302, EPI_ISL_720303, EPI_ISL_720304, EPI_ISL_720307, EPI_ISL_720309, EPI_ISL_720310, EPI_ISL_720311, EPI_ISL_720312, EPI_ISL_720314, EPI_ISL_720316, EPI_ISL_720317, EPI_ISL_720318, EPI_ISL_720320, EPI_ISL_720321, EPI_ISL_720324, EPI_ISL_720325, EPI_ISL_720328, EPI_ISL_720329, EPI_ISL_720330, EPI_ISL_720334, EPI_ISL_720336, EPI_ISL_720338, EPI_ISL_720341, EPI_ISL_720342, EPI_ISL_720343, EPI_ISL_720344, EPI_ISL_720348, EPI_ISL_720352, EPI_ISL_720355, EPI_ISL_720357, EPI_ISL_720360, EPI_ISL_720364, EPI_ISL_720365, EPI_ISL_720366, EPI_ISL_720370, EPI_ISL_720376, EPI_ISL_720377, EPI_ISL_720378, EPI_ISL_720379, EPI_ISL_720380, EPI_ISL_720382, EPI_ISL_720383, EPI_ISL_720384, EPI_ISL_720386, EPI_ISL_720388, EPI_ISL_720389, EPI_ISL_720392, EPI_ISL_720393, EPI_ISL_720395, EPI_ISL_720397, EPI_ISL_720398, EPI_ISL_720399, EPI_ISL_720400, EPI_ISL_720402, EPI_ISL_720403, EPI_ISL_720404, EPI_ISL_720405, EPI_ISL_720407, EPI_ISL_720410, EPI_ISL_720411, EPI_ISL_720417, EPI_ISL_720419, EPI_ISL_720423, EPI_ISL_720424, EPI_ISL_720426, EPI_ISL_720427, EPI_ISL_720428, EPI_ISL_720429, EPI_ISL_720430, EPI_ISL_720432, EPI_ISL_720433, EPI_ISL_720435, EPI_ISL_720438, EPI_ISL_720439, EPI_ISL_720440, EPI_ISL_720441, EPI_ISL_720443, EPI_ISL_720444, EPI_ISL_720445, EPI_ISL_720446, EPI_ISL_720447, EPI_ISL_720449, EPI_ISL_720450, EPI_ISL_720452, EPI_ISL_720455, EPI_ISL_720456, EPI_ISL_720457, EPI_ISL_720458, EPI_ISL_720459, EPI_ISL_720462, EPI_ISL_720464, EPI_ISL_720465, EPI_ISL_720468, EPI_ISL_720470, EPI_ISL_720471, EPI_ISL_720473, EPI_ISL_720474, EPI_ISL_720476, EPI_ISL_720477, EPI_ISL_720479, EPI_ISL_720480, EPI_ISL_720481, EPI_ISL_720484, EPI_ISL_720485, EPI_ISL_720487, EPI_ISL_720488, EPI_ISL_720491, EPI_ISL_720492, EPI_ISL_720495, EPI_ISL_720496, EPI_ISL_720497, EPI_ISL_720500, EPI_ISL_720501, EPI_ISL_720503, EPI_ISL_720505, EPI_ISL_720506, EPI_ISL_720507, EPI_ISL_720508, EPI_ISL_720509, EPI_ISL_720510, EPI_ISL_720511, EPI_ISL_720513, EPI_ISL_720515, EPI_ISL_720516, EPI_ISL_720517, EPI_ISL_720518, EPI_ISL_720520, EPI_ISL_720523, EPI_ISL_720526, EPI_ISL_720528, EPI_ISL_720530, EPI_ISL_720531, EPI_ISL_720534, EPI_ISL_720535, EPI_ISL_720536, EPI_ISL_720538, EPI_ISL_720540, EPI_ISL_720542, EPI_ISL_720544, EPI_ISL_720546, EPI_ISL_720547, EPI_ISL_720549, EPI_ISL_720552, EPI_ISL_720553, EPI_ISL_720554, EPI_ISL_720555, EPI_ISL_720556, EPI_ISL_720557, EPI_ISL_720560, EPI_ISL_720562, EPI_ISL_720564, EPI_ISL_720566, EPI_ISL_720567, EPI_ISL_720568, EPI_ISL_720569, EPI_ISL_720570, EPI_ISL_720575, EPI_ISL_720582, EPI_ISL_720583, EPI_ISL_720586, EPI_ISL_720587, EPI_ISL_720589, EPI_ISL_720590, EPI_ISL_720591, EPI_ISL_720592, EPI_ISL_720593, EPI_ISL_720595, EPI_ISL_720596, EPI_ISL_720598, EPI_ISL_720599, EPI_ISL_720600, EPI_ISL_720601, EPI_ISL_720603, EPI_ISL_720604, EPI_ISL_720606, EPI_ISL_720608, EPI_ISL_720610, EPI_ISL_720611, EPI_ISL_720612, EPI_ISL_720613, EPI_ISL_720615, EPI_ISL_720616, EPI_ISL_720617, EPI_ISL_720618, EPI_ISL_720620, EPI_ISL_720624, EPI_ISL_720625, EPI_ISL_720626, EPI_ISL_720628, EPI_ISL_720632, EPI_ISL_720633, EPI_ISL_720635, EPI_ISL_720636, EPI_ISL_720639, EPI_ISL_720640, EPI_ISL_720642, EPI_ISL_720643, EPI_ISL_720644, EPI_ISL_720645, EPI_ISL_720648, EPI_ISL_720650, EPI_ISL_720653, EPI_ISL_720655, EPI_ISL_720656, EPI_ISL_720657, EPI_ISL_720658, EPI_ISL_720660, EPI_ISL_720662, EPI_ISL_720665, EPI_ISL_720666, EPI_ISL_720667, EPI_ISL_720673, EPI_ISL_721528, EPI_ISL_721531, EPI_ISL_731253, EPI_ISL_767239, EPI_ISL_783069, EPI_ISL_783070, EPI_ISL_783073, EPI_ISL_783074, EPI_ISL_783076, EPI_ISL_783079, EPI_ISL_783080, EPI_ISL_783081, EPI_ISL_783082, EPI_ISL_783084, EPI_ISL_783086, EPI_ISL_783088, EPI_ISL_783089, EPI_ISL_783091, EPI_ISL_783092, EPI_ISL_783093, EPI_ISL_783094, EPI_ISL_783095, EPI_ISL_783096, EPI_ISL_783097, EPI_ISL_783098, EPI_ISL_783100, EPI_ISL_783101, EPI_ISL_783102, EPI_ISL_783103, EPI_ISL_783104, EPI_ISL_783107, EPI_ISL_783108, EPI_ISL_783109, EPI_ISL_783111, EPI_ISL_783115, EPI_ISL_783116, EPI_ISL_783117, EPI_ISL_783118, EPI_ISL_783119, EPI_ISL_783120, EPI_ISL_783121, EPI_ISL_783122, EPI_ISL_783123, EPI_ISL_783125, EPI_ISL_783126, EPI_ISL_783127, EPI_ISL_783128, EPI_ISL_783130, EPI_ISL_783131, EPI_ISL_783133, EPI_ISL_783134, EPI_ISL_783135, EPI_ISL_783137, EPI_ISL_783138, EPI_ISL_783139, EPI_ISL_783140, EPI_ISL_783141, EPI_ISL_783143, EPI_ISL_783144, EPI_ISL_783145, EPI_ISL_783146, EPI_ISL_783147, EPI_ISL_783151, EPI_ISL_783152, EPI_ISL_783156, EPI_ISL_783160, EPI_ISL_783163, EPI_ISL_783164, EPI_ISL_783166, EPI_ISL_783167, EPI_ISL_783169, EPI_ISL_783172, EPI_ISL_783173, EPI_ISL_783175, EPI_ISL_783176, EPI_ISL_783180, EPI_ISL_783181, EPI_ISL_783182, EPI_ISL_783183, EPI_ISL_783185, EPI_ISL_783186, EPI_ISL_783187, EPI_ISL_783190, EPI_ISL_783192, EPI_ISL_783193, EPI_ISL_783194, EPI_ISL_783197, EPI_ISL_783199, EPI_ISL_783202, EPI_ISL_783203, EPI_ISL_783204, EPI_ISL_783205, EPI_ISL_783208, EPI_ISL_783209, EPI_ISL_783210, EPI_ISL_783212, EPI_ISL_783213, EPI_ISL_783214, EPI_ISL_783216, EPI_ISL_783217, EPI_ISL_783218, EPI_ISL_783219, EPI_ISL_783220, EPI_ISL_783222, EPI_ISL_783223, EPI_ISL_783227, EPI_ISL_783228, EPI_ISL_783229, EPI_ISL_783232, EPI_ISL_783233, EPI_ISL_783234, EPI_ISL_783236, EPI_ISL_783239, EPI_ISL_783242, EPI_ISL_783244, EPI_ISL_783248, EPI_ISL_783249, EPI_ISL_783251, EPI_ISL_783252, EPI_ISL_783254, EPI_ISL_783256, EPI_ISL_783258, EPI_ISL_783259, EPI_ISL_783260, EPI_ISL_783261, EPI_ISL_783262, EPI_ISL_783264, EPI_ISL_783265, EPI_ISL_783266, EPI_ISL_783271, EPI_ISL_783272, EPI_ISL_783273, EPI_ISL_783274, EPI_ISL_783275, EPI_ISL_783276, EPI_ISL_783277, EPI_ISL_783278, EPI_ISL_783280, EPI_ISL_783282, EPI_ISL_783285, EPI_ISL_783286, EPI_ISL_783287, EPI_ISL_783288, EPI_ISL_783289, EPI_ISL_783290, EPI_ISL_783292, EPI_ISL_783294, EPI_ISL_783295, EPI_ISL_783297, EPI_ISL_783299, EPI_ISL_783300, EPI_ISL_783301, EPI_ISL_783302, EPI_ISL_783304, EPI_ISL_783305, EPI_ISL_783307, EPI_ISL_783309, EPI_ISL_783310, EPI_ISL_783313, EPI_ISL_783315, EPI_ISL_783318, EPI_ISL_783319, EPI_ISL_783320, EPI_ISL_783323, EPI_ISL_783325, EPI_ISL_783326, EPI_ISL_783327, EPI_ISL_783328, EPI_ISL_783331, EPI_ISL_783333, EPI_ISL_783335, EPI_ISL_783338, EPI_ISL_783341, EPI_ISL_783343, EPI_ISL_783346, EPI_ISL_783349, EPI_ISL_783350, EPI_ISL_783352, EPI_ISL_783355, EPI_ISL_783356, EPI_ISL_783357, EPI_ISL_783358, EPI_ISL_783361, EPI_ISL_783362, EPI_ISL_783363, EPI_ISL_783364, EPI_ISL_783366, EPI_ISL_801182, EPI_ISL_801193, EPI_ISL_801194, EPI_ISL_801197, EPI_ISL_801199, EPI_ISL_801200, EPI_ISL_801204, EPI_ISL_801208, EPI_ISL_801210, EPI_ISL_801212, EPI_ISL_801219, EPI_ISL_801223, EPI_ISL_801225

| | | | |
|---|---|---|---|
| see above | Lighthouse Lab in Milton Keynes | Wellcome Sanger Institute for the COVID-19 Genomics UK (COG-UK) Consortium | Cordelia Langford; David K. Jackson; Dominic Kwiatkowski; Ewan Harrison; Ian Johnston; John Sillitoe on behalf of the Wellcome Sanger Institute COVID-19 Surveillance Team; Roberto Amato; Sonia Goncalves; The Lighthouse Lab in Milton Keynes and Alex Alderton |

EPI_ISL_679946, EPI_ISL_679947, EPI_ISL_679948, EPI_ISL_679949, EPI_ISL_679950, EPI_ISL_679951, EPI_ISL_679952, EPI_ISL_679953, EPI_ISL_679954, EPI_ISL_679955, EPI_ISL_679956, EPI_ISL_679957, EPI_ISL_679958, EPI_ISL_679959, EPI_ISL_679960, EPI_ISL_679961, EPI_ISL_679962, EPI_ISL_679963, EPI_ISL_679964, EPI_ISL_679965, EPI_ISL_679966, EPI_ISL_725540, EPI_ISL_725541, EPI_ISL_725542, EPI_ISL_725543, EPI_ISL_725544, EPI_ISL_725545, EPI_ISL_725546, EPI_ISL_725547, EPI_ISL_725548, EPI_ISL_725549, EPI_ISL_725550, EPI_ISL_725551, EPI_ISL_725552, EPI_ISL_725553, EPI_ISL_725554, EPI_ISL_725555, EPI_ISL_725556, EPI_ISL_725557, EPI_ISL_725558, EPI_ISL_725559, EPI_ISL_725560, EPI_ISL_725561, EPI_ISL_725562, EPI_ISL_725563, EPI_ISL_725564, EPI_ISL_725565, EPI_ISL_725566, EPI_ISL_725567, EPI_ISL_725568, EPI_ISL_725569, EPI_ISL_725570, EPI_ISL_725571, EPI_ISL_725572, EPI_ISL_725573, EPI_ISL_725574, EPI_ISL_725575, EPI_ISL_725576, EPI_ISL_725577, EPI_ISL_725578, EPI_ISL_725579, EPI_ISL_725580, EPI_ISL_725581, EPI_ISL_725582, EPI_ISL_725583, EPI_ISL_725584, EPI_ISL_725585, EPI_ISL_725586, EPI_ISL_725587, EPI_ISL_725588, EPI_ISL_725589, EPI_ISL_725590, EPI_ISL_725591, EPI_ISL_725592, EPI_ISL_725593, EPI_ISL_725594, EPI_ISL_725595, EPI_ISL_819478, EPI_ISL_819479, EPI_ISL_819480, EPI_ISL_819481, EPI_ISL_819482, EPI_ISL_1475495

| | | | |
|---|---|---|---|
| see above | Lincolnshire Hospitals and DeepSeq Nottingham | COVID-19 Genomics UK (COG-UK) Consortium | Christopher Moore; Fei Sang; Joeseph Chappell; Johnny Debebe; Jonathan Ball; Matthew Carlisle; Matthew Loose; Nadine Holmes; Nichola Duckworth; Patrick McClure; Sarah Walsh; Tim Sloan; Victoria Wright |

EPI_ISL_679393, EPI_ISL_679394, EPI_ISL_679395, EPI_ISL_679396, EPI_ISL_679397, EPI_ISL_679398, EPI_ISL_679399, EPI_ISL_679400, EPI_ISL_679401, EPI_ISL_679402, EPI_ISL_679403, EPI_ISL_679404, EPI_ISL_679405, EPI_ISL_702471, EPI_ISL_702481, EPI_ISL_702492, EPI_ISL_702510, EPI_ISL_702523, EPI_ISL_702574, EPI_ISL_702577, EPI_ISL_702605, EPI_ISL_702615, EPI_ISL_702623, EPI_ISL_702626, EPI_ISL_702631, EPI_ISL_702639, EPI_ISL_702644, EPI_ISL_702649, EPI_ISL_702674, EPI_ISL_702677, EPI_ISL_702718, EPI_ISL_702801, EPI_ISL_702806, EPI_ISL_702826, EPI_ISL_702845, EPI_ISL_702866, EPI_ISL_702871, EPI_ISL_702878, EPI_ISL_702884, EPI_ISL_702894, EPI_ISL_702907, EPI_ISL_702932, EPI_ISL_702950, EPI_ISL_702956, EPI_ISL_702969, EPI_ISL_702996, EPI_ISL_703023, EPI_ISL_703088, EPI_ISL_703090, EPI_ISL_703102, EPI_ISL_703110, EPI_ISL_703147, EPI_ISL_703162, EPI_ISL_703165, EPI_ISL_703167, EPI_ISL_703170, EPI_ISL_703176, EPI_ISL_703182, EPI_ISL_703184, EPI_ISL_703206, EPI_ISL_703208, EPI_ISL_703216, EPI_ISL_703261, EPI_ISL_703275, EPI_ISL_703289, EPI_ISL_703305, EPI_ISL_703313, EPI_ISL_703324, EPI_ISL_703348, EPI_ISL_703353, EPI_ISL_703356, EPI_ISL_703361, EPI_ISL_703380, EPI_ISL_703421, EPI_ISL_703427, EPI_ISL_703481, EPI_ISL_703527, EPI_ISL_703690, EPI_ISL_703695, EPI_ISL_703734, EPI_ISL_703735, EPI_ISL_703738, EPI_ISL_703744, EPI_ISL_703759, EPI_ISL_703762, EPI_ISL_703763, EPI_ISL_703770, EPI_ISL_703807, EPI_ISL_703815, EPI_ISL_703818, EPI_ISL_703821, EPI_ISL_703823, EPI_ISL_703847, EPI_ISL_703876, EPI_ISL_703897, EPI_ISL_703913, EPI_ISL_703916, EPI_ISL_703971, EPI_ISL_703977, EPI_ISL_704003, EPI_ISL_704006, EPI_ISL_704009, EPI_ISL_704131, EPI_ISL_704134, EPI_ISL_704137, EPI_ISL_704153, EPI_ISL_704156, EPI_ISL_704195, EPI_ISL_704260, EPI_ISL_704562, EPI_ISL_704895, EPI_ISL_704897, EPI_ISL_704900, EPI_ISL_704903, EPI_ISL_704905, EPI_ISL_704945, EPI_ISL_704968, EPI_ISL_704973, EPI_ISL_704983, EPI_ISL_704989, EPI_ISL_705005, EPI_ISL_705024, EPI_ISL_705030, EPI_ISL_705038, EPI_ISL_705043, EPI_ISL_705054, EPI_ISL_705056, EPI_ISL_705059, EPI_ISL_705083, EPI_ISL_705121, EPI_ISL_705150, EPI_ISL_705176, EPI_ISL_705193, EPI_ISL_705203, EPI_ISL_705206, EPI_ISL_705208, EPI_ISL_705241, EPI_ISL_705242, EPI_ISL_705246, EPI_ISL_705248, EPI_ISL_705269, EPI_ISL_705274, EPI_ISL_705286, EPI_ISL_705287, EPI_ISL_705299, EPI_ISL_705303, EPI_ISL_705313, EPI_ISL_705315, EPI_ISL_705321, EPI_ISL_705329, EPI_ISL_705334, EPI_ISL_705336, EPI_ISL_705349, EPI_ISL_705358, EPI_ISL_705372, EPI_ISL_705377, EPI_ISL_705385, EPI_ISL_705386, EPI_ISL_705387, EPI_ISL_705388, EPI_ISL_705398, EPI_ISL_705400, EPI_ISL_705471, EPI_ISL_705809, EPI_ISL_705810, EPI_ISL_705811, EPI_ISL_705812, EPI_ISL_705813, EPI_ISL_705814, EPI_ISL_705815, EPI_ISL_705816, EPI_ISL_705817, EPI_ISL_705818, EPI_ISL_705819, EPI_ISL_705820, EPI_ISL_705821, EPI_ISL_705822, EPI_ISL_705823, EPI_ISL_705824, EPI_ISL_705825, EPI_ISL_705826, EPI_ISL_705827, EPI_ISL_705828, EPI_ISL_705829, EPI_ISL_705830, EPI_ISL_705831, EPI_ISL_705832, EPI_ISL_705833, EPI_ISL_705834, EPI_ISL_705835, EPI_ISL_705836, EPI_ISL_705837, EPI_ISL_705838, EPI_ISL_705839, EPI_ISL_705840, EPI_ISL_705841, EPI_ISL_705842, EPI_ISL_705843, EPI_ISL_705844, EPI_ISL_705845, EPI_ISL_705846, EPI_ISL_705847, EPI_ISL_705848, EPI_ISL_705849, EPI_ISL_705850, EPI_ISL_705851, EPI_ISL_705852, EPI_ISL_705853, EPI_ISL_705854, EPI_ISL_705855, EPI_ISL_705856, EPI_ISL_705857, EPI_ISL_705858, EPI_ISL_705859, EPI_ISL_705860, EPI_ISL_705861, EPI_ISL_705862, EPI_ISL_705863, EPI_ISL_705864, EPI_ISL_705923, EPI_ISL_705924, EPI_ISL_705925, EPI_ISL_705926, EPI_ISL_705927, EPI_ISL_705940, EPI_ISL_724396, EPI_ISL_724397, EPI_ISL_724398, EPI_ISL_724399, EPI_ISL_724400, EPI_ISL_724401, EPI_ISL_724402, EPI_ISL_724403, EPI_ISL_724404, EPI_ISL_724405, EPI_ISL_724406, EPI_ISL_724407, EPI_ISL_724408, EPI_ISL_724409, EPI_ISL_724410, EPI_ISL_724411, EPI_ISL_724412, EPI_ISL_724413, EPI_ISL_724414, EPI_ISL_724415, EPI_ISL_724416, EPI_ISL_724417, EPI_ISL_724418, EPI_ISL_724419, EPI_ISL_724420, EPI_ISL_724421, EPI_ISL_724422, EPI_ISL_724423, EPI_ISL_724424, EPI_ISL_724425, EPI_ISL_724426, EPI_ISL_724427, EPI_ISL_724428, EPI_ISL_724429, EPI_ISL_724430, EPI_ISL_724431, EPI_ISL_724432, EPI_ISL_724433, EPI_ISL_724434, EPI_ISL_724435, EPI_ISL_724436, EPI_ISL_724437, EPI_ISL_724438, EPI_ISL_724439, EPI_ISL_724440, EPI_ISL_724441, EPI_ISL_724442, EPI_ISL_724443, EPI_ISL_724444, EPI_ISL_724445, EPI_ISL_724446, EPI_ISL_724447, EPI_ISL_724448, EPI_ISL_724449, EPI_ISL_724450, EPI_ISL_724451, EPI_ISL_724452, EPI_ISL_724453, EPI_ISL_724454, EPI_ISL_724455, EPI_ISL_724456, EPI_ISL_724457, EPI_ISL_724458, EPI_ISL_724459, EPI_ISL_724460, EPI_ISL_724461, EPI_ISL_724464, EPI_ISL_724465, EPI_ISL_724466, EPI_ISL_724467, EPI_ISL_724468, EPI_ISL_724469, EPI_ISL_724470, EPI_ISL_724471, EPI_ISL_724472, EPI_ISL_724473, EPI_ISL_724474, EPI_ISL_724475, EPI_ISL_724476, EPI_ISL_724477, EPI_ISL_724478, EPI_ISL_724479, EPI_ISL_724480, EPI_ISL_724481, EPI_ISL_724482, EPI_ISL_724483, EPI_ISL_724484, EPI_ISL_724485, EPI_ISL_724486, EPI_ISL_724487, EPI_ISL_724488, EPI_ISL_724489, EPI_ISL_724490, EPI_ISL_724491, EPI_ISL_724492, EPI_ISL_724493, EPI_ISL_724494, EPI_ISL_724495, EPI_ISL_724496, EPI_ISL_724499, EPI_ISL_724500, EPI_ISL_724501, EPI_ISL_724502, EPI_ISL_724503, EPI_ISL_724504, EPI_ISL_724505, EPI_ISL_724506, EPI_ISL_724507, EPI_ISL_724509, EPI_ISL_724510, EPI_ISL_724511, EPI_ISL_724512, EPI_ISL_724513, EPI_ISL_724514, EPI_ISL_724515, EPI_ISL_724516, EPI_ISL_724518, EPI_ISL_724523, EPI_ISL_724524, EPI_ISL_813751, EPI_ISL_865174, EPI_ISL_1104320, EPI_ISL_1177824, EPI_ISL_1177825, EPI_ISL_1177826, EPI_ISL_1177827, EPI_ISL_1177828, EPI_ISL_1177829, EPI_ISL_1177830, EPI_ISL_1636817, EPI_ISL_1636818, EPI_ISL_1636819

| | | | |
|---|---|---|---|
| see above | Liverpool Clinical Laboratories | COVID-19 Genomics UK (COG-UK) Consortium | A Alrezaihi; Alessandro Gerada; Alistair Darby; Angela Cowell; Anita Lucaci; Anu Chawla; Cassie Olateju; Catherine Hartley; Charlotte Nelson; Ecaterina Vamos; Elaine O'Toole; Eleanor G Bentley; Ghada T Shawli; Hermione Webster; Isabel García-Dorival; James Johnson; James P Stewart; Jenifer Manson; Joanne Watts; Jones Benjamin; Jordan J Clark; Julian Hiscox; L Luu; Lucille Rainbow; M Almsaud; Margaret Hughes; Mark Whitehead; Matthew Gemmell; Miren Iturriza-Gomara; Muhannad Alruwaili; N.P Randle; Neil Swainston; PKF Gilmore; Parul Sharma; Rebekah Penrice-Randal; Richard Eccles; Richard Gregory; Sam Haldenby; Steve Paterson; Stuart D Armstrong; Trevor Ian Robinson; Ximeng Han |

EPI_ISL_702495, EPI_ISL_702539, EPI_ISL_702747, EPI_ISL_702803, EPI_ISL_702815, EPI_ISL_702827, EPI_ISL_703010, EPI_ISL_703013, EPI_ISL_703105, EPI_ISL_703377, EPI_ISL_703484, EPI_ISL_703487, EPI_ISL_703545, EPI_ISL_703704, EPI_ISL_704198, EPI_ISL_704422, EPI_ISL_704425, EPI_ISL_704572, EPI_ISL_704573, EPI_ISL_704574, EPI_ISL_704575, EPI_ISL_704576, EPI_ISL_704976, EPI_ISL_705010, EPI_ISL_705067, EPI_ISL_705201, EPI_ISL_705207, EPI_ISL_705214, EPI_ISL_705273, EPI_ISL_705282, EPI_ISL_705343, EPI_ISL_705356, EPI_ISL_705364, EPI_ISL_705486, EPI_ISL_705494, EPI_ISL_705495, EPI_ISL_705496, EPI_ISL_706312, EPI_ISL_706317, EPI_ISL_706318, EPI_ISL_706319, EPI_ISL_706320, EPI_ISL_706321, EPI_ISL_706322, EPI_ISL_706323, EPI_ISL_706324, EPI_ISL_706325, EPI_ISL_706326, EPI_ISL_706327, EPI_ISL_706328, EPI_ISL_706329, EPI_ISL_706330, EPI_ISL_706331, EPI_ISL_706349, EPI_ISL_706350, EPI_ISL_706351, EPI_ISL_706352, EPI_ISL_724063, EPI_ISL_724064, EPI_ISL_724065, EPI_ISL_724066, EPI_ISL_724067, EPI_ISL_724068, EPI_ISL_724069, EPI_ISL_724070, EPI_ISL_724071, EPI_ISL_724072, EPI_ISL_724073, EPI_ISL_724074, EPI_ISL_724075, EPI_ISL_724076, EPI_ISL_724077, EPI_ISL_724078, EPI_ISL_724079, EPI_ISL_724080, EPI_ISL_724081, EPI_ISL_724082, EPI_ISL_724083, EPI_ISL_724084, EPI_ISL_724085, EPI_ISL_724086, EPI_ISL_724087, EPI_ISL_724088, EPI_ISL_724089, EPI_ISL_724090, EPI_ISL_724091, EPI_ISL_724092, EPI_ISL_724093, EPI_ISL_724094, EPI_ISL_724095, EPI_ISL_724096, EPI_ISL_724097, EPI_ISL_724098, EPI_ISL_724099, EPI_ISL_724100, EPI_ISL_724101, EPI_ISL_724102, EPI_ISL_724103, EPI_ISL_724104, EPI_ISL_724105, EPI_ISL_724106, EPI_ISL_724107, EPI_ISL_724108, EPI_ISL_724109, EPI_ISL_724110, EPI_ISL_724111, EPI_ISL_724112, EPI_ISL_724113, EPI_ISL_724114, EPI_ISL_724115, EPI_ISL_724116, EPI_ISL_724117, EPI_ISL_724118, EPI_ISL_724119, EPI_ISL_724120, EPI_ISL_724121, EPI_ISL_724122, EPI_ISL_724123, EPI_ISL_724855, EPI_ISL_724856, EPI_ISL_724857, EPI_ISL_724858, EPI_ISL_724859, EPI_ISL_724860, EPI_ISL_724861, EPI_ISL_724862, EPI_ISL_724863, EPI_ISL_724864, EPI_ISL_724865, EPI_ISL_724866, EPI_ISL_724867, EPI_ISL_724868, EPI_ISL_724869, EPI_ISL_724870, EPI_ISL_724871, EPI_ISL_724872, EPI_ISL_724873, EPI_ISL_724874, EPI_ISL_724875, EPI_ISL_724876, EPI_ISL_724879, EPI_ISL_724880, EPI_ISL_724881, EPI_ISL_724924, EPI_ISL_724925, EPI_ISL_724926, EPI_ISL_724927, EPI_ISL_724928, EPI_ISL_724929, EPI_ISL_724930, EPI_ISL_724931, EPI_ISL_724932, EPI_ISL_724933, EPI_ISL_724934, EPI_ISL_724935, EPI_ISL_724936, EPI_ISL_724937, EPI_ISL_724938, EPI_ISL_724939, EPI_ISL_724940, EPI_ISL_724941, EPI_ISL_724942, EPI_ISL_724943, EPI_ISL_724944, EPI_ISL_724945, EPI_ISL_724946, EPI_ISL_724947, EPI_ISL_724948, EPI_ISL_724949, EPI_ISL_724950, EPI_ISL_724951, EPI_ISL_724952, EPI_ISL_724953, EPI_ISL_724954, EPI_ISL_724955, EPI_ISL_724956, EPI_ISL_724957, EPI_ISL_724958, EPI_ISL_724959, EPI_ISL_724960, EPI_ISL_724961, EPI_ISL_724962, EPI_ISL_724970, EPI_ISL_725012, EPI_ISL_725013, EPI_ISL_725014, EPI_ISL_725015, EPI_ISL_725016, EPI_ISL_725017, EPI_ISL_725018, EPI_ISL_725019, EPI_ISL_741439, EPI_ISL_741441, EPI_ISL_741442, EPI_ISL_741443, EPI_ISL_741444, EPI_ISL_741445, EPI_ISL_741446, EPI_ISL_741447, EPI_ISL_741448, EPI_ISL_741449, EPI_ISL_741450, EPI_ISL_741451, EPI_ISL_741452

| | | | |
|---|---|---|---|
| see above | Northumbria University / South Tees Hospitals NHS Foundation Trust / North Cumbria Integrated Care NHS Foundation Trust / North Tees and Hartlepool NHS Foundation Trust / Newcastle Hospitals NHS Foundation Trust | COVID-19 Genomics UK (COG-UK) Consortium | Andrew Nelson; Brendan Payne; Clive Graham; Darren L Smith; Debra Padgett; Edward Barton; Emma Swindells; Garren Scott; Gary Black; Gary Eltringham; Giles S Holt; Greg R Young; Jane Greenaway; Jennifer Collins; John Allan; Joshua Loh; Lynn Dover; Matthew Bashton; Mohammad A Tariq; Paul Baker; Sarah Essex; Steve Liggett; Wen C Yew; Yusri Taha |

EPI_ISL_841531, EPI_ISL_841559, EPI_ISL_841560, EPI_ISL_841561, EPI_ISL_841563, EPI_ISL_841564, EPI_ISL_841565, EPI_ISL_841566, EPI_ISL_841568, EPI_ISL_841569, EPI_ISL_841570, EPI_ISL_841571, EPI_ISL_951788

| | | | |
|---|---|---|---|
| see above | Originating lab: Wales Specialist Virology Centre Sequencing lab: Pathogen Genomics Unit | Public Health Wales Microbiology Cardiff Wales Specialist Virology Centre | Alec Birchley; Alexander Adams; Amy Gaskin; Angela Marchbank; Bree Gatica-Wilcox; Catherine Moore; Jason Coombes; Joanne Watkins; Joel Southgate; Johnathan Evans; Laura Gifford; Lauren Gilbert; Lee Graham; Malorie Perry; Matthew Bull; Nicole Pacchiarini; Sally Corden; Sara Kumziene-Summerhayes; Sara Rey; Sarah Taylor; Simon Cottrell; Sophie Jones; Tom Connor |

EPI_ISL_840188, EPI_ISL_840189, EPI_ISL_840190, EPI_ISL_840191, EPI_ISL_840192, EPI_ISL_840193, EPI_ISL_840204, EPI_ISL_840205, EPI_ISL_840206, EPI_ISL_840207, EPI_ISL_840210, EPI_ISL_840211, EPI_ISL_840212, EPI_ISL_840213, EPI_ISL_840214, EPI_ISL_840215, EPI_ISL_840216, EPI_ISL_840217, EPI_ISL_840220, EPI_ISL_840221, EPI_ISL_1833035, EPI_ISL_1833036, EPI_ISL_1833037, EPI_ISL_1833038, EPI_ISL_1833039, EPI_ISL_1833040, EPI_ISL_1833041, EPI_ISL_1833042

| | | | |
|---|---|---|---|
| see above | Oxford Viromics, NDM, University of Oxford; Oxford University Hospitals; Basingstoke and North Hampshire Hospital | COVID-19 Genomics UK (COG-UK) Consortium | Alex Mobbs; Amy Trebes; Anita Justice; Catrin Moore; Christophe Fraser; David Bonsall; David Buck; Emma Wise; George Macintyre; Jessica Lynch; John Todd; Mariateresa de Cesare; Matilde Mori; Monique Andersson; Nathan Moore; Nick Cortes; Robert Shaw; Stephen Kidd; Tanya Golubchik; Timothy Peto |

EPI_ISL_665268, EPI_ISL_665295, EPI_ISL_665316, EPI_ISL_665336, EPI_ISL_665362, EPI_ISL_665369, EPI_ISL_665377, EPI_ISL_665380, EPI_ISL_665381, EPI_ISL_665423, EPI_ISL_665429, EPI_ISL_665440, EPI_ISL_665450, EPI_ISL_665453, EPI_ISL_665463, EPI_ISL_665465, EPI_ISL_665475, EPI_ISL_665508, EPI_ISL_665518, EPI_ISL_665519, EPI_ISL_665534, EPI_ISL_665574, EPI_ISL_665594, EPI_ISL_665596, EPI_ISL_665599, EPI_ISL_665603, EPI_ISL_665639, EPI_ISL_665665, EPI_ISL_665668, EPI_ISL_665678, EPI_ISL_665689, EPI_ISL_665692, EPI_ISL_665697, EPI_ISL_665712, EPI_ISL_665714, EPI_ISL_665730, EPI_ISL_665732, EPI_ISL_665737, EPI_ISL_665749, EPI_ISL_665876, EPI_ISL_665897, EPI_ISL_702716, EPI_ISL_702861, EPI_ISL_702875, EPI_ISL_702882, EPI_ISL_702902, EPI_ISL_702925, EPI_ISL_702941, EPI_ISL_702944, EPI_ISL_703099, EPI_ISL_703113, EPI_ISL_703187, EPI_ISL_703200, EPI_ISL_703211, EPI_ISL_703264, EPI_ISL_703424, EPI_ISL_703590, EPI_ISL_703616, EPI_ISL_703714, EPI_ISL_703733, EPI_ISL_703736, EPI_ISL_703764, EPI_ISL_703769, EPI_ISL_703800, EPI_ISL_703802, EPI_ISL_703805, EPI_ISL_703905, EPI_ISL_703932, EPI_ISL_703945, EPI_ISL_703947, EPI_ISL_703963, EPI_ISL_703979, EPI_ISL_703998, EPI_ISL_704054, EPI_ISL_704073, EPI_ISL_704100, EPI_ISL_704123, EPI_ISL_704139, EPI_ISL_704150, EPI_ISL_704158, EPI_ISL_704174, EPI_ISL_704181, EPI_ISL_704236, EPI_ISL_704241, EPI_ISL_704580, EPI_ISL_704598, EPI_ISL_704599, EPI_ISL_704619, EPI_ISL_704620, EPI_ISL_704621, EPI_ISL_704622, EPI_ISL_704623, EPI_ISL_704624, EPI_ISL_704625, EPI_ISL_704626, EPI_ISL_704627, EPI_ISL_704628, EPI_ISL_704629, EPI_ISL_704630, EPI_ISL_704631, EPI_ISL_704632, EPI_ISL_704633, EPI_ISL_704634, EPI_ISL_704635, EPI_ISL_704719, EPI_ISL_704722, EPI_ISL_704725, EPI_ISL_704749, EPI_ISL_704752, EPI_ISL_704754, EPI_ISL_704762, EPI_ISL_704765, EPI_ISL_704768, EPI_ISL_704770, EPI_ISL_704772, EPI_ISL_704775, EPI_ISL_704777, EPI_ISL_704786, EPI_ISL_704797, EPI_ISL_704799, EPI_ISL_704802, EPI_ISL_704820, EPI_ISL_704823, EPI_ISL_704826, EPI_ISL_704831, EPI_ISL_704842, EPI_ISL_704847, EPI_ISL_704858, EPI_ISL_704892, EPI_ISL_704911, EPI_ISL_704914, EPI_ISL_704916, EPI_ISL_705211, EPI_ISL_705228, EPI_ISL_705262, EPI_ISL_705332, EPI_ISL_705375, EPI_ISL_705422, EPI_ISL_705432, EPI_ISL_705491, EPI_ISL_705567, EPI_ISL_705568, EPI_ISL_705569, EPI_ISL_705570, EPI_ISL_705571, EPI_ISL_705572, EPI_ISL_705573, EPI_ISL_705574, EPI_ISL_705575, EPI_ISL_705576, EPI_ISL_705577, EPI_ISL_705578, EPI_ISL_705579, EPI_ISL_705580, EPI_ISL_705581, EPI_ISL_705582, EPI_ISL_705583, EPI_ISL_705584, EPI_ISL_705585, EPI_ISL_705586, EPI_ISL_705587, EPI_ISL_705588, EPI_ISL_705589, EPI_ISL_705590, EPI_ISL_705591, EPI_ISL_705592, EPI_ISL_705593, EPI_ISL_705594, EPI_ISL_705595, EPI_ISL_705596, EPI_ISL_705597, EPI_ISL_705598, EPI_ISL_705599, EPI_ISL_705600, EPI_ISL_705601, EPI_ISL_705602, EPI_ISL_705603, EPI_ISL_705604, EPI_ISL_705605, EPI_ISL_705606, EPI_ISL_705607, EPI_ISL_705608, EPI_ISL_705609, EPI_ISL_705610, EPI_ISL_705611, EPI_ISL_705612, EPI_ISL_706361, EPI_ISL_706362, EPI_ISL_706363, EPI_ISL_706364, EPI_ISL_706365, EPI_ISL_706366, EPI_ISL_706384, EPI_ISL_706385, EPI_ISL_706386, EPI_ISL_706387, EPI_ISL_706388, EPI_ISL_706389, EPI_ISL_706390, EPI_ISL_706391, EPI_ISL_706392, EPI_ISL_706393, EPI_ISL_706394, EPI_ISL_706395, EPI_ISL_706396, EPI_ISL_706402, EPI_ISL_706408, EPI_ISL_706414, EPI_ISL_724124, EPI_ISL_724127, EPI_ISL_724128, EPI_ISL_724129, EPI_ISL_724130, EPI_ISL_724131, EPI_ISL_724132, EPI_ISL_724133, EPI_ISL_724134, EPI_ISL_724135, EPI_ISL_724136, EPI_ISL_724137, EPI_ISL_724138, EPI_ISL_724139, EPI_ISL_724140, EPI_ISL_724141, EPI_ISL_724142, EPI_ISL_724143, EPI_ISL_724144, EPI_ISL_724145, EPI_ISL_724146, EPI_ISL_724147, EPI_ISL_724148, EPI_ISL_724149, EPI_ISL_724150, EPI_ISL_724151, EPI_ISL_724152, EPI_ISL_724153, EPI_ISL_724154, EPI_ISL_724155, EPI_ISL_724156, EPI_ISL_724157, EPI_ISL_724158, EPI_ISL_724159, EPI_ISL_724160, EPI_ISL_724161, EPI_ISL_724162, EPI_ISL_724163, EPI_ISL_724164, EPI_ISL_724165, EPI_ISL_724166, EPI_ISL_724167, EPI_ISL_724168, EPI_ISL_724169, EPI_ISL_724170, EPI_ISL_724171, EPI_ISL_724172, EPI_ISL_724173, EPI_ISL_724174, EPI_ISL_724175, EPI_ISL_724176, EPI_ISL_724177, EPI_ISL_724178, EPI_ISL_724179, EPI_ISL_724180, EPI_ISL_724181, EPI_ISL_724182, EPI_ISL_724183, EPI_ISL_724184, EPI_ISL_724185, EPI_ISL_724186, EPI_ISL_724187, EPI_ISL_724189, EPI_ISL_724190, EPI_ISL_724191, EPI_ISL_724192, EPI_ISL_724193, EPI_ISL_724194, EPI_ISL_724195, EPI_ISL_724196, EPI_ISL_724197, EPI_ISL_724198, EPI_ISL_724199, EPI_ISL_724200, EPI_ISL_724201, EPI_ISL_724202, EPI_ISL_724203, EPI_ISL_724204, EPI_ISL_724205, EPI_ISL_724206, EPI_ISL_724207, EPI_ISL_724208, EPI_ISL_724209, EPI_ISL_725160, EPI_ISL_725174, EPI_ISL_725178, EPI_ISL_725183, EPI_ISL_725186, EPI_ISL_725188, EPI_ISL_725189, EPI_ISL_725192, EPI_ISL_725194, EPI_ISL_725199, EPI_ISL_725200, EPI_ISL_725201, EPI_ISL_725203, EPI_ISL_725205, EPI_ISL_725206, EPI_ISL_725207, EPI_ISL_725208, EPI_ISL_725209, EPI_ISL_725210, EPI_ISL_725213, EPI_ISL_725215, EPI_ISL_725217, EPI_ISL_725219, EPI_ISL_725220, EPI_ISL_725221, EPI_ISL_725223, EPI_ISL_725224, EPI_ISL_725225, EPI_ISL_725226, EPI_ISL_725229, EPI_ISL_725232, EPI_ISL_725234, EPI_ISL_725235, EPI_ISL_725236, EPI_ISL_725240, EPI_ISL_725241, EPI_ISL_725243, EPI_ISL_725244, EPI_ISL_725251, EPI_ISL_725252, EPI_ISL_725253, EPI_ISL_725259, EPI_ISL_725260, EPI_ISL_725262, EPI_ISL_725281, EPI_ISL_725284, EPI_ISL_725288, EPI_ISL_725370, EPI_ISL_725425, EPI_ISL_725513, EPI_ISL_741516, EPI_ISL_866822, EPI_ISL_998829, EPI_ISL_998834, EPI_ISL_998835, EPI_ISL_1832930, EPI_ISL_1832943, EPI_ISL_1832945, EPI_ISL_1832953, EPI_ISL_1832954, EPI_ISL_1832958, EPI_ISL_1832968

| | | | |
|---|---|---|---|
| see above | Quadram Institute Bioscience | COVID-19 Genomics UK (COG-UK) Consortium | Alexander J Trotter; Alison E. Mather; Alp Aydin; Ana P. Tedim; Anastasia Kolyva; Andrew Bell; Andrew J. Page; Claire Stuart; Dave J. Baker; Gemma L. Kay; John Wain; Justin O'Grady; Leonardo de Oliveira Martins; Lizzie Meadows; Maria Diaz; Mark Webber; Muhammed Yasir; Nabil-Fareed Alikhan; Ngozi Elumogo; Nicholas M. Thomson; Rachael Stanley; Rachel Gilroy; Reenesh Prakash; Samir Dervisevic; Samuel Bloomfield; Steven Rudder; Thanh Le-Viet |

EPI_ISL_665290, EPI_ISL_665291, EPI_ISL_665344, EPI_ISL_665345, EPI_ISL_665365, EPI_ISL_665366, EPI_ISL_665367, EPI_ISL_665415, EPI_ISL_665421, EPI_ISL_665424, EPI_ISL_665430, EPI_ISL_665452, EPI_ISL_665510, EPI_ISL_665512, EPI_ISL_665513, EPI_ISL_665528, EPI_ISL_665544, EPI_ISL_665545, EPI_ISL_665562, EPI_ISL_665601, EPI_ISL_665610, EPI_ISL_665612, EPI_ISL_665623, EPI_ISL_665724, EPI_ISL_665740, EPI_ISL_665789, EPI_ISL_665906, EPI_ISL_665907, EPI_ISL_665908, EPI_ISL_665909, EPI_ISL_665910, EPI_ISL_665911, EPI_ISL_665912, EPI_ISL_665913, EPI_ISL_665914, EPI_ISL_665915, EPI_ISL_665916, EPI_ISL_665917, EPI_ISL_665918, EPI_ISL_665919, EPI_ISL_665920, EPI_ISL_665921, EPI_ISL_665922, EPI_ISL_665923, EPI_ISL_665924, EPI_ISL_665925, EPI_ISL_665926, EPI_ISL_665927, EPI_ISL_665928, EPI_ISL_665929, EPI_ISL_665930, EPI_ISL_665931, EPI_ISL_665932, EPI_ISL_665933, EPI_ISL_665934, EPI_ISL_665935, EPI_ISL_665936, EPI_ISL_665937, EPI_ISL_665938, EPI_ISL_665939, EPI_ISL_665943, EPI_ISL_665944, EPI_ISL_665945, EPI_ISL_679249, EPI_ISL_679250, EPI_ISL_679251, EPI_ISL_679252, EPI_ISL_679253, EPI_ISL_679254, EPI_ISL_679255, EPI_ISL_679256, EPI_ISL_679257, EPI_ISL_679258, EPI_ISL_679259, EPI_ISL_679260, EPI_ISL_679261, EPI_ISL_679262, EPI_ISL_679263, EPI_ISL_679264, EPI_ISL_679265, EPI_ISL_679266, EPI_ISL_679267, EPI_ISL_679268, EPI_ISL_679269, EPI_ISL_679270, EPI_ISL_679271, EPI_ISL_679272, EPI_ISL_679273, EPI_ISL_679274, EPI_ISL_679275, EPI_ISL_679276, EPI_ISL_679277, EPI_ISL_679278, EPI_ISL_679279, EPI_ISL_679280, EPI_ISL_679281, EPI_ISL_679282, EPI_ISL_679283, EPI_ISL_679284, EPI_ISL_679285, EPI_ISL_679286, EPI_ISL_679287, EPI_ISL_679288, EPI_ISL_679289, EPI_ISL_679290, EPI_ISL_679291, EPI_ISL_679292, EPI_ISL_679293, EPI_ISL_679294, EPI_ISL_679295, EPI_ISL_679296, EPI_ISL_679297, EPI_ISL_679298, EPI_ISL_679299, EPI_ISL_679300, EPI_ISL_679301, EPI_ISL_679302, EPI_ISL_679303, EPI_ISL_679304, EPI_ISL_679305, EPI_ISL_679306, EPI_ISL_679307, EPI_ISL_679309, EPI_ISL_679310, EPI_ISL_679311, EPI_ISL_679312, EPI_ISL_679313, EPI_ISL_679314, EPI_ISL_679933, EPI_ISL_679934, EPI_ISL_679935, EPI_ISL_679936, EPI_ISL_679937, EPI_ISL_679938, EPI_ISL_679939, EPI_ISL_679940, EPI_ISL_679941, EPI_ISL_679942, EPI_ISL_679943, EPI_ISL_702846, EPI_ISL_702879, EPI_ISL_702889, EPI_ISL_703124, EPI_ISL_703720, EPI_ISL_703775, EPI_ISL_703776, EPI_ISL_703789, EPI_ISL_703792, EPI_ISL_703794, EPI_ISL_703853, EPI_ISL_705094, EPI_ISL_705223, EPI_ISL_705487, EPI_ISL_705613, EPI_ISL_705614, EPI_ISL_706420, EPI_ISL_706421, EPI_ISL_706422, EPI_ISL_706423, EPI_ISL_706424, EPI_ISL_706425, EPI_ISL_706426, EPI_ISL_706427, EPI_ISL_706431, EPI_ISL_706432, EPI_ISL_706433, EPI_ISL_706434, EPI_ISL_725519, EPI_ISL_725520, EPI_ISL_725521, EPI_ISL_725522, EPI_ISL_725523, EPI_ISL_725524, EPI_ISL_725525, EPI_ISL_725526, EPI_ISL_725527, EPI_ISL_741610, EPI_ISL_819464, EPI_ISL_838344, EPI_ISL_838345, EPI_ISL_838346, EPI_ISL_838347, EPI_ISL_838350, EPI_ISL_950602, EPI_ISL_950603, EPI_ISL_950613, EPI_ISL_950614, EPI_ISL_950617, EPI_ISL_950618, EPI_ISL_950619, EPI_ISL_950620, EPI_ISL_950621, EPI_ISL_950622, EPI_ISL_950623, EPI_ISL_950624, EPI_ISL_950625, EPI_ISL_950626, EPI_ISL_950627, EPI_ISL_950628, EPI_ISL_950629, EPI_ISL_950630, EPI_ISL_950631, EPI_ISL_950632, EPI_ISL_950633, EPI_ISL_950634, EPI_ISL_950635

| | | | |
|---|---|---|---|
| see above | Queens Medical Centre, Clinical Microbiology Department / DeepSeq Nottingham | COVID-19 Genomics UK (COG-UK) Consortium | Christopher Moore; Fei Sang; Gemma Clark; Hannah Howson-Wells; Johnny Debebe; Jonathan Ball; Joseph Chappell; Manjinder Khakh; Matthew Carlisle; Matthew Loose; Michelle M Lister; Nadine Holmes; Patrick McClure; Theocharis Tsoleridis; Vicki M Fleming; Victoria Wright; Wendy Smith |
| EPI_ISL_680440, EPI_ISL_742174 | Regional Virus Laboratory, Belfast Health and Social Care Trust | COVID-19 Genomics UK (COG-UK) Consortium | Alison Watt; Ciara Cox; Conall McCaughey; David Simpson; Derek Fairley; James McKenna; Mairead Connor; Susan Feeney; Tanya Curran; Zoltan Molnar |

EPI_ISL_660794, EPI_ISL_661175, EPI_ISL_661176, EPI_ISL_676511, EPI_ISL_678727, EPI_ISL_678729, EPI_ISL_678730, EPI_ISL_678731, EPI_ISL_678732, EPI_ISL_678733, EPI_ISL_678734, EPI_ISL_678735, EPI_ISL_678736, EPI_ISL_678737, EPI_ISL_678738, EPI_ISL_678739, EPI_ISL_678740, EPI_ISL_678742, EPI_ISL_678743, EPI_ISL_678744, EPI_ISL_678745, EPI_ISL_678746, EPI_ISL_678747, EPI_ISL_678748, EPI_ISL_678749, EPI_ISL_678750, EPI_ISL_678752, EPI_ISL_678753, EPI_ISL_678754, EPI_ISL_678755, EPI_ISL_678756, EPI_ISL_678757, EPI_ISL_678758, EPI_ISL_678759, EPI_ISL_678761, EPI_ISL_678762, EPI_ISL_678763, EPI_ISL_678764, EPI_ISL_678765, EPI_ISL_678766, EPI_ISL_678767, EPI_ISL_678768, EPI_ISL_678769, EPI_ISL_678770, EPI_ISL_678771, EPI_ISL_678772, EPI_ISL_678773, EPI_ISL_678774, EPI_ISL_678775, EPI_ISL_678776, EPI_ISL_678777, EPI_ISL_678778, EPI_ISL_678779, EPI_ISL_678780, EPI_ISL_678781, EPI_ISL_678782, EPI_ISL_678783, EPI_ISL_678784, EPI_ISL_678785, EPI_ISL_678786, EPI_ISL_678828, EPI_ISL_683453, EPI_ISL_683459, EPI_ISL_683460, EPI_ISL_683462, EPI_ISL_683463, EPI_ISL_683464, EPI_ISL_683465, EPI_ISL_683466, EPI_ISL_683467, EPI_ISL_683468, EPI_ISL_683469, EPI_ISL_683470, EPI_ISL_683471, EPI_ISL_683472, EPI_ISL_683473, EPI_ISL_683474, EPI_ISL_683475, EPI_ISL_683476, EPI_ISL_683478, EPI_ISL_683482, EPI_ISL_683483, EPI_ISL_683484, EPI_ISL_683489, EPI_ISL_683490, EPI_ISL_683546, EPI_ISL_683547, EPI_ISL_683548, EPI_ISL_683549, EPI_ISL_683550, EPI_ISL_683551, EPI_ISL_683552, EPI_ISL_683553, EPI_ISL_683554, EPI_ISL_683555, EPI_ISL_683556, EPI_ISL_683557, EPI_ISL_683558, EPI_ISL_683559, EPI_ISL_683560, EPI_ISL_683561, EPI_ISL_683562, EPI_ISL_683563, EPI_ISL_683564, EPI_ISL_683565, EPI_ISL_683566, EPI_ISL_683567, EPI_ISL_683568, EPI_ISL_683569, EPI_ISL_683570, EPI_ISL_683571, EPI_ISL_683572, EPI_ISL_683573, EPI_ISL_683574, EPI_ISL_683575, EPI_ISL_683576, EPI_ISL_683577, EPI_ISL_683578, EPI_ISL_683579, EPI_ISL_683580, EPI_ISL_683581, EPI_ISL_683582, EPI_ISL_683583, EPI_ISL_683584, EPI_ISL_683585, EPI_ISL_683586, EPI_ISL_683587, EPI_ISL_683588, EPI_ISL_683589, EPI_ISL_683590, EPI_ISL_683591, EPI_ISL_686611, EPI_ISL_693406, EPI_ISL_693468

| | | | |
|---|---|---|---|
| see above | Respiratory Virus Unit, Microbiology Services Colindale, Public Health England | COVID-19 Genomics UK (COG-UK) Consortium | PHE Covid Sequencing Team |

EPI_ISL_722971, EPI_ISL_722972, EPI_ISL_722973, EPI_ISL_722975, EPI_ISL_723016, EPI_ISL_723017, EPI_ISL_723018, EPI_ISL_723019, EPI_ISL_723020, EPI_ISL_723021, EPI_ISL_747492, EPI_ISL_754257, EPI_ISL_754287, EPI_ISL_754294, EPI_ISL_754295, EPI_ISL_754296, EPI_ISL_754297, EPI_ISL_754332, EPI_ISL_754333, EPI_ISL_754334, EPI_ISL_754343, EPI_ISL_766071, EPI_ISL_766087, EPI_ISL_766115, EPI_ISL_766116, EPI_ISL_766233, EPI_ISL_766235, EPI_ISL_766274, EPI_ISL_766275, EPI_ISL_766276, EPI_ISL_766277, EPI_ISL_766278, EPI_ISL_769866, EPI_ISL_825552, EPI_ISL_826281

| | | | |
|---|---|---|---|
| see above | Respiratory Virus Unit, National Infection Service, Public Health England | COVID-19 Genomics UK (COG-UK) Consortium | PHE Covid Sequencing Team |

EPI_ISL_679406, EPI_ISL_679407, EPI_ISL_679416, EPI_ISL_679417, EPI_ISL_679418, EPI_ISL_679419, EPI_ISL_679420, EPI_ISL_679421, EPI_ISL_679422, EPI_ISL_679423, EPI_ISL_679424, EPI_ISL_679425, EPI_ISL_679426, EPI_ISL_679427, EPI_ISL_679428, EPI_ISL_679429, EPI_ISL_679432, EPI_ISL_679433, EPI_ISL_679435, EPI_ISL_679436, EPI_ISL_679437, EPI_ISL_679438, EPI_ISL_679439, EPI_ISL_679440, EPI_ISL_679441, EPI_ISL_679442, EPI_ISL_679443, EPI_ISL_679444, EPI_ISL_679445, EPI_ISL_679446, EPI_ISL_702507, EPI_ISL_702602, EPI_ISL_702669, EPI_ISL_703222, EPI_ISL_704052, EPI_ISL_704102, EPI_ISL_704666, EPI_ISL_705247, EPI_ISL_705472, EPI_ISL_705942, EPI_ISL_705943, EPI_ISL_705944, EPI_ISL_705945, EPI_ISL_705946, EPI_ISL_705947, EPI_ISL_705948, EPI_ISL_705952, EPI_ISL_705953, EPI_ISL_705954, EPI_ISL_705955, EPI_ISL_705956, EPI_ISL_920355, EPI_ISL_920364, EPI_ISL_920482, EPI_ISL_920491, EPI_ISL_920509, EPI_ISL_920517, EPI_ISL_1104868, EPI_ISL_1104878, EPI_ISL_1104884, EPI_ISL_1104890, EPI_ISL_1104891, EPI_ISL_1104898, EPI_ISL_1104914, EPI_ISL_1104928, EPI_ISL_1104935, EPI_ISL_1104941, EPI_ISL_1104947, EPI_ISL_1105058

| | | | |
|---|---|---|---|
| see above | University College London Hospital | COVID-19 Genomics UK (COG-UK) Consortium | Catherine Houlihan; Dan Frampton; Judith Heaney; Matthew Byott; Moira Spyer and Eleni Nastouli; Stuart Kirk |

EPI_ISL_679549, EPI_ISL_679581, EPI_ISL_679582, EPI_ISL_679583, EPI_ISL_679585, EPI_ISL_679586, EPI_ISL_679588, EPI_ISL_679589, EPI_ISL_679590, EPI_ISL_679591, EPI_ISL_679592, EPI_ISL_679593, EPI_ISL_679594, EPI_ISL_679595, EPI_ISL_679596, EPI_ISL_679597, EPI_ISL_679598, EPI_ISL_679599, EPI_ISL_679600, EPI_ISL_679601, EPI_ISL_679602, EPI_ISL_679603, EPI_ISL_679604, EPI_ISL_679605, EPI_ISL_679606, EPI_ISL_679607, EPI_ISL_679608, EPI_ISL_679609, EPI_ISL_679610, EPI_ISL_679611, EPI_ISL_679612, EPI_ISL_679613, EPI_ISL_679614, EPI_ISL_679616, EPI_ISL_679617, EPI_ISL_679619, EPI_ISL_679620, EPI_ISL_702544, EPI_ISL_702610, EPI_ISL_702731, EPI_ISL_702999, EPI_ISL_703385, EPI_ISL_703865, EPI_ISL_704025, EPI_ISL_704036, EPI_ISL_704179, EPI_ISL_704955, EPI_ISL_704965, EPI_ISL_704997, EPI_ISL_704999, EPI_ISL_705072, EPI_ISL_705209, EPI_ISL_705215, EPI_ISL_705216, EPI_ISL_705278, EPI_ISL_705310, EPI_ISL_705342, EPI_ISL_705963, EPI_ISL_705967, EPI_ISL_705968, EPI_ISL_705970, EPI_ISL_724541, EPI_ISL_724542, EPI_ISL_724543, EPI_ISL_724544, EPI_ISL_724545, EPI_ISL_724546, EPI_ISL_724547, EPI_ISL_724548, EPI_ISL_724549, EPI_ISL_724550, EPI_ISL_724551, EPI_ISL_724552, EPI_ISL_724553, EPI_ISL_724554, EPI_ISL_724555, EPI_ISL_724556, EPI_ISL_724557, EPI_ISL_724558, EPI_ISL_724559, EPI_ISL_724560, EPI_ISL_724561, EPI_ISL_724562, EPI_ISL_724563, EPI_ISL_724564, EPI_ISL_724565, EPI_ISL_724566, EPI_ISL_724567, EPI_ISL_724568, EPI_ISL_724569, EPI_ISL_724570, EPI_ISL_724571, EPI_ISL_724572, EPI_ISL_724573, EPI_ISL_724574, EPI_ISL_724575, EPI_ISL_724576, EPI_ISL_724577, EPI_ISL_724578, EPI_ISL_724579, EPI_ISL_724580, EPI_ISL_724581, EPI_ISL_724582, EPI_ISL_724583, EPI_ISL_724584, EPI_ISL_724585, EPI_ISL_724586, EPI_ISL_724587, EPI_ISL_724588, EPI_ISL_724589, EPI_ISL_724590, EPI_ISL_724591, EPI_ISL_724592, EPI_ISL_724595, EPI_ISL_724596, EPI_ISL_819462, EPI_ISL_819463, EPI_ISL_819465, EPI_ISL_819466, EPI_ISL_819467, EPI_ISL_819468, EPI_ISL_819469, EPI_ISL_819470, EPI_ISL_819471, EPI_ISL_819472, EPI_ISL_819473, EPI_ISL_819474, EPI_ISL_819475, EPI_ISL_819477, EPI_ISL_865699, EPI_ISL_865700, EPI_ISL_865701, EPI_ISL_865702, EPI_ISL_865703, EPI_ISL_865704, EPI_ISL_865705, EPI_ISL_1386878, EPI_ISL_1474800

| | | | |
|---|---|---|---|
| see above | University College London, Great Ormond Street Hospital for Children NHS Foundation Trust, Imperial College Healthcare NHS Trust | COVID-19 Genomics UK (COG-UK) Consortium | Alison Holmes; Charlotte Williams; Helena Tutill; Jacqueline Findlay; James Price; Judith Breuer; Julianne Brown; Kathryn Harris; Leysa Forrest; Mark Kristiansen; Paola Niola; Paola Resende Silva; Patricia Dyal; Paul Randell; Rachel Williams; Samuel Weeks; Sergi Castellano; Sunando Roy; Tony Brooks; Yasmin Panchbhaya |
| EPI_ISL_1104292, EPI_ISL_1177799, EPI_ISL_2125907 | University Hospitals Of Leicester NHS Trust and DeepSeq Nottingham | COVID-19 Genomics UK (COG-UK) Consortium | Christopher Holmes; Christopher Moore; Fei Sang; Joeseph Chappell; Johnny Debebe; Jonathan Ball; Julian Tang; Karlie Fallon; Matthew Carlisle; Matthew Loose; Nadine Holmes; Patrick McClure; Paul Bird; Thomas Helmer; Victoria Wright |

EPI_ISL_813045, EPI_ISL_813046, EPI_ISL_813047, EPI_ISL_813048, EPI_ISL_813049, EPI_ISL_813050, EPI_ISL_813051, EPI_ISL_813053, EPI_ISL_813054, EPI_ISL_813055, EPI_ISL_813056, EPI_ISL_813057, EPI_ISL_813058, EPI_ISL_813059, EPI_ISL_918565, EPI_ISL_918566, EPI_ISL_918567, EPI_ISL_918568, EPI_ISL_918569, EPI_ISL_918570, EPI_ISL_918571, EPI_ISL_918572, EPI_ISL_918573, EPI_ISL_918574, EPI_ISL_918575, EPI_ISL_918576, EPI_ISL_918577, EPI_ISL_918585, EPI_ISL_918586, EPI_ISL_918587, EPI_ISL_918588, EPI_ISL_918589, EPI_ISL_918590, EPI_ISL_918591, EPI_ISL_918592, EPI_ISL_918593, EPI_ISL_918594, EPI_ISL_918595, EPI_ISL_918596, EPI_ISL_918597, EPI_ISL_918598, EPI_ISL_918666, EPI_ISL_918668, EPI_ISL_918669, EPI_ISL_918673, EPI_ISL_918682, EPI_ISL_918683

| | | | |
|---|---|---|---|
| see above | University of Birmingham | COVID-19 Genomics UK (COG-UK) Consortium | Alex Richter; Andrew Bosworth; Andrew Bosworth. Queen Elizabeth Hospital: Anna Casey; Andrew D Beggs PHE Heartlands Lab: Husam Osman; Charlotte Poxon; Institute of Microbiology; Joanne Stockton; Josh Quick; Kasun Wanigasooriya; Mike Kidd; Nicholas Loman. University of Birmingham Testing Laboratory: Celina M Whalley; Oliver Pickles; Radoslaw Poplawski; Samuel Nicholls; University of Birmingham: Claire McMurray; Will Rowe |

EPI_ISL_702562, EPI_ISL_702790, EPI_ISL_702844, EPI_ISL_702994, EPI_ISL_705099, EPI_ISL_705134, EPI_ISL_705237, EPI_ISL_705251, EPI_ISL_705254, EPI_ISL_705373, EPI_ISL_705383, EPI_ISL_724262, EPI_ISL_724263, EPI_ISL_724264, EPI_ISL_724265, EPI_ISL_724266, EPI_ISL_996589, EPI_ISL_997315, EPI_ISL_1247534, EPI_ISL_1247535, EPI_ISL_1247538, EPI_ISL_1247539, EPI_ISL_1247544, EPI_ISL_1247547, EPI_ISL_1545813

| | | | |
|---|---|---|---|
| see above | University of Exeter | COVID-19 Genomics UK (COG-UK) Consortium | Aaron Jeffries; Audrey Farbos; Ben Temperton; Jane Masoli; Joanna Warwick-Dugdale; Michelle Michelsen; Robyn Manley; Stephen Michell |

EPI_ISL_680461, EPI_ISL_680462, EPI_ISL_680463, EPI_ISL_680464, EPI_ISL_680465, EPI_ISL_680466, EPI_ISL_680467, EPI_ISL_680468, EPI_ISL_680486, EPI_ISL_680487, EPI_ISL_680488, EPI_ISL_680489, EPI_ISL_680490, EPI_ISL_680491, EPI_ISL_680492, EPI_ISL_680493, EPI_ISL_680494, EPI_ISL_680555, EPI_ISL_703572, EPI_ISL_703813, EPI_ISL_704559, EPI_ISL_705118, EPI_ISL_705734, EPI_ISL_705735, EPI_ISL_705736, EPI_ISL_705737, EPI_ISL_705738, EPI_ISL_705739, EPI_ISL_705740, EPI_ISL_705742, EPI_ISL_705743, EPI_ISL_705744, EPI_ISL_705745, EPI_ISL_705747, EPI_ISL_705776, EPI_ISL_705804, EPI_ISL_724222, EPI_ISL_724223, EPI_ISL_724224, EPI_ISL_764360, EPI_ISL_838281, EPI_ISL_838282, EPI_ISL_919347

| | | | |
|---|---|---|---|
| see above | Virology Department, Royal Infirmary of Edinburgh, NHS Lothian / School of Biological Sciences, University of Edinburgh / Institute of Genetics and Molecular Medicine, University of Edinburgh | COVID-19 Genomics UK (COG-UK) Consortium | Balcaza C; Colquhoun R; Dewar R; Gallagher M; Hill V; Jackson B; McCrone JT; McHugh M; O'Toole Á; Rambaut A; Rooke S; Scher E; Templeton K; Williams TC; Yu X |

EPI_ISL_680109, EPI_ISL_680110, EPI_ISL_680111, EPI_ISL_680112, EPI_ISL_680113, EPI_ISL_680114, EPI_ISL_680115, EPI_ISL_680118, EPI_ISL_680121, EPI_ISL_680128, EPI_ISL_680134, EPI_ISL_680135, EPI_ISL_680136, EPI_ISL_680137, EPI_ISL_680141, EPI_ISL_680142, EPI_ISL_680144, EPI_ISL_680146, EPI_ISL_680147, EPI_ISL_680148, EPI_ISL_680150, EPI_ISL_680154, EPI_ISL_680155, EPI_ISL_680158, EPI_ISL_680168, EPI_ISL_680170, EPI_ISL_680171, EPI_ISL_680178, EPI_ISL_680181, EPI_ISL_680183, EPI_ISL_680186, EPI_ISL_680187, EPI_ISL_680190, EPI_ISL_680192, EPI_ISL_680194, EPI_ISL_702476, EPI_ISL_702518, EPI_ISL_702531, EPI_ISL_702705, EPI_ISL_702761, EPI_ISL_702776, EPI_ISL_702798, EPI_ISL_702829, EPI_ISL_702830, EPI_ISL_702833, EPI_ISL_702843, EPI_ISL_702880, EPI_ISL_702964, EPI_ISL_703018, EPI_ISL_703020, EPI_ISL_703026, EPI_ISL_703029, EPI_ISL_703040, EPI_ISL_703056, EPI_ISL_703505, EPI_ISL_703569, EPI_ISL_703575, EPI_ISL_703579, EPI_ISL_703582, EPI_ISL_703585, EPI_ISL_703587, EPI_ISL_703593, EPI_ISL_703601, EPI_ISL_703606, EPI_ISL_703636, EPI_ISL_703639, EPI_ISL_703647, EPI_ISL_703655, EPI_ISL_703658, EPI_ISL_703684, EPI_ISL_703723, EPI_ISL_703894, EPI_ISL_704190, EPI_ISL_704600, EPI_ISL_704601, EPI_ISL_704602, EPI_ISL_704603, EPI_ISL_704604, EPI_ISL_704605, EPI_ISL_704636, EPI_ISL_704637, EPI_ISL_704638, EPI_ISL_704639, EPI_ISL_704828, EPI_ISL_704960, EPI_ISL_705088, EPI_ISL_705163, EPI_ISL_705195, EPI_ISL_705270, EPI_ISL_705272, EPI_ISL_705317, EPI_ISL_705319, EPI_ISL_705335, EPI_ISL_705371, EPI_ISL_705405, EPI_ISL_705615, EPI_ISL_705616, EPI_ISL_705617, EPI_ISL_705618, EPI_ISL_705619, EPI_ISL_705620, EPI_ISL_705621, EPI_ISL_705622, EPI_ISL_705623, EPI_ISL_705624, EPI_ISL_705625, EPI_ISL_705626, EPI_ISL_705627, EPI_ISL_705628, EPI_ISL_706983, EPI_ISL_706987, EPI_ISL_706989, EPI_ISL_706992, EPI_ISL_706993, EPI_ISL_706995, EPI_ISL_706996, EPI_ISL_706997, EPI_ISL_706998, EPI_ISL_706999, EPI_ISL_707000, EPI_ISL_707001, EPI_ISL_707002, EPI_ISL_707003, EPI_ISL_707005, EPI_ISL_707006, EPI_ISL_727868, EPI_ISL_727908, EPI_ISL_727917, EPI_ISL_727931, EPI_ISL_727934, EPI_ISL_727940, EPI_ISL_727967, EPI_ISL_727968, EPI_ISL_727969, EPI_ISL_727976, EPI_ISL_727978, EPI_ISL_727981, EPI_ISL_727996, EPI_ISL_816324

| | | | |
|---|---|---|---|
| see above | Virology Department, Sheffield Teaching Hospitals NHS Foundation Trust/Department of Infection, Immunity and Cardiovascular Disease, The Medical School, University of Sheffield | COVID-19 Genomics UK (COG-UK) Consortium | Adri Angyal; Alex Keeley; Benjamin Lindsey; Cariad Evans; Danielle Groves; Dave Partridge; Katie Johnson; Laura Carrilero; Luke Green; Matthew Parker; Matthew Wyles; Mehmet Yavuz; Mohammad Raza; Nikki Smith; Paul Parsons; Rachel Tucker; Rebecca Brown; Thushan de Silva |

EPI_ISL_702713, EPI_ISL_703074, EPI_ISL_703746, EPI_ISL_703761, EPI_ISL_703955, EPI_ISL_704001, EPI_ISL_704111, EPI_ISL_704584, EPI_ISL_705244, EPI_ISL_705435, EPI_ISL_705441, EPI_ISL_706445, EPI_ISL_706459, EPI_ISL_706460, EPI_ISL_706461, EPI_ISL_706462, EPI_ISL_706463, EPI_ISL_706576, EPI_ISL_706577, EPI_ISL_706578, EPI_ISL_706579, EPI_ISL_706580, EPI_ISL_706581, EPI_ISL_706582, EPI_ISL_706583, EPI_ISL_706584, EPI_ISL_706585, EPI_ISL_706586, EPI_ISL_706587, EPI_ISL_706588, EPI_ISL_706602, EPI_ISL_706610, EPI_ISL_706611, EPI_ISL_706612, EPI_ISL_706646, EPI_ISL_706647, EPI_ISL_706652, EPI_ISL_706653, EPI_ISL_706654, EPI_ISL_706655, EPI_ISL_706656, EPI_ISL_706657, EPI_ISL_706839, EPI_ISL_706853, EPI_ISL_706854, EPI_ISL_706855, EPI_ISL_706856, EPI_ISL_706857, EPI_ISL_706858, EPI_ISL_706859, EPI_ISL_706860, EPI_ISL_706861, EPI_ISL_706862, EPI_ISL_706863, EPI_ISL_725672, EPI_ISL_725673, EPI_ISL_725676, EPI_ISL_725677, EPI_ISL_725678, EPI_ISL_725679, EPI_ISL_725680, EPI_ISL_725681, EPI_ISL_725682, EPI_ISL_725683, EPI_ISL_725684, EPI_ISL_725685, EPI_ISL_725686, EPI_ISL_725687, EPI_ISL_725688, EPI_ISL_725713, EPI_ISL_725714, EPI_ISL_725716, EPI_ISL_725717, EPI_ISL_725718, EPI_ISL_725719, EPI_ISL_725720, EPI_ISL_725721, EPI_ISL_725722, EPI_ISL_725723, EPI_ISL_725724, EPI_ISL_725725, EPI_ISL_725727, EPI_ISL_725728, EPI_ISL_725729, EPI_ISL_725730, EPI_ISL_725731, EPI_ISL_725732, EPI_ISL_725733, EPI_ISL_725734, EPI_ISL_725735, EPI_ISL_725736, EPI_ISL_725737, EPI_ISL_725738, EPI_ISL_725739, EPI_ISL_725740, EPI_ISL_725741, EPI_ISL_725742, EPI_ISL_725744, EPI_ISL_725745, EPI_ISL_725746, EPI_ISL_725747, EPI_ISL_725749, EPI_ISL_725750, EPI_ISL_725751, EPI_ISL_725753, EPI_ISL_725754, EPI_ISL_725755, EPI_ISL_725756, EPI_ISL_725757, EPI_ISL_725758, EPI_ISL_725759, EPI_ISL_725760, EPI_ISL_725761, EPI_ISL_725762, EPI_ISL_725763, EPI_ISL_725764, EPI_ISL_725765, EPI_ISL_725766, EPI_ISL_725768, EPI_ISL_725769, EPI_ISL_725771, EPI_ISL_725772, EPI_ISL_725773, EPI_ISL_725774, EPI_ISL_725776, EPI_ISL_725777, EPI_ISL_725778, EPI_ISL_725780, EPI_ISL_725781, EPI_ISL_725782, EPI_ISL_725784, EPI_ISL_725785, EPI_ISL_725786, EPI_ISL_725787, EPI_ISL_725788, EPI_ISL_725789, EPI_ISL_725790, EPI_ISL_725791, EPI_ISL_725792, EPI_ISL_725793, EPI_ISL_725795, EPI_ISL_725796, EPI_ISL_725797, EPI_ISL_725798, EPI_ISL_725799, EPI_ISL_725800, EPI_ISL_725982, EPI_ISL_725983, EPI_ISL_725984, EPI_ISL_725997, EPI_ISL_726000, EPI_ISL_726001, EPI_ISL_726002, EPI_ISL_726003, EPI_ISL_726004, EPI_ISL_726006, EPI_ISL_726007, EPI_ISL_726008, EPI_ISL_726009, EPI_ISL_726012, EPI_ISL_726013, EPI_ISL_726014, EPI_ISL_726015, EPI_ISL_726016, EPI_ISL_726022, EPI_ISL_726023, EPI_ISL_726024, EPI_ISL_726025, EPI_ISL_726026, EPI_ISL_726027, EPI_ISL_726028, EPI_ISL_726029, EPI_ISL_726030, EPI_ISL_726031, EPI_ISL_726032, EPI_ISL_726053, EPI_ISL_726084, EPI_ISL_726085, EPI_ISL_726097, EPI_ISL_726098, EPI_ISL_726100, EPI_ISL_726101, EPI_ISL_726102, EPI_ISL_726103, EPI_ISL_726104, EPI_ISL_726105, EPI_ISL_726106, EPI_ISL_726107, EPI_ISL_726108, EPI_ISL_726109, EPI_ISL_726110, EPI_ISL_726111, EPI_ISL_726112, EPI_ISL_726113, EPI_ISL_726114, EPI_ISL_726115, EPI_ISL_726116, EPI_ISL_726117, EPI_ISL_726118, EPI_ISL_726119, EPI_ISL_726120, EPI_ISL_726121, EPI_ISL_726122, EPI_ISL_726123, EPI_ISL_726124, EPI_ISL_726125, EPI_ISL_726126, EPI_ISL_726127, EPI_ISL_726128, EPI_ISL_726129, EPI_ISL_726130, EPI_ISL_726131, EPI_ISL_726132, EPI_ISL_726133, EPI_ISL_726134, EPI_ISL_726135, EPI_ISL_726136, EPI_ISL_726137, EPI_ISL_726138, EPI_ISL_726139, EPI_ISL_726140, EPI_ISL_726141, EPI_ISL_726142, EPI_ISL_726143, EPI_ISL_726144, EPI_ISL_726145, EPI_ISL_726146, EPI_ISL_726147, EPI_ISL_726148, EPI_ISL_726149, EPI_ISL_726150, EPI_ISL_726151, EPI_ISL_726152, EPI_ISL_726153, EPI_ISL_726154, EPI_ISL_726155, EPI_ISL_726156, EPI_ISL_726157, EPI_ISL_726158, EPI_ISL_726159, EPI_ISL_726160, EPI_ISL_726161, EPI_ISL_726162, EPI_ISL_726163, EPI_ISL_726164, EPI_ISL_726165, EPI_ISL_726166, EPI_ISL_726167, EPI_ISL_726168, EPI_ISL_726169, EPI_ISL_726170, EPI_ISL_726171, EPI_ISL_726172, EPI_ISL_726173, EPI_ISL_726174, EPI_ISL_726175, EPI_ISL_726176, EPI_ISL_726177, EPI_ISL_726178, EPI_ISL_726179, EPI_ISL_726180, EPI_ISL_726181, EPI_ISL_726182, EPI_ISL_726183, EPI_ISL_726184, EPI_ISL_726185, EPI_ISL_726186, EPI_ISL_726187, EPI_ISL_726188, EPI_ISL_726189, EPI_ISL_726190, EPI_ISL_726191, EPI_ISL_726192, EPI_ISL_726193, EPI_ISL_726194, EPI_ISL_726195, EPI_ISL_726196, EPI_ISL_726197, EPI_ISL_726198, EPI_ISL_726199, EPI_ISL_726200, EPI_ISL_726201, EPI_ISL_726202, EPI_ISL_726203, EPI_ISL_726204, EPI_ISL_726205, EPI_ISL_726206, EPI_ISL_726207,

EPI_ISL_726208, EPI_ISL_726209, EPI_ISL_726210, EPI_ISL_726211, EPI_ISL_726212, EPI_ISL_726214, EPI_ISL_726215, EPI_ISL_726216, EPI_ISL_726217, EPI_ISL_726218, EPI_ISL_726219, EPI_ISL_726220, EPI_ISL_726221, EPI_ISL_726222, EPI_ISL_726223, EPI_ISL_726224, EPI_ISL_726225, EPI_ISL_726226, EPI_ISL_726227, EPI_ISL_726228, EPI_ISL_726229, EPI_ISL_726230, EPI_ISL_726231, EPI_ISL_726232, EPI_ISL_726233, EPI_ISL_726234, EPI_ISL_726235, EPI_ISL_726236, EPI_ISL_726237, EPI_ISL_726238, EPI_ISL_726239, EPI_ISL_726240, EPI_ISL_726241, EPI_ISL_726242, EPI_ISL_726243, EPI_ISL_726244, EPI_ISL_726245, EPI_ISL_726246, EPI_ISL_726247, EPI_ISL_726248, EPI_ISL_726249, EPI_ISL_726250, EPI_ISL_726251, EPI_ISL_726252, EPI_ISL_726255, EPI_ISL_726256, EPI_ISL_726257, EPI_ISL_726258, EPI_ISL_726259, EPI_ISL_726260, EPI_ISL_726261, EPI_ISL_726262, EPI_ISL_726263, EPI_ISL_726264, EPI_ISL_726265, EPI_ISL_726266, EPI_ISL_726267, EPI_ISL_726268, EPI_ISL_726269, EPI_ISL_726270, EPI_ISL_726271, EPI_ISL_726272, EPI_ISL_726273, EPI_ISL_726274, EPI_ISL_726275, EPI_ISL_726276, EPI_ISL_726277, EPI_ISL_726278, EPI_ISL_726279, EPI_ISL_726280, EPI_ISL_726281, EPI_ISL_726282, EPI_ISL_726283, EPI_ISL_726284, EPI_ISL_726285, EPI_ISL_726286, EPI_ISL_726287, EPI_ISL_726288, EPI_ISL_726289, EPI_ISL_726290, EPI_ISL_726291, EPI_ISL_726292, EPI_ISL_726293, EPI_ISL_726294, EPI_ISL_726295, EPI_ISL_726296, EPI_ISL_726297, EPI_ISL_726298, EPI_ISL_726299, EPI_ISL_726300, EPI_ISL_726301, EPI_ISL_726302, EPI_ISL_726303, EPI_ISL_726304, EPI_ISL_726305, EPI_ISL_726306, EPI_ISL_726307, EPI_ISL_726308, EPI_ISL_726309, EPI_ISL_726310, EPI_ISL_726311, EPI_ISL_726312, EPI_ISL_726313, EPI_ISL_726314, EPI_ISL_726315, EPI_ISL_726316, EPI_ISL_726317, EPI_ISL_726318, EPI_ISL_726319, EPI_ISL_726320, EPI_ISL_726321, EPI_ISL_726322, EPI_ISL_726323, EPI_ISL_726324, EPI_ISL_726325, EPI_ISL_726326, EPI_ISL_726327, EPI_ISL_726328, EPI_ISL_726329, EPI_ISL_726330, EPI_ISL_726331, EPI_ISL_726332, EPI_ISL_726333, EPI_ISL_726334, EPI_ISL_726335, EPI_ISL_726336, EPI_ISL_726337, EPI_ISL_726338, EPI_ISL_726339, EPI_ISL_726340, EPI_ISL_726341, EPI_ISL_726342, EPI_ISL_726343, EPI_ISL_726344, EPI_ISL_726345, EPI_ISL_726346, EPI_ISL_726347, EPI_ISL_726348, EPI_ISL_726349, EPI_ISL_726350, EPI_ISL_726351, EPI_ISL_726352, EPI_ISL_726353, EPI_ISL_726354, EPI_ISL_726355, EPI_ISL_726356, EPI_ISL_726357, EPI_ISL_726358, EPI_ISL_726359, EPI_ISL_726360, EPI_ISL_726361, EPI_ISL_726362, EPI_ISL_726363, EPI_ISL_726364, EPI_ISL_726365, EPI_ISL_726366, EPI_ISL_726367, EPI_ISL_726368, EPI_ISL_726369, EPI_ISL_726370, EPI_ISL_726371, EPI_ISL_726372, EPI_ISL_726373, EPI_ISL_726374, EPI_ISL_726375, EPI_ISL_726376, EPI_ISL_726377, EPI_ISL_726378, EPI_ISL_726379, EPI_ISL_726380, EPI_ISL_726381, EPI_ISL_726382, EPI_ISL_726383, EPI_ISL_726384, EPI_ISL_726385, EPI_ISL_726386, EPI_ISL_726387, EPI_ISL_726388, EPI_ISL_726389, EPI_ISL_726390, EPI_ISL_726391, EPI_ISL_726392, EPI_ISL_726393, EPI_ISL_726394, EPI_ISL_726395, EPI_ISL_726396, EPI_ISL_726397, EPI_ISL_726398, EPI_ISL_726399, EPI_ISL_726400, EPI_ISL_726401, EPI_ISL_726402, EPI_ISL_726403, EPI_ISL_726404, EPI_ISL_726405, EPI_ISL_726406, EPI_ISL_726407, EPI_ISL_726408, EPI_ISL_726409, EPI_ISL_726410, EPI_ISL_726411, EPI_ISL_726412, EPI_ISL_726413, EPI_ISL_726414, EPI_ISL_726415, EPI_ISL_726416, EPI_ISL_726417, EPI_ISL_726418, EPI_ISL_726419, EPI_ISL_726420, EPI_ISL_726421, EPI_ISL_726422, EPI_ISL_726423, EPI_ISL_726424, EPI_ISL_726425, EPI_ISL_726426, EPI_ISL_726427, EPI_ISL_726428, EPI_ISL_726429, EPI_ISL_726430, EPI_ISL_726431, EPI_ISL_726432, EPI_ISL_726433, EPI_ISL_726434, EPI_ISL_726435, EPI_ISL_726436, EPI_ISL_726437, EPI_ISL_726438, EPI_ISL_726439, EPI_ISL_726440, EPI_ISL_726441, EPI_ISL_726442, EPI_ISL_726443, EPI_ISL_726444, EPI_ISL_726445, EPI_ISL_726446, EPI_ISL_726447, EPI_ISL_726448, EPI_ISL_726449, EPI_ISL_726450, EPI_ISL_726451, EPI_ISL_726452, EPI_ISL_726453, EPI_ISL_726454, EPI_ISL_726455, EPI_ISL_726456, EPI_ISL_726457, EPI_ISL_726458, EPI_ISL_726459, EPI_ISL_726460, EPI_ISL_726461, EPI_ISL_726462, EPI_ISL_726463, EPI_ISL_726464, EPI_ISL_726465, EPI_ISL_726466, EPI_ISL_726467, EPI_ISL_726468, EPI_ISL_726469, EPI_ISL_726470, EPI_ISL_726471, EPI_ISL_726472, EPI_ISL_726473, EPI_ISL_726474, EPI_ISL_726475, EPI_ISL_726476, EPI_ISL_726477, EPI_ISL_726478, EPI_ISL_726479, EPI_ISL_726480, EPI_ISL_726481, EPI_ISL_726482, EPI_ISL_726483, EPI_ISL_726484, EPI_ISL_726485, EPI_ISL_726486, EPI_ISL_726487, EPI_ISL_726488, EPI_ISL_726489, EPI_ISL_726490, EPI_ISL_726491, EPI_ISL_726492, EPI_ISL_726493, EPI_ISL_726494, EPI_ISL_726495, EPI_ISL_726496, EPI_ISL_726497, EPI_ISL_726498, EPI_ISL_726499, EPI_ISL_726500, EPI_ISL_726501, EPI_ISL_726502, EPI_ISL_726503, EPI_ISL_726504, EPI_ISL_726505, EPI_ISL_726506, EPI_ISL_726507, EPI_ISL_726508, EPI_ISL_726509, EPI_ISL_726510, EPI_ISL_726511, EPI_ISL_726512, EPI_ISL_726513, EPI_ISL_726514, EPI_ISL_726515, EPI_ISL_726516, EPI_ISL_726517, EPI_ISL_726518, EPI_ISL_726519, EPI_ISL_726520, EPI_ISL_726521, EPI_ISL_726522, EPI_ISL_726523, EPI_ISL_726524, EPI_ISL_726525, EPI_ISL_726526, EPI_ISL_726527, EPI_ISL_726528, EPI_ISL_726529, EPI_ISL_726530, EPI_ISL_726531, EPI_ISL_726532, EPI_ISL_726533, EPI_ISL_726534, EPI_ISL_726535, EPI_ISL_726536, EPI_ISL_726537, EPI_ISL_726538, EPI_ISL_726539, EPI_ISL_726540, EPI_ISL_726541, EPI_ISL_726542, EPI_ISL_726543, EPI_ISL_726544, EPI_ISL_726545, EPI_ISL_726546, EPI_ISL_726547, EPI_ISL_726548, EPI_ISL_726549, EPI_ISL_726550, EPI_ISL_726551, EPI_ISL_726552, EPI_ISL_726553, EPI_ISL_726554, EPI_ISL_726555, EPI_ISL_726556, EPI_ISL_726557, EPI_ISL_726558, EPI_ISL_726559, EPI_ISL_726560, EPI_ISL_726561, EPI_ISL_726562, EPI_ISL_726563, EPI_ISL_726564, EPI_ISL_726565, EPI_ISL_726566, EPI_ISL_726567, EPI_ISL_726568, EPI_ISL_726569, EPI_ISL_726570, EPI_ISL_726571, EPI_ISL_726572, EPI_ISL_726573, EPI_ISL_726574, EPI_ISL_726575, EPI_ISL_726576, EPI_ISL_726577, EPI_ISL_726578, EPI_ISL_726579, EPI_ISL_726580, EPI_ISL_726581, EPI_ISL_726582, EPI_ISL_726583, EPI_ISL_726584, EPI_ISL_726585, EPI_ISL_726586, EPI_ISL_726587, EPI_ISL_726588, EPI_ISL_726589, EPI_ISL_726590, EPI_ISL_726591, EPI_ISL_726592, EPI_ISL_726593, EPI_ISL_726594, EPI_ISL_726595, EPI_ISL_726596, EPI_ISL_726597, EPI_ISL_726598, EPI_ISL_726599, EPI_ISL_726600, EPI_ISL_726601, EPI_ISL_726602, EPI_ISL_726603, EPI_ISL_726604, EPI_ISL_726605, EPI_ISL_726606, EPI_ISL_726607, EPI_ISL_726608, EPI_ISL_726609, EPI_ISL_726610, EPI_ISL_726611, EPI_ISL_726612, EPI_ISL_726613, EPI_ISL_726614, EPI_ISL_726615, EPI_ISL_726616, EPI_ISL_726617, EPI_ISL_726618, EPI_ISL_726619, EPI_ISL_726620, EPI_ISL_726621, EPI_ISL_726622, EPI_ISL_726623, EPI_ISL_726624, EPI_ISL_726625, EPI_ISL_726626, EPI_ISL_726627, EPI_ISL_726628, EPI_ISL_726629, EPI_ISL_726630, EPI_ISL_726631, EPI_ISL_726632, EPI_ISL_726633, EPI_ISL_726634, EPI_ISL_726635, EPI_ISL_726636, EPI_ISL_726637, EPI_ISL_726638, EPI_ISL_726639, EPI_ISL_726640, EPI_ISL_726641, EPI_ISL_726642, EPI_ISL_726643, EPI_ISL_726644, EPI_ISL_726645, EPI_ISL_726646, EPI_ISL_726647, EPI_ISL_726648, EPI_ISL_726649, EPI_ISL_726650, EPI_ISL_726651, EPI_ISL_726652, EPI_ISL_726653, EPI_ISL_726654, EPI_ISL_726655, EPI_ISL_726656, EPI_ISL_726657, EPI_ISL_726658, EPI_ISL_726659, EPI_ISL_726660, EPI_ISL_726661, EPI_ISL_726662, EPI_ISL_726663, EPI_ISL_726664, EPI_ISL_726665, EPI_ISL_726666, EPI_ISL_726667, EPI_ISL_726668, EPI_ISL_726670, EPI_ISL_726671, EPI_ISL_726672, EPI_ISL_726673, EPI_ISL_726674, EPI_ISL_726675, EPI_ISL_726676, EPI_ISL_726677, EPI_ISL_726678, EPI_ISL_726679, EPI_ISL_726680, EPI_ISL_726681, EPI_ISL_726682, EPI_ISL_726683, EPI_ISL_726684, EPI_ISL_726685, EPI_ISL_726686, EPI_ISL_726687, EPI_ISL_726688, EPI_ISL_726689, EPI_ISL_726690, EPI_ISL_726691, EPI_ISL_726692, EPI_ISL_726693, EPI_ISL_726694, EPI_ISL_726695, EPI_ISL_726696, EPI_ISL_726697, EPI_ISL_726698, EPI_ISL_726699, EPI_ISL_726700, EPI_ISL_726701, EPI_ISL_726702, EPI_ISL_726703, EPI_ISL_726704, EPI_ISL_726705, EPI_ISL_726706, EPI_ISL_726707, EPI_ISL_726708, EPI_ISL_726709, EPI_ISL_726710, EPI_ISL_726711, EPI_ISL_726712, EPI_ISL_726713, EPI_ISL_726714, EPI_ISL_726715, EPI_ISL_726716, EPI_ISL_726717, EPI_ISL_726718, EPI_ISL_726719, EPI_ISL_726720, EPI_ISL_726721, EPI_ISL_726723, EPI_ISL_726724, EPI_ISL_726725, EPI_ISL_726726, EPI_ISL_726727, EPI_ISL_726728, EPI_ISL_726729, EPI_ISL_726730, EPI_ISL_726731, EPI_ISL_726732, EPI_ISL_726733, EPI_ISL_726734, EPI_ISL_726735, EPI_ISL_726736, EPI_ISL_726737, EPI_ISL_726738, EPI_ISL_726739, EPI_ISL_726740, EPI_ISL_726741, EPI_ISL_726742, EPI_ISL_726743, EPI_ISL_726744, EPI_ISL_726745, EPI_ISL_726746, EPI_ISL_726747, EPI_ISL_726748, EPI_ISL_726749, EPI_ISL_726750, EPI_ISL_726751, EPI_ISL_726752, EPI_ISL_726753, EPI_ISL_726754, EPI_ISL_726755, EPI_ISL_726756, EPI_ISL_726757, EPI_ISL_726758, EPI_ISL_726759, EPI_ISL_726760, EPI_ISL_726761, EPI_ISL_726762, EPI_ISL_726763, EPI_ISL_726764, EPI_ISL_726765, EPI_ISL_726766, EPI_ISL_726767, EPI_ISL_726768, EPI_ISL_726769, EPI_ISL_726770, EPI_ISL_726771, EPI_ISL_726772, EPI_ISL_726773, EPI_ISL_726774, EPI_ISL_726775, EPI_ISL_726776, EPI_ISL_726777, EPI_ISL_726778, EPI_ISL_726779, EPI_ISL_726780, EPI_ISL_726781, EPI_ISL_726782, EPI_ISL_726783, EPI_ISL_726784, EPI_ISL_726785, EPI_ISL_726786, EPI_ISL_726787, EPI_ISL_726788, EPI_ISL_726789, EPI_ISL_726790, EPI_ISL_726791, EPI_ISL_726792, EPI_ISL_726793, EPI_ISL_726794, EPI_ISL_726795, EPI_ISL_726796, EPI_ISL_726797, EPI_ISL_726798, EPI_ISL_726799, EPI_ISL_726800, EPI_ISL_726801, EPI_ISL_726802, EPI_ISL_726803, EPI_ISL_726804, EPI_ISL_726805, EPI_ISL_726806, EPI_ISL_726807, EPI_ISL_726808, EPI_ISL_726809, EPI_ISL_726810, EPI_ISL_726811, EPI_ISL_726812, EPI_ISL_726813, EPI_ISL_726814, EPI_ISL_726815, EPI_ISL_726816, EPI_ISL_726817, EPI_ISL_726818, EPI_ISL_726819, EPI_ISL_726820, EPI_ISL_726821, EPI_ISL_726822, EPI_ISL_726823, EPI_ISL_726824, EPI_ISL_726825, EPI_ISL_726826, EPI_ISL_726827, EPI_ISL_726828, EPI_ISL_726829, EPI_ISL_726830, EPI_ISL_726831, EPI_ISL_726832, EPI_ISL_726833, EPI_ISL_726834, EPI_ISL_726835, EPI_ISL_726837, EPI_ISL_726839, EPI_ISL_726865, EPI_ISL_726866, EPI_ISL_726867, EPI_ISL_726868, EPI_ISL_726869, EPI_ISL_726870, EPI_ISL_726871, EPI_ISL_726872, EPI_ISL_726873, EPI_ISL_726874, EPI_ISL_726875, EPI_ISL_726876, EPI_ISL_726877, EPI_ISL_726878, EPI_ISL_726879, EPI_ISL_726880, EPI_ISL_726881, EPI_ISL_726882, EPI_ISL_726883, EPI_ISL_726884, EPI_ISL_726885, EPI_ISL_726886, EPI_ISL_726887, EPI_ISL_726888, EPI_ISL_726889, EPI_ISL_726890, EPI_ISL_726891, EPI_ISL_726892, EPI_ISL_726893, EPI_ISL_726894, EPI_ISL_726895, EPI_ISL_726896, EPI_ISL_726897, EPI_ISL_726898, EPI_ISL_726899, EPI_ISL_726900, EPI_ISL_726901, EPI_ISL_726902, EPI_ISL_726903, EPI_ISL_726904, EPI_ISL_726905, EPI_ISL_726906, EPI_ISL_726907, EPI_ISL_726908, EPI_ISL_726909, EPI_ISL_726910, EPI_ISL_726911, EPI_ISL_726912, EPI_ISL_726913, EPI_ISL_726914, EPI_ISL_726915, EPI_ISL_726916, EPI_ISL_726917, EPI_ISL_726918, EPI_ISL_726919, EPI_ISL_726920, EPI_ISL_726921, EPI_ISL_726922, EPI_ISL_726923, EPI_ISL_726924, EPI_ISL_726925, EPI_ISL_726926, EPI_ISL_726927, EPI_ISL_726928, EPI_ISL_726929, EPI_ISL_726930, EPI_ISL_726931, EPI_ISL_726932, EPI_ISL_726933, EPI_ISL_726934, EPI_ISL_726935, EPI_ISL_726936, EPI_ISL_726937, EPI_ISL_726938, EPI_ISL_726939, EPI_ISL_726940, EPI_ISL_726941, EPI_ISL_726942, EPI_ISL_726943, EPI_ISL_726944, EPI_ISL_726945, EPI_ISL_726946, EPI_ISL_726947, EPI_ISL_726948, EPI_ISL_726949, EPI_ISL_726950, EPI_ISL_726951, EPI_ISL_726952, EPI_ISL_726953, EPI_ISL_726954, EPI_ISL_726955, EPI_ISL_726956, EPI_ISL_726957, EPI_ISL_726958, EPI_ISL_726959, EPI_ISL_727103, EPI_ISL_727104, EPI_ISL_727105, EPI_ISL_727106, EPI_ISL_727107, EPI_ISL_727108, EPI_ISL_727109, EPI_ISL_727110, EPI_ISL_727111, EPI_ISL_727112, EPI_ISL_727113, EPI_ISL_727114, EPI_ISL_727115, EPI_ISL_727116, EPI_ISL_727117, EPI_ISL_727124, EPI_ISL_727125, EPI_ISL_727126, EPI_ISL_727127, EPI_ISL_727128, EPI_ISL_727129, EPI_ISL_727130, EPI_ISL_727131, EPI_ISL_727132, EPI_ISL_727133, EPI_ISL_727134, EPI_ISL_727135, EPI_ISL_727136, EPI_ISL_727137, EPI_ISL_727138, EPI_ISL_727181, EPI_ISL_727182, EPI_ISL_727183, EPI_ISL_727184, EPI_ISL_727185, EPI_ISL_727186, EPI_ISL_727188, EPI_ISL_727189, EPI_ISL_727190, EPI_ISL_727191, EPI_ISL_727192, EPI_ISL_727193, EPI_ISL_727195, EPI_ISL_727196, EPI_ISL_727197, EPI_ISL_727198, EPI_ISL_727199, EPI_ISL_727200, EPI_ISL_727201, EPI_ISL_727202, EPI_ISL_727203, EPI_ISL_727204, EPI_ISL_727206, EPI_ISL_727207, EPI_ISL_727208, EPI_ISL_727210, EPI_ISL_727212, EPI_ISL_727409, EPI_ISL_727412, EPI_ISL_727413, EPI_ISL_727414, EPI_ISL_727415, EPI_ISL_727416, EPI_ISL_727432, EPI_ISL_727433, EPI_ISL_727434, EPI_ISL_727435, EPI_ISL_727436, EPI_ISL_727437, EPI_ISL_727438, EPI_ISL_727439, EPI_ISL_727440, EPI_ISL_727441, EPI_ISL_727442, EPI_ISL_727443, EPI_ISL_727444, EPI_ISL_727445, EPI_ISL_727446, EPI_ISL_727447, EPI_ISL_727448, EPI_ISL_727449, EPI_ISL_727450, EPI_ISL_727451, EPI_ISL_727452, EPI_ISL_727453, EPI_ISL_727454, EPI_ISL_727455, EPI_ISL_727456, EPI_ISL_727457, EPI_ISL_727458, EPI_ISL_727459, EPI_ISL_727460, EPI_ISL_727461, EPI_ISL_727462, EPI_ISL_727463, EPI_ISL_727464, EPI_ISL_727465, EPI_ISL_727466, EPI_ISL_727467, EPI_ISL_727468, EPI_ISL_727469, EPI_ISL_727470, EPI_ISL_727471, EPI_ISL_727472, EPI_ISL_727473, EPI_ISL_727474, EPI_ISL_727475, EPI_ISL_727476, EPI_ISL_727477, EPI_ISL_727478, EPI_ISL_727479, EPI_ISL_727480, EPI_ISL_727481, EPI_ISL_727482, EPI_ISL_742541, EPI_ISL_742542, EPI_ISL_742543, EPI_ISL_742544, EPI_ISL_742547, EPI_ISL_742548, EPI_ISL_742549, EPI_ISL_742561, EPI_ISL_742563, EPI_ISL_742564, EPI_ISL_742585, EPI_ISL_742588, EPI_ISL_742589, EPI_ISL_742590, EPI_ISL_742591, EPI_ISL_742592, EPI_ISL_742593, EPI_ISL_742594, EPI_ISL_742595, EPI_ISL_742596, EPI_ISL_742597, EPI_ISL_742598, EPI_ISL_742599, EPI_ISL_742600, EPI_ISL_742601, EPI_ISL_742602, EPI_ISL_742603, EPI_ISL_742604, EPI_ISL_742605, EPI_ISL_742606, EPI_ISL_742607, EPI_ISL_742608, EPI_ISL_742609, EPI_ISL_742610, EPI_ISL_742611, EPI_ISL_742612, EPI_ISL_742613, EPI_ISL_742614, EPI_ISL_742615, EPI_ISL_742616, EPI_ISL_742617, EPI_ISL_742618, EPI_ISL_742619, EPI_ISL_742620, EPI_ISL_742621, EPI_ISL_742622, EPI_ISL_742623, EPI_ISL_742624, EPI_ISL_742625, EPI_ISL_742626, EPI_ISL_742627, EPI_ISL_742628, EPI_ISL_742629, EPI_ISL_742630, EPI_ISL_742631, EPI_ISL_742632, EPI_ISL_742633, EPI_ISL_742634, EPI_ISL_742635, EPI_ISL_742636, EPI_ISL_742840, EPI_ISL_742880, EPI_ISL_742881, EPI_ISL_742882, EPI_ISL_742883, EPI_ISL_742884, EPI_ISL_742885, EPI_ISL_742886, EPI_ISL_742887, EPI_ISL_742888, EPI_ISL_742889, EPI_ISL_743117, EPI_ISL_743126, EPI_ISL_743161, EPI_ISL_743592, EPI_ISL_743593, EPI_ISL_743594, EPI_ISL_743595, EPI_ISL_743606, EPI_ISL_743607, EPI_ISL_743608, EPI_ISL_743609, EPI_ISL_743610, EPI_ISL_743611, EPI_ISL_743612, EPI_ISL_743613, EPI_ISL_743614, EPI_ISL_743615, EPI_ISL_743616, EPI_ISL_743617, EPI_ISL_743618, EPI_ISL_743619, EPI_ISL_743620, EPI_ISL_743621, EPI_ISL_743622, EPI_ISL_743623, EPI_ISL_743624, EPI_ISL_743625, EPI_ISL_743626, EPI_ISL_743627, EPI_ISL_743628, EPI_ISL_743629, EPI_ISL_743630, EPI_ISL_743631, EPI_ISL_743632, EPI_ISL_743633, EPI_ISL_743634, EPI_ISL_743636, EPI_ISL_814276, EPI_ISL_814321, EPI_ISL_814343, EPI_ISL_814416, EPI_ISL_814695, EPI_ISL_814764, EPI_ISL_814765, EPI_ISL_814766, EPI_ISL_814767, EPI_ISL_814768, EPI_ISL_814769, EPI_ISL_814770, EPI_ISL_814771, EPI_ISL_814772, EPI_ISL_814773, EPI_ISL_814774, EPI_ISL_814775, EPI_ISL_814776, EPI_ISL_814777, EPI_ISL_814778, EPI_ISL_814779, EPI_ISL_814780, EPI_ISL_814781, EPI_ISL_814782, EPI_ISL_814783, EPI_ISL_814784, EPI_ISL_814785, EPI_ISL_814786, EPI_ISL_814787, EPI_ISL_814788, EPI_ISL_814796, EPI_ISL_814797

| | | | |
|---|---|---|---|
| see above | Wales Specialist Virology Centre Sequencing lab: Pathogen Genomics Unit | COVID-19 Genomics UK (COG-UK) Consortium | Alec Birchley; Alexander Adams; Amy Gaskin; Angela Marchbank; Bree Gatica-Wilcox; Catherine Moore; Jason Coombes; Joanne Watkins; Joel Southgate; Johnathan Evans; Laura Gifford; Lauren Gilbert; Lee Graham; Malorie Perry; Matthew Bull; Nicole Pacchiarini; Sally Corden; Sara Kumziene-Summerhayes; Sara Rey; Sarah Taylor; Simon Cottrell; Sophie Jones; Tom Connor |

EPI_ISL_680452, EPI_ISL_680453, EPI_ISL_680454, EPI_ISL_702659, EPI_ISL_702771, EPI_ISL_702893, EPI_ISL_703141, EPI_ISL_703155, EPI_ISL_703240, EPI_ISL_703243, EPI_ISL_703246, EPI_ISL_703717, EPI_ISL_703719, EPI_ISL_703743, EPI_ISL_703750, EPI_ISL_703751, EPI_ISL_703752, EPI_ISL_703753, EPI_ISL_703754, EPI_ISL_703756, EPI_ISL_703773, EPI_ISL_703842, EPI_ISL_703845, EPI_ISL_703921, EPI_ISL_703924, EPI_ISL_703968, EPI_ISL_704118, EPI_ISL_704184, EPI_ISL_704228, EPI_ISL_704809, EPI_ISL_705300, EPI_ISL_705399, EPI_ISL_705490, EPI_ISL_705649, EPI_ISL_705665, EPI_ISL_705666, EPI_ISL_705667, EPI_ISL_705668, EPI_ISL_705669, EPI_ISL_705670, EPI_ISL_705671, EPI_ISL_705672, EPI_ISL_705673, EPI_ISL_705674, EPI_ISL_705675, EPI_ISL_705676, EPI_ISL_705677, EPI_ISL_705678, EPI_ISL_705679, EPI_ISL_705680, EPI_ISL_705681, EPI_ISL_705682, EPI_ISL_705683, EPI_ISL_705684, EPI_ISL_705685, EPI_ISL_705686, EPI_ISL_705687, EPI_ISL_705688, EPI_ISL_705689, EPI_ISL_705690, EPI_ISL_705691, EPI_ISL_705692, EPI_ISL_705693, EPI_ISL_705694, EPI_ISL_705695, EPI_ISL_705696, EPI_ISL_705697, EPI_ISL_705698, EPI_ISL_705699, EPI_ISL_705700, EPI_ISL_705701, EPI_ISL_705702, EPI_ISL_705703, EPI_ISL_705704, EPI_ISL_705705, EPI_ISL_705706, EPI_ISL_705707, EPI_ISL_705708, EPI_ISL_705709, EPI_ISL_705710, EPI_ISL_705718, EPI_ISL_705720, EPI_ISL_705727, EPI_ISL_705729, EPI_ISL_705731, EPI_ISL_724213, EPI_ISL_838055, EPI_ISL_838056, EPI_ISL_838057, EPI_ISL_838058, EPI_ISL_838060, EPI_ISL_838061, EPI_ISL_838062, EPI_ISL_838063, EPI_ISL_919239, EPI_ISL_919270, EPI_ISL_919272, EPI_ISL_919273, EPI_ISL_919274, EPI_ISL_919275

| | | | |
|---|---|---|---|
| see above | West of Scotland Specialist Virology Centre, NHSGGC / MRC-University of Glasgow Centre for Virus Research | COVID-19 Genomics UK (COG-UK) Consortium | Alasdair MacLean; Alice Broos; Ana da Silva Filipe; Antonia Ho; Daniel Mair; David L Robertson; Emma Thomson; Guy Mollett; James Shepherd; Jenna Nichols; Joseph Hughes; Kathy Li; Kathy Smollett; Kyriaki Nomikou; Lily Tong; Matthew Holden; Natasha Johnson; Rachel Blacow; Richard Orton; Rory Gunson; Sarah McDonald; Sharif Shaaban; Sreenu Vattipally; Stephen Carmichael |

We gratefully acknowledge the following Authors from the Originating laboratories responsible for obtaining the specimens, as well as the Submitting laboratories where the genome data were generated and shared via GISAID, on which this research is based.

All Submitters of data may be contacted directly via www.gisaid.org

Authors are sorted alphabetically.

| Accession ID | Originating Laboratory | Submitting Laboratory | Authors |
|---|---|---|---|
| EPI_ISL_842966, EPI_ISL_842982, EPI_ISL_842983, EPI_ISL_843006, EPI_ISL_843008, EPI_ISL_843009, EPI_ISL_843010, EPI_ISL_843011, EPI_ISL_843012, EPI_ISL_843021, EPI_ISL_843022, EPI_ISL_843023, EPI_ISL_843024, EPI_ISL_843025, EPI_ISL_843026, EPI_ISL_843027, EPI_ISL_843028, EPI_ISL_843029, EPI_ISL_843030, EPI_ISL_843031, EPI_ISL_843032, EPI_ISL_843033, EPI_ISL_843034, EPI_ISL_843035, EPI_ISL_843036, EPI_ISL_843037, EPI_ISL_843038, EPI_ISL_843039, EPI_ISL_843040, EPI_ISL_843042, EPI_ISL_843043, EPI_ISL_843044, EPI_ISL_843045, EPI_ISL_843046, EPI_ISL_843047, EPI_ISL_843048, EPI_ISL_843049, EPI_ISL_843050, EPI_ISL_843051, EPI_ISL_843052, EPI_ISL_843053, EPI_ISL_843054, EPI_ISL_843055, EPI_ISL_843056, EPI_ISL_843057, EPI_ISL_843058, EPI_ISL_843059, EPI_ISL_843060, EPI_ISL_843061, EPI_ISL_843062, EPI_ISL_843063, EPI_ISL_843064, EPI_ISL_843065, EPI_ISL_843066, EPI_ISL_843067, EPI_ISL_843068, EPI_ISL_843069, EPI_ISL_843070, EPI_ISL_843088, EPI_ISL_1636931, EPI_ISL_1636932, EPI_ISL_1636933, EPI_ISL_1636934, EPI_ISL_1636935, EPI_ISL_1636936, EPI_ISL_1636938, EPI_ISL_1636939, EPI_ISL_1636940, EPI_ISL_1636941, EPI_ISL_1636942, EPI_ISL_1636943, EPI_ISL_1636944, EPI_ISL_1636945, EPI_ISL_1636946, EPI_ISL_1636966, EPI_ISL_1636967, EPI_ISL_1636988 | | | |
| see above | Barts Health NHS Trust | COVID-19 Genomics UK (COG-UK) Consortium | BROAD; Beatrix; CUTINO-MOGUEL; Claire; David; Dola; HARRINGTON; KELE; KULASEGARAN-SHYLINI; Maria-Teresa; OWOYEMI; Raghavendran; SHYLINI |
| EPI_ISL_816861, EPI_ISL_816862, EPI_ISL_816863, EPI_ISL_816864, EPI_ISL_816865, EPI_ISL_816866, EPI_ISL_816867, EPI_ISL_816868, EPI_ISL_816869, EPI_ISL_816870, EPI_ISL_816871, EPI_ISL_816872, EPI_ISL_816873, EPI_ISL_816874, EPI_ISL_816875, EPI_ISL_816876, EPI_ISL_816877, EPI_ISL_816878, EPI_ISL_816879, EPI_ISL_816880, EPI_ISL_816881, EPI_ISL_816882, EPI_ISL_816883, EPI_ISL_816884, EPI_ISL_816885, EPI_ISL_816886, EPI_ISL_816887, EPI_ISL_816888, EPI_ISL_816889, EPI_ISL_816890, EPI_ISL_816891, EPI_ISL_816893, EPI_ISL_816895, EPI_ISL_816900, EPI_ISL_816901, EPI_ISL_816902, EPI_ISL_816904, EPI_ISL_816906, EPI_ISL_816909, EPI_ISL_816910, EPI_ISL_816911, EPI_ISL_816913, EPI_ISL_816915, EPI_ISL_816916, EPI_ISL_816918, EPI_ISL_816920, EPI_ISL_816923, EPI_ISL_816924, EPI_ISL_816925, EPI_ISL_816958, EPI_ISL_816960, EPI_ISL_816962, EPI_ISL_816967, EPI_ISL_816968, EPI_ISL_816969, EPI_ISL_816971, EPI_ISL_816973, EPI_ISL_816976, EPI_ISL_816977, EPI_ISL_816978, EPI_ISL_816980, EPI_ISL_816982, EPI_ISL_816983, EPI_ISL_816985, EPI_ISL_816987, EPI_ISL_816990, EPI_ISL_816991, EPI_ISL_816992, EPI_ISL_842355, EPI_ISL_842357, EPI_ISL_842359, EPI_ISL_842364, EPI_ISL_842365, EPI_ISL_842366, EPI_ISL_842368, EPI_ISL_842370, EPI_ISL_842373, EPI_ISL_842374, EPI_ISL_842375, EPI_ISL_842377, EPI_ISL_842379, EPI_ISL_842380, EPI_ISL_842382, EPI_ISL_842384, EPI_ISL_842387, EPI_ISL_842388, EPI_ISL_842389, EPI_ISL_868843, EPI_ISL_868844, EPI_ISL_868845, EPI_ISL_868848, EPI_ISL_868850, EPI_ISL_868851, EPI_ISL_868853, EPI_ISL_868854, EPI_ISL_868855, EPI_ISL_868856, EPI_ISL_868859, EPI_ISL_868860, EPI_ISL_868861, EPI_ISL_868863, EPI_ISL_868864, EPI_ISL_868865, EPI_ISL_868866, EPI_ISL_868867, EPI_ISL_868868, EPI_ISL_868869, EPI_ISL_868872, EPI_ISL_868873, EPI_ISL_868875, EPI_ISL_868950, EPI_ISL_868951, EPI_ISL_868952, EPI_ISL_868953, EPI_ISL_868954, EPI_ISL_868955, EPI_ISL_868956, EPI_ISL_868957, EPI_ISL_868958, EPI_ISL_868959, EPI_ISL_868960, EPI_ISL_868961, EPI_ISL_868962, EPI_ISL_868963, EPI_ISL_868964, EPI_ISL_868965, EPI_ISL_868966, EPI_ISL_868967, EPI_ISL_868968, EPI_ISL_868969, EPI_ISL_868970, EPI_ISL_868971, EPI_ISL_868972, EPI_ISL_868973, EPI_ISL_868974, EPI_ISL_868975, EPI_ISL_868976, EPI_ISL_868977, EPI_ISL_868978, EPI_ISL_868979, EPI_ISL_868980, EPI_ISL_868981, EPI_ISL_868982, EPI_ISL_868983, EPI_ISL_868984, EPI_ISL_868987, EPI_ISL_868993, EPI_ISL_868995, EPI_ISL_868996, EPI_ISL_868999, EPI_ISL_869000, EPI_ISL_869001, EPI_ISL_869005, EPI_ISL_869006, EPI_ISL_869008 | | | |
| see above | Bioinformatics and Biostatistics Lab, Advanced Sequencing Facility | COVID-19 Genomics UK (COG-UK) Consortium | Aengus Stewart; Chelsea Sawyer; Harshil Patel; Jerome Nicod; Laura Cubitt; Margaret Crawford |
| EPI_ISL_702870, EPI_ISL_727713, EPI_ISL_727714, EPI_ISL_727715, EPI_ISL_727716, EPI_ISL_727717, EPI_ISL_727718, EPI_ISL_727719, EPI_ISL_727720, EPI_ISL_727721, EPI_ISL_727722, EPI_ISL_727723, EPI_ISL_741734, EPI_ISL_741736, EPI_ISL_741737, EPI_ISL_741750, EPI_ISL_741751, EPI_ISL_741752, EPI_ISL_741754, EPI_ISL_741756, EPI_ISL_841767, EPI_ISL_867948, EPI_ISL_867949, EPI_ISL_867952, EPI_ISL_867953, EPI_ISL_923217, EPI_ISL_923219, EPI_ISL_923221, EPI_ISL_923222, EPI_ISL_1000274, EPI_ISL_1000278, EPI_ISL_1107754, EPI_ISL_1730847, EPI_ISL_1979794, EPI_ISL_1979985, EPI_ISL_1979987, EPI_ISL_1979988, EPI_ISL_1979990, EPI_ISL_1979991, EPI_ISL_1979992, EPI_ISL_1979994, EPI_ISL_1979996, EPI_ISL_1979997, EPI_ISL_2127960 | | | |
| see above | Centre for Enzyme Innovation, University of Portsmouth / Translational Research Laboratory, Portsmouth Hospitals NHS Trust | COVID-19 Genomics UK (COG-UK) Consortium | Allyson Lloyd; Angela Beckett; Anoop Chauhan; Christopher Fearn; Ethan Butcher; Garry Scarlett; Kate Cook; Katie Loveson; Kelly Bicknell; Robert Impey; Salman Goudarzi; Samuel Robson; Sarah Wyllie; Scott Elliott; Sharon Glaysher; Yann Bourgeois |
| EPI_ISL_723863, EPI_ISL_723864, EPI_ISL_723865, EPI_ISL_723871, EPI_ISL_723885, EPI_ISL_723886, EPI_ISL_723887, EPI_ISL_723888, EPI_ISL_723889, EPI_ISL_723890, EPI_ISL_723891, EPI_ISL_723892, EPI_ISL_723893, EPI_ISL_723894, EPI_ISL_723895, EPI_ISL_723897, EPI_ISL_723898, EPI_ISL_723899, EPI_ISL_723901, EPI_ISL_723902, EPI_ISL_723903, EPI_ISL_723904, EPI_ISL_723905, EPI_ISL_723906, EPI_ISL_723907, EPI_ISL_723908, EPI_ISL_723909, EPI_ISL_723910, EPI_ISL_723911, EPI_ISL_723912, EPI_ISL_723913, EPI_ISL_723914, EPI_ISL_723915, EPI_ISL_723916, EPI_ISL_723919, EPI_ISL_723920, EPI_ISL_723921, EPI_ISL_723933, EPI_ISL_723935, EPI_ISL_723936, EPI_ISL_723947, EPI_ISL_723948, EPI_ISL_740955, EPI_ISL_740956, EPI_ISL_740957, EPI_ISL_740958, EPI_ISL_740959, EPI_ISL_740960, EPI_ISL_740961, EPI_ISL_740962, EPI_ISL_740963, EPI_ISL_740964, EPI_ISL_740965, EPI_ISL_740966, EPI_ISL_740967, EPI_ISL_740968, EPI_ISL_740969, EPI_ISL_740970, EPI_ISL_740972, EPI_ISL_740973, EPI_ISL_740974, EPI_ISL_740975, EPI_ISL_740976, EPI_ISL_740977, EPI_ISL_740978, EPI_ISL_740979, EPI_ISL_740980, EPI_ISL_740981, EPI_ISL_740982, EPI_ISL_740983, EPI_ISL_740984, EPI_ISL_740985, EPI_ISL_740990, EPI_ISL_740991, EPI_ISL_740992, EPI_ISL_740993, EPI_ISL_740994, EPI_ISL_763370, EPI_ISL_763375, EPI_ISL_763438, EPI_ISL_763569, EPI_ISL_763593, EPI_ISL_763641, EPI_ISL_763648, EPI_ISL_763792, EPI_ISL_763793, EPI_ISL_763794, EPI_ISL_763801, EPI_ISL_763815, EPI_ISL_763848, EPI_ISL_763872, EPI_ISL_764159, EPI_ISL_764171, EPI_ISL_764174, EPI_ISL_764176, EPI_ISL_764177, EPI_ISL_764191, EPI_ISL_764209, EPI_ISL_764281, EPI_ISL_764284, EPI_ISL_764285, EPI_ISL_764286, EPI_ISL_764287, EPI_ISL_764288, EPI_ISL_764289, EPI_ISL_764290, EPI_ISL_764291, EPI_ISL_764292, EPI_ISL_764293, EPI_ISL_764294, EPI_ISL_764295, EPI_ISL_764296, EPI_ISL_764297, EPI_ISL_764298, EPI_ISL_764299, EPI_ISL_764300, EPI_ISL_764301, EPI_ISL_764303, EPI_ISL_764304, EPI_ISL_764305, EPI_ISL_764307, EPI_ISL_764309, EPI_ISL_764312, EPI_ISL_764313, EPI_ISL_764314, EPI_ISL_764316, EPI_ISL_764317, EPI_ISL_764319, EPI_ISL_764322, EPI_ISL_764323, EPI_ISL_764324, EPI_ISL_764325, EPI_ISL_764326, EPI_ISL_864782, EPI_ISL_864783, EPI_ISL_864868, EPI_ISL_864870, EPI_ISL_864888, EPI_ISL_864890, EPI_ISL_864892, EPI_ISL_864894 | | | |
| see above | Department of Pathology, University of Cambridge | COVID-19 Genomics UK (COG-UK) Consortium | Aminu S. Jahun; Grant Hall; Ian Goodfellow; Iliana Georgana; Malte Pinckert; Martin D. Curran; Myra Hosmillo; Surendra Parmar; Yasmin Chaudhry |
| EPI_ISL_708841, EPI_ISL_708842, EPI_ISL_708845, EPI_ISL_708846, EPI_ISL_708851, EPI_ISL_708852, EPI_ISL_708853, EPI_ISL_708854, EPI_ISL_708859, EPI_ISL_708860, EPI_ISL_708861, EPI_ISL_708863, EPI_ISL_708864, EPI_ISL_708865, EPI_ISL_708866, EPI_ISL_708867, EPI_ISL_708868, EPI_ISL_708870, EPI_ISL_708872, EPI_ISL_708873, EPI_ISL_708874, EPI_ISL_708875, EPI_ISL_708876, EPI_ISL_708877, EPI_ISL_708878, EPI_ISL_708880, EPI_ISL_708882, EPI_ISL_708885, EPI_ISL_708886, EPI_ISL_708887, EPI_ISL_708888, EPI_ISL_708891, EPI_ISL_708892, EPI_ISL_708894, EPI_ISL_708895, EPI_ISL_708899, EPI_ISL_708902, EPI_ISL_708903, EPI_ISL_708907, EPI_ISL_708909, EPI_ISL_708911, EPI_ISL_708914, EPI_ISL_708915, EPI_ISL_708917, EPI_ISL_708918, EPI_ISL_708919, EPI_ISL_708923, EPI_ISL_708924, EPI_ISL_708926, EPI_ISL_708927, EPI_ISL_708929, EPI_ISL_708931, EPI_ISL_708932, EPI_ISL_708934, EPI_ISL_708935, EPI_ISL_708936, EPI_ISL_708937, EPI_ISL_708938, EPI_ISL_708939, EPI_ISL_708940, EPI_ISL_708942, EPI_ISL_708943, EPI_ISL_708946, EPI_ISL_708947, EPI_ISL_708949, EPI_ISL_708950, EPI_ISL_708952, EPI_ISL_708954, EPI_ISL_708955, EPI_ISL_708956, EPI_ISL_708959, EPI_ISL_708960, EPI_ISL_708961, EPI_ISL_708962, EPI_ISL_708965, EPI_ISL_708966, EPI_ISL_708967, EPI_ISL_708968, EPI_ISL_708970, EPI_ISL_708971, EPI_ISL_708972, EPI_ISL_708973, EPI_ISL_708974, EPI_ISL_708976, EPI_ISL_708977, EPI_ISL_708978, EPI_ISL_708980, EPI_ISL_708981, EPI_ISL_708982, EPI_ISL_708987, EPI_ISL_708988, EPI_ISL_709295, EPI_ISL_709296, EPI_ISL_709297, EPI_ISL_709298, EPI_ISL_709299, EPI_ISL_709300, EPI_ISL_709302, EPI_ISL_709303, EPI_ISL_709304, EPI_ISL_709305, EPI_ISL_709306, EPI_ISL_709308, EPI_ISL_709309, EPI_ISL_709310, EPI_ISL_709313, EPI_ISL_709315, EPI_ISL_709316, EPI_ISL_709317, EPI_ISL_709318, EPI_ISL_709319, EPI_ISL_709321, EPI_ISL_709322, EPI_ISL_709323, EPI_ISL_709325, EPI_ISL_709326, EPI_ISL_709328, EPI_ISL_709329, EPI_ISL_709330, EPI_ISL_709331, EPI_ISL_709333, EPI_ISL_709335, EPI_ISL_709337, EPI_ISL_709338, EPI_ISL_709339, EPI_ISL_709340, EPI_ISL_709342, EPI_ISL_709344, EPI_ISL_709346, EPI_ISL_709347, EPI_ISL_709348, EPI_ISL_709349, EPI_ISL_709350, EPI_ISL_709351, EPI_ISL_709352, EPI_ISL_709355, EPI_ISL_709356, EPI_ISL_709357, EPI_ISL_709361, EPI_ISL_709362, EPI_ISL_709363, EPI_ISL_709368, EPI_ISL_709369, EPI_ISL_709370, EPI_ISL_709371, EPI_ISL_709372, EPI_ISL_709374, EPI_ISL_709375, EPI_ISL_709377, EPI_ISL_709379, EPI_ISL_709381, EPI_ISL_709382, EPI_ISL_709385, EPI_ISL_709386, EPI_ISL_709387, EPI_ISL_709389, EPI_ISL_709390, EPI_ISL_709391, EPI_ISL_709393, EPI_ISL_709397, EPI_ISL_709398, EPI_ISL_709400, EPI_ISL_709401, EPI_ISL_709402, EPI_ISL_709405, EPI_ISL_709408, EPI_ISL_709410, EPI_ISL_709413, EPI_ISL_709414, EPI_ISL_709415, EPI_ISL_709416, EPI_ISL_709418, EPI_ISL_709420, EPI_ISL_709421, EPI_ISL_709425, EPI_ISL_709427, EPI_ISL_709428, EPI_ISL_709430, EPI_ISL_709431, EPI_ISL_709432, EPI_ISL_709433, EPI_ISL_709434, EPI_ISL_709435, EPI_ISL_709437, EPI_ISL_709442, EPI_ISL_709444, EPI_ISL_709445, EPI_ISL_709447, EPI_ISL_709448, EPI_ISL_709449, EPI_ISL_709451, EPI_ISL_709452, EPI_ISL_709455, EPI_ISL_709460, EPI_ISL_709461, EPI_ISL_709463, EPI_ISL_709467, EPI_ISL_709468, EPI_ISL_709470, EPI_ISL_709471, EPI_ISL_709473, EPI_ISL_709475, EPI_ISL_709476, EPI_ISL_709477, EPI_ISL_709478, EPI_ISL_709479, EPI_ISL_709480, EPI_ISL_709484, EPI_ISL_709485, EPI_ISL_709488, EPI_ISL_709489, EPI_ISL_709491, EPI_ISL_709492, EPI_ISL_709499, EPI_ISL_709501, EPI_ISL_709504, EPI_ISL_709505, EPI_ISL_709506, EPI_ISL_709507, EPI_ISL_709508, EPI_ISL_709512, EPI_ISL_709513, EPI_ISL_709514, EPI_ISL_709516, EPI_ISL_709519, EPI_ISL_709520, EPI_ISL_709521, EPI_ISL_709523, EPI_ISL_709526, EPI_ISL_709529, EPI_ISL_709531, EPI_ISL_709532, EPI_ISL_709534, EPI_ISL_709535, EPI_ISL_709536, EPI_ISL_709538, EPI_ISL_709544, EPI_ISL_709545, EPI_ISL_709546, EPI_ISL_709547, EPI_ISL_709548, EPI_ISL_709550, EPI_ISL_709551, EPI_ISL_709552, EPI_ISL_709553, EPI_ISL_709554, EPI_ISL_709559, EPI_ISL_709560, EPI_ISL_709562, EPI_ISL_709563, EPI_ISL_709564, EPI_ISL_709566, EPI_ISL_710614, EPI_ISL_710616, EPI_ISL_710617, EPI_ISL_710618, EPI_ISL_710620, EPI_ISL_710625, EPI_ISL_710628, EPI_ISL_710631, EPI_ISL_710632, EPI_ISL_710636, EPI_ISL_710638, EPI_ISL_710645, EPI_ISL_710647, EPI_ISL_710648, EPI_ISL_710656, EPI_ISL_710661, EPI_ISL_710662, EPI_ISL_710663, EPI_ISL_710665, EPI_ISL_710671, EPI_ISL_710676, EPI_ISL_710682, EPI_ISL_710689, EPI_ISL_710693, EPI_ISL_710694, EPI_ISL_710698, EPI_ISL_720683, EPI_ISL_720685, EPI_ISL_720686, EPI_ISL_720688, EPI_ISL_720694, EPI_ISL_720695, EPI_ISL_720697, EPI_ISL_720702, EPI_ISL_720709, EPI_ISL_720711, EPI_ISL_720713, EPI_ISL_720715, EPI_ISL_720722, EPI_ISL_720734, EPI_ISL_720741, EPI_ISL_720745, EPI_ISL_720750, EPI_ISL_720756, EPI_ISL_720762, EPI_ISL_720764, EPI_ISL_720765, EPI_ISL_720770, EPI_ISL_720773, EPI_ISL_720779, EPI_ISL_720780, EPI_ISL_720790, EPI_ISL_720796, EPI_ISL_720810, EPI_ISL_720814, EPI_ISL_720816, EPI_ISL_720823, EPI_ISL_720825, EPI_ISL_720827, EPI_ISL_720836, EPI_ISL_720842, EPI_ISL_720845, EPI_ISL_720846, EPI_ISL_720848, EPI_ISL_720849, EPI_ISL_720853, EPI_ISL_720857, EPI_ISL_720858, EPI_ISL_720864, EPI_ISL_720869, EPI_ISL_720872, EPI_ISL_720875, EPI_ISL_720880, EPI_ISL_720886, EPI_ISL_720891, EPI_ISL_720893, EPI_ISL_720894, EPI_ISL_720898, EPI_ISL_720900, EPI_ISL_720907, EPI_ISL_721517, EPI_ISL_721518, EPI_ISL_721520, EPI_ISL_721522, EPI_ISL_721523, EPI_ISL_721525, EPI_ISL_730654, EPI_ISL_730660, EPI_ISL_730661, EPI_ISL_730662, EPI_ISL_730663, EPI_ISL_730665, EPI_ISL_730666, EPI_ISL_730669, EPI_ISL_730670, EPI_ISL_730671, EPI_ISL_730672, EPI_ISL_730674, EPI_ISL_730677, EPI_ISL_730679, EPI_ISL_730681, EPI_ISL_730684, EPI_ISL_730687, EPI_ISL_730688, EPI_ISL_730689, EPI_ISL_730690, EPI_ISL_730691, EPI_ISL_730693, EPI_ISL_730694, EPI_ISL_730698, EPI_ISL_730700, EPI_ISL_730701, EPI_ISL_730703, EPI_ISL_730706, EPI_ISL_730707, EPI_ISL_730708, EPI_ISL_730713, EPI_ISL_730714, EPI_ISL_730715, EPI_ISL_730716, EPI_ISL_730717, EPI_ISL_730719, EPI_ISL_730720, EPI_ISL_730721, EPI_ISL_730724, EPI_ISL_730725, EPI_ISL_730726, EPI_ISL_730727, EPI_ISL_730728, EPI_ISL_730730, EPI_ISL_730735, EPI_ISL_730736, EPI_ISL_730738, EPI_ISL_730739, EPI_ISL_730740, EPI_ISL_730741, EPI_ISL_730744, EPI_ISL_730745, EPI_ISL_730746, EPI_ISL_730747, EPI_ISL_730748, EPI_ISL_730749, EPI_ISL_730755, EPI_ISL_730756, EPI_ISL_730757, EPI_ISL_730760, EPI_ISL_730761, EPI_ISL_730762, EPI_ISL_730763, EPI_ISL_730764, EPI_ISL_730766, EPI_ISL_730767, EPI_ISL_730768, EPI_ISL_730769, EPI_ISL_730770, EPI_ISL_730772, EPI_ISL_730775, EPI_ISL_730776, EPI_ISL_730779, EPI_ISL_730781, EPI_ISL_730782, EPI_ISL_730783, EPI_ISL_730787, EPI_ISL_730790, EPI_ISL_730791, EPI_ISL_730797, EPI_ISL_730798, EPI_ISL_730799, EPI_ISL_730800, EPI_ISL_730807, EPI_ISL_730810, EPI_ISL_730811, EPI_ISL_730812, EPI_ISL_730816, EPI_ISL_730817, EPI_ISL_730820, EPI_ISL_730821, EPI_ISL_730822, EPI_ISL_730827, EPI_ISL_730835, EPI_ISL_730836, EPI_ISL_730837, EPI_ISL_730838, EPI_ISL_730841, EPI_ISL_730842, EPI_ISL_730843, EPI_ISL_730851, EPI_ISL_730853, EPI_ISL_730856, EPI_ISL_730857, EPI_ISL_730863, EPI_ISL_730866, EPI_ISL_730868, EPI_ISL_730871, EPI_ISL_730872, EPI_ISL_730874, EPI_ISL_730879, EPI_ISL_730880, EPI_ISL_730882, EPI_ISL_730883, EPI_ISL_730884, EPI_ISL_730885, EPI_ISL_730888, EPI_ISL_730891, EPI_ISL_730892, EPI_ISL_730895, EPI_ISL_730896, EPI_ISL_730897, EPI_ISL_730899, EPI_ISL_730901, EPI_ISL_730903, EPI_ISL_730905, EPI_ISL_730906, EPI_ISL_730907, EPI_ISL_730909, EPI_ISL_730912, EPI_ISL_730915, EPI_ISL_730916, EPI_ISL_730921, EPI_ISL_730924, EPI_ISL_730929, EPI_ISL_730935, EPI_ISL_730939, EPI_ISL_730948, EPI_ISL_730963, EPI_ISL_730966, EPI_ISL_730967, EPI_ISL_730969, EPI_ISL_730970, EPI_ISL_730971, EPI_ISL_730973, EPI_ISL_730980, EPI_ISL_730987, EPI_ISL_730989, EPI_ISL_730994, EPI_ISL_731000, EPI_ISL_731006, EPI_ISL_731017, EPI_ISL_731019, EPI_ISL_731020, EPI_ISL_731024, EPI_ISL_731026, EPI_ISL_731027, EPI_ISL_731033, EPI_ISL_731035, EPI_ISL_731040, EPI_ISL_731045, EPI_ISL_731050, EPI_ISL_731052, EPI_ISL_731054, EPI_ISL_731055, EPI_ISL_731057, EPI_ISL_731059, EPI_ISL_731065, EPI_ISL_731066, EPI_ISL_731074, EPI_ISL_731076, EPI_ISL_731079, EPI_ISL_731080, EPI_ISL_731082, EPI_ISL_731083, EPI_ISL_731086, EPI_ISL_731087, EPI_ISL_731088, EPI_ISL_731094, EPI_ISL_731096, EPI_ISL_731104, EPI_ISL_731106, EPI_ISL_731107, EPI_ISL_731110, EPI_ISL_731116, EPI_ISL_731119, EPI_ISL_731120, EPI_ISL_731123, EPI_ISL_731126, EPI_ISL_731129, EPI_ISL_731130, EPI_ISL_731131, EPI_ISL_731133, EPI_ISL_731135, EPI_ISL_731137, EPI_ISL_731141, EPI_ISL_731143, EPI_ISL_731147, EPI_ISL_731153, EPI_ISL_731154, EPI_ISL_731157, EPI_ISL_731158, EPI_ISL_731160, EPI_ISL_731161, EPI_ISL_731169, EPI_ISL_731171, EPI_ISL_731181, EPI_ISL_731183, EPI_ISL_731188, EPI_ISL_731196, EPI_ISL_731199, EPI_ISL_731200, EPI_ISL_731204, EPI_ISL_731206, EPI_ISL_731214, EPI_ISL_731215, EPI_ISL_731233, EPI_ISL_731235, EPI_ISL_731240, EPI_ISL_731247, EPI_ISL_731248, EPI_ISL_731254, EPI_ISL_731260, EPI_ISL_731263, EPI_ISL_731265, EPI_ISL_731266, EPI_ISL_731268, EPI_ISL_731269, EPI_ISL_731270, EPI_ISL_731272, EPI_ISL_731274, EPI_ISL_731275, EPI_ISL_731276, EPI_ISL_731277, EPI_ISL_731278, EPI_ISL_731279, EPI_ISL_731285, EPI_ISL_731288, EPI_ISL_731289, EPI_ISL_731292, EPI_ISL_731294, EPI_ISL_731296, EPI_ISL_731298, EPI_ISL_731300, EPI_ISL_731301, EPI_ISL_731302, EPI_ISL_731303, EPI_ISL_731304, EPI_ISL_731307, EPI_ISL_731311, EPI_ISL_731312, EPI_ISL_731313, EPI_ISL_731315, EPI_ISL_731316, EPI_ISL_731317, EPI_ISL_731318, EPI_ISL_731323, EPI_ISL_731324, EPI_ISL_731325, EPI_ISL_731328, EPI_ISL_731329, EPI_ISL_731331, EPI_ISL_731335, EPI_ISL_731336, EPI_ISL_731339, EPI_ISL_731342, EPI_ISL_731343, EPI_ISL_731344, EPI_ISL_731348, EPI_ISL_731350, EPI_ISL_731351, EPI_ISL_731352, EPI_ISL_731356, EPI_ISL_731358, EPI_ISL_731359, EPI_ISL_731360, EPI_ISL_731362, EPI_ISL_731363, EPI_ISL_731364, EPI_ISL_731365, EPI_ISL_731366, EPI_ISL_731369, EPI_ISL_731370, EPI_ISL_731371, EPI_ISL_731374, EPI_ISL_731375, EPI_ISL_731377, EPI_ISL_731381, EPI_ISL_731384, EPI_ISL_731385, EPI_ISL_731387, EPI_ISL_731388, EPI_ISL_731389, EPI_ISL_731391, EPI_ISL_731394, EPI_ISL_731395, EPI_ISL_731399, EPI_ISL_731409, EPI_ISL_731410, EPI_ISL_731412, EPI_ISL_731413, EPI_ISL_731415, EPI_ISL_731416, EPI_ISL_731418, EPI_ISL_731419, EPI_ISL_731422, EPI_ISL_731423, EPI_ISL_731424, EPI_ISL_731427, EPI_ISL_731428, EPI_ISL_731430, EPI_ISL_731434, EPI_ISL_731436, EPI_ISL_731439, EPI_ISL_731440, EPI_ISL_731441, EPI_ISL_731442, EPI_ISL_731443, EPI_ISL_731445, EPI_ISL_731446, EPI_ISL_731448, EPI_ISL_731451, EPI_ISL_731455, EPI_ISL_731457, EPI_ISL_731460, EPI_ISL_731461, EPI_ISL_731462, EPI_ISL_731463, EPI_ISL_731464, EPI_ISL_731465, EPI_ISL_731467, EPI_ISL_731468, EPI_ISL_731470, EPI_ISL_731473, EPI_ISL_731474, EPI_ISL_731475, EPI_ISL_731478, EPI_ISL_731480, EPI_ISL_731481, EPI_ISL_731483, EPI_ISL_731487, EPI_ISL_731489, EPI_ISL_731490, EPI_ISL_731492, EPI_ISL_731495, EPI_ISL_731498, EPI_ISL_731500, EPI_ISL_731502, EPI_ISL_731503, EPI_ISL_731504, EPI_ISL_731506, EPI_ISL_731509, EPI_ISL_731510, EPI_ISL_731511, EPI_ISL_731512, EPI_ISL_731514, EPI_ISL_731515, EPI_ISL_731516, EPI_ISL_731518, EPI_ISL_731520, EPI_ISL_731521, EPI_ISL_731524, EPI_ISL_731526, EPI_ISL_731527, EPI_ISL_731528, EPI_ISL_731529, EPI_ISL_731530, EPI_ISL_731531, EPI_ISL_731532, EPI_ISL_731533, EPI_ISL_731534, EPI_ISL_731538, EPI_ISL_731539, EPI_ISL_731540, EPI_ISL_731542, EPI_ISL_731543, EPI_ISL_731545, EPI_ISL_731546, EPI_ISL_731547, EPI_ISL_731549, EPI_ISL_731555, EPI_ISL_731565, EPI_ISL_731570, EPI_ISL_731571, EPI_ISL_731576, EPI_ISL_731580, EPI_ISL_731584, EPI_ISL_731585, EPI_ISL_731588, EPI_ISL_731592, EPI_ISL_731599, EPI_ISL_731611, EPI_ISL_731624, EPI_ISL_731626, EPI_ISL_731629, EPI_ISL_731632, EPI_ISL_731635, EPI_ISL_731639, EPI_ISL_731640, EPI_ISL_731641, EPI_ISL_731644, EPI_ISL_731647, EPI_ISL_731648, EPI_ISL_731649, EPI_ISL_731655, EPI_ISL_731656, EPI_ISL_731664, EPI_ISL_731665, EPI_ISL_731667, EPI_ISL_731671, EPI_ISL_731680, EPI_ISL_731684, EPI_ISL_731686, EPI_ISL_731688, EPI_ISL_731690, EPI_ISL_731691, EPI_ISL_731692, EPI_ISL_731693, EPI_ISL_731697, EPI_ISL_731701, EPI_ISL_731705, EPI_ISL_731707, EPI_ISL_731709, EPI_ISL_731711, EPI_ISL_731714, EPI_ISL_731719, EPI_ISL_731721, EPI_ISL_731722, EPI_ISL_731724, EPI_ISL_731731, EPI_ISL_731736, EPI_ISL_731737, EPI_ISL_731763, EPI_ISL_731767, EPI_ISL_731770, EPI_ISL_731775, EPI_ISL_731785, EPI_ISL_731787, EPI_ISL_731788, EPI_ISL_731792, EPI_ISL_731795, EPI_ISL_731796, EPI_ISL_731799, EPI_ISL_731800, EPI_ISL_731806, EPI_ISL_731808, EPI_ISL_731810, EPI_ISL_731819, EPI_ISL_731824, EPI_ISL_731827, EPI_ISL_731836, EPI_ISL_731838, EPI_ISL_731840, EPI_ISL_731841, EPI_ISL_731842, EPI_ISL_731846, EPI_ISL_731850, EPI_ISL_731858, EPI_ISL_731863, EPI_ISL_731868, EPI_ISL_731874, EPI_ISL_731875, EPI_ISL_731877, EPI_ISL_731881, EPI_ISL_731884, EPI_ISL_731885, EPI_ISL_731887, EPI_ISL_731888, EPI_ISL_733598, EPI_ISL_733603, EPI_ISL_733605, EPI_ISL_733609, EPI_ISL_733611, EPI_ISL_733613, EPI_ISL_733618, EPI_ISL_733621, EPI_ISL_733623, EPI_ISL_733625, EPI_ISL_733635, EPI_ISL_733638, EPI_ISL_733639, EPI_ISL_733641, EPI_ISL_733642, EPI_ISL_733643, EPI_ISL_733647, EPI_ISL_733653, EPI_ISL_733654, EPI_ISL_733659, EPI_ISL_733682, EPI_ISL_733683, EPI_ISL_733685, EPI_ISL_733693, EPI_ISL_733697, EPI_ISL_733699, EPI_ISL_733701, EPI_ISL_733703, EPI_ISL_733706, EPI_ISL_733710, EPI_ISL_733712, EPI_ISL_733718, EPI_ISL_733725, EPI_ISL_733731, EPI_ISL_733734, EPI_ISL_733737, EPI_ISL_733742, EPI_ISL_733754, EPI_ISL_733755, EPI_ISL_733759, EPI_ISL_733762, EPI_ISL_733766, EPI_ISL_733769, EPI_ISL_733777, EPI_ISL_733783, EPI_ISL_733785, EPI_ISL_733794, EPI_ISL_733795, EPI_ISL_733797, EPI_ISL_733803, EPI_ISL_733806, EPI_ISL_733807, EPI_ISL_733808, EPI_ISL_733815, EPI_ISL_733818, EPI_ISL_733819, EPI_ISL_733830, EPI_ISL_733833, EPI_ISL_733837, EPI_ISL_733843, EPI_ISL_733844, EPI_ISL_733848, EPI_ISL_733850, EPI_ISL_733859, EPI_ISL_733867, EPI_ISL_733877, EPI_ISL_733879, EPI_ISL_733880, EPI_ISL_733882, EPI_ISL_733885, EPI_ISL_733897, EPI_ISL_733901, EPI_ISL_733912, EPI_ISL_733913, EPI_ISL_736758, EPI_ISL_736772, EPI_ISL_736773, EPI_ISL_760846, EPI_ISL_760860, EPI_ISL_760878, EPI_ISL_760888, EPI_ISL_760891, EPI_ISL_760929, EPI_ISL_760949, EPI_ISL_760951, EPI_ISL_760952, EPI_ISL_760978, EPI_ISL_760982, EPI_ISL_761001, EPI_ISL_761013, EPI_ISL_761042, EPI_ISL_761046, EPI_ISL_761061, EPI_ISL_767151, EPI_ISL_767152, EPI_ISL_767153, EPI_ISL_767154, EPI_ISL_767155, EPI_ISL_767156, EPI_ISL_767157, EPI_ISL_767158, EPI_ISL_767159, EPI_ISL_767160, EPI_ISL_767161, EPI_ISL_767162, EPI_ISL_767163, EPI_ISL_767164, EPI_ISL_767165, EPI_ISL_767166, EPI_ISL_767167, EPI_ISL_767168, EPI_ISL_767169, EPI_ISL_767170, EPI_ISL_767171, EPI_ISL_767172, EPI_ISL_767173, EPI_ISL_767174, EPI_ISL_767175, EPI_ISL_767176, EPI_ISL_767177, EPI_ISL_767179, EPI_ISL_767180, EPI_ISL_767181, EPI_ISL_767182, EPI_ISL_767183, EPI_ISL_767184, EPI_ISL_767185, EPI_ISL_767186, EPI_ISL_767187, EPI_ISL_767188, EPI_ISL_767189, EPI_ISL_767190, EPI_ISL_767191, EPI_ISL_767192, EPI_ISL_767193, EPI_ISL_767194, EPI_ISL_767195, EPI_ISL_767196, EPI_ISL_767197, EPI_ISL_767198, EPI_ISL_767199, EPI_ISL_767200, EPI_ISL_767201, EPI_ISL_767202, EPI_ISL_767203, EPI_ISL_767204, EPI_ISL_767205, EPI_ISL_767206, EPI_ISL_767207, EPI_ISL_767208, EPI_ISL_767209, EPI_ISL_767210, EPI_ISL_767211, EPI_ISL_767212, EPI_ISL_767213, EPI_ISL_767214, EPI_ISL_767215, EPI_ISL_767221, EPI_ISL_767230, EPI_ISL_767240, EPI_ISL_767241, EPI_ISL_767242, EPI_ISL_767243, EPI_ISL_767244, EPI_ISL_767245, EPI_ISL_767246, EPI_ISL_767247, EPI_ISL_767248, EPI_ISL_767249, EPI_ISL_767250, EPI_ISL_767251, EPI_ISL_767252, EPI_ISL_767253, EPI_ISL_767254, EPI_ISL_767255, EPI_ISL_767256, EPI_ISL_767257, EPI_ISL_767258, EPI_ISL_767259, EPI_ISL_767260, EPI_ISL_767261, EPI_ISL_767262, EPI_ISL_767263, EPI_ISL_767264, EPI_ISL_767265, EPI_ISL_767266, EPI_ISL_767267, EPI_ISL_767268, EPI_ISL_767269, EPI_ISL_767270, EPI_ISL_767271, EPI_ISL_767272, EPI_ISL_767273, EPI_ISL_767274, EPI_ISL_767275, EPI_ISL_767276, EPI_ISL_767278, EPI_ISL_767284, EPI_ISL_767291, EPI_ISL_767292, EPI_ISL_767293, EPI_ISL_767294, EPI_ISL_767295, EPI_ISL_767296, EPI_ISL_767297, EPI_ISL_767298, EPI_ISL_767299, EPI_ISL_767300, EPI_ISL_767301, EPI_ISL_767302, EPI_ISL_767303, EPI_ISL_767304, EPI_ISL_767305, EPI_ISL_767306, EPI_ISL_767307, EPI_ISL_767308, EPI_ISL_767309, EPI_ISL_767310, EPI_ISL_767311, EPI_ISL_767312, EPI_ISL_767313, EPI_ISL_767314, EPI_ISL_767315, EPI_ISL_767316, EPI_ISL_767317, EPI_ISL_767318, EPI_ISL_777169, EPI_ISL_782745, EPI_ISL_782747, EPI_ISL_782748, EPI_ISL_782758, EPI_ISL_782768, EPI_ISL_782771, EPI_ISL_782774, EPI_ISL_782783, EPI_ISL_782784, EPI_ISL_782787, EPI_ISL_782791, EPI_ISL_782792, EPI_ISL_782795, EPI_ISL_782801, EPI_ISL_782805, EPI_ISL_782806, EPI_ISL_782808, EPI_ISL_782813, EPI_ISL_782820, EPI_ISL_782825, EPI_ISL_782827, EPI_ISL_782832, EPI_ISL_782838, EPI_ISL_782841, EPI_ISL_782844, EPI_ISL_782849, EPI_ISL_782855, EPI_ISL_782856, EPI_ISL_782869, EPI_ISL_782872, EPI_ISL_782873, EPI_ISL_782877, EPI_ISL_782878, EPI_ISL_782892, EPI_ISL_782893, EPI_ISL_782894, EPI_ISL_782897, EPI_ISL_782899, EPI_ISL_782902, EPI_ISL_782903, EPI_ISL_782904, EPI_ISL_782905, EPI_ISL_782911, EPI_ISL_782918, EPI_ISL_782919, EPI_ISL_782924, EPI_ISL_782926, EPI_ISL_782936, EPI_ISL_782945, EPI_ISL_782956, EPI_ISL_782963, EPI_ISL_782964, EPI_ISL_782967, EPI_ISL_782969, EPI_ISL_782970, EPI_ISL_782976, EPI_ISL_782988, EPI_ISL_782993, EPI_ISL_782994, EPI_ISL_782997, EPI_ISL_783000, EPI_ISL_783006, EPI_ISL_783007, EPI_ISL_783008, EPI_ISL_783014, EPI_ISL_783015, EPI_ISL_783026, EPI_ISL_783030, EPI_ISL_783032, EPI_ISL_783033, EPI_ISL_783034, EPI_ISL_783043, EPI_ISL_783045, EPI_ISL_783046, EPI_ISL_783047, EPI_ISL_783052, EPI_ISL_783056, EPI_ISL_783068, EPI_ISL_822093, EPI_ISL_835474, EPI_ISL_859533, EPI_ISL_859534, EPI_ISL_859535, EPI_ISL_859536, EPI_ISL_859537, EPI_ISL_859538, EPI_ISL_859539, EPI_ISL_882333 | | | |
| see above | Lighthouse Lab in Alderley Park | Wellcome Sanger Institute for the COVID-19 Genomics UK (COG-UK) Consortium | Cordelia Langford; David K. Jackson; Dominic Kwiatkowski; Ewan Harrison; Ian Johnston; Jacquelyn Wynn; John Sillitoe on behalf of the Wellcome Sanger Institute COVID-19 Surveillance Team; Mairead Hyland; Roberto Amato; Sonia Goncalves; The Lighthouse Lab in Alderley Park and Alex Alderton |
| EPI_ISL_702025, EPI_ISL_702027, EPI_ISL_702028, EPI_ISL_702031, EPI_ISL_702032, EPI_ISL_702033, EPI_ISL_702034, EPI_ISL_702035, EPI_ISL_702036, EPI_ISL_702037, EPI_ISL_702041, EPI_ISL_702042, EPI_ISL_702043, EPI_ISL_702044, EPI_ISL_702045, EPI_ISL_702046, EPI_ISL_702048, EPI_ISL_702049, EPI_ISL_702050, EPI_ISL_702054, EPI_ISL_702055, EPI_ISL_702056, EPI_ISL_702057, EPI_ISL_702061, EPI_ISL_702062, EPI_ISL_702063, EPI_ISL_702068, EPI_ISL_702069, EPI_ISL_702071, EPI_ISL_702072, EPI_ISL_702075, EPI_ISL_702076, EPI_ISL_702077, EPI_ISL_702078, EPI_ISL_702080, EPI_ISL_702081, EPI_ISL_702082, EPI_ISL_702085, EPI_ISL_702087, EPI_ISL_702090, EPI_ISL_702091, EPI_ISL_702093, EPI_ISL_702094, EPI_ISL_702098, EPI_ISL_702099, EPI_ISL_702100, EPI_ISL_702105, EPI_ISL_702106, EPI_ISL_702109, EPI_ISL_702112, EPI_ISL_702114, EPI_ISL_702115, EPI_ISL_702116, EPI_ISL_702117, EPI_ISL_702119, EPI_ISL_702120, EPI_ISL_702121, EPI_ISL_702122, EPI_ISL_702123, EPI_ISL_702124, EPI_ISL_702126, EPI_ISL_702130, EPI_ISL_702132, EPI_ISL_702138, EPI_ISL_702139, EPI_ISL_702140, EPI_ISL_702141, EPI_ISL_702144, EPI_ISL_702145, EPI_ISL_702147, EPI_ISL_702148, EPI_ISL_702151, EPI_ISL_702156, EPI_ISL_702158, EPI_ISL_702159, EPI_ISL_702162, EPI_ISL_702163, EPI_ISL_702164, EPI_ISL_702165, EPI_ISL_702166, EPI_ISL_702169, EPI_ISL_702170, EPI_ISL_702173, EPI_ISL_702174, EPI_ISL_702175, EPI_ISL_702180, EPI_ISL_702182, EPI_ISL_702183, EPI_ISL_702188, EPI_ISL_702192, EPI_ISL_702193, EPI_ISL_702195, EPI_ISL_702196, EPI_ISL_702197, EPI_ISL_702199, EPI_ISL_702200, EPI_ISL_702202, EPI_ISL_702204, EPI_ISL_702207, EPI_ISL_702210, EPI_ISL_702212, EPI_ISL_702214, EPI_ISL_702215, EPI_ISL_702217, EPI_ISL_702218, EPI_ISL_702219, EPI_ISL_702220, EPI_ISL_702221, EPI_ISL_702224, EPI_ISL_702226, EPI_ISL_702227, EPI_ISL_702228, EPI_ISL_702229, EPI_ISL_702231, EPI_ISL_702233, EPI_ISL_702234, EPI_ISL_702235, EPI_ISL_702238, EPI_ISL_702239, EPI_ISL_702241, EPI_ISL_702246, EPI_ISL_702247, EPI_ISL_702248, EPI_ISL_702249, EPI_ISL_702253, EPI_ISL_702254, EPI_ISL_702255, EPI_ISL_702257, EPI_ISL_702258, EPI_ISL_702262, EPI_ISL_702272, EPI_ISL_702273, EPI_ISL_702274, EPI_ISL_702275, EPI_ISL_702276, EPI_ISL_702278, EPI_ISL_702279, EPI_ISL_702280, EPI_ISL_702281, EPI_ISL_702284, EPI_ISL_702288, EPI_ISL_702290, EPI_ISL_702292, EPI_ISL_702293, EPI_ISL_702296, EPI_ISL_702298, EPI_ISL_702299, EPI_ISL_702300, EPI_ISL_702301, EPI_ISL_702303, EPI_ISL_702305, EPI_ISL_702307, EPI_ISL_702308, EPI_ISL_702309, EPI_ISL_702311, EPI_ISL_702313, EPI_ISL_702314, EPI_ISL_702315, EPI_ISL_702316, EPI_ISL_702320, EPI_ISL_702327, EPI_ISL_702328, EPI_ISL_702333, EPI_ISL_702335, EPI_ISL_702336, EPI_ISL_702337, EPI_ISL_702341, EPI_ISL_702344, EPI_ISL_702352, EPI_ISL_702353, EPI_ISL_702358, EPI_ISL_702359, EPI_ISL_702360, EPI_ISL_702361, EPI_ISL_702368, EPI_ISL_702369, EPI_ISL_708989, EPI_ISL_708990, EPI_ISL_708992, EPI_ISL_708993, EPI_ISL_708994, EPI_ISL_708995, EPI_ISL_708996, EPI_ISL_708997, EPI_ISL_708999, EPI_ISL_709000, EPI_ISL_709001, EPI_ISL_709002, EPI_ISL_709003, EPI_ISL_709006, EPI_ISL_709007, EPI_ISL_709008, EPI_ISL_709010, EPI_ISL_709011, EPI_ISL_709014, EPI_ISL_709015, EPI_ISL_709016, EPI_ISL_709017, EPI_ISL_709018, EPI_ISL_709019, EPI_ISL_709020, EPI_ISL_709021, EPI_ISL_709023, EPI_ISL_709024, EPI_ISL_709025, EPI_ISL_709026, EPI_ISL_709027, EPI_ISL_709028, EPI_ISL_709029, EPI_ISL_709030, EPI_ISL_709032, EPI_ISL_709033, EPI_ISL_709034, EPI_ISL_709035, EPI_ISL_709037, EPI_ISL_709038, EPI_ISL_709039, EPI_ISL_709041, EPI_ISL_709042, EPI_ISL_709043, EPI_ISL_709045, EPI_ISL_709046, EPI_ISL_709047, EPI_ISL_709049, EPI_ISL_709051, EPI_ISL_709052, EPI_ISL_709053, EPI_ISL_709054, EPI_ISL_709055, EPI_ISL_709057, EPI_ISL_709059, EPI_ISL_709061, EPI_ISL_709062, EPI_ISL_709063, EPI_ISL_709064, EPI_ISL_709066, EPI_ISL_709067, EPI_ISL_709068, EPI_ISL_709069, EPI_ISL_709070, EPI_ISL_709071, EPI_ISL_709072, EPI_ISL_709073, EPI_ISL_709074, EPI_ISL_709075, EPI_ISL_709076, EPI_ISL_709080, EPI_ISL_709081, EPI_ISL_709083, EPI_ISL_709084, EPI_ISL_709085, EPI_ISL_709086, EPI_ISL_709087, EPI_ISL_709088, EPI_ISL_709089, EPI_ISL_709090, EPI_ISL_709092, EPI_ISL_709093, EPI_ISL_709094, EPI_ISL_709095, EPI_ISL_709096, EPI_ISL_709097, EPI_ISL_709099, EPI_ISL_709100, EPI_ISL_709101, EPI_ISL_709102, EPI_ISL_709103, EPI_ISL_709105, EPI_ISL_709106, EPI_ISL_709111, EPI_ISL_709112, EPI_ISL_709113, EPI_ISL_709114, EPI_ISL_709115, EPI_ISL_709116, EPI_ISL_709117, EPI_ISL_709118, EPI_ISL_709120, EPI_ISL_709121, EPI_ISL_709122, EPI_ISL_709123, EPI_ISL_709124, EPI_ISL_709125, EPI_ISL_709127, EPI_ISL_709128, EPI_ISL_709129, EPI_ISL_709130, EPI_ISL_709134, EPI_ISL_709136, EPI_ISL_709137, EPI_ISL_709138, EPI_ISL_709143, EPI_ISL_709145, EPI_ISL_709146, EPI_ISL_709147, EPI_ISL_709149, EPI_ISL_709152, EPI_ISL_709153, EPI_ISL_709154, EPI_ISL_709155, EPI_ISL_709156, EPI_ISL_709157, EPI_ISL_709158, EPI_ISL_709162, EPI_ISL_709163, EPI_ISL_709164, EPI_ISL_709165, EPI_ISL_709166, EPI_ISL_709167, EPI_ISL_709168, EPI_ISL_709169, EPI_ISL_709172, EPI_ISL_709173, EPI_ISL_709175, EPI_ISL_709176, EPI_ISL_709177, EPI_ISL_709178, EPI_ISL_709179, EPI_ISL_709180, EPI_ISL_709181, EPI_ISL_709184, EPI_ISL_709185, EPI_ISL_709186, EPI_ISL_709187, EPI_ISL_709188, EPI_ISL_709192, EPI_ISL_709193, EPI_ISL_709194, EPI_ISL_709195, EPI_ISL_709196, EPI_ISL_709197, EPI_ISL_709198, EPI_ISL_709200, EPI_ISL_709203, EPI_ISL_709205, EPI_ISL_709206, EPI_ISL_709207, EPI_ISL_709208, EPI_ISL_709209, EPI_ISL_709210, EPI_ISL_709211, EPI_ISL_709212, EPI_ISL_709214, EPI_ISL_709216, EPI_ISL_709217, EPI_ISL_709218, EPI_ISL_709219, EPI_ISL_709220, EPI_ISL_709222, EPI_ISL_709223, EPI_ISL_709224, EPI_ISL_709225, EPI_ISL_709226, EPI_ISL_709227, EPI_ISL_709228, EPI_ISL_709229, EPI_ISL_709230, EPI_ISL_709232, EPI_ISL_709233, EPI_ISL_709237, EPI_ISL_709238, EPI_ISL_709240, EPI_ISL_709241, EPI_ISL_709244, EPI_ISL_709246, EPI_ISL_709247, EPI_ISL_709249, EPI_ISL_709250, EPI_ISL_709251, EPI_ISL_709252, EPI_ISL_709253, EPI_ISL_709254, EPI_ISL_709255, EPI_ISL_709256, EPI_ISL_709257, EPI_ISL_709258, EPI_ISL_709259, EPI_ISL_709260, | | | |

| | | | |
|---|---|---|---|
| EPI_ISL_709261, EPI_ISL_709262, EPI_ISL_709263, EPI_ISL_709265, EPI_ISL_709266, EPI_ISL_709267, EPI_ISL_709268, EPI_ISL_709269, EPI_ISL_709270, EPI_ISL_709271, EPI_ISL_709272, EPI_ISL_709273, EPI_ISL_709275, EPI_ISL_709276, EPI_ISL_709278, EPI_ISL_709279, EPI_ISL_709280, EPI_ISL_709281, EPI_ISL_709284, EPI_ISL_709286, EPI_ISL_709287, EPI_ISL_709289, EPI_ISL_709293, EPI_ISL_709294, EPI_ISL_709804, EPI_ISL_709805, EPI_ISL_709807, EPI_ISL_709808, EPI_ISL_709810, EPI_ISL_709813, EPI_ISL_709814, EPI_ISL_709817, EPI_ISL_709821, EPI_ISL_709822, EPI_ISL_709825, EPI_ISL_709827, EPI_ISL_709829, EPI_ISL_709836, EPI_ISL_709837, EPI_ISL_709842, EPI_ISL_709843, EPI_ISL_709844, EPI_ISL_709846, EPI_ISL_709850, EPI_ISL_709854, EPI_ISL_709864, EPI_ISL_709865, EPI_ISL_709867, EPI_ISL_709869, EPI_ISL_709870, EPI_ISL_709873, EPI_ISL_709876, EPI_ISL_709877, EPI_ISL_709878, EPI_ISL_709880, EPI_ISL_733596, EPI_ISL_733597, EPI_ISL_733599, EPI_ISL_733601, EPI_ISL_733602, EPI_ISL_733604, EPI_ISL_733606, EPI_ISL_733608, EPI_ISL_733610, EPI_ISL_733612, EPI_ISL_733614, EPI_ISL_733615, EPI_ISL_733619, EPI_ISL_733620, EPI_ISL_733622, EPI_ISL_733624, EPI_ISL_733626, EPI_ISL_733627, EPI_ISL_733629, EPI_ISL_733631, EPI_ISL_733632, EPI_ISL_733633, EPI_ISL_733634, EPI_ISL_733637, EPI_ISL_733640, EPI_ISL_733646, EPI_ISL_733649, EPI_ISL_733650, EPI_ISL_733651, EPI_ISL_733655, EPI_ISL_733656, EPI_ISL_733660, EPI_ISL_733661, EPI_ISL_733662, EPI_ISL_733663, EPI_ISL_733665, EPI_ISL_733667, EPI_ISL_733668, EPI_ISL_733670, EPI_ISL_733671, EPI_ISL_733672, EPI_ISL_733674, EPI_ISL_733675, EPI_ISL_733676, EPI_ISL_733677, EPI_ISL_733680, EPI_ISL_733681, EPI_ISL_733684, EPI_ISL_733688, EPI_ISL_733689, EPI_ISL_733690, EPI_ISL_733691, EPI_ISL_733694, EPI_ISL_733695, EPI_ISL_733696, EPI_ISL_733698, EPI_ISL_733700, EPI_ISL_733702, EPI_ISL_733704, EPI_ISL_733705, EPI_ISL_733708, EPI_ISL_733709, EPI_ISL_733714, EPI_ISL_733715, EPI_ISL_733716, EPI_ISL_733717, EPI_ISL_733719, EPI_ISL_733720, EPI_ISL_733721, EPI_ISL_733722, EPI_ISL_733723, EPI_ISL_733724, EPI_ISL_733726, EPI_ISL_733727, EPI_ISL_733728, EPI_ISL_733729, EPI_ISL_733730, EPI_ISL_733732, EPI_ISL_733733, EPI_ISL_733736, EPI_ISL_733738, EPI_ISL_733740, EPI_ISL_733741, EPI_ISL_733744, EPI_ISL_733745, EPI_ISL_733750, EPI_ISL_733751, EPI_ISL_733752, EPI_ISL_733753, EPI_ISL_733758, EPI_ISL_733763, EPI_ISL_733765, EPI_ISL_733767, EPI_ISL_733770, EPI_ISL_733772, EPI_ISL_733773, EPI_ISL_733774, EPI_ISL_733775, EPI_ISL_733778, EPI_ISL_733779, EPI_ISL_733782, EPI_ISL_733784, EPI_ISL_733786, EPI_ISL_733787, EPI_ISL_733788, EPI_ISL_733790, EPI_ISL_733791, EPI_ISL_733792, EPI_ISL_733793, EPI_ISL_733799, EPI_ISL_733800, EPI_ISL_733802, EPI_ISL_733804, EPI_ISL_733805, EPI_ISL_733809, EPI_ISL_733810, EPI_ISL_733812, EPI_ISL_733814, EPI_ISL_733816, EPI_ISL_733817, EPI_ISL_733821, EPI_ISL_733822, EPI_ISL_733823, EPI_ISL_733824, EPI_ISL_733827, EPI_ISL_733831, EPI_ISL_733834, EPI_ISL_733835, EPI_ISL_733839, EPI_ISL_733840, EPI_ISL_733841, EPI_ISL_733846, EPI_ISL_733847, EPI_ISL_733849, EPI_ISL_733852, EPI_ISL_733853, EPI_ISL_733854, EPI_ISL_733856, EPI_ISL_733857, EPI_ISL_733860, EPI_ISL_733862, EPI_ISL_733866, EPI_ISL_733868, EPI_ISL_733869, EPI_ISL_733870, EPI_ISL_733871, EPI_ISL_733874, EPI_ISL_733876, EPI_ISL_733878, EPI_ISL_733881, EPI_ISL_733883, EPI_ISL_733886, EPI_ISL_733888, EPI_ISL_733892, EPI_ISL_733895, EPI_ISL_733896, EPI_ISL_733898, EPI_ISL_733899, EPI_ISL_733906, EPI_ISL_733907, EPI_ISL_733908, EPI_ISL_733909, EPI_ISL_733910, EPI_ISL_733911, EPI_ISL_733915, EPI_ISL_760813, EPI_ISL_760822, EPI_ISL_760833, EPI_ISL_760858, EPI_ISL_760859, EPI_ISL_760894, EPI_ISL_760903, EPI_ISL_760920, EPI_ISL_760922, EPI_ISL_760932, EPI_ISL_760936, EPI_ISL_760939, EPI_ISL_760969, EPI_ISL_760976, EPI_ISL_760988, EPI_ISL_761017, EPI_ISL_761021, EPI_ISL_761057, EPI_ISL_761074, EPI_ISL_761075, EPI_ISL_777201, EPI_ISL_777323, EPI_ISL_782739, EPI_ISL_782746, EPI_ISL_782757, EPI_ISL_782759, EPI_ISL_782761, EPI_ISL_782763, EPI_ISL_782764, EPI_ISL_782769, EPI_ISL_782776, EPI_ISL_782777, EPI_ISL_782780, EPI_ISL_782788, EPI_ISL_782796, EPI_ISL_782797, EPI_ISL_782800, EPI_ISL_782803, EPI_ISL_782809, EPI_ISL_782810, EPI_ISL_782814, EPI_ISL_782816, EPI_ISL_782817, EPI_ISL_782818, EPI_ISL_782822, EPI_ISL_782830, EPI_ISL_782834, EPI_ISL_782840, EPI_ISL_782842, EPI_ISL_782846, EPI_ISL_782850, EPI_ISL_782853, EPI_ISL_782854, EPI_ISL_782857, EPI_ISL_782860, EPI_ISL_782862, EPI_ISL_782866, EPI_ISL_782867, EPI_ISL_782868, EPI_ISL_782880, EPI_ISL_782882, EPI_ISL_782883, EPI_ISL_782885, EPI_ISL_782886, EPI_ISL_782887, EPI_ISL_782889, EPI_ISL_782890, EPI_ISL_782907, EPI_ISL_782908, EPI_ISL_782909, EPI_ISL_782910, EPI_ISL_782912, EPI_ISL_782913, EPI_ISL_782920, EPI_ISL_782923, EPI_ISL_782928, EPI_ISL_782929, EPI_ISL_782930, EPI_ISL_782932, EPI_ISL_782937, EPI_ISL_782941, EPI_ISL_782942, EPI_ISL_782947, EPI_ISL_782954, EPI_ISL_782959, EPI_ISL_782960, EPI_ISL_782966, EPI_ISL_782971, EPI_ISL_782974, EPI_ISL_782979, EPI_ISL_782983, EPI_ISL_782984, EPI_ISL_782985, EPI_ISL_782987, EPI_ISL_782991, EPI_ISL_782996, EPI_ISL_782998, EPI_ISL_783012, EPI_ISL_783019, EPI_ISL_783023, EPI_ISL_783028, EPI_ISL_783031, EPI_ISL_783036, EPI_ISL_783037, EPI_ISL_783038, EPI_ISL_783042, EPI_ISL_783044, EPI_ISL_783051, EPI_ISL_783055, EPI_ISL_783057, EPI_ISL_783066, EPI_ISL_800089, EPI_ISL_800157, EPI_ISL_800203, EPI_ISL_800586, EPI_ISL_800587, EPI_ISL_800589, EPI_ISL_800590, EPI_ISL_800592, EPI_ISL_800597, EPI_ISL_800600, EPI_ISL_800606, EPI_ISL_800609, EPI_ISL_800610, EPI_ISL_800615, EPI_ISL_800622, EPI_ISL_800626, EPI_ISL_800637, EPI_ISL_800641, EPI_ISL_800645, EPI_ISL_800647, EPI_ISL_800648, EPI_ISL_800649, EPI_ISL_800658, EPI_ISL_800663, EPI_ISL_800665, EPI_ISL_800669, EPI_ISL_800672, EPI_ISL_800679, EPI_ISL_800680, EPI_ISL_800682, EPI_ISL_800683, EPI_ISL_800686, EPI_ISL_800687, EPI_ISL_800690, EPI_ISL_800702, EPI_ISL_800707, EPI_ISL_800708, EPI_ISL_800709, EPI_ISL_800712, EPI_ISL_800714, EPI_ISL_800716, EPI_ISL_800717, EPI_ISL_800720, EPI_ISL_800721, EPI_ISL_800724, EPI_ISL_800730, EPI_ISL_800732, EPI_ISL_800733, EPI_ISL_800734, EPI_ISL_800737, EPI_ISL_800744, EPI_ISL_800746, EPI_ISL_800755, EPI_ISL_800761, EPI_ISL_800763, EPI_ISL_800768, EPI_ISL_800770, EPI_ISL_800771, EPI_ISL_800776, EPI_ISL_800784, EPI_ISL_800791, EPI_ISL_800792, EPI_ISL_800793, EPI_ISL_800795, EPI_ISL_800798, EPI_ISL_800802, EPI_ISL_800803, EPI_ISL_800806, EPI_ISL_800808, EPI_ISL_800810, EPI_ISL_800816, EPI_ISL_800817, EPI_ISL_800819, EPI_ISL_800828, EPI_ISL_800832, EPI_ISL_800839, EPI_ISL_800842, EPI_ISL_800853, EPI_ISL_800854, EPI_ISL_800855, EPI_ISL_800859, EPI_ISL_800865, EPI_ISL_800870, EPI_ISL_800872, EPI_ISL_800873, EPI_ISL_800876, EPI_ISL_800883 | | | |
| see above | Lighthouse Lab in Cambridge | Wellcome Sanger Institute for the COVID-19 Genomics UK (COG-UK) Consortium | Cordelia Langford; David K. Jackson; Dominic Kwiatkowski; Ewan Harrison; Ian Johnston; John Sillitoe on behalf of the Wellcome Sanger Institute COVID-19 Surveillance Team; Rob Howes; Roberto Amato; Sonia Goncalves; The Lighthouse Lab in Cambridge and Alex Alderton |
| EPI_ISL_708855, EPI_ISL_708884, EPI_ISL_708890, EPI_ISL_708893, EPI_ISL_708904, EPI_ISL_708905, EPI_ISL_708906, EPI_ISL_708910, EPI_ISL_708921, EPI_ISL_708922, EPI_ISL_708925, EPI_ISL_708933, EPI_ISL_708944, EPI_ISL_708953, EPI_ISL_708957, EPI_ISL_708985, EPI_ISL_708991, EPI_ISL_708998, EPI_ISL_709004, EPI_ISL_709005, EPI_ISL_709009, EPI_ISL_709013, EPI_ISL_709022, EPI_ISL_709031, EPI_ISL_709036, EPI_ISL_709040, EPI_ISL_709048, EPI_ISL_709050, EPI_ISL_709056, EPI_ISL_709058, EPI_ISL_709060, EPI_ISL_709065, EPI_ISL_709077, EPI_ISL_709078, EPI_ISL_709079, EPI_ISL_709082, EPI_ISL_709091, EPI_ISL_709098, EPI_ISL_709104, EPI_ISL_709107, EPI_ISL_709108, EPI_ISL_709109, EPI_ISL_709110, EPI_ISL_709119, EPI_ISL_709126, EPI_ISL_709131, EPI_ISL_709132, EPI_ISL_709133, EPI_ISL_709135, EPI_ISL_709139, EPI_ISL_709140, EPI_ISL_709141, EPI_ISL_709142, EPI_ISL_709144, EPI_ISL_709148, EPI_ISL_709150, EPI_ISL_709151, EPI_ISL_709159, EPI_ISL_709160, EPI_ISL_709161, EPI_ISL_709170, EPI_ISL_709171, EPI_ISL_709174, EPI_ISL_709182, EPI_ISL_709183, EPI_ISL_709189, EPI_ISL_709190, EPI_ISL_709191, EPI_ISL_709199, EPI_ISL_709201, EPI_ISL_709202, EPI_ISL_709215, EPI_ISL_709231, EPI_ISL_709234, EPI_ISL_709235, EPI_ISL_709236, EPI_ISL_709239, EPI_ISL_709242, EPI_ISL_709243, EPI_ISL_709245, EPI_ISL_709248, EPI_ISL_709264, EPI_ISL_709274, EPI_ISL_709277, EPI_ISL_709282, EPI_ISL_709283, EPI_ISL_709285, EPI_ISL_709288, EPI_ISL_709290, EPI_ISL_709291, EPI_ISL_709292, EPI_ISL_709301, EPI_ISL_709307, EPI_ISL_709311, EPI_ISL_709312, EPI_ISL_709314, EPI_ISL_709320, EPI_ISL_709324, EPI_ISL_709327, EPI_ISL_709332, EPI_ISL_709334, EPI_ISL_709336, EPI_ISL_709341, EPI_ISL_709343, EPI_ISL_709345, EPI_ISL_709353, EPI_ISL_709354, EPI_ISL_709358, EPI_ISL_709359, EPI_ISL_709360, EPI_ISL_709364, EPI_ISL_709365, EPI_ISL_709366, EPI_ISL_709367, EPI_ISL_709373, EPI_ISL_709376, EPI_ISL_709378, EPI_ISL_709380, EPI_ISL_709383, EPI_ISL_709384, EPI_ISL_709388, EPI_ISL_709392, EPI_ISL_709394, EPI_ISL_709395, EPI_ISL_709396, EPI_ISL_709399, EPI_ISL_709403, EPI_ISL_709404, EPI_ISL_709406, EPI_ISL_709407, EPI_ISL_709409, EPI_ISL_709411, EPI_ISL_709412, EPI_ISL_709417, EPI_ISL_709419, EPI_ISL_709422, EPI_ISL_709423, EPI_ISL_709424, EPI_ISL_709426, EPI_ISL_709429, EPI_ISL_709436, EPI_ISL_709438, EPI_ISL_709439, EPI_ISL_709440, EPI_ISL_709441, EPI_ISL_709443, EPI_ISL_709446, EPI_ISL_709450, EPI_ISL_709453, EPI_ISL_709454, EPI_ISL_709456, EPI_ISL_709457, EPI_ISL_709458, EPI_ISL_709459, EPI_ISL_709462, EPI_ISL_709464, EPI_ISL_709465, EPI_ISL_709466, EPI_ISL_709469, EPI_ISL_709472, EPI_ISL_709474, EPI_ISL_709481, EPI_ISL_709482, EPI_ISL_709483, EPI_ISL_709486, EPI_ISL_709487, EPI_ISL_709490, EPI_ISL_709493, EPI_ISL_709494, EPI_ISL_709495, EPI_ISL_709496, EPI_ISL_709497, EPI_ISL_709498, EPI_ISL_709500, EPI_ISL_709502, EPI_ISL_709503, EPI_ISL_709509, EPI_ISL_709510, EPI_ISL_709511, EPI_ISL_709515, EPI_ISL_709517, EPI_ISL_709518, EPI_ISL_709522, EPI_ISL_709524, EPI_ISL_709525, EPI_ISL_709527, EPI_ISL_709528, EPI_ISL_709530, EPI_ISL_709533, EPI_ISL_709537, EPI_ISL_709539, EPI_ISL_709540, EPI_ISL_709541, EPI_ISL_709549, EPI_ISL_709555, EPI_ISL_709556, EPI_ISL_709557, EPI_ISL_709558, EPI_ISL_709561, EPI_ISL_709565, EPI_ISL_709800, EPI_ISL_709806, EPI_ISL_709809, EPI_ISL_709811, EPI_ISL_709812, EPI_ISL_709816, EPI_ISL_709818, EPI_ISL_709820, EPI_ISL_709826, EPI_ISL_709831, EPI_ISL_709832, EPI_ISL_709834, EPI_ISL_709835, EPI_ISL_709840, EPI_ISL_709845, EPI_ISL_709848, EPI_ISL_709849, EPI_ISL_709851, EPI_ISL_709855, EPI_ISL_709857, EPI_ISL_709858, EPI_ISL_709859, EPI_ISL_709861, EPI_ISL_709862, EPI_ISL_709875, EPI_ISL_710615, EPI_ISL_710619, EPI_ISL_710621, EPI_ISL_710622, EPI_ISL_710623, EPI_ISL_710624, EPI_ISL_710626, EPI_ISL_710627, EPI_ISL_710629, EPI_ISL_710630, EPI_ISL_710633, EPI_ISL_710634, EPI_ISL_710635, EPI_ISL_710637, EPI_ISL_710639, EPI_ISL_710640, EPI_ISL_710641, EPI_ISL_710642, EPI_ISL_710643, EPI_ISL_710644, EPI_ISL_710646, EPI_ISL_710649, EPI_ISL_710650, EPI_ISL_710651, EPI_ISL_710652, EPI_ISL_710653, EPI_ISL_710654, EPI_ISL_710655, EPI_ISL_710657, EPI_ISL_710658, EPI_ISL_710659, EPI_ISL_710660, EPI_ISL_710664, EPI_ISL_710666, EPI_ISL_710667, EPI_ISL_710668, EPI_ISL_710669, EPI_ISL_710670, EPI_ISL_710672, EPI_ISL_710673, EPI_ISL_710674, EPI_ISL_710675, EPI_ISL_710677, EPI_ISL_710678, EPI_ISL_710679, EPI_ISL_710680, EPI_ISL_710681, EPI_ISL_710683, EPI_ISL_710684, EPI_ISL_710685, EPI_ISL_710686, EPI_ISL_710687, EPI_ISL_710688, EPI_ISL_710690, EPI_ISL_710691, EPI_ISL_710692, EPI_ISL_710695, EPI_ISL_710696, EPI_ISL_710697, EPI_ISL_720668, EPI_ISL_720672, EPI_ISL_720684, EPI_ISL_720704, EPI_ISL_720726, EPI_ISL_720728, EPI_ISL_720732, EPI_ISL_720733, EPI_ISL_720738, EPI_ISL_720740, EPI_ISL_720749, EPI_ISL_720752, EPI_ISL_720755, EPI_ISL_720761, EPI_ISL_720767, EPI_ISL_720776, EPI_ISL_720786, EPI_ISL_720787, EPI_ISL_720792, EPI_ISL_720802, EPI_ISL_720805, EPI_ISL_720811, EPI_ISL_720819, EPI_ISL_720831, EPI_ISL_720834, EPI_ISL_720837, EPI_ISL_720841, EPI_ISL_720850, EPI_ISL_720856, EPI_ISL_720867, EPI_ISL_720870, EPI_ISL_720877, EPI_ISL_720889, EPI_ISL_720897, EPI_ISL_720904, EPI_ISL_721519, EPI_ISL_721521, EPI_ISL_721524, EPI_ISL_730723, EPI_ISL_730808, EPI_ISL_731261, EPI_ISL_731262, EPI_ISL_731264, EPI_ISL_731283, EPI_ISL_731284, EPI_ISL_731290, EPI_ISL_731293, EPI_ISL_731299, EPI_ISL_731306, EPI_ISL_731333, EPI_ISL_731334, EPI_ISL_731349, EPI_ISL_731353, EPI_ISL_731378, EPI_ISL_731393, EPI_ISL_731396, EPI_ISL_731398, EPI_ISL_731400, EPI_ISL_731405, EPI_ISL_731421, EPI_ISL_731433, EPI_ISL_731438, EPI_ISL_731456, EPI_ISL_731459, EPI_ISL_731466, EPI_ISL_731479, EPI_ISL_731522, EPI_ISL_731535, EPI_ISL_731541, EPI_ISL_733928, EPI_ISL_733950, EPI_ISL_734041, EPI_ISL_734044, EPI_ISL_734093, EPI_ISL_734117, EPI_ISL_734131, EPI_ISL_736458, EPI_ISL_736477, EPI_ISL_757226, EPI_ISL_757227, EPI_ISL_757229, EPI_ISL_757231, EPI_ISL_757232, EPI_ISL_757233, EPI_ISL_757234, EPI_ISL_757235, EPI_ISL_757236, EPI_ISL_757237, EPI_ISL_757238, EPI_ISL_757239, EPI_ISL_757240, EPI_ISL_757241, EPI_ISL_757242, EPI_ISL_757243, EPI_ISL_757244, EPI_ISL_757245, EPI_ISL_757246, EPI_ISL_757247, EPI_ISL_757248, EPI_ISL_757249, EPI_ISL_757250, EPI_ISL_757251, EPI_ISL_757252, EPI_ISL_757253, EPI_ISL_757254, EPI_ISL_757255, EPI_ISL_757256, EPI_ISL_757257, EPI_ISL_757258, EPI_ISL_757259, EPI_ISL_757260, EPI_ISL_757261, EPI_ISL_757262, EPI_ISL_757263, EPI_ISL_757264, EPI_ISL_757265, EPI_ISL_757266, EPI_ISL_757267, EPI_ISL_757268, EPI_ISL_757269, EPI_ISL_757270, EPI_ISL_757271, EPI_ISL_757272, EPI_ISL_760866, EPI_ISL_760872, EPI_ISL_760876, EPI_ISL_760986, EPI_ISL_761043, EPI_ISL_761047, EPI_ISL_761067, EPI_ISL_777013, EPI_ISL_777081, EPI_ISL_777101, EPI_ISL_777142, EPI_ISL_777192, EPI_ISL_782486, EPI_ISL_782494, EPI_ISL_782500, EPI_ISL_782504, EPI_ISL_782512, EPI_ISL_782536, EPI_ISL_782538, EPI_ISL_782568, EPI_ISL_782580, EPI_ISL_782584, EPI_ISL_782596, EPI_ISL_782605, EPI_ISL_782615, EPI_ISL_782618, EPI_ISL_782624, EPI_ISL_782631, EPI_ISL_782633, EPI_ISL_782674, EPI_ISL_782679, EPI_ISL_782689, EPI_ISL_782710, EPI_ISL_800591, EPI_ISL_800594, EPI_ISL_800596, EPI_ISL_800601, EPI_ISL_800602, EPI_ISL_800603, EPI_ISL_800604, EPI_ISL_800605, EPI_ISL_800607, EPI_ISL_800608, EPI_ISL_800611, EPI_ISL_800612, EPI_ISL_800614, EPI_ISL_800616, EPI_ISL_800618, EPI_ISL_800620, EPI_ISL_800621, EPI_ISL_800625, EPI_ISL_800627, EPI_ISL_800628, EPI_ISL_800629, EPI_ISL_800630, EPI_ISL_800631, EPI_ISL_800633, EPI_ISL_800634, EPI_ISL_800635, EPI_ISL_800638, EPI_ISL_800639, EPI_ISL_800640, EPI_ISL_800643, EPI_ISL_800646, EPI_ISL_800651, EPI_ISL_800653, EPI_ISL_800654, EPI_ISL_800655, EPI_ISL_800656, EPI_ISL_800659, EPI_ISL_800662, EPI_ISL_800666, EPI_ISL_800667, EPI_ISL_800668, EPI_ISL_800670, EPI_ISL_800671, EPI_ISL_800675, EPI_ISL_800685, EPI_ISL_800688, EPI_ISL_800689, EPI_ISL_800691, EPI_ISL_800692, EPI_ISL_800693, EPI_ISL_800694, EPI_ISL_800695, EPI_ISL_800696, EPI_ISL_800697, EPI_ISL_800699, EPI_ISL_800700, EPI_ISL_800703, EPI_ISL_800704, EPI_ISL_800705, EPI_ISL_800706, EPI_ISL_800715, EPI_ISL_800719, EPI_ISL_800722, EPI_ISL_800723, EPI_ISL_800725, EPI_ISL_800726, EPI_ISL_800727, EPI_ISL_800728, EPI_ISL_800731, EPI_ISL_800735, EPI_ISL_800736, EPI_ISL_800739, EPI_ISL_800742, EPI_ISL_800745, EPI_ISL_800748, EPI_ISL_800749, EPI_ISL_800753, EPI_ISL_800754, EPI_ISL_800759, EPI_ISL_800760, EPI_ISL_800765, EPI_ISL_800766, EPI_ISL_800767, EPI_ISL_800772, EPI_ISL_800775, EPI_ISL_800779, EPI_ISL_800780, EPI_ISL_800781, EPI_ISL_800785, EPI_ISL_800786, EPI_ISL_800787, EPI_ISL_800790, EPI_ISL_800794, EPI_ISL_800796, EPI_ISL_800797, EPI_ISL_800799, EPI_ISL_800800, EPI_ISL_800801, EPI_ISL_800804, EPI_ISL_800805, EPI_ISL_800809, EPI_ISL_800812, EPI_ISL_800813, EPI_ISL_800815, EPI_ISL_800818, EPI_ISL_800821, EPI_ISL_800823, EPI_ISL_800825, EPI_ISL_800826, EPI_ISL_800827, EPI_ISL_800829, EPI_ISL_800833, EPI_ISL_800835, EPI_ISL_800838, EPI_ISL_800841, EPI_ISL_800843, EPI_ISL_800847, EPI_ISL_800848, EPI_ISL_800850, EPI_ISL_800851, EPI_ISL_800852, EPI_ISL_800856, EPI_ISL_800858, EPI_ISL_800860, EPI_ISL_800861, EPI_ISL_800862, EPI_ISL_800869, EPI_ISL_800874, EPI_ISL_800875, EPI_ISL_800880, EPI_ISL_800882, EPI_ISL_800884, EPI_ISL_859541, EPI_ISL_859542, EPI_ISL_859543, EPI_ISL_859544, EPI_ISL_859545, EPI_ISL_859546, EPI_ISL_859547, EPI_ISL_859548, EPI_ISL_859549, EPI_ISL_859550, EPI_ISL_859551, EPI_ISL_859552, EPI_ISL_933493, EPI_ISL_933494, EPI_ISL_989989, EPI_ISL_989990, EPI_ISL_989993, EPI_ISL_1115111 | | | |
| see above | Lighthouse Lab in Glasgow | Wellcome Sanger Institute for the COVID-19 Genomics UK (COG-UK) Consortium | Anna Dominiczak and Alex Alderton; Carol Clugston; Cordelia Langford; David Gray; David K. Jackson; Dominic Kwiatkowski; Ewan Harrison; Harper VanSteenhouse; Ian Johnston; Jeffrey Barrett; John Sillitoe on behalf of the Wellcome Sanger Institute COVID-19 Surveillance Team; Roberto Amato; Sonia Goncalves; Yumi Kasai |
| EPI_ISL_708844, EPI_ISL_708847, EPI_ISL_708848, EPI_ISL_708849, EPI_ISL_708850, EPI_ISL_708856, EPI_ISL_708857, EPI_ISL_708858, EPI_ISL_708862, EPI_ISL_708869, EPI_ISL_708871, EPI_ISL_708879, EPI_ISL_708881, EPI_ISL_708883, EPI_ISL_708889, EPI_ISL_708896, EPI_ISL_708897, EPI_ISL_708898, EPI_ISL_708900, EPI_ISL_708901, EPI_ISL_708908, EPI_ISL_708912, EPI_ISL_708913, EPI_ISL_708916, EPI_ISL_708920, EPI_ISL_708928, EPI_ISL_708930, EPI_ISL_708941, EPI_ISL_708945, EPI_ISL_708948, EPI_ISL_708951, EPI_ISL_708958, EPI_ISL_708963, EPI_ISL_708964, EPI_ISL_708969, EPI_ISL_708975, EPI_ISL_708979, EPI_ISL_708983, EPI_ISL_708984, EPI_ISL_708986, EPI_ISL_709801, EPI_ISL_709802, EPI_ISL_709803, EPI_ISL_709815, EPI_ISL_709819, EPI_ISL_709823, EPI_ISL_709824, EPI_ISL_709828, EPI_ISL_709830, EPI_ISL_709833, EPI_ISL_709838, EPI_ISL_709839, EPI_ISL_709841, EPI_ISL_709847, EPI_ISL_709852, EPI_ISL_709853, EPI_ISL_709856, EPI_ISL_709860, EPI_ISL_709863, EPI_ISL_709866, EPI_ISL_709868, EPI_ISL_709871, EPI_ISL_709872, EPI_ISL_709874, EPI_ISL_709879, EPI_ISL_709881, EPI_ISL_720669, EPI_ISL_720671, EPI_ISL_720674, EPI_ISL_720675, EPI_ISL_720676, EPI_ISL_720677, EPI_ISL_720678, EPI_ISL_720679, EPI_ISL_720680, EPI_ISL_720681, EPI_ISL_720682, EPI_ISL_720687, EPI_ISL_720689, EPI_ISL_720690, EPI_ISL_720691, EPI_ISL_720692, EPI_ISL_720693, EPI_ISL_720696, EPI_ISL_720698, EPI_ISL_720699, EPI_ISL_720700, EPI_ISL_720701, EPI_ISL_720703, EPI_ISL_720705, EPI_ISL_720706, EPI_ISL_720707, EPI_ISL_720708, EPI_ISL_720710, EPI_ISL_720712, EPI_ISL_720714, EPI_ISL_720716, EPI_ISL_720717, EPI_ISL_720718, EPI_ISL_720719, EPI_ISL_720720, EPI_ISL_720721, EPI_ISL_720723, EPI_ISL_720724, EPI_ISL_720725, EPI_ISL_720727, EPI_ISL_720729, EPI_ISL_720730, EPI_ISL_720731, EPI_ISL_720735, EPI_ISL_720736, EPI_ISL_720737, EPI_ISL_720739, EPI_ISL_720742, EPI_ISL_720743, EPI_ISL_720744, EPI_ISL_720746, EPI_ISL_720747, EPI_ISL_720748, EPI_ISL_720751, EPI_ISL_720753, EPI_ISL_720754, EPI_ISL_720757, EPI_ISL_720758, EPI_ISL_720759, EPI_ISL_720760, EPI_ISL_720763, EPI_ISL_720766, EPI_ISL_720768, EPI_ISL_720769, EPI_ISL_720771, EPI_ISL_720772, EPI_ISL_720774, EPI_ISL_720775, EPI_ISL_720777, EPI_ISL_720778, EPI_ISL_720781, EPI_ISL_720782, EPI_ISL_720783, EPI_ISL_720784, EPI_ISL_720785, EPI_ISL_720788, EPI_ISL_720789, EPI_ISL_720791, EPI_ISL_720793, EPI_ISL_720794, EPI_ISL_720795, EPI_ISL_720797, EPI_ISL_720798, EPI_ISL_720799, EPI_ISL_720800, EPI_ISL_720801, EPI_ISL_720803, EPI_ISL_720804, EPI_ISL_720806, EPI_ISL_720807, EPI_ISL_720808, EPI_ISL_720809, EPI_ISL_720812, EPI_ISL_720813, EPI_ISL_720815, EPI_ISL_720817, EPI_ISL_720818, EPI_ISL_720820, EPI_ISL_720821, EPI_ISL_720822, EPI_ISL_720824, EPI_ISL_720826, EPI_ISL_720828, EPI_ISL_720829, EPI_ISL_720830, EPI_ISL_720832, EPI_ISL_720833, EPI_ISL_720835, EPI_ISL_720838, EPI_ISL_720839, EPI_ISL_720840, EPI_ISL_720843, EPI_ISL_720844, EPI_ISL_720847, EPI_ISL_720851, EPI_ISL_720852, EPI_ISL_720854, EPI_ISL_720855, EPI_ISL_720859, EPI_ISL_720860, EPI_ISL_720861, EPI_ISL_720862, EPI_ISL_720863, EPI_ISL_720865, EPI_ISL_720866, EPI_ISL_720868, EPI_ISL_720871, EPI_ISL_720873, EPI_ISL_720874, EPI_ISL_720876, EPI_ISL_720878, EPI_ISL_720879, EPI_ISL_720881, EPI_ISL_720882, EPI_ISL_720883, EPI_ISL_720884, EPI_ISL_720887, EPI_ISL_720888, EPI_ISL_720890, EPI_ISL_720892, EPI_ISL_720895, EPI_ISL_720896, EPI_ISL_720899, EPI_ISL_720901, EPI_ISL_720902, EPI_ISL_720903, EPI_ISL_720905, EPI_ISL_720906, EPI_ISL_720908, EPI_ISL_721526, EPI_ISL_721527, EPI_ISL_721529, EPI_ISL_721530, EPI_ISL_721532, EPI_ISL_731891, EPI_ISL_731892, EPI_ISL_731893, EPI_ISL_733600, EPI_ISL_733607, EPI_ISL_733616, EPI_ISL_733617, EPI_ISL_733628, EPI_ISL_733630, EPI_ISL_733636, EPI_ISL_733644, EPI_ISL_733645, EPI_ISL_733648, EPI_ISL_733652, EPI_ISL_733657, EPI_ISL_733658, EPI_ISL_733664, EPI_ISL_733666, EPI_ISL_733669, EPI_ISL_733673, EPI_ISL_733678, EPI_ISL_733679, EPI_ISL_733686, EPI_ISL_733687, EPI_ISL_733692, EPI_ISL_733707, EPI_ISL_733711, EPI_ISL_733713, EPI_ISL_733735, EPI_ISL_733739, EPI_ISL_733743, EPI_ISL_733746, EPI_ISL_733747, EPI_ISL_733748, EPI_ISL_733749, EPI_ISL_733756, EPI_ISL_733757, EPI_ISL_733760, EPI_ISL_733761, EPI_ISL_733764, EPI_ISL_733768, EPI_ISL_733771, EPI_ISL_733776, EPI_ISL_733780, EPI_ISL_733781, EPI_ISL_733789, EPI_ISL_733796, EPI_ISL_733798, EPI_ISL_733801, EPI_ISL_733811, EPI_ISL_733813, EPI_ISL_733820, EPI_ISL_733825, EPI_ISL_733826, EPI_ISL_733828, EPI_ISL_733829, EPI_ISL_733832, EPI_ISL_733836, EPI_ISL_733838, EPI_ISL_733842, EPI_ISL_733845, EPI_ISL_733851, EPI_ISL_733855, EPI_ISL_733858, EPI_ISL_733861, EPI_ISL_733863, EPI_ISL_733864, EPI_ISL_733865, EPI_ISL_733872, EPI_ISL_733873, EPI_ISL_733875, EPI_ISL_733884, EPI_ISL_733887, EPI_ISL_733889, EPI_ISL_733890, EPI_ISL_733891, EPI_ISL_733893, EPI_ISL_733894, EPI_ISL_733900, EPI_ISL_733902, EPI_ISL_733903, EPI_ISL_733904, EPI_ISL_733905, EPI_ISL_733914, EPI_ISL_736202, EPI_ISL_736240, EPI_ISL_736252, EPI_ISL_736264, EPI_ISL_736274, EPI_ISL_736289, EPI_ISL_736292, EPI_ISL_736293, EPI_ISL_736303, EPI_ISL_736349, EPI_ISL_736369, EPI_ISL_736380, EPI_ISL_736392, EPI_ISL_736408, EPI_ISL_736449, EPI_ISL_736469, EPI_ISL_736547, EPI_ISL_736686, EPI_ISL_736694, EPI_ISL_736749, EPI_ISL_756765, EPI_ISL_756860, EPI_ISL_756865, EPI_ISL_756876, EPI_ISL_756925, EPI_ISL_757014, EPI_ISL_760809, EPI_ISL_760810, EPI_ISL_760844, EPI_ISL_760863, EPI_ISL_760871, EPI_ISL_760873, EPI_ISL_760881, EPI_ISL_760896, EPI_ISL_760904, EPI_ISL_760905, EPI_ISL_760943, EPI_ISL_760954, EPI_ISL_760959, EPI_ISL_760961, EPI_ISL_760963, EPI_ISL_760975, EPI_ISL_760983, EPI_ISL_760987, EPI_ISL_761002, EPI_ISL_761023, EPI_ISL_761039, EPI_ISL_761050, EPI_ISL_761079, EPI_ISL_767090, EPI_ISL_767134, EPI_ISL_769545, EPI_ISL_769584, EPI_ISL_769590, EPI_ISL_769597, EPI_ISL_769646, EPI_ISL_769715, EPI_ISL_769744, EPI_ISL_769747, EPI_ISL_769764, EPI_ISL_769776, EPI_ISL_769845, EPI_ISL_769852, EPI_ISL_769857, EPI_ISL_777039, EPI_ISL_777056, EPI_ISL_777085, EPI_ISL_777102, EPI_ISL_777108, EPI_ISL_777145, EPI_ISL_777295, EPI_ISL_777313, EPI_ISL_782734, EPI_ISL_782735, EPI_ISL_782736, EPI_ISL_782738, EPI_ISL_782740, EPI_ISL_782741, EPI_ISL_782742, EPI_ISL_782743, EPI_ISL_782744, EPI_ISL_782749, EPI_ISL_782750, EPI_ISL_782751, EPI_ISL_782752, EPI_ISL_782753, EPI_ISL_782754, EPI_ISL_782755, EPI_ISL_782756, EPI_ISL_782760, EPI_ISL_782762, EPI_ISL_782765, EPI_ISL_782766, EPI_ISL_782767, EPI_ISL_782770, EPI_ISL_782772, EPI_ISL_782773, EPI_ISL_782775, EPI_ISL_782778, EPI_ISL_782779, EPI_ISL_782781, EPI_ISL_782782, EPI_ISL_782785, EPI_ISL_782786, EPI_ISL_782789, EPI_ISL_782790, EPI_ISL_782793, EPI_ISL_782794, EPI_ISL_782798, EPI_ISL_782799, EPI_ISL_782802, EPI_ISL_782804, EPI_ISL_782807, EPI_ISL_782811, EPI_ISL_782812, EPI_ISL_782815, EPI_ISL_782819, EPI_ISL_782821, EPI_ISL_782823, EPI_ISL_782824, EPI_ISL_782826, EPI_ISL_782828, EPI_ISL_782829, EPI_ISL_782831, EPI_ISL_782833, EPI_ISL_782835, EPI_ISL_782836, EPI_ISL_782837, EPI_ISL_782839, EPI_ISL_782845, EPI_ISL_782847, EPI_ISL_782848, EPI_ISL_782851, EPI_ISL_782852, EPI_ISL_782858, EPI_ISL_782859, EPI_ISL_782861, EPI_ISL_782863, EPI_ISL_782864, EPI_ISL_782865, EPI_ISL_782870, EPI_ISL_782871, EPI_ISL_782874, EPI_ISL_782875, EPI_ISL_782876, EPI_ISL_782879, EPI_ISL_782881, EPI_ISL_782884, EPI_ISL_782888, EPI_ISL_782891, EPI_ISL_782895, EPI_ISL_782896, EPI_ISL_782898, EPI_ISL_782900, EPI_ISL_782901, EPI_ISL_782906, EPI_ISL_782914, EPI_ISL_782915, EPI_ISL_782916, EPI_ISL_782917, EPI_ISL_782921, EPI_ISL_782922, EPI_ISL_782925, EPI_ISL_782927, EPI_ISL_782933, EPI_ISL_782934, EPI_ISL_782935, EPI_ISL_782938, EPI_ISL_782939, EPI_ISL_782940, EPI_ISL_782943, EPI_ISL_782944, EPI_ISL_782948, EPI_ISL_782949, EPI_ISL_782950, EPI_ISL_782951, EPI_ISL_782952, EPI_ISL_782953, EPI_ISL_782955, EPI_ISL_782957, EPI_ISL_782958, EPI_ISL_782961, EPI_ISL_782962, EPI_ISL_782965, EPI_ISL_782968, EPI_ISL_782972, EPI_ISL_782973, EPI_ISL_782975, EPI_ISL_782977, EPI_ISL_782978, EPI_ISL_782980, EPI_ISL_782981, EPI_ISL_782982, EPI_ISL_782986, EPI_ISL_782989, EPI_ISL_782990, EPI_ISL_782992, EPI_ISL_782995, EPI_ISL_782999, EPI_ISL_783001, EPI_ISL_783003, EPI_ISL_783004, EPI_ISL_783005, EPI_ISL_783006, EPI_ISL_783009, EPI_ISL_783010, EPI_ISL_783011, EPI_ISL_783013, EPI_ISL_783016, EPI_ISL_783017, EPI_ISL_783018, EPI_ISL_783020, EPI_ISL_783021, EPI_ISL_783022, EPI_ISL_783024, EPI_ISL_783025, EPI_ISL_783027, EPI_ISL_783029, EPI_ISL_783035, EPI_ISL_783039, EPI_ISL_783040, EPI_ISL_783041, EPI_ISL_783048, EPI_ISL_783049, EPI_ISL_783050, EPI_ISL_783053, EPI_ISL_783054, EPI_ISL_783058, EPI_ISL_783059, EPI_ISL_783060, EPI_ISL_783061, EPI_ISL_783062, EPI_ISL_783063, EPI_ISL_783064, EPI_ISL_783065, EPI_ISL_783067, EPI_ISL_800583, EPI_ISL_800584, EPI_ISL_800585, EPI_ISL_800588, EPI_ISL_800593, EPI_ISL_800595, EPI_ISL_800598, EPI_ISL_800599, EPI_ISL_800613, EPI_ISL_800617, EPI_ISL_800619, EPI_ISL_800623, EPI_ISL_800624, EPI_ISL_800632, EPI_ISL_800636, EPI_ISL_800642, EPI_ISL_800644, EPI_ISL_800650, EPI_ISL_800652, EPI_ISL_800657, EPI_ISL_800660, EPI_ISL_800661, EPI_ISL_800664, EPI_ISL_800673, EPI_ISL_800674, EPI_ISL_800676, EPI_ISL_800677, EPI_ISL_800678, EPI_ISL_800681, EPI_ISL_800684, EPI_ISL_800698, EPI_ISL_800701, EPI_ISL_800710, EPI_ISL_800711, EPI_ISL_800713, EPI_ISL_800718, EPI_ISL_800729, EPI_ISL_800738, EPI_ISL_800740, EPI_ISL_800741, EPI_ISL_800743, EPI_ISL_800747, EPI_ISL_800750, EPI_ISL_800751, EPI_ISL_800752, EPI_ISL_800756, EPI_ISL_800757, EPI_ISL_800758, EPI_ISL_800762, EPI_ISL_800764, EPI_ISL_800769, EPI_ISL_800773, EPI_ISL_800774, EPI_ISL_800777, EPI_ISL_800778, EPI_ISL_800782, EPI_ISL_800783, EPI_ISL_800788, EPI_ISL_800789, EPI_ISL_800807, EPI_ISL_800811, EPI_ISL_800814, EPI_ISL_800820, EPI_ISL_800822, EPI_ISL_800824, EPI_ISL_800830, EPI_ISL_800831, EPI_ISL_800834, EPI_ISL_800836, EPI_ISL_800837, EPI_ISL_800840, EPI_ISL_800844, EPI_ISL_800845, EPI_ISL_800846, EPI_ISL_800849, EPI_ISL_800857, EPI_ISL_800863, EPI_ISL_800864, EPI_ISL_800866, EPI_ISL_800867, EPI_ISL_800868, EPI_ISL_800871, EPI_ISL_800877, EPI_ISL_800878, EPI_ISL_800879, EPI_ISL_800881, EPI_ISL_801180, EPI_ISL_801181, EPI_ISL_859469, EPI_ISL_859540, EPI_ISL_859553, EPI_ISL_933496, EPI_ISL_933497, EPI_ISL_933498, EPI_ISL_933499, EPI_ISL_933500, EPI_ISL_933501, EPI_ISL_933502, EPI_ISL_933503, EPI_ISL_933504, EPI_ISL_933505, EPI_ISL_933506, EPI_ISL_933507, EPI_ISL_933508, EPI_ISL_933509 | | | |
| see above | Lighthouse Lab in Milton Keynes | Wellcome Sanger Institute for the COVID-19 Genomics UK (COG-UK) Consortium | Cordelia Langford; David K. Jackson; Dominic Kwiatkowski; Ewan Harrison; Ian Johnston; John Sillitoe on behalf of the Wellcome Sanger Institute COVID-19 Surveillance Team; Roberto Amato; Sonia Goncalves; The Lighthouse Lab in Milton Keynes and Alex Alderton |
| EPI_ISL_725596, EPI_ISL_725597, EPI_ISL_725598, EPI_ISL_725599, EPI_ISL_725600, EPI_ISL_725601, EPI_ISL_725602, EPI_ISL_725603, EPI_ISL_725604, EPI_ISL_819487, EPI_ISL_819488, EPI_ISL_819489, EPI_ISL_819493, EPI_ISL_819499, EPI_ISL_819503, EPI_ISL_819504, EPI_ISL_819506, EPI_ISL_819507, EPI_ISL_819520, EPI_ISL_819521, EPI_ISL_819522, EPI_ISL_819523, EPI_ISL_819529, EPI_ISL_819532, EPI_ISL_840017, EPI_ISL_840018, EPI_ISL_840019, EPI_ISL_840020, EPI_ISL_840021, EPI_ISL_840022, EPI_ISL_840023, EPI_ISL_840024, EPI_ISL_840025, EPI_ISL_840026, EPI_ISL_840027, EPI_ISL_840028, EPI_ISL_840029, EPI_ISL_840030, EPI_ISL_840031, EPI_ISL_840032, EPI_ISL_840033, EPI_ISL_840034, EPI_ISL_840035, EPI_ISL_840036, EPI_ISL_840037, EPI_ISL_840038, EPI_ISL_840039, EPI_ISL_840040 | | | |
| see above | Lincolnshire Hospitals and DeepSeq Nottingham | COVID-19 Genomics UK (COG-UK) Consortium | Christopher Moore; Fei Sang; Joeseph Chappell; Johnny Debebe; Jonathan Ball; Matthew Carlisle; Matthew Loose; Nadine Holmes; Nichola Duckworth; Patrick McClure; Sarah Walsh; Tim Sloan; Victoria Wright |
| EPI_ISL_702469, EPI_ISL_702484, EPI_ISL_702515, EPI_ISL_702558, EPI_ISL_702853, EPI_ISL_702872, EPI_ISL_702874, EPI_ISL_702974, EPI_ISL_703043, EPI_ISL_703292, EPI_ISL_703310, EPI_ISL_703409, EPI_ISL_703611, EPI_ISL_703850, EPI_ISL_704939, EPI_ISL_704942, EPI_ISL_704991, EPI_ISL_705021, EPI_ISL_705032, EPI_ISL_705046, EPI_ISL_705048, EPI_ISL_705051, EPI_ISL_705062, EPI_ISL_705075, EPI_ISL_705108, EPI_ISL_705126, EPI_ISL_705156, EPI_ISL_705158, EPI_ISL_705161, EPI_ISL_705174, EPI_ISL_705221, EPI_ISL_705222, EPI_ISL_705235, EPI_ISL_705245, EPI_ISL_705258, EPI_ISL_705267, EPI_ISL_705291, EPI_ISL_705292, EPI_ISL_705304, EPI_ISL_705311, EPI_ISL_705324, EPI_ISL_705345, EPI_ISL_705347, EPI_ISL_705350, EPI_ISL_705352, EPI_ISL_705389, EPI_ISL_705394, EPI_ISL_705403, EPI_ISL_705921, EPI_ISL_705922, EPI_ISL_705928, EPI_ISL_705941, EPI_ISL_724347, EPI_ISL_724348, EPI_ISL_724349, EPI_ISL_724350, EPI_ISL_724351, EPI_ISL_724352, EPI_ISL_724353, EPI_ISL_724354, EPI_ISL_724355, EPI_ISL_724356, EPI_ISL_724357, EPI_ISL_724358, EPI_ISL_724359, EPI_ISL_724360, EPI_ISL_724361, EPI_ISL_724362, EPI_ISL_724363, EPI_ISL_724364, EPI_ISL_724365, EPI_ISL_724366, EPI_ISL_724367, EPI_ISL_724368, EPI_ISL_724369, EPI_ISL_724370, EPI_ISL_724372, EPI_ISL_724373, EPI_ISL_724374, EPI_ISL_724375, EPI_ISL_724376, EPI_ISL_724377, EPI_ISL_724378, EPI_ISL_724379, EPI_ISL_724380, EPI_ISL_724381, EPI_ISL_724382, EPI_ISL_724383, EPI_ISL_724384, EPI_ISL_724385, EPI_ISL_724386, EPI_ISL_724387, EPI_ISL_724388, EPI_ISL_724389, EPI_ISL_724390, EPI_ISL_724391, EPI_ISL_724392, EPI_ISL_724393, EPI_ISL_724394, EPI_ISL_724395, EPI_ISL_724462, EPI_ISL_724463, EPI_ISL_724497, EPI_ISL_724498, EPI_ISL_724517, EPI_ISL_724519, EPI_ISL_724520, EPI_ISL_724521, EPI_ISL_724522, EPI_ISL_813677, EPI_ISL_813678, EPI_ISL_813679, EPI_ISL_813680, EPI_ISL_813681, EPI_ISL_813682, EPI_ISL_813683, EPI_ISL_813684, EPI_ISL_813685, EPI_ISL_813686, EPI_ISL_813687, EPI_ISL_813688, EPI_ISL_813689, EPI_ISL_813690, EPI_ISL_813691, EPI_ISL_813692, EPI_ISL_813693, EPI_ISL_813694, EPI_ISL_813695, EPI_ISL_813696, EPI_ISL_813697, EPI_ISL_813698, EPI_ISL_813699, EPI_ISL_813700, EPI_ISL_813701, EPI_ISL_813702, EPI_ISL_813703, EPI_ISL_813704, EPI_ISL_813705, EPI_ISL_813706, EPI_ISL_813707, EPI_ISL_813708, EPI_ISL_813709, EPI_ISL_813710, EPI_ISL_813711, EPI_ISL_813712, EPI_ISL_813713, EPI_ISL_813714, EPI_ISL_813715, EPI_ISL_813716, EPI_ISL_813717, EPI_ISL_813718, EPI_ISL_813719, EPI_ISL_813720, EPI_ISL_813721, EPI_ISL_813722, EPI_ISL_813723, EPI_ISL_813724, EPI_ISL_813725, EPI_ISL_813726, EPI_ISL_813727, EPI_ISL_813728, EPI_ISL_813729, EPI_ISL_813730, EPI_ISL_813731, EPI_ISL_813752, EPI_ISL_813753, EPI_ISL_813754, EPI_ISL_813755, EPI_ISL_813756, EPI_ISL_813757, EPI_ISL_813758, EPI_ISL_813759, EPI_ISL_813760, EPI_ISL_813761, EPI_ISL_813762, EPI_ISL_813763, EPI_ISL_838351, EPI_ISL_838353, EPI_ISL_838354, EPI_ISL_838355, EPI_ISL_838356, EPI_ISL_838357, EPI_ISL_838358, EPI_ISL_838359, EPI_ISL_838360, EPI_ISL_838361, EPI_ISL_838362, EPI_ISL_838363, EPI_ISL_838364, EPI_ISL_838365, EPI_ISL_838366, EPI_ISL_838367, EPI_ISL_838368, EPI_ISL_838369, EPI_ISL_838370, EPI_ISL_838371, EPI_ISL_838372, EPI_ISL_838373, EPI_ISL_838374, EPI_ISL_838375, EPI_ISL_838376, EPI_ISL_838377, EPI_ISL_838378, EPI_ISL_838379, EPI_ISL_838380, EPI_ISL_838381, EPI_ISL_838382, EPI_ISL_838383, EPI_ISL_838384, EPI_ISL_838385, EPI_ISL_838386, EPI_ISL_838387, EPI_ISL_838388, EPI_ISL_838389, EPI_ISL_838390, EPI_ISL_838391, EPI_ISL_838392, EPI_ISL_838393, EPI_ISL_838394, EPI_ISL_838395, EPI_ISL_838396, EPI_ISL_838397, EPI_ISL_838398, EPI_ISL_838399, EPI_ISL_838400, EPI_ISL_838401, EPI_ISL_838402, EPI_ISL_838403, EPI_ISL_838404, EPI_ISL_838405, EPI_ISL_838406, EPI_ISL_838407, EPI_ISL_838408, EPI_ISL_838409, EPI_ISL_838410, EPI_ISL_838411, EPI_ISL_838412, EPI_ISL_838413, EPI_ISL_838414, EPI_ISL_838415, EPI_ISL_838416, EPI_ISL_838417, EPI_ISL_838418, EPI_ISL_838419, EPI_ISL_838420, EPI_ISL_838421, EPI_ISL_838422, EPI_ISL_838423, EPI_ISL_838424, EPI_ISL_838425, EPI_ISL_838426, EPI_ISL_838427, EPI_ISL_838428, EPI_ISL_838429, EPI_ISL_838430, EPI_ISL_838431, EPI_ISL_838432, EPI_ISL_838433, EPI_ISL_838434, EPI_ISL_838435, EPI_ISL_838436, EPI_ISL_838437, EPI_ISL_838438, EPI_ISL_838439, EPI_ISL_838440, EPI_ISL_838441, EPI_ISL_838442, EPI_ISL_838443, EPI_ISL_838444, EPI_ISL_838445, EPI_ISL_838446, EPI_ISL_838447, EPI_ISL_838448, EPI_ISL_838449, EPI_ISL_838450, EPI_ISL_838451, EPI_ISL_838452, EPI_ISL_838453, EPI_ISL_838454, EPI_ISL_838455, EPI_ISL_838456, EPI_ISL_838457, EPI_ISL_838541, EPI_ISL_838585, EPI_ISL_838664, EPI_ISL_865207, EPI_ISL_865209, EPI_ISL_865211, EPI_ISL_865212, EPI_ISL_865213, EPI_ISL_865214, EPI_ISL_865215, EPI_ISL_865216, EPI_ISL_865261, EPI_ISL_865264, EPI_ISL_865265, EPI_ISL_865268, EPI_ISL_919445, EPI_ISL_919458, EPI_ISL_1177831, EPI_ISL_1177832, EPI_ISL_1636820, EPI_ISL_1636821, EPI_ISL_1636822, EPI_ISL_1636823, EPI_ISL_1636824 | | | |
| see above | Liverpool Clinical Laboratories | COVID-19 Genomics UK (COG-UK) Consortium | A Alrezaihi; Alessandro Gerada; Alistair Darby; Angela Cowell; Anita Lucaci; Anu Chawla; Cassie Olateju; Catherine Hartley; Charlotte Nelson; Ecaterina Vamos; Elaine O'Toole; Eleanor G Bentley; Ghada T Shawli; Hermione Webster; Isabel García-Dorival; James Johnson; James P Stewart; Jenifer Manson; Joanne Watts; Jones Benjamin; Jordan J Clark; Julian Hiscox; L Luu; Lucille Rainbow; M Almsaud; Margaret Hughes; Mark Whitehead; Matthew Gemmell; Miren Iturriza-Gomara; Muhannad Alruwaili; N.P Randle; Neil Swainston; PKF Gilmore; Parul Sharma; Rebekah Penrice-Randal; Richard Eccles; Richard Gregory; Sam Haldenby; Steve Paterson; Stuart D Armstrong; Trevor Ian Robinson; Ximeng Han |
| EPI_ISL_702520, EPI_ISL_703096, EPI_ISL_703727, EPI_ISL_704262, EPI_ISL_704564, EPI_ISL_705145, EPI_ISL_705357, EPI_ISL_705457, EPI_ISL_706280, EPI_ISL_706281, EPI_ISL_706282, EPI_ISL_706284, EPI_ISL_706285, EPI_ISL_724837, EPI_ISL_724841, EPI_ISL_724842, EPI_ISL_724845, EPI_ISL_724848, EPI_ISL_724849, EPI_ISL_724850, EPI_ISL_724851, EPI_ISL_724852, EPI_ISL_724853, EPI_ISL_724854, EPI_ISL_724877, EPI_ISL_724878, EPI_ISL_724882, EPI_ISL_724883, EPI_ISL_724884, EPI_ISL_724885, EPI_ISL_724886, EPI_ISL_724887, EPI_ISL_724888, EPI_ISL_724889, EPI_ISL_724890, EPI_ISL_724891, EPI_ISL_724892, EPI_ISL_724893, EPI_ISL_724894, EPI_ISL_724895, EPI_ISL_724896, EPI_ISL_724897, EPI_ISL_724898, EPI_ISL_724899, EPI_ISL_724900, EPI_ISL_724901, EPI_ISL_724902, EPI_ISL_724903, EPI_ISL_724904, EPI_ISL_724905, EPI_ISL_724906, EPI_ISL_724907, EPI_ISL_724908, EPI_ISL_724909, EPI_ISL_724910, EPI_ISL_724911, EPI_ISL_724912, EPI_ISL_724913, EPI_ISL_724914, EPI_ISL_724915, EPI_ISL_724916, EPI_ISL_724917, EPI_ISL_724918, EPI_ISL_724919, EPI_ISL_724920, EPI_ISL_724921, EPI_ISL_724922, EPI_ISL_724923, EPI_ISL_725020, EPI_ISL_725021, EPI_ISL_725022, EPI_ISL_725023, EPI_ISL_725024, EPI_ISL_725025, EPI_ISL_725026, EPI_ISL_725027, EPI_ISL_725028, EPI_ISL_741425, EPI_ISL_741426, EPI_ISL_741427, EPI_ISL_741428, EPI_ISL_741429, EPI_ISL_741430, EPI_ISL_741431, EPI_ISL_741432, EPI_ISL_741433, EPI_ISL_741434, EPI_ISL_741435, EPI_ISL_741436, EPI_ISL_741437, EPI_ISL_741438, EPI_ISL_741440, EPI_ISL_819568, EPI_ISL_819569, EPI_ISL_819570, EPI_ISL_819571, EPI_ISL_819572, EPI_ISL_819573, EPI_ISL_819574, EPI_ISL_839520, EPI_ISL_839521, EPI_ISL_839522, EPI_ISL_839523, EPI_ISL_839524, EPI_ISL_839525, EPI_ISL_839526, EPI_ISL_839527, EPI_ISL_839528, EPI_ISL_839529, EPI_ISL_839530, EPI_ISL_839531, EPI_ISL_839532, EPI_ISL_839533, EPI_ISL_839543, EPI_ISL_839735, EPI_ISL_839736, EPI_ISL_839737, EPI_ISL_839738, EPI_ISL_839739, EPI_ISL_839740, EPI_ISL_839741, EPI_ISL_839742, EPI_ISL_839743, EPI_ISL_839744, EPI_ISL_839745, EPI_ISL_839746, EPI_ISL_839747, EPI_ISL_839748, EPI_ISL_866427, EPI_ISL_866428, EPI_ISL_866429, EPI_ISL_866430, EPI_ISL_866431, EPI_ISL_866433, EPI_ISL_866434, EPI_ISL_866435, EPI_ISL_866436, EPI_ISL_866437, EPI_ISL_866438, EPI_ISL_866439, EPI_ISL_866440, EPI_ISL_866442, EPI_ISL_866443, EPI_ISL_866444, | | | |

| | | | |
|---|---|---|---|
| EPI_ISL_866445, EPI_ISL_866446, EPI_ISL_866447, EPI_ISL_866448, EPI_ISL_866449, EPI_ISL_866450, EPI_ISL_866451, EPI_ISL_866452, EPI_ISL_866453, EPI_ISL_866454, EPI_ISL_866455, EPI_ISL_866456, EPI_ISL_950459, EPI_ISL_1637201, EPI_ISL_1637202, EPI_ISL_1637206, EPI_ISL_1637207, EPI_ISL_1637208, EPI_ISL_1637209, EPI_ISL_1637210, EPI_ISL_1637211 | | | |
| see above | Northumbria University / South Tees Hospitals NHS Foundation Trust / North Cumbria Integrated Care NHS Foundation Trust / North Tees and Hartlepool NHS Foundation Trust / Newcastle Hospitals NHS Foundation Trust | COVID-19 Genomics UK (COG-UK) Consortium | Andrew Nelson; Brendan Payne; Clive Graham; Darren L Smith; Debra Padgett; Edward Barton; Emma Swindells; Garren Scott; Gary Black; Gary Eltringham; Giles S Holt; Greg R Young; Jane Greenaway; Jennifer Collins; John Allan; Joshua Loh; Lynn Dover; Matthew Bashton; Mohammad A Tariq; Paul Baker; Sarah Essex; Steve Liggett; Wen C Yew; Yusri Taha |
| EPI_ISL_841356, EPI_ISL_841357, EPI_ISL_841503, EPI_ISL_841504, EPI_ISL_841507, EPI_ISL_841529, EPI_ISL_841562, EPI_ISL_841572, EPI_ISL_841573, EPI_ISL_841574, EPI_ISL_841575, EPI_ISL_841576, EPI_ISL_841577, EPI_ISL_841578, EPI_ISL_841579, EPI_ISL_867198, EPI_ISL_867199, EPI_ISL_867200, EPI_ISL_867201, EPI_ISL_867271, EPI_ISL_867272, EPI_ISL_867273, EPI_ISL_867274, EPI_ISL_867275, EPI_ISL_867276, EPI_ISL_867277, EPI_ISL_867278, EPI_ISL_867279, EPI_ISL_867280, EPI_ISL_867281, EPI_ISL_867282, EPI_ISL_867283, EPI_ISL_867284, EPI_ISL_867285, EPI_ISL_867427 | | | |
| see above | Originating lab: Wales Specialist Virology Centre Sequencing lab: Pathogen Genomics Unit | Public Health Wales Microbiology Cardiff Wales Specialist Virology Centre | Alec Birchley; Alexander Adams; Amy Gaskin; Angela Marchbank; Bree Gatica-Wilcox; Catherine Moore; Jason Coombes; Joanne Watkins; Joel Southgate; Johnathan Evans; Laura Gifford; Lauren Gilbert; Lee Graham; Malorie Perry; Matthew Bull; Nicole Pacchiarini; Sally Corden; Sara Kumziene-Summerhayes; Sara Rey; Sarah Taylor; Simon Cottrell; Sophie Jones; Tom Connor |
| EPI_ISL_763934, EPI_ISL_763935, EPI_ISL_763940, EPI_ISL_763941, EPI_ISL_763945, EPI_ISL_764066, EPI_ISL_764087, EPI_ISL_764532, EPI_ISL_764533, EPI_ISL_764538, EPI_ISL_764539, EPI_ISL_840209, EPI_ISL_840222, EPI_ISL_840223, EPI_ISL_840224, EPI_ISL_840226, EPI_ISL_840249, EPI_ISL_1833043, EPI_ISL_1833046, EPI_ISL_1833047, EPI_ISL_1833049 | | | |
| see above | Oxford Viromics, NDM, University of Oxford; Oxford University Hospitals; Basingstoke and North Hampshire Hospital | COVID-19 Genomics UK (COG-UK) Consortium | Alex Mobbs; Amy Trebes; Anita Justice; Catrin Moore; Christophe Fraser; David Bonsall; David Buck; Emma Wise; George Macintyre; Jessica Lynch; John Todd; Mariateresa de Cesare; Matilde Mori; Monique Andersson; Nathan Moore; Nick Cortes; Robert Shaw; Stephen Kidd; Tanya Golubchik; Timothy Peto |
| EPI_ISL_702478, EPI_ISL_702652, EPI_ISL_702694, EPI_ISL_702700, EPI_ISL_702749, EPI_ISL_702825, EPI_ISL_702840, EPI_ISL_702873, EPI_ISL_703295, EPI_ISL_703974, EPI_ISL_703982, EPI_ISL_704070, EPI_ISL_704142, EPI_ISL_704145, EPI_ISL_704148, EPI_ISL_704176, EPI_ISL_704217, EPI_ISL_704233, EPI_ISL_704238, EPI_ISL_704609, EPI_ISL_704709, EPI_ISL_704711, EPI_ISL_704714, EPI_ISL_704716, EPI_ISL_704728, EPI_ISL_704731, EPI_ISL_704734, EPI_ISL_704736, EPI_ISL_704739, EPI_ISL_704741, EPI_ISL_704744, EPI_ISL_704746, EPI_ISL_704757, EPI_ISL_704759, EPI_ISL_704788, EPI_ISL_704791, EPI_ISL_704794, EPI_ISL_704850, EPI_ISL_704853, EPI_ISL_704855, EPI_ISL_704887, EPI_ISL_704890, EPI_ISL_704908, EPI_ISL_704921, EPI_ISL_704923, EPI_ISL_704926, EPI_ISL_704929, EPI_ISL_705091, EPI_ISL_705115, EPI_ISL_705255, EPI_ISL_705256, EPI_ISL_705261, EPI_ISL_705401, EPI_ISL_705431, EPI_ISL_706356, EPI_ISL_706357, EPI_ISL_706358, EPI_ISL_706359, EPI_ISL_706360, EPI_ISL_706367, EPI_ISL_706368, EPI_ISL_706369, EPI_ISL_706370, EPI_ISL_706371, EPI_ISL_706372, EPI_ISL_706373, EPI_ISL_706374, EPI_ISL_706375, EPI_ISL_706376, EPI_ISL_706377, EPI_ISL_706378, EPI_ISL_706379, EPI_ISL_706380, EPI_ISL_706381, EPI_ISL_706382, EPI_ISL_706397, EPI_ISL_706398, EPI_ISL_706399, EPI_ISL_706400, EPI_ISL_706401, EPI_ISL_706403, EPI_ISL_706404, EPI_ISL_706405, EPI_ISL_706406, EPI_ISL_706407, EPI_ISL_706409, EPI_ISL_706410, EPI_ISL_706411, EPI_ISL_706413, EPI_ISL_706415, EPI_ISL_706416, EPI_ISL_706417, EPI_ISL_706418, EPI_ISL_706419, EPI_ISL_725029, EPI_ISL_725030, EPI_ISL_725031, EPI_ISL_725032, EPI_ISL_725036, EPI_ISL_725037, EPI_ISL_725039, EPI_ISL_725040, EPI_ISL_725042, EPI_ISL_725043, EPI_ISL_725044, EPI_ISL_725045, EPI_ISL_725046, EPI_ISL_725047, EPI_ISL_725048, EPI_ISL_725049, EPI_ISL_725050, EPI_ISL_725051, EPI_ISL_725052, EPI_ISL_725053, EPI_ISL_725054, EPI_ISL_725056, EPI_ISL_725057, EPI_ISL_725058, EPI_ISL_725059, EPI_ISL_725060, EPI_ISL_725061, EPI_ISL_725106, EPI_ISL_725107, EPI_ISL_725108, EPI_ISL_725109, EPI_ISL_725110, EPI_ISL_725111, EPI_ISL_725138, EPI_ISL_725139, EPI_ISL_725140, EPI_ISL_725141, EPI_ISL_725142, EPI_ISL_725143, EPI_ISL_725144, EPI_ISL_725145, EPI_ISL_725146, EPI_ISL_725147, EPI_ISL_725148, EPI_ISL_725149, EPI_ISL_725150, EPI_ISL_725152, EPI_ISL_725153, EPI_ISL_725154, EPI_ISL_725156, EPI_ISL_725157, EPI_ISL_725158, EPI_ISL_725275, EPI_ISL_725302, EPI_ISL_725303, EPI_ISL_725304, EPI_ISL_725305, EPI_ISL_725306, EPI_ISL_725307, EPI_ISL_725308, EPI_ISL_725309, EPI_ISL_725310, EPI_ISL_725311, EPI_ISL_725312, EPI_ISL_725313, EPI_ISL_725314, EPI_ISL_725315, EPI_ISL_725316, EPI_ISL_725317, EPI_ISL_725318, EPI_ISL_725319, EPI_ISL_725320, EPI_ISL_725321, EPI_ISL_725322, EPI_ISL_725323, EPI_ISL_725324, EPI_ISL_725325, EPI_ISL_725326, EPI_ISL_725327, EPI_ISL_725328, EPI_ISL_725329, EPI_ISL_725330, EPI_ISL_725331, EPI_ISL_725332, EPI_ISL_725333, EPI_ISL_725334, EPI_ISL_725335, EPI_ISL_725336, EPI_ISL_725337, EPI_ISL_725338, EPI_ISL_725339, EPI_ISL_725340, EPI_ISL_725341, EPI_ISL_725342, EPI_ISL_725365, EPI_ISL_725371, EPI_ISL_725372, EPI_ISL_725373, EPI_ISL_725374, EPI_ISL_725375, EPI_ISL_725376, EPI_ISL_725377, EPI_ISL_725378, EPI_ISL_725379, EPI_ISL_725380, EPI_ISL_725381, EPI_ISL_725382, EPI_ISL_725383, EPI_ISL_725384, EPI_ISL_725385, EPI_ISL_725386, EPI_ISL_725387, EPI_ISL_725388, EPI_ISL_725389, EPI_ISL_725390, EPI_ISL_725391, EPI_ISL_725392, EPI_ISL_725393, EPI_ISL_725394, EPI_ISL_725395, EPI_ISL_725396, EPI_ISL_725397, EPI_ISL_725398, EPI_ISL_725399, EPI_ISL_725400, EPI_ISL_725401, EPI_ISL_725402, EPI_ISL_725403, EPI_ISL_725404, EPI_ISL_725405, EPI_ISL_725407, EPI_ISL_725408, EPI_ISL_725409, EPI_ISL_725410, EPI_ISL_725411, EPI_ISL_725412, EPI_ISL_725413, EPI_ISL_725414, EPI_ISL_725415, EPI_ISL_725416, EPI_ISL_725418, EPI_ISL_725421, EPI_ISL_725426, EPI_ISL_725427, EPI_ISL_725429, EPI_ISL_725431, EPI_ISL_725432, EPI_ISL_725434, EPI_ISL_725435, EPI_ISL_725436, EPI_ISL_725437, EPI_ISL_725439, EPI_ISL_725440, EPI_ISL_725441, EPI_ISL_725442, EPI_ISL_725443, EPI_ISL_725444, EPI_ISL_725445, EPI_ISL_725446, EPI_ISL_725447, EPI_ISL_725448, EPI_ISL_725449, EPI_ISL_725450, EPI_ISL_725451, EPI_ISL_725452, EPI_ISL_725453, EPI_ISL_725454, EPI_ISL_725455, EPI_ISL_725456, EPI_ISL_725457, EPI_ISL_725460, EPI_ISL_725461, EPI_ISL_725462, EPI_ISL_725465, EPI_ISL_725470, EPI_ISL_725473, EPI_ISL_725474, EPI_ISL_725477, EPI_ISL_725478, EPI_ISL_725479, EPI_ISL_725482, EPI_ISL_725483, EPI_ISL_725484, EPI_ISL_725485, EPI_ISL_725486, EPI_ISL_725488, EPI_ISL_725489, EPI_ISL_725490, EPI_ISL_725491, EPI_ISL_725496, EPI_ISL_725497, EPI_ISL_725498, EPI_ISL_725499, EPI_ISL_725500, EPI_ISL_725501, EPI_ISL_725502, EPI_ISL_725503, EPI_ISL_725504, EPI_ISL_725505, EPI_ISL_725506, EPI_ISL_725507, EPI_ISL_725508, EPI_ISL_725509, EPI_ISL_725510, EPI_ISL_725511, EPI_ISL_725512, EPI_ISL_725514, EPI_ISL_725515, EPI_ISL_725516, EPI_ISL_725517, EPI_ISL_725518, EPI_ISL_741544, EPI_ISL_866560, EPI_ISL_866579, EPI_ISL_866580, EPI_ISL_866583, EPI_ISL_866585, EPI_ISL_866804, EPI_ISL_866809, EPI_ISL_866810, EPI_ISL_866811, EPI_ISL_866813, EPI_ISL_866816, EPI_ISL_866823, EPI_ISL_866827, EPI_ISL_866829, EPI_ISL_950573, EPI_ISL_950577, EPI_ISL_950581, EPI_ISL_950587, EPI_ISL_950596, EPI_ISL_998821, EPI_ISL_998822, EPI_ISL_998823, EPI_ISL_998825, EPI_ISL_998827, EPI_ISL_998828, EPI_ISL_1051438, EPI_ISL_1051517 | | | |
| see above | Quadram Institute Bioscience | COVID-19 Genomics UK (COG-UK) Consortium | Alexander J Trotter; Alison E. Mather; Alp Aydin; Ana P. Tedim; Anastasia Kolyva; Andrew Bell; Andrew J. Page; Claire Stuart; Dave J. Baker; Gemma L. Kay; John Wain; Justin O'Grady; Leonardo de Oliveira Martins; Lizzie Meadows; Maria Diaz; Mark Webber; Muhammed Yasir; Nabil-Fareed Alikhan; Ngozi Elumogo; Nicholas M. Thomson; Rachael Stanley; Rachel Gilroy; Reenesh Prakash; Samir Dervisevic; Samuel Bloomfield; Steven Rudder; Thanh Le-Viet |
| EPI_ISL_679944, EPI_ISL_679945, EPI_ISL_702841, EPI_ISL_704582, EPI_ISL_704583, EPI_ISL_704780, EPI_ISL_705213, EPI_ISL_705380, EPI_ISL_706428, EPI_ISL_706429, EPI_ISL_706430, EPI_ISL_706435, EPI_ISL_706436, EPI_ISL_706437, EPI_ISL_725528, EPI_ISL_725529, EPI_ISL_725530, EPI_ISL_725531, EPI_ISL_725532, EPI_ISL_725533, EPI_ISL_725534, EPI_ISL_725535, EPI_ISL_725536, EPI_ISL_725537, EPI_ISL_725538, EPI_ISL_725539, EPI_ISL_725605, EPI_ISL_725606, EPI_ISL_725607, EPI_ISL_725608, EPI_ISL_725609, EPI_ISL_725610, EPI_ISL_725611, EPI_ISL_725612, EPI_ISL_725613, EPI_ISL_725614, EPI_ISL_725615, EPI_ISL_725616, EPI_ISL_725617, EPI_ISL_725618, EPI_ISL_725619, EPI_ISL_725620, EPI_ISL_725621, EPI_ISL_725622, EPI_ISL_725623, EPI_ISL_725624, EPI_ISL_725625, EPI_ISL_725626, EPI_ISL_725627, EPI_ISL_725628, EPI_ISL_725629, EPI_ISL_725630, EPI_ISL_725631, EPI_ISL_725632, EPI_ISL_725633, EPI_ISL_725634, EPI_ISL_725635, EPI_ISL_725636, EPI_ISL_725637, EPI_ISL_725638, EPI_ISL_725639, EPI_ISL_725640, EPI_ISL_725641, EPI_ISL_725642, EPI_ISL_741611, EPI_ISL_741612, EPI_ISL_741613, EPI_ISL_819558, EPI_ISL_838348, EPI_ISL_839987, EPI_ISL_839988, EPI_ISL_839989, EPI_ISL_949650 | | | |
| see above | Queens Medical Centre, Clinical Microbiology Department / DeepSeq Nottingham | COVID-19 Genomics UK (COG-UK) Consortium | Christopher Moore; Fei Sang; Gemma Clark; Hannah Howson-Wells; Johnny Debebe; Jonathan Ball; Joseph Chappell; Manjinder Khakh; Matthew Carlisle; Matthew Loose; Michelle M Lister; Nadine Holmes; Patrick McClure; Theocharis Tsoleridis; Vicki M Fleming; Victoria Wright; Wendy Smith |
| EPI_ISL_678830 | Respiratory Virus Unit, Microbiology Services Colindale, Public Health England | COVID-19 Genomics UK (COG-UK) Consortium | PHE Covid Sequencing Team |
| EPI_ISL_700624, EPI_ISL_700628, EPI_ISL_700637, EPI_ISL_700651, EPI_ISL_700658, EPI_ISL_700659, EPI_ISL_700665, EPI_ISL_700686, EPI_ISL_700687, EPI_ISL_700688, EPI_ISL_700689, EPI_ISL_700690, EPI_ISL_700691, EPI_ISL_700692, EPI_ISL_700693, EPI_ISL_700694, EPI_ISL_700695, EPI_ISL_700696, EPI_ISL_700697, EPI_ISL_700698, EPI_ISL_707703, EPI_ISL_707704, EPI_ISL_707705, EPI_ISL_709949, EPI_ISL_709958, EPI_ISL_722967, EPI_ISL_722968, EPI_ISL_722969, EPI_ISL_722970, EPI_ISL_722974, EPI_ISL_722976, EPI_ISL_722977, EPI_ISL_722978, EPI_ISL_722979, EPI_ISL_722980, EPI_ISL_722981, EPI_ISL_722982, EPI_ISL_722983, EPI_ISL_722984, EPI_ISL_722985, EPI_ISL_722986, EPI_ISL_722987, EPI_ISL_722988, EPI_ISL_722989, EPI_ISL_722990, EPI_ISL_722991, EPI_ISL_722992, EPI_ISL_722993, EPI_ISL_722994, EPI_ISL_722995, EPI_ISL_722996, EPI_ISL_722997, EPI_ISL_722998, EPI_ISL_723001, EPI_ISL_723002, EPI_ISL_723003, EPI_ISL_723004, EPI_ISL_723005, EPI_ISL_723008, EPI_ISL_723009, EPI_ISL_723010, EPI_ISL_723011, EPI_ISL_723012, EPI_ISL_723013, EPI_ISL_723014, EPI_ISL_723022, EPI_ISL_723023, EPI_ISL_723024, EPI_ISL_723025, EPI_ISL_723026, EPI_ISL_723027, EPI_ISL_723028, EPI_ISL_723029, EPI_ISL_723030, EPI_ISL_723031, EPI_ISL_723032, EPI_ISL_723033, EPI_ISL_723034, EPI_ISL_723035, EPI_ISL_723036, EPI_ISL_723037, EPI_ISL_723038, EPI_ISL_723039, EPI_ISL_723040, EPI_ISL_723041, EPI_ISL_723042, EPI_ISL_723043, EPI_ISL_733589, EPI_ISL_733590, EPI_ISL_733591, EPI_ISL_733592, EPI_ISL_733593, EPI_ISL_747490, EPI_ISL_747491, EPI_ISL_747493, EPI_ISL_747494, EPI_ISL_747495, EPI_ISL_747496, EPI_ISL_747497, EPI_ISL_754242, EPI_ISL_754243, EPI_ISL_754244, EPI_ISL_754247, EPI_ISL_754248, EPI_ISL_754249, EPI_ISL_754250, EPI_ISL_754251, EPI_ISL_754252, EPI_ISL_754253, EPI_ISL_754256, EPI_ISL_754258, EPI_ISL_754259, EPI_ISL_754260, EPI_ISL_754261, EPI_ISL_754262, EPI_ISL_754263, EPI_ISL_754264, EPI_ISL_754265, EPI_ISL_754274, EPI_ISL_754275, EPI_ISL_754276, EPI_ISL_754286, EPI_ISL_754288, EPI_ISL_754290, EPI_ISL_754291, EPI_ISL_754292, EPI_ISL_754293, EPI_ISL_754298, EPI_ISL_754299, EPI_ISL_754300, EPI_ISL_754301, EPI_ISL_754303, EPI_ISL_754304, EPI_ISL_754306, EPI_ISL_754307, EPI_ISL_754308, EPI_ISL_754309, EPI_ISL_754310, EPI_ISL_754311, EPI_ISL_754335, EPI_ISL_754336, EPI_ISL_754337, EPI_ISL_754338, EPI_ISL_754339, EPI_ISL_754340, EPI_ISL_754341, EPI_ISL_754342, EPI_ISL_754344, EPI_ISL_754345, EPI_ISL_754346, EPI_ISL_754347, EPI_ISL_754348, EPI_ISL_754349, EPI_ISL_754350, EPI_ISL_754351, EPI_ISL_754352, EPI_ISL_754353, EPI_ISL_754354, EPI_ISL_754355, EPI_ISL_754356, EPI_ISL_754357, EPI_ISL_754358, EPI_ISL_754359, EPI_ISL_754360, EPI_ISL_754361, EPI_ISL_766064, EPI_ISL_766069, EPI_ISL_766090, EPI_ISL_766096, EPI_ISL_766123, EPI_ISL_766124, EPI_ISL_766125, EPI_ISL_766126, EPI_ISL_766129, EPI_ISL_766130, EPI_ISL_766213, EPI_ISL_766231, EPI_ISL_766239, EPI_ISL_766246, EPI_ISL_766247, EPI_ISL_766279, EPI_ISL_766280, EPI_ISL_766281, EPI_ISL_766282, EPI_ISL_766283, EPI_ISL_766284, EPI_ISL_766285, EPI_ISL_766286, EPI_ISL_766287, EPI_ISL_766289, EPI_ISL_766291, EPI_ISL_766292, EPI_ISL_766401, EPI_ISL_766506, EPI_ISL_766507, EPI_ISL_766510, EPI_ISL_766511, EPI_ISL_766512, EPI_ISL_766513, EPI_ISL_766514, EPI_ISL_766515, EPI_ISL_766516, EPI_ISL_766558, EPI_ISL_766559, EPI_ISL_766561, EPI_ISL_766567, EPI_ISL_766842, EPI_ISL_766853, EPI_ISL_768800, EPI_ISL_768801, EPI_ISL_768802, EPI_ISL_768803, EPI_ISL_769867, EPI_ISL_769868, EPI_ISL_769870, EPI_ISL_769878, EPI_ISL_804226, EPI_ISL_804265, EPI_ISL_836979, EPI_ISL_837083, EPI_ISL_837100, EPI_ISL_837107, EPI_ISL_837108, EPI_ISL_837109, EPI_ISL_837110, EPI_ISL_837111, EPI_ISL_837112, EPI_ISL_837185, EPI_ISL_837198, EPI_ISL_837199, EPI_ISL_846588, EPI_ISL_1296546 | | | |
| see above | Respiratory Virus Unit, National Infection Service, Public Health England | COVID-19 Genomics UK (COG-UK) Consortium | PHE Covid Sequencing Team |
| EPI_ISL_702862, EPI_ISL_702961, EPI_ISL_703340, EPI_ISL_703369, EPI_ISL_703886, EPI_ISL_703966, EPI_ISL_704041, EPI_ISL_704105, EPI_ISL_704607, EPI_ISL_704608, EPI_ISL_704669, EPI_ISL_704671, EPI_ISL_704674, EPI_ISL_705378, EPI_ISL_705950, EPI_ISL_705951, EPI_ISL_705957, EPI_ISL_705958, EPI_ISL_705959, EPI_ISL_705960, EPI_ISL_842653, EPI_ISL_842654, EPI_ISL_842656, EPI_ISL_842657, EPI_ISL_842659, EPI_ISL_842660, EPI_ISL_842661, EPI_ISL_842662, EPI_ISL_842663, EPI_ISL_842664, EPI_ISL_842665, EPI_ISL_842666, EPI_ISL_842667, EPI_ISL_842668, EPI_ISL_842669, EPI_ISL_842696, EPI_ISL_842697, EPI_ISL_842766, EPI_ISL_842767, EPI_ISL_842783, EPI_ISL_842790, EPI_ISL_866055, EPI_ISL_866056, EPI_ISL_866057, EPI_ISL_866058, EPI_ISL_866059, EPI_ISL_866060, EPI_ISL_866061, EPI_ISL_866062, EPI_ISL_866063, EPI_ISL_866064, EPI_ISL_920187, EPI_ISL_920223, EPI_ISL_920377, EPI_ISL_920504, EPI_ISL_1104834, EPI_ISL_1104882, EPI_ISL_1104888, EPI_ISL_1104893, EPI_ISL_1104954, EPI_ISL_1105056, EPI_ISL_1105148 | | | |
| see above | University College London Hospital | COVID-19 Genomics UK (COG-UK) Consortium | Catherine Houlihan; Dan Frampton; Judith Heaney; Matthew Byott; Moira Spyer and Eleni Nastouli; Stuart Kirk |
| EPI_ISL_679615, EPI_ISL_679618, EPI_ISL_702661, EPI_ISL_702824, EPI_ISL_702910, EPI_ISL_703069, EPI_ISL_703107, EPI_ISL_703740, EPI_ISL_704963, EPI_ISL_705007, EPI_ISL_705078, EPI_ISL_705132, EPI_ISL_705184, EPI_ISL_705232, EPI_ISL_705283, EPI_ISL_705285, EPI_ISL_705297, EPI_ISL_705331, EPI_ISL_705337, EPI_ISL_705392, EPI_ISL_705396, EPI_ISL_724597, EPI_ISL_724598, EPI_ISL_724599, EPI_ISL_724600, EPI_ISL_724601, EPI_ISL_724602, EPI_ISL_724603, EPI_ISL_724604, EPI_ISL_724605, EPI_ISL_741187, EPI_ISL_741207, EPI_ISL_741255, EPI_ISL_741256, EPI_ISL_741257, EPI_ISL_741258, EPI_ISL_741264, EPI_ISL_741265, EPI_ISL_741266, EPI_ISL_741267, EPI_ISL_741268, EPI_ISL_741269, EPI_ISL_741271, EPI_ISL_741272, EPI_ISL_741273, EPI_ISL_741274, EPI_ISL_741276, EPI_ISL_741278, EPI_ISL_741279, EPI_ISL_741280, EPI_ISL_741281, EPI_ISL_741282, EPI_ISL_741283, EPI_ISL_741284, EPI_ISL_741285, EPI_ISL_741286, EPI_ISL_741288, EPI_ISL_741289, EPI_ISL_741290, EPI_ISL_741291, EPI_ISL_741292, EPI_ISL_741293, EPI_ISL_741300, EPI_ISL_741303, EPI_ISL_741304, EPI_ISL_741305, EPI_ISL_741306, EPI_ISL_741307, EPI_ISL_741308, EPI_ISL_741312, EPI_ISL_741313, EPI_ISL_741314, EPI_ISL_764238, EPI_ISL_819483, EPI_ISL_819484, EPI_ISL_819485, EPI_ISL_819486, EPI_ISL_819490, EPI_ISL_819491, EPI_ISL_819492, EPI_ISL_819494, EPI_ISL_819495, EPI_ISL_819496, EPI_ISL_819497, EPI_ISL_819498, EPI_ISL_819500, EPI_ISL_819501, EPI_ISL_819502, EPI_ISL_819505, EPI_ISL_819508, EPI_ISL_819509, EPI_ISL_819510, EPI_ISL_819511, EPI_ISL_819512, EPI_ISL_819513, EPI_ISL_819514, EPI_ISL_819515, EPI_ISL_819516, EPI_ISL_819517, EPI_ISL_819518, EPI_ISL_819519, EPI_ISL_819524, EPI_ISL_819526, EPI_ISL_819527, EPI_ISL_819528, EPI_ISL_819530, EPI_ISL_819531, EPI_ISL_819533, EPI_ISL_819534, EPI_ISL_819535, EPI_ISL_819536, EPI_ISL_819537, EPI_ISL_819538, EPI_ISL_819539, EPI_ISL_819540, EPI_ISL_819541, EPI_ISL_819542, EPI_ISL_819543, EPI_ISL_819544, EPI_ISL_819545, EPI_ISL_819546, EPI_ISL_819547, EPI_ISL_819548, EPI_ISL_819549, EPI_ISL_819550, EPI_ISL_819551, EPI_ISL_819552, EPI_ISL_819553, EPI_ISL_819554, EPI_ISL_819555, EPI_ISL_819556, EPI_ISL_819557, EPI_ISL_819559, EPI_ISL_819560, EPI_ISL_819561, EPI_ISL_819562, EPI_ISL_819563, EPI_ISL_819564, EPI_ISL_819565, EPI_ISL_819566, EPI_ISL_819567, EPI_ISL_839331, EPI_ISL_839332, EPI_ISL_839333, EPI_ISL_839334, EPI_ISL_839335, EPI_ISL_839336, EPI_ISL_839338, EPI_ISL_839339, EPI_ISL_839340, EPI_ISL_839341, EPI_ISL_839358, EPI_ISL_839359, EPI_ISL_865706, EPI_ISL_865707, EPI_ISL_865708, EPI_ISL_865709, EPI_ISL_865710, EPI_ISL_865711, EPI_ISL_865712, EPI_ISL_865713, EPI_ISL_865714, EPI_ISL_865715, EPI_ISL_865716, EPI_ISL_865717, EPI_ISL_866334, EPI_ISL_866335, EPI_ISL_866336, EPI_ISL_866337, EPI_ISL_866338, EPI_ISL_866339, EPI_ISL_866340, EPI_ISL_866341, EPI_ISL_866342, EPI_ISL_866343, EPI_ISL_866344, EPI_ISL_866345, EPI_ISL_949787, EPI_ISL_949925, EPI_ISL_998006, EPI_ISL_998007, EPI_ISL_998009, EPI_ISL_998010, EPI_ISL_1048048, EPI_ISL_1104573, EPI_ISL_1104574, EPI_ISL_1104575, EPI_ISL_1104576, EPI_ISL_1104577, EPI_ISL_1104578, EPI_ISL_1104665, EPI_ISL_1104666, EPI_ISL_1104667 | | | |
| see above | University College London, Great Ormond Street Hospital for Children NHS Foundation Trust, Imperial College Healthcare NHS Trust | COVID-19 Genomics UK (COG-UK) Consortium | Alison Holmes; Charlotte Williams; Helena Tutill; Jacqueline Findlay; James Price; Judith Breuer; Julianne Brown; Kathryn Harris; Leysa Forrest; Mark Kristiansen; Paola Niola; Paola Resende Silva; Patricia Dyal; Paul Randell; Rachel Williams; Samuel Weeks; Sergi Castellano; Sunando Roy; Tony Brooks; Yasmin Panchbhaya |
| EPI_ISL_1047916, EPI_ISL_1047917, EPI_ISL_1047918, EPI_ISL_1047919, EPI_ISL_1047954, EPI_ISL_2125919 | University Hospitals Of Leicester NHS Trust and DeepSeq Nottingham | COVID-19 Genomics UK (COG-UK) Consortium | Christopher Holmes; Christopher Moore; Fei Sang; Joeseph Chappell; Johnny Debebe; Jonathan Ball; Julian Tang; Karlie Fallon; Matthew Carlisle; Matthew Loose; Nadine Holmes; Patrick McClure; Paul Bird; Thomas Helmer; Victoria Wright |
| EPI_ISL_813030, EPI_ISL_813031, EPI_ISL_813032, EPI_ISL_813033, EPI_ISL_813034, EPI_ISL_813035, EPI_ISL_813036, EPI_ISL_813071, EPI_ISL_918578, EPI_ISL_918600, EPI_ISL_918601, EPI_ISL_918602, EPI_ISL_918604, EPI_ISL_918605, EPI_ISL_918606, EPI_ISL_918607, EPI_ISL_918608, EPI_ISL_918609, EPI_ISL_918610, EPI_ISL_918611, EPI_ISL_918612, EPI_ISL_918613, EPI_ISL_918614, EPI_ISL_918615, EPI_ISL_918616, EPI_ISL_918617, EPI_ISL_918618, EPI_ISL_918619, EPI_ISL_918620, EPI_ISL_918621, EPI_ISL_918622, EPI_ISL_918623, EPI_ISL_918624, EPI_ISL_918625, EPI_ISL_918626, EPI_ISL_918627, EPI_ISL_918628, EPI_ISL_918629, EPI_ISL_918630, EPI_ISL_918631, EPI_ISL_918632, EPI_ISL_918633, EPI_ISL_918634, EPI_ISL_918635, EPI_ISL_918636, EPI_ISL_918637, EPI_ISL_918638, EPI_ISL_918639, EPI_ISL_918640, EPI_ISL_918641, EPI_ISL_918642, EPI_ISL_918643, EPI_ISL_918644, EPI_ISL_918645, EPI_ISL_918646, EPI_ISL_918648, EPI_ISL_918649, EPI_ISL_918650, EPI_ISL_918651, EPI_ISL_918652, EPI_ISL_918653, EPI_ISL_918654, EPI_ISL_918655, EPI_ISL_918656, EPI_ISL_918657, EPI_ISL_918658, EPI_ISL_918659, EPI_ISL_918660, EPI_ISL_918661, EPI_ISL_918662, EPI_ISL_918663, EPI_ISL_918664, EPI_ISL_918667, EPI_ISL_918670, EPI_ISL_918671, EPI_ISL_918672, EPI_ISL_918674, EPI_ISL_918675, EPI_ISL_918676, EPI_ISL_918677, EPI_ISL_918678, EPI_ISL_918679, EPI_ISL_918680, EPI_ISL_918681, EPI_ISL_918684, EPI_ISL_918685, EPI_ISL_949391, EPI_ISL_949392, EPI_ISL_949393, EPI_ISL_949394, EPI_ISL_949395, EPI_ISL_949399, EPI_ISL_949400, EPI_ISL_949401, EPI_ISL_949402 | | | |
| see above | University of Birmingham | COVID-19 Genomics UK (COG-UK) Consortium | Alex Richter; Andrew Bosworth; Andrew Bosworth. Queen Elizabeth Hospital: Anna Casey; Andrew D Beggs PHE Heartlands Lab: Husam Osman; Charlotte Poxon; Institute of Microbiology; Joanne Stockton; Josh Quick; Kasun Wanigasooriya; Mike Kidd; Nicholas Loman. University of Birmingham Testing Laboratory: Celina M Whalley; Oliver Pickles; Radoslaw Poplawski; Samuel Nicholls; University of Birmingham: Claire McMurray; Will Rowe |
| EPI_ISL_724267, EPI_ISL_724268, EPI_ISL_724276, EPI_ISL_724277, EPI_ISL_724278, EPI_ISL_724279, EPI_ISL_724280, EPI_ISL_724281, EPI_ISL_724282, EPI_ISL_724283, EPI_ISL_724284, EPI_ISL_724285, EPI_ISL_724286, EPI_ISL_724287, EPI_ISL_724288, EPI_ISL_724289, EPI_ISL_724290, EPI_ISL_724291, EPI_ISL_724292, EPI_ISL_724293, EPI_ISL_724294, EPI_ISL_724295, EPI_ISL_724296, EPI_ISL_724297, EPI_ISL_724299, EPI_ISL_724300, EPI_ISL_724301, EPI_ISL_724302, EPI_ISL_724303, EPI_ISL_724304, EPI_ISL_724305, EPI_ISL_724306, EPI_ISL_724307, EPI_ISL_724308, EPI_ISL_724309, EPI_ISL_724310, EPI_ISL_724311, EPI_ISL_724312, EPI_ISL_724313, EPI_ISL_724314, EPI_ISL_724315, EPI_ISL_724316, EPI_ISL_724317, EPI_ISL_724318, EPI_ISL_724319, EPI_ISL_724320, EPI_ISL_724321, EPI_ISL_724322, EPI_ISL_724323, EPI_ISL_996587, EPI_ISL_996588, EPI_ISL_997316, EPI_ISL_997317, EPI_ISL_1103803, EPI_ISL_1247541, EPI_ISL_1545811, EPI_ISL_1545814 | | | |
| see above | University of Exeter | COVID-19 Genomics UK (COG-UK) Consortium | Aaron Jeffries; Audrey Farbos; Ben Temperton; Jane Masoli; Joanna Warwick-Dugdale; Michelle Michelsen; Robyn Manley; Stephen Michell |
| EPI_ISL_1730123 | Virology Department, Royal Infirmary of Edinburgh, NHS Lothian / School of Biological Sciences, University of Edinburgh | COVID-19 Genomics UK (COG-UK) Consortium | Colquhoun R; Cotton S; Dewar R; Gallagher A; Hill V; Jackson B; Maloney D; McCrone JT; McHugh M; O'Toole Á; Rambaut A; Rooke S; Scher E; Templeton K; Yu X |
| EPI_ISL_703414, EPI_ISL_705492, EPI_ISL_705746, EPI_ISL_705748, EPI_ISL_705749, EPI_ISL_705750, EPI_ISL_705774, EPI_ISL_705775, EPI_ISL_705777, EPI_ISL_705780, EPI_ISL_705781, EPI_ISL_705782, EPI_ISL_705783, EPI_ISL_705784, EPI_ISL_705785, EPI_ISL_705786, EPI_ISL_705787, EPI_ISL_705788, EPI_ISL_705789, EPI_ISL_705790, EPI_ISL_705791, EPI_ISL_705792, EPI_ISL_705793, EPI_ISL_705794, EPI_ISL_724225, EPI_ISL_724226, EPI_ISL_724227, EPI_ISL_724228, EPI_ISL_724229, EPI_ISL_724230, EPI_ISL_724231, EPI_ISL_724232, EPI_ISL_724233, EPI_ISL_724234, EPI_ISL_724235, EPI_ISL_724236, EPI_ISL_724237, EPI_ISL_724238, EPI_ISL_724239, EPI_ISL_724240, EPI_ISL_724241, EPI_ISL_724242, EPI_ISL_724243, EPI_ISL_724244, EPI_ISL_724246, EPI_ISL_724247, EPI_ISL_742175, EPI_ISL_742176, EPI_ISL_742180, EPI_ISL_763511, EPI_ISL_763649, EPI_ISL_763903, EPI_ISL_764357, EPI_ISL_764358, EPI_ISL_814365, EPI_ISL_814533, EPI_ISL_814535, EPI_ISL_814665, EPI_ISL_814666, EPI_ISL_814667, EPI_ISL_814668, EPI_ISL_814669, EPI_ISL_814670, EPI_ISL_814671, EPI_ISL_814672, EPI_ISL_814673, EPI_ISL_814674, EPI_ISL_814675, EPI_ISL_814676, EPI_ISL_814677, EPI_ISL_814680, EPI_ISL_838214, EPI_ISL_838215, EPI_ISL_838216, EPI_ISL_838217, EPI_ISL_838218, EPI_ISL_838219, EPI_ISL_838283, EPI_ISL_865069, EPI_ISL_865070, EPI_ISL_865071, EPI_ISL_865153, EPI_ISL_865154, EPI_ISL_865155, EPI_ISL_865156, EPI_ISL_865157, EPI_ISL_865158, EPI_ISL_865159, EPI_ISL_865160, EPI_ISL_865161, EPI_ISL_919308, EPI_ISL_919309, EPI_ISL_919310, EPI_ISL_919348, EPI_ISL_919349, EPI_ISL_919350, EPI_ISL_919351, EPI_ISL_919352, EPI_ISL_919353, EPI_ISL_919354, EPI_ISL_919355, EPI_ISL_919356, EPI_ISL_919357, EPI_ISL_919358, EPI_ISL_919359, EPI_ISL_919360, EPI_ISL_919361, EPI_ISL_919362, EPI_ISL_919363, EPI_ISL_919364, EPI_ISL_919382, EPI_ISL_919383, EPI_ISL_919384, EPI_ISL_919385, EPI_ISL_919386, EPI_ISL_919387, EPI_ISL_919402, EPI_ISL_919403 | | | |
| see above | Virology Department, Royal Infirmary of Edinburgh, NHS Lothian / School of Biological Sciences, University of Edinburgh / Institute of Genetics and Molecular Medicine, University of Edinburgh | COVID-19 Genomics UK (COG-UK) Consortium | Balcaza C; Colquhoun R; Dewar R; Gallagher M; Hill V; Jackson B; McCrone JT; McHugh M; O'Toole Á; Rambaut A; Rooke S; Scher E; Templeton K; Williams TC; Yu X |
| EPI_ISL_702487, EPI_ISL_702550, EPI_ISL_702564, EPI_ISL_702579, EPI_ISL_702607, EPI_ISL_702633, EPI_ISL_702682, EPI_ISL_702842, EPI_ISL_702852, EPI_ISL_702881, EPI_ISL_703038, EPI_ISL_703133, EPI_ISL_703400, EPI_ISL_703449, EPI_ISL_703604, EPI_ISL_703644, EPI_ISL_703649, EPI_ISL_703660, EPI_ISL_703671, EPI_ISL_703676, EPI_ISL_703765, EPI_ISL_703766, EPI_ISL_704978, EPI_ISL_705026, EPI_ISL_705096, EPI_ISL_705129, EPI_ISL_705153, EPI_ISL_705204, EPI_ISL_705210, EPI_ISL_705220, EPI_ISL_705230, EPI_ISL_705233, EPI_ISL_705288, EPI_ISL_705351, EPI_ISL_705421, EPI_ISL_706975, EPI_ISL_706976, EPI_ISL_706977, EPI_ISL_706978, EPI_ISL_706979, EPI_ISL_706980, EPI_ISL_706981, EPI_ISL_706982, EPI_ISL_706984, EPI_ISL_706985, EPI_ISL_706986, EPI_ISL_706990, EPI_ISL_707004, EPI_ISL_727863, EPI_ISL_727869, EPI_ISL_727871, EPI_ISL_727875, EPI_ISL_727883, EPI_ISL_727885, EPI_ISL_727888, EPI_ISL_727892, EPI_ISL_727903, EPI_ISL_727905, EPI_ISL_727906, EPI_ISL_727914, EPI_ISL_727918, EPI_ISL_727920, EPI_ISL_727922, EPI_ISL_727923, EPI_ISL_727929, EPI_ISL_727936, EPI_ISL_727939, EPI_ISL_727941, EPI_ISL_727943, EPI_ISL_727947, EPI_ISL_727951, EPI_ISL_727957, EPI_ISL_727964, EPI_ISL_727966, EPI_ISL_727970, EPI_ISL_727971, EPI_ISL_727975, EPI_ISL_727982, EPI_ISL_727983, EPI_ISL_727985, EPI_ISL_727986, EPI_ISL_727987, EPI_ISL_727988, EPI_ISL_741939, EPI_ISL_741940, EPI_ISL_741941, EPI_ISL_741944, EPI_ISL_741946, EPI_ISL_741947, EPI_ISL_741949, EPI_ISL_741952, EPI_ISL_741955, EPI_ISL_741957, EPI_ISL_741959, EPI_ISL_741963, EPI_ISL_741968, EPI_ISL_741970, EPI_ISL_741972, EPI_ISL_741973, EPI_ISL_741979, EPI_ISL_741980, EPI_ISL_741981, EPI_ISL_741985, EPI_ISL_741987, EPI_ISL_741988, EPI_ISL_741992, EPI_ISL_741993, EPI_ISL_741994, EPI_ISL_741995, EPI_ISL_741996, EPI_ISL_741997, EPI_ISL_741998, EPI_ISL_742001, EPI_ISL_742002, EPI_ISL_742003, EPI_ISL_742005, EPI_ISL_742006, EPI_ISL_742008, EPI_ISL_742011, EPI_ISL_742012, EPI_ISL_742013, EPI_ISL_742014, EPI_ISL_742016, EPI_ISL_742019, EPI_ISL_742021, EPI_ISL_742024, EPI_ISL_742026, EPI_ISL_742027, EPI_ISL_742029, EPI_ISL_742032, EPI_ISL_742035, EPI_ISL_742036, EPI_ISL_742037, EPI_ISL_742038, EPI_ISL_742040, EPI_ISL_742042, EPI_ISL_742043, EPI_ISL_742045, EPI_ISL_742051, EPI_ISL_742053, EPI_ISL_742056, EPI_ISL_742057, EPI_ISL_742058, EPI_ISL_742061, EPI_ISL_742069, EPI_ISL_742070, EPI_ISL_742073, EPI_ISL_742078, EPI_ISL_742081, EPI_ISL_742085, EPI_ISL_742087, EPI_ISL_742088, EPI_ISL_742089, EPI_ISL_742091, EPI_ISL_742093, EPI_ISL_742095, EPI_ISL_742096, EPI_ISL_742097, EPI_ISL_742098, EPI_ISL_742101, EPI_ISL_742102, EPI_ISL_742104, EPI_ISL_742110, EPI_ISL_816238, EPI_ISL_816241, EPI_ISL_816263, EPI_ISL_816270, EPI_ISL_816279, EPI_ISL_816295, EPI_ISL_816330, EPI_ISL_816342, EPI_ISL_816356, EPI_ISL_816370, EPI_ISL_816393, EPI_ISL_816397, EPI_ISL_816403, EPI_ISL_816428, EPI_ISL_816443, EPI_ISL_816445, EPI_ISL_816468, EPI_ISL_816473, EPI_ISL_816476, EPI_ISL_816484, EPI_ISL_816485, EPI_ISL_816489, EPI_ISL_816499, EPI_ISL_816512, EPI_ISL_816516, EPI_ISL_816522, EPI_ISL_816530, EPI_ISL_816532, EPI_ISL_816545, EPI_ISL_816551, EPI_ISL_816570, EPI_ISL_816602, EPI_ISL_816627, EPI_ISL_816643, EPI_ISL_868427, EPI_ISL_924141 | | | |
| see above | Virology Department, Sheffield Teaching Hospitals NHS Foundation | COVID-19 Genomics UK | Adri Angyal; Alex Keeley; Benjamin Lindsey; Cariad Evans; Danielle Groves; Dave Partridge; Katie Johnson; Laura Carrilero; Luke Green; Matthew Parker; Matthew Wyles; Mehmet Yavuz; Mohammad Raza; Nikki Smith; Paul Parsons; Rachel Tucker; |

| | | | |
|---|---|---|---|
| | Trust/Department of Infection, Immunity and Cardiovascular Disease, The Medical School, University of Sheffield | (COG-UK) Consortium | Rebecca Brown; Thushan de Silva |
| EPI_ISL_725671, EPI_ISL_725674, EPI_ISL_725675, EPI_ISL_725689, EPI_ISL_725690, EPI_ISL_725691, EPI_ISL_725692, EPI_ISL_725693, EPI_ISL_725694, EPI_ISL_725695, EPI_ISL_725696, EPI_ISL_725697, EPI_ISL_725698, EPI_ISL_725699, EPI_ISL_725700, EPI_ISL_725701, EPI_ISL_725702, EPI_ISL_725703, EPI_ISL_725704, EPI_ISL_725705, EPI_ISL_725706, EPI_ISL_725707, EPI_ISL_725708, EPI_ISL_725709, EPI_ISL_725710, EPI_ISL_725711, EPI_ISL_725712, EPI_ISL_725715, EPI_ISL_725726, EPI_ISL_725743, EPI_ISL_725748, EPI_ISL_725752, EPI_ISL_725767, EPI_ISL_725770, EPI_ISL_725775, EPI_ISL_725779, EPI_ISL_725783, EPI_ISL_725794, EPI_ISL_725801, EPI_ISL_725802, EPI_ISL_725803, EPI_ISL_725804, EPI_ISL_725805, EPI_ISL_725806, EPI_ISL_725807, EPI_ISL_725808, EPI_ISL_725809, EPI_ISL_725810, EPI_ISL_725811, EPI_ISL_725812, EPI_ISL_725813, EPI_ISL_725814, EPI_ISL_725815, EPI_ISL_725816, EPI_ISL_725817, EPI_ISL_725818, EPI_ISL_725819, EPI_ISL_725820, EPI_ISL_725821, EPI_ISL_725822, EPI_ISL_725823, EPI_ISL_725824, EPI_ISL_725825, EPI_ISL_725826, EPI_ISL_725827, EPI_ISL_725828, EPI_ISL_725829, EPI_ISL_725830, EPI_ISL_725831, EPI_ISL_725832, EPI_ISL_725833, EPI_ISL_725834, EPI_ISL_725835, EPI_ISL_725836, EPI_ISL_725837, EPI_ISL_725838, EPI_ISL_725839, EPI_ISL_725840, EPI_ISL_725841, EPI_ISL_725842, EPI_ISL_725843, EPI_ISL_725844, EPI_ISL_725845, EPI_ISL_725846, EPI_ISL_725847, EPI_ISL_725848, EPI_ISL_725849, EPI_ISL_725850, EPI_ISL_725851, EPI_ISL_725852, EPI_ISL_725853, EPI_ISL_725854, EPI_ISL_725855, EPI_ISL_725856, EPI_ISL_725857, EPI_ISL_725858, EPI_ISL_725859, EPI_ISL_725860, EPI_ISL_725861, EPI_ISL_725862, EPI_ISL_725863, EPI_ISL_725864, EPI_ISL_725865, EPI_ISL_725866, EPI_ISL_725867, EPI_ISL_725868, EPI_ISL_725869, EPI_ISL_725870, EPI_ISL_725871, EPI_ISL_725872, EPI_ISL_725873, EPI_ISL_725874, EPI_ISL_725875, EPI_ISL_725876, EPI_ISL_725877, EPI_ISL_725878, EPI_ISL_725879, EPI_ISL_725880, EPI_ISL_725881, EPI_ISL_725882, EPI_ISL_725883, EPI_ISL_725884, EPI_ISL_725885, EPI_ISL_725886, EPI_ISL_725887, EPI_ISL_725888, EPI_ISL_725889, EPI_ISL_725890, EPI_ISL_725891, EPI_ISL_725892, EPI_ISL_725893, EPI_ISL_725894, EPI_ISL_725895, EPI_ISL_725896, EPI_ISL_725897, EPI_ISL_725898, EPI_ISL_725899, EPI_ISL_725900, EPI_ISL_725901, EPI_ISL_725902, EPI_ISL_725903, EPI_ISL_725904, EPI_ISL_725905, EPI_ISL_725906, EPI_ISL_725907, EPI_ISL_725908, EPI_ISL_725909, EPI_ISL_725910, EPI_ISL_725911, EPI_ISL_725912, EPI_ISL_725913, EPI_ISL_725914, EPI_ISL_725915, EPI_ISL_725916, EPI_ISL_725917, EPI_ISL_725918, EPI_ISL_725919, EPI_ISL_725920, EPI_ISL_725921, EPI_ISL_725922, EPI_ISL_725923, EPI_ISL_725924, EPI_ISL_725925, EPI_ISL_725926, EPI_ISL_725927, EPI_ISL_725928, EPI_ISL_725929, EPI_ISL_725930, EPI_ISL_725931, EPI_ISL_725932, EPI_ISL_725933, EPI_ISL_725934, EPI_ISL_725935, EPI_ISL_725936, EPI_ISL_725937, EPI_ISL_725938, EPI_ISL_725939, EPI_ISL_725940, EPI_ISL_725941, EPI_ISL_725942, EPI_ISL_725943, EPI_ISL_725944, EPI_ISL_725945, EPI_ISL_725946, EPI_ISL_725947, EPI_ISL_725948, EPI_ISL_725949, EPI_ISL_725950, EPI_ISL_725951, EPI_ISL_725952, EPI_ISL_725953, EPI_ISL_725954, EPI_ISL_725955, EPI_ISL_725956, EPI_ISL_725957, EPI_ISL_725958, EPI_ISL_725959, EPI_ISL_725960, EPI_ISL_725961, EPI_ISL_725962, EPI_ISL_725963, EPI_ISL_725964, EPI_ISL_725965, EPI_ISL_725966, EPI_ISL_725967, EPI_ISL_725968, EPI_ISL_725969, EPI_ISL_725970, EPI_ISL_725971, EPI_ISL_725972, EPI_ISL_725973, EPI_ISL_725974, EPI_ISL_725975, EPI_ISL_725976, EPI_ISL_725977, EPI_ISL_725978, EPI_ISL_725979, EPI_ISL_725980, EPI_ISL_725981, EPI_ISL_726836, EPI_ISL_726838, EPI_ISL_726840, EPI_ISL_726841, EPI_ISL_726842, EPI_ISL_726843, EPI_ISL_726844, EPI_ISL_726845, EPI_ISL_726846, EPI_ISL_726847, EPI_ISL_726848, EPI_ISL_726849, EPI_ISL_726850, EPI_ISL_726851, EPI_ISL_726852, EPI_ISL_726853, EPI_ISL_726854, EPI_ISL_726855, EPI_ISL_726856, EPI_ISL_726857, EPI_ISL_726858, EPI_ISL_726859, EPI_ISL_726860, EPI_ISL_726861, EPI_ISL_726862, EPI_ISL_726864, EPI_ISL_726960, EPI_ISL_726961, EPI_ISL_726962, EPI_ISL_726963, EPI_ISL_726964, EPI_ISL_726965, EPI_ISL_726966, EPI_ISL_726967, EPI_ISL_726968, EPI_ISL_726969, EPI_ISL_726970, EPI_ISL_726971, EPI_ISL_726972, EPI_ISL_726973, EPI_ISL_726974, EPI_ISL_726975, EPI_ISL_726976, EPI_ISL_726977, EPI_ISL_726978, EPI_ISL_726979, EPI_ISL_726980, EPI_ISL_726981, EPI_ISL_726982, EPI_ISL_726983, EPI_ISL_726984, EPI_ISL_726985, EPI_ISL_726986, EPI_ISL_726987, EPI_ISL_726988, EPI_ISL_726989, EPI_ISL_726990, EPI_ISL_726991, EPI_ISL_726992, EPI_ISL_726993, EPI_ISL_726994, EPI_ISL_726995, EPI_ISL_726996, EPI_ISL_726997, EPI_ISL_726998, EPI_ISL_726999, EPI_ISL_727000, EPI_ISL_727001, EPI_ISL_727002, EPI_ISL_727003, EPI_ISL_727004, EPI_ISL_727005, EPI_ISL_727006, EPI_ISL_727007, EPI_ISL_727008, EPI_ISL_727009, EPI_ISL_727010, EPI_ISL_727011, EPI_ISL_727012, EPI_ISL_727013, EPI_ISL_727014, EPI_ISL_727015, EPI_ISL_727016, EPI_ISL_727017, EPI_ISL_727018, EPI_ISL_727019, EPI_ISL_727020, EPI_ISL_727021, EPI_ISL_727022, EPI_ISL_727023, EPI_ISL_727024, EPI_ISL_727025, EPI_ISL_727026, EPI_ISL_727027, EPI_ISL_727028, EPI_ISL_727029, EPI_ISL_727030, EPI_ISL_727031, EPI_ISL_727032, EPI_ISL_727033, EPI_ISL_727034, EPI_ISL_727035, EPI_ISL_727036, EPI_ISL_727037, EPI_ISL_727038, EPI_ISL_727039, EPI_ISL_727040, EPI_ISL_727041, EPI_ISL_727042, EPI_ISL_727043, EPI_ISL_727044, EPI_ISL_727045, EPI_ISL_727046, EPI_ISL_727047, EPI_ISL_727048, EPI_ISL_727049, EPI_ISL_727050, EPI_ISL_727051, EPI_ISL_727052, EPI_ISL_727053, EPI_ISL_727054, EPI_ISL_727055, EPI_ISL_727056, EPI_ISL_727057, EPI_ISL_727058, EPI_ISL_727059, EPI_ISL_727060, EPI_ISL_727061, EPI_ISL_727062, EPI_ISL_727063, EPI_ISL_727064, EPI_ISL_727065, EPI_ISL_727066, EPI_ISL_727067, EPI_ISL_727068, EPI_ISL_727069, EPI_ISL_727070, EPI_ISL_727071, EPI_ISL_727072, EPI_ISL_727073, EPI_ISL_727074, EPI_ISL_727075, EPI_ISL_727076, EPI_ISL_727077, EPI_ISL_727078, EPI_ISL_727079, EPI_ISL_727080, EPI_ISL_727081, EPI_ISL_727082, EPI_ISL_727083, EPI_ISL_727084, EPI_ISL_727085, EPI_ISL_727086, EPI_ISL_727087, EPI_ISL_727088, EPI_ISL_727089, EPI_ISL_727090, EPI_ISL_727091, EPI_ISL_727092, EPI_ISL_727093, EPI_ISL_727094, EPI_ISL_727095, EPI_ISL_727096, EPI_ISL_727097, EPI_ISL_727098, EPI_ISL_727099, EPI_ISL_727100, EPI_ISL_727101, EPI_ISL_727102, EPI_ISL_727118, EPI_ISL_727119, EPI_ISL_727120, EPI_ISL_727121, EPI_ISL_727122, EPI_ISL_727123, EPI_ISL_727139, EPI_ISL_727140, EPI_ISL_727141, EPI_ISL_727142, EPI_ISL_727143, EPI_ISL_727144, EPI_ISL_727145, EPI_ISL_727146, EPI_ISL_727147, EPI_ISL_727148, EPI_ISL_727149, EPI_ISL_727150, EPI_ISL_727151, EPI_ISL_727152, EPI_ISL_727153, EPI_ISL_727154, EPI_ISL_727155, EPI_ISL_727156, EPI_ISL_727157, EPI_ISL_727158, EPI_ISL_727159, EPI_ISL_727160, EPI_ISL_727161, EPI_ISL_727162, EPI_ISL_727163, EPI_ISL_727164, EPI_ISL_727165, EPI_ISL_727166, EPI_ISL_727167, EPI_ISL_727168, EPI_ISL_727169, EPI_ISL_727170, EPI_ISL_727171, EPI_ISL_727172, EPI_ISL_727173, EPI_ISL_727174, EPI_ISL_727175, EPI_ISL_727176, EPI_ISL_727177, EPI_ISL_727178, EPI_ISL_727179, EPI_ISL_727187, EPI_ISL_727194, EPI_ISL_727205, EPI_ISL_727209, EPI_ISL_727211, EPI_ISL_727213, EPI_ISL_727214, EPI_ISL_727215, EPI_ISL_727216, EPI_ISL_727217, EPI_ISL_727218, EPI_ISL_727219, EPI_ISL_727220, EPI_ISL_727221, EPI_ISL_727222, EPI_ISL_727223, EPI_ISL_727224, EPI_ISL_727225, EPI_ISL_727226, EPI_ISL_727227, EPI_ISL_727228, EPI_ISL_727229, EPI_ISL_727230, EPI_ISL_727231, EPI_ISL_727232, EPI_ISL_727233, EPI_ISL_727234, EPI_ISL_727235, EPI_ISL_727236, EPI_ISL_727237, EPI_ISL_727238, EPI_ISL_727239, EPI_ISL_727240, EPI_ISL_727241, EPI_ISL_727242, EPI_ISL_727243, EPI_ISL_727244, EPI_ISL_727245, EPI_ISL_727246, EPI_ISL_727247, EPI_ISL_727248, EPI_ISL_727249, EPI_ISL_727250, EPI_ISL_727251, EPI_ISL_727252, EPI_ISL_727253, EPI_ISL_727254, EPI_ISL_727255, EPI_ISL_727256, EPI_ISL_727257, EPI_ISL_727258, EPI_ISL_727259, EPI_ISL_727260, EPI_ISL_727261, EPI_ISL_727262, EPI_ISL_727263, EPI_ISL_727264, EPI_ISL_727265, EPI_ISL_727266, EPI_ISL_727267, EPI_ISL_727268, EPI_ISL_727269, EPI_ISL_727270, EPI_ISL_727271, EPI_ISL_727272, EPI_ISL_727273, EPI_ISL_727274, EPI_ISL_727275, EPI_ISL_727276, EPI_ISL_727277, EPI_ISL_727278, EPI_ISL_727279, EPI_ISL_727280, EPI_ISL_727281, EPI_ISL_727282, EPI_ISL_727283, EPI_ISL_727284, EPI_ISL_727285, EPI_ISL_727286, EPI_ISL_727287, EPI_ISL_727288, EPI_ISL_727289, EPI_ISL_727290, EPI_ISL_727291, EPI_ISL_727292, EPI_ISL_727293, EPI_ISL_727294, EPI_ISL_727295, EPI_ISL_727296, EPI_ISL_727297, EPI_ISL_727298, EPI_ISL_727299, EPI_ISL_727300, EPI_ISL_727301, EPI_ISL_727302, EPI_ISL_727303, EPI_ISL_727304, EPI_ISL_727305, EPI_ISL_727306, EPI_ISL_727307, EPI_ISL_727308, EPI_ISL_727309, EPI_ISL_727310, EPI_ISL_727311, EPI_ISL_727312, EPI_ISL_727313, EPI_ISL_727314, EPI_ISL_727315, EPI_ISL_727316, EPI_ISL_727317, EPI_ISL_727318, EPI_ISL_727319, EPI_ISL_727320, EPI_ISL_727321, EPI_ISL_727322, EPI_ISL_727323, EPI_ISL_727324, EPI_ISL_727325, EPI_ISL_727326, EPI_ISL_727327, EPI_ISL_727328, EPI_ISL_727329, EPI_ISL_727330, EPI_ISL_727331, EPI_ISL_727332, EPI_ISL_727333, EPI_ISL_727334, EPI_ISL_727335, EPI_ISL_727336, EPI_ISL_727337, EPI_ISL_727338, EPI_ISL_727339, EPI_ISL_727340, EPI_ISL_727341, EPI_ISL_727342, EPI_ISL_727343, EPI_ISL_727344, EPI_ISL_727345, EPI_ISL_727346, EPI_ISL_727347, EPI_ISL_727348, EPI_ISL_727349, EPI_ISL_727350, EPI_ISL_727351, EPI_ISL_727352, EPI_ISL_727353, EPI_ISL_727354, EPI_ISL_727355, EPI_ISL_727356, EPI_ISL_727357, EPI_ISL_727358, EPI_ISL_727359, EPI_ISL_727360, EPI_ISL_727361, EPI_ISL_727362, EPI_ISL_727363, EPI_ISL_727364, EPI_ISL_727365, EPI_ISL_727366, EPI_ISL_727367, EPI_ISL_727368, EPI_ISL_727369, EPI_ISL_727370, EPI_ISL_727371, EPI_ISL_727372, EPI_ISL_727373, EPI_ISL_727374, EPI_ISL_727375, EPI_ISL_727376, EPI_ISL_727377, EPI_ISL_727378, EPI_ISL_727379, EPI_ISL_727380, EPI_ISL_727381, EPI_ISL_727382, EPI_ISL_727383, EPI_ISL_727384, EPI_ISL_727385, EPI_ISL_727386, EPI_ISL_727387, EPI_ISL_727388, EPI_ISL_727389, EPI_ISL_727390, EPI_ISL_727391, EPI_ISL_727392, EPI_ISL_727393, EPI_ISL_727394, EPI_ISL_727395, EPI_ISL_727396, EPI_ISL_727397, EPI_ISL_727398, EPI_ISL_727399, EPI_ISL_727400, EPI_ISL_727401, EPI_ISL_727402, EPI_ISL_727403, EPI_ISL_727404, EPI_ISL_727405, EPI_ISL_727406, EPI_ISL_727407, EPI_ISL_727408, EPI_ISL_727410, EPI_ISL_727411, EPI_ISL_727417, EPI_ISL_727418, EPI_ISL_727419, EPI_ISL_727420, EPI_ISL_727421, EPI_ISL_727422, EPI_ISL_727423, EPI_ISL_727424, EPI_ISL_727425, EPI_ISL_727426, EPI_ISL_727427, EPI_ISL_727428, EPI_ISL_727429, EPI_ISL_727430, EPI_ISL_727431, EPI_ISL_727483, EPI_ISL_727484, EPI_ISL_727485, EPI_ISL_727486, EPI_ISL_727487, EPI_ISL_727488, EPI_ISL_727489, EPI_ISL_727490, EPI_ISL_727491, EPI_ISL_727492, EPI_ISL_727493, EPI_ISL_727494, EPI_ISL_727495, EPI_ISL_727496, EPI_ISL_727497, EPI_ISL_727498, EPI_ISL_727499, EPI_ISL_727500, EPI_ISL_727501, EPI_ISL_727502, EPI_ISL_727503, EPI_ISL_727504, EPI_ISL_727505, EPI_ISL_727506, EPI_ISL_727507, EPI_ISL_727508, EPI_ISL_727509, EPI_ISL_727510, EPI_ISL_727511, EPI_ISL_727512, EPI_ISL_727513, EPI_ISL_727514, EPI_ISL_727515, EPI_ISL_727516, EPI_ISL_727517, EPI_ISL_727518, EPI_ISL_727519, EPI_ISL_727520, EPI_ISL_727521, EPI_ISL_727522, EPI_ISL_727523, EPI_ISL_727524, EPI_ISL_727525, EPI_ISL_727526, EPI_ISL_727527, EPI_ISL_727528, EPI_ISL_727529, EPI_ISL_727530, EPI_ISL_727531, EPI_ISL_727532, EPI_ISL_727533, EPI_ISL_727534, EPI_ISL_727535, EPI_ISL_727536, EPI_ISL_727537, EPI_ISL_727538, EPI_ISL_727539, EPI_ISL_727540, EPI_ISL_727541, EPI_ISL_727542, EPI_ISL_727543, EPI_ISL_727544, EPI_ISL_727545, EPI_ISL_727546, EPI_ISL_727547, EPI_ISL_727548, EPI_ISL_727549, EPI_ISL_727550, EPI_ISL_727551, EPI_ISL_727552, EPI_ISL_727553, EPI_ISL_727554, EPI_ISL_727555, EPI_ISL_727556, EPI_ISL_727557, EPI_ISL_727558, EPI_ISL_727559, EPI_ISL_727560, EPI_ISL_727561, EPI_ISL_727562, EPI_ISL_727563, EPI_ISL_727564, EPI_ISL_727565, EPI_ISL_727566, EPI_ISL_727567, EPI_ISL_727568, EPI_ISL_727569, EPI_ISL_727570, EPI_ISL_727571, EPI_ISL_727572, EPI_ISL_727573, EPI_ISL_727574, EPI_ISL_727575, EPI_ISL_727576, EPI_ISL_727577, EPI_ISL_727578, EPI_ISL_727579, EPI_ISL_727580, EPI_ISL_727581, EPI_ISL_727582, EPI_ISL_727583, EPI_ISL_727584, EPI_ISL_727585, EPI_ISL_727586, EPI_ISL_727587, EPI_ISL_727588, EPI_ISL_727589, EPI_ISL_727590, EPI_ISL_727591, EPI_ISL_727592, EPI_ISL_727593, EPI_ISL_727594, EPI_ISL_727595, EPI_ISL_727596, EPI_ISL_727597, EPI_ISL_727598, EPI_ISL_727599, EPI_ISL_727600, EPI_ISL_727601, EPI_ISL_727602, EPI_ISL_727603, EPI_ISL_727604, EPI_ISL_727605, EPI_ISL_727606, EPI_ISL_727607, EPI_ISL_727608, EPI_ISL_727609, EPI_ISL_727610, EPI_ISL_727611, EPI_ISL_727612, EPI_ISL_727613, EPI_ISL_727614, EPI_ISL_727615, EPI_ISL_727616, EPI_ISL_727617, EPI_ISL_727618, EPI_ISL_727619, EPI_ISL_727620, EPI_ISL_727621, EPI_ISL_727622, EPI_ISL_727623, EPI_ISL_727624, EPI_ISL_727625, EPI_ISL_727626, EPI_ISL_727627, EPI_ISL_727628, EPI_ISL_727629, EPI_ISL_727630, EPI_ISL_727631, EPI_ISL_727632, EPI_ISL_727633, EPI_ISL_727634, EPI_ISL_727635, EPI_ISL_727636, EPI_ISL_727637, EPI_ISL_727638, EPI_ISL_727639, EPI_ISL_727640, EPI_ISL_727641, EPI_ISL_727642, EPI_ISL_727643, EPI_ISL_727644, EPI_ISL_727645, EPI_ISL_727646, EPI_ISL_727647, EPI_ISL_727648, EPI_ISL_727649, EPI_ISL_727650, EPI_ISL_727651, EPI_ISL_727652, EPI_ISL_727653, EPI_ISL_727654, EPI_ISL_727655, EPI_ISL_727656, EPI_ISL_727657, EPI_ISL_727658, EPI_ISL_727659, EPI_ISL_727660, EPI_ISL_727661, EPI_ISL_727662, EPI_ISL_727663, EPI_ISL_727664, EPI_ISL_727665, EPI_ISL_727666, EPI_ISL_727667, EPI_ISL_727668, EPI_ISL_727669, EPI_ISL_727670, EPI_ISL_727671, EPI_ISL_727672, EPI_ISL_727673, EPI_ISL_727674, EPI_ISL_727675, EPI_ISL_727676, EPI_ISL_727677, EPI_ISL_727678, EPI_ISL_727679, EPI_ISL_727680, EPI_ISL_727681, EPI_ISL_727682, EPI_ISL_727683, EPI_ISL_727684, EPI_ISL_727685, EPI_ISL_727686, EPI_ISL_727687, EPI_ISL_727688, EPI_ISL_727689, EPI_ISL_727690, EPI_ISL_727691, EPI_ISL_727692, EPI_ISL_727693, EPI_ISL_727694, EPI_ISL_727695, EPI_ISL_727696, EPI_ISL_742262, EPI_ISL_742263, EPI_ISL_742264, EPI_ISL_742265, EPI_ISL_742266, EPI_ISL_742267, EPI_ISL_742268, EPI_ISL_742269, EPI_ISL_742270, EPI_ISL_742271, EPI_ISL_742272, EPI_ISL_742273, EPI_ISL_742274, EPI_ISL_742275, EPI_ISL_742277, EPI_ISL_742286, EPI_ISL_742287, EPI_ISL_742288, EPI_ISL_742289, EPI_ISL_742290, EPI_ISL_742291, EPI_ISL_742292, EPI_ISL_742293, EPI_ISL_742294, EPI_ISL_742295, EPI_ISL_742296, EPI_ISL_742297, EPI_ISL_742298, EPI_ISL_742299, EPI_ISL_742351, EPI_ISL_742352, EPI_ISL_742353, EPI_ISL_742354, EPI_ISL_742355, EPI_ISL_742356, EPI_ISL_742357, EPI_ISL_742358, EPI_ISL_742359, EPI_ISL_742360, EPI_ISL_742361, EPI_ISL_742362, EPI_ISL_742363, EPI_ISL_742364, EPI_ISL_742365, EPI_ISL_742366, EPI_ISL_742367, EPI_ISL_742368, EPI_ISL_742369, EPI_ISL_742370, EPI_ISL_742371, EPI_ISL_742372, EPI_ISL_742373, EPI_ISL_742374, EPI_ISL_742375, EPI_ISL_742376, EPI_ISL_742377, EPI_ISL_742378, EPI_ISL_742379, EPI_ISL_742380, EPI_ISL_742381, EPI_ISL_742382, EPI_ISL_742383, EPI_ISL_742384, EPI_ISL_742385, EPI_ISL_742386, EPI_ISL_742387, EPI_ISL_742388, EPI_ISL_742389, EPI_ISL_742390, EPI_ISL_742391, EPI_ISL_742392, EPI_ISL_742393, EPI_ISL_742394, EPI_ISL_742395, EPI_ISL_742396, EPI_ISL_742397, EPI_ISL_742398, EPI_ISL_742399, EPI_ISL_742400, EPI_ISL_742401, EPI_ISL_742402, EPI_ISL_742403, EPI_ISL_742404, EPI_ISL_742405, EPI_ISL_742406, EPI_ISL_742407, EPI_ISL_742408, EPI_ISL_742409, EPI_ISL_742410, EPI_ISL_742411, EPI_ISL_742412, EPI_ISL_742413, EPI_ISL_742414, EPI_ISL_742415, EPI_ISL_742416, EPI_ISL_742417, EPI_ISL_742418, EPI_ISL_742419, EPI_ISL_742420, EPI_ISL_742421, EPI_ISL_742422, EPI_ISL_742423, EPI_ISL_742424, EPI_ISL_742425, EPI_ISL_742426, EPI_ISL_742427, EPI_ISL_742428, EPI_ISL_742429, EPI_ISL_742430, EPI_ISL_742431, EPI_ISL_742432, EPI_ISL_742433, EPI_ISL_742434, EPI_ISL_742435, EPI_ISL_742436, EPI_ISL_742437, EPI_ISL_742438, EPI_ISL_742439, EPI_ISL_742440, EPI_ISL_742441, EPI_ISL_742442, EPI_ISL_742443, EPI_ISL_742444, EPI_ISL_742445, EPI_ISL_742446, EPI_ISL_742447, EPI_ISL_742448, EPI_ISL_742449, EPI_ISL_742450, EPI_ISL_742451, EPI_ISL_742452, EPI_ISL_742453, EPI_ISL_742454, EPI_ISL_742455, EPI_ISL_742456, EPI_ISL_742457, EPI_ISL_742458, EPI_ISL_742459, EPI_ISL_742460, EPI_ISL_742461, EPI_ISL_742462, EPI_ISL_742463, EPI_ISL_742464, EPI_ISL_742465, EPI_ISL_742466, EPI_ISL_742467, EPI_ISL_742468, EPI_ISL_742469, EPI_ISL_742470, EPI_ISL_742471, EPI_ISL_742472, EPI_ISL_742473, EPI_ISL_742474, EPI_ISL_742475, EPI_ISL_742476, EPI_ISL_742477, EPI_ISL_742478, EPI_ISL_742479, EPI_ISL_742480, EPI_ISL_742481, EPI_ISL_742482, EPI_ISL_742483, EPI_ISL_742484, EPI_ISL_742485, EPI_ISL_742486, EPI_ISL_742487, EPI_ISL_742488, EPI_ISL_742489, EPI_ISL_742490, EPI_ISL_742491, EPI_ISL_742492, EPI_ISL_742493, EPI_ISL_742494, EPI_ISL_742495, EPI_ISL_742496, EPI_ISL_742497, EPI_ISL_742498, EPI_ISL_742499, EPI_ISL_742500, EPI_ISL_742501, EPI_ISL_742502, EPI_ISL_742503, EPI_ISL_742504, EPI_ISL_742505, EPI_ISL_742506, EPI_ISL_742507, EPI_ISL_742508, EPI_ISL_742509, EPI_ISL_742510, EPI_ISL_742511, EPI_ISL_742512, EPI_ISL_742513, EPI_ISL_742514, EPI_ISL_742515, EPI_ISL_742516, EPI_ISL_742517, EPI_ISL_742518, EPI_ISL_742519, EPI_ISL_742520, EPI_ISL_742521, EPI_ISL_742522, EPI_ISL_742523, EPI_ISL_742524, EPI_ISL_742525, EPI_ISL_742526, EPI_ISL_742527, EPI_ISL_742528, EPI_ISL_742529, EPI_ISL_742531, EPI_ISL_742532, EPI_ISL_742533, EPI_ISL_742534, EPI_ISL_742535, EPI_ISL_742536, EPI_ISL_742537, EPI_ISL_742538, EPI_ISL_742539, EPI_ISL_742540, EPI_ISL_742545, EPI_ISL_742550, EPI_ISL_742551, EPI_ISL_742552, EPI_ISL_742553, EPI_ISL_742554, EPI_ISL_742565, EPI_ISL_742637, EPI_ISL_742638, EPI_ISL_742639, EPI_ISL_742640, EPI_ISL_742641, EPI_ISL_742642, EPI_ISL_742643, EPI_ISL_742644, EPI_ISL_742645, EPI_ISL_742646, EPI_ISL_742649, EPI_ISL_742651, EPI_ISL_742653, EPI_ISL_742654, EPI_ISL_742655, EPI_ISL_742656, EPI_ISL_742657, EPI_ISL_742658, EPI_ISL_742659, EPI_ISL_742660, EPI_ISL_742661, EPI_ISL_742662, EPI_ISL_742663, EPI_ISL_742664, EPI_ISL_742665, EPI_ISL_742666, EPI_ISL_742667, EPI_ISL_742668, EPI_ISL_742669, EPI_ISL_742670, EPI_ISL_742671, EPI_ISL_742672, EPI_ISL_742673, EPI_ISL_742674, EPI_ISL_742675, EPI_ISL_742676, EPI_ISL_742677, EPI_ISL_742678, EPI_ISL_742679, EPI_ISL_742683, EPI_ISL_742684, EPI_ISL_742685, EPI_ISL_742686, EPI_ISL_742687, EPI_ISL_742688, EPI_ISL_742689, EPI_ISL_742690, EPI_ISL_742691, EPI_ISL_742694, EPI_ISL_742695, EPI_ISL_742696, EPI_ISL_742697, EPI_ISL_742700, EPI_ISL_742701, EPI_ISL_742702, EPI_ISL_742704, EPI_ISL_742705, EPI_ISL_742707, EPI_ISL_742709, EPI_ISL_742710, EPI_ISL_742713, EPI_ISL_742714, EPI_ISL_742716, EPI_ISL_742717, EPI_ISL_742718, EPI_ISL_742719, EPI_ISL_742720, EPI_ISL_742838, EPI_ISL_742839, EPI_ISL_742841, EPI_ISL_742842, EPI_ISL_742843, EPI_ISL_742844, EPI_ISL_742845, EPI_ISL_742846, EPI_ISL_742847, EPI_ISL_742848, EPI_ISL_742849, EPI_ISL_742850, EPI_ISL_742851, EPI_ISL_742852, EPI_ISL_742853, EPI_ISL_742854, EPI_ISL_742855, EPI_ISL_742856, EPI_ISL_742857, EPI_ISL_742858, EPI_ISL_742859, EPI_ISL_742860, EPI_ISL_742861, EPI_ISL_742862, EPI_ISL_742863, EPI_ISL_742864, EPI_ISL_742865, EPI_ISL_742866, EPI_ISL_742867, EPI_ISL_742868, EPI_ISL_742869, EPI_ISL_742870, EPI_ISL_742871, EPI_ISL_742872, EPI_ISL_742873, EPI_ISL_742874, EPI_ISL_742875, EPI_ISL_742876, EPI_ISL_742877, EPI_ISL_742878, EPI_ISL_742879, EPI_ISL_742890, EPI_ISL_742891, EPI_ISL_742892, EPI_ISL_742893, EPI_ISL_742894, EPI_ISL_742895, EPI_ISL_742896, EPI_ISL_742897, EPI_ISL_742898, EPI_ISL_742899, EPI_ISL_742900, EPI_ISL_742901, EPI_ISL_742902, EPI_ISL_742903, EPI_ISL_742904, EPI_ISL_742905, EPI_ISL_742906, EPI_ISL_742907, EPI_ISL_742908, EPI_ISL_742909, EPI_ISL_742910, EPI_ISL_742911, EPI_ISL_742912, EPI_ISL_742913, EPI_ISL_742914, EPI_ISL_742915, EPI_ISL_742916, EPI_ISL_742917, EPI_ISL_742918, EPI_ISL_742919, EPI_ISL_742920, EPI_ISL_742921, EPI_ISL_742922, EPI_ISL_742923, EPI_ISL_742924, EPI_ISL_742925, EPI_ISL_742926, EPI_ISL_742927, EPI_ISL_742928, EPI_ISL_742929, EPI_ISL_742930, EPI_ISL_742931, EPI_ISL_742932, EPI_ISL_742933, EPI_ISL_742934, EPI_ISL_742935, EPI_ISL_742936, EPI_ISL_742937, EPI_ISL_742938, EPI_ISL_742939, EPI_ISL_742940, EPI_ISL_742941, EPI_ISL_742942, EPI_ISL_742943, EPI_ISL_742944, EPI_ISL_742945, EPI_ISL_742946, EPI_ISL_742947, EPI_ISL_742948, EPI_ISL_742949, EPI_ISL_742950, EPI_ISL_742951, EPI_ISL_742952, EPI_ISL_742953, EPI_ISL_742954, EPI_ISL_742955, EPI_ISL_742956, EPI_ISL_742957, EPI_ISL_742958, EPI_ISL_742959, EPI_ISL_742960, EPI_ISL_742961, EPI_ISL_742962, EPI_ISL_742963, EPI_ISL_742964, EPI_ISL_742965, EPI_ISL_742966, EPI_ISL_742967, EPI_ISL_742968, EPI_ISL_742969, EPI_ISL_742970, EPI_ISL_742971, EPI_ISL_742972, EPI_ISL_742973, EPI_ISL_742974, EPI_ISL_742975, EPI_ISL_742976, EPI_ISL_742977, EPI_ISL_742978, EPI_ISL_742979, EPI_ISL_742980, EPI_ISL_742981, EPI_ISL_742982, EPI_ISL_742983, EPI_ISL_742984, EPI_ISL_742985, EPI_ISL_742986, EPI_ISL_742987, EPI_ISL_742988, EPI_ISL_742989, EPI_ISL_742990, EPI_ISL_742991, EPI_ISL_742992, EPI_ISL_742993, EPI_ISL_742994, EPI_ISL_742995, EPI_ISL_742996, EPI_ISL_742997, EPI_ISL_742998, EPI_ISL_742999, EPI_ISL_743000, EPI_ISL_743001, EPI_ISL_743002, EPI_ISL_743003, EPI_ISL_743004, EPI_ISL_743005, EPI_ISL_743006, EPI_ISL_743007, EPI_ISL_743008, EPI_ISL_743009, EPI_ISL_743010, EPI_ISL_743011, EPI_ISL_743012, EPI_ISL_743013, EPI_ISL_743014, EPI_ISL_743015, EPI_ISL_743016, EPI_ISL_743017, EPI_ISL_743018, EPI_ISL_743019, EPI_ISL_743020, EPI_ISL_743021, EPI_ISL_743022, EPI_ISL_743023, EPI_ISL_743024, EPI_ISL_743025, EPI_ISL_743026, EPI_ISL_743027, EPI_ISL_743028, EPI_ISL_743029, EPI_ISL_743030, EPI_ISL_743031, EPI_ISL_743032, EPI_ISL_743033, EPI_ISL_743034, EPI_ISL_743035, EPI_ISL_743036, EPI_ISL_743037, EPI_ISL_743038, EPI_ISL_743039, EPI_ISL_743040, EPI_ISL_743041, EPI_ISL_743042, EPI_ISL_743043, EPI_ISL_743044, EPI_ISL_743045, EPI_ISL_743046, EPI_ISL_743047, EPI_ISL_743048, EPI_ISL_743049, EPI_ISL_743050, EPI_ISL_743051, EPI_ISL_743052, EPI_ISL_743053, EPI_ISL_743054, EPI_ISL_743055, EPI_ISL_743056, EPI_ISL_743057, EPI_ISL_743058, EPI_ISL_743059, EPI_ISL_743060, EPI_ISL_743061, EPI_ISL_743062, EPI_ISL_743063, EPI_ISL_743064, EPI_ISL_743065, EPI_ISL_743066, EPI_ISL_743067, EPI_ISL_743068, EPI_ISL_743069, EPI_ISL_743070, EPI_ISL_743071, EPI_ISL_743072, EPI_ISL_743073, EPI_ISL_743074, EPI_ISL_743075, EPI_ISL_743076, EPI_ISL_743077, EPI_ISL_743078, EPI_ISL_743079, EPI_ISL_743080, EPI_ISL_743081, EPI_ISL_743082, EPI_ISL_743083, EPI_ISL_743084, EPI_ISL_743085, EPI_ISL_743086, EPI_ISL_743087, EPI_ISL_743088, EPI_ISL_743089, EPI_ISL_743090, EPI_ISL_743091, EPI_ISL_743092, EPI_ISL_743093, EPI_ISL_743094, EPI_ISL_743095, EPI_ISL_743096, EPI_ISL_743097, EPI_ISL_743098, EPI_ISL_743099, EPI_ISL_743100, EPI_ISL_743101, EPI_ISL_743102, EPI_ISL_743103, EPI_ISL_743104, EPI_ISL_743105, EPI_ISL_743106, EPI_ISL_743107, EPI_ISL_743108, EPI_ISL_743109, EPI_ISL_743110, EPI_ISL_743111, EPI_ISL_743112, EPI_ISL_743113, EPI_ISL_743114, EPI_ISL_743115, EPI_ISL_743116, EPI_ISL_743118, EPI_ISL_743119, EPI_ISL_743120, EPI_ISL_743121, EPI_ISL_743122, EPI_ISL_743123, EPI_ISL_743124, EPI_ISL_743125, EPI_ISL_743128, EPI_ISL_743129, EPI_ISL_743130, EPI_ISL_743131, EPI_ISL_743132, EPI_ISL_743133, EPI_ISL_743134, EPI_ISL_743135, EPI_ISL_743136, EPI_ISL_743137, EPI_ISL_743138, EPI_ISL_743139, EPI_ISL_743140, EPI_ISL_743141, EPI_ISL_743142, EPI_ISL_743143, EPI_ISL_743144, EPI_ISL_743145, EPI_ISL_743159, EPI_ISL_743160, EPI_ISL_743162, EPI_ISL_743163, EPI_ISL_743164, EPI_ISL_743165, EPI_ISL_743166, EPI_ISL_743167, EPI_ISL_743168, EPI_ISL_743169, EPI_ISL_743170, EPI_ISL_743171, EPI_ISL_743172, EPI_ISL_743173, EPI_ISL_743174, EPI_ISL_743175, EPI_ISL_743176, EPI_ISL_743177, EPI_ISL_743178, EPI_ISL_743179, EPI_ISL_743180, EPI_ISL_743181, EPI_ISL_743182, EPI_ISL_743183, EPI_ISL_743184, EPI_ISL_743185, EPI_ISL_743186, EPI_ISL_743187, EPI_ISL_743188, EPI_ISL_743189, EPI_ISL_743190, EPI_ISL_743191, EPI_ISL_743192, EPI_ISL_743193, EPI_ISL_743194, EPI_ISL_743195, EPI_ISL_743196, EPI_ISL_743197, EPI_ISL_743198, EPI_ISL_743199, EPI_ISL_743200, EPI_ISL_743201, EPI_ISL_743202, EPI_ISL_743203, EPI_ISL_743204, EPI_ISL_743205, EPI_ISL_743206, EPI_ISL_743207, EPI_ISL_743208, EPI_ISL_743209, EPI_ISL_743210, EPI_ISL_743211, EPI_ISL_743212, EPI_ISL_743213, EPI_ISL_743214, EPI_ISL_743215, EPI_ISL_743216, EPI_ISL_743217, EPI_ISL_743218, EPI_ISL_743219, EPI_ISL_743220, EPI_ISL_743221, EPI_ISL_743222, EPI_ISL_743223, EPI_ISL_743224, EPI_ISL_743225, EPI_ISL_743226, EPI_ISL_743227, EPI_ISL_743228, EPI_ISL_743229, EPI_ISL_743230, EPI_ISL_743231, EPI_ISL_743232, EPI_ISL_743233, EPI_ISL_743234, EPI_ISL_743235, EPI_ISL_743236, EPI_ISL_743237, EPI_ISL_743238, EPI_ISL_743239, EPI_ISL_743240, EPI_ISL_743241, EPI_ISL_743242, EPI_ISL_743243, EPI_ISL_743244, EPI_ISL_743245, EPI_ISL_743246, EPI_ISL_743247, EPI_ISL_743248, EPI_ISL_743249, EPI_ISL_743250, EPI_ISL_743251, EPI_ISL_743252, EPI_ISL_743253, EPI_ISL_743254, EPI_ISL_743255, EPI_ISL_743256, EPI_ISL_743257, EPI_ISL_743258, EPI_ISL_743259, EPI_ISL_743260, EPI_ISL_743261, EPI_ISL_743262, EPI_ISL_743263, EPI_ISL_743264, EPI_ISL_743265, EPI_ISL_743266, EPI_ISL_743267, EPI_ISL_743268, EPI_ISL_743269, EPI_ISL_743270, EPI_ISL_743271, EPI_ISL_743272, EPI_ISL_743273, EPI_ISL_743274, EPI_ISL_743275, EPI_ISL_743276, EPI_ISL_743277, EPI_ISL_743278, EPI_ISL_743279, EPI_ISL_743280, EPI_ISL_743281, EPI_ISL_743282, EPI_ISL_743283, EPI_ISL_743284, EPI_ISL_743285, EPI_ISL_743286, EPI_ISL_743287, EPI_ISL_743288, EPI_ISL_743289, EPI_ISL_743290, EPI_ISL_743291, EPI_ISL_743292, EPI_ISL_743293, EPI_ISL_743294, EPI_ISL_743295, EPI_ISL_743296, EPI_ISL_743297, EPI_ISL_743298, EPI_ISL_743299, EPI_ISL_743300, EPI_ISL_743301, EPI_ISL_743302, EPI_ISL_743303, EPI_ISL_743304, EPI_ISL_743305, EPI_ISL_743306, EPI_ISL_743307, EPI_ISL_743308, EPI_ISL_743309, EPI_ISL_743310, EPI_ISL_743311, EPI_ISL_743312, EPI_ISL_743313, EPI_ISL_743314, EPI_ISL_743315, EPI_ISL_743316, EPI_ISL_743317, EPI_ISL_743318, EPI_ISL_743319, EPI_ISL_743320, EPI_ISL_743323, EPI_ISL_743324, EPI_ISL_743325, EPI_ISL_743327, EPI_ISL_743328, EPI_ISL_743329, EPI_ISL_743330, EPI_ISL_743331, EPI_ISL_743332, EPI_ISL_743333, EPI_ISL_743334, EPI_ISL_743335, EPI_ISL_743336, EPI_ISL_743337, EPI_ISL_743338, EPI_ISL_743339, EPI_ISL_743340, EPI_ISL_743342, EPI_ISL_743343, EPI_ISL_743344, EPI_ISL_743345, EPI_ISL_743346, EPI_ISL_743347, EPI_ISL_743348, EPI_ISL_743349, EPI_ISL_743350, EPI_ISL_743351, EPI_ISL_743353, EPI_ISL_743354, EPI_ISL_743355, EPI_ISL_743356, EPI_ISL_743357, EPI_ISL_743358, EPI_ISL_743359, EPI_ISL_743360, EPI_ISL_743361, EPI_ISL_743362, EPI_ISL_743363, EPI_ISL_743364, EPI_ISL_743380, EPI_ISL_743381, EPI_ISL_743382, EPI_ISL_743383, EPI_ISL_743384, EPI_ISL_743385, EPI_ISL_743386, EPI_ISL_743387, EPI_ISL_743388, EPI_ISL_743389, EPI_ISL_743390, EPI_ISL_743391, EPI_ISL_743392, EPI_ISL_743393, EPI_ISL_743394, EPI_ISL_743395, EPI_ISL_743396, EPI_ISL_743397, EPI_ISL_743398, EPI_ISL_743399, EPI_ISL_743400, EPI_ISL_743401, EPI_ISL_743402, EPI_ISL_743403, EPI_ISL_743404, EPI_ISL_743405, EPI_ISL_743406, EPI_ISL_743407, EPI_ISL_743408, EPI_ISL_743409, EPI_ISL_743410, EPI_ISL_743411, EPI_ISL_743412, EPI_ISL_743413, EPI_ISL_743414, EPI_ISL_743415, EPI_ISL_743416, EPI_ISL_743417, EPI_ISL_743418, EPI_ISL_743419, EPI_ISL_743420, EPI_ISL_743421, EPI_ISL_743422, EPI_ISL_743423, EPI_ISL_743424, EPI_ISL_743432, EPI_ISL_743435, EPI_ISL_743436, EPI_ISL_743437, EPI_ISL_743438, EPI_ISL_743439, EPI_ISL_743440, EPI_ISL_743441, EPI_ISL_743442, EPI_ISL_743443, EPI_ISL_743444, EPI_ISL_743450, EPI_ISL_743451, EPI_ISL_743452, EPI_ISL_743453, EPI_ISL_743454, EPI_ISL_743455, EPI_ISL_743456, EPI_ISL_743457, EPI_ISL_743458, EPI_ISL_743459, EPI_ISL_743460, EPI_ISL_743461, EPI_ISL_743462, EPI_ISL_743463, EPI_ISL_743464, EPI_ISL_743465, EPI_ISL_743466, EPI_ISL_743467, EPI_ISL_743468, EPI_ISL_743469, EPI_ISL_743470, EPI_ISL_743471, EPI_ISL_743472, EPI_ISL_743473, EPI_ISL_743474, EPI_ISL_743475, EPI_ISL_743476, EPI_ISL_743477, EPI_ISL_743478, EPI_ISL_743479, EPI_ISL_743480, EPI_ISL_743481, EPI_ISL_743482, EPI_ISL_743483, EPI_ISL_743484, EPI_ISL_743485, EPI_ISL_743486, EPI_ISL_743487, EPI_ISL_743488, EPI_ISL_743489, EPI_ISL_743490, EPI_ISL_743491, EPI_ISL_743492, EPI_ISL_743493, EPI_ISL_743494, EPI_ISL_743495, EPI_ISL_743496, EPI_ISL_743497, EPI_ISL_743498, EPI_ISL_743499, EPI_ISL_743500, EPI_ISL_743501, EPI_ISL_743502, EPI_ISL_743503, EPI_ISL_743504, EPI_ISL_743505, EPI_ISL_743506, EPI_ISL_743507, EPI_ISL_743508, EPI_ISL_743510, EPI_ISL_743568, EPI_ISL_743580, EPI_ISL_743635, EPI_ISL_743637, EPI_ISL_743638, EPI_ISL_743639, EPI_ISL_743640, EPI_ISL_743642, EPI_ISL_743643, EPI_ISL_743644, EPI_ISL_743645, EPI_ISL_743646, EPI_ISL_743647, EPI_ISL_743648, EPI_ISL_743649, EPI_ISL_743650, EPI_ISL_743651, EPI_ISL_743652, EPI_ISL_743653, EPI_ISL_743654, EPI_ISL_743655, EPI_ISL_743656, EPI_ISL_743657, EPI_ISL_743658, EPI_ISL_743659, EPI_ISL_743660, EPI_ISL_743661, EPI_ISL_743665, EPI_ISL_743666, EPI_ISL_743667, EPI_ISL_743668, EPI_ISL_743669, EPI_ISL_743670, EPI_ISL_743671, EPI_ISL_743672, EPI_ISL_743673, EPI_ISL_743674, EPI_ISL_743676, EPI_ISL_743677, EPI_ISL_743679, EPI_ISL_743901, EPI_ISL_743943, EPI_ISL_743945, EPI_ISL_743946, EPI_ISL_743947, EPI_ISL_743948, EPI_ISL_743949, EPI_ISL_743950, EPI_ISL_743951, EPI_ISL_743953, EPI_ISL_743954, EPI_ISL_743955, EPI_ISL_743956, EPI_ISL_743957, EPI_ISL_763392, EPI_ISL_763409, EPI_ISL_763416, EPI_ISL_763420, EPI_ISL_763422, EPI_ISL_763423, EPI_ISL_763454, EPI_ISL_763462, EPI_ISL_763467, EPI_ISL_763470, EPI_ISL_763479, EPI_ISL_763480, EPI_ISL_763481, EPI_ISL_763489, EPI_ISL_763492, EPI_ISL_763565, EPI_ISL_763572, EPI_ISL_763590, EPI_ISL_763658, EPI_ISL_763721, EPI_ISL_763725, EPI_ISL_763750, EPI_ISL_763760, EPI_ISL_763782, EPI_ISL_763821, EPI_ISL_764064, EPI_ISL_764133, EPI_ISL_764138, EPI_ISL_764155, EPI_ISL_764157, EPI_ISL_764192, EPI_ISL_764200, EPI_ISL_764201, EPI_ISL_764235, EPI_ISL_764255, EPI_ISL_764582, EPI_ISL_764746, EPI_ISL_764747, EPI_ISL_764753, EPI_ISL_764754, EPI_ISL_764755, EPI_ISL_764766, EPI_ISL_764767, EPI_ISL_764769, EPI_ISL_764770, EPI_ISL_764771, EPI_ISL_764772, EPI_ISL_764773, EPI_ISL_764774, EPI_ISL_764775, EPI_ISL_764776, EPI_ISL_764777, EPI_ISL_764778, EPI_ISL_764779, EPI_ISL_764780, EPI_ISL_764781, EPI_ISL_764782, EPI_ISL_764783, EPI_ISL_764784, EPI_ISL_764785, EPI_ISL_764786, EPI_ISL_764787, EPI_ISL_764788, EPI_ISL_764789, EPI_ISL_764790, EPI_ISL_764791, EPI_ISL_764792, EPI_ISL_764793, EPI_ISL_764794, EPI_ISL_764795, EPI_ISL_764796, EPI_ISL_764797, EPI_ISL_764798, EPI_ISL_764799, EPI_ISL_764800, EPI_ISL_764801, EPI_ISL_764802, EPI_ISL_764803, EPI_ISL_764804, EPI_ISL_814297, EPI_ISL_814298, EPI_ISL_814299, EPI_ISL_814307, EPI_ISL_814316, EPI_ISL_814322, EPI_ISL_814324, EPI_ISL_814327, EPI_ISL_814346, EPI_ISL_814362, EPI_ISL_814390, EPI_ISL_814391, EPI_ISL_814393, EPI_ISL_814399, EPI_ISL_814409, EPI_ISL_814472, EPI_ISL_814514, EPI_ISL_814516, EPI_ISL_814545, EPI_ISL_814559, EPI_ISL_814562, EPI_ISL_814563, EPI_ISL_814564, EPI_ISL_814565, EPI_ISL_814576, EPI_ISL_814577, EPI_ISL_814603, EPI_ISL_814607, EPI_ISL_814696, EPI_ISL_814697, EPI_ISL_814698, EPI_ISL_814699, EPI_ISL_814700, EPI_ISL_814701, EPI_ISL_814705, EPI_ISL_814706, EPI_ISL_814707, EPI_ISL_814708, EPI_ISL_814709, EPI_ISL_814710, EPI_ISL_814711, EPI_ISL_814712, EPI_ISL_814713, EPI_ISL_814714, EPI_ISL_814715, EPI_ISL_814716, EPI_ISL_814754, EPI_ISL_814755, EPI_ISL_814789, EPI_ISL_814790, EPI_ISL_814791, EPI_ISL_814792, EPI_ISL_814793, EPI_ISL_814794, EPI_ISL_814795, EPI_ISL_814800, EPI_ISL_814801, EPI_ISL_814802, EPI_ISL_814803, EPI_ISL_814804, EPI_ISL_814805, EPI_ISL_814806, EPI_ISL_814807, EPI_ISL_814808, EPI_ISL_814809, EPI_ISL_814810, EPI_ISL_814811, EPI_ISL_814812, EPI_ISL_814813, EPI_ISL_814814, EPI_ISL_814815, EPI_ISL_814816, EPI_ISL_814817, EPI_ISL_814818, EPI_ISL_814819, EPI_ISL_814820, EPI_ISL_814821, EPI_ISL_814822, EPI_ISL_814823, EPI_ISL_814824, EPI_ISL_814825, EPI_ISL_814826, EPI_ISL_814827, EPI_ISL_814828, EPI_ISL_814829, EPI_ISL_814830, EPI_ISL_814831, EPI_ISL_814832, EPI_ISL_814833, EPI_ISL_814834, EPI_ISL_814835, EPI_ISL_814837, EPI_ISL_814838, EPI_ISL_814839, EPI_ISL_814840, EPI_ISL_814841, EPI_ISL_814842, EPI_ISL_814843, EPI_ISL_814844, EPI_ISL_814845, EPI_ISL_814846, EPI_ISL_814847, EPI_ISL_814848, EPI_ISL_814849, EPI_ISL_814850, EPI_ISL_814851, EPI_ISL_814852, EPI_ISL_814853, EPI_ISL_814854, EPI_ISL_814855, EPI_ISL_814856, EPI_ISL_814857, EPI_ISL_814858, EPI_ISL_814859, EPI_ISL_815088, EPI_ISL_815089, EPI_ISL_815091, EPI_ISL_815092, EPI_ISL_822900, EPI_ISL_823206, EPI_ISL_823207 | | | |
| see above | Wales Specialist Virology Centre Sequencing lab: Pathogen Genomics Unit | COVID-19 Genomics UK (COG-UK) Consortium | Alec Birchley; Alexander Adams; Amy Gaskin; Angela Marchbank; Bree Gatica-Wilcox; Catherine Moore; Jason Coombes; Joanne Watkins; Joel Southgate; Johnathan Evans; Laura Gifford; Lauren Gilbert; Lee Graham; Malorie Perry; Matthew Bull; Nicole Pacchiarini; Sally Corden; Sara Kumziene-Summerhayes; Sara Rey; Sarah Taylor; Simon Cottrell; Sophie Jones; Tom Connor |
| EPI_ISL_922892, EPI_ISL_922893, EPI_ISL_922901, EPI_ISL_922967, EPI_ISL_922968, EPI_ISL_922969 | Wales Specialist Virology Centre Sequencing lab: Pathogen Genomics Unit | Public Health Wales Microbiology Cardiff Wales Specialist Virology Centre | Alec Birchley; Alexander Adams; Amy Gaskin; Angela Marchbank; Bree Gatica-Wilcox; Catherine Moore; Jason Coombes; Joanne Watkins; Joel Southgate; Johnathan Evans; Laura Gifford; Lauren Gilbert; Lee Graham; Malorie Perry; Matthew Bull; Nicole Pacchiarini; Sally Corden; Sara Kumziene-Summerhayes; Sara Rey; Sarah Taylor; Simon Cottrell; Sophie Jones; Tom Connor |
| EPI_ISL_680455, EPI_ISL_680456, EPI_ISL_680457, EPI_ISL_680458, EPI_ISL_680459, EPI_ISL_680460, EPI_ISL_703136, EPI_ISL_703233, EPI_ISL_703430, EPI_ISL_703737, EPI_ISL_703757, EPI_ISL_703758, EPI_ISL_703927, EPI_ISL_703929, EPI_ISL_704596, EPI_ISL_704597, EPI_ISL_704804, EPI_ISL_704807, EPI_ISL_705002, EPI_ISL_705253, EPI_ISL_705296, EPI_ISL_705712, EPI_ISL_705713, EPI_ISL_705714, EPI_ISL_705715, EPI_ISL_705716, EPI_ISL_705717, EPI_ISL_705721, EPI_ISL_705722, EPI_ISL_705723, EPI_ISL_705724, EPI_ISL_705725, EPI_ISL_705726, EPI_ISL_705728, EPI_ISL_705732, EPI_ISL_705733, EPI_ISL_724210, EPI_ISL_724211, EPI_ISL_724214, EPI_ISL_724215, EPI_ISL_724216, EPI_ISL_724217, EPI_ISL_724218, EPI_ISL_724219, EPI_ISL_724220, EPI_ISL_724221, EPI_ISL_814590, EPI_ISL_814591, EPI_ISL_814592, EPI_ISL_814593, EPI_ISL_838059, EPI_ISL_838064, EPI_ISL_838065, EPI_ISL_838066, EPI_ISL_838067, EPI_ISL_838068, EPI_ISL_838069, EPI_ISL_838070, EPI_ISL_838071, EPI_ISL_838072, EPI_ISL_838073, EPI_ISL_838074, EPI_ISL_838075, EPI_ISL_838076, EPI_ISL_838077, EPI_ISL_838078, EPI_ISL_838079, EPI_ISL_838080, EPI_ISL_838081, EPI_ISL_838082, EPI_ISL_838083, EPI_ISL_838084, EPI_ISL_838085, EPI_ISL_838086, EPI_ISL_838087, EPI_ISL_838088, EPI_ISL_838089, EPI_ISL_838090, EPI_ISL_838091, EPI_ISL_838092, EPI_ISL_838093, EPI_ISL_838094, EPI_ISL_838095, EPI_ISL_838096, EPI_ISL_838097, EPI_ISL_838098, EPI_ISL_838099, EPI_ISL_838100, EPI_ISL_838101, EPI_ISL_838102, EPI_ISL_838103, EPI_ISL_838104, EPI_ISL_838105, EPI_ISL_838106, EPI_ISL_838107, EPI_ISL_838108, EPI_ISL_838109, EPI_ISL_838110, EPI_ISL_838111, EPI_ISL_838112, EPI_ISL_838113, EPI_ISL_838114, EPI_ISL_838115, EPI_ISL_838209, EPI_ISL_838210, EPI_ISL_838211, EPI_ISL_838212, EPI_ISL_919234, EPI_ISL_919269, EPI_ISL_919271, EPI_ISL_949631, EPI_ISL_949633, EPI_ISL_949635, EPI_ISL_949636 | | | |
| see above | West of Scotland Specialist Virology Centre, NHSGGC / MRC-University of Glasgow Centre for Virus Research | COVID-19 Genomics UK (COG-UK) Consortium | Alasdair MacLean; Alice Broos; Ana da Silva Filipe; Antonia Ho; Daniel Mair; David L Robertson; Emma Thomson; Guy Mollett; James Shepherd; Jenna Nichols; Joseph Hughes; Kathy Li; Kathy Smollett; Kyriaki Nomikou; Lily Tong; Matthew Holden; Natasha Johnson; Rachel Blacow; Richard Orton; Rory Gunson; Sarah McDonald; Sharif Shaaban; Sreenu Vattipally; Stephen Carmichael |

We gratefully acknowledge the following Authors from the Originating laboratories responsible for obtaining the specimens, as well as the Submitting laboratories where the genome data were generated and shared via GISAID, on which this research is based.

All Submitters of data may be contacted directly via www.gisaid.org

Authors are sorted alphabetically.

| Accession ID | Originating Laboratory | Submitting Laboratory | Authors |
|---|---|---|---|
| EPI_ISL_843071, EPI_ISL_843072, EPI_ISL_843073, EPI_ISL_843074, EPI_ISL_843075, EPI_ISL_843076, EPI_ISL_843078, EPI_ISL_843089, EPI_ISL_1636891, EPI_ISL_1636968, EPI_ISL_1636969, EPI_ISL_1636970, EPI_ISL_1636971, EPI_ISL_1636972, EPI_ISL_1636973, EPI_ISL_1636974, EPI_ISL_1636975, EPI_ISL_1636976, EPI_ISL_1636977, EPI_ISL_1636978, EPI_ISL_1636979, EPI_ISL_1636980, EPI_ISL_1636981, EPI_ISL_1636982, EPI_ISL_1636983, EPI_ISL_1636984, EPI_ISL_1636989, EPI_ISL_1636990, EPI_ISL_1636991, EPI_ISL_1636992, EPI_ISL_1636993 | | | |
| see above | Barts Health NHS Trust | COVID-19 Genomics UK (COG-UK) Consortium | BROAD; Beatrix; CUTINO-MOGUEL; Claire; David; Dola; HARRINGTON; KELE; KULASEGARAN-SHYLINI; Maria-Teresa; OWOYEMI; Raghavendran; SHYLINI |
| EPI_ISL_816894, EPI_ISL_816896, EPI_ISL_816899, EPI_ISL_816905, EPI_ISL_816907, EPI_ISL_816908, EPI_ISL_816912, EPI_ISL_816914, EPI_ISL_816917, EPI_ISL_816919, EPI_ISL_816921, EPI_ISL_816922, EPI_ISL_816926, EPI_ISL_816927, EPI_ISL_816928, EPI_ISL_816929, EPI_ISL_816930, EPI_ISL_816931, EPI_ISL_816932, EPI_ISL_816933, EPI_ISL_816934, EPI_ISL_816935, EPI_ISL_816936, EPI_ISL_816937, EPI_ISL_816938, EPI_ISL_816939, EPI_ISL_816940, EPI_ISL_816941, EPI_ISL_816942, EPI_ISL_816943, EPI_ISL_816944, EPI_ISL_816945, EPI_ISL_816946, EPI_ISL_816947, EPI_ISL_816948, EPI_ISL_816949, EPI_ISL_816950, EPI_ISL_816951, EPI_ISL_816952, EPI_ISL_816953, EPI_ISL_816954, EPI_ISL_816955, EPI_ISL_816956, EPI_ISL_816957, EPI_ISL_816961, EPI_ISL_816963, EPI_ISL_816966, EPI_ISL_816972, EPI_ISL_816974, EPI_ISL_816975, EPI_ISL_816979, EPI_ISL_816981, EPI_ISL_816984, EPI_ISL_816986, EPI_ISL_816988, EPI_ISL_816989, EPI_ISL_816993, EPI_ISL_816994, EPI_ISL_816995, EPI_ISL_816996, EPI_ISL_816997, EPI_ISL_816998, EPI_ISL_816999, EPI_ISL_817000, EPI_ISL_817001, EPI_ISL_817002, EPI_ISL_817003, EPI_ISL_817004, EPI_ISL_817005, EPI_ISL_817006, EPI_ISL_817007, EPI_ISL_817008, EPI_ISL_817009, EPI_ISL_817010, EPI_ISL_817011, EPI_ISL_817012, EPI_ISL_817013, EPI_ISL_817014, EPI_ISL_817015, EPI_ISL_817016, EPI_ISL_817017, EPI_ISL_817018, EPI_ISL_817019, EPI_ISL_817020, EPI_ISL_817021, EPI_ISL_817022, EPI_ISL_817023, EPI_ISL_817024, EPI_ISL_817025, EPI_ISL_817027, EPI_ISL_817028, EPI_ISL_817030, EPI_ISL_817031, EPI_ISL_817033, EPI_ISL_817034, EPI_ISL_817035, EPI_ISL_817036, EPI_ISL_817037, EPI_ISL_817041, EPI_ISL_817042, EPI_ISL_817043, EPI_ISL_817045, EPI_ISL_817046, EPI_ISL_817049, EPI_ISL_817060, EPI_ISL_817064, EPI_ISL_817065, EPI_ISL_842358, EPI_ISL_842360, EPI_ISL_842363, EPI_ISL_842369, EPI_ISL_842371, EPI_ISL_842372, EPI_ISL_842376, EPI_ISL_842378, EPI_ISL_842381, EPI_ISL_842383, EPI_ISL_842385, EPI_ISL_842386, EPI_ISL_842390, EPI_ISL_842391, EPI_ISL_842392, EPI_ISL_842393, EPI_ISL_842394, EPI_ISL_842395, EPI_ISL_842396, EPI_ISL_842397, EPI_ISL_842398, EPI_ISL_842399, EPI_ISL_842400, EPI_ISL_842401, EPI_ISL_842402, EPI_ISL_842403, EPI_ISL_842404, EPI_ISL_842405, EPI_ISL_842406, EPI_ISL_842407, EPI_ISL_842408, EPI_ISL_842409, EPI_ISL_842410, EPI_ISL_842411, EPI_ISL_842412, EPI_ISL_842413, EPI_ISL_842414, EPI_ISL_842415, EPI_ISL_842416, EPI_ISL_842417, EPI_ISL_842418, EPI_ISL_842419, EPI_ISL_842420, EPI_ISL_842421 | | | |
| see above | Bioinformatics and Biostatistics Lab, Advanced Sequencing Facility | COVID-19 Genomics UK (COG-UK) Consortium | Aengus Stewart; Chelsea Sawyer; Harshil Patel; Jerome Nicod; Laura Cubitt; Margaret Crawford |
| EPI_ISL_741658, EPI_ISL_741675, EPI_ISL_741693, EPI_ISL_741697, EPI_ISL_741730, EPI_ISL_741731, EPI_ISL_741732, EPI_ISL_741733, EPI_ISL_741747, EPI_ISL_741748, EPI_ISL_741749, EPI_ISL_763607, EPI_ISL_763611, EPI_ISL_841822, EPI_ISL_841852 | | | |
| see above | Centre for Enzyme Innovation, University of Portsmouth / Translational Research Laboratory, Portsmouth Hospitals NHS Trust | COVID-19 Genomics UK (COG-UK) Consortium | Allyson Lloyd; Angela Beckett; Anoop Chauhan; Ethan Butcher; Garry Scarlett; Kelly Bicknell; Robert Impey; Samuel Robson; Sarah Wyllie; Scott Elliott; Sharon Glaysher; Yann Bourgeois |
| EPI_ISL_723922, EPI_ISL_723923, EPI_ISL_723924, EPI_ISL_723925, EPI_ISL_723926, EPI_ISL_723927, EPI_ISL_723928, EPI_ISL_723929, EPI_ISL_723930, EPI_ISL_723931, EPI_ISL_723932, EPI_ISL_723934, EPI_ISL_723937, EPI_ISL_723938, EPI_ISL_723939, EPI_ISL_723940, EPI_ISL_723941, EPI_ISL_723942, EPI_ISL_723943, EPI_ISL_723944, EPI_ISL_723945, EPI_ISL_723946, EPI_ISL_740995, EPI_ISL_740996, EPI_ISL_740997, EPI_ISL_740998, EPI_ISL_740999, EPI_ISL_741000, EPI_ISL_741001, EPI_ISL_741002, EPI_ISL_741004, EPI_ISL_741007, EPI_ISL_741008, EPI_ISL_741009, EPI_ISL_741010, EPI_ISL_741011, EPI_ISL_741012, EPI_ISL_763526, EPI_ISL_763542, EPI_ISL_763637, EPI_ISL_763638, EPI_ISL_763796, EPI_ISL_763849, EPI_ISL_763874, EPI_ISL_763875, EPI_ISL_763876, EPI_ISL_763877, EPI_ISL_763878, EPI_ISL_763879, EPI_ISL_763880, EPI_ISL_763881, EPI_ISL_764162, EPI_ISL_764202, EPI_ISL_764276, EPI_ISL_764329, EPI_ISL_764330, EPI_ISL_764331, EPI_ISL_764332, EPI_ISL_764333, EPI_ISL_764334, EPI_ISL_764335, EPI_ISL_764336, EPI_ISL_764337, EPI_ISL_764338, EPI_ISL_764339, EPI_ISL_764340, EPI_ISL_764341, EPI_ISL_764342, EPI_ISL_813189, EPI_ISL_813225, EPI_ISL_813226, EPI_ISL_813239, EPI_ISL_813243, EPI_ISL_813252, EPI_ISL_813255, EPI_ISL_813262, EPI_ISL_813264, EPI_ISL_813274, EPI_ISL_813275, EPI_ISL_813276, EPI_ISL_813277, EPI_ISL_813279, EPI_ISL_813286, EPI_ISL_813288, EPI_ISL_813290, EPI_ISL_813292, EPI_ISL_864786, EPI_ISL_864788, EPI_ISL_864793, EPI_ISL_864796, EPI_ISL_864802, EPI_ISL_864804, EPI_ISL_864806, EPI_ISL_864808, EPI_ISL_864852, EPI_ISL_864874, EPI_ISL_864882 | | | |
| see above | Department of Pathology, University of Cambridge | COVID-19 Genomics UK (COG-UK) Consortium | Aminu S. Jahun; Grant Hall; Ian Goodfellow; Iliana Georgana; Malte Pinckert; Martin D. Curran; Myra Hosmillo; Surendra Parmar; Yasmin Chaudhry |
| EPI_ISL_813655 | Kettering General Hospital | COVID-19 Genomics UK (COG-UK) Consortium | Christopher Moore; Fei Sang; Johnny Debebe; Jonathan Ball; Joseph Chappell; Matthew Carlisle; Matthew Loose; Nadine Holmes; Patrick McClure; Theocharis Tsoleridis; Victoria Wright |
| EPI_ISL_718629, EPI_ISL_718631, EPI_ISL_718633, EPI_ISL_718635, EPI_ISL_718637, EPI_ISL_718639, EPI_ISL_718640, EPI_ISL_718641, EPI_ISL_718643, EPI_ISL_718644, EPI_ISL_718647, EPI_ISL_718650, EPI_ISL_718653, EPI_ISL_718657, EPI_ISL_718658, EPI_ISL_718662, EPI_ISL_718663, EPI_ISL_718664, EPI_ISL_718665, EPI_ISL_718666, EPI_ISL_718670, EPI_ISL_718672, EPI_ISL_718674, EPI_ISL_718675, EPI_ISL_718676, EPI_ISL_718678, EPI_ISL_718682, EPI_ISL_718686, EPI_ISL_718687, EPI_ISL_718691, EPI_ISL_718694, EPI_ISL_718695, EPI_ISL_718700, EPI_ISL_718705, EPI_ISL_718707, EPI_ISL_718710, EPI_ISL_718711, EPI_ISL_718712, EPI_ISL_718713, EPI_ISL_718714, EPI_ISL_718715, EPI_ISL_718716, EPI_ISL_718718, EPI_ISL_718719, EPI_ISL_718721, EPI_ISL_718723, EPI_ISL_718724, EPI_ISL_718725, EPI_ISL_718728, EPI_ISL_718729, EPI_ISL_718735, EPI_ISL_718738, EPI_ISL_718739, EPI_ISL_718740, EPI_ISL_718742, EPI_ISL_718744, EPI_ISL_718745, EPI_ISL_718746, EPI_ISL_718747, EPI_ISL_718750, EPI_ISL_718754, EPI_ISL_718755, EPI_ISL_718759, EPI_ISL_718761, EPI_ISL_718762, EPI_ISL_718764, EPI_ISL_718765, EPI_ISL_718766, EPI_ISL_718767, EPI_ISL_718769, EPI_ISL_718772, EPI_ISL_718773, EPI_ISL_718774, EPI_ISL_718777, EPI_ISL_718779, EPI_ISL_718780, EPI_ISL_718781, EPI_ISL_718782, EPI_ISL_718783, EPI_ISL_718787, EPI_ISL_718789, EPI_ISL_718791, EPI_ISL_718795, EPI_ISL_718796, EPI_ISL_718798, EPI_ISL_718799, EPI_ISL_718801, EPI_ISL_718802, EPI_ISL_718803, EPI_ISL_718806, EPI_ISL_718807, EPI_ISL_718808, EPI_ISL_718811, EPI_ISL_718812, EPI_ISL_718813, EPI_ISL_718819, EPI_ISL_718821, EPI_ISL_718823, EPI_ISL_718827, EPI_ISL_718828, EPI_ISL_718829, EPI_ISL_718830, EPI_ISL_718831, EPI_ISL_718832, EPI_ISL_718833, EPI_ISL_718837, EPI_ISL_718838, EPI_ISL_718839, EPI_ISL_718841, EPI_ISL_718844, EPI_ISL_718847, EPI_ISL_718849, EPI_ISL_718850, EPI_ISL_718853, EPI_ISL_718854, EPI_ISL_718858, EPI_ISL_718859, EPI_ISL_718864, EPI_ISL_718865, EPI_ISL_718866, EPI_ISL_718870, EPI_ISL_718871, EPI_ISL_718872, EPI_ISL_718873, EPI_ISL_718877, EPI_ISL_718878, EPI_ISL_718880, EPI_ISL_718881, EPI_ISL_718885, EPI_ISL_718886, EPI_ISL_718887, EPI_ISL_718888, EPI_ISL_718889, EPI_ISL_718890, EPI_ISL_718891, EPI_ISL_718894, EPI_ISL_718895, EPI_ISL_718901, EPI_ISL_718902, EPI_ISL_718903, EPI_ISL_718907, EPI_ISL_718908, EPI_ISL_718911, EPI_ISL_718914, EPI_ISL_718915, EPI_ISL_718917, EPI_ISL_718919, EPI_ISL_718922, EPI_ISL_718923, EPI_ISL_718926, EPI_ISL_718929, EPI_ISL_718930, EPI_ISL_718932, EPI_ISL_718935, EPI_ISL_718936, EPI_ISL_718939, EPI_ISL_718943, EPI_ISL_718945, EPI_ISL_718950, EPI_ISL_718952, EPI_ISL_718953, EPI_ISL_718955, EPI_ISL_718957, EPI_ISL_718959, EPI_ISL_718962, EPI_ISL_718963, EPI_ISL_718964, EPI_ISL_718966, EPI_ISL_718967, EPI_ISL_718970, EPI_ISL_718974, EPI_ISL_718975, EPI_ISL_718976, EPI_ISL_718977, EPI_ISL_718978, EPI_ISL_718979, EPI_ISL_730655, EPI_ISL_730658, EPI_ISL_730667, EPI_ISL_730668, EPI_ISL_730680, EPI_ISL_730682, EPI_ISL_730683, EPI_ISL_730686, EPI_ISL_730692, EPI_ISL_730695, EPI_ISL_730696, EPI_ISL_730699, EPI_ISL_730702, EPI_ISL_730704, EPI_ISL_730711, EPI_ISL_730712, EPI_ISL_730722, EPI_ISL_730729, EPI_ISL_730731, EPI_ISL_730732, EPI_ISL_730733, EPI_ISL_730734, EPI_ISL_730742, EPI_ISL_730743, EPI_ISL_730750, EPI_ISL_730751, EPI_ISL_730753, EPI_ISL_730754, EPI_ISL_730758, EPI_ISL_730771, EPI_ISL_730773, EPI_ISL_730774, EPI_ISL_730784, EPI_ISL_730786, EPI_ISL_730788, EPI_ISL_730794, EPI_ISL_730814, EPI_ISL_730818, EPI_ISL_730819, EPI_ISL_730823, EPI_ISL_730825, EPI_ISL_730826, EPI_ISL_730829, EPI_ISL_730830, EPI_ISL_730834, EPI_ISL_730839, EPI_ISL_730844, EPI_ISL_730845, EPI_ISL_730848, EPI_ISL_730852, EPI_ISL_730854, EPI_ISL_730855, EPI_ISL_730859, EPI_ISL_730861, EPI_ISL_730865, EPI_ISL_730870, EPI_ISL_730873, EPI_ISL_730878, EPI_ISL_730881, EPI_ISL_730886, EPI_ISL_730890, EPI_ISL_730893, EPI_ISL_730900, EPI_ISL_730902, EPI_ISL_730904, EPI_ISL_730914, EPI_ISL_730917, EPI_ISL_730918, EPI_ISL_730919, EPI_ISL_730920, EPI_ISL_730922, EPI_ISL_730923, EPI_ISL_730925, EPI_ISL_730926, EPI_ISL_730927, EPI_ISL_730928, EPI_ISL_730930, EPI_ISL_730931, EPI_ISL_730932, EPI_ISL_730933, EPI_ISL_730934, EPI_ISL_730936, EPI_ISL_730937, EPI_ISL_730938, EPI_ISL_730940, EPI_ISL_730941, EPI_ISL_730942, EPI_ISL_730943, EPI_ISL_730944, EPI_ISL_730945, EPI_ISL_730946, EPI_ISL_730947, EPI_ISL_730949, EPI_ISL_730950, EPI_ISL_730951, EPI_ISL_730952, EPI_ISL_730953, EPI_ISL_730954, EPI_ISL_730955, EPI_ISL_730956, EPI_ISL_730957, EPI_ISL_730958, EPI_ISL_730960, EPI_ISL_730961, EPI_ISL_730962, EPI_ISL_730963, EPI_ISL_730964, EPI_ISL_730965, EPI_ISL_730968, EPI_ISL_730972, EPI_ISL_730974, EPI_ISL_730975, EPI_ISL_730976, EPI_ISL_730977, EPI_ISL_730978, EPI_ISL_730979, EPI_ISL_730981, EPI_ISL_730982, EPI_ISL_730983, EPI_ISL_730984, EPI_ISL_730985, EPI_ISL_730986, EPI_ISL_730988, EPI_ISL_730990, EPI_ISL_730991, EPI_ISL_730992, EPI_ISL_730993, EPI_ISL_730995, EPI_ISL_730996, EPI_ISL_730997, EPI_ISL_730998, EPI_ISL_730999, EPI_ISL_731001, EPI_ISL_731002, EPI_ISL_731003, EPI_ISL_731004, EPI_ISL_731005, EPI_ISL_731007, EPI_ISL_731008, EPI_ISL_731009, EPI_ISL_731010, EPI_ISL_731011, EPI_ISL_731012, EPI_ISL_731013, EPI_ISL_731014, EPI_ISL_731015, EPI_ISL_731018, EPI_ISL_731021, EPI_ISL_731022, EPI_ISL_731023, EPI_ISL_731025, EPI_ISL_731028, EPI_ISL_731029, EPI_ISL_731030, EPI_ISL_731031, EPI_ISL_731032, EPI_ISL_731034, EPI_ISL_731036, EPI_ISL_731037, EPI_ISL_731038, EPI_ISL_731039, EPI_ISL_731041, EPI_ISL_731042, EPI_ISL_731043, EPI_ISL_731044, EPI_ISL_731046, EPI_ISL_731047, EPI_ISL_731048, EPI_ISL_731049, EPI_ISL_731051, EPI_ISL_731053, EPI_ISL_731056, EPI_ISL_731058, EPI_ISL_731060, EPI_ISL_731061, EPI_ISL_731062, EPI_ISL_731063, EPI_ISL_731064, EPI_ISL_731067, EPI_ISL_731068, EPI_ISL_731069, EPI_ISL_731070, EPI_ISL_731071, EPI_ISL_731072, EPI_ISL_731073, EPI_ISL_731075, EPI_ISL_731077, EPI_ISL_731078, EPI_ISL_731081, EPI_ISL_731084, EPI_ISL_731085, EPI_ISL_731089, EPI_ISL_731090, EPI_ISL_731091, EPI_ISL_731092, EPI_ISL_731093, EPI_ISL_731095, EPI_ISL_731097, EPI_ISL_731098, EPI_ISL_731099, EPI_ISL_731100, EPI_ISL_731101, EPI_ISL_731102, EPI_ISL_731103, EPI_ISL_731105, EPI_ISL_731108, EPI_ISL_731109, EPI_ISL_731111, EPI_ISL_731112, EPI_ISL_731113, EPI_ISL_731114, EPI_ISL_731115, EPI_ISL_731117, EPI_ISL_731118, EPI_ISL_731121, EPI_ISL_731122, EPI_ISL_731124, EPI_ISL_731125, EPI_ISL_731127, EPI_ISL_731128, EPI_ISL_731132, EPI_ISL_731134, EPI_ISL_731136, EPI_ISL_731138, EPI_ISL_731139, EPI_ISL_731140, EPI_ISL_731142, EPI_ISL_731144, EPI_ISL_731145, EPI_ISL_731146, EPI_ISL_731148, EPI_ISL_731149, EPI_ISL_731150, EPI_ISL_731151, EPI_ISL_731152, EPI_ISL_731156, EPI_ISL_731159, EPI_ISL_731162, EPI_ISL_731163, EPI_ISL_731164, EPI_ISL_731165, EPI_ISL_731166, EPI_ISL_731167, EPI_ISL_731168, EPI_ISL_731170, EPI_ISL_731172, EPI_ISL_731173, EPI_ISL_731174, EPI_ISL_731175, EPI_ISL_731176, EPI_ISL_731177, EPI_ISL_731178, EPI_ISL_731179, EPI_ISL_731180, EPI_ISL_731182, EPI_ISL_731184, EPI_ISL_731185, EPI_ISL_731186, EPI_ISL_731187, EPI_ISL_731189, EPI_ISL_731190, EPI_ISL_731191, EPI_ISL_731192, EPI_ISL_731193, EPI_ISL_731194, EPI_ISL_731195, EPI_ISL_731197, EPI_ISL_731198, EPI_ISL_731201, EPI_ISL_731202, EPI_ISL_731203, EPI_ISL_731205, EPI_ISL_731207, EPI_ISL_731208, EPI_ISL_731209, EPI_ISL_731210, EPI_ISL_731211, EPI_ISL_731212, EPI_ISL_731213, EPI_ISL_731216, EPI_ISL_731217, EPI_ISL_731218, EPI_ISL_731219, EPI_ISL_731220, EPI_ISL_731221, EPI_ISL_731222, EPI_ISL_731223, EPI_ISL_731224, EPI_ISL_731225, EPI_ISL_731226, EPI_ISL_731227, EPI_ISL_731228, EPI_ISL_731229, EPI_ISL_731230, EPI_ISL_731231, EPI_ISL_731232, EPI_ISL_731234, EPI_ISL_731236, EPI_ISL_731237, EPI_ISL_731238, EPI_ISL_731239, EPI_ISL_731241, EPI_ISL_731242, EPI_ISL_731243, EPI_ISL_731244, EPI_ISL_731245, EPI_ISL_731246, EPI_ISL_731249, EPI_ISL_731250, EPI_ISL_731552, EPI_ISL_731553, EPI_ISL_731554, EPI_ISL_731556, EPI_ISL_731557, EPI_ISL_731558, EPI_ISL_731559, EPI_ISL_731560, EPI_ISL_731561, EPI_ISL_731562, EPI_ISL_731563, EPI_ISL_731564, EPI_ISL_731566, EPI_ISL_731567, EPI_ISL_731568, EPI_ISL_731569, EPI_ISL_731572, EPI_ISL_731573, EPI_ISL_731574, EPI_ISL_731575, EPI_ISL_731577, EPI_ISL_731578, EPI_ISL_731579, EPI_ISL_731581, EPI_ISL_731582, EPI_ISL_731583, EPI_ISL_731586, EPI_ISL_731587, EPI_ISL_731589, EPI_ISL_731590, EPI_ISL_731591, EPI_ISL_731593, EPI_ISL_731594, EPI_ISL_731595, EPI_ISL_731596, EPI_ISL_731597, EPI_ISL_731598, EPI_ISL_731600, EPI_ISL_731601, EPI_ISL_731602, EPI_ISL_731603, EPI_ISL_731604, EPI_ISL_731605, EPI_ISL_731606, EPI_ISL_731607, EPI_ISL_731608, EPI_ISL_731609, EPI_ISL_731610, EPI_ISL_731612, EPI_ISL_731613, EPI_ISL_731614, EPI_ISL_731615, EPI_ISL_731616, EPI_ISL_731617, EPI_ISL_731618, EPI_ISL_731619, EPI_ISL_731620, EPI_ISL_731621, EPI_ISL_731622, EPI_ISL_731623, EPI_ISL_731625, EPI_ISL_731627, EPI_ISL_731628, EPI_ISL_731630, EPI_ISL_731631, EPI_ISL_731633, EPI_ISL_731634, EPI_ISL_731636, EPI_ISL_731637, EPI_ISL_731638, EPI_ISL_731642, EPI_ISL_731643, EPI_ISL_731645, EPI_ISL_731646, EPI_ISL_731650, EPI_ISL_731651, EPI_ISL_731652, EPI_ISL_731653, EPI_ISL_731654, EPI_ISL_731657, EPI_ISL_731658, EPI_ISL_731659, EPI_ISL_731660, EPI_ISL_731661, EPI_ISL_731662, EPI_ISL_731663, EPI_ISL_731666, EPI_ISL_731668, EPI_ISL_731669, EPI_ISL_731670, EPI_ISL_731672, EPI_ISL_731673, EPI_ISL_731674, EPI_ISL_731675, EPI_ISL_731676, EPI_ISL_731677, EPI_ISL_731678, EPI_ISL_731679, EPI_ISL_731681, EPI_ISL_731682, EPI_ISL_731683, EPI_ISL_731685, EPI_ISL_731687, EPI_ISL_731689, EPI_ISL_731694, EPI_ISL_731695, EPI_ISL_731696, EPI_ISL_731698, EPI_ISL_731699, EPI_ISL_731700, EPI_ISL_731702, EPI_ISL_731703, EPI_ISL_731704, EPI_ISL_731706, EPI_ISL_731708, EPI_ISL_731710, EPI_ISL_731712, EPI_ISL_731713, EPI_ISL_731715, EPI_ISL_731716, EPI_ISL_731717, EPI_ISL_731718, EPI_ISL_731720, EPI_ISL_731723, EPI_ISL_731725, EPI_ISL_731726, EPI_ISL_731727, EPI_ISL_731729, EPI_ISL_731730, EPI_ISL_731732, EPI_ISL_731733, EPI_ISL_731734, EPI_ISL_731738, EPI_ISL_731739, EPI_ISL_731740, EPI_ISL_731741, EPI_ISL_731742, EPI_ISL_731743, EPI_ISL_731744, EPI_ISL_731745, EPI_ISL_731746, EPI_ISL_731747, EPI_ISL_731748, EPI_ISL_731749, EPI_ISL_731750, EPI_ISL_731751, EPI_ISL_731752, EPI_ISL_731753, EPI_ISL_731754, EPI_ISL_731755, EPI_ISL_731756, EPI_ISL_731757, EPI_ISL_731758, EPI_ISL_731759, EPI_ISL_731760, EPI_ISL_731761, EPI_ISL_731762, EPI_ISL_731764, EPI_ISL_731765, EPI_ISL_731766, EPI_ISL_731768, EPI_ISL_731769, EPI_ISL_731771, EPI_ISL_731772, EPI_ISL_731773, EPI_ISL_731774, EPI_ISL_731776, EPI_ISL_731777, EPI_ISL_731778, EPI_ISL_731779, EPI_ISL_731780, EPI_ISL_731781, EPI_ISL_731782, EPI_ISL_731783, EPI_ISL_731784, EPI_ISL_731786, EPI_ISL_731789, EPI_ISL_731790, EPI_ISL_731791, EPI_ISL_731793, EPI_ISL_731794, EPI_ISL_731797, EPI_ISL_731798, EPI_ISL_731801, EPI_ISL_731802, EPI_ISL_731803, EPI_ISL_731804, EPI_ISL_731805, EPI_ISL_731807, EPI_ISL_731809, EPI_ISL_731811, EPI_ISL_731812, EPI_ISL_731813, EPI_ISL_731814, EPI_ISL_731815, EPI_ISL_731816, EPI_ISL_731817, EPI_ISL_731818, EPI_ISL_731820, EPI_ISL_731821, EPI_ISL_731822, EPI_ISL_731823, EPI_ISL_731825, EPI_ISL_731826, EPI_ISL_731828, EPI_ISL_731829, EPI_ISL_731830, EPI_ISL_731831, EPI_ISL_731832, EPI_ISL_731833, EPI_ISL_731834, EPI_ISL_731835, EPI_ISL_731837, EPI_ISL_731839, EPI_ISL_731843, EPI_ISL_731844, EPI_ISL_731845, EPI_ISL_731847, EPI_ISL_731848, EPI_ISL_731849, EPI_ISL_731851, EPI_ISL_731852, EPI_ISL_731853, EPI_ISL_731855, EPI_ISL_731856, EPI_ISL_731857, EPI_ISL_731859, EPI_ISL_731860, EPI_ISL_731861, EPI_ISL_731862, EPI_ISL_731864, EPI_ISL_731865, EPI_ISL_731866, EPI_ISL_731867, EPI_ISL_731869, EPI_ISL_731870, EPI_ISL_731871, EPI_ISL_731872, EPI_ISL_731873, EPI_ISL_731876, EPI_ISL_731878, EPI_ISL_731879, EPI_ISL_731880, EPI_ISL_731882, EPI_ISL_731883, EPI_ISL_731886, EPI_ISL_731890, EPI_ISL_735860, EPI_ISL_735862, EPI_ISL_735863, EPI_ISL_735865, EPI_ISL_735866, EPI_ISL_735867, EPI_ISL_735868, EPI_ISL_735869, EPI_ISL_735870, EPI_ISL_735871, EPI_ISL_735872, EPI_ISL_735873, EPI_ISL_735874, EPI_ISL_735875, EPI_ISL_735876, EPI_ISL_735877, EPI_ISL_735879, EPI_ISL_735881, EPI_ISL_735882, EPI_ISL_735883, EPI_ISL_735884, EPI_ISL_735885, EPI_ISL_735887, EPI_ISL_735888, EPI_ISL_735889, EPI_ISL_735890, EPI_ISL_735891, EPI_ISL_735892, EPI_ISL_735893, EPI_ISL_735895, EPI_ISL_735896, EPI_ISL_735897, EPI_ISL_735898, EPI_ISL_735899, EPI_ISL_735900, EPI_ISL_735901, EPI_ISL_735902, EPI_ISL_735903, EPI_ISL_735904, EPI_ISL_735905, EPI_ISL_735906, EPI_ISL_735907, EPI_ISL_735908, EPI_ISL_735910, EPI_ISL_735911, EPI_ISL_735912, EPI_ISL_735913, EPI_ISL_735914, EPI_ISL_735915, EPI_ISL_735916, EPI_ISL_735917, EPI_ISL_735918, EPI_ISL_735919, EPI_ISL_735921, EPI_ISL_735922, EPI_ISL_735923, EPI_ISL_735924, EPI_ISL_735925, EPI_ISL_735926, EPI_ISL_735928, EPI_ISL_735929, EPI_ISL_735930, EPI_ISL_735931, EPI_ISL_735932, EPI_ISL_735933, EPI_ISL_735934, EPI_ISL_735935, EPI_ISL_735936, EPI_ISL_735938, EPI_ISL_735940, EPI_ISL_735941, EPI_ISL_735942, EPI_ISL_735943, EPI_ISL_735944, EPI_ISL_735946, EPI_ISL_735947, EPI_ISL_735948, EPI_ISL_735950, EPI_ISL_735951, EPI_ISL_735953, EPI_ISL_735954, EPI_ISL_735955, EPI_ISL_735957, EPI_ISL_735958, EPI_ISL_735959, EPI_ISL_735960, EPI_ISL_735962, EPI_ISL_735963, EPI_ISL_735964, EPI_ISL_735966, EPI_ISL_735967, EPI_ISL_735969, EPI_ISL_735970, EPI_ISL_735971, EPI_ISL_735974, EPI_ISL_735975, EPI_ISL_735976, EPI_ISL_735977, EPI_ISL_735979, EPI_ISL_735980, EPI_ISL_735981, EPI_ISL_735983, EPI_ISL_735984, EPI_ISL_735985, EPI_ISL_735986, EPI_ISL_735988, EPI_ISL_735989, EPI_ISL_735990, EPI_ISL_735991, EPI_ISL_735992, EPI_ISL_735994, EPI_ISL_735997, EPI_ISL_735998, EPI_ISL_735999, EPI_ISL_736000, EPI_ISL_736002, EPI_ISL_736004, EPI_ISL_736005, EPI_ISL_736006, EPI_ISL_736008, EPI_ISL_736009, EPI_ISL_736010, EPI_ISL_736011, EPI_ISL_736012, EPI_ISL_736013, EPI_ISL_736014, EPI_ISL_736016, EPI_ISL_736020, EPI_ISL_736021, EPI_ISL_736022, EPI_ISL_736023, EPI_ISL_736026, EPI_ISL_736028, EPI_ISL_736029, EPI_ISL_736030, EPI_ISL_736031, EPI_ISL_736032, EPI_ISL_736034, EPI_ISL_736035, EPI_ISL_736036, EPI_ISL_736037, EPI_ISL_736039, EPI_ISL_736040, EPI_ISL_736042, EPI_ISL_736043, EPI_ISL_736044, EPI_ISL_736045, EPI_ISL_736046, EPI_ISL_736047, EPI_ISL_736048, EPI_ISL_736049, EPI_ISL_736050, EPI_ISL_736051, EPI_ISL_736053, EPI_ISL_736054, EPI_ISL_736055, EPI_ISL_736056, EPI_ISL_736058, EPI_ISL_736060, EPI_ISL_736061, EPI_ISL_736062, EPI_ISL_736063, EPI_ISL_736065, EPI_ISL_736067, EPI_ISL_736068, EPI_ISL_736069, EPI_ISL_736070, EPI_ISL_736071, EPI_ISL_736073, EPI_ISL_736074, EPI_ISL_736075, EPI_ISL_736076, EPI_ISL_736077, EPI_ISL_736078, EPI_ISL_736079, EPI_ISL_736080, EPI_ISL_736081, EPI_ISL_736082, EPI_ISL_736083, EPI_ISL_736084, EPI_ISL_736085, EPI_ISL_736086, EPI_ISL_736087, EPI_ISL_736088, EPI_ISL_736089, EPI_ISL_736093, EPI_ISL_736094, EPI_ISL_736095, EPI_ISL_736097, EPI_ISL_736098, EPI_ISL_736103, EPI_ISL_736105, EPI_ISL_736106, EPI_ISL_736108, EPI_ISL_736109, EPI_ISL_736110, EPI_ISL_736111, EPI_ISL_736112, EPI_ISL_736113, EPI_ISL_736114, EPI_ISL_736116, EPI_ISL_736117, EPI_ISL_736118, EPI_ISL_736119, EPI_ISL_736120, EPI_ISL_736121, EPI_ISL_736122, EPI_ISL_736123, EPI_ISL_736125, EPI_ISL_736126, EPI_ISL_736127, EPI_ISL_736128, EPI_ISL_736129, EPI_ISL_736133, EPI_ISL_736134, EPI_ISL_736136, EPI_ISL_736137, EPI_ISL_736138, EPI_ISL_736139, EPI_ISL_736140, EPI_ISL_736141, EPI_ISL_736142, EPI_ISL_736144, EPI_ISL_736145, EPI_ISL_736147, EPI_ISL_736148, EPI_ISL_736150, EPI_ISL_736152, EPI_ISL_736153, EPI_ISL_736154, EPI_ISL_736155, EPI_ISL_736156, EPI_ISL_736157, EPI_ISL_736158, EPI_ISL_736160, EPI_ISL_736161, EPI_ISL_736163, EPI_ISL_736164, EPI_ISL_736165, EPI_ISL_736166, EPI_ISL_736170, EPI_ISL_736171, EPI_ISL_736172, EPI_ISL_736173, EPI_ISL_736174, EPI_ISL_736176, EPI_ISL_736177, EPI_ISL_736178, EPI_ISL_736179, EPI_ISL_736180, EPI_ISL_736181, EPI_ISL_736185, EPI_ISL_736186, EPI_ISL_736187, EPI_ISL_736188, EPI_ISL_736189, EPI_ISL_736192, EPI_ISL_736193, EPI_ISL_736194, EPI_ISL_736195, EPI_ISL_736196, EPI_ISL_736197, EPI_ISL_736198, EPI_ISL_736199, EPI_ISL_736200, EPI_ISL_736481, EPI_ISL_736482, EPI_ISL_736484, EPI_ISL_736485, EPI_ISL_736486, EPI_ISL_736493, EPI_ISL_736494, EPI_ISL_736495, EPI_ISL_736496, EPI_ISL_736498, EPI_ISL_736499, EPI_ISL_736500, EPI_ISL_736505, EPI_ISL_736506, EPI_ISL_736507, EPI_ISL_736509, EPI_ISL_736510, EPI_ISL_736511, EPI_ISL_736513, EPI_ISL_736516, EPI_ISL_736517, EPI_ISL_736518, EPI_ISL_736520, EPI_ISL_736521, EPI_ISL_736522, EPI_ISL_736524, EPI_ISL_736526, EPI_ISL_736530, EPI_ISL_736531, EPI_ISL_736534, EPI_ISL_736535, EPI_ISL_736537, EPI_ISL_736539, EPI_ISL_736540, EPI_ISL_736541, EPI_ISL_736544, EPI_ISL_736545, EPI_ISL_736546, EPI_ISL_736548, EPI_ISL_736553, EPI_ISL_736554, EPI_ISL_736555, EPI_ISL_736557, EPI_ISL_736559, EPI_ISL_736560, EPI_ISL_736561, EPI_ISL_736562, EPI_ISL_736564, EPI_ISL_736566, EPI_ISL_736568, EPI_ISL_736569, EPI_ISL_736570, EPI_ISL_736572, EPI_ISL_736574, EPI_ISL_736575, EPI_ISL_736576, EPI_ISL_736577, EPI_ISL_736579, EPI_ISL_736581, EPI_ISL_736583, EPI_ISL_736584, EPI_ISL_736585, EPI_ISL_736587, EPI_ISL_736589, EPI_ISL_736591, EPI_ISL_736592, EPI_ISL_736595, EPI_ISL_736596, EPI_ISL_736599, EPI_ISL_736600, EPI_ISL_736603, EPI_ISL_736604, EPI_ISL_736606, EPI_ISL_736607, EPI_ISL_736611, EPI_ISL_736612, EPI_ISL_736613, EPI_ISL_736614, EPI_ISL_736615, EPI_ISL_736616, EPI_ISL_736619, EPI_ISL_736621, EPI_ISL_736623, EPI_ISL_736625, EPI_ISL_736626, EPI_ISL_736629, EPI_ISL_736632, EPI_ISL_736634, EPI_ISL_736635, EPI_ISL_736636, EPI_ISL_736638, EPI_ISL_736639, EPI_ISL_736640, EPI_ISL_736641, EPI_ISL_736642, EPI_ISL_736644, EPI_ISL_736645, EPI_ISL_736647, EPI_ISL_736649, EPI_ISL_736651, EPI_ISL_736652, EPI_ISL_736653, EPI_ISL_736654, EPI_ISL_736655, EPI_ISL_736656, EPI_ISL_736658, EPI_ISL_736660, EPI_ISL_736661, EPI_ISL_736662, EPI_ISL_736663, EPI_ISL_736664, EPI_ISL_736665, EPI_ISL_736666, EPI_ISL_736667, EPI_ISL_736669, EPI_ISL_736670, EPI_ISL_736671, EPI_ISL_736673, EPI_ISL_736674, EPI_ISL_736676, EPI_ISL_736679, EPI_ISL_736680, EPI_ISL_736684, EPI_ISL_736687, EPI_ISL_736688, EPI_ISL_736690, EPI_ISL_736692, EPI_ISL_736698, EPI_ISL_736701, EPI_ISL_736702, EPI_ISL_736704, EPI_ISL_736705, EPI_ISL_736706, EPI_ISL_736707, EPI_ISL_736708, EPI_ISL_736709, EPI_ISL_736710, EPI_ISL_736711, EPI_ISL_736714, EPI_ISL_736715, EPI_ISL_736718, EPI_ISL_736720, EPI_ISL_736722, EPI_ISL_736726, EPI_ISL_736729, EPI_ISL_736730, EPI_ISL_736732, EPI_ISL_736735, EPI_ISL_736739, EPI_ISL_736741, EPI_ISL_736742, EPI_ISL_736747, EPI_ISL_736748, EPI_ISL_736750, EPI_ISL_736751, EPI_ISL_736753, EPI_ISL_736755, EPI_ISL_736756, EPI_ISL_736759, EPI_ISL_736760, EPI_ISL_736761, EPI_ISL_736762, EPI_ISL_736763, EPI_ISL_736764, EPI_ISL_736765, EPI_ISL_736766, EPI_ISL_736767, EPI_ISL_736768, EPI_ISL_736769, EPI_ISL_736770, EPI_ISL_736771, EPI_ISL_756470, EPI_ISL_760567, EPI_ISL_760600, EPI_ISL_760649, EPI_ISL_760924, EPI_ISL_760989, EPI_ISL_761096, EPI_ISL_761727, EPI_ISL_767120, EPI_ISL_767121, EPI_ISL_767122, EPI_ISL_767123, EPI_ISL_767124, EPI_ISL_767125, EPI_ISL_767126, EPI_ISL_767127, EPI_ISL_767128, EPI_ISL_767129, EPI_ISL_767130, EPI_ISL_767131, EPI_ISL_767132, EPI_ISL_767133, EPI_ISL_767135, EPI_ISL_767136, EPI_ISL_767137, EPI_ISL_767139, EPI_ISL_767140, EPI_ISL_767141, EPI_ISL_767142, EPI_ISL_767143, EPI_ISL_767144, EPI_ISL_767145, EPI_ISL_767146, EPI_ISL_767147, EPI_ISL_767148, EPI_ISL_767149, EPI_ISL_767150, EPI_ISL_767178, EPI_ISL_767216, EPI_ISL_767217, EPI_ISL_767218, EPI_ISL_767219, EPI_ISL_767220, EPI_ISL_767222, EPI_ISL_767223, EPI_ISL_767224, EPI_ISL_767225, EPI_ISL_767226, EPI_ISL_767227, EPI_ISL_767228, EPI_ISL_767229, EPI_ISL_767231, EPI_ISL_767232, EPI_ISL_767233, EPI_ISL_767235, EPI_ISL_767236, EPI_ISL_767237, EPI_ISL_767238, EPI_ISL_767277, EPI_ISL_767279, EPI_ISL_767280, EPI_ISL_767281, EPI_ISL_767282, EPI_ISL_767283, EPI_ISL_767285, EPI_ISL_767286, EPI_ISL_767287, EPI_ISL_767288, EPI_ISL_767289, EPI_ISL_767290, EPI_ISL_777012, EPI_ISL_777015, EPI_ISL_777033, EPI_ISL_777042, EPI_ISL_777044, EPI_ISL_777046, EPI_ISL_777053, EPI_ISL_777057, EPI_ISL_777063, EPI_ISL_777067, EPI_ISL_777070, EPI_ISL_777071, EPI_ISL_777078, EPI_ISL_777083, EPI_ISL_777091, EPI_ISL_777097, EPI_ISL_777106, EPI_ISL_777114, EPI_ISL_777128, EPI_ISL_777129, EPI_ISL_777132, EPI_ISL_777153, EPI_ISL_777155, EPI_ISL_777167, EPI_ISL_777177, EPI_ISL_777182, EPI_ISL_777183, EPI_ISL_777190, EPI_ISL_777193, EPI_ISL_777197, EPI_ISL_777202, EPI_ISL_777211, EPI_ISL_777212, EPI_ISL_777213, EPI_ISL_777221, EPI_ISL_777226, EPI_ISL_777228, EPI_ISL_777240, EPI_ISL_777243, EPI_ISL_777244, EPI_ISL_777267, EPI_ISL_777270, EPI_ISL_777273, EPI_ISL_777278, EPI_ISL_777279, EPI_ISL_777282, EPI_ISL_777285, EPI_ISL_777286, EPI_ISL_777292, EPI_ISL_777311, EPI_ISL_777314, EPI_ISL_777333, EPI_ISL_777380, EPI_ISL_777430, EPI_ISL_777520, EPI_ISL_777524, EPI_ISL_777592, EPI_ISL_777650, EPI_ISL_777669, EPI_ISL_778305, EPI_ISL_778309, EPI_ISL_778311, EPI_ISL_778315, EPI_ISL_778327, EPI_ISL_778328, EPI_ISL_778331, EPI_ISL_778333, EPI_ISL_778335, EPI_ISL_778342, EPI_ISL_778346, EPI_ISL_778347, EPI_ISL_778348, EPI_ISL_778353, EPI_ISL_778355, EPI_ISL_778357, EPI_ISL_778358, EPI_ISL_778360, EPI_ISL_778365, EPI_ISL_778368, EPI_ISL_778374, EPI_ISL_778380, EPI_ISL_778382, EPI_ISL_778385, EPI_ISL_778390, EPI_ISL_778392, EPI_ISL_778396, EPI_ISL_778399, EPI_ISL_778401, EPI_ISL_778404, EPI_ISL_778408, EPI_ISL_778410, EPI_ISL_778412, EPI_ISL_778415, EPI_ISL_778416, EPI_ISL_778417, EPI_ISL_778426, EPI_ISL_778430, EPI_ISL_778432, EPI_ISL_778444, EPI_ISL_778448, EPI_ISL_778454, EPI_ISL_778455, EPI_ISL_778471, EPI_ISL_778472, EPI_ISL_778473, EPI_ISL_778478, EPI_ISL_778484, EPI_ISL_778487, EPI_ISL_778488, EPI_ISL_778490, EPI_ISL_778491, EPI_ISL_778494, EPI_ISL_778500, EPI_ISL_778510, EPI_ISL_778512, EPI_ISL_778517, EPI_ISL_778519, EPI_ISL_778524, EPI_ISL_778528, EPI_ISL_778529, EPI_ISL_778531, EPI_ISL_778539, EPI_ISL_778543, EPI_ISL_778547, EPI_ISL_778556, EPI_ISL_778558, EPI_ISL_778562, EPI_ISL_778564, EPI_ISL_778569, EPI_ISL_778571, EPI_ISL_778578, EPI_ISL_778584, EPI_ISL_778593, EPI_ISL_778598, EPI_ISL_778599, EPI_ISL_778601, EPI_ISL_778605, EPI_ISL_778607, EPI_ISL_778612, EPI_ISL_778614, EPI_ISL_778615, EPI_ISL_778616, EPI_ISL_778618, EPI_ISL_778621, EPI_ISL_778623, EPI_ISL_778627, EPI_ISL_778628, EPI_ISL_778630, EPI_ISL_778635, EPI_ISL_778638, EPI_ISL_820129, EPI_ISL_820130, EPI_ISL_820132, EPI_ISL_820133, EPI_ISL_820135, EPI_ISL_820137, EPI_ISL_820139, EPI_ISL_820140, EPI_ISL_820142, EPI_ISL_820143, EPI_ISL_820145, EPI_ISL_820148, EPI_ISL_820153, EPI_ISL_820155, EPI_ISL_820157, EPI_ISL_820158, EPI_ISL_820160, EPI_ISL_820162, EPI_ISL_820163, EPI_ISL_820165, EPI_ISL_820166, EPI_ISL_820168, EPI_ISL_820170, EPI_ISL_820173, EPI_ISL_820174, EPI_ISL_820179, EPI_ISL_820181, EPI_ISL_820185, EPI_ISL_820187, EPI_ISL_820189, EPI_ISL_820192, EPI_ISL_820194, EPI_ISL_820195, EPI_ISL_820198, EPI_ISL_820201, EPI_ISL_820203, EPI_ISL_820205, EPI_ISL_820207, EPI_ISL_820208, EPI_ISL_820210, EPI_ISL_820214, EPI_ISL_820217, EPI_ISL_820219, EPI_ISL_820222, EPI_ISL_820224, EPI_ISL_820228, EPI_ISL_820230, EPI_ISL_820232, EPI_ISL_820235, EPI_ISL_820237, EPI_ISL_820239, EPI_ISL_820240, EPI_ISL_820242, EPI_ISL_820245, EPI_ISL_820247, EPI_ISL_820248, EPI_ISL_820250, EPI_ISL_820252, EPI_ISL_820254, EPI_ISL_820255, EPI_ISL_820257, EPI_ISL_820258, EPI_ISL_820263, EPI_ISL_820265, EPI_ISL_820267, EPI_ISL_820269, EPI_ISL_820274, EPI_ISL_820275, EPI_ISL_820277, EPI_ISL_820278, EPI_ISL_820280, EPI_ISL_820281, EPI_ISL_820285, EPI_ISL_820286, EPI_ISL_820288, EPI_ISL_820290, EPI_ISL_820291, EPI_ISL_820293, EPI_ISL_820296, EPI_ISL_820298, EPI_ISL_820299, EPI_ISL_820301, EPI_ISL_820305, EPI_ISL_820307, EPI_ISL_820308, EPI_ISL_820310, EPI_ISL_820313, EPI_ISL_820315, EPI_ISL_820317, EPI_ISL_820322, EPI_ISL_820324, EPI_ISL_820325, EPI_ISL_820327, EPI_ISL_820329, EPI_ISL_820330, EPI_ISL_820332, EPI_ISL_820334, EPI_ISL_820336, EPI_ISL_820338, EPI_ISL_820339, EPI_ISL_820341, EPI_ISL_820342, EPI_ISL_820344, EPI_ISL_820346, EPI_ISL_820348, EPI_ISL_820351, EPI_ISL_820352, EPI_ISL_820354, EPI_ISL_820357, EPI_ISL_820359, EPI_ISL_820361, EPI_ISL_820362, EPI_ISL_820364, EPI_ISL_820366, EPI_ISL_820367, EPI_ISL_820369, EPI_ISL_820370, EPI_ISL_820372, EPI_ISL_820374, EPI_ISL_820375, EPI_ISL_820377, EPI_ISL_820379, EPI_ISL_820380, EPI_ISL_820382, EPI_ISL_820389, EPI_ISL_820390, EPI_ISL_820392, EPI_ISL_820393, EPI_ISL_820395, EPI_ISL_820398, EPI_ISL_820400, EPI_ISL_820401, EPI_ISL_820403, EPI_ISL_820405, EPI_ISL_820406, EPI_ISL_820408, EPI_ISL_820411, EPI_ISL_820413, EPI_ISL_820414, EPI_ISL_820416, EPI_ISL_820419, EPI_ISL_820422, EPI_ISL_820424, EPI_ISL_820425, EPI_ISL_820427, EPI_ISL_820429, EPI_ISL_820430, EPI_ISL_820432, EPI_ISL_820434, EPI_ISL_820435, EPI_ISL_820438, EPI_ISL_820440, EPI_ISL_820442, EPI_ISL_820443, EPI_ISL_820445, EPI_ISL_820447, EPI_ISL_820450, EPI_ISL_820452, EPI_ISL_820453, EPI_ISL_820455, EPI_ISL_820457, EPI_ISL_820458, EPI_ISL_820460, EPI_ISL_820463, EPI_ISL_820466, EPI_ISL_820469, EPI_ISL_820471, EPI_ISL_820473, EPI_ISL_820474, EPI_ISL_820476, EPI_ISL_820482, EPI_ISL_820486, EPI_ISL_820487, EPI_ISL_820490, EPI_ISL_820492, EPI_ISL_820494, EPI_ISL_820495, EPI_ISL_820497, EPI_ISL_820499, EPI_ISL_820500, EPI_ISL_820502, EPI_ISL_820503, EPI_ISL_820505, EPI_ISL_820508, EPI_ISL_820515, EPI_ISL_820517, EPI_ISL_820520, EPI_ISL_820522, EPI_ISL_820523, EPI_ISL_820525, EPI_ISL_820527, EPI_ISL_820529, EPI_ISL_820530, EPI_ISL_820532, EPI_ISL_820534, EPI_ISL_820535, EPI_ISL_820537, EPI_ISL_820538, EPI_ISL_820540, EPI_ISL_820542, EPI_ISL_820544, EPI_ISL_820545, EPI_ISL_820548, EPI_ISL_820550, EPI_ISL_820551, EPI_ISL_820553, EPI_ISL_820555, EPI_ISL_820556, EPI_ISL_820559, EPI_ISL_820561, EPI_ISL_820562, EPI_ISL_820564, EPI_ISL_820566, EPI_ISL_820567, EPI_ISL_820569, EPI_ISL_820571, EPI_ISL_820572, EPI_ISL_820574, EPI_ISL_820576, EPI_ISL_820579, EPI_ISL_820581, EPI_ISL_820582, EPI_ISL_820584, EPI_ISL_820585, EPI_ISL_820587, EPI_ISL_820589, EPI_ISL_820592, EPI_ISL_820594, EPI_ISL_820595, EPI_ISL_820599, EPI_ISL_820600, EPI_ISL_820602, EPI_ISL_820604, EPI_ISL_820606, EPI_ISL_820608, EPI_ISL_820611, EPI_ISL_820612, EPI_ISL_820613, EPI_ISL_820615, EPI_ISL_820616, EPI_ISL_820619, EPI_ISL_820621, EPI_ISL_820622, EPI_ISL_820623, EPI_ISL_820624, EPI_ISL_820625, EPI_ISL_820626, EPI_ISL_820627, EPI_ISL_820629, EPI_ISL_820630, EPI_ISL_820631, EPI_ISL_820632, EPI_ISL_820635, EPI_ISL_820636, EPI_ISL_820637, EPI_ISL_820638, EPI_ISL_820640, EPI_ISL_820642, EPI_ISL_820643, EPI_ISL_820646, EPI_ISL_826464, EPI_ISL_826465, EPI_ISL_858408, EPI_ISL_858409, EPI_ISL_858411, EPI_ISL_858413, EPI_ISL_858414, EPI_ISL_858416, EPI_ISL_858418, EPI_ISL_858422, EPI_ISL_858994, EPI_ISL_858998, EPI_ISL_859001, EPI_ISL_859004, EPI_ISL_859005, EPI_ISL_859008, EPI_ISL_859015, EPI_ISL_859018, EPI_ISL_859020, EPI_ISL_859021, EPI_ISL_859024, EPI_ISL_859401, EPI_ISL_892215, EPI_ISL_892216, EPI_ISL_945240, | | | |

EPI_ISL_945289

see above | Lighthouse Lab in Alderley Park | Wellcome Sanger Institute for the COVID-19 Genomics UK (COG-UK) Consortium | Cordelia Langford; David K. Jackson; Dominic Kwiatkowski; Ewan Harrison; Ian Johnston; Jacquelyn Wynn; John Sillitoe on behalf of the Wellcome Sanger Institute COVID-19 Surveillance Team; Mairead Hyland; Roberto Amato; Sonia Goncalves; The Lighthouse Lab in Alderley Park and Alex Alderton

EPI_ISL_718318, EPI_ISL_718319, EPI_ISL_718320, EPI_ISL_718321, EPI_ISL_718322, EPI_ISL_718323, EPI_ISL_718324, EPI_ISL_718325, EPI_ISL_718326, EPI_ISL_718327, EPI_ISL_718328, EPI_ISL_718329, EPI_ISL_718330, EPI_ISL_718331, EPI_ISL_718332, EPI_ISL_718333, EPI_ISL_718334, EPI_ISL_718335, EPI_ISL_718336, EPI_ISL_718337, EPI_ISL_718338, EPI_ISL_718339, EPI_ISL_718340, EPI_ISL_718341,
EPI_ISL_718342, EPI_ISL_718343, EPI_ISL_718344, EPI_ISL_718345, EPI_ISL_718346, EPI_ISL_718347, EPI_ISL_718348, EPI_ISL_718349, EPI_ISL_718350, EPI_ISL_718351, EPI_ISL_718352, EPI_ISL_718353, EPI_ISL_718354, EPI_ISL_718355, EPI_ISL_718356, EPI_ISL_718357, EPI_ISL_718358, EPI_ISL_718359, EPI_ISL_718360, EPI_ISL_718361, EPI_ISL_718362, EPI_ISL_718363, EPI_ISL_718364, EPI_ISL_718365,
EPI_ISL_718366, EPI_ISL_718367, EPI_ISL_718368, EPI_ISL_718369, EPI_ISL_718370, EPI_ISL_718371, EPI_ISL_718372, EPI_ISL_718373, EPI_ISL_718374, EPI_ISL_718375, EPI_ISL_718376, EPI_ISL_718377, EPI_ISL_718378, EPI_ISL_718379, EPI_ISL_718380, EPI_ISL_718381, EPI_ISL_718382, EPI_ISL_718383, EPI_ISL_718384, EPI_ISL_718385, EPI_ISL_718386, EPI_ISL_718387, EPI_ISL_718388, EPI_ISL_718389,
EPI_ISL_718390, EPI_ISL_718391, EPI_ISL_718392, EPI_ISL_718393, EPI_ISL_718394, EPI_ISL_718395, EPI_ISL_718396, EPI_ISL_718397, EPI_ISL_718398, EPI_ISL_718399, EPI_ISL_718400, EPI_ISL_718401, EPI_ISL_718402, EPI_ISL_718403, EPI_ISL_718404, EPI_ISL_718405, EPI_ISL_718406, EPI_ISL_718407, EPI_ISL_718408, EPI_ISL_718409, EPI_ISL_718410, EPI_ISL_718411, EPI_ISL_718412, EPI_ISL_718413,
EPI_ISL_718414, EPI_ISL_718415, EPI_ISL_718416, EPI_ISL_718417, EPI_ISL_718418, EPI_ISL_718419, EPI_ISL_718420, EPI_ISL_718421, EPI_ISL_718422, EPI_ISL_718423, EPI_ISL_718424, EPI_ISL_718425, EPI_ISL_718426, EPI_ISL_718427, EPI_ISL_718428, EPI_ISL_718429, EPI_ISL_718430, EPI_ISL_718431, EPI_ISL_718432, EPI_ISL_718433, EPI_ISL_718434, EPI_ISL_718435, EPI_ISL_718436, EPI_ISL_718437,
EPI_ISL_718438, EPI_ISL_718439, EPI_ISL_718440, EPI_ISL_718441, EPI_ISL_718442, EPI_ISL_718443, EPI_ISL_718444, EPI_ISL_718445, EPI_ISL_718446, EPI_ISL_718447, EPI_ISL_718448, EPI_ISL_718449, EPI_ISL_718450, EPI_ISL_718451, EPI_ISL_718452, EPI_ISL_718453, EPI_ISL_718454, EPI_ISL_718455, EPI_ISL_718456, EPI_ISL_718457, EPI_ISL_718458, EPI_ISL_718459, EPI_ISL_718460, EPI_ISL_718461,
EPI_ISL_718462, EPI_ISL_718463, EPI_ISL_718464, EPI_ISL_718465, EPI_ISL_718466, EPI_ISL_718467, EPI_ISL_718468, EPI_ISL_718469, EPI_ISL_718470, EPI_ISL_718471, EPI_ISL_718472, EPI_ISL_718473, EPI_ISL_718474, EPI_ISL_718475, EPI_ISL_718476, EPI_ISL_718477, EPI_ISL_718478, EPI_ISL_718479, EPI_ISL_718480, EPI_ISL_718481, EPI_ISL_718482, EPI_ISL_718483, EPI_ISL_718484, EPI_ISL_718485,
EPI_ISL_718486, EPI_ISL_718487, EPI_ISL_718488, EPI_ISL_718489, EPI_ISL_718490, EPI_ISL_718491, EPI_ISL_718492, EPI_ISL_718493, EPI_ISL_718494, EPI_ISL_718495, EPI_ISL_718496, EPI_ISL_718497, EPI_ISL_718498, EPI_ISL_718499, EPI_ISL_718500, EPI_ISL_718501, EPI_ISL_718502, EPI_ISL_718503, EPI_ISL_718504, EPI_ISL_718505, EPI_ISL_718506, EPI_ISL_718507, EPI_ISL_718508, EPI_ISL_718509,
EPI_ISL_718510, EPI_ISL_718511, EPI_ISL_718512, EPI_ISL_718513, EPI_ISL_718514, EPI_ISL_718515, EPI_ISL_718516, EPI_ISL_718517, EPI_ISL_718518, EPI_ISL_718519, EPI_ISL_718520, EPI_ISL_718521, EPI_ISL_718522, EPI_ISL_718523, EPI_ISL_718524, EPI_ISL_718525, EPI_ISL_718526, EPI_ISL_718527, EPI_ISL_718528, EPI_ISL_718529, EPI_ISL_718530, EPI_ISL_718531, EPI_ISL_718532, EPI_ISL_718533,
EPI_ISL_718534, EPI_ISL_718535, EPI_ISL_718536, EPI_ISL_718537, EPI_ISL_718538, EPI_ISL_718539, EPI_ISL_718540, EPI_ISL_718541, EPI_ISL_718542, EPI_ISL_718543, EPI_ISL_718544, EPI_ISL_718545, EPI_ISL_718546, EPI_ISL_718547, EPI_ISL_718548, EPI_ISL_718549, EPI_ISL_718550, EPI_ISL_718551, EPI_ISL_718552, EPI_ISL_718553, EPI_ISL_718554, EPI_ISL_718555, EPI_ISL_718556, EPI_ISL_718557,
EPI_ISL_718558, EPI_ISL_718559, EPI_ISL_718560, EPI_ISL_718561, EPI_ISL_718562, EPI_ISL_718563, EPI_ISL_718564, EPI_ISL_718565, EPI_ISL_718566, EPI_ISL_718567, EPI_ISL_718568, EPI_ISL_718569, EPI_ISL_718570, EPI_ISL_718571, EPI_ISL_718572, EPI_ISL_718573, EPI_ISL_718574, EPI_ISL_718575, EPI_ISL_718576, EPI_ISL_718577, EPI_ISL_718578, EPI_ISL_718579, EPI_ISL_718580, EPI_ISL_718581,
EPI_ISL_718582, EPI_ISL_718583, EPI_ISL_718584, EPI_ISL_718585, EPI_ISL_718586, EPI_ISL_718587, EPI_ISL_718588, EPI_ISL_718589, EPI_ISL_718590, EPI_ISL_718591, EPI_ISL_718592, EPI_ISL_718593, EPI_ISL_718594, EPI_ISL_718595, EPI_ISL_718596, EPI_ISL_718597, EPI_ISL_718598, EPI_ISL_718599, EPI_ISL_718600, EPI_ISL_718601, EPI_ISL_718602, EPI_ISL_718603, EPI_ISL_718604, EPI_ISL_718605,
EPI_ISL_718606, EPI_ISL_718607, EPI_ISL_718608, EPI_ISL_718609, EPI_ISL_718610, EPI_ISL_718611, EPI_ISL_718612, EPI_ISL_718613, EPI_ISL_718614, EPI_ISL_718615, EPI_ISL_718616, EPI_ISL_718617, EPI_ISL_718618, EPI_ISL_718619, EPI_ISL_718620, EPI_ISL_718621, EPI_ISL_718622, EPI_ISL_718623, EPI_ISL_718624, EPI_ISL_718625, EPI_ISL_718626, EPI_ISL_718627, EPI_ISL_718628, EPI_ISL_718630,
EPI_ISL_718632, EPI_ISL_718634, EPI_ISL_718636, EPI_ISL_718638, EPI_ISL_718642, EPI_ISL_718645, EPI_ISL_718646, EPI_ISL_718648, EPI_ISL_718649, EPI_ISL_718651, EPI_ISL_718652, EPI_ISL_718654, EPI_ISL_718655, EPI_ISL_718656, EPI_ISL_718659, EPI_ISL_718660, EPI_ISL_718661, EPI_ISL_718667, EPI_ISL_718668, EPI_ISL_718669, EPI_ISL_718671, EPI_ISL_718673, EPI_ISL_718677, EPI_ISL_718679,
EPI_ISL_718680, EPI_ISL_718681, EPI_ISL_718683, EPI_ISL_718684, EPI_ISL_718685, EPI_ISL_718688, EPI_ISL_718689, EPI_ISL_718690, EPI_ISL_718692, EPI_ISL_718693, EPI_ISL_718696, EPI_ISL_718697, EPI_ISL_718698, EPI_ISL_718699, EPI_ISL_718701, EPI_ISL_718702, EPI_ISL_718703, EPI_ISL_718704, EPI_ISL_718706, EPI_ISL_718708, EPI_ISL_718709, EPI_ISL_718717, EPI_ISL_718720, EPI_ISL_718722,
EPI_ISL_718726, EPI_ISL_718727, EPI_ISL_718730, EPI_ISL_718731, EPI_ISL_718732, EPI_ISL_718733, EPI_ISL_718734, EPI_ISL_718736, EPI_ISL_718737, EPI_ISL_718741, EPI_ISL_718743, EPI_ISL_718748, EPI_ISL_718749, EPI_ISL_718751, EPI_ISL_718752, EPI_ISL_718753, EPI_ISL_718756, EPI_ISL_718757, EPI_ISL_718758, EPI_ISL_718760, EPI_ISL_718763, EPI_ISL_718768, EPI_ISL_718770, EPI_ISL_718771,
EPI_ISL_718775, EPI_ISL_718776, EPI_ISL_718778, EPI_ISL_718784, EPI_ISL_718785, EPI_ISL_718786, EPI_ISL_718788, EPI_ISL_718790, EPI_ISL_718792, EPI_ISL_718793, EPI_ISL_718794, EPI_ISL_718797, EPI_ISL_718800, EPI_ISL_718804, EPI_ISL_718805, EPI_ISL_718809, EPI_ISL_718810, EPI_ISL_718814, EPI_ISL_718815, EPI_ISL_718816, EPI_ISL_718817, EPI_ISL_718818, EPI_ISL_718820, EPI_ISL_718822,
EPI_ISL_718824, EPI_ISL_718825, EPI_ISL_718826, EPI_ISL_718834, EPI_ISL_718835, EPI_ISL_718836, EPI_ISL_718840, EPI_ISL_718842, EPI_ISL_718843, EPI_ISL_718845, EPI_ISL_718846, EPI_ISL_718848, EPI_ISL_718851, EPI_ISL_718852, EPI_ISL_718855, EPI_ISL_718856, EPI_ISL_718857, EPI_ISL_718860, EPI_ISL_718861, EPI_ISL_718862, EPI_ISL_718863, EPI_ISL_718867, EPI_ISL_718868, EPI_ISL_718869,
EPI_ISL_718874, EPI_ISL_718875, EPI_ISL_718876, EPI_ISL_718879, EPI_ISL_718882, EPI_ISL_718883, EPI_ISL_718884, EPI_ISL_718892, EPI_ISL_718893, EPI_ISL_718896, EPI_ISL_718897, EPI_ISL_718898, EPI_ISL_718899, EPI_ISL_718900, EPI_ISL_718904, EPI_ISL_718905, EPI_ISL_718906, EPI_ISL_718909, EPI_ISL_718910, EPI_ISL_718912, EPI_ISL_718913, EPI_ISL_718916, EPI_ISL_718918, EPI_ISL_718920,
EPI_ISL_718921, EPI_ISL_718924, EPI_ISL_718925, EPI_ISL_718927, EPI_ISL_718928, EPI_ISL_718931, EPI_ISL_718933, EPI_ISL_718934, EPI_ISL_718937, EPI_ISL_718938, EPI_ISL_718940, EPI_ISL_718941, EPI_ISL_718942, EPI_ISL_718944, EPI_ISL_718946, EPI_ISL_718947, EPI_ISL_718948, EPI_ISL_718949, EPI_ISL_718951, EPI_ISL_718954, EPI_ISL_718956, EPI_ISL_718958, EPI_ISL_718960, EPI_ISL_718961,
EPI_ISL_718965, EPI_ISL_718968, EPI_ISL_718969, EPI_ISL_718971, EPI_ISL_718972, EPI_ISL_718973, EPI_ISL_718980, EPI_ISL_718981, EPI_ISL_718982, EPI_ISL_718983, EPI_ISL_718984, EPI_ISL_718985, EPI_ISL_718986, EPI_ISL_718987, EPI_ISL_718988, EPI_ISL_718989, EPI_ISL_718990, EPI_ISL_718991, EPI_ISL_718992, EPI_ISL_718993, EPI_ISL_718994, EPI_ISL_718995, EPI_ISL_718996, EPI_ISL_718997,
EPI_ISL_718998, EPI_ISL_718999, EPI_ISL_719000, EPI_ISL_719001, EPI_ISL_719002, EPI_ISL_719003, EPI_ISL_719004, EPI_ISL_719005, EPI_ISL_719006, EPI_ISL_719007, EPI_ISL_719008, EPI_ISL_719009, EPI_ISL_719010, EPI_ISL_719011, EPI_ISL_719012, EPI_ISL_719013, EPI_ISL_719014, EPI_ISL_719015, EPI_ISL_719016, EPI_ISL_719017, EPI_ISL_719018, EPI_ISL_719019, EPI_ISL_719020, EPI_ISL_719021,
EPI_ISL_719022, EPI_ISL_719023, EPI_ISL_719024, EPI_ISL_719025, EPI_ISL_719026, EPI_ISL_719027, EPI_ISL_719028, EPI_ISL_719029, EPI_ISL_719030, EPI_ISL_719031, EPI_ISL_719032, EPI_ISL_719033, EPI_ISL_719034, EPI_ISL_719035, EPI_ISL_719036, EPI_ISL_719037, EPI_ISL_719038, EPI_ISL_719039, EPI_ISL_719040, EPI_ISL_719041, EPI_ISL_719042, EPI_ISL_719043, EPI_ISL_719044, EPI_ISL_719045,
EPI_ISL_719046, EPI_ISL_719047, EPI_ISL_719048, EPI_ISL_719049, EPI_ISL_719050, EPI_ISL_719051, EPI_ISL_719052, EPI_ISL_719053, EPI_ISL_719054, EPI_ISL_719055, EPI_ISL_719056, EPI_ISL_719057, EPI_ISL_719058, EPI_ISL_719059, EPI_ISL_719060, EPI_ISL_719061, EPI_ISL_719062, EPI_ISL_719063, EPI_ISL_719064, EPI_ISL_719065, EPI_ISL_719066, EPI_ISL_719067, EPI_ISL_719068, EPI_ISL_719069,
EPI_ISL_719070, EPI_ISL_719071, EPI_ISL_719072, EPI_ISL_719073, EPI_ISL_719074, EPI_ISL_719075, EPI_ISL_719076, EPI_ISL_719077, EPI_ISL_719078, EPI_ISL_719079, EPI_ISL_719080, EPI_ISL_719081, EPI_ISL_719082, EPI_ISL_719083, EPI_ISL_719084, EPI_ISL_719085, EPI_ISL_719086, EPI_ISL_719087, EPI_ISL_719088, EPI_ISL_719089, EPI_ISL_719090, EPI_ISL_719091, EPI_ISL_719092, EPI_ISL_719093,
EPI_ISL_719094, EPI_ISL_719095, EPI_ISL_719096, EPI_ISL_719097, EPI_ISL_719098, EPI_ISL_719099, EPI_ISL_719100, EPI_ISL_719101, EPI_ISL_719102, EPI_ISL_719103, EPI_ISL_719104, EPI_ISL_719105, EPI_ISL_719106, EPI_ISL_719107, EPI_ISL_719108, EPI_ISL_719109, EPI_ISL_719110, EPI_ISL_719111, EPI_ISL_719112, EPI_ISL_719113, EPI_ISL_719114, EPI_ISL_719115, EPI_ISL_719116, EPI_ISL_719117,
EPI_ISL_719118, EPI_ISL_719119, EPI_ISL_719120, EPI_ISL_719121, EPI_ISL_719122, EPI_ISL_719123, EPI_ISL_719124, EPI_ISL_719125, EPI_ISL_719126, EPI_ISL_719127, EPI_ISL_719128, EPI_ISL_719129, EPI_ISL_719130, EPI_ISL_719131, EPI_ISL_719132, EPI_ISL_719133, EPI_ISL_719134, EPI_ISL_719135, EPI_ISL_719136, EPI_ISL_719137, EPI_ISL_719138, EPI_ISL_719139, EPI_ISL_719140, EPI_ISL_719141,
EPI_ISL_719142, EPI_ISL_719143, EPI_ISL_719144, EPI_ISL_719145, EPI_ISL_719146, EPI_ISL_719147, EPI_ISL_719148, EPI_ISL_719149, EPI_ISL_719150, EPI_ISL_719151, EPI_ISL_719152, EPI_ISL_719153, EPI_ISL_719154, EPI_ISL_719155, EPI_ISL_719156, EPI_ISL_719157, EPI_ISL_719158, EPI_ISL_719159, EPI_ISL_719160, EPI_ISL_719161, EPI_ISL_719162, EPI_ISL_719163, EPI_ISL_719164, EPI_ISL_719165,
EPI_ISL_719166, EPI_ISL_719167, EPI_ISL_719168, EPI_ISL_719169, EPI_ISL_719170, EPI_ISL_719171, EPI_ISL_719172, EPI_ISL_719173, EPI_ISL_719174, EPI_ISL_719175, EPI_ISL_719176, EPI_ISL_719177, EPI_ISL_719178, EPI_ISL_719179, EPI_ISL_719180, EPI_ISL_719181, EPI_ISL_719182, EPI_ISL_719183, EPI_ISL_719184, EPI_ISL_719185, EPI_ISL_719186, EPI_ISL_719187, EPI_ISL_719188, EPI_ISL_719189,
EPI_ISL_719190, EPI_ISL_719191, EPI_ISL_719192, EPI_ISL_719193, EPI_ISL_719194, EPI_ISL_719195, EPI_ISL_719196, EPI_ISL_719197, EPI_ISL_719198, EPI_ISL_719199, EPI_ISL_719200, EPI_ISL_719201, EPI_ISL_719202, EPI_ISL_719203, EPI_ISL_719204, EPI_ISL_719205, EPI_ISL_719206, EPI_ISL_719207, EPI_ISL_719208, EPI_ISL_719209, EPI_ISL_719210, EPI_ISL_719211, EPI_ISL_719212, EPI_ISL_719213,
EPI_ISL_719214, EPI_ISL_719215, EPI_ISL_719216, EPI_ISL_719217, EPI_ISL_719218, EPI_ISL_719219, EPI_ISL_719220, EPI_ISL_719221, EPI_ISL_719222, EPI_ISL_719223, EPI_ISL_719224, EPI_ISL_719225, EPI_ISL_719226, EPI_ISL_719227, EPI_ISL_719228, EPI_ISL_719229, EPI_ISL_719230, EPI_ISL_719231, EPI_ISL_719232, EPI_ISL_719233, EPI_ISL_719234, EPI_ISL_719235, EPI_ISL_719236, EPI_ISL_719237,
EPI_ISL_719238, EPI_ISL_719239, EPI_ISL_719240, EPI_ISL_719241, EPI_ISL_719242, EPI_ISL_719243, EPI_ISL_719244, EPI_ISL_719245, EPI_ISL_719246, EPI_ISL_719247, EPI_ISL_719248, EPI_ISL_719249, EPI_ISL_719250, EPI_ISL_719251, EPI_ISL_719252, EPI_ISL_719253, EPI_ISL_719254, EPI_ISL_719255, EPI_ISL_719256, EPI_ISL_719257, EPI_ISL_719258, EPI_ISL_719259, EPI_ISL_719260, EPI_ISL_719261,
EPI_ISL_719262, EPI_ISL_719263, EPI_ISL_719264, EPI_ISL_719265, EPI_ISL_719266, EPI_ISL_719267, EPI_ISL_719268, EPI_ISL_719269, EPI_ISL_719270, EPI_ISL_719271, EPI_ISL_719272, EPI_ISL_719273, EPI_ISL_719274, EPI_ISL_719275, EPI_ISL_719276, EPI_ISL_719277, EPI_ISL_719278, EPI_ISL_719279, EPI_ISL_719280, EPI_ISL_719281, EPI_ISL_719282, EPI_ISL_719283, EPI_ISL_719284, EPI_ISL_719285,
EPI_ISL_719286, EPI_ISL_719287, EPI_ISL_719288, EPI_ISL_719289, EPI_ISL_719290, EPI_ISL_719291, EPI_ISL_719292, EPI_ISL_719293, EPI_ISL_719294, EPI_ISL_719295, EPI_ISL_719296, EPI_ISL_719297, EPI_ISL_719298, EPI_ISL_719299, EPI_ISL_719300, EPI_ISL_719301, EPI_ISL_719302, EPI_ISL_719303, EPI_ISL_719304, EPI_ISL_719305, EPI_ISL_719306, EPI_ISL_719307, EPI_ISL_719308, EPI_ISL_719309,
EPI_ISL_719310, EPI_ISL_719311, EPI_ISL_719312, EPI_ISL_719313, EPI_ISL_719314, EPI_ISL_719315, EPI_ISL_719316, EPI_ISL_719317, EPI_ISL_719318, EPI_ISL_719319, EPI_ISL_719320, EPI_ISL_719321, EPI_ISL_719322, EPI_ISL_719323, EPI_ISL_719324, EPI_ISL_719325, EPI_ISL_719326, EPI_ISL_719327, EPI_ISL_719328, EPI_ISL_719329, EPI_ISL_719330, EPI_ISL_719331, EPI_ISL_719332, EPI_ISL_719333,
EPI_ISL_719334, EPI_ISL_719335, EPI_ISL_719336, EPI_ISL_719337, EPI_ISL_719338, EPI_ISL_719339, EPI_ISL_719340, EPI_ISL_719341, EPI_ISL_719342, EPI_ISL_719343, EPI_ISL_719344, EPI_ISL_719345, EPI_ISL_719346, EPI_ISL_719347, EPI_ISL_719348, EPI_ISL_719349, EPI_ISL_719350, EPI_ISL_719351, EPI_ISL_719352, EPI_ISL_719353, EPI_ISL_719354, EPI_ISL_719355, EPI_ISL_719356, EPI_ISL_719357,
EPI_ISL_719358, EPI_ISL_719359, EPI_ISL_719360, EPI_ISL_719361, EPI_ISL_719362, EPI_ISL_719363, EPI_ISL_719364, EPI_ISL_719365, EPI_ISL_719366, EPI_ISL_719367, EPI_ISL_719368, EPI_ISL_719369, EPI_ISL_719370, EPI_ISL_719371, EPI_ISL_719372, EPI_ISL_719373, EPI_ISL_719374, EPI_ISL_719375, EPI_ISL_719376, EPI_ISL_719377, EPI_ISL_719378, EPI_ISL_719379, EPI_ISL_719380, EPI_ISL_719381,
EPI_ISL_719382, EPI_ISL_719383, EPI_ISL_719384, EPI_ISL_719385, EPI_ISL_719386, EPI_ISL_719387, EPI_ISL_719388, EPI_ISL_719389, EPI_ISL_719390, EPI_ISL_719391, EPI_ISL_719392, EPI_ISL_719393, EPI_ISL_719394, EPI_ISL_719395, EPI_ISL_719396, EPI_ISL_719397, EPI_ISL_719398, EPI_ISL_719399, EPI_ISL_719400, EPI_ISL_719401, EPI_ISL_719402, EPI_ISL_719403, EPI_ISL_719404, EPI_ISL_719405,
EPI_ISL_719406, EPI_ISL_719407, EPI_ISL_719408, EPI_ISL_719409, EPI_ISL_719410, EPI_ISL_719411, EPI_ISL_719412, EPI_ISL_719413, EPI_ISL_719414, EPI_ISL_719415, EPI_ISL_719416, EPI_ISL_719417, EPI_ISL_719418, EPI_ISL_719419, EPI_ISL_719420, EPI_ISL_719421, EPI_ISL_719422, EPI_ISL_719423, EPI_ISL_719424, EPI_ISL_719425, EPI_ISL_719426, EPI_ISL_719427, EPI_ISL_719428, EPI_ISL_719429,
EPI_ISL_719430, EPI_ISL_719431, EPI_ISL_719432, EPI_ISL_719433, EPI_ISL_719434, EPI_ISL_719435, EPI_ISL_719436, EPI_ISL_719437, EPI_ISL_719438, EPI_ISL_719439, EPI_ISL_719440, EPI_ISL_719441, EPI_ISL_719442, EPI_ISL_719443, EPI_ISL_719444, EPI_ISL_719445, EPI_ISL_719446, EPI_ISL_719447, EPI_ISL_719448, EPI_ISL_719449, EPI_ISL_719450, EPI_ISL_719451, EPI_ISL_719452, EPI_ISL_719453,
EPI_ISL_719454, EPI_ISL_719455, EPI_ISL_719456, EPI_ISL_719457, EPI_ISL_719458, EPI_ISL_719459, EPI_ISL_719460, EPI_ISL_719461, EPI_ISL_719462, EPI_ISL_719463, EPI_ISL_719464, EPI_ISL_719465, EPI_ISL_719466, EPI_ISL_719467, EPI_ISL_719468, EPI_ISL_719469, EPI_ISL_719470, EPI_ISL_719471, EPI_ISL_719472, EPI_ISL_719473, EPI_ISL_719474, EPI_ISL_719475, EPI_ISL_719476, EPI_ISL_719477,
EPI_ISL_719478, EPI_ISL_719479, EPI_ISL_719480, EPI_ISL_719481, EPI_ISL_719482, EPI_ISL_719483, EPI_ISL_719484, EPI_ISL_719485, EPI_ISL_719486, EPI_ISL_719487, EPI_ISL_719488, EPI_ISL_719489, EPI_ISL_719490, EPI_ISL_719491, EPI_ISL_719492, EPI_ISL_719493, EPI_ISL_719494, EPI_ISL_719495, EPI_ISL_719496, EPI_ISL_719497, EPI_ISL_719498, EPI_ISL_719499, EPI_ISL_719500, EPI_ISL_719501,
EPI_ISL_719502, EPI_ISL_719503, EPI_ISL_719504, EPI_ISL_719505, EPI_ISL_719506, EPI_ISL_719507, EPI_ISL_719508, EPI_ISL_719509, EPI_ISL_719510, EPI_ISL_719511, EPI_ISL_719512, EPI_ISL_719513, EPI_ISL_719514, EPI_ISL_719515, EPI_ISL_719516, EPI_ISL_719517, EPI_ISL_719518, EPI_ISL_719519, EPI_ISL_719520, EPI_ISL_719521, EPI_ISL_719522, EPI_ISL_719523, EPI_ISL_719524, EPI_ISL_719525,
EPI_ISL_719526, EPI_ISL_719527, EPI_ISL_719528, EPI_ISL_719529, EPI_ISL_719530, EPI_ISL_719531, EPI_ISL_719532, EPI_ISL_719533, EPI_ISL_719534, EPI_ISL_719535, EPI_ISL_719536, EPI_ISL_719537, EPI_ISL_719538, EPI_ISL_719539, EPI_ISL_719540, EPI_ISL_719541, EPI_ISL_719542, EPI_ISL_719543, EPI_ISL_719544, EPI_ISL_719545, EPI_ISL_719546, EPI_ISL_719547, EPI_ISL_719548, EPI_ISL_719549,
EPI_ISL_719550, EPI_ISL_719551, EPI_ISL_719552, EPI_ISL_719553, EPI_ISL_719554, EPI_ISL_719555, EPI_ISL_719556, EPI_ISL_719557, EPI_ISL_719558, EPI_ISL_719559, EPI_ISL_719560, EPI_ISL_719561, EPI_ISL_719562, EPI_ISL_719563, EPI_ISL_719564, EPI_ISL_719565, EPI_ISL_719566, EPI_ISL_719567, EPI_ISL_719568, EPI_ISL_719569, EPI_ISL_719570, EPI_ISL_719571, EPI_ISL_719572, EPI_ISL_719573,
EPI_ISL_719574, EPI_ISL_719575, EPI_ISL_719576, EPI_ISL_719577, EPI_ISL_719578, EPI_ISL_719579, EPI_ISL_719580, EPI_ISL_719581, EPI_ISL_719582, EPI_ISL_719583, EPI_ISL_719584, EPI_ISL_719585, EPI_ISL_719586, EPI_ISL_719587, EPI_ISL_719588, EPI_ISL_719589, EPI_ISL_719590, EPI_ISL_719591, EPI_ISL_719592, EPI_ISL_719593, EPI_ISL_719594, EPI_ISL_719595, EPI_ISL_719596, EPI_ISL_719597,
EPI_ISL_719598, EPI_ISL_719599, EPI_ISL_719600, EPI_ISL_719601, EPI_ISL_719602, EPI_ISL_719603, EPI_ISL_719604, EPI_ISL_728299, EPI_ISL_757119, EPI_ISL_767138, EPI_ISL_769107, EPI_ISL_777133, EPI_ISL_777173, EPI_ISL_777181, EPI_ISL_777218, EPI_ISL_777275, EPI_ISL_777289, EPI_ISL_777293, EPI_ISL_777320, EPI_ISL_781551, EPI_ISL_781571, EPI_ISL_781625, EPI_ISL_781671, EPI_ISL_781690,
EPI_ISL_781710, EPI_ISL_781824, EPI_ISL_781863, EPI_ISL_799936, EPI_ISL_799971, EPI_ISL_799972, EPI_ISL_799993, EPI_ISL_799994, EPI_ISL_800054, EPI_ISL_800100, EPI_ISL_800139, EPI_ISL_800192, EPI_ISL_800225, EPI_ISL_800244

see above | Lighthouse Lab in Cambridge | Wellcome Sanger Institute for the COVID-19 Genomics UK (COG-UK) Consortium | Cordelia Langford; David K. Jackson; Dominic Kwiatkowski; Ewan Harrison; Ian Johnston; John Sillitoe on behalf of the Wellcome Sanger Institute COVID-19 Surveillance Team; Rob Howes; Roberto Amato; Sonia Goncalves; The Lighthouse Lab in Cambridge and Alex Alderton

EPI_ISL_730656, EPI_ISL_730657, EPI_ISL_730659, EPI_ISL_730664, EPI_ISL_730673, EPI_ISL_730675, EPI_ISL_730676, EPI_ISL_730678, EPI_ISL_730685, EPI_ISL_730697, EPI_ISL_730705, EPI_ISL_730709, EPI_ISL_730710, EPI_ISL_730718, EPI_ISL_730737, EPI_ISL_730752, EPI_ISL_730759, EPI_ISL_730765, EPI_ISL_730777, EPI_ISL_730778, EPI_ISL_730780, EPI_ISL_730785, EPI_ISL_730789, EPI_ISL_730793,
EPI_ISL_730795, EPI_ISL_730796, EPI_ISL_730801, EPI_ISL_730802, EPI_ISL_730803, EPI_ISL_730804, EPI_ISL_730805, EPI_ISL_730806, EPI_ISL_730809, EPI_ISL_730813, EPI_ISL_730815, EPI_ISL_730824, EPI_ISL_730828, EPI_ISL_730831, EPI_ISL_730832, EPI_ISL_730833, EPI_ISL_730840, EPI_ISL_730846, EPI_ISL_730847, EPI_ISL_730849, EPI_ISL_730850, EPI_ISL_730858, EPI_ISL_730860, EPI_ISL_730862,
EPI_ISL_730864, EPI_ISL_730867, EPI_ISL_730869, EPI_ISL_730875, EPI_ISL_730876, EPI_ISL_730877, EPI_ISL_730887, EPI_ISL_730889, EPI_ISL_730894, EPI_ISL_730898, EPI_ISL_730908, EPI_ISL_730910, EPI_ISL_730911, EPI_ISL_730913, EPI_ISL_731255, EPI_ISL_731256, EPI_ISL_731257, EPI_ISL_731258, EPI_ISL_731259, EPI_ISL_731267, EPI_ISL_731271, EPI_ISL_731273, EPI_ISL_731280, EPI_ISL_731281,
EPI_ISL_731282, EPI_ISL_731286, EPI_ISL_731287, EPI_ISL_731291, EPI_ISL_731295, EPI_ISL_731297, EPI_ISL_731305, EPI_ISL_731308, EPI_ISL_731309, EPI_ISL_731310, EPI_ISL_731314, EPI_ISL_731319, EPI_ISL_731320, EPI_ISL_731321, EPI_ISL_731322, EPI_ISL_731326, EPI_ISL_731327, EPI_ISL_731330, EPI_ISL_731332, EPI_ISL_731337, EPI_ISL_731338, EPI_ISL_731340, EPI_ISL_731341, EPI_ISL_731345,
EPI_ISL_731346, EPI_ISL_731354, EPI_ISL_731355, EPI_ISL_731357, EPI_ISL_731361, EPI_ISL_731367, EPI_ISL_731368, EPI_ISL_731372, EPI_ISL_731373, EPI_ISL_731376, EPI_ISL_731379, EPI_ISL_731380, EPI_ISL_731382, EPI_ISL_731383, EPI_ISL_731386, EPI_ISL_731390, EPI_ISL_731392, EPI_ISL_731397, EPI_ISL_731401, EPI_ISL_731402, EPI_ISL_731403, EPI_ISL_731404, EPI_ISL_731406, EPI_ISL_731407,
EPI_ISL_731408, EPI_ISL_731411, EPI_ISL_731414, EPI_ISL_731417, EPI_ISL_731420, EPI_ISL_731425, EPI_ISL_731426, EPI_ISL_731429, EPI_ISL_731431, EPI_ISL_731432, EPI_ISL_731435, EPI_ISL_731437, EPI_ISL_731444, EPI_ISL_731447, EPI_ISL_731449, EPI_ISL_731450, EPI_ISL_731452, EPI_ISL_731453, EPI_ISL_731454, EPI_ISL_731458, EPI_ISL_731469, EPI_ISL_731471, EPI_ISL_731472, EPI_ISL_731476,
EPI_ISL_731477, EPI_ISL_731482, EPI_ISL_731484, EPI_ISL_731485, EPI_ISL_731486, EPI_ISL_731488, EPI_ISL_731491, EPI_ISL_731493, EPI_ISL_731494, EPI_ISL_731496, EPI_ISL_731497, EPI_ISL_731499, EPI_ISL_731501, EPI_ISL_731505, EPI_ISL_731507, EPI_ISL_731508, EPI_ISL_731513, EPI_ISL_731517, EPI_ISL_731519, EPI_ISL_731523, EPI_ISL_731536, EPI_ISL_731537, EPI_ISL_731544, EPI_ISL_731548,
EPI_ISL_731550, EPI_ISL_731551, EPI_ISL_733916, EPI_ISL_733917, EPI_ISL_733918, EPI_ISL_733919, EPI_ISL_733920, EPI_ISL_733921, EPI_ISL_733922, EPI_ISL_733923, EPI_ISL_733924, EPI_ISL_733925, EPI_ISL_733926, EPI_ISL_733927, EPI_ISL_733929, EPI_ISL_733930, EPI_ISL_733931, EPI_ISL_733932, EPI_ISL_733933, EPI_ISL_733934, EPI_ISL_733935, EPI_ISL_733936, EPI_ISL_733937, EPI_ISL_733938,
EPI_ISL_733939, EPI_ISL_733940, EPI_ISL_733941, EPI_ISL_733942, EPI_ISL_733943, EPI_ISL_733944, EPI_ISL_733945, EPI_ISL_733946, EPI_ISL_733947, EPI_ISL_733948, EPI_ISL_733949, EPI_ISL_733951, EPI_ISL_733952, EPI_ISL_733953, EPI_ISL_733954, EPI_ISL_733955, EPI_ISL_733956, EPI_ISL_733957, EPI_ISL_733958, EPI_ISL_733959, EPI_ISL_733960, EPI_ISL_733961, EPI_ISL_733962, EPI_ISL_733963,
EPI_ISL_733964, EPI_ISL_733965, EPI_ISL_733966, EPI_ISL_733967, EPI_ISL_733968, EPI_ISL_733969, EPI_ISL_733970, EPI_ISL_733971, EPI_ISL_733972, EPI_ISL_733973, EPI_ISL_733974, EPI_ISL_733975, EPI_ISL_733976, EPI_ISL_733977, EPI_ISL_733978, EPI_ISL_733979, EPI_ISL_733980, EPI_ISL_733981, EPI_ISL_733982, EPI_ISL_733983, EPI_ISL_733984, EPI_ISL_733985, EPI_ISL_733986, EPI_ISL_733987,
EPI_ISL_733988, EPI_ISL_733989, EPI_ISL_733990, EPI_ISL_733991, EPI_ISL_733992, EPI_ISL_733993, EPI_ISL_733994, EPI_ISL_733995, EPI_ISL_733996, EPI_ISL_733997, EPI_ISL_733998, EPI_ISL_733999, EPI_ISL_734000, EPI_ISL_734001, EPI_ISL_734002, EPI_ISL_734003, EPI_ISL_734004, EPI_ISL_734005, EPI_ISL_734006, EPI_ISL_734007, EPI_ISL_734008, EPI_ISL_734009, EPI_ISL_734010, EPI_ISL_734011,
EPI_ISL_734012, EPI_ISL_734013, EPI_ISL_734014, EPI_ISL_734015, EPI_ISL_734016, EPI_ISL_734017, EPI_ISL_734018, EPI_ISL_734019, EPI_ISL_734020, EPI_ISL_734021, EPI_ISL_734022, EPI_ISL_734023, EPI_ISL_734024, EPI_ISL_734025, EPI_ISL_734026, EPI_ISL_734027, EPI_ISL_734028, EPI_ISL_734029, EPI_ISL_734030, EPI_ISL_734031, EPI_ISL_734032, EPI_ISL_734033, EPI_ISL_734034, EPI_ISL_734035,
EPI_ISL_734036, EPI_ISL_734037, EPI_ISL_734038, EPI_ISL_734039, EPI_ISL_734040, EPI_ISL_734042, EPI_ISL_734043, EPI_ISL_734045, EPI_ISL_734046, EPI_ISL_734047, EPI_ISL_734048, EPI_ISL_734049, EPI_ISL_734050, EPI_ISL_734051, EPI_ISL_734052, EPI_ISL_734053, EPI_ISL_734054, EPI_ISL_734055, EPI_ISL_734056, EPI_ISL_734057, EPI_ISL_734058, EPI_ISL_734059, EPI_ISL_734060, EPI_ISL_734061,
EPI_ISL_734062, EPI_ISL_734063, EPI_ISL_734064, EPI_ISL_734065, EPI_ISL_734066, EPI_ISL_734067, EPI_ISL_734068, EPI_ISL_734069, EPI_ISL_734070, EPI_ISL_734071, EPI_ISL_734072, EPI_ISL_734073, EPI_ISL_734074, EPI_ISL_734075, EPI_ISL_734076, EPI_ISL_734077, EPI_ISL_734078, EPI_ISL_734079, EPI_ISL_734080, EPI_ISL_734081, EPI_ISL_734082, EPI_ISL_734083, EPI_ISL_734084, EPI_ISL_734085,
EPI_ISL_734086, EPI_ISL_734087, EPI_ISL_734088, EPI_ISL_734089, EPI_ISL_734090, EPI_ISL_734091, EPI_ISL_734092, EPI_ISL_734094, EPI_ISL_734095, EPI_ISL_734096, EPI_ISL_734097, EPI_ISL_734098, EPI_ISL_734099, EPI_ISL_734100, EPI_ISL_734101, EPI_ISL_734102, EPI_ISL_734103, EPI_ISL_734104, EPI_ISL_734105, EPI_ISL_734106, EPI_ISL_734107, EPI_ISL_734108, EPI_ISL_734109, EPI_ISL_734110,
EPI_ISL_734111, EPI_ISL_734112, EPI_ISL_734113, EPI_ISL_734114, EPI_ISL_734115, EPI_ISL_734116, EPI_ISL_734118, EPI_ISL_734119, EPI_ISL_734120, EPI_ISL_734121, EPI_ISL_734122, EPI_ISL_734123, EPI_ISL_734124, EPI_ISL_734125, EPI_ISL_734126, EPI_ISL_734127, EPI_ISL_734128, EPI_ISL_734129, EPI_ISL_734130, EPI_ISL_734132, EPI_ISL_734133, EPI_ISL_734134, EPI_ISL_734135, EPI_ISL_734136,
EPI_ISL_734137, EPI_ISL_734138, EPI_ISL_734139, EPI_ISL_734140, EPI_ISL_734141, EPI_ISL_734142, EPI_ISL_734143, EPI_ISL_734144, EPI_ISL_734145, EPI_ISL_734146, EPI_ISL_734147, EPI_ISL_734148, EPI_ISL_734149, EPI_ISL_734150, EPI_ISL_734151, EPI_ISL_734152, EPI_ISL_734153, EPI_ISL_734154, EPI_ISL_734155, EPI_ISL_734156, EPI_ISL_734157, EPI_ISL_734158, EPI_ISL_734159, EPI_ISL_734160,
EPI_ISL_734161, EPI_ISL_734162, EPI_ISL_734163, EPI_ISL_734164, EPI_ISL_734165, EPI_ISL_735861, EPI_ISL_735864, EPI_ISL_735878, EPI_ISL_735880, EPI_ISL_735886, EPI_ISL_735894, EPI_ISL_735909, EPI_ISL_735920, EPI_ISL_735927, EPI_ISL_735937, EPI_ISL_735939, EPI_ISL_735945, EPI_ISL_735949, EPI_ISL_735952, EPI_ISL_735956, EPI_ISL_735961, EPI_ISL_735965, EPI_ISL_735968, EPI_ISL_735972,
EPI_ISL_735973, EPI_ISL_735978, EPI_ISL_735982, EPI_ISL_735987, EPI_ISL_735993, EPI_ISL_735995, EPI_ISL_735996, EPI_ISL_736001, EPI_ISL_736003, EPI_ISL_736007, EPI_ISL_736015, EPI_ISL_736017, EPI_ISL_736018, EPI_ISL_736019, EPI_ISL_736024, EPI_ISL_736025, EPI_ISL_736027, EPI_ISL_736033, EPI_ISL_736038, EPI_ISL_736041, EPI_ISL_736052, EPI_ISL_736057, EPI_ISL_736059, EPI_ISL_736064,
EPI_ISL_736066, EPI_ISL_736072, EPI_ISL_736090, EPI_ISL_736091, EPI_ISL_736092, EPI_ISL_736096, EPI_ISL_736099, EPI_ISL_736100, EPI_ISL_736101, EPI_ISL_736102, EPI_ISL_736104, EPI_ISL_736107, EPI_ISL_736115, EPI_ISL_736124, EPI_ISL_736130, EPI_ISL_736131, EPI_ISL_736132, EPI_ISL_736135, EPI_ISL_736143, EPI_ISL_736146, EPI_ISL_736149, EPI_ISL_736151, EPI_ISL_736159, EPI_ISL_736162,
EPI_ISL_736167, EPI_ISL_736168, EPI_ISL_736169, EPI_ISL_736175, EPI_ISL_736182, EPI_ISL_736183, EPI_ISL_736184, EPI_ISL_736190, EPI_ISL_736191, EPI_ISL_736204, EPI_ISL_736205, EPI_ISL_736206, EPI_ISL_736209, EPI_ISL_736212, EPI_ISL_736220, EPI_ISL_736221, EPI_ISL_736222, EPI_ISL_736223, EPI_ISL_736231, EPI_ISL_736234, EPI_ISL_736235, EPI_ISL_736236, EPI_ISL_736237, EPI_ISL_736238,
EPI_ISL_736239, EPI_ISL_736241, EPI_ISL_736242, EPI_ISL_736243, EPI_ISL_736245, EPI_ISL_736248, EPI_ISL_736250, EPI_ISL_736254, EPI_ISL_736255, EPI_ISL_736257, EPI_ISL_736258, EPI_ISL_736261, EPI_ISL_736266, EPI_ISL_736269, EPI_ISL_736271, EPI_ISL_736275, EPI_ISL_736277, EPI_ISL_736278, EPI_ISL_736284, EPI_ISL_736287, EPI_ISL_736290, EPI_ISL_736301, EPI_ISL_736304, EPI_ISL_736306,
EPI_ISL_736309, EPI_ISL_736311, EPI_ISL_736312, EPI_ISL_736313, EPI_ISL_736315, EPI_ISL_736316, EPI_ISL_736318, EPI_ISL_736320, EPI_ISL_736321, EPI_ISL_736323, EPI_ISL_736325, EPI_ISL_736327, EPI_ISL_736330, EPI_ISL_736331, EPI_ISL_736332, EPI_ISL_736334, EPI_ISL_736343, EPI_ISL_736346, EPI_ISL_736347, EPI_ISL_736350, EPI_ISL_736352, EPI_ISL_736355, EPI_ISL_736356, EPI_ISL_736358,
EPI_ISL_736362, EPI_ISL_736367, EPI_ISL_736370, EPI_ISL_736371, EPI_ISL_736373, EPI_ISL_736376, EPI_ISL_736378, EPI_ISL_736382, EPI_ISL_736383, EPI_ISL_736386, EPI_ISL_736388, EPI_ISL_736389, EPI_ISL_736390, EPI_ISL_736391, EPI_ISL_736394, EPI_ISL_736398, EPI_ISL_736399, EPI_ISL_736400, EPI_ISL_736404, EPI_ISL_736411, EPI_ISL_736412, EPI_ISL_736414, EPI_ISL_736416, EPI_ISL_736417,
EPI_ISL_736419, EPI_ISL_736423, EPI_ISL_736425, EPI_ISL_736426, EPI_ISL_736428, EPI_ISL_736429, EPI_ISL_736430, EPI_ISL_736431, EPI_ISL_736437, EPI_ISL_736438, EPI_ISL_736442, EPI_ISL_736444, EPI_ISL_736445, EPI_ISL_736451, EPI_ISL_736455, EPI_ISL_736456, EPI_ISL_736459, EPI_ISL_736460, EPI_ISL_736461, EPI_ISL_736464, EPI_ISL_736466, EPI_ISL_736468, EPI_ISL_736470, EPI_ISL_736471,
EPI_ISL_736474, EPI_ISL_736476, EPI_ISL_736478, EPI_ISL_736479, EPI_ISL_736483, EPI_ISL_736488, EPI_ISL_736489, EPI_ISL_736490, EPI_ISL_736491, EPI_ISL_736492, EPI_ISL_736503, EPI_ISL_736512, EPI_ISL_736519, EPI_ISL_736529, EPI_ISL_736532, EPI_ISL_736533, EPI_ISL_736556, EPI_ISL_736567, EPI_ISL_736573, EPI_ISL_736580, EPI_ISL_736582, EPI_ISL_736605, EPI_ISL_736620, EPI_ISL_736624,
EPI_ISL_736627, EPI_ISL_736628, EPI_ISL_736631, EPI_ISL_736633, EPI_ISL_736646, EPI_ISL_736650, EPI_ISL_736657, EPI_ISL_736659, EPI_ISL_736668, EPI_ISL_736672, EPI_ISL_736678, EPI_ISL_736685, EPI_ISL_736689, EPI_ISL_736691, EPI_ISL_736695, EPI_ISL_736700, EPI_ISL_736721, EPI_ISL_736734, EPI_ISL_736736, EPI_ISL_736737, EPI_ISL_736743, EPI_ISL_736745, EPI_ISL_736752, EPI_ISL_736754,
EPI_ISL_736774, EPI_ISL_757034, EPI_ISL_757035, EPI_ISL_757036, EPI_ISL_757037, EPI_ISL_757038, EPI_ISL_757039, EPI_ISL_757040, EPI_ISL_757041, EPI_ISL_757042, EPI_ISL_757043, EPI_ISL_757044, EPI_ISL_757045, EPI_ISL_757046, EPI_ISL_757047, EPI_ISL_757048, EPI_ISL_757049, EPI_ISL_757050, EPI_ISL_757051, EPI_ISL_757052, EPI_ISL_757053, EPI_ISL_757054, EPI_ISL_757055, EPI_ISL_757056,
EPI_ISL_757057, EPI_ISL_757058, EPI_ISL_757059, EPI_ISL_757060, EPI_ISL_757061, EPI_ISL_757062, EPI_ISL_757063, EPI_ISL_757064, EPI_ISL_757065, EPI_ISL_757066, EPI_ISL_757067, EPI_ISL_757068, EPI_ISL_757069, EPI_ISL_757070, EPI_ISL_757071, EPI_ISL_757072, EPI_ISL_757073, EPI_ISL_757074, EPI_ISL_757075, EPI_ISL_757076, EPI_ISL_757077, EPI_ISL_757078, EPI_ISL_757079, EPI_ISL_757080,
EPI_ISL_757081, EPI_ISL_757082, EPI_ISL_757083, EPI_ISL_757084, EPI_ISL_757085, EPI_ISL_757086, EPI_ISL_757087, EPI_ISL_757088, EPI_ISL_757089, EPI_ISL_757090, EPI_ISL_757091, EPI_ISL_757092, EPI_ISL_757093, EPI_ISL_757094, EPI_ISL_757095, EPI_ISL_757096, EPI_ISL_757097, EPI_ISL_757098, EPI_ISL_757099, EPI_ISL_757100, EPI_ISL_757101, EPI_ISL_757102, EPI_ISL_757103, EPI_ISL_757104,
EPI_ISL_757105, EPI_ISL_757106, EPI_ISL_757107, EPI_ISL_757108, EPI_ISL_757109, EPI_ISL_757110, EPI_ISL_757111, EPI_ISL_757112, EPI_ISL_757113, EPI_ISL_757114, EPI_ISL_757115, EPI_ISL_757116, EPI_ISL_757117, EPI_ISL_757118, EPI_ISL_757120, EPI_ISL_757121, EPI_ISL_757122, EPI_ISL_757123, EPI_ISL_757124, EPI_ISL_757125, EPI_ISL_757126, EPI_ISL_757127, EPI_ISL_757128, EPI_ISL_757129,
EPI_ISL_757130, EPI_ISL_757131, EPI_ISL_757132, EPI_ISL_757133, EPI_ISL_757134, EPI_ISL_757135, EPI_ISL_757136, EPI_ISL_757137, EPI_ISL_757138, EPI_ISL_757139, EPI_ISL_757140, EPI_ISL_757141, EPI_ISL_757142, EPI_ISL_757143, EPI_ISL_757144, EPI_ISL_757145, EPI_ISL_757146, EPI_ISL_757147, EPI_ISL_757148, EPI_ISL_757149, EPI_ISL_757150, EPI_ISL_757151, EPI_ISL_757152, EPI_ISL_757153,
EPI_ISL_757154, EPI_ISL_757155, EPI_ISL_757156, EPI_ISL_757157, EPI_ISL_757158, EPI_ISL_757159, EPI_ISL_757160, EPI_ISL_757161, EPI_ISL_757162, EPI_ISL_757163, EPI_ISL_757164, EPI_ISL_757165, EPI_ISL_757166, EPI_ISL_757167, EPI_ISL_757168, EPI_ISL_757169, EPI_ISL_757170, EPI_ISL_757171, EPI_ISL_757172, EPI_ISL_757173, EPI_ISL_757174, EPI_ISL_757175, EPI_ISL_757176, EPI_ISL_757177,
EPI_ISL_757178, EPI_ISL_757179, EPI_ISL_757180, EPI_ISL_757181, EPI_ISL_757182, EPI_ISL_757183, EPI_ISL_757184, EPI_ISL_757185, EPI_ISL_757186, EPI_ISL_757225, EPI_ISL_757228, EPI_ISL_757230, EPI_ISL_760847, EPI_ISL_760977, EPI_ISL_760980, EPI_ISL_760984, EPI_ISL_761018, EPI_ISL_762062, EPI_ISL_762156, EPI_ISL_762979, EPI_ISL_769223, EPI_ISL_769224, EPI_ISL_769225, EPI_ISL_769229,
EPI_ISL_769233, EPI_ISL_769236, EPI_ISL_769243, EPI_ISL_769244, EPI_ISL_769245, EPI_ISL_769246, EPI_ISL_769249, EPI_ISL_769250, EPI_ISL_769252, EPI_ISL_769254, EPI_ISL_769255, EPI_ISL_769257, EPI_ISL_769259, EPI_ISL_769260, EPI_ISL_769261, EPI_ISL_769266, EPI_ISL_769267, EPI_ISL_769273, EPI_ISL_769275, EPI_ISL_769276, EPI_ISL_769280, EPI_ISL_769281, EPI_ISL_769282, EPI_ISL_769284,
EPI_ISL_769286, EPI_ISL_769288, EPI_ISL_769289, EPI_ISL_769290, EPI_ISL_769291, EPI_ISL_769294, EPI_ISL_769297, EPI_ISL_769306, EPI_ISL_769308, EPI_ISL_769310, EPI_ISL_769312, EPI_ISL_769314, EPI_ISL_769316, EPI_ISL_769319, EPI_ISL_769320, EPI_ISL_769321, EPI_ISL_769322, EPI_ISL_769323, EPI_ISL_769325, EPI_ISL_769327, EPI_ISL_769329, EPI_ISL_769330, EPI_ISL_769334, EPI_ISL_769335,
EPI_ISL_769341, EPI_ISL_769343, EPI_ISL_769345, EPI_ISL_769348, EPI_ISL_769349, EPI_ISL_769352, EPI_ISL_769353, EPI_ISL_769355, EPI_ISL_769360, EPI_ISL_769361, EPI_ISL_769365, EPI_ISL_769367, EPI_ISL_769370, EPI_ISL_769374, EPI_ISL_769380, EPI_ISL_769381, EPI_ISL_769383, EPI_ISL_769384, EPI_ISL_769385, EPI_ISL_769387, EPI_ISL_769388, EPI_ISL_769389, EPI_ISL_769390, EPI_ISL_769391,
EPI_ISL_769392, EPI_ISL_769394, EPI_ISL_769396, EPI_ISL_769399, EPI_ISL_769401, EPI_ISL_769402, EPI_ISL_769404, EPI_ISL_769405, EPI_ISL_769406, EPI_ISL_769412, EPI_ISL_769413, EPI_ISL_769416, EPI_ISL_769417, EPI_ISL_769418, EPI_ISL_769419, EPI_ISL_769420, EPI_ISL_769421, EPI_ISL_769422, EPI_ISL_769427, EPI_ISL_769428, EPI_ISL_769430, EPI_ISL_769431, EPI_ISL_769434, EPI_ISL_769440,
EPI_ISL_769441, EPI_ISL_769442, EPI_ISL_769445, EPI_ISL_769446, EPI_ISL_769449, EPI_ISL_769450, EPI_ISL_769452, EPI_ISL_769454, EPI_ISL_769457, EPI_ISL_769458, EPI_ISL_769459, EPI_ISL_769460, EPI_ISL_769462, EPI_ISL_769466, EPI_ISL_769468, EPI_ISL_769469, EPI_ISL_769470, EPI_ISL_769471, EPI_ISL_769473, EPI_ISL_769474, EPI_ISL_769475, EPI_ISL_769476, EPI_ISL_769481, EPI_ISL_769482,
EPI_ISL_769485, EPI_ISL_769489, EPI_ISL_769497, EPI_ISL_769499, EPI_ISL_769500, EPI_ISL_769501, EPI_ISL_769502, EPI_ISL_769503, EPI_ISL_769504, EPI_ISL_769506, EPI_ISL_769511, EPI_ISL_769512, EPI_ISL_769513, EPI_ISL_769517, EPI_ISL_769518, EPI_ISL_769519, EPI_ISL_769522, EPI_ISL_769524, EPI_ISL_769525, EPI_ISL_769528, EPI_ISL_769530, EPI_ISL_769533, EPI_ISL_769535, EPI_ISL_769539,
EPI_ISL_769540, EPI_ISL_769542, EPI_ISL_769543, EPI_ISL_769551, EPI_ISL_769556, EPI_ISL_769567, EPI_ISL_769568, EPI_ISL_769577, EPI_ISL_769583, EPI_ISL_769589, EPI_ISL_769594, EPI_ISL_769606, EPI_ISL_769607, EPI_ISL_769609, EPI_ISL_769611, EPI_ISL_769612, EPI_ISL_769613, EPI_ISL_769614, EPI_ISL_769617, EPI_ISL_769619, EPI_ISL_769625, EPI_ISL_769641, EPI_ISL_769644, EPI_ISL_769648,
EPI_ISL_769652, EPI_ISL_769655, EPI_ISL_769661, EPI_ISL_769662, EPI_ISL_769666, EPI_ISL_769667, EPI_ISL_769677, EPI_ISL_769687, EPI_ISL_769689, EPI_ISL_769691, EPI_ISL_769703, EPI_ISL_769705, EPI_ISL_769711, EPI_ISL_769713, EPI_ISL_769714, EPI_ISL_769716, EPI_ISL_769720, EPI_ISL_769724, EPI_ISL_769735, EPI_ISL_769741, EPI_ISL_769743, EPI_ISL_769751, EPI_ISL_769752, EPI_ISL_769753,
EPI_ISL_769758, EPI_ISL_769759, EPI_ISL_769761, EPI_ISL_769768, EPI_ISL_769773, EPI_ISL_769787, EPI_ISL_769788, EPI_ISL_769794, EPI_ISL_769802, EPI_ISL_769806, EPI_ISL_769813, EPI_ISL_769815, EPI_ISL_769818, EPI_ISL_769826, EPI_ISL_769834, EPI_ISL_769836, EPI_ISL_769837, EPI_ISL_769842, EPI_ISL_769850, EPI_ISL_769861, EPI_ISL_769862, EPI_ISL_777023, EPI_ISL_777058, EPI_ISL_777065,
EPI_ISL_777148, EPI_ISL_777149, EPI_ISL_777255, EPI_ISL_777284, EPI_ISL_778297, EPI_ISL_778299, EPI_ISL_778301, EPI_ISL_778302, EPI_ISL_778303, EPI_ISL_778322, EPI_ISL_778324, EPI_ISL_778325, EPI_ISL_778330, EPI_ISL_778332, EPI_ISL_778334, EPI_ISL_778336, EPI_ISL_778337, EPI_ISL_778345, EPI_ISL_778352, EPI_ISL_778356, EPI_ISL_778359, EPI_ISL_778361, EPI_ISL_778362, EPI_ISL_778364,
EPI_ISL_778372, EPI_ISL_778373, EPI_ISL_778375, EPI_ISL_778376, EPI_ISL_778377, EPI_ISL_778378, EPI_ISL_778397, EPI_ISL_778400, EPI_ISL_778403, EPI_ISL_778407, EPI_ISL_778411, EPI_ISL_778419, EPI_ISL_778420, EPI_ISL_778421, EPI_ISL_778422, EPI_ISL_778423, EPI_ISL_778424, EPI_ISL_778428, EPI_ISL_778433, EPI_ISL_778450, EPI_ISL_778451, EPI_ISL_778459, EPI_ISL_778461, EPI_ISL_778462,
EPI_ISL_778463, EPI_ISL_778465, EPI_ISL_778496, EPI_ISL_778497, EPI_ISL_778499, EPI_ISL_778504, EPI_ISL_778506, EPI_ISL_778516, EPI_ISL_778522, EPI_ISL_778525, EPI_ISL_778527, EPI_ISL_778530, EPI_ISL_778540, EPI_ISL_778541, EPI_ISL_778548, EPI_ISL_778551, EPI_ISL_778553, EPI_ISL_778560, EPI_ISL_778568, EPI_ISL_778570, EPI_ISL_778577, EPI_ISL_778586, EPI_ISL_778594, EPI_ISL_778617,
EPI_ISL_778619, EPI_ISL_778620, EPI_ISL_778629, EPI_ISL_778631, EPI_ISL_782482, EPI_ISL_782483, EPI_ISL_782484, EPI_ISL_782485, EPI_ISL_782487, EPI_ISL_782488, EPI_ISL_782489, EPI_ISL_782490, EPI_ISL_782491, EPI_ISL_782492, EPI_ISL_782493, EPI_ISL_782495, EPI_ISL_782496, EPI_ISL_782497, EPI_ISL_782498, EPI_ISL_782499, EPI_ISL_782501, EPI_ISL_782502, EPI_ISL_782503, EPI_ISL_782505,
EPI_ISL_782506, EPI_ISL_782507, EPI_ISL_782508, EPI_ISL_782509, EPI_ISL_782510, EPI_ISL_782511, EPI_ISL_782513, EPI_ISL_782514, EPI_ISL_782515, EPI_ISL_782516, EPI_ISL_782517, EPI_ISL_782518, EPI_ISL_782519, EPI_ISL_782520, EPI_ISL_782521, EPI_ISL_782522, EPI_ISL_782523, EPI_ISL_782524, EPI_ISL_782525, EPI_ISL_782526, EPI_ISL_782527, EPI_ISL_782528, EPI_ISL_782529, EPI_ISL_782530,
EPI_ISL_782531, EPI_ISL_782532, EPI_ISL_782533, EPI_ISL_782534, EPI_ISL_782535, EPI_ISL_782537, EPI_ISL_782539, EPI_ISL_782540, EPI_ISL_782541, EPI_ISL_782542, EPI_ISL_782543, EPI_ISL_782544, EPI_ISL_782545, EPI_ISL_782546, EPI_ISL_782547, EPI_ISL_782548, EPI_ISL_782549, EPI_ISL_782550, EPI_ISL_782551, EPI_ISL_782552, EPI_ISL_782553, EPI_ISL_782554, EPI_ISL_782555, EPI_ISL_782556,
EPI_ISL_782557, EPI_ISL_782558, EPI_ISL_782559, EPI_ISL_782560, EPI_ISL_782561, EPI_ISL_782562, EPI_ISL_782563, EPI_ISL_782564, EPI_ISL_782565, EPI_ISL_782566, EPI_ISL_782567, EPI_ISL_782569, EPI_ISL_782570, EPI_ISL_782571, EPI_ISL_782572, EPI_ISL_782573, EPI_ISL_782574, EPI_ISL_782575, EPI_ISL_782576, EPI_ISL_782577, EPI_ISL_782578, EPI_ISL_782579, EPI_ISL_782581, EPI_ISL_782582,
EPI_ISL_782583, EPI_ISL_782585, EPI_ISL_782586, EPI_ISL_782587, EPI_ISL_782588, EPI_ISL_782589, EPI_ISL_782590, EPI_ISL_782591, EPI_ISL_782592, EPI_ISL_782593, EPI_ISL_782594, EPI_ISL_782595, EPI_ISL_782597, EPI_ISL_782598, EPI_ISL_782599, EPI_ISL_782600, EPI_ISL_782601, EPI_ISL_782602, EPI_ISL_782603, EPI_ISL_782604, EPI_ISL_782606, EPI_ISL_782607, EPI_ISL_782608, EPI_ISL_782609,
EPI_ISL_782610, EPI_ISL_782611, EPI_ISL_782612, EPI_ISL_782613, EPI_ISL_782614, EPI_ISL_782616, EPI_ISL_782617, EPI_ISL_782619, EPI_ISL_782620, EPI_ISL_782621, EPI_ISL_782622, EPI_ISL_782623, EPI_ISL_782625, EPI_ISL_782626, EPI_ISL_782627, EPI_ISL_782628, EPI_ISL_782629, EPI_ISL_782630, EPI_ISL_782632, EPI_ISL_782634, EPI_ISL_782635, EPI_ISL_782636, EPI_ISL_782637, EPI_ISL_782638,
EPI_ISL_782639, EPI_ISL_782640, EPI_ISL_782641, EPI_ISL_782642, EPI_ISL_782643, EPI_ISL_782644, EPI_ISL_782645, EPI_ISL_782646, EPI_ISL_782647, EPI_ISL_782648, EPI_ISL_782649, EPI_ISL_782650, EPI_ISL_782651, EPI_ISL_782652, EPI_ISL_782653, EPI_ISL_782654, EPI_ISL_782655, EPI_ISL_782656, EPI_ISL_782657, EPI_ISL_782658, EPI_ISL_782659, EPI_ISL_782660, EPI_ISL_782661, EPI_ISL_782662,
EPI_ISL_782663, EPI_ISL_782664, EPI_ISL_782665, EPI_ISL_782666, EPI_ISL_782667, EPI_ISL_782668, EPI_ISL_782669, EPI_ISL_782670, EPI_ISL_782671, EPI_ISL_782672, EPI_ISL_782673, EPI_ISL_782675, EPI_ISL_782676, EPI_ISL_782677, EPI_ISL_782678, EPI_ISL_782680, EPI_ISL_782681, EPI_ISL_782682, EPI_ISL_782683, EPI_ISL_782684, EPI_ISL_782685, EPI_ISL_782686, EPI_ISL_782687, EPI_ISL_782688,

EPI_ISL_782690, EPI_ISL_782691, EPI_ISL_782692, EPI_ISL_782693, EPI_ISL_782694, EPI_ISL_782695, EPI_ISL_782696, EPI_ISL_782697, EPI_ISL_782698, EPI_ISL_782699, EPI_ISL_782700, EPI_ISL_782701, EPI_ISL_782702, EPI_ISL_782703, EPI_ISL_782704, EPI_ISL_782705, EPI_ISL_782706, EPI_ISL_782707, EPI_ISL_782708, EPI_ISL_782709, EPI_ISL_782711, EPI_ISL_782712, EPI_ISL_782713, EPI_ISL_782714, EPI_ISL_782715, EPI_ISL_782716, EPI_ISL_782717, EPI_ISL_782718, EPI_ISL_782719, EPI_ISL_782720, EPI_ISL_782721, EPI_ISL_782722, EPI_ISL_782723, EPI_ISL_782724, EPI_ISL_782725, EPI_ISL_782726, EPI_ISL_782727, EPI_ISL_782728, EPI_ISL_782729, EPI_ISL_782730, EPI_ISL_782731, EPI_ISL_782732, EPI_ISL_782733, EPI_ISL_801179, EPI_ISL_820147, EPI_ISL_820150, EPI_ISL_820152, EPI_ISL_820171, EPI_ISL_820176, EPI_ISL_820177, EPI_ISL_820184, EPI_ISL_820197, EPI_ISL_820212, EPI_ISL_820215, EPI_ISL_820220, EPI_ISL_820225, EPI_ISL_820227, EPI_ISL_820233, EPI_ISL_820243, EPI_ISL_820260, EPI_ISL_820262, EPI_ISL_820270, EPI_ISL_820272, EPI_ISL_820283, EPI_ISL_820295, EPI_ISL_820312, EPI_ISL_820319, EPI_ISL_820320, EPI_ISL_820384, EPI_ISL_820386, EPI_ISL_820387, EPI_ISL_820409, EPI_ISL_820417, EPI_ISL_820421, EPI_ISL_820437, EPI_ISL_820448, EPI_ISL_820461, EPI_ISL_820465, EPI_ISL_820468, EPI_ISL_820477, EPI_ISL_820479, EPI_ISL_820484, EPI_ISL_820489, EPI_ISL_820505, EPI_ISL_820507, EPI_ISL_820510, EPI_ISL_820512, EPI_ISL_820518, EPI_ISL_820547, EPI_ISL_820558, EPI_ISL_820577, EPI_ISL_820590, EPI_ISL_820609, EPI_ISL_820613, EPI_ISL_820617, EPI_ISL_820618, EPI_ISL_820620, EPI_ISL_820628, EPI_ISL_820634, EPI_ISL_820639, EPI_ISL_820641, EPI_ISL_820645, EPI_ISL_820647, EPI_ISL_858412, EPI_ISL_859456, EPI_ISL_859457, EPI_ISL_859458, EPI_ISL_859459, EPI_ISL_859461, EPI_ISL_859462, EPI_ISL_859463, EPI_ISL_859464, EPI_ISL_859465, EPI_ISL_859466, EPI_ISL_859467, EPI_ISL_859468, EPI_ISL_859470, EPI_ISL_859471, EPI_ISL_859472, EPI_ISL_859473, EPI_ISL_859474, EPI_ISL_859475, EPI_ISL_859476, EPI_ISL_859477, EPI_ISL_859478, EPI_ISL_859479, EPI_ISL_859480, EPI_ISL_859481, EPI_ISL_859482, EPI_ISL_859483, EPI_ISL_859484, EPI_ISL_859485, EPI_ISL_859486, EPI_ISL_859487, EPI_ISL_859488, EPI_ISL_859489, EPI_ISL_859490, EPI_ISL_859491, EPI_ISL_859492, EPI_ISL_859493, EPI_ISL_859494, EPI_ISL_859495, EPI_ISL_859496, EPI_ISL_859497, EPI_ISL_859498, EPI_ISL_859499, EPI_ISL_859500, EPI_ISL_859501, EPI_ISL_859502, EPI_ISL_859503, EPI_ISL_859504, EPI_ISL_859505, EPI_ISL_859506, EPI_ISL_859507, EPI_ISL_859508, EPI_ISL_859509, EPI_ISL_859510, EPI_ISL_859511, EPI_ISL_859512, EPI_ISL_859513, EPI_ISL_859514, EPI_ISL_859515, EPI_ISL_859516, EPI_ISL_859517, EPI_ISL_859518, EPI_ISL_859519, EPI_ISL_859520, EPI_ISL_859521, EPI_ISL_859522, EPI_ISL_859523, EPI_ISL_859524, EPI_ISL_859525, EPI_ISL_859526, EPI_ISL_859527, EPI_ISL_859528, EPI_ISL_859529, EPI_ISL_859530, EPI_ISL_859531, EPI_ISL_859532, EPI_ISL_933492, EPI_ISL_945255, EPI_ISL_989983, EPI_ISL_989984, EPI_ISL_989985, EPI_ISL_989988, EPI_ISL_989991, EPI_ISL_989992, EPI_ISL_989994, EPI_ISL_989995, EPI_ISL_989997

| | | | |
|---|---|---|---|
| see above | Lighthouse Lab in Glasgow | Wellcome Sanger Institute for the COVID-19 Genomics UK (COG-UK) Consortium | Anna Dominiczak and Alex Alderton; Carol Clugston; Cordelia Langford; David Gray; David K. Jackson; Dominic Kwiatkowski; Ewan Harrison; Harper VanSteenhouse; Ian Johnston; John Sillitoe on behalf of the Wellcome Sanger Institute COVID-19 Surveillance Team; Roberto Amato; Sonia Goncalves; Yumi Kasai |

EPI_ISL_735512, EPI_ISL_735513, EPI_ISL_735514, EPI_ISL_735515, EPI_ISL_735516, EPI_ISL_735517, EPI_ISL_735518, EPI_ISL_735519, EPI_ISL_735520, EPI_ISL_735521, EPI_ISL_735522, EPI_ISL_735523, EPI_ISL_735524, EPI_ISL_735525, EPI_ISL_735526, EPI_ISL_735527, EPI_ISL_735528, EPI_ISL_735529, EPI_ISL_735530, EPI_ISL_735531, EPI_ISL_735532, EPI_ISL_735533, EPI_ISL_735534, EPI_ISL_735535, EPI_ISL_735536, EPI_ISL_735537, EPI_ISL_735538, EPI_ISL_735539, EPI_ISL_735540, EPI_ISL_735541, EPI_ISL_735542, EPI_ISL_735543, EPI_ISL_735544, EPI_ISL_735545, EPI_ISL_735546, EPI_ISL_735547, EPI_ISL_735548, EPI_ISL_735549, EPI_ISL_735550, EPI_ISL_735551, EPI_ISL_735552, EPI_ISL_735553, EPI_ISL_735554, EPI_ISL_735555, EPI_ISL_735556, EPI_ISL_735557, EPI_ISL_735558, EPI_ISL_735559, EPI_ISL_735560, EPI_ISL_735561, EPI_ISL_735562, EPI_ISL_735563, EPI_ISL_735564, EPI_ISL_735565, EPI_ISL_735566, EPI_ISL_735567, EPI_ISL_735568, EPI_ISL_735569, EPI_ISL_735570, EPI_ISL_735571, EPI_ISL_735572, EPI_ISL_735573, EPI_ISL_735574, EPI_ISL_735575, EPI_ISL_735576, EPI_ISL_735577, EPI_ISL_735578, EPI_ISL_735579, EPI_ISL_735580, EPI_ISL_735581, EPI_ISL_735582, EPI_ISL_735583, EPI_ISL_735584, EPI_ISL_735585, EPI_ISL_735586, EPI_ISL_735587, EPI_ISL_735588, EPI_ISL_735589, EPI_ISL_735590, EPI_ISL_735591, EPI_ISL_735592, EPI_ISL_735593, EPI_ISL_735594, EPI_ISL_735595, EPI_ISL_735596, EPI_ISL_735597, EPI_ISL_735598, EPI_ISL_735599, EPI_ISL_735600, EPI_ISL_735601, EPI_ISL_735602, EPI_ISL_735603, EPI_ISL_735604, EPI_ISL_735605, EPI_ISL_735606, EPI_ISL_735607, EPI_ISL_735608, EPI_ISL_735609, EPI_ISL_735610, EPI_ISL_735611, EPI_ISL_735612, EPI_ISL_735613, EPI_ISL_735614, EPI_ISL_735615, EPI_ISL_735616, EPI_ISL_735617, EPI_ISL_735618, EPI_ISL_735619, EPI_ISL_735620, EPI_ISL_735621, EPI_ISL_735622, EPI_ISL_735623, EPI_ISL_735624, EPI_ISL_735625, EPI_ISL_735626, EPI_ISL_735627, EPI_ISL_735628, EPI_ISL_735629, EPI_ISL_735630, EPI_ISL_735631, EPI_ISL_735632, EPI_ISL_735633, EPI_ISL_735634, EPI_ISL_735635, EPI_ISL_735636, EPI_ISL_735637, EPI_ISL_735638, EPI_ISL_735639, EPI_ISL_735640, EPI_ISL_735641, EPI_ISL_735642, EPI_ISL_735643, EPI_ISL_735644, EPI_ISL_735645, EPI_ISL_735646, EPI_ISL_735647, EPI_ISL_735648, EPI_ISL_735649, EPI_ISL_735650, EPI_ISL_735651, EPI_ISL_735652, EPI_ISL_735653, EPI_ISL_735654, EPI_ISL_735655, EPI_ISL_735656, EPI_ISL_735657, EPI_ISL_735658, EPI_ISL_735659, EPI_ISL_735660, EPI_ISL_735661, EPI_ISL_735662, EPI_ISL_735663, EPI_ISL_735664, EPI_ISL_735665, EPI_ISL_735666, EPI_ISL_735667, EPI_ISL_735668, EPI_ISL_735669, EPI_ISL_735670, EPI_ISL_735671, EPI_ISL_735672, EPI_ISL_735673, EPI_ISL_735674, EPI_ISL_735675, EPI_ISL_735676, EPI_ISL_735677, EPI_ISL_735678, EPI_ISL_735679, EPI_ISL_735680, EPI_ISL_735681, EPI_ISL_735682, EPI_ISL_735683, EPI_ISL_735684, EPI_ISL_735685, EPI_ISL_735686, EPI_ISL_735687, EPI_ISL_735688, EPI_ISL_735689, EPI_ISL_735690, EPI_ISL_735691, EPI_ISL_735692, EPI_ISL_735693, EPI_ISL_735694, EPI_ISL_735695, EPI_ISL_735696, EPI_ISL_735697, EPI_ISL_735698, EPI_ISL_735699, EPI_ISL_735700, EPI_ISL_735701, EPI_ISL_735702, EPI_ISL_735703, EPI_ISL_735704, EPI_ISL_735705, EPI_ISL_735706, EPI_ISL_735707, EPI_ISL_735708, EPI_ISL_735709, EPI_ISL_735710, EPI_ISL_735711, EPI_ISL_735712, EPI_ISL_735713, EPI_ISL_735714, EPI_ISL_735715, EPI_ISL_735716, EPI_ISL_735717, EPI_ISL_735718, EPI_ISL_735719, EPI_ISL_735720, EPI_ISL_735721, EPI_ISL_735722, EPI_ISL_735723, EPI_ISL_735724, EPI_ISL_735725, EPI_ISL_735726, EPI_ISL_735727, EPI_ISL_735728, EPI_ISL_735729, EPI_ISL_735730, EPI_ISL_735731, EPI_ISL_735732, EPI_ISL_735733, EPI_ISL_735734, EPI_ISL_735735, EPI_ISL_735736, EPI_ISL_735737, EPI_ISL_735738, EPI_ISL_735739, EPI_ISL_735740, EPI_ISL_735741, EPI_ISL_735742, EPI_ISL_735743, EPI_ISL_735744, EPI_ISL_735745, EPI_ISL_735746, EPI_ISL_735747, EPI_ISL_735748, EPI_ISL_735749, EPI_ISL_735750, EPI_ISL_735751, EPI_ISL_735752, EPI_ISL_735753, EPI_ISL_735754, EPI_ISL_735755, EPI_ISL_735756, EPI_ISL_735757, EPI_ISL_735758, EPI_ISL_735759, EPI_ISL_735760, EPI_ISL_735761, EPI_ISL_735762, EPI_ISL_735763, EPI_ISL_735764, EPI_ISL_735765, EPI_ISL_735766, EPI_ISL_735767, EPI_ISL_735768, EPI_ISL_735769, EPI_ISL_735770, EPI_ISL_735771, EPI_ISL_735772, EPI_ISL_735773, EPI_ISL_735774, EPI_ISL_735775, EPI_ISL_735776, EPI_ISL_735777, EPI_ISL_735778, EPI_ISL_735779, EPI_ISL_735780, EPI_ISL_735781, EPI_ISL_735782, EPI_ISL_735783, EPI_ISL_735784, EPI_ISL_735785, EPI_ISL_735786, EPI_ISL_735787, EPI_ISL_735788, EPI_ISL_735789, EPI_ISL_735790, EPI_ISL_735791, EPI_ISL_735792, EPI_ISL_735793, EPI_ISL_735794, EPI_ISL_735795, EPI_ISL_735796, EPI_ISL_735797, EPI_ISL_735798, EPI_ISL_735799, EPI_ISL_735800, EPI_ISL_735801, EPI_ISL_735802, EPI_ISL_735803, EPI_ISL_735804, EPI_ISL_735805, EPI_ISL_735806, EPI_ISL_735807, EPI_ISL_735808, EPI_ISL_735809, EPI_ISL_735810, EPI_ISL_735811, EPI_ISL_735812, EPI_ISL_735813, EPI_ISL_735814, EPI_ISL_735815, EPI_ISL_735816, EPI_ISL_735817, EPI_ISL_735818, EPI_ISL_735819, EPI_ISL_735820, EPI_ISL_735821, EPI_ISL_735822, EPI_ISL_735823, EPI_ISL_735824, EPI_ISL_735825, EPI_ISL_735826, EPI_ISL_735827, EPI_ISL_735828, EPI_ISL_735829, EPI_ISL_735830, EPI_ISL_735831, EPI_ISL_735832, EPI_ISL_735833, EPI_ISL_735834, EPI_ISL_735835, EPI_ISL_735836, EPI_ISL_735837, EPI_ISL_735838, EPI_ISL_735839, EPI_ISL_735840, EPI_ISL_735841, EPI_ISL_735842, EPI_ISL_735843, EPI_ISL_735844, EPI_ISL_735845, EPI_ISL_735846, EPI_ISL_735847, EPI_ISL_735848, EPI_ISL_735849, EPI_ISL_735850, EPI_ISL_735851, EPI_ISL_735852, EPI_ISL_735853, EPI_ISL_735854, EPI_ISL_735855, EPI_ISL_735856, EPI_ISL_735857, EPI_ISL_735858, EPI_ISL_735859, EPI_ISL_736201, EPI_ISL_736203, EPI_ISL_736207, EPI_ISL_736208, EPI_ISL_736210, EPI_ISL_736211, EPI_ISL_736213, EPI_ISL_736214, EPI_ISL_736215, EPI_ISL_736216, EPI_ISL_736217, EPI_ISL_736218, EPI_ISL_736219, EPI_ISL_736224, EPI_ISL_736225, EPI_ISL_736226, EPI_ISL_736227, EPI_ISL_736228, EPI_ISL_736229, EPI_ISL_736230, EPI_ISL_736232, EPI_ISL_736233, EPI_ISL_736244, EPI_ISL_736246, EPI_ISL_736247, EPI_ISL_736249, EPI_ISL_736251, EPI_ISL_736253, EPI_ISL_736256, EPI_ISL_736259, EPI_ISL_736260, EPI_ISL_736262, EPI_ISL_736263, EPI_ISL_736265, EPI_ISL_736267, EPI_ISL_736268, EPI_ISL_736270, EPI_ISL_736272, EPI_ISL_736273, EPI_ISL_736276, EPI_ISL_736279, EPI_ISL_736280, EPI_ISL_736281, EPI_ISL_736282, EPI_ISL_736283, EPI_ISL_736285, EPI_ISL_736286, EPI_ISL_736288, EPI_ISL_736291, EPI_ISL_736294, EPI_ISL_736295, EPI_ISL_736296, EPI_ISL_736297, EPI_ISL_736298, EPI_ISL_736299, EPI_ISL_736300, EPI_ISL_736302, EPI_ISL_736305, EPI_ISL_736307, EPI_ISL_736308, EPI_ISL_736310, EPI_ISL_736314, EPI_ISL_736317, EPI_ISL_736319, EPI_ISL_736322, EPI_ISL_736324, EPI_ISL_736326, EPI_ISL_736328, EPI_ISL_736329, EPI_ISL_736333, EPI_ISL_736335, EPI_ISL_736336, EPI_ISL_736337, EPI_ISL_736338, EPI_ISL_736339, EPI_ISL_736340, EPI_ISL_736341, EPI_ISL_736342, EPI_ISL_736344, EPI_ISL_736345, EPI_ISL_736348, EPI_ISL_736351, EPI_ISL_736353, EPI_ISL_736354, EPI_ISL_736357, EPI_ISL_736359, EPI_ISL_736360, EPI_ISL_736361, EPI_ISL_736363, EPI_ISL_736364, EPI_ISL_736365, EPI_ISL_736366, EPI_ISL_736368, EPI_ISL_736372, EPI_ISL_736374, EPI_ISL_736375, EPI_ISL_736377, EPI_ISL_736379, EPI_ISL_736381, EPI_ISL_736384, EPI_ISL_736385, EPI_ISL_736387, EPI_ISL_736393, EPI_ISL_736395, EPI_ISL_736396, EPI_ISL_736397, EPI_ISL_736401, EPI_ISL_736402, EPI_ISL_736403, EPI_ISL_736405, EPI_ISL_736406, EPI_ISL_736407, EPI_ISL_736409, EPI_ISL_736410, EPI_ISL_736413, EPI_ISL_736415, EPI_ISL_736418, EPI_ISL_736420, EPI_ISL_736421, EPI_ISL_736422, EPI_ISL_736424, EPI_ISL_736427, EPI_ISL_736432, EPI_ISL_736433, EPI_ISL_736434, EPI_ISL_736435, EPI_ISL_736436, EPI_ISL_736439, EPI_ISL_736440, EPI_ISL_736441, EPI_ISL_736443, EPI_ISL_736446, EPI_ISL_736447, EPI_ISL_736448, EPI_ISL_736450, EPI_ISL_736452, EPI_ISL_736453, EPI_ISL_736454, EPI_ISL_736457, EPI_ISL_736462, EPI_ISL_736463, EPI_ISL_736465, EPI_ISL_736467, EPI_ISL_736472, EPI_ISL_736473, EPI_ISL_736475, EPI_ISL_736480, EPI_ISL_736487, EPI_ISL_736497, EPI_ISL_736501, EPI_ISL_736502, EPI_ISL_736504, EPI_ISL_736508, EPI_ISL_736514, EPI_ISL_736515, EPI_ISL_736523, EPI_ISL_736525, EPI_ISL_736527, EPI_ISL_736528, EPI_ISL_736536, EPI_ISL_736538, EPI_ISL_736542, EPI_ISL_736549, EPI_ISL_736550, EPI_ISL_736551, EPI_ISL_736552, EPI_ISL_736558, EPI_ISL_736563, EPI_ISL_736565, EPI_ISL_736571, EPI_ISL_736578, EPI_ISL_736586, EPI_ISL_736588, EPI_ISL_736590, EPI_ISL_736593, EPI_ISL_736594, EPI_ISL_736597, EPI_ISL_736598, EPI_ISL_736601, EPI_ISL_736602, EPI_ISL_736608, EPI_ISL_736609, EPI_ISL_736610, EPI_ISL_736617, EPI_ISL_736618, EPI_ISL_736622, EPI_ISL_736630, EPI_ISL_736637, EPI_ISL_736643, EPI_ISL_736648, EPI_ISL_736675, EPI_ISL_736677, EPI_ISL_736681, EPI_ISL_736682, EPI_ISL_736683, EPI_ISL_736693, EPI_ISL_736696, EPI_ISL_736697, EPI_ISL_736699, EPI_ISL_736703, EPI_ISL_736712, EPI_ISL_736713, EPI_ISL_736716, EPI_ISL_736719, EPI_ISL_736723, EPI_ISL_736724, EPI_ISL_736725, EPI_ISL_736727, EPI_ISL_736728, EPI_ISL_736731, EPI_ISL_736733, EPI_ISL_736738, EPI_ISL_736740, EPI_ISL_736744, EPI_ISL_736746, EPI_ISL_736757, EPI_ISL_756707, EPI_ISL_756753, EPI_ISL_756772, EPI_ISL_756804, EPI_ISL_756808, EPI_ISL_756823, EPI_ISL_756846, EPI_ISL_756872, EPI_ISL_756887, EPI_ISL_756890, EPI_ISL_756903, EPI_ISL_756917, EPI_ISL_756929, EPI_ISL_756946, EPI_ISL_756989, EPI_ISL_756999, EPI_ISL_757020, EPI_ISL_760842, EPI_ISL_761059, EPI_ISL_762237, EPI_ISL_762281, EPI_ISL_762399, EPI_ISL_762493, EPI_ISL_762494, EPI_ISL_769226, EPI_ISL_769227, EPI_ISL_769228, EPI_ISL_769230, EPI_ISL_769231, EPI_ISL_769232, EPI_ISL_769234, EPI_ISL_769235, EPI_ISL_769237, EPI_ISL_769238, EPI_ISL_769239, EPI_ISL_769240, EPI_ISL_769241, EPI_ISL_769242, EPI_ISL_769247, EPI_ISL_769248, EPI_ISL_769251, EPI_ISL_769253, EPI_ISL_769256, EPI_ISL_769258, EPI_ISL_769262, EPI_ISL_769263, EPI_ISL_769264, EPI_ISL_769265, EPI_ISL_769268, EPI_ISL_769269, EPI_ISL_769270, EPI_ISL_769271, EPI_ISL_769272, EPI_ISL_769274, EPI_ISL_769277, EPI_ISL_769278, EPI_ISL_769279, EPI_ISL_769283, EPI_ISL_769285, EPI_ISL_769287, EPI_ISL_769292, EPI_ISL_769293, EPI_ISL_769295, EPI_ISL_769296, EPI_ISL_769298, EPI_ISL_769299, EPI_ISL_769300, EPI_ISL_769301, EPI_ISL_769302, EPI_ISL_769303, EPI_ISL_769304, EPI_ISL_769305, EPI_ISL_769307, EPI_ISL_769309, EPI_ISL_769311, EPI_ISL_769313, EPI_ISL_769315, EPI_ISL_769317, EPI_ISL_769318, EPI_ISL_769324, EPI_ISL_769326, EPI_ISL_769328, EPI_ISL_769331, EPI_ISL_769332, EPI_ISL_769333, EPI_ISL_769336, EPI_ISL_769337, EPI_ISL_769338, EPI_ISL_769339, EPI_ISL_769340, EPI_ISL_769342, EPI_ISL_769344, EPI_ISL_769346, EPI_ISL_769347, EPI_ISL_769350, EPI_ISL_769351, EPI_ISL_769354, EPI_ISL_769356, EPI_ISL_769357, EPI_ISL_769358, EPI_ISL_769359, EPI_ISL_769362, EPI_ISL_769363, EPI_ISL_769364, EPI_ISL_769366, EPI_ISL_769368, EPI_ISL_769369, EPI_ISL_769371, EPI_ISL_769372, EPI_ISL_769373, EPI_ISL_769375, EPI_ISL_769376, EPI_ISL_769377, EPI_ISL_769378, EPI_ISL_769379, EPI_ISL_769382, EPI_ISL_769386, EPI_ISL_769393, EPI_ISL_769395, EPI_ISL_769397, EPI_ISL_769398, EPI_ISL_769400, EPI_ISL_769403, EPI_ISL_769407, EPI_ISL_769408, EPI_ISL_769409, EPI_ISL_769410, EPI_ISL_769411, EPI_ISL_769414, EPI_ISL_769415, EPI_ISL_769423, EPI_ISL_769424, EPI_ISL_769425, EPI_ISL_769426, EPI_ISL_769429, EPI_ISL_769432, EPI_ISL_769433, EPI_ISL_769435, EPI_ISL_769436, EPI_ISL_769437, EPI_ISL_769438, EPI_ISL_769439, EPI_ISL_769443, EPI_ISL_769444, EPI_ISL_769447, EPI_ISL_769448, EPI_ISL_769451, EPI_ISL_769453, EPI_ISL_769455, EPI_ISL_769456, EPI_ISL_769461, EPI_ISL_769463, EPI_ISL_769464, EPI_ISL_769465, EPI_ISL_769467, EPI_ISL_769472, EPI_ISL_769477, EPI_ISL_769478, EPI_ISL_769479, EPI_ISL_769480, EPI_ISL_769483, EPI_ISL_769484, EPI_ISL_769486, EPI_ISL_769487, EPI_ISL_769488, EPI_ISL_769490, EPI_ISL_769491, EPI_ISL_769492, EPI_ISL_769493, EPI_ISL_769494, EPI_ISL_769495, EPI_ISL_769496, EPI_ISL_769498, EPI_ISL_769505, EPI_ISL_769507, EPI_ISL_769508, EPI_ISL_769509, EPI_ISL_769510, EPI_ISL_769514, EPI_ISL_769515, EPI_ISL_769516, EPI_ISL_769520, EPI_ISL_769521, EPI_ISL_769523, EPI_ISL_769526, EPI_ISL_769527, EPI_ISL_769529, EPI_ISL_769531, EPI_ISL_769532, EPI_ISL_769534, EPI_ISL_769536, EPI_ISL_769537, EPI_ISL_769538, EPI_ISL_769541, EPI_ISL_769544, EPI_ISL_769546, EPI_ISL_769547, EPI_ISL_769548, EPI_ISL_769549, EPI_ISL_769550, EPI_ISL_769552, EPI_ISL_769553, EPI_ISL_769554, EPI_ISL_769555, EPI_ISL_769557, EPI_ISL_769558, EPI_ISL_769559, EPI_ISL_769560, EPI_ISL_769561, EPI_ISL_769562, EPI_ISL_769563, EPI_ISL_769564, EPI_ISL_769565, EPI_ISL_769566, EPI_ISL_769569, EPI_ISL_769570, EPI_ISL_769571, EPI_ISL_769572, EPI_ISL_769573, EPI_ISL_769574, EPI_ISL_769575, EPI_ISL_769576, EPI_ISL_769578, EPI_ISL_769579, EPI_ISL_769580, EPI_ISL_769581, EPI_ISL_769582, EPI_ISL_769585, EPI_ISL_769586, EPI_ISL_769587, EPI_ISL_769588, EPI_ISL_769591, EPI_ISL_769592, EPI_ISL_769593, EPI_ISL_769595, EPI_ISL_769596, EPI_ISL_769598, EPI_ISL_769599, EPI_ISL_769600, EPI_ISL_769601, EPI_ISL_769602, EPI_ISL_769603, EPI_ISL_769604, EPI_ISL_769605, EPI_ISL_769608, EPI_ISL_769610, EPI_ISL_769615, EPI_ISL_769616, EPI_ISL_769618, EPI_ISL_769620, EPI_ISL_769621, EPI_ISL_769622, EPI_ISL_769623, EPI_ISL_769624, EPI_ISL_769626, EPI_ISL_769627, EPI_ISL_769628, EPI_ISL_769629, EPI_ISL_769630, EPI_ISL_769631, EPI_ISL_769632, EPI_ISL_769633, EPI_ISL_769634, EPI_ISL_769635, EPI_ISL_769636, EPI_ISL_769637, EPI_ISL_769638, EPI_ISL_769639, EPI_ISL_769640, EPI_ISL_769642, EPI_ISL_769643, EPI_ISL_769645, EPI_ISL_769647, EPI_ISL_769649, EPI_ISL_769650, EPI_ISL_769651, EPI_ISL_769653, EPI_ISL_769654, EPI_ISL_769656, EPI_ISL_769657, EPI_ISL_769658, EPI_ISL_769659, EPI_ISL_769660, EPI_ISL_769663, EPI_ISL_769664, EPI_ISL_769665, EPI_ISL_769668, EPI_ISL_769669, EPI_ISL_769670, EPI_ISL_769671, EPI_ISL_769672, EPI_ISL_769673, EPI_ISL_769674, EPI_ISL_769675, EPI_ISL_769676, EPI_ISL_769678, EPI_ISL_769679, EPI_ISL_769680, EPI_ISL_769681, EPI_ISL_769682, EPI_ISL_769683, EPI_ISL_769684, EPI_ISL_769685, EPI_ISL_769686, EPI_ISL_769688, EPI_ISL_769690, EPI_ISL_769692, EPI_ISL_769693, EPI_ISL_769694, EPI_ISL_769695, EPI_ISL_769696, EPI_ISL_769697, EPI_ISL_769698, EPI_ISL_769699, EPI_ISL_769700, EPI_ISL_769701, EPI_ISL_769702, EPI_ISL_769704, EPI_ISL_769706, EPI_ISL_769707, EPI_ISL_769708, EPI_ISL_769709, EPI_ISL_769710, EPI_ISL_769712, EPI_ISL_769717, EPI_ISL_769718, EPI_ISL_769719, EPI_ISL_769721, EPI_ISL_769722, EPI_ISL_769723, EPI_ISL_769725, EPI_ISL_769726, EPI_ISL_769727, EPI_ISL_769728, EPI_ISL_769729, EPI_ISL_769730, EPI_ISL_769731, EPI_ISL_769732, EPI_ISL_769733, EPI_ISL_769734, EPI_ISL_769736, EPI_ISL_769737, EPI_ISL_769738, EPI_ISL_769739, EPI_ISL_769740, EPI_ISL_769742, EPI_ISL_769745, EPI_ISL_769746, EPI_ISL_769748, EPI_ISL_769749, EPI_ISL_769750, EPI_ISL_769754, EPI_ISL_769755, EPI_ISL_769756, EPI_ISL_769757, EPI_ISL_769760, EPI_ISL_769762, EPI_ISL_769763, EPI_ISL_769765, EPI_ISL_769766, EPI_ISL_769767, EPI_ISL_769769, EPI_ISL_769770, EPI_ISL_769771, EPI_ISL_769772, EPI_ISL_769774, EPI_ISL_769775, EPI_ISL_769777, EPI_ISL_769778, EPI_ISL_769779, EPI_ISL_769780, EPI_ISL_769781, EPI_ISL_769782, EPI_ISL_769783, EPI_ISL_769784, EPI_ISL_769785, EPI_ISL_769786, EPI_ISL_769789, EPI_ISL_769790, EPI_ISL_769791, EPI_ISL_769792, EPI_ISL_769793, EPI_ISL_769795, EPI_ISL_769796, EPI_ISL_769797, EPI_ISL_769798, EPI_ISL_769799, EPI_ISL_769800, EPI_ISL_769801, EPI_ISL_769803, EPI_ISL_769804, EPI_ISL_769805, EPI_ISL_769807, EPI_ISL_769808, EPI_ISL_769809, EPI_ISL_769810, EPI_ISL_769811, EPI_ISL_769812, EPI_ISL_769814, EPI_ISL_769816, EPI_ISL_769817, EPI_ISL_769819, EPI_ISL_769820, EPI_ISL_769821, EPI_ISL_769822, EPI_ISL_769823, EPI_ISL_769824, EPI_ISL_769825, EPI_ISL_769827, EPI_ISL_769828, EPI_ISL_769829, EPI_ISL_769830, EPI_ISL_769831, EPI_ISL_769832, EPI_ISL_769833, EPI_ISL_769835, EPI_ISL_769838, EPI_ISL_769839, EPI_ISL_769840, EPI_ISL_769841, EPI_ISL_769843, EPI_ISL_769844, EPI_ISL_769846, EPI_ISL_769847, EPI_ISL_769848, EPI_ISL_769849, EPI_ISL_769851, EPI_ISL_769853, EPI_ISL_769854, EPI_ISL_769855, EPI_ISL_769856, EPI_ISL_769858, EPI_ISL_769859, EPI_ISL_769860, EPI_ISL_777064, EPI_ISL_777080, EPI_ISL_777084, EPI_ISL_777104, EPI_ISL_777141, EPI_ISL_777158, EPI_ISL_777184, EPI_ISL_777187, EPI_ISL_777200, EPI_ISL_777207, EPI_ISL_777224, EPI_ISL_777229, EPI_ISL_777245, EPI_ISL_777269, EPI_ISL_777303, EPI_ISL_777304, EPI_ISL_777310, EPI_ISL_778298, EPI_ISL_778300, EPI_ISL_778304, EPI_ISL_778306, EPI_ISL_778307, EPI_ISL_778308, EPI_ISL_778310, EPI_ISL_778312, EPI_ISL_778313, EPI_ISL_778314, EPI_ISL_778316, EPI_ISL_778317, EPI_ISL_778318, EPI_ISL_778319, EPI_ISL_778320, EPI_ISL_778321, EPI_ISL_778323, EPI_ISL_778326, EPI_ISL_778329, EPI_ISL_778338, EPI_ISL_778339, EPI_ISL_778340, EPI_ISL_778341, EPI_ISL_778343, EPI_ISL_778344, EPI_ISL_778349, EPI_ISL_778350, EPI_ISL_778351, EPI_ISL_778354, EPI_ISL_778363, EPI_ISL_778366, EPI_ISL_778367, EPI_ISL_778369, EPI_ISL_778370, EPI_ISL_778371, EPI_ISL_778379, EPI_ISL_778381, EPI_ISL_778383, EPI_ISL_778384, EPI_ISL_778386, EPI_ISL_778387, EPI_ISL_778388, EPI_ISL_778389, EPI_ISL_778391, EPI_ISL_778393, EPI_ISL_778394, EPI_ISL_778395, EPI_ISL_778398, EPI_ISL_778402, EPI_ISL_778405, EPI_ISL_778406, EPI_ISL_778409, EPI_ISL_778413, EPI_ISL_778414, EPI_ISL_778418, EPI_ISL_778425, EPI_ISL_778427, EPI_ISL_778429, EPI_ISL_778431, EPI_ISL_778434, EPI_ISL_778435, EPI_ISL_778436, EPI_ISL_778437, EPI_ISL_778438, EPI_ISL_778439, EPI_ISL_778440, EPI_ISL_778441, EPI_ISL_778442, EPI_ISL_778443, EPI_ISL_778446, EPI_ISL_778449, EPI_ISL_778452, EPI_ISL_778453, EPI_ISL_778456, EPI_ISL_778458, EPI_ISL_778460, EPI_ISL_778464, EPI_ISL_778466, EPI_ISL_778467, EPI_ISL_778468, EPI_ISL_778469, EPI_ISL_778470, EPI_ISL_778474, EPI_ISL_778475, EPI_ISL_778476, EPI_ISL_778477, EPI_ISL_778479, EPI_ISL_778480, EPI_ISL_778481, EPI_ISL_778483, EPI_ISL_778485, EPI_ISL_778486, EPI_ISL_778489, EPI_ISL_778492, EPI_ISL_778493, EPI_ISL_778495, EPI_ISL_778498, EPI_ISL_778501, EPI_ISL_778502, EPI_ISL_778503, EPI_ISL_778505, EPI_ISL_778507, EPI_ISL_778508, EPI_ISL_778509, EPI_ISL_778511, EPI_ISL_778513, EPI_ISL_778514, EPI_ISL_778515, EPI_ISL_778518, EPI_ISL_778520, EPI_ISL_778521, EPI_ISL_778523, EPI_ISL_778526, EPI_ISL_778532, EPI_ISL_778533, EPI_ISL_778535, EPI_ISL_778536, EPI_ISL_778537, EPI_ISL_778538, EPI_ISL_778542, EPI_ISL_778544, EPI_ISL_778545, EPI_ISL_778546, EPI_ISL_778549, EPI_ISL_778550, EPI_ISL_778552, EPI_ISL_778554, EPI_ISL_778555, EPI_ISL_778557, EPI_ISL_778559, EPI_ISL_778561, EPI_ISL_778563, EPI_ISL_778565, EPI_ISL_778566, EPI_ISL_778567, EPI_ISL_778572, EPI_ISL_778573, EPI_ISL_778574, EPI_ISL_778575, EPI_ISL_778576, EPI_ISL_778580, EPI_ISL_778581, EPI_ISL_778582, EPI_ISL_778583, EPI_ISL_778585, EPI_ISL_778587, EPI_ISL_778588, EPI_ISL_778589, EPI_ISL_778590, EPI_ISL_778591, EPI_ISL_778592, EPI_ISL_778595, EPI_ISL_778596, EPI_ISL_778597, EPI_ISL_778600, EPI_ISL_778602, EPI_ISL_778603, EPI_ISL_778604, EPI_ISL_778606, EPI_ISL_778608, EPI_ISL_778609, EPI_ISL_778610, EPI_ISL_778611, EPI_ISL_778613, EPI_ISL_778622, EPI_ISL_778624, EPI_ISL_778625, EPI_ISL_778626, EPI_ISL_778632, EPI_ISL_778633, EPI_ISL_778634, EPI_ISL_778636, EPI_ISL_778637, EPI_ISL_778639, EPI_ISL_778640, EPI_ISL_778641, EPI_ISL_778642, EPI_ISL_778643, EPI_ISL_933482, EPI_ISL_933483, EPI_ISL_933484, EPI_ISL_933485, EPI_ISL_933486, EPI_ISL_933487, EPI_ISL_933488, EPI_ISL_933489, EPI_ISL_933490, EPI_ISL_933491, EPI_ISL_945140, EPI_ISL_945338

| | | | |
|---|---|---|---|
| see above | Lighthouse Lab in Milton Keynes | Wellcome Sanger Institute for the COVID-19 Genomics UK (COG-UK) Consortium | Cordelia Langford; David K. Jackson; Dominic Kwiatkowski; Ewan Harrison; Ian Johnston; John Sillitoe on behalf of the Wellcome Sanger Institute COVID-19 Surveillance Team; Roberto Amato; Sonia Goncalves; The Lighthouse Lab in Milton Keynes and Alex Alderton |

EPI_ISL_840041, EPI_ISL_840042, EPI_ISL_840043, EPI_ISL_840044, EPI_ISL_840045, EPI_ISL_840046, EPI_ISL_840047, EPI_ISL_840048, EPI_ISL_840049, EPI_ISL_840050, EPI_ISL_840146, EPI_ISL_840147, EPI_ISL_840148, EPI_ISL_840149, EPI_ISL_840150, EPI_ISL_840151, EPI_ISL_840152, EPI_ISL_840153, EPI_ISL_840154, EPI_ISL_840155, EPI_ISL_840156, EPI_ISL_840157, EPI_ISL_840158, EPI_ISL_840159, EPI_ISL_840160, EPI_ISL_840169, EPI_ISL_950698, EPI_ISL_950699, EPI_ISL_950700, EPI_ISL_950701

| | | | |
|---|---|---|---|
| see above | Lincolnshire Hospitals and DeepSeq Nottingham | COVID-19 Genomics UK (COG-UK) Consortium | Christopher Moore; Fei Sang; Joeseph Chappell; Johnny Debebe; Jonathan Ball; Matthew Carlisle; Matthew Loose; Nadine Holmes; Nichola Duckworth; Patrick McClure; Sarah Walsh; Tim Sloan; Victoria Wright |

EPI_ISL_724525, EPI_ISL_724526, EPI_ISL_724527, EPI_ISL_724528, EPI_ISL_724529, EPI_ISL_724530, EPI_ISL_724531, EPI_ISL_724532, EPI_ISL_724533, EPI_ISL_724534, EPI_ISL_724535, EPI_ISL_724536, EPI_ISL_724537, EPI_ISL_724538, EPI_ISL_724539, EPI_ISL_813732, EPI_ISL_813733, EPI_ISL_813734, EPI_ISL_813735, EPI_ISL_813736, EPI_ISL_813737, EPI_ISL_813738, EPI_ISL_813739, EPI_ISL_813740, EPI_ISL_813741, EPI_ISL_813742, EPI_ISL_813743, EPI_ISL_813744, EPI_ISL_813745, EPI_ISL_813746, EPI_ISL_813747, EPI_ISL_813748, EPI_ISL_813749, EPI_ISL_813750, EPI_ISL_813764, EPI_ISL_813765, EPI_ISL_813766, EPI_ISL_813767, EPI_ISL_813768, EPI_ISL_813769, EPI_ISL_813770, EPI_ISL_813771, EPI_ISL_813772, EPI_ISL_814285, EPI_ISL_814286, EPI_ISL_814552, EPI_ISL_838352, EPI_ISL_838458, EPI_ISL_838459, EPI_ISL_838460, EPI_ISL_838461, EPI_ISL_838462, EPI_ISL_838463, EPI_ISL_838464, EPI_ISL_838465, EPI_ISL_838466, EPI_ISL_838467, EPI_ISL_838468, EPI_ISL_838469, EPI_ISL_838470, EPI_ISL_838471, EPI_ISL_838472, EPI_ISL_838473, EPI_ISL_838475, EPI_ISL_838476, EPI_ISL_838477, EPI_ISL_838478, EPI_ISL_838479, EPI_ISL_838480, EPI_ISL_838481, EPI_ISL_838482, EPI_ISL_838483, EPI_ISL_838484, EPI_ISL_838485, EPI_ISL_838486, EPI_ISL_838487, EPI_ISL_838488, EPI_ISL_838489, EPI_ISL_838490, EPI_ISL_838491, EPI_ISL_838492, EPI_ISL_838493, EPI_ISL_838494, EPI_ISL_838495, EPI_ISL_838496, EPI_ISL_838497, EPI_ISL_838498, EPI_ISL_838500, EPI_ISL_838506, EPI_ISL_838508, EPI_ISL_838509, EPI_ISL_838517, EPI_ISL_838518, EPI_ISL_838519, EPI_ISL_838521, EPI_ISL_838523, EPI_ISL_838665, EPI_ISL_838672, EPI_ISL_865208, EPI_ISL_865210, EPI_ISL_865255, EPI_ISL_865256, EPI_ISL_865257, EPI_ISL_865258, EPI_ISL_865259, EPI_ISL_865260, EPI_ISL_865262, EPI_ISL_865263, EPI_ISL_865266, EPI_ISL_865267, EPI_ISL_865269, EPI_ISL_865412, EPI_ISL_865413, EPI_ISL_865415, EPI_ISL_865416, EPI_ISL_865417, EPI_ISL_865418, EPI_ISL_865419, EPI_ISL_865428, EPI_ISL_865432, EPI_ISL_865436, EPI_ISL_997376, EPI_ISL_1177833

| | | | |
|---|---|---|---|
| see above | Liverpool Clinical Laboratories | COVID-19 Genomics UK (COG-UK) Consortium | A Alrezaihi; Alessandro Gerada; Alistair Darby; Angela Cowell; Anita Lucaci; Anu Chawla; Cassie Olateju; Catherine Hartley; Charlotte Nelson; Ecaterina Vamos; Elaine O'Toole; Eleanor G Bentley; Ghada T Shawli; Hermione Webster; Isabel García-Dorival; James Johnson; James P Stewart; Jenifer Manson; Joanne Watts; Jones Benjamin; Jordan J Clark; Julian Hiscox; L Luu; Lucille Rainbow; M Almsaud; Margaret Hughes; Mark Whitehead; Matthew Gemmell; Miren Iturriza-Gomara; Muhannad Alruwaili; N.P Randle; Neil Swainston; PKF Gilmore; Parul Sharma; Rebekah Penrice-Randal; Richard Eccles; Richard Gregory; Sam Haldenby; Steve Paterson; Stuart D Armstrong; Trevor Ian Robinson; Ximeng Han |

EPI_ISL_819575, EPI_ISL_819576, EPI_ISL_839485, EPI_ISL_839486, EPI_ISL_839487, EPI_ISL_839488, EPI_ISL_839489, EPI_ISL_839490, EPI_ISL_839491, EPI_ISL_839534, EPI_ISL_839535, EPI_ISL_839536, EPI_ISL_839537, EPI_ISL_839538, EPI_ISL_839539, EPI_ISL_839540, EPI_ISL_839541, EPI_ISL_839542, EPI_ISL_839544, EPI_ISL_839545, EPI_ISL_839546, EPI_ISL_839547, EPI_ISL_839548, EPI_ISL_839549, EPI_ISL_839550, EPI_ISL_839551, EPI_ISL_839552, EPI_ISL_839553, EPI_ISL_839554, EPI_ISL_839555, EPI_ISL_839556, EPI_ISL_839557, EPI_ISL_839558, EPI_ISL_839559, EPI_ISL_839560, EPI_ISL_839602, EPI_ISL_839662, EPI_ISL_839749, EPI_ISL_839750, EPI_ISL_839751, EPI_ISL_839752, EPI_ISL_839753, EPI_ISL_839754, EPI_ISL_839755, EPI_ISL_839756, EPI_ISL_839757, EPI_ISL_839758, EPI_ISL_839759, EPI_ISL_839760, EPI_ISL_839761, EPI_ISL_839762, EPI_ISL_839763, EPI_ISL_839764, EPI_ISL_839765, EPI_ISL_839766, EPI_ISL_839767, EPI_ISL_839768, EPI_ISL_839769, EPI_ISL_839770, EPI_ISL_839771, EPI_ISL_839772, EPI_ISL_839773, EPI_ISL_839774, EPI_ISL_839775, EPI_ISL_839776, EPI_ISL_839777, EPI_ISL_839778, EPI_ISL_839779, EPI_ISL_839780, EPI_ISL_839781, EPI_ISL_839782, EPI_ISL_839783, EPI_ISL_839784, EPI_ISL_839785, EPI_ISL_839786, EPI_ISL_839787, EPI_ISL_839788, EPI_ISL_839789, EPI_ISL_839790, EPI_ISL_839791, EPI_ISL_839792, EPI_ISL_839793, EPI_ISL_839794, EPI_ISL_839795, EPI_ISL_839796, EPI_ISL_839797, EPI_ISL_839798, EPI_ISL_839799, EPI_ISL_839800, EPI_ISL_839801, EPI_ISL_839802, EPI_ISL_839803, EPI_ISL_839804, EPI_ISL_839805, EPI_ISL_839806, EPI_ISL_839807, EPI_ISL_839808, EPI_ISL_839809, EPI_ISL_839810, EPI_ISL_839811, EPI_ISL_839812, EPI_ISL_839813, EPI_ISL_839814, EPI_ISL_839815, EPI_ISL_866400, EPI_ISL_866401, EPI_ISL_866402, EPI_ISL_866403, EPI_ISL_866404, EPI_ISL_866405, EPI_ISL_866406, EPI_ISL_866407, EPI_ISL_866408, EPI_ISL_866409, EPI_ISL_866410, EPI_ISL_866411, EPI_ISL_866412, EPI_ISL_866413, EPI_ISL_866414, EPI_ISL_866415, EPI_ISL_866416, EPI_ISL_866417, EPI_ISL_866418, EPI_ISL_866420, EPI_ISL_866421, EPI_ISL_866422, EPI_ISL_866423, EPI_ISL_866424, EPI_ISL_866425, EPI_ISL_866426, EPI_ISL_866457, EPI_ISL_866458, EPI_ISL_866459, EPI_ISL_866460, EPI_ISL_866461, EPI_ISL_866462, EPI_ISL_866463, EPI_ISL_866464, EPI_ISL_866465, EPI_ISL_866469, EPI_ISL_866470, EPI_ISL_866471, EPI_ISL_866472, EPI_ISL_866473, EPI_ISL_866474, EPI_ISL_866475, EPI_ISL_866476, EPI_ISL_866477, EPI_ISL_866478, EPI_ISL_866479, EPI_ISL_866480, EPI_ISL_998704, EPI_ISL_998713, EPI_ISL_1637197, EPI_ISL_1637198

| | | | |
|---|---|---|---|
| see above | Northumbria University / South Tees Hospitals NHS Foundation Trust / North Cumbria Integrated Care NHS Foundation Trust / North Tees and Hartlepool NHS Foundation Trust / Newcastle Hospitals NHS Foundation Trust | COVID-19 Genomics UK (COG-UK) Consortium | Andrew Nelson; Brendan Payne; Clive Graham; Darren L Smith; Debra Padgett; Edward Barton; Emma Swindells; Garren Scott; Gary Black; Gary Eltringham; Giles S Holt; Greg R Young; Jane Greenaway; Jennifer Collins; John Allan; Joshua Loh; Lynn Dover; Matthew Bashton; Mohammad A Tariq; Paul Baker; Sarah Essex; Steve Liggett; Wen C Yew; Yusri Taha |

EPI_ISL_867286, EPI_ISL_867287, EPI_ISL_867288, EPI_ISL_867289, EPI_ISL_867290, EPI_ISL_867291, EPI_ISL_867292, EPI_ISL_867293, EPI_ISL_867294, EPI_ISL_867295, EPI_ISL_867296, EPI_ISL_867297, EPI_ISL_867499

| | | | |
|---|---|---|---|
| see above | Originating lab: Wales Specialist Virology Centre Sequencing lab: Pathogen Genomics Unit | Public Health Wales Microbiology Cardiff Wales Specialist Virology Centre | Alec Birchley; Alexander Adams; Amy Gaskin; Angela Marchbank; Bree Gatica-Wilcox; Catherine Moore; Jason Coombes; Joanne Watkins; Joel Southgate; Johnathan Evans; Laura Gifford; Lauren Gilbert; Lee Graham; Malorie Perry; Matthew Bull; Nicole Pacchiarini; Sally Corden; Sara Kumziene-Summerhayes; Sara Rey; Sarah Taylor; Simon Cottrell; Sophie Jones; Tom Connor |

EPI_ISL_763841, EPI_ISL_763844, EPI_ISL_763859, EPI_ISL_763860, EPI_ISL_763861, EPI_ISL_763862, EPI_ISL_763930, EPI_ISL_763931, EPI_ISL_763932, EPI_ISL_763933, EPI_ISL_763936, EPI_ISL_763937, EPI_ISL_763938, EPI_ISL_763939, EPI_ISL_763942, EPI_ISL_763943, EPI_ISL_763944, EPI_ISL_763946, EPI_ISL_763947, EPI_ISL_763948, EPI_ISL_763949, EPI_ISL_763951, EPI_ISL_763952, EPI_ISL_763959, EPI_ISL_763960, EPI_ISL_763966, EPI_ISL_764077, EPI_ISL_764078, EPI_ISL_764079, EPI_ISL_764086, EPI_ISL_764088, EPI_ISL_764090, EPI_ISL_764117, EPI_ISL_764250, EPI_ISL_764264, EPI_ISL_764265, EPI_ISL_764271, EPI_ISL_764525, EPI_ISL_764528, EPI_ISL_764529, EPI_ISL_764534, EPI_ISL_764535, EPI_ISL_764536, EPI_ISL_764537, EPI_ISL_764540, EPI_ISL_764541, EPI_ISL_764542, EPI_ISL_764543, EPI_ISL_764544, EPI_ISL_764545, EPI_ISL_764547, EPI_ISL_764548, EPI_ISL_764552, EPI_ISL_764556, EPI_ISL_764557, EPI_ISL_764564, EPI_ISL_764565, EPI_ISL_764567, EPI_ISL_764569, EPI_ISL_764570, EPI_ISL_764572, EPI_ISL_819578, EPI_ISL_840250, EPI_ISL_840252, EPI_ISL_840253, EPI_ISL_840254, EPI_ISL_840259, EPI_ISL_840265, EPI_ISL_840281, EPI_ISL_840286

| | | | |
|---|---|---|---|
| see above | Oxford Viromics, NDM, University of Oxford; Oxford University Hospitals; Basingstoke and North Hampshire Hospital | COVID-19 Genomics UK (COG-UK) Consortium | Alex Mobbs; Amy Trebes; Anita Justice; Catrin Moore; Christophe Fraser; David Bonsall; David Buck; Emma Wise; George Macintyre; Jessica Lynch; John Todd; Mariateresa de Cesare; Matilde Mori; Monique Andersson; Nathan Moore; Nick Cortes; Robert Shaw; Stephen Kidd; Tanya Golubchik; Timothy Peto |

EPI_ISL_725033, EPI_ISL_725034, EPI_ISL_725035, EPI_ISL_725038, EPI_ISL_725041, EPI_ISL_725055, EPI_ISL_725062, EPI_ISL_725063, EPI_ISL_725064, EPI_ISL_725065, EPI_ISL_725066, EPI_ISL_725067, EPI_ISL_725068, EPI_ISL_725069, EPI_ISL_725070, EPI_ISL_725071, EPI_ISL_725072, EPI_ISL_725073, EPI_ISL_725074, EPI_ISL_725075, EPI_ISL_725076, EPI_ISL_725077, EPI_ISL_725078, EPI_ISL_725079, EPI_ISL_725080, EPI_ISL_725081, EPI_ISL_725082, EPI_ISL_725083, EPI_ISL_725084, EPI_ISL_725085, EPI_ISL_725086, EPI_ISL_725087, EPI_ISL_725088, EPI_ISL_725089, EPI_ISL_725090, EPI_ISL_725091, EPI_ISL_725092, EPI_ISL_725093, EPI_ISL_725094, EPI_ISL_725095, EPI_ISL_725096, EPI_ISL_725097, EPI_ISL_725098, EPI_ISL_725099, EPI_ISL_725100, EPI_ISL_725101, EPI_ISL_725102, EPI_ISL_725103, EPI_ISL_725104, EPI_ISL_725105, EPI_ISL_725151, EPI_ISL_725155, EPI_ISL_725296, EPI_ISL_725297, EPI_ISL_725298, EPI_ISL_725299, EPI_ISL_725300, EPI_ISL_725301, EPI_ISL_725343, EPI_ISL_725344, EPI_ISL_725346, EPI_ISL_725347, EPI_ISL_725348, EPI_ISL_725349, EPI_ISL_725350, EPI_ISL_725351, EPI_ISL_725352, EPI_ISL_725353, EPI_ISL_725354, EPI_ISL_725355, EPI_ISL_725356, EPI_ISL_725357, EPI_ISL_725358, EPI_ISL_725359, EPI_ISL_725360, EPI_ISL_725361, EPI_ISL_725362, EPI_ISL_725363, EPI_ISL_725364, EPI_ISL_725366, EPI_ISL_725367, EPI_ISL_725368, EPI_ISL_725369, EPI_ISL_725417, EPI_ISL_725419, EPI_ISL_725420, EPI_ISL_725422, EPI_ISL_725423, EPI_ISL_725424, EPI_ISL_725428, EPI_ISL_725430, EPI_ISL_725433, EPI_ISL_725438, EPI_ISL_725458, EPI_ISL_725459, EPI_ISL_725463, EPI_ISL_725464, EPI_ISL_725466, EPI_ISL_725467, EPI_ISL_725468, EPI_ISL_725469, EPI_ISL_725471, EPI_ISL_725472, EPI_ISL_725475, EPI_ISL_725476, EPI_ISL_725480, EPI_ISL_725481, EPI_ISL_725487, EPI_ISL_725492, EPI_ISL_725493, EPI_ISL_725494, EPI_ISL_725495, EPI_ISL_741453, EPI_ISL_741454, EPI_ISL_741455, EPI_ISL_741456, EPI_ISL_741457, EPI_ISL_741458, EPI_ISL_741459, EPI_ISL_741460, EPI_ISL_741461, EPI_ISL_741465, EPI_ISL_741470, EPI_ISL_741471, EPI_ISL_741473, EPI_ISL_741476, EPI_ISL_741477, EPI_ISL_741478, EPI_ISL_741480, EPI_ISL_741481, EPI_ISL_741483, EPI_ISL_741484, EPI_ISL_741485, EPI_ISL_741532, EPI_ISL_741583, EPI_ISL_741584, EPI_ISL_741585, EPI_ISL_741586, EPI_ISL_741587, EPI_ISL_741592, EPI_ISL_741594, EPI_ISL_741595, EPI_ISL_741596, EPI_ISL_741597, EPI_ISL_741598, EPI_ISL_741599, EPI_ISL_741600, EPI_ISL_741602, EPI_ISL_741603, EPI_ISL_741604, EPI_ISL_741605, EPI_ISL_741606, EPI_ISL_741607, EPI_ISL_741608, EPI_ISL_866581, EPI_ISL_866587, EPI_ISL_866806, EPI_ISL_866815, EPI_ISL_866817, EPI_ISL_866830

| | | | |
|---|---|---|---|
| see above | Quadram Institute Bioscience | COVID-19 Genomics UK (COG-UK) Consortium | Alexander J Trotter; Alison E. Mather; Alp Aydin; Ana P. Tedim; Anastasia Kolyva; Andrew Bell; Andrew J. Page; Claire Stuart; Dave J. Baker; Gemma L. Kay; John Wain; Justin O'Grady; Leonardo de Oliveira Martins; Lizzie Meadows; Maria Diaz; Mark Webber; Muhammed Yasir; Nabil-Fareed Alikhan; Ngozi Elumogo; Nicholas M. Thomson; Rachael Stanley; Rachel Gilroy; Reenesh Prakash; Samir Dervisevic; Samuel Bloomfield; Steven Rudder; Thanh Le-Viet |
| EPI_ISL_725643, EPI_ISL_725644, EPI_ISL_725645, EPI_ISL_725646, EPI_ISL_725647, EPI_ISL_725648, EPI_ISL_725649, EPI_ISL_725650, EPI_ISL_725651, EPI_ISL_725652, EPI_ISL_725653, EPI_ISL_725654, EPI_ISL_725655, EPI_ISL_725656, EPI_ISL_725657, EPI_ISL_725658, EPI_ISL_725659, EPI_ISL_725660, EPI_ISL_725661, EPI_ISL_725662, EPI_ISL_725663, EPI_ISL_725664, EPI_ISL_725665, EPI_ISL_725666, EPI_ISL_725667, EPI_ISL_725668, EPI_ISL_725669, EPI_ISL_741614, EPI_ISL_741615, EPI_ISL_741616, EPI_ISL_741617, EPI_ISL_741618, EPI_ISL_741619, EPI_ISL_741620, EPI_ISL_741621, EPI_ISL_741622, EPI_ISL_741623, EPI_ISL_741624, EPI_ISL_819577, EPI_ISL_866896, EPI_ISL_866897, EPI_ISL_866898, EPI_ISL_866899, EPI_ISL_866900, EPI_ISL_949651 | | | |
| see above | Queens Medical Centre, Clinical Microbiology Department / DeepSeq Nottingham | COVID-19 Genomics UK (COG-UK) Consortium | Christopher Moore; Fei Sang; Gemma Clark; Hannah Howson-Wells; Johnny Debebe; Jonathan Ball; Joseph Chappell; Manjinder Khakh; Matthew Carlisle; Matthew Loose; Michelle M Lister; Nadine Holmes; Patrick McClure; Theocharis Tsoleridis; Vicki M Fleming; Victoria Wright; Wendy Smith |
| EPI_ISL_722999, EPI_ISL_723000, EPI_ISL_723006, EPI_ISL_723007, EPI_ISL_723015, EPI_ISL_723044, EPI_ISL_728343, EPI_ISL_747498, EPI_ISL_747499, EPI_ISL_747500, EPI_ISL_747501, EPI_ISL_747502, EPI_ISL_747503, EPI_ISL_747504, EPI_ISL_747505, EPI_ISL_747506, EPI_ISL_747507, EPI_ISL_747508, EPI_ISL_747509, EPI_ISL_747510, EPI_ISL_747511, EPI_ISL_747512, EPI_ISL_747513, EPI_ISL_747514, EPI_ISL_747515, EPI_ISL_747516, EPI_ISL_747517, EPI_ISL_747518, EPI_ISL_747519, EPI_ISL_766070, EPI_ISL_766072, EPI_ISL_766073, EPI_ISL_766079, EPI_ISL_766080, EPI_ISL_766081, EPI_ISL_766084, EPI_ISL_766097, EPI_ISL_766102, EPI_ISL_766105, EPI_ISL_766106, EPI_ISL_766107, EPI_ISL_766109, EPI_ISL_766110, EPI_ISL_766117, EPI_ISL_766118, EPI_ISL_766119, EPI_ISL_766120, EPI_ISL_766121, EPI_ISL_766127, EPI_ISL_766128, EPI_ISL_766131, EPI_ISL_766132, EPI_ISL_766133, EPI_ISL_766134, EPI_ISL_766135, EPI_ISL_766136, EPI_ISL_766137, EPI_ISL_766138, EPI_ISL_766139, EPI_ISL_766140, EPI_ISL_766141, EPI_ISL_766144, EPI_ISL_766158, EPI_ISL_766202, EPI_ISL_766203, EPI_ISL_766224, EPI_ISL_766225, EPI_ISL_766226, EPI_ISL_766230, EPI_ISL_766237, EPI_ISL_766238, EPI_ISL_766248, EPI_ISL_766249, EPI_ISL_766250, EPI_ISL_766260, EPI_ISL_766261, EPI_ISL_766288, EPI_ISL_766290, EPI_ISL_766293, EPI_ISL_766294, EPI_ISL_766295, EPI_ISL_766296, EPI_ISL_766297, EPI_ISL_766298, EPI_ISL_766299, EPI_ISL_766300, EPI_ISL_766301, EPI_ISL_766302, EPI_ISL_766303, EPI_ISL_766304, EPI_ISL_766305, EPI_ISL_766306, EPI_ISL_766307, EPI_ISL_766308, EPI_ISL_766309, EPI_ISL_766311, EPI_ISL_766312, EPI_ISL_766313, EPI_ISL_766315, EPI_ISL_766317, EPI_ISL_766395, EPI_ISL_766396, EPI_ISL_766399, EPI_ISL_766400, EPI_ISL_766403, EPI_ISL_766405, EPI_ISL_766407, EPI_ISL_766409, EPI_ISL_766410, EPI_ISL_766411, EPI_ISL_766425, EPI_ISL_766463, EPI_ISL_766508, EPI_ISL_766555, EPI_ISL_766562, EPI_ISL_766563, EPI_ISL_766564, EPI_ISL_768804, EPI_ISL_768805, EPI_ISL_768806, EPI_ISL_768807, EPI_ISL_768808, EPI_ISL_768809, EPI_ISL_768810, EPI_ISL_768811, EPI_ISL_768812, EPI_ISL_768813, EPI_ISL_768814, EPI_ISL_769869, EPI_ISL_769877, EPI_ISL_791184, EPI_ISL_791185, EPI_ISL_791186, EPI_ISL_791208, EPI_ISL_791220, EPI_ISL_836980, EPI_ISL_837076, EPI_ISL_837113, EPI_ISL_837114, EPI_ISL_837186, EPI_ISL_846591, EPI_ISL_860690, EPI_ISL_862251 | | | |
| see above | Respiratory Virus Unit, National Infection Service, Public Health England | COVID-19 Genomics UK (COG-UK) Consortium | PHE Covid Sequencing Team |
| EPI_ISL_1833813 | St George's University Hospitals NHS Foundation Trust | COVID-19 Genomics UK (COG-UK) Consortium | Adam Witney; Cassie Pope; Irene Monahan; Joshua Taylor; Ken Laing; NgeeKeong Tan |
| EPI_ISL_842670, EPI_ISL_842671, EPI_ISL_842672, EPI_ISL_842673, EPI_ISL_842674, EPI_ISL_842675, EPI_ISL_842677, EPI_ISL_842678, EPI_ISL_842679, EPI_ISL_842680, EPI_ISL_842681, EPI_ISL_842693, EPI_ISL_842694, EPI_ISL_842712, EPI_ISL_842713, EPI_ISL_842714, EPI_ISL_842715, EPI_ISL_842716, EPI_ISL_842717, EPI_ISL_842718, EPI_ISL_842720, EPI_ISL_842729, EPI_ISL_842731, EPI_ISL_842732, EPI_ISL_842733, EPI_ISL_842734, EPI_ISL_842735, EPI_ISL_842736, EPI_ISL_842737, EPI_ISL_842738, EPI_ISL_842739, EPI_ISL_842740, EPI_ISL_842741, EPI_ISL_842742, EPI_ISL_842743, EPI_ISL_842744, EPI_ISL_842745, EPI_ISL_842746, EPI_ISL_842747, EPI_ISL_842748, EPI_ISL_842749, EPI_ISL_842750, EPI_ISL_842751, EPI_ISL_842752, EPI_ISL_842753, EPI_ISL_842754, EPI_ISL_842755, EPI_ISL_842756, EPI_ISL_842757, EPI_ISL_842758, EPI_ISL_842759, EPI_ISL_842760, EPI_ISL_842761, EPI_ISL_842762, EPI_ISL_842763, EPI_ISL_842764, EPI_ISL_842765, EPI_ISL_842768, EPI_ISL_842769, EPI_ISL_842770, EPI_ISL_842771, EPI_ISL_842772, EPI_ISL_842773, EPI_ISL_842774, EPI_ISL_842775, EPI_ISL_842776, EPI_ISL_842777, EPI_ISL_842778, EPI_ISL_842779, EPI_ISL_842780, EPI_ISL_842781, EPI_ISL_842782, EPI_ISL_842784, EPI_ISL_842785, EPI_ISL_842786, EPI_ISL_842787, EPI_ISL_842788, EPI_ISL_842789, EPI_ISL_842791, EPI_ISL_866065, EPI_ISL_866066, EPI_ISL_866067, EPI_ISL_866106, EPI_ISL_866107, EPI_ISL_866108, EPI_ISL_866138, EPI_ISL_920199, EPI_ISL_920216, EPI_ISL_920267, EPI_ISL_920277, EPI_ISL_920322, EPI_ISL_920331, EPI_ISL_920365, EPI_ISL_920372, EPI_ISL_920378, EPI_ISL_920459, EPI_ISL_920460, EPI_ISL_920500, EPI_ISL_920503, EPI_ISL_920511, EPI_ISL_920516, EPI_ISL_920576, EPI_ISL_920595, EPI_ISL_1104875, EPI_ISL_1104912, EPI_ISL_1105126, EPI_ISL_1105141, EPI_ISL_1248498, EPI_ISL_1637074 | | | |
| see above | University College London Hospital | COVID-19 Genomics UK (COG-UK) Consortium | Catherine Houlihan; Dan Frampton; Dr Catherine Houlihan; Dr Daniel Frampton; Dr Judith Heaney; Dr Moira Spyer; Dr Paul Grant and Dr Eleni Nastouli; Judith Heaney; Matthew Byott; Moira Spyer and Eleni Nastouli; Stuart Kirk |
| EPI_ISL_724593, EPI_ISL_724606, EPI_ISL_724607, EPI_ISL_724609, EPI_ISL_741186, EPI_ISL_741188, EPI_ISL_741189, EPI_ISL_741190, EPI_ISL_741191, EPI_ISL_741192, EPI_ISL_741193, EPI_ISL_741194, EPI_ISL_741195, EPI_ISL_741196, EPI_ISL_741197, EPI_ISL_741198, EPI_ISL_741199, EPI_ISL_741200, EPI_ISL_741201, EPI_ISL_741202, EPI_ISL_741203, EPI_ISL_741204, EPI_ISL_741205, EPI_ISL_741206, EPI_ISL_741208, EPI_ISL_741209, EPI_ISL_741210, EPI_ISL_741211, EPI_ISL_741212, EPI_ISL_741213, EPI_ISL_741214, EPI_ISL_741215, EPI_ISL_741216, EPI_ISL_741217, EPI_ISL_741218, EPI_ISL_741219, EPI_ISL_741220, EPI_ISL_741221, EPI_ISL_741222, EPI_ISL_741223, EPI_ISL_741224, EPI_ISL_741225, EPI_ISL_741226, EPI_ISL_741227, EPI_ISL_741228, EPI_ISL_741229, EPI_ISL_741230, EPI_ISL_741231, EPI_ISL_741232, EPI_ISL_741233, EPI_ISL_741234, EPI_ISL_741235, EPI_ISL_741236, EPI_ISL_741237, EPI_ISL_741238, EPI_ISL_741239, EPI_ISL_741240, EPI_ISL_741241, EPI_ISL_741242, EPI_ISL_741243, EPI_ISL_741244, EPI_ISL_741245, EPI_ISL_741246, EPI_ISL_741247, EPI_ISL_741248, EPI_ISL_741249, EPI_ISL_741250, EPI_ISL_741251, EPI_ISL_741252, EPI_ISL_741253, EPI_ISL_741254, EPI_ISL_741259, EPI_ISL_741260, EPI_ISL_741261, EPI_ISL_741262, EPI_ISL_741263, EPI_ISL_741270, EPI_ISL_741275, EPI_ISL_741277, EPI_ISL_741287, EPI_ISL_741294, EPI_ISL_741296, EPI_ISL_741297, EPI_ISL_741298, EPI_ISL_741299, EPI_ISL_741301, EPI_ISL_741302, EPI_ISL_741315, EPI_ISL_741316, EPI_ISL_741317, EPI_ISL_741318, EPI_ISL_741319, EPI_ISL_741320, EPI_ISL_741321, EPI_ISL_741322, EPI_ISL_741323, EPI_ISL_741324, EPI_ISL_741325, EPI_ISL_741328, EPI_ISL_763381, EPI_ISL_838673, EPI_ISL_838674, EPI_ISL_838675, EPI_ISL_838676, EPI_ISL_838677, EPI_ISL_838678, EPI_ISL_838679, EPI_ISL_838680, EPI_ISL_838681, EPI_ISL_838682, EPI_ISL_838683, EPI_ISL_838684, EPI_ISL_838685, EPI_ISL_838686, EPI_ISL_838690, EPI_ISL_838694, EPI_ISL_838700, EPI_ISL_838701, EPI_ISL_838702, EPI_ISL_838703, EPI_ISL_838704, EPI_ISL_838716, EPI_ISL_838719, EPI_ISL_838720, EPI_ISL_838722, EPI_ISL_838728, EPI_ISL_838729, EPI_ISL_865718, EPI_ISL_865719, EPI_ISL_949786, EPI_ISL_998008, EPI_ISL_1104662, EPI_ISL_1104663, EPI_ISL_1104664, EPI_ISL_1104668, EPI_ISL_1104669, EPI_ISL_1104670, EPI_ISL_1104671, EPI_ISL_1104672, EPI_ISL_1104673, EPI_ISL_1104674, EPI_ISL_1104675, EPI_ISL_1104676, EPI_ISL_1104677, EPI_ISL_1104678, EPI_ISL_1104679, EPI_ISL_1104680, EPI_ISL_1104681, EPI_ISL_1637129, EPI_ISL_1730359, EPI_ISL_1730360, EPI_ISL_1730391 | | | |
| see above | University College London, Great Ormond Street Hospital for Children NHS Foundation Trust, Imperial College Healthcare NHS Trust | COVID-19 Genomics UK (COG-UK) Consortium | Alison Holmes; Charlotte Williams; Helena Tutill; Jacqueline Findlay; James Price; Judith Breuer; Julianne Brown; Kathryn Harris; Leysa Forrest; Mark Kristiansen; Paola Niola; Paola Resende Silva; Patricia Dyal; Paul Randell; Rachel Williams; Samuel Weeks; Sergi Castellano; Sunando Roy; Tony Brooks; Yasmin Panchbhaya |
| EPI_ISL_1047920, EPI_ISL_1047942, EPI_ISL_1047943, EPI_ISL_1047945, EPI_ISL_1047956, EPI_ISL_1047957, EPI_ISL_1047958, EPI_ISL_2125918, EPI_ISL_2125923 | | | |
| see above | University Hospitals Of Leicester NHS Trust and DeepSeq Nottingham | COVID-19 Genomics UK (COG-UK) Consortium | Christopher Holmes; Christopher Moore; Fei Sang; Joeseph Chappell; Johnny Debebe; Jonathan Ball; Julian Tang; Karlie Fallon; Matthew Carlisle; Matthew Loose; Nadine Holmes; Patrick McClure; Paul Bird; Thomas Helmer; Victoria Wright |
| EPI_ISL_813037, EPI_ISL_813038, EPI_ISL_813039, EPI_ISL_813040, EPI_ISL_813041, EPI_ISL_813042, EPI_ISL_813043, EPI_ISL_813044, EPI_ISL_813063, EPI_ISL_813064, EPI_ISL_813065, EPI_ISL_813066, EPI_ISL_813067, EPI_ISL_813068, EPI_ISL_813069, EPI_ISL_813070, EPI_ISL_813072, EPI_ISL_813073, EPI_ISL_813074, EPI_ISL_813075, EPI_ISL_813076, EPI_ISL_813077, EPI_ISL_813078, EPI_ISL_813079, EPI_ISL_813080, EPI_ISL_813081, EPI_ISL_813082, EPI_ISL_813083, EPI_ISL_813084, EPI_ISL_813085, EPI_ISL_813086, EPI_ISL_813088, EPI_ISL_813089, EPI_ISL_813090, EPI_ISL_918579, EPI_ISL_918580, EPI_ISL_918581, EPI_ISL_918582, EPI_ISL_918599, EPI_ISL_918603, EPI_ISL_949396, EPI_ISL_949397, EPI_ISL_949398, EPI_ISL_949403, EPI_ISL_949404 | | | |
| see above | University of Birmingham | COVID-19 Genomics UK (COG-UK) Consortium | Alex Richter; Andrew Bosworth; Andrew Bosworth. Queen Elizabeth Hospital: Anna Casey; Andrew D Beggs PHE Heartlands Lab: Husam Osman; Charlotte Poxon; Institute of Microbiology; Joanne Stockton; Josh Quick; Kasun Wanigasooriya; Mike Kidd; Nicholas Loman. University of Birmingham Testing Laboratory: Celina M Whalley; Oliver Pickles; Radoslaw Poplawski; Samuel Nicholls; University of Birmingham: Claire McMurray; Will Rowe |
| EPI_ISL_763382, EPI_ISL_763384, EPI_ISL_763385, EPI_ISL_763390, EPI_ISL_763391, EPI_ISL_763432, EPI_ISL_763453, EPI_ISL_763457, EPI_ISL_763459, EPI_ISL_763460, EPI_ISL_763527, EPI_ISL_763528, EPI_ISL_763529, EPI_ISL_763543, EPI_ISL_763553, EPI_ISL_763554, EPI_ISL_763555, EPI_ISL_763556, EPI_ISL_763561, EPI_ISL_763562, EPI_ISL_763563, EPI_ISL_763564, EPI_ISL_763669, EPI_ISL_763696, EPI_ISL_763697, EPI_ISL_763701, EPI_ISL_763703, EPI_ISL_763704, EPI_ISL_763705, EPI_ISL_763706, EPI_ISL_763707, EPI_ISL_763708, EPI_ISL_763709, EPI_ISL_763710, EPI_ISL_763711, EPI_ISL_763763, EPI_ISL_763907, EPI_ISL_764132, EPI_ISL_764134, EPI_ISL_764135, EPI_ISL_764137, EPI_ISL_764165, EPI_ISL_764166, EPI_ISL_764170, EPI_ISL_764178, EPI_ISL_764181, EPI_ISL_764182, EPI_ISL_764183, EPI_ISL_764184, EPI_ISL_764197, EPI_ISL_764211, EPI_ISL_764217, EPI_ISL_764218, EPI_ISL_764219, EPI_ISL_764220, EPI_ISL_764223, EPI_ISL_764224, EPI_ISL_764240, EPI_ISL_764373, EPI_ISL_764374, EPI_ISL_764375, EPI_ISL_764376, EPI_ISL_764377, EPI_ISL_996586 | | | |
| see above | University of Exeter | COVID-19 Genomics UK (COG-UK) Consortium | Aaron Jeffries; Audrey Farbos; Ben Temperton; Jane Masoli; Joanna Warwick-Dugdale; Michelle Michelsen; Robyn Manley; Stephen Michell |
| EPI_ISL_997113, EPI_ISL_1047856, EPI_ISL_1104251, EPI_ISL_1104252, EPI_ISL_1104253, EPI_ISL_1104254, EPI_ISL_2125887 | | | |
| see above | Virology Department, Royal Infirmary of Edinburgh, NHS Lothian / School of Biological Sciences, University of Edinburgh | COVID-19 Genomics UK (COG-UK) Consortium | Colquhoun R; Cotton S; Dewar R; Gallagher A; Hill V; Jackson B; Maloney D; McCrone JT; McHugh M; O'Toole Á; Rambaut A; Rooke S; Scher E; Templeton K; Yu X |
| EPI_ISL_724245, EPI_ISL_724248, EPI_ISL_724249, EPI_ISL_724250, EPI_ISL_724251, EPI_ISL_724252, EPI_ISL_742177, EPI_ISL_742178, EPI_ISL_742179, EPI_ISL_742181, EPI_ISL_742182, EPI_ISL_742183, EPI_ISL_742184, EPI_ISL_742185, EPI_ISL_742186, EPI_ISL_742187, EPI_ISL_742188, EPI_ISL_742189, EPI_ISL_742190, EPI_ISL_742191, EPI_ISL_742192, EPI_ISL_742193, EPI_ISL_742194, EPI_ISL_742195, EPI_ISL_742196, EPI_ISL_742197, EPI_ISL_742198, EPI_ISL_763732, EPI_ISL_764061, EPI_ISL_764069, EPI_ISL_764070, EPI_ISL_764071, EPI_ISL_764072, EPI_ISL_764147, EPI_ISL_764198, EPI_ISL_764359, EPI_ISL_838220, EPI_ISL_838221, EPI_ISL_838222, EPI_ISL_838223, EPI_ISL_838224, EPI_ISL_838226, EPI_ISL_838234, EPI_ISL_838284, EPI_ISL_838285, EPI_ISL_838286, EPI_ISL_838287, EPI_ISL_838288, EPI_ISL_919311, EPI_ISL_919312, EPI_ISL_919313, EPI_ISL_919365, EPI_ISL_919366, EPI_ISL_919367, EPI_ISL_919368, EPI_ISL_919369, EPI_ISL_919370, EPI_ISL_919380, EPI_ISL_919381, EPI_ISL_919388, EPI_ISL_919389, EPI_ISL_919390, EPI_ISL_919391, EPI_ISL_919392, EPI_ISL_919393, EPI_ISL_919394, EPI_ISL_919395, EPI_ISL_919396, EPI_ISL_919397, EPI_ISL_919398 | | | |
| see above | Virology Department, Royal Infirmary of Edinburgh, NHS Lothian / School of Biological Sciences, University of Edinburgh / Institute of Genetics and Molecular Medicine, University of Edinburgh | COVID-19 Genomics UK (COG-UK) Consortium | Balcaza C; Colquhoun R; Dewar R; Gallagher M; Hill V; Jackson B; McCrone JT; McHugh M; O'Toole Á; Rambaut A; Rooke S; Scher E; Templeton K; Williams TC; Yu X |
| EPI_ISL_727862, EPI_ISL_727864, EPI_ISL_727865, EPI_ISL_727866, EPI_ISL_727867, EPI_ISL_727870, EPI_ISL_727872, EPI_ISL_727873, EPI_ISL_727876, EPI_ISL_727878, EPI_ISL_727881, EPI_ISL_727884, EPI_ISL_727886, EPI_ISL_727889, EPI_ISL_727893, EPI_ISL_727894, EPI_ISL_727898, EPI_ISL_727899, EPI_ISL_727900, EPI_ISL_727902, EPI_ISL_727904, EPI_ISL_727907, EPI_ISL_727910, EPI_ISL_727912, EPI_ISL_727913, EPI_ISL_727916, EPI_ISL_727919, EPI_ISL_727921, EPI_ISL_727924, EPI_ISL_727925, EPI_ISL_727926, EPI_ISL_727928, EPI_ISL_727930, EPI_ISL_727933, EPI_ISL_727935, EPI_ISL_727937, EPI_ISL_727938, EPI_ISL_727944, EPI_ISL_727945, EPI_ISL_727946, EPI_ISL_727952, EPI_ISL_727953, EPI_ISL_727958, EPI_ISL_727959, EPI_ISL_727962, EPI_ISL_727965, EPI_ISL_727974, EPI_ISL_727977, EPI_ISL_727984, EPI_ISL_727989, EPI_ISL_741945, EPI_ISL_741966, EPI_ISL_742009, EPI_ISL_742017, EPI_ISL_742022, EPI_ISL_742023, EPI_ISL_742028, EPI_ISL_742044, EPI_ISL_742047, EPI_ISL_742050, EPI_ISL_742052, EPI_ISL_742055, EPI_ISL_742059, EPI_ISL_742064, EPI_ISL_742065, EPI_ISL_742066, EPI_ISL_742068, EPI_ISL_742071, EPI_ISL_742075, EPI_ISL_742099, EPI_ISL_742103, EPI_ISL_742106, EPI_ISL_742114, EPI_ISL_763634, EPI_ISL_763652, EPI_ISL_763740, EPI_ISL_764104, EPI_ISL_764108, EPI_ISL_764109, EPI_ISL_765149, EPI_ISL_765179, EPI_ISL_816234, EPI_ISL_816249, EPI_ISL_816250, EPI_ISL_816254, EPI_ISL_816257, EPI_ISL_816265, EPI_ISL_816266, EPI_ISL_816269, EPI_ISL_816278, EPI_ISL_816298, EPI_ISL_816300, EPI_ISL_816301, EPI_ISL_816306, EPI_ISL_816325, EPI_ISL_816329, EPI_ISL_816346, EPI_ISL_816349, EPI_ISL_816354, EPI_ISL_816360, EPI_ISL_816375, EPI_ISL_816388, EPI_ISL_816410, EPI_ISL_816419, EPI_ISL_816420, EPI_ISL_816422, EPI_ISL_816440, EPI_ISL_816454, EPI_ISL_816459, EPI_ISL_816464, EPI_ISL_816465, EPI_ISL_816466, EPI_ISL_816482, EPI_ISL_816483, EPI_ISL_816490, EPI_ISL_816515, EPI_ISL_816538, EPI_ISL_816541, EPI_ISL_816544, EPI_ISL_816553, EPI_ISL_816556, EPI_ISL_816568, EPI_ISL_816569, EPI_ISL_816579, EPI_ISL_816587, EPI_ISL_816593, EPI_ISL_816594, EPI_ISL_816595, EPI_ISL_816598, EPI_ISL_816604, EPI_ISL_816610, EPI_ISL_816612, EPI_ISL_816615, EPI_ISL_816628, EPI_ISL_816640, EPI_ISL_816654, EPI_ISL_816656 | | | |
| see above | Virology Department, Sheffield Teaching Hospitals NHS Foundation Trust/Department of Infection, Immunity and Cardiovascular Disease, The Medical School, University of Sheffield | COVID-19 Genomics UK (COG-UK) Consortium | Adri Angyal; Alex Keeley; Benjamin Lindsey; Cariad Evans; Danielle Groves; Dave Partridge; Katie Johnson; Laura Carrilero; Luke Green; Matthew Parker; Matthew Wyles; Mehmet Yavuz; Mohammad Raza; Nikki Smith; Paul Parsons; Rachel Tucker; Rebecca Brown; Thushan de Silva |
| EPI_ISL_742546, EPI_ISL_742555, EPI_ISL_742556, EPI_ISL_742557, EPI_ISL_742558, EPI_ISL_742560, EPI_ISL_742562, EPI_ISL_742566, EPI_ISL_742567, EPI_ISL_742568, EPI_ISL_742569, EPI_ISL_742570, EPI_ISL_742571, EPI_ISL_742572, EPI_ISL_742573, EPI_ISL_742574, EPI_ISL_742575, EPI_ISL_742576, EPI_ISL_742577, EPI_ISL_742578, EPI_ISL_742579, EPI_ISL_742580, EPI_ISL_742581, EPI_ISL_742582, EPI_ISL_742583, EPI_ISL_742584, EPI_ISL_742647, EPI_ISL_742648, EPI_ISL_742650, EPI_ISL_742652, EPI_ISL_742680, EPI_ISL_742681, EPI_ISL_742682, EPI_ISL_742692, EPI_ISL_742693, EPI_ISL_742698, EPI_ISL_742699, EPI_ISL_742703, EPI_ISL_742706, EPI_ISL_742708, EPI_ISL_742711, EPI_ISL_742712, EPI_ISL_742715, EPI_ISL_742721, EPI_ISL_742722, EPI_ISL_742723, EPI_ISL_742724, EPI_ISL_742725, EPI_ISL_742726, EPI_ISL_742727, EPI_ISL_742728, EPI_ISL_742729, EPI_ISL_742730, EPI_ISL_742731, EPI_ISL_742732, EPI_ISL_742733, EPI_ISL_742734, EPI_ISL_742735, EPI_ISL_742736, EPI_ISL_742737, EPI_ISL_742738, EPI_ISL_742739, EPI_ISL_742740, EPI_ISL_742741, EPI_ISL_742742, EPI_ISL_742743, EPI_ISL_742744, EPI_ISL_742745, EPI_ISL_742746, EPI_ISL_742747, EPI_ISL_742748, EPI_ISL_742749, EPI_ISL_742750, EPI_ISL_742751, EPI_ISL_742752, EPI_ISL_742753, EPI_ISL_742754, EPI_ISL_742755, EPI_ISL_742756, EPI_ISL_742757, EPI_ISL_742758, EPI_ISL_742759, EPI_ISL_742760, EPI_ISL_742761, EPI_ISL_742762, EPI_ISL_742763, EPI_ISL_742764, EPI_ISL_742765, EPI_ISL_742766, EPI_ISL_742767, EPI_ISL_742768, EPI_ISL_742769, EPI_ISL_742770, EPI_ISL_742771, EPI_ISL_742772, EPI_ISL_742773, EPI_ISL_742774, EPI_ISL_742775, EPI_ISL_742776, EPI_ISL_742777, EPI_ISL_742778, EPI_ISL_742779, EPI_ISL_742780, EPI_ISL_742781, EPI_ISL_742782, EPI_ISL_742783, EPI_ISL_742784, EPI_ISL_742785, EPI_ISL_742786, EPI_ISL_742787, EPI_ISL_742788, EPI_ISL_742789, EPI_ISL_742790, EPI_ISL_742791, EPI_ISL_742792, EPI_ISL_742793, EPI_ISL_742794, EPI_ISL_742795, EPI_ISL_742796, EPI_ISL_742797, EPI_ISL_742798, EPI_ISL_742799, EPI_ISL_742800, EPI_ISL_742801, EPI_ISL_742802, EPI_ISL_742803, EPI_ISL_742804, EPI_ISL_742805, EPI_ISL_742806, EPI_ISL_742807, EPI_ISL_742808, EPI_ISL_742809, EPI_ISL_742810, EPI_ISL_742811, EPI_ISL_742812, EPI_ISL_742813, EPI_ISL_742814, EPI_ISL_742815, EPI_ISL_742816, EPI_ISL_742817, EPI_ISL_742818, EPI_ISL_742819, EPI_ISL_742820, EPI_ISL_742821, EPI_ISL_742822, EPI_ISL_742823, EPI_ISL_742824, EPI_ISL_742825, EPI_ISL_742826, EPI_ISL_742827, EPI_ISL_742828, EPI_ISL_742829, EPI_ISL_742830, EPI_ISL_742831, EPI_ISL_742832, EPI_ISL_742833, EPI_ISL_742834, EPI_ISL_742835, EPI_ISL_742836, EPI_ISL_742837, EPI_ISL_743127, EPI_ISL_743146, EPI_ISL_743147, EPI_ISL_743148, EPI_ISL_743149, EPI_ISL_743150, EPI_ISL_743151, EPI_ISL_743152, EPI_ISL_743153, EPI_ISL_743154, EPI_ISL_743155, EPI_ISL_743156, EPI_ISL_743157, EPI_ISL_743158, EPI_ISL_743321, EPI_ISL_743322, EPI_ISL_743326, EPI_ISL_743341, EPI_ISL_743352, EPI_ISL_743365, EPI_ISL_743366, EPI_ISL_743367, EPI_ISL_743368, EPI_ISL_743369, EPI_ISL_743370, EPI_ISL_743371, EPI_ISL_743372, EPI_ISL_743373, EPI_ISL_743374, EPI_ISL_743375, EPI_ISL_743376, EPI_ISL_743377, EPI_ISL_743378, EPI_ISL_743379, EPI_ISL_743425, EPI_ISL_743426, EPI_ISL_743427, EPI_ISL_743428, EPI_ISL_743429, EPI_ISL_743430, EPI_ISL_743431, EPI_ISL_743433, EPI_ISL_743434, EPI_ISL_743445, EPI_ISL_743446, EPI_ISL_743447, EPI_ISL_743448, EPI_ISL_743449, EPI_ISL_743509, EPI_ISL_743511, EPI_ISL_743512, EPI_ISL_743513, EPI_ISL_743514, EPI_ISL_743515, EPI_ISL_743516, EPI_ISL_743517, EPI_ISL_743518, EPI_ISL_743519, EPI_ISL_743520, EPI_ISL_743521, EPI_ISL_743522, EPI_ISL_743523, EPI_ISL_743524, EPI_ISL_743525, EPI_ISL_743526, EPI_ISL_743527, EPI_ISL_743528, EPI_ISL_743529, EPI_ISL_743530, EPI_ISL_743531, EPI_ISL_743532, EPI_ISL_743533, EPI_ISL_743534, EPI_ISL_743535, EPI_ISL_743536, EPI_ISL_743537, EPI_ISL_743538, EPI_ISL_743539, EPI_ISL_743540, EPI_ISL_743541, EPI_ISL_743542, EPI_ISL_743543, EPI_ISL_743544, EPI_ISL_743545, EPI_ISL_743546, EPI_ISL_743547, EPI_ISL_743548, EPI_ISL_743549, EPI_ISL_743550, EPI_ISL_743551, EPI_ISL_743552, EPI_ISL_743553, EPI_ISL_743554, EPI_ISL_743555, EPI_ISL_743556, EPI_ISL_743557, EPI_ISL_743558, EPI_ISL_743559, EPI_ISL_743560, EPI_ISL_743561, EPI_ISL_743562, EPI_ISL_743563, EPI_ISL_743564, EPI_ISL_743565, EPI_ISL_743566, EPI_ISL_743567, EPI_ISL_743569, EPI_ISL_743570, EPI_ISL_743571, EPI_ISL_743572, EPI_ISL_743573, EPI_ISL_743574, EPI_ISL_743575, EPI_ISL_743576, EPI_ISL_743577, EPI_ISL_743578, EPI_ISL_743579, EPI_ISL_743581, EPI_ISL_743582, EPI_ISL_743583, EPI_ISL_743584, EPI_ISL_743585, EPI_ISL_743586, EPI_ISL_743587, EPI_ISL_743588, EPI_ISL_743589, EPI_ISL_743590, EPI_ISL_743591, EPI_ISL_743596, EPI_ISL_743597, EPI_ISL_743598, EPI_ISL_743599, EPI_ISL_743600, EPI_ISL_743601, EPI_ISL_743602, EPI_ISL_743603, EPI_ISL_743604, EPI_ISL_743605, EPI_ISL_743641, EPI_ISL_743662, EPI_ISL_743663, EPI_ISL_743664, EPI_ISL_743675, EPI_ISL_743678, EPI_ISL_743680, EPI_ISL_743681, EPI_ISL_743682, EPI_ISL_743683, EPI_ISL_743684, EPI_ISL_743685, EPI_ISL_743686, EPI_ISL_743687, EPI_ISL_743688, EPI_ISL_743689, EPI_ISL_743690, EPI_ISL_743691, EPI_ISL_743692, EPI_ISL_743693, EPI_ISL_743694, EPI_ISL_743695, EPI_ISL_743696, EPI_ISL_743697, EPI_ISL_743698, EPI_ISL_743699, EPI_ISL_743700, EPI_ISL_743701, EPI_ISL_743702, EPI_ISL_743703, EPI_ISL_743704, EPI_ISL_743705, EPI_ISL_743706, EPI_ISL_743707, EPI_ISL_743708, EPI_ISL_743709, EPI_ISL_743710, EPI_ISL_743711, EPI_ISL_743712, EPI_ISL_743713, EPI_ISL_743714, EPI_ISL_743715, EPI_ISL_743716, EPI_ISL_743717, EPI_ISL_743718, EPI_ISL_743719, EPI_ISL_743720, EPI_ISL_743721, EPI_ISL_743722, EPI_ISL_743723, EPI_ISL_743724, EPI_ISL_743725, EPI_ISL_743726, EPI_ISL_743727, EPI_ISL_743728, EPI_ISL_743729, EPI_ISL_743730, EPI_ISL_743731, EPI_ISL_743732, EPI_ISL_743733, EPI_ISL_743734, EPI_ISL_743735, EPI_ISL_743736, EPI_ISL_743737, EPI_ISL_743738, EPI_ISL_743739, EPI_ISL_743740, EPI_ISL_743741, EPI_ISL_743742, EPI_ISL_743743, EPI_ISL_743744, EPI_ISL_743745, EPI_ISL_743746, EPI_ISL_743747, EPI_ISL_743748, EPI_ISL_743749, EPI_ISL_743750, EPI_ISL_743751, EPI_ISL_743752, EPI_ISL_743753, EPI_ISL_743754, EPI_ISL_743755, EPI_ISL_743756, EPI_ISL_743757, EPI_ISL_743758, EPI_ISL_743759, EPI_ISL_743760, EPI_ISL_743761, EPI_ISL_743762, EPI_ISL_743763, EPI_ISL_743764, EPI_ISL_743765, EPI_ISL_743766, EPI_ISL_743767, EPI_ISL_743768, EPI_ISL_743769, EPI_ISL_743770, EPI_ISL_743771, EPI_ISL_743772, EPI_ISL_743773, EPI_ISL_743774, EPI_ISL_743775, EPI_ISL_743776, EPI_ISL_743777, EPI_ISL_743778, EPI_ISL_743779, EPI_ISL_743780, EPI_ISL_743781, EPI_ISL_743782, EPI_ISL_743783, EPI_ISL_743784, EPI_ISL_743785, EPI_ISL_743786, EPI_ISL_743787, EPI_ISL_743788, EPI_ISL_743789, EPI_ISL_743790, EPI_ISL_743791, EPI_ISL_743792, EPI_ISL_743793, EPI_ISL_743794, EPI_ISL_743795, EPI_ISL_743796, EPI_ISL_743797, EPI_ISL_743798, EPI_ISL_743799, EPI_ISL_743800, EPI_ISL_743801, EPI_ISL_743802, EPI_ISL_743803, EPI_ISL_743804, EPI_ISL_743805, EPI_ISL_743806, EPI_ISL_743807, EPI_ISL_743808, EPI_ISL_743809, EPI_ISL_743810, EPI_ISL_743811, EPI_ISL_743812, EPI_ISL_743813, EPI_ISL_743814, EPI_ISL_743815, EPI_ISL_743816, EPI_ISL_743817, EPI_ISL_743818, EPI_ISL_743819, EPI_ISL_743820, EPI_ISL_743821, EPI_ISL_743822, EPI_ISL_743823, EPI_ISL_743824, EPI_ISL_743825, EPI_ISL_743826, EPI_ISL_743827, EPI_ISL_743828, EPI_ISL_743829, EPI_ISL_743830, EPI_ISL_743831, EPI_ISL_743832, EPI_ISL_743833, EPI_ISL_743834, EPI_ISL_743835, EPI_ISL_743836, EPI_ISL_743837, EPI_ISL_743838, EPI_ISL_743839, EPI_ISL_743840, EPI_ISL_743841, EPI_ISL_743842, EPI_ISL_743843, EPI_ISL_743844, EPI_ISL_743845, EPI_ISL_743846, EPI_ISL_743847, EPI_ISL_743848, EPI_ISL_743849, EPI_ISL_743850, EPI_ISL_743851, EPI_ISL_743852, EPI_ISL_743853, EPI_ISL_743854, EPI_ISL_743855, EPI_ISL_743856, EPI_ISL_743857, EPI_ISL_743858, EPI_ISL_743859, EPI_ISL_743860, EPI_ISL_743861, EPI_ISL_743862, EPI_ISL_743863, EPI_ISL_743864, EPI_ISL_743865, EPI_ISL_743866, EPI_ISL_743867, EPI_ISL_743868, EPI_ISL_743869, EPI_ISL_743870, EPI_ISL_743871, EPI_ISL_743872, EPI_ISL_743873, EPI_ISL_743874, EPI_ISL_743875, EPI_ISL_743876, EPI_ISL_743877, EPI_ISL_743879, EPI_ISL_743880, EPI_ISL_743881, EPI_ISL_743882, EPI_ISL_743883, EPI_ISL_743884, EPI_ISL_743885, EPI_ISL_743886, EPI_ISL_743887, EPI_ISL_743888, EPI_ISL_743889, EPI_ISL_743890, EPI_ISL_743891, EPI_ISL_743892, EPI_ISL_743893, EPI_ISL_743894, EPI_ISL_743895, EPI_ISL_743896, EPI_ISL_743897, EPI_ISL_743898, EPI_ISL_743899, EPI_ISL_743900, EPI_ISL_743902, EPI_ISL_743903, EPI_ISL_743904, EPI_ISL_743937, EPI_ISL_743938, EPI_ISL_743939, EPI_ISL_743942, EPI_ISL_743944, EPI_ISL_743952, EPI_ISL_743958, EPI_ISL_743959, EPI_ISL_743960, EPI_ISL_743961, EPI_ISL_743962, EPI_ISL_743963, EPI_ISL_743964, EPI_ISL_743965, EPI_ISL_743966, EPI_ISL_743967, EPI_ISL_743968, EPI_ISL_743969, EPI_ISL_743970, EPI_ISL_743971, EPI_ISL_743972, EPI_ISL_743973, EPI_ISL_743974, EPI_ISL_743975, EPI_ISL_743976, EPI_ISL_743977, EPI_ISL_743978, EPI_ISL_743979, EPI_ISL_743980, EPI_ISL_743981, EPI_ISL_743982, EPI_ISL_743983, EPI_ISL_743984, EPI_ISL_743985, EPI_ISL_743986, EPI_ISL_743987, EPI_ISL_743988, EPI_ISL_743989, EPI_ISL_743990, EPI_ISL_743991, EPI_ISL_743992, EPI_ISL_743993, EPI_ISL_743994, EPI_ISL_743995, EPI_ISL_743996, EPI_ISL_743997, EPI_ISL_743998, EPI_ISL_743999, EPI_ISL_744000, EPI_ISL_744001, EPI_ISL_744002, EPI_ISL_744003, EPI_ISL_744004, EPI_ISL_744005, EPI_ISL_744006, EPI_ISL_744007, EPI_ISL_744008, EPI_ISL_744009, EPI_ISL_744010, EPI_ISL_744011, EPI_ISL_744012, EPI_ISL_744013, EPI_ISL_744014, EPI_ISL_744015, EPI_ISL_744016, EPI_ISL_744017, EPI_ISL_744018, EPI_ISL_744019, EPI_ISL_744020, EPI_ISL_744021, EPI_ISL_744022, EPI_ISL_744023, EPI_ISL_744024, EPI_ISL_744025, EPI_ISL_744026, EPI_ISL_744027, EPI_ISL_744028, EPI_ISL_744029, EPI_ISL_744030, EPI_ISL_744031, EPI_ISL_744032, EPI_ISL_744033, EPI_ISL_744034, EPI_ISL_744035, EPI_ISL_744036, EPI_ISL_744037, EPI_ISL_744038, EPI_ISL_744039, EPI_ISL_744040, EPI_ISL_744041, EPI_ISL_744042, EPI_ISL_744043, EPI_ISL_744044, EPI_ISL_744045, EPI_ISL_744046, EPI_ISL_744047, EPI_ISL_744048, EPI_ISL_744049, EPI_ISL_744050, EPI_ISL_744051, EPI_ISL_744052, EPI_ISL_744053, EPI_ISL_744054, EPI_ISL_744055, EPI_ISL_744056, EPI_ISL_744057, EPI_ISL_744058, EPI_ISL_744059, EPI_ISL_744060, EPI_ISL_744061, EPI_ISL_744062, EPI_ISL_744063, EPI_ISL_744064, EPI_ISL_744065, EPI_ISL_744066, EPI_ISL_744068, EPI_ISL_744069, EPI_ISL_744070, EPI_ISL_744071, EPI_ISL_744072, EPI_ISL_744073, EPI_ISL_744074, EPI_ISL_744075, EPI_ISL_744076, EPI_ISL_744077, EPI_ISL_744078, EPI_ISL_744079, EPI_ISL_744080, EPI_ISL_744081, EPI_ISL_744082, EPI_ISL_744083, EPI_ISL_744084, EPI_ISL_744086, EPI_ISL_744087, EPI_ISL_744088, EPI_ISL_744089, EPI_ISL_744090, EPI_ISL_744091, EPI_ISL_744092, EPI_ISL_744093, EPI_ISL_744094, EPI_ISL_744095, EPI_ISL_744096, EPI_ISL_744097, EPI_ISL_744099, EPI_ISL_744100, EPI_ISL_744102, EPI_ISL_744106, EPI_ISL_744108, EPI_ISL_744109, EPI_ISL_744110, EPI_ISL_744113, EPI_ISL_744114, EPI_ISL_763413, EPI_ISL_763472, EPI_ISL_763484, EPI_ISL_763487, EPI_ISL_763493, EPI_ISL_763497, EPI_ISL_763519, EPI_ISL_763522, EPI_ISL_763539, EPI_ISL_763566, EPI_ISL_763568, EPI_ISL_763570, EPI_ISL_763571, EPI_ISL_763574, EPI_ISL_763575, EPI_ISL_763576, EPI_ISL_763584, EPI_ISL_763656, EPI_ISL_763664, EPI_ISL_763668, EPI_ISL_763674, EPI_ISL_763686, EPI_ISL_763724, EPI_ISL_763738, EPI_ISL_763747, EPI_ISL_763764, EPI_ISL_763768, EPI_ISL_763771, EPI_ISL_763776, EPI_ISL_763778, EPI_ISL_763786, EPI_ISL_763787, EPI_ISL_763820, EPI_ISL_763976, EPI_ISL_763977, EPI_ISL_763978, EPI_ISL_763979, EPI_ISL_763980, EPI_ISL_763981, EPI_ISL_763982, EPI_ISL_763989, EPI_ISL_764035, EPI_ISL_764150, EPI_ISL_764156, EPI_ISL_764161, EPI_ISL_764186, EPI_ISL_764203, EPI_ISL_764229, EPI_ISL_764232, EPI_ISL_764234, EPI_ISL_764266, EPI_ISL_764267, EPI_ISL_764573, EPI_ISL_764574, EPI_ISL_764575, EPI_ISL_764576, EPI_ISL_764581, EPI_ISL_764585, EPI_ISL_764599, EPI_ISL_764600, EPI_ISL_764601, EPI_ISL_764602, EPI_ISL_764603, EPI_ISL_764604, EPI_ISL_764605, EPI_ISL_764606, EPI_ISL_764607, EPI_ISL_764608, EPI_ISL_764609, EPI_ISL_764610, EPI_ISL_764611, EPI_ISL_764612, EPI_ISL_764613, EPI_ISL_764614, EPI_ISL_764615, EPI_ISL_764616, EPI_ISL_764617, EPI_ISL_764618, EPI_ISL_764619, EPI_ISL_764620, EPI_ISL_764621, EPI_ISL_764622, EPI_ISL_764623, EPI_ISL_764625, EPI_ISL_764626, EPI_ISL_764627, EPI_ISL_764628, EPI_ISL_764629, EPI_ISL_764630, EPI_ISL_764631, EPI_ISL_764632, EPI_ISL_764633, EPI_ISL_764634, EPI_ISL_764635, EPI_ISL_764636, EPI_ISL_764637, EPI_ISL_764638, EPI_ISL_764639, EPI_ISL_764640, EPI_ISL_764641, EPI_ISL_764642, EPI_ISL_764643, EPI_ISL_764644, EPI_ISL_764645, EPI_ISL_764646, EPI_ISL_764647, EPI_ISL_764648, EPI_ISL_764649, EPI_ISL_764650, EPI_ISL_764651, EPI_ISL_764652, EPI_ISL_764653, EPI_ISL_764654, EPI_ISL_764655, EPI_ISL_764656, EPI_ISL_764739, EPI_ISL_764740, EPI_ISL_764741, EPI_ISL_764748, EPI_ISL_764749, EPI_ISL_764750, EPI_ISL_764751, EPI_ISL_764752, EPI_ISL_764756, EPI_ISL_764768, EPI_ISL_764805, EPI_ISL_764806, EPI_ISL_764807, EPI_ISL_764808, EPI_ISL_764809, EPI_ISL_764810, EPI_ISL_764811, EPI_ISL_764812, EPI_ISL_764813, EPI_ISL_764814, EPI_ISL_764815, EPI_ISL_764819, EPI_ISL_764820, EPI_ISL_764821, EPI_ISL_764822, EPI_ISL_764823, EPI_ISL_764825, EPI_ISL_764828, EPI_ISL_764829, EPI_ISL_764952, EPI_ISL_764953, EPI_ISL_764954, EPI_ISL_764955, EPI_ISL_764956, EPI_ISL_764957, EPI_ISL_764958, EPI_ISL_764959, EPI_ISL_764960, EPI_ISL_764961, EPI_ISL_764962, EPI_ISL_764963, EPI_ISL_764964, EPI_ISL_764965, EPI_ISL_764966, EPI_ISL_764967, EPI_ISL_764968, EPI_ISL_764969, EPI_ISL_764970, EPI_ISL_764972, EPI_ISL_764973, EPI_ISL_764976, EPI_ISL_764977, EPI_ISL_764978, EPI_ISL_764979, EPI_ISL_814277, EPI_ISL_814278, EPI_ISL_814282, EPI_ISL_814295, EPI_ISL_814320, EPI_ISL_814326, EPI_ISL_814328, EPI_ISL_814356, EPI_ISL_814367, EPI_ISL_814369, EPI_ISL_814372, EPI_ISL_814377, EPI_ISL_814386, EPI_ISL_814387, EPI_ISL_814400, EPI_ISL_814404, EPI_ISL_814405, EPI_ISL_814408, EPI_ISL_814411, EPI_ISL_814413, EPI_ISL_814415, EPI_ISL_814468, EPI_ISL_814473, EPI_ISL_814474, EPI_ISL_814475, EPI_ISL_814476, EPI_ISL_814477, EPI_ISL_814478, EPI_ISL_814479, EPI_ISL_814492, EPI_ISL_814529, EPI_ISL_814531, EPI_ISL_814536, EPI_ISL_814551, EPI_ISL_814561, EPI_ISL_814573, EPI_ISL_814578, EPI_ISL_814584, EPI_ISL_814595, EPI_ISL_814606, EPI_ISL_814702, EPI_ISL_814703, EPI_ISL_814704, EPI_ISL_814717, EPI_ISL_814718, EPI_ISL_814719, EPI_ISL_814720, EPI_ISL_814721, EPI_ISL_814722, EPI_ISL_814723, EPI_ISL_814724, EPI_ISL_814725, EPI_ISL_814726, EPI_ISL_814727, EPI_ISL_814728, EPI_ISL_814729, EPI_ISL_814730, EPI_ISL_814731, EPI_ISL_814732, EPI_ISL_814733, EPI_ISL_814734, EPI_ISL_814735, EPI_ISL_814736, | | | |

| | | | |
|---|---|---|---|
| EPI_ISL_814737, EPI_ISL_814738, EPI_ISL_814739, EPI_ISL_814740, EPI_ISL_814753, EPI_ISL_814798, EPI_ISL_814799, EPI_ISL_814860, EPI_ISL_814861, EPI_ISL_814862, EPI_ISL_814863, EPI_ISL_814864, EPI_ISL_814865, EPI_ISL_814866, EPI_ISL_814867, EPI_ISL_814868, EPI_ISL_814869, EPI_ISL_814870, EPI_ISL_814871, EPI_ISL_814873, EPI_ISL_814874, EPI_ISL_814875, EPI_ISL_814876, EPI_ISL_814877, EPI_ISL_814878, EPI_ISL_814879, EPI_ISL_814880, EPI_ISL_814881, EPI_ISL_814882, EPI_ISL_814883, EPI_ISL_814884, EPI_ISL_814885, EPI_ISL_814886, EPI_ISL_814887, EPI_ISL_814888, EPI_ISL_814889, EPI_ISL_814890, EPI_ISL_814891, EPI_ISL_814892, EPI_ISL_814893, EPI_ISL_814894, EPI_ISL_814895, EPI_ISL_814896, EPI_ISL_814897, EPI_ISL_814898, EPI_ISL_814899, EPI_ISL_814902, EPI_ISL_814903, EPI_ISL_814904, EPI_ISL_814905, EPI_ISL_814906, EPI_ISL_814907, EPI_ISL_814908, EPI_ISL_814909, EPI_ISL_814910, EPI_ISL_814911, EPI_ISL_814912, EPI_ISL_814913, EPI_ISL_814914, EPI_ISL_814915, EPI_ISL_814916, EPI_ISL_814917, EPI_ISL_814918, EPI_ISL_814919, EPI_ISL_814920, EPI_ISL_814921, EPI_ISL_814922, EPI_ISL_814923, EPI_ISL_814924, EPI_ISL_815035, EPI_ISL_815036, EPI_ISL_815037, EPI_ISL_815052, EPI_ISL_815053, EPI_ISL_815054, EPI_ISL_815055, EPI_ISL_815056, EPI_ISL_815057, EPI_ISL_815058, EPI_ISL_815060, EPI_ISL_815093, EPI_ISL_815094, EPI_ISL_815095, EPI_ISL_815096, EPI_ISL_815097, EPI_ISL_815098, EPI_ISL_815099, EPI_ISL_815100, EPI_ISL_815101, EPI_ISL_815102, EPI_ISL_815103, EPI_ISL_815104, EPI_ISL_815105, EPI_ISL_815106, EPI_ISL_815107, EPI_ISL_815109, EPI_ISL_815110, EPI_ISL_815249, EPI_ISL_815250, EPI_ISL_815251, EPI_ISL_815252, EPI_ISL_815253, EPI_ISL_822842, EPI_ISL_822848, EPI_ISL_822849, EPI_ISL_822850, EPI_ISL_822853, EPI_ISL_822863, EPI_ISL_822868, EPI_ISL_822869, EPI_ISL_822872, EPI_ISL_822916, EPI_ISL_822927, EPI_ISL_822936, EPI_ISL_822937, EPI_ISL_822977, EPI_ISL_823004, EPI_ISL_823008, EPI_ISL_823012, EPI_ISL_823014, EPI_ISL_823015, EPI_ISL_823021, EPI_ISL_823022, EPI_ISL_823023, EPI_ISL_823024, EPI_ISL_823056, EPI_ISL_823057, EPI_ISL_823058, EPI_ISL_823059, EPI_ISL_823060, EPI_ISL_823061, EPI_ISL_823062, EPI_ISL_823063, EPI_ISL_823129, EPI_ISL_823131, EPI_ISL_823137, EPI_ISL_823143, EPI_ISL_823156, EPI_ISL_823162, EPI_ISL_823170, EPI_ISL_823171, EPI_ISL_823175, EPI_ISL_823205, EPI_ISL_823208, EPI_ISL_823209, EPI_ISL_823210, EPI_ISL_823211, EPI_ISL_823212, EPI_ISL_823213, EPI_ISL_823214, EPI_ISL_823215, EPI_ISL_823216, EPI_ISL_823217, EPI_ISL_823218, EPI_ISL_823219, EPI_ISL_823220, EPI_ISL_823221, EPI_ISL_823222, EPI_ISL_823223, EPI_ISL_823224, EPI_ISL_823225, EPI_ISL_823226, EPI_ISL_823227, EPI_ISL_823228, EPI_ISL_823229, EPI_ISL_823230, EPI_ISL_823397, EPI_ISL_823398, EPI_ISL_823399, EPI_ISL_823400, EPI_ISL_823401, EPI_ISL_823569, EPI_ISL_823570, EPI_ISL_823571, EPI_ISL_823572, EPI_ISL_823573, EPI_ISL_823574, EPI_ISL_823575, EPI_ISL_823576, EPI_ISL_823577, EPI_ISL_823578, EPI_ISL_823579, EPI_ISL_823580, EPI_ISL_823581, EPI_ISL_823582, EPI_ISL_823583, EPI_ISL_823584, EPI_ISL_823585, EPI_ISL_823586, EPI_ISL_823587, EPI_ISL_823588, EPI_ISL_823589, EPI_ISL_823590, EPI_ISL_823591, EPI_ISL_823592, EPI_ISL_823593, EPI_ISL_823594, EPI_ISL_823595, EPI_ISL_823596, EPI_ISL_823597, EPI_ISL_823598, EPI_ISL_823599, EPI_ISL_823600, EPI_ISL_823601, EPI_ISL_823602, EPI_ISL_823603, EPI_ISL_823604, EPI_ISL_823605, EPI_ISL_823606, EPI_ISL_823607, EPI_ISL_823608, EPI_ISL_823609, EPI_ISL_823610, EPI_ISL_823611, EPI_ISL_823612, EPI_ISL_823613, EPI_ISL_823614, EPI_ISL_823615, EPI_ISL_823616, EPI_ISL_823617, EPI_ISL_823618, EPI_ISL_823619, EPI_ISL_823620, EPI_ISL_823621, EPI_ISL_823622, EPI_ISL_823623, EPI_ISL_823624, EPI_ISL_823625, EPI_ISL_823626, EPI_ISL_823627, EPI_ISL_823628, EPI_ISL_823629, EPI_ISL_823630, EPI_ISL_823631, EPI_ISL_823632, EPI_ISL_823633 | | | |
| see above | Wales Specialist Virology Centre Sequencing lab: Pathogen Genomics Unit | COVID-19 Genomics UK (COG-UK) Consortium | Alec Birchley; Alexander Adams; Amy Gaskin; Angela Marchbank; Bree Gatica-Wilcox; Catherine Moore; Jason Coombes; Joanne Watkins; Joel Southgate; Johnathan Evans; Laura Gifford; Lauren Gilbert; Lee Graham; Malorie Perry; Matthew Bull; Nicole Pacchiarini; Sally Corden; Sara Kumziene-Summerhayes; Sara Rey; Sarah Taylor; Simon Cottrell; Sophie Jones; Tom Connor |
| EPI_ISL_840835, EPI_ISL_922894 | Wales Specialist Virology Centre Sequencing lab: Pathogen Genomics Unit | Public Health Wales Microbiology Cardiff Wales Specialist Virology Centre | Alec Birchley; Alexander Adams; Amy Gaskin; Angela Marchbank; Bree Gatica-Wilcox; Catherine Moore; Jason Coombes; Joanne Watkins; Joel Southgate; Johnathan Evans; Laura Gifford; Lauren Gilbert; Lee Graham; Malorie Perry; Matthew Bull; Nicole Pacchiarini; Sally Corden; Sara Kumziene-Summerhayes; Sara Rey; Sarah Taylor; Simon Cottrell; Sophie Jones; Tom Connor |
| EPI_ISL_814659, EPI_ISL_838129, EPI_ISL_919232, EPI_ISL_919233, EPI_ISL_919235, EPI_ISL_919236, EPI_ISL_919237, EPI_ISL_919238, EPI_ISL_919240, EPI_ISL_919244, EPI_ISL_919252, EPI_ISL_919259, EPI_ISL_919261, EPI_ISL_919266, EPI_ISL_919267 | | | |
| see above | West of Scotland Specialist Virology Centre, NHSGGC / MRC-University of Glasgow Centre for Virus Research | COVID-19 Genomics UK (COG-UK) Consortium | Alasdair MacLean; Alice Broos; Ana da Silva Filipe; Antonia Ho; Daniel Mair; David L Robertson; Emma Thomson; Guy Mollett; James Shepherd; Jenna Nichols; Joseph Hughes; Kathy Li; Kathy Smollett; Kyriaki Nomikou; Lily Tong; Matthew Holden; Natasha Johnson; Rachel Blacow; Richard Orton; Rory Gunson; Sarah McDonald; Sharif Shaaban; Sreenu Vattipally; Stephen Carmichael |

We gratefully acknowledge the following Authors from the Originating laboratories responsible for obtaining the specimens, as well as the Submitting laboratories where the genome data were generated and shared via GISAID, on which this research is based.

All Submitters of data may be contacted directly via www.gisaid.org

Authors are sorted alphabetically.

| Accession ID | Originating Laboratory | Submitting Laboratory | Authors |
|---|---|---|---|
| EPI_ISL_843090, EPI_ISL_843091, EPI_ISL_843092, EPI_ISL_843105, EPI_ISL_843106, EPI_ISL_843107, EPI_ISL_843108, EPI_ISL_843109, EPI_ISL_843110, EPI_ISL_843116, EPI_ISL_843117, EPI_ISL_843118, EPI_ISL_843119, EPI_ISL_843120, EPI_ISL_865492, EPI_ISL_1636892, EPI_ISL_1636937, EPI_ISL_1636947, EPI_ISL_1636948, EPI_ISL_1636949, EPI_ISL_1636950, EPI_ISL_1636951, EPI_ISL_1636952, EPI_ISL_1636953, EPI_ISL_1636954, EPI_ISL_1636955, EPI_ISL_1636994, EPI_ISL_1636995, EPI_ISL_1636996, EPI_ISL_1636997 | | | |
| see above | Barts Health NHS Trust | COVID-19 Genomics UK (COG-UK) Consortium | BROAD; Beatrix; CUTINO-MOGUEL; Claire; David; Dola; HARRINGTON; KELE; KULASEGARAN-SHYLINI; Maria-Teresa; OWOYEMI; Raghavendran; SHYLINI |
| EPI_ISL_814287, EPI_ISL_814288, EPI_ISL_814289, EPI_ISL_814290, EPI_ISL_814292, EPI_ISL_814351, EPI_ISL_814357, EPI_ISL_814359, EPI_ISL_814360, EPI_ISL_814361, EPI_ISL_814417, EPI_ISL_814418, EPI_ISL_814419, EPI_ISL_814421, EPI_ISL_814424, EPI_ISL_814425, EPI_ISL_814426, EPI_ISL_814427, EPI_ISL_814428, EPI_ISL_814430, EPI_ISL_814431, EPI_ISL_814432, EPI_ISL_814435, EPI_ISL_814436, EPI_ISL_814437, EPI_ISL_814438, EPI_ISL_814517, EPI_ISL_814518, EPI_ISL_814519, EPI_ISL_814520, EPI_ISL_814521, EPI_ISL_814522, EPI_ISL_814524, EPI_ISL_814525, EPI_ISL_814530, EPI_ISL_814532, EPI_ISL_814538, EPI_ISL_814539, EPI_ISL_814543, EPI_ISL_814544, EPI_ISL_814548, EPI_ISL_814610, EPI_ISL_814611, EPI_ISL_814612, EPI_ISL_814613, EPI_ISL_814614, EPI_ISL_814615, EPI_ISL_814616, EPI_ISL_814617, EPI_ISL_814618, EPI_ISL_814619, EPI_ISL_817026, EPI_ISL_817029, EPI_ISL_817032, EPI_ISL_817038, EPI_ISL_817040, EPI_ISL_817047, EPI_ISL_817048, EPI_ISL_817050, EPI_ISL_817051, EPI_ISL_817052, EPI_ISL_817053, EPI_ISL_817054, EPI_ISL_817055, EPI_ISL_817056, EPI_ISL_817057, EPI_ISL_817058, EPI_ISL_817059, EPI_ISL_817061, EPI_ISL_817062, EPI_ISL_817063, EPI_ISL_817066, EPI_ISL_817067, EPI_ISL_817069, EPI_ISL_817070, EPI_ISL_817071, EPI_ISL_817072, EPI_ISL_817073, EPI_ISL_817074, EPI_ISL_817075, EPI_ISL_817076, EPI_ISL_817078, EPI_ISL_817079, EPI_ISL_817080, EPI_ISL_817081, EPI_ISL_817082, EPI_ISL_817083, EPI_ISL_817084, EPI_ISL_817085, EPI_ISL_817086, EPI_ISL_817087, EPI_ISL_817088, EPI_ISL_817089, EPI_ISL_817090, EPI_ISL_817091, EPI_ISL_817092, EPI_ISL_817093, EPI_ISL_817094, EPI_ISL_817095, EPI_ISL_817096, EPI_ISL_817097, EPI_ISL_817098, EPI_ISL_817099, EPI_ISL_817100, EPI_ISL_817101, EPI_ISL_817102, EPI_ISL_817103, EPI_ISL_817104, EPI_ISL_817105, EPI_ISL_817106, EPI_ISL_817107, EPI_ISL_817108, EPI_ISL_817109, EPI_ISL_817110, EPI_ISL_817111, EPI_ISL_817112, EPI_ISL_817113, EPI_ISL_817114, EPI_ISL_817115, EPI_ISL_817116, EPI_ISL_817117, EPI_ISL_817118, EPI_ISL_817119, EPI_ISL_817120, EPI_ISL_817121, EPI_ISL_817122, EPI_ISL_869047, EPI_ISL_924463, EPI_ISL_953118, EPI_ISL_953126, EPI_ISL_953129, EPI_ISL_953132, EPI_ISL_953133, EPI_ISL_953135, EPI_ISL_953138, EPI_ISL_953143, EPI_ISL_1476741, EPI_ISL_1476746, EPI_ISL_1476747, EPI_ISL_1476759, EPI_ISL_1476855, EPI_ISL_1476856, EPI_ISL_1476858, EPI_ISL_1476860, EPI_ISL_1476867, EPI_ISL_1476872, EPI_ISL_1476874, EPI_ISL_1476878, EPI_ISL_1476880, EPI_ISL_1476895, EPI_ISL_1476896, EPI_ISL_1476897, EPI_ISL_1476899, EPI_ISL_1476900, EPI_ISL_1638111, EPI_ISL_1638125, EPI_ISL_1638129, EPI_ISL_1638133, EPI_ISL_1638159, EPI_ISL_1638161, EPI_ISL_1638167, EPI_ISL_1638173, EPI_ISL_1638185, EPI_ISL_1638186 | | | |
| see above | Bioinformatics and Biostatistics Lab, Advanced Sequencing Facility | COVID-19 Genomics UK (COG-UK) Consortium | Aengus Stewart; Chelsea Sawyer; Harshil Patel; Jerome Nicod; Laura Cubitt; Margaret Crawford |
| EPI_ISL_763396, EPI_ISL_763407, EPI_ISL_763425, EPI_ISL_763451, EPI_ISL_763465, EPI_ISL_763471, EPI_ISL_763476, EPI_ISL_763495, EPI_ISL_763505, EPI_ISL_763512, EPI_ISL_763516, EPI_ISL_763521, EPI_ISL_763541, EPI_ISL_763550, EPI_ISL_763595, EPI_ISL_763609, EPI_ISL_763610, EPI_ISL_763612, EPI_ISL_763617, EPI_ISL_763624, EPI_ISL_763659, EPI_ISL_763660, EPI_ISL_763661, EPI_ISL_763672, EPI_ISL_763676, EPI_ISL_763685, EPI_ISL_763690, EPI_ISL_763693, EPI_ISL_763734, EPI_ISL_763755, EPI_ISL_763791, EPI_ISL_763798, EPI_ISL_763799, EPI_ISL_763800, EPI_ISL_763804, EPI_ISL_763805, EPI_ISL_763806, EPI_ISL_763823, EPI_ISL_763824, EPI_ISL_763825, EPI_ISL_763827, EPI_ISL_763828, EPI_ISL_763829, EPI_ISL_763830, EPI_ISL_763831, EPI_ISL_763832, EPI_ISL_763833, EPI_ISL_763834, EPI_ISL_763835, EPI_ISL_763836, EPI_ISL_763865, EPI_ISL_764007, EPI_ISL_764008, EPI_ISL_764009, EPI_ISL_764010, EPI_ISL_764011, EPI_ISL_764012, EPI_ISL_764013, EPI_ISL_764014, EPI_ISL_764015, EPI_ISL_764016, EPI_ISL_764017, EPI_ISL_764018, EPI_ISL_764019, EPI_ISL_764020, EPI_ISL_764021, EPI_ISL_764022, EPI_ISL_764023, EPI_ISL_764024, EPI_ISL_764025, EPI_ISL_764026, EPI_ISL_764027, EPI_ISL_764028, EPI_ISL_764029, EPI_ISL_764030, EPI_ISL_764036, EPI_ISL_764081, EPI_ISL_764094, EPI_ISL_764095, EPI_ISL_764096, EPI_ISL_764101, EPI_ISL_764102, EPI_ISL_764118, EPI_ISL_764119, EPI_ISL_764120, EPI_ISL_764121, EPI_ISL_764122, EPI_ISL_764123, EPI_ISL_764124, EPI_ISL_764125, EPI_ISL_764136, EPI_ISL_764169, EPI_ISL_764172, EPI_ISL_764193, EPI_ISL_765112, EPI_ISL_765113, EPI_ISL_765114, EPI_ISL_765115, EPI_ISL_765116, EPI_ISL_765119, EPI_ISL_765120, EPI_ISL_765121, EPI_ISL_765122, EPI_ISL_765123, EPI_ISL_765124, EPI_ISL_765125, EPI_ISL_765126, EPI_ISL_765127, EPI_ISL_765128, EPI_ISL_765129, EPI_ISL_765130, EPI_ISL_765131, EPI_ISL_765132, EPI_ISL_765133, EPI_ISL_765134, EPI_ISL_765135, EPI_ISL_765136, EPI_ISL_765137, EPI_ISL_765138, EPI_ISL_765139, EPI_ISL_765140, EPI_ISL_765141, EPI_ISL_765142, EPI_ISL_765144, EPI_ISL_765145, EPI_ISL_765146, EPI_ISL_765147, EPI_ISL_765148, EPI_ISL_816208, EPI_ISL_816209, EPI_ISL_841754, EPI_ISL_841755, EPI_ISL_841756, EPI_ISL_841757, EPI_ISL_841758, EPI_ISL_841759, EPI_ISL_841769, EPI_ISL_841770, EPI_ISL_841771, EPI_ISL_841772, EPI_ISL_841773, EPI_ISL_841774, EPI_ISL_841775, EPI_ISL_841776, EPI_ISL_841777, EPI_ISL_841778, EPI_ISL_841779, EPI_ISL_841780, EPI_ISL_841781, EPI_ISL_841782, EPI_ISL_841783, EPI_ISL_841784, EPI_ISL_841785, EPI_ISL_841786, EPI_ISL_841787, EPI_ISL_841788, EPI_ISL_841789, EPI_ISL_841790, EPI_ISL_841791, EPI_ISL_841792, EPI_ISL_841793, EPI_ISL_841794, EPI_ISL_841795, EPI_ISL_841796, EPI_ISL_841797, EPI_ISL_841798, EPI_ISL_841799, EPI_ISL_841800, EPI_ISL_841801, EPI_ISL_841802, EPI_ISL_841803, EPI_ISL_841804, EPI_ISL_841805, EPI_ISL_841806, EPI_ISL_841807, EPI_ISL_841808, EPI_ISL_841809, EPI_ISL_841810, EPI_ISL_841811, EPI_ISL_841812, EPI_ISL_841813, EPI_ISL_841814, EPI_ISL_841815, EPI_ISL_841816, EPI_ISL_841817, EPI_ISL_841818, EPI_ISL_841819, EPI_ISL_841821, EPI_ISL_841823, EPI_ISL_841824, EPI_ISL_841825, EPI_ISL_841826, EPI_ISL_841827, EPI_ISL_841828, EPI_ISL_841829, EPI_ISL_841830, EPI_ISL_841831, EPI_ISL_841832, EPI_ISL_841833, EPI_ISL_841834, EPI_ISL_841835, EPI_ISL_841836, EPI_ISL_841837, EPI_ISL_841838, EPI_ISL_841839, EPI_ISL_841840, EPI_ISL_841841, EPI_ISL_841842, EPI_ISL_841843, EPI_ISL_841844, EPI_ISL_841845, EPI_ISL_841848, EPI_ISL_841849, EPI_ISL_841850, EPI_ISL_841851, EPI_ISL_841853, EPI_ISL_841854, EPI_ISL_841855, EPI_ISL_841856, EPI_ISL_841857, EPI_ISL_841858, EPI_ISL_841859, EPI_ISL_841860, EPI_ISL_841861, EPI_ISL_841862, EPI_ISL_841863, EPI_ISL_841864, EPI_ISL_841865, EPI_ISL_841866, EPI_ISL_841867, EPI_ISL_841868, EPI_ISL_841869, EPI_ISL_841870, EPI_ISL_842012, EPI_ISL_923234, EPI_ISL_1000235, EPI_ISL_1107543, EPI_ISL_1107699, EPI_ISL_1107749, EPI_ISL_1107752, EPI_ISL_1107753, EPI_ISL_1179794, EPI_ISL_1730820, EPI_ISL_1730850, EPI_ISL_2127922, EPI_ISL_2127924, EPI_ISL_2127926, EPI_ISL_2127928, EPI_ISL_2127930, EPI_ISL_2127963, EPI_ISL_2127964 | | | |
| see above | Centre for Enzyme Innovation, University of Portsmouth / Translational Research Laboratory, Portsmouth Hospitals NHS Trust | COVID-19 Genomics UK (COG-UK) Consortium | Allyson Lloyd; Angela Beckett; Anoop Chauhan; Christopher Fearn; Ethan Butcher; Garry Scarlett; Kate Cook; Katie Loveson; Kelly Bicknell; Robert Impey; Salman Goudarzi; Samuel Robson; Sarah Wyllie; Scott Elliott; Sharon Glaysher; Yann Bourgeois |
| EPI_ISL_741005, EPI_ISL_741006, EPI_ISL_741013, EPI_ISL_741014, EPI_ISL_741015, EPI_ISL_741016, EPI_ISL_741017, EPI_ISL_763448, EPI_ISL_813172, EPI_ISL_813196, EPI_ISL_813232, EPI_ISL_813233, EPI_ISL_813234, EPI_ISL_813235, EPI_ISL_813237, EPI_ISL_813238, EPI_ISL_813240, EPI_ISL_813241, EPI_ISL_813242, EPI_ISL_813246, EPI_ISL_813253, EPI_ISL_813254, EPI_ISL_813259, EPI_ISL_813260, EPI_ISL_813261, EPI_ISL_813272, EPI_ISL_813273, EPI_ISL_813282, EPI_ISL_813284, EPI_ISL_813289, EPI_ISL_813297, EPI_ISL_813298, EPI_ISL_813299, EPI_ISL_813300, EPI_ISL_813301, EPI_ISL_813302, EPI_ISL_813303, EPI_ISL_813304, EPI_ISL_813307, EPI_ISL_813308, EPI_ISL_813309, EPI_ISL_813310, EPI_ISL_813311, EPI_ISL_813312, EPI_ISL_813313, EPI_ISL_813314, EPI_ISL_813315, EPI_ISL_813316, EPI_ISL_813318, EPI_ISL_813319, EPI_ISL_813321, EPI_ISL_813322, EPI_ISL_813324, EPI_ISL_813327, EPI_ISL_813329, EPI_ISL_813330, EPI_ISL_813333, EPI_ISL_813334, EPI_ISL_813335, EPI_ISL_813336, EPI_ISL_813337, EPI_ISL_813338, EPI_ISL_813340, EPI_ISL_813341, EPI_ISL_813342, EPI_ISL_813343, EPI_ISL_813345, EPI_ISL_813347, EPI_ISL_813348, EPI_ISL_813349, EPI_ISL_813350, EPI_ISL_813351, EPI_ISL_813352, EPI_ISL_813353, EPI_ISL_813354, EPI_ISL_813355, EPI_ISL_837837, EPI_ISL_837838, EPI_ISL_837839, EPI_ISL_837840, EPI_ISL_837841, EPI_ISL_837842, EPI_ISL_864833, EPI_ISL_864837, EPI_ISL_864839, EPI_ISL_864847, EPI_ISL_864849, EPI_ISL_864851, EPI_ISL_864854 | | | |
| see above | Department of Pathology, University of Cambridge | COVID-19 Genomics UK (COG-UK) Consortium | Aminu S. Jahun; Grant Hall; Ian Goodfellow; Iliana Georgana; Malte Pinckert; Martin D. Curran; Myra Hosmillo; Surendra Parmar; Yasmin Chaudhry |
| EPI_ISL_813656 | Kettering General Hospital | COVID-19 Genomics UK (COG-UK) Consortium | Christopher Moore; Fei Sang; Johnny Debebe; Jonathan Ball; Joseph Chappell; Matthew Carlisle; Matthew Loose; Nadine Holmes; Patrick McClure; Theocharis Tsoleridis; Victoria Wright |
| EPI_ISL_756378, EPI_ISL_756381, EPI_ISL_756382, EPI_ISL_756383, EPI_ISL_756385, EPI_ISL_756387, EPI_ISL_756388, EPI_ISL_756390, EPI_ISL_756391, EPI_ISL_756394, EPI_ISL_756396, EPI_ISL_756397, EPI_ISL_756398, EPI_ISL_756399, EPI_ISL_756400, EPI_ISL_756402, EPI_ISL_756404, EPI_ISL_756405, EPI_ISL_756406, EPI_ISL_756407, EPI_ISL_756408, EPI_ISL_756410, EPI_ISL_756411, EPI_ISL_756412, EPI_ISL_756414, EPI_ISL_756421, EPI_ISL_756423, EPI_ISL_756425, EPI_ISL_756427, EPI_ISL_756428, EPI_ISL_756430, EPI_ISL_756433, EPI_ISL_756435, EPI_ISL_756436, EPI_ISL_756437, EPI_ISL_756439, EPI_ISL_756440, EPI_ISL_756441, EPI_ISL_756443, EPI_ISL_756445, EPI_ISL_756447, EPI_ISL_756449, EPI_ISL_756450, EPI_ISL_756451, EPI_ISL_756452, EPI_ISL_756455, EPI_ISL_756457, EPI_ISL_756458, EPI_ISL_756459, EPI_ISL_756461, EPI_ISL_756462, EPI_ISL_756463, EPI_ISL_756464, EPI_ISL_756465, EPI_ISL_756466, EPI_ISL_756468, EPI_ISL_756471, EPI_ISL_756472, EPI_ISL_756473, EPI_ISL_756476, EPI_ISL_756477, EPI_ISL_756479, EPI_ISL_756482, EPI_ISL_756483, EPI_ISL_756488, EPI_ISL_756489, EPI_ISL_756493, EPI_ISL_756494, EPI_ISL_756495, EPI_ISL_756503, EPI_ISL_756504, EPI_ISL_756509, EPI_ISL_756510, EPI_ISL_756514, EPI_ISL_756515, EPI_ISL_756516, EPI_ISL_756520, EPI_ISL_756523, EPI_ISL_756524, EPI_ISL_756526, EPI_ISL_756527, EPI_ISL_756530, EPI_ISL_756531, EPI_ISL_756532, EPI_ISL_756533, EPI_ISL_756536, EPI_ISL_756537, EPI_ISL_756538, EPI_ISL_756539, EPI_ISL_756541, EPI_ISL_756544, EPI_ISL_756546, EPI_ISL_756547, EPI_ISL_756548, EPI_ISL_756550, EPI_ISL_756551, EPI_ISL_756552, EPI_ISL_756555, EPI_ISL_756556, EPI_ISL_756557, EPI_ISL_756560, EPI_ISL_756561, EPI_ISL_756562, EPI_ISL_756563, EPI_ISL_756565, EPI_ISL_756566, EPI_ISL_756569, EPI_ISL_756571, EPI_ISL_756573, EPI_ISL_756574, EPI_ISL_756575, EPI_ISL_756578, EPI_ISL_756580, EPI_ISL_756581, EPI_ISL_756583, EPI_ISL_756587, EPI_ISL_756592, EPI_ISL_756593, EPI_ISL_756596, EPI_ISL_756598, EPI_ISL_756599, EPI_ISL_756600, EPI_ISL_756602, EPI_ISL_756603, EPI_ISL_756609, EPI_ISL_756612, EPI_ISL_756616, EPI_ISL_756618, EPI_ISL_756619, EPI_ISL_756620, EPI_ISL_756624, EPI_ISL_756626, EPI_ISL_756632, EPI_ISL_756634, EPI_ISL_756635, EPI_ISL_756636, EPI_ISL_756638, EPI_ISL_756639, EPI_ISL_756640, EPI_ISL_756641, EPI_ISL_756644, EPI_ISL_756645, EPI_ISL_756646, EPI_ISL_756647, EPI_ISL_756648, EPI_ISL_756650, EPI_ISL_756651, EPI_ISL_756652, EPI_ISL_756654, EPI_ISL_756656, EPI_ISL_756657, EPI_ISL_756658, EPI_ISL_756661, EPI_ISL_756662, EPI_ISL_756663, EPI_ISL_756664, EPI_ISL_756669, EPI_ISL_756670, EPI_ISL_756673, EPI_ISL_756681, EPI_ISL_756682, EPI_ISL_756683, EPI_ISL_756684, EPI_ISL_756685, EPI_ISL_756686, EPI_ISL_756687, EPI_ISL_756688, EPI_ISL_760249, EPI_ISL_760259, EPI_ISL_760261, EPI_ISL_760263, EPI_ISL_760266, EPI_ISL_760267, EPI_ISL_760268, EPI_ISL_760273, EPI_ISL_760276, EPI_ISL_760278, EPI_ISL_760281, EPI_ISL_760284, EPI_ISL_760285, EPI_ISL_760286, EPI_ISL_760288, EPI_ISL_760293, EPI_ISL_760295, EPI_ISL_760296, EPI_ISL_760301, EPI_ISL_760305, EPI_ISL_760306, EPI_ISL_760311, EPI_ISL_760326, EPI_ISL_760333, EPI_ISL_760341, EPI_ISL_760344, EPI_ISL_760346, EPI_ISL_760350, EPI_ISL_760358, EPI_ISL_760359, EPI_ISL_760360, EPI_ISL_760364, EPI_ISL_760369, EPI_ISL_760370, EPI_ISL_760371, EPI_ISL_760372, EPI_ISL_760381, EPI_ISL_760386, EPI_ISL_760387, EPI_ISL_760389, EPI_ISL_760390, EPI_ISL_760392, EPI_ISL_760397, EPI_ISL_760405, EPI_ISL_760408, EPI_ISL_760415, EPI_ISL_760418, EPI_ISL_760419, EPI_ISL_760421, EPI_ISL_760424, EPI_ISL_760430, EPI_ISL_760432, EPI_ISL_760434, EPI_ISL_760436, EPI_ISL_760441, EPI_ISL_760442, EPI_ISL_760445, EPI_ISL_760448, EPI_ISL_760450, EPI_ISL_760460, EPI_ISL_760461, EPI_ISL_760471, EPI_ISL_760474, EPI_ISL_760475, EPI_ISL_760485, EPI_ISL_760486, EPI_ISL_760491, EPI_ISL_760495, EPI_ISL_760498, EPI_ISL_760500, EPI_ISL_760501, EPI_ISL_760508, EPI_ISL_760509, EPI_ISL_760512, EPI_ISL_760516, EPI_ISL_760521, EPI_ISL_760524, EPI_ISL_760530, EPI_ISL_760531, EPI_ISL_760534, EPI_ISL_760535, EPI_ISL_760538, EPI_ISL_760543, EPI_ISL_760544, EPI_ISL_760545, EPI_ISL_760546, EPI_ISL_760549, EPI_ISL_760551, EPI_ISL_760552, EPI_ISL_760555, EPI_ISL_760557, EPI_ISL_760558, EPI_ISL_760559, EPI_ISL_760561, EPI_ISL_760562, EPI_ISL_760564, EPI_ISL_760565, EPI_ISL_760570, EPI_ISL_760572, EPI_ISL_760574, EPI_ISL_760582, EPI_ISL_760586, EPI_ISL_760587, EPI_ISL_760588, EPI_ISL_760589, EPI_ISL_760591, EPI_ISL_760593, EPI_ISL_760595, EPI_ISL_760596, EPI_ISL_760597, EPI_ISL_760599, EPI_ISL_760601, EPI_ISL_760603, EPI_ISL_760608, EPI_ISL_760609, EPI_ISL_760612, EPI_ISL_760613, EPI_ISL_760615, EPI_ISL_760617, EPI_ISL_760623, EPI_ISL_760624, EPI_ISL_760625, EPI_ISL_760627, EPI_ISL_760628, EPI_ISL_760633, EPI_ISL_760637, EPI_ISL_760639, EPI_ISL_760640, EPI_ISL_760644, EPI_ISL_760647, EPI_ISL_760650, EPI_ISL_760652, EPI_ISL_760654, EPI_ISL_760656, EPI_ISL_760660, EPI_ISL_760661, EPI_ISL_760663, EPI_ISL_760664, EPI_ISL_760665, EPI_ISL_760666, EPI_ISL_760667, EPI_ISL_760670, EPI_ISL_760677, EPI_ISL_760679, EPI_ISL_760680, EPI_ISL_760684, EPI_ISL_760685, EPI_ISL_760688, EPI_ISL_760691, EPI_ISL_760693, EPI_ISL_760697, EPI_ISL_760699, EPI_ISL_760701, EPI_ISL_760702, EPI_ISL_760705, EPI_ISL_760708, EPI_ISL_760710, EPI_ISL_760711, EPI_ISL_760714, EPI_ISL_760715, EPI_ISL_760719, EPI_ISL_760721, EPI_ISL_760722, EPI_ISL_760723, EPI_ISL_760724, EPI_ISL_760726, EPI_ISL_760727, EPI_ISL_760728, EPI_ISL_760733, EPI_ISL_760734, EPI_ISL_760736, EPI_ISL_760737, EPI_ISL_760738, EPI_ISL_760745, EPI_ISL_760748, EPI_ISL_760752, EPI_ISL_760753, EPI_ISL_760755, EPI_ISL_760756, EPI_ISL_760759, EPI_ISL_760761, EPI_ISL_760762, EPI_ISL_760763, EPI_ISL_760769, EPI_ISL_760776, EPI_ISL_760780, EPI_ISL_760783, EPI_ISL_760784, EPI_ISL_760785, EPI_ISL_760786, EPI_ISL_760788, EPI_ISL_760790, EPI_ISL_760792, EPI_ISL_760794, EPI_ISL_760795, EPI_ISL_760799, EPI_ISL_760802, EPI_ISL_760804, EPI_ISL_760806, EPI_ISL_760808, EPI_ISL_760812, EPI_ISL_760817, EPI_ISL_760825, EPI_ISL_760826, EPI_ISL_760832, EPI_ISL_760838, EPI_ISL_760839, EPI_ISL_760852, EPI_ISL_760853, EPI_ISL_760857, EPI_ISL_760861, EPI_ISL_760862, EPI_ISL_760864, EPI_ISL_760865, EPI_ISL_760867, EPI_ISL_760868, EPI_ISL_760874, EPI_ISL_760884, EPI_ISL_760887, EPI_ISL_760889, EPI_ISL_760890, EPI_ISL_760892, EPI_ISL_760897, EPI_ISL_760898, EPI_ISL_760900, EPI_ISL_760906, EPI_ISL_760908, EPI_ISL_760912, EPI_ISL_760915, EPI_ISL_760916, EPI_ISL_760918, EPI_ISL_760921, EPI_ISL_760925, EPI_ISL_760928, EPI_ISL_760931, EPI_ISL_760933, EPI_ISL_760937, EPI_ISL_760938, EPI_ISL_760941, EPI_ISL_760942, EPI_ISL_760945, EPI_ISL_760948, EPI_ISL_760950, EPI_ISL_760960, EPI_ISL_760962, EPI_ISL_760967, EPI_ISL_760968, EPI_ISL_760971, EPI_ISL_760979, EPI_ISL_760994, EPI_ISL_761000, EPI_ISL_761008, EPI_ISL_761011, EPI_ISL_761022, EPI_ISL_761025, EPI_ISL_761027, EPI_ISL_761029, EPI_ISL_761036, EPI_ISL_761040, EPI_ISL_761048, EPI_ISL_761049, EPI_ISL_761052, EPI_ISL_761054, EPI_ISL_761056, EPI_ISL_761060, EPI_ISL_761064, EPI_ISL_761065, EPI_ISL_761066, EPI_ISL_761069, EPI_ISL_761080, EPI_ISL_761081, EPI_ISL_761082, EPI_ISL_761083, EPI_ISL_761084, EPI_ISL_761085, EPI_ISL_761086, EPI_ISL_761087, EPI_ISL_761088, EPI_ISL_761089, EPI_ISL_761090, EPI_ISL_761091, EPI_ISL_761092, EPI_ISL_761093, EPI_ISL_761097, EPI_ISL_761098, EPI_ISL_761099, EPI_ISL_761100, EPI_ISL_761101, EPI_ISL_761102, EPI_ISL_761103, EPI_ISL_761104, EPI_ISL_761105, EPI_ISL_761106, EPI_ISL_761107, EPI_ISL_761108, EPI_ISL_761109, EPI_ISL_761110, EPI_ISL_761111, EPI_ISL_761112, EPI_ISL_761115, EPI_ISL_761117, EPI_ISL_761119, EPI_ISL_761122, EPI_ISL_761123, EPI_ISL_761124, EPI_ISL_761126, EPI_ISL_761127, EPI_ISL_761129, EPI_ISL_761130, EPI_ISL_761132, EPI_ISL_761135, EPI_ISL_761136, EPI_ISL_761137, EPI_ISL_761138, EPI_ISL_761139, EPI_ISL_761140, EPI_ISL_761141, EPI_ISL_761142, EPI_ISL_761144, EPI_ISL_761146, EPI_ISL_761147, EPI_ISL_761148, EPI_ISL_761149, EPI_ISL_761150, EPI_ISL_761151, EPI_ISL_761152, EPI_ISL_761153, EPI_ISL_761155, EPI_ISL_761156, EPI_ISL_761159, EPI_ISL_761160, EPI_ISL_761161, EPI_ISL_761163, EPI_ISL_761164, EPI_ISL_761165, EPI_ISL_761166, EPI_ISL_761167, EPI_ISL_761169, EPI_ISL_761170, EPI_ISL_761171, EPI_ISL_761173, EPI_ISL_761174, EPI_ISL_761175, EPI_ISL_761177, EPI_ISL_761178, EPI_ISL_761179, EPI_ISL_761180, EPI_ISL_761181, EPI_ISL_761182, EPI_ISL_761183, EPI_ISL_761184, EPI_ISL_761186, EPI_ISL_761188, EPI_ISL_761189, EPI_ISL_761190, EPI_ISL_761192, EPI_ISL_761195, EPI_ISL_761196, EPI_ISL_761198, EPI_ISL_761199, EPI_ISL_761200, EPI_ISL_761201, EPI_ISL_761202, EPI_ISL_761203, EPI_ISL_761205, EPI_ISL_761206, EPI_ISL_761208, EPI_ISL_761210, EPI_ISL_761211, EPI_ISL_761212, EPI_ISL_761213, EPI_ISL_761214, EPI_ISL_761215, EPI_ISL_761216, EPI_ISL_761217, EPI_ISL_761218, EPI_ISL_761219, EPI_ISL_761220, EPI_ISL_761222, EPI_ISL_761223, EPI_ISL_761224, EPI_ISL_761226, EPI_ISL_761230, EPI_ISL_761231, EPI_ISL_761232, EPI_ISL_761233, EPI_ISL_761234, EPI_ISL_761235, EPI_ISL_761236, EPI_ISL_761237, EPI_ISL_761238, EPI_ISL_761240, EPI_ISL_761242, EPI_ISL_761243, EPI_ISL_761244, EPI_ISL_761245, EPI_ISL_761246, EPI_ISL_761247, EPI_ISL_761248, EPI_ISL_761249, EPI_ISL_761250, EPI_ISL_761251, EPI_ISL_761253, EPI_ISL_761254, EPI_ISL_761256, EPI_ISL_761258, EPI_ISL_761260, EPI_ISL_761261, EPI_ISL_761262, EPI_ISL_761263, EPI_ISL_761264, EPI_ISL_761265, EPI_ISL_761267, EPI_ISL_761268, EPI_ISL_761269, EPI_ISL_761270, EPI_ISL_761271, EPI_ISL_761272, EPI_ISL_761273, EPI_ISL_761274, EPI_ISL_761275, EPI_ISL_761277, EPI_ISL_761279, EPI_ISL_761281, EPI_ISL_761282, EPI_ISL_761284, EPI_ISL_761285, EPI_ISL_761286, EPI_ISL_761288, EPI_ISL_761290, EPI_ISL_761291, EPI_ISL_761294, EPI_ISL_761295, EPI_ISL_761297, EPI_ISL_761299, EPI_ISL_761300, EPI_ISL_761301, EPI_ISL_761304, EPI_ISL_761305, EPI_ISL_761306, EPI_ISL_761308, EPI_ISL_761309, EPI_ISL_761310, EPI_ISL_761311, EPI_ISL_761312, EPI_ISL_761314, EPI_ISL_761315, EPI_ISL_761316, EPI_ISL_761317, EPI_ISL_761318, EPI_ISL_761319, EPI_ISL_761320, EPI_ISL_761321, EPI_ISL_761322, EPI_ISL_761323, EPI_ISL_761324, EPI_ISL_761325, EPI_ISL_761327, EPI_ISL_761328, EPI_ISL_761329, EPI_ISL_761330, EPI_ISL_761332, EPI_ISL_761333, EPI_ISL_761334, EPI_ISL_761335, EPI_ISL_761336, EPI_ISL_761337, EPI_ISL_761340, EPI_ISL_761341, EPI_ISL_761342, EPI_ISL_761343, EPI_ISL_761344, EPI_ISL_761345, EPI_ISL_761346, EPI_ISL_761347, EPI_ISL_761349, EPI_ISL_761350, EPI_ISL_761352, EPI_ISL_761354, EPI_ISL_761356, EPI_ISL_761665, EPI_ISL_761666, EPI_ISL_761667, EPI_ISL_761668, EPI_ISL_761669, EPI_ISL_761670, EPI_ISL_761671, EPI_ISL_761672, EPI_ISL_761673, EPI_ISL_761674, EPI_ISL_761675, EPI_ISL_761676, EPI_ISL_761677, EPI_ISL_761678, EPI_ISL_761679, EPI_ISL_761680, EPI_ISL_761681, EPI_ISL_761682, EPI_ISL_761683, EPI_ISL_761684, EPI_ISL_761685, EPI_ISL_761686, EPI_ISL_761687, EPI_ISL_761688, EPI_ISL_761689, EPI_ISL_761690, EPI_ISL_761691, EPI_ISL_761692, EPI_ISL_761693, EPI_ISL_761694, EPI_ISL_761695, EPI_ISL_761696, EPI_ISL_761697, EPI_ISL_761698, EPI_ISL_761699, EPI_ISL_761700, EPI_ISL_761701, EPI_ISL_761702, EPI_ISL_761703, EPI_ISL_761704, EPI_ISL_761705, EPI_ISL_761706, EPI_ISL_761707, EPI_ISL_761708, EPI_ISL_761709, EPI_ISL_761710, EPI_ISL_761711, EPI_ISL_761712, EPI_ISL_761713, EPI_ISL_761714, EPI_ISL_761715, EPI_ISL_761716, EPI_ISL_761717, EPI_ISL_761718, EPI_ISL_761719, EPI_ISL_761720, EPI_ISL_761721, EPI_ISL_761722, EPI_ISL_761723, EPI_ISL_761724, EPI_ISL_761725, EPI_ISL_761726, EPI_ISL_761728, EPI_ISL_761729, EPI_ISL_761730, EPI_ISL_761731, EPI_ISL_761732, EPI_ISL_761733, EPI_ISL_761734, EPI_ISL_761735, EPI_ISL_761736, EPI_ISL_761737, EPI_ISL_761738, EPI_ISL_761739, EPI_ISL_761740, EPI_ISL_761741, EPI_ISL_761742, EPI_ISL_761743, EPI_ISL_761744, EPI_ISL_761745, EPI_ISL_761746, EPI_ISL_761747, EPI_ISL_761748, EPI_ISL_761749, EPI_ISL_761750, EPI_ISL_761751, EPI_ISL_761752, EPI_ISL_761753, EPI_ISL_761754, EPI_ISL_761755, EPI_ISL_761756, EPI_ISL_761757, EPI_ISL_761758, EPI_ISL_761759, EPI_ISL_761760, EPI_ISL_761761, EPI_ISL_761762, EPI_ISL_761763, EPI_ISL_761764, EPI_ISL_761765, EPI_ISL_761766, EPI_ISL_761767, EPI_ISL_761768, EPI_ISL_761769, EPI_ISL_761770, EPI_ISL_761771, EPI_ISL_761772, EPI_ISL_761773, EPI_ISL_761774, EPI_ISL_761775, EPI_ISL_761776, EPI_ISL_761777, EPI_ISL_761778, EPI_ISL_761779, EPI_ISL_761780, EPI_ISL_761781, EPI_ISL_761782, EPI_ISL_761783, EPI_ISL_761784, EPI_ISL_761785, EPI_ISL_761786, EPI_ISL_761787, EPI_ISL_761788, EPI_ISL_761789, EPI_ISL_761790, EPI_ISL_761791, EPI_ISL_761792, EPI_ISL_761793, EPI_ISL_761794, EPI_ISL_761795, EPI_ISL_761796, EPI_ISL_761797, EPI_ISL_761798, EPI_ISL_761799, EPI_ISL_761800, EPI_ISL_761801, EPI_ISL_761802, EPI_ISL_761803, EPI_ISL_761804, EPI_ISL_761805, EPI_ISL_761806, EPI_ISL_761807, EPI_ISL_761808, EPI_ISL_761809, EPI_ISL_761810, EPI_ISL_761811, EPI_ISL_761812, EPI_ISL_761813, EPI_ISL_761814, EPI_ISL_761815, EPI_ISL_761816, EPI_ISL_761817, EPI_ISL_761818, EPI_ISL_761819, EPI_ISL_761820, EPI_ISL_761821, EPI_ISL_761822, EPI_ISL_761823, EPI_ISL_761824, EPI_ISL_761825, EPI_ISL_761826, EPI_ISL_761827, EPI_ISL_761828, EPI_ISL_761829, EPI_ISL_761830, EPI_ISL_761831, EPI_ISL_761832, EPI_ISL_761833, EPI_ISL_761834, EPI_ISL_761835, EPI_ISL_761836, EPI_ISL_761837, EPI_ISL_761838, EPI_ISL_761839, EPI_ISL_761840, EPI_ISL_761841, EPI_ISL_761842, EPI_ISL_761843, EPI_ISL_761844, EPI_ISL_761845, EPI_ISL_761846, EPI_ISL_761847, EPI_ISL_761848, EPI_ISL_761849, EPI_ISL_761850, EPI_ISL_761851, EPI_ISL_761852, EPI_ISL_761853, EPI_ISL_761854, EPI_ISL_761855, EPI_ISL_761856, EPI_ISL_761857, EPI_ISL_761858, EPI_ISL_761859, EPI_ISL_761860, EPI_ISL_761861, EPI_ISL_761862, EPI_ISL_761863, EPI_ISL_761864, EPI_ISL_761865, EPI_ISL_761866, EPI_ISL_761867, EPI_ISL_761868, EPI_ISL_761869, EPI_ISL_761870, EPI_ISL_761871, EPI_ISL_761872, EPI_ISL_761873, EPI_ISL_761874, EPI_ISL_761875, EPI_ISL_761876, EPI_ISL_761877, EPI_ISL_761878, EPI_ISL_761879, EPI_ISL_761880, EPI_ISL_761881, EPI_ISL_761882, EPI_ISL_761883, EPI_ISL_761884, EPI_ISL_761885, EPI_ISL_761886, EPI_ISL_761887, EPI_ISL_761888, EPI_ISL_761889, EPI_ISL_761890, EPI_ISL_761891, EPI_ISL_761893, EPI_ISL_761894, EPI_ISL_761895, EPI_ISL_761896, EPI_ISL_761897, EPI_ISL_761898, EPI_ISL_761899, EPI_ISL_761900, EPI_ISL_761901, EPI_ISL_761902, EPI_ISL_761903, EPI_ISL_761904, EPI_ISL_761905, EPI_ISL_761906, EPI_ISL_761907, EPI_ISL_761908, EPI_ISL_761909, EPI_ISL_761910, EPI_ISL_761911, EPI_ISL_761912, EPI_ISL_761913, EPI_ISL_761914, EPI_ISL_761915, EPI_ISL_761916, EPI_ISL_761917, EPI_ISL_761918, EPI_ISL_761919, EPI_ISL_761920, EPI_ISL_762547, EPI_ISL_762554, EPI_ISL_762557, EPI_ISL_762558, EPI_ISL_762562, EPI_ISL_762564, EPI_ISL_762581, EPI_ISL_762588, EPI_ISL_762599, EPI_ISL_762605, EPI_ISL_762606, EPI_ISL_762610, EPI_ISL_762617, EPI_ISL_762620, EPI_ISL_762625, EPI_ISL_762629, EPI_ISL_762634, EPI_ISL_762638, EPI_ISL_762642, EPI_ISL_762643, EPI_ISL_762647, EPI_ISL_762655, EPI_ISL_762659, EPI_ISL_762666, EPI_ISL_762671, EPI_ISL_762672, EPI_ISL_762674, EPI_ISL_762683, EPI_ISL_762686, EPI_ISL_762691, EPI_ISL_762692, EPI_ISL_762693, EPI_ISL_762702, EPI_ISL_762703, EPI_ISL_762706, EPI_ISL_762707, EPI_ISL_762709, EPI_ISL_762710, EPI_ISL_762712, EPI_ISL_762714, EPI_ISL_762720, EPI_ISL_762722, EPI_ISL_762724, EPI_ISL_762730, EPI_ISL_762733, EPI_ISL_762735, EPI_ISL_762750, EPI_ISL_762754, EPI_ISL_762756, EPI_ISL_762763, EPI_ISL_762764, EPI_ISL_762771, EPI_ISL_762777, EPI_ISL_762786, EPI_ISL_762789, EPI_ISL_762791, EPI_ISL_762793, EPI_ISL_762796, EPI_ISL_762798, EPI_ISL_762799, EPI_ISL_762812, EPI_ISL_762815, EPI_ISL_762819, EPI_ISL_762821, EPI_ISL_762823, EPI_ISL_762828, EPI_ISL_762830, EPI_ISL_762832, EPI_ISL_762835, EPI_ISL_762839, EPI_ISL_762841, EPI_ISL_762843, EPI_ISL_762844, EPI_ISL_762845, EPI_ISL_762847, EPI_ISL_762849, EPI_ISL_762862, EPI_ISL_762869, EPI_ISL_762878, EPI_ISL_762879, EPI_ISL_762883, EPI_ISL_762886, EPI_ISL_762887, EPI_ISL_762890, EPI_ISL_762894, EPI_ISL_762900, EPI_ISL_762901, EPI_ISL_762902, EPI_ISL_762906, EPI_ISL_762910, EPI_ISL_762911, EPI_ISL_762913, EPI_ISL_762916, EPI_ISL_762919, EPI_ISL_762921, EPI_ISL_762922, EPI_ISL_762926, EPI_ISL_762931, EPI_ISL_762932, EPI_ISL_762933, EPI_ISL_762935, EPI_ISL_762938, EPI_ISL_762939, EPI_ISL_762947, EPI_ISL_762949, EPI_ISL_762951, EPI_ISL_762952, EPI_ISL_762955, EPI_ISL_762957, EPI_ISL_762964, EPI_ISL_762970, EPI_ISL_762971, EPI_ISL_762973, EPI_ISL_762974, EPI_ISL_762975, EPI_ISL_762977, EPI_ISL_762988, EPI_ISL_762989, EPI_ISL_767089, EPI_ISL_767091, EPI_ISL_767093, EPI_ISL_767094, EPI_ISL_767095, EPI_ISL_767097, EPI_ISL_767102, EPI_ISL_767103, EPI_ISL_767104, EPI_ISL_767105, EPI_ISL_767106, EPI_ISL_767116, EPI_ISL_767119, EPI_ISL_768841, EPI_ISL_768842, EPI_ISL_768843, EPI_ISL_768844, EPI_ISL_768898, EPI_ISL_768899, EPI_ISL_768900, EPI_ISL_777011, EPI_ISL_777014, EPI_ISL_777017, EPI_ISL_777018, EPI_ISL_777020, EPI_ISL_777024, EPI_ISL_777025, EPI_ISL_777026, EPI_ISL_777027, EPI_ISL_777028, EPI_ISL_777030, EPI_ISL_777032, EPI_ISL_777035, EPI_ISL_777036, EPI_ISL_777041, EPI_ISL_777043, EPI_ISL_777045, EPI_ISL_777048, EPI_ISL_777049, EPI_ISL_777050, EPI_ISL_777051, EPI_ISL_777052, EPI_ISL_777054, EPI_ISL_777060, EPI_ISL_777069, EPI_ISL_777072, EPI_ISL_777074, EPI_ISL_777075, EPI_ISL_777076, EPI_ISL_777077, EPI_ISL_777079, EPI_ISL_777086, EPI_ISL_777087, EPI_ISL_777088, EPI_ISL_777090, EPI_ISL_777092, EPI_ISL_777093, EPI_ISL_777094, EPI_ISL_777096, EPI_ISL_777103, EPI_ISL_777107, EPI_ISL_777109, EPI_ISL_777110, EPI_ISL_777111, EPI_ISL_777113, EPI_ISL_777116, EPI_ISL_777119, EPI_ISL_777123, EPI_ISL_777125, EPI_ISL_777126, EPI_ISL_777134, EPI_ISL_777135, EPI_ISL_777136, EPI_ISL_777137, EPI_ISL_777138, EPI_ISL_777139, EPI_ISL_777143, EPI_ISL_777144, EPI_ISL_777147, EPI_ISL_777150, EPI_ISL_777156, EPI_ISL_777157, EPI_ISL_777159, EPI_ISL_777163, EPI_ISL_777165, EPI_ISL_777166, EPI_ISL_777168, EPI_ISL_777170, EPI_ISL_777171, EPI_ISL_777172, EPI_ISL_777174, EPI_ISL_777175, EPI_ISL_777178, EPI_ISL_777179, EPI_ISL_777185, EPI_ISL_777186, EPI_ISL_777188, EPI_ISL_777194, EPI_ISL_777195, EPI_ISL_777198, EPI_ISL_777199, EPI_ISL_777203, EPI_ISL_777204, EPI_ISL_777206, EPI_ISL_777210, EPI_ISL_777214, EPI_ISL_777216, EPI_ISL_777217, EPI_ISL_777220, EPI_ISL_777231, EPI_ISL_777234, EPI_ISL_777237, EPI_ISL_777239, EPI_ISL_777247, EPI_ISL_777248, EPI_ISL_777249, EPI_ISL_777251, EPI_ISL_777254, EPI_ISL_777256, EPI_ISL_777257, EPI_ISL_777258, EPI_ISL_777259, EPI_ISL_777260, EPI_ISL_777262, EPI_ISL_777263, EPI_ISL_777264, EPI_ISL_777265, EPI_ISL_777266, EPI_ISL_777271, EPI_ISL_777272, EPI_ISL_777274, EPI_ISL_777276, EPI_ISL_777277, EPI_ISL_777280, EPI_ISL_777281, EPI_ISL_777287, EPI_ISL_777288, EPI_ISL_777291, EPI_ISL_777294, EPI_ISL_777296, EPI_ISL_777297, EPI_ISL_777299, EPI_ISL_777307, EPI_ISL_777308, EPI_ISL_777309, EPI_ISL_777312, EPI_ISL_777315, EPI_ISL_777317, EPI_ISL_777318, EPI_ISL_777319, EPI_ISL_777326, EPI_ISL_777327, EPI_ISL_777336, EPI_ISL_777338, EPI_ISL_777349, EPI_ISL_777375, EPI_ISL_777386, EPI_ISL_777389, EPI_ISL_777394, EPI_ISL_777411, EPI_ISL_777412, EPI_ISL_777416, EPI_ISL_777421, EPI_ISL_777424, EPI_ISL_777425, EPI_ISL_777427, EPI_ISL_777435, EPI_ISL_777437, EPI_ISL_777440, EPI_ISL_777448, EPI_ISL_777451, EPI_ISL_777455, EPI_ISL_777458, EPI_ISL_777465, EPI_ISL_777467, EPI_ISL_777470, EPI_ISL_777477, EPI_ISL_777479, EPI_ISL_777481, EPI_ISL_777489, EPI_ISL_777491, EPI_ISL_777495, EPI_ISL_777502, EPI_ISL_777509, EPI_ISL_777515, EPI_ISL_777522, EPI_ISL_777525, EPI_ISL_777529, EPI_ISL_777530, EPI_ISL_777532, EPI_ISL_777534, EPI_ISL_777535, EPI_ISL_777537, EPI_ISL_777538, EPI_ISL_777539, EPI_ISL_777543, EPI_ISL_777548, EPI_ISL_777550, EPI_ISL_777552, EPI_ISL_777558, EPI_ISL_777561, EPI_ISL_777563, EPI_ISL_777567, EPI_ISL_777568, EPI_ISL_777572, EPI_ISL_777573, EPI_ISL_777576, EPI_ISL_777580, EPI_ISL_777582, EPI_ISL_777584, EPI_ISL_777587, EPI_ISL_777589, EPI_ISL_777591, EPI_ISL_777593, EPI_ISL_777595, EPI_ISL_777597, EPI_ISL_777605, EPI_ISL_777621, EPI_ISL_777627, EPI_ISL_777629, EPI_ISL_777633, EPI_ISL_777634, EPI_ISL_777638, EPI_ISL_777639, EPI_ISL_777644, EPI_ISL_777649, EPI_ISL_777651, EPI_ISL_777655, EPI_ISL_777659, EPI_ISL_777672, EPI_ISL_778016, EPI_ISL_778018, EPI_ISL_778021, EPI_ISL_778029, EPI_ISL_778033, EPI_ISL_778035, EPI_ISL_778040, EPI_ISL_778043, EPI_ISL_778044, EPI_ISL_778047, EPI_ISL_778048, EPI_ISL_778050, EPI_ISL_778051, EPI_ISL_778052, EPI_ISL_778055, EPI_ISL_778059, EPI_ISL_778064, EPI_ISL_778065, EPI_ISL_778069, EPI_ISL_778070, EPI_ISL_778074, EPI_ISL_778083, EPI_ISL_778089, EPI_ISL_778101, EPI_ISL_778104, EPI_ISL_778108, EPI_ISL_778113, EPI_ISL_778116, EPI_ISL_778121, EPI_ISL_778128, EPI_ISL_778132, EPI_ISL_778140, EPI_ISL_778144, EPI_ISL_778163, EPI_ISL_778164, EPI_ISL_778165, EPI_ISL_778166, EPI_ISL_778183, EPI_ISL_778187, EPI_ISL_778190, EPI_ISL_778191, EPI_ISL_778195, EPI_ISL_778196, EPI_ISL_778201, EPI_ISL_778207, EPI_ISL_778212, EPI_ISL_778225, EPI_ISL_778228, EPI_ISL_778230, EPI_ISL_778233, EPI_ISL_778234, EPI_ISL_778238, EPI_ISL_778239, EPI_ISL_778240, EPI_ISL_778246, EPI_ISL_778249, EPI_ISL_778256, EPI_ISL_778259, EPI_ISL_778261, EPI_ISL_778264, EPI_ISL_778266, EPI_ISL_778270, EPI_ISL_778272, EPI_ISL_778282, EPI_ISL_778287, EPI_ISL_778288, EPI_ISL_778290, EPI_ISL_778292, EPI_ISL_778295, EPI_ISL_781201, EPI_ISL_781204, EPI_ISL_781205, EPI_ISL_781259, EPI_ISL_781294, EPI_ISL_781301, EPI_ISL_781307, EPI_ISL_781336, EPI_ISL_781366, EPI_ISL_781374, EPI_ISL_781375, EPI_ISL_781408, EPI_ISL_781433, EPI_ISL_781457, EPI_ISL_781469, EPI_ISL_781495, EPI_ISL_781500, EPI_ISL_781872, EPI_ISL_781874, EPI_ISL_781877, EPI_ISL_781879, EPI_ISL_781881, EPI_ISL_781883, EPI_ISL_781884, EPI_ISL_781887, EPI_ISL_781889, EPI_ISL_781892, EPI_ISL_781893, EPI_ISL_781894, EPI_ISL_781896, EPI_ISL_781899, EPI_ISL_781900, EPI_ISL_781901, EPI_ISL_781902, EPI_ISL_781904, EPI_ISL_781905, EPI_ISL_781906, EPI_ISL_781911, EPI_ISL_781912, EPI_ISL_781917, EPI_ISL_781923, EPI_ISL_781926, EPI_ISL_781927, EPI_ISL_781929, EPI_ISL_781931, EPI_ISL_781932, EPI_ISL_781934, EPI_ISL_781935, EPI_ISL_781936, EPI_ISL_781937, EPI_ISL_781939, EPI_ISL_781943, EPI_ISL_781945, EPI_ISL_781949, EPI_ISL_781951, EPI_ISL_781954, EPI_ISL_781957, EPI_ISL_781958, EPI_ISL_781959, EPI_ISL_781961, EPI_ISL_781962, EPI_ISL_781963, EPI_ISL_781964, EPI_ISL_781965, EPI_ISL_781966, EPI_ISL_781969, EPI_ISL_781970, EPI_ISL_781972, EPI_ISL_781974, EPI_ISL_781975, EPI_ISL_781976, EPI_ISL_781981, EPI_ISL_781983, EPI_ISL_781985, EPI_ISL_781986, EPI_ISL_781989, EPI_ISL_781991, EPI_ISL_781993, EPI_ISL_781995, EPI_ISL_781996, EPI_ISL_782002, EPI_ISL_782005, EPI_ISL_782006, EPI_ISL_782007, EPI_ISL_782010, EPI_ISL_782011, EPI_ISL_782012, EPI_ISL_782013, EPI_ISL_782016, EPI_ISL_782019, EPI_ISL_782020, EPI_ISL_782022, EPI_ISL_782023, EPI_ISL_782024, EPI_ISL_782025, EPI_ISL_782029, EPI_ISL_782033, EPI_ISL_782034, EPI_ISL_782041, EPI_ISL_782043, EPI_ISL_782045, EPI_ISL_782047, EPI_ISL_782048, EPI_ISL_782050, EPI_ISL_782053, EPI_ISL_782057, EPI_ISL_782060, EPI_ISL_782063, EPI_ISL_782068, EPI_ISL_782070, EPI_ISL_782071, EPI_ISL_782072, EPI_ISL_782076, EPI_ISL_782078, EPI_ISL_782082, EPI_ISL_782083, EPI_ISL_782085, EPI_ISL_782086, EPI_ISL_782089, EPI_ISL_782090, EPI_ISL_782091, EPI_ISL_782095, EPI_ISL_782096, EPI_ISL_782097, EPI_ISL_782099, EPI_ISL_782100, EPI_ISL_782102, EPI_ISL_782104, EPI_ISL_782108, EPI_ISL_782109, EPI_ISL_782110, EPI_ISL_782111, EPI_ISL_782117, EPI_ISL_782119, EPI_ISL_782120, EPI_ISL_782122, EPI_ISL_782123, EPI_ISL_782124, EPI_ISL_782126, EPI_ISL_782132, EPI_ISL_782135, EPI_ISL_782139, EPI_ISL_782140, EPI_ISL_782141, EPI_ISL_782142, EPI_ISL_782148, | | | |

EPI_ISL_782149, EPI_ISL_782150, EPI_ISL_782184, EPI_ISL_782220, EPI_ISL_782243, EPI_ISL_782249, EPI_ISL_782259, EPI_ISL_782267, EPI_ISL_782307, EPI_ISL_782315, EPI_ISL_782358, EPI_ISL_782362, EPI_ISL_782402, EPI_ISL_782412, EPI_ISL_782435, EPI_ISL_782478, EPI_ISL_799637, EPI_ISL_799638, EPI_ISL_799683, EPI_ISL_799706, EPI_ISL_799749, EPI_ISL_799750, EPI_ISL_799796, EPI_ISL_799818, EPI_ISL_799840, EPI_ISL_799868, EPI_ISL_799877, EPI_ISL_799891, EPI_ISL_799898, EPI_ISL_799908, EPI_ISL_799922, EPI_ISL_799925, EPI_ISL_800575, EPI_ISL_800577, EPI_ISL_800578, EPI_ISL_800579, EPI_ISL_851989, EPI_ISL_852074, EPI_ISL_852083, EPI_ISL_857891, EPI_ISL_858902, EPI_ISL_858905, EPI_ISL_858907, EPI_ISL_858909, EPI_ISL_858913, EPI_ISL_858914, EPI_ISL_858915, EPI_ISL_858917, EPI_ISL_858918, EPI_ISL_858923, EPI_ISL_858924, EPI_ISL_858925, EPI_ISL_858926, EPI_ISL_858929, EPI_ISL_858931, EPI_ISL_858932, EPI_ISL_858933, EPI_ISL_858934, EPI_ISL_858938, EPI_ISL_858939, EPI_ISL_858940, EPI_ISL_858941, EPI_ISL_858942, EPI_ISL_858944, EPI_ISL_858946, EPI_ISL_858947, EPI_ISL_858948, EPI_ISL_858949, EPI_ISL_858950, EPI_ISL_858951, EPI_ISL_858952, EPI_ISL_858954, EPI_ISL_858955, EPI_ISL_858956, EPI_ISL_858957, EPI_ISL_858959, EPI_ISL_858961, EPI_ISL_858962, EPI_ISL_858963, EPI_ISL_858964, EPI_ISL_858965, EPI_ISL_858966, EPI_ISL_858967, EPI_ISL_858968, EPI_ISL_858971, EPI_ISL_858972, EPI_ISL_858973, EPI_ISL_858976, EPI_ISL_858977, EPI_ISL_858978, EPI_ISL_858984, EPI_ISL_858992, EPI_ISL_858993, EPI_ISL_858995, EPI_ISL_858996, EPI_ISL_858999, EPI_ISL_859000, EPI_ISL_859002, EPI_ISL_859006, EPI_ISL_859007, EPI_ISL_859009, EPI_ISL_859010, EPI_ISL_859011, EPI_ISL_859013, EPI_ISL_859014, EPI_ISL_859016, EPI_ISL_859017, EPI_ISL_859019, EPI_ISL_859022, EPI_ISL_859023, EPI_ISL_859025, EPI_ISL_859026, EPI_ISL_859027, EPI_ISL_859028, EPI_ISL_859029, EPI_ISL_859035, EPI_ISL_859043, EPI_ISL_859044, EPI_ISL_859049, EPI_ISL_859051, EPI_ISL_859056, EPI_ISL_859061, EPI_ISL_859063, EPI_ISL_859066, EPI_ISL_859067, EPI_ISL_859070, EPI_ISL_859073, EPI_ISL_859075, EPI_ISL_859076, EPI_ISL_859080, EPI_ISL_859082, EPI_ISL_859085, EPI_ISL_859086, EPI_ISL_859089, EPI_ISL_859090, EPI_ISL_859091, EPI_ISL_859092, EPI_ISL_859095, EPI_ISL_859096, EPI_ISL_859097, EPI_ISL_859099, EPI_ISL_859101, EPI_ISL_859103, EPI_ISL_859104, EPI_ISL_859107, EPI_ISL_859109, EPI_ISL_859110, EPI_ISL_859111, EPI_ISL_859113, EPI_ISL_859115, EPI_ISL_859117, EPI_ISL_859121, EPI_ISL_859122, EPI_ISL_859124, EPI_ISL_859126, EPI_ISL_859128, EPI_ISL_859129, EPI_ISL_859132, EPI_ISL_859134, EPI_ISL_859135, EPI_ISL_859136, EPI_ISL_859138, EPI_ISL_859141, EPI_ISL_859142, EPI_ISL_859143, EPI_ISL_859144, EPI_ISL_859145, EPI_ISL_859146, EPI_ISL_859147, EPI_ISL_859148, EPI_ISL_859154, EPI_ISL_859161, EPI_ISL_859165, EPI_ISL_859179, EPI_ISL_859181, EPI_ISL_859186, EPI_ISL_859194, EPI_ISL_859198, EPI_ISL_859201, EPI_ISL_859202, EPI_ISL_859203, EPI_ISL_859204, EPI_ISL_859206, EPI_ISL_859207, EPI_ISL_859208, EPI_ISL_859209, EPI_ISL_859210, EPI_ISL_859212, EPI_ISL_859213, EPI_ISL_859215, EPI_ISL_859216, EPI_ISL_859217, EPI_ISL_859220, EPI_ISL_859221, EPI_ISL_859222, EPI_ISL_859223, EPI_ISL_859224, EPI_ISL_859225, EPI_ISL_859226, EPI_ISL_859227, EPI_ISL_859228, EPI_ISL_859229, EPI_ISL_859230, EPI_ISL_859231, EPI_ISL_859233, EPI_ISL_859235, EPI_ISL_859237, EPI_ISL_859238, EPI_ISL_859239, EPI_ISL_859240, EPI_ISL_859241, EPI_ISL_859242, EPI_ISL_859243, EPI_ISL_859244, EPI_ISL_859245, EPI_ISL_859246, EPI_ISL_859247, EPI_ISL_859248, EPI_ISL_859249, EPI_ISL_859250, EPI_ISL_859251, EPI_ISL_859252, EPI_ISL_859257, EPI_ISL_859258, EPI_ISL_859259, EPI_ISL_859260, EPI_ISL_859261, EPI_ISL_859262, EPI_ISL_859264, EPI_ISL_859266, EPI_ISL_859267, EPI_ISL_859269, EPI_ISL_859271, EPI_ISL_859272, EPI_ISL_859273, EPI_ISL_859274, EPI_ISL_859275, EPI_ISL_859276, EPI_ISL_859277, EPI_ISL_859278, EPI_ISL_859279, EPI_ISL_859281, EPI_ISL_859283, EPI_ISL_859284, EPI_ISL_859285, EPI_ISL_859286, EPI_ISL_859287, EPI_ISL_859289, EPI_ISL_859290, EPI_ISL_859291, EPI_ISL_859292, EPI_ISL_859293, EPI_ISL_859296, EPI_ISL_859298, EPI_ISL_859299, EPI_ISL_859300, EPI_ISL_859302, EPI_ISL_859305, EPI_ISL_859307, EPI_ISL_859308, EPI_ISL_859310, EPI_ISL_859314, EPI_ISL_859315, EPI_ISL_859328, EPI_ISL_859329, EPI_ISL_859330, EPI_ISL_859331, EPI_ISL_859332, EPI_ISL_859333, EPI_ISL_859334, EPI_ISL_859335, EPI_ISL_859336, EPI_ISL_859337, EPI_ISL_859338, EPI_ISL_859339, EPI_ISL_859340, EPI_ISL_859341, EPI_ISL_859342, EPI_ISL_859343, EPI_ISL_859344, EPI_ISL_859345, EPI_ISL_859346, EPI_ISL_859347, EPI_ISL_859348, EPI_ISL_859349, EPI_ISL_859350, EPI_ISL_859351, EPI_ISL_859352, EPI_ISL_859353, EPI_ISL_859354, EPI_ISL_859355, EPI_ISL_859356, EPI_ISL_859357, EPI_ISL_859358, EPI_ISL_859359, EPI_ISL_859360, EPI_ISL_859361, EPI_ISL_859362, EPI_ISL_859363, EPI_ISL_859364, EPI_ISL_859365, EPI_ISL_859366, EPI_ISL_859367, EPI_ISL_859368, EPI_ISL_859369, EPI_ISL_859370, EPI_ISL_859371, EPI_ISL_859372, EPI_ISL_859373, EPI_ISL_859374, EPI_ISL_859375, EPI_ISL_859376, EPI_ISL_859377, EPI_ISL_859378, EPI_ISL_859379, EPI_ISL_859380, EPI_ISL_859381, EPI_ISL_859382, EPI_ISL_859383, EPI_ISL_859384, EPI_ISL_859385, EPI_ISL_859386, EPI_ISL_859387, EPI_ISL_859388, EPI_ISL_859389, EPI_ISL_859390, EPI_ISL_859391, EPI_ISL_859392, EPI_ISL_859393, EPI_ISL_859394, EPI_ISL_859395, EPI_ISL_859396, EPI_ISL_859397, EPI_ISL_859398, EPI_ISL_859399, EPI_ISL_859400, EPI_ISL_859402, EPI_ISL_859403, EPI_ISL_859404, EPI_ISL_859405, EPI_ISL_859406, EPI_ISL_859407, EPI_ISL_859408, EPI_ISL_859409, EPI_ISL_859410, EPI_ISL_859411, EPI_ISL_859412, EPI_ISL_859413, EPI_ISL_859414, EPI_ISL_859415, EPI_ISL_859416, EPI_ISL_859417, EPI_ISL_859418, EPI_ISL_859419, EPI_ISL_859420, EPI_ISL_859421, EPI_ISL_859423, EPI_ISL_859424, EPI_ISL_859425, EPI_ISL_859427, EPI_ISL_859428, EPI_ISL_859429, EPI_ISL_859430, EPI_ISL_859431, EPI_ISL_859432, EPI_ISL_859433, EPI_ISL_859434, EPI_ISL_859436, EPI_ISL_859437, EPI_ISL_859438, EPI_ISL_859439, EPI_ISL_859440, EPI_ISL_859441, EPI_ISL_859442, EPI_ISL_859443, EPI_ISL_859444, EPI_ISL_859445, EPI_ISL_859446, EPI_ISL_859447, EPI_ISL_859448, EPI_ISL_859449, EPI_ISL_859450, EPI_ISL_859451, EPI_ISL_859452, EPI_ISL_859453, EPI_ISL_859454, EPI_ISL_859455, EPI_ISL_892213, EPI_ISL_945088, EPI_ISL_945099, EPI_ISL_945113, EPI_ISL_945117, EPI_ISL_945160, EPI_ISL_945201, EPI_ISL_945204, EPI_ISL_945252, EPI_ISL_945282, EPI_ISL_945306, EPI_ISL_945310, EPI_ISL_945323, EPI_ISL_945343, EPI_ISL_945361

| | | | |
|---|---|---|---|
| see above | Lighthouse Lab in Alderley Park | Wellcome Sanger Institute for the COVID-19 Genomics UK (COG-UK) Consortium | Cordelia Langford; David K. Jackson; Dominic Kwiatkowski; Ewan Harrison; Ian Johnston; Jacquelyn Wynn; John Sillitoe on behalf of the Wellcome Sanger Institute COVID-19 Surveillance Team; Mairead Hyland; Roberto Amato; Sonia Goncalves; The Lighthouse Lab in Alderley Park and Alex Alderton |

EPI_ISL_756379, EPI_ISL_756380, EPI_ISL_756384, EPI_ISL_756386, EPI_ISL_756389, EPI_ISL_756392, EPI_ISL_756393, EPI_ISL_756395, EPI_ISL_756403, EPI_ISL_756409, EPI_ISL_756413, EPI_ISL_756415, EPI_ISL_756416, EPI_ISL_756417, EPI_ISL_756418, EPI_ISL_756419, EPI_ISL_756420, EPI_ISL_756422, EPI_ISL_756424, EPI_ISL_756426, EPI_ISL_756429, EPI_ISL_756431, EPI_ISL_756432, EPI_ISL_756434, EPI_ISL_756438, EPI_ISL_756442, EPI_ISL_756444, EPI_ISL_756446, EPI_ISL_756448, EPI_ISL_756453, EPI_ISL_756454, EPI_ISL_756456, EPI_ISL_756460, EPI_ISL_756467, EPI_ISL_756469, EPI_ISL_756474, EPI_ISL_756475, EPI_ISL_756478, EPI_ISL_756480, EPI_ISL_756481, EPI_ISL_756484, EPI_ISL_756485, EPI_ISL_756486, EPI_ISL_756487, EPI_ISL_756490, EPI_ISL_756491, EPI_ISL_756492, EPI_ISL_756496, EPI_ISL_756497, EPI_ISL_756498, EPI_ISL_756499, EPI_ISL_756500, EPI_ISL_756501, EPI_ISL_756502, EPI_ISL_756505, EPI_ISL_756506, EPI_ISL_756507, EPI_ISL_756508, EPI_ISL_756511, EPI_ISL_756512, EPI_ISL_756513, EPI_ISL_756517, EPI_ISL_756518, EPI_ISL_756519, EPI_ISL_756521, EPI_ISL_756522, EPI_ISL_756525, EPI_ISL_756528, EPI_ISL_756529, EPI_ISL_756534, EPI_ISL_756535, EPI_ISL_756540, EPI_ISL_756542, EPI_ISL_756543, EPI_ISL_756545, EPI_ISL_756549, EPI_ISL_756553, EPI_ISL_756554, EPI_ISL_756558, EPI_ISL_756559, EPI_ISL_756564, EPI_ISL_756567, EPI_ISL_756568, EPI_ISL_756570, EPI_ISL_756572, EPI_ISL_756576, EPI_ISL_756577, EPI_ISL_756579, EPI_ISL_756582, EPI_ISL_756584, EPI_ISL_756585, EPI_ISL_756586, EPI_ISL_756588, EPI_ISL_756589, EPI_ISL_756590, EPI_ISL_756591, EPI_ISL_756594, EPI_ISL_756595, EPI_ISL_756597, EPI_ISL_756601, EPI_ISL_756604, EPI_ISL_756605, EPI_ISL_756606, EPI_ISL_756607, EPI_ISL_756608, EPI_ISL_756610, EPI_ISL_756611, EPI_ISL_756613, EPI_ISL_756614, EPI_ISL_756615, EPI_ISL_756617, EPI_ISL_756621, EPI_ISL_756622, EPI_ISL_756623, EPI_ISL_756625, EPI_ISL_756627, EPI_ISL_756628, EPI_ISL_756629, EPI_ISL_756630, EPI_ISL_756631, EPI_ISL_756633, EPI_ISL_756637, EPI_ISL_756642, EPI_ISL_756643, EPI_ISL_756649, EPI_ISL_756653, EPI_ISL_756655, EPI_ISL_756659, EPI_ISL_756660, EPI_ISL_756665, EPI_ISL_756666, EPI_ISL_756667, EPI_ISL_756668, EPI_ISL_756671, EPI_ISL_756672, EPI_ISL_756674, EPI_ISL_756675, EPI_ISL_756676, EPI_ISL_756677, EPI_ISL_756678, EPI_ISL_756679, EPI_ISL_756680, EPI_ISL_760883, EPI_ISL_761922, EPI_ISL_761927, EPI_ISL_761929, EPI_ISL_761935, EPI_ISL_761937, EPI_ISL_761939, EPI_ISL_761946, EPI_ISL_761952, EPI_ISL_761953, EPI_ISL_761955, EPI_ISL_761956, EPI_ISL_761961, EPI_ISL_761967, EPI_ISL_761968, EPI_ISL_761970, EPI_ISL_761979, EPI_ISL_761981, EPI_ISL_761994, EPI_ISL_761995, EPI_ISL_761997, EPI_ISL_761999, EPI_ISL_762007, EPI_ISL_762008, EPI_ISL_762009, EPI_ISL_762014, EPI_ISL_762015, EPI_ISL_762016, EPI_ISL_762018, EPI_ISL_762019, EPI_ISL_762023, EPI_ISL_762024, EPI_ISL_762025, EPI_ISL_762026, EPI_ISL_762027, EPI_ISL_762028, EPI_ISL_762029, EPI_ISL_762030, EPI_ISL_762036, EPI_ISL_762037, EPI_ISL_762038, EPI_ISL_762041, EPI_ISL_762042, EPI_ISL_762043, EPI_ISL_762045, EPI_ISL_762050, EPI_ISL_762057, EPI_ISL_762059, EPI_ISL_762060, EPI_ISL_762061, EPI_ISL_762063, EPI_ISL_762064, EPI_ISL_762065, EPI_ISL_762066, EPI_ISL_762067, EPI_ISL_762069, EPI_ISL_762072, EPI_ISL_762073, EPI_ISL_762074, EPI_ISL_762075, EPI_ISL_762078, EPI_ISL_762085, EPI_ISL_762087, EPI_ISL_762089, EPI_ISL_762090, EPI_ISL_762091, EPI_ISL_762096, EPI_ISL_762097, EPI_ISL_762098, EPI_ISL_762103, EPI_ISL_762109, EPI_ISL_762112, EPI_ISL_762113, EPI_ISL_762114, EPI_ISL_762117, EPI_ISL_762118, EPI_ISL_762122, EPI_ISL_762124, EPI_ISL_762125, EPI_ISL_762126, EPI_ISL_762131, EPI_ISL_762133, EPI_ISL_762135, EPI_ISL_762136, EPI_ISL_762137, EPI_ISL_762138, EPI_ISL_762139, EPI_ISL_762140, EPI_ISL_762142, EPI_ISL_762146, EPI_ISL_762147, EPI_ISL_762148, EPI_ISL_762152, EPI_ISL_762153, EPI_ISL_762155, EPI_ISL_762163, EPI_ISL_762166, EPI_ISL_762173, EPI_ISL_762175, EPI_ISL_762178, EPI_ISL_762181, EPI_ISL_762183, EPI_ISL_762186, EPI_ISL_762187, EPI_ISL_762189, EPI_ISL_762192, EPI_ISL_762193, EPI_ISL_762195, EPI_ISL_762546, EPI_ISL_762549, EPI_ISL_762550, EPI_ISL_762551, EPI_ISL_762552, EPI_ISL_762553, EPI_ISL_762559, EPI_ISL_762563, EPI_ISL_762565, EPI_ISL_762566, EPI_ISL_762568, EPI_ISL_762569, EPI_ISL_762570, EPI_ISL_762571, EPI_ISL_762572, EPI_ISL_762575, EPI_ISL_762577, EPI_ISL_762578, EPI_ISL_762579, EPI_ISL_762580, EPI_ISL_762582, EPI_ISL_762584, EPI_ISL_762585, EPI_ISL_762587, EPI_ISL_762590, EPI_ISL_762591, EPI_ISL_762592, EPI_ISL_762594, EPI_ISL_762595, EPI_ISL_762596, EPI_ISL_762598, EPI_ISL_762600, EPI_ISL_762601, EPI_ISL_762603, EPI_ISL_762607, EPI_ISL_762608, EPI_ISL_762611, EPI_ISL_762612, EPI_ISL_762613, EPI_ISL_762615, EPI_ISL_762616, EPI_ISL_762619, EPI_ISL_762621, EPI_ISL_762623, EPI_ISL_762624, EPI_ISL_762626, EPI_ISL_762627, EPI_ISL_762628, EPI_ISL_762632, EPI_ISL_762635, EPI_ISL_762636, EPI_ISL_762637, EPI_ISL_762640, EPI_ISL_762641, EPI_ISL_762645, EPI_ISL_762649, EPI_ISL_762650, EPI_ISL_762651, EPI_ISL_762658, EPI_ISL_762660, EPI_ISL_762664, EPI_ISL_762665, EPI_ISL_762667, EPI_ISL_762668, EPI_ISL_762670, EPI_ISL_762673, EPI_ISL_762675, EPI_ISL_762677, EPI_ISL_762679, EPI_ISL_762680, EPI_ISL_762681, EPI_ISL_762682, EPI_ISL_762684, EPI_ISL_762685, EPI_ISL_762687, EPI_ISL_762688, EPI_ISL_762689, EPI_ISL_762695, EPI_ISL_762696, EPI_ISL_762697, EPI_ISL_762699, EPI_ISL_762700, EPI_ISL_762705, EPI_ISL_762713, EPI_ISL_762715, EPI_ISL_762716, EPI_ISL_762717, EPI_ISL_762718, EPI_ISL_762719, EPI_ISL_762721, EPI_ISL_762723, EPI_ISL_762725, EPI_ISL_762727, EPI_ISL_762729, EPI_ISL_762736, EPI_ISL_762737, EPI_ISL_762738, EPI_ISL_762740, EPI_ISL_762741, EPI_ISL_762742, EPI_ISL_762745, EPI_ISL_762747, EPI_ISL_762748, EPI_ISL_762751, EPI_ISL_762752, EPI_ISL_762753, EPI_ISL_762755, EPI_ISL_762757, EPI_ISL_762762, EPI_ISL_762765, EPI_ISL_762767, EPI_ISL_762768, EPI_ISL_762769, EPI_ISL_762772, EPI_ISL_762774, EPI_ISL_762778, EPI_ISL_762780, EPI_ISL_762781, EPI_ISL_762782, EPI_ISL_762783, EPI_ISL_762784, EPI_ISL_762787, EPI_ISL_762788, EPI_ISL_762790, EPI_ISL_762792, EPI_ISL_762795, EPI_ISL_762797, EPI_ISL_762800, EPI_ISL_762803, EPI_ISL_762804, EPI_ISL_762805, EPI_ISL_762806, EPI_ISL_762807, EPI_ISL_762808, EPI_ISL_762810, EPI_ISL_762811, EPI_ISL_762817, EPI_ISL_762818, EPI_ISL_762820, EPI_ISL_762822, EPI_ISL_762825, EPI_ISL_762827, EPI_ISL_762829, EPI_ISL_762831, EPI_ISL_762833, EPI_ISL_762834, EPI_ISL_762836, EPI_ISL_762838, EPI_ISL_762840, EPI_ISL_762842, EPI_ISL_762848, EPI_ISL_762852, EPI_ISL_762855, EPI_ISL_762857, EPI_ISL_762858, EPI_ISL_767115, EPI_ISL_768845, EPI_ISL_768901, EPI_ISL_768902, EPI_ISL_768903, EPI_ISL_768904, EPI_ISL_768905, EPI_ISL_768906, EPI_ISL_768907, EPI_ISL_768908, EPI_ISL_768909, EPI_ISL_768910, EPI_ISL_768911, EPI_ISL_768912, EPI_ISL_768913, EPI_ISL_768915, EPI_ISL_768916, EPI_ISL_768917, EPI_ISL_768918, EPI_ISL_768919, EPI_ISL_768920, EPI_ISL_768921, EPI_ISL_768922, EPI_ISL_768923, EPI_ISL_768924, EPI_ISL_768925, EPI_ISL_768926, EPI_ISL_768927, EPI_ISL_768928, EPI_ISL_768929, EPI_ISL_768930, EPI_ISL_768931, EPI_ISL_768932, EPI_ISL_768933, EPI_ISL_768934, EPI_ISL_768935, EPI_ISL_768936, EPI_ISL_768937, EPI_ISL_768938, EPI_ISL_768939, EPI_ISL_768940, EPI_ISL_768941, EPI_ISL_768942, EPI_ISL_768943, EPI_ISL_768944, EPI_ISL_768945, EPI_ISL_768946, EPI_ISL_768947, EPI_ISL_768948, EPI_ISL_768949, EPI_ISL_768950, EPI_ISL_768951, EPI_ISL_768952, EPI_ISL_768953, EPI_ISL_768954, EPI_ISL_768955, EPI_ISL_768956, EPI_ISL_768957, EPI_ISL_768958, EPI_ISL_768959, EPI_ISL_768960, EPI_ISL_768961, EPI_ISL_768962, EPI_ISL_768963, EPI_ISL_768964, EPI_ISL_768965, EPI_ISL_768966, EPI_ISL_768967, EPI_ISL_768968, EPI_ISL_768969, EPI_ISL_768970, EPI_ISL_768971, EPI_ISL_768972, EPI_ISL_768973, EPI_ISL_768974, EPI_ISL_768975, EPI_ISL_768976, EPI_ISL_768977, EPI_ISL_768978, EPI_ISL_768979, EPI_ISL_768980, EPI_ISL_768981, EPI_ISL_768982, EPI_ISL_768983, EPI_ISL_768984, EPI_ISL_768985, EPI_ISL_768986, EPI_ISL_768987, EPI_ISL_768988, EPI_ISL_768989, EPI_ISL_768990, EPI_ISL_768991, EPI_ISL_768992, EPI_ISL_768993, EPI_ISL_768994, EPI_ISL_768995, EPI_ISL_768996, EPI_ISL_768997, EPI_ISL_768998, EPI_ISL_768999, EPI_ISL_769000, EPI_ISL_769002, EPI_ISL_769003, EPI_ISL_769004, EPI_ISL_769005, EPI_ISL_769006, EPI_ISL_769007, EPI_ISL_769008, EPI_ISL_769009, EPI_ISL_769011, EPI_ISL_769012, EPI_ISL_769013, EPI_ISL_769014, EPI_ISL_769015, EPI_ISL_769016, EPI_ISL_769017, EPI_ISL_769018, EPI_ISL_769019, EPI_ISL_769020, EPI_ISL_769021, EPI_ISL_769022, EPI_ISL_769023, EPI_ISL_769024, EPI_ISL_769025, EPI_ISL_769026, EPI_ISL_769027, EPI_ISL_769028, EPI_ISL_769029, EPI_ISL_769030, EPI_ISL_769031, EPI_ISL_769032, EPI_ISL_769033, EPI_ISL_769034, EPI_ISL_769035, EPI_ISL_769036, EPI_ISL_769037, EPI_ISL_769038, EPI_ISL_769039, EPI_ISL_769040, EPI_ISL_769041, EPI_ISL_769042, EPI_ISL_769043, EPI_ISL_769044, EPI_ISL_769045, EPI_ISL_769046, EPI_ISL_769047, EPI_ISL_769048, EPI_ISL_769049, EPI_ISL_769050, EPI_ISL_769051, EPI_ISL_769052, EPI_ISL_769053, EPI_ISL_769054, EPI_ISL_769055, EPI_ISL_769056, EPI_ISL_769057, EPI_ISL_769058, EPI_ISL_769059, EPI_ISL_769060, EPI_ISL_769061, EPI_ISL_769062, EPI_ISL_769063, EPI_ISL_769064, EPI_ISL_769065, EPI_ISL_769066, EPI_ISL_769067, EPI_ISL_769068, EPI_ISL_769069, EPI_ISL_769070, EPI_ISL_769071, EPI_ISL_769072, EPI_ISL_769073, EPI_ISL_769074, EPI_ISL_769075, EPI_ISL_769076, EPI_ISL_769077, EPI_ISL_769078, EPI_ISL_769079, EPI_ISL_769080, EPI_ISL_769081, EPI_ISL_769082, EPI_ISL_769083, EPI_ISL_769084, EPI_ISL_769085, EPI_ISL_769086, EPI_ISL_769087, EPI_ISL_769088, EPI_ISL_769089, EPI_ISL_769090, EPI_ISL_769091, EPI_ISL_769092, EPI_ISL_769093, EPI_ISL_769094, EPI_ISL_769095, EPI_ISL_769096, EPI_ISL_769097, EPI_ISL_769098, EPI_ISL_769099, EPI_ISL_769100, EPI_ISL_769101, EPI_ISL_769102, EPI_ISL_769103, EPI_ISL_769104, EPI_ISL_769105, EPI_ISL_769106, EPI_ISL_769108, EPI_ISL_769109, EPI_ISL_769110, EPI_ISL_769111, EPI_ISL_769112, EPI_ISL_769113, EPI_ISL_769114, EPI_ISL_769115, EPI_ISL_769116, EPI_ISL_769117, EPI_ISL_769118, EPI_ISL_769119, EPI_ISL_769120, EPI_ISL_769121, EPI_ISL_769122, EPI_ISL_769123, EPI_ISL_769124, EPI_ISL_769125, EPI_ISL_769126, EPI_ISL_769127, EPI_ISL_769128, EPI_ISL_769129, EPI_ISL_769130, EPI_ISL_769131, EPI_ISL_769132, EPI_ISL_769133, EPI_ISL_769134, EPI_ISL_769135, EPI_ISL_769136, EPI_ISL_769137, EPI_ISL_769138, EPI_ISL_769139, EPI_ISL_769140, EPI_ISL_769141, EPI_ISL_769142, EPI_ISL_769143, EPI_ISL_769144, EPI_ISL_769145, EPI_ISL_769146, EPI_ISL_769147, EPI_ISL_769148, EPI_ISL_769149, EPI_ISL_769150, EPI_ISL_769151, EPI_ISL_769152, EPI_ISL_769153, EPI_ISL_769154, EPI_ISL_769155, EPI_ISL_769156, EPI_ISL_769157, EPI_ISL_769158, EPI_ISL_769159, EPI_ISL_769160, EPI_ISL_769161, EPI_ISL_769162, EPI_ISL_769163, EPI_ISL_769164, EPI_ISL_769165, EPI_ISL_769166, EPI_ISL_769167, EPI_ISL_769168, EPI_ISL_769169, EPI_ISL_769170, EPI_ISL_769171, EPI_ISL_769172, EPI_ISL_769173, EPI_ISL_769174, EPI_ISL_769175, EPI_ISL_769176, EPI_ISL_769177, EPI_ISL_769178, EPI_ISL_769179, EPI_ISL_769180, EPI_ISL_769181, EPI_ISL_769182, EPI_ISL_769183, EPI_ISL_769184, EPI_ISL_769185, EPI_ISL_769186, EPI_ISL_769187, EPI_ISL_769188, EPI_ISL_769189, EPI_ISL_769190, EPI_ISL_769191, EPI_ISL_769192, EPI_ISL_769193, EPI_ISL_769194, EPI_ISL_769195, EPI_ISL_769196, EPI_ISL_769197, EPI_ISL_769198, EPI_ISL_769199, EPI_ISL_769200, EPI_ISL_769201, EPI_ISL_769202, EPI_ISL_769203, EPI_ISL_769204, EPI_ISL_769205, EPI_ISL_769206, EPI_ISL_769207, EPI_ISL_769208, EPI_ISL_769209, EPI_ISL_769210, EPI_ISL_769211, EPI_ISL_769212, EPI_ISL_769213, EPI_ISL_769214, EPI_ISL_769215, EPI_ISL_769216, EPI_ISL_769217, EPI_ISL_769218, EPI_ISL_769219, EPI_ISL_769220, EPI_ISL_769221, EPI_ISL_769222, EPI_ISL_777328, EPI_ISL_777329, EPI_ISL_777330, EPI_ISL_777331, EPI_ISL_777332, EPI_ISL_777334, EPI_ISL_777339, EPI_ISL_777342, EPI_ISL_777343, EPI_ISL_777344, EPI_ISL_777345, EPI_ISL_777347, EPI_ISL_777348, EPI_ISL_777350, EPI_ISL_777351, EPI_ISL_777352, EPI_ISL_777354, EPI_ISL_777355, EPI_ISL_777357, EPI_ISL_777359, EPI_ISL_777360, EPI_ISL_777362, EPI_ISL_777365, EPI_ISL_777366, EPI_ISL_777367, EPI_ISL_777368, EPI_ISL_777370, EPI_ISL_777371, EPI_ISL_777374, EPI_ISL_777378, EPI_ISL_777379, EPI_ISL_777381, EPI_ISL_777384, EPI_ISL_777387, EPI_ISL_777388, EPI_ISL_777390, EPI_ISL_777391, EPI_ISL_777396, EPI_ISL_777398, EPI_ISL_777400, EPI_ISL_777401, EPI_ISL_777403, EPI_ISL_777404, EPI_ISL_777405, EPI_ISL_777406, EPI_ISL_777408, EPI_ISL_777409, EPI_ISL_777413, EPI_ISL_777414, EPI_ISL_777415, EPI_ISL_777418, EPI_ISL_777419, EPI_ISL_777422, EPI_ISL_777426, EPI_ISL_777428, EPI_ISL_777429, EPI_ISL_777431, EPI_ISL_777433, EPI_ISL_777434, EPI_ISL_777439, EPI_ISL_777441, EPI_ISL_777442, EPI_ISL_777445, EPI_ISL_777446, EPI_ISL_777450, EPI_ISL_777452, EPI_ISL_777453, EPI_ISL_777454, EPI_ISL_777456, EPI_ISL_777457, EPI_ISL_777460, EPI_ISL_777463, EPI_ISL_777464, EPI_ISL_777468, EPI_ISL_777469, EPI_ISL_777472, EPI_ISL_777473, EPI_ISL_777476, EPI_ISL_777478, EPI_ISL_777480, EPI_ISL_777482, EPI_ISL_777483, EPI_ISL_777485, EPI_ISL_777486, EPI_ISL_777488, EPI_ISL_777492, EPI_ISL_777494, EPI_ISL_777499, EPI_ISL_777500, EPI_ISL_777501, EPI_ISL_777503, EPI_ISL_777507, EPI_ISL_777508, EPI_ISL_777511, EPI_ISL_777512, EPI_ISL_777513, EPI_ISL_777516, EPI_ISL_777517, EPI_ISL_777519, EPI_ISL_777521, EPI_ISL_777527, EPI_ISL_777528, EPI_ISL_777536, EPI_ISL_777540, EPI_ISL_777541, EPI_ISL_777542, EPI_ISL_777544, EPI_ISL_777545, EPI_ISL_777546, EPI_ISL_777547, EPI_ISL_777549, EPI_ISL_777551, EPI_ISL_777555, EPI_ISL_777556, EPI_ISL_777559, EPI_ISL_777564, EPI_ISL_777565, EPI_ISL_777566, EPI_ISL_777570, EPI_ISL_777571, EPI_ISL_777575, EPI_ISL_777577, EPI_ISL_777578, EPI_ISL_777581, EPI_ISL_777585, EPI_ISL_777586, EPI_ISL_777588, EPI_ISL_777590, EPI_ISL_777594, EPI_ISL_777596, EPI_ISL_777599, EPI_ISL_777602, EPI_ISL_777603, EPI_ISL_777604, EPI_ISL_777606, EPI_ISL_777607, EPI_ISL_777608, EPI_ISL_777609, EPI_ISL_777611, EPI_ISL_777613, EPI_ISL_777614, EPI_ISL_777615, EPI_ISL_777616, EPI_ISL_777617, EPI_ISL_777618, EPI_ISL_777619, EPI_ISL_777620, EPI_ISL_777624, EPI_ISL_777625, EPI_ISL_777626, EPI_ISL_777631, EPI_ISL_777632, EPI_ISL_777635, EPI_ISL_777636, EPI_ISL_777637, EPI_ISL_777640, EPI_ISL_777642, EPI_ISL_777643, EPI_ISL_777645, EPI_ISL_777647, EPI_ISL_777648, EPI_ISL_777656, EPI_ISL_777661, EPI_ISL_777664, EPI_ISL_777665, EPI_ISL_777666, EPI_ISL_777668, EPI_ISL_777671, EPI_ISL_777673, EPI_ISL_777674, EPI_ISL_777675, EPI_ISL_777676, EPI_ISL_777677, EPI_ISL_777678, EPI_ISL_777679, EPI_ISL_777680, EPI_ISL_777681, EPI_ISL_777682, EPI_ISL_777683, EPI_ISL_777684, EPI_ISL_777685, EPI_ISL_777686, EPI_ISL_777687, EPI_ISL_777688, EPI_ISL_777689, EPI_ISL_777690, EPI_ISL_777691, EPI_ISL_777692, EPI_ISL_777693, EPI_ISL_777694, EPI_ISL_777695, EPI_ISL_777696, EPI_ISL_777697, EPI_ISL_777698, EPI_ISL_777699, EPI_ISL_777700, EPI_ISL_777701, EPI_ISL_777702, EPI_ISL_777703, EPI_ISL_777704, EPI_ISL_777705, EPI_ISL_777706, EPI_ISL_777707, EPI_ISL_777708, EPI_ISL_777709, EPI_ISL_777710, EPI_ISL_777711, EPI_ISL_777712, EPI_ISL_777713, EPI_ISL_777714, EPI_ISL_777715, EPI_ISL_777716, EPI_ISL_777717, EPI_ISL_777718, EPI_ISL_777719, EPI_ISL_777720, EPI_ISL_777721, EPI_ISL_777722, EPI_ISL_777723, EPI_ISL_777724, EPI_ISL_777725, EPI_ISL_777726, EPI_ISL_777727, EPI_ISL_777728, EPI_ISL_777729, EPI_ISL_777730, EPI_ISL_777731, EPI_ISL_777732, EPI_ISL_777733, EPI_ISL_777734, EPI_ISL_777735, EPI_ISL_777736, EPI_ISL_777737, EPI_ISL_777738, EPI_ISL_777739, EPI_ISL_777740, EPI_ISL_777741, EPI_ISL_777742, EPI_ISL_777743, EPI_ISL_777744, EPI_ISL_777745, EPI_ISL_777746, EPI_ISL_777747, EPI_ISL_777748, EPI_ISL_777749, EPI_ISL_777750, EPI_ISL_777751, EPI_ISL_777752, EPI_ISL_777753, EPI_ISL_777754, EPI_ISL_777755, EPI_ISL_777756, EPI_ISL_777757, EPI_ISL_777758, EPI_ISL_777759, EPI_ISL_777760, EPI_ISL_777761, EPI_ISL_777762, EPI_ISL_777763, EPI_ISL_777764, EPI_ISL_777765, EPI_ISL_777766, EPI_ISL_777767, EPI_ISL_777768, EPI_ISL_777769, EPI_ISL_777770, EPI_ISL_777771, EPI_ISL_777772, EPI_ISL_777773, EPI_ISL_777774, EPI_ISL_777775, EPI_ISL_777776, EPI_ISL_777777, EPI_ISL_777778, EPI_ISL_777779, EPI_ISL_777780, EPI_ISL_777781, EPI_ISL_777782, EPI_ISL_777783, EPI_ISL_777784, EPI_ISL_777785, EPI_ISL_777786, EPI_ISL_777787, EPI_ISL_777788, EPI_ISL_777789, EPI_ISL_777790, EPI_ISL_777791, EPI_ISL_777792, EPI_ISL_777793, EPI_ISL_777794, EPI_ISL_777795, EPI_ISL_777796, EPI_ISL_777797, EPI_ISL_777798, EPI_ISL_777799, EPI_ISL_777800, EPI_ISL_777801, EPI_ISL_777802, EPI_ISL_777803, EPI_ISL_777804, EPI_ISL_777805, EPI_ISL_777806, EPI_ISL_777807, EPI_ISL_777808, EPI_ISL_777809, EPI_ISL_777810, EPI_ISL_777811, EPI_ISL_777812, EPI_ISL_777813, EPI_ISL_777814, EPI_ISL_777815, EPI_ISL_777816, EPI_ISL_777817, EPI_ISL_777818, EPI_ISL_777819, EPI_ISL_777820, EPI_ISL_777821, EPI_ISL_777822, EPI_ISL_777823, EPI_ISL_777824, EPI_ISL_777825, EPI_ISL_777826, EPI_ISL_777827, EPI_ISL_777828, EPI_ISL_777829, EPI_ISL_777830, EPI_ISL_777831, EPI_ISL_777832, EPI_ISL_777833, EPI_ISL_777834, EPI_ISL_777835, EPI_ISL_777836, EPI_ISL_777837, EPI_ISL_777838, EPI_ISL_777839, EPI_ISL_777840, EPI_ISL_777841, EPI_ISL_777842, EPI_ISL_777843, EPI_ISL_777844, EPI_ISL_777845, EPI_ISL_777846, EPI_ISL_777847, EPI_ISL_777848, EPI_ISL_777849, EPI_ISL_777850, EPI_ISL_777851, EPI_ISL_777852, EPI_ISL_777853, EPI_ISL_777854, EPI_ISL_777855, EPI_ISL_777856, EPI_ISL_777857, EPI_ISL_777858, EPI_ISL_777859, EPI_ISL_777860, EPI_ISL_777861, EPI_ISL_777862, EPI_ISL_777863, EPI_ISL_777864, EPI_ISL_777865, EPI_ISL_777866, EPI_ISL_777867, EPI_ISL_777868, EPI_ISL_777869, EPI_ISL_777870, EPI_ISL_777871, EPI_ISL_777872, EPI_ISL_777873, EPI_ISL_777874, EPI_ISL_777875, EPI_ISL_777876, EPI_ISL_777877, EPI_ISL_777878, EPI_ISL_777879, EPI_ISL_777880, EPI_ISL_777881, EPI_ISL_777882, EPI_ISL_777883, EPI_ISL_777884, EPI_ISL_777885, EPI_ISL_777886, EPI_ISL_777887, EPI_ISL_777888, EPI_ISL_777889, EPI_ISL_777890, EPI_ISL_777891, EPI_ISL_777892, EPI_ISL_777893, EPI_ISL_777894, EPI_ISL_777895, EPI_ISL_777896, EPI_ISL_777897, EPI_ISL_777898, EPI_ISL_777899, EPI_ISL_777900, EPI_ISL_777901, EPI_ISL_777903, EPI_ISL_777904, EPI_ISL_777905, EPI_ISL_777906, EPI_ISL_777907, EPI_ISL_777908, EPI_ISL_777909, EPI_ISL_777910, EPI_ISL_777911, EPI_ISL_777912, EPI_ISL_777913, EPI_ISL_777914, EPI_ISL_777915, EPI_ISL_777916, EPI_ISL_777917, EPI_ISL_777918, EPI_ISL_777919, EPI_ISL_777920, EPI_ISL_777921, EPI_ISL_777922, EPI_ISL_777923, EPI_ISL_777924, EPI_ISL_777925, EPI_ISL_777926, EPI_ISL_777927, EPI_ISL_777928, EPI_ISL_777929, EPI_ISL_777930, EPI_ISL_777931, EPI_ISL_777932, EPI_ISL_777934, EPI_ISL_777935, EPI_ISL_777936, EPI_ISL_777937, EPI_ISL_777938, EPI_ISL_777939, EPI_ISL_777940, EPI_ISL_777941, EPI_ISL_777942, EPI_ISL_777943, EPI_ISL_777944, EPI_ISL_777946, EPI_ISL_777947, EPI_ISL_777948, EPI_ISL_777949, EPI_ISL_777950, EPI_ISL_777951, EPI_ISL_777952, EPI_ISL_777953, EPI_ISL_777954, EPI_ISL_777955, EPI_ISL_777956, EPI_ISL_777957, EPI_ISL_777958, EPI_ISL_777959, EPI_ISL_777960, EPI_ISL_777961, EPI_ISL_777962, EPI_ISL_777963, EPI_ISL_777964, EPI_ISL_777965, EPI_ISL_777966, EPI_ISL_777967, EPI_ISL_777968, EPI_ISL_777969, EPI_ISL_777970, EPI_ISL_777971, EPI_ISL_777972, EPI_ISL_777973, EPI_ISL_777974, EPI_ISL_777975, EPI_ISL_777976, EPI_ISL_777977, EPI_ISL_777979, EPI_ISL_777980, EPI_ISL_777981, EPI_ISL_777982, EPI_ISL_777983, EPI_ISL_777984, EPI_ISL_777986, EPI_ISL_777987, EPI_ISL_777988, EPI_ISL_777989, EPI_ISL_777990, EPI_ISL_777991, EPI_ISL_777992, EPI_ISL_777993, EPI_ISL_777995, EPI_ISL_777996, EPI_ISL_777997, EPI_ISL_777998, EPI_ISL_777999, EPI_ISL_778000, EPI_ISL_778001, EPI_ISL_778002, EPI_ISL_778005, EPI_ISL_778006, EPI_ISL_778010, EPI_ISL_778011, EPI_ISL_778012, EPI_ISL_778013, EPI_ISL_778015, EPI_ISL_778017, EPI_ISL_778019, EPI_ISL_778020, EPI_ISL_778022, EPI_ISL_778023, EPI_ISL_778025, EPI_ISL_778027, EPI_ISL_778028, EPI_ISL_778030, EPI_ISL_778032, EPI_ISL_778037, EPI_ISL_778038, EPI_ISL_778039, EPI_ISL_778041, EPI_ISL_778045, EPI_ISL_778049, EPI_ISL_778054, EPI_ISL_778056, EPI_ISL_778057, EPI_ISL_778061, EPI_ISL_778063, EPI_ISL_778066, EPI_ISL_778067, EPI_ISL_778071, EPI_ISL_778072, EPI_ISL_778075, EPI_ISL_778078, EPI_ISL_778079, EPI_ISL_778080, EPI_ISL_778081, EPI_ISL_778084, EPI_ISL_778085, EPI_ISL_778086, EPI_ISL_778087, EPI_ISL_778091, EPI_ISL_778094, EPI_ISL_778095, EPI_ISL_778096, EPI_ISL_778097, EPI_ISL_778099, EPI_ISL_778100, EPI_ISL_778103, EPI_ISL_778105, EPI_ISL_778109, EPI_ISL_778110, EPI_ISL_778112, EPI_ISL_778114, EPI_ISL_778115, EPI_ISL_778118, EPI_ISL_778120, EPI_ISL_778123, EPI_ISL_778124, EPI_ISL_778125, EPI_ISL_778126, EPI_ISL_778130, EPI_ISL_778134, EPI_ISL_778135, EPI_ISL_778137, EPI_ISL_778138, EPI_ISL_778139, EPI_ISL_778141, EPI_ISL_778142, EPI_ISL_778146, EPI_ISL_778147, EPI_ISL_778148, EPI_ISL_778150, EPI_ISL_778152, EPI_ISL_778157, EPI_ISL_778158, EPI_ISL_778160, EPI_ISL_778161, EPI_ISL_778169, EPI_ISL_778170, EPI_ISL_778172, EPI_ISL_778173, EPI_ISL_778174, EPI_ISL_778175, EPI_ISL_778177, EPI_ISL_778178, EPI_ISL_778179, EPI_ISL_778180, EPI_ISL_778181, EPI_ISL_778182, EPI_ISL_778185, EPI_ISL_778186, EPI_ISL_778188, EPI_ISL_778189, EPI_ISL_778193, EPI_ISL_778194, EPI_ISL_778197, EPI_ISL_778198, EPI_ISL_778199, EPI_ISL_778200, EPI_ISL_778204, EPI_ISL_778205, EPI_ISL_778206, EPI_ISL_778208, EPI_ISL_778209, EPI_ISL_778210, EPI_ISL_778211, EPI_ISL_778213, EPI_ISL_778214, EPI_ISL_778217, EPI_ISL_778218, EPI_ISL_778220, EPI_ISL_778221, EPI_ISL_778222, EPI_ISL_778224, EPI_ISL_778226, EPI_ISL_778227, EPI_ISL_778231, EPI_ISL_778232, EPI_ISL_778235, EPI_ISL_778237, EPI_ISL_778241, EPI_ISL_778242, EPI_ISL_778243, EPI_ISL_778245, EPI_ISL_778252, EPI_ISL_778253, EPI_ISL_778254, EPI_ISL_778255, EPI_ISL_778257, EPI_ISL_778262, EPI_ISL_778265, EPI_ISL_778267, EPI_ISL_778268, EPI_ISL_778271, EPI_ISL_778273, EPI_ISL_778275, EPI_ISL_778277, EPI_ISL_778278, EPI_ISL_778280, EPI_ISL_778284, EPI_ISL_778285, EPI_ISL_778291, EPI_ISL_778293, EPI_ISL_778294, EPI_ISL_778296, EPI_ISL_781181, EPI_ISL_781185, EPI_ISL_781188, EPI_ISL_781192, EPI_ISL_781199, EPI_ISL_781221, EPI_ISL_781223, EPI_ISL_781224, EPI_ISL_781260, EPI_ISL_781263, EPI_ISL_781279, EPI_ISL_781281, EPI_ISL_781287, EPI_ISL_781299, EPI_ISL_781306, EPI_ISL_781308, EPI_ISL_781327, EPI_ISL_781332, EPI_ISL_781340, EPI_ISL_781342, EPI_ISL_781345, EPI_ISL_781358, EPI_ISL_781364, EPI_ISL_781368, EPI_ISL_781376, EPI_ISL_781377, EPI_ISL_781379, EPI_ISL_781385, EPI_ISL_781387, EPI_ISL_781396, EPI_ISL_781402, EPI_ISL_781403, EPI_ISL_781407, EPI_ISL_781409, EPI_ISL_781413, EPI_ISL_781426, EPI_ISL_781431, EPI_ISL_781432, EPI_ISL_781436, EPI_ISL_781438, EPI_ISL_781440, EPI_ISL_781441, EPI_ISL_781442, EPI_ISL_781466, EPI_ISL_781471, EPI_ISL_781472, EPI_ISL_781480, EPI_ISL_781488, EPI_ISL_781490, EPI_ISL_781494, EPI_ISL_781498, EPI_ISL_781512, EPI_ISL_781513, EPI_ISL_781514, EPI_ISL_781519, EPI_ISL_781523, EPI_ISL_781524, EPI_ISL_781528, EPI_ISL_781532, EPI_ISL_781538, EPI_ISL_781542, EPI_ISL_781545, EPI_ISL_781546, EPI_ISL_781553, EPI_ISL_781557, EPI_ISL_781565, EPI_ISL_781566, EPI_ISL_781581, EPI_ISL_781587, EPI_ISL_781590, EPI_ISL_781591, EPI_ISL_781593, EPI_ISL_781595, EPI_ISL_781603, EPI_ISL_781606, EPI_ISL_781610, EPI_ISL_781620, EPI_ISL_781622, EPI_ISL_781637, EPI_ISL_781638, EPI_ISL_781642, EPI_ISL_781645, EPI_ISL_781654, EPI_ISL_781656, EPI_ISL_781666, EPI_ISL_781678, EPI_ISL_781685, EPI_ISL_781691, EPI_ISL_781698, EPI_ISL_781703, EPI_ISL_781705, EPI_ISL_781706, EPI_ISL_781707, EPI_ISL_781711, EPI_ISL_781712, EPI_ISL_781713, EPI_ISL_781715, EPI_ISL_781716, EPI_ISL_781728, EPI_ISL_781729, EPI_ISL_781734, EPI_ISL_781736, EPI_ISL_781741, EPI_ISL_781745, EPI_ISL_781758, EPI_ISL_781759, EPI_ISL_781760, EPI_ISL_781762, EPI_ISL_781764, EPI_ISL_781766, EPI_ISL_781771, EPI_ISL_781772, EPI_ISL_781780, EPI_ISL_781794, EPI_ISL_781797, EPI_ISL_781805, EPI_ISL_781808, EPI_ISL_781809, EPI_ISL_781815, EPI_ISL_781818, EPI_ISL_781822, EPI_ISL_781828, EPI_ISL_781838, EPI_ISL_781842, EPI_ISL_781843, EPI_ISL_781844, EPI_ISL_781846, EPI_ISL_781848, EPI_ISL_781867, EPI_ISL_945054, EPI_ISL_945085, EPI_ISL_945089, EPI_ISL_945102, EPI_ISL_945109, EPI_ISL_945170, EPI_ISL_945188, EPI_ISL_945202, EPI_ISL_945247, EPI_ISL_945249, EPI_ISL_945305, EPI_ISL_945320, EPI_ISL_945350, EPI_ISL_945367

| | | | |
|---|---|---|---|
| see above | Lighthouse Lab in Cambridge | Wellcome Sanger Institute for the COVID-19 Genomics UK (COG-UK) Consortium | Cordelia Langford; David K. Jackson; Dominic Kwiatkowski; Ewan Harrison; Ian Johnston; John Sillitoe on behalf of the Wellcome Sanger Institute COVID-19 Surveillance Team; Rob Howes; Roberto Amato; Sonia Goncalves; The Lighthouse Lab in Cambridge and Alex Alderton |

EPI_ISL_731347, EPI_ISL_756690, EPI_ISL_756695, EPI_ISL_756696, EPI_ISL_756702, EPI_ISL_756705, EPI_ISL_756706, EPI_ISL_756709, EPI_ISL_756711, EPI_ISL_756716, EPI_ISL_756723, EPI_ISL_756729, EPI_ISL_756737, EPI_ISL_756744, EPI_ISL_756745, EPI_ISL_756748, EPI_ISL_756750, EPI_ISL_756761, EPI_ISL_756773, EPI_ISL_756775, EPI_ISL_756780, EPI_ISL_756784, EPI_ISL_756789, EPI_ISL_756795, EPI_ISL_756800, EPI_ISL_756803, EPI_ISL_756805, EPI_ISL_756824, EPI_ISL_756835, EPI_ISL_756840, EPI_ISL_756849, EPI_ISL_756850, EPI_ISL_756852, EPI_ISL_756861, EPI_ISL_756867, EPI_ISL_756877, EPI_ISL_756878, EPI_ISL_756879, EPI_ISL_756881, EPI_ISL_756885, EPI_ISL_756886, EPI_ISL_756888, EPI_ISL_756889, EPI_ISL_756893, EPI_ISL_756894, EPI_ISL_756895, EPI_ISL_756897, EPI_ISL_756900, EPI_ISL_756909, EPI_ISL_756910, EPI_ISL_756912, EPI_ISL_756918, EPI_ISL_756920, EPI_ISL_756923, EPI_ISL_756928, EPI_ISL_756931, EPI_ISL_756935, EPI_ISL_756940, EPI_ISL_756947, EPI_ISL_756951, EPI_ISL_756952, EPI_ISL_756955, EPI_ISL_756959, EPI_ISL_756969, EPI_ISL_756970, EPI_ISL_756971, EPI_ISL_756973, EPI_ISL_756977, EPI_ISL_756982, EPI_ISL_756998, EPI_ISL_757005, EPI_ISL_757007, EPI_ISL_757008, EPI_ISL_757009, EPI_ISL_757011, EPI_ISL_757012, EPI_ISL_757017, EPI_ISL_757018, EPI_ISL_757021, EPI_ISL_757027, EPI_ISL_757028, EPI_ISL_757032, EPI_ISL_760277, EPI_ISL_760280, EPI_ISL_760282, EPI_ISL_760287, EPI_ISL_760303, EPI_ISL_760313, EPI_ISL_760315, EPI_ISL_760317, EPI_ISL_760365, EPI_ISL_760374, EPI_ISL_760377, EPI_ISL_760380, EPI_ISL_760414, EPI_ISL_760433, EPI_ISL_760446, EPI_ISL_760511, EPI_ISL_760542, EPI_ISL_760560, EPI_ISL_760598, EPI_ISL_760638, EPI_ISL_760657, EPI_ISL_760658, EPI_ISL_760669, EPI_ISL_760673, EPI_ISL_760686, EPI_ISL_760692, EPI_ISL_760720, EPI_ISL_760739, EPI_ISL_760740, EPI_ISL_760741, EPI_ISL_760750, EPI_ISL_760782, EPI_ISL_760789, EPI_ISL_760796, EPI_ISL_760811, EPI_ISL_760814, EPI_ISL_760815, EPI_ISL_760816, EPI_ISL_760818, EPI_ISL_760819, EPI_ISL_760820, EPI_ISL_760821, EPI_ISL_760823, EPI_ISL_760824, EPI_ISL_760827, EPI_ISL_760828, EPI_ISL_760829, EPI_ISL_760830, EPI_ISL_760831, EPI_ISL_760834, EPI_ISL_760835, EPI_ISL_760836, EPI_ISL_760837, EPI_ISL_760840, EPI_ISL_760841, EPI_ISL_760843, EPI_ISL_760845, EPI_ISL_760848, EPI_ISL_760849, EPI_ISL_760850, EPI_ISL_760851, EPI_ISL_760854, EPI_ISL_760855, EPI_ISL_760856, EPI_ISL_760869, EPI_ISL_760870, EPI_ISL_760877, EPI_ISL_760879, EPI_ISL_760880, EPI_ISL_760882, EPI_ISL_760885, EPI_ISL_760886, EPI_ISL_760893, EPI_ISL_760895, EPI_ISL_760899, EPI_ISL_760901, EPI_ISL_760902, EPI_ISL_760907, EPI_ISL_760909, EPI_ISL_760910, EPI_ISL_760913, EPI_ISL_760914, EPI_ISL_760917, EPI_ISL_760919, EPI_ISL_760923, EPI_ISL_760926, EPI_ISL_760927, EPI_ISL_760930, EPI_ISL_760934, EPI_ISL_760935, EPI_ISL_760940, EPI_ISL_760944, EPI_ISL_760946, EPI_ISL_760947, EPI_ISL_760953, EPI_ISL_760955, EPI_ISL_760956, EPI_ISL_760957, EPI_ISL_760958, EPI_ISL_760964, EPI_ISL_760965, EPI_ISL_760966, EPI_ISL_760970, EPI_ISL_760972, EPI_ISL_760973, EPI_ISL_760974, EPI_ISL_760981, EPI_ISL_760985, EPI_ISL_760990, EPI_ISL_760991, EPI_ISL_760992, EPI_ISL_760993, EPI_ISL_760995, EPI_ISL_760996, EPI_ISL_760997, EPI_ISL_760998, EPI_ISL_760999, EPI_ISL_761003, EPI_ISL_761004, EPI_ISL_761005, EPI_ISL_761006, EPI_ISL_761007, EPI_ISL_761009, EPI_ISL_761010, EPI_ISL_761012, EPI_ISL_761014, EPI_ISL_761015, EPI_ISL_761016, EPI_ISL_761019, EPI_ISL_761020, EPI_ISL_761024, EPI_ISL_761026, EPI_ISL_761028, EPI_ISL_761030, EPI_ISL_761031, EPI_ISL_761032, EPI_ISL_761033, EPI_ISL_761034, EPI_ISL_761035, EPI_ISL_761037, EPI_ISL_761038, EPI_ISL_761041, EPI_ISL_761044, EPI_ISL_761045, EPI_ISL_761051, EPI_ISL_761053, EPI_ISL_761055, EPI_ISL_761058, EPI_ISL_761062, EPI_ISL_761063, EPI_ISL_761068, EPI_ISL_761070, EPI_ISL_761071, EPI_ISL_761072, EPI_ISL_761073, EPI_ISL_761076, EPI_ISL_761077, EPI_ISL_761078, EPI_ISL_761094, EPI_ISL_761095, EPI_ISL_761113, EPI_ISL_761114, EPI_ISL_761116, EPI_ISL_761118, EPI_ISL_761120, EPI_ISL_761121, EPI_ISL_761125, EPI_ISL_761128, EPI_ISL_761131, EPI_ISL_761133, EPI_ISL_761134, EPI_ISL_761143, EPI_ISL_761145, EPI_ISL_761154, EPI_ISL_761157, EPI_ISL_761158, EPI_ISL_761162, EPI_ISL_761168, EPI_ISL_761172, EPI_ISL_761176, EPI_ISL_761187, EPI_ISL_761191, EPI_ISL_761193, EPI_ISL_761194, EPI_ISL_761197, EPI_ISL_761204, EPI_ISL_761207, EPI_ISL_761209, EPI_ISL_761221, EPI_ISL_761225, EPI_ISL_761227, EPI_ISL_761228, EPI_ISL_761229, EPI_ISL_761239, EPI_ISL_761241, EPI_ISL_761252, EPI_ISL_761255, EPI_ISL_761257, EPI_ISL_761259, EPI_ISL_761266, EPI_ISL_761276, EPI_ISL_761278, EPI_ISL_761280, EPI_ISL_761283, EPI_ISL_761287, EPI_ISL_761289, EPI_ISL_761292, EPI_ISL_761293, EPI_ISL_761296, EPI_ISL_761298, EPI_ISL_761302, EPI_ISL_761303, EPI_ISL_761307, EPI_ISL_761313, EPI_ISL_761326, EPI_ISL_761331, EPI_ISL_761338, EPI_ISL_761339, EPI_ISL_761348, EPI_ISL_761351, EPI_ISL_761353, EPI_ISL_761355, EPI_ISL_761921, EPI_ISL_761923, EPI_ISL_761924, EPI_ISL_761925, EPI_ISL_761926, EPI_ISL_761928, EPI_ISL_761930, EPI_ISL_761931, EPI_ISL_761932, EPI_ISL_761933, EPI_ISL_761934, EPI_ISL_761936, EPI_ISL_761938, EPI_ISL_761940, EPI_ISL_761941, EPI_ISL_761942, EPI_ISL_761943, EPI_ISL_761944, EPI_ISL_761945, EPI_ISL_761947, EPI_ISL_761948, EPI_ISL_761949, EPI_ISL_761950, EPI_ISL_761951, EPI_ISL_761954, EPI_ISL_761957, EPI_ISL_761958, EPI_ISL_761959, EPI_ISL_761960, EPI_ISL_761962, EPI_ISL_761963, EPI_ISL_761964, EPI_ISL_761965, EPI_ISL_761966, EPI_ISL_761969, EPI_ISL_761971, EPI_ISL_761972, EPI_ISL_761973, EPI_ISL_761974, EPI_ISL_761975, EPI_ISL_761976, EPI_ISL_761977, EPI_ISL_761978, EPI_ISL_761980, EPI_ISL_761982, EPI_ISL_761983, EPI_ISL_761984, EPI_ISL_761985, EPI_ISL_761986, EPI_ISL_761987, EPI_ISL_761988, EPI_ISL_761989, EPI_ISL_761990, EPI_ISL_761991, EPI_ISL_761992, EPI_ISL_761993, EPI_ISL_761996, EPI_ISL_761998, EPI_ISL_762000, EPI_ISL_762001, EPI_ISL_762002, EPI_ISL_762003, EPI_ISL_762004, EPI_ISL_762005, EPI_ISL_762006, EPI_ISL_762010, EPI_ISL_762011, EPI_ISL_762012, EPI_ISL_762013, EPI_ISL_762017, EPI_ISL_762020, EPI_ISL_762021, EPI_ISL_762022, EPI_ISL_762031, EPI_ISL_762032, EPI_ISL_762033, EPI_ISL_762034, EPI_ISL_762035, EPI_ISL_762039, EPI_ISL_762040, EPI_ISL_762044, EPI_ISL_762046, EPI_ISL_762047, EPI_ISL_762048, EPI_ISL_762049, EPI_ISL_762051, EPI_ISL_762052, EPI_ISL_762053, EPI_ISL_762054, EPI_ISL_762055, EPI_ISL_762056, EPI_ISL_762058, EPI_ISL_762068, EPI_ISL_762070, EPI_ISL_762071, EPI_ISL_762076, EPI_ISL_762077, EPI_ISL_762079, EPI_ISL_762080, EPI_ISL_762081, EPI_ISL_762082, EPI_ISL_762083, EPI_ISL_762084, EPI_ISL_762086, EPI_ISL_762088, EPI_ISL_762092, EPI_ISL_762093, EPI_ISL_762094, EPI_ISL_762095, EPI_ISL_762099, EPI_ISL_762100, EPI_ISL_762101, EPI_ISL_762102, EPI_ISL_762104, EPI_ISL_762105, EPI_ISL_762106, EPI_ISL_762107, EPI_ISL_762108, EPI_ISL_762110, EPI_ISL_762111, EPI_ISL_762115, EPI_ISL_762116, EPI_ISL_762119, EPI_ISL_762120, EPI_ISL_762121, EPI_ISL_762123, EPI_ISL_762127, EPI_ISL_762128, EPI_ISL_762129, EPI_ISL_762130, EPI_ISL_762132, EPI_ISL_762134, EPI_ISL_762141, EPI_ISL_762143, EPI_ISL_762144, EPI_ISL_762145, EPI_ISL_762149, EPI_ISL_762150, EPI_ISL_762151, EPI_ISL_762154, EPI_ISL_762157, EPI_ISL_762158, EPI_ISL_762159, EPI_ISL_762160, EPI_ISL_762161, EPI_ISL_762162, EPI_ISL_762164, EPI_ISL_762165, EPI_ISL_762167, EPI_ISL_762168, EPI_ISL_762169, EPI_ISL_762170, EPI_ISL_762171, EPI_ISL_762172, EPI_ISL_762174, EPI_ISL_762176, EPI_ISL_762177, EPI_ISL_762179, EPI_ISL_762180, EPI_ISL_762182, EPI_ISL_762184, EPI_ISL_762185, EPI_ISL_762188, EPI_ISL_762190, EPI_ISL_762191, EPI_ISL_762194, EPI_ISL_762196, EPI_ISL_762197, EPI_ISL_762548, EPI_ISL_762555, EPI_ISL_762556, EPI_ISL_762560, EPI_ISL_762561, EPI_ISL_762567, EPI_ISL_762573, EPI_ISL_762574, EPI_ISL_762576, EPI_ISL_762583, EPI_ISL_762586, EPI_ISL_762589, EPI_ISL_762597, EPI_ISL_762602, EPI_ISL_762604, EPI_ISL_762609, EPI_ISL_762614, EPI_ISL_762618, EPI_ISL_762622, EPI_ISL_762630, EPI_ISL_762631, EPI_ISL_762633, EPI_ISL_762639, EPI_ISL_762644, EPI_ISL_762646, EPI_ISL_762648, EPI_ISL_762652, EPI_ISL_762653, EPI_ISL_762654,

EPI_ISL_762656, EPI_ISL_762657, EPI_ISL_762661, EPI_ISL_762662, EPI_ISL_762663, EPI_ISL_762669, EPI_ISL_762676, EPI_ISL_762678, EPI_ISL_762690, EPI_ISL_762694, EPI_ISL_762698, EPI_ISL_762701, EPI_ISL_762704, EPI_ISL_762708, EPI_ISL_762711, EPI_ISL_762726, EPI_ISL_762728, EPI_ISL_762731, EPI_ISL_762732, EPI_ISL_762734, EPI_ISL_762739, EPI_ISL_762743, EPI_ISL_762744, EPI_ISL_762746, EPI_ISL_762749, EPI_ISL_762758, EPI_ISL_762759, EPI_ISL_762760, EPI_ISL_762761, EPI_ISL_762766, EPI_ISL_762770, EPI_ISL_762773, EPI_ISL_762775, EPI_ISL_762776, EPI_ISL_762779, EPI_ISL_762785, EPI_ISL_762794, EPI_ISL_762801, EPI_ISL_762802, EPI_ISL_762809, EPI_ISL_762813, EPI_ISL_762814, EPI_ISL_762816, EPI_ISL_762824, EPI_ISL_762826, EPI_ISL_762837, EPI_ISL_762846, EPI_ISL_762850, EPI_ISL_762851, EPI_ISL_762853, EPI_ISL_762854, EPI_ISL_762856, EPI_ISL_762859, EPI_ISL_762860, EPI_ISL_762861, EPI_ISL_762863, EPI_ISL_762865, EPI_ISL_762866, EPI_ISL_762868, EPI_ISL_762870, EPI_ISL_762871, EPI_ISL_762872, EPI_ISL_762873, EPI_ISL_762874, EPI_ISL_762876, EPI_ISL_762877, EPI_ISL_762881, EPI_ISL_762882, EPI_ISL_762885, EPI_ISL_762888, EPI_ISL_762889, EPI_ISL_762892, EPI_ISL_762893, EPI_ISL_762897, EPI_ISL_762898, EPI_ISL_762899, EPI_ISL_762904, EPI_ISL_762905, EPI_ISL_762907, EPI_ISL_762909, EPI_ISL_762915, EPI_ISL_762918, EPI_ISL_762923, EPI_ISL_762924, EPI_ISL_762925, EPI_ISL_762927, EPI_ISL_762928, EPI_ISL_762929, EPI_ISL_762930, EPI_ISL_762934, EPI_ISL_762936, EPI_ISL_762940, EPI_ISL_762941, EPI_ISL_762942, EPI_ISL_762944, EPI_ISL_762946, EPI_ISL_762948, EPI_ISL_762950, EPI_ISL_762953, EPI_ISL_762954, EPI_ISL_762956, EPI_ISL_762958, EPI_ISL_762959, EPI_ISL_762960, EPI_ISL_762961, EPI_ISL_762962, EPI_ISL_762966, EPI_ISL_762969, EPI_ISL_762972, EPI_ISL_762980, EPI_ISL_762982, EPI_ISL_762983, EPI_ISL_762984, EPI_ISL_762985, EPI_ISL_762986, EPI_ISL_762987, EPI_ISL_767092, EPI_ISL_767113, EPI_ISL_767114, EPI_ISL_767117, EPI_ISL_767118, EPI_ISL_777016, EPI_ISL_777019, EPI_ISL_777021, EPI_ISL_777022, EPI_ISL_777029, EPI_ISL_777031, EPI_ISL_777034, EPI_ISL_777037, EPI_ISL_777038, EPI_ISL_777040, EPI_ISL_777047, EPI_ISL_777055, EPI_ISL_777059, EPI_ISL_777061, EPI_ISL_777062, EPI_ISL_777066, EPI_ISL_777068, EPI_ISL_777073, EPI_ISL_777082, EPI_ISL_777089, EPI_ISL_777095, EPI_ISL_777098, EPI_ISL_777099, EPI_ISL_777100, EPI_ISL_777105, EPI_ISL_777112, EPI_ISL_777115, EPI_ISL_777117, EPI_ISL_777118, EPI_ISL_777120, EPI_ISL_777121, EPI_ISL_777122, EPI_ISL_777124, EPI_ISL_777127, EPI_ISL_777130, EPI_ISL_777131, EPI_ISL_777140, EPI_ISL_777146, EPI_ISL_777151, EPI_ISL_777152, EPI_ISL_777154, EPI_ISL_777160, EPI_ISL_777161, EPI_ISL_777162, EPI_ISL_777164, EPI_ISL_777176, EPI_ISL_777180, EPI_ISL_777189, EPI_ISL_777191, EPI_ISL_777196, EPI_ISL_777205, EPI_ISL_777208, EPI_ISL_777209, EPI_ISL_777215, EPI_ISL_777219, EPI_ISL_777222, EPI_ISL_777223, EPI_ISL_777225, EPI_ISL_777227, EPI_ISL_777230, EPI_ISL_777232, EPI_ISL_777233, EPI_ISL_777235, EPI_ISL_777236, EPI_ISL_777238, EPI_ISL_777241, EPI_ISL_777242, EPI_ISL_777246, EPI_ISL_777250, EPI_ISL_777252, EPI_ISL_777253, EPI_ISL_777261, EPI_ISL_777268, EPI_ISL_777283, EPI_ISL_777290, EPI_ISL_777298, EPI_ISL_777300, EPI_ISL_777301, EPI_ISL_777302, EPI_ISL_777305, EPI_ISL_777306, EPI_ISL_777316, EPI_ISL_777321, EPI_ISL_777322, EPI_ISL_777324, EPI_ISL_777325, EPI_ISL_777335, EPI_ISL_777337, EPI_ISL_777340, EPI_ISL_777341, EPI_ISL_777346, EPI_ISL_777353, EPI_ISL_777356, EPI_ISL_777358, EPI_ISL_777361, EPI_ISL_777363, EPI_ISL_777364, EPI_ISL_777369, EPI_ISL_777372, EPI_ISL_777373, EPI_ISL_777376, EPI_ISL_777377, EPI_ISL_777382, EPI_ISL_777383, EPI_ISL_777385, EPI_ISL_777392, EPI_ISL_777393, EPI_ISL_777395, EPI_ISL_777397, EPI_ISL_777399, EPI_ISL_777402, EPI_ISL_777407, EPI_ISL_777410, EPI_ISL_777417, EPI_ISL_777420, EPI_ISL_777423, EPI_ISL_777432, EPI_ISL_777436, EPI_ISL_777438, EPI_ISL_777443, EPI_ISL_777444, EPI_ISL_777447, EPI_ISL_777449, EPI_ISL_777459, EPI_ISL_777461, EPI_ISL_777462, EPI_ISL_777466, EPI_ISL_777471, EPI_ISL_777474, EPI_ISL_777475, EPI_ISL_777484, EPI_ISL_777487, EPI_ISL_777493, EPI_ISL_777496, EPI_ISL_777497, EPI_ISL_777498, EPI_ISL_777504, EPI_ISL_777505, EPI_ISL_777506, EPI_ISL_777510, EPI_ISL_777514, EPI_ISL_777518, EPI_ISL_777523, EPI_ISL_777526, EPI_ISL_777531, EPI_ISL_777533, EPI_ISL_777554, EPI_ISL_777557, EPI_ISL_777560, EPI_ISL_777562, EPI_ISL_777569, EPI_ISL_777574, EPI_ISL_777579, EPI_ISL_777583, EPI_ISL_777600, EPI_ISL_777601, EPI_ISL_777610, EPI_ISL_777612, EPI_ISL_777622, EPI_ISL_777623, EPI_ISL_777628, EPI_ISL_777630, EPI_ISL_777641, EPI_ISL_777646, EPI_ISL_777653, EPI_ISL_777654, EPI_ISL_777657, EPI_ISL_777658, EPI_ISL_777660, EPI_ISL_777662, EPI_ISL_777663, EPI_ISL_777667, EPI_ISL_777670, EPI_ISL_778098, EPI_ISL_778155, EPI_ISL_778184, EPI_ISL_778281, EPI_ISL_778447, EPI_ISL_778457, EPI_ISL_778482, EPI_ISL_778534, EPI_ISL_778579, EPI_ISL_781982, EPI_ISL_782133, EPI_ISL_782327, EPI_ISL_820182, EPI_ISL_820190, EPI_ISL_820200, EPI_ISL_820303, EPI_ISL_820349, EPI_ISL_820356, EPI_ISL_820397, EPI_ISL_820481, EPI_ISL_820513, EPI_ISL_820597, EPI_ISL_820633, EPI_ISL_858410, EPI_ISL_858415, EPI_ISL_858417, EPI_ISL_858419, EPI_ISL_858420, EPI_ISL_858421, EPI_ISL_858423, EPI_ISL_858424, EPI_ISL_858911, EPI_ISL_858997, EPI_ISL_859003, EPI_ISL_859012, EPI_ISL_859030, EPI_ISL_859031, EPI_ISL_859032, EPI_ISL_859033, EPI_ISL_859034, EPI_ISL_859036, EPI_ISL_859037, EPI_ISL_859038, EPI_ISL_859039, EPI_ISL_859040, EPI_ISL_859041, EPI_ISL_859042, EPI_ISL_859045, EPI_ISL_859046, EPI_ISL_859047, EPI_ISL_859048, EPI_ISL_859050, EPI_ISL_859052, EPI_ISL_859053, EPI_ISL_859054, EPI_ISL_859055, EPI_ISL_859057, EPI_ISL_859058, EPI_ISL_859059, EPI_ISL_859060, EPI_ISL_859062, EPI_ISL_859064, EPI_ISL_859065, EPI_ISL_859068, EPI_ISL_859069, EPI_ISL_859071, EPI_ISL_859072, EPI_ISL_859074, EPI_ISL_859078, EPI_ISL_859079, EPI_ISL_859081, EPI_ISL_859083, EPI_ISL_859084, EPI_ISL_859087, EPI_ISL_859088, EPI_ISL_859093, EPI_ISL_859094, EPI_ISL_859098, EPI_ISL_859100, EPI_ISL_859102, EPI_ISL_859105, EPI_ISL_859106, EPI_ISL_859108, EPI_ISL_859112, EPI_ISL_859114, EPI_ISL_859116, EPI_ISL_859118, EPI_ISL_859119, EPI_ISL_859120, EPI_ISL_859123, EPI_ISL_859125, EPI_ISL_859127, EPI_ISL_859130, EPI_ISL_859131, EPI_ISL_859137, EPI_ISL_859139, EPI_ISL_859140, EPI_ISL_859320, EPI_ISL_859321, EPI_ISL_859322, EPI_ISL_859323, EPI_ISL_859324, EPI_ISL_859325, EPI_ISL_859326, EPI_ISL_859435, EPI_ISL_859460, EPI_ISL_945073, EPI_ISL_945209, EPI_ISL_945357, EPI_ISL_989986, EPI_ISL_989987

| | | | |
|---|---|---|---|
| see above | Lighthouse Lab in Glasgow | Wellcome Sanger Institute for the COVID-19 Genomics UK (COG-UK) Consortium | Anna Dominiczak and Alex Alderton; Carol Clugston; Cordelia Langford; David Gray; David K. Jackson; Dominic Kwiatkowski; Ewan Harrison; Harper VanSteenhouse; Ian Johnston; John Sillitoe on behalf of the Wellcome Sanger Institute COVID-19 Surveillance Team; Roberto Amato; Sonia Goncalves; Yumi Kasai |

EPI_ISL_756689, EPI_ISL_756691, EPI_ISL_756692, EPI_ISL_756693, EPI_ISL_756694, EPI_ISL_756697, EPI_ISL_756698, EPI_ISL_756699, EPI_ISL_756700, EPI_ISL_756701, EPI_ISL_756703, EPI_ISL_756704, EPI_ISL_756708, EPI_ISL_756710, EPI_ISL_756712, EPI_ISL_756713, EPI_ISL_756714, EPI_ISL_756715, EPI_ISL_756717, EPI_ISL_756718, EPI_ISL_756719, EPI_ISL_756720, EPI_ISL_756721, EPI_ISL_756722, EPI_ISL_756724, EPI_ISL_756725, EPI_ISL_756726, EPI_ISL_756727, EPI_ISL_756730, EPI_ISL_756731, EPI_ISL_756732, EPI_ISL_756733, EPI_ISL_756734, EPI_ISL_756735, EPI_ISL_756736, EPI_ISL_756738, EPI_ISL_756739, EPI_ISL_756740, EPI_ISL_756741, EPI_ISL_756742, EPI_ISL_756743, EPI_ISL_756746, EPI_ISL_756747, EPI_ISL_756749, EPI_ISL_756751, EPI_ISL_756752, EPI_ISL_756754, EPI_ISL_756755, EPI_ISL_756756, EPI_ISL_756757, EPI_ISL_756758, EPI_ISL_756759, EPI_ISL_756760, EPI_ISL_756762, EPI_ISL_756763, EPI_ISL_756764, EPI_ISL_756766, EPI_ISL_756767, EPI_ISL_756768, EPI_ISL_756769, EPI_ISL_756770, EPI_ISL_756771, EPI_ISL_756774, EPI_ISL_756776, EPI_ISL_756777, EPI_ISL_756778, EPI_ISL_756779, EPI_ISL_756781, EPI_ISL_756782, EPI_ISL_756783, EPI_ISL_756785, EPI_ISL_756786, EPI_ISL_756787, EPI_ISL_756788, EPI_ISL_756790, EPI_ISL_756791, EPI_ISL_756792, EPI_ISL_756793, EPI_ISL_756794, EPI_ISL_756796, EPI_ISL_756797, EPI_ISL_756798, EPI_ISL_756799, EPI_ISL_756801, EPI_ISL_756802, EPI_ISL_756806, EPI_ISL_756807, EPI_ISL_756809, EPI_ISL_756810, EPI_ISL_756811, EPI_ISL_756812, EPI_ISL_756813, EPI_ISL_756814, EPI_ISL_756815, EPI_ISL_756816, EPI_ISL_756817, EPI_ISL_756818, EPI_ISL_756819, EPI_ISL_756820, EPI_ISL_756821, EPI_ISL_756822, EPI_ISL_756825, EPI_ISL_756826, EPI_ISL_756827, EPI_ISL_756828, EPI_ISL_756829, EPI_ISL_756830, EPI_ISL_756831, EPI_ISL_756832, EPI_ISL_756833, EPI_ISL_756834, EPI_ISL_756836, EPI_ISL_756837, EPI_ISL_756838, EPI_ISL_756839, EPI_ISL_756841, EPI_ISL_756842, EPI_ISL_756843, EPI_ISL_756844, EPI_ISL_756845, EPI_ISL_756848, EPI_ISL_756851, EPI_ISL_756853, EPI_ISL_756854, EPI_ISL_756855, EPI_ISL_756856, EPI_ISL_756857, EPI_ISL_756858, EPI_ISL_756859, EPI_ISL_756862, EPI_ISL_756863, EPI_ISL_756864, EPI_ISL_756866, EPI_ISL_756868, EPI_ISL_756869, EPI_ISL_756870, EPI_ISL_756871, EPI_ISL_756873, EPI_ISL_756874, EPI_ISL_756875, EPI_ISL_756880, EPI_ISL_756882, EPI_ISL_756883, EPI_ISL_756884, EPI_ISL_756891, EPI_ISL_756892, EPI_ISL_756896, EPI_ISL_756898, EPI_ISL_756899, EPI_ISL_756901, EPI_ISL_756902, EPI_ISL_756904, EPI_ISL_756905, EPI_ISL_756906, EPI_ISL_756907, EPI_ISL_756908, EPI_ISL_756911, EPI_ISL_756913, EPI_ISL_756914, EPI_ISL_756915, EPI_ISL_756916, EPI_ISL_756919, EPI_ISL_756921, EPI_ISL_756922, EPI_ISL_756924, EPI_ISL_756926, EPI_ISL_756927, EPI_ISL_756930, EPI_ISL_756932, EPI_ISL_756933, EPI_ISL_756934, EPI_ISL_756936, EPI_ISL_756937, EPI_ISL_756938, EPI_ISL_756939, EPI_ISL_756941, EPI_ISL_756942, EPI_ISL_756943, EPI_ISL_756944, EPI_ISL_756945, EPI_ISL_756948, EPI_ISL_756949, EPI_ISL_756950, EPI_ISL_756954, EPI_ISL_756956, EPI_ISL_756957, EPI_ISL_756958, EPI_ISL_756960, EPI_ISL_756961, EPI_ISL_756962, EPI_ISL_756963, EPI_ISL_756964, EPI_ISL_756965, EPI_ISL_756966, EPI_ISL_756967, EPI_ISL_756968, EPI_ISL_756972, EPI_ISL_756974, EPI_ISL_756975, EPI_ISL_756976, EPI_ISL_756978, EPI_ISL_756979, EPI_ISL_756980, EPI_ISL_756981, EPI_ISL_756983, EPI_ISL_756984, EPI_ISL_756985, EPI_ISL_756986, EPI_ISL_756987, EPI_ISL_756988, EPI_ISL_756990, EPI_ISL_756991, EPI_ISL_756992, EPI_ISL_756993, EPI_ISL_756994, EPI_ISL_756995, EPI_ISL_756996, EPI_ISL_756997, EPI_ISL_757000, EPI_ISL_757001, EPI_ISL_757002, EPI_ISL_757003, EPI_ISL_757004, EPI_ISL_757006, EPI_ISL_757010, EPI_ISL_757013, EPI_ISL_757015, EPI_ISL_757016, EPI_ISL_757019, EPI_ISL_757022, EPI_ISL_757023, EPI_ISL_757024, EPI_ISL_757025, EPI_ISL_757026, EPI_ISL_757029, EPI_ISL_757030, EPI_ISL_757031, EPI_ISL_757033, EPI_ISL_760698, EPI_ISL_760801, EPI_ISL_761357, EPI_ISL_761358, EPI_ISL_761359, EPI_ISL_761360, EPI_ISL_761361, EPI_ISL_761362, EPI_ISL_761363, EPI_ISL_761364, EPI_ISL_761365, EPI_ISL_761366, EPI_ISL_761367, EPI_ISL_761368, EPI_ISL_761369, EPI_ISL_761370, EPI_ISL_761371, EPI_ISL_761372, EPI_ISL_761373, EPI_ISL_761374, EPI_ISL_761375, EPI_ISL_761376, EPI_ISL_761377, EPI_ISL_761378, EPI_ISL_761379, EPI_ISL_761380, EPI_ISL_761381, EPI_ISL_761382, EPI_ISL_761383, EPI_ISL_761384, EPI_ISL_761385, EPI_ISL_761386, EPI_ISL_761387, EPI_ISL_761388, EPI_ISL_761389, EPI_ISL_761390, EPI_ISL_761391, EPI_ISL_761392, EPI_ISL_761393, EPI_ISL_761394, EPI_ISL_761395, EPI_ISL_761396, EPI_ISL_761397, EPI_ISL_761398, EPI_ISL_761399, EPI_ISL_761400, EPI_ISL_761401, EPI_ISL_761402, EPI_ISL_761403, EPI_ISL_761404, EPI_ISL_761405, EPI_ISL_761406, EPI_ISL_761407, EPI_ISL_761408, EPI_ISL_761409, EPI_ISL_761410, EPI_ISL_761411, EPI_ISL_761412, EPI_ISL_761413, EPI_ISL_761414, EPI_ISL_761415, EPI_ISL_761416, EPI_ISL_761417, EPI_ISL_761418, EPI_ISL_761419, EPI_ISL_761420, EPI_ISL_761421, EPI_ISL_761422, EPI_ISL_761423, EPI_ISL_761424, EPI_ISL_761425, EPI_ISL_761426, EPI_ISL_761427, EPI_ISL_761428, EPI_ISL_761429, EPI_ISL_761430, EPI_ISL_761431, EPI_ISL_761432, EPI_ISL_761433, EPI_ISL_761434, EPI_ISL_761435, EPI_ISL_761436, EPI_ISL_761437, EPI_ISL_761438, EPI_ISL_761439, EPI_ISL_761440, EPI_ISL_761441, EPI_ISL_761442, EPI_ISL_761443, EPI_ISL_761444, EPI_ISL_761445, EPI_ISL_761446, EPI_ISL_761447, EPI_ISL_761448, EPI_ISL_761449, EPI_ISL_761450, EPI_ISL_761451, EPI_ISL_761452, EPI_ISL_761453, EPI_ISL_761454, EPI_ISL_761455, EPI_ISL_761456, EPI_ISL_761457, EPI_ISL_761458, EPI_ISL_761459, EPI_ISL_761460, EPI_ISL_761461, EPI_ISL_761462, EPI_ISL_761463, EPI_ISL_761464, EPI_ISL_761465, EPI_ISL_761466, EPI_ISL_761467, EPI_ISL_761468, EPI_ISL_761469, EPI_ISL_761470, EPI_ISL_761471, EPI_ISL_761472, EPI_ISL_761473, EPI_ISL_761474, EPI_ISL_761475, EPI_ISL_761476, EPI_ISL_761477, EPI_ISL_761478, EPI_ISL_761479, EPI_ISL_761480, EPI_ISL_761481, EPI_ISL_761482, EPI_ISL_761483, EPI_ISL_761484, EPI_ISL_761485, EPI_ISL_761486, EPI_ISL_761487, EPI_ISL_761488, EPI_ISL_761489, EPI_ISL_761490, EPI_ISL_761491, EPI_ISL_761492, EPI_ISL_761493, EPI_ISL_761494, EPI_ISL_761495, EPI_ISL_761496, EPI_ISL_761497, EPI_ISL_761498, EPI_ISL_761499, EPI_ISL_761500, EPI_ISL_761501, EPI_ISL_761502, EPI_ISL_761503, EPI_ISL_761504, EPI_ISL_761505, EPI_ISL_761506, EPI_ISL_761507, EPI_ISL_761508, EPI_ISL_761509, EPI_ISL_761510, EPI_ISL_761511, EPI_ISL_761512, EPI_ISL_761513, EPI_ISL_761514, EPI_ISL_761515, EPI_ISL_761516, EPI_ISL_761517, EPI_ISL_761518, EPI_ISL_761519, EPI_ISL_761520, EPI_ISL_761521, EPI_ISL_761522, EPI_ISL_761523, EPI_ISL_761524, EPI_ISL_761525, EPI_ISL_761526, EPI_ISL_761527, EPI_ISL_761528, EPI_ISL_761529, EPI_ISL_761530, EPI_ISL_761531, EPI_ISL_761532, EPI_ISL_761533, EPI_ISL_761534, EPI_ISL_761535, EPI_ISL_761536, EPI_ISL_761537, EPI_ISL_761538, EPI_ISL_761539, EPI_ISL_761540, EPI_ISL_761541, EPI_ISL_761542, EPI_ISL_761543, EPI_ISL_761544, EPI_ISL_761545, EPI_ISL_761546, EPI_ISL_761547, EPI_ISL_761548, EPI_ISL_761549, EPI_ISL_761550, EPI_ISL_761551, EPI_ISL_761552, EPI_ISL_761553, EPI_ISL_761554, EPI_ISL_761555, EPI_ISL_761556, EPI_ISL_761557, EPI_ISL_761558, EPI_ISL_761559, EPI_ISL_761560, EPI_ISL_761561, EPI_ISL_761562, EPI_ISL_761563, EPI_ISL_761564, EPI_ISL_761565, EPI_ISL_761566, EPI_ISL_761567, EPI_ISL_761568, EPI_ISL_761569, EPI_ISL_761570, EPI_ISL_761571, EPI_ISL_761572, EPI_ISL_761573, EPI_ISL_761574, EPI_ISL_761575, EPI_ISL_761576, EPI_ISL_761577, EPI_ISL_761578, EPI_ISL_761579, EPI_ISL_761580, EPI_ISL_761581, EPI_ISL_761582, EPI_ISL_761583, EPI_ISL_761584, EPI_ISL_761585, EPI_ISL_761586, EPI_ISL_761587, EPI_ISL_761588, EPI_ISL_761589, EPI_ISL_761590, EPI_ISL_761591, EPI_ISL_761592, EPI_ISL_761593, EPI_ISL_761594, EPI_ISL_761595, EPI_ISL_761596, EPI_ISL_761597, EPI_ISL_761598, EPI_ISL_761599, EPI_ISL_761600, EPI_ISL_761601, EPI_ISL_761602, EPI_ISL_761603, EPI_ISL_761604, EPI_ISL_761605, EPI_ISL_761606, EPI_ISL_761607, EPI_ISL_761608, EPI_ISL_761609, EPI_ISL_761610, EPI_ISL_761611, EPI_ISL_761612, EPI_ISL_761613, EPI_ISL_761614, EPI_ISL_761615, EPI_ISL_761616, EPI_ISL_761617, EPI_ISL_761618, EPI_ISL_761619, EPI_ISL_761620, EPI_ISL_761621, EPI_ISL_761622, EPI_ISL_761624, EPI_ISL_761625, EPI_ISL_761626, EPI_ISL_761627, EPI_ISL_761628, EPI_ISL_761629, EPI_ISL_761630, EPI_ISL_761631, EPI_ISL_761632, EPI_ISL_761633, EPI_ISL_761634, EPI_ISL_761635, EPI_ISL_761636, EPI_ISL_761637, EPI_ISL_761638, EPI_ISL_761639, EPI_ISL_761640, EPI_ISL_761641, EPI_ISL_761642, EPI_ISL_761643, EPI_ISL_761644, EPI_ISL_761645, EPI_ISL_761646, EPI_ISL_761647, EPI_ISL_761648, EPI_ISL_761649, EPI_ISL_761650, EPI_ISL_761651, EPI_ISL_761652, EPI_ISL_761653, EPI_ISL_761654, EPI_ISL_761655, EPI_ISL_761656, EPI_ISL_761657, EPI_ISL_761658, EPI_ISL_761659, EPI_ISL_761660, EPI_ISL_761661, EPI_ISL_761662, EPI_ISL_761663, EPI_ISL_761664, EPI_ISL_762198, EPI_ISL_762199, EPI_ISL_762200, EPI_ISL_762201, EPI_ISL_762202, EPI_ISL_762203, EPI_ISL_762204, EPI_ISL_762205, EPI_ISL_762206, EPI_ISL_762207, EPI_ISL_762208, EPI_ISL_762209, EPI_ISL_762210, EPI_ISL_762211, EPI_ISL_762212, EPI_ISL_762213, EPI_ISL_762214, EPI_ISL_762215, EPI_ISL_762216, EPI_ISL_762217, EPI_ISL_762218, EPI_ISL_762219, EPI_ISL_762220, EPI_ISL_762221, EPI_ISL_762222, EPI_ISL_762223, EPI_ISL_762224, EPI_ISL_762225, EPI_ISL_762226, EPI_ISL_762227, EPI_ISL_762228, EPI_ISL_762229, EPI_ISL_762230, EPI_ISL_762231, EPI_ISL_762232, EPI_ISL_762233, EPI_ISL_762234, EPI_ISL_762235, EPI_ISL_762236, EPI_ISL_762238, EPI_ISL_762239, EPI_ISL_762240, EPI_ISL_762241, EPI_ISL_762242, EPI_ISL_762243, EPI_ISL_762244, EPI_ISL_762245, EPI_ISL_762246, EPI_ISL_762247, EPI_ISL_762248, EPI_ISL_762249, EPI_ISL_762250, EPI_ISL_762251, EPI_ISL_762252, EPI_ISL_762254, EPI_ISL_762255, EPI_ISL_762256, EPI_ISL_762257, EPI_ISL_762258, EPI_ISL_762259, EPI_ISL_762260, EPI_ISL_762261, EPI_ISL_762262, EPI_ISL_762263, EPI_ISL_762264, EPI_ISL_762265, EPI_ISL_762266, EPI_ISL_762267, EPI_ISL_762268, EPI_ISL_762269, EPI_ISL_762270, EPI_ISL_762271, EPI_ISL_762272, EPI_ISL_762273, EPI_ISL_762274, EPI_ISL_762275, EPI_ISL_762276, EPI_ISL_762277, EPI_ISL_762278, EPI_ISL_762279, EPI_ISL_762280, EPI_ISL_762282, EPI_ISL_762283, EPI_ISL_762284, EPI_ISL_762285, EPI_ISL_762286, EPI_ISL_762287, EPI_ISL_762288, EPI_ISL_762289, EPI_ISL_762290, EPI_ISL_762291, EPI_ISL_762292, EPI_ISL_762293, EPI_ISL_762294, EPI_ISL_762295, EPI_ISL_762296, EPI_ISL_762297, EPI_ISL_762298, EPI_ISL_762299, EPI_ISL_762300, EPI_ISL_762301, EPI_ISL_762302, EPI_ISL_762303, EPI_ISL_762304, EPI_ISL_762305, EPI_ISL_762306, EPI_ISL_762307, EPI_ISL_762308, EPI_ISL_762309, EPI_ISL_762310, EPI_ISL_762311, EPI_ISL_762312, EPI_ISL_762313, EPI_ISL_762314, EPI_ISL_762315, EPI_ISL_762316, EPI_ISL_762317, EPI_ISL_762318, EPI_ISL_762319, EPI_ISL_762320, EPI_ISL_762321, EPI_ISL_762322, EPI_ISL_762323, EPI_ISL_762324, EPI_ISL_762325, EPI_ISL_762326, EPI_ISL_762327, EPI_ISL_762328, EPI_ISL_762329, EPI_ISL_762330, EPI_ISL_762331, EPI_ISL_762332, EPI_ISL_762333, EPI_ISL_762334, EPI_ISL_762335, EPI_ISL_762336, EPI_ISL_762337, EPI_ISL_762338, EPI_ISL_762339, EPI_ISL_762340, EPI_ISL_762341, EPI_ISL_762342, EPI_ISL_762343, EPI_ISL_762344, EPI_ISL_762345, EPI_ISL_762346, EPI_ISL_762347, EPI_ISL_762348, EPI_ISL_762349, EPI_ISL_762350, EPI_ISL_762351, EPI_ISL_762352, EPI_ISL_762353, EPI_ISL_762354, EPI_ISL_762355, EPI_ISL_762356, EPI_ISL_762357, EPI_ISL_762358, EPI_ISL_762360, EPI_ISL_762361, EPI_ISL_762362, EPI_ISL_762363, EPI_ISL_762364, EPI_ISL_762365, EPI_ISL_762366, EPI_ISL_762367, EPI_ISL_762368, EPI_ISL_762369, EPI_ISL_762370, EPI_ISL_762371, EPI_ISL_762372, EPI_ISL_762373, EPI_ISL_762374, EPI_ISL_762375, EPI_ISL_762376, EPI_ISL_762377, EPI_ISL_762378, EPI_ISL_762379, EPI_ISL_762380, EPI_ISL_762381, EPI_ISL_762382, EPI_ISL_762383, EPI_ISL_762384, EPI_ISL_762385, EPI_ISL_762386, EPI_ISL_762387, EPI_ISL_762388, EPI_ISL_762389, EPI_ISL_762390, EPI_ISL_762391, EPI_ISL_762392, EPI_ISL_762393, EPI_ISL_762394, EPI_ISL_762395, EPI_ISL_762396, EPI_ISL_762397, EPI_ISL_762398, EPI_ISL_762400, EPI_ISL_762401, EPI_ISL_762402, EPI_ISL_762403, EPI_ISL_762404, EPI_ISL_762405, EPI_ISL_762406, EPI_ISL_762407, EPI_ISL_762408, EPI_ISL_762409, EPI_ISL_762410, EPI_ISL_762411, EPI_ISL_762412, EPI_ISL_762413, EPI_ISL_762414, EPI_ISL_762415, EPI_ISL_762416, EPI_ISL_762417, EPI_ISL_762418, EPI_ISL_762419, EPI_ISL_762420, EPI_ISL_762421, EPI_ISL_762422, EPI_ISL_762423, EPI_ISL_762424, EPI_ISL_762425, EPI_ISL_762426, EPI_ISL_762427, EPI_ISL_762428, EPI_ISL_762429, EPI_ISL_762430, EPI_ISL_762431, EPI_ISL_762432, EPI_ISL_762433, EPI_ISL_762434, EPI_ISL_762435, EPI_ISL_762436, EPI_ISL_762437, EPI_ISL_762438, EPI_ISL_762439, EPI_ISL_762440, EPI_ISL_762441, EPI_ISL_762442, EPI_ISL_762443, EPI_ISL_762444, EPI_ISL_762445, EPI_ISL_762446, EPI_ISL_762447, EPI_ISL_762448, EPI_ISL_762449, EPI_ISL_762450, EPI_ISL_762451, EPI_ISL_762452, EPI_ISL_762453, EPI_ISL_762454, EPI_ISL_762455, EPI_ISL_762456, EPI_ISL_762457, EPI_ISL_762458, EPI_ISL_762459, EPI_ISL_762460, EPI_ISL_762461, EPI_ISL_762462, EPI_ISL_762463, EPI_ISL_762464, EPI_ISL_762465, EPI_ISL_762466, EPI_ISL_762467, EPI_ISL_762468, EPI_ISL_762469, EPI_ISL_762470, EPI_ISL_762471, EPI_ISL_762472, EPI_ISL_762473, EPI_ISL_762474, EPI_ISL_762475, EPI_ISL_762476, EPI_ISL_762477, EPI_ISL_762478, EPI_ISL_762479, EPI_ISL_762480, EPI_ISL_762481, EPI_ISL_762482, EPI_ISL_762483, EPI_ISL_762484, EPI_ISL_762485, EPI_ISL_762486, EPI_ISL_762487, EPI_ISL_762488, EPI_ISL_762489, EPI_ISL_762490, EPI_ISL_762491, EPI_ISL_762492, EPI_ISL_762495, EPI_ISL_762496, EPI_ISL_762497, EPI_ISL_762498, EPI_ISL_762499, EPI_ISL_762500, EPI_ISL_762501, EPI_ISL_762502, EPI_ISL_762504, EPI_ISL_762505, EPI_ISL_762506, EPI_ISL_762507, EPI_ISL_762508, EPI_ISL_762509, EPI_ISL_762510, EPI_ISL_762511, EPI_ISL_762512, EPI_ISL_762513, EPI_ISL_762514, EPI_ISL_762515, EPI_ISL_762516, EPI_ISL_762517, EPI_ISL_762518, EPI_ISL_762519, EPI_ISL_762520, EPI_ISL_762521, EPI_ISL_762522, EPI_ISL_762523, EPI_ISL_762524, EPI_ISL_762525, EPI_ISL_762526, EPI_ISL_762527, EPI_ISL_762528, EPI_ISL_762529, EPI_ISL_762530, EPI_ISL_762531, EPI_ISL_762532, EPI_ISL_762533, EPI_ISL_762534, EPI_ISL_762535, EPI_ISL_762536, EPI_ISL_762537, EPI_ISL_762538, EPI_ISL_762539, EPI_ISL_762540, EPI_ISL_762541, EPI_ISL_762542, EPI_ISL_762543, EPI_ISL_762544, EPI_ISL_762545, EPI_ISL_762864, EPI_ISL_762867, EPI_ISL_762875, EPI_ISL_762880, EPI_ISL_762884, EPI_ISL_762891, EPI_ISL_762895, EPI_ISL_762896, EPI_ISL_762903, EPI_ISL_762908, EPI_ISL_762912, EPI_ISL_762914, EPI_ISL_762917, EPI_ISL_762920, EPI_ISL_762937, EPI_ISL_762943, EPI_ISL_762945, EPI_ISL_762963, EPI_ISL_762965, EPI_ISL_762967, EPI_ISL_762968, EPI_ISL_762976, EPI_ISL_762978, EPI_ISL_762981, EPI_ISL_767098, EPI_ISL_767099, EPI_ISL_767100, EPI_ISL_767101, EPI_ISL_767107, EPI_ISL_767108, EPI_ISL_767109, EPI_ISL_767110, EPI_ISL_767111, EPI_ISL_767112, EPI_ISL_768846, EPI_ISL_768847, EPI_ISL_768848, EPI_ISL_768849, EPI_ISL_768850, EPI_ISL_768851, EPI_ISL_768852, EPI_ISL_768853, EPI_ISL_768854, EPI_ISL_768855, EPI_ISL_768856, EPI_ISL_768857, EPI_ISL_768858, EPI_ISL_768859, EPI_ISL_768860, EPI_ISL_768861, EPI_ISL_768862, EPI_ISL_768863, EPI_ISL_768864, EPI_ISL_768865, EPI_ISL_768866, EPI_ISL_768867, EPI_ISL_768868, EPI_ISL_768869, EPI_ISL_768870, EPI_ISL_768871, EPI_ISL_768872, EPI_ISL_768873, EPI_ISL_768874, EPI_ISL_768876, EPI_ISL_768877, EPI_ISL_768878, EPI_ISL_768879, EPI_ISL_768880, EPI_ISL_768881, EPI_ISL_768882, EPI_ISL_768883, EPI_ISL_768884, EPI_ISL_768885, EPI_ISL_768886, EPI_ISL_768887, EPI_ISL_768888, EPI_ISL_768889, EPI_ISL_768890, EPI_ISL_768891, EPI_ISL_768892, EPI_ISL_768893, EPI_ISL_768894, EPI_ISL_768895, EPI_ISL_768896, EPI_ISL_768897, EPI_ISL_781179, EPI_ISL_781180, EPI_ISL_781184, EPI_ISL_781193, EPI_ISL_781197, EPI_ISL_781198, EPI_ISL_781208, EPI_ISL_781211, EPI_ISL_781212, EPI_ISL_781213, EPI_ISL_781217, EPI_ISL_781227, EPI_ISL_781228, EPI_ISL_781230, EPI_ISL_781231, EPI_ISL_781233, EPI_ISL_781236, EPI_ISL_781238, EPI_ISL_781240, EPI_ISL_781242, EPI_ISL_781243, EPI_ISL_781245, EPI_ISL_781246, EPI_ISL_781252, EPI_ISL_781253, EPI_ISL_781254, EPI_ISL_781257, EPI_ISL_781264, EPI_ISL_781265, EPI_ISL_781266, EPI_ISL_781270, EPI_ISL_781271, EPI_ISL_781274, EPI_ISL_781275, EPI_ISL_781277, EPI_ISL_781283, EPI_ISL_781284, EPI_ISL_781285, EPI_ISL_781289, EPI_ISL_781293, EPI_ISL_781296, EPI_ISL_781302, EPI_ISL_781303, EPI_ISL_781305, EPI_ISL_781311, EPI_ISL_781314, EPI_ISL_781317, EPI_ISL_781321, EPI_ISL_781322, EPI_ISL_781326, EPI_ISL_781328, EPI_ISL_781334, EPI_ISL_781338, EPI_ISL_781339, EPI_ISL_781344, EPI_ISL_781346, EPI_ISL_781347, EPI_ISL_781351, EPI_ISL_781353, EPI_ISL_781361, EPI_ISL_781362, EPI_ISL_781365, EPI_ISL_781369, EPI_ISL_781370, EPI_ISL_781373, EPI_ISL_781382, EPI_ISL_781383, EPI_ISL_781386, EPI_ISL_781388, EPI_ISL_781391, EPI_ISL_781397, EPI_ISL_781399, EPI_ISL_781410, EPI_ISL_781411, EPI_ISL_781414, EPI_ISL_781416, EPI_ISL_781417, EPI_ISL_781419, EPI_ISL_781420, EPI_ISL_781421, EPI_ISL_781422, EPI_ISL_781423, EPI_ISL_781425, EPI_ISL_781427, EPI_ISL_781428, EPI_ISL_781430, EPI_ISL_781445, EPI_ISL_781449, EPI_ISL_781450, EPI_ISL_781452, EPI_ISL_781453, EPI_ISL_781454, EPI_ISL_781459, EPI_ISL_781462, EPI_ISL_781467, EPI_ISL_781470, EPI_ISL_781474, EPI_ISL_781477, EPI_ISL_781479, EPI_ISL_781482, EPI_ISL_781486, EPI_ISL_781487, EPI_ISL_781491, EPI_ISL_781492, EPI_ISL_781493, EPI_ISL_781496, EPI_ISL_781501, EPI_ISL_781503, EPI_ISL_781504, EPI_ISL_781507, EPI_ISL_781509, EPI_ISL_781511, EPI_ISL_781522, EPI_ISL_781527, EPI_ISL_781529, EPI_ISL_781536, EPI_ISL_781537, EPI_ISL_781539, EPI_ISL_781541, EPI_ISL_781544, EPI_ISL_781547, EPI_ISL_781548, EPI_ISL_781554, EPI_ISL_781555, EPI_ISL_781556, EPI_ISL_781558, EPI_ISL_781567, EPI_ISL_781576, EPI_ISL_781578, EPI_ISL_781579, EPI_ISL_781582, EPI_ISL_781597, EPI_ISL_781598, EPI_ISL_781599, EPI_ISL_781605, EPI_ISL_781609, EPI_ISL_781611, EPI_ISL_781615, EPI_ISL_781617, EPI_ISL_781618, EPI_ISL_781619, EPI_ISL_781621, EPI_ISL_781623, EPI_ISL_781624, EPI_ISL_781627, EPI_ISL_781636, EPI_ISL_781649, EPI_ISL_781651, EPI_ISL_781652, EPI_ISL_781655, EPI_ISL_781663, EPI_ISL_781665, EPI_ISL_781669, EPI_ISL_781674, EPI_ISL_781677, EPI_ISL_781680, EPI_ISL_781681, EPI_ISL_781684, EPI_ISL_781686, EPI_ISL_781695, EPI_ISL_781699, EPI_ISL_781701, EPI_ISL_781720, EPI_ISL_781721, EPI_ISL_781724, EPI_ISL_781725, EPI_ISL_781730, EPI_ISL_781732, EPI_ISL_781735, EPI_ISL_781742, EPI_ISL_781743, EPI_ISL_781746, EPI_ISL_781747, EPI_ISL_781749, EPI_ISL_781753, EPI_ISL_781754, EPI_ISL_781763, EPI_ISL_781767, EPI_ISL_781770, EPI_ISL_781773, EPI_ISL_781777, EPI_ISL_781783, EPI_ISL_781784, EPI_ISL_781785, EPI_ISL_781788, EPI_ISL_781789, EPI_ISL_781795, EPI_ISL_781798, EPI_ISL_781801, EPI_ISL_781807, EPI_ISL_781832, EPI_ISL_781834, EPI_ISL_781837, EPI_ISL_781839, EPI_ISL_781840, EPI_ISL_781841, EPI_ISL_781851, EPI_ISL_781854, EPI_ISL_781858, EPI_ISL_781862, EPI_ISL_781864, EPI_ISL_781869, EPI_ISL_781873, EPI_ISL_781876, EPI_ISL_781878, EPI_ISL_781880, EPI_ISL_781885, EPI_ISL_781888, EPI_ISL_781891, EPI_ISL_781897, EPI_ISL_781898, EPI_ISL_781903, EPI_ISL_781907, EPI_ISL_781908, EPI_ISL_781909, EPI_ISL_781914, EPI_ISL_781915, EPI_ISL_781916, EPI_ISL_781920, EPI_ISL_781921, EPI_ISL_781924, EPI_ISL_781925, EPI_ISL_781938, EPI_ISL_781940, EPI_ISL_781941, EPI_ISL_781942, EPI_ISL_781944, EPI_ISL_781946, EPI_ISL_781947, EPI_ISL_781948, EPI_ISL_781950, EPI_ISL_781952, EPI_ISL_781955, EPI_ISL_781968, EPI_ISL_781973, EPI_ISL_781977, EPI_ISL_781979, EPI_ISL_781984, EPI_ISL_781987, EPI_ISL_781988, EPI_ISL_781994, EPI_ISL_781998, EPI_ISL_781999, EPI_ISL_782000, EPI_ISL_782009, EPI_ISL_782015, EPI_ISL_782017, EPI_ISL_782021, EPI_ISL_782026, EPI_ISL_782028, EPI_ISL_782030, EPI_ISL_782031, EPI_ISL_782032, EPI_ISL_782035, EPI_ISL_782037, EPI_ISL_782040, EPI_ISL_782042, EPI_ISL_782044, EPI_ISL_782046, EPI_ISL_782049, EPI_ISL_782051, EPI_ISL_782052, EPI_ISL_782054, EPI_ISL_782055, EPI_ISL_782058, EPI_ISL_782061, EPI_ISL_782069, EPI_ISL_782073, EPI_ISL_782077, EPI_ISL_782080, EPI_ISL_782081, EPI_ISL_782087, EPI_ISL_782088, EPI_ISL_782092, EPI_ISL_782093, EPI_ISL_782094, EPI_ISL_782098, EPI_ISL_782103, EPI_ISL_782105, EPI_ISL_782106, EPI_ISL_782113, EPI_ISL_782118, EPI_ISL_782125, EPI_ISL_782129, EPI_ISL_782130, EPI_ISL_782131, EPI_ISL_782134, EPI_ISL_782136, EPI_ISL_782137, EPI_ISL_782143, EPI_ISL_782144, EPI_ISL_782147, EPI_ISL_799430, EPI_ISL_799465, EPI_ISL_799607, EPI_ISL_799710, EPI_ISL_799740, EPI_ISL_799900, EPI_ISL_800581, EPI_ISL_800582, EPI_ISL_859327, EPI_ISL_933481, EPI_ISL_945111, EPI_ISL_945119, EPI_ISL_945171, EPI_ISL_945184, EPI_ISL_945223, EPI_ISL_945269, EPI_ISL_945270, EPI_ISL_945293, EPI_ISL_945299, EPI_ISL_945307, EPI_ISL_945340, EPI_ISL_945342

| | | | |
|---|---|---|---|
| see above | Lighthouse Lab in Milton Keynes | Wellcome Sanger Institute for the COVID-19 Genomics UK (COG-UK) Consortium | Cordelia Langford; David K. Jackson; Dominic Kwiatkowski; Ewan Harrison; Ian Johnston; John Sillitoe on behalf of the Wellcome Sanger Institute COVID-19 Surveillance Team; Roberto Amato; Sonia Goncalves; The Lighthouse Lab in Milton Keynes and Alex Alderton |

EPI_ISL_840062, EPI_ISL_840063, EPI_ISL_840064, EPI_ISL_840065, EPI_ISL_840066, EPI_ISL_840067, EPI_ISL_840068, EPI_ISL_840069, EPI_ISL_840070, EPI_ISL_840071, EPI_ISL_840072, EPI_ISL_840073, EPI_ISL_840074, EPI_ISL_840075, EPI_ISL_840076, EPI_ISL_840077, EPI_ISL_840078, EPI_ISL_840079, EPI_ISL_840080, EPI_ISL_840081, EPI_ISL_840161, EPI_ISL_840162, EPI_ISL_840163, EPI_ISL_840164, EPI_ISL_840165, EPI_ISL_840166, EPI_ISL_840167, EPI_ISL_840168, EPI_ISL_840170, EPI_ISL_840171, EPI_ISL_840172, EPI_ISL_840173, EPI_ISL_840174, EPI_ISL_840175, EPI_ISL_840176, EPI_ISL_840177, EPI_ISL_840178, EPI_ISL_840179, EPI_ISL_840180, EPI_ISL_840181, EPI_ISL_840182, EPI_ISL_840183, EPI_ISL_840184, EPI_ISL_840185, EPI_ISL_950702, EPI_ISL_950703, EPI_ISL_950704, EPI_ISL_950705

| | | | |
|---|---|---|---|
| see above | Lincolnshire Hospitals and DeepSeq Nottingham | COVID-19 Genomics UK (COG-UK) Consortium | Christopher Moore; Fei Sang; Joeseph Chappell; Johnny Debebe; Jonathan Ball; Matthew Carlisle; Matthew Loose; Nadine Holmes; Nichola Duckworth; Patrick McClure; Sarah Walsh; Tim Sloan; Victoria Wright |

EPI_ISL_813803, EPI_ISL_813805, EPI_ISL_813807, EPI_ISL_813808, EPI_ISL_813809, EPI_ISL_814293, EPI_ISL_838499, EPI_ISL_838501, EPI_ISL_838502, EPI_ISL_838503, EPI_ISL_838504, EPI_ISL_838505, EPI_ISL_838507, EPI_ISL_838510, EPI_ISL_838511, EPI_ISL_838512, EPI_ISL_838513, EPI_ISL_838516, EPI_ISL_838520, EPI_ISL_838522, EPI_ISL_838524, EPI_ISL_838525, EPI_ISL_838526, EPI_ISL_838527, EPI_ISL_838528, EPI_ISL_838529, EPI_ISL_838530, EPI_ISL_838531, EPI_ISL_838532, EPI_ISL_838533, EPI_ISL_838534, EPI_ISL_838535, EPI_ISL_838536, EPI_ISL_838537, EPI_ISL_838538, EPI_ISL_838539, EPI_ISL_838540, EPI_ISL_838542, EPI_ISL_838544, EPI_ISL_838545, EPI_ISL_838546, EPI_ISL_838547, EPI_ISL_838548, EPI_ISL_838549, EPI_ISL_838550, EPI_ISL_838551, EPI_ISL_838552, EPI_ISL_838553, EPI_ISL_838554, EPI_ISL_838555, EPI_ISL_838556, EPI_ISL_838557, EPI_ISL_838558, EPI_ISL_838560, EPI_ISL_838561, EPI_ISL_838562, EPI_ISL_838563, EPI_ISL_838564, EPI_ISL_838565, EPI_ISL_838566, EPI_ISL_838567, EPI_ISL_838568, EPI_ISL_838569, EPI_ISL_838570, EPI_ISL_838578, EPI_ISL_838579, EPI_ISL_838646, EPI_ISL_838666, EPI_ISL_865176, EPI_ISL_865239, EPI_ISL_865241, EPI_ISL_865242, EPI_ISL_865243, EPI_ISL_865244, EPI_ISL_865245, EPI_ISL_865247, EPI_ISL_865250, EPI_ISL_865251, EPI_ISL_865252, EPI_ISL_865253, EPI_ISL_865254, EPI_ISL_865278, EPI_ISL_865411, EPI_ISL_865414, EPI_ISL_865420, EPI_ISL_865422, EPI_ISL_865427, EPI_ISL_865433, EPI_ISL_865437, EPI_ISL_865448, EPI_ISL_865459, EPI_ISL_865460, EPI_ISL_865461, EPI_ISL_865462, EPI_ISL_865463, EPI_ISL_865464, EPI_ISL_865465, EPI_ISL_865466, EPI_ISL_919446, EPI_ISL_919447, EPI_ISL_1177842, EPI_ISL_1177848, EPI_ISL_1177849, EPI_ISL_1177853, EPI_ISL_1177855, EPI_ISL_1177873, EPI_ISL_1545984, EPI_ISL_1545985

| | | | |
|---|---|---|---|
| see above | Liverpool Clinical Laboratories | COVID-19 Genomics UK (COG-UK) Consortium | A Alrezaihi; Alessandro Gerada; Alistair Darby; Angela Cowell; Anita Lucaci; Anu Chawla; Cassie Olateju; Catherine Hartley; Charlotte Nelson; Ecaterina Vamos; Elaine O'Toole; Eleanor G Bentley; Ghada T Shawli; Hermione Webster; Isabel García-Dorival; James Johnson; James P Stewart; Jenifer Manson; Joanne Watts; Jones Benjamin; Jordan J Clark; Julian Hiscox; L Luu; Lucille Rainbow; M Almsaud; Margaret Hughes; Mark Whitehead; Matthew Gemmell; Miren Iturriza-Gomara; Muhannad Alruwaili; N.P Randle; Neil Swainston; PKF Gilmore; Parul Sharma; Rebekah Penrice-Randal; Richard Eccles; Richard Gregory; Sam Haldenby; Steve Paterson; Stuart D Armstrong; Trevor Ian Robinson; Ximeng Han |

EPI_ISL_839484, EPI_ISL_839493, EPI_ISL_839494, EPI_ISL_839497, EPI_ISL_839500, EPI_ISL_839504, EPI_ISL_839507, EPI_ISL_839508, EPI_ISL_839509, EPI_ISL_839510, EPI_ISL_839511, EPI_ISL_839516, EPI_ISL_839517, EPI_ISL_839518, EPI_ISL_839561, EPI_ISL_839562, EPI_ISL_839563, EPI_ISL_839564, EPI_ISL_839565, EPI_ISL_839566, EPI_ISL_839567, EPI_ISL_839568, EPI_ISL_839569, EPI_ISL_839570, EPI_ISL_839571, EPI_ISL_839572, EPI_ISL_839573, EPI_ISL_839574, EPI_ISL_839575, EPI_ISL_839576, EPI_ISL_839577, EPI_ISL_839578, EPI_ISL_839579, EPI_ISL_839580, EPI_ISL_839581, EPI_ISL_839582, EPI_ISL_839583, EPI_ISL_839584, EPI_ISL_839585, EPI_ISL_839586, EPI_ISL_839587, EPI_ISL_839588, EPI_ISL_839589, EPI_ISL_839590, EPI_ISL_839591, EPI_ISL_839592, EPI_ISL_839599, EPI_ISL_839600, EPI_ISL_839601, EPI_ISL_839603, EPI_ISL_839615, EPI_ISL_839624, EPI_ISL_839631, EPI_ISL_839632, EPI_ISL_839633, EPI_ISL_839634, EPI_ISL_839635, EPI_ISL_839636, EPI_ISL_839637, EPI_ISL_839638, EPI_ISL_839639, EPI_ISL_839640, EPI_ISL_839642, EPI_ISL_839643, EPI_ISL_839644, EPI_ISL_839645, EPI_ISL_839646, EPI_ISL_839647, EPI_ISL_839648, EPI_ISL_839651, EPI_ISL_839653, EPI_ISL_839658, EPI_ISL_839660, EPI_ISL_839661, EPI_ISL_839663, EPI_ISL_839664, EPI_ISL_839665, EPI_ISL_839666, EPI_ISL_839731, EPI_ISL_866419, EPI_ISL_866441, EPI_ISL_866466, EPI_ISL_866467, EPI_ISL_866468, EPI_ISL_866481, EPI_ISL_866482, EPI_ISL_866483, EPI_ISL_866484, EPI_ISL_866485, EPI_ISL_921137, EPI_ISL_921138, EPI_ISL_921139, EPI_ISL_921140, EPI_ISL_921141, EPI_ISL_921142, EPI_ISL_921143, EPI_ISL_921144, EPI_ISL_921145, EPI_ISL_921146, EPI_ISL_921147, EPI_ISL_921148, EPI_ISL_921149, EPI_ISL_921150, EPI_ISL_921151, EPI_ISL_921152, EPI_ISL_921153, EPI_ISL_921154, EPI_ISL_921155, EPI_ISL_921156, EPI_ISL_921157, EPI_ISL_921158, EPI_ISL_921159, EPI_ISL_921160, EPI_ISL_921161, EPI_ISL_921162, EPI_ISL_921163, EPI_ISL_921164, EPI_ISL_921165, EPI_ISL_921166, EPI_ISL_921167, EPI_ISL_921168, EPI_ISL_921169, EPI_ISL_921170, EPI_ISL_921171, EPI_ISL_921172, EPI_ISL_921173, EPI_ISL_921174, EPI_ISL_921175, EPI_ISL_921176, EPI_ISL_921177, EPI_ISL_921178, EPI_ISL_921179, EPI_ISL_921180, EPI_ISL_921181, EPI_ISL_921182, EPI_ISL_921183, EPI_ISL_921184, EPI_ISL_921187, EPI_ISL_921190, EPI_ISL_921191, EPI_ISL_921192, EPI_ISL_921193, EPI_ISL_921194, EPI_ISL_921195, EPI_ISL_921344, EPI_ISL_921345, EPI_ISL_921346, EPI_ISL_921347, EPI_ISL_921348, EPI_ISL_921351, EPI_ISL_921352, EPI_ISL_921353, EPI_ISL_921354, EPI_ISL_921355, EPI_ISL_921356, EPI_ISL_921358, EPI_ISL_921359, EPI_ISL_921361, EPI_ISL_921682, EPI_ISL_921683, EPI_ISL_921684, EPI_ISL_921685, EPI_ISL_921686, EPI_ISL_921687, EPI_ISL_921688, EPI_ISL_921689, EPI_ISL_921690, EPI_ISL_921691, EPI_ISL_921692, EPI_ISL_921699, EPI_ISL_921700, EPI_ISL_921701, EPI_ISL_921702, EPI_ISL_921703, EPI_ISL_921704, EPI_ISL_921705, EPI_ISL_921706, EPI_ISL_921707, EPI_ISL_921708, EPI_ISL_921709, EPI_ISL_921710, EPI_ISL_921711, EPI_ISL_921712, EPI_ISL_921713, EPI_ISL_921714, EPI_ISL_921715, EPI_ISL_921716, EPI_ISL_921717, EPI_ISL_921718, EPI_ISL_921719, EPI_ISL_921720, EPI_ISL_921721, EPI_ISL_921722, EPI_ISL_921723, EPI_ISL_921724, EPI_ISL_921725, EPI_ISL_921752, EPI_ISL_921753, EPI_ISL_921754, EPI_ISL_921755, EPI_ISL_921756, EPI_ISL_921757, EPI_ISL_921758, EPI_ISL_998587, EPI_ISL_998592, EPI_ISL_998598, EPI_ISL_998616, EPI_ISL_998624, EPI_ISL_998626, EPI_ISL_998630, EPI_ISL_998633, EPI_ISL_998636, EPI_ISL_998639, EPI_ISL_998644, EPI_ISL_998651, EPI_ISL_998676, EPI_ISL_998677, EPI_ISL_998678, EPI_ISL_998679, EPI_ISL_998680, EPI_ISL_998681, EPI_ISL_998682, EPI_ISL_998683, EPI_ISL_998684, EPI_ISL_998685, EPI_ISL_998686, EPI_ISL_998687, EPI_ISL_998688, EPI_ISL_998689, EPI_ISL_998690, EPI_ISL_998691, EPI_ISL_998692, EPI_ISL_998693, EPI_ISL_998694, EPI_ISL_998695, EPI_ISL_998696, EPI_ISL_998697, EPI_ISL_998698, EPI_ISL_998699, EPI_ISL_998700, EPI_ISL_998701, EPI_ISL_998702, EPI_ISL_998703, EPI_ISL_998704, EPI_ISL_998705, EPI_ISL_998706, EPI_ISL_998707, EPI_ISL_998708, EPI_ISL_998709, EPI_ISL_998710, EPI_ISL_998711, EPI_ISL_998712, EPI_ISL_998714, EPI_ISL_998715, EPI_ISL_998716, EPI_ISL_998717, EPI_ISL_998718, EPI_ISL_998729

| | | | |
|---|---|---|---|
| see above | Northumbria University / South Tees Hospitals NHS Foundation Trust / North Cumbria Integrated Care NHS Foundation Trust / North Tees and Hartlepool NHS Foundation Trust / Newcastle Hospitals NHS Foundation Trust | COVID-19 Genomics UK (COG-UK) Consortium | Andrew Nelson; Brendan Payne; Clive Graham; Darren L Smith; Debra Padgett; Edward Barton; Emma Swindells; Garren Scott; Gary Black; Gary Eltringham; Giles S Holt; Greg R Young; Jane Greenaway; Jennifer Collins; John Allan; Joshua Loh; Lynn Dover; Matthew Bashton; Mohammad A Tariq; Paul Baker; Sarah Essex; Steve Liggett; Wen C Yew; Yusri Taha |

EPI_ISL_840712, EPI_ISL_840713, EPI_ISL_840714, EPI_ISL_840716, EPI_ISL_840719, EPI_ISL_840720, EPI_ISL_840723, EPI_ISL_840725, EPI_ISL_840733, EPI_ISL_840734, EPI_ISL_841505, EPI_ISL_867500, EPI_ISL_867501, EPI_ISL_951751

| | | | |
|---|---|---|---|
| see above | Originating lab: Wales Specialist Virology Centre Sequencing lab: Pathogen | Public Health Wales | Alec Birchley; Alexander Adams; Amy Gaskin; Angela Marchbank; Bree Gatica-Wilcox; Catherine Moore; Jason Coombes; Joanne Watkins; Joel Southgate; Johnathan Evans; Laura Gifford; Lauren Gilbert; Lee Graham; Malorie Perry; Matthew Bull; Nicole |

| | | | |
|---|---|---|---|
| | Genomics Unit | Microbiology Cardiff Wales Specialist Virology Centre | Pacchiarini; Sally Corden; Sara Kumziene-Summerhayes; Sara Rey; Sarah Taylor; Simon Cottrell; Sophie Jones; Tom Connor |
| EPI_ISL_763838, EPI_ISL_763845, EPI_ISL_763846, EPI_ISL_763863, EPI_ISL_763928, EPI_ISL_763929, EPI_ISL_763950, EPI_ISL_763953, EPI_ISL_763954, EPI_ISL_763955, EPI_ISL_763956, EPI_ISL_763957, EPI_ISL_763958, EPI_ISL_763961, EPI_ISL_763962, EPI_ISL_763963, EPI_ISL_763964, EPI_ISL_763965, EPI_ISL_763967, EPI_ISL_763968, EPI_ISL_763969, EPI_ISL_763970, EPI_ISL_763971, EPI_ISL_763972, EPI_ISL_763973, EPI_ISL_763974, EPI_ISL_764067, EPI_ISL_764068, EPI_ISL_764085, EPI_ISL_764089, EPI_ISL_764091, EPI_ISL_764092, EPI_ISL_764093, EPI_ISL_764243, EPI_ISL_764526, EPI_ISL_764527, EPI_ISL_764530, EPI_ISL_764531, EPI_ISL_764546, EPI_ISL_764549, EPI_ISL_764550, EPI_ISL_764551, EPI_ISL_764553, EPI_ISL_764554, EPI_ISL_764555, EPI_ISL_764558, EPI_ISL_764559, EPI_ISL_764560, EPI_ISL_764561, EPI_ISL_764562, EPI_ISL_764563, EPI_ISL_764566, EPI_ISL_764568, EPI_ISL_764571, EPI_ISL_814466, EPI_ISL_814467, EPI_ISL_814572, EPI_ISL_814594, EPI_ISL_814604, EPI_ISL_814605, EPI_ISL_814688, EPI_ISL_814689, EPI_ISL_814690, EPI_ISL_814691, EPI_ISL_814692, EPI_ISL_814693, EPI_ISL_814694, EPI_ISL_819585, EPI_ISL_819586, EPI_ISL_819587, EPI_ISL_819588, EPI_ISL_819589, EPI_ISL_819590, EPI_ISL_819591, EPI_ISL_819592, EPI_ISL_819593, EPI_ISL_819605, EPI_ISL_819606, EPI_ISL_819607, EPI_ISL_819608, EPI_ISL_819612, EPI_ISL_819613, EPI_ISL_819614, EPI_ISL_819615, EPI_ISL_819616, EPI_ISL_819617, EPI_ISL_819618, EPI_ISL_819619, EPI_ISL_819620, EPI_ISL_819621, EPI_ISL_819622, EPI_ISL_819623, EPI_ISL_819624, EPI_ISL_819625, EPI_ISL_819626, EPI_ISL_819627, EPI_ISL_819628, EPI_ISL_819629, EPI_ISL_819630, EPI_ISL_819631, EPI_ISL_819632, EPI_ISL_819633, EPI_ISL_819634, EPI_ISL_819635, EPI_ISL_819636, EPI_ISL_819637, EPI_ISL_819638, EPI_ISL_819639, EPI_ISL_819640, EPI_ISL_819641, EPI_ISL_819642, EPI_ISL_819643, EPI_ISL_819644, EPI_ISL_819645, EPI_ISL_819646, EPI_ISL_819653, EPI_ISL_819654, EPI_ISL_819655, EPI_ISL_819656, EPI_ISL_819657, EPI_ISL_819658, EPI_ISL_819659, EPI_ISL_819660, EPI_ISL_819661, EPI_ISL_819662, EPI_ISL_819663, EPI_ISL_819664, EPI_ISL_819665, EPI_ISL_819666, EPI_ISL_819667, EPI_ISL_819668, EPI_ISL_819669, EPI_ISL_819670, EPI_ISL_819675, EPI_ISL_819676, EPI_ISL_819677, EPI_ISL_819678, EPI_ISL_819679, EPI_ISL_819680, EPI_ISL_819681, EPI_ISL_819682, EPI_ISL_819683, EPI_ISL_819684, EPI_ISL_819685, EPI_ISL_819689, EPI_ISL_819690, EPI_ISL_819691, EPI_ISL_819692, EPI_ISL_819693, EPI_ISL_819694, EPI_ISL_819695, EPI_ISL_819696, EPI_ISL_819697, EPI_ISL_819698, EPI_ISL_819699, EPI_ISL_819700, EPI_ISL_819701, EPI_ISL_819702, EPI_ISL_819703, EPI_ISL_819704, EPI_ISL_819705, EPI_ISL_819706, EPI_ISL_819707, EPI_ISL_819708, EPI_ISL_819709, EPI_ISL_819710, EPI_ISL_819711, EPI_ISL_819712, EPI_ISL_819713, EPI_ISL_819714, EPI_ISL_819715, EPI_ISL_819716, EPI_ISL_819717, EPI_ISL_819718, EPI_ISL_819719, EPI_ISL_819720, EPI_ISL_819721, EPI_ISL_819722, EPI_ISL_819723, EPI_ISL_819732, EPI_ISL_819733, EPI_ISL_819734, EPI_ISL_819735, EPI_ISL_819736, EPI_ISL_819737, EPI_ISL_819738, EPI_ISL_819739, EPI_ISL_819740, EPI_ISL_819741, EPI_ISL_819742, EPI_ISL_819743, EPI_ISL_819744, EPI_ISL_819745, EPI_ISL_819746, EPI_ISL_819747, EPI_ISL_819748, EPI_ISL_819749, EPI_ISL_819750, EPI_ISL_819751, EPI_ISL_819752, EPI_ISL_819753, EPI_ISL_819754, EPI_ISL_819755, EPI_ISL_819756, EPI_ISL_819759, EPI_ISL_819760, EPI_ISL_819761, EPI_ISL_819762, EPI_ISL_819764, EPI_ISL_819765, EPI_ISL_819766, EPI_ISL_819767, EPI_ISL_819768, EPI_ISL_819769, EPI_ISL_819770, EPI_ISL_819771, EPI_ISL_819772, EPI_ISL_819773, EPI_ISL_819774, EPI_ISL_819775, EPI_ISL_819776, EPI_ISL_819777, EPI_ISL_819778, EPI_ISL_819779, EPI_ISL_819780, EPI_ISL_819781, EPI_ISL_819782, EPI_ISL_819783, EPI_ISL_819784, EPI_ISL_819785, EPI_ISL_819786, EPI_ISL_819787, EPI_ISL_819788, EPI_ISL_819789, EPI_ISL_819790, EPI_ISL_819791, EPI_ISL_819792, EPI_ISL_819793, EPI_ISL_819794, EPI_ISL_819796, EPI_ISL_819797, EPI_ISL_819798, EPI_ISL_819803, EPI_ISL_819804, EPI_ISL_819805, EPI_ISL_819806, EPI_ISL_819807, EPI_ISL_819808, EPI_ISL_819809, EPI_ISL_819810, EPI_ISL_819811, EPI_ISL_819812, EPI_ISL_819813, EPI_ISL_819814, EPI_ISL_819815, EPI_ISL_819816, EPI_ISL_819817, EPI_ISL_819818, EPI_ISL_819819, EPI_ISL_819820, EPI_ISL_819822, EPI_ISL_819823, EPI_ISL_819824, EPI_ISL_819825, EPI_ISL_819826, EPI_ISL_819827, EPI_ISL_819835, EPI_ISL_819836, EPI_ISL_819837, EPI_ISL_819838, EPI_ISL_819839, EPI_ISL_819840, EPI_ISL_819841, EPI_ISL_819842, EPI_ISL_819843, EPI_ISL_819844, EPI_ISL_819845, EPI_ISL_819846, EPI_ISL_819847, EPI_ISL_819848, EPI_ISL_819849, EPI_ISL_819850, EPI_ISL_819852, EPI_ISL_819853, EPI_ISL_819854, EPI_ISL_819855, EPI_ISL_819856, EPI_ISL_819857, EPI_ISL_819858, EPI_ISL_819859, EPI_ISL_819860, EPI_ISL_819861, EPI_ISL_819862, EPI_ISL_819863, EPI_ISL_819866, EPI_ISL_819868, EPI_ISL_819869, EPI_ISL_819870, EPI_ISL_819871, EPI_ISL_819872, EPI_ISL_840256, EPI_ISL_840257, EPI_ISL_840260, EPI_ISL_840261, EPI_ISL_840262, EPI_ISL_840263, EPI_ISL_840264, EPI_ISL_840266, EPI_ISL_840267, EPI_ISL_840269, EPI_ISL_840270, EPI_ISL_840271, EPI_ISL_840272, EPI_ISL_840274, EPI_ISL_840276, EPI_ISL_840278, EPI_ISL_840279, EPI_ISL_840280, EPI_ISL_840282, EPI_ISL_840283, EPI_ISL_840284, EPI_ISL_840299, EPI_ISL_840300, EPI_ISL_840310, EPI_ISL_840312, EPI_ISL_840349, EPI_ISL_840350, EPI_ISL_840351, EPI_ISL_840353, EPI_ISL_840357, EPI_ISL_840358, EPI_ISL_840359, EPI_ISL_840360, EPI_ISL_840361, EPI_ISL_840362, EPI_ISL_840363, EPI_ISL_867035, EPI_ISL_867175, EPI_ISL_867178, EPI_ISL_867182, EPI_ISL_950780, EPI_ISL_950797, EPI_ISL_950800, EPI_ISL_950801, EPI_ISL_950900, EPI_ISL_950901, EPI_ISL_950903, EPI_ISL_950904, EPI_ISL_950905, EPI_ISL_950908, EPI_ISL_950909, EPI_ISL_950910, EPI_ISL_950911, EPI_ISL_950912, EPI_ISL_950913, EPI_ISL_950914, EPI_ISL_950915, EPI_ISL_950916, EPI_ISL_950917, EPI_ISL_950918, EPI_ISL_950921, EPI_ISL_1833051, EPI_ISL_1833052 | | | |
| see above | Oxford Viromics, NDM, University of Oxford; Oxford University Hospitals; Basingstoke and North Hampshire Hospital | COVID-19 Genomics UK (COG-UK) Consortium | Alex Mobbs; Amy Trebes; Anita Justice; Catrin Moore; Christophe Fraser; David Bonsall; David Buck; Emma Wise; George Macintyre; Jessica Lynch; John Todd; Mariateresa de Cesare; Matilde Mori; Monique Andersson; Nathan Moore; Nick Cortes; Robert Shaw; Stephen Kidd; Tanya Golubchik; Timothy Peto |
| EPI_ISL_741462, EPI_ISL_741463, EPI_ISL_741464, EPI_ISL_741466, EPI_ISL_741467, EPI_ISL_741468, EPI_ISL_741469, EPI_ISL_741472, EPI_ISL_741474, EPI_ISL_741475, EPI_ISL_741479, EPI_ISL_741482, EPI_ISL_741486, EPI_ISL_741487, EPI_ISL_741488, EPI_ISL_741489, EPI_ISL_741490, EPI_ISL_741491, EPI_ISL_741492, EPI_ISL_741493, EPI_ISL_741494, EPI_ISL_741495, EPI_ISL_741496, EPI_ISL_741497, EPI_ISL_741498, EPI_ISL_741499, EPI_ISL_741500, EPI_ISL_741501, EPI_ISL_741502, EPI_ISL_741503, EPI_ISL_741504, EPI_ISL_741505, EPI_ISL_741506, EPI_ISL_741507, EPI_ISL_741508, EPI_ISL_741509, EPI_ISL_741510, EPI_ISL_741511, EPI_ISL_741512, EPI_ISL_741513, EPI_ISL_741514, EPI_ISL_741515, EPI_ISL_741517, EPI_ISL_741518, EPI_ISL_741519, EPI_ISL_741520, EPI_ISL_741521, EPI_ISL_741522, EPI_ISL_741523, EPI_ISL_741524, EPI_ISL_741525, EPI_ISL_741526, EPI_ISL_741527, EPI_ISL_741528, EPI_ISL_741530, EPI_ISL_741531, EPI_ISL_741533, EPI_ISL_741534, EPI_ISL_741535, EPI_ISL_741536, EPI_ISL_741537, EPI_ISL_741538, EPI_ISL_741539, EPI_ISL_741540, EPI_ISL_741541, EPI_ISL_741542, EPI_ISL_741543, EPI_ISL_741545, EPI_ISL_741546, EPI_ISL_741547, EPI_ISL_741548, EPI_ISL_741549, EPI_ISL_741550, EPI_ISL_741551, EPI_ISL_741553, EPI_ISL_741554, EPI_ISL_741555, EPI_ISL_741556, EPI_ISL_741557, EPI_ISL_741558, EPI_ISL_741559, EPI_ISL_741560, EPI_ISL_741561, EPI_ISL_741562, EPI_ISL_741563, EPI_ISL_741564, EPI_ISL_741565, EPI_ISL_741566, EPI_ISL_741568, EPI_ISL_741569, EPI_ISL_741570, EPI_ISL_741572, EPI_ISL_741573, EPI_ISL_741574, EPI_ISL_741575, EPI_ISL_741576, EPI_ISL_741577, EPI_ISL_741578, EPI_ISL_741579, EPI_ISL_741580, EPI_ISL_741581, EPI_ISL_741582, EPI_ISL_741588, EPI_ISL_741589, EPI_ISL_741590, EPI_ISL_741591, EPI_ISL_741593, EPI_ISL_741601, EPI_ISL_763372, EPI_ISL_763483, EPI_ISL_763490, EPI_ISL_763502, EPI_ISL_763662, EPI_ISL_763683, EPI_ISL_763684, EPI_ISL_763689, EPI_ISL_763727, EPI_ISL_763781, EPI_ISL_763851, EPI_ISL_763852, EPI_ISL_763908, EPI_ISL_763909, EPI_ISL_763910, EPI_ISL_763911, EPI_ISL_763912, EPI_ISL_763913, EPI_ISL_763914, EPI_ISL_763915, EPI_ISL_763916, EPI_ISL_763917, EPI_ISL_763918, EPI_ISL_763919, EPI_ISL_763920, EPI_ISL_763921, EPI_ISL_763922, EPI_ISL_763923, EPI_ISL_764116, EPI_ISL_764126, EPI_ISL_764194, EPI_ISL_764205, EPI_ISL_764207, EPI_ISL_764237, EPI_ISL_764270, EPI_ISL_764445, EPI_ISL_764446, EPI_ISL_764447, EPI_ISL_764448, EPI_ISL_764449, EPI_ISL_764450, EPI_ISL_764451, EPI_ISL_764452, EPI_ISL_764453, EPI_ISL_764454, EPI_ISL_764455, EPI_ISL_764456, EPI_ISL_764457, EPI_ISL_764458, EPI_ISL_764459, EPI_ISL_764460, EPI_ISL_764461, EPI_ISL_764462, EPI_ISL_764463, EPI_ISL_764464, EPI_ISL_764465, EPI_ISL_764466, EPI_ISL_764467, EPI_ISL_764468, EPI_ISL_764469, EPI_ISL_764470, EPI_ISL_764471, EPI_ISL_764472, EPI_ISL_764473, EPI_ISL_764474, EPI_ISL_764475, EPI_ISL_764476, EPI_ISL_764477, EPI_ISL_764478, EPI_ISL_764479, EPI_ISL_764480, EPI_ISL_764481, EPI_ISL_764482, EPI_ISL_764483, EPI_ISL_764484, EPI_ISL_764489, EPI_ISL_764492, EPI_ISL_866577, EPI_ISL_866578, EPI_ISL_866584, EPI_ISL_866586, EPI_ISL_866802, EPI_ISL_866807, EPI_ISL_866808, EPI_ISL_866812, EPI_ISL_866814, EPI_ISL_866828, EPI_ISL_950592 | | | |
| see above | Quadram Institute Bioscience | COVID-19 Genomics UK (COG-UK) Consortium | Alexander J Trotter; Alison E. Mather; Alp Aydin; Ana P. Tedim; Anastasia Kolyva; Andrew Bell; Andrew J. Page; Claire Stuart; Dave J. Baker; Gemma L. Kay; John Wain; Justin O'Grady; Leonardo de Oliveira Martins; Lizzie Meadows; Maria Diaz; Mark Webber; Muhammed Yasir; Nabil-Fareed Alikhan; Ngozi Elumogo; Nicholas M. Thomson; Rachael Stanley; Rachel Gilroy; Reenesh Prakash; Samir Dervisevic; Samuel Bloomfield; Steven Rudder; Thanh Le-Viet |
| EPI_ISL_741625, EPI_ISL_741626, EPI_ISL_741627, EPI_ISL_741628, EPI_ISL_741629, EPI_ISL_741630, EPI_ISL_741631, EPI_ISL_741632, EPI_ISL_741633, EPI_ISL_741634, EPI_ISL_741635, EPI_ISL_741636, EPI_ISL_741637, EPI_ISL_741638, EPI_ISL_741639, EPI_ISL_741640, EPI_ISL_741641, EPI_ISL_741642, EPI_ISL_741643, EPI_ISL_741644, EPI_ISL_741645, EPI_ISL_741646, EPI_ISL_741647, EPI_ISL_741648, EPI_ISL_741649, EPI_ISL_741650, EPI_ISL_741651, EPI_ISL_741652, EPI_ISL_741653, EPI_ISL_741654, EPI_ISL_741655, EPI_ISL_764493, EPI_ISL_764494, EPI_ISL_819579, EPI_ISL_819580, EPI_ISL_819581, EPI_ISL_819582, EPI_ISL_819583, EPI_ISL_819584, EPI_ISL_819594, EPI_ISL_819595, EPI_ISL_819596, EPI_ISL_819597, EPI_ISL_819598, EPI_ISL_819599, EPI_ISL_819600, EPI_ISL_819601, EPI_ISL_819602, EPI_ISL_819603, EPI_ISL_819604, EPI_ISL_819609, EPI_ISL_819610, EPI_ISL_819611, EPI_ISL_819647, EPI_ISL_819648, EPI_ISL_819649, EPI_ISL_819650, EPI_ISL_819651, EPI_ISL_819652, EPI_ISL_819671, EPI_ISL_819672, EPI_ISL_819673, EPI_ISL_819674, EPI_ISL_819686, EPI_ISL_819687, EPI_ISL_819688, EPI_ISL_819724, EPI_ISL_819725, EPI_ISL_819726, EPI_ISL_819727, EPI_ISL_819728, EPI_ISL_819729, EPI_ISL_819730, EPI_ISL_819731, EPI_ISL_819757, EPI_ISL_819758, EPI_ISL_819763, EPI_ISL_819795, EPI_ISL_819799, EPI_ISL_819800, EPI_ISL_819801, EPI_ISL_819802, EPI_ISL_819821, EPI_ISL_819828, EPI_ISL_819829, EPI_ISL_819830, EPI_ISL_819831, EPI_ISL_819832, EPI_ISL_819833, EPI_ISL_819834, EPI_ISL_819851, EPI_ISL_819864, EPI_ISL_819865, EPI_ISL_819867, EPI_ISL_866891, EPI_ISL_866892, EPI_ISL_866893, EPI_ISL_866894, EPI_ISL_866895, EPI_ISL_866904, EPI_ISL_866990, EPI_ISL_866991, EPI_ISL_949652, EPI_ISL_949653, EPI_ISL_949654 | | | |
| see above | Queens Medical Centre, Clinical Microbiology Department / DeepSeq Nottingham | COVID-19 Genomics UK (COG-UK) Consortium | Christopher Moore; Fei Sang; Gemma Clark; Hannah Howson-Wells; Johnny Debebe; Jonathan Ball; Joseph Chappell; Manjinder Khakh; Matthew Carlisle; Matthew Loose; Michelle M Lister; Nadine Holmes; Patrick McClure; Theocharis Tsoleridis; Vicki M Fleming; Victoria Wright; Wendy Smith |
| EPI_ISL_921037, EPI_ISL_998201, EPI_ISL_998202, EPI_ISL_998203, EPI_ISL_998204, EPI_ISL_998205, EPI_ISL_998206, EPI_ISL_998207, EPI_ISL_998208, EPI_ISL_998209, EPI_ISL_998210 | | | |
| see above | Regional Virus Laboratory, Belfast Health and Social Care Trust | COVID-19 Genomics UK (COG-UK) Consortium | Alison Watt; Ciara Cox; Conall McCaughey; David Simpson; Derek Fairley; James McKenna; Mairead Connor; Susan Feeney; Tanya Curran; Zoltan Molnar |
| EPI_ISL_754245, EPI_ISL_754254, EPI_ISL_754267, EPI_ISL_754268, EPI_ISL_754269, EPI_ISL_754272, EPI_ISL_754273, EPI_ISL_754277, EPI_ISL_754278, EPI_ISL_754282, EPI_ISL_754283, EPI_ISL_754284, EPI_ISL_754289, EPI_ISL_754312, EPI_ISL_754313, EPI_ISL_754315, EPI_ISL_754317, EPI_ISL_754319, EPI_ISL_754320, EPI_ISL_754321, EPI_ISL_754322, EPI_ISL_754323, EPI_ISL_754324, EPI_ISL_754325, EPI_ISL_754326, EPI_ISL_754327, EPI_ISL_754328, EPI_ISL_754329, EPI_ISL_754331, EPI_ISL_754362, EPI_ISL_754363, EPI_ISL_754365, EPI_ISL_754366, EPI_ISL_754367, EPI_ISL_754368, EPI_ISL_754369, EPI_ISL_754370, EPI_ISL_754371, EPI_ISL_754372, EPI_ISL_754373, EPI_ISL_754374, EPI_ISL_754375, EPI_ISL_754377, EPI_ISL_754378, EPI_ISL_754379, EPI_ISL_754381, EPI_ISL_754382, EPI_ISL_754383, EPI_ISL_754384, EPI_ISL_754385, EPI_ISL_754386, EPI_ISL_754387, EPI_ISL_756135, EPI_ISL_756136, EPI_ISL_756137, EPI_ISL_756138, EPI_ISL_756139, EPI_ISL_756140, EPI_ISL_756141, EPI_ISL_756142, EPI_ISL_756143, EPI_ISL_756144, EPI_ISL_756145, EPI_ISL_756146, EPI_ISL_756147, EPI_ISL_756148, EPI_ISL_756149, EPI_ISL_756150, EPI_ISL_756151, EPI_ISL_756152, EPI_ISL_756153, EPI_ISL_756154, EPI_ISL_756155, EPI_ISL_756156, EPI_ISL_756157, EPI_ISL_756158, EPI_ISL_756159, EPI_ISL_756160, EPI_ISL_756161, EPI_ISL_756162, EPI_ISL_756163, EPI_ISL_762995, EPI_ISL_762996, EPI_ISL_766060, EPI_ISL_766061, EPI_ISL_766062, EPI_ISL_766065, EPI_ISL_766074, EPI_ISL_766075, EPI_ISL_766078, EPI_ISL_766082, EPI_ISL_766085, EPI_ISL_766086, EPI_ISL_766088, EPI_ISL_766089, EPI_ISL_766093, EPI_ISL_766099, EPI_ISL_766100, EPI_ISL_766104, EPI_ISL_766108, EPI_ISL_766111, EPI_ISL_766122, EPI_ISL_766142, EPI_ISL_766143, EPI_ISL_766145, EPI_ISL_766146, EPI_ISL_766147, EPI_ISL_766148, EPI_ISL_766149, EPI_ISL_766150, EPI_ISL_766151, EPI_ISL_766152, EPI_ISL_766153, EPI_ISL_766154, EPI_ISL_766155, EPI_ISL_766156, EPI_ISL_766157, EPI_ISL_766159, EPI_ISL_766160, EPI_ISL_766161, EPI_ISL_766162, EPI_ISL_766163, EPI_ISL_766164, EPI_ISL_766165, EPI_ISL_766166, EPI_ISL_766167, EPI_ISL_766168, EPI_ISL_766169, EPI_ISL_766170, EPI_ISL_766171, EPI_ISL_766172, EPI_ISL_766173, EPI_ISL_766174, EPI_ISL_766175, EPI_ISL_766176, EPI_ISL_766177, EPI_ISL_766178, EPI_ISL_766179, EPI_ISL_766180, EPI_ISL_766181, EPI_ISL_766182, EPI_ISL_766183, EPI_ISL_766184, EPI_ISL_766185, EPI_ISL_766186, EPI_ISL_766187, EPI_ISL_766188, EPI_ISL_766189, EPI_ISL_766190, EPI_ISL_766191, EPI_ISL_766192, EPI_ISL_766193, EPI_ISL_766194, EPI_ISL_766195, EPI_ISL_766196, EPI_ISL_766197, EPI_ISL_766198, EPI_ISL_766199, EPI_ISL_766200, EPI_ISL_766201, EPI_ISL_766204, EPI_ISL_766205, EPI_ISL_766215, EPI_ISL_766216, EPI_ISL_766217, EPI_ISL_766223, EPI_ISL_766227, EPI_ISL_766228, EPI_ISL_766229, EPI_ISL_766234, EPI_ISL_766236, EPI_ISL_766243, EPI_ISL_766244, EPI_ISL_766251, EPI_ISL_766252, EPI_ISL_766253, EPI_ISL_766254, EPI_ISL_766257, EPI_ISL_766258, EPI_ISL_766310, EPI_ISL_766314, EPI_ISL_766316, EPI_ISL_766397, EPI_ISL_766398, EPI_ISL_766402, EPI_ISL_766404, EPI_ISL_766406, EPI_ISL_766408, EPI_ISL_766412, EPI_ISL_766413, EPI_ISL_766414, EPI_ISL_766415, EPI_ISL_766416, EPI_ISL_766417, EPI_ISL_766418, EPI_ISL_766419, EPI_ISL_766420, EPI_ISL_766421, EPI_ISL_766422, EPI_ISL_766423, EPI_ISL_766424, EPI_ISL_766426, EPI_ISL_766427, EPI_ISL_766428, EPI_ISL_766429, EPI_ISL_766430, EPI_ISL_766431, EPI_ISL_766432, EPI_ISL_766433, EPI_ISL_766434, EPI_ISL_766435, EPI_ISL_766436, EPI_ISL_766437, EPI_ISL_766438, EPI_ISL_766439, EPI_ISL_766440, EPI_ISL_766441, EPI_ISL_766442, EPI_ISL_766443, EPI_ISL_766444, EPI_ISL_766445, EPI_ISL_766446, EPI_ISL_766447, EPI_ISL_766448, EPI_ISL_766449, EPI_ISL_766450, EPI_ISL_766451, EPI_ISL_766452, EPI_ISL_766453, EPI_ISL_766454, EPI_ISL_766455, EPI_ISL_766456, EPI_ISL_766457, EPI_ISL_766458, EPI_ISL_766459, EPI_ISL_766460, EPI_ISL_766461, EPI_ISL_766462, EPI_ISL_766464, EPI_ISL_766465, EPI_ISL_766466, EPI_ISL_766467, EPI_ISL_766468, EPI_ISL_766469, EPI_ISL_766470, EPI_ISL_766471, EPI_ISL_766472, EPI_ISL_766473, EPI_ISL_766474, EPI_ISL_766475, EPI_ISL_766476, EPI_ISL_766477, EPI_ISL_766478, EPI_ISL_766479, EPI_ISL_766480, EPI_ISL_766481, EPI_ISL_766482, EPI_ISL_766483, EPI_ISL_766484, EPI_ISL_766485, EPI_ISL_766486, EPI_ISL_766487, EPI_ISL_766488, EPI_ISL_766489, EPI_ISL_766490, EPI_ISL_766491, EPI_ISL_766492, EPI_ISL_766493, EPI_ISL_766494, EPI_ISL_766495, EPI_ISL_766496, EPI_ISL_766497, EPI_ISL_766498, EPI_ISL_766499, EPI_ISL_766500, EPI_ISL_766501, EPI_ISL_766502, EPI_ISL_766503, EPI_ISL_766504, EPI_ISL_766517, EPI_ISL_766519, EPI_ISL_766520, EPI_ISL_766521, EPI_ISL_766553, EPI_ISL_766554, EPI_ISL_766560, EPI_ISL_766565, EPI_ISL_766566, EPI_ISL_766859, EPI_ISL_766860, EPI_ISL_769873, EPI_ISL_791183, EPI_ISL_791187, EPI_ISL_791188, EPI_ISL_791189, EPI_ISL_791204, EPI_ISL_791205, EPI_ISL_791206, EPI_ISL_791207, EPI_ISL_791210, EPI_ISL_791212, EPI_ISL_791213, EPI_ISL_791214, EPI_ISL_791215, EPI_ISL_791216, EPI_ISL_791217, EPI_ISL_791218, EPI_ISL_791219, EPI_ISL_791221, EPI_ISL_804214, EPI_ISL_804215, EPI_ISL_804227, EPI_ISL_804228, EPI_ISL_804229, EPI_ISL_804230, EPI_ISL_804231, EPI_ISL_804254, EPI_ISL_804266, EPI_ISL_804267, EPI_ISL_804268, EPI_ISL_804269, EPI_ISL_804270, EPI_ISL_804271, EPI_ISL_836981, EPI_ISL_836982, EPI_ISL_836983, EPI_ISL_836984, EPI_ISL_836985, EPI_ISL_836986, EPI_ISL_836987, EPI_ISL_836988, EPI_ISL_837066, EPI_ISL_837068, EPI_ISL_837071, EPI_ISL_837074, EPI_ISL_837082, EPI_ISL_837084, EPI_ISL_837085, EPI_ISL_837090, EPI_ISL_837097, EPI_ISL_837098, EPI_ISL_837099, EPI_ISL_837101, EPI_ISL_837102, EPI_ISL_837106, EPI_ISL_837115, EPI_ISL_837116, EPI_ISL_837117, EPI_ISL_837118, EPI_ISL_837119, EPI_ISL_837187, EPI_ISL_837195, EPI_ISL_837200, EPI_ISL_837201, EPI_ISL_837202, EPI_ISL_837203, EPI_ISL_837204, EPI_ISL_837205, EPI_ISL_837206, EPI_ISL_837207, EPI_ISL_837208, EPI_ISL_846589, EPI_ISL_846590, EPI_ISL_846593, EPI_ISL_862190, EPI_ISL_1296543, EPI_ISL_1296544 | | | |
| see above | Respiratory Virus Unit, National Infection Service, Public Health England | COVID-19 Genomics UK (COG-UK) Consortium | PHE Covid Sequencing Team |
| EPI_ISL_1833410 | St George's University Hospitals NHS Foundation Trust | COVID-19 Genomics UK (COG-UK) Consortium | Adam Witney; Cassie Pope; Irene Monahan; Joshua Taylor; Ken Laing; NgeeKeong Tan |
| EPI_ISL_842676, EPI_ISL_842682, EPI_ISL_842683, EPI_ISL_842684, EPI_ISL_842685, EPI_ISL_842686, EPI_ISL_842687, EPI_ISL_842688, EPI_ISL_842689, EPI_ISL_842690, EPI_ISL_842691, EPI_ISL_842692, EPI_ISL_842695, EPI_ISL_842698, EPI_ISL_842699, EPI_ISL_842700, EPI_ISL_842701, EPI_ISL_842702, EPI_ISL_842703, EPI_ISL_842704, EPI_ISL_842705, EPI_ISL_842706, EPI_ISL_842707, EPI_ISL_842708, EPI_ISL_842709, EPI_ISL_842710, EPI_ISL_842711, EPI_ISL_842719, EPI_ISL_842721, EPI_ISL_842722, EPI_ISL_842723, EPI_ISL_842724, EPI_ISL_842725, EPI_ISL_842726, EPI_ISL_842727, EPI_ISL_842728, EPI_ISL_842730, EPI_ISL_866049, EPI_ISL_866050, EPI_ISL_866051, EPI_ISL_866068, EPI_ISL_866069, EPI_ISL_866070, EPI_ISL_866116, EPI_ISL_866118, EPI_ISL_866119, EPI_ISL_866121, EPI_ISL_866132, EPI_ISL_866133, EPI_ISL_866136, EPI_ISL_866148, EPI_ISL_866149, EPI_ISL_866151, EPI_ISL_866164, EPI_ISL_866179, EPI_ISL_920176, EPI_ISL_920178, EPI_ISL_920185, EPI_ISL_920220, EPI_ISL_920221, EPI_ISL_920240, EPI_ISL_920242, EPI_ISL_920243, EPI_ISL_920248, EPI_ISL_920249, EPI_ISL_920250, EPI_ISL_920251, EPI_ISL_920256, EPI_ISL_920257, EPI_ISL_920258, EPI_ISL_920259, EPI_ISL_920260, EPI_ISL_920269, EPI_ISL_920270, EPI_ISL_920279, EPI_ISL_920280, EPI_ISL_920281, EPI_ISL_920290, EPI_ISL_920291, EPI_ISL_920292, EPI_ISL_920301, EPI_ISL_920302, EPI_ISL_920303, EPI_ISL_920321, EPI_ISL_920356, EPI_ISL_920373, EPI_ISL_920461, EPI_ISL_920468, EPI_ISL_920469, EPI_ISL_920474, EPI_ISL_920476, EPI_ISL_920477, EPI_ISL_920478, EPI_ISL_920479, EPI_ISL_920483, EPI_ISL_920485, EPI_ISL_920486, EPI_ISL_920487, EPI_ISL_920492, EPI_ISL_920494, EPI_ISL_920495, EPI_ISL_920502, EPI_ISL_920505, EPI_ISL_920508, EPI_ISL_920510, EPI_ISL_920513, EPI_ISL_920520, EPI_ISL_920521, EPI_ISL_920526, EPI_ISL_920527, EPI_ISL_920538, EPI_ISL_920539, EPI_ISL_920540, EPI_ISL_920547, EPI_ISL_920548, EPI_ISL_920549, EPI_ISL_920550, EPI_ISL_920551, EPI_ISL_920558, EPI_ISL_920559, EPI_ISL_920560, EPI_ISL_920568, EPI_ISL_920575, EPI_ISL_920582, EPI_ISL_920584, EPI_ISL_920585, EPI_ISL_920594, EPI_ISL_920596, EPI_ISL_920603, EPI_ISL_920612, EPI_ISL_920613, EPI_ISL_920620, EPI_ISL_920621, EPI_ISL_920631, EPI_ISL_920636, EPI_ISL_920637, EPI_ISL_920647, EPI_ISL_920659, EPI_ISL_920660, EPI_ISL_920661, EPI_ISL_920666, EPI_ISL_920667, EPI_ISL_920674, EPI_ISL_920676, EPI_ISL_920688, EPI_ISL_920696, EPI_ISL_920709, EPI_ISL_920710, EPI_ISL_920717, EPI_ISL_920719, EPI_ISL_920720, EPI_ISL_920721, EPI_ISL_920729, EPI_ISL_920730, EPI_ISL_920735, EPI_ISL_920736, EPI_ISL_920737, EPI_ISL_920744, EPI_ISL_920745, EPI_ISL_920746, EPI_ISL_920753, EPI_ISL_920754, EPI_ISL_920762, EPI_ISL_920767, EPI_ISL_920768, EPI_ISL_920773, EPI_ISL_920774, EPI_ISL_1104800, EPI_ISL_1104904, EPI_ISL_1104921, EPI_ISL_1104926, EPI_ISL_1104980, EPI_ISL_1105109, EPI_ISL_1105121, EPI_ISL_1248370, EPI_ISL_1248371, EPI_ISL_1248381, EPI_ISL_1248392, EPI_ISL_1248393, EPI_ISL_1248404, EPI_ISL_1248415, EPI_ISL_1248426, EPI_ISL_1248437, EPI_ISL_1637061 | | | |
| see above | University College London Hospital | COVID-19 Genomics UK (COG-UK) Consortium | Catherine Houlihan; Dan Frampton; Dr Catherine Houlihan; Dr Daniel Frampton; Dr Judith Heaney; Dr Moira Spyer; Dr Paul Grant and Dr Eleni Nastouli; Judith Heaney; Matthew Byott; Moira Spyer and Eleni Nastouli; Stuart Kirk |
| EPI_ISL_724608, EPI_ISL_741309, EPI_ISL_741310, EPI_ISL_741311, EPI_ISL_741326, EPI_ISL_741327, EPI_ISL_741329, EPI_ISL_741330, EPI_ISL_741331, EPI_ISL_741332, EPI_ISL_741333, EPI_ISL_741334, EPI_ISL_741335, EPI_ISL_741336, EPI_ISL_741337, EPI_ISL_741338, EPI_ISL_741339, EPI_ISL_741340, EPI_ISL_741341, EPI_ISL_741342, EPI_ISL_741343, EPI_ISL_741344, EPI_ISL_741345, EPI_ISL_741346, EPI_ISL_763377, EPI_ISL_763393, EPI_ISL_763397, EPI_ISL_763398, EPI_ISL_763399, EPI_ISL_763404, EPI_ISL_763442, EPI_ISL_763455, EPI_ISL_763468, EPI_ISL_763485, EPI_ISL_763486, EPI_ISL_763491, EPI_ISL_763500, EPI_ISL_763510, EPI_ISL_763520, EPI_ISL_763540, EPI_ISL_763544, EPI_ISL_763545, EPI_ISL_763551, EPI_ISL_763552, EPI_ISL_763567, EPI_ISL_763578, EPI_ISL_763582, EPI_ISL_763628, EPI_ISL_763640, EPI_ISL_763643, EPI_ISL_763644, EPI_ISL_763682, EPI_ISL_763687, EPI_ISL_763700, EPI_ISL_763720, EPI_ISL_763728, EPI_ISL_763730, EPI_ISL_763745, EPI_ISL_763756, EPI_ISL_763757, EPI_ISL_763767, EPI_ISL_763772, EPI_ISL_763784, EPI_ISL_763785, EPI_ISL_763809, EPI_ISL_763810, EPI_ISL_763811, EPI_ISL_763813, EPI_ISL_763826, EPI_ISL_764037, EPI_ISL_764038, EPI_ISL_764039, EPI_ISL_764040, EPI_ISL_764041, EPI_ISL_764042, EPI_ISL_764043, EPI_ISL_764044, EPI_ISL_764045, EPI_ISL_764046, EPI_ISL_764047, EPI_ISL_764048, EPI_ISL_764049, EPI_ISL_764050, EPI_ISL_764051, EPI_ISL_764052, EPI_ISL_764053, EPI_ISL_764054, EPI_ISL_764055, EPI_ISL_764056, EPI_ISL_764057, EPI_ISL_764058, EPI_ISL_764059, EPI_ISL_764060, EPI_ISL_764128, EPI_ISL_764131, EPI_ISL_764146, EPI_ISL_764160, EPI_ISL_764175, EPI_ISL_764228, EPI_ISL_764241, EPI_ISL_764383, EPI_ISL_764384, EPI_ISL_764385, EPI_ISL_764386, EPI_ISL_813832, EPI_ISL_813833, EPI_ISL_813834, EPI_ISL_813835, EPI_ISL_813836, EPI_ISL_813837, EPI_ISL_813838, EPI_ISL_813839, EPI_ISL_813840, EPI_ISL_813841, EPI_ISL_813842, EPI_ISL_813843, EPI_ISL_813844, EPI_ISL_813847, EPI_ISL_813866, EPI_ISL_813880, EPI_ISL_813894, EPI_ISL_813895, EPI_ISL_813907, EPI_ISL_813908, EPI_ISL_813909, EPI_ISL_813910, EPI_ISL_813911, EPI_ISL_813912, EPI_ISL_813914, EPI_ISL_813915, EPI_ISL_813916, EPI_ISL_813917, EPI_ISL_813918, EPI_ISL_813919, EPI_ISL_813920, EPI_ISL_813921, EPI_ISL_813922, EPI_ISL_813923, EPI_ISL_813924, EPI_ISL_813925, EPI_ISL_813926, EPI_ISL_813927, EPI_ISL_813928, EPI_ISL_813929, EPI_ISL_813931, EPI_ISL_838687, EPI_ISL_838688, EPI_ISL_838689, EPI_ISL_838691, EPI_ISL_838692, EPI_ISL_838693, EPI_ISL_838695, EPI_ISL_838696, EPI_ISL_838697, EPI_ISL_838698, EPI_ISL_838699, EPI_ISL_838705, EPI_ISL_838706, EPI_ISL_838707, EPI_ISL_838708, EPI_ISL_838709, EPI_ISL_838710, EPI_ISL_838711, EPI_ISL_838712, EPI_ISL_838713, EPI_ISL_838715, EPI_ISL_838717, EPI_ISL_838718, EPI_ISL_838721, EPI_ISL_838723, EPI_ISL_838724, EPI_ISL_838725, EPI_ISL_838726, EPI_ISL_838727, EPI_ISL_838730, EPI_ISL_838731, EPI_ISL_838732, EPI_ISL_838733, EPI_ISL_838739, EPI_ISL_838744, EPI_ISL_839337, EPI_ISL_839342, EPI_ISL_839343, EPI_ISL_839344, EPI_ISL_839345, EPI_ISL_839346, EPI_ISL_839347, EPI_ISL_839348, EPI_ISL_839349, EPI_ISL_839350, EPI_ISL_839351, EPI_ISL_839352, EPI_ISL_839353, EPI_ISL_839354, EPI_ISL_839355, EPI_ISL_839356, EPI_ISL_839357, EPI_ISL_865687, EPI_ISL_865690, EPI_ISL_865720, EPI_ISL_865721, EPI_ISL_865722, EPI_ISL_865723, EPI_ISL_865724, EPI_ISL_865725, EPI_ISL_865726, EPI_ISL_865727, EPI_ISL_865728, EPI_ISL_865729, EPI_ISL_865730, EPI_ISL_865731, EPI_ISL_865732, EPI_ISL_865733, EPI_ISL_865734, EPI_ISL_865735, EPI_ISL_865736, EPI_ISL_865737, EPI_ISL_865738, EPI_ISL_865740, EPI_ISL_865741, EPI_ISL_865743, EPI_ISL_865745, EPI_ISL_865746, EPI_ISL_865747, EPI_ISL_919725, EPI_ISL_919726, EPI_ISL_919727, EPI_ISL_919737, EPI_ISL_919750, EPI_ISL_919751, EPI_ISL_919752, EPI_ISL_919753, EPI_ISL_919754, EPI_ISL_949785, EPI_ISL_950166, EPI_ISL_997571, EPI_ISL_998011, EPI_ISL_1048049, EPI_ISL_1309096, EPI_ISL_1386879, EPI_ISL_1474865, EPI_ISL_1637105, EPI_ISL_1637106, EPI_ISL_1637107, EPI_ISL_1730339, EPI_ISL_1730357, EPI_ISL_1730358, EPI_ISL_1730392 | | | |
| see above | University College London, Great Ormond Street Hospital for Children NHS Foundation Trust, Imperial College Healthcare NHS Trust | COVID-19 Genomics UK (COG-UK) Consortium | Alison Holmes; Charlotte Williams; Helena Tutill; Jacqueline Findlay; James Price; Judith Breuer; Julianne Brown; Kathryn Harris; Leysa Forrest; Mark Kristiansen; Paola Niola; Paola Resende Silva; Patricia Dyal; Paul Randell; Rachel Williams; Samuel Weeks; Sergi Castellano; Sunando Roy; Tony Brooks; Yasmin Panchbhaya |
| EPI_ISL_1047914, EPI_ISL_1047915, EPI_ISL_1047921, EPI_ISL_1047922, EPI_ISL_1047923, EPI_ISL_1047924, EPI_ISL_1047925, EPI_ISL_1047926, EPI_ISL_1047927, EPI_ISL_1047928, EPI_ISL_1047929, EPI_ISL_1047931, EPI_ISL_1047938, EPI_ISL_1047939, EPI_ISL_1047940, EPI_ISL_1047941, EPI_ISL_1047955, EPI_ISL_1050352, EPI_ISL_1050353, EPI_ISL_2125934 | | | |
| see above | University Hospitals Of Leicester NHS Trust and DeepSeq Nottingham | COVID-19 Genomics UK (COG-UK) Consortium | Christopher Holmes; Christopher Moore; Fei Sang; Joeseph Chappell; Johnny Debebe; Jonathan Ball; Julian Tang; Karlie Fallon; Matthew Carlisle; Matthew Loose; Nadine Holmes; Patrick McClure; Paul Bird; Thomas Helmer; Victoria Wright |
| EPI_ISL_813060, EPI_ISL_813061, EPI_ISL_813062, EPI_ISL_813087, EPI_ISL_813144, EPI_ISL_813145, EPI_ISL_813146, EPI_ISL_813147, EPI_ISL_813148, EPI_ISL_813149, EPI_ISL_813150, EPI_ISL_813151, EPI_ISL_813152, EPI_ISL_813153, EPI_ISL_918828, EPI_ISL_918829, EPI_ISL_918830, EPI_ISL_918831, EPI_ISL_918832, EPI_ISL_918833, EPI_ISL_918834, EPI_ISL_918835, EPI_ISL_918836, EPI_ISL_918837, EPI_ISL_918838, EPI_ISL_918839, EPI_ISL_918840, EPI_ISL_918841, EPI_ISL_918846, EPI_ISL_918847, EPI_ISL_918848, EPI_ISL_918849, EPI_ISL_918957, EPI_ISL_918969, EPI_ISL_918970, EPI_ISL_918971, EPI_ISL_918972 | | | |
| see above | University of Birmingham | COVID-19 Genomics UK (COG-UK) Consortium | Alex Richter; Andrew Bosworth; Andrew Bosworth. Queen Elizabeth Hospital: Anna Casey; Andrew D Beggs PHE Heartlands Lab: Husam Osman; Charlotte Poxon; Institute of Microbiology; Joanne Stockton; Josh Quick; Kasun Wanigasooriya; Mike Kidd; Nicholas Loman. University of Birmingham Testing Laboratory: Celina M Whalley; Oliver Pickles; Radoslaw Poplawski; Samuel Nicholls; University of Birmingham: Claire McMurray; Will Rowe |
| EPI_ISL_763376, EPI_ISL_763383, EPI_ISL_763387, EPI_ISL_763412, EPI_ISL_763414, EPI_ISL_763431, EPI_ISL_763433, EPI_ISL_763434, EPI_ISL_763435, EPI_ISL_763436, EPI_ISL_763437, EPI_ISL_763452, EPI_ISL_763461, EPI_ISL_763496, EPI_ISL_763530, EPI_ISL_763531, EPI_ISL_763532, EPI_ISL_763533, EPI_ISL_763534, EPI_ISL_763535, EPI_ISL_763536, EPI_ISL_763558, EPI_ISL_763559, EPI_ISL_763629, EPI_ISL_763630, EPI_ISL_763631, EPI_ISL_763670, EPI_ISL_763698, EPI_ISL_763699, EPI_ISL_763712, EPI_ISL_763713, EPI_ISL_763714, EPI_ISL_763715, EPI_ISL_763716, EPI_ISL_763717, EPI_ISL_763818, EPI_ISL_764084, EPI_ISL_764129, EPI_ISL_764139, EPI_ISL_764158, EPI_ISL_764163, EPI_ISL_764179, EPI_ISL_764185, EPI_ISL_764196, EPI_ISL_764214, EPI_ISL_764215, EPI_ISL_764225, EPI_ISL_764378, EPI_ISL_764379, EPI_ISL_764380, EPI_ISL_764381, EPI_ISL_764382, EPI_ISL_813357, EPI_ISL_813358, EPI_ISL_813359, EPI_ISL_813360, EPI_ISL_813361, EPI_ISL_813362, EPI_ISL_813363, EPI_ISL_813364, EPI_ISL_813365, EPI_ISL_813366, EPI_ISL_813367, EPI_ISL_813368, EPI_ISL_813369, EPI_ISL_813370, EPI_ISL_813371, EPI_ISL_813372, EPI_ISL_813373, EPI_ISL_813374, EPI_ISL_813375, EPI_ISL_813376, EPI_ISL_813377, EPI_ISL_813378, EPI_ISL_813379, EPI_ISL_813380, EPI_ISL_813381, EPI_ISL_813382, EPI_ISL_813383, EPI_ISL_813384, EPI_ISL_813385, EPI_ISL_813386, EPI_ISL_813387, EPI_ISL_813388, EPI_ISL_813389, EPI_ISL_813390, EPI_ISL_813391, EPI_ISL_813392, EPI_ISL_813393, EPI_ISL_813394, EPI_ISL_813395, EPI_ISL_813396, EPI_ISL_813397, EPI_ISL_813398, EPI_ISL_813399, EPI_ISL_813400, EPI_ISL_813401, EPI_ISL_813402, EPI_ISL_813403, EPI_ISL_813404, EPI_ISL_813405, EPI_ISL_813406, EPI_ISL_813407, EPI_ISL_813408, EPI_ISL_813409, EPI_ISL_813410, EPI_ISL_813411, EPI_ISL_813412, EPI_ISL_813413, EPI_ISL_813414, EPI_ISL_813415, EPI_ISL_813416, EPI_ISL_813417, EPI_ISL_813418, EPI_ISL_813419, EPI_ISL_813420, EPI_ISL_813421, EPI_ISL_813422, EPI_ISL_813423, EPI_ISL_813424, EPI_ISL_813425, EPI_ISL_813426, EPI_ISL_813427, EPI_ISL_813428, EPI_ISL_813429, EPI_ISL_813430, EPI_ISL_813431, EPI_ISL_813432, EPI_ISL_813433, EPI_ISL_813434, EPI_ISL_813435, EPI_ISL_813436, EPI_ISL_813437, EPI_ISL_813438, EPI_ISL_813439, EPI_ISL_813440, EPI_ISL_813441, EPI_ISL_813442, EPI_ISL_813443, EPI_ISL_813444, EPI_ISL_813445, EPI_ISL_813446, EPI_ISL_813447, EPI_ISL_813448, EPI_ISL_813449, EPI_ISL_813450, EPI_ISL_813451, EPI_ISL_813452, EPI_ISL_813457, EPI_ISL_813458, EPI_ISL_813459, EPI_ISL_813460, EPI_ISL_813461, EPI_ISL_813462, EPI_ISL_813463, EPI_ISL_813464, EPI_ISL_813465, EPI_ISL_813466, EPI_ISL_996576, EPI_ISL_996582, EPI_ISL_996585, EPI_ISL_996596, EPI_ISL_997318, EPI_ISL_1103877 | | | |
| see above | University of Exeter | COVID-19 Genomics UK (COG-UK) Consortium | Aaron Jeffries; Audrey Farbos; Ben Temperton; Jane Masoli; Joanna Warwick-Dugdale; Michelle Michelsen; Robyn Manley; Stephen Michell |
| EPI_ISL_997114, EPI_ISL_997115, EPI_ISL_997116, EPI_ISL_997123, EPI_ISL_997124, EPI_ISL_1104244, EPI_ISL_1104245, EPI_ISL_1104246, EPI_ISL_1104247, EPI_ISL_1104248, EPI_ISL_1104249, EPI_ISL_1104250, EPI_ISL_1104255, EPI_ISL_1104258 | | | |

| | | | |
|---|---|---|---|
| see above | Virology Department, Royal Infirmary of Edinburgh, NHS Lothian / School of Biological Sciences, University of Edinburgh | COVID-19 Genomics UK (COG-UK) Consortium | Colquhoun R; Cotton S; Dewar R; Hill V; Jackson B; McCrone JT; McHugh M; O'Toole Á; Rambaut A; Rooke S; Scher E; Templeton K; Yu X |

EPI_ISL_742199, EPI_ISL_742200, EPI_ISL_742201, EPI_ISL_742203, EPI_ISL_742204, EPI_ISL_742205, EPI_ISL_742206, EPI_ISL_742207, EPI_ISL_742208, EPI_ISL_742209, EPI_ISL_742210, EPI_ISL_742211, EPI_ISL_742212, EPI_ISL_742213, EPI_ISL_742214, EPI_ISL_742215, EPI_ISL_742216, EPI_ISL_742217, EPI_ISL_742218, EPI_ISL_742219, EPI_ISL_742220, EPI_ISL_742221, EPI_ISL_742222, EPI_ISL_742223, EPI_ISL_742224, EPI_ISL_742225, EPI_ISL_742226, EPI_ISL_742227, EPI_ISL_742228, EPI_ISL_742229, EPI_ISL_742230, EPI_ISL_742231, EPI_ISL_742232, EPI_ISL_742233, EPI_ISL_742234, EPI_ISL_742235, EPI_ISL_742236, EPI_ISL_742237, EPI_ISL_742238, EPI_ISL_742239, EPI_ISL_742240, EPI_ISL_742241, EPI_ISL_742242, EPI_ISL_763647, EPI_ISL_763892, EPI_ISL_763893, EPI_ISL_763894, EPI_ISL_763895, EPI_ISL_763896, EPI_ISL_763897, EPI_ISL_763898, EPI_ISL_763899, EPI_ISL_763900, EPI_ISL_763901, EPI_ISL_763902, EPI_ISL_763904, EPI_ISL_763905, EPI_ISL_763906, EPI_ISL_764073, EPI_ISL_764074, EPI_ISL_764075, EPI_ISL_764083, EPI_ISL_814460, EPI_ISL_814461, EPI_ISL_814462, EPI_ISL_814463, EPI_ISL_814464, EPI_ISL_814465, EPI_ISL_814679, EPI_ISL_838225, EPI_ISL_838227, EPI_ISL_838228, EPI_ISL_838229, EPI_ISL_838230, EPI_ISL_838231, EPI_ISL_838232, EPI_ISL_838233, EPI_ISL_838235, EPI_ISL_838236, EPI_ISL_838237, EPI_ISL_838238, EPI_ISL_838239, EPI_ISL_838275, EPI_ISL_838280, EPI_ISL_838289, EPI_ISL_865072, EPI_ISL_919314, EPI_ISL_919315, EPI_ISL_919316, EPI_ISL_919317, EPI_ISL_919318, EPI_ISL_919319, EPI_ISL_919320, EPI_ISL_919371, EPI_ISL_919372, EPI_ISL_919373, EPI_ISL_919374, EPI_ISL_919375, EPI_ISL_919376, EPI_ISL_919377, EPI_ISL_919399, EPI_ISL_919400, EPI_ISL_919401

| | | | |
|---|---|---|---|
| see above | Virology Department, Royal Infirmary of Edinburgh, NHS Lothian / School of Biological Sciences, University of Edinburgh / Institute of Genetics and Molecular Medicine, University of Edinburgh | COVID-19 Genomics UK (COG-UK) Consortium | Balcaza C; Colquhoun R; Dewar R; Gallagher M; Hill V; Jackson B; McCrone JT; McHugh M; O'Toole Á; Rambaut A; Rooke S; Scher E; Templeton K; Williams TC; Yu X |

EPI_ISL_727861, EPI_ISL_727874, EPI_ISL_727877, EPI_ISL_727879, EPI_ISL_727880, EPI_ISL_727882, EPI_ISL_727887, EPI_ISL_727890, EPI_ISL_727891, EPI_ISL_727896, EPI_ISL_727897, EPI_ISL_727901, EPI_ISL_727909, EPI_ISL_727911, EPI_ISL_727915, EPI_ISL_727927, EPI_ISL_727932, EPI_ISL_727942, EPI_ISL_727948, EPI_ISL_727949, EPI_ISL_727950, EPI_ISL_727954, EPI_ISL_727955, EPI_ISL_727956, EPI_ISL_727960, EPI_ISL_727961, EPI_ISL_727963, EPI_ISL_727972, EPI_ISL_727973, EPI_ISL_727979, EPI_ISL_727980, EPI_ISL_741936, EPI_ISL_741937, EPI_ISL_741942, EPI_ISL_741943, EPI_ISL_741948, EPI_ISL_741950, EPI_ISL_741951, EPI_ISL_741953, EPI_ISL_741954, EPI_ISL_741956, EPI_ISL_741958, EPI_ISL_741960, EPI_ISL_741961, EPI_ISL_741962, EPI_ISL_741964, EPI_ISL_741965, EPI_ISL_741967, EPI_ISL_741969, EPI_ISL_741971, EPI_ISL_741974, EPI_ISL_741975, EPI_ISL_741976, EPI_ISL_741977, EPI_ISL_741978, EPI_ISL_741983, EPI_ISL_741984, EPI_ISL_741986, EPI_ISL_741989, EPI_ISL_741990, EPI_ISL_741991, EPI_ISL_741999, EPI_ISL_742000, EPI_ISL_742004, EPI_ISL_742007, EPI_ISL_742010, EPI_ISL_742015, EPI_ISL_742018, EPI_ISL_742020, EPI_ISL_742025, EPI_ISL_742030, EPI_ISL_742031, EPI_ISL_742033, EPI_ISL_742034, EPI_ISL_742039, EPI_ISL_742041, EPI_ISL_742046, EPI_ISL_742048, EPI_ISL_742049, EPI_ISL_742054, EPI_ISL_742060, EPI_ISL_742062, EPI_ISL_742063, EPI_ISL_742067, EPI_ISL_742072, EPI_ISL_742074, EPI_ISL_742076, EPI_ISL_742077, EPI_ISL_742079, EPI_ISL_742080, EPI_ISL_742082, EPI_ISL_742083, EPI_ISL_742084, EPI_ISL_742086, EPI_ISL_742090, EPI_ISL_742092, EPI_ISL_742094, EPI_ISL_742100, EPI_ISL_742105, EPI_ISL_742107, EPI_ISL_742108, EPI_ISL_742109, EPI_ISL_742111, EPI_ISL_742112, EPI_ISL_742113, EPI_ISL_742115, EPI_ISL_742116, EPI_ISL_763365, EPI_ISL_763402, EPI_ISL_763403, EPI_ISL_763419, EPI_ISL_763427, EPI_ISL_763428, EPI_ISL_763429, EPI_ISL_763501, EPI_ISL_763506, EPI_ISL_763507, EPI_ISL_763588, EPI_ISL_763635, EPI_ISL_763639, EPI_ISL_763645, EPI_ISL_763651, EPI_ISL_763653, EPI_ISL_763654, EPI_ISL_763677, EPI_ISL_763741, EPI_ISL_763742, EPI_ISL_763743, EPI_ISL_763744, EPI_ISL_763746, EPI_ISL_763868, EPI_ISL_763869, EPI_ISL_763871, EPI_ISL_764032, EPI_ISL_764033, EPI_ISL_764076, EPI_ISL_764100, EPI_ISL_764107, EPI_ISL_764111, EPI_ISL_764226, EPI_ISL_764227, EPI_ISL_764269, EPI_ISL_765152, EPI_ISL_765153, EPI_ISL_765156, EPI_ISL_765157, EPI_ISL_765158, EPI_ISL_765159, EPI_ISL_765160, EPI_ISL_765161, EPI_ISL_765162, EPI_ISL_765165, EPI_ISL_765166, EPI_ISL_765167, EPI_ISL_765168, EPI_ISL_765175, EPI_ISL_765176, EPI_ISL_765177, EPI_ISL_765178, EPI_ISL_765180, EPI_ISL_765181, EPI_ISL_765182, EPI_ISL_765183, EPI_ISL_765185, EPI_ISL_765189, EPI_ISL_765192, EPI_ISL_765193, EPI_ISL_765194, EPI_ISL_765198, EPI_ISL_765199, EPI_ISL_765200, EPI_ISL_765201, EPI_ISL_765207, EPI_ISL_816240, EPI_ISL_816245, EPI_ISL_816259, EPI_ISL_816272, EPI_ISL_816274, EPI_ISL_816282, EPI_ISL_816304, EPI_ISL_816344, EPI_ISL_816347, EPI_ISL_816385, EPI_ISL_816400, EPI_ISL_816404, EPI_ISL_816418, EPI_ISL_816446, EPI_ISL_816460, EPI_ISL_816481, EPI_ISL_816486, EPI_ISL_816487, EPI_ISL_816488, EPI_ISL_816504, EPI_ISL_816511, EPI_ISL_816514, EPI_ISL_816528, EPI_ISL_816529, EPI_ISL_816535, EPI_ISL_816547, EPI_ISL_816554, EPI_ISL_816567, EPI_ISL_816580, EPI_ISL_816617, EPI_ISL_816626, EPI_ISL_816630, EPI_ISL_816644, EPI_ISL_816650, EPI_ISL_842313, EPI_ISL_842317, EPI_ISL_924268, EPI_ISL_924316

| | | | |
|---|---|---|---|
| see above | Virology Department, Sheffield Teaching Hospitals NHS Foundation Trust/Department of Infection, Immunity and Cardiovascular Disease, The Medical School, University of Sheffield | COVID-19 Genomics UK (COG-UK) Consortium | Adri Angyal; Alex Keeley; Benjamin Lindsey; Cariad Evans; Danielle Groves; Dave Partridge; Katie Johnson; Laura Carrilero; Luke Green; Matthew Parker; Matthew Wyles; Mehmet Yavuz; Mohammad Raza; Nikki Smith; Paul Parsons; Rachel Tucker; Rebecca Brown; Thushan de Silva |

EPI_ISL_743878, EPI_ISL_743905, EPI_ISL_743906, EPI_ISL_743907, EPI_ISL_743908, EPI_ISL_743909, EPI_ISL_743910, EPI_ISL_743911, EPI_ISL_743912, EPI_ISL_743913, EPI_ISL_743914, EPI_ISL_743915, EPI_ISL_743916, EPI_ISL_743917, EPI_ISL_743918, EPI_ISL_743919, EPI_ISL_743920, EPI_ISL_743921, EPI_ISL_743922, EPI_ISL_743923, EPI_ISL_743924, EPI_ISL_743925, EPI_ISL_743926, EPI_ISL_743927, EPI_ISL_743928, EPI_ISL_743929, EPI_ISL_743930, EPI_ISL_743931, EPI_ISL_743932, EPI_ISL_743933, EPI_ISL_743934, EPI_ISL_743935, EPI_ISL_743936, EPI_ISL_743940, EPI_ISL_743941, EPI_ISL_744067, EPI_ISL_744085, EPI_ISL_744098, EPI_ISL_744101, EPI_ISL_744103, EPI_ISL_744104, EPI_ISL_744105, EPI_ISL_744107, EPI_ISL_744111, EPI_ISL_744112, EPI_ISL_744115, EPI_ISL_744116, EPI_ISL_744117, EPI_ISL_744118, EPI_ISL_744119, EPI_ISL_744120, EPI_ISL_744121, EPI_ISL_744122, EPI_ISL_744123, EPI_ISL_744124, EPI_ISL_744125, EPI_ISL_744126, EPI_ISL_744127, EPI_ISL_744128, EPI_ISL_744129, EPI_ISL_744130, EPI_ISL_744131, EPI_ISL_744132, EPI_ISL_744133, EPI_ISL_744134, EPI_ISL_763371, EPI_ISL_763374, EPI_ISL_763378, EPI_ISL_763410, EPI_ISL_763411, EPI_ISL_763418, EPI_ISL_763441, EPI_ISL_763443, EPI_ISL_763445, EPI_ISL_763458, EPI_ISL_763473, EPI_ISL_763474, EPI_ISL_763488, EPI_ISL_763494, EPI_ISL_763509, EPI_ISL_763525, EPI_ISL_763538, EPI_ISL_763546, EPI_ISL_763557, EPI_ISL_763577, EPI_ISL_763579, EPI_ISL_763581, EPI_ISL_763585, EPI_ISL_763589, EPI_ISL_763591, EPI_ISL_763594, EPI_ISL_763597, EPI_ISL_763599, EPI_ISL_763633, EPI_ISL_763657, EPI_ISL_763671, EPI_ISL_763679, EPI_ISL_763680, EPI_ISL_763692, EPI_ISL_763718, EPI_ISL_763722, EPI_ISL_763733, EPI_ISL_763735, EPI_ISL_763739, EPI_ISL_763748, EPI_ISL_763751, EPI_ISL_763752, EPI_ISL_763753, EPI_ISL_763754, EPI_ISL_763758, EPI_ISL_763759, EPI_ISL_763761, EPI_ISL_763762, EPI_ISL_763765, EPI_ISL_763766, EPI_ISL_763770, EPI_ISL_763773, EPI_ISL_763774, EPI_ISL_763775, EPI_ISL_763779, EPI_ISL_763783, EPI_ISL_763788, EPI_ISL_763814, EPI_ISL_763822, EPI_ISL_763864, EPI_ISL_763975, EPI_ISL_763983, EPI_ISL_763984, EPI_ISL_763985, EPI_ISL_763986, EPI_ISL_763987, EPI_ISL_763988, EPI_ISL_763990, EPI_ISL_763991, EPI_ISL_763992, EPI_ISL_763993, EPI_ISL_763994, EPI_ISL_763995, EPI_ISL_763996, EPI_ISL_763997, EPI_ISL_763998, EPI_ISL_763999, EPI_ISL_764000, EPI_ISL_764001, EPI_ISL_764002, EPI_ISL_764003, EPI_ISL_764004, EPI_ISL_764005, EPI_ISL_764034, EPI_ISL_764062, EPI_ISL_764063, EPI_ISL_764065, EPI_ISL_764080, EPI_ISL_764142, EPI_ISL_764168, EPI_ISL_764180, EPI_ISL_764199, EPI_ISL_764212, EPI_ISL_764222, EPI_ISL_764242, EPI_ISL_764244, EPI_ISL_764245, EPI_ISL_764246, EPI_ISL_764247, EPI_ISL_764248, EPI_ISL_764249, EPI_ISL_764251, EPI_ISL_764252, EPI_ISL_764253, EPI_ISL_764254, EPI_ISL_764256, EPI_ISL_764257, EPI_ISL_764258, EPI_ISL_764259, EPI_ISL_764260, EPI_ISL_764268, EPI_ISL_764273, EPI_ISL_764274, EPI_ISL_764275, EPI_ISL_764277, EPI_ISL_764282, EPI_ISL_764283, EPI_ISL_764577, EPI_ISL_764578, EPI_ISL_764579, EPI_ISL_764580, EPI_ISL_764583, EPI_ISL_764584, EPI_ISL_764586, EPI_ISL_764587, EPI_ISL_764588, EPI_ISL_764589, EPI_ISL_764590, EPI_ISL_764591, EPI_ISL_764592, EPI_ISL_764593, EPI_ISL_764594, EPI_ISL_764595, EPI_ISL_764596, EPI_ISL_764597, EPI_ISL_764598, EPI_ISL_764624, EPI_ISL_764657, EPI_ISL_764658, EPI_ISL_764659, EPI_ISL_764660, EPI_ISL_764661, EPI_ISL_764662, EPI_ISL_764663, EPI_ISL_764664, EPI_ISL_764665, EPI_ISL_764666, EPI_ISL_764667, EPI_ISL_764668, EPI_ISL_764669, EPI_ISL_764670, EPI_ISL_764671, EPI_ISL_764672, EPI_ISL_764673, EPI_ISL_764674, EPI_ISL_764675, EPI_ISL_764676, EPI_ISL_764677, EPI_ISL_764678, EPI_ISL_764679, EPI_ISL_764680, EPI_ISL_764681, EPI_ISL_764682, EPI_ISL_764683, EPI_ISL_764684, EPI_ISL_764685, EPI_ISL_764686, EPI_ISL_764687, EPI_ISL_764688, EPI_ISL_764689, EPI_ISL_764690, EPI_ISL_764691, EPI_ISL_764692, EPI_ISL_764693, EPI_ISL_764694, EPI_ISL_764695, EPI_ISL_764696, EPI_ISL_764697, EPI_ISL_764698, EPI_ISL_764699, EPI_ISL_764700, EPI_ISL_764701, EPI_ISL_764702, EPI_ISL_764703, EPI_ISL_764704, EPI_ISL_764705, EPI_ISL_764706, EPI_ISL_764707, EPI_ISL_764708, EPI_ISL_764709, EPI_ISL_764710, EPI_ISL_764711, EPI_ISL_764712, EPI_ISL_764713, EPI_ISL_764714, EPI_ISL_764715, EPI_ISL_764716, EPI_ISL_764717, EPI_ISL_764718, EPI_ISL_764719, EPI_ISL_764720, EPI_ISL_764721, EPI_ISL_764722, EPI_ISL_764724, EPI_ISL_764725, EPI_ISL_764726, EPI_ISL_764727, EPI_ISL_764728, EPI_ISL_764729, EPI_ISL_764730, EPI_ISL_764731, EPI_ISL_764732, EPI_ISL_764733, EPI_ISL_764734, EPI_ISL_764735, EPI_ISL_764736, EPI_ISL_764737, EPI_ISL_764738, EPI_ISL_764742, EPI_ISL_764743, EPI_ISL_764744, EPI_ISL_764745, EPI_ISL_764757, EPI_ISL_764758, EPI_ISL_764759, EPI_ISL_764760, EPI_ISL_764761, EPI_ISL_764762, EPI_ISL_764763, EPI_ISL_764764, EPI_ISL_764765, EPI_ISL_764816, EPI_ISL_764817, EPI_ISL_764818, EPI_ISL_764824, EPI_ISL_764826, EPI_ISL_764827, EPI_ISL_764830, EPI_ISL_764831, EPI_ISL_764832, EPI_ISL_764833, EPI_ISL_764834, EPI_ISL_764835, EPI_ISL_764836, EPI_ISL_764837, EPI_ISL_764838, EPI_ISL_764839, EPI_ISL_764840, EPI_ISL_764841, EPI_ISL_764842, EPI_ISL_764844, EPI_ISL_764845, EPI_ISL_764846, EPI_ISL_764847, EPI_ISL_764848, EPI_ISL_764849, EPI_ISL_764850, EPI_ISL_764851, EPI_ISL_764852, EPI_ISL_764853, EPI_ISL_764854, EPI_ISL_764855, EPI_ISL_764856, EPI_ISL_764857, EPI_ISL_764858, EPI_ISL_764859, EPI_ISL_764860, EPI_ISL_764861, EPI_ISL_764862, EPI_ISL_764863, EPI_ISL_764864, EPI_ISL_764865, EPI_ISL_764866, EPI_ISL_764867, EPI_ISL_764868, EPI_ISL_764869, EPI_ISL_764870, EPI_ISL_764871, EPI_ISL_764872, EPI_ISL_764873, EPI_ISL_764874, EPI_ISL_764875, EPI_ISL_764876, EPI_ISL_764877, EPI_ISL_764878, EPI_ISL_764879, EPI_ISL_764880, EPI_ISL_764881, EPI_ISL_764882, EPI_ISL_764883, EPI_ISL_764884, EPI_ISL_764885, EPI_ISL_764886, EPI_ISL_764887, EPI_ISL_764888, EPI_ISL_764889, EPI_ISL_764890, EPI_ISL_764891, EPI_ISL_764892, EPI_ISL_764893, EPI_ISL_764894, EPI_ISL_764895, EPI_ISL_764896, EPI_ISL_764897, EPI_ISL_764898, EPI_ISL_764899, EPI_ISL_764900, EPI_ISL_764901, EPI_ISL_764902, EPI_ISL_764903, EPI_ISL_764904, EPI_ISL_764905, EPI_ISL_764906, EPI_ISL_764908, EPI_ISL_764909, EPI_ISL_764910, EPI_ISL_764911, EPI_ISL_764912, EPI_ISL_764913, EPI_ISL_764914, EPI_ISL_764915, EPI_ISL_764916, EPI_ISL_764917, EPI_ISL_764918, EPI_ISL_764919, EPI_ISL_764920, EPI_ISL_764921, EPI_ISL_764922, EPI_ISL_764923, EPI_ISL_764924, EPI_ISL_764925, EPI_ISL_764926, EPI_ISL_764927, EPI_ISL_764928, EPI_ISL_764929, EPI_ISL_764930, EPI_ISL_764931, EPI_ISL_764932, EPI_ISL_764933, EPI_ISL_764934, EPI_ISL_764935, EPI_ISL_764936, EPI_ISL_764937, EPI_ISL_764938, EPI_ISL_764939, EPI_ISL_764940, EPI_ISL_764941, EPI_ISL_764942, EPI_ISL_764943, EPI_ISL_764944, EPI_ISL_764945, EPI_ISL_764946, EPI_ISL_764947, EPI_ISL_764949, EPI_ISL_764950, EPI_ISL_764951, EPI_ISL_764971, EPI_ISL_764974, EPI_ISL_764980, EPI_ISL_764981, EPI_ISL_764982, EPI_ISL_764983, EPI_ISL_764984, EPI_ISL_764985, EPI_ISL_764986, EPI_ISL_764987, EPI_ISL_764988, EPI_ISL_764989, EPI_ISL_764990, EPI_ISL_764991, EPI_ISL_764992, EPI_ISL_764993, EPI_ISL_764994, EPI_ISL_764995, EPI_ISL_764996, EPI_ISL_764997, EPI_ISL_764998, EPI_ISL_764999, EPI_ISL_765000, EPI_ISL_765001, EPI_ISL_765002, EPI_ISL_765003, EPI_ISL_765004, EPI_ISL_765005, EPI_ISL_765006, EPI_ISL_765007, EPI_ISL_765008, EPI_ISL_765009, EPI_ISL_765010, EPI_ISL_765011, EPI_ISL_765012, EPI_ISL_765014, EPI_ISL_765015, EPI_ISL_765016, EPI_ISL_765017, EPI_ISL_765018, EPI_ISL_765019, EPI_ISL_765020, EPI_ISL_765021, EPI_ISL_765022, EPI_ISL_765023, EPI_ISL_765024, EPI_ISL_765026, EPI_ISL_765027, EPI_ISL_765028, EPI_ISL_765029, EPI_ISL_765030, EPI_ISL_765031, EPI_ISL_765032, EPI_ISL_765033, EPI_ISL_765034, EPI_ISL_765035, EPI_ISL_765036, EPI_ISL_765037, EPI_ISL_765038, EPI_ISL_765039, EPI_ISL_765040, EPI_ISL_765041, EPI_ISL_765042, EPI_ISL_765043, EPI_ISL_765044, EPI_ISL_765045, EPI_ISL_765046, EPI_ISL_765047, EPI_ISL_765048, EPI_ISL_765049, EPI_ISL_765050, EPI_ISL_765051, EPI_ISL_765052, EPI_ISL_765053, EPI_ISL_765054, EPI_ISL_765055, EPI_ISL_765056, EPI_ISL_765057, EPI_ISL_765058, EPI_ISL_765059, EPI_ISL_765060, EPI_ISL_765061, EPI_ISL_765062, EPI_ISL_765063, EPI_ISL_765064, EPI_ISL_765065, EPI_ISL_765066, EPI_ISL_765067, EPI_ISL_765068, EPI_ISL_765069, EPI_ISL_765070, EPI_ISL_765071, EPI_ISL_765072, EPI_ISL_765073, EPI_ISL_765074, EPI_ISL_765075, EPI_ISL_765076, EPI_ISL_765077, EPI_ISL_765078, EPI_ISL_765079, EPI_ISL_765080, EPI_ISL_765081, EPI_ISL_765082, EPI_ISL_765083, EPI_ISL_765084, EPI_ISL_765085, EPI_ISL_765086, EPI_ISL_765087, EPI_ISL_765088, EPI_ISL_765089, EPI_ISL_765090, EPI_ISL_765091, EPI_ISL_765092, EPI_ISL_765093, EPI_ISL_765094, EPI_ISL_765095, EPI_ISL_765096, EPI_ISL_765097, EPI_ISL_765098, EPI_ISL_765099, EPI_ISL_765100, EPI_ISL_765101, EPI_ISL_765102, EPI_ISL_765103, EPI_ISL_765104, EPI_ISL_765105, EPI_ISL_765106, EPI_ISL_765107, EPI_ISL_765108, EPI_ISL_765109, EPI_ISL_765110, EPI_ISL_765111, EPI_ISL_814274, EPI_ISL_814279, EPI_ISL_814284, EPI_ISL_814294, EPI_ISL_814296, EPI_ISL_814300, EPI_ISL_814302, EPI_ISL_814303, EPI_ISL_814304, EPI_ISL_814306, EPI_ISL_814309, EPI_ISL_814310, EPI_ISL_814313, EPI_ISL_814315, EPI_ISL_814317, EPI_ISL_814319, EPI_ISL_814323, EPI_ISL_814329, EPI_ISL_814331, EPI_ISL_814334, EPI_ISL_814337, EPI_ISL_814340, EPI_ISL_814341, EPI_ISL_814349, EPI_ISL_814352, EPI_ISL_814353, EPI_ISL_814354, EPI_ISL_814363, EPI_ISL_814366, EPI_ISL_814368, EPI_ISL_814370, EPI_ISL_814371, EPI_ISL_814373, EPI_ISL_814375, EPI_ISL_814378, EPI_ISL_814380, EPI_ISL_814383, EPI_ISL_814384, EPI_ISL_814389, EPI_ISL_814392, EPI_ISL_814394, EPI_ISL_814395, EPI_ISL_814396, EPI_ISL_814397, EPI_ISL_814401, EPI_ISL_814402, EPI_ISL_814403, EPI_ISL_814406, EPI_ISL_814407, EPI_ISL_814410, EPI_ISL_814414, EPI_ISL_814422, EPI_ISL_814480, EPI_ISL_814481, EPI_ISL_814482, EPI_ISL_814483, EPI_ISL_814484, EPI_ISL_814485, EPI_ISL_814486, EPI_ISL_814493, EPI_ISL_814494, EPI_ISL_814495, EPI_ISL_814496, EPI_ISL_814497, EPI_ISL_814498, EPI_ISL_814499, EPI_ISL_814500, EPI_ISL_814501, EPI_ISL_814511, EPI_ISL_814512, EPI_ISL_814513, EPI_ISL_814523, EPI_ISL_814526, EPI_ISL_814537, EPI_ISL_814540, EPI_ISL_814542, EPI_ISL_814550, EPI_ISL_814553, EPI_ISL_814555, EPI_ISL_814556, EPI_ISL_814558, EPI_ISL_814566, EPI_ISL_814579, EPI_ISL_814581, EPI_ISL_814583, EPI_ISL_814587, EPI_ISL_814597, EPI_ISL_814741, EPI_ISL_814742, EPI_ISL_814743, EPI_ISL_814744, EPI_ISL_814745, EPI_ISL_814746, EPI_ISL_814747, EPI_ISL_814748, EPI_ISL_814749, EPI_ISL_814750, EPI_ISL_814751, EPI_ISL_814752, EPI_ISL_814756, EPI_ISL_814872, EPI_ISL_814900, EPI_ISL_814901, EPI_ISL_814925, EPI_ISL_814926, EPI_ISL_814927, EPI_ISL_814928, EPI_ISL_814929, EPI_ISL_814930, EPI_ISL_814931, EPI_ISL_814932, EPI_ISL_814933, EPI_ISL_814934, EPI_ISL_814935, EPI_ISL_814936, EPI_ISL_814937, EPI_ISL_814938, EPI_ISL_814939, EPI_ISL_814940, EPI_ISL_814941, EPI_ISL_814942, EPI_ISL_814943, EPI_ISL_814944, EPI_ISL_814945, EPI_ISL_814946, EPI_ISL_814947, EPI_ISL_814948, EPI_ISL_814949, EPI_ISL_814950, EPI_ISL_814951, EPI_ISL_814952, EPI_ISL_814953, EPI_ISL_814954, EPI_ISL_814955, EPI_ISL_814956, EPI_ISL_814957, EPI_ISL_814958, EPI_ISL_814959, EPI_ISL_814960, EPI_ISL_814961, EPI_ISL_814962, EPI_ISL_814963, EPI_ISL_814964, EPI_ISL_814965, EPI_ISL_814966, EPI_ISL_814967, EPI_ISL_814968, EPI_ISL_814969, EPI_ISL_814970, EPI_ISL_814971, EPI_ISL_814972, EPI_ISL_814985, EPI_ISL_814986, EPI_ISL_814987, EPI_ISL_814989, EPI_ISL_814990, EPI_ISL_814991, EPI_ISL_814992, EPI_ISL_814993, EPI_ISL_814994, EPI_ISL_814995, EPI_ISL_814996, EPI_ISL_814997, EPI_ISL_814998, EPI_ISL_814999, EPI_ISL_815000, EPI_ISL_815001, EPI_ISL_815002, EPI_ISL_815003, EPI_ISL_815004, EPI_ISL_815005, EPI_ISL_815006, EPI_ISL_815007, EPI_ISL_815008, EPI_ISL_815009, EPI_ISL_815010, EPI_ISL_815011, EPI_ISL_815012, EPI_ISL_815013, EPI_ISL_815014, EPI_ISL_815015, EPI_ISL_815016, EPI_ISL_815033, EPI_ISL_815034, EPI_ISL_815038, EPI_ISL_815039, EPI_ISL_815040, EPI_ISL_815041, EPI_ISL_815042, EPI_ISL_815044, EPI_ISL_815048, EPI_ISL_815049, EPI_ISL_815050, EPI_ISL_815059, EPI_ISL_815061, EPI_ISL_815072, EPI_ISL_815073, EPI_ISL_815074, EPI_ISL_815075, EPI_ISL_815090, EPI_ISL_815108, EPI_ISL_815111, EPI_ISL_815112, EPI_ISL_815113, EPI_ISL_815114, EPI_ISL_815115, EPI_ISL_815116, EPI_ISL_815117, EPI_ISL_815118, EPI_ISL_815119, EPI_ISL_815120, EPI_ISL_815121, EPI_ISL_815122, EPI_ISL_815123, EPI_ISL_815124, EPI_ISL_815125, EPI_ISL_815126, EPI_ISL_815127, EPI_ISL_815128, EPI_ISL_815129, EPI_ISL_815130, EPI_ISL_815131, EPI_ISL_815132, EPI_ISL_815133, EPI_ISL_815134, EPI_ISL_815135, EPI_ISL_815136, EPI_ISL_815138, EPI_ISL_815139, EPI_ISL_815140, EPI_ISL_815141, EPI_ISL_815142, EPI_ISL_815143, EPI_ISL_815145, EPI_ISL_815147, EPI_ISL_815148, EPI_ISL_815149, EPI_ISL_815151, EPI_ISL_815152, EPI_ISL_815153, EPI_ISL_815154, EPI_ISL_815155, EPI_ISL_815156, EPI_ISL_815157, EPI_ISL_815158, EPI_ISL_815159, EPI_ISL_815160, EPI_ISL_815161, EPI_ISL_815162, EPI_ISL_815163, EPI_ISL_815164, EPI_ISL_815165, EPI_ISL_815166, EPI_ISL_815167, EPI_ISL_815168, EPI_ISL_815169, EPI_ISL_815170, EPI_ISL_815172, EPI_ISL_815173, EPI_ISL_815174, EPI_ISL_815175, EPI_ISL_815176, EPI_ISL_815177, EPI_ISL_815178, EPI_ISL_815179, EPI_ISL_815180, EPI_ISL_815181, EPI_ISL_815182, EPI_ISL_815183, EPI_ISL_815184, EPI_ISL_815244, EPI_ISL_815245, EPI_ISL_815246, EPI_ISL_815247, EPI_ISL_815248, EPI_ISL_815254, EPI_ISL_822346, EPI_ISL_822347, EPI_ISL_822348, EPI_ISL_822349, EPI_ISL_822350, EPI_ISL_822351, EPI_ISL_822352, EPI_ISL_822353, EPI_ISL_822354, EPI_ISL_822355, EPI_ISL_822356, EPI_ISL_822357, EPI_ISL_822358, EPI_ISL_822359, EPI_ISL_822360, EPI_ISL_822361, EPI_ISL_822362, EPI_ISL_822363, EPI_ISL_822364, EPI_ISL_822365, EPI_ISL_822840, EPI_ISL_822846, EPI_ISL_822847, EPI_ISL_822851, EPI_ISL_822852, EPI_ISL_822854, EPI_ISL_822855, EPI_ISL_822856, EPI_ISL_822857, EPI_ISL_822858, EPI_ISL_822860, EPI_ISL_822861, EPI_ISL_822862, EPI_ISL_822864, EPI_ISL_822866, EPI_ISL_822870, EPI_ISL_822871, EPI_ISL_822873, EPI_ISL_822876, EPI_ISL_822877, EPI_ISL_822878, EPI_ISL_822883, EPI_ISL_822888, EPI_ISL_822890, EPI_ISL_822891, EPI_ISL_822893, EPI_ISL_822896, EPI_ISL_822897, EPI_ISL_822899, EPI_ISL_822901, EPI_ISL_822902, EPI_ISL_822904, EPI_ISL_822906, EPI_ISL_822907, EPI_ISL_822908, EPI_ISL_822909, EPI_ISL_822910, EPI_ISL_822911, EPI_ISL_822912, EPI_ISL_822913, EPI_ISL_822915, EPI_ISL_822917, EPI_ISL_822918, EPI_ISL_822919, EPI_ISL_822924, EPI_ISL_822926, EPI_ISL_822928, EPI_ISL_822929, EPI_ISL_822930, EPI_ISL_822935, EPI_ISL_822938, EPI_ISL_822939, EPI_ISL_822942, EPI_ISL_822943, EPI_ISL_822945, EPI_ISL_822946, EPI_ISL_822948, EPI_ISL_822956, EPI_ISL_822959, EPI_ISL_822960, EPI_ISL_822964, EPI_ISL_822965, EPI_ISL_822966, EPI_ISL_822967, EPI_ISL_822968, EPI_ISL_822969, EPI_ISL_822970, EPI_ISL_822973, EPI_ISL_822976, EPI_ISL_822979, EPI_ISL_822980, EPI_ISL_822981, EPI_ISL_822982, EPI_ISL_822983, EPI_ISL_822984, EPI_ISL_822985, EPI_ISL_822986, EPI_ISL_822987, EPI_ISL_822988, EPI_ISL_822989, EPI_ISL_822990, EPI_ISL_822991, EPI_ISL_822992, EPI_ISL_822993, EPI_ISL_822994, EPI_ISL_822995, EPI_ISL_822996, EPI_ISL_822997, EPI_ISL_822999, EPI_ISL_823000, EPI_ISL_823001, EPI_ISL_823006, EPI_ISL_823007, EPI_ISL_823009, EPI_ISL_823010, EPI_ISL_823016, EPI_ISL_823020, EPI_ISL_823025, EPI_ISL_823026, EPI_ISL_823027, EPI_ISL_823028, EPI_ISL_823029, EPI_ISL_823030, EPI_ISL_823031, EPI_ISL_823032, EPI_ISL_823033, EPI_ISL_823034, EPI_ISL_823035, EPI_ISL_823036, EPI_ISL_823037, EPI_ISL_823038, EPI_ISL_823039, EPI_ISL_823040, EPI_ISL_823041, EPI_ISL_823042, EPI_ISL_823043, EPI_ISL_823044, EPI_ISL_823045, EPI_ISL_823046, EPI_ISL_823048, EPI_ISL_823049, EPI_ISL_823050, EPI_ISL_823051, EPI_ISL_823064, EPI_ISL_823109, EPI_ISL_823111, EPI_ISL_823114, EPI_ISL_823116, EPI_ISL_823118, EPI_ISL_823120, EPI_ISL_823122, EPI_ISL_823125, EPI_ISL_823126, EPI_ISL_823127, EPI_ISL_823128, EPI_ISL_823130, EPI_ISL_823134, EPI_ISL_823138, EPI_ISL_823139, EPI_ISL_823142, EPI_ISL_823144, EPI_ISL_823145, EPI_ISL_823146, EPI_ISL_823157, EPI_ISL_823163, EPI_ISL_823164, EPI_ISL_823165, EPI_ISL_823166, EPI_ISL_823167, EPI_ISL_823168, EPI_ISL_823174, EPI_ISL_823176, EPI_ISL_823177, EPI_ISL_823178, EPI_ISL_823179, EPI_ISL_823180, EPI_ISL_823181, EPI_ISL_823182, EPI_ISL_823183, EPI_ISL_823184, EPI_ISL_823185, EPI_ISL_823186, EPI_ISL_823187, EPI_ISL_823188, EPI_ISL_823189, EPI_ISL_823190, EPI_ISL_823191, EPI_ISL_823192, EPI_ISL_823193, EPI_ISL_823194, EPI_ISL_823195, EPI_ISL_823196, EPI_ISL_823197, EPI_ISL_823198, EPI_ISL_823199, EPI_ISL_823200, EPI_ISL_823201, EPI_ISL_823202, EPI_ISL_823203, EPI_ISL_823231, EPI_ISL_823232, EPI_ISL_823233, EPI_ISL_823234, EPI_ISL_823236, EPI_ISL_823237, EPI_ISL_823238, EPI_ISL_823239, EPI_ISL_823240, EPI_ISL_823241, EPI_ISL_823242, EPI_ISL_823243, EPI_ISL_823244, EPI_ISL_823245, EPI_ISL_823246, EPI_ISL_823247, EPI_ISL_823248, EPI_ISL_823249, EPI_ISL_823250, EPI_ISL_823251, EPI_ISL_823252, EPI_ISL_823253, EPI_ISL_823254, EPI_ISL_823255, EPI_ISL_823256, EPI_ISL_823257, EPI_ISL_823258, EPI_ISL_823259, EPI_ISL_823260, EPI_ISL_823261, EPI_ISL_823262, EPI_ISL_823263, EPI_ISL_823264, EPI_ISL_823265, EPI_ISL_823266, EPI_ISL_823267, EPI_ISL_823268, EPI_ISL_823269, EPI_ISL_823270, EPI_ISL_823271, EPI_ISL_823272, EPI_ISL_823273, EPI_ISL_823274, EPI_ISL_823275, EPI_ISL_823276, EPI_ISL_823277, EPI_ISL_823278, EPI_ISL_823280, EPI_ISL_823281, EPI_ISL_823282, EPI_ISL_823283, EPI_ISL_823284, EPI_ISL_823285, EPI_ISL_823286, EPI_ISL_823287, EPI_ISL_823288, EPI_ISL_823289, EPI_ISL_823290, EPI_ISL_823291, EPI_ISL_823292, EPI_ISL_823293, EPI_ISL_823294, EPI_ISL_823295, EPI_ISL_823296, EPI_ISL_823297, EPI_ISL_823298, EPI_ISL_823299, EPI_ISL_823300, EPI_ISL_823301, EPI_ISL_823302, EPI_ISL_823303, EPI_ISL_823304, EPI_ISL_823305, EPI_ISL_823306, EPI_ISL_823307, EPI_ISL_823308, EPI_ISL_823309, EPI_ISL_823310, EPI_ISL_823311, EPI_ISL_823312, EPI_ISL_823313, EPI_ISL_823314, EPI_ISL_823315, EPI_ISL_823316, EPI_ISL_823317, EPI_ISL_823318, EPI_ISL_823319, EPI_ISL_823320, EPI_ISL_823321, EPI_ISL_823322, EPI_ISL_823323, EPI_ISL_823324, EPI_ISL_823325, EPI_ISL_823326, EPI_ISL_823327, EPI_ISL_823328, EPI_ISL_823329, EPI_ISL_823330, EPI_ISL_823331, EPI_ISL_823332, EPI_ISL_823333, EPI_ISL_823334, EPI_ISL_823335, EPI_ISL_823336, EPI_ISL_823337, EPI_ISL_823338, EPI_ISL_823339, EPI_ISL_823340, EPI_ISL_823341, EPI_ISL_823342, EPI_ISL_823343, EPI_ISL_823344, EPI_ISL_823345, EPI_ISL_823346, EPI_ISL_823347, EPI_ISL_823348, EPI_ISL_823349, EPI_ISL_823350, EPI_ISL_823351, EPI_ISL_823352, EPI_ISL_823353, EPI_ISL_823354, EPI_ISL_823355, EPI_ISL_823356, EPI_ISL_823357, EPI_ISL_823358, EPI_ISL_823359, EPI_ISL_823360, EPI_ISL_823361, EPI_ISL_823362, EPI_ISL_823363, EPI_ISL_823364, EPI_ISL_823365, EPI_ISL_823366, EPI_ISL_823370, EPI_ISL_823371, EPI_ISL_823379, EPI_ISL_823384, EPI_ISL_823385, EPI_ISL_823386, EPI_ISL_823387, EPI_ISL_823388, EPI_ISL_823389, EPI_ISL_823402, EPI_ISL_823403, EPI_ISL_823404, EPI_ISL_823405, EPI_ISL_823406, EPI_ISL_823407, EPI_ISL_823408, EPI_ISL_823409, EPI_ISL_823410, EPI_ISL_823411, EPI_ISL_823412, EPI_ISL_823413, EPI_ISL_823414, EPI_ISL_823415, EPI_ISL_823416, EPI_ISL_823417, EPI_ISL_823418, EPI_ISL_823419, EPI_ISL_823420, EPI_ISL_823421, EPI_ISL_823422, EPI_ISL_823423, EPI_ISL_823424, EPI_ISL_823425, EPI_ISL_823426, EPI_ISL_823427, EPI_ISL_823428, EPI_ISL_823429, EPI_ISL_823430, EPI_ISL_823431, EPI_ISL_823432, EPI_ISL_823433, EPI_ISL_823434, EPI_ISL_823435, EPI_ISL_823436, EPI_ISL_823437, EPI_ISL_823438, EPI_ISL_823439, EPI_ISL_823440, EPI_ISL_823441, EPI_ISL_823442, EPI_ISL_823443, EPI_ISL_823444, EPI_ISL_823445, EPI_ISL_823446, EPI_ISL_823447, EPI_ISL_823449, EPI_ISL_823450, EPI_ISL_823451, EPI_ISL_823452, EPI_ISL_823453, EPI_ISL_823454, EPI_ISL_823455, EPI_ISL_823456, EPI_ISL_823457, EPI_ISL_823459, EPI_ISL_823460, EPI_ISL_823461, EPI_ISL_823462, EPI_ISL_823463, EPI_ISL_823464, EPI_ISL_823465, EPI_ISL_823466, EPI_ISL_823467, EPI_ISL_823468, EPI_ISL_823469, EPI_ISL_823470, EPI_ISL_823471, EPI_ISL_823472, EPI_ISL_823473, EPI_ISL_823474, EPI_ISL_823475, EPI_ISL_823476, EPI_ISL_823477, EPI_ISL_823478, EPI_ISL_823479, EPI_ISL_823480, EPI_ISL_823481, EPI_ISL_823482, EPI_ISL_823483, EPI_ISL_823484, EPI_ISL_823485, EPI_ISL_823486, EPI_ISL_823487, EPI_ISL_823488, EPI_ISL_823489, EPI_ISL_823490, EPI_ISL_823491, EPI_ISL_823492, EPI_ISL_823493, EPI_ISL_823494, EPI_ISL_823495, EPI_ISL_823496, EPI_ISL_823497, EPI_ISL_823498, EPI_ISL_823499, EPI_ISL_823500, EPI_ISL_823501, EPI_ISL_823502, EPI_ISL_823503, EPI_ISL_823504, EPI_ISL_823505, EPI_ISL_823506, EPI_ISL_823654, EPI_ISL_823655, EPI_ISL_823656, EPI_ISL_823658, EPI_ISL_823806

| | | | |
|---|---|---|---|
| see above | Wales Specialist Virology Centre Sequencing lab: Pathogen Genomics Unit | COVID-19 Genomics UK (COG-UK) Consortium | Alec Birchley; Alexander Adams; Amy Gaskin; Angela Marchbank; Bree Gatica-Wilcox; Catherine Moore; Jason Coombes; Joanne Watkins; Joel Southgate; Johnathan Evans; Laura Gifford; Lauren Gilbert; Lee Graham; Malorie Perry; Matthew Bull; Nicole Pacchiarini; Sally Corden; Sara Kumziene-Summerhayes; Sara Rey; Sarah Taylor; Simon Cottrell; Sophie Jones; Tom Connor |
| EPI_ISL_840836, EPI_ISL_840885, EPI_ISL_840886, EPI_ISL_840887, EPI_ISL_840888 | Wales Specialist Virology Centre Sequencing lab: Pathogen Genomics Unit | Public Health Wales Microbiology Cardiff Wales Specialist Virology Centre | Alec Birchley; Alexander Adams; Amy Gaskin; Angela Marchbank; Bree Gatica-Wilcox; Catherine Moore; Jason Coombes; Joanne Watkins; Joel Southgate; Johnathan Evans; Laura Gifford; Lauren Gilbert; Lee Graham; Malorie Perry; Matthew Bull; Nicole Pacchiarini; Sally Corden; Sara Kumziene-Summerhayes; Sara Rey; Sarah Taylor; Simon Cottrell; Sophie Jones; Tom Connor |

EPI_ISL_814312, EPI_ISL_814339, EPI_ISL_814344, EPI_ISL_814345, EPI_ISL_814348, EPI_ISL_814355, EPI_ISL_814439, EPI_ISL_814440, EPI_ISL_814441, EPI_ISL_814442, EPI_ISL_814443, EPI_ISL_814444, EPI_ISL_814445, EPI_ISL_814446, EPI_ISL_814447, EPI_ISL_814448, EPI_ISL_814449, EPI_ISL_814450, EPI_ISL_814451, EPI_ISL_814452, EPI_ISL_814453, EPI_ISL_814455, EPI_ISL_814457, EPI_ISL_814458, EPI_ISL_814547, EPI_ISL_814549, EPI_ISL_814568, EPI_ISL_814609, EPI_ISL_814620, EPI_ISL_814621, EPI_ISL_814622, EPI_ISL_814624, EPI_ISL_814626, EPI_ISL_814627, EPI_ISL_814628, EPI_ISL_814629, EPI_ISL_814630, EPI_ISL_814631, EPI_ISL_814632, EPI_ISL_814633, EPI_ISL_814634, EPI_ISL_814636, EPI_ISL_814638, EPI_ISL_814639, EPI_ISL_814642, EPI_ISL_814643, EPI_ISL_814644, EPI_ISL_814645, EPI_ISL_814646, EPI_ISL_814647, EPI_ISL_814648, EPI_ISL_814649, EPI_ISL_814650, EPI_ISL_814651, EPI_ISL_814652, EPI_ISL_814653, EPI_ISL_814654, EPI_ISL_814655, EPI_ISL_814656, EPI_ISL_814657, EPI_ISL_814658, EPI_ISL_814660, EPI_ISL_814661, EPI_ISL_814662, EPI_ISL_814664, EPI_ISL_838116, EPI_ISL_838117, EPI_ISL_838118, EPI_ISL_838119, EPI_ISL_838120, EPI_ISL_838121, EPI_ISL_838122, EPI_ISL_838123, EPI_ISL_838124, EPI_ISL_838125, EPI_ISL_838126, EPI_ISL_838127, EPI_ISL_838128, EPI_ISL_838130, EPI_ISL_838131, EPI_ISL_838132, EPI_ISL_838133, EPI_ISL_838134, EPI_ISL_838135, EPI_ISL_864960, EPI_ISL_919241, EPI_ISL_919242, EPI_ISL_919246, EPI_ISL_919249, EPI_ISL_919251, EPI_ISL_919258, EPI_ISL_919262, EPI_ISL_919263, EPI_ISL_919265, EPI_ISL_919268

| | | | |
|---|---|---|---|
| see above | West of Scotland Specialist Virology Centre, NHSGGC / MRC-University of Glasgow Centre for Virus Research | COVID-19 Genomics UK (COG-UK) Consortium | Alasdair MacLean; Alice Broos; Ana da Silva Filipe; Antonia Ho; Daniel Mair; David L Robertson; Emma Thomson; Guy Mollett; James Shepherd; Jenna Nichols; Joseph Hughes; Kathy Li; Kathy Smollett; Kyriaki Nomikou; Lily Tong; Matthew Holden; Natasha Johnson; Rachel Blacow; Richard Orton; Rory Gunson; Sarah McDonald; Sharif Shaaban; Sreenu Vattipally; Stephen Carmichael |

We gratefully acknowledge the following Authors from the Originating laboratories responsible for obtaining the specimens, as well as the Submitting laboratories where the genome data were generated and shared via GISAID, on which this research is based.

All Submitters of data may be contacted directly via www.gisaid.org

Authors are sorted alphabetically.

| Accession ID | Originating Laboratory | Submitting Laboratory | Authors |
|---|---|---|---|
| EPI_ISL_843077, EPI_ISL_843080, EPI_ISL_843081, EPI_ISL_843082, EPI_ISL_843083, EPI_ISL_843084, EPI_ISL_843085, EPI_ISL_843086, EPI_ISL_843087, EPI_ISL_843093, EPI_ISL_843094, EPI_ISL_843095, EPI_ISL_843096, EPI_ISL_843097, EPI_ISL_843098, EPI_ISL_843099, EPI_ISL_843100, EPI_ISL_843101, EPI_ISL_843102, EPI_ISL_843103, EPI_ISL_843104, EPI_ISL_843111, EPI_ISL_843112, EPI_ISL_843113, EPI_ISL_843114, EPI_ISL_843115, EPI_ISL_843121, EPI_ISL_843122, EPI_ISL_843123, EPI_ISL_843124, EPI_ISL_843125, EPI_ISL_843126, EPI_ISL_843127, EPI_ISL_843128, EPI_ISL_843129, EPI_ISL_843130, EPI_ISL_843131, EPI_ISL_843132, EPI_ISL_843133, EPI_ISL_843134, EPI_ISL_843135, EPI_ISL_843136, EPI_ISL_843137, EPI_ISL_843138, EPI_ISL_843139, EPI_ISL_843140, EPI_ISL_843141, EPI_ISL_843142, EPI_ISL_843143, EPI_ISL_843144, EPI_ISL_843145, EPI_ISL_843146, EPI_ISL_865488, EPI_ISL_865489, EPI_ISL_865490, EPI_ISL_865491, EPI_ISL_865492, EPI_ISL_865493, EPI_ISL_865494, EPI_ISL_865495, EPI_ISL_865496, EPI_ISL_865497, EPI_ISL_865501, EPI_ISL_865503, EPI_ISL_865504, EPI_ISL_865505, EPI_ISL_865506, EPI_ISL_865507, EPI_ISL_865508, EPI_ISL_865509, EPI_ISL_865510, EPI_ISL_865511, EPI_ISL_865512, EPI_ISL_865513, EPI_ISL_865514, EPI_ISL_865515, EPI_ISL_865516, EPI_ISL_865517, EPI_ISL_865518, EPI_ISL_865519, EPI_ISL_865520, EPI_ISL_865521, EPI_ISL_865522, EPI_ISL_865523, EPI_ISL_865524, EPI_ISL_865525, EPI_ISL_865526, EPI_ISL_865543, EPI_ISL_865544, EPI_ISL_865545, EPI_ISL_865559, EPI_ISL_865560, EPI_ISL_865563, EPI_ISL_865564, EPI_ISL_865565, EPI_ISL_865566, EPI_ISL_865576, EPI_ISL_865577, EPI_ISL_865578, EPI_ISL_949761, EPI_ISL_1474801, EPI_ISL_1636893, EPI_ISL_1636956, EPI_ISL_1636957, EPI_ISL_1636958, EPI_ISL_1636959, EPI_ISL_1636960, EPI_ISL_1730305, EPI_ISL_1730306, EPI_ISL_1975078, EPI_ISL_1975080, EPI_ISL_1975082, EPI_ISL_1975083, EPI_ISL_1975084, EPI_ISL_1975086, EPI_ISL_1975087, EPI_ISL_1975089, EPI_ISL_1975091, EPI_ISL_1975092, EPI_ISL_1975094, EPI_ISL_1975096, EPI_ISL_1975098, EPI_ISL_2126015, EPI_ISL_2126017, EPI_ISL_2126038, EPI_ISL_2126040, EPI_ISL_2126042, EPI_ISL_2126044, EPI_ISL_2126046, EPI_ISL_2126047, EPI_ISL_2126049, EPI_ISL_2126051, EPI_ISL_2126053, EPI_ISL_2126055 | | | |
| see above | Barts Health NHS Trust | COVID-19 Genomics UK (COG-UK) Consortium | BROAD; Beatrix; CUTINO-MOGUEL; Claire; David; Dola; HARRINGTON; KELE; KULASEGARAN-SHYLINI; Maria-Teresa; OWOYEMI; Raghavendran; SHYLINI |
| EPI_ISL_814420, EPI_ISL_817044, EPI_ISL_817068, EPI_ISL_817077, EPI_ISL_842514, EPI_ISL_842515, EPI_ISL_842516, EPI_ISL_842517, EPI_ISL_842518, EPI_ISL_842519, EPI_ISL_842520, EPI_ISL_842521, EPI_ISL_842522, EPI_ISL_842523, EPI_ISL_842524, EPI_ISL_842525, EPI_ISL_842526, EPI_ISL_842527, EPI_ISL_842528, EPI_ISL_842529, EPI_ISL_842530, EPI_ISL_842531, EPI_ISL_842532, EPI_ISL_842533, EPI_ISL_842534, EPI_ISL_842535, EPI_ISL_842536, EPI_ISL_842537, EPI_ISL_842538, EPI_ISL_842539, EPI_ISL_842540, EPI_ISL_842541, EPI_ISL_842542, EPI_ISL_842543, EPI_ISL_842544, EPI_ISL_842545, EPI_ISL_842546, EPI_ISL_842547, EPI_ISL_842548, EPI_ISL_842549, EPI_ISL_842550, EPI_ISL_842551, EPI_ISL_842552, EPI_ISL_842553, EPI_ISL_842554, EPI_ISL_842555, EPI_ISL_842556, EPI_ISL_842557, EPI_ISL_842558, EPI_ISL_842559, EPI_ISL_842560, EPI_ISL_842561, EPI_ISL_842562, EPI_ISL_842563, EPI_ISL_842564, EPI_ISL_842565, EPI_ISL_842566, EPI_ISL_842567, EPI_ISL_842568, EPI_ISL_842569, EPI_ISL_842570, EPI_ISL_842571, EPI_ISL_842572, EPI_ISL_842573, EPI_ISL_842574, EPI_ISL_842575, EPI_ISL_842576, EPI_ISL_842577, EPI_ISL_842578, EPI_ISL_842579, EPI_ISL_842580, EPI_ISL_842581, EPI_ISL_842582, EPI_ISL_842584, EPI_ISL_842595, EPI_ISL_953119, EPI_ISL_953120, EPI_ISL_953121, EPI_ISL_953122, EPI_ISL_953123, EPI_ISL_953124, EPI_ISL_953125, EPI_ISL_953127, EPI_ISL_953128, EPI_ISL_953130, EPI_ISL_953131, EPI_ISL_953136, EPI_ISL_953137, EPI_ISL_953139, EPI_ISL_953140, EPI_ISL_953142, EPI_ISL_953144, EPI_ISL_953145, EPI_ISL_953146, EPI_ISL_953147, EPI_ISL_953148, EPI_ISL_953149, EPI_ISL_953150, EPI_ISL_953151, EPI_ISL_953152, EPI_ISL_953153, EPI_ISL_953154, EPI_ISL_953156, EPI_ISL_953157, EPI_ISL_953158, EPI_ISL_953159, EPI_ISL_953160, EPI_ISL_953161, EPI_ISL_953162, EPI_ISL_953163, EPI_ISL_953164, EPI_ISL_953165, EPI_ISL_953166, EPI_ISL_953167, EPI_ISL_953169, EPI_ISL_953170, EPI_ISL_953171, EPI_ISL_953172, EPI_ISL_953173, EPI_ISL_1476857, EPI_ISL_1476892, EPI_ISL_1638110, EPI_ISL_1638120, EPI_ISL_1638122, EPI_ISL_1638137, EPI_ISL_1638141, EPI_ISL_1638178, EPI_ISL_1638197 | | | |
| see above | Bioinformatics and Biostatistics Lab, Advanced Sequencing Facility | COVID-19 Genomics UK (COG-UK) Consortium | Aengus Stewart; Chelsea Sawyer; Harshil Patel; Jerome Nicod; Laura Cubitt; Margaret Crawford |
| EPI_ISL_763606, EPI_ISL_763608, EPI_ISL_816153, EPI_ISL_816172, EPI_ISL_816174, EPI_ISL_816194, EPI_ISL_816195, EPI_ISL_816196, EPI_ISL_816197, EPI_ISL_816198, EPI_ISL_816199, EPI_ISL_816200, EPI_ISL_816201, EPI_ISL_816202, EPI_ISL_816203, EPI_ISL_816204, EPI_ISL_816205, EPI_ISL_816206, EPI_ISL_816207, EPI_ISL_816224, EPI_ISL_816225, EPI_ISL_841846, EPI_ISL_841847, EPI_ISL_842010, EPI_ISL_842011, EPI_ISL_842013, EPI_ISL_923212, EPI_ISL_923218, EPI_ISL_923220, EPI_ISL_923227, EPI_ISL_923229, EPI_ISL_923230, EPI_ISL_923231, EPI_ISL_923232, EPI_ISL_923233, EPI_ISL_923235, EPI_ISL_923237, EPI_ISL_923238, EPI_ISL_923239, EPI_ISL_923240, EPI_ISL_923243, EPI_ISL_923245, EPI_ISL_923246, EPI_ISL_923247, EPI_ISL_923248, EPI_ISL_923249, EPI_ISL_923250, EPI_ISL_923251, EPI_ISL_923258, EPI_ISL_923267, EPI_ISL_923268, EPI_ISL_952817, EPI_ISL_952818, EPI_ISL_1000261, EPI_ISL_1000268, EPI_ISL_1107698, EPI_ISL_1107751, EPI_ISL_1637981, EPI_ISL_1638000, EPI_ISL_1730851, EPI_ISL_1730852, EPI_ISL_1730854, EPI_ISL_1730855, EPI_ISL_1730856, EPI_ISL_1730857, EPI_ISL_1730858, EPI_ISL_1730859, EPI_ISL_1730860, EPI_ISL_1979897, EPI_ISL_1979899, EPI_ISL_1979900, EPI_ISL_1979902, EPI_ISL_1979903, EPI_ISL_1979905, EPI_ISL_1979908, EPI_ISL_2127966, EPI_ISL_2127967, EPI_ISL_2127969, EPI_ISL_2127971, EPI_ISL_2127972, EPI_ISL_2127975, EPI_ISL_2127977, EPI_ISL_2127979, EPI_ISL_2127981, EPI_ISL_2127983, EPI_ISL_2127985, EPI_ISL_2127986, EPI_ISL_2127987, EPI_ISL_2127988, EPI_ISL_2127990, EPI_ISL_2127991, EPI_ISL_2127994, EPI_ISL_2127995, EPI_ISL_2127997, EPI_ISL_2128053, EPI_ISL_2128055, EPI_ISL_2128056, EPI_ISL_2128058, EPI_ISL_2128060, EPI_ISL_2128062, EPI_ISL_2128064, EPI_ISL_2128065, EPI_ISL_2128067, EPI_ISL_2128069, EPI_ISL_2128071, EPI_ISL_2128073, EPI_ISL_2128075, EPI_ISL_2128077, EPI_ISL_2128079, EPI_ISL_2128080, EPI_ISL_2128082, EPI_ISL_2128084, EPI_ISL_2128086, EPI_ISL_2128087, EPI_ISL_2128089, EPI_ISL_2128091, EPI_ISL_2128093, EPI_ISL_2128094, EPI_ISL_2128095, EPI_ISL_2128097, EPI_ISL_2128099, EPI_ISL_2128101, EPI_ISL_2128103, EPI_ISL_2128104, EPI_ISL_2128106, EPI_ISL_2128108, EPI_ISL_2128110, EPI_ISL_2128112, EPI_ISL_2128113, EPI_ISL_2128115, EPI_ISL_2128117, EPI_ISL_2128119, EPI_ISL_2128121, EPI_ISL_2128123, EPI_ISL_2128124, EPI_ISL_2128126, EPI_ISL_2128127, EPI_ISL_2128128, EPI_ISL_2128130, EPI_ISL_2128131, EPI_ISL_2128132, EPI_ISL_2128134, EPI_ISL_2128136, EPI_ISL_2128137, EPI_ISL_2128139, EPI_ISL_2128141, EPI_ISL_2128143, EPI_ISL_2128145, EPI_ISL_2128147, EPI_ISL_2128149, EPI_ISL_2128150, EPI_ISL_2128173, EPI_ISL_2128174, EPI_ISL_2128186, EPI_ISL_2128188, EPI_ISL_2128190, EPI_ISL_2128194, EPI_ISL_2128196, EPI_ISL_2128198, EPI_ISL_2128199, EPI_ISL_2128210, EPI_ISL_2128214, EPI_ISL_2128216, EPI_ISL_2128218, EPI_ISL_2128219 | | | |
| see above | Centre for Enzyme Innovation, University of Portsmouth / Translational Research Laboratory, Portsmouth Hospitals NHS Trust | COVID-19 Genomics UK (COG-UK) Consortium | Allyson Lloyd; Angela Beckett; Anoop Chauhan; Christopher Fearn; Ethan Butcher; Garry Scarlett; Kate Cook; Katie Loveson; Kelly Bicknell; Robert Impey; Salman Goudarzi; Samuel Robson; Sarah Wyllie; Scott Elliott; Sharon Glaysher; Yann Bourgeois |
| EPI_ISL_763379, EPI_ISL_763380, EPI_ISL_763464, EPI_ISL_763469, EPI_ISL_763508, EPI_ISL_763518, EPI_ISL_763592, EPI_ISL_763666, EPI_ISL_763688, EPI_ISL_763702, EPI_ISL_763816, EPI_ISL_763839, EPI_ISL_763843, EPI_ISL_763850, EPI_ISL_763883, EPI_ISL_763884, EPI_ISL_763885, EPI_ISL_763886, EPI_ISL_763887, EPI_ISL_763888, EPI_ISL_763889, EPI_ISL_763890, EPI_ISL_763891, EPI_ISL_764141, EPI_ISL_764164, EPI_ISL_764236, EPI_ISL_764343, EPI_ISL_764344, EPI_ISL_764345, EPI_ISL_764347, EPI_ISL_764348, EPI_ISL_764349, EPI_ISL_764350, EPI_ISL_764351, EPI_ISL_764352, EPI_ISL_764353, EPI_ISL_764354, EPI_ISL_764356, EPI_ISL_813166, EPI_ISL_813167, EPI_ISL_813168, EPI_ISL_813169, EPI_ISL_813170, EPI_ISL_813171, EPI_ISL_813173, EPI_ISL_813175, EPI_ISL_813176, EPI_ISL_813177, EPI_ISL_813178, EPI_ISL_813179, EPI_ISL_813180, EPI_ISL_813181, EPI_ISL_813182, EPI_ISL_813183, EPI_ISL_813184, EPI_ISL_813185, EPI_ISL_813186, EPI_ISL_813187, EPI_ISL_813190, EPI_ISL_813191, EPI_ISL_813192, EPI_ISL_813193, EPI_ISL_813194, EPI_ISL_813227, EPI_ISL_813230, EPI_ISL_813231, EPI_ISL_813245, EPI_ISL_813249, EPI_ISL_813250, EPI_ISL_813251, EPI_ISL_813256, EPI_ISL_813257, EPI_ISL_813258, EPI_ISL_813265, EPI_ISL_813266, EPI_ISL_813267, EPI_ISL_813268, EPI_ISL_813269, EPI_ISL_813278, EPI_ISL_813281, EPI_ISL_813283, EPI_ISL_813285, EPI_ISL_813287, EPI_ISL_813293, EPI_ISL_813294, EPI_ISL_813305, EPI_ISL_813306, EPI_ISL_813317, EPI_ISL_813320, EPI_ISL_813323, EPI_ISL_837879, EPI_ISL_837882, EPI_ISL_837889, EPI_ISL_837891, EPI_ISL_837892, EPI_ISL_837893, EPI_ISL_837895, EPI_ISL_837898, EPI_ISL_837901, EPI_ISL_837902 | | | |
| see above | Department of Pathology, University of Cambridge | COVID-19 Genomics UK (COG-UK) Consortium | Aminu S. Jahun; Grant Hall; Ian Goodfellow; Iliana Georgana; Malte Pinckert; Martin D. Curran; Myra Hosmillo; Surendra Parmar; Yasmin Chaudhry |
| EPI_ISL_761185, EPI_ISL_761892, EPI_ISL_781186, EPI_ISL_781194, EPI_ISL_781195, EPI_ISL_781202, EPI_ISL_781206, EPI_ISL_781207, EPI_ISL_781209, EPI_ISL_781210, EPI_ISL_781214, EPI_ISL_781216, EPI_ISL_781219, EPI_ISL_781226, EPI_ISL_781230, EPI_ISL_781234, EPI_ISL_781235, EPI_ISL_781250, EPI_ISL_781256, EPI_ISL_781262, EPI_ISL_781273, EPI_ISL_781280, EPI_ISL_781282, EPI_ISL_781288, EPI_ISL_781291, EPI_ISL_781292, EPI_ISL_781295, EPI_ISL_781297, EPI_ISL_781313, EPI_ISL_781315, EPI_ISL_781318, EPI_ISL_781323, EPI_ISL_781324, EPI_ISL_781330, EPI_ISL_781331, EPI_ISL_781337, EPI_ISL_781348, EPI_ISL_781349, EPI_ISL_781352, EPI_ISL_781356, EPI_ISL_781372, EPI_ISL_781378, EPI_ISL_781389, EPI_ISL_781400, EPI_ISL_781405, EPI_ISL_781406, EPI_ISL_781412, EPI_ISL_781424, EPI_ISL_781435, EPI_ISL_781437, EPI_ISL_781443, EPI_ISL_781446, EPI_ISL_781447, EPI_ISL_781451, EPI_ISL_781456, EPI_ISL_781458, EPI_ISL_781463, EPI_ISL_781464, EPI_ISL_781465, EPI_ISL_781473, EPI_ISL_781475, EPI_ISL_781478, EPI_ISL_781499, EPI_ISL_781502, EPI_ISL_781505, EPI_ISL_781508, EPI_ISL_781510, EPI_ISL_781515, EPI_ISL_781516, EPI_ISL_782161, EPI_ISL_782165, EPI_ISL_782171, EPI_ISL_782197, EPI_ISL_782201, EPI_ISL_782202, EPI_ISL_782207, EPI_ISL_782211, EPI_ISL_782212, EPI_ISL_782224, EPI_ISL_782226, EPI_ISL_782235, EPI_ISL_782242, EPI_ISL_782245, EPI_ISL_782250, EPI_ISL_782252, EPI_ISL_782256, EPI_ISL_782264, EPI_ISL_782268, EPI_ISL_782271, EPI_ISL_782274, EPI_ISL_782278, EPI_ISL_782282, EPI_ISL_782287, EPI_ISL_782292, EPI_ISL_782309, EPI_ISL_782313, EPI_ISL_782314, EPI_ISL_782317, EPI_ISL_782325, EPI_ISL_782326, EPI_ISL_782328, EPI_ISL_782335, EPI_ISL_782345, EPI_ISL_782352, EPI_ISL_782355, EPI_ISL_782361, EPI_ISL_782369, EPI_ISL_782371, EPI_ISL_782372, EPI_ISL_782377, EPI_ISL_782380, EPI_ISL_782388, EPI_ISL_782390, EPI_ISL_782393, EPI_ISL_782409, EPI_ISL_782420, EPI_ISL_782423, EPI_ISL_782425, EPI_ISL_782426, EPI_ISL_782439, EPI_ISL_782451, EPI_ISL_782454, EPI_ISL_782461, EPI_ISL_782466, EPI_ISL_782471, EPI_ISL_782474, EPI_ISL_796795, EPI_ISL_796997, EPI_ISL_797087, EPI_ISL_797999, EPI_ISL_798002, EPI_ISL_798007, EPI_ISL_798009, EPI_ISL_798011, EPI_ISL_798013, EPI_ISL_798015, EPI_ISL_798025, EPI_ISL_798027, EPI_ISL_798029, EPI_ISL_798035, EPI_ISL_798054, EPI_ISL_798061, EPI_ISL_798062, EPI_ISL_798064, EPI_ISL_798066, EPI_ISL_798068, EPI_ISL_798073, EPI_ISL_798083, EPI_ISL_798084, EPI_ISL_798096, EPI_ISL_798101, EPI_ISL_798103, EPI_ISL_798106, EPI_ISL_798108, EPI_ISL_798119, EPI_ISL_798122, EPI_ISL_798123, EPI_ISL_798124, EPI_ISL_798125, EPI_ISL_798127, EPI_ISL_798131, EPI_ISL_798133, EPI_ISL_798134, EPI_ISL_798139, EPI_ISL_798152, EPI_ISL_798154, EPI_ISL_798169, EPI_ISL_798170, EPI_ISL_798173, EPI_ISL_798174, EPI_ISL_798175, EPI_ISL_798176, EPI_ISL_798181, EPI_ISL_798182, EPI_ISL_798202, EPI_ISL_798204, EPI_ISL_798208, EPI_ISL_798220, EPI_ISL_798221, EPI_ISL_798222, EPI_ISL_798225, EPI_ISL_798229, EPI_ISL_798234, EPI_ISL_798235, EPI_ISL_798236, EPI_ISL_798239, EPI_ISL_798241, EPI_ISL_798242, EPI_ISL_798245, EPI_ISL_798246, EPI_ISL_798248, EPI_ISL_798250, EPI_ISL_798255, EPI_ISL_798268, EPI_ISL_798274, EPI_ISL_798275, EPI_ISL_798277, EPI_ISL_798280, EPI_ISL_798282, EPI_ISL_798283, EPI_ISL_798284, EPI_ISL_798285, EPI_ISL_798292, EPI_ISL_798295, EPI_ISL_798296, EPI_ISL_798299, EPI_ISL_798305, EPI_ISL_798310, EPI_ISL_798314, EPI_ISL_798321, EPI_ISL_798323, EPI_ISL_798324, EPI_ISL_798328, EPI_ISL_798329, EPI_ISL_798334, EPI_ISL_798336, EPI_ISL_798341, EPI_ISL_798345, EPI_ISL_798346, EPI_ISL_798349, EPI_ISL_798356, EPI_ISL_798362, EPI_ISL_798368, EPI_ISL_798369, EPI_ISL_798370, EPI_ISL_798378, EPI_ISL_798379, EPI_ISL_798384, EPI_ISL_798386, EPI_ISL_798388, EPI_ISL_798389, EPI_ISL_798396, EPI_ISL_798399, EPI_ISL_798401, EPI_ISL_798404, EPI_ISL_798405, EPI_ISL_798413, EPI_ISL_798416, EPI_ISL_798417, EPI_ISL_798431, EPI_ISL_798436, EPI_ISL_798444, EPI_ISL_798455, EPI_ISL_798465, EPI_ISL_798468, EPI_ISL_798475, EPI_ISL_798479, EPI_ISL_798483, EPI_ISL_798492, EPI_ISL_798493, EPI_ISL_798500, EPI_ISL_798502, EPI_ISL_798508, EPI_ISL_798510, EPI_ISL_798513, EPI_ISL_798516, EPI_ISL_798517, EPI_ISL_798519, EPI_ISL_798524, EPI_ISL_798531, EPI_ISL_798534, EPI_ISL_798539, EPI_ISL_798541, EPI_ISL_798542, EPI_ISL_798544, EPI_ISL_798548, EPI_ISL_798550, EPI_ISL_798555, EPI_ISL_798561, EPI_ISL_798564, EPI_ISL_798591, EPI_ISL_798593, EPI_ISL_798598, EPI_ISL_798601, EPI_ISL_798607, EPI_ISL_798609, EPI_ISL_798612, EPI_ISL_798615, EPI_ISL_798622, EPI_ISL_798631, EPI_ISL_798632, EPI_ISL_798633, EPI_ISL_798635, EPI_ISL_798637, EPI_ISL_798639, EPI_ISL_798640, EPI_ISL_798641, EPI_ISL_798643, EPI_ISL_798652, EPI_ISL_798659, EPI_ISL_798661, EPI_ISL_798665, EPI_ISL_798667, EPI_ISL_798668, EPI_ISL_798669, EPI_ISL_798671, EPI_ISL_798676, EPI_ISL_798684, EPI_ISL_798688, EPI_ISL_798689, EPI_ISL_798690, EPI_ISL_798692, EPI_ISL_798695, EPI_ISL_798704, EPI_ISL_798705, EPI_ISL_798709, EPI_ISL_798719, EPI_ISL_798722, EPI_ISL_798727, EPI_ISL_798729, EPI_ISL_798735, EPI_ISL_798742, EPI_ISL_798744, EPI_ISL_798749, EPI_ISL_798768, EPI_ISL_798771, EPI_ISL_798784, EPI_ISL_798791, EPI_ISL_798793, EPI_ISL_798798, EPI_ISL_798804, EPI_ISL_798811, EPI_ISL_798814, EPI_ISL_798819, EPI_ISL_798824, EPI_ISL_798826, EPI_ISL_798833, EPI_ISL_798845, EPI_ISL_798850, EPI_ISL_798852, EPI_ISL_798853, EPI_ISL_798869, EPI_ISL_798872, EPI_ISL_798878, EPI_ISL_798880, EPI_ISL_798884, EPI_ISL_798885, EPI_ISL_798888, EPI_ISL_798891, EPI_ISL_798898, EPI_ISL_798902, EPI_ISL_798905, EPI_ISL_798906, EPI_ISL_798914, EPI_ISL_798921, EPI_ISL_798924, EPI_ISL_798926, EPI_ISL_798928, EPI_ISL_798934, EPI_ISL_798935, EPI_ISL_798939, EPI_ISL_798941, EPI_ISL_798951, EPI_ISL_798952, EPI_ISL_798957, EPI_ISL_798962, EPI_ISL_798963, EPI_ISL_798964, EPI_ISL_798968, EPI_ISL_798969, EPI_ISL_798984, EPI_ISL_798986, EPI_ISL_798988, EPI_ISL_798991, EPI_ISL_799000, EPI_ISL_799003, EPI_ISL_799004, EPI_ISL_799005, EPI_ISL_799010, EPI_ISL_799021, EPI_ISL_799022, EPI_ISL_799025, EPI_ISL_799026, EPI_ISL_799029, EPI_ISL_799040, EPI_ISL_799045, EPI_ISL_799047, EPI_ISL_799053, EPI_ISL_799055, EPI_ISL_799057, EPI_ISL_799059, EPI_ISL_799069, EPI_ISL_799070, EPI_ISL_799079, EPI_ISL_799085, EPI_ISL_799086, EPI_ISL_799099, EPI_ISL_799103, EPI_ISL_799104, EPI_ISL_799105, EPI_ISL_799106, EPI_ISL_799111, EPI_ISL_799113, EPI_ISL_799114, EPI_ISL_799122, EPI_ISL_799124, EPI_ISL_799137, EPI_ISL_799138, EPI_ISL_799139, EPI_ISL_799143, EPI_ISL_799145, EPI_ISL_799149, EPI_ISL_799154, EPI_ISL_799160, EPI_ISL_799161, EPI_ISL_799165, EPI_ISL_799166, EPI_ISL_799170, EPI_ISL_799171, EPI_ISL_799177, EPI_ISL_799178, EPI_ISL_799179, EPI_ISL_799182, EPI_ISL_799187, EPI_ISL_799188, EPI_ISL_799191, EPI_ISL_799198, EPI_ISL_799200, EPI_ISL_799202, EPI_ISL_799212, EPI_ISL_799213, EPI_ISL_799214, EPI_ISL_799219, EPI_ISL_799227, EPI_ISL_799228, EPI_ISL_799229, EPI_ISL_799231, EPI_ISL_799233, EPI_ISL_799234, EPI_ISL_799235, EPI_ISL_799237, EPI_ISL_799244, EPI_ISL_799248, EPI_ISL_799252, EPI_ISL_799257, EPI_ISL_799261, EPI_ISL_799265, EPI_ISL_799269, EPI_ISL_799271, EPI_ISL_799276, EPI_ISL_799278, EPI_ISL_799281, EPI_ISL_799644, EPI_ISL_799652, EPI_ISL_799655, EPI_ISL_799662, EPI_ISL_799665, EPI_ISL_799678, EPI_ISL_799687, EPI_ISL_799691, EPI_ISL_799693, EPI_ISL_799702, EPI_ISL_799711, EPI_ISL_799732, EPI_ISL_799734, EPI_ISL_799744, EPI_ISL_799747, EPI_ISL_799751, EPI_ISL_799754, EPI_ISL_799760, EPI_ISL_799767, EPI_ISL_799784, EPI_ISL_799787, EPI_ISL_799797, EPI_ISL_799799, EPI_ISL_799807, EPI_ISL_799809, EPI_ISL_799812, EPI_ISL_799814, EPI_ISL_799821, EPI_ISL_799822, EPI_ISL_799823, EPI_ISL_799831, EPI_ISL_799832, EPI_ISL_799841, EPI_ISL_799844, EPI_ISL_799846, EPI_ISL_799859, EPI_ISL_799881, EPI_ISL_799897, EPI_ISL_799899, EPI_ISL_799902, EPI_ISL_799914, EPI_ISL_799920, EPI_ISL_799924, EPI_ISL_799930, EPI_ISL_799933, EPI_ISL_799938, EPI_ISL_799940, EPI_ISL_799941, EPI_ISL_799942, EPI_ISL_799943, EPI_ISL_799948, EPI_ISL_799956, EPI_ISL_799961, EPI_ISL_799964, EPI_ISL_799966, EPI_ISL_799967, EPI_ISL_799970, EPI_ISL_799975, EPI_ISL_799977, EPI_ISL_799978, EPI_ISL_799979, EPI_ISL_799982, EPI_ISL_799983, EPI_ISL_799984, EPI_ISL_799989, EPI_ISL_799991, EPI_ISL_799995, EPI_ISL_799998, EPI_ISL_799999, EPI_ISL_800002, EPI_ISL_800003, EPI_ISL_800009, EPI_ISL_800011, EPI_ISL_800013, EPI_ISL_800014, EPI_ISL_800016, EPI_ISL_800017, EPI_ISL_800019, EPI_ISL_800020, EPI_ISL_800021, EPI_ISL_800022, EPI_ISL_800023, EPI_ISL_800025, EPI_ISL_800031, EPI_ISL_800033, EPI_ISL_800035, EPI_ISL_800036, EPI_ISL_800039, EPI_ISL_800040, EPI_ISL_800041, EPI_ISL_800042, EPI_ISL_800043, EPI_ISL_800044, EPI_ISL_800046, EPI_ISL_800049, EPI_ISL_800050, EPI_ISL_800051, EPI_ISL_800053, EPI_ISL_800058, EPI_ISL_800059, EPI_ISL_800060, EPI_ISL_800061, EPI_ISL_800062, EPI_ISL_800063, EPI_ISL_800066, EPI_ISL_800067, EPI_ISL_800069, EPI_ISL_800073, EPI_ISL_800074, EPI_ISL_800075, EPI_ISL_800076, EPI_ISL_800080, EPI_ISL_800081, EPI_ISL_800087, EPI_ISL_800090, EPI_ISL_800091, EPI_ISL_800094, EPI_ISL_800097, EPI_ISL_800099, EPI_ISL_800101, EPI_ISL_800104, EPI_ISL_800105, EPI_ISL_800107, EPI_ISL_800108, EPI_ISL_800109, EPI_ISL_800110, EPI_ISL_800112, EPI_ISL_800118, EPI_ISL_800126, EPI_ISL_800127, EPI_ISL_800128, EPI_ISL_800129, EPI_ISL_800130, EPI_ISL_800132, EPI_ISL_800134, EPI_ISL_800135, EPI_ISL_800136, EPI_ISL_800140, EPI_ISL_800143, EPI_ISL_800144, EPI_ISL_800145, EPI_ISL_800146, EPI_ISL_800148, EPI_ISL_800150, EPI_ISL_800152, EPI_ISL_800154, EPI_ISL_800155, EPI_ISL_800159, EPI_ISL_800160, EPI_ISL_800162, EPI_ISL_800163, EPI_ISL_800165, EPI_ISL_800169, EPI_ISL_800170, EPI_ISL_800171, EPI_ISL_800179, EPI_ISL_800181, EPI_ISL_800184, EPI_ISL_800185, EPI_ISL_800187, EPI_ISL_800193, EPI_ISL_800194, EPI_ISL_800197, EPI_ISL_800198, EPI_ISL_800199, EPI_ISL_800202, EPI_ISL_800205, EPI_ISL_800206, EPI_ISL_800207, EPI_ISL_800208, EPI_ISL_800209, EPI_ISL_800212, EPI_ISL_800213, EPI_ISL_800215, EPI_ISL_800216, EPI_ISL_800219, EPI_ISL_800222, EPI_ISL_800223, EPI_ISL_800224, EPI_ISL_800226, EPI_ISL_800227, EPI_ISL_800229, EPI_ISL_800230, EPI_ISL_800231, EPI_ISL_800232, EPI_ISL_800239, EPI_ISL_800240, EPI_ISL_800242, EPI_ISL_800243, EPI_ISL_800246, EPI_ISL_800247, EPI_ISL_800248, EPI_ISL_800249, EPI_ISL_800262, EPI_ISL_800267, EPI_ISL_800271, EPI_ISL_800274, EPI_ISL_800280, EPI_ISL_800281, EPI_ISL_800282, EPI_ISL_800286, EPI_ISL_800299, EPI_ISL_800301, EPI_ISL_800303, EPI_ISL_800308, EPI_ISL_800317, EPI_ISL_800338, EPI_ISL_800342, EPI_ISL_800347, EPI_ISL_800348, EPI_ISL_800350, EPI_ISL_800351, EPI_ISL_800354, EPI_ISL_800355, EPI_ISL_800359, EPI_ISL_800360, EPI_ISL_800362, EPI_ISL_800365, EPI_ISL_800367, EPI_ISL_800370, EPI_ISL_800376, EPI_ISL_800384, EPI_ISL_800392, EPI_ISL_800395, EPI_ISL_800407, EPI_ISL_800420, EPI_ISL_800428, EPI_ISL_800441, EPI_ISL_800444, EPI_ISL_800446, EPI_ISL_800452, EPI_ISL_800453, EPI_ISL_800454, EPI_ISL_800456, EPI_ISL_800457, EPI_ISL_800460, EPI_ISL_800468, EPI_ISL_800474, EPI_ISL_800481, EPI_ISL_800485, EPI_ISL_800494, EPI_ISL_800495, EPI_ISL_800501, EPI_ISL_800502, EPI_ISL_800506, EPI_ISL_800509, EPI_ISL_800516, EPI_ISL_800518, EPI_ISL_800524, EPI_ISL_800525, EPI_ISL_800526, EPI_ISL_800528, EPI_ISL_800531, EPI_ISL_800542, EPI_ISL_800555, EPI_ISL_800558, EPI_ISL_800566, EPI_ISL_800580, EPI_ISL_822291, EPI_ISL_822294, EPI_ISL_822297, EPI_ISL_822298, EPI_ISL_822300, EPI_ISL_822303, EPI_ISL_822308, EPI_ISL_834343, EPI_ISL_858701, EPI_ISL_858702, EPI_ISL_858703, EPI_ISL_858704, EPI_ISL_858705, EPI_ISL_858706, EPI_ISL_858707, EPI_ISL_858708, EPI_ISL_858709, EPI_ISL_858710, EPI_ISL_858713, EPI_ISL_858714, EPI_ISL_858719, EPI_ISL_858720, EPI_ISL_858725, EPI_ISL_858732, EPI_ISL_858737, EPI_ISL_858739, EPI_ISL_858748, EPI_ISL_858752, EPI_ISL_858753, EPI_ISL_858779, EPI_ISL_858780, EPI_ISL_858784, EPI_ISL_858785, EPI_ISL_858786, EPI_ISL_858787, EPI_ISL_858788, EPI_ISL_858789, EPI_ISL_858790, EPI_ISL_858791, EPI_ISL_858809, EPI_ISL_858811, EPI_ISL_858812, EPI_ISL_858813, EPI_ISL_858814, EPI_ISL_858816, EPI_ISL_858817, EPI_ISL_858818, EPI_ISL_858819, EPI_ISL_858820, EPI_ISL_858821, EPI_ISL_858822, EPI_ISL_858823, EPI_ISL_858824, EPI_ISL_858825, EPI_ISL_858826, EPI_ISL_858827, EPI_ISL_858828, EPI_ISL_858829, EPI_ISL_858831, EPI_ISL_858832, EPI_ISL_858833, EPI_ISL_858834, EPI_ISL_858835, EPI_ISL_858836, EPI_ISL_858837, EPI_ISL_858838, EPI_ISL_858839, EPI_ISL_858840, EPI_ISL_858841, EPI_ISL_858845, EPI_ISL_858846, EPI_ISL_858847, EPI_ISL_858848, EPI_ISL_858849, EPI_ISL_858850, EPI_ISL_858851, EPI_ISL_858852, EPI_ISL_858853, EPI_ISL_858854, EPI_ISL_858855, EPI_ISL_858856, EPI_ISL_858857, EPI_ISL_858858, EPI_ISL_858861, EPI_ISL_858862, EPI_ISL_858864, EPI_ISL_858866, EPI_ISL_858868, EPI_ISL_858869, EPI_ISL_858870, EPI_ISL_858871, EPI_ISL_858872, EPI_ISL_858873, EPI_ISL_858875, EPI_ISL_858876, EPI_ISL_858877, EPI_ISL_858878, EPI_ISL_858879, EPI_ISL_858880, EPI_ISL_858881, EPI_ISL_858882, EPI_ISL_858883, EPI_ISL_858884, EPI_ISL_858886, EPI_ISL_858887, EPI_ISL_858888, EPI_ISL_858889, EPI_ISL_858890, EPI_ISL_858891, EPI_ISL_858892, EPI_ISL_858893, EPI_ISL_858894, EPI_ISL_858895, EPI_ISL_858896, EPI_ISL_858897, EPI_ISL_858898, EPI_ISL_858899, EPI_ISL_858979, EPI_ISL_858980, EPI_ISL_858981, EPI_ISL_858982, EPI_ISL_858983, EPI_ISL_858985, EPI_ISL_858986, EPI_ISL_858987, EPI_ISL_858988, EPI_ISL_858989, EPI_ISL_858991, EPI_ISL_882314, EPI_ISL_882590, EPI_ISL_892212, EPI_ISL_945235, EPI_ISL_945335 | | | |
| see above | Lighthouse Lab in Alderley Park | Wellcome Sanger Institute for the COVID-19 Genomics UK (COG-UK) Consortium | Cordelia Langford; David K. Jackson; Dominic Kwiatkowski; Ewan Harrison; Ian Johnston; Jacquelyn Wynn; John Sillitoe on behalf of the Wellcome Sanger Institute COVID-19 Surveillance Team; Mairead Hyland; Roberto Amato; Sonia Goncalves; The Lighthouse Lab in Alderley Park and Alex Alderton |
| EPI_ISL_777985, EPI_ISL_781183, EPI_ISL_781187, EPI_ISL_781189, EPI_ISL_781190, EPI_ISL_781220, EPI_ISL_781248, EPI_ISL_781249, EPI_ISL_781251, EPI_ISL_781258, EPI_ISL_781261, EPI_ISL_781269, EPI_ISL_781319, EPI_ISL_781325, EPI_ISL_781333, EPI_ISL_781341, EPI_ISL_781354, EPI_ISL_781355, EPI_ISL_781359, EPI_ISL_781360, EPI_ISL_781367, EPI_ISL_781371, EPI_ISL_781392, EPI_ISL_781401, EPI_ISL_781434, EPI_ISL_781448, EPI_ISL_781481, EPI_ISL_781483, EPI_ISL_781489, EPI_ISL_781497, EPI_ISL_781506, EPI_ISL_781517, EPI_ISL_781518, EPI_ISL_781520, EPI_ISL_781521, EPI_ISL_781525, EPI_ISL_781526, EPI_ISL_781530, EPI_ISL_781531, EPI_ISL_781533, EPI_ISL_781534, EPI_ISL_781535, EPI_ISL_781540, EPI_ISL_781549, EPI_ISL_781550, EPI_ISL_781559, EPI_ISL_781560, EPI_ISL_781561, EPI_ISL_781562, EPI_ISL_781563, EPI_ISL_781564, EPI_ISL_781568, EPI_ISL_781569, EPI_ISL_781570, EPI_ISL_781572, EPI_ISL_781573, EPI_ISL_781574, EPI_ISL_781575, EPI_ISL_781577, EPI_ISL_781580, EPI_ISL_781583, EPI_ISL_781584, EPI_ISL_781585, EPI_ISL_781586, EPI_ISL_781588, EPI_ISL_781589, EPI_ISL_781592, EPI_ISL_781594, EPI_ISL_781596, EPI_ISL_781600, EPI_ISL_781601, EPI_ISL_781602, EPI_ISL_781604, EPI_ISL_781607, EPI_ISL_781608, EPI_ISL_781612, EPI_ISL_781613, EPI_ISL_781614, EPI_ISL_781616, EPI_ISL_781626, EPI_ISL_781628, EPI_ISL_781629, EPI_ISL_781630, EPI_ISL_781631, EPI_ISL_781633, EPI_ISL_781634, EPI_ISL_781635, EPI_ISL_781639, EPI_ISL_781640, EPI_ISL_781641, EPI_ISL_781643, EPI_ISL_781644, EPI_ISL_781646, EPI_ISL_781647, EPI_ISL_781648, EPI_ISL_781650, EPI_ISL_781653, EPI_ISL_781657, EPI_ISL_781658, EPI_ISL_781659, EPI_ISL_781660, EPI_ISL_781661, EPI_ISL_781662, EPI_ISL_781664, EPI_ISL_781667, EPI_ISL_781668, EPI_ISL_781670, EPI_ISL_781672, EPI_ISL_781673, EPI_ISL_781675, EPI_ISL_781676, EPI_ISL_781679, EPI_ISL_781682, EPI_ISL_781683, EPI_ISL_781687, EPI_ISL_781688, EPI_ISL_781689, EPI_ISL_781692, EPI_ISL_781693, EPI_ISL_781694, EPI_ISL_781696, EPI_ISL_781697, EPI_ISL_781700, EPI_ISL_781702, EPI_ISL_781704, EPI_ISL_781708, EPI_ISL_781709, EPI_ISL_781714, EPI_ISL_781717, EPI_ISL_781718, EPI_ISL_781719, EPI_ISL_781722, EPI_ISL_781723, EPI_ISL_781726, EPI_ISL_781727, EPI_ISL_781731, EPI_ISL_781733, EPI_ISL_781737, EPI_ISL_781738, EPI_ISL_781739, EPI_ISL_781740, EPI_ISL_781744, EPI_ISL_781748, EPI_ISL_781750, EPI_ISL_781751, EPI_ISL_781752, EPI_ISL_781755, EPI_ISL_781756, EPI_ISL_781757, EPI_ISL_781761, EPI_ISL_781765, EPI_ISL_781768, EPI_ISL_781769, EPI_ISL_781774, EPI_ISL_781775, EPI_ISL_781776, EPI_ISL_781778, EPI_ISL_781779, EPI_ISL_781781, EPI_ISL_781782, EPI_ISL_781786, EPI_ISL_781787, EPI_ISL_781790, EPI_ISL_781791, EPI_ISL_781792, EPI_ISL_781793, EPI_ISL_781796, EPI_ISL_781799, EPI_ISL_781800, EPI_ISL_781802, EPI_ISL_781803, EPI_ISL_781804, EPI_ISL_781806, EPI_ISL_781810, EPI_ISL_781811, EPI_ISL_781812, EPI_ISL_781813, EPI_ISL_781814, EPI_ISL_781816, EPI_ISL_781817, EPI_ISL_781819, EPI_ISL_781820, EPI_ISL_781821, EPI_ISL_781823, EPI_ISL_781825, EPI_ISL_781826, EPI_ISL_781827, EPI_ISL_781829, EPI_ISL_781830, EPI_ISL_781831, EPI_ISL_781833, EPI_ISL_781835, EPI_ISL_781836, EPI_ISL_781845, EPI_ISL_781847, EPI_ISL_781849, EPI_ISL_781850, EPI_ISL_781852, EPI_ISL_781853, EPI_ISL_781855, EPI_ISL_781856, EPI_ISL_781857, EPI_ISL_781859, EPI_ISL_781860, EPI_ISL_781861, EPI_ISL_781865, EPI_ISL_781866, EPI_ISL_781868, EPI_ISL_797400, EPI_ISL_797403, EPI_ISL_797434, EPI_ISL_797446, EPI_ISL_797465, EPI_ISL_797481, EPI_ISL_797483, EPI_ISL_797492, EPI_ISL_797497, EPI_ISL_797498, EPI_ISL_797502, EPI_ISL_797505, EPI_ISL_797510, EPI_ISL_797511, EPI_ISL_797532, EPI_ISL_797537, EPI_ISL_797548, EPI_ISL_797566, EPI_ISL_797578, EPI_ISL_797579, EPI_ISL_797580, EPI_ISL_797590, EPI_ISL_797595, EPI_ISL_797597, EPI_ISL_797600, EPI_ISL_797625, EPI_ISL_797631, EPI_ISL_797643, EPI_ISL_797645, EPI_ISL_797646, EPI_ISL_797647, EPI_ISL_797648, EPI_ISL_797652, EPI_ISL_797657, EPI_ISL_797659, EPI_ISL_797664, EPI_ISL_797668, EPI_ISL_797676, EPI_ISL_797688, EPI_ISL_797690, EPI_ISL_797692, EPI_ISL_797694, EPI_ISL_797705, EPI_ISL_798000, EPI_ISL_798003, EPI_ISL_798018, EPI_ISL_798032, EPI_ISL_798038, EPI_ISL_798040, EPI_ISL_798046, EPI_ISL_798048, EPI_ISL_798053, EPI_ISL_798055, EPI_ISL_798058, EPI_ISL_798060, EPI_ISL_798069, EPI_ISL_798072, EPI_ISL_798076, EPI_ISL_798077, EPI_ISL_798078, EPI_ISL_798079, EPI_ISL_798086, EPI_ISL_798088, EPI_ISL_798093, EPI_ISL_798094, EPI_ISL_798095, EPI_ISL_798097, EPI_ISL_798105, EPI_ISL_798110, EPI_ISL_798111, EPI_ISL_798112, EPI_ISL_798113, EPI_ISL_798115, EPI_ISL_798117, EPI_ISL_798121, EPI_ISL_798126, EPI_ISL_798128, EPI_ISL_798130, EPI_ISL_798132, EPI_ISL_798137, EPI_ISL_798140, EPI_ISL_798143, EPI_ISL_798145, EPI_ISL_798148, EPI_ISL_798149, EPI_ISL_798151, EPI_ISL_798153, EPI_ISL_798160, EPI_ISL_798161, EPI_ISL_798166, EPI_ISL_798167, EPI_ISL_798168, EPI_ISL_798183, EPI_ISL_798185, EPI_ISL_798186, EPI_ISL_798193, EPI_ISL_798197, EPI_ISL_798199, EPI_ISL_798201, EPI_ISL_798203, EPI_ISL_798211, EPI_ISL_798213, EPI_ISL_798214, EPI_ISL_798216, EPI_ISL_798217, EPI_ISL_798218, EPI_ISL_798224, EPI_ISL_798230, EPI_ISL_798243, EPI_ISL_798244, EPI_ISL_798251, EPI_ISL_798252, EPI_ISL_798253, EPI_ISL_798254, EPI_ISL_798256, EPI_ISL_798258, EPI_ISL_798266, EPI_ISL_798270, EPI_ISL_798272, EPI_ISL_798273, EPI_ISL_798281, EPI_ISL_798288, EPI_ISL_798289, EPI_ISL_798294, EPI_ISL_798298, EPI_ISL_798300, EPI_ISL_798307, EPI_ISL_798311, EPI_ISL_798312, EPI_ISL_798315, EPI_ISL_798316, EPI_ISL_798319, EPI_ISL_798322, EPI_ISL_798326, EPI_ISL_798327, EPI_ISL_798664, EPI_ISL_798673, EPI_ISL_798674, EPI_ISL_798677, EPI_ISL_798678, EPI_ISL_798679, EPI_ISL_798681, EPI_ISL_798682, EPI_ISL_798683, EPI_ISL_798685, EPI_ISL_798686, EPI_ISL_798687, EPI_ISL_798694, EPI_ISL_798697, EPI_ISL_798698, EPI_ISL_798699, EPI_ISL_798700, EPI_ISL_798703, EPI_ISL_798706, EPI_ISL_798708, EPI_ISL_798711, EPI_ISL_798713, EPI_ISL_798714, EPI_ISL_798715, EPI_ISL_798717, EPI_ISL_798718, EPI_ISL_798723, EPI_ISL_798726, EPI_ISL_798730, EPI_ISL_798731, EPI_ISL_798732, EPI_ISL_798733, EPI_ISL_798734, EPI_ISL_798736, EPI_ISL_798737, EPI_ISL_798743, EPI_ISL_798745, EPI_ISL_798746, EPI_ISL_798747, EPI_ISL_798748, EPI_ISL_798751, EPI_ISL_798753, EPI_ISL_798754, EPI_ISL_798756, EPI_ISL_798757, EPI_ISL_798758, EPI_ISL_798759, EPI_ISL_798760, EPI_ISL_798761, EPI_ISL_798762, EPI_ISL_798763, EPI_ISL_798765, EPI_ISL_798766, EPI_ISL_798772, EPI_ISL_798773, EPI_ISL_798775, EPI_ISL_798778, EPI_ISL_798780, EPI_ISL_798782, EPI_ISL_798783, EPI_ISL_798785, EPI_ISL_798786, EPI_ISL_798787, EPI_ISL_798788, EPI_ISL_798789, EPI_ISL_798790, EPI_ISL_798792, EPI_ISL_798794, EPI_ISL_798795, EPI_ISL_798796, EPI_ISL_798797, EPI_ISL_798799, EPI_ISL_798800, EPI_ISL_798802, EPI_ISL_798805, EPI_ISL_798806, EPI_ISL_798807, EPI_ISL_798810, EPI_ISL_798813, EPI_ISL_798815, EPI_ISL_798816, EPI_ISL_798818, EPI_ISL_798820, EPI_ISL_798825, EPI_ISL_798827, EPI_ISL_798828, EPI_ISL_798829, EPI_ISL_798831, EPI_ISL_798834, EPI_ISL_798835, EPI_ISL_798836, EPI_ISL_798839, EPI_ISL_798840, EPI_ISL_798841, EPI_ISL_798842, EPI_ISL_798843, EPI_ISL_798846, EPI_ISL_798847, EPI_ISL_798848, EPI_ISL_798849, EPI_ISL_798854, EPI_ISL_798857, EPI_ISL_798858, EPI_ISL_798859, EPI_ISL_798860, EPI_ISL_798861, EPI_ISL_798862, EPI_ISL_798865, EPI_ISL_798866, EPI_ISL_798867, EPI_ISL_798868, EPI_ISL_798871, EPI_ISL_798873, EPI_ISL_798877, EPI_ISL_798881, EPI_ISL_798883, EPI_ISL_798886, EPI_ISL_798887, EPI_ISL_798890, EPI_ISL_798893, EPI_ISL_798894, EPI_ISL_798895, EPI_ISL_798896, EPI_ISL_798897, EPI_ISL_798899, EPI_ISL_798900, EPI_ISL_798901, EPI_ISL_798903, EPI_ISL_798904, EPI_ISL_798907, EPI_ISL_798908, EPI_ISL_798909, EPI_ISL_798912, EPI_ISL_798913, EPI_ISL_798916, EPI_ISL_798917, EPI_ISL_798918, EPI_ISL_798920, EPI_ISL_798923, EPI_ISL_798927, EPI_ISL_798929, EPI_ISL_798931, EPI_ISL_798932, EPI_ISL_798937, EPI_ISL_798938, EPI_ISL_798940, EPI_ISL_798943, EPI_ISL_798944, EPI_ISL_798945, EPI_ISL_798946, EPI_ISL_798947, EPI_ISL_798949, EPI_ISL_798953, EPI_ISL_798954, EPI_ISL_798956, EPI_ISL_798958, EPI_ISL_798959, EPI_ISL_798960, EPI_ISL_798965, EPI_ISL_798971, EPI_ISL_798974, EPI_ISL_798976, EPI_ISL_798978, EPI_ISL_798980, EPI_ISL_798982, EPI_ISL_798985, EPI_ISL_798989, EPI_ISL_798990, EPI_ISL_798992, EPI_ISL_798993, EPI_ISL_798997, EPI_ISL_798998, EPI_ISL_798999, EPI_ISL_799001, EPI_ISL_799002, EPI_ISL_799007, EPI_ISL_799008, EPI_ISL_799013, EPI_ISL_799017, EPI_ISL_799018, EPI_ISL_799020, EPI_ISL_799031, EPI_ISL_799032, EPI_ISL_799035, | | | |

EPI_ISL_799036, EPI_ISL_799037, EPI_ISL_799039, EPI_ISL_799048, EPI_ISL_799052, EPI_ISL_799058, EPI_ISL_799061, EPI_ISL_799074, EPI_ISL_799076, EPI_ISL_799078, EPI_ISL_799083, EPI_ISL_799087, EPI_ISL_799088, EPI_ISL_799091, EPI_ISL_799092, EPI_ISL_799093, EPI_ISL_799094, EPI_ISL_799095, EPI_ISL_799097, EPI_ISL_799109, EPI_ISL_799110, EPI_ISL_799116, EPI_ISL_799117, EPI_ISL_799119,
EPI_ISL_799121, EPI_ISL_799128, EPI_ISL_799133, EPI_ISL_799135, EPI_ISL_799136, EPI_ISL_799141, EPI_ISL_799142, EPI_ISL_799146, EPI_ISL_799148, EPI_ISL_799152, EPI_ISL_799153, EPI_ISL_799159, EPI_ISL_799168, EPI_ISL_799173, EPI_ISL_799174, EPI_ISL_799181, EPI_ISL_799184, EPI_ISL_799185, EPI_ISL_799186, EPI_ISL_799195, EPI_ISL_799206, EPI_ISL_799208, EPI_ISL_799210, EPI_ISL_799211,
EPI_ISL_799217, EPI_ISL_799220, EPI_ISL_799223, EPI_ISL_799225, EPI_ISL_799226, EPI_ISL_799238, EPI_ISL_799239, EPI_ISL_799241, EPI_ISL_799242, EPI_ISL_799247, EPI_ISL_799251, EPI_ISL_799253, EPI_ISL_799254, EPI_ISL_799259, EPI_ISL_799266, EPI_ISL_799284, EPI_ISL_799291, EPI_ISL_799292, EPI_ISL_799293, EPI_ISL_799295, EPI_ISL_799935, EPI_ISL_799939, EPI_ISL_799944, EPI_ISL_799947,
EPI_ISL_799950, EPI_ISL_799952, EPI_ISL_799953, EPI_ISL_799955, EPI_ISL_799957, EPI_ISL_799958, EPI_ISL_799959, EPI_ISL_799960, EPI_ISL_799962, EPI_ISL_799963, EPI_ISL_799974, EPI_ISL_799988, EPI_ISL_799996, EPI_ISL_800004, EPI_ISL_800006, EPI_ISL_800007, EPI_ISL_800008, EPI_ISL_800012, EPI_ISL_800018, EPI_ISL_800024, EPI_ISL_800026, EPI_ISL_800028, EPI_ISL_800037, EPI_ISL_800045,
EPI_ISL_800047, EPI_ISL_800055, EPI_ISL_800065, EPI_ISL_800072, EPI_ISL_800077, EPI_ISL_800082, EPI_ISL_800083, EPI_ISL_800085, EPI_ISL_800088, EPI_ISL_800092, EPI_ISL_800093, EPI_ISL_800095, EPI_ISL_800096, EPI_ISL_800098, EPI_ISL_800102, EPI_ISL_800103, EPI_ISL_800114, EPI_ISL_800115, EPI_ISL_800116, EPI_ISL_800119, EPI_ISL_800121, EPI_ISL_800123, EPI_ISL_800125, EPI_ISL_800133,
EPI_ISL_800137, EPI_ISL_800149, EPI_ISL_800153, EPI_ISL_800156, EPI_ISL_800166, EPI_ISL_800167, EPI_ISL_800168, EPI_ISL_800172, EPI_ISL_800173, EPI_ISL_800177, EPI_ISL_800178, EPI_ISL_800182, EPI_ISL_800195, EPI_ISL_800196, EPI_ISL_800211, EPI_ISL_800214, EPI_ISL_800218, EPI_ISL_800228, EPI_ISL_800233, EPI_ISL_800238, EPI_ISL_800250, EPI_ISL_800252, EPI_ISL_800254, EPI_ISL_800255,
EPI_ISL_800256, EPI_ISL_800257, EPI_ISL_800258, EPI_ISL_800259, EPI_ISL_800260, EPI_ISL_800261, EPI_ISL_800263, EPI_ISL_800264, EPI_ISL_800265, EPI_ISL_800266, EPI_ISL_800268, EPI_ISL_800269, EPI_ISL_800270, EPI_ISL_800272, EPI_ISL_800273, EPI_ISL_800275, EPI_ISL_800276, EPI_ISL_800277, EPI_ISL_800278, EPI_ISL_800279, EPI_ISL_800283, EPI_ISL_800284, EPI_ISL_800285, EPI_ISL_800287,
EPI_ISL_800288, EPI_ISL_800289, EPI_ISL_800290, EPI_ISL_800291, EPI_ISL_800292, EPI_ISL_800293, EPI_ISL_800294, EPI_ISL_800295, EPI_ISL_800296, EPI_ISL_800297, EPI_ISL_800298, EPI_ISL_800300, EPI_ISL_800302, EPI_ISL_800304, EPI_ISL_800305, EPI_ISL_800306, EPI_ISL_800307, EPI_ISL_800309, EPI_ISL_800310, EPI_ISL_800311, EPI_ISL_800312, EPI_ISL_800313, EPI_ISL_800314, EPI_ISL_800315,
EPI_ISL_800316, EPI_ISL_800318, EPI_ISL_800319, EPI_ISL_800320, EPI_ISL_800321, EPI_ISL_800322, EPI_ISL_800323, EPI_ISL_800324, EPI_ISL_800325, EPI_ISL_800326, EPI_ISL_800327, EPI_ISL_800328, EPI_ISL_800329, EPI_ISL_800330, EPI_ISL_800331, EPI_ISL_800332, EPI_ISL_800333, EPI_ISL_800334, EPI_ISL_800335, EPI_ISL_800336, EPI_ISL_800337, EPI_ISL_800339, EPI_ISL_800341, EPI_ISL_800343,
EPI_ISL_800344, EPI_ISL_800345, EPI_ISL_800346, EPI_ISL_800349, EPI_ISL_800352, EPI_ISL_800353, EPI_ISL_800356, EPI_ISL_800357, EPI_ISL_800358, EPI_ISL_800361, EPI_ISL_800363, EPI_ISL_800364, EPI_ISL_800366, EPI_ISL_800368, EPI_ISL_800369, EPI_ISL_800371, EPI_ISL_800372, EPI_ISL_800373, EPI_ISL_800374, EPI_ISL_800375, EPI_ISL_800377, EPI_ISL_800378, EPI_ISL_800379, EPI_ISL_800380,
EPI_ISL_800381, EPI_ISL_800382, EPI_ISL_800383, EPI_ISL_800385, EPI_ISL_800386, EPI_ISL_800387, EPI_ISL_800388, EPI_ISL_800389, EPI_ISL_800390, EPI_ISL_800391, EPI_ISL_800393, EPI_ISL_800394, EPI_ISL_800396, EPI_ISL_800397, EPI_ISL_800398, EPI_ISL_800399, EPI_ISL_800400, EPI_ISL_800401, EPI_ISL_800402, EPI_ISL_800403, EPI_ISL_800404, EPI_ISL_800405, EPI_ISL_800406, EPI_ISL_800408,
EPI_ISL_800409, EPI_ISL_800410, EPI_ISL_800411, EPI_ISL_800412, EPI_ISL_800413, EPI_ISL_800414, EPI_ISL_800415, EPI_ISL_800416, EPI_ISL_800417, EPI_ISL_800418, EPI_ISL_800419, EPI_ISL_800421, EPI_ISL_800422, EPI_ISL_800423, EPI_ISL_800424, EPI_ISL_800425, EPI_ISL_800426, EPI_ISL_800427, EPI_ISL_800429, EPI_ISL_800430, EPI_ISL_800431, EPI_ISL_800432, EPI_ISL_800433, EPI_ISL_800434,
EPI_ISL_800435, EPI_ISL_800436, EPI_ISL_800437, EPI_ISL_800438, EPI_ISL_800439, EPI_ISL_800440, EPI_ISL_800442, EPI_ISL_800443, EPI_ISL_800445, EPI_ISL_800447, EPI_ISL_800448, EPI_ISL_800449, EPI_ISL_800450, EPI_ISL_800451, EPI_ISL_800455, EPI_ISL_800458, EPI_ISL_800459, EPI_ISL_800461, EPI_ISL_800462, EPI_ISL_800463, EPI_ISL_800464, EPI_ISL_800465, EPI_ISL_800466, EPI_ISL_800467,
EPI_ISL_800469, EPI_ISL_800470, EPI_ISL_800471, EPI_ISL_800472, EPI_ISL_800473, EPI_ISL_800475, EPI_ISL_800476, EPI_ISL_800477, EPI_ISL_800478, EPI_ISL_800479, EPI_ISL_800480, EPI_ISL_800482, EPI_ISL_800483, EPI_ISL_800484, EPI_ISL_800486, EPI_ISL_800487, EPI_ISL_800488, EPI_ISL_800489, EPI_ISL_800490, EPI_ISL_800491, EPI_ISL_800492, EPI_ISL_800493, EPI_ISL_800496, EPI_ISL_800497,
EPI_ISL_800498, EPI_ISL_800499, EPI_ISL_800500, EPI_ISL_800503, EPI_ISL_800504, EPI_ISL_800505, EPI_ISL_800507, EPI_ISL_800508, EPI_ISL_800510, EPI_ISL_800511, EPI_ISL_800512, EPI_ISL_800513, EPI_ISL_800514, EPI_ISL_800515, EPI_ISL_800517, EPI_ISL_800519, EPI_ISL_800520, EPI_ISL_800521, EPI_ISL_800522, EPI_ISL_800523, EPI_ISL_800527, EPI_ISL_800529, EPI_ISL_800530, EPI_ISL_800532,
EPI_ISL_800533, EPI_ISL_800534, EPI_ISL_800535, EPI_ISL_800536, EPI_ISL_800537, EPI_ISL_800538, EPI_ISL_800539, EPI_ISL_800540, EPI_ISL_800541, EPI_ISL_800543, EPI_ISL_800544, EPI_ISL_800545, EPI_ISL_800546, EPI_ISL_800547, EPI_ISL_800548, EPI_ISL_800549, EPI_ISL_800550, EPI_ISL_800551, EPI_ISL_800552, EPI_ISL_800553, EPI_ISL_800554, EPI_ISL_800556, EPI_ISL_800557, EPI_ISL_800559,
EPI_ISL_800560, EPI_ISL_800561, EPI_ISL_800562, EPI_ISL_800563, EPI_ISL_800564, EPI_ISL_800565, EPI_ISL_800567, EPI_ISL_800568, EPI_ISL_800569, EPI_ISL_800570, EPI_ISL_800571, EPI_ISL_800572, EPI_ISL_800573, EPI_ISL_800574, EPI_ISL_821667, EPI_ISL_821672, EPI_ISL_821677, EPI_ISL_821733, EPI_ISL_821787, EPI_ISL_821790, EPI_ISL_821827, EPI_ISL_821877, EPI_ISL_821889, EPI_ISL_821934,
EPI_ISL_821943, EPI_ISL_821959, EPI_ISL_822299, EPI_ISL_822301, EPI_ISL_945051, EPI_ISL_945226

| | | | |
|---|---|---|---|
| see above | Lighthouse Lab in Cambridge | Wellcome Sanger Institute for the COVID-19 Genomics UK (COG-UK) Consortium | Cordelia Langford; David K. Jackson; Dominic Kwiatkowski; Ewan Harrison; Ian Johnston; John Sillitoe on behalf of the Wellcome Sanger Institute COVID-19 Surveillance Team; Rob Howes; Roberto Amato; Sonia Goncalves; The Lighthouse Lab in Cambridge and Alex Alderton |

EPI_ISL_760250, EPI_ISL_760251, EPI_ISL_760252, EPI_ISL_760253, EPI_ISL_760254, EPI_ISL_760255, EPI_ISL_760256, EPI_ISL_760257, EPI_ISL_760258, EPI_ISL_760260, EPI_ISL_760262, EPI_ISL_760264, EPI_ISL_760265, EPI_ISL_760269, EPI_ISL_760270, EPI_ISL_760271, EPI_ISL_760272, EPI_ISL_760274, EPI_ISL_760275, EPI_ISL_760279, EPI_ISL_760283, EPI_ISL_760289, EPI_ISL_760290, EPI_ISL_760291,
EPI_ISL_760292, EPI_ISL_760294, EPI_ISL_760297, EPI_ISL_760298, EPI_ISL_760299, EPI_ISL_760300, EPI_ISL_760302, EPI_ISL_760304, EPI_ISL_760307, EPI_ISL_760308, EPI_ISL_760309, EPI_ISL_760310, EPI_ISL_760312, EPI_ISL_760314, EPI_ISL_760316, EPI_ISL_760318, EPI_ISL_760319, EPI_ISL_760320, EPI_ISL_760321, EPI_ISL_760322, EPI_ISL_760323, EPI_ISL_760324, EPI_ISL_760325, EPI_ISL_760327,
EPI_ISL_760328, EPI_ISL_760329, EPI_ISL_760330, EPI_ISL_760331, EPI_ISL_760332, EPI_ISL_760334, EPI_ISL_760335, EPI_ISL_760336, EPI_ISL_760337, EPI_ISL_760338, EPI_ISL_760339, EPI_ISL_760340, EPI_ISL_760342, EPI_ISL_760343, EPI_ISL_760345, EPI_ISL_760347, EPI_ISL_760348, EPI_ISL_760349, EPI_ISL_760351, EPI_ISL_760352, EPI_ISL_760353, EPI_ISL_760354, EPI_ISL_760355, EPI_ISL_760356,
EPI_ISL_760357, EPI_ISL_760361, EPI_ISL_760362, EPI_ISL_760363, EPI_ISL_760366, EPI_ISL_760367, EPI_ISL_760368, EPI_ISL_760373, EPI_ISL_760375, EPI_ISL_760376, EPI_ISL_760378, EPI_ISL_760379, EPI_ISL_760382, EPI_ISL_760383, EPI_ISL_760384, EPI_ISL_760385, EPI_ISL_760388, EPI_ISL_760391, EPI_ISL_760393, EPI_ISL_760394, EPI_ISL_760395, EPI_ISL_760396, EPI_ISL_760398, EPI_ISL_760399,
EPI_ISL_760400, EPI_ISL_760401, EPI_ISL_760402, EPI_ISL_760403, EPI_ISL_760404, EPI_ISL_760406, EPI_ISL_760407, EPI_ISL_760409, EPI_ISL_760410, EPI_ISL_760411, EPI_ISL_760412, EPI_ISL_760413, EPI_ISL_760416, EPI_ISL_760417, EPI_ISL_760420, EPI_ISL_760422, EPI_ISL_760423, EPI_ISL_760425, EPI_ISL_760426, EPI_ISL_760427, EPI_ISL_760428, EPI_ISL_760429, EPI_ISL_760431, EPI_ISL_760435,
EPI_ISL_760437, EPI_ISL_760438, EPI_ISL_760439, EPI_ISL_760440, EPI_ISL_760443, EPI_ISL_760444, EPI_ISL_760447, EPI_ISL_760449, EPI_ISL_760451, EPI_ISL_760452, EPI_ISL_760453, EPI_ISL_760454, EPI_ISL_760455, EPI_ISL_760456, EPI_ISL_760457, EPI_ISL_760458, EPI_ISL_760459, EPI_ISL_760462, EPI_ISL_760463, EPI_ISL_760464, EPI_ISL_760465, EPI_ISL_760466, EPI_ISL_760467, EPI_ISL_760468,
EPI_ISL_760469, EPI_ISL_760470, EPI_ISL_760472, EPI_ISL_760473, EPI_ISL_760476, EPI_ISL_760477, EPI_ISL_760478, EPI_ISL_760479, EPI_ISL_760480, EPI_ISL_760481, EPI_ISL_760482, EPI_ISL_760483, EPI_ISL_760484, EPI_ISL_760487, EPI_ISL_760488, EPI_ISL_760489, EPI_ISL_760492, EPI_ISL_760493, EPI_ISL_760494, EPI_ISL_760496, EPI_ISL_760497, EPI_ISL_760499, EPI_ISL_760502, EPI_ISL_760503,
EPI_ISL_760504, EPI_ISL_760505, EPI_ISL_760506, EPI_ISL_760507, EPI_ISL_760510, EPI_ISL_760513, EPI_ISL_760514, EPI_ISL_760515, EPI_ISL_760517, EPI_ISL_760518, EPI_ISL_760519, EPI_ISL_760520, EPI_ISL_760522, EPI_ISL_760523, EPI_ISL_760525, EPI_ISL_760526, EPI_ISL_760527, EPI_ISL_760528, EPI_ISL_760529, EPI_ISL_760532, EPI_ISL_760533, EPI_ISL_760536, EPI_ISL_760537, EPI_ISL_760539,
EPI_ISL_760540, EPI_ISL_760541, EPI_ISL_760547, EPI_ISL_760548, EPI_ISL_760550, EPI_ISL_760553, EPI_ISL_760554, EPI_ISL_760556, EPI_ISL_760563, EPI_ISL_760566, EPI_ISL_760568, EPI_ISL_760569, EPI_ISL_760571, EPI_ISL_760573, EPI_ISL_760575, EPI_ISL_760576, EPI_ISL_760577, EPI_ISL_760578, EPI_ISL_760579, EPI_ISL_760580, EPI_ISL_760581, EPI_ISL_760583, EPI_ISL_760584, EPI_ISL_760585,
EPI_ISL_760590, EPI_ISL_760592, EPI_ISL_760594, EPI_ISL_760602, EPI_ISL_760604, EPI_ISL_760605, EPI_ISL_760606, EPI_ISL_760607, EPI_ISL_760610, EPI_ISL_760611, EPI_ISL_760614, EPI_ISL_760616, EPI_ISL_760618, EPI_ISL_760619, EPI_ISL_760620, EPI_ISL_760621, EPI_ISL_760622, EPI_ISL_760626, EPI_ISL_760629, EPI_ISL_760630, EPI_ISL_760631, EPI_ISL_760632, EPI_ISL_760634, EPI_ISL_760635,
EPI_ISL_760636, EPI_ISL_760641, EPI_ISL_760642, EPI_ISL_760643, EPI_ISL_760645, EPI_ISL_760646, EPI_ISL_760648, EPI_ISL_760651, EPI_ISL_760653, EPI_ISL_760655, EPI_ISL_760659, EPI_ISL_760662, EPI_ISL_760668, EPI_ISL_760671, EPI_ISL_760672, EPI_ISL_760674, EPI_ISL_760675, EPI_ISL_760676, EPI_ISL_760678, EPI_ISL_760681, EPI_ISL_760682, EPI_ISL_760683, EPI_ISL_760687, EPI_ISL_760689,
EPI_ISL_760690, EPI_ISL_760694, EPI_ISL_760695, EPI_ISL_760696, EPI_ISL_760700, EPI_ISL_760703, EPI_ISL_760704, EPI_ISL_760706, EPI_ISL_760707, EPI_ISL_760709, EPI_ISL_760712, EPI_ISL_760713, EPI_ISL_760716, EPI_ISL_760717, EPI_ISL_760718, EPI_ISL_760725, EPI_ISL_760729, EPI_ISL_760730, EPI_ISL_760731, EPI_ISL_760732, EPI_ISL_760735, EPI_ISL_760742, EPI_ISL_760743, EPI_ISL_760744,
EPI_ISL_760746, EPI_ISL_760747, EPI_ISL_760749, EPI_ISL_760751, EPI_ISL_760754, EPI_ISL_760757, EPI_ISL_760758, EPI_ISL_760760, EPI_ISL_760764, EPI_ISL_760765, EPI_ISL_760766, EPI_ISL_760767, EPI_ISL_760768, EPI_ISL_760770, EPI_ISL_760771, EPI_ISL_760772, EPI_ISL_760773, EPI_ISL_760774, EPI_ISL_760775, EPI_ISL_760777, EPI_ISL_760778, EPI_ISL_760779, EPI_ISL_760781, EPI_ISL_760787,
EPI_ISL_760791, EPI_ISL_760793, EPI_ISL_760797, EPI_ISL_760798, EPI_ISL_760800, EPI_ISL_760803, EPI_ISL_760805, EPI_ISL_760807, EPI_ISL_767096, EPI_ISL_778003, EPI_ISL_778004, EPI_ISL_778007, EPI_ISL_778008, EPI_ISL_778009, EPI_ISL_778014, EPI_ISL_778024, EPI_ISL_778026, EPI_ISL_778031, EPI_ISL_778034, EPI_ISL_778036, EPI_ISL_778042, EPI_ISL_778046, EPI_ISL_778053, EPI_ISL_778058,
EPI_ISL_778060, EPI_ISL_778062, EPI_ISL_778068, EPI_ISL_778073, EPI_ISL_778076, EPI_ISL_778077, EPI_ISL_778082, EPI_ISL_778088, EPI_ISL_778090, EPI_ISL_778092, EPI_ISL_778093, EPI_ISL_778102, EPI_ISL_778106, EPI_ISL_778107, EPI_ISL_778111, EPI_ISL_778117, EPI_ISL_778119, EPI_ISL_778122, EPI_ISL_778127, EPI_ISL_778129, EPI_ISL_778131, EPI_ISL_778133, EPI_ISL_778136, EPI_ISL_778145,
EPI_ISL_778149, EPI_ISL_778151, EPI_ISL_778153, EPI_ISL_778154, EPI_ISL_778156, EPI_ISL_778159, EPI_ISL_778167, EPI_ISL_778168, EPI_ISL_778176, EPI_ISL_778192, EPI_ISL_778202, EPI_ISL_778203, EPI_ISL_778215, EPI_ISL_778216, EPI_ISL_778219, EPI_ISL_778229, EPI_ISL_778236, EPI_ISL_778244, EPI_ISL_778247, EPI_ISL_778248, EPI_ISL_778250, EPI_ISL_778258, EPI_ISL_778260, EPI_ISL_778263,
EPI_ISL_778269, EPI_ISL_778274, EPI_ISL_778276, EPI_ISL_778279, EPI_ISL_778283, EPI_ISL_778286, EPI_ISL_778289, EPI_ISL_781870, EPI_ISL_781871, EPI_ISL_781875, EPI_ISL_781882, EPI_ISL_781886, EPI_ISL_781890, EPI_ISL_781895, EPI_ISL_781910, EPI_ISL_781913, EPI_ISL_781918, EPI_ISL_781919, EPI_ISL_781922, EPI_ISL_781928, EPI_ISL_781930, EPI_ISL_781933, EPI_ISL_781953, EPI_ISL_781956,
EPI_ISL_781960, EPI_ISL_781967, EPI_ISL_781971, EPI_ISL_781978, EPI_ISL_781980, EPI_ISL_781990, EPI_ISL_781992, EPI_ISL_781997, EPI_ISL_782001, EPI_ISL_782003, EPI_ISL_782004, EPI_ISL_782008, EPI_ISL_782014, EPI_ISL_782018, EPI_ISL_782027, EPI_ISL_782036, EPI_ISL_782038, EPI_ISL_782039, EPI_ISL_782056, EPI_ISL_782059, EPI_ISL_782062, EPI_ISL_782064, EPI_ISL_782065, EPI_ISL_782066,
EPI_ISL_782067, EPI_ISL_782074, EPI_ISL_782075, EPI_ISL_782079, EPI_ISL_782084, EPI_ISL_782101, EPI_ISL_782107, EPI_ISL_782112, EPI_ISL_782114, EPI_ISL_782115, EPI_ISL_782116, EPI_ISL_782121, EPI_ISL_782127, EPI_ISL_782128, EPI_ISL_782138, EPI_ISL_782145, EPI_ISL_782146, EPI_ISL_782151, EPI_ISL_782152, EPI_ISL_782153, EPI_ISL_782154, EPI_ISL_782155, EPI_ISL_782156, EPI_ISL_782157,
EPI_ISL_782158, EPI_ISL_782159, EPI_ISL_782160, EPI_ISL_782162, EPI_ISL_782163, EPI_ISL_782164, EPI_ISL_782166, EPI_ISL_782167, EPI_ISL_782168, EPI_ISL_782169, EPI_ISL_782170, EPI_ISL_782172, EPI_ISL_782173, EPI_ISL_782174, EPI_ISL_782175, EPI_ISL_782176, EPI_ISL_782177, EPI_ISL_782178, EPI_ISL_782179, EPI_ISL_782180, EPI_ISL_782181, EPI_ISL_782182, EPI_ISL_782183, EPI_ISL_782185,
EPI_ISL_782186, EPI_ISL_782187, EPI_ISL_782188, EPI_ISL_782189, EPI_ISL_782190, EPI_ISL_782191, EPI_ISL_782192, EPI_ISL_782193, EPI_ISL_782194, EPI_ISL_782195, EPI_ISL_782196, EPI_ISL_782198, EPI_ISL_782199, EPI_ISL_782200, EPI_ISL_782203, EPI_ISL_782204, EPI_ISL_782205, EPI_ISL_782206, EPI_ISL_782208, EPI_ISL_782209, EPI_ISL_782210, EPI_ISL_782213, EPI_ISL_782214, EPI_ISL_782215,
EPI_ISL_782216, EPI_ISL_782217, EPI_ISL_782218, EPI_ISL_782219, EPI_ISL_782221, EPI_ISL_782222, EPI_ISL_782223, EPI_ISL_782225, EPI_ISL_782227, EPI_ISL_782228, EPI_ISL_782229, EPI_ISL_782230, EPI_ISL_782231, EPI_ISL_782232, EPI_ISL_782233, EPI_ISL_782234, EPI_ISL_782236, EPI_ISL_782237, EPI_ISL_782238, EPI_ISL_782239, EPI_ISL_782240, EPI_ISL_782241, EPI_ISL_782244, EPI_ISL_782246,
EPI_ISL_782247, EPI_ISL_782248, EPI_ISL_782251, EPI_ISL_782253, EPI_ISL_782254, EPI_ISL_782255, EPI_ISL_782257, EPI_ISL_782258, EPI_ISL_782260, EPI_ISL_782261, EPI_ISL_782262, EPI_ISL_782263, EPI_ISL_782265, EPI_ISL_782266, EPI_ISL_782269, EPI_ISL_782270, EPI_ISL_782272, EPI_ISL_782273, EPI_ISL_782275, EPI_ISL_782276, EPI_ISL_782277, EPI_ISL_782279, EPI_ISL_782280, EPI_ISL_782281,
EPI_ISL_782283, EPI_ISL_782284, EPI_ISL_782285, EPI_ISL_782286, EPI_ISL_782288, EPI_ISL_782289, EPI_ISL_782290, EPI_ISL_782291, EPI_ISL_782293, EPI_ISL_782294, EPI_ISL_782295, EPI_ISL_782296, EPI_ISL_782297, EPI_ISL_782298, EPI_ISL_782299, EPI_ISL_782300, EPI_ISL_782301, EPI_ISL_782302, EPI_ISL_782303, EPI_ISL_782304, EPI_ISL_782305, EPI_ISL_782306, EPI_ISL_782308, EPI_ISL_782310,
EPI_ISL_782311, EPI_ISL_782312, EPI_ISL_782316, EPI_ISL_782318, EPI_ISL_782319, EPI_ISL_782320, EPI_ISL_782321, EPI_ISL_782322, EPI_ISL_782323, EPI_ISL_782324, EPI_ISL_782329, EPI_ISL_782330, EPI_ISL_782331, EPI_ISL_782332, EPI_ISL_782333, EPI_ISL_782334, EPI_ISL_782336, EPI_ISL_782337, EPI_ISL_782338, EPI_ISL_782339, EPI_ISL_782340, EPI_ISL_782341, EPI_ISL_782342, EPI_ISL_782343,
EPI_ISL_782344, EPI_ISL_782346, EPI_ISL_782347, EPI_ISL_782348, EPI_ISL_782349, EPI_ISL_782350, EPI_ISL_782351, EPI_ISL_782353, EPI_ISL_782354, EPI_ISL_782356, EPI_ISL_782357, EPI_ISL_782359, EPI_ISL_782360, EPI_ISL_782363, EPI_ISL_782364, EPI_ISL_782365, EPI_ISL_782366, EPI_ISL_782367, EPI_ISL_782368, EPI_ISL_782370, EPI_ISL_782373, EPI_ISL_782374, EPI_ISL_782375, EPI_ISL_782376,
EPI_ISL_782378, EPI_ISL_782379, EPI_ISL_782381, EPI_ISL_782382, EPI_ISL_782383, EPI_ISL_782384, EPI_ISL_782385, EPI_ISL_782386, EPI_ISL_782387, EPI_ISL_782389, EPI_ISL_782391, EPI_ISL_782392, EPI_ISL_782394, EPI_ISL_782395, EPI_ISL_782396, EPI_ISL_782397, EPI_ISL_782398, EPI_ISL_782399, EPI_ISL_782400, EPI_ISL_782401, EPI_ISL_782403, EPI_ISL_782404, EPI_ISL_782405, EPI_ISL_782406,
EPI_ISL_782407, EPI_ISL_782408, EPI_ISL_782410, EPI_ISL_782411, EPI_ISL_782413, EPI_ISL_782414, EPI_ISL_782415, EPI_ISL_782416, EPI_ISL_782417, EPI_ISL_782418, EPI_ISL_782419, EPI_ISL_782421, EPI_ISL_782422, EPI_ISL_782424, EPI_ISL_782427, EPI_ISL_782428, EPI_ISL_782429, EPI_ISL_782430, EPI_ISL_782431, EPI_ISL_782432, EPI_ISL_782433, EPI_ISL_782434, EPI_ISL_782436, EPI_ISL_782437,
EPI_ISL_782438, EPI_ISL_782440, EPI_ISL_782441, EPI_ISL_782442, EPI_ISL_782443, EPI_ISL_782444, EPI_ISL_782445, EPI_ISL_782446, EPI_ISL_782447, EPI_ISL_782448, EPI_ISL_782449, EPI_ISL_782450, EPI_ISL_782452, EPI_ISL_782453, EPI_ISL_782455, EPI_ISL_782456, EPI_ISL_782457, EPI_ISL_782458, EPI_ISL_782459, EPI_ISL_782460, EPI_ISL_782462, EPI_ISL_782463, EPI_ISL_782464, EPI_ISL_782465,
EPI_ISL_782467, EPI_ISL_782468, EPI_ISL_782469, EPI_ISL_782470, EPI_ISL_782472, EPI_ISL_782473, EPI_ISL_782475, EPI_ISL_782476, EPI_ISL_782477, EPI_ISL_782479, EPI_ISL_782480, EPI_ISL_782481, EPI_ISL_797739, EPI_ISL_797766, EPI_ISL_797775, EPI_ISL_797789, EPI_ISL_797797, EPI_ISL_797817, EPI_ISL_797825, EPI_ISL_797829, EPI_ISL_797830, EPI_ISL_797831, EPI_ISL_797838, EPI_ISL_797850,
EPI_ISL_797851, EPI_ISL_797853, EPI_ISL_797868, EPI_ISL_797884, EPI_ISL_797885, EPI_ISL_797903, EPI_ISL_797916, EPI_ISL_797921, EPI_ISL_797937, EPI_ISL_797952, EPI_ISL_797965, EPI_ISL_797985, EPI_ISL_797994, EPI_ISL_798017, EPI_ISL_798022, EPI_ISL_798043, EPI_ISL_798051, EPI_ISL_798080, EPI_ISL_798107, EPI_ISL_798141, EPI_ISL_798159, EPI_ISL_798171, EPI_ISL_798178, EPI_ISL_798184,
EPI_ISL_798192, EPI_ISL_798223, EPI_ISL_798231, EPI_ISL_798249, EPI_ISL_798261, EPI_ISL_798271, EPI_ISL_798278, EPI_ISL_798287, EPI_ISL_798308, EPI_ISL_798325, EPI_ISL_798331, EPI_ISL_798332, EPI_ISL_799297, EPI_ISL_799298, EPI_ISL_799300, EPI_ISL_799301, EPI_ISL_799302, EPI_ISL_799304, EPI_ISL_799307, EPI_ISL_799309, EPI_ISL_799310, EPI_ISL_799312, EPI_ISL_799313, EPI_ISL_799315,
EPI_ISL_799316, EPI_ISL_799318, EPI_ISL_799321, EPI_ISL_799322, EPI_ISL_799324, EPI_ISL_799326, EPI_ISL_799328, EPI_ISL_799329, EPI_ISL_799330, EPI_ISL_799331, EPI_ISL_799332, EPI_ISL_799335, EPI_ISL_799336, EPI_ISL_799337, EPI_ISL_799338, EPI_ISL_799339, EPI_ISL_799340, EPI_ISL_799341, EPI_ISL_799342, EPI_ISL_799343, EPI_ISL_799344, EPI_ISL_799345, EPI_ISL_799349, EPI_ISL_799355,
EPI_ISL_799358, EPI_ISL_799359, EPI_ISL_799361, EPI_ISL_799365, EPI_ISL_799367, EPI_ISL_799369, EPI_ISL_799371, EPI_ISL_799374, EPI_ISL_799377, EPI_ISL_799379, EPI_ISL_799381, EPI_ISL_799382, EPI_ISL_799383, EPI_ISL_799385, EPI_ISL_799389, EPI_ISL_799391, EPI_ISL_799393, EPI_ISL_799396, EPI_ISL_799398, EPI_ISL_799399, EPI_ISL_799401, EPI_ISL_799402, EPI_ISL_799405, EPI_ISL_799407,
EPI_ISL_799408, EPI_ISL_799409, EPI_ISL_799414, EPI_ISL_799417, EPI_ISL_799418, EPI_ISL_799419, EPI_ISL_799421, EPI_ISL_799422, EPI_ISL_799423, EPI_ISL_799424, EPI_ISL_799425, EPI_ISL_799426, EPI_ISL_799428, EPI_ISL_799429, EPI_ISL_799431, EPI_ISL_799433, EPI_ISL_799435, EPI_ISL_799438, EPI_ISL_799442, EPI_ISL_799444, EPI_ISL_799449, EPI_ISL_799450, EPI_ISL_799451, EPI_ISL_799452,
EPI_ISL_799453, EPI_ISL_799455, EPI_ISL_799456, EPI_ISL_799457, EPI_ISL_799458, EPI_ISL_799459, EPI_ISL_799460, EPI_ISL_799461, EPI_ISL_799462, EPI_ISL_799463, EPI_ISL_799464, EPI_ISL_799471, EPI_ISL_799472, EPI_ISL_799474, EPI_ISL_799475, EPI_ISL_799478, EPI_ISL_799480, EPI_ISL_799481, EPI_ISL_799483, EPI_ISL_799484, EPI_ISL_799486, EPI_ISL_799489, EPI_ISL_799491, EPI_ISL_799493,
EPI_ISL_799495, EPI_ISL_799499, EPI_ISL_799503, EPI_ISL_799504, EPI_ISL_799507, EPI_ISL_799510, EPI_ISL_799511, EPI_ISL_799513, EPI_ISL_799516, EPI_ISL_799517, EPI_ISL_799518, EPI_ISL_799521, EPI_ISL_799524, EPI_ISL_799525, EPI_ISL_799526, EPI_ISL_799527, EPI_ISL_799530, EPI_ISL_799532, EPI_ISL_799537, EPI_ISL_799539, EPI_ISL_799541, EPI_ISL_799542, EPI_ISL_799543, EPI_ISL_799544,
EPI_ISL_799548, EPI_ISL_799549, EPI_ISL_799552, EPI_ISL_799553, EPI_ISL_799554, EPI_ISL_799556, EPI_ISL_799557, EPI_ISL_799558, EPI_ISL_799559, EPI_ISL_799560, EPI_ISL_799561, EPI_ISL_799564, EPI_ISL_799566, EPI_ISL_799568, EPI_ISL_799569, EPI_ISL_799571, EPI_ISL_799574, EPI_ISL_799576, EPI_ISL_799581, EPI_ISL_799584, EPI_ISL_799585, EPI_ISL_799586, EPI_ISL_799589, EPI_ISL_799591,
EPI_ISL_799592, EPI_ISL_799593, EPI_ISL_799594, EPI_ISL_799596, EPI_ISL_799599, EPI_ISL_799601, EPI_ISL_799605, EPI_ISL_799606, EPI_ISL_799609, EPI_ISL_799612, EPI_ISL_799613, EPI_ISL_799615, EPI_ISL_799619, EPI_ISL_799622, EPI_ISL_799630, EPI_ISL_799634, EPI_ISL_799641, EPI_ISL_799648, EPI_ISL_799649, EPI_ISL_799650, EPI_ISL_799659, EPI_ISL_799661, EPI_ISL_799664, EPI_ISL_799669,
EPI_ISL_799676, EPI_ISL_799679, EPI_ISL_799681, EPI_ISL_799682, EPI_ISL_799686, EPI_ISL_799699, EPI_ISL_799707, EPI_ISL_799708, EPI_ISL_799713, EPI_ISL_799718, EPI_ISL_799722, EPI_ISL_799726, EPI_ISL_799729, EPI_ISL_799730, EPI_ISL_799731, EPI_ISL_799735, EPI_ISL_799737, EPI_ISL_799739, EPI_ISL_799748, EPI_ISL_799758, EPI_ISL_799759, EPI_ISL_799761, EPI_ISL_799763, EPI_ISL_799765,
EPI_ISL_799768, EPI_ISL_799775, EPI_ISL_799776, EPI_ISL_799780, EPI_ISL_799781, EPI_ISL_799783, EPI_ISL_799786, EPI_ISL_799791, EPI_ISL_799792, EPI_ISL_799801, EPI_ISL_799805, EPI_ISL_799811, EPI_ISL_799819, EPI_ISL_799825, EPI_ISL_799827, EPI_ISL_799830, EPI_ISL_799838, EPI_ISL_799839, EPI_ISL_799843, EPI_ISL_799855, EPI_ISL_799861, EPI_ISL_799864, EPI_ISL_799865, EPI_ISL_799866,
EPI_ISL_799869, EPI_ISL_799871, EPI_ISL_799875, EPI_ISL_799883, EPI_ISL_799888, EPI_ISL_799889, EPI_ISL_799892, EPI_ISL_799893, EPI_ISL_799894, EPI_ISL_799901, EPI_ISL_799903, EPI_ISL_799906, EPI_ISL_799910, EPI_ISL_799912, EPI_ISL_799916, EPI_ISL_799917, EPI_ISL_799918, EPI_ISL_799919, EPI_ISL_799932, EPI_ISL_799937, EPI_ISL_799945, EPI_ISL_799946, EPI_ISL_799949, EPI_ISL_799951,
EPI_ISL_799954, EPI_ISL_799965, EPI_ISL_799968, EPI_ISL_799969, EPI_ISL_799973, EPI_ISL_799976, EPI_ISL_799980, EPI_ISL_799981, EPI_ISL_799985, EPI_ISL_799986, EPI_ISL_799987, EPI_ISL_799990, EPI_ISL_799992, EPI_ISL_799997, EPI_ISL_800000, EPI_ISL_800001, EPI_ISL_800005, EPI_ISL_800010, EPI_ISL_800015, EPI_ISL_800027, EPI_ISL_800029, EPI_ISL_800030, EPI_ISL_800032, EPI_ISL_800034,
EPI_ISL_800038, EPI_ISL_800048, EPI_ISL_800052, EPI_ISL_800056, EPI_ISL_800057, EPI_ISL_800064, EPI_ISL_800068, EPI_ISL_800070, EPI_ISL_800071, EPI_ISL_800078, EPI_ISL_800079, EPI_ISL_800084, EPI_ISL_800086, EPI_ISL_800106, EPI_ISL_800111, EPI_ISL_800113, EPI_ISL_800117, EPI_ISL_800120, EPI_ISL_800122, EPI_ISL_800124, EPI_ISL_800131, EPI_ISL_800138, EPI_ISL_800141, EPI_ISL_800142,
EPI_ISL_800147, EPI_ISL_800151, EPI_ISL_800158, EPI_ISL_800161, EPI_ISL_800164, EPI_ISL_800174, EPI_ISL_800175, EPI_ISL_800176, EPI_ISL_800180, EPI_ISL_800183, EPI_ISL_800186, EPI_ISL_800188, EPI_ISL_800189, EPI_ISL_800190, EPI_ISL_800191, EPI_ISL_800200, EPI_ISL_800201, EPI_ISL_800204, EPI_ISL_800210, EPI_ISL_800217, EPI_ISL_800221, EPI_ISL_800234, EPI_ISL_800235, EPI_ISL_800236,
EPI_ISL_800237, EPI_ISL_800241, EPI_ISL_800245, EPI_ISL_800251, EPI_ISL_800253, EPI_ISL_800576, EPI_ISL_822307, EPI_ISL_834578, EPI_ISL_858590, EPI_ISL_858597, EPI_ISL_858619, EPI_ISL_858778, EPI_ISL_858782, EPI_ISL_858793, EPI_ISL_858794, EPI_ISL_858796, EPI_ISL_858797, EPI_ISL_858801, EPI_ISL_858802, EPI_ISL_858806, EPI_ISL_858810, EPI_ISL_858815, EPI_ISL_858830, EPI_ISL_858842,
EPI_ISL_858843, EPI_ISL_858844, EPI_ISL_858859, EPI_ISL_858860, EPI_ISL_858863, EPI_ISL_858865, EPI_ISL_858867, EPI_ISL_858874, EPI_ISL_858900, EPI_ISL_858901, EPI_ISL_858903, EPI_ISL_858904, EPI_ISL_858906, EPI_ISL_858908, EPI_ISL_858910, EPI_ISL_858912, EPI_ISL_858916, EPI_ISL_858919, EPI_ISL_858920, EPI_ISL_858921, EPI_ISL_858922, EPI_ISL_858927, EPI_ISL_858928, EPI_ISL_858930,
EPI_ISL_858935, EPI_ISL_858936, EPI_ISL_858937, EPI_ISL_858943, EPI_ISL_858945, EPI_ISL_858953, EPI_ISL_858958, EPI_ISL_858960, EPI_ISL_858969, EPI_ISL_858970, EPI_ISL_858974, EPI_ISL_858975, EPI_ISL_858990, EPI_ISL_859077, EPI_ISL_859149, EPI_ISL_859150, EPI_ISL_859151, EPI_ISL_859152, EPI_ISL_859153, EPI_ISL_859155, EPI_ISL_859156, EPI_ISL_859157, EPI_ISL_859158, EPI_ISL_859159,
EPI_ISL_859160, EPI_ISL_859162, EPI_ISL_859163, EPI_ISL_859164, EPI_ISL_859166, EPI_ISL_859167, EPI_ISL_859168, EPI_ISL_859169, EPI_ISL_859170, EPI_ISL_859171, EPI_ISL_859172, EPI_ISL_859173, EPI_ISL_859174, EPI_ISL_859175, EPI_ISL_859176, EPI_ISL_859177, EPI_ISL_859178, EPI_ISL_859180, EPI_ISL_859182, EPI_ISL_859183, EPI_ISL_859184, EPI_ISL_859185, EPI_ISL_859187, EPI_ISL_859188,
EPI_ISL_859189, EPI_ISL_859190, EPI_ISL_859191, EPI_ISL_859192, EPI_ISL_859193, EPI_ISL_859195, EPI_ISL_859196, EPI_ISL_859197, EPI_ISL_859199, EPI_ISL_859200, EPI_ISL_859205, EPI_ISL_859211, EPI_ISL_859214, EPI_ISL_859218, EPI_ISL_859219, EPI_ISL_859232, EPI_ISL_859234, EPI_ISL_859236, EPI_ISL_859253, EPI_ISL_859254, EPI_ISL_859255, EPI_ISL_859256, EPI_ISL_859263, EPI_ISL_859265,
EPI_ISL_859268, EPI_ISL_859270, EPI_ISL_859280, EPI_ISL_859282, EPI_ISL_859288, EPI_ISL_859294, EPI_ISL_859295, EPI_ISL_859297, EPI_ISL_859301, EPI_ISL_859303, EPI_ISL_859304, EPI_ISL_859306, EPI_ISL_859309, EPI_ISL_859311, EPI_ISL_859312, EPI_ISL_859313, EPI_ISL_859316, EPI_ISL_859317, EPI_ISL_859318, EPI_ISL_859319, EPI_ISL_892214, EPI_ISL_945098, EPI_ISL_945123, EPI_ISL_945126,
EPI_ISL_945136, EPI_ISL_945234, EPI_ISL_945351, EPI_ISL_989996

| | | | |
|---|---|---|---|
| see above | Lighthouse Lab in Glasgow | Wellcome Sanger Institute for the COVID-19 Genomics UK (COG-UK) Consortium | Anna Dominiczak and Alex Alderton; Carol Clugston; Cordelia Langford; David Gray; David K. Jackson; Dominic Kwiatkowski; Ewan Harrison; Harper VanSteenhouse; Ian Johnston; John Sillitoe on behalf of the Wellcome Sanger Institute COVID-19 Surveillance Team; Roberto Amato; Sonia Goncalves; Yumi Kasai |

EPI_ISL_781178, EPI_ISL_781182, EPI_ISL_781191, EPI_ISL_781196, EPI_ISL_781200, EPI_ISL_781203, EPI_ISL_781215, EPI_ISL_781218, EPI_ISL_781222, EPI_ISL_781225, EPI_ISL_781229, EPI_ISL_781232, EPI_ISL_781237, EPI_ISL_781239, EPI_ISL_781244, EPI_ISL_781247, EPI_ISL_781255, EPI_ISL_781267, EPI_ISL_781268, EPI_ISL_781272, EPI_ISL_781276, EPI_ISL_781278, EPI_ISL_781286, EPI_ISL_781290,
EPI_ISL_781298, EPI_ISL_781300, EPI_ISL_781309, EPI_ISL_781310, EPI_ISL_781312, EPI_ISL_781316, EPI_ISL_781320, EPI_ISL_781329, EPI_ISL_781335, EPI_ISL_781343, EPI_ISL_781350, EPI_ISL_781357, EPI_ISL_781363, EPI_ISL_781380, EPI_ISL_781381, EPI_ISL_781384, EPI_ISL_781390, EPI_ISL_781393, EPI_ISL_781394, EPI_ISL_781395, EPI_ISL_781398, EPI_ISL_781404, EPI_ISL_781415, EPI_ISL_781418,
EPI_ISL_781429, EPI_ISL_781439, EPI_ISL_781444, EPI_ISL_781455, EPI_ISL_781460, EPI_ISL_781461, EPI_ISL_781468, EPI_ISL_781476, EPI_ISL_781484, EPI_ISL_781485, EPI_ISL_797119, EPI_ISL_797122, EPI_ISL_797123, EPI_ISL_797124, EPI_ISL_797127, EPI_ISL_797130, EPI_ISL_797131, EPI_ISL_797132, EPI_ISL_797135, EPI_ISL_797136, EPI_ISL_797137, EPI_ISL_797140, EPI_ISL_797145, EPI_ISL_797147,
EPI_ISL_797150, EPI_ISL_797152, EPI_ISL_797157, EPI_ISL_797158, EPI_ISL_797159, EPI_ISL_797162, EPI_ISL_797166, EPI_ISL_797167, EPI_ISL_797169, EPI_ISL_797170, EPI_ISL_797172, EPI_ISL_797174, EPI_ISL_797180, EPI_ISL_797181, EPI_ISL_797182, EPI_ISL_797183, EPI_ISL_797184, EPI_ISL_797186, EPI_ISL_797188, EPI_ISL_797189, EPI_ISL_797191, EPI_ISL_797192, EPI_ISL_797193, EPI_ISL_797195,
EPI_ISL_797196, EPI_ISL_797197, EPI_ISL_797199, EPI_ISL_797200, EPI_ISL_797201, EPI_ISL_797206, EPI_ISL_797208, EPI_ISL_797209, EPI_ISL_797211, EPI_ISL_797213, EPI_ISL_797214, EPI_ISL_797215, EPI_ISL_797217, EPI_ISL_797219, EPI_ISL_797221, EPI_ISL_797222, EPI_ISL_797223, EPI_ISL_797224, EPI_ISL_797227, EPI_ISL_797230, EPI_ISL_797233, EPI_ISL_797235, EPI_ISL_797236, EPI_ISL_797237,
EPI_ISL_797238, EPI_ISL_797241, EPI_ISL_797244, EPI_ISL_797245, EPI_ISL_797249, EPI_ISL_797250, EPI_ISL_797251, EPI_ISL_797255, EPI_ISL_797257, EPI_ISL_797260, EPI_ISL_797262, EPI_ISL_797263, EPI_ISL_797265, EPI_ISL_797267, EPI_ISL_797268, EPI_ISL_797270, EPI_ISL_797271, EPI_ISL_797273, EPI_ISL_797274, EPI_ISL_797275, EPI_ISL_797276, EPI_ISL_797278, EPI_ISL_797279, EPI_ISL_797280,
EPI_ISL_797282, EPI_ISL_797283, EPI_ISL_797284, EPI_ISL_797285, EPI_ISL_797287, EPI_ISL_797288, EPI_ISL_797291, EPI_ISL_797292, EPI_ISL_797293, EPI_ISL_797294, EPI_ISL_797297, EPI_ISL_797298, EPI_ISL_797299, EPI_ISL_797300, EPI_ISL_797302, EPI_ISL_797303, EPI_ISL_797304, EPI_ISL_797306, EPI_ISL_797307, EPI_ISL_797308, EPI_ISL_797309, EPI_ISL_797310, EPI_ISL_797311, EPI_ISL_797312,
EPI_ISL_797314, EPI_ISL_797316, EPI_ISL_797320, EPI_ISL_797321, EPI_ISL_797322, EPI_ISL_797324, EPI_ISL_797328, EPI_ISL_797329, EPI_ISL_797330, EPI_ISL_797331, EPI_ISL_797333, EPI_ISL_797334, EPI_ISL_797336, EPI_ISL_797340, EPI_ISL_797341, EPI_ISL_797343, EPI_ISL_797344, EPI_ISL_797347, EPI_ISL_797348, EPI_ISL_797351, EPI_ISL_797352, EPI_ISL_797353, EPI_ISL_797355, EPI_ISL_797356,
EPI_ISL_797358, EPI_ISL_797359, EPI_ISL_797360, EPI_ISL_797361, EPI_ISL_797364, EPI_ISL_797368, EPI_ISL_797369, EPI_ISL_797370, EPI_ISL_797371, EPI_ISL_797372, EPI_ISL_797373, EPI_ISL_797374, EPI_ISL_797375, EPI_ISL_797377, EPI_ISL_797378, EPI_ISL_797381, EPI_ISL_797382, EPI_ISL_797383, EPI_ISL_797384, EPI_ISL_797386, EPI_ISL_797387, EPI_ISL_797388, EPI_ISL_797392, EPI_ISL_797393,
EPI_ISL_797394, EPI_ISL_797396, EPI_ISL_798333, EPI_ISL_798335, EPI_ISL_798337, EPI_ISL_798338, EPI_ISL_798339, EPI_ISL_798340, EPI_ISL_798342, EPI_ISL_798343, EPI_ISL_798344, EPI_ISL_798347, EPI_ISL_798348, EPI_ISL_798350, EPI_ISL_798351, EPI_ISL_798352, EPI_ISL_798353, EPI_ISL_798354, EPI_ISL_798355, EPI_ISL_798357, EPI_ISL_798358, EPI_ISL_798359, EPI_ISL_798360, EPI_ISL_798361,
EPI_ISL_798363, EPI_ISL_798364, EPI_ISL_798365, EPI_ISL_798366, EPI_ISL_798367, EPI_ISL_798371, EPI_ISL_798372, EPI_ISL_798373, EPI_ISL_798374, EPI_ISL_798375, EPI_ISL_798376, EPI_ISL_798377, EPI_ISL_798380, EPI_ISL_798381, EPI_ISL_798382, EPI_ISL_798383, EPI_ISL_798385, EPI_ISL_798387, EPI_ISL_798390, EPI_ISL_798391, EPI_ISL_798392, EPI_ISL_798393, EPI_ISL_798394, EPI_ISL_798395,
EPI_ISL_798397, EPI_ISL_798398, EPI_ISL_798400, EPI_ISL_798402, EPI_ISL_798403, EPI_ISL_798406, EPI_ISL_798407, EPI_ISL_798408, EPI_ISL_798409, EPI_ISL_798410, EPI_ISL_798411, EPI_ISL_798412, EPI_ISL_798414, EPI_ISL_798415, EPI_ISL_798418, EPI_ISL_798419, EPI_ISL_798420, EPI_ISL_798421, EPI_ISL_798422, EPI_ISL_798423, EPI_ISL_798424, EPI_ISL_798425, EPI_ISL_798426, EPI_ISL_798427,
EPI_ISL_798428, EPI_ISL_798429, EPI_ISL_798430, EPI_ISL_798432, EPI_ISL_798433, EPI_ISL_798434, EPI_ISL_798435, EPI_ISL_798437, EPI_ISL_798439, EPI_ISL_798440, EPI_ISL_798441, EPI_ISL_798442, EPI_ISL_798443, EPI_ISL_798445, EPI_ISL_798446, EPI_ISL_798447, EPI_ISL_798448, EPI_ISL_798449, EPI_ISL_798450, EPI_ISL_798451, EPI_ISL_798452, EPI_ISL_798453, EPI_ISL_798454, EPI_ISL_798456,
EPI_ISL_798457, EPI_ISL_798458, EPI_ISL_798459, EPI_ISL_798460, EPI_ISL_798461, EPI_ISL_798462, EPI_ISL_798463, EPI_ISL_798464, EPI_ISL_798466, EPI_ISL_798467, EPI_ISL_798469, EPI_ISL_798470, EPI_ISL_798471, EPI_ISL_798472, EPI_ISL_798473, EPI_ISL_798474, EPI_ISL_798476, EPI_ISL_798477, EPI_ISL_798478, EPI_ISL_798480, EPI_ISL_798481, EPI_ISL_798482, EPI_ISL_798484, EPI_ISL_798485,
EPI_ISL_798486, EPI_ISL_798488, EPI_ISL_798489, EPI_ISL_798490, EPI_ISL_798491, EPI_ISL_798494, EPI_ISL_798495, EPI_ISL_798496, EPI_ISL_798497, EPI_ISL_798498, EPI_ISL_798499, EPI_ISL_798501, EPI_ISL_798503, EPI_ISL_798504, EPI_ISL_798505, EPI_ISL_798506, EPI_ISL_798507, EPI_ISL_798509, EPI_ISL_798511, EPI_ISL_798512, EPI_ISL_798514, EPI_ISL_798515, EPI_ISL_798518, EPI_ISL_798520,
EPI_ISL_798521, EPI_ISL_798522, EPI_ISL_798523, EPI_ISL_798525, EPI_ISL_798526, EPI_ISL_798527, EPI_ISL_798528, EPI_ISL_798529, EPI_ISL_798530, EPI_ISL_798532, EPI_ISL_798533, EPI_ISL_798535, EPI_ISL_798536, EPI_ISL_798537, EPI_ISL_798538, EPI_ISL_798540, EPI_ISL_798543, EPI_ISL_798545, EPI_ISL_798546, EPI_ISL_798547, EPI_ISL_798549, EPI_ISL_798552, EPI_ISL_798553, EPI_ISL_798554,
EPI_ISL_798556, EPI_ISL_798557, EPI_ISL_798558, EPI_ISL_798559, EPI_ISL_798560, EPI_ISL_798562, EPI_ISL_798563, EPI_ISL_798565, EPI_ISL_798566, EPI_ISL_798567, EPI_ISL_798568, EPI_ISL_798569, EPI_ISL_798570, EPI_ISL_798571, EPI_ISL_798572, EPI_ISL_798573, EPI_ISL_798574, EPI_ISL_798575, EPI_ISL_798576, EPI_ISL_798577, EPI_ISL_798578, EPI_ISL_798580, EPI_ISL_798581, EPI_ISL_798582,
EPI_ISL_798583, EPI_ISL_798584, EPI_ISL_798585, EPI_ISL_798586, EPI_ISL_798587, EPI_ISL_798588, EPI_ISL_798589, EPI_ISL_798590, EPI_ISL_798592, EPI_ISL_798594, EPI_ISL_798595, EPI_ISL_798596, EPI_ISL_798597, EPI_ISL_798599, EPI_ISL_798600, EPI_ISL_798602, EPI_ISL_798603, EPI_ISL_798604, EPI_ISL_798605, EPI_ISL_798606, EPI_ISL_798608, EPI_ISL_798610, EPI_ISL_798611, EPI_ISL_798613,
EPI_ISL_798614, EPI_ISL_798616, EPI_ISL_798617, EPI_ISL_798618, EPI_ISL_798619, EPI_ISL_798620, EPI_ISL_798621, EPI_ISL_798623, EPI_ISL_798624, EPI_ISL_798625, EPI_ISL_798626, EPI_ISL_798627, EPI_ISL_798628, EPI_ISL_798629, EPI_ISL_798630, EPI_ISL_798634, EPI_ISL_798636, EPI_ISL_798638, EPI_ISL_798642, EPI_ISL_798644, EPI_ISL_798645, EPI_ISL_798646, EPI_ISL_798647, EPI_ISL_798648,
EPI_ISL_798649, EPI_ISL_798650, EPI_ISL_798651, EPI_ISL_798653, EPI_ISL_798654, EPI_ISL_798655, EPI_ISL_798656, EPI_ISL_798657, EPI_ISL_798658, EPI_ISL_798660, EPI_ISL_798662, EPI_ISL_798663, EPI_ISL_798666, EPI_ISL_798670, EPI_ISL_798672, EPI_ISL_798675, EPI_ISL_798680, EPI_ISL_798691, EPI_ISL_798693, EPI_ISL_798696, EPI_ISL_798701, EPI_ISL_798702, EPI_ISL_798707, EPI_ISL_798710,
EPI_ISL_798712, EPI_ISL_798716, EPI_ISL_798720, EPI_ISL_798721, EPI_ISL_798724, EPI_ISL_798725, EPI_ISL_798728, EPI_ISL_798738, EPI_ISL_798739, EPI_ISL_798740, EPI_ISL_798741, EPI_ISL_798750, EPI_ISL_798752, EPI_ISL_798755, EPI_ISL_798764, EPI_ISL_798767, EPI_ISL_798769, EPI_ISL_798770, EPI_ISL_798774, EPI_ISL_798776, EPI_ISL_798777, EPI_ISL_798779, EPI_ISL_798781, EPI_ISL_798801,
EPI_ISL_798803, EPI_ISL_798808, EPI_ISL_798809, EPI_ISL_798812, EPI_ISL_798817, EPI_ISL_798821, EPI_ISL_798822, EPI_ISL_798823, EPI_ISL_798830, EPI_ISL_798832, EPI_ISL_798837, EPI_ISL_798838, EPI_ISL_798844, EPI_ISL_798851, EPI_ISL_798855, EPI_ISL_798856, EPI_ISL_798863, EPI_ISL_798864, EPI_ISL_798870, EPI_ISL_798874, EPI_ISL_798875, EPI_ISL_798876, EPI_ISL_798879, EPI_ISL_798882,
EPI_ISL_798889, EPI_ISL_798892, EPI_ISL_798910, EPI_ISL_798911, EPI_ISL_798915, EPI_ISL_798919, EPI_ISL_798922, EPI_ISL_798925, EPI_ISL_798930, EPI_ISL_798933, EPI_ISL_798936, EPI_ISL_798942, EPI_ISL_798948, EPI_ISL_798950, EPI_ISL_798955, EPI_ISL_798961, EPI_ISL_798966, EPI_ISL_798967, EPI_ISL_798970, EPI_ISL_798972, EPI_ISL_798973, EPI_ISL_798975, EPI_ISL_798977, EPI_ISL_798979,
EPI_ISL_798981, EPI_ISL_798983, EPI_ISL_798987, EPI_ISL_798994, EPI_ISL_798995, EPI_ISL_798996, EPI_ISL_799296, EPI_ISL_799299, EPI_ISL_799303, EPI_ISL_799305, EPI_ISL_799306, EPI_ISL_799308, EPI_ISL_799311, EPI_ISL_799314, EPI_ISL_799317, EPI_ISL_799319, EPI_ISL_799320, EPI_ISL_799323, EPI_ISL_799325, EPI_ISL_799327, EPI_ISL_799333, EPI_ISL_799334, EPI_ISL_799346, EPI_ISL_799347,
EPI_ISL_799348, EPI_ISL_799350, EPI_ISL_799351, EPI_ISL_799352, EPI_ISL_799353, EPI_ISL_799354, EPI_ISL_799356, EPI_ISL_799357, EPI_ISL_799360, EPI_ISL_799362, EPI_ISL_799363, EPI_ISL_799364, EPI_ISL_799366, EPI_ISL_799368, EPI_ISL_799370, EPI_ISL_799372, EPI_ISL_799373, EPI_ISL_799375, EPI_ISL_799376, EPI_ISL_799378, EPI_ISL_799380, EPI_ISL_799384, EPI_ISL_799386, EPI_ISL_799387,
EPI_ISL_799388, EPI_ISL_799390, EPI_ISL_799392, EPI_ISL_799394, EPI_ISL_799395, EPI_ISL_799397, EPI_ISL_799400, EPI_ISL_799403, EPI_ISL_799404, EPI_ISL_799406, EPI_ISL_799410, EPI_ISL_799411, EPI_ISL_799412, EPI_ISL_799413, EPI_ISL_799415, EPI_ISL_799416, EPI_ISL_799420, EPI_ISL_799427, EPI_ISL_799432, EPI_ISL_799434, EPI_ISL_799436, EPI_ISL_799437, EPI_ISL_799439, EPI_ISL_799440,
EPI_ISL_799441, EPI_ISL_799443, EPI_ISL_799445, EPI_ISL_799446, EPI_ISL_799447, EPI_ISL_799448, EPI_ISL_799454, EPI_ISL_799466, EPI_ISL_799467, EPI_ISL_799468, EPI_ISL_799469, EPI_ISL_799470, EPI_ISL_799473, EPI_ISL_799476, EPI_ISL_799477, EPI_ISL_799479, EPI_ISL_799482, EPI_ISL_799485, EPI_ISL_799487, EPI_ISL_799488, EPI_ISL_799490, EPI_ISL_799492, EPI_ISL_799494, EPI_ISL_799496,
EPI_ISL_799497, EPI_ISL_799498, EPI_ISL_799500, EPI_ISL_799501, EPI_ISL_799502, EPI_ISL_799505, EPI_ISL_799506, EPI_ISL_799508, EPI_ISL_799509, EPI_ISL_799512, EPI_ISL_799514, EPI_ISL_799515, EPI_ISL_799519, EPI_ISL_799520, EPI_ISL_799522, EPI_ISL_799523, EPI_ISL_799528, EPI_ISL_799529, EPI_ISL_799531, EPI_ISL_799533, EPI_ISL_799534, EPI_ISL_799535, EPI_ISL_799536, EPI_ISL_799538,
EPI_ISL_799540, EPI_ISL_799545, EPI_ISL_799546, EPI_ISL_799547, EPI_ISL_799550, EPI_ISL_799551, EPI_ISL_799555, EPI_ISL_799562, EPI_ISL_799563, EPI_ISL_799565, EPI_ISL_799567, EPI_ISL_799570, EPI_ISL_799572, EPI_ISL_799573, EPI_ISL_799575, EPI_ISL_799577, EPI_ISL_799578, EPI_ISL_799579, EPI_ISL_799580, EPI_ISL_799582, EPI_ISL_799583, EPI_ISL_799587, EPI_ISL_799588, EPI_ISL_799590,
EPI_ISL_799595, EPI_ISL_799597, EPI_ISL_799598, EPI_ISL_799600, EPI_ISL_799602, EPI_ISL_799603, EPI_ISL_799604, EPI_ISL_799608, EPI_ISL_799610, EPI_ISL_799611, EPI_ISL_799614, EPI_ISL_799616, EPI_ISL_799617, EPI_ISL_799618, EPI_ISL_799620, EPI_ISL_799621, EPI_ISL_799623, EPI_ISL_799624, EPI_ISL_799625, EPI_ISL_799626, EPI_ISL_799627, EPI_ISL_799628, EPI_ISL_799629, EPI_ISL_799631,
EPI_ISL_799632, EPI_ISL_799633, EPI_ISL_799635, EPI_ISL_799636, EPI_ISL_799639, EPI_ISL_799640, EPI_ISL_799642, EPI_ISL_799643, EPI_ISL_799645, EPI_ISL_799646, EPI_ISL_799647, EPI_ISL_799651, EPI_ISL_799653, EPI_ISL_799654, EPI_ISL_799656, EPI_ISL_799657, EPI_ISL_799658, EPI_ISL_799660, EPI_ISL_799663, EPI_ISL_799666, EPI_ISL_799667, EPI_ISL_799668, EPI_ISL_799670, EPI_ISL_799671,
EPI_ISL_799672, EPI_ISL_799673, EPI_ISL_799674, EPI_ISL_799675, EPI_ISL_799677, EPI_ISL_799680, EPI_ISL_799684, EPI_ISL_799685, EPI_ISL_799688, EPI_ISL_799690, EPI_ISL_799692, EPI_ISL_799694, EPI_ISL_799695, EPI_ISL_799696, EPI_ISL_799697, EPI_ISL_799698, EPI_ISL_799700, EPI_ISL_799701, EPI_ISL_799703, EPI_ISL_799704, EPI_ISL_799705, EPI_ISL_799709, EPI_ISL_799712, EPI_ISL_799714,
EPI_ISL_799715, EPI_ISL_799716, EPI_ISL_799717, EPI_ISL_799719, EPI_ISL_799720, EPI_ISL_799721, EPI_ISL_799723, EPI_ISL_799724, EPI_ISL_799725, EPI_ISL_799727, EPI_ISL_799728, EPI_ISL_799733, EPI_ISL_799736, EPI_ISL_799738, EPI_ISL_799741, EPI_ISL_799742, EPI_ISL_799743, EPI_ISL_799745, EPI_ISL_799746, EPI_ISL_799752, EPI_ISL_799753, EPI_ISL_799755, EPI_ISL_799756, EPI_ISL_799757,

EPI_ISL_799762, EPI_ISL_799764, EPI_ISL_799766, EPI_ISL_799769, EPI_ISL_799770, EPI_ISL_799771, EPI_ISL_799772, EPI_ISL_799773, EPI_ISL_799774, EPI_ISL_799777, EPI_ISL_799778, EPI_ISL_799779, EPI_ISL_799782, EPI_ISL_799785, EPI_ISL_799788, EPI_ISL_799789, EPI_ISL_799790, EPI_ISL_799793, EPI_ISL_799794, EPI_ISL_799795, EPI_ISL_799798, EPI_ISL_799800, EPI_ISL_799802, EPI_ISL_799803,
EPI_ISL_799804, EPI_ISL_799806, EPI_ISL_799808, EPI_ISL_799810, EPI_ISL_799813, EPI_ISL_799815, EPI_ISL_799816, EPI_ISL_799817, EPI_ISL_799820, EPI_ISL_799824, EPI_ISL_799826, EPI_ISL_799828, EPI_ISL_799829, EPI_ISL_799833, EPI_ISL_799834, EPI_ISL_799835, EPI_ISL_799836, EPI_ISL_799837, EPI_ISL_799842, EPI_ISL_799845, EPI_ISL_799847, EPI_ISL_799848, EPI_ISL_799849, EPI_ISL_799850,
EPI_ISL_799851, EPI_ISL_799852, EPI_ISL_799853, EPI_ISL_799854, EPI_ISL_799856, EPI_ISL_799857, EPI_ISL_799858, EPI_ISL_799860, EPI_ISL_799862, EPI_ISL_799863, EPI_ISL_799867, EPI_ISL_799870, EPI_ISL_799872, EPI_ISL_799873, EPI_ISL_799874, EPI_ISL_799876, EPI_ISL_799878, EPI_ISL_799879, EPI_ISL_799880, EPI_ISL_799882, EPI_ISL_799884, EPI_ISL_799885, EPI_ISL_799886, EPI_ISL_799887,
EPI_ISL_799890, EPI_ISL_799895, EPI_ISL_799896, EPI_ISL_799904, EPI_ISL_799905, EPI_ISL_799907, EPI_ISL_799909, EPI_ISL_799911, EPI_ISL_799913, EPI_ISL_799915, EPI_ISL_799921, EPI_ISL_799923, EPI_ISL_799926, EPI_ISL_799927, EPI_ISL_799928, EPI_ISL_799929, EPI_ISL_799931, EPI_ISL_799934, EPI_ISL_811809, EPI_ISL_811810, EPI_ISL_811813, EPI_ISL_811815, EPI_ISL_811816, EPI_ISL_811820,
EPI_ISL_811822, EPI_ISL_811829, EPI_ISL_811838, EPI_ISL_811842, EPI_ISL_811844, EPI_ISL_811863, EPI_ISL_811864, EPI_ISL_811866, EPI_ISL_811872, EPI_ISL_811876, EPI_ISL_811882, EPI_ISL_811895, EPI_ISL_811901, EPI_ISL_811903, EPI_ISL_811904, EPI_ISL_811906, EPI_ISL_811908, EPI_ISL_811911, EPI_ISL_811920, EPI_ISL_811926, EPI_ISL_811933, EPI_ISL_811934, EPI_ISL_811940, EPI_ISL_811947,
EPI_ISL_811961, EPI_ISL_811963, EPI_ISL_811964, EPI_ISL_811966, EPI_ISL_811980, EPI_ISL_811982, EPI_ISL_811992, EPI_ISL_811995, EPI_ISL_812001, EPI_ISL_812004, EPI_ISL_812005, EPI_ISL_812006, EPI_ISL_812008, EPI_ISL_812011, EPI_ISL_812013, EPI_ISL_812016, EPI_ISL_812023, EPI_ISL_812029, EPI_ISL_812031, EPI_ISL_812032, EPI_ISL_812034, EPI_ISL_812039, EPI_ISL_812043, EPI_ISL_812044,
EPI_ISL_812053, EPI_ISL_812056, EPI_ISL_812057, EPI_ISL_812060, EPI_ISL_812065, EPI_ISL_812071, EPI_ISL_812072, EPI_ISL_812081, EPI_ISL_812082, EPI_ISL_812087, EPI_ISL_812089, EPI_ISL_812095, EPI_ISL_812107, EPI_ISL_812108, EPI_ISL_812110, EPI_ISL_812111, EPI_ISL_812112, EPI_ISL_821629, EPI_ISL_821630, EPI_ISL_821631, EPI_ISL_821633, EPI_ISL_821634, EPI_ISL_821635, EPI_ISL_821636,
EPI_ISL_821637, EPI_ISL_821641, EPI_ISL_821648, EPI_ISL_821653, EPI_ISL_822305, EPI_ISL_822306, EPI_ISL_858792, EPI_ISL_858795, EPI_ISL_858798, EPI_ISL_858799, EPI_ISL_858800, EPI_ISL_858803, EPI_ISL_858804, EPI_ISL_858805, EPI_ISL_858807, EPI_ISL_858808, EPI_ISL_933426, EPI_ISL_933427, EPI_ISL_933428, EPI_ISL_933429, EPI_ISL_933430, EPI_ISL_933431, EPI_ISL_933432, EPI_ISL_933433,
EPI_ISL_933434, EPI_ISL_933435, EPI_ISL_933436, EPI_ISL_933437, EPI_ISL_933438, EPI_ISL_933439, EPI_ISL_933440, EPI_ISL_933447, EPI_ISL_933453, EPI_ISL_933456, EPI_ISL_933463, EPI_ISL_933467, EPI_ISL_933469, EPI_ISL_933472, EPI_ISL_933474, EPI_ISL_933475, EPI_ISL_933479, EPI_ISL_933480, EPI_ISL_945052, EPI_ISL_945138, EPI_ISL_945195, EPI_ISL_945224, EPI_ISL_945231, EPI_ISL_945358,
EPI_ISL_945360

| | | | |
|---|---|---|---|
| see above | Lighthouse Lab in Milton Keynes | Wellcome Sanger Institute for the COVID-19 Genomics UK (COG-UK) Consortium | Cordelia Langford; David K. Jackson; Dominic Kwiatkowski; Ewan Harrison; Ian Johnston; John Sillitoe on behalf of the Wellcome Sanger Institute COVID-19 Surveillance Team; Roberto Amato; Sonia Goncalves; The Lighthouse Lab in Milton Keynes and Alex Alderton |

EPI_ISL_840082, EPI_ISL_840083, EPI_ISL_840084, EPI_ISL_840085, EPI_ISL_840086, EPI_ISL_840087, EPI_ISL_840088, EPI_ISL_840089, EPI_ISL_840090, EPI_ISL_840091, EPI_ISL_840092, EPI_ISL_840093, EPI_ISL_840094, EPI_ISL_840095, EPI_ISL_840096, EPI_ISL_840097, EPI_ISL_840098, EPI_ISL_840099, EPI_ISL_840100, EPI_ISL_840101, EPI_ISL_840102, EPI_ISL_840103, EPI_ISL_840104, EPI_ISL_840105,
EPI_ISL_840106, EPI_ISL_840107, EPI_ISL_840111, EPI_ISL_950706, EPI_ISL_950707, EPI_ISL_950708, EPI_ISL_950709, EPI_ISL_998921, EPI_ISL_998922, EPI_ISL_998923, EPI_ISL_998924, EPI_ISL_998925, EPI_ISL_998926, EPI_ISL_998927

| | | | |
|---|---|---|---|
| see above | Lincolnshire Hospitals and DeepSeq Nottingham | COVID-19 Genomics UK (COG-UK) Consortium | Christopher Moore; Fei Sang; Joseph Chappell; Johnny Debebe; Jonathan Ball; Matthew Carlisle; Matthew Loose; Nadine Holmes; Nichola Duckworth; Patrick McClure; Sarah Walsh; Tim Sloan; Victoria Wright |

EPI_ISL_813773, EPI_ISL_813774, EPI_ISL_813775, EPI_ISL_813776, EPI_ISL_813777, EPI_ISL_813778, EPI_ISL_813779, EPI_ISL_813780, EPI_ISL_813781, EPI_ISL_813782, EPI_ISL_813783, EPI_ISL_813784, EPI_ISL_813785, EPI_ISL_813786, EPI_ISL_813787, EPI_ISL_813788, EPI_ISL_813789, EPI_ISL_813790, EPI_ISL_813791, EPI_ISL_813792, EPI_ISL_813793, EPI_ISL_813794, EPI_ISL_813795, EPI_ISL_813796,
EPI_ISL_813797, EPI_ISL_813798, EPI_ISL_813799, EPI_ISL_813800, EPI_ISL_813801, EPI_ISL_813802, EPI_ISL_813804, EPI_ISL_813806, EPI_ISL_814557, EPI_ISL_838571, EPI_ISL_838572, EPI_ISL_838573, EPI_ISL_838574, EPI_ISL_838575, EPI_ISL_838576, EPI_ISL_838577, EPI_ISL_865217, EPI_ISL_865218, EPI_ISL_865219, EPI_ISL_865220, EPI_ISL_865221, EPI_ISL_865222, EPI_ISL_865223, EPI_ISL_865224,
EPI_ISL_865225, EPI_ISL_865226, EPI_ISL_865227, EPI_ISL_865228, EPI_ISL_865229, EPI_ISL_865230, EPI_ISL_865231, EPI_ISL_865232, EPI_ISL_865233, EPI_ISL_865234, EPI_ISL_865235, EPI_ISL_865236, EPI_ISL_865237, EPI_ISL_865238, EPI_ISL_865240, EPI_ISL_865246, EPI_ISL_865248, EPI_ISL_865249, EPI_ISL_865373, EPI_ISL_865374, EPI_ISL_865376, EPI_ISL_865391, EPI_ISL_865392, EPI_ISL_865393,
EPI_ISL_865394, EPI_ISL_865395, EPI_ISL_865396, EPI_ISL_865397, EPI_ISL_865398, EPI_ISL_865399, EPI_ISL_865400, EPI_ISL_865401, EPI_ISL_865402, EPI_ISL_865403, EPI_ISL_865404, EPI_ISL_865405, EPI_ISL_865406, EPI_ISL_865407, EPI_ISL_865408, EPI_ISL_865409, EPI_ISL_865410, EPI_ISL_865421, EPI_ISL_865423, EPI_ISL_865424, EPI_ISL_865425, EPI_ISL_865426, EPI_ISL_865429, EPI_ISL_865431,
EPI_ISL_865434, EPI_ISL_865435, EPI_ISL_865438, EPI_ISL_865439, EPI_ISL_865440, EPI_ISL_865441, EPI_ISL_865442, EPI_ISL_865443, EPI_ISL_865444, EPI_ISL_865445, EPI_ISL_865446, EPI_ISL_865447, EPI_ISL_865451, EPI_ISL_865452, EPI_ISL_865453, EPI_ISL_865454, EPI_ISL_865455, EPI_ISL_865457, EPI_ISL_865458, EPI_ISL_919456, EPI_ISL_1247934

| | | | |
|---|---|---|---|
| see above | Liverpool Clinical Laboratories | COVID-19 Genomics UK (COG-UK) Consortium | A Alrezaihi; Alessandro Gerada; Alistair Darby; Angela Cowell; Anita Lucaci; Anu Chawla; Cassie Olateju; Catherine Hartley; Charlotte Nelson; Ecaterina Vamos; Elaine O'Toole; Eleanor G Bentley; Ghada T Shawli; Hermione Webster; Isabel García-Dorival; James Johnson; James P Stewart; Jenifer Manson; Joanne Watts; Jones Benjamin; Jordan J Clark; Julian Hiscox; L Luu; Lucille Rainbow; M Almsaud; Margaret Hughes; Mark Whitehead; Matthew Gemmell; Miren Iturriza-Gomara; Muhannad Alruwaili; N.P Randle; Neil Swainston; PKF Gilmore; Parul Sharma; Rebekah Penrice-Randal; Richard Eccles; Richard Gregory; Sam Haldenby; Steve Paterson; Stuart D Armstrong; Trevor Ian Robinson; Ximeng Han |

EPI_ISL_839607, EPI_ISL_839608, EPI_ISL_839609, EPI_ISL_839610, EPI_ISL_839611, EPI_ISL_839612, EPI_ISL_839613, EPI_ISL_839614, EPI_ISL_839616, EPI_ISL_839617, EPI_ISL_839618, EPI_ISL_839619, EPI_ISL_839620, EPI_ISL_839621, EPI_ISL_839622, EPI_ISL_839623, EPI_ISL_839625, EPI_ISL_839626, EPI_ISL_839627, EPI_ISL_839628, EPI_ISL_839629, EPI_ISL_839630, EPI_ISL_839641, EPI_ISL_839649,
EPI_ISL_839650, EPI_ISL_839652, EPI_ISL_839654, EPI_ISL_839655, EPI_ISL_839656, EPI_ISL_839657, EPI_ISL_839659, EPI_ISL_839667, EPI_ISL_839668, EPI_ISL_839669, EPI_ISL_839670, EPI_ISL_839671, EPI_ISL_839672, EPI_ISL_839673, EPI_ISL_839674, EPI_ISL_839675, EPI_ISL_839676, EPI_ISL_839677, EPI_ISL_839678, EPI_ISL_839679, EPI_ISL_839680, EPI_ISL_839681, EPI_ISL_839682, EPI_ISL_839683,
EPI_ISL_839684, EPI_ISL_839685, EPI_ISL_839686, EPI_ISL_839687, EPI_ISL_839688, EPI_ISL_839689, EPI_ISL_839690, EPI_ISL_839691, EPI_ISL_839692, EPI_ISL_839693, EPI_ISL_839694, EPI_ISL_839695, EPI_ISL_839696, EPI_ISL_839697, EPI_ISL_839698, EPI_ISL_839699, EPI_ISL_839700, EPI_ISL_839701, EPI_ISL_839702, EPI_ISL_839703, EPI_ISL_839704, EPI_ISL_839705, EPI_ISL_839706, EPI_ISL_839707,
EPI_ISL_839708, EPI_ISL_839709, EPI_ISL_839816, EPI_ISL_839817, EPI_ISL_839818, EPI_ISL_839819, EPI_ISL_839820, EPI_ISL_839821, EPI_ISL_839822, EPI_ISL_921185, EPI_ISL_921186, EPI_ISL_921188, EPI_ISL_921189, EPI_ISL_921196, EPI_ISL_921197, EPI_ISL_921198, EPI_ISL_921199, EPI_ISL_921200, EPI_ISL_921201, EPI_ISL_921202, EPI_ISL_921203, EPI_ISL_921204, EPI_ISL_921205, EPI_ISL_921206,
EPI_ISL_921207, EPI_ISL_921209, EPI_ISL_921210, EPI_ISL_921211, EPI_ISL_921213, EPI_ISL_921214, EPI_ISL_921215, EPI_ISL_921216, EPI_ISL_921217, EPI_ISL_921218, EPI_ISL_921219, EPI_ISL_921220, EPI_ISL_921221, EPI_ISL_921223, EPI_ISL_921224, EPI_ISL_921225, EPI_ISL_921226, EPI_ISL_921227, EPI_ISL_921229, EPI_ISL_921230, EPI_ISL_921231, EPI_ISL_921232, EPI_ISL_921233,
EPI_ISL_921234, EPI_ISL_921235, EPI_ISL_921236, EPI_ISL_921237, EPI_ISL_921238, EPI_ISL_921239, EPI_ISL_921240, EPI_ISL_921241, EPI_ISL_921242, EPI_ISL_921321, EPI_ISL_921322, EPI_ISL_921323, EPI_ISL_921324, EPI_ISL_921325, EPI_ISL_921326, EPI_ISL_921327, EPI_ISL_921328, EPI_ISL_921329, EPI_ISL_921330, EPI_ISL_921331, EPI_ISL_921332, EPI_ISL_921333, EPI_ISL_921334, EPI_ISL_921335,
EPI_ISL_921336, EPI_ISL_921337, EPI_ISL_921338, EPI_ISL_921339, EPI_ISL_921340, EPI_ISL_921341, EPI_ISL_921342, EPI_ISL_921343, EPI_ISL_921349, EPI_ISL_921350, EPI_ISL_921357, EPI_ISL_921360, EPI_ISL_921362, EPI_ISL_921363, EPI_ISL_921364, EPI_ISL_921365, EPI_ISL_921366, EPI_ISL_921367, EPI_ISL_921368, EPI_ISL_921369, EPI_ISL_921370, EPI_ISL_921372, EPI_ISL_921373, EPI_ISL_921374,
EPI_ISL_921375, EPI_ISL_921376, EPI_ISL_921377, EPI_ISL_921378, EPI_ISL_921663, EPI_ISL_921664, EPI_ISL_921665, EPI_ISL_921666, EPI_ISL_921667, EPI_ISL_921668, EPI_ISL_921669, EPI_ISL_921670, EPI_ISL_921671, EPI_ISL_921672, EPI_ISL_921673, EPI_ISL_921674, EPI_ISL_921675, EPI_ISL_921677, EPI_ISL_921678, EPI_ISL_921679, EPI_ISL_921680, EPI_ISL_921681, EPI_ISL_921693, EPI_ISL_921694,
EPI_ISL_921695, EPI_ISL_921696, EPI_ISL_921697, EPI_ISL_921698, EPI_ISL_921726, EPI_ISL_921727, EPI_ISL_921728, EPI_ISL_921729, EPI_ISL_921730, EPI_ISL_921731, EPI_ISL_921732, EPI_ISL_921733, EPI_ISL_921734, EPI_ISL_921735, EPI_ISL_921736, EPI_ISL_921737, EPI_ISL_921738, EPI_ISL_921739, EPI_ISL_921740, EPI_ISL_921741, EPI_ISL_921742, EPI_ISL_921743, EPI_ISL_921744, EPI_ISL_921745,
EPI_ISL_921746, EPI_ISL_921747, EPI_ISL_921748, EPI_ISL_921749, EPI_ISL_921750, EPI_ISL_921751, EPI_ISL_921759, EPI_ISL_921760, EPI_ISL_921761, EPI_ISL_921762, EPI_ISL_921763, EPI_ISL_921764, EPI_ISL_921765, EPI_ISL_921766, EPI_ISL_921767, EPI_ISL_921768, EPI_ISL_921769, EPI_ISL_921770, EPI_ISL_921771, EPI_ISL_921772, EPI_ISL_921773, EPI_ISL_921774, EPI_ISL_921775, EPI_ISL_921776,
EPI_ISL_921777, EPI_ISL_921778, EPI_ISL_921779, EPI_ISL_921780, EPI_ISL_921781, EPI_ISL_921782, EPI_ISL_950240, EPI_ISL_950436, EPI_ISL_950438, EPI_ISL_950439, EPI_ISL_950440, EPI_ISL_950441, EPI_ISL_950442, EPI_ISL_950443, EPI_ISL_998588, EPI_ISL_998589, EPI_ISL_998590, EPI_ISL_998591, EPI_ISL_998593, EPI_ISL_998594, EPI_ISL_998595, EPI_ISL_998596, EPI_ISL_998597, EPI_ISL_998599,
EPI_ISL_998600, EPI_ISL_998601, EPI_ISL_998602, EPI_ISL_998603, EPI_ISL_998604, EPI_ISL_998605, EPI_ISL_998606, EPI_ISL_998607, EPI_ISL_998608, EPI_ISL_998609, EPI_ISL_998610, EPI_ISL_998611, EPI_ISL_998612, EPI_ISL_998613, EPI_ISL_998614, EPI_ISL_998615, EPI_ISL_998617, EPI_ISL_998618, EPI_ISL_998619, EPI_ISL_998620, EPI_ISL_998621, EPI_ISL_998622, EPI_ISL_998623, EPI_ISL_998625,
EPI_ISL_998627, EPI_ISL_998629, EPI_ISL_998631, EPI_ISL_998632, EPI_ISL_998634, EPI_ISL_998635, EPI_ISL_998637, EPI_ISL_998638, EPI_ISL_998640, EPI_ISL_998641, EPI_ISL_998642, EPI_ISL_998643, EPI_ISL_998645, EPI_ISL_998646, EPI_ISL_998647, EPI_ISL_998648, EPI_ISL_998649, EPI_ISL_998650, EPI_ISL_998652, EPI_ISL_998653, EPI_ISL_998654, EPI_ISL_998655, EPI_ISL_998656, EPI_ISL_998657,
EPI_ISL_998658, EPI_ISL_998659, EPI_ISL_998660, EPI_ISL_998661, EPI_ISL_998662, EPI_ISL_998663, EPI_ISL_998664, EPI_ISL_998665, EPI_ISL_998666, EPI_ISL_998667, EPI_ISL_998668, EPI_ISL_998669, EPI_ISL_998670, EPI_ISL_998671, EPI_ISL_998672, EPI_ISL_998673, EPI_ISL_998674, EPI_ISL_998675, EPI_ISL_998719, EPI_ISL_998720, EPI_ISL_998721, EPI_ISL_998722, EPI_ISL_998723, EPI_ISL_998724,
EPI_ISL_998725, EPI_ISL_998726, EPI_ISL_998727, EPI_ISL_998728, EPI_ISL_998730, EPI_ISL_998731, EPI_ISL_998732, EPI_ISL_998733, EPI_ISL_998734, EPI_ISL_998735, EPI_ISL_998736, EPI_ISL_1050916, EPI_ISL_1050917, EPI_ISL_1050918, EPI_ISL_1050919, EPI_ISL_1051341

| | | | |
|---|---|---|---|
| see above | Northumbria University / South Tees Hospitals NHS Foundation Trust / North Cumbria Integrated Care NHS Foundation Trust / North Tees and Hartlepool NHS Foundation Trust / Newcastle Hospitals NHS Foundation Trust | COVID-19 Genomics UK (COG-UK) Consortium | Andrew Nelson; Brendan Payne; Clive Graham; Darren L Smith; Debra Padgett; Edward Barton; Emma Swindells; Garren Scott; Gary Black; Gary Eltringham; Giles S Holt; Greg R Young; Jane Greenaway; Jennifer Collins; John Allan; Joshua Loh; Lynn Dover; Matthew Bashton; Mohammad A Tariq; Paul Baker; Sarah Essex; Steve Liggett; Wen C Yew; Yusri Taha |

EPI_ISL_840370, EPI_ISL_840374, EPI_ISL_840375, EPI_ISL_840376, EPI_ISL_840377, EPI_ISL_840378, EPI_ISL_840380, EPI_ISL_840381, EPI_ISL_840382, EPI_ISL_840383, EPI_ISL_840384, EPI_ISL_840385, EPI_ISL_840386, EPI_ISL_840387, EPI_ISL_840388, EPI_ISL_840389, EPI_ISL_840398, EPI_ISL_840400, EPI_ISL_840401, EPI_ISL_840402, EPI_ISL_840403, EPI_ISL_840404, EPI_ISL_840405, EPI_ISL_840406,
EPI_ISL_840407, EPI_ISL_840408, EPI_ISL_840409, EPI_ISL_840410, EPI_ISL_840411, EPI_ISL_840412, EPI_ISL_840413, EPI_ISL_840414, EPI_ISL_840415, EPI_ISL_840416, EPI_ISL_840417, EPI_ISL_840418, EPI_ISL_840419, EPI_ISL_840420, EPI_ISL_840421, EPI_ISL_840422, EPI_ISL_840423, EPI_ISL_840424, EPI_ISL_840425, EPI_ISL_840426, EPI_ISL_840427, EPI_ISL_840428, EPI_ISL_840429, EPI_ISL_840430,
EPI_ISL_840431, EPI_ISL_840432, EPI_ISL_840433, EPI_ISL_840434, EPI_ISL_840435, EPI_ISL_840436, EPI_ISL_840437, EPI_ISL_840438, EPI_ISL_840439, EPI_ISL_840440, EPI_ISL_840441, EPI_ISL_840442, EPI_ISL_840443, EPI_ISL_840444, EPI_ISL_840445, EPI_ISL_840446, EPI_ISL_840447, EPI_ISL_840448, EPI_ISL_840449, EPI_ISL_840450, EPI_ISL_840451, EPI_ISL_840452, EPI_ISL_840453, EPI_ISL_840454,
EPI_ISL_840455, EPI_ISL_840456, EPI_ISL_840457, EPI_ISL_840458, EPI_ISL_840459, EPI_ISL_840460, EPI_ISL_840461, EPI_ISL_840462, EPI_ISL_840463, EPI_ISL_840464, EPI_ISL_840465, EPI_ISL_840466, EPI_ISL_840467, EPI_ISL_840468, EPI_ISL_840469, EPI_ISL_840470, EPI_ISL_840471, EPI_ISL_840472, EPI_ISL_840473, EPI_ISL_840474, EPI_ISL_840475, EPI_ISL_840476, EPI_ISL_840477, EPI_ISL_840478,
EPI_ISL_840479, EPI_ISL_840480, EPI_ISL_840481, EPI_ISL_840482, EPI_ISL_840483, EPI_ISL_840484, EPI_ISL_840485, EPI_ISL_840486, EPI_ISL_840487, EPI_ISL_840488, EPI_ISL_840489, EPI_ISL_840490, EPI_ISL_840491, EPI_ISL_840492, EPI_ISL_840493, EPI_ISL_840494, EPI_ISL_840495, EPI_ISL_840496, EPI_ISL_840497, EPI_ISL_840498, EPI_ISL_840499, EPI_ISL_840500, EPI_ISL_840501, EPI_ISL_840502,
EPI_ISL_840503, EPI_ISL_840505, EPI_ISL_840506, EPI_ISL_840507, EPI_ISL_840509, EPI_ISL_840510, EPI_ISL_840513, EPI_ISL_840514, EPI_ISL_840516, EPI_ISL_840517, EPI_ISL_840518, EPI_ISL_840519, EPI_ISL_840520, EPI_ISL_840521, EPI_ISL_840522, EPI_ISL_840523, EPI_ISL_840524, EPI_ISL_840525, EPI_ISL_840526, EPI_ISL_840527, EPI_ISL_840528, EPI_ISL_840529, EPI_ISL_840530, EPI_ISL_840531,
EPI_ISL_840532, EPI_ISL_840533, EPI_ISL_840534, EPI_ISL_840535, EPI_ISL_840536, EPI_ISL_840537, EPI_ISL_840538, EPI_ISL_840539, EPI_ISL_840540, EPI_ISL_840541, EPI_ISL_840542, EPI_ISL_840543, EPI_ISL_840544, EPI_ISL_840545, EPI_ISL_840546, EPI_ISL_840547, EPI_ISL_840548, EPI_ISL_840549, EPI_ISL_840550, EPI_ISL_840551, EPI_ISL_840553, EPI_ISL_840555, EPI_ISL_840556, EPI_ISL_840557,
EPI_ISL_840558, EPI_ISL_840559, EPI_ISL_840560, EPI_ISL_840561, EPI_ISL_840562, EPI_ISL_840563, EPI_ISL_840564, EPI_ISL_840565, EPI_ISL_840566, EPI_ISL_840567, EPI_ISL_840568, EPI_ISL_840569, EPI_ISL_840571, EPI_ISL_840572, EPI_ISL_840573, EPI_ISL_840574, EPI_ISL_840575, EPI_ISL_840576, EPI_ISL_840577, EPI_ISL_840578, EPI_ISL_840581, EPI_ISL_840582, EPI_ISL_840583, EPI_ISL_840585,
EPI_ISL_840587, EPI_ISL_840588, EPI_ISL_840589, EPI_ISL_840591, EPI_ISL_840592, EPI_ISL_840593, EPI_ISL_840594, EPI_ISL_840595, EPI_ISL_840599, EPI_ISL_840662, EPI_ISL_840667, EPI_ISL_840668, EPI_ISL_840669, EPI_ISL_840670, EPI_ISL_840705, EPI_ISL_840706, EPI_ISL_840707, EPI_ISL_840708, EPI_ISL_840709, EPI_ISL_840710, EPI_ISL_840728, EPI_ISL_840729, EPI_ISL_840730,
EPI_ISL_840737, EPI_ISL_840738, EPI_ISL_840739, EPI_ISL_840740, EPI_ISL_840742, EPI_ISL_840743, EPI_ISL_840744, EPI_ISL_840745, EPI_ISL_840746, EPI_ISL_840749, EPI_ISL_840750, EPI_ISL_840751, EPI_ISL_840752, EPI_ISL_840754, EPI_ISL_840756, EPI_ISL_840757, EPI_ISL_840758, EPI_ISL_840759, EPI_ISL_840760, EPI_ISL_840761, EPI_ISL_840762, EPI_ISL_840763, EPI_ISL_840764, EPI_ISL_840765,
EPI_ISL_840766, EPI_ISL_840767, EPI_ISL_840769, EPI_ISL_867218, EPI_ISL_951750, EPI_ISL_951789, EPI_ISL_951790, EPI_ISL_951791, EPI_ISL_951792, EPI_ISL_951793, EPI_ISL_951794, EPI_ISL_951795, EPI_ISL_951796, EPI_ISL_951797, EPI_ISL_951798, EPI_ISL_951799, EPI_ISL_951800, EPI_ISL_951801, EPI_ISL_951802, EPI_ISL_951803, EPI_ISL_951804, EPI_ISL_951805, EPI_ISL_951806, EPI_ISL_951807,
EPI_ISL_951808, EPI_ISL_951809, EPI_ISL_951810, EPI_ISL_951811, EPI_ISL_951812, EPI_ISL_951813, EPI_ISL_951814

| | | | |
|---|---|---|---|
| see above | Originating lab: Wales Specialist Virology Centre Sequencing lab: Pathogen Genomics Unit | Public Health Wales Microbiology Cardiff Wales Specialist Virology Centre | Alec Birchley; Alexander Adams; Amy Gaskin; Angela Marchbank; Bree Gatica-Wilcox; Catherine Moore; Jason Coombes; Joanne Watkins; Joel Southgate; Johnathan Evans; Laura Gifford; Lauren Gilbert; Lee Graham; Malorie Perry; Matthew Bull; Nicole Pacchiarini; Sally Corden; Sara Kumziene-Summerhayes; Sara Rey; Sarah Taylor; Simon Cottrell; Sophie Jones; Tom Connor |

EPI_ISL_840268, EPI_ISL_840288, EPI_ISL_840289, EPI_ISL_840290, EPI_ISL_840291, EPI_ISL_840292, EPI_ISL_840294, EPI_ISL_840295, EPI_ISL_840296, EPI_ISL_840297, EPI_ISL_840298, EPI_ISL_840301, EPI_ISL_840302, EPI_ISL_840303, EPI_ISL_840304, EPI_ISL_840305, EPI_ISL_840306, EPI_ISL_840307, EPI_ISL_840308, EPI_ISL_840309, EPI_ISL_840311, EPI_ISL_840313, EPI_ISL_840314, EPI_ISL_840315,
EPI_ISL_840316, EPI_ISL_840317, EPI_ISL_840318, EPI_ISL_840319, EPI_ISL_840320, EPI_ISL_840321, EPI_ISL_840322, EPI_ISL_840323, EPI_ISL_840324, EPI_ISL_840325, EPI_ISL_840326, EPI_ISL_840327, EPI_ISL_840328, EPI_ISL_840329, EPI_ISL_840330, EPI_ISL_840331, EPI_ISL_840332, EPI_ISL_840333, EPI_ISL_840334, EPI_ISL_840335, EPI_ISL_840336, EPI_ISL_840338, EPI_ISL_840339, EPI_ISL_840340,
EPI_ISL_840341, EPI_ISL_840342, EPI_ISL_840343, EPI_ISL_840344, EPI_ISL_840345, EPI_ISL_840348, EPI_ISL_840365, EPI_ISL_840366, EPI_ISL_867037, EPI_ISL_867038, EPI_ISL_867047, EPI_ISL_867051, EPI_ISL_867055, EPI_ISL_867172, EPI_ISL_867173, EPI_ISL_867174, EPI_ISL_867177, EPI_ISL_867179, EPI_ISL_867183, EPI_ISL_867184, EPI_ISL_867185, EPI_ISL_922172, EPI_ISL_922173, EPI_ISL_922253,
EPI_ISL_922268, EPI_ISL_922269, EPI_ISL_922270, EPI_ISL_922273, EPI_ISL_922276, EPI_ISL_922281, EPI_ISL_922282, EPI_ISL_922307, EPI_ISL_922312, EPI_ISL_922313, EPI_ISL_922314, EPI_ISL_922316, EPI_ISL_922319, EPI_ISL_950763, EPI_ISL_950764, EPI_ISL_950765, EPI_ISL_950766, EPI_ISL_950767, EPI_ISL_950769, EPI_ISL_950772, EPI_ISL_950773, EPI_ISL_950774, EPI_ISL_950775,
EPI_ISL_950776, EPI_ISL_950777, EPI_ISL_950778, EPI_ISL_950779, EPI_ISL_950781, EPI_ISL_950783, EPI_ISL_950784, EPI_ISL_950785, EPI_ISL_950786, EPI_ISL_950788, EPI_ISL_950789, EPI_ISL_950790, EPI_ISL_950791, EPI_ISL_950793, EPI_ISL_950794, EPI_ISL_950795, EPI_ISL_950796, EPI_ISL_950798, EPI_ISL_950799, EPI_ISL_950802, EPI_ISL_950803, EPI_ISL_950804, EPI_ISL_950805, EPI_ISL_950806,
EPI_ISL_950807, EPI_ISL_950808, EPI_ISL_950809, EPI_ISL_950810, EPI_ISL_950811, EPI_ISL_950812, EPI_ISL_950814, EPI_ISL_950815, EPI_ISL_950816, EPI_ISL_950817, EPI_ISL_950820, EPI_ISL_950821, EPI_ISL_950822, EPI_ISL_950823, EPI_ISL_950825, EPI_ISL_950826, EPI_ISL_950827, EPI_ISL_950828, EPI_ISL_950831, EPI_ISL_950832, EPI_ISL_950833, EPI_ISL_950834, EPI_ISL_950835, EPI_ISL_950836,
EPI_ISL_950837, EPI_ISL_950838, EPI_ISL_950840, EPI_ISL_950841, EPI_ISL_950842, EPI_ISL_950843, EPI_ISL_950845, EPI_ISL_950846, EPI_ISL_950848, EPI_ISL_950849, EPI_ISL_950850, EPI_ISL_950851, EPI_ISL_950852, EPI_ISL_950853, EPI_ISL_950854, EPI_ISL_950855, EPI_ISL_950856, EPI_ISL_950857, EPI_ISL_950858, EPI_ISL_950859, EPI_ISL_950861, EPI_ISL_950862, EPI_ISL_950863, EPI_ISL_950864,
EPI_ISL_950865, EPI_ISL_950868, EPI_ISL_950869, EPI_ISL_950870, EPI_ISL_950871, EPI_ISL_950873, EPI_ISL_950874, EPI_ISL_950875, EPI_ISL_950876, EPI_ISL_950877, EPI_ISL_950879, EPI_ISL_950880, EPI_ISL_950881, EPI_ISL_950882, EPI_ISL_950884, EPI_ISL_950885, EPI_ISL_950886, EPI_ISL_950887, EPI_ISL_950889, EPI_ISL_950890, EPI_ISL_950891, EPI_ISL_950892, EPI_ISL_950893, EPI_ISL_950894,
EPI_ISL_950895, EPI_ISL_950896, EPI_ISL_950897, EPI_ISL_950898, EPI_ISL_950899, EPI_ISL_950920, EPI_ISL_950922, EPI_ISL_950923, EPI_ISL_950924, EPI_ISL_950925, EPI_ISL_950926, EPI_ISL_950927, EPI_ISL_950928, EPI_ISL_950929, EPI_ISL_950931, EPI_ISL_950933, EPI_ISL_950934, EPI_ISL_950935, EPI_ISL_950936, EPI_ISL_950938, EPI_ISL_950939, EPI_ISL_950940, EPI_ISL_950941, EPI_ISL_950943,
EPI_ISL_950944, EPI_ISL_950946, EPI_ISL_950947, EPI_ISL_950948, EPI_ISL_950949, EPI_ISL_950950, EPI_ISL_950951, EPI_ISL_950952, EPI_ISL_950955, EPI_ISL_950961, EPI_ISL_950963, EPI_ISL_950964, EPI_ISL_950966, EPI_ISL_950968, EPI_ISL_950970, EPI_ISL_950971, EPI_ISL_950973, EPI_ISL_950974, EPI_ISL_950975, EPI_ISL_950976, EPI_ISL_950977, EPI_ISL_950984, EPI_ISL_950997,
EPI_ISL_951005, EPI_ISL_951006, EPI_ISL_951007, EPI_ISL_951012, EPI_ISL_951015, EPI_ISL_951343, EPI_ISL_1309735, EPI_ISL_1309736, EPI_ISL_1309737, EPI_ISL_1309738, EPI_ISL_1833050, EPI_ISL_1833055, EPI_ISL_1833060, EPI_ISL_1833061, EPI_ISL_2127632

| | | | |
|---|---|---|---|
| see above | Oxford Viromics, NDM, University of Oxford; Oxford University Hospitals; Basingstoke and North Hampshire Hospital | COVID-19 Genomics UK (COG-UK) Consortium | Alex Mobbs; Amy Trebes; Anita Justice; Catrin Moore; Christophe Fraser; David Bonsall; David Buck; Emma Wise; George Macintyre; Jessica Lynch; John Todd; Mariateresa de Cesare; Matilde Mori; Monique Andersson; Nathan Moore; Nick Cortes; Robert Shaw; Stephen Kidd; Tanya Golubchik; Timothy Peto |

EPI_ISL_763406, EPI_ISL_763456, EPI_ISL_763466, EPI_ISL_763478, EPI_ISL_763537, EPI_ISL_763549, EPI_ISL_763560, EPI_ISL_763587, EPI_ISL_763655, EPI_ISL_763665, EPI_ISL_763681, EPI_ISL_763737, EPI_ISL_763749, EPI_ISL_763777, EPI_ISL_763780, EPI_ISL_763797, EPI_ISL_763853, EPI_ISL_763924, EPI_ISL_764113, EPI_ISL_764114, EPI_ISL_764115, EPI_ISL_764130, EPI_ISL_764485, EPI_ISL_764486,
EPI_ISL_764487, EPI_ISL_764488, EPI_ISL_764490, EPI_ISL_764491, EPI_ISL_819370, EPI_ISL_819430, EPI_ISL_819437, EPI_ISL_819439, EPI_ISL_819877, EPI_ISL_819885, EPI_ISL_819886, EPI_ISL_819887, EPI_ISL_819890, EPI_ISL_819891, EPI_ISL_819894, EPI_ISL_819895, EPI_ISL_819896, EPI_ISL_819897, EPI_ISL_819898, EPI_ISL_819899, EPI_ISL_819900, EPI_ISL_819901, EPI_ISL_819902, EPI_ISL_819903,
EPI_ISL_819904, EPI_ISL_819905, EPI_ISL_819907, EPI_ISL_819908, EPI_ISL_819909, EPI_ISL_819910, EPI_ISL_819911, EPI_ISL_819912, EPI_ISL_819913, EPI_ISL_819914, EPI_ISL_819915, EPI_ISL_819916, EPI_ISL_819917, EPI_ISL_819918, EPI_ISL_819928, EPI_ISL_819931, EPI_ISL_819932, EPI_ISL_819933, EPI_ISL_819934, EPI_ISL_819935, EPI_ISL_819945, EPI_ISL_819953, EPI_ISL_819956,
EPI_ISL_819959, EPI_ISL_819960, EPI_ISL_819963, EPI_ISL_819970, EPI_ISL_819971, EPI_ISL_819973, EPI_ISL_819974, EPI_ISL_819976, EPI_ISL_819977, EPI_ISL_819990, EPI_ISL_819996, EPI_ISL_820001, EPI_ISL_820002, EPI_ISL_820003, EPI_ISL_820004, EPI_ISL_820005, EPI_ISL_820011, EPI_ISL_820015, EPI_ISL_820021, EPI_ISL_820022, EPI_ISL_820023, EPI_ISL_820025,
EPI_ISL_820026, EPI_ISL_820027, EPI_ISL_820028, EPI_ISL_820029, EPI_ISL_820030, EPI_ISL_820041, EPI_ISL_820042, EPI_ISL_820043, EPI_ISL_820044, EPI_ISL_820055, EPI_ISL_820056, EPI_ISL_820062, EPI_ISL_820067, EPI_ISL_820068, EPI_ISL_820069, EPI_ISL_820070, EPI_ISL_820071, EPI_ISL_820072, EPI_ISL_820073, EPI_ISL_820074, EPI_ISL_820075, EPI_ISL_820076, EPI_ISL_820077, EPI_ISL_820078,
EPI_ISL_820079, EPI_ISL_820080, EPI_ISL_820082, EPI_ISL_820088, EPI_ISL_820090, EPI_ISL_820091, EPI_ISL_820092, EPI_ISL_820093, EPI_ISL_820096, EPI_ISL_820097, EPI_ISL_820098, EPI_ISL_820102, EPI_ISL_820103, EPI_ISL_820104, EPI_ISL_820105, EPI_ISL_820106, EPI_ISL_820107, EPI_ISL_820108, EPI_ISL_820110, EPI_ISL_820112, EPI_ISL_820119, EPI_ISL_820124, EPI_ISL_820126,
EPI_ISL_820131, EPI_ISL_820134, EPI_ISL_820136, EPI_ISL_820141, EPI_ISL_820144, EPI_ISL_820146, EPI_ISL_820149, EPI_ISL_820151, EPI_ISL_820154, EPI_ISL_820156, EPI_ISL_820159, EPI_ISL_820161, EPI_ISL_820164, EPI_ISL_820167, EPI_ISL_820169, EPI_ISL_820172, EPI_ISL_820175, EPI_ISL_820178, EPI_ISL_820180, EPI_ISL_820183, EPI_ISL_820186, EPI_ISL_820188, EPI_ISL_820191, EPI_ISL_820193,
EPI_ISL_820196, EPI_ISL_820199, EPI_ISL_820202, EPI_ISL_820221, EPI_ISL_820223, EPI_ISL_820231, EPI_ISL_820234, EPI_ISL_820238, EPI_ISL_820241, EPI_ISL_820244, EPI_ISL_820253, EPI_ISL_820261, EPI_ISL_820279, EPI_ISL_820284, EPI_ISL_820287, EPI_ISL_820289, EPI_ISL_820292, EPI_ISL_820294, EPI_ISL_820300, EPI_ISL_820302, EPI_ISL_820304, EPI_ISL_820306, EPI_ISL_820311, EPI_ISL_820323,
EPI_ISL_866582, EPI_ISL_866824, EPI_ISL_921783, EPI_ISL_921784, EPI_ISL_921785, EPI_ISL_921789, EPI_ISL_921790, EPI_ISL_921791, EPI_ISL_921792, EPI_ISL_921794, EPI_ISL_921795, EPI_ISL_921796, EPI_ISL_921797, EPI_ISL_921798, EPI_ISL_921799, EPI_ISL_921801, EPI_ISL_921802, EPI_ISL_921803, EPI_ISL_921804, EPI_ISL_921806, EPI_ISL_921807, EPI_ISL_921811, EPI_ISL_921812, EPI_ISL_921814,
EPI_ISL_921815, EPI_ISL_921816, EPI_ISL_921817, EPI_ISL_921818, EPI_ISL_921820, EPI_ISL_921821, EPI_ISL_921822, EPI_ISL_921823, EPI_ISL_921824, EPI_ISL_921826, EPI_ISL_921827, EPI_ISL_921828, EPI_ISL_921829, EPI_ISL_921830, EPI_ISL_921831, EPI_ISL_921832, EPI_ISL_921833, EPI_ISL_921834, EPI_ISL_921835, EPI_ISL_921836, EPI_ISL_921837, EPI_ISL_921838, EPI_ISL_921839, EPI_ISL_921840,
EPI_ISL_921841, EPI_ISL_921842, EPI_ISL_921843, EPI_ISL_921844, EPI_ISL_921845, EPI_ISL_921846, EPI_ISL_921847, EPI_ISL_921848, EPI_ISL_921849, EPI_ISL_921850, EPI_ISL_950544, EPI_ISL_950545, EPI_ISL_950546, EPI_ISL_950547, EPI_ISL_950548, EPI_ISL_950549, EPI_ISL_950550, EPI_ISL_950552, EPI_ISL_950553, EPI_ISL_950554, EPI_ISL_950556, EPI_ISL_950557, EPI_ISL_950558, EPI_ISL_950559,
EPI_ISL_950560, EPI_ISL_950561, EPI_ISL_950562, EPI_ISL_950572, EPI_ISL_950574, EPI_ISL_950575, EPI_ISL_950576, EPI_ISL_950578, EPI_ISL_950579, EPI_ISL_950580, EPI_ISL_950583, EPI_ISL_950588, EPI_ISL_950589, EPI_ISL_950590, EPI_ISL_950591, EPI_ISL_950593, EPI_ISL_950595, EPI_ISL_950597, EPI_ISL_950598, EPI_ISL_950599, EPI_ISL_950600,
EPI_ISL_1051466, EPI_ISL_1051523

| | | | |
|---|---|---|---|
| see above | Quadram Institute Bioscience | COVID-19 Genomics UK (COG-UK) Consortium | Alexander J Trotter; Alison E. Mather; Alp Aydin; Ana P. Tedim; Anastasia Kolyva; Andrew Bell; Andrew J. Page; Claire Stuart; Dave J. Baker; Gemma L. Kay; John Wain; Justin O'Grady; Leonardo de Oliveira Martins; Lizzie Meadows; Maria Diaz; Mark Webber; Muhammed Yasir; Nabil-Fareed Alikhan; Ngozi Elumogo; Nicholas M. Thomson; Rachael Stanley; Rachel Gilroy; Reenesh Prakash; Samir Dervisevic; Samuel Bloomfield; Steven Rudder; Thanh Le-Viet |

EPI_ISL_741656, EPI_ISL_741657, EPI_ISL_763389, EPI_ISL_763444, EPI_ISL_763449, EPI_ISL_763547, EPI_ISL_763580, EPI_ISL_763650, EPI_ISL_763663, EPI_ISL_763673, EPI_ISL_763675, EPI_ISL_763678, EPI_ISL_763726, EPI_ISL_763819, EPI_ISL_763854, EPI_ISL_763855, EPI_ISL_763856, EPI_ISL_763857, EPI_ISL_763858, EPI_ISL_763925, EPI_ISL_763926, EPI_ISL_763927, EPI_ISL_764189, EPI_ISL_764190,
EPI_ISL_764230, EPI_ISL_764233, EPI_ISL_764495, EPI_ISL_764496, EPI_ISL_764497, EPI_ISL_764498, EPI_ISL_764499, EPI_ISL_764500, EPI_ISL_764501, EPI_ISL_764502, EPI_ISL_764503, EPI_ISL_764504, EPI_ISL_764505, EPI_ISL_764506, EPI_ISL_764507, EPI_ISL_764508, EPI_ISL_764509, EPI_ISL_764510, EPI_ISL_764511, EPI_ISL_764512, EPI_ISL_764513, EPI_ISL_764514, EPI_ISL_764515, EPI_ISL_764516,
EPI_ISL_764517, EPI_ISL_764518, EPI_ISL_764519, EPI_ISL_764520, EPI_ISL_764521, EPI_ISL_764522, EPI_ISL_764523, EPI_ISL_764524, EPI_ISL_819942, EPI_ISL_820006, EPI_ISL_820297, EPI_ISL_839990, EPI_ISL_839991, EPI_ISL_839992, EPI_ISL_839993, EPI_ISL_839994, EPI_ISL_866890

| | | | |
|---|---|---|---|
| see above | Queens Medical Centre, Clinical Microbiology Department / DeepSeq Nottingham | COVID-19 Genomics UK (COG-UK) Consortium | Christopher Moore; Fei Sang; Gemma Clark; Hannah Howson-Wells; Johnny Debebe; Jonathan Ball; Joseph Chappell; Manjinder Khakh; Matthew Carlisle; Matthew Loose; Michelle M Lister; Nadine Holmes; Patrick McClure; Theocharis Tsoleridis; Vicki M Fleming; Victoria Wright; Wendy Smith |

EPI_ISL_843151, EPI_ISL_843152, EPI_ISL_843153, EPI_ISL_843154, EPI_ISL_843163, EPI_ISL_843167, EPI_ISL_843168, EPI_ISL_843169, EPI_ISL_921090

| | | | |
|---|---|---|---|
| see above | Regional Virus Laboratory, Belfast Health and Social Care Trust | COVID-19 Genomics UK (COG-UK) Consortium | Alison Watt; Ciara Cox; Conall McCaughey; David Simpson; Derek Fairley; James McKenna; Mairead Connor; Susan Feeney; Tanya Curran; Zoltan Molnar |

EPI_ISL_754246, EPI_ISL_754255, EPI_ISL_754270, EPI_ISL_754271, EPI_ISL_754279, EPI_ISL_754280, EPI_ISL_754281, EPI_ISL_754285, EPI_ISL_754302, EPI_ISL_754314, EPI_ISL_754316, EPI_ISL_754318, EPI_ISL_754330, EPI_ISL_754364, EPI_ISL_754376, EPI_ISL_754380, EPI_ISL_766059, EPI_ISL_766063, EPI_ISL_766068, EPI_ISL_766092, EPI_ISL_766113, EPI_ISL_766114, EPI_ISL_766206, EPI_ISL_766207,
EPI_ISL_766208, EPI_ISL_766209, EPI_ISL_766210, EPI_ISL_766211, EPI_ISL_766212, EPI_ISL_766214, EPI_ISL_766218, EPI_ISL_766220, EPI_ISL_766232, EPI_ISL_766241, EPI_ISL_766245, EPI_ISL_766505, EPI_ISL_766518, EPI_ISL_766522, EPI_ISL_769874, EPI_ISL_769875, EPI_ISL_769876, EPI_ISL_770439, EPI_ISL_770440, EPI_ISL_770441, EPI_ISL_770462, EPI_ISL_770463, EPI_ISL_770464, EPI_ISL_770465,
EPI_ISL_770466, EPI_ISL_770467, EPI_ISL_770468, EPI_ISL_770469, EPI_ISL_791190, EPI_ISL_791191, EPI_ISL_791192, EPI_ISL_791211, EPI_ISL_804218, EPI_ISL_804219, EPI_ISL_804220, EPI_ISL_804221, EPI_ISL_804223, EPI_ISL_804232, EPI_ISL_804233, EPI_ISL_804234, EPI_ISL_804235, EPI_ISL_804236, EPI_ISL_804237, EPI_ISL_804238, EPI_ISL_804239, EPI_ISL_804240, EPI_ISL_804241, EPI_ISL_804250,
EPI_ISL_804253, EPI_ISL_804255, EPI_ISL_804256, EPI_ISL_804257, EPI_ISL_804272, EPI_ISL_804273, EPI_ISL_804274, EPI_ISL_804275, EPI_ISL_804277, EPI_ISL_804278, EPI_ISL_804279, EPI_ISL_804294, EPI_ISL_804295, EPI_ISL_804296, EPI_ISL_804297, EPI_ISL_804298, EPI_ISL_804299, EPI_ISL_804300, EPI_ISL_804301, EPI_ISL_804302, EPI_ISL_804303, EPI_ISL_804304, EPI_ISL_804305, EPI_ISL_804306,
EPI_ISL_804307, EPI_ISL_804308, EPI_ISL_804309, EPI_ISL_804310, EPI_ISL_804311, EPI_ISL_804312, EPI_ISL_804313, EPI_ISL_804314, EPI_ISL_804315, EPI_ISL_804316, EPI_ISL_804317, EPI_ISL_804318, EPI_ISL_804319, EPI_ISL_804320, EPI_ISL_804321, EPI_ISL_804322, EPI_ISL_804323, EPI_ISL_804324, EPI_ISL_804325, EPI_ISL_804326, EPI_ISL_804327, EPI_ISL_804328, EPI_ISL_804329, EPI_ISL_804330,
EPI_ISL_804331, EPI_ISL_804332, EPI_ISL_804333, EPI_ISL_804334, EPI_ISL_804335, EPI_ISL_804336, EPI_ISL_804337, EPI_ISL_804338, EPI_ISL_804339, EPI_ISL_804340, EPI_ISL_804341, EPI_ISL_804342, EPI_ISL_804343, EPI_ISL_804344, EPI_ISL_804345, EPI_ISL_804346, EPI_ISL_804347, EPI_ISL_804348, EPI_ISL_804349, EPI_ISL_804350, EPI_ISL_804351, EPI_ISL_804352, EPI_ISL_804353, EPI_ISL_804354,
EPI_ISL_804355, EPI_ISL_804356, EPI_ISL_804357, EPI_ISL_804358, EPI_ISL_804359, EPI_ISL_804360, EPI_ISL_804361, EPI_ISL_804362, EPI_ISL_804363, EPI_ISL_804364, EPI_ISL_804365, EPI_ISL_804366, EPI_ISL_804367, EPI_ISL_804368, EPI_ISL_804812, EPI_ISL_811120, EPI_ISL_811122, EPI_ISL_825498, EPI_ISL_825501, EPI_ISL_825502, EPI_ISL_825503, EPI_ISL_825504, EPI_ISL_825505, EPI_ISL_825506,

| | | | |
|---|---|---|---|
| EPI_ISL_825507, EPI_ISL_825508, EPI_ISL_825509, EPI_ISL_825510, EPI_ISL_825511, EPI_ISL_825512, EPI_ISL_825513, EPI_ISL_825514, EPI_ISL_825515, EPI_ISL_825516, EPI_ISL_825517, EPI_ISL_825518, EPI_ISL_825519, EPI_ISL_825520, EPI_ISL_825521, EPI_ISL_825522, EPI_ISL_825523, EPI_ISL_825524, EPI_ISL_825525, EPI_ISL_825526, EPI_ISL_825527, EPI_ISL_825528, EPI_ISL_825529, EPI_ISL_825530, EPI_ISL_825531, EPI_ISL_825532, EPI_ISL_825533, EPI_ISL_825546, EPI_ISL_825547, EPI_ISL_825554, EPI_ISL_825555, EPI_ISL_825556, EPI_ISL_825557, EPI_ISL_825558, EPI_ISL_825559, EPI_ISL_825560, EPI_ISL_825561, EPI_ISL_825562, EPI_ISL_825563, EPI_ISL_825564, EPI_ISL_825565, EPI_ISL_825566, EPI_ISL_825567, EPI_ISL_825572, EPI_ISL_825605, EPI_ISL_825606, EPI_ISL_825607, EPI_ISL_825608, EPI_ISL_825609, EPI_ISL_836989, EPI_ISL_836990, EPI_ISL_836991, EPI_ISL_836992, EPI_ISL_836993, EPI_ISL_836994, EPI_ISL_836995, EPI_ISL_836996, EPI_ISL_836997, EPI_ISL_836998, EPI_ISL_836999, EPI_ISL_837000, EPI_ISL_837001, EPI_ISL_837002, EPI_ISL_837003, EPI_ISL_837004, EPI_ISL_837005, EPI_ISL_837006, EPI_ISL_837007, EPI_ISL_837008, EPI_ISL_837009, EPI_ISL_837010, EPI_ISL_837011, EPI_ISL_837012, EPI_ISL_837013, EPI_ISL_837014, EPI_ISL_837015, EPI_ISL_837016, EPI_ISL_837017, EPI_ISL_837018, EPI_ISL_837019, EPI_ISL_837020, EPI_ISL_837021, EPI_ISL_837022, EPI_ISL_837023, EPI_ISL_837024, EPI_ISL_837025, EPI_ISL_837026, EPI_ISL_837027, EPI_ISL_837028, EPI_ISL_837029, EPI_ISL_837030, EPI_ISL_837031, EPI_ISL_837032, EPI_ISL_837033, EPI_ISL_837034, EPI_ISL_837035, EPI_ISL_837036, EPI_ISL_837037, EPI_ISL_837038, EPI_ISL_837039, EPI_ISL_837040, EPI_ISL_837061, EPI_ISL_837062, EPI_ISL_837072, EPI_ISL_837075, EPI_ISL_837079, EPI_ISL_837081, EPI_ISL_837088, EPI_ISL_837089, EPI_ISL_837091, EPI_ISL_837095, EPI_ISL_837103, EPI_ISL_837104, EPI_ISL_837105, EPI_ISL_837120, EPI_ISL_837121, EPI_ISL_837122, EPI_ISL_837123, EPI_ISL_837124, EPI_ISL_837125, EPI_ISL_837126, EPI_ISL_837127, EPI_ISL_837128, EPI_ISL_837129, EPI_ISL_837130, EPI_ISL_837131, EPI_ISL_837132, EPI_ISL_837133, EPI_ISL_837134, EPI_ISL_837189, EPI_ISL_837209, EPI_ISL_837210, EPI_ISL_837211, EPI_ISL_837212, EPI_ISL_837213, EPI_ISL_837214, EPI_ISL_837215, EPI_ISL_837216, EPI_ISL_837217, EPI_ISL_837218, EPI_ISL_837219, EPI_ISL_837220, EPI_ISL_837221, EPI_ISL_837222, EPI_ISL_837223, EPI_ISL_837224, EPI_ISL_837225, EPI_ISL_846592, EPI_ISL_846594, EPI_ISL_855576, EPI_ISL_855580, EPI_ISL_855583, EPI_ISL_855585, EPI_ISL_855586, EPI_ISL_855587, EPI_ISL_860635, EPI_ISL_860636, EPI_ISL_860637, EPI_ISL_860638, EPI_ISL_860639, EPI_ISL_860640, EPI_ISL_860641, EPI_ISL_860687 | | | |
| see above | Respiratory Virus Unit, National Infection Service, Public Health England | COVID-19 Genomics UK (COG-UK) Consortium | PHE Covid Sequencing Team |
| EPI_ISL_1833394, EPI_ISL_1833395, EPI_ISL_1833396, EPI_ISL_1833399, EPI_ISL_1833400, EPI_ISL_1833401, EPI_ISL_1833402, EPI_ISL_1833403, EPI_ISL_1833428 | | | |
| see above | St George's University Hospitals NHS Foundation Trust | COVID-19 Genomics UK (COG-UK) Consortium | Adam Witney; Cassie Pope; Irene Monahan; Joshua Taylor; Ken Laing; NgeeKeong Tan |
| EPI_ISL_866052, EPI_ISL_866123, EPI_ISL_866124, EPI_ISL_866128, EPI_ISL_866129, EPI_ISL_866130, EPI_ISL_866131, EPI_ISL_866137, EPI_ISL_866139, EPI_ISL_866141, EPI_ISL_866145, EPI_ISL_866183, EPI_ISL_920189, EPI_ISL_920192, EPI_ISL_920282, EPI_ISL_920319, EPI_ISL_920320, EPI_ISL_920354, EPI_ISL_920370, EPI_ISL_920462, EPI_ISL_920470, EPI_ISL_920471, EPI_ISL_920475, EPI_ISL_920484, EPI_ISL_920493, EPI_ISL_920496, EPI_ISL_920506, EPI_ISL_920512, EPI_ISL_920515, EPI_ISL_920518, EPI_ISL_920519, EPI_ISL_920528, EPI_ISL_920537, EPI_ISL_920567, EPI_ISL_920569, EPI_ISL_920574, EPI_ISL_920583, EPI_ISL_920629, EPI_ISL_920630, EPI_ISL_920646, EPI_ISL_920653, EPI_ISL_920680, EPI_ISL_920697, EPI_ISL_920706, EPI_ISL_1104830, EPI_ISL_1104876, EPI_ISL_1104894, EPI_ISL_1104903, EPI_ISL_1104913, EPI_ISL_1104922, EPI_ISL_1104927, EPI_ISL_1104932, EPI_ISL_1104951, EPI_ISL_1104958, EPI_ISL_1104963, EPI_ISL_1104969, EPI_ISL_1104977, EPI_ISL_1104979, EPI_ISL_1104983, EPI_ISL_1104984, EPI_ISL_1104987, EPI_ISL_1104989, EPI_ISL_1104993, EPI_ISL_1104996, EPI_ISL_1105002, EPI_ISL_1105003, EPI_ISL_1105006, EPI_ISL_1105007, EPI_ISL_1105012, EPI_ISL_1105016, EPI_ISL_1105019, EPI_ISL_1105020, EPI_ISL_1105021, EPI_ISL_1105024, EPI_ISL_1105031, EPI_ISL_1105032, EPI_ISL_1105036, EPI_ISL_1105037, EPI_ISL_1105099, EPI_ISL_1105103, EPI_ISL_1105104, EPI_ISL_1105116, EPI_ISL_1105117, EPI_ISL_1105122, EPI_ISL_1105123, EPI_ISL_1105127, EPI_ISL_1105128, EPI_ISL_1105132, EPI_ISL_1105133, EPI_ISL_1105140, EPI_ISL_1105143, EPI_ISL_1105261, EPI_ISL_1105282, EPI_ISL_1105294, EPI_ISL_1105296, EPI_ISL_1248368, EPI_ISL_1248369, EPI_ISL_1248380, EPI_ISL_1248391, EPI_ISL_1248399, EPI_ISL_1248403, EPI_ISL_1248425, EPI_ISL_1248436, EPI_ISL_1248450, EPI_ISL_1248455, EPI_ISL_1248456, EPI_ISL_1248460, EPI_ISL_1248461, EPI_ISL_1248464, EPI_ISL_1248468, EPI_ISL_1248472, EPI_ISL_1248475, EPI_ISL_1248479, EPI_ISL_1248483, EPI_ISL_1248489, EPI_ISL_1248490, EPI_ISL_1637020, EPI_ISL_1637021, EPI_ISL_1637022, EPI_ISL_1637023, EPI_ISL_1637024, EPI_ISL_1637025, EPI_ISL_1637026, EPI_ISL_1637027, EPI_ISL_1637053, EPI_ISL_1637055, EPI_ISL_1637057, EPI_ISL_1637062, EPI_ISL_1637063, EPI_ISL_1637064, EPI_ISL_1637066, EPI_ISL_1637069, EPI_ISL_1637070, EPI_ISL_1637073, EPI_ISL_1637080 | | | |
| see above | University College London Hospital | COVID-19 Genomics UK (COG-UK) Consortium | Catherine Houlihan; Dan Frampton; Dr Catherine Houlihan; Dr Daniel Frampton; Dr Judith Heaney; Dr Moira Spyer; Dr Paul Grant and Dr Eleni Nastouli; Judith Heaney; Matthew Byott; Moira Spyer and Eleni Nastouli; Stuart Kirk |
| EPI_ISL_764188, EPI_ISL_813845, EPI_ISL_813846, EPI_ISL_813848, EPI_ISL_813849, EPI_ISL_813850, EPI_ISL_813851, EPI_ISL_813852, EPI_ISL_813853, EPI_ISL_813854, EPI_ISL_813855, EPI_ISL_813856, EPI_ISL_813857, EPI_ISL_813858, EPI_ISL_813859, EPI_ISL_813860, EPI_ISL_813861, EPI_ISL_813862, EPI_ISL_813863, EPI_ISL_813864, EPI_ISL_813865, EPI_ISL_813867, EPI_ISL_813868, EPI_ISL_813869, EPI_ISL_813870, EPI_ISL_813871, EPI_ISL_813872, EPI_ISL_813873, EPI_ISL_813874, EPI_ISL_813875, EPI_ISL_813876, EPI_ISL_813877, EPI_ISL_813878, EPI_ISL_813879, EPI_ISL_813881, EPI_ISL_813883, EPI_ISL_813884, EPI_ISL_813885, EPI_ISL_813886, EPI_ISL_813887, EPI_ISL_813888, EPI_ISL_813889, EPI_ISL_813890, EPI_ISL_813891, EPI_ISL_813892, EPI_ISL_813893, EPI_ISL_813896, EPI_ISL_813897, EPI_ISL_813898, EPI_ISL_813899, EPI_ISL_813900, EPI_ISL_813901, EPI_ISL_813902, EPI_ISL_813903, EPI_ISL_813904, EPI_ISL_813905, EPI_ISL_813906, EPI_ISL_813913, EPI_ISL_813930, EPI_ISL_813932, EPI_ISL_813933, EPI_ISL_813934, EPI_ISL_813935, EPI_ISL_813936, EPI_ISL_813937, EPI_ISL_813938, EPI_ISL_813939, EPI_ISL_813940, EPI_ISL_813941, EPI_ISL_813942, EPI_ISL_813943, EPI_ISL_813944, EPI_ISL_813945, EPI_ISL_813946, EPI_ISL_813947, EPI_ISL_813948, EPI_ISL_813949, EPI_ISL_813950, EPI_ISL_813951, EPI_ISL_813952, EPI_ISL_813953, EPI_ISL_813954, EPI_ISL_813955, EPI_ISL_813956, EPI_ISL_813957, EPI_ISL_813959, EPI_ISL_813960, EPI_ISL_813961, EPI_ISL_813962, EPI_ISL_813963, EPI_ISL_813964, EPI_ISL_813965, EPI_ISL_813966, EPI_ISL_813967, EPI_ISL_813968, EPI_ISL_813969, EPI_ISL_813970, EPI_ISL_819873, EPI_ISL_819874, EPI_ISL_819875, EPI_ISL_819876, EPI_ISL_819878, EPI_ISL_819879, EPI_ISL_819880, EPI_ISL_819881, EPI_ISL_819882, EPI_ISL_819883, EPI_ISL_819884, EPI_ISL_819888, EPI_ISL_819889, EPI_ISL_819892, EPI_ISL_819893, EPI_ISL_819919, EPI_ISL_819920, EPI_ISL_819921, EPI_ISL_819922, EPI_ISL_819923, EPI_ISL_819924, EPI_ISL_819925, EPI_ISL_819926, EPI_ISL_819927, EPI_ISL_819929, EPI_ISL_819930, EPI_ISL_819936, EPI_ISL_819937, EPI_ISL_819938, EPI_ISL_819939, EPI_ISL_819940, EPI_ISL_819941, EPI_ISL_819943, EPI_ISL_819944, EPI_ISL_819946, EPI_ISL_819947, EPI_ISL_819948, EPI_ISL_819949, EPI_ISL_819950, EPI_ISL_819951, EPI_ISL_819952, EPI_ISL_819954, EPI_ISL_819955, EPI_ISL_819957, EPI_ISL_819958, EPI_ISL_819961, EPI_ISL_819962, EPI_ISL_819964, EPI_ISL_819965, EPI_ISL_819966, EPI_ISL_819967, EPI_ISL_819968, EPI_ISL_819969, EPI_ISL_819978, EPI_ISL_819979, EPI_ISL_819980, EPI_ISL_819981, EPI_ISL_819982, EPI_ISL_819983, EPI_ISL_819984, EPI_ISL_819985, EPI_ISL_819986, EPI_ISL_819987, EPI_ISL_819988, EPI_ISL_819989, EPI_ISL_819991, EPI_ISL_819992, EPI_ISL_819993, EPI_ISL_819994, EPI_ISL_819995, EPI_ISL_819997, EPI_ISL_819998, EPI_ISL_819999, EPI_ISL_820000, EPI_ISL_820007, EPI_ISL_820008, EPI_ISL_820009, EPI_ISL_820010, EPI_ISL_820012, EPI_ISL_820013, EPI_ISL_820014, EPI_ISL_820016, EPI_ISL_820017, EPI_ISL_820018, EPI_ISL_820019, EPI_ISL_820020, EPI_ISL_820024, EPI_ISL_820031, EPI_ISL_820032, EPI_ISL_820033, EPI_ISL_820034, EPI_ISL_820035, EPI_ISL_820036, EPI_ISL_820037, EPI_ISL_820038, EPI_ISL_820039, EPI_ISL_820040, EPI_ISL_820045, EPI_ISL_820046, EPI_ISL_820047, EPI_ISL_820048, EPI_ISL_820049, EPI_ISL_820050, EPI_ISL_820051, EPI_ISL_820052, EPI_ISL_820053, EPI_ISL_820054, EPI_ISL_820057, EPI_ISL_820058, EPI_ISL_820059, EPI_ISL_820060, EPI_ISL_820061, EPI_ISL_820063, EPI_ISL_820064, EPI_ISL_820065, EPI_ISL_820066, EPI_ISL_820081, EPI_ISL_820083, EPI_ISL_820084, EPI_ISL_820085, EPI_ISL_820086, EPI_ISL_820087, EPI_ISL_820089, EPI_ISL_820094, EPI_ISL_820099, EPI_ISL_820100, EPI_ISL_820101, EPI_ISL_820109, EPI_ISL_820111, EPI_ISL_820113, EPI_ISL_820114, EPI_ISL_820115, EPI_ISL_820116, EPI_ISL_820117, EPI_ISL_820118, EPI_ISL_820120, EPI_ISL_820121, EPI_ISL_820122, EPI_ISL_820123, EPI_ISL_820125, EPI_ISL_820127, EPI_ISL_820128, EPI_ISL_820138, EPI_ISL_820204, EPI_ISL_820206, EPI_ISL_820209, EPI_ISL_820211, EPI_ISL_820213, EPI_ISL_820216, EPI_ISL_820218, EPI_ISL_820226, EPI_ISL_820229, EPI_ISL_820236, EPI_ISL_820246, EPI_ISL_820249, EPI_ISL_820251, EPI_ISL_820256, EPI_ISL_820259, EPI_ISL_820264, EPI_ISL_820266, EPI_ISL_820268, EPI_ISL_820271, EPI_ISL_820273, EPI_ISL_820276, EPI_ISL_820282, EPI_ISL_820309, EPI_ISL_820314, EPI_ISL_820316, EPI_ISL_820318, EPI_ISL_820321, EPI_ISL_838734, EPI_ISL_838735, EPI_ISL_838736, EPI_ISL_838737, EPI_ISL_838738, EPI_ISL_838740, EPI_ISL_838741, EPI_ISL_838742, EPI_ISL_838743, EPI_ISL_838745, EPI_ISL_838746, EPI_ISL_838747, EPI_ISL_838748, EPI_ISL_838749, EPI_ISL_838750, EPI_ISL_838751, EPI_ISL_838752, EPI_ISL_838753, EPI_ISL_838754, EPI_ISL_838755, EPI_ISL_838756, EPI_ISL_838757, EPI_ISL_838758, EPI_ISL_838759, EPI_ISL_838760, EPI_ISL_838761, EPI_ISL_838762, EPI_ISL_838763, EPI_ISL_838764, EPI_ISL_838765, EPI_ISL_838766, EPI_ISL_838772, EPI_ISL_838774, EPI_ISL_838779, EPI_ISL_838780, EPI_ISL_838781, EPI_ISL_838782, EPI_ISL_838783, EPI_ISL_838784, EPI_ISL_838785, EPI_ISL_838786, EPI_ISL_838787, EPI_ISL_838788, EPI_ISL_838789, EPI_ISL_838790, EPI_ISL_838791, EPI_ISL_838792, EPI_ISL_838793, EPI_ISL_838794, EPI_ISL_838795, EPI_ISL_838796, EPI_ISL_838797, EPI_ISL_839309, EPI_ISL_839311, EPI_ISL_839312, EPI_ISL_839313, EPI_ISL_839314, EPI_ISL_839315, EPI_ISL_839318, EPI_ISL_839319, EPI_ISL_839320, EPI_ISL_839321, EPI_ISL_839322, EPI_ISL_839323, EPI_ISL_839324, EPI_ISL_839325, EPI_ISL_839326, EPI_ISL_839327, EPI_ISL_839328, EPI_ISL_839329, EPI_ISL_865688, EPI_ISL_865689, EPI_ISL_865739, EPI_ISL_865742, EPI_ISL_865744, EPI_ISL_865748, EPI_ISL_865749, EPI_ISL_865750, EPI_ISL_865751, EPI_ISL_865752, EPI_ISL_865753, EPI_ISL_865754, EPI_ISL_865755, EPI_ISL_865756, EPI_ISL_865757, EPI_ISL_865758, EPI_ISL_865759, EPI_ISL_865760, EPI_ISL_865761, EPI_ISL_865762, EPI_ISL_865763, EPI_ISL_865764, EPI_ISL_865765, EPI_ISL_865766, EPI_ISL_865767, EPI_ISL_865768, EPI_ISL_865769, EPI_ISL_865770, EPI_ISL_865771, EPI_ISL_865772, EPI_ISL_865773, EPI_ISL_865774, EPI_ISL_865775, EPI_ISL_865776, EPI_ISL_865777, EPI_ISL_865778, EPI_ISL_865779, EPI_ISL_865780, EPI_ISL_865781, EPI_ISL_865782, EPI_ISL_865783, EPI_ISL_865784, EPI_ISL_865785, EPI_ISL_865786, EPI_ISL_865787, EPI_ISL_865788, EPI_ISL_865789, EPI_ISL_865790, EPI_ISL_865791, EPI_ISL_865792, EPI_ISL_865793, EPI_ISL_865794, EPI_ISL_865795, EPI_ISL_865796, EPI_ISL_865797, EPI_ISL_865798, EPI_ISL_865799, EPI_ISL_865800, EPI_ISL_865801, EPI_ISL_865802, EPI_ISL_865803, EPI_ISL_865804, EPI_ISL_865805, EPI_ISL_865806, EPI_ISL_865807, EPI_ISL_865808, EPI_ISL_865809, EPI_ISL_865810, EPI_ISL_865811, EPI_ISL_865813, EPI_ISL_865814, EPI_ISL_865815, EPI_ISL_865818, EPI_ISL_865837, EPI_ISL_919728, EPI_ISL_919729, EPI_ISL_919730, EPI_ISL_919731, EPI_ISL_919732, EPI_ISL_919733, EPI_ISL_919734, EPI_ISL_919735, EPI_ISL_919736, EPI_ISL_919738, EPI_ISL_919739, EPI_ISL_919740, EPI_ISL_919741, EPI_ISL_919742, EPI_ISL_919743, EPI_ISL_919744, EPI_ISL_919745, EPI_ISL_919746, EPI_ISL_919747, EPI_ISL_919748, EPI_ISL_919749, EPI_ISL_919755, EPI_ISL_919756, EPI_ISL_919757, EPI_ISL_919758, EPI_ISL_919759, EPI_ISL_919760, EPI_ISL_919761, EPI_ISL_919762, EPI_ISL_919763, EPI_ISL_919764, EPI_ISL_919765, EPI_ISL_919766, EPI_ISL_919767, EPI_ISL_919768, EPI_ISL_919769, EPI_ISL_919770, EPI_ISL_919771, EPI_ISL_919772, EPI_ISL_919773, EPI_ISL_919774, EPI_ISL_919775, EPI_ISL_920841, EPI_ISL_920854, EPI_ISL_920855, EPI_ISL_949747, EPI_ISL_950105, EPI_ISL_950138, EPI_ISL_950145, EPI_ISL_950168, EPI_ISL_997572, EPI_ISL_997573, EPI_ISL_997574, EPI_ISL_997575, EPI_ISL_997576, EPI_ISL_997577, EPI_ISL_997578, EPI_ISL_1308971, EPI_ISL_1308972, EPI_ISL_1308973, EPI_ISL_1308974, EPI_ISL_1308975, EPI_ISL_1308976, EPI_ISL_1308977, EPI_ISL_1308978, EPI_ISL_1309028, EPI_ISL_1309029, EPI_ISL_1309030, EPI_ISL_1309031, EPI_ISL_1309033, EPI_ISL_1309034, EPI_ISL_1309035, EPI_ISL_1309036, EPI_ISL_1309047, EPI_ISL_1309048, EPI_ISL_1309068, EPI_ISL_1309119, EPI_ISL_1309125, EPI_ISL_1386831, EPI_ISL_1386832, EPI_ISL_1386833, EPI_ISL_1386834, EPI_ISL_1386835, EPI_ISL_1386836, EPI_ISL_1386860, EPI_ISL_1386861, EPI_ISL_1386877, EPI_ISL_1386880, EPI_ISL_1386884, EPI_ISL_1386887, EPI_ISL_1386888, EPI_ISL_1386891, EPI_ISL_1386897, EPI_ISL_1386898, EPI_ISL_1386902, EPI_ISL_1386905, EPI_ISL_1474836, EPI_ISL_1474855, EPI_ISL_1474858, EPI_ISL_1637108, EPI_ISL_1637112, EPI_ISL_1637122, EPI_ISL_1730355, EPI_ISL_1730356 | | | |
| see above | University College London, Great Ormond Street Hospital for Children NHS Foundation Trust, Imperial College Healthcare NHS Trust | COVID-19 Genomics UK (COG-UK) Consortium | Alison Holmes; Charlotte Williams; Helena Tutill; Jacqueline Findlay; James Price; Judith Breuer; Julianne Brown; Kathryn Harris; Leysa Forrest; Mark Kristiansen; Paola Niola; Paola Resende Silva; Patricia Dyal; Paul Randell; Rachel Williams; Samuel Weeks; Sergi Castellano; Sunando Roy; Tony Brooks; Yasmin Panchbhaya |
| EPI_ISL_1047930, EPI_ISL_1047932, EPI_ISL_1047933, EPI_ISL_1047935, EPI_ISL_1047946, EPI_ISL_1047947, EPI_ISL_1047948, EPI_ISL_1047949, EPI_ISL_1047950, EPI_ISL_1047951, EPI_ISL_1050354, EPI_ISL_2125932 | | | |
| see above | University Hospitals Of Leicester NHS Trust and DeepSeq Nottingham | COVID-19 Genomics UK (COG-UK) Consortium | Christopher Holmes; Christopher Moore; Fei Sang; Joeseph Chappell; Johnny Debebe; Jonathan Ball; Julian Tang; Karlie Fallon; Matthew Carlisle; Matthew Loose; Nadine Holmes; Patrick McClure; Paul Bird; Thomas Helmer; Victoria Wright |
| EPI_ISL_813094, EPI_ISL_813095, EPI_ISL_813096, EPI_ISL_813097, EPI_ISL_813098, EPI_ISL_813099, EPI_ISL_813100, EPI_ISL_813101, EPI_ISL_813102, EPI_ISL_813103, EPI_ISL_813104, EPI_ISL_813105, EPI_ISL_813106, EPI_ISL_813107, EPI_ISL_813108, EPI_ISL_813109, EPI_ISL_813110, EPI_ISL_813111, EPI_ISL_813112, EPI_ISL_813113, EPI_ISL_813114, EPI_ISL_813115, EPI_ISL_813117, EPI_ISL_813118, EPI_ISL_813119, EPI_ISL_813120, EPI_ISL_813121, EPI_ISL_813122, EPI_ISL_813123, EPI_ISL_813124, EPI_ISL_813125, EPI_ISL_813126, EPI_ISL_813127, EPI_ISL_813128, EPI_ISL_813129, EPI_ISL_813130, EPI_ISL_813131, EPI_ISL_813132, EPI_ISL_813133, EPI_ISL_813134, EPI_ISL_813135, EPI_ISL_813136, EPI_ISL_813137, EPI_ISL_813138, EPI_ISL_813139, EPI_ISL_813140, EPI_ISL_813141, EPI_ISL_813142, EPI_ISL_813143, EPI_ISL_918758, EPI_ISL_918759, EPI_ISL_918850, EPI_ISL_918851, EPI_ISL_918852, EPI_ISL_918853, EPI_ISL_918854, EPI_ISL_918855, EPI_ISL_918858, EPI_ISL_918955, EPI_ISL_918956 | | | |
| see above | University of Birmingham | COVID-19 Genomics UK (COG-UK) Consortium | Alex Richter; Andrew Bosworth; Andrew Bosworth. Queen Elizabeth Hospital: Anna Casey; Andrew D Beggs PHE Heartlands Lab: Husam Osman; Charlotte Poxon; Institute of Microbiology; Joanne Stockton; Josh Quick; Kasun Wanigasooriya; Mike Kidd; Nicholas Loman. University of Birmingham Testing Laboratory: Celina M Whalley; Oliver Pickles; Radoslaw Poplawski; Samuel Nicholls; University of Birmingham: Claire McMurray; Will Rowe |
| EPI_ISL_813453, EPI_ISL_813454, EPI_ISL_813455, EPI_ISL_813456, EPI_ISL_813467, EPI_ISL_813468, EPI_ISL_813469, EPI_ISL_813470, EPI_ISL_813471, EPI_ISL_813472, EPI_ISL_813473, EPI_ISL_813474, EPI_ISL_813475, EPI_ISL_813476, EPI_ISL_813477, EPI_ISL_813478, EPI_ISL_813479, EPI_ISL_813480, EPI_ISL_813481, EPI_ISL_813482, EPI_ISL_813483, EPI_ISL_813484, EPI_ISL_813485, EPI_ISL_813486, EPI_ISL_813487, EPI_ISL_813488, EPI_ISL_813489, EPI_ISL_813490, EPI_ISL_813491, EPI_ISL_813492, EPI_ISL_813493, EPI_ISL_813494, EPI_ISL_813495, EPI_ISL_813496, EPI_ISL_813497, EPI_ISL_813498, EPI_ISL_813499, EPI_ISL_813500, EPI_ISL_813501, EPI_ISL_813502, EPI_ISL_813503, EPI_ISL_813504, EPI_ISL_813505, EPI_ISL_813506, EPI_ISL_813507, EPI_ISL_813508, EPI_ISL_813509, EPI_ISL_813510, EPI_ISL_813511, EPI_ISL_813512, EPI_ISL_813513, EPI_ISL_813514, EPI_ISL_813515, EPI_ISL_813516, EPI_ISL_813517, EPI_ISL_813518, EPI_ISL_813519, EPI_ISL_813520, EPI_ISL_813521, EPI_ISL_813522, EPI_ISL_813523, EPI_ISL_813524, EPI_ISL_813525, EPI_ISL_813526, EPI_ISL_813527, EPI_ISL_813528, EPI_ISL_813529, EPI_ISL_813530, EPI_ISL_813531, EPI_ISL_813532, EPI_ISL_813533, EPI_ISL_813534, EPI_ISL_813535, EPI_ISL_813536, EPI_ISL_813537, EPI_ISL_813538, EPI_ISL_813539, EPI_ISL_813540, EPI_ISL_813541, EPI_ISL_813542, EPI_ISL_813543, EPI_ISL_813544, EPI_ISL_813545, EPI_ISL_813546, EPI_ISL_813547, EPI_ISL_813548, EPI_ISL_813549, EPI_ISL_813550, EPI_ISL_813551, EPI_ISL_813552, EPI_ISL_813553, EPI_ISL_813554, EPI_ISL_813555, EPI_ISL_813556, EPI_ISL_813557, EPI_ISL_813558, EPI_ISL_813559, EPI_ISL_813560, EPI_ISL_813561, EPI_ISL_813562, EPI_ISL_813564, EPI_ISL_813565, EPI_ISL_813566, EPI_ISL_813567, EPI_ISL_813568, EPI_ISL_813569, EPI_ISL_813570, EPI_ISL_813571, EPI_ISL_813572, EPI_ISL_813573, EPI_ISL_813574, EPI_ISL_813575, EPI_ISL_813576, EPI_ISL_813577, EPI_ISL_813578, EPI_ISL_813579, EPI_ISL_813580, EPI_ISL_813581, EPI_ISL_813582, EPI_ISL_813583, EPI_ISL_813584, EPI_ISL_813585, EPI_ISL_813586, EPI_ISL_813587, EPI_ISL_813588, EPI_ISL_813589, EPI_ISL_813590, EPI_ISL_813591, EPI_ISL_813592, EPI_ISL_813593, EPI_ISL_813594, EPI_ISL_813595, EPI_ISL_813596, EPI_ISL_813597, EPI_ISL_813598, EPI_ISL_813600, EPI_ISL_813601, EPI_ISL_813602, EPI_ISL_813603, EPI_ISL_813604, EPI_ISL_813605, EPI_ISL_813606, EPI_ISL_813607, EPI_ISL_813609, EPI_ISL_813610, EPI_ISL_813611, EPI_ISL_813612, EPI_ISL_813613, EPI_ISL_813614, EPI_ISL_813615, EPI_ISL_813616, EPI_ISL_813617, EPI_ISL_813618, EPI_ISL_813619, EPI_ISL_813620, EPI_ISL_813621, EPI_ISL_813622, EPI_ISL_813624, EPI_ISL_813637, EPI_ISL_996485, EPI_ISL_996569, EPI_ISL_996591, EPI_ISL_997288, EPI_ISL_997289, EPI_ISL_997290, EPI_ISL_997291, EPI_ISL_997292, EPI_ISL_997293, EPI_ISL_997294, EPI_ISL_997295, EPI_ISL_997296, EPI_ISL_997297, EPI_ISL_997298, EPI_ISL_997299, EPI_ISL_997300, EPI_ISL_997301, EPI_ISL_997302, EPI_ISL_997303, EPI_ISL_997304, EPI_ISL_997305, EPI_ISL_997306, EPI_ISL_997319, EPI_ISL_997340, EPI_ISL_997341, EPI_ISL_997342, EPI_ISL_997343, EPI_ISL_997344, EPI_ISL_997345, EPI_ISL_997346, EPI_ISL_997348, EPI_ISL_997349, EPI_ISL_997350, EPI_ISL_997351, EPI_ISL_997352, EPI_ISL_997353, EPI_ISL_997354, EPI_ISL_997355, EPI_ISL_997356, EPI_ISL_997357, EPI_ISL_997358, EPI_ISL_997359, EPI_ISL_997360, EPI_ISL_997361, EPI_ISL_997362, EPI_ISL_997363, EPI_ISL_997364, EPI_ISL_997365, EPI_ISL_1308596, EPI_ISL_1545822 | | | |
| see above | University of Exeter | COVID-19 Genomics UK (COG-UK) Consortium | Aaron Jeffries; Audrey Farbos; Ben Temperton; Jane Masoli; Joanna Warwick-Dugdale; Michelle Michelsen; Robyn Manley; Stephen Michell |
| EPI_ISL_997117, EPI_ISL_997122, EPI_ISL_1308860 | Virology Department, Royal Infirmary of Edinburgh, NHS Lothian / School of Biological Sciences, University of Edinburgh | COVID-19 Genomics UK (COG-UK) Consortium | Colquhoun R; Cotton S; Dewar R; Hill V; Jackson B; McCrone JT; McHugh M; O'Toole Á; Rambaut A; Rooke S; Scher E; Templeton K; Yu X |
| EPI_ISL_763447, EPI_ISL_763548, EPI_ISL_763694, EPI_ISL_763695, EPI_ISL_763817, EPI_ISL_764195, EPI_ISL_764221, EPI_ISL_764278, EPI_ISL_764279, EPI_ISL_764361, EPI_ISL_764362, EPI_ISL_764363, EPI_ISL_764364, EPI_ISL_764365, EPI_ISL_764366, EPI_ISL_764367, EPI_ISL_764368, EPI_ISL_764369, EPI_ISL_764370, EPI_ISL_764371, EPI_ISL_764372, EPI_ISL_838240, EPI_ISL_838241, EPI_ISL_838242, EPI_ISL_838243, EPI_ISL_838244, EPI_ISL_838245, EPI_ISL_838246, EPI_ISL_838247, EPI_ISL_838248, EPI_ISL_838249, EPI_ISL_838250, EPI_ISL_838251, EPI_ISL_838252, EPI_ISL_838253, EPI_ISL_838254, EPI_ISL_838255, EPI_ISL_838256, EPI_ISL_838257, EPI_ISL_838258, EPI_ISL_838259, EPI_ISL_838260, EPI_ISL_838261, EPI_ISL_838262, EPI_ISL_838263, EPI_ISL_838264, EPI_ISL_838265, EPI_ISL_838266, EPI_ISL_838267, EPI_ISL_865079, EPI_ISL_865083, EPI_ISL_865084, EPI_ISL_865085, EPI_ISL_865086, EPI_ISL_865088, EPI_ISL_865090, EPI_ISL_865091, EPI_ISL_865142, EPI_ISL_865146, EPI_ISL_865147, EPI_ISL_865148, EPI_ISL_919343, EPI_ISL_919344, EPI_ISL_919378, EPI_ISL_919379, EPI_ISL_919406 | | | |
| see above | Virology Department, Royal Infirmary of Edinburgh, NHS Lothian / School of Biological Sciences, University of Edinburgh / Institute of Genetics and Molecular Medicine, University of Edinburgh | COVID-19 Genomics UK (COG-UK) Consortium | Balcaza C; Colquhoun R; Dewar R; Gallagher M; Hill V; Jackson B; McCrone JT; McHugh M; O'Toole Á; Rambaut A; Rooke S; Scher E; Templeton K; Williams TC; Yu X |
| EPI_ISL_741938, EPI_ISL_741982, EPI_ISL_763368, EPI_ISL_763369, EPI_ISL_763394, EPI_ISL_763415, EPI_ISL_763424, EPI_ISL_763439, EPI_ISL_763499, EPI_ISL_763504, EPI_ISL_763514, EPI_ISL_763632, EPI_ISL_763636, EPI_ISL_763731, EPI_ISL_763736, EPI_ISL_763842, EPI_ISL_763866, EPI_ISL_763867, EPI_ISL_763870, EPI_ISL_764099, EPI_ISL_764103, EPI_ISL_764105, EPI_ISL_764106, EPI_ISL_764110, EPI_ISL_764112, EPI_ISL_764144, EPI_ISL_764145, EPI_ISL_764149, EPI_ISL_764153, EPI_ISL_764167, EPI_ISL_764210, EPI_ISL_764213, EPI_ISL_764216, EPI_ISL_764231, EPI_ISL_764239, EPI_ISL_765150, EPI_ISL_765151, EPI_ISL_765154, EPI_ISL_765155, EPI_ISL_765163, EPI_ISL_765164, EPI_ISL_765169, EPI_ISL_765170, EPI_ISL_765171, EPI_ISL_765172, EPI_ISL_765173, EPI_ISL_765174, EPI_ISL_765184, EPI_ISL_765186, EPI_ISL_765187, EPI_ISL_765190, EPI_ISL_765191, EPI_ISL_765195, EPI_ISL_765196, EPI_ISL_765197, EPI_ISL_765202, EPI_ISL_765203, EPI_ISL_765204, EPI_ISL_765205, EPI_ISL_765206, EPI_ISL_765208, EPI_ISL_816230, EPI_ISL_816231, EPI_ISL_816236, EPI_ISL_816239, EPI_ISL_816242, EPI_ISL_816247, EPI_ISL_816252, EPI_ISL_816255, EPI_ISL_816262, EPI_ISL_816268, EPI_ISL_816271, EPI_ISL_816273, EPI_ISL_816280, EPI_ISL_816283, EPI_ISL_816288, EPI_ISL_816289, EPI_ISL_816290, EPI_ISL_816292, EPI_ISL_816302, EPI_ISL_816308, EPI_ISL_816310, EPI_ISL_816313, EPI_ISL_816316, EPI_ISL_816320, EPI_ISL_816321, EPI_ISL_816332, EPI_ISL_816335, EPI_ISL_816337, EPI_ISL_816339, EPI_ISL_816340, EPI_ISL_816343, EPI_ISL_816345, EPI_ISL_816350, EPI_ISL_816351, EPI_ISL_816352, EPI_ISL_816357, EPI_ISL_816359, EPI_ISL_816361, EPI_ISL_816365, EPI_ISL_816366, EPI_ISL_816372, EPI_ISL_816374, EPI_ISL_816379, EPI_ISL_816380, EPI_ISL_816381, EPI_ISL_816382, EPI_ISL_816384, EPI_ISL_816386, EPI_ISL_816392, EPI_ISL_816398, EPI_ISL_816401, EPI_ISL_816411, EPI_ISL_816412, EPI_ISL_816414, EPI_ISL_816417, EPI_ISL_816421, EPI_ISL_816425, EPI_ISL_816429, EPI_ISL_816430, EPI_ISL_816435, EPI_ISL_816447, EPI_ISL_816456, EPI_ISL_816457, EPI_ISL_816461, EPI_ISL_816467, EPI_ISL_816474, EPI_ISL_816478, EPI_ISL_816480, EPI_ISL_816491, EPI_ISL_816497, EPI_ISL_816500, EPI_ISL_816503, EPI_ISL_816505, EPI_ISL_816507, EPI_ISL_816513, EPI_ISL_816521, EPI_ISL_816531, EPI_ISL_816533, EPI_ISL_816534, EPI_ISL_816537, EPI_ISL_816540, EPI_ISL_816542, EPI_ISL_816548, EPI_ISL_816549, EPI_ISL_816550, EPI_ISL_816557, EPI_ISL_816558, EPI_ISL_816560, EPI_ISL_816562, EPI_ISL_816563, EPI_ISL_816575, EPI_ISL_816577, EPI_ISL_816578, EPI_ISL_816585, EPI_ISL_816586, EPI_ISL_816589, EPI_ISL_816592, EPI_ISL_816599, EPI_ISL_816600, EPI_ISL_816605, EPI_ISL_816606, EPI_ISL_816607, EPI_ISL_816609, EPI_ISL_816616, EPI_ISL_816619, EPI_ISL_816620, EPI_ISL_816621, EPI_ISL_816625, EPI_ISL_816631, EPI_ISL_816635, EPI_ISL_816637, EPI_ISL_816638, EPI_ISL_816647, EPI_ISL_816651, EPI_ISL_842214, EPI_ISL_842217, EPI_ISL_842218, EPI_ISL_842228, EPI_ISL_842230, EPI_ISL_842232, EPI_ISL_842233, EPI_ISL_842235, EPI_ISL_842248, EPI_ISL_842254, EPI_ISL_842255, EPI_ISL_842256, EPI_ISL_842263, EPI_ISL_842264, EPI_ISL_842266, EPI_ISL_842270, EPI_ISL_842272, EPI_ISL_842281, EPI_ISL_842282, EPI_ISL_842291, EPI_ISL_842292, EPI_ISL_842295, EPI_ISL_842297, EPI_ISL_842306, EPI_ISL_842316, EPI_ISL_842318, EPI_ISL_842319, EPI_ISL_842325, EPI_ISL_842335, EPI_ISL_868372, EPI_ISL_868411, EPI_ISL_868414, EPI_ISL_868419, EPI_ISL_868422, EPI_ISL_868436, EPI_ISL_868438, EPI_ISL_868452, EPI_ISL_868475, EPI_ISL_868483, EPI_ISL_868512, EPI_ISL_868559, EPI_ISL_868570, EPI_ISL_868576, EPI_ISL_868577, EPI_ISL_868614, EPI_ISL_868628, EPI_ISL_868635, EPI_ISL_868645, EPI_ISL_868649, EPI_ISL_868669, EPI_ISL_868673, EPI_ISL_924323, EPI_ISL_924357 | | | |
| see above | Virology Department, Sheffield Teaching Hospitals NHS Foundation Trust/Department of Infection, Immunity and Cardiovascular Disease, The Medical School, University of Sheffield | COVID-19 Genomics UK (COG-UK) Consortium | Adri Angyal; Alex Keeley; Benjamin Lindsey; Cariad Evans; Danielle Groves; Dave Partridge; Katie Johnson; Laura Carrilero; Luke Green; Matthew Parker; Matthew Wyles; Mehmet Yavuz; Mohammad Raza; Nikki Smith; Paul Parsons; Rachel Tucker; Rebecca Brown; Thushan de Silva |
| EPI_ISL_814275, EPI_ISL_814280, EPI_ISL_814281, EPI_ISL_814283, EPI_ISL_814291, EPI_ISL_814301, EPI_ISL_814305, EPI_ISL_814308, EPI_ISL_814311, EPI_ISL_814314, EPI_ISL_814318, EPI_ISL_814325, EPI_ISL_814330, EPI_ISL_814332, EPI_ISL_814336, EPI_ISL_814338, EPI_ISL_814342, EPI_ISL_814347, EPI_ISL_814350, EPI_ISL_814364, EPI_ISL_814374, EPI_ISL_814376, EPI_ISL_814379, EPI_ISL_814381, EPI_ISL_814382, EPI_ISL_814385, EPI_ISL_814388, EPI_ISL_814398, EPI_ISL_814412, EPI_ISL_814423, EPI_ISL_814469, EPI_ISL_814470, EPI_ISL_814471, EPI_ISL_814487, EPI_ISL_814488, EPI_ISL_814489, EPI_ISL_814490, EPI_ISL_814491, EPI_ISL_814502, EPI_ISL_814503, EPI_ISL_814504, EPI_ISL_814505, EPI_ISL_814506, EPI_ISL_814507, EPI_ISL_814508, EPI_ISL_814509, EPI_ISL_814510, EPI_ISL_814515, EPI_ISL_814527, EPI_ISL_814528, EPI_ISL_814541, EPI_ISL_814554, EPI_ISL_814567, EPI_ISL_814574, EPI_ISL_814575, EPI_ISL_814580, EPI_ISL_814582, EPI_ISL_814585, EPI_ISL_814586, EPI_ISL_814588, EPI_ISL_814589, EPI_ISL_814596, EPI_ISL_814598, EPI_ISL_814599, EPI_ISL_814600, EPI_ISL_814601, EPI_ISL_814602, EPI_ISL_814757, EPI_ISL_814758, EPI_ISL_814759, EPI_ISL_814760, EPI_ISL_814761, EPI_ISL_814762, EPI_ISL_814763, EPI_ISL_814973, EPI_ISL_814974, EPI_ISL_814975, EPI_ISL_814976, EPI_ISL_814977, EPI_ISL_814978, EPI_ISL_814979, EPI_ISL_814980, EPI_ISL_814981, EPI_ISL_814982, EPI_ISL_814983, EPI_ISL_814984, EPI_ISL_814988, EPI_ISL_815017, EPI_ISL_815018, EPI_ISL_815019, EPI_ISL_815020, EPI_ISL_815021, EPI_ISL_815022, EPI_ISL_815023, EPI_ISL_815024, EPI_ISL_815025, EPI_ISL_815026, EPI_ISL_815027, EPI_ISL_815028, EPI_ISL_815029, EPI_ISL_815030, EPI_ISL_815031, EPI_ISL_815032, EPI_ISL_815045, EPI_ISL_815046, EPI_ISL_815047, EPI_ISL_815051, EPI_ISL_815062, EPI_ISL_815063, EPI_ISL_815064, EPI_ISL_815065, EPI_ISL_815066, EPI_ISL_815067, EPI_ISL_815068, EPI_ISL_815069, EPI_ISL_815070, EPI_ISL_815071, EPI_ISL_815076, EPI_ISL_815077, EPI_ISL_815078, EPI_ISL_815079, EPI_ISL_815080, EPI_ISL_815081, EPI_ISL_815082, EPI_ISL_815083, EPI_ISL_815084, EPI_ISL_815085, EPI_ISL_815086, EPI_ISL_815087, EPI_ISL_815137, EPI_ISL_815144, EPI_ISL_815146, EPI_ISL_815150, EPI_ISL_815171, EPI_ISL_815185, EPI_ISL_815186, EPI_ISL_815187, EPI_ISL_815188, EPI_ISL_815189, EPI_ISL_815190, EPI_ISL_815191, EPI_ISL_815192, EPI_ISL_815193, EPI_ISL_815194, EPI_ISL_815195, EPI_ISL_815196, EPI_ISL_815197, EPI_ISL_815198, EPI_ISL_815199, EPI_ISL_815200, EPI_ISL_815201, EPI_ISL_815202, EPI_ISL_815203, EPI_ISL_815204, EPI_ISL_815205, EPI_ISL_815207, EPI_ISL_815208, EPI_ISL_815209, EPI_ISL_815210, EPI_ISL_815211, EPI_ISL_815212, EPI_ISL_815213, EPI_ISL_815214, EPI_ISL_815215, EPI_ISL_815216, EPI_ISL_815217, EPI_ISL_815218, EPI_ISL_815219, EPI_ISL_815220, EPI_ISL_815221, EPI_ISL_815222, EPI_ISL_815223, EPI_ISL_815224, EPI_ISL_815225, EPI_ISL_815226, EPI_ISL_815227, EPI_ISL_815228, EPI_ISL_815229, EPI_ISL_815230, EPI_ISL_815231, EPI_ISL_815232, EPI_ISL_815233, EPI_ISL_815234, EPI_ISL_815235, EPI_ISL_815236, EPI_ISL_815237, EPI_ISL_815238, EPI_ISL_815239, EPI_ISL_815240, EPI_ISL_815241, EPI_ISL_815242, EPI_ISL_815243, EPI_ISL_822366, EPI_ISL_822367, EPI_ISL_822368, EPI_ISL_822369, EPI_ISL_822370, EPI_ISL_822371, EPI_ISL_822372, EPI_ISL_822373, EPI_ISL_822374, EPI_ISL_822375, EPI_ISL_822376, EPI_ISL_822377, EPI_ISL_822378, EPI_ISL_822379, EPI_ISL_822380, EPI_ISL_822381, EPI_ISL_822382, EPI_ISL_822383, EPI_ISL_822384, EPI_ISL_822385, EPI_ISL_822386, EPI_ISL_822387, EPI_ISL_822388, EPI_ISL_822389, EPI_ISL_822390, EPI_ISL_822391, EPI_ISL_822392, EPI_ISL_822393, EPI_ISL_822394, EPI_ISL_822395, EPI_ISL_822396, EPI_ISL_822397, EPI_ISL_822398, EPI_ISL_822399, EPI_ISL_822400, EPI_ISL_822401, EPI_ISL_822402, EPI_ISL_822403, EPI_ISL_822404, EPI_ISL_822405, EPI_ISL_822406, EPI_ISL_822407, EPI_ISL_822408, EPI_ISL_822409, EPI_ISL_822410, EPI_ISL_822411, EPI_ISL_822412, EPI_ISL_822413, EPI_ISL_822414, EPI_ISL_822415, EPI_ISL_822416, EPI_ISL_822417, EPI_ISL_822418, EPI_ISL_822419, EPI_ISL_822420, EPI_ISL_822421, EPI_ISL_822422, EPI_ISL_822423, EPI_ISL_822424, EPI_ISL_822425, EPI_ISL_822426, EPI_ISL_822427, EPI_ISL_822428, EPI_ISL_822429, EPI_ISL_822430, EPI_ISL_822431, EPI_ISL_822432, EPI_ISL_822433, EPI_ISL_822434, EPI_ISL_822435, EPI_ISL_822436, EPI_ISL_822437, EPI_ISL_822438, EPI_ISL_822439, EPI_ISL_822440, EPI_ISL_822441, EPI_ISL_822442, EPI_ISL_822443, EPI_ISL_822444, EPI_ISL_822445, EPI_ISL_822446, EPI_ISL_822447, EPI_ISL_822448, EPI_ISL_822449, EPI_ISL_822450, EPI_ISL_822451, EPI_ISL_822452, EPI_ISL_822453, EPI_ISL_822454, EPI_ISL_822455, EPI_ISL_822456, EPI_ISL_822457, EPI_ISL_822458, EPI_ISL_822459, EPI_ISL_822460, EPI_ISL_822461, EPI_ISL_822462, EPI_ISL_822463, EPI_ISL_822464, EPI_ISL_822466, EPI_ISL_822467, EPI_ISL_822468, EPI_ISL_822469, EPI_ISL_822470, EPI_ISL_822471, EPI_ISL_822472, EPI_ISL_822473, EPI_ISL_822474, EPI_ISL_822475, EPI_ISL_822476, EPI_ISL_822477, EPI_ISL_822478, EPI_ISL_822479, EPI_ISL_822480, EPI_ISL_822481, EPI_ISL_822482, EPI_ISL_822483, EPI_ISL_822484, EPI_ISL_822485, EPI_ISL_822486, EPI_ISL_822487, EPI_ISL_822488, EPI_ISL_822489, EPI_ISL_822490, EPI_ISL_822491, EPI_ISL_822492, EPI_ISL_822493, EPI_ISL_822494, EPI_ISL_822495, EPI_ISL_822496, EPI_ISL_822497, EPI_ISL_822498, EPI_ISL_822499, EPI_ISL_822500, EPI_ISL_822501, EPI_ISL_822502, EPI_ISL_822503, EPI_ISL_822504, EPI_ISL_822505, EPI_ISL_822506, EPI_ISL_822507, EPI_ISL_822508, EPI_ISL_822509, EPI_ISL_822510, EPI_ISL_822511, EPI_ISL_822512, EPI_ISL_822513, EPI_ISL_822514, EPI_ISL_822515, EPI_ISL_822516, EPI_ISL_822517, EPI_ISL_822518, EPI_ISL_822519, EPI_ISL_822520, EPI_ISL_822521, EPI_ISL_822522, EPI_ISL_822523, EPI_ISL_822524, EPI_ISL_822525, EPI_ISL_822526, EPI_ISL_822527, EPI_ISL_822528, EPI_ISL_822529, EPI_ISL_822530, EPI_ISL_822531, EPI_ISL_822532, EPI_ISL_822533, EPI_ISL_822534, EPI_ISL_822535, EPI_ISL_822536, EPI_ISL_822537, EPI_ISL_822538, EPI_ISL_822539, EPI_ISL_822540, EPI_ISL_822541, EPI_ISL_822542, EPI_ISL_822543, EPI_ISL_822544, EPI_ISL_822545, EPI_ISL_822546, EPI_ISL_822547, EPI_ISL_822548, EPI_ISL_822549, EPI_ISL_822550, EPI_ISL_822551, EPI_ISL_822552, EPI_ISL_822553, EPI_ISL_822554, EPI_ISL_822555, EPI_ISL_822556, EPI_ISL_822557, EPI_ISL_822558, EPI_ISL_822559, EPI_ISL_822560, EPI_ISL_822561, EPI_ISL_822562, EPI_ISL_822563, EPI_ISL_822564, EPI_ISL_822565, EPI_ISL_822566, EPI_ISL_822567, EPI_ISL_822568, EPI_ISL_822569, EPI_ISL_822570, EPI_ISL_822571, EPI_ISL_822572, EPI_ISL_822573, EPI_ISL_822574, EPI_ISL_822575, EPI_ISL_822576, EPI_ISL_822577, EPI_ISL_822578, EPI_ISL_822579, EPI_ISL_822580, | | | |

EPI_ISL_822581, EPI_ISL_822582, EPI_ISL_822583, EPI_ISL_822584, EPI_ISL_822585, EPI_ISL_822586, EPI_ISL_822587, EPI_ISL_822588, EPI_ISL_822589, EPI_ISL_822590, EPI_ISL_822591, EPI_ISL_822592, EPI_ISL_822593, EPI_ISL_822594, EPI_ISL_822595, EPI_ISL_822596, EPI_ISL_822597, EPI_ISL_822598, EPI_ISL_822599, EPI_ISL_822600, EPI_ISL_822601, EPI_ISL_822602, EPI_ISL_822603, EPI_ISL_822604, EPI_ISL_822605, EPI_ISL_822606, EPI_ISL_822607, EPI_ISL_822608, EPI_ISL_822609, EPI_ISL_822610, EPI_ISL_822611, EPI_ISL_822612, EPI_ISL_822613, EPI_ISL_822614, EPI_ISL_822615, EPI_ISL_822616, EPI_ISL_822617, EPI_ISL_822618, EPI_ISL_822619, EPI_ISL_822620, EPI_ISL_822621, EPI_ISL_822622, EPI_ISL_822623, EPI_ISL_822624, EPI_ISL_822625, EPI_ISL_822626, EPI_ISL_822627, EPI_ISL_822628, EPI_ISL_822629, EPI_ISL_822630, EPI_ISL_822631, EPI_ISL_822632, EPI_ISL_822633, EPI_ISL_822634, EPI_ISL_822635, EPI_ISL_822636, EPI_ISL_822637, EPI_ISL_822638, EPI_ISL_822639, EPI_ISL_822640, EPI_ISL_822641, EPI_ISL_822642, EPI_ISL_822643, EPI_ISL_822644, EPI_ISL_822645, EPI_ISL_822646, EPI_ISL_822647, EPI_ISL_822648, EPI_ISL_822649, EPI_ISL_822650, EPI_ISL_822651, EPI_ISL_822652, EPI_ISL_822653, EPI_ISL_822654, EPI_ISL_822655, EPI_ISL_822656, EPI_ISL_822657, EPI_ISL_822658, EPI_ISL_822838, EPI_ISL_822839, EPI_ISL_822841, EPI_ISL_822843, EPI_ISL_822845, EPI_ISL_822865, EPI_ISL_822867, EPI_ISL_822874, EPI_ISL_822875, EPI_ISL_822879, EPI_ISL_822881, EPI_ISL_822882, EPI_ISL_822884, EPI_ISL_822885, EPI_ISL_822886, EPI_ISL_822887, EPI_ISL_822889, EPI_ISL_822892, EPI_ISL_822894, EPI_ISL_822895, EPI_ISL_822898, EPI_ISL_822903, EPI_ISL_822905, EPI_ISL_822914, EPI_ISL_822920, EPI_ISL_822921, EPI_ISL_822922, EPI_ISL_822923, EPI_ISL_822931, EPI_ISL_822932, EPI_ISL_822933, EPI_ISL_822941, EPI_ISL_822944, EPI_ISL_822949, EPI_ISL_822950, EPI_ISL_822953, EPI_ISL_822954, EPI_ISL_822958, EPI_ISL_822961, EPI_ISL_822962, EPI_ISL_822963, EPI_ISL_822972, EPI_ISL_822974, EPI_ISL_822975, EPI_ISL_822978, EPI_ISL_822998, EPI_ISL_823002, EPI_ISL_823005, EPI_ISL_823013, EPI_ISL_823017, EPI_ISL_823018, EPI_ISL_823019, EPI_ISL_823047, EPI_ISL_823052, EPI_ISL_823053, EPI_ISL_823054, EPI_ISL_823055, EPI_ISL_823065, EPI_ISL_823067, EPI_ISL_823068, EPI_ISL_823069, EPI_ISL_823070, EPI_ISL_823071, EPI_ISL_823072, EPI_ISL_823073, EPI_ISL_823074, EPI_ISL_823075, EPI_ISL_823076, EPI_ISL_823077, EPI_ISL_823078, EPI_ISL_823079, EPI_ISL_823080, EPI_ISL_823081, EPI_ISL_823082, EPI_ISL_823083, EPI_ISL_823084, EPI_ISL_823085, EPI_ISL_823086, EPI_ISL_823087, EPI_ISL_823088, EPI_ISL_823089, EPI_ISL_823090, EPI_ISL_823091, EPI_ISL_823092, EPI_ISL_823093, EPI_ISL_823094, EPI_ISL_823095, EPI_ISL_823096, EPI_ISL_823097, EPI_ISL_823104, EPI_ISL_823105, EPI_ISL_823106, EPI_ISL_823107, EPI_ISL_823108, EPI_ISL_823110, EPI_ISL_823113, EPI_ISL_823115, EPI_ISL_823117, EPI_ISL_823119, EPI_ISL_823121, EPI_ISL_823124, EPI_ISL_823132, EPI_ISL_823133, EPI_ISL_823135, EPI_ISL_823136, EPI_ISL_823140, EPI_ISL_823141, EPI_ISL_823147, EPI_ISL_823148, EPI_ISL_823150, EPI_ISL_823151, EPI_ISL_823153, EPI_ISL_823158, EPI_ISL_823159, EPI_ISL_823160, EPI_ISL_823161, EPI_ISL_823169, EPI_ISL_823172, EPI_ISL_823173, EPI_ISL_823204, EPI_ISL_823367, EPI_ISL_823368, EPI_ISL_823369, EPI_ISL_823372, EPI_ISL_823373, EPI_ISL_823374, EPI_ISL_823375, EPI_ISL_823376, EPI_ISL_823377, EPI_ISL_823378, EPI_ISL_823380, EPI_ISL_823381, EPI_ISL_823382, EPI_ISL_823383, EPI_ISL_823390, EPI_ISL_823391, EPI_ISL_823392, EPI_ISL_823393, EPI_ISL_823394, EPI_ISL_823395, EPI_ISL_823396, EPI_ISL_823458, EPI_ISL_823507, EPI_ISL_823508, EPI_ISL_823509, EPI_ISL_823510, EPI_ISL_823511, EPI_ISL_823512, EPI_ISL_823513, EPI_ISL_823514, EPI_ISL_823515, EPI_ISL_823516, EPI_ISL_823517, EPI_ISL_823518, EPI_ISL_823519, EPI_ISL_823520, EPI_ISL_823521, EPI_ISL_823522, EPI_ISL_823523, EPI_ISL_823524, EPI_ISL_823525, EPI_ISL_823526, EPI_ISL_823527, EPI_ISL_823528, EPI_ISL_823529, EPI_ISL_823530, EPI_ISL_823531, EPI_ISL_823532, EPI_ISL_823533, EPI_ISL_823534, EPI_ISL_823535, EPI_ISL_823536, EPI_ISL_823537, EPI_ISL_823538, EPI_ISL_823539, EPI_ISL_823540, EPI_ISL_823541, EPI_ISL_823542, EPI_ISL_823543, EPI_ISL_823544, EPI_ISL_823545, EPI_ISL_823546, EPI_ISL_823547, EPI_ISL_823548, EPI_ISL_823549, EPI_ISL_823550, EPI_ISL_823551, EPI_ISL_823552, EPI_ISL_823553, EPI_ISL_823554, EPI_ISL_823555, EPI_ISL_823556, EPI_ISL_823557, EPI_ISL_823558, EPI_ISL_823559, EPI_ISL_823560, EPI_ISL_823561, EPI_ISL_823562, EPI_ISL_823563, EPI_ISL_823564, EPI_ISL_823565, EPI_ISL_823566, EPI_ISL_823567, EPI_ISL_823568, EPI_ISL_823634, EPI_ISL_823635, EPI_ISL_823636, EPI_ISL_823637, EPI_ISL_823638, EPI_ISL_823639, EPI_ISL_823640, EPI_ISL_823641, EPI_ISL_823642, EPI_ISL_823643, EPI_ISL_823644, EPI_ISL_823645, EPI_ISL_823650, EPI_ISL_823651, EPI_ISL_823652, EPI_ISL_823653, EPI_ISL_823657, EPI_ISL_823659, EPI_ISL_823660, EPI_ISL_823661, EPI_ISL_823662, EPI_ISL_823663, EPI_ISL_823664, EPI_ISL_823665, EPI_ISL_823666, EPI_ISL_823667, EPI_ISL_823668, EPI_ISL_823669, EPI_ISL_823670, EPI_ISL_823671, EPI_ISL_823672, EPI_ISL_823673, EPI_ISL_823674, EPI_ISL_823675, EPI_ISL_823676, EPI_ISL_823677, EPI_ISL_823678, EPI_ISL_823679, EPI_ISL_823680, EPI_ISL_823681, EPI_ISL_823682, EPI_ISL_823683, EPI_ISL_823684, EPI_ISL_823685, EPI_ISL_823686, EPI_ISL_823687, EPI_ISL_823688, EPI_ISL_823689, EPI_ISL_823690, EPI_ISL_823691, EPI_ISL_823692, EPI_ISL_823693, EPI_ISL_823694, EPI_ISL_823695, EPI_ISL_823696, EPI_ISL_823697, EPI_ISL_823698, EPI_ISL_823699, EPI_ISL_823700, EPI_ISL_823701, EPI_ISL_823702, EPI_ISL_823703, EPI_ISL_823704, EPI_ISL_823705, EPI_ISL_823706, EPI_ISL_823707, EPI_ISL_823708, EPI_ISL_823709, EPI_ISL_823710, EPI_ISL_823711, EPI_ISL_823712, EPI_ISL_823713, EPI_ISL_823714, EPI_ISL_823715, EPI_ISL_823716, EPI_ISL_823717, EPI_ISL_823718, EPI_ISL_823719, EPI_ISL_823720, EPI_ISL_823721, EPI_ISL_823722, EPI_ISL_823723, EPI_ISL_823724, EPI_ISL_823725, EPI_ISL_823726, EPI_ISL_823727, EPI_ISL_823728, EPI_ISL_823729, EPI_ISL_823730, EPI_ISL_823731, EPI_ISL_823732, EPI_ISL_823733, EPI_ISL_823734, EPI_ISL_823735, EPI_ISL_823736, EPI_ISL_823737, EPI_ISL_823738, EPI_ISL_823739, EPI_ISL_823740, EPI_ISL_823741, EPI_ISL_823742, EPI_ISL_823743, EPI_ISL_823744, EPI_ISL_823745, EPI_ISL_823746, EPI_ISL_823747, EPI_ISL_823748, EPI_ISL_823749, EPI_ISL_823750, EPI_ISL_823751, EPI_ISL_823752, EPI_ISL_823753, EPI_ISL_823754, EPI_ISL_823755, EPI_ISL_823756, EPI_ISL_823757, EPI_ISL_823758, EPI_ISL_823759, EPI_ISL_823760, EPI_ISL_823761, EPI_ISL_823762, EPI_ISL_823763, EPI_ISL_823764, EPI_ISL_823765, EPI_ISL_823766, EPI_ISL_823767, EPI_ISL_823768, EPI_ISL_823769, EPI_ISL_823770, EPI_ISL_823771, EPI_ISL_823772, EPI_ISL_823773, EPI_ISL_823774, EPI_ISL_823775, EPI_ISL_823776, EPI_ISL_823777, EPI_ISL_823778, EPI_ISL_823779, EPI_ISL_823796, EPI_ISL_823797, EPI_ISL_823798, EPI_ISL_823799, EPI_ISL_823800, EPI_ISL_823801, EPI_ISL_823802, EPI_ISL_823803, EPI_ISL_823804, EPI_ISL_823805, EPI_ISL_823807, EPI_ISL_823808, EPI_ISL_823809, EPI_ISL_823810, EPI_ISL_823811, EPI_ISL_823812

| | | | |
|---|---|---|---|
| see above | Wales Specialist Virology Centre Sequencing lab: Pathogen Genomics Unit | COVID-19 Genomics UK (COG-UK) Consortium | Alec Birchley; Alexander Adams; Amy Gaskin; Angela Marchbank; Bree Gatica-Wilcox; Catherine Moore; Jason Coombes; Joanne Watkins; Joel Southgate; Johnathan Evans; Laura Gifford; Lauren Gilbert; Lee Graham; Malorie Perry; Matthew Bull; Nicole Pacchiarini; Sally Corden; Sara Kumziene-Summerhayes; Sara Rey; Sarah Taylor; Simon Cottrell; Sophie Jones; Tom Connor |

EPI_ISL_840844, EPI_ISL_840890, EPI_ISL_840891, EPI_ISL_840893, EPI_ISL_840894, EPI_ISL_841065, EPI_ISL_841066

| | | | |
|---|---|---|---|
| see above | Wales Specialist Virology Centre Sequencing lab: Pathogen Genomics Unit | Public Health Wales Microbiology Cardiff Wales Specialist Virology Centre | Alec Birchley; Alexander Adams; Amy Gaskin; Angela Marchbank; Bree Gatica-Wilcox; Catherine Moore; Jason Coombes; Joanne Watkins; Joel Southgate; Johnathan Evans; Laura Gifford; Lauren Gilbert; Lee Graham; Malorie Perry; Matthew Bull; Nicole Pacchiarini; Sally Corden; Sara Kumziene-Summerhayes; Sara Rey; Sarah Taylor; Simon Cottrell; Sophie Jones; Tom Connor |

EPI_ISL_814335, EPI_ISL_814454, EPI_ISL_814456, EPI_ISL_814459, EPI_ISL_814608, EPI_ISL_814623, EPI_ISL_814625, EPI_ISL_814635, EPI_ISL_814637, EPI_ISL_814640, EPI_ISL_814641, EPI_ISL_814663, EPI_ISL_838136, EPI_ISL_838137, EPI_ISL_838138, EPI_ISL_838139, EPI_ISL_838140, EPI_ISL_838141, EPI_ISL_838142, EPI_ISL_838143, EPI_ISL_838144, EPI_ISL_838145, EPI_ISL_838146, EPI_ISL_838147, EPI_ISL_838148, EPI_ISL_838149, EPI_ISL_838150, EPI_ISL_838151, EPI_ISL_838152, EPI_ISL_838153, EPI_ISL_838154, EPI_ISL_838155, EPI_ISL_838156, EPI_ISL_838157, EPI_ISL_838158, EPI_ISL_838159, EPI_ISL_838160, EPI_ISL_838161, EPI_ISL_838162, EPI_ISL_838163, EPI_ISL_838164, EPI_ISL_838165, EPI_ISL_838166, EPI_ISL_838167, EPI_ISL_838168, EPI_ISL_838169, EPI_ISL_838170, EPI_ISL_838171, EPI_ISL_838172, EPI_ISL_838173, EPI_ISL_838174, EPI_ISL_838175, EPI_ISL_838176, EPI_ISL_838177, EPI_ISL_838178, EPI_ISL_838179, EPI_ISL_838180, EPI_ISL_838181, EPI_ISL_838182, EPI_ISL_838183, EPI_ISL_838184, EPI_ISL_838185, EPI_ISL_838186, EPI_ISL_838187, EPI_ISL_838188, EPI_ISL_838189, EPI_ISL_838190, EPI_ISL_838191, EPI_ISL_838192, EPI_ISL_838193, EPI_ISL_838194, EPI_ISL_838195, EPI_ISL_838196, EPI_ISL_838197, EPI_ISL_838198, EPI_ISL_838199, EPI_ISL_838200, EPI_ISL_838201, EPI_ISL_838202, EPI_ISL_838203, EPI_ISL_838204, EPI_ISL_838205, EPI_ISL_838206, EPI_ISL_838207, EPI_ISL_838208, EPI_ISL_838213, EPI_ISL_919243, EPI_ISL_919245, EPI_ISL_919247, EPI_ISL_919248, EPI_ISL_919250, EPI_ISL_919255, EPI_ISL_919256, EPI_ISL_919260, EPI_ISL_919276, EPI_ISL_997051

| | | | |
|---|---|---|---|
| see above | West of Scotland Specialist Virology Centre, NHSGGC / MRC-University of Glasgow Centre for Virus Research | COVID-19 Genomics UK (COG-UK) Consortium | Alasdair MacLean; Alice Broos; Ana da Silva Filipe; Antonia Ho; Daniel Mair; David L Robertson; Emma Thomson; Guy Mollett; James Shepherd; Jenna Nichols; Joseph Hughes; Kathy Li; Kathy Smollett; Kyriaki Nomikou; Lily Tong; Matthew Holden; Natasha Johnson; Rachel Blacow; Richard Orton; Rory Gunson; Sarah McDonald; Sharif Shaaban; Sreenu Vattipally; Stephen Carmichael |

We gratefully acknowledge the following Authors from the Originating laboratories responsible for obtaining the specimens, as well as the Submitting laboratories where the genome data were generated and shared via GISAID, on which this research is based.

All Submitters of data may be contacted directly via www.gisaid.org

Authors are sorted alphabetically.

| Accession ID | Originating Laboratory | Submitting Laboratory | Authors |
|---|---|---|---|
| EPI_ISL_843147, EPI_ISL_843148, EPI_ISL_843149, EPI_ISL_843150, EPI_ISL_865498, EPI_ISL_865502, EPI_ISL_865527, EPI_ISL_865528, EPI_ISL_865529, EPI_ISL_865530, EPI_ISL_865531, EPI_ISL_865532, EPI_ISL_865533, EPI_ISL_865534, EPI_ISL_865535, EPI_ISL_865536, EPI_ISL_865537, EPI_ISL_865538, EPI_ISL_865539, EPI_ISL_865540, EPI_ISL_865541, EPI_ISL_865542, EPI_ISL_865546, EPI_ISL_865547, EPI_ISL_865548, EPI_ISL_865549, EPI_ISL_865550, EPI_ISL_865551, EPI_ISL_865552, EPI_ISL_865553, EPI_ISL_865554, EPI_ISL_865555, EPI_ISL_865556, EPI_ISL_865557, EPI_ISL_865558, EPI_ISL_865561, EPI_ISL_865562, EPI_ISL_865567, EPI_ISL_865568, EPI_ISL_865569, EPI_ISL_865570, EPI_ISL_865572, EPI_ISL_865573, EPI_ISL_865579, EPI_ISL_865580, EPI_ISL_865581, EPI_ISL_865582, EPI_ISL_865583, EPI_ISL_919781, EPI_ISL_919782, EPI_ISL_919783, EPI_ISL_919784, EPI_ISL_919785, EPI_ISL_919786, EPI_ISL_919787, EPI_ISL_919788, EPI_ISL_919789, EPI_ISL_919790, EPI_ISL_919791, EPI_ISL_919792, EPI_ISL_919793, EPI_ISL_919794, EPI_ISL_919795, EPI_ISL_919796, EPI_ISL_1048067, EPI_ISL_2126057, EPI_ISL_2126058, EPI_ISL_2126061 | | | |
| see above | Barts Health NHS Trust | COVID-19 Genomics UK (COG-UK) Consortium | BROAD; Beatrix; CUTINO-MOGUEL; Claire; David; Dola; HARRINGTON; KELE; KULASEGARAN-SHYLINI; Maria-Teresa; OWOYEMI; Raghavendran; SHYLINI |
| EPI_ISL_814358, EPI_ISL_814429, EPI_ISL_814433, EPI_ISL_814434, EPI_ISL_816825, EPI_ISL_816826, EPI_ISL_816827, EPI_ISL_816828, EPI_ISL_816829, EPI_ISL_816830, EPI_ISL_816831, EPI_ISL_816832, EPI_ISL_816833, EPI_ISL_816834, EPI_ISL_816835, EPI_ISL_816836, EPI_ISL_816837, EPI_ISL_816838, EPI_ISL_816839, EPI_ISL_816840, EPI_ISL_816841, EPI_ISL_816842, EPI_ISL_816843, EPI_ISL_816844, EPI_ISL_816845, EPI_ISL_816846, EPI_ISL_816847, EPI_ISL_816848, EPI_ISL_816849, EPI_ISL_816850, EPI_ISL_816851, EPI_ISL_816852, EPI_ISL_816853, EPI_ISL_816854, EPI_ISL_816855, EPI_ISL_816856, EPI_ISL_816857, EPI_ISL_816858, EPI_ISL_816859, EPI_ISL_816860, EPI_ISL_816892, EPI_ISL_816897, EPI_ISL_816898, EPI_ISL_816903, EPI_ISL_816959, EPI_ISL_816964, EPI_ISL_816965, EPI_ISL_816970, EPI_ISL_817039, EPI_ISL_842356, EPI_ISL_842361, EPI_ISL_842362, EPI_ISL_842367, EPI_ISL_842422, EPI_ISL_842423, EPI_ISL_842424, EPI_ISL_842425, EPI_ISL_842426, EPI_ISL_842427, EPI_ISL_842428, EPI_ISL_842429, EPI_ISL_842430, EPI_ISL_842431, EPI_ISL_842432, EPI_ISL_842433, EPI_ISL_842434, EPI_ISL_842435, EPI_ISL_842436, EPI_ISL_842437, EPI_ISL_842438, EPI_ISL_842439, EPI_ISL_842440, EPI_ISL_842441, EPI_ISL_842442, EPI_ISL_842443, EPI_ISL_842444, EPI_ISL_842445, EPI_ISL_842447, EPI_ISL_842448, EPI_ISL_842449, EPI_ISL_842450, EPI_ISL_842451, EPI_ISL_842452, EPI_ISL_842453, EPI_ISL_842454, EPI_ISL_842455, EPI_ISL_842456, EPI_ISL_842457, EPI_ISL_842458, EPI_ISL_842459, EPI_ISL_842460, EPI_ISL_842461, EPI_ISL_842462, EPI_ISL_842463, EPI_ISL_842464, EPI_ISL_842465, EPI_ISL_842466, EPI_ISL_842467, EPI_ISL_842468, EPI_ISL_842469, EPI_ISL_842470, EPI_ISL_842472, EPI_ISL_842473, EPI_ISL_842474, EPI_ISL_842475, EPI_ISL_842476, EPI_ISL_842477, EPI_ISL_842478, EPI_ISL_842479, EPI_ISL_842480, EPI_ISL_842481, EPI_ISL_842482, EPI_ISL_842483, EPI_ISL_842484, EPI_ISL_842485, EPI_ISL_842486, EPI_ISL_842487, EPI_ISL_842488, EPI_ISL_842489, EPI_ISL_842490, EPI_ISL_842491, EPI_ISL_842492, EPI_ISL_842493, EPI_ISL_842494, EPI_ISL_842496, EPI_ISL_842497, EPI_ISL_842498, EPI_ISL_842499, EPI_ISL_842500, EPI_ISL_842501, EPI_ISL_842502, EPI_ISL_842503, EPI_ISL_842504, EPI_ISL_842505, EPI_ISL_842506, EPI_ISL_842507, EPI_ISL_842508, EPI_ISL_842509, EPI_ISL_842510, EPI_ISL_842511, EPI_ISL_842513, EPI_ISL_842583, EPI_ISL_842585, EPI_ISL_842586, EPI_ISL_842587, EPI_ISL_842588, EPI_ISL_842589, EPI_ISL_842590, EPI_ISL_842591, EPI_ISL_842592, EPI_ISL_842593, EPI_ISL_842594, EPI_ISL_842596, EPI_ISL_842598, EPI_ISL_842599, EPI_ISL_842600, EPI_ISL_842601, EPI_ISL_842602, EPI_ISL_842603, EPI_ISL_842604, EPI_ISL_842605, EPI_ISL_842606, EPI_ISL_842607, EPI_ISL_842608, EPI_ISL_868716, EPI_ISL_868717, EPI_ISL_868718, EPI_ISL_868719, EPI_ISL_868720, EPI_ISL_868721, EPI_ISL_868722, EPI_ISL_868723, EPI_ISL_868724, EPI_ISL_868725, EPI_ISL_868726, EPI_ISL_868727, EPI_ISL_868728, EPI_ISL_868729, EPI_ISL_868730, EPI_ISL_868731, EPI_ISL_868732, EPI_ISL_868733, EPI_ISL_868734, EPI_ISL_868735, EPI_ISL_868736, EPI_ISL_868737, EPI_ISL_868738, EPI_ISL_868739, EPI_ISL_868740, EPI_ISL_868741, EPI_ISL_868742, EPI_ISL_868743, EPI_ISL_868744, EPI_ISL_868745, EPI_ISL_868746, EPI_ISL_868747, EPI_ISL_868748, EPI_ISL_868749, EPI_ISL_868750, EPI_ISL_868751, EPI_ISL_868752, EPI_ISL_868753, EPI_ISL_868754, EPI_ISL_868755, EPI_ISL_868756, EPI_ISL_868757, EPI_ISL_868842, EPI_ISL_868846, EPI_ISL_868847, EPI_ISL_868849, EPI_ISL_868852, EPI_ISL_868857, EPI_ISL_868858, EPI_ISL_868862, EPI_ISL_868870, EPI_ISL_868871, EPI_ISL_868874, EPI_ISL_868876, EPI_ISL_868877, EPI_ISL_868901, EPI_ISL_868902, EPI_ISL_868903, EPI_ISL_868904, EPI_ISL_868905, EPI_ISL_868906, EPI_ISL_868907, EPI_ISL_868908, EPI_ISL_868909, EPI_ISL_868910, EPI_ISL_868911, EPI_ISL_868912, EPI_ISL_868913, EPI_ISL_868914, EPI_ISL_868915, EPI_ISL_868916, EPI_ISL_868917, EPI_ISL_868918, EPI_ISL_868919, EPI_ISL_868920, EPI_ISL_868921, EPI_ISL_868922, EPI_ISL_868923, EPI_ISL_868924, EPI_ISL_868925, EPI_ISL_868926, EPI_ISL_868927, EPI_ISL_868928, EPI_ISL_868929, EPI_ISL_868930, EPI_ISL_868931, EPI_ISL_868932, EPI_ISL_868933, EPI_ISL_868934, EPI_ISL_868935, EPI_ISL_868936, EPI_ISL_868937, EPI_ISL_869014, EPI_ISL_869015, EPI_ISL_869016, EPI_ISL_869017, EPI_ISL_869018, EPI_ISL_869019, EPI_ISL_869020, EPI_ISL_869021, EPI_ISL_869022, EPI_ISL_869023, EPI_ISL_869024, EPI_ISL_869025, EPI_ISL_869027, EPI_ISL_869028, EPI_ISL_869029, EPI_ISL_869030, EPI_ISL_869031, EPI_ISL_869032, EPI_ISL_869033, EPI_ISL_869034, EPI_ISL_869035, EPI_ISL_869036, EPI_ISL_869037, EPI_ISL_869038, EPI_ISL_869039, EPI_ISL_869040, EPI_ISL_869041, EPI_ISL_869042, EPI_ISL_869043, EPI_ISL_869044, EPI_ISL_869045, EPI_ISL_869046, EPI_ISL_869048, EPI_ISL_869049, EPI_ISL_869050, EPI_ISL_869051, EPI_ISL_869052, EPI_ISL_869053, EPI_ISL_869054, EPI_ISL_869055, EPI_ISL_869056, EPI_ISL_869057, EPI_ISL_869058, EPI_ISL_869059, EPI_ISL_869060, EPI_ISL_869061, EPI_ISL_869062, EPI_ISL_869063, EPI_ISL_869064, EPI_ISL_869065, EPI_ISL_869066, EPI_ISL_869067, EPI_ISL_869068, EPI_ISL_869069, EPI_ISL_869070, EPI_ISL_869071, EPI_ISL_869072, EPI_ISL_869073, EPI_ISL_869074, EPI_ISL_869075, EPI_ISL_869076, EPI_ISL_869077, EPI_ISL_869078, EPI_ISL_924430, EPI_ISL_924431, EPI_ISL_924432, EPI_ISL_924433, EPI_ISL_924434, EPI_ISL_924435, EPI_ISL_924436, EPI_ISL_924437, EPI_ISL_924438, EPI_ISL_924439, EPI_ISL_924440, EPI_ISL_924441, EPI_ISL_924443, EPI_ISL_924444, EPI_ISL_924445, EPI_ISL_924446, EPI_ISL_924447, EPI_ISL_924448, EPI_ISL_924449, EPI_ISL_924450, EPI_ISL_924451, EPI_ISL_924452, EPI_ISL_924453, EPI_ISL_924454, EPI_ISL_924455, EPI_ISL_924456, EPI_ISL_924457, EPI_ISL_924458, EPI_ISL_924459, EPI_ISL_924460, EPI_ISL_924461, EPI_ISL_924462, EPI_ISL_924464, EPI_ISL_924465, EPI_ISL_924466, EPI_ISL_924467, EPI_ISL_924468, EPI_ISL_924469, EPI_ISL_924470, EPI_ISL_924471, EPI_ISL_924472, EPI_ISL_924473, EPI_ISL_924474, EPI_ISL_924475, EPI_ISL_924476, EPI_ISL_924477, EPI_ISL_924478, EPI_ISL_924479, EPI_ISL_924480, EPI_ISL_924481, EPI_ISL_924482, EPI_ISL_924483, EPI_ISL_924484, EPI_ISL_924485, EPI_ISL_924486, EPI_ISL_924487, EPI_ISL_924488, EPI_ISL_924489, EPI_ISL_924490, EPI_ISL_924491, EPI_ISL_924492, EPI_ISL_924493, EPI_ISL_924494, EPI_ISL_924495, EPI_ISL_924497, EPI_ISL_924498, EPI_ISL_924500, EPI_ISL_924501, EPI_ISL_924502, EPI_ISL_924503, EPI_ISL_924504, EPI_ISL_924505, EPI_ISL_924506, EPI_ISL_924507, EPI_ISL_924508, EPI_ISL_924509, EPI_ISL_924510, EPI_ISL_924511, EPI_ISL_924512, EPI_ISL_924513, EPI_ISL_924514, EPI_ISL_924515, EPI_ISL_924516, EPI_ISL_924517, EPI_ISL_924518, EPI_ISL_924519, EPI_ISL_924520, EPI_ISL_924521, EPI_ISL_924522, EPI_ISL_924523, EPI_ISL_924524, EPI_ISL_924525, EPI_ISL_924526, EPI_ISL_924527, EPI_ISL_924528, EPI_ISL_924529, EPI_ISL_924530, EPI_ISL_924531, EPI_ISL_924532, EPI_ISL_924533, EPI_ISL_924534, EPI_ISL_924535, EPI_ISL_924537, EPI_ISL_924538, EPI_ISL_924539, EPI_ISL_924540, EPI_ISL_924542, EPI_ISL_924544, EPI_ISL_924545, EPI_ISL_924547, EPI_ISL_924548, EPI_ISL_924549, EPI_ISL_924550, EPI_ISL_924551, EPI_ISL_924552, EPI_ISL_924553, EPI_ISL_924554, EPI_ISL_924555, EPI_ISL_924556, EPI_ISL_924557, EPI_ISL_924558, EPI_ISL_924559, EPI_ISL_924560, EPI_ISL_924561, EPI_ISL_924562, EPI_ISL_924563, EPI_ISL_924564, EPI_ISL_924565, EPI_ISL_924566, EPI_ISL_924567, EPI_ISL_924568, EPI_ISL_924569, EPI_ISL_924570, EPI_ISL_924571, EPI_ISL_924572, EPI_ISL_924573, EPI_ISL_924574, EPI_ISL_924575, EPI_ISL_924576, EPI_ISL_924577, EPI_ISL_924578, EPI_ISL_924579, EPI_ISL_924580, EPI_ISL_924581, EPI_ISL_924582, EPI_ISL_924583, EPI_ISL_924584, EPI_ISL_924588, EPI_ISL_924591, EPI_ISL_924599, EPI_ISL_924602, EPI_ISL_1638206 | | | |
| see above | Bioinformatics and Biostatistics Lab, Advanced Sequencing Facility | COVID-19 Genomics UK (COG-UK) Consortium | Aengus Stewart; Chelsea Sawyer; Harshil Patel; Jerome Nicod; Laura Cubitt; Margaret Crawford |
| EPI_ISL_816212, EPI_ISL_816213, EPI_ISL_816214, EPI_ISL_816215, EPI_ISL_816217, EPI_ISL_816218, EPI_ISL_816220, EPI_ISL_816221, EPI_ISL_816222, EPI_ISL_816223, EPI_ISL_841978, EPI_ISL_842004, EPI_ISL_842014, EPI_ISL_842015, EPI_ISL_867982, EPI_ISL_867983, EPI_ISL_867984, EPI_ISL_867985, EPI_ISL_867986, EPI_ISL_867987, EPI_ISL_867988, EPI_ISL_867989, EPI_ISL_867991, EPI_ISL_867992, EPI_ISL_867994, EPI_ISL_867995, EPI_ISL_867996, EPI_ISL_867997, EPI_ISL_867998, EPI_ISL_868000, EPI_ISL_868002, EPI_ISL_868003, EPI_ISL_868004, EPI_ISL_868005, EPI_ISL_868006, EPI_ISL_868007, EPI_ISL_868009, EPI_ISL_868010, EPI_ISL_868011, EPI_ISL_868012, EPI_ISL_868013, EPI_ISL_868014, EPI_ISL_868015, EPI_ISL_868016, EPI_ISL_868017, EPI_ISL_868018, EPI_ISL_868019, EPI_ISL_868020, EPI_ISL_868021, EPI_ISL_868022, EPI_ISL_868024, EPI_ISL_868025, EPI_ISL_868026, EPI_ISL_868027, EPI_ISL_868028, EPI_ISL_868029, EPI_ISL_868030, EPI_ISL_868031, EPI_ISL_868032, EPI_ISL_868033, EPI_ISL_868035, EPI_ISL_868036, EPI_ISL_868037, EPI_ISL_868038, EPI_ISL_868039, EPI_ISL_868040, EPI_ISL_868041, EPI_ISL_868042, EPI_ISL_868043, EPI_ISL_868044, EPI_ISL_868045, EPI_ISL_868046, EPI_ISL_868047, EPI_ISL_868048, EPI_ISL_868049, EPI_ISL_868050, EPI_ISL_868051, EPI_ISL_868052, EPI_ISL_868053, EPI_ISL_868054, EPI_ISL_868055, EPI_ISL_868056, EPI_ISL_868057, EPI_ISL_868058, EPI_ISL_868059, EPI_ISL_868060, EPI_ISL_868061, EPI_ISL_868062, EPI_ISL_868063, EPI_ISL_868064, EPI_ISL_868065, EPI_ISL_868066, EPI_ISL_868067, EPI_ISL_868068, EPI_ISL_868069, EPI_ISL_868070, EPI_ISL_868071, EPI_ISL_868072, EPI_ISL_868073, EPI_ISL_868074, EPI_ISL_868075, EPI_ISL_868076, EPI_ISL_868077, EPI_ISL_868078, EPI_ISL_868079, EPI_ISL_868080, EPI_ISL_868081, EPI_ISL_868082, EPI_ISL_868083, EPI_ISL_868084, EPI_ISL_868085, EPI_ISL_868086, EPI_ISL_868087, EPI_ISL_868088, EPI_ISL_868089, EPI_ISL_868090, EPI_ISL_868091, EPI_ISL_868092, EPI_ISL_868093, EPI_ISL_868094, EPI_ISL_868095, EPI_ISL_868096, EPI_ISL_868097, EPI_ISL_868098, EPI_ISL_868099, EPI_ISL_868100, EPI_ISL_868101, EPI_ISL_868103, EPI_ISL_868104, EPI_ISL_868105, EPI_ISL_868106, EPI_ISL_868107, EPI_ISL_868108, EPI_ISL_868109, EPI_ISL_868110, EPI_ISL_868111, EPI_ISL_868112, EPI_ISL_868113, EPI_ISL_868114, EPI_ISL_868115, EPI_ISL_868116, EPI_ISL_868117, EPI_ISL_868118, EPI_ISL_868119, EPI_ISL_868120, EPI_ISL_868121, EPI_ISL_868122, EPI_ISL_868123, EPI_ISL_868124, EPI_ISL_868125, EPI_ISL_868126, EPI_ISL_868127, EPI_ISL_868128, EPI_ISL_868129, EPI_ISL_868130, EPI_ISL_868131, EPI_ISL_868132, EPI_ISL_868133, EPI_ISL_868134, EPI_ISL_868135, EPI_ISL_868136, EPI_ISL_868137, EPI_ISL_868138, EPI_ISL_868139, EPI_ISL_868140, EPI_ISL_868259, EPI_ISL_868260, EPI_ISL_868261, EPI_ISL_868262, EPI_ISL_868263, EPI_ISL_868264, EPI_ISL_868265, EPI_ISL_868266, EPI_ISL_868267, EPI_ISL_868268, EPI_ISL_868269, EPI_ISL_868270, EPI_ISL_868271, EPI_ISL_868272, EPI_ISL_868273, EPI_ISL_868274, EPI_ISL_868275, EPI_ISL_868276, EPI_ISL_868277, EPI_ISL_868278, EPI_ISL_868279, EPI_ISL_868280, EPI_ISL_868281, EPI_ISL_868282, EPI_ISL_868283, EPI_ISL_868284, EPI_ISL_868285, EPI_ISL_868286, EPI_ISL_868287, EPI_ISL_868288, EPI_ISL_868289, EPI_ISL_868290, EPI_ISL_868291, EPI_ISL_868292, EPI_ISL_868293, EPI_ISL_868294, EPI_ISL_868295, EPI_ISL_868296, EPI_ISL_868297, EPI_ISL_868298, EPI_ISL_868299, EPI_ISL_868300, EPI_ISL_868301, EPI_ISL_868302, EPI_ISL_868303, EPI_ISL_868304, EPI_ISL_868305, EPI_ISL_868306, EPI_ISL_868307, EPI_ISL_868308, EPI_ISL_868318, EPI_ISL_868320, EPI_ISL_923242, EPI_ISL_923244, EPI_ISL_923253, EPI_ISL_923265, EPI_ISL_923295, EPI_ISL_923425, EPI_ISL_923426, EPI_ISL_923427, EPI_ISL_923428, EPI_ISL_923429, EPI_ISL_923431, EPI_ISL_923432, EPI_ISL_923433, EPI_ISL_923434, EPI_ISL_923435, EPI_ISL_923437, EPI_ISL_923439, EPI_ISL_923440, EPI_ISL_923441, EPI_ISL_923442, EPI_ISL_923444, EPI_ISL_923445, EPI_ISL_923446, EPI_ISL_923447, EPI_ISL_923448, EPI_ISL_923449, EPI_ISL_923450, EPI_ISL_923451, EPI_ISL_923452, EPI_ISL_923453, EPI_ISL_923454, EPI_ISL_923455, EPI_ISL_923456, EPI_ISL_923457, EPI_ISL_923458, EPI_ISL_923459, EPI_ISL_923460, EPI_ISL_923461, EPI_ISL_923462, EPI_ISL_923463, EPI_ISL_923464, EPI_ISL_923465, EPI_ISL_923466, EPI_ISL_923467, EPI_ISL_923468, EPI_ISL_923469, EPI_ISL_923470, EPI_ISL_923471, EPI_ISL_923472, EPI_ISL_923473, EPI_ISL_923474, EPI_ISL_923475, EPI_ISL_923476, EPI_ISL_923477, EPI_ISL_923478, EPI_ISL_923479, EPI_ISL_923480, EPI_ISL_923481, EPI_ISL_923482, EPI_ISL_923483, EPI_ISL_923484, EPI_ISL_923485, EPI_ISL_923486, EPI_ISL_923487, EPI_ISL_923488, EPI_ISL_923489, EPI_ISL_923490, EPI_ISL_923491, EPI_ISL_923492, EPI_ISL_923493, EPI_ISL_923494, EPI_ISL_923495, EPI_ISL_923496, EPI_ISL_923497, EPI_ISL_923498, EPI_ISL_923499, EPI_ISL_923500, EPI_ISL_923501, EPI_ISL_923502, EPI_ISL_923503, EPI_ISL_923504, EPI_ISL_923505, EPI_ISL_923506, EPI_ISL_923507, EPI_ISL_923508, EPI_ISL_923509, EPI_ISL_923510, EPI_ISL_923511, EPI_ISL_923512, EPI_ISL_923513, EPI_ISL_923514, EPI_ISL_923515, EPI_ISL_923516, EPI_ISL_923517, EPI_ISL_923518, EPI_ISL_923519, EPI_ISL_923520, EPI_ISL_923521, EPI_ISL_923522, EPI_ISL_923523, EPI_ISL_923524, EPI_ISL_923525, EPI_ISL_923526, EPI_ISL_923527, EPI_ISL_923528, EPI_ISL_923529, EPI_ISL_923530, EPI_ISL_923531, EPI_ISL_923532, EPI_ISL_923533, EPI_ISL_923534, EPI_ISL_923535, EPI_ISL_923536, EPI_ISL_923537, EPI_ISL_923538, EPI_ISL_923540, EPI_ISL_923541, EPI_ISL_923542, EPI_ISL_923543, EPI_ISL_923544, EPI_ISL_923545, EPI_ISL_923546, EPI_ISL_923547, EPI_ISL_923548, EPI_ISL_923549, EPI_ISL_923550, EPI_ISL_923551, EPI_ISL_923552, EPI_ISL_923553, EPI_ISL_923554, EPI_ISL_923555, EPI_ISL_923556, EPI_ISL_923557, EPI_ISL_923558, EPI_ISL_923559, EPI_ISL_923560, EPI_ISL_923561, EPI_ISL_923562, EPI_ISL_923563, EPI_ISL_923564, EPI_ISL_923565, EPI_ISL_923566, EPI_ISL_923567, EPI_ISL_923568, EPI_ISL_923569, EPI_ISL_923570, EPI_ISL_923571, EPI_ISL_923572, EPI_ISL_923573, EPI_ISL_923574, EPI_ISL_923575, EPI_ISL_923576, EPI_ISL_923578, EPI_ISL_923579, EPI_ISL_923581, EPI_ISL_923582, EPI_ISL_923583, EPI_ISL_923584, EPI_ISL_923585, EPI_ISL_923586, EPI_ISL_923587, EPI_ISL_923599, EPI_ISL_923600, EPI_ISL_923601, EPI_ISL_923602, EPI_ISL_923603, EPI_ISL_923604, EPI_ISL_923606, EPI_ISL_923607, EPI_ISL_923608, EPI_ISL_923609, EPI_ISL_923610, EPI_ISL_923612, EPI_ISL_923614, EPI_ISL_923615, EPI_ISL_923616, EPI_ISL_923617, EPI_ISL_923618, EPI_ISL_923619, EPI_ISL_923620, EPI_ISL_923621, EPI_ISL_923622, EPI_ISL_923623, EPI_ISL_923624, EPI_ISL_923625, EPI_ISL_923626, EPI_ISL_923627, EPI_ISL_923629, EPI_ISL_923630, EPI_ISL_923631, EPI_ISL_923632, EPI_ISL_923633, EPI_ISL_923634, EPI_ISL_923635, EPI_ISL_923636, EPI_ISL_923637, EPI_ISL_923638, EPI_ISL_923639, EPI_ISL_923640, EPI_ISL_923641, EPI_ISL_923642, EPI_ISL_923643, EPI_ISL_923644, EPI_ISL_923645, EPI_ISL_923646, EPI_ISL_923647, EPI_ISL_923648, EPI_ISL_923649, EPI_ISL_923650, EPI_ISL_923651, EPI_ISL_923652, EPI_ISL_923653, EPI_ISL_923654, EPI_ISL_923655, EPI_ISL_923656, EPI_ISL_923657, EPI_ISL_923658, EPI_ISL_923659, EPI_ISL_923661, EPI_ISL_923662, EPI_ISL_952430, EPI_ISL_952435, EPI_ISL_952500, EPI_ISL_952501, EPI_ISL_952502, EPI_ISL_952503, EPI_ISL_952504, EPI_ISL_952505, EPI_ISL_952506, EPI_ISL_952507, EPI_ISL_952508, EPI_ISL_952509, EPI_ISL_952510, EPI_ISL_952511, EPI_ISL_952512, EPI_ISL_952513, EPI_ISL_952514, EPI_ISL_952515, EPI_ISL_952516, EPI_ISL_952517, EPI_ISL_952518, EPI_ISL_952519, EPI_ISL_952520, EPI_ISL_952603, EPI_ISL_952604, EPI_ISL_952605, EPI_ISL_952606, EPI_ISL_952607, EPI_ISL_952608, EPI_ISL_952609, EPI_ISL_952610, EPI_ISL_952611, EPI_ISL_952612, EPI_ISL_952613, EPI_ISL_952614, EPI_ISL_952615, EPI_ISL_952616, EPI_ISL_952617, EPI_ISL_952618, EPI_ISL_952619, EPI_ISL_952620, EPI_ISL_952621, EPI_ISL_952622, EPI_ISL_952623, EPI_ISL_952625, EPI_ISL_952626, EPI_ISL_952627, EPI_ISL_952628, EPI_ISL_952629, EPI_ISL_952630, EPI_ISL_952631, EPI_ISL_952632, EPI_ISL_952633, EPI_ISL_952634, EPI_ISL_952635, EPI_ISL_952637, EPI_ISL_952638, EPI_ISL_952639, EPI_ISL_952640, EPI_ISL_952641, EPI_ISL_952642, EPI_ISL_952643, EPI_ISL_952644, EPI_ISL_952645, EPI_ISL_952646, EPI_ISL_952647, EPI_ISL_952648, EPI_ISL_952649, EPI_ISL_952650, EPI_ISL_952651, EPI_ISL_952652, EPI_ISL_952653, EPI_ISL_952654, EPI_ISL_952655, EPI_ISL_952656, EPI_ISL_952657, EPI_ISL_952658, EPI_ISL_952659, EPI_ISL_952660, EPI_ISL_952661, EPI_ISL_952662, EPI_ISL_952663, EPI_ISL_952664, EPI_ISL_952665, EPI_ISL_952666, EPI_ISL_952667, EPI_ISL_952668, EPI_ISL_952669, EPI_ISL_952670, EPI_ISL_952671, EPI_ISL_952672, EPI_ISL_952674, EPI_ISL_952676, EPI_ISL_952678, EPI_ISL_952679, EPI_ISL_952681, EPI_ISL_952682, EPI_ISL_952683, EPI_ISL_952684, EPI_ISL_952685, EPI_ISL_952686, EPI_ISL_952687, EPI_ISL_952688, EPI_ISL_952690, EPI_ISL_952691, EPI_ISL_952692, EPI_ISL_952693, EPI_ISL_952694, EPI_ISL_952695, EPI_ISL_952696, EPI_ISL_952697, EPI_ISL_952698, EPI_ISL_952699, EPI_ISL_952700, EPI_ISL_952701, EPI_ISL_952702, EPI_ISL_952703, EPI_ISL_952704, EPI_ISL_952705, EPI_ISL_952706, EPI_ISL_952707, EPI_ISL_952708, EPI_ISL_952709, EPI_ISL_952710, EPI_ISL_952711, EPI_ISL_952712, EPI_ISL_952713, EPI_ISL_952714, EPI_ISL_952715, EPI_ISL_952716, EPI_ISL_952717, EPI_ISL_952718, EPI_ISL_952719, EPI_ISL_952720, EPI_ISL_952721, EPI_ISL_952722, EPI_ISL_952723, EPI_ISL_952724, EPI_ISL_952725, EPI_ISL_952726, EPI_ISL_952727, EPI_ISL_952728, EPI_ISL_952729, EPI_ISL_952730, EPI_ISL_952731, EPI_ISL_952732, EPI_ISL_952733, EPI_ISL_952734, EPI_ISL_952735, EPI_ISL_952736, EPI_ISL_952737, EPI_ISL_952738, EPI_ISL_952739, EPI_ISL_952740, EPI_ISL_952741, EPI_ISL_952742, EPI_ISL_952743, EPI_ISL_952744, EPI_ISL_952745, EPI_ISL_952746, EPI_ISL_952747, EPI_ISL_952748, EPI_ISL_952749, EPI_ISL_952750, EPI_ISL_952751, EPI_ISL_952752, EPI_ISL_952753, EPI_ISL_952754, EPI_ISL_952755, EPI_ISL_952756, EPI_ISL_952757, EPI_ISL_952758, EPI_ISL_952759, EPI_ISL_952760, EPI_ISL_952761, EPI_ISL_952762, EPI_ISL_952763, EPI_ISL_952764, EPI_ISL_952765, EPI_ISL_952766, EPI_ISL_952767, EPI_ISL_952768, EPI_ISL_952769, EPI_ISL_952770, EPI_ISL_952771, EPI_ISL_952772, EPI_ISL_952773, EPI_ISL_952774, EPI_ISL_952775, EPI_ISL_952776, EPI_ISL_1000237, EPI_ISL_1000238, EPI_ISL_1000239, EPI_ISL_1000255, EPI_ISL_1000264, EPI_ISL_1000266, EPI_ISL_1000275, EPI_ISL_1000276, EPI_ISL_1000279, EPI_ISL_1000283, EPI_ISL_1000284, EPI_ISL_1000285, EPI_ISL_1000286, EPI_ISL_1000287, EPI_ISL_1000325, EPI_ISL_1000326, EPI_ISL_1000327, EPI_ISL_1000328, EPI_ISL_1000329, EPI_ISL_1000330, EPI_ISL_1000331, EPI_ISL_1000332, EPI_ISL_1000333, EPI_ISL_1000334, EPI_ISL_1000335, EPI_ISL_1000336, EPI_ISL_1000337, EPI_ISL_1000338, EPI_ISL_1000339, EPI_ISL_1000340, EPI_ISL_1000341, EPI_ISL_1000342, EPI_ISL_1000343, EPI_ISL_1000344, EPI_ISL_1000345, EPI_ISL_1000346, EPI_ISL_1000347, EPI_ISL_1000348, EPI_ISL_1000349, EPI_ISL_1000350, EPI_ISL_1000351, EPI_ISL_1000352, EPI_ISL_1000353, EPI_ISL_1000354, EPI_ISL_1000355, EPI_ISL_1000356, EPI_ISL_1000357, EPI_ISL_1000358, EPI_ISL_1000359, EPI_ISL_1000360, EPI_ISL_1000361, EPI_ISL_1000362, EPI_ISL_1000363, EPI_ISL_1000364, EPI_ISL_1000365, EPI_ISL_1000366, EPI_ISL_1000367, EPI_ISL_1000368, EPI_ISL_1000369, EPI_ISL_1000370, EPI_ISL_1000371, EPI_ISL_1000372, EPI_ISL_1000373, EPI_ISL_1000374, EPI_ISL_1000375, EPI_ISL_1000376, EPI_ISL_1000377, EPI_ISL_1000378, EPI_ISL_1000379, EPI_ISL_1000380, EPI_ISL_1000381, EPI_ISL_1000382, EPI_ISL_1000383, EPI_ISL_1000384, EPI_ISL_1000385, EPI_ISL_1000386, EPI_ISL_1000387, EPI_ISL_1000388, EPI_ISL_1000389, EPI_ISL_1000390, EPI_ISL_1000391, EPI_ISL_1000392, EPI_ISL_1000393, EPI_ISL_1000394, EPI_ISL_1000395, EPI_ISL_1000396, EPI_ISL_1000397, EPI_ISL_1000398, EPI_ISL_1000399, EPI_ISL_1000400, EPI_ISL_1000401, EPI_ISL_1000402, EPI_ISL_1000403, EPI_ISL_1000478, EPI_ISL_1000479, EPI_ISL_1000480, EPI_ISL_1000481, EPI_ISL_1000484, EPI_ISL_1000485, EPI_ISL_1000486, EPI_ISL_1000487, EPI_ISL_1000488, EPI_ISL_1000489, EPI_ISL_1000490, EPI_ISL_1000491, EPI_ISL_1000492, EPI_ISL_1000493, EPI_ISL_1000494, EPI_ISL_1000495, EPI_ISL_1000496, EPI_ISL_1000497, EPI_ISL_1000498, EPI_ISL_1000499, EPI_ISL_1000500, EPI_ISL_1000501, EPI_ISL_1000502, EPI_ISL_1000503, EPI_ISL_1000504, EPI_ISL_1000505, EPI_ISL_1000506, EPI_ISL_1000507, EPI_ISL_1000508, EPI_ISL_1000509, EPI_ISL_1000510, EPI_ISL_1000511, EPI_ISL_1000512, EPI_ISL_1000513, EPI_ISL_1000514, EPI_ISL_1000515, EPI_ISL_1000516, EPI_ISL_1000517, EPI_ISL_1000518, EPI_ISL_1000519, EPI_ISL_1000520, EPI_ISL_1000521, EPI_ISL_1000522, EPI_ISL_1000523, EPI_ISL_1000524, EPI_ISL_1000526, EPI_ISL_1000527, EPI_ISL_1000528, EPI_ISL_1000529, EPI_ISL_1000530, EPI_ISL_1000531, EPI_ISL_1000532, EPI_ISL_1000533, EPI_ISL_1000534, EPI_ISL_1000535, EPI_ISL_1000536, EPI_ISL_1000537, EPI_ISL_1000538, EPI_ISL_1000539, EPI_ISL_1000540, EPI_ISL_1000541, EPI_ISL_1000543, EPI_ISL_1000544, EPI_ISL_1000545, EPI_ISL_1000547, EPI_ISL_1000548, EPI_ISL_1000549, EPI_ISL_1000550, EPI_ISL_1000551, EPI_ISL_1000552, EPI_ISL_1000553, EPI_ISL_1000554, EPI_ISL_1000555, EPI_ISL_1000556, EPI_ISL_1000557, EPI_ISL_1000558, EPI_ISL_1000559, EPI_ISL_1000560, EPI_ISL_1000561, EPI_ISL_1000562, EPI_ISL_1000563, EPI_ISL_1000564, EPI_ISL_1000565, EPI_ISL_1000566, EPI_ISL_1000567, EPI_ISL_1000568, EPI_ISL_1000569, EPI_ISL_1000570, EPI_ISL_1000571, EPI_ISL_1000572, EPI_ISL_1000574, EPI_ISL_1000575, EPI_ISL_1000577, EPI_ISL_1000578, EPI_ISL_1000579, EPI_ISL_1000580, EPI_ISL_1000581, EPI_ISL_1000582, EPI_ISL_1000583, EPI_ISL_1000584, EPI_ISL_1000585, EPI_ISL_1000586, EPI_ISL_1000587, EPI_ISL_1000588, EPI_ISL_1000589, EPI_ISL_1000590, EPI_ISL_1000591, EPI_ISL_1000592, EPI_ISL_1000593, EPI_ISL_1000594, EPI_ISL_1053754, EPI_ISL_1053755, EPI_ISL_1053756, EPI_ISL_1053757, EPI_ISL_1053758, EPI_ISL_1053759, EPI_ISL_1053760, EPI_ISL_1053761, EPI_ISL_1053762, EPI_ISL_1053763, EPI_ISL_1053764, EPI_ISL_1053765, EPI_ISL_1053766, EPI_ISL_1053767, EPI_ISL_1053768, EPI_ISL_1053769, EPI_ISL_1053770, EPI_ISL_1053771, EPI_ISL_1053772, EPI_ISL_1053773, EPI_ISL_1053774, EPI_ISL_1053775, EPI_ISL_1053776, EPI_ISL_1053777, EPI_ISL_1053778, EPI_ISL_1053779, EPI_ISL_1053780, EPI_ISL_1053781, EPI_ISL_1053782, EPI_ISL_1053783, EPI_ISL_1053784, EPI_ISL_1053785, EPI_ISL_1053786, EPI_ISL_1053787, EPI_ISL_1053788, EPI_ISL_1053789, EPI_ISL_1053790, EPI_ISL_1053791, EPI_ISL_1053792, EPI_ISL_1053793, EPI_ISL_1053794, EPI_ISL_1053795, EPI_ISL_1053796, EPI_ISL_1053797, EPI_ISL_1053798, EPI_ISL_1053799, EPI_ISL_1053800, EPI_ISL_1053801, EPI_ISL_1053802, EPI_ISL_1053803, EPI_ISL_1053804, EPI_ISL_1053805, EPI_ISL_1053806, EPI_ISL_1053807, EPI_ISL_1053808, EPI_ISL_1053809, EPI_ISL_1053810, EPI_ISL_1053811, EPI_ISL_1053812, EPI_ISL_1053813, EPI_ISL_1053814, EPI_ISL_1053815, EPI_ISL_1053816, EPI_ISL_1053903, EPI_ISL_1053904, EPI_ISL_1053905, EPI_ISL_1053908, EPI_ISL_1053909, EPI_ISL_1053910, EPI_ISL_1053911, EPI_ISL_1053912, EPI_ISL_1053913, EPI_ISL_1053914, EPI_ISL_1053917, EPI_ISL_1053918, EPI_ISL_1053919, EPI_ISL_1053920, EPI_ISL_1053921, EPI_ISL_1053924, EPI_ISL_1053925, EPI_ISL_1053928, EPI_ISL_1053929, EPI_ISL_1053930, EPI_ISL_1053931, EPI_ISL_1107544, EPI_ISL_1107545, EPI_ISL_1107546, EPI_ISL_1107547, EPI_ISL_1107548, EPI_ISL_1107549, EPI_ISL_1107550, EPI_ISL_1107551, EPI_ISL_1107552, EPI_ISL_1107553, EPI_ISL_1107554, EPI_ISL_1107555, EPI_ISL_1107556, EPI_ISL_1107557, EPI_ISL_1107558, EPI_ISL_1107559, EPI_ISL_1107560, EPI_ISL_1107561, EPI_ISL_1107562, EPI_ISL_1107563, EPI_ISL_1107564, EPI_ISL_1107565, EPI_ISL_1107566, EPI_ISL_1107567, EPI_ISL_1107568, EPI_ISL_1107569, EPI_ISL_1107570, EPI_ISL_1107571, EPI_ISL_1107572, EPI_ISL_1107573, EPI_ISL_1107574, EPI_ISL_1107575, EPI_ISL_1107576, EPI_ISL_1107577, EPI_ISL_1107578, EPI_ISL_1107579, EPI_ISL_1107580, EPI_ISL_1107581, EPI_ISL_1107582, EPI_ISL_1107583, EPI_ISL_1107584, EPI_ISL_1107585, EPI_ISL_1107588, EPI_ISL_1107589, EPI_ISL_1107590, EPI_ISL_1107591, EPI_ISL_1107592, EPI_ISL_1107593, EPI_ISL_1107594, EPI_ISL_1107595, EPI_ISL_1107596, EPI_ISL_1107597, EPI_ISL_1107598, EPI_ISL_1107599, EPI_ISL_1107600, EPI_ISL_1107601, EPI_ISL_1107602, EPI_ISL_1107603, EPI_ISL_1107604, EPI_ISL_1107605, EPI_ISL_1107606, EPI_ISL_1107607, EPI_ISL_1107608, EPI_ISL_1107609, EPI_ISL_1107610, EPI_ISL_1107611, EPI_ISL_1107612, EPI_ISL_1107614, EPI_ISL_1107615, EPI_ISL_1107616, EPI_ISL_1107617, EPI_ISL_1107618, EPI_ISL_1107619, EPI_ISL_1107620, EPI_ISL_1107621, EPI_ISL_1107622, EPI_ISL_1107623, EPI_ISL_1107624, EPI_ISL_1107625, EPI_ISL_1107626, EPI_ISL_1107627, EPI_ISL_1107628, EPI_ISL_1107629, EPI_ISL_1107630, EPI_ISL_1107700, EPI_ISL_1107701, EPI_ISL_1107702, EPI_ISL_1107703, EPI_ISL_1107704, EPI_ISL_1107705, EPI_ISL_1107706, EPI_ISL_1107707, EPI_ISL_1107709, EPI_ISL_1107710, EPI_ISL_1107711, EPI_ISL_1107712, EPI_ISL_1179680, EPI_ISL_1179733, EPI_ISL_1179734, EPI_ISL_1179736, EPI_ISL_1179763, EPI_ISL_1179779, EPI_ISL_1179781, EPI_ISL_1179786, EPI_ISL_1179799, EPI_ISL_1179801, EPI_ISL_1179805, EPI_ISL_1179807, EPI_ISL_1179810, EPI_ISL_1179826, EPI_ISL_1179835, EPI_ISL_1179840, EPI_ISL_1179841, EPI_ISL_1179842, EPI_ISL_1179844, EPI_ISL_1179845, EPI_ISL_1179846, EPI_ISL_1179847, EPI_ISL_1179848, EPI_ISL_1179849, EPI_ISL_1179850, EPI_ISL_1179851, EPI_ISL_1249888, EPI_ISL_1249889, EPI_ISL_1249890, EPI_ISL_1249891, EPI_ISL_1249892, EPI_ISL_1249893, EPI_ISL_1249894, EPI_ISL_1249895, EPI_ISL_1249896, EPI_ISL_1249897, EPI_ISL_1249898, EPI_ISL_1249899, EPI_ISL_1249900, EPI_ISL_1249901, EPI_ISL_1249902, EPI_ISL_1249903, EPI_ISL_1249904, EPI_ISL_1249905, EPI_ISL_1249906, EPI_ISL_1249907, EPI_ISL_1249908, EPI_ISL_1249909, EPI_ISL_1249910, EPI_ISL_1249911, EPI_ISL_1249912, EPI_ISL_1249913, EPI_ISL_1249914, EPI_ISL_1249915, EPI_ISL_1249916, EPI_ISL_1249917, EPI_ISL_1249918, EPI_ISL_1249920, EPI_ISL_1249921, EPI_ISL_1249922, EPI_ISL_1249923, EPI_ISL_1249924, EPI_ISL_1249925, EPI_ISL_1249926, EPI_ISL_1249927, EPI_ISL_1249928, EPI_ISL_1249929, EPI_ISL_1249930, EPI_ISL_1249931, EPI_ISL_1249932, EPI_ISL_1249933, EPI_ISL_1249934, EPI_ISL_1249935, EPI_ISL_1249936, EPI_ISL_1249937, EPI_ISL_1249938, EPI_ISL_1249939, EPI_ISL_1249940, EPI_ISL_1249941, EPI_ISL_1249942, EPI_ISL_1249943, EPI_ISL_1249944, EPI_ISL_1249945, EPI_ISL_1249946, EPI_ISL_1249947, EPI_ISL_1249948, EPI_ISL_1249949, EPI_ISL_1249951, EPI_ISL_1249952, EPI_ISL_1249953, EPI_ISL_1249954, EPI_ISL_1249955, EPI_ISL_1249956, EPI_ISL_1249957, EPI_ISL_1249958, EPI_ISL_1249960, EPI_ISL_1249961, EPI_ISL_1249962, EPI_ISL_1249963, EPI_ISL_1249964, EPI_ISL_1249965, EPI_ISL_1249966, EPI_ISL_1250031, EPI_ISL_1250032, EPI_ISL_1250033, EPI_ISL_1250034, EPI_ISL_1250035, EPI_ISL_1250036, EPI_ISL_1250037, EPI_ISL_1250038, EPI_ISL_1250039, EPI_ISL_1250040, EPI_ISL_1250041, EPI_ISL_1250042, EPI_ISL_1250043, EPI_ISL_1250044, EPI_ISL_1250045, EPI_ISL_1250046, EPI_ISL_1250047, EPI_ISL_1250048, EPI_ISL_1250049, EPI_ISL_1250050, EPI_ISL_1250051, EPI_ISL_1250052, EPI_ISL_1250053, EPI_ISL_1250054, EPI_ISL_1250055, EPI_ISL_1250057, EPI_ISL_1250058, EPI_ISL_1250059, EPI_ISL_1250060, EPI_ISL_1250062, EPI_ISL_1250063, EPI_ISL_1250064, EPI_ISL_1250065, EPI_ISL_1250066, EPI_ISL_1250067, EPI_ISL_1250068, EPI_ISL_1250069, EPI_ISL_1250070, EPI_ISL_1250071, EPI_ISL_1250072, EPI_ISL_1250073, EPI_ISL_1250074, EPI_ISL_1250075, EPI_ISL_1250076, EPI_ISL_1250077, EPI_ISL_1250078, EPI_ISL_1250079, EPI_ISL_1250080, EPI_ISL_1250081, EPI_ISL_1250082, EPI_ISL_1250083, EPI_ISL_1250084, EPI_ISL_1250085, EPI_ISL_1250086, EPI_ISL_1250087, EPI_ISL_1250088, EPI_ISL_1250089, EPI_ISL_1250090, EPI_ISL_1250091, EPI_ISL_1250092, EPI_ISL_1250093, EPI_ISL_1250096, EPI_ISL_1250097, EPI_ISL_1250098, EPI_ISL_1250099, EPI_ISL_1250100, EPI_ISL_1250101, EPI_ISL_1250102, EPI_ISL_1250103, EPI_ISL_1250104, EPI_ISL_1250106, EPI_ISL_1250107, EPI_ISL_1250108, EPI_ISL_1250120, EPI_ISL_1250121, EPI_ISL_1250122, EPI_ISL_1310289, EPI_ISL_1310290, EPI_ISL_1310327, EPI_ISL_1310362, EPI_ISL_1310363, EPI_ISL_1310364, EPI_ISL_1310365, EPI_ISL_1310403, EPI_ISL_1310404, EPI_ISL_1310405, EPI_ISL_1310406, EPI_ISL_1310407, EPI_ISL_1310408, EPI_ISL_1310409, EPI_ISL_1310410, EPI_ISL_1310411, EPI_ISL_1310412, EPI_ISL_1310413, EPI_ISL_1310414, EPI_ISL_1310415, EPI_ISL_1310416, EPI_ISL_1310417, EPI_ISL_1310418, EPI_ISL_1310419, EPI_ISL_1310420, EPI_ISL_1310421, EPI_ISL_1310422, EPI_ISL_1310423, EPI_ISL_1310424, EPI_ISL_1310425, EPI_ISL_1310426, EPI_ISL_1310427, EPI_ISL_1310428, EPI_ISL_1310429, EPI_ISL_1310430, EPI_ISL_1310431, EPI_ISL_1310432, EPI_ISL_1310433, EPI_ISL_1310434, EPI_ISL_1310435, EPI_ISL_1310436, EPI_ISL_1310437, EPI_ISL_1388654, EPI_ISL_1388656, EPI_ISL_1388658, EPI_ISL_1388741, EPI_ISL_1388762, EPI_ISL_1388811, EPI_ISL_1388814, EPI_ISL_1388816, EPI_ISL_1388822, EPI_ISL_1388824, EPI_ISL_1388826, EPI_ISL_1388828, EPI_ISL_1388831, EPI_ISL_1388832, EPI_ISL_1388835, EPI_ISL_1388837, EPI_ISL_1388839, EPI_ISL_1388841, EPI_ISL_1388843, EPI_ISL_1388846, EPI_ISL_1388848, EPI_ISL_1388849, EPI_ISL_1388851, EPI_ISL_1388853, EPI_ISL_1388855, EPI_ISL_1388857, EPI_ISL_1388859, EPI_ISL_1388861, EPI_ISL_1476566, EPI_ISL_1476567, EPI_ISL_1476568, EPI_ISL_1476569, EPI_ISL_1476570, EPI_ISL_1476571, EPI_ISL_1476572, EPI_ISL_1476573, EPI_ISL_1476574, EPI_ISL_1476575, EPI_ISL_1476576, EPI_ISL_1476577, EPI_ISL_1476578, EPI_ISL_1476579, EPI_ISL_1476580, EPI_ISL_1476581, EPI_ISL_1476582, EPI_ISL_1476583, EPI_ISL_1476584, EPI_ISL_1476585, EPI_ISL_1476586, EPI_ISL_1476587, EPI_ISL_1476588, EPI_ISL_1476589, EPI_ISL_1476590, EPI_ISL_1476591, EPI_ISL_1476592, EPI_ISL_1476593, EPI_ISL_1476594, EPI_ISL_1476595, EPI_ISL_1476596, EPI_ISL_1476597, EPI_ISL_1476598, EPI_ISL_1476599, EPI_ISL_1476600, EPI_ISL_1476601, EPI_ISL_1476602, EPI_ISL_1476603, EPI_ISL_1476604, EPI_ISL_1476605, EPI_ISL_1476606, EPI_ISL_1476607, EPI_ISL_1476608, EPI_ISL_1476609, EPI_ISL_1476610, EPI_ISL_1476611, EPI_ISL_1476612, EPI_ISL_1476613, EPI_ISL_1476614, EPI_ISL_1476615, EPI_ISL_1476616, EPI_ISL_1476617, EPI_ISL_1476618, EPI_ISL_1476619, EPI_ISL_1476620, EPI_ISL_1476621, EPI_ISL_1476622, EPI_ISL_1476623, EPI_ISL_1476624, EPI_ISL_1476625, EPI_ISL_1476630, EPI_ISL_1476631, EPI_ISL_1476632, EPI_ISL_1476633, EPI_ISL_1476634, EPI_ISL_1476635, EPI_ISL_1476636, EPI_ISL_1476637, EPI_ISL_1476638, EPI_ISL_1476639, EPI_ISL_1476640, EPI_ISL_1476641, EPI_ISL_1476642, EPI_ISL_1476643, EPI_ISL_1476644, EPI_ISL_1476645, EPI_ISL_1476646, EPI_ISL_1476647, EPI_ISL_1476648, EPI_ISL_1476649, EPI_ISL_1476651, EPI_ISL_1476652, EPI_ISL_1476653, EPI_ISL_1476654, EPI_ISL_1476655, EPI_ISL_1476656, EPI_ISL_1476657, EPI_ISL_1476658, EPI_ISL_1476659, EPI_ISL_1476660, EPI_ISL_1476661, EPI_ISL_1476662, EPI_ISL_1476663, EPI_ISL_1476664, EPI_ISL_1476666, EPI_ISL_1476667, EPI_ISL_1476668, EPI_ISL_1476669, EPI_ISL_1476670, EPI_ISL_1476671, EPI_ISL_1547228, EPI_ISL_1547229, EPI_ISL_1547259, EPI_ISL_1547260, EPI_ISL_1547261, EPI_ISL_1547262, EPI_ISL_1547263, EPI_ISL_1547264, EPI_ISL_1547265, EPI_ISL_1547266, EPI_ISL_1547267, EPI_ISL_1547268, EPI_ISL_1547269, EPI_ISL_1547270, EPI_ISL_1547271, EPI_ISL_1547272, EPI_ISL_1547273, EPI_ISL_1547274, EPI_ISL_1547275, EPI_ISL_1547276, EPI_ISL_1547277, EPI_ISL_1547278, EPI_ISL_1547279, EPI_ISL_1547280, EPI_ISL_1547281, EPI_ISL_1547282, EPI_ISL_1547283, EPI_ISL_1547284, EPI_ISL_1547288, EPI_ISL_1547289, EPI_ISL_1547290, EPI_ISL_1547291, EPI_ISL_1547294, EPI_ISL_1547295, EPI_ISL_1547296, EPI_ISL_1547297, EPI_ISL_1547298, EPI_ISL_1547299, EPI_ISL_1547301, EPI_ISL_1547302, EPI_ISL_1547303, EPI_ISL_1637986, EPI_ISL_1638002, EPI_ISL_1730821, EPI_ISL_1730831, EPI_ISL_1730832, EPI_ISL_1730862, EPI_ISL_1730863, EPI_ISL_1730911, EPI_ISL_1833260, EPI_ISL_1833261, EPI_ISL_1833262, EPI_ISL_1833263, EPI_ISL_1833264, EPI_ISL_1833265, EPI_ISL_1833266, EPI_ISL_1833267, EPI_ISL_1833268, EPI_ISL_1833270, EPI_ISL_1833271, EPI_ISL_1833272, EPI_ISL_1833273, EPI_ISL_1833274, EPI_ISL_1833275, EPI_ISL_1833276, EPI_ISL_1833277, EPI_ISL_1833289, EPI_ISL_1833290, EPI_ISL_1833291, EPI_ISL_1833292, EPI_ISL_1833293, EPI_ISL_1833295, EPI_ISL_1833298, EPI_ISL_1833299, EPI_ISL_1833300, EPI_ISL_1833301, EPI_ISL_1833302, EPI_ISL_1833304, EPI_ISL_1833305, EPI_ISL_1833306, EPI_ISL_1833307, EPI_ISL_1833308, EPI_ISL_1833309, EPI_ISL_1833310, EPI_ISL_1833311, EPI_ISL_1833312, EPI_ISL_1833313, EPI_ISL_1833314, EPI_ISL_1833315, EPI_ISL_1833316, EPI_ISL_1833317, EPI_ISL_1833318, EPI_ISL_1833319, EPI_ISL_1833320, EPI_ISL_1833321, EPI_ISL_1833322, EPI_ISL_1833323, EPI_ISL_1833324, EPI_ISL_1833325, EPI_ISL_1833326, EPI_ISL_1833327, EPI_ISL_1833328, EPI_ISL_1833329, EPI_ISL_1833330, EPI_ISL_1833331, EPI_ISL_1833332, EPI_ISL_1833333, EPI_ISL_1833334, EPI_ISL_1833335, EPI_ISL_1833336, EPI_ISL_1833337, EPI_ISL_1833338, EPI_ISL_1833342, EPI_ISL_1833343, EPI_ISL_1833344, EPI_ISL_1980125, EPI_ISL_1980166, EPI_ISL_1980167, EPI_ISL_1980169, EPI_ISL_1980207, EPI_ISL_1980209, EPI_ISL_2127918, EPI_ISL_2127931, EPI_ISL_2127932, EPI_ISL_2127934, EPI_ISL_2127936, EPI_ISL_2127938, EPI_ISL_2127940, EPI_ISL_2127942, EPI_ISL_2127945, EPI_ISL_2127948, EPI_ISL_2127950, EPI_ISL_2127952, EPI_ISL_2127999, EPI_ISL_2128001, EPI_ISL_2128003, EPI_ISL_2128005, EPI_ISL_2128007, EPI_ISL_2128009, EPI_ISL_2128011, EPI_ISL_2128013, EPI_ISL_2128015, EPI_ISL_2128035, EPI_ISL_2128042, EPI_ISL_2128044, EPI_ISL_2128046, EPI_ISL_2128047, EPI_ISL_2128151, EPI_ISL_2128153, EPI_ISL_2128155, EPI_ISL_2128157, EPI_ISL_2128158, EPI_ISL_2128160, EPI_ISL_2128162, EPI_ISL_2128164, EPI_ISL_2128165, EPI_ISL_2128167, EPI_ISL_2128169, EPI_ISL_2128171 | | | |
| see above | Centre for Enzyme Innovation, University of Portsmouth / Translational Research Laboratory, Portsmouth Hospitals NHS Trust | COVID-19 Genomics UK (COG-UK) Consortium | Allyson Lloyd; Angela Beckett; Anoop Chauhan; Christopher Fearn; Ethan Butcher; Garry Scarlett; Kate Cook; Katie Loveson; Kelly Bicknell; Robert Impey; Salman Goudarzi; Samuel Robson; Sarah Wyllie; Scott Elliott; Sharon Glaysher; Yann Bourgeois |

EPI_ISL_813198, EPI_ISL_813199, EPI_ISL_813201, EPI_ISL_813202, EPI_ISL_813203, EPI_ISL_813204, EPI_ISL_813205, EPI_ISL_813206, EPI_ISL_813207, EPI_ISL_813209, EPI_ISL_813210, EPI_ISL_813211, EPI_ISL_813212, EPI_ISL_813213, EPI_ISL_813214, EPI_ISL_813215, EPI_ISL_813216, EPI_ISL_813217, EPI_ISL_813219, EPI_ISL_813220, EPI_ISL_813221, EPI_ISL_813222, EPI_ISL_813223, EPI_ISL_813224, EPI_ISL_837849, EPI_ISL_837851, EPI_ISL_837853, EPI_ISL_837855, EPI_ISL_837858, EPI_ISL_837860, EPI_ISL_837862, EPI_ISL_837864, EPI_ISL_837866, EPI_ISL_837867, EPI_ISL_837868, EPI_ISL_837870, EPI_ISL_837872, EPI_ISL_837873, EPI_ISL_837874, EPI_ISL_837875, EPI_ISL_837878, EPI_ISL_837880, EPI_ISL_837881, EPI_ISL_837883, EPI_ISL_837884, EPI_ISL_837885, EPI_ISL_837888, EPI_ISL_837890, EPI_ISL_837894, EPI_ISL_837897, EPI_ISL_837899, EPI_ISL_837900, EPI_ISL_837926, EPI_ISL_837928, EPI_ISL_864738, EPI_ISL_864739, EPI_ISL_864740, EPI_ISL_864741, EPI_ISL_864742, EPI_ISL_864743, EPI_ISL_864744, EPI_ISL_864745, EPI_ISL_864746, EPI_ISL_864747, EPI_ISL_864749, EPI_ISL_864750, EPI_ISL_864752, EPI_ISL_864753, EPI_ISL_864757, EPI_ISL_864759, EPI_ISL_864761, EPI_ISL_864763, EPI_ISL_864765, EPI_ISL_864767, EPI_ISL_864769, EPI_ISL_864770, EPI_ISL_864772, EPI_ISL_864785, EPI_ISL_864787, EPI_ISL_864790, EPI_ISL_864795, EPI_ISL_864801, EPI_ISL_864812, EPI_ISL_864816, EPI_ISL_864817, EPI_ISL_864820, EPI_ISL_864822, EPI_ISL_864823, EPI_ISL_864824, EPI_ISL_864825, EPI_ISL_864826, EPI_ISL_864827, EPI_ISL_864830, EPI_ISL_864831, EPI_ISL_864836, EPI_ISL_864848, EPI_ISL_864853, EPI_ISL_864855, EPI_ISL_864858, EPI_ISL_864860, EPI_ISL_864869, EPI_ISL_864871, EPI_ISL_864873, EPI_ISL_864877, EPI_ISL_864879, EPI_ISL_864881, EPI_ISL_864885, EPI_ISL_864887, EPI_ISL_864891, EPI_ISL_864895, EPI_ISL_864917, EPI_ISL_864918, EPI_ISL_864925, EPI_ISL_864947, EPI_ISL_864948, EPI_ISL_918984, EPI_ISL_918997, EPI_ISL_919005, EPI_ISL_919018, EPI_ISL_919019, EPI_ISL_919020, EPI_ISL_919021, EPI_ISL_919022, EPI_ISL_919023, EPI_ISL_919024, EPI_ISL_919025, EPI_ISL_919026, EPI_ISL_919027, EPI_ISL_919028, EPI_ISL_919030, EPI_ISL_919031, EPI_ISL_919032, EPI_ISL_919033, EPI_ISL_919034, EPI_ISL_919035, EPI_ISL_919036, EPI_ISL_919038, EPI_ISL_919040, EPI_ISL_919042, EPI_ISL_919053, EPI_ISL_919054, EPI_ISL_919055, EPI_ISL_919058, EPI_ISL_919062, EPI_ISL_919063, EPI_ISL_919067, EPI_ISL_919070, EPI_ISL_919074, EPI_ISL_919075, EPI_ISL_919079, EPI_ISL_919080, EPI_ISL_919082, EPI_ISL_919084, EPI_ISL_919087, EPI_ISL_919091, EPI_ISL_919092, EPI_ISL_919093, EPI_ISL_919095, EPI_ISL_919097, EPI_ISL_919099, EPI_ISL_919101, EPI_ISL_919102, EPI_ISL_919103, EPI_ISL_919106, EPI_ISL_919107, EPI_ISL_919114, EPI_ISL_919119, EPI_ISL_919134, EPI_ISL_919136, EPI_ISL_919139, EPI_ISL_919143, EPI_ISL_949430, EPI_ISL_949433, EPI_ISL_949479, EPI_ISL_949485, EPI_ISL_949487, EPI_ISL_949490, EPI_ISL_949500, EPI_ISL_949502, EPI_ISL_949508, EPI_ISL_949510, EPI_ISL_949512, EPI_ISL_949514, EPI_ISL_949516, EPI_ISL_949518, EPI_ISL_949519, EPI_ISL_949520, EPI_ISL_949521, EPI_ISL_949541, EPI_ISL_996627, EPI_ISL_996630, EPI_ISL_996672, EPI_ISL_996674, EPI_ISL_996678, EPI_ISL_996681, EPI_ISL_996683, EPI_ISL_996684, EPI_ISL_996685, EPI_ISL_996689, EPI_ISL_996690, EPI_ISL_996694, EPI_ISL_996695, EPI_ISL_996696, EPI_ISL_996704, EPI_ISL_996707, EPI_ISL_996708, EPI_ISL_996712, EPI_ISL_996719, EPI_ISL_996726, EPI_ISL_996728, EPI_ISL_996730, EPI_ISL_996735, EPI_ISL_996737, EPI_ISL_996741, EPI_ISL_996742, EPI_ISL_996778, EPI_ISL_996791, EPI_ISL_996807, EPI_ISL_996834, EPI_ISL_996852, EPI_ISL_996853, EPI_ISL_1047071, EPI_ISL_1047078, EPI_ISL_1047079, EPI_ISL_1047085, EPI_ISL_1047089, EPI_ISL_1047099, EPI_ISL_1103878, EPI_ISL_1103879, EPI_ISL_1103880, EPI_ISL_1103881, EPI_ISL_1103882, EPI_ISL_1103886, EPI_ISL_1103887, EPI_ISL_1103889, EPI_ISL_1177643, EPI_ISL_1177644, EPI_ISL_1177645, EPI_ISL_1177646, EPI_ISL_1177648, EPI_ISL_1177650, EPI_ISL_1177676, EPI_ISL_1247572, EPI_ISL_1247575, EPI_ISL_1247577, EPI_ISL_1247579, EPI_ISL_1247602, EPI_ISL_1247604, EPI_ISL_1247606, EPI_ISL_1247607, EPI_ISL_1247609, EPI_ISL_1247623, EPI_ISL_1247630, EPI_ISL_1308698, EPI_ISL_1386576, EPI_ISL_1386582, EPI_ISL_1386584, EPI_ISL_1386585, EPI_ISL_1386587, EPI_ISL_1545846, EPI_ISL_1730069, EPI_ISL_1832203, EPI_ISL_1832204, EPI_ISL_1832206, EPI_ISL_1832208, EPI_ISL_1832209, EPI_ISL_1832211, EPI_ISL_1832212, EPI_ISL_1832213, EPI_ISL_1832215, EPI_ISL_1832217, EPI_ISL_1974787, EPI_ISL_1974790, EPI_ISL_1974792, EPI_ISL_1974794, EPI_ISL_1974797

| | | | |
|---|---|---|---|
| see above | Department of Pathology, University of Cambridge | COVID-19 Genomics UK (COG-UK) Consortium | Aminu S. Jahun; Grant Hall; Ian Goodfellow; Iliana Georgana; Malte Pinckert; Martin D. Curran; Myra Hosmillo; Rhys Izu; Rhys Izuagbe; Surendra Parmar; William L. Hamilton; Yasmin Chaudhry |

EPI_ISL_796784, EPI_ISL_796785, EPI_ISL_796786, EPI_ISL_796787, EPI_ISL_796788, EPI_ISL_796789, EPI_ISL_796790, EPI_ISL_796791, EPI_ISL_796792, EPI_ISL_796793, EPI_ISL_796794, EPI_ISL_796796, EPI_ISL_796797, EPI_ISL_796798, EPI_ISL_796799, EPI_ISL_796800, EPI_ISL_796801, EPI_ISL_796802, EPI_ISL_796803, EPI_ISL_796804, EPI_ISL_796805, EPI_ISL_796806, EPI_ISL_796807, EPI_ISL_796808, EPI_ISL_796809, EPI_ISL_796810, EPI_ISL_796811, EPI_ISL_796812, EPI_ISL_796813, EPI_ISL_796814, EPI_ISL_796815, EPI_ISL_796816, EPI_ISL_796817, EPI_ISL_796818, EPI_ISL_796819, EPI_ISL_796820, EPI_ISL_796821, EPI_ISL_796822, EPI_ISL_796823, EPI_ISL_796824, EPI_ISL_796825, EPI_ISL_796826, EPI_ISL_796827, EPI_ISL_796828, EPI_ISL_796829, EPI_ISL_796830, EPI_ISL_796831, EPI_ISL_796832, EPI_ISL_796833, EPI_ISL_796834, EPI_ISL_796835, EPI_ISL_796836, EPI_ISL_796837, EPI_ISL_796838, EPI_ISL_796839, EPI_ISL_796840, EPI_ISL_796841, EPI_ISL_796842, EPI_ISL_796843, EPI_ISL_796844, EPI_ISL_796845, EPI_ISL_796846, EPI_ISL_796847, EPI_ISL_796848, EPI_ISL_796849, EPI_ISL_796850, EPI_ISL_796851, EPI_ISL_796852, EPI_ISL_796853, EPI_ISL_796854, EPI_ISL_796855, EPI_ISL_796856, EPI_ISL_796857, EPI_ISL_796858, EPI_ISL_796859, EPI_ISL_796860, EPI_ISL_796861, EPI_ISL_796862, EPI_ISL_796863, EPI_ISL_796864, EPI_ISL_796865, EPI_ISL_796866, EPI_ISL_796867, EPI_ISL_796868, EPI_ISL_796869, EPI_ISL_796870, EPI_ISL_796871, EPI_ISL_796872, EPI_ISL_796873, EPI_ISL_796874, EPI_ISL_796875, EPI_ISL_796876, EPI_ISL_796877, EPI_ISL_796878, EPI_ISL_796879, EPI_ISL_796880, EPI_ISL_796881, EPI_ISL_796882, EPI_ISL_796883, EPI_ISL_796884, EPI_ISL_796885, EPI_ISL_796886, EPI_ISL_796887, EPI_ISL_796888, EPI_ISL_796889, EPI_ISL_796890, EPI_ISL_796891, EPI_ISL_796892, EPI_ISL_796893, EPI_ISL_796894, EPI_ISL_796895, EPI_ISL_796896, EPI_ISL_796897, EPI_ISL_796898, EPI_ISL_796899, EPI_ISL_796900, EPI_ISL_796901, EPI_ISL_796902, EPI_ISL_796903, EPI_ISL_796904, EPI_ISL_796905, EPI_ISL_796906, EPI_ISL_796907, EPI_ISL_796908, EPI_ISL_796909, EPI_ISL_796910, EPI_ISL_796911, EPI_ISL_796912, EPI_ISL_796913, EPI_ISL_796914, EPI_ISL_796915, EPI_ISL_796916, EPI_ISL_796917, EPI_ISL_796918, EPI_ISL_796919, EPI_ISL_796920, EPI_ISL_796921, EPI_ISL_796922, EPI_ISL_796923, EPI_ISL_796924, EPI_ISL_796926, EPI_ISL_796927, EPI_ISL_796928, EPI_ISL_796929, EPI_ISL_796930, EPI_ISL_796931, EPI_ISL_796932, EPI_ISL_796933, EPI_ISL_796934, EPI_ISL_796935, EPI_ISL_796936, EPI_ISL_796937, EPI_ISL_796938, EPI_ISL_796939, EPI_ISL_796940, EPI_ISL_796941, EPI_ISL_796942, EPI_ISL_796943, EPI_ISL_796944, EPI_ISL_796945, EPI_ISL_796946, EPI_ISL_796947, EPI_ISL_796948, EPI_ISL_796949, EPI_ISL_796950, EPI_ISL_796951, EPI_ISL_796952, EPI_ISL_796953, EPI_ISL_796954, EPI_ISL_796955, EPI_ISL_796956, EPI_ISL_796957, EPI_ISL_796958, EPI_ISL_796959, EPI_ISL_796960, EPI_ISL_796961, EPI_ISL_796962, EPI_ISL_796963, EPI_ISL_796964, EPI_ISL_796965, EPI_ISL_796966, EPI_ISL_796967, EPI_ISL_796968, EPI_ISL_796969, EPI_ISL_796970, EPI_ISL_796971, EPI_ISL_796972, EPI_ISL_796973, EPI_ISL_796974, EPI_ISL_796975, EPI_ISL_796976, EPI_ISL_796977, EPI_ISL_796978, EPI_ISL_796979, EPI_ISL_796980, EPI_ISL_796981, EPI_ISL_796982, EPI_ISL_796983, EPI_ISL_796984, EPI_ISL_796985, EPI_ISL_796986, EPI_ISL_796987, EPI_ISL_796988, EPI_ISL_796989, EPI_ISL_796990, EPI_ISL_796991, EPI_ISL_796992, EPI_ISL_796993, EPI_ISL_796994, EPI_ISL_796995, EPI_ISL_796996, EPI_ISL_796998, EPI_ISL_796999, EPI_ISL_797000, EPI_ISL_797001, EPI_ISL_797002, EPI_ISL_797003, EPI_ISL_797004, EPI_ISL_797005, EPI_ISL_797006, EPI_ISL_797007, EPI_ISL_797008, EPI_ISL_797009, EPI_ISL_797010, EPI_ISL_797011, EPI_ISL_797012, EPI_ISL_797013, EPI_ISL_797014, EPI_ISL_797015, EPI_ISL_797016, EPI_ISL_797017, EPI_ISL_797018, EPI_ISL_797019, EPI_ISL_797020, EPI_ISL_797021, EPI_ISL_797022, EPI_ISL_797023, EPI_ISL_797024, EPI_ISL_797025, EPI_ISL_797026, EPI_ISL_797027, EPI_ISL_797028, EPI_ISL_797029, EPI_ISL_797030, EPI_ISL_797031, EPI_ISL_797032, EPI_ISL_797033, EPI_ISL_797034, EPI_ISL_797035, EPI_ISL_797036, EPI_ISL_797037, EPI_ISL_797038, EPI_ISL_797039, EPI_ISL_797040, EPI_ISL_797041, EPI_ISL_797042, EPI_ISL_797043, EPI_ISL_797044, EPI_ISL_797045, EPI_ISL_797046, EPI_ISL_797047, EPI_ISL_797048, EPI_ISL_797049, EPI_ISL_797050, EPI_ISL_797051, EPI_ISL_797052, EPI_ISL_797053, EPI_ISL_797054, EPI_ISL_797055, EPI_ISL_797056, EPI_ISL_797057, EPI_ISL_797058, EPI_ISL_797059, EPI_ISL_797060, EPI_ISL_797061, EPI_ISL_797062, EPI_ISL_797063, EPI_ISL_797064, EPI_ISL_797065, EPI_ISL_797066, EPI_ISL_797067, EPI_ISL_797068, EPI_ISL_797069, EPI_ISL_797070, EPI_ISL_797071, EPI_ISL_797072, EPI_ISL_797073, EPI_ISL_797074, EPI_ISL_797075, EPI_ISL_797076, EPI_ISL_797077, EPI_ISL_797078, EPI_ISL_797079, EPI_ISL_797080, EPI_ISL_797081, EPI_ISL_797082, EPI_ISL_797083, EPI_ISL_797084, EPI_ISL_797085, EPI_ISL_797086, EPI_ISL_797088, EPI_ISL_797089, EPI_ISL_797090, EPI_ISL_797091, EPI_ISL_797092, EPI_ISL_797093, EPI_ISL_797094, EPI_ISL_797095, EPI_ISL_797096, EPI_ISL_797097, EPI_ISL_797098, EPI_ISL_797099, EPI_ISL_797100, EPI_ISL_797101, EPI_ISL_797102, EPI_ISL_797103, EPI_ISL_797104, EPI_ISL_797105, EPI_ISL_797106, EPI_ISL_797107, EPI_ISL_797108, EPI_ISL_797109, EPI_ISL_797110, EPI_ISL_797111, EPI_ISL_797112, EPI_ISL_797113, EPI_ISL_797114, EPI_ISL_797115, EPI_ISL_797116, EPI_ISL_797117, EPI_ISL_797399, EPI_ISL_797402, EPI_ISL_797409, EPI_ISL_797411, EPI_ISL_797413, EPI_ISL_797416, EPI_ISL_797420, EPI_ISL_797421, EPI_ISL_797424, EPI_ISL_797428, EPI_ISL_797430, EPI_ISL_797435, EPI_ISL_797438, EPI_ISL_797441, EPI_ISL_797456, EPI_ISL_797461, EPI_ISL_797462, EPI_ISL_797468, EPI_ISL_797475, EPI_ISL_797480, EPI_ISL_797484, EPI_ISL_797489, EPI_ISL_797491, EPI_ISL_797499, EPI_ISL_797504, EPI_ISL_797509, EPI_ISL_797512, EPI_ISL_797533, EPI_ISL_797535, EPI_ISL_797540, EPI_ISL_797541, EPI_ISL_797543, EPI_ISL_797547, EPI_ISL_797549, EPI_ISL_797553, EPI_ISL_797559, EPI_ISL_797560, EPI_ISL_797568, EPI_ISL_797575, EPI_ISL_797576, EPI_ISL_797586, EPI_ISL_797587, EPI_ISL_797599, EPI_ISL_797605, EPI_ISL_797606, EPI_ISL_797610, EPI_ISL_797613, EPI_ISL_797616, EPI_ISL_797619, EPI_ISL_797635, EPI_ISL_797637, EPI_ISL_797641, EPI_ISL_797642, EPI_ISL_797644, EPI_ISL_797654, EPI_ISL_797660, EPI_ISL_797663, EPI_ISL_797666, EPI_ISL_797673, EPI_ISL_797686, EPI_ISL_797699, EPI_ISL_797701, EPI_ISL_797709, EPI_ISL_797712, EPI_ISL_797714, EPI_ISL_797715, EPI_ISL_797717, EPI_ISL_821003, EPI_ISL_821005, EPI_ISL_821010, EPI_ISL_821011, EPI_ISL_821012, EPI_ISL_821013, EPI_ISL_821018, EPI_ISL_821020, EPI_ISL_821021, EPI_ISL_821022, EPI_ISL_821025, EPI_ISL_821026, EPI_ISL_821028, EPI_ISL_821030, EPI_ISL_821035, EPI_ISL_821036, EPI_ISL_821038, EPI_ISL_821039, EPI_ISL_821042, EPI_ISL_821046, EPI_ISL_821047, EPI_ISL_821050, EPI_ISL_821055, EPI_ISL_821060, EPI_ISL_821063, EPI_ISL_821066, EPI_ISL_821073, EPI_ISL_821074, EPI_ISL_821078, EPI_ISL_821082, EPI_ISL_821084, EPI_ISL_821085, EPI_ISL_821090, EPI_ISL_821091, EPI_ISL_821093, EPI_ISL_821095, EPI_ISL_821099, EPI_ISL_821102, EPI_ISL_821104, EPI_ISL_821109, EPI_ISL_821110, EPI_ISL_821111, EPI_ISL_821117, EPI_ISL_821124, EPI_ISL_821125, EPI_ISL_821126, EPI_ISL_821127, EPI_ISL_821128, EPI_ISL_821131, EPI_ISL_821133, EPI_ISL_821134, EPI_ISL_821136, EPI_ISL_821137, EPI_ISL_821140, EPI_ISL_821144, EPI_ISL_821151, EPI_ISL_821152, EPI_ISL_821156, EPI_ISL_821158, EPI_ISL_821159, EPI_ISL_821163, EPI_ISL_821165, EPI_ISL_821166, EPI_ISL_821169, EPI_ISL_821170, EPI_ISL_821172, EPI_ISL_821173, EPI_ISL_821174, EPI_ISL_821175, EPI_ISL_821178, EPI_ISL_821180, EPI_ISL_821181, EPI_ISL_821187, EPI_ISL_821190, EPI_ISL_821191, EPI_ISL_821194, EPI_ISL_821197, EPI_ISL_821199, EPI_ISL_821200, EPI_ISL_821201, EPI_ISL_821206, EPI_ISL_821208, EPI_ISL_821214, EPI_ISL_821215, EPI_ISL_821220, EPI_ISL_821222, EPI_ISL_821223, EPI_ISL_821228, EPI_ISL_821230, EPI_ISL_821231, EPI_ISL_821235, EPI_ISL_821243, EPI_ISL_821244, EPI_ISL_821248, EPI_ISL_821250, EPI_ISL_821251, EPI_ISL_821253, EPI_ISL_821254, EPI_ISL_821255, EPI_ISL_821261, EPI_ISL_821263, EPI_ISL_821264, EPI_ISL_821270, EPI_ISL_821271, EPI_ISL_821285, EPI_ISL_821292, EPI_ISL_821294, EPI_ISL_821298, EPI_ISL_821303, EPI_ISL_821308, EPI_ISL_821318, EPI_ISL_821327, EPI_ISL_821329, EPI_ISL_821335, EPI_ISL_821347, EPI_ISL_821370, EPI_ISL_821371, EPI_ISL_821382, EPI_ISL_821384, EPI_ISL_821412, EPI_ISL_821414, EPI_ISL_821415, EPI_ISL_821433, EPI_ISL_821436, EPI_ISL_821437, EPI_ISL_821446, EPI_ISL_821456, EPI_ISL_821469, EPI_ISL_821482, EPI_ISL_821483, EPI_ISL_821490, EPI_ISL_821493, EPI_ISL_821496, EPI_ISL_821500, EPI_ISL_821503, EPI_ISL_821525, EPI_ISL_821526, EPI_ISL_821528, EPI_ISL_821534, EPI_ISL_821550, EPI_ISL_821551, EPI_ISL_821556, EPI_ISL_821600, EPI_ISL_821606, EPI_ISL_821617, EPI_ISL_821626, EPI_ISL_821627, EPI_ISL_821628, EPI_ISL_821971, EPI_ISL_821972, EPI_ISL_821973, EPI_ISL_821974, EPI_ISL_821976, EPI_ISL_821977, EPI_ISL_821978, EPI_ISL_821979, EPI_ISL_821980, EPI_ISL_821981, EPI_ISL_821982, EPI_ISL_821983, EPI_ISL_821984, EPI_ISL_821985, EPI_ISL_821986, EPI_ISL_821989, EPI_ISL_821990, EPI_ISL_821991, EPI_ISL_821992, EPI_ISL_821994, EPI_ISL_821996, EPI_ISL_821997, EPI_ISL_821998, EPI_ISL_821999, EPI_ISL_822000, EPI_ISL_822002, EPI_ISL_822003, EPI_ISL_822007, EPI_ISL_822008, EPI_ISL_822009, EPI_ISL_822010, EPI_ISL_822012, EPI_ISL_822013, EPI_ISL_822014, EPI_ISL_822015, EPI_ISL_822016, EPI_ISL_822017, EPI_ISL_822018, EPI_ISL_822019, EPI_ISL_822020, EPI_ISL_822021, EPI_ISL_822024, EPI_ISL_822025, EPI_ISL_822026, EPI_ISL_822027, EPI_ISL_822028, EPI_ISL_822030, EPI_ISL_822031, EPI_ISL_822032, EPI_ISL_822034, EPI_ISL_822036, EPI_ISL_822037, EPI_ISL_822038, EPI_ISL_822039, EPI_ISL_822042, EPI_ISL_822043, EPI_ISL_822046, EPI_ISL_822047, EPI_ISL_822049, EPI_ISL_822050, EPI_ISL_822052, EPI_ISL_822054, EPI_ISL_822055, EPI_ISL_822056, EPI_ISL_822057, EPI_ISL_822058, EPI_ISL_822060, EPI_ISL_822061, EPI_ISL_822062, EPI_ISL_822064, EPI_ISL_822066, EPI_ISL_822068, EPI_ISL_822070, EPI_ISL_822071, EPI_ISL_822073, EPI_ISL_822074, EPI_ISL_822076, EPI_ISL_822077, EPI_ISL_822079, EPI_ISL_822081, EPI_ISL_822082, EPI_ISL_822084, EPI_ISL_822085, EPI_ISL_822087, EPI_ISL_822088, EPI_ISL_822089, EPI_ISL_822090, EPI_ISL_822091, EPI_ISL_822092, EPI_ISL_822094, EPI_ISL_822095, EPI_ISL_822096, EPI_ISL_822097, EPI_ISL_822100, EPI_ISL_822101, EPI_ISL_822102, EPI_ISL_822103, EPI_ISL_822104, EPI_ISL_822105, EPI_ISL_822106, EPI_ISL_822107, EPI_ISL_822108, EPI_ISL_822109, EPI_ISL_822111, EPI_ISL_822112, EPI_ISL_822114, EPI_ISL_822115, EPI_ISL_822116, EPI_ISL_822118, EPI_ISL_822120, EPI_ISL_822121, EPI_ISL_822123, EPI_ISL_822124, EPI_ISL_822125, EPI_ISL_822126, EPI_ISL_822128, EPI_ISL_822129, EPI_ISL_822131, EPI_ISL_822132, EPI_ISL_822133, EPI_ISL_822134, EPI_ISL_822135, EPI_ISL_822136, EPI_ISL_822137, EPI_ISL_822138, EPI_ISL_822139, EPI_ISL_822141, EPI_ISL_822142, EPI_ISL_822143, EPI_ISL_822144, EPI_ISL_822145, EPI_ISL_822146, EPI_ISL_822147, EPI_ISL_822149, EPI_ISL_822150, EPI_ISL_822151, EPI_ISL_822152, EPI_ISL_822154, EPI_ISL_822155, EPI_ISL_822157, EPI_ISL_822160, EPI_ISL_822161, EPI_ISL_822162, EPI_ISL_822163, EPI_ISL_822164, EPI_ISL_822166, EPI_ISL_822167, EPI_ISL_822168, EPI_ISL_822171, EPI_ISL_822172, EPI_ISL_822173, EPI_ISL_822174, EPI_ISL_822175, EPI_ISL_822176, EPI_ISL_822177, EPI_ISL_822179, EPI_ISL_822180, EPI_ISL_822181, EPI_ISL_822183, EPI_ISL_822184, EPI_ISL_822185, EPI_ISL_822186, EPI_ISL_822187, EPI_ISL_822188, EPI_ISL_822189, EPI_ISL_822190, EPI_ISL_822191, EPI_ISL_822192, EPI_ISL_822194, EPI_ISL_822195, EPI_ISL_822196, EPI_ISL_822197, EPI_ISL_822198, EPI_ISL_822199, EPI_ISL_822200, EPI_ISL_822202, EPI_ISL_822203, EPI_ISL_822204, EPI_ISL_822205, EPI_ISL_822206, EPI_ISL_822207, EPI_ISL_822208, EPI_ISL_822210, EPI_ISL_822211, EPI_ISL_822212, EPI_ISL_822213, EPI_ISL_822215, EPI_ISL_822216, EPI_ISL_822217, EPI_ISL_822218, EPI_ISL_822219, EPI_ISL_822220, EPI_ISL_822221, EPI_ISL_822222, EPI_ISL_822224, EPI_ISL_822225, EPI_ISL_822226, EPI_ISL_822227, EPI_ISL_822228, EPI_ISL_822229, EPI_ISL_822231, EPI_ISL_822233, EPI_ISL_822234, EPI_ISL_822235, EPI_ISL_822236, EPI_ISL_822238, EPI_ISL_822239, EPI_ISL_822241, EPI_ISL_822246, EPI_ISL_822248, EPI_ISL_822250, EPI_ISL_822251, EPI_ISL_822252, EPI_ISL_822254, EPI_ISL_822255, EPI_ISL_822257, EPI_ISL_822258, EPI_ISL_822259, EPI_ISL_822261, EPI_ISL_822262, EPI_ISL_822263, EPI_ISL_822266, EPI_ISL_822269, EPI_ISL_822270, EPI_ISL_822271, EPI_ISL_822273, EPI_ISL_822274, EPI_ISL_822276, EPI_ISL_822277, EPI_ISL_822278, EPI_ISL_822279, EPI_ISL_822280, EPI_ISL_822283, EPI_ISL_822284, EPI_ISL_822285, EPI_ISL_822286, EPI_ISL_822287, EPI_ISL_822289, EPI_ISL_822290, EPI_ISL_826467, EPI_ISL_826472, EPI_ISL_826476, EPI_ISL_834146, EPI_ISL_834211, EPI_ISL_834212, EPI_ISL_834223, EPI_ISL_834226, EPI_ISL_834228, EPI_ISL_834230, EPI_ISL_834231, EPI_ISL_834233, EPI_ISL_834235, EPI_ISL_834236, EPI_ISL_834237, EPI_ISL_834246, EPI_ISL_834248, EPI_ISL_834250, EPI_ISL_834256, EPI_ISL_834263, EPI_ISL_834264, EPI_ISL_834271, EPI_ISL_834274, EPI_ISL_834280, EPI_ISL_834283, EPI_ISL_834288, EPI_ISL_834290, EPI_ISL_834293, EPI_ISL_834298, EPI_ISL_834299, EPI_ISL_834311, EPI_ISL_834312, EPI_ISL_834313, EPI_ISL_834344, EPI_ISL_834352, EPI_ISL_834353, EPI_ISL_834360, EPI_ISL_834368, EPI_ISL_834370, EPI_ISL_834373, EPI_ISL_834376, EPI_ISL_834377, EPI_ISL_834378, EPI_ISL_834380, EPI_ISL_834381, EPI_ISL_834382, EPI_ISL_834388, EPI_ISL_834402, EPI_ISL_834404, EPI_ISL_834407, EPI_ISL_834409, EPI_ISL_834417, EPI_ISL_834423, EPI_ISL_834427, EPI_ISL_834430, EPI_ISL_834436, EPI_ISL_834440, EPI_ISL_834443, EPI_ISL_834445, EPI_ISL_834448, EPI_ISL_834449, EPI_ISL_834456, EPI_ISL_834459, EPI_ISL_834460, EPI_ISL_834466, EPI_ISL_834467, EPI_ISL_834468, EPI_ISL_834484, EPI_ISL_834485, EPI_ISL_834488, EPI_ISL_834492, EPI_ISL_834494, EPI_ISL_834497, EPI_ISL_834499, EPI_ISL_834500, EPI_ISL_834503, EPI_ISL_834507, EPI_ISL_834511, EPI_ISL_834517, EPI_ISL_834519, EPI_ISL_834524, EPI_ISL_834529, EPI_ISL_834541, EPI_ISL_834545, EPI_ISL_834804, EPI_ISL_834814, EPI_ISL_834823, EPI_ISL_834825, EPI_ISL_834826, EPI_ISL_834833, EPI_ISL_834834, EPI_ISL_834837, EPI_ISL_834848, EPI_ISL_834865, EPI_ISL_834867, EPI_ISL_834870, EPI_ISL_834874, EPI_ISL_834875, EPI_ISL_834886, EPI_ISL_834889, EPI_ISL_834901, EPI_ISL_834903, EPI_ISL_834909, EPI_ISL_834910, EPI_ISL_834920, EPI_ISL_834921, EPI_ISL_834925, EPI_ISL_834931, EPI_ISL_834933, EPI_ISL_834947, EPI_ISL_834949, EPI_ISL_834959, EPI_ISL_834963, EPI_ISL_834972, EPI_ISL_834979, EPI_ISL_834981, EPI_ISL_834982, EPI_ISL_834983, EPI_ISL_834989, EPI_ISL_834991, EPI_ISL_835001, EPI_ISL_835004, EPI_ISL_835008, EPI_ISL_835017, EPI_ISL_835031, EPI_ISL_835038, EPI_ISL_835043, EPI_ISL_835047, EPI_ISL_835055, EPI_ISL_835058, EPI_ISL_835060, EPI_ISL_835067, EPI_ISL_835091, EPI_ISL_835094, EPI_ISL_835112, EPI_ISL_835114, EPI_ISL_835115, EPI_ISL_835127, EPI_ISL_835130, EPI_ISL_835132, EPI_ISL_835140, EPI_ISL_835483, EPI_ISL_835492, EPI_ISL_835495, EPI_ISL_835504, EPI_ISL_835532, EPI_ISL_835546, EPI_ISL_835550, EPI_ISL_835562, EPI_ISL_835615, EPI_ISL_835639, EPI_ISL_835655, EPI_ISL_835680, EPI_ISL_835698, EPI_ISL_846526, EPI_ISL_846536, EPI_ISL_852191, EPI_ISL_858573, EPI_ISL_858575, EPI_ISL_858577, EPI_ISL_858578, EPI_ISL_858580, EPI_ISL_858582, EPI_ISL_858644, EPI_ISL_858645, EPI_ISL_858647, EPI_ISL_858648, EPI_ISL_858649, EPI_ISL_858653, EPI_ISL_858654, EPI_ISL_858655, EPI_ISL_858662, EPI_ISL_858663, EPI_ISL_858666, EPI_ISL_858667, EPI_ISL_858669, EPI_ISL_858670, EPI_ISL_858671, EPI_ISL_858672, EPI_ISL_858673, EPI_ISL_858674, EPI_ISL_858675, EPI_ISL_858676, EPI_ISL_858677, EPI_ISL_858678, EPI_ISL_858679, EPI_ISL_858680, EPI_ISL_858681, EPI_ISL_858682, EPI_ISL_858683, EPI_ISL_858684, EPI_ISL_858685, EPI_ISL_858686, EPI_ISL_858687, EPI_ISL_858688, EPI_ISL_858689, EPI_ISL_858690, EPI_ISL_858691, EPI_ISL_858692, EPI_ISL_858693, EPI_ISL_858694, EPI_ISL_858695, EPI_ISL_858696, EPI_ISL_858697, EPI_ISL_858698, EPI_ISL_858699, EPI_ISL_858700, EPI_ISL_882303, EPI_ISL_882305, EPI_ISL_882307, EPI_ISL_882308, EPI_ISL_882309, EPI_ISL_882316, EPI_ISL_882319, EPI_ISL_882320, EPI_ISL_882321, EPI_ISL_882323, EPI_ISL_882327, EPI_ISL_882332, EPI_ISL_882339, EPI_ISL_882341, EPI_ISL_882343, EPI_ISL_882344, EPI_ISL_882352, EPI_ISL_882360, EPI_ISL_882361, EPI_ISL_882363, EPI_ISL_882364, EPI_ISL_882365, EPI_ISL_882371, EPI_ISL_882372, EPI_ISL_882375, EPI_ISL_882376, EPI_ISL_882377, EPI_ISL_882379, EPI_ISL_882382, EPI_ISL_882385, EPI_ISL_882388, EPI_ISL_882390, EPI_ISL_882393, EPI_ISL_882397, EPI_ISL_882398, EPI_ISL_882402, EPI_ISL_882404, EPI_ISL_882409, EPI_ISL_882413, EPI_ISL_882415, EPI_ISL_882418, EPI_ISL_882420, EPI_ISL_882421, EPI_ISL_882427, EPI_ISL_882428, EPI_ISL_882429, EPI_ISL_882434, EPI_ISL_882437, EPI_ISL_882438, EPI_ISL_882440, EPI_ISL_882441, EPI_ISL_882442, EPI_ISL_882445, EPI_ISL_882452, EPI_ISL_882471, EPI_ISL_882473, EPI_ISL_882475, EPI_ISL_882476, EPI_ISL_882480, EPI_ISL_882481, EPI_ISL_882482, EPI_ISL_882484, EPI_ISL_882491, EPI_ISL_882494, EPI_ISL_882505, EPI_ISL_882506, EPI_ISL_882512, EPI_ISL_882518, EPI_ISL_882520, EPI_ISL_882524, EPI_ISL_882525, EPI_ISL_882534, EPI_ISL_882537, EPI_ISL_882538, EPI_ISL_882539, EPI_ISL_882542, EPI_ISL_882544, EPI_ISL_882548, EPI_ISL_882549, EPI_ISL_882550, EPI_ISL_882559, EPI_ISL_882562, EPI_ISL_882563, EPI_ISL_882564, EPI_ISL_882565, EPI_ISL_882566, EPI_ISL_882568, EPI_ISL_882571, EPI_ISL_882576, EPI_ISL_882579, EPI_ISL_882580, EPI_ISL_882585, EPI_ISL_882586, EPI_ISL_882588, EPI_ISL_882589, EPI_ISL_882593, EPI_ISL_882595, EPI_ISL_882600, EPI_ISL_882601, EPI_ISL_882602

| | | | |
|---|---|---|---|
| see above | Lighthouse Lab in Alderley Park | Wellcome Sanger Institute for the COVID-19 Genomics UK (COG-UK) Consortium | Cordelia Langford; David K. Jackson; Dominic Kwiatkowski; Ewan Harrison; Ian Johnston; Jacquelyn Wynn; John Sillitoe on behalf of the Wellcome Sanger Institute COVID-19 Surveillance Team; Mairead Hyland; Roberto Amato; Sonia Goncalves; The Lighthouse Lab in Alderley Park and Alex Alderton |

EPI_ISL_797397, EPI_ISL_797398, EPI_ISL_797401, EPI_ISL_797404, EPI_ISL_797406, EPI_ISL_797407, EPI_ISL_797410, EPI_ISL_797412, EPI_ISL_797414, EPI_ISL_797415, EPI_ISL_797417, EPI_ISL_797418, EPI_ISL_797419, EPI_ISL_797422, EPI_ISL_797423, EPI_ISL_797425, EPI_ISL_797426, EPI_ISL_797427, EPI_ISL_797429, EPI_ISL_797431, EPI_ISL_797433, EPI_ISL_797436, EPI_ISL_797437, EPI_ISL_797439, EPI_ISL_797440, EPI_ISL_797442, EPI_ISL_797443, EPI_ISL_797444, EPI_ISL_797447, EPI_ISL_797448, EPI_ISL_797449, EPI_ISL_797450, EPI_ISL_797452, EPI_ISL_797453, EPI_ISL_797454, EPI_ISL_797455, EPI_ISL_797457, EPI_ISL_797458, EPI_ISL_797460, EPI_ISL_797463, EPI_ISL_797464, EPI_ISL_797466, EPI_ISL_797467, EPI_ISL_797469, EPI_ISL_797470, EPI_ISL_797471, EPI_ISL_797472, EPI_ISL_797473, EPI_ISL_797474, EPI_ISL_797476, EPI_ISL_797477, EPI_ISL_797478, EPI_ISL_797479, EPI_ISL_797482, EPI_ISL_797485, EPI_ISL_797486, EPI_ISL_797487, EPI_ISL_797488, EPI_ISL_797490, EPI_ISL_797493, EPI_ISL_797494, EPI_ISL_797495, EPI_ISL_797496, EPI_ISL_797501, EPI_ISL_797503, EPI_ISL_797505, EPI_ISL_797506, EPI_ISL_797507, EPI_ISL_797508, EPI_ISL_797513, EPI_ISL_797515, EPI_ISL_797516, EPI_ISL_797517, EPI_ISL_797518, EPI_ISL_797519, EPI_ISL_797520, EPI_ISL_797521, EPI_ISL_797522, EPI_ISL_797523, EPI_ISL_797524, EPI_ISL_797525, EPI_ISL_797526, EPI_ISL_797527, EPI_ISL_797528, EPI_ISL_797529, EPI_ISL_797530, EPI_ISL_797534, EPI_ISL_797536, EPI_ISL_797538, EPI_ISL_797539, EPI_ISL_797542, EPI_ISL_797544, EPI_ISL_797545, EPI_ISL_797546, EPI_ISL_797550, EPI_ISL_797551, EPI_ISL_797552, EPI_ISL_797555, EPI_ISL_797556, EPI_ISL_797557, EPI_ISL_797558, EPI_ISL_797561, EPI_ISL_797562, EPI_ISL_797565, EPI_ISL_797567, EPI_ISL_797569, EPI_ISL_797570, EPI_ISL_797571, EPI_ISL_797572, EPI_ISL_797573, EPI_ISL_797574, EPI_ISL_797577, EPI_ISL_797581, EPI_ISL_797582, EPI_ISL_797583, EPI_ISL_797584, EPI_ISL_797585, EPI_ISL_797589, EPI_ISL_797592, EPI_ISL_797594, EPI_ISL_797596, EPI_ISL_797598, EPI_ISL_797601, EPI_ISL_797602, EPI_ISL_797603, EPI_ISL_797604, EPI_ISL_797607, EPI_ISL_797608, EPI_ISL_797609, EPI_ISL_797611, EPI_ISL_797612, EPI_ISL_797614, EPI_ISL_797617, EPI_ISL_797618, EPI_ISL_797621, EPI_ISL_797622, EPI_ISL_797623, EPI_ISL_797626, EPI_ISL_797628, EPI_ISL_797629, EPI_ISL_797630, EPI_ISL_797633, EPI_ISL_797634, EPI_ISL_797636, EPI_ISL_797638, EPI_ISL_797639, EPI_ISL_797640, EPI_ISL_797649, EPI_ISL_797650, EPI_ISL_797651, EPI_ISL_797653, EPI_ISL_797655, EPI_ISL_797656, EPI_ISL_797658, EPI_ISL_797661, EPI_ISL_797662, EPI_ISL_797665, EPI_ISL_797667, EPI_ISL_797669, EPI_ISL_797670, EPI_ISL_797671, EPI_ISL_797672, EPI_ISL_797674, EPI_ISL_797675, EPI_ISL_797679, EPI_ISL_797680, EPI_ISL_797681, EPI_ISL_797682, EPI_ISL_797683, EPI_ISL_797684, EPI_ISL_797687, EPI_ISL_797689, EPI_ISL_797691, EPI_ISL_797693, EPI_ISL_797695, EPI_ISL_797696, EPI_ISL_797697, EPI_ISL_797700, EPI_ISL_797702, EPI_ISL_797703, EPI_ISL_797706, EPI_ISL_797707, EPI_ISL_797708, EPI_ISL_797710, EPI_ISL_797711, EPI_ISL_797718, EPI_ISL_811595, EPI_ISL_811681, EPI_ISL_811713, EPI_ISL_821278, EPI_ISL_821286, EPI_ISL_821295, EPI_ISL_821311, EPI_ISL_821316, EPI_ISL_821320, EPI_ISL_821339, EPI_ISL_821360, EPI_ISL_821368, EPI_ISL_821374, EPI_ISL_821376, EPI_ISL_821392, EPI_ISL_821393, EPI_ISL_821399, EPI_ISL_821421, EPI_ISL_821427, EPI_ISL_821441, EPI_ISL_821444, EPI_ISL_821447, EPI_ISL_821454, EPI_ISL_821463, EPI_ISL_821473, EPI_ISL_821479, EPI_ISL_821485, EPI_ISL_821492, EPI_ISL_821506, EPI_ISL_821513, EPI_ISL_821544, EPI_ISL_821546, EPI_ISL_821549, EPI_ISL_821563, EPI_ISL_821564, EPI_ISL_821577, EPI_ISL_821579, EPI_ISL_821583, EPI_ISL_821590, EPI_ISL_821602, EPI_ISL_821604, EPI_ISL_821605, EPI_ISL_821613, EPI_ISL_821661, EPI_ISL_821662, EPI_ISL_821663, EPI_ISL_821664, EPI_ISL_821665, EPI_ISL_821666, EPI_ISL_821668, EPI_ISL_821669, EPI_ISL_821670, EPI_ISL_821671, EPI_ISL_821673, EPI_ISL_821674, EPI_ISL_821676, EPI_ISL_821678, EPI_ISL_821680, EPI_ISL_821681, EPI_ISL_821682, EPI_ISL_821683, EPI_ISL_821684, EPI_ISL_821685, EPI_ISL_821686, EPI_ISL_821687, EPI_ISL_821688, EPI_ISL_821689, EPI_ISL_821690, EPI_ISL_821691, EPI_ISL_821692, EPI_ISL_821693, EPI_ISL_821694, EPI_ISL_821695, EPI_ISL_821696, EPI_ISL_821697, EPI_ISL_821698, EPI_ISL_821699, EPI_ISL_821700, EPI_ISL_821701, EPI_ISL_821702, EPI_ISL_821703, EPI_ISL_821704, EPI_ISL_821705, EPI_ISL_821706, EPI_ISL_821707, EPI_ISL_821708, EPI_ISL_821709, EPI_ISL_821710, EPI_ISL_821711, EPI_ISL_821712, EPI_ISL_821713, EPI_ISL_821714, EPI_ISL_821715, EPI_ISL_821716, EPI_ISL_821717, EPI_ISL_821718, EPI_ISL_821719, EPI_ISL_821720, EPI_ISL_821721, EPI_ISL_821722, EPI_ISL_821723, EPI_ISL_821724, EPI_ISL_821725, EPI_ISL_821726, EPI_ISL_821727, EPI_ISL_821728, EPI_ISL_821729, EPI_ISL_821730, EPI_ISL_821731, EPI_ISL_821732, EPI_ISL_821734, EPI_ISL_821735, EPI_ISL_821736, EPI_ISL_821737, EPI_ISL_821738, EPI_ISL_821739, EPI_ISL_821740, EPI_ISL_821741, EPI_ISL_821742, EPI_ISL_821743, EPI_ISL_821745, EPI_ISL_821746, EPI_ISL_821747, EPI_ISL_821748, EPI_ISL_821749, EPI_ISL_821750, EPI_ISL_821751, EPI_ISL_821752, EPI_ISL_821753, EPI_ISL_821754, EPI_ISL_821755, EPI_ISL_821756, EPI_ISL_821757, EPI_ISL_821758, EPI_ISL_821759, EPI_ISL_821760, EPI_ISL_821761, EPI_ISL_821762, EPI_ISL_821763, EPI_ISL_821764, EPI_ISL_821765, EPI_ISL_821766, EPI_ISL_821767, EPI_ISL_821768, EPI_ISL_821769, EPI_ISL_821770, EPI_ISL_821771, EPI_ISL_821772, EPI_ISL_821773, EPI_ISL_821774, EPI_ISL_821775, EPI_ISL_821776, EPI_ISL_821777, EPI_ISL_821778, EPI_ISL_821779, EPI_ISL_821780, EPI_ISL_821781, EPI_ISL_821782, EPI_ISL_821783, EPI_ISL_821784, EPI_ISL_821785, EPI_ISL_821786, EPI_ISL_821788, EPI_ISL_821789, EPI_ISL_821791, EPI_ISL_821792, EPI_ISL_821793, EPI_ISL_821794, EPI_ISL_821795, EPI_ISL_821796, EPI_ISL_821797, EPI_ISL_821798, EPI_ISL_821799, EPI_ISL_821800, EPI_ISL_821801, EPI_ISL_821802, EPI_ISL_821803, EPI_ISL_821804, EPI_ISL_821805, EPI_ISL_821806, EPI_ISL_821807, EPI_ISL_821808, EPI_ISL_821809, EPI_ISL_821810, EPI_ISL_821811, EPI_ISL_821812, EPI_ISL_821813, EPI_ISL_821814, EPI_ISL_821815, EPI_ISL_821816, EPI_ISL_821817, EPI_ISL_821818, EPI_ISL_821819, EPI_ISL_821820, EPI_ISL_821821, EPI_ISL_821822, EPI_ISL_821823, EPI_ISL_821824, EPI_ISL_821825, EPI_ISL_821826, EPI_ISL_821828, EPI_ISL_821829, EPI_ISL_821830, EPI_ISL_821831, EPI_ISL_821832, EPI_ISL_821833, EPI_ISL_821834, EPI_ISL_821835, EPI_ISL_821836, EPI_ISL_821837, EPI_ISL_821838, EPI_ISL_821839, EPI_ISL_821840, EPI_ISL_821841, EPI_ISL_821842, EPI_ISL_821843, EPI_ISL_821844, EPI_ISL_821845, EPI_ISL_821846, EPI_ISL_821847, EPI_ISL_821848, EPI_ISL_821849, EPI_ISL_821850, EPI_ISL_821851, EPI_ISL_821852, EPI_ISL_821853, EPI_ISL_821854, EPI_ISL_821855, EPI_ISL_821856, EPI_ISL_821857, EPI_ISL_821858, EPI_ISL_821859, EPI_ISL_821860, EPI_ISL_821861, EPI_ISL_821862, EPI_ISL_821863, EPI_ISL_821864, EPI_ISL_821865, EPI_ISL_821866, EPI_ISL_821867, EPI_ISL_821868, EPI_ISL_821869, EPI_ISL_821870, EPI_ISL_821871, EPI_ISL_821872, EPI_ISL_821873, EPI_ISL_821874, EPI_ISL_821875, EPI_ISL_821876, EPI_ISL_821878, EPI_ISL_821879, EPI_ISL_821880, EPI_ISL_821881, EPI_ISL_821882, EPI_ISL_821883, EPI_ISL_821885, EPI_ISL_821886, EPI_ISL_821887, EPI_ISL_821888, EPI_ISL_821890, EPI_ISL_821891, EPI_ISL_821892, EPI_ISL_821893, EPI_ISL_821894, EPI_ISL_821895, EPI_ISL_821896, EPI_ISL_821897, EPI_ISL_821898, EPI_ISL_821899, EPI_ISL_821900, EPI_ISL_821901, EPI_ISL_821902, EPI_ISL_821903, EPI_ISL_821904, EPI_ISL_821905, EPI_ISL_821906, EPI_ISL_821907, EPI_ISL_821908, EPI_ISL_821909, EPI_ISL_821910, EPI_ISL_821911, EPI_ISL_821912, EPI_ISL_821913, EPI_ISL_821914, EPI_ISL_821915, EPI_ISL_821916, EPI_ISL_821917, EPI_ISL_821918, EPI_ISL_821919, EPI_ISL_821920, EPI_ISL_821921, EPI_ISL_821922, EPI_ISL_821923, EPI_ISL_821924, EPI_ISL_821925, EPI_ISL_821926, EPI_ISL_821927, EPI_ISL_821928, EPI_ISL_821929, EPI_ISL_821930, EPI_ISL_821931, EPI_ISL_821932, EPI_ISL_821933, EPI_ISL_821935, EPI_ISL_821936, EPI_ISL_821937, EPI_ISL_821938, EPI_ISL_821939, EPI_ISL_821940, EPI_ISL_821941, EPI_ISL_821942, EPI_ISL_821944, EPI_ISL_821945, EPI_ISL_821946, EPI_ISL_821947, EPI_ISL_821948, EPI_ISL_821949, EPI_ISL_821950, EPI_ISL_821951, EPI_ISL_821952, EPI_ISL_821953, EPI_ISL_821954, EPI_ISL_821955, EPI_ISL_821956, EPI_ISL_821957, EPI_ISL_821958, EPI_ISL_821960, EPI_ISL_821961, EPI_ISL_821962, EPI_ISL_821963, EPI_ISL_821964, EPI_ISL_821965, EPI_ISL_821966, EPI_ISL_821968, EPI_ISL_821969, EPI_ISL_821970, EPI_ISL_826473, EPI_ISL_826475, EPI_ISL_945068

| | | | |
|---|---|---|---|
| see above | Lighthouse Lab in Cambridge | Wellcome Sanger Institute for the COVID-19 Genomics UK (COG-UK) Consortium | Cordelia Langford; David K. Jackson; Dominic Kwiatkowski; Ewan Harrison; Ian Johnston; John Sillitoe on behalf of the Wellcome Sanger Institute COVID-19 Surveillance Team; Rob Howes; Roberto Amato; Sonia Goncalves; The Lighthouse Lab in Cambridge and Alex Alderton |

EPI_ISL_797719, EPI_ISL_797720, EPI_ISL_797721, EPI_ISL_797722, EPI_ISL_797723, EPI_ISL_797724, EPI_ISL_797725, EPI_ISL_797726, EPI_ISL_797727, EPI_ISL_797728, EPI_ISL_797729, EPI_ISL_797730, EPI_ISL_797731, EPI_ISL_797732, EPI_ISL_797733, EPI_ISL_797734, EPI_ISL_797735, EPI_ISL_797736, EPI_ISL_797737, EPI_ISL_797738, EPI_ISL_797740, EPI_ISL_797741, EPI_ISL_797742, EPI_ISL_797743, EPI_ISL_797744, EPI_ISL_797745, EPI_ISL_797746, EPI_ISL_797747, EPI_ISL_797748, EPI_ISL_797749, EPI_ISL_797750, EPI_ISL_797751, EPI_ISL_797752, EPI_ISL_797753, EPI_ISL_797754, EPI_ISL_797755, EPI_ISL_797756, EPI_ISL_797757, EPI_ISL_797758, EPI_ISL_797759, EPI_ISL_797760, EPI_ISL_797761, EPI_ISL_797762, EPI_ISL_797763, EPI_ISL_797764, EPI_ISL_797765, EPI_ISL_797767, EPI_ISL_797768, EPI_ISL_797769, EPI_ISL_797770, EPI_ISL_797771, EPI_ISL_797772, EPI_ISL_797774, EPI_ISL_797776, EPI_ISL_797777, EPI_ISL_797778, EPI_ISL_797779, EPI_ISL_797780, EPI_ISL_797781, EPI_ISL_797782, EPI_ISL_797783, EPI_ISL_797785, EPI_ISL_797786, EPI_ISL_797787, EPI_ISL_797788, EPI_ISL_797790, EPI_ISL_797791, EPI_ISL_797792, EPI_ISL_797794, EPI_ISL_797795, EPI_ISL_797796, EPI_ISL_797798, EPI_ISL_797799, EPI_ISL_797800, EPI_ISL_797801, EPI_ISL_797802, EPI_ISL_797803, EPI_ISL_797804, EPI_ISL_797805, EPI_ISL_797806, EPI_ISL_797807, EPI_ISL_797808, EPI_ISL_797809, EPI_ISL_797810, EPI_ISL_797811, EPI_ISL_797812, EPI_ISL_797813, EPI_ISL_797814, EPI_ISL_797815, EPI_ISL_797816, EPI_ISL_797818, EPI_ISL_797819, EPI_ISL_797820, EPI_ISL_797821, EPI_ISL_797822, EPI_ISL_797823, EPI_ISL_797824, EPI_ISL_797826, EPI_ISL_797827, EPI_ISL_797828, EPI_ISL_797832, EPI_ISL_797833, EPI_ISL_797834, EPI_ISL_797835, EPI_ISL_797836, EPI_ISL_797837, EPI_ISL_797839, EPI_ISL_797840, EPI_ISL_797841, EPI_ISL_797842, EPI_ISL_797843, EPI_ISL_797844, EPI_ISL_797845, EPI_ISL_797846, EPI_ISL_797848, EPI_ISL_797849, EPI_ISL_797852, EPI_ISL_797854, EPI_ISL_797855, EPI_ISL_797856, EPI_ISL_797857, EPI_ISL_797858, EPI_ISL_797859, EPI_ISL_797860, EPI_ISL_797861, EPI_ISL_797862, EPI_ISL_797863, EPI_ISL_797864, EPI_ISL_797865, EPI_ISL_797866, EPI_ISL_797869, EPI_ISL_797871, EPI_ISL_797872, EPI_ISL_797873, EPI_ISL_797874, EPI_ISL_797875, EPI_ISL_797876, EPI_ISL_797877, EPI_ISL_797878, EPI_ISL_797879, EPI_ISL_797880, EPI_ISL_797881, EPI_ISL_797882, EPI_ISL_797883, EPI_ISL_797886, EPI_ISL_797887, EPI_ISL_797888, EPI_ISL_797889, EPI_ISL_797890, EPI_ISL_797891, EPI_ISL_797892, EPI_ISL_797893, EPI_ISL_797894, EPI_ISL_797895, EPI_ISL_797896, EPI_ISL_797897, EPI_ISL_797898, EPI_ISL_797899, EPI_ISL_797900, EPI_ISL_797901, EPI_ISL_797902, EPI_ISL_797904, EPI_ISL_797905, EPI_ISL_797906, EPI_ISL_797907, EPI_ISL_797908, EPI_ISL_797909, EPI_ISL_797910, EPI_ISL_797911, EPI_ISL_797912, EPI_ISL_797913, EPI_ISL_797915, EPI_ISL_797917, EPI_ISL_797918, EPI_ISL_797919, EPI_ISL_797920, EPI_ISL_797922, EPI_ISL_797923, EPI_ISL_797924, EPI_ISL_797925, EPI_ISL_797926, EPI_ISL_797927, EPI_ISL_797928, EPI_ISL_797930, EPI_ISL_797931, EPI_ISL_797932, EPI_ISL_797933, EPI_ISL_797934, EPI_ISL_797935, EPI_ISL_797936, EPI_ISL_797938, EPI_ISL_797939, EPI_ISL_797940, EPI_ISL_797941, EPI_ISL_797942, EPI_ISL_797943, EPI_ISL_797944, EPI_ISL_797946, EPI_ISL_797947, EPI_ISL_797948, EPI_ISL_797949, EPI_ISL_797950, EPI_ISL_797951, EPI_ISL_797953, EPI_ISL_797954, EPI_ISL_797955, EPI_ISL_797956, EPI_ISL_797957, EPI_ISL_797958, EPI_ISL_797959, EPI_ISL_797960, EPI_ISL_797962, EPI_ISL_797963, EPI_ISL_797964, EPI_ISL_797966, EPI_ISL_797967, EPI_ISL_797968, EPI_ISL_797969, EPI_ISL_797971, EPI_ISL_797972, EPI_ISL_797973, EPI_ISL_797974, EPI_ISL_797975, EPI_ISL_797976, EPI_ISL_797977, EPI_ISL_797978, EPI_ISL_797979, EPI_ISL_797981, EPI_ISL_797982, EPI_ISL_797983, EPI_ISL_797984, EPI_ISL_797986, EPI_ISL_797987, EPI_ISL_797988, EPI_ISL_797989, EPI_ISL_797990, EPI_ISL_797991, EPI_ISL_797992, EPI_ISL_797993, EPI_ISL_797995, EPI_ISL_797997, EPI_ISL_797998, EPI_ISL_798001, EPI_ISL_798004, EPI_ISL_798005, EPI_ISL_798006, EPI_ISL_798008, EPI_ISL_798010, EPI_ISL_798012, EPI_ISL_798014, EPI_ISL_798016, EPI_ISL_798019, EPI_ISL_798020, EPI_ISL_798021, EPI_ISL_798023, EPI_ISL_798024, EPI_ISL_798026, EPI_ISL_798028, EPI_ISL_798030, EPI_ISL_798031, EPI_ISL_798033, EPI_ISL_798034, EPI_ISL_798036, EPI_ISL_798037, EPI_ISL_798039, EPI_ISL_798041, EPI_ISL_798042, EPI_ISL_798044, EPI_ISL_798045, EPI_ISL_798047, EPI_ISL_798049, EPI_ISL_798050, EPI_ISL_798052, EPI_ISL_798056, EPI_ISL_798057, EPI_ISL_798059, EPI_ISL_798063, EPI_ISL_798065, EPI_ISL_798067, EPI_ISL_798070, EPI_ISL_798071, EPI_ISL_798074, EPI_ISL_798075, EPI_ISL_798081, EPI_ISL_798082, EPI_ISL_798085, EPI_ISL_798087, EPI_ISL_798089, EPI_ISL_798090, EPI_ISL_798091, EPI_ISL_798092, EPI_ISL_798098, EPI_ISL_798099, EPI_ISL_798100, EPI_ISL_798102, EPI_ISL_798104, EPI_ISL_798109, EPI_ISL_798114, EPI_ISL_798116, EPI_ISL_798118, EPI_ISL_798120, EPI_ISL_798129, EPI_ISL_798135, EPI_ISL_798136, EPI_ISL_798138, EPI_ISL_798142, EPI_ISL_798144, EPI_ISL_798146, EPI_ISL_798147, EPI_ISL_798150, EPI_ISL_798155, EPI_ISL_798156, EPI_ISL_798157, EPI_ISL_798158, EPI_ISL_798162, EPI_ISL_798163, EPI_ISL_798164, EPI_ISL_798165, EPI_ISL_798172, EPI_ISL_798177, EPI_ISL_798179, EPI_ISL_798180, EPI_ISL_798187, EPI_ISL_798188, EPI_ISL_798189, EPI_ISL_798190, EPI_ISL_798191, EPI_ISL_798194, EPI_ISL_798195, EPI_ISL_798196, EPI_ISL_798198, EPI_ISL_798200, EPI_ISL_798205, EPI_ISL_798206, EPI_ISL_798207, EPI_ISL_798209, EPI_ISL_798210, EPI_ISL_798212, EPI_ISL_798215, EPI_ISL_798219, EPI_ISL_798226, EPI_ISL_798227, EPI_ISL_798228, EPI_ISL_798232, EPI_ISL_798233, EPI_ISL_798237, EPI_ISL_798238, EPI_ISL_798240, EPI_ISL_798247, EPI_ISL_798257, EPI_ISL_798259, EPI_ISL_798260, EPI_ISL_798262, EPI_ISL_798263, EPI_ISL_798264, EPI_ISL_798265, EPI_ISL_798267, EPI_ISL_798269, EPI_ISL_798276, EPI_ISL_798279, EPI_ISL_798286, EPI_ISL_798290, EPI_ISL_798291, EPI_ISL_798293, EPI_ISL_798297, EPI_ISL_798301, EPI_ISL_798302, EPI_ISL_798303, EPI_ISL_798304, EPI_ISL_798306, EPI_ISL_798309, EPI_ISL_798313, EPI_ISL_798317, EPI_ISL_798318, EPI_ISL_798320, EPI_ISL_798330, EPI_ISL_799006, EPI_ISL_799009, EPI_ISL_799011, EPI_ISL_799012, EPI_ISL_799014, EPI_ISL_799015, EPI_ISL_799016, EPI_ISL_799019, EPI_ISL_799024, EPI_ISL_799027, EPI_ISL_799028, EPI_ISL_799030, EPI_ISL_799033, EPI_ISL_799034, EPI_ISL_799038, EPI_ISL_799041, EPI_ISL_799042, EPI_ISL_799043, EPI_ISL_799044, EPI_ISL_799049, EPI_ISL_799050, EPI_ISL_799051, EPI_ISL_799054, EPI_ISL_799056, EPI_ISL_799060, EPI_ISL_799062, EPI_ISL_799063, EPI_ISL_799064, EPI_ISL_799065, EPI_ISL_799066, EPI_ISL_799067, EPI_ISL_799068, EPI_ISL_799071, EPI_ISL_799072, EPI_ISL_799073, EPI_ISL_799075, EPI_ISL_799077, EPI_ISL_799080, EPI_ISL_799081, EPI_ISL_799082, EPI_ISL_799084, EPI_ISL_799089, EPI_ISL_799090, EPI_ISL_799096, EPI_ISL_799098, EPI_ISL_799100, EPI_ISL_799101, EPI_ISL_799102, EPI_ISL_799107, EPI_ISL_799108, EPI_ISL_799112, EPI_ISL_799115, EPI_ISL_799118, EPI_ISL_799120, EPI_ISL_799123, EPI_ISL_799125, EPI_ISL_799126, EPI_ISL_799127, EPI_ISL_799129, EPI_ISL_799130, EPI_ISL_799131, EPI_ISL_799132, EPI_ISL_799134, EPI_ISL_799140, EPI_ISL_799144, EPI_ISL_799147, EPI_ISL_799150, EPI_ISL_799151, EPI_ISL_799155, EPI_ISL_799156, EPI_ISL_799157, EPI_ISL_799158, EPI_ISL_799162, EPI_ISL_799163, EPI_ISL_799164, EPI_ISL_799167,

EPI_ISL_799169, EPI_ISL_799172, EPI_ISL_799175, EPI_ISL_799176, EPI_ISL_799180, EPI_ISL_799183, EPI_ISL_799189, EPI_ISL_799190, EPI_ISL_799192, EPI_ISL_799193, EPI_ISL_799194, EPI_ISL_799196, EPI_ISL_799197, EPI_ISL_799201, EPI_ISL_799203, EPI_ISL_799204, EPI_ISL_799205, EPI_ISL_799207, EPI_ISL_799209, EPI_ISL_799215, EPI_ISL_799216, EPI_ISL_799218, EPI_ISL_799221, EPI_ISL_799222, EPI_ISL_799224, EPI_ISL_799230, EPI_ISL_799232, EPI_ISL_799236, EPI_ISL_799240, EPI_ISL_799243, EPI_ISL_799245, EPI_ISL_799246, EPI_ISL_799249, EPI_ISL_799250, EPI_ISL_799255, EPI_ISL_799256, EPI_ISL_799258, EPI_ISL_799260, EPI_ISL_799262, EPI_ISL_799263, EPI_ISL_799264, EPI_ISL_799267, EPI_ISL_799268, EPI_ISL_799270, EPI_ISL_799272, EPI_ISL_799273, EPI_ISL_799274, EPI_ISL_799275, EPI_ISL_799277, EPI_ISL_799279, EPI_ISL_799280, EPI_ISL_799282, EPI_ISL_799283, EPI_ISL_799285, EPI_ISL_799286, EPI_ISL_799287, EPI_ISL_799288, EPI_ISL_799289, EPI_ISL_799290, EPI_ISL_799294, EPI_ISL_821659, EPI_ISL_821660, EPI_ISL_822292, EPI_ISL_822293, EPI_ISL_822295, EPI_ISL_822296, EPI_ISL_822302, EPI_ISL_822304, EPI_ISL_858274, EPI_ISL_858313, EPI_ISL_858583, EPI_ISL_858584, EPI_ISL_858586, EPI_ISL_858589, EPI_ISL_858591, EPI_ISL_858593, EPI_ISL_858595, EPI_ISL_858596, EPI_ISL_858599, EPI_ISL_858602, EPI_ISL_858603, EPI_ISL_858605, EPI_ISL_858606, EPI_ISL_858611, EPI_ISL_858614, EPI_ISL_858617, EPI_ISL_858620, EPI_ISL_858621, EPI_ISL_858622, EPI_ISL_858624, EPI_ISL_858625, EPI_ISL_858629, EPI_ISL_858631, EPI_ISL_858632, EPI_ISL_858633, EPI_ISL_858634, EPI_ISL_858636, EPI_ISL_858639, EPI_ISL_858711, EPI_ISL_858712, EPI_ISL_858715, EPI_ISL_858716, EPI_ISL_858717, EPI_ISL_858718, EPI_ISL_858721, EPI_ISL_858722, EPI_ISL_858723, EPI_ISL_858724, EPI_ISL_858726, EPI_ISL_858727, EPI_ISL_858728, EPI_ISL_858729, EPI_ISL_858730, EPI_ISL_858731, EPI_ISL_858733, EPI_ISL_858734, EPI_ISL_858735, EPI_ISL_858736, EPI_ISL_858738, EPI_ISL_858740, EPI_ISL_858741, EPI_ISL_858742, EPI_ISL_858743, EPI_ISL_858744, EPI_ISL_858745, EPI_ISL_858746, EPI_ISL_858747, EPI_ISL_858749, EPI_ISL_858750, EPI_ISL_858751, EPI_ISL_858754, EPI_ISL_858755, EPI_ISL_858756, EPI_ISL_858757, EPI_ISL_858758, EPI_ISL_858759, EPI_ISL_858760, EPI_ISL_858761, EPI_ISL_858762, EPI_ISL_858763, EPI_ISL_858764, EPI_ISL_858765, EPI_ISL_858766, EPI_ISL_858767, EPI_ISL_858768, EPI_ISL_858769, EPI_ISL_858770, EPI_ISL_858771, EPI_ISL_858772, EPI_ISL_858773, EPI_ISL_858774, EPI_ISL_858775, EPI_ISL_858776, EPI_ISL_858777, EPI_ISL_858781, EPI_ISL_858783, EPI_ISL_863532, EPI_ISL_863590, EPI_ISL_863600, EPI_ISL_863631, EPI_ISL_863635, EPI_ISL_863639, EPI_ISL_863645, EPI_ISL_863651, EPI_ISL_863707, EPI_ISL_863721, EPI_ISL_863729, EPI_ISL_863746, EPI_ISL_863803, EPI_ISL_863811, EPI_ISL_863830, EPI_ISL_863841, EPI_ISL_863853, EPI_ISL_945065, EPI_ISL_945301

| | | | |
|---|---|---|---|
| see above | Lighthouse Lab in Glasgow | Wellcome Sanger Institute for the COVID-19 Genomics UK (COG-UK) Consortium | Anna Dominiczak and Alex Alderton; Carol Clugston; Cordelia Langford; David Gray; David K. Jackson; Dominic Kwiatkowski; Ewan Harrison; Harper VanSteenhouse; Ian Johnston; John Sillitoe on behalf of the Wellcome Sanger Institute COVID-19 Surveillance Team; Roberto Amato; Sonia Goncalves; Yumi Kasai |

EPI_ISL_865000, EPI_ISL_865001, EPI_ISL_865002, EPI_ISL_865003, EPI_ISL_865004, EPI_ISL_865005, EPI_ISL_865006, EPI_ISL_865008

| | | | |
|---|---|---|---|
| see above | Lighthouse Lab in Glasgow / MRC-University of Glasgow Centre for Virus Research | COVID-19 Genomics UK (COG-UK) Consortium | Alasdair MacLean; Alice Broos; Ana da Silva Filipe; Anna Dominiczak; Antonia Ho; Carol Clugston; Daniel Mair; David Gray; David L Robertson; Emma Thomson; Harper VanSteenhouse; James Shepherd; Jenna Nichols; Joseph Hughes; Kathy Li; Kathy Smollett; Kyriaki Nomikou; Lily Tong; Matthew Holden; Natasha Johnson; Richard Orton; Rory Gunson; Sarah McDonald; Sharif Shaaban; Sreenu Vattipally; Stephen Carmichael; Yumi Kasai |

EPI_ISL_797118, EPI_ISL_797120, EPI_ISL_797121, EPI_ISL_797125, EPI_ISL_797126, EPI_ISL_797128, EPI_ISL_797129, EPI_ISL_797133, EPI_ISL_797134, EPI_ISL_797138, EPI_ISL_797139, EPI_ISL_797141, EPI_ISL_797142, EPI_ISL_797143, EPI_ISL_797144, EPI_ISL_797148, EPI_ISL_797149, EPI_ISL_797151, EPI_ISL_797153, EPI_ISL_797154, EPI_ISL_797155, EPI_ISL_797156, EPI_ISL_797160, EPI_ISL_797161, EPI_ISL_797163, EPI_ISL_797164, EPI_ISL_797165, EPI_ISL_797168, EPI_ISL_797171, EPI_ISL_797173, EPI_ISL_797175, EPI_ISL_797176, EPI_ISL_797177, EPI_ISL_797178, EPI_ISL_797179, EPI_ISL_797185, EPI_ISL_797187, EPI_ISL_797190, EPI_ISL_797194, EPI_ISL_797198, EPI_ISL_797202, EPI_ISL_797203, EPI_ISL_797204, EPI_ISL_797205, EPI_ISL_797207, EPI_ISL_797210, EPI_ISL_797212, EPI_ISL_797216, EPI_ISL_797218, EPI_ISL_797220, EPI_ISL_797225, EPI_ISL_797226, EPI_ISL_797228, EPI_ISL_797229, EPI_ISL_797231, EPI_ISL_797232, EPI_ISL_797234, EPI_ISL_797239, EPI_ISL_797240, EPI_ISL_797242, EPI_ISL_797243, EPI_ISL_797246, EPI_ISL_797247, EPI_ISL_797248, EPI_ISL_797252, EPI_ISL_797253, EPI_ISL_797254, EPI_ISL_797256, EPI_ISL_797258, EPI_ISL_797259, EPI_ISL_797261, EPI_ISL_797264, EPI_ISL_797266, EPI_ISL_797269, EPI_ISL_797272, EPI_ISL_797277, EPI_ISL_797281, EPI_ISL_797286, EPI_ISL_797289, EPI_ISL_797290, EPI_ISL_797295, EPI_ISL_797296, EPI_ISL_797301, EPI_ISL_797305, EPI_ISL_797313, EPI_ISL_797315, EPI_ISL_797317, EPI_ISL_797318, EPI_ISL_797319, EPI_ISL_797323, EPI_ISL_797325, EPI_ISL_797326, EPI_ISL_797332, EPI_ISL_797335, EPI_ISL_797337, EPI_ISL_797338, EPI_ISL_797339, EPI_ISL_797342, EPI_ISL_797345, EPI_ISL_797346, EPI_ISL_797349, EPI_ISL_797357, EPI_ISL_797362, EPI_ISL_797363, EPI_ISL_797365, EPI_ISL_797366, EPI_ISL_797367, EPI_ISL_797376, EPI_ISL_797379, EPI_ISL_797380, EPI_ISL_797385, EPI_ISL_797389, EPI_ISL_797390, EPI_ISL_797391, EPI_ISL_797395, EPI_ISL_811207, EPI_ISL_811211, EPI_ISL_811214, EPI_ISL_811215, EPI_ISL_811217, EPI_ISL_811229, EPI_ISL_811234, EPI_ISL_811236, EPI_ISL_811241, EPI_ISL_811247, EPI_ISL_811248, EPI_ISL_811262, EPI_ISL_811275, EPI_ISL_811280, EPI_ISL_811284, EPI_ISL_811293, EPI_ISL_811309, EPI_ISL_811310, EPI_ISL_811312, EPI_ISL_811321, EPI_ISL_811325, EPI_ISL_811335, EPI_ISL_811344, EPI_ISL_811345, EPI_ISL_811350, EPI_ISL_811353, EPI_ISL_811362, EPI_ISL_811366, EPI_ISL_811377, EPI_ISL_811380, EPI_ISL_811381, EPI_ISL_811382, EPI_ISL_811385, EPI_ISL_811389, EPI_ISL_811392, EPI_ISL_811394, EPI_ISL_811395, EPI_ISL_811400, EPI_ISL_811404, EPI_ISL_811406, EPI_ISL_811410, EPI_ISL_811414, EPI_ISL_811420, EPI_ISL_811422, EPI_ISL_811433, EPI_ISL_811436, EPI_ISL_811454, EPI_ISL_811472, EPI_ISL_811478, EPI_ISL_811482, EPI_ISL_811490, EPI_ISL_811494, EPI_ISL_811500, EPI_ISL_811502, EPI_ISL_811516, EPI_ISL_811519, EPI_ISL_811524, EPI_ISL_811530, EPI_ISL_811538, EPI_ISL_811548, EPI_ISL_811563, EPI_ISL_811565, EPI_ISL_811573, EPI_ISL_811580, EPI_ISL_811584, EPI_ISL_811594, EPI_ISL_811606, EPI_ISL_811610, EPI_ISL_811617, EPI_ISL_811618, EPI_ISL_811620, EPI_ISL_811621, EPI_ISL_811626, EPI_ISL_811627, EPI_ISL_811634, EPI_ISL_811645, EPI_ISL_811650, EPI_ISL_811651, EPI_ISL_811660, EPI_ISL_811661, EPI_ISL_811664, EPI_ISL_811666, EPI_ISL_811669, EPI_ISL_811670, EPI_ISL_811671, EPI_ISL_811672, EPI_ISL_811677, EPI_ISL_811678, EPI_ISL_811688, EPI_ISL_811698, EPI_ISL_811701, EPI_ISL_811702, EPI_ISL_811703, EPI_ISL_811708, EPI_ISL_811720, EPI_ISL_811726, EPI_ISL_811727, EPI_ISL_811736, EPI_ISL_811740, EPI_ISL_811750, EPI_ISL_811753, EPI_ISL_811756, EPI_ISL_811759, EPI_ISL_811767, EPI_ISL_811776, EPI_ISL_811785, EPI_ISL_811802, EPI_ISL_811808, EPI_ISL_811811, EPI_ISL_811812, EPI_ISL_811814, EPI_ISL_811817, EPI_ISL_811818, EPI_ISL_811819, EPI_ISL_811821, EPI_ISL_811823, EPI_ISL_811824, EPI_ISL_811825, EPI_ISL_811826, EPI_ISL_811827, EPI_ISL_811828, EPI_ISL_811830, EPI_ISL_811831, EPI_ISL_811832, EPI_ISL_811833, EPI_ISL_811834, EPI_ISL_811835, EPI_ISL_811836, EPI_ISL_811837, EPI_ISL_811839, EPI_ISL_811840, EPI_ISL_811841, EPI_ISL_811843, EPI_ISL_811845, EPI_ISL_811846, EPI_ISL_811847, EPI_ISL_811848, EPI_ISL_811849, EPI_ISL_811850, EPI_ISL_811851, EPI_ISL_811852, EPI_ISL_811853, EPI_ISL_811854, EPI_ISL_811855, EPI_ISL_811856, EPI_ISL_811857, EPI_ISL_811858, EPI_ISL_811859, EPI_ISL_811860, EPI_ISL_811861, EPI_ISL_811862, EPI_ISL_811865, EPI_ISL_811867, EPI_ISL_811868, EPI_ISL_811869, EPI_ISL_811870, EPI_ISL_811871, EPI_ISL_811873, EPI_ISL_811874, EPI_ISL_811875, EPI_ISL_811877, EPI_ISL_811878, EPI_ISL_811879, EPI_ISL_811880, EPI_ISL_811881, EPI_ISL_811883, EPI_ISL_811884, EPI_ISL_811885, EPI_ISL_811886, EPI_ISL_811887, EPI_ISL_811888, EPI_ISL_811889, EPI_ISL_811890, EPI_ISL_811891, EPI_ISL_811892, EPI_ISL_811893, EPI_ISL_811894, EPI_ISL_811896, EPI_ISL_811897, EPI_ISL_811898, EPI_ISL_811899, EPI_ISL_811900, EPI_ISL_811902, EPI_ISL_811905, EPI_ISL_811907, EPI_ISL_811909, EPI_ISL_811910, EPI_ISL_811912, EPI_ISL_811913, EPI_ISL_811914, EPI_ISL_811915, EPI_ISL_811916, EPI_ISL_811917, EPI_ISL_811918, EPI_ISL_811919, EPI_ISL_811921, EPI_ISL_811922, EPI_ISL_811923, EPI_ISL_811924, EPI_ISL_811925, EPI_ISL_811927, EPI_ISL_811928, EPI_ISL_811929, EPI_ISL_811930, EPI_ISL_811931, EPI_ISL_811932, EPI_ISL_811935, EPI_ISL_811936, EPI_ISL_811937, EPI_ISL_811938, EPI_ISL_811939, EPI_ISL_811941, EPI_ISL_811942, EPI_ISL_811943, EPI_ISL_811944, EPI_ISL_811945, EPI_ISL_811946, EPI_ISL_811948, EPI_ISL_811949, EPI_ISL_811950, EPI_ISL_811951, EPI_ISL_811952, EPI_ISL_811953, EPI_ISL_811954, EPI_ISL_811955, EPI_ISL_811956, EPI_ISL_811957, EPI_ISL_811958, EPI_ISL_811959, EPI_ISL_811960, EPI_ISL_811962, EPI_ISL_811965, EPI_ISL_811967, EPI_ISL_811968, EPI_ISL_811969, EPI_ISL_811970, EPI_ISL_811971, EPI_ISL_811972, EPI_ISL_811973, EPI_ISL_811974, EPI_ISL_811975, EPI_ISL_811976, EPI_ISL_811977, EPI_ISL_811978, EPI_ISL_811979, EPI_ISL_811981, EPI_ISL_811983, EPI_ISL_811984, EPI_ISL_811985, EPI_ISL_811986, EPI_ISL_811987, EPI_ISL_811988, EPI_ISL_811989, EPI_ISL_811990, EPI_ISL_811991, EPI_ISL_811993, EPI_ISL_811994, EPI_ISL_811996, EPI_ISL_811997, EPI_ISL_811998, EPI_ISL_811999, EPI_ISL_812000, EPI_ISL_812002, EPI_ISL_812003, EPI_ISL_812007, EPI_ISL_812009, EPI_ISL_812010, EPI_ISL_812012, EPI_ISL_812014, EPI_ISL_812015, EPI_ISL_812017, EPI_ISL_812018, EPI_ISL_812019, EPI_ISL_812020, EPI_ISL_812021, EPI_ISL_812024, EPI_ISL_812025, EPI_ISL_812026, EPI_ISL_812027, EPI_ISL_812028, EPI_ISL_812030, EPI_ISL_812033, EPI_ISL_812035, EPI_ISL_812036, EPI_ISL_812037, EPI_ISL_812038, EPI_ISL_812040, EPI_ISL_812041, EPI_ISL_812042, EPI_ISL_812045, EPI_ISL_812046, EPI_ISL_812047, EPI_ISL_812048, EPI_ISL_812049, EPI_ISL_812050, EPI_ISL_812051, EPI_ISL_812052, EPI_ISL_812054, EPI_ISL_812055, EPI_ISL_812058, EPI_ISL_812059, EPI_ISL_812061, EPI_ISL_812062, EPI_ISL_812063, EPI_ISL_812064, EPI_ISL_812066, EPI_ISL_812067, EPI_ISL_812068, EPI_ISL_812069, EPI_ISL_812070, EPI_ISL_812073, EPI_ISL_812074, EPI_ISL_812075, EPI_ISL_812076, EPI_ISL_812077, EPI_ISL_812078, EPI_ISL_812079, EPI_ISL_812080, EPI_ISL_812083, EPI_ISL_812084, EPI_ISL_812085, EPI_ISL_812086, EPI_ISL_812088, EPI_ISL_812090, EPI_ISL_812091, EPI_ISL_812092, EPI_ISL_812093, EPI_ISL_812094, EPI_ISL_812096, EPI_ISL_812097, EPI_ISL_812098, EPI_ISL_812099, EPI_ISL_812100, EPI_ISL_812101, EPI_ISL_812102, EPI_ISL_812103, EPI_ISL_812104, EPI_ISL_812105, EPI_ISL_812106, EPI_ISL_812109, EPI_ISL_812113, EPI_ISL_821275, EPI_ISL_821277, EPI_ISL_821280, EPI_ISL_821287, EPI_ISL_821291, EPI_ISL_821293, EPI_ISL_821299, EPI_ISL_821300, EPI_ISL_821302, EPI_ISL_821304, EPI_ISL_821305, EPI_ISL_821307, EPI_ISL_821312, EPI_ISL_821313, EPI_ISL_821315, EPI_ISL_821321, EPI_ISL_821322, EPI_ISL_821325, EPI_ISL_821326, EPI_ISL_821330, EPI_ISL_821332, EPI_ISL_821337, EPI_ISL_821345, EPI_ISL_821349, EPI_ISL_821350, EPI_ISL_821355, EPI_ISL_821356, EPI_ISL_821357, EPI_ISL_821361, EPI_ISL_821364, EPI_ISL_821369, EPI_ISL_821385, EPI_ISL_821396, EPI_ISL_821406, EPI_ISL_821407, EPI_ISL_821413, EPI_ISL_821417, EPI_ISL_821420, EPI_ISL_821422, EPI_ISL_821423, EPI_ISL_821424, EPI_ISL_821428, EPI_ISL_821429, EPI_ISL_821430, EPI_ISL_821438, EPI_ISL_821450, EPI_ISL_821452, EPI_ISL_821455, EPI_ISL_821457, EPI_ISL_821458, EPI_ISL_821464, EPI_ISL_821467, EPI_ISL_821471, EPI_ISL_821481, EPI_ISL_821487, EPI_ISL_821489, EPI_ISL_821494, EPI_ISL_821495, EPI_ISL_821499, EPI_ISL_821505, EPI_ISL_821508, EPI_ISL_821512, EPI_ISL_821530, EPI_ISL_821531, EPI_ISL_821532, EPI_ISL_821535, EPI_ISL_821554, EPI_ISL_821557, EPI_ISL_821567, EPI_ISL_821578, EPI_ISL_821581, EPI_ISL_821587, EPI_ISL_821589, EPI_ISL_821591, EPI_ISL_821594, EPI_ISL_821598, EPI_ISL_821610, EPI_ISL_821611, EPI_ISL_821615, EPI_ISL_821619, EPI_ISL_821632, EPI_ISL_821638, EPI_ISL_821639, EPI_ISL_821640, EPI_ISL_821642, EPI_ISL_821643, EPI_ISL_821644, EPI_ISL_821645, EPI_ISL_821646, EPI_ISL_821647, EPI_ISL_821649, EPI_ISL_821650, EPI_ISL_821651, EPI_ISL_821652, EPI_ISL_821654, EPI_ISL_821655, EPI_ISL_821656, EPI_ISL_821657, EPI_ISL_821658, EPI_ISL_852017, EPI_ISL_933441, EPI_ISL_933442, EPI_ISL_933443, EPI_ISL_933444, EPI_ISL_933445, EPI_ISL_933446, EPI_ISL_933448, EPI_ISL_933449, EPI_ISL_933450, EPI_ISL_933451, EPI_ISL_933452, EPI_ISL_933454, EPI_ISL_933455, EPI_ISL_933457, EPI_ISL_933458, EPI_ISL_933459, EPI_ISL_933460, EPI_ISL_933461, EPI_ISL_933462, EPI_ISL_933464, EPI_ISL_933465, EPI_ISL_933466, EPI_ISL_933468, EPI_ISL_933470, EPI_ISL_933471, EPI_ISL_933473, EPI_ISL_933476, EPI_ISL_933477, EPI_ISL_933478, EPI_ISL_945273, EPI_ISL_945339

| | | | |
|---|---|---|---|
| see above | Lighthouse Lab in Milton Keynes | Wellcome Sanger Institute for the COVID-19 Genomics UK (COG-UK) Consortium | Cordelia Langford; David K. Jackson; Dominic Kwiatkowski; Ewan Harrison; Ian Johnston; John Sillitoe on behalf of the Wellcome Sanger Institute COVID-19 Surveillance Team; Roberto Amato; Sonia Goncalves; The Lighthouse Lab in Milton Keynes and Alex Alderton |

EPI_ISL_840108, EPI_ISL_840109, EPI_ISL_840110, EPI_ISL_840112, EPI_ISL_840113, EPI_ISL_840114, EPI_ISL_840115, EPI_ISL_840116, EPI_ISL_840117, EPI_ISL_840118, EPI_ISL_840119, EPI_ISL_840120, EPI_ISL_840121, EPI_ISL_840122, EPI_ISL_840123, EPI_ISL_840124, EPI_ISL_840126, EPI_ISL_866992, EPI_ISL_866993, EPI_ISL_866994, EPI_ISL_866995, EPI_ISL_866996, EPI_ISL_866997, EPI_ISL_866998, EPI_ISL_866999, EPI_ISL_867000, EPI_ISL_867001, EPI_ISL_867002, EPI_ISL_867003, EPI_ISL_867004, EPI_ISL_867005, EPI_ISL_867006, EPI_ISL_867007, EPI_ISL_867008, EPI_ISL_867009, EPI_ISL_867010, EPI_ISL_867011, EPI_ISL_867012, EPI_ISL_867013, EPI_ISL_867014, EPI_ISL_867015, EPI_ISL_867016, EPI_ISL_867017, EPI_ISL_950710, EPI_ISL_950711, EPI_ISL_998928, EPI_ISL_998929, EPI_ISL_998930, EPI_ISL_998931, EPI_ISL_998932, EPI_ISL_998933, EPI_ISL_998934

| | | | |
|---|---|---|---|
| see above | Lincolnshire Hospitals and DeepSeq Nottingham | COVID-19 Genomics UK (COG-UK) Consortium | Christopher Moore; Fei Sang; Joeseph Chappell; Johnny Debebe; Jonathan Ball; Matthew Carlisle; Matthew Loose; Nadine Holmes; Nichola Duckworth; Patrick McClure; Sarah Walsh; Tim Sloan; Victoria Wright |

EPI_ISL_813810, EPI_ISL_813811, EPI_ISL_813812, EPI_ISL_813813, EPI_ISL_813814, EPI_ISL_813815, EPI_ISL_838580, EPI_ISL_838581, EPI_ISL_838582, EPI_ISL_838583, EPI_ISL_838584, EPI_ISL_838589, EPI_ISL_838590, EPI_ISL_838591, EPI_ISL_838594, EPI_ISL_838595, EPI_ISL_838596, EPI_ISL_838598, EPI_ISL_838601, EPI_ISL_838604, EPI_ISL_838606, EPI_ISL_838607, EPI_ISL_838608, EPI_ISL_838609, EPI_ISL_838610, EPI_ISL_838611, EPI_ISL_838612, EPI_ISL_838614, EPI_ISL_838615, EPI_ISL_838616, EPI_ISL_838618, EPI_ISL_838621, EPI_ISL_838622, EPI_ISL_838623, EPI_ISL_838624, EPI_ISL_838629, EPI_ISL_838630, EPI_ISL_838631, EPI_ISL_838632, EPI_ISL_838633, EPI_ISL_838634, EPI_ISL_838635, EPI_ISL_838636, EPI_ISL_838637, EPI_ISL_838639, EPI_ISL_838640, EPI_ISL_838641, EPI_ISL_838642, EPI_ISL_838643, EPI_ISL_838644, EPI_ISL_838647, EPI_ISL_838648, EPI_ISL_838649, EPI_ISL_838650, EPI_ISL_838651, EPI_ISL_838652, EPI_ISL_838653, EPI_ISL_838654, EPI_ISL_838656, EPI_ISL_838657, EPI_ISL_838658, EPI_ISL_838659, EPI_ISL_838660, EPI_ISL_838661, EPI_ISL_838663, EPI_ISL_838667, EPI_ISL_838668, EPI_ISL_865177, EPI_ISL_865354, EPI_ISL_865355, EPI_ISL_865356, EPI_ISL_865357, EPI_ISL_865358, EPI_ISL_865359, EPI_ISL_865360, EPI_ISL_865361, EPI_ISL_865362, EPI_ISL_865363, EPI_ISL_865364, EPI_ISL_865367, EPI_ISL_865368, EPI_ISL_865371, EPI_ISL_865372, EPI_ISL_865375, EPI_ISL_865377, EPI_ISL_865378, EPI_ISL_865379, EPI_ISL_865380, EPI_ISL_865381, EPI_ISL_865382, EPI_ISL_865386, EPI_ISL_865389, EPI_ISL_919452, EPI_ISL_919453, EPI_ISL_919476, EPI_ISL_1474655, EPI_ISL_1545986, EPI_ISL_1545987, EPI_ISL_1545988, EPI_ISL_1545989

| | | | |
|---|---|---|---|
| see above | Liverpool Clinical Laboratories | COVID-19 Genomics UK (COG-UK) Consortium | A Alrezaihi; Alessandro Gerada; Alistair Darby; Angela Cowell; Anita Lucaci; Anu Chawla; Cassie Olateju; Catherine Hartley; Charlotte Nelson; Ecaterina Vamos; Elaine O'Toole; Eleanor G Bentley; Ghada T Shawli; Hermione Webster; Isabel García-Dorival; James Johnson; James P Stewart; Jenifer Manson; Joanne Watts; Jones Benjamin; Jordan J Clark; Julian Hiscox; L Luu; Lucille Rainbow; M Almsaud; Margaret Hughes; Mark Whitehead; Matthew Gemmell; Miren Iturriza-Gomara; Muhannad Alruwaili; N.P Randle; Neil Swainston; PKF Gilmore; Parul Sharma; Rebekah Penrice-Randal; Richard Eccles; Richard Gregory; Sam Haldenby; Steve Paterson; Stuart D Armstrong; Trevor Ian Robinson; Ximeng Han |

EPI_ISL_839710, EPI_ISL_839711, EPI_ISL_839712, EPI_ISL_839713, EPI_ISL_839714, EPI_ISL_839715, EPI_ISL_839716, EPI_ISL_839717, EPI_ISL_839718, EPI_ISL_839719, EPI_ISL_839720, EPI_ISL_839721, EPI_ISL_839722, EPI_ISL_839723, EPI_ISL_839724, EPI_ISL_839725, EPI_ISL_839726, EPI_ISL_839727, EPI_ISL_839728, EPI_ISL_839729, EPI_ISL_839730, EPI_ISL_866486, EPI_ISL_866487, EPI_ISL_866488, EPI_ISL_866489, EPI_ISL_866490, EPI_ISL_866491, EPI_ISL_866492, EPI_ISL_866493, EPI_ISL_866494, EPI_ISL_866495, EPI_ISL_866496, EPI_ISL_866497, EPI_ISL_866498, EPI_ISL_866499, EPI_ISL_866500, EPI_ISL_866501, EPI_ISL_866502, EPI_ISL_866503, EPI_ISL_866504, EPI_ISL_866505, EPI_ISL_866506, EPI_ISL_866507, EPI_ISL_866508, EPI_ISL_866509, EPI_ISL_866510, EPI_ISL_866511, EPI_ISL_866512, EPI_ISL_866513, EPI_ISL_866514, EPI_ISL_866515, EPI_ISL_866516, EPI_ISL_866517, EPI_ISL_866518, EPI_ISL_866519, EPI_ISL_866520, EPI_ISL_866521, EPI_ISL_866522, EPI_ISL_866523, EPI_ISL_866524, EPI_ISL_866525, EPI_ISL_866526, EPI_ISL_866535, EPI_ISL_921249, EPI_ISL_921253, EPI_ISL_921254, EPI_ISL_921255, EPI_ISL_921258, EPI_ISL_921259, EPI_ISL_921260, EPI_ISL_921261, EPI_ISL_921262, EPI_ISL_921264, EPI_ISL_921265, EPI_ISL_921268, EPI_ISL_921271, EPI_ISL_921272, EPI_ISL_921273, EPI_ISL_921274, EPI_ISL_921275, EPI_ISL_921276, EPI_ISL_921278, EPI_ISL_921280, EPI_ISL_921281, EPI_ISL_921282, EPI_ISL_921283, EPI_ISL_921284, EPI_ISL_921286, EPI_ISL_921288, EPI_ISL_921295, EPI_ISL_921297, EPI_ISL_921298, EPI_ISL_921301, EPI_ISL_921303, EPI_ISL_921305, EPI_ISL_921313, EPI_ISL_921314, EPI_ISL_921317, EPI_ISL_921318, EPI_ISL_921371, EPI_ISL_921379, EPI_ISL_921380, EPI_ISL_921381, EPI_ISL_921382, EPI_ISL_921383, EPI_ISL_921384, EPI_ISL_921385, EPI_ISL_921386, EPI_ISL_921387, EPI_ISL_921388, EPI_ISL_921389, EPI_ISL_921390, EPI_ISL_921391, EPI_ISL_921392, EPI_ISL_921393, EPI_ISL_921394, EPI_ISL_921395, EPI_ISL_921396, EPI_ISL_921397, EPI_ISL_921398, EPI_ISL_921399, EPI_ISL_921400, EPI_ISL_921401, EPI_ISL_921402, EPI_ISL_921403, EPI_ISL_921404, EPI_ISL_921405, EPI_ISL_921406, EPI_ISL_921414, EPI_ISL_921415, EPI_ISL_921416, EPI_ISL_921417, EPI_ISL_921418, EPI_ISL_921420, EPI_ISL_921421, EPI_ISL_921422, EPI_ISL_921423, EPI_ISL_921424, EPI_ISL_921425, EPI_ISL_921426, EPI_ISL_921432, EPI_ISL_921433, EPI_ISL_950233, EPI_ISL_950234, EPI_ISL_950235, EPI_ISL_950236, EPI_ISL_950237, EPI_ISL_950238, EPI_ISL_950239, EPI_ISL_950241, EPI_ISL_950242, EPI_ISL_950243, EPI_ISL_950244, EPI_ISL_950245, EPI_ISL_950246, EPI_ISL_950247, EPI_ISL_950248, EPI_ISL_950249, EPI_ISL_950250, EPI_ISL_950251, EPI_ISL_950252, EPI_ISL_950253, EPI_ISL_950256, EPI_ISL_950271, EPI_ISL_950273, EPI_ISL_950274, EPI_ISL_950275, EPI_ISL_950276, EPI_ISL_950277, EPI_ISL_950278, EPI_ISL_950279, EPI_ISL_950281, EPI_ISL_950286, EPI_ISL_950287, EPI_ISL_950295, EPI_ISL_950296, EPI_ISL_950299, EPI_ISL_950300, EPI_ISL_950301, EPI_ISL_950302, EPI_ISL_950303, EPI_ISL_950313, EPI_ISL_950314, EPI_ISL_950315, EPI_ISL_950316, EPI_ISL_950318, EPI_ISL_950319, EPI_ISL_950320, EPI_ISL_950322, EPI_ISL_950351, EPI_ISL_950352, EPI_ISL_950353, EPI_ISL_950354, EPI_ISL_950355, EPI_ISL_950356, EPI_ISL_950357, EPI_ISL_950358, EPI_ISL_950359, EPI_ISL_950360, EPI_ISL_950361, EPI_ISL_950362, EPI_ISL_950363, EPI_ISL_950364, EPI_ISL_950365, EPI_ISL_950366, EPI_ISL_950367, EPI_ISL_950368, EPI_ISL_950369, EPI_ISL_950370, EPI_ISL_950371, EPI_ISL_950372, EPI_ISL_950374, EPI_ISL_950375, EPI_ISL_950377, EPI_ISL_950378, EPI_ISL_950380, EPI_ISL_950381, EPI_ISL_950383, EPI_ISL_950384, EPI_ISL_950385, EPI_ISL_950386, EPI_ISL_950387, EPI_ISL_950388, EPI_ISL_950389, EPI_ISL_950390, EPI_ISL_950416, EPI_ISL_950417, EPI_ISL_950418, EPI_ISL_950419, EPI_ISL_950420, EPI_ISL_950421, EPI_ISL_950422, EPI_ISL_950423, EPI_ISL_950437, EPI_ISL_950444, EPI_ISL_950445, EPI_ISL_950446, EPI_ISL_950447, EPI_ISL_950448, EPI_ISL_950449, EPI_ISL_950450, EPI_ISL_950451, EPI_ISL_950452, EPI_ISL_950453, EPI_ISL_950454, EPI_ISL_950455, EPI_ISL_950456, EPI_ISL_950457, EPI_ISL_950458, EPI_ISL_998315, EPI_ISL_998322, EPI_ISL_998461, EPI_ISL_998462, EPI_ISL_998463, EPI_ISL_998492, EPI_ISL_998505, EPI_ISL_998506, EPI_ISL_998517, EPI_ISL_998520, EPI_ISL_998525, EPI_ISL_998526, EPI_ISL_998536, EPI_ISL_998537, EPI_ISL_998538, EPI_ISL_998539, EPI_ISL_998540, EPI_ISL_998541, EPI_ISL_998545, EPI_ISL_998547, EPI_ISL_998548, EPI_ISL_998549, EPI_ISL_998550, EPI_ISL_998551, EPI_ISL_1050920, EPI_ISL_1050921, EPI_ISL_1050923, EPI_ISL_1050924, EPI_ISL_1050926, EPI_ISL_1050927, EPI_ISL_1050929, EPI_ISL_1050930, EPI_ISL_1050932, EPI_ISL_1050933, EPI_ISL_1050934, EPI_ISL_1050935, EPI_ISL_1050936, EPI_ISL_1050937, EPI_ISL_1050938, EPI_ISL_1050939, EPI_ISL_1050940, EPI_ISL_1050942, EPI_ISL_1050947, EPI_ISL_1050948, EPI_ISL_1050967, EPI_ISL_1050971, EPI_ISL_1050972, EPI_ISL_1050973, EPI_ISL_1050975, EPI_ISL_1050976, EPI_ISL_1050977, EPI_ISL_1050978, EPI_ISL_1050979, EPI_ISL_1050980, EPI_ISL_1050981, EPI_ISL_1050982, EPI_ISL_1050983, EPI_ISL_1050984, EPI_ISL_1050985, EPI_ISL_1050986, EPI_ISL_1050987, EPI_ISL_1050988, EPI_ISL_1050989, EPI_ISL_1050996, EPI_ISL_1050997, EPI_ISL_1050998, EPI_ISL_1050999, EPI_ISL_1051000, EPI_ISL_1051006, EPI_ISL_1051018, EPI_ISL_1051020, EPI_ISL_1051021, EPI_ISL_1051028, EPI_ISL_1051029, EPI_ISL_1051030, EPI_ISL_1051039, EPI_ISL_1105657, EPI_ISL_1105711, EPI_ISL_1105712, EPI_ISL_1105716, EPI_ISL_1105717, EPI_ISL_1105720, EPI_ISL_1105724, EPI_ISL_1105725, EPI_ISL_1105727, EPI_ISL_1105728, EPI_ISL_1309434, EPI_ISL_1309436, EPI_ISL_1309437, EPI_ISL_1309438, EPI_ISL_1309439, EPI_ISL_1309440, EPI_ISL_1309441, EPI_ISL_1309442, EPI_ISL_1309443, EPI_ISL_1309444, EPI_ISL_1309447, EPI_ISL_1309448, EPI_ISL_1309449, EPI_ISL_1309457, EPI_ISL_1309458, EPI_ISL_1309479, EPI_ISL_1309486, EPI_ISL_1309487, EPI_ISL_1309556, EPI_ISL_1309557, EPI_ISL_1309558, EPI_ISL_1309562, EPI_ISL_1309563, EPI_ISL_1309564, EPI_ISL_1309567, EPI_ISL_1309568, EPI_ISL_1309569, EPI_ISL_1309570, EPI_ISL_1309572, EPI_ISL_1309573, EPI_ISL_1309574, EPI_ISL_1309575, EPI_ISL_1309576, EPI_ISL_1309577, EPI_ISL_1309578, EPI_ISL_1309579, EPI_ISL_1309580, EPI_ISL_1309581, EPI_ISL_1309582, EPI_ISL_1309583, EPI_ISL_1309584, EPI_ISL_1309585, EPI_ISL_1309586, EPI_ISL_1309587, EPI_ISL_1309588, EPI_ISL_1309589, EPI_ISL_1309590, EPI_ISL_1309591, EPI_ISL_1309592, EPI_ISL_1309593, EPI_ISL_1309594, EPI_ISL_1309595, EPI_ISL_1309596, EPI_ISL_1309599, EPI_ISL_1309600, EPI_ISL_1309601, EPI_ISL_1309602, EPI_ISL_1309603, EPI_ISL_1309604, EPI_ISL_1309605, EPI_ISL_1309606, EPI_ISL_1309607, EPI_ISL_1309609, EPI_ISL_1309611, EPI_ISL_1309634, EPI_ISL_1387053, EPI_ISL_1387055, EPI_ISL_1387056, EPI_ISL_1387057, EPI_ISL_1387058, EPI_ISL_1387059, EPI_ISL_1387060, EPI_ISL_1387061, EPI_ISL_1387076, EPI_ISL_1387086, EPI_ISL_1387087, EPI_ISL_1387089, EPI_ISL_1387092, EPI_ISL_1387093, EPI_ISL_1387101, EPI_ISL_1387102, EPI_ISL_1387103, EPI_ISL_1387104, EPI_ISL_1387105, EPI_ISL_1387106, EPI_ISL_1387107, EPI_ISL_1387109, EPI_ISL_1387110, EPI_ISL_1387111, EPI_ISL_1387112, EPI_ISL_1475190, EPI_ISL_1475194, EPI_ISL_1475195, EPI_ISL_1475197, EPI_ISL_1475198, EPI_ISL_1475199, EPI_ISL_1475200, EPI_ISL_1475201, EPI_ISL_1475202, EPI_ISL_1475203, EPI_ISL_1475205, EPI_ISL_1475223, EPI_ISL_1475224, EPI_ISL_1475225, EPI_ISL_1475226, EPI_ISL_1475227, EPI_ISL_1475228, EPI_ISL_1475229, EPI_ISL_1475230, EPI_ISL_1475231, EPI_ISL_1475232, EPI_ISL_1475233, EPI_ISL_1475234, EPI_ISL_1475235, EPI_ISL_1475236, EPI_ISL_1475237, EPI_ISL_1475238, EPI_ISL_1475240, EPI_ISL_1475241, EPI_ISL_1475242, EPI_ISL_1475243, EPI_ISL_1475244, EPI_ISL_1475245, EPI_ISL_1475246, EPI_ISL_1475247, EPI_ISL_1475248, EPI_ISL_1475249, EPI_ISL_1475250, EPI_ISL_1475251, EPI_ISL_1475252, EPI_ISL_1475253, EPI_ISL_1475254, EPI_ISL_1475255, EPI_ISL_1475256, EPI_ISL_1475257, EPI_ISL_1475258, EPI_ISL_1475259, EPI_ISL_1475260, EPI_ISL_1475261, EPI_ISL_1475262, EPI_ISL_1475263, EPI_ISL_1475264, EPI_ISL_1475265, EPI_ISL_1475266, EPI_ISL_1475267, EPI_ISL_1475268, EPI_ISL_1475269, EPI_ISL_1475270, EPI_ISL_1475271, EPI_ISL_1475272, EPI_ISL_1475273, EPI_ISL_1475274, EPI_ISL_1475275, EPI_ISL_1475276, EPI_ISL_1475277, EPI_ISL_1475278, EPI_ISL_1475279, EPI_ISL_1475280, EPI_ISL_1475281, EPI_ISL_1475282, EPI_ISL_1475283, EPI_ISL_1475284, EPI_ISL_1475285, EPI_ISL_1475286, EPI_ISL_1475287, EPI_ISL_1475288, EPI_ISL_1475289, EPI_ISL_1475290, EPI_ISL_1475291, EPI_ISL_1475292, EPI_ISL_1475293, EPI_ISL_1475294, EPI_ISL_1475295, EPI_ISL_1475296, EPI_ISL_1475297, EPI_ISL_1475298, EPI_ISL_1475299, EPI_ISL_1475300, EPI_ISL_1475301, EPI_ISL_1475302, EPI_ISL_1475303, EPI_ISL_1475304, EPI_ISL_1475305, EPI_ISL_1475387, EPI_ISL_1730401, EPI_ISL_1978386, EPI_ISL_1978398, EPI_ISL_1978400, EPI_ISL_1978410, EPI_ISL_1978422, EPI_ISL_1978427, EPI_ISL_1978429, EPI_ISL_1978443, EPI_ISL_1978453, EPI_ISL_1978454, EPI_ISL_1978458, EPI_ISL_1978464, EPI_ISL_1978522, EPI_ISL_1978523, EPI_ISL_1978549, EPI_ISL_1978550, EPI_ISL_1978558, EPI_ISL_1978569, EPI_ISL_1978576, EPI_ISL_1978596, EPI_ISL_1978642, EPI_ISL_1978653, EPI_ISL_1978658, EPI_ISL_1978667, EPI_ISL_1978707, EPI_ISL_1978716, EPI_ISL_1978731, EPI_ISL_1978748, EPI_ISL_1978749, EPI_ISL_1978751, EPI_ISL_1978752, EPI_ISL_1978754, EPI_ISL_1978755, EPI_ISL_1978757, EPI_ISL_1978759, EPI_ISL_1978760, EPI_ISL_1978762, EPI_ISL_1978763, EPI_ISL_1978765, EPI_ISL_1978767, EPI_ISL_1978768, EPI_ISL_1978770, EPI_ISL_1978771, EPI_ISL_1978773, EPI_ISL_1978775, EPI_ISL_1978776, EPI_ISL_1978778, EPI_ISL_1978780, EPI_ISL_1978781, EPI_ISL_1978783, EPI_ISL_1978784, EPI_ISL_1978786, EPI_ISL_1978788, EPI_ISL_1978790, EPI_ISL_1978791, EPI_ISL_1978793, EPI_ISL_1978795, EPI_ISL_1978796, EPI_ISL_1978798, EPI_ISL_1978799, EPI_ISL_1978801, EPI_ISL_1978803, EPI_ISL_1978804, EPI_ISL_1978806, EPI_ISL_1978808, EPI_ISL_1978810, EPI_ISL_1978811, EPI_ISL_1978813, EPI_ISL_1978815, EPI_ISL_1978817, EPI_ISL_1978818, EPI_ISL_1978820, EPI_ISL_1978822, EPI_ISL_1978823, EPI_ISL_1978825, EPI_ISL_1978826, EPI_ISL_1978828, EPI_ISL_1978830, EPI_ISL_1978832, EPI_ISL_1978833, EPI_ISL_1978834, EPI_ISL_1978836, EPI_ISL_1978838, EPI_ISL_1978840, EPI_ISL_1978841, EPI_ISL_1978843, EPI_ISL_1978844, EPI_ISL_1978846, EPI_ISL_1978847, EPI_ISL_1978849, EPI_ISL_1978851, EPI_ISL_1978853, EPI_ISL_1978854, EPI_ISL_1978856, EPI_ISL_1978857, EPI_ISL_1978859, EPI_ISL_1978861, EPI_ISL_1978863, EPI_ISL_1978864, EPI_ISL_1978866, EPI_ISL_1978868, EPI_ISL_1978869, EPI_ISL_1978871, EPI_ISL_1978873, EPI_ISL_1978874, EPI_ISL_1978876, EPI_ISL_1978878, EPI_ISL_1978879, EPI_ISL_1978881, EPI_ISL_1978883, EPI_ISL_1978884, EPI_ISL_1978886, EPI_ISL_1978887, EPI_ISL_2126253, EPI_ISL_2126256, EPI_ISL_2126386, EPI_ISL_2126639, EPI_ISL_2126686, EPI_ISL_2126718, EPI_ISL_2126726, EPI_ISL_2126728, EPI_ISL_2126730, EPI_ISL_2126800, EPI_ISL_2126802, EPI_ISL_2126803, EPI_ISL_2126805, EPI_ISL_2126807, EPI_ISL_2126809, EPI_ISL_2126811, EPI_ISL_2126813, EPI_ISL_2126815, EPI_ISL_2126817, EPI_ISL_2126819, EPI_ISL_2126820, EPI_ISL_2126822, EPI_ISL_2126824, EPI_ISL_2126825, EPI_ISL_2126827, EPI_ISL_2126829, EPI_ISL_2126831, EPI_ISL_2126833, EPI_ISL_2126835, EPI_ISL_2126837, EPI_ISL_2126838, EPI_ISL_2126840, EPI_ISL_2126842, EPI_ISL_2126844, EPI_ISL_2126846, EPI_ISL_2126849, EPI_ISL_2126850, EPI_ISL_2126853, EPI_ISL_2126854, EPI_ISL_2126856, EPI_ISL_2126859, EPI_ISL_2126861, EPI_ISL_2126863, EPI_ISL_2126865, EPI_ISL_2126867, EPI_ISL_2126869, EPI_ISL_2126871, EPI_ISL_2126873, EPI_ISL_2126875, EPI_ISL_2126877, EPI_ISL_2126879, EPI_ISL_2126881, EPI_ISL_2126882, EPI_ISL_2126884, EPI_ISL_2126886, EPI_ISL_2126888, EPI_ISL_2126890, EPI_ISL_2126892, EPI_ISL_2126893, EPI_ISL_2126896, EPI_ISL_2126898, EPI_ISL_2126899, EPI_ISL_2126901, EPI_ISL_2126903, EPI_ISL_2126905, EPI_ISL_2126907, EPI_ISL_2126909, EPI_ISL_2126911, EPI_ISL_2126913, EPI_ISL_2126915, EPI_ISL_2126917, EPI_ISL_2126922, EPI_ISL_2126924, EPI_ISL_2126926, EPI_ISL_2126928, EPI_ISL_2126930, EPI_ISL_2126931, EPI_ISL_2126933, EPI_ISL_2126936, EPI_ISL_2126938, EPI_ISL_2126940, EPI_ISL_2126942, EPI_ISL_2126944, EPI_ISL_2126946, EPI_ISL_2126948, EPI_ISL_2126950, EPI_ISL_2126952, EPI_ISL_2126954, EPI_ISL_2126956, EPI_ISL_2126958, EPI_ISL_2126960, EPI_ISL_2126962, EPI_ISL_2126964, EPI_ISL_2126966, EPI_ISL_2126968, EPI_ISL_2126970, EPI_ISL_2126972, EPI_ISL_2126973, EPI_ISL_2126975, EPI_ISL_2126976, EPI_ISL_2126977, EPI_ISL_2126979, EPI_ISL_2126981, EPI_ISL_2126982, EPI_ISL_2126983, EPI_ISL_2126984, EPI_ISL_2126986, EPI_ISL_2126988, EPI_ISL_2126990, EPI_ISL_2126992, EPI_ISL_2126994, EPI_ISL_2126996, EPI_ISL_2126998, EPI_ISL_2127000, EPI_ISL_2127001, EPI_ISL_2127003, EPI_ISL_2127005, EPI_ISL_2127007, EPI_ISL_2127009, EPI_ISL_2127011, EPI_ISL_2127012, EPI_ISL_2127013, EPI_ISL_2127015, EPI_ISL_2127016, EPI_ISL_2127018, EPI_ISL_2127019, EPI_ISL_2127021, EPI_ISL_2127023, EPI_ISL_2127024, EPI_ISL_2127026, EPI_ISL_2127029, EPI_ISL_2127031, EPI_ISL_2127032, EPI_ISL_2127034, EPI_ISL_2127036, EPI_ISL_2127038, EPI_ISL_2127041, EPI_ISL_2127042, EPI_ISL_2127044, EPI_ISL_2127055, EPI_ISL_2127057, EPI_ISL_2127059, EPI_ISL_2127063, EPI_ISL_2127065, EPI_ISL_2127066, EPI_ISL_2127068, EPI_ISL_2127070, EPI_ISL_2127073, EPI_ISL_2127074, EPI_ISL_2127077, EPI_ISL_2127079, EPI_ISL_2127081, EPI_ISL_2127082, EPI_ISL_2127084, EPI_ISL_2127086, EPI_ISL_2127087, EPI_ISL_2127090, EPI_ISL_2127092, EPI_ISL_2127093, EPI_ISL_2127095, EPI_ISL_2127097, EPI_ISL_2127099, EPI_ISL_2127100, EPI_ISL_2127102, EPI_ISL_2127104, EPI_ISL_2127105, EPI_ISL_2127108, EPI_ISL_2127109, EPI_ISL_2127111, EPI_ISL_2127113, EPI_ISL_2127115, EPI_ISL_2127116, EPI_ISL_2127117, EPI_ISL_2127118, EPI_ISL_2127120, EPI_ISL_2127122, EPI_ISL_2127124, EPI_ISL_2127126, EPI_ISL_2127128, EPI_ISL_2127130, EPI_ISL_2127131, EPI_ISL_2127133, EPI_ISL_2127135, EPI_ISL_2127137, EPI_ISL_2127139, EPI_ISL_2127141, EPI_ISL_2127142, EPI_ISL_2127144, EPI_ISL_2127146, EPI_ISL_2127147, EPI_ISL_2127149, EPI_ISL_2127151, EPI_ISL_2127153, EPI_ISL_2127154, EPI_ISL_2127156, EPI_ISL_2127158, EPI_ISL_2127160, EPI_ISL_2127162, EPI_ISL_2127164, EPI_ISL_2127165, EPI_ISL_2127167, EPI_ISL_2127169, EPI_ISL_2127171, EPI_ISL_2127173, EPI_ISL_2127175, EPI_ISL_2127177, EPI_ISL_2127179, EPI_ISL_2127181, EPI_ISL_2127183, EPI_ISL_2127184, EPI_ISL_2127187, EPI_ISL_2127189, EPI_ISL_2127191, EPI_ISL_2127193, EPI_ISL_2127195, EPI_ISL_2127197, EPI_ISL_2127199, EPI_ISL_2127201, EPI_ISL_2127202, EPI_ISL_2127205, EPI_ISL_2127206, EPI_ISL_2127208, EPI_ISL_2127210, EPI_ISL_2127212, EPI_ISL_2127214, EPI_ISL_2127217, EPI_ISL_2127220, EPI_ISL_2127222, EPI_ISL_2127224, EPI_ISL_2127226, EPI_ISL_2127228, EPI_ISL_2127230, EPI_ISL_2127232, EPI_ISL_2127233, EPI_ISL_2127235, EPI_ISL_2127237, EPI_ISL_2127239, EPI_ISL_2127241, EPI_ISL_2127243, EPI_ISL_2127244, EPI_ISL_2127247, EPI_ISL_2127249, EPI_ISL_2127250, EPI_ISL_2127252, EPI_ISL_2127254, EPI_ISL_2127257, EPI_ISL_2127259, EPI_ISL_2127261, EPI_ISL_2127263, EPI_ISL_2127265, EPI_ISL_2127266, EPI_ISL_2127268, EPI_ISL_2127270, EPI_ISL_2127272, EPI_ISL_2127274, EPI_ISL_2127276, EPI_ISL_2127278, EPI_ISL_2127280, EPI_ISL_2127281, EPI_ISL_2127284, EPI_ISL_2127285, EPI_ISL_2127287, EPI_ISL_2127289, EPI_ISL_2127291, EPI_ISL_2127292, EPI_ISL_2127294, EPI_ISL_2127296, EPI_ISL_2127298, EPI_ISL_2127300, EPI_ISL_2127307, EPI_ISL_2127308, EPI_ISL_2127310, EPI_ISL_2127312, EPI_ISL_2127314, EPI_ISL_2127316, EPI_ISL_2127318, EPI_ISL_2127320, EPI_ISL_2127322, EPI_ISL_2127324, EPI_ISL_2127326, EPI_ISL_2127328, EPI_ISL_2127330, EPI_ISL_2127332, EPI_ISL_2127334, EPI_ISL_2127336, EPI_ISL_2127338, EPI_ISL_2127340, EPI_ISL_2127342, EPI_ISL_2127344, EPI_ISL_2127346, EPI_ISL_2127348, EPI_ISL_2127350, EPI_ISL_2127351, EPI_ISL_2127353, EPI_ISL_2127355, EPI_ISL_2127357, EPI_ISL_2127359, EPI_ISL_2127360, EPI_ISL_2127362, EPI_ISL_2127365, EPI_ISL_2127366, EPI_ISL_2127367, EPI_ISL_2127368, EPI_ISL_2127369, EPI_ISL_2127371, EPI_ISL_2127373, EPI_ISL_2127375, EPI_ISL_2127377, EPI_ISL_2127378, EPI_ISL_2127380, EPI_ISL_2127382, EPI_ISL_2127386, EPI_ISL_2127388, EPI_ISL_2127390, EPI_ISL_2127391, EPI_ISL_2127393, EPI_ISL_2127394, EPI_ISL_2127396, EPI_ISL_2127398, EPI_ISL_2127400, EPI_ISL_2127402, EPI_ISL_2127404, EPI_ISL_2127406, EPI_ISL_2127407, EPI_ISL_2127409, EPI_ISL_2127410, EPI_ISL_2127413, EPI_ISL_2127415, EPI_ISL_2127417, EPI_ISL_2127418, EPI_ISL_2127420, EPI_ISL_2127422, EPI_ISL_2127424, EPI_ISL_2127426, EPI_ISL_2127428, EPI_ISL_2127430, EPI_ISL_2127432, EPI_ISL_2127435, EPI_ISL_2127437, EPI_ISL_2127438, EPI_ISL_2127440, EPI_ISL_2127442, EPI_ISL_2127444, EPI_ISL_2127446, EPI_ISL_2127448, EPI_ISL_2127449, EPI_ISL_2127451, EPI_ISL_2127453, EPI_ISL_2127455, EPI_ISL_2127456, EPI_ISL_2127458, EPI_ISL_2127460, EPI_ISL_2127462, EPI_ISL_2127465, EPI_ISL_2127467, EPI_ISL_2127469, EPI_ISL_2127471, EPI_ISL_2127473, EPI_ISL_2127476, EPI_ISL_2127478, EPI_ISL_2127480, EPI_ISL_2127482, EPI_ISL_2127483, EPI_ISL_2127485, EPI_ISL_2127488, EPI_ISL_2127490, EPI_ISL_2127491, EPI_ISL_2127493, EPI_ISL_2127495, EPI_ISL_2127497, EPI_ISL_2127499, EPI_ISL_2127501, EPI_ISL_2127503, EPI_ISL_2127505, EPI_ISL_2127507, EPI_ISL_2127509, EPI_ISL_2127511, EPI_ISL_2127513, EPI_ISL_2127514, EPI_ISL_2127516, EPI_ISL_2127518, EPI_ISL_2127520, EPI_ISL_2127522, EPI_ISL_2127524, EPI_ISL_2127529, EPI_ISL_2127530, EPI_ISL_2127532, EPI_ISL_2127545, EPI_ISL_2127597, EPI_ISL_2127599

| | | | |
|---|---|---|---|
| see above | Northumbria University / South Tees Hospitals NHS Foundation Trust / North Cumbria Integrated Care NHS Foundation Trust / North Tees and Hartlepool NHS Foundation Trust / Newcastle Hospitals NHS Foundation Trust | COVID-19 Genomics UK (COG-UK) Consortium | Andrew Nelson; Brendan Payne; Clive Graham; Darren L Smith; Debra Padgett; Edward Barton; Emma Swindells; Garren Scott; Gary Black; Gary Eltringham; Giles S Holt; Greg R Young; Jane Greenaway; Jennifer Collins; John Allan; Joshua Loh; Lynn Dover; Matthew Bashton; Mohammad A Tariq; Paul Baker; Sarah Essex; Steve Liggett; Wen C Yew; Yusri Taha |

| | | | |
|---|---|---|---|
| EPI_ISL_840371, EPI_ISL_840372, EPI_ISL_840390, EPI_ISL_840391, EPI_ISL_840392, EPI_ISL_840393, EPI_ISL_840394, EPI_ISL_840395, EPI_ISL_840396, EPI_ISL_840397, EPI_ISL_840399, EPI_ISL_840600, EPI_ISL_840603, EPI_ISL_840604, EPI_ISL_840606, EPI_ISL_840609, EPI_ISL_840610, EPI_ISL_840611, EPI_ISL_840612, EPI_ISL_840615, EPI_ISL_840617, EPI_ISL_840618, EPI_ISL_840619, EPI_ISL_840620, EPI_ISL_840621, EPI_ISL_840622, EPI_ISL_840623, EPI_ISL_840624, EPI_ISL_840625, EPI_ISL_840626, EPI_ISL_840627, EPI_ISL_840628, EPI_ISL_840629, EPI_ISL_840630, EPI_ISL_840631, EPI_ISL_840632, EPI_ISL_840633, EPI_ISL_840634, EPI_ISL_840635, EPI_ISL_840636, EPI_ISL_840638, EPI_ISL_840639, EPI_ISL_840640, EPI_ISL_840641, EPI_ISL_840643, EPI_ISL_840644, EPI_ISL_840645, EPI_ISL_840646, EPI_ISL_840647, EPI_ISL_840648, EPI_ISL_840649, EPI_ISL_840651, EPI_ISL_840652, EPI_ISL_840653, EPI_ISL_840655, EPI_ISL_840656, EPI_ISL_840657, EPI_ISL_840658, EPI_ISL_840671, EPI_ISL_840672, EPI_ISL_840673, EPI_ISL_840674, EPI_ISL_840675, EPI_ISL_840676, EPI_ISL_840695, EPI_ISL_840696, EPI_ISL_840697, EPI_ISL_840698, EPI_ISL_840699, EPI_ISL_840700, EPI_ISL_840701, EPI_ISL_840702, EPI_ISL_840703, EPI_ISL_840774, EPI_ISL_840775, EPI_ISL_840776, EPI_ISL_840777, EPI_ISL_840778, EPI_ISL_840779, EPI_ISL_840781, EPI_ISL_840782, EPI_ISL_840783, EPI_ISL_840785, EPI_ISL_840787, EPI_ISL_840790, EPI_ISL_840794, EPI_ISL_840795, EPI_ISL_840796, EPI_ISL_840798, EPI_ISL_840799, EPI_ISL_840801, EPI_ISL_840802, EPI_ISL_840803, EPI_ISL_840804, EPI_ISL_840805, EPI_ISL_840806, EPI_ISL_840808, EPI_ISL_840810, EPI_ISL_840811, EPI_ISL_840812, EPI_ISL_840813, EPI_ISL_840814, EPI_ISL_840815, EPI_ISL_841321, EPI_ISL_841355, EPI_ISL_841359, EPI_ISL_841360, EPI_ISL_841361, EPI_ISL_841362, EPI_ISL_841363, EPI_ISL_841364, EPI_ISL_841365, EPI_ISL_841366, EPI_ISL_841367, EPI_ISL_841368, EPI_ISL_841369, EPI_ISL_841370, EPI_ISL_841508, EPI_ISL_841509, EPI_ISL_841521, EPI_ISL_841522, EPI_ISL_841580, EPI_ISL_841581, EPI_ISL_841584, EPI_ISL_841587, EPI_ISL_841591, EPI_ISL_841608, EPI_ISL_867216, EPI_ISL_867217, EPI_ISL_867431, EPI_ISL_867437, EPI_ISL_867438, EPI_ISL_867439, EPI_ISL_867440, EPI_ISL_867441, EPI_ISL_867442, EPI_ISL_867443, EPI_ISL_867486, EPI_ISL_1053526, EPI_ISL_1106644, EPI_ISL_1476517, EPI_ISL_1476519 | | | |
| see above | Originating lab: Wales Specialist Virology Centre Sequencing lab: Pathogen Genomics Unit | Public Health Wales Microbiology Cardiff Wales Specialist Virology Centre | Alec Birchley; Alexander Adams; Amy Gaskin; Angela Marchbank; Bree Gatica-Wilcox; Catherine Moore; Jason Coombes; Joanne Watkins; Joel Southgate; Johnathan Evans; Laura Gifford; Lauren Gilbert; Lee Graham; Malorie Perry; Matthew Bull; Nicole Pacchiarini; Sally Corden; Sara Kumziene-Summerhayes; Sara Rey; Sarah Taylor; Simon Cottrell; Sophie Jones; Tom Connor |
| EPI_ISL_867039, EPI_ISL_867040, EPI_ISL_867041, EPI_ISL_867044, EPI_ISL_867045, EPI_ISL_867046, EPI_ISL_867048, EPI_ISL_867049, EPI_ISL_867050, EPI_ISL_867052, EPI_ISL_867053, EPI_ISL_867054, EPI_ISL_867056, EPI_ISL_867057, EPI_ISL_867058, EPI_ISL_867059, EPI_ISL_867060, EPI_ISL_867061, EPI_ISL_867062, EPI_ISL_867063, EPI_ISL_867064, EPI_ISL_867065, EPI_ISL_867066, EPI_ISL_867067, EPI_ISL_867068, EPI_ISL_867069, EPI_ISL_867071, EPI_ISL_867072, EPI_ISL_867073, EPI_ISL_867074, EPI_ISL_867075, EPI_ISL_867077, EPI_ISL_867078, EPI_ISL_867079, EPI_ISL_867080, EPI_ISL_867081, EPI_ISL_867082, EPI_ISL_867084, EPI_ISL_867087, EPI_ISL_867088, EPI_ISL_867089, EPI_ISL_867090, EPI_ISL_867093, EPI_ISL_867094, EPI_ISL_867095, EPI_ISL_867096, EPI_ISL_867097, EPI_ISL_867098, EPI_ISL_867130, EPI_ISL_867155, EPI_ISL_867159, EPI_ISL_867161, EPI_ISL_867162, EPI_ISL_867168, EPI_ISL_867169, EPI_ISL_867170, EPI_ISL_867171, EPI_ISL_867181, EPI_ISL_922241, EPI_ISL_922260, EPI_ISL_922261, EPI_ISL_922262, EPI_ISL_922263, EPI_ISL_922264, EPI_ISL_922267, EPI_ISL_922271, EPI_ISL_922272, EPI_ISL_922274, EPI_ISL_922275, EPI_ISL_922277, EPI_ISL_922279, EPI_ISL_922287, EPI_ISL_922288, EPI_ISL_922289, EPI_ISL_922290, EPI_ISL_922291, EPI_ISL_922292, EPI_ISL_922293, EPI_ISL_922296, EPI_ISL_922297, EPI_ISL_922299, EPI_ISL_922300, EPI_ISL_922302, EPI_ISL_922305, EPI_ISL_950953, EPI_ISL_950954, EPI_ISL_950956, EPI_ISL_950957, EPI_ISL_950958, EPI_ISL_950959, EPI_ISL_950965, EPI_ISL_950979, EPI_ISL_950980, EPI_ISL_950981, EPI_ISL_950982, EPI_ISL_950983, EPI_ISL_950985, EPI_ISL_950987, EPI_ISL_950988, EPI_ISL_950989, EPI_ISL_950990, EPI_ISL_950991, EPI_ISL_950992, EPI_ISL_950993, EPI_ISL_950994, EPI_ISL_950995, EPI_ISL_950996, EPI_ISL_950999, EPI_ISL_951000, EPI_ISL_951002, EPI_ISL_951003, EPI_ISL_951004, EPI_ISL_951008, EPI_ISL_951009, EPI_ISL_951010, EPI_ISL_951011, EPI_ISL_951013, EPI_ISL_951014, EPI_ISL_951016, EPI_ISL_951018, EPI_ISL_951020, EPI_ISL_951021, EPI_ISL_951023, EPI_ISL_951024, EPI_ISL_951025, EPI_ISL_951026, EPI_ISL_951027, EPI_ISL_951028, EPI_ISL_951029, EPI_ISL_951030, EPI_ISL_951031, EPI_ISL_951032, EPI_ISL_951033, EPI_ISL_951034, EPI_ISL_951036, EPI_ISL_951037, EPI_ISL_951038, EPI_ISL_951042, EPI_ISL_951043, EPI_ISL_951044, EPI_ISL_951045, EPI_ISL_951046, EPI_ISL_951049, EPI_ISL_951050, EPI_ISL_951052, EPI_ISL_951053, EPI_ISL_951054, EPI_ISL_951055, EPI_ISL_951056, EPI_ISL_951057, EPI_ISL_951058, EPI_ISL_951059, EPI_ISL_951061, EPI_ISL_951062, EPI_ISL_951064, EPI_ISL_951066, EPI_ISL_951079, EPI_ISL_951080, EPI_ISL_951081, EPI_ISL_951083, EPI_ISL_951094, EPI_ISL_951097, EPI_ISL_951098, EPI_ISL_951163, EPI_ISL_951164, EPI_ISL_951165, EPI_ISL_951166, EPI_ISL_951169, EPI_ISL_951170, EPI_ISL_951171, EPI_ISL_951249, EPI_ISL_998991, EPI_ISL_2127630, EPI_ISL_2127635 | | | |
| see above | Oxford Viromics, NDM, University of Oxford; Oxford University Hospitals; Basingstoke and North Hampshire Hospital | COVID-19 Genomics UK (COG-UK) Consortium | Alex Mobbs; Amy Trebes; Anita Justice; Catrin Moore; Christophe Fraser; David Bonsall; David Buck; Emma Wise; George Macintyre; Jessica Lynch; John Todd; Mariateresa de Cesare; Matilde Mori; Monique Andersson; Nathan Moore; Nick Cortes; Robert Shaw; Stephen Kidd; Tanya Golubchik; Timothy Peto |
| EPI_ISL_763769, EPI_ISL_819378, EPI_ISL_819387, EPI_ISL_819388, EPI_ISL_819389, EPI_ISL_819390, EPI_ISL_819398, EPI_ISL_819399, EPI_ISL_819400, EPI_ISL_819401, EPI_ISL_819402, EPI_ISL_819403, EPI_ISL_819404, EPI_ISL_819428, EPI_ISL_819431, EPI_ISL_819433, EPI_ISL_819438, EPI_ISL_820353, EPI_ISL_820355, EPI_ISL_820358, EPI_ISL_820360, EPI_ISL_820363, EPI_ISL_820365, EPI_ISL_820368, EPI_ISL_820371, EPI_ISL_820378, EPI_ISL_820388, EPI_ISL_820391, EPI_ISL_820399, EPI_ISL_820404, EPI_ISL_820407, EPI_ISL_820410, EPI_ISL_820412, EPI_ISL_820415, EPI_ISL_820426, EPI_ISL_820436, EPI_ISL_820439, EPI_ISL_820441, EPI_ISL_820444, EPI_ISL_820446, EPI_ISL_820449, EPI_ISL_820451, EPI_ISL_820454, EPI_ISL_820456, EPI_ISL_820459, EPI_ISL_820462, EPI_ISL_820464, EPI_ISL_820467, EPI_ISL_820470, EPI_ISL_820472, EPI_ISL_820475, EPI_ISL_820478, EPI_ISL_820480, EPI_ISL_820483, EPI_ISL_820485, EPI_ISL_820488, EPI_ISL_820490, EPI_ISL_820493, EPI_ISL_820496, EPI_ISL_820498, EPI_ISL_820501, EPI_ISL_820504, EPI_ISL_820506, EPI_ISL_820509, EPI_ISL_820511, EPI_ISL_820514, EPI_ISL_820516, EPI_ISL_820519, EPI_ISL_820521, EPI_ISL_820524, EPI_ISL_820526, EPI_ISL_820528, EPI_ISL_820531, EPI_ISL_820533, EPI_ISL_820536, EPI_ISL_820539, EPI_ISL_820541, EPI_ISL_820543, EPI_ISL_820546, EPI_ISL_820549, EPI_ISL_820552, EPI_ISL_820554, EPI_ISL_820557, EPI_ISL_820560, EPI_ISL_820563, EPI_ISL_820565, EPI_ISL_820568, EPI_ISL_820570, EPI_ISL_866601, EPI_ISL_921786, EPI_ISL_921787, EPI_ISL_921788, EPI_ISL_921793, EPI_ISL_921800, EPI_ISL_921805, EPI_ISL_921808, EPI_ISL_921809, EPI_ISL_921810, EPI_ISL_921813, EPI_ISL_921819, EPI_ISL_921825, EPI_ISL_955055, EPI_ISL_996082, EPI_ISL_996083, EPI_ISL_996084, EPI_ISL_996085, EPI_ISL_996086, EPI_ISL_996087, EPI_ISL_996088, EPI_ISL_996089, EPI_ISL_996090, EPI_ISL_996091, EPI_ISL_996092, EPI_ISL_996093, EPI_ISL_996094, EPI_ISL_996095, EPI_ISL_996096, EPI_ISL_996097, EPI_ISL_996098, EPI_ISL_996099, EPI_ISL_996100, EPI_ISL_996101, EPI_ISL_996102, EPI_ISL_996103, EPI_ISL_996104, EPI_ISL_996105, EPI_ISL_996106, EPI_ISL_996107, EPI_ISL_996108, EPI_ISL_996109, EPI_ISL_996110, EPI_ISL_996111, EPI_ISL_996112, EPI_ISL_996113, EPI_ISL_996114, EPI_ISL_996115, EPI_ISL_996116, EPI_ISL_996117, EPI_ISL_996118, EPI_ISL_996119, EPI_ISL_996120, EPI_ISL_996121, EPI_ISL_996122, EPI_ISL_996123, EPI_ISL_996124, EPI_ISL_996125, EPI_ISL_996126, EPI_ISL_996127, EPI_ISL_996128, EPI_ISL_996129, EPI_ISL_996130, EPI_ISL_996131, EPI_ISL_996132, EPI_ISL_996133, EPI_ISL_996134, EPI_ISL_996135, EPI_ISL_996136, EPI_ISL_996137, EPI_ISL_996138, EPI_ISL_996139, EPI_ISL_996140, EPI_ISL_996141, EPI_ISL_996142, EPI_ISL_996143, EPI_ISL_996144, EPI_ISL_996145, EPI_ISL_996146, EPI_ISL_996147, EPI_ISL_996148, EPI_ISL_996149, EPI_ISL_996150, EPI_ISL_996151, EPI_ISL_996152, EPI_ISL_996153, EPI_ISL_996154, EPI_ISL_996155, EPI_ISL_996156, EPI_ISL_996157, EPI_ISL_996158, EPI_ISL_996159, EPI_ISL_996160, EPI_ISL_996161, EPI_ISL_996162, EPI_ISL_996163, EPI_ISL_996164, EPI_ISL_996165, EPI_ISL_996166, EPI_ISL_996167, EPI_ISL_996168, EPI_ISL_996169, EPI_ISL_996170, EPI_ISL_996171, EPI_ISL_996172, EPI_ISL_996173, EPI_ISL_996174, EPI_ISL_996175, EPI_ISL_996176, EPI_ISL_996177, EPI_ISL_996178, EPI_ISL_996179, EPI_ISL_996180, EPI_ISL_996181, EPI_ISL_996182, EPI_ISL_996183, EPI_ISL_996184, EPI_ISL_996185, EPI_ISL_996186, EPI_ISL_996187, EPI_ISL_996188, EPI_ISL_996189, EPI_ISL_996190, EPI_ISL_996191, EPI_ISL_996192, EPI_ISL_996193, EPI_ISL_996194, EPI_ISL_996195, EPI_ISL_996196, EPI_ISL_996197, EPI_ISL_996198, EPI_ISL_996199, EPI_ISL_996200, EPI_ISL_996201, EPI_ISL_996202, EPI_ISL_996203, EPI_ISL_996204, EPI_ISL_996205, EPI_ISL_996206, EPI_ISL_996207, EPI_ISL_996208, EPI_ISL_996209, EPI_ISL_996210, EPI_ISL_996211, EPI_ISL_996212, EPI_ISL_996213, EPI_ISL_996214, EPI_ISL_996215, EPI_ISL_996216, EPI_ISL_996217, EPI_ISL_996218, EPI_ISL_996219, EPI_ISL_996220, EPI_ISL_996221, EPI_ISL_996222, EPI_ISL_996223, EPI_ISL_996224, EPI_ISL_996225, EPI_ISL_996226, EPI_ISL_996227, EPI_ISL_996228, EPI_ISL_996229, EPI_ISL_996230, EPI_ISL_996231, EPI_ISL_996232, EPI_ISL_996233, EPI_ISL_996234, EPI_ISL_996235, EPI_ISL_996236, EPI_ISL_996237, EPI_ISL_996238, EPI_ISL_996239, EPI_ISL_996240, EPI_ISL_996241, EPI_ISL_996242, EPI_ISL_996243, EPI_ISL_996244, EPI_ISL_996245, EPI_ISL_996246, EPI_ISL_996247, EPI_ISL_996248, EPI_ISL_996249, EPI_ISL_996250, EPI_ISL_996251, EPI_ISL_996252, EPI_ISL_996253, EPI_ISL_996254, EPI_ISL_996255, EPI_ISL_996256, EPI_ISL_996257, EPI_ISL_996258, EPI_ISL_996259, EPI_ISL_996260, EPI_ISL_996261, EPI_ISL_996262, EPI_ISL_996263, EPI_ISL_996264, EPI_ISL_996265, EPI_ISL_996266, EPI_ISL_996267, EPI_ISL_996268, EPI_ISL_996269, EPI_ISL_996270, EPI_ISL_996271, EPI_ISL_996272, EPI_ISL_996273, EPI_ISL_996274, EPI_ISL_996275, EPI_ISL_996276, EPI_ISL_996277, EPI_ISL_996278, EPI_ISL_996279, EPI_ISL_996280, EPI_ISL_996281, EPI_ISL_996282, EPI_ISL_996283, EPI_ISL_996284, EPI_ISL_996285, EPI_ISL_996286, EPI_ISL_996287, EPI_ISL_996288, EPI_ISL_996289, EPI_ISL_996290, EPI_ISL_996291, EPI_ISL_996292, EPI_ISL_996293, EPI_ISL_996294, EPI_ISL_996295, EPI_ISL_996296, EPI_ISL_996297, EPI_ISL_996298, EPI_ISL_996299, EPI_ISL_996300, EPI_ISL_996301, EPI_ISL_996302, EPI_ISL_996303, EPI_ISL_996304, EPI_ISL_996305, EPI_ISL_996306, EPI_ISL_996307, EPI_ISL_996308, EPI_ISL_996309, EPI_ISL_996310, EPI_ISL_996311, EPI_ISL_996312, EPI_ISL_996313, EPI_ISL_996314, EPI_ISL_996315, EPI_ISL_996316, EPI_ISL_996317, EPI_ISL_996318, EPI_ISL_996319, EPI_ISL_996320, EPI_ISL_996321, EPI_ISL_996322, EPI_ISL_996323, EPI_ISL_996324, EPI_ISL_996325, EPI_ISL_996326, EPI_ISL_996327, EPI_ISL_996328, EPI_ISL_996329, EPI_ISL_996330, EPI_ISL_996331, EPI_ISL_996332, EPI_ISL_996333, EPI_ISL_996334, EPI_ISL_996335, EPI_ISL_996336, EPI_ISL_996337, EPI_ISL_996338, EPI_ISL_996339, EPI_ISL_996340, EPI_ISL_996341, EPI_ISL_996342, EPI_ISL_996343, EPI_ISL_996344, EPI_ISL_996345, EPI_ISL_996346, EPI_ISL_996347, EPI_ISL_996348, EPI_ISL_996349, EPI_ISL_996350, EPI_ISL_996351, EPI_ISL_996352, EPI_ISL_996353, EPI_ISL_996354, EPI_ISL_996355, EPI_ISL_996356, EPI_ISL_996357, EPI_ISL_996358, EPI_ISL_996359, EPI_ISL_996360, EPI_ISL_996361, EPI_ISL_996362, EPI_ISL_996363, EPI_ISL_996364, EPI_ISL_996365, EPI_ISL_996366, EPI_ISL_996367, EPI_ISL_996368, EPI_ISL_996369, EPI_ISL_996370, EPI_ISL_996371, EPI_ISL_996372, EPI_ISL_996373, EPI_ISL_996374, EPI_ISL_996375, EPI_ISL_996376, EPI_ISL_996377, EPI_ISL_996378, EPI_ISL_996379, EPI_ISL_996380, EPI_ISL_996381, EPI_ISL_996382, EPI_ISL_996383, EPI_ISL_996384, EPI_ISL_996385, EPI_ISL_996386, EPI_ISL_996387, EPI_ISL_996388, EPI_ISL_1051440, EPI_ISL_1051446, EPI_ISL_1051470, EPI_ISL_1051498, EPI_ISL_1051505, EPI_ISL_1051508, EPI_ISL_1051525, EPI_ISL_1051526, EPI_ISL_1051529, EPI_ISL_1247369, EPI_ISL_1247370, EPI_ISL_1247371, EPI_ISL_1247374, EPI_ISL_1247375, EPI_ISL_1247377, EPI_ISL_1247378, EPI_ISL_1247379, EPI_ISL_1247380, EPI_ISL_1247382, EPI_ISL_1247383, EPI_ISL_1247384, EPI_ISL_1247385, EPI_ISL_1247387, EPI_ISL_1247388, EPI_ISL_1247389, EPI_ISL_1247390, EPI_ISL_1247392, EPI_ISL_1247393, EPI_ISL_1247396, EPI_ISL_1247397, EPI_ISL_1247398, EPI_ISL_1247399, EPI_ISL_1247400, EPI_ISL_1247401, EPI_ISL_1247402, EPI_ISL_1247403, EPI_ISL_1247404, EPI_ISL_1386412, EPI_ISL_1386414, EPI_ISL_1386415, EPI_ISL_1386420, EPI_ISL_1386421, EPI_ISL_1386422, EPI_ISL_1386423, EPI_ISL_1386425, EPI_ISL_1386428, EPI_ISL_1386429, EPI_ISL_1386430, EPI_ISL_1386431, EPI_ISL_1386432, EPI_ISL_1386435, EPI_ISL_1386436, EPI_ISL_1386437, EPI_ISL_1386438, EPI_ISL_1386442, EPI_ISL_1386444, EPI_ISL_1386445, EPI_ISL_1386446, EPI_ISL_1386447, EPI_ISL_1386448, EPI_ISL_1386449, EPI_ISL_1386452, EPI_ISL_1386454, EPI_ISL_1386457, EPI_ISL_1386459, EPI_ISL_1386462, EPI_ISL_1386463, EPI_ISL_1386465, EPI_ISL_1386466, EPI_ISL_1386467, EPI_ISL_1386469, EPI_ISL_1386470, EPI_ISL_1386471, EPI_ISL_1386474, EPI_ISL_1386475, EPI_ISL_1831944, EPI_ISL_1831946, EPI_ISL_1831948, EPI_ISL_1831950, EPI_ISL_1831952, EPI_ISL_1831954, EPI_ISL_1831956, EPI_ISL_1831958, EPI_ISL_1831960, EPI_ISL_1831962, EPI_ISL_1831964, EPI_ISL_1831966, EPI_ISL_1831968, EPI_ISL_1831971, EPI_ISL_1831973, EPI_ISL_1831975, EPI_ISL_1831977, EPI_ISL_1831979, EPI_ISL_1831981, EPI_ISL_1831983, EPI_ISL_1831985, EPI_ISL_1831987, EPI_ISL_1831988, EPI_ISL_1831991, EPI_ISL_1831993, EPI_ISL_1831995, EPI_ISL_1831997, EPI_ISL_1831999, EPI_ISL_1832001, EPI_ISL_1832003, EPI_ISL_1832005, EPI_ISL_1832007, EPI_ISL_1832009, EPI_ISL_1832011, EPI_ISL_1832013, EPI_ISL_1832015, EPI_ISL_1832017, EPI_ISL_1832019, EPI_ISL_1832021, EPI_ISL_1832023, EPI_ISL_1832025, EPI_ISL_1832027, EPI_ISL_1832029, EPI_ISL_1832031, EPI_ISL_1832034, EPI_ISL_1832036, EPI_ISL_1832038, EPI_ISL_1832040, EPI_ISL_1832042, EPI_ISL_1832044, EPI_ISL_1832046, EPI_ISL_1832049, EPI_ISL_1832051, EPI_ISL_1832053, EPI_ISL_1832055, EPI_ISL_1832057, EPI_ISL_1832059, EPI_ISL_1832061, EPI_ISL_1832063, EPI_ISL_1832065, EPI_ISL_1832067, EPI_ISL_1832069, EPI_ISL_1832071, EPI_ISL_1974709, EPI_ISL_1974711, EPI_ISL_1974713, EPI_ISL_1974715, EPI_ISL_1974716, EPI_ISL_1974720, EPI_ISL_1978894, EPI_ISL_1978896, EPI_ISL_1978898 | | | |
| see above | Quadram Institute Bioscience | COVID-19 Genomics UK (COG-UK) Consortium | Alexander J Trotter; Alison E. Mather; Alp Aydin; Ana P. Tedim; Anastasia Kolyva; Andrew Bell; Andrew J. Page; Claire Stuart; Dave J. Baker; Gemma L. Kay; John Wain; Justin O'Grady; Leonardo de Oliveira Martins; Lizzie Meadows; Maria Diaz; Mark Webber; Muhammed Yasir; Nabil-Fareed Alikhan; Ngozi Elumogo; Nicholas M. Thomson; Rachael Stanley; Rachel Gilroy; Reenesh Prakash; Samir Dervisevic; Samuel Bloomfield; Steven Rudder; Thanh Le-Viet |
| EPI_ISL_820394, EPI_ISL_820402, EPI_ISL_820418, EPI_ISL_820423, EPI_ISL_820428, EPI_ISL_820431, EPI_ISL_820433, EPI_ISL_820560, EPI_ISL_866905 | | | |
| see above | Queens Medical Centre, Clinical Microbiology Department / DeepSeq Nottingham | COVID-19 Genomics UK (COG-UK) Consortium | Christopher Moore; Fei Sang; Gemma Clark; Hannah Howson-Wells; Johnny Debebe; Jonathan Ball; Joseph Chappell; Manjinder Khakh; Matthew Carlisle; Matthew Loose; Michelle M Lister; Nadine Holmes; Patrick McClure; Theocharis Tsoleridis; Vicki M Fleming; Victoria Wright; Wendy Smith |
| EPI_ISL_843155, EPI_ISL_843156, EPI_ISL_843157, EPI_ISL_843158, EPI_ISL_843159, EPI_ISL_843160, EPI_ISL_843161, EPI_ISL_843164, EPI_ISL_843165, EPI_ISL_843166, EPI_ISL_866346, EPI_ISL_866347, EPI_ISL_866348, EPI_ISL_866349, EPI_ISL_866350, EPI_ISL_866351, EPI_ISL_866352, EPI_ISL_866364, EPI_ISL_866365, EPI_ISL_866372, EPI_ISL_921022, EPI_ISL_921027, EPI_ISL_921028, EPI_ISL_921029, EPI_ISL_921030, EPI_ISL_921031, EPI_ISL_921032, EPI_ISL_921033, EPI_ISL_921034, EPI_ISL_921035, EPI_ISL_921036, EPI_ISL_921038, EPI_ISL_921039, EPI_ISL_921041, EPI_ISL_921042, EPI_ISL_921043, EPI_ISL_921044, EPI_ISL_921045, EPI_ISL_921046, EPI_ISL_921072, EPI_ISL_921073, EPI_ISL_921074, EPI_ISL_921075, EPI_ISL_921076, EPI_ISL_921077, EPI_ISL_921085, EPI_ISL_921087, EPI_ISL_921088, EPI_ISL_921089, EPI_ISL_921118, EPI_ISL_921119, EPI_ISL_921120, EPI_ISL_921133, EPI_ISL_921135, EPI_ISL_921136, EPI_ISL_998138, EPI_ISL_998139, EPI_ISL_998141, EPI_ISL_998142, EPI_ISL_998143, EPI_ISL_998305, EPI_ISL_998307, EPI_ISL_998308, EPI_ISL_1475188 | | | |
| see above | Regional Virus Laboratory, Belfast Health and Social Care Trust | COVID-19 Genomics UK (COG-UK) Consortium | Alison Watt; Ciara Cox; Conall McCaughey; David Simpson; Derek Fairley; James McKenna; Mairead Connor; Susan Feeney; Tanya Curran; Zoltan Molnar |
| EPI_ISL_766221, EPI_ISL_791193, EPI_ISL_791195, EPI_ISL_791196, EPI_ISL_791199, EPI_ISL_791200, EPI_ISL_791201, EPI_ISL_791202, EPI_ISL_791203, EPI_ISL_791209, EPI_ISL_791222, EPI_ISL_791249, EPI_ISL_791250, EPI_ISL_791251, EPI_ISL_791252, EPI_ISL_791253, EPI_ISL_791269, EPI_ISL_791270, EPI_ISL_791271, EPI_ISL_791272, EPI_ISL_791273, EPI_ISL_791274, EPI_ISL_791275, EPI_ISL_791276, EPI_ISL_791277, EPI_ISL_791278, EPI_ISL_804222, EPI_ISL_804224, EPI_ISL_804242, EPI_ISL_804243, EPI_ISL_804244, EPI_ISL_804245, EPI_ISL_804246, EPI_ISL_804247, EPI_ISL_804248, EPI_ISL_804251, EPI_ISL_804252, EPI_ISL_804260, EPI_ISL_804280, EPI_ISL_804281, EPI_ISL_804282, EPI_ISL_804283, EPI_ISL_804284, EPI_ISL_804285, EPI_ISL_804286, EPI_ISL_804287, EPI_ISL_804288, EPI_ISL_804289, EPI_ISL_804290, EPI_ISL_804291, EPI_ISL_804292, EPI_ISL_804293, EPI_ISL_811040, EPI_ISL_811041, EPI_ISL_811042, EPI_ISL_811043, EPI_ISL_811044, EPI_ISL_811045, EPI_ISL_811046, EPI_ISL_811047, EPI_ISL_811048, EPI_ISL_811049, EPI_ISL_811050, EPI_ISL_811051, EPI_ISL_811052, EPI_ISL_811053, EPI_ISL_811054, EPI_ISL_811055, EPI_ISL_811056, EPI_ISL_811057, EPI_ISL_811058, EPI_ISL_811059, EPI_ISL_811060, EPI_ISL_811061, EPI_ISL_811062, EPI_ISL_811063, EPI_ISL_811064, EPI_ISL_811065, EPI_ISL_811066, EPI_ISL_811067, EPI_ISL_811068, EPI_ISL_811069, EPI_ISL_811070, EPI_ISL_811071, EPI_ISL_811072, EPI_ISL_811073, EPI_ISL_811074, EPI_ISL_811075, EPI_ISL_811076, EPI_ISL_811077, EPI_ISL_811078, EPI_ISL_811079, EPI_ISL_811080, EPI_ISL_811081, EPI_ISL_811082, EPI_ISL_811083, EPI_ISL_811084, EPI_ISL_811085, EPI_ISL_811086, EPI_ISL_811087, EPI_ISL_811088, EPI_ISL_811089, EPI_ISL_811090, EPI_ISL_811091, EPI_ISL_811092, EPI_ISL_811093, EPI_ISL_811094, EPI_ISL_811095, EPI_ISL_811096, EPI_ISL_811097, EPI_ISL_811098, EPI_ISL_811099, EPI_ISL_811100, EPI_ISL_811101, EPI_ISL_811102, EPI_ISL_811103, EPI_ISL_811104, EPI_ISL_811105, EPI_ISL_811106, EPI_ISL_811115, EPI_ISL_811117, EPI_ISL_811118, EPI_ISL_811119, EPI_ISL_825499, EPI_ISL_825500, EPI_ISL_825534, EPI_ISL_825535, EPI_ISL_825536, EPI_ISL_825537, EPI_ISL_825538, EPI_ISL_825539, EPI_ISL_825540, EPI_ISL_825541, EPI_ISL_825542, EPI_ISL_825543, EPI_ISL_825544, EPI_ISL_825545, EPI_ISL_825568, EPI_ISL_825569, EPI_ISL_825570, EPI_ISL_825610, EPI_ISL_825611, EPI_ISL_825612, EPI_ISL_825613, EPI_ISL_825614, EPI_ISL_825615, EPI_ISL_837041, EPI_ISL_837042, EPI_ISL_837043, EPI_ISL_837044, EPI_ISL_837045, EPI_ISL_837046, EPI_ISL_837047, EPI_ISL_837048, EPI_ISL_837049, EPI_ISL_837050, EPI_ISL_837051, EPI_ISL_837052, EPI_ISL_837064, EPI_ISL_837067, EPI_ISL_837070, EPI_ISL_837077, EPI_ISL_837080, EPI_ISL_837086, EPI_ISL_837092, EPI_ISL_837093, EPI_ISL_837094, EPI_ISL_837096, EPI_ISL_837135, EPI_ISL_837136, EPI_ISL_837137, EPI_ISL_837138, EPI_ISL_837139, EPI_ISL_837140, EPI_ISL_837141, EPI_ISL_837142, EPI_ISL_837143, EPI_ISL_837144, EPI_ISL_837145, EPI_ISL_837146, EPI_ISL_837147, EPI_ISL_837148, EPI_ISL_837149, EPI_ISL_837150, EPI_ISL_837151, EPI_ISL_837152, EPI_ISL_837153, EPI_ISL_837154, EPI_ISL_837155, EPI_ISL_837156, EPI_ISL_837157, EPI_ISL_837158, EPI_ISL_837159, EPI_ISL_837160, EPI_ISL_837161, EPI_ISL_837162, EPI_ISL_837163, EPI_ISL_837164, EPI_ISL_837165, EPI_ISL_837166, EPI_ISL_837167, EPI_ISL_837168, EPI_ISL_837169, EPI_ISL_837170, EPI_ISL_837171, EPI_ISL_837172, EPI_ISL_837173, EPI_ISL_837174, EPI_ISL_837175, EPI_ISL_837184, EPI_ISL_837188, EPI_ISL_837190, EPI_ISL_837191, EPI_ISL_837192, EPI_ISL_837193, EPI_ISL_837194, EPI_ISL_837226, EPI_ISL_837227, EPI_ISL_837228, EPI_ISL_837229, EPI_ISL_837230, EPI_ISL_837231, EPI_ISL_837232, EPI_ISL_837233, EPI_ISL_837234, EPI_ISL_837235, EPI_ISL_837236, EPI_ISL_837237, EPI_ISL_837238, EPI_ISL_837239, EPI_ISL_860642, EPI_ISL_860643, EPI_ISL_860644, EPI_ISL_860645, EPI_ISL_860646, EPI_ISL_860647, EPI_ISL_860648, EPI_ISL_860649, EPI_ISL_860650, EPI_ISL_860651, EPI_ISL_860653, EPI_ISL_860654, EPI_ISL_860655, EPI_ISL_860656, EPI_ISL_860657, EPI_ISL_860658, EPI_ISL_860659, EPI_ISL_860660, EPI_ISL_860661, EPI_ISL_860662, EPI_ISL_860663, EPI_ISL_860664, EPI_ISL_860665, EPI_ISL_860782, EPI_ISL_1296494, EPI_ISL_1296500 | | | |
| see above | Respiratory Virus Unit, National Infection Service, Public Health England | COVID-19 Genomics UK (COG-UK) Consortium | PHE Covid Sequencing Team |
| EPI_ISL_1833388, EPI_ISL_1833389, EPI_ISL_1833390, EPI_ISL_1833391, EPI_ISL_1833392, EPI_ISL_1833393, EPI_ISL_1833404, EPI_ISL_1833413, EPI_ISL_1833414, EPI_ISL_1833419, EPI_ISL_1833420, EPI_ISL_1833421, EPI_ISL_1833425, EPI_ISL_1833426, EPI_ISL_1833427, EPI_ISL_1833429, EPI_ISL_1833430, EPI_ISL_1833435, EPI_ISL_1833437, EPI_ISL_1833440, EPI_ISL_1833441, EPI_ISL_1833442, EPI_ISL_1833445, EPI_ISL_1833466, EPI_ISL_1833505, EPI_ISL_1833508 | | | |
| see above | St George's University Hospitals NHS Foundation Trust | COVID-19 Genomics UK (COG-UK) Consortium | Adam Witney; Cassie Pope; Irene Monahan; Joshua Taylor; Ken Laing; NgeeKeong Tan |
| EPI_ISL_866071, EPI_ISL_866072, EPI_ISL_866073, EPI_ISL_866074, EPI_ISL_866115, EPI_ISL_866122, EPI_ISL_866127, EPI_ISL_866135, EPI_ISL_866140, EPI_ISL_866162, EPI_ISL_866168, EPI_ISL_920344, EPI_ISL_920457, EPI_ISL_920499, EPI_ISL_1104931, EPI_ISL_1104937, EPI_ISL_1104938, EPI_ISL_1104949, EPI_ISL_1104972, EPI_ISL_1104973, EPI_ISL_1104975, EPI_ISL_1104976, EPI_ISL_1104978, EPI_ISL_1104981, EPI_ISL_1104982, EPI_ISL_1104986, EPI_ISL_1104988, EPI_ISL_1104990, EPI_ISL_1104991, EPI_ISL_1104992, EPI_ISL_1104997, EPI_ISL_1104998, EPI_ISL_1104999, EPI_ISL_1105000, EPI_ISL_1105001, EPI_ISL_1105005, EPI_ISL_1105009, EPI_ISL_1105010, EPI_ISL_1105011, EPI_ISL_1105018, EPI_ISL_1105026, EPI_ISL_1105027, EPI_ISL_1105034, EPI_ISL_1105035, EPI_ISL_1105069, EPI_ISL_1105169, EPI_ISL_1105255, EPI_ISL_1105283, EPI_ISL_1105301, EPI_ISL_1105302, EPI_ISL_1105310, EPI_ISL_1248377, EPI_ISL_1248379, EPI_ISL_1248387, EPI_ISL_1248398, EPI_ISL_1248410, EPI_ISL_1248421, EPI_ISL_1248431, EPI_ISL_1248435, EPI_ISL_1248443, EPI_ISL_1248448, EPI_ISL_1248491, EPI_ISL_1637028, EPI_ISL_1637029, EPI_ISL_1637030, EPI_ISL_1637054, EPI_ISL_1637058, EPI_ISL_1637059, EPI_ISL_1637060, EPI_ISL_1637065, EPI_ISL_1637081, EPI_ISL_1637082, EPI_ISL_1637089 | | | |
| see above | University College London Hospital | COVID-19 Genomics UK (COG-UK) Consortium | Catherine Houlihan; Dan Frampton; Dr Catherine Houlihan; Dr Daniel Frampton; Dr Judith Heaney; Dr Moira Spyer; Dr Paul Grant and Dr Eleni Nastouli; Judith Heaney; Matthew Byott; Moira Spyer and Eleni Nastouli; Stuart Kirk |
| EPI_ISL_813958, EPI_ISL_820326, EPI_ISL_820328, EPI_ISL_820331, EPI_ISL_820333, EPI_ISL_820335, EPI_ISL_820337, EPI_ISL_820340, EPI_ISL_820343, EPI_ISL_820345, EPI_ISL_820347, EPI_ISL_820350, EPI_ISL_820373, EPI_ISL_820376, EPI_ISL_820381, EPI_ISL_820383, EPI_ISL_820385, EPI_ISL_820396, EPI_ISL_820420, EPI_ISL_838767, EPI_ISL_838768, EPI_ISL_838769, EPI_ISL_838770, EPI_ISL_838771, EPI_ISL_838773, EPI_ISL_838775, EPI_ISL_838776, EPI_ISL_838777, EPI_ISL_838778, EPI_ISL_838798, EPI_ISL_838799, EPI_ISL_838800, EPI_ISL_838801, EPI_ISL_838802, EPI_ISL_838803, EPI_ISL_838804, EPI_ISL_838805, EPI_ISL_838806, EPI_ISL_838808, EPI_ISL_838809, EPI_ISL_838810, EPI_ISL_838811, EPI_ISL_838812, EPI_ISL_838813, EPI_ISL_838814, EPI_ISL_838815, EPI_ISL_838816, EPI_ISL_838817, EPI_ISL_838818, EPI_ISL_838819, EPI_ISL_838820, EPI_ISL_838821, EPI_ISL_838822, EPI_ISL_838823, EPI_ISL_838824, EPI_ISL_838825, EPI_ISL_838826, EPI_ISL_838827, EPI_ISL_838828, EPI_ISL_838829, EPI_ISL_838830, EPI_ISL_838831, EPI_ISL_838832, EPI_ISL_838833, EPI_ISL_838834, EPI_ISL_838838, EPI_ISL_838839, EPI_ISL_838840, EPI_ISL_838841, EPI_ISL_839041, EPI_ISL_839042, EPI_ISL_839043, EPI_ISL_839054, EPI_ISL_839060, EPI_ISL_839063, EPI_ISL_839064, EPI_ISL_839065, EPI_ISL_839066, EPI_ISL_839067, EPI_ISL_839068, EPI_ISL_839069, EPI_ISL_839074, EPI_ISL_839075, EPI_ISL_839080, EPI_ISL_839081, EPI_ISL_839082, EPI_ISL_839083, EPI_ISL_839084, EPI_ISL_839085, EPI_ISL_839086, EPI_ISL_839087, EPI_ISL_839088, EPI_ISL_839089, EPI_ISL_839090, EPI_ISL_839091, EPI_ISL_839092, EPI_ISL_839093, EPI_ISL_839094, EPI_ISL_839095, EPI_ISL_839096, EPI_ISL_839097, EPI_ISL_839098, EPI_ISL_839099, EPI_ISL_839100, EPI_ISL_839101, EPI_ISL_839102, EPI_ISL_839103, EPI_ISL_839104, EPI_ISL_839105, EPI_ISL_839106, EPI_ISL_839107, EPI_ISL_839108, EPI_ISL_839109, EPI_ISL_839110, EPI_ISL_839111, EPI_ISL_839112, EPI_ISL_839113, EPI_ISL_839114, EPI_ISL_839116, EPI_ISL_839209, EPI_ISL_839215, EPI_ISL_839218, EPI_ISL_839219, EPI_ISL_839220, EPI_ISL_839221, EPI_ISL_839222, EPI_ISL_839223, EPI_ISL_839224, EPI_ISL_839225, EPI_ISL_839226, EPI_ISL_839227, EPI_ISL_839232, EPI_ISL_839305, EPI_ISL_839306, EPI_ISL_839307, EPI_ISL_839308, EPI_ISL_839310, EPI_ISL_839316, EPI_ISL_839317, EPI_ISL_839330, EPI_ISL_865682, EPI_ISL_865691, EPI_ISL_865692, EPI_ISL_865693, EPI_ISL_865694, EPI_ISL_865812, EPI_ISL_865816, EPI_ISL_865817, EPI_ISL_865819, EPI_ISL_865820, EPI_ISL_865821, EPI_ISL_865822, EPI_ISL_865823, EPI_ISL_865824, EPI_ISL_865825, EPI_ISL_865826, EPI_ISL_865827, EPI_ISL_865828, EPI_ISL_865829, EPI_ISL_865830, EPI_ISL_865831, EPI_ISL_865832, EPI_ISL_865833, EPI_ISL_865834, EPI_ISL_865835, EPI_ISL_865836, EPI_ISL_865838, EPI_ISL_865839, EPI_ISL_865840, EPI_ISL_865841, EPI_ISL_865842, EPI_ISL_865843, EPI_ISL_865844, EPI_ISL_865845, EPI_ISL_865846, EPI_ISL_919961, EPI_ISL_919964, EPI_ISL_919966, EPI_ISL_920842, EPI_ISL_920843, EPI_ISL_920844, EPI_ISL_920845, EPI_ISL_920846, EPI_ISL_920847, EPI_ISL_949959, EPI_ISL_950121, EPI_ISL_950131, EPI_ISL_950132, EPI_ISL_950137, EPI_ISL_950139, EPI_ISL_950140, EPI_ISL_950141, EPI_ISL_950143, EPI_ISL_950144, EPI_ISL_950149, EPI_ISL_950167, EPI_ISL_997579, EPI_ISL_997580, EPI_ISL_997581, EPI_ISL_997582, EPI_ISL_997583, EPI_ISL_997584, EPI_ISL_997585, EPI_ISL_997586, EPI_ISL_997587, EPI_ISL_997588, EPI_ISL_997589, EPI_ISL_997590, EPI_ISL_997591, EPI_ISL_997592, EPI_ISL_997593, EPI_ISL_997594, EPI_ISL_997595, EPI_ISL_997596, EPI_ISL_997597, EPI_ISL_997598, EPI_ISL_997599, EPI_ISL_997600, EPI_ISL_997601, EPI_ISL_997602, EPI_ISL_997603, EPI_ISL_997604, EPI_ISL_997605, EPI_ISL_997606, EPI_ISL_997607, EPI_ISL_997608, EPI_ISL_997609, EPI_ISL_997610, EPI_ISL_997611, EPI_ISL_997612, EPI_ISL_997613, EPI_ISL_1048050, EPI_ISL_1104579, EPI_ISL_1104580, EPI_ISL_1104581, EPI_ISL_1104582, EPI_ISL_1104583, EPI_ISL_1104584, EPI_ISL_1104585, EPI_ISL_1104586, EPI_ISL_1104587, EPI_ISL_1104588, EPI_ISL_1104589, EPI_ISL_1104590, EPI_ISL_1104591, EPI_ISL_1104592, EPI_ISL_1104593, EPI_ISL_1104594, EPI_ISL_1104595, EPI_ISL_1104596, EPI_ISL_1104597, EPI_ISL_1104598, EPI_ISL_1104599, EPI_ISL_1105450, EPI_ISL_1247997, EPI_ISL_1247998, EPI_ISL_1247999, EPI_ISL_1248001, EPI_ISL_1248002, EPI_ISL_1248003, EPI_ISL_1248004, EPI_ISL_1248005, EPI_ISL_1248006, EPI_ISL_1248007, EPI_ISL_1248008, EPI_ISL_1248009, EPI_ISL_1248010, EPI_ISL_1248011, EPI_ISL_1248012, EPI_ISL_1248013, EPI_ISL_1308979, EPI_ISL_1308980, EPI_ISL_1308989, EPI_ISL_1309005, EPI_ISL_1309007, EPI_ISL_1309008, EPI_ISL_1309009, EPI_ISL_1309010, EPI_ISL_1309011, EPI_ISL_1309012, EPI_ISL_1309013, EPI_ISL_1309014, EPI_ISL_1309015, EPI_ISL_1309016, EPI_ISL_1309017, EPI_ISL_1309018, EPI_ISL_1309019, EPI_ISL_1309020, EPI_ISL_1309021, EPI_ISL_1309032, EPI_ISL_1309041, EPI_ISL_1309044, EPI_ISL_1309046, EPI_ISL_1309049, EPI_ISL_1309054, EPI_ISL_1309092, EPI_ISL_1309093, EPI_ISL_1309094, EPI_ISL_1309121, EPI_ISL_1309124, EPI_ISL_1386901, EPI_ISL_1386909, EPI_ISL_1474837, EPI_ISL_1474848, EPI_ISL_1474850, EPI_ISL_1637111, EPI_ISL_1637118, EPI_ISL_1637119, EPI_ISL_1637120, EPI_ISL_1637124, EPI_ISL_1637125, EPI_ISL_1730393 | | | |
| see above | University College London, Great Ormond Street Hospital for Children NHS Foundation Trust, Imperial College Healthcare NHS Trust | COVID-19 Genomics UK (COG-UK) Consortium | Alison Holmes; Charlotte Williams; Helena Tutill; Jacqueline Findlay; James Price; Judith Breuer; Julianne Brown; Kathryn Harris; Leysa Forrest; Mark Kristiansen; Paola Niola; Paola Resende Silva; Patricia Dyal; Paul Randell; Rachel Williams; Samuel Weeks; Sergi Castellano; Sunando Roy; Tony Brooks; Yasmin Panchbhaya |
| EPI_ISL_1047934, EPI_ISL_1047936, EPI_ISL_1047937 | University Hospitals Of Leicester NHS Trust and DeepSeq Nottingham | COVID-19 Genomics UK (COG-UK) Consortium | Christopher Holmes; Christopher Moore; Fei Sang; Joeseph Chappell; Johnny Debebe; Jonathan Ball; Julian Tang; Karlie Fallon; Matthew Carlisle; Matthew Loose; Nadine Holmes; Patrick McClure; Paul Bird; Thomas Helmer; Victoria Wright |
| EPI_ISL_813091, EPI_ISL_813092, EPI_ISL_813093, EPI_ISL_813116, EPI_ISL_918748, EPI_ISL_918749, EPI_ISL_918750, EPI_ISL_918751, EPI_ISL_918752, EPI_ISL_918753, EPI_ISL_918754, EPI_ISL_918755, EPI_ISL_918756, EPI_ISL_918757, EPI_ISL_918760, EPI_ISL_918761, EPI_ISL_918762, EPI_ISL_918763, EPI_ISL_918940, EPI_ISL_918941, EPI_ISL_918942, EPI_ISL_918943, EPI_ISL_918944, EPI_ISL_918945, EPI_ISL_918946, EPI_ISL_918947, EPI_ISL_918948, EPI_ISL_918949, EPI_ISL_918950, EPI_ISL_918951, EPI_ISL_918952, EPI_ISL_918953, EPI_ISL_918954, EPI_ISL_918958, EPI_ISL_918959, EPI_ISL_918960, EPI_ISL_918961, EPI_ISL_918962, EPI_ISL_918963, EPI_ISL_918964, EPI_ISL_918965, EPI_ISL_918966, EPI_ISL_918967, EPI_ISL_918968, EPI_ISL_996424, EPI_ISL_996441, EPI_ISL_1047015, EPI_ISL_1047065, EPI_ISL_1247405, EPI_ISL_1247406, EPI_ISL_1247407, EPI_ISL_1247408, EPI_ISL_1247409, EPI_ISL_1247410, EPI_ISL_1247411, EPI_ISL_1247412, EPI_ISL_1247413, EPI_ISL_1247414, EPI_ISL_1247416, EPI_ISL_1247417, EPI_ISL_1247418, EPI_ISL_1247419, EPI_ISL_1247420, EPI_ISL_1247421, EPI_ISL_1247422, EPI_ISL_1247423, EPI_ISL_1247424, EPI_ISL_1247425, EPI_ISL_1247426, EPI_ISL_1247427, EPI_ISL_1247428, EPI_ISL_1247429, EPI_ISL_1247431, EPI_ISL_1247432, EPI_ISL_1308448, EPI_ISL_1308449, EPI_ISL_1308450, EPI_ISL_1308452, EPI_ISL_1308453, EPI_ISL_1308454, EPI_ISL_1308455, EPI_ISL_1308456, EPI_ISL_1308457, EPI_ISL_1308458, EPI_ISL_1308459, EPI_ISL_1308460, EPI_ISL_1308461, EPI_ISL_1308462, EPI_ISL_1308463, EPI_ISL_1308464, EPI_ISL_1308465, EPI_ISL_1308466, EPI_ISL_1308467, EPI_ISL_1308468, EPI_ISL_1308469, EPI_ISL_1308470, EPI_ISL_1308480, EPI_ISL_1386523, EPI_ISL_1386529, EPI_ISL_1386534, EPI_ISL_1386535, EPI_ISL_1386536, EPI_ISL_1386537, EPI_ISL_1386539, EPI_ISL_1474397, EPI_ISL_1474441, EPI_ISL_1474443, EPI_ISL_1474445, EPI_ISL_1474455, EPI_ISL_1474463, EPI_ISL_1832093, EPI_ISL_1832111, EPI_ISL_1832116 | | | |
| see above | University of Birmingham | COVID-19 Genomics UK (COG-UK) Consortium | Alex Richter; Andrew Bosworth; Andrew Bosworth. Queen Elizabeth Hospital: Anna Casey; Andrew D Beggs PHE Heartlands Lab: Husam Osman; Charlotte Poxon; Institute of Microbiology: Joanne Stockton; Josh Quick; Kasun Wanigasooriya; Mike Kidd; Nicholas Loman. University of Birmingham Testing Laboratory: Celina M Whalley; Oliver Pickles; Radoslaw Poplawski; Samuel Nicholls; University of Birmingham: Claire McMurray; Will Rowe |

| | | | |
|---|---|---|---|
| EPI_ISL_813608, EPI_ISL_813623, EPI_ISL_813625, EPI_ISL_813626, EPI_ISL_813627, EPI_ISL_813628, EPI_ISL_813629, EPI_ISL_813630, EPI_ISL_813631, EPI_ISL_813632, EPI_ISL_813633, EPI_ISL_813634, EPI_ISL_813635, EPI_ISL_813636, EPI_ISL_813638, EPI_ISL_813639, EPI_ISL_813640, EPI_ISL_813641, EPI_ISL_813642, EPI_ISL_813643, EPI_ISL_813644, EPI_ISL_813645, EPI_ISL_813646, EPI_ISL_813647, EPI_ISL_813648, EPI_ISL_813649, EPI_ISL_813650, EPI_ISL_813651, EPI_ISL_813652, EPI_ISL_813653, EPI_ISL_813654, EPI_ISL_838330, EPI_ISL_838332, EPI_ISL_838333, EPI_ISL_838338, EPI_ISL_919407, EPI_ISL_919408, EPI_ISL_919409, EPI_ISL_919410, EPI_ISL_919411, EPI_ISL_919412, EPI_ISL_919413, EPI_ISL_919414, EPI_ISL_919415, EPI_ISL_919416, EPI_ISL_919417, EPI_ISL_919418, EPI_ISL_919419, EPI_ISL_919420, EPI_ISL_919421, EPI_ISL_919422, EPI_ISL_919423, EPI_ISL_919424, EPI_ISL_919425, EPI_ISL_919426, EPI_ISL_919427, EPI_ISL_919428, EPI_ISL_919429, EPI_ISL_919430, EPI_ISL_919431, EPI_ISL_919432, EPI_ISL_919433, EPI_ISL_919434, EPI_ISL_919435, EPI_ISL_919436, EPI_ISL_919437, EPI_ISL_919438, EPI_ISL_919439, EPI_ISL_919440, EPI_ISL_919441, EPI_ISL_919442, EPI_ISL_919443, EPI_ISL_919444, EPI_ISL_996483, EPI_ISL_996490, EPI_ISL_996492, EPI_ISL_996493, EPI_ISL_996494, EPI_ISL_996495, EPI_ISL_996496, EPI_ISL_996497, EPI_ISL_996498, EPI_ISL_996499, EPI_ISL_996506, EPI_ISL_996507, EPI_ISL_996509, EPI_ISL_996510, EPI_ISL_996512, EPI_ISL_996513, EPI_ISL_997209, EPI_ISL_997210, EPI_ISL_997211, EPI_ISL_997212, EPI_ISL_997213, EPI_ISL_997214, EPI_ISL_997215, EPI_ISL_997217, EPI_ISL_997218, EPI_ISL_997219, EPI_ISL_997220, EPI_ISL_997221, EPI_ISL_997222, EPI_ISL_997223, EPI_ISL_997224, EPI_ISL_997225, EPI_ISL_997226, EPI_ISL_997227, EPI_ISL_997228, EPI_ISL_997229, EPI_ISL_997230, EPI_ISL_997231, EPI_ISL_997232, EPI_ISL_997233, EPI_ISL_997234, EPI_ISL_997235, EPI_ISL_997236, EPI_ISL_997237, EPI_ISL_997238, EPI_ISL_997240, EPI_ISL_997241, EPI_ISL_997242, EPI_ISL_997243, EPI_ISL_997244, EPI_ISL_997245, EPI_ISL_997246, EPI_ISL_997247, EPI_ISL_997248, EPI_ISL_997249, EPI_ISL_997285, EPI_ISL_997347, EPI_ISL_997366, EPI_ISL_1050245, EPI_ISL_1050246, EPI_ISL_1050247, EPI_ISL_1050248, EPI_ISL_1050249, EPI_ISL_1050250, EPI_ISL_1050251, EPI_ISL_1050252, EPI_ISL_1050253, EPI_ISL_1050254, EPI_ISL_1050255, EPI_ISL_1050256, EPI_ISL_1050257, EPI_ISL_1050258, EPI_ISL_1050259, EPI_ISL_1050260, EPI_ISL_1050261, EPI_ISL_1050262, EPI_ISL_1050263, EPI_ISL_1050264, EPI_ISL_1050265, EPI_ISL_1050266, EPI_ISL_1050267, EPI_ISL_1050268, EPI_ISL_1050269, EPI_ISL_1050270, EPI_ISL_1050271, EPI_ISL_1050272, EPI_ISL_1050273, EPI_ISL_1050274, EPI_ISL_1050275, EPI_ISL_1050276, EPI_ISL_1050277, EPI_ISL_1050278, EPI_ISL_1050279, EPI_ISL_1050280, EPI_ISL_1050281, EPI_ISL_1050282, EPI_ISL_1050283, EPI_ISL_1050284, EPI_ISL_1050285, EPI_ISL_1050286, EPI_ISL_1050287, EPI_ISL_1050288, EPI_ISL_1050289, EPI_ISL_1050290, EPI_ISL_1050291, EPI_ISL_1050292, EPI_ISL_1050307, EPI_ISL_1050308, EPI_ISL_1050309, EPI_ISL_1050310, EPI_ISL_1050311, EPI_ISL_1050312, EPI_ISL_1050313, EPI_ISL_1050314, EPI_ISL_1050315, EPI_ISL_1050316, EPI_ISL_1050317, EPI_ISL_1050318, EPI_ISL_1050319, EPI_ISL_1050320, EPI_ISL_1050321, EPI_ISL_1050322, EPI_ISL_1050325, EPI_ISL_1050348, EPI_ISL_1050349, EPI_ISL_1050350, EPI_ISL_1177785, EPI_ISL_1308580, EPI_ISL_1308597, EPI_ISL_1308599, EPI_ISL_1474644, EPI_ISL_1545819, EPI_ISL_1545943, EPI_ISL_1545945, EPI_ISL_1545960, EPI_ISL_1545961, EPI_ISL_1545962, EPI_ISL_1545963, EPI_ISL_1545964, EPI_ISL_1545965, EPI_ISL_1545966, EPI_ISL_1545967, EPI_ISL_1545968, EPI_ISL_1545970, EPI_ISL_1545971, EPI_ISL_1545972, EPI_ISL_1730136, EPI_ISL_1730137, EPI_ISL_1730138, EPI_ISL_1730139, EPI_ISL_1730140, EPI_ISL_1730141, EPI_ISL_1730145, EPI_ISL_1730146, EPI_ISL_1730147, EPI_ISL_1730148, EPI_ISL_1730156, EPI_ISL_1730157, EPI_ISL_1730158, EPI_ISL_1730159, EPI_ISL_1730160, EPI_ISL_1730173, EPI_ISL_1730174, EPI_ISL_2125760, EPI_ISL_2125761, EPI_ISL_2125763, EPI_ISL_2125765, EPI_ISL_2125767, EPI_ISL_2125768, EPI_ISL_2125770, EPI_ISL_2125772, EPI_ISL_2125774, EPI_ISL_2125776, EPI_ISL_2125780, EPI_ISL_2125782, EPI_ISL_2125784, EPI_ISL_2125786, EPI_ISL_2125793, EPI_ISL_2125798, EPI_ISL_2125799, EPI_ISL_2125803, EPI_ISL_2125804, EPI_ISL_2125806, EPI_ISL_2125808, EPI_ISL_2125810, EPI_ISL_2125813, EPI_ISL_2125815, EPI_ISL_2125817, EPI_ISL_2125818, EPI_ISL_2125820, EPI_ISL_2125822, EPI_ISL_2125824, EPI_ISL_2125826, EPI_ISL_2125828, EPI_ISL_2125830, EPI_ISL_2125831, EPI_ISL_2125833, EPI_ISL_2125835, EPI_ISL_2125837, EPI_ISL_2125839, EPI_ISL_2125841, EPI_ISL_2125843, EPI_ISL_2125844 | | | |
| see above | University of Exeter | COVID-19 Genomics UK (COG-UK) Consortium | Aaron Jeffries; Audrey Farbos; Ben Temperton; Jane Masoli; Joanna Warwick-Dugdale; Michelle Michelsen; Robyn Manley; Stephen Michell |
| EPI_ISL_997118, EPI_ISL_997119, EPI_ISL_1308832, EPI_ISL_1308833, EPI_ISL_1308834, EPI_ISL_1308835 | Virology Department, Royal Infirmary of Edinburgh, NHS Lothian / School of Biological Sciences, University of Edinburgh | COVID-19 Genomics UK (COG-UK) Consortium | Colquhoun R; Cotton S; Dewar R; Hill V; Jackson B; McCrone JT; McHugh M; O'Toole Á; Rambaut A; Rooke S; Scher E; Templeton K; Yu X |
| EPI_ISL_838268, EPI_ISL_838269, EPI_ISL_838270, EPI_ISL_838271, EPI_ISL_838272, EPI_ISL_838273, EPI_ISL_838274, EPI_ISL_838276, EPI_ISL_838277, EPI_ISL_838278, EPI_ISL_838279, EPI_ISL_865073, EPI_ISL_865080, EPI_ISL_865081, EPI_ISL_865082, EPI_ISL_865087, EPI_ISL_919321, EPI_ISL_919322, EPI_ISL_919323, EPI_ISL_919324, EPI_ISL_919325, EPI_ISL_919326, EPI_ISL_919327, EPI_ISL_919328, EPI_ISL_919329, EPI_ISL_919330, EPI_ISL_919331, EPI_ISL_919332, EPI_ISL_919333, EPI_ISL_919334, EPI_ISL_919335, EPI_ISL_919336 | | | |
| see above | Virology Department, Royal Infirmary of Edinburgh, NHS Lothian / School of Biological Sciences, University of Edinburgh / Institute of Genetics and Molecular Medicine, University of Edinburgh | COVID-19 Genomics UK (COG-UK) Consortium | Balcaza C; Colquhoun R; Dewar R; Gallagher M; Hill V; Jackson B; McCrone JT; McHugh M; O'Toole Á; Rambaut A; Rooke S; Scher E; Templeton K; Williams TC; Yu X |
| EPI_ISL_816226, EPI_ISL_816227, EPI_ISL_816228, EPI_ISL_816229, EPI_ISL_816232, EPI_ISL_816233, EPI_ISL_816235, EPI_ISL_816243, EPI_ISL_816246, EPI_ISL_816248, EPI_ISL_816256, EPI_ISL_816261, EPI_ISL_816275, EPI_ISL_816276, EPI_ISL_816277, EPI_ISL_816285, EPI_ISL_816291, EPI_ISL_816293, EPI_ISL_816294, EPI_ISL_816296, EPI_ISL_816297, EPI_ISL_816305, EPI_ISL_816311, EPI_ISL_816314, EPI_ISL_816318, EPI_ISL_816319, EPI_ISL_816323, EPI_ISL_816326, EPI_ISL_816327, EPI_ISL_816331, EPI_ISL_816348, EPI_ISL_816353, EPI_ISL_816355, EPI_ISL_816364, EPI_ISL_816368, EPI_ISL_816383, EPI_ISL_816390, EPI_ISL_816396, EPI_ISL_816405, EPI_ISL_816406, EPI_ISL_816407, EPI_ISL_816409, EPI_ISL_816415, EPI_ISL_816423, EPI_ISL_816426, EPI_ISL_816427, EPI_ISL_816431, EPI_ISL_816434, EPI_ISL_816436, EPI_ISL_816437, EPI_ISL_816438, EPI_ISL_816441, EPI_ISL_816442, EPI_ISL_816444, EPI_ISL_816448, EPI_ISL_816449, EPI_ISL_816458, EPI_ISL_816462, EPI_ISL_816469, EPI_ISL_816472, EPI_ISL_816475, EPI_ISL_816477, EPI_ISL_816492, EPI_ISL_816494, EPI_ISL_816495, EPI_ISL_816496, EPI_ISL_816501, EPI_ISL_816508, EPI_ISL_816517, EPI_ISL_816525, EPI_ISL_816536, EPI_ISL_816539, EPI_ISL_816546, EPI_ISL_816555, EPI_ISL_816561, EPI_ISL_816566, EPI_ISL_816571, EPI_ISL_816576, EPI_ISL_816581, EPI_ISL_816582, EPI_ISL_816584, EPI_ISL_816601, EPI_ISL_816608, EPI_ISL_816611, EPI_ISL_816613, EPI_ISL_816629, EPI_ISL_816632, EPI_ISL_816633, EPI_ISL_816634, EPI_ISL_816641, EPI_ISL_816642, EPI_ISL_816646, EPI_ISL_842209, EPI_ISL_842265, EPI_ISL_842273, EPI_ISL_842293, EPI_ISL_842312, EPI_ISL_842338, EPI_ISL_842341, EPI_ISL_868420, EPI_ISL_868423, EPI_ISL_868485, EPI_ISL_868629, EPI_ISL_924094, EPI_ISL_924168, EPI_ISL_924213, EPI_ISL_924237, EPI_ISL_924250, EPI_ISL_924280, EPI_ISL_924283, EPI_ISL_924402 | | | |
| see above | Virology Department, Sheffield Teaching Hospitals NHS Foundation Trust/Department of Infection, Immunity and Cardiovascular Disease, The Medical School, University of Sheffield | COVID-19 Genomics UK (COG-UK) Consortium | Adri Angyal; Alex Keeley; Benjamin Lindsey; Cariad Evans; Danielle Groves; Dave Partridge; Katie Johnson; Laura Carrilero; Luke Green; Matthew Parker; Matthew Wyles; Mehmet Yavuz; Mohammad Raza; Nikki Smith; Paul Parsons; Rachel Tucker; Rebecca Brown; Thushan de Silva |
| EPI_ISL_822659, EPI_ISL_822660, EPI_ISL_822661, EPI_ISL_822662, EPI_ISL_822663, EPI_ISL_822664, EPI_ISL_822665, EPI_ISL_822666, EPI_ISL_822667, EPI_ISL_822668, EPI_ISL_822669, EPI_ISL_822670, EPI_ISL_822671, EPI_ISL_822672, EPI_ISL_822673, EPI_ISL_822674, EPI_ISL_822675, EPI_ISL_822676, EPI_ISL_822677, EPI_ISL_822678, EPI_ISL_822679, EPI_ISL_822680, EPI_ISL_822681, EPI_ISL_822682, EPI_ISL_822683, EPI_ISL_822684, EPI_ISL_822685, EPI_ISL_822686, EPI_ISL_822687, EPI_ISL_822688, EPI_ISL_822689, EPI_ISL_822690, EPI_ISL_822691, EPI_ISL_822692, EPI_ISL_822693, EPI_ISL_822694, EPI_ISL_822695, EPI_ISL_822696, EPI_ISL_822697, EPI_ISL_822698, EPI_ISL_822699, EPI_ISL_822700, EPI_ISL_822701, EPI_ISL_822702, EPI_ISL_822703, EPI_ISL_822704, EPI_ISL_822705, EPI_ISL_822706, EPI_ISL_822707, EPI_ISL_822708, EPI_ISL_822709, EPI_ISL_822710, EPI_ISL_822711, EPI_ISL_822712, EPI_ISL_822713, EPI_ISL_822714, EPI_ISL_822715, EPI_ISL_822716, EPI_ISL_822717, EPI_ISL_822718, EPI_ISL_822719, EPI_ISL_822720, EPI_ISL_822721, EPI_ISL_822722, EPI_ISL_822723, EPI_ISL_822724, EPI_ISL_822725, EPI_ISL_822726, EPI_ISL_822727, EPI_ISL_822728, EPI_ISL_822729, EPI_ISL_822730, EPI_ISL_822731, EPI_ISL_822732, EPI_ISL_822733, EPI_ISL_822734, EPI_ISL_822735, EPI_ISL_822736, EPI_ISL_822737, EPI_ISL_822738, EPI_ISL_822739, EPI_ISL_822740, EPI_ISL_822741, EPI_ISL_822742, EPI_ISL_822743, EPI_ISL_822744, EPI_ISL_822745, EPI_ISL_822746, EPI_ISL_822747, EPI_ISL_822748, EPI_ISL_822749, EPI_ISL_822750, EPI_ISL_822751, EPI_ISL_822752, EPI_ISL_822753, EPI_ISL_822754, EPI_ISL_822755, EPI_ISL_822756, EPI_ISL_822757, EPI_ISL_822758, EPI_ISL_822759, EPI_ISL_822760, EPI_ISL_822761, EPI_ISL_822762, EPI_ISL_822763, EPI_ISL_822764, EPI_ISL_822765, EPI_ISL_822766, EPI_ISL_822767, EPI_ISL_822768, EPI_ISL_822769, EPI_ISL_822770, EPI_ISL_822771, EPI_ISL_822772, EPI_ISL_822773, EPI_ISL_822774, EPI_ISL_822775, EPI_ISL_822776, EPI_ISL_822777, EPI_ISL_822778, EPI_ISL_822779, EPI_ISL_822780, EPI_ISL_822781, EPI_ISL_822782, EPI_ISL_822783, EPI_ISL_822784, EPI_ISL_822785, EPI_ISL_822786, EPI_ISL_822787, EPI_ISL_822788, EPI_ISL_822789, EPI_ISL_822790, EPI_ISL_822791, EPI_ISL_822792, EPI_ISL_822793, EPI_ISL_822794, EPI_ISL_822795, EPI_ISL_822796, EPI_ISL_822797, EPI_ISL_822798, EPI_ISL_822799, EPI_ISL_822800, EPI_ISL_822801, EPI_ISL_822802, EPI_ISL_822803, EPI_ISL_822804, EPI_ISL_822805, EPI_ISL_822806, EPI_ISL_822807, EPI_ISL_822808, EPI_ISL_822809, EPI_ISL_822810, EPI_ISL_822811, EPI_ISL_822812, EPI_ISL_822813, EPI_ISL_822814, EPI_ISL_822815, EPI_ISL_822844, EPI_ISL_822880, EPI_ISL_822925, EPI_ISL_822940, EPI_ISL_822947, EPI_ISL_822951, EPI_ISL_822952, EPI_ISL_822955, EPI_ISL_822957, EPI_ISL_822971, EPI_ISL_823003, EPI_ISL_823066, EPI_ISL_823098, EPI_ISL_823099, EPI_ISL_823100, EPI_ISL_823101, EPI_ISL_823102, EPI_ISL_823103, EPI_ISL_823112, EPI_ISL_823149, EPI_ISL_823152, EPI_ISL_823154, EPI_ISL_823155, EPI_ISL_823646, EPI_ISL_823647, EPI_ISL_823648, EPI_ISL_823649, EPI_ISL_823780, EPI_ISL_823781, EPI_ISL_823782, EPI_ISL_823783, EPI_ISL_823784, EPI_ISL_823785, EPI_ISL_823786, EPI_ISL_823787, EPI_ISL_823788, EPI_ISL_823789, EPI_ISL_823790, EPI_ISL_823791, EPI_ISL_823792, EPI_ISL_823793, EPI_ISL_823794, EPI_ISL_823795 | | | |
| see above | Wales Specialist Virology Centre Sequencing lab: Pathogen Genomics Unit | COVID-19 Genomics UK (COG-UK) Consortium | Alec Birchley; Alexander Adams; Amy Gaskin; Angela Marchbank; Bree Gatica-Wilcox; Catherine Moore; Jason Coombes; Joanne Watkins; Joel Southgate; Johnathan Evans; Laura Gifford; Lauren Gilbert; Lee Graham; Malorie Perry; Matthew Bull; Nicole Pacchiarini; Sally Corden; Sara Kumziene-Summerhayes; Sara Rey; Sarah Taylor; Simon Cottrell; Sophie Jones; Tom Connor |
| EPI_ISL_840816, EPI_ISL_840819, EPI_ISL_840821, EPI_ISL_840837, EPI_ISL_840839, EPI_ISL_840841, EPI_ISL_840895, EPI_ISL_840897, EPI_ISL_840898, EPI_ISL_840899, EPI_ISL_840900, EPI_ISL_840916, EPI_ISL_840917, EPI_ISL_840918, EPI_ISL_840919, EPI_ISL_841057, EPI_ISL_841058, EPI_ISL_841083, EPI_ISL_841087, EPI_ISL_841088, EPI_ISL_841089, EPI_ISL_841090, EPI_ISL_841091, EPI_ISL_841092, EPI_ISL_841093, EPI_ISL_841094, EPI_ISL_841095, EPI_ISL_841096, EPI_ISL_841097, EPI_ISL_841099, EPI_ISL_841100, EPI_ISL_841101, EPI_ISL_841102, EPI_ISL_841103, EPI_ISL_841104, EPI_ISL_841105, EPI_ISL_841106, EPI_ISL_841107, EPI_ISL_841109, EPI_ISL_841110, EPI_ISL_841111, EPI_ISL_841112, EPI_ISL_841113, EPI_ISL_841114, EPI_ISL_841115, EPI_ISL_841117, EPI_ISL_841118, EPI_ISL_841119, EPI_ISL_841123, EPI_ISL_841302, EPI_ISL_841303, EPI_ISL_841304, EPI_ISL_841305, EPI_ISL_1000023 | | | |
| see above | Wales Specialist Virology Centre Sequencing lab: Pathogen Genomics Unit | Public Health Wales Microbiology Cardiff Wales Specialist Virology Centre | Alec Birchley; Alexander Adams; Amy Gaskin; Angela Marchbank; Bree Gatica-Wilcox; Catherine Moore; Jason Coombes; Joanne Watkins; Joel Southgate; Johnathan Evans; Laura Gifford; Lauren Gilbert; Lee Graham; Malorie Perry; Matthew Bull; Nicole Pacchiarini; Sally Corden; Sara Kumziene-Summerhayes; Sara Rey; Sarah Taylor; Simon Cottrell; Sophie Jones; Tom Connor |
| EPI_ISL_864961, EPI_ISL_864962, EPI_ISL_864963, EPI_ISL_864964, EPI_ISL_864965, EPI_ISL_864966, EPI_ISL_864967, EPI_ISL_864968, EPI_ISL_864969, EPI_ISL_864970, EPI_ISL_864971, EPI_ISL_864972, EPI_ISL_864973, EPI_ISL_864974, EPI_ISL_864975, EPI_ISL_864976, EPI_ISL_864977, EPI_ISL_864978, EPI_ISL_864979, EPI_ISL_864980, EPI_ISL_864981, EPI_ISL_864982, EPI_ISL_864983, EPI_ISL_864992, EPI_ISL_864993, EPI_ISL_919257 | | | |
| see above | West of Scotland Specialist Virology Centre, NHSGGC / MRC-University of Glasgow Centre for Virus Research | COVID-19 Genomics UK (COG-UK) Consortium | Alasdair MacLean; Alice Broos; Ana da Silva Filipe; Antonia Ho; Daniel Mair; David L Robertson; Emma Thomson; Guy Mollett; James Shepherd; Jenna Nichols; Joseph Hughes; Kathy Li; Kathy Smollett; Kyriaki Nomikou; Lily Tong; Matthew Holden; Natasha Johnson; Rachel Blacow; Richard Orton; Rory Gunson; Sarah McDonald; Sharif Shaaban; Sreenu Vattipally; Stephen Carmichael |

We gratefully acknowledge the following Authors from the Originating laboratories responsible for obtaining the specimens, as well as the Submitting laboratories where the genome data were generated and shared via GISAID, on which this research is based.

All Submitters of data may be contacted directly via www.gisaid.org

Authors are sorted alphabetically.

| Accession ID | Originating Laboratory | Submitting Laboratory | Authors |
|---|---|---|---|
| EPI_ISL_865499, EPI_ISL_865500, EPI_ISL_865571, EPI_ISL_865574, EPI_ISL_865575, EPI_ISL_919779, EPI_ISL_919780, EPI_ISL_919797, EPI_ISL_919798, EPI_ISL_919799, EPI_ISL_919800, EPI_ISL_919801, EPI_ISL_919802, EPI_ISL_919803, EPI_ISL_919804, EPI_ISL_919805, EPI_ISL_919806, EPI_ISL_919807, EPI_ISL_919808, EPI_ISL_919809, EPI_ISL_919810, EPI_ISL_919811, EPI_ISL_919812, EPI_ISL_919813, EPI_ISL_919814, EPI_ISL_919815, EPI_ISL_919816, EPI_ISL_919817, EPI_ISL_919818, EPI_ISL_919819, EPI_ISL_919820, EPI_ISL_919821, EPI_ISL_919822, EPI_ISL_919823, EPI_ISL_919824, EPI_ISL_919825, EPI_ISL_919826, EPI_ISL_919827, EPI_ISL_919828, EPI_ISL_919829, EPI_ISL_919830, EPI_ISL_919831, EPI_ISL_919832, EPI_ISL_919833, EPI_ISL_919834, EPI_ISL_919835, EPI_ISL_919836, EPI_ISL_919837, EPI_ISL_919838, EPI_ISL_919839, EPI_ISL_919840, EPI_ISL_919842, EPI_ISL_919843, EPI_ISL_919844, EPI_ISL_919845, EPI_ISL_919846, EPI_ISL_919847, EPI_ISL_919848, EPI_ISL_919849, EPI_ISL_919850, EPI_ISL_919851, EPI_ISL_949748, EPI_ISL_949754, EPI_ISL_949755, EPI_ISL_949756, EPI_ISL_949767, EPI_ISL_949769, EPI_ISL_949775, EPI_ISL_949784, EPI_ISL_2126036, EPI_ISL_2126064, EPI_ISL_2126066 | | | |
| see above | Barts Health NHS Trust | COVID-19 Genomics UK (COG-UK) Consortium | BROAD; Beatrix; CUTINO-MOGUEL; Claire; David; Dola; HARRINGTON; KELE; KULASEGARAN-SHYLINI; Maria-Teresa; OWOYEMI; Raghavendran |
| EPI_ISL_924585, EPI_ISL_924586, EPI_ISL_924587, EPI_ISL_924589, EPI_ISL_924590, EPI_ISL_924592, EPI_ISL_924593, EPI_ISL_924594, EPI_ISL_924595, EPI_ISL_924596, EPI_ISL_924597, EPI_ISL_924598, EPI_ISL_924600, EPI_ISL_924601, EPI_ISL_924603, EPI_ISL_924604, EPI_ISL_924622, EPI_ISL_924623, EPI_ISL_924625, EPI_ISL_924626, EPI_ISL_924627, EPI_ISL_924628, EPI_ISL_924629, EPI_ISL_924630, EPI_ISL_924631, EPI_ISL_924632, EPI_ISL_924633, EPI_ISL_924634, EPI_ISL_924635, EPI_ISL_924636, EPI_ISL_924637, EPI_ISL_924638, EPI_ISL_924639, EPI_ISL_924640, EPI_ISL_924641, EPI_ISL_924642, EPI_ISL_924644, EPI_ISL_924645, EPI_ISL_924646, EPI_ISL_924647, EPI_ISL_924648, EPI_ISL_924649, EPI_ISL_924650, EPI_ISL_924651, EPI_ISL_924723, EPI_ISL_924726, EPI_ISL_924730, EPI_ISL_924732, EPI_ISL_924734, EPI_ISL_924735, EPI_ISL_924737, EPI_ISL_924738, EPI_ISL_924739, EPI_ISL_924741, EPI_ISL_924743, EPI_ISL_924745, EPI_ISL_924747, EPI_ISL_924757, EPI_ISL_924758, EPI_ISL_924759, EPI_ISL_924760, EPI_ISL_924761, EPI_ISL_924762, EPI_ISL_924763, EPI_ISL_924765, EPI_ISL_924766, EPI_ISL_924768, EPI_ISL_924771, EPI_ISL_924772, EPI_ISL_924773, EPI_ISL_924775, EPI_ISL_924776, EPI_ISL_924777, EPI_ISL_924778, EPI_ISL_924779, EPI_ISL_924780, EPI_ISL_924781, EPI_ISL_924782, EPI_ISL_924893, EPI_ISL_924894, EPI_ISL_924896, EPI_ISL_924900, EPI_ISL_924906, EPI_ISL_953174, EPI_ISL_953175, EPI_ISL_953176, EPI_ISL_953178, EPI_ISL_953179, EPI_ISL_953183, EPI_ISL_953184, EPI_ISL_953186, EPI_ISL_953188, EPI_ISL_953189, EPI_ISL_953190, EPI_ISL_953191, EPI_ISL_953192, EPI_ISL_953194, EPI_ISL_953195, EPI_ISL_953196, EPI_ISL_953197, EPI_ISL_953198, EPI_ISL_953200, EPI_ISL_953201, EPI_ISL_953202, EPI_ISL_953203, EPI_ISL_953204, EPI_ISL_953206, EPI_ISL_953208, EPI_ISL_953209, EPI_ISL_953210, EPI_ISL_953211, EPI_ISL_953212, EPI_ISL_953213, EPI_ISL_953214, EPI_ISL_953215, EPI_ISL_953218, EPI_ISL_953220, EPI_ISL_953221, EPI_ISL_953223, EPI_ISL_953224, EPI_ISL_953225, EPI_ISL_953227, EPI_ISL_953228, EPI_ISL_953229, EPI_ISL_953234, EPI_ISL_953235, EPI_ISL_953237, EPI_ISL_953242, EPI_ISL_953260, EPI_ISL_953273, EPI_ISL_953274, EPI_ISL_953278, EPI_ISL_1054760 | | | |
| see above | Bioinformatics and Biostatistics Lab, Advanced Sequencing Facility | COVID-19 Genomics UK (COG-UK) Consortium | Aengus Stewart; Chelsea Sawyer; Harshil Patel; Jerome Nicod; Laura Cubitt; Margaret Crawford |
| EPI_ISL_816210, EPI_ISL_816211, EPI_ISL_816216, EPI_ISL_816219, EPI_ISL_841922, EPI_ISL_841927, EPI_ISL_841945, EPI_ISL_841960, EPI_ISL_841961, EPI_ISL_842008, EPI_ISL_867990, EPI_ISL_868001, EPI_ISL_868008, EPI_ISL_868309, EPI_ISL_868310, EPI_ISL_868311, EPI_ISL_868312, EPI_ISL_868313, EPI_ISL_868314, EPI_ISL_868315, EPI_ISL_868316, EPI_ISL_868317, EPI_ISL_868319, EPI_ISL_868321, EPI_ISL_868322, EPI_ISL_868324, EPI_ISL_868325, EPI_ISL_868326, EPI_ISL_868327, EPI_ISL_868328, EPI_ISL_868343, EPI_ISL_868344, EPI_ISL_868345, EPI_ISL_868346, EPI_ISL_868347, EPI_ISL_868348, EPI_ISL_868349, EPI_ISL_868350, EPI_ISL_868351, EPI_ISL_868352, EPI_ISL_923241, EPI_ISL_923252, EPI_ISL_923254, EPI_ISL_923255, EPI_ISL_923256, EPI_ISL_923257, EPI_ISL_923261, EPI_ISL_923263, EPI_ISL_923266, EPI_ISL_923379, EPI_ISL_923381, EPI_ISL_923443, EPI_ISL_923577, EPI_ISL_923580, EPI_ISL_923663, EPI_ISL_952407, EPI_ISL_952408, EPI_ISL_952420, EPI_ISL_952423, EPI_ISL_952785, EPI_ISL_952813, EPI_ISL_952814, EPI_ISL_952815, EPI_ISL_1179836, EPI_ISL_1179837, EPI_ISL_1179843, EPI_ISL_1730833, EPI_ISL_1730834, EPI_ISL_1730835, EPI_ISL_1730836, EPI_ISL_1730837, EPI_ISL_1730838, EPI_ISL_1730839, EPI_ISL_1730840, EPI_ISL_1730882, EPI_ISL_1730883, EPI_ISL_1730884, EPI_ISL_1730885, EPI_ISL_1730886, EPI_ISL_1730887, EPI_ISL_1730888, EPI_ISL_1979881, EPI_ISL_1979882, EPI_ISL_1979884, EPI_ISL_1979886, EPI_ISL_1979887, EPI_ISL_1979889, EPI_ISL_1979891, EPI_ISL_1979892, EPI_ISL_1979894, EPI_ISL_1979895, EPI_ISL_1979977, EPI_ISL_1979978, EPI_ISL_1979980, EPI_ISL_1979982, EPI_ISL_1979984, EPI_ISL_1980081, EPI_ISL_1980083, EPI_ISL_1980085, EPI_ISL_1980086, EPI_ISL_1980088, EPI_ISL_1980089, EPI_ISL_1980090, EPI_ISL_1980091, EPI_ISL_1980093, EPI_ISL_1980094, EPI_ISL_1980096, EPI_ISL_1980097, EPI_ISL_1980099, EPI_ISL_1980100, EPI_ISL_1980102, EPI_ISL_1980104, EPI_ISL_1980106, EPI_ISL_1980107, EPI_ISL_2127920, EPI_ISL_2127954, EPI_ISL_2127956, EPI_ISL_2128032, EPI_ISL_2128037, EPI_ISL_2128038, EPI_ISL_2128049, EPI_ISL_2128051, EPI_ISL_2128192, EPI_ISL_2128201, EPI_ISL_2128203, EPI_ISL_2128205, EPI_ISL_2128207, EPI_ISL_2128208, EPI_ISL_2128222, EPI_ISL_2128224 | | | |
| see above | Centre for Enzyme Innovation, University of Portsmouth / Translational Research Laboratory, Portsmouth Hospitals NHS Trust | COVID-19 Genomics UK (COG-UK) Consortium | Allyson Lloyd; Angela Beckett; Anoop Chauhan; Christopher Fearn; Ethan Butcher; Garry Scarlett; Kate Cook; Katie Loveson; Kelly Bicknell; Robert Impey; Salman Goudarzi; Samuel Robson; Sarah Wyllie; Scott Elliott; Sharon Glaysher; Yann Bourgeois |
| EPI_ISL_837843, EPI_ISL_837844, EPI_ISL_837845, EPI_ISL_837846, EPI_ISL_837847, EPI_ISL_837848, EPI_ISL_837850, EPI_ISL_837852, EPI_ISL_837854, EPI_ISL_837856, EPI_ISL_837857, EPI_ISL_837859, EPI_ISL_837861, EPI_ISL_837863, EPI_ISL_837865, EPI_ISL_837869, EPI_ISL_837871, EPI_ISL_837903, EPI_ISL_837904, EPI_ISL_837905, EPI_ISL_837906, EPI_ISL_837907, EPI_ISL_837908, EPI_ISL_837909, EPI_ISL_837910, EPI_ISL_837911, EPI_ISL_837912, EPI_ISL_837913, EPI_ISL_837915, EPI_ISL_837916, EPI_ISL_837917, EPI_ISL_837918, EPI_ISL_837919, EPI_ISL_837920, EPI_ISL_837921, EPI_ISL_837922, EPI_ISL_837923, EPI_ISL_837924, EPI_ISL_837925, EPI_ISL_837927, EPI_ISL_837929, EPI_ISL_837930, EPI_ISL_837931, EPI_ISL_837932, EPI_ISL_837933, EPI_ISL_837934, EPI_ISL_837935, EPI_ISL_837936, EPI_ISL_837937, EPI_ISL_837938, EPI_ISL_837939, EPI_ISL_837940, EPI_ISL_837941, EPI_ISL_837942, EPI_ISL_837943, EPI_ISL_837944, EPI_ISL_837945, EPI_ISL_837946, EPI_ISL_837947, EPI_ISL_837948, EPI_ISL_837949, EPI_ISL_837950, EPI_ISL_837951, EPI_ISL_837952, EPI_ISL_837953, EPI_ISL_837954, EPI_ISL_837955, EPI_ISL_837956, EPI_ISL_837958, EPI_ISL_837960, EPI_ISL_837962, EPI_ISL_837963, EPI_ISL_837964, EPI_ISL_837965, EPI_ISL_837966, EPI_ISL_837967, EPI_ISL_837968, EPI_ISL_837969, EPI_ISL_837970, EPI_ISL_837971, EPI_ISL_864748, EPI_ISL_864751, EPI_ISL_864754, EPI_ISL_864755, EPI_ISL_864756, EPI_ISL_864758, EPI_ISL_864760, EPI_ISL_864762, EPI_ISL_864764, EPI_ISL_864766, EPI_ISL_864768, EPI_ISL_864771, EPI_ISL_864773, EPI_ISL_864774, EPI_ISL_864775, EPI_ISL_864776, EPI_ISL_864777, EPI_ISL_864778, EPI_ISL_864779, EPI_ISL_864780, EPI_ISL_864797, EPI_ISL_864799, EPI_ISL_864803, EPI_ISL_864805, EPI_ISL_864807, EPI_ISL_864809, EPI_ISL_864811, EPI_ISL_864813, EPI_ISL_864815, EPI_ISL_864818, EPI_ISL_864819, EPI_ISL_864821, EPI_ISL_864828, EPI_ISL_864829, EPI_ISL_864832, EPI_ISL_864834, EPI_ISL_864838, EPI_ISL_864846, EPI_ISL_864850, EPI_ISL_864856, EPI_ISL_864875, EPI_ISL_864883, EPI_ISL_864889 | | | |
| see above | Department of Pathology, University of Cambridge | COVID-19 Genomics UK (COG-UK) Consortium | Aminu S. Jahun; Grant Hall; Ian Goodfellow; Iliana Georgana; Malte Pinckert; Martin D. Curran; Myra Hosmillo; Surendra Parmar; Yasmin Chaudhry |
| EPI_ISL_821002, EPI_ISL_821004, EPI_ISL_821006, EPI_ISL_821007, EPI_ISL_821008, EPI_ISL_821009, EPI_ISL_821014, EPI_ISL_821015, EPI_ISL_821016, EPI_ISL_821017, EPI_ISL_821019, EPI_ISL_821023, EPI_ISL_821024, EPI_ISL_821027, EPI_ISL_821029, EPI_ISL_821031, EPI_ISL_821033, EPI_ISL_821034, EPI_ISL_821040, EPI_ISL_821041, EPI_ISL_821043, EPI_ISL_821044, EPI_ISL_821045, EPI_ISL_821048, EPI_ISL_821049, EPI_ISL_821051, EPI_ISL_821052, EPI_ISL_821053, EPI_ISL_821054, EPI_ISL_821057, EPI_ISL_821058, EPI_ISL_821059, EPI_ISL_821064, EPI_ISL_821065, EPI_ISL_821067, EPI_ISL_821070, EPI_ISL_821071, EPI_ISL_821072, EPI_ISL_821075, EPI_ISL_821076, EPI_ISL_821079, EPI_ISL_821080, EPI_ISL_821081, EPI_ISL_821083, EPI_ISL_821086, EPI_ISL_821087, EPI_ISL_821088, EPI_ISL_821089, EPI_ISL_821092, EPI_ISL_821094, EPI_ISL_821096, EPI_ISL_821097, EPI_ISL_821100, EPI_ISL_821101, EPI_ISL_821103, EPI_ISL_821106, EPI_ISL_821107, EPI_ISL_821108, EPI_ISL_821112, EPI_ISL_821113, EPI_ISL_821114, EPI_ISL_821115, EPI_ISL_821118, EPI_ISL_821119, EPI_ISL_821120, EPI_ISL_821121, EPI_ISL_821122, EPI_ISL_821123, EPI_ISL_821129, EPI_ISL_821130, EPI_ISL_821132, EPI_ISL_821135, EPI_ISL_821138, EPI_ISL_821139, EPI_ISL_821141, EPI_ISL_821143, EPI_ISL_821145, EPI_ISL_821146, EPI_ISL_821147, EPI_ISL_821148, EPI_ISL_821153, EPI_ISL_821154, EPI_ISL_821155, EPI_ISL_821157, EPI_ISL_821161, EPI_ISL_821162, EPI_ISL_821164, EPI_ISL_821167, EPI_ISL_821168, EPI_ISL_821171, EPI_ISL_821176, EPI_ISL_821177, EPI_ISL_821179, EPI_ISL_821182, EPI_ISL_821183, EPI_ISL_821184, EPI_ISL_821185, EPI_ISL_821186, EPI_ISL_821188, EPI_ISL_821189, EPI_ISL_821192, EPI_ISL_821193, EPI_ISL_821196, EPI_ISL_821202, EPI_ISL_821203, EPI_ISL_821204, EPI_ISL_821205, EPI_ISL_821209, EPI_ISL_821210, EPI_ISL_821211, EPI_ISL_821212, EPI_ISL_821213, EPI_ISL_821216, EPI_ISL_821217, EPI_ISL_821218, EPI_ISL_821219, EPI_ISL_821221, EPI_ISL_821224, EPI_ISL_821226, EPI_ISL_821229, EPI_ISL_821232, EPI_ISL_821233, EPI_ISL_821234, EPI_ISL_821236, EPI_ISL_821237, EPI_ISL_821238, EPI_ISL_821239, EPI_ISL_821240, EPI_ISL_821242, EPI_ISL_821246, EPI_ISL_821247, EPI_ISL_821252, EPI_ISL_821256, EPI_ISL_821259, EPI_ISL_821260, EPI_ISL_821262, EPI_ISL_821265, EPI_ISL_821267, EPI_ISL_821268, EPI_ISL_821272, EPI_ISL_821273, EPI_ISL_821283, EPI_ISL_821288, EPI_ISL_821314, EPI_ISL_821351, EPI_ISL_821366, EPI_ISL_821372, EPI_ISL_821389, EPI_ISL_821390, EPI_ISL_821398, EPI_ISL_821401, EPI_ISL_821403, EPI_ISL_821418, EPI_ISL_821419, EPI_ISL_821425, EPI_ISL_821426, EPI_ISL_821440, EPI_ISL_821451, EPI_ISL_821453, EPI_ISL_821459, EPI_ISL_821466, EPI_ISL_821470, EPI_ISL_821476, EPI_ISL_821497, EPI_ISL_821502, EPI_ISL_821511, EPI_ISL_821518, EPI_ISL_821533, EPI_ISL_821538, EPI_ISL_821548, EPI_ISL_821552, EPI_ISL_821553, EPI_ISL_821558, EPI_ISL_821566, EPI_ISL_821584, EPI_ISL_821585, EPI_ISL_821586, EPI_ISL_821599, EPI_ISL_821607, EPI_ISL_821614, EPI_ISL_821987, EPI_ISL_821988, EPI_ISL_822065, EPI_ISL_822080, EPI_ISL_822086, EPI_ISL_822193, EPI_ISL_822209, EPI_ISL_826468, EPI_ISL_826469, EPI_ISL_826470, EPI_ISL_826471, EPI_ISL_826474, EPI_ISL_826478, EPI_ISL_833933, EPI_ISL_833934, EPI_ISL_833935, EPI_ISL_833936, EPI_ISL_833937, EPI_ISL_833938, EPI_ISL_833939, EPI_ISL_833940, EPI_ISL_833941, EPI_ISL_833942, EPI_ISL_833943, EPI_ISL_833944, EPI_ISL_833945, EPI_ISL_833946, EPI_ISL_833947, EPI_ISL_833948, EPI_ISL_833949, EPI_ISL_833950, EPI_ISL_833951, EPI_ISL_833952, EPI_ISL_833953, EPI_ISL_833954, EPI_ISL_833955, EPI_ISL_833956, EPI_ISL_833957, EPI_ISL_833958, EPI_ISL_833959, EPI_ISL_833960, EPI_ISL_833961, EPI_ISL_833962, EPI_ISL_833963, EPI_ISL_833964, EPI_ISL_833965, EPI_ISL_833966, EPI_ISL_833967, EPI_ISL_833968, EPI_ISL_833969, EPI_ISL_833970, EPI_ISL_833971, EPI_ISL_833972, EPI_ISL_833973, EPI_ISL_833974, EPI_ISL_833975, EPI_ISL_833976, EPI_ISL_833977, EPI_ISL_833978, EPI_ISL_833979, EPI_ISL_833980, EPI_ISL_833981, EPI_ISL_833982, EPI_ISL_833983, EPI_ISL_833984, EPI_ISL_833985, EPI_ISL_833986, EPI_ISL_833987, EPI_ISL_833988, EPI_ISL_833989, EPI_ISL_833990, EPI_ISL_833991, EPI_ISL_833992, EPI_ISL_833993, EPI_ISL_833994, EPI_ISL_833995, EPI_ISL_833996, EPI_ISL_833997, EPI_ISL_833999, EPI_ISL_834000, EPI_ISL_834001, EPI_ISL_834002, EPI_ISL_834003, EPI_ISL_834004, EPI_ISL_834005, EPI_ISL_834006, EPI_ISL_834007, EPI_ISL_834008, EPI_ISL_834009, EPI_ISL_834010, EPI_ISL_834011, EPI_ISL_834012, EPI_ISL_834013, EPI_ISL_834014, EPI_ISL_834015, EPI_ISL_834016, EPI_ISL_834017, EPI_ISL_834018, EPI_ISL_834019, EPI_ISL_834020, EPI_ISL_834021, EPI_ISL_834022, EPI_ISL_834023, EPI_ISL_834024, EPI_ISL_834025, EPI_ISL_834026, EPI_ISL_834027, EPI_ISL_834028, EPI_ISL_834029, EPI_ISL_834030, EPI_ISL_834031, EPI_ISL_834032, EPI_ISL_834033, EPI_ISL_834034, EPI_ISL_834035, EPI_ISL_834036, EPI_ISL_834037, EPI_ISL_834038, EPI_ISL_834039, EPI_ISL_834040, EPI_ISL_834041, EPI_ISL_834042, EPI_ISL_834043, EPI_ISL_834044, EPI_ISL_834045, EPI_ISL_834047, EPI_ISL_834048, EPI_ISL_834049, EPI_ISL_834050, EPI_ISL_834051, EPI_ISL_834052, EPI_ISL_834053, EPI_ISL_834054, EPI_ISL_834055, EPI_ISL_834056, EPI_ISL_834057, EPI_ISL_834058, EPI_ISL_834059, EPI_ISL_834060, EPI_ISL_834061, EPI_ISL_834062, EPI_ISL_834063, EPI_ISL_834064, EPI_ISL_834065, EPI_ISL_834066, EPI_ISL_834067, EPI_ISL_834068, EPI_ISL_834069, EPI_ISL_834070, EPI_ISL_834071, EPI_ISL_834072, EPI_ISL_834073, EPI_ISL_834074, EPI_ISL_834075, EPI_ISL_834076, EPI_ISL_834077, EPI_ISL_834078, EPI_ISL_834079, EPI_ISL_834080, EPI_ISL_834081, EPI_ISL_834082, EPI_ISL_834083, EPI_ISL_834084, EPI_ISL_834085, EPI_ISL_834086, EPI_ISL_834087, EPI_ISL_834088, EPI_ISL_834089, EPI_ISL_834090, EPI_ISL_834091, EPI_ISL_834092, EPI_ISL_834093, EPI_ISL_834094, EPI_ISL_834095, EPI_ISL_834096, EPI_ISL_834097, EPI_ISL_834098, EPI_ISL_834099, EPI_ISL_834100, EPI_ISL_834101, EPI_ISL_834102, EPI_ISL_834103, EPI_ISL_834104, EPI_ISL_834105, EPI_ISL_834106, EPI_ISL_834107, EPI_ISL_834108, EPI_ISL_834109, EPI_ISL_834110, EPI_ISL_834111, EPI_ISL_834112, EPI_ISL_834113, EPI_ISL_834114, EPI_ISL_834115, EPI_ISL_834116, EPI_ISL_834117, EPI_ISL_834118, EPI_ISL_834119, EPI_ISL_834120, EPI_ISL_834121, EPI_ISL_834122, EPI_ISL_834123, EPI_ISL_834124, EPI_ISL_834125, EPI_ISL_834126, EPI_ISL_834127, EPI_ISL_834128, EPI_ISL_834129, EPI_ISL_834130, EPI_ISL_834131, EPI_ISL_834132, EPI_ISL_834133, EPI_ISL_834134, EPI_ISL_834135, EPI_ISL_834136, EPI_ISL_834137, EPI_ISL_834138, EPI_ISL_834139, EPI_ISL_834140, EPI_ISL_834141, EPI_ISL_834142, EPI_ISL_834143, EPI_ISL_834144, EPI_ISL_834145, EPI_ISL_834147, EPI_ISL_834148, EPI_ISL_834149, EPI_ISL_834151, EPI_ISL_834152, EPI_ISL_834153, EPI_ISL_834154, EPI_ISL_834155, EPI_ISL_834156, EPI_ISL_834157, EPI_ISL_834158, EPI_ISL_834159, EPI_ISL_834160, EPI_ISL_834161, EPI_ISL_834162, EPI_ISL_834163, EPI_ISL_834164, EPI_ISL_834165, EPI_ISL_834166, EPI_ISL_834167, EPI_ISL_834168, EPI_ISL_834169, EPI_ISL_834170, EPI_ISL_834171, EPI_ISL_834172, EPI_ISL_834173, EPI_ISL_834174, EPI_ISL_834175, EPI_ISL_834176, EPI_ISL_834177, EPI_ISL_834178, EPI_ISL_834179, EPI_ISL_834180, EPI_ISL_834181, EPI_ISL_834182, EPI_ISL_834183, EPI_ISL_834184, EPI_ISL_834185, EPI_ISL_834186, EPI_ISL_834187, EPI_ISL_834188, EPI_ISL_834189, EPI_ISL_834190, EPI_ISL_834191, EPI_ISL_834192, EPI_ISL_834193, EPI_ISL_834195, EPI_ISL_834196, EPI_ISL_834197, EPI_ISL_834198, EPI_ISL_834199, EPI_ISL_834200, EPI_ISL_834201, EPI_ISL_834202, EPI_ISL_834203, EPI_ISL_834204, EPI_ISL_834205, EPI_ISL_834206, EPI_ISL_834207, EPI_ISL_834208, EPI_ISL_834209, EPI_ISL_834210, EPI_ISL_834213, EPI_ISL_834214, EPI_ISL_834215, EPI_ISL_834216, EPI_ISL_834217, EPI_ISL_834218, EPI_ISL_834219, EPI_ISL_834220, EPI_ISL_834221, EPI_ISL_834222, EPI_ISL_834224, EPI_ISL_834225, EPI_ISL_834227, EPI_ISL_834229, EPI_ISL_834232, EPI_ISL_834234, EPI_ISL_834238, EPI_ISL_834239, EPI_ISL_834240, EPI_ISL_834241, EPI_ISL_834242, EPI_ISL_834243, EPI_ISL_834244, EPI_ISL_834245, EPI_ISL_834247, EPI_ISL_834249, EPI_ISL_834251, EPI_ISL_834252, EPI_ISL_834253, EPI_ISL_834254, EPI_ISL_834255, EPI_ISL_834257, EPI_ISL_834258, EPI_ISL_834259, EPI_ISL_834260, EPI_ISL_834261, EPI_ISL_834262, EPI_ISL_834265, EPI_ISL_834266, EPI_ISL_834267, EPI_ISL_834268, EPI_ISL_834269, EPI_ISL_834270, EPI_ISL_834272, EPI_ISL_834273, EPI_ISL_834275, EPI_ISL_834276, EPI_ISL_834277, EPI_ISL_834278, EPI_ISL_834279, EPI_ISL_834281, EPI_ISL_834282, EPI_ISL_834284, EPI_ISL_834285, EPI_ISL_834286, EPI_ISL_834287, EPI_ISL_834289, EPI_ISL_834291, EPI_ISL_834292, EPI_ISL_834294, EPI_ISL_834295, EPI_ISL_834296, EPI_ISL_834297, EPI_ISL_834300, EPI_ISL_834301, EPI_ISL_834302, EPI_ISL_834303, EPI_ISL_834304, EPI_ISL_834305, EPI_ISL_834306, EPI_ISL_834307, EPI_ISL_834308, EPI_ISL_834309, EPI_ISL_834310, EPI_ISL_834314, EPI_ISL_834315, EPI_ISL_834316, EPI_ISL_834317, EPI_ISL_834318, EPI_ISL_834319, EPI_ISL_834320, EPI_ISL_834321, EPI_ISL_834322, EPI_ISL_834323, EPI_ISL_834324, EPI_ISL_834325, EPI_ISL_834326, EPI_ISL_834327, EPI_ISL_834328, EPI_ISL_834329, EPI_ISL_834330, EPI_ISL_834331, EPI_ISL_834332, EPI_ISL_834333, EPI_ISL_834334, EPI_ISL_834335, EPI_ISL_834336, EPI_ISL_834337, EPI_ISL_834338, EPI_ISL_834339, EPI_ISL_834340, EPI_ISL_834341, EPI_ISL_834342, EPI_ISL_834345, EPI_ISL_834346, EPI_ISL_834347, EPI_ISL_834348, EPI_ISL_834349, EPI_ISL_834350, EPI_ISL_834351, EPI_ISL_834354, EPI_ISL_834355, EPI_ISL_834356, EPI_ISL_834357, EPI_ISL_834358, EPI_ISL_834359, EPI_ISL_834361, EPI_ISL_834362, EPI_ISL_834363, EPI_ISL_834364, EPI_ISL_834365, EPI_ISL_834366, EPI_ISL_834367, EPI_ISL_834369, EPI_ISL_834371, EPI_ISL_834372, EPI_ISL_834374, EPI_ISL_834375, EPI_ISL_834379, EPI_ISL_834383, EPI_ISL_834384, EPI_ISL_834385, EPI_ISL_834386, EPI_ISL_834387, EPI_ISL_834389, EPI_ISL_834390, EPI_ISL_834391, EPI_ISL_834392, EPI_ISL_834393, EPI_ISL_834394, EPI_ISL_834395, EPI_ISL_834396, EPI_ISL_834397, EPI_ISL_834398, EPI_ISL_834399, EPI_ISL_834400, EPI_ISL_834401, EPI_ISL_834403, EPI_ISL_834405, EPI_ISL_834406, EPI_ISL_834408, EPI_ISL_834410, EPI_ISL_834411, EPI_ISL_834412, EPI_ISL_834413, EPI_ISL_834414, EPI_ISL_834415, EPI_ISL_834416, EPI_ISL_834418, EPI_ISL_834419, EPI_ISL_834420, EPI_ISL_834421, EPI_ISL_834422, EPI_ISL_834424, EPI_ISL_834425, EPI_ISL_834426, EPI_ISL_834428, EPI_ISL_834429, EPI_ISL_834431, EPI_ISL_834432, EPI_ISL_834434, EPI_ISL_834435, EPI_ISL_834437, EPI_ISL_834438, EPI_ISL_834439, EPI_ISL_834441, EPI_ISL_834442, EPI_ISL_834444, EPI_ISL_834446, EPI_ISL_834447, EPI_ISL_834450, EPI_ISL_834451, EPI_ISL_834452, EPI_ISL_834453, EPI_ISL_834454, EPI_ISL_834455, EPI_ISL_834457, EPI_ISL_834458, EPI_ISL_834461, EPI_ISL_834462, EPI_ISL_834463, EPI_ISL_834464, EPI_ISL_834465, EPI_ISL_834469, EPI_ISL_834470, EPI_ISL_834471, EPI_ISL_834472, EPI_ISL_834473, EPI_ISL_834474, EPI_ISL_834475, EPI_ISL_834476, EPI_ISL_834477, EPI_ISL_834478, EPI_ISL_834479, EPI_ISL_834480, EPI_ISL_834481, EPI_ISL_834482, EPI_ISL_834483, EPI_ISL_834487, EPI_ISL_834489, EPI_ISL_834490, EPI_ISL_834491, EPI_ISL_834493, EPI_ISL_834495, EPI_ISL_834496, EPI_ISL_834498, EPI_ISL_834501, EPI_ISL_834502, EPI_ISL_834504, EPI_ISL_834505, EPI_ISL_834506, EPI_ISL_834508, EPI_ISL_834509, EPI_ISL_834510, EPI_ISL_834512, EPI_ISL_834513, EPI_ISL_834514, EPI_ISL_834515, EPI_ISL_834516, EPI_ISL_834518, EPI_ISL_834520, EPI_ISL_834521, EPI_ISL_834522, EPI_ISL_834523, EPI_ISL_834525, EPI_ISL_834526, EPI_ISL_834527, EPI_ISL_834528, EPI_ISL_834530, EPI_ISL_834531, EPI_ISL_834532, EPI_ISL_834533, EPI_ISL_834534, EPI_ISL_834535, EPI_ISL_834536, EPI_ISL_834537, EPI_ISL_834538, EPI_ISL_834539, EPI_ISL_834540, EPI_ISL_834542, EPI_ISL_834543, EPI_ISL_834544, EPI_ISL_834546, EPI_ISL_834547, EPI_ISL_834548, EPI_ISL_834549, EPI_ISL_834550, EPI_ISL_834551, EPI_ISL_834552, EPI_ISL_834798, EPI_ISL_834799, EPI_ISL_834800, EPI_ISL_834801, EPI_ISL_834802, EPI_ISL_834803, EPI_ISL_834805, EPI_ISL_834806, EPI_ISL_834807, EPI_ISL_834808, EPI_ISL_834809, EPI_ISL_834810, EPI_ISL_834811, EPI_ISL_834812, EPI_ISL_834813, EPI_ISL_834815, EPI_ISL_834816, EPI_ISL_834817, EPI_ISL_834818, EPI_ISL_834819, EPI_ISL_834820, EPI_ISL_834821, EPI_ISL_834822, EPI_ISL_834824, EPI_ISL_834827, EPI_ISL_834828, EPI_ISL_834829, EPI_ISL_834830, EPI_ISL_834831, EPI_ISL_834832, EPI_ISL_834835, EPI_ISL_834836, EPI_ISL_834838, EPI_ISL_834839, EPI_ISL_834840, EPI_ISL_834841, EPI_ISL_834842, EPI_ISL_834843, EPI_ISL_834844, EPI_ISL_834846, EPI_ISL_834847, EPI_ISL_834849, EPI_ISL_834850, EPI_ISL_834851, EPI_ISL_834852, EPI_ISL_834853, EPI_ISL_834854, EPI_ISL_834855, EPI_ISL_834856, EPI_ISL_834857, EPI_ISL_834858, EPI_ISL_834859, EPI_ISL_834860, EPI_ISL_834861, EPI_ISL_834862, EPI_ISL_834863, EPI_ISL_834864, EPI_ISL_834866, EPI_ISL_834868, EPI_ISL_834869, EPI_ISL_834871, EPI_ISL_834872, EPI_ISL_834873, EPI_ISL_834876, EPI_ISL_834877, EPI_ISL_834878, EPI_ISL_834879, EPI_ISL_834880, EPI_ISL_834881, EPI_ISL_834882, EPI_ISL_834883, EPI_ISL_834884, EPI_ISL_834885, EPI_ISL_834887, EPI_ISL_834888, EPI_ISL_834890, EPI_ISL_834891, EPI_ISL_834892, EPI_ISL_834893, EPI_ISL_834894, EPI_ISL_834895, EPI_ISL_834896, EPI_ISL_834897, EPI_ISL_834898, EPI_ISL_834899, EPI_ISL_834900, EPI_ISL_834902, EPI_ISL_834904, EPI_ISL_834905, EPI_ISL_834906, EPI_ISL_834907, EPI_ISL_834908, EPI_ISL_834911, EPI_ISL_834912, EPI_ISL_834913, EPI_ISL_834914, EPI_ISL_834915, EPI_ISL_834916, EPI_ISL_834917, EPI_ISL_834918, EPI_ISL_834919, EPI_ISL_834922, EPI_ISL_834923, EPI_ISL_834924, EPI_ISL_834926, EPI_ISL_834927, EPI_ISL_834928, EPI_ISL_834929, EPI_ISL_834930, EPI_ISL_834932, EPI_ISL_834934, EPI_ISL_834935, EPI_ISL_834936, EPI_ISL_834937, EPI_ISL_834938, EPI_ISL_834939, EPI_ISL_834940, EPI_ISL_834941, EPI_ISL_834942, EPI_ISL_834943, EPI_ISL_834944, EPI_ISL_834945, EPI_ISL_834946, EPI_ISL_834948, EPI_ISL_834950, EPI_ISL_834951, EPI_ISL_834952, EPI_ISL_834953, EPI_ISL_834954, EPI_ISL_834955, EPI_ISL_834956, EPI_ISL_834957, EPI_ISL_834958, EPI_ISL_834960, EPI_ISL_834961, EPI_ISL_834962, EPI_ISL_834964, EPI_ISL_834965, EPI_ISL_834966, EPI_ISL_834967, EPI_ISL_834968, EPI_ISL_834969, EPI_ISL_834970, EPI_ISL_834971, EPI_ISL_834973, EPI_ISL_834974, EPI_ISL_834975, EPI_ISL_834976, EPI_ISL_834977, EPI_ISL_834978, EPI_ISL_834980, EPI_ISL_834984, EPI_ISL_834985, EPI_ISL_834986, EPI_ISL_834987, EPI_ISL_834988, EPI_ISL_834990, EPI_ISL_834992, EPI_ISL_834993, EPI_ISL_834994, EPI_ISL_834995, EPI_ISL_834996, EPI_ISL_834997, EPI_ISL_834998, EPI_ISL_834999, EPI_ISL_835000, EPI_ISL_835002, EPI_ISL_835003, EPI_ISL_835005, EPI_ISL_835006, EPI_ISL_835007, EPI_ISL_835009, EPI_ISL_835010, EPI_ISL_835011, EPI_ISL_835012, EPI_ISL_835013, EPI_ISL_835014, EPI_ISL_835015, EPI_ISL_835016, EPI_ISL_835018, EPI_ISL_835019, EPI_ISL_835020, EPI_ISL_835021, EPI_ISL_835022, EPI_ISL_835023, EPI_ISL_835024, EPI_ISL_835025, EPI_ISL_835026, EPI_ISL_835027, EPI_ISL_835028, EPI_ISL_835029, EPI_ISL_835030, EPI_ISL_835032, EPI_ISL_835033, EPI_ISL_835034, EPI_ISL_835036, EPI_ISL_835037, EPI_ISL_835040, EPI_ISL_835042, EPI_ISL_835044, EPI_ISL_835045, EPI_ISL_835046, EPI_ISL_835048, EPI_ISL_835049, EPI_ISL_835050, EPI_ISL_835051, EPI_ISL_835052, EPI_ISL_835053, EPI_ISL_835054, EPI_ISL_835056, EPI_ISL_835057, EPI_ISL_835059, EPI_ISL_835061, EPI_ISL_835062, EPI_ISL_835063, EPI_ISL_835064, EPI_ISL_835065, EPI_ISL_835066, EPI_ISL_835068, EPI_ISL_835069, EPI_ISL_835070, EPI_ISL_835071, EPI_ISL_835072, EPI_ISL_835073, EPI_ISL_835074, EPI_ISL_835075, EPI_ISL_835076, EPI_ISL_835077, EPI_ISL_835078, EPI_ISL_835079, EPI_ISL_835080, EPI_ISL_835081, EPI_ISL_835082, EPI_ISL_835083, EPI_ISL_835084, EPI_ISL_835085, EPI_ISL_835086, EPI_ISL_835087, EPI_ISL_835088, EPI_ISL_835089, EPI_ISL_835090, EPI_ISL_835092, EPI_ISL_835093, EPI_ISL_835095, EPI_ISL_835096, EPI_ISL_835097, EPI_ISL_835098, EPI_ISL_835099, EPI_ISL_835100, EPI_ISL_835101, EPI_ISL_835102, EPI_ISL_835103, EPI_ISL_835104, EPI_ISL_835105, EPI_ISL_835106, EPI_ISL_835107, EPI_ISL_835108, EPI_ISL_835109, EPI_ISL_835110, EPI_ISL_835111, EPI_ISL_835113, EPI_ISL_835116, EPI_ISL_835117, EPI_ISL_835118, EPI_ISL_835119, EPI_ISL_835120, EPI_ISL_835121, EPI_ISL_835122, EPI_ISL_835123, EPI_ISL_835124, EPI_ISL_835125, EPI_ISL_835126, EPI_ISL_835128, EPI_ISL_835129, EPI_ISL_835131, EPI_ISL_835133, EPI_ISL_835134, EPI_ISL_835135, EPI_ISL_835136, EPI_ISL_835137, EPI_ISL_835138, EPI_ISL_835139, EPI_ISL_835141, EPI_ISL_835142, EPI_ISL_835426, EPI_ISL_835427, EPI_ISL_835428, EPI_ISL_835429, EPI_ISL_835430, EPI_ISL_835431, EPI_ISL_835432, EPI_ISL_835434, EPI_ISL_835435, EPI_ISL_835436, EPI_ISL_835437, EPI_ISL_835438, EPI_ISL_835440, EPI_ISL_835441, EPI_ISL_835442, EPI_ISL_835443, EPI_ISL_835444, EPI_ISL_835445, EPI_ISL_835446, EPI_ISL_835449, EPI_ISL_835450, EPI_ISL_835451, EPI_ISL_835452, EPI_ISL_835453, EPI_ISL_835455, EPI_ISL_835456, EPI_ISL_835458, EPI_ISL_835459, EPI_ISL_835460, EPI_ISL_835461, EPI_ISL_835462, EPI_ISL_835463, EPI_ISL_835466, EPI_ISL_835467, EPI_ISL_835468, EPI_ISL_835469, EPI_ISL_835471, EPI_ISL_835472, EPI_ISL_835473, EPI_ISL_835476, EPI_ISL_835477, EPI_ISL_835478, EPI_ISL_835479, EPI_ISL_835480, EPI_ISL_835482, EPI_ISL_835484, EPI_ISL_835485, EPI_ISL_835489, EPI_ISL_835490, EPI_ISL_835491, EPI_ISL_835494, EPI_ISL_835496, EPI_ISL_835498, EPI_ISL_835501, EPI_ISL_835508, EPI_ISL_835509, EPI_ISL_835510, EPI_ISL_835511, EPI_ISL_835512, EPI_ISL_835513, EPI_ISL_835514, EPI_ISL_835516, EPI_ISL_835517, EPI_ISL_835519, EPI_ISL_835520, EPI_ISL_835521, EPI_ISL_835522, EPI_ISL_835523, EPI_ISL_835524, EPI_ISL_835525, EPI_ISL_835526, EPI_ISL_835528, EPI_ISL_835529, EPI_ISL_835534, EPI_ISL_835536, EPI_ISL_835537, EPI_ISL_835538, EPI_ISL_835542, EPI_ISL_835543, EPI_ISL_835544, EPI_ISL_835545, EPI_ISL_835549, EPI_ISL_835551, EPI_ISL_835552, EPI_ISL_835554, EPI_ISL_835556, EPI_ISL_835557, EPI_ISL_835558, EPI_ISL_835559, EPI_ISL_835560, EPI_ISL_835561, EPI_ISL_835563, EPI_ISL_835565, EPI_ISL_835566, EPI_ISL_835567, EPI_ISL_835568, EPI_ISL_835569, EPI_ISL_835571, EPI_ISL_835572, EPI_ISL_835575, EPI_ISL_835578, EPI_ISL_835580, EPI_ISL_835581, EPI_ISL_835583, EPI_ISL_835584, EPI_ISL_835585, EPI_ISL_835586, EPI_ISL_835587, EPI_ISL_835588, EPI_ISL_835591, EPI_ISL_835593, EPI_ISL_835594, EPI_ISL_835595, EPI_ISL_835596, EPI_ISL_835598, EPI_ISL_835599, EPI_ISL_835601, EPI_ISL_835602, EPI_ISL_835604, EPI_ISL_835605, EPI_ISL_835606, EPI_ISL_835607, EPI_ISL_835608, EPI_ISL_835609, EPI_ISL_835611, EPI_ISL_835612, EPI_ISL_835613, EPI_ISL_835617, EPI_ISL_835619, EPI_ISL_835620, EPI_ISL_835621, EPI_ISL_835624, EPI_ISL_835625, EPI_ISL_835626, EPI_ISL_835627, EPI_ISL_835628, EPI_ISL_835629, EPI_ISL_835630, EPI_ISL_835632, EPI_ISL_835633, EPI_ISL_835634, EPI_ISL_835635, EPI_ISL_835636, EPI_ISL_835637, EPI_ISL_835638, EPI_ISL_835640, EPI_ISL_835641, EPI_ISL_835642, EPI_ISL_835644, EPI_ISL_835645, EPI_ISL_835648, EPI_ISL_835649, EPI_ISL_835650, EPI_ISL_835652, EPI_ISL_835653, EPI_ISL_835654, EPI_ISL_835656, EPI_ISL_835657, EPI_ISL_835658, EPI_ISL_835660, EPI_ISL_835661, EPI_ISL_835662, EPI_ISL_835663, EPI_ISL_835665, EPI_ISL_835666, EPI_ISL_835667, EPI_ISL_835668, EPI_ISL_835669, EPI_ISL_835670, EPI_ISL_835672, EPI_ISL_835674, EPI_ISL_835675, EPI_ISL_835676, EPI_ISL_835679, EPI_ISL_835681, EPI_ISL_835684, EPI_ISL_835685, EPI_ISL_835689, EPI_ISL_835690, EPI_ISL_835691, EPI_ISL_835693, EPI_ISL_835694, EPI_ISL_835696, EPI_ISL_835697, EPI_ISL_835699, EPI_ISL_835700, EPI_ISL_835702, EPI_ISL_835704, EPI_ISL_835705, EPI_ISL_835706, EPI_ISL_835708, EPI_ISL_835709, EPI_ISL_836754, EPI_ISL_836765, EPI_ISL_836766, EPI_ISL_836769, EPI_ISL_836777, EPI_ISL_836781, EPI_ISL_836788, EPI_ISL_836792, EPI_ISL_836795, EPI_ISL_836797, EPI_ISL_836802, EPI_ISL_836803, EPI_ISL_836806, EPI_ISL_836807, EPI_ISL_836811, EPI_ISL_836813, EPI_ISL_836814, EPI_ISL_836817, EPI_ISL_836819, EPI_ISL_836822, EPI_ISL_836836, EPI_ISL_836837, EPI_ISL_836840, EPI_ISL_836842, EPI_ISL_836843, EPI_ISL_836848, EPI_ISL_836851, EPI_ISL_836858, EPI_ISL_836859, EPI_ISL_836862, EPI_ISL_836863, EPI_ISL_836866, EPI_ISL_836867, EPI_ISL_836876, EPI_ISL_836877, EPI_ISL_836878, EPI_ISL_836883, EPI_ISL_836887, EPI_ISL_836890, EPI_ISL_836891, EPI_ISL_836897, EPI_ISL_836903, EPI_ISL_836906, EPI_ISL_836909, EPI_ISL_836910, EPI_ISL_836913, EPI_ISL_836914, EPI_ISL_836920, EPI_ISL_836921, EPI_ISL_836922, EPI_ISL_836926, EPI_ISL_836928, EPI_ISL_836935, EPI_ISL_836936, EPI_ISL_836940, EPI_ISL_836946, EPI_ISL_836947, EPI_ISL_836948, EPI_ISL_836955, EPI_ISL_836962, EPI_ISL_836975, EPI_ISL_836976, EPI_ISL_846319, EPI_ISL_846400, EPI_ISL_846510, EPI_ISL_846511, EPI_ISL_846513, EPI_ISL_846514, EPI_ISL_846515, EPI_ISL_846516, EPI_ISL_846517, EPI_ISL_846518, EPI_ISL_846519, EPI_ISL_846520, EPI_ISL_846521, EPI_ISL_846522, EPI_ISL_846523, EPI_ISL_846524, EPI_ISL_846525, EPI_ISL_846537, EPI_ISL_851750, EPI_ISL_851964, EPI_ISL_852047, EPI_ISL_852225, EPI_ISL_857879, EPI_ISL_857890, EPI_ISL_858064, EPI_ISL_858065, EPI_ISL_858066, EPI_ISL_858067, EPI_ISL_858068, EPI_ISL_858069, EPI_ISL_858070, EPI_ISL_858071, EPI_ISL_858072, EPI_ISL_858073, EPI_ISL_858074, EPI_ISL_858075, EPI_ISL_858076, EPI_ISL_858077, EPI_ISL_858078, EPI_ISL_858079, EPI_ISL_858080, EPI_ISL_858081, EPI_ISL_858082, EPI_ISL_858083, EPI_ISL_858084, EPI_ISL_858085, EPI_ISL_858086, EPI_ISL_858087, EPI_ISL_858088, EPI_ISL_858089, EPI_ISL_858090, EPI_ISL_858091, EPI_ISL_858092, EPI_ISL_858093, EPI_ISL_858094, EPI_ISL_858095, EPI_ISL_858096, EPI_ISL_858097, EPI_ISL_858098, EPI_ISL_858099, EPI_ISL_858100, EPI_ISL_858101, EPI_ISL_858102, EPI_ISL_858103, EPI_ISL_858104, EPI_ISL_858105, EPI_ISL_858106, EPI_ISL_858107, EPI_ISL_858108, EPI_ISL_858109, EPI_ISL_858110, EPI_ISL_858111, EPI_ISL_858112, EPI_ISL_858113, EPI_ISL_858114, EPI_ISL_858115, EPI_ISL_858116, EPI_ISL_858117, EPI_ISL_858118, EPI_ISL_858119, EPI_ISL_858120, EPI_ISL_858121, EPI_ISL_858122, EPI_ISL_858123, EPI_ISL_858124, EPI_ISL_858125, EPI_ISL_858126, EPI_ISL_858127, EPI_ISL_858128, EPI_ISL_858129, EPI_ISL_858130, EPI_ISL_858131, EPI_ISL_858132, EPI_ISL_858133, EPI_ISL_858134, EPI_ISL_858135, EPI_ISL_858136, EPI_ISL_858137, EPI_ISL_858138, EPI_ISL_858139, EPI_ISL_858140, EPI_ISL_858141, EPI_ISL_858142, EPI_ISL_858143, EPI_ISL_858144, EPI_ISL_858145, EPI_ISL_858146, EPI_ISL_858147, EPI_ISL_858148, EPI_ISL_858149, EPI_ISL_858150, EPI_ISL_858151, EPI_ISL_858152, EPI_ISL_858153, EPI_ISL_858154, EPI_ISL_858155, EPI_ISL_858156, EPI_ISL_858157, EPI_ISL_858207, EPI_ISL_858208, EPI_ISL_858209, EPI_ISL_858210, EPI_ISL_858211, EPI_ISL_858212, EPI_ISL_858213, EPI_ISL_858214, EPI_ISL_858215, EPI_ISL_858216, EPI_ISL_858217, EPI_ISL_858218, EPI_ISL_858219, EPI_ISL_858220, EPI_ISL_858221, EPI_ISL_858222, EPI_ISL_858269, EPI_ISL_858270, EPI_ISL_858287, EPI_ISL_858296, EPI_ISL_858306, EPI_ISL_858307, EPI_ISL_858308, EPI_ISL_858309, EPI_ISL_858311, EPI_ISL_858324, EPI_ISL_858325, EPI_ISL_858326, EPI_ISL_858328, EPI_ISL_858329, EPI_ISL_858332, EPI_ISL_858334, EPI_ISL_858343, EPI_ISL_858354, EPI_ISL_858357, EPI_ISL_858368, EPI_ISL_858370, EPI_ISL_858375, EPI_ISL_858381, EPI_ISL_858382, EPI_ISL_858387, EPI_ISL_858388, EPI_ISL_858389, EPI_ISL_858393, EPI_ISL_858397, EPI_ISL_858400, EPI_ISL_858403, EPI_ISL_858516, EPI_ISL_858521, EPI_ISL_858523, EPI_ISL_858531, EPI_ISL_858557, EPI_ISL_858563, EPI_ISL_858572, EPI_ISL_858574, EPI_ISL_858576, EPI_ISL_858581, EPI_ISL_864039, EPI_ISL_874170, EPI_ISL_874281, EPI_ISL_882301, EPI_ISL_882302, EPI_ISL_882304, EPI_ISL_882306, EPI_ISL_882310, EPI_ISL_882311, EPI_ISL_882312, EPI_ISL_882313, EPI_ISL_882315, EPI_ISL_882317, EPI_ISL_882318, EPI_ISL_882322, EPI_ISL_882324, EPI_ISL_882325, EPI_ISL_882326, EPI_ISL_882328, EPI_ISL_882329, EPI_ISL_882330, EPI_ISL_882331, EPI_ISL_882334, EPI_ISL_882335, EPI_ISL_882336, EPI_ISL_882337, EPI_ISL_882338, EPI_ISL_882340, EPI_ISL_882342, EPI_ISL_882345, EPI_ISL_882346, EPI_ISL_882347, EPI_ISL_882348, EPI_ISL_882349, EPI_ISL_882350, EPI_ISL_882351, EPI_ISL_882353, EPI_ISL_882354, EPI_ISL_882355, EPI_ISL_882356, EPI_ISL_882357, EPI_ISL_882358, EPI_ISL_882359, EPI_ISL_882362, EPI_ISL_882366, EPI_ISL_882367, EPI_ISL_882368, EPI_ISL_882370, EPI_ISL_882373, EPI_ISL_882374, EPI_ISL_882378, EPI_ISL_882380, EPI_ISL_882381, EPI_ISL_882383, EPI_ISL_882384, EPI_ISL_882386, EPI_ISL_882387, EPI_ISL_882389, EPI_ISL_882392, EPI_ISL_882394, EPI_ISL_882395, EPI_ISL_882396, EPI_ISL_882399, EPI_ISL_882400, EPI_ISL_882401, EPI_ISL_882403, EPI_ISL_882405, EPI_ISL_882406, EPI_ISL_882407, EPI_ISL_882408, EPI_ISL_882410, EPI_ISL_882411, EPI_ISL_882412, EPI_ISL_882414, EPI_ISL_882416, EPI_ISL_882417, EPI_ISL_882419, EPI_ISL_882422, EPI_ISL_882423, EPI_ISL_882424, EPI_ISL_882425, EPI_ISL_882426, EPI_ISL_882430, EPI_ISL_882431, EPI_ISL_882432, EPI_ISL_882433, EPI_ISL_882435, EPI_ISL_882436, EPI_ISL_882439, EPI_ISL_882443, EPI_ISL_882444, EPI_ISL_882446, EPI_ISL_882447, EPI_ISL_882448, EPI_ISL_882449, EPI_ISL_882450, EPI_ISL_882451, EPI_ISL_882453, EPI_ISL_882454, EPI_ISL_882455, EPI_ISL_882456, EPI_ISL_882457, EPI_ISL_882458, EPI_ISL_882459, EPI_ISL_882460, EPI_ISL_882461, EPI_ISL_882462, EPI_ISL_882463, EPI_ISL_882464, EPI_ISL_882465, | | | |

EPI_ISL_882466, EPI_ISL_882467, EPI_ISL_882468, EPI_ISL_882469, EPI_ISL_882470, EPI_ISL_882472, EPI_ISL_882474, EPI_ISL_882477, EPI_ISL_882478, EPI_ISL_882479, EPI_ISL_882483, EPI_ISL_882485, EPI_ISL_882486, EPI_ISL_882487, EPI_ISL_882488, EPI_ISL_882489, EPI_ISL_882490, EPI_ISL_882492, EPI_ISL_882493, EPI_ISL_882495, EPI_ISL_882496, EPI_ISL_882497, EPI_ISL_882498, EPI_ISL_882499, EPI_ISL_882500, EPI_ISL_882501, EPI_ISL_882502, EPI_ISL_882503, EPI_ISL_882504, EPI_ISL_882507, EPI_ISL_882508, EPI_ISL_882509, EPI_ISL_882510, EPI_ISL_882511, EPI_ISL_882513, EPI_ISL_882514, EPI_ISL_882515, EPI_ISL_882516, EPI_ISL_882517, EPI_ISL_882519, EPI_ISL_882521, EPI_ISL_882522, EPI_ISL_882523, EPI_ISL_882526, EPI_ISL_882527, EPI_ISL_882528, EPI_ISL_882529, EPI_ISL_882530, EPI_ISL_882531, EPI_ISL_882532, EPI_ISL_882533, EPI_ISL_882535, EPI_ISL_882536, EPI_ISL_882540, EPI_ISL_882541, EPI_ISL_882543, EPI_ISL_882545, EPI_ISL_882546, EPI_ISL_882547, EPI_ISL_882551, EPI_ISL_882552, EPI_ISL_882553, EPI_ISL_882554, EPI_ISL_882555, EPI_ISL_882556, EPI_ISL_882557, EPI_ISL_882558, EPI_ISL_882560, EPI_ISL_882561, EPI_ISL_882567, EPI_ISL_882569, EPI_ISL_882570, EPI_ISL_882572, EPI_ISL_882573, EPI_ISL_882574, EPI_ISL_882575, EPI_ISL_882577, EPI_ISL_882578, EPI_ISL_882581, EPI_ISL_882582, EPI_ISL_882583, EPI_ISL_882584, EPI_ISL_882587, EPI_ISL_882591, EPI_ISL_882592, EPI_ISL_882594, EPI_ISL_882596, EPI_ISL_882597, EPI_ISL_882598, EPI_ISL_882599, EPI_ISL_882603, EPI_ISL_882604, EPI_ISL_892059, EPI_ISL_892060, EPI_ISL_892061, EPI_ISL_892062, EPI_ISL_892063, EPI_ISL_892064, EPI_ISL_892065, EPI_ISL_892066, EPI_ISL_892207, EPI_ISL_892208, EPI_ISL_892209, EPI_ISL_931818, EPI_ISL_946746, EPI_ISL_947649, EPI_ISL_963106, EPI_ISL_963980, EPI_ISL_964107, EPI_ISL_964219, EPI_ISL_964234, EPI_ISL_985788, EPI_ISL_994651, EPI_ISL_1719437

| | | | |
|---|---|---|---|
| see above | Lighthouse Lab in Alderley Park | Wellcome Sanger Institute for the COVID-19 Genomics UK (COG-UK) Consortium | Cordelia Langford; David K. Jackson; Dominic Kwiatkowski; Ewan Harrison; Ian Johnston; Jacquelyn Wynn; Jeffrey Barrett; John Sillitoe on behalf of the Wellcome Sanger Institute COVID-19 Surveillance Team; Mairead Hyland; Roberto Amato; Sonia Goncalves; The Lighthouse Lab in Alderley Park and Alex Alderton |

EPI_ISL_811457, EPI_ISL_811458, EPI_ISL_811459, EPI_ISL_811460, EPI_ISL_811461, EPI_ISL_811462, EPI_ISL_811463, EPI_ISL_811464, EPI_ISL_811465, EPI_ISL_811466, EPI_ISL_811467, EPI_ISL_811468, EPI_ISL_811470, EPI_ISL_811471, EPI_ISL_811473, EPI_ISL_811474, EPI_ISL_811475, EPI_ISL_811476, EPI_ISL_811477, EPI_ISL_811479, EPI_ISL_811480, EPI_ISL_811481, EPI_ISL_811483, EPI_ISL_811485, EPI_ISL_811486, EPI_ISL_811488, EPI_ISL_811489, EPI_ISL_811491, EPI_ISL_811492, EPI_ISL_811493, EPI_ISL_811495, EPI_ISL_811496, EPI_ISL_811497, EPI_ISL_811499, EPI_ISL_811501, EPI_ISL_811503, EPI_ISL_811504, EPI_ISL_811505, EPI_ISL_811506, EPI_ISL_811507, EPI_ISL_811508, EPI_ISL_811509, EPI_ISL_811510, EPI_ISL_811512, EPI_ISL_811513, EPI_ISL_811514, EPI_ISL_811515, EPI_ISL_811517, EPI_ISL_811518, EPI_ISL_811520, EPI_ISL_811521, EPI_ISL_811522, EPI_ISL_811523, EPI_ISL_811526, EPI_ISL_811527, EPI_ISL_811528, EPI_ISL_811529, EPI_ISL_811531, EPI_ISL_811532, EPI_ISL_811533, EPI_ISL_811534, EPI_ISL_811535, EPI_ISL_811536, EPI_ISL_811537, EPI_ISL_811539, EPI_ISL_811540, EPI_ISL_811542, EPI_ISL_811543, EPI_ISL_811544, EPI_ISL_811546, EPI_ISL_811547, EPI_ISL_811550, EPI_ISL_811551, EPI_ISL_811552, EPI_ISL_811553, EPI_ISL_811554, EPI_ISL_811556, EPI_ISL_811557, EPI_ISL_811558, EPI_ISL_811560, EPI_ISL_811561, EPI_ISL_811562, EPI_ISL_811564, EPI_ISL_811566, EPI_ISL_811567, EPI_ISL_811568, EPI_ISL_811570, EPI_ISL_811571, EPI_ISL_811572, EPI_ISL_811574, EPI_ISL_811575, EPI_ISL_811576, EPI_ISL_811577, EPI_ISL_811578, EPI_ISL_811579, EPI_ISL_811581, EPI_ISL_811582, EPI_ISL_811583, EPI_ISL_811585, EPI_ISL_811586, EPI_ISL_811587, EPI_ISL_811588, EPI_ISL_811589, EPI_ISL_811590, EPI_ISL_811592, EPI_ISL_811593, EPI_ISL_811596, EPI_ISL_811597, EPI_ISL_811598, EPI_ISL_811599, EPI_ISL_811600, EPI_ISL_811601, EPI_ISL_811602, EPI_ISL_811604, EPI_ISL_811605, EPI_ISL_811607, EPI_ISL_811608, EPI_ISL_811609, EPI_ISL_811612, EPI_ISL_811613, EPI_ISL_811614, EPI_ISL_811615, EPI_ISL_811616, EPI_ISL_811619, EPI_ISL_811623, EPI_ISL_811624, EPI_ISL_811628, EPI_ISL_811629, EPI_ISL_811630, EPI_ISL_811631, EPI_ISL_811632, EPI_ISL_811633, EPI_ISL_811635, EPI_ISL_811636, EPI_ISL_811637, EPI_ISL_811638, EPI_ISL_811639, EPI_ISL_811640, EPI_ISL_811641, EPI_ISL_811642, EPI_ISL_811643, EPI_ISL_811646, EPI_ISL_811647, EPI_ISL_811648, EPI_ISL_811649, EPI_ISL_811652, EPI_ISL_811653, EPI_ISL_811654, EPI_ISL_811655, EPI_ISL_811656, EPI_ISL_811658, EPI_ISL_811659, EPI_ISL_811662, EPI_ISL_811663, EPI_ISL_811665, EPI_ISL_811667, EPI_ISL_811668, EPI_ISL_811674, EPI_ISL_811675, EPI_ISL_811676, EPI_ISL_811679, EPI_ISL_811680, EPI_ISL_811683, EPI_ISL_811684, EPI_ISL_811685, EPI_ISL_811686, EPI_ISL_811687, EPI_ISL_811689, EPI_ISL_811690, EPI_ISL_811691, EPI_ISL_811692, EPI_ISL_811693, EPI_ISL_811694, EPI_ISL_811695, EPI_ISL_811697, EPI_ISL_811699, EPI_ISL_811700, EPI_ISL_811704, EPI_ISL_811705, EPI_ISL_811706, EPI_ISL_811707, EPI_ISL_811709, EPI_ISL_811710, EPI_ISL_811711, EPI_ISL_811712, EPI_ISL_811714, EPI_ISL_811715, EPI_ISL_811716, EPI_ISL_811717, EPI_ISL_811718, EPI_ISL_811719, EPI_ISL_811721, EPI_ISL_811722, EPI_ISL_811723, EPI_ISL_811724, EPI_ISL_811725, EPI_ISL_811728, EPI_ISL_811729, EPI_ISL_811730, EPI_ISL_811731, EPI_ISL_811732, EPI_ISL_811733, EPI_ISL_811734, EPI_ISL_811735, EPI_ISL_811737, EPI_ISL_811738, EPI_ISL_811739, EPI_ISL_811742, EPI_ISL_811743, EPI_ISL_811744, EPI_ISL_811745, EPI_ISL_811746, EPI_ISL_811747, EPI_ISL_811748, EPI_ISL_811749, EPI_ISL_811751, EPI_ISL_811752, EPI_ISL_811755, EPI_ISL_811757, EPI_ISL_811758, EPI_ISL_811760, EPI_ISL_811761, EPI_ISL_811763, EPI_ISL_811764, EPI_ISL_811765, EPI_ISL_811766, EPI_ISL_811768, EPI_ISL_811769, EPI_ISL_811770, EPI_ISL_811771, EPI_ISL_811772, EPI_ISL_811773, EPI_ISL_811774, EPI_ISL_811775, EPI_ISL_811777, EPI_ISL_811778, EPI_ISL_811779, EPI_ISL_811780, EPI_ISL_811781, EPI_ISL_811782, EPI_ISL_811783, EPI_ISL_811784, EPI_ISL_811786, EPI_ISL_811788, EPI_ISL_811789, EPI_ISL_811790, EPI_ISL_811791, EPI_ISL_811792, EPI_ISL_811793, EPI_ISL_811794, EPI_ISL_811795, EPI_ISL_811796, EPI_ISL_811797, EPI_ISL_811798, EPI_ISL_811799, EPI_ISL_811800, EPI_ISL_811801, EPI_ISL_811803, EPI_ISL_811804, EPI_ISL_811805, EPI_ISL_811806, EPI_ISL_821276, EPI_ISL_821279, EPI_ISL_821284, EPI_ISL_821289, EPI_ISL_821290, EPI_ISL_821309, EPI_ISL_821310, EPI_ISL_821333, EPI_ISL_821338, EPI_ISL_821342, EPI_ISL_821343, EPI_ISL_821348, EPI_ISL_821354, EPI_ISL_821363, EPI_ISL_821365, EPI_ISL_821367, EPI_ISL_821375, EPI_ISL_821386, EPI_ISL_821394, EPI_ISL_821404, EPI_ISL_821408, EPI_ISL_821411, EPI_ISL_821416, EPI_ISL_821435, EPI_ISL_821439, EPI_ISL_821484, EPI_ISL_821488, EPI_ISL_821491, EPI_ISL_821510, EPI_ISL_821516, EPI_ISL_821519, EPI_ISL_821523, EPI_ISL_821529, EPI_ISL_821547, EPI_ISL_821555, EPI_ISL_821565, EPI_ISL_821568, EPI_ISL_821570, EPI_ISL_821576, EPI_ISL_821588, EPI_ISL_821592, EPI_ISL_821593, EPI_ISL_821596, EPI_ISL_821608, EPI_ISL_821616, EPI_ISL_821618, EPI_ISL_821620, EPI_ISL_821622, EPI_ISL_821623, EPI_ISL_821624, EPI_ISL_821967, EPI_ISL_833626, EPI_ISL_833630, EPI_ISL_833631, EPI_ISL_833654, EPI_ISL_833662, EPI_ISL_833671, EPI_ISL_833679, EPI_ISL_833680, EPI_ISL_833683, EPI_ISL_833707, EPI_ISL_833746, EPI_ISL_833770, EPI_ISL_833833, EPI_ISL_833842, EPI_ISL_833844, EPI_ISL_833849, EPI_ISL_833855, EPI_ISL_833859, EPI_ISL_833860, EPI_ISL_833896, EPI_ISL_833903, EPI_ISL_833909, EPI_ISL_833910, EPI_ISL_836421, EPI_ISL_836422, EPI_ISL_836423, EPI_ISL_836424, EPI_ISL_836425, EPI_ISL_836426, EPI_ISL_836427, EPI_ISL_836428, EPI_ISL_836429, EPI_ISL_836430, EPI_ISL_836431, EPI_ISL_836432, EPI_ISL_836433, EPI_ISL_836434, EPI_ISL_836435, EPI_ISL_836436, EPI_ISL_836437, EPI_ISL_836438, EPI_ISL_836439, EPI_ISL_836440, EPI_ISL_836441, EPI_ISL_836442, EPI_ISL_836443, EPI_ISL_836444, EPI_ISL_836445, EPI_ISL_836446, EPI_ISL_836447, EPI_ISL_836448, EPI_ISL_836449, EPI_ISL_836450, EPI_ISL_836451, EPI_ISL_836452, EPI_ISL_836453, EPI_ISL_836454, EPI_ISL_836455, EPI_ISL_836456, EPI_ISL_836457, EPI_ISL_836458, EPI_ISL_836459, EPI_ISL_836460, EPI_ISL_836461, EPI_ISL_836462, EPI_ISL_836463, EPI_ISL_836464, EPI_ISL_836465, EPI_ISL_836466, EPI_ISL_836467, EPI_ISL_836468, EPI_ISL_836469, EPI_ISL_836470, EPI_ISL_836471, EPI_ISL_836472, EPI_ISL_836473, EPI_ISL_836474, EPI_ISL_836475, EPI_ISL_836476, EPI_ISL_836477, EPI_ISL_836478, EPI_ISL_836479, EPI_ISL_836480, EPI_ISL_836481, EPI_ISL_836482, EPI_ISL_836483, EPI_ISL_836484, EPI_ISL_836485, EPI_ISL_836486, EPI_ISL_836487, EPI_ISL_836488, EPI_ISL_836489, EPI_ISL_836490, EPI_ISL_836491, EPI_ISL_836492, EPI_ISL_836493, EPI_ISL_836494, EPI_ISL_836495, EPI_ISL_836496, EPI_ISL_836497, EPI_ISL_836498, EPI_ISL_836499, EPI_ISL_836500, EPI_ISL_836501, EPI_ISL_836503, EPI_ISL_836504, EPI_ISL_836505, EPI_ISL_836506, EPI_ISL_836507, EPI_ISL_836508, EPI_ISL_836509, EPI_ISL_836510, EPI_ISL_836511, EPI_ISL_836512, EPI_ISL_836513, EPI_ISL_836514, EPI_ISL_836515, EPI_ISL_836516, EPI_ISL_836517, EPI_ISL_836518, EPI_ISL_836519, EPI_ISL_836520, EPI_ISL_836521, EPI_ISL_836522, EPI_ISL_836523, EPI_ISL_836524, EPI_ISL_836525, EPI_ISL_836526, EPI_ISL_836527, EPI_ISL_836528, EPI_ISL_836529, EPI_ISL_836530, EPI_ISL_836531, EPI_ISL_836532, EPI_ISL_836533, EPI_ISL_836534, EPI_ISL_836535, EPI_ISL_836536, EPI_ISL_836537, EPI_ISL_836538, EPI_ISL_836539, EPI_ISL_836540, EPI_ISL_836541, EPI_ISL_836542, EPI_ISL_836543, EPI_ISL_836544, EPI_ISL_836545, EPI_ISL_836546, EPI_ISL_836547, EPI_ISL_836548, EPI_ISL_836549, EPI_ISL_836550, EPI_ISL_836551, EPI_ISL_836552, EPI_ISL_836553, EPI_ISL_836554, EPI_ISL_836555, EPI_ISL_836556, EPI_ISL_836557, EPI_ISL_836558, EPI_ISL_836559, EPI_ISL_836560, EPI_ISL_836561, EPI_ISL_836562, EPI_ISL_836563, EPI_ISL_836564, EPI_ISL_836565, EPI_ISL_836566, EPI_ISL_836567, EPI_ISL_836568, EPI_ISL_836569, EPI_ISL_836570, EPI_ISL_836571, EPI_ISL_836572, EPI_ISL_836573, EPI_ISL_836574, EPI_ISL_836575, EPI_ISL_836576, EPI_ISL_836577, EPI_ISL_836578, EPI_ISL_836579, EPI_ISL_836580, EPI_ISL_836581, EPI_ISL_836582, EPI_ISL_836583, EPI_ISL_836584, EPI_ISL_836585, EPI_ISL_836586, EPI_ISL_836587, EPI_ISL_836588, EPI_ISL_836589, EPI_ISL_836590, EPI_ISL_836591, EPI_ISL_836592, EPI_ISL_836593, EPI_ISL_836594, EPI_ISL_836595, EPI_ISL_836596, EPI_ISL_836597, EPI_ISL_836598, EPI_ISL_836599, EPI_ISL_836600, EPI_ISL_836601, EPI_ISL_836602, EPI_ISL_836603, EPI_ISL_836604, EPI_ISL_836605, EPI_ISL_836606, EPI_ISL_836607, EPI_ISL_836608, EPI_ISL_836609, EPI_ISL_836610, EPI_ISL_836611, EPI_ISL_836612, EPI_ISL_836613, EPI_ISL_836614, EPI_ISL_836615, EPI_ISL_836616, EPI_ISL_836617, EPI_ISL_836618, EPI_ISL_836619, EPI_ISL_836620, EPI_ISL_836621, EPI_ISL_836622, EPI_ISL_836623, EPI_ISL_836624, EPI_ISL_836625, EPI_ISL_836626, EPI_ISL_836627, EPI_ISL_836628, EPI_ISL_836629, EPI_ISL_836630, EPI_ISL_836631, EPI_ISL_836632, EPI_ISL_836633, EPI_ISL_836634, EPI_ISL_836635, EPI_ISL_836636, EPI_ISL_836637, EPI_ISL_836638, EPI_ISL_836639, EPI_ISL_836640, EPI_ISL_836641, EPI_ISL_836642, EPI_ISL_836643, EPI_ISL_836644, EPI_ISL_836645, EPI_ISL_836646, EPI_ISL_836647, EPI_ISL_836648, EPI_ISL_836649, EPI_ISL_836650, EPI_ISL_836651, EPI_ISL_836652, EPI_ISL_836653, EPI_ISL_836654, EPI_ISL_836655, EPI_ISL_836656, EPI_ISL_836657, EPI_ISL_836658, EPI_ISL_836659, EPI_ISL_836660, EPI_ISL_836661, EPI_ISL_836662, EPI_ISL_836663, EPI_ISL_836664, EPI_ISL_836665, EPI_ISL_836666, EPI_ISL_836667, EPI_ISL_836668, EPI_ISL_836669, EPI_ISL_836670, EPI_ISL_836671, EPI_ISL_836672, EPI_ISL_836673, EPI_ISL_836674, EPI_ISL_836675, EPI_ISL_836676, EPI_ISL_836677, EPI_ISL_836678, EPI_ISL_836679, EPI_ISL_836680, EPI_ISL_836681, EPI_ISL_836682, EPI_ISL_836683, EPI_ISL_836684, EPI_ISL_836685, EPI_ISL_836686, EPI_ISL_836687, EPI_ISL_836688, EPI_ISL_836689, EPI_ISL_836690, EPI_ISL_836691, EPI_ISL_836692, EPI_ISL_836693, EPI_ISL_836694, EPI_ISL_836695, EPI_ISL_836696, EPI_ISL_836697, EPI_ISL_836698, EPI_ISL_836699, EPI_ISL_836700, EPI_ISL_836701, EPI_ISL_836702, EPI_ISL_836703, EPI_ISL_836704, EPI_ISL_836705, EPI_ISL_836706, EPI_ISL_836707, EPI_ISL_836708, EPI_ISL_836709, EPI_ISL_836710, EPI_ISL_836711, EPI_ISL_836712, EPI_ISL_836713, EPI_ISL_836714, EPI_ISL_836715, EPI_ISL_836716, EPI_ISL_836717, EPI_ISL_836718, EPI_ISL_836719, EPI_ISL_836720, EPI_ISL_836721, EPI_ISL_836722, EPI_ISL_836723, EPI_ISL_836724, EPI_ISL_836725, EPI_ISL_836726, EPI_ISL_836727, EPI_ISL_836728, EPI_ISL_836729, EPI_ISL_836730, EPI_ISL_836731, EPI_ISL_836732, EPI_ISL_836733, EPI_ISL_836734, EPI_ISL_836735, EPI_ISL_836736, EPI_ISL_836737, EPI_ISL_836738, EPI_ISL_836739, EPI_ISL_836740, EPI_ISL_836741, EPI_ISL_836742, EPI_ISL_836743, EPI_ISL_836744, EPI_ISL_836745, EPI_ISL_836746, EPI_ISL_836747, EPI_ISL_845973, EPI_ISL_846022, EPI_ISL_846070, EPI_ISL_851079, EPI_ISL_851133, EPI_ISL_851170, EPI_ISL_851251, EPI_ISL_851269, EPI_ISL_851273, EPI_ISL_851298, EPI_ISL_851343, EPI_ISL_857546, EPI_ISL_857553, EPI_ISL_857562, EPI_ISL_857584, EPI_ISL_857599, EPI_ISL_857604, EPI_ISL_857642, EPI_ISL_857645, EPI_ISL_857654, EPI_ISL_857682, EPI_ISL_857759, EPI_ISL_857818, EPI_ISL_857819, EPI_ISL_857823, EPI_ISL_857852, EPI_ISL_945063, EPI_ISL_945067, EPI_ISL_945070, EPI_ISL_945076, EPI_ISL_945083, EPI_ISL_945100, EPI_ISL_945103, EPI_ISL_945106, EPI_ISL_945107, EPI_ISL_945116, EPI_ISL_945131, EPI_ISL_945139, EPI_ISL_945148, EPI_ISL_945157, EPI_ISL_945166, EPI_ISL_945176, EPI_ISL_945180, EPI_ISL_945186, EPI_ISL_945191, EPI_ISL_945192, EPI_ISL_945199, EPI_ISL_945203, EPI_ISL_945222, EPI_ISL_945232, EPI_ISL_945237, EPI_ISL_945257, EPI_ISL_945261, EPI_ISL_945268, EPI_ISL_945271, EPI_ISL_945321, EPI_ISL_945341, EPI_ISL_945346, EPI_ISL_945356, EPI_ISL_945365, EPI_ISL_1012629

| | | | |
|---|---|---|---|
| see above | Lighthouse Lab in Cambridge | Wellcome Sanger Institute for the COVID-19 Genomics UK (COG-UK) Consortium | Cordelia Langford; David K. Jackson; Dominic Kwiatkowski; Ewan Harrison; Ian Johnston; John Sillitoe on behalf of the Wellcome Sanger Institute COVID-19 Surveillance Team; Rob Howes; Roberto Amato; Sonia Goncalves; The Lighthouse Lab in Cambridge and Alex Alderton |

EPI_ISL_797405, EPI_ISL_797408, EPI_ISL_797432, EPI_ISL_797445, EPI_ISL_797451, EPI_ISL_797459, EPI_ISL_797500, EPI_ISL_797514, EPI_ISL_797531, EPI_ISL_797554, EPI_ISL_797563, EPI_ISL_797564, EPI_ISL_797615, EPI_ISL_797620, EPI_ISL_797627, EPI_ISL_797632, EPI_ISL_797677, EPI_ISL_797678, EPI_ISL_797685, EPI_ISL_797704, EPI_ISL_797713, EPI_ISL_797716, EPI_ISL_797847, EPI_ISL_797867, EPI_ISL_797870, EPI_ISL_797914, EPI_ISL_797929, EPI_ISL_797945, EPI_ISL_797961, EPI_ISL_797970, EPI_ISL_797980, EPI_ISL_797996, EPI_ISL_811194, EPI_ISL_811195, EPI_ISL_811196, EPI_ISL_811198, EPI_ISL_811199, EPI_ISL_811200, EPI_ISL_811201, EPI_ISL_811202, EPI_ISL_811203, EPI_ISL_811204, EPI_ISL_811206, EPI_ISL_811208, EPI_ISL_811209, EPI_ISL_811210, EPI_ISL_811213, EPI_ISL_811216, EPI_ISL_811218, EPI_ISL_811219, EPI_ISL_811220, EPI_ISL_811221, EPI_ISL_811223, EPI_ISL_811224, EPI_ISL_811225, EPI_ISL_811226, EPI_ISL_811227, EPI_ISL_811228, EPI_ISL_811230, EPI_ISL_811231, EPI_ISL_811232, EPI_ISL_811233, EPI_ISL_811235, EPI_ISL_811237, EPI_ISL_811238, EPI_ISL_811239, EPI_ISL_811240, EPI_ISL_811242, EPI_ISL_811243, EPI_ISL_811244, EPI_ISL_811246, EPI_ISL_811249, EPI_ISL_811250, EPI_ISL_811252, EPI_ISL_811253, EPI_ISL_811254, EPI_ISL_811256, EPI_ISL_811257, EPI_ISL_811258, EPI_ISL_811259, EPI_ISL_811260, EPI_ISL_811261, EPI_ISL_811263, EPI_ISL_811264, EPI_ISL_811265, EPI_ISL_811266, EPI_ISL_811267, EPI_ISL_811268, EPI_ISL_811270, EPI_ISL_811271, EPI_ISL_811272, EPI_ISL_811273, EPI_ISL_811274, EPI_ISL_811276, EPI_ISL_811277, EPI_ISL_811278, EPI_ISL_811279, EPI_ISL_811281, EPI_ISL_811283, EPI_ISL_811286, EPI_ISL_811287, EPI_ISL_811288, EPI_ISL_811289, EPI_ISL_811290, EPI_ISL_811291, EPI_ISL_811292, EPI_ISL_811294, EPI_ISL_811295, EPI_ISL_811296, EPI_ISL_811297, EPI_ISL_811298, EPI_ISL_811299, EPI_ISL_811301, EPI_ISL_811303, EPI_ISL_811305, EPI_ISL_811308, EPI_ISL_811313, EPI_ISL_811314, EPI_ISL_811316, EPI_ISL_811317, EPI_ISL_811318, EPI_ISL_811319, EPI_ISL_811320, EPI_ISL_811323, EPI_ISL_811324, EPI_ISL_811326, EPI_ISL_811327, EPI_ISL_811328, EPI_ISL_811330, EPI_ISL_811331, EPI_ISL_811333, EPI_ISL_811337, EPI_ISL_811338, EPI_ISL_811340, EPI_ISL_811341, EPI_ISL_811343, EPI_ISL_811346, EPI_ISL_811347, EPI_ISL_811348, EPI_ISL_811349, EPI_ISL_811351, EPI_ISL_811352, EPI_ISL_811354, EPI_ISL_811355, EPI_ISL_811356, EPI_ISL_811357, EPI_ISL_811360, EPI_ISL_811361, EPI_ISL_811363, EPI_ISL_811364, EPI_ISL_811365, EPI_ISL_811370, EPI_ISL_811371, EPI_ISL_811372, EPI_ISL_811374, EPI_ISL_811375, EPI_ISL_811376, EPI_ISL_811379, EPI_ISL_811384, EPI_ISL_811386, EPI_ISL_811387, EPI_ISL_811388, EPI_ISL_811390, EPI_ISL_811391, EPI_ISL_811393, EPI_ISL_811396, EPI_ISL_811398, EPI_ISL_811399, EPI_ISL_811401, EPI_ISL_811402, EPI_ISL_811403, EPI_ISL_811405, EPI_ISL_811407, EPI_ISL_811408, EPI_ISL_811411, EPI_ISL_811412, EPI_ISL_811413, EPI_ISL_811415, EPI_ISL_811416, EPI_ISL_811417, EPI_ISL_811418, EPI_ISL_811419, EPI_ISL_811421, EPI_ISL_811423, EPI_ISL_811424, EPI_ISL_811425, EPI_ISL_811426, EPI_ISL_811427, EPI_ISL_811428, EPI_ISL_811432, EPI_ISL_811434, EPI_ISL_811435, EPI_ISL_811436, EPI_ISL_811437, EPI_ISL_811438, EPI_ISL_811439, EPI_ISL_811440, EPI_ISL_811441, EPI_ISL_811443, EPI_ISL_811446, EPI_ISL_811447, EPI_ISL_811449, EPI_ISL_811450, EPI_ISL_811451, EPI_ISL_811453, EPI_ISL_821032, EPI_ISL_821037, EPI_ISL_821056, EPI_ISL_821061, EPI_ISL_821062, EPI_ISL_821069, EPI_ISL_821077, EPI_ISL_821098, EPI_ISL_821105, EPI_ISL_821142, EPI_ISL_821150, EPI_ISL_821160, EPI_ISL_821195, EPI_ISL_821198, EPI_ISL_821207, EPI_ISL_821225, EPI_ISL_821227, EPI_ISL_821241, EPI_ISL_821245, EPI_ISL_821249, EPI_ISL_821257, EPI_ISL_821258, EPI_ISL_821266, EPI_ISL_821269, EPI_ISL_821621, EPI_ISL_821975, EPI_ISL_821993, EPI_ISL_821995, EPI_ISL_822001, EPI_ISL_822004, EPI_ISL_822005, EPI_ISL_822006, EPI_ISL_822011, EPI_ISL_822022, EPI_ISL_822023, EPI_ISL_822029, EPI_ISL_822033, EPI_ISL_822035, EPI_ISL_822040, EPI_ISL_822041, EPI_ISL_822044, EPI_ISL_822048, EPI_ISL_822051, EPI_ISL_822053, EPI_ISL_822059, EPI_ISL_822063, EPI_ISL_822067, EPI_ISL_822069, EPI_ISL_822072, EPI_ISL_822075, EPI_ISL_822078, EPI_ISL_822083, EPI_ISL_822099, EPI_ISL_822113, EPI_ISL_822117, EPI_ISL_822119, EPI_ISL_822122, EPI_ISL_822127, EPI_ISL_822130, EPI_ISL_822140, EPI_ISL_822148, EPI_ISL_822153, EPI_ISL_822156, EPI_ISL_822158, EPI_ISL_822159, EPI_ISL_822165, EPI_ISL_822169, EPI_ISL_822170, EPI_ISL_822178, EPI_ISL_822182, EPI_ISL_822201, EPI_ISL_822214, EPI_ISL_822223, EPI_ISL_822230, EPI_ISL_822237, EPI_ISL_822240, EPI_ISL_822242, EPI_ISL_822243, EPI_ISL_822244, EPI_ISL_822245, EPI_ISL_822247, EPI_ISL_822249, EPI_ISL_822253, EPI_ISL_822256, EPI_ISL_822260, EPI_ISL_822264, EPI_ISL_822265, EPI_ISL_822267, EPI_ISL_822268, EPI_ISL_822272, EPI_ISL_822275, EPI_ISL_822281, EPI_ISL_822282, EPI_ISL_822288, EPI_ISL_826477, EPI_ISL_826479, EPI_ISL_834553, EPI_ISL_834554, EPI_ISL_834555, EPI_ISL_834556, EPI_ISL_834557, EPI_ISL_834558, EPI_ISL_834559, EPI_ISL_834560, EPI_ISL_834561, EPI_ISL_834562, EPI_ISL_834563, EPI_ISL_834564, EPI_ISL_834565, EPI_ISL_834566, EPI_ISL_834567, EPI_ISL_834568, EPI_ISL_834569, EPI_ISL_834570, EPI_ISL_834571, EPI_ISL_834572, EPI_ISL_834573, EPI_ISL_834574, EPI_ISL_834575, EPI_ISL_834576, EPI_ISL_834577, EPI_ISL_834579, EPI_ISL_834580, EPI_ISL_834581, EPI_ISL_834582, EPI_ISL_834583, EPI_ISL_834584, EPI_ISL_834585, EPI_ISL_834586, EPI_ISL_834587, EPI_ISL_834588, EPI_ISL_834589, EPI_ISL_834590, EPI_ISL_834591, EPI_ISL_834592, EPI_ISL_834593, EPI_ISL_834594, EPI_ISL_834595, EPI_ISL_834597, EPI_ISL_834598, EPI_ISL_834599, EPI_ISL_834600, EPI_ISL_834601, EPI_ISL_834602, EPI_ISL_834603, EPI_ISL_834604, EPI_ISL_834605, EPI_ISL_834606, EPI_ISL_834607, EPI_ISL_834608, EPI_ISL_834609, EPI_ISL_834610, EPI_ISL_834611, EPI_ISL_834612, EPI_ISL_834613, EPI_ISL_834614, EPI_ISL_834615, EPI_ISL_834616, EPI_ISL_834617, EPI_ISL_834618, EPI_ISL_834619, EPI_ISL_834620, EPI_ISL_834621, EPI_ISL_834622, EPI_ISL_834623, EPI_ISL_834624, EPI_ISL_834625, EPI_ISL_834626, EPI_ISL_834627, EPI_ISL_834628, EPI_ISL_834629, EPI_ISL_834630, EPI_ISL_834631, EPI_ISL_834632, EPI_ISL_834633, EPI_ISL_834634, EPI_ISL_834635, EPI_ISL_834636, EPI_ISL_834637, EPI_ISL_834638, EPI_ISL_834639, EPI_ISL_834640, EPI_ISL_834641, EPI_ISL_834642, EPI_ISL_834643, EPI_ISL_834644, EPI_ISL_834645, EPI_ISL_834646, EPI_ISL_834647, EPI_ISL_834648, EPI_ISL_834649, EPI_ISL_834650, EPI_ISL_834651, EPI_ISL_834652, EPI_ISL_834653, EPI_ISL_834654, EPI_ISL_834655, EPI_ISL_834656, EPI_ISL_834657, EPI_ISL_834658, EPI_ISL_834659, EPI_ISL_834660, EPI_ISL_834661, EPI_ISL_834662, EPI_ISL_834663, EPI_ISL_834664, EPI_ISL_834665, EPI_ISL_834666, EPI_ISL_834667, EPI_ISL_834668, EPI_ISL_834669, EPI_ISL_834670, EPI_ISL_834671, EPI_ISL_834672, EPI_ISL_834673, EPI_ISL_834674, EPI_ISL_834675, EPI_ISL_834676, EPI_ISL_834677, EPI_ISL_834678, EPI_ISL_834679, EPI_ISL_834680, EPI_ISL_834681, EPI_ISL_834682, EPI_ISL_834683, EPI_ISL_834684, EPI_ISL_834685, EPI_ISL_834686, EPI_ISL_834687, EPI_ISL_834688, EPI_ISL_834689, EPI_ISL_834690, EPI_ISL_834691, EPI_ISL_834693, EPI_ISL_834694, EPI_ISL_834695, EPI_ISL_834696, EPI_ISL_834697, EPI_ISL_834698, EPI_ISL_834699, EPI_ISL_834700, EPI_ISL_834701, EPI_ISL_834702, EPI_ISL_834703, EPI_ISL_834704, EPI_ISL_834705, EPI_ISL_834706, EPI_ISL_834707, EPI_ISL_834708, EPI_ISL_834709, EPI_ISL_834710, EPI_ISL_834711, EPI_ISL_834712, EPI_ISL_834713, EPI_ISL_834714, EPI_ISL_834715, EPI_ISL_834716, EPI_ISL_834717, EPI_ISL_834718, EPI_ISL_834719, EPI_ISL_834720, EPI_ISL_834721, EPI_ISL_834722, EPI_ISL_834723, EPI_ISL_834724, EPI_ISL_834725, EPI_ISL_834726, EPI_ISL_834727, EPI_ISL_834728, EPI_ISL_834729, EPI_ISL_834730, EPI_ISL_834731, EPI_ISL_834732, EPI_ISL_834733, EPI_ISL_834734, EPI_ISL_834735, EPI_ISL_834736, EPI_ISL_834737, EPI_ISL_834738, EPI_ISL_834739, EPI_ISL_834740, EPI_ISL_834741, EPI_ISL_834742, EPI_ISL_834743, EPI_ISL_834744, EPI_ISL_834745, EPI_ISL_834746, EPI_ISL_834747, EPI_ISL_834748, EPI_ISL_834749, EPI_ISL_834750, EPI_ISL_834751, EPI_ISL_834752, EPI_ISL_834753, EPI_ISL_834754, EPI_ISL_834755, EPI_ISL_834756, EPI_ISL_834757, EPI_ISL_834758, EPI_ISL_834759, EPI_ISL_834760, EPI_ISL_834761, EPI_ISL_834762, EPI_ISL_834763, EPI_ISL_834764, EPI_ISL_834765, EPI_ISL_834766, EPI_ISL_834767, EPI_ISL_834768, EPI_ISL_834769, EPI_ISL_834770, EPI_ISL_834771, EPI_ISL_834772, EPI_ISL_834773, EPI_ISL_834774, EPI_ISL_834775, EPI_ISL_834776, EPI_ISL_834777, EPI_ISL_834778, EPI_ISL_834779, EPI_ISL_834780, EPI_ISL_834781, EPI_ISL_834782, EPI_ISL_834783, EPI_ISL_834784, EPI_ISL_834785, EPI_ISL_834786, EPI_ISL_834787, EPI_ISL_834788, EPI_ISL_834789, EPI_ISL_834790, EPI_ISL_834791, EPI_ISL_834792, EPI_ISL_834793, EPI_ISL_834794, EPI_ISL_834795, EPI_ISL_834796, EPI_ISL_834797, EPI_ISL_835143, EPI_ISL_835144, EPI_ISL_835145, EPI_ISL_835146, EPI_ISL_835147, EPI_ISL_835148, EPI_ISL_835149, EPI_ISL_835150, EPI_ISL_835151, EPI_ISL_835152, EPI_ISL_835153, EPI_ISL_835154, EPI_ISL_835155, EPI_ISL_835156, EPI_ISL_835157, EPI_ISL_835158, EPI_ISL_835159, EPI_ISL_835160, EPI_ISL_835161, EPI_ISL_835162, EPI_ISL_835163, EPI_ISL_835164, EPI_ISL_835165, EPI_ISL_835166, EPI_ISL_835167, EPI_ISL_835168, EPI_ISL_835169, EPI_ISL_835170, EPI_ISL_835171, EPI_ISL_835172, EPI_ISL_835173, EPI_ISL_835175, EPI_ISL_835176, EPI_ISL_835177, EPI_ISL_835178, EPI_ISL_835179, EPI_ISL_835180, EPI_ISL_835181, EPI_ISL_835182, EPI_ISL_835183, EPI_ISL_835184, EPI_ISL_835185, EPI_ISL_835186, EPI_ISL_835187, EPI_ISL_835188, EPI_ISL_835189, EPI_ISL_835190, EPI_ISL_835191, EPI_ISL_835192, EPI_ISL_835193, EPI_ISL_835194, EPI_ISL_835195, EPI_ISL_835196, EPI_ISL_835197, EPI_ISL_835198, EPI_ISL_835199, EPI_ISL_835200, EPI_ISL_835201, EPI_ISL_835202, EPI_ISL_835203, EPI_ISL_835204, EPI_ISL_835205, EPI_ISL_835206, EPI_ISL_835207, EPI_ISL_835208, EPI_ISL_835209, EPI_ISL_835210, EPI_ISL_835211, EPI_ISL_835212, EPI_ISL_835213, EPI_ISL_835214, EPI_ISL_835215, EPI_ISL_835216, EPI_ISL_835217, EPI_ISL_835218, EPI_ISL_835219, EPI_ISL_835220, EPI_ISL_835221, EPI_ISL_835222, EPI_ISL_835223, EPI_ISL_835224, EPI_ISL_835226, EPI_ISL_835227, EPI_ISL_835228, EPI_ISL_835229, EPI_ISL_835230, EPI_ISL_835231, EPI_ISL_835232, EPI_ISL_835233, EPI_ISL_835234, EPI_ISL_835236, EPI_ISL_835237, EPI_ISL_835238, EPI_ISL_835239, EPI_ISL_835240, EPI_ISL_835241, EPI_ISL_835242, EPI_ISL_835243, EPI_ISL_835244, EPI_ISL_835245, EPI_ISL_835246, EPI_ISL_835247, EPI_ISL_835248, EPI_ISL_835249, EPI_ISL_835250, EPI_ISL_835251, EPI_ISL_835252, EPI_ISL_835253, EPI_ISL_835254, EPI_ISL_835255, EPI_ISL_835256, EPI_ISL_835257, EPI_ISL_835258, EPI_ISL_835259, EPI_ISL_835260, EPI_ISL_835261, EPI_ISL_835262, EPI_ISL_835263, EPI_ISL_835264, EPI_ISL_835265, EPI_ISL_835266, EPI_ISL_835267, EPI_ISL_835268, EPI_ISL_835269, EPI_ISL_835270, EPI_ISL_835271, EPI_ISL_835272, EPI_ISL_835273, EPI_ISL_835274, EPI_ISL_835275, EPI_ISL_835276, EPI_ISL_835277, EPI_ISL_835278, EPI_ISL_835279, EPI_ISL_835280, EPI_ISL_835281, EPI_ISL_835282, EPI_ISL_835283, EPI_ISL_835284, EPI_ISL_835285, EPI_ISL_835286, EPI_ISL_835287, EPI_ISL_835288, EPI_ISL_835289, EPI_ISL_835290, EPI_ISL_835291, EPI_ISL_835292, EPI_ISL_835294, EPI_ISL_835295, EPI_ISL_835296, EPI_ISL_835297, EPI_ISL_835298, EPI_ISL_835299, EPI_ISL_835300, EPI_ISL_835301, EPI_ISL_835302, EPI_ISL_835303, EPI_ISL_835304, EPI_ISL_835305, EPI_ISL_835306, EPI_ISL_835307, EPI_ISL_835308, EPI_ISL_835309, EPI_ISL_835310, EPI_ISL_835311, EPI_ISL_835312, EPI_ISL_835313, EPI_ISL_835314, EPI_ISL_835315, EPI_ISL_835316, EPI_ISL_835317, EPI_ISL_835318, EPI_ISL_835319, EPI_ISL_835320, EPI_ISL_835321, EPI_ISL_835322, EPI_ISL_835323, EPI_ISL_835324, EPI_ISL_835325, EPI_ISL_835326, EPI_ISL_835327, EPI_ISL_835328, EPI_ISL_835329, EPI_ISL_835330, EPI_ISL_835331, EPI_ISL_835332, EPI_ISL_835333, EPI_ISL_835334, EPI_ISL_835335, EPI_ISL_835337, EPI_ISL_835338, EPI_ISL_835339, EPI_ISL_835340, EPI_ISL_835341, EPI_ISL_835342, EPI_ISL_835343, EPI_ISL_835344, EPI_ISL_835345, EPI_ISL_835346, EPI_ISL_835347, EPI_ISL_835348, EPI_ISL_835349, EPI_ISL_835350, EPI_ISL_835351, EPI_ISL_835352, EPI_ISL_835353, EPI_ISL_835354, EPI_ISL_835355, EPI_ISL_835356, EPI_ISL_835357, EPI_ISL_835358, EPI_ISL_835359, EPI_ISL_835360, EPI_ISL_835361, EPI_ISL_835362, EPI_ISL_835363, EPI_ISL_835364, EPI_ISL_835365, EPI_ISL_835366, EPI_ISL_835367, EPI_ISL_835368, EPI_ISL_835369, EPI_ISL_835370, EPI_ISL_835371, EPI_ISL_835372, EPI_ISL_835373, EPI_ISL_835374, EPI_ISL_835375, EPI_ISL_835376, EPI_ISL_835377, EPI_ISL_835378, EPI_ISL_835379, EPI_ISL_835380, EPI_ISL_835381, EPI_ISL_835382, EPI_ISL_835383, EPI_ISL_835384, EPI_ISL_835385, EPI_ISL_835386, EPI_ISL_835387, EPI_ISL_835388, EPI_ISL_835389, EPI_ISL_835390, EPI_ISL_835391, EPI_ISL_835392, EPI_ISL_835393, EPI_ISL_835394, EPI_ISL_835395, EPI_ISL_835396, EPI_ISL_835397, EPI_ISL_835398, EPI_ISL_835399, EPI_ISL_835400, EPI_ISL_835401, EPI_ISL_835402, EPI_ISL_835403, EPI_ISL_835404, EPI_ISL_835405, EPI_ISL_835406, EPI_ISL_835407, EPI_ISL_835408, EPI_ISL_835409, EPI_ISL_835410, EPI_ISL_835411, EPI_ISL_835412, EPI_ISL_835413, EPI_ISL_835414, EPI_ISL_835415, EPI_ISL_835416, EPI_ISL_835417, EPI_ISL_835418, EPI_ISL_835419, EPI_ISL_835420, EPI_ISL_835421, EPI_ISL_835422, EPI_ISL_835423, EPI_ISL_835424, EPI_ISL_835425, EPI_ISL_835433, EPI_ISL_835439, EPI_ISL_835447, EPI_ISL_835448, EPI_ISL_835454, EPI_ISL_835457, EPI_ISL_835464, EPI_ISL_835465, EPI_ISL_835470, EPI_ISL_835475, EPI_ISL_835481, EPI_ISL_835486, EPI_ISL_835487, EPI_ISL_835488, EPI_ISL_835493, EPI_ISL_835497, EPI_ISL_835499, EPI_ISL_835500, EPI_ISL_835502, EPI_ISL_835503, EPI_ISL_835505, EPI_ISL_835506, EPI_ISL_835507, EPI_ISL_835515, EPI_ISL_835518, EPI_ISL_835527, EPI_ISL_835530, EPI_ISL_835531, EPI_ISL_835533, EPI_ISL_835535, EPI_ISL_835539, EPI_ISL_835540, EPI_ISL_835541, EPI_ISL_835547, EPI_ISL_835548, EPI_ISL_835553, EPI_ISL_835555, EPI_ISL_835564, EPI_ISL_835570, EPI_ISL_835573, EPI_ISL_835574, EPI_ISL_835576, EPI_ISL_835577, EPI_ISL_835579, EPI_ISL_835582, EPI_ISL_835589, EPI_ISL_835590, EPI_ISL_835592, EPI_ISL_835597, EPI_ISL_835600, EPI_ISL_835603, EPI_ISL_835610, EPI_ISL_835614, EPI_ISL_835616, EPI_ISL_835618, EPI_ISL_835622, EPI_ISL_835623, EPI_ISL_835631, EPI_ISL_835643, EPI_ISL_835646, EPI_ISL_835647, EPI_ISL_835651, EPI_ISL_835659, EPI_ISL_835664, EPI_ISL_835671, EPI_ISL_835673, EPI_ISL_835677, EPI_ISL_835678, EPI_ISL_835682, EPI_ISL_835683, EPI_ISL_835686, EPI_ISL_835687, EPI_ISL_835688, EPI_ISL_835692, EPI_ISL_835695, EPI_ISL_835701, EPI_ISL_835703, EPI_ISL_835707, EPI_ISL_836748, EPI_ISL_836749, EPI_ISL_836750, EPI_ISL_836751, EPI_ISL_836752, EPI_ISL_836753, EPI_ISL_836755, EPI_ISL_836756, EPI_ISL_836757, EPI_ISL_836758, EPI_ISL_836759, EPI_ISL_836760, EPI_ISL_836761, EPI_ISL_836762, EPI_ISL_836763, EPI_ISL_836764, EPI_ISL_836767, EPI_ISL_836768, EPI_ISL_836770, EPI_ISL_836771, EPI_ISL_836773, EPI_ISL_836774, EPI_ISL_836775, EPI_ISL_836776, EPI_ISL_836778, EPI_ISL_836779, EPI_ISL_836780, EPI_ISL_836782, EPI_ISL_836783, EPI_ISL_836784, EPI_ISL_836785, EPI_ISL_836786, EPI_ISL_836787, EPI_ISL_836789, EPI_ISL_836790, EPI_ISL_836791, EPI_ISL_836793, EPI_ISL_836794, EPI_ISL_836796, EPI_ISL_836798, EPI_ISL_836799, EPI_ISL_836800, EPI_ISL_836801, EPI_ISL_836804, EPI_ISL_836805, EPI_ISL_836808, EPI_ISL_836809, EPI_ISL_836810, EPI_ISL_836812, EPI_ISL_836815, EPI_ISL_836816, EPI_ISL_836818, EPI_ISL_836820, EPI_ISL_836821, EPI_ISL_836823, EPI_ISL_836824, EPI_ISL_836825, EPI_ISL_836826, EPI_ISL_836827, EPI_ISL_836828, EPI_ISL_836829, EPI_ISL_836830, EPI_ISL_836831, EPI_ISL_836832, EPI_ISL_836833, EPI_ISL_836834, EPI_ISL_836835, EPI_ISL_836838, EPI_ISL_836839, EPI_ISL_836841, EPI_ISL_836844, EPI_ISL_836845, EPI_ISL_836846, EPI_ISL_836847, EPI_ISL_836849, EPI_ISL_836850, EPI_ISL_836852, EPI_ISL_836853, EPI_ISL_836854, EPI_ISL_836855, EPI_ISL_836856, EPI_ISL_836857, EPI_ISL_836860, EPI_ISL_836861, EPI_ISL_836864, EPI_ISL_836865, EPI_ISL_836868, EPI_ISL_836869, EPI_ISL_836870, EPI_ISL_836871, EPI_ISL_836872, EPI_ISL_836874, EPI_ISL_836876, EPI_ISL_836879, EPI_ISL_836880, EPI_ISL_836881, EPI_ISL_836882, EPI_ISL_836884, EPI_ISL_836885, EPI_ISL_836886, EPI_ISL_836888, EPI_ISL_836889, EPI_ISL_836892, EPI_ISL_836893, EPI_ISL_836894, EPI_ISL_836895, EPI_ISL_836896, EPI_ISL_836898, EPI_ISL_836899, EPI_ISL_836900, EPI_ISL_836901, EPI_ISL_836902, EPI_ISL_836904, EPI_ISL_836905, EPI_ISL_836907, EPI_ISL_836908, EPI_ISL_836911, EPI_ISL_836912, EPI_ISL_836915, EPI_ISL_836916, EPI_ISL_836917, EPI_ISL_836918, EPI_ISL_836919, EPI_ISL_836923, EPI_ISL_836924, EPI_ISL_836925, EPI_ISL_836927, EPI_ISL_836930, EPI_ISL_836931, EPI_ISL_836932, EPI_ISL_836933, EPI_ISL_836934, EPI_ISL_836937, EPI_ISL_836938, EPI_ISL_836939, EPI_ISL_836941, EPI_ISL_836942, EPI_ISL_836943, EPI_ISL_836944, EPI_ISL_836945, EPI_ISL_836949, EPI_ISL_836950, EPI_ISL_836951, EPI_ISL_836952, EPI_ISL_836953, EPI_ISL_836954, EPI_ISL_836956, EPI_ISL_836957, EPI_ISL_836958, EPI_ISL_836959, EPI_ISL_836960, EPI_ISL_836961, EPI_ISL_836963, EPI_ISL_836964, EPI_ISL_836965, EPI_ISL_836966, EPI_ISL_836967, EPI_ISL_836968, EPI_ISL_836969, EPI_ISL_836970, EPI_ISL_836971, EPI_ISL_836972, EPI_ISL_836973, EPI_ISL_836974, EPI_ISL_846527, EPI_ISL_846528, EPI_ISL_846529, EPI_ISL_846530, EPI_ISL_846531, EPI_ISL_846532, EPI_ISL_846533, EPI_ISL_846534, EPI_ISL_846535, EPI_ISL_846538, EPI_ISL_846539, EPI_ISL_851455, EPI_ISL_851467, EPI_ISL_851559, EPI_ISL_851594, EPI_ISL_851614, EPI_ISL_851648, EPI_ISL_851657, EPI_ISL_851954, EPI_ISL_852256, EPI_ISL_852261, EPI_ISL_852262, EPI_ISL_852263, EPI_ISL_852265, EPI_ISL_852266, EPI_ISL_852267, EPI_ISL_852270, EPI_ISL_852271, EPI_ISL_852272, EPI_ISL_852273, EPI_ISL_852274, EPI_ISL_852277, EPI_ISL_852278, EPI_ISL_852280, EPI_ISL_852282, EPI_ISL_852284, EPI_ISL_852290, EPI_ISL_852291, EPI_ISL_852294, EPI_ISL_852296, EPI_ISL_852297, EPI_ISL_852298, EPI_ISL_852299, EPI_ISL_852302, EPI_ISL_852304, EPI_ISL_852308, EPI_ISL_852309, EPI_ISL_852311, EPI_ISL_852315, EPI_ISL_852317, EPI_ISL_852318, EPI_ISL_852320, EPI_ISL_852323, EPI_ISL_852324, EPI_ISL_852327, EPI_ISL_852329, EPI_ISL_852330, EPI_ISL_852331, EPI_ISL_852332, EPI_ISL_852333, EPI_ISL_852336, EPI_ISL_852337, EPI_ISL_852341, EPI_ISL_852342, EPI_ISL_852346, EPI_ISL_852349, EPI_ISL_852351, EPI_ISL_852352, EPI_ISL_852354, EPI_ISL_852355, EPI_ISL_852356, EPI_ISL_852357, EPI_ISL_852359, EPI_ISL_852362, EPI_ISL_852363, EPI_ISL_852365, EPI_ISL_852366, EPI_ISL_852371, EPI_ISL_852372, EPI_ISL_852373, EPI_ISL_852376, EPI_ISL_852377, EPI_ISL_852379, EPI_ISL_852380, EPI_ISL_852381, EPI_ISL_852382, EPI_ISL_852383, EPI_ISL_852384, EPI_ISL_852385, EPI_ISL_852386, EPI_ISL_852387, EPI_ISL_852388, EPI_ISL_852390, EPI_ISL_852392, EPI_ISL_852394, EPI_ISL_852396, EPI_ISL_852399, EPI_ISL_852400, EPI_ISL_852401, EPI_ISL_852402, EPI_ISL_852403, EPI_ISL_852405, EPI_ISL_852408, EPI_ISL_852409, EPI_ISL_852410, EPI_ISL_852411, EPI_ISL_852418, EPI_ISL_852419, EPI_ISL_852423, EPI_ISL_852427, EPI_ISL_852428, EPI_ISL_852430, EPI_ISL_852431, EPI_ISL_852432, EPI_ISL_852434, EPI_ISL_852436, EPI_ISL_852437, EPI_ISL_852441, EPI_ISL_852442, EPI_ISL_852443, EPI_ISL_852449, EPI_ISL_852450, EPI_ISL_852452, EPI_ISL_852453, EPI_ISL_852455, EPI_ISL_852456, EPI_ISL_852457, EPI_ISL_852458, EPI_ISL_852461, EPI_ISL_852464, EPI_ISL_852465, EPI_ISL_852466, EPI_ISL_852468, EPI_ISL_852469, EPI_ISL_852470, EPI_ISL_852473, EPI_ISL_852477, EPI_ISL_852478, EPI_ISL_852480, EPI_ISL_852482, EPI_ISL_852483, EPI_ISL_852486, EPI_ISL_852488, EPI_ISL_852489, EPI_ISL_852490, EPI_ISL_852492, EPI_ISL_852493, EPI_ISL_852494, EPI_ISL_852495, EPI_ISL_852496, EPI_ISL_852499, EPI_ISL_852501, EPI_ISL_852502, EPI_ISL_852505, EPI_ISL_852507, EPI_ISL_852511, EPI_ISL_852512, EPI_ISL_852513, EPI_ISL_852515, EPI_ISL_852516, EPI_ISL_852518, EPI_ISL_852521, EPI_ISL_852523, EPI_ISL_852526, EPI_ISL_852528, EPI_ISL_852538, EPI_ISL_852539, EPI_ISL_852542, EPI_ISL_852543, EPI_ISL_852545, EPI_ISL_852546, EPI_ISL_852547, EPI_ISL_852551, EPI_ISL_852552, EPI_ISL_852553, EPI_ISL_852554, EPI_ISL_852556, EPI_ISL_858014, EPI_ISL_858020, EPI_ISL_858028, EPI_ISL_858029, EPI_ISL_858037, EPI_ISL_858158, EPI_ISL_858159, EPI_ISL_858160, EPI_ISL_858161, EPI_ISL_858162, EPI_ISL_858163, EPI_ISL_858164, EPI_ISL_858165, EPI_ISL_858166, EPI_ISL_858167, EPI_ISL_858168, EPI_ISL_858169, EPI_ISL_858170, EPI_ISL_858171, EPI_ISL_858172, EPI_ISL_858173, EPI_ISL_858174, EPI_ISL_858175, EPI_ISL_858176, EPI_ISL_858177, EPI_ISL_858178, EPI_ISL_858179, EPI_ISL_858180, EPI_ISL_858181, EPI_ISL_858182, EPI_ISL_858183, EPI_ISL_858184, EPI_ISL_858185, EPI_ISL_858186, EPI_ISL_858187, EPI_ISL_858188, EPI_ISL_858189, EPI_ISL_858190, EPI_ISL_858191, EPI_ISL_858192, EPI_ISL_858193, EPI_ISL_858194, EPI_ISL_858195, EPI_ISL_858196, EPI_ISL_858197, EPI_ISL_858198, EPI_ISL_858199, EPI_ISL_858200, EPI_ISL_858201, EPI_ISL_858202, EPI_ISL_858203, EPI_ISL_858204, EPI_ISL_858205, EPI_ISL_858206, EPI_ISL_858223, EPI_ISL_858224, EPI_ISL_858225, EPI_ISL_858226, EPI_ISL_858227, EPI_ISL_858228, EPI_ISL_858229, EPI_ISL_858230, EPI_ISL_858231, EPI_ISL_858232, EPI_ISL_858233, EPI_ISL_858234, EPI_ISL_858235, EPI_ISL_858236, EPI_ISL_858237, EPI_ISL_858238, EPI_ISL_858239, EPI_ISL_858240, EPI_ISL_858241, EPI_ISL_858242, EPI_ISL_858243, EPI_ISL_858244, EPI_ISL_858245, EPI_ISL_858246, EPI_ISL_858247, EPI_ISL_858248, EPI_ISL_858249, EPI_ISL_858250, EPI_ISL_858251, EPI_ISL_858252, EPI_ISL_858253, EPI_ISL_858254, EPI_ISL_858255, EPI_ISL_858256, EPI_ISL_858257, EPI_ISL_858258, EPI_ISL_858259, EPI_ISL_858260, EPI_ISL_858261, EPI_ISL_858262, EPI_ISL_858263, EPI_ISL_858264, EPI_ISL_858265, EPI_ISL_858266, EPI_ISL_858267, EPI_ISL_858271, EPI_ISL_858272, EPI_ISL_858273, EPI_ISL_858275, EPI_ISL_858276, EPI_ISL_858277, EPI_ISL_858278, EPI_ISL_858279, EPI_ISL_858280, EPI_ISL_858281, EPI_ISL_858282, EPI_ISL_858283, EPI_ISL_858284, EPI_ISL_858285, EPI_ISL_858286, EPI_ISL_858288, EPI_ISL_858289, EPI_ISL_858290, EPI_ISL_858291, EPI_ISL_858292, EPI_ISL_858293, EPI_ISL_858294, EPI_ISL_858295, EPI_ISL_858297, EPI_ISL_858298, EPI_ISL_858299, EPI_ISL_858300, EPI_ISL_858301, EPI_ISL_858302, EPI_ISL_858303, EPI_ISL_858304, EPI_ISL_858305, EPI_ISL_858310, EPI_ISL_858312, EPI_ISL_858314, EPI_ISL_858315, EPI_ISL_858317, EPI_ISL_858318, EPI_ISL_858319, EPI_ISL_858320, EPI_ISL_858321, EPI_ISL_858322, EPI_ISL_858323, EPI_ISL_858327, EPI_ISL_858330, EPI_ISL_858331, EPI_ISL_858333, EPI_ISL_858335, EPI_ISL_858336, EPI_ISL_858337, EPI_ISL_858338, EPI_ISL_858339, EPI_ISL_858340, EPI_ISL_858341, EPI_ISL_858342, EPI_ISL_858344, EPI_ISL_858345, EPI_ISL_858346, EPI_ISL_858347, EPI_ISL_858348, EPI_ISL_858349, EPI_ISL_858350, EPI_ISL_858351, EPI_ISL_858352, EPI_ISL_858353, EPI_ISL_858355, EPI_ISL_858356, EPI_ISL_858358, EPI_ISL_858359, EPI_ISL_858360, EPI_ISL_858361, EPI_ISL_858362, EPI_ISL_858363, EPI_ISL_858364, EPI_ISL_858365, EPI_ISL_858366, EPI_ISL_858367, EPI_ISL_858369, EPI_ISL_858371, EPI_ISL_858372, EPI_ISL_858373, EPI_ISL_858374, EPI_ISL_858376, EPI_ISL_858377, EPI_ISL_858379, EPI_ISL_858380, EPI_ISL_858381, EPI_ISL_858384, EPI_ISL_858385, EPI_ISL_858386, EPI_ISL_858390, EPI_ISL_858391, EPI_ISL_858392, EPI_ISL_858394, EPI_ISL_858395, EPI_ISL_858396, EPI_ISL_858398, EPI_ISL_858399, EPI_ISL_858401, EPI_ISL_858402, EPI_ISL_858404, EPI_ISL_858405, EPI_ISL_858406, EPI_ISL_858407, EPI_ISL_858425, EPI_ISL_858426, EPI_ISL_858427, EPI_ISL_858428, EPI_ISL_858429, EPI_ISL_858430, EPI_ISL_858431, EPI_ISL_858432, EPI_ISL_858433, EPI_ISL_858434, EPI_ISL_858435, EPI_ISL_858436, EPI_ISL_858437, EPI_ISL_858438, EPI_ISL_858439, EPI_ISL_858440, EPI_ISL_858441, EPI_ISL_858442, EPI_ISL_858443, EPI_ISL_858444, EPI_ISL_858445, EPI_ISL_858446, EPI_ISL_858447, EPI_ISL_858448, EPI_ISL_858449, EPI_ISL_858450, EPI_ISL_858451, EPI_ISL_858452, EPI_ISL_858453, EPI_ISL_858454, EPI_ISL_858455, EPI_ISL_858456, EPI_ISL_858457, EPI_ISL_858458, EPI_ISL_858459, EPI_ISL_858460, EPI_ISL_858461, EPI_ISL_858462, EPI_ISL_858463, EPI_ISL_858464, EPI_ISL_858465, EPI_ISL_858466, EPI_ISL_858467, EPI_ISL_858468, EPI_ISL_858469, EPI_ISL_858470, EPI_ISL_858471, EPI_ISL_858472, EPI_ISL_858473, EPI_ISL_858474, EPI_ISL_858475, EPI_ISL_858476, EPI_ISL_858477, EPI_ISL_858478, EPI_ISL_858479, EPI_ISL_858480, EPI_ISL_858481, EPI_ISL_858482, EPI_ISL_858483, EPI_ISL_858484, EPI_ISL_858485, EPI_ISL_858486, EPI_ISL_858487, EPI_ISL_858488, EPI_ISL_858489, EPI_ISL_858490, EPI_ISL_858491, EPI_ISL_858492, EPI_ISL_858493, EPI_ISL_858494, EPI_ISL_858495, EPI_ISL_858496, EPI_ISL_858497, EPI_ISL_858498, EPI_ISL_858499, EPI_ISL_858500, EPI_ISL_858501, EPI_ISL_858502, EPI_ISL_858503, EPI_ISL_858504, EPI_ISL_858505, EPI_ISL_858506, EPI_ISL_858507, EPI_ISL_858508, EPI_ISL_858509, EPI_ISL_858510, EPI_ISL_858511, EPI_ISL_858512, EPI_ISL_858513, EPI_ISL_858514, EPI_ISL_858515, EPI_ISL_858517, EPI_ISL_858518, EPI_ISL_858519, EPI_ISL_858520, EPI_ISL_858522, EPI_ISL_858524, EPI_ISL_858525, EPI_ISL_858526, EPI_ISL_858527, EPI_ISL_858528, EPI_ISL_858529, EPI_ISL_858530, EPI_ISL_858532, EPI_ISL_858533, EPI_ISL_858534, EPI_ISL_858535, EPI_ISL_858536, EPI_ISL_858537, EPI_ISL_858538, EPI_ISL_858539, EPI_ISL_858540, EPI_ISL_858541, EPI_ISL_858542, EPI_ISL_858543, EPI_ISL_858544, EPI_ISL_858545, EPI_ISL_858546, EPI_ISL_858547, EPI_ISL_858548, EPI_ISL_858550, EPI_ISL_858551, EPI_ISL_858552, EPI_ISL_858553, EPI_ISL_858555, EPI_ISL_858556, EPI_ISL_858558, EPI_ISL_858559, EPI_ISL_858560, EPI_ISL_858561, EPI_ISL_858562, EPI_ISL_858565, EPI_ISL_858566, EPI_ISL_858567, EPI_ISL_858568, EPI_ISL_858569, EPI_ISL_858570, EPI_ISL_858571, EPI_ISL_858585, EPI_ISL_858587, EPI_ISL_858588, EPI_ISL_858592, EPI_ISL_858594, EPI_ISL_858598, EPI_ISL_858600, EPI_ISL_858601, EPI_ISL_858604, EPI_ISL_858607,

EPI_ISL_858608, EPI_ISL_858609, EPI_ISL_858610, EPI_ISL_858612, EPI_ISL_858613, EPI_ISL_858615, EPI_ISL_858616, EPI_ISL_858618, EPI_ISL_858623, EPI_ISL_858626, EPI_ISL_858627, EPI_ISL_858628, EPI_ISL_858630, EPI_ISL_858635, EPI_ISL_858637, EPI_ISL_858638, EPI_ISL_858640, EPI_ISL_858641, EPI_ISL_858642, EPI_ISL_858643, EPI_ISL_858646, EPI_ISL_858650, EPI_ISL_858651, EPI_ISL_858652, EPI_ISL_858656, EPI_ISL_858657, EPI_ISL_858658, EPI_ISL_858659, EPI_ISL_858660, EPI_ISL_858661, EPI_ISL_858664, EPI_ISL_858665, EPI_ISL_858668, EPI_ISL_864543, EPI_ISL_892210, EPI_ISL_892211, EPI_ISL_932832, EPI_ISL_933425, EPI_ISL_945144, EPI_ISL_945162, EPI_ISL_986354

| | | | |
|---|---|---|---|
| see above | Lighthouse Lab in Glasgow | Wellcome Sanger Institute for the COVID-19 Genomics UK (COG-UK) Consortium | Anna Dominiczak and Alex Alderton; Carol Clugston; Cordelia Langford; David Gray; David K. Jackson; Dominic Kwiatkowski; Ewan Harrison; Harper VanSteenhouse; Ian Johnston; John Sillitoe on behalf of the Wellcome Sanger Institute COVID-19 Surveillance Team; John Sillitoe on behalf of the Wellcome Sanger Institute COVID-19 Surveillance Team (http://www.sanger.ac.uk/covid-team); Roberto Amato; Sonia Goncalves; Yumi Kasai |
| EPI_ISL_865007, EPI_ISL_865009, EPI_ISL_865010, EPI_ISL_865011, EPI_ISL_865012 | Lighthouse Lab in Glasgow / MRC-University of Glasgow Centre for Virus Research | COVID-19 Genomics UK (COG-UK) Consortium | Alasdair MacLean; Alice Broos; Ana da Silva Filipe; Anna Dominiczak; Antonia Ho; Carol Clugston; Daniel Mair; David Gray; David L Robertson; Emma Thomson; Harper VanSteenhouse; James Shepherd; Jenna Nichols; Joseph Hughes; Kathy Li; Kathy Smollett; Kyriaki Nomikou; Lily Tong; Matthew Holden; Natasha Johnson; Richard Orton; Rory Gunson; Sarah McDonald; Sharif Shaaban; Sreenu Vattipally; Stephen Carmichael; Yumi Kasai |

EPI_ISL_811197, EPI_ISL_811205, EPI_ISL_811212, EPI_ISL_811245, EPI_ISL_811251, EPI_ISL_811255, EPI_ISL_811269, EPI_ISL_811282, EPI_ISL_811285, EPI_ISL_811300, EPI_ISL_811302, EPI_ISL_811304, EPI_ISL_811306, EPI_ISL_811307, EPI_ISL_811311, EPI_ISL_811315, EPI_ISL_811322, EPI_ISL_811329, EPI_ISL_811332, EPI_ISL_811334, EPI_ISL_811336, EPI_ISL_811339, EPI_ISL_811342, EPI_ISL_811358, EPI_ISL_811359, EPI_ISL_811367, EPI_ISL_811368, EPI_ISL_811369, EPI_ISL_811373, EPI_ISL_811383, EPI_ISL_811397, EPI_ISL_811409, EPI_ISL_811429, EPI_ISL_811430, EPI_ISL_811431, EPI_ISL_811444, EPI_ISL_811445, EPI_ISL_811452, EPI_ISL_811455, EPI_ISL_811456, EPI_ISL_811469, EPI_ISL_811484, EPI_ISL_811487, EPI_ISL_811498, EPI_ISL_811511, EPI_ISL_811525, EPI_ISL_811541, EPI_ISL_811545, EPI_ISL_811549, EPI_ISL_811556, EPI_ISL_811559, EPI_ISL_811569, EPI_ISL_811591, EPI_ISL_811603, EPI_ISL_811611, EPI_ISL_811622, EPI_ISL_811625, EPI_ISL_811644, EPI_ISL_811657, EPI_ISL_811673, EPI_ISL_811696, EPI_ISL_811741, EPI_ISL_811754, EPI_ISL_811762, EPI_ISL_811787, EPI_ISL_811807, EPI_ISL_820648, EPI_ISL_820649, EPI_ISL_820650, EPI_ISL_820651, EPI_ISL_820652, EPI_ISL_820653, EPI_ISL_820654, EPI_ISL_820655, EPI_ISL_820656, EPI_ISL_820657, EPI_ISL_820658, EPI_ISL_820659, EPI_ISL_820660, EPI_ISL_820661, EPI_ISL_820662, EPI_ISL_820663, EPI_ISL_820664, EPI_ISL_820665, EPI_ISL_820666, EPI_ISL_820667, EPI_ISL_820668, EPI_ISL_820669, EPI_ISL_820670, EPI_ISL_820671, EPI_ISL_820672, EPI_ISL_820673, EPI_ISL_820674, EPI_ISL_820675, EPI_ISL_820676, EPI_ISL_820677, EPI_ISL_820678, EPI_ISL_820679, EPI_ISL_820680, EPI_ISL_820681, EPI_ISL_820682, EPI_ISL_820683, EPI_ISL_820684, EPI_ISL_820685, EPI_ISL_820686, EPI_ISL_820687, EPI_ISL_820688, EPI_ISL_820689, EPI_ISL_820690, EPI_ISL_820691, EPI_ISL_820692, EPI_ISL_820693, EPI_ISL_820694, EPI_ISL_820695, EPI_ISL_820696, EPI_ISL_820697, EPI_ISL_820698, EPI_ISL_820699, EPI_ISL_820700, EPI_ISL_820701, EPI_ISL_820702, EPI_ISL_820703, EPI_ISL_820704, EPI_ISL_820705, EPI_ISL_820706, EPI_ISL_820707, EPI_ISL_820708, EPI_ISL_820709, EPI_ISL_820710, EPI_ISL_820711, EPI_ISL_820712, EPI_ISL_820713, EPI_ISL_820714, EPI_ISL_820715, EPI_ISL_820716, EPI_ISL_820717, EPI_ISL_820718, EPI_ISL_820719, EPI_ISL_820720, EPI_ISL_820721, EPI_ISL_820722, EPI_ISL_820723, EPI_ISL_820724, EPI_ISL_820725, EPI_ISL_820726, EPI_ISL_820727, EPI_ISL_820728, EPI_ISL_820729, EPI_ISL_820730, EPI_ISL_820731, EPI_ISL_820732, EPI_ISL_820733, EPI_ISL_820734, EPI_ISL_820735, EPI_ISL_820736, EPI_ISL_820737, EPI_ISL_820738, EPI_ISL_820739, EPI_ISL_820740, EPI_ISL_820741, EPI_ISL_820742, EPI_ISL_820743, EPI_ISL_820744, EPI_ISL_820745, EPI_ISL_820746, EPI_ISL_820747, EPI_ISL_820748, EPI_ISL_820749, EPI_ISL_820750, EPI_ISL_820751, EPI_ISL_820752, EPI_ISL_820753, EPI_ISL_820754, EPI_ISL_820755, EPI_ISL_820756, EPI_ISL_820757, EPI_ISL_820758, EPI_ISL_820759, EPI_ISL_820760, EPI_ISL_820761, EPI_ISL_820762, EPI_ISL_820763, EPI_ISL_820764, EPI_ISL_820765, EPI_ISL_820766, EPI_ISL_820767, EPI_ISL_820768, EPI_ISL_820769, EPI_ISL_820770, EPI_ISL_820771, EPI_ISL_820772, EPI_ISL_820773, EPI_ISL_820774, EPI_ISL_820775, EPI_ISL_820776, EPI_ISL_820777, EPI_ISL_820778, EPI_ISL_820779, EPI_ISL_820780, EPI_ISL_820781, EPI_ISL_820782, EPI_ISL_820783, EPI_ISL_820784, EPI_ISL_820785, EPI_ISL_820786, EPI_ISL_820787, EPI_ISL_820788, EPI_ISL_820789, EPI_ISL_820790, EPI_ISL_820791, EPI_ISL_820792, EPI_ISL_820793, EPI_ISL_820794, EPI_ISL_820795, EPI_ISL_820796, EPI_ISL_820797, EPI_ISL_820798, EPI_ISL_820799, EPI_ISL_820800, EPI_ISL_820801, EPI_ISL_820802, EPI_ISL_820803, EPI_ISL_820804, EPI_ISL_820805, EPI_ISL_820806, EPI_ISL_820807, EPI_ISL_820808, EPI_ISL_820809, EPI_ISL_820810, EPI_ISL_820811, EPI_ISL_820812, EPI_ISL_820813, EPI_ISL_820814, EPI_ISL_820815, EPI_ISL_820816, EPI_ISL_820817, EPI_ISL_820818, EPI_ISL_820819, EPI_ISL_820820, EPI_ISL_820821, EPI_ISL_820822, EPI_ISL_820823, EPI_ISL_820824, EPI_ISL_820825, EPI_ISL_820826, EPI_ISL_820827, EPI_ISL_820828, EPI_ISL_820829, EPI_ISL_820830, EPI_ISL_820831, EPI_ISL_820832, EPI_ISL_820833, EPI_ISL_820834, EPI_ISL_820835, EPI_ISL_820836, EPI_ISL_820837, EPI_ISL_820838, EPI_ISL_820839, EPI_ISL_820840, EPI_ISL_820841, EPI_ISL_820842, EPI_ISL_820843, EPI_ISL_820844, EPI_ISL_820845, EPI_ISL_820846, EPI_ISL_820847, EPI_ISL_820848, EPI_ISL_820849, EPI_ISL_820850, EPI_ISL_820851, EPI_ISL_820852, EPI_ISL_820853, EPI_ISL_820854, EPI_ISL_820855, EPI_ISL_820856, EPI_ISL_820857, EPI_ISL_820858, EPI_ISL_820859, EPI_ISL_820860, EPI_ISL_820861, EPI_ISL_820862, EPI_ISL_820863, EPI_ISL_820864, EPI_ISL_820865, EPI_ISL_820866, EPI_ISL_820867, EPI_ISL_820868, EPI_ISL_820869, EPI_ISL_820870, EPI_ISL_820871, EPI_ISL_820872, EPI_ISL_820873, EPI_ISL_820874, EPI_ISL_820875, EPI_ISL_820876, EPI_ISL_820877, EPI_ISL_820878, EPI_ISL_820879, EPI_ISL_820880, EPI_ISL_820881, EPI_ISL_820882, EPI_ISL_820883, EPI_ISL_820884, EPI_ISL_820885, EPI_ISL_820886, EPI_ISL_820887, EPI_ISL_820888, EPI_ISL_820889, EPI_ISL_820890, EPI_ISL_820891, EPI_ISL_820892, EPI_ISL_820893, EPI_ISL_820894, EPI_ISL_820895, EPI_ISL_820896, EPI_ISL_820897, EPI_ISL_820898, EPI_ISL_820899, EPI_ISL_820900, EPI_ISL_820901, EPI_ISL_820902, EPI_ISL_820903, EPI_ISL_820904, EPI_ISL_820905, EPI_ISL_820906, EPI_ISL_820907, EPI_ISL_820908, EPI_ISL_820909, EPI_ISL_820910, EPI_ISL_820911, EPI_ISL_820912, EPI_ISL_820913, EPI_ISL_820914, EPI_ISL_820915, EPI_ISL_820916, EPI_ISL_820917, EPI_ISL_820918, EPI_ISL_820919, EPI_ISL_820920, EPI_ISL_820921, EPI_ISL_820922, EPI_ISL_820923, EPI_ISL_820924, EPI_ISL_820925, EPI_ISL_820926, EPI_ISL_820927, EPI_ISL_820928, EPI_ISL_820929, EPI_ISL_820930, EPI_ISL_820931, EPI_ISL_820932, EPI_ISL_820933, EPI_ISL_820934, EPI_ISL_820935, EPI_ISL_820936, EPI_ISL_820937, EPI_ISL_820938, EPI_ISL_820939, EPI_ISL_820940, EPI_ISL_820941, EPI_ISL_820942, EPI_ISL_820943, EPI_ISL_820944, EPI_ISL_820945, EPI_ISL_820946, EPI_ISL_820947, EPI_ISL_820948, EPI_ISL_820949, EPI_ISL_820950, EPI_ISL_820951, EPI_ISL_820952, EPI_ISL_820953, EPI_ISL_820954, EPI_ISL_820955, EPI_ISL_820956, EPI_ISL_820957, EPI_ISL_820958, EPI_ISL_820959, EPI_ISL_820960, EPI_ISL_820961, EPI_ISL_820962, EPI_ISL_820963, EPI_ISL_820964, EPI_ISL_820965, EPI_ISL_820966, EPI_ISL_820967, EPI_ISL_820968, EPI_ISL_820969, EPI_ISL_820970, EPI_ISL_820971, EPI_ISL_820972, EPI_ISL_820973, EPI_ISL_820974, EPI_ISL_820975, EPI_ISL_820976, EPI_ISL_820977, EPI_ISL_820978, EPI_ISL_820979, EPI_ISL_820980, EPI_ISL_820981, EPI_ISL_820982, EPI_ISL_820983, EPI_ISL_820984, EPI_ISL_820985, EPI_ISL_820986, EPI_ISL_820987, EPI_ISL_820988, EPI_ISL_820989, EPI_ISL_820990, EPI_ISL_820991, EPI_ISL_820992, EPI_ISL_820993, EPI_ISL_820994, EPI_ISL_820995, EPI_ISL_820996, EPI_ISL_820997, EPI_ISL_820998, EPI_ISL_820999, EPI_ISL_821000, EPI_ISL_821001, EPI_ISL_821274, EPI_ISL_821281, EPI_ISL_821282, EPI_ISL_821296, EPI_ISL_821297, EPI_ISL_821301, EPI_ISL_821306, EPI_ISL_821317, EPI_ISL_821319, EPI_ISL_821323, EPI_ISL_821324, EPI_ISL_821328, EPI_ISL_821331, EPI_ISL_821334, EPI_ISL_821336, EPI_ISL_821340, EPI_ISL_821341, EPI_ISL_821344, EPI_ISL_821346, EPI_ISL_821352, EPI_ISL_821353, EPI_ISL_821358, EPI_ISL_821359, EPI_ISL_821362, EPI_ISL_821373, EPI_ISL_821377, EPI_ISL_821378, EPI_ISL_821379, EPI_ISL_821380, EPI_ISL_821381, EPI_ISL_821383, EPI_ISL_821387, EPI_ISL_821388, EPI_ISL_821391, EPI_ISL_821395, EPI_ISL_821397, EPI_ISL_821400, EPI_ISL_821402, EPI_ISL_821405, EPI_ISL_821409, EPI_ISL_821410, EPI_ISL_821431, EPI_ISL_821432, EPI_ISL_821434, EPI_ISL_821443, EPI_ISL_821445, EPI_ISL_821448, EPI_ISL_821449, EPI_ISL_821460, EPI_ISL_821461, EPI_ISL_821462, EPI_ISL_821468, EPI_ISL_821472, EPI_ISL_821474, EPI_ISL_821475, EPI_ISL_821477, EPI_ISL_821478, EPI_ISL_821480, EPI_ISL_821486, EPI_ISL_821498, EPI_ISL_821501, EPI_ISL_821504, EPI_ISL_821507, EPI_ISL_821509, EPI_ISL_821514, EPI_ISL_821515, EPI_ISL_821517, EPI_ISL_821520, EPI_ISL_821521, EPI_ISL_821522, EPI_ISL_821524, EPI_ISL_821527, EPI_ISL_821536, EPI_ISL_821537, EPI_ISL_821539, EPI_ISL_821540, EPI_ISL_821541, EPI_ISL_821542, EPI_ISL_821543, EPI_ISL_821545, EPI_ISL_821559, EPI_ISL_821560, EPI_ISL_821561, EPI_ISL_821562, EPI_ISL_821569, EPI_ISL_821571, EPI_ISL_821572, EPI_ISL_821573, EPI_ISL_821574, EPI_ISL_821575, EPI_ISL_821580, EPI_ISL_821582, EPI_ISL_821595, EPI_ISL_821597, EPI_ISL_821601, EPI_ISL_821603, EPI_ISL_821609, EPI_ISL_821612, EPI_ISL_821625, EPI_ISL_826466, EPI_ISL_833586, EPI_ISL_833590, EPI_ISL_833593, EPI_ISL_833596, EPI_ISL_833597, EPI_ISL_833599, EPI_ISL_833602, EPI_ISL_833604, EPI_ISL_833606, EPI_ISL_833607, EPI_ISL_833611, EPI_ISL_833612, EPI_ISL_833614, EPI_ISL_833615, EPI_ISL_833617, EPI_ISL_833618, EPI_ISL_833619, EPI_ISL_833620, EPI_ISL_833621, EPI_ISL_833629, EPI_ISL_833632, EPI_ISL_833634, EPI_ISL_833643, EPI_ISL_833647, EPI_ISL_833648, EPI_ISL_833652, EPI_ISL_833656, EPI_ISL_833658, EPI_ISL_833667, EPI_ISL_833676, EPI_ISL_833678, EPI_ISL_833685, EPI_ISL_833693, EPI_ISL_833700, EPI_ISL_833712, EPI_ISL_833716, EPI_ISL_833719, EPI_ISL_833721, EPI_ISL_833725, EPI_ISL_833726, EPI_ISL_833734, EPI_ISL_833735, EPI_ISL_833736, EPI_ISL_833738, EPI_ISL_833743, EPI_ISL_833755, EPI_ISL_833756, EPI_ISL_833759, EPI_ISL_833766, EPI_ISL_833767, EPI_ISL_833768, EPI_ISL_833779, EPI_ISL_833780, EPI_ISL_833789, EPI_ISL_833791, EPI_ISL_833795, EPI_ISL_833796, EPI_ISL_833797, EPI_ISL_833801, EPI_ISL_833803, EPI_ISL_833804, EPI_ISL_833809, EPI_ISL_833814, EPI_ISL_833817, EPI_ISL_833818, EPI_ISL_833821, EPI_ISL_833823, EPI_ISL_833830, EPI_ISL_833834, EPI_ISL_833835, EPI_ISL_833841, EPI_ISL_833845, EPI_ISL_833846, EPI_ISL_833848, EPI_ISL_833856, EPI_ISL_833861, EPI_ISL_833865, EPI_ISL_833868, EPI_ISL_833872, EPI_ISL_833879, EPI_ISL_833881, EPI_ISL_833884, EPI_ISL_833889, EPI_ISL_833890, EPI_ISL_833894, EPI_ISL_833895, EPI_ISL_833901, EPI_ISL_833908, EPI_ISL_833916, EPI_ISL_833922, EPI_ISL_833924, EPI_ISL_833930, EPI_ISL_833931, EPI_ISL_835710, EPI_ISL_835711, EPI_ISL_835712, EPI_ISL_835713, EPI_ISL_835714, EPI_ISL_835715, EPI_ISL_835716, EPI_ISL_835717, EPI_ISL_835718, EPI_ISL_835719, EPI_ISL_835720, EPI_ISL_835721, EPI_ISL_835722, EPI_ISL_835723, EPI_ISL_835724, EPI_ISL_835725, EPI_ISL_835726, EPI_ISL_835727, EPI_ISL_835728, EPI_ISL_835729, EPI_ISL_835730, EPI_ISL_835731, EPI_ISL_835732, EPI_ISL_835733, EPI_ISL_835734, EPI_ISL_835735, EPI_ISL_835736, EPI_ISL_835737, EPI_ISL_835738, EPI_ISL_835739, EPI_ISL_835740, EPI_ISL_835741, EPI_ISL_835742, EPI_ISL_835743, EPI_ISL_835744, EPI_ISL_835745, EPI_ISL_835746, EPI_ISL_835747, EPI_ISL_835748, EPI_ISL_835749, EPI_ISL_835750, EPI_ISL_835751, EPI_ISL_835752, EPI_ISL_835753, EPI_ISL_835754, EPI_ISL_835755, EPI_ISL_835756, EPI_ISL_835757, EPI_ISL_835758, EPI_ISL_835759, EPI_ISL_835760, EPI_ISL_835761, EPI_ISL_835762, EPI_ISL_835763, EPI_ISL_835764, EPI_ISL_835765, EPI_ISL_835766, EPI_ISL_835767, EPI_ISL_835768, EPI_ISL_835769, EPI_ISL_835770, EPI_ISL_835771, EPI_ISL_835772, EPI_ISL_835773, EPI_ISL_835774, EPI_ISL_835775, EPI_ISL_835776, EPI_ISL_835777, EPI_ISL_835778, EPI_ISL_835779, EPI_ISL_835780, EPI_ISL_835781, EPI_ISL_835782, EPI_ISL_835783, EPI_ISL_835784, EPI_ISL_835785, EPI_ISL_835786, EPI_ISL_835787, EPI_ISL_835788, EPI_ISL_835789, EPI_ISL_835790, EPI_ISL_835791, EPI_ISL_835792, EPI_ISL_835793, EPI_ISL_835794, EPI_ISL_835795, EPI_ISL_835796, EPI_ISL_835797, EPI_ISL_835798, EPI_ISL_835799, EPI_ISL_835800, EPI_ISL_835801, EPI_ISL_835802, EPI_ISL_835803, EPI_ISL_835804, EPI_ISL_835805, EPI_ISL_835806, EPI_ISL_835807, EPI_ISL_835808, EPI_ISL_835809, EPI_ISL_835810, EPI_ISL_835811, EPI_ISL_835812, EPI_ISL_835813, EPI_ISL_835814, EPI_ISL_835815, EPI_ISL_835816, EPI_ISL_835817, EPI_ISL_835818, EPI_ISL_835819, EPI_ISL_835820, EPI_ISL_835821, EPI_ISL_835822, EPI_ISL_835823, EPI_ISL_835824, EPI_ISL_835825, EPI_ISL_835826, EPI_ISL_835827, EPI_ISL_835828, EPI_ISL_835829, EPI_ISL_835830, EPI_ISL_835831, EPI_ISL_835832, EPI_ISL_835833, EPI_ISL_835834, EPI_ISL_835835, EPI_ISL_835836, EPI_ISL_835837, EPI_ISL_835838, EPI_ISL_835839, EPI_ISL_835840, EPI_ISL_835841, EPI_ISL_835842, EPI_ISL_835843, EPI_ISL_835844, EPI_ISL_835845, EPI_ISL_835846, EPI_ISL_835847, EPI_ISL_835848, EPI_ISL_835849, EPI_ISL_835850, EPI_ISL_835851, EPI_ISL_835852, EPI_ISL_835853, EPI_ISL_835854, EPI_ISL_835855, EPI_ISL_835856, EPI_ISL_835857, EPI_ISL_835858, EPI_ISL_835859, EPI_ISL_835860, EPI_ISL_835861, EPI_ISL_835862, EPI_ISL_835863, EPI_ISL_835864, EPI_ISL_835865, EPI_ISL_835866, EPI_ISL_835867, EPI_ISL_835868, EPI_ISL_835869, EPI_ISL_835870, EPI_ISL_835871, EPI_ISL_835872, EPI_ISL_835873, EPI_ISL_835874, EPI_ISL_835875, EPI_ISL_835876, EPI_ISL_835877, EPI_ISL_835878, EPI_ISL_835879, EPI_ISL_835880, EPI_ISL_835881, EPI_ISL_835882, EPI_ISL_835883, EPI_ISL_835884, EPI_ISL_835885, EPI_ISL_835886, EPI_ISL_835887, EPI_ISL_835888, EPI_ISL_835889, EPI_ISL_835890, EPI_ISL_835891, EPI_ISL_835892, EPI_ISL_835893, EPI_ISL_835894, EPI_ISL_835895, EPI_ISL_835896, EPI_ISL_835897, EPI_ISL_835898, EPI_ISL_835899, EPI_ISL_835900, EPI_ISL_835901, EPI_ISL_835902, EPI_ISL_835903, EPI_ISL_835904, EPI_ISL_835905, EPI_ISL_835906, EPI_ISL_835907, EPI_ISL_835908, EPI_ISL_835909, EPI_ISL_835910, EPI_ISL_835911, EPI_ISL_835912, EPI_ISL_835913, EPI_ISL_835914, EPI_ISL_835915, EPI_ISL_835916, EPI_ISL_835917, EPI_ISL_835918, EPI_ISL_835919, EPI_ISL_835920, EPI_ISL_835921, EPI_ISL_835922, EPI_ISL_835923, EPI_ISL_835924, EPI_ISL_835925, EPI_ISL_835926, EPI_ISL_835927, EPI_ISL_835928, EPI_ISL_835929, EPI_ISL_835930, EPI_ISL_835931, EPI_ISL_835932, EPI_ISL_835933, EPI_ISL_835934, EPI_ISL_835935, EPI_ISL_835936, EPI_ISL_835937, EPI_ISL_835938, EPI_ISL_835939, EPI_ISL_835940, EPI_ISL_835941, EPI_ISL_835942, EPI_ISL_835943, EPI_ISL_835944, EPI_ISL_835945, EPI_ISL_835946, EPI_ISL_835947, EPI_ISL_835948, EPI_ISL_835949, EPI_ISL_835950, EPI_ISL_835951, EPI_ISL_835952, EPI_ISL_835953, EPI_ISL_835954, EPI_ISL_835955, EPI_ISL_835956, EPI_ISL_835957, EPI_ISL_835958, EPI_ISL_835959, EPI_ISL_835960, EPI_ISL_835961, EPI_ISL_835962, EPI_ISL_835963, EPI_ISL_835964, EPI_ISL_835965, EPI_ISL_835966, EPI_ISL_835967, EPI_ISL_835968, EPI_ISL_835969, EPI_ISL_835970, EPI_ISL_835971, EPI_ISL_835972, EPI_ISL_835973, EPI_ISL_835974, EPI_ISL_835975, EPI_ISL_835976, EPI_ISL_835977, EPI_ISL_835978, EPI_ISL_835979, EPI_ISL_835980, EPI_ISL_835981, EPI_ISL_835982, EPI_ISL_835983, EPI_ISL_835984, EPI_ISL_835985, EPI_ISL_835986, EPI_ISL_835987, EPI_ISL_835988, EPI_ISL_835989, EPI_ISL_835990, EPI_ISL_835991, EPI_ISL_835992, EPI_ISL_835993, EPI_ISL_835994, EPI_ISL_835995, EPI_ISL_835996, EPI_ISL_835997, EPI_ISL_835998, EPI_ISL_835999, EPI_ISL_836000, EPI_ISL_836001, EPI_ISL_836002, EPI_ISL_836003, EPI_ISL_836004, EPI_ISL_836005, EPI_ISL_836006, EPI_ISL_836007, EPI_ISL_836008, EPI_ISL_836009, EPI_ISL_836010, EPI_ISL_836011, EPI_ISL_836012, EPI_ISL_836013, EPI_ISL_836014, EPI_ISL_836015, EPI_ISL_836016, EPI_ISL_836017, EPI_ISL_836018, EPI_ISL_836019, EPI_ISL_836020, EPI_ISL_836021, EPI_ISL_836022, EPI_ISL_836023, EPI_ISL_836024, EPI_ISL_836025, EPI_ISL_836026, EPI_ISL_836027, EPI_ISL_836028, EPI_ISL_836029, EPI_ISL_836030, EPI_ISL_836031, EPI_ISL_836032, EPI_ISL_836033, EPI_ISL_836034, EPI_ISL_836035, EPI_ISL_836036, EPI_ISL_836037, EPI_ISL_836038, EPI_ISL_836039, EPI_ISL_836040, EPI_ISL_836041, EPI_ISL_836042, EPI_ISL_836043, EPI_ISL_836044, EPI_ISL_836045, EPI_ISL_836046, EPI_ISL_836047, EPI_ISL_836048, EPI_ISL_836049, EPI_ISL_836050, EPI_ISL_836051, EPI_ISL_836052, EPI_ISL_836053, EPI_ISL_836054, EPI_ISL_836055, EPI_ISL_836056, EPI_ISL_836057, EPI_ISL_836058, EPI_ISL_836059, EPI_ISL_836060, EPI_ISL_836061, EPI_ISL_836062, EPI_ISL_836063, EPI_ISL_836064, EPI_ISL_836065, EPI_ISL_836066, EPI_ISL_836067, EPI_ISL_836068, EPI_ISL_836069, EPI_ISL_836070, EPI_ISL_836071, EPI_ISL_836072, EPI_ISL_836073, EPI_ISL_836074, EPI_ISL_836075, EPI_ISL_836076, EPI_ISL_836077, EPI_ISL_836078, EPI_ISL_836079, EPI_ISL_836080, EPI_ISL_836081, EPI_ISL_836082, EPI_ISL_836083, EPI_ISL_836084, EPI_ISL_836085, EPI_ISL_836086, EPI_ISL_836087, EPI_ISL_836088, EPI_ISL_836089, EPI_ISL_836090, EPI_ISL_836091, EPI_ISL_836092, EPI_ISL_836093, EPI_ISL_836094, EPI_ISL_836095, EPI_ISL_836096, EPI_ISL_836097, EPI_ISL_836098, EPI_ISL_836099, EPI_ISL_836100, EPI_ISL_836101, EPI_ISL_836102, EPI_ISL_836103, EPI_ISL_836104, EPI_ISL_836105, EPI_ISL_836106, EPI_ISL_836107, EPI_ISL_836108, EPI_ISL_836109, EPI_ISL_836110, EPI_ISL_836111, EPI_ISL_836112, EPI_ISL_836113, EPI_ISL_836114, EPI_ISL_836115, EPI_ISL_836116, EPI_ISL_836117, EPI_ISL_836118, EPI_ISL_836119, EPI_ISL_836120, EPI_ISL_836121, EPI_ISL_836122, EPI_ISL_836123, EPI_ISL_836124, EPI_ISL_836125, EPI_ISL_836126, EPI_ISL_836127, EPI_ISL_836128, EPI_ISL_836129, EPI_ISL_836130, EPI_ISL_836131, EPI_ISL_836132, EPI_ISL_836133, EPI_ISL_836134, EPI_ISL_836135, EPI_ISL_836136, EPI_ISL_836137, EPI_ISL_836138, EPI_ISL_836139, EPI_ISL_836140, EPI_ISL_836141, EPI_ISL_836142, EPI_ISL_836144, EPI_ISL_836145, EPI_ISL_836146, EPI_ISL_836147, EPI_ISL_836148, EPI_ISL_836149, EPI_ISL_836150, EPI_ISL_836151, EPI_ISL_836152, EPI_ISL_836153, EPI_ISL_836154, EPI_ISL_836155, EPI_ISL_836156, EPI_ISL_836157, EPI_ISL_836158, EPI_ISL_836159, EPI_ISL_836160, EPI_ISL_836161, EPI_ISL_836162, EPI_ISL_836163, EPI_ISL_836164, EPI_ISL_836165, EPI_ISL_836166, EPI_ISL_836167, EPI_ISL_836168, EPI_ISL_836169, EPI_ISL_836170, EPI_ISL_836171, EPI_ISL_836172, EPI_ISL_836173, EPI_ISL_836174, EPI_ISL_836175, EPI_ISL_836176, EPI_ISL_836177, EPI_ISL_836178, EPI_ISL_836179, EPI_ISL_836180, EPI_ISL_836181, EPI_ISL_836182, EPI_ISL_836183, EPI_ISL_836184, EPI_ISL_836185, EPI_ISL_836186, EPI_ISL_836187, EPI_ISL_836188, EPI_ISL_836189, EPI_ISL_836190, EPI_ISL_836191, EPI_ISL_836192, EPI_ISL_836193, EPI_ISL_836194, EPI_ISL_836195, EPI_ISL_836196, EPI_ISL_836197, EPI_ISL_836198, EPI_ISL_836199, EPI_ISL_836200, EPI_ISL_836201, EPI_ISL_836202, EPI_ISL_836203, EPI_ISL_836204, EPI_ISL_836205, EPI_ISL_836206, EPI_ISL_836207, EPI_ISL_836208, EPI_ISL_836209, EPI_ISL_836210, EPI_ISL_836211, EPI_ISL_836212, EPI_ISL_836213, EPI_ISL_836214, EPI_ISL_836215, EPI_ISL_836216, EPI_ISL_836217, EPI_ISL_836218, EPI_ISL_836219, EPI_ISL_836220, EPI_ISL_836221, EPI_ISL_836222, EPI_ISL_836223, EPI_ISL_836224, EPI_ISL_836225, EPI_ISL_836226, EPI_ISL_836227, EPI_ISL_836228, EPI_ISL_836229, EPI_ISL_836230, EPI_ISL_836231, EPI_ISL_836232, EPI_ISL_836233, EPI_ISL_836234, EPI_ISL_836235, EPI_ISL_836236, EPI_ISL_836237, EPI_ISL_836238, EPI_ISL_836239, EPI_ISL_836240, EPI_ISL_836241, EPI_ISL_836242, EPI_ISL_836243, EPI_ISL_836244, EPI_ISL_836245, EPI_ISL_836246, EPI_ISL_836247, EPI_ISL_836248, EPI_ISL_836249, EPI_ISL_836250, EPI_ISL_836251, EPI_ISL_836252, EPI_ISL_836253, EPI_ISL_836254, EPI_ISL_836255, EPI_ISL_836256, EPI_ISL_836257, EPI_ISL_836258, EPI_ISL_836259, EPI_ISL_836260, EPI_ISL_836261, EPI_ISL_836262, EPI_ISL_836263, EPI_ISL_836264, EPI_ISL_836265, EPI_ISL_836266, EPI_ISL_836267, EPI_ISL_836268, EPI_ISL_836269, EPI_ISL_836270, EPI_ISL_836271, EPI_ISL_836272, EPI_ISL_836273, EPI_ISL_836274, EPI_ISL_836275, EPI_ISL_836276, EPI_ISL_836277, EPI_ISL_836278, EPI_ISL_836279, EPI_ISL_836280, EPI_ISL_836281, EPI_ISL_836282, EPI_ISL_836283, EPI_ISL_836284, EPI_ISL_836285, EPI_ISL_836286, EPI_ISL_836287, EPI_ISL_836288, EPI_ISL_836289, EPI_ISL_836290, EPI_ISL_836291, EPI_ISL_836292, EPI_ISL_836293, EPI_ISL_836294, EPI_ISL_836295, EPI_ISL_836296, EPI_ISL_836297, EPI_ISL_836298, EPI_ISL_836299, EPI_ISL_836300, EPI_ISL_836301, EPI_ISL_836302, EPI_ISL_836303, EPI_ISL_836304, EPI_ISL_836305, EPI_ISL_836306, EPI_ISL_836307, EPI_ISL_836308, EPI_ISL_836309, EPI_ISL_836310, EPI_ISL_836311, EPI_ISL_836312, EPI_ISL_836313, EPI_ISL_836314, EPI_ISL_836315, EPI_ISL_836316, EPI_ISL_836317, EPI_ISL_836318, EPI_ISL_836319, EPI_ISL_836320, EPI_ISL_836321, EPI_ISL_836322, EPI_ISL_836323, EPI_ISL_836324, EPI_ISL_836325, EPI_ISL_836326, EPI_ISL_836327, EPI_ISL_836328, EPI_ISL_836329, EPI_ISL_836330, EPI_ISL_836331, EPI_ISL_836332, EPI_ISL_836333, EPI_ISL_836334, EPI_ISL_836335, EPI_ISL_836336, EPI_ISL_836337, EPI_ISL_836338, EPI_ISL_836339, EPI_ISL_836340, EPI_ISL_836341, EPI_ISL_836342, EPI_ISL_836343, EPI_ISL_836344, EPI_ISL_836345, EPI_ISL_836346, EPI_ISL_836347, EPI_ISL_836348, EPI_ISL_836349, EPI_ISL_836350, EPI_ISL_836351, EPI_ISL_836352, EPI_ISL_836353, EPI_ISL_836354, EPI_ISL_836355, EPI_ISL_836356, EPI_ISL_836357, EPI_ISL_836358, EPI_ISL_836359, EPI_ISL_836360, EPI_ISL_836361, EPI_ISL_836362, EPI_ISL_836363, EPI_ISL_836364, EPI_ISL_836365, EPI_ISL_836366, EPI_ISL_836367, EPI_ISL_836368, EPI_ISL_836369, EPI_ISL_836370, EPI_ISL_836371, EPI_ISL_836372, EPI_ISL_836373, EPI_ISL_836374, EPI_ISL_836375, EPI_ISL_836376, EPI_ISL_836377, EPI_ISL_836378, EPI_ISL_836379, EPI_ISL_836380, EPI_ISL_836381, EPI_ISL_836382, EPI_ISL_836383, EPI_ISL_836384, EPI_ISL_836385, EPI_ISL_836386, EPI_ISL_836387, EPI_ISL_836388, EPI_ISL_836389, EPI_ISL_836390, EPI_ISL_836391, EPI_ISL_836392, EPI_ISL_836393, EPI_ISL_836394, EPI_ISL_836395, EPI_ISL_836396, EPI_ISL_836397, EPI_ISL_836398, EPI_ISL_836399, EPI_ISL_836400, EPI_ISL_836401, EPI_ISL_836402, EPI_ISL_836403, EPI_ISL_836404, EPI_ISL_836405, EPI_ISL_836406, EPI_ISL_836407, EPI_ISL_836408, EPI_ISL_836409, EPI_ISL_836410, EPI_ISL_836411, EPI_ISL_836412, EPI_ISL_836413, EPI_ISL_836414, EPI_ISL_836415, EPI_ISL_836416, EPI_ISL_836417, EPI_ISL_836418, EPI_ISL_836419, EPI_ISL_836420, EPI_ISL_852002, EPI_ISL_852190, EPI_ISL_852206, EPI_ISL_858268, EPI_ISL_863534, EPI_ISL_863535, EPI_ISL_863536, EPI_ISL_863541, EPI_ISL_863545, EPI_ISL_863548, EPI_ISL_863550, EPI_ISL_863554, EPI_ISL_863559, EPI_ISL_863563, EPI_ISL_863564, EPI_ISL_863568, EPI_ISL_863569, EPI_ISL_863571, EPI_ISL_863573, EPI_ISL_863575, EPI_ISL_863585, EPI_ISL_863596, EPI_ISL_863597, EPI_ISL_863598, EPI_ISL_863602, EPI_ISL_863607, EPI_ISL_863611, EPI_ISL_863613, EPI_ISL_863615, EPI_ISL_863617, EPI_ISL_863620, EPI_ISL_863624, EPI_ISL_863632, EPI_ISL_863634, EPI_ISL_863637, EPI_ISL_863638, EPI_ISL_863642, EPI_ISL_863646, EPI_ISL_863653, EPI_ISL_863657, EPI_ISL_863660, EPI_ISL_863669, EPI_ISL_863677, EPI_ISL_863678, EPI_ISL_863679, EPI_ISL_863681, EPI_ISL_863696, EPI_ISL_863699, EPI_ISL_863700, EPI_ISL_863705, EPI_ISL_863709, EPI_ISL_863711, EPI_ISL_863723, EPI_ISL_863725, EPI_ISL_863726, EPI_ISL_863733, EPI_ISL_863734, EPI_ISL_863741, EPI_ISL_863742, EPI_ISL_863747, EPI_ISL_863759, EPI_ISL_863761, EPI_ISL_863766, EPI_ISL_863773, EPI_ISL_863776, EPI_ISL_863781, EPI_ISL_863784, EPI_ISL_863785, EPI_ISL_863788, EPI_ISL_863789, EPI_ISL_863792, EPI_ISL_863793, EPI_ISL_863795, EPI_ISL_863796, EPI_ISL_863797, EPI_ISL_863802, EPI_ISL_863804, EPI_ISL_863806, EPI_ISL_863809, EPI_ISL_863810, EPI_ISL_863812, EPI_ISL_863838, EPI_ISL_863842, EPI_ISL_863845, EPI_ISL_863847, EPI_ISL_863848, EPI_ISL_863850, EPI_ISL_863851, EPI_ISL_863856, EPI_ISL_863858, EPI_ISL_863860, EPI_ISL_863863, EPI_ISL_863865, EPI_ISL_864204, EPI_ISL_864213, EPI_ISL_864216, EPI_ISL_864217, EPI_ISL_864239, EPI_ISL_864244, EPI_ISL_864255, EPI_ISL_864262, EPI_ISL_864266, EPI_ISL_864274, EPI_ISL_864281, EPI_ISL_864291, EPI_ISL_864297, EPI_ISL_864305, EPI_ISL_864309, EPI_ISL_864311, EPI_ISL_864317, EPI_ISL_864321, EPI_ISL_864324, EPI_ISL_864336, EPI_ISL_864356, EPI_ISL_864361, EPI_ISL_864365, EPI_ISL_864368, EPI_ISL_864383, EPI_ISL_864384, EPI_ISL_864396, EPI_ISL_864399, EPI_ISL_864400, EPI_ISL_864402, EPI_ISL_864403, EPI_ISL_864409, EPI_ISL_864411, EPI_ISL_864414, EPI_ISL_864422, EPI_ISL_864439, EPI_ISL_864440, EPI_ISL_864447, EPI_ISL_864452, EPI_ISL_864461, EPI_ISL_864469, EPI_ISL_864472, EPI_ISL_864475, EPI_ISL_864484, EPI_ISL_864493, EPI_ISL_864515, EPI_ISL_864516, EPI_ISL_864520, EPI_ISL_864527, EPI_ISL_864528, EPI_ISL_864529, EPI_ISL_864534, EPI_ISL_864539, EPI_ISL_864540, EPI_ISL_864542, EPI_ISL_945050, EPI_ISL_945053, EPI_ISL_945055, EPI_ISL_945057, EPI_ISL_945058, EPI_ISL_945059, EPI_ISL_945060, EPI_ISL_945062, EPI_ISL_945064, EPI_ISL_945066, EPI_ISL_945069, EPI_ISL_945071, EPI_ISL_945074, EPI_ISL_945075, EPI_ISL_945077, EPI_ISL_945078, EPI_ISL_945079, EPI_ISL_945080, EPI_ISL_945081, EPI_ISL_945082, EPI_ISL_945086, EPI_ISL_945087, EPI_ISL_945090, EPI_ISL_945091, EPI_ISL_945092, EPI_ISL_945093, EPI_ISL_945094, EPI_ISL_945096, EPI_ISL_945097, EPI_ISL_945101, EPI_ISL_945104, EPI_ISL_945105, EPI_ISL_945108, EPI_ISL_945110, EPI_ISL_945112, EPI_ISL_945118, EPI_ISL_945120, EPI_ISL_945121, EPI_ISL_945124, EPI_ISL_945127, EPI_ISL_945128, EPI_ISL_945129, EPI_ISL_945132, EPI_ISL_945133, EPI_ISL_945134, EPI_ISL_945135, EPI_ISL_945137, EPI_ISL_945141, EPI_ISL_945145, EPI_ISL_945149, EPI_ISL_945150, EPI_ISL_945151, EPI_ISL_945153, EPI_ISL_945154, EPI_ISL_945156, EPI_ISL_945158, EPI_ISL_945159, EPI_ISL_945161, EPI_ISL_945164, EPI_ISL_945165, EPI_ISL_945167, EPI_ISL_945168, EPI_ISL_945172, EPI_ISL_945173, EPI_ISL_945174, EPI_ISL_945175, EPI_ISL_945177, EPI_ISL_945178, EPI_ISL_945179, EPI_ISL_945182, EPI_ISL_945183, EPI_ISL_945185, EPI_ISL_945187, EPI_ISL_945189, EPI_ISL_945193, EPI_ISL_945194, EPI_ISL_945196, EPI_ISL_945198, EPI_ISL_945200, EPI_ISL_945205, EPI_ISL_945206, EPI_ISL_945210, EPI_ISL_945211, EPI_ISL_945213, EPI_ISL_945214, EPI_ISL_945215, EPI_ISL_945216, EPI_ISL_945218, EPI_ISL_945219, EPI_ISL_945220, EPI_ISL_945225, EPI_ISL_945227, EPI_ISL_945228, EPI_ISL_945229, EPI_ISL_945230, EPI_ISL_945233, EPI_ISL_945239, EPI_ISL_945242, EPI_ISL_945245, EPI_ISL_945246, EPI_ISL_945251, EPI_ISL_945253, EPI_ISL_945256, EPI_ISL_945258, EPI_ISL_945259, EPI_ISL_945260, EPI_ISL_945262, EPI_ISL_945263, EPI_ISL_945264, EPI_ISL_945265, EPI_ISL_945266, EPI_ISL_945267, EPI_ISL_945272, EPI_ISL_945274, EPI_ISL_945276, EPI_ISL_945277, EPI_ISL_945278, EPI_ISL_945280, EPI_ISL_945281, EPI_ISL_945283, EPI_ISL_945284, EPI_ISL_945286, EPI_ISL_945287, EPI_ISL_945290, EPI_ISL_945291, EPI_ISL_945292, EPI_ISL_945294, EPI_ISL_945295, EPI_ISL_945296, EPI_ISL_945298, EPI_ISL_945300, EPI_ISL_945302, EPI_ISL_945303, EPI_ISL_945308, EPI_ISL_945312, EPI_ISL_945313, EPI_ISL_945314, EPI_ISL_945315, EPI_ISL_945316, EPI_ISL_945317, EPI_ISL_945318, EPI_ISL_945322, EPI_ISL_945325, EPI_ISL_945326, EPI_ISL_945327, EPI_ISL_945328, EPI_ISL_945329, EPI_ISL_945330, EPI_ISL_945331, EPI_ISL_945333, EPI_ISL_945336, EPI_ISL_945344, EPI_ISL_945348, EPI_ISL_945349, EPI_ISL_945352, EPI_ISL_945353, EPI_ISL_945354, EPI_ISL_945355, EPI_ISL_945359, EPI_ISL_945362, EPI_ISL_945368, EPI_ISL_945369, EPI_ISL_945370, EPI_ISL_945371, EPI_ISL_945372, EPI_ISL_945373, EPI_ISL_958564, EPI_ISL_958565, EPI_ISL_993805, EPI_ISL_993914, EPI_ISL_993916, EPI_ISL_993955, EPI_ISL_993962, EPI_ISL_993991, EPI_ISL_994034, EPI_ISL_994056, EPI_ISL_994652, EPI_ISL_1985381

| | | | |
|---|---|---|---|
| see above | Lighthouse Lab in Milton Keynes | Wellcome Sanger Institute for the COVID-19 Genomics UK (COG-UK) Consortium | Cordelia Langford; David K. Jackson; Dominic Kwiatkowski; Ewan Harrison; Ian Johnston; Jeffrey Barrett; John Sillitoe on behalf of the Wellcome Sanger Institute COVID-19 Surveillance Team; John Sillitoe on behalf of the Wellcome Sanger Institute COVID-19 Surveillance Team (http://www.sanger.ac.uk/covid-team); Roberto Amato; Sonia Goncalves; The Lighthouse Lab in Milton Keynes and Alex Alderton |

EPI_ISL_840125, EPI_ISL_840127, EPI_ISL_840128, EPI_ISL_840129, EPI_ISL_840130, EPI_ISL_840131, EPI_ISL_840132, EPI_ISL_840133, EPI_ISL_840134, EPI_ISL_840135, EPI_ISL_840136, EPI_ISL_840137, EPI_ISL_840138, EPI_ISL_840139, EPI_ISL_840140, EPI_ISL_840141, EPI_ISL_840142, EPI_ISL_840143, EPI_ISL_840144, EPI_ISL_840145, EPI_ISL_867018, EPI_ISL_867019, EPI_ISL_867020, EPI_ISL_867021, EPI_ISL_867022, EPI_ISL_867023, EPI_ISL_867024, EPI_ISL_867025, EPI_ISL_867026, EPI_ISL_867027, EPI_ISL_867028, EPI_ISL_867029, EPI_ISL_867030, EPI_ISL_867031, EPI_ISL_867032, EPI_ISL_867033, EPI_ISL_867034, EPI_ISL_922041, EPI_ISL_922042, EPI_ISL_922043, EPI_ISL_922044, EPI_ISL_922045, EPI_ISL_922046, EPI_ISL_922047, EPI_ISL_922048, EPI_ISL_922049, EPI_ISL_922050, EPI_ISL_922051, EPI_ISL_922052, EPI_ISL_922053, EPI_ISL_922054, EPI_ISL_922055, EPI_ISL_922056, EPI_ISL_922057, EPI_ISL_922058, EPI_ISL_922059, EPI_ISL_922061, EPI_ISL_922062, EPI_ISL_950712, EPI_ISL_998935, EPI_ISL_998936

| | | | |
|---|---|---|---|
| see above | Lincolnshire Hospitals and DeepSeq Nottingham | COVID-19 Genomics UK (COG-UK) Consortium | Christopher Moore; Fei Sang; Joeseph Chappell; Johnny Debebe; Jonathan Ball; Matthew Carlisle; Matthew Loose; Nadine Holmes; Nichola Duckworth; Patrick McClure; Sarah Walsh; Tim Sloan; Victoria Wright |

EPI_ISL_813816, EPI_ISL_813817, EPI_ISL_813818, EPI_ISL_813819, EPI_ISL_813820, EPI_ISL_813821, EPI_ISL_813822, EPI_ISL_813823, EPI_ISL_813824, EPI_ISL_813825, EPI_ISL_813826, EPI_ISL_813827, EPI_ISL_813828, EPI_ISL_813829, EPI_ISL_813830, EPI_ISL_813831, EPI_ISL_838586, EPI_ISL_838587, EPI_ISL_838592, EPI_ISL_838593, EPI_ISL_838597, EPI_ISL_838600, EPI_ISL_838602, EPI_ISL_838603, EPI_ISL_838605, EPI_ISL_838613, EPI_ISL_838620, EPI_ISL_838628, EPI_ISL_838655, EPI_ISL_838669, EPI_ISL_838670, EPI_ISL_838671, EPI_ISL_865178, EPI_ISL_865179, EPI_ISL_865181, EPI_ISL_865182, EPI_ISL_865183, EPI_ISL_865184, EPI_ISL_865185, EPI_ISL_865186, EPI_ISL_865187, EPI_ISL_865188, EPI_ISL_865189, EPI_ISL_865190, EPI_ISL_865191, EPI_ISL_865192, EPI_ISL_865193, EPI_ISL_865194, EPI_ISL_865196, EPI_ISL_865197, EPI_ISL_865198, EPI_ISL_865199, EPI_ISL_865200, EPI_ISL_865201, EPI_ISL_865202, EPI_ISL_865203, EPI_ISL_865204, EPI_ISL_865205, EPI_ISL_865206, EPI_ISL_865331, EPI_ISL_865332, EPI_ISL_865333, EPI_ISL_865334, EPI_ISL_865335, EPI_ISL_865336, EPI_ISL_865337, EPI_ISL_865338, EPI_ISL_865339, EPI_ISL_865340, EPI_ISL_865365, EPI_ISL_865366, EPI_ISL_865369, EPI_ISL_865370, EPI_ISL_865383, EPI_ISL_865384, EPI_ISL_865385, EPI_ISL_865387, EPI_ISL_865388, EPI_ISL_865390, EPI_ISL_865450, EPI_ISL_865478, EPI_ISL_865479, EPI_ISL_865480, EPI_ISL_865481, EPI_ISL_865482, EPI_ISL_865483, EPI_ISL_865484, EPI_ISL_919449, EPI_ISL_919450, EPI_ISL_919451, EPI_ISL_919454, EPI_ISL_919455, EPI_ISL_919459, EPI_ISL_919460, EPI_ISL_919461, EPI_ISL_1104572, EPI_ISL_1177834, EPI_ISL_1177835, EPI_ISL_1177836, EPI_ISL_1177837, EPI_ISL_1177838, EPI_ISL_1474654, EPI_ISL_1474656, EPI_ISL_1474657, EPI_ISL_1474658, EPI_ISL_1474659, EPI_ISL_1474660, EPI_ISL_1474661

| | | | |
|---|---|---|---|
| see above | Liverpool Clinical Laboratories | COVID-19 Genomics UK (COG-UK) Consortium | A Alrezaihi; Alessandro Gerada; Alistair Darby; Angela Cowell; Anita Lucaci; Anu Chawla; Cassie Olateju; Catherine Hartley; Charlotte Nelson; Ecaterina Vamos; Elaine O'Toole; Eleanor G Bentley; Ghada T Shawli; Hermione Webster; Isabel García-Dorival; James Johnson; James P Stewart; Jenifer Manson; Joanne Watts; Jones Benjamin; Jordan J Clark; Julian Hiscox; L Luu; Lucille Rainbow; M Almsaud; Margaret Hughes; Mark Whitehead; Matthew Gemmell; Miren Iturriza-Gomara; Muhannad Alruwaili; N.P Randle; Neil Swainston; PKF Gilmore; Parul Sharma; Rebekah Penrice-Randal; Richard Eccles; Richard Gregory; Sam Haldenby; Steve Paterson; Stuart D Armstrong; Trevor Ian Robinson; Ximeng Han |

EPI_ISL_839733, EPI_ISL_839734, EPI_ISL_866527, EPI_ISL_866528, EPI_ISL_866529, EPI_ISL_866530, EPI_ISL_866531, EPI_ISL_866532, EPI_ISL_866533, EPI_ISL_866534, EPI_ISL_866536, EPI_ISL_866537, EPI_ISL_866538, EPI_ISL_866539, EPI_ISL_866540, EPI_ISL_921243, EPI_ISL_921244, EPI_ISL_921245, EPI_ISL_921246, EPI_ISL_921247, EPI_ISL_921248, EPI_ISL_921250, EPI_ISL_921251, EPI_ISL_921277, EPI_ISL_921279, EPI_ISL_921285, EPI_ISL_921287, EPI_ISL_921289, EPI_ISL_921290, EPI_ISL_921291, EPI_ISL_921292, EPI_ISL_921293, EPI_ISL_921294, EPI_ISL_921296, EPI_ISL_921299, EPI_ISL_921302, EPI_ISL_921304, EPI_ISL_921306, EPI_ISL_921307, EPI_ISL_921308, EPI_ISL_921309, EPI_ISL_921310, EPI_ISL_921311, EPI_ISL_921312, EPI_ISL_921315, EPI_ISL_921316, EPI_ISL_921319, EPI_ISL_921320, EPI_ISL_921407, EPI_ISL_921408, EPI_ISL_921409, EPI_ISL_921410, EPI_ISL_921411, EPI_ISL_921412, EPI_ISL_921413, EPI_ISL_921419, EPI_ISL_921427, EPI_ISL_921429, EPI_ISL_921430, EPI_ISL_921431, EPI_ISL_921434, EPI_ISL_921435, EPI_ISL_921436, EPI_ISL_921437, EPI_ISL_921438, EPI_ISL_921439, EPI_ISL_921440, EPI_ISL_921441, EPI_ISL_921442, EPI_ISL_921443, EPI_ISL_921444, EPI_ISL_921445, EPI_ISL_921446, EPI_ISL_921447, EPI_ISL_921448, EPI_ISL_921449, EPI_ISL_921450, EPI_ISL_921451, EPI_ISL_921452, EPI_ISL_921453, EPI_ISL_921454, EPI_ISL_921455, EPI_ISL_921456, EPI_ISL_921457, EPI_ISL_921458, EPI_ISL_921459, EPI_ISL_921460, EPI_ISL_921461, EPI_ISL_921462, EPI_ISL_921463, EPI_ISL_921464, EPI_ISL_921465, EPI_ISL_921466, EPI_ISL_921467, EPI_ISL_921468, EPI_ISL_921469, EPI_ISL_921470, EPI_ISL_921471, EPI_ISL_921472, EPI_ISL_921473, EPI_ISL_921474, EPI_ISL_950254, EPI_ISL_950255, EPI_ISL_950257, EPI_ISL_950258, EPI_ISL_950259, EPI_ISL_950260, EPI_ISL_950261, EPI_ISL_950262, EPI_ISL_950263, EPI_ISL_950264, EPI_ISL_950265, EPI_ISL_950266, EPI_ISL_950267, EPI_ISL_950268, EPI_ISL_950269, EPI_ISL_950270, EPI_ISL_950288, EPI_ISL_950460, EPI_ISL_950461, EPI_ISL_950462, EPI_ISL_950463, EPI_ISL_950464, EPI_ISL_950465, EPI_ISL_950466, EPI_ISL_950467, EPI_ISL_950468, EPI_ISL_950469, EPI_ISL_950470, EPI_ISL_950471, EPI_ISL_950472, EPI_ISL_950473, EPI_ISL_950474, EPI_ISL_950475, EPI_ISL_950476, EPI_ISL_950477, EPI_ISL_950478, EPI_ISL_950479, EPI_ISL_950480, EPI_ISL_950481, EPI_ISL_950482, EPI_ISL_950483, EPI_ISL_950484, EPI_ISL_950485, EPI_ISL_950486, EPI_ISL_950487, EPI_ISL_950488, EPI_ISL_950489, EPI_ISL_950490, EPI_ISL_950491, EPI_ISL_950492, EPI_ISL_950493, EPI_ISL_998368, EPI_ISL_998369, EPI_ISL_998370, EPI_ISL_998372, EPI_ISL_998737, EPI_ISL_998738, EPI_ISL_998739, EPI_ISL_998740, EPI_ISL_998741, EPI_ISL_998742, EPI_ISL_998743, EPI_ISL_998744, EPI_ISL_998745, EPI_ISL_998746, EPI_ISL_998747, EPI_ISL_998748, EPI_ISL_998750, EPI_ISL_998751, EPI_ISL_998752, EPI_ISL_998753, EPI_ISL_998754, EPI_ISL_998755, EPI_ISL_998756, EPI_ISL_998757, EPI_ISL_998758, EPI_ISL_998759, EPI_ISL_998760, EPI_ISL_998761, EPI_ISL_998762, EPI_ISL_998763, EPI_ISL_998764, EPI_ISL_998765, EPI_ISL_998766, EPI_ISL_998767, EPI_ISL_998768, EPI_ISL_998769, EPI_ISL_998770, EPI_ISL_998771, EPI_ISL_1050922, EPI_ISL_1050925, EPI_ISL_1050928, EPI_ISL_1050931, EPI_ISL_1050957, EPI_ISL_1050958, EPI_ISL_1050959, EPI_ISL_1051040, EPI_ISL_1051041, EPI_ISL_1051043, EPI_ISL_1051045, EPI_ISL_1051048, EPI_ISL_1051050, EPI_ISL_1051052, EPI_ISL_1051054, EPI_ISL_1051056, EPI_ISL_1051058, EPI_ISL_1051060, EPI_ISL_1051062, EPI_ISL_1051063, EPI_ISL_1051065, EPI_ISL_1051067, EPI_ISL_1051073, EPI_ISL_1051075, EPI_ISL_1051080, EPI_ISL_1051082, EPI_ISL_1051089, EPI_ISL_1051093, EPI_ISL_1051118, EPI_ISL_1051119, EPI_ISL_1051342, EPI_ISL_1051343, EPI_ISL_1051344, EPI_ISL_1105706, EPI_ISL_1105708, EPI_ISL_1105709, EPI_ISL_1105772, EPI_ISL_1105773, EPI_ISL_1105774, EPI_ISL_1105775, EPI_ISL_1105776, EPI_ISL_1105777, EPI_ISL_1105778, EPI_ISL_1105779, EPI_ISL_1105780, EPI_ISL_1105781, EPI_ISL_1105782, EPI_ISL_1105783, EPI_ISL_1105784, EPI_ISL_1105785, EPI_ISL_1105786, EPI_ISL_1105787, EPI_ISL_1105788, EPI_ISL_1105789, EPI_ISL_1105790, EPI_ISL_1105791, EPI_ISL_1105792, EPI_ISL_1105793, EPI_ISL_1105794, EPI_ISL_1105795, EPI_ISL_1105796, EPI_ISL_1105797, EPI_ISL_1105798, EPI_ISL_1105799, EPI_ISL_1105800, EPI_ISL_1105801, EPI_ISL_1105802,

| | | | |
|---|---|---|---|
| EPI_ISL_1105803, EPI_ISL_1105804, EPI_ISL_1105805, EPI_ISL_1105806, EPI_ISL_1105807, EPI_ISL_1105808, EPI_ISL_1105809, EPI_ISL_1105810, EPI_ISL_1105811, EPI_ISL_1178334, EPI_ISL_1178335, EPI_ISL_1178339, EPI_ISL_1178340, EPI_ISL_1178341, EPI_ISL_1178342, EPI_ISL_1178344, EPI_ISL_1178345, EPI_ISL_1178346, EPI_ISL_1178347, EPI_ISL_1309550, EPI_ISL_1309551, EPI_ISL_1309552, EPI_ISL_1309553, EPI_ISL_1309554, EPI_ISL_1309555, EPI_ISL_1309559, EPI_ISL_1309560, EPI_ISL_1309561, EPI_ISL_1309565, EPI_ISL_1309566, EPI_ISL_1309571, EPI_ISL_1309597, EPI_ISL_1309598, EPI_ISL_1309608, EPI_ISL_1309610 | | | |
| see above | Northumbria University / South Tees Hospitals NHS Foundation Trust / North Cumbria Integrated Care NHS Foundation Trust / North Tees and Hartlepool NHS Foundation Trust / Newcastle Hospitals NHS Foundation Trust | COVID-19 Genomics UK (COG-UK) Consortium | Andrew Nelson; Brendan Payne; Clive Graham; Darren L Smith; Debra Padgett; Edward Barton; Emma Swindells; Garren Scott; Gary Black; Gary Eltringham; Giles S Holt; Greg R Young; Jane Greenaway; Jennifer Collins; John Allan; Joshua Loh; Lynn Dover; Matthew Bashton; Mohammad A Tariq; Paul Baker; Sarah Essex; Steve Liggett; Wen C Yew; Yusri Taha |
| EPI_ISL_840650, EPI_ISL_840660, EPI_ISL_840661, EPI_ISL_840663, EPI_ISL_840664, EPI_ISL_840666, EPI_ISL_840677, EPI_ISL_840678, EPI_ISL_840679, EPI_ISL_840680, EPI_ISL_840681, EPI_ISL_840682, EPI_ISL_840683, EPI_ISL_840684, EPI_ISL_840685, EPI_ISL_840686, EPI_ISL_840687, EPI_ISL_840688, EPI_ISL_840689, EPI_ISL_840690, EPI_ISL_840691, EPI_ISL_840692, EPI_ISL_840693, EPI_ISL_840694, EPI_ISL_841318, EPI_ISL_841319, EPI_ISL_841320, EPI_ISL_841323, EPI_ISL_841332, EPI_ISL_841333, EPI_ISL_841334, EPI_ISL_841335, EPI_ISL_841336, EPI_ISL_841337, EPI_ISL_841338, EPI_ISL_841341, EPI_ISL_841342, EPI_ISL_841358, EPI_ISL_841371, EPI_ISL_841372, EPI_ISL_841374, EPI_ISL_841375, EPI_ISL_841376, EPI_ISL_841377, EPI_ISL_841378, EPI_ISL_841379, EPI_ISL_841380, EPI_ISL_841381, EPI_ISL_841382, EPI_ISL_841383, EPI_ISL_841384, EPI_ISL_841385, EPI_ISL_841386, EPI_ISL_841387, EPI_ISL_841388, EPI_ISL_841389, EPI_ISL_841390, EPI_ISL_841391, EPI_ISL_841392, EPI_ISL_841393, EPI_ISL_841394, EPI_ISL_841395, EPI_ISL_841396, EPI_ISL_841397, EPI_ISL_841399, EPI_ISL_841400, EPI_ISL_841401, EPI_ISL_841402, EPI_ISL_841403, EPI_ISL_841404, EPI_ISL_841405, EPI_ISL_841406, EPI_ISL_841413, EPI_ISL_841414, EPI_ISL_841416, EPI_ISL_841418, EPI_ISL_841419, EPI_ISL_841420, EPI_ISL_841421, EPI_ISL_841422, EPI_ISL_841423, EPI_ISL_841424, EPI_ISL_841425, EPI_ISL_841428, EPI_ISL_841429, EPI_ISL_841432, EPI_ISL_841435, EPI_ISL_841436, EPI_ISL_841498, EPI_ISL_841506, EPI_ISL_841510, EPI_ISL_841511, EPI_ISL_841512, EPI_ISL_841513, EPI_ISL_841514, EPI_ISL_841515, EPI_ISL_841548, EPI_ISL_841586, EPI_ISL_841588, EPI_ISL_841590, EPI_ISL_841594, EPI_ISL_841595, EPI_ISL_841596, EPI_ISL_841597, EPI_ISL_841598, EPI_ISL_841599, EPI_ISL_841600, EPI_ISL_841601, EPI_ISL_841602, EPI_ISL_841603, EPI_ISL_841604, EPI_ISL_841605, EPI_ISL_841606, EPI_ISL_841607, EPI_ISL_841610, EPI_ISL_841611, EPI_ISL_841613, EPI_ISL_841615, EPI_ISL_841616, EPI_ISL_867186, EPI_ISL_867203, EPI_ISL_867204, EPI_ISL_867205, EPI_ISL_867206, EPI_ISL_867207, EPI_ISL_867208, EPI_ISL_867209, EPI_ISL_867210, EPI_ISL_867432, EPI_ISL_867433, EPI_ISL_867434, EPI_ISL_867456, EPI_ISL_867503, EPI_ISL_867512, EPI_ISL_867751, EPI_ISL_867758, EPI_ISL_1476506, EPI_ISL_1476507, EPI_ISL_1476508, EPI_ISL_1476509, EPI_ISL_1476510, EPI_ISL_1476511, EPI_ISL_1476512, EPI_ISL_1476513, EPI_ISL_1476518, EPI_ISL_1476520, EPI_ISL_1476521, EPI_ISL_1476526, EPI_ISL_1476529, EPI_ISL_1476530, EPI_ISL_1476531, EPI_ISL_1476532, EPI_ISL_1476533, EPI_ISL_1476536, EPI_ISL_1476537, EPI_ISL_1476538, EPI_ISL_1476539, EPI_ISL_1476541, EPI_ISL_1476542, EPI_ISL_1476543, EPI_ISL_1476547, EPI_ISL_1546812, EPI_ISL_1546813, EPI_ISL_1546818, EPI_ISL_1546821, EPI_ISL_1546822, EPI_ISL_1546823, EPI_ISL_1546917, EPI_ISL_1546918, EPI_ISL_1546919, EPI_ISL_1546923, EPI_ISL_1546924, EPI_ISL_1546925, EPI_ISL_1546926, EPI_ISL_1546927, EPI_ISL_1546928, EPI_ISL_1546930, EPI_ISL_1546931, EPI_ISL_1546932, EPI_ISL_1546933, EPI_ISL_1546934, EPI_ISL_1546935, EPI_ISL_1546936, EPI_ISL_1546937, EPI_ISL_1546938, EPI_ISL_1546939, EPI_ISL_1546940, EPI_ISL_1546941, EPI_ISL_1546942, EPI_ISL_1546943, EPI_ISL_1546944, EPI_ISL_1546945, EPI_ISL_1546946 | | | |
| see above | Originating lab: Wales Specialist Virology Centre Sequencing lab: Pathogen Genomics Unit | Public Health Wales Microbiology Cardiff Wales Specialist Virology Centre | Alec Birchley; Alexander Adams; Amy Gaskin; Angela Marchbank; Bree Gatica-Wilcox; Catherine Moore; Jason Coombes; Joanne Watkins; Joel Southgate; Johnathan Evans; Laura Gifford; Lauren Gilbert; Lee Graham; Malorie Perry; Matthew Bull; Nicole Pacchiarini; Sally Corden; Sara Kumziene-Summerhayes; Sara Rey; Sarah Taylor; Simon Cottrell; Sophie Jones; Tom Connor |
| EPI_ISL_867091, EPI_ISL_867092, EPI_ISL_867099, EPI_ISL_867100, EPI_ISL_867101, EPI_ISL_867103, EPI_ISL_867105, EPI_ISL_867106, EPI_ISL_867107, EPI_ISL_867108, EPI_ISL_867110, EPI_ISL_867111, EPI_ISL_867112, EPI_ISL_867113, EPI_ISL_867114, EPI_ISL_867118, EPI_ISL_867119, EPI_ISL_867120, EPI_ISL_867121, EPI_ISL_867123, EPI_ISL_867125, EPI_ISL_867127, EPI_ISL_867128, EPI_ISL_867129, EPI_ISL_867131, EPI_ISL_867132, EPI_ISL_867133, EPI_ISL_867134, EPI_ISL_867137, EPI_ISL_867138, EPI_ISL_867139, EPI_ISL_867140, EPI_ISL_867141, EPI_ISL_867142, EPI_ISL_867144, EPI_ISL_867145, EPI_ISL_867147, EPI_ISL_867148, EPI_ISL_867150, EPI_ISL_867151, EPI_ISL_867152, EPI_ISL_867163, EPI_ISL_867164, EPI_ISL_867165, EPI_ISL_867166, EPI_ISL_867167, EPI_ISL_922174, EPI_ISL_922175, EPI_ISL_922177, EPI_ISL_922178, EPI_ISL_922180, EPI_ISL_922182, EPI_ISL_922183, EPI_ISL_922184, EPI_ISL_922185, EPI_ISL_922188, EPI_ISL_922189, EPI_ISL_922190, EPI_ISL_922191, EPI_ISL_922192, EPI_ISL_922198, EPI_ISL_922199, EPI_ISL_922201, EPI_ISL_922242, EPI_ISL_922244, EPI_ISL_922246, EPI_ISL_922249, EPI_ISL_922252, EPI_ISL_922254, EPI_ISL_922255, EPI_ISL_922283, EPI_ISL_922284, EPI_ISL_922285, EPI_ISL_922286, EPI_ISL_922304, EPI_ISL_922308, EPI_ISL_922309, EPI_ISL_922310, EPI_ISL_922311, EPI_ISL_922317, EPI_ISL_922321, EPI_ISL_922323, EPI_ISL_922324, EPI_ISL_922325, EPI_ISL_922326, EPI_ISL_922327, EPI_ISL_922328, EPI_ISL_922329, EPI_ISL_922330, EPI_ISL_922331, EPI_ISL_922332, EPI_ISL_922333, EPI_ISL_922334, EPI_ISL_922335, EPI_ISL_922336, EPI_ISL_922337, EPI_ISL_922338, EPI_ISL_922339, EPI_ISL_922340, EPI_ISL_922341, EPI_ISL_922342, EPI_ISL_922343, EPI_ISL_922345, EPI_ISL_951068, EPI_ISL_951069, EPI_ISL_951070, EPI_ISL_951071, EPI_ISL_951072, EPI_ISL_951073, EPI_ISL_951074, EPI_ISL_951075, EPI_ISL_951077, EPI_ISL_951078, EPI_ISL_951082, EPI_ISL_951084, EPI_ISL_951085, EPI_ISL_951087, EPI_ISL_951088, EPI_ISL_951089, EPI_ISL_951090, EPI_ISL_951091, EPI_ISL_951093, EPI_ISL_951101, EPI_ISL_951102, EPI_ISL_951103, EPI_ISL_951104, EPI_ISL_951105, EPI_ISL_951106, EPI_ISL_951107, EPI_ISL_951108, EPI_ISL_951109, EPI_ISL_951110, EPI_ISL_951111, EPI_ISL_951112, EPI_ISL_951114, EPI_ISL_951115, EPI_ISL_951116, EPI_ISL_951117, EPI_ISL_951118, EPI_ISL_951119, EPI_ISL_951120, EPI_ISL_951121, EPI_ISL_951122, EPI_ISL_951125, EPI_ISL_951126, EPI_ISL_951131, EPI_ISL_951132, EPI_ISL_951134, EPI_ISL_951135, EPI_ISL_951136, EPI_ISL_951137, EPI_ISL_951138, EPI_ISL_951139, EPI_ISL_951141, EPI_ISL_951143, EPI_ISL_951144, EPI_ISL_951145, EPI_ISL_951147, EPI_ISL_951148, EPI_ISL_951149, EPI_ISL_951152, EPI_ISL_951153, EPI_ISL_951154, EPI_ISL_951155, EPI_ISL_951158, EPI_ISL_951159, EPI_ISL_951160, EPI_ISL_951161, EPI_ISL_951162, EPI_ISL_951172, EPI_ISL_951173, EPI_ISL_951174, EPI_ISL_951175, EPI_ISL_951176, EPI_ISL_951177, EPI_ISL_951178, EPI_ISL_951180, EPI_ISL_951181, EPI_ISL_951182, EPI_ISL_951183, EPI_ISL_951184, EPI_ISL_951185, EPI_ISL_951186, EPI_ISL_951187, EPI_ISL_951188, EPI_ISL_951189, EPI_ISL_951191, EPI_ISL_951192, EPI_ISL_951193, EPI_ISL_951195, EPI_ISL_951196, EPI_ISL_951197, EPI_ISL_951198, EPI_ISL_951199, EPI_ISL_951200, EPI_ISL_951201, EPI_ISL_951202, EPI_ISL_951204, EPI_ISL_951205, EPI_ISL_951206, EPI_ISL_951207, EPI_ISL_951209, EPI_ISL_951210, EPI_ISL_951211, EPI_ISL_951212, EPI_ISL_951213, EPI_ISL_951214, EPI_ISL_951215, EPI_ISL_951216, EPI_ISL_951218, EPI_ISL_951220, EPI_ISL_951221, EPI_ISL_951223, EPI_ISL_951224, EPI_ISL_951225, EPI_ISL_951226, EPI_ISL_951227, EPI_ISL_951228, EPI_ISL_951229, EPI_ISL_951230, EPI_ISL_951233, EPI_ISL_951234, EPI_ISL_951235, EPI_ISL_951242, EPI_ISL_951319, EPI_ISL_951320, EPI_ISL_951323, EPI_ISL_951324, EPI_ISL_951326, EPI_ISL_951327, EPI_ISL_951328, EPI_ISL_951329, EPI_ISL_951331, EPI_ISL_951333, EPI_ISL_951334, EPI_ISL_951335, EPI_ISL_951336, EPI_ISL_951337, EPI_ISL_951338, EPI_ISL_951339, EPI_ISL_951340, EPI_ISL_951341, EPI_ISL_951342, EPI_ISL_951344, EPI_ISL_951381, EPI_ISL_951382, EPI_ISL_951384, EPI_ISL_951386, EPI_ISL_951397, EPI_ISL_951398, EPI_ISL_951400, EPI_ISL_951401, EPI_ISL_951402, EPI_ISL_951410, EPI_ISL_998986, EPI_ISL_998987, EPI_ISL_998988, EPI_ISL_998989, EPI_ISL_998992, EPI_ISL_998996, EPI_ISL_998998, EPI_ISL_999001, EPI_ISL_999002, EPI_ISL_999004, EPI_ISL_999005, EPI_ISL_999006, EPI_ISL_999007, EPI_ISL_999008, EPI_ISL_999009, EPI_ISL_999010, EPI_ISL_999012, EPI_ISL_999013, EPI_ISL_999014, EPI_ISL_999015, EPI_ISL_999017, EPI_ISL_999018, EPI_ISL_999019, EPI_ISL_999020, EPI_ISL_999021, EPI_ISL_999023, EPI_ISL_999024, EPI_ISL_999025, EPI_ISL_999027, EPI_ISL_999028, EPI_ISL_999029, EPI_ISL_999031, EPI_ISL_999051, EPI_ISL_999052, EPI_ISL_999053, EPI_ISL_999054, EPI_ISL_999055, EPI_ISL_999056, EPI_ISL_999057, EPI_ISL_999058, EPI_ISL_999213, EPI_ISL_999215, EPI_ISL_999216, EPI_ISL_999348, EPI_ISL_999349, EPI_ISL_999350, EPI_ISL_999351, EPI_ISL_999352, EPI_ISL_999353, EPI_ISL_999355, EPI_ISL_999356, EPI_ISL_999360, EPI_ISL_999361, EPI_ISL_999362, EPI_ISL_999386, EPI_ISL_999418, EPI_ISL_999424, EPI_ISL_999454, EPI_ISL_999459, EPI_ISL_999465, EPI_ISL_999466, EPI_ISL_999470, EPI_ISL_999509, EPI_ISL_1051598, EPI_ISL_1051876, EPI_ISL_1178724, EPI_ISL_1249234, EPI_ISL_1249235, EPI_ISL_1249237, EPI_ISL_1309739, EPI_ISL_1309748, EPI_ISL_1387236, EPI_ISL_1833057, EPI_ISL_1833059, EPI_ISL_1833105, EPI_ISL_2127634 | | | |
| see above | Oxford Viromics, NDM, University of Oxford; Oxford University Hospitals; Basingstoke and North Hampshire Hospital | COVID-19 Genomics UK (COG-UK) Consortium | Alex Mobbs; Amy Trebes; Anita Justice; Catrin Moore; Christophe Fraser; David Bonsall; David Buck; Emma Wise; George Macintyre; Jessica Lynch; John Todd; Mariateresa de Cesare; Matilde Mori; Monique Andersson; Nathan Moore; Nick Cortes; Robert Shaw; Stephen Kidd; Tanya Golubchik; Timothy Peto |
| EPI_ISL_819366, EPI_ISL_819375, EPI_ISL_819377, EPI_ISL_819384, EPI_ISL_819391, EPI_ISL_819392, EPI_ISL_819393, EPI_ISL_819394, EPI_ISL_819395, EPI_ISL_819405, EPI_ISL_819406, EPI_ISL_819407, EPI_ISL_819408, EPI_ISL_819409, EPI_ISL_819410, EPI_ISL_819411, EPI_ISL_819412, EPI_ISL_819413, EPI_ISL_819414, EPI_ISL_819415, EPI_ISL_819416, EPI_ISL_819417, EPI_ISL_819418, EPI_ISL_819419, EPI_ISL_819420, EPI_ISL_819421, EPI_ISL_819424, EPI_ISL_819425, EPI_ISL_819427, EPI_ISL_819429, EPI_ISL_819440, EPI_ISL_819441, EPI_ISL_819442, EPI_ISL_819443, EPI_ISL_819444, EPI_ISL_819445, EPI_ISL_819446, EPI_ISL_839823, EPI_ISL_839824, EPI_ISL_839825, EPI_ISL_839826, EPI_ISL_839827, EPI_ISL_839828, EPI_ISL_839829, EPI_ISL_839830, EPI_ISL_839831, EPI_ISL_839832, EPI_ISL_839833, EPI_ISL_839834, EPI_ISL_839835, EPI_ISL_839836, EPI_ISL_839837, EPI_ISL_839838, EPI_ISL_839839, EPI_ISL_839840, EPI_ISL_839841, EPI_ISL_839842, EPI_ISL_839843, EPI_ISL_839844, EPI_ISL_839845, EPI_ISL_839846, EPI_ISL_839847, EPI_ISL_839848, EPI_ISL_839849, EPI_ISL_839850, EPI_ISL_839851, EPI_ISL_839852, EPI_ISL_839853, EPI_ISL_839854, EPI_ISL_839855, EPI_ISL_839856, EPI_ISL_839857, EPI_ISL_839858, EPI_ISL_839859, EPI_ISL_839860, EPI_ISL_839861, EPI_ISL_839862, EPI_ISL_839863, EPI_ISL_839864, EPI_ISL_839865, EPI_ISL_839866, EPI_ISL_839867, EPI_ISL_839868, EPI_ISL_839869, EPI_ISL_839870, EPI_ISL_839871, EPI_ISL_839872, EPI_ISL_839873, EPI_ISL_839874, EPI_ISL_839875, EPI_ISL_839876, EPI_ISL_839877, EPI_ISL_839878, EPI_ISL_839879, EPI_ISL_839880, EPI_ISL_839881, EPI_ISL_839882, EPI_ISL_839883, EPI_ISL_839884, EPI_ISL_839885, EPI_ISL_839886, EPI_ISL_839887, EPI_ISL_839888, EPI_ISL_839889, EPI_ISL_839890, EPI_ISL_839891, EPI_ISL_839892, EPI_ISL_839893, EPI_ISL_839894, EPI_ISL_839895, EPI_ISL_839896, EPI_ISL_839897, EPI_ISL_839898, EPI_ISL_839899, EPI_ISL_839900, EPI_ISL_839901, EPI_ISL_839902, EPI_ISL_839903, EPI_ISL_839904, EPI_ISL_839905, EPI_ISL_839906, EPI_ISL_839907, EPI_ISL_839908, EPI_ISL_839909, EPI_ISL_839910, EPI_ISL_839911, EPI_ISL_839912, EPI_ISL_839913, EPI_ISL_839914, EPI_ISL_839915, EPI_ISL_839916, EPI_ISL_839917, EPI_ISL_839918, EPI_ISL_839919, EPI_ISL_839920, EPI_ISL_839921, EPI_ISL_839922, EPI_ISL_839923, EPI_ISL_839924, EPI_ISL_839925, EPI_ISL_839926, EPI_ISL_839927, EPI_ISL_839928, EPI_ISL_839929, EPI_ISL_839930, EPI_ISL_839931, EPI_ISL_839932, EPI_ISL_839933, EPI_ISL_839934, EPI_ISL_839935, EPI_ISL_839936, EPI_ISL_839937, EPI_ISL_839938, EPI_ISL_839939, EPI_ISL_839940, EPI_ISL_839941, EPI_ISL_839942, EPI_ISL_839943, EPI_ISL_839944, EPI_ISL_839945, EPI_ISL_839946, EPI_ISL_839947, EPI_ISL_839948, EPI_ISL_839949, EPI_ISL_839950, EPI_ISL_839951, EPI_ISL_839952, EPI_ISL_839953, EPI_ISL_839954, EPI_ISL_839955, EPI_ISL_839956, EPI_ISL_839957, EPI_ISL_839958, EPI_ISL_839959, EPI_ISL_839960, EPI_ISL_839961, EPI_ISL_839962, EPI_ISL_839963, EPI_ISL_839964, EPI_ISL_839965, EPI_ISL_839966, EPI_ISL_839967, EPI_ISL_839968, EPI_ISL_839969, EPI_ISL_839970, EPI_ISL_839971, EPI_ISL_839972, EPI_ISL_839973, EPI_ISL_839974, EPI_ISL_839975, EPI_ISL_839976, EPI_ISL_839977, EPI_ISL_839978, EPI_ISL_839979, EPI_ISL_839980, EPI_ISL_866588, EPI_ISL_866589, EPI_ISL_866590, EPI_ISL_866591, EPI_ISL_866592, EPI_ISL_866593, EPI_ISL_866594, EPI_ISL_866595, EPI_ISL_866596, EPI_ISL_866597, EPI_ISL_866598, EPI_ISL_866599, EPI_ISL_866600, EPI_ISL_866602, EPI_ISL_950551, EPI_ISL_950582, EPI_ISL_950585, EPI_ISL_1051429, EPI_ISL_1051432, EPI_ISL_1051434, EPI_ISL_1051437, EPI_ISL_1051439, EPI_ISL_1051442, EPI_ISL_1051444, EPI_ISL_1051448, EPI_ISL_1051453, EPI_ISL_1051457, EPI_ISL_1051458, EPI_ISL_1051460, EPI_ISL_1051461, EPI_ISL_1051463, EPI_ISL_1051464, EPI_ISL_1051472, EPI_ISL_1051480, EPI_ISL_1051483, EPI_ISL_1051487, EPI_ISL_1051492, EPI_ISL_1051496, EPI_ISL_1051499, EPI_ISL_1051502, EPI_ISL_1051509, EPI_ISL_1051511, EPI_ISL_1051512, EPI_ISL_1051513, EPI_ISL_1051518, EPI_ISL_1051524, EPI_ISL_1051527, EPI_ISL_1051528 | | | |
| see above | Quadram Institute Bioscience | COVID-19 Genomics UK (COG-UK) Consortium | Alexander J Trotter; Alison E. Mather; Alp Aydin; Ana P. Tedim; Anastasia Kolyva; Andrew Bell; Andrew J. Page; Claire Stuart; Dave J. Baker; Gemma L. Kay; John Wain; Justin O'Grady; Leonardo de Oliveira Martins; Lizzie Meadows; Maria Diaz; Mark Webber; Muhammed Yasir; Nabil-Fareed Alikhan; Ngozi Elumogo; Nicholas M. Thomson; Rachael Stanley; Rachel Gilroy; Reenesh Prakash; Samir Dervisevic; Samuel Bloomfield; Steven Rudder; Thanh Le-Viet |
| EPI_ISL_820573, EPI_ISL_820575, EPI_ISL_820578, EPI_ISL_820580, EPI_ISL_820583, EPI_ISL_820586, EPI_ISL_820588, EPI_ISL_820591, EPI_ISL_820593, EPI_ISL_820596, EPI_ISL_820598, EPI_ISL_820601, EPI_ISL_820603, EPI_ISL_820605, EPI_ISL_820607, EPI_ISL_820610, EPI_ISL_839995, EPI_ISL_839996, EPI_ISL_839997, EPI_ISL_839998, EPI_ISL_839999, EPI_ISL_840000, EPI_ISL_840001, EPI_ISL_840002, EPI_ISL_840003, EPI_ISL_840004, EPI_ISL_840005, EPI_ISL_840006, EPI_ISL_840007, EPI_ISL_840008, EPI_ISL_866906, EPI_ISL_866907 | | | |
| see above | Queens Medical Centre, Clinical Microbiology Department / DeepSeq Nottingham | COVID-19 Genomics UK (COG-UK) Consortium | Christopher Moore; Fei Sang; Gemma Clark; Hannah Howson-Wells; Johnny Debebe; Jonathan Ball; Joseph Chappell; Manjinder Khakh; Matthew Carlisle; Matthew Loose; Michelle M Lister; Nadine Holmes; Patrick McClure; Theocharis Tsoleridis; Vicki M Fleming; Victoria Wright; Wendy Smith |
| EPI_ISL_866353, EPI_ISL_866354, EPI_ISL_866355, EPI_ISL_866357, EPI_ISL_866358, EPI_ISL_866359, EPI_ISL_866360, EPI_ISL_866361, EPI_ISL_866362, EPI_ISL_866367, EPI_ISL_866370, EPI_ISL_866373, EPI_ISL_866374, EPI_ISL_866375, EPI_ISL_866376, EPI_ISL_866377, EPI_ISL_866378, EPI_ISL_866379, EPI_ISL_866380, EPI_ISL_866381, EPI_ISL_866382, EPI_ISL_866384, EPI_ISL_866385, EPI_ISL_866386, EPI_ISL_866387, EPI_ISL_866388, EPI_ISL_866389, EPI_ISL_866390, EPI_ISL_866391, EPI_ISL_866392, EPI_ISL_866395, EPI_ISL_866396, EPI_ISL_866397, EPI_ISL_866398, EPI_ISL_866399, EPI_ISL_920980, EPI_ISL_921019, EPI_ISL_921020, EPI_ISL_921021, EPI_ISL_921023, EPI_ISL_921024, EPI_ISL_921026, EPI_ISL_921040, EPI_ISL_921047, EPI_ISL_921048, EPI_ISL_921049, EPI_ISL_921050, EPI_ISL_921051, EPI_ISL_921052, EPI_ISL_921053, EPI_ISL_921054, EPI_ISL_921055, EPI_ISL_921056, EPI_ISL_921057, EPI_ISL_921058, EPI_ISL_921059, EPI_ISL_921060, EPI_ISL_921061, EPI_ISL_921062, EPI_ISL_921063, EPI_ISL_921064, EPI_ISL_921065, EPI_ISL_921066, EPI_ISL_921067, EPI_ISL_921068, EPI_ISL_921069, EPI_ISL_921070, EPI_ISL_921071, EPI_ISL_921080, EPI_ISL_921081, EPI_ISL_921082, EPI_ISL_921083, EPI_ISL_921084, EPI_ISL_921091, EPI_ISL_921092, EPI_ISL_921093, EPI_ISL_921094, EPI_ISL_921095, EPI_ISL_921098, EPI_ISL_921099, EPI_ISL_921100, EPI_ISL_921102, EPI_ISL_921103, EPI_ISL_921104, EPI_ISL_921106, EPI_ISL_921107, EPI_ISL_921108, EPI_ISL_921109, EPI_ISL_921110, EPI_ISL_921111, EPI_ISL_921112, EPI_ISL_921114, EPI_ISL_921115, EPI_ISL_921116, EPI_ISL_921117, EPI_ISL_921123, EPI_ISL_921124, EPI_ISL_921125, EPI_ISL_921126, EPI_ISL_921127, EPI_ISL_921129, EPI_ISL_921130, EPI_ISL_921131, EPI_ISL_921134, EPI_ISL_998239, EPI_ISL_1474958, EPI_ISL_1474959, EPI_ISL_1474960, EPI_ISL_1474963, EPI_ISL_1474964, EPI_ISL_1474995, EPI_ISL_1475062, EPI_ISL_1475063, EPI_ISL_1475064, EPI_ISL_1475065, EPI_ISL_1475066, EPI_ISL_1475067, EPI_ISL_1475068, EPI_ISL_1475069, EPI_ISL_1475070, EPI_ISL_1475071, EPI_ISL_1475072, EPI_ISL_1475073, EPI_ISL_1475074, EPI_ISL_1475075, EPI_ISL_1475076, EPI_ISL_1475077, EPI_ISL_1475078, EPI_ISL_1475079, EPI_ISL_1475081, EPI_ISL_1475083 | | | |
| see above | Regional Virus Laboratory, Belfast Health and Social Care Trust | COVID-19 Genomics UK (COG-UK) Consortium | Alison Watt; Ciara Cox; Conall McCaughey; David Simpson; Derek Fairley; James McKenna; Mairead Connor; Susan Feeney; Tanya Curran; Zoltan Molnar |
| EPI_ISL_768815, EPI_ISL_768816, EPI_ISL_791194, EPI_ISL_791197, EPI_ISL_791198, EPI_ISL_791223, EPI_ISL_791224, EPI_ISL_791225, EPI_ISL_791226, EPI_ISL_791227, EPI_ISL_791228, EPI_ISL_791229, EPI_ISL_791230, EPI_ISL_791231, EPI_ISL_791232, EPI_ISL_791233, EPI_ISL_791234, EPI_ISL_791235, EPI_ISL_791236, EPI_ISL_791237, EPI_ISL_791238, EPI_ISL_791239, EPI_ISL_791240, EPI_ISL_791241, EPI_ISL_791242, EPI_ISL_791243, EPI_ISL_791244, EPI_ISL_791245, EPI_ISL_791246, EPI_ISL_791247, EPI_ISL_791248, EPI_ISL_791254, EPI_ISL_791255, EPI_ISL_791256, EPI_ISL_791257, EPI_ISL_791258, EPI_ISL_791259, EPI_ISL_791260, EPI_ISL_791261, EPI_ISL_791262, EPI_ISL_791263, EPI_ISL_791264, EPI_ISL_791265, EPI_ISL_791266, EPI_ISL_791267, EPI_ISL_791268, EPI_ISL_804217, EPI_ISL_804225, EPI_ISL_804249, EPI_ISL_804259, EPI_ISL_804276, EPI_ISL_811107, EPI_ISL_811108, EPI_ISL_811109, EPI_ISL_811110, EPI_ISL_811111, EPI_ISL_811112, EPI_ISL_811113, EPI_ISL_811114, EPI_ISL_811116, EPI_ISL_811121, EPI_ISL_837240, EPI_ISL_837241, EPI_ISL_837242, EPI_ISL_846595, EPI_ISL_846596, EPI_ISL_855574, EPI_ISL_855575, EPI_ISL_855577, EPI_ISL_855578, EPI_ISL_855579, EPI_ISL_855581, EPI_ISL_855582, EPI_ISL_855584, EPI_ISL_855588, EPI_ISL_855589, EPI_ISL_855590, EPI_ISL_855591, EPI_ISL_855592, EPI_ISL_855593, EPI_ISL_855594, EPI_ISL_855596, EPI_ISL_855597, EPI_ISL_855598, EPI_ISL_855599, EPI_ISL_855600, EPI_ISL_855601, EPI_ISL_855606, EPI_ISL_855607, EPI_ISL_855608, EPI_ISL_855609, EPI_ISL_856703, EPI_ISL_856704, EPI_ISL_856705, EPI_ISL_856706, EPI_ISL_860666, EPI_ISL_860667, EPI_ISL_860668, EPI_ISL_860669, EPI_ISL_860670, EPI_ISL_860671, EPI_ISL_860672, EPI_ISL_860673, EPI_ISL_860674, EPI_ISL_860675, EPI_ISL_860676, EPI_ISL_860678, EPI_ISL_860679, EPI_ISL_860680, EPI_ISL_860681, EPI_ISL_860682, EPI_ISL_860683, EPI_ISL_860684, EPI_ISL_1296492, EPI_ISL_1296493, EPI_ISL_1296495, EPI_ISL_1296496, EPI_ISL_1296497, EPI_ISL_1296498, EPI_ISL_1296499, EPI_ISL_1296501, EPI_ISL_1296502, EPI_ISL_1296503, EPI_ISL_1296504, EPI_ISL_1296508, EPI_ISL_1296542 | | | |
| see above | Respiratory Virus Unit, National Infection Service, Public Health England | COVID-19 Genomics UK (COG-UK) Consortium | PHE Covid Sequencing Team |
| EPI_ISL_1833415, EPI_ISL_1833417, EPI_ISL_1833431, EPI_ISL_1833434, EPI_ISL_1833436, EPI_ISL_1833446, EPI_ISL_1833448, EPI_ISL_1833449, EPI_ISL_1833450, EPI_ISL_1833451, EPI_ISL_1833452, EPI_ISL_1833453, EPI_ISL_1833454, EPI_ISL_1833455, EPI_ISL_1833457, EPI_ISL_1833460, EPI_ISL_1833461, EPI_ISL_1833462, EPI_ISL_1833464, EPI_ISL_1833468, EPI_ISL_1833469, EPI_ISL_1833470, EPI_ISL_1833473, EPI_ISL_1833474, EPI_ISL_1833475, EPI_ISL_1833476, EPI_ISL_1833477, EPI_ISL_1833478, EPI_ISL_1833479, EPI_ISL_1833480 | | | |
| see above | St George's University Hospitals NHS Foundation Trust | COVID-19 Genomics UK (COG-UK) Consortium | Adam Witney; Cassie Pope; Irene Monahan; Joshua Taylor; Ken Laing; NgeeKeong Tan |
| EPI_ISL_866075, EPI_ISL_866076, EPI_ISL_866077, EPI_ISL_866078, EPI_ISL_866079, EPI_ISL_866109, EPI_ISL_866110, EPI_ISL_866111, EPI_ISL_866147, EPI_ISL_920179, EPI_ISL_920288, EPI_ISL_920297, EPI_ISL_920311, EPI_ISL_920689, EPI_ISL_920711, EPI_ISL_1104974, EPI_ISL_1105008, EPI_ISL_1105017, EPI_ISL_1105025, EPI_ISL_1105033, EPI_ISL_1105042, EPI_ISL_1105164, EPI_ISL_1105177, EPI_ISL_1105193, EPI_ISL_1105208, EPI_ISL_1105257, EPI_ISL_1105260, EPI_ISL_1105267, EPI_ISL_1105274, EPI_ISL_1105275, EPI_ISL_1105281, EPI_ISL_1105287, EPI_ISL_1105288, EPI_ISL_1105293, EPI_ISL_1248367, EPI_ISL_1248376, EPI_ISL_1248378, EPI_ISL_1248386, EPI_ISL_1248388, EPI_ISL_1248390, EPI_ISL_1248400, EPI_ISL_1248401, EPI_ISL_1248402, EPI_ISL_1248405, EPI_ISL_1248411, EPI_ISL_1248412, EPI_ISL_1248413, EPI_ISL_1248414, EPI_ISL_1248416, EPI_ISL_1248420, EPI_ISL_1248422, EPI_ISL_1248423, EPI_ISL_1248424, EPI_ISL_1248427, EPI_ISL_1248430, EPI_ISL_1248432, EPI_ISL_1248433, EPI_ISL_1248434, EPI_ISL_1248442, EPI_ISL_1248444, EPI_ISL_1248445, EPI_ISL_1248459, EPI_ISL_1248486, EPI_ISL_1248499, EPI_ISL_1637068, EPI_ISL_1637071, EPI_ISL_1637072, EPI_ISL_1637075, EPI_ISL_1637076, EPI_ISL_1637077, EPI_ISL_1637078, EPI_ISL_1637079, EPI_ISL_1637083, EPI_ISL_1637084, EPI_ISL_1637087, EPI_ISL_1637088 | | | |
| see above | University College London Hospital | COVID-19 Genomics UK (COG-UK) Consortium | Catherine Houlihan; Dan Frampton; Dr Catherine Houlihan; Dr Daniel Frampton; Dr Judith Heaney; Dr Moira Spyer; Dr Paul Grant and Dr Eleni Nastouli; Judith Heaney; Matthew Byott; Moira Spyer and Eleni Nastouli; Stuart Kirk |
| EPI_ISL_838807, EPI_ISL_838835, EPI_ISL_838836, EPI_ISL_838837, EPI_ISL_838842, EPI_ISL_838843, EPI_ISL_838844, EPI_ISL_838845, EPI_ISL_838846, EPI_ISL_838847, EPI_ISL_838848, EPI_ISL_838849, EPI_ISL_838850, EPI_ISL_838851, EPI_ISL_838852, EPI_ISL_838853, EPI_ISL_838854, EPI_ISL_838855, EPI_ISL_838856, EPI_ISL_838857, EPI_ISL_838858, EPI_ISL_838859, EPI_ISL_838860, EPI_ISL_838861, EPI_ISL_838862, EPI_ISL_838863, EPI_ISL_838864, EPI_ISL_838865, EPI_ISL_838866, EPI_ISL_838867, EPI_ISL_838868, EPI_ISL_838869, EPI_ISL_838870, EPI_ISL_838871, EPI_ISL_838872, EPI_ISL_838873, EPI_ISL_838874, EPI_ISL_838875, EPI_ISL_838876, EPI_ISL_838877, EPI_ISL_838878, EPI_ISL_838879, EPI_ISL_838880, EPI_ISL_838881, EPI_ISL_838882, EPI_ISL_838883, EPI_ISL_838884, EPI_ISL_838885, EPI_ISL_838886, EPI_ISL_838887, EPI_ISL_838888, EPI_ISL_838889, EPI_ISL_838890, EPI_ISL_838891, EPI_ISL_838892, EPI_ISL_838893, EPI_ISL_838894, EPI_ISL_838895, EPI_ISL_838896, EPI_ISL_838897, EPI_ISL_838898, EPI_ISL_838899, EPI_ISL_838900, EPI_ISL_838901, EPI_ISL_838902, EPI_ISL_838903, EPI_ISL_838904, EPI_ISL_838905, EPI_ISL_838906, EPI_ISL_838907, EPI_ISL_838908, EPI_ISL_838909, EPI_ISL_838910, EPI_ISL_838911, EPI_ISL_838912, EPI_ISL_838913, EPI_ISL_838914, EPI_ISL_838915, EPI_ISL_838916, EPI_ISL_838917, EPI_ISL_838918, EPI_ISL_838919, EPI_ISL_838920, EPI_ISL_838921, EPI_ISL_838922, EPI_ISL_838923, EPI_ISL_838924, EPI_ISL_838925, EPI_ISL_838926, EPI_ISL_838927, EPI_ISL_838928, EPI_ISL_838929, EPI_ISL_838930, EPI_ISL_838931, EPI_ISL_838932, EPI_ISL_838933, EPI_ISL_838934, EPI_ISL_838935, EPI_ISL_838936, EPI_ISL_838950, EPI_ISL_838952, EPI_ISL_838953, EPI_ISL_838954, EPI_ISL_838955, EPI_ISL_838956, EPI_ISL_838957, EPI_ISL_839026, EPI_ISL_839027, EPI_ISL_839028, EPI_ISL_839030, EPI_ISL_839031, EPI_ISL_839032, EPI_ISL_839033, EPI_ISL_839034, EPI_ISL_839035, EPI_ISL_839036, EPI_ISL_839037, EPI_ISL_839038, EPI_ISL_839039, EPI_ISL_839040, EPI_ISL_839044, EPI_ISL_839045, EPI_ISL_839046, EPI_ISL_839047, EPI_ISL_839048, EPI_ISL_839049, EPI_ISL_839050, EPI_ISL_839051, EPI_ISL_839052, EPI_ISL_839053, EPI_ISL_839055, EPI_ISL_839056, EPI_ISL_839057, EPI_ISL_839058, EPI_ISL_839059, EPI_ISL_839061, EPI_ISL_839062, EPI_ISL_839070, EPI_ISL_839071, EPI_ISL_839072, EPI_ISL_839073, EPI_ISL_839076, EPI_ISL_839077, EPI_ISL_839078, EPI_ISL_839079, EPI_ISL_839115, EPI_ISL_839117, EPI_ISL_839118, EPI_ISL_839119, EPI_ISL_839120, EPI_ISL_839121, EPI_ISL_839122, EPI_ISL_839123, EPI_ISL_839124, EPI_ISL_839125, EPI_ISL_839135, EPI_ISL_839137, EPI_ISL_839138, EPI_ISL_839139, EPI_ISL_839140, EPI_ISL_839141, EPI_ISL_839142, EPI_ISL_839143, EPI_ISL_839144, EPI_ISL_839146, EPI_ISL_839147, EPI_ISL_839148, EPI_ISL_839149, EPI_ISL_839150, EPI_ISL_839151, EPI_ISL_839152, EPI_ISL_839153, EPI_ISL_839154, EPI_ISL_839155, EPI_ISL_839156, EPI_ISL_839157, EPI_ISL_839158, EPI_ISL_839159, EPI_ISL_839160, EPI_ISL_839161, EPI_ISL_839162, EPI_ISL_839163, EPI_ISL_839164, EPI_ISL_839165, EPI_ISL_839166, EPI_ISL_839167, EPI_ISL_839168, EPI_ISL_839169, EPI_ISL_839170, EPI_ISL_839171, EPI_ISL_839172, EPI_ISL_839173, EPI_ISL_839174, EPI_ISL_839175, EPI_ISL_839176, EPI_ISL_839177, EPI_ISL_839178, EPI_ISL_839179, EPI_ISL_839180, EPI_ISL_839181, EPI_ISL_839182, EPI_ISL_839183, EPI_ISL_839184, EPI_ISL_839185, EPI_ISL_839186, EPI_ISL_839187, EPI_ISL_839188, EPI_ISL_839189, EPI_ISL_839190, EPI_ISL_839191, EPI_ISL_839192, EPI_ISL_839193, EPI_ISL_839194, EPI_ISL_839195, EPI_ISL_839196, EPI_ISL_839197, EPI_ISL_839198, EPI_ISL_839199, EPI_ISL_839200, EPI_ISL_839201, EPI_ISL_839202, EPI_ISL_839203, EPI_ISL_839204, EPI_ISL_839205, EPI_ISL_839206, EPI_ISL_839207, EPI_ISL_839208, EPI_ISL_839210, EPI_ISL_839211, EPI_ISL_839212, EPI_ISL_839213, EPI_ISL_839214, EPI_ISL_839216, EPI_ISL_839217, EPI_ISL_839228, EPI_ISL_839229, EPI_ISL_839230, EPI_ISL_839231, EPI_ISL_839250, EPI_ISL_839252, EPI_ISL_839260, EPI_ISL_839264, EPI_ISL_839265, EPI_ISL_839273, EPI_ISL_866187, EPI_ISL_866188, EPI_ISL_866189, EPI_ISL_866190, EPI_ISL_866191, EPI_ISL_866192, EPI_ISL_866193, EPI_ISL_919962, EPI_ISL_919963, EPI_ISL_919965, EPI_ISL_919967, EPI_ISL_919968, EPI_ISL_919969, EPI_ISL_919970, EPI_ISL_919971, EPI_ISL_919972, EPI_ISL_919973, EPI_ISL_919974, EPI_ISL_919975, EPI_ISL_919976, EPI_ISL_919977, EPI_ISL_919978, EPI_ISL_919979, EPI_ISL_919980, EPI_ISL_919981, EPI_ISL_919982, EPI_ISL_919983, EPI_ISL_919984, EPI_ISL_919985, EPI_ISL_919986, EPI_ISL_919987, EPI_ISL_919988, EPI_ISL_919989, EPI_ISL_919990, EPI_ISL_919992, EPI_ISL_919993, EPI_ISL_919994, EPI_ISL_919996, EPI_ISL_919997, EPI_ISL_919998, EPI_ISL_919999, EPI_ISL_920040, EPI_ISL_920041, EPI_ISL_920046, EPI_ISL_920047, EPI_ISL_920048, EPI_ISL_920839, EPI_ISL_920840, EPI_ISL_920848, EPI_ISL_920849, EPI_ISL_920850, EPI_ISL_920856, EPI_ISL_920857, EPI_ISL_920858, EPI_ISL_920859, EPI_ISL_950048, EPI_ISL_950068, EPI_ISL_950073, EPI_ISL_950076, EPI_ISL_950078, EPI_ISL_950081, EPI_ISL_950082, EPI_ISL_950086, EPI_ISL_950089, EPI_ISL_950091, EPI_ISL_950093, EPI_ISL_950094, EPI_ISL_950095, EPI_ISL_950096, EPI_ISL_950099, EPI_ISL_950103, EPI_ISL_950106, EPI_ISL_950111, EPI_ISL_950113, EPI_ISL_950116, EPI_ISL_950118, EPI_ISL_950122, EPI_ISL_950123, EPI_ISL_950124, EPI_ISL_950127, EPI_ISL_950130, EPI_ISL_950133, EPI_ISL_950134, EPI_ISL_950135, EPI_ISL_950136, EPI_ISL_950142, EPI_ISL_950146, EPI_ISL_950147, EPI_ISL_950148, EPI_ISL_950151, EPI_ISL_950152, EPI_ISL_950153, EPI_ISL_950154, EPI_ISL_950165, EPI_ISL_997614, EPI_ISL_1050434, EPI_ISL_1050435, EPI_ISL_1050436, EPI_ISL_1050498, EPI_ISL_1104600, EPI_ISL_1104601, EPI_ISL_1104602, EPI_ISL_1104603, EPI_ISL_1104604, EPI_ISL_1104605, EPI_ISL_1104606, EPI_ISL_1104607, EPI_ISL_1104608, EPI_ISL_1104609, EPI_ISL_1104610, EPI_ISL_1247995, EPI_ISL_1247996, EPI_ISL_1248000, EPI_ISL_1248014, EPI_ISL_1248015, EPI_ISL_1248016, EPI_ISL_1248017, EPI_ISL_1248018, EPI_ISL_1248019, EPI_ISL_1248020, EPI_ISL_1248021, EPI_ISL_1248022, EPI_ISL_1248023, EPI_ISL_1248024, EPI_ISL_1248025, EPI_ISL_1248026, EPI_ISL_1248027, EPI_ISL_1248028, EPI_ISL_1248029, EPI_ISL_1248030, EPI_ISL_1248031, EPI_ISL_1248032, EPI_ISL_1248033, EPI_ISL_1248034, EPI_ISL_1248035, EPI_ISL_1248036, EPI_ISL_1248037, EPI_ISL_1248038, EPI_ISL_1248039, EPI_ISL_1248040, EPI_ISL_1248041, EPI_ISL_1248042, EPI_ISL_1248043, EPI_ISL_1248044, EPI_ISL_1248045, EPI_ISL_1248046, EPI_ISL_1248047, EPI_ISL_1248048, EPI_ISL_1248049, EPI_ISL_1248050, EPI_ISL_1248051, EPI_ISL_1248052, EPI_ISL_1248053, EPI_ISL_1248054, EPI_ISL_1248055, EPI_ISL_1248056, EPI_ISL_1248066, EPI_ISL_1248067, EPI_ISL_1308981, EPI_ISL_1308982, EPI_ISL_1308983, EPI_ISL_1308984, EPI_ISL_1308985, EPI_ISL_1308986, EPI_ISL_1308987, EPI_ISL_1308988, EPI_ISL_1308990, EPI_ISL_1308991, EPI_ISL_1308992, EPI_ISL_1308993, EPI_ISL_1308994, EPI_ISL_1308995, EPI_ISL_1308996, EPI_ISL_1308997, EPI_ISL_1308998, EPI_ISL_1308999, EPI_ISL_1309000, EPI_ISL_1309001, EPI_ISL_1309002, EPI_ISL_1309003, EPI_ISL_1309004, EPI_ISL_1309006, EPI_ISL_1309022, EPI_ISL_1309023, EPI_ISL_1309024, EPI_ISL_1309025, EPI_ISL_1309053, EPI_ISL_1309079, EPI_ISL_1309084, EPI_ISL_1309095, EPI_ISL_1309100, EPI_ISL_1309115, EPI_ISL_1309116, EPI_ISL_1309117, EPI_ISL_1309118, EPI_ISL_1309127, EPI_ISL_1386862, EPI_ISL_1386885, EPI_ISL_1386890, EPI_ISL_1386892, EPI_ISL_1386893, EPI_ISL_1386894, EPI_ISL_1386899, EPI_ISL_1386903, EPI_ISL_1386904, EPI_ISL_1386906, EPI_ISL_1386907, EPI_ISL_1386908, EPI_ISL_1386910, EPI_ISL_1386911, EPI_ISL_1474849, EPI_ISL_1474851, EPI_ISL_1474853, EPI_ISL_1637114, EPI_ISL_1637116, EPI_ISL_1637121, EPI_ISL_1637123, EPI_ISL_1637146, EPI_ISL_1730361 | | | |
| see above | University College London, Great Ormond Street Hospital for Children NHS Foundation Trust, Imperial College Healthcare NHS Trust | COVID-19 Genomics UK (COG-UK) Consortium | Alison Holmes; Charlotte Williams; Helena Tutill; Jacqueline Findlay; James Price; Judith Breuer; Julianne Brown; Kathryn Harris; Leysa Forrest; Mark Kristiansen; Paola Niola; Paola Resende Silva; Patricia Dyal; Paul Randell; Rachel Williams; Samuel Weeks; Sergi Castellano; Sunando Roy; Tony Brooks; Yasmin Panchbhaya |
| EPI_ISL_1047952, EPI_ISL_2125925 | University Hospitals Of Leicester NHS Trust and DeepSeq Nottingham | COVID-19 Genomics UK (COG-UK) Consortium | Christopher Holmes; Christopher Moore; Fei Sang; Joeseph Chappell; Johnny Debebe; Jonathan Ball; Julian Tang; Karlie Fallon; Matthew Carlisle; Matthew Loose; Nadine Holmes; Patrick McClure; Paul Bird; Thomas Helmer; Victoria Wright |

| | | | |
|---|---|---|---|
| EPI_ISL_918584, EPI_ISL_918702, EPI_ISL_918704, EPI_ISL_918705, EPI_ISL_918706, EPI_ISL_918707, EPI_ISL_918712, EPI_ISL_918714, EPI_ISL_918718, EPI_ISL_918720, EPI_ISL_918721, EPI_ISL_918722, EPI_ISL_918723, EPI_ISL_918724, EPI_ISL_918725, EPI_ISL_918726, EPI_ISL_918727, EPI_ISL_918728, EPI_ISL_918729, EPI_ISL_918730, EPI_ISL_918731, EPI_ISL_918735, EPI_ISL_918736, EPI_ISL_918737, EPI_ISL_918738, EPI_ISL_918739, EPI_ISL_918740, EPI_ISL_918741, EPI_ISL_918742, EPI_ISL_918743, EPI_ISL_918744, EPI_ISL_918745, EPI_ISL_918746, EPI_ISL_918747, EPI_ISL_918901, EPI_ISL_918902, EPI_ISL_918903, EPI_ISL_918904, EPI_ISL_918905, EPI_ISL_918906, EPI_ISL_918907, EPI_ISL_918908, EPI_ISL_918909, EPI_ISL_918910, EPI_ISL_918911, EPI_ISL_949406, EPI_ISL_996433, EPI_ISL_996434, EPI_ISL_996435, EPI_ISL_996437, EPI_ISL_996438, EPI_ISL_996454, EPI_ISL_1046999, EPI_ISL_1047064, EPI_ISL_1047066, EPI_ISL_1474497, EPI_ISL_2125451 | | | |
| see above | University of Birmingham | COVID-19 Genomics UK (COG-UK) Consortium | Alex Richter; Andrew Bosworth; Andrew Bosworth. Queen Elizabeth Hospital: Anna Casey; Andrew D Beggs PHE Heartlands Lab: Husam Osman; Charlotte Poxon; Institute of Microbiology; Joanne Stockton; Josh Quick; Kasun Wanigasooriya; Mike Kidd; Nicholas Loman. University of Birmingham Testing Laboratory: Celina M Whalley; Oliver Pickles; Radoslaw Poplawski; Samuel Nicholls; University of Birmingham: Claire McMurray; Will Rowe |
| EPI_ISL_838331, EPI_ISL_838334, EPI_ISL_838335, EPI_ISL_838336, EPI_ISL_838337, EPI_ISL_838339, EPI_ISL_838340, EPI_ISL_838341, EPI_ISL_996482, EPI_ISL_996484, EPI_ISL_996486, EPI_ISL_996487, EPI_ISL_996488, EPI_ISL_996489, EPI_ISL_996491, EPI_ISL_996500, EPI_ISL_996501, EPI_ISL_996502, EPI_ISL_996503, EPI_ISL_996504, EPI_ISL_996505, EPI_ISL_996508, EPI_ISL_996511, EPI_ISL_996527, EPI_ISL_996528, EPI_ISL_996529, EPI_ISL_996570, EPI_ISL_997323, EPI_ISL_997324, EPI_ISL_997325, EPI_ISL_1103805, EPI_ISL_1308593, EPI_ISL_1308594, EPI_ISL_1308595, EPI_ISL_1308598, EPI_ISL_1474558 | | | |
| see above | University of Exeter | COVID-19 Genomics UK (COG-UK) Consortium | Aaron Jeffries; Audrey Farbos; Ben Temperton; Jane Masoli; Joanna Warwick-Dugdale; Michelle Michelsen; Robyn Manley; Stephen Michell |
| EPI_ISL_997072, EPI_ISL_997073, EPI_ISL_997120, EPI_ISL_997161, EPI_ISL_997162, EPI_ISL_1104234, EPI_ISL_1104237, EPI_ISL_1104239, EPI_ISL_1104240, EPI_ISL_1104256, EPI_ISL_1308845, EPI_ISL_1308846, EPI_ISL_1308848, EPI_ISL_1308849 | | | |
| see above | Virology Department, Royal Infirmary of Edinburgh, NHS Lothian / School of Biological Sciences, University of Edinburgh | COVID-19 Genomics UK (COG-UK) Consortium | Colquhoun R; Cotton S; Dewar R; Hill V; Jackson B; McCrone JT; McHugh M; O'Toole Á; Rambaut A; Rooke S; Scher E; Templeton K; Yu X |
| EPI_ISL_838290, EPI_ISL_838291, EPI_ISL_838292, EPI_ISL_838293, EPI_ISL_838294, EPI_ISL_838295, EPI_ISL_838296, EPI_ISL_838297, EPI_ISL_838298, EPI_ISL_838299, EPI_ISL_838300, EPI_ISL_838301, EPI_ISL_838302, EPI_ISL_838303, EPI_ISL_838304, EPI_ISL_838305, EPI_ISL_838306, EPI_ISL_838307, EPI_ISL_838308, EPI_ISL_838309, EPI_ISL_838310, EPI_ISL_838311, EPI_ISL_838312, EPI_ISL_865074, EPI_ISL_865075, EPI_ISL_865076, EPI_ISL_865077, EPI_ISL_865078, EPI_ISL_865094, EPI_ISL_865095, EPI_ISL_865096, EPI_ISL_865097, EPI_ISL_865098, EPI_ISL_865099, EPI_ISL_865100, EPI_ISL_865101, EPI_ISL_865102, EPI_ISL_865103, EPI_ISL_865104, EPI_ISL_865131, EPI_ISL_865149, EPI_ISL_865150, EPI_ISL_865151, EPI_ISL_865152 | | | |
| see above | Virology Department, Royal Infirmary of Edinburgh, NHS Lothian / School of Biological Sciences, University of Edinburgh / Institute of Genetics and Molecular Medicine, University of Edinburgh | COVID-19 Genomics UK (COG-UK) Consortium | Balcaza C; Colquhoun R; Dewar R; Gallagher M; Hill V; Jackson B; McCrone JT; McHugh M; O'Toole Á; Rambaut A; Rooke S; Scher E; Templeton K; Williams TC; Yu X |
| EPI_ISL_816237, EPI_ISL_816244, EPI_ISL_816251, EPI_ISL_816253, EPI_ISL_816258, EPI_ISL_816260, EPI_ISL_816264, EPI_ISL_816267, EPI_ISL_816281, EPI_ISL_816284, EPI_ISL_816286, EPI_ISL_816287, EPI_ISL_816299, EPI_ISL_816303, EPI_ISL_816307, EPI_ISL_816309, EPI_ISL_816312, EPI_ISL_816315, EPI_ISL_816322, EPI_ISL_816328, EPI_ISL_816333, EPI_ISL_816334, EPI_ISL_816336, EPI_ISL_816338, EPI_ISL_816341, EPI_ISL_816358, EPI_ISL_816363, EPI_ISL_816367, EPI_ISL_816369, EPI_ISL_816371, EPI_ISL_816373, EPI_ISL_816376, EPI_ISL_816377, EPI_ISL_816378, EPI_ISL_816387, EPI_ISL_816389, EPI_ISL_816391, EPI_ISL_816394, EPI_ISL_816395, EPI_ISL_816399, EPI_ISL_816402, EPI_ISL_816408, EPI_ISL_816413, EPI_ISL_816416, EPI_ISL_816424, EPI_ISL_816433, EPI_ISL_816439, EPI_ISL_816450, EPI_ISL_816451, EPI_ISL_816452, EPI_ISL_816455, EPI_ISL_816463, EPI_ISL_816470, EPI_ISL_816471, EPI_ISL_816479, EPI_ISL_816493, EPI_ISL_816498, EPI_ISL_816502, EPI_ISL_816506, EPI_ISL_816509, EPI_ISL_816518, EPI_ISL_816519, EPI_ISL_816520, EPI_ISL_816523, EPI_ISL_816524, EPI_ISL_816526, EPI_ISL_816527, EPI_ISL_816543, EPI_ISL_816552, EPI_ISL_816559, EPI_ISL_816564, EPI_ISL_816565, EPI_ISL_816572, EPI_ISL_816573, EPI_ISL_816583, EPI_ISL_816588, EPI_ISL_816590, EPI_ISL_816591, EPI_ISL_816596, EPI_ISL_816603, EPI_ISL_816614, EPI_ISL_816618, EPI_ISL_816622, EPI_ISL_816623, EPI_ISL_816624, EPI_ISL_816636, EPI_ISL_816639, EPI_ISL_816648, EPI_ISL_816649, EPI_ISL_816652, EPI_ISL_816653, EPI_ISL_816655, EPI_ISL_842202, EPI_ISL_842205, EPI_ISL_842206, EPI_ISL_842208, EPI_ISL_842215, EPI_ISL_842216, EPI_ISL_842220, EPI_ISL_842221, EPI_ISL_842224, EPI_ISL_842226, EPI_ISL_842231, EPI_ISL_842239, EPI_ISL_842240, EPI_ISL_842241, EPI_ISL_842244, EPI_ISL_842245, EPI_ISL_842246, EPI_ISL_842251, EPI_ISL_842258, EPI_ISL_842260, EPI_ISL_842261, EPI_ISL_842262, EPI_ISL_842268, EPI_ISL_842279, EPI_ISL_842280, EPI_ISL_842284, EPI_ISL_842285, EPI_ISL_842286, EPI_ISL_842287, EPI_ISL_842288, EPI_ISL_842289, EPI_ISL_842294, EPI_ISL_842299, EPI_ISL_842300, EPI_ISL_842301, EPI_ISL_842303, EPI_ISL_842305, EPI_ISL_842311, EPI_ISL_842315, EPI_ISL_842322, EPI_ISL_842324, EPI_ISL_842329, EPI_ISL_842330, EPI_ISL_842334, EPI_ISL_842340, EPI_ISL_842345, EPI_ISL_842346, EPI_ISL_868357, EPI_ISL_868370, EPI_ISL_868390, EPI_ISL_868400, EPI_ISL_868406, EPI_ISL_868416, EPI_ISL_868431, EPI_ISL_868453, EPI_ISL_868455, EPI_ISL_868468, EPI_ISL_868470, EPI_ISL_868479, EPI_ISL_868481, EPI_ISL_868498, EPI_ISL_868499, EPI_ISL_868516, EPI_ISL_868518, EPI_ISL_868531, EPI_ISL_868543, EPI_ISL_868545, EPI_ISL_868561, EPI_ISL_868565, EPI_ISL_868572, EPI_ISL_868590, EPI_ISL_868598, EPI_ISL_868607, EPI_ISL_868609, EPI_ISL_868620, EPI_ISL_868627, EPI_ISL_868659, EPI_ISL_868664, EPI_ISL_868665, EPI_ISL_868675, EPI_ISL_868683, EPI_ISL_868699, EPI_ISL_868704, EPI_ISL_924079, EPI_ISL_924107, EPI_ISL_924132, EPI_ISL_924157, EPI_ISL_924292, EPI_ISL_924333, EPI_ISL_924342, EPI_ISL_924383, EPI_ISL_924387, EPI_ISL_924400, EPI_ISL_924412, EPI_ISL_1000747 | | | |
| see above | Virology Department, Sheffield Teaching Hospitals NHS Foundation Trust/Department of Infection, Immunity and Cardiovascular Disease, The Medical School, University of Sheffield | COVID-19 Genomics UK (COG-UK) Consortium | Adri Angyal; Alex Keeley; Benjamin Lindsey; Cariad Evans; Danielle Groves; Dave Partridge; Katie Johnson; Laura Carrilero; Luke Green; Matthew Parker; Matthew Wyles; Mehmet Yavuz; Mohammad Raza; Nikki Smith; Paul Parsons; Rachel Tucker; Rebecca Brown; Thushan de Silva |
| EPI_ISL_822816, EPI_ISL_822817, EPI_ISL_822818, EPI_ISL_822819, EPI_ISL_822820, EPI_ISL_822821, EPI_ISL_822822, EPI_ISL_822823, EPI_ISL_822824, EPI_ISL_822825, EPI_ISL_822826, EPI_ISL_822827, EPI_ISL_822828, EPI_ISL_822829, EPI_ISL_822830, EPI_ISL_822831, EPI_ISL_822832, EPI_ISL_822833, EPI_ISL_822834, EPI_ISL_822835, EPI_ISL_822836 | | | |
| see above | Wales Specialist Virology Centre Sequencing lab: Pathogen Genomics Unit | COVID-19 Genomics UK (COG-UK) Consortium | Alec Birchley; Alexander Adams; Amy Gaskin; Angela Marchbank; Bree Gatica-Wilcox; Catherine Moore; Jason Coombes; Joanne Watkins; Joel Southgate; Johnathan Evans; Laura Gifford; Lauren Gilbert; Lee Graham; Malorie Perry; Matthew Bull; Nicole Pacchiarini; Sally Corden; Sara Kumziene-Summerhayes; Sara Rey; Sarah Taylor; Simon Cottrell; Sophie Jones; Tom Connor |
| EPI_ISL_840820, EPI_ISL_840822, EPI_ISL_840824, EPI_ISL_840826, EPI_ISL_840827, EPI_ISL_840840, EPI_ISL_840842, EPI_ISL_840843, EPI_ISL_840845, EPI_ISL_840846, EPI_ISL_840847, EPI_ISL_840848, EPI_ISL_840849, EPI_ISL_840852, EPI_ISL_840853, EPI_ISL_840855, EPI_ISL_840856, EPI_ISL_840857, EPI_ISL_840858, EPI_ISL_840859, EPI_ISL_840861, EPI_ISL_840862, EPI_ISL_840863, EPI_ISL_840864, EPI_ISL_840865, EPI_ISL_840866, EPI_ISL_840867, EPI_ISL_840869, EPI_ISL_840870, EPI_ISL_840871, EPI_ISL_840872, EPI_ISL_840874, EPI_ISL_840875, EPI_ISL_840876, EPI_ISL_840877, EPI_ISL_840878, EPI_ISL_840880, EPI_ISL_840881, EPI_ISL_840882, EPI_ISL_840883, EPI_ISL_840884, EPI_ISL_840907, EPI_ISL_840908, EPI_ISL_840922, EPI_ISL_840925, EPI_ISL_840926, EPI_ISL_840928, EPI_ISL_840929, EPI_ISL_840930, EPI_ISL_840932, EPI_ISL_840933, EPI_ISL_840934, EPI_ISL_840935, EPI_ISL_840936, EPI_ISL_840942, EPI_ISL_840944, EPI_ISL_840945, EPI_ISL_840947, EPI_ISL_840951, EPI_ISL_840952, EPI_ISL_840954, EPI_ISL_840955, EPI_ISL_840956, EPI_ISL_840957, EPI_ISL_840958, EPI_ISL_840959, EPI_ISL_840960, EPI_ISL_840961, EPI_ISL_840962, EPI_ISL_840963, EPI_ISL_840964, EPI_ISL_840965, EPI_ISL_840966, EPI_ISL_840967, EPI_ISL_840968, EPI_ISL_840969, EPI_ISL_840970, EPI_ISL_840971, EPI_ISL_840974, EPI_ISL_840975, EPI_ISL_840976, EPI_ISL_840977, EPI_ISL_840978, EPI_ISL_840979, EPI_ISL_840981, EPI_ISL_840982, EPI_ISL_840983, EPI_ISL_840984, EPI_ISL_840985, EPI_ISL_840989, EPI_ISL_840990, EPI_ISL_840991, EPI_ISL_840992, EPI_ISL_840993, EPI_ISL_840994, EPI_ISL_840995, EPI_ISL_840996, EPI_ISL_840997, EPI_ISL_840998, EPI_ISL_841000, EPI_ISL_841002, EPI_ISL_841004, EPI_ISL_841008, EPI_ISL_841010, EPI_ISL_841012, EPI_ISL_841013, EPI_ISL_841014, EPI_ISL_841015, EPI_ISL_841016, EPI_ISL_841017, EPI_ISL_841018, EPI_ISL_841019, EPI_ISL_841020, EPI_ISL_841021, EPI_ISL_841027, EPI_ISL_841029, EPI_ISL_841030, EPI_ISL_841031, EPI_ISL_841032, EPI_ISL_841036, EPI_ISL_841037, EPI_ISL_841038, EPI_ISL_841040, EPI_ISL_841041, EPI_ISL_841042, EPI_ISL_841043, EPI_ISL_841044, EPI_ISL_841045, EPI_ISL_841046, EPI_ISL_841047, EPI_ISL_841049, EPI_ISL_841050, EPI_ISL_841051, EPI_ISL_841052, EPI_ISL_841053, EPI_ISL_841054, EPI_ISL_841055, EPI_ISL_841056, EPI_ISL_841059, EPI_ISL_841060, EPI_ISL_841062, EPI_ISL_841063, EPI_ISL_841064, EPI_ISL_841067, EPI_ISL_841068, EPI_ISL_841069, EPI_ISL_841070, EPI_ISL_841071, EPI_ISL_841072, EPI_ISL_841073, EPI_ISL_841074, EPI_ISL_841075, EPI_ISL_841076, EPI_ISL_841077, EPI_ISL_841078, EPI_ISL_841079, EPI_ISL_841080, EPI_ISL_841081, EPI_ISL_841086, EPI_ISL_841098, EPI_ISL_841108, EPI_ISL_841116, EPI_ISL_841120, EPI_ISL_841121, EPI_ISL_841122, EPI_ISL_841124, EPI_ISL_841125, EPI_ISL_841126, EPI_ISL_841127, EPI_ISL_841128, EPI_ISL_841129, EPI_ISL_841130, EPI_ISL_841131, EPI_ISL_841133, EPI_ISL_841134, EPI_ISL_841137, EPI_ISL_841138, EPI_ISL_841139, EPI_ISL_841140, EPI_ISL_841141, EPI_ISL_841142, EPI_ISL_841143, EPI_ISL_841144, EPI_ISL_841145, EPI_ISL_841146, EPI_ISL_841147, EPI_ISL_841149, EPI_ISL_841151, EPI_ISL_841152, EPI_ISL_841153, EPI_ISL_841154, EPI_ISL_841155, EPI_ISL_841156, EPI_ISL_841157, EPI_ISL_841158, EPI_ISL_841159, EPI_ISL_841161, EPI_ISL_841162, EPI_ISL_841163, EPI_ISL_841164, EPI_ISL_841165, EPI_ISL_841166, EPI_ISL_841167, EPI_ISL_841168, EPI_ISL_841169, EPI_ISL_841170, EPI_ISL_841172, EPI_ISL_841174, EPI_ISL_841175, EPI_ISL_841176, EPI_ISL_841178, EPI_ISL_841179, EPI_ISL_841182, EPI_ISL_841183, EPI_ISL_841184, EPI_ISL_841185, EPI_ISL_841186, EPI_ISL_841191, EPI_ISL_841192, EPI_ISL_841193, EPI_ISL_841194, EPI_ISL_841195, EPI_ISL_841196, EPI_ISL_841197, EPI_ISL_841200, EPI_ISL_841201, EPI_ISL_841202, EPI_ISL_841203, EPI_ISL_841204, EPI_ISL_841205, EPI_ISL_841206, EPI_ISL_841207, EPI_ISL_841208, EPI_ISL_841209, EPI_ISL_841210, EPI_ISL_841213, EPI_ISL_841214, EPI_ISL_841216, EPI_ISL_841217, EPI_ISL_841220, EPI_ISL_841221, EPI_ISL_841222, EPI_ISL_841223, EPI_ISL_841224, EPI_ISL_841226, EPI_ISL_841227, EPI_ISL_841229, EPI_ISL_841230, EPI_ISL_841231, EPI_ISL_841232, EPI_ISL_841233, EPI_ISL_841234, EPI_ISL_841235, EPI_ISL_841236, EPI_ISL_841237, EPI_ISL_841238, EPI_ISL_841240, EPI_ISL_841242, EPI_ISL_841244, EPI_ISL_841245, EPI_ISL_841246, EPI_ISL_841247, EPI_ISL_841248, EPI_ISL_841250, EPI_ISL_841251, EPI_ISL_841252, EPI_ISL_841253, EPI_ISL_841255, EPI_ISL_841256, EPI_ISL_841257, EPI_ISL_841258, EPI_ISL_841259, EPI_ISL_841261, EPI_ISL_841262, EPI_ISL_841263, EPI_ISL_841264, EPI_ISL_841265, EPI_ISL_841266, EPI_ISL_841267, EPI_ISL_841268, EPI_ISL_841269, EPI_ISL_841270, EPI_ISL_841271, EPI_ISL_841273, EPI_ISL_841275, EPI_ISL_841276, EPI_ISL_841280, EPI_ISL_841281, EPI_ISL_841288, EPI_ISL_841292, EPI_ISL_841293, EPI_ISL_841296, EPI_ISL_841297, EPI_ISL_841299, EPI_ISL_841300, EPI_ISL_841306, EPI_ISL_841307, EPI_ISL_841308, EPI_ISL_841309, EPI_ISL_841310, EPI_ISL_841311, EPI_ISL_841312, EPI_ISL_841313, EPI_ISL_841314, EPI_ISL_841315, EPI_ISL_841316, EPI_ISL_1546806, EPI_ISL_1833128, EPI_ISL_1833129, EPI_ISL_1833131, EPI_ISL_1833133, EPI_ISL_1833134, EPI_ISL_1833136, EPI_ISL_1833138, EPI_ISL_1833140, EPI_ISL_1833141, EPI_ISL_1833142, EPI_ISL_1833143, EPI_ISL_1833145, EPI_ISL_1833146, EPI_ISL_1833147, EPI_ISL_1833149, EPI_ISL_1833150, EPI_ISL_1833151, EPI_ISL_1833152 | | | |
| see above | Wales Specialist Virology Centre Sequencing lab: Pathogen Genomics Unit | Public Health Wales Microbiology Cardiff Wales Specialist Virology Centre | Alec Birchley; Alexander Adams; Amy Gaskin; Angela Marchbank; Bree Gatica-Wilcox; Catherine Moore; Jason Coombes; Joanne Watkins; Joel Southgate; Johnathan Evans; Laura Gifford; Lauren Gilbert; Lee Graham; Malorie Perry; Matthew Bull; Nicole Pacchiarini; Sally Corden; Sara Kumziene-Summerhayes; Sara Rey; Sarah Taylor; Simon Cottrell; Sophie Jones; Tom Connor |
| EPI_ISL_864984, EPI_ISL_864985, EPI_ISL_864986, EPI_ISL_864987, EPI_ISL_864988, EPI_ISL_864989, EPI_ISL_864990, EPI_ISL_864991, EPI_ISL_864994, EPI_ISL_864995, EPI_ISL_864996, EPI_ISL_864997, EPI_ISL_864998, EPI_ISL_864999, EPI_ISL_865013, EPI_ISL_865014, EPI_ISL_865041, EPI_ISL_865042, EPI_ISL_865043, EPI_ISL_865044, EPI_ISL_865045, EPI_ISL_865046, EPI_ISL_865047, EPI_ISL_865048, EPI_ISL_865049, EPI_ISL_865051, EPI_ISL_865054 | | | |
| see above | West of Scotland Specialist Virology Centre, NHSGGC / MRC-University of Glasgow Centre for Virus Research | COVID-19 Genomics UK (COG-UK) Consortium | Alasdair MacLean; Alice Broos; Ana da Silva Filipe; Antonia Ho; Daniel Mair; David L Robertson; Emma Thomson; Guy Mollett; James Shepherd; Jenna Nichols; Joseph Hughes; Kathy Li; Kathy Smollett; Kyriaki Nomikou; Lily Tong; Matthew Holden; Natasha Johnson; Rachel Blacow; Richard Orton; Rory Gunson; Sarah McDonald; Sharif Shaaban; Sreenu Vattipally; Stephen Carmichael |

We gratefully acknowledge the following Authors from the Originating laboratories responsible for obtaining the specimens, as well as the Submitting laboratories where the genome data were generated and shared via GISAID, on which this research is based.

All Submitters of data may be contacted directly via www.gisaid.org

Authors are sorted alphabetically.

| Accession ID | Originating Laboratory | Submitting Laboratory | Authors |
|---|---|---|---|
| EPI_ISL_919841, EPI_ISL_919852, EPI_ISL_949750, EPI_ISL_949757, EPI_ISL_949758, EPI_ISL_949759, EPI_ISL_949760, EPI_ISL_949762, EPI_ISL_949763, EPI_ISL_949764, EPI_ISL_949765, EPI_ISL_949766, EPI_ISL_949768, EPI_ISL_949771, EPI_ISL_949772, EPI_ISL_949773, EPI_ISL_949774, EPI_ISL_949776, EPI_ISL_949779, EPI_ISL_949781, EPI_ISL_949782, EPI_ISL_997629, EPI_ISL_997675, EPI_ISL_997676, EPI_ISL_1048056, EPI_ISL_1048096, EPI_ISL_1730307, EPI_ISL_1832541, EPI_ISL_2126068, EPI_ISL_2126070, EPI_ISL_2126072, EPI_ISL_2126074, EPI_ISL_2126076, EPI_ISL_2126078, EPI_ISL_2126080, EPI_ISL_2126082, EPI_ISL_2126084 | | | |
| see above | Barts Health NHS Trust | COVID-19 Genomics UK (COG-UK) Consortium | BROAD; Beatrix; CUTINO-MOGUEL; Claire; David; Dola; HARRINGTON; KELE; KULASEGARAN-SHYLINI; Maria-Teresa; OWOYEMI; Raghavendran |
| EPI_ISL_924724, EPI_ISL_924725, EPI_ISL_924727, EPI_ISL_924728, EPI_ISL_924729, EPI_ISL_924731, EPI_ISL_924733, EPI_ISL_924736, EPI_ISL_924740, EPI_ISL_924742, EPI_ISL_924744, EPI_ISL_924746, EPI_ISL_924748, EPI_ISL_924749, EPI_ISL_924750, EPI_ISL_924752, EPI_ISL_924753, EPI_ISL_924754, EPI_ISL_924755, EPI_ISL_924756, EPI_ISL_924764, EPI_ISL_924769, EPI_ISL_924770, EPI_ISL_924774, EPI_ISL_924794, EPI_ISL_924795, EPI_ISL_924797, EPI_ISL_924798, EPI_ISL_924800, EPI_ISL_924805, EPI_ISL_924807, EPI_ISL_924808, EPI_ISL_924809, EPI_ISL_924816, EPI_ISL_924818, EPI_ISL_924819, EPI_ISL_924827, EPI_ISL_924828, EPI_ISL_924832, EPI_ISL_924836, EPI_ISL_924838, EPI_ISL_924839, EPI_ISL_924846, EPI_ISL_924848, EPI_ISL_924849, EPI_ISL_924850, EPI_ISL_924851, EPI_ISL_924855, EPI_ISL_924860, EPI_ISL_924863, EPI_ISL_924864, EPI_ISL_924865, EPI_ISL_924867, EPI_ISL_924871, EPI_ISL_924877, EPI_ISL_924879, EPI_ISL_924880, EPI_ISL_924883, EPI_ISL_924886, EPI_ISL_924887, EPI_ISL_924890, EPI_ISL_924891, EPI_ISL_924892, EPI_ISL_924895, EPI_ISL_924897, EPI_ISL_924899, EPI_ISL_924901, EPI_ISL_924902, EPI_ISL_924904, EPI_ISL_924905, EPI_ISL_924907, EPI_ISL_924908, EPI_ISL_924909, EPI_ISL_924910, EPI_ISL_924911, EPI_ISL_924912, EPI_ISL_924913, EPI_ISL_924914, EPI_ISL_924915, EPI_ISL_924916, EPI_ISL_924917, EPI_ISL_924918, EPI_ISL_924919, EPI_ISL_924921, EPI_ISL_924922, EPI_ISL_924923, EPI_ISL_924924, EPI_ISL_924925, EPI_ISL_924926, EPI_ISL_924927, EPI_ISL_924928, EPI_ISL_924929, EPI_ISL_924930, EPI_ISL_924931, EPI_ISL_924933, EPI_ISL_924935, EPI_ISL_924936, EPI_ISL_924937, EPI_ISL_924938, EPI_ISL_924939, EPI_ISL_924940, EPI_ISL_924941, EPI_ISL_924942, EPI_ISL_924943, EPI_ISL_924944, EPI_ISL_924945, EPI_ISL_924946, EPI_ISL_924947, EPI_ISL_924950, EPI_ISL_924975, EPI_ISL_924980, EPI_ISL_953177, EPI_ISL_953180, EPI_ISL_953181, EPI_ISL_953182, EPI_ISL_953185, EPI_ISL_953187, EPI_ISL_953193, EPI_ISL_953199, EPI_ISL_953205, EPI_ISL_953207, EPI_ISL_953216, EPI_ISL_953217, EPI_ISL_953219, EPI_ISL_953222, EPI_ISL_953226, EPI_ISL_953230, EPI_ISL_953231, EPI_ISL_953232, EPI_ISL_953233, EPI_ISL_953236, EPI_ISL_953238, EPI_ISL_953239, EPI_ISL_1000881, EPI_ISL_1000883, EPI_ISL_1000885, EPI_ISL_1000886, EPI_ISL_1000887, EPI_ISL_1000892, EPI_ISL_1000893, EPI_ISL_1000898, EPI_ISL_1000900, EPI_ISL_1000903, EPI_ISL_1000905, EPI_ISL_1000907, EPI_ISL_1000908, EPI_ISL_1054779, EPI_ISL_1054824, EPI_ISL_1180108, EPI_ISL_1180126 | | | |
| see above | Bioinformatics and Biostatistics Lab, Advanced Sequencing Facility | COVID-19 Genomics UK (COG-UK) Consortium | Aengus Stewart; Chelsea Sawyer; Harshil Patel; Jerome Nicod; Laura Cubitt; Margaret Crawford |
| EPI_ISL_841921, EPI_ISL_868141, EPI_ISL_868142, EPI_ISL_868143, EPI_ISL_868144, EPI_ISL_868145, EPI_ISL_868146, EPI_ISL_868147, EPI_ISL_868148, EPI_ISL_868149, EPI_ISL_868150, EPI_ISL_868151, EPI_ISL_868152, EPI_ISL_868153, EPI_ISL_868154, EPI_ISL_868161, EPI_ISL_868162, EPI_ISL_868163, EPI_ISL_868164, EPI_ISL_868165, EPI_ISL_868166, EPI_ISL_868167, EPI_ISL_868168, EPI_ISL_868169, EPI_ISL_868170, EPI_ISL_868171, EPI_ISL_868172, EPI_ISL_868173, EPI_ISL_868174, EPI_ISL_868177, EPI_ISL_868178, EPI_ISL_868179, EPI_ISL_868181, EPI_ISL_868182, EPI_ISL_868183, EPI_ISL_868184, EPI_ISL_868185, EPI_ISL_868186, EPI_ISL_868187, EPI_ISL_868188, EPI_ISL_868189, EPI_ISL_868190, EPI_ISL_868191, EPI_ISL_868192, EPI_ISL_868193, EPI_ISL_868194, EPI_ISL_868196, EPI_ISL_868197, EPI_ISL_868198, EPI_ISL_868199, EPI_ISL_868200, EPI_ISL_868201, EPI_ISL_868202, EPI_ISL_868203, EPI_ISL_868204, EPI_ISL_868217, EPI_ISL_868218, EPI_ISL_868323, EPI_ISL_868329, EPI_ISL_868330, EPI_ISL_868331, EPI_ISL_868332, EPI_ISL_868333, EPI_ISL_868334, EPI_ISL_868335, EPI_ISL_868336, EPI_ISL_868337, EPI_ISL_868338, EPI_ISL_868339, EPI_ISL_868340, EPI_ISL_868341, EPI_ISL_868342, EPI_ISL_868353, EPI_ISL_868354, EPI_ISL_868355, EPI_ISL_923260, EPI_ISL_923262, EPI_ISL_923275, EPI_ISL_923279, EPI_ISL_923284, EPI_ISL_923293, EPI_ISL_923296, EPI_ISL_923300, EPI_ISL_923301, EPI_ISL_923305, EPI_ISL_923306, EPI_ISL_923372, EPI_ISL_923373, EPI_ISL_923374, EPI_ISL_923375, EPI_ISL_923376, EPI_ISL_923377, EPI_ISL_923378, EPI_ISL_923380, EPI_ISL_923382, EPI_ISL_923383, EPI_ISL_923384, EPI_ISL_923385, EPI_ISL_923386, EPI_ISL_923387, EPI_ISL_923388, EPI_ISL_923389, EPI_ISL_923390, EPI_ISL_923391, EPI_ISL_923392, EPI_ISL_923393, EPI_ISL_923394, EPI_ISL_923395, EPI_ISL_923396, EPI_ISL_923397, EPI_ISL_923398, EPI_ISL_923399, EPI_ISL_923400, EPI_ISL_923401, EPI_ISL_923402, EPI_ISL_923403, EPI_ISL_923404, EPI_ISL_923405, EPI_ISL_923406, EPI_ISL_923407, EPI_ISL_923408, EPI_ISL_923409, EPI_ISL_923410, EPI_ISL_923411, EPI_ISL_923412, EPI_ISL_923413, EPI_ISL_923430, EPI_ISL_923539, EPI_ISL_923588, EPI_ISL_923593, EPI_ISL_923605, EPI_ISL_923611, EPI_ISL_923613, EPI_ISL_923671, EPI_ISL_923678, EPI_ISL_923679, EPI_ISL_923684, EPI_ISL_952381, EPI_ISL_952384, EPI_ISL_952388, EPI_ISL_952396, EPI_ISL_952400, EPI_ISL_952401, EPI_ISL_952402, EPI_ISL_952406, EPI_ISL_952413, EPI_ISL_952415, EPI_ISL_952419, EPI_ISL_952421, EPI_ISL_952425, EPI_ISL_952426, EPI_ISL_952434, EPI_ISL_952559, EPI_ISL_952560, EPI_ISL_952561, EPI_ISL_952562, EPI_ISL_952563, EPI_ISL_952564, EPI_ISL_952565, EPI_ISL_952566, EPI_ISL_952567, EPI_ISL_952568, EPI_ISL_952777, EPI_ISL_952781, EPI_ISL_952782, EPI_ISL_952784, EPI_ISL_952786, EPI_ISL_952787, EPI_ISL_952788, EPI_ISL_952790, EPI_ISL_952791, EPI_ISL_952792, EPI_ISL_952796, EPI_ISL_952797, EPI_ISL_952800, EPI_ISL_952801, EPI_ISL_952802, EPI_ISL_952803, EPI_ISL_952805, EPI_ISL_952806, EPI_ISL_952807, EPI_ISL_952808, EPI_ISL_952809, EPI_ISL_952810, EPI_ISL_952811, EPI_ISL_952812, EPI_ISL_952816, EPI_ISL_952819, EPI_ISL_1000244, EPI_ISL_1000245, EPI_ISL_1000246, EPI_ISL_1000258, EPI_ISL_1000270, EPI_ISL_1000272, EPI_ISL_1107587, EPI_ISL_1107631, EPI_ISL_1179796, EPI_ISL_1179822, EPI_ISL_1179823, EPI_ISL_1179830, EPI_ISL_1179838, EPI_ISL_1249959, EPI_ISL_1476665, EPI_ISL_1547230, EPI_ISL_1547305, EPI_ISL_1730864, EPI_ISL_1730866, EPI_ISL_1730867, EPI_ISL_1979800, EPI_ISL_1979802, EPI_ISL_1979803, EPI_ISL_1979804, EPI_ISL_1979806, EPI_ISL_1979808, EPI_ISL_1979809, EPI_ISL_1979811, EPI_ISL_1979812, EPI_ISL_1979814, EPI_ISL_1979816, EPI_ISL_1979818, EPI_ISL_1979820, EPI_ISL_1979821, EPI_ISL_1979823, EPI_ISL_1979825, EPI_ISL_1979826, EPI_ISL_1979828, EPI_ISL_1979830, EPI_ISL_1979831, EPI_ISL_1979833, EPI_ISL_1979834, EPI_ISL_1979836, EPI_ISL_1979838, EPI_ISL_1979840, EPI_ISL_1979841, EPI_ISL_1979843, EPI_ISL_1979844, EPI_ISL_1979910, EPI_ISL_1979912, EPI_ISL_1979914, EPI_ISL_1979915, EPI_ISL_1979917, EPI_ISL_1979919, EPI_ISL_1979921, EPI_ISL_1979922, EPI_ISL_1979924, EPI_ISL_1979925, EPI_ISL_1979927, EPI_ISL_1979928, EPI_ISL_1979930, EPI_ISL_1979932, EPI_ISL_1979933, EPI_ISL_1979935, EPI_ISL_1979937, EPI_ISL_1979938, EPI_ISL_1979940, EPI_ISL_1979942, EPI_ISL_1979943, EPI_ISL_1979945, EPI_ISL_1979947, EPI_ISL_2128018, EPI_ISL_2128020, EPI_ISL_2128023, EPI_ISL_2128025, EPI_ISL_2128027, EPI_ISL_2128040, EPI_ISL_2128184 | | | |
| see above | Centre for Enzyme Innovation, University of Portsmouth / Translational Research Laboratory, Portsmouth Hospitals NHS Trust | COVID-19 Genomics UK (COG-UK) Consortium | Allyson Lloyd; Angela Beckett; Anoop Chauhan; Christopher Fearn; Ethan Butcher; Garry Scarlett; Kate Cook; Katie Loveson; Kelly Bicknell; Robert Impey; Salman Goudarzi; Samuel Robson; Sarah Wyllie; Scott Elliott; Sharon Glaysher; Yann Bourgeois |
| EPI_ISL_837957, EPI_ISL_837959, EPI_ISL_837961, EPI_ISL_837973, EPI_ISL_837974, EPI_ISL_837975, EPI_ISL_837976, EPI_ISL_837977, EPI_ISL_837978, EPI_ISL_837979, EPI_ISL_837980, EPI_ISL_837981, EPI_ISL_837982, EPI_ISL_837983, EPI_ISL_837984, EPI_ISL_837985, EPI_ISL_837986, EPI_ISL_837987, EPI_ISL_837988, EPI_ISL_837990, EPI_ISL_837991, EPI_ISL_837993, EPI_ISL_837997, EPI_ISL_838000, EPI_ISL_838001, EPI_ISL_838003, EPI_ISL_838004, EPI_ISL_838005, EPI_ISL_838006, EPI_ISL_838007, EPI_ISL_838008, EPI_ISL_838009, EPI_ISL_838010, EPI_ISL_838011, EPI_ISL_838012, EPI_ISL_838013, EPI_ISL_838014, EPI_ISL_838016, EPI_ISL_838019, EPI_ISL_838020, EPI_ISL_838023, EPI_ISL_838024, EPI_ISL_838026, EPI_ISL_838027, EPI_ISL_838028, EPI_ISL_838029, EPI_ISL_838030, EPI_ISL_838031, EPI_ISL_838032, EPI_ISL_838033, EPI_ISL_838036, EPI_ISL_838039, EPI_ISL_838040, EPI_ISL_838041, EPI_ISL_838042, EPI_ISL_838043, EPI_ISL_838044, EPI_ISL_838045, EPI_ISL_838046, EPI_ISL_838047, EPI_ISL_838049, EPI_ISL_838050, EPI_ISL_838051, EPI_ISL_838052, EPI_ISL_838053, EPI_ISL_838054, EPI_ISL_864810, EPI_ISL_864814, EPI_ISL_864893, EPI_ISL_864926, EPI_ISL_919011, EPI_ISL_919012, EPI_ISL_919017, EPI_ISL_919029, EPI_ISL_919037, EPI_ISL_919039, EPI_ISL_919041, EPI_ISL_919043, EPI_ISL_919044, EPI_ISL_919045, EPI_ISL_919046, EPI_ISL_919047, EPI_ISL_919048, EPI_ISL_919049, EPI_ISL_919050, EPI_ISL_919051, EPI_ISL_919052, EPI_ISL_919056, EPI_ISL_919071, EPI_ISL_919073, EPI_ISL_919077, EPI_ISL_919081, EPI_ISL_919089, EPI_ISL_919104, EPI_ISL_919105, EPI_ISL_920894, EPI_ISL_920895, EPI_ISL_920896, EPI_ISL_920897, EPI_ISL_920898, EPI_ISL_920899, EPI_ISL_920900, EPI_ISL_923842, EPI_ISL_1247594 | | | |
| see above | Department of Pathology, University of Cambridge | COVID-19 Genomics UK (COG-UK) Consortium | Aminu S. Jahun; Grant Hall; Ian Goodfellow; Iliana Georgana; Malte Pinckert; Martin D. Curran; Myra Hosmillo; Rhys Izu; Rhys Izuagbe; Surendra Parmar; William L. Hamilton; Yasmin Chaudhry |
| EPI_ISL_834150, EPI_ISL_835041, EPI_ISL_846200, EPI_ISL_846201, EPI_ISL_846202, EPI_ISL_846203, EPI_ISL_846204, EPI_ISL_846205, EPI_ISL_846206, EPI_ISL_846207, EPI_ISL_846208, EPI_ISL_846209, EPI_ISL_846210, EPI_ISL_846211, EPI_ISL_846212, EPI_ISL_846213, EPI_ISL_846214, EPI_ISL_846215, EPI_ISL_846216, EPI_ISL_846217, EPI_ISL_846218, EPI_ISL_846219, EPI_ISL_846220, EPI_ISL_846221, EPI_ISL_846222, EPI_ISL_846223, EPI_ISL_846224, EPI_ISL_846225, EPI_ISL_846226, EPI_ISL_846227, EPI_ISL_846228, EPI_ISL_846229, EPI_ISL_846230, EPI_ISL_846231, EPI_ISL_846232, EPI_ISL_846233, EPI_ISL_846234, EPI_ISL_846235, EPI_ISL_846236, EPI_ISL_846237, EPI_ISL_846238, EPI_ISL_846239, EPI_ISL_846240, EPI_ISL_846241, EPI_ISL_846242, EPI_ISL_846243, EPI_ISL_846244, EPI_ISL_846245, EPI_ISL_846246, EPI_ISL_846247, EPI_ISL_846248, EPI_ISL_846249, EPI_ISL_846250, EPI_ISL_846251, EPI_ISL_846252, EPI_ISL_846253, EPI_ISL_846254, EPI_ISL_846255, EPI_ISL_846256, EPI_ISL_846257, EPI_ISL_846258, EPI_ISL_846259, EPI_ISL_846260, EPI_ISL_846261, EPI_ISL_846262, EPI_ISL_846263, EPI_ISL_846264, EPI_ISL_846265, EPI_ISL_846266, EPI_ISL_846267, EPI_ISL_846268, EPI_ISL_846269, EPI_ISL_846270, EPI_ISL_846271, EPI_ISL_846272, EPI_ISL_846273, EPI_ISL_846274, EPI_ISL_846275, EPI_ISL_846276, EPI_ISL_846277, EPI_ISL_846278, EPI_ISL_846279, EPI_ISL_846280, EPI_ISL_846281, EPI_ISL_846282, EPI_ISL_846283, EPI_ISL_846284, EPI_ISL_846285, EPI_ISL_846286, EPI_ISL_846287, EPI_ISL_846288, EPI_ISL_846289, EPI_ISL_846290, EPI_ISL_846291, EPI_ISL_846292, EPI_ISL_846293, EPI_ISL_846294, EPI_ISL_846295, EPI_ISL_846296, EPI_ISL_846297, EPI_ISL_846298, EPI_ISL_846299, EPI_ISL_846300, EPI_ISL_846301, EPI_ISL_846302, EPI_ISL_846303, EPI_ISL_846304, EPI_ISL_846305, EPI_ISL_846306, EPI_ISL_846307, EPI_ISL_846308, EPI_ISL_846309, EPI_ISL_846310, EPI_ISL_846311, EPI_ISL_846312, EPI_ISL_846313, EPI_ISL_846314, EPI_ISL_846315, EPI_ISL_846316, EPI_ISL_846317, EPI_ISL_846318, EPI_ISL_846320, EPI_ISL_846321, EPI_ISL_846322, EPI_ISL_846323, EPI_ISL_846324, EPI_ISL_846325, EPI_ISL_846326, EPI_ISL_846327, EPI_ISL_846328, EPI_ISL_846329, EPI_ISL_846330, EPI_ISL_846331, EPI_ISL_846332, EPI_ISL_846333, EPI_ISL_846334, EPI_ISL_846335, EPI_ISL_846336, EPI_ISL_846337, EPI_ISL_846338, EPI_ISL_846339, EPI_ISL_846340, EPI_ISL_846341, EPI_ISL_846342, EPI_ISL_846343, EPI_ISL_846344, EPI_ISL_846345, EPI_ISL_846346, EPI_ISL_846347, EPI_ISL_846348, EPI_ISL_846349, EPI_ISL_846350, EPI_ISL_846351, EPI_ISL_846352, EPI_ISL_846353, EPI_ISL_846354, EPI_ISL_846355, EPI_ISL_846356, EPI_ISL_846357, EPI_ISL_846358, EPI_ISL_846359, EPI_ISL_846360, EPI_ISL_846361, EPI_ISL_846362, EPI_ISL_846363, EPI_ISL_846364, EPI_ISL_846365, EPI_ISL_846366, EPI_ISL_846367, EPI_ISL_846368, EPI_ISL_846369, EPI_ISL_846370, EPI_ISL_846371, EPI_ISL_846372, EPI_ISL_846373, EPI_ISL_846374, EPI_ISL_846375, EPI_ISL_846376, EPI_ISL_846377, EPI_ISL_846378, EPI_ISL_846379, EPI_ISL_846380, EPI_ISL_846381, EPI_ISL_846382, EPI_ISL_846383, EPI_ISL_846384, EPI_ISL_846385, EPI_ISL_846386, EPI_ISL_846387, EPI_ISL_846388, EPI_ISL_846389, EPI_ISL_846390, EPI_ISL_846391, EPI_ISL_846392, EPI_ISL_846393, EPI_ISL_846394, EPI_ISL_846395, EPI_ISL_846396, EPI_ISL_846397, EPI_ISL_846398, EPI_ISL_846399, EPI_ISL_846401, EPI_ISL_846402, EPI_ISL_846403, EPI_ISL_846404, EPI_ISL_846405, EPI_ISL_846406, EPI_ISL_846407, EPI_ISL_846408, EPI_ISL_846409, EPI_ISL_846410, EPI_ISL_846411, EPI_ISL_846412, EPI_ISL_846413, EPI_ISL_846414, EPI_ISL_846415, EPI_ISL_846416, EPI_ISL_846417, EPI_ISL_846418, EPI_ISL_846419, EPI_ISL_846420, EPI_ISL_846421, EPI_ISL_846422, EPI_ISL_846423, EPI_ISL_846424, EPI_ISL_846425, EPI_ISL_846426, EPI_ISL_846427, EPI_ISL_846428, EPI_ISL_846429, EPI_ISL_846430, EPI_ISL_846431, EPI_ISL_846432, EPI_ISL_846433, EPI_ISL_846434, EPI_ISL_846435, EPI_ISL_846436, EPI_ISL_846437, EPI_ISL_846438, EPI_ISL_846439, EPI_ISL_846440, EPI_ISL_846441, EPI_ISL_846442, EPI_ISL_846443, EPI_ISL_846444, EPI_ISL_846445, EPI_ISL_846446, EPI_ISL_846447, EPI_ISL_846448, EPI_ISL_846449, EPI_ISL_846450, EPI_ISL_846451, EPI_ISL_846452, EPI_ISL_846453, EPI_ISL_846454, EPI_ISL_846455, EPI_ISL_846456, EPI_ISL_846457, EPI_ISL_846458, EPI_ISL_846459, EPI_ISL_846460, EPI_ISL_846461, EPI_ISL_846462, EPI_ISL_846463, EPI_ISL_846464, EPI_ISL_846465, EPI_ISL_846466, EPI_ISL_846467, EPI_ISL_846468, EPI_ISL_846469, EPI_ISL_846470, EPI_ISL_846471, EPI_ISL_846472, EPI_ISL_846473, EPI_ISL_846474, EPI_ISL_846475, EPI_ISL_846476, EPI_ISL_846477, EPI_ISL_846478, EPI_ISL_846479, EPI_ISL_846480, EPI_ISL_846481, EPI_ISL_846482, EPI_ISL_846483, EPI_ISL_846484, EPI_ISL_846485, EPI_ISL_846486, EPI_ISL_846487, EPI_ISL_846488, EPI_ISL_846489, EPI_ISL_846490, EPI_ISL_846491, EPI_ISL_846492, EPI_ISL_846493, EPI_ISL_846494, EPI_ISL_846495, EPI_ISL_846496, EPI_ISL_846497, EPI_ISL_846498, EPI_ISL_846499, EPI_ISL_846500, EPI_ISL_846501, EPI_ISL_846502, EPI_ISL_846503, EPI_ISL_846504, EPI_ISL_846505, EPI_ISL_846506, EPI_ISL_846507, EPI_ISL_846508, EPI_ISL_846512, EPI_ISL_851673, EPI_ISL_851675, EPI_ISL_851677, EPI_ISL_851678, EPI_ISL_851679, EPI_ISL_851680, EPI_ISL_851682, EPI_ISL_851683, EPI_ISL_851684, EPI_ISL_851688, EPI_ISL_851691, EPI_ISL_851692, EPI_ISL_851694, EPI_ISL_851695, EPI_ISL_851696, EPI_ISL_851697, EPI_ISL_851698, EPI_ISL_851699, EPI_ISL_851700, EPI_ISL_851702, EPI_ISL_851703, EPI_ISL_851704, EPI_ISL_851705, EPI_ISL_851706, EPI_ISL_851707, EPI_ISL_851708, EPI_ISL_851709, EPI_ISL_851710, EPI_ISL_851711, EPI_ISL_851712, EPI_ISL_851713, EPI_ISL_851715, EPI_ISL_851716, EPI_ISL_851717, EPI_ISL_851718, EPI_ISL_851719, EPI_ISL_851721, EPI_ISL_851722, EPI_ISL_851723, EPI_ISL_851724, EPI_ISL_851726, EPI_ISL_851728, EPI_ISL_851730, EPI_ISL_851731, EPI_ISL_851732, EPI_ISL_851733, EPI_ISL_851735, EPI_ISL_851736, EPI_ISL_851737, EPI_ISL_851739, EPI_ISL_851740, EPI_ISL_851741, EPI_ISL_851742, EPI_ISL_851744, EPI_ISL_851745, EPI_ISL_851746, EPI_ISL_851748, EPI_ISL_851751, EPI_ISL_851752, EPI_ISL_851753, EPI_ISL_851755, EPI_ISL_851756, EPI_ISL_851758, EPI_ISL_851759, EPI_ISL_851761, EPI_ISL_851762, EPI_ISL_851764, EPI_ISL_851765, EPI_ISL_851766, EPI_ISL_851768, EPI_ISL_851769, EPI_ISL_851771, EPI_ISL_851773, EPI_ISL_851775, EPI_ISL_851776, EPI_ISL_851777, EPI_ISL_851778, EPI_ISL_851779, EPI_ISL_851782, EPI_ISL_851783, EPI_ISL_851784, EPI_ISL_851786, EPI_ISL_851787, EPI_ISL_851788, EPI_ISL_851789, EPI_ISL_851790, EPI_ISL_851791, EPI_ISL_851793, EPI_ISL_851795, EPI_ISL_851800, EPI_ISL_851801, EPI_ISL_851802, EPI_ISL_851804, EPI_ISL_851806, EPI_ISL_851808, EPI_ISL_851809, EPI_ISL_851810, EPI_ISL_851811, EPI_ISL_851813, EPI_ISL_851815, EPI_ISL_851816, EPI_ISL_851817, EPI_ISL_851818, EPI_ISL_851819, EPI_ISL_851820, EPI_ISL_851821, EPI_ISL_851822, EPI_ISL_851823, EPI_ISL_851824, EPI_ISL_851825, EPI_ISL_851826, EPI_ISL_851827, EPI_ISL_851828, EPI_ISL_851830, EPI_ISL_851831, EPI_ISL_851832, EPI_ISL_851833, EPI_ISL_851834, EPI_ISL_851835, EPI_ISL_851836, EPI_ISL_851838, EPI_ISL_851839, EPI_ISL_851841, EPI_ISL_851842, EPI_ISL_851843, EPI_ISL_851844, EPI_ISL_851845, EPI_ISL_851848, EPI_ISL_851849, EPI_ISL_851850, EPI_ISL_851852, EPI_ISL_851853, EPI_ISL_851854, EPI_ISL_851855, EPI_ISL_851858, EPI_ISL_851860, EPI_ISL_851861, EPI_ISL_851862, EPI_ISL_851867, EPI_ISL_851868, EPI_ISL_851869, EPI_ISL_851872, EPI_ISL_851873, EPI_ISL_851874, EPI_ISL_851875, EPI_ISL_851878, EPI_ISL_851879, EPI_ISL_851880, EPI_ISL_851881, EPI_ISL_851882, EPI_ISL_851883, EPI_ISL_851884, EPI_ISL_851886, EPI_ISL_851887, EPI_ISL_851889, EPI_ISL_851890, EPI_ISL_851893, EPI_ISL_851894, EPI_ISL_851896, EPI_ISL_851897, EPI_ISL_851899, EPI_ISL_851900, EPI_ISL_851902, EPI_ISL_851903, EPI_ISL_851904, EPI_ISL_851907, EPI_ISL_851908, EPI_ISL_851909, EPI_ISL_851910, EPI_ISL_851911, EPI_ISL_851912, EPI_ISL_851913, EPI_ISL_851914, EPI_ISL_851915, EPI_ISL_851917, EPI_ISL_851918, EPI_ISL_851919, EPI_ISL_851921, EPI_ISL_851923, EPI_ISL_851924, EPI_ISL_851925, EPI_ISL_851926, EPI_ISL_851927, EPI_ISL_851928, EPI_ISL_851930, EPI_ISL_851931, EPI_ISL_851932, EPI_ISL_851933, EPI_ISL_851935, EPI_ISL_851937, EPI_ISL_851938, EPI_ISL_851939, EPI_ISL_851941, EPI_ISL_851942, EPI_ISL_851943, EPI_ISL_851944, EPI_ISL_851945, EPI_ISL_851946, EPI_ISL_851947, EPI_ISL_851948, EPI_ISL_851949, EPI_ISL_851951, EPI_ISL_851952, EPI_ISL_851953, EPI_ISL_852025, EPI_ISL_852091, EPI_ISL_852107, EPI_ISL_852183, EPI_ISL_852214, EPI_ISL_857874, EPI_ISL_857875, EPI_ISL_857877, EPI_ISL_857880, EPI_ISL_857882, EPI_ISL_857883, EPI_ISL_857884, EPI_ISL_857886, EPI_ISL_857960, EPI_ISL_857961, EPI_ISL_857962, EPI_ISL_857963, EPI_ISL_857964, EPI_ISL_857965, EPI_ISL_857966, EPI_ISL_857967, EPI_ISL_857968, EPI_ISL_857969, EPI_ISL_857970, EPI_ISL_857971, EPI_ISL_857972, EPI_ISL_857973, EPI_ISL_857974, EPI_ISL_857975, EPI_ISL_857976, EPI_ISL_857977, EPI_ISL_857978, EPI_ISL_857979, EPI_ISL_857980, EPI_ISL_857981, EPI_ISL_857982, EPI_ISL_857983, EPI_ISL_857984, EPI_ISL_857985, EPI_ISL_857986, EPI_ISL_857987, EPI_ISL_857988, EPI_ISL_857989, EPI_ISL_857990, EPI_ISL_857991, EPI_ISL_857992, EPI_ISL_857993, EPI_ISL_857994, EPI_ISL_857995, EPI_ISL_857996, EPI_ISL_857997, EPI_ISL_857998, EPI_ISL_857999, EPI_ISL_858000, EPI_ISL_858001, EPI_ISL_858002, EPI_ISL_858003, EPI_ISL_858004, EPI_ISL_858005, EPI_ISL_858006, EPI_ISL_858007, EPI_ISL_858008, EPI_ISL_858009, EPI_ISL_858010, EPI_ISL_862886, EPI_ISL_862887, EPI_ISL_862888, EPI_ISL_862889, EPI_ISL_862890, EPI_ISL_862891, EPI_ISL_862892, EPI_ISL_862893, EPI_ISL_862894, EPI_ISL_862895, EPI_ISL_862896, EPI_ISL_862897, EPI_ISL_862898, EPI_ISL_862899, EPI_ISL_862900, EPI_ISL_862901, EPI_ISL_862902, EPI_ISL_862903, EPI_ISL_862904, EPI_ISL_862905, EPI_ISL_862906, EPI_ISL_862907, EPI_ISL_862908, EPI_ISL_862909, EPI_ISL_862910, EPI_ISL_862911, EPI_ISL_862912, EPI_ISL_862913, EPI_ISL_862914, EPI_ISL_862915, EPI_ISL_862916, EPI_ISL_862917, EPI_ISL_862918, EPI_ISL_862919, EPI_ISL_862920, EPI_ISL_862921, EPI_ISL_862922, EPI_ISL_862923, EPI_ISL_862924, EPI_ISL_862925, EPI_ISL_862926, EPI_ISL_862927, EPI_ISL_862928, EPI_ISL_862929, EPI_ISL_862930, EPI_ISL_862931, EPI_ISL_862932, EPI_ISL_862933, EPI_ISL_862934, EPI_ISL_862935, EPI_ISL_862936, EPI_ISL_862937, EPI_ISL_862938, EPI_ISL_862939, EPI_ISL_862940, EPI_ISL_862941, EPI_ISL_862942, EPI_ISL_862943, EPI_ISL_862944, EPI_ISL_862945, EPI_ISL_862946, EPI_ISL_862947, EPI_ISL_862948, EPI_ISL_862949, EPI_ISL_862950, EPI_ISL_862951, EPI_ISL_862952, EPI_ISL_862953, EPI_ISL_862954, EPI_ISL_862955, EPI_ISL_862956, EPI_ISL_862957, EPI_ISL_862958, EPI_ISL_862959, EPI_ISL_862960, EPI_ISL_862961, EPI_ISL_862962, EPI_ISL_862963, EPI_ISL_862964, EPI_ISL_862965, EPI_ISL_862966, EPI_ISL_862967, EPI_ISL_862968, EPI_ISL_862969, EPI_ISL_862970, EPI_ISL_862971, EPI_ISL_862972, EPI_ISL_862973, EPI_ISL_862974, EPI_ISL_862975, EPI_ISL_862976, EPI_ISL_862977, EPI_ISL_862978, EPI_ISL_862979, EPI_ISL_862980, EPI_ISL_862981, EPI_ISL_862982, EPI_ISL_862983, EPI_ISL_862984, EPI_ISL_862985, EPI_ISL_862986, EPI_ISL_862987, EPI_ISL_862988, EPI_ISL_862989, EPI_ISL_862990, EPI_ISL_862991, EPI_ISL_862992, EPI_ISL_862993, EPI_ISL_862994, EPI_ISL_862995, EPI_ISL_862996, EPI_ISL_862997, EPI_ISL_862998, EPI_ISL_862999, EPI_ISL_863000, EPI_ISL_863001, EPI_ISL_863002, EPI_ISL_863003, EPI_ISL_863004, EPI_ISL_863005, EPI_ISL_863006, EPI_ISL_863007, EPI_ISL_863008, EPI_ISL_863009, EPI_ISL_863010, EPI_ISL_863011, EPI_ISL_863012, EPI_ISL_863013, EPI_ISL_863014, EPI_ISL_863015, EPI_ISL_863016, EPI_ISL_863017, EPI_ISL_863018, EPI_ISL_863019, EPI_ISL_863020, EPI_ISL_863021, EPI_ISL_863022, EPI_ISL_863023, EPI_ISL_863024, EPI_ISL_863025, EPI_ISL_863026, EPI_ISL_863027, EPI_ISL_863028, EPI_ISL_863029, EPI_ISL_863030, EPI_ISL_863031, EPI_ISL_863032, EPI_ISL_863033, EPI_ISL_863034, EPI_ISL_863035, EPI_ISL_863036, EPI_ISL_863037, EPI_ISL_863038, EPI_ISL_863039, EPI_ISL_863040, EPI_ISL_863041, EPI_ISL_863042, EPI_ISL_863043, EPI_ISL_863044, EPI_ISL_863045, EPI_ISL_863046, EPI_ISL_863047, EPI_ISL_863048, EPI_ISL_863049, EPI_ISL_863050, EPI_ISL_863051, EPI_ISL_863052, EPI_ISL_863053, EPI_ISL_863054, EPI_ISL_863055, EPI_ISL_863056, EPI_ISL_863057, EPI_ISL_863058, EPI_ISL_863059, EPI_ISL_863060, EPI_ISL_863061, EPI_ISL_863062, EPI_ISL_863063, EPI_ISL_863064, EPI_ISL_863065, EPI_ISL_863066, EPI_ISL_863067, EPI_ISL_863068, EPI_ISL_863069, EPI_ISL_863070, EPI_ISL_863071, EPI_ISL_863072, EPI_ISL_863073, EPI_ISL_863074, EPI_ISL_863075, EPI_ISL_863076, EPI_ISL_863077, EPI_ISL_863078, EPI_ISL_863079, EPI_ISL_863080, EPI_ISL_863081, EPI_ISL_863082, EPI_ISL_863083, EPI_ISL_863084, EPI_ISL_863085, EPI_ISL_863086, EPI_ISL_863087, EPI_ISL_863088, EPI_ISL_863089, EPI_ISL_863090, EPI_ISL_863091, EPI_ISL_863092, EPI_ISL_863093, EPI_ISL_863094, EPI_ISL_863095, EPI_ISL_863096, EPI_ISL_863097, EPI_ISL_863098, EPI_ISL_863099, EPI_ISL_863100, EPI_ISL_863101, EPI_ISL_863102, EPI_ISL_863103, EPI_ISL_863104, EPI_ISL_863105, EPI_ISL_863106, EPI_ISL_863107, EPI_ISL_863108, EPI_ISL_863109, EPI_ISL_863110, EPI_ISL_863111, EPI_ISL_863112, EPI_ISL_863113, EPI_ISL_863114, EPI_ISL_863115, EPI_ISL_863116, EPI_ISL_863117, EPI_ISL_863118, EPI_ISL_863119, EPI_ISL_863120, EPI_ISL_863121, EPI_ISL_863122, EPI_ISL_863123, EPI_ISL_863124, EPI_ISL_863125, EPI_ISL_863126, EPI_ISL_863127, EPI_ISL_863128, EPI_ISL_863129, EPI_ISL_863130, EPI_ISL_863131, EPI_ISL_863132, EPI_ISL_863133, EPI_ISL_863134, EPI_ISL_863135, EPI_ISL_863136, EPI_ISL_863137, EPI_ISL_863138, EPI_ISL_863139, EPI_ISL_863140, EPI_ISL_863141, EPI_ISL_863142, EPI_ISL_863143, EPI_ISL_863144, EPI_ISL_863145, EPI_ISL_863146, EPI_ISL_863147, EPI_ISL_863148, EPI_ISL_863149, EPI_ISL_863150, EPI_ISL_863151, EPI_ISL_863152, EPI_ISL_863153, EPI_ISL_863154, EPI_ISL_863155, EPI_ISL_863156, EPI_ISL_863157, EPI_ISL_863158, EPI_ISL_863159, EPI_ISL_863160, EPI_ISL_863161, EPI_ISL_863162, EPI_ISL_863163, EPI_ISL_863164, EPI_ISL_863165, EPI_ISL_863166, EPI_ISL_863167, EPI_ISL_863168, EPI_ISL_863169, EPI_ISL_863170, EPI_ISL_863171, EPI_ISL_863172, EPI_ISL_863173, EPI_ISL_863174, EPI_ISL_863175, EPI_ISL_863176, EPI_ISL_863177, EPI_ISL_863178, EPI_ISL_863179, EPI_ISL_863180, EPI_ISL_863181, EPI_ISL_863182, EPI_ISL_863183, EPI_ISL_863184, EPI_ISL_863185, EPI_ISL_863186, EPI_ISL_863187, EPI_ISL_863188, EPI_ISL_863189, EPI_ISL_863190, EPI_ISL_863191, EPI_ISL_863192, EPI_ISL_863193, EPI_ISL_863194, EPI_ISL_863195, EPI_ISL_863196, EPI_ISL_863197, EPI_ISL_863198, EPI_ISL_863199, EPI_ISL_863200, EPI_ISL_863201, EPI_ISL_863202, EPI_ISL_863203, EPI_ISL_863204, EPI_ISL_863205, EPI_ISL_863206, EPI_ISL_863207, EPI_ISL_863208, EPI_ISL_863209, EPI_ISL_863210, EPI_ISL_863211, EPI_ISL_863212, EPI_ISL_863213, EPI_ISL_863214, EPI_ISL_863215, EPI_ISL_863216, EPI_ISL_863217, EPI_ISL_863218, EPI_ISL_863219, EPI_ISL_863220, EPI_ISL_863221, EPI_ISL_863222, EPI_ISL_863223, EPI_ISL_863224, EPI_ISL_863225, EPI_ISL_863226, EPI_ISL_863227, EPI_ISL_863228, EPI_ISL_863229, EPI_ISL_863230, EPI_ISL_863231, EPI_ISL_863232, EPI_ISL_863233, EPI_ISL_863234, EPI_ISL_863235, EPI_ISL_863236, EPI_ISL_863237, EPI_ISL_863238, EPI_ISL_863239, EPI_ISL_863240, EPI_ISL_863241, EPI_ISL_863242, EPI_ISL_863243, EPI_ISL_863244, EPI_ISL_863334, EPI_ISL_863396, EPI_ISL_863401, EPI_ISL_863403, EPI_ISL_863424, EPI_ISL_863432, EPI_ISL_863467, EPI_ISL_863514, EPI_ISL_863529, EPI_ISL_863873, EPI_ISL_863946, EPI_ISL_864004, EPI_ISL_864064, EPI_ISL_864094, EPI_ISL_864110, EPI_ISL_864142, EPI_ISL_864145, EPI_ISL_864178, EPI_ISL_864226, EPI_ISL_864267, EPI_ISL_864308, EPI_ISL_873961, EPI_ISL_873962, EPI_ISL_873963, EPI_ISL_873966, EPI_ISL_873967, EPI_ISL_873968, EPI_ISL_873969, EPI_ISL_873970, EPI_ISL_873971, EPI_ISL_873972, EPI_ISL_873973, EPI_ISL_873975, EPI_ISL_873977, EPI_ISL_873978, EPI_ISL_873979, EPI_ISL_873980, EPI_ISL_873981, EPI_ISL_873982, EPI_ISL_873983, EPI_ISL_873985, EPI_ISL_873986, EPI_ISL_873988, EPI_ISL_873989, EPI_ISL_873991, EPI_ISL_873992, EPI_ISL_873993, EPI_ISL_873994, EPI_ISL_873995, EPI_ISL_873996, EPI_ISL_873997, EPI_ISL_873998, EPI_ISL_874000, EPI_ISL_874001, EPI_ISL_874002, EPI_ISL_874003, EPI_ISL_874005, EPI_ISL_874006, EPI_ISL_874008, EPI_ISL_874011, EPI_ISL_874013, EPI_ISL_874015, EPI_ISL_874016, EPI_ISL_874018, EPI_ISL_874019, EPI_ISL_874020, EPI_ISL_874023, EPI_ISL_874024, EPI_ISL_874025, EPI_ISL_874026, EPI_ISL_874027, EPI_ISL_874028, EPI_ISL_874029, EPI_ISL_874030, EPI_ISL_874031, EPI_ISL_874033, EPI_ISL_874034, EPI_ISL_874035, EPI_ISL_874036, EPI_ISL_874040, EPI_ISL_874041, EPI_ISL_874042, EPI_ISL_874043, EPI_ISL_874044, EPI_ISL_874045, EPI_ISL_874046, EPI_ISL_874048, EPI_ISL_874049, EPI_ISL_874050, EPI_ISL_874051, EPI_ISL_874052, EPI_ISL_874053, EPI_ISL_874054, EPI_ISL_874056, EPI_ISL_874057, EPI_ISL_874058, EPI_ISL_874059, EPI_ISL_874060, EPI_ISL_874063, EPI_ISL_874065, EPI_ISL_874066, EPI_ISL_874067, EPI_ISL_874068, EPI_ISL_874069, EPI_ISL_874070, EPI_ISL_874072, EPI_ISL_874073, EPI_ISL_874074, EPI_ISL_874075, EPI_ISL_874077, EPI_ISL_874078, EPI_ISL_874079, EPI_ISL_874080, EPI_ISL_874081, EPI_ISL_874082, EPI_ISL_874083, EPI_ISL_874084, EPI_ISL_874086, EPI_ISL_874087, EPI_ISL_874088, EPI_ISL_874089, EPI_ISL_874091, EPI_ISL_874093, EPI_ISL_874094, EPI_ISL_874095, EPI_ISL_874098, EPI_ISL_874099, EPI_ISL_874100, EPI_ISL_874101, EPI_ISL_874102, EPI_ISL_874103, EPI_ISL_874104, EPI_ISL_874105, EPI_ISL_874106, EPI_ISL_874107, EPI_ISL_874108, EPI_ISL_874109, EPI_ISL_874110, EPI_ISL_874111, EPI_ISL_874112, EPI_ISL_874113, EPI_ISL_874114, EPI_ISL_874116, EPI_ISL_874117, EPI_ISL_874119, EPI_ISL_874120, EPI_ISL_874121, EPI_ISL_874122, EPI_ISL_874123, EPI_ISL_874124, EPI_ISL_874125, EPI_ISL_874126, EPI_ISL_874127, EPI_ISL_874128, EPI_ISL_874129, EPI_ISL_874131, EPI_ISL_874132, EPI_ISL_874133, EPI_ISL_874134, EPI_ISL_874135, EPI_ISL_874136, EPI_ISL_874137, EPI_ISL_874138, EPI_ISL_874139, EPI_ISL_874140, EPI_ISL_874142, EPI_ISL_874144, EPI_ISL_874145, EPI_ISL_874146, EPI_ISL_874147, EPI_ISL_874149, EPI_ISL_874150, EPI_ISL_874151, EPI_ISL_874152, EPI_ISL_874155, EPI_ISL_874156, EPI_ISL_874158, EPI_ISL_874159, EPI_ISL_874161, EPI_ISL_874162, EPI_ISL_874166, EPI_ISL_874167, EPI_ISL_874168, EPI_ISL_874171, EPI_ISL_874172, EPI_ISL_874173, EPI_ISL_874174, EPI_ISL_874175, EPI_ISL_874176, EPI_ISL_874177, EPI_ISL_874178, EPI_ISL_874179, EPI_ISL_874181, EPI_ISL_874182, EPI_ISL_874183, EPI_ISL_874184, EPI_ISL_874185, EPI_ISL_874186, EPI_ISL_874187, EPI_ISL_874189, EPI_ISL_874190, EPI_ISL_874192, EPI_ISL_874194, EPI_ISL_874196, EPI_ISL_874200, EPI_ISL_874201, EPI_ISL_874203, EPI_ISL_874205, EPI_ISL_874206, EPI_ISL_874208, EPI_ISL_874210, EPI_ISL_874212, EPI_ISL_874213, EPI_ISL_874214, EPI_ISL_874216, EPI_ISL_874217, EPI_ISL_874218, EPI_ISL_874219, EPI_ISL_874220, EPI_ISL_874221, EPI_ISL_874222, EPI_ISL_874223, EPI_ISL_874224, EPI_ISL_874225, EPI_ISL_874226, EPI_ISL_874227, EPI_ISL_874228, EPI_ISL_874229, EPI_ISL_874230, EPI_ISL_874231, EPI_ISL_874232, EPI_ISL_874233, EPI_ISL_874234, EPI_ISL_874235, EPI_ISL_874236, EPI_ISL_874237, EPI_ISL_874239, EPI_ISL_874241, EPI_ISL_874242, EPI_ISL_874243, EPI_ISL_874244, EPI_ISL_874245, EPI_ISL_874247, EPI_ISL_874249, EPI_ISL_874250, EPI_ISL_874251, EPI_ISL_874252, EPI_ISL_874253, EPI_ISL_874255, EPI_ISL_874256, EPI_ISL_874257, EPI_ISL_874259, EPI_ISL_874260, EPI_ISL_874261, EPI_ISL_874262, EPI_ISL_874264, EPI_ISL_874265, EPI_ISL_874267, EPI_ISL_874269, EPI_ISL_874270, EPI_ISL_874271, EPI_ISL_874272, EPI_ISL_874273, EPI_ISL_874274, EPI_ISL_874276, EPI_ISL_874277, EPI_ISL_874279, EPI_ISL_874280, EPI_ISL_874282, EPI_ISL_874284, EPI_ISL_874285, EPI_ISL_874286, EPI_ISL_874287, EPI_ISL_874288, EPI_ISL_874289, EPI_ISL_874291, EPI_ISL_874292, EPI_ISL_874294, EPI_ISL_874295, EPI_ISL_874296, EPI_ISL_874298, EPI_ISL_874299, EPI_ISL_874300, EPI_ISL_874302, EPI_ISL_874303, EPI_ISL_874304, EPI_ISL_874305, EPI_ISL_874306, EPI_ISL_874307, EPI_ISL_874308, EPI_ISL_874309, EPI_ISL_874310, EPI_ISL_874315, EPI_ISL_874379, EPI_ISL_874401, EPI_ISL_874404, EPI_ISL_874443, EPI_ISL_874448, EPI_ISL_874469, EPI_ISL_874482, EPI_ISL_874499, EPI_ISL_874502, EPI_ISL_874523, EPI_ISL_874527, EPI_ISL_874528, EPI_ISL_874530, EPI_ISL_874543, EPI_ISL_874558, EPI_ISL_874624, EPI_ISL_874759, EPI_ISL_874816, EPI_ISL_874853, EPI_ISL_874938, EPI_ISL_874956, EPI_ISL_874966, EPI_ISL_877913, EPI_ISL_877955, EPI_ISL_878844, EPI_ISL_879274, EPI_ISL_881277, EPI_ISL_881281, EPI_ISL_881283, EPI_ISL_881289, EPI_ISL_881290, EPI_ISL_881291, EPI_ISL_881293, EPI_ISL_881298, EPI_ISL_881306, EPI_ISL_881307, EPI_ISL_881309, EPI_ISL_881310, EPI_ISL_881315, EPI_ISL_881318, EPI_ISL_881326, EPI_ISL_881341, EPI_ISL_881359, EPI_ISL_881386, EPI_ISL_881388, EPI_ISL_881414, EPI_ISL_881429, EPI_ISL_881455, EPI_ISL_881461, EPI_ISL_881487, EPI_ISL_881494, EPI_ISL_881495, EPI_ISL_881509, EPI_ISL_881518, EPI_ISL_881521, EPI_ISL_881522, EPI_ISL_881531, EPI_ISL_881540, EPI_ISL_881543, EPI_ISL_881546, EPI_ISL_881550, EPI_ISL_881567, EPI_ISL_881570, EPI_ISL_881571, EPI_ISL_881576, EPI_ISL_881580, EPI_ISL_881582, EPI_ISL_881586, EPI_ISL_881588, EPI_ISL_881598, EPI_ISL_881600, EPI_ISL_881607, EPI_ISL_881612, EPI_ISL_881623, EPI_ISL_881624, EPI_ISL_881625, EPI_ISL_881626, EPI_ISL_881627, EPI_ISL_881628, EPI_ISL_881629, EPI_ISL_881630, EPI_ISL_881631, EPI_ISL_881632, EPI_ISL_881633, EPI_ISL_881634, EPI_ISL_881635, EPI_ISL_881636, EPI_ISL_881637, EPI_ISL_881638, EPI_ISL_881639, EPI_ISL_881640, EPI_ISL_881641, EPI_ISL_881642, EPI_ISL_881643, EPI_ISL_881644, EPI_ISL_881645, EPI_ISL_881646, EPI_ISL_881647, EPI_ISL_881648, EPI_ISL_881649, EPI_ISL_881650, EPI_ISL_881651, EPI_ISL_881652, EPI_ISL_881653, EPI_ISL_881654, EPI_ISL_881655, EPI_ISL_881656, EPI_ISL_881657, EPI_ISL_881658, EPI_ISL_881659, EPI_ISL_881660, EPI_ISL_881661, EPI_ISL_881662, EPI_ISL_881663, EPI_ISL_881664, EPI_ISL_881665, EPI_ISL_881666, EPI_ISL_881667, EPI_ISL_881669, EPI_ISL_881670, EPI_ISL_881671, EPI_ISL_881672, EPI_ISL_881673, EPI_ISL_881674, EPI_ISL_881675, EPI_ISL_881676, EPI_ISL_881677, EPI_ISL_881678, EPI_ISL_881679, EPI_ISL_881680, EPI_ISL_881681, EPI_ISL_881682, EPI_ISL_881683, EPI_ISL_881684, EPI_ISL_881685, EPI_ISL_881686, EPI_ISL_881687, EPI_ISL_881688, EPI_ISL_881689, EPI_ISL_881690, EPI_ISL_881691, EPI_ISL_881692, EPI_ISL_881693, EPI_ISL_881694, EPI_ISL_881695, EPI_ISL_881696, EPI_ISL_881697, EPI_ISL_881698, EPI_ISL_881699, EPI_ISL_881700, EPI_ISL_881701, EPI_ISL_881702, EPI_ISL_881703, EPI_ISL_881704, EPI_ISL_881705, EPI_ISL_881706, EPI_ISL_881707, EPI_ISL_881708, EPI_ISL_881709, EPI_ISL_881710, EPI_ISL_881711, EPI_ISL_881712, EPI_ISL_881713, EPI_ISL_881714, EPI_ISL_881715, EPI_ISL_881716, EPI_ISL_881717, EPI_ISL_881718, EPI_ISL_881719, EPI_ISL_881720, EPI_ISL_881721, EPI_ISL_881722, EPI_ISL_881723, EPI_ISL_881724, EPI_ISL_881725, EPI_ISL_881726, EPI_ISL_881727, EPI_ISL_881728, EPI_ISL_881729, EPI_ISL_881730, EPI_ISL_881731, EPI_ISL_881732, EPI_ISL_881733, EPI_ISL_881734, EPI_ISL_881735, EPI_ISL_881736, EPI_ISL_881737, EPI_ISL_881738, EPI_ISL_881739, EPI_ISL_881740, EPI_ISL_881741, EPI_ISL_881742, EPI_ISL_881743, EPI_ISL_881744, EPI_ISL_881745, EPI_ISL_881746, EPI_ISL_881747, EPI_ISL_881748, EPI_ISL_881749, EPI_ISL_881750, EPI_ISL_881751, EPI_ISL_881752, EPI_ISL_881753, EPI_ISL_881754, EPI_ISL_881755, EPI_ISL_881756, EPI_ISL_881757, EPI_ISL_881758, EPI_ISL_881759, EPI_ISL_881760, EPI_ISL_881761, EPI_ISL_881762, EPI_ISL_881763, EPI_ISL_881764, EPI_ISL_881765, EPI_ISL_881766, EPI_ISL_881767, EPI_ISL_881768, EPI_ISL_881769, EPI_ISL_881770, EPI_ISL_881771, EPI_ISL_881772, EPI_ISL_881773, EPI_ISL_881774, EPI_ISL_881775, EPI_ISL_881776, EPI_ISL_881777, EPI_ISL_881778, EPI_ISL_881779, EPI_ISL_881780, EPI_ISL_881781, EPI_ISL_881782, EPI_ISL_881783, EPI_ISL_881784, | | | |

EPI_ISL_881785, EPI_ISL_881786, EPI_ISL_881787, EPI_ISL_881788, EPI_ISL_881789, EPI_ISL_881790, EPI_ISL_881791, EPI_ISL_881792, EPI_ISL_881793, EPI_ISL_881794, EPI_ISL_881795, EPI_ISL_881796, EPI_ISL_881797, EPI_ISL_881798, EPI_ISL_881799, EPI_ISL_881800, EPI_ISL_881801, EPI_ISL_881802, EPI_ISL_881803, EPI_ISL_881804, EPI_ISL_881805, EPI_ISL_881806, EPI_ISL_881807, EPI_ISL_881808,
EPI_ISL_881809, EPI_ISL_881810, EPI_ISL_881811, EPI_ISL_881812, EPI_ISL_881813, EPI_ISL_881814, EPI_ISL_881815, EPI_ISL_881816, EPI_ISL_881817, EPI_ISL_881818, EPI_ISL_881819, EPI_ISL_881820, EPI_ISL_881821, EPI_ISL_881822, EPI_ISL_881823, EPI_ISL_881824, EPI_ISL_881825, EPI_ISL_881826, EPI_ISL_881827, EPI_ISL_881828, EPI_ISL_881829, EPI_ISL_881830, EPI_ISL_881831, EPI_ISL_881832,
EPI_ISL_881833, EPI_ISL_881834, EPI_ISL_881835, EPI_ISL_881836, EPI_ISL_881837, EPI_ISL_881838, EPI_ISL_881839, EPI_ISL_881840, EPI_ISL_881841, EPI_ISL_881842, EPI_ISL_881843, EPI_ISL_881844, EPI_ISL_881845, EPI_ISL_881846, EPI_ISL_881847, EPI_ISL_881848, EPI_ISL_881849, EPI_ISL_881850, EPI_ISL_881851, EPI_ISL_881852, EPI_ISL_881853, EPI_ISL_881854, EPI_ISL_881855, EPI_ISL_881856,
EPI_ISL_881857, EPI_ISL_881858, EPI_ISL_881859, EPI_ISL_881860, EPI_ISL_881861, EPI_ISL_881862, EPI_ISL_881863, EPI_ISL_881864, EPI_ISL_881865, EPI_ISL_881866, EPI_ISL_881867, EPI_ISL_881868, EPI_ISL_881869, EPI_ISL_881870, EPI_ISL_881871, EPI_ISL_881872, EPI_ISL_881873, EPI_ISL_881874, EPI_ISL_881875, EPI_ISL_881876, EPI_ISL_881877, EPI_ISL_881878, EPI_ISL_881879, EPI_ISL_881880,
EPI_ISL_881881, EPI_ISL_881882, EPI_ISL_881883, EPI_ISL_881884, EPI_ISL_881885, EPI_ISL_881886, EPI_ISL_881887, EPI_ISL_881888, EPI_ISL_881889, EPI_ISL_881890, EPI_ISL_881891, EPI_ISL_881892, EPI_ISL_881893, EPI_ISL_881894, EPI_ISL_881895, EPI_ISL_881896, EPI_ISL_881897, EPI_ISL_881898, EPI_ISL_881899, EPI_ISL_881900, EPI_ISL_881901, EPI_ISL_881902, EPI_ISL_881903, EPI_ISL_881904,
EPI_ISL_881905, EPI_ISL_881906, EPI_ISL_881907, EPI_ISL_881908, EPI_ISL_881909, EPI_ISL_881910, EPI_ISL_881911, EPI_ISL_881912, EPI_ISL_881913, EPI_ISL_881914, EPI_ISL_881915, EPI_ISL_881916, EPI_ISL_881917, EPI_ISL_881918, EPI_ISL_881919, EPI_ISL_881920, EPI_ISL_881921, EPI_ISL_881922, EPI_ISL_881923, EPI_ISL_881924, EPI_ISL_881925, EPI_ISL_881926, EPI_ISL_881927, EPI_ISL_881928,
EPI_ISL_881929, EPI_ISL_881930, EPI_ISL_881931, EPI_ISL_881932, EPI_ISL_881933, EPI_ISL_881934, EPI_ISL_881935, EPI_ISL_881936, EPI_ISL_881937, EPI_ISL_881938, EPI_ISL_881939, EPI_ISL_881940, EPI_ISL_881941, EPI_ISL_881942, EPI_ISL_881943, EPI_ISL_881944, EPI_ISL_881945, EPI_ISL_881946, EPI_ISL_881947, EPI_ISL_881948, EPI_ISL_881949, EPI_ISL_881950, EPI_ISL_881951, EPI_ISL_881952,
EPI_ISL_881953, EPI_ISL_881954, EPI_ISL_881955, EPI_ISL_881956, EPI_ISL_881957, EPI_ISL_881958, EPI_ISL_881959, EPI_ISL_881960, EPI_ISL_881961, EPI_ISL_881962, EPI_ISL_881963, EPI_ISL_881964, EPI_ISL_881965, EPI_ISL_881966, EPI_ISL_881967, EPI_ISL_881968, EPI_ISL_881969, EPI_ISL_881970, EPI_ISL_881971, EPI_ISL_881972, EPI_ISL_881973, EPI_ISL_881974, EPI_ISL_881975, EPI_ISL_881976,
EPI_ISL_881977, EPI_ISL_881978, EPI_ISL_881979, EPI_ISL_881980, EPI_ISL_881981, EPI_ISL_881982, EPI_ISL_881983, EPI_ISL_881984, EPI_ISL_881985, EPI_ISL_881986, EPI_ISL_881987, EPI_ISL_881988, EPI_ISL_881989, EPI_ISL_881990, EPI_ISL_881991, EPI_ISL_881992, EPI_ISL_881993, EPI_ISL_881994, EPI_ISL_881995, EPI_ISL_881996, EPI_ISL_881997, EPI_ISL_881998, EPI_ISL_881999, EPI_ISL_882000,
EPI_ISL_882001, EPI_ISL_882002, EPI_ISL_882003, EPI_ISL_882004, EPI_ISL_882005, EPI_ISL_882006, EPI_ISL_882007, EPI_ISL_882008, EPI_ISL_882009, EPI_ISL_882010, EPI_ISL_882011, EPI_ISL_882012, EPI_ISL_882013, EPI_ISL_882014, EPI_ISL_882015, EPI_ISL_882016, EPI_ISL_882017, EPI_ISL_882018, EPI_ISL_882019, EPI_ISL_882020, EPI_ISL_882021, EPI_ISL_882022, EPI_ISL_882023, EPI_ISL_882024,
EPI_ISL_882025, EPI_ISL_882026, EPI_ISL_882027, EPI_ISL_882028, EPI_ISL_882029, EPI_ISL_882030, EPI_ISL_882031, EPI_ISL_882032, EPI_ISL_882033, EPI_ISL_882034, EPI_ISL_882035, EPI_ISL_882036, EPI_ISL_882037, EPI_ISL_882038, EPI_ISL_882039, EPI_ISL_882040, EPI_ISL_882041, EPI_ISL_882042, EPI_ISL_882043, EPI_ISL_882044, EPI_ISL_882045, EPI_ISL_882046, EPI_ISL_882047, EPI_ISL_882048,
EPI_ISL_882049, EPI_ISL_882050, EPI_ISL_882051, EPI_ISL_882052, EPI_ISL_882053, EPI_ISL_882054, EPI_ISL_882055, EPI_ISL_882056, EPI_ISL_882057, EPI_ISL_882058, EPI_ISL_882059, EPI_ISL_882060, EPI_ISL_882061, EPI_ISL_882062, EPI_ISL_882063, EPI_ISL_882064, EPI_ISL_882065, EPI_ISL_882066, EPI_ISL_882067, EPI_ISL_882068, EPI_ISL_882069, EPI_ISL_882070, EPI_ISL_882071, EPI_ISL_882072,
EPI_ISL_882073, EPI_ISL_882074, EPI_ISL_882075, EPI_ISL_882076, EPI_ISL_882077, EPI_ISL_882078, EPI_ISL_882079, EPI_ISL_882080, EPI_ISL_882081, EPI_ISL_882082, EPI_ISL_882083, EPI_ISL_882084, EPI_ISL_882085, EPI_ISL_882086, EPI_ISL_882087, EPI_ISL_882088, EPI_ISL_882089, EPI_ISL_882090, EPI_ISL_882091, EPI_ISL_882092, EPI_ISL_882093, EPI_ISL_882094, EPI_ISL_882095, EPI_ISL_882096,
EPI_ISL_882097, EPI_ISL_882098, EPI_ISL_882099, EPI_ISL_882100, EPI_ISL_882101, EPI_ISL_882102, EPI_ISL_882103, EPI_ISL_882104, EPI_ISL_882105, EPI_ISL_882106, EPI_ISL_882107, EPI_ISL_882108, EPI_ISL_882109, EPI_ISL_882110, EPI_ISL_882111, EPI_ISL_882112, EPI_ISL_882113, EPI_ISL_882114, EPI_ISL_882115, EPI_ISL_882116, EPI_ISL_882117, EPI_ISL_882118, EPI_ISL_882119, EPI_ISL_882120,
EPI_ISL_882121, EPI_ISL_882122, EPI_ISL_882123, EPI_ISL_882124, EPI_ISL_882125, EPI_ISL_882126, EPI_ISL_882127, EPI_ISL_882128, EPI_ISL_882129, EPI_ISL_882130, EPI_ISL_882131, EPI_ISL_882132, EPI_ISL_882133, EPI_ISL_882134, EPI_ISL_882135, EPI_ISL_882136, EPI_ISL_882137, EPI_ISL_882138, EPI_ISL_882139, EPI_ISL_882140, EPI_ISL_882141, EPI_ISL_882142, EPI_ISL_882143, EPI_ISL_882144,
EPI_ISL_882145, EPI_ISL_882146, EPI_ISL_882147, EPI_ISL_882148, EPI_ISL_882149, EPI_ISL_882150, EPI_ISL_882151, EPI_ISL_882152, EPI_ISL_882153, EPI_ISL_882154, EPI_ISL_882155, EPI_ISL_882156, EPI_ISL_882157, EPI_ISL_882158, EPI_ISL_882159, EPI_ISL_882160, EPI_ISL_882161, EPI_ISL_882162, EPI_ISL_882163, EPI_ISL_882164, EPI_ISL_882165, EPI_ISL_882166, EPI_ISL_882167, EPI_ISL_882168,
EPI_ISL_882169, EPI_ISL_882170, EPI_ISL_882171, EPI_ISL_882172, EPI_ISL_882173, EPI_ISL_882174, EPI_ISL_882175, EPI_ISL_882176, EPI_ISL_882177, EPI_ISL_882178, EPI_ISL_882179, EPI_ISL_882180, EPI_ISL_882181, EPI_ISL_882182, EPI_ISL_882183, EPI_ISL_882184, EPI_ISL_882185, EPI_ISL_882186, EPI_ISL_882187, EPI_ISL_882188, EPI_ISL_882189, EPI_ISL_882190, EPI_ISL_882191, EPI_ISL_882192,
EPI_ISL_882193, EPI_ISL_882194, EPI_ISL_882195, EPI_ISL_882196, EPI_ISL_882197, EPI_ISL_882198, EPI_ISL_882199, EPI_ISL_882200, EPI_ISL_882201, EPI_ISL_882202, EPI_ISL_882203, EPI_ISL_882204, EPI_ISL_882205, EPI_ISL_882206, EPI_ISL_882207, EPI_ISL_882208, EPI_ISL_882209, EPI_ISL_882210, EPI_ISL_882211, EPI_ISL_882212, EPI_ISL_882213, EPI_ISL_882214, EPI_ISL_882215, EPI_ISL_882216,
EPI_ISL_882217, EPI_ISL_882218, EPI_ISL_882219, EPI_ISL_882220, EPI_ISL_882221, EPI_ISL_882222, EPI_ISL_882223, EPI_ISL_882224, EPI_ISL_882225, EPI_ISL_882226, EPI_ISL_882227, EPI_ISL_882228, EPI_ISL_882229, EPI_ISL_882230, EPI_ISL_882231, EPI_ISL_882232, EPI_ISL_882233, EPI_ISL_882234, EPI_ISL_882235, EPI_ISL_882236, EPI_ISL_882237, EPI_ISL_882238, EPI_ISL_882239, EPI_ISL_882240,
EPI_ISL_882241, EPI_ISL_882242, EPI_ISL_882243, EPI_ISL_882244, EPI_ISL_882245, EPI_ISL_882246, EPI_ISL_882247, EPI_ISL_882248, EPI_ISL_882249, EPI_ISL_882250, EPI_ISL_882251, EPI_ISL_882252, EPI_ISL_882253, EPI_ISL_882254, EPI_ISL_882255, EPI_ISL_882256, EPI_ISL_882257, EPI_ISL_882258, EPI_ISL_882259, EPI_ISL_882260, EPI_ISL_882261, EPI_ISL_882262, EPI_ISL_882263, EPI_ISL_882264,
EPI_ISL_882265, EPI_ISL_882266, EPI_ISL_882267, EPI_ISL_882268, EPI_ISL_882269, EPI_ISL_882270, EPI_ISL_882271, EPI_ISL_882272, EPI_ISL_882273, EPI_ISL_882274, EPI_ISL_882275, EPI_ISL_882276, EPI_ISL_882277, EPI_ISL_882278, EPI_ISL_882279, EPI_ISL_882280, EPI_ISL_882281, EPI_ISL_882282, EPI_ISL_882283, EPI_ISL_882284, EPI_ISL_882285, EPI_ISL_882286, EPI_ISL_882287, EPI_ISL_882288,
EPI_ISL_882289, EPI_ISL_882290, EPI_ISL_882291, EPI_ISL_882292, EPI_ISL_882293, EPI_ISL_882294, EPI_ISL_882295, EPI_ISL_882296, EPI_ISL_882297, EPI_ISL_882298, EPI_ISL_882299, EPI_ISL_885240, EPI_ISL_885327, EPI_ISL_885409, EPI_ISL_885514, EPI_ISL_892050, EPI_ISL_892051, EPI_ISL_892052, EPI_ISL_892053, EPI_ISL_892054, EPI_ISL_892055, EPI_ISL_892057, EPI_ISL_892058, EPI_ISL_892067,
EPI_ISL_892068, EPI_ISL_892069, EPI_ISL_892070, EPI_ISL_892071, EPI_ISL_892072, EPI_ISL_892073, EPI_ISL_892074, EPI_ISL_892075, EPI_ISL_892076, EPI_ISL_892077, EPI_ISL_892078, EPI_ISL_892079, EPI_ISL_892080, EPI_ISL_892081, EPI_ISL_892082, EPI_ISL_892083, EPI_ISL_892084, EPI_ISL_892085, EPI_ISL_892086, EPI_ISL_892087, EPI_ISL_892088, EPI_ISL_892089, EPI_ISL_892090, EPI_ISL_892091,
EPI_ISL_892092, EPI_ISL_892093, EPI_ISL_892094, EPI_ISL_892095, EPI_ISL_892096, EPI_ISL_892097, EPI_ISL_892098, EPI_ISL_892099, EPI_ISL_892100, EPI_ISL_892101, EPI_ISL_892102, EPI_ISL_892103, EPI_ISL_892104, EPI_ISL_892105, EPI_ISL_892106, EPI_ISL_892107, EPI_ISL_892108, EPI_ISL_892109, EPI_ISL_892110, EPI_ISL_892111, EPI_ISL_892112, EPI_ISL_892113, EPI_ISL_892114, EPI_ISL_892115,
EPI_ISL_892116, EPI_ISL_892117, EPI_ISL_892118, EPI_ISL_892119, EPI_ISL_892120, EPI_ISL_892121, EPI_ISL_892122, EPI_ISL_892123, EPI_ISL_892124, EPI_ISL_892125, EPI_ISL_892126, EPI_ISL_892179, EPI_ISL_892189, EPI_ISL_892190, EPI_ISL_892191, EPI_ISL_892192, EPI_ISL_892193, EPI_ISL_892194, EPI_ISL_892195, EPI_ISL_892196, EPI_ISL_892197, EPI_ISL_892206, EPI_ISL_909595, EPI_ISL_909596,
EPI_ISL_909597, EPI_ISL_909598, EPI_ISL_909599, EPI_ISL_909600, EPI_ISL_909601, EPI_ISL_909602, EPI_ISL_909603, EPI_ISL_909604, EPI_ISL_909605, EPI_ISL_909606, EPI_ISL_909607, EPI_ISL_909608, EPI_ISL_909609, EPI_ISL_909610, EPI_ISL_909611, EPI_ISL_909612, EPI_ISL_909613, EPI_ISL_909614, EPI_ISL_909615, EPI_ISL_909616, EPI_ISL_909617, EPI_ISL_909618, EPI_ISL_909619, EPI_ISL_909620,
EPI_ISL_909621, EPI_ISL_909622, EPI_ISL_909623, EPI_ISL_909624, EPI_ISL_909625, EPI_ISL_909626, EPI_ISL_909627, EPI_ISL_909628, EPI_ISL_909629, EPI_ISL_909630, EPI_ISL_909631, EPI_ISL_909632, EPI_ISL_909633, EPI_ISL_909634, EPI_ISL_909635, EPI_ISL_909636, EPI_ISL_909637, EPI_ISL_909638, EPI_ISL_909639, EPI_ISL_909640, EPI_ISL_909641, EPI_ISL_909642, EPI_ISL_909643, EPI_ISL_909644,
EPI_ISL_909645, EPI_ISL_909646, EPI_ISL_909647, EPI_ISL_909648, EPI_ISL_909649, EPI_ISL_909650, EPI_ISL_909651, EPI_ISL_912226, EPI_ISL_916630, EPI_ISL_916637, EPI_ISL_916651, EPI_ISL_916732, EPI_ISL_916746, EPI_ISL_916766, EPI_ISL_916775, EPI_ISL_916779, EPI_ISL_916849, EPI_ISL_916919, EPI_ISL_916934, EPI_ISL_916950, EPI_ISL_917011, EPI_ISL_917060, EPI_ISL_917095, EPI_ISL_917121,
EPI_ISL_917792, EPI_ISL_937818, EPI_ISL_946701, EPI_ISL_946780, EPI_ISL_963061, EPI_ISL_963296, EPI_ISL_963955, EPI_ISL_990545, EPI_ISL_994649, EPI_ISL_994650, EPI_ISL_1346482

| | | | |
|---|---|---|---|
| see above | Lighthouse Lab in Alderley Park | Wellcome Sanger Institute for the COVID-19 Genomics UK (COG-UK) Consortium | Cordelia Langford; David K. Jackson; Dominic Kwiatkowski; Ewan Harrison; Ian Johnston; Jacquelyn Wynn; Jeffrey Barrett; John Sillitoe on behalf of the Wellcome Sanger Institute COVID-19 Surveillance Team; John Sillitoe on behalf of the Wellcome Sanger Institute COVID-19 Surveillance Team (http://www.sanger.ac.uk/covid-team); Mairead Hyland; Roberto Amato; Sonia Goncalves; The Lighthouse Lab in Alderley Park and Alex Alderton |

EPI_ISL_833585, EPI_ISL_833587, EPI_ISL_833588, EPI_ISL_833589, EPI_ISL_833591, EPI_ISL_833592, EPI_ISL_833594, EPI_ISL_833595, EPI_ISL_833598, EPI_ISL_833600, EPI_ISL_833601, EPI_ISL_833603, EPI_ISL_833605, EPI_ISL_833608, EPI_ISL_833609, EPI_ISL_833610, EPI_ISL_833613, EPI_ISL_833616, EPI_ISL_833622, EPI_ISL_833623, EPI_ISL_833624, EPI_ISL_833625, EPI_ISL_833627, EPI_ISL_833628,
EPI_ISL_833633, EPI_ISL_833635, EPI_ISL_833636, EPI_ISL_833637, EPI_ISL_833638, EPI_ISL_833639, EPI_ISL_833640, EPI_ISL_833641, EPI_ISL_833642, EPI_ISL_833644, EPI_ISL_833645, EPI_ISL_833646, EPI_ISL_833649, EPI_ISL_833650, EPI_ISL_833651, EPI_ISL_833653, EPI_ISL_833655, EPI_ISL_833657, EPI_ISL_833659, EPI_ISL_833660, EPI_ISL_833661, EPI_ISL_833663, EPI_ISL_833664, EPI_ISL_833665,
EPI_ISL_833666, EPI_ISL_833668, EPI_ISL_833669, EPI_ISL_833670, EPI_ISL_833672, EPI_ISL_833673, EPI_ISL_833674, EPI_ISL_833675, EPI_ISL_833677, EPI_ISL_833681, EPI_ISL_833682, EPI_ISL_833684, EPI_ISL_833686, EPI_ISL_833687, EPI_ISL_833688, EPI_ISL_833689, EPI_ISL_833690, EPI_ISL_833691, EPI_ISL_833692, EPI_ISL_833694, EPI_ISL_833695, EPI_ISL_833696, EPI_ISL_833697, EPI_ISL_833698,
EPI_ISL_833699, EPI_ISL_833701, EPI_ISL_833702, EPI_ISL_833703, EPI_ISL_833704, EPI_ISL_833705, EPI_ISL_833706, EPI_ISL_833708, EPI_ISL_833709, EPI_ISL_833710, EPI_ISL_833711, EPI_ISL_833713, EPI_ISL_833714, EPI_ISL_833715, EPI_ISL_833717, EPI_ISL_833718, EPI_ISL_833720, EPI_ISL_833722, EPI_ISL_833723, EPI_ISL_833724, EPI_ISL_833727, EPI_ISL_833728, EPI_ISL_833729, EPI_ISL_833730,
EPI_ISL_833731, EPI_ISL_833732, EPI_ISL_833733, EPI_ISL_833737, EPI_ISL_833739, EPI_ISL_833740, EPI_ISL_833741, EPI_ISL_833742, EPI_ISL_833744, EPI_ISL_833745, EPI_ISL_833747, EPI_ISL_833748, EPI_ISL_833749, EPI_ISL_833750, EPI_ISL_833751, EPI_ISL_833752, EPI_ISL_833753, EPI_ISL_833754, EPI_ISL_833757, EPI_ISL_833758, EPI_ISL_833760, EPI_ISL_833761, EPI_ISL_833762, EPI_ISL_833763,
EPI_ISL_833764, EPI_ISL_833765, EPI_ISL_833769, EPI_ISL_833771, EPI_ISL_833772, EPI_ISL_833773, EPI_ISL_833774, EPI_ISL_833775, EPI_ISL_833776, EPI_ISL_833777, EPI_ISL_833778, EPI_ISL_833781, EPI_ISL_833782, EPI_ISL_833783, EPI_ISL_833784, EPI_ISL_833785, EPI_ISL_833786, EPI_ISL_833787, EPI_ISL_833788, EPI_ISL_833790, EPI_ISL_833792, EPI_ISL_833793, EPI_ISL_833794, EPI_ISL_833798,
EPI_ISL_833799, EPI_ISL_833800, EPI_ISL_833802, EPI_ISL_833805, EPI_ISL_833806, EPI_ISL_833807, EPI_ISL_833808, EPI_ISL_833810, EPI_ISL_833811, EPI_ISL_833812, EPI_ISL_833813, EPI_ISL_833815, EPI_ISL_833816, EPI_ISL_833819, EPI_ISL_833820, EPI_ISL_833822, EPI_ISL_833824, EPI_ISL_833825, EPI_ISL_833826, EPI_ISL_833827, EPI_ISL_833828, EPI_ISL_833829, EPI_ISL_833831, EPI_ISL_833832,
EPI_ISL_833836, EPI_ISL_833837, EPI_ISL_833838, EPI_ISL_833839, EPI_ISL_833840, EPI_ISL_833843, EPI_ISL_833847, EPI_ISL_833850, EPI_ISL_833851, EPI_ISL_833852, EPI_ISL_833853, EPI_ISL_833854, EPI_ISL_833857, EPI_ISL_833858, EPI_ISL_833862, EPI_ISL_833863, EPI_ISL_833864, EPI_ISL_833866, EPI_ISL_833867, EPI_ISL_833869, EPI_ISL_833870, EPI_ISL_833871, EPI_ISL_833873, EPI_ISL_833874,
EPI_ISL_833875, EPI_ISL_833876, EPI_ISL_833877, EPI_ISL_833878, EPI_ISL_833880, EPI_ISL_833882, EPI_ISL_833883, EPI_ISL_833885, EPI_ISL_833886, EPI_ISL_833887, EPI_ISL_833888, EPI_ISL_833891, EPI_ISL_833892, EPI_ISL_833893, EPI_ISL_833897, EPI_ISL_833898, EPI_ISL_833899, EPI_ISL_833900, EPI_ISL_833902, EPI_ISL_833904, EPI_ISL_833905, EPI_ISL_833906, EPI_ISL_833907, EPI_ISL_833911,
EPI_ISL_833912, EPI_ISL_833913, EPI_ISL_833914, EPI_ISL_833915, EPI_ISL_833917, EPI_ISL_833918, EPI_ISL_833919, EPI_ISL_833920, EPI_ISL_833921, EPI_ISL_833923, EPI_ISL_833925, EPI_ISL_833926, EPI_ISL_833927, EPI_ISL_833928, EPI_ISL_833929, EPI_ISL_833932, EPI_ISL_845893, EPI_ISL_845894, EPI_ISL_845895, EPI_ISL_845896, EPI_ISL_845897, EPI_ISL_845898, EPI_ISL_845899, EPI_ISL_845900,
EPI_ISL_845901, EPI_ISL_845902, EPI_ISL_845903, EPI_ISL_845904, EPI_ISL_845905, EPI_ISL_845906, EPI_ISL_845907, EPI_ISL_845908, EPI_ISL_845909, EPI_ISL_845910, EPI_ISL_845911, EPI_ISL_845912, EPI_ISL_845913, EPI_ISL_845914, EPI_ISL_845915, EPI_ISL_845916, EPI_ISL_845917, EPI_ISL_845918, EPI_ISL_845919, EPI_ISL_845920, EPI_ISL_845921, EPI_ISL_845922, EPI_ISL_845923, EPI_ISL_845924,
EPI_ISL_845925, EPI_ISL_845926, EPI_ISL_845927, EPI_ISL_845928, EPI_ISL_845929, EPI_ISL_845930, EPI_ISL_845931, EPI_ISL_845932, EPI_ISL_845933, EPI_ISL_845934, EPI_ISL_845935, EPI_ISL_845936, EPI_ISL_845937, EPI_ISL_845938, EPI_ISL_845939, EPI_ISL_845940, EPI_ISL_845941, EPI_ISL_845942, EPI_ISL_845943, EPI_ISL_845944, EPI_ISL_845945, EPI_ISL_845946, EPI_ISL_845947, EPI_ISL_845948,
EPI_ISL_845949, EPI_ISL_845950, EPI_ISL_845951, EPI_ISL_845952, EPI_ISL_845953, EPI_ISL_845954, EPI_ISL_845955, EPI_ISL_845956, EPI_ISL_845957, EPI_ISL_845958, EPI_ISL_845959, EPI_ISL_845960, EPI_ISL_845961, EPI_ISL_845962, EPI_ISL_845963, EPI_ISL_845964, EPI_ISL_845965, EPI_ISL_845966, EPI_ISL_845967, EPI_ISL_845968, EPI_ISL_845969, EPI_ISL_845970, EPI_ISL_845971, EPI_ISL_845972,
EPI_ISL_845974, EPI_ISL_845975, EPI_ISL_845976, EPI_ISL_845977, EPI_ISL_845978, EPI_ISL_845979, EPI_ISL_845980, EPI_ISL_845981, EPI_ISL_845982, EPI_ISL_845983, EPI_ISL_845984, EPI_ISL_845985, EPI_ISL_845986, EPI_ISL_845987, EPI_ISL_845988, EPI_ISL_845989, EPI_ISL_845990, EPI_ISL_845991, EPI_ISL_845992, EPI_ISL_845993, EPI_ISL_845994, EPI_ISL_845995, EPI_ISL_845996, EPI_ISL_845997,
EPI_ISL_845998, EPI_ISL_845999, EPI_ISL_846000, EPI_ISL_846001, EPI_ISL_846002, EPI_ISL_846003, EPI_ISL_846004, EPI_ISL_846005, EPI_ISL_846006, EPI_ISL_846007, EPI_ISL_846008, EPI_ISL_846009, EPI_ISL_846010, EPI_ISL_846011, EPI_ISL_846012, EPI_ISL_846013, EPI_ISL_846014, EPI_ISL_846015, EPI_ISL_846016, EPI_ISL_846017, EPI_ISL_846018, EPI_ISL_846019, EPI_ISL_846020, EPI_ISL_846021,
EPI_ISL_846023, EPI_ISL_846024, EPI_ISL_846025, EPI_ISL_846026, EPI_ISL_846027, EPI_ISL_846028, EPI_ISL_846029, EPI_ISL_846030, EPI_ISL_846031, EPI_ISL_846032, EPI_ISL_846033, EPI_ISL_846034, EPI_ISL_846035, EPI_ISL_846036, EPI_ISL_846037, EPI_ISL_846038, EPI_ISL_846039, EPI_ISL_846040, EPI_ISL_846041, EPI_ISL_846042, EPI_ISL_846043, EPI_ISL_846044, EPI_ISL_846045, EPI_ISL_846046,
EPI_ISL_846047, EPI_ISL_846048, EPI_ISL_846049, EPI_ISL_846050, EPI_ISL_846051, EPI_ISL_846052, EPI_ISL_846053, EPI_ISL_846054, EPI_ISL_846055, EPI_ISL_846056, EPI_ISL_846057, EPI_ISL_846058, EPI_ISL_846059, EPI_ISL_846060, EPI_ISL_846061, EPI_ISL_846062, EPI_ISL_846063, EPI_ISL_846064, EPI_ISL_846065, EPI_ISL_846066, EPI_ISL_846067, EPI_ISL_846068, EPI_ISL_846069, EPI_ISL_846071,
EPI_ISL_846072, EPI_ISL_846073, EPI_ISL_846074, EPI_ISL_846075, EPI_ISL_846076, EPI_ISL_846077, EPI_ISL_846078, EPI_ISL_846079, EPI_ISL_846080, EPI_ISL_846081, EPI_ISL_846082, EPI_ISL_846083, EPI_ISL_846084, EPI_ISL_846085, EPI_ISL_846086, EPI_ISL_846087, EPI_ISL_846088, EPI_ISL_846089, EPI_ISL_846090, EPI_ISL_846091, EPI_ISL_846092, EPI_ISL_846093, EPI_ISL_846094, EPI_ISL_846095,
EPI_ISL_846096, EPI_ISL_846097, EPI_ISL_846098, EPI_ISL_846099, EPI_ISL_846100, EPI_ISL_846101, EPI_ISL_846102, EPI_ISL_846103, EPI_ISL_846104, EPI_ISL_846105, EPI_ISL_846106, EPI_ISL_846107, EPI_ISL_846108, EPI_ISL_846109, EPI_ISL_846110, EPI_ISL_846111, EPI_ISL_846112, EPI_ISL_846113, EPI_ISL_846114, EPI_ISL_846115, EPI_ISL_846116, EPI_ISL_846117, EPI_ISL_846118, EPI_ISL_846119,
EPI_ISL_846120, EPI_ISL_846121, EPI_ISL_846122, EPI_ISL_846123, EPI_ISL_846124, EPI_ISL_846125, EPI_ISL_846126, EPI_ISL_846127, EPI_ISL_846128, EPI_ISL_846129, EPI_ISL_846130, EPI_ISL_846131, EPI_ISL_846132, EPI_ISL_846133, EPI_ISL_846134, EPI_ISL_846135, EPI_ISL_846136, EPI_ISL_846137, EPI_ISL_846138, EPI_ISL_846139, EPI_ISL_846140, EPI_ISL_846141, EPI_ISL_846142, EPI_ISL_846143,
EPI_ISL_846144, EPI_ISL_846145, EPI_ISL_846146, EPI_ISL_846147, EPI_ISL_846148, EPI_ISL_846149, EPI_ISL_846150, EPI_ISL_846151, EPI_ISL_846152, EPI_ISL_846153, EPI_ISL_846154, EPI_ISL_846155, EPI_ISL_846156, EPI_ISL_846157, EPI_ISL_846158, EPI_ISL_846159, EPI_ISL_846160, EPI_ISL_846161, EPI_ISL_846162, EPI_ISL_846163, EPI_ISL_846164, EPI_ISL_846165, EPI_ISL_846166, EPI_ISL_846167,
EPI_ISL_846168, EPI_ISL_846169, EPI_ISL_846170, EPI_ISL_846172, EPI_ISL_846173, EPI_ISL_846174, EPI_ISL_846175, EPI_ISL_846176, EPI_ISL_846177, EPI_ISL_846178, EPI_ISL_846179, EPI_ISL_846180, EPI_ISL_846181, EPI_ISL_846182, EPI_ISL_846183, EPI_ISL_846184, EPI_ISL_846185, EPI_ISL_846186, EPI_ISL_846187, EPI_ISL_846188, EPI_ISL_846189, EPI_ISL_846190, EPI_ISL_846191, EPI_ISL_846192,
EPI_ISL_846193, EPI_ISL_846194, EPI_ISL_846195, EPI_ISL_846196, EPI_ISL_846197, EPI_ISL_846198, EPI_ISL_846199, EPI_ISL_846509, EPI_ISL_851062, EPI_ISL_851063, EPI_ISL_851064, EPI_ISL_851065, EPI_ISL_851066, EPI_ISL_851067, EPI_ISL_851068, EPI_ISL_851069, EPI_ISL_851071, EPI_ISL_851072, EPI_ISL_851073, EPI_ISL_851074, EPI_ISL_851075, EPI_ISL_851076, EPI_ISL_851078, EPI_ISL_851080,
EPI_ISL_851083, EPI_ISL_851084, EPI_ISL_851086, EPI_ISL_851087, EPI_ISL_851088, EPI_ISL_851089, EPI_ISL_851090, EPI_ISL_851091, EPI_ISL_851092, EPI_ISL_851093, EPI_ISL_851096, EPI_ISL_851097, EPI_ISL_851098, EPI_ISL_851099, EPI_ISL_851100, EPI_ISL_851103, EPI_ISL_851105, EPI_ISL_851106, EPI_ISL_851108, EPI_ISL_851109, EPI_ISL_851110, EPI_ISL_851111, EPI_ISL_851112, EPI_ISL_851113,
EPI_ISL_851114, EPI_ISL_851115, EPI_ISL_851117, EPI_ISL_851118, EPI_ISL_851120, EPI_ISL_851121, EPI_ISL_851122, EPI_ISL_851123, EPI_ISL_851127, EPI_ISL_851128, EPI_ISL_851129, EPI_ISL_851131, EPI_ISL_851132, EPI_ISL_851136, EPI_ISL_851138, EPI_ISL_851139, EPI_ISL_851140, EPI_ISL_851141, EPI_ISL_851143, EPI_ISL_851144, EPI_ISL_851145, EPI_ISL_851149, EPI_ISL_851150, EPI_ISL_851151,
EPI_ISL_851152, EPI_ISL_851153, EPI_ISL_851155, EPI_ISL_851156, EPI_ISL_851157, EPI_ISL_851158, EPI_ISL_851159, EPI_ISL_851160, EPI_ISL_851161, EPI_ISL_851162, EPI_ISL_851163, EPI_ISL_851164, EPI_ISL_851165, EPI_ISL_851166, EPI_ISL_851168, EPI_ISL_851169, EPI_ISL_851171, EPI_ISL_851172, EPI_ISL_851173, EPI_ISL_851174, EPI_ISL_851175, EPI_ISL_851176, EPI_ISL_851177, EPI_ISL_851179,
EPI_ISL_851181, EPI_ISL_851183, EPI_ISL_851184, EPI_ISL_851185, EPI_ISL_851186, EPI_ISL_851188, EPI_ISL_851189, EPI_ISL_851191, EPI_ISL_851192, EPI_ISL_851193, EPI_ISL_851194, EPI_ISL_851195, EPI_ISL_851197, EPI_ISL_851198, EPI_ISL_851199, EPI_ISL_851200, EPI_ISL_851201, EPI_ISL_851202, EPI_ISL_851203, EPI_ISL_851204, EPI_ISL_851205, EPI_ISL_851208, EPI_ISL_851209, EPI_ISL_851210,
EPI_ISL_851211, EPI_ISL_851212, EPI_ISL_851213, EPI_ISL_851214, EPI_ISL_851216, EPI_ISL_851219, EPI_ISL_851220, EPI_ISL_851221, EPI_ISL_851222, EPI_ISL_851223, EPI_ISL_851224, EPI_ISL_851226, EPI_ISL_851227, EPI_ISL_851228, EPI_ISL_851230, EPI_ISL_851231, EPI_ISL_851232, EPI_ISL_851233, EPI_ISL_851234, EPI_ISL_851235, EPI_ISL_851238, EPI_ISL_851239, EPI_ISL_851240, EPI_ISL_851242,
EPI_ISL_851243, EPI_ISL_851244, EPI_ISL_851245, EPI_ISL_851247, EPI_ISL_851248, EPI_ISL_851249, EPI_ISL_851250, EPI_ISL_851254, EPI_ISL_851255, EPI_ISL_851256, EPI_ISL_851260, EPI_ISL_851261, EPI_ISL_851262, EPI_ISL_851263, EPI_ISL_851265, EPI_ISL_851266, EPI_ISL_851267, EPI_ISL_851268, EPI_ISL_851271, EPI_ISL_851272, EPI_ISL_851274, EPI_ISL_851275, EPI_ISL_851277, EPI_ISL_851278,
EPI_ISL_851279, EPI_ISL_851280, EPI_ISL_851281, EPI_ISL_851282, EPI_ISL_851283, EPI_ISL_851284, EPI_ISL_851286, EPI_ISL_851287, EPI_ISL_851288, EPI_ISL_851289, EPI_ISL_851290, EPI_ISL_851291, EPI_ISL_851292, EPI_ISL_851293, EPI_ISL_851294, EPI_ISL_851295, EPI_ISL_851297, EPI_ISL_851299, EPI_ISL_851300, EPI_ISL_851302, EPI_ISL_851303, EPI_ISL_851304, EPI_ISL_851308, EPI_ISL_851309,
EPI_ISL_851311, EPI_ISL_851312, EPI_ISL_851313, EPI_ISL_851314, EPI_ISL_851315, EPI_ISL_851316, EPI_ISL_851317, EPI_ISL_851318, EPI_ISL_851319, EPI_ISL_851320, EPI_ISL_851321, EPI_ISL_851322, EPI_ISL_851323, EPI_ISL_851324, EPI_ISL_851326, EPI_ISL_851328, EPI_ISL_851330, EPI_ISL_851332, EPI_ISL_851333, EPI_ISL_851334, EPI_ISL_851336, EPI_ISL_851337, EPI_ISL_851338, EPI_ISL_851339,
EPI_ISL_851340, EPI_ISL_851342, EPI_ISL_851344, EPI_ISL_851345, EPI_ISL_851347, EPI_ISL_851348, EPI_ISL_851349, EPI_ISL_851350, EPI_ISL_851352, EPI_ISL_851353, EPI_ISL_851354, EPI_ISL_851355, EPI_ISL_851356, EPI_ISL_851357, EPI_ISL_851359, EPI_ISL_851360, EPI_ISL_851361, EPI_ISL_851362, EPI_ISL_851363, EPI_ISL_851364, EPI_ISL_851366, EPI_ISL_851368, EPI_ISL_851369, EPI_ISL_851370,
EPI_ISL_851372, EPI_ISL_851373, EPI_ISL_851375, EPI_ISL_851376, EPI_ISL_851377, EPI_ISL_851378, EPI_ISL_851379, EPI_ISL_851381, EPI_ISL_857542, EPI_ISL_857543, EPI_ISL_857544, EPI_ISL_857545, EPI_ISL_857547, EPI_ISL_857548, EPI_ISL_857549, EPI_ISL_857550, EPI_ISL_857551, EPI_ISL_857552, EPI_ISL_857554, EPI_ISL_857555, EPI_ISL_857556, EPI_ISL_857557, EPI_ISL_857558, EPI_ISL_857559,
EPI_ISL_857560, EPI_ISL_857561, EPI_ISL_857563, EPI_ISL_857564, EPI_ISL_857565, EPI_ISL_857566, EPI_ISL_857567, EPI_ISL_857568, EPI_ISL_857569, EPI_ISL_857570, EPI_ISL_857571, EPI_ISL_857572, EPI_ISL_857573, EPI_ISL_857574, EPI_ISL_857575, EPI_ISL_857576, EPI_ISL_857577, EPI_ISL_857578, EPI_ISL_857579, EPI_ISL_857580, EPI_ISL_857581, EPI_ISL_857582, EPI_ISL_857583, EPI_ISL_857585,
EPI_ISL_857586, EPI_ISL_857587, EPI_ISL_857588, EPI_ISL_857589, EPI_ISL_857590, EPI_ISL_857591, EPI_ISL_857592, EPI_ISL_857593, EPI_ISL_857594, EPI_ISL_857595, EPI_ISL_857596, EPI_ISL_857597, EPI_ISL_857598, EPI_ISL_857600, EPI_ISL_857601, EPI_ISL_857602, EPI_ISL_857603, EPI_ISL_857605, EPI_ISL_857606, EPI_ISL_857607, EPI_ISL_857608, EPI_ISL_857609, EPI_ISL_857610, EPI_ISL_857611,
EPI_ISL_857612, EPI_ISL_857613, EPI_ISL_857614, EPI_ISL_857615, EPI_ISL_857616, EPI_ISL_857617, EPI_ISL_857618, EPI_ISL_857619, EPI_ISL_857620, EPI_ISL_857621, EPI_ISL_857622, EPI_ISL_857623, EPI_ISL_857624, EPI_ISL_857625, EPI_ISL_857626, EPI_ISL_857627, EPI_ISL_857628, EPI_ISL_857629, EPI_ISL_857630, EPI_ISL_857631, EPI_ISL_857632, EPI_ISL_857633, EPI_ISL_857634, EPI_ISL_857635,
EPI_ISL_857636, EPI_ISL_857637, EPI_ISL_857638, EPI_ISL_857639, EPI_ISL_857640, EPI_ISL_857641, EPI_ISL_857643, EPI_ISL_857644, EPI_ISL_857646, EPI_ISL_857647, EPI_ISL_857648, EPI_ISL_857649, EPI_ISL_857650, EPI_ISL_857651, EPI_ISL_857652, EPI_ISL_857653, EPI_ISL_857655, EPI_ISL_857656, EPI_ISL_857657, EPI_ISL_857658, EPI_ISL_857659, EPI_ISL_857660, EPI_ISL_857661, EPI_ISL_857662,
EPI_ISL_857663, EPI_ISL_857664, EPI_ISL_857665, EPI_ISL_857666, EPI_ISL_857667, EPI_ISL_857668, EPI_ISL_857669, EPI_ISL_857670, EPI_ISL_857671, EPI_ISL_857672, EPI_ISL_857673, EPI_ISL_857674, EPI_ISL_857675, EPI_ISL_857676, EPI_ISL_857677, EPI_ISL_857678, EPI_ISL_857679, EPI_ISL_857680, EPI_ISL_857681, EPI_ISL_857683, EPI_ISL_857684, EPI_ISL_857685, EPI_ISL_857686, EPI_ISL_857687,
EPI_ISL_857688, EPI_ISL_857689, EPI_ISL_857690, EPI_ISL_857691, EPI_ISL_857692, EPI_ISL_857693, EPI_ISL_857694, EPI_ISL_857695, EPI_ISL_857696, EPI_ISL_857697, EPI_ISL_857698, EPI_ISL_857699, EPI_ISL_857700, EPI_ISL_857701, EPI_ISL_857702, EPI_ISL_857703, EPI_ISL_857704, EPI_ISL_857705, EPI_ISL_857706, EPI_ISL_857707, EPI_ISL_857708, EPI_ISL_857709, EPI_ISL_857710, EPI_ISL_857711,
EPI_ISL_857712, EPI_ISL_857713, EPI_ISL_857714, EPI_ISL_857715, EPI_ISL_857716, EPI_ISL_857717, EPI_ISL_857718, EPI_ISL_857719, EPI_ISL_857720, EPI_ISL_857721, EPI_ISL_857722, EPI_ISL_857723, EPI_ISL_857724, EPI_ISL_857725, EPI_ISL_857726, EPI_ISL_857727, EPI_ISL_857728, EPI_ISL_857729, EPI_ISL_857730, EPI_ISL_857731, EPI_ISL_857732, EPI_ISL_857733, EPI_ISL_857734, EPI_ISL_857735,
EPI_ISL_857736, EPI_ISL_857737, EPI_ISL_857738, EPI_ISL_857739, EPI_ISL_857740, EPI_ISL_857741, EPI_ISL_857742, EPI_ISL_857743, EPI_ISL_857744, EPI_ISL_857745, EPI_ISL_857746, EPI_ISL_857747, EPI_ISL_857748, EPI_ISL_857749, EPI_ISL_857750, EPI_ISL_857751, EPI_ISL_857752, EPI_ISL_857753, EPI_ISL_857754, EPI_ISL_857755, EPI_ISL_857756, EPI_ISL_857757, EPI_ISL_857758, EPI_ISL_857760,
EPI_ISL_857761, EPI_ISL_857762, EPI_ISL_857763, EPI_ISL_857764, EPI_ISL_857765, EPI_ISL_857766, EPI_ISL_857767, EPI_ISL_857768, EPI_ISL_857769, EPI_ISL_857770, EPI_ISL_857771, EPI_ISL_857772, EPI_ISL_857773, EPI_ISL_857774, EPI_ISL_857775, EPI_ISL_857776, EPI_ISL_857777, EPI_ISL_857778, EPI_ISL_857779, EPI_ISL_857780, EPI_ISL_857781, EPI_ISL_857782, EPI_ISL_857783, EPI_ISL_857784,
EPI_ISL_857785, EPI_ISL_857786, EPI_ISL_857787, EPI_ISL_857788, EPI_ISL_857789, EPI_ISL_857790, EPI_ISL_857791, EPI_ISL_857792, EPI_ISL_857793, EPI_ISL_857794, EPI_ISL_857795, EPI_ISL_857796, EPI_ISL_857797, EPI_ISL_857798, EPI_ISL_857799, EPI_ISL_857800, EPI_ISL_857801, EPI_ISL_857802, EPI_ISL_857803, EPI_ISL_857804, EPI_ISL_857805, EPI_ISL_857806, EPI_ISL_857807, EPI_ISL_857808,
EPI_ISL_857809, EPI_ISL_857810, EPI_ISL_857811, EPI_ISL_857812, EPI_ISL_857813, EPI_ISL_857814, EPI_ISL_857815, EPI_ISL_857816, EPI_ISL_857817, EPI_ISL_857820, EPI_ISL_857821, EPI_ISL_857822, EPI_ISL_857824, EPI_ISL_857825, EPI_ISL_857826, EPI_ISL_857827, EPI_ISL_857828, EPI_ISL_857829, EPI_ISL_857830, EPI_ISL_857831, EPI_ISL_857832, EPI_ISL_857833, EPI_ISL_857834, EPI_ISL_857835,
EPI_ISL_857836, EPI_ISL_857837, EPI_ISL_857838, EPI_ISL_857839, EPI_ISL_857840, EPI_ISL_857841, EPI_ISL_857842, EPI_ISL_857843, EPI_ISL_857844, EPI_ISL_857845, EPI_ISL_857846, EPI_ISL_857847, EPI_ISL_857848, EPI_ISL_857849, EPI_ISL_857850, EPI_ISL_857851, EPI_ISL_857853, EPI_ISL_857854, EPI_ISL_857856, EPI_ISL_857857, EPI_ISL_857858, EPI_ISL_857859, EPI_ISL_857860, EPI_ISL_857861,
EPI_ISL_857862, EPI_ISL_857863, EPI_ISL_857864, EPI_ISL_857865, EPI_ISL_857866, EPI_ISL_857867, EPI_ISL_857868, EPI_ISL_857869, EPI_ISL_873282, EPI_ISL_873295, EPI_ISL_873365, EPI_ISL_873368, EPI_ISL_873418, EPI_ISL_873432, EPI_ISL_873489, EPI_ISL_873513, EPI_ISL_873561, EPI_ISL_873611, EPI_ISL_873613, EPI_ISL_873624, EPI_ISL_873746, EPI_ISL_873750, EPI_ISL_873752, EPI_ISL_873825,
EPI_ISL_873882, EPI_ISL_879820, EPI_ISL_880038, EPI_ISL_881619, EPI_ISL_945049, EPI_ISL_945056, EPI_ISL_945061, EPI_ISL_945072, EPI_ISL_945084, EPI_ISL_945095, EPI_ISL_945114, EPI_ISL_945115, EPI_ISL_945122, EPI_ISL_945125, EPI_ISL_945130, EPI_ISL_945142, EPI_ISL_945143, EPI_ISL_945146, EPI_ISL_945147, EPI_ISL_945152, EPI_ISL_945155, EPI_ISL_945163, EPI_ISL_945169, EPI_ISL_945181,
EPI_ISL_945190, EPI_ISL_945197, EPI_ISL_945207, EPI_ISL_945208, EPI_ISL_945212, EPI_ISL_945217, EPI_ISL_945221, EPI_ISL_945236, EPI_ISL_945238, EPI_ISL_945241, EPI_ISL_945243, EPI_ISL_945244, EPI_ISL_945248, EPI_ISL_945250, EPI_ISL_945254, EPI_ISL_945275, EPI_ISL_945279, EPI_ISL_945285, EPI_ISL_945288, EPI_ISL_945297, EPI_ISL_945304, EPI_ISL_945309, EPI_ISL_945311, EPI_ISL_945319,
EPI_ISL_945324, EPI_ISL_945332, EPI_ISL_945334, EPI_ISL_945337, EPI_ISL_945345, EPI_ISL_945347, EPI_ISL_945363, EPI_ISL_945364, EPI_ISL_945366, EPI_ISL_1005947, EPI_ISL_1012376, EPI_ISL_1012388, EPI_ISL_1018924

| | | | |
|---|---|---|---|
| see above | Lighthouse Lab in Cambridge | Wellcome Sanger Institute for the COVID-19 Genomics UK (COG-UK) Consortium | Cordelia Langford; David K. Jackson; Dominic Kwiatkowski; Ewan Harrison; Ian Johnston; Jeffrey Barrett; John Sillitoe on behalf of the Wellcome Sanger Institute COVID-19 Surveillance Team; Rob Howes; Roberto Amato; Sonia Goncalves; The Lighthouse Lab in Cambridge and Alex Alderton |

EPI_ISL_851070, EPI_ISL_851077, EPI_ISL_851081, EPI_ISL_851082, EPI_ISL_851085, EPI_ISL_851094, EPI_ISL_851095, EPI_ISL_851101, EPI_ISL_851102, EPI_ISL_851104, EPI_ISL_851107, EPI_ISL_851116, EPI_ISL_851119, EPI_ISL_851124, EPI_ISL_851125, EPI_ISL_851126, EPI_ISL_851130, EPI_ISL_851134, EPI_ISL_851135, EPI_ISL_851137, EPI_ISL_851142, EPI_ISL_851146, EPI_ISL_851147, EPI_ISL_851148,
EPI_ISL_851154, EPI_ISL_851167, EPI_ISL_851178, EPI_ISL_851180, EPI_ISL_851182, EPI_ISL_851187, EPI_ISL_851190, EPI_ISL_851196, EPI_ISL_851206, EPI_ISL_851207, EPI_ISL_851215, EPI_ISL_851217, EPI_ISL_851218, EPI_ISL_851225, EPI_ISL_851229, EPI_ISL_851236, EPI_ISL_851237, EPI_ISL_851241, EPI_ISL_851246, EPI_ISL_851252, EPI_ISL_851253, EPI_ISL_851257, EPI_ISL_851258, EPI_ISL_851259,
EPI_ISL_851264, EPI_ISL_851270, EPI_ISL_851276, EPI_ISL_851285, EPI_ISL_851296, EPI_ISL_851301, EPI_ISL_851305, EPI_ISL_851306, EPI_ISL_851307, EPI_ISL_851310, EPI_ISL_851325, EPI_ISL_851327, EPI_ISL_851329, EPI_ISL_851331, EPI_ISL_851335, EPI_ISL_851341, EPI_ISL_851346, EPI_ISL_851351, EPI_ISL_851358, EPI_ISL_851365, EPI_ISL_851367, EPI_ISL_851371, EPI_ISL_851374, EPI_ISL_851380,
EPI_ISL_851382, EPI_ISL_851383, EPI_ISL_851384, EPI_ISL_851385, EPI_ISL_851386, EPI_ISL_851387, EPI_ISL_851388, EPI_ISL_851389, EPI_ISL_851390, EPI_ISL_851391, EPI_ISL_851392, EPI_ISL_851393, EPI_ISL_851394, EPI_ISL_851395, EPI_ISL_851396, EPI_ISL_851397, EPI_ISL_851398, EPI_ISL_851399, EPI_ISL_851400, EPI_ISL_851401, EPI_ISL_851402, EPI_ISL_851403, EPI_ISL_851404, EPI_ISL_851405,
EPI_ISL_851406, EPI_ISL_851407, EPI_ISL_851408, EPI_ISL_851409, EPI_ISL_851410, EPI_ISL_851411, EPI_ISL_851412, EPI_ISL_851413, EPI_ISL_851414, EPI_ISL_851415, EPI_ISL_851416, EPI_ISL_851417, EPI_ISL_851418, EPI_ISL_851419, EPI_ISL_851420, EPI_ISL_851421, EPI_ISL_851422, EPI_ISL_851423, EPI_ISL_851424, EPI_ISL_851425, EPI_ISL_851426, EPI_ISL_851427, EPI_ISL_851428, EPI_ISL_851429,
EPI_ISL_851430, EPI_ISL_851431, EPI_ISL_851432, EPI_ISL_851433, EPI_ISL_851434, EPI_ISL_851435, EPI_ISL_851436, EPI_ISL_851437, EPI_ISL_851438, EPI_ISL_851439, EPI_ISL_851440, EPI_ISL_851441, EPI_ISL_851442, EPI_ISL_851443, EPI_ISL_851444, EPI_ISL_851445, EPI_ISL_851446, EPI_ISL_851447, EPI_ISL_851448, EPI_ISL_851449, EPI_ISL_851450, EPI_ISL_851451, EPI_ISL_851452, EPI_ISL_851453,
EPI_ISL_851454, EPI_ISL_851456, EPI_ISL_851457, EPI_ISL_851458, EPI_ISL_851459, EPI_ISL_851460, EPI_ISL_851462, EPI_ISL_851463, EPI_ISL_851464, EPI_ISL_851465, EPI_ISL_851466, EPI_ISL_851468, EPI_ISL_851469, EPI_ISL_851470, EPI_ISL_851471, EPI_ISL_851472, EPI_ISL_851473, EPI_ISL_851474, EPI_ISL_851475, EPI_ISL_851476, EPI_ISL_851477, EPI_ISL_851478, EPI_ISL_851479, EPI_ISL_851480,
EPI_ISL_851481, EPI_ISL_851482, EPI_ISL_851483, EPI_ISL_851484, EPI_ISL_851485, EPI_ISL_851486, EPI_ISL_851487, EPI_ISL_851488, EPI_ISL_851489, EPI_ISL_851490, EPI_ISL_851491, EPI_ISL_851492, EPI_ISL_851493, EPI_ISL_851494, EPI_ISL_851495, EPI_ISL_851496, EPI_ISL_851497, EPI_ISL_851498, EPI_ISL_851499, EPI_ISL_851500, EPI_ISL_851501, EPI_ISL_851502, EPI_ISL_851503, EPI_ISL_851504,
EPI_ISL_851505, EPI_ISL_851506, EPI_ISL_851507, EPI_ISL_851508, EPI_ISL_851509, EPI_ISL_851510, EPI_ISL_851511, EPI_ISL_851512, EPI_ISL_851513, EPI_ISL_851514, EPI_ISL_851515, EPI_ISL_851516, EPI_ISL_851517, EPI_ISL_851518, EPI_ISL_851520, EPI_ISL_851521, EPI_ISL_851522, EPI_ISL_851523, EPI_ISL_851524, EPI_ISL_851525, EPI_ISL_851526, EPI_ISL_851527, EPI_ISL_851528, EPI_ISL_851529,
EPI_ISL_851530, EPI_ISL_851531, EPI_ISL_851532, EPI_ISL_851533, EPI_ISL_851534, EPI_ISL_851535, EPI_ISL_851536, EPI_ISL_851537, EPI_ISL_851538, EPI_ISL_851539, EPI_ISL_851540, EPI_ISL_851541, EPI_ISL_851542, EPI_ISL_851543, EPI_ISL_851544, EPI_ISL_851545, EPI_ISL_851546, EPI_ISL_851547, EPI_ISL_851548, EPI_ISL_851549, EPI_ISL_851550, EPI_ISL_851551, EPI_ISL_851552, EPI_ISL_851553,
EPI_ISL_851554, EPI_ISL_851555, EPI_ISL_851556, EPI_ISL_851557, EPI_ISL_851558, EPI_ISL_851560, EPI_ISL_851561, EPI_ISL_851562, EPI_ISL_851563, EPI_ISL_851564, EPI_ISL_851565, EPI_ISL_851566, EPI_ISL_851567, EPI_ISL_851568, EPI_ISL_851569, EPI_ISL_851570, EPI_ISL_851571, EPI_ISL_851572, EPI_ISL_851573, EPI_ISL_851574, EPI_ISL_851575, EPI_ISL_851576, EPI_ISL_851577, EPI_ISL_851578,
EPI_ISL_851579, EPI_ISL_851580, EPI_ISL_851581, EPI_ISL_851582, EPI_ISL_851583, EPI_ISL_851584, EPI_ISL_851585, EPI_ISL_851586, EPI_ISL_851587, EPI_ISL_851588, EPI_ISL_851589, EPI_ISL_851590, EPI_ISL_851591, EPI_ISL_851592, EPI_ISL_851593, EPI_ISL_851595, EPI_ISL_851596, EPI_ISL_851597, EPI_ISL_851598, EPI_ISL_851599, EPI_ISL_851600, EPI_ISL_851601, EPI_ISL_851602, EPI_ISL_851603,
EPI_ISL_851604, EPI_ISL_851605, EPI_ISL_851606, EPI_ISL_851607, EPI_ISL_851608, EPI_ISL_851609, EPI_ISL_851610, EPI_ISL_851611, EPI_ISL_851612, EPI_ISL_851613, EPI_ISL_851615, EPI_ISL_851616, EPI_ISL_851617, EPI_ISL_851618, EPI_ISL_851619, EPI_ISL_851620, EPI_ISL_851621, EPI_ISL_851622, EPI_ISL_851623, EPI_ISL_851624, EPI_ISL_851625, EPI_ISL_851626, EPI_ISL_851627, EPI_ISL_851629,
EPI_ISL_851630, EPI_ISL_851631, EPI_ISL_851632, EPI_ISL_851633, EPI_ISL_851634, EPI_ISL_851635, EPI_ISL_851636, EPI_ISL_851637, EPI_ISL_851638, EPI_ISL_851639, EPI_ISL_851640, EPI_ISL_851641, EPI_ISL_851642, EPI_ISL_851643, EPI_ISL_851644, EPI_ISL_851645, EPI_ISL_851646, EPI_ISL_851647, EPI_ISL_851649, EPI_ISL_851650, EPI_ISL_851651, EPI_ISL_851652, EPI_ISL_851653, EPI_ISL_851654,
EPI_ISL_851655, EPI_ISL_851656, EPI_ISL_851658, EPI_ISL_851659, EPI_ISL_851660, EPI_ISL_851661, EPI_ISL_851662, EPI_ISL_851663, EPI_ISL_851664, EPI_ISL_851665, EPI_ISL_851666, EPI_ISL_851667, EPI_ISL_851668, EPI_ISL_851669, EPI_ISL_851670, EPI_ISL_851671, EPI_ISL_851672, EPI_ISL_851674, EPI_ISL_851676, EPI_ISL_851681, EPI_ISL_851685, EPI_ISL_851686, EPI_ISL_851687, EPI_ISL_851689,
EPI_ISL_851690, EPI_ISL_851693, EPI_ISL_851701, EPI_ISL_851714, EPI_ISL_851720, EPI_ISL_851725, EPI_ISL_851727, EPI_ISL_851729, EPI_ISL_851734, EPI_ISL_851738, EPI_ISL_851743, EPI_ISL_851747, EPI_ISL_851749, EPI_ISL_851754, EPI_ISL_851757, EPI_ISL_851760, EPI_ISL_851763, EPI_ISL_851767, EPI_ISL_851770, EPI_ISL_851772, EPI_ISL_851774, EPI_ISL_851780, EPI_ISL_851781, EPI_ISL_851785,
EPI_ISL_851792, EPI_ISL_851794, EPI_ISL_851796, EPI_ISL_851797, EPI_ISL_851798, EPI_ISL_851799, EPI_ISL_851803, EPI_ISL_851805, EPI_ISL_851807, EPI_ISL_851812, EPI_ISL_851814, EPI_ISL_851829, EPI_ISL_851837, EPI_ISL_851840, EPI_ISL_851846, EPI_ISL_851847, EPI_ISL_851851, EPI_ISL_851856, EPI_ISL_851857, EPI_ISL_851859, EPI_ISL_851863, EPI_ISL_851864, EPI_ISL_851865, EPI_ISL_851866,
EPI_ISL_851870, EPI_ISL_851871, EPI_ISL_851876, EPI_ISL_851877, EPI_ISL_851885, EPI_ISL_851888, EPI_ISL_851891, EPI_ISL_851892, EPI_ISL_851895, EPI_ISL_851898, EPI_ISL_851901, EPI_ISL_851905, EPI_ISL_851906, EPI_ISL_851916, EPI_ISL_851920, EPI_ISL_851922, EPI_ISL_851929, EPI_ISL_851934, EPI_ISL_851936, EPI_ISL_851940, EPI_ISL_851950, EPI_ISL_851955, EPI_ISL_851956, EPI_ISL_852257,
EPI_ISL_852258, EPI_ISL_852259, EPI_ISL_852260, EPI_ISL_852264, EPI_ISL_852268, EPI_ISL_852269, EPI_ISL_852275, EPI_ISL_852276, EPI_ISL_852279, EPI_ISL_852281, EPI_ISL_852283, EPI_ISL_852285, EPI_ISL_852286, EPI_ISL_852287, EPI_ISL_852288, EPI_ISL_852289, EPI_ISL_852292, EPI_ISL_852293, EPI_ISL_852295, EPI_ISL_852300, EPI_ISL_852301, EPI_ISL_852303, EPI_ISL_852305, EPI_ISL_852306,
EPI_ISL_852307, EPI_ISL_852310, EPI_ISL_852312, EPI_ISL_852313, EPI_ISL_852314, EPI_ISL_852316, EPI_ISL_852319, EPI_ISL_852321, EPI_ISL_852322, EPI_ISL_852325, EPI_ISL_852326, EPI_ISL_852328, EPI_ISL_852334, EPI_ISL_852335, EPI_ISL_852338, EPI_ISL_852339, EPI_ISL_852340, EPI_ISL_852343, EPI_ISL_852344, EPI_ISL_852345, EPI_ISL_852347, EPI_ISL_852348, EPI_ISL_852350, EPI_ISL_852353,
EPI_ISL_852358, EPI_ISL_852360, EPI_ISL_852361, EPI_ISL_852364, EPI_ISL_852367, EPI_ISL_852368, EPI_ISL_852369, EPI_ISL_852370, EPI_ISL_852374, EPI_ISL_852375, EPI_ISL_852378, EPI_ISL_852389, EPI_ISL_852391, EPI_ISL_852393, EPI_ISL_852395, EPI_ISL_852397, EPI_ISL_852398, EPI_ISL_852404, EPI_ISL_852406, EPI_ISL_852407, EPI_ISL_852412, EPI_ISL_852413, EPI_ISL_852414, EPI_ISL_852415,
EPI_ISL_852416, EPI_ISL_852417, EPI_ISL_852420, EPI_ISL_852421, EPI_ISL_852422, EPI_ISL_852424, EPI_ISL_852425, EPI_ISL_852426, EPI_ISL_852429, EPI_ISL_852433, EPI_ISL_852435, EPI_ISL_852438, EPI_ISL_852439, EPI_ISL_852440, EPI_ISL_852444, EPI_ISL_852445, EPI_ISL_852446, EPI_ISL_852447, EPI_ISL_852448, EPI_ISL_852451, EPI_ISL_852454, EPI_ISL_852459, EPI_ISL_852460, EPI_ISL_852462,
EPI_ISL_852463, EPI_ISL_852467, EPI_ISL_852471, EPI_ISL_852472, EPI_ISL_852474, EPI_ISL_852475, EPI_ISL_852476, EPI_ISL_852479, EPI_ISL_852481, EPI_ISL_852484, EPI_ISL_852485, EPI_ISL_852487, EPI_ISL_852491, EPI_ISL_852497, EPI_ISL_852498, EPI_ISL_852500, EPI_ISL_852503, EPI_ISL_852504, EPI_ISL_852506, EPI_ISL_852508, EPI_ISL_852509, EPI_ISL_852510, EPI_ISL_852514, EPI_ISL_852517,
EPI_ISL_852519, EPI_ISL_852520, EPI_ISL_852522, EPI_ISL_852524, EPI_ISL_852527, EPI_ISL_852529, EPI_ISL_852530, EPI_ISL_852531, EPI_ISL_852532, EPI_ISL_852533, EPI_ISL_852534, EPI_ISL_852535, EPI_ISL_852536, EPI_ISL_852537, EPI_ISL_852540, EPI_ISL_852541, EPI_ISL_852544, EPI_ISL_852548, EPI_ISL_852549, EPI_ISL_852550, EPI_ISL_852555, EPI_ISL_852557, EPI_ISL_852558, EPI_ISL_852559,

| | | | |
|---|---|---|---|
| EPI_ISL_852560, EPI_ISL_852561, EPI_ISL_852562, EPI_ISL_852563, EPI_ISL_857870, EPI_ISL_857871, EPI_ISL_857872, EPI_ISL_857873, EPI_ISL_857876, EPI_ISL_857878, EPI_ISL_857881, EPI_ISL_857885, EPI_ISL_857887, EPI_ISL_857888, EPI_ISL_857889, EPI_ISL_857892, EPI_ISL_857893, EPI_ISL_857894, EPI_ISL_857895, EPI_ISL_857896, EPI_ISL_857897, EPI_ISL_857898, EPI_ISL_857899, EPI_ISL_857900, EPI_ISL_857901, EPI_ISL_857902, EPI_ISL_857903, EPI_ISL_857904, EPI_ISL_857905, EPI_ISL_857906, EPI_ISL_857907, EPI_ISL_857908, EPI_ISL_857909, EPI_ISL_857910, EPI_ISL_857911, EPI_ISL_857912, EPI_ISL_857913, EPI_ISL_857914, EPI_ISL_857915, EPI_ISL_857916, EPI_ISL_857917, EPI_ISL_857918, EPI_ISL_857919, EPI_ISL_857920, EPI_ISL_857921, EPI_ISL_857922, EPI_ISL_857923, EPI_ISL_857924, EPI_ISL_857925, EPI_ISL_857926, EPI_ISL_857927, EPI_ISL_857928, EPI_ISL_857929, EPI_ISL_857930, EPI_ISL_857931, EPI_ISL_857932, EPI_ISL_857933, EPI_ISL_857934, EPI_ISL_857935, EPI_ISL_857936, EPI_ISL_857937, EPI_ISL_857938, EPI_ISL_857939, EPI_ISL_857940, EPI_ISL_857941, EPI_ISL_857942, EPI_ISL_857943, EPI_ISL_857944, EPI_ISL_857945, EPI_ISL_857946, EPI_ISL_857947, EPI_ISL_857948, EPI_ISL_857949, EPI_ISL_857950, EPI_ISL_857951, EPI_ISL_857952, EPI_ISL_857953, EPI_ISL_857954, EPI_ISL_857955, EPI_ISL_857956, EPI_ISL_857957, EPI_ISL_857958, EPI_ISL_857959, EPI_ISL_858011, EPI_ISL_858012, EPI_ISL_858013, EPI_ISL_858015, EPI_ISL_858016, EPI_ISL_858017, EPI_ISL_858018, EPI_ISL_858019, EPI_ISL_858021, EPI_ISL_858022, EPI_ISL_858023, EPI_ISL_858024, EPI_ISL_858025, EPI_ISL_858026, EPI_ISL_858027, EPI_ISL_858030, EPI_ISL_858031, EPI_ISL_858032, EPI_ISL_858033, EPI_ISL_858034, EPI_ISL_858035, EPI_ISL_858036, EPI_ISL_858038, EPI_ISL_858039, EPI_ISL_858040, EPI_ISL_858041, EPI_ISL_858042, EPI_ISL_858043, EPI_ISL_858044, EPI_ISL_858045, EPI_ISL_858046, EPI_ISL_858047, EPI_ISL_858048, EPI_ISL_858049, EPI_ISL_858050, EPI_ISL_858051, EPI_ISL_858052, EPI_ISL_858053, EPI_ISL_858054, EPI_ISL_858055, EPI_ISL_858056, EPI_ISL_858057, EPI_ISL_858058, EPI_ISL_858059, EPI_ISL_858060, EPI_ISL_858061, EPI_ISL_858062, EPI_ISL_858063, EPI_ISL_858316, EPI_ISL_863431, EPI_ISL_907210, EPI_ISL_907750 | | | |
| see above | Lighthouse Lab in Glasgow | Wellcome Sanger Institute for the COVID-19 Genomics UK (COG-UK) Consortium | Anna Dominiczak and Alex Alderton; Carol Clugston; Cordelia Langford; David Gray; David K. Jackson; Dominic Kwiatkowski; Ewan Harrison; Harper VanSteenhouse; Ian Johnston; John Sillitoe on behalf of the Wellcome Sanger Institute COVID-19 Surveillance Team; Roberto Amato; Sonia Goncalves; Yumi Kasai |
| EPI_ISL_865068 | Lighthouse Lab in Glasgow / MRC-University of Glasgow Centre for Virus Research | COVID-19 Genomics UK (COG-UK) Consortium | Alasdair MacLean; Alice Broos; Ana da Silva Filipe; Anna Dominiczak; Antonia Ho; Carol Clugston; Daniel Mair; David Gray; David L Robertson; Emma Thomson; Harper VanSteenhouse; James Shepherd; Jenna Nichols; Joseph Hughes; Kathy Li; Kathy Smollett; Kyriaki Nomikou; Lily Tong; Matthew Holden; Natasha Johnson; Richard Orton; Rory Gunson; Sarah McDonald; Sharif Shaaban; Sreenu Vattipally; Stephen Carmichael; Yumi Kasai |
| EPI_ISL_851957, EPI_ISL_851958, EPI_ISL_851959, EPI_ISL_851960, EPI_ISL_851961, EPI_ISL_851962, EPI_ISL_851963, EPI_ISL_851965, EPI_ISL_851966, EPI_ISL_851967, EPI_ISL_851968, EPI_ISL_851969, EPI_ISL_851970, EPI_ISL_851971, EPI_ISL_851972, EPI_ISL_851973, EPI_ISL_851974, EPI_ISL_851975, EPI_ISL_851976, EPI_ISL_851977, EPI_ISL_851978, EPI_ISL_851979, EPI_ISL_851980, EPI_ISL_851981, EPI_ISL_851982, EPI_ISL_851983, EPI_ISL_851984, EPI_ISL_851985, EPI_ISL_851986, EPI_ISL_851987, EPI_ISL_851988, EPI_ISL_851990, EPI_ISL_851991, EPI_ISL_851992, EPI_ISL_851993, EPI_ISL_851994, EPI_ISL_851995, EPI_ISL_851996, EPI_ISL_851997, EPI_ISL_851998, EPI_ISL_851999, EPI_ISL_852000, EPI_ISL_852001, EPI_ISL_852003, EPI_ISL_852004, EPI_ISL_852005, EPI_ISL_852006, EPI_ISL_852007, EPI_ISL_852008, EPI_ISL_852009, EPI_ISL_852010, EPI_ISL_852011, EPI_ISL_852012, EPI_ISL_852013, EPI_ISL_852015, EPI_ISL_852016, EPI_ISL_852018, EPI_ISL_852019, EPI_ISL_852020, EPI_ISL_852021, EPI_ISL_852022, EPI_ISL_852023, EPI_ISL_852024, EPI_ISL_852026, EPI_ISL_852027, EPI_ISL_852028, EPI_ISL_852029, EPI_ISL_852030, EPI_ISL_852031, EPI_ISL_852032, EPI_ISL_852033, EPI_ISL_852034, EPI_ISL_852035, EPI_ISL_852036, EPI_ISL_852037, EPI_ISL_852038, EPI_ISL_852039, EPI_ISL_852040, EPI_ISL_852041, EPI_ISL_852042, EPI_ISL_852043, EPI_ISL_852044, EPI_ISL_852045, EPI_ISL_852046, EPI_ISL_852048, EPI_ISL_852049, EPI_ISL_852050, EPI_ISL_852051, EPI_ISL_852052, EPI_ISL_852053, EPI_ISL_852054, EPI_ISL_852055, EPI_ISL_852056, EPI_ISL_852057, EPI_ISL_852058, EPI_ISL_852059, EPI_ISL_852060, EPI_ISL_852061, EPI_ISL_852062, EPI_ISL_852063, EPI_ISL_852064, EPI_ISL_852065, EPI_ISL_852066, EPI_ISL_852067, EPI_ISL_852068, EPI_ISL_852069, EPI_ISL_852070, EPI_ISL_852071, EPI_ISL_852072, EPI_ISL_852073, EPI_ISL_852075, EPI_ISL_852076, EPI_ISL_852077, EPI_ISL_852078, EPI_ISL_852079, EPI_ISL_852080, EPI_ISL_852081, EPI_ISL_852082, EPI_ISL_852084, EPI_ISL_852085, EPI_ISL_852086, EPI_ISL_852087, EPI_ISL_852088, EPI_ISL_852089, EPI_ISL_852090, EPI_ISL_852092, EPI_ISL_852093, EPI_ISL_852094, EPI_ISL_852095, EPI_ISL_852096, EPI_ISL_852097, EPI_ISL_852098, EPI_ISL_852099, EPI_ISL_852100, EPI_ISL_852101, EPI_ISL_852102, EPI_ISL_852103, EPI_ISL_852104, EPI_ISL_852105, EPI_ISL_852106, EPI_ISL_852108, EPI_ISL_852109, EPI_ISL_852110, EPI_ISL_852111, EPI_ISL_852112, EPI_ISL_852113, EPI_ISL_852114, EPI_ISL_852115, EPI_ISL_852116, EPI_ISL_852117, EPI_ISL_852118, EPI_ISL_852119, EPI_ISL_852120, EPI_ISL_852121, EPI_ISL_852122, EPI_ISL_852123, EPI_ISL_852124, EPI_ISL_852125, EPI_ISL_852126, EPI_ISL_852127, EPI_ISL_852128, EPI_ISL_852129, EPI_ISL_852130, EPI_ISL_852131, EPI_ISL_852132, EPI_ISL_852133, EPI_ISL_852134, EPI_ISL_852135, EPI_ISL_852136, EPI_ISL_852137, EPI_ISL_852138, EPI_ISL_852139, EPI_ISL_852140, EPI_ISL_852141, EPI_ISL_852142, EPI_ISL_852143, EPI_ISL_852144, EPI_ISL_852145, EPI_ISL_852146, EPI_ISL_852147, EPI_ISL_852148, EPI_ISL_852149, EPI_ISL_852150, EPI_ISL_852151, EPI_ISL_852152, EPI_ISL_852153, EPI_ISL_852154, EPI_ISL_852155, EPI_ISL_852156, EPI_ISL_852157, EPI_ISL_852158, EPI_ISL_852159, EPI_ISL_852160, EPI_ISL_852161, EPI_ISL_852162, EPI_ISL_852163, EPI_ISL_852164, EPI_ISL_852165, EPI_ISL_852166, EPI_ISL_852167, EPI_ISL_852168, EPI_ISL_852169, EPI_ISL_852170, EPI_ISL_852171, EPI_ISL_852172, EPI_ISL_852173, EPI_ISL_852174, EPI_ISL_852175, EPI_ISL_852176, EPI_ISL_852177, EPI_ISL_852178, EPI_ISL_852180, EPI_ISL_852181, EPI_ISL_852182, EPI_ISL_852184, EPI_ISL_852185, EPI_ISL_852186, EPI_ISL_852187, EPI_ISL_852188, EPI_ISL_852189, EPI_ISL_852192, EPI_ISL_852193, EPI_ISL_852194, EPI_ISL_852195, EPI_ISL_852196, EPI_ISL_852197, EPI_ISL_852198, EPI_ISL_852199, EPI_ISL_852200, EPI_ISL_852201, EPI_ISL_852202, EPI_ISL_852203, EPI_ISL_852204, EPI_ISL_852205, EPI_ISL_852207, EPI_ISL_852208, EPI_ISL_852209, EPI_ISL_852210, EPI_ISL_852211, EPI_ISL_852212, EPI_ISL_852213, EPI_ISL_852215, EPI_ISL_852216, EPI_ISL_852217, EPI_ISL_852218, EPI_ISL_852219, EPI_ISL_852220, EPI_ISL_852221, EPI_ISL_852222, EPI_ISL_852223, EPI_ISL_852224, EPI_ISL_852226, EPI_ISL_852227, EPI_ISL_852228, EPI_ISL_852229, EPI_ISL_852230, EPI_ISL_852231, EPI_ISL_852232, EPI_ISL_852233, EPI_ISL_852234, EPI_ISL_852235, EPI_ISL_852236, EPI_ISL_852237, EPI_ISL_852238, EPI_ISL_852239, EPI_ISL_852240, EPI_ISL_852241, EPI_ISL_852242, EPI_ISL_852243, EPI_ISL_852244, EPI_ISL_852245, EPI_ISL_852246, EPI_ISL_852247, EPI_ISL_852248, EPI_ISL_852249, EPI_ISL_852250, EPI_ISL_852251, EPI_ISL_852252, EPI_ISL_852253, EPI_ISL_852254, EPI_ISL_852255, EPI_ISL_863531, EPI_ISL_863533, EPI_ISL_863537, EPI_ISL_863539, EPI_ISL_863540, EPI_ISL_863542, EPI_ISL_863543, EPI_ISL_863544, EPI_ISL_863546, EPI_ISL_863547, EPI_ISL_863549, EPI_ISL_863551, EPI_ISL_863552, EPI_ISL_863553, EPI_ISL_863555, EPI_ISL_863556, EPI_ISL_863557, EPI_ISL_863558, EPI_ISL_863560, EPI_ISL_863561, EPI_ISL_863562, EPI_ISL_863565, EPI_ISL_863566, EPI_ISL_863567, EPI_ISL_863570, EPI_ISL_863572, EPI_ISL_863574, EPI_ISL_863576, EPI_ISL_863577, EPI_ISL_863578, EPI_ISL_863579, EPI_ISL_863580, EPI_ISL_863581, EPI_ISL_863582, EPI_ISL_863583, EPI_ISL_863584, EPI_ISL_863586, EPI_ISL_863587, EPI_ISL_863588, EPI_ISL_863589, EPI_ISL_863591, EPI_ISL_863592, EPI_ISL_863593, EPI_ISL_863594, EPI_ISL_863595, EPI_ISL_863599, EPI_ISL_863601, EPI_ISL_863603, EPI_ISL_863604, EPI_ISL_863605, EPI_ISL_863606, EPI_ISL_863608, EPI_ISL_863609, EPI_ISL_863610, EPI_ISL_863612, EPI_ISL_863614, EPI_ISL_863616, EPI_ISL_863618, EPI_ISL_863619, EPI_ISL_863621, EPI_ISL_863622, EPI_ISL_863623, EPI_ISL_863625, EPI_ISL_863626, EPI_ISL_863627, EPI_ISL_863628, EPI_ISL_863629, EPI_ISL_863630, EPI_ISL_863633, EPI_ISL_863636, EPI_ISL_863640, EPI_ISL_863641, EPI_ISL_863643, EPI_ISL_863644, EPI_ISL_863647, EPI_ISL_863648, EPI_ISL_863649, EPI_ISL_863650, EPI_ISL_863652, EPI_ISL_863654, EPI_ISL_863655, EPI_ISL_863656, EPI_ISL_863658, EPI_ISL_863659, EPI_ISL_863661, EPI_ISL_863662, EPI_ISL_863663, EPI_ISL_863664, EPI_ISL_863665, EPI_ISL_863666, EPI_ISL_863667, EPI_ISL_863668, EPI_ISL_863670, EPI_ISL_863671, EPI_ISL_863672, EPI_ISL_863673, EPI_ISL_863674, EPI_ISL_863675, EPI_ISL_863676, EPI_ISL_863680, EPI_ISL_863682, EPI_ISL_863683, EPI_ISL_863684, EPI_ISL_863685, EPI_ISL_863686, EPI_ISL_863687, EPI_ISL_863688, EPI_ISL_863689, EPI_ISL_863690, EPI_ISL_863691, EPI_ISL_863692, EPI_ISL_863693, EPI_ISL_863694, EPI_ISL_863695, EPI_ISL_863697, EPI_ISL_863698, EPI_ISL_863701, EPI_ISL_863702, EPI_ISL_863703, EPI_ISL_863704, EPI_ISL_863706, EPI_ISL_863708, EPI_ISL_863710, EPI_ISL_863712, EPI_ISL_863713, EPI_ISL_863714, EPI_ISL_863716, EPI_ISL_863717, EPI_ISL_863718, EPI_ISL_863719, EPI_ISL_863720, EPI_ISL_863722, EPI_ISL_863724, EPI_ISL_863727, EPI_ISL_863728, EPI_ISL_863730, EPI_ISL_863731, EPI_ISL_863735, EPI_ISL_863736, EPI_ISL_863737, EPI_ISL_863738, EPI_ISL_863739, EPI_ISL_863740, EPI_ISL_863743, EPI_ISL_863744, EPI_ISL_863745, EPI_ISL_863748, EPI_ISL_863749, EPI_ISL_863750, EPI_ISL_863751, EPI_ISL_863752, EPI_ISL_863753, EPI_ISL_863754, EPI_ISL_863755, EPI_ISL_863756, EPI_ISL_863757, EPI_ISL_863758, EPI_ISL_863760, EPI_ISL_863763, EPI_ISL_863764, EPI_ISL_863765, EPI_ISL_863767, EPI_ISL_863768, EPI_ISL_863769, EPI_ISL_863770, EPI_ISL_863771, EPI_ISL_863772, EPI_ISL_863774, EPI_ISL_863775, EPI_ISL_863777, EPI_ISL_863778, EPI_ISL_863779, EPI_ISL_863780, EPI_ISL_863782, EPI_ISL_863783, EPI_ISL_863786, EPI_ISL_863787, EPI_ISL_863791, EPI_ISL_863794, EPI_ISL_863798, EPI_ISL_863799, EPI_ISL_863800, EPI_ISL_863801, EPI_ISL_863805, EPI_ISL_863807, EPI_ISL_863808, EPI_ISL_863813, EPI_ISL_863814, EPI_ISL_863815, EPI_ISL_863816, EPI_ISL_863817, EPI_ISL_863818, EPI_ISL_863819, EPI_ISL_863820, EPI_ISL_863821, EPI_ISL_863822, EPI_ISL_863823, EPI_ISL_863824, EPI_ISL_863825, EPI_ISL_863826, EPI_ISL_863827, EPI_ISL_863828, EPI_ISL_863829, EPI_ISL_863831, EPI_ISL_863832, EPI_ISL_863833, EPI_ISL_863834, EPI_ISL_863835, EPI_ISL_863836, EPI_ISL_863837, EPI_ISL_863839, EPI_ISL_863840, EPI_ISL_863843, EPI_ISL_863844, EPI_ISL_863846, EPI_ISL_863849, EPI_ISL_863852, EPI_ISL_863854, EPI_ISL_863855, EPI_ISL_863857, EPI_ISL_863859, EPI_ISL_863861, EPI_ISL_863862, EPI_ISL_863864, EPI_ISL_863866, EPI_ISL_863867, EPI_ISL_863868, EPI_ISL_863899, EPI_ISL_863901, EPI_ISL_863912, EPI_ISL_863913, EPI_ISL_863921, EPI_ISL_863922, EPI_ISL_863956, EPI_ISL_863969, EPI_ISL_863996, EPI_ISL_864005, EPI_ISL_864009, EPI_ISL_864026, EPI_ISL_864036, EPI_ISL_864043, EPI_ISL_864098, EPI_ISL_864124, EPI_ISL_864133, EPI_ISL_864165, EPI_ISL_864169, EPI_ISL_864173, EPI_ISL_864175, EPI_ISL_864176, EPI_ISL_864191, EPI_ISL_864193, EPI_ISL_864203, EPI_ISL_864205, EPI_ISL_864206, EPI_ISL_864207, EPI_ISL_864208, EPI_ISL_864209, EPI_ISL_864210, EPI_ISL_864211, EPI_ISL_864212, EPI_ISL_864214, EPI_ISL_864215, EPI_ISL_864218, EPI_ISL_864219, EPI_ISL_864220, EPI_ISL_864221, EPI_ISL_864222, EPI_ISL_864223, EPI_ISL_864224, EPI_ISL_864225, EPI_ISL_864227, EPI_ISL_864228, EPI_ISL_864229, EPI_ISL_864230, EPI_ISL_864231, EPI_ISL_864232, EPI_ISL_864233, EPI_ISL_864234, EPI_ISL_864235, EPI_ISL_864236, EPI_ISL_864237, EPI_ISL_864238, EPI_ISL_864240, EPI_ISL_864241, EPI_ISL_864242, EPI_ISL_864243, EPI_ISL_864245, EPI_ISL_864246, EPI_ISL_864247, EPI_ISL_864248, EPI_ISL_864249, EPI_ISL_864250, EPI_ISL_864251, EPI_ISL_864252, EPI_ISL_864253, EPI_ISL_864254, EPI_ISL_864256, EPI_ISL_864257, EPI_ISL_864258, EPI_ISL_864259, EPI_ISL_864260, EPI_ISL_864261, EPI_ISL_864263, EPI_ISL_864264, EPI_ISL_864265, EPI_ISL_864268, EPI_ISL_864269, EPI_ISL_864270, EPI_ISL_864271, EPI_ISL_864272, EPI_ISL_864273, EPI_ISL_864275, EPI_ISL_864276, EPI_ISL_864277, EPI_ISL_864278, EPI_ISL_864279, EPI_ISL_864280, EPI_ISL_864282, EPI_ISL_864283, EPI_ISL_864284, EPI_ISL_864285, EPI_ISL_864286, EPI_ISL_864287, EPI_ISL_864288, EPI_ISL_864289, EPI_ISL_864290, EPI_ISL_864292, EPI_ISL_864293, EPI_ISL_864294, EPI_ISL_864295, EPI_ISL_864296, EPI_ISL_864298, EPI_ISL_864299, EPI_ISL_864300, EPI_ISL_864301, EPI_ISL_864302, EPI_ISL_864303, EPI_ISL_864304, EPI_ISL_864306, EPI_ISL_864307, EPI_ISL_864310, EPI_ISL_864312, EPI_ISL_864313, EPI_ISL_864314, EPI_ISL_864315, EPI_ISL_864316, EPI_ISL_864318, EPI_ISL_864319, EPI_ISL_864320, EPI_ISL_864322, EPI_ISL_864323, EPI_ISL_864325, EPI_ISL_864326, EPI_ISL_864327, EPI_ISL_864328, EPI_ISL_864329, EPI_ISL_864330, EPI_ISL_864331, EPI_ISL_864332, EPI_ISL_864333, EPI_ISL_864334, EPI_ISL_864335, EPI_ISL_864337, EPI_ISL_864338, EPI_ISL_864339, EPI_ISL_864340, EPI_ISL_864341, EPI_ISL_864342, EPI_ISL_864343, EPI_ISL_864344, EPI_ISL_864345, EPI_ISL_864346, EPI_ISL_864347, EPI_ISL_864348, EPI_ISL_864349, EPI_ISL_864350, EPI_ISL_864351, EPI_ISL_864352, EPI_ISL_864353, EPI_ISL_864354, EPI_ISL_864355, EPI_ISL_864357, EPI_ISL_864358, EPI_ISL_864359, EPI_ISL_864360, EPI_ISL_864362, EPI_ISL_864363, EPI_ISL_864364, EPI_ISL_864366, EPI_ISL_864367, EPI_ISL_864369, EPI_ISL_864370, EPI_ISL_864371, EPI_ISL_864372, EPI_ISL_864373, EPI_ISL_864374, EPI_ISL_864375, EPI_ISL_864376, EPI_ISL_864377, EPI_ISL_864378, EPI_ISL_864379, EPI_ISL_864380, EPI_ISL_864381, EPI_ISL_864382, EPI_ISL_864385, EPI_ISL_864386, EPI_ISL_864387, EPI_ISL_864388, EPI_ISL_864389, EPI_ISL_864390, EPI_ISL_864391, EPI_ISL_864392, EPI_ISL_864393, EPI_ISL_864394, EPI_ISL_864395, EPI_ISL_864397, EPI_ISL_864398, EPI_ISL_864401, EPI_ISL_864404, EPI_ISL_864405, EPI_ISL_864406, EPI_ISL_864407, EPI_ISL_864408, EPI_ISL_864410, EPI_ISL_864412, EPI_ISL_864413, EPI_ISL_864415, EPI_ISL_864416, EPI_ISL_864417, EPI_ISL_864418, EPI_ISL_864419, EPI_ISL_864420, EPI_ISL_864421, EPI_ISL_864423, EPI_ISL_864424, EPI_ISL_864425, EPI_ISL_864426, EPI_ISL_864427, EPI_ISL_864428, EPI_ISL_864429, EPI_ISL_864430, EPI_ISL_864431, EPI_ISL_864432, EPI_ISL_864433, EPI_ISL_864434, EPI_ISL_864435, EPI_ISL_864436, EPI_ISL_864437, EPI_ISL_864438, EPI_ISL_864441, EPI_ISL_864442, EPI_ISL_864443, EPI_ISL_864444, EPI_ISL_864445, EPI_ISL_864446, EPI_ISL_864448, EPI_ISL_864449, EPI_ISL_864450, EPI_ISL_864451, EPI_ISL_864453, EPI_ISL_864454, EPI_ISL_864455, EPI_ISL_864456, EPI_ISL_864457, EPI_ISL_864458, EPI_ISL_864459, EPI_ISL_864460, EPI_ISL_864462, EPI_ISL_864463, EPI_ISL_864464, EPI_ISL_864465, EPI_ISL_864466, EPI_ISL_864467, EPI_ISL_864468, EPI_ISL_864470, EPI_ISL_864471, EPI_ISL_864473, EPI_ISL_864474, EPI_ISL_864476, EPI_ISL_864477, EPI_ISL_864478, EPI_ISL_864479, EPI_ISL_864480, EPI_ISL_864481, EPI_ISL_864482, EPI_ISL_864483, EPI_ISL_864485, EPI_ISL_864486, EPI_ISL_864487, EPI_ISL_864488, EPI_ISL_864489, EPI_ISL_864490, EPI_ISL_864491, EPI_ISL_864492, EPI_ISL_864494, EPI_ISL_864495, EPI_ISL_864496, EPI_ISL_864497, EPI_ISL_864498, EPI_ISL_864499, EPI_ISL_864500, EPI_ISL_864501, EPI_ISL_864502, EPI_ISL_864503, EPI_ISL_864504, EPI_ISL_864505, EPI_ISL_864506, EPI_ISL_864507, EPI_ISL_864508, EPI_ISL_864509, EPI_ISL_864510, EPI_ISL_864511, EPI_ISL_864512, EPI_ISL_864513, EPI_ISL_864514, EPI_ISL_864517, EPI_ISL_864518, EPI_ISL_864519, EPI_ISL_864521, EPI_ISL_864522, EPI_ISL_864523, EPI_ISL_864524, EPI_ISL_864525, EPI_ISL_864526, EPI_ISL_864530, EPI_ISL_864531, EPI_ISL_864532, EPI_ISL_864533, EPI_ISL_864535, EPI_ISL_864536, EPI_ISL_864537, EPI_ISL_864538, EPI_ISL_864541, EPI_ISL_873279, EPI_ISL_873280, EPI_ISL_873284, EPI_ISL_873285, EPI_ISL_873287, EPI_ISL_873288, EPI_ISL_873289, EPI_ISL_873290, EPI_ISL_873293, EPI_ISL_873296, EPI_ISL_873299, EPI_ISL_873300, EPI_ISL_873301, EPI_ISL_873303, EPI_ISL_873305, EPI_ISL_873307, EPI_ISL_873308, EPI_ISL_873309, EPI_ISL_873312, EPI_ISL_873314, EPI_ISL_873315, EPI_ISL_873317, EPI_ISL_873319, EPI_ISL_873322, EPI_ISL_873323, EPI_ISL_873327, EPI_ISL_873328, EPI_ISL_873329, EPI_ISL_873331, EPI_ISL_873332, EPI_ISL_873333, EPI_ISL_873335, EPI_ISL_873338, EPI_ISL_873340, EPI_ISL_873344, EPI_ISL_873345, EPI_ISL_873346, EPI_ISL_873349, EPI_ISL_873351, EPI_ISL_873352, EPI_ISL_873353, EPI_ISL_873355, EPI_ISL_873356, EPI_ISL_873358, EPI_ISL_873360, EPI_ISL_873363, EPI_ISL_873366, EPI_ISL_873367, EPI_ISL_873372, EPI_ISL_873373, EPI_ISL_873375, EPI_ISL_873377, EPI_ISL_873379, EPI_ISL_873380, EPI_ISL_873382, EPI_ISL_873383, EPI_ISL_873384, EPI_ISL_873385, EPI_ISL_873387, EPI_ISL_873390, EPI_ISL_873391, EPI_ISL_873392, EPI_ISL_873393, EPI_ISL_873394, EPI_ISL_873395, EPI_ISL_873397, EPI_ISL_873398, EPI_ISL_873400, EPI_ISL_873401, EPI_ISL_873402, EPI_ISL_873403, EPI_ISL_873404, EPI_ISL_873405, EPI_ISL_873406, EPI_ISL_873407, EPI_ISL_873408, EPI_ISL_873409, EPI_ISL_873410, EPI_ISL_873417, EPI_ISL_873419, EPI_ISL_873421, EPI_ISL_873423, EPI_ISL_873424, EPI_ISL_873425, EPI_ISL_873427, EPI_ISL_873434, EPI_ISL_873436, EPI_ISL_873437, EPI_ISL_873440, EPI_ISL_873441, EPI_ISL_873442, EPI_ISL_873446, EPI_ISL_873447, EPI_ISL_873448, EPI_ISL_873449, EPI_ISL_873452, EPI_ISL_873453, EPI_ISL_873456, EPI_ISL_873458, EPI_ISL_873460, EPI_ISL_873461, EPI_ISL_873463, EPI_ISL_873464, EPI_ISL_873465, EPI_ISL_873467, EPI_ISL_873468, EPI_ISL_873469, EPI_ISL_873470, EPI_ISL_873471, EPI_ISL_873472, EPI_ISL_873474, EPI_ISL_873475, EPI_ISL_873477, EPI_ISL_873478, EPI_ISL_873479, EPI_ISL_873480, EPI_ISL_873483, EPI_ISL_873484, EPI_ISL_873485, EPI_ISL_873486, EPI_ISL_873488, EPI_ISL_873490, EPI_ISL_873491, EPI_ISL_873498, EPI_ISL_873502, EPI_ISL_873505, EPI_ISL_873506, EPI_ISL_873507, EPI_ISL_873508, EPI_ISL_873511, EPI_ISL_873512, EPI_ISL_873514, EPI_ISL_873516, EPI_ISL_873517, EPI_ISL_873519, EPI_ISL_873521, EPI_ISL_873522, EPI_ISL_873523, EPI_ISL_873524, EPI_ISL_873525, EPI_ISL_873526, EPI_ISL_873527, EPI_ISL_873528, EPI_ISL_873529, EPI_ISL_873530, EPI_ISL_873533, EPI_ISL_873534, EPI_ISL_873535, EPI_ISL_873536, EPI_ISL_873538, EPI_ISL_873539, EPI_ISL_873540, EPI_ISL_873541, EPI_ISL_873542, EPI_ISL_873543, EPI_ISL_873544, EPI_ISL_873545, EPI_ISL_873546, EPI_ISL_873547, EPI_ISL_873548, EPI_ISL_873549, EPI_ISL_873550, EPI_ISL_873553, EPI_ISL_873554, EPI_ISL_873555, EPI_ISL_873556, EPI_ISL_873557, EPI_ISL_873558, EPI_ISL_873559, EPI_ISL_873560, EPI_ISL_873562, EPI_ISL_873563, EPI_ISL_873564, EPI_ISL_873565, EPI_ISL_873567, EPI_ISL_873568, EPI_ISL_873569, EPI_ISL_873570, EPI_ISL_873572, EPI_ISL_873575, EPI_ISL_873577, EPI_ISL_873578, EPI_ISL_873579, EPI_ISL_873580, EPI_ISL_873581, EPI_ISL_873582, EPI_ISL_873583, EPI_ISL_873584, EPI_ISL_873586, EPI_ISL_873587, EPI_ISL_873588, EPI_ISL_873589, EPI_ISL_873590, EPI_ISL_873591, EPI_ISL_873592, EPI_ISL_873593, EPI_ISL_873594, EPI_ISL_873595, EPI_ISL_873597, EPI_ISL_873598, EPI_ISL_873599, EPI_ISL_873601, EPI_ISL_873602, EPI_ISL_873606, EPI_ISL_873608, EPI_ISL_873609, EPI_ISL_873610, EPI_ISL_873612, EPI_ISL_873615, EPI_ISL_873616, EPI_ISL_873617, EPI_ISL_873618, EPI_ISL_873619, EPI_ISL_873620, EPI_ISL_873622, EPI_ISL_873623, EPI_ISL_873626, EPI_ISL_873627, EPI_ISL_873628, EPI_ISL_873631, EPI_ISL_873637, EPI_ISL_873641, EPI_ISL_873654, EPI_ISL_873656, EPI_ISL_873657, EPI_ISL_873660, EPI_ISL_873669, EPI_ISL_873672, EPI_ISL_873704, EPI_ISL_873724, EPI_ISL_873725, EPI_ISL_873726, EPI_ISL_873729, EPI_ISL_873730, EPI_ISL_873734, EPI_ISL_873741, EPI_ISL_873743, EPI_ISL_873744, EPI_ISL_873749, EPI_ISL_873760, EPI_ISL_873764, EPI_ISL_873769, EPI_ISL_873786, EPI_ISL_873794, EPI_ISL_873806, EPI_ISL_873809, EPI_ISL_873812, EPI_ISL_873817, EPI_ISL_873833, EPI_ISL_873834, EPI_ISL_873847, EPI_ISL_873850, EPI_ISL_873857, EPI_ISL_873859, EPI_ISL_873865, EPI_ISL_873870, EPI_ISL_873873, EPI_ISL_873875, EPI_ISL_873880, EPI_ISL_873884, EPI_ISL_873887, EPI_ISL_873900, EPI_ISL_873904, EPI_ISL_873905, EPI_ISL_873910, EPI_ISL_873913, EPI_ISL_873916, EPI_ISL_873923, EPI_ISL_873924, EPI_ISL_873925, EPI_ISL_873928, EPI_ISL_873934, EPI_ISL_873935, EPI_ISL_873937, EPI_ISL_873943, EPI_ISL_873947, EPI_ISL_873948, EPI_ISL_873954, EPI_ISL_873960, EPI_ISL_874990, EPI_ISL_874991, EPI_ISL_874992, EPI_ISL_874993, EPI_ISL_874994, EPI_ISL_874995, EPI_ISL_874996, EPI_ISL_874997, EPI_ISL_874998, EPI_ISL_874999, EPI_ISL_875000, EPI_ISL_875001, EPI_ISL_875002, EPI_ISL_875003, EPI_ISL_875004, EPI_ISL_875005, EPI_ISL_875006, EPI_ISL_875007, EPI_ISL_875008, EPI_ISL_875009, EPI_ISL_875010, EPI_ISL_875011, EPI_ISL_875012, EPI_ISL_875013, EPI_ISL_875014, EPI_ISL_875015, EPI_ISL_875016, EPI_ISL_875017, EPI_ISL_875018, EPI_ISL_875019, EPI_ISL_875020, EPI_ISL_875021, EPI_ISL_875022, EPI_ISL_875023, EPI_ISL_875024, EPI_ISL_875025, EPI_ISL_875026, EPI_ISL_875027, EPI_ISL_875028, EPI_ISL_875029, EPI_ISL_875030, EPI_ISL_875031, EPI_ISL_875032, EPI_ISL_875033, EPI_ISL_875034, EPI_ISL_875035, EPI_ISL_875036, EPI_ISL_875037, EPI_ISL_875038, EPI_ISL_875039, EPI_ISL_875040, EPI_ISL_875041, EPI_ISL_875042, EPI_ISL_875043, EPI_ISL_875044, EPI_ISL_875045, EPI_ISL_875046, EPI_ISL_875047, EPI_ISL_875048, EPI_ISL_875049, EPI_ISL_875050, EPI_ISL_875051, EPI_ISL_875052, EPI_ISL_875053, EPI_ISL_875054, EPI_ISL_875055, EPI_ISL_875056, EPI_ISL_875057, EPI_ISL_875058, EPI_ISL_875059, EPI_ISL_875060, EPI_ISL_875061, EPI_ISL_875062, EPI_ISL_875063, EPI_ISL_875064, EPI_ISL_875065, EPI_ISL_875066, EPI_ISL_875067, EPI_ISL_875068, EPI_ISL_875069, EPI_ISL_875070, EPI_ISL_875071, EPI_ISL_875072, EPI_ISL_875073, EPI_ISL_875074, EPI_ISL_875075, EPI_ISL_875076, EPI_ISL_875077, EPI_ISL_875078, EPI_ISL_875079, EPI_ISL_875080, EPI_ISL_875081, EPI_ISL_875082, EPI_ISL_875083, EPI_ISL_875084, EPI_ISL_875085, EPI_ISL_875086, EPI_ISL_875087, EPI_ISL_875088, EPI_ISL_875089, EPI_ISL_875090, EPI_ISL_875091, EPI_ISL_875092, EPI_ISL_875093, EPI_ISL_875094, EPI_ISL_875095, EPI_ISL_875096, EPI_ISL_875097, EPI_ISL_875098, EPI_ISL_875099, EPI_ISL_875100, EPI_ISL_875101, EPI_ISL_875102, EPI_ISL_875103, EPI_ISL_875104, EPI_ISL_875105, EPI_ISL_875106, EPI_ISL_875107, EPI_ISL_875108, EPI_ISL_875109, EPI_ISL_875110, EPI_ISL_875111, EPI_ISL_875112, EPI_ISL_875113, EPI_ISL_875114, EPI_ISL_875115, EPI_ISL_875116, EPI_ISL_875117, EPI_ISL_875118, EPI_ISL_875119, EPI_ISL_875120, EPI_ISL_875121, EPI_ISL_875122, EPI_ISL_875123, EPI_ISL_875124, EPI_ISL_875125, EPI_ISL_875126, EPI_ISL_875127, EPI_ISL_875128, EPI_ISL_875129, EPI_ISL_875130, EPI_ISL_875131, EPI_ISL_875132, EPI_ISL_875133, EPI_ISL_875134, EPI_ISL_875135, EPI_ISL_875136, EPI_ISL_875137, EPI_ISL_875138, EPI_ISL_875139, EPI_ISL_875140, EPI_ISL_875141, EPI_ISL_875142, EPI_ISL_875143, EPI_ISL_875144, EPI_ISL_875145, EPI_ISL_875146, EPI_ISL_875147, EPI_ISL_875148, EPI_ISL_875149, EPI_ISL_875150, EPI_ISL_875151, EPI_ISL_875152, EPI_ISL_875153, EPI_ISL_875154, EPI_ISL_875155, EPI_ISL_875156, EPI_ISL_875157, EPI_ISL_875158, EPI_ISL_875159, EPI_ISL_875160, EPI_ISL_875161, EPI_ISL_875162, EPI_ISL_875163, EPI_ISL_875164, EPI_ISL_875165, EPI_ISL_875166, EPI_ISL_875167, EPI_ISL_875168, EPI_ISL_875169, EPI_ISL_875170, EPI_ISL_875171, EPI_ISL_875172, EPI_ISL_875173, EPI_ISL_875174, EPI_ISL_875175, EPI_ISL_875176, EPI_ISL_875177, EPI_ISL_875178, EPI_ISL_875179, EPI_ISL_875180, EPI_ISL_875181, EPI_ISL_875182, EPI_ISL_875183, EPI_ISL_875184, EPI_ISL_875185, EPI_ISL_875186, EPI_ISL_875187, EPI_ISL_875188, EPI_ISL_875189, EPI_ISL_875190, EPI_ISL_875191, EPI_ISL_875192, EPI_ISL_875193, EPI_ISL_875194, EPI_ISL_875195, EPI_ISL_875196, EPI_ISL_875197, EPI_ISL_875198, EPI_ISL_875199, EPI_ISL_875200, EPI_ISL_875201, EPI_ISL_875202, EPI_ISL_875203, EPI_ISL_875204, EPI_ISL_875205, EPI_ISL_875206, EPI_ISL_875207, EPI_ISL_875208, EPI_ISL_875209, EPI_ISL_875210, EPI_ISL_875211, EPI_ISL_875212, EPI_ISL_875213, EPI_ISL_875214, EPI_ISL_875215, EPI_ISL_875216, EPI_ISL_875217, EPI_ISL_875218, EPI_ISL_875219, EPI_ISL_875220, EPI_ISL_875221, EPI_ISL_875222, EPI_ISL_875223, EPI_ISL_875224, EPI_ISL_875225, EPI_ISL_875226, EPI_ISL_875227, EPI_ISL_875228, EPI_ISL_875229, EPI_ISL_875230, EPI_ISL_875231, EPI_ISL_875232, EPI_ISL_875233, EPI_ISL_875234, EPI_ISL_875235, EPI_ISL_875236, EPI_ISL_875237, EPI_ISL_875238, EPI_ISL_875239, EPI_ISL_875240, EPI_ISL_875241, EPI_ISL_875242, EPI_ISL_875243, EPI_ISL_875244, EPI_ISL_875245, EPI_ISL_875246, EPI_ISL_875247, EPI_ISL_875248, EPI_ISL_875249, EPI_ISL_875250, EPI_ISL_875251, EPI_ISL_875252, EPI_ISL_875253, EPI_ISL_875254, EPI_ISL_875255, EPI_ISL_875256, EPI_ISL_875257, EPI_ISL_875258, EPI_ISL_875259, EPI_ISL_875260, EPI_ISL_875261, EPI_ISL_875262, EPI_ISL_875263, EPI_ISL_875264, EPI_ISL_875265, EPI_ISL_875266, EPI_ISL_875267, EPI_ISL_875268, EPI_ISL_875269, EPI_ISL_875270, EPI_ISL_875271, EPI_ISL_875272, EPI_ISL_875273, EPI_ISL_875274, EPI_ISL_875275, EPI_ISL_875276, EPI_ISL_875277, EPI_ISL_875278, EPI_ISL_875279, EPI_ISL_875280, EPI_ISL_875281, EPI_ISL_875282, EPI_ISL_875283, EPI_ISL_875284, EPI_ISL_875285, EPI_ISL_875286, EPI_ISL_875287, EPI_ISL_875288, EPI_ISL_875289, EPI_ISL_875290, EPI_ISL_875291, EPI_ISL_875292, EPI_ISL_875293, EPI_ISL_875294, EPI_ISL_875295, EPI_ISL_875296, EPI_ISL_875297, EPI_ISL_875298, EPI_ISL_875299, EPI_ISL_875300, EPI_ISL_875301, EPI_ISL_875302, EPI_ISL_875303, EPI_ISL_875304, EPI_ISL_875305, EPI_ISL_875306, EPI_ISL_875307, EPI_ISL_875308, EPI_ISL_875309, EPI_ISL_875310, EPI_ISL_875311, EPI_ISL_875312, EPI_ISL_875313, EPI_ISL_875314, EPI_ISL_875315, EPI_ISL_875316, EPI_ISL_875317, EPI_ISL_875318, EPI_ISL_875319, EPI_ISL_875320, EPI_ISL_875321, EPI_ISL_875322, EPI_ISL_875323, EPI_ISL_875324, EPI_ISL_875325, EPI_ISL_875326, EPI_ISL_875327, EPI_ISL_875328, EPI_ISL_875329, EPI_ISL_875330, EPI_ISL_875331, EPI_ISL_875332, EPI_ISL_875333, EPI_ISL_875334, EPI_ISL_875335, EPI_ISL_875336, EPI_ISL_875337, EPI_ISL_875338, EPI_ISL_875339, EPI_ISL_875340, EPI_ISL_875341, EPI_ISL_878769, EPI_ISL_878833, EPI_ISL_880586, EPI_ISL_880590, EPI_ISL_880591, EPI_ISL_880595, EPI_ISL_880596, EPI_ISL_880599, EPI_ISL_880601, EPI_ISL_880602, EPI_ISL_880606, EPI_ISL_880608, EPI_ISL_880609, EPI_ISL_880610, EPI_ISL_880611, EPI_ISL_880613, EPI_ISL_880615, EPI_ISL_880616, EPI_ISL_880618, EPI_ISL_880619, EPI_ISL_880620, EPI_ISL_880623, EPI_ISL_880626, EPI_ISL_880627, EPI_ISL_880629, EPI_ISL_880637, EPI_ISL_880639, EPI_ISL_880641, EPI_ISL_880642, EPI_ISL_880647, EPI_ISL_880651, EPI_ISL_880653, EPI_ISL_880654, EPI_ISL_880655, EPI_ISL_880657, EPI_ISL_880660, EPI_ISL_880662, EPI_ISL_880665, EPI_ISL_880667, EPI_ISL_880668, EPI_ISL_880669, EPI_ISL_880672, EPI_ISL_880673, EPI_ISL_880674, EPI_ISL_880675, EPI_ISL_880677, EPI_ISL_880679, EPI_ISL_880682, EPI_ISL_880695, EPI_ISL_880701, EPI_ISL_880705, EPI_ISL_880706, EPI_ISL_880708, EPI_ISL_880709, EPI_ISL_880710, EPI_ISL_880713, EPI_ISL_880717, EPI_ISL_880718, EPI_ISL_880721, EPI_ISL_880722, EPI_ISL_880724, EPI_ISL_880725, EPI_ISL_880726, EPI_ISL_880727, EPI_ISL_880729, EPI_ISL_880736, EPI_ISL_880737, EPI_ISL_880738, EPI_ISL_880739, EPI_ISL_880740, EPI_ISL_880745, EPI_ISL_880746, EPI_ISL_880751, EPI_ISL_880752, EPI_ISL_880753, EPI_ISL_880755, EPI_ISL_880759, EPI_ISL_880762, EPI_ISL_880763, EPI_ISL_880766, EPI_ISL_880769, EPI_ISL_880774, EPI_ISL_880775, EPI_ISL_880776, EPI_ISL_880779, EPI_ISL_880781, EPI_ISL_880785, EPI_ISL_880786, EPI_ISL_880789, EPI_ISL_880791, EPI_ISL_880792, EPI_ISL_880797, EPI_ISL_880798, EPI_ISL_880799, EPI_ISL_880800, EPI_ISL_880803, EPI_ISL_880809, EPI_ISL_880812, EPI_ISL_880813, EPI_ISL_880817, EPI_ISL_880818, EPI_ISL_880819, EPI_ISL_880822, EPI_ISL_880824, EPI_ISL_880826, EPI_ISL_880829, EPI_ISL_880830, EPI_ISL_880831, EPI_ISL_880833, EPI_ISL_880835, EPI_ISL_880836, EPI_ISL_880838, EPI_ISL_880839, EPI_ISL_880840, EPI_ISL_880841, EPI_ISL_880843, EPI_ISL_880847, EPI_ISL_880853, EPI_ISL_880854, EPI_ISL_880856, EPI_ISL_880857, EPI_ISL_880858, EPI_ISL_880862, EPI_ISL_880866, EPI_ISL_880868, EPI_ISL_880870, EPI_ISL_880878, EPI_ISL_880879, EPI_ISL_880883, EPI_ISL_880884, EPI_ISL_880886, EPI_ISL_880889, EPI_ISL_880894, EPI_ISL_880895, EPI_ISL_880898, EPI_ISL_880900, EPI_ISL_880901, EPI_ISL_880903, EPI_ISL_880908, EPI_ISL_880909, EPI_ISL_880910, EPI_ISL_880911, EPI_ISL_880914, EPI_ISL_880915, EPI_ISL_880916, EPI_ISL_880921, EPI_ISL_880925, EPI_ISL_880933, EPI_ISL_880934, EPI_ISL_880935, EPI_ISL_881618, EPI_ISL_881621, EPI_ISL_882610, EPI_ISL_959273, EPI_ISL_987284, EPI_ISL_987285, EPI_ISL_987286, EPI_ISL_987297, EPI_ISL_987298, EPI_ISL_987305, EPI_ISL_987329, EPI_ISL_987338, EPI_ISL_987342, EPI_ISL_987343, EPI_ISL_987346, EPI_ISL_987351, EPI_ISL_987352, EPI_ISL_987353, EPI_ISL_987358, EPI_ISL_987364, EPI_ISL_987366, EPI_ISL_987371, EPI_ISL_987374, EPI_ISL_987386, EPI_ISL_987388, EPI_ISL_987396, EPI_ISL_987400, EPI_ISL_987403, EPI_ISL_987423, EPI_ISL_987431, EPI_ISL_987434, EPI_ISL_987442, EPI_ISL_987447, EPI_ISL_987459, EPI_ISL_987460, EPI_ISL_987463, EPI_ISL_987466, EPI_ISL_987468, EPI_ISL_987473, EPI_ISL_987477, EPI_ISL_987487, EPI_ISL_987492, EPI_ISL_987494, EPI_ISL_987495, EPI_ISL_987500, EPI_ISL_987504, EPI_ISL_987512, EPI_ISL_987515, EPI_ISL_987528, EPI_ISL_987531, EPI_ISL_987533, EPI_ISL_987537, EPI_ISL_987538, EPI_ISL_987539, EPI_ISL_987543, EPI_ISL_987551, EPI_ISL_987558, EPI_ISL_987563, EPI_ISL_987564, EPI_ISL_987565, EPI_ISL_987569, EPI_ISL_987580, EPI_ISL_987582, EPI_ISL_987586, EPI_ISL_987588, EPI_ISL_987592, EPI_ISL_987600, EPI_ISL_987601, EPI_ISL_987608, EPI_ISL_987618, EPI_ISL_987626, EPI_ISL_987630, EPI_ISL_987632, EPI_ISL_987634, EPI_ISL_987635, EPI_ISL_1043207, EPI_ISL_1346483 | | | |
| see above | Lighthouse Lab in Milton Keynes | Wellcome Sanger Institute for the COVID-19 Genomics UK (COG-UK) Consortium | Cordelia Langford; David K. Jackson; Dominic Kwiatkowski; Ewan Harrison; Ian Johnston; Jeffrey Barrett; John Sillitoe on behalf of the Wellcome Sanger Institute COVID-19 Surveillance Team; John Sillitoe on behalf of the Wellcome Sanger Institute COVID-19 Surveillance Team (http://www.sanger.ac.uk/covid-team); Roberto Amato; Sonia Goncalves; The Lighthouse Lab in Milton Keynes and Alex Alderton |
| EPI_ISL_922060, EPI_ISL_922063, EPI_ISL_922064, EPI_ISL_922065, EPI_ISL_922066, EPI_ISL_922067, EPI_ISL_922068, EPI_ISL_922069, EPI_ISL_922070, EPI_ISL_922071, EPI_ISL_922072, EPI_ISL_922073, EPI_ISL_922074, EPI_ISL_922075, EPI_ISL_922077, EPI_ISL_922078, EPI_ISL_922079, EPI_ISL_922080, EPI_ISL_922081, EPI_ISL_922082, EPI_ISL_922083, EPI_ISL_922084, EPI_ISL_922085, EPI_ISL_922086, EPI_ISL_922087, EPI_ISL_922088, EPI_ISL_922089, EPI_ISL_922090, EPI_ISL_922091, EPI_ISL_922092, EPI_ISL_922093, EPI_ISL_922094, EPI_ISL_922095, EPI_ISL_922096, EPI_ISL_922097, EPI_ISL_922098, EPI_ISL_922099, EPI_ISL_922100, EPI_ISL_922101, EPI_ISL_922102, EPI_ISL_922103, EPI_ISL_922104, EPI_ISL_922105, EPI_ISL_922106, EPI_ISL_922107, EPI_ISL_922108, EPI_ISL_922109, EPI_ISL_922110, EPI_ISL_922111, EPI_ISL_922112, EPI_ISL_922115, EPI_ISL_922116, EPI_ISL_950713, EPI_ISL_998937, EPI_ISL_998938, EPI_ISL_998939, EPI_ISL_998940, EPI_ISL_998941, EPI_ISL_998942, EPI_ISL_998943, EPI_ISL_998944 | | | |
| see above | Lincolnshire Hospitals and DeepSeq Nottingham | COVID-19 Genomics UK (COG-UK) Consortium | Christopher Moore; Fei Sang; Joeseph Chappell; Johnny Debebe; Jonathan Ball; Matthew Carlisle; Matthew Loose; Nadine Holmes; Nichola Duckworth; Patrick McClure; Sarah Walsh; Tim Sloan; Victoria Wright |
| EPI_ISL_865270, EPI_ISL_865271, EPI_ISL_865272, EPI_ISL_865273, EPI_ISL_865274, EPI_ISL_865275, EPI_ISL_865276, EPI_ISL_865277, EPI_ISL_865280, EPI_ISL_865281, EPI_ISL_865282, EPI_ISL_865284, EPI_ISL_865285, EPI_ISL_865286, EPI_ISL_865287, EPI_ISL_865288, EPI_ISL_865289, EPI_ISL_865290, EPI_ISL_865291, EPI_ISL_865292, EPI_ISL_865293, EPI_ISL_865294, EPI_ISL_865295, EPI_ISL_865296, EPI_ISL_865297, EPI_ISL_865298, EPI_ISL_865300, EPI_ISL_865301, EPI_ISL_865302, EPI_ISL_865303, EPI_ISL_865304, EPI_ISL_865305, EPI_ISL_865306, EPI_ISL_865307, EPI_ISL_865308, EPI_ISL_865309, EPI_ISL_865310, EPI_ISL_865311, EPI_ISL_865312, EPI_ISL_865313, EPI_ISL_865314, EPI_ISL_865315, EPI_ISL_865317, EPI_ISL_865318, EPI_ISL_865319, EPI_ISL_865320, EPI_ISL_865321, EPI_ISL_865322, EPI_ISL_865324, EPI_ISL_865326, EPI_ISL_865327, EPI_ISL_865328, EPI_ISL_865341, EPI_ISL_865342, EPI_ISL_865343, EPI_ISL_865344, EPI_ISL_865345, EPI_ISL_865346, EPI_ISL_865347, EPI_ISL_865348, EPI_ISL_865349, EPI_ISL_865350, EPI_ISL_865351, EPI_ISL_865353, EPI_ISL_865449, EPI_ISL_919462, EPI_ISL_919463, EPI_ISL_919465, EPI_ISL_919467, EPI_ISL_919468, EPI_ISL_919469, EPI_ISL_919470, EPI_ISL_919471, EPI_ISL_919472, EPI_ISL_919473, EPI_ISL_919474, EPI_ISL_919475, EPI_ISL_919478, EPI_ISL_919515, EPI_ISL_919517, EPI_ISL_919591, EPI_ISL_919593, EPI_ISL_919594, EPI_ISL_919595, EPI_ISL_919644, EPI_ISL_997377, EPI_ISL_1104346, EPI_ISL_1104347, EPI_ISL_1104348, EPI_ISL_1104359, EPI_ISL_1104360, EPI_ISL_1104361, EPI_ISL_1104380, EPI_ISL_1104381, EPI_ISL_1104382, EPI_ISL_1104383, EPI_ISL_1104384, EPI_ISL_1177839, EPI_ISL_1177840, EPI_ISL_1177841, EPI_ISL_1177843, EPI_ISL_1177844, EPI_ISL_1177845, EPI_ISL_1177846, EPI_ISL_1177847, EPI_ISL_1177850, EPI_ISL_1177851, EPI_ISL_1177852, EPI_ISL_1177854, EPI_ISL_1177856, EPI_ISL_1177857, EPI_ISL_1177858, EPI_ISL_1177859, EPI_ISL_1177860, EPI_ISL_1177861, EPI_ISL_1177862, EPI_ISL_1177863, EPI_ISL_1177864, EPI_ISL_1177865, EPI_ISL_1177866, EPI_ISL_1177867, EPI_ISL_1177868, EPI_ISL_1177869, EPI_ISL_1177870, EPI_ISL_1177871, EPI_ISL_1177872, EPI_ISL_1177874, EPI_ISL_1177875, EPI_ISL_1177876, EPI_ISL_1177877, EPI_ISL_1545990, EPI_ISL_1545991, EPI_ISL_1545992, EPI_ISL_1545993, EPI_ISL_1545994, EPI_ISL_1545995, EPI_ISL_1545996, EPI_ISL_1545998, EPI_ISL_1545999, EPI_ISL_1546000, EPI_ISL_1546001, EPI_ISL_1832341 | | | |
| see above | Liverpool Clinical Laboratories | COVID-19 Genomics UK (COG-UK) Consortium | A Alrezaihi; Alessandro Gerada; Alistair Darby; Angela Cowell; Anita Lucaci; Anu Chawla; Cassie Olateju; Catherine Hartley; Charlotte Nelson; Ecaterina Vamos; Elaine O'Toole; Eleanor G Bentley; Ghada T Shawli; Hermione Webster; Isabel García-Dorival; James Johnson; James P Stewart; Jenifer Manson; Joanne Watts; Jones Benjamin; Jordan J Clark; Julian Hiscox; L Luu; Lucille Rainbow; M Almsaud; Margaret Hughes; Mark Whitehead; Matthew Gemmell; Miren Iturriza-Gomara; Muhannad Alruwaili; N.P Randle; Neil Swainston; PKF Gilmore; Parul Sharma; Rebekah Penrice-Randal; Richard Eccles; Richard Gregory; Sam Haldenby; Steve Paterson; Stuart D Armstrong; Trevor Ian Robinson; Ximeng Han |
| EPI_ISL_921300, EPI_ISL_921475, EPI_ISL_921476, EPI_ISL_921477, EPI_ISL_921478, EPI_ISL_921479, EPI_ISL_921480, EPI_ISL_921481, EPI_ISL_921482, EPI_ISL_921483, EPI_ISL_921484, EPI_ISL_921488, EPI_ISL_921489, EPI_ISL_921490, EPI_ISL_921491, EPI_ISL_921492, EPI_ISL_921493, EPI_ISL_921494, EPI_ISL_921495, EPI_ISL_921496, EPI_ISL_921497, EPI_ISL_921498, EPI_ISL_921499, EPI_ISL_921500, EPI_ISL_921501, EPI_ISL_921503, EPI_ISL_921504, EPI_ISL_921505, EPI_ISL_921506, EPI_ISL_921507, EPI_ISL_921508, EPI_ISL_921509, EPI_ISL_921510, EPI_ISL_921511, EPI_ISL_921512, EPI_ISL_921513, EPI_ISL_921514, EPI_ISL_921515, EPI_ISL_921516, EPI_ISL_921517, EPI_ISL_921518, EPI_ISL_921519, EPI_ISL_921520, EPI_ISL_921541, EPI_ISL_950272, EPI_ISL_950280, EPI_ISL_950282, EPI_ISL_950283, EPI_ISL_950284, EPI_ISL_950285, EPI_ISL_950289, EPI_ISL_950290, EPI_ISL_950291, EPI_ISL_950292, EPI_ISL_950293, EPI_ISL_950297, EPI_ISL_998371, EPI_ISL_998749, EPI_ISL_1051069, EPI_ISL_1051071, EPI_ISL_1051077, EPI_ISL_1051084, EPI_ISL_1051087, EPI_ISL_1051091, EPI_ISL_1051095, EPI_ISL_1051097, EPI_ISL_1051099, EPI_ISL_1051101, EPI_ISL_1051103, EPI_ISL_1051105, EPI_ISL_1051107, EPI_ISL_1051110, EPI_ISL_1051112, EPI_ISL_1051114, EPI_ISL_1051116, EPI_ISL_1051117, EPI_ISL_1051120, EPI_ISL_1051121, EPI_ISL_1051122, EPI_ISL_1051123, EPI_ISL_1051124, EPI_ISL_1051125, EPI_ISL_1051126, EPI_ISL_1051127, EPI_ISL_1051128, EPI_ISL_1051129, EPI_ISL_1051130, EPI_ISL_1051131, EPI_ISL_1051132, EPI_ISL_1051133, EPI_ISL_1051134, EPI_ISL_1051135, EPI_ISL_1051136, EPI_ISL_1051137, EPI_ISL_1051138, EPI_ISL_1051139, EPI_ISL_1051140, EPI_ISL_1051141, EPI_ISL_1051142, EPI_ISL_1051143, EPI_ISL_1051144, EPI_ISL_1051145, EPI_ISL_1051146, EPI_ISL_1051147, EPI_ISL_1051148, EPI_ISL_1051149, EPI_ISL_1051150, EPI_ISL_1051151, EPI_ISL_1051152, EPI_ISL_1051153, EPI_ISL_1051154, EPI_ISL_1051155, EPI_ISL_1051156, EPI_ISL_1051157, EPI_ISL_1051158, EPI_ISL_1051159, EPI_ISL_1051160, EPI_ISL_1051161, EPI_ISL_1051162, EPI_ISL_1051163, EPI_ISL_1051164, EPI_ISL_1051165, EPI_ISL_1051166, EPI_ISL_1051167, EPI_ISL_1051168, EPI_ISL_1051169, EPI_ISL_1051170, EPI_ISL_1051171, EPI_ISL_1051172, EPI_ISL_1051173, EPI_ISL_1051174, EPI_ISL_1051175, EPI_ISL_1051177, EPI_ISL_1051178, EPI_ISL_1051179, EPI_ISL_1051180, EPI_ISL_1051181, EPI_ISL_1051182, EPI_ISL_1051183, EPI_ISL_1051184, EPI_ISL_1051185, EPI_ISL_1051186, EPI_ISL_1051187, EPI_ISL_1051188, EPI_ISL_1051189, EPI_ISL_1051190, EPI_ISL_1051191, EPI_ISL_1051192, EPI_ISL_1051193, EPI_ISL_1051194, EPI_ISL_1051195, EPI_ISL_1051196, EPI_ISL_1051197, EPI_ISL_1051198, EPI_ISL_1051199, EPI_ISL_1051200, EPI_ISL_1051201, EPI_ISL_1051202, EPI_ISL_1051203, | | | |

| | | | |
|---|---|---|---|
| EPI_ISL_1051204, EPI_ISL_1051205, EPI_ISL_1051206, EPI_ISL_1051207, EPI_ISL_1051208, EPI_ISL_1051209, EPI_ISL_1051210, EPI_ISL_1051211, EPI_ISL_1051212, EPI_ISL_1051214, EPI_ISL_1051217, EPI_ISL_1051220, EPI_ISL_1051221, EPI_ISL_1051345, EPI_ISL_1051346, EPI_ISL_1051347, EPI_ISL_1051348, EPI_ISL_1051349, EPI_ISL_1051350, EPI_ISL_1051351, EPI_ISL_1051352, EPI_ISL_1051353, EPI_ISL_1051354, EPI_ISL_1051355, EPI_ISL_1051356, EPI_ISL_1051357, EPI_ISL_1051358, EPI_ISL_1051359, EPI_ISL_1051360, EPI_ISL_1051361, EPI_ISL_1051362, EPI_ISL_1051363, EPI_ISL_1051364, EPI_ISL_1051365, EPI_ISL_1051366, EPI_ISL_1051367, EPI_ISL_1051368, EPI_ISL_1051369, EPI_ISL_1051370, EPI_ISL_1051371, EPI_ISL_1051372, EPI_ISL_1051373, EPI_ISL_1051374, EPI_ISL_1051375, EPI_ISL_1051376, EPI_ISL_1051377, EPI_ISL_1051378, EPI_ISL_1051379, EPI_ISL_1051380, EPI_ISL_1051381, EPI_ISL_1051382, EPI_ISL_1051383, EPI_ISL_1051384, EPI_ISL_1051385, EPI_ISL_1051386, EPI_ISL_1051387, EPI_ISL_1051388, EPI_ISL_1051389, EPI_ISL_1051390, EPI_ISL_1051391, EPI_ISL_1051392, EPI_ISL_1051393, EPI_ISL_1051394, EPI_ISL_1051395, EPI_ISL_1051396, EPI_ISL_1051397, EPI_ISL_1051398, EPI_ISL_1051399, EPI_ISL_1051400, EPI_ISL_1051401, EPI_ISL_1105658, EPI_ISL_1105659, EPI_ISL_1105660, EPI_ISL_1105661, EPI_ISL_1105662, EPI_ISL_1105663, EPI_ISL_1105664, EPI_ISL_1105665, EPI_ISL_1105666, EPI_ISL_1105667, EPI_ISL_1105668, EPI_ISL_1105669, EPI_ISL_1105670, EPI_ISL_1105671, EPI_ISL_1105672, EPI_ISL_1105673, EPI_ISL_1105674, EPI_ISL_1105675, EPI_ISL_1105676, EPI_ISL_1105677, EPI_ISL_1105678, EPI_ISL_1105679, EPI_ISL_1105680, EPI_ISL_1105681, EPI_ISL_1105682, EPI_ISL_1105683, EPI_ISL_1105684, EPI_ISL_1105685, EPI_ISL_1105686, EPI_ISL_1105687, EPI_ISL_1105688, EPI_ISL_1105689, EPI_ISL_1105690, EPI_ISL_1105691, EPI_ISL_1105692, EPI_ISL_1105693, EPI_ISL_1105694, EPI_ISL_1105695, EPI_ISL_1105696, EPI_ISL_1105697, EPI_ISL_1105698, EPI_ISL_1105699, EPI_ISL_1105700, EPI_ISL_1105701, EPI_ISL_1105702, EPI_ISL_1105703, EPI_ISL_1105704, EPI_ISL_1105705, EPI_ISL_1105707, EPI_ISL_1178326, EPI_ISL_1178327, EPI_ISL_1178328, EPI_ISL_1178329, EPI_ISL_1178330, EPI_ISL_1178331, EPI_ISL_1178332, EPI_ISL_1178333, EPI_ISL_1178336, EPI_ISL_1178337, EPI_ISL_1178338, EPI_ISL_1178348, EPI_ISL_1178353, EPI_ISL_1178356, EPI_ISL_1178357, EPI_ISL_1178359, EPI_ISL_1178361, EPI_ISL_1178362, EPI_ISL_1178363, EPI_ISL_1178364, EPI_ISL_1178365, EPI_ISL_1178366, EPI_ISL_1178367, EPI_ISL_1178368, EPI_ISL_1178369, EPI_ISL_1178370, EPI_ISL_1178371, EPI_ISL_1178372, EPI_ISL_1178373, EPI_ISL_1178374, EPI_ISL_1178375, EPI_ISL_1178376, EPI_ISL_1178378, EPI_ISL_1178379, EPI_ISL_1178380, EPI_ISL_1178381, EPI_ISL_1178382, EPI_ISL_1178383, EPI_ISL_1178384, EPI_ISL_1178385, EPI_ISL_1178386, EPI_ISL_1178387, EPI_ISL_1178388, EPI_ISL_1178389, EPI_ISL_1178390, EPI_ISL_1178391, EPI_ISL_1178392, EPI_ISL_1178393, EPI_ISL_1178394, EPI_ISL_1178395, EPI_ISL_1178396, EPI_ISL_1178397, EPI_ISL_1178398, EPI_ISL_1178400, EPI_ISL_1178401, EPI_ISL_1178402, EPI_ISL_1178403, EPI_ISL_1178404, EPI_ISL_1178405, EPI_ISL_1178406, EPI_ISL_1178417, EPI_ISL_1178418, EPI_ISL_1178419, EPI_ISL_1178420, EPI_ISL_1178421, EPI_ISL_1387095 | | | |
| see above | Northumbria University / South Tees Hospitals NHS Foundation Trust / North Cumbria Integrated Care NHS Foundation Trust / North Tees and Hartlepool NHS Foundation Trust / Newcastle Hospitals NHS Foundation Trust | COVID-19 Genomics UK (COG-UK) Consortium | Andrew Nelson; Brendan Payne; Clive Graham; Darren L Smith; Debra Padgett; Edward Barton; Emma Swindells; Garren Scott; Gary Black; Gary Eltringham; Giles S Holt; Greg R Young; Jane Greenaway; Jennifer Collins; John Allan; Joshua Loh; Lynn Dover; Matthew Bashton; Mohammad A Tariq; Paul Baker; Sarah Essex; Steve Liggett; Wen C Yew; Yusri Taha |
| EPI_ISL_841317, EPI_ISL_841322, EPI_ISL_841324, EPI_ISL_841326, EPI_ISL_841327, EPI_ISL_841328, EPI_ISL_841330, EPI_ISL_841331, EPI_ISL_841339, EPI_ISL_841340, EPI_ISL_841343, EPI_ISL_841344, EPI_ISL_841345, EPI_ISL_841346, EPI_ISL_841347, EPI_ISL_841348, EPI_ISL_841349, EPI_ISL_841350, EPI_ISL_841351, EPI_ISL_841352, EPI_ISL_841353, EPI_ISL_841354, EPI_ISL_841417, EPI_ISL_841426, EPI_ISL_841427, EPI_ISL_841430, EPI_ISL_841431, EPI_ISL_841434, EPI_ISL_841437, EPI_ISL_841440, EPI_ISL_841443, EPI_ISL_841444, EPI_ISL_841445, EPI_ISL_841446, EPI_ISL_841448, EPI_ISL_841451, EPI_ISL_841453, EPI_ISL_841454, EPI_ISL_841455, EPI_ISL_841456, EPI_ISL_841457, EPI_ISL_841458, EPI_ISL_841460, EPI_ISL_841461, EPI_ISL_841462, EPI_ISL_841464, EPI_ISL_841465, EPI_ISL_841467, EPI_ISL_841468, EPI_ISL_841469, EPI_ISL_841470, EPI_ISL_841472, EPI_ISL_841473, EPI_ISL_841474, EPI_ISL_841475, EPI_ISL_841476, EPI_ISL_841477, EPI_ISL_841478, EPI_ISL_841480, EPI_ISL_841481, EPI_ISL_841484, EPI_ISL_841487, EPI_ISL_841488, EPI_ISL_841489, EPI_ISL_841492, EPI_ISL_841493, EPI_ISL_841494, EPI_ISL_841495, EPI_ISL_841496, EPI_ISL_841497, EPI_ISL_841502, EPI_ISL_841516, EPI_ISL_841517, EPI_ISL_841518, EPI_ISL_841519, EPI_ISL_841520, EPI_ISL_841523, EPI_ISL_841524, EPI_ISL_841525, EPI_ISL_841526, EPI_ISL_841527, EPI_ISL_841528, EPI_ISL_841530, EPI_ISL_841532, EPI_ISL_841533, EPI_ISL_841534, EPI_ISL_841535, EPI_ISL_841536, EPI_ISL_841537, EPI_ISL_841538, EPI_ISL_841539, EPI_ISL_841540, EPI_ISL_841541, EPI_ISL_841542, EPI_ISL_841543, EPI_ISL_841544, EPI_ISL_841545, EPI_ISL_841546, EPI_ISL_841547, EPI_ISL_841549, EPI_ISL_841550, EPI_ISL_841551, EPI_ISL_841552, EPI_ISL_841553, EPI_ISL_841554, EPI_ISL_841555, EPI_ISL_841556, EPI_ISL_841557, EPI_ISL_841558, EPI_ISL_841614, EPI_ISL_841621, EPI_ISL_841624, EPI_ISL_841625, EPI_ISL_841628, EPI_ISL_841629, EPI_ISL_841631, EPI_ISL_841632, EPI_ISL_841642, EPI_ISL_841644, EPI_ISL_841645, EPI_ISL_841659, EPI_ISL_841661, EPI_ISL_841665, EPI_ISL_841667, EPI_ISL_841669, EPI_ISL_841670, EPI_ISL_841673, EPI_ISL_841676, EPI_ISL_841677, EPI_ISL_841679, EPI_ISL_841684, EPI_ISL_841685, EPI_ISL_841686, EPI_ISL_841687, EPI_ISL_841688, EPI_ISL_841689, EPI_ISL_841690, EPI_ISL_841694, EPI_ISL_841695, EPI_ISL_841700, EPI_ISL_841702, EPI_ISL_841704, EPI_ISL_841705, EPI_ISL_841706, EPI_ISL_841707, EPI_ISL_841709, EPI_ISL_841710, EPI_ISL_841711, EPI_ISL_841712, EPI_ISL_841713, EPI_ISL_841714, EPI_ISL_841715, EPI_ISL_841716, EPI_ISL_841717, EPI_ISL_841718, EPI_ISL_841719, EPI_ISL_841720, EPI_ISL_841721, EPI_ISL_841722, EPI_ISL_841723, EPI_ISL_841724, EPI_ISL_841725, EPI_ISL_841726, EPI_ISL_841727, EPI_ISL_841728, EPI_ISL_841729, EPI_ISL_841730, EPI_ISL_841733, EPI_ISL_841734, EPI_ISL_841735, EPI_ISL_841736, EPI_ISL_841738, EPI_ISL_841739, EPI_ISL_841740, EPI_ISL_841741, EPI_ISL_841742, EPI_ISL_841743, EPI_ISL_841744, EPI_ISL_841745, EPI_ISL_841746, EPI_ISL_841747, EPI_ISL_841748, EPI_ISL_841749, EPI_ISL_841750, EPI_ISL_841751, EPI_ISL_841752, EPI_ISL_841753, EPI_ISL_867190, EPI_ISL_867191, EPI_ISL_867192, EPI_ISL_867193, EPI_ISL_867194, EPI_ISL_867195, EPI_ISL_867196, EPI_ISL_867197, EPI_ISL_867202, EPI_ISL_867211, EPI_ISL_867212, EPI_ISL_867213, EPI_ISL_867214, EPI_ISL_867215, EPI_ISL_867219, EPI_ISL_867220, EPI_ISL_867221, EPI_ISL_867222, EPI_ISL_867223, EPI_ISL_867224, EPI_ISL_867225, EPI_ISL_867226, EPI_ISL_867227, EPI_ISL_867228, EPI_ISL_867229, EPI_ISL_867230, EPI_ISL_867231, EPI_ISL_867232, EPI_ISL_867233, EPI_ISL_867237, EPI_ISL_867238, EPI_ISL_867239, EPI_ISL_867240, EPI_ISL_867241, EPI_ISL_867242, EPI_ISL_867243, EPI_ISL_867244, EPI_ISL_867245, EPI_ISL_867246, EPI_ISL_867247, EPI_ISL_867248, EPI_ISL_867249, EPI_ISL_867250, EPI_ISL_867251, EPI_ISL_867252, EPI_ISL_867253, EPI_ISL_867254, EPI_ISL_867255, EPI_ISL_867256, EPI_ISL_867257, EPI_ISL_867258, EPI_ISL_867261, EPI_ISL_867269, EPI_ISL_867270, EPI_ISL_867298, EPI_ISL_867299, EPI_ISL_867300, EPI_ISL_867301, EPI_ISL_867305, EPI_ISL_867306, EPI_ISL_867307, EPI_ISL_867308, EPI_ISL_867309, EPI_ISL_867310, EPI_ISL_867311, EPI_ISL_867312, EPI_ISL_867313, EPI_ISL_867314, EPI_ISL_867315, EPI_ISL_867317, EPI_ISL_867318, EPI_ISL_867319, EPI_ISL_867320, EPI_ISL_867321, EPI_ISL_867322, EPI_ISL_867323, EPI_ISL_867324, EPI_ISL_867325, EPI_ISL_867326, EPI_ISL_867327, EPI_ISL_867328, EPI_ISL_867329, EPI_ISL_867330, EPI_ISL_867331, EPI_ISL_867332, EPI_ISL_867333, EPI_ISL_867334, EPI_ISL_867335, EPI_ISL_867336, EPI_ISL_867337, EPI_ISL_867338, EPI_ISL_867339, EPI_ISL_867340, EPI_ISL_867341, EPI_ISL_867342, EPI_ISL_867343, EPI_ISL_867344, EPI_ISL_867345, EPI_ISL_867346, EPI_ISL_867348, EPI_ISL_867349, EPI_ISL_867350, EPI_ISL_867351, EPI_ISL_867352, EPI_ISL_867353, EPI_ISL_867354, EPI_ISL_867355, EPI_ISL_867357, EPI_ISL_867358, EPI_ISL_867359, EPI_ISL_867360, EPI_ISL_867361, EPI_ISL_867362, EPI_ISL_867364, EPI_ISL_867365, EPI_ISL_867366, EPI_ISL_867367, EPI_ISL_867368, EPI_ISL_867369, EPI_ISL_867370, EPI_ISL_867371, EPI_ISL_867372, EPI_ISL_867373, EPI_ISL_867374, EPI_ISL_867375, EPI_ISL_867377, EPI_ISL_867378, EPI_ISL_867379, EPI_ISL_867381, EPI_ISL_867382, EPI_ISL_867383, EPI_ISL_867384, EPI_ISL_867385, EPI_ISL_867386, EPI_ISL_867387, EPI_ISL_867388, EPI_ISL_867390, EPI_ISL_867391, EPI_ISL_867392, EPI_ISL_867393, EPI_ISL_867394, EPI_ISL_867395, EPI_ISL_867396, EPI_ISL_867397, EPI_ISL_867398, EPI_ISL_867399, EPI_ISL_867400, EPI_ISL_867401, EPI_ISL_867403, EPI_ISL_867404, EPI_ISL_867405, EPI_ISL_867416, EPI_ISL_867417, EPI_ISL_867419, EPI_ISL_867424, EPI_ISL_867429, EPI_ISL_867430, EPI_ISL_867450, EPI_ISL_867451, EPI_ISL_867457, EPI_ISL_867458, EPI_ISL_867459, EPI_ISL_867491, EPI_ISL_867492, EPI_ISL_867494, EPI_ISL_867497, EPI_ISL_867498, EPI_ISL_867506, EPI_ISL_867509, EPI_ISL_867510, EPI_ISL_867511, EPI_ISL_867515, EPI_ISL_867518, EPI_ISL_867519, EPI_ISL_867520, EPI_ISL_867521, EPI_ISL_867522, EPI_ISL_867523, EPI_ISL_867525, EPI_ISL_867536, EPI_ISL_867538, EPI_ISL_867539, EPI_ISL_867540, EPI_ISL_867541, EPI_ISL_867542, EPI_ISL_867543, EPI_ISL_867547, EPI_ISL_867550, EPI_ISL_867552, EPI_ISL_867553, EPI_ISL_867556, EPI_ISL_867557, EPI_ISL_867561, EPI_ISL_867562, EPI_ISL_867567, EPI_ISL_867572, EPI_ISL_867573, EPI_ISL_867574, EPI_ISL_867575, EPI_ISL_867579, EPI_ISL_867582, EPI_ISL_867584, EPI_ISL_867586, EPI_ISL_867587, EPI_ISL_867588, EPI_ISL_867589, EPI_ISL_867590, EPI_ISL_867704, EPI_ISL_867705, EPI_ISL_867706, EPI_ISL_867708, EPI_ISL_867716, EPI_ISL_867717, EPI_ISL_867718, EPI_ISL_867719, EPI_ISL_867720, EPI_ISL_867721, EPI_ISL_867722, EPI_ISL_867723, EPI_ISL_867724, EPI_ISL_867725, EPI_ISL_867726, EPI_ISL_867728, EPI_ISL_867730, EPI_ISL_867731, EPI_ISL_867737, EPI_ISL_867738, EPI_ISL_867739, EPI_ISL_867740, EPI_ISL_867741, EPI_ISL_867759, EPI_ISL_867762, EPI_ISL_867763, EPI_ISL_867764, EPI_ISL_867766, EPI_ISL_867768, EPI_ISL_867771, EPI_ISL_867773, EPI_ISL_867776, EPI_ISL_867779, EPI_ISL_867795, EPI_ISL_867796, EPI_ISL_867797, EPI_ISL_867800, EPI_ISL_867801, EPI_ISL_867802, EPI_ISL_867804, EPI_ISL_867805, EPI_ISL_867806, EPI_ISL_867807, EPI_ISL_867808, EPI_ISL_867809, EPI_ISL_867810, EPI_ISL_867811, EPI_ISL_867812, EPI_ISL_867815, EPI_ISL_867816, EPI_ISL_867819, EPI_ISL_867821, EPI_ISL_867823, EPI_ISL_867824, EPI_ISL_867825, EPI_ISL_867828, EPI_ISL_867830, EPI_ISL_867832, EPI_ISL_867833, EPI_ISL_867834, EPI_ISL_867837, EPI_ISL_867838, EPI_ISL_867839, EPI_ISL_867841, EPI_ISL_867842, EPI_ISL_867843, EPI_ISL_867844, EPI_ISL_867845, EPI_ISL_867847, EPI_ISL_867848, EPI_ISL_867849, EPI_ISL_867850, EPI_ISL_867851, EPI_ISL_867853, EPI_ISL_867855, EPI_ISL_867856, EPI_ISL_867859, EPI_ISL_867860, EPI_ISL_867861, EPI_ISL_867862, EPI_ISL_867863, EPI_ISL_867868, EPI_ISL_867873, EPI_ISL_867875, EPI_ISL_867876, EPI_ISL_867878, EPI_ISL_867879, EPI_ISL_867880, EPI_ISL_867881, EPI_ISL_867883, EPI_ISL_867884, EPI_ISL_867885, EPI_ISL_867889, EPI_ISL_867890, EPI_ISL_867893, EPI_ISL_867895, EPI_ISL_867899, EPI_ISL_867900, EPI_ISL_867901, EPI_ISL_867904, EPI_ISL_867906, EPI_ISL_867911, EPI_ISL_867912, EPI_ISL_867913, EPI_ISL_867914, EPI_ISL_867915, EPI_ISL_867916, EPI_ISL_867918, EPI_ISL_867921, EPI_ISL_999515, EPI_ISL_1476085, EPI_ISL_1476095, EPI_ISL_1476101, EPI_ISL_1476236, EPI_ISL_1476237, EPI_ISL_1476238, EPI_ISL_1476239, EPI_ISL_1476240, EPI_ISL_1476241, EPI_ISL_1476242, EPI_ISL_1476247, EPI_ISL_1476522, EPI_ISL_1476523, EPI_ISL_1476524, EPI_ISL_1476525, EPI_ISL_1476527, EPI_ISL_1476534, EPI_ISL_1476535, EPI_ISL_1476540, EPI_ISL_1476544, EPI_ISL_1476545, EPI_ISL_1476546, EPI_ISL_1476548, EPI_ISL_1546802, EPI_ISL_1546803, EPI_ISL_1546804, EPI_ISL_1546805, EPI_ISL_1546808, EPI_ISL_1546809, EPI_ISL_1546810, EPI_ISL_1546811, EPI_ISL_1546814, EPI_ISL_1546817, EPI_ISL_1546820, EPI_ISL_1546912, EPI_ISL_1546929 | | | |
| see above | Originating lab: Wales Specialist Virology Centre Sequencing lab: Pathogen Genomics Unit | Public Health Wales Microbiology Cardiff Wales Specialist Virology Centre | Alec Birchley; Alexander Adams; Amy Gaskin; Angela Marchbank; Bree Gatica-Wilcox; Catherine Moore; Jason Coombes; Joanne Watkins; Joel Southgate; Johnathan Evans; Laura Gifford; Lauren Gilbert; Lee Graham; Malorie Perry; Matthew Bull; Nicole Pacchiarini; Sally Corden; Sara Kumziene-Summerhayes; Sara Rey; Sarah Taylor; Simon Cottrell; Sophie Jones; Tom Connor |
| EPI_ISL_922194, EPI_ISL_922197, EPI_ISL_922200, EPI_ISL_922203, EPI_ISL_922206, EPI_ISL_922207, EPI_ISL_922208, EPI_ISL_922209, EPI_ISL_922211, EPI_ISL_922212, EPI_ISL_922213, EPI_ISL_922214, EPI_ISL_922215, EPI_ISL_922216, EPI_ISL_922217, EPI_ISL_922218, EPI_ISL_922219, EPI_ISL_922220, EPI_ISL_922223, EPI_ISL_922224, EPI_ISL_922225, EPI_ISL_922226, EPI_ISL_922227, EPI_ISL_922228, EPI_ISL_922229, EPI_ISL_922230, EPI_ISL_922237, EPI_ISL_922258, EPI_ISL_922259, EPI_ISL_922294, EPI_ISL_922295, EPI_ISL_922346, EPI_ISL_922347, EPI_ISL_922348, EPI_ISL_922349, EPI_ISL_951231, EPI_ISL_951232, EPI_ISL_951237, EPI_ISL_951238, EPI_ISL_951239, EPI_ISL_951241, EPI_ISL_951245, EPI_ISL_951246, EPI_ISL_951247, EPI_ISL_951248, EPI_ISL_951252, EPI_ISL_951254, EPI_ISL_951255, EPI_ISL_951256, EPI_ISL_951258, EPI_ISL_951264, EPI_ISL_951265, EPI_ISL_951267, EPI_ISL_951268, EPI_ISL_951269, EPI_ISL_951270, EPI_ISL_951271, EPI_ISL_951272, EPI_ISL_951273, EPI_ISL_951274, EPI_ISL_951275, EPI_ISL_951276, EPI_ISL_951277, EPI_ISL_951281, EPI_ISL_951282, EPI_ISL_951283, EPI_ISL_951286, EPI_ISL_951287, EPI_ISL_951288, EPI_ISL_951289, EPI_ISL_951290, EPI_ISL_951291, EPI_ISL_951293, EPI_ISL_951294, EPI_ISL_951297, EPI_ISL_951299, EPI_ISL_951300, EPI_ISL_951301, EPI_ISL_951304, EPI_ISL_951307, EPI_ISL_951308, EPI_ISL_951309, EPI_ISL_951316, EPI_ISL_951317, EPI_ISL_951345, EPI_ISL_951346, EPI_ISL_951347, EPI_ISL_951348, EPI_ISL_951349, EPI_ISL_951350, EPI_ISL_951351, EPI_ISL_951352, EPI_ISL_951353, EPI_ISL_951354, EPI_ISL_951356, EPI_ISL_951362, EPI_ISL_951363, EPI_ISL_951364, EPI_ISL_951366, EPI_ISL_951367, EPI_ISL_951376, EPI_ISL_951377, EPI_ISL_951389, EPI_ISL_951391, EPI_ISL_951392, EPI_ISL_951393, EPI_ISL_951395, EPI_ISL_951396, EPI_ISL_951404, EPI_ISL_951405, EPI_ISL_951406, EPI_ISL_951407, EPI_ISL_951408, EPI_ISL_951409, EPI_ISL_951412, EPI_ISL_951413, EPI_ISL_951414, EPI_ISL_951415, EPI_ISL_951417, EPI_ISL_951426, EPI_ISL_951427, EPI_ISL_951429, EPI_ISL_951439, EPI_ISL_999033, EPI_ISL_999034, EPI_ISL_999035, EPI_ISL_999036, EPI_ISL_999037, EPI_ISL_999038, EPI_ISL_999039, EPI_ISL_999040, EPI_ISL_999041, EPI_ISL_999042, EPI_ISL_999044, EPI_ISL_999045, EPI_ISL_999048, EPI_ISL_999049, EPI_ISL_999050, EPI_ISL_999059, EPI_ISL_999060, EPI_ISL_999061, EPI_ISL_999062, EPI_ISL_999063, EPI_ISL_999064, EPI_ISL_999065, EPI_ISL_999066, EPI_ISL_999067, EPI_ISL_999076, EPI_ISL_999077, EPI_ISL_999080, EPI_ISL_999088, EPI_ISL_999094, EPI_ISL_999115, EPI_ISL_999128, EPI_ISL_999129, EPI_ISL_999133, EPI_ISL_999199, EPI_ISL_999211, EPI_ISL_999212, EPI_ISL_999364, EPI_ISL_999365, EPI_ISL_999368, EPI_ISL_999369, EPI_ISL_999370, EPI_ISL_999371, EPI_ISL_999372, EPI_ISL_999373, EPI_ISL_999377, EPI_ISL_999378, EPI_ISL_999383, EPI_ISL_999384, EPI_ISL_999388, EPI_ISL_999390, EPI_ISL_999391, EPI_ISL_999392, EPI_ISL_999393, EPI_ISL_999396, EPI_ISL_999399, EPI_ISL_999403, EPI_ISL_999419, EPI_ISL_999420, EPI_ISL_999423, EPI_ISL_999428, EPI_ISL_999431, EPI_ISL_999432, EPI_ISL_999433, EPI_ISL_999434, EPI_ISL_999440, EPI_ISL_999446, EPI_ISL_999447, EPI_ISL_999449, EPI_ISL_999452, EPI_ISL_999455, EPI_ISL_999456, EPI_ISL_999457, EPI_ISL_999458, EPI_ISL_999460, EPI_ISL_999471, EPI_ISL_999472, EPI_ISL_999482, EPI_ISL_999512, EPI_ISL_999513, EPI_ISL_999514, EPI_ISL_1051789, EPI_ISL_1051791, EPI_ISL_1051792, EPI_ISL_1051810, EPI_ISL_1051811, EPI_ISL_1051812, EPI_ISL_1051828, EPI_ISL_1178721, EPI_ISL_1178822, EPI_ISL_1178847, EPI_ISL_1249226, EPI_ISL_1309745, EPI_ISL_1309746, EPI_ISL_1309747, EPI_ISL_1833053, EPI_ISL_1833054, EPI_ISL_1833079, EPI_ISL_1833088, EPI_ISL_1833090, EPI_ISL_1833100, EPI_ISL_1833103, EPI_ISL_1833109 | | | |
| see above | Oxford Viromics, NDM, University of Oxford; Oxford University Hospitals; Basingstoke and North Hampshire Hospital | COVID-19 Genomics UK (COG-UK) Consortium | Alex Mobbs; Amy Trebes; Anita Justice; Catrin Moore; Christophe Fraser; David Bonsall; David Buck; Emma Wise; George Macintyre; Jessica Lynch; John Todd; Mariateresa de Cesare; Matilde Mori; Monique Andersson; Nathan Moore; Nick Cortes; Robert Shaw; Stephen Kidd; Tanya Golubchik; Timothy Peto |
| EPI_ISL_866542, EPI_ISL_866543, EPI_ISL_866544, EPI_ISL_866545, EPI_ISL_866546, EPI_ISL_866547, EPI_ISL_866549, EPI_ISL_866550, EPI_ISL_866551, EPI_ISL_866552, EPI_ISL_866553, EPI_ISL_866554, EPI_ISL_866555, EPI_ISL_866557, EPI_ISL_866558, EPI_ISL_866559, EPI_ISL_866561, EPI_ISL_866562, EPI_ISL_866563, EPI_ISL_866564, EPI_ISL_866565, EPI_ISL_866567, EPI_ISL_866568, EPI_ISL_866570, EPI_ISL_866571, EPI_ISL_866572, EPI_ISL_866573, EPI_ISL_866574, EPI_ISL_866575, EPI_ISL_866576, EPI_ISL_866603, EPI_ISL_866604, EPI_ISL_866605, EPI_ISL_866606, EPI_ISL_866607, EPI_ISL_866608, EPI_ISL_866609, EPI_ISL_866610, EPI_ISL_866611, EPI_ISL_866612, EPI_ISL_866613, EPI_ISL_866614, EPI_ISL_866615, EPI_ISL_866616, EPI_ISL_866617, EPI_ISL_866618, EPI_ISL_866620, EPI_ISL_866621, EPI_ISL_866622, EPI_ISL_866623, EPI_ISL_866624, EPI_ISL_866625, EPI_ISL_866626, EPI_ISL_866627, EPI_ISL_866628, EPI_ISL_866629, EPI_ISL_866630, EPI_ISL_866631, EPI_ISL_866632, EPI_ISL_866633, EPI_ISL_866634, EPI_ISL_866635, EPI_ISL_866636, EPI_ISL_866637, EPI_ISL_866638, EPI_ISL_866639, EPI_ISL_866640, EPI_ISL_866641, EPI_ISL_866642, EPI_ISL_866643, EPI_ISL_866644, EPI_ISL_866645, EPI_ISL_866646, EPI_ISL_866647, EPI_ISL_866648, EPI_ISL_866649, EPI_ISL_866650, EPI_ISL_866651, EPI_ISL_866652, EPI_ISL_866653, EPI_ISL_866654, EPI_ISL_866655, EPI_ISL_866656, EPI_ISL_866657, EPI_ISL_866658, EPI_ISL_866659, EPI_ISL_866660, EPI_ISL_866661, EPI_ISL_866662, EPI_ISL_866663, EPI_ISL_866664, EPI_ISL_866665, EPI_ISL_866666, EPI_ISL_866667, EPI_ISL_866668, EPI_ISL_866669, EPI_ISL_866670, EPI_ISL_866671, EPI_ISL_866672, EPI_ISL_866673, EPI_ISL_866674, EPI_ISL_866675, EPI_ISL_866676, EPI_ISL_866677, EPI_ISL_866678, EPI_ISL_866679, EPI_ISL_866680, EPI_ISL_866681, EPI_ISL_866682, EPI_ISL_866683, EPI_ISL_866684, EPI_ISL_866685, EPI_ISL_866686, EPI_ISL_866687, EPI_ISL_866688, EPI_ISL_866689, EPI_ISL_866690, EPI_ISL_866691, EPI_ISL_866692, EPI_ISL_866693, EPI_ISL_866694, EPI_ISL_866695, EPI_ISL_866696, EPI_ISL_866697, EPI_ISL_866698, EPI_ISL_866699, EPI_ISL_866700, EPI_ISL_866701, EPI_ISL_866702, EPI_ISL_866703, EPI_ISL_866704, EPI_ISL_866705, EPI_ISL_866706, EPI_ISL_866707, EPI_ISL_866708, EPI_ISL_866709, EPI_ISL_866710, EPI_ISL_866711, EPI_ISL_866712, EPI_ISL_866713, EPI_ISL_866714, EPI_ISL_866715, EPI_ISL_866716, EPI_ISL_866717, EPI_ISL_866718, EPI_ISL_866719, EPI_ISL_866720, EPI_ISL_866721, EPI_ISL_866722, EPI_ISL_866723, EPI_ISL_866724, EPI_ISL_866725, EPI_ISL_866726, EPI_ISL_866727, EPI_ISL_866728, EPI_ISL_866729, EPI_ISL_866730, EPI_ISL_866731, EPI_ISL_866732, EPI_ISL_866733, EPI_ISL_866734, EPI_ISL_866735, EPI_ISL_866736, EPI_ISL_866737, EPI_ISL_866738, EPI_ISL_866739, EPI_ISL_866740, EPI_ISL_866741, EPI_ISL_866742, EPI_ISL_866743, EPI_ISL_866744, EPI_ISL_866745, EPI_ISL_866746, EPI_ISL_866747, EPI_ISL_866758, EPI_ISL_866759, EPI_ISL_866761, EPI_ISL_866762, EPI_ISL_866765, EPI_ISL_866766, EPI_ISL_866768, EPI_ISL_866769, EPI_ISL_866771, EPI_ISL_866772, EPI_ISL_866773, EPI_ISL_866776, EPI_ISL_866777, EPI_ISL_866778, EPI_ISL_866779, EPI_ISL_866780, EPI_ISL_866782, EPI_ISL_866783, EPI_ISL_866784, EPI_ISL_866785, EPI_ISL_866786, EPI_ISL_866787, EPI_ISL_866790, EPI_ISL_866791, EPI_ISL_866792, EPI_ISL_866794, EPI_ISL_866795, EPI_ISL_866796, EPI_ISL_866797, EPI_ISL_866798, EPI_ISL_866799, EPI_ISL_866800, EPI_ISL_866831, EPI_ISL_866832, EPI_ISL_866833, EPI_ISL_866834, EPI_ISL_998830, EPI_ISL_998832, EPI_ISL_998836, EPI_ISL_1051450, EPI_ISL_1178430, EPI_ISL_1178431, EPI_ISL_1178432, EPI_ISL_1178442, EPI_ISL_1178443, EPI_ISL_1178446, EPI_ISL_1178454, EPI_ISL_1178476, EPI_ISL_1249006, EPI_ISL_1249055, EPI_ISL_1249059, EPI_ISL_1832940, EPI_ISL_1832949, EPI_ISL_1832950, EPI_ISL_1832952, EPI_ISL_1832959, EPI_ISL_1832960, EPI_ISL_1832962, EPI_ISL_1832964, EPI_ISL_1832965, EPI_ISL_1832966, EPI_ISL_1832967, EPI_ISL_1832980 | | | |
| see above | Quadram Institute Bioscience | COVID-19 Genomics UK (COG-UK) Consortium | Alexander J Trotter; Alison E. Mather; Alp Aydin; Ana P. Tedim; Anastasia Kolyva; Andrew Bell; Andrew J. Page; Claire Stuart; Dave J. Baker; Gemma L. Kay; John Wain; Justin O'Grady; Leonardo de Oliveira Martins; Lizzie Meadows; Maria Diaz; Mark Webber; Muhammed Yasir; Nabil-Fareed Alikhan; Ngozi Elumogo; Nicholas M. Thomson; Rachael Stanley; Rachel Gilroy; Reenesh Prakash; Samir Dervisevic; Samuel Bloomfield; Steven Rudder; Thanh Le-Viet |
| EPI_ISL_840009, EPI_ISL_840010, EPI_ISL_840011, EPI_ISL_840012, EPI_ISL_840013, EPI_ISL_840014, EPI_ISL_840015, EPI_ISL_840016, EPI_ISL_840051, EPI_ISL_840052, EPI_ISL_840053, EPI_ISL_840054, EPI_ISL_840055, EPI_ISL_840056, EPI_ISL_840057, EPI_ISL_840058, EPI_ISL_840059, EPI_ISL_840060, EPI_ISL_840061, EPI_ISL_866908, EPI_ISL_866909, EPI_ISL_866910, EPI_ISL_866911, EPI_ISL_866912, EPI_ISL_866913, EPI_ISL_866914, EPI_ISL_866915, EPI_ISL_866916, EPI_ISL_866917, EPI_ISL_866918, EPI_ISL_866919, EPI_ISL_866920 | | | |
| see above | Queens Medical Centre, Clinical Microbiology Department / DeepSeq Nottingham | COVID-19 Genomics UK (COG-UK) Consortium | Christopher Moore; Fei Sang; Gemma Clark; Hannah Howson-Wells; Johnny Debebe; Jonathan Ball; Joseph Chappell; Manjinder Khakh; Matthew Carlisle; Matthew Loose; Michelle M Lister; Nadine Holmes; Patrick McClure; Theocharis Tsoleridis; Vicki M Fleming; Victoria Wright; Wendy Smith |
| EPI_ISL_866366, EPI_ISL_866393, EPI_ISL_866394, EPI_ISL_920996, EPI_ISL_920997, EPI_ISL_920998, EPI_ISL_920999, EPI_ISL_921000, EPI_ISL_921001, EPI_ISL_921002, EPI_ISL_921003, EPI_ISL_921004, EPI_ISL_921005, EPI_ISL_921006, EPI_ISL_921007, EPI_ISL_921008, EPI_ISL_921009, EPI_ISL_921010, EPI_ISL_921018, EPI_ISL_998136, EPI_ISL_998140, EPI_ISL_998144, EPI_ISL_998154, EPI_ISL_998158, EPI_ISL_998159, EPI_ISL_998163, EPI_ISL_1474961, EPI_ISL_1474962, EPI_ISL_1474965, EPI_ISL_1474966, EPI_ISL_1474967, EPI_ISL_1474968, EPI_ISL_1474969, EPI_ISL_1474970, EPI_ISL_1474971, EPI_ISL_1474972, EPI_ISL_1474974, EPI_ISL_1474975, EPI_ISL_1474976, EPI_ISL_1474977, EPI_ISL_1474978, EPI_ISL_1474979, EPI_ISL_1474980, EPI_ISL_1474981, EPI_ISL_1474982, EPI_ISL_1474984, EPI_ISL_1474985, EPI_ISL_1474991, EPI_ISL_1474992, EPI_ISL_1474993, EPI_ISL_1474994, EPI_ISL_1475004, EPI_ISL_1475005, EPI_ISL_1475006, EPI_ISL_1475082 | | | |
| see above | Regional Virus Laboratory, Belfast Health and Social Care Trust | COVID-19 Genomics UK (COG-UK) Consortium | Alison Watt; Ciara Cox; Conall McCaughey; David Simpson; Derek Fairley; James McKenna; Mairead Connor; Susan Feeney; Tanya Curran; Zoltan Molnar |
| EPI_ISL_804369, EPI_ISL_811123, EPI_ISL_811124, EPI_ISL_811125, EPI_ISL_811126, EPI_ISL_811150, EPI_ISL_825491, EPI_ISL_825550, EPI_ISL_825551, EPI_ISL_825573, EPI_ISL_825574, EPI_ISL_825616, EPI_ISL_826282, EPI_ISL_837055, EPI_ISL_837056, EPI_ISL_837057, EPI_ISL_837059, EPI_ISL_837060, EPI_ISL_837069, EPI_ISL_837176, EPI_ISL_837177, EPI_ISL_837178, EPI_ISL_837179, EPI_ISL_837180, EPI_ISL_837181, EPI_ISL_837182, EPI_ISL_837243, EPI_ISL_837244, EPI_ISL_846597, EPI_ISL_846598, EPI_ISL_855595, EPI_ISL_855602, EPI_ISL_855605, EPI_ISL_860688, EPI_ISL_1296505, EPI_ISL_1296506, EPI_ISL_1296507, EPI_ISL_1296509, EPI_ISL_1296510, EPI_ISL_1296511, EPI_ISL_1296512, EPI_ISL_1296513, EPI_ISL_1296514, EPI_ISL_1296515, EPI_ISL_1296516, EPI_ISL_1296517, EPI_ISL_1296518, EPI_ISL_1296519, EPI_ISL_1296520, EPI_ISL_1296521, EPI_ISL_1296522, EPI_ISL_1296523, EPI_ISL_1296525, EPI_ISL_1296526, EPI_ISL_1296527, EPI_ISL_1296530, EPI_ISL_1296531, EPI_ISL_1296532, EPI_ISL_1296536, EPI_ISL_1296537, EPI_ISL_1296549 | | | |
| see above | Respiratory Virus Unit, National Infection Service, Public Health England | COVID-19 Genomics UK (COG-UK) Consortium | PHE Covid Sequencing Team |
| EPI_ISL_1833472, EPI_ISL_1833481, EPI_ISL_1833482, EPI_ISL_1833483, EPI_ISL_1833486, EPI_ISL_1833487, EPI_ISL_1833489, EPI_ISL_1833490, EPI_ISL_1833492, EPI_ISL_1833493, EPI_ISL_1833495, EPI_ISL_1833496, EPI_ISL_1833497, EPI_ISL_1833498, EPI_ISL_1833499, EPI_ISL_1833500, EPI_ISL_1833502, EPI_ISL_1833510, EPI_ISL_1833511, EPI_ISL_1833512, EPI_ISL_1833513, EPI_ISL_1833514, EPI_ISL_1833516, EPI_ISL_1833518, EPI_ISL_1833519, EPI_ISL_1833520, EPI_ISL_1833521, EPI_ISL_1833522, EPI_ISL_1833523, EPI_ISL_1833524, EPI_ISL_1833525, EPI_ISL_1833527, EPI_ISL_1833528, EPI_ISL_1833529, EPI_ISL_1833530, EPI_ISL_1833537, EPI_ISL_1833538, EPI_ISL_1833540, EPI_ISL_1833541, EPI_ISL_1833542, EPI_ISL_1833544, EPI_ISL_1833545, EPI_ISL_1833546, EPI_ISL_1833547 | | | |
| see above | St George's University Hospitals NHS Foundation Trust | COVID-19 Genomics UK (COG-UK) Consortium | Adam Witney; Cassie Pope; Irene Monahan; Joshua Taylor; Ken Laing; NgeeKeong Tan |
| EPI_ISL_866080, EPI_ISL_866081, EPI_ISL_866082, EPI_ISL_866083, EPI_ISL_866084, EPI_ISL_866085, EPI_ISL_866086, EPI_ISL_866087, EPI_ISL_866088, EPI_ISL_866089, EPI_ISL_866090, EPI_ISL_866091, EPI_ISL_866092, EPI_ISL_866093, EPI_ISL_866094, EPI_ISL_866095, EPI_ISL_866096, EPI_ISL_866097, EPI_ISL_866098, EPI_ISL_866099, EPI_ISL_866100, EPI_ISL_866101, EPI_ISL_866102, EPI_ISL_866103, EPI_ISL_866104, EPI_ISL_866105, EPI_ISL_866112, EPI_ISL_866113, EPI_ISL_866114, EPI_ISL_866117, EPI_ISL_866120, EPI_ISL_866125, EPI_ISL_866126, EPI_ISL_866134, EPI_ISL_866142, EPI_ISL_866143, EPI_ISL_866146, EPI_ISL_866152, EPI_ISL_866154, EPI_ISL_866157, EPI_ISL_866160, EPI_ISL_866171, EPI_ISL_866174, EPI_ISL_866178, EPI_ISL_866180, EPI_ISL_866186, EPI_ISL_920188, EPI_ISL_920191, EPI_ISL_920193, EPI_ISL_920241, EPI_ISL_920298, EPI_ISL_920299, EPI_ISL_920309, EPI_ISL_920310, EPI_ISL_920318, EPI_ISL_920327, EPI_ISL_920328, EPI_ISL_920329, EPI_ISL_920330, EPI_ISL_920337, EPI_ISL_920338, EPI_ISL_920339, EPI_ISL_920345, EPI_ISL_920353, EPI_ISL_920357, EPI_ISL_920362, EPI_ISL_920363, EPI_ISL_920371, EPI_ISL_920458, EPI_ISL_920466, EPI_ISL_920467, EPI_ISL_920501, EPI_ISL_1105184, EPI_ISL_1105258, EPI_ISL_1105265, EPI_ISL_1105284, EPI_ISL_1105297, EPI_ISL_1105303, EPI_ISL_1248374, EPI_ISL_1248375, EPI_ISL_1248384, EPI_ISL_1248385, EPI_ISL_1248389, EPI_ISL_1248395, EPI_ISL_1248396, EPI_ISL_1248397, EPI_ISL_1248407, EPI_ISL_1248408, EPI_ISL_1248409, EPI_ISL_1248418, EPI_ISL_1248419, EPI_ISL_1248428, EPI_ISL_1248429, EPI_ISL_1248440, EPI_ISL_1248441, EPI_ISL_1248467, EPI_ISL_1248487, EPI_ISL_1248495 | | | |
| see above | University College London Hospital | COVID-19 Genomics UK (COG-UK) Consortium | Catherine Houlihan; Dan Frampton; Dr Catherine Houlihan; Dr Daniel Frampton; Dr Judith Heaney; Dr Moira Spyer; Dr Paul Grant and Dr Eleni Nastouli; Judith Heaney; Matthew Byott; Moira Spyer and Eleni Nastouli; Stuart Kirk |
| EPI_ISL_838937, EPI_ISL_838938, EPI_ISL_838939, EPI_ISL_838940, EPI_ISL_838941, EPI_ISL_838942, EPI_ISL_838943, EPI_ISL_838944, EPI_ISL_838945, EPI_ISL_838946, EPI_ISL_838947, EPI_ISL_838948, EPI_ISL_838949, EPI_ISL_838951, EPI_ISL_838958, EPI_ISL_838959, EPI_ISL_838960, EPI_ISL_838961, EPI_ISL_838962, EPI_ISL_838963, EPI_ISL_838964, EPI_ISL_838965, EPI_ISL_838966, EPI_ISL_838967, EPI_ISL_838968, EPI_ISL_838969, EPI_ISL_838970, EPI_ISL_838971, EPI_ISL_838972, EPI_ISL_838973, EPI_ISL_838974, EPI_ISL_838975, EPI_ISL_838976, EPI_ISL_838977, EPI_ISL_838978, EPI_ISL_838979, EPI_ISL_838980, EPI_ISL_838981, EPI_ISL_838982, EPI_ISL_838983, EPI_ISL_838984, EPI_ISL_838985, EPI_ISL_838986, EPI_ISL_838987, EPI_ISL_838988, EPI_ISL_838989, EPI_ISL_838990, EPI_ISL_838991, EPI_ISL_838992, EPI_ISL_838993, EPI_ISL_838994, EPI_ISL_838995, EPI_ISL_838996, EPI_ISL_838997, EPI_ISL_838998, EPI_ISL_838999, EPI_ISL_839000, EPI_ISL_839001, EPI_ISL_839002, EPI_ISL_839003, EPI_ISL_839004, EPI_ISL_839005, EPI_ISL_839007, EPI_ISL_839008, EPI_ISL_839009, EPI_ISL_839010, EPI_ISL_839011, EPI_ISL_839012, EPI_ISL_839013, EPI_ISL_839014, EPI_ISL_839015, EPI_ISL_839016, EPI_ISL_839017, EPI_ISL_839018, EPI_ISL_839019, EPI_ISL_839021, EPI_ISL_839022, EPI_ISL_839023, EPI_ISL_839024, EPI_ISL_839025, EPI_ISL_839029, EPI_ISL_839126, EPI_ISL_839127, EPI_ISL_839128, EPI_ISL_839129, EPI_ISL_839130, EPI_ISL_839131, EPI_ISL_839132, EPI_ISL_839133, EPI_ISL_839134, EPI_ISL_839136, EPI_ISL_839145, EPI_ISL_839233, EPI_ISL_839234, EPI_ISL_839235, EPI_ISL_839236, EPI_ISL_839237, EPI_ISL_839238, EPI_ISL_839239, EPI_ISL_839240, EPI_ISL_839241, EPI_ISL_839242, EPI_ISL_839243, EPI_ISL_839244, EPI_ISL_839245, EPI_ISL_839246, EPI_ISL_839247, EPI_ISL_839248, EPI_ISL_839249, EPI_ISL_839251, EPI_ISL_839253, EPI_ISL_839254, EPI_ISL_839255, EPI_ISL_839256, EPI_ISL_839257, EPI_ISL_839258, EPI_ISL_839259, EPI_ISL_839261, EPI_ISL_839262, EPI_ISL_839263, EPI_ISL_839266, EPI_ISL_839267, EPI_ISL_839268, EPI_ISL_839269, EPI_ISL_839270, EPI_ISL_839271, EPI_ISL_839272, EPI_ISL_839274, EPI_ISL_839275, EPI_ISL_839276, EPI_ISL_839277, EPI_ISL_839278, EPI_ISL_839279, EPI_ISL_839280, EPI_ISL_839281, EPI_ISL_839282, EPI_ISL_839283, EPI_ISL_839284, EPI_ISL_839285, EPI_ISL_839286, EPI_ISL_839287, EPI_ISL_839288, EPI_ISL_839289, EPI_ISL_839290, EPI_ISL_839291, EPI_ISL_839292, EPI_ISL_839293, EPI_ISL_839294, EPI_ISL_839295, EPI_ISL_839296, EPI_ISL_839297, EPI_ISL_839298, EPI_ISL_839299, EPI_ISL_839300, EPI_ISL_839301, EPI_ISL_839302, EPI_ISL_839303, EPI_ISL_839304, EPI_ISL_865847, EPI_ISL_866194, EPI_ISL_866195, EPI_ISL_866196, EPI_ISL_866197, EPI_ISL_866199, EPI_ISL_866200, EPI_ISL_866202, EPI_ISL_866203, EPI_ISL_866204, EPI_ISL_866205, EPI_ISL_866206, EPI_ISL_866207, EPI_ISL_866208, EPI_ISL_866209, EPI_ISL_866210, EPI_ISL_866211, EPI_ISL_866212, EPI_ISL_866213, EPI_ISL_866214, EPI_ISL_866216, EPI_ISL_866218, EPI_ISL_866220, EPI_ISL_866222, EPI_ISL_866223, EPI_ISL_866224, EPI_ISL_866225, EPI_ISL_866226, EPI_ISL_866227, EPI_ISL_866228, EPI_ISL_866229, EPI_ISL_866230, EPI_ISL_866232, EPI_ISL_866233, EPI_ISL_866234, EPI_ISL_866235, EPI_ISL_866236, EPI_ISL_866237, EPI_ISL_866238, EPI_ISL_866239, EPI_ISL_866240, EPI_ISL_866241, EPI_ISL_866242, EPI_ISL_866243, EPI_ISL_866244, EPI_ISL_866245, EPI_ISL_866246, EPI_ISL_866247, EPI_ISL_866248, EPI_ISL_866249, EPI_ISL_866250, EPI_ISL_866251, EPI_ISL_866252, EPI_ISL_866253, EPI_ISL_866254, EPI_ISL_866255, EPI_ISL_866256, EPI_ISL_866257, EPI_ISL_866258, EPI_ISL_866259, EPI_ISL_866260, EPI_ISL_866261, EPI_ISL_919995, EPI_ISL_920000, EPI_ISL_920001, EPI_ISL_920002, EPI_ISL_920003, EPI_ISL_920004, EPI_ISL_920005, EPI_ISL_920006, EPI_ISL_920007, EPI_ISL_920008, EPI_ISL_920009, EPI_ISL_920010, EPI_ISL_920011, EPI_ISL_920012, EPI_ISL_920013, EPI_ISL_920014, EPI_ISL_920015, EPI_ISL_920016, EPI_ISL_920017, EPI_ISL_920018, EPI_ISL_920019, EPI_ISL_920020, EPI_ISL_920021, EPI_ISL_920022, EPI_ISL_920023, EPI_ISL_920024, EPI_ISL_920025, EPI_ISL_920027, EPI_ISL_920028, EPI_ISL_920029, EPI_ISL_920034, EPI_ISL_920042, EPI_ISL_920049, EPI_ISL_920050, EPI_ISL_920051, EPI_ISL_920851, EPI_ISL_920852, EPI_ISL_920853, EPI_ISL_920860, EPI_ISL_920861, EPI_ISL_920862, EPI_ISL_920863, EPI_ISL_920864, EPI_ISL_920865, EPI_ISL_920866, EPI_ISL_920867, EPI_ISL_920868, EPI_ISL_920869, EPI_ISL_920870, EPI_ISL_920871, EPI_ISL_920872, EPI_ISL_920873, EPI_ISL_920874, EPI_ISL_920875, EPI_ISL_920876, EPI_ISL_920877, EPI_ISL_920878, EPI_ISL_920879, EPI_ISL_920880, EPI_ISL_920881, EPI_ISL_949788, EPI_ISL_949789, EPI_ISL_949790, EPI_ISL_949927, EPI_ISL_950069, EPI_ISL_950070, EPI_ISL_950071, EPI_ISL_950072, EPI_ISL_950074, EPI_ISL_950075, EPI_ISL_950077, EPI_ISL_950079, EPI_ISL_950080, EPI_ISL_950083, EPI_ISL_950084, EPI_ISL_950085, EPI_ISL_950087, EPI_ISL_950088, EPI_ISL_950090, EPI_ISL_950092, EPI_ISL_950097, EPI_ISL_950098, EPI_ISL_950100, EPI_ISL_950101, EPI_ISL_950102, EPI_ISL_950104, EPI_ISL_950107, EPI_ISL_950108, EPI_ISL_950109, EPI_ISL_950110, EPI_ISL_950112, EPI_ISL_950114, EPI_ISL_950115, EPI_ISL_950117, EPI_ISL_950119, EPI_ISL_950120, EPI_ISL_950125, EPI_ISL_950126, EPI_ISL_950128, EPI_ISL_950129, EPI_ISL_950150, EPI_ISL_950155, EPI_ISL_950162, EPI_ISL_950163, EPI_ISL_950164, EPI_ISL_950169, EPI_ISL_997839, EPI_ISL_1050431, EPI_ISL_1050432, EPI_ISL_1050433, EPI_ISL_1050437, EPI_ISL_1050438, EPI_ISL_1050449, EPI_ISL_1050452, EPI_ISL_1050453, EPI_ISL_1050454, EPI_ISL_1050455, EPI_ISL_1050456, EPI_ISL_1050463, EPI_ISL_1050467, EPI_ISL_1050468, EPI_ISL_1050469, EPI_ISL_1050470, EPI_ISL_1050471, EPI_ISL_1050472, EPI_ISL_1050473, EPI_ISL_1050474, EPI_ISL_1050475, EPI_ISL_1050476, EPI_ISL_1050477, EPI_ISL_1050478, EPI_ISL_1050479, EPI_ISL_1050480, EPI_ISL_1050481, EPI_ISL_1050482, EPI_ISL_1050483, EPI_ISL_1050484, | | | |

| | | | |
|---|---|---|---|
| EPI_ISL_1050485, EPI_ISL_1050486, EPI_ISL_1050487, EPI_ISL_1050488, EPI_ISL_1050489, EPI_ISL_1050490, EPI_ISL_1050491, EPI_ISL_1050492, EPI_ISL_1050493, EPI_ISL_1050494, EPI_ISL_1050495, EPI_ISL_1050496, EPI_ISL_1050497, EPI_ISL_1050499, EPI_ISL_1050500, EPI_ISL_1050501, EPI_ISL_1050503, EPI_ISL_1050504, EPI_ISL_1050505, EPI_ISL_1050506, EPI_ISL_1050507, EPI_ISL_1050509, EPI_ISL_1050512, EPI_ISL_1050513, EPI_ISL_1050514, EPI_ISL_1050515, EPI_ISL_1050518, EPI_ISL_1050522, EPI_ISL_1050523, EPI_ISL_1248057, EPI_ISL_1248058, EPI_ISL_1248059, EPI_ISL_1248060, EPI_ISL_1248061, EPI_ISL_1248062, EPI_ISL_1248063, EPI_ISL_1248064, EPI_ISL_1248065, EPI_ISL_1248068, EPI_ISL_1248069, EPI_ISL_1248070, EPI_ISL_1248071, EPI_ISL_1248072, EPI_ISL_1248073, EPI_ISL_1248074, EPI_ISL_1248075, EPI_ISL_1248076, EPI_ISL_1248077, EPI_ISL_1248078, EPI_ISL_1248079, EPI_ISL_1248080, EPI_ISL_1248081, EPI_ISL_1248082, EPI_ISL_1248083, EPI_ISL_1248084, EPI_ISL_1248085, EPI_ISL_1248086, EPI_ISL_1248087, EPI_ISL_1248088, EPI_ISL_1248089, EPI_ISL_1248091, EPI_ISL_1248092, EPI_ISL_1248093, EPI_ISL_1248094, EPI_ISL_1248095, EPI_ISL_1248096, EPI_ISL_1248097, EPI_ISL_1248098, EPI_ISL_1248099, EPI_ISL_1248100, EPI_ISL_1248101, EPI_ISL_1248102, EPI_ISL_1248103, EPI_ISL_1248104, EPI_ISL_1248105, EPI_ISL_1248106, EPI_ISL_1248113, EPI_ISL_1248117, EPI_ISL_1248118, EPI_ISL_1248119, EPI_ISL_1248120, EPI_ISL_1248121, EPI_ISL_1309037, EPI_ISL_1309045, EPI_ISL_1309050, EPI_ISL_1309057, EPI_ISL_1309064, EPI_ISL_1309065, EPI_ISL_1309069, EPI_ISL_1309075, EPI_ISL_1309083, EPI_ISL_1309086, EPI_ISL_1309087, EPI_ISL_1309088, EPI_ISL_1309098, EPI_ISL_1309102, EPI_ISL_1309107, EPI_ISL_1309111, EPI_ISL_1386858, EPI_ISL_1386876, EPI_ISL_1386881, EPI_ISL_1386886, EPI_ISL_1386889, EPI_ISL_1386896, EPI_ISL_1386912, EPI_ISL_1386913, EPI_ISL_1637109, EPI_ISL_1730336, EPI_ISL_1730394, EPI_ISL_1832593 | | | |
| see above | University College London, Great Ormond Street Hospital for Children NHS Foundation Trust, Imperial College Healthcare NHS Trust | COVID-19 Genomics UK (COG-UK) Consortium | Alison Holmes; Charlotte Williams; Helena Tutill; Jacqueline Findlay; James Price; Judith Breuer; Julianne Brown; Kathryn Harris; Leysa Forrest; Mark Kristiansen; Paola Niola; Paola Resende Silva; Patricia Dyal; Paul Randell; Rachel Williams; Samuel Weeks; Sergi Castellano; Sunando Roy; Tony Brooks; Yasmin Panchbhaya |
| EPI_ISL_1104305, EPI_ISL_2125926 | University Hospitals Of Leicester NHS Trust and DeepSeq Nottingham | COVID-19 Genomics UK (COG-UK) Consortium | Christopher Holmes; Christopher Moore; Fei Sang; Joeseph Chappell; Johnny Debebe; Jonathan Ball; Julian Tang; Karlie Fallon; Matthew Carlisle; Matthew Loose; Nadine Holmes; Patrick McClure; Paul Bird; Thomas Helmer; Victoria Wright |
| EPI_ISL_918686, EPI_ISL_918687, EPI_ISL_918688, EPI_ISL_918689, EPI_ISL_918690, EPI_ISL_918691, EPI_ISL_918692, EPI_ISL_918693, EPI_ISL_918694, EPI_ISL_918695, EPI_ISL_918696, EPI_ISL_918697, EPI_ISL_918698, EPI_ISL_918699, EPI_ISL_918700, EPI_ISL_918701, EPI_ISL_918703, EPI_ISL_918708, EPI_ISL_918709, EPI_ISL_918710, EPI_ISL_918713, EPI_ISL_918715, EPI_ISL_918716, EPI_ISL_918717, EPI_ISL_918719, EPI_ISL_918732, EPI_ISL_918733, EPI_ISL_918734, EPI_ISL_918859, EPI_ISL_918860, EPI_ISL_918861, EPI_ISL_918862, EPI_ISL_918863, EPI_ISL_918864, EPI_ISL_918866, EPI_ISL_918867, EPI_ISL_918868, EPI_ISL_918870, EPI_ISL_918871, EPI_ISL_918872, EPI_ISL_918873, EPI_ISL_918879, EPI_ISL_918880, EPI_ISL_918881, EPI_ISL_918884, EPI_ISL_918885, EPI_ISL_918886, EPI_ISL_918887, EPI_ISL_918888, EPI_ISL_918889, EPI_ISL_918890, EPI_ISL_918891, EPI_ISL_918892, EPI_ISL_918893, EPI_ISL_918894, EPI_ISL_918895, EPI_ISL_918896, EPI_ISL_918897, EPI_ISL_918898, EPI_ISL_918899, EPI_ISL_918914, EPI_ISL_918915, EPI_ISL_918916, EPI_ISL_918917, EPI_ISL_918918, EPI_ISL_918919, EPI_ISL_918920, EPI_ISL_918921, EPI_ISL_918922, EPI_ISL_918923, EPI_ISL_918924, EPI_ISL_918925, EPI_ISL_918926, EPI_ISL_918927, EPI_ISL_918928, EPI_ISL_918929, EPI_ISL_918930, EPI_ISL_918931, EPI_ISL_918932, EPI_ISL_918933, EPI_ISL_918934, EPI_ISL_918935, EPI_ISL_918936, EPI_ISL_918937, EPI_ISL_918938, EPI_ISL_918939, EPI_ISL_918973, EPI_ISL_918974, EPI_ISL_918975, EPI_ISL_918976, EPI_ISL_949411, EPI_ISL_996427, EPI_ISL_996428, EPI_ISL_996429, EPI_ISL_996430, EPI_ISL_996436, EPI_ISL_1046997, EPI_ISL_1046998, EPI_ISL_1047067 | | | |
| see above | University of Birmingham | COVID-19 Genomics UK (COG-UK) Consortium | Alex Richter; Andrew Bosworth; Andrew Bosworth. Queen Elizabeth Hospital: Anna Casey; Andrew D Beggs PHE Heartlands Lab: Husam Osman; Charlotte Poxon; Institute of Microbiology; Joanne Stockton; Josh Quick; Kasun Wanigasooriya; Mike Kidd; Nicholas Loman. University of Birmingham Testing Laboratory: Celina M Whalley; Oliver Pickles; Radoslaw Poplawski; Samuel Nicholls; University of Birmingham: Claire McMurray; Will Rowe |
| EPI_ISL_838315, EPI_ISL_996514, EPI_ISL_996515, EPI_ISL_996516, EPI_ISL_996517, EPI_ISL_996518, EPI_ISL_996519, EPI_ISL_996520, EPI_ISL_996522, EPI_ISL_996523, EPI_ISL_996524, EPI_ISL_996525, EPI_ISL_996526, EPI_ISL_996531, EPI_ISL_996532, EPI_ISL_996533, EPI_ISL_996534, EPI_ISL_996535, EPI_ISL_996536, EPI_ISL_996537, EPI_ISL_996571, EPI_ISL_996580, EPI_ISL_996581, EPI_ISL_996583, EPI_ISL_996584, EPI_ISL_996592, EPI_ISL_997326, EPI_ISL_997327, EPI_ISL_1103806, EPI_ISL_1308583, EPI_ISL_1308584, EPI_ISL_1545815, EPI_ISL_1545816 | | | |
| see above | University of Exeter | COVID-19 Genomics UK (COG-UK) Consortium | Aaron Jeffries; Audrey Farbos; Ben Temperton; Jane Masoli; Joanna Warwick-Dugdale; Michelle Michelsen; Robyn Manley; Stephen Michell |
| EPI_ISL_997074, EPI_ISL_1047855, EPI_ISL_1047858, EPI_ISL_1047859, EPI_ISL_1104197, EPI_ISL_1104220, EPI_ISL_1104221, EPI_ISL_1104231, EPI_ISL_1104235, EPI_ISL_1104236, EPI_ISL_1104242, EPI_ISL_1104243, EPI_ISL_1308703, EPI_ISL_1308704, EPI_ISL_1308705, EPI_ISL_1308838, EPI_ISL_1308839, EPI_ISL_1308842, EPI_ISL_1308843 | | | |
| see above | Virology Department, Royal Infirmary of Edinburgh, NHS Lothian / School of Biological Sciences, University of Edinburgh | COVID-19 Genomics UK (COG-UK) Consortium | Colquhoun R; Cotton S; Dewar R; Hill V; Jackson B; McCrone JT; McHugh M; O'Toole Á; Rambaut A; Rooke S; Scher E; Templeton K; Yu X |
| EPI_ISL_865089, EPI_ISL_865111, EPI_ISL_865112, EPI_ISL_865113, EPI_ISL_865114, EPI_ISL_865115, EPI_ISL_865116, EPI_ISL_865117, EPI_ISL_865118, EPI_ISL_865119, EPI_ISL_865120, EPI_ISL_865121, EPI_ISL_865122, EPI_ISL_865123, EPI_ISL_865124, EPI_ISL_865125, EPI_ISL_865126, EPI_ISL_865127, EPI_ISL_865128, EPI_ISL_865129, EPI_ISL_865130, EPI_ISL_865132, EPI_ISL_865133, EPI_ISL_865134, EPI_ISL_865135, EPI_ISL_865140, EPI_ISL_865143, EPI_ISL_865145, EPI_ISL_919337, EPI_ISL_919338 | | | |
| see above | Virology Department, Royal Infirmary of Edinburgh, NHS Lothian / School of Biological Sciences, University of Edinburgh / Institute of Genetics and Molecular Medicine, University of Edinburgh | COVID-19 Genomics UK (COG-UK) Consortium | Balcaza C; Colquhoun R; Dewar R; Gallagher M; Hill V; Jackson B; McCrone JT; McHugh M; O'Toole Á; Rambaut A; Rooke S; Scher E; Templeton K; Williams TC; Yu X |
| EPI_ISL_842201, EPI_ISL_842203, EPI_ISL_842207, EPI_ISL_842210, EPI_ISL_842213, EPI_ISL_842219, EPI_ISL_842222, EPI_ISL_842223, EPI_ISL_842225, EPI_ISL_842227, EPI_ISL_842229, EPI_ISL_842234, EPI_ISL_842237, EPI_ISL_842242, EPI_ISL_842243, EPI_ISL_842247, EPI_ISL_842249, EPI_ISL_842252, EPI_ISL_842253, EPI_ISL_842257, EPI_ISL_842259, EPI_ISL_842267, EPI_ISL_842269, EPI_ISL_842271, EPI_ISL_842275, EPI_ISL_842276, EPI_ISL_842277, EPI_ISL_842290, EPI_ISL_842296, EPI_ISL_842298, EPI_ISL_842302, EPI_ISL_842307, EPI_ISL_842308, EPI_ISL_842309, EPI_ISL_842310, EPI_ISL_842314, EPI_ISL_842320, EPI_ISL_842321, EPI_ISL_842323, EPI_ISL_842326, EPI_ISL_842328, EPI_ISL_842331, EPI_ISL_842332, EPI_ISL_842333, EPI_ISL_842336, EPI_ISL_842337, EPI_ISL_842343, EPI_ISL_842344, EPI_ISL_868358, EPI_ISL_868364, EPI_ISL_868373, EPI_ISL_868381, EPI_ISL_868382, EPI_ISL_868386, EPI_ISL_868388, EPI_ISL_868392, EPI_ISL_868395, EPI_ISL_868399, EPI_ISL_868401, EPI_ISL_868402, EPI_ISL_868408, EPI_ISL_868409, EPI_ISL_868413, EPI_ISL_868415, EPI_ISL_868428, EPI_ISL_868429, EPI_ISL_868430, EPI_ISL_868432, EPI_ISL_868434, EPI_ISL_868442, EPI_ISL_868444, EPI_ISL_868445, EPI_ISL_868446, EPI_ISL_868448, EPI_ISL_868450, EPI_ISL_868454, EPI_ISL_868459, EPI_ISL_868464, EPI_ISL_868465, EPI_ISL_868467, EPI_ISL_868469, EPI_ISL_868471, EPI_ISL_868474, EPI_ISL_868478, EPI_ISL_868480, EPI_ISL_868486, EPI_ISL_868487, EPI_ISL_868491, EPI_ISL_868495, EPI_ISL_868502, EPI_ISL_868503, EPI_ISL_868507, EPI_ISL_868508, EPI_ISL_868513, EPI_ISL_868515, EPI_ISL_868521, EPI_ISL_868522, EPI_ISL_868524, EPI_ISL_868525, EPI_ISL_868533, EPI_ISL_868535, EPI_ISL_868537, EPI_ISL_868541, EPI_ISL_868542, EPI_ISL_868547, EPI_ISL_868552, EPI_ISL_868556, EPI_ISL_868557, EPI_ISL_868560, EPI_ISL_868564, EPI_ISL_868566, EPI_ISL_868573, EPI_ISL_868588, EPI_ISL_868593, EPI_ISL_868594, EPI_ISL_868597, EPI_ISL_868599, EPI_ISL_868601, EPI_ISL_868603, EPI_ISL_868608, EPI_ISL_868612, EPI_ISL_868613, EPI_ISL_868621, EPI_ISL_868622, EPI_ISL_868625, EPI_ISL_868630, EPI_ISL_868631, EPI_ISL_868638, EPI_ISL_868643, EPI_ISL_868644, EPI_ISL_868648, EPI_ISL_868651, EPI_ISL_868654, EPI_ISL_868655, EPI_ISL_868656, EPI_ISL_868660, EPI_ISL_868674, EPI_ISL_868677, EPI_ISL_868678, EPI_ISL_868680, EPI_ISL_868681, EPI_ISL_868684, EPI_ISL_868688, EPI_ISL_868692, EPI_ISL_868693, EPI_ISL_868700, EPI_ISL_868701, EPI_ISL_868715, EPI_ISL_924091, EPI_ISL_924182, EPI_ISL_924270, EPI_ISL_924285, EPI_ISL_924369, EPI_ISL_1000665 | | | |
| see above | Virology Department, Sheffield Teaching Hospitals NHS Foundation Trust/Department of Infection, Immunity and Cardiovascular Disease, The Medical School, University of Sheffield | COVID-19 Genomics UK (COG-UK) Consortium | Adri Angyal; Alex Keeley; Benjamin Lindsey; Cariad Evans; Danielle Groves; Dave Partridge; Katie Johnson; Laura Carrilero; Luke Green; Matthew Parker; Matthew Wyles; Mehmet Yavuz; Mohammad Raza; Nikki Smith; Paul Parsons; Rachel Tucker; Rebecca Brown; Thushan de Silva |
| EPI_ISL_841039, EPI_ISL_841148, EPI_ISL_841150, EPI_ISL_841277, EPI_ISL_841278, EPI_ISL_841283, EPI_ISL_841284, EPI_ISL_841287, EPI_ISL_841290, EPI_ISL_841291, EPI_ISL_841294, EPI_ISL_841295, EPI_ISL_841298, EPI_ISL_922361, EPI_ISL_922364, EPI_ISL_922380, EPI_ISL_922387, EPI_ISL_922388, EPI_ISL_922393, EPI_ISL_922397, EPI_ISL_922558, EPI_ISL_922588, EPI_ISL_922589, EPI_ISL_922644, EPI_ISL_999985, EPI_ISL_1476080, EPI_ISL_1476089, EPI_ISL_1476194, EPI_ISL_1476198, EPI_ISL_1476203, EPI_ISL_1476528, EPI_ISL_1546807, EPI_ISL_1546815, EPI_ISL_1546819 | | | |
| see above | Wales Specialist Virology Centre Sequencing lab: Pathogen Genomics Unit | Public Health Wales Microbiology Cardiff Wales Specialist Virology Centre | Alec Birchley; Alexander Adams; Amy Gaskin; Angela Marchbank; Bree Gatica-Wilcox; Catherine Moore; Jason Coombes; Joanne Watkins; Joel Southgate; Johnathan Evans; Laura Gifford; Lauren Gilbert; Lee Graham; Malorie Perry; Matthew Bull; Nicole Pacchiarini; Sally Corden; Sara Kumziene-Summerhayes; Sara Rey; Sarah Taylor; Simon Cottrell; Sophie Jones; Tom Connor |
| EPI_ISL_865015, EPI_ISL_865016, EPI_ISL_865017, EPI_ISL_865018, EPI_ISL_865019, EPI_ISL_865020, EPI_ISL_865021, EPI_ISL_865022, EPI_ISL_865023, EPI_ISL_865024, EPI_ISL_865025, EPI_ISL_865026, EPI_ISL_865027, EPI_ISL_865028, EPI_ISL_865029, EPI_ISL_865030, EPI_ISL_865031, EPI_ISL_865032, EPI_ISL_865033, EPI_ISL_865034, EPI_ISL_865035, EPI_ISL_865036, EPI_ISL_865037, EPI_ISL_865038, EPI_ISL_865039, EPI_ISL_865040, EPI_ISL_865050, EPI_ISL_865052, EPI_ISL_865053, EPI_ISL_865055, EPI_ISL_865056, EPI_ISL_865057, EPI_ISL_865058, EPI_ISL_865059, EPI_ISL_865060, EPI_ISL_865061, EPI_ISL_865062, EPI_ISL_865063, EPI_ISL_865064, EPI_ISL_865065, EPI_ISL_865066, EPI_ISL_865067, EPI_ISL_919190, EPI_ISL_919191, EPI_ISL_919192, EPI_ISL_919193, EPI_ISL_919194, EPI_ISL_919195, EPI_ISL_919196, EPI_ISL_919197, EPI_ISL_1050233, EPI_ISL_1050234, EPI_ISL_1050235, EPI_ISL_1050236, EPI_ISL_1050237, EPI_ISL_1050238 | | | |
| see above | West of Scotland Specialist Virology Centre, NHSGGC / MRC-University of Glasgow Centre for Virus Research | COVID-19 Genomics UK (COG-UK) Consortium | Alasdair MacLean; Alice Broos; Ana da Silva Filipe; Antonia Ho; Daniel Mair; David L Robertson; Emma Thomson; Guy Mollett; James Shepherd; Jenna Nichols; Joseph Hughes; Kathy Li; Kathy Smollett; Kyriaki Nomikou; Lily Tong; Matthew Holden; Natasha Johnson; Rachel Blacow; Richard Orton; Rory Gunson; Sarah McDonald; Sharif Shaaban; Sreenu Vattipally; Stephen Carmichael |

We gratefully acknowledge the following Authors from the Originating laboratories responsible for obtaining the specimens, as well as the Submitting laboratories where the genome data were generated and shared via GISAID, on which this research is based.

All Submitters of data may be contacted directly via www.gisaid.org

Authors are sorted alphabetically.

| Accession ID | Originating Laboratory | Submitting Laboratory | Authors |
|---|---|---|---|
| EPI_ISL_919853, EPI_ISL_949749, EPI_ISL_949777, EPI_ISL_949778, EPI_ISL_949780, EPI_ISL_949783, EPI_ISL_997616, EPI_ISL_997617, EPI_ISL_997618, EPI_ISL_997619, EPI_ISL_997620, EPI_ISL_997621, EPI_ISL_997622, EPI_ISL_997623, EPI_ISL_997624, EPI_ISL_997630, EPI_ISL_997631, EPI_ISL_997632, EPI_ISL_997633, EPI_ISL_997634, EPI_ISL_997635, EPI_ISL_997636, EPI_ISL_997637, EPI_ISL_997638, EPI_ISL_997639, EPI_ISL_997640, EPI_ISL_997641, EPI_ISL_997643, EPI_ISL_997644, EPI_ISL_1048071, EPI_ISL_1048088, EPI_ISL_1177999, EPI_ISL_2126086, EPI_ISL_2126088, EPI_ISL_2126090, EPI_ISL_2126092, EPI_ISL_2126094, EPI_ISL_2126096, EPI_ISL_2126097, EPI_ISL_2126099, EPI_ISL_2126101, EPI_ISL_2126103, EPI_ISL_2126105, EPI_ISL_2126107, EPI_ISL_2126109 | | | |
| see above | Barts Health NHS Trust | COVID-19 Genomics UK (COG-UK) Consortium | BROAD; Beatrix; CUTINO-MOGUEL; Claire; David; Dola; HARRINGTON; KELE; KULASEGARAN-SHYLINI; Maria-Teresa; OWOYEMI; Raghavendran |
| EPI_ISL_924796, EPI_ISL_924801, EPI_ISL_924802, EPI_ISL_924803, EPI_ISL_924804, EPI_ISL_924806, EPI_ISL_924810, EPI_ISL_924811, EPI_ISL_924812, EPI_ISL_924813, EPI_ISL_924814, EPI_ISL_924815, EPI_ISL_924817, EPI_ISL_924820, EPI_ISL_924821, EPI_ISL_924822, EPI_ISL_924823, EPI_ISL_924824, EPI_ISL_924825, EPI_ISL_924826, EPI_ISL_924830, EPI_ISL_924834, EPI_ISL_924841, EPI_ISL_924842, EPI_ISL_924845, EPI_ISL_924857, EPI_ISL_924859, EPI_ISL_924948, EPI_ISL_924949, EPI_ISL_924952, EPI_ISL_924953, EPI_ISL_924954, EPI_ISL_924956, EPI_ISL_924957, EPI_ISL_924958, EPI_ISL_924959, EPI_ISL_924960, EPI_ISL_924961, EPI_ISL_924962, EPI_ISL_924963, EPI_ISL_924964, EPI_ISL_924965, EPI_ISL_924966, EPI_ISL_924967, EPI_ISL_924968, EPI_ISL_924969, EPI_ISL_924970, EPI_ISL_924971, EPI_ISL_924972, EPI_ISL_924973, EPI_ISL_924974, EPI_ISL_924976, EPI_ISL_924977, EPI_ISL_924978, EPI_ISL_924979, EPI_ISL_924981, EPI_ISL_924982, EPI_ISL_924983, EPI_ISL_924984, EPI_ISL_924985, EPI_ISL_924986, EPI_ISL_924988, EPI_ISL_924989, EPI_ISL_924990, EPI_ISL_924991, EPI_ISL_924992, EPI_ISL_924993, EPI_ISL_924994, EPI_ISL_924995, EPI_ISL_924996, EPI_ISL_924997, EPI_ISL_924998, EPI_ISL_924999, EPI_ISL_925000, EPI_ISL_925001, EPI_ISL_925002, EPI_ISL_925003, EPI_ISL_925004, EPI_ISL_925005, EPI_ISL_925006, EPI_ISL_925007, EPI_ISL_925008, EPI_ISL_925009, EPI_ISL_925010, EPI_ISL_925011, EPI_ISL_925012, EPI_ISL_925013, EPI_ISL_953024, EPI_ISL_953025, EPI_ISL_953026, EPI_ISL_953027, EPI_ISL_953028, EPI_ISL_953029, EPI_ISL_953030, EPI_ISL_953031, EPI_ISL_953032, EPI_ISL_953033, EPI_ISL_953034, EPI_ISL_953035, EPI_ISL_953036, EPI_ISL_953037, EPI_ISL_953038, EPI_ISL_953039, EPI_ISL_953040, EPI_ISL_953041, EPI_ISL_953042, EPI_ISL_953043, EPI_ISL_953044, EPI_ISL_953045, EPI_ISL_953046, EPI_ISL_953047, EPI_ISL_953048, EPI_ISL_953049, EPI_ISL_953050, EPI_ISL_953051, EPI_ISL_953052, EPI_ISL_953053, EPI_ISL_953054, EPI_ISL_953055, EPI_ISL_953056, EPI_ISL_953057, EPI_ISL_953058, EPI_ISL_953059, EPI_ISL_953060, EPI_ISL_953061, EPI_ISL_953062, EPI_ISL_953063, EPI_ISL_953064, EPI_ISL_953065, EPI_ISL_953066, EPI_ISL_953067, EPI_ISL_953068, EPI_ISL_953069, EPI_ISL_953070, EPI_ISL_953071, EPI_ISL_953072, EPI_ISL_953073, EPI_ISL_953074, EPI_ISL_953075, EPI_ISL_953076, EPI_ISL_953077, EPI_ISL_953078, EPI_ISL_953079, EPI_ISL_953080, EPI_ISL_953081, EPI_ISL_953082, EPI_ISL_953085, EPI_ISL_953086, EPI_ISL_953087, EPI_ISL_953088, EPI_ISL_953090, EPI_ISL_953091, EPI_ISL_953092, EPI_ISL_953093, EPI_ISL_953094, EPI_ISL_953095, EPI_ISL_953100, EPI_ISL_1000811, EPI_ISL_1000884, EPI_ISL_1000889, EPI_ISL_1000890, EPI_ISL_1000899, EPI_ISL_1000901, EPI_ISL_1000904, EPI_ISL_1000912, EPI_ISL_1000936, EPI_ISL_1000938, EPI_ISL_1000939, EPI_ISL_1000941, EPI_ISL_1000942, EPI_ISL_1000944, EPI_ISL_1000946, EPI_ISL_1000948, EPI_ISL_1000949, EPI_ISL_1000950, EPI_ISL_1000951, EPI_ISL_1000952, EPI_ISL_1000953, EPI_ISL_1000954, EPI_ISL_1000955, EPI_ISL_1000956, EPI_ISL_1000957, EPI_ISL_1000958, EPI_ISL_1000959, EPI_ISL_1000961, EPI_ISL_1000962, EPI_ISL_1000965, EPI_ISL_1000966, EPI_ISL_1054792, EPI_ISL_1054794, EPI_ISL_1054796, EPI_ISL_1054798, EPI_ISL_1054801, EPI_ISL_1054803, EPI_ISL_1054804, EPI_ISL_1054805, EPI_ISL_1054806, EPI_ISL_1054807, EPI_ISL_1054809, EPI_ISL_1054810, EPI_ISL_1054811, EPI_ISL_1054812, EPI_ISL_1054813, EPI_ISL_1054814, EPI_ISL_1054816, EPI_ISL_1054817, EPI_ISL_1054819, EPI_ISL_1054821, EPI_ISL_1638087 | | | |
| see above | Bioinformatics and Biostatistics Lab, Advanced Sequencing Facility | COVID-19 Genomics UK (COG-UK) Consortium | Aengus Stewart; Chelsea Sawyer; Harshil Patel; Jerome Nicod; Laura Cubitt; Margaret Crawford |
| EPI_ISL_868034, EPI_ISL_868102, EPI_ISL_868155, EPI_ISL_868156, EPI_ISL_868157, EPI_ISL_868158, EPI_ISL_868159, EPI_ISL_868160, EPI_ISL_868175, EPI_ISL_868176, EPI_ISL_868180, EPI_ISL_868195, EPI_ISL_868205, EPI_ISL_868206, EPI_ISL_868207, EPI_ISL_868208, EPI_ISL_868209, EPI_ISL_868210, EPI_ISL_868211, EPI_ISL_868212, EPI_ISL_868213, EPI_ISL_868214, EPI_ISL_868215, EPI_ISL_868216, EPI_ISL_868219, EPI_ISL_868220, EPI_ISL_868221, EPI_ISL_868222, EPI_ISL_868223, EPI_ISL_868224, EPI_ISL_868225, EPI_ISL_868227, EPI_ISL_868228, EPI_ISL_868229, EPI_ISL_868230, EPI_ISL_868231, EPI_ISL_868232, EPI_ISL_868233, EPI_ISL_868234, EPI_ISL_868235, EPI_ISL_868236, EPI_ISL_868237, EPI_ISL_868238, EPI_ISL_868239, EPI_ISL_868240, EPI_ISL_923276, EPI_ISL_923280, EPI_ISL_923281, EPI_ISL_923287, EPI_ISL_923290, EPI_ISL_923291, EPI_ISL_923292, EPI_ISL_923294, EPI_ISL_923298, EPI_ISL_923302, EPI_ISL_923303, EPI_ISL_923304, EPI_ISL_923307, EPI_ISL_923308, EPI_ISL_923309, EPI_ISL_923310, EPI_ISL_923311, EPI_ISL_923312, EPI_ISL_923313, EPI_ISL_923314, EPI_ISL_923317, EPI_ISL_923355, EPI_ISL_923356, EPI_ISL_923359, EPI_ISL_923360, EPI_ISL_923361, EPI_ISL_923362, EPI_ISL_923364, EPI_ISL_923365, EPI_ISL_923368, EPI_ISL_923370, EPI_ISL_923371, EPI_ISL_923414, EPI_ISL_923416, EPI_ISL_923419, EPI_ISL_923438, EPI_ISL_923589, EPI_ISL_923590, EPI_ISL_923591, EPI_ISL_923592, EPI_ISL_923594, EPI_ISL_923628, EPI_ISL_923660, EPI_ISL_923664, EPI_ISL_923665, EPI_ISL_923666, EPI_ISL_923667, EPI_ISL_923668, EPI_ISL_923669, EPI_ISL_923670, EPI_ISL_923672, EPI_ISL_923673, EPI_ISL_923674, EPI_ISL_923675, EPI_ISL_923676, EPI_ISL_923677, EPI_ISL_923681, EPI_ISL_923682, EPI_ISL_923683, EPI_ISL_952377, EPI_ISL_952378, EPI_ISL_952379, EPI_ISL_952382, EPI_ISL_952383, EPI_ISL_952385, EPI_ISL_952389, EPI_ISL_952392, EPI_ISL_952394, EPI_ISL_952397, EPI_ISL_952398, EPI_ISL_952409, EPI_ISL_952411, EPI_ISL_952414, EPI_ISL_952418, EPI_ISL_952422, EPI_ISL_952424, EPI_ISL_952427, EPI_ISL_952429, EPI_ISL_952433, EPI_ISL_952471, EPI_ISL_952472, EPI_ISL_952473, EPI_ISL_952474, EPI_ISL_952475, EPI_ISL_952476, EPI_ISL_952526, EPI_ISL_952527, EPI_ISL_952569, EPI_ISL_952570, EPI_ISL_952571, EPI_ISL_952572, EPI_ISL_952573, EPI_ISL_952574, EPI_ISL_952575, EPI_ISL_952576, EPI_ISL_952577, EPI_ISL_952578, EPI_ISL_952579, EPI_ISL_952580, EPI_ISL_952581, EPI_ISL_952582, EPI_ISL_952583, EPI_ISL_952584, EPI_ISL_952585, EPI_ISL_952586, EPI_ISL_952587, EPI_ISL_952588, EPI_ISL_952589, EPI_ISL_952590, EPI_ISL_952591, EPI_ISL_952592, EPI_ISL_952593, EPI_ISL_952594, EPI_ISL_952595, EPI_ISL_952596, EPI_ISL_952597, EPI_ISL_952598, EPI_ISL_952599, EPI_ISL_952600, EPI_ISL_952601, EPI_ISL_952602, EPI_ISL_952624, EPI_ISL_952778, EPI_ISL_952779, EPI_ISL_952780, EPI_ISL_952789, EPI_ISL_952793, EPI_ISL_952794, EPI_ISL_952795, EPI_ISL_952798, EPI_ISL_952799, EPI_ISL_952804, EPI_ISL_952854, EPI_ISL_1000247, EPI_ISL_1000417, EPI_ISL_1000418, EPI_ISL_1000419, EPI_ISL_1000420, EPI_ISL_1000421, EPI_ISL_1000422, EPI_ISL_1000423, EPI_ISL_1000424, EPI_ISL_1000425, EPI_ISL_1000426, EPI_ISL_1000427, EPI_ISL_1000428, EPI_ISL_1000429, EPI_ISL_1000430, EPI_ISL_1000431, EPI_ISL_1000432, EPI_ISL_1000433, EPI_ISL_1000434, EPI_ISL_1000435, EPI_ISL_1000436, EPI_ISL_1000437, EPI_ISL_1000438, EPI_ISL_1000439, EPI_ISL_1000440, EPI_ISL_1000441, EPI_ISL_1000442, EPI_ISL_1000443, EPI_ISL_1000444, EPI_ISL_1000445, EPI_ISL_1000448, EPI_ISL_1000449, EPI_ISL_1053817, EPI_ISL_1053818, EPI_ISL_1053819, EPI_ISL_1107750, EPI_ISL_1179792, EPI_ISL_1179825, EPI_ISL_1476563, EPI_ISL_1547231, EPI_ISL_1637982, EPI_ISL_1730869, EPI_ISL_1730870, EPI_ISL_1730871, EPI_ISL_1730872, EPI_ISL_1979846, EPI_ISL_1979848, EPI_ISL_1979849, EPI_ISL_1979851, EPI_ISL_1979853, EPI_ISL_1979854, EPI_ISL_1979856, EPI_ISL_1979857, EPI_ISL_1979859, EPI_ISL_1979860, EPI_ISL_1979862, EPI_ISL_1979949, EPI_ISL_1979950, EPI_ISL_1979952, EPI_ISL_1979954, EPI_ISL_1979955, EPI_ISL_1979956, EPI_ISL_1979958, EPI_ISL_1979960, EPI_ISL_1979962, EPI_ISL_1980023, EPI_ISL_1980025, EPI_ISL_1980027, EPI_ISL_1980028, EPI_ISL_1980030, EPI_ISL_1980031, EPI_ISL_1980033, EPI_ISL_1980035, EPI_ISL_1980037, EPI_ISL_1980038, EPI_ISL_1980040, EPI_ISL_1980042, EPI_ISL_1980044, EPI_ISL_1980046, EPI_ISL_1980047, EPI_ISL_1980049, EPI_ISL_1980050, EPI_ISL_1980052, EPI_ISL_1980054 | | | |
| see above | Centre for Enzyme Innovation, University of Portsmouth / Translational Research Laboratory, Portsmouth Hospitals NHS Trust | COVID-19 Genomics UK (COG-UK) Consortium | Allyson Lloyd; Angela Beckett; Anoop Chauhan; Christopher Fearn; Ethan Butcher; Garry Scarlett; Kate Cook; Katie Loveson; Kelly Bicknell; Robert Impey; Salman Goudarzi; Samuel Robson; Sarah Wyllie; Scott Elliott; Sharon Glaysher; Yann Bourgeois |
| EPI_ISL_838015, EPI_ISL_838017, EPI_ISL_838018, EPI_ISL_838021, EPI_ISL_838022, EPI_ISL_838025, EPI_ISL_838035, EPI_ISL_838038, EPI_ISL_838048, EPI_ISL_864896, EPI_ISL_864897, EPI_ISL_864898, EPI_ISL_864899, EPI_ISL_864900, EPI_ISL_864901, EPI_ISL_864902, EPI_ISL_864903, EPI_ISL_864904, EPI_ISL_864905, EPI_ISL_864906, EPI_ISL_864907, EPI_ISL_864908, EPI_ISL_864909, EPI_ISL_864910, EPI_ISL_864911, EPI_ISL_864912, EPI_ISL_864913, EPI_ISL_864914, EPI_ISL_864915, EPI_ISL_864916, EPI_ISL_864919, EPI_ISL_864920, EPI_ISL_864921, EPI_ISL_864922, EPI_ISL_864923, EPI_ISL_864924, EPI_ISL_864927, EPI_ISL_864928, EPI_ISL_864930, EPI_ISL_864931, EPI_ISL_864933, EPI_ISL_864934, EPI_ISL_864935, EPI_ISL_864937, EPI_ISL_864938, EPI_ISL_864939, EPI_ISL_864940, EPI_ISL_864941, EPI_ISL_864942, EPI_ISL_864943, EPI_ISL_864944, EPI_ISL_864946, EPI_ISL_864949, EPI_ISL_864950, EPI_ISL_864951, EPI_ISL_864952, EPI_ISL_864953, EPI_ISL_864954, EPI_ISL_864955, EPI_ISL_864956, EPI_ISL_864957, EPI_ISL_864958, EPI_ISL_864959, EPI_ISL_919057, EPI_ISL_919059, EPI_ISL_919060, EPI_ISL_919061, EPI_ISL_919064, EPI_ISL_919065, EPI_ISL_919066, EPI_ISL_919068, EPI_ISL_919069, EPI_ISL_919072, EPI_ISL_919076, EPI_ISL_919078, EPI_ISL_919083, EPI_ISL_919085, EPI_ISL_919086, EPI_ISL_919088, EPI_ISL_919090, EPI_ISL_919094, EPI_ISL_919096, EPI_ISL_919098, EPI_ISL_919100, EPI_ISL_920901, EPI_ISL_920902, EPI_ISL_920903, EPI_ISL_920904, EPI_ISL_920905, EPI_ISL_920906, EPI_ISL_920907, EPI_ISL_920908, EPI_ISL_920909, EPI_ISL_920910, EPI_ISL_920911, EPI_ISL_920912, EPI_ISL_920913, EPI_ISL_920914, EPI_ISL_920915, EPI_ISL_920916, EPI_ISL_920917, EPI_ISL_920918, EPI_ISL_920919, EPI_ISL_920920, EPI_ISL_920921, EPI_ISL_920922, EPI_ISL_920923, EPI_ISL_920924, EPI_ISL_920925, EPI_ISL_920926, EPI_ISL_920927, EPI_ISL_920928, EPI_ISL_920929, EPI_ISL_920930, EPI_ISL_920931, EPI_ISL_920932, EPI_ISL_920933, EPI_ISL_920934, EPI_ISL_920935, EPI_ISL_920936, EPI_ISL_920937, EPI_ISL_920938, EPI_ISL_920939, EPI_ISL_920940, EPI_ISL_920941, EPI_ISL_920942, EPI_ISL_920943, EPI_ISL_920944, EPI_ISL_920945, EPI_ISL_920946, EPI_ISL_920947, EPI_ISL_920948, EPI_ISL_920949, EPI_ISL_920950, EPI_ISL_920951, EPI_ISL_920952, EPI_ISL_920953, EPI_ISL_920954, EPI_ISL_920955, EPI_ISL_920956, EPI_ISL_920957, EPI_ISL_920958, EPI_ISL_920959, EPI_ISL_920960, EPI_ISL_920961, EPI_ISL_920962, EPI_ISL_920963, EPI_ISL_920964, EPI_ISL_920965, EPI_ISL_920966, EPI_ISL_920967, EPI_ISL_920968, EPI_ISL_920969, EPI_ISL_920970, EPI_ISL_920971, EPI_ISL_920972, EPI_ISL_920973, EPI_ISL_920974, EPI_ISL_923685, EPI_ISL_923686, EPI_ISL_923687, EPI_ISL_923689, EPI_ISL_923690, EPI_ISL_923691, EPI_ISL_923692, EPI_ISL_923693, EPI_ISL_923694, EPI_ISL_923695, EPI_ISL_923696, EPI_ISL_923697, EPI_ISL_923698, EPI_ISL_923699, EPI_ISL_923700, EPI_ISL_923701, EPI_ISL_923702, EPI_ISL_923703, EPI_ISL_923704, EPI_ISL_923705, EPI_ISL_923706, EPI_ISL_923707, EPI_ISL_923708, EPI_ISL_923709, EPI_ISL_923710, EPI_ISL_923711, EPI_ISL_923712, EPI_ISL_923713, EPI_ISL_923714, EPI_ISL_923715, EPI_ISL_923716, EPI_ISL_923717, EPI_ISL_923718, EPI_ISL_923719, EPI_ISL_923720, EPI_ISL_923721, EPI_ISL_923722, EPI_ISL_923723, EPI_ISL_923724, EPI_ISL_923725, EPI_ISL_923726, EPI_ISL_923727, EPI_ISL_923728, EPI_ISL_923729, EPI_ISL_923730, EPI_ISL_923731, EPI_ISL_923732, EPI_ISL_923733, EPI_ISL_923734, EPI_ISL_923735, EPI_ISL_923736, EPI_ISL_923737, EPI_ISL_923738, EPI_ISL_923739, EPI_ISL_923740, EPI_ISL_923741, EPI_ISL_923742, EPI_ISL_923743, EPI_ISL_923744, EPI_ISL_923745, EPI_ISL_923746, EPI_ISL_923747, EPI_ISL_923748, EPI_ISL_923749, EPI_ISL_923750, EPI_ISL_923751, EPI_ISL_923752, EPI_ISL_923755, EPI_ISL_923757, EPI_ISL_923761, EPI_ISL_923762, EPI_ISL_923765, EPI_ISL_923766, EPI_ISL_923767, EPI_ISL_923768, EPI_ISL_923769, EPI_ISL_923770, EPI_ISL_923771, EPI_ISL_923772, EPI_ISL_923773, EPI_ISL_923774, EPI_ISL_923775, EPI_ISL_923776, EPI_ISL_923777, EPI_ISL_923778, EPI_ISL_923795, EPI_ISL_923796, EPI_ISL_923797, EPI_ISL_923798, EPI_ISL_923799, EPI_ISL_923800, EPI_ISL_923801, EPI_ISL_923802, EPI_ISL_923803, EPI_ISL_923804, EPI_ISL_923805, EPI_ISL_923806, EPI_ISL_923807, EPI_ISL_923808, EPI_ISL_923809, EPI_ISL_923810, EPI_ISL_923811, EPI_ISL_923812, EPI_ISL_923813, EPI_ISL_923814, EPI_ISL_923815, EPI_ISL_923816, EPI_ISL_923817, EPI_ISL_923818, EPI_ISL_923819, EPI_ISL_923843, EPI_ISL_923844, EPI_ISL_923845, EPI_ISL_923846, EPI_ISL_923847, EPI_ISL_923848, EPI_ISL_923849, EPI_ISL_923850, EPI_ISL_923851, EPI_ISL_923852, EPI_ISL_923853, EPI_ISL_923854, EPI_ISL_923855, EPI_ISL_923856, EPI_ISL_923857, EPI_ISL_923858, EPI_ISL_923859, EPI_ISL_923860, EPI_ISL_923861, EPI_ISL_923862, EPI_ISL_923863, EPI_ISL_923864, EPI_ISL_923865, EPI_ISL_923866, EPI_ISL_923867, EPI_ISL_923868, EPI_ISL_923869, EPI_ISL_923870, EPI_ISL_923871, EPI_ISL_923872, EPI_ISL_923873, EPI_ISL_923874, EPI_ISL_923904, EPI_ISL_923905, EPI_ISL_923906, EPI_ISL_923907, EPI_ISL_923908, EPI_ISL_923909, EPI_ISL_923910, EPI_ISL_923911, EPI_ISL_923912, EPI_ISL_923913, EPI_ISL_923914, EPI_ISL_923915, EPI_ISL_923916, EPI_ISL_923917, EPI_ISL_923918, EPI_ISL_923919, EPI_ISL_923920, EPI_ISL_923921, EPI_ISL_923922, EPI_ISL_923923, EPI_ISL_923924, EPI_ISL_923925, EPI_ISL_923926, EPI_ISL_923927, EPI_ISL_923928, EPI_ISL_923929, EPI_ISL_923930, EPI_ISL_923931, EPI_ISL_923932, EPI_ISL_923933, EPI_ISL_923934, EPI_ISL_923935, EPI_ISL_923936, EPI_ISL_923937, EPI_ISL_923938, EPI_ISL_923939, EPI_ISL_923940, EPI_ISL_923941, EPI_ISL_923942, EPI_ISL_923943, EPI_ISL_923944, EPI_ISL_923945, EPI_ISL_923946, EPI_ISL_923947, EPI_ISL_923948, EPI_ISL_923949, EPI_ISL_923950, EPI_ISL_923951, EPI_ISL_923952, EPI_ISL_923953, EPI_ISL_923954, EPI_ISL_923955, EPI_ISL_923956, EPI_ISL_923957, EPI_ISL_923958, EPI_ISL_923959, EPI_ISL_923960, EPI_ISL_923961, EPI_ISL_923962, EPI_ISL_923963, EPI_ISL_923964, EPI_ISL_923965, EPI_ISL_923966, EPI_ISL_923967, EPI_ISL_923968, EPI_ISL_923969, EPI_ISL_923970, EPI_ISL_923972, EPI_ISL_923973, EPI_ISL_923976, EPI_ISL_923977, EPI_ISL_923978, EPI_ISL_923979, EPI_ISL_923980, EPI_ISL_923981, EPI_ISL_923982, EPI_ISL_923983, EPI_ISL_923984, EPI_ISL_923985, EPI_ISL_923986, EPI_ISL_923987, EPI_ISL_923988, EPI_ISL_923989, EPI_ISL_923990, EPI_ISL_923991, EPI_ISL_923992, EPI_ISL_923993, EPI_ISL_923994, EPI_ISL_923995, EPI_ISL_923996, EPI_ISL_923997, EPI_ISL_923998, EPI_ISL_923999, EPI_ISL_924000, EPI_ISL_924001, EPI_ISL_924002, EPI_ISL_924003, EPI_ISL_924004, EPI_ISL_924005, EPI_ISL_924006, EPI_ISL_924007, EPI_ISL_924008, EPI_ISL_924009, EPI_ISL_924010, EPI_ISL_924011, EPI_ISL_924012, EPI_ISL_924013, EPI_ISL_924014, EPI_ISL_924015, EPI_ISL_924016, EPI_ISL_924017, EPI_ISL_924018, EPI_ISL_924019, EPI_ISL_924020, EPI_ISL_924021, EPI_ISL_924022, EPI_ISL_924023, EPI_ISL_924024, EPI_ISL_924025, EPI_ISL_924026, EPI_ISL_924029, EPI_ISL_924030, EPI_ISL_924031, EPI_ISL_924032, EPI_ISL_924033, EPI_ISL_924034, EPI_ISL_924035, EPI_ISL_924036, EPI_ISL_924037, EPI_ISL_924038, EPI_ISL_924039, EPI_ISL_924040, EPI_ISL_924041, EPI_ISL_924042, EPI_ISL_924043, EPI_ISL_924044, EPI_ISL_924045, EPI_ISL_924046, EPI_ISL_924047, EPI_ISL_924048, EPI_ISL_924049, EPI_ISL_924050, EPI_ISL_924051, EPI_ISL_924052, EPI_ISL_924053, EPI_ISL_924054, EPI_ISL_924055, EPI_ISL_924056, EPI_ISL_924057, EPI_ISL_924058, EPI_ISL_924059, EPI_ISL_924060, EPI_ISL_924061, EPI_ISL_924062, EPI_ISL_924063, EPI_ISL_924064, EPI_ISL_924065, EPI_ISL_924066, EPI_ISL_924067, EPI_ISL_924068, EPI_ISL_924069, EPI_ISL_924070, EPI_ISL_924071, EPI_ISL_924072, EPI_ISL_924073, EPI_ISL_924074, EPI_ISL_924075, EPI_ISL_924076, EPI_ISL_949256, EPI_ISL_949257, EPI_ISL_949258, EPI_ISL_949259, EPI_ISL_949260, EPI_ISL_949261, EPI_ISL_949262, EPI_ISL_949263, EPI_ISL_949264, EPI_ISL_949265, EPI_ISL_949266, EPI_ISL_949267, EPI_ISL_949268, EPI_ISL_949269, EPI_ISL_949270, EPI_ISL_949271, EPI_ISL_949272, EPI_ISL_949273, EPI_ISL_949274, EPI_ISL_949275, EPI_ISL_949276, EPI_ISL_949277, EPI_ISL_949278, EPI_ISL_949279, EPI_ISL_949280, EPI_ISL_949281, EPI_ISL_949282, EPI_ISL_949283, EPI_ISL_949284, EPI_ISL_949285, EPI_ISL_949286, EPI_ISL_949287, EPI_ISL_949288, EPI_ISL_949289, EPI_ISL_949290, EPI_ISL_949291, EPI_ISL_949292, EPI_ISL_949293, EPI_ISL_949294, EPI_ISL_949295, EPI_ISL_949296, EPI_ISL_949297, EPI_ISL_949298, EPI_ISL_949311, EPI_ISL_949315, EPI_ISL_949316, EPI_ISL_949318, EPI_ISL_949319, EPI_ISL_949320, EPI_ISL_949321, EPI_ISL_949323, EPI_ISL_949324, EPI_ISL_949325, EPI_ISL_949326, EPI_ISL_949492, EPI_ISL_949494, EPI_ISL_949496, EPI_ISL_949498, EPI_ISL_949504, EPI_ISL_949506, EPI_ISL_952896, EPI_ISL_952897, EPI_ISL_952898, EPI_ISL_952899, EPI_ISL_952900, EPI_ISL_952901, EPI_ISL_952902, EPI_ISL_952903, EPI_ISL_952904, EPI_ISL_952905, EPI_ISL_952906, EPI_ISL_952907, EPI_ISL_952908, EPI_ISL_952909, EPI_ISL_952910, EPI_ISL_952911, EPI_ISL_952912, EPI_ISL_952913, EPI_ISL_952932, EPI_ISL_996066, EPI_ISL_996067, EPI_ISL_996068, EPI_ISL_996069, EPI_ISL_996070, EPI_ISL_996071, EPI_ISL_996072, EPI_ISL_996073, EPI_ISL_996074, EPI_ISL_996075, EPI_ISL_996076, EPI_ISL_996077, EPI_ISL_996078, EPI_ISL_996079, EPI_ISL_996080, EPI_ISL_996081, EPI_ISL_1000631, EPI_ISL_1000632, EPI_ISL_1000633, EPI_ISL_1000634, EPI_ISL_1000635, EPI_ISL_1000636, EPI_ISL_1000637, EPI_ISL_1000638, EPI_ISL_1000639, EPI_ISL_1000640, EPI_ISL_1247571, EPI_ISL_1247573, EPI_ISL_1247574, EPI_ISL_1247576, EPI_ISL_1247578, EPI_ISL_1247580, EPI_ISL_1247581, EPI_ISL_1247582, EPI_ISL_1247583, EPI_ISL_1247584, EPI_ISL_1247585, EPI_ISL_1247586, EPI_ISL_1247587, EPI_ISL_1247588, EPI_ISL_1247589, EPI_ISL_1247590, EPI_ISL_1247591, EPI_ISL_1247592, EPI_ISL_1247595, EPI_ISL_1247596, EPI_ISL_1247597, EPI_ISL_1247598, EPI_ISL_1247599 | | | |
| see above | Department of Pathology, University of Cambridge | COVID-19 Genomics UK (COG-UK) Consortium | Aminu S. Jahun; Grant Hall; Ian Goodfellow; Iliana Georgana; Malte Pinckert; Martin D. Curran; Myra Hosmillo; Rhys Izu; Rhys Izuagbe; Surendra Parmar; William L. Hamilton; Yasmin Chaudhry |
| EPI_ISL_863251, EPI_ISL_863254, EPI_ISL_863258, EPI_ISL_863268, EPI_ISL_863270, EPI_ISL_863271, EPI_ISL_863272, EPI_ISL_863274, EPI_ISL_863275, EPI_ISL_863280, EPI_ISL_863281, EPI_ISL_863289, EPI_ISL_863290, EPI_ISL_863299, EPI_ISL_863302, EPI_ISL_863306, EPI_ISL_863310, EPI_ISL_863311, EPI_ISL_863318, EPI_ISL_863319, EPI_ISL_863326, EPI_ISL_863338, EPI_ISL_863342, EPI_ISL_863343, EPI_ISL_863346, EPI_ISL_863356, EPI_ISL_863357, EPI_ISL_863360, EPI_ISL_863362, EPI_ISL_863363, EPI_ISL_863364, EPI_ISL_863377, EPI_ISL_863379, EPI_ISL_863383, EPI_ISL_863387, EPI_ISL_863395, EPI_ISL_863397, EPI_ISL_863404, EPI_ISL_863406, EPI_ISL_863408, EPI_ISL_863411, EPI_ISL_863414, EPI_ISL_863416, EPI_ISL_863420, EPI_ISL_863421, EPI_ISL_863423, EPI_ISL_863426, EPI_ISL_863434, EPI_ISL_863437, EPI_ISL_863443, EPI_ISL_863446, EPI_ISL_863447, EPI_ISL_863448, EPI_ISL_863454, EPI_ISL_863461, EPI_ISL_863470, EPI_ISL_863477, EPI_ISL_863479, EPI_ISL_863484, EPI_ISL_863491, EPI_ISL_863503, EPI_ISL_863504, EPI_ISL_863506, EPI_ISL_863515, EPI_ISL_863516, EPI_ISL_863519, EPI_ISL_863520, EPI_ISL_863524, EPI_ISL_863527, EPI_ISL_863869, EPI_ISL_863870, EPI_ISL_863871, EPI_ISL_863872, EPI_ISL_863874, EPI_ISL_863875, EPI_ISL_863876, EPI_ISL_863877, EPI_ISL_863878, EPI_ISL_863879, EPI_ISL_863880, EPI_ISL_863881, EPI_ISL_863882, EPI_ISL_863883, EPI_ISL_863884, EPI_ISL_863885, EPI_ISL_863886, EPI_ISL_863887, EPI_ISL_863888, EPI_ISL_863889, EPI_ISL_863890, EPI_ISL_863891, EPI_ISL_863892, EPI_ISL_863893, EPI_ISL_863894, EPI_ISL_863895, EPI_ISL_863896, EPI_ISL_863897, EPI_ISL_863898, EPI_ISL_863900, EPI_ISL_863902, EPI_ISL_863903, EPI_ISL_863904, EPI_ISL_863905, EPI_ISL_863906, EPI_ISL_863907, EPI_ISL_863908, EPI_ISL_863909, EPI_ISL_863910, EPI_ISL_863911, EPI_ISL_863914, EPI_ISL_863915, EPI_ISL_863916, EPI_ISL_863917, EPI_ISL_863918, EPI_ISL_863919, EPI_ISL_863920, EPI_ISL_863923, EPI_ISL_863924, EPI_ISL_863925, EPI_ISL_863926, EPI_ISL_863927, EPI_ISL_863928, EPI_ISL_863929, EPI_ISL_863930, EPI_ISL_863931, EPI_ISL_863932, EPI_ISL_863933, EPI_ISL_863934, EPI_ISL_863935, EPI_ISL_863936, EPI_ISL_863937, EPI_ISL_863938, EPI_ISL_863939, EPI_ISL_863940, EPI_ISL_863941, EPI_ISL_863942, EPI_ISL_863943, EPI_ISL_863944, EPI_ISL_863945, EPI_ISL_863947, EPI_ISL_863948, EPI_ISL_863949, EPI_ISL_863950, EPI_ISL_863951, EPI_ISL_863952, EPI_ISL_863953, EPI_ISL_863954, EPI_ISL_863955, EPI_ISL_863957, EPI_ISL_863958, EPI_ISL_863959, EPI_ISL_863960, EPI_ISL_863961, EPI_ISL_863962, EPI_ISL_863963, EPI_ISL_863964, EPI_ISL_863965, EPI_ISL_863966, EPI_ISL_863967, EPI_ISL_863968, EPI_ISL_863970, EPI_ISL_863971, EPI_ISL_863972, EPI_ISL_863973, EPI_ISL_863974, EPI_ISL_863975, EPI_ISL_863976, EPI_ISL_863977, EPI_ISL_863978, EPI_ISL_863979, EPI_ISL_863980, EPI_ISL_863981, EPI_ISL_863982, EPI_ISL_863983, EPI_ISL_863984, EPI_ISL_863985, EPI_ISL_863986, EPI_ISL_863987, EPI_ISL_863988, EPI_ISL_863989, EPI_ISL_863990, EPI_ISL_863991, EPI_ISL_863992, EPI_ISL_863993, EPI_ISL_863994, EPI_ISL_863995, EPI_ISL_863997, EPI_ISL_863998, EPI_ISL_863999, EPI_ISL_864000, EPI_ISL_864001, EPI_ISL_864002, EPI_ISL_864003, EPI_ISL_864006, EPI_ISL_864007, EPI_ISL_864008, EPI_ISL_864010, EPI_ISL_864011, EPI_ISL_864012, EPI_ISL_864013, EPI_ISL_864014, EPI_ISL_864015, EPI_ISL_864016, EPI_ISL_864017, EPI_ISL_864018, EPI_ISL_864019, EPI_ISL_864020, EPI_ISL_864021, EPI_ISL_864022, EPI_ISL_864023, EPI_ISL_864024, EPI_ISL_864025, EPI_ISL_864027, EPI_ISL_864028, EPI_ISL_864029, EPI_ISL_864030, EPI_ISL_864031, EPI_ISL_864032, EPI_ISL_864033, EPI_ISL_864034, EPI_ISL_864035, EPI_ISL_864037, EPI_ISL_864038, EPI_ISL_864040, EPI_ISL_864041, EPI_ISL_864042, EPI_ISL_864044, EPI_ISL_864045, EPI_ISL_864046, EPI_ISL_864047, EPI_ISL_864048, EPI_ISL_864049, EPI_ISL_864050, EPI_ISL_864051, EPI_ISL_864052, EPI_ISL_864053, EPI_ISL_864054, EPI_ISL_864055, EPI_ISL_864056, EPI_ISL_864057, EPI_ISL_864058, EPI_ISL_864059, EPI_ISL_864060, EPI_ISL_864061, EPI_ISL_864062, EPI_ISL_864063, EPI_ISL_864065, EPI_ISL_864066, EPI_ISL_864067, EPI_ISL_864068, EPI_ISL_864069, EPI_ISL_864070, EPI_ISL_864071, EPI_ISL_864073, EPI_ISL_864074, EPI_ISL_864075, EPI_ISL_864076, EPI_ISL_864077, EPI_ISL_864078, EPI_ISL_864079, EPI_ISL_864080, EPI_ISL_864081, EPI_ISL_864082, EPI_ISL_864083, EPI_ISL_864084, EPI_ISL_864085, EPI_ISL_864086, EPI_ISL_864087, EPI_ISL_864088, EPI_ISL_864089, EPI_ISL_864090, EPI_ISL_864091, EPI_ISL_864092, EPI_ISL_864093, EPI_ISL_864095, EPI_ISL_864096, EPI_ISL_864097, EPI_ISL_864099, EPI_ISL_864100, EPI_ISL_864101, EPI_ISL_864102, EPI_ISL_864103, EPI_ISL_864104, EPI_ISL_864105, EPI_ISL_864106, EPI_ISL_864107, EPI_ISL_864108, EPI_ISL_864109, EPI_ISL_864111, EPI_ISL_864112, EPI_ISL_864113, EPI_ISL_864114, EPI_ISL_864115, EPI_ISL_864116, EPI_ISL_864117, EPI_ISL_864118, EPI_ISL_864119, EPI_ISL_864120, EPI_ISL_864121, EPI_ISL_864122, EPI_ISL_864123, EPI_ISL_864125, EPI_ISL_864126, EPI_ISL_864127, EPI_ISL_864128, EPI_ISL_864129, EPI_ISL_864130, EPI_ISL_864131, EPI_ISL_864132, EPI_ISL_864134, EPI_ISL_864135, EPI_ISL_864136, EPI_ISL_864137, EPI_ISL_864138, EPI_ISL_864139, EPI_ISL_864140, EPI_ISL_864141, EPI_ISL_864143, EPI_ISL_864144, EPI_ISL_864146, EPI_ISL_864147, EPI_ISL_864148, EPI_ISL_864149, EPI_ISL_864150, EPI_ISL_864151, EPI_ISL_864152, EPI_ISL_864153, EPI_ISL_864154, EPI_ISL_864155, EPI_ISL_864156, EPI_ISL_864157, EPI_ISL_864158, EPI_ISL_864159, EPI_ISL_864160, EPI_ISL_864161, EPI_ISL_864162, EPI_ISL_864163, EPI_ISL_864164, EPI_ISL_864166, EPI_ISL_864167, EPI_ISL_864168, EPI_ISL_864170, EPI_ISL_864171, EPI_ISL_864172, EPI_ISL_864174, EPI_ISL_864177, EPI_ISL_864179, EPI_ISL_864180, EPI_ISL_864181, EPI_ISL_864182, EPI_ISL_864183, EPI_ISL_864184, EPI_ISL_864185, EPI_ISL_864186, EPI_ISL_864187, EPI_ISL_864188, EPI_ISL_864189, EPI_ISL_864190, EPI_ISL_864192, EPI_ISL_864194, EPI_ISL_864195, EPI_ISL_864196, EPI_ISL_864197, EPI_ISL_864198, EPI_ISL_864199, EPI_ISL_864200, EPI_ISL_864201, EPI_ISL_864202, EPI_ISL_873964, EPI_ISL_873965, EPI_ISL_873974, EPI_ISL_873976, EPI_ISL_873984, EPI_ISL_873987, EPI_ISL_873990, EPI_ISL_873999, EPI_ISL_874004, EPI_ISL_874007, EPI_ISL_874009, EPI_ISL_874010, EPI_ISL_874012, EPI_ISL_874014, EPI_ISL_874017, EPI_ISL_874021, EPI_ISL_874022, EPI_ISL_874032, EPI_ISL_874037, EPI_ISL_874038, EPI_ISL_874039, EPI_ISL_874047, EPI_ISL_874055, EPI_ISL_874061, EPI_ISL_874062, EPI_ISL_874064, EPI_ISL_874071, EPI_ISL_874076, EPI_ISL_874085, EPI_ISL_874090, EPI_ISL_874092, EPI_ISL_874096, EPI_ISL_874097, EPI_ISL_874115, EPI_ISL_874118, EPI_ISL_874130, EPI_ISL_874141, EPI_ISL_874143, EPI_ISL_874148, EPI_ISL_874153, EPI_ISL_874154, EPI_ISL_874157, EPI_ISL_874160, EPI_ISL_874163, EPI_ISL_874164, EPI_ISL_874165, EPI_ISL_874169, EPI_ISL_874180, EPI_ISL_874188, EPI_ISL_874191, EPI_ISL_874193, EPI_ISL_874195, EPI_ISL_874197, EPI_ISL_874198, EPI_ISL_874199, EPI_ISL_874202, EPI_ISL_874204, EPI_ISL_874207, EPI_ISL_874209, EPI_ISL_874211, EPI_ISL_874215, EPI_ISL_874238, EPI_ISL_874240, EPI_ISL_874246, EPI_ISL_874248, EPI_ISL_874254, EPI_ISL_874258, EPI_ISL_874263, EPI_ISL_874266, EPI_ISL_874268, EPI_ISL_874275, EPI_ISL_874278, EPI_ISL_874283, EPI_ISL_874290, EPI_ISL_874293, EPI_ISL_874297, EPI_ISL_874301, EPI_ISL_874311, EPI_ISL_874312, EPI_ISL_874313, EPI_ISL_874314, EPI_ISL_874316, EPI_ISL_874317, EPI_ISL_874318, EPI_ISL_874319, EPI_ISL_874320, EPI_ISL_874321, EPI_ISL_874322, EPI_ISL_874323, EPI_ISL_874324, EPI_ISL_874325, EPI_ISL_874326, EPI_ISL_874327, EPI_ISL_874328, EPI_ISL_874329, EPI_ISL_874330, EPI_ISL_874331, EPI_ISL_874332, EPI_ISL_874333, EPI_ISL_874334, EPI_ISL_874335, EPI_ISL_874336, EPI_ISL_874337, EPI_ISL_874338, EPI_ISL_874339, EPI_ISL_874340, EPI_ISL_874341, EPI_ISL_874342, EPI_ISL_874343, EPI_ISL_874344, EPI_ISL_874345, EPI_ISL_874346, EPI_ISL_874347, EPI_ISL_874348, EPI_ISL_874349, EPI_ISL_874350, EPI_ISL_874351, EPI_ISL_874352, EPI_ISL_874353, EPI_ISL_874354, EPI_ISL_874355, EPI_ISL_874356, EPI_ISL_874357, EPI_ISL_874358, EPI_ISL_874359, EPI_ISL_874360, EPI_ISL_874361, EPI_ISL_874362, EPI_ISL_874363, EPI_ISL_874364, EPI_ISL_874365, EPI_ISL_874366, EPI_ISL_874367, EPI_ISL_874368, EPI_ISL_874369, EPI_ISL_874370, EPI_ISL_874371, EPI_ISL_874372, EPI_ISL_874373, EPI_ISL_874374, EPI_ISL_874375, EPI_ISL_874376, EPI_ISL_874377, EPI_ISL_874378, EPI_ISL_874380, EPI_ISL_874381, EPI_ISL_874382, EPI_ISL_874383, EPI_ISL_874384, EPI_ISL_874385, EPI_ISL_874386, EPI_ISL_874387, EPI_ISL_874388, EPI_ISL_874389, EPI_ISL_874390, EPI_ISL_874391, EPI_ISL_874392, EPI_ISL_874393, EPI_ISL_874394, EPI_ISL_874395, EPI_ISL_874396, EPI_ISL_874397, EPI_ISL_874398, EPI_ISL_874399, EPI_ISL_874400, EPI_ISL_874402, EPI_ISL_874403, EPI_ISL_874405, EPI_ISL_874406, EPI_ISL_874407, EPI_ISL_874408, EPI_ISL_874409, EPI_ISL_874410, EPI_ISL_874411, EPI_ISL_874412, EPI_ISL_874413, EPI_ISL_874414, EPI_ISL_874415, EPI_ISL_874416, EPI_ISL_874417, EPI_ISL_874418, EPI_ISL_874419, EPI_ISL_874420, EPI_ISL_874421, EPI_ISL_874422, EPI_ISL_874423, EPI_ISL_874424, EPI_ISL_874425, EPI_ISL_874426, EPI_ISL_874427, EPI_ISL_874428, EPI_ISL_874429, EPI_ISL_874430, EPI_ISL_874431, EPI_ISL_874432, EPI_ISL_874433, EPI_ISL_874434, EPI_ISL_874435, EPI_ISL_874436, EPI_ISL_874437, EPI_ISL_874438, EPI_ISL_874439, EPI_ISL_874440, EPI_ISL_874441, EPI_ISL_874442, EPI_ISL_874444, EPI_ISL_874445, EPI_ISL_874446, EPI_ISL_874447, EPI_ISL_874449, EPI_ISL_874450, EPI_ISL_874451, EPI_ISL_874452, EPI_ISL_874453, EPI_ISL_874454, EPI_ISL_874455, EPI_ISL_874456, EPI_ISL_874457, EPI_ISL_874458, EPI_ISL_874459, EPI_ISL_874460, EPI_ISL_874461, EPI_ISL_874462, EPI_ISL_874463, EPI_ISL_874464, EPI_ISL_874465, EPI_ISL_874466, EPI_ISL_874467, EPI_ISL_874468, EPI_ISL_874470, EPI_ISL_874471, EPI_ISL_874472, EPI_ISL_874473, EPI_ISL_874474, EPI_ISL_874475, EPI_ISL_874476, EPI_ISL_874477, EPI_ISL_874478, EPI_ISL_874479, EPI_ISL_874480, EPI_ISL_874481, EPI_ISL_874483, EPI_ISL_874484, EPI_ISL_874485, EPI_ISL_874486, EPI_ISL_874487, EPI_ISL_874488, EPI_ISL_874489, EPI_ISL_874490, EPI_ISL_874491, EPI_ISL_874492, EPI_ISL_874493, EPI_ISL_874494, EPI_ISL_874495, EPI_ISL_874496, EPI_ISL_874497, EPI_ISL_874498, EPI_ISL_874500, EPI_ISL_874501, EPI_ISL_874503, EPI_ISL_874504, EPI_ISL_874505, EPI_ISL_874506, EPI_ISL_874507, EPI_ISL_874508, EPI_ISL_874509, EPI_ISL_874510, EPI_ISL_874511, EPI_ISL_874512, EPI_ISL_874513, EPI_ISL_874514, EPI_ISL_874515, EPI_ISL_874516, EPI_ISL_874517, EPI_ISL_874518, EPI_ISL_874519, EPI_ISL_874520, EPI_ISL_874521, EPI_ISL_874522, EPI_ISL_874524, EPI_ISL_874525, EPI_ISL_874526, EPI_ISL_874529, EPI_ISL_874531, EPI_ISL_874532, EPI_ISL_874533, EPI_ISL_874534, EPI_ISL_874535, EPI_ISL_874536, EPI_ISL_874537, EPI_ISL_874538, EPI_ISL_874539, EPI_ISL_874540, EPI_ISL_874541, EPI_ISL_874542, EPI_ISL_874544, EPI_ISL_874545, EPI_ISL_874546, EPI_ISL_874547, EPI_ISL_874548, EPI_ISL_874549, EPI_ISL_874550, EPI_ISL_874551, EPI_ISL_874552, EPI_ISL_874553, EPI_ISL_874554, EPI_ISL_874555, EPI_ISL_874556, EPI_ISL_874557, EPI_ISL_874559, EPI_ISL_874560, EPI_ISL_874561, EPI_ISL_874562, EPI_ISL_874563, EPI_ISL_874564, EPI_ISL_874565, EPI_ISL_874566, EPI_ISL_874567, EPI_ISL_874568, EPI_ISL_874569, EPI_ISL_874570, EPI_ISL_874571, EPI_ISL_874572, EPI_ISL_874573, EPI_ISL_874574, EPI_ISL_874575, EPI_ISL_874576, EPI_ISL_874577, EPI_ISL_874578, EPI_ISL_874579, EPI_ISL_874580, EPI_ISL_874581, EPI_ISL_874582, EPI_ISL_874583, EPI_ISL_874584, EPI_ISL_874585, EPI_ISL_874586, EPI_ISL_874587, EPI_ISL_874588, EPI_ISL_874589, EPI_ISL_874590, EPI_ISL_874591, EPI_ISL_874592, EPI_ISL_874593, EPI_ISL_874594, EPI_ISL_874595, EPI_ISL_874596, EPI_ISL_874597, EPI_ISL_874598, EPI_ISL_874599, EPI_ISL_874600, EPI_ISL_874601, EPI_ISL_874602, EPI_ISL_874603, EPI_ISL_874604, EPI_ISL_874605, EPI_ISL_874606, EPI_ISL_874607, EPI_ISL_874608, EPI_ISL_874609, EPI_ISL_874610, EPI_ISL_874611, EPI_ISL_874612, EPI_ISL_874613, EPI_ISL_874614, EPI_ISL_874615, EPI_ISL_874616, EPI_ISL_874617, EPI_ISL_874618, EPI_ISL_874619, EPI_ISL_874620, EPI_ISL_874621, EPI_ISL_874622, EPI_ISL_874623, EPI_ISL_874625, EPI_ISL_874626, EPI_ISL_874627, EPI_ISL_874628, EPI_ISL_874629, EPI_ISL_874630, EPI_ISL_874631, EPI_ISL_874632, EPI_ISL_874633, EPI_ISL_874634, EPI_ISL_874635, EPI_ISL_874636, EPI_ISL_874637, EPI_ISL_874638, EPI_ISL_874639, EPI_ISL_874640, EPI_ISL_874641, EPI_ISL_874642, EPI_ISL_874643, EPI_ISL_874644, EPI_ISL_874645, EPI_ISL_874646, EPI_ISL_874647, EPI_ISL_874648, EPI_ISL_874649, EPI_ISL_874650, EPI_ISL_874651, EPI_ISL_874652, EPI_ISL_874653, EPI_ISL_874654, EPI_ISL_874655, EPI_ISL_874657, EPI_ISL_874658, EPI_ISL_874659, EPI_ISL_874660, EPI_ISL_874661, EPI_ISL_874662, EPI_ISL_874663, EPI_ISL_874664, EPI_ISL_874665, EPI_ISL_874666, EPI_ISL_874667, EPI_ISL_874668, EPI_ISL_874669, EPI_ISL_874670, EPI_ISL_874671, EPI_ISL_874672, EPI_ISL_874673, EPI_ISL_874674, EPI_ISL_874675, EPI_ISL_874677, EPI_ISL_874678, EPI_ISL_874679, EPI_ISL_874680, EPI_ISL_874681, EPI_ISL_874682, EPI_ISL_874683, EPI_ISL_874684, EPI_ISL_874685, EPI_ISL_874686, EPI_ISL_874687, EPI_ISL_874688, EPI_ISL_874689, EPI_ISL_874690, EPI_ISL_874691, EPI_ISL_874692, EPI_ISL_874693, EPI_ISL_874694, EPI_ISL_874695, EPI_ISL_874696, EPI_ISL_874697, EPI_ISL_874698, EPI_ISL_874699, EPI_ISL_874700, EPI_ISL_874701, EPI_ISL_874702, EPI_ISL_874703, EPI_ISL_874704, EPI_ISL_874705, EPI_ISL_874706, EPI_ISL_874707, EPI_ISL_874708, EPI_ISL_874709, EPI_ISL_874710, EPI_ISL_874711, EPI_ISL_874712, EPI_ISL_874713, EPI_ISL_874714, EPI_ISL_874715, EPI_ISL_874716, EPI_ISL_874717, EPI_ISL_874718, EPI_ISL_874719, EPI_ISL_874720, EPI_ISL_874721, EPI_ISL_874722, EPI_ISL_874723, EPI_ISL_874724, EPI_ISL_874725, EPI_ISL_874726, EPI_ISL_874727, EPI_ISL_874728, EPI_ISL_874729, EPI_ISL_874730, EPI_ISL_874731, EPI_ISL_874732, EPI_ISL_874734, EPI_ISL_874735, EPI_ISL_874738, EPI_ISL_874740, EPI_ISL_874741, EPI_ISL_874742, EPI_ISL_874743, EPI_ISL_874744, EPI_ISL_874745, EPI_ISL_874746, EPI_ISL_874747, EPI_ISL_874748, EPI_ISL_874749, EPI_ISL_874750, EPI_ISL_874751, EPI_ISL_874752, EPI_ISL_874754, EPI_ISL_874755, EPI_ISL_874756, EPI_ISL_874757, EPI_ISL_874758, EPI_ISL_874760, EPI_ISL_874761, EPI_ISL_874762, EPI_ISL_874763, EPI_ISL_874764, EPI_ISL_874765, EPI_ISL_874766, EPI_ISL_874767, EPI_ISL_874768, EPI_ISL_874769, EPI_ISL_874770, EPI_ISL_874771, EPI_ISL_874772, EPI_ISL_874773, EPI_ISL_874774, EPI_ISL_874776, EPI_ISL_874777, EPI_ISL_874778, EPI_ISL_874779, EPI_ISL_874780, EPI_ISL_874781, EPI_ISL_874782, EPI_ISL_874783, EPI_ISL_874784, EPI_ISL_874785, EPI_ISL_874786, EPI_ISL_874787, EPI_ISL_874788, EPI_ISL_874789, EPI_ISL_874790, EPI_ISL_874791, EPI_ISL_874792, EPI_ISL_874793, EPI_ISL_874794, EPI_ISL_874795, EPI_ISL_874796, EPI_ISL_874797, EPI_ISL_874799, EPI_ISL_874800, EPI_ISL_874801, EPI_ISL_874802, EPI_ISL_874803, EPI_ISL_874804, EPI_ISL_874805, EPI_ISL_874807, EPI_ISL_874809, EPI_ISL_874810, EPI_ISL_874811, EPI_ISL_874812, EPI_ISL_874813, EPI_ISL_874814, EPI_ISL_874815, EPI_ISL_874817, EPI_ISL_874818, EPI_ISL_874819, EPI_ISL_874820, EPI_ISL_874821, EPI_ISL_874822, EPI_ISL_874823, EPI_ISL_874824, EPI_ISL_874825, EPI_ISL_874826, EPI_ISL_874827, EPI_ISL_874828, EPI_ISL_874829, EPI_ISL_874830, EPI_ISL_874831, | | | |

EPI_ISL_874832, EPI_ISL_874833, EPI_ISL_874834, EPI_ISL_874835, EPI_ISL_874836, EPI_ISL_874837, EPI_ISL_874838, EPI_ISL_874839, EPI_ISL_874840, EPI_ISL_874841, EPI_ISL_874842, EPI_ISL_874843, EPI_ISL_874844, EPI_ISL_874845, EPI_ISL_874846, EPI_ISL_874847, EPI_ISL_874848, EPI_ISL_874849, EPI_ISL_874850, EPI_ISL_874851, EPI_ISL_874852, EPI_ISL_874854, EPI_ISL_874855, EPI_ISL_874856, EPI_ISL_874857, EPI_ISL_874858, EPI_ISL_874859, EPI_ISL_874860, EPI_ISL_874861, EPI_ISL_874862, EPI_ISL_874864, EPI_ISL_874865, EPI_ISL_874866, EPI_ISL_874867, EPI_ISL_874868, EPI_ISL_874869, EPI_ISL_874870, EPI_ISL_874871, EPI_ISL_874872, EPI_ISL_874873, EPI_ISL_874874, EPI_ISL_874875, EPI_ISL_874876, EPI_ISL_874877, EPI_ISL_874878, EPI_ISL_874879, EPI_ISL_874880, EPI_ISL_874881, EPI_ISL_874882, EPI_ISL_874883, EPI_ISL_874884, EPI_ISL_874885, EPI_ISL_874886, EPI_ISL_874887, EPI_ISL_874888, EPI_ISL_874889, EPI_ISL_874890, EPI_ISL_874891, EPI_ISL_874892, EPI_ISL_874893, EPI_ISL_874894, EPI_ISL_874895, EPI_ISL_874897, EPI_ISL_874899, EPI_ISL_874900, EPI_ISL_874901, EPI_ISL_874902, EPI_ISL_874903, EPI_ISL_874904, EPI_ISL_874906, EPI_ISL_874907, EPI_ISL_874908, EPI_ISL_874909, EPI_ISL_874910, EPI_ISL_874911, EPI_ISL_874912, EPI_ISL_874913, EPI_ISL_874914, EPI_ISL_874915, EPI_ISL_874916, EPI_ISL_874917, EPI_ISL_874918, EPI_ISL_874919, EPI_ISL_874920, EPI_ISL_874921, EPI_ISL_874923, EPI_ISL_874924, EPI_ISL_874925, EPI_ISL_874926, EPI_ISL_874927, EPI_ISL_874928, EPI_ISL_874929, EPI_ISL_874930, EPI_ISL_874931, EPI_ISL_874932, EPI_ISL_874933, EPI_ISL_874934, EPI_ISL_874936, EPI_ISL_874937, EPI_ISL_874939, EPI_ISL_874940, EPI_ISL_874941, EPI_ISL_874942, EPI_ISL_874944, EPI_ISL_874945, EPI_ISL_874946, EPI_ISL_874947, EPI_ISL_874948, EPI_ISL_874949, EPI_ISL_874951, EPI_ISL_874952, EPI_ISL_874953, EPI_ISL_874954, EPI_ISL_874955, EPI_ISL_874957, EPI_ISL_874958, EPI_ISL_874959, EPI_ISL_874960, EPI_ISL_874961, EPI_ISL_874962, EPI_ISL_874963, EPI_ISL_874964, EPI_ISL_874968, EPI_ISL_874969, EPI_ISL_874970, EPI_ISL_874971, EPI_ISL_874972, EPI_ISL_874973, EPI_ISL_874974, EPI_ISL_874975, EPI_ISL_874976, EPI_ISL_874977, EPI_ISL_874978, EPI_ISL_874979, EPI_ISL_874980, EPI_ISL_874981, EPI_ISL_874982, EPI_ISL_874983, EPI_ISL_874984, EPI_ISL_874985, EPI_ISL_874986, EPI_ISL_874987, EPI_ISL_874988, EPI_ISL_874989, EPI_ISL_877771, EPI_ISL_877781, EPI_ISL_877787, EPI_ISL_877788, EPI_ISL_877796, EPI_ISL_877797, EPI_ISL_877800, EPI_ISL_877802, EPI_ISL_877804, EPI_ISL_877807, EPI_ISL_877809, EPI_ISL_877821, EPI_ISL_877828, EPI_ISL_877833, EPI_ISL_877844, EPI_ISL_877881, EPI_ISL_877884, EPI_ISL_877885, EPI_ISL_877889, EPI_ISL_877899, EPI_ISL_877903, EPI_ISL_877907, EPI_ISL_877918, EPI_ISL_877922, EPI_ISL_877924, EPI_ISL_877930, EPI_ISL_877938, EPI_ISL_877940, EPI_ISL_877948, EPI_ISL_877953, EPI_ISL_877964, EPI_ISL_877969, EPI_ISL_877972, EPI_ISL_877975, EPI_ISL_877981, EPI_ISL_877989, EPI_ISL_877995, EPI_ISL_877997, EPI_ISL_878011, EPI_ISL_878013, EPI_ISL_878021, EPI_ISL_878025, EPI_ISL_878026, EPI_ISL_878032, EPI_ISL_878038, EPI_ISL_878040, EPI_ISL_878041, EPI_ISL_878042, EPI_ISL_878044, EPI_ISL_878053, EPI_ISL_878054, EPI_ISL_878062, EPI_ISL_878064, EPI_ISL_878067, EPI_ISL_878078, EPI_ISL_878083, EPI_ISL_878088, EPI_ISL_878089, EPI_ISL_878098, EPI_ISL_878102, EPI_ISL_878105, EPI_ISL_878108, EPI_ISL_878111, EPI_ISL_878112, EPI_ISL_878113, EPI_ISL_878114, EPI_ISL_878115, EPI_ISL_878116, EPI_ISL_878117, EPI_ISL_878118, EPI_ISL_878119, EPI_ISL_878120, EPI_ISL_878121, EPI_ISL_878122, EPI_ISL_878123, EPI_ISL_878124, EPI_ISL_878125, EPI_ISL_878126, EPI_ISL_878127, EPI_ISL_878128, EPI_ISL_878129, EPI_ISL_878130, EPI_ISL_878131, EPI_ISL_878132, EPI_ISL_878133, EPI_ISL_878135, EPI_ISL_878136, EPI_ISL_878137, EPI_ISL_878138, EPI_ISL_878139, EPI_ISL_878140, EPI_ISL_878141, EPI_ISL_878142, EPI_ISL_878143, EPI_ISL_878144, EPI_ISL_878145, EPI_ISL_878146, EPI_ISL_878147, EPI_ISL_878148, EPI_ISL_878149, EPI_ISL_878150, EPI_ISL_878151, EPI_ISL_878153, EPI_ISL_878154, EPI_ISL_878156, EPI_ISL_878160, EPI_ISL_878161, EPI_ISL_878163, EPI_ISL_878164, EPI_ISL_878166, EPI_ISL_878169, EPI_ISL_878171, EPI_ISL_878173, EPI_ISL_878174, EPI_ISL_878176, EPI_ISL_878177, EPI_ISL_878179, EPI_ISL_878181, EPI_ISL_878182, EPI_ISL_878184, EPI_ISL_878185, EPI_ISL_878187, EPI_ISL_878188, EPI_ISL_878190, EPI_ISL_878192, EPI_ISL_878193, EPI_ISL_878195, EPI_ISL_878196, EPI_ISL_878198, EPI_ISL_878200, EPI_ISL_878201, EPI_ISL_878204, EPI_ISL_878206, EPI_ISL_878208, EPI_ISL_878210, EPI_ISL_878213, EPI_ISL_878214, EPI_ISL_878216, EPI_ISL_878218, EPI_ISL_878219, EPI_ISL_878220, EPI_ISL_878222, EPI_ISL_878224, EPI_ISL_878225, EPI_ISL_878227, EPI_ISL_878228, EPI_ISL_878230, EPI_ISL_878231, EPI_ISL_878233, EPI_ISL_878235, EPI_ISL_878236, EPI_ISL_878240, EPI_ISL_878241, EPI_ISL_878243, EPI_ISL_878244, EPI_ISL_878246, EPI_ISL_878247, EPI_ISL_878249, EPI_ISL_878251, EPI_ISL_878252, EPI_ISL_878254, EPI_ISL_878255, EPI_ISL_878257, EPI_ISL_878259, EPI_ISL_878261, EPI_ISL_878262, EPI_ISL_878265, EPI_ISL_878267, EPI_ISL_878269, EPI_ISL_878270, EPI_ISL_878272, EPI_ISL_878273, EPI_ISL_878275, EPI_ISL_878277, EPI_ISL_878278, EPI_ISL_878280, EPI_ISL_878282, EPI_ISL_878283, EPI_ISL_878285, EPI_ISL_878286, EPI_ISL_878288, EPI_ISL_878289, EPI_ISL_878291, EPI_ISL_878293, EPI_ISL_878294, EPI_ISL_878296, EPI_ISL_878297, EPI_ISL_878299, EPI_ISL_878301, EPI_ISL_878302, EPI_ISL_878304, EPI_ISL_878306, EPI_ISL_878307, EPI_ISL_878309, EPI_ISL_878310, EPI_ISL_878312, EPI_ISL_878313, EPI_ISL_878315, EPI_ISL_878317, EPI_ISL_878318, EPI_ISL_878320, EPI_ISL_878321, EPI_ISL_878323, EPI_ISL_878324, EPI_ISL_878326, EPI_ISL_878328, EPI_ISL_878331, EPI_ISL_878332, EPI_ISL_878334, EPI_ISL_878335, EPI_ISL_878337, EPI_ISL_878339, EPI_ISL_878340, EPI_ISL_878409, EPI_ISL_878410, EPI_ISL_878412, EPI_ISL_878414, EPI_ISL_878415, EPI_ISL_878417, EPI_ISL_878419, EPI_ISL_878420, EPI_ISL_878422, EPI_ISL_878424, EPI_ISL_878425, EPI_ISL_878427, EPI_ISL_878428, EPI_ISL_878430, EPI_ISL_878432, EPI_ISL_878435, EPI_ISL_878436, EPI_ISL_878438, EPI_ISL_878439, EPI_ISL_878441, EPI_ISL_878442, EPI_ISL_878444, EPI_ISL_878446, EPI_ISL_878449, EPI_ISL_878451, EPI_ISL_878453, EPI_ISL_878454, EPI_ISL_878456, EPI_ISL_878458, EPI_ISL_878459, EPI_ISL_878461, EPI_ISL_878462, EPI_ISL_878464, EPI_ISL_878466, EPI_ISL_878467, EPI_ISL_878469, EPI_ISL_878471, EPI_ISL_878473, EPI_ISL_878475, EPI_ISL_878476, EPI_ISL_878478, EPI_ISL_878481, EPI_ISL_878482, EPI_ISL_878484, EPI_ISL_878486, EPI_ISL_878487, EPI_ISL_878489, EPI_ISL_878490, EPI_ISL_878493, EPI_ISL_878494, EPI_ISL_878496, EPI_ISL_878497, EPI_ISL_878499, EPI_ISL_878501, EPI_ISL_878502, EPI_ISL_878504, EPI_ISL_878505, EPI_ISL_878507, EPI_ISL_878508, EPI_ISL_878510, EPI_ISL_878512, EPI_ISL_878513, EPI_ISL_878515, EPI_ISL_878517, EPI_ISL_878518, EPI_ISL_878520, EPI_ISL_878521, EPI_ISL_878523, EPI_ISL_878525, EPI_ISL_878527, EPI_ISL_878528, EPI_ISL_878530, EPI_ISL_878532, EPI_ISL_878533, EPI_ISL_878535, EPI_ISL_878538, EPI_ISL_878540, EPI_ISL_878542, EPI_ISL_878543, EPI_ISL_878546, EPI_ISL_878548, EPI_ISL_878550, EPI_ISL_878551, EPI_ISL_878555, EPI_ISL_878556, EPI_ISL_878558, EPI_ISL_878559, EPI_ISL_878561, EPI_ISL_878564, EPI_ISL_878567, EPI_ISL_878569, EPI_ISL_878570, EPI_ISL_878572, EPI_ISL_878573, EPI_ISL_878575, EPI_ISL_878577, EPI_ISL_878580, EPI_ISL_878582, EPI_ISL_878583, EPI_ISL_878585, EPI_ISL_878586, EPI_ISL_878588, EPI_ISL_878590, EPI_ISL_878591, EPI_ISL_878593, EPI_ISL_878594, EPI_ISL_878596, EPI_ISL_878598, EPI_ISL_878599, EPI_ISL_878601, EPI_ISL_878602, EPI_ISL_878604, EPI_ISL_878606, EPI_ISL_878607, EPI_ISL_878609, EPI_ISL_878612, EPI_ISL_878614, EPI_ISL_878615, EPI_ISL_878617, EPI_ISL_878618, EPI_ISL_878620, EPI_ISL_878621, EPI_ISL_878625, EPI_ISL_878626, EPI_ISL_878628, EPI_ISL_878630, EPI_ISL_878631, EPI_ISL_878633, EPI_ISL_878634, EPI_ISL_878636, EPI_ISL_878638, EPI_ISL_878639, EPI_ISL_878641, EPI_ISL_878642, EPI_ISL_878644, EPI_ISL_878646, EPI_ISL_878647, EPI_ISL_878649, EPI_ISL_878650, EPI_ISL_878652, EPI_ISL_878654, EPI_ISL_878655, EPI_ISL_878657, EPI_ISL_878659, EPI_ISL_878660, EPI_ISL_878662, EPI_ISL_878663, EPI_ISL_878665, EPI_ISL_878667, EPI_ISL_878668, EPI_ISL_878670, EPI_ISL_878672, EPI_ISL_878673, EPI_ISL_878675, EPI_ISL_878676, EPI_ISL_878678, EPI_ISL_878680, EPI_ISL_878681, EPI_ISL_878683, EPI_ISL_878685, EPI_ISL_878686, EPI_ISL_878688, EPI_ISL_878689, EPI_ISL_878691, EPI_ISL_878692, EPI_ISL_878694, EPI_ISL_878696, EPI_ISL_878698, EPI_ISL_878701, EPI_ISL_878703, EPI_ISL_878704, EPI_ISL_878706, EPI_ISL_878709, EPI_ISL_878711, EPI_ISL_878712, EPI_ISL_878714, EPI_ISL_878715, EPI_ISL_878718, EPI_ISL_878720, EPI_ISL_878722, EPI_ISL_878723, EPI_ISL_878725, EPI_ISL_878727, EPI_ISL_878730, EPI_ISL_878790, EPI_ISL_878805, EPI_ISL_878819, EPI_ISL_878825, EPI_ISL_878856, EPI_ISL_878876, EPI_ISL_878886, EPI_ISL_878913, EPI_ISL_878915, EPI_ISL_878959, EPI_ISL_878966, EPI_ISL_878993, EPI_ISL_878999, EPI_ISL_879007, EPI_ISL_879040, EPI_ISL_879056, EPI_ISL_879070, EPI_ISL_879073, EPI_ISL_879098, EPI_ISL_879100, EPI_ISL_879109, EPI_ISL_879110, EPI_ISL_879116, EPI_ISL_879117, EPI_ISL_879118, EPI_ISL_879120, EPI_ISL_879121, EPI_ISL_879124, EPI_ISL_879134, EPI_ISL_879138, EPI_ISL_879145, EPI_ISL_879149, EPI_ISL_879154, EPI_ISL_879161, EPI_ISL_879164, EPI_ISL_879167, EPI_ISL_879169, EPI_ISL_879171, EPI_ISL_879173, EPI_ISL_879185, EPI_ISL_879187, EPI_ISL_879193, EPI_ISL_879195, EPI_ISL_879201, EPI_ISL_879206, EPI_ISL_879207, EPI_ISL_879214, EPI_ISL_879220, EPI_ISL_879226, EPI_ISL_879227, EPI_ISL_879230, EPI_ISL_879234, EPI_ISL_879235, EPI_ISL_879239, EPI_ISL_879241, EPI_ISL_879243, EPI_ISL_879247, EPI_ISL_879248, EPI_ISL_879249, EPI_ISL_879253, EPI_ISL_879254, EPI_ISL_879256, EPI_ISL_879258, EPI_ISL_879262, EPI_ISL_879268, EPI_ISL_879270, EPI_ISL_879278, EPI_ISL_879284, EPI_ISL_879286, EPI_ISL_879290, EPI_ISL_879305, EPI_ISL_879314, EPI_ISL_879318, EPI_ISL_879322, EPI_ISL_879329, EPI_ISL_879331, EPI_ISL_879333, EPI_ISL_879338, EPI_ISL_879339, EPI_ISL_879341, EPI_ISL_879343, EPI_ISL_879344, EPI_ISL_879348, EPI_ISL_879354, EPI_ISL_879356, EPI_ISL_879358, EPI_ISL_879363, EPI_ISL_879374, EPI_ISL_879375, EPI_ISL_879377, EPI_ISL_879382, EPI_ISL_879384, EPI_ISL_879393, EPI_ISL_879416, EPI_ISL_879417, EPI_ISL_879433, EPI_ISL_879434, EPI_ISL_879451, EPI_ISL_879452, EPI_ISL_879460, EPI_ISL_879462, EPI_ISL_879466, EPI_ISL_879473, EPI_ISL_879475, EPI_ISL_879480, EPI_ISL_879486, EPI_ISL_879487, EPI_ISL_879504, EPI_ISL_879510, EPI_ISL_879519, EPI_ISL_879520, EPI_ISL_879524, EPI_ISL_879546, EPI_ISL_879547, EPI_ISL_879550, EPI_ISL_879556, EPI_ISL_879563, EPI_ISL_879565, EPI_ISL_879596, EPI_ISL_879614, EPI_ISL_879617, EPI_ISL_879629, EPI_ISL_879631, EPI_ISL_879646, EPI_ISL_879666, EPI_ISL_879672, EPI_ISL_879692, EPI_ISL_879693, EPI_ISL_879700, EPI_ISL_879714, EPI_ISL_879718, EPI_ISL_879719, EPI_ISL_879721, EPI_ISL_879726, EPI_ISL_879728, EPI_ISL_879736, EPI_ISL_879737, EPI_ISL_879740, EPI_ISL_879742, EPI_ISL_879744, EPI_ISL_879747, EPI_ISL_879748, EPI_ISL_879750, EPI_ISL_879751, EPI_ISL_879752, EPI_ISL_879753, EPI_ISL_879755, EPI_ISL_879756, EPI_ISL_879757, EPI_ISL_879758, EPI_ISL_879759, EPI_ISL_879760, EPI_ISL_879761, EPI_ISL_879762, EPI_ISL_879765, EPI_ISL_879766, EPI_ISL_879767, EPI_ISL_879775, EPI_ISL_879778, EPI_ISL_879780, EPI_ISL_879783, EPI_ISL_879784, EPI_ISL_879785, EPI_ISL_879787, EPI_ISL_879789, EPI_ISL_879791, EPI_ISL_879792, EPI_ISL_879793, EPI_ISL_879794, EPI_ISL_879800, EPI_ISL_879801, EPI_ISL_879802, EPI_ISL_879803, EPI_ISL_879808, EPI_ISL_879810, EPI_ISL_879811, EPI_ISL_879813, EPI_ISL_879814, EPI_ISL_879817, EPI_ISL_879819, EPI_ISL_879821, EPI_ISL_879822, EPI_ISL_879825, EPI_ISL_879826, EPI_ISL_879827, EPI_ISL_879829, EPI_ISL_879831, EPI_ISL_879836, EPI_ISL_879837, EPI_ISL_879838, EPI_ISL_879841, EPI_ISL_879842, EPI_ISL_879843, EPI_ISL_879844, EPI_ISL_879846, EPI_ISL_879849, EPI_ISL_879851, EPI_ISL_879854, EPI_ISL_879855, EPI_ISL_879856, EPI_ISL_879858, EPI_ISL_879859, EPI_ISL_879863, EPI_ISL_879865, EPI_ISL_879866, EPI_ISL_879868, EPI_ISL_879869, EPI_ISL_879870, EPI_ISL_879871, EPI_ISL_879872, EPI_ISL_879873, EPI_ISL_879875, EPI_ISL_879878, EPI_ISL_879879, EPI_ISL_879882, EPI_ISL_879884, EPI_ISL_879885, EPI_ISL_879886, EPI_ISL_879887, EPI_ISL_879888, EPI_ISL_879894, EPI_ISL_879895, EPI_ISL_879897, EPI_ISL_879898, EPI_ISL_879901, EPI_ISL_879903, EPI_ISL_879905, EPI_ISL_879906, EPI_ISL_879907, EPI_ISL_879912, EPI_ISL_879913, EPI_ISL_879914, EPI_ISL_879915, EPI_ISL_879916, EPI_ISL_879918, EPI_ISL_879919, EPI_ISL_879922, EPI_ISL_879923, EPI_ISL_879925, EPI_ISL_879927, EPI_ISL_879931, EPI_ISL_879933, EPI_ISL_879936, EPI_ISL_879938, EPI_ISL_879948, EPI_ISL_879949, EPI_ISL_879955, EPI_ISL_879957, EPI_ISL_879965, EPI_ISL_879966, EPI_ISL_879968, EPI_ISL_879970, EPI_ISL_879971, EPI_ISL_879974, EPI_ISL_879976, EPI_ISL_879978, EPI_ISL_879979, EPI_ISL_879981, EPI_ISL_879984, EPI_ISL_879986, EPI_ISL_879988, EPI_ISL_879991, EPI_ISL_879997, EPI_ISL_879999, EPI_ISL_880002, EPI_ISL_880007, EPI_ISL_880008, EPI_ISL_880010, EPI_ISL_880016, EPI_ISL_880018, EPI_ISL_880021, EPI_ISL_880023, EPI_ISL_880028, EPI_ISL_880030, EPI_ISL_880031, EPI_ISL_880036, EPI_ISL_880041, EPI_ISL_880043, EPI_ISL_880052, EPI_ISL_880055, EPI_ISL_880057, EPI_ISL_880062, EPI_ISL_880064, EPI_ISL_880067, EPI_ISL_880072, EPI_ISL_880083, EPI_ISL_880084, EPI_ISL_880086, EPI_ISL_880088, EPI_ISL_880089, EPI_ISL_880091, EPI_ISL_880093, EPI_ISL_880094, EPI_ISL_880096, EPI_ISL_880097, EPI_ISL_880099, EPI_ISL_880100, EPI_ISL_880102, EPI_ISL_880104, EPI_ISL_880105, EPI_ISL_880107, EPI_ISL_880108, EPI_ISL_880110, EPI_ISL_880112, EPI_ISL_880113, EPI_ISL_880115, EPI_ISL_880116, EPI_ISL_880118, EPI_ISL_880120, EPI_ISL_880121, EPI_ISL_880123, EPI_ISL_880124, EPI_ISL_880126, EPI_ISL_880127, EPI_ISL_880129, EPI_ISL_880131, EPI_ISL_880132, EPI_ISL_880134, EPI_ISL_880136, EPI_ISL_880137, EPI_ISL_880139, EPI_ISL_880210, EPI_ISL_880211, EPI_ISL_880213, EPI_ISL_880214, EPI_ISL_880216, EPI_ISL_880217, EPI_ISL_880219, EPI_ISL_880221, EPI_ISL_880222, EPI_ISL_880224, EPI_ISL_880226, EPI_ISL_880227, EPI_ISL_880229, EPI_ISL_880230, EPI_ISL_880232, EPI_ISL_880234, EPI_ISL_880235, EPI_ISL_880237, EPI_ISL_880238, EPI_ISL_880240, EPI_ISL_880242, EPI_ISL_880243, EPI_ISL_880245, EPI_ISL_880247, EPI_ISL_880248, EPI_ISL_880250, EPI_ISL_880252, EPI_ISL_880254, EPI_ISL_880255, EPI_ISL_880257, EPI_ISL_880258, EPI_ISL_880260, EPI_ISL_880262, EPI_ISL_880264, EPI_ISL_880265, EPI_ISL_880267, EPI_ISL_880269, EPI_ISL_880270, EPI_ISL_880272, EPI_ISL_880273, EPI_ISL_880275, EPI_ISL_880276, EPI_ISL_880278, EPI_ISL_880280, EPI_ISL_880281, EPI_ISL_880283, EPI_ISL_880285, EPI_ISL_880286, EPI_ISL_880288, EPI_ISL_880289, EPI_ISL_880291, EPI_ISL_880293, EPI_ISL_880295, EPI_ISL_880296, EPI_ISL_880298, EPI_ISL_880299, EPI_ISL_880301, EPI_ISL_880302, EPI_ISL_880304, EPI_ISL_880306, EPI_ISL_880307, EPI_ISL_880309, EPI_ISL_880311, EPI_ISL_880312, EPI_ISL_880314, EPI_ISL_880317, EPI_ISL_880318, EPI_ISL_880320, EPI_ISL_880322, EPI_ISL_880323, EPI_ISL_880325, EPI_ISL_880327, EPI_ISL_880328, EPI_ISL_880330, EPI_ISL_880331, EPI_ISL_880332, EPI_ISL_880333, EPI_ISL_880334, EPI_ISL_880335, EPI_ISL_880336, EPI_ISL_880337, EPI_ISL_880338, EPI_ISL_880339, EPI_ISL_880340, EPI_ISL_880341, EPI_ISL_880342, EPI_ISL_880343, EPI_ISL_880344, EPI_ISL_880345, EPI_ISL_880346, EPI_ISL_880347, EPI_ISL_880348, EPI_ISL_880349, EPI_ISL_880350, EPI_ISL_880351, EPI_ISL_880352, EPI_ISL_880353, EPI_ISL_880354, EPI_ISL_880355, EPI_ISL_880356, EPI_ISL_880357, EPI_ISL_880358, EPI_ISL_880359, EPI_ISL_880360, EPI_ISL_880361, EPI_ISL_880362, EPI_ISL_880363, EPI_ISL_880364, EPI_ISL_880365, EPI_ISL_880366, EPI_ISL_880367, EPI_ISL_880368, EPI_ISL_880369, EPI_ISL_880370, EPI_ISL_880371, EPI_ISL_880372, EPI_ISL_880373, EPI_ISL_880374, EPI_ISL_880375, EPI_ISL_880376, EPI_ISL_880377, EPI_ISL_880378, EPI_ISL_880379, EPI_ISL_880380, EPI_ISL_880381, EPI_ISL_880382, EPI_ISL_880383, EPI_ISL_880384, EPI_ISL_880385, EPI_ISL_880386, EPI_ISL_880387, EPI_ISL_880388, EPI_ISL_880389, EPI_ISL_880390, EPI_ISL_880391, EPI_ISL_880392, EPI_ISL_880393, EPI_ISL_880394, EPI_ISL_880395, EPI_ISL_880396, EPI_ISL_880397, EPI_ISL_880398, EPI_ISL_880399, EPI_ISL_880400, EPI_ISL_880401, EPI_ISL_880402, EPI_ISL_880403, EPI_ISL_880404, EPI_ISL_880405, EPI_ISL_880406, EPI_ISL_880407, EPI_ISL_880408, EPI_ISL_880409, EPI_ISL_880410, EPI_ISL_880411, EPI_ISL_880412, EPI_ISL_880413, EPI_ISL_880414, EPI_ISL_880415, EPI_ISL_880416, EPI_ISL_880417, EPI_ISL_880418, EPI_ISL_880419, EPI_ISL_880420, EPI_ISL_880421, EPI_ISL_880422, EPI_ISL_880423, EPI_ISL_880424, EPI_ISL_880425, EPI_ISL_880426, EPI_ISL_880427, EPI_ISL_880428, EPI_ISL_880429, EPI_ISL_880430, EPI_ISL_880431, EPI_ISL_880432, EPI_ISL_880433, EPI_ISL_880434, EPI_ISL_880435, EPI_ISL_880436, EPI_ISL_880437, EPI_ISL_880438, EPI_ISL_880439, EPI_ISL_880440, EPI_ISL_880441, EPI_ISL_880442, EPI_ISL_880443, EPI_ISL_880444, EPI_ISL_880445, EPI_ISL_880446, EPI_ISL_880447, EPI_ISL_880448, EPI_ISL_880449, EPI_ISL_880450, EPI_ISL_880451, EPI_ISL_880452, EPI_ISL_880453, EPI_ISL_880454, EPI_ISL_880455, EPI_ISL_880456, EPI_ISL_880457, EPI_ISL_880458, EPI_ISL_880459, EPI_ISL_880460, EPI_ISL_880461, EPI_ISL_880462, EPI_ISL_880463, EPI_ISL_880464, EPI_ISL_880465, EPI_ISL_880466, EPI_ISL_880467, EPI_ISL_880468, EPI_ISL_880469, EPI_ISL_880470, EPI_ISL_880471, EPI_ISL_880472, EPI_ISL_880473, EPI_ISL_880474, EPI_ISL_880475, EPI_ISL_880476, EPI_ISL_880477, EPI_ISL_880478, EPI_ISL_880479, EPI_ISL_880480, EPI_ISL_880481, EPI_ISL_880482, EPI_ISL_880483, EPI_ISL_880484, EPI_ISL_880485, EPI_ISL_880486, EPI_ISL_880487, EPI_ISL_880488, EPI_ISL_880489, EPI_ISL_880490, EPI_ISL_880491, EPI_ISL_880492, EPI_ISL_880493, EPI_ISL_880494, EPI_ISL_880495, EPI_ISL_880496, EPI_ISL_880497, EPI_ISL_880498, EPI_ISL_880499, EPI_ISL_880500, EPI_ISL_880501, EPI_ISL_880502, EPI_ISL_880503, EPI_ISL_880504, EPI_ISL_880505, EPI_ISL_880506, EPI_ISL_880507, EPI_ISL_880508, EPI_ISL_880509, EPI_ISL_880510, EPI_ISL_880511, EPI_ISL_880512, EPI_ISL_880513, EPI_ISL_880514, EPI_ISL_880515, EPI_ISL_880516, EPI_ISL_880517, EPI_ISL_880518, EPI_ISL_880519, EPI_ISL_880520, EPI_ISL_880521, EPI_ISL_880522, EPI_ISL_880523, EPI_ISL_880524, EPI_ISL_880525, EPI_ISL_880526, EPI_ISL_880527, EPI_ISL_880528, EPI_ISL_880529, EPI_ISL_880530, EPI_ISL_880531, EPI_ISL_880532, EPI_ISL_880533, EPI_ISL_880534, EPI_ISL_880535, EPI_ISL_880536, EPI_ISL_880537, EPI_ISL_880538, EPI_ISL_880539, EPI_ISL_880540, EPI_ISL_880541, EPI_ISL_880542, EPI_ISL_880543, EPI_ISL_880544, EPI_ISL_880545, EPI_ISL_880546, EPI_ISL_880547, EPI_ISL_880548, EPI_ISL_880549, EPI_ISL_880550, EPI_ISL_880551, EPI_ISL_880552, EPI_ISL_880553, EPI_ISL_880554, EPI_ISL_880555, EPI_ISL_880556, EPI_ISL_880557, EPI_ISL_880558, EPI_ISL_880559, EPI_ISL_880560, EPI_ISL_880562, EPI_ISL_880563, EPI_ISL_880564, EPI_ISL_880565, EPI_ISL_880566, EPI_ISL_880567, EPI_ISL_880568, EPI_ISL_880569, EPI_ISL_880570, EPI_ISL_880571, EPI_ISL_880572, EPI_ISL_880573, EPI_ISL_880574, EPI_ISL_880575, EPI_ISL_880576, EPI_ISL_880577, EPI_ISL_880578, EPI_ISL_880579, EPI_ISL_880580, EPI_ISL_880581, EPI_ISL_880582, EPI_ISL_880583, EPI_ISL_880936, EPI_ISL_880937, EPI_ISL_880938, EPI_ISL_880939, EPI_ISL_880940, EPI_ISL_880941, EPI_ISL_880942, EPI_ISL_880943, EPI_ISL_880944, EPI_ISL_880945, EPI_ISL_880946, EPI_ISL_880947, EPI_ISL_880948, EPI_ISL_880949, EPI_ISL_880950, EPI_ISL_880951, EPI_ISL_880952, EPI_ISL_880953, EPI_ISL_880954, EPI_ISL_880955, EPI_ISL_880956, EPI_ISL_880957, EPI_ISL_880958, EPI_ISL_880959, EPI_ISL_880960, EPI_ISL_880961, EPI_ISL_880962, EPI_ISL_880963, EPI_ISL_880964, EPI_ISL_880965, EPI_ISL_880966, EPI_ISL_880967, EPI_ISL_880968, EPI_ISL_880969, EPI_ISL_880970, EPI_ISL_880971, EPI_ISL_880972, EPI_ISL_880973, EPI_ISL_880974, EPI_ISL_880975, EPI_ISL_880976, EPI_ISL_880977, EPI_ISL_880978, EPI_ISL_880979, EPI_ISL_880980, EPI_ISL_880981, EPI_ISL_880982, EPI_ISL_880983, EPI_ISL_880984, EPI_ISL_880985, EPI_ISL_880986, EPI_ISL_880987, EPI_ISL_880988, EPI_ISL_880989, EPI_ISL_880990, EPI_ISL_880991, EPI_ISL_880992, EPI_ISL_880993, EPI_ISL_880994, EPI_ISL_880995, EPI_ISL_880996, EPI_ISL_880997, EPI_ISL_880998, EPI_ISL_880999, EPI_ISL_881000, EPI_ISL_881001, EPI_ISL_881002, EPI_ISL_881003, EPI_ISL_881004, EPI_ISL_881005, EPI_ISL_881006, EPI_ISL_881007, EPI_ISL_881008, EPI_ISL_881009, EPI_ISL_881010, EPI_ISL_881011, EPI_ISL_881012, EPI_ISL_881013, EPI_ISL_881014, EPI_ISL_881015, EPI_ISL_881016, EPI_ISL_881017, EPI_ISL_881018, EPI_ISL_881019, EPI_ISL_881020, EPI_ISL_881021, EPI_ISL_881022, EPI_ISL_881023, EPI_ISL_881024, EPI_ISL_881025, EPI_ISL_881026, EPI_ISL_881027, EPI_ISL_881028, EPI_ISL_881029, EPI_ISL_881030, EPI_ISL_881031, EPI_ISL_881032, EPI_ISL_881033, EPI_ISL_881034, EPI_ISL_881035, EPI_ISL_881036, EPI_ISL_881037, EPI_ISL_881038, EPI_ISL_881039, EPI_ISL_881040, EPI_ISL_881041, EPI_ISL_881042, EPI_ISL_881043, EPI_ISL_881044, EPI_ISL_881045, EPI_ISL_881046, EPI_ISL_881047, EPI_ISL_881048, EPI_ISL_881049, EPI_ISL_881050, EPI_ISL_881051, EPI_ISL_881052, EPI_ISL_881053, EPI_ISL_881054, EPI_ISL_881055, EPI_ISL_881056, EPI_ISL_881057, EPI_ISL_881058, EPI_ISL_881059, EPI_ISL_881060, EPI_ISL_881061, EPI_ISL_881062, EPI_ISL_881063, EPI_ISL_881064, EPI_ISL_881065, EPI_ISL_881066, EPI_ISL_881067, EPI_ISL_881068, EPI_ISL_881069, EPI_ISL_881070, EPI_ISL_881071, EPI_ISL_881072, EPI_ISL_881073, EPI_ISL_881074, EPI_ISL_881075, EPI_ISL_881076, EPI_ISL_881077, EPI_ISL_881078, EPI_ISL_881079, EPI_ISL_881080, EPI_ISL_881081, EPI_ISL_881082, EPI_ISL_881083, EPI_ISL_881084, EPI_ISL_881085, EPI_ISL_881086, EPI_ISL_881087, EPI_ISL_881088, EPI_ISL_881089, EPI_ISL_881090, EPI_ISL_881091, EPI_ISL_881092, EPI_ISL_881093, EPI_ISL_881094, EPI_ISL_881095, EPI_ISL_881096, EPI_ISL_881097, EPI_ISL_881098, EPI_ISL_881099, EPI_ISL_881100, EPI_ISL_881101, EPI_ISL_881102, EPI_ISL_881103, EPI_ISL_881104, EPI_ISL_881105, EPI_ISL_881106, EPI_ISL_881107, EPI_ISL_881108, EPI_ISL_881109, EPI_ISL_881110, EPI_ISL_881111, EPI_ISL_881112, EPI_ISL_881113, EPI_ISL_881114, EPI_ISL_881115, EPI_ISL_881116, EPI_ISL_881117, EPI_ISL_881118, EPI_ISL_881119, EPI_ISL_881120, EPI_ISL_881121, EPI_ISL_881122, EPI_ISL_881123, EPI_ISL_881124, EPI_ISL_881125, EPI_ISL_881126, EPI_ISL_881127, EPI_ISL_881128, EPI_ISL_881129, EPI_ISL_881130, EPI_ISL_881131, EPI_ISL_881132, EPI_ISL_881133, EPI_ISL_881134, EPI_ISL_881135, EPI_ISL_881136, EPI_ISL_881137, EPI_ISL_881138, EPI_ISL_881139, EPI_ISL_881140, EPI_ISL_881141, EPI_ISL_881142, EPI_ISL_881143, EPI_ISL_881144, EPI_ISL_881145, EPI_ISL_881146, EPI_ISL_881147, EPI_ISL_881148, EPI_ISL_881149, EPI_ISL_881150, EPI_ISL_881151, EPI_ISL_881152, EPI_ISL_881153, EPI_ISL_881154, EPI_ISL_881155, EPI_ISL_881156, EPI_ISL_881157, EPI_ISL_881158, EPI_ISL_881159, EPI_ISL_881160, EPI_ISL_881161, EPI_ISL_881162, EPI_ISL_881163, EPI_ISL_881164, EPI_ISL_881165, EPI_ISL_881166, EPI_ISL_881167, EPI_ISL_881168, EPI_ISL_881169, EPI_ISL_881170, EPI_ISL_881171, EPI_ISL_881172, EPI_ISL_881173, EPI_ISL_881174, EPI_ISL_881175, EPI_ISL_881176, EPI_ISL_881177, EPI_ISL_881178, EPI_ISL_881179, EPI_ISL_881180, EPI_ISL_881181, EPI_ISL_881182, EPI_ISL_881183, EPI_ISL_881184, EPI_ISL_881185, EPI_ISL_881186, EPI_ISL_881187, EPI_ISL_881188, EPI_ISL_881189, EPI_ISL_881190, EPI_ISL_881191, EPI_ISL_881192, EPI_ISL_881193, EPI_ISL_881194, EPI_ISL_881195, EPI_ISL_881196, EPI_ISL_881197, EPI_ISL_881198, EPI_ISL_881199, EPI_ISL_881200, EPI_ISL_881201, EPI_ISL_881202, EPI_ISL_881203, EPI_ISL_881204, EPI_ISL_881205, EPI_ISL_881206, EPI_ISL_881207, EPI_ISL_881208, EPI_ISL_881209, EPI_ISL_881210, EPI_ISL_881211, EPI_ISL_881212, EPI_ISL_881213, EPI_ISL_881214, EPI_ISL_881215, EPI_ISL_881216, EPI_ISL_881217, EPI_ISL_881218, EPI_ISL_881219, EPI_ISL_881220, EPI_ISL_881221, EPI_ISL_881222, EPI_ISL_881223, EPI_ISL_881224, EPI_ISL_881225, EPI_ISL_881226, EPI_ISL_881227, EPI_ISL_881228, EPI_ISL_881229, EPI_ISL_881230, EPI_ISL_881231, EPI_ISL_881232, EPI_ISL_881233, EPI_ISL_881234, EPI_ISL_881235, EPI_ISL_881236, EPI_ISL_881237, EPI_ISL_881238, EPI_ISL_881239, EPI_ISL_881240, EPI_ISL_881241, EPI_ISL_881242, EPI_ISL_881243, EPI_ISL_881244, EPI_ISL_881245, EPI_ISL_881246, EPI_ISL_881247, EPI_ISL_881248, EPI_ISL_881249, EPI_ISL_881250, EPI_ISL_881251, EPI_ISL_881252, EPI_ISL_881253, EPI_ISL_881254, EPI_ISL_881255, EPI_ISL_881256, EPI_ISL_881257, EPI_ISL_881258, EPI_ISL_881259, EPI_ISL_881260, EPI_ISL_881261, EPI_ISL_881262, EPI_ISL_881263, EPI_ISL_881264, EPI_ISL_881265, EPI_ISL_881266, EPI_ISL_881267, EPI_ISL_881268, EPI_ISL_881269, EPI_ISL_881270, EPI_ISL_881271, EPI_ISL_881272, EPI_ISL_881273, EPI_ISL_881274, EPI_ISL_881275, EPI_ISL_881276, EPI_ISL_881278, EPI_ISL_881279, EPI_ISL_881280, EPI_ISL_881282, EPI_ISL_881284, EPI_ISL_881285, EPI_ISL_881286, EPI_ISL_881287, EPI_ISL_881288, EPI_ISL_881292, EPI_ISL_881294, EPI_ISL_881295, EPI_ISL_881296, EPI_ISL_881297, EPI_ISL_881299, EPI_ISL_881300, EPI_ISL_881301, EPI_ISL_881302, EPI_ISL_881303, EPI_ISL_881304, EPI_ISL_881305, EPI_ISL_881308, EPI_ISL_881311, EPI_ISL_881312, EPI_ISL_881313, EPI_ISL_881314, EPI_ISL_881316, EPI_ISL_881317, EPI_ISL_881319, EPI_ISL_881320, EPI_ISL_881321, EPI_ISL_881322, EPI_ISL_881324, EPI_ISL_881325, EPI_ISL_881327, EPI_ISL_881328, EPI_ISL_881329, EPI_ISL_881330, EPI_ISL_881331, EPI_ISL_881332, EPI_ISL_881333, EPI_ISL_881334, EPI_ISL_881335, EPI_ISL_881336, EPI_ISL_881337, EPI_ISL_881338, EPI_ISL_881339, EPI_ISL_881340, EPI_ISL_881342, EPI_ISL_881343, EPI_ISL_881344, EPI_ISL_881345, EPI_ISL_881346, EPI_ISL_881347, EPI_ISL_881348, EPI_ISL_881349, EPI_ISL_881350, EPI_ISL_881351, EPI_ISL_881352, EPI_ISL_881353, EPI_ISL_881354, EPI_ISL_881355, EPI_ISL_881356, EPI_ISL_881357, EPI_ISL_881358, EPI_ISL_881360, EPI_ISL_881361, EPI_ISL_881362, EPI_ISL_881363, EPI_ISL_881364, EPI_ISL_881365, EPI_ISL_881366, EPI_ISL_881367, EPI_ISL_881368, EPI_ISL_881369, EPI_ISL_881370, EPI_ISL_881371, EPI_ISL_881372, EPI_ISL_881373, EPI_ISL_881374, EPI_ISL_881375, EPI_ISL_881376, EPI_ISL_881377, EPI_ISL_881378, EPI_ISL_881379, EPI_ISL_881380, EPI_ISL_881381, EPI_ISL_881382, EPI_ISL_881383, EPI_ISL_881384, EPI_ISL_881385, EPI_ISL_881387, EPI_ISL_881389, EPI_ISL_881390, EPI_ISL_881391, EPI_ISL_881392, EPI_ISL_881393, EPI_ISL_881394, EPI_ISL_881395, EPI_ISL_881396, EPI_ISL_881397, EPI_ISL_881398, EPI_ISL_881399, EPI_ISL_881400, EPI_ISL_881401, EPI_ISL_881402, EPI_ISL_881403, EPI_ISL_881404, EPI_ISL_881405, EPI_ISL_881406, EPI_ISL_881407, EPI_ISL_881408, EPI_ISL_881409, EPI_ISL_881410, EPI_ISL_881411, EPI_ISL_881412, EPI_ISL_881413, EPI_ISL_881415, EPI_ISL_881416, EPI_ISL_881417, EPI_ISL_881418, EPI_ISL_881419, EPI_ISL_881420, EPI_ISL_881421, EPI_ISL_881423, EPI_ISL_881424, EPI_ISL_881425, EPI_ISL_881426, EPI_ISL_881427, EPI_ISL_881428, EPI_ISL_881430, EPI_ISL_881431, EPI_ISL_881432, EPI_ISL_881433, EPI_ISL_881434, EPI_ISL_881435, EPI_ISL_881436, EPI_ISL_881437, EPI_ISL_881438, EPI_ISL_881439, EPI_ISL_881440, EPI_ISL_881441, EPI_ISL_881442, EPI_ISL_881443, EPI_ISL_881444, EPI_ISL_881445, EPI_ISL_881446, EPI_ISL_881447, EPI_ISL_881448, EPI_ISL_881449, EPI_ISL_881450, EPI_ISL_881451, EPI_ISL_881452, EPI_ISL_881453, EPI_ISL_881454, EPI_ISL_881456, EPI_ISL_881457, EPI_ISL_881458, EPI_ISL_881459, EPI_ISL_881460, EPI_ISL_881462, EPI_ISL_881463, EPI_ISL_881464, EPI_ISL_881465, EPI_ISL_881466, EPI_ISL_881467, EPI_ISL_881468, EPI_ISL_881469, EPI_ISL_881470, EPI_ISL_881471, EPI_ISL_881472, EPI_ISL_881473, EPI_ISL_881474, EPI_ISL_881475, EPI_ISL_881476, EPI_ISL_881477, EPI_ISL_881478, EPI_ISL_881479, EPI_ISL_881480, EPI_ISL_881481, EPI_ISL_881482, EPI_ISL_881483, EPI_ISL_881484, EPI_ISL_881485, EPI_ISL_881486, EPI_ISL_881488, EPI_ISL_881489, EPI_ISL_881490, EPI_ISL_881491, EPI_ISL_881492, EPI_ISL_881493, EPI_ISL_881496, EPI_ISL_881497, EPI_ISL_881498, EPI_ISL_881499, EPI_ISL_881500, EPI_ISL_881501, EPI_ISL_881502, EPI_ISL_881503, EPI_ISL_881504, EPI_ISL_881505, EPI_ISL_881506, EPI_ISL_881507, EPI_ISL_881508, EPI_ISL_881510, EPI_ISL_881511, EPI_ISL_881512, EPI_ISL_881513, EPI_ISL_881514, EPI_ISL_881515, EPI_ISL_881516, EPI_ISL_881517, EPI_ISL_881519, EPI_ISL_881520, EPI_ISL_881523, EPI_ISL_881524, EPI_ISL_881525, EPI_ISL_881526, EPI_ISL_881527, EPI_ISL_881528, EPI_ISL_881529, EPI_ISL_881530, EPI_ISL_881532, EPI_ISL_881533, EPI_ISL_881534, EPI_ISL_881535, EPI_ISL_881536, EPI_ISL_881537, EPI_ISL_881538, EPI_ISL_881539, EPI_ISL_881541, EPI_ISL_881542, EPI_ISL_881544, EPI_ISL_881545, EPI_ISL_881547, EPI_ISL_881548, EPI_ISL_881549, EPI_ISL_881551, EPI_ISL_881552, EPI_ISL_881553, EPI_ISL_881554, EPI_ISL_881555, EPI_ISL_881556, EPI_ISL_881557, EPI_ISL_881558, EPI_ISL_881559, EPI_ISL_881560, EPI_ISL_881561, EPI_ISL_881562, EPI_ISL_881563, EPI_ISL_881564, EPI_ISL_881565, EPI_ISL_881566, EPI_ISL_881568, EPI_ISL_881569, EPI_ISL_881572, EPI_ISL_881573, EPI_ISL_881574, EPI_ISL_881575, EPI_ISL_881577, EPI_ISL_881578, EPI_ISL_881579, EPI_ISL_881581, EPI_ISL_881583, EPI_ISL_881584, EPI_ISL_881585, EPI_ISL_881587, EPI_ISL_881589, EPI_ISL_881590, EPI_ISL_881591, EPI_ISL_881592, EPI_ISL_881593, EPI_ISL_881594, EPI_ISL_881595, EPI_ISL_881596, EPI_ISL_881597, EPI_ISL_881599, EPI_ISL_881601, EPI_ISL_881602, EPI_ISL_881603, EPI_ISL_881604, EPI_ISL_881605, EPI_ISL_881606, EPI_ISL_881608, EPI_ISL_881609, EPI_ISL_881610, EPI_ISL_881611, EPI_ISL_881613, EPI_ISL_881614, EPI_ISL_881615, EPI_ISL_881616, EPI_ISL_881617, EPI_ISL_882300, EPI_ISL_882605, EPI_ISL_882606, EPI_ISL_882607, EPI_ISL_882608, EPI_ISL_885160, EPI_ISL_885163, EPI_ISL_885165, EPI_ISL_885178, EPI_ISL_885180, EPI_ISL_885183, EPI_ISL_885187, EPI_ISL_885189, EPI_ISL_885197, EPI_ISL_885204, EPI_ISL_885209, EPI_ISL_885211, EPI_ISL_885213, EPI_ISL_885214, EPI_ISL_885215, EPI_ISL_885234, EPI_ISL_885238, EPI_ISL_885244, EPI_ISL_885248, EPI_ISL_885250, EPI_ISL_885264, EPI_ISL_885273, EPI_ISL_885274, EPI_ISL_885275, EPI_ISL_885277, EPI_ISL_885288, EPI_ISL_885289, EPI_ISL_885291, EPI_ISL_885295, EPI_ISL_885297, EPI_ISL_885299, EPI_ISL_885302, EPI_ISL_885305, EPI_ISL_885307, EPI_ISL_885311, EPI_ISL_885317, EPI_ISL_885319, EPI_ISL_885321, EPI_ISL_885323, EPI_ISL_885326, EPI_ISL_885335, EPI_ISL_885337, EPI_ISL_885349, EPI_ISL_885352, EPI_ISL_885353, EPI_ISL_885359, EPI_ISL_885361, EPI_ISL_885364, EPI_ISL_885365, EPI_ISL_885367, EPI_ISL_885369, EPI_ISL_885371, EPI_ISL_885374, EPI_ISL_885376, EPI_ISL_885388, EPI_ISL_885390, EPI_ISL_885392, EPI_ISL_885396, EPI_ISL_885397, EPI_ISL_885402, EPI_ISL_885411, EPI_ISL_885413, EPI_ISL_885419, EPI_ISL_885422, EPI_ISL_885427, EPI_ISL_885431, EPI_ISL_885452, EPI_ISL_885455, EPI_ISL_885458, EPI_ISL_885459, EPI_ISL_885461, EPI_ISL_885463, EPI_ISL_885471, EPI_ISL_885472, EPI_ISL_885473, EPI_ISL_885476, EPI_ISL_885478, EPI_ISL_885479, EPI_ISL_885482, EPI_ISL_885484, EPI_ISL_885499, EPI_ISL_885500, EPI_ISL_885503, EPI_ISL_885513, EPI_ISL_885541, EPI_ISL_885543, EPI_ISL_885557, EPI_ISL_885580, EPI_ISL_885586, EPI_ISL_885587, EPI_ISL_885589, EPI_ISL_885597, EPI_ISL_885619, EPI_ISL_885630, EPI_ISL_885637, EPI_ISL_885640, EPI_ISL_885649, EPI_ISL_885655, EPI_ISL_885661, EPI_ISL_885680, EPI_ISL_885683, EPI_ISL_885690, EPI_ISL_885705, EPI_ISL_885725, EPI_ISL_885754, EPI_ISL_885758, EPI_ISL_885762, EPI_ISL_885766, EPI_ISL_885813, EPI_ISL_885828, EPI_ISL_885860, EPI_ISL_885867, EPI_ISL_885880, EPI_ISL_885889, EPI_ISL_885912, EPI_ISL_885916, EPI_ISL_885918, EPI_ISL_885935, EPI_ISL_885954, EPI_ISL_885958, EPI_ISL_885962, EPI_ISL_885966, EPI_ISL_886009, EPI_ISL_886016, EPI_ISL_886039, EPI_ISL_886056, EPI_ISL_886057, EPI_ISL_886103, EPI_ISL_886121, EPI_ISL_886136, EPI_ISL_886142, EPI_ISL_886143, EPI_ISL_891899, EPI_ISL_891901, EPI_ISL_891903, EPI_ISL_891905, EPI_ISL_891908, EPI_ISL_891909, EPI_ISL_891914, EPI_ISL_891918, EPI_ISL_891920, EPI_ISL_891921, EPI_ISL_891923, EPI_ISL_891924, EPI_ISL_891925, EPI_ISL_891926, EPI_ISL_891927, EPI_ISL_891928, EPI_ISL_891929, EPI_ISL_891930, EPI_ISL_891931, EPI_ISL_891932, EPI_ISL_891934, EPI_ISL_891935, EPI_ISL_891936, EPI_ISL_891937, EPI_ISL_891938, EPI_ISL_891939, EPI_ISL_891940, EPI_ISL_891941, EPI_ISL_891942, EPI_ISL_891943, EPI_ISL_891944, EPI_ISL_891945, EPI_ISL_891946, EPI_ISL_891949, EPI_ISL_891950, EPI_ISL_891953, EPI_ISL_891954, EPI_ISL_891957, EPI_ISL_891958, EPI_ISL_891959, EPI_ISL_891962, EPI_ISL_891963, EPI_ISL_891967, EPI_ISL_891969, EPI_ISL_891970, EPI_ISL_891972, EPI_ISL_891974, EPI_ISL_891975, EPI_ISL_891976, EPI_ISL_891977, EPI_ISL_891978, EPI_ISL_891979, EPI_ISL_891980, EPI_ISL_891981, EPI_ISL_891982, EPI_ISL_891983, EPI_ISL_891984, EPI_ISL_891985, EPI_ISL_891986, EPI_ISL_891987, EPI_ISL_891988, EPI_ISL_891989, EPI_ISL_891990, EPI_ISL_891991, EPI_ISL_891992, EPI_ISL_891993, EPI_ISL_891994, EPI_ISL_891995, EPI_ISL_891996, EPI_ISL_891997, EPI_ISL_891998, EPI_ISL_891999, EPI_ISL_892000, EPI_ISL_892001, EPI_ISL_892002, EPI_ISL_892003, EPI_ISL_892004, EPI_ISL_892005, EPI_ISL_892006, EPI_ISL_892007, EPI_ISL_892008, EPI_ISL_892009, EPI_ISL_892012, EPI_ISL_892013, EPI_ISL_892014, EPI_ISL_892015, EPI_ISL_892016, EPI_ISL_892017, EPI_ISL_892018, EPI_ISL_892019, EPI_ISL_892020, EPI_ISL_892021, EPI_ISL_892022, EPI_ISL_892023, EPI_ISL_892024, EPI_ISL_892025, EPI_ISL_892026, EPI_ISL_892027, EPI_ISL_892028, EPI_ISL_892029, EPI_ISL_892030, EPI_ISL_892031, EPI_ISL_892032, EPI_ISL_892033, EPI_ISL_892034, EPI_ISL_892035, EPI_ISL_892036, EPI_ISL_892037, EPI_ISL_892038, EPI_ISL_892039, EPI_ISL_892040, EPI_ISL_892041, EPI_ISL_892042, EPI_ISL_892043, EPI_ISL_892044, EPI_ISL_892045, EPI_ISL_892046, EPI_ISL_892047, EPI_ISL_892048, EPI_ISL_892049, EPI_ISL_892056, EPI_ISL_892127, EPI_ISL_892128, EPI_ISL_892129, EPI_ISL_892130, EPI_ISL_892131, EPI_ISL_892132, EPI_ISL_892133, EPI_ISL_892134, EPI_ISL_892135, EPI_ISL_892136, EPI_ISL_892137, EPI_ISL_892138, EPI_ISL_892139, EPI_ISL_892140, EPI_ISL_892141, EPI_ISL_892142, EPI_ISL_892143, EPI_ISL_892144, EPI_ISL_892145, EPI_ISL_892146, EPI_ISL_892147, EPI_ISL_892148, EPI_ISL_892149, EPI_ISL_892150, EPI_ISL_892151, EPI_ISL_892152, EPI_ISL_892153, EPI_ISL_892154, EPI_ISL_892155, EPI_ISL_892156, EPI_ISL_892157, EPI_ISL_892158, EPI_ISL_892159, EPI_ISL_892160, EPI_ISL_892161, EPI_ISL_892162, EPI_ISL_892163, EPI_ISL_892164, EPI_ISL_892165, EPI_ISL_892166, EPI_ISL_892167, EPI_ISL_892168, EPI_ISL_892169, EPI_ISL_892170, EPI_ISL_892171, EPI_ISL_892172, EPI_ISL_892173, EPI_ISL_892174, EPI_ISL_892175, EPI_ISL_892176, EPI_ISL_892177, EPI_ISL_892178, EPI_ISL_892180, EPI_ISL_892181, EPI_ISL_892182, EPI_ISL_892183, EPI_ISL_892184, EPI_ISL_892185, EPI_ISL_892186, EPI_ISL_892187, EPI_ISL_892188, EPI_ISL_892198, EPI_ISL_892199, EPI_ISL_892200, EPI_ISL_892202, EPI_ISL_892203, EPI_ISL_892204, EPI_ISL_892205, EPI_ISL_909594, EPI_ISL_916622, EPI_ISL_916623, EPI_ISL_916624, EPI_ISL_916627, EPI_ISL_916628, EPI_ISL_916629, EPI_ISL_916631, EPI_ISL_916632, EPI_ISL_916634, EPI_ISL_916635, EPI_ISL_916636, EPI_ISL_916638, EPI_ISL_916639, EPI_ISL_916640, EPI_ISL_916641, EPI_ISL_916642, EPI_ISL_916643, EPI_ISL_916644, EPI_ISL_916646, EPI_ISL_916647, EPI_ISL_916650, EPI_ISL_916652, EPI_ISL_916653, EPI_ISL_916654, EPI_ISL_916655, EPI_ISL_916656, EPI_ISL_916657, EPI_ISL_916658, EPI_ISL_916659, EPI_ISL_916660, EPI_ISL_916663, EPI_ISL_916664, EPI_ISL_916665, EPI_ISL_916666, EPI_ISL_916667, EPI_ISL_916669, EPI_ISL_916670, EPI_ISL_916671, EPI_ISL_916672, EPI_ISL_916673, EPI_ISL_916674, EPI_ISL_916676, EPI_ISL_916677, EPI_ISL_916678, EPI_ISL_916679, EPI_ISL_916680, EPI_ISL_916681, EPI_ISL_916683, EPI_ISL_916685, EPI_ISL_916686, EPI_ISL_916687, EPI_ISL_916689, EPI_ISL_916690, EPI_ISL_916691, EPI_ISL_916692, EPI_ISL_916693, EPI_ISL_916695, EPI_ISL_916697, EPI_ISL_916698, EPI_ISL_916699, EPI_ISL_916700, EPI_ISL_916701, EPI_ISL_916702, EPI_ISL_916703, EPI_ISL_916706, EPI_ISL_916707, EPI_ISL_916708, EPI_ISL_916709, EPI_ISL_916710, EPI_ISL_916712, EPI_ISL_916714, EPI_ISL_916715, EPI_ISL_916717, EPI_ISL_916718, EPI_ISL_916719, EPI_ISL_916720, EPI_ISL_916721, EPI_ISL_916722, EPI_ISL_916723, EPI_ISL_916724, EPI_ISL_916725, EPI_ISL_916726, EPI_ISL_916727, EPI_ISL_916729, EPI_ISL_916731, EPI_ISL_916733, EPI_ISL_916734, EPI_ISL_916735, EPI_ISL_916737, EPI_ISL_916739, EPI_ISL_916740, EPI_ISL_916741, EPI_ISL_916742, EPI_ISL_916743, EPI_ISL_916744, EPI_ISL_916745, EPI_ISL_916747, EPI_ISL_916748, EPI_ISL_916749, EPI_ISL_916750, EPI_ISL_916751, EPI_ISL_916752, EPI_ISL_916753, EPI_ISL_916754, EPI_ISL_916755, EPI_ISL_916756, EPI_ISL_916757, EPI_ISL_916759, EPI_ISL_916760, EPI_ISL_916762, EPI_ISL_916763, EPI_ISL_916764, EPI_ISL_916765, EPI_ISL_916767, EPI_ISL_916768, EPI_ISL_916769, EPI_ISL_916770, EPI_ISL_916772, EPI_ISL_916773, EPI_ISL_916774, EPI_ISL_916776, EPI_ISL_916777, EPI_ISL_916778, EPI_ISL_916780, EPI_ISL_916781, EPI_ISL_916783, EPI_ISL_916785, EPI_ISL_916786, EPI_ISL_916787, EPI_ISL_916788, EPI_ISL_916789, EPI_ISL_916790, EPI_ISL_916791, EPI_ISL_916792, EPI_ISL_916794, EPI_ISL_916795, EPI_ISL_916796, EPI_ISL_916797, EPI_ISL_916798, EPI_ISL_916799, EPI_ISL_916801, EPI_ISL_916803, EPI_ISL_916804, EPI_ISL_916805, EPI_ISL_916806, EPI_ISL_916807, EPI_ISL_916808, EPI_ISL_916811, EPI_ISL_916813, EPI_ISL_916814, EPI_ISL_916816, EPI_ISL_916817, EPI_ISL_916818, EPI_ISL_916819, EPI_ISL_916822, EPI_ISL_916823, EPI_ISL_916824, EPI_ISL_916826, EPI_ISL_916827, EPI_ISL_916828, EPI_ISL_916829, EPI_ISL_916830, EPI_ISL_916831, EPI_ISL_916832, EPI_ISL_916835, EPI_ISL_916836, EPI_ISL_916837, EPI_ISL_916839, EPI_ISL_916841, EPI_ISL_916843, EPI_ISL_916844, EPI_ISL_916846, EPI_ISL_916848, EPI_ISL_916851, EPI_ISL_916852, EPI_ISL_916853, EPI_ISL_916854, EPI_ISL_916856, EPI_ISL_916857, EPI_ISL_916858, EPI_ISL_916859, EPI_ISL_916860, EPI_ISL_916861, EPI_ISL_916862, EPI_ISL_916863, EPI_ISL_916864, EPI_ISL_916865, EPI_ISL_916866, EPI_ISL_916867, EPI_ISL_916868, EPI_ISL_916870, EPI_ISL_916871, EPI_ISL_916872, EPI_ISL_916873, EPI_ISL_916874, EPI_ISL_916875, EPI_ISL_916876, EPI_ISL_916877, EPI_ISL_916878, EPI_ISL_916880, EPI_ISL_916881, EPI_ISL_916883, EPI_ISL_916884, EPI_ISL_916885, EPI_ISL_916886, EPI_ISL_916888, EPI_ISL_916889, EPI_ISL_916890, EPI_ISL_916892, EPI_ISL_916893, EPI_ISL_916895, EPI_ISL_916897, EPI_ISL_916901, EPI_ISL_916902, EPI_ISL_916903, EPI_ISL_916904, EPI_ISL_916906, EPI_ISL_916910, EPI_ISL_916913, EPI_ISL_916914, EPI_ISL_916915, EPI_ISL_916917, EPI_ISL_916920, EPI_ISL_916921, EPI_ISL_916922, EPI_ISL_916924, EPI_ISL_916925, EPI_ISL_916926, EPI_ISL_916927, EPI_ISL_916928, EPI_ISL_916929, EPI_ISL_916930, EPI_ISL_916931, EPI_ISL_916932, EPI_ISL_916933, EPI_ISL_916935, EPI_ISL_916936, EPI_ISL_916937, EPI_ISL_916938, EPI_ISL_916939, EPI_ISL_916941, EPI_ISL_916943, EPI_ISL_916944, EPI_ISL_916947, EPI_ISL_916952, EPI_ISL_916955, EPI_ISL_916957, EPI_ISL_916958, EPI_ISL_916959, EPI_ISL_916960, EPI_ISL_916962, EPI_ISL_916963, EPI_ISL_916964, EPI_ISL_916965, EPI_ISL_916966, EPI_ISL_916969, EPI_ISL_916971, EPI_ISL_916972, EPI_ISL_916973, EPI_ISL_916975, EPI_ISL_916978, EPI_ISL_916980, EPI_ISL_916982, EPI_ISL_916983, EPI_ISL_916986, EPI_ISL_916988, EPI_ISL_916989, EPI_ISL_916990, EPI_ISL_916992, EPI_ISL_916993, EPI_ISL_916994, EPI_ISL_916996, EPI_ISL_916997, EPI_ISL_917000, EPI_ISL_917001, EPI_ISL_917003, EPI_ISL_917004, EPI_ISL_917005, EPI_ISL_917007, EPI_ISL_917008, EPI_ISL_917010, EPI_ISL_917015, EPI_ISL_917016, EPI_ISL_917017, EPI_ISL_917018, EPI_ISL_917020, EPI_ISL_917023, EPI_ISL_917027, EPI_ISL_917028, EPI_ISL_917030, EPI_ISL_917033, EPI_ISL_917034, EPI_ISL_917035, EPI_ISL_917037, EPI_ISL_917039, EPI_ISL_917040, EPI_ISL_917041, EPI_ISL_917043, EPI_ISL_917044, EPI_ISL_917045, EPI_ISL_917046, EPI_ISL_917048, EPI_ISL_917050, EPI_ISL_917051, EPI_ISL_917052, EPI_ISL_917056, EPI_ISL_917057, EPI_ISL_917058, EPI_ISL_917059, EPI_ISL_917061, EPI_ISL_917062, EPI_ISL_917064, EPI_ISL_917065, EPI_ISL_917066, EPI_ISL_917068, EPI_ISL_917069, EPI_ISL_917072, EPI_ISL_917073, EPI_ISL_917078, EPI_ISL_917080, EPI_ISL_917084, EPI_ISL_917085, EPI_ISL_917087, EPI_ISL_917089, EPI_ISL_917090, EPI_ISL_917092, EPI_ISL_917094, EPI_ISL_917097, EPI_ISL_917098, EPI_ISL_917100, EPI_ISL_917101, EPI_ISL_917102, EPI_ISL_917103, EPI_ISL_917106, EPI_ISL_917108, EPI_ISL_917109, EPI_ISL_917111, EPI_ISL_917113, EPI_ISL_917115, EPI_ISL_917117, EPI_ISL_917118, EPI_ISL_917119, EPI_ISL_917122, EPI_ISL_917123, EPI_ISL_917124, EPI_ISL_917126, EPI_ISL_917127, EPI_ISL_917130, EPI_ISL_917131, EPI_ISL_917132, EPI_ISL_917134, EPI_ISL_917135, EPI_ISL_917136, EPI_ISL_917138, EPI_ISL_917140, EPI_ISL_917142, EPI_ISL_917146, EPI_ISL_917147, EPI_ISL_917150, EPI_ISL_917152, EPI_ISL_917153, EPI_ISL_917154, EPI_ISL_917155, EPI_ISL_917157, EPI_ISL_917158, EPI_ISL_917162, EPI_ISL_917164, EPI_ISL_917165, EPI_ISL_917167, EPI_ISL_918060, EPI_ISL_918061, EPI_ISL_939608, EPI_ISL_963321, EPI_ISL_991916, EPI_ISL_991917, EPI_ISL_991918, EPI_ISL_991919, EPI_ISL_991920, EPI_ISL_991921, EPI_ISL_991922, EPI_ISL_991923, EPI_ISL_991924, EPI_ISL_991925, EPI_ISL_991926, EPI_ISL_991927, EPI_ISL_991928, EPI_ISL_991929, EPI_ISL_991930, EPI_ISL_991931, EPI_ISL_991932, EPI_ISL_991933, EPI_ISL_991934, EPI_ISL_991935, EPI_ISL_991936, EPI_ISL_991937, EPI_ISL_991938, EPI_ISL_991939, EPI_ISL_991940, EPI_ISL_991941, EPI_ISL_991942, EPI_ISL_991943, EPI_ISL_991944, EPI_ISL_991945, EPI_ISL_991946, EPI_ISL_991947, EPI_ISL_991948, EPI_ISL_991949, EPI_ISL_991950, EPI_ISL_991951, EPI_ISL_991952, EPI_ISL_991953, EPI_ISL_991954, EPI_ISL_991955, EPI_ISL_991956, EPI_ISL_991957, EPI_ISL_991958, EPI_ISL_991959, EPI_ISL_991960, EPI_ISL_991961, EPI_ISL_991962, EPI_ISL_992647, EPI_ISL_993111, EPI_ISL_993149, EPI_ISL_993198, EPI_ISL_993239, EPI_ISL_993362, EPI_ISL_993401, EPI_ISL_994288, EPI_ISL_994297, EPI_ISL_994338, EPI_ISL_994339, EPI_ISL_994343, EPI_ISL_994357, EPI_ISL_994360, EPI_ISL_994361, EPI_ISL_994386, EPI_ISL_994389, EPI_ISL_994391, EPI_ISL_994402, EPI_ISL_994407, EPI_ISL_994411, EPI_ISL_994413, EPI_ISL_994416, EPI_ISL_994419, EPI_ISL_994424, EPI_ISL_994446, EPI_ISL_994448, EPI_ISL_994456, EPI_ISL_994464, EPI_ISL_994467, EPI_ISL_994468, EPI_ISL_994473, EPI_ISL_994478, EPI_ISL_994481, EPI_ISL_994482, EPI_ISL_994484, EPI_ISL_994488, EPI_ISL_994496, EPI_ISL_994507, EPI_ISL_994513, EPI_ISL_994514, EPI_ISL_994515, EPI_ISL_994529, EPI_ISL_994538, EPI_ISL_994543, EPI_ISL_994590, EPI_ISL_994615, EPI_ISL_994619, EPI_ISL_994622, EPI_ISL_994623, EPI_ISL_994624, EPI_ISL_994626, EPI_ISL_994629, EPI_ISL_994630, EPI_ISL_994633, EPI_ISL_994634, EPI_ISL_994635, EPI_ISL_994636, EPI_ISL_994637, EPI_ISL_994639,

EPI_ISL_994641, EPI_ISL_994643, EPI_ISL_994646, EPI_ISL_1007580, EPI_ISL_1007581, EPI_ISL_1007582, EPI_ISL_1101110

see above | Lighthouse Lab in Alderley Park | Wellcome Sanger Institute for the COVID-19 Genomics UK (COG-UK) Consortium | Cordelia Langford; David K. Jackson; Dominic Kwiatkowski; Ewan Harrison; Ian Johnston; Jacquelyn Wynn; Jeffrey Barrett; John Sillitoe on behalf of the Wellcome Sanger Institute COVID-19 Surveillance Team; John Sillitoe on behalf of the Wellcome Sanger Institute COVID-19 Surveillance Team (http://www.sanger.ac.uk/covid-team); Mairead Hyland; Roberto Amato; Sonia Goncalves; The Lighthouse Lab in Alderley Park and Alex Alderton

EPI_ISL_857855, EPI_ISL_873283, EPI_ISL_873286, EPI_ISL_873291, EPI_ISL_873292, EPI_ISL_873297, EPI_ISL_873302, EPI_ISL_873304, EPI_ISL_873306, EPI_ISL_873310, EPI_ISL_873313, EPI_ISL_873316, EPI_ISL_873318, EPI_ISL_873320, EPI_ISL_873324, EPI_ISL_873325, EPI_ISL_873326, EPI_ISL_873334, EPI_ISL_873337, EPI_ISL_873339, EPI_ISL_873341, EPI_ISL_873342, EPI_ISL_873343, EPI_ISL_873347, EPI_ISL_873348, EPI_ISL_873350, EPI_ISL_873357, EPI_ISL_873359, EPI_ISL_873362, EPI_ISL_873364, EPI_ISL_873369, EPI_ISL_873370, EPI_ISL_873371, EPI_ISL_873374, EPI_ISL_873381, EPI_ISL_873388, EPI_ISL_873399, EPI_ISL_873412, EPI_ISL_873416, EPI_ISL_873422, EPI_ISL_873426, EPI_ISL_873431, EPI_ISL_873433, EPI_ISL_873451, EPI_ISL_873454, EPI_ISL_873457, EPI_ISL_873459, EPI_ISL_873476, EPI_ISL_873481, EPI_ISL_873482, EPI_ISL_873492, EPI_ISL_873494, EPI_ISL_873495, EPI_ISL_873497, EPI_ISL_873499, EPI_ISL_873500, EPI_ISL_873501, EPI_ISL_873510, EPI_ISL_873518, EPI_ISL_873532, EPI_ISL_873537, EPI_ISL_873552, EPI_ISL_873566, EPI_ISL_873571, EPI_ISL_873573, EPI_ISL_873574, EPI_ISL_873576, EPI_ISL_873605, EPI_ISL_873614, EPI_ISL_873621, EPI_ISL_873625, EPI_ISL_873629, EPI_ISL_873630, EPI_ISL_873632, EPI_ISL_873633, EPI_ISL_873634, EPI_ISL_873635, EPI_ISL_873636, EPI_ISL_873638, EPI_ISL_873639, EPI_ISL_873640, EPI_ISL_873642, EPI_ISL_873643, EPI_ISL_873644, EPI_ISL_873646, EPI_ISL_873647, EPI_ISL_873649, EPI_ISL_873650, EPI_ISL_873651, EPI_ISL_873653, EPI_ISL_873655, EPI_ISL_873658, EPI_ISL_873659, EPI_ISL_873661, EPI_ISL_873662, EPI_ISL_873663, EPI_ISL_873664, EPI_ISL_873665, EPI_ISL_873666, EPI_ISL_873667, EPI_ISL_873668, EPI_ISL_873670, EPI_ISL_873671, EPI_ISL_873673, EPI_ISL_873674, EPI_ISL_873675, EPI_ISL_873676, EPI_ISL_873677, EPI_ISL_873678, EPI_ISL_873680, EPI_ISL_873681, EPI_ISL_873682, EPI_ISL_873683, EPI_ISL_873685, EPI_ISL_873686, EPI_ISL_873687, EPI_ISL_873688, EPI_ISL_873689, EPI_ISL_873690, EPI_ISL_873691, EPI_ISL_873692, EPI_ISL_873693, EPI_ISL_873694, EPI_ISL_873695, EPI_ISL_873696, EPI_ISL_873697, EPI_ISL_873698, EPI_ISL_873699, EPI_ISL_873700, EPI_ISL_873701, EPI_ISL_873702, EPI_ISL_873703, EPI_ISL_873705, EPI_ISL_873706, EPI_ISL_873707, EPI_ISL_873708, EPI_ISL_873709, EPI_ISL_873710, EPI_ISL_873711, EPI_ISL_873712, EPI_ISL_873713, EPI_ISL_873714, EPI_ISL_873715, EPI_ISL_873717, EPI_ISL_873718, EPI_ISL_873719, EPI_ISL_873720, EPI_ISL_873721, EPI_ISL_873722, EPI_ISL_873723, EPI_ISL_873727, EPI_ISL_873728, EPI_ISL_873731, EPI_ISL_873732, EPI_ISL_873733, EPI_ISL_873735, EPI_ISL_873736, EPI_ISL_873737, EPI_ISL_873738, EPI_ISL_873739, EPI_ISL_873740, EPI_ISL_873742, EPI_ISL_873745, EPI_ISL_873747, EPI_ISL_873751, EPI_ISL_873753, EPI_ISL_873754, EPI_ISL_873756, EPI_ISL_873757, EPI_ISL_873758, EPI_ISL_873759, EPI_ISL_873761, EPI_ISL_873762, EPI_ISL_873763, EPI_ISL_873765, EPI_ISL_873766, EPI_ISL_873767, EPI_ISL_873768, EPI_ISL_873770, EPI_ISL_873771, EPI_ISL_873772, EPI_ISL_873773, EPI_ISL_873774, EPI_ISL_873775, EPI_ISL_873776, EPI_ISL_873777, EPI_ISL_873778, EPI_ISL_873779, EPI_ISL_873780, EPI_ISL_873781, EPI_ISL_873782, EPI_ISL_873784, EPI_ISL_873785, EPI_ISL_873787, EPI_ISL_873788, EPI_ISL_873789, EPI_ISL_873790, EPI_ISL_873791, EPI_ISL_873792, EPI_ISL_873793, EPI_ISL_873795, EPI_ISL_873796, EPI_ISL_873798, EPI_ISL_873799, EPI_ISL_873800, EPI_ISL_873801, EPI_ISL_873802, EPI_ISL_873803, EPI_ISL_873804, EPI_ISL_873805, EPI_ISL_873807, EPI_ISL_873808, EPI_ISL_873810, EPI_ISL_873811, EPI_ISL_873813, EPI_ISL_873814, EPI_ISL_873815, EPI_ISL_873816, EPI_ISL_873818, EPI_ISL_873819, EPI_ISL_873820, EPI_ISL_873821, EPI_ISL_873823, EPI_ISL_873824, EPI_ISL_873826, EPI_ISL_873828, EPI_ISL_873830, EPI_ISL_873831, EPI_ISL_873832, EPI_ISL_873835, EPI_ISL_873836, EPI_ISL_873837, EPI_ISL_873838, EPI_ISL_873839, EPI_ISL_873841, EPI_ISL_873843, EPI_ISL_873844, EPI_ISL_873845, EPI_ISL_873846, EPI_ISL_873849, EPI_ISL_873851, EPI_ISL_873852, EPI_ISL_873854, EPI_ISL_873855, EPI_ISL_873856, EPI_ISL_873858, EPI_ISL_873861, EPI_ISL_873863, EPI_ISL_873864, EPI_ISL_873866, EPI_ISL_873867, EPI_ISL_873868, EPI_ISL_873869, EPI_ISL_873871, EPI_ISL_873872, EPI_ISL_873874, EPI_ISL_873876, EPI_ISL_873877, EPI_ISL_873878, EPI_ISL_873879, EPI_ISL_873883, EPI_ISL_873885, EPI_ISL_873888, EPI_ISL_873889, EPI_ISL_873890, EPI_ISL_873891, EPI_ISL_873892, EPI_ISL_873893, EPI_ISL_873894, EPI_ISL_873895, EPI_ISL_873896, EPI_ISL_873897, EPI_ISL_873898, EPI_ISL_873899, EPI_ISL_873901, EPI_ISL_873902, EPI_ISL_873903, EPI_ISL_873906, EPI_ISL_873907, EPI_ISL_873908, EPI_ISL_873909, EPI_ISL_873911, EPI_ISL_873912, EPI_ISL_873914, EPI_ISL_873915, EPI_ISL_873917, EPI_ISL_873918, EPI_ISL_873919, EPI_ISL_873920, EPI_ISL_873922, EPI_ISL_873927, EPI_ISL_873929, EPI_ISL_873930, EPI_ISL_873931, EPI_ISL_873932, EPI_ISL_873933, EPI_ISL_873936, EPI_ISL_873938, EPI_ISL_873939, EPI_ISL_873940, EPI_ISL_873941, EPI_ISL_873944, EPI_ISL_873945, EPI_ISL_873946, EPI_ISL_873949, EPI_ISL_873951, EPI_ISL_873952, EPI_ISL_873953, EPI_ISL_873955, EPI_ISL_873957, EPI_ISL_873958, EPI_ISL_873959, EPI_ISL_878731, EPI_ISL_878740, EPI_ISL_878758, EPI_ISL_878761, EPI_ISL_878766, EPI_ISL_878770, EPI_ISL_878796, EPI_ISL_878801, EPI_ISL_878802, EPI_ISL_878803, EPI_ISL_878810, EPI_ISL_878815, EPI_ISL_878830, EPI_ISL_878831, EPI_ISL_878832, EPI_ISL_878834, EPI_ISL_878839, EPI_ISL_878841, EPI_ISL_878847, EPI_ISL_878850, EPI_ISL_878857, EPI_ISL_878862, EPI_ISL_878863, EPI_ISL_878864, EPI_ISL_878869, EPI_ISL_878870, EPI_ISL_878871, EPI_ISL_878889, EPI_ISL_878894, EPI_ISL_878899, EPI_ISL_878903, EPI_ISL_878904, EPI_ISL_878905, EPI_ISL_878912, EPI_ISL_878924, EPI_ISL_878931, EPI_ISL_878935, EPI_ISL_878950, EPI_ISL_878964, EPI_ISL_878975, EPI_ISL_878980, EPI_ISL_878989, EPI_ISL_878991, EPI_ISL_878995, EPI_ISL_878997, EPI_ISL_878998, EPI_ISL_879011, EPI_ISL_879012, EPI_ISL_879022, EPI_ISL_879029, EPI_ISL_879031, EPI_ISL_879045, EPI_ISL_879051, EPI_ISL_879054, EPI_ISL_879058, EPI_ISL_879062, EPI_ISL_879068, EPI_ISL_879069, EPI_ISL_879071, EPI_ISL_879074, EPI_ISL_879076, EPI_ISL_879078, EPI_ISL_879082, EPI_ISL_879095, EPI_ISL_879381, EPI_ISL_879383, EPI_ISL_879385, EPI_ISL_879387, EPI_ISL_879389, EPI_ISL_879390, EPI_ISL_879391, EPI_ISL_879394, EPI_ISL_879398, EPI_ISL_879400, EPI_ISL_879401, EPI_ISL_879402, EPI_ISL_879403, EPI_ISL_879406, EPI_ISL_879407, EPI_ISL_879408, EPI_ISL_879410, EPI_ISL_879411, EPI_ISL_879414, EPI_ISL_879419, EPI_ISL_879421, EPI_ISL_879422, EPI_ISL_879423, EPI_ISL_879428, EPI_ISL_879430, EPI_ISL_879431, EPI_ISL_879432, EPI_ISL_879435, EPI_ISL_879436, EPI_ISL_879437, EPI_ISL_879438, EPI_ISL_879439, EPI_ISL_879440, EPI_ISL_879441, EPI_ISL_879443, EPI_ISL_879445, EPI_ISL_879453, EPI_ISL_879454, EPI_ISL_879463, EPI_ISL_879465, EPI_ISL_879468, EPI_ISL_879469, EPI_ISL_879470, EPI_ISL_879472, EPI_ISL_879474, EPI_ISL_879478, EPI_ISL_879481, EPI_ISL_879482, EPI_ISL_879484, EPI_ISL_879485, EPI_ISL_879489, EPI_ISL_879490, EPI_ISL_879491, EPI_ISL_879494, EPI_ISL_879496, EPI_ISL_879498, EPI_ISL_879500, EPI_ISL_879503, EPI_ISL_879506, EPI_ISL_879507, EPI_ISL_879508, EPI_ISL_879509, EPI_ISL_879511, EPI_ISL_879513, EPI_ISL_879516, EPI_ISL_879517, EPI_ISL_879523, EPI_ISL_879525, EPI_ISL_879527, EPI_ISL_879528, EPI_ISL_879529, EPI_ISL_879530, EPI_ISL_879531, EPI_ISL_879537, EPI_ISL_879543, EPI_ISL_879544, EPI_ISL_879545, EPI_ISL_879551, EPI_ISL_879557, EPI_ISL_879558, EPI_ISL_879559, EPI_ISL_879562, EPI_ISL_879567, EPI_ISL_879568, EPI_ISL_879569, EPI_ISL_879570, EPI_ISL_879573, EPI_ISL_879574, EPI_ISL_879577, EPI_ISL_879579, EPI_ISL_879580, EPI_ISL_879583, EPI_ISL_879587, EPI_ISL_879588, EPI_ISL_879590, EPI_ISL_879595, EPI_ISL_879598, EPI_ISL_879599, EPI_ISL_879605, EPI_ISL_879606, EPI_ISL_879607, EPI_ISL_879609, EPI_ISL_879610, EPI_ISL_879612, EPI_ISL_879615, EPI_ISL_879620, EPI_ISL_879622, EPI_ISL_879624, EPI_ISL_879625, EPI_ISL_879626, EPI_ISL_879630, EPI_ISL_879632, EPI_ISL_879633, EPI_ISL_879634, EPI_ISL_879635, EPI_ISL_879636, EPI_ISL_879637, EPI_ISL_879639, EPI_ISL_879640, EPI_ISL_879644, EPI_ISL_879645, EPI_ISL_879648, EPI_ISL_879650, EPI_ISL_879651, EPI_ISL_879658, EPI_ISL_879659, EPI_ISL_879660, EPI_ISL_879667, EPI_ISL_879668, EPI_ISL_879669, EPI_ISL_879670, EPI_ISL_879673, EPI_ISL_879676, EPI_ISL_879677, EPI_ISL_879680, EPI_ISL_879684, EPI_ISL_879685, EPI_ISL_879688, EPI_ISL_879690, EPI_ISL_879691, EPI_ISL_879694, EPI_ISL_879696, EPI_ISL_879697, EPI_ISL_879698, EPI_ISL_879699, EPI_ISL_879701, EPI_ISL_879704, EPI_ISL_879705, EPI_ISL_879706, EPI_ISL_879708, EPI_ISL_879709, EPI_ISL_879716, EPI_ISL_879720, EPI_ISL_879722, EPI_ISL_879723, EPI_ISL_879724, EPI_ISL_879725, EPI_ISL_879727, EPI_ISL_879729, EPI_ISL_879730, EPI_ISL_879731, EPI_ISL_879732, EPI_ISL_879733, EPI_ISL_879734, EPI_ISL_879735, EPI_ISL_879738, EPI_ISL_879739, EPI_ISL_879741, EPI_ISL_879743, EPI_ISL_879745, EPI_ISL_879746, EPI_ISL_879749, EPI_ISL_879754, EPI_ISL_879763, EPI_ISL_879764, EPI_ISL_879768, EPI_ISL_879769, EPI_ISL_879770, EPI_ISL_879771, EPI_ISL_879772, EPI_ISL_879773, EPI_ISL_879774, EPI_ISL_879776, EPI_ISL_879777, EPI_ISL_879779, EPI_ISL_879781, EPI_ISL_879782, EPI_ISL_879786, EPI_ISL_879788, EPI_ISL_879795, EPI_ISL_879796, EPI_ISL_879797, EPI_ISL_879798, EPI_ISL_879799, EPI_ISL_879804, EPI_ISL_879805, EPI_ISL_879806, EPI_ISL_879807, EPI_ISL_879809, EPI_ISL_879812, EPI_ISL_879815, EPI_ISL_879816, EPI_ISL_879818, EPI_ISL_879823, EPI_ISL_879824, EPI_ISL_879828, EPI_ISL_879830, EPI_ISL_879832, EPI_ISL_879833, EPI_ISL_879834, EPI_ISL_879835, EPI_ISL_879839, EPI_ISL_879840, EPI_ISL_879845, EPI_ISL_879847, EPI_ISL_879848, EPI_ISL_879850, EPI_ISL_879853, EPI_ISL_879857, EPI_ISL_879860, EPI_ISL_879861, EPI_ISL_879862, EPI_ISL_879864, EPI_ISL_879867, EPI_ISL_879874, EPI_ISL_879876, EPI_ISL_879877, EPI_ISL_879880, EPI_ISL_879881, EPI_ISL_879883, EPI_ISL_879889, EPI_ISL_879890, EPI_ISL_879891, EPI_ISL_879892, EPI_ISL_879893, EPI_ISL_879899, EPI_ISL_879900, EPI_ISL_879902, EPI_ISL_879904, EPI_ISL_879908, EPI_ISL_879909, EPI_ISL_879910, EPI_ISL_879911, EPI_ISL_879917, EPI_ISL_879920, EPI_ISL_879921, EPI_ISL_879928, EPI_ISL_879930, EPI_ISL_879935, EPI_ISL_879940, EPI_ISL_879941, EPI_ISL_879943, EPI_ISL_879944, EPI_ISL_879946, EPI_ISL_879951, EPI_ISL_879952, EPI_ISL_879954, EPI_ISL_879958, EPI_ISL_879960, EPI_ISL_879962, EPI_ISL_879963, EPI_ISL_879973, EPI_ISL_879983, EPI_ISL_879989, EPI_ISL_879992, EPI_ISL_879994, EPI_ISL_879996, EPI_ISL_880001, EPI_ISL_880004, EPI_ISL_880005, EPI_ISL_880012, EPI_ISL_880013, EPI_ISL_880015, EPI_ISL_880020, EPI_ISL_880026, EPI_ISL_880033, EPI_ISL_880035, EPI_ISL_880039, EPI_ISL_880044, EPI_ISL_880046, EPI_ISL_880047, EPI_ISL_880049, EPI_ISL_880051, EPI_ISL_880054, EPI_ISL_880059, EPI_ISL_880060, EPI_ISL_880065, EPI_ISL_880068, EPI_ISL_880070, EPI_ISL_880073, EPI_ISL_880075, EPI_ISL_880076, EPI_ISL_880078, EPI_ISL_880080, EPI_ISL_880081, EPI_ISL_881620, EPI_ISL_881622, EPI_ISL_987283, EPI_ISL_987287, EPI_ISL_987290, EPI_ISL_987291, EPI_ISL_987299, EPI_ISL_987303, EPI_ISL_987310, EPI_ISL_987312, EPI_ISL_987319, EPI_ISL_987322, EPI_ISL_987326, EPI_ISL_987328, EPI_ISL_987332, EPI_ISL_987339, EPI_ISL_987340, EPI_ISL_987357, EPI_ISL_987369, EPI_ISL_987378, EPI_ISL_987380, EPI_ISL_987381, EPI_ISL_987382, EPI_ISL_987384, EPI_ISL_987389, EPI_ISL_987393, EPI_ISL_987398, EPI_ISL_987399, EPI_ISL_987402, EPI_ISL_987406, EPI_ISL_987411, EPI_ISL_987414, EPI_ISL_987415, EPI_ISL_987418, EPI_ISL_987419, EPI_ISL_987424, EPI_ISL_987426, EPI_ISL_987427, EPI_ISL_987428, EPI_ISL_987429, EPI_ISL_987435, EPI_ISL_987441, EPI_ISL_987443, EPI_ISL_987445, EPI_ISL_987446, EPI_ISL_987452, EPI_ISL_987453, EPI_ISL_987455, EPI_ISL_987464, EPI_ISL_987465, EPI_ISL_987474, EPI_ISL_987475, EPI_ISL_987479, EPI_ISL_987484, EPI_ISL_987488, EPI_ISL_987498, EPI_ISL_987499, EPI_ISL_987506, EPI_ISL_987509, EPI_ISL_987510, EPI_ISL_987514, EPI_ISL_987530, EPI_ISL_987547, EPI_ISL_987557, EPI_ISL_987560, EPI_ISL_987567, EPI_ISL_987573, EPI_ISL_987575, EPI_ISL_987577, EPI_ISL_987587, EPI_ISL_987589, EPI_ISL_987591, EPI_ISL_987594, EPI_ISL_987597, EPI_ISL_987598, EPI_ISL_987602, EPI_ISL_987609, EPI_ISL_987610, EPI_ISL_987612, EPI_ISL_987622, EPI_ISL_987624, EPI_ISL_987625, EPI_ISL_987627, EPI_ISL_987628, EPI_ISL_987631, EPI_ISL_987633, EPI_ISL_987637, EPI_ISL_993736, EPI_ISL_993738, EPI_ISL_993739, EPI_ISL_993740, EPI_ISL_993741, EPI_ISL_993742, EPI_ISL_993744, EPI_ISL_993746, EPI_ISL_993747, EPI_ISL_993750, EPI_ISL_993754, EPI_ISL_993756, EPI_ISL_993757, EPI_ISL_993758, EPI_ISL_993761, EPI_ISL_993762, EPI_ISL_993766, EPI_ISL_993770, EPI_ISL_993771, EPI_ISL_993774, EPI_ISL_993776, EPI_ISL_993778, EPI_ISL_993779, EPI_ISL_993782, EPI_ISL_993784, EPI_ISL_993791, EPI_ISL_993792, EPI_ISL_993794, EPI_ISL_993797, EPI_ISL_993798, EPI_ISL_993802, EPI_ISL_993803, EPI_ISL_993804, EPI_ISL_993807, EPI_ISL_993808, EPI_ISL_993809, EPI_ISL_993810, EPI_ISL_993811, EPI_ISL_993813, EPI_ISL_993814, EPI_ISL_993816, EPI_ISL_993819, EPI_ISL_993821, EPI_ISL_993824, EPI_ISL_993827, EPI_ISL_993830, EPI_ISL_993832, EPI_ISL_993836, EPI_ISL_993837, EPI_ISL_993838, EPI_ISL_993843, EPI_ISL_993847, EPI_ISL_993849, EPI_ISL_993850, EPI_ISL_993853, EPI_ISL_993854, EPI_ISL_993855, EPI_ISL_993856, EPI_ISL_993859, EPI_ISL_993863, EPI_ISL_993864, EPI_ISL_993865, EPI_ISL_993866, EPI_ISL_993867, EPI_ISL_993868, EPI_ISL_993869, EPI_ISL_993871, EPI_ISL_993872, EPI_ISL_993873, EPI_ISL_993878, EPI_ISL_993879, EPI_ISL_993881, EPI_ISL_993887, EPI_ISL_993891, EPI_ISL_993893, EPI_ISL_993894, EPI_ISL_993895, EPI_ISL_993900, EPI_ISL_993903, EPI_ISL_993906, EPI_ISL_993909, EPI_ISL_993911, EPI_ISL_993912, EPI_ISL_993921, EPI_ISL_993922, EPI_ISL_993926, EPI_ISL_993927, EPI_ISL_993937, EPI_ISL_993942, EPI_ISL_993945, EPI_ISL_993946, EPI_ISL_993949, EPI_ISL_993954, EPI_ISL_993956, EPI_ISL_993957, EPI_ISL_993960, EPI_ISL_993961, EPI_ISL_993963, EPI_ISL_993966, EPI_ISL_993968, EPI_ISL_993969, EPI_ISL_993972, EPI_ISL_993973, EPI_ISL_993975, EPI_ISL_993976, EPI_ISL_993979, EPI_ISL_993982, EPI_ISL_993983, EPI_ISL_993984, EPI_ISL_993987, EPI_ISL_993992, EPI_ISL_993994, EPI_ISL_993996, EPI_ISL_993997, EPI_ISL_994000, EPI_ISL_994001, EPI_ISL_994004, EPI_ISL_994006, EPI_ISL_994007, EPI_ISL_994008, EPI_ISL_994010, EPI_ISL_994011, EPI_ISL_994014, EPI_ISL_994015, EPI_ISL_994018, EPI_ISL_994019, EPI_ISL_994020, EPI_ISL_994022, EPI_ISL_994024, EPI_ISL_994025, EPI_ISL_994028, EPI_ISL_994031, EPI_ISL_994032, EPI_ISL_994033, EPI_ISL_994040, EPI_ISL_994042, EPI_ISL_994044, EPI_ISL_994045, EPI_ISL_994046, EPI_ISL_994048, EPI_ISL_994050, EPI_ISL_994052, EPI_ISL_994053, EPI_ISL_994055, EPI_ISL_994058, EPI_ISL_994060, EPI_ISL_994061, EPI_ISL_994064, EPI_ISL_994066, EPI_ISL_1007433

see above | Lighthouse Lab in Cambridge | Wellcome Sanger Institute for the COVID-19 Genomics UK (COG-UK) Consortium | Cordelia Langford; David K. Jackson; Dominic Kwiatkowski; Ewan Harrison; Ian Johnston; John Sillitoe on behalf of the Wellcome Sanger Institute COVID-19 Surveillance Team; John Sillitoe on behalf of the Wellcome Sanger Institute COVID-19 Surveillance Team (http://www.sanger.ac.uk/covid-team); Rob Howes; Roberto Amato; Sonia Goncalves; The Lighthouse Lab in Cambridge and Alex Alderton

EPI_ISL_863245, EPI_ISL_863246, EPI_ISL_863247, EPI_ISL_863248, EPI_ISL_863249, EPI_ISL_863250, EPI_ISL_863252, EPI_ISL_863253, EPI_ISL_863255, EPI_ISL_863256, EPI_ISL_863257, EPI_ISL_863259, EPI_ISL_863260, EPI_ISL_863261, EPI_ISL_863262, EPI_ISL_863263, EPI_ISL_863264, EPI_ISL_863265, EPI_ISL_863266, EPI_ISL_863267, EPI_ISL_863269, EPI_ISL_863273, EPI_ISL_863276, EPI_ISL_863277, EPI_ISL_863278, EPI_ISL_863279, EPI_ISL_863282, EPI_ISL_863283, EPI_ISL_863284, EPI_ISL_863285, EPI_ISL_863286, EPI_ISL_863287, EPI_ISL_863288, EPI_ISL_863291, EPI_ISL_863292, EPI_ISL_863293, EPI_ISL_863294, EPI_ISL_863295, EPI_ISL_863296, EPI_ISL_863297, EPI_ISL_863298, EPI_ISL_863300, EPI_ISL_863301, EPI_ISL_863303, EPI_ISL_863304, EPI_ISL_863305, EPI_ISL_863307, EPI_ISL_863308, EPI_ISL_863309, EPI_ISL_863310, EPI_ISL_863312, EPI_ISL_863313, EPI_ISL_863315, EPI_ISL_863316, EPI_ISL_863317, EPI_ISL_863320, EPI_ISL_863321, EPI_ISL_863322, EPI_ISL_863323, EPI_ISL_863324, EPI_ISL_863325, EPI_ISL_863327, EPI_ISL_863328, EPI_ISL_863329, EPI_ISL_863330, EPI_ISL_863331, EPI_ISL_863332, EPI_ISL_863333, EPI_ISL_863335, EPI_ISL_863336, EPI_ISL_863337, EPI_ISL_863339, EPI_ISL_863340, EPI_ISL_863341, EPI_ISL_863344, EPI_ISL_863345, EPI_ISL_863347, EPI_ISL_863348, EPI_ISL_863349, EPI_ISL_863350, EPI_ISL_863351, EPI_ISL_863352, EPI_ISL_863353, EPI_ISL_863354, EPI_ISL_863355, EPI_ISL_863358, EPI_ISL_863359, EPI_ISL_863361, EPI_ISL_863365, EPI_ISL_863366, EPI_ISL_863367, EPI_ISL_863368, EPI_ISL_863369, EPI_ISL_863370, EPI_ISL_863371, EPI_ISL_863372, EPI_ISL_863373, EPI_ISL_863374, EPI_ISL_863375, EPI_ISL_863376, EPI_ISL_863378, EPI_ISL_863380, EPI_ISL_863381, EPI_ISL_863382, EPI_ISL_863384, EPI_ISL_863385, EPI_ISL_863386, EPI_ISL_863388, EPI_ISL_863389, EPI_ISL_863390, EPI_ISL_863391, EPI_ISL_863392, EPI_ISL_863393, EPI_ISL_863394, EPI_ISL_863398, EPI_ISL_863399, EPI_ISL_863400, EPI_ISL_863402, EPI_ISL_863405, EPI_ISL_863407, EPI_ISL_863409, EPI_ISL_863410, EPI_ISL_863412, EPI_ISL_863413, EPI_ISL_863415, EPI_ISL_863417, EPI_ISL_863418, EPI_ISL_863419, EPI_ISL_863422, EPI_ISL_863425, EPI_ISL_863427, EPI_ISL_863428, EPI_ISL_863429, EPI_ISL_863430, EPI_ISL_863433, EPI_ISL_863435, EPI_ISL_863436, EPI_ISL_863438, EPI_ISL_863439, EPI_ISL_863440, EPI_ISL_863441, EPI_ISL_863442, EPI_ISL_863444, EPI_ISL_863445, EPI_ISL_863449, EPI_ISL_863450, EPI_ISL_863451, EPI_ISL_863452, EPI_ISL_863455, EPI_ISL_863456, EPI_ISL_863457, EPI_ISL_863458, EPI_ISL_863459, EPI_ISL_863460, EPI_ISL_863462, EPI_ISL_863463, EPI_ISL_863464, EPI_ISL_863465, EPI_ISL_863466, EPI_ISL_863468, EPI_ISL_863469, EPI_ISL_863471, EPI_ISL_863472, EPI_ISL_863473, EPI_ISL_863474, EPI_ISL_863475, EPI_ISL_863476, EPI_ISL_863478, EPI_ISL_863480, EPI_ISL_863481, EPI_ISL_863482, EPI_ISL_863483, EPI_ISL_863485, EPI_ISL_863486, EPI_ISL_863487, EPI_ISL_863488, EPI_ISL_863489, EPI_ISL_863490, EPI_ISL_863492, EPI_ISL_863493, EPI_ISL_863494, EPI_ISL_863495, EPI_ISL_863496, EPI_ISL_863497, EPI_ISL_863498, EPI_ISL_863499, EPI_ISL_863500, EPI_ISL_863501, EPI_ISL_863502, EPI_ISL_863505, EPI_ISL_863507, EPI_ISL_863508, EPI_ISL_863509, EPI_ISL_863510, EPI_ISL_863511, EPI_ISL_863512, EPI_ISL_863513, EPI_ISL_863517, EPI_ISL_863518, EPI_ISL_863521, EPI_ISL_863522, EPI_ISL_863523, EPI_ISL_863525, EPI_ISL_863526, EPI_ISL_863528, EPI_ISL_863530, EPI_ISL_874656, EPI_ISL_874676, EPI_ISL_874733, EPI_ISL_874736, EPI_ISL_874737, EPI_ISL_874739, EPI_ISL_874753, EPI_ISL_874775, EPI_ISL_874798, EPI_ISL_874806, EPI_ISL_874808, EPI_ISL_874863, EPI_ISL_874896, EPI_ISL_874898, EPI_ISL_874905, EPI_ISL_874922, EPI_ISL_874935, EPI_ISL_874943, EPI_ISL_874950, EPI_ISL_874965, EPI_ISL_874967, EPI_ISL_882609, EPI_ISL_891700, EPI_ISL_892201, EPI_ISL_916625, EPI_ISL_916626, EPI_ISL_916633, EPI_ISL_916645, EPI_ISL_916648, EPI_ISL_916649, EPI_ISL_916661, EPI_ISL_916662, EPI_ISL_916668, EPI_ISL_916675, EPI_ISL_916682, EPI_ISL_916684, EPI_ISL_916688, EPI_ISL_916694, EPI_ISL_916696, EPI_ISL_916704, EPI_ISL_916705, EPI_ISL_916711, EPI_ISL_916713, EPI_ISL_916716, EPI_ISL_916728, EPI_ISL_916730, EPI_ISL_916736, EPI_ISL_916738, EPI_ISL_916758, EPI_ISL_916761, EPI_ISL_916771, EPI_ISL_916782, EPI_ISL_916784, EPI_ISL_916793, EPI_ISL_916800, EPI_ISL_916802, EPI_ISL_916809, EPI_ISL_916810, EPI_ISL_916812, EPI_ISL_916815, EPI_ISL_916820, EPI_ISL_916821, EPI_ISL_916825, EPI_ISL_916833, EPI_ISL_916834, EPI_ISL_916838, EPI_ISL_916840, EPI_ISL_916842, EPI_ISL_916845, EPI_ISL_916847, EPI_ISL_916850, EPI_ISL_916855, EPI_ISL_916869, EPI_ISL_916879, EPI_ISL_916882, EPI_ISL_916887, EPI_ISL_916891, EPI_ISL_916894, EPI_ISL_916896, EPI_ISL_916898, EPI_ISL_916899, EPI_ISL_916900, EPI_ISL_916905, EPI_ISL_916907, EPI_ISL_916908, EPI_ISL_916909, EPI_ISL_916911, EPI_ISL_916912, EPI_ISL_916916, EPI_ISL_916918, EPI_ISL_916923, EPI_ISL_916940, EPI_ISL_916942, EPI_ISL_916945, EPI_ISL_916946, EPI_ISL_916948, EPI_ISL_916949, EPI_ISL_916951, EPI_ISL_916953, EPI_ISL_916954, EPI_ISL_916956, EPI_ISL_916961, EPI_ISL_916967, EPI_ISL_916968, EPI_ISL_916970, EPI_ISL_916974, EPI_ISL_916976, EPI_ISL_916977, EPI_ISL_916979, EPI_ISL_916981, EPI_ISL_916984, EPI_ISL_916985, EPI_ISL_916987, EPI_ISL_916991, EPI_ISL_916998, EPI_ISL_916999, EPI_ISL_917002, EPI_ISL_917009, EPI_ISL_917012, EPI_ISL_917013, EPI_ISL_917014, EPI_ISL_917019, EPI_ISL_917021, EPI_ISL_917022, EPI_ISL_917024, EPI_ISL_917025, EPI_ISL_917026, EPI_ISL_917029, EPI_ISL_917031, EPI_ISL_917032, EPI_ISL_917036, EPI_ISL_917038, EPI_ISL_917042, EPI_ISL_917047, EPI_ISL_917049, EPI_ISL_917053, EPI_ISL_917054, EPI_ISL_917055, EPI_ISL_917063, EPI_ISL_917067, EPI_ISL_917070, EPI_ISL_917071, EPI_ISL_917074, EPI_ISL_917075, EPI_ISL_917076, EPI_ISL_917077, EPI_ISL_917079, EPI_ISL_917081, EPI_ISL_917082, EPI_ISL_917083, EPI_ISL_917086, EPI_ISL_917088, EPI_ISL_917091, EPI_ISL_917093, EPI_ISL_917096, EPI_ISL_917099, EPI_ISL_917104, EPI_ISL_917105, EPI_ISL_917107, EPI_ISL_917110, EPI_ISL_917112, EPI_ISL_917114, EPI_ISL_917116, EPI_ISL_917120, EPI_ISL_917125, EPI_ISL_917128, EPI_ISL_917129, EPI_ISL_917133, EPI_ISL_917139, EPI_ISL_917141, EPI_ISL_917143, EPI_ISL_917144, EPI_ISL_917145, EPI_ISL_917148, EPI_ISL_917149, EPI_ISL_917151, EPI_ISL_917156, EPI_ISL_917159, EPI_ISL_917160, EPI_ISL_917161, EPI_ISL_917163, EPI_ISL_917166, EPI_ISL_917975, EPI_ISL_918062, EPI_ISL_918063, EPI_ISL_918064, EPI_ISL_918065, EPI_ISL_918066, EPI_ISL_918067, EPI_ISL_918068, EPI_ISL_918069, EPI_ISL_918070, EPI_ISL_918071, EPI_ISL_918072, EPI_ISL_918073, EPI_ISL_918074, EPI_ISL_918075, EPI_ISL_918076, EPI_ISL_918077, EPI_ISL_918078, EPI_ISL_918079, EPI_ISL_918080, EPI_ISL_918081, EPI_ISL_918082, EPI_ISL_918083, EPI_ISL_918084, EPI_ISL_918085, EPI_ISL_918086, EPI_ISL_918087, EPI_ISL_918088, EPI_ISL_918089, EPI_ISL_918090, EPI_ISL_918091, EPI_ISL_918092, EPI_ISL_918093, EPI_ISL_918094, EPI_ISL_918095, EPI_ISL_918096, EPI_ISL_918097, EPI_ISL_918098, EPI_ISL_918099, EPI_ISL_918100, EPI_ISL_918101, EPI_ISL_918102, EPI_ISL_918103, EPI_ISL_918104, EPI_ISL_918105, EPI_ISL_918106, EPI_ISL_918107, EPI_ISL_918108, EPI_ISL_918109, EPI_ISL_918110, EPI_ISL_918111, EPI_ISL_918112, EPI_ISL_918113, EPI_ISL_918114, EPI_ISL_918115, EPI_ISL_918116, EPI_ISL_918117, EPI_ISL_918118, EPI_ISL_918119, EPI_ISL_918120, EPI_ISL_918121, EPI_ISL_918122, EPI_ISL_918123, EPI_ISL_918124, EPI_ISL_948967, EPI_ISL_948968, EPI_ISL_948969, EPI_ISL_948970, EPI_ISL_948971, EPI_ISL_948972, EPI_ISL_948974, EPI_ISL_948976, EPI_ISL_948977, EPI_ISL_948979, EPI_ISL_948980, EPI_ISL_948982, EPI_ISL_948983, EPI_ISL_948985, EPI_ISL_948987, EPI_ISL_948988, EPI_ISL_948990, EPI_ISL_948992, EPI_ISL_948993, EPI_ISL_948995, EPI_ISL_948996, EPI_ISL_948998, EPI_ISL_948999, EPI_ISL_949001, EPI_ISL_949002, EPI_ISL_949004, EPI_ISL_949006, EPI_ISL_949007, EPI_ISL_949009, EPI_ISL_949010, EPI_ISL_949012, EPI_ISL_949014, EPI_ISL_949015, EPI_ISL_949017, EPI_ISL_949018, EPI_ISL_949020, EPI_ISL_949021, EPI_ISL_949023, EPI_ISL_949025, EPI_ISL_949026, EPI_ISL_949027, EPI_ISL_949029, EPI_ISL_949030, EPI_ISL_949031, EPI_ISL_949032, EPI_ISL_949033, EPI_ISL_949034, EPI_ISL_949035, EPI_ISL_949036, EPI_ISL_949037, EPI_ISL_949038, EPI_ISL_949039, EPI_ISL_949040, EPI_ISL_949041, EPI_ISL_949042, EPI_ISL_949043, EPI_ISL_949044, EPI_ISL_949045, EPI_ISL_949046, EPI_ISL_949047, EPI_ISL_949048, EPI_ISL_949049, EPI_ISL_949050, EPI_ISL_949051, EPI_ISL_949052, EPI_ISL_949053, EPI_ISL_949054, EPI_ISL_949055, EPI_ISL_949056, EPI_ISL_949057, EPI_ISL_949058, EPI_ISL_949060, EPI_ISL_949061, EPI_ISL_949062, EPI_ISL_949063, EPI_ISL_949064, EPI_ISL_949065, EPI_ISL_949066, EPI_ISL_987300, EPI_ISL_987311, EPI_ISL_987317, EPI_ISL_987335, EPI_ISL_987373, EPI_ISL_987390, EPI_ISL_987395, EPI_ISL_987420, EPI_ISL_987422, EPI_ISL_987454, EPI_ISL_987461, EPI_ISL_987481, EPI_ISL_987491, EPI_ISL_987501, EPI_ISL_987507, EPI_ISL_987549, EPI_ISL_987552, EPI_ISL_987555, EPI_ISL_987556, EPI_ISL_987570, EPI_ISL_987572, EPI_ISL_987574, EPI_ISL_987614, EPI_ISL_1019835

see above | Lighthouse Lab in Glasgow | Wellcome Sanger Institute for the COVID-19 Genomics UK (COG-UK) Consortium | Anna Dominiczak and Alex Alderton; Carol Clugston; Cordelia Langford; David Gray; David K. Jackson; Dominic Kwiatkowski; Ewan Harrison; Harper VanSteenhouse; Ian Johnston; Jeffrey Barrett; John Sillitoe on behalf of the Wellcome Sanger Institute COVID-19 Surveillance Team; John Sillitoe on behalf of the Wellcome Sanger Institute COVID-19 Surveillance Team (http://www.sanger.ac.uk/covid-team); Roberto Amato; Sonia Goncalves; Yumi Kasai

EPI_ISL_852014, EPI_ISL_873281, EPI_ISL_873294, EPI_ISL_873298, EPI_ISL_873311, EPI_ISL_873321, EPI_ISL_873330, EPI_ISL_873336, EPI_ISL_873354, EPI_ISL_873361, EPI_ISL_873376, EPI_ISL_873378, EPI_ISL_873386, EPI_ISL_873389, EPI_ISL_873396, EPI_ISL_873411, EPI_ISL_873413, EPI_ISL_873414, EPI_ISL_873415, EPI_ISL_873420, EPI_ISL_873429, EPI_ISL_873430, EPI_ISL_873435, EPI_ISL_873438, EPI_ISL_873439, EPI_ISL_873444, EPI_ISL_873445, EPI_ISL_873450, EPI_ISL_873455, EPI_ISL_873462, EPI_ISL_873466, EPI_ISL_873473, EPI_ISL_873487, EPI_ISL_873493, EPI_ISL_873496, EPI_ISL_873503, EPI_ISL_873504, EPI_ISL_873509, EPI_ISL_873515, EPI_ISL_873520, EPI_ISL_873531, EPI_ISL_873551, EPI_ISL_873585, EPI_ISL_873596, EPI_ISL_873600, EPI_ISL_873603, EPI_ISL_873604, EPI_ISL_873607, EPI_ISL_873645, EPI_ISL_873652, EPI_ISL_873679, EPI_ISL_873716, EPI_ISL_873748, EPI_ISL_873755, EPI_ISL_873783, EPI_ISL_873797, EPI_ISL_873822, EPI_ISL_873827, EPI_ISL_873829, EPI_ISL_873840, EPI_ISL_873842, EPI_ISL_873853, EPI_ISL_873860, EPI_ISL_873862, EPI_ISL_873881, EPI_ISL_873886, EPI_ISL_873921, EPI_ISL_873926, EPI_ISL_873942, EPI_ISL_873950, EPI_ISL_873956, EPI_ISL_878751, EPI_ISL_878756, EPI_ISL_878759, EPI_ISL_878772, EPI_ISL_878775, EPI_ISL_878778, EPI_ISL_878780, EPI_ISL_878782, EPI_ISL_878792, EPI_ISL_878793, EPI_ISL_878798, EPI_ISL_878800, EPI_ISL_878806, EPI_ISL_878807, EPI_ISL_878814, EPI_ISL_878824, EPI_ISL_878842, EPI_ISL_878845, EPI_ISL_878846, EPI_ISL_878848, EPI_ISL_878851, EPI_ISL_878852, EPI_ISL_878854, EPI_ISL_878858, EPI_ISL_878860, EPI_ISL_878861, EPI_ISL_878874, EPI_ISL_878879, EPI_ISL_878880, EPI_ISL_878882, EPI_ISL_878884, EPI_ISL_878892, EPI_ISL_878896, EPI_ISL_878902, EPI_ISL_878906, EPI_ISL_878918, EPI_ISL_878934, EPI_ISL_878941, EPI_ISL_878942, EPI_ISL_878943, EPI_ISL_878944, EPI_ISL_878945, EPI_ISL_878951, EPI_ISL_878952, EPI_ISL_878960, EPI_ISL_878962, EPI_ISL_878967, EPI_ISL_878971, EPI_ISL_878972, EPI_ISL_878982, EPI_ISL_878990, EPI_ISL_879001, EPI_ISL_879009, EPI_ISL_879013, EPI_ISL_879014, EPI_ISL_879016, EPI_ISL_879019, EPI_ISL_879021, EPI_ISL_879023, EPI_ISL_879025, EPI_ISL_879028, EPI_ISL_879032, EPI_ISL_879034, EPI_ISL_879037, EPI_ISL_879038, EPI_ISL_879042, EPI_ISL_879043, EPI_ISL_879044, EPI_ISL_879047, EPI_ISL_879049, EPI_ISL_879050, EPI_ISL_879052, EPI_ISL_879055, EPI_ISL_879057, EPI_ISL_879063, EPI_ISL_879064, EPI_ISL_879065, EPI_ISL_879075, EPI_ISL_879086, EPI_ISL_879087, EPI_ISL_879089, EPI_ISL_879093, EPI_ISL_879094, EPI_ISL_879096, EPI_ISL_879378, EPI_ISL_879379, EPI_ISL_879380, EPI_ISL_879388, EPI_ISL_879395, EPI_ISL_879396, EPI_ISL_879397, EPI_ISL_879399, EPI_ISL_879404, EPI_ISL_879409, EPI_ISL_879412, EPI_ISL_879413, EPI_ISL_879424, EPI_ISL_879425, EPI_ISL_879426, EPI_ISL_879429, EPI_ISL_879442, EPI_ISL_879448, EPI_ISL_879449, EPI_ISL_879450, EPI_ISL_879455, EPI_ISL_879457, EPI_ISL_879458, EPI_ISL_879459, EPI_ISL_879461, EPI_ISL_879464, EPI_ISL_879467, EPI_ISL_879471, EPI_ISL_879477, EPI_ISL_879479, EPI_ISL_879492, EPI_ISL_879495, EPI_ISL_879499, EPI_ISL_879502, EPI_ISL_879514, EPI_ISL_879526, EPI_ISL_879532, EPI_ISL_879538, EPI_ISL_879539, EPI_ISL_879540, EPI_ISL_879548, EPI_ISL_879549, EPI_ISL_879552, EPI_ISL_879553, EPI_ISL_879554, EPI_ISL_879560, EPI_ISL_879566, EPI_ISL_879571, EPI_ISL_879572, EPI_ISL_879575, EPI_ISL_879576, EPI_ISL_879581, EPI_ISL_879585, EPI_ISL_879586, EPI_ISL_879591, EPI_ISL_879592, EPI_ISL_879594, EPI_ISL_879597, EPI_ISL_879600, EPI_ISL_879601, EPI_ISL_879608, EPI_ISL_879611, EPI_ISL_879613, EPI_ISL_879618, EPI_ISL_879621, EPI_ISL_879623, EPI_ISL_879628, EPI_ISL_879642, EPI_ISL_879643, EPI_ISL_879653, EPI_ISL_879654, EPI_ISL_879655, EPI_ISL_879656, EPI_ISL_879663, EPI_ISL_879664, EPI_ISL_879675, EPI_ISL_879678, EPI_ISL_879679, EPI_ISL_879681, EPI_ISL_879686, EPI_ISL_879689, EPI_ISL_879703, EPI_ISL_879707, EPI_ISL_879710, EPI_ISL_879711, EPI_ISL_879712, EPI_ISL_879713, EPI_ISL_879715, EPI_ISL_879717, EPI_ISL_880584, EPI_ISL_880585, EPI_ISL_880587, EPI_ISL_880588, EPI_ISL_880589, EPI_ISL_880592, EPI_ISL_880593, EPI_ISL_880594, EPI_ISL_880597, EPI_ISL_880598, EPI_ISL_880600, EPI_ISL_880603, EPI_ISL_880604, EPI_ISL_880605, EPI_ISL_880607, EPI_ISL_880612, EPI_ISL_880614, EPI_ISL_880617, EPI_ISL_880621, EPI_ISL_880622, EPI_ISL_880624, EPI_ISL_880625, EPI_ISL_880628, EPI_ISL_880630, EPI_ISL_880631, EPI_ISL_880632, EPI_ISL_880633, EPI_ISL_880634, EPI_ISL_880635, EPI_ISL_880636, EPI_ISL_880638, EPI_ISL_880640, EPI_ISL_880643, EPI_ISL_880644, EPI_ISL_880645, EPI_ISL_880646, EPI_ISL_880648, EPI_ISL_880649, EPI_ISL_880650, EPI_ISL_880652, EPI_ISL_880656, EPI_ISL_880658, EPI_ISL_880659, EPI_ISL_880661, EPI_ISL_880663, EPI_ISL_880664, EPI_ISL_880666, EPI_ISL_880670, EPI_ISL_880671, EPI_ISL_880676, EPI_ISL_880678, EPI_ISL_880680, EPI_ISL_880681, EPI_ISL_880683, EPI_ISL_880684, EPI_ISL_880685, EPI_ISL_880686, EPI_ISL_880687, EPI_ISL_880688, EPI_ISL_880689, EPI_ISL_880690, EPI_ISL_880691, EPI_ISL_880692, EPI_ISL_880693, EPI_ISL_880694, EPI_ISL_880696, EPI_ISL_880697, EPI_ISL_880698, EPI_ISL_880699, EPI_ISL_880700, EPI_ISL_880702, EPI_ISL_880704, EPI_ISL_880707, EPI_ISL_880711, EPI_ISL_880712, EPI_ISL_880714, EPI_ISL_880715, EPI_ISL_880716, EPI_ISL_880719, EPI_ISL_880720, EPI_ISL_880723, EPI_ISL_880728, EPI_ISL_880730, EPI_ISL_880731, EPI_ISL_880732, EPI_ISL_880733, EPI_ISL_880734, EPI_ISL_880735, EPI_ISL_880741, EPI_ISL_880743, EPI_ISL_880744, EPI_ISL_880747, EPI_ISL_880748, EPI_ISL_880749, EPI_ISL_880750, EPI_ISL_880754, EPI_ISL_880756, EPI_ISL_880757, EPI_ISL_880758, EPI_ISL_880760, EPI_ISL_880761, EPI_ISL_880764, EPI_ISL_880765, EPI_ISL_880767, EPI_ISL_880768, EPI_ISL_880770, EPI_ISL_880771, EPI_ISL_880772, EPI_ISL_880773, EPI_ISL_880777, EPI_ISL_880778, EPI_ISL_880780, EPI_ISL_880782, EPI_ISL_880783, EPI_ISL_880784, EPI_ISL_880787, EPI_ISL_880788, EPI_ISL_880790, EPI_ISL_880793, EPI_ISL_880794, EPI_ISL_880795, EPI_ISL_880796, EPI_ISL_880801, EPI_ISL_880802, EPI_ISL_880804, EPI_ISL_880805, EPI_ISL_880806, EPI_ISL_880807, EPI_ISL_880808, EPI_ISL_880810, EPI_ISL_880811, EPI_ISL_880814, EPI_ISL_880815, EPI_ISL_880816, EPI_ISL_880820, EPI_ISL_880821, EPI_ISL_880823, EPI_ISL_880825, EPI_ISL_880827, EPI_ISL_880828, EPI_ISL_880832, EPI_ISL_880834, EPI_ISL_880837, EPI_ISL_880842, EPI_ISL_880844, EPI_ISL_880845, EPI_ISL_880846, EPI_ISL_880848, EPI_ISL_880849, EPI_ISL_880850, EPI_ISL_880851, EPI_ISL_880852, EPI_ISL_880855, EPI_ISL_880859, EPI_ISL_880860, EPI_ISL_880861, EPI_ISL_880863, EPI_ISL_880864, EPI_ISL_880865, EPI_ISL_880867, EPI_ISL_880869, EPI_ISL_880871, EPI_ISL_880872, EPI_ISL_880873, EPI_ISL_880874, EPI_ISL_880875, EPI_ISL_880876, EPI_ISL_880877, EPI_ISL_880880, EPI_ISL_880881, EPI_ISL_880882, EPI_ISL_880885, EPI_ISL_880887, EPI_ISL_880888, EPI_ISL_880890, EPI_ISL_880891, EPI_ISL_880893, EPI_ISL_880896, EPI_ISL_880897, EPI_ISL_880899, EPI_ISL_880902, EPI_ISL_880904, EPI_ISL_880905, EPI_ISL_880906, EPI_ISL_880907, EPI_ISL_880912, EPI_ISL_880913, EPI_ISL_880917, EPI_ISL_880918, EPI_ISL_880919, EPI_ISL_880920, EPI_ISL_880922, EPI_ISL_880923, EPI_ISL_880924, EPI_ISL_880926, EPI_ISL_880927, EPI_ISL_880928, EPI_ISL_880929, EPI_ISL_880930, EPI_ISL_880931, EPI_ISL_880932, EPI_ISL_908934, EPI_ISL_908936, EPI_ISL_908943, EPI_ISL_908960, EPI_ISL_908966, EPI_ISL_908969, EPI_ISL_908971, EPI_ISL_908972, EPI_ISL_908973, EPI_ISL_908975, EPI_ISL_908976, EPI_ISL_908980, EPI_ISL_908981, EPI_ISL_908984, EPI_ISL_908994, EPI_ISL_909000, EPI_ISL_909001, EPI_ISL_909013, EPI_ISL_909020, EPI_ISL_909025, EPI_ISL_909028, EPI_ISL_909032, EPI_ISL_909033, EPI_ISL_909039, EPI_ISL_909042, EPI_ISL_909045, EPI_ISL_909055, EPI_ISL_909064, EPI_ISL_909073, EPI_ISL_909074, EPI_ISL_909077, EPI_ISL_909092, EPI_ISL_909098, EPI_ISL_909100, EPI_ISL_909104, EPI_ISL_909112, EPI_ISL_909115, EPI_ISL_909126, EPI_ISL_909134, EPI_ISL_909135, EPI_ISL_909138, EPI_ISL_909140, EPI_ISL_909141, EPI_ISL_909142, EPI_ISL_909143, EPI_ISL_909151, EPI_ISL_909161, EPI_ISL_909163, EPI_ISL_909164, EPI_ISL_909168, EPI_ISL_909175, EPI_ISL_909183, EPI_ISL_909188, EPI_ISL_909190, EPI_ISL_909196, EPI_ISL_909199, EPI_ISL_909209, EPI_ISL_909213, EPI_ISL_909221, EPI_ISL_909225, EPI_ISL_909229, EPI_ISL_909232, EPI_ISL_909239, EPI_ISL_909244, EPI_ISL_909249, EPI_ISL_909253, EPI_ISL_909258, EPI_ISL_909264, EPI_ISL_909404, EPI_ISL_909495, EPI_ISL_912225, EPI_ISL_987330, EPI_ISL_987331, EPI_ISL_987334, EPI_ISL_987345, EPI_ISL_987355, EPI_ISL_987356, EPI_ISL_987377, EPI_ISL_987438, EPI_ISL_987444, EPI_ISL_987457, EPI_ISL_987516, EPI_ISL_987517, EPI_ISL_987522, EPI_ISL_987544, EPI_ISL_987548, EPI_ISL_987585, EPI_ISL_987593, EPI_ISL_987636, EPI_ISL_993085, EPI_ISL_993087, EPI_ISL_993088, EPI_ISL_993090, EPI_ISL_993092, EPI_ISL_993093, EPI_ISL_993097, EPI_ISL_993102, EPI_ISL_993103, EPI_ISL_993107, EPI_ISL_993115, EPI_ISL_993120, EPI_ISL_993121, EPI_ISL_993124, EPI_ISL_993126, EPI_ISL_993130, EPI_ISL_993139, EPI_ISL_993142, EPI_ISL_993144, EPI_ISL_993145, EPI_ISL_993147, EPI_ISL_993148, EPI_ISL_993150, EPI_ISL_993151, EPI_ISL_993156, EPI_ISL_993159, EPI_ISL_993167, EPI_ISL_993170, EPI_ISL_993172, EPI_ISL_993173, EPI_ISL_993175, EPI_ISL_993179, EPI_ISL_993181, EPI_ISL_993186, EPI_ISL_993188, EPI_ISL_993191, EPI_ISL_993193, EPI_ISL_993195, EPI_ISL_993200, EPI_ISL_993204, EPI_ISL_993210, EPI_ISL_993214, EPI_ISL_993215, EPI_ISL_993218, EPI_ISL_993220, EPI_ISL_993226, EPI_ISL_993228, EPI_ISL_993229, EPI_ISL_993230, EPI_ISL_993243, EPI_ISL_993246, EPI_ISL_993250, EPI_ISL_993253, EPI_ISL_993254, EPI_ISL_993256, EPI_ISL_993259, EPI_ISL_993260, EPI_ISL_993262, EPI_ISL_993265, EPI_ISL_993267, EPI_ISL_993268, EPI_ISL_993269, EPI_ISL_993271, EPI_ISL_993272, EPI_ISL_993274, EPI_ISL_993276, EPI_ISL_993278, EPI_ISL_993281, EPI_ISL_993284, EPI_ISL_993286, EPI_ISL_993288, EPI_ISL_993289, EPI_ISL_993290, EPI_ISL_993295, EPI_ISL_993298, EPI_ISL_993299, EPI_ISL_993303, EPI_ISL_993304, EPI_ISL_993307, EPI_ISL_993308, EPI_ISL_993314, EPI_ISL_993318, EPI_ISL_993319, EPI_ISL_993322, EPI_ISL_993323, EPI_ISL_993330, EPI_ISL_993332, EPI_ISL_993333, EPI_ISL_993339, EPI_ISL_993347, EPI_ISL_993349, EPI_ISL_993355, EPI_ISL_993356, EPI_ISL_993357, EPI_ISL_993358, EPI_ISL_993359, EPI_ISL_993366, EPI_ISL_993372, EPI_ISL_993373, EPI_ISL_993375, EPI_ISL_993376, EPI_ISL_993380, EPI_ISL_993382, EPI_ISL_993387, EPI_ISL_993428, EPI_ISL_993429, EPI_ISL_993445, EPI_ISL_993460, EPI_ISL_993470, EPI_ISL_993502, EPI_ISL_993525, EPI_ISL_993559, EPI_ISL_993569, EPI_ISL_993577, EPI_ISL_993587, EPI_ISL_993602, EPI_ISL_993615, EPI_ISL_993618, EPI_ISL_993624, EPI_ISL_993625, EPI_ISL_993648, EPI_ISL_993649, EPI_ISL_993662, EPI_ISL_993666, EPI_ISL_993672, EPI_ISL_993675, EPI_ISL_993702, EPI_ISL_993703, EPI_ISL_993733, EPI_ISL_993737, EPI_ISL_993745, EPI_ISL_993749, EPI_ISL_993751, EPI_ISL_993752, EPI_ISL_993753, EPI_ISL_993755, EPI_ISL_993759, EPI_ISL_993760, EPI_ISL_993763, EPI_ISL_993764, EPI_ISL_993765, EPI_ISL_993767, EPI_ISL_993768, EPI_ISL_993769, EPI_ISL_993772, EPI_ISL_993773, EPI_ISL_993775, EPI_ISL_993777, EPI_ISL_993780, EPI_ISL_993781, EPI_ISL_993783, EPI_ISL_993785, EPI_ISL_993786, EPI_ISL_993787, EPI_ISL_993788, EPI_ISL_993789, EPI_ISL_993795, EPI_ISL_993796, EPI_ISL_993799, EPI_ISL_993800, EPI_ISL_993801, EPI_ISL_993805, EPI_ISL_993806, EPI_ISL_993812, EPI_ISL_993815, EPI_ISL_993818, EPI_ISL_993820, EPI_ISL_993822, EPI_ISL_993823, EPI_ISL_993825, EPI_ISL_993826, EPI_ISL_993828, EPI_ISL_993829, EPI_ISL_993831, EPI_ISL_993834, EPI_ISL_993835, EPI_ISL_993839, EPI_ISL_993840, EPI_ISL_993841, EPI_ISL_993844, EPI_ISL_993845, EPI_ISL_993846, EPI_ISL_993848, EPI_ISL_993851, EPI_ISL_993852, EPI_ISL_993857, EPI_ISL_993858, EPI_ISL_993860, EPI_ISL_993861, EPI_ISL_993862, EPI_ISL_993870, EPI_ISL_993874, EPI_ISL_993875, EPI_ISL_993876, EPI_ISL_993877, EPI_ISL_993880, EPI_ISL_993882, EPI_ISL_993883, EPI_ISL_993884, EPI_ISL_993885, EPI_ISL_993886, EPI_ISL_993888, EPI_ISL_993889, EPI_ISL_993891, EPI_ISL_993896, EPI_ISL_993897, EPI_ISL_993898, EPI_ISL_993899, EPI_ISL_993901, EPI_ISL_993902, EPI_ISL_993904, EPI_ISL_993905, EPI_ISL_993908, EPI_ISL_993910, EPI_ISL_993913, EPI_ISL_993917, EPI_ISL_993918, EPI_ISL_993919, EPI_ISL_993923, EPI_ISL_993924, EPI_ISL_993925, EPI_ISL_993928, EPI_ISL_993929, EPI_ISL_993931, EPI_ISL_993932, EPI_ISL_993933, EPI_ISL_993934, EPI_ISL_993935, EPI_ISL_993936, EPI_ISL_993938, EPI_ISL_993939, EPI_ISL_993940, EPI_ISL_993941, EPI_ISL_993943, EPI_ISL_993944, EPI_ISL_993947, EPI_ISL_993948, EPI_ISL_993950, EPI_ISL_993951, EPI_ISL_993952, EPI_ISL_993953, EPI_ISL_993958, EPI_ISL_993959, EPI_ISL_993964, EPI_ISL_993965, EPI_ISL_993967, EPI_ISL_993970, EPI_ISL_993971, EPI_ISL_993974, EPI_ISL_993977, EPI_ISL_993978, EPI_ISL_993980, EPI_ISL_993981, EPI_ISL_993985, EPI_ISL_993986, EPI_ISL_993988, EPI_ISL_993989, EPI_ISL_993990, EPI_ISL_993991, EPI_ISL_993993, EPI_ISL_993995, EPI_ISL_993998, EPI_ISL_993999, EPI_ISL_994002, EPI_ISL_994005, EPI_ISL_994009, EPI_ISL_994012, EPI_ISL_994013, EPI_ISL_994016, EPI_ISL_994017, EPI_ISL_994021, EPI_ISL_994023, EPI_ISL_994026, EPI_ISL_994027, EPI_ISL_994029, EPI_ISL_994030, EPI_ISL_994035, EPI_ISL_994036, EPI_ISL_994037, EPI_ISL_994038, EPI_ISL_994039, EPI_ISL_994041, EPI_ISL_994043, EPI_ISL_994047, EPI_ISL_994049, EPI_ISL_994051, EPI_ISL_994054, EPI_ISL_994057, EPI_ISL_994062, EPI_ISL_994063, EPI_ISL_994065, EPI_ISL_1007575 see above — Lighthouse Lab in Milton Keynes — Wellcome Sanger Institute for the COVID-19 Genomics UK (COG-UK) Consortium — Cordelia Langford; David K. Jackson; Dominic Kwiatkowski; Ewan Harrison; Ian Johnston; John Sillitoe on behalf of the Wellcome Sanger Institute COVID-19 Surveillance Team; John Sillitoe on behalf of the Wellcome Sanger Institute COVID-19 Surveillance Team (http://www.sanger.ac.uk/covid-team); Roberto Amato; Sonia Goncalves; The Lighthouse Lab in Milton Keynes and Alex Alderton

EPI_ISL_922113, EPI_ISL_922114, EPI_ISL_922117, EPI_ISL_922118, EPI_ISL_922119, EPI_ISL_922120, EPI_ISL_922121, EPI_ISL_922122, EPI_ISL_922123, EPI_ISL_922124, EPI_ISL_922125, EPI_ISL_922126, EPI_ISL_922127, EPI_ISL_922128, EPI_ISL_922129, EPI_ISL_922130, EPI_ISL_922131, EPI_ISL_922132, EPI_ISL_922133, EPI_ISL_922134, EPI_ISL_922135, EPI_ISL_922136, EPI_ISL_922137, EPI_ISL_922138, EPI_ISL_922139, EPI_ISL_922140, EPI_ISL_922141, EPI_ISL_922142, EPI_ISL_922143, EPI_ISL_922144, EPI_ISL_922145, EPI_ISL_922146, EPI_ISL_922148, EPI_ISL_922157, EPI_ISL_950714, EPI_ISL_998945, EPI_ISL_998946, EPI_ISL_998947, EPI_ISL_998948, EPI_ISL_998949, EPI_ISL_998950, EPI_ISL_998951, EPI_ISL_998952, EPI_ISL_998953, EPI_ISL_998954, EPI_ISL_998955, EPI_ISL_998956

see above — Lincolnshire Hospitals and DeepSeq Nottingham — COVID-19 Genomics UK (COG-UK) Consortium — Christopher Moore; Fei Sang; Joeseph Chappell; Johnny Debebe; Jonathan Ball; Matthew Carlisle; Matthew Loose; Nadine Holmes; Nichola Duckworth; Patrick McClure; Sarah Walsh; Tim Sloan; Victoria Wright

EPI_ISL_865472, EPI_ISL_865474, EPI_ISL_865475, EPI_ISL_865476, EPI_ISL_865477, EPI_ISL_919479, EPI_ISL_919480, EPI_ISL_919481, EPI_ISL_919482, EPI_ISL_919483, EPI_ISL_919484, EPI_ISL_919485, EPI_ISL_919486, EPI_ISL_919487, EPI_ISL_919516, EPI_ISL_919524, EPI_ISL_919526, EPI_ISL_919527, EPI_ISL_919528, EPI_ISL_919529, EPI_ISL_919531, EPI_ISL_919537, EPI_ISL_919538, EPI_ISL_919539, EPI_ISL_919540, EPI_ISL_919541, EPI_ISL_919544, EPI_ISL_919545, EPI_ISL_919546, EPI_ISL_919549, EPI_ISL_919551, EPI_ISL_919552, EPI_ISL_919553, EPI_ISL_919554, EPI_ISL_919555, EPI_ISL_919558, EPI_ISL_919559, EPI_ISL_919560, EPI_ISL_919561, EPI_ISL_919565, EPI_ISL_919566, EPI_ISL_919567, EPI_ISL_919569, EPI_ISL_919580, EPI_ISL_919581, EPI_ISL_919583, EPI_ISL_919584, EPI_ISL_919586, EPI_ISL_919592, EPI_ISL_919596, EPI_ISL_919597, EPI_ISL_919598, EPI_ISL_919599, EPI_ISL_919600, EPI_ISL_919601, EPI_ISL_919602, EPI_ISL_919632, EPI_ISL_919634, EPI_ISL_919637, EPI_ISL_919641, EPI_ISL_919642, EPI_ISL_919643, EPI_ISL_919645, EPI_ISL_919646, EPI_ISL_919647, EPI_ISL_919648, EPI_ISL_919650, EPI_ISL_919651, EPI_ISL_919652, EPI_ISL_919653, EPI_ISL_919655, EPI_ISL_919656, EPI_ISL_919657, EPI_ISL_919659, EPI_ISL_919660, EPI_ISL_919661, EPI_ISL_919662, EPI_ISL_997378, EPI_ISL_997384, EPI_ISL_997385, EPI_ISL_997386, EPI_ISL_997387, EPI_ISL_997388, EPI_ISL_997389, EPI_ISL_997390, EPI_ISL_997391, EPI_ISL_997392, EPI_ISL_1104321, EPI_ISL_1104333, EPI_ISL_1104334, EPI_ISL_1104335, EPI_ISL_1104336, EPI_ISL_1104362, EPI_ISL_1104367, EPI_ISL_1104368, EPI_ISL_1104388, EPI_ISL_1177878

see above — Liverpool Clinical Laboratories — COVID-19 Genomics UK (COG-UK) Consortium — A Alrezaihi; Alessandro Gerada; Alistair Darby; Angela Cowell; Anita Lucaci; Anu Chawla; Cassie Olateju; Catherine Hartley; Charlotte Nelson; Ecaterina Vamos; Elaine O'Toole; Eleanor G Bentley; Ghada T Shawli; Hermione Webster; Isabel García-Dorival; James Johnson; James P Stewart; Jenifer Manson; Joanne Watts; Jones Benjamin; Jordan J Clark; Julian Hiscox; L Luu; Lucille Rainbow; M Almsaud; Margaret Hughes; Mark Whitehead; Matthew Gemmell; Miren Iturriza-Gomara; Muhannad Alruwaili; N.P Randle; Neil Swainston; PKF Gilmore; Parul Sharma; Rebekah Penrice-Randal; Richard Eccles; Richard Gregory; Sam Haldenby; Steve Paterson; Stuart D Armstrong; Trevor Ian Robinson; Ximeng Han

EPI_ISL_921428, EPI_ISL_921485, EPI_ISL_921486, EPI_ISL_921487, EPI_ISL_921502, EPI_ISL_921521, EPI_ISL_921522, EPI_ISL_921523, EPI_ISL_921524, EPI_ISL_921525, EPI_ISL_921526, EPI_ISL_921527, EPI_ISL_921528, EPI_ISL_921529, EPI_ISL_921530, EPI_ISL_921531, EPI_ISL_921532, EPI_ISL_921533, EPI_ISL_921534, EPI_ISL_921535, EPI_ISL_921536, EPI_ISL_921537, EPI_ISL_921538, EPI_ISL_921539, EPI_ISL_921540, EPI_ISL_921542, EPI_ISL_921543, EPI_ISL_921544, EPI_ISL_921545, EPI_ISL_921546, EPI_ISL_921547, EPI_ISL_921548, EPI_ISL_921549, EPI_ISL_921550, EPI_ISL_921551, EPI_ISL_921552, EPI_ISL_921553, EPI_ISL_921554, EPI_ISL_921555, EPI_ISL_921556, EPI_ISL_921557, EPI_ISL_921558, EPI_ISL_921559, EPI_ISL_921560, EPI_ISL_921561, EPI_ISL_921562, EPI_ISL_921563, EPI_ISL_921564, EPI_ISL_921565, EPI_ISL_921566, EPI_ISL_921567, EPI_ISL_921568, EPI_ISL_921569, EPI_ISL_921570, EPI_ISL_921571, EPI_ISL_921572, EPI_ISL_921573, EPI_ISL_921574, EPI_ISL_921575, EPI_ISL_921576, EPI_ISL_921577, EPI_ISL_921578, EPI_ISL_921579, EPI_ISL_921580, EPI_ISL_921581, EPI_ISL_921582, EPI_ISL_921583, EPI_ISL_921584, EPI_ISL_921585, EPI_ISL_950294, EPI_ISL_950298, EPI_ISL_950304, EPI_ISL_950305, EPI_ISL_950306, EPI_ISL_950307, EPI_ISL_950308, EPI_ISL_950309, EPI_ISL_950310, EPI_ISL_950311, EPI_ISL_950312, EPI_ISL_950317, EPI_ISL_950321, EPI_ISL_950323, EPI_ISL_950324, EPI_ISL_950325, EPI_ISL_950326, EPI_ISL_950327, EPI_ISL_950328, EPI_ISL_950329, EPI_ISL_950330, EPI_ISL_950331, EPI_ISL_950332, EPI_ISL_950333, EPI_ISL_950334, EPI_ISL_950335, EPI_ISL_950336, EPI_ISL_998313, EPI_ISL_998314, EPI_ISL_998316, EPI_ISL_998317, EPI_ISL_998318, EPI_ISL_998319, EPI_ISL_998320, EPI_ISL_998321, EPI_ISL_998323, EPI_ISL_998324, EPI_ISL_998325, EPI_ISL_998326, EPI_ISL_998327, EPI_ISL_998328, EPI_ISL_998329, EPI_ISL_998330, EPI_ISL_998331, EPI_ISL_998332, EPI_ISL_998333, EPI_ISL_998334, EPI_ISL_998335, EPI_ISL_998336, EPI_ISL_998337, EPI_ISL_998338, EPI_ISL_998339, EPI_ISL_998340, EPI_ISL_998341, EPI_ISL_998346, EPI_ISL_998351, EPI_ISL_998352, EPI_ISL_998353, EPI_ISL_998354, EPI_ISL_998355, EPI_ISL_998356, EPI_ISL_998357, EPI_ISL_998358, EPI_ISL_998359, EPI_ISL_998360, EPI_ISL_998361, EPI_ISL_998362, EPI_ISL_998363, EPI_ISL_998365, EPI_ISL_998366, EPI_ISL_998367, EPI_ISL_998373, EPI_ISL_998374, EPI_ISL_998375, EPI_ISL_998376, EPI_ISL_998377, EPI_ISL_998378, EPI_ISL_998379, EPI_ISL_1050949, EPI_ISL_1050951, EPI_ISL_1050952, EPI_ISL_1050953, EPI_ISL_1050954, EPI_ISL_1050955, EPI_ISL_1050956, EPI_ISL_1050960, EPI_ISL_1050992, EPI_ISL_1051213, EPI_ISL_1051215, EPI_ISL_1051216, EPI_ISL_1051218, EPI_ISL_1051219, EPI_ISL_1051411, EPI_ISL_1178262, EPI_ISL_1178343, EPI_ISL_1178349, EPI_ISL_1178350, EPI_ISL_1178351, EPI_ISL_1178352, EPI_ISL_1178354, EPI_ISL_1178355, EPI_ISL_1178358, EPI_ISL_1178360, EPI_ISL_1178407, EPI_ISL_1178408, EPI_ISL_1178409, EPI_ISL_1178410, EPI_ISL_1178411, EPI_ISL_1178412, EPI_ISL_1178413, EPI_ISL_1178414, EPI_ISL_1178415, EPI_ISL_1178416, EPI_ISL_1475239

see above — Northumbria University / South Tees Hospitals NHS Foundation Trust / North Cumbria Integrated Care NHS Foundation Trust / North Tees and Hartlepool NHS Foundation Trust / Newcastle Hospitals NHS Foundation Trust — COVID-19 Genomics UK (COG-UK) Consortium — Andrew Nelson; Brendan Payne; Clive Graham; Darren L Smith; Debra Padgett; Edward Barton; Emma Swindells; Garren Scott; Gary Black; Gary Eltringham; Giles S Holt; Greg R Young; Jane Greenaway; Jennifer Collins; John Allan; Joshua Loh; Lynn Dover; Matthew Bashton; Mohammad A Tariq; Paul Baker; Sarah Essex; Steve Liggett; Wen C Yew; Yusri Taha

EPI_ISL_867234, EPI_ISL_867235, EPI_ISL_867236, EPI_ISL_867259, EPI_ISL_867260, EPI_ISL_867262, EPI_ISL_867263, EPI_ISL_867264, EPI_ISL_867265, EPI_ISL_867266, EPI_ISL_867267, EPI_ISL_867268, EPI_ISL_867407, EPI_ISL_867408, EPI_ISL_867409, EPI_ISL_867410, EPI_ISL_867411, EPI_ISL_867412, EPI_ISL_867413, EPI_ISL_867414, EPI_ISL_867415, EPI_ISL_867420, EPI_ISL_867421, EPI_ISL_867422, EPI_ISL_867423, EPI_ISL_867425, EPI_ISL_867426, EPI_ISL_867428, EPI_ISL_867436, EPI_ISL_867444, EPI_ISL_867445, EPI_ISL_867446, EPI_ISL_867447, EPI_ISL_867448, EPI_ISL_867449, EPI_ISL_867452, EPI_ISL_867453, EPI_ISL_867454, EPI_ISL_867455, EPI_ISL_867460, EPI_ISL_867462, EPI_ISL_867463, EPI_ISL_867464, EPI_ISL_867465, EPI_ISL_867466, EPI_ISL_867467, EPI_ISL_867468, EPI_ISL_867469, EPI_ISL_867470, EPI_ISL_867471, EPI_ISL_867472, EPI_ISL_867473, EPI_ISL_867474, EPI_ISL_867475, EPI_ISL_867476, EPI_ISL_867477, EPI_ISL_867478, EPI_ISL_867479, EPI_ISL_867480, EPI_ISL_867481, EPI_ISL_867482, EPI_ISL_867483, EPI_ISL_867484, EPI_ISL_867485, EPI_ISL_867487, EPI_ISL_867488, EPI_ISL_867489, EPI_ISL_867495, EPI_ISL_867496, EPI_ISL_867502, EPI_ISL_867549, EPI_ISL_867576, EPI_ISL_867577, EPI_ISL_867585, EPI_ISL_867591, EPI_ISL_867592, EPI_ISL_867594, EPI_ISL_867595, EPI_ISL_867596, EPI_ISL_867597, EPI_ISL_867599, EPI_ISL_867600, EPI_ISL_867601, EPI_ISL_867602, EPI_ISL_867605, EPI_ISL_867607, EPI_ISL_867608, EPI_ISL_867610, EPI_ISL_867611, EPI_ISL_867613, EPI_ISL_867616, EPI_ISL_867617, EPI_ISL_867618, EPI_ISL_867620, EPI_ISL_867621, EPI_ISL_867623, EPI_ISL_867624, EPI_ISL_867628, EPI_ISL_867629, EPI_ISL_867630, EPI_ISL_867631, EPI_ISL_867633, EPI_ISL_867634, EPI_ISL_867635, EPI_ISL_867636, EPI_ISL_867637, EPI_ISL_867639, EPI_ISL_867640, EPI_ISL_867641, EPI_ISL_867642, EPI_ISL_867643, EPI_ISL_867644, EPI_ISL_867647, EPI_ISL_867649, EPI_ISL_867650, EPI_ISL_867651, EPI_ISL_867652, EPI_ISL_867653, EPI_ISL_867654, EPI_ISL_867655, EPI_ISL_867656, EPI_ISL_867657, EPI_ISL_867658, EPI_ISL_867659, EPI_ISL_867660, EPI_ISL_867661, EPI_ISL_867662, EPI_ISL_867663, EPI_ISL_867664, EPI_ISL_867665, EPI_ISL_867667, EPI_ISL_867668, EPI_ISL_867670, EPI_ISL_867671, EPI_ISL_867672, EPI_ISL_867673, EPI_ISL_867674, EPI_ISL_867675, EPI_ISL_867676, EPI_ISL_867677, EPI_ISL_867678, EPI_ISL_867679, EPI_ISL_867680, EPI_ISL_867681, EPI_ISL_867682, EPI_ISL_867683, EPI_ISL_867684, EPI_ISL_867686, EPI_ISL_867688, EPI_ISL_867689, EPI_ISL_867690, EPI_ISL_867692, EPI_ISL_867696, EPI_ISL_867697, EPI_ISL_867698, EPI_ISL_867699, EPI_ISL_867700, EPI_ISL_867701, EPI_ISL_867727, EPI_ISL_867732, EPI_ISL_867733, EPI_ISL_867734, EPI_ISL_867735, EPI_ISL_867909, EPI_ISL_867917, EPI_ISL_867922, EPI_ISL_867923, EPI_ISL_867924, EPI_ISL_867926, EPI_ISL_867927, EPI_ISL_867928, EPI_ISL_867929, EPI_ISL_951629, EPI_ISL_951816, EPI_ISL_1476078, EPI_ISL_1476079, EPI_ISL_1476098, EPI_ISL_1476099, EPI_ISL_1476102, EPI_ISL_1476176, EPI_ISL_1476182, EPI_ISL_1476185, EPI_ISL_1476189, EPI_ISL_1476192, EPI_ISL_1476193, EPI_ISL_1476195, EPI_ISL_1476196, EPI_ISL_1476197, EPI_ISL_1476201, EPI_ISL_1476204, EPI_ISL_1476256

see above — Originating lab: Wales Specialist Virology Centre Sequencing lab: Pathogen Genomics Unit — Public Health Wales Microbiology Cardiff Wales Specialist Virology Centre — Alec Birchley; Alexander Adams; Amy Gaskin; Angela Marchbank; Bree Gatica-Wilcox; Catherine Moore; Jason Coombes; Joanne Watkins; Joel Southgate; Johnathan Evans; Laura Gifford; Lauren Gilbert; Lee Graham; Malorie Perry; Matthew Bull; Nicole Pacchiarini; Sally Corden; Sara Kumziene-Summerhayes; Sara Rey; Sarah Taylor; Simon Cottrell; Sophie Jones; Tom Connor

EPI_ISL_922231, EPI_ISL_922232, EPI_ISL_922233, EPI_ISL_922234, EPI_ISL_922238, EPI_ISL_922239, EPI_ISL_922240, EPI_ISL_951305, EPI_ISL_951311, EPI_ISL_951313, EPI_ISL_951318, EPI_ISL_951361, EPI_ISL_951374, EPI_ISL_951375, EPI_ISL_951378, EPI_ISL_951379, EPI_ISL_951380, EPI_ISL_951383, EPI_ISL_951387, EPI_ISL_951388, EPI_ISL_951416, EPI_ISL_951419, EPI_ISL_951420, EPI_ISL_951421, EPI_ISL_951423, EPI_ISL_951424, EPI_ISL_951425, EPI_ISL_951428, EPI_ISL_951435, EPI_ISL_951436, EPI_ISL_951437, EPI_ISL_951438, EPI_ISL_951442, EPI_ISL_951444, EPI_ISL_951445, EPI_ISL_951446, EPI_ISL_951447, EPI_ISL_951449, EPI_ISL_951450, EPI_ISL_951451, EPI_ISL_951452, EPI_ISL_951453, EPI_ISL_951456, EPI_ISL_951457, EPI_ISL_951458, EPI_ISL_951459, EPI_ISL_999068, EPI_ISL_999069, EPI_ISL_999071, EPI_ISL_999078, EPI_ISL_999081, EPI_ISL_999083, EPI_ISL_999084, EPI_ISL_999085, EPI_ISL_999086, EPI_ISL_999087, EPI_ISL_999089, EPI_ISL_999092, EPI_ISL_999093, EPI_ISL_999095, EPI_ISL_999099, EPI_ISL_999100, EPI_ISL_999101, EPI_ISL_999102, EPI_ISL_999103, EPI_ISL_999104, EPI_ISL_999105, EPI_ISL_999106, EPI_ISL_999108, EPI_ISL_999109, EPI_ISL_999111, EPI_ISL_999112, EPI_ISL_999113, EPI_ISL_999114, EPI_ISL_999116, EPI_ISL_999119, EPI_ISL_999120, EPI_ISL_999121, EPI_ISL_999122, EPI_ISL_999123, EPI_ISL_999124, EPI_ISL_999125, EPI_ISL_999126, EPI_ISL_999127, EPI_ISL_999132, EPI_ISL_999134, EPI_ISL_999136, EPI_ISL_999137, EPI_ISL_999138, EPI_ISL_999139, EPI_ISL_999141, EPI_ISL_999142, EPI_ISL_999143, EPI_ISL_999144, EPI_ISL_999145, EPI_ISL_999146, EPI_ISL_999148, EPI_ISL_999150, EPI_ISL_999151, EPI_ISL_999152, EPI_ISL_999153, EPI_ISL_999154, EPI_ISL_999156, EPI_ISL_999161, EPI_ISL_999169, EPI_ISL_999204, EPI_ISL_999205, EPI_ISL_999207, EPI_ISL_999208, EPI_ISL_999210, EPI_ISL_999223, EPI_ISL_999374, EPI_ISL_999379, EPI_ISL_999381, EPI_ISL_999382, EPI_ISL_999385, EPI_ISL_999394, EPI_ISL_999395, EPI_ISL_999397, EPI_ISL_999401, EPI_ISL_999402, EPI_ISL_999404, EPI_ISL_999405, EPI_ISL_999406, EPI_ISL_999407, EPI_ISL_999427, EPI_ISL_999436, EPI_ISL_999462, EPI_ISL_999463, EPI_ISL_999479, EPI_ISL_999481, EPI_ISL_999506, EPI_ISL_999507, EPI_ISL_1051788, EPI_ISL_1051790, EPI_ISL_1051794, EPI_ISL_1051797, EPI_ISL_1051798, EPI_ISL_1051799, EPI_ISL_1051800, EPI_ISL_1051801, EPI_ISL_1051802, EPI_ISL_1051803, EPI_ISL_1051805, EPI_ISL_1051806, EPI_ISL_1051807, EPI_ISL_1051808, EPI_ISL_1051809, EPI_ISL_1051813, EPI_ISL_1051814, EPI_ISL_1051816, EPI_ISL_1051822, EPI_ISL_1051824, EPI_ISL_1051825, EPI_ISL_1051826, EPI_ISL_1051827, EPI_ISL_1051829, EPI_ISL_1051832, EPI_ISL_1051838, EPI_ISL_1051840, EPI_ISL_1051841, EPI_ISL_1051843, EPI_ISL_1051844, EPI_ISL_1051845, EPI_ISL_1051849, EPI_ISL_1051851, EPI_ISL_1051852, EPI_ISL_1051853, EPI_ISL_1051854, EPI_ISL_1051855, EPI_ISL_1051856, EPI_ISL_1051858, EPI_ISL_1051873, EPI_ISL_1051874, EPI_ISL_1178720, EPI_ISL_1178726, EPI_ISL_1178746, EPI_ISL_1178749, EPI_ISL_1178777, EPI_ISL_1178778, EPI_ISL_1178783, EPI_ISL_1178801, EPI_ISL_1178858, EPI_ISL_1249242, EPI_ISL_1249244, EPI_ISL_1833078, EPI_ISL_1833081, EPI_ISL_1833082, EPI_ISL_1833083, EPI_ISL_1833085, EPI_ISL_1833087, EPI_ISL_1833095, EPI_ISL_1833096, EPI_ISL_1833101, EPI_ISL_1833102, EPI_ISL_1833106

see above — Oxford Viromics, NDM, University of Oxford; Oxford University Hospitals; Basingstoke and North Hampshire Hospital — COVID-19 Genomics UK (COG-UK) Consortium — Alex Mobbs; Amy Trebes; Anita Justice; Catrin Moore; Christophe Fraser; David Bonsall; David Buck; Emma Wise; George Macintyre; Jessica Lynch; John Todd; Mariateresa de Cesare; Matilde Mori; Monique Andersson; Nathan Moore; Nick Cortes; Robert Shaw; Stephen Kidd; Tanya Golubchik; Timothy Peto

EPI_ISL_866541, EPI_ISL_866548, EPI_ISL_866556, EPI_ISL_866566, EPI_ISL_866569, EPI_ISL_866748, EPI_ISL_866749, EPI_ISL_866750, EPI_ISL_866751, EPI_ISL_866752, EPI_ISL_866753, EPI_ISL_866754, EPI_ISL_866755, EPI_ISL_866756, EPI_ISL_866757, EPI_ISL_866760, EPI_ISL_866763, EPI_ISL_866764, EPI_ISL_866767, EPI_ISL_866770, EPI_ISL_866774, EPI_ISL_866775, EPI_ISL_866781, EPI_ISL_866788, EPI_ISL_866789, EPI_ISL_866801, EPI_ISL_866835, EPI_ISL_866836, EPI_ISL_866837, EPI_ISL_866838, EPI_ISL_866839, EPI_ISL_866840, EPI_ISL_866841, EPI_ISL_866842, EPI_ISL_866843, EPI_ISL_866844, EPI_ISL_866845, EPI_ISL_866846, EPI_ISL_866847, EPI_ISL_866848, EPI_ISL_866849, EPI_ISL_866850, EPI_ISL_866851, EPI_ISL_866852, EPI_ISL_866853, EPI_ISL_866854, EPI_ISL_866855, EPI_ISL_866856, EPI_ISL_866857, EPI_ISL_866858, EPI_ISL_866859, EPI_ISL_866860, EPI_ISL_866861, EPI_ISL_866862, EPI_ISL_866863, EPI_ISL_866864, EPI_ISL_866865, EPI_ISL_866866, EPI_ISL_866867, EPI_ISL_866868, EPI_ISL_866869, EPI_ISL_866870, EPI_ISL_866871, EPI_ISL_866872, EPI_ISL_866873, EPI_ISL_866874, EPI_ISL_866875, EPI_ISL_866876, EPI_ISL_866877, EPI_ISL_866878, EPI_ISL_866879, EPI_ISL_866880, EPI_ISL_866881, EPI_ISL_866882, EPI_ISL_866883, EPI_ISL_866884, EPI_ISL_866885, EPI_ISL_866886, EPI_ISL_866887, EPI_ISL_866888, EPI_ISL_1051427, EPI_ISL_1051430, EPI_ISL_1051447, EPI_ISL_1051449, EPI_ISL_1051454, EPI_ISL_1051456, EPI_ISL_1051469, EPI_ISL_1051474, EPI_ISL_1051477, EPI_ISL_1051478, EPI_ISL_1051493, EPI_ISL_1051494, EPI_ISL_1051495, EPI_ISL_1051497, EPI_ISL_1051500, EPI_ISL_1051515, EPI_ISL_1248990, EPI_ISL_1249057, EPI_ISL_1832928, EPI_ISL_1832929, EPI_ISL_1832931, EPI_ISL_1832932, EPI_ISL_1832933, EPI_ISL_1832934, EPI_ISL_1832935, EPI_ISL_1832936, EPI_ISL_1832971, EPI_ISL_1832972, EPI_ISL_1832974, EPI_ISL_1832975, EPI_ISL_1832978, EPI_ISL_1832979, EPI_ISL_1832982, EPI_ISL_1832983, EPI_ISL_1832984, EPI_ISL_1832985, EPI_ISL_1832987, EPI_ISL_1832995, EPI_ISL_1832997, EPI_ISL_1832998, EPI_ISL_1833000, EPI_ISL_1833001

see above — Quadram Institute Bioscience — COVID-19 Genomics UK (COG-UK) Consortium — Alexander J Trotter; Alison E. Mather; Alp Aydin; Ana P. Tedim; Anastasia Kolyva; Andrew Bell; Andrew J. Page; Claire Stuart; Dave J. Baker; Gemma L. Kay; John Wain; Justin O'Grady; Leonardo de Oliveira Martins; Lizzie Meadows; Maria Diaz; Mark Webber; Muhammed Yasir; Nabil-Fareed Alikhan; Ngozi Elumogo; Nicholas M. Thomson; Rachael Stanley; Rachel Gilroy; Reenesh Prakash; Samir Dervisevic; Samuel Bloomfield; Steven Rudder; Thanh Le-Viet

EPI_ISL_866921, EPI_ISL_866922, EPI_ISL_866923, EPI_ISL_866924, EPI_ISL_866925, EPI_ISL_866926, EPI_ISL_866927, EPI_ISL_866928, EPI_ISL_866929, EPI_ISL_866930, EPI_ISL_866931, EPI_ISL_866932, EPI_ISL_866933, EPI_ISL_866934, EPI_ISL_866935, EPI_ISL_866936, EPI_ISL_866937, EPI_ISL_866938, EPI_ISL_866939, EPI_ISL_866940, EPI_ISL_866941, EPI_ISL_866942, EPI_ISL_866943, EPI_ISL_866944, EPI_ISL_866945, EPI_ISL_866946, EPI_ISL_866947, EPI_ISL_866948, EPI_ISL_866949, EPI_ISL_866950, EPI_ISL_866971, EPI_ISL_866972, EPI_ISL_866973, EPI_ISL_866974, EPI_ISL_866975, EPI_ISL_866976, EPI_ISL_866977, EPI_ISL_866978, EPI_ISL_866979, EPI_ISL_866980, EPI_ISL_866981, EPI_ISL_866982, EPI_ISL_866983, EPI_ISL_866984, EPI_ISL_866986, EPI_ISL_866987, EPI_ISL_866988, EPI_ISL_866989, EPI_ISL_1178590

see above — Queens Medical Centre, Clinical Microbiology Department / DeepSeq Nottingham — COVID-19 Genomics UK (COG-UK) Consortium — Christopher Moore; Emily Park; Fei Sang; Gemma Clark; Hannah Howson-Wells; Jack Hill; Jayasree Dey; Johnny Debebe; Jonathan Ball; Joseph Chappell; Manjinder Khakh; Matthew Carlisle; Matthew Loose; Michelle M Lister; Nadine Holmes; Patrick McClure; Theocharis Tsoleridis; Timothy Byaruhanga; Vicki M Fleming; Victoria Wright; Wendy Smith

EPI_ISL_920976, EPI_ISL_920977, EPI_ISL_920978, EPI_ISL_920979, EPI_ISL_920981, EPI_ISL_920982, EPI_ISL_920983, EPI_ISL_920984, EPI_ISL_920985, EPI_ISL_920986, EPI_ISL_920987, EPI_ISL_920988, EPI_ISL_920989, EPI_ISL_920990, EPI_ISL_920991, EPI_ISL_920992, EPI_ISL_920993, EPI_ISL_920994, EPI_ISL_920995, EPI_ISL_921011, EPI_ISL_921012, EPI_ISL_921013, EPI_ISL_921014, EPI_ISL_921015, EPI_ISL_921016, EPI_ISL_921017, EPI_ISL_998150, EPI_ISL_998151, EPI_ISL_998152, EPI_ISL_998153, EPI_ISL_998155, EPI_ISL_998156, EPI_ISL_998157, EPI_ISL_998160, EPI_ISL_998161, EPI_ISL_998162, EPI_ISL_998171, EPI_ISL_998172, EPI_ISL_998174, EPI_ISL_998189, EPI_ISL_998190, EPI_ISL_998191, EPI_ISL_998192, EPI_ISL_998193, EPI_ISL_998194, EPI_ISL_998195, EPI_ISL_998196, EPI_ISL_998198, EPI_ISL_998200, EPI_ISL_998218, EPI_ISL_998219, EPI_ISL_998220, EPI_ISL_998221, EPI_ISL_998222, EPI_ISL_998240, EPI_ISL_998241, EPI_ISL_998243, EPI_ISL_998244, EPI_ISL_998245, EPI_ISL_998246, EPI_ISL_998247, EPI_ISL_1474986, EPI_ISL_1474987, EPI_ISL_1474988, EPI_ISL_1474989, EPI_ISL_1474990, EPI_ISL_1475003

see above — Regional Virus Laboratory, Belfast Health and Social Care Trust — COVID-19 Genomics UK (COG-UK) Consortium — Alison Watt; Ciara Cox; Conall McCaughey; David Simpson; Derek Fairley; James McKenna; Mairead Connor; Susan Feeney; Tanya Curran; Zoltan Molnar

EPI_ISL_837058, EPI_ISL_837063, EPI_ISL_837065, EPI_ISL_837073, EPI_ISL_837087, EPI_ISL_837183, EPI_ISL_837196, EPI_ISL_837197, EPI_ISL_837245, EPI_ISL_837246, EPI_ISL_846774, EPI_ISL_855603, EPI_ISL_860686, EPI_ISL_860691, EPI_ISL_860694, EPI_ISL_860695, EPI_ISL_860709, EPI_ISL_862193, EPI_ISL_862259, EPI_ISL_1296524, EPI_ISL_1296528, EPI_ISL_1296529, EPI_ISL_1296533, EPI_ISL_1296534, EPI_ISL_1296535, EPI_ISL_1296538, EPI_ISL_1296547, EPI_ISL_1296548, EPI_ISL_1296551, EPI_ISL_1296571

see above — Respiratory Virus Unit, National Infection Service, Public Health England — COVID-19 Genomics UK (COG-UK) Consortium — PHE Covid Sequencing Team

EPI_ISL_1833532, EPI_ISL_1833533, EPI_ISL_1833534, EPI_ISL_1833535, EPI_ISL_1833536, EPI_ISL_1833549, EPI_ISL_1833550, EPI_ISL_1833551, EPI_ISL_1833552, EPI_ISL_1833553, EPI_ISL_1833554, EPI_ISL_1833555, EPI_ISL_1833556, EPI_ISL_1833557, EPI_ISL_1833558, EPI_ISL_1833559, EPI_ISL_1833561, EPI_ISL_1833562, EPI_ISL_1833563, EPI_ISL_1833564, EPI_ISL_1833565, EPI_ISL_1833566, EPI_ISL_1833567, EPI_ISL_1833568, EPI_ISL_1833569, EPI_ISL_1833570, EPI_ISL_1833571, EPI_ISL_1833572, EPI_ISL_1833573, EPI_ISL_1833574, EPI_ISL_1833575, EPI_ISL_1833576, EPI_ISL_1833577, EPI_ISL_1833578, EPI_ISL_1833579, EPI_ISL_1833580, EPI_ISL_1833581, EPI_ISL_1833582, EPI_ISL_1833583, EPI_ISL_1833584, EPI_ISL_1833585, EPI_ISL_1833586, EPI_ISL_1833587, EPI_ISL_1833588, EPI_ISL_1833589, EPI_ISL_1833591, EPI_ISL_1833592, EPI_ISL_1833596, EPI_ISL_1833597, EPI_ISL_1833598, EPI_ISL_1833599, EPI_ISL_1833604

see above — St George's University Hospitals NHS Foundation Trust — COVID-19 Genomics UK (COG-UK) Consortium — Adam Witney; Cassie Pope; Irene Monahan; Joshua Taylor; Ken Laing; NgeeKeong Tan

EPI_ISL_866144, EPI_ISL_866150, EPI_ISL_866153, EPI_ISL_866155, EPI_ISL_866156, EPI_ISL_866158, EPI_ISL_866159, EPI_ISL_866161, EPI_ISL_866163, EPI_ISL_866165, EPI_ISL_866166, EPI_ISL_866167, EPI_ISL_866169, EPI_ISL_866170, EPI_ISL_866172, EPI_ISL_866173, EPI_ISL_866175, EPI_ISL_866176, EPI_ISL_866177, EPI_ISL_866181, EPI_ISL_866182, EPI_ISL_866184, EPI_ISL_866185, EPI_ISL_920174, EPI_ISL_920175, EPI_ISL_920177, EPI_ISL_920180, EPI_ISL_920181, EPI_ISL_920182, EPI_ISL_920183, EPI_ISL_920184, EPI_ISL_920186, EPI_ISL_920190, EPI_ISL_920194, EPI_ISL_920195, EPI_ISL_920196, EPI_ISL_920197, EPI_ISL_920198, EPI_ISL_920200, EPI_ISL_920201, EPI_ISL_920202, EPI_ISL_920203, EPI_ISL_920204, EPI_ISL_920205, EPI_ISL_920206, EPI_ISL_920207, EPI_ISL_920208, EPI_ISL_920210, EPI_ISL_920211, EPI_ISL_920212, EPI_ISL_920214, EPI_ISL_920215, EPI_ISL_920217, EPI_ISL_920218, EPI_ISL_920219, EPI_ISL_920222, EPI_ISL_920224, EPI_ISL_920225, EPI_ISL_920226, EPI_ISL_920227, EPI_ISL_920228, EPI_ISL_920229, EPI_ISL_920230, EPI_ISL_920231, EPI_ISL_920232, EPI_ISL_920233, EPI_ISL_920234, EPI_ISL_920235, EPI_ISL_920236, EPI_ISL_920237, EPI_ISL_920238, EPI_ISL_920239, EPI_ISL_920244, EPI_ISL_920245, EPI_ISL_920246, EPI_ISL_920247, EPI_ISL_920252, EPI_ISL_920253, EPI_ISL_920254, EPI_ISL_920255, EPI_ISL_920261, EPI_ISL_920262, EPI_ISL_920263, EPI_ISL_920264, EPI_ISL_920265, EPI_ISL_920266, EPI_ISL_920271, EPI_ISL_920272, EPI_ISL_920273, EPI_ISL_920274, EPI_ISL_920275, EPI_ISL_920276, EPI_ISL_920283, EPI_ISL_920284, EPI_ISL_920285, EPI_ISL_920286, EPI_ISL_920287, EPI_ISL_920293, EPI_ISL_920294, EPI_ISL_920295, EPI_ISL_920296, EPI_ISL_920304, EPI_ISL_920305, EPI_ISL_920306, EPI_ISL_920307, EPI_ISL_920308, EPI_ISL_920315, EPI_ISL_920316, EPI_ISL_920326, EPI_ISL_920334, EPI_ISL_920335, EPI_ISL_920343, EPI_ISL_920350, EPI_ISL_920351, EPI_ISL_920358, EPI_ISL_920360, EPI_ISL_920367, EPI_ISL_920368, EPI_ISL_920700, EPI_ISL_1105149, EPI_ISL_1105157, EPI_ISL_1105266, EPI_ISL_1105272, EPI_ISL_1105273, EPI_ISL_1105289, EPI_ISL_1105333, EPI_ISL_1248372, EPI_ISL_1248373, EPI_ISL_1248383, EPI_ISL_1248394, EPI_ISL_1248417, EPI_ISL_1248438, EPI_ISL_1248439, EPI_ISL_1248482

see above — University College London Hospital — COVID-19 Genomics UK (COG-UK) Consortium — Catherine Houlihan; Dan Frampton; Dr Catherine Houlihan; Dr Daniel Frampton; Dr Judith Heaney; Dr Moira Spyer; Dr Paul Grant and Dr Eleni Nastouli; Judith Heaney; Matthew Byott; Moira Spyer and Eleni Nastouli; Stuart Kirk

EPI_ISL_839006, EPI_ISL_839020, EPI_ISL_865485, EPI_ISL_865486, EPI_ISL_865487, EPI_ISL_865584, EPI_ISL_865585, EPI_ISL_865586, EPI_ISL_865587, EPI_ISL_865588, EPI_ISL_865589, EPI_ISL_865590, EPI_ISL_865591, EPI_ISL_865592, EPI_ISL_865593, EPI_ISL_865594, EPI_ISL_865595, EPI_ISL_865596, EPI_ISL_865597, EPI_ISL_865598, EPI_ISL_865599, EPI_ISL_865600, EPI_ISL_865601, EPI_ISL_865602, EPI_ISL_865603, EPI_ISL_865604, EPI_ISL_865644, EPI_ISL_865848, EPI_ISL_865849, EPI_ISL_865850, EPI_ISL_865851, EPI_ISL_865852, EPI_ISL_865853, EPI_ISL_865854, EPI_ISL_865855, EPI_ISL_865856, EPI_ISL_865857, EPI_ISL_865858, EPI_ISL_865859, EPI_ISL_865860, EPI_ISL_865861, EPI_ISL_865862, EPI_ISL_865863, EPI_ISL_865864, EPI_ISL_865865, EPI_ISL_865866, EPI_ISL_865867, EPI_ISL_865868, EPI_ISL_865869, EPI_ISL_865870, EPI_ISL_865871, EPI_ISL_865872, EPI_ISL_865873, EPI_ISL_865874, EPI_ISL_865875, EPI_ISL_865876, EPI_ISL_865877, EPI_ISL_865878, EPI_ISL_865879, EPI_ISL_865880, EPI_ISL_865881, EPI_ISL_865882, EPI_ISL_865883, EPI_ISL_865891, EPI_ISL_865895, EPI_ISL_865897, EPI_ISL_865901, EPI_ISL_865905, EPI_ISL_865906, EPI_ISL_865909, EPI_ISL_865910, EPI_ISL_865911, EPI_ISL_865912, EPI_ISL_865913, EPI_ISL_865914, EPI_ISL_865915, EPI_ISL_865916, EPI_ISL_865918, EPI_ISL_865919, EPI_ISL_865920, EPI_ISL_865921, EPI_ISL_865922, EPI_ISL_865923, EPI_ISL_865924, EPI_ISL_865925, EPI_ISL_865926, EPI_ISL_865927, EPI_ISL_865928, EPI_ISL_865929, EPI_ISL_865930, EPI_ISL_865931, EPI_ISL_865932, EPI_ISL_865933, EPI_ISL_865934, EPI_ISL_865935, EPI_ISL_865938, EPI_ISL_865939, EPI_ISL_865940, EPI_ISL_865941, EPI_ISL_865942, EPI_ISL_865943, EPI_ISL_865944, EPI_ISL_865945, EPI_ISL_865947, EPI_ISL_865948, EPI_ISL_865949, EPI_ISL_865950, EPI_ISL_865952, EPI_ISL_865953, EPI_ISL_865954, EPI_ISL_865955, EPI_ISL_865956, EPI_ISL_865957, EPI_ISL_865958, EPI_ISL_865959, EPI_ISL_865960, EPI_ISL_865961, EPI_ISL_865962, EPI_ISL_865963, EPI_ISL_865964, EPI_ISL_865965, EPI_ISL_865966, EPI_ISL_865968, EPI_ISL_865977, EPI_ISL_865978, EPI_ISL_865986, EPI_ISL_865987, EPI_ISL_865994, EPI_ISL_866000, EPI_ISL_866001, EPI_ISL_866007, EPI_ISL_866009, EPI_ISL_866011, EPI_ISL_866012, EPI_ISL_866198, EPI_ISL_866201, EPI_ISL_866215, EPI_ISL_866217, EPI_ISL_866219, EPI_ISL_866221, EPI_ISL_866231, EPI_ISL_866262, EPI_ISL_866263, EPI_ISL_866264, EPI_ISL_866265, EPI_ISL_866266, EPI_ISL_866267, EPI_ISL_866268, EPI_ISL_866269, EPI_ISL_866270, EPI_ISL_866271, EPI_ISL_866272, EPI_ISL_866273, EPI_ISL_866274, EPI_ISL_866275, EPI_ISL_866276, EPI_ISL_866277, EPI_ISL_866278, EPI_ISL_866279, EPI_ISL_866280, EPI_ISL_866281, EPI_ISL_866282, EPI_ISL_866283, EPI_ISL_866284, EPI_ISL_866285, EPI_ISL_866286, EPI_ISL_866287, EPI_ISL_866288, EPI_ISL_866289, EPI_ISL_866290, EPI_ISL_866291, EPI_ISL_866292, EPI_ISL_866293, EPI_ISL_866294, EPI_ISL_866295, EPI_ISL_866296, EPI_ISL_866297, EPI_ISL_866299, EPI_ISL_866300, EPI_ISL_866301, EPI_ISL_866302, EPI_ISL_866303, EPI_ISL_866304, EPI_ISL_866305, EPI_ISL_866307, EPI_ISL_866308, EPI_ISL_866309, EPI_ISL_866310, EPI_ISL_866313, EPI_ISL_866314, EPI_ISL_866315, EPI_ISL_866316, EPI_ISL_866317, EPI_ISL_866318, EPI_ISL_866319, EPI_ISL_920026, EPI_ISL_920030, EPI_ISL_920031, EPI_ISL_920032, EPI_ISL_920033, EPI_ISL_920035, EPI_ISL_920036, EPI_ISL_920037, EPI_ISL_920038, EPI_ISL_920039, EPI_ISL_920043, EPI_ISL_920044, EPI_ISL_920045, EPI_ISL_920052, EPI_ISL_920053, EPI_ISL_920054, EPI_ISL_920055, EPI_ISL_920056, EPI_ISL_920057, EPI_ISL_920058, EPI_ISL_920079, EPI_ISL_920080, EPI_ISL_920882, EPI_ISL_920883, EPI_ISL_920884, EPI_ISL_920885, EPI_ISL_920886, EPI_ISL_920887, EPI_ISL_920888, EPI_ISL_920889, EPI_ISL_920890, EPI_ISL_920891, EPI_ISL_920892, EPI_ISL_920893, EPI_ISL_950159, EPI_ISL_950211, EPI_ISL_950212, EPI_ISL_950213, EPI_ISL_950214, EPI_ISL_950215, EPI_ISL_950216, EPI_ISL_950217, EPI_ISL_950218, EPI_ISL_950219, EPI_ISL_950220, EPI_ISL_950221, EPI_ISL_950222, EPI_ISL_950223, EPI_ISL_950224, EPI_ISL_950225, EPI_ISL_950226, EPI_ISL_950227, EPI_ISL_950228, EPI_ISL_950229, EPI_ISL_950230, EPI_ISL_950231, EPI_ISL_997677, EPI_ISL_997840, EPI_ISL_997841, EPI_ISL_997842, EPI_ISL_997995, EPI_ISL_997996, EPI_ISL_998002, EPI_ISL_998003, EPI_ISL_998004, EPI_ISL_998005, EPI_ISL_1048051, EPI_ISL_1048052, EPI_ISL_1048136, EPI_ISL_1048137, EPI_ISL_1050357, EPI_ISL_1050439, EPI_ISL_1050440, EPI_ISL_1050441, EPI_ISL_1050442, EPI_ISL_1050443, EPI_ISL_1050444, EPI_ISL_1050445, EPI_ISL_1050446, EPI_ISL_1050447, EPI_ISL_1050448, EPI_ISL_1050450, EPI_ISL_1050451, EPI_ISL_1050457, EPI_ISL_1050458, EPI_ISL_1050459, EPI_ISL_1050460, EPI_ISL_1050461, EPI_ISL_1050462, EPI_ISL_1050464, EPI_ISL_1050465, EPI_ISL_1050466, EPI_ISL_1050508, EPI_ISL_1050510, EPI_ISL_1050511, EPI_ISL_1050516, EPI_ISL_1050517, EPI_ISL_1050519, EPI_ISL_1050520, EPI_ISL_1050521, EPI_ISL_1050524, EPI_ISL_1050525, EPI_ISL_1178009, EPI_ISL_1248107, EPI_ISL_1248108, EPI_ISL_1248109, EPI_ISL_1248110, EPI_ISL_1248111, EPI_ISL_1248112, EPI_ISL_1248114, EPI_ISL_1248115, EPI_ISL_1248116, EPI_ISL_1248122, EPI_ISL_1248123, EPI_ISL_1248124, EPI_ISL_1248125, EPI_ISL_1248126, EPI_ISL_1248127, EPI_ISL_1248128, EPI_ISL_1248129, EPI_ISL_1248130, EPI_ISL_1248131, EPI_ISL_1248132, EPI_ISL_1248133, EPI_ISL_1248134, EPI_ISL_1248135, EPI_ISL_1248136, EPI_ISL_1248137, EPI_ISL_1248138, EPI_ISL_1248139, EPI_ISL_1248140, EPI_ISL_1248141, EPI_ISL_1248142, EPI_ISL_1248143, EPI_ISL_1248144, EPI_ISL_1248145, EPI_ISL_1248146, EPI_ISL_1248147, EPI_ISL_1248148, EPI_ISL_1248151, EPI_ISL_1248152, EPI_ISL_1248156, EPI_ISL_1248158, EPI_ISL_1248164, EPI_ISL_1248165, EPI_ISL_1248173, EPI_ISL_1248174, EPI_ISL_1248175, EPI_ISL_1248181, EPI_ISL_1248182, EPI_ISL_1248184, EPI_ISL_1248191, EPI_ISL_1248193, EPI_ISL_1248203, EPI_ISL_1248205, EPI_ISL_1309051, EPI_ISL_1309070, EPI_ISL_1309077, EPI_ISL_1309078, EPI_ISL_1309085, EPI_ISL_1309090, EPI_ISL_1309097, EPI_ISL_1309099, EPI_ISL_1309103, EPI_ISL_1309109, EPI_ISL_1309113, EPI_ISL_1386855, EPI_ISL_1386856, EPI_ISL_1386859, EPI_ISL_1386873, EPI_ISL_1386895, EPI_ISL_1474857, EPI_ISL_1637117, EPI_ISL_1637127, EPI_ISL_1730337, EPI_ISL_1730395

| | | | |
|---|---|---|---|
| see above | University College London, Great Ormond Street Hospital for Children NHS Foundation Trust, Imperial College Healthcare NHS Trust | COVID-19 Genomics UK (COG-UK) Consortium | Alison Holmes; Charlotte Williams; Helena Tutill; Jacqueline Findlay; James Price; Judith Breuer; Julianne Brown; Kathryn Harris; Leysa Forrest; Mark Kristiansen; Paola Niola; Paola Resende Silva; Patricia Dyal; Paul Randell; Rachel Williams; Samuel Weeks; Sergi Castellano; Sunando Roy; Tony Brooks; Yasmin Panchbhaya |
| EPI_ISL_1104291, EPI_ISL_1104293, EPI_ISL_1104296, EPI_ISL_2125914 | University Hospitals Of Leicester NHS Trust and DeepSeq Nottingham | COVID-19 Genomics UK (COG-UK) Consortium | Christopher Holmes; Christopher Moore; Fei Sang; Joeseph Chappell; Johnny Debebe; Jonathan Ball; Julian Tang; Karlie Fallon; Matthew Carlisle; Matthew Loose; Nadine Holmes; Patrick McClure; Paul Bird; Thomas Helmer; Victoria Wright |
| EPI_ISL_918764, EPI_ISL_918765, EPI_ISL_918766, EPI_ISL_918767, EPI_ISL_918768, EPI_ISL_918769, EPI_ISL_918770, EPI_ISL_918771, EPI_ISL_918772, EPI_ISL_918773, EPI_ISL_918774, EPI_ISL_918776, EPI_ISL_918777, EPI_ISL_918778, EPI_ISL_918780, EPI_ISL_918781, EPI_ISL_918782, EPI_ISL_918783, EPI_ISL_918784, EPI_ISL_918785, EPI_ISL_918786, EPI_ISL_918787, EPI_ISL_918788, EPI_ISL_918789, EPI_ISL_918790, EPI_ISL_918791, EPI_ISL_918792, EPI_ISL_918793, EPI_ISL_918794, EPI_ISL_918795, EPI_ISL_918796, EPI_ISL_918797, EPI_ISL_918798, EPI_ISL_918799, EPI_ISL_918800, EPI_ISL_918801, EPI_ISL_918802, EPI_ISL_918803, EPI_ISL_918804, EPI_ISL_918805, EPI_ISL_918806, EPI_ISL_918807, EPI_ISL_918808, EPI_ISL_918809, EPI_ISL_918810, EPI_ISL_918811, EPI_ISL_918812, EPI_ISL_918813, EPI_ISL_918814, EPI_ISL_918815, EPI_ISL_918816, EPI_ISL_918817, EPI_ISL_918818, EPI_ISL_918819, EPI_ISL_918820, EPI_ISL_918821, EPI_ISL_918822, EPI_ISL_918823, EPI_ISL_918824, EPI_ISL_918825, EPI_ISL_918826, EPI_ISL_918827, EPI_ISL_918842, EPI_ISL_918843, EPI_ISL_918844, EPI_ISL_918845, EPI_ISL_918856, EPI_ISL_918869, EPI_ISL_918874, EPI_ISL_918875, EPI_ISL_918876, EPI_ISL_918877, EPI_ISL_918878, EPI_ISL_918882, EPI_ISL_918912, EPI_ISL_918913, EPI_ISL_918977, EPI_ISL_918978, EPI_ISL_918979, EPI_ISL_918980, EPI_ISL_918981, EPI_ISL_918982, EPI_ISL_918983, EPI_ISL_949417, EPI_ISL_949420, EPI_ISL_949421, EPI_ISL_996422, EPI_ISL_996423, EPI_ISL_996431, EPI_ISL_996432, EPI_ISL_996449, EPI_ISL_996453, EPI_ISL_996455, EPI_ISL_996459, EPI_ISL_1046996, EPI_ISL_1047060, EPI_ISL_1047063, EPI_ISL_1047068, EPI_ISL_1474382, EPI_ISL_1474383, EPI_ISL_1474385, EPI_ISL_1474386, EPI_ISL_1474387, EPI_ISL_1474388, EPI_ISL_1474389, EPI_ISL_1474390, EPI_ISL_1474391, EPI_ISL_1474392, EPI_ISL_1474393, EPI_ISL_1474394, EPI_ISL_1474395 | | | |
| see above | University of Birmingham | COVID-19 Genomics UK (COG-UK) Consortium | Alex Richter; Andrew Bosworth; Andrew Bosworth. Queen Elizabeth Hospital: Anna Casey; Andrew D Beggs PHE Heartlands Lab: Husam Osman; Charlotte Poxon; Institute of Microbiology; Joanne Stockton; Josh Quick; Kasun Wanigasooriya; Mike Kidd; Nicholas Loman. University of Birmingham Testing Laboratory: Celina M Whalley; Oliver Pickles; Radoslaw Poplawski; Samuel Nicholls; University of Birmingham: Claire McMurray; Will Rowe |
| EPI_ISL_996579, EPI_ISL_996594, EPI_ISL_997328, EPI_ISL_997329, EPI_ISL_997330, EPI_ISL_997331, EPI_ISL_997332, EPI_ISL_997337, EPI_ISL_1308578, EPI_ISL_1308587, EPI_ISL_1308589, EPI_ISL_1308590, EPI_ISL_1308591 | | | |
| see above | University of Exeter | COVID-19 Genomics UK (COG-UK) Consortium | Aaron Jeffries; Audrey Farbos; Ben Temperton; Jane Masoli; Joanna Warwick-Dugdale; Michelle Michelsen; Robyn Manley; Stephen Michell |
| EPI_ISL_997075, EPI_ISL_997076, EPI_ISL_997159, EPI_ISL_997160, EPI_ISL_997163, EPI_ISL_1104218, EPI_ISL_1104232, EPI_ISL_1104238, EPI_ISL_1104241, EPI_ISL_1104257, EPI_ISL_1247720, EPI_ISL_1247724, EPI_ISL_1474578, EPI_ISL_1474579, EPI_ISL_1474580, EPI_ISL_1636712, EPI_ISL_2125729 | | | |
| see above | Virology Department, Royal Infirmary of Edinburgh, NHS Lothian / School of Biological Sciences, University of Edinburgh | COVID-19 Genomics UK (COG-UK) Consortium | Colquhoun R; Cotton S; Dewar R; Gallagher A; Hill V; Jackson B; Maloney D; McCrone JT; McHugh M; O'Toole Á; Rambaut A; Rooke S; Scher E; Templeton K; Yu X |
| EPI_ISL_865092, EPI_ISL_865093, EPI_ISL_865105, EPI_ISL_865106, EPI_ISL_865107, EPI_ISL_865108, EPI_ISL_865109, EPI_ISL_865110, EPI_ISL_865136, EPI_ISL_865137, EPI_ISL_865138, EPI_ISL_865139, EPI_ISL_865141, EPI_ISL_865144, EPI_ISL_919339, EPI_ISL_919340, EPI_ISL_919341, EPI_ISL_919342, EPI_ISL_949640, EPI_ISL_949643, EPI_ISL_949644 | | | |
| see above | Virology Department, Royal Infirmary of Edinburgh, NHS Lothian / School of Biological Sciences, University of Edinburgh / Institute of Genetics and Molecular Medicine, University of Edinburgh | COVID-19 Genomics UK (COG-UK) Consortium | Balcaza C; Colquhoun R; Dewar R; Gallagher M; Hill V; Jackson B; McCrone JT; McHugh M; O'Toole Á; Rambaut A; Rooke S; Scher E; Templeton K; Williams TC; Yu X |
| EPI_ISL_868361, EPI_ISL_868362, EPI_ISL_868365, EPI_ISL_868367, EPI_ISL_868368, EPI_ISL_868378, EPI_ISL_868379, EPI_ISL_868385, EPI_ISL_868387, EPI_ISL_868389, EPI_ISL_868391, EPI_ISL_868397, EPI_ISL_868398, EPI_ISL_868407, EPI_ISL_868424, EPI_ISL_868425, EPI_ISL_868437, EPI_ISL_868439, EPI_ISL_868440, EPI_ISL_868451, EPI_ISL_868476, EPI_ISL_868482, EPI_ISL_868484, EPI_ISL_868492, EPI_ISL_868493, EPI_ISL_868496, EPI_ISL_868500, EPI_ISL_868501, EPI_ISL_868517, EPI_ISL_868523, EPI_ISL_868527, EPI_ISL_868532, EPI_ISL_868536, EPI_ISL_868546, EPI_ISL_868562, EPI_ISL_868569, EPI_ISL_868578, EPI_ISL_868579, EPI_ISL_868581, EPI_ISL_868582, EPI_ISL_868584, EPI_ISL_868592, EPI_ISL_868602, EPI_ISL_868618, EPI_ISL_868623, EPI_ISL_868626, EPI_ISL_868632, EPI_ISL_868634, EPI_ISL_868639, EPI_ISL_868642, EPI_ISL_868661, EPI_ISL_868662, EPI_ISL_868666, EPI_ISL_868679, EPI_ISL_868690, EPI_ISL_868696, EPI_ISL_868698, EPI_ISL_868703, EPI_ISL_868705, EPI_ISL_868708, EPI_ISL_924083, EPI_ISL_924096, EPI_ISL_924145, EPI_ISL_924150, EPI_ISL_924177, EPI_ISL_924181, EPI_ISL_924220, EPI_ISL_924224, EPI_ISL_924230, EPI_ISL_924233, EPI_ISL_924239, EPI_ISL_924242, EPI_ISL_924244, EPI_ISL_924247, EPI_ISL_924278, EPI_ISL_924295, EPI_ISL_924303, EPI_ISL_924325, EPI_ISL_924326, EPI_ISL_924354, EPI_ISL_924399, EPI_ISL_924408 | | | |
| see above | Virology Department, Sheffield Teaching Hospitals NHS Foundation Trust/Department of Infection, Immunity and Cardiovascular Disease, The Medical School, University of Sheffield | COVID-19 Genomics UK (COG-UK) Consortium | Adri Angyal; Alex Keeley; Benjamin Lindsey; Cariad Evans; Danielle Groves; Dave Partridge; Katie Johnson; Laura Carrilero; Luke Green; Matthew Parker; Matthew Wyles; Mehmet Yavuz; Mohammad Raza; Nikki Smith; Paul Parsons; Rachel Tucker; Rebecca Brown; Thushan de Silva |
| EPI_ISL_922359, EPI_ISL_922360, EPI_ISL_922362, EPI_ISL_922363, EPI_ISL_922365, EPI_ISL_922367, EPI_ISL_922368, EPI_ISL_922369, EPI_ISL_922370, EPI_ISL_922372, EPI_ISL_922373, EPI_ISL_922384, EPI_ISL_922392, EPI_ISL_922395, EPI_ISL_922396, EPI_ISL_922398, EPI_ISL_922399, EPI_ISL_922401, EPI_ISL_922402, EPI_ISL_922403, EPI_ISL_922404, EPI_ISL_922406, EPI_ISL_922407, EPI_ISL_922408, EPI_ISL_922409, EPI_ISL_922410, EPI_ISL_922411, EPI_ISL_922413, EPI_ISL_922415, EPI_ISL_922416, EPI_ISL_922417, EPI_ISL_922418, EPI_ISL_922419, EPI_ISL_922420, EPI_ISL_922421, EPI_ISL_922423, EPI_ISL_922425, EPI_ISL_922428, EPI_ISL_922430, EPI_ISL_922432, EPI_ISL_922433, EPI_ISL_922434, EPI_ISL_922435, EPI_ISL_922436, EPI_ISL_922437, EPI_ISL_922439, EPI_ISL_922440, EPI_ISL_922442, EPI_ISL_922443, EPI_ISL_922444, EPI_ISL_922445, EPI_ISL_922446, EPI_ISL_922448, EPI_ISL_922452, EPI_ISL_922455, EPI_ISL_922457, EPI_ISL_922458, EPI_ISL_922459, EPI_ISL_922460, EPI_ISL_922462, EPI_ISL_922463, EPI_ISL_922464, EPI_ISL_922465, EPI_ISL_922466, EPI_ISL_922467, EPI_ISL_922468, EPI_ISL_922469, EPI_ISL_922470, EPI_ISL_922473, EPI_ISL_922474, EPI_ISL_922481, EPI_ISL_922483, EPI_ISL_922485, EPI_ISL_922488, EPI_ISL_922491, EPI_ISL_922494, EPI_ISL_922495, EPI_ISL_922497, EPI_ISL_922498, EPI_ISL_922499, EPI_ISL_922500, EPI_ISL_922502, EPI_ISL_922504, EPI_ISL_922505, EPI_ISL_922507, EPI_ISL_922510, EPI_ISL_922511, EPI_ISL_922515, EPI_ISL_922516, EPI_ISL_922553, EPI_ISL_922559, EPI_ISL_922560, EPI_ISL_922563, EPI_ISL_922565, EPI_ISL_922566, EPI_ISL_922567, EPI_ISL_922568, EPI_ISL_922570, EPI_ISL_922571, EPI_ISL_922572, EPI_ISL_922574, EPI_ISL_922580, EPI_ISL_922581, EPI_ISL_922582, EPI_ISL_922590, EPI_ISL_922591, EPI_ISL_922592, EPI_ISL_922593, EPI_ISL_922594, EPI_ISL_922595, EPI_ISL_922596, EPI_ISL_922597, EPI_ISL_922598, EPI_ISL_922599, EPI_ISL_922600, EPI_ISL_922601, EPI_ISL_922602, EPI_ISL_922603, EPI_ISL_922604, EPI_ISL_922605, EPI_ISL_922606, EPI_ISL_922607, EPI_ISL_922626, EPI_ISL_922627, EPI_ISL_922628, EPI_ISL_922629, EPI_ISL_922633, EPI_ISL_922634, EPI_ISL_922643, EPI_ISL_922645, EPI_ISL_922646, EPI_ISL_922647, EPI_ISL_922648, EPI_ISL_922649, EPI_ISL_922650, EPI_ISL_922651, EPI_ISL_922652, EPI_ISL_922653, EPI_ISL_922654, EPI_ISL_922655, EPI_ISL_922656, EPI_ISL_922659, EPI_ISL_922660, EPI_ISL_922663, EPI_ISL_922664, EPI_ISL_922665, EPI_ISL_922666, EPI_ISL_922667, EPI_ISL_922668, EPI_ISL_922669, EPI_ISL_922672, EPI_ISL_922673, EPI_ISL_922674, EPI_ISL_922675, EPI_ISL_922676, EPI_ISL_922677, EPI_ISL_922678, EPI_ISL_922679, EPI_ISL_922680, EPI_ISL_922681, EPI_ISL_922682, EPI_ISL_922683, EPI_ISL_922684, EPI_ISL_922685, EPI_ISL_922686, EPI_ISL_922687, EPI_ISL_922688, EPI_ISL_922689, EPI_ISL_922690, EPI_ISL_922692, EPI_ISL_922693, EPI_ISL_922695, EPI_ISL_922696, EPI_ISL_922697, EPI_ISL_922698, EPI_ISL_922699, EPI_ISL_922700, EPI_ISL_922701, EPI_ISL_922702, EPI_ISL_922703, EPI_ISL_922704, EPI_ISL_922705, EPI_ISL_922706, EPI_ISL_922707, EPI_ISL_922708, EPI_ISL_922709, EPI_ISL_922710, EPI_ISL_922712, EPI_ISL_922713, EPI_ISL_922719, EPI_ISL_922721, EPI_ISL_922722, EPI_ISL_922723, EPI_ISL_922724, EPI_ISL_922726, EPI_ISL_922727, EPI_ISL_922732, EPI_ISL_922734, EPI_ISL_922741, EPI_ISL_922743, EPI_ISL_922895, EPI_ISL_922903, EPI_ISL_922904, EPI_ISL_922905, EPI_ISL_922906, EPI_ISL_922907, EPI_ISL_922908, EPI_ISL_922909, EPI_ISL_922910, EPI_ISL_922912, EPI_ISL_922913, EPI_ISL_922914, EPI_ISL_922923, EPI_ISL_922925, EPI_ISL_922926, EPI_ISL_922927, EPI_ISL_922928, EPI_ISL_922929, EPI_ISL_922930, EPI_ISL_922931, EPI_ISL_922932, EPI_ISL_922965, EPI_ISL_922970, EPI_ISL_922974, EPI_ISL_922975, EPI_ISL_922976, EPI_ISL_922977, EPI_ISL_922978, EPI_ISL_922979, EPI_ISL_922980, EPI_ISL_922981, EPI_ISL_922982, EPI_ISL_922983, EPI_ISL_922984, EPI_ISL_922985, EPI_ISL_922986, EPI_ISL_922987, EPI_ISL_922988, EPI_ISL_922989, EPI_ISL_922990, EPI_ISL_922991, EPI_ISL_922992, EPI_ISL_922993, EPI_ISL_922994, EPI_ISL_922995, EPI_ISL_922996, EPI_ISL_922997, EPI_ISL_922998, EPI_ISL_922999, EPI_ISL_923000, EPI_ISL_923001, EPI_ISL_923002, EPI_ISL_923003, EPI_ISL_923004, EPI_ISL_923005, EPI_ISL_923006, EPI_ISL_923007, EPI_ISL_923008, EPI_ISL_923009, EPI_ISL_923010, EPI_ISL_923011, EPI_ISL_923013, EPI_ISL_923014, EPI_ISL_923015, EPI_ISL_923016, EPI_ISL_923017, EPI_ISL_923018, EPI_ISL_923019, EPI_ISL_923020, EPI_ISL_923021, EPI_ISL_923022, EPI_ISL_923023, EPI_ISL_923025, EPI_ISL_923026, EPI_ISL_923027, EPI_ISL_1476077, EPI_ISL_1476081, EPI_ISL_1476082, EPI_ISL_1476083, EPI_ISL_1476084, EPI_ISL_1476086, EPI_ISL_1476087, EPI_ISL_1476088, EPI_ISL_1476090, EPI_ISL_1476091, EPI_ISL_1476092, EPI_ISL_1476093, EPI_ISL_1476094, EPI_ISL_1476096, EPI_ISL_1476097, EPI_ISL_1476100, EPI_ISL_1476186, EPI_ISL_1476187, EPI_ISL_1476188, EPI_ISL_1476191, EPI_ISL_1476200, EPI_ISL_1476202, EPI_ISL_1833130, EPI_ISL_1833139 | | | |
| see above | Wales Specialist Virology Centre Sequencing lab: Pathogen Genomics Unit | Public Health Wales Microbiology Cardiff Wales Specialist Virology Centre | Alec Birchley; Alexander Adams; Amy Gaskin; Angela Marchbank; Bree Gatica-Wilcox; Catherine Moore; Jason Coombes; Joanne Watkins; Joel Southgate; Johnathan Evans; Laura Gifford; Lauren Gilbert; Lee Graham; Malorie Perry; Matthew Bull; Nicole Pacchiarini; Sally Corden; Sara Kumziene-Summerhayes; Sara Rey; Sarah Taylor; Simon Cottrell; Sophie Jones; Tom Connor |
| EPI_ISL_919198, EPI_ISL_919199, EPI_ISL_919200, EPI_ISL_919201, EPI_ISL_919202, EPI_ISL_919203, EPI_ISL_919204, EPI_ISL_919205, EPI_ISL_919206, EPI_ISL_919207, EPI_ISL_919209, EPI_ISL_919210, EPI_ISL_919212, EPI_ISL_1047839, EPI_ISL_1047840 | | | |
| see above | West of Scotland Specialist Virology Centre, NHSGGC / MRC-University of Glasgow Centre for Virus Research | COVID-19 Genomics UK (COG-UK) Consortium | Alasdair MacLean; Alice Broos; Ana da Silva Filipe; Antonia Ho; Daniel Mair; David L Robertson; Emma Thomson; Guy Mollett; James Shepherd; Jenna Nichols; Joseph Hughes; Kathy Li; Kathy Smollett; Kyriaki Nomikou; Lily Tong; Matthew Holden; Natasha Johnson; Rachel Blacow; Richard Orton; Rory Gunson; Sarah McDonald; Sharif Shaaban; Sreenu Vattipally; Stephen Carmichael |

We gratefully acknowledge the following Authors from the Originating laboratories responsible for obtaining the specimens, as well as the Submitting laboratories where the genome data were generated and shared via GISAID, on which this research is based.

All Submitters of data may be contacted directly via www.gisaid.org

Authors are sorted alphabetically.

| Accession ID | Originating Laboratory | Submitting Laboratory | Authors |
|---|---|---|---|
| EPI_ISL_919854, EPI_ISL_919855, EPI_ISL_997625, EPI_ISL_997626, EPI_ISL_997627, EPI_ISL_997628, EPI_ISL_997642, EPI_ISL_997645, EPI_ISL_997646, EPI_ISL_997647, EPI_ISL_997648, EPI_ISL_997649, EPI_ISL_997650, EPI_ISL_997651, EPI_ISL_1048090, EPI_ISL_1048094, EPI_ISL_2126110, EPI_ISL_2126111, EPI_ISL_2126113, EPI_ISL_2126127 | | | |
| see above | Barts Health NHS Trust | COVID-19 Genomics UK (COG-UK) Consortium | BROAD; Beatrix; CUTINO-MOGUEL; Claire; David; Dola; HARRINGTON; KELE; KULASEGARAN-SHYLINI; Maria-Teresa; OWOYEMI; Raghavendran |
| EPI_ISL_924652, EPI_ISL_924654, EPI_ISL_924655, EPI_ISL_924656, EPI_ISL_924658, EPI_ISL_924659, EPI_ISL_924660, EPI_ISL_924662, EPI_ISL_924663, EPI_ISL_924665, EPI_ISL_924670, EPI_ISL_924673, EPI_ISL_924678, EPI_ISL_924679, EPI_ISL_924680, EPI_ISL_924681, EPI_ISL_924682, EPI_ISL_924686, EPI_ISL_924687, EPI_ISL_924703, EPI_ISL_924710, EPI_ISL_924711, EPI_ISL_924829, EPI_ISL_924831, EPI_ISL_924833, EPI_ISL_924835, EPI_ISL_924837, EPI_ISL_924840, EPI_ISL_924843, EPI_ISL_924844, EPI_ISL_924847, EPI_ISL_924852, EPI_ISL_924854, EPI_ISL_924856, EPI_ISL_924858, EPI_ISL_924861, EPI_ISL_924862, EPI_ISL_924866, EPI_ISL_924868, EPI_ISL_924869, EPI_ISL_924870, EPI_ISL_924872, EPI_ISL_924873, EPI_ISL_924874, EPI_ISL_924875, EPI_ISL_924878, EPI_ISL_924881, EPI_ISL_924882, EPI_ISL_924884, EPI_ISL_924885, EPI_ISL_924888, EPI_ISL_924889, EPI_ISL_953083, EPI_ISL_953089, EPI_ISL_953096, EPI_ISL_953097, EPI_ISL_953098, EPI_ISL_953099, EPI_ISL_953101, EPI_ISL_953102, EPI_ISL_953103, EPI_ISL_953104, EPI_ISL_953105, EPI_ISL_953106, EPI_ISL_953107, EPI_ISL_953108, EPI_ISL_953109, EPI_ISL_953111, EPI_ISL_953113, EPI_ISL_953114, EPI_ISL_953115, EPI_ISL_953116, EPI_ISL_953117, EPI_ISL_1000888, EPI_ISL_1000895, EPI_ISL_1000896, EPI_ISL_1000897, EPI_ISL_1000902, EPI_ISL_1000906, EPI_ISL_1000913, EPI_ISL_1000937, EPI_ISL_1000943, EPI_ISL_1000945, EPI_ISL_1000947, EPI_ISL_1054710, EPI_ISL_1054713, EPI_ISL_1054714, EPI_ISL_1054719, EPI_ISL_1054720, EPI_ISL_1054723, EPI_ISL_1054729, EPI_ISL_1054793, EPI_ISL_1054822, EPI_ISL_1638235, EPI_ISL_1638245 | | | |
| see above | Bioinformatics and Biostatistics Lab, Advanced Sequencing Facility | COVID-19 Genomics UK (COG-UK) Consortium | Aengus Stewart; Chelsea Sawyer; Harshil Patel; Jerome Nicod; Laura Cubitt; Margaret Crawford |
| EPI_ISL_1832313, EPI_ISL_1832315, EPI_ISL_1832317, EPI_ISL_1832320, EPI_ISL_1832323, EPI_ISL_1832326, EPI_ISL_1832330, EPI_ISL_1832332 | | | |
| see above | Centre for Clinical Infection and Diagnostics Research and Genomics Innovation Unit, Guy's and St. Thomas' NHS Trust | COVID-19 Genomics UK (COG-UK) Consortium | Ali Raza Awan; Chloe Fisher; Jonathan Edgeworth; Luke Snell; Penny Cliff; Rahul Batra |
| EPI_ISL_867993, EPI_ISL_867999, EPI_ISL_868226, EPI_ISL_868241, EPI_ISL_868242, EPI_ISL_868243, EPI_ISL_868244, EPI_ISL_868245, EPI_ISL_868246, EPI_ISL_868247, EPI_ISL_868248, EPI_ISL_868249, EPI_ISL_868250, EPI_ISL_868251, EPI_ISL_868252, EPI_ISL_868253, EPI_ISL_868254, EPI_ISL_868255, EPI_ISL_868256, EPI_ISL_868257, EPI_ISL_868258, EPI_ISL_923315, EPI_ISL_923316, EPI_ISL_923318, EPI_ISL_923319, EPI_ISL_923320, EPI_ISL_923321, EPI_ISL_923322, EPI_ISL_923323, EPI_ISL_923324, EPI_ISL_923325, EPI_ISL_923326, EPI_ISL_923327, EPI_ISL_923329, EPI_ISL_923331, EPI_ISL_923332, EPI_ISL_923333, EPI_ISL_923334, EPI_ISL_923335, EPI_ISL_923336, EPI_ISL_923337, EPI_ISL_923338, EPI_ISL_923339, EPI_ISL_923340, EPI_ISL_923341, EPI_ISL_923342, EPI_ISL_923343, EPI_ISL_923344, EPI_ISL_923346, EPI_ISL_923347, EPI_ISL_923348, EPI_ISL_923350, EPI_ISL_923352, EPI_ISL_923353, EPI_ISL_923354, EPI_ISL_923357, EPI_ISL_923358, EPI_ISL_923363, EPI_ISL_923366, EPI_ISL_923367, EPI_ISL_923369, EPI_ISL_923415, EPI_ISL_923417, EPI_ISL_923418, EPI_ISL_923420, EPI_ISL_923421, EPI_ISL_923436, EPI_ISL_923595, EPI_ISL_923596, EPI_ISL_923597, EPI_ISL_923598, EPI_ISL_923680, EPI_ISL_952386, EPI_ISL_952387, EPI_ISL_952393, EPI_ISL_952405, EPI_ISL_952410, EPI_ISL_952412, EPI_ISL_952416, EPI_ISL_952432, EPI_ISL_952489, EPI_ISL_952490, EPI_ISL_952491, EPI_ISL_952521, EPI_ISL_952522, EPI_ISL_952528, EPI_ISL_952529, EPI_ISL_952538, EPI_ISL_952554, EPI_ISL_952636, EPI_ISL_952673, EPI_ISL_952675, EPI_ISL_952677, EPI_ISL_952680, EPI_ISL_952689, EPI_ISL_952783, EPI_ISL_952820, EPI_ISL_952821, EPI_ISL_952830, EPI_ISL_952831, EPI_ISL_952832, EPI_ISL_952833, EPI_ISL_952834, EPI_ISL_952835, EPI_ISL_952842, EPI_ISL_952843, EPI_ISL_952844, EPI_ISL_952845, EPI_ISL_952846, EPI_ISL_952847, EPI_ISL_952848, EPI_ISL_952849, EPI_ISL_952850, EPI_ISL_952852, EPI_ISL_952853, EPI_ISL_1000256, EPI_ISL_1000404, EPI_ISL_1000405, EPI_ISL_1000406, EPI_ISL_1000407, EPI_ISL_1000408, EPI_ISL_1000409, EPI_ISL_1000410, EPI_ISL_1000411, EPI_ISL_1000412, EPI_ISL_1000413, EPI_ISL_1000414, EPI_ISL_1000415, EPI_ISL_1000446, EPI_ISL_1000447, EPI_ISL_1000450, EPI_ISL_1000451, EPI_ISL_1000452, EPI_ISL_1000453, EPI_ISL_1000472, EPI_ISL_1000473, EPI_ISL_1000474, EPI_ISL_1000475, EPI_ISL_1000476, EPI_ISL_1053820, EPI_ISL_1053822, EPI_ISL_1053824, EPI_ISL_1053861, EPI_ISL_1053862, EPI_ISL_1053863, EPI_ISL_1053864, EPI_ISL_1053865, EPI_ISL_1053866, EPI_ISL_1053867, EPI_ISL_1053868, EPI_ISL_1053869, EPI_ISL_1053870, EPI_ISL_1053871, EPI_ISL_1053872, EPI_ISL_1053873, EPI_ISL_1053874, EPI_ISL_1053875, EPI_ISL_1053876, EPI_ISL_1179834, EPI_ISL_1179839, EPI_ISL_1310348, EPI_ISL_1310350, EPI_ISL_1730873, EPI_ISL_1730874, EPI_ISL_1730875, EPI_ISL_1979963, EPI_ISL_1979965, EPI_ISL_1979967, EPI_ISL_1980055, EPI_ISL_1980057, EPI_ISL_1980059, EPI_ISL_1980061, EPI_ISL_1980062, EPI_ISL_1980064, EPI_ISL_1980065, EPI_ISL_1980066, EPI_ISL_1980067, EPI_ISL_1980068, EPI_ISL_1980069, EPI_ISL_1980070, EPI_ISL_2128028, EPI_ISL_2128031 | | | |
| see above | Centre for Enzyme Innovation, University of Portsmouth / Translational Research Laboratory, Portsmouth Hospitals NHS Trust | COVID-19 Genomics UK (COG-UK) Consortium | Allyson Lloyd; Angela Beckett; Anoop Chauhan; Christopher Fearn; Ethan Butcher; Garry Scarlett; Kate Cook; Katie Loveson; Kelly Bicknell; Robert Impey; Salman Goudarzi; Samuel Robson; Sarah Wyllie; Scott Elliott; Sharon Glaysher; Yann Bourgeois |
| EPI_ISL_864929, EPI_ISL_864932, EPI_ISL_864936, EPI_ISL_864945, EPI_ISL_918985, EPI_ISL_918986, EPI_ISL_918987, EPI_ISL_918988, EPI_ISL_918989, EPI_ISL_918990, EPI_ISL_918991, EPI_ISL_918992, EPI_ISL_918993, EPI_ISL_918994, EPI_ISL_918995, EPI_ISL_918996, EPI_ISL_918998, EPI_ISL_918999, EPI_ISL_919000, EPI_ISL_919001, EPI_ISL_919002, EPI_ISL_919003, EPI_ISL_919004, EPI_ISL_919006, EPI_ISL_919008, EPI_ISL_919009, EPI_ISL_919010, EPI_ISL_919013, EPI_ISL_919014, EPI_ISL_919015, EPI_ISL_919016, EPI_ISL_919115, EPI_ISL_919148, EPI_ISL_923688, EPI_ISL_923753, EPI_ISL_923754, EPI_ISL_923756, EPI_ISL_923758, EPI_ISL_923759, EPI_ISL_923760, EPI_ISL_923763, EPI_ISL_923764, EPI_ISL_923779, EPI_ISL_923780, EPI_ISL_923781, EPI_ISL_923782, EPI_ISL_923783, EPI_ISL_923784, EPI_ISL_923785, EPI_ISL_923786, EPI_ISL_923787, EPI_ISL_923788, EPI_ISL_923789, EPI_ISL_923790, EPI_ISL_923791, EPI_ISL_923792, EPI_ISL_923793, EPI_ISL_923794, EPI_ISL_923820, EPI_ISL_923821, EPI_ISL_923822, EPI_ISL_923823, EPI_ISL_923824, EPI_ISL_923825, EPI_ISL_923826, EPI_ISL_923827, EPI_ISL_923828, EPI_ISL_923829, EPI_ISL_923830, EPI_ISL_923831, EPI_ISL_923832, EPI_ISL_923833, EPI_ISL_923834, EPI_ISL_923835, EPI_ISL_923836, EPI_ISL_923837, EPI_ISL_923838, EPI_ISL_923839, EPI_ISL_923840, EPI_ISL_923841, EPI_ISL_923875, EPI_ISL_923876, EPI_ISL_923877, EPI_ISL_923878, EPI_ISL_923879, EPI_ISL_923880, EPI_ISL_923881, EPI_ISL_923882, EPI_ISL_923883, EPI_ISL_923884, EPI_ISL_923885, EPI_ISL_923886, EPI_ISL_923887, EPI_ISL_923888, EPI_ISL_923889, EPI_ISL_923890, EPI_ISL_923891, EPI_ISL_923892, EPI_ISL_923893, EPI_ISL_923894, EPI_ISL_923895, EPI_ISL_923896, EPI_ISL_923897, EPI_ISL_923898, EPI_ISL_923899, EPI_ISL_923900, EPI_ISL_923901, EPI_ISL_923902, EPI_ISL_923903, EPI_ISL_923971, EPI_ISL_923974, EPI_ISL_923975, EPI_ISL_924027, EPI_ISL_924028, EPI_ISL_949299, EPI_ISL_949300, EPI_ISL_949301, EPI_ISL_949302, EPI_ISL_949303, EPI_ISL_949304, EPI_ISL_949305, EPI_ISL_949306, EPI_ISL_949307, EPI_ISL_949308, EPI_ISL_949309, EPI_ISL_949310, EPI_ISL_949312, EPI_ISL_949313, EPI_ISL_949314, EPI_ISL_949317, EPI_ISL_949322, EPI_ISL_949428, EPI_ISL_949442, EPI_ISL_949443, EPI_ISL_949444, EPI_ISL_949448, EPI_ISL_949478, EPI_ISL_949480, EPI_ISL_949481, EPI_ISL_949482, EPI_ISL_949483, EPI_ISL_949484, EPI_ISL_949486, EPI_ISL_949488, EPI_ISL_949489, EPI_ISL_949491, EPI_ISL_949493, EPI_ISL_949495, EPI_ISL_949497, EPI_ISL_949499, EPI_ISL_949501, EPI_ISL_949503, EPI_ISL_949505, EPI_ISL_949507, EPI_ISL_949509, EPI_ISL_949511, EPI_ISL_949513, EPI_ISL_949515, EPI_ISL_949517, EPI_ISL_952858, EPI_ISL_952859, EPI_ISL_952860, EPI_ISL_952861, EPI_ISL_952862, EPI_ISL_952863, EPI_ISL_952864, EPI_ISL_952865, EPI_ISL_952866, EPI_ISL_952867, EPI_ISL_952868, EPI_ISL_952869, EPI_ISL_952870, EPI_ISL_952871, EPI_ISL_952872, EPI_ISL_952873, EPI_ISL_952874, EPI_ISL_952875, EPI_ISL_952876, EPI_ISL_952877, EPI_ISL_952878, EPI_ISL_952879, EPI_ISL_952880, EPI_ISL_952881, EPI_ISL_952882, EPI_ISL_952883, EPI_ISL_952884, EPI_ISL_952885, EPI_ISL_952886, EPI_ISL_952887, EPI_ISL_952888, EPI_ISL_952889, EPI_ISL_952890, EPI_ISL_952891, EPI_ISL_952892, EPI_ISL_952893, EPI_ISL_952894, EPI_ISL_952895, EPI_ISL_952914, EPI_ISL_952915, EPI_ISL_952916, EPI_ISL_952917, EPI_ISL_952918, EPI_ISL_952919, EPI_ISL_952920, EPI_ISL_952921, EPI_ISL_952922, EPI_ISL_952923, EPI_ISL_952924, EPI_ISL_952925, EPI_ISL_952926, EPI_ISL_952927, EPI_ISL_952928, EPI_ISL_952929, EPI_ISL_952930, EPI_ISL_952931, EPI_ISL_952933, EPI_ISL_952934, EPI_ISL_952935, EPI_ISL_952936, EPI_ISL_952937, EPI_ISL_952938, EPI_ISL_952939, EPI_ISL_952940, EPI_ISL_952941, EPI_ISL_996700, EPI_ISL_996702, EPI_ISL_996706, EPI_ISL_996710, EPI_ISL_996714, EPI_ISL_996716, EPI_ISL_996718, EPI_ISL_996721, EPI_ISL_996723, EPI_ISL_1047069 | | | |
| see above | Department of Pathology, University of Cambridge | COVID-19 Genomics UK (COG-UK) Consortium | Aminu S. Jahun; Grant Hall; Ian Goodfellow; Iliana Georgana; Malte Pinckert; Martin D. Curran; Myra Hosmillo; Rhys Izu; Rhys Izuagbe; Surendra Parmar; William L. Hamilton; Yasmin Chaudhry |
| EPI_ISL_877770, EPI_ISL_877772, EPI_ISL_877773, EPI_ISL_877774, EPI_ISL_877775, EPI_ISL_877776, EPI_ISL_877777, EPI_ISL_877778, EPI_ISL_877779, EPI_ISL_877780, EPI_ISL_877782, EPI_ISL_877783, EPI_ISL_877784, EPI_ISL_877785, EPI_ISL_877786, EPI_ISL_877789, EPI_ISL_877790, EPI_ISL_877791, EPI_ISL_877792, EPI_ISL_877793, EPI_ISL_877794, EPI_ISL_877795, EPI_ISL_877798, EPI_ISL_877799, EPI_ISL_877801, EPI_ISL_877803, EPI_ISL_877805, EPI_ISL_877806, EPI_ISL_877808, EPI_ISL_877810, EPI_ISL_877811, EPI_ISL_877812, EPI_ISL_877813, EPI_ISL_877814, EPI_ISL_877815, EPI_ISL_877816, EPI_ISL_877817, EPI_ISL_877818, EPI_ISL_877819, EPI_ISL_877820, EPI_ISL_877822, EPI_ISL_877823, EPI_ISL_877824, EPI_ISL_877825, EPI_ISL_877826, EPI_ISL_877827, EPI_ISL_877829, EPI_ISL_877830, EPI_ISL_877831, EPI_ISL_877832, EPI_ISL_877834, EPI_ISL_877835, EPI_ISL_877836, EPI_ISL_877837, EPI_ISL_877838, EPI_ISL_877839, EPI_ISL_877840, EPI_ISL_877841, EPI_ISL_877842, EPI_ISL_877843, EPI_ISL_877845, EPI_ISL_877846, EPI_ISL_877847, EPI_ISL_877848, EPI_ISL_877849, EPI_ISL_877850, EPI_ISL_877851, EPI_ISL_877852, EPI_ISL_877853, EPI_ISL_877854, EPI_ISL_877855, EPI_ISL_877856, EPI_ISL_877857, EPI_ISL_877858, EPI_ISL_877859, EPI_ISL_877860, EPI_ISL_877861, EPI_ISL_877862, EPI_ISL_877863, EPI_ISL_877864, EPI_ISL_877865, EPI_ISL_877866, EPI_ISL_877867, EPI_ISL_877868, EPI_ISL_877869, EPI_ISL_877870, EPI_ISL_877871, EPI_ISL_877872, EPI_ISL_877873, EPI_ISL_877874, EPI_ISL_877875, EPI_ISL_877876, EPI_ISL_877877, EPI_ISL_877878, EPI_ISL_877879, EPI_ISL_877880, EPI_ISL_877882, EPI_ISL_877883, EPI_ISL_877886, EPI_ISL_877887, EPI_ISL_877888, EPI_ISL_877890, EPI_ISL_877891, EPI_ISL_877892, EPI_ISL_877893, EPI_ISL_877894, EPI_ISL_877895, EPI_ISL_877896, EPI_ISL_877897, EPI_ISL_877898, EPI_ISL_877900, EPI_ISL_877901, EPI_ISL_877902, EPI_ISL_877904, EPI_ISL_877905, EPI_ISL_877906, EPI_ISL_877908, EPI_ISL_877909, EPI_ISL_877910, EPI_ISL_877911, EPI_ISL_877912, EPI_ISL_877914, EPI_ISL_877915, EPI_ISL_877916, EPI_ISL_877917, EPI_ISL_877919, EPI_ISL_877920, EPI_ISL_877921, EPI_ISL_877923, EPI_ISL_877925, EPI_ISL_877926, EPI_ISL_877927, EPI_ISL_877928, EPI_ISL_877929, EPI_ISL_877931, EPI_ISL_877932, EPI_ISL_877933, EPI_ISL_877934, EPI_ISL_877935, EPI_ISL_877936, EPI_ISL_877937, EPI_ISL_877939, EPI_ISL_877941, EPI_ISL_877942, EPI_ISL_877943, EPI_ISL_877944, EPI_ISL_877945, EPI_ISL_877946, EPI_ISL_877947, EPI_ISL_877949, EPI_ISL_877950, EPI_ISL_877951, EPI_ISL_877952, EPI_ISL_877954, EPI_ISL_877956, EPI_ISL_877957, EPI_ISL_877958, EPI_ISL_877959, EPI_ISL_877960, EPI_ISL_877961, EPI_ISL_877962, EPI_ISL_877963, EPI_ISL_877965, EPI_ISL_877966, EPI_ISL_877967, EPI_ISL_877968, EPI_ISL_877970, EPI_ISL_877971, EPI_ISL_877973, EPI_ISL_877974, EPI_ISL_877976, EPI_ISL_877977, EPI_ISL_877978, EPI_ISL_877979, EPI_ISL_877980, EPI_ISL_877982, EPI_ISL_877983, EPI_ISL_877984, EPI_ISL_877985, EPI_ISL_877986, EPI_ISL_877987, EPI_ISL_877988, EPI_ISL_877990, EPI_ISL_877991, EPI_ISL_877992, EPI_ISL_877993, EPI_ISL_877994, EPI_ISL_877996, EPI_ISL_877998, EPI_ISL_877999, EPI_ISL_878000, EPI_ISL_878001, EPI_ISL_878002, EPI_ISL_878003, EPI_ISL_878004, EPI_ISL_878005, EPI_ISL_878006, EPI_ISL_878007, EPI_ISL_878008, EPI_ISL_878009, EPI_ISL_878010, EPI_ISL_878012, EPI_ISL_878014, EPI_ISL_878015, EPI_ISL_878016, EPI_ISL_878017, EPI_ISL_878018, EPI_ISL_878019, EPI_ISL_878020, EPI_ISL_878022, EPI_ISL_878023, EPI_ISL_878024, EPI_ISL_878027, EPI_ISL_878028, EPI_ISL_878029, EPI_ISL_878030, EPI_ISL_878031, EPI_ISL_878033, EPI_ISL_878034, EPI_ISL_878035, EPI_ISL_878036, EPI_ISL_878037, EPI_ISL_878039, EPI_ISL_878043, EPI_ISL_878045, EPI_ISL_878046, EPI_ISL_878047, EPI_ISL_878048, EPI_ISL_878049, EPI_ISL_878050, EPI_ISL_878051, EPI_ISL_878052, EPI_ISL_878055, EPI_ISL_878056, EPI_ISL_878057, EPI_ISL_878058, EPI_ISL_878059, EPI_ISL_878060, EPI_ISL_878061, EPI_ISL_878063, EPI_ISL_878065, EPI_ISL_878066, EPI_ISL_878068, EPI_ISL_878069, EPI_ISL_878070, EPI_ISL_878071, EPI_ISL_878072, EPI_ISL_878073, EPI_ISL_878074, EPI_ISL_878075, EPI_ISL_878076, EPI_ISL_878077, EPI_ISL_878079, EPI_ISL_878080, EPI_ISL_878081, EPI_ISL_878082, EPI_ISL_878084, EPI_ISL_878085, EPI_ISL_878086, EPI_ISL_878087, EPI_ISL_878090, EPI_ISL_878091, EPI_ISL_878092, EPI_ISL_878093, EPI_ISL_878094, EPI_ISL_878095, EPI_ISL_878096, EPI_ISL_878097, EPI_ISL_878099, EPI_ISL_878100, EPI_ISL_878101, EPI_ISL_878103, EPI_ISL_878104, EPI_ISL_878106, EPI_ISL_878107, EPI_ISL_878109, EPI_ISL_878110, EPI_ISL_878134, EPI_ISL_878158, EPI_ISL_878168, EPI_ISL_878203, EPI_ISL_878211, EPI_ISL_878238, EPI_ISL_878329, EPI_ISL_878433, EPI_ISL_878447, EPI_ISL_878479, EPI_ISL_878536, EPI_ISL_878545, EPI_ISL_878553, EPI_ISL_878562, EPI_ISL_878566, EPI_ISL_878578, EPI_ISL_878610, EPI_ISL_878623, EPI_ISL_878699, EPI_ISL_878708, EPI_ISL_878717, EPI_ISL_878741, EPI_ISL_878746, EPI_ISL_878754, EPI_ISL_878777, EPI_ISL_878785, EPI_ISL_878804, EPI_ISL_878808, EPI_ISL_878809, EPI_ISL_878812, EPI_ISL_878817, EPI_ISL_878820, EPI_ISL_878822, EPI_ISL_878835, EPI_ISL_878836, EPI_ISL_878840, EPI_ISL_878843, EPI_ISL_878849, EPI_ISL_878853, EPI_ISL_878859, EPI_ISL_878865, EPI_ISL_878866, EPI_ISL_878868, EPI_ISL_878875, EPI_ISL_878878, EPI_ISL_878883, EPI_ISL_878887, EPI_ISL_878888, EPI_ISL_878891, EPI_ISL_878909, EPI_ISL_878914, EPI_ISL_878917, EPI_ISL_878920, EPI_ISL_878925, EPI_ISL_878926, EPI_ISL_878927, EPI_ISL_878929, EPI_ISL_878932, EPI_ISL_878936, EPI_ISL_878938, EPI_ISL_878946, EPI_ISL_878947, EPI_ISL_878955, EPI_ISL_878958, EPI_ISL_878961, EPI_ISL_878965, EPI_ISL_878970, EPI_ISL_878979, EPI_ISL_878981, EPI_ISL_878983, EPI_ISL_878985, EPI_ISL_878992, EPI_ISL_878994, EPI_ISL_879000, EPI_ISL_879003, EPI_ISL_879008, EPI_ISL_879015, EPI_ISL_879018, EPI_ISL_879026, EPI_ISL_879033, EPI_ISL_879061, EPI_ISL_879066, EPI_ISL_879072, EPI_ISL_879077, EPI_ISL_879079, EPI_ISL_879081, EPI_ISL_879083, EPI_ISL_879088, EPI_ISL_879091, EPI_ISL_879097, EPI_ISL_879099, EPI_ISL_879101, EPI_ISL_879103, EPI_ISL_879108, EPI_ISL_879111, EPI_ISL_879113, EPI_ISL_879114, EPI_ISL_879125, EPI_ISL_879126, EPI_ISL_879128, EPI_ISL_879129, EPI_ISL_879130, EPI_ISL_879131, EPI_ISL_879132, EPI_ISL_879137, EPI_ISL_879140, EPI_ISL_879144, EPI_ISL_879146, EPI_ISL_879147, EPI_ISL_879151, EPI_ISL_879152, EPI_ISL_879157, EPI_ISL_879159, EPI_ISL_879160, EPI_ISL_879162, EPI_ISL_879181, EPI_ISL_879188, EPI_ISL_879191, EPI_ISL_879194, EPI_ISL_879199, EPI_ISL_879202, EPI_ISL_879203, EPI_ISL_879212, EPI_ISL_879218, EPI_ISL_879219, EPI_ISL_879224, EPI_ISL_879225, EPI_ISL_879229, EPI_ISL_879231, EPI_ISL_879237, EPI_ISL_879244, EPI_ISL_879250, EPI_ISL_879252, EPI_ISL_879259, EPI_ISL_879263, EPI_ISL_879265, EPI_ISL_879266, EPI_ISL_879271, EPI_ISL_879273, EPI_ISL_879276, EPI_ISL_879277, EPI_ISL_879279, EPI_ISL_879296, EPI_ISL_879297, EPI_ISL_879298, EPI_ISL_879299, EPI_ISL_879300, EPI_ISL_879302, EPI_ISL_879303, EPI_ISL_879306, EPI_ISL_879307, EPI_ISL_879308, EPI_ISL_879313, EPI_ISL_879316, EPI_ISL_879323, EPI_ISL_879324, EPI_ISL_879340, EPI_ISL_879342, EPI_ISL_879345, EPI_ISL_879346, EPI_ISL_879347, EPI_ISL_879351, EPI_ISL_879357, EPI_ISL_879359, EPI_ISL_879360, EPI_ISL_879361, EPI_ISL_879362, EPI_ISL_879367, EPI_ISL_879371, EPI_ISL_879386, EPI_ISL_879392, EPI_ISL_879415, EPI_ISL_879418, EPI_ISL_879420, EPI_ISL_879427, EPI_ISL_879444, EPI_ISL_879446, EPI_ISL_879456, EPI_ISL_879476, EPI_ISL_879483, EPI_ISL_879493, EPI_ISL_879497, EPI_ISL_879501, EPI_ISL_879505, EPI_ISL_879515, EPI_ISL_879518, EPI_ISL_879522, EPI_ISL_879534, EPI_ISL_879535, EPI_ISL_879542, EPI_ISL_879564, EPI_ISL_879578, EPI_ISL_879582, EPI_ISL_879589, EPI_ISL_879593, EPI_ISL_879602, EPI_ISL_879603, EPI_ISL_879604, EPI_ISL_879616, EPI_ISL_879619, EPI_ISL_879627, EPI_ISL_879641, EPI_ISL_879647, EPI_ISL_879649, EPI_ISL_879665, EPI_ISL_879674, EPI_ISL_879682, EPI_ISL_879683, EPI_ISL_879687, EPI_ISL_879695, EPI_ISL_879702, EPI_ISL_885155, EPI_ISL_885156, EPI_ISL_885157, EPI_ISL_885158, EPI_ISL_885159, EPI_ISL_885161, EPI_ISL_885162, EPI_ISL_885164, EPI_ISL_885166, EPI_ISL_885167, EPI_ISL_885168, EPI_ISL_885169, EPI_ISL_885170, EPI_ISL_885171, EPI_ISL_885172, EPI_ISL_885173, EPI_ISL_885174, EPI_ISL_885175, EPI_ISL_885176, EPI_ISL_885177, EPI_ISL_885179, EPI_ISL_885181, EPI_ISL_885182, EPI_ISL_885184, EPI_ISL_885185, EPI_ISL_885186, EPI_ISL_885188, EPI_ISL_885190, EPI_ISL_885191, EPI_ISL_885192, EPI_ISL_885193, EPI_ISL_885194, EPI_ISL_885195, EPI_ISL_885196, EPI_ISL_885198, EPI_ISL_885199, EPI_ISL_885200, EPI_ISL_885201, EPI_ISL_885202, EPI_ISL_885203, EPI_ISL_885205, EPI_ISL_885206, EPI_ISL_885207, EPI_ISL_885208, EPI_ISL_885210, EPI_ISL_885212, EPI_ISL_885216, EPI_ISL_885217, EPI_ISL_885218, EPI_ISL_885219, EPI_ISL_885220, EPI_ISL_885221, EPI_ISL_885222, EPI_ISL_885223, EPI_ISL_885224, EPI_ISL_885225, EPI_ISL_885226, EPI_ISL_885227, EPI_ISL_885228, EPI_ISL_885229, EPI_ISL_885230, EPI_ISL_885231, EPI_ISL_885232, EPI_ISL_885233, EPI_ISL_885235, EPI_ISL_885236, EPI_ISL_885237, EPI_ISL_885239, EPI_ISL_885241, EPI_ISL_885242, EPI_ISL_885243, EPI_ISL_885245, EPI_ISL_885246, EPI_ISL_885247, EPI_ISL_885249, EPI_ISL_885251, EPI_ISL_885252, EPI_ISL_885253, EPI_ISL_885254, EPI_ISL_885255, EPI_ISL_885256, EPI_ISL_885257, EPI_ISL_885258, EPI_ISL_885259, EPI_ISL_885260, EPI_ISL_885261, EPI_ISL_885262, EPI_ISL_885263, EPI_ISL_885265, EPI_ISL_885266, EPI_ISL_885267, EPI_ISL_885268, EPI_ISL_885269, EPI_ISL_885270, EPI_ISL_885271, EPI_ISL_885272, EPI_ISL_885276, EPI_ISL_885278, EPI_ISL_885279, EPI_ISL_885280, EPI_ISL_885281, EPI_ISL_885282, EPI_ISL_885283, EPI_ISL_885284, EPI_ISL_885285, EPI_ISL_885286, EPI_ISL_885287, EPI_ISL_885290, EPI_ISL_885292, EPI_ISL_885293, EPI_ISL_885294, EPI_ISL_885296, EPI_ISL_885298, EPI_ISL_885300, EPI_ISL_885301, EPI_ISL_885303, EPI_ISL_885304, EPI_ISL_885306, EPI_ISL_885308, EPI_ISL_885309, EPI_ISL_885310, EPI_ISL_885312, EPI_ISL_885313, EPI_ISL_885314, EPI_ISL_885315, EPI_ISL_885316, EPI_ISL_885318, EPI_ISL_885320, EPI_ISL_885322, EPI_ISL_885324, EPI_ISL_885325, EPI_ISL_885328, EPI_ISL_885329, EPI_ISL_885330, EPI_ISL_885331, EPI_ISL_885332, EPI_ISL_885333, EPI_ISL_885334, EPI_ISL_885336, EPI_ISL_885338, EPI_ISL_885339, EPI_ISL_885340, EPI_ISL_885341, EPI_ISL_885342, EPI_ISL_885343, EPI_ISL_885344, EPI_ISL_885345, EPI_ISL_885346, EPI_ISL_885347, EPI_ISL_885348, EPI_ISL_885350, EPI_ISL_885351, EPI_ISL_885354, EPI_ISL_885355, EPI_ISL_885356, EPI_ISL_885357, EPI_ISL_885358, EPI_ISL_885360, EPI_ISL_885362, EPI_ISL_885363, EPI_ISL_885366, EPI_ISL_885368, EPI_ISL_885370, EPI_ISL_885372, EPI_ISL_885373, EPI_ISL_885375, EPI_ISL_885377, EPI_ISL_885378, EPI_ISL_885379, EPI_ISL_885380, EPI_ISL_885381, EPI_ISL_885382, EPI_ISL_885383, EPI_ISL_885384, EPI_ISL_885385, EPI_ISL_885386, EPI_ISL_885387, EPI_ISL_885389, EPI_ISL_885391, EPI_ISL_885393, EPI_ISL_885394, EPI_ISL_885395, EPI_ISL_885398, EPI_ISL_885399, EPI_ISL_885400, EPI_ISL_885401, EPI_ISL_885403, EPI_ISL_885404, EPI_ISL_885405, EPI_ISL_885406, EPI_ISL_885407, EPI_ISL_885408, EPI_ISL_885410, EPI_ISL_885412, EPI_ISL_885414, EPI_ISL_885415, EPI_ISL_885416, EPI_ISL_885417, EPI_ISL_885418, EPI_ISL_885420, EPI_ISL_885421, EPI_ISL_885423, EPI_ISL_885424, EPI_ISL_885425, EPI_ISL_885426, EPI_ISL_885428, EPI_ISL_885429, EPI_ISL_885430, EPI_ISL_885432, EPI_ISL_885433, EPI_ISL_885435, EPI_ISL_885436, EPI_ISL_885437, EPI_ISL_885438, EPI_ISL_885439, EPI_ISL_885440, EPI_ISL_885441, EPI_ISL_885442, EPI_ISL_885443, EPI_ISL_885444, EPI_ISL_885445, EPI_ISL_885446, EPI_ISL_885447, EPI_ISL_885448, EPI_ISL_885449, EPI_ISL_885450, EPI_ISL_885451, EPI_ISL_885453, EPI_ISL_885454, EPI_ISL_885456, EPI_ISL_885457, EPI_ISL_885460, EPI_ISL_885462, EPI_ISL_885464, EPI_ISL_885465, EPI_ISL_885466, EPI_ISL_885467, EPI_ISL_885468, EPI_ISL_885469, EPI_ISL_885470, EPI_ISL_885474, EPI_ISL_885475, EPI_ISL_885477, EPI_ISL_885480, EPI_ISL_885481, EPI_ISL_885483, EPI_ISL_885485, EPI_ISL_885486, EPI_ISL_885487, EPI_ISL_885488, EPI_ISL_885489, EPI_ISL_885490, EPI_ISL_885491, EPI_ISL_885492, EPI_ISL_885493, EPI_ISL_885494, EPI_ISL_885495, EPI_ISL_885496, EPI_ISL_885497, EPI_ISL_885498, EPI_ISL_885501, EPI_ISL_885502, EPI_ISL_885504, EPI_ISL_885505, EPI_ISL_885506, EPI_ISL_885507, EPI_ISL_885508, EPI_ISL_885509, EPI_ISL_885510, EPI_ISL_885511, EPI_ISL_885512, EPI_ISL_885515, EPI_ISL_885516, EPI_ISL_885517, EPI_ISL_885518, EPI_ISL_885519, EPI_ISL_885520, EPI_ISL_885521, EPI_ISL_885522, EPI_ISL_885523, EPI_ISL_885524, EPI_ISL_885525, EPI_ISL_885526, EPI_ISL_885527, EPI_ISL_885528, EPI_ISL_885529, EPI_ISL_885530, EPI_ISL_885531, EPI_ISL_885532, EPI_ISL_885533, EPI_ISL_885534, EPI_ISL_885535, EPI_ISL_885536, EPI_ISL_885537, EPI_ISL_885538, EPI_ISL_885539, EPI_ISL_885540, EPI_ISL_885542, EPI_ISL_885544, EPI_ISL_885545, EPI_ISL_885546, EPI_ISL_885547, EPI_ISL_885548, EPI_ISL_885549, EPI_ISL_885550, EPI_ISL_885551, EPI_ISL_885552, EPI_ISL_885553, EPI_ISL_885554, EPI_ISL_885555, EPI_ISL_885556, EPI_ISL_885558, EPI_ISL_885559, EPI_ISL_885560, EPI_ISL_885561, EPI_ISL_885562, EPI_ISL_885563, EPI_ISL_885564, EPI_ISL_885565, EPI_ISL_885566, EPI_ISL_885567, EPI_ISL_885568, EPI_ISL_885569, EPI_ISL_885570, EPI_ISL_885571, EPI_ISL_885572, EPI_ISL_885573, EPI_ISL_885574, EPI_ISL_885575, EPI_ISL_885576, EPI_ISL_885577, EPI_ISL_885578, EPI_ISL_885579, EPI_ISL_885581, EPI_ISL_885582, EPI_ISL_885583, EPI_ISL_885584, EPI_ISL_885585, EPI_ISL_885588, EPI_ISL_885590, EPI_ISL_885591, EPI_ISL_885592, EPI_ISL_885593, EPI_ISL_885594, EPI_ISL_885595, EPI_ISL_885596, EPI_ISL_885598, EPI_ISL_885599, EPI_ISL_885600, EPI_ISL_885601, EPI_ISL_885602, EPI_ISL_885603, EPI_ISL_885604, EPI_ISL_885605, EPI_ISL_885606, EPI_ISL_885607, EPI_ISL_885608, EPI_ISL_885609, EPI_ISL_885610, EPI_ISL_885611, EPI_ISL_885612, EPI_ISL_885613, EPI_ISL_885614, EPI_ISL_885615, EPI_ISL_885616, EPI_ISL_885617, EPI_ISL_885618, EPI_ISL_885620, EPI_ISL_885621, EPI_ISL_885622, EPI_ISL_885623, EPI_ISL_885624, EPI_ISL_885625, EPI_ISL_885626, EPI_ISL_885627, EPI_ISL_885628, EPI_ISL_885629, EPI_ISL_885631, EPI_ISL_885632, EPI_ISL_885633, EPI_ISL_885634, EPI_ISL_885635, EPI_ISL_885636, EPI_ISL_885638, EPI_ISL_885639, EPI_ISL_885641, EPI_ISL_885642, EPI_ISL_885643, EPI_ISL_885644, EPI_ISL_885645, EPI_ISL_885646, EPI_ISL_885647, EPI_ISL_885648, EPI_ISL_885650, EPI_ISL_885651, EPI_ISL_885652, EPI_ISL_885653, EPI_ISL_885654, EPI_ISL_885656, EPI_ISL_885657, EPI_ISL_885658, EPI_ISL_885659, EPI_ISL_885660, EPI_ISL_885662, EPI_ISL_885663, EPI_ISL_885664, EPI_ISL_885665, EPI_ISL_885666, EPI_ISL_885667, EPI_ISL_885668, EPI_ISL_885669, EPI_ISL_885670, EPI_ISL_885671, EPI_ISL_885672, EPI_ISL_885673, EPI_ISL_885674, EPI_ISL_885675, EPI_ISL_885676, EPI_ISL_885677, EPI_ISL_885678, EPI_ISL_885679, EPI_ISL_885681, EPI_ISL_885682, EPI_ISL_885684, EPI_ISL_885685, EPI_ISL_885686, EPI_ISL_885687, EPI_ISL_885688, EPI_ISL_885689, EPI_ISL_885691, EPI_ISL_885692, EPI_ISL_885693, EPI_ISL_885694, EPI_ISL_885695, EPI_ISL_885696, EPI_ISL_885697, EPI_ISL_885698, EPI_ISL_885699, EPI_ISL_885700, EPI_ISL_885701, EPI_ISL_885702, EPI_ISL_885703, EPI_ISL_885704, EPI_ISL_885706, EPI_ISL_885707, EPI_ISL_885708, EPI_ISL_885709, EPI_ISL_885710, EPI_ISL_885711, EPI_ISL_885712, EPI_ISL_885713, EPI_ISL_885714, EPI_ISL_885715, EPI_ISL_885716, EPI_ISL_885717, EPI_ISL_885718, EPI_ISL_885719, EPI_ISL_885720, EPI_ISL_885721, EPI_ISL_885722, EPI_ISL_885723, EPI_ISL_885724, EPI_ISL_885726, EPI_ISL_885727, EPI_ISL_885728, EPI_ISL_885729, EPI_ISL_885730, EPI_ISL_885731, EPI_ISL_885732, EPI_ISL_885733, EPI_ISL_885734, EPI_ISL_885735, EPI_ISL_885736, EPI_ISL_885737, EPI_ISL_885738, EPI_ISL_885739, EPI_ISL_885740, EPI_ISL_885741, EPI_ISL_885742, EPI_ISL_885743, EPI_ISL_885744, EPI_ISL_885745, EPI_ISL_885746, EPI_ISL_885747, EPI_ISL_885748, EPI_ISL_885749, EPI_ISL_885750, EPI_ISL_885751, EPI_ISL_885752, EPI_ISL_885753, EPI_ISL_885755, EPI_ISL_885756, EPI_ISL_885757, EPI_ISL_885759, EPI_ISL_885760, EPI_ISL_885761, EPI_ISL_885763, EPI_ISL_885764, EPI_ISL_885765, EPI_ISL_885767, EPI_ISL_885768, EPI_ISL_885769, EPI_ISL_885770, EPI_ISL_885771, EPI_ISL_885772, EPI_ISL_885773, EPI_ISL_885774, EPI_ISL_885775, EPI_ISL_885776, EPI_ISL_885777, EPI_ISL_885778, EPI_ISL_885779, EPI_ISL_885780, EPI_ISL_885781, EPI_ISL_885782, EPI_ISL_885783, EPI_ISL_885784, EPI_ISL_885785, EPI_ISL_885786, EPI_ISL_885787, EPI_ISL_885788, EPI_ISL_885789, EPI_ISL_885790, EPI_ISL_885791, EPI_ISL_885792, EPI_ISL_885793, EPI_ISL_885794, EPI_ISL_885795, EPI_ISL_885796, EPI_ISL_885797, EPI_ISL_885798, EPI_ISL_885799, EPI_ISL_885800, EPI_ISL_885801, EPI_ISL_885802, EPI_ISL_885803, EPI_ISL_885804, EPI_ISL_885805, EPI_ISL_885806, EPI_ISL_885807, EPI_ISL_885808, EPI_ISL_885809, EPI_ISL_885810, EPI_ISL_885811, EPI_ISL_885812, EPI_ISL_885814, EPI_ISL_885815, EPI_ISL_885816, EPI_ISL_885817, EPI_ISL_885818, EPI_ISL_885819, EPI_ISL_885820, EPI_ISL_885821, EPI_ISL_885822, EPI_ISL_885823, EPI_ISL_885824, EPI_ISL_885825, EPI_ISL_885826, EPI_ISL_885827, EPI_ISL_885829, EPI_ISL_885830, EPI_ISL_885831, EPI_ISL_885832, EPI_ISL_885833, EPI_ISL_885834, EPI_ISL_885835, EPI_ISL_885836, EPI_ISL_885837, EPI_ISL_885838, EPI_ISL_885839, EPI_ISL_885840, EPI_ISL_885841, EPI_ISL_885842, EPI_ISL_885843, EPI_ISL_885844, EPI_ISL_885845, EPI_ISL_885846, EPI_ISL_885847, EPI_ISL_885848, EPI_ISL_885849, EPI_ISL_885850, EPI_ISL_885851, EPI_ISL_885852, EPI_ISL_885853, EPI_ISL_885854, EPI_ISL_885856, EPI_ISL_885857, EPI_ISL_885858, EPI_ISL_885859, EPI_ISL_885861, EPI_ISL_885862, EPI_ISL_885863, EPI_ISL_885864, EPI_ISL_885865, EPI_ISL_885866, EPI_ISL_885868, EPI_ISL_885869, EPI_ISL_885870, EPI_ISL_885871, EPI_ISL_885872, EPI_ISL_885873, EPI_ISL_885874, EPI_ISL_885875, EPI_ISL_885876, EPI_ISL_885877, EPI_ISL_885878, EPI_ISL_885879, EPI_ISL_885881, EPI_ISL_885882, EPI_ISL_885883, EPI_ISL_885884, EPI_ISL_885885, EPI_ISL_885886, EPI_ISL_885887, EPI_ISL_885888, EPI_ISL_885890, EPI_ISL_885891, EPI_ISL_885892, EPI_ISL_885893, EPI_ISL_885894, EPI_ISL_885895, EPI_ISL_885896, EPI_ISL_885897, EPI_ISL_885898, EPI_ISL_885899, EPI_ISL_885900, EPI_ISL_885901, EPI_ISL_885902, EPI_ISL_885903, EPI_ISL_885904, EPI_ISL_885905, EPI_ISL_885906, EPI_ISL_885907, EPI_ISL_885908, EPI_ISL_885909, EPI_ISL_885910, EPI_ISL_885911, EPI_ISL_885913, EPI_ISL_885914, EPI_ISL_885915, EPI_ISL_885917, EPI_ISL_885919, EPI_ISL_885920, EPI_ISL_885921, EPI_ISL_885922, EPI_ISL_885923, EPI_ISL_885924, EPI_ISL_885925, EPI_ISL_885926, EPI_ISL_885927, EPI_ISL_885928, EPI_ISL_885929, EPI_ISL_885930, EPI_ISL_885931, EPI_ISL_885932, EPI_ISL_885933, EPI_ISL_885934, EPI_ISL_885936, EPI_ISL_885937, EPI_ISL_885938, EPI_ISL_885939, EPI_ISL_885940, EPI_ISL_885941, EPI_ISL_885942, EPI_ISL_885943, EPI_ISL_885944, EPI_ISL_885945, EPI_ISL_885946, EPI_ISL_885947, EPI_ISL_885948, EPI_ISL_885949, EPI_ISL_885950, EPI_ISL_885951, EPI_ISL_885952, EPI_ISL_885953, EPI_ISL_885955, EPI_ISL_885956, EPI_ISL_885957, EPI_ISL_885959, EPI_ISL_885960, EPI_ISL_885961, EPI_ISL_885963, EPI_ISL_885964, EPI_ISL_885965, EPI_ISL_885967, EPI_ISL_885968, EPI_ISL_885969, EPI_ISL_885970, EPI_ISL_885971, EPI_ISL_885972, EPI_ISL_885973, EPI_ISL_885974, EPI_ISL_885975, EPI_ISL_885976, EPI_ISL_885977, EPI_ISL_885978, EPI_ISL_885979, EPI_ISL_885980, EPI_ISL_885981, EPI_ISL_885982, EPI_ISL_885983, EPI_ISL_885984, EPI_ISL_885985, EPI_ISL_885986, EPI_ISL_885987, EPI_ISL_885988, EPI_ISL_885989, EPI_ISL_885990, EPI_ISL_885991, EPI_ISL_885992, EPI_ISL_885993, EPI_ISL_885994, EPI_ISL_885995, EPI_ISL_885996, EPI_ISL_885997, EPI_ISL_885998, EPI_ISL_885999, EPI_ISL_886000, EPI_ISL_886001, EPI_ISL_886002, EPI_ISL_886003, EPI_ISL_886004, EPI_ISL_886005, EPI_ISL_886006, EPI_ISL_886007, EPI_ISL_886008, EPI_ISL_886010, EPI_ISL_886011, EPI_ISL_886012, EPI_ISL_886013, EPI_ISL_886014, EPI_ISL_886015, EPI_ISL_886017, EPI_ISL_886018, EPI_ISL_886019, EPI_ISL_886020, EPI_ISL_886021, EPI_ISL_886022, EPI_ISL_886023, EPI_ISL_886024, EPI_ISL_886025, EPI_ISL_886026, EPI_ISL_886027, EPI_ISL_886028, EPI_ISL_886029, EPI_ISL_886030, EPI_ISL_886031, EPI_ISL_886032, EPI_ISL_886033, EPI_ISL_886034, EPI_ISL_886035, EPI_ISL_886036, EPI_ISL_886037, EPI_ISL_886038, EPI_ISL_886040, EPI_ISL_886041, EPI_ISL_886042, EPI_ISL_886043, EPI_ISL_886044, EPI_ISL_886045, EPI_ISL_886046, EPI_ISL_886047, EPI_ISL_886048, EPI_ISL_886049, EPI_ISL_886050, EPI_ISL_886051, EPI_ISL_886052, EPI_ISL_886053, EPI_ISL_886054, EPI_ISL_886055, EPI_ISL_886058, EPI_ISL_886059, EPI_ISL_886060, EPI_ISL_886061, EPI_ISL_886062, EPI_ISL_886063, EPI_ISL_886064, EPI_ISL_886065, EPI_ISL_886066, EPI_ISL_886067, EPI_ISL_886068, EPI_ISL_886069, EPI_ISL_886070, EPI_ISL_886071, EPI_ISL_886072, EPI_ISL_886073, EPI_ISL_886074, EPI_ISL_886075, EPI_ISL_886076, EPI_ISL_886077, EPI_ISL_886078, EPI_ISL_886079, EPI_ISL_886080, EPI_ISL_886081, EPI_ISL_886082, EPI_ISL_886083, EPI_ISL_886084, EPI_ISL_886085, EPI_ISL_886086, EPI_ISL_886087, EPI_ISL_886088, EPI_ISL_886089, EPI_ISL_886090, EPI_ISL_886091, EPI_ISL_886092, EPI_ISL_886093, EPI_ISL_886094, EPI_ISL_886095, EPI_ISL_886096, EPI_ISL_886097, EPI_ISL_886098, EPI_ISL_886099, EPI_ISL_886100, EPI_ISL_886101, EPI_ISL_886102, EPI_ISL_886104, EPI_ISL_886105, EPI_ISL_886106, EPI_ISL_886107, EPI_ISL_886108, EPI_ISL_886109, EPI_ISL_886110, EPI_ISL_886111, EPI_ISL_886112, EPI_ISL_886113, EPI_ISL_886114, EPI_ISL_886115, EPI_ISL_886116, EPI_ISL_886117, EPI_ISL_886118, EPI_ISL_886119, EPI_ISL_886120, EPI_ISL_886122, EPI_ISL_886123, EPI_ISL_886124, EPI_ISL_886125, EPI_ISL_886126, EPI_ISL_886127, EPI_ISL_886128, EPI_ISL_886129, EPI_ISL_886130, EPI_ISL_886131, EPI_ISL_886132, EPI_ISL_886133, EPI_ISL_886134, EPI_ISL_886135, EPI_ISL_886137, EPI_ISL_886138, EPI_ISL_886139, EPI_ISL_886140, EPI_ISL_886141, EPI_ISL_886144, EPI_ISL_886145, EPI_ISL_886146, EPI_ISL_886147, EPI_ISL_886148, EPI_ISL_886149, EPI_ISL_886150, EPI_ISL_886151, EPI_ISL_886152, EPI_ISL_886153, EPI_ISL_886154, EPI_ISL_886155, EPI_ISL_886156, EPI_ISL_891278, EPI_ISL_891280, EPI_ISL_891285, EPI_ISL_891295, EPI_ISL_891299, EPI_ISL_891305, EPI_ISL_891311, EPI_ISL_891312, EPI_ISL_891313, EPI_ISL_891317, EPI_ISL_891318, EPI_ISL_891319, EPI_ISL_891323, EPI_ISL_891324, EPI_ISL_891326, EPI_ISL_891334, EPI_ISL_891337, EPI_ISL_891340, EPI_ISL_891342, EPI_ISL_891344, EPI_ISL_891349, EPI_ISL_891352, EPI_ISL_891357, EPI_ISL_891360, EPI_ISL_891364, EPI_ISL_891371, EPI_ISL_891374, EPI_ISL_891376, EPI_ISL_891377, EPI_ISL_891378, EPI_ISL_891383, EPI_ISL_891384, EPI_ISL_891385, EPI_ISL_891386, EPI_ISL_891392, EPI_ISL_891400, EPI_ISL_891401, EPI_ISL_891403, EPI_ISL_891404, EPI_ISL_891411, EPI_ISL_891416, EPI_ISL_891417, EPI_ISL_891419, EPI_ISL_891421, EPI_ISL_891423, EPI_ISL_891438, EPI_ISL_891444, EPI_ISL_891448, EPI_ISL_891451, EPI_ISL_891454, EPI_ISL_891455, EPI_ISL_891458, EPI_ISL_891459, EPI_ISL_891460, EPI_ISL_891463, EPI_ISL_891478, EPI_ISL_891484, EPI_ISL_891485, EPI_ISL_891489, EPI_ISL_891490, EPI_ISL_891496, EPI_ISL_891499, EPI_ISL_891503, EPI_ISL_891507, | | | |

EPI_ISL_891511, EPI_ISL_891514, EPI_ISL_891520, EPI_ISL_891522, EPI_ISL_891525, EPI_ISL_891527, EPI_ISL_891528, EPI_ISL_891529, EPI_ISL_891531, EPI_ISL_891536, EPI_ISL_891544, EPI_ISL_891545, EPI_ISL_891547, EPI_ISL_891548, EPI_ISL_891555, EPI_ISL_891560, EPI_ISL_891563, EPI_ISL_891564, EPI_ISL_891569, EPI_ISL_891572, EPI_ISL_891582, EPI_ISL_891586, EPI_ISL_891596, EPI_ISL_891599,
EPI_ISL_891600, EPI_ISL_891602, EPI_ISL_891606, EPI_ISL_891615, EPI_ISL_891616, EPI_ISL_891623, EPI_ISL_891625, EPI_ISL_891629, EPI_ISL_891630, EPI_ISL_891633, EPI_ISL_891634, EPI_ISL_891640, EPI_ISL_891643, EPI_ISL_891647, EPI_ISL_891650, EPI_ISL_891659, EPI_ISL_891661, EPI_ISL_891662, EPI_ISL_891670, EPI_ISL_891672, EPI_ISL_891673, EPI_ISL_891677, EPI_ISL_891680, EPI_ISL_891681,
EPI_ISL_891683, EPI_ISL_891687, EPI_ISL_891704, EPI_ISL_891710, EPI_ISL_891713, EPI_ISL_891714, EPI_ISL_891718, EPI_ISL_891719, EPI_ISL_891723, EPI_ISL_891724, EPI_ISL_891726, EPI_ISL_891729, EPI_ISL_891735, EPI_ISL_891736, EPI_ISL_891738, EPI_ISL_891742, EPI_ISL_891746, EPI_ISL_891751, EPI_ISL_891753, EPI_ISL_891757, EPI_ISL_891760, EPI_ISL_891762, EPI_ISL_891765, EPI_ISL_891766,
EPI_ISL_891773, EPI_ISL_891774, EPI_ISL_891775, EPI_ISL_891776, EPI_ISL_891777, EPI_ISL_891780, EPI_ISL_891786, EPI_ISL_891787, EPI_ISL_891788, EPI_ISL_891789, EPI_ISL_891794, EPI_ISL_891804, EPI_ISL_891807, EPI_ISL_891811, EPI_ISL_891813, EPI_ISL_891822, EPI_ISL_891824, EPI_ISL_891833, EPI_ISL_891838, EPI_ISL_891839, EPI_ISL_891843, EPI_ISL_891847, EPI_ISL_891848, EPI_ISL_891850,
EPI_ISL_891856, EPI_ISL_891858, EPI_ISL_891860, EPI_ISL_891863, EPI_ISL_891864, EPI_ISL_891866, EPI_ISL_891867, EPI_ISL_891870, EPI_ISL_891878, EPI_ISL_891887, EPI_ISL_891895, EPI_ISL_891900, EPI_ISL_891902, EPI_ISL_891904, EPI_ISL_891906, EPI_ISL_891907, EPI_ISL_891910, EPI_ISL_891911, EPI_ISL_891912, EPI_ISL_891913, EPI_ISL_891915, EPI_ISL_891916, EPI_ISL_891917, EPI_ISL_891919,
EPI_ISL_891922, EPI_ISL_891947, EPI_ISL_891948, EPI_ISL_891951, EPI_ISL_891952, EPI_ISL_891955, EPI_ISL_891956, EPI_ISL_891960, EPI_ISL_891961, EPI_ISL_891964, EPI_ISL_891965, EPI_ISL_891966, EPI_ISL_891968, EPI_ISL_891971, EPI_ISL_892010, EPI_ISL_892011, EPI_ISL_907479, EPI_ISL_907492, EPI_ISL_907636, EPI_ISL_907700, EPI_ISL_907701, EPI_ISL_907704, EPI_ISL_907709, EPI_ISL_907710,
EPI_ISL_907711, EPI_ISL_907712, EPI_ISL_907713, EPI_ISL_907714, EPI_ISL_907715, EPI_ISL_907717, EPI_ISL_907718, EPI_ISL_907719, EPI_ISL_907720, EPI_ISL_907724, EPI_ISL_907725, EPI_ISL_907726, EPI_ISL_907727, EPI_ISL_907728, EPI_ISL_907729, EPI_ISL_907730, EPI_ISL_907732, EPI_ISL_907733, EPI_ISL_907734, EPI_ISL_907737, EPI_ISL_907739, EPI_ISL_907740, EPI_ISL_907741, EPI_ISL_907742,
EPI_ISL_907743, EPI_ISL_907744, EPI_ISL_907746, EPI_ISL_907748, EPI_ISL_907751, EPI_ISL_907752, EPI_ISL_907753, EPI_ISL_907754, EPI_ISL_907758, EPI_ISL_907759, EPI_ISL_907761, EPI_ISL_907762, EPI_ISL_907764, EPI_ISL_907766, EPI_ISL_907768, EPI_ISL_907769, EPI_ISL_907770, EPI_ISL_907774, EPI_ISL_907775, EPI_ISL_907777, EPI_ISL_907779, EPI_ISL_907780, EPI_ISL_907781, EPI_ISL_907782,
EPI_ISL_907783, EPI_ISL_907784, EPI_ISL_907785, EPI_ISL_907790, EPI_ISL_907791, EPI_ISL_907792, EPI_ISL_907798, EPI_ISL_907799, EPI_ISL_907803, EPI_ISL_907804, EPI_ISL_907806, EPI_ISL_907807, EPI_ISL_907808, EPI_ISL_907810, EPI_ISL_907815, EPI_ISL_907816, EPI_ISL_907818, EPI_ISL_907819, EPI_ISL_907822, EPI_ISL_907823, EPI_ISL_907825, EPI_ISL_907827, EPI_ISL_907828, EPI_ISL_907829,
EPI_ISL_907830, EPI_ISL_907831, EPI_ISL_907836, EPI_ISL_907839, EPI_ISL_907840, EPI_ISL_907841, EPI_ISL_907842, EPI_ISL_907843, EPI_ISL_907844, EPI_ISL_907845, EPI_ISL_907846, EPI_ISL_907848, EPI_ISL_907849, EPI_ISL_907850, EPI_ISL_907853, EPI_ISL_907856, EPI_ISL_907857, EPI_ISL_907858, EPI_ISL_907859, EPI_ISL_907861, EPI_ISL_907862, EPI_ISL_907864, EPI_ISL_907866, EPI_ISL_907869,
EPI_ISL_907870, EPI_ISL_907871, EPI_ISL_907873, EPI_ISL_907876, EPI_ISL_907880, EPI_ISL_907883, EPI_ISL_907884, EPI_ISL_907885, EPI_ISL_907886, EPI_ISL_907887, EPI_ISL_907888, EPI_ISL_907892, EPI_ISL_907896, EPI_ISL_907897, EPI_ISL_907900, EPI_ISL_907901, EPI_ISL_907904, EPI_ISL_907905, EPI_ISL_907906, EPI_ISL_907907, EPI_ISL_907908, EPI_ISL_907909, EPI_ISL_907910, EPI_ISL_907913,
EPI_ISL_907916, EPI_ISL_907917, EPI_ISL_907918, EPI_ISL_907919, EPI_ISL_907920, EPI_ISL_907922, EPI_ISL_907924, EPI_ISL_907927, EPI_ISL_907928, EPI_ISL_907929, EPI_ISL_907930, EPI_ISL_907932, EPI_ISL_907933, EPI_ISL_907935, EPI_ISL_907937, EPI_ISL_907938, EPI_ISL_907939, EPI_ISL_907941, EPI_ISL_907942, EPI_ISL_907947, EPI_ISL_907950, EPI_ISL_907951, EPI_ISL_907952, EPI_ISL_907954,
EPI_ISL_907955, EPI_ISL_907957, EPI_ISL_907958, EPI_ISL_907959, EPI_ISL_907961, EPI_ISL_907962, EPI_ISL_907963, EPI_ISL_907964, EPI_ISL_907965, EPI_ISL_907966, EPI_ISL_907968, EPI_ISL_907971, EPI_ISL_907977, EPI_ISL_907978, EPI_ISL_907980, EPI_ISL_907983, EPI_ISL_907985, EPI_ISL_907986, EPI_ISL_907990, EPI_ISL_907991, EPI_ISL_907992, EPI_ISL_907993, EPI_ISL_907996, EPI_ISL_907997,
EPI_ISL_907999, EPI_ISL_908000, EPI_ISL_908001, EPI_ISL_908004, EPI_ISL_908005, EPI_ISL_908006, EPI_ISL_908008, EPI_ISL_908012, EPI_ISL_908014, EPI_ISL_908015, EPI_ISL_908018, EPI_ISL_908019, EPI_ISL_908020, EPI_ISL_908023, EPI_ISL_908024, EPI_ISL_908029, EPI_ISL_908031, EPI_ISL_908032, EPI_ISL_908268, EPI_ISL_908271, EPI_ISL_908272, EPI_ISL_908273, EPI_ISL_908276, EPI_ISL_908278,
EPI_ISL_908280, EPI_ISL_908281, EPI_ISL_908283, EPI_ISL_908286, EPI_ISL_908287, EPI_ISL_908288, EPI_ISL_908294, EPI_ISL_908297, EPI_ISL_908299, EPI_ISL_908300, EPI_ISL_908304, EPI_ISL_908305, EPI_ISL_908311, EPI_ISL_908312, EPI_ISL_908313, EPI_ISL_908314, EPI_ISL_908318, EPI_ISL_908319, EPI_ISL_908320, EPI_ISL_908324, EPI_ISL_908325, EPI_ISL_908327, EPI_ISL_908328, EPI_ISL_908330,
EPI_ISL_908331, EPI_ISL_908332, EPI_ISL_908336, EPI_ISL_908339, EPI_ISL_908342, EPI_ISL_908345, EPI_ISL_908346, EPI_ISL_908347, EPI_ISL_908349, EPI_ISL_908350, EPI_ISL_908352, EPI_ISL_908354, EPI_ISL_908355, EPI_ISL_908356, EPI_ISL_908358, EPI_ISL_908359, EPI_ISL_908361, EPI_ISL_908362, EPI_ISL_908363, EPI_ISL_908364, EPI_ISL_908368, EPI_ISL_908370, EPI_ISL_908371, EPI_ISL_908373,
EPI_ISL_908374, EPI_ISL_908375, EPI_ISL_908376, EPI_ISL_908377, EPI_ISL_908378, EPI_ISL_908379, EPI_ISL_908381, EPI_ISL_908382, EPI_ISL_908386, EPI_ISL_908388, EPI_ISL_908390, EPI_ISL_908391, EPI_ISL_908392, EPI_ISL_908395, EPI_ISL_908396, EPI_ISL_908398, EPI_ISL_908399, EPI_ISL_908400, EPI_ISL_908404, EPI_ISL_908406, EPI_ISL_908409, EPI_ISL_908410, EPI_ISL_908414, EPI_ISL_908417,
EPI_ISL_908418, EPI_ISL_908420, EPI_ISL_908423, EPI_ISL_908424, EPI_ISL_908425, EPI_ISL_908426, EPI_ISL_908427, EPI_ISL_908428, EPI_ISL_908430, EPI_ISL_908432, EPI_ISL_908434, EPI_ISL_908436, EPI_ISL_908437, EPI_ISL_908438, EPI_ISL_908440, EPI_ISL_908441, EPI_ISL_908445, EPI_ISL_908448, EPI_ISL_908449, EPI_ISL_908450, EPI_ISL_908454, EPI_ISL_908457, EPI_ISL_908458, EPI_ISL_908459,
EPI_ISL_908460, EPI_ISL_908461, EPI_ISL_908463, EPI_ISL_908464, EPI_ISL_908465, EPI_ISL_908466, EPI_ISL_908468, EPI_ISL_908469, EPI_ISL_908472, EPI_ISL_908473, EPI_ISL_908474, EPI_ISL_908475, EPI_ISL_908476, EPI_ISL_908477, EPI_ISL_908480, EPI_ISL_908482, EPI_ISL_908483, EPI_ISL_908484, EPI_ISL_908485, EPI_ISL_908487, EPI_ISL_908490, EPI_ISL_908491, EPI_ISL_908492, EPI_ISL_908493,
EPI_ISL_908497, EPI_ISL_908498, EPI_ISL_908499, EPI_ISL_908501, EPI_ISL_908502, EPI_ISL_908503, EPI_ISL_908504, EPI_ISL_908505, EPI_ISL_908510, EPI_ISL_908513, EPI_ISL_908514, EPI_ISL_908515, EPI_ISL_908517, EPI_ISL_908523, EPI_ISL_908531, EPI_ISL_908532, EPI_ISL_908533, EPI_ISL_908537, EPI_ISL_908539, EPI_ISL_908540, EPI_ISL_908541, EPI_ISL_908543, EPI_ISL_908544, EPI_ISL_908545,
EPI_ISL_908548, EPI_ISL_908549, EPI_ISL_908552, EPI_ISL_908554, EPI_ISL_908555, EPI_ISL_908556, EPI_ISL_908562, EPI_ISL_908563, EPI_ISL_908565, EPI_ISL_908566, EPI_ISL_908567, EPI_ISL_908569, EPI_ISL_908570, EPI_ISL_908572, EPI_ISL_908574, EPI_ISL_908575, EPI_ISL_908577, EPI_ISL_908578, EPI_ISL_908579, EPI_ISL_908583, EPI_ISL_908584, EPI_ISL_908586, EPI_ISL_908587, EPI_ISL_908588,
EPI_ISL_908592, EPI_ISL_908593, EPI_ISL_908594, EPI_ISL_909260, EPI_ISL_909270, EPI_ISL_909273, EPI_ISL_909275, EPI_ISL_909281, EPI_ISL_909285, EPI_ISL_909292, EPI_ISL_909294, EPI_ISL_909297, EPI_ISL_909302, EPI_ISL_909309, EPI_ISL_909311, EPI_ISL_909312, EPI_ISL_909314, EPI_ISL_909316, EPI_ISL_909318, EPI_ISL_909321, EPI_ISL_909322, EPI_ISL_909326, EPI_ISL_909328, EPI_ISL_909334,
EPI_ISL_909339, EPI_ISL_909341, EPI_ISL_909343, EPI_ISL_909344, EPI_ISL_909358, EPI_ISL_909359, EPI_ISL_909364, EPI_ISL_909366, EPI_ISL_909367, EPI_ISL_909368, EPI_ISL_909370, EPI_ISL_909372, EPI_ISL_909373, EPI_ISL_909380, EPI_ISL_909381, EPI_ISL_909385, EPI_ISL_909388, EPI_ISL_909389, EPI_ISL_909397, EPI_ISL_909401, EPI_ISL_909409, EPI_ISL_909414, EPI_ISL_909416, EPI_ISL_909417,
EPI_ISL_909421, EPI_ISL_909424, EPI_ISL_909425, EPI_ISL_909426, EPI_ISL_909440, EPI_ISL_909446, EPI_ISL_909447, EPI_ISL_909450, EPI_ISL_909453, EPI_ISL_909456, EPI_ISL_909457, EPI_ISL_909465, EPI_ISL_909467, EPI_ISL_909471, EPI_ISL_909476, EPI_ISL_909478, EPI_ISL_909482, EPI_ISL_909487, EPI_ISL_909488, EPI_ISL_909503, EPI_ISL_909504, EPI_ISL_909513, EPI_ISL_909518, EPI_ISL_909526,
EPI_ISL_909531, EPI_ISL_909536, EPI_ISL_909539, EPI_ISL_909545, EPI_ISL_909546, EPI_ISL_909552, EPI_ISL_909553, EPI_ISL_909561, EPI_ISL_909564, EPI_ISL_909571, EPI_ISL_909572, EPI_ISL_909573, EPI_ISL_909575, EPI_ISL_909577, EPI_ISL_909578, EPI_ISL_909580, EPI_ISL_909581, EPI_ISL_909586, EPI_ISL_912220, EPI_ISL_912221, EPI_ISL_912222, EPI_ISL_912223, EPI_ISL_917214, EPI_ISL_917501,
EPI_ISL_987327, EPI_ISL_987333, EPI_ISL_987336, EPI_ISL_987410, EPI_ISL_987448, EPI_ISL_987469, EPI_ISL_987524, EPI_ISL_987534, EPI_ISL_987584, EPI_ISL_987620, EPI_ISL_992001, EPI_ISL_992009, EPI_ISL_992013, EPI_ISL_992083, EPI_ISL_992092, EPI_ISL_992125, EPI_ISL_992166, EPI_ISL_992172, EPI_ISL_992226, EPI_ISL_992258, EPI_ISL_992261, EPI_ISL_992283, EPI_ISL_992292, EPI_ISL_992305,
EPI_ISL_992360, EPI_ISL_992362, EPI_ISL_992410, EPI_ISL_992437, EPI_ISL_992439, EPI_ISL_992449, EPI_ISL_992460, EPI_ISL_992463, EPI_ISL_992466, EPI_ISL_992484, EPI_ISL_992486, EPI_ISL_992501, EPI_ISL_992547, EPI_ISL_992549, EPI_ISL_992643, EPI_ISL_992665, EPI_ISL_992694, EPI_ISL_992701, EPI_ISL_992710, EPI_ISL_992722, EPI_ISL_992729, EPI_ISL_992743, EPI_ISL_992800, EPI_ISL_992858,
EPI_ISL_992864, EPI_ISL_992866, EPI_ISL_992930, EPI_ISL_992985, EPI_ISL_992989, EPI_ISL_993012, EPI_ISL_993015, EPI_ISL_993017, EPI_ISL_993022, EPI_ISL_993043, EPI_ISL_993044, EPI_ISL_993045, EPI_ISL_993046, EPI_ISL_993047, EPI_ISL_993048, EPI_ISL_993049, EPI_ISL_993050, EPI_ISL_993051, EPI_ISL_993052, EPI_ISL_993053, EPI_ISL_993054, EPI_ISL_993055, EPI_ISL_993057, EPI_ISL_993058,
EPI_ISL_993059, EPI_ISL_993060, EPI_ISL_993061, EPI_ISL_993062, EPI_ISL_993063, EPI_ISL_993064, EPI_ISL_993065, EPI_ISL_993066, EPI_ISL_993067, EPI_ISL_993068, EPI_ISL_993069, EPI_ISL_993070, EPI_ISL_993071, EPI_ISL_993072, EPI_ISL_993073, EPI_ISL_993074, EPI_ISL_993075, EPI_ISL_993076, EPI_ISL_993077, EPI_ISL_993078, EPI_ISL_993079, EPI_ISL_993080, EPI_ISL_993081, EPI_ISL_993082,
EPI_ISL_993084, EPI_ISL_993096, EPI_ISL_993098, EPI_ISL_993100, EPI_ISL_993105, EPI_ISL_993106, EPI_ISL_993109, EPI_ISL_993110, EPI_ISL_993113, EPI_ISL_993114, EPI_ISL_993119, EPI_ISL_993127, EPI_ISL_993133, EPI_ISL_993140, EPI_ISL_993146, EPI_ISL_993152, EPI_ISL_993153, EPI_ISL_993154, EPI_ISL_993155, EPI_ISL_993157, EPI_ISL_993161, EPI_ISL_993162, EPI_ISL_993165, EPI_ISL_993168,
EPI_ISL_993171, EPI_ISL_993176, EPI_ISL_993184, EPI_ISL_993187, EPI_ISL_993194, EPI_ISL_993199, EPI_ISL_993202, EPI_ISL_993205, EPI_ISL_993206, EPI_ISL_993208, EPI_ISL_993211, EPI_ISL_993213, EPI_ISL_993221, EPI_ISL_993225, EPI_ISL_993227, EPI_ISL_993244, EPI_ISL_993245, EPI_ISL_993248, EPI_ISL_993255, EPI_ISL_993257, EPI_ISL_993258, EPI_ISL_993266, EPI_ISL_993282, EPI_ISL_993291,
EPI_ISL_993292, EPI_ISL_993297, EPI_ISL_993306, EPI_ISL_993309, EPI_ISL_993311, EPI_ISL_993313, EPI_ISL_993316, EPI_ISL_993317, EPI_ISL_993321, EPI_ISL_993325, EPI_ISL_993327, EPI_ISL_993334, EPI_ISL_993338, EPI_ISL_993341, EPI_ISL_993348, EPI_ISL_993353, EPI_ISL_993364, EPI_ISL_993369, EPI_ISL_993370, EPI_ISL_993377, EPI_ISL_993383, EPI_ISL_993385, EPI_ISL_993386, EPI_ISL_993390,
EPI_ISL_993391, EPI_ISL_993393, EPI_ISL_993397, EPI_ISL_993398, EPI_ISL_993399, EPI_ISL_993400, EPI_ISL_993402, EPI_ISL_993403, EPI_ISL_993404, EPI_ISL_993405, EPI_ISL_993406, EPI_ISL_993407, EPI_ISL_993408, EPI_ISL_993409, EPI_ISL_993410, EPI_ISL_993411, EPI_ISL_993412, EPI_ISL_994289, EPI_ISL_994291, EPI_ISL_994292, EPI_ISL_994293, EPI_ISL_994298, EPI_ISL_994303, EPI_ISL_994304,
EPI_ISL_994306, EPI_ISL_994307, EPI_ISL_994308, EPI_ISL_994309, EPI_ISL_994313, EPI_ISL_994314, EPI_ISL_994318, EPI_ISL_994320, EPI_ISL_994321, EPI_ISL_994322, EPI_ISL_994323, EPI_ISL_994324, EPI_ISL_994325, EPI_ISL_994329, EPI_ISL_994330, EPI_ISL_994335, EPI_ISL_994340, EPI_ISL_994342, EPI_ISL_994345, EPI_ISL_994346, EPI_ISL_994348, EPI_ISL_994351, EPI_ISL_994352, EPI_ISL_994354,
EPI_ISL_994355, EPI_ISL_994358, EPI_ISL_994364, EPI_ISL_994368, EPI_ISL_994369, EPI_ISL_994370, EPI_ISL_994375, EPI_ISL_994376, EPI_ISL_994379, EPI_ISL_994385, EPI_ISL_994387, EPI_ISL_994388, EPI_ISL_994392, EPI_ISL_994393, EPI_ISL_994394, EPI_ISL_994396, EPI_ISL_994399, EPI_ISL_994400, EPI_ISL_994404, EPI_ISL_994405, EPI_ISL_994412, EPI_ISL_994414, EPI_ISL_994415, EPI_ISL_994417,
EPI_ISL_994418, EPI_ISL_994422, EPI_ISL_994423, EPI_ISL_994426, EPI_ISL_994428, EPI_ISL_994430, EPI_ISL_994433, EPI_ISL_994438, EPI_ISL_994442, EPI_ISL_994445, EPI_ISL_994449, EPI_ISL_994451, EPI_ISL_994453, EPI_ISL_994454, EPI_ISL_994455, EPI_ISL_994460, EPI_ISL_994461, EPI_ISL_994462, EPI_ISL_994463, EPI_ISL_994465, EPI_ISL_994466, EPI_ISL_994471, EPI_ISL_994474, EPI_ISL_994475,
EPI_ISL_994476, EPI_ISL_994479, EPI_ISL_994483, EPI_ISL_994485, EPI_ISL_994489, EPI_ISL_994491, EPI_ISL_994492, EPI_ISL_994493, EPI_ISL_994494, EPI_ISL_994495, EPI_ISL_994498, EPI_ISL_994499, EPI_ISL_994500, EPI_ISL_994505, EPI_ISL_994506, EPI_ISL_994512, EPI_ISL_994519, EPI_ISL_994520, EPI_ISL_994522, EPI_ISL_994525, EPI_ISL_994526, EPI_ISL_994527, EPI_ISL_994531, EPI_ISL_994532,
EPI_ISL_994534, EPI_ISL_994535, EPI_ISL_994539, EPI_ISL_994540, EPI_ISL_994541, EPI_ISL_994545, EPI_ISL_994547, EPI_ISL_994548, EPI_ISL_994549, EPI_ISL_994550, EPI_ISL_994551, EPI_ISL_994552, EPI_ISL_994553, EPI_ISL_994554, EPI_ISL_994555, EPI_ISL_994556, EPI_ISL_994557, EPI_ISL_994558, EPI_ISL_994559, EPI_ISL_994560, EPI_ISL_994561, EPI_ISL_994562, EPI_ISL_994563, EPI_ISL_994564,
EPI_ISL_994565, EPI_ISL_994566, EPI_ISL_994567, EPI_ISL_994568, EPI_ISL_994569, EPI_ISL_994570, EPI_ISL_994571, EPI_ISL_994572, EPI_ISL_994573, EPI_ISL_994574, EPI_ISL_994575, EPI_ISL_994576, EPI_ISL_994577, EPI_ISL_994578, EPI_ISL_994579, EPI_ISL_994580, EPI_ISL_994581, EPI_ISL_994582, EPI_ISL_994583, EPI_ISL_994584, EPI_ISL_994585, EPI_ISL_994586, EPI_ISL_994587, EPI_ISL_994588,
EPI_ISL_994589, EPI_ISL_994591, EPI_ISL_994592, EPI_ISL_994593, EPI_ISL_994594, EPI_ISL_994595, EPI_ISL_994596, EPI_ISL_994597, EPI_ISL_994598, EPI_ISL_994599, EPI_ISL_994600, EPI_ISL_994601, EPI_ISL_994602, EPI_ISL_994603, EPI_ISL_994604, EPI_ISL_994605, EPI_ISL_994606, EPI_ISL_994607, EPI_ISL_994608, EPI_ISL_994609, EPI_ISL_994611, EPI_ISL_994612, EPI_ISL_994613, EPI_ISL_994614,
EPI_ISL_994616, EPI_ISL_994617, EPI_ISL_994618, EPI_ISL_994620, EPI_ISL_994621, EPI_ISL_994625, EPI_ISL_994627, EPI_ISL_994628, EPI_ISL_994631, EPI_ISL_994632, EPI_ISL_994638, EPI_ISL_994640, EPI_ISL_994642, EPI_ISL_994644, EPI_ISL_994645, EPI_ISL_994647, EPI_ISL_994648, EPI_ISL_1007574, EPI_ISL_1012831, EPI_ISL_1012833, EPI_ISL_1012834, EPI_ISL_1012844, EPI_ISL_1012849,
EPI_ISL_1012857, EPI_ISL_1012858, EPI_ISL_1012859, EPI_ISL_1012860, EPI_ISL_1012861, EPI_ISL_1012863, EPI_ISL_1012864, EPI_ISL_1012866, EPI_ISL_1012867, EPI_ISL_1012868, EPI_ISL_1012869, EPI_ISL_1012870, EPI_ISL_1012871, EPI_ISL_1012872, EPI_ISL_1190728

see above | Lighthouse Lab in Alderley Park | Wellcome Sanger Institute for the COVID-19 Genomics UK (COG-UK) Consortium | Cordelia Langford; David K. Jackson; Dominic Kwiatkowski; Ewan Harrison; Ian Johnston; Jacquelyn Wynn; Jeffrey Barrett; John Sillitoe on behalf of the Wellcome Sanger Institute COVID-19 Surveillance Team; John Sillitoe on behalf of the Wellcome Sanger Institute COVID-19 Surveillance Team (http://www.sanger.ac.uk/covid-team); Mairead Hyland; Roberto Amato; Sonia Goncalves; The Lighthouse Lab in Alderley Park and Alex Alderton

EPI_ISL_878728, EPI_ISL_878733, EPI_ISL_878735, EPI_ISL_878737, EPI_ISL_878738, EPI_ISL_878743, EPI_ISL_878745, EPI_ISL_878748, EPI_ISL_878749, EPI_ISL_878753, EPI_ISL_878762, EPI_ISL_878764, EPI_ISL_878767, EPI_ISL_878774, EPI_ISL_878783, EPI_ISL_878787, EPI_ISL_878788, EPI_ISL_878795, EPI_ISL_878799, EPI_ISL_878811, EPI_ISL_878813, EPI_ISL_878816, EPI_ISL_878818, EPI_ISL_878821,
EPI_ISL_878823, EPI_ISL_878826, EPI_ISL_878827, EPI_ISL_878828, EPI_ISL_878829, EPI_ISL_878837, EPI_ISL_878838, EPI_ISL_878855, EPI_ISL_878867, EPI_ISL_878872, EPI_ISL_878873, EPI_ISL_878877, EPI_ISL_878881, EPI_ISL_878885, EPI_ISL_878890, EPI_ISL_878893, EPI_ISL_878895, EPI_ISL_878897, EPI_ISL_878898, EPI_ISL_878900, EPI_ISL_878901, EPI_ISL_878907, EPI_ISL_878908, EPI_ISL_878910,
EPI_ISL_878911, EPI_ISL_878916, EPI_ISL_878921, EPI_ISL_878922, EPI_ISL_878923, EPI_ISL_878928, EPI_ISL_878930, EPI_ISL_878933, EPI_ISL_878937, EPI_ISL_878939, EPI_ISL_878940, EPI_ISL_878948, EPI_ISL_878949, EPI_ISL_878953, EPI_ISL_878954, EPI_ISL_878956, EPI_ISL_878957, EPI_ISL_878963, EPI_ISL_878968, EPI_ISL_878969, EPI_ISL_878973, EPI_ISL_878974, EPI_ISL_878976, EPI_ISL_878977,
EPI_ISL_878978, EPI_ISL_878984, EPI_ISL_878986, EPI_ISL_878987, EPI_ISL_878988, EPI_ISL_878996, EPI_ISL_879002, EPI_ISL_879004, EPI_ISL_879005, EPI_ISL_879006, EPI_ISL_879010, EPI_ISL_879017, EPI_ISL_879020, EPI_ISL_879024, EPI_ISL_879027, EPI_ISL_879030, EPI_ISL_879035, EPI_ISL_879036, EPI_ISL_879039, EPI_ISL_879041, EPI_ISL_879046, EPI_ISL_879048, EPI_ISL_879053, EPI_ISL_879059,
EPI_ISL_879060, EPI_ISL_879067, EPI_ISL_879080, EPI_ISL_879084, EPI_ISL_879085, EPI_ISL_879090, EPI_ISL_879102, EPI_ISL_879104, EPI_ISL_879105, EPI_ISL_879106, EPI_ISL_879107, EPI_ISL_879112, EPI_ISL_879115, EPI_ISL_879119, EPI_ISL_879122, EPI_ISL_879123, EPI_ISL_879127, EPI_ISL_879133, EPI_ISL_879135, EPI_ISL_879136, EPI_ISL_879139, EPI_ISL_879141, EPI_ISL_879142, EPI_ISL_879143,
EPI_ISL_879148, EPI_ISL_879150, EPI_ISL_879153, EPI_ISL_879155, EPI_ISL_879156, EPI_ISL_879158, EPI_ISL_879163, EPI_ISL_879165, EPI_ISL_879166, EPI_ISL_879168, EPI_ISL_879170, EPI_ISL_879172, EPI_ISL_879174, EPI_ISL_879175, EPI_ISL_879176, EPI_ISL_879177, EPI_ISL_879178, EPI_ISL_879179, EPI_ISL_879180, EPI_ISL_879182, EPI_ISL_879183, EPI_ISL_879184, EPI_ISL_879186, EPI_ISL_879189,
EPI_ISL_879190, EPI_ISL_879192, EPI_ISL_879196, EPI_ISL_879197, EPI_ISL_879198, EPI_ISL_879200, EPI_ISL_879204, EPI_ISL_879205, EPI_ISL_879208, EPI_ISL_879209, EPI_ISL_879210, EPI_ISL_879211, EPI_ISL_879213, EPI_ISL_879215, EPI_ISL_879216, EPI_ISL_879217, EPI_ISL_879221, EPI_ISL_879222, EPI_ISL_879223, EPI_ISL_879228, EPI_ISL_879232, EPI_ISL_879233, EPI_ISL_879236, EPI_ISL_879238,
EPI_ISL_879240, EPI_ISL_879242, EPI_ISL_879245, EPI_ISL_879246, EPI_ISL_879251, EPI_ISL_879255, EPI_ISL_879257, EPI_ISL_879260, EPI_ISL_879261, EPI_ISL_879264, EPI_ISL_879267, EPI_ISL_879269, EPI_ISL_879272, EPI_ISL_879275, EPI_ISL_879280, EPI_ISL_879281, EPI_ISL_879282, EPI_ISL_879283, EPI_ISL_879285, EPI_ISL_879287, EPI_ISL_879288, EPI_ISL_879289, EPI_ISL_879291, EPI_ISL_879292,
EPI_ISL_879293, EPI_ISL_879294, EPI_ISL_879295, EPI_ISL_879301, EPI_ISL_879304, EPI_ISL_879309, EPI_ISL_879310, EPI_ISL_879311, EPI_ISL_879312, EPI_ISL_879315, EPI_ISL_879317, EPI_ISL_879319, EPI_ISL_879320, EPI_ISL_879321, EPI_ISL_879325, EPI_ISL_879326, EPI_ISL_879327, EPI_ISL_879328, EPI_ISL_879330, EPI_ISL_879332, EPI_ISL_879334, EPI_ISL_879335, EPI_ISL_879336, EPI_ISL_879337,
EPI_ISL_879349, EPI_ISL_879350, EPI_ISL_879352, EPI_ISL_879353, EPI_ISL_879355, EPI_ISL_879364, EPI_ISL_879365, EPI_ISL_879366, EPI_ISL_879368, EPI_ISL_879369, EPI_ISL_879370, EPI_ISL_879372, EPI_ISL_879373, EPI_ISL_879376, EPI_ISL_879405, EPI_ISL_879447, EPI_ISL_879488, EPI_ISL_879512, EPI_ISL_879521, EPI_ISL_879533, EPI_ISL_879536, EPI_ISL_879541, EPI_ISL_879638, EPI_ISL_879657,
EPI_ISL_879661, EPI_ISL_879662, EPI_ISL_879671, EPI_ISL_891273, EPI_ISL_891274, EPI_ISL_891275, EPI_ISL_891276, EPI_ISL_891277, EPI_ISL_891282, EPI_ISL_891284, EPI_ISL_891287, EPI_ISL_891289, EPI_ISL_891290, EPI_ISL_891291, EPI_ISL_891292, EPI_ISL_891293, EPI_ISL_891294, EPI_ISL_891296, EPI_ISL_891300, EPI_ISL_891301, EPI_ISL_891302, EPI_ISL_891303, EPI_ISL_891306, EPI_ISL_891307,
EPI_ISL_891308, EPI_ISL_891309, EPI_ISL_891310, EPI_ISL_891314, EPI_ISL_891316, EPI_ISL_891321, EPI_ISL_891322, EPI_ISL_891325, EPI_ISL_891328, EPI_ISL_891329, EPI_ISL_891331, EPI_ISL_891332, EPI_ISL_891333, EPI_ISL_891336, EPI_ISL_891339, EPI_ISL_891341, EPI_ISL_891343, EPI_ISL_891346, EPI_ISL_891347, EPI_ISL_891348, EPI_ISL_891350, EPI_ISL_891351, EPI_ISL_891354, EPI_ISL_891355,
EPI_ISL_891356, EPI_ISL_891359, EPI_ISL_891362, EPI_ISL_891363, EPI_ISL_891366, EPI_ISL_891367, EPI_ISL_891368, EPI_ISL_891369, EPI_ISL_891370, EPI_ISL_891373, EPI_ISL_891379, EPI_ISL_891381, EPI_ISL_891387, EPI_ISL_891388, EPI_ISL_891389, EPI_ISL_891390, EPI_ISL_891394, EPI_ISL_891395, EPI_ISL_891397, EPI_ISL_891398, EPI_ISL_891399, EPI_ISL_891406, EPI_ISL_891407, EPI_ISL_891408,
EPI_ISL_891409, EPI_ISL_891412, EPI_ISL_891413, EPI_ISL_891414, EPI_ISL_891418, EPI_ISL_891422, EPI_ISL_891425, EPI_ISL_891427, EPI_ISL_891428, EPI_ISL_891429, EPI_ISL_891431, EPI_ISL_891432, EPI_ISL_891433, EPI_ISL_891434, EPI_ISL_891436, EPI_ISL_891437, EPI_ISL_891439, EPI_ISL_891441, EPI_ISL_891442, EPI_ISL_891443, EPI_ISL_891447, EPI_ISL_891449, EPI_ISL_891450, EPI_ISL_891452,
EPI_ISL_891457, EPI_ISL_891461, EPI_ISL_891466, EPI_ISL_891467, EPI_ISL_891468, EPI_ISL_891470, EPI_ISL_891471, EPI_ISL_891472, EPI_ISL_891474, EPI_ISL_891475, EPI_ISL_891476, EPI_ISL_891477, EPI_ISL_891480, EPI_ISL_891481, EPI_ISL_891482, EPI_ISL_891483, EPI_ISL_891486, EPI_ISL_891488, EPI_ISL_891492, EPI_ISL_891493, EPI_ISL_891494, EPI_ISL_891498, EPI_ISL_891500, EPI_ISL_891501,
EPI_ISL_891504, EPI_ISL_891506, EPI_ISL_891510, EPI_ISL_891513, EPI_ISL_891515, EPI_ISL_891517, EPI_ISL_891518, EPI_ISL_891519, EPI_ISL_891521, EPI_ISL_891523, EPI_ISL_891524, EPI_ISL_891526, EPI_ISL_891530, EPI_ISL_891533, EPI_ISL_891534, EPI_ISL_891535, EPI_ISL_891537, EPI_ISL_891540, EPI_ISL_891541, EPI_ISL_891543, EPI_ISL_891546, EPI_ISL_891550, EPI_ISL_891551, EPI_ISL_891552,
EPI_ISL_891553, EPI_ISL_891554, EPI_ISL_891556, EPI_ISL_891557, EPI_ISL_891558, EPI_ISL_891561, EPI_ISL_891565, EPI_ISL_891566, EPI_ISL_891567, EPI_ISL_891570, EPI_ISL_891573, EPI_ISL_891575, EPI_ISL_891576, EPI_ISL_891577, EPI_ISL_891578, EPI_ISL_891579, EPI_ISL_891580, EPI_ISL_891581, EPI_ISL_891583, EPI_ISL_891584, EPI_ISL_891589, EPI_ISL_891590, EPI_ISL_891591, EPI_ISL_891592,
EPI_ISL_891593, EPI_ISL_891594, EPI_ISL_891595, EPI_ISL_891597, EPI_ISL_891598, EPI_ISL_891601, EPI_ISL_891607, EPI_ISL_891608, EPI_ISL_891614, EPI_ISL_891617, EPI_ISL_891619, EPI_ISL_891620, EPI_ISL_891621, EPI_ISL_891627, EPI_ISL_891631, EPI_ISL_891632, EPI_ISL_891635, EPI_ISL_891637, EPI_ISL_891638, EPI_ISL_891641, EPI_ISL_891642, EPI_ISL_891644, EPI_ISL_891645, EPI_ISL_891646,
EPI_ISL_891648, EPI_ISL_891649, EPI_ISL_891651, EPI_ISL_891652, EPI_ISL_891653, EPI_ISL_891654, EPI_ISL_891655, EPI_ISL_891657, EPI_ISL_891660, EPI_ISL_891663, EPI_ISL_891665, EPI_ISL_891666, EPI_ISL_891667, EPI_ISL_891669, EPI_ISL_891671, EPI_ISL_891674, EPI_ISL_891675, EPI_ISL_891676, EPI_ISL_891678, EPI_ISL_891679, EPI_ISL_891682, EPI_ISL_891684, EPI_ISL_891686, EPI_ISL_891688,
EPI_ISL_891690, EPI_ISL_891691, EPI_ISL_891692, EPI_ISL_891693, EPI_ISL_891695, EPI_ISL_891696, EPI_ISL_891697, EPI_ISL_891698, EPI_ISL_891699, EPI_ISL_891701, EPI_ISL_891702, EPI_ISL_891703, EPI_ISL_891706, EPI_ISL_891707, EPI_ISL_891708, EPI_ISL_891709, EPI_ISL_891712, EPI_ISL_891715, EPI_ISL_891716, EPI_ISL_891720, EPI_ISL_891728, EPI_ISL_891730, EPI_ISL_891731, EPI_ISL_891732,
EPI_ISL_891733, EPI_ISL_891734, EPI_ISL_891737, EPI_ISL_891739, EPI_ISL_891743, EPI_ISL_891744, EPI_ISL_891745, EPI_ISL_891748, EPI_ISL_891750, EPI_ISL_891752, EPI_ISL_891754, EPI_ISL_891755, EPI_ISL_891756, EPI_ISL_891758, EPI_ISL_891759, EPI_ISL_891763, EPI_ISL_891764, EPI_ISL_891767, EPI_ISL_891768, EPI_ISL_891770, EPI_ISL_891778, EPI_ISL_891779, EPI_ISL_891781, EPI_ISL_891782,
EPI_ISL_891783, EPI_ISL_891784, EPI_ISL_891785, EPI_ISL_891792, EPI_ISL_891795, EPI_ISL_891796, EPI_ISL_891797, EPI_ISL_891798, EPI_ISL_891799, EPI_ISL_891800, EPI_ISL_891801, EPI_ISL_891803, EPI_ISL_891805, EPI_ISL_891806, EPI_ISL_891808, EPI_ISL_891810, EPI_ISL_891812, EPI_ISL_891814, EPI_ISL_891815, EPI_ISL_891816, EPI_ISL_891817, EPI_ISL_891818, EPI_ISL_891820, EPI_ISL_891825,
EPI_ISL_891827, EPI_ISL_891829, EPI_ISL_891830, EPI_ISL_891831, EPI_ISL_891832, EPI_ISL_891835, EPI_ISL_891837, EPI_ISL_891840, EPI_ISL_891841, EPI_ISL_891842, EPI_ISL_891844, EPI_ISL_891849, EPI_ISL_891854, EPI_ISL_891859, EPI_ISL_891862, EPI_ISL_891868, EPI_ISL_891871, EPI_ISL_891872, EPI_ISL_891874, EPI_ISL_891876, EPI_ISL_891877, EPI_ISL_891879, EPI_ISL_891880, EPI_ISL_891882,
EPI_ISL_891883, EPI_ISL_891886, EPI_ISL_891888, EPI_ISL_891889, EPI_ISL_891890, EPI_ISL_891891, EPI_ISL_891892, EPI_ISL_891893, EPI_ISL_891894, EPI_ISL_891896, EPI_ISL_891897, EPI_ISL_891898, EPI_ISL_907119, EPI_ISL_907133, EPI_ISL_907136, EPI_ISL_907139, EPI_ISL_907144, EPI_ISL_907148, EPI_ISL_907152, EPI_ISL_907155, EPI_ISL_907160, EPI_ISL_907164, EPI_ISL_907167, EPI_ISL_907173,
EPI_ISL_907179, EPI_ISL_907182, EPI_ISL_907186, EPI_ISL_907187, EPI_ISL_907190, EPI_ISL_907193, EPI_ISL_907195, EPI_ISL_907200, EPI_ISL_907206, EPI_ISL_907209, EPI_ISL_907212, EPI_ISL_907214, EPI_ISL_907219, EPI_ISL_907221, EPI_ISL_907223, EPI_ISL_907224, EPI_ISL_907225, EPI_ISL_907238, EPI_ISL_907241, EPI_ISL_907250, EPI_ISL_907254, EPI_ISL_907256, EPI_ISL_907260, EPI_ISL_907263,
EPI_ISL_907264, EPI_ISL_907267, EPI_ISL_907269, EPI_ISL_907279, EPI_ISL_907284, EPI_ISL_907287, EPI_ISL_907288, EPI_ISL_907292, EPI_ISL_907300, EPI_ISL_907304, EPI_ISL_907308, EPI_ISL_907315, EPI_ISL_907318, EPI_ISL_907321, EPI_ISL_907322, EPI_ISL_907323, EPI_ISL_907325, EPI_ISL_907328, EPI_ISL_907336, EPI_ISL_907341, EPI_ISL_907347, EPI_ISL_907351, EPI_ISL_907352, EPI_ISL_907353,
EPI_ISL_907355, EPI_ISL_907362, EPI_ISL_907363, EPI_ISL_907367, EPI_ISL_908035, EPI_ISL_908037, EPI_ISL_908038, EPI_ISL_908039, EPI_ISL_908040, EPI_ISL_908041, EPI_ISL_908042, EPI_ISL_908043, EPI_ISL_908044, EPI_ISL_908045, EPI_ISL_908046, EPI_ISL_908047, EPI_ISL_908048, EPI_ISL_908049, EPI_ISL_908050, EPI_ISL_908052, EPI_ISL_908053, EPI_ISL_908055, EPI_ISL_908057, EPI_ISL_908058,
EPI_ISL_908062, EPI_ISL_908063, EPI_ISL_908065, EPI_ISL_908066, EPI_ISL_908067, EPI_ISL_908068, EPI_ISL_908069, EPI_ISL_908071, EPI_ISL_908076, EPI_ISL_908077, EPI_ISL_908078, EPI_ISL_908079, EPI_ISL_908080, EPI_ISL_908082, EPI_ISL_908083, EPI_ISL_908088, EPI_ISL_908090, EPI_ISL_908091, EPI_ISL_908093, EPI_ISL_908095, EPI_ISL_908098, EPI_ISL_908099, EPI_ISL_908100, EPI_ISL_908101,
EPI_ISL_908102, EPI_ISL_908103, EPI_ISL_908105, EPI_ISL_908106, EPI_ISL_908107, EPI_ISL_908108, EPI_ISL_908109, EPI_ISL_908110, EPI_ISL_908111, EPI_ISL_908112, EPI_ISL_908114, EPI_ISL_908115, EPI_ISL_908117, EPI_ISL_908118, EPI_ISL_908121, EPI_ISL_908123, EPI_ISL_908124, EPI_ISL_908125, EPI_ISL_908127, EPI_ISL_908128, EPI_ISL_908129, EPI_ISL_908130, EPI_ISL_908132, EPI_ISL_908133,
EPI_ISL_908134, EPI_ISL_908135, EPI_ISL_908136, EPI_ISL_908137, EPI_ISL_908138, EPI_ISL_908140, EPI_ISL_908141, EPI_ISL_908142, EPI_ISL_908143, EPI_ISL_908144, EPI_ISL_908145, EPI_ISL_908146, EPI_ISL_908147, EPI_ISL_908148, EPI_ISL_908149, EPI_ISL_908150, EPI_ISL_908153, EPI_ISL_908154, EPI_ISL_908155, EPI_ISL_908156, EPI_ISL_908157, EPI_ISL_908158, EPI_ISL_908159, EPI_ISL_908160,
EPI_ISL_908161, EPI_ISL_908162, EPI_ISL_908163, EPI_ISL_908164, EPI_ISL_908165, EPI_ISL_908166, EPI_ISL_908167, EPI_ISL_908169, EPI_ISL_908170, EPI_ISL_908172, EPI_ISL_908173, EPI_ISL_908175, EPI_ISL_908176, EPI_ISL_908178, EPI_ISL_908180, EPI_ISL_908182, EPI_ISL_908184, EPI_ISL_908185, EPI_ISL_908186, EPI_ISL_908188, EPI_ISL_908189, EPI_ISL_908190, EPI_ISL_908191, EPI_ISL_908192,
EPI_ISL_908193, EPI_ISL_908195, EPI_ISL_908196, EPI_ISL_908197, EPI_ISL_908198, EPI_ISL_908199, EPI_ISL_908201, EPI_ISL_908202, EPI_ISL_908203, EPI_ISL_908204, EPI_ISL_908205, EPI_ISL_908207, EPI_ISL_908209, EPI_ISL_908210, EPI_ISL_908211, EPI_ISL_908212, EPI_ISL_908213, EPI_ISL_908215, EPI_ISL_908216, EPI_ISL_908217, EPI_ISL_908219, EPI_ISL_908220, EPI_ISL_908221, EPI_ISL_908222,
EPI_ISL_908223, EPI_ISL_908224, EPI_ISL_908225, EPI_ISL_908226, EPI_ISL_908227, EPI_ISL_908228, EPI_ISL_908230, EPI_ISL_908232, EPI_ISL_908233, EPI_ISL_908234, EPI_ISL_908235, EPI_ISL_908236, EPI_ISL_908239, EPI_ISL_908240, EPI_ISL_908244, EPI_ISL_908245, EPI_ISL_908246, EPI_ISL_908247, EPI_ISL_908248, EPI_ISL_908249, EPI_ISL_908250, EPI_ISL_908251, EPI_ISL_908253, EPI_ISL_908255,
EPI_ISL_908257, EPI_ISL_908258, EPI_ISL_908259, EPI_ISL_908264, EPI_ISL_908267, EPI_ISL_908597, EPI_ISL_908598, EPI_ISL_908599, EPI_ISL_908600, EPI_ISL_908601, EPI_ISL_908602, EPI_ISL_908603, EPI_ISL_908604, EPI_ISL_908605, EPI_ISL_908606, EPI_ISL_908607, EPI_ISL_908608, EPI_ISL_908609, EPI_ISL_908610, EPI_ISL_908611, EPI_ISL_908612, EPI_ISL_908613, EPI_ISL_908614, EPI_ISL_908615,
EPI_ISL_908616, EPI_ISL_908617, EPI_ISL_908618, EPI_ISL_908619, EPI_ISL_908620, EPI_ISL_908621, EPI_ISL_908622, EPI_ISL_908623, EPI_ISL_908624, EPI_ISL_908625, EPI_ISL_908626, EPI_ISL_908627, EPI_ISL_908628, EPI_ISL_908629, EPI_ISL_908630, EPI_ISL_908631, EPI_ISL_908632, EPI_ISL_908633, EPI_ISL_908634, EPI_ISL_908635, EPI_ISL_908636, EPI_ISL_908637, EPI_ISL_908638, EPI_ISL_908639,
EPI_ISL_908640, EPI_ISL_908641, EPI_ISL_908642, EPI_ISL_908643, EPI_ISL_908644, EPI_ISL_908645, EPI_ISL_908646, EPI_ISL_908647, EPI_ISL_908648, EPI_ISL_908649, EPI_ISL_908650, EPI_ISL_908651, EPI_ISL_908652, EPI_ISL_908653, EPI_ISL_908654, EPI_ISL_908655, EPI_ISL_908656, EPI_ISL_908657, EPI_ISL_908658, EPI_ISL_908659, EPI_ISL_908660, EPI_ISL_908661, EPI_ISL_908662, EPI_ISL_908663,
EPI_ISL_908664, EPI_ISL_908665, EPI_ISL_908666, EPI_ISL_908667, EPI_ISL_908668, EPI_ISL_908669, EPI_ISL_908670, EPI_ISL_908671, EPI_ISL_908672, EPI_ISL_908673, EPI_ISL_908674, EPI_ISL_908675, EPI_ISL_908676, EPI_ISL_908677, EPI_ISL_908678, EPI_ISL_908679, EPI_ISL_908680, EPI_ISL_908681, EPI_ISL_908682, EPI_ISL_908683, EPI_ISL_908684, EPI_ISL_908685, EPI_ISL_908686, EPI_ISL_908687,
EPI_ISL_908688, EPI_ISL_908689, EPI_ISL_908690, EPI_ISL_908691, EPI_ISL_908692, EPI_ISL_908693, EPI_ISL_908694, EPI_ISL_908695, EPI_ISL_908696, EPI_ISL_908697, EPI_ISL_908698, EPI_ISL_908699, EPI_ISL_908700, EPI_ISL_908701, EPI_ISL_908702, EPI_ISL_908703, EPI_ISL_908704, EPI_ISL_908705, EPI_ISL_908706, EPI_ISL_908707, EPI_ISL_908708, EPI_ISL_908709, EPI_ISL_908710, EPI_ISL_908711,
EPI_ISL_908712, EPI_ISL_908713, EPI_ISL_908714, EPI_ISL_908715, EPI_ISL_908716, EPI_ISL_908717, EPI_ISL_908718, EPI_ISL_908719, EPI_ISL_908720, EPI_ISL_908721, EPI_ISL_908722, EPI_ISL_908723, EPI_ISL_908724, EPI_ISL_908725, EPI_ISL_908726, EPI_ISL_908727, EPI_ISL_908728, EPI_ISL_908729, EPI_ISL_908730, EPI_ISL_908731, EPI_ISL_908732, EPI_ISL_908733, EPI_ISL_908734, EPI_ISL_908735,
EPI_ISL_908736, EPI_ISL_908737, EPI_ISL_908738, EPI_ISL_908739, EPI_ISL_908740, EPI_ISL_908741, EPI_ISL_908742, EPI_ISL_908743, EPI_ISL_908744, EPI_ISL_908745, EPI_ISL_908746, EPI_ISL_908747, EPI_ISL_908748, EPI_ISL_908749, EPI_ISL_908750, EPI_ISL_908751, EPI_ISL_908752, EPI_ISL_908753, EPI_ISL_908754, EPI_ISL_908755, EPI_ISL_908756, EPI_ISL_908757, EPI_ISL_908758, EPI_ISL_908759,
EPI_ISL_908760, EPI_ISL_908762, EPI_ISL_908763, EPI_ISL_908764, EPI_ISL_908765, EPI_ISL_908766, EPI_ISL_908767, EPI_ISL_908768, EPI_ISL_908769, EPI_ISL_908770, EPI_ISL_908771, EPI_ISL_908772, EPI_ISL_908773, EPI_ISL_908774, EPI_ISL_908775, EPI_ISL_908776, EPI_ISL_908777, EPI_ISL_908778, EPI_ISL_908779, EPI_ISL_908780, EPI_ISL_908781, EPI_ISL_908782, EPI_ISL_908783, EPI_ISL_908784,
EPI_ISL_908785, EPI_ISL_908786, EPI_ISL_908787, EPI_ISL_908788, EPI_ISL_908789, EPI_ISL_908790, EPI_ISL_908791, EPI_ISL_908792, EPI_ISL_908793, EPI_ISL_908794, EPI_ISL_908795, EPI_ISL_908796, EPI_ISL_908797, EPI_ISL_908798, EPI_ISL_908799, EPI_ISL_908800, EPI_ISL_908801, EPI_ISL_908802, EPI_ISL_908803, EPI_ISL_908804, EPI_ISL_908805, EPI_ISL_908806, EPI_ISL_908807, EPI_ISL_908808,
EPI_ISL_908809, EPI_ISL_908810, EPI_ISL_908811, EPI_ISL_908812, EPI_ISL_908813, EPI_ISL_908814, EPI_ISL_908815, EPI_ISL_908816, EPI_ISL_908817, EPI_ISL_908818, EPI_ISL_908819, EPI_ISL_908820, EPI_ISL_908821, EPI_ISL_908822, EPI_ISL_908823, EPI_ISL_908824, EPI_ISL_908825, EPI_ISL_908826, EPI_ISL_908827, EPI_ISL_908828, EPI_ISL_908829, EPI_ISL_908830, EPI_ISL_908831, EPI_ISL_908832,
EPI_ISL_908833, EPI_ISL_908834, EPI_ISL_908835, EPI_ISL_908836, EPI_ISL_908837, EPI_ISL_908838, EPI_ISL_908839, EPI_ISL_908840, EPI_ISL_908841, EPI_ISL_908842, EPI_ISL_908843, EPI_ISL_908844, EPI_ISL_908845, EPI_ISL_908846, EPI_ISL_908847, EPI_ISL_908848, EPI_ISL_908849, EPI_ISL_908850, EPI_ISL_908851, EPI_ISL_908852, EPI_ISL_908853, EPI_ISL_908854, EPI_ISL_908855, EPI_ISL_908856,
EPI_ISL_908857, EPI_ISL_908858, EPI_ISL_908859, EPI_ISL_908860, EPI_ISL_908861, EPI_ISL_908862, EPI_ISL_908863, EPI_ISL_908864, EPI_ISL_908865, EPI_ISL_908866, EPI_ISL_908867, EPI_ISL_908868, EPI_ISL_908869, EPI_ISL_908870, EPI_ISL_908871, EPI_ISL_908872, EPI_ISL_908873, EPI_ISL_908874, EPI_ISL_908875, EPI_ISL_908876, EPI_ISL_908877, EPI_ISL_908878, EPI_ISL_908879, EPI_ISL_908880,
EPI_ISL_908881, EPI_ISL_908882, EPI_ISL_908883, EPI_ISL_908884, EPI_ISL_908885, EPI_ISL_908886, EPI_ISL_908887, EPI_ISL_908888, EPI_ISL_908889, EPI_ISL_908890, EPI_ISL_908891, EPI_ISL_908892, EPI_ISL_908893, EPI_ISL_908894, EPI_ISL_908895, EPI_ISL_908896, EPI_ISL_908897, EPI_ISL_908898, EPI_ISL_908899, EPI_ISL_908900, EPI_ISL_908901, EPI_ISL_908902, EPI_ISL_908903, EPI_ISL_908904,
EPI_ISL_908905, EPI_ISL_908906, EPI_ISL_908907, EPI_ISL_908908, EPI_ISL_908909, EPI_ISL_908910, EPI_ISL_908911, EPI_ISL_908912, EPI_ISL_908913, EPI_ISL_908914, EPI_ISL_908915, EPI_ISL_908916, EPI_ISL_908917, EPI_ISL_908918, EPI_ISL_908919, EPI_ISL_908920, EPI_ISL_908921, EPI_ISL_908922, EPI_ISL_908923, EPI_ISL_908924, EPI_ISL_908925, EPI_ISL_908926, EPI_ISL_908927, EPI_ISL_908928,
EPI_ISL_908929, EPI_ISL_908930, EPI_ISL_908931, EPI_ISL_908933, EPI_ISL_908935, EPI_ISL_908942, EPI_ISL_908946, EPI_ISL_908949, EPI_ISL_908951, EPI_ISL_908952, EPI_ISL_908955, EPI_ISL_908957, EPI_ISL_908959, EPI_ISL_908961, EPI_ISL_908963, EPI_ISL_908967, EPI_ISL_908968, EPI_ISL_908977, EPI_ISL_908978, EPI_ISL_908979, EPI_ISL_908983, EPI_ISL_908985, EPI_ISL_908990, EPI_ISL_908991,
EPI_ISL_908992, EPI_ISL_908998, EPI_ISL_908999, EPI_ISL_909006, EPI_ISL_909007, EPI_ISL_909009, EPI_ISL_909011, EPI_ISL_909015, EPI_ISL_909019, EPI_ISL_909035, EPI_ISL_909036, EPI_ISL_909040, EPI_ISL_909046, EPI_ISL_909047, EPI_ISL_909048, EPI_ISL_909049, EPI_ISL_909050, EPI_ISL_909052, EPI_ISL_909060, EPI_ISL_909066, EPI_ISL_909067, EPI_ISL_909071, EPI_ISL_909072, EPI_ISL_909075,
EPI_ISL_909079, EPI_ISL_909080, EPI_ISL_909081, EPI_ISL_909088, EPI_ISL_909093, EPI_ISL_909095, EPI_ISL_909096, EPI_ISL_909097, EPI_ISL_909102, EPI_ISL_909103, EPI_ISL_909105, EPI_ISL_909109, EPI_ISL_909119, EPI_ISL_909122, EPI_ISL_909128, EPI_ISL_909144, EPI_ISL_909145, EPI_ISL_909148, EPI_ISL_909155, EPI_ISL_909156, EPI_ISL_909157, EPI_ISL_909173, EPI_ISL_909178, EPI_ISL_909180,
EPI_ISL_909182, EPI_ISL_909187, EPI_ISL_909189, EPI_ISL_909192, EPI_ISL_909193, EPI_ISL_909202, EPI_ISL_909203, EPI_ISL_909204, EPI_ISL_909217, EPI_ISL_909220, EPI_ISL_909223, EPI_ISL_909230, EPI_ISL_909234, EPI_ISL_909238, EPI_ISL_909240, EPI_ISL_909243, EPI_ISL_909251, EPI_ISL_909254, EPI_ISL_912214, EPI_ISL_916191, EPI_ISL_916192, EPI_ISL_916196, EPI_ISL_916210, EPI_ISL_916211,
EPI_ISL_916212, EPI_ISL_916229, EPI_ISL_916231, EPI_ISL_916237, EPI_ISL_916242, EPI_ISL_916243, EPI_ISL_916247, EPI_ISL_916249, EPI_ISL_916252, EPI_ISL_916260, EPI_ISL_916262, EPI_ISL_916269, EPI_ISL_916270, EPI_ISL_916271, EPI_ISL_916274, EPI_ISL_916277, EPI_ISL_916280, EPI_ISL_916292, EPI_ISL_916297, EPI_ISL_916304, EPI_ISL_916313, EPI_ISL_916331, EPI_ISL_916334, EPI_ISL_916346,
EPI_ISL_916348, EPI_ISL_916354, EPI_ISL_916355, EPI_ISL_916362, EPI_ISL_916363, EPI_ISL_916376, EPI_ISL_916382, EPI_ISL_916391, EPI_ISL_916392, EPI_ISL_916414, EPI_ISL_916416, EPI_ISL_916420, EPI_ISL_916427, EPI_ISL_916435, EPI_ISL_916444, EPI_ISL_916445, EPI_ISL_916450, EPI_ISL_916461, EPI_ISL_916470, EPI_ISL_916475, EPI_ISL_916495, EPI_ISL_916496, EPI_ISL_916497, EPI_ISL_933113,
EPI_ISL_933123, EPI_ISL_933124, EPI_ISL_933127, EPI_ISL_933130, EPI_ISL_933134, EPI_ISL_933136, EPI_ISL_933141, EPI_ISL_933143, EPI_ISL_933149, EPI_ISL_933150, EPI_ISL_933154, EPI_ISL_933158, EPI_ISL_933166, EPI_ISL_933167, EPI_ISL_933170, EPI_ISL_933171, EPI_ISL_933181, EPI_ISL_933187, EPI_ISL_933188, EPI_ISL_933196, EPI_ISL_933199, EPI_ISL_933200, EPI_ISL_933207, EPI_ISL_933213,
EPI_ISL_933214, EPI_ISL_933215, EPI_ISL_933221, EPI_ISL_933226, EPI_ISL_933227, EPI_ISL_933228, EPI_ISL_933233, EPI_ISL_933239, EPI_ISL_933243, EPI_ISL_933256, EPI_ISL_933259, EPI_ISL_933265, EPI_ISL_933266, EPI_ISL_933271, EPI_ISL_933272, EPI_ISL_933273, EPI_ISL_933277, EPI_ISL_933282, EPI_ISL_933283, EPI_ISL_933289, EPI_ISL_933293, EPI_ISL_933298, EPI_ISL_933303, EPI_ISL_933321,
EPI_ISL_933325, EPI_ISL_933326, EPI_ISL_933336, EPI_ISL_933340, EPI_ISL_933341, EPI_ISL_933343, EPI_ISL_933344, EPI_ISL_933345, EPI_ISL_933346, EPI_ISL_933348, EPI_ISL_933350, EPI_ISL_933354, EPI_ISL_933358, EPI_ISL_933359, EPI_ISL_933364, EPI_ISL_933371, EPI_ISL_933375, EPI_ISL_933382, EPI_ISL_933386, EPI_ISL_933387, EPI_ISL_933388, EPI_ISL_933393, EPI_ISL_933396, EPI_ISL_933397,
EPI_ISL_933400, EPI_ISL_933401, EPI_ISL_933410, EPI_ISL_933412, EPI_ISL_933414, EPI_ISL_933417, EPI_ISL_993415, EPI_ISL_993418, EPI_ISL_993420, EPI_ISL_993424, EPI_ISL_993430, EPI_ISL_993431, EPI_ISL_993435, EPI_ISL_993437, EPI_ISL_993439, EPI_ISL_993440, EPI_ISL_993448, EPI_ISL_993449, EPI_ISL_993455, EPI_ISL_993457, EPI_ISL_993465, EPI_ISL_993469, EPI_ISL_993477, EPI_ISL_993487,
EPI_ISL_993496, EPI_ISL_993504, EPI_ISL_993507, EPI_ISL_993519, EPI_ISL_993522, EPI_ISL_993523, EPI_ISL_993532, EPI_ISL_993538, EPI_ISL_993542, EPI_ISL_993543, EPI_ISL_993546, EPI_ISL_993549, EPI_ISL_993550, EPI_ISL_993560, EPI_ISL_993562, EPI_ISL_993571, EPI_ISL_993581, EPI_ISL_993582, EPI_ISL_993584, EPI_ISL_993589, EPI_ISL_993593, EPI_ISL_993595, EPI_ISL_993598, EPI_ISL_993599,
EPI_ISL_993616, EPI_ISL_993619, EPI_ISL_993620, EPI_ISL_993621, EPI_ISL_993634, EPI_ISL_993638, EPI_ISL_993639, EPI_ISL_993640, EPI_ISL_993641, EPI_ISL_993654, EPI_ISL_993656, EPI_ISL_993661, EPI_ISL_993682, EPI_ISL_993683, EPI_ISL_993684, EPI_ISL_993690, EPI_ISL_993697, EPI_ISL_993698, EPI_ISL_993704, EPI_ISL_993706, EPI_ISL_993718, EPI_ISL_993720, EPI_ISL_993722, EPI_ISL_993743,
EPI_ISL_993793, EPI_ISL_993817, EPI_ISL_993823, EPI_ISL_993833, EPI_ISL_993890, EPI_ISL_993907, EPI_ISL_993915, EPI_ISL_993920, EPI_ISL_994003, EPI_ISL_994059, EPI_ISL_994287, EPI_ISL_994290, EPI_ISL_994294, EPI_ISL_994295, EPI_ISL_994296, EPI_ISL_994299, EPI_ISL_994300, EPI_ISL_994310, EPI_ISL_994315, EPI_ISL_994319, EPI_ISL_994327, EPI_ISL_994331, EPI_ISL_994334, EPI_ISL_994341,
EPI_ISL_994349, EPI_ISL_994350, EPI_ISL_994356, EPI_ISL_994359, EPI_ISL_994362, EPI_ISL_994363, EPI_ISL_994372, EPI_ISL_994373, EPI_ISL_994374, EPI_ISL_994382, EPI_ISL_994383, EPI_ISL_994390, EPI_ISL_994395, EPI_ISL_994397, EPI_ISL_994401, EPI_ISL_994403, EPI_ISL_994408, EPI_ISL_994409, EPI_ISL_994410, EPI_ISL_994420, EPI_ISL_994421, EPI_ISL_994425, EPI_ISL_994427, EPI_ISL_994429,
EPI_ISL_994431, EPI_ISL_994432, EPI_ISL_994434, EPI_ISL_994435, EPI_ISL_994436, EPI_ISL_994440, EPI_ISL_994441, EPI_ISL_994444, EPI_ISL_994447, EPI_ISL_994452, EPI_ISL_994457, EPI_ISL_994458, EPI_ISL_994459, EPI_ISL_994470, EPI_ISL_994472, EPI_ISL_994477, EPI_ISL_994480, EPI_ISL_994497, EPI_ISL_994501, EPI_ISL_994504, EPI_ISL_994508, EPI_ISL_994511, EPI_ISL_994516, EPI_ISL_994517,
EPI_ISL_994521, EPI_ISL_994524, EPI_ISL_994530, EPI_ISL_994536, EPI_ISL_994544, EPI_ISL_994610, EPI_ISL_1007583

see above | Lighthouse Lab in Cambridge | Wellcome Sanger Institute | Cordelia Langford; David K. Jackson; Dominic Kwiatkowski; Ewan Harrison; Ian Johnston; John Sillitoe on behalf of the Wellcome Sanger Institute COVID-19 Surveillance Team; John Sillitoe on behalf of the Wellcome Sanger Institute COVID-19 Surveillance for the COVID-19 Genomics UK (COG-UK) Consortium | Team (http://www.sanger.ac.uk/covid-team); Rob Howes; Roberto Amato; Sonia Goncalves; The Lighthouse Lab in Cambridge and Alex Alderton

EPI_ISL_891281, EPI_ISL_891283, EPI_ISL_891286, EPI_ISL_891288, EPI_ISL_891297, EPI_ISL_891298, EPI_ISL_891304, EPI_ISL_891320, EPI_ISL_891327, EPI_ISL_891330, EPI_ISL_891335, EPI_ISL_891338, EPI_ISL_891345, EPI_ISL_891361, EPI_ISL_891372, EPI_ISL_891380, EPI_ISL_891391, EPI_ISL_891393, EPI_ISL_891396, EPI_ISL_891410, EPI_ISL_891415, EPI_ISL_891420, EPI_ISL_891426, EPI_ISL_891435, EPI_ISL_891445, EPI_ISL_891446, EPI_ISL_891453, EPI_ISL_891462, EPI_ISL_891464, EPI_ISL_891465, EPI_ISL_891473, EPI_ISL_891491, EPI_ISL_891495, EPI_ISL_891497, EPI_ISL_891502, EPI_ISL_891505, EPI_ISL_891508, EPI_ISL_891509, EPI_ISL_891516, EPI_ISL_891539, EPI_ISL_891549, EPI_ISL_891562, EPI_ISL_891568, EPI_ISL_891571, EPI_ISL_891574, EPI_ISL_891585, EPI_ISL_891587, EPI_ISL_891588, EPI_ISL_891604, EPI_ISL_891605, EPI_ISL_891610, EPI_ISL_891612, EPI_ISL_891618, EPI_ISL_891624, EPI_ISL_891626, EPI_ISL_891628, EPI_ISL_891636, EPI_ISL_891656, EPI_ISL_891658, EPI_ISL_891689, EPI_ISL_891705, EPI_ISL_891717, EPI_ISL_891721, EPI_ISL_891722, EPI_ISL_891725, EPI_ISL_891727, EPI_ISL_891740, EPI_ISL_891741, EPI_ISL_891761, EPI_ISL_891771, EPI_ISL_891772, EPI_ISL_891790, EPI_ISL_891791, EPI_ISL_891793, EPI_ISL_891802, EPI_ISL_891809, EPI_ISL_891828, EPI_ISL_891836, EPI_ISL_891846, EPI_ISL_891851, EPI_ISL_891852, EPI_ISL_891853, EPI_ISL_891855, EPI_ISL_891857, EPI_ISL_891861, EPI_ISL_891865, EPI_ISL_891869, EPI_ISL_891873, EPI_ISL_891875, EPI_ISL_891881, EPI_ISL_891884, EPI_ISL_891885, EPI_ISL_907788, EPI_ISL_908036, EPI_ISL_908051, EPI_ISL_908054, EPI_ISL_908056, EPI_ISL_908059, EPI_ISL_908060, EPI_ISL_908061, EPI_ISL_908064, EPI_ISL_908070, EPI_ISL_908072, EPI_ISL_908073, EPI_ISL_908074, EPI_ISL_908075, EPI_ISL_908081, EPI_ISL_908084, EPI_ISL_908085, EPI_ISL_908086, EPI_ISL_908087, EPI_ISL_908089, EPI_ISL_908092, EPI_ISL_908094, EPI_ISL_908096, EPI_ISL_908097, EPI_ISL_908104, EPI_ISL_908113, EPI_ISL_908116, EPI_ISL_908119, EPI_ISL_908120, EPI_ISL_908122, EPI_ISL_908126, EPI_ISL_908131, EPI_ISL_908139, EPI_ISL_908151, EPI_ISL_908152, EPI_ISL_908168, EPI_ISL_908171, EPI_ISL_908174, EPI_ISL_908177, EPI_ISL_908179, EPI_ISL_908181, EPI_ISL_908183, EPI_ISL_908187, EPI_ISL_908194, EPI_ISL_908200, EPI_ISL_908206, EPI_ISL_908208, EPI_ISL_908214, EPI_ISL_908218, EPI_ISL_908229, EPI_ISL_908231, EPI_ISL_908237, EPI_ISL_908238, EPI_ISL_908241, EPI_ISL_908242, EPI_ISL_908243, EPI_ISL_908252, EPI_ISL_908254, EPI_ISL_908256, EPI_ISL_908260, EPI_ISL_908261, EPI_ISL_908262, EPI_ISL_908263, EPI_ISL_908265, EPI_ISL_908298, EPI_ISL_908419, EPI_ISL_908945, EPI_ISL_908953, EPI_ISL_908958, EPI_ISL_908962, EPI_ISL_908965, EPI_ISL_908974, EPI_ISL_908982, EPI_ISL_908986, EPI_ISL_908989, EPI_ISL_908993, EPI_ISL_909002, EPI_ISL_909004, EPI_ISL_909005, EPI_ISL_909008, EPI_ISL_909010, EPI_ISL_909017, EPI_ISL_909021, EPI_ISL_909024, EPI_ISL_909029, EPI_ISL_909030, EPI_ISL_909034, EPI_ISL_909038, EPI_ISL_909053, EPI_ISL_909056, EPI_ISL_909057, EPI_ISL_909058, EPI_ISL_909061, EPI_ISL_909063, EPI_ISL_909068, EPI_ISL_909069, EPI_ISL_909083, EPI_ISL_909085, EPI_ISL_909086, EPI_ISL_909087, EPI_ISL_909089, EPI_ISL_909090, EPI_ISL_909094, EPI_ISL_909101, EPI_ISL_909106, EPI_ISL_909111, EPI_ISL_909113, EPI_ISL_909114, EPI_ISL_909116, EPI_ISL_909117, EPI_ISL_909118, EPI_ISL_909121, EPI_ISL_909131, EPI_ISL_909150, EPI_ISL_909158, EPI_ISL_909162, EPI_ISL_909165, EPI_ISL_909169, EPI_ISL_909170, EPI_ISL_909171, EPI_ISL_909176, EPI_ISL_909177, EPI_ISL_909184, EPI_ISL_909194, EPI_ISL_909200, EPI_ISL_909206, EPI_ISL_909208, EPI_ISL_909210, EPI_ISL_909211, EPI_ISL_909218, EPI_ISL_909219, EPI_ISL_909227, EPI_ISL_909233, EPI_ISL_909235, EPI_ISL_909236, EPI_ISL_909237, EPI_ISL_909242, EPI_ISL_909255, EPI_ISL_909257, EPI_ISL_909327, EPI_ISL_909351, EPI_ISL_909356, EPI_ISL_909369, EPI_ISL_909375, EPI_ISL_909382, EPI_ISL_909390, EPI_ISL_909405, EPI_ISL_909435, EPI_ISL_909497, EPI_ISL_909508, EPI_ISL_909538, EPI_ISL_909549, EPI_ISL_909568, EPI_ISL_909588, EPI_ISL_909590, EPI_ISL_917849, EPI_ISL_917850, EPI_ISL_917851, EPI_ISL_917852, EPI_ISL_917853, EPI_ISL_917854, EPI_ISL_917855, EPI_ISL_917856, EPI_ISL_917857, EPI_ISL_917858, EPI_ISL_917859, EPI_ISL_917860, EPI_ISL_917861, EPI_ISL_917862, EPI_ISL_917863, EPI_ISL_917864, EPI_ISL_917865, EPI_ISL_917866, EPI_ISL_917867, EPI_ISL_917868, EPI_ISL_917869, EPI_ISL_917870, EPI_ISL_917871, EPI_ISL_917872, EPI_ISL_917873, EPI_ISL_917874, EPI_ISL_917875, EPI_ISL_917876, EPI_ISL_917877, EPI_ISL_917878, EPI_ISL_917879, EPI_ISL_917880, EPI_ISL_917881, EPI_ISL_917882, EPI_ISL_917883, EPI_ISL_917884, EPI_ISL_917885, EPI_ISL_917886, EPI_ISL_917887, EPI_ISL_917888, EPI_ISL_917889, EPI_ISL_917890, EPI_ISL_917891, EPI_ISL_917892, EPI_ISL_917893, EPI_ISL_917894, EPI_ISL_917895, EPI_ISL_917896, EPI_ISL_917897, EPI_ISL_917898, EPI_ISL_917899, EPI_ISL_917900, EPI_ISL_917901, EPI_ISL_917902, EPI_ISL_917903, EPI_ISL_917904, EPI_ISL_917905, EPI_ISL_917906, EPI_ISL_917907, EPI_ISL_917908, EPI_ISL_917909, EPI_ISL_917910, EPI_ISL_917911, EPI_ISL_917912, EPI_ISL_917913, EPI_ISL_917914, EPI_ISL_917915, EPI_ISL_917916, EPI_ISL_917917, EPI_ISL_917918, EPI_ISL_917919, EPI_ISL_917920, EPI_ISL_917921, EPI_ISL_917922, EPI_ISL_917923, EPI_ISL_917924, EPI_ISL_917925, EPI_ISL_917926, EPI_ISL_917927, EPI_ISL_917928, EPI_ISL_917929, EPI_ISL_917930, EPI_ISL_917931, EPI_ISL_917932, EPI_ISL_917933, EPI_ISL_917934, EPI_ISL_917935, EPI_ISL_917937, EPI_ISL_917939, EPI_ISL_917940, EPI_ISL_917942, EPI_ISL_917956, EPI_ISL_917957, EPI_ISL_917958, EPI_ISL_917959, EPI_ISL_917960, EPI_ISL_917961, EPI_ISL_917962, EPI_ISL_917963, EPI_ISL_917964, EPI_ISL_917965, EPI_ISL_917966, EPI_ISL_917967, EPI_ISL_917968, EPI_ISL_917969, EPI_ISL_917970, EPI_ISL_917971, EPI_ISL_917972, EPI_ISL_917973, EPI_ISL_917974, EPI_ISL_917976, EPI_ISL_917977, EPI_ISL_917979, EPI_ISL_917980, EPI_ISL_917981, EPI_ISL_917982, EPI_ISL_917983, EPI_ISL_917984, EPI_ISL_918020, EPI_ISL_918021, EPI_ISL_918022, EPI_ISL_918023, EPI_ISL_918024, EPI_ISL_918025, EPI_ISL_918026, EPI_ISL_918027, EPI_ISL_918028, EPI_ISL_918029, EPI_ISL_918030, EPI_ISL_918031, EPI_ISL_918032, EPI_ISL_918033, EPI_ISL_918034, EPI_ISL_918035, EPI_ISL_918036, EPI_ISL_918037, EPI_ISL_918038, EPI_ISL_987282, EPI_ISL_987294, EPI_ISL_987308, EPI_ISL_987314, EPI_ISL_987316, EPI_ISL_987320, EPI_ISL_987323, EPI_ISL_987324, EPI_ISL_987337, EPI_ISL_987341, EPI_ISL_987347, EPI_ISL_987349, EPI_ISL_987350, EPI_ISL_987354, EPI_ISL_987359, EPI_ISL_987360, EPI_ISL_987362, EPI_ISL_987363, EPI_ISL_987370, EPI_ISL_987375, EPI_ISL_987376, EPI_ISL_987385, EPI_ISL_987391, EPI_ISL_987394, EPI_ISL_987404, EPI_ISL_987405, EPI_ISL_987433, EPI_ISL_987436, EPI_ISL_987437, EPI_ISL_987439, EPI_ISL_987440, EPI_ISL_987450, EPI_ISL_987470, EPI_ISL_987471, EPI_ISL_987483, EPI_ISL_987485, EPI_ISL_987486, EPI_ISL_987496, EPI_ISL_987497, EPI_ISL_987502, EPI_ISL_987503, EPI_ISL_987511, EPI_ISL_987529, EPI_ISL_987532, EPI_ISL_987535, EPI_ISL_987536, EPI_ISL_987550, EPI_ISL_987568, EPI_ISL_987571, EPI_ISL_987576, EPI_ISL_987590, EPI_ISL_987616, EPI_ISL_987619, EPI_ISL_987621, EPI_ISL_987629, EPI_ISL_994067, EPI_ISL_994068, EPI_ISL_994069, EPI_ISL_994070, EPI_ISL_994071, EPI_ISL_994072, EPI_ISL_994073, EPI_ISL_994074, EPI_ISL_994075, EPI_ISL_994076, EPI_ISL_994077, EPI_ISL_994078, EPI_ISL_994079, EPI_ISL_994080, EPI_ISL_994081, EPI_ISL_994082, EPI_ISL_994083, EPI_ISL_994084, EPI_ISL_994085, EPI_ISL_994086, EPI_ISL_994087, EPI_ISL_994088, EPI_ISL_994089, EPI_ISL_994090, EPI_ISL_994091, EPI_ISL_994092, EPI_ISL_994093, EPI_ISL_994094, EPI_ISL_994095, EPI_ISL_994096, EPI_ISL_994097, EPI_ISL_994098, EPI_ISL_994099, EPI_ISL_994100, EPI_ISL_994101, EPI_ISL_994102, EPI_ISL_994103, EPI_ISL_994104, EPI_ISL_994105, EPI_ISL_994106, EPI_ISL_994107, EPI_ISL_994108, EPI_ISL_994109, EPI_ISL_994110, EPI_ISL_994111, EPI_ISL_994112, EPI_ISL_994113, EPI_ISL_994114, EPI_ISL_994115, EPI_ISL_994116, EPI_ISL_994117, EPI_ISL_994118, EPI_ISL_994119, EPI_ISL_994120, EPI_ISL_994121, EPI_ISL_994122, EPI_ISL_994123, EPI_ISL_994124, EPI_ISL_994125, EPI_ISL_994126, EPI_ISL_994127, EPI_ISL_994128, EPI_ISL_994129, EPI_ISL_994130, EPI_ISL_994131, EPI_ISL_994132, EPI_ISL_994133, EPI_ISL_994134, EPI_ISL_994135, EPI_ISL_994136, EPI_ISL_994137, EPI_ISL_994138, EPI_ISL_994139, EPI_ISL_994140, EPI_ISL_994141, EPI_ISL_994142, EPI_ISL_994143, EPI_ISL_994144, EPI_ISL_994145, EPI_ISL_994146, EPI_ISL_994147, EPI_ISL_994148, EPI_ISL_994149, EPI_ISL_994150, EPI_ISL_994151, EPI_ISL_994152, EPI_ISL_994153, EPI_ISL_994154, EPI_ISL_994155, EPI_ISL_994156, EPI_ISL_994157, EPI_ISL_994158, EPI_ISL_994159, EPI_ISL_994160, EPI_ISL_994161, EPI_ISL_994162, EPI_ISL_994163, EPI_ISL_994164, EPI_ISL_994165, EPI_ISL_994166, EPI_ISL_994167, EPI_ISL_994168, EPI_ISL_994169, EPI_ISL_994170, EPI_ISL_994171, EPI_ISL_994172, EPI_ISL_994173, EPI_ISL_994174, EPI_ISL_994175, EPI_ISL_994176, EPI_ISL_994177, EPI_ISL_994178, EPI_ISL_994179, EPI_ISL_994180, EPI_ISL_994181, EPI_ISL_994182, EPI_ISL_994183, EPI_ISL_994184, EPI_ISL_994185, EPI_ISL_994186, EPI_ISL_994187, EPI_ISL_994188, EPI_ISL_994189, EPI_ISL_994190, EPI_ISL_994191, EPI_ISL_994192, EPI_ISL_994193, EPI_ISL_994194, EPI_ISL_994195, EPI_ISL_994196, EPI_ISL_994197, EPI_ISL_994198, EPI_ISL_994199, EPI_ISL_994200, EPI_ISL_994201, EPI_ISL_994202, EPI_ISL_994203, EPI_ISL_994204, EPI_ISL_994205, EPI_ISL_994206, EPI_ISL_994207, EPI_ISL_994208, EPI_ISL_994209, EPI_ISL_994210, EPI_ISL_994211, EPI_ISL_994212, EPI_ISL_994213, EPI_ISL_994214, EPI_ISL_994215, EPI_ISL_994216, EPI_ISL_994217, EPI_ISL_994218, EPI_ISL_994219, EPI_ISL_994220, EPI_ISL_994221, EPI_ISL_994222, EPI_ISL_994223, EPI_ISL_994224, EPI_ISL_994225, EPI_ISL_994226, EPI_ISL_994227, EPI_ISL_994228, EPI_ISL_994229, EPI_ISL_994230, EPI_ISL_994231, EPI_ISL_994232, EPI_ISL_994233, EPI_ISL_994234, EPI_ISL_994235, EPI_ISL_994236, EPI_ISL_994237, EPI_ISL_994238, EPI_ISL_994239, EPI_ISL_994240, EPI_ISL_994241, EPI_ISL_994242, EPI_ISL_994243, EPI_ISL_994244, EPI_ISL_994245, EPI_ISL_994246, EPI_ISL_994247, EPI_ISL_994248, EPI_ISL_994249, EPI_ISL_994250, EPI_ISL_994251, EPI_ISL_994252, EPI_ISL_994253, EPI_ISL_994254, EPI_ISL_994255, EPI_ISL_994256, EPI_ISL_994257, EPI_ISL_994258, EPI_ISL_994259, EPI_ISL_994260, EPI_ISL_994261, EPI_ISL_994262, EPI_ISL_994263, EPI_ISL_994264, EPI_ISL_994265, EPI_ISL_994266, EPI_ISL_994267, EPI_ISL_994268, EPI_ISL_994269, EPI_ISL_994270, EPI_ISL_994271, EPI_ISL_994272, EPI_ISL_994273, EPI_ISL_994274, EPI_ISL_994275, EPI_ISL_994276, EPI_ISL_994277, EPI_ISL_994278, EPI_ISL_994279, EPI_ISL_994280, EPI_ISL_994281, EPI_ISL_994282, EPI_ISL_994283, EPI_ISL_994284, EPI_ISL_994285, EPI_ISL_994286, EPI_ISL_994301, EPI_ISL_994302, EPI_ISL_994305, EPI_ISL_994311, EPI_ISL_994312, EPI_ISL_994316, EPI_ISL_994317, EPI_ISL_994326, EPI_ISL_994332, EPI_ISL_994333, EPI_ISL_994336, EPI_ISL_994337, EPI_ISL_994344, EPI_ISL_994347, EPI_ISL_994353, EPI_ISL_994365, EPI_ISL_994366, EPI_ISL_994367, EPI_ISL_994371, EPI_ISL_994377, EPI_ISL_994378, EPI_ISL_994380, EPI_ISL_994381, EPI_ISL_994384, EPI_ISL_994398, EPI_ISL_994406, EPI_ISL_994437, EPI_ISL_994439, EPI_ISL_994443, EPI_ISL_994450, EPI_ISL_994469, EPI_ISL_994486, EPI_ISL_994487, EPI_ISL_994490, EPI_ISL_994502, EPI_ISL_994503, EPI_ISL_994509, EPI_ISL_994510, EPI_ISL_994518, EPI_ISL_994523, EPI_ISL_994528, EPI_ISL_994533, EPI_ISL_994537, EPI_ISL_994542, EPI_ISL_994546, EPI_ISL_1007434, EPI_ISL_1007576, EPI_ISL_1007577, EPI_ISL_1007578, EPI_ISL_1007579, EPI_ISL_1012865, EPI_ISL_1019583, EPI_ISL_1019584, EPI_ISL_1019585, EPI_ISL_1019816, EPI_ISL_1019817, EPI_ISL_1019819, EPI_ISL_1019821, EPI_ISL_1019822, EPI_ISL_1019824, EPI_ISL_1019826, EPI_ISL_1019827, EPI_ISL_1019829, EPI_ISL_1019831, EPI_ISL_1019832, EPI_ISL_1019834, EPI_ISL_1019837, EPI_ISL_1019839, EPI_ISL_1019840, EPI_ISL_1019842, EPI_ISL_1019844, EPI_ISL_1019845, EPI_ISL_1019847, EPI_ISL_1019848, EPI_ISL_1019850, EPI_ISL_1019852, EPI_ISL_1019854, EPI_ISL_1019856, EPI_ISL_1019857, EPI_ISL_1019859, EPI_ISL_1019860, EPI_ISL_1019862, EPI_ISL_1019864, EPI_ISL_1019865, EPI_ISL_1019867, EPI_ISL_1019868, EPI_ISL_1019870, EPI_ISL_1019871, EPI_ISL_1019873, EPI_ISL_1019874, EPI_ISL_1019876, EPI_ISL_1019878, EPI_ISL_1019879, EPI_ISL_1019881, EPI_ISL_1019883, EPI_ISL_1019884, EPI_ISL_1019886, EPI_ISL_1019887, EPI_ISL_1019889, EPI_ISL_1019891, EPI_ISL_1019892, EPI_ISL_1019894, EPI_ISL_1019895, EPI_ISL_1019896, EPI_ISL_1019898, EPI_ISL_1019899, EPI_ISL_1019900, EPI_ISL_1019901, EPI_ISL_1019902, EPI_ISL_1019903, EPI_ISL_1019904, EPI_ISL_1019905, EPI_ISL_1019906, EPI_ISL_1019907, EPI_ISL_1019908, EPI_ISL_1019909, EPI_ISL_1019910, EPI_ISL_1019911, EPI_ISL_1019912, EPI_ISL_1019913, EPI_ISL_1019914, EPI_ISL_1019915, EPI_ISL_1019916, EPI_ISL_1019917, EPI_ISL_1019918, EPI_ISL_1019919, EPI_ISL_1019920, EPI_ISL_1019921, EPI_ISL_1019922, EPI_ISL_1019923, EPI_ISL_1019924, EPI_ISL_1019925, EPI_ISL_1019926, EPI_ISL_1019927, EPI_ISL_1019928, EPI_ISL_1019929, EPI_ISL_1019930, EPI_ISL_1019931, EPI_ISL_1019932, EPI_ISL_1019933, EPI_ISL_1019934, EPI_ISL_1019935, EPI_ISL_1019936, EPI_ISL_1019937, EPI_ISL_1019938, EPI_ISL_1019939, EPI_ISL_1019940, EPI_ISL_1019941, EPI_ISL_1019942, EPI_ISL_1019943, EPI_ISL_1019944, EPI_ISL_1019945, EPI_ISL_1019946, EPI_ISL_1019947, EPI_ISL_1019948, EPI_ISL_1019949, EPI_ISL_1019950, EPI_ISL_1019951, EPI_ISL_1019952, EPI_ISL_1019953, EPI_ISL_1019954, EPI_ISL_1019955, EPI_ISL_1019956, EPI_ISL_1019957, EPI_ISL_1019958, EPI_ISL_1019959, EPI_ISL_1019960, EPI_ISL_1019961, EPI_ISL_1019962, EPI_ISL_1019963, EPI_ISL_1019964, EPI_ISL_1019965, EPI_ISL_1019966, EPI_ISL_1019967, EPI_ISL_1019968, EPI_ISL_1019969, EPI_ISL_1019970, EPI_ISL_1019971, EPI_ISL_1019972, EPI_ISL_1019973, EPI_ISL_1019974, EPI_ISL_1019975, EPI_ISL_1019976, EPI_ISL_1019977, EPI_ISL_1019978, EPI_ISL_1019979, EPI_ISL_1019980, EPI_ISL_1019981, EPI_ISL_1019982, EPI_ISL_1019983, EPI_ISL_1019984, EPI_ISL_1019985, EPI_ISL_1019986, EPI_ISL_1019987, EPI_ISL_1019988, EPI_ISL_1019989, EPI_ISL_1019990, EPI_ISL_1019991, EPI_ISL_1019992, EPI_ISL_1019993, EPI_ISL_1019994, EPI_ISL_1019995, EPI_ISL_1019996, EPI_ISL_1019997, EPI_ISL_1019998, EPI_ISL_1019999, EPI_ISL_1020000, EPI_ISL_1020001, EPI_ISL_1020002, EPI_ISL_1020003, EPI_ISL_1020004, EPI_ISL_1020005, EPI_ISL_1020006, EPI_ISL_1020007, EPI_ISL_1020008, EPI_ISL_1020009, EPI_ISL_1020010, EPI_ISL_1020011, EPI_ISL_1020012, EPI_ISL_1020013, EPI_ISL_1020014, EPI_ISL_1020015, EPI_ISL_1069731, EPI_ISL_1175965

see above | Lighthouse Lab in Glasgow | Wellcome Sanger Institute for the COVID-19 Genomics UK (COG-UK) Consortium | Anna Dominiczak and Alex Alderton; Carol Clugston; Cordelia Langford; David Gray; David K. Jackson; Dominic Kwiatkowski; Ewan Harrison; Harper VanSteenhouse; Ian Johnston; Jeffrey Barrett; John Sillitoe on behalf of the Wellcome Sanger Institute COVID-19 Surveillance Team; John Sillitoe on behalf of the Wellcome Sanger Institute COVID-19 Surveillance Team (http://www.sanger.ac.uk/covid-team); Roberto Amato; Sonia Goncalves; Yumi Kasai

EPI_ISL_1050239, EPI_ISL_1104104, EPI_ISL_1104105, EPI_ISL_1104106 | Lighthouse Lab in Glasgow / MRC-University of Glasgow Centre for Virus Research | COVID-19 Genomics UK (COG-UK) Consortium | Alasdair MacLean; Alice Broos; Ana da Silva Filipe; Anna Dominiczak; Antonia Ho; Carol Clugston; Daniel Mair; David Gray; David L Robertson; Emma Thomson; Harper VanSteenhouse; James Shepherd; Jenna Nichols; Joseph Hughes; Kathy Li; Kathy Smollett; Kyriaki Nomikou; Lily Tong; Matthew Holden; Natasha Johnson; Richard Orton; Rory Gunson; Sarah McDonald; Sharif Shaaban; Sreenu Vattipally; Stephen Carmichael; Yumi Kasai

EPI_ISL_908932, EPI_ISL_908937, EPI_ISL_908938, EPI_ISL_908939, EPI_ISL_908940, EPI_ISL_908941, EPI_ISL_908944, EPI_ISL_908947, EPI_ISL_908948, EPI_ISL_908950, EPI_ISL_908954, EPI_ISL_908956, EPI_ISL_908964, EPI_ISL_908970, EPI_ISL_908987, EPI_ISL_908988, EPI_ISL_908995, EPI_ISL_908996, EPI_ISL_908997, EPI_ISL_909003, EPI_ISL_909014, EPI_ISL_909016, EPI_ISL_909018, EPI_ISL_909022, EPI_ISL_909023, EPI_ISL_909026, EPI_ISL_909027, EPI_ISL_909031, EPI_ISL_909037, EPI_ISL_909041, EPI_ISL_909043, EPI_ISL_909044, EPI_ISL_909051, EPI_ISL_909054, EPI_ISL_909059, EPI_ISL_909062, EPI_ISL_909065, EPI_ISL_909070, EPI_ISL_909078, EPI_ISL_909082, EPI_ISL_909084, EPI_ISL_909091, EPI_ISL_909099, EPI_ISL_909107, EPI_ISL_909108, EPI_ISL_909110, EPI_ISL_909120, EPI_ISL_909123, EPI_ISL_909124, EPI_ISL_909125, EPI_ISL_909127, EPI_ISL_909129, EPI_ISL_909130, EPI_ISL_909132, EPI_ISL_909133, EPI_ISL_909136, EPI_ISL_909137, EPI_ISL_909139, EPI_ISL_909146, EPI_ISL_909147, EPI_ISL_909149, EPI_ISL_909152, EPI_ISL_909153, EPI_ISL_909154, EPI_ISL_909159, EPI_ISL_909160, EPI_ISL_909166, EPI_ISL_909167, EPI_ISL_909172, EPI_ISL_909174, EPI_ISL_909179, EPI_ISL_909181, EPI_ISL_909185, EPI_ISL_909186, EPI_ISL_909191, EPI_ISL_909195, EPI_ISL_909197, EPI_ISL_909198, EPI_ISL_909201, EPI_ISL_909205, EPI_ISL_909207, EPI_ISL_909212, EPI_ISL_909214, EPI_ISL_909215, EPI_ISL_909216, EPI_ISL_909222, EPI_ISL_909224, EPI_ISL_909226, EPI_ISL_909228, EPI_ISL_909231, EPI_ISL_909241, EPI_ISL_909245, EPI_ISL_909246, EPI_ISL_909247, EPI_ISL_909248, EPI_ISL_909250, EPI_ISL_909252, EPI_ISL_909256, EPI_ISL_909259, EPI_ISL_909261, EPI_ISL_909263, EPI_ISL_909265, EPI_ISL_909266, EPI_ISL_909267, EPI_ISL_909268, EPI_ISL_909269, EPI_ISL_909271, EPI_ISL_909274, EPI_ISL_909276, EPI_ISL_909277, EPI_ISL_909279, EPI_ISL_909280, EPI_ISL_909282, EPI_ISL_909284, EPI_ISL_909286, EPI_ISL_909287, EPI_ISL_909288, EPI_ISL_909290, EPI_ISL_909291, EPI_ISL_909293, EPI_ISL_909295, EPI_ISL_909299, EPI_ISL_909300, EPI_ISL_909301, EPI_ISL_909305, EPI_ISL_909307, EPI_ISL_909308, EPI_ISL_909310, EPI_ISL_909313, EPI_ISL_909315, EPI_ISL_909319, EPI_ISL_909320, EPI_ISL_909323, EPI_ISL_909324, EPI_ISL_909329, EPI_ISL_909330, EPI_ISL_909331, EPI_ISL_909332, EPI_ISL_909335, EPI_ISL_909336, EPI_ISL_909337, EPI_ISL_909338, EPI_ISL_909340, EPI_ISL_909345, EPI_ISL_909346, EPI_ISL_909347, EPI_ISL_909350, EPI_ISL_909352, EPI_ISL_909354, EPI_ISL_909355, EPI_ISL_909357, EPI_ISL_909360, EPI_ISL_909362, EPI_ISL_909363, EPI_ISL_909365, EPI_ISL_909377, EPI_ISL_909378, EPI_ISL_909383, EPI_ISL_909384, EPI_ISL_909386, EPI_ISL_909394, EPI_ISL_909395, EPI_ISL_909396, EPI_ISL_909398, EPI_ISL_909399, EPI_ISL_909400, EPI_ISL_909402, EPI_ISL_909411, EPI_ISL_909412, EPI_ISL_909413, EPI_ISL_909415, EPI_ISL_909418, EPI_ISL_909419, EPI_ISL_909420, EPI_ISL_909423, EPI_ISL_909427, EPI_ISL_909430, EPI_ISL_909431, EPI_ISL_909432, EPI_ISL_909433, EPI_ISL_909434, EPI_ISL_909436, EPI_ISL_909437, EPI_ISL_909439, EPI_ISL_909441, EPI_ISL_909442, EPI_ISL_909443, EPI_ISL_909444, EPI_ISL_909445, EPI_ISL_909448, EPI_ISL_909449, EPI_ISL_909451, EPI_ISL_909452, EPI_ISL_909454, EPI_ISL_909455, EPI_ISL_909458, EPI_ISL_909459, EPI_ISL_909460, EPI_ISL_909461, EPI_ISL_909462, EPI_ISL_909463, EPI_ISL_909464, EPI_ISL_909466, EPI_ISL_909469, EPI_ISL_909470, EPI_ISL_909472, EPI_ISL_909473, EPI_ISL_909474, EPI_ISL_909475, EPI_ISL_909477, EPI_ISL_909479, EPI_ISL_909480, EPI_ISL_909483, EPI_ISL_909484, EPI_ISL_909485, EPI_ISL_909486, EPI_ISL_909489, EPI_ISL_909491, EPI_ISL_909492, EPI_ISL_909493, EPI_ISL_909494, EPI_ISL_909498, EPI_ISL_909499, EPI_ISL_909500, EPI_ISL_909501, EPI_ISL_909502, EPI_ISL_909505, EPI_ISL_909506, EPI_ISL_909507, EPI_ISL_909509, EPI_ISL_909510, EPI_ISL_909511, EPI_ISL_909514, EPI_ISL_909515, EPI_ISL_909516, EPI_ISL_909519, EPI_ISL_909520, EPI_ISL_909521, EPI_ISL_909523, EPI_ISL_909527, EPI_ISL_909529, EPI_ISL_909530, EPI_ISL_909533, EPI_ISL_909535, EPI_ISL_909540, EPI_ISL_909541, EPI_ISL_909542, EPI_ISL_909543, EPI_ISL_909547, EPI_ISL_909548, EPI_ISL_909550, EPI_ISL_909551, EPI_ISL_909555, EPI_ISL_909556, EPI_ISL_909558, EPI_ISL_909559, EPI_ISL_909560, EPI_ISL_909562, EPI_ISL_909563, EPI_ISL_909565, EPI_ISL_909566, EPI_ISL_909567, EPI_ISL_909569, EPI_ISL_909574, EPI_ISL_909576, EPI_ISL_909579, EPI_ISL_909584, EPI_ISL_909585, EPI_ISL_909587, EPI_ISL_909591, EPI_ISL_909592, EPI_ISL_912224, EPI_ISL_932257, EPI_ISL_932263, EPI_ISL_932276, EPI_ISL_932294, EPI_ISL_932319, EPI_ISL_932329, EPI_ISL_932358, EPI_ISL_932366, EPI_ISL_932372, EPI_ISL_932380, EPI_ISL_932391, EPI_ISL_932395, EPI_ISL_932403, EPI_ISL_932417, EPI_ISL_932456, EPI_ISL_932505, EPI_ISL_932507, EPI_ISL_932542, EPI_ISL_932563, EPI_ISL_932575, EPI_ISL_932627, EPI_ISL_932631, EPI_ISL_932756, EPI_ISL_932793, EPI_ISL_932874, EPI_ISL_932886, EPI_ISL_932895, EPI_ISL_932913, EPI_ISL_932974, EPI_ISL_933022, EPI_ISL_933061, EPI_ISL_938057, EPI_ISL_938120, EPI_ISL_938151, EPI_ISL_991351, EPI_ISL_991355, EPI_ISL_991358, EPI_ISL_991359, EPI_ISL_991367, EPI_ISL_991368, EPI_ISL_991370, EPI_ISL_991377, EPI_ISL_991379, EPI_ISL_991381, EPI_ISL_991382, EPI_ISL_991383, EPI_ISL_991385, EPI_ISL_991386, EPI_ISL_991387, EPI_ISL_991389, EPI_ISL_991395, EPI_ISL_991397, EPI_ISL_991399, EPI_ISL_991403, EPI_ISL_991409, EPI_ISL_991412, EPI_ISL_991417, EPI_ISL_991419, EPI_ISL_991422, EPI_ISL_991428, EPI_ISL_991432, EPI_ISL_991434, EPI_ISL_991435, EPI_ISL_991437, EPI_ISL_991443, EPI_ISL_991446, EPI_ISL_991448, EPI_ISL_991450, EPI_ISL_991451, EPI_ISL_991453, EPI_ISL_991455, EPI_ISL_991458, EPI_ISL_991460, EPI_ISL_991464, EPI_ISL_991465, EPI_ISL_991467, EPI_ISL_991470, EPI_ISL_991475, EPI_ISL_991476, EPI_ISL_991478, EPI_ISL_991482, EPI_ISL_991486, EPI_ISL_991489, EPI_ISL_991491, EPI_ISL_991493, EPI_ISL_991494, EPI_ISL_991496, EPI_ISL_991498, EPI_ISL_991499, EPI_ISL_991500, EPI_ISL_991503, EPI_ISL_991506, EPI_ISL_991515, EPI_ISL_991517, EPI_ISL_991525, EPI_ISL_991527, EPI_ISL_991528, EPI_ISL_991530, EPI_ISL_991531, EPI_ISL_991532, EPI_ISL_991534, EPI_ISL_991536, EPI_ISL_991537, EPI_ISL_991540, EPI_ISL_991543, EPI_ISL_991544, EPI_ISL_991546, EPI_ISL_991547, EPI_ISL_991548, EPI_ISL_991549, EPI_ISL_991551, EPI_ISL_991553, EPI_ISL_991556, EPI_ISL_991558, EPI_ISL_991561, EPI_ISL_993083, EPI_ISL_993086, EPI_ISL_993089, EPI_ISL_993091, EPI_ISL_993094, EPI_ISL_993095, EPI_ISL_993099, EPI_ISL_993101, EPI_ISL_993104, EPI_ISL_993108, EPI_ISL_993112, EPI_ISL_993117, EPI_ISL_993118, EPI_ISL_993122, EPI_ISL_993123, EPI_ISL_993125, EPI_ISL_993128, EPI_ISL_993129, EPI_ISL_993131, EPI_ISL_993132, EPI_ISL_993134, EPI_ISL_993135, EPI_ISL_993136, EPI_ISL_993137, EPI_ISL_993138, EPI_ISL_993141, EPI_ISL_993143, EPI_ISL_993158, EPI_ISL_993160, EPI_ISL_993163, EPI_ISL_993164, EPI_ISL_993166, EPI_ISL_993169, EPI_ISL_993174, EPI_ISL_993177, EPI_ISL_993178, EPI_ISL_993180, EPI_ISL_993182, EPI_ISL_993183, EPI_ISL_993185, EPI_ISL_993189, EPI_ISL_993190, EPI_ISL_993192, EPI_ISL_993196, EPI_ISL_993197, EPI_ISL_993201, EPI_ISL_993203, EPI_ISL_993207, EPI_ISL_993209, EPI_ISL_993212, EPI_ISL_993216, EPI_ISL_993217, EPI_ISL_993219, EPI_ISL_993222, EPI_ISL_993223, EPI_ISL_993224, EPI_ISL_993231, EPI_ISL_993232, EPI_ISL_993233, EPI_ISL_993234, EPI_ISL_993235, EPI_ISL_993236, EPI_ISL_993237, EPI_ISL_993238, EPI_ISL_993240, EPI_ISL_993241, EPI_ISL_993242, EPI_ISL_993247, EPI_ISL_993249, EPI_ISL_993251, EPI_ISL_993252, EPI_ISL_993261, EPI_ISL_993263, EPI_ISL_993264, EPI_ISL_993270, EPI_ISL_993273, EPI_ISL_993275, EPI_ISL_993277, EPI_ISL_993279, EPI_ISL_993280, EPI_ISL_993283, EPI_ISL_993285, EPI_ISL_993287, EPI_ISL_993293, EPI_ISL_993294, EPI_ISL_993296, EPI_ISL_993300, EPI_ISL_993301, EPI_ISL_993302, EPI_ISL_993305, EPI_ISL_993310, EPI_ISL_993312, EPI_ISL_993315, EPI_ISL_993320, EPI_ISL_993324, EPI_ISL_993326, EPI_ISL_993328, EPI_ISL_993329, EPI_ISL_993331, EPI_ISL_993335, EPI_ISL_993336, EPI_ISL_993337, EPI_ISL_993340, EPI_ISL_993342, EPI_ISL_993343, EPI_ISL_993344, EPI_ISL_993345, EPI_ISL_993346, EPI_ISL_993350, EPI_ISL_993351, EPI_ISL_993352, EPI_ISL_993354, EPI_ISL_993360, EPI_ISL_993361, EPI_ISL_993363, EPI_ISL_993365, EPI_ISL_993367, EPI_ISL_993368, EPI_ISL_993371, EPI_ISL_993374, EPI_ISL_993378, EPI_ISL_993379, EPI_ISL_993381, EPI_ISL_993384, EPI_ISL_993388, EPI_ISL_993389, EPI_ISL_993392, EPI_ISL_993394, EPI_ISL_993395, EPI_ISL_993396, EPI_ISL_993413, EPI_ISL_993414, EPI_ISL_993416, EPI_ISL_993417, EPI_ISL_993419, EPI_ISL_993421, EPI_ISL_993422, EPI_ISL_993423, EPI_ISL_993425, EPI_ISL_993426, EPI_ISL_993427, EPI_ISL_993432, EPI_ISL_993433, EPI_ISL_993434, EPI_ISL_993436, EPI_ISL_993438, EPI_ISL_993441, EPI_ISL_993442, EPI_ISL_993443, EPI_ISL_993444, EPI_ISL_993446, EPI_ISL_993447, EPI_ISL_993450, EPI_ISL_993451, EPI_ISL_993452, EPI_ISL_993453, EPI_ISL_993454, EPI_ISL_993456, EPI_ISL_993458, EPI_ISL_993459, EPI_ISL_993461, EPI_ISL_993462, EPI_ISL_993463, EPI_ISL_993464, EPI_ISL_993466, EPI_ISL_993467, EPI_ISL_993468, EPI_ISL_993471, EPI_ISL_993472, EPI_ISL_993473, EPI_ISL_993474, EPI_ISL_993475, EPI_ISL_993476, EPI_ISL_993478, EPI_ISL_993479, EPI_ISL_993480, EPI_ISL_993481, EPI_ISL_993482, EPI_ISL_993483, EPI_ISL_993484, EPI_ISL_993485, EPI_ISL_993486, EPI_ISL_993488, EPI_ISL_993489, EPI_ISL_993490, EPI_ISL_993491, EPI_ISL_993492, EPI_ISL_993493, EPI_ISL_993494, EPI_ISL_993495, EPI_ISL_993497, EPI_ISL_993498, EPI_ISL_993499, EPI_ISL_993500, EPI_ISL_993501, EPI_ISL_993503, EPI_ISL_993505, EPI_ISL_993506, EPI_ISL_993508, EPI_ISL_993509, EPI_ISL_993510, EPI_ISL_993511, EPI_ISL_993512, EPI_ISL_993513, EPI_ISL_993514, EPI_ISL_993515, EPI_ISL_993516, EPI_ISL_993517, EPI_ISL_993518, EPI_ISL_993520, EPI_ISL_993521, EPI_ISL_993524, EPI_ISL_993526, EPI_ISL_993527, EPI_ISL_993528, EPI_ISL_993529, EPI_ISL_993530, EPI_ISL_993531, EPI_ISL_993533, EPI_ISL_993534, EPI_ISL_993535, EPI_ISL_993536, EPI_ISL_993537, EPI_ISL_993539, EPI_ISL_993540, EPI_ISL_993541, EPI_ISL_993544, EPI_ISL_993545, EPI_ISL_993547, EPI_ISL_993548, EPI_ISL_993551, EPI_ISL_993552, EPI_ISL_993553, EPI_ISL_993554, EPI_ISL_993555, EPI_ISL_993556, EPI_ISL_993557, EPI_ISL_993558, EPI_ISL_993561, EPI_ISL_993563, EPI_ISL_993564, EPI_ISL_993565, EPI_ISL_993566, EPI_ISL_993567, EPI_ISL_993568, EPI_ISL_993570, EPI_ISL_993572, EPI_ISL_993573, EPI_ISL_993574, EPI_ISL_993575, EPI_ISL_993576, EPI_ISL_993578, EPI_ISL_993579, EPI_ISL_993580, EPI_ISL_993583, EPI_ISL_993585, EPI_ISL_993586, EPI_ISL_993588, EPI_ISL_993590, EPI_ISL_993591, EPI_ISL_993592, EPI_ISL_993594, EPI_ISL_993596, EPI_ISL_993597, EPI_ISL_993600, EPI_ISL_993601, EPI_ISL_993603, EPI_ISL_993604, EPI_ISL_993605, EPI_ISL_993606, EPI_ISL_993607, EPI_ISL_993608, EPI_ISL_993609, EPI_ISL_993610, EPI_ISL_993611, EPI_ISL_993612, EPI_ISL_993613, EPI_ISL_993614, EPI_ISL_993617, EPI_ISL_993622, EPI_ISL_993623, EPI_ISL_993626, EPI_ISL_993627, EPI_ISL_993628, EPI_ISL_993629, EPI_ISL_993631, EPI_ISL_993632, EPI_ISL_993633, EPI_ISL_993635, EPI_ISL_993636, EPI_ISL_993637, EPI_ISL_993642, EPI_ISL_993643, EPI_ISL_993644, EPI_ISL_993645, EPI_ISL_993646, EPI_ISL_993647, EPI_ISL_993650, EPI_ISL_993651, EPI_ISL_993652, EPI_ISL_993653, EPI_ISL_993655, EPI_ISL_993657, EPI_ISL_993658, EPI_ISL_993659, EPI_ISL_993660, EPI_ISL_993663, EPI_ISL_993664, EPI_ISL_993665, EPI_ISL_993667, EPI_ISL_993668, EPI_ISL_993669, EPI_ISL_993670, EPI_ISL_993671, EPI_ISL_993673, EPI_ISL_993674, EPI_ISL_993676, EPI_ISL_993677, EPI_ISL_993678, EPI_ISL_993679, EPI_ISL_993680, EPI_ISL_993681, EPI_ISL_993685, EPI_ISL_993686, EPI_ISL_993687, EPI_ISL_993688, EPI_ISL_993689, EPI_ISL_993691, EPI_ISL_993692, EPI_ISL_993693, EPI_ISL_993694, EPI_ISL_993695, EPI_ISL_993696, EPI_ISL_993699, EPI_ISL_993700, EPI_ISL_993701, EPI_ISL_993705, EPI_ISL_993707, EPI_ISL_993708, EPI_ISL_993709, EPI_ISL_993710, EPI_ISL_993711, EPI_ISL_993712, EPI_ISL_993713, EPI_ISL_993714, EPI_ISL_993715, EPI_ISL_993716, EPI_ISL_993717, EPI_ISL_993719, EPI_ISL_993721, EPI_ISL_993723, EPI_ISL_993724, EPI_ISL_993725, EPI_ISL_993726, EPI_ISL_993727, EPI_ISL_993728, EPI_ISL_993729, EPI_ISL_993730, EPI_ISL_993731, EPI_ISL_993732, EPI_ISL_993734, EPI_ISL_993735, EPI_ISL_1012862

see above | Lighthouse Lab in Milton Keynes | Wellcome Sanger Institute for the COVID-19 Genomics UK (COG-UK) Consortium | Cordelia Langford; David K. Jackson; Dominic Kwiatkowski; Ewan Harrison; Ian Johnston; John Sillitoe on behalf of the Wellcome Sanger Institute COVID-19 Surveillance Team; John Sillitoe on behalf of the Wellcome Sanger Institute COVID-19 Surveillance Team (http://www.sanger.ac.uk/covid-team); Roberto Amato; Sonia Goncalves; The Lighthouse Lab in Milton Keynes and Alex Alderton

EPI_ISL_922147, EPI_ISL_922149, EPI_ISL_922150, EPI_ISL_922151, EPI_ISL_922152, EPI_ISL_922153, EPI_ISL_922154, EPI_ISL_922155, EPI_ISL_922156, EPI_ISL_922158, EPI_ISL_922159, EPI_ISL_922160, EPI_ISL_922161, EPI_ISL_922162, EPI_ISL_922163, EPI_ISL_922164, EPI_ISL_922165, EPI_ISL_922166, EPI_ISL_922167, EPI_ISL_922168, EPI_ISL_950715, EPI_ISL_950716, EPI_ISL_998957, EPI_ISL_998958, EPI_ISL_998959, EPI_ISL_998960, EPI_ISL_998961, EPI_ISL_998962, EPI_ISL_998963, EPI_ISL_998964, EPI_ISL_998965, EPI_ISL_998966, EPI_ISL_998967, EPI_ISL_998968, EPI_ISL_998969, EPI_ISL_998970, EPI_ISL_998971

see above | Lincolnshire Hospitals and DeepSeq Nottingham | COVID-19 Genomics UK (COG-UK) Consortium | Christopher Moore; Fei Sang; Joeseph Chappell; Johnny Debebe; Jonathan Ball; Matthew Carlisle; Matthew Loose; Nadine Holmes; Nichola Duckworth; Patrick McClure; Sarah Walsh; Tim Sloan; Victoria Wright

EPI_ISL_865467, EPI_ISL_865468, EPI_ISL_865469, EPI_ISL_865470, EPI_ISL_865471, EPI_ISL_919488, EPI_ISL_919489, EPI_ISL_919490, EPI_ISL_919491, EPI_ISL_919492, EPI_ISL_919493, EPI_ISL_919494, EPI_ISL_919495, EPI_ISL_919496, EPI_ISL_919497, EPI_ISL_919498, EPI_ISL_919499, EPI_ISL_919500, EPI_ISL_919501, EPI_ISL_919502, EPI_ISL_919503, EPI_ISL_919504, EPI_ISL_919505, EPI_ISL_919507, EPI_ISL_919508, EPI_ISL_919509, EPI_ISL_919510, EPI_ISL_919512, EPI_ISL_919514, EPI_ISL_919518, EPI_ISL_919525, EPI_ISL_919530, EPI_ISL_919532, EPI_ISL_919533, EPI_ISL_919535, EPI_ISL_919536, EPI_ISL_919542, EPI_ISL_919543, EPI_ISL_919547, EPI_ISL_919548, EPI_ISL_919550, EPI_ISL_919556, EPI_ISL_919557, EPI_ISL_919562, EPI_ISL_919563, EPI_ISL_919564, EPI_ISL_919568, EPI_ISL_919570, EPI_ISL_919571, EPI_ISL_919572, EPI_ISL_919573, EPI_ISL_919574, EPI_ISL_919575, EPI_ISL_919576, EPI_ISL_919577, EPI_ISL_919578, EPI_ISL_919579, EPI_ISL_919582, EPI_ISL_919585, EPI_ISL_919587, EPI_ISL_919588, EPI_ISL_919589, EPI_ISL_919590, EPI_ISL_919604, EPI_ISL_919605, EPI_ISL_919606, EPI_ISL_919607, EPI_ISL_919608, EPI_ISL_919609, EPI_ISL_919611, EPI_ISL_919612, EPI_ISL_919613, EPI_ISL_919614, EPI_ISL_919615, EPI_ISL_919617, EPI_ISL_919618, EPI_ISL_919619, EPI_ISL_919635, EPI_ISL_919638, EPI_ISL_919639, EPI_ISL_919649, EPI_ISL_919654, EPI_ISL_919691, EPI_ISL_997405, EPI_ISL_997412, EPI_ISL_997413, EPI_ISL_997414, EPI_ISL_997415, EPI_ISL_997416, EPI_ISL_997417, EPI_ISL_1048023, EPI_ISL_1048024, EPI_ISL_1048025, EPI_ISL_1104337, EPI_ISL_1104338, EPI_ISL_1104340, EPI_ISL_1104342, EPI_ISL_1104345, EPI_ISL_1104349, EPI_ISL_1104350, EPI_ISL_1104352, EPI_ISL_1104353, EPI_ISL_1104355, EPI_ISL_1104358, EPI_ISL_1104364, EPI_ISL_1104365, EPI_ISL_1104366, EPI_ISL_1177879, EPI_ISL_1177880, EPI_ISL_1177881, EPI_ISL_1177882, EPI_ISL_1177883, EPI_ISL_1177884, EPI_ISL_1177885, EPI_ISL_1177886, EPI_ISL_1177887, EPI_ISL_1177905, EPI_ISL_1177906, EPI_ISL_1247767, EPI_ISL_1247775, EPI_ISL_1247776, EPI_ISL_1247781, EPI_ISL_1247784, EPI_ISL_1247790, EPI_ISL_1247809, EPI_ISL_1247814, EPI_ISL_1475758, EPI_ISL_1546002, EPI_ISL_1546004, EPI_ISL_1546006, EPI_ISL_1546007

see above | Liverpool Clinical Laboratories | COVID-19 Genomics UK (COG-UK) Consortium | A Alrezaihi; Alessandro Gerada; Alistair Darby; Angela Cowell; Anita Lucaci; Anu Chawla; Cassie Olateju; Catherine Hartley; Charlotte Nelson; Ecaterina Vamos; Elaine O'Toole; Eleanor G Bentley; Ghada T Shawli; Hermione Webster; Isabel García-Dorival; James Johnson; James P Stewart; Jenifer Manson; Joanne Watts; Jones Benjamin; Jordan J Clark; Julian Hiscox; L Luu; Lucille Rainbow; M Almsaud; Margaret Hughes; Mark Whitehead; Matthew Gemmell; Miren Iturriza-Gomara; Muhannad Alruwaili; N.P Randle; Neil Swainston; PKF Gilmore; Parul Sharma; Rebekah Penrice-Randal; Richard Eccles; Richard Gregory; Sam Haldenby; Steve Paterson; Stuart D Armstrong; Trevor Ian Robinson; Ximeng Han

EPI_ISL_921586, EPI_ISL_921587, EPI_ISL_921588, EPI_ISL_921589, EPI_ISL_921590, EPI_ISL_921591, EPI_ISL_921592, EPI_ISL_921593, EPI_ISL_921594, EPI_ISL_921595, EPI_ISL_921596, EPI_ISL_921597, EPI_ISL_921598, EPI_ISL_921599, EPI_ISL_921600, EPI_ISL_921601, EPI_ISL_921602, EPI_ISL_921603, EPI_ISL_921604, EPI_ISL_921605, EPI_ISL_921606, EPI_ISL_921607, EPI_ISL_921608, EPI_ISL_921609, EPI_ISL_921610, EPI_ISL_921611, EPI_ISL_921612, EPI_ISL_921613, EPI_ISL_921614, EPI_ISL_921615, EPI_ISL_921616, EPI_ISL_921617, EPI_ISL_921618, EPI_ISL_921619, EPI_ISL_921620, EPI_ISL_921621, EPI_ISL_921622, EPI_ISL_921623, EPI_ISL_921624, EPI_ISL_921625, EPI_ISL_921626, EPI_ISL_921627, EPI_ISL_921628, EPI_ISL_921629, EPI_ISL_921630, EPI_ISL_950339, EPI_ISL_950340, EPI_ISL_950341, EPI_ISL_950343, EPI_ISL_950344, EPI_ISL_950345, EPI_ISL_950346, EPI_ISL_998342, EPI_ISL_998343, EPI_ISL_998344, EPI_ISL_998345, EPI_ISL_998347, EPI_ISL_998348, EPI_ISL_998349, EPI_ISL_998350, EPI_ISL_998364, EPI_ISL_998380, EPI_ISL_998381, EPI_ISL_998382, EPI_ISL_998383, EPI_ISL_998384, EPI_ISL_998385, EPI_ISL_998386, EPI_ISL_998387, EPI_ISL_998388, EPI_ISL_998389, EPI_ISL_998390, EPI_ISL_998391, EPI_ISL_998392, EPI_ISL_998393, EPI_ISL_998394, EPI_ISL_998395, EPI_ISL_998396, EPI_ISL_998397, EPI_ISL_998398, EPI_ISL_998399, EPI_ISL_998400, EPI_ISL_998401, EPI_ISL_998402, EPI_ISL_998403, EPI_ISL_998404, EPI_ISL_998405, EPI_ISL_998406, EPI_ISL_998407, EPI_ISL_998408, EPI_ISL_998409, EPI_ISL_998410, EPI_ISL_998411, EPI_ISL_998412, EPI_ISL_998413, EPI_ISL_998441, EPI_ISL_998442, EPI_ISL_998443, EPI_ISL_998444, EPI_ISL_998447, EPI_ISL_998475, EPI_ISL_1050941, EPI_ISL_1050943, EPI_ISL_1050944, EPI_ISL_1050945, EPI_ISL_1050946, EPI_ISL_1050961, EPI_ISL_1050962, EPI_ISL_1050963, EPI_ISL_1050964, EPI_ISL_1050965, EPI_ISL_1050966, EPI_ISL_1050968, EPI_ISL_1050969, EPI_ISL_1050970, EPI_ISL_1050974, EPI_ISL_1050990, EPI_ISL_1050991, EPI_ISL_1051412, EPI_ISL_1178223, EPI_ISL_1178224, EPI_ISL_1178225, EPI_ISL_1178226, EPI_ISL_1178227, EPI_ISL_1178228, EPI_ISL_1178229, EPI_ISL_1178230, EPI_ISL_1178231, EPI_ISL_1178233, EPI_ISL_1178234, EPI_ISL_1178235, EPI_ISL_1178236, EPI_ISL_1178238, EPI_ISL_1178239, EPI_ISL_1178241, EPI_ISL_1178242, EPI_ISL_1178243, EPI_ISL_1178245, EPI_ISL_1178246, EPI_ISL_1178248, EPI_ISL_1178249, EPI_ISL_1178250, EPI_ISL_1178251, EPI_ISL_1178252, EPI_ISL_1178254, EPI_ISL_1178255, EPI_ISL_1178256, EPI_ISL_1178257, EPI_ISL_1178259, EPI_ISL_1178260, EPI_ISL_1178261, EPI_ISL_1178263, EPI_ISL_1178264, EPI_ISL_1178265, EPI_ISL_1178266, EPI_ISL_1178268, EPI_ISL_1178270, EPI_ISL_1178271, EPI_ISL_1178272, EPI_ISL_1178273, EPI_ISL_1178274, EPI_ISL_1178275, EPI_ISL_1178276, EPI_ISL_1178277, EPI_ISL_1178278, EPI_ISL_1178279, EPI_ISL_1178280, EPI_ISL_1178281, EPI_ISL_1178282, EPI_ISL_1178283, EPI_ISL_1178284, EPI_ISL_1178286, EPI_ISL_1178291, EPI_ISL_1178292, EPI_ISL_1178293, EPI_ISL_1178294, EPI_ISL_1178295, EPI_ISL_1178296, EPI_ISL_1178297, EPI_ISL_1178298, EPI_ISL_1178299, EPI_ISL_1178300, EPI_ISL_1178301, EPI_ISL_1178302, EPI_ISL_1178303, EPI_ISL_1178304, EPI_ISL_1178305, EPI_ISL_1178306, EPI_ISL_1178307, EPI_ISL_1178308, EPI_ISL_1178309, EPI_ISL_1178310, EPI_ISL_1178311, EPI_ISL_1178312, EPI_ISL_1178313, EPI_ISL_1178315, EPI_ISL_1178316, EPI_ISL_1178317, EPI_ISL_1178318, EPI_ISL_1178319, EPI_ISL_1178320, EPI_ISL_1178321, EPI_ISL_1178322, EPI_ISL_1178323, EPI_ISL_1178377

see above | Northumbria University / South Tees Hospitals NHS Foundation Trust / North | COVID-19 Genomics UK | Andrew Nelson; Brendan Payne; Clive Graham; Darren L Smith; Debra Padgett; Edward Barton; Emma Swindells; Garren Scott; Gary Black; Gary Eltringham; Giles S Holt; Greg R Young; Jane Greenaway; Jennifer Collins; John Allan; Joshua Loh; Lynn Dover;

| | | | |
|---|---|---|---|
| | Cumbria Integrated Care NHS Foundation Trust / North Tees and Hartlepool NHS Foundation Trust / Newcastle Hospitals NHS Foundation Trust | (COG-UK) Consortium | Matthew Bashton; Mohammad A Tariq; Paul Baker; Sarah Essex; Steve Liggett; Wen C Yew; Yusri Taha |
| EPI_ISL_951590, EPI_ISL_951591, EPI_ISL_951592, EPI_ISL_951593, EPI_ISL_951632, EPI_ISL_951633, EPI_ISL_951634, EPI_ISL_951635, EPI_ISL_951637, EPI_ISL_951648, EPI_ISL_951740, EPI_ISL_951756, EPI_ISL_951819, EPI_ISL_952187, EPI_ISL_952188, EPI_ISL_952189, EPI_ISL_952190, EPI_ISL_1052497, EPI_ISL_1106643, EPI_ISL_1178925, EPI_ISL_1249600, EPI_ISL_1476177, EPI_ISL_1476178, EPI_ISL_1476179, EPI_ISL_1476184, EPI_ISL_1476199, EPI_ISL_1476248, EPI_ISL_1476255, EPI_ISL_1476461, EPI_ISL_1476469, EPI_ISL_1546816, EPI_ISL_1546886, EPI_ISL_1546887, EPI_ISL_1546888, EPI_ISL_1546889, EPI_ISL_1546890, EPI_ISL_1546891, EPI_ISL_1546892, EPI_ISL_1546893, EPI_ISL_1546894, EPI_ISL_1546895, EPI_ISL_1546896, EPI_ISL_1546897, EPI_ISL_1546898, EPI_ISL_1546899, EPI_ISL_1546900, EPI_ISL_1546901, EPI_ISL_1546902, EPI_ISL_1546903, EPI_ISL_1546904, EPI_ISL_1546905, EPI_ISL_1546906, EPI_ISL_1546907, EPI_ISL_1546908, EPI_ISL_1546909, EPI_ISL_1546910, EPI_ISL_1546911, EPI_ISL_1546913, EPI_ISL_1546914, EPI_ISL_1546915, EPI_ISL_1546916, EPI_ISL_1546921, EPI_ISL_1547007, EPI_ISL_1547009, EPI_ISL_1547011, EPI_ISL_1547012, EPI_ISL_1547018, EPI_ISL_1547022, EPI_ISL_1547028 | | | |
| see above | Originating lab: Wales Specialist Virology Centre Sequencing lab: Pathogen Genomics Unit | Public Health Wales Microbiology Cardiff Wales Specialist Virology Centre | Alec Birchley; Alexander Adams; Amy Gaskin; Angela Marchbank; Bree Gatica-Wilcox; Catherine Moore; Jason Coombes; Joanne Watkins; Joel Southgate; Johnathan Evans; Laura Gifford; Lauren Gilbert; Lee Graham; Malorie Perry; Matthew Bull; Nicole Pacchiarini; Sally Corden; Sara Kumziene-Summerhayes; Sara Rey; Sarah Taylor; Simon Cottrell; Sophie Jones; Tom Connor |
| EPI_ISL_951359, EPI_ISL_951360, EPI_ISL_951368, EPI_ISL_951369, EPI_ISL_951370, EPI_ISL_951371, EPI_ISL_951372, EPI_ISL_951373, EPI_ISL_951461, EPI_ISL_951462, EPI_ISL_951463, EPI_ISL_951464, EPI_ISL_951466, EPI_ISL_951468, EPI_ISL_951469, EPI_ISL_951470, EPI_ISL_951471, EPI_ISL_951475, EPI_ISL_951476, EPI_ISL_951477, EPI_ISL_951478, EPI_ISL_951479, EPI_ISL_951480, EPI_ISL_951481, EPI_ISL_951482, EPI_ISL_951484, EPI_ISL_951485, EPI_ISL_951489, EPI_ISL_999135, EPI_ISL_999149, EPI_ISL_999157, EPI_ISL_999158, EPI_ISL_999159, EPI_ISL_999160, EPI_ISL_999162, EPI_ISL_999163, EPI_ISL_999164, EPI_ISL_999165, EPI_ISL_999166, EPI_ISL_999167, EPI_ISL_999168, EPI_ISL_999170, EPI_ISL_999173, EPI_ISL_999174, EPI_ISL_999175, EPI_ISL_999177, EPI_ISL_999178, EPI_ISL_999179, EPI_ISL_999180, EPI_ISL_999181, EPI_ISL_999182, EPI_ISL_999183, EPI_ISL_999184, EPI_ISL_999196, EPI_ISL_999198, EPI_ISL_999200, EPI_ISL_999201, EPI_ISL_999202, EPI_ISL_999203, EPI_ISL_999219, EPI_ISL_999222, EPI_ISL_999225, EPI_ISL_999226, EPI_ISL_999290, EPI_ISL_999291, EPI_ISL_999292, EPI_ISL_999408, EPI_ISL_999409, EPI_ISL_999410, EPI_ISL_999411, EPI_ISL_999412, EPI_ISL_999413, EPI_ISL_999414, EPI_ISL_999416, EPI_ISL_999425, EPI_ISL_999426, EPI_ISL_999437, EPI_ISL_999438, EPI_ISL_999443, EPI_ISL_999444, EPI_ISL_999445, EPI_ISL_999453, EPI_ISL_999467, EPI_ISL_999468, EPI_ISL_999469, EPI_ISL_999473, EPI_ISL_999474, EPI_ISL_999475, EPI_ISL_999476, EPI_ISL_999478, EPI_ISL_999496, EPI_ISL_999497, EPI_ISL_999498, EPI_ISL_999499, EPI_ISL_999500, EPI_ISL_999501, EPI_ISL_999502, EPI_ISL_999503, EPI_ISL_999504, EPI_ISL_1051595, EPI_ISL_1051830, EPI_ISL_1051834, EPI_ISL_1051837, EPI_ISL_1051859, EPI_ISL_1051863, EPI_ISL_1051864, EPI_ISL_1051865, EPI_ISL_1051866, EPI_ISL_1051867, EPI_ISL_1051868, EPI_ISL_1051869, EPI_ISL_1051872, EPI_ISL_1051879, EPI_ISL_1051888, EPI_ISL_1051889, EPI_ISL_1178705, EPI_ISL_1178706, EPI_ISL_1178707, EPI_ISL_1178711, EPI_ISL_1178714, EPI_ISL_1178716, EPI_ISL_1178718, EPI_ISL_1178737, EPI_ISL_1178739, EPI_ISL_1178752, EPI_ISL_1178761, EPI_ISL_1178762, EPI_ISL_1178764, EPI_ISL_1178767, EPI_ISL_1178771, EPI_ISL_1178785, EPI_ISL_1178787, EPI_ISL_1178789, EPI_ISL_1178790, EPI_ISL_1178791, EPI_ISL_1178793, EPI_ISL_1178794, EPI_ISL_1178795, EPI_ISL_1178798, EPI_ISL_1178799, EPI_ISL_1178800, EPI_ISL_1178803, EPI_ISL_1178806, EPI_ISL_1178811, EPI_ISL_1178812, EPI_ISL_1178813, EPI_ISL_1178814, EPI_ISL_1178815, EPI_ISL_1178828, EPI_ISL_1178830, EPI_ISL_1178835, EPI_ISL_1178837, EPI_ISL_1178839, EPI_ISL_1178840, EPI_ISL_1178842, EPI_ISL_1178844, EPI_ISL_1178849, EPI_ISL_1178850, EPI_ISL_1178855, EPI_ISL_1178859, EPI_ISL_1178860, EPI_ISL_1178864, EPI_ISL_1178865, EPI_ISL_1178866, EPI_ISL_1178869, EPI_ISL_1309740, EPI_ISL_1730530, EPI_ISL_1730531, EPI_ISL_1833062, EPI_ISL_1833084 | | | |
| see above | Oxford Viromics, NDM, University of Oxford; Oxford University Hospitals; Basingstoke and North Hampshire Hospital | COVID-19 Genomics UK (COG-UK) Consortium | Alex Mobbs; Amy Trebes; Anita Justice; Catrin Moore; Christophe Fraser; David Bonsall; David Buck; Emma Wise; George Macintyre; Jessica Lynch; John Todd; Mariateresa de Cesare; Matilde Mori; Monique Andersson; Nathan Moore; Nick Cortes; Robert Shaw; Stephen Kidd; Tanya Golubchik; Timothy Peto |
| EPI_ISL_921854, EPI_ISL_921855, EPI_ISL_921856, EPI_ISL_921860, EPI_ISL_921862, EPI_ISL_921869, EPI_ISL_921871, EPI_ISL_921873, EPI_ISL_921878, EPI_ISL_921879, EPI_ISL_921880, EPI_ISL_921881, EPI_ISL_921882, EPI_ISL_921883, EPI_ISL_921884, EPI_ISL_921885, EPI_ISL_921886, EPI_ISL_921887, EPI_ISL_921888, EPI_ISL_921889, EPI_ISL_921890, EPI_ISL_921891, EPI_ISL_921892, EPI_ISL_921893, EPI_ISL_921894, EPI_ISL_921895, EPI_ISL_921896, EPI_ISL_921897, EPI_ISL_921898, EPI_ISL_921899, EPI_ISL_921900, EPI_ISL_921901, EPI_ISL_921902, EPI_ISL_921903, EPI_ISL_921905, EPI_ISL_921906, EPI_ISL_921907, EPI_ISL_921908, EPI_ISL_921911, EPI_ISL_921915, EPI_ISL_921916, EPI_ISL_921918, EPI_ISL_921919, EPI_ISL_921920, EPI_ISL_921921, EPI_ISL_921922, EPI_ISL_921924, EPI_ISL_921925, EPI_ISL_921926, EPI_ISL_921927, EPI_ISL_921928, EPI_ISL_921929, EPI_ISL_921930, EPI_ISL_921932, EPI_ISL_921936, EPI_ISL_921937, EPI_ISL_921939, EPI_ISL_921941, EPI_ISL_921943, EPI_ISL_921944, EPI_ISL_921945, EPI_ISL_921946, EPI_ISL_921948, EPI_ISL_921949, EPI_ISL_921950, EPI_ISL_921951, EPI_ISL_921954, EPI_ISL_921955, EPI_ISL_921956, EPI_ISL_921957, EPI_ISL_921958, EPI_ISL_921959, EPI_ISL_921960, EPI_ISL_921961, EPI_ISL_921962, EPI_ISL_921963, EPI_ISL_921964, EPI_ISL_921965, EPI_ISL_921967, EPI_ISL_921970, EPI_ISL_921971, EPI_ISL_921972, EPI_ISL_921973, EPI_ISL_921974, EPI_ISL_921975, EPI_ISL_921976, EPI_ISL_921977, EPI_ISL_921979, EPI_ISL_921980, EPI_ISL_950563, EPI_ISL_950564, EPI_ISL_950565, EPI_ISL_950566, EPI_ISL_950567, EPI_ISL_950568, EPI_ISL_950569, EPI_ISL_950570, EPI_ISL_950571, EPI_ISL_1051428, EPI_ISL_1051443, EPI_ISL_1051445, EPI_ISL_1051451, EPI_ISL_1051455, EPI_ISL_1051467, EPI_ISL_1051471, EPI_ISL_1051482, EPI_ISL_1051486, EPI_ISL_1051488, EPI_ISL_1051491, EPI_ISL_1178465, EPI_ISL_1178471, EPI_ISL_1248982, EPI_ISL_1248988, EPI_ISL_1248993, EPI_ISL_1248996, EPI_ISL_1249002, EPI_ISL_1832941, EPI_ISL_1832942, EPI_ISL_1832944, EPI_ISL_1832946, EPI_ISL_1832947, EPI_ISL_1832951, EPI_ISL_1832955, EPI_ISL_1832956, EPI_ISL_1832957, EPI_ISL_1832961, EPI_ISL_1832963, EPI_ISL_1832969, EPI_ISL_1832970, EPI_ISL_1832976, EPI_ISL_1832988, EPI_ISL_1832989, EPI_ISL_1832990, EPI_ISL_1832991, EPI_ISL_1832992, EPI_ISL_1832993, EPI_ISL_1832994, EPI_ISL_1832996 | | | |
| see above | Quadram Institute Bioscience | COVID-19 Genomics UK (COG-UK) Consortium | Alexander J Trotter; Alison E. Mather; Alp Aydin; Ana P. Tedim; Anastasia Kolyva; Andrew Bell; Andrew J. Page; Claire Stuart; Dave J. Baker; Gemma L. Kay; John Wain; Justin O'Grady; Leonardo de Oliveira Martins; Lizzie Meadows; Maria Diaz; Mark Webber; Muhammed Yasir; Nabil-Fareed Alikhan; Ngozi Elumogo; Nicholas M. Thomson; Rachael Stanley; Rachel Gilroy; Reenesh Prakash; Samir Dervisevic; Samuel Bloomfield; Steven Rudder; Thanh Le-Viet |
| EPI_ISL_866951, EPI_ISL_866952, EPI_ISL_866953, EPI_ISL_866954, EPI_ISL_866955, EPI_ISL_866956, EPI_ISL_866957, EPI_ISL_866958, EPI_ISL_866959, EPI_ISL_866960, EPI_ISL_866961, EPI_ISL_866962, EPI_ISL_866963, EPI_ISL_866964, EPI_ISL_866965, EPI_ISL_866966, EPI_ISL_866967, EPI_ISL_866968, EPI_ISL_866969, EPI_ISL_866970, EPI_ISL_921981, EPI_ISL_921982, EPI_ISL_921983, EPI_ISL_921984, EPI_ISL_921985, EPI_ISL_921986, EPI_ISL_921987, EPI_ISL_921988, EPI_ISL_921989, EPI_ISL_921990, EPI_ISL_921991, EPI_ISL_921992, EPI_ISL_921993, EPI_ISL_921994, EPI_ISL_921995, EPI_ISL_921996, EPI_ISL_921997, EPI_ISL_921998, EPI_ISL_1178589 | | | |
| see above | Queens Medical Centre, Clinical Microbiology Department / DeepSeq Nottingham | COVID-19 Genomics UK (COG-UK) Consortium | Christopher Moore; Emily Park; Fei Sang; Gemma Clark; Hannah Howson-Wells; Jack Hill; Jayasree Dey; Johnny Debebe; Jonathan Ball; Joseph Chappell; Manjinder Khakh; Matthew Carlisle; Matthew Loose; Michelle M Lister; Nadine Holmes; Patrick McClure; Theocharis Tsoleridis; Timothy Byaruhanga; Vicki M Fleming; Victoria Wright; Wendy Smith |
| EPI_ISL_998145, EPI_ISL_998146, EPI_ISL_998148, EPI_ISL_998149, EPI_ISL_998164, EPI_ISL_998165, EPI_ISL_998166, EPI_ISL_998168, EPI_ISL_998169, EPI_ISL_998170, EPI_ISL_998173, EPI_ISL_998180, EPI_ISL_998197, EPI_ISL_998199, EPI_ISL_998212, EPI_ISL_998213, EPI_ISL_998214, EPI_ISL_998215, EPI_ISL_998216, EPI_ISL_998217, EPI_ISL_998222, EPI_ISL_998223, EPI_ISL_998224, EPI_ISL_998248, EPI_ISL_998249, EPI_ISL_998250, EPI_ISL_998251, EPI_ISL_998252, EPI_ISL_998253, EPI_ISL_998254, EPI_ISL_998255, EPI_ISL_998257, EPI_ISL_998258, EPI_ISL_998259, EPI_ISL_998260, EPI_ISL_998261, EPI_ISL_998262, EPI_ISL_998263, EPI_ISL_998265, EPI_ISL_998266, EPI_ISL_998309, EPI_ISL_998310, EPI_ISL_998311, EPI_ISL_998312, EPI_ISL_1475018, EPI_ISL_1475021, EPI_ISL_1475092, EPI_ISL_1475093, EPI_ISL_1475094, EPI_ISL_1475095, EPI_ISL_1475096, EPI_ISL_1475097, EPI_ISL_1475098, EPI_ISL_1475099, EPI_ISL_1475100, EPI_ISL_1475101, EPI_ISL_1475103, EPI_ISL_1475104, EPI_ISL_1475105, EPI_ISL_1475107, EPI_ISL_1475108, EPI_ISL_1475109, EPI_ISL_1475110, EPI_ISL_1475111, EPI_ISL_1475112, EPI_ISL_1475115, EPI_ISL_1475116, EPI_ISL_1475117 | | | |
| see above | Regional Virus Laboratory, Belfast Health and Social Care Trust | COVID-19 Genomics UK (COG-UK) Consortium | Alison Watt; Ciara Cox; Conall McCaughey; David Simpson; Derek Fairley; James McKenna; Mairead Connor; Susan Feeney; Tanya Curran; Zoltan Molnar |
| EPI_ISL_855604, EPI_ISL_860692, EPI_ISL_860693, EPI_ISL_860696, EPI_ISL_860697, EPI_ISL_860698, EPI_ISL_860699, EPI_ISL_860700, EPI_ISL_860701, EPI_ISL_860702, EPI_ISL_860704, EPI_ISL_860705, EPI_ISL_860706, EPI_ISL_860707, EPI_ISL_860708, EPI_ISL_860783, EPI_ISL_862048, EPI_ISL_862188, EPI_ISL_862189, EPI_ISL_862191, EPI_ISL_862192, EPI_ISL_862194, EPI_ISL_862195, EPI_ISL_862196, EPI_ISL_862197, EPI_ISL_862198, EPI_ISL_862199, EPI_ISL_862200, EPI_ISL_862201, EPI_ISL_862202, EPI_ISL_862203, EPI_ISL_862204, EPI_ISL_862205, EPI_ISL_862206, EPI_ISL_862207, EPI_ISL_862208, EPI_ISL_862209, EPI_ISL_862210, EPI_ISL_862211, EPI_ISL_862212, EPI_ISL_862213, EPI_ISL_862214, EPI_ISL_862215, EPI_ISL_862216, EPI_ISL_862217, EPI_ISL_862218, EPI_ISL_862219, EPI_ISL_862220, EPI_ISL_862221, EPI_ISL_862222, EPI_ISL_862223, EPI_ISL_862224, EPI_ISL_862225, EPI_ISL_862226, EPI_ISL_862227, EPI_ISL_862228, EPI_ISL_862229, EPI_ISL_862230, EPI_ISL_862231, EPI_ISL_862232, EPI_ISL_862233, EPI_ISL_862234, EPI_ISL_862235, EPI_ISL_862236, EPI_ISL_862237, EPI_ISL_862238, EPI_ISL_862239, EPI_ISL_862240, EPI_ISL_862241, EPI_ISL_862242, EPI_ISL_862243, EPI_ISL_862244, EPI_ISL_862245, EPI_ISL_862246, EPI_ISL_862247, EPI_ISL_862248, EPI_ISL_862249, EPI_ISL_862250, EPI_ISL_862252, EPI_ISL_862253, EPI_ISL_862254, EPI_ISL_862255, EPI_ISL_862256, EPI_ISL_862257, EPI_ISL_862258, EPI_ISL_862260, EPI_ISL_862261, EPI_ISL_862262, EPI_ISL_862263, EPI_ISL_862264, EPI_ISL_862265, EPI_ISL_862266, EPI_ISL_866432, EPI_ISL_866619, EPI_ISL_866793, EPI_ISL_866985, EPI_ISL_867156, EPI_ISL_1296539, EPI_ISL_1296540, EPI_ISL_1296541, EPI_ISL_1296545, EPI_ISL_1296550, EPI_ISL_1296552, EPI_ISL_1296553, EPI_ISL_1296554, EPI_ISL_1296555, EPI_ISL_1296556, EPI_ISL_1296557, EPI_ISL_1296558, EPI_ISL_1296559, EPI_ISL_1296560, EPI_ISL_1296561, EPI_ISL_1296562 | | | |
| see above | Respiratory Virus Unit, National Infection Service, Public Health England | COVID-19 Genomics UK (COG-UK) Consortium | PHE Covid Sequencing Team |
| EPI_ISL_1833590, EPI_ISL_1833593, EPI_ISL_1833594, EPI_ISL_1833595, EPI_ISL_1833600, EPI_ISL_1833601, EPI_ISL_1833602, EPI_ISL_1833603, EPI_ISL_1833605, EPI_ISL_1833606, EPI_ISL_1833607, EPI_ISL_1833608, EPI_ISL_1833609, EPI_ISL_1833610, EPI_ISL_1833612, EPI_ISL_1833613, EPI_ISL_1833614, EPI_ISL_1833615, EPI_ISL_1833791, EPI_ISL_1833808, EPI_ISL_1833827 | | | |
| see above | St George's University Hospitals NHS Foundation Trust | COVID-19 Genomics UK (COG-UK) Consortium | Adam Witney; Cassie Pope; Irene Monahan; Joshua Taylor; Ken Laing; NgeeKeong Tan |
| EPI_ISL_920289, EPI_ISL_920300, EPI_ISL_920312, EPI_ISL_920313, EPI_ISL_920314, EPI_ISL_920317, EPI_ISL_920323, EPI_ISL_920324, EPI_ISL_920325, EPI_ISL_920332, EPI_ISL_920333, EPI_ISL_920336, EPI_ISL_920340, EPI_ISL_920341, EPI_ISL_920342, EPI_ISL_920346, EPI_ISL_920347, EPI_ISL_920348, EPI_ISL_920349, EPI_ISL_920352, EPI_ISL_920359, EPI_ISL_920361, EPI_ISL_920366, EPI_ISL_920369, EPI_ISL_920374, EPI_ISL_920375, EPI_ISL_920379, EPI_ISL_920380, EPI_ISL_920381, EPI_ISL_920382, EPI_ISL_920383, EPI_ISL_920384, EPI_ISL_920385, EPI_ISL_920386, EPI_ISL_920387, EPI_ISL_920388, EPI_ISL_920389, EPI_ISL_920390, EPI_ISL_920391, EPI_ISL_920392, EPI_ISL_920393, EPI_ISL_920394, EPI_ISL_920395, EPI_ISL_920396, EPI_ISL_920397, EPI_ISL_920398, EPI_ISL_920399, EPI_ISL_920400, EPI_ISL_920401, EPI_ISL_920402, EPI_ISL_920403, EPI_ISL_920404, EPI_ISL_920405, EPI_ISL_920406, EPI_ISL_920407, EPI_ISL_920408, EPI_ISL_920409, EPI_ISL_920411, EPI_ISL_920412, EPI_ISL_920413, EPI_ISL_920414, EPI_ISL_920415, EPI_ISL_920416, EPI_ISL_920417, EPI_ISL_920418, EPI_ISL_920419, EPI_ISL_920420, EPI_ISL_920421, EPI_ISL_920422, EPI_ISL_920423, EPI_ISL_920425, EPI_ISL_920426, EPI_ISL_920428, EPI_ISL_920429, EPI_ISL_920430, EPI_ISL_920431, EPI_ISL_920432, EPI_ISL_920433, EPI_ISL_920434, EPI_ISL_920435, EPI_ISL_920436, EPI_ISL_920438, EPI_ISL_920439, EPI_ISL_920441, EPI_ISL_920442, EPI_ISL_920443, EPI_ISL_920444, EPI_ISL_920445, EPI_ISL_920446, EPI_ISL_920448, EPI_ISL_920449, EPI_ISL_920450, EPI_ISL_920451, EPI_ISL_920452, EPI_ISL_920453, EPI_ISL_920456, EPI_ISL_920464, EPI_ISL_920465, EPI_ISL_920473, EPI_ISL_920481, EPI_ISL_920489, EPI_ISL_920490, EPI_ISL_920497, EPI_ISL_920498, EPI_ISL_920507, EPI_ISL_920514, EPI_ISL_920627, EPI_ISL_920670, EPI_ISL_920683, EPI_ISL_920684, EPI_ISL_920691, EPI_ISL_920705, EPI_ISL_920714, EPI_ISL_920722, EPI_ISL_920738, EPI_ISL_920739, EPI_ISL_920747, EPI_ISL_920748, EPI_ISL_920755, EPI_ISL_920770, EPI_ISL_920775, EPI_ISL_1104773, EPI_ISL_1104781, EPI_ISL_1104782, EPI_ISL_1104783, EPI_ISL_1104787, EPI_ISL_1104789, EPI_ISL_1104794, EPI_ISL_1104796, EPI_ISL_1104802, EPI_ISL_1104810, EPI_ISL_1104818, EPI_ISL_1104819, EPI_ISL_1104824, EPI_ISL_1104825, EPI_ISL_1104826, EPI_ISL_1105045, EPI_ISL_1105051, EPI_ISL_1105060, EPI_ISL_1105065, EPI_ISL_1105072, EPI_ISL_1105085, EPI_ISL_1105092, EPI_ISL_1105165, EPI_ISL_1105199, EPI_ISL_1105200, EPI_ISL_1105264, EPI_ISL_1105268, EPI_ISL_1105276, EPI_ISL_1105280 | | | |
| see above | University College London Hospital | COVID-19 Genomics UK (COG-UK) Consortium | Catherine Houlihan; Dan Frampton; Judith Heaney; Matthew Byott; Moira Spyer and Eleni Nastouli; Stuart Kirk |
| EPI_ISL_865605, EPI_ISL_865606, EPI_ISL_865607, EPI_ISL_865608, EPI_ISL_865609, EPI_ISL_865610, EPI_ISL_865611, EPI_ISL_865612, EPI_ISL_865613, EPI_ISL_865619, EPI_ISL_865620, EPI_ISL_865621, EPI_ISL_865622, EPI_ISL_865624, EPI_ISL_865625, EPI_ISL_865626, EPI_ISL_865627, EPI_ISL_865628, EPI_ISL_865629, EPI_ISL_865631, EPI_ISL_865632, EPI_ISL_865633, EPI_ISL_865634, EPI_ISL_865635, EPI_ISL_865636, EPI_ISL_865637, EPI_ISL_865638, EPI_ISL_865640, EPI_ISL_865641, EPI_ISL_865642, EPI_ISL_865643, EPI_ISL_865645, EPI_ISL_865646, EPI_ISL_865647, EPI_ISL_865648, EPI_ISL_865649, EPI_ISL_865650, EPI_ISL_865651, EPI_ISL_865652, EPI_ISL_865653, EPI_ISL_865654, EPI_ISL_865655, EPI_ISL_865656, EPI_ISL_865657, EPI_ISL_865658, EPI_ISL_865659, EPI_ISL_865660, EPI_ISL_865661, EPI_ISL_865662, EPI_ISL_865663, EPI_ISL_865664, EPI_ISL_865665, EPI_ISL_865666, EPI_ISL_865667, EPI_ISL_865668, EPI_ISL_865669, EPI_ISL_865670, EPI_ISL_865671, EPI_ISL_865672, EPI_ISL_865673, EPI_ISL_865674, EPI_ISL_865675, EPI_ISL_865676, EPI_ISL_865677, EPI_ISL_865678, EPI_ISL_865679, EPI_ISL_865680, EPI_ISL_865681, EPI_ISL_865884, EPI_ISL_865885, EPI_ISL_865886, EPI_ISL_865887, EPI_ISL_865888, EPI_ISL_865889, EPI_ISL_865890, EPI_ISL_865902, EPI_ISL_865904, EPI_ISL_865907, EPI_ISL_865908, EPI_ISL_865917, EPI_ISL_865920, EPI_ISL_865936, EPI_ISL_865937, EPI_ISL_865946, EPI_ISL_865951, EPI_ISL_865967, EPI_ISL_865969, EPI_ISL_865970, EPI_ISL_865971, EPI_ISL_865972, EPI_ISL_865973, EPI_ISL_865974, EPI_ISL_865975, EPI_ISL_865976, EPI_ISL_865979, EPI_ISL_865980, EPI_ISL_865981, EPI_ISL_865982, EPI_ISL_865983, EPI_ISL_865984, EPI_ISL_865985, EPI_ISL_865988, EPI_ISL_865989, EPI_ISL_865990, EPI_ISL_865991, EPI_ISL_865992, EPI_ISL_865993, EPI_ISL_865995, EPI_ISL_865996, EPI_ISL_865997, EPI_ISL_865998, EPI_ISL_865999, EPI_ISL_866002, EPI_ISL_866003, EPI_ISL_866004, EPI_ISL_866005, EPI_ISL_866006, EPI_ISL_866008, EPI_ISL_866010, EPI_ISL_866013, EPI_ISL_866014, EPI_ISL_866015, EPI_ISL_866016, EPI_ISL_866017, EPI_ISL_866018, EPI_ISL_866019, EPI_ISL_866020, EPI_ISL_866022, EPI_ISL_866023, EPI_ISL_866025, EPI_ISL_866026, EPI_ISL_866029, EPI_ISL_866038, EPI_ISL_866040, EPI_ISL_866043, EPI_ISL_866044, EPI_ISL_866298, EPI_ISL_866306, EPI_ISL_866311, EPI_ISL_866312, EPI_ISL_866320, EPI_ISL_866321, EPI_ISL_866322, EPI_ISL_866323, EPI_ISL_866324, EPI_ISL_866325, EPI_ISL_866326, EPI_ISL_866327, EPI_ISL_866328, EPI_ISL_866329, EPI_ISL_866330, EPI_ISL_866331, EPI_ISL_866332, EPI_ISL_866333, EPI_ISL_920059, EPI_ISL_920060, EPI_ISL_920061, EPI_ISL_920062, EPI_ISL_920063, EPI_ISL_920064, EPI_ISL_920065, EPI_ISL_920066, EPI_ISL_920068, EPI_ISL_920071, EPI_ISL_920072, EPI_ISL_920782, EPI_ISL_920783, EPI_ISL_920784, EPI_ISL_920785, EPI_ISL_920786, EPI_ISL_920787, EPI_ISL_920788, EPI_ISL_920789, EPI_ISL_920790, EPI_ISL_920791, EPI_ISL_920792, EPI_ISL_920793, EPI_ISL_920794, EPI_ISL_920795, EPI_ISL_920796, EPI_ISL_920797, EPI_ISL_920798, EPI_ISL_920799, EPI_ISL_920800, EPI_ISL_920801, EPI_ISL_920802, EPI_ISL_920803, EPI_ISL_920804, EPI_ISL_920805, EPI_ISL_920806, EPI_ISL_920807, EPI_ISL_920808, EPI_ISL_920809, EPI_ISL_920811, EPI_ISL_920812, EPI_ISL_920813, EPI_ISL_920814, EPI_ISL_920815, EPI_ISL_920816, EPI_ISL_920817, EPI_ISL_920831, EPI_ISL_920832, EPI_ISL_949791, EPI_ISL_949905, EPI_ISL_949979, EPI_ISL_950156, EPI_ISL_950157, EPI_ISL_950158, EPI_ISL_950160, EPI_ISL_950161, EPI_ISL_997678, EPI_ISL_997680, EPI_ISL_997681, EPI_ISL_997682, EPI_ISL_997997, EPI_ISL_997998, EPI_ISL_997999, EPI_ISL_998000, EPI_ISL_998001, EPI_ISL_1048138, EPI_ISL_1178010, EPI_ISL_1248149, EPI_ISL_1248150, EPI_ISL_1248153, EPI_ISL_1248154, EPI_ISL_1248155, EPI_ISL_1248157, EPI_ISL_1248159, EPI_ISL_1248163, EPI_ISL_1248166, EPI_ISL_1248168, EPI_ISL_1248172, EPI_ISL_1248176, EPI_ISL_1248178, EPI_ISL_1248179, EPI_ISL_1248183, EPI_ISL_1248185, EPI_ISL_1248186, EPI_ISL_1248187, EPI_ISL_1248192, EPI_ISL_1248194, EPI_ISL_1248195, EPI_ISL_1248196, EPI_ISL_1248201, EPI_ISL_1248202, EPI_ISL_1248204, EPI_ISL_1248206, EPI_ISL_1248230, EPI_ISL_1248231, EPI_ISL_1248235, EPI_ISL_1248236, EPI_ISL_1248237, EPI_ISL_1248240, EPI_ISL_1248244, EPI_ISL_1248247, EPI_ISL_1309080, EPI_ISL_1309082, EPI_ISL_1309112, EPI_ISL_1309114, EPI_ISL_1386857, EPI_ISL_1386875, EPI_ISL_1637153, EPI_ISL_1730396, EPI_ISL_1730397, EPI_ISL_1975182 | | | |
| see above | University College London, Great Ormond Street Hospital for Children NHS Foundation Trust, Imperial College Healthcare NHS Trust | COVID-19 Genomics UK (COG-UK) Consortium | Alison Holmes; Charlotte Williams; Helena Tutill; Jacqueline Findlay; James Price; Judith Breuer; Julianne Brown; Kathryn Harris; Leysa Forrest; Mark Kristiansen; Paola Niola; Paola Resende Silva; Patricia Dyal; Paul Randell; Rachel Williams; Samuel Weeks; Sergi Castellano; Sunando Roy; Tony Brooks; Yasmin Panchbhaya |
| EPI_ISL_1047944, EPI_ISL_1104295, EPI_ISL_1104306 | University Hospitals Of Leicester NHS Trust and DeepSeq Nottingham | COVID-19 Genomics UK (COG-UK) Consortium | Christopher Holmes; Christopher Moore; Fei Sang; Joeseph Chappell; Johnny Debebe; Jonathan Ball; Julian Tang; Karlie Fallon; Matthew Carlisle; Matthew Loose; Nadine Holmes; Patrick McClure; Paul Bird; Thomas Helmer; Victoria Wright |
| EPI_ISL_949407, EPI_ISL_949408, EPI_ISL_949418, EPI_ISL_949419, EPI_ISL_949422, EPI_ISL_949423, EPI_ISL_949424, EPI_ISL_949425, EPI_ISL_949426, EPI_ISL_996389, EPI_ISL_996390, EPI_ISL_996391, EPI_ISL_996392, EPI_ISL_996393, EPI_ISL_996394, EPI_ISL_996397, EPI_ISL_996398, EPI_ISL_996399, EPI_ISL_996400, EPI_ISL_996425, EPI_ISL_996426, EPI_ISL_996447, EPI_ISL_996450, EPI_ISL_996451, EPI_ISL_1046985, EPI_ISL_1046986, EPI_ISL_1046987, EPI_ISL_1046992, EPI_ISL_1046993, EPI_ISL_1046994 | | | |
| see above | University of Birmingham | COVID-19 Genomics UK (COG-UK) Consortium | Alex Richter; Andrew Bosworth; Andrew Bosworth. Queen Elizabeth Hospital: Anna Casey; Andrew D Beggs PHE Heartlands Lab: Husam Osman; Charlotte Poxon; Institute of Microbiology; Joanne Stockton; Josh Quick; Kasun Wanigasooriya; Mike Kidd; Nicholas Loman. University of Birmingham Testing Laboratory: Celina M Whalley; Oliver Pickles; Radoslaw Poplawski; Samuel Nicholls; University of Birmingham: Claire McMurray; Will Rowe |
| EPI_ISL_865163, EPI_ISL_996572, EPI_ISL_996573, EPI_ISL_996574, EPI_ISL_996575, EPI_ISL_996590, EPI_ISL_996593, EPI_ISL_1308564, EPI_ISL_1308585, EPI_ISL_1308586, EPI_ISL_1308592 | | | |
| see above | University of Exeter | COVID-19 Genomics UK (COG-UK) Consortium | Aaron Jeffries; Audrey Farbos; Ben Temperton; Jane Masoli; Joanna Warwick-Dugdale; Michelle Michelsen; Robyn Manley; Stephen Michell |
| EPI_ISL_997077, EPI_ISL_997078, EPI_ISL_997079, EPI_ISL_997151, EPI_ISL_997152, EPI_ISL_997153, EPI_ISL_997154, EPI_ISL_997155, EPI_ISL_997156, EPI_ISL_997157, EPI_ISL_997158, EPI_ISL_1047857, EPI_ISL_1104217, EPI_ISL_1177692, EPI_ISL_1177704, EPI_ISL_1177705, EPI_ISL_1177706, EPI_ISL_1308716, EPI_ISL_1308847, EPI_ISL_1474581 | | | |
| see above | Virology Department, Royal Infirmary of Edinburgh, NHS Lothian / School of Biological Sciences, University of Edinburgh | COVID-19 Genomics UK (COG-UK) Consortium | Colquhoun R; Cotton S; Dewar R; Hill V; Jackson B; McCrone JT; McHugh M; O'Toole Á; Rambaut A; Rooke S; Scher E; Templeton K; Yu X |
| EPI_ISL_919345, EPI_ISL_919346, EPI_ISL_949637, EPI_ISL_949638, EPI_ISL_949639, EPI_ISL_949641 | Virology Department, Royal Infirmary of Edinburgh, NHS Lothian / School of Biological Sciences, University of Edinburgh / Institute of Genetics and Molecular Medicine, University of Edinburgh | COVID-19 Genomics UK (COG-UK) Consortium | Balcaza C; Colquhoun R; Dewar R; Gallagher M; Hill V; Jackson B; McCrone JT; McHugh M; O'Toole Á; Rambaut A; Rooke S; Scher E; Templeton K; Williams TC; Yu X |
| EPI_ISL_868363, EPI_ISL_868366, EPI_ISL_868369, EPI_ISL_868371, EPI_ISL_868374, EPI_ISL_868375, EPI_ISL_868376, EPI_ISL_868380, EPI_ISL_868383, EPI_ISL_868384, EPI_ISL_868394, EPI_ISL_868396, EPI_ISL_868404, EPI_ISL_868405, EPI_ISL_868410, EPI_ISL_868412, EPI_ISL_868417, EPI_ISL_868421, EPI_ISL_868426, EPI_ISL_868435, EPI_ISL_868447, EPI_ISL_868456, EPI_ISL_868457, EPI_ISL_868458, EPI_ISL_868460, EPI_ISL_868461, EPI_ISL_868462, EPI_ISL_868463, EPI_ISL_868473, EPI_ISL_868477, EPI_ISL_868489, EPI_ISL_868490, EPI_ISL_868494, EPI_ISL_868497, EPI_ISL_868504, EPI_ISL_868506, EPI_ISL_868509, EPI_ISL_868510, EPI_ISL_868519, EPI_ISL_868528, EPI_ISL_868529, EPI_ISL_868544, EPI_ISL_868550, EPI_ISL_868551, EPI_ISL_868553, EPI_ISL_868555, EPI_ISL_868563, EPI_ISL_868568, EPI_ISL_868571, EPI_ISL_868580, EPI_ISL_868583, EPI_ISL_868585, EPI_ISL_868587, EPI_ISL_868591, EPI_ISL_868595, EPI_ISL_868596, EPI_ISL_868600, EPI_ISL_868604, EPI_ISL_868605, EPI_ISL_868610, EPI_ISL_868611, EPI_ISL_868616, EPI_ISL_868619, EPI_ISL_868624, EPI_ISL_868633, EPI_ISL_868636, EPI_ISL_868637, EPI_ISL_868640, EPI_ISL_868641, EPI_ISL_868646, EPI_ISL_868650, EPI_ISL_868652, EPI_ISL_868657, EPI_ISL_868667, EPI_ISL_868668, EPI_ISL_868672, EPI_ISL_868676, EPI_ISL_868682, EPI_ISL_868685, EPI_ISL_868687, EPI_ISL_868689, EPI_ISL_868691, EPI_ISL_868694, EPI_ISL_868695, EPI_ISL_868697, EPI_ISL_868706, EPI_ISL_868707, EPI_ISL_868709, EPI_ISL_868711, EPI_ISL_868712, EPI_ISL_868714, EPI_ISL_924098, EPI_ISL_924102, EPI_ISL_924106, EPI_ISL_924114, EPI_ISL_924121, EPI_ISL_924123, EPI_ISL_924159, EPI_ISL_924169, EPI_ISL_924170, EPI_ISL_924174, EPI_ISL_924189, EPI_ISL_924199, EPI_ISL_924204, EPI_ISL_924206, EPI_ISL_924207, EPI_ISL_924208, EPI_ISL_924218, EPI_ISL_924253, EPI_ISL_924255, EPI_ISL_924256, EPI_ISL_924266, EPI_ISL_924284, EPI_ISL_924291, EPI_ISL_924299, EPI_ISL_924307, EPI_ISL_924310, EPI_ISL_924311, EPI_ISL_924312, EPI_ISL_924314, EPI_ISL_924319, EPI_ISL_924324, EPI_ISL_924337, EPI_ISL_924343, EPI_ISL_924345, EPI_ISL_924348, EPI_ISL_924358, EPI_ISL_924360, EPI_ISL_924363, EPI_ISL_924364, EPI_ISL_924380, EPI_ISL_924386, EPI_ISL_924388, EPI_ISL_924394, EPI_ISL_924413, EPI_ISL_1000702, EPI_ISL_1000705, EPI_ISL_1000738, EPI_ISL_1054625, EPI_ISL_1054630, EPI_ISL_1054681, EPI_ISL_1054704, EPI_ISL_1180081 | | | |
| see above | Virology Department, Sheffield Teaching Hospitals NHS Foundation Trust/Department of Infection, Immunity and Cardiovascular Disease, The Medical School, University of Sheffield | COVID-19 Genomics UK (COG-UK) Consortium | Adri Angyal; Alex Keeley; Benjamin Lindsey; Cariad Evans; Danielle Groves; Dave Partridge; Katie Johnson; Laura Carrilero; Luke Green; Matthew Parker; Matthew Wyles; Mehmet Yavuz; Mohammad Raza; Nikki Smith; Paul Parsons; Rachel Tucker; Rebecca Brown; Thushan de Silva |
| EPI_ISL_922353, EPI_ISL_922356, EPI_ISL_922357, EPI_ISL_922358, EPI_ISL_922371, EPI_ISL_922512, EPI_ISL_922513, EPI_ISL_922514, EPI_ISL_922518, EPI_ISL_922520, EPI_ISL_922521, EPI_ISL_922522, EPI_ISL_922530, EPI_ISL_922536, EPI_ISL_922537, EPI_ISL_922538, EPI_ISL_922542, EPI_ISL_922545, EPI_ISL_922546, EPI_ISL_922552, EPI_ISL_922555, EPI_ISL_922556, EPI_ISL_922557, EPI_ISL_922561, EPI_ISL_922562, EPI_ISL_922564, EPI_ISL_922569, EPI_ISL_922575, EPI_ISL_922576, EPI_ISL_922577, EPI_ISL_922578, EPI_ISL_922579, EPI_ISL_922583, EPI_ISL_922584, EPI_ISL_922585, EPI_ISL_922586, EPI_ISL_922587, EPI_ISL_922608, EPI_ISL_922609, EPI_ISL_922610, EPI_ISL_922611, EPI_ISL_922612, EPI_ISL_922613, EPI_ISL_922614, EPI_ISL_922615, EPI_ISL_922616, EPI_ISL_922617, EPI_ISL_922618, EPI_ISL_922619, EPI_ISL_922620, EPI_ISL_922621, EPI_ISL_922622, EPI_ISL_922623, EPI_ISL_922624, EPI_ISL_922625, EPI_ISL_922630, EPI_ISL_922631, EPI_ISL_922632, EPI_ISL_922635, EPI_ISL_922636, EPI_ISL_922714, EPI_ISL_922716, EPI_ISL_922717, EPI_ISL_922728, EPI_ISL_922731, EPI_ISL_922733, EPI_ISL_922735, EPI_ISL_922736, EPI_ISL_922737, EPI_ISL_922739, EPI_ISL_922740, EPI_ISL_922742, EPI_ISL_922744, EPI_ISL_922746, EPI_ISL_922747, EPI_ISL_922748, EPI_ISL_922749, EPI_ISL_922751, EPI_ISL_922752, EPI_ISL_922753, EPI_ISL_922754, EPI_ISL_922755, EPI_ISL_922756, EPI_ISL_922757, EPI_ISL_922758, EPI_ISL_922759, EPI_ISL_922760, EPI_ISL_922761, EPI_ISL_922762, EPI_ISL_922763, EPI_ISL_922764, EPI_ISL_922765, EPI_ISL_922766, EPI_ISL_922767, EPI_ISL_922768, EPI_ISL_922769, | | | |

EPI_ISL_922770, EPI_ISL_922771, EPI_ISL_922772, EPI_ISL_922773, EPI_ISL_922774, EPI_ISL_922775, EPI_ISL_922776, EPI_ISL_922777, EPI_ISL_922778, EPI_ISL_922779, EPI_ISL_922781, EPI_ISL_922782, EPI_ISL_922783, EPI_ISL_922784, EPI_ISL_922785, EPI_ISL_922786, EPI_ISL_922787, EPI_ISL_922788, EPI_ISL_922789, EPI_ISL_922790, EPI_ISL_922791, EPI_ISL_922792, EPI_ISL_922793, EPI_ISL_922794, EPI_ISL_922795, EPI_ISL_922796, EPI_ISL_922798, EPI_ISL_922799, EPI_ISL_922800, EPI_ISL_922804, EPI_ISL_922806, EPI_ISL_922808, EPI_ISL_922810, EPI_ISL_922811, EPI_ISL_922812, EPI_ISL_922813, EPI_ISL_922814, EPI_ISL_922815, EPI_ISL_922816, EPI_ISL_922817, EPI_ISL_922819, EPI_ISL_922820, EPI_ISL_922821, EPI_ISL_922822, EPI_ISL_922823, EPI_ISL_922824, EPI_ISL_922826, EPI_ISL_922827, EPI_ISL_922828, EPI_ISL_922829, EPI_ISL_922830, EPI_ISL_922831, EPI_ISL_922832, EPI_ISL_922833, EPI_ISL_922834, EPI_ISL_922839, EPI_ISL_922840, EPI_ISL_922842, EPI_ISL_922843, EPI_ISL_922844, EPI_ISL_922845, EPI_ISL_922847, EPI_ISL_922848, EPI_ISL_922850, EPI_ISL_922851, EPI_ISL_922854, EPI_ISL_922857, EPI_ISL_922859, EPI_ISL_922860, EPI_ISL_922861, EPI_ISL_922863, EPI_ISL_922864, EPI_ISL_922865, EPI_ISL_922866, EPI_ISL_922867, EPI_ISL_922868, EPI_ISL_922869, EPI_ISL_922870, EPI_ISL_922871, EPI_ISL_922872, EPI_ISL_922873, EPI_ISL_922874, EPI_ISL_922875, EPI_ISL_922876, EPI_ISL_922877, EPI_ISL_922878, EPI_ISL_922879, EPI_ISL_922880, EPI_ISL_922881, EPI_ISL_922885, EPI_ISL_922886, EPI_ISL_922888, EPI_ISL_922889, EPI_ISL_922890, EPI_ISL_922896, EPI_ISL_922897, EPI_ISL_922898, EPI_ISL_922899, EPI_ISL_922900, EPI_ISL_922911, EPI_ISL_922915, EPI_ISL_922916, EPI_ISL_922917, EPI_ISL_922918, EPI_ISL_922919, EPI_ISL_922920, EPI_ISL_922921, EPI_ISL_922922, EPI_ISL_922933, EPI_ISL_922934, EPI_ISL_922935, EPI_ISL_922936, EPI_ISL_922937, EPI_ISL_922939, EPI_ISL_922956, EPI_ISL_922962, EPI_ISL_922963, EPI_ISL_922966, EPI_ISL_922971, EPI_ISL_923028, EPI_ISL_923045, EPI_ISL_923046, EPI_ISL_923048, EPI_ISL_923049, EPI_ISL_923050, EPI_ISL_923052, EPI_ISL_923053, EPI_ISL_923054, EPI_ISL_923055, EPI_ISL_923056, EPI_ISL_923057, EPI_ISL_923058, EPI_ISL_923059, EPI_ISL_923060, EPI_ISL_923061, EPI_ISL_923062, EPI_ISL_923063, EPI_ISL_923064, EPI_ISL_923065, EPI_ISL_923066, EPI_ISL_923067, EPI_ISL_923068, EPI_ISL_923069, EPI_ISL_923070, EPI_ISL_923071, EPI_ISL_923073, EPI_ISL_923074, EPI_ISL_923075, EPI_ISL_923076, EPI_ISL_923084, EPI_ISL_923088, EPI_ISL_923089, EPI_ISL_923091, EPI_ISL_923092, EPI_ISL_923093, EPI_ISL_923095, EPI_ISL_923096, EPI_ISL_923097, EPI_ISL_923098, EPI_ISL_923099, EPI_ISL_923100, EPI_ISL_923103, EPI_ISL_923104, EPI_ISL_923105, EPI_ISL_923106, EPI_ISL_923107, EPI_ISL_923108, EPI_ISL_923109, EPI_ISL_923111, EPI_ISL_923112, EPI_ISL_923115, EPI_ISL_923116, EPI_ISL_923118, EPI_ISL_923121, EPI_ISL_923123, EPI_ISL_923125, EPI_ISL_923126, EPI_ISL_923127, EPI_ISL_923129, EPI_ISL_923130, EPI_ISL_923131, EPI_ISL_923133, EPI_ISL_923134, EPI_ISL_923135, EPI_ISL_923137, EPI_ISL_923138, EPI_ISL_923139, EPI_ISL_923140, EPI_ISL_923141, EPI_ISL_923144, EPI_ISL_923145, EPI_ISL_923148, EPI_ISL_923149, EPI_ISL_923150, EPI_ISL_923151, EPI_ISL_923152, EPI_ISL_923154, EPI_ISL_923157, EPI_ISL_923161, EPI_ISL_923163, EPI_ISL_923164, EPI_ISL_923165, EPI_ISL_923166, EPI_ISL_923167, EPI_ISL_1178889, EPI_ISL_1178923, EPI_ISL_1178924, EPI_ISL_1476449, EPI_ISL_1476459, EPI_ISL_1476462, EPI_ISL_1476463, EPI_ISL_1476468, EPI_ISL_1476470, EPI_ISL_1476473, EPI_ISL_1476475

| | | | |
|---|---|---|---|
| see above | Wales Specialist Virology Centre Sequencing lab: Pathogen Genomics Unit | Public Health Wales Microbiology Cardiff Wales Specialist Virology Centre | Alec Birchley; Alexander Adams; Amy Gaskin; Angela Marchbank; Bree Gatica-Wilcox; Catherine Moore; Jason Coombes; Joanne Watkins; Joel Southgate; Johnathan Evans; Laura Gifford; Lauren Gilbert; Lee Graham; Malorie Perry; Matthew Bull; Nicole Pacchiarini; Sally Corden; Sara Kumziene-Summerhayes; Sara Rey; Sarah Taylor; Simon Cottrell; Sophie Jones; Tom Connor |

EPI_ISL_919208, EPI_ISL_919211, EPI_ISL_919213, EPI_ISL_919214, EPI_ISL_919215, EPI_ISL_919216, EPI_ISL_919217, EPI_ISL_919218, EPI_ISL_919219, EPI_ISL_919220, EPI_ISL_919221, EPI_ISL_919222, EPI_ISL_919223, EPI_ISL_919224, EPI_ISL_919225, EPI_ISL_919226, EPI_ISL_919227, EPI_ISL_919228, EPI_ISL_919229, EPI_ISL_919230, EPI_ISL_919231, EPI_ISL_919277, EPI_ISL_919278, EPI_ISL_919279, EPI_ISL_919280, EPI_ISL_949612, EPI_ISL_997061, EPI_ISL_1047838, EPI_ISL_1050221

| | | | |
|---|---|---|---|
| see above | West of Scotland Specialist Virology Centre, NHSGGC / MRC-University of Glasgow Centre for Virus Research | COVID-19 Genomics UK (COG-UK) Consortium | Alasdair MacLean; Alice Broos; Ana da Silva Filipe; Antonia Ho; Daniel Mair; David L Robertson; Emma Thomson; Guy Mollett; James Shepherd; Jenna Nichols; Joseph Hughes; Kathy Li; Kathy Smollett; Kyriaki Nomikou; Lily Tong; Matthew Holden; Natasha Johnson; Rachel Blacow; Richard Orton; Rory Gunson; Sarah McDonald; Sharif Shaaban; Sreenu Vattipally; Stephen Carmichael |

We gratefully acknowledge the following Authors from the Originating laboratories responsible for obtaining the specimens, as well as the Submitting laboratories where the genome data were generated and shared via GISAID, on which this research is based.

All Submitters of data may be contacted directly via www.gisaid.org

Authors are sorted alphabetically.

| Accession ID | Originating Laboratory | Submitting Laboratory | Authors |
|---|---|---|---|
| EPI_ISL_1391410 | Barts Health NHS Trust | Barts Health NHS TRust | BROAD; Beatrix; CUTINO-MOGUEL; Claire; David; Dola; Esin; HARRINGTON; KARAA; KELE; KULASEGARAN-SHYLINI; Maria-Teresa; OWOYEMI; Raghavendran |
| EPI_ISL_1265369, EPI_ISL_1391409, EPI_ISL_1391411, EPI_ISL_1391412 | Barts Health NHS Trust | Barts Health NHS Trust | BROAD; Beatrix; CUTINO-MOGUEL; Claire; David; Dola; Esin; HARRINGTON; KARAA; KELE; KULASEGARAN-SHYLINI; Maria-Teresa; OWOYEMI; Raghavendran |
| EPI_ISL_949751, EPI_ISL_949752, EPI_ISL_949753, EPI_ISL_1048078, EPI_ISL_1048083, EPI_ISL_1048095 | Barts Health NHS Trust | COVID-19 Genomics UK (COG-UK) Consortium | BROAD; Beatrix; CUTINO-MOGUEL; Claire; David; Dola; HARRINGTON; KELE; KULASEGARAN-SHYLINI; Maria-Teresa; OWOYEMI; Raghavendran |
| EPI_ISL_924653, EPI_ISL_924657, EPI_ISL_924661, EPI_ISL_924664, EPI_ISL_924666, EPI_ISL_924667, EPI_ISL_924668, EPI_ISL_924669, EPI_ISL_924672, EPI_ISL_924676, EPI_ISL_924677, EPI_ISL_924683, EPI_ISL_924684, EPI_ISL_924685, EPI_ISL_924688, EPI_ISL_924689, EPI_ISL_924690, EPI_ISL_924691, EPI_ISL_924692, EPI_ISL_924693, EPI_ISL_924694, EPI_ISL_924695, EPI_ISL_924696, EPI_ISL_924697, EPI_ISL_924698, EPI_ISL_924699, EPI_ISL_924700, EPI_ISL_924701, EPI_ISL_924702, EPI_ISL_924704, EPI_ISL_924705, EPI_ISL_924706, EPI_ISL_924707, EPI_ISL_924708, EPI_ISL_924709, EPI_ISL_924712, EPI_ISL_924713, EPI_ISL_924714, EPI_ISL_924715, EPI_ISL_924716, EPI_ISL_924717, EPI_ISL_924718, EPI_ISL_924719, EPI_ISL_924720, EPI_ISL_924721, EPI_ISL_924722, EPI_ISL_1000795, EPI_ISL_1000796, EPI_ISL_1000797, EPI_ISL_1000798, EPI_ISL_1000799, EPI_ISL_1000800, EPI_ISL_1000801, EPI_ISL_1000802, EPI_ISL_1000803, EPI_ISL_1000804, EPI_ISL_1000805, EPI_ISL_1000807, EPI_ISL_1000809, EPI_ISL_1000810, EPI_ISL_1000812, EPI_ISL_1000813, EPI_ISL_1000814, EPI_ISL_1000815, EPI_ISL_1000817, EPI_ISL_1000818, EPI_ISL_1000820, EPI_ISL_1000821, EPI_ISL_1000823, EPI_ISL_1000824, EPI_ISL_1000825, EPI_ISL_1000827, EPI_ISL_1000828, EPI_ISL_1000829, EPI_ISL_1000830, EPI_ISL_1000831, EPI_ISL_1000891, EPI_ISL_1000894, EPI_ISL_1000909, EPI_ISL_1000911, EPI_ISL_1000963, EPI_ISL_1054711, EPI_ISL_1054712, EPI_ISL_1054715, EPI_ISL_1054716, EPI_ISL_1054717, EPI_ISL_1054718, EPI_ISL_1054721, EPI_ISL_1054722, EPI_ISL_1054724, EPI_ISL_1054725, EPI_ISL_1054726, EPI_ISL_1054727, EPI_ISL_1054728, EPI_ISL_1054730, EPI_ISL_1054731, EPI_ISL_1054732, EPI_ISL_1054733, EPI_ISL_1054734, EPI_ISL_1054735, EPI_ISL_1054736, EPI_ISL_1054737, EPI_ISL_1054738, EPI_ISL_1054739, EPI_ISL_1054740, EPI_ISL_1054741, EPI_ISL_1054742, EPI_ISL_1054743, EPI_ISL_1054744, EPI_ISL_1054745, EPI_ISL_1054746, EPI_ISL_1054747, EPI_ISL_1054748, EPI_ISL_1054749, EPI_ISL_1054750, EPI_ISL_1054751, EPI_ISL_1054752, EPI_ISL_1054753, EPI_ISL_1054754, EPI_ISL_1054755, EPI_ISL_1054818 | | | |
| see above | Bioinformatics and Biostatistics Lab, Advanced Sequencing Facility | COVID-19 Genomics UK (COG-UK) Consortium | Aengus Stewart; Chelsea Sawyer; Harshil Patel; Jerome Nicod; Laura Cubitt; Margaret Crawford |
| EPI_ISL_1832314, EPI_ISL_1832316, EPI_ISL_1832319, EPI_ISL_1832321, EPI_ISL_1832325, EPI_ISL_1832329, EPI_ISL_1832331, EPI_ISL_1832334 | | | |
| see above | Centre for Clinical Infection and Diagnostics Research and Genomics Innovation Unit, Guy's and St. Thomas' NHS Trust | COVID-19 Genomics UK (COG-UK) Consortium | Ali Raza Awan; Chloe Fisher; Jonathan Edgeworth; Luke Snell; Penny Cliff; Rahul Batra |
| EPI_ISL_868023, EPI_ISL_923328, EPI_ISL_923330, EPI_ISL_923345, EPI_ISL_923351, EPI_ISL_923422, EPI_ISL_923423, EPI_ISL_923424, EPI_ISL_952380, EPI_ISL_952391, EPI_ISL_952395, EPI_ISL_952417, EPI_ISL_952437, EPI_ISL_952438, EPI_ISL_952439, EPI_ISL_952440, EPI_ISL_952441, EPI_ISL_952442, EPI_ISL_952443, EPI_ISL_952444, EPI_ISL_952445, EPI_ISL_952446, EPI_ISL_952447, EPI_ISL_952448, EPI_ISL_952449, EPI_ISL_952450, EPI_ISL_952451, EPI_ISL_952452, EPI_ISL_952455, EPI_ISL_952457, EPI_ISL_952458, EPI_ISL_952461, EPI_ISL_952463, EPI_ISL_952468, EPI_ISL_952492, EPI_ISL_952493, EPI_ISL_952494, EPI_ISL_952495, EPI_ISL_952496, EPI_ISL_952497, EPI_ISL_952498, EPI_ISL_952499, EPI_ISL_952523, EPI_ISL_952524, EPI_ISL_952525, EPI_ISL_952530, EPI_ISL_952531, EPI_ISL_952532, EPI_ISL_952533, EPI_ISL_952534, EPI_ISL_952535, EPI_ISL_952536, EPI_ISL_952537, EPI_ISL_952539, EPI_ISL_952540, EPI_ISL_952541, EPI_ISL_952542, EPI_ISL_952543, EPI_ISL_952544, EPI_ISL_952545, EPI_ISL_952546, EPI_ISL_952547, EPI_ISL_952548, EPI_ISL_952549, EPI_ISL_952550, EPI_ISL_952551, EPI_ISL_952552, EPI_ISL_952553, EPI_ISL_952555, EPI_ISL_952556, EPI_ISL_952557, EPI_ISL_952822, EPI_ISL_952823, EPI_ISL_952824, EPI_ISL_952825, EPI_ISL_952826, EPI_ISL_952827, EPI_ISL_952828, EPI_ISL_952829, EPI_ISL_952836, EPI_ISL_952837, EPI_ISL_952838, EPI_ISL_952839, EPI_ISL_952840, EPI_ISL_952841, EPI_ISL_952851, EPI_ISL_952855, EPI_ISL_952856, EPI_ISL_952857, EPI_ISL_1000248, EPI_ISL_1000249, EPI_ISL_1000250, EPI_ISL_1000251, EPI_ISL_1000252, EPI_ISL_1000253, EPI_ISL_1000254, EPI_ISL_1000259, EPI_ISL_1000263, EPI_ISL_1000454, EPI_ISL_1000455, EPI_ISL_1000626, EPI_ISL_1053825, EPI_ISL_1053826, EPI_ISL_1053827, EPI_ISL_1053828, EPI_ISL_1053829, EPI_ISL_1053830, EPI_ISL_1053831, EPI_ISL_1053832, EPI_ISL_1053833, EPI_ISL_1053834, EPI_ISL_1053835, EPI_ISL_1053836, EPI_ISL_1053837, EPI_ISL_1053838, EPI_ISL_1053839, EPI_ISL_1053840, EPI_ISL_1053841, EPI_ISL_1053842, EPI_ISL_1053843, EPI_ISL_1053877, EPI_ISL_1053878, EPI_ISL_1053879, EPI_ISL_1053880, EPI_ISL_1053881, EPI_ISL_1053882, EPI_ISL_1053883, EPI_ISL_1053884, EPI_ISL_1053885, EPI_ISL_1053886, EPI_ISL_1053887, EPI_ISL_1053888, EPI_ISL_1053889, EPI_ISL_1053890, EPI_ISL_1053891, EPI_ISL_1053934, EPI_ISL_1053935, EPI_ISL_1053936, EPI_ISL_1053937, EPI_ISL_1053976, EPI_ISL_1107633, EPI_ISL_1107637, EPI_ISL_1107697, EPI_ISL_1179832, EPI_ISL_1637977, EPI_ISL_1979970, EPI_ISL_1979971, EPI_ISL_1980071, EPI_ISL_1980073, EPI_ISL_1980075, EPI_ISL_1980077, EPI_ISL_1980078, EPI_ISL_1980080 | | | |
| see above | Centre for Enzyme Innovation, University of Portsmouth / Translational Research Laboratory, Portsmouth Hospitals NHS Trust | COVID-19 Genomics UK (COG-UK) Consortium | Allyson Lloyd; Angela Beckett; Anoop Chauhan; Christopher Fearn; Ethan Butcher; Garry Scarlett; Kate Cook; Katie Loveson; Kelly Bicknell; Robert Impey; Salman Goudarzi; Samuel Robson; Sarah Wyllie; Scott Elliott; Sharon Glaysher; Yann Bourgeois |
| EPI_ISL_919007, EPI_ISL_919108, EPI_ISL_919109, EPI_ISL_919110, EPI_ISL_919111, EPI_ISL_919112, EPI_ISL_919113, EPI_ISL_919116, EPI_ISL_919117, EPI_ISL_919118, EPI_ISL_919120, EPI_ISL_919121, EPI_ISL_919122, EPI_ISL_919123, EPI_ISL_919124, EPI_ISL_919125, EPI_ISL_919126, EPI_ISL_919127, EPI_ISL_919128, EPI_ISL_919129, EPI_ISL_919130, EPI_ISL_919131, EPI_ISL_919132, EPI_ISL_919133, EPI_ISL_919135, EPI_ISL_919137, EPI_ISL_919138, EPI_ISL_919140, EPI_ISL_919141, EPI_ISL_919142, EPI_ISL_919144, EPI_ISL_919145, EPI_ISL_919146, EPI_ISL_919147, EPI_ISL_919178, EPI_ISL_949427, EPI_ISL_949429, EPI_ISL_949431, EPI_ISL_949432, EPI_ISL_949434, EPI_ISL_949435, EPI_ISL_949436, EPI_ISL_949437, EPI_ISL_949438, EPI_ISL_949439, EPI_ISL_949440, EPI_ISL_949441, EPI_ISL_949445, EPI_ISL_949446, EPI_ISL_949447, EPI_ISL_949449, EPI_ISL_949450, EPI_ISL_949451, EPI_ISL_949452, EPI_ISL_949453, EPI_ISL_949454, EPI_ISL_949455, EPI_ISL_949456, EPI_ISL_949457, EPI_ISL_949458, EPI_ISL_949459, EPI_ISL_949460, EPI_ISL_949461, EPI_ISL_949466, EPI_ISL_949470, EPI_ISL_949471, EPI_ISL_949472, EPI_ISL_949474, EPI_ISL_949522, EPI_ISL_949523, EPI_ISL_996598, EPI_ISL_996599, EPI_ISL_996600, EPI_ISL_996601, EPI_ISL_996602, EPI_ISL_996603, EPI_ISL_996676, EPI_ISL_996680, EPI_ISL_996682, EPI_ISL_996686, EPI_ISL_996688, EPI_ISL_996692, EPI_ISL_996698, EPI_ISL_996725, EPI_ISL_996727, EPI_ISL_996729, EPI_ISL_996731, EPI_ISL_996733, EPI_ISL_996738, EPI_ISL_996739, EPI_ISL_996740, EPI_ISL_1047184, EPI_ISL_1047187, EPI_ISL_1047191, EPI_ISL_1047331, EPI_ISL_1047511, EPI_ISL_1247603 | | | |
| see above | Department of Pathology, University of Cambridge | COVID-19 Genomics UK (COG-UK) Consortium | Aminu S. Jahun; Ian Goodfellow; Iliana Georgana; Martin D. Curran; Myra Hosmillo; Rhys Izu; Rhys Izuagbe; Surendra Parmar; William L. Hamilton; Yasmin Chaudhry |
| EPI_ISL_907371, EPI_ISL_907372, EPI_ISL_907373, EPI_ISL_907374, EPI_ISL_907375, EPI_ISL_907376, EPI_ISL_907377, EPI_ISL_907380, EPI_ISL_907382, EPI_ISL_907383, EPI_ISL_907387, EPI_ISL_907388, EPI_ISL_907389, EPI_ISL_907390, EPI_ISL_907391, EPI_ISL_907392, EPI_ISL_907396, EPI_ISL_907399, EPI_ISL_907400, EPI_ISL_907403, EPI_ISL_907404, EPI_ISL_907406, EPI_ISL_907407, EPI_ISL_907409, EPI_ISL_907410, EPI_ISL_907411, EPI_ISL_907412, EPI_ISL_907413, EPI_ISL_907414, EPI_ISL_907416, EPI_ISL_907417, EPI_ISL_907422, EPI_ISL_907424, EPI_ISL_907427, EPI_ISL_907428, EPI_ISL_907429, EPI_ISL_907432, EPI_ISL_907437, EPI_ISL_907438, EPI_ISL_907440, EPI_ISL_907441, EPI_ISL_907443, EPI_ISL_907444, EPI_ISL_907446, EPI_ISL_907447, EPI_ISL_907448, EPI_ISL_907449, EPI_ISL_907450, EPI_ISL_907452, EPI_ISL_907457, EPI_ISL_907458, EPI_ISL_907459, EPI_ISL_907460, EPI_ISL_907462, EPI_ISL_907463, EPI_ISL_907464, EPI_ISL_907466, EPI_ISL_907467, EPI_ISL_907468, EPI_ISL_907470, EPI_ISL_907471, EPI_ISL_907477, EPI_ISL_907478, EPI_ISL_907480, EPI_ISL_907482, EPI_ISL_907484, EPI_ISL_907485, EPI_ISL_907489, EPI_ISL_907491, EPI_ISL_907493, EPI_ISL_907496, EPI_ISL_907497, EPI_ISL_907500, EPI_ISL_907503, EPI_ISL_907505, EPI_ISL_907510, EPI_ISL_907511, EPI_ISL_907512, EPI_ISL_907516, EPI_ISL_907517, EPI_ISL_907518, EPI_ISL_907519, EPI_ISL_907521, EPI_ISL_907522, EPI_ISL_907523, EPI_ISL_907526, EPI_ISL_907528, EPI_ISL_907529, EPI_ISL_907532, EPI_ISL_907533, EPI_ISL_907536, EPI_ISL_907537, EPI_ISL_907538, EPI_ISL_907542, EPI_ISL_907543, EPI_ISL_907546, EPI_ISL_907549, EPI_ISL_907551, EPI_ISL_907552, EPI_ISL_907557, EPI_ISL_907559, EPI_ISL_907561, EPI_ISL_907563, EPI_ISL_907567, EPI_ISL_907568, EPI_ISL_907571, EPI_ISL_907573, EPI_ISL_907576, EPI_ISL_907578, EPI_ISL_907579, EPI_ISL_907580, EPI_ISL_907582, EPI_ISL_907583, EPI_ISL_907584, EPI_ISL_907587, EPI_ISL_907589, EPI_ISL_907590, EPI_ISL_907594, EPI_ISL_907596, EPI_ISL_907598, EPI_ISL_907600, EPI_ISL_907601, EPI_ISL_907605, EPI_ISL_907606, EPI_ISL_907607, EPI_ISL_907610, EPI_ISL_907611, EPI_ISL_907612, EPI_ISL_907615, EPI_ISL_907616, EPI_ISL_907618, EPI_ISL_907621, EPI_ISL_907622, EPI_ISL_907623, EPI_ISL_907626, EPI_ISL_907628, EPI_ISL_907630, EPI_ISL_907631, EPI_ISL_907640, EPI_ISL_907641, EPI_ISL_907642, EPI_ISL_907645, EPI_ISL_907647, EPI_ISL_907648, EPI_ISL_907651, EPI_ISL_907652, EPI_ISL_907654, EPI_ISL_907658, EPI_ISL_907659, EPI_ISL_907661, EPI_ISL_907664, EPI_ISL_907667, EPI_ISL_907672, EPI_ISL_907673, EPI_ISL_907675, EPI_ISL_907676, EPI_ISL_907677, EPI_ISL_907680, EPI_ISL_907684, EPI_ISL_907686, EPI_ISL_907689, EPI_ISL_907690, EPI_ISL_907692, EPI_ISL_907693, EPI_ISL_907695, EPI_ISL_907696, EPI_ISL_907697, EPI_ISL_907699, EPI_ISL_907838, EPI_ISL_908269, EPI_ISL_908270, EPI_ISL_908274, EPI_ISL_908277, EPI_ISL_908279, EPI_ISL_908282, EPI_ISL_908285, EPI_ISL_908292, EPI_ISL_908293, EPI_ISL_908296, EPI_ISL_908301, EPI_ISL_908306, EPI_ISL_908309, EPI_ISL_908310, EPI_ISL_908315, EPI_ISL_908316, EPI_ISL_908322, EPI_ISL_908333, EPI_ISL_908341, EPI_ISL_908343, EPI_ISL_908344, EPI_ISL_908351, EPI_ISL_908367, EPI_ISL_908369, EPI_ISL_908372, EPI_ISL_908384, EPI_ISL_908385, EPI_ISL_908389, EPI_ISL_908397, EPI_ISL_908401, EPI_ISL_908402, EPI_ISL_908407, EPI_ISL_908408, EPI_ISL_908413, EPI_ISL_908415, EPI_ISL_908435, EPI_ISL_908439, EPI_ISL_908446, EPI_ISL_908452, EPI_ISL_908453, EPI_ISL_908456, EPI_ISL_908467, EPI_ISL_908478, EPI_ISL_908479, EPI_ISL_908494, EPI_ISL_908506, EPI_ISL_908511, EPI_ISL_908518, EPI_ISL_908519, EPI_ISL_908520, EPI_ISL_908524, EPI_ISL_908526, EPI_ISL_908527, EPI_ISL_908528, EPI_ISL_908529, EPI_ISL_908534, EPI_ISL_908535, EPI_ISL_908542, EPI_ISL_908546, EPI_ISL_908550, EPI_ISL_908553, EPI_ISL_908560, EPI_ISL_908561, EPI_ISL_908571, EPI_ISL_908576, EPI_ISL_908580, EPI_ISL_908581, EPI_ISL_908585, EPI_ISL_908590, EPI_ISL_908596, EPI_ISL_912215, EPI_ISL_912216, EPI_ISL_912217, EPI_ISL_912218, EPI_ISL_916184, EPI_ISL_916186, EPI_ISL_916195, EPI_ISL_916197, EPI_ISL_916198, EPI_ISL_916199, EPI_ISL_916204, EPI_ISL_916206, EPI_ISL_916214, EPI_ISL_916215, EPI_ISL_916217, EPI_ISL_916218, EPI_ISL_916219, EPI_ISL_916220, EPI_ISL_916221, EPI_ISL_916223, EPI_ISL_916224, EPI_ISL_916230, EPI_ISL_916235, EPI_ISL_916239, EPI_ISL_916240, EPI_ISL_916251, EPI_ISL_916253, EPI_ISL_916255, EPI_ISL_916257, EPI_ISL_916259, EPI_ISL_916265, EPI_ISL_916267, EPI_ISL_916272, EPI_ISL_916282, EPI_ISL_916286, EPI_ISL_916289, EPI_ISL_916293, EPI_ISL_916294, EPI_ISL_916298, EPI_ISL_916300, EPI_ISL_916302, EPI_ISL_916309, EPI_ISL_916311, EPI_ISL_916317, EPI_ISL_916319, EPI_ISL_916324, EPI_ISL_916326, EPI_ISL_916330, EPI_ISL_916337, EPI_ISL_916351, EPI_ISL_916353, EPI_ISL_916356, EPI_ISL_916359, EPI_ISL_916361, EPI_ISL_916385, EPI_ISL_916386, EPI_ISL_916387, EPI_ISL_916389, EPI_ISL_916393, EPI_ISL_916395, EPI_ISL_916396, EPI_ISL_916398, EPI_ISL_916402, EPI_ISL_916404, EPI_ISL_916418, EPI_ISL_916422, EPI_ISL_916423, EPI_ISL_916424, EPI_ISL_916426, EPI_ISL_916429, EPI_ISL_916448, EPI_ISL_916452, EPI_ISL_916465, EPI_ISL_916466, EPI_ISL_916468, EPI_ISL_916469, EPI_ISL_916471, EPI_ISL_916474, EPI_ISL_916482, EPI_ISL_916483, EPI_ISL_916488, EPI_ISL_916490, EPI_ISL_916498, EPI_ISL_916501, EPI_ISL_917168, EPI_ISL_917169, EPI_ISL_917170, EPI_ISL_917171, EPI_ISL_917172, EPI_ISL_917173, EPI_ISL_917174, EPI_ISL_917175, EPI_ISL_917176, EPI_ISL_917177, EPI_ISL_917178, EPI_ISL_917179, EPI_ISL_917180, EPI_ISL_917181, EPI_ISL_917182, EPI_ISL_917183, EPI_ISL_917184, EPI_ISL_917185, EPI_ISL_917186, EPI_ISL_917187, EPI_ISL_917188, EPI_ISL_917189, EPI_ISL_917190, EPI_ISL_917191, EPI_ISL_917192, EPI_ISL_917193, EPI_ISL_917194, EPI_ISL_917195, EPI_ISL_917196, EPI_ISL_917197, EPI_ISL_917198, EPI_ISL_917199, EPI_ISL_917200, EPI_ISL_917201, EPI_ISL_917202, EPI_ISL_917203, EPI_ISL_917204, EPI_ISL_917205, EPI_ISL_917206, EPI_ISL_917207, EPI_ISL_917208, EPI_ISL_917209, EPI_ISL_917210, EPI_ISL_917211, EPI_ISL_917212, EPI_ISL_917213, EPI_ISL_917215, EPI_ISL_917216, EPI_ISL_917217, EPI_ISL_917218, EPI_ISL_917219, EPI_ISL_917220, EPI_ISL_917221, EPI_ISL_917222, EPI_ISL_917223, EPI_ISL_917224, EPI_ISL_917225, EPI_ISL_917226, EPI_ISL_917227, EPI_ISL_917228, EPI_ISL_917229, EPI_ISL_917230, EPI_ISL_917231, EPI_ISL_917232, EPI_ISL_917234, EPI_ISL_917235, EPI_ISL_917236, EPI_ISL_917237, EPI_ISL_917238, EPI_ISL_917239, EPI_ISL_917240, EPI_ISL_917241, EPI_ISL_917242, EPI_ISL_917243, EPI_ISL_917244, EPI_ISL_917245, EPI_ISL_917246, EPI_ISL_917247, EPI_ISL_917248, EPI_ISL_917249, EPI_ISL_917250, EPI_ISL_917251, EPI_ISL_917252, EPI_ISL_917253, EPI_ISL_917254, EPI_ISL_917255, EPI_ISL_917256, EPI_ISL_917257, EPI_ISL_917258, EPI_ISL_917259, EPI_ISL_917260, EPI_ISL_917261, EPI_ISL_917262, EPI_ISL_917263, EPI_ISL_917264, EPI_ISL_917265, EPI_ISL_917266, EPI_ISL_917267, EPI_ISL_917268, EPI_ISL_917269, EPI_ISL_917270, EPI_ISL_917271, EPI_ISL_917272, EPI_ISL_917273, EPI_ISL_917274, EPI_ISL_917275, EPI_ISL_917276, EPI_ISL_917277, EPI_ISL_917278, EPI_ISL_917279, EPI_ISL_917280, EPI_ISL_917281, EPI_ISL_917282, EPI_ISL_917283, EPI_ISL_917284, EPI_ISL_917285, EPI_ISL_917286, EPI_ISL_917287, EPI_ISL_917288, EPI_ISL_917289, EPI_ISL_917290, EPI_ISL_917291, EPI_ISL_917292, EPI_ISL_917293, EPI_ISL_917294, EPI_ISL_917295, EPI_ISL_917296, EPI_ISL_917297, EPI_ISL_917298, EPI_ISL_917299, EPI_ISL_917300, EPI_ISL_917301, EPI_ISL_917302, EPI_ISL_917303, EPI_ISL_917304, EPI_ISL_917305, EPI_ISL_917306, EPI_ISL_917307, EPI_ISL_917308, EPI_ISL_917309, EPI_ISL_917310, EPI_ISL_917311, EPI_ISL_917312, EPI_ISL_917313, EPI_ISL_917314, EPI_ISL_917315, EPI_ISL_917316, EPI_ISL_917317, EPI_ISL_917318, EPI_ISL_917319, EPI_ISL_917320, EPI_ISL_917321, EPI_ISL_917322, EPI_ISL_917323, EPI_ISL_917324, EPI_ISL_917325, EPI_ISL_917326, EPI_ISL_917327, EPI_ISL_917328, EPI_ISL_917329, EPI_ISL_917330, EPI_ISL_917331, EPI_ISL_917332, EPI_ISL_917333, EPI_ISL_917334, EPI_ISL_917335, EPI_ISL_917336, EPI_ISL_917337, EPI_ISL_917338, EPI_ISL_917339, EPI_ISL_917340, EPI_ISL_917341, EPI_ISL_917342, EPI_ISL_917343, EPI_ISL_917344, EPI_ISL_917345, EPI_ISL_917346, EPI_ISL_917347, EPI_ISL_917348, EPI_ISL_917349, EPI_ISL_917350, EPI_ISL_917351, EPI_ISL_917352, EPI_ISL_917353, EPI_ISL_917354, EPI_ISL_917355, EPI_ISL_917356, EPI_ISL_917357, EPI_ISL_917358, EPI_ISL_917359, EPI_ISL_917360, EPI_ISL_917361, EPI_ISL_917362, EPI_ISL_917363, EPI_ISL_917364, EPI_ISL_917365, EPI_ISL_917366, EPI_ISL_917367, EPI_ISL_917368, EPI_ISL_917369, EPI_ISL_917370, EPI_ISL_917371, EPI_ISL_917372, EPI_ISL_917373, EPI_ISL_917374, EPI_ISL_917375, EPI_ISL_917376, EPI_ISL_917377, EPI_ISL_917378, EPI_ISL_917379, EPI_ISL_917380, EPI_ISL_917381, EPI_ISL_917382, EPI_ISL_917383, EPI_ISL_917384, EPI_ISL_917385, EPI_ISL_917386, EPI_ISL_917387, EPI_ISL_917388, EPI_ISL_917389, EPI_ISL_917390, EPI_ISL_917391, EPI_ISL_917392, EPI_ISL_917393, EPI_ISL_917394, EPI_ISL_917395, EPI_ISL_917396, EPI_ISL_917397, EPI_ISL_917398, EPI_ISL_917399, EPI_ISL_917400, EPI_ISL_917401, EPI_ISL_917402, EPI_ISL_917403, EPI_ISL_917404, EPI_ISL_917405, EPI_ISL_917406, EPI_ISL_917407, EPI_ISL_917408, EPI_ISL_917409, EPI_ISL_917410, EPI_ISL_917411, EPI_ISL_917412, EPI_ISL_917413, EPI_ISL_917414, EPI_ISL_917415, EPI_ISL_917416, EPI_ISL_917417, EPI_ISL_917418, EPI_ISL_917419, EPI_ISL_917420, EPI_ISL_917421, EPI_ISL_917422, EPI_ISL_917423, EPI_ISL_917424, EPI_ISL_917425, EPI_ISL_917426, EPI_ISL_917427, EPI_ISL_917428, EPI_ISL_917429, EPI_ISL_917430, EPI_ISL_917431, EPI_ISL_917432, EPI_ISL_917433, EPI_ISL_917434, EPI_ISL_917435, EPI_ISL_917436, EPI_ISL_917437, EPI_ISL_917438, EPI_ISL_917439, EPI_ISL_917440, EPI_ISL_917441, EPI_ISL_917442, EPI_ISL_917443, EPI_ISL_917444, EPI_ISL_917445, EPI_ISL_917446, EPI_ISL_917447, EPI_ISL_917448, EPI_ISL_917449, EPI_ISL_917450, EPI_ISL_917451, EPI_ISL_917452, EPI_ISL_917453, EPI_ISL_917454, EPI_ISL_917455, EPI_ISL_917456, EPI_ISL_917457, EPI_ISL_917458, EPI_ISL_917459, EPI_ISL_917460, EPI_ISL_917461, EPI_ISL_917462, EPI_ISL_917463, EPI_ISL_917464, EPI_ISL_917465, EPI_ISL_917466, EPI_ISL_917467, EPI_ISL_917468, EPI_ISL_917469, EPI_ISL_917470, EPI_ISL_917471, EPI_ISL_917472, EPI_ISL_917473, EPI_ISL_917474, EPI_ISL_917475, EPI_ISL_917476, EPI_ISL_917477, EPI_ISL_917478, EPI_ISL_917479, EPI_ISL_917480, EPI_ISL_917481, EPI_ISL_917482, EPI_ISL_917483, EPI_ISL_917484, EPI_ISL_917485, EPI_ISL_917486, EPI_ISL_917487, EPI_ISL_917488, EPI_ISL_917489, EPI_ISL_917490, EPI_ISL_917491, EPI_ISL_917492, EPI_ISL_917493, EPI_ISL_917494, EPI_ISL_917495, EPI_ISL_917496, EPI_ISL_917497, EPI_ISL_917498, EPI_ISL_917499, EPI_ISL_917500, EPI_ISL_917502, EPI_ISL_917503, EPI_ISL_917504, EPI_ISL_917505, EPI_ISL_917506, EPI_ISL_917507, EPI_ISL_917508, EPI_ISL_917509, EPI_ISL_917510, EPI_ISL_917511, EPI_ISL_917512, EPI_ISL_917513, EPI_ISL_917514, EPI_ISL_917515, EPI_ISL_917516, EPI_ISL_917517, EPI_ISL_917518, EPI_ISL_917519, EPI_ISL_917520, EPI_ISL_917521, EPI_ISL_917522, EPI_ISL_917523, EPI_ISL_917524, EPI_ISL_917525, EPI_ISL_917526, EPI_ISL_917527, EPI_ISL_917528, EPI_ISL_917529, EPI_ISL_917530, EPI_ISL_917531, EPI_ISL_917532, EPI_ISL_917533, EPI_ISL_917534, EPI_ISL_917535, EPI_ISL_917536, EPI_ISL_917537, EPI_ISL_917538, EPI_ISL_917539, EPI_ISL_917540, EPI_ISL_917541, EPI_ISL_917542, EPI_ISL_917543, EPI_ISL_917544, EPI_ISL_917545, EPI_ISL_917546, EPI_ISL_917547, EPI_ISL_917548, EPI_ISL_917549, EPI_ISL_917550, EPI_ISL_917551, EPI_ISL_917552, EPI_ISL_917553, EPI_ISL_917554, EPI_ISL_917555, EPI_ISL_917556, EPI_ISL_917557, EPI_ISL_917558, EPI_ISL_917559, EPI_ISL_917560, EPI_ISL_917561, EPI_ISL_917562, EPI_ISL_917563, EPI_ISL_917564, EPI_ISL_917565, EPI_ISL_917566, EPI_ISL_917567, EPI_ISL_917568, EPI_ISL_917569, EPI_ISL_917570, EPI_ISL_917571, EPI_ISL_917572, EPI_ISL_917573, EPI_ISL_917574, EPI_ISL_917575, EPI_ISL_917576, EPI_ISL_917577, EPI_ISL_917578, EPI_ISL_917579, EPI_ISL_917580, EPI_ISL_917581, EPI_ISL_917582, EPI_ISL_917583, EPI_ISL_917584, EPI_ISL_917585, EPI_ISL_917586, EPI_ISL_917587, EPI_ISL_917588, EPI_ISL_917589, EPI_ISL_917590, EPI_ISL_917591, EPI_ISL_917592, EPI_ISL_917593, EPI_ISL_917594, EPI_ISL_917595, EPI_ISL_917596, EPI_ISL_917597, EPI_ISL_917598, EPI_ISL_917599, EPI_ISL_917600, EPI_ISL_917601, EPI_ISL_917602, EPI_ISL_917603, EPI_ISL_917604, EPI_ISL_917605, EPI_ISL_917606, EPI_ISL_917607, EPI_ISL_917608, EPI_ISL_917609, EPI_ISL_917610, EPI_ISL_917611, EPI_ISL_917612, EPI_ISL_917613, EPI_ISL_917614, EPI_ISL_917615, EPI_ISL_917616, EPI_ISL_917617, EPI_ISL_917618, EPI_ISL_917619, EPI_ISL_917620, EPI_ISL_917621, EPI_ISL_917622, EPI_ISL_917623, EPI_ISL_917624, EPI_ISL_917625, EPI_ISL_917626, EPI_ISL_917627, EPI_ISL_917628, EPI_ISL_917629, EPI_ISL_917630, EPI_ISL_917631, EPI_ISL_917632, EPI_ISL_917633, EPI_ISL_917634, EPI_ISL_917635, EPI_ISL_917636, EPI_ISL_917637, EPI_ISL_917638, EPI_ISL_917639, EPI_ISL_917640, EPI_ISL_917641, EPI_ISL_917642, EPI_ISL_917643, EPI_ISL_917644, EPI_ISL_917645, EPI_ISL_917646, EPI_ISL_917647, EPI_ISL_917648, EPI_ISL_917649, EPI_ISL_917650, EPI_ISL_917651, EPI_ISL_917652, EPI_ISL_917653, EPI_ISL_917654, EPI_ISL_917655, EPI_ISL_917656, EPI_ISL_917657, EPI_ISL_917658, EPI_ISL_917659, EPI_ISL_917660, EPI_ISL_917661, EPI_ISL_917662, EPI_ISL_917663, EPI_ISL_917664, EPI_ISL_917665, EPI_ISL_917666, EPI_ISL_917667, EPI_ISL_917668, EPI_ISL_917669, EPI_ISL_917670, EPI_ISL_917671, EPI_ISL_917672, EPI_ISL_917673, EPI_ISL_917674, EPI_ISL_917675, EPI_ISL_917676, EPI_ISL_917677, EPI_ISL_917678, EPI_ISL_917679, EPI_ISL_917680, EPI_ISL_917681, EPI_ISL_917682, EPI_ISL_917683, EPI_ISL_917684, EPI_ISL_917685, EPI_ISL_917686, EPI_ISL_917687, EPI_ISL_917688, EPI_ISL_917689, EPI_ISL_917690, EPI_ISL_917691, EPI_ISL_917692, EPI_ISL_917693, EPI_ISL_917694, EPI_ISL_917695, EPI_ISL_917696, EPI_ISL_917697, EPI_ISL_917698, EPI_ISL_917699, EPI_ISL_917700, EPI_ISL_917701, EPI_ISL_917702, EPI_ISL_917703, EPI_ISL_917704, EPI_ISL_917705, EPI_ISL_917706, EPI_ISL_917707, EPI_ISL_917708, EPI_ISL_917709, EPI_ISL_917710, EPI_ISL_917711, EPI_ISL_917712, EPI_ISL_917713, EPI_ISL_917714, EPI_ISL_917715, EPI_ISL_917716, EPI_ISL_917717, EPI_ISL_917718, EPI_ISL_917719, EPI_ISL_917720, EPI_ISL_917721, EPI_ISL_917722, EPI_ISL_917723, EPI_ISL_917724, EPI_ISL_917725, EPI_ISL_917726, EPI_ISL_917727, EPI_ISL_917728, EPI_ISL_917729, EPI_ISL_917730, EPI_ISL_917731, EPI_ISL_917732, EPI_ISL_917733, EPI_ISL_917734, EPI_ISL_917735, EPI_ISL_917736, EPI_ISL_917737, EPI_ISL_917738, EPI_ISL_917739, EPI_ISL_917740, EPI_ISL_917741, EPI_ISL_917742, EPI_ISL_917743, EPI_ISL_917744, EPI_ISL_917745, EPI_ISL_917746, EPI_ISL_917747, EPI_ISL_917748, EPI_ISL_917749, EPI_ISL_917750, EPI_ISL_917751, EPI_ISL_917752, EPI_ISL_917753, EPI_ISL_917754, EPI_ISL_917755, EPI_ISL_917756, EPI_ISL_917757, EPI_ISL_917758, EPI_ISL_917759, EPI_ISL_917760, EPI_ISL_917761, EPI_ISL_917762, EPI_ISL_917763, EPI_ISL_917764, EPI_ISL_917765, EPI_ISL_917766, EPI_ISL_917767, EPI_ISL_917768, EPI_ISL_917769, EPI_ISL_917770, EPI_ISL_917771, EPI_ISL_917772, EPI_ISL_917773, EPI_ISL_917774, EPI_ISL_917775, EPI_ISL_917776, EPI_ISL_917777, EPI_ISL_917778, EPI_ISL_917779, EPI_ISL_917780, EPI_ISL_917781, EPI_ISL_917782, EPI_ISL_917783, EPI_ISL_917784, EPI_ISL_917785, EPI_ISL_917786, EPI_ISL_917787, EPI_ISL_917788, EPI_ISL_917789, EPI_ISL_917790, EPI_ISL_917791, EPI_ISL_917793, EPI_ISL_917794, EPI_ISL_917795, EPI_ISL_917796, EPI_ISL_917797, EPI_ISL_917798, EPI_ISL_917799, EPI_ISL_917800, EPI_ISL_917801, EPI_ISL_917802, EPI_ISL_917803, EPI_ISL_917804, EPI_ISL_917805, EPI_ISL_917806, EPI_ISL_917807, EPI_ISL_917808, EPI_ISL_917809, EPI_ISL_917810, EPI_ISL_917811, EPI_ISL_917812, EPI_ISL_917813, EPI_ISL_917814, EPI_ISL_917815, EPI_ISL_917816, EPI_ISL_917817, EPI_ISL_917818, EPI_ISL_917819, EPI_ISL_917820, EPI_ISL_917821, EPI_ISL_917822, EPI_ISL_917823, EPI_ISL_917824, EPI_ISL_917825, EPI_ISL_917826, EPI_ISL_917827, EPI_ISL_917828, EPI_ISL_917829, EPI_ISL_917830, EPI_ISL_917831, EPI_ISL_917832, EPI_ISL_917833, EPI_ISL_917834, EPI_ISL_917835, EPI_ISL_917836, EPI_ISL_917837, EPI_ISL_917838, EPI_ISL_917839, EPI_ISL_917840, EPI_ISL_917841, EPI_ISL_917842, EPI_ISL_917843, EPI_ISL_931589, EPI_ISL_931595, EPI_ISL_931690, EPI_ISL_931726, EPI_ISL_931822, EPI_ISL_931827, EPI_ISL_931874, EPI_ISL_931918, EPI_ISL_931921, EPI_ISL_931937, EPI_ISL_931939, EPI_ISL_931943, EPI_ISL_931953, EPI_ISL_931957, EPI_ISL_931959, EPI_ISL_931979, EPI_ISL_932017, EPI_ISL_932039, EPI_ISL_932046, EPI_ISL_932057, EPI_ISL_932061, EPI_ISL_932068, EPI_ISL_932108, EPI_ISL_932114, EPI_ISL_932157, EPI_ISL_932201, EPI_ISL_932222, EPI_ISL_932231, EPI_ISL_932240, EPI_ISL_932241, EPI_ISL_932254, EPI_ISL_932261, EPI_ISL_932264, EPI_ISL_932268, EPI_ISL_932273, EPI_ISL_932274, EPI_ISL_932284, EPI_ISL_932285, EPI_ISL_932287, EPI_ISL_932290, EPI_ISL_932292, EPI_ISL_932297, EPI_ISL_932300, EPI_ISL_932303, EPI_ISL_932310, EPI_ISL_932313, EPI_ISL_932325, EPI_ISL_932334, EPI_ISL_932345, EPI_ISL_932348, EPI_ISL_932357, EPI_ISL_932371, EPI_ISL_932376, EPI_ISL_932388, EPI_ISL_932393, EPI_ISL_932425, EPI_ISL_932433, EPI_ISL_932436, EPI_ISL_932440, EPI_ISL_932444, EPI_ISL_932454, EPI_ISL_932455, EPI_ISL_932457, EPI_ISL_932458, EPI_ISL_932463, EPI_ISL_932470, EPI_ISL_932473, EPI_ISL_932475, EPI_ISL_932479, EPI_ISL_932480, EPI_ISL_932486, EPI_ISL_932489, EPI_ISL_932502, EPI_ISL_932503, EPI_ISL_932511, EPI_ISL_932515, EPI_ISL_932517, EPI_ISL_932522, EPI_ISL_932530, EPI_ISL_932532, EPI_ISL_932539, EPI_ISL_932543, EPI_ISL_932552, EPI_ISL_932567, EPI_ISL_932599, EPI_ISL_932607, EPI_ISL_932620, EPI_ISL_932640, EPI_ISL_932645, EPI_ISL_932647, EPI_ISL_932661, EPI_ISL_932668, EPI_ISL_932670, EPI_ISL_932673, EPI_ISL_932690, EPI_ISL_932696, EPI_ISL_932704, EPI_ISL_932709, EPI_ISL_932725, EPI_ISL_932734, EPI_ISL_932743, EPI_ISL_932749, EPI_ISL_932753, EPI_ISL_932760, EPI_ISL_932768, EPI_ISL_932791, EPI_ISL_932800, EPI_ISL_932814, EPI_ISL_932817, EPI_ISL_932818, EPI_ISL_932834, EPI_ISL_932835, EPI_ISL_932840, EPI_ISL_932844, EPI_ISL_932862, EPI_ISL_932866, EPI_ISL_932867, EPI_ISL_932868, EPI_ISL_932876, EPI_ISL_932878, EPI_ISL_932881, EPI_ISL_932882, EPI_ISL_932883, EPI_ISL_932889, EPI_ISL_932890, EPI_ISL_932891, EPI_ISL_932893, EPI_ISL_932900, EPI_ISL_932901, EPI_ISL_932902, EPI_ISL_932906, EPI_ISL_932907, EPI_ISL_932911, EPI_ISL_932912, EPI_ISL_932922, EPI_ISL_932923, EPI_ISL_932924, EPI_ISL_932927, EPI_ISL_932935, EPI_ISL_932937, EPI_ISL_932938, EPI_ISL_932940, EPI_ISL_932942, EPI_ISL_932943, EPI_ISL_932947, EPI_ISL_932952, EPI_ISL_932956, EPI_ISL_932964, EPI_ISL_932972, EPI_ISL_932973, EPI_ISL_932975, EPI_ISL_932976, EPI_ISL_932979, EPI_ISL_932981, EPI_ISL_932982, EPI_ISL_932984, EPI_ISL_932988, EPI_ISL_932990, EPI_ISL_932993, EPI_ISL_933001, EPI_ISL_933003, EPI_ISL_933010, EPI_ISL_933013, EPI_ISL_933015, EPI_ISL_933024, EPI_ISL_933025, EPI_ISL_933026, EPI_ISL_933029, EPI_ISL_933031, EPI_ISL_933041, EPI_ISL_933045, EPI_ISL_933049, EPI_ISL_933052, EPI_ISL_933059, EPI_ISL_933062, EPI_ISL_933064, EPI_ISL_933066, EPI_ISL_933083, EPI_ISL_933084, EPI_ISL_933086, EPI_ISL_933087, EPI_ISL_933089, EPI_ISL_933090, EPI_ISL_933092, EPI_ISL_933093, EPI_ISL_933094, EPI_ISL_933095, EPI_ISL_933097, EPI_ISL_933098, EPI_ISL_933102, EPI_ISL_933103, EPI_ISL_933104, EPI_ISL_933105, EPI_ISL_933106, EPI_ISL_933111, EPI_ISL_933112, EPI_ISL_937549, EPI_ISL_937550, EPI_ISL_937551, EPI_ISL_937553, EPI_ISL_937554, EPI_ISL_937557, EPI_ISL_937558, EPI_ISL_937559, EPI_ISL_937562, EPI_ISL_937564, EPI_ISL_937568, EPI_ISL_937569, EPI_ISL_937570, EPI_ISL_937571, EPI_ISL_937573, EPI_ISL_937576, EPI_ISL_937578, EPI_ISL_937580, EPI_ISL_937581, EPI_ISL_937583, EPI_ISL_937586, EPI_ISL_937587, EPI_ISL_937588, EPI_ISL_937590, EPI_ISL_937592, EPI_ISL_937594, EPI_ISL_937595, EPI_ISL_937596, EPI_ISL_937599, EPI_ISL_937601, EPI_ISL_937604, EPI_ISL_937605, EPI_ISL_937609, EPI_ISL_937610, EPI_ISL_937612, EPI_ISL_937614, EPI_ISL_937616, EPI_ISL_937618, EPI_ISL_937620, EPI_ISL_937621, EPI_ISL_937623, EPI_ISL_937627, EPI_ISL_937630, EPI_ISL_937634, EPI_ISL_937636, EPI_ISL_937638, EPI_ISL_937639, EPI_ISL_937640, EPI_ISL_937641, EPI_ISL_937643, EPI_ISL_937648, EPI_ISL_937649, EPI_ISL_937650, EPI_ISL_937651, EPI_ISL_937653, EPI_ISL_937658, EPI_ISL_937663, EPI_ISL_937664, EPI_ISL_937665, EPI_ISL_937666, EPI_ISL_937668, EPI_ISL_937669, EPI_ISL_937670, EPI_ISL_937672, EPI_ISL_937674, EPI_ISL_937675, EPI_ISL_937680, EPI_ISL_937682, EPI_ISL_937685, EPI_ISL_937687, EPI_ISL_937688, EPI_ISL_937689, EPI_ISL_937693, EPI_ISL_937697, EPI_ISL_937698, EPI_ISL_937700, EPI_ISL_937703, EPI_ISL_937704, EPI_ISL_937707, EPI_ISL_937709, EPI_ISL_937710, EPI_ISL_937711, EPI_ISL_937712, EPI_ISL_937714, EPI_ISL_937715, EPI_ISL_937716, EPI_ISL_937717, EPI_ISL_937720, EPI_ISL_937721, EPI_ISL_937724, EPI_ISL_937726, EPI_ISL_937730, EPI_ISL_937734, EPI_ISL_937737, EPI_ISL_937738, EPI_ISL_937741, EPI_ISL_937742, EPI_ISL_937744, EPI_ISL_937745, EPI_ISL_937746, EPI_ISL_937748, EPI_ISL_937749, EPI_ISL_937751, EPI_ISL_937753, EPI_ISL_937761, EPI_ISL_937762, EPI_ISL_937763, EPI_ISL_937765, EPI_ISL_937766, EPI_ISL_937767, EPI_ISL_937768, EPI_ISL_937769, EPI_ISL_937770, EPI_ISL_937777, EPI_ISL_937780, EPI_ISL_937781, EPI_ISL_937784, EPI_ISL_937787, EPI_ISL_937792, EPI_ISL_937794, EPI_ISL_937798, EPI_ISL_937799, EPI_ISL_937800, EPI_ISL_937801, EPI_ISL_937802, EPI_ISL_937803, EPI_ISL_937805, EPI_ISL_937806, EPI_ISL_937808, EPI_ISL_937811, EPI_ISL_937815, EPI_ISL_937817, EPI_ISL_937820, EPI_ISL_937822, EPI_ISL_937824, EPI_ISL_937826, EPI_ISL_937829, EPI_ISL_937830, EPI_ISL_937831, EPI_ISL_937835, EPI_ISL_937837, EPI_ISL_937839, EPI_ISL_937840, EPI_ISL_937843, EPI_ISL_937849, EPI_ISL_937850, EPI_ISL_937852, EPI_ISL_937853, EPI_ISL_937854, EPI_ISL_937860, EPI_ISL_937862, EPI_ISL_937863, EPI_ISL_937864, EPI_ISL_937865, EPI_ISL_937867, EPI_ISL_937872, EPI_ISL_937873, EPI_ISL_937874, EPI_ISL_937875, EPI_ISL_937877, EPI_ISL_937881, EPI_ISL_937883, EPI_ISL_938940, EPI_ISL_939604, EPI_ISL_939607, EPI_ISL_946421, EPI_ISL_946427, EPI_ISL_946445, EPI_ISL_946454, EPI_ISL_946471, EPI_ISL_946493, EPI_ISL_946547, EPI_ISL_946580, EPI_ISL_946585, EPI_ISL_946646, EPI_ISL_946672, EPI_ISL_946956, EPI_ISL_946963, EPI_ISL_946992, EPI_ISL_947406, EPI_ISL_947407, EPI_ISL_947408, EPI_ISL_947409, EPI_ISL_947410, EPI_ISL_947411, EPI_ISL_947412, EPI_ISL_947413, EPI_ISL_947414, EPI_ISL_947415, EPI_ISL_947416, EPI_ISL_947417, EPI_ISL_947418, EPI_ISL_947419, EPI_ISL_947420, EPI_ISL_947421, EPI_ISL_947422, EPI_ISL_947423, EPI_ISL_947424, EPI_ISL_947425, EPI_ISL_947426, EPI_ISL_947427, EPI_ISL_947428, EPI_ISL_947429, EPI_ISL_947430, EPI_ISL_947431, EPI_ISL_947432, EPI_ISL_947433, EPI_ISL_947434, EPI_ISL_947435, EPI_ISL_947436, EPI_ISL_947437, EPI_ISL_947438, EPI_ISL_947439, EPI_ISL_947440, EPI_ISL_947441, EPI_ISL_947442, EPI_ISL_947443, EPI_ISL_947444, EPI_ISL_947445, EPI_ISL_947446, EPI_ISL_947447, EPI_ISL_947448, EPI_ISL_947449, EPI_ISL_947450, EPI_ISL_947451, EPI_ISL_947452, EPI_ISL_947453, EPI_ISL_947454, EPI_ISL_947455, EPI_ISL_947456, EPI_ISL_947457, EPI_ISL_947458, EPI_ISL_947459, EPI_ISL_947460, EPI_ISL_947461, EPI_ISL_947462, EPI_ISL_947463, EPI_ISL_947464, EPI_ISL_947465, EPI_ISL_947466, EPI_ISL_947467, EPI_ISL_947468, EPI_ISL_947469, EPI_ISL_947470, EPI_ISL_947471, EPI_ISL_947472, EPI_ISL_947473, EPI_ISL_947474, EPI_ISL_947475, EPI_ISL_947476, EPI_ISL_947477, EPI_ISL_947478, EPI_ISL_947479, EPI_ISL_947480, EPI_ISL_947481, EPI_ISL_947482, EPI_ISL_947483, EPI_ISL_947484, EPI_ISL_947485, EPI_ISL_947486, EPI_ISL_947487, EPI_ISL_947488, EPI_ISL_947489, EPI_ISL_947490, EPI_ISL_947491, | | | |

EPI_ISL_947492, EPI_ISL_947493, EPI_ISL_947494, EPI_ISL_947495, EPI_ISL_947496, EPI_ISL_947497, EPI_ISL_947498, EPI_ISL_947499, EPI_ISL_947500, EPI_ISL_947501, EPI_ISL_947502, EPI_ISL_947503, EPI_ISL_947504, EPI_ISL_947505, EPI_ISL_947506, EPI_ISL_947507, EPI_ISL_947508, EPI_ISL_947509, EPI_ISL_947510, EPI_ISL_947511, EPI_ISL_947512, EPI_ISL_947513, EPI_ISL_947514, EPI_ISL_947515,
EPI_ISL_947516, EPI_ISL_947517, EPI_ISL_947518, EPI_ISL_947519, EPI_ISL_947520, EPI_ISL_947521, EPI_ISL_947522, EPI_ISL_947523, EPI_ISL_947524, EPI_ISL_947525, EPI_ISL_947526, EPI_ISL_947527, EPI_ISL_947528, EPI_ISL_947529, EPI_ISL_947530, EPI_ISL_947531, EPI_ISL_947532, EPI_ISL_947533, EPI_ISL_947534, EPI_ISL_947535, EPI_ISL_947536, EPI_ISL_947537, EPI_ISL_947538, EPI_ISL_947539,
EPI_ISL_947540, EPI_ISL_947541, EPI_ISL_947542, EPI_ISL_947543, EPI_ISL_947544, EPI_ISL_947545, EPI_ISL_947546, EPI_ISL_947547, EPI_ISL_947548, EPI_ISL_947549, EPI_ISL_947550, EPI_ISL_947551, EPI_ISL_947552, EPI_ISL_947553, EPI_ISL_947554, EPI_ISL_947555, EPI_ISL_947556, EPI_ISL_947557, EPI_ISL_947558, EPI_ISL_947559, EPI_ISL_947560, EPI_ISL_947561, EPI_ISL_947562, EPI_ISL_947563,
EPI_ISL_947564, EPI_ISL_947565, EPI_ISL_947566, EPI_ISL_947567, EPI_ISL_947568, EPI_ISL_947569, EPI_ISL_947570, EPI_ISL_947571, EPI_ISL_947572, EPI_ISL_947573, EPI_ISL_947574, EPI_ISL_947575, EPI_ISL_947576, EPI_ISL_947577, EPI_ISL_947578, EPI_ISL_947579, EPI_ISL_947580, EPI_ISL_947581, EPI_ISL_947582, EPI_ISL_947583, EPI_ISL_947584, EPI_ISL_947585, EPI_ISL_947586, EPI_ISL_947587,
EPI_ISL_947588, EPI_ISL_947589, EPI_ISL_947590, EPI_ISL_947591, EPI_ISL_947592, EPI_ISL_947593, EPI_ISL_947594, EPI_ISL_947595, EPI_ISL_947596, EPI_ISL_947597, EPI_ISL_947598, EPI_ISL_947599, EPI_ISL_947600, EPI_ISL_947601, EPI_ISL_947602, EPI_ISL_947603, EPI_ISL_947604, EPI_ISL_947605, EPI_ISL_947606, EPI_ISL_947607, EPI_ISL_947608, EPI_ISL_947609, EPI_ISL_947610, EPI_ISL_947611,
EPI_ISL_947612, EPI_ISL_947613, EPI_ISL_947614, EPI_ISL_947615, EPI_ISL_947616, EPI_ISL_947617, EPI_ISL_947618, EPI_ISL_947619, EPI_ISL_947620, EPI_ISL_947621, EPI_ISL_947622, EPI_ISL_947623, EPI_ISL_947624, EPI_ISL_947625, EPI_ISL_947626, EPI_ISL_947627, EPI_ISL_947628, EPI_ISL_947629, EPI_ISL_947630, EPI_ISL_947631, EPI_ISL_947632, EPI_ISL_947633, EPI_ISL_947634, EPI_ISL_947635,
EPI_ISL_947636, EPI_ISL_947637, EPI_ISL_947638, EPI_ISL_947639, EPI_ISL_947640, EPI_ISL_947641, EPI_ISL_947642, EPI_ISL_947643, EPI_ISL_947644, EPI_ISL_947645, EPI_ISL_947646, EPI_ISL_947647, EPI_ISL_947648, EPI_ISL_947650, EPI_ISL_947651, EPI_ISL_947652, EPI_ISL_947653, EPI_ISL_947654, EPI_ISL_947655, EPI_ISL_947656, EPI_ISL_947657, EPI_ISL_947658, EPI_ISL_947659, EPI_ISL_947660,
EPI_ISL_947661, EPI_ISL_947662, EPI_ISL_947663, EPI_ISL_947664, EPI_ISL_947665, EPI_ISL_947666, EPI_ISL_947667, EPI_ISL_947668, EPI_ISL_947669, EPI_ISL_947670, EPI_ISL_947671, EPI_ISL_947672, EPI_ISL_947673, EPI_ISL_947674, EPI_ISL_947675, EPI_ISL_947676, EPI_ISL_947677, EPI_ISL_947679, EPI_ISL_947680, EPI_ISL_947681, EPI_ISL_947682, EPI_ISL_947683, EPI_ISL_947684, EPI_ISL_947685,
EPI_ISL_947686, EPI_ISL_947687, EPI_ISL_947688, EPI_ISL_947689, EPI_ISL_947690, EPI_ISL_947691, EPI_ISL_947692, EPI_ISL_947693, EPI_ISL_947694, EPI_ISL_947695, EPI_ISL_947696, EPI_ISL_947697, EPI_ISL_947698, EPI_ISL_947699, EPI_ISL_947700, EPI_ISL_947701, EPI_ISL_947702, EPI_ISL_947703, EPI_ISL_947704, EPI_ISL_947705, EPI_ISL_947706, EPI_ISL_947707, EPI_ISL_947708, EPI_ISL_947709,
EPI_ISL_947710, EPI_ISL_947711, EPI_ISL_947712, EPI_ISL_947713, EPI_ISL_947714, EPI_ISL_947715, EPI_ISL_947716, EPI_ISL_947717, EPI_ISL_947718, EPI_ISL_947719, EPI_ISL_947720, EPI_ISL_947721, EPI_ISL_947722, EPI_ISL_947723, EPI_ISL_947724, EPI_ISL_947725, EPI_ISL_947726, EPI_ISL_947727, EPI_ISL_947728, EPI_ISL_947729, EPI_ISL_947730, EPI_ISL_947731, EPI_ISL_947732, EPI_ISL_947733,
EPI_ISL_947734, EPI_ISL_947735, EPI_ISL_947736, EPI_ISL_947737, EPI_ISL_947738, EPI_ISL_947739, EPI_ISL_947740, EPI_ISL_947741, EPI_ISL_947742, EPI_ISL_947743, EPI_ISL_947744, EPI_ISL_947745, EPI_ISL_947746, EPI_ISL_947747, EPI_ISL_947748, EPI_ISL_947749, EPI_ISL_947750, EPI_ISL_947751, EPI_ISL_947752, EPI_ISL_947753, EPI_ISL_947754, EPI_ISL_947755, EPI_ISL_947756, EPI_ISL_947757,
EPI_ISL_947758, EPI_ISL_947759, EPI_ISL_947760, EPI_ISL_947761, EPI_ISL_947762, EPI_ISL_947763, EPI_ISL_947764, EPI_ISL_947765, EPI_ISL_947766, EPI_ISL_947767, EPI_ISL_947768, EPI_ISL_947769, EPI_ISL_947770, EPI_ISL_947771, EPI_ISL_947772, EPI_ISL_947773, EPI_ISL_947774, EPI_ISL_947775, EPI_ISL_947776, EPI_ISL_947777, EPI_ISL_947778, EPI_ISL_947779, EPI_ISL_947780, EPI_ISL_947781,
EPI_ISL_947782, EPI_ISL_947783, EPI_ISL_947784, EPI_ISL_947785, EPI_ISL_947786, EPI_ISL_947787, EPI_ISL_947788, EPI_ISL_947789, EPI_ISL_947790, EPI_ISL_947791, EPI_ISL_947792, EPI_ISL_947793, EPI_ISL_947794, EPI_ISL_947795, EPI_ISL_947796, EPI_ISL_947797, EPI_ISL_947798, EPI_ISL_947799, EPI_ISL_947800, EPI_ISL_947801, EPI_ISL_947802, EPI_ISL_947803, EPI_ISL_947804, EPI_ISL_947805,
EPI_ISL_947806, EPI_ISL_947807, EPI_ISL_947808, EPI_ISL_947809, EPI_ISL_947810, EPI_ISL_947811, EPI_ISL_947812, EPI_ISL_947813, EPI_ISL_947814, EPI_ISL_947815, EPI_ISL_947816, EPI_ISL_947817, EPI_ISL_947818, EPI_ISL_947819, EPI_ISL_947820, EPI_ISL_947821, EPI_ISL_947822, EPI_ISL_947823, EPI_ISL_947824, EPI_ISL_947825, EPI_ISL_947826, EPI_ISL_947827, EPI_ISL_947828, EPI_ISL_947829,
EPI_ISL_947830, EPI_ISL_947831, EPI_ISL_947832, EPI_ISL_947833, EPI_ISL_947834, EPI_ISL_947835, EPI_ISL_947836, EPI_ISL_947837, EPI_ISL_947838, EPI_ISL_947839, EPI_ISL_947840, EPI_ISL_947841, EPI_ISL_947842, EPI_ISL_947843, EPI_ISL_947844, EPI_ISL_947845, EPI_ISL_947846, EPI_ISL_947847, EPI_ISL_947848, EPI_ISL_947849, EPI_ISL_947850, EPI_ISL_947851, EPI_ISL_947852, EPI_ISL_947853,
EPI_ISL_947854, EPI_ISL_947855, EPI_ISL_947856, EPI_ISL_947857, EPI_ISL_947858, EPI_ISL_947859, EPI_ISL_947860, EPI_ISL_947861, EPI_ISL_947862, EPI_ISL_947863, EPI_ISL_947864, EPI_ISL_947865, EPI_ISL_947866, EPI_ISL_947867, EPI_ISL_947868, EPI_ISL_947869, EPI_ISL_947870, EPI_ISL_947871, EPI_ISL_947872, EPI_ISL_947873, EPI_ISL_947874, EPI_ISL_947875, EPI_ISL_947876, EPI_ISL_947877,
EPI_ISL_947878, EPI_ISL_947879, EPI_ISL_947880, EPI_ISL_947881, EPI_ISL_947882, EPI_ISL_947883, EPI_ISL_947884, EPI_ISL_947885, EPI_ISL_947886, EPI_ISL_947887, EPI_ISL_947888, EPI_ISL_947889, EPI_ISL_947890, EPI_ISL_947891, EPI_ISL_947892, EPI_ISL_947893, EPI_ISL_947894, EPI_ISL_947895, EPI_ISL_947896, EPI_ISL_947897, EPI_ISL_947898, EPI_ISL_947899, EPI_ISL_947900, EPI_ISL_947901,
EPI_ISL_947902, EPI_ISL_947903, EPI_ISL_947904, EPI_ISL_947905, EPI_ISL_947906, EPI_ISL_947907, EPI_ISL_947908, EPI_ISL_947909, EPI_ISL_947910, EPI_ISL_947911, EPI_ISL_947912, EPI_ISL_947913, EPI_ISL_947914, EPI_ISL_947915, EPI_ISL_947916, EPI_ISL_947917, EPI_ISL_947918, EPI_ISL_947919, EPI_ISL_947920, EPI_ISL_947921, EPI_ISL_947922, EPI_ISL_947923, EPI_ISL_947924, EPI_ISL_947925,
EPI_ISL_947926, EPI_ISL_947927, EPI_ISL_947928, EPI_ISL_947929, EPI_ISL_947930, EPI_ISL_947931, EPI_ISL_947932, EPI_ISL_947933, EPI_ISL_947934, EPI_ISL_947935, EPI_ISL_947936, EPI_ISL_947937, EPI_ISL_947938, EPI_ISL_947939, EPI_ISL_947940, EPI_ISL_947941, EPI_ISL_947942, EPI_ISL_947943, EPI_ISL_947944, EPI_ISL_947945, EPI_ISL_947946, EPI_ISL_947947, EPI_ISL_947948, EPI_ISL_947949,
EPI_ISL_947950, EPI_ISL_947951, EPI_ISL_947952, EPI_ISL_947953, EPI_ISL_947954, EPI_ISL_947955, EPI_ISL_947956, EPI_ISL_947957, EPI_ISL_947958, EPI_ISL_947959, EPI_ISL_947960, EPI_ISL_947961, EPI_ISL_947962, EPI_ISL_947963, EPI_ISL_947964, EPI_ISL_947965, EPI_ISL_947966, EPI_ISL_947967, EPI_ISL_947968, EPI_ISL_947969, EPI_ISL_947970, EPI_ISL_947971, EPI_ISL_947972, EPI_ISL_947973,
EPI_ISL_947974, EPI_ISL_947975, EPI_ISL_947976, EPI_ISL_947977, EPI_ISL_947978, EPI_ISL_947979, EPI_ISL_947980, EPI_ISL_947981, EPI_ISL_947982, EPI_ISL_947983, EPI_ISL_947984, EPI_ISL_947985, EPI_ISL_947986, EPI_ISL_947987, EPI_ISL_947988, EPI_ISL_947989, EPI_ISL_947990, EPI_ISL_947991, EPI_ISL_947992, EPI_ISL_947993, EPI_ISL_947994, EPI_ISL_947995, EPI_ISL_947996, EPI_ISL_947997,
EPI_ISL_947998, EPI_ISL_947999, EPI_ISL_948000, EPI_ISL_948001, EPI_ISL_948002, EPI_ISL_948003, EPI_ISL_948004, EPI_ISL_948005, EPI_ISL_948006, EPI_ISL_948007, EPI_ISL_948008, EPI_ISL_948009, EPI_ISL_948010, EPI_ISL_948011, EPI_ISL_948012, EPI_ISL_948013, EPI_ISL_948014, EPI_ISL_948015, EPI_ISL_948016, EPI_ISL_948017, EPI_ISL_948018, EPI_ISL_948019, EPI_ISL_948020, EPI_ISL_948021,
EPI_ISL_948022, EPI_ISL_948023, EPI_ISL_948024, EPI_ISL_948025, EPI_ISL_948026, EPI_ISL_948027, EPI_ISL_948028, EPI_ISL_948029, EPI_ISL_948030, EPI_ISL_948031, EPI_ISL_948032, EPI_ISL_948033, EPI_ISL_948034, EPI_ISL_948035, EPI_ISL_948036, EPI_ISL_948037, EPI_ISL_948038, EPI_ISL_948039, EPI_ISL_948040, EPI_ISL_948041, EPI_ISL_948042, EPI_ISL_948043, EPI_ISL_948044, EPI_ISL_948045,
EPI_ISL_948046, EPI_ISL_948047, EPI_ISL_948048, EPI_ISL_948049, EPI_ISL_948050, EPI_ISL_948051, EPI_ISL_948052, EPI_ISL_948053, EPI_ISL_948054, EPI_ISL_948055, EPI_ISL_948056, EPI_ISL_948057, EPI_ISL_948058, EPI_ISL_948059, EPI_ISL_948060, EPI_ISL_948061, EPI_ISL_948062, EPI_ISL_948063, EPI_ISL_948064, EPI_ISL_948065, EPI_ISL_948066, EPI_ISL_948067, EPI_ISL_948068, EPI_ISL_948069,
EPI_ISL_948070, EPI_ISL_948071, EPI_ISL_948072, EPI_ISL_948073, EPI_ISL_948074, EPI_ISL_948075, EPI_ISL_948076, EPI_ISL_948077, EPI_ISL_948078, EPI_ISL_948079, EPI_ISL_948080, EPI_ISL_948081, EPI_ISL_948082, EPI_ISL_948083, EPI_ISL_948084, EPI_ISL_948085, EPI_ISL_948086, EPI_ISL_948087, EPI_ISL_948088, EPI_ISL_948089, EPI_ISL_948090, EPI_ISL_948091, EPI_ISL_948092, EPI_ISL_948093,
EPI_ISL_948094, EPI_ISL_948095, EPI_ISL_948096, EPI_ISL_948097, EPI_ISL_948098, EPI_ISL_948449, EPI_ISL_948450, EPI_ISL_948451, EPI_ISL_948452, EPI_ISL_948453, EPI_ISL_948454, EPI_ISL_948455, EPI_ISL_948456, EPI_ISL_948457, EPI_ISL_948458, EPI_ISL_948459, EPI_ISL_948460, EPI_ISL_948461, EPI_ISL_948462, EPI_ISL_948463, EPI_ISL_948464, EPI_ISL_948465, EPI_ISL_948466,
EPI_ISL_948467, EPI_ISL_948468, EPI_ISL_948469, EPI_ISL_948470, EPI_ISL_948471, EPI_ISL_948472, EPI_ISL_948473, EPI_ISL_948474, EPI_ISL_948475, EPI_ISL_948476, EPI_ISL_948477, EPI_ISL_948478, EPI_ISL_948479, EPI_ISL_948480, EPI_ISL_948481, EPI_ISL_948482, EPI_ISL_948483, EPI_ISL_948484, EPI_ISL_948485, EPI_ISL_948486, EPI_ISL_948487, EPI_ISL_948488, EPI_ISL_948489, EPI_ISL_948490,
EPI_ISL_948491, EPI_ISL_948492, EPI_ISL_948493, EPI_ISL_948494, EPI_ISL_948495, EPI_ISL_948496, EPI_ISL_948497, EPI_ISL_948498, EPI_ISL_948499, EPI_ISL_948500, EPI_ISL_948501, EPI_ISL_948502, EPI_ISL_948503, EPI_ISL_948504, EPI_ISL_948505, EPI_ISL_948506, EPI_ISL_948507, EPI_ISL_948508, EPI_ISL_948509, EPI_ISL_948510, EPI_ISL_948511, EPI_ISL_948512, EPI_ISL_948513, EPI_ISL_948514,
EPI_ISL_948515, EPI_ISL_948516, EPI_ISL_948517, EPI_ISL_948518, EPI_ISL_948519, EPI_ISL_948520, EPI_ISL_948521, EPI_ISL_948522, EPI_ISL_948523, EPI_ISL_948524, EPI_ISL_948525, EPI_ISL_948526, EPI_ISL_948527, EPI_ISL_948528, EPI_ISL_948529, EPI_ISL_948530, EPI_ISL_948531, EPI_ISL_948532, EPI_ISL_948533, EPI_ISL_948534, EPI_ISL_948535, EPI_ISL_948536, EPI_ISL_948537, EPI_ISL_948538,
EPI_ISL_948539, EPI_ISL_948540, EPI_ISL_948541, EPI_ISL_948542, EPI_ISL_948543, EPI_ISL_948544, EPI_ISL_948545, EPI_ISL_948546, EPI_ISL_948547, EPI_ISL_948548, EPI_ISL_948549, EPI_ISL_948550, EPI_ISL_948551, EPI_ISL_948552, EPI_ISL_948553, EPI_ISL_948554, EPI_ISL_948555, EPI_ISL_948556, EPI_ISL_948557, EPI_ISL_948558, EPI_ISL_948559, EPI_ISL_948560, EPI_ISL_948561, EPI_ISL_948562,
EPI_ISL_948563, EPI_ISL_948564, EPI_ISL_948565, EPI_ISL_948566, EPI_ISL_948567, EPI_ISL_948568, EPI_ISL_948569, EPI_ISL_948570, EPI_ISL_948571, EPI_ISL_948572, EPI_ISL_948573, EPI_ISL_948574, EPI_ISL_948575, EPI_ISL_948576, EPI_ISL_948577, EPI_ISL_948578, EPI_ISL_948579, EPI_ISL_948580, EPI_ISL_948581, EPI_ISL_948582, EPI_ISL_948583, EPI_ISL_948584, EPI_ISL_948585, EPI_ISL_948586,
EPI_ISL_948587, EPI_ISL_948588, EPI_ISL_948589, EPI_ISL_948590, EPI_ISL_948591, EPI_ISL_948592, EPI_ISL_948593, EPI_ISL_948594, EPI_ISL_948595, EPI_ISL_948596, EPI_ISL_948597, EPI_ISL_948598, EPI_ISL_948599, EPI_ISL_948600, EPI_ISL_948601, EPI_ISL_948602, EPI_ISL_948603, EPI_ISL_948604, EPI_ISL_948605, EPI_ISL_948606, EPI_ISL_948607, EPI_ISL_948608, EPI_ISL_948609, EPI_ISL_948610,
EPI_ISL_948611, EPI_ISL_948612, EPI_ISL_948613, EPI_ISL_948614, EPI_ISL_948615, EPI_ISL_948616, EPI_ISL_948617, EPI_ISL_948618, EPI_ISL_948619, EPI_ISL_948620, EPI_ISL_948621, EPI_ISL_948622, EPI_ISL_948623, EPI_ISL_948624, EPI_ISL_948625, EPI_ISL_948626, EPI_ISL_948627, EPI_ISL_948628, EPI_ISL_948629, EPI_ISL_948630, EPI_ISL_948631, EPI_ISL_948632, EPI_ISL_948633, EPI_ISL_948634,
EPI_ISL_948635, EPI_ISL_948636, EPI_ISL_948637, EPI_ISL_948638, EPI_ISL_948639, EPI_ISL_948640, EPI_ISL_948641, EPI_ISL_948642, EPI_ISL_948643, EPI_ISL_948644, EPI_ISL_948645, EPI_ISL_948646, EPI_ISL_948647, EPI_ISL_948648, EPI_ISL_948649, EPI_ISL_948650, EPI_ISL_948651, EPI_ISL_948652, EPI_ISL_948653, EPI_ISL_948654, EPI_ISL_948655, EPI_ISL_948656, EPI_ISL_948657, EPI_ISL_948658,
EPI_ISL_948659, EPI_ISL_948660, EPI_ISL_948661, EPI_ISL_948662, EPI_ISL_948663, EPI_ISL_948664, EPI_ISL_948665, EPI_ISL_948666, EPI_ISL_948667, EPI_ISL_948668, EPI_ISL_948669, EPI_ISL_948670, EPI_ISL_948671, EPI_ISL_948672, EPI_ISL_948673, EPI_ISL_948674, EPI_ISL_948675, EPI_ISL_948676, EPI_ISL_948677, EPI_ISL_948678, EPI_ISL_948679, EPI_ISL_948680, EPI_ISL_948681, EPI_ISL_948682,
EPI_ISL_948683, EPI_ISL_948684, EPI_ISL_948685, EPI_ISL_948686, EPI_ISL_948687, EPI_ISL_948688, EPI_ISL_948689, EPI_ISL_948690, EPI_ISL_948691, EPI_ISL_948692, EPI_ISL_948693, EPI_ISL_948694, EPI_ISL_948695, EPI_ISL_948696, EPI_ISL_948697, EPI_ISL_948698, EPI_ISL_948699, EPI_ISL_948700, EPI_ISL_948701, EPI_ISL_948702, EPI_ISL_948703, EPI_ISL_948704, EPI_ISL_948705, EPI_ISL_948706,
EPI_ISL_948707, EPI_ISL_948708, EPI_ISL_948709, EPI_ISL_948710, EPI_ISL_948711, EPI_ISL_948712, EPI_ISL_948713, EPI_ISL_948714, EPI_ISL_948715, EPI_ISL_948716, EPI_ISL_948717, EPI_ISL_948718, EPI_ISL_948719, EPI_ISL_948720, EPI_ISL_948721, EPI_ISL_948722, EPI_ISL_948723, EPI_ISL_948724, EPI_ISL_948725, EPI_ISL_948726, EPI_ISL_948727, EPI_ISL_948728, EPI_ISL_948729, EPI_ISL_948730,
EPI_ISL_948731, EPI_ISL_948732, EPI_ISL_948733, EPI_ISL_948734, EPI_ISL_948735, EPI_ISL_948736, EPI_ISL_948737, EPI_ISL_948738, EPI_ISL_948739, EPI_ISL_948740, EPI_ISL_948741, EPI_ISL_948742, EPI_ISL_948743, EPI_ISL_948744, EPI_ISL_948745, EPI_ISL_948746, EPI_ISL_948747, EPI_ISL_948748, EPI_ISL_948749, EPI_ISL_948750, EPI_ISL_948751, EPI_ISL_948752, EPI_ISL_948753, EPI_ISL_948754,
EPI_ISL_948755, EPI_ISL_948756, EPI_ISL_948757, EPI_ISL_948758, EPI_ISL_948759, EPI_ISL_948760, EPI_ISL_948761, EPI_ISL_948762, EPI_ISL_948763, EPI_ISL_948764, EPI_ISL_948765, EPI_ISL_948766, EPI_ISL_948767, EPI_ISL_948768, EPI_ISL_948769, EPI_ISL_948770, EPI_ISL_948771, EPI_ISL_948772, EPI_ISL_948773, EPI_ISL_948774, EPI_ISL_948775, EPI_ISL_948776, EPI_ISL_948777, EPI_ISL_948778,
EPI_ISL_948779, EPI_ISL_948780, EPI_ISL_948781, EPI_ISL_948782, EPI_ISL_948783, EPI_ISL_948784, EPI_ISL_948785, EPI_ISL_948786, EPI_ISL_990003, EPI_ISL_990043, EPI_ISL_990044, EPI_ISL_990045, EPI_ISL_990046, EPI_ISL_990047, EPI_ISL_990048, EPI_ISL_990049, EPI_ISL_990050, EPI_ISL_990051, EPI_ISL_990052, EPI_ISL_990053, EPI_ISL_990054, EPI_ISL_990055, EPI_ISL_990056, EPI_ISL_990057,
EPI_ISL_990286, EPI_ISL_990435, EPI_ISL_990500, EPI_ISL_990502, EPI_ISL_990510, EPI_ISL_990557, EPI_ISL_990571, EPI_ISL_990576, EPI_ISL_990579, EPI_ISL_990616, EPI_ISL_990618, EPI_ISL_990652, EPI_ISL_990658, EPI_ISL_990662, EPI_ISL_990664, EPI_ISL_990680, EPI_ISL_990712, EPI_ISL_990726, EPI_ISL_990745, EPI_ISL_990779, EPI_ISL_990881, EPI_ISL_991174, EPI_ISL_991175, EPI_ISL_991176,
EPI_ISL_991177, EPI_ISL_991179, EPI_ISL_991180, EPI_ISL_991181, EPI_ISL_991182, EPI_ISL_991183, EPI_ISL_991184, EPI_ISL_991185, EPI_ISL_991186, EPI_ISL_991187, EPI_ISL_991188, EPI_ISL_991189, EPI_ISL_991190, EPI_ISL_991191, EPI_ISL_991192, EPI_ISL_991193, EPI_ISL_991194, EPI_ISL_991195, EPI_ISL_991196, EPI_ISL_991197, EPI_ISL_991198, EPI_ISL_991199, EPI_ISL_991200, EPI_ISL_991201,
EPI_ISL_991202, EPI_ISL_991203, EPI_ISL_991204, EPI_ISL_991205, EPI_ISL_991206, EPI_ISL_991207, EPI_ISL_991208, EPI_ISL_991209, EPI_ISL_991210, EPI_ISL_991211, EPI_ISL_991212, EPI_ISL_991213, EPI_ISL_991214, EPI_ISL_991215, EPI_ISL_991216, EPI_ISL_991217, EPI_ISL_991218, EPI_ISL_991219, EPI_ISL_991220, EPI_ISL_991221, EPI_ISL_991269, EPI_ISL_991272, EPI_ISL_991274, EPI_ISL_991291,
EPI_ISL_991292, EPI_ISL_991293, EPI_ISL_991294, EPI_ISL_991295, EPI_ISL_991300, EPI_ISL_991302, EPI_ISL_991306, EPI_ISL_991566, EPI_ISL_991567, EPI_ISL_991568, EPI_ISL_991569, EPI_ISL_991570, EPI_ISL_991571, EPI_ISL_991572, EPI_ISL_991573, EPI_ISL_991574, EPI_ISL_991575, EPI_ISL_991576, EPI_ISL_991577, EPI_ISL_991578, EPI_ISL_991579, EPI_ISL_991580, EPI_ISL_991581, EPI_ISL_991582,
EPI_ISL_991583, EPI_ISL_991584, EPI_ISL_991585, EPI_ISL_991586, EPI_ISL_991587, EPI_ISL_991588, EPI_ISL_991589, EPI_ISL_991590, EPI_ISL_991591, EPI_ISL_991592, EPI_ISL_991593, EPI_ISL_991594, EPI_ISL_991596, EPI_ISL_991597, EPI_ISL_991598, EPI_ISL_991599, EPI_ISL_991600, EPI_ISL_991601, EPI_ISL_991602, EPI_ISL_991603, EPI_ISL_991604, EPI_ISL_991605, EPI_ISL_991606, EPI_ISL_991607,
EPI_ISL_991608, EPI_ISL_991609, EPI_ISL_991610, EPI_ISL_991611, EPI_ISL_991612, EPI_ISL_991613, EPI_ISL_991614, EPI_ISL_991615, EPI_ISL_991616, EPI_ISL_991617, EPI_ISL_991618, EPI_ISL_991619, EPI_ISL_991620, EPI_ISL_991621, EPI_ISL_991622, EPI_ISL_991623, EPI_ISL_991624, EPI_ISL_991625, EPI_ISL_991626, EPI_ISL_991627, EPI_ISL_991628, EPI_ISL_991630, EPI_ISL_991631, EPI_ISL_991632,
EPI_ISL_991633, EPI_ISL_991634, EPI_ISL_991635, EPI_ISL_991636, EPI_ISL_991638, EPI_ISL_991639, EPI_ISL_991640, EPI_ISL_991641, EPI_ISL_991642, EPI_ISL_991643, EPI_ISL_991644, EPI_ISL_991645, EPI_ISL_991646, EPI_ISL_991647, EPI_ISL_991648, EPI_ISL_991649, EPI_ISL_991650, EPI_ISL_991651, EPI_ISL_991652, EPI_ISL_991654, EPI_ISL_991655, EPI_ISL_991656, EPI_ISL_991657, EPI_ISL_991658,
EPI_ISL_991659, EPI_ISL_991660, EPI_ISL_991661, EPI_ISL_991662, EPI_ISL_991663, EPI_ISL_991664, EPI_ISL_991665, EPI_ISL_991666, EPI_ISL_991667, EPI_ISL_991668, EPI_ISL_991669, EPI_ISL_991670, EPI_ISL_991671, EPI_ISL_991672, EPI_ISL_991673, EPI_ISL_991674, EPI_ISL_991675, EPI_ISL_991676, EPI_ISL_991677, EPI_ISL_991678, EPI_ISL_991679, EPI_ISL_991681, EPI_ISL_991682, EPI_ISL_991683,
EPI_ISL_991684, EPI_ISL_991685, EPI_ISL_991686, EPI_ISL_991687, EPI_ISL_991688, EPI_ISL_991689, EPI_ISL_991690, EPI_ISL_991691, EPI_ISL_991693, EPI_ISL_991695, EPI_ISL_991696, EPI_ISL_991697, EPI_ISL_991698, EPI_ISL_991699, EPI_ISL_991700, EPI_ISL_991701, EPI_ISL_991702, EPI_ISL_991704, EPI_ISL_991705, EPI_ISL_991706, EPI_ISL_991707, EPI_ISL_991708, EPI_ISL_991709, EPI_ISL_991710,
EPI_ISL_991711, EPI_ISL_991712, EPI_ISL_991713, EPI_ISL_991714, EPI_ISL_991716, EPI_ISL_991717, EPI_ISL_991718, EPI_ISL_991719, EPI_ISL_991720, EPI_ISL_991721, EPI_ISL_991722, EPI_ISL_991723, EPI_ISL_991724, EPI_ISL_991725, EPI_ISL_991726, EPI_ISL_991727, EPI_ISL_991728, EPI_ISL_991729, EPI_ISL_991730, EPI_ISL_991731, EPI_ISL_991732, EPI_ISL_991733, EPI_ISL_991734, EPI_ISL_991735,
EPI_ISL_991736, EPI_ISL_991737, EPI_ISL_991738, EPI_ISL_991739, EPI_ISL_991740, EPI_ISL_991741, EPI_ISL_991742, EPI_ISL_991743, EPI_ISL_991744, EPI_ISL_991745, EPI_ISL_991746, EPI_ISL_991747, EPI_ISL_991748, EPI_ISL_991749, EPI_ISL_991750, EPI_ISL_991751, EPI_ISL_991752, EPI_ISL_991753, EPI_ISL_991754, EPI_ISL_991756, EPI_ISL_991757, EPI_ISL_991758, EPI_ISL_991759, EPI_ISL_991760,
EPI_ISL_991761, EPI_ISL_991762, EPI_ISL_991763, EPI_ISL_991764, EPI_ISL_991765, EPI_ISL_991766, EPI_ISL_991767, EPI_ISL_991768, EPI_ISL_991769, EPI_ISL_991770, EPI_ISL_991771, EPI_ISL_991772, EPI_ISL_991773, EPI_ISL_991774, EPI_ISL_991775, EPI_ISL_991776, EPI_ISL_991777, EPI_ISL_991778, EPI_ISL_991779, EPI_ISL_991780, EPI_ISL_991782, EPI_ISL_991783, EPI_ISL_991784, EPI_ISL_991785,
EPI_ISL_991786, EPI_ISL_991787, EPI_ISL_991788, EPI_ISL_991789, EPI_ISL_991790, EPI_ISL_991791, EPI_ISL_991792, EPI_ISL_991794, EPI_ISL_991795, EPI_ISL_991796, EPI_ISL_991797, EPI_ISL_991798, EPI_ISL_991799, EPI_ISL_991800, EPI_ISL_991801, EPI_ISL_991802, EPI_ISL_991803, EPI_ISL_991804, EPI_ISL_991805, EPI_ISL_991806, EPI_ISL_991807, EPI_ISL_991808, EPI_ISL_991809, EPI_ISL_991810,
EPI_ISL_991811, EPI_ISL_991812, EPI_ISL_991813, EPI_ISL_991814, EPI_ISL_991815, EPI_ISL_991816, EPI_ISL_991818, EPI_ISL_991819, EPI_ISL_991820, EPI_ISL_991821, EPI_ISL_991822, EPI_ISL_991823, EPI_ISL_991824, EPI_ISL_991825, EPI_ISL_991827, EPI_ISL_991828, EPI_ISL_991829, EPI_ISL_991830, EPI_ISL_991831, EPI_ISL_991832, EPI_ISL_991833, EPI_ISL_991835, EPI_ISL_991836, EPI_ISL_991837,
EPI_ISL_991838, EPI_ISL_991839, EPI_ISL_991840, EPI_ISL_991841, EPI_ISL_991842, EPI_ISL_991843, EPI_ISL_991844, EPI_ISL_991845, EPI_ISL_991847, EPI_ISL_991849, EPI_ISL_991850, EPI_ISL_991851, EPI_ISL_991852, EPI_ISL_991853, EPI_ISL_991854, EPI_ISL_991855, EPI_ISL_991856, EPI_ISL_991857, EPI_ISL_991858, EPI_ISL_991859, EPI_ISL_991861, EPI_ISL_991862, EPI_ISL_991863, EPI_ISL_991864,
EPI_ISL_991865, EPI_ISL_991866, EPI_ISL_991867, EPI_ISL_991868, EPI_ISL_991869, EPI_ISL_991870, EPI_ISL_991871, EPI_ISL_991872, EPI_ISL_991874, EPI_ISL_991875, EPI_ISL_991876, EPI_ISL_991877, EPI_ISL_991878, EPI_ISL_991879, EPI_ISL_991880, EPI_ISL_991881, EPI_ISL_991882, EPI_ISL_991883, EPI_ISL_991884, EPI_ISL_991885, EPI_ISL_991886, EPI_ISL_991887, EPI_ISL_991888, EPI_ISL_991889,
EPI_ISL_991890, EPI_ISL_991891, EPI_ISL_991892, EPI_ISL_991893, EPI_ISL_991894, EPI_ISL_991895, EPI_ISL_991896, EPI_ISL_991897, EPI_ISL_991898, EPI_ISL_991899, EPI_ISL_991901, EPI_ISL_991902, EPI_ISL_991903, EPI_ISL_991905, EPI_ISL_991906, EPI_ISL_991907, EPI_ISL_991908, EPI_ISL_991909, EPI_ISL_991910, EPI_ISL_991911, EPI_ISL_991912, EPI_ISL_991913, EPI_ISL_991914, EPI_ISL_991915,
EPI_ISL_991964, EPI_ISL_991965, EPI_ISL_991966, EPI_ISL_991967, EPI_ISL_991968, EPI_ISL_991969, EPI_ISL_991970, EPI_ISL_991971, EPI_ISL_991972, EPI_ISL_991973, EPI_ISL_991974, EPI_ISL_991975, EPI_ISL_991976, EPI_ISL_991977, EPI_ISL_991978, EPI_ISL_991979, EPI_ISL_991980, EPI_ISL_991981, EPI_ISL_991982, EPI_ISL_991983, EPI_ISL_991984, EPI_ISL_991985, EPI_ISL_991986, EPI_ISL_991987,
EPI_ISL_991988, EPI_ISL_991989, EPI_ISL_991990, EPI_ISL_991991, EPI_ISL_991992, EPI_ISL_991993, EPI_ISL_991994, EPI_ISL_991995, EPI_ISL_991996, EPI_ISL_991997, EPI_ISL_991998, EPI_ISL_991999, EPI_ISL_992000, EPI_ISL_992002, EPI_ISL_992003, EPI_ISL_992004, EPI_ISL_992005, EPI_ISL_992006, EPI_ISL_992007, EPI_ISL_992008, EPI_ISL_992010, EPI_ISL_992011, EPI_ISL_992012, EPI_ISL_992014,
EPI_ISL_992015, EPI_ISL_992016, EPI_ISL_992017, EPI_ISL_992018, EPI_ISL_992019, EPI_ISL_992020, EPI_ISL_992021, EPI_ISL_992022, EPI_ISL_992023, EPI_ISL_992024, EPI_ISL_992025, EPI_ISL_992026, EPI_ISL_992027, EPI_ISL_992028, EPI_ISL_992029, EPI_ISL_992030, EPI_ISL_992031, EPI_ISL_992032, EPI_ISL_992033, EPI_ISL_992034, EPI_ISL_992035, EPI_ISL_992036, EPI_ISL_992037, EPI_ISL_992038,
EPI_ISL_992039, EPI_ISL_992040, EPI_ISL_992041, EPI_ISL_992042, EPI_ISL_992044, EPI_ISL_992046, EPI_ISL_992048, EPI_ISL_992050, EPI_ISL_992053, EPI_ISL_992054, EPI_ISL_992055, EPI_ISL_992057, EPI_ISL_992058, EPI_ISL_992059, EPI_ISL_992061, EPI_ISL_992062, EPI_ISL_992063, EPI_ISL_992064, EPI_ISL_992066, EPI_ISL_992067, EPI_ISL_992069, EPI_ISL_992071, EPI_ISL_992073, EPI_ISL_992074,
EPI_ISL_992076, EPI_ISL_992077, EPI_ISL_992078, EPI_ISL_992079, EPI_ISL_992080, EPI_ISL_992081, EPI_ISL_992082, EPI_ISL_992085, EPI_ISL_992087, EPI_ISL_992089, EPI_ISL_992090, EPI_ISL_992091, EPI_ISL_992097, EPI_ISL_992099, EPI_ISL_992100, EPI_ISL_992101, EPI_ISL_992103, EPI_ISL_992105, EPI_ISL_992108, EPI_ISL_992109, EPI_ISL_992111, EPI_ISL_992116, EPI_ISL_992117, EPI_ISL_992118,
EPI_ISL_992119, EPI_ISL_992122, EPI_ISL_992127, EPI_ISL_992128, EPI_ISL_992130, EPI_ISL_992131, EPI_ISL_992135, EPI_ISL_992138, EPI_ISL_992143, EPI_ISL_992144, EPI_ISL_992146, EPI_ISL_992147, EPI_ISL_992149, EPI_ISL_992151, EPI_ISL_992152, EPI_ISL_992153, EPI_ISL_992155, EPI_ISL_992156, EPI_ISL_992159, EPI_ISL_992161, EPI_ISL_992164, EPI_ISL_992173, EPI_ISL_992175, EPI_ISL_992177,
EPI_ISL_992178, EPI_ISL_992179, EPI_ISL_992180, EPI_ISL_992181, EPI_ISL_992183, EPI_ISL_992184, EPI_ISL_992187, EPI_ISL_992188, EPI_ISL_992189, EPI_ISL_992196, EPI_ISL_992197, EPI_ISL_992198, EPI_ISL_992204, EPI_ISL_992205, EPI_ISL_992206, EPI_ISL_992207, EPI_ISL_992209, EPI_ISL_992211, EPI_ISL_992214, EPI_ISL_992215, EPI_ISL_992218, EPI_ISL_992219, EPI_ISL_992220, EPI_ISL_992221,
EPI_ISL_992225, EPI_ISL_992229, EPI_ISL_992234, EPI_ISL_992236, EPI_ISL_992237, EPI_ISL_992240, EPI_ISL_992241, EPI_ISL_992243, EPI_ISL_992246, EPI_ISL_992250, EPI_ISL_992252, EPI_ISL_992260, EPI_ISL_992262, EPI_ISL_992265, EPI_ISL_992267, EPI_ISL_992269, EPI_ISL_992271, EPI_ISL_992272, EPI_ISL_992273, EPI_ISL_992275, EPI_ISL_992276, EPI_ISL_992279, EPI_ISL_992282, EPI_ISL_992286,
EPI_ISL_992287, EPI_ISL_992288, EPI_ISL_992289, EPI_ISL_992291, EPI_ISL_992294, EPI_ISL_992295, EPI_ISL_992301, EPI_ISL_992302, EPI_ISL_992303, EPI_ISL_992304, EPI_ISL_992306, EPI_ISL_992309, EPI_ISL_992310, EPI_ISL_992311, EPI_ISL_992312, EPI_ISL_992314, EPI_ISL_992315, EPI_ISL_992317, EPI_ISL_992318, EPI_ISL_992321, EPI_ISL_992322, EPI_ISL_992323, EPI_ISL_992329, EPI_ISL_992330,
EPI_ISL_992331, EPI_ISL_992332, EPI_ISL_992335, EPI_ISL_992336, EPI_ISL_992337, EPI_ISL_992338, EPI_ISL_992344, EPI_ISL_992345, EPI_ISL_992348, EPI_ISL_992349, EPI_ISL_992350, EPI_ISL_992351, EPI_ISL_992352, EPI_ISL_992353, EPI_ISL_992354, EPI_ISL_992355, EPI_ISL_992356, EPI_ISL_992357, EPI_ISL_992358, EPI_ISL_992359, EPI_ISL_992361, EPI_ISL_992363, EPI_ISL_992364, EPI_ISL_992365,
EPI_ISL_992366, EPI_ISL_992367, EPI_ISL_992368, EPI_ISL_992369, EPI_ISL_992370, EPI_ISL_992371, EPI_ISL_992372, EPI_ISL_992373, EPI_ISL_992374, EPI_ISL_992375, EPI_ISL_992376, EPI_ISL_992377, EPI_ISL_992378, EPI_ISL_992379, EPI_ISL_992380, EPI_ISL_992381, EPI_ISL_992382, EPI_ISL_992383, EPI_ISL_992384, EPI_ISL_992385, EPI_ISL_992386, EPI_ISL_992387, EPI_ISL_992388, EPI_ISL_992389,
EPI_ISL_992390, EPI_ISL_992391, EPI_ISL_992392, EPI_ISL_992393, EPI_ISL_992394, EPI_ISL_992395, EPI_ISL_992396, EPI_ISL_992397, EPI_ISL_992398, EPI_ISL_992399, EPI_ISL_992400, EPI_ISL_992401, EPI_ISL_992402, EPI_ISL_992403, EPI_ISL_992404, EPI_ISL_992405, EPI_ISL_992406, EPI_ISL_992407, EPI_ISL_992408, EPI_ISL_992409, EPI_ISL_992411, EPI_ISL_992412, EPI_ISL_992413, EPI_ISL_992414,
EPI_ISL_992415, EPI_ISL_992416, EPI_ISL_992417, EPI_ISL_992418, EPI_ISL_992419, EPI_ISL_992420, EPI_ISL_992421, EPI_ISL_992422, EPI_ISL_992423, EPI_ISL_992424, EPI_ISL_992425, EPI_ISL_992426, EPI_ISL_992427, EPI_ISL_992428, EPI_ISL_992429, EPI_ISL_992430, EPI_ISL_992431, EPI_ISL_992432, EPI_ISL_992433, EPI_ISL_992434, EPI_ISL_992435, EPI_ISL_992436, EPI_ISL_992438, EPI_ISL_992440,
EPI_ISL_992441, EPI_ISL_992442, EPI_ISL_992443, EPI_ISL_992444, EPI_ISL_992445, EPI_ISL_992446, EPI_ISL_992447, EPI_ISL_992448, EPI_ISL_992450, EPI_ISL_992451, EPI_ISL_992452, EPI_ISL_992453, EPI_ISL_992454, EPI_ISL_992455, EPI_ISL_992456, EPI_ISL_992457, EPI_ISL_992458, EPI_ISL_992459, EPI_ISL_992461, EPI_ISL_992462, EPI_ISL_992464, EPI_ISL_992465, EPI_ISL_992467, EPI_ISL_992468,
EPI_ISL_992469, EPI_ISL_992470, EPI_ISL_992471, EPI_ISL_992472, EPI_ISL_992473, EPI_ISL_992474, EPI_ISL_992475, EPI_ISL_992476, EPI_ISL_992477, EPI_ISL_992478, EPI_ISL_992479, EPI_ISL_992480, EPI_ISL_992481, EPI_ISL_992482, EPI_ISL_992483, EPI_ISL_992485, EPI_ISL_992487, EPI_ISL_992488, EPI_ISL_992489, EPI_ISL_992490, EPI_ISL_992491, EPI_ISL_992492, EPI_ISL_992493, EPI_ISL_992494,
EPI_ISL_992495, EPI_ISL_992496, EPI_ISL_992497, EPI_ISL_992498, EPI_ISL_992499, EPI_ISL_992500, EPI_ISL_992502, EPI_ISL_992503, EPI_ISL_992504, EPI_ISL_992505, EPI_ISL_992506, EPI_ISL_992507, EPI_ISL_992508, EPI_ISL_992509, EPI_ISL_992510, EPI_ISL_992511, EPI_ISL_992512, EPI_ISL_992513, EPI_ISL_992514, EPI_ISL_992515, EPI_ISL_992516, EPI_ISL_992517, EPI_ISL_992518, EPI_ISL_992519,
EPI_ISL_992520, EPI_ISL_992521, EPI_ISL_992522, EPI_ISL_992523, EPI_ISL_992524, EPI_ISL_992525, EPI_ISL_992526, EPI_ISL_992527, EPI_ISL_992528, EPI_ISL_992529, EPI_ISL_992530, EPI_ISL_992531, EPI_ISL_992532, EPI_ISL_992533, EPI_ISL_992534, EPI_ISL_992535, EPI_ISL_992536, EPI_ISL_992537, EPI_ISL_992538, EPI_ISL_992539, EPI_ISL_992540, EPI_ISL_992541, EPI_ISL_992542, EPI_ISL_992543,
EPI_ISL_992544, EPI_ISL_992545, EPI_ISL_992546, EPI_ISL_992548, EPI_ISL_992550, EPI_ISL_992551, EPI_ISL_992552, EPI_ISL_992553, EPI_ISL_992554, EPI_ISL_992555, EPI_ISL_992556, EPI_ISL_992557, EPI_ISL_992558, EPI_ISL_992559, EPI_ISL_992560, EPI_ISL_992561, EPI_ISL_992562, EPI_ISL_992563, EPI_ISL_992564, EPI_ISL_992565, EPI_ISL_992566, EPI_ISL_992567, EPI_ISL_992568, EPI_ISL_992569,
EPI_ISL_992570, EPI_ISL_992571, EPI_ISL_992572, EPI_ISL_992573, EPI_ISL_992574, EPI_ISL_992575, EPI_ISL_992576, EPI_ISL_992577, EPI_ISL_992578, EPI_ISL_992579, EPI_ISL_992580, EPI_ISL_992581, EPI_ISL_992582, EPI_ISL_992583, EPI_ISL_992584, EPI_ISL_992585, EPI_ISL_992586, EPI_ISL_992587, EPI_ISL_992588, EPI_ISL_992589, EPI_ISL_992590, EPI_ISL_992591, EPI_ISL_992592, EPI_ISL_992593,
EPI_ISL_992594, EPI_ISL_992595, EPI_ISL_992596, EPI_ISL_992597, EPI_ISL_992598, EPI_ISL_992599, EPI_ISL_992600, EPI_ISL_992601, EPI_ISL_992602, EPI_ISL_992603, EPI_ISL_992604, EPI_ISL_992605, EPI_ISL_992606, EPI_ISL_992607, EPI_ISL_992608, EPI_ISL_992609, EPI_ISL_992610, EPI_ISL_992611, EPI_ISL_992612, EPI_ISL_992613, EPI_ISL_992614, EPI_ISL_992615, EPI_ISL_992616, EPI_ISL_992617,
EPI_ISL_992618, EPI_ISL_992619, EPI_ISL_992620, EPI_ISL_992621, EPI_ISL_992622, EPI_ISL_992623, EPI_ISL_992624, EPI_ISL_992625, EPI_ISL_992626, EPI_ISL_992627, EPI_ISL_992628, EPI_ISL_992629, EPI_ISL_992630, EPI_ISL_992631, EPI_ISL_992632, EPI_ISL_992633, EPI_ISL_992634, EPI_ISL_992635, EPI_ISL_992636, EPI_ISL_992637, EPI_ISL_992638, EPI_ISL_992639, EPI_ISL_992640, EPI_ISL_992641,
EPI_ISL_992642, EPI_ISL_992644, EPI_ISL_992645, EPI_ISL_992646, EPI_ISL_992648, EPI_ISL_992649, EPI_ISL_992650, EPI_ISL_992651, EPI_ISL_992652, EPI_ISL_992653, EPI_ISL_992654, EPI_ISL_992655, EPI_ISL_992656, EPI_ISL_992657, EPI_ISL_992658, EPI_ISL_992659, EPI_ISL_992660, EPI_ISL_992661, EPI_ISL_992662, EPI_ISL_992663, EPI_ISL_992664, EPI_ISL_992666, EPI_ISL_992667, EPI_ISL_992668,
EPI_ISL_992669, EPI_ISL_992670, EPI_ISL_992671, EPI_ISL_992672, EPI_ISL_992673, EPI_ISL_992674, EPI_ISL_992675, EPI_ISL_992676, EPI_ISL_992677, EPI_ISL_992678, EPI_ISL_992679, EPI_ISL_992680, EPI_ISL_992681, EPI_ISL_992682, EPI_ISL_992683, EPI_ISL_992684, EPI_ISL_992685, EPI_ISL_992686, EPI_ISL_992687, EPI_ISL_992688, EPI_ISL_992689, EPI_ISL_992690, EPI_ISL_992691, EPI_ISL_992692,
EPI_ISL_992693, EPI_ISL_992695, EPI_ISL_992696, EPI_ISL_992697, EPI_ISL_992698, EPI_ISL_992699, EPI_ISL_992700, EPI_ISL_992702, EPI_ISL_992703, EPI_ISL_992704, EPI_ISL_992705, EPI_ISL_992706, EPI_ISL_992707, EPI_ISL_992708, EPI_ISL_992709, EPI_ISL_992711, EPI_ISL_992712, EPI_ISL_992713, EPI_ISL_992714, EPI_ISL_992715, EPI_ISL_992716, EPI_ISL_992717, EPI_ISL_992718, EPI_ISL_992719,
EPI_ISL_992720, EPI_ISL_992721, EPI_ISL_992723, EPI_ISL_992724, EPI_ISL_992725, EPI_ISL_992726, EPI_ISL_992727, EPI_ISL_992728, EPI_ISL_992730, EPI_ISL_992731, EPI_ISL_992732, EPI_ISL_992733, EPI_ISL_992734, EPI_ISL_992735, EPI_ISL_992736, EPI_ISL_992737, EPI_ISL_992738, EPI_ISL_992739, EPI_ISL_992740, EPI_ISL_992741, EPI_ISL_992742, EPI_ISL_992744, EPI_ISL_992745, EPI_ISL_992746,
EPI_ISL_992747, EPI_ISL_992748, EPI_ISL_992749, EPI_ISL_992750, EPI_ISL_992751, EPI_ISL_992752, EPI_ISL_992753, EPI_ISL_992754, EPI_ISL_992755, EPI_ISL_992756, EPI_ISL_992757, EPI_ISL_992758, EPI_ISL_992759, EPI_ISL_992760, EPI_ISL_992761, EPI_ISL_992762, EPI_ISL_992763, EPI_ISL_992764, EPI_ISL_992765, EPI_ISL_992766, EPI_ISL_992767, EPI_ISL_992768, EPI_ISL_992769, EPI_ISL_992770,
EPI_ISL_992771, EPI_ISL_992772, EPI_ISL_992773, EPI_ISL_992774, EPI_ISL_992775, EPI_ISL_992776, EPI_ISL_992777, EPI_ISL_992778, EPI_ISL_992779, EPI_ISL_992780, EPI_ISL_992781, EPI_ISL_992782, EPI_ISL_992783, EPI_ISL_992784, EPI_ISL_992785, EPI_ISL_992786, EPI_ISL_992787, EPI_ISL_992788, EPI_ISL_992789, EPI_ISL_992790, EPI_ISL_992791, EPI_ISL_992792, EPI_ISL_992793, EPI_ISL_992794,
EPI_ISL_992795, EPI_ISL_992796, EPI_ISL_992797, EPI_ISL_992798, EPI_ISL_992799, EPI_ISL_992801, EPI_ISL_992802, EPI_ISL_992803, EPI_ISL_992804, EPI_ISL_992805, EPI_ISL_992806, EPI_ISL_992807, EPI_ISL_992808, EPI_ISL_992809, EPI_ISL_992810, EPI_ISL_992811, EPI_ISL_992812, EPI_ISL_992813, EPI_ISL_992814, EPI_ISL_992815, EPI_ISL_992816, EPI_ISL_992817, EPI_ISL_992818, EPI_ISL_992819,
EPI_ISL_992820, EPI_ISL_992821, EPI_ISL_992822, EPI_ISL_992823, EPI_ISL_992824, EPI_ISL_992825, EPI_ISL_992826, EPI_ISL_992827, EPI_ISL_992828, EPI_ISL_992829, EPI_ISL_992830, EPI_ISL_992831, EPI_ISL_992832, EPI_ISL_992833, EPI_ISL_992834, EPI_ISL_992835, EPI_ISL_992836, EPI_ISL_992837, EPI_ISL_992838, EPI_ISL_992839, EPI_ISL_992840, EPI_ISL_992841, EPI_ISL_992842, EPI_ISL_992843,
EPI_ISL_992844, EPI_ISL_992845, EPI_ISL_992846, EPI_ISL_992847, EPI_ISL_992848, EPI_ISL_992849, EPI_ISL_992850, EPI_ISL_992851, EPI_ISL_992852, EPI_ISL_992853, EPI_ISL_992854, EPI_ISL_992855, EPI_ISL_992856, EPI_ISL_992857, EPI_ISL_992859, EPI_ISL_992860, EPI_ISL_992861, EPI_ISL_992862, EPI_ISL_992863, EPI_ISL_992865, EPI_ISL_992867, EPI_ISL_992868, EPI_ISL_992869, EPI_ISL_992870,
EPI_ISL_992871, EPI_ISL_992872, EPI_ISL_992873, EPI_ISL_992874, EPI_ISL_992875, EPI_ISL_992876, EPI_ISL_992877, EPI_ISL_992878, EPI_ISL_992879, EPI_ISL_992880, EPI_ISL_992881, EPI_ISL_992882, EPI_ISL_992883, EPI_ISL_992884, EPI_ISL_992885, EPI_ISL_992886, EPI_ISL_992887, EPI_ISL_992888, EPI_ISL_992889, EPI_ISL_992890, EPI_ISL_992891, EPI_ISL_992892, EPI_ISL_992893, EPI_ISL_992894,
EPI_ISL_992895, EPI_ISL_992896, EPI_ISL_992897, EPI_ISL_992898, EPI_ISL_992899, EPI_ISL_992900, EPI_ISL_992901, EPI_ISL_992902, EPI_ISL_992903, EPI_ISL_992904, EPI_ISL_992905, EPI_ISL_992906, EPI_ISL_992907, EPI_ISL_992908, EPI_ISL_992909, EPI_ISL_992910, EPI_ISL_992911, EPI_ISL_992912, EPI_ISL_992913, EPI_ISL_992914, EPI_ISL_992915, EPI_ISL_992916, EPI_ISL_992917, EPI_ISL_992918,
EPI_ISL_992919, EPI_ISL_992920, EPI_ISL_992921, EPI_ISL_992922, EPI_ISL_992923, EPI_ISL_992924, EPI_ISL_992925, EPI_ISL_992926, EPI_ISL_992927, EPI_ISL_992928, EPI_ISL_992929, EPI_ISL_992931, EPI_ISL_992932, EPI_ISL_992933, EPI_ISL_992934, EPI_ISL_992935, EPI_ISL_992936, EPI_ISL_992937, EPI_ISL_992938, EPI_ISL_992939, EPI_ISL_992940, EPI_ISL_992941, EPI_ISL_992942, EPI_ISL_992943,
EPI_ISL_992944, EPI_ISL_992945, EPI_ISL_992946, EPI_ISL_992947, EPI_ISL_992948, EPI_ISL_992949, EPI_ISL_992950, EPI_ISL_992951, EPI_ISL_992952, EPI_ISL_992953, EPI_ISL_992954, EPI_ISL_992955, EPI_ISL_992957, EPI_ISL_992958, EPI_ISL_992959, EPI_ISL_992960, EPI_ISL_992961, EPI_ISL_992962, EPI_ISL_992963, EPI_ISL_992964, EPI_ISL_992965, EPI_ISL_992966, EPI_ISL_992967, EPI_ISL_992968,
EPI_ISL_992969, EPI_ISL_992970, EPI_ISL_992971, EPI_ISL_992972, EPI_ISL_992973, EPI_ISL_992974, EPI_ISL_992975, EPI_ISL_992976, EPI_ISL_992977, EPI_ISL_992978, EPI_ISL_992979, EPI_ISL_992980, EPI_ISL_992981, EPI_ISL_992982, EPI_ISL_992983, EPI_ISL_992984, EPI_ISL_992986, EPI_ISL_992987, EPI_ISL_992988, EPI_ISL_992990, EPI_ISL_992991, EPI_ISL_992992, EPI_ISL_992993, EPI_ISL_992994,
EPI_ISL_992995, EPI_ISL_992996, EPI_ISL_992997, EPI_ISL_992998, EPI_ISL_992999, EPI_ISL_993000, EPI_ISL_993001, EPI_ISL_993002, EPI_ISL_993003, EPI_ISL_993005, EPI_ISL_993006, EPI_ISL_993007, EPI_ISL_993008, EPI_ISL_993009, EPI_ISL_993010, EPI_ISL_993011, EPI_ISL_993013, EPI_ISL_993014, EPI_ISL_993016, EPI_ISL_993018, EPI_ISL_993019, EPI_ISL_993020, EPI_ISL_993021, EPI_ISL_993023,
EPI_ISL_993024, EPI_ISL_993025, EPI_ISL_993026, EPI_ISL_993027, EPI_ISL_993028, EPI_ISL_993029, EPI_ISL_993030, EPI_ISL_993031, EPI_ISL_993032, EPI_ISL_993033, EPI_ISL_993034, EPI_ISL_993035, EPI_ISL_993036, EPI_ISL_993037, EPI_ISL_993038, EPI_ISL_993039, EPI_ISL_993040, EPI_ISL_993041, EPI_ISL_993042, EPI_ISL_1007564, EPI_ISL_1007565, EPI_ISL_1007566, EPI_ISL_1007567,
EPI_ISL_1007568, EPI_ISL_1007569, EPI_ISL_1007570, EPI_ISL_1007571, EPI_ISL_1007572, EPI_ISL_1007573, EPI_ISL_1012754, EPI_ISL_1012762, EPI_ISL_1012787, EPI_ISL_1012788, EPI_ISL_1012789, EPI_ISL_1012790, EPI_ISL_1012791, EPI_ISL_1012792, EPI_ISL_1012793, EPI_ISL_1012794, EPI_ISL_1012795, EPI_ISL_1012796, EPI_ISL_1012797, EPI_ISL_1012798, EPI_ISL_1012799, EPI_ISL_1012800,
EPI_ISL_1012801, EPI_ISL_1012802, EPI_ISL_1012803, EPI_ISL_1012804, EPI_ISL_1012805, EPI_ISL_1012806, EPI_ISL_1012807, EPI_ISL_1012809, EPI_ISL_1012810, EPI_ISL_1012811, EPI_ISL_1012812, EPI_ISL_1012813, EPI_ISL_1012814, EPI_ISL_1012815, EPI_ISL_1012816, EPI_ISL_1012817, EPI_ISL_1012818, EPI_ISL_1012819, EPI_ISL_1012820, EPI_ISL_1012821, EPI_ISL_1012822, EPI_ISL_1012823,
EPI_ISL_1012824, EPI_ISL_1012825, EPI_ISL_1012826, EPI_ISL_1012827, EPI_ISL_1012828, EPI_ISL_1012829, EPI_ISL_1012830, EPI_ISL_1012832, EPI_ISL_1012835, EPI_ISL_1012836, EPI_ISL_1012837, EPI_ISL_1012838, EPI_ISL_1012839, EPI_ISL_1012840, EPI_ISL_1012841, EPI_ISL_1012842, EPI_ISL_1012843, EPI_ISL_1012845, EPI_ISL_1012846, EPI_ISL_1012847, EPI_ISL_1012848, EPI_ISL_1012850,
EPI_ISL_1012851, EPI_ISL_1012852, EPI_ISL_1012853, EPI_ISL_1012854, EPI_ISL_1012855, EPI_ISL_1012856, EPI_ISL_1043901, EPI_ISL_1057732, EPI_ISL_1057745, EPI_ISL_1189117

see above | Lighthouse Lab in Alderley Park | Wellcome Sanger Institute for the COVID-19 Genomics UK (COG-UK) Consortium | Cordelia Langford; David K. Jackson; Dominic Kwiatkowski; Ewan Harrison; Ian Johnston; Jacquelyn Wynn; Jeffrey Barrett; John Sillitoe on behalf of the Wellcome Sanger Institute COVID-19 Surveillance Team; John Sillitoe on behalf of the Wellcome Sanger Institute COVID-19 Surveillance Team (http://www.sanger.ac.uk/covid-team); Mairead Hyland; Roberto Amato; Sonia Goncalves; The Lighthouse Lab in Alderley Park and Alex Alderton

EPI_ISL_907147, EPI_ISL_907163, EPI_ISL_907168, EPI_ISL_907192, EPI_ISL_907194, EPI_ISL_907222, EPI_ISL_907273, EPI_ISL_907281, EPI_ISL_907303, EPI_ISL_907348, EPI_ISL_907350, EPI_ISL_907359, EPI_ISL_907364, EPI_ISL_907369, EPI_ISL_908761, EPI_ISL_916180, EPI_ISL_916181, EPI_ISL_916182, EPI_ISL_916183, EPI_ISL_916185, EPI_ISL_916187, EPI_ISL_916188, EPI_ISL_916189, EPI_ISL_916190,
EPI_ISL_916193, EPI_ISL_916194, EPI_ISL_916200, EPI_ISL_916201, EPI_ISL_916202, EPI_ISL_916203, EPI_ISL_916205, EPI_ISL_916207, EPI_ISL_916208, EPI_ISL_916209, EPI_ISL_916213, EPI_ISL_916216, EPI_ISL_916222, EPI_ISL_916225, EPI_ISL_916226, EPI_ISL_916227, EPI_ISL_916228, EPI_ISL_916232, EPI_ISL_916233, EPI_ISL_916234, EPI_ISL_916236, EPI_ISL_916238, EPI_ISL_916241, EPI_ISL_916244,
EPI_ISL_916245, EPI_ISL_916246, EPI_ISL_916248, EPI_ISL_916250, EPI_ISL_916254, EPI_ISL_916256, EPI_ISL_916258, EPI_ISL_916261, EPI_ISL_916263, EPI_ISL_916264, EPI_ISL_916266, EPI_ISL_916268, EPI_ISL_916273, EPI_ISL_916275, EPI_ISL_916276, EPI_ISL_916278, EPI_ISL_916279, EPI_ISL_916281, EPI_ISL_916283, EPI_ISL_916284, EPI_ISL_916285, EPI_ISL_916287, EPI_ISL_916288, EPI_ISL_916290,
EPI_ISL_916291, EPI_ISL_916295, EPI_ISL_916296, EPI_ISL_916299, EPI_ISL_916301, EPI_ISL_916303, EPI_ISL_916305, EPI_ISL_916306, EPI_ISL_916307, EPI_ISL_916308, EPI_ISL_916310, EPI_ISL_916312, EPI_ISL_916314, EPI_ISL_916315, EPI_ISL_916316, EPI_ISL_916318, EPI_ISL_916320, EPI_ISL_916321, EPI_ISL_916322, EPI_ISL_916323, EPI_ISL_916325, EPI_ISL_916327, EPI_ISL_916328, EPI_ISL_916329,
EPI_ISL_916332, EPI_ISL_916333, EPI_ISL_916335, EPI_ISL_916336, EPI_ISL_916338, EPI_ISL_916339, EPI_ISL_916340, EPI_ISL_916341, EPI_ISL_916342, EPI_ISL_916343, EPI_ISL_916344, EPI_ISL_916345, EPI_ISL_916347, EPI_ISL_916349, EPI_ISL_916350, EPI_ISL_916352, EPI_ISL_916357, EPI_ISL_916358, EPI_ISL_916360, EPI_ISL_916364, EPI_ISL_916365, EPI_ISL_916366, EPI_ISL_916367, EPI_ISL_916368,
EPI_ISL_916369, EPI_ISL_916370, EPI_ISL_916371, EPI_ISL_916372, EPI_ISL_916373, EPI_ISL_916374, EPI_ISL_916375, EPI_ISL_916377, EPI_ISL_916378, EPI_ISL_916379, EPI_ISL_916380, EPI_ISL_916381, EPI_ISL_916383, EPI_ISL_916384, EPI_ISL_916388, EPI_ISL_916390, EPI_ISL_916394, EPI_ISL_916397, EPI_ISL_916399, EPI_ISL_916400, EPI_ISL_916401, EPI_ISL_916403, EPI_ISL_916405, EPI_ISL_916406,
EPI_ISL_916407, EPI_ISL_916408, EPI_ISL_916409, EPI_ISL_916410, EPI_ISL_916411, EPI_ISL_916412, EPI_ISL_916413, EPI_ISL_916415, EPI_ISL_916417, EPI_ISL_916419, EPI_ISL_916421, EPI_ISL_916425, EPI_ISL_916428, EPI_ISL_916430, EPI_ISL_916431, EPI_ISL_916432, EPI_ISL_916433, EPI_ISL_916434, EPI_ISL_916436, EPI_ISL_916437, EPI_ISL_916438, EPI_ISL_916439, EPI_ISL_916440, EPI_ISL_916441,
EPI_ISL_916442, EPI_ISL_916443, EPI_ISL_916446, EPI_ISL_916447, EPI_ISL_916449, EPI_ISL_916451, EPI_ISL_916453, EPI_ISL_916454, EPI_ISL_916455, EPI_ISL_916456, EPI_ISL_916457, EPI_ISL_916458, EPI_ISL_916459, EPI_ISL_916460, EPI_ISL_916462, EPI_ISL_916463, EPI_ISL_916464, EPI_ISL_916467, EPI_ISL_916472, EPI_ISL_916473, EPI_ISL_916476, EPI_ISL_916477, EPI_ISL_916478, EPI_ISL_916479,

EPI_ISL_916480, EPI_ISL_916481, EPI_ISL_916484, EPI_ISL_916485, EPI_ISL_916486, EPI_ISL_916487, EPI_ISL_916489, EPI_ISL_916491, EPI_ISL_916492, EPI_ISL_916493, EPI_ISL_916494, EPI_ISL_916499, EPI_ISL_916500, EPI_ISL_916502, EPI_ISL_932576, EPI_ISL_932581, EPI_ISL_932596, EPI_ISL_932598, EPI_ISL_932603, EPI_ISL_932611, EPI_ISL_932613, EPI_ISL_932614, EPI_ISL_932619, EPI_ISL_932621,
EPI_ISL_932644, EPI_ISL_932646, EPI_ISL_932648, EPI_ISL_932651, EPI_ISL_932652, EPI_ISL_932655, EPI_ISL_932664, EPI_ISL_932666, EPI_ISL_932669, EPI_ISL_932671, EPI_ISL_932674, EPI_ISL_932677, EPI_ISL_932678, EPI_ISL_932680, EPI_ISL_932686, EPI_ISL_932687, EPI_ISL_932689, EPI_ISL_932691, EPI_ISL_932693, EPI_ISL_932703, EPI_ISL_932706, EPI_ISL_932710, EPI_ISL_932712, EPI_ISL_932715,
EPI_ISL_932718, EPI_ISL_932721, EPI_ISL_932722, EPI_ISL_932728, EPI_ISL_932737, EPI_ISL_932739, EPI_ISL_932742, EPI_ISL_932754, EPI_ISL_932755, EPI_ISL_932758, EPI_ISL_932770, EPI_ISL_932771, EPI_ISL_932776, EPI_ISL_932778, EPI_ISL_932782, EPI_ISL_932790, EPI_ISL_932794, EPI_ISL_932796, EPI_ISL_932805, EPI_ISL_932808, EPI_ISL_932812, EPI_ISL_932816, EPI_ISL_932820, EPI_ISL_932822,
EPI_ISL_932827, EPI_ISL_932829, EPI_ISL_932830, EPI_ISL_932836, EPI_ISL_932838, EPI_ISL_932839, EPI_ISL_932842, EPI_ISL_932843, EPI_ISL_933114, EPI_ISL_933115, EPI_ISL_933116, EPI_ISL_933117, EPI_ISL_933118, EPI_ISL_933119, EPI_ISL_933120, EPI_ISL_933121, EPI_ISL_933122, EPI_ISL_933125, EPI_ISL_933126, EPI_ISL_933128, EPI_ISL_933129, EPI_ISL_933131, EPI_ISL_933132, EPI_ISL_933133,
EPI_ISL_933135, EPI_ISL_933137, EPI_ISL_933138, EPI_ISL_933139, EPI_ISL_933142, EPI_ISL_933144, EPI_ISL_933145, EPI_ISL_933146, EPI_ISL_933147, EPI_ISL_933148, EPI_ISL_933151, EPI_ISL_933152, EPI_ISL_933153, EPI_ISL_933155, EPI_ISL_933156, EPI_ISL_933157, EPI_ISL_933159, EPI_ISL_933160, EPI_ISL_933161, EPI_ISL_933162, EPI_ISL_933163, EPI_ISL_933164, EPI_ISL_933165, EPI_ISL_933168,
EPI_ISL_933169, EPI_ISL_933172, EPI_ISL_933173, EPI_ISL_933174, EPI_ISL_933175, EPI_ISL_933176, EPI_ISL_933177, EPI_ISL_933178, EPI_ISL_933179, EPI_ISL_933180, EPI_ISL_933182, EPI_ISL_933183, EPI_ISL_933184, EPI_ISL_933185, EPI_ISL_933186, EPI_ISL_933189, EPI_ISL_933190, EPI_ISL_933191, EPI_ISL_933192, EPI_ISL_933193, EPI_ISL_933194, EPI_ISL_933195, EPI_ISL_933197, EPI_ISL_933198,
EPI_ISL_933201, EPI_ISL_933202, EPI_ISL_933203, EPI_ISL_933204, EPI_ISL_933205, EPI_ISL_933206, EPI_ISL_933208, EPI_ISL_933209, EPI_ISL_933210, EPI_ISL_933211, EPI_ISL_933212, EPI_ISL_933216, EPI_ISL_933217, EPI_ISL_933218, EPI_ISL_933219, EPI_ISL_933220, EPI_ISL_933222, EPI_ISL_933223, EPI_ISL_933224, EPI_ISL_933225, EPI_ISL_933229, EPI_ISL_933230, EPI_ISL_933231, EPI_ISL_933232,
EPI_ISL_933234, EPI_ISL_933235, EPI_ISL_933236, EPI_ISL_933237, EPI_ISL_933238, EPI_ISL_933240, EPI_ISL_933241, EPI_ISL_933242, EPI_ISL_933244, EPI_ISL_933245, EPI_ISL_933246, EPI_ISL_933247, EPI_ISL_933248, EPI_ISL_933249, EPI_ISL_933250, EPI_ISL_933251, EPI_ISL_933252, EPI_ISL_933253, EPI_ISL_933254, EPI_ISL_933255, EPI_ISL_933257, EPI_ISL_933258, EPI_ISL_933260, EPI_ISL_933261,
EPI_ISL_933262, EPI_ISL_933263, EPI_ISL_933264, EPI_ISL_933267, EPI_ISL_933268, EPI_ISL_933269, EPI_ISL_933270, EPI_ISL_933274, EPI_ISL_933275, EPI_ISL_933276, EPI_ISL_933278, EPI_ISL_933279, EPI_ISL_933280, EPI_ISL_933281, EPI_ISL_933284, EPI_ISL_933285, EPI_ISL_933286, EPI_ISL_933287, EPI_ISL_933288, EPI_ISL_933290, EPI_ISL_933291, EPI_ISL_933292, EPI_ISL_933294, EPI_ISL_933295,
EPI_ISL_933296, EPI_ISL_933297, EPI_ISL_933299, EPI_ISL_933300, EPI_ISL_933301, EPI_ISL_933302, EPI_ISL_933304, EPI_ISL_933305, EPI_ISL_933306, EPI_ISL_933307, EPI_ISL_933308, EPI_ISL_933309, EPI_ISL_933310, EPI_ISL_933311, EPI_ISL_933312, EPI_ISL_933313, EPI_ISL_933314, EPI_ISL_933315, EPI_ISL_933316, EPI_ISL_933317, EPI_ISL_933318, EPI_ISL_933319, EPI_ISL_933320, EPI_ISL_933322,
EPI_ISL_933323, EPI_ISL_933324, EPI_ISL_933327, EPI_ISL_933328, EPI_ISL_933329, EPI_ISL_933330, EPI_ISL_933331, EPI_ISL_933332, EPI_ISL_933333, EPI_ISL_933334, EPI_ISL_933335, EPI_ISL_933337, EPI_ISL_933338, EPI_ISL_933339, EPI_ISL_933342, EPI_ISL_933347, EPI_ISL_933349, EPI_ISL_933351, EPI_ISL_933352, EPI_ISL_933353, EPI_ISL_933355, EPI_ISL_933356, EPI_ISL_933357, EPI_ISL_933360,
EPI_ISL_933361, EPI_ISL_933362, EPI_ISL_933363, EPI_ISL_933365, EPI_ISL_933366, EPI_ISL_933367, EPI_ISL_933368, EPI_ISL_933369, EPI_ISL_933370, EPI_ISL_933372, EPI_ISL_933373, EPI_ISL_933374, EPI_ISL_933376, EPI_ISL_933377, EPI_ISL_933378, EPI_ISL_933379, EPI_ISL_933380, EPI_ISL_933381, EPI_ISL_933383, EPI_ISL_933384, EPI_ISL_933385, EPI_ISL_933389, EPI_ISL_933390, EPI_ISL_933391,
EPI_ISL_933392, EPI_ISL_933394, EPI_ISL_933395, EPI_ISL_933398, EPI_ISL_933399, EPI_ISL_933402, EPI_ISL_933403, EPI_ISL_933404, EPI_ISL_933405, EPI_ISL_933406, EPI_ISL_933407, EPI_ISL_933408, EPI_ISL_933409, EPI_ISL_933411, EPI_ISL_933413, EPI_ISL_933415, EPI_ISL_933416, EPI_ISL_933418, EPI_ISL_933419, EPI_ISL_933420, EPI_ISL_933421, EPI_ISL_933422, EPI_ISL_933423, EPI_ISL_933424,
EPI_ISL_938234, EPI_ISL_938235, EPI_ISL_938236, EPI_ISL_938237, EPI_ISL_938238, EPI_ISL_938239, EPI_ISL_938240, EPI_ISL_938241, EPI_ISL_938242, EPI_ISL_938243, EPI_ISL_938244, EPI_ISL_938245, EPI_ISL_938246, EPI_ISL_938247, EPI_ISL_938248, EPI_ISL_938249, EPI_ISL_938250, EPI_ISL_938251, EPI_ISL_938252, EPI_ISL_938253, EPI_ISL_938254, EPI_ISL_938255, EPI_ISL_938256, EPI_ISL_938257,
EPI_ISL_938258, EPI_ISL_938259, EPI_ISL_938260, EPI_ISL_938261, EPI_ISL_938262, EPI_ISL_938263, EPI_ISL_938264, EPI_ISL_938265, EPI_ISL_938266, EPI_ISL_938267, EPI_ISL_938268, EPI_ISL_938269, EPI_ISL_938270, EPI_ISL_938271, EPI_ISL_938272, EPI_ISL_938273, EPI_ISL_938274, EPI_ISL_938275, EPI_ISL_938276, EPI_ISL_938277, EPI_ISL_938278, EPI_ISL_938279, EPI_ISL_938280, EPI_ISL_938281,
EPI_ISL_938282, EPI_ISL_938283, EPI_ISL_938284, EPI_ISL_938285, EPI_ISL_938286, EPI_ISL_938287, EPI_ISL_938288, EPI_ISL_938289, EPI_ISL_938290, EPI_ISL_938291, EPI_ISL_938292, EPI_ISL_938293, EPI_ISL_938294, EPI_ISL_938295, EPI_ISL_938296, EPI_ISL_938297, EPI_ISL_938298, EPI_ISL_938299, EPI_ISL_938300, EPI_ISL_938301, EPI_ISL_938302, EPI_ISL_938303, EPI_ISL_938304, EPI_ISL_938305,
EPI_ISL_938306, EPI_ISL_938307, EPI_ISL_938308, EPI_ISL_938309, EPI_ISL_938310, EPI_ISL_938311, EPI_ISL_938312, EPI_ISL_938313, EPI_ISL_938314, EPI_ISL_938315, EPI_ISL_938316, EPI_ISL_938317, EPI_ISL_938318, EPI_ISL_938319, EPI_ISL_938320, EPI_ISL_938321, EPI_ISL_938322, EPI_ISL_938323, EPI_ISL_938324, EPI_ISL_938325, EPI_ISL_938326, EPI_ISL_938327, EPI_ISL_938328, EPI_ISL_938329,
EPI_ISL_938330, EPI_ISL_938331, EPI_ISL_938332, EPI_ISL_938333, EPI_ISL_938334, EPI_ISL_938335, EPI_ISL_938336, EPI_ISL_938337, EPI_ISL_938338, EPI_ISL_938339, EPI_ISL_938340, EPI_ISL_938341, EPI_ISL_938342, EPI_ISL_938343, EPI_ISL_938344, EPI_ISL_938345, EPI_ISL_938346, EPI_ISL_938347, EPI_ISL_938348, EPI_ISL_938349, EPI_ISL_938350, EPI_ISL_938351, EPI_ISL_938352, EPI_ISL_938353,
EPI_ISL_938354, EPI_ISL_938355, EPI_ISL_938356, EPI_ISL_938357, EPI_ISL_938358, EPI_ISL_938359, EPI_ISL_938360, EPI_ISL_938361, EPI_ISL_938362, EPI_ISL_938363, EPI_ISL_938364, EPI_ISL_938365, EPI_ISL_938366, EPI_ISL_938367, EPI_ISL_938368, EPI_ISL_938369, EPI_ISL_938370, EPI_ISL_938371, EPI_ISL_938372, EPI_ISL_938373, EPI_ISL_938374, EPI_ISL_938375, EPI_ISL_938376, EPI_ISL_938377,
EPI_ISL_938378, EPI_ISL_938379, EPI_ISL_938380, EPI_ISL_938381, EPI_ISL_938382, EPI_ISL_938383, EPI_ISL_938384, EPI_ISL_938385, EPI_ISL_938386, EPI_ISL_938387, EPI_ISL_938388, EPI_ISL_938389, EPI_ISL_938390, EPI_ISL_938391, EPI_ISL_938392, EPI_ISL_938393, EPI_ISL_938394, EPI_ISL_938395, EPI_ISL_938396, EPI_ISL_938397, EPI_ISL_938398, EPI_ISL_938399, EPI_ISL_938400, EPI_ISL_938401,
EPI_ISL_938402, EPI_ISL_938403, EPI_ISL_938404, EPI_ISL_938405, EPI_ISL_938406, EPI_ISL_938407, EPI_ISL_938408, EPI_ISL_938409, EPI_ISL_938410, EPI_ISL_938411, EPI_ISL_938412, EPI_ISL_938413, EPI_ISL_938414, EPI_ISL_938415, EPI_ISL_938416, EPI_ISL_938417, EPI_ISL_938418, EPI_ISL_938419, EPI_ISL_938420, EPI_ISL_938421, EPI_ISL_938422, EPI_ISL_938423, EPI_ISL_938424, EPI_ISL_938425,
EPI_ISL_938426, EPI_ISL_938427, EPI_ISL_938428, EPI_ISL_938429, EPI_ISL_938430, EPI_ISL_938431, EPI_ISL_938432, EPI_ISL_938433, EPI_ISL_938434, EPI_ISL_938435, EPI_ISL_938436, EPI_ISL_938437, EPI_ISL_938438, EPI_ISL_938439, EPI_ISL_938440, EPI_ISL_938441, EPI_ISL_938442, EPI_ISL_938443, EPI_ISL_938444, EPI_ISL_938445, EPI_ISL_938446, EPI_ISL_938447, EPI_ISL_938448, EPI_ISL_938449,
EPI_ISL_938450, EPI_ISL_938451, EPI_ISL_938452, EPI_ISL_938453, EPI_ISL_938454, EPI_ISL_938455, EPI_ISL_938456, EPI_ISL_938457, EPI_ISL_938458, EPI_ISL_938459, EPI_ISL_938460, EPI_ISL_938461, EPI_ISL_938462, EPI_ISL_938463, EPI_ISL_938464, EPI_ISL_938465, EPI_ISL_938466, EPI_ISL_938467, EPI_ISL_938468, EPI_ISL_938469, EPI_ISL_938470, EPI_ISL_938471, EPI_ISL_938472, EPI_ISL_938473,
EPI_ISL_938474, EPI_ISL_938475, EPI_ISL_938476, EPI_ISL_938477, EPI_ISL_938478, EPI_ISL_938479, EPI_ISL_938480, EPI_ISL_938481, EPI_ISL_938482, EPI_ISL_938483, EPI_ISL_938484, EPI_ISL_938485, EPI_ISL_938486, EPI_ISL_938487, EPI_ISL_938488, EPI_ISL_938489, EPI_ISL_938490, EPI_ISL_938491, EPI_ISL_938492, EPI_ISL_938493, EPI_ISL_938494, EPI_ISL_938495, EPI_ISL_938496, EPI_ISL_938497,
EPI_ISL_938498, EPI_ISL_938499, EPI_ISL_938500, EPI_ISL_938501, EPI_ISL_938502, EPI_ISL_938503, EPI_ISL_938504, EPI_ISL_938505, EPI_ISL_938506, EPI_ISL_938507, EPI_ISL_938508, EPI_ISL_938509, EPI_ISL_938510, EPI_ISL_938511, EPI_ISL_938512, EPI_ISL_938513, EPI_ISL_938514, EPI_ISL_938515, EPI_ISL_938516, EPI_ISL_938517, EPI_ISL_938518, EPI_ISL_938519, EPI_ISL_938520, EPI_ISL_938521,
EPI_ISL_938522, EPI_ISL_938523, EPI_ISL_938524, EPI_ISL_938525, EPI_ISL_938526, EPI_ISL_938527, EPI_ISL_938528, EPI_ISL_938529, EPI_ISL_938530, EPI_ISL_938531, EPI_ISL_938532, EPI_ISL_938533, EPI_ISL_938534, EPI_ISL_938535, EPI_ISL_938536, EPI_ISL_938537, EPI_ISL_938538, EPI_ISL_938539, EPI_ISL_938540, EPI_ISL_938541, EPI_ISL_938542, EPI_ISL_938543, EPI_ISL_938544, EPI_ISL_938545,
EPI_ISL_938546, EPI_ISL_938547, EPI_ISL_938548, EPI_ISL_938549, EPI_ISL_938550, EPI_ISL_938551, EPI_ISL_938552, EPI_ISL_938553, EPI_ISL_938554, EPI_ISL_938555, EPI_ISL_938556, EPI_ISL_938557, EPI_ISL_938558, EPI_ISL_938559, EPI_ISL_938560, EPI_ISL_938561, EPI_ISL_938562, EPI_ISL_938563, EPI_ISL_938564, EPI_ISL_938565, EPI_ISL_938566, EPI_ISL_938567, EPI_ISL_938568, EPI_ISL_938569,
EPI_ISL_938570, EPI_ISL_938571, EPI_ISL_938572, EPI_ISL_938573, EPI_ISL_938574, EPI_ISL_938575, EPI_ISL_938576, EPI_ISL_938577, EPI_ISL_938578, EPI_ISL_948101, EPI_ISL_948116, EPI_ISL_948125, EPI_ISL_948128, EPI_ISL_948133, EPI_ISL_948150, EPI_ISL_948155, EPI_ISL_948159, EPI_ISL_948160, EPI_ISL_948177, EPI_ISL_948178, EPI_ISL_948185, EPI_ISL_948186, EPI_ISL_948194, EPI_ISL_948204,
EPI_ISL_948210, EPI_ISL_948215, EPI_ISL_948221, EPI_ISL_948231, EPI_ISL_948266, EPI_ISL_948289, EPI_ISL_948292, EPI_ISL_948294, EPI_ISL_948330, EPI_ISL_948368, EPI_ISL_948374, EPI_ISL_948376, EPI_ISL_948388, EPI_ISL_948407, EPI_ISL_987606, EPI_ISL_988214, EPI_ISL_988226, EPI_ISL_988228, EPI_ISL_988232, EPI_ISL_988247, EPI_ISL_988253, EPI_ISL_988255, EPI_ISL_988259, EPI_ISL_988264,
EPI_ISL_988268, EPI_ISL_988274, EPI_ISL_988280, EPI_ISL_988285, EPI_ISL_988293, EPI_ISL_988298, EPI_ISL_988300, EPI_ISL_988302, EPI_ISL_988304, EPI_ISL_988315, EPI_ISL_988323, EPI_ISL_988324, EPI_ISL_988333, EPI_ISL_988339, EPI_ISL_988340, EPI_ISL_988343, EPI_ISL_988355, EPI_ISL_988359, EPI_ISL_988381, EPI_ISL_988390, EPI_ISL_988397, EPI_ISL_988409, EPI_ISL_988420, EPI_ISL_988429,
EPI_ISL_988433, EPI_ISL_988435, EPI_ISL_988437, EPI_ISL_988442, EPI_ISL_988451, EPI_ISL_988456, EPI_ISL_988457, EPI_ISL_988465, EPI_ISL_988467, EPI_ISL_988475, EPI_ISL_988477, EPI_ISL_988478, EPI_ISL_988481, EPI_ISL_1007525

| | | | |
|---|---|---|---|
| see above | Lighthouse Lab in Cambridge | Wellcome Sanger Institute for the COVID-19 Genomics UK (COG-UK) Consortium | Cordelia Langford; David K. Jackson; Dominic Kwiatkowski; Ewan Harrison; Ian Johnston; John Sillitoe on behalf of the Wellcome Sanger Institute COVID-19 Surveillance Team; John Sillitoe on behalf of the Wellcome Sanger Institute COVID-19 Surveillance Team (http://www.sanger.ac.uk/covid-team); Rob Howes; Roberto Amato; Sonia Goncalves; The Lighthouse Lab in Cambridge and Alex Alderton |

EPI_ISL_891279, EPI_ISL_891315, EPI_ISL_891353, EPI_ISL_891358, EPI_ISL_891365, EPI_ISL_891375, EPI_ISL_891382, EPI_ISL_891402, EPI_ISL_891405, EPI_ISL_891424, EPI_ISL_891430, EPI_ISL_891440, EPI_ISL_891456, EPI_ISL_891469, EPI_ISL_891479, EPI_ISL_891487, EPI_ISL_891512, EPI_ISL_891532, EPI_ISL_891538, EPI_ISL_891542, EPI_ISL_891559, EPI_ISL_891603, EPI_ISL_891609, EPI_ISL_891611,
EPI_ISL_891613, EPI_ISL_891622, EPI_ISL_891639, EPI_ISL_891664, EPI_ISL_891668, EPI_ISL_891694, EPI_ISL_891711, EPI_ISL_891747, EPI_ISL_891749, EPI_ISL_891769, EPI_ISL_891819, EPI_ISL_891821, EPI_ISL_891823, EPI_ISL_891826, EPI_ISL_891834, EPI_ISL_891845, EPI_ISL_907120, EPI_ISL_907121, EPI_ISL_907122, EPI_ISL_907123, EPI_ISL_907124, EPI_ISL_907125, EPI_ISL_907126, EPI_ISL_907127,
EPI_ISL_907128, EPI_ISL_907129, EPI_ISL_907130, EPI_ISL_907131, EPI_ISL_907132, EPI_ISL_907134, EPI_ISL_907135, EPI_ISL_907137, EPI_ISL_907138, EPI_ISL_907140, EPI_ISL_907141, EPI_ISL_907142, EPI_ISL_907143, EPI_ISL_907145, EPI_ISL_907146, EPI_ISL_907149, EPI_ISL_907150, EPI_ISL_907151, EPI_ISL_907153, EPI_ISL_907154, EPI_ISL_907156, EPI_ISL_907157, EPI_ISL_907158, EPI_ISL_907159,
EPI_ISL_907161, EPI_ISL_907162, EPI_ISL_907165, EPI_ISL_907166, EPI_ISL_907169, EPI_ISL_907170, EPI_ISL_907171, EPI_ISL_907172, EPI_ISL_907174, EPI_ISL_907175, EPI_ISL_907176, EPI_ISL_907177, EPI_ISL_907178, EPI_ISL_907180, EPI_ISL_907181, EPI_ISL_907183, EPI_ISL_907184, EPI_ISL_907185, EPI_ISL_907188, EPI_ISL_907189, EPI_ISL_907191, EPI_ISL_907196, EPI_ISL_907197, EPI_ISL_907198,
EPI_ISL_907199, EPI_ISL_907201, EPI_ISL_907202, EPI_ISL_907203, EPI_ISL_907204, EPI_ISL_907205, EPI_ISL_907207, EPI_ISL_907208, EPI_ISL_907211, EPI_ISL_907213, EPI_ISL_907215, EPI_ISL_907216, EPI_ISL_907217, EPI_ISL_907218, EPI_ISL_907220, EPI_ISL_907226, EPI_ISL_907227, EPI_ISL_907228, EPI_ISL_907229, EPI_ISL_907230, EPI_ISL_907231, EPI_ISL_907232, EPI_ISL_907233, EPI_ISL_907234,
EPI_ISL_907235, EPI_ISL_907236, EPI_ISL_907239, EPI_ISL_907240, EPI_ISL_907242, EPI_ISL_907243, EPI_ISL_907244, EPI_ISL_907245, EPI_ISL_907246, EPI_ISL_907247, EPI_ISL_907248, EPI_ISL_907249, EPI_ISL_907251, EPI_ISL_907252, EPI_ISL_907253, EPI_ISL_907255, EPI_ISL_907257, EPI_ISL_907258, EPI_ISL_907259, EPI_ISL_907261, EPI_ISL_907262, EPI_ISL_907265, EPI_ISL_907266, EPI_ISL_907268,
EPI_ISL_907270, EPI_ISL_907271, EPI_ISL_907272, EPI_ISL_907274, EPI_ISL_907275, EPI_ISL_907276, EPI_ISL_907277, EPI_ISL_907278, EPI_ISL_907280, EPI_ISL_907282, EPI_ISL_907283, EPI_ISL_907285, EPI_ISL_907286, EPI_ISL_907289, EPI_ISL_907290, EPI_ISL_907291, EPI_ISL_907293, EPI_ISL_907294, EPI_ISL_907295, EPI_ISL_907296, EPI_ISL_907297, EPI_ISL_907298, EPI_ISL_907299, EPI_ISL_907301,
EPI_ISL_907302, EPI_ISL_907305, EPI_ISL_907306, EPI_ISL_907307, EPI_ISL_907309, EPI_ISL_907310, EPI_ISL_907311, EPI_ISL_907312, EPI_ISL_907313, EPI_ISL_907314, EPI_ISL_907316, EPI_ISL_907317, EPI_ISL_907319, EPI_ISL_907320, EPI_ISL_907324, EPI_ISL_907326, EPI_ISL_907327, EPI_ISL_907329, EPI_ISL_907330, EPI_ISL_907331, EPI_ISL_907332, EPI_ISL_907333, EPI_ISL_907334, EPI_ISL_907335,
EPI_ISL_907337, EPI_ISL_907338, EPI_ISL_907339, EPI_ISL_907340, EPI_ISL_907342, EPI_ISL_907343, EPI_ISL_907344, EPI_ISL_907345, EPI_ISL_907346, EPI_ISL_907349, EPI_ISL_907354, EPI_ISL_907356, EPI_ISL_907357, EPI_ISL_907358, EPI_ISL_907360, EPI_ISL_907361, EPI_ISL_907365, EPI_ISL_907366, EPI_ISL_907368, EPI_ISL_907370, EPI_ISL_907378, EPI_ISL_907379, EPI_ISL_907381, EPI_ISL_907384,
EPI_ISL_907385, EPI_ISL_907386, EPI_ISL_907393, EPI_ISL_907394, EPI_ISL_907395, EPI_ISL_907397, EPI_ISL_907398, EPI_ISL_907401, EPI_ISL_907402, EPI_ISL_907405, EPI_ISL_907408, EPI_ISL_907415, EPI_ISL_907418, EPI_ISL_907419, EPI_ISL_907420, EPI_ISL_907421, EPI_ISL_907423, EPI_ISL_907425, EPI_ISL_907426, EPI_ISL_907430, EPI_ISL_907431, EPI_ISL_907433, EPI_ISL_907434, EPI_ISL_907435,
EPI_ISL_907436, EPI_ISL_907439, EPI_ISL_907442, EPI_ISL_907445, EPI_ISL_907451, EPI_ISL_907453, EPI_ISL_907454, EPI_ISL_907455, EPI_ISL_907456, EPI_ISL_907461, EPI_ISL_907465, EPI_ISL_907469, EPI_ISL_907472, EPI_ISL_907473, EPI_ISL_907474, EPI_ISL_907475, EPI_ISL_907476, EPI_ISL_907481, EPI_ISL_907483, EPI_ISL_907486, EPI_ISL_907487, EPI_ISL_907488, EPI_ISL_907490, EPI_ISL_907494,
EPI_ISL_907495, EPI_ISL_907498, EPI_ISL_907499, EPI_ISL_907501, EPI_ISL_907502, EPI_ISL_907504, EPI_ISL_907506, EPI_ISL_907507, EPI_ISL_907508, EPI_ISL_907509, EPI_ISL_907513, EPI_ISL_907514, EPI_ISL_907515, EPI_ISL_907520, EPI_ISL_907524, EPI_ISL_907525, EPI_ISL_907527, EPI_ISL_907530, EPI_ISL_907531, EPI_ISL_907534, EPI_ISL_907535, EPI_ISL_907539, EPI_ISL_907540, EPI_ISL_907541,
EPI_ISL_907544, EPI_ISL_907545, EPI_ISL_907547, EPI_ISL_907548, EPI_ISL_907550, EPI_ISL_907553, EPI_ISL_907554, EPI_ISL_907555, EPI_ISL_907556, EPI_ISL_907558, EPI_ISL_907560, EPI_ISL_907562, EPI_ISL_907564, EPI_ISL_907565, EPI_ISL_907566, EPI_ISL_907569, EPI_ISL_907570, EPI_ISL_907572, EPI_ISL_907574, EPI_ISL_907575, EPI_ISL_907577, EPI_ISL_907581, EPI_ISL_907585, EPI_ISL_907586,
EPI_ISL_907588, EPI_ISL_907591, EPI_ISL_907592, EPI_ISL_907593, EPI_ISL_907595, EPI_ISL_907597, EPI_ISL_907599, EPI_ISL_907602, EPI_ISL_907603, EPI_ISL_907604, EPI_ISL_907608, EPI_ISL_907609, EPI_ISL_907613, EPI_ISL_907614, EPI_ISL_907617, EPI_ISL_907619, EPI_ISL_907620, EPI_ISL_907624, EPI_ISL_907625, EPI_ISL_907627, EPI_ISL_907629, EPI_ISL_907632, EPI_ISL_907633, EPI_ISL_907634,
EPI_ISL_907635, EPI_ISL_907637, EPI_ISL_907638, EPI_ISL_907639, EPI_ISL_907643, EPI_ISL_907644, EPI_ISL_907646, EPI_ISL_907649, EPI_ISL_907650, EPI_ISL_907653, EPI_ISL_907655, EPI_ISL_907656, EPI_ISL_907657, EPI_ISL_907660, EPI_ISL_907662, EPI_ISL_907663, EPI_ISL_907665, EPI_ISL_907666, EPI_ISL_907668, EPI_ISL_907669, EPI_ISL_907670, EPI_ISL_907671, EPI_ISL_907674, EPI_ISL_907678,
EPI_ISL_907679, EPI_ISL_907681, EPI_ISL_907682, EPI_ISL_907683, EPI_ISL_907685, EPI_ISL_907687, EPI_ISL_907688, EPI_ISL_907691, EPI_ISL_907698, EPI_ISL_907702, EPI_ISL_907703, EPI_ISL_907705, EPI_ISL_907706, EPI_ISL_907707, EPI_ISL_907708, EPI_ISL_907716, EPI_ISL_907721, EPI_ISL_907722, EPI_ISL_907723, EPI_ISL_907731, EPI_ISL_907735, EPI_ISL_907736, EPI_ISL_907738, EPI_ISL_907745,
EPI_ISL_907747, EPI_ISL_907749, EPI_ISL_907755, EPI_ISL_907756, EPI_ISL_907757, EPI_ISL_907760, EPI_ISL_907763, EPI_ISL_907765, EPI_ISL_907767, EPI_ISL_907771, EPI_ISL_907772, EPI_ISL_907773, EPI_ISL_907776, EPI_ISL_907778, EPI_ISL_907786, EPI_ISL_907787, EPI_ISL_907789, EPI_ISL_907793, EPI_ISL_907794, EPI_ISL_907795, EPI_ISL_907796, EPI_ISL_907797, EPI_ISL_907800, EPI_ISL_907801,
EPI_ISL_907802, EPI_ISL_907805, EPI_ISL_907809, EPI_ISL_907811, EPI_ISL_907812, EPI_ISL_907813, EPI_ISL_907814, EPI_ISL_907817, EPI_ISL_907820, EPI_ISL_907821, EPI_ISL_907824, EPI_ISL_907826, EPI_ISL_907832, EPI_ISL_907833, EPI_ISL_907834, EPI_ISL_907835, EPI_ISL_907837, EPI_ISL_907847, EPI_ISL_907851, EPI_ISL_907852, EPI_ISL_907854, EPI_ISL_907855, EPI_ISL_907860, EPI_ISL_907863,
EPI_ISL_907865, EPI_ISL_907867, EPI_ISL_907868, EPI_ISL_907872, EPI_ISL_907874, EPI_ISL_907875, EPI_ISL_907877, EPI_ISL_907878, EPI_ISL_907879, EPI_ISL_907881, EPI_ISL_907882, EPI_ISL_907889, EPI_ISL_907890, EPI_ISL_907891, EPI_ISL_907893, EPI_ISL_907894, EPI_ISL_907895, EPI_ISL_907898, EPI_ISL_907899, EPI_ISL_907902, EPI_ISL_907903, EPI_ISL_907911, EPI_ISL_907912, EPI_ISL_907914,
EPI_ISL_907915, EPI_ISL_907921, EPI_ISL_907923, EPI_ISL_907925, EPI_ISL_907926, EPI_ISL_907931, EPI_ISL_907934, EPI_ISL_907936, EPI_ISL_907940, EPI_ISL_907943, EPI_ISL_907944, EPI_ISL_907945, EPI_ISL_907946, EPI_ISL_907948, EPI_ISL_907949, EPI_ISL_907953, EPI_ISL_907956, EPI_ISL_907960, EPI_ISL_907967, EPI_ISL_907969, EPI_ISL_907970, EPI_ISL_907972, EPI_ISL_907973, EPI_ISL_907974,
EPI_ISL_907975, EPI_ISL_907976, EPI_ISL_907979, EPI_ISL_907981, EPI_ISL_907982, EPI_ISL_907984, EPI_ISL_907987, EPI_ISL_907988, EPI_ISL_907989, EPI_ISL_907994, EPI_ISL_907995, EPI_ISL_907998, EPI_ISL_908002, EPI_ISL_908003, EPI_ISL_908007, EPI_ISL_908009, EPI_ISL_908010, EPI_ISL_908011, EPI_ISL_908013, EPI_ISL_908016, EPI_ISL_908017, EPI_ISL_908021, EPI_ISL_908022, EPI_ISL_908025,
EPI_ISL_908026, EPI_ISL_908027, EPI_ISL_908028, EPI_ISL_908030, EPI_ISL_908033, EPI_ISL_908034, EPI_ISL_908275, EPI_ISL_908284, EPI_ISL_908289, EPI_ISL_908290, EPI_ISL_908291, EPI_ISL_908295, EPI_ISL_908302, EPI_ISL_908303, EPI_ISL_908307, EPI_ISL_908308, EPI_ISL_908317, EPI_ISL_908321, EPI_ISL_908323, EPI_ISL_908326, EPI_ISL_908329, EPI_ISL_908334, EPI_ISL_908335, EPI_ISL_908337,
EPI_ISL_908338, EPI_ISL_908340, EPI_ISL_908348, EPI_ISL_908353, EPI_ISL_908357, EPI_ISL_908360, EPI_ISL_908365, EPI_ISL_908366, EPI_ISL_908380, EPI_ISL_908383, EPI_ISL_908387, EPI_ISL_908393, EPI_ISL_908394, EPI_ISL_908403, EPI_ISL_908405, EPI_ISL_908411, EPI_ISL_908412, EPI_ISL_908416, EPI_ISL_908421, EPI_ISL_908422, EPI_ISL_908429, EPI_ISL_908431, EPI_ISL_908433, EPI_ISL_908442,
EPI_ISL_908443, EPI_ISL_908444, EPI_ISL_908447, EPI_ISL_908451, EPI_ISL_908455, EPI_ISL_908462, EPI_ISL_908470, EPI_ISL_908471, EPI_ISL_908481, EPI_ISL_908486, EPI_ISL_908488, EPI_ISL_908489, EPI_ISL_908495, EPI_ISL_908496, EPI_ISL_908500, EPI_ISL_908507, EPI_ISL_908508, EPI_ISL_908509, EPI_ISL_908512, EPI_ISL_908516, EPI_ISL_908521, EPI_ISL_908522, EPI_ISL_908525, EPI_ISL_908530,
EPI_ISL_908536, EPI_ISL_908538, EPI_ISL_908547, EPI_ISL_908551, EPI_ISL_908557, EPI_ISL_908558, EPI_ISL_908559, EPI_ISL_908564, EPI_ISL_908568, EPI_ISL_908573, EPI_ISL_908582, EPI_ISL_908589, EPI_ISL_908591, EPI_ISL_908595, EPI_ISL_909262, EPI_ISL_909272, EPI_ISL_909278, EPI_ISL_909283, EPI_ISL_909289, EPI_ISL_909296, EPI_ISL_909298, EPI_ISL_909303, EPI_ISL_909304, EPI_ISL_909306,
EPI_ISL_909317, EPI_ISL_909325, EPI_ISL_909333, EPI_ISL_909342, EPI_ISL_909348, EPI_ISL_909349, EPI_ISL_909353, EPI_ISL_909361, EPI_ISL_909371, EPI_ISL_909374, EPI_ISL_909376, EPI_ISL_909379, EPI_ISL_909387, EPI_ISL_909391, EPI_ISL_909392, EPI_ISL_909393, EPI_ISL_909403, EPI_ISL_909406, EPI_ISL_909407, EPI_ISL_909408, EPI_ISL_909410, EPI_ISL_909422, EPI_ISL_909428, EPI_ISL_909429,
EPI_ISL_909438, EPI_ISL_909468, EPI_ISL_909481, EPI_ISL_909490, EPI_ISL_909496, EPI_ISL_909512, EPI_ISL_909522, EPI_ISL_909524, EPI_ISL_909525, EPI_ISL_909528, EPI_ISL_909532, EPI_ISL_909534, EPI_ISL_909537, EPI_ISL_909544, EPI_ISL_909554, EPI_ISL_909557, EPI_ISL_909570, EPI_ISL_909582, EPI_ISL_909583, EPI_ISL_909589, EPI_ISL_909593, EPI_ISL_912212, EPI_ISL_912213, EPI_ISL_912219,
EPI_ISL_916503, EPI_ISL_916504, EPI_ISL_916505, EPI_ISL_916506, EPI_ISL_916507, EPI_ISL_916508, EPI_ISL_916509, EPI_ISL_916510, EPI_ISL_916511, EPI_ISL_916512, EPI_ISL_916513, EPI_ISL_916514, EPI_ISL_916515, EPI_ISL_916516, EPI_ISL_916517, EPI_ISL_916518, EPI_ISL_916519, EPI_ISL_916520, EPI_ISL_916521, EPI_ISL_916522, EPI_ISL_916523, EPI_ISL_916524, EPI_ISL_916525, EPI_ISL_916526,
EPI_ISL_916527, EPI_ISL_916528, EPI_ISL_916529, EPI_ISL_916530, EPI_ISL_916531, EPI_ISL_916532, EPI_ISL_916533, EPI_ISL_916534, EPI_ISL_916535, EPI_ISL_916536, EPI_ISL_916537, EPI_ISL_916538, EPI_ISL_916539, EPI_ISL_916540, EPI_ISL_916541, EPI_ISL_916542, EPI_ISL_916543, EPI_ISL_916544, EPI_ISL_916545, EPI_ISL_916546, EPI_ISL_916547, EPI_ISL_916548, EPI_ISL_916549, EPI_ISL_916550,
EPI_ISL_916551, EPI_ISL_916552, EPI_ISL_916553, EPI_ISL_916554, EPI_ISL_916555, EPI_ISL_916556, EPI_ISL_916557, EPI_ISL_916558, EPI_ISL_916559, EPI_ISL_916560, EPI_ISL_916561, EPI_ISL_916562, EPI_ISL_916563, EPI_ISL_916564, EPI_ISL_916565, EPI_ISL_916566, EPI_ISL_916567, EPI_ISL_916568, EPI_ISL_916569, EPI_ISL_916570, EPI_ISL_916571, EPI_ISL_916572, EPI_ISL_916573, EPI_ISL_916574,
EPI_ISL_916575, EPI_ISL_916576, EPI_ISL_916577, EPI_ISL_916578, EPI_ISL_916579, EPI_ISL_916580, EPI_ISL_916581, EPI_ISL_916582, EPI_ISL_916583, EPI_ISL_916584, EPI_ISL_916585, EPI_ISL_916586, EPI_ISL_916587, EPI_ISL_916588, EPI_ISL_916589, EPI_ISL_916590, EPI_ISL_916591, EPI_ISL_916592, EPI_ISL_916593, EPI_ISL_916594, EPI_ISL_916595, EPI_ISL_916596, EPI_ISL_916597, EPI_ISL_916598,
EPI_ISL_916599, EPI_ISL_916600, EPI_ISL_916601, EPI_ISL_916602, EPI_ISL_916603, EPI_ISL_916604, EPI_ISL_916605, EPI_ISL_916606, EPI_ISL_916607, EPI_ISL_916608, EPI_ISL_916609, EPI_ISL_916610, EPI_ISL_916611, EPI_ISL_916612, EPI_ISL_916613, EPI_ISL_916614, EPI_ISL_916615, EPI_ISL_916616, EPI_ISL_916617, EPI_ISL_916618, EPI_ISL_916619, EPI_ISL_916620, EPI_ISL_916621, EPI_ISL_917844,
EPI_ISL_917845, EPI_ISL_917846, EPI_ISL_917847, EPI_ISL_917848, EPI_ISL_917985, EPI_ISL_917986, EPI_ISL_917987, EPI_ISL_917988, EPI_ISL_917989, EPI_ISL_917990, EPI_ISL_917991, EPI_ISL_917992, EPI_ISL_917993, EPI_ISL_917994, EPI_ISL_917995, EPI_ISL_917996, EPI_ISL_917997, EPI_ISL_917998, EPI_ISL_917999, EPI_ISL_918000, EPI_ISL_918001, EPI_ISL_918002, EPI_ISL_918003, EPI_ISL_918004,
EPI_ISL_918005, EPI_ISL_918006, EPI_ISL_918007, EPI_ISL_918008, EPI_ISL_918009, EPI_ISL_918010, EPI_ISL_918011, EPI_ISL_918012, EPI_ISL_918013, EPI_ISL_918014, EPI_ISL_918015, EPI_ISL_918016, EPI_ISL_918017, EPI_ISL_918018, EPI_ISL_918019, EPI_ISL_918039, EPI_ISL_918040, EPI_ISL_918041, EPI_ISL_918042, EPI_ISL_918043, EPI_ISL_918044, EPI_ISL_918045, EPI_ISL_918046, EPI_ISL_918047,
EPI_ISL_918048, EPI_ISL_918049, EPI_ISL_918050, EPI_ISL_918051, EPI_ISL_918052, EPI_ISL_918053, EPI_ISL_918054, EPI_ISL_918055, EPI_ISL_918056, EPI_ISL_918057, EPI_ISL_918058, EPI_ISL_918059, EPI_ISL_932251, EPI_ISL_932252, EPI_ISL_932255, EPI_ISL_932256, EPI_ISL_932265, EPI_ISL_932266, EPI_ISL_932267, EPI_ISL_932269, EPI_ISL_932270, EPI_ISL_932271, EPI_ISL_932272, EPI_ISL_932275,
EPI_ISL_932277, EPI_ISL_932279, EPI_ISL_932280, EPI_ISL_932281, EPI_ISL_932282, EPI_ISL_932286, EPI_ISL_932289, EPI_ISL_932291, EPI_ISL_932293, EPI_ISL_932295, EPI_ISL_932296, EPI_ISL_932299, EPI_ISL_932301, EPI_ISL_932302, EPI_ISL_932304, EPI_ISL_932305, EPI_ISL_932308, EPI_ISL_932309, EPI_ISL_932311, EPI_ISL_932312, EPI_ISL_932314, EPI_ISL_932316, EPI_ISL_932317, EPI_ISL_932318,
EPI_ISL_932321, EPI_ISL_932322, EPI_ISL_932323, EPI_ISL_932326, EPI_ISL_932328, EPI_ISL_932330, EPI_ISL_932331, EPI_ISL_932333, EPI_ISL_932335, EPI_ISL_932336, EPI_ISL_932337, EPI_ISL_932338, EPI_ISL_932341, EPI_ISL_932342, EPI_ISL_932344, EPI_ISL_932349, EPI_ISL_932351, EPI_ISL_932352, EPI_ISL_932353, EPI_ISL_932355, EPI_ISL_932356, EPI_ISL_932359, EPI_ISL_932360, EPI_ISL_932361,
EPI_ISL_932362, EPI_ISL_932367, EPI_ISL_932369, EPI_ISL_932370, EPI_ISL_932373, EPI_ISL_932379, EPI_ISL_932381, EPI_ISL_932382, EPI_ISL_932384, EPI_ISL_932386, EPI_ISL_932389, EPI_ISL_932390, EPI_ISL_932394, EPI_ISL_932397, EPI_ISL_932398, EPI_ISL_932401, EPI_ISL_932404, EPI_ISL_932405, EPI_ISL_932407, EPI_ISL_932409, EPI_ISL_932410, EPI_ISL_932412, EPI_ISL_932415, EPI_ISL_932416,
EPI_ISL_932418, EPI_ISL_932420, EPI_ISL_932421, EPI_ISL_932422, EPI_ISL_932423, EPI_ISL_932424, EPI_ISL_932427, EPI_ISL_932431, EPI_ISL_932432, EPI_ISL_932435, EPI_ISL_932438, EPI_ISL_932448, EPI_ISL_932449, EPI_ISL_932451, EPI_ISL_932452, EPI_ISL_932454, EPI_ISL_932459, EPI_ISL_932461, EPI_ISL_932462, EPI_ISL_932464, EPI_ISL_932465, EPI_ISL_932466, EPI_ISL_932467, EPI_ISL_932482,
EPI_ISL_932487, EPI_ISL_932488, EPI_ISL_932490, EPI_ISL_932493, EPI_ISL_932494, EPI_ISL_932497, EPI_ISL_932498, EPI_ISL_932499, EPI_ISL_932500, EPI_ISL_932501, EPI_ISL_932513, EPI_ISL_932518, EPI_ISL_932523, EPI_ISL_932524, EPI_ISL_932526, EPI_ISL_932531, EPI_ISL_932533, EPI_ISL_932534, EPI_ISL_932535, EPI_ISL_932536, EPI_ISL_932537, EPI_ISL_932538, EPI_ISL_932540, EPI_ISL_932541,
EPI_ISL_932545, EPI_ISL_932547, EPI_ISL_932554, EPI_ISL_932556, EPI_ISL_932557, EPI_ISL_932558, EPI_ISL_932562, EPI_ISL_932564, EPI_ISL_932566, EPI_ISL_932568, EPI_ISL_932588, EPI_ISL_932606, EPI_ISL_932705, EPI_ISL_932711, EPI_ISL_932732, EPI_ISL_932746, EPI_ISL_932764, EPI_ISL_932781, EPI_ISL_932807, EPI_ISL_932831, EPI_ISL_944844, EPI_ISL_947208, EPI_ISL_947350, EPI_ISL_947352,
EPI_ISL_948377, EPI_ISL_948432, EPI_ISL_948829, EPI_ISL_948830, EPI_ISL_948834, EPI_ISL_948845, EPI_ISL_948848, EPI_ISL_948945, EPI_ISL_948946, EPI_ISL_948947, EPI_ISL_948948, EPI_ISL_948949, EPI_ISL_948950, EPI_ISL_948951, EPI_ISL_948952, EPI_ISL_948953, EPI_ISL_948954, EPI_ISL_948955, EPI_ISL_948956, EPI_ISL_948957, EPI_ISL_948958, EPI_ISL_948959, EPI_ISL_948960, EPI_ISL_948961,
EPI_ISL_948962, EPI_ISL_948963, EPI_ISL_948964, EPI_ISL_948965, EPI_ISL_948966, EPI_ISL_957101, EPI_ISL_991173, EPI_ISL_991354, EPI_ISL_991361, EPI_ISL_991366, EPI_ISL_991371, EPI_ISL_991380, EPI_ISL_991392, EPI_ISL_991394, EPI_ISL_991398, EPI_ISL_991400, EPI_ISL_991405, EPI_ISL_991415, EPI_ISL_991418, EPI_ISL_991423, EPI_ISL_991425, EPI_ISL_991440, EPI_ISL_991447, EPI_ISL_991454,
EPI_ISL_991463, EPI_ISL_991477, EPI_ISL_991479, EPI_ISL_991481, EPI_ISL_991483, EPI_ISL_991501, EPI_ISL_991510, EPI_ISL_991511, EPI_ISL_991523, EPI_ISL_991533, EPI_ISL_991541, EPI_ISL_991904, EPI_ISL_992043, EPI_ISL_992045, EPI_ISL_992047, EPI_ISL_992049, EPI_ISL_992051, EPI_ISL_992052, EPI_ISL_992056, EPI_ISL_992060, EPI_ISL_992065, EPI_ISL_992068, EPI_ISL_992070, EPI_ISL_992072,
EPI_ISL_992075, EPI_ISL_992084, EPI_ISL_992086, EPI_ISL_992088, EPI_ISL_992093, EPI_ISL_992094, EPI_ISL_992095, EPI_ISL_992096, EPI_ISL_992098, EPI_ISL_992102, EPI_ISL_992104, EPI_ISL_992106, EPI_ISL_992107, EPI_ISL_992110, EPI_ISL_992112, EPI_ISL_992113, EPI_ISL_992114, EPI_ISL_992115, EPI_ISL_992120, EPI_ISL_992121, EPI_ISL_992123, EPI_ISL_992124, EPI_ISL_992126, EPI_ISL_992129,
EPI_ISL_992132, EPI_ISL_992133, EPI_ISL_992134, EPI_ISL_992136, EPI_ISL_992137, EPI_ISL_992139, EPI_ISL_992140, EPI_ISL_992141, EPI_ISL_992142, EPI_ISL_992145, EPI_ISL_992148, EPI_ISL_992150, EPI_ISL_992154, EPI_ISL_992157, EPI_ISL_992158, EPI_ISL_992160, EPI_ISL_992162, EPI_ISL_992163, EPI_ISL_992165, EPI_ISL_992167, EPI_ISL_992168, EPI_ISL_992169, EPI_ISL_992170, EPI_ISL_992171,
EPI_ISL_992174, EPI_ISL_992176, EPI_ISL_992182, EPI_ISL_992185, EPI_ISL_992186, EPI_ISL_992190, EPI_ISL_992191, EPI_ISL_992192, EPI_ISL_992193, EPI_ISL_992194, EPI_ISL_992195, EPI_ISL_992199, EPI_ISL_992200, EPI_ISL_992201, EPI_ISL_992202, EPI_ISL_992203, EPI_ISL_992208, EPI_ISL_992210, EPI_ISL_992212, EPI_ISL_992213, EPI_ISL_992216, EPI_ISL_992217, EPI_ISL_992222, EPI_ISL_992223,
EPI_ISL_992224, EPI_ISL_992227, EPI_ISL_992228, EPI_ISL_992230, EPI_ISL_992231, EPI_ISL_992232, EPI_ISL_992233, EPI_ISL_992235, EPI_ISL_992238, EPI_ISL_992239, EPI_ISL_992242, EPI_ISL_992244, EPI_ISL_992245, EPI_ISL_992247, EPI_ISL_992248, EPI_ISL_992249, EPI_ISL_992251, EPI_ISL_992253, EPI_ISL_992254, EPI_ISL_992255, EPI_ISL_992256, EPI_ISL_992257, EPI_ISL_992259, EPI_ISL_992263,
EPI_ISL_992264, EPI_ISL_992266, EPI_ISL_992268, EPI_ISL_992270, EPI_ISL_992274, EPI_ISL_992277, EPI_ISL_992278, EPI_ISL_992280, EPI_ISL_992281, EPI_ISL_992284, EPI_ISL_992285, EPI_ISL_992290, EPI_ISL_992293, EPI_ISL_992296, EPI_ISL_992297, EPI_ISL_992298, EPI_ISL_992299, EPI_ISL_992300, EPI_ISL_992307, EPI_ISL_992308, EPI_ISL_992313, EPI_ISL_992316, EPI_ISL_992319, EPI_ISL_992320,
EPI_ISL_992324, EPI_ISL_992325, EPI_ISL_992326, EPI_ISL_992327, EPI_ISL_992328, EPI_ISL_992333, EPI_ISL_992334, EPI_ISL_992339, EPI_ISL_992340, EPI_ISL_992341, EPI_ISL_992342, EPI_ISL_992343, EPI_ISL_992346, EPI_ISL_992347, EPI_ISL_1012808, EPI_ISL_1019608, EPI_ISL_1019615, EPI_ISL_1019628, EPI_ISL_1019641, EPI_ISL_1019666, EPI_ISL_1019667, EPI_ISL_1019690, EPI_ISL_1019706,
EPI_ISL_1019711, EPI_ISL_1019730, EPI_ISL_1019732, EPI_ISL_1019741, EPI_ISL_1019744, EPI_ISL_1019747, EPI_ISL_1019754, EPI_ISL_1019781, EPI_ISL_1019782, EPI_ISL_1019784, EPI_ISL_1019786, EPI_ISL_1019787, EPI_ISL_1019789, EPI_ISL_1019791, EPI_ISL_1019792, EPI_ISL_1019794, EPI_ISL_1019796, EPI_ISL_1019797, EPI_ISL_1019799, EPI_ISL_1019801, EPI_ISL_1019802, EPI_ISL_1019804,
EPI_ISL_1019806, EPI_ISL_1019808, EPI_ISL_1019809, EPI_ISL_1019811, EPI_ISL_1019812, EPI_ISL_1019814, EPI_ISL_1365323

| | | | |
|---|---|---|---|
| see above | Lighthouse Lab in Glasgow | Wellcome Sanger Institute for the COVID-19 Genomics UK (COG-UK) Consortium | Anna Dominiczak and Alex Alderton; Carol Clugston; Cordelia Langford; David Gray; David K. Jackson; Dominic Kwiatkowski; Ewan Harrison; Harper VanSteenhouse; Ian Johnston; Jeffrey Barrett; John Sillitoe on behalf of the Wellcome Sanger Institute COVID-19 Surveillance Team; John Sillitoe on behalf of the Wellcome Sanger Institute COVID-19 Surveillance Team (http://www.sanger.ac.uk/covid-team); Roberto Amato; Sonia Goncalves; Yumi Kasai |

EPI_ISL_1050224, EPI_ISL_1050226, EPI_ISL_1050228, EPI_ISL_1050229, EPI_ISL_1050230, EPI_ISL_1050231, EPI_ISL_1050232, EPI_ISL_1104107, EPI_ISL_1104108, EPI_ISL_1104109, EPI_ISL_1104110, EPI_ISL_1104111, EPI_ISL_1104112, EPI_ISL_1104113, EPI_ISL_1104114, EPI_ISL_1104115, EPI_ISL_1104116, EPI_ISL_1104117, EPI_ISL_1104118, EPI_ISL_1104119, EPI_ISL_1104120, EPI_ISL_1104121,
EPI_ISL_1104122

| | | | |
|---|---|---|---|
| see above | Lighthouse Lab in Glasgow / MRC-University of Glasgow Centre for Virus Research | COVID-19 Genomics UK (COG-UK) Consortium | Alasdair MacLean; Alice Broos; Ana da Silva Filipe; Anna Dominiczak; Antonia Ho; Carol Clugston; Daniel Mair; David Gray; David L Robertson; Emma Thomson; Harper VanSteenhouse; James Shepherd; Jenna Nichols; Joseph Hughes; Kathy Li; Kathy Smollett; Kyriaki Nomikou; Lily Tong; Matthew Holden; Natasha Johnson; Richard Orton; Rory Gunson; Sarah McDonald; Sharif Shaaban; Sreenu Vattipally; Stephen Carmichael; Yumi Kasai |

EPI_ISL_882611, EPI_ISL_932253, EPI_ISL_932258, EPI_ISL_932259, EPI_ISL_932260, EPI_ISL_932262, EPI_ISL_932278, EPI_ISL_932283, EPI_ISL_932288, EPI_ISL_932298, EPI_ISL_932306, EPI_ISL_932307, EPI_ISL_932315, EPI_ISL_932320, EPI_ISL_932324, EPI_ISL_932327, EPI_ISL_932332, EPI_ISL_932339, EPI_ISL_932340, EPI_ISL_932343, EPI_ISL_932346, EPI_ISL_932347, EPI_ISL_932350, EPI_ISL_932354,
EPI_ISL_932363, EPI_ISL_932364, EPI_ISL_932365, EPI_ISL_932368, EPI_ISL_932374, EPI_ISL_932375, EPI_ISL_932377, EPI_ISL_932378, EPI_ISL_932383, EPI_ISL_932385, EPI_ISL_932387, EPI_ISL_932392, EPI_ISL_932396, EPI_ISL_932399, EPI_ISL_932400, EPI_ISL_932402, EPI_ISL_932406, EPI_ISL_932408, EPI_ISL_932411, EPI_ISL_932413, EPI_ISL_932414, EPI_ISL_932419, EPI_ISL_932426, EPI_ISL_932428,
EPI_ISL_932429, EPI_ISL_932430, EPI_ISL_932434, EPI_ISL_932437, EPI_ISL_932439, EPI_ISL_932441, EPI_ISL_932442, EPI_ISL_932443, EPI_ISL_932445, EPI_ISL_932446, EPI_ISL_932447, EPI_ISL_932450, EPI_ISL_932460, EPI_ISL_932468, EPI_ISL_932469, EPI_ISL_932471, EPI_ISL_932472, EPI_ISL_932474, EPI_ISL_932476, EPI_ISL_932477, EPI_ISL_932478, EPI_ISL_932481, EPI_ISL_932483, EPI_ISL_932484,
EPI_ISL_932485, EPI_ISL_932491, EPI_ISL_932492, EPI_ISL_932495, EPI_ISL_932496, EPI_ISL_932504, EPI_ISL_932506, EPI_ISL_932508, EPI_ISL_932509, EPI_ISL_932510, EPI_ISL_932514, EPI_ISL_932516, EPI_ISL_932519, EPI_ISL_932520, EPI_ISL_932521, EPI_ISL_932525, EPI_ISL_932527, EPI_ISL_932528, EPI_ISL_932529, EPI_ISL_932544, EPI_ISL_932546, EPI_ISL_932548, EPI_ISL_932549, EPI_ISL_932550,
EPI_ISL_932551, EPI_ISL_932553, EPI_ISL_932555, EPI_ISL_932559, EPI_ISL_932560, EPI_ISL_932561, EPI_ISL_932565, EPI_ISL_932569, EPI_ISL_932570, EPI_ISL_932571, EPI_ISL_932572, EPI_ISL_932573, EPI_ISL_932574, EPI_ISL_932577, EPI_ISL_932578, EPI_ISL_932579, EPI_ISL_932580, EPI_ISL_932582, EPI_ISL_932583, EPI_ISL_932584, EPI_ISL_932585, EPI_ISL_932586, EPI_ISL_932589, EPI_ISL_932590,
EPI_ISL_932591, EPI_ISL_932592, EPI_ISL_932593, EPI_ISL_932594, EPI_ISL_932595, EPI_ISL_932597, EPI_ISL_932600, EPI_ISL_932601, EPI_ISL_932602, EPI_ISL_932604, EPI_ISL_932605, EPI_ISL_932608, EPI_ISL_932609, EPI_ISL_932610, EPI_ISL_932612, EPI_ISL_932615, EPI_ISL_932616, EPI_ISL_932617, EPI_ISL_932618, EPI_ISL_932622, EPI_ISL_932623, EPI_ISL_932624, EPI_ISL_932625, EPI_ISL_932626,
EPI_ISL_932628, EPI_ISL_932629, EPI_ISL_932630, EPI_ISL_932632, EPI_ISL_932633, EPI_ISL_932634, EPI_ISL_932635, EPI_ISL_932636, EPI_ISL_932637, EPI_ISL_932638, EPI_ISL_932639, EPI_ISL_932641, EPI_ISL_932642, EPI_ISL_932643, EPI_ISL_932649, EPI_ISL_932650, EPI_ISL_932653, EPI_ISL_932654, EPI_ISL_932656, EPI_ISL_932657, EPI_ISL_932658, EPI_ISL_932659, EPI_ISL_932660, EPI_ISL_932662,
EPI_ISL_932663, EPI_ISL_932665, EPI_ISL_932667, EPI_ISL_932672, EPI_ISL_932675, EPI_ISL_932676, EPI_ISL_932679, EPI_ISL_932681, EPI_ISL_932682, EPI_ISL_932683, EPI_ISL_932684, EPI_ISL_932685, EPI_ISL_932688, EPI_ISL_932692, EPI_ISL_932694, EPI_ISL_932695, EPI_ISL_932697, EPI_ISL_932698, EPI_ISL_932699, EPI_ISL_932700, EPI_ISL_932701, EPI_ISL_932702, EPI_ISL_932707, EPI_ISL_932708,
EPI_ISL_932713, EPI_ISL_932714, EPI_ISL_932716, EPI_ISL_932717, EPI_ISL_932719, EPI_ISL_932720, EPI_ISL_932723, EPI_ISL_932724, EPI_ISL_932726, EPI_ISL_932727, EPI_ISL_932729, EPI_ISL_932730, EPI_ISL_932731, EPI_ISL_932733, EPI_ISL_932735, EPI_ISL_932736, EPI_ISL_932738, EPI_ISL_932740, EPI_ISL_932741, EPI_ISL_932744, EPI_ISL_932745, EPI_ISL_932747, EPI_ISL_932748, EPI_ISL_932750,
EPI_ISL_932751, EPI_ISL_932752, EPI_ISL_932757, EPI_ISL_932759, EPI_ISL_932761, EPI_ISL_932762, EPI_ISL_932763, EPI_ISL_932765, EPI_ISL_932766, EPI_ISL_932767, EPI_ISL_932769, EPI_ISL_932772, EPI_ISL_932773, EPI_ISL_932774, EPI_ISL_932775, EPI_ISL_932777, EPI_ISL_932779, EPI_ISL_932780, EPI_ISL_932783, EPI_ISL_932784, EPI_ISL_932785, EPI_ISL_932786, EPI_ISL_932787, EPI_ISL_932788,
EPI_ISL_932789, EPI_ISL_932792, EPI_ISL_932795, EPI_ISL_932797, EPI_ISL_932798, EPI_ISL_932799, EPI_ISL_932801, EPI_ISL_932802, EPI_ISL_932803, EPI_ISL_932804, EPI_ISL_932806, EPI_ISL_932809, EPI_ISL_932810, EPI_ISL_932811, EPI_ISL_932813, EPI_ISL_932815, EPI_ISL_932819, EPI_ISL_932821, EPI_ISL_932823, EPI_ISL_932824, EPI_ISL_932825, EPI_ISL_932826, EPI_ISL_932828, EPI_ISL_932833,
EPI_ISL_932837, EPI_ISL_932841, EPI_ISL_932845, EPI_ISL_932846, EPI_ISL_932847, EPI_ISL_932848, EPI_ISL_932849, EPI_ISL_932851, EPI_ISL_932852, EPI_ISL_932854, EPI_ISL_932861, EPI_ISL_932869, EPI_ISL_932872, EPI_ISL_932873, EPI_ISL_932875, EPI_ISL_932877, EPI_ISL_932880, EPI_ISL_932887, EPI_ISL_932888, EPI_ISL_932892, EPI_ISL_932898, EPI_ISL_932904, EPI_ISL_932909, EPI_ISL_932910,
EPI_ISL_932916, EPI_ISL_932926, EPI_ISL_932928, EPI_ISL_932933, EPI_ISL_932934, EPI_ISL_932941, EPI_ISL_932945, EPI_ISL_932946, EPI_ISL_932950, EPI_ISL_932951, EPI_ISL_932953, EPI_ISL_932954, EPI_ISL_932959, EPI_ISL_932960, EPI_ISL_932961, EPI_ISL_932962, EPI_ISL_932963, EPI_ISL_932968, EPI_ISL_932969, EPI_ISL_932971, EPI_ISL_932977, EPI_ISL_932978, EPI_ISL_932985, EPI_ISL_932989,
EPI_ISL_932992, EPI_ISL_932997, EPI_ISL_932998, EPI_ISL_932999, EPI_ISL_933005, EPI_ISL_933008, EPI_ISL_933012, EPI_ISL_933014, EPI_ISL_933016, EPI_ISL_933019, EPI_ISL_933023, EPI_ISL_933027, EPI_ISL_933030, EPI_ISL_933032, EPI_ISL_933034, EPI_ISL_933039, EPI_ISL_933043, EPI_ISL_933046, EPI_ISL_933047, EPI_ISL_933048, EPI_ISL_933055, EPI_ISL_933057, EPI_ISL_933060, EPI_ISL_933063,
EPI_ISL_933068, EPI_ISL_933071, EPI_ISL_933075, EPI_ISL_933076, EPI_ISL_933077, EPI_ISL_933078, EPI_ISL_933081, EPI_ISL_933082, EPI_ISL_933085, EPI_ISL_933088, EPI_ISL_933091, EPI_ISL_933101, EPI_ISL_933107, EPI_ISL_933108, EPI_ISL_937886, EPI_ISL_937887, EPI_ISL_937888, EPI_ISL_937889, EPI_ISL_937890, EPI_ISL_937891, EPI_ISL_937892, EPI_ISL_937893, EPI_ISL_937894, EPI_ISL_937895,

| | | | |
|---|---|---|---|
| EPI_ISL_937896, EPI_ISL_937897, EPI_ISL_937898, EPI_ISL_937899, EPI_ISL_937900, EPI_ISL_937901, EPI_ISL_937902, EPI_ISL_937903, EPI_ISL_937904, EPI_ISL_937905, EPI_ISL_937906, EPI_ISL_937907, EPI_ISL_937908, EPI_ISL_937909, EPI_ISL_937910, EPI_ISL_937911, EPI_ISL_937912, EPI_ISL_937913, EPI_ISL_937914, EPI_ISL_937915, EPI_ISL_937916, EPI_ISL_937917, EPI_ISL_937918, EPI_ISL_937919, EPI_ISL_937920, EPI_ISL_937921, EPI_ISL_937922, EPI_ISL_937923, EPI_ISL_937924, EPI_ISL_937925, EPI_ISL_937926, EPI_ISL_937927, EPI_ISL_937928, EPI_ISL_937929, EPI_ISL_937930, EPI_ISL_937931, EPI_ISL_937932, EPI_ISL_937933, EPI_ISL_937934, EPI_ISL_937935, EPI_ISL_937936, EPI_ISL_937937, EPI_ISL_937938, EPI_ISL_937939, EPI_ISL_937940, EPI_ISL_937941, EPI_ISL_937942, EPI_ISL_937943, EPI_ISL_937944, EPI_ISL_937945, EPI_ISL_937946, EPI_ISL_937947, EPI_ISL_937948, EPI_ISL_937949, EPI_ISL_937950, EPI_ISL_937951, EPI_ISL_937952, EPI_ISL_937953, EPI_ISL_937954, EPI_ISL_937955, EPI_ISL_937956, EPI_ISL_937957, EPI_ISL_937958, EPI_ISL_937959, EPI_ISL_937960, EPI_ISL_937961, EPI_ISL_937962, EPI_ISL_937963, EPI_ISL_937964, EPI_ISL_937965, EPI_ISL_937966, EPI_ISL_937967, EPI_ISL_937968, EPI_ISL_937969, EPI_ISL_937970, EPI_ISL_937971, EPI_ISL_937972, EPI_ISL_937973, EPI_ISL_937974, EPI_ISL_937975, EPI_ISL_937976, EPI_ISL_937977, EPI_ISL_937978, EPI_ISL_937979, EPI_ISL_937980, EPI_ISL_937981, EPI_ISL_937982, EPI_ISL_937983, EPI_ISL_937984, EPI_ISL_937985, EPI_ISL_937986, EPI_ISL_937987, EPI_ISL_937988, EPI_ISL_937989, EPI_ISL_937990, EPI_ISL_937991, EPI_ISL_937992, EPI_ISL_937993, EPI_ISL_937994, EPI_ISL_937995, EPI_ISL_937996, EPI_ISL_937997, EPI_ISL_937998, EPI_ISL_937999, EPI_ISL_938000, EPI_ISL_938001, EPI_ISL_938002, EPI_ISL_938003, EPI_ISL_938004, EPI_ISL_938005, EPI_ISL_938006, EPI_ISL_938007, EPI_ISL_938008, EPI_ISL_938009, EPI_ISL_938010, EPI_ISL_938011, EPI_ISL_938012, EPI_ISL_938013, EPI_ISL_938014, EPI_ISL_938015, EPI_ISL_938016, EPI_ISL_938017, EPI_ISL_938018, EPI_ISL_938019, EPI_ISL_938020, EPI_ISL_938021, EPI_ISL_938022, EPI_ISL_938023, EPI_ISL_938024, EPI_ISL_938025, EPI_ISL_938026, EPI_ISL_938027, EPI_ISL_938028, EPI_ISL_938029, EPI_ISL_938030, EPI_ISL_938031, EPI_ISL_938032, EPI_ISL_938033, EPI_ISL_938034, EPI_ISL_938035, EPI_ISL_938036, EPI_ISL_938037, EPI_ISL_938038, EPI_ISL_938039, EPI_ISL_938040, EPI_ISL_938041, EPI_ISL_938042, EPI_ISL_938043, EPI_ISL_938044, EPI_ISL_938045, EPI_ISL_938046, EPI_ISL_938047, EPI_ISL_938048, EPI_ISL_938049, EPI_ISL_938050, EPI_ISL_938051, EPI_ISL_938052, EPI_ISL_938053, EPI_ISL_938054, EPI_ISL_938055, EPI_ISL_938056, EPI_ISL_938058, EPI_ISL_938059, EPI_ISL_938060, EPI_ISL_938061, EPI_ISL_938062, EPI_ISL_938063, EPI_ISL_938064, EPI_ISL_938065, EPI_ISL_938066, EPI_ISL_938067, EPI_ISL_938068, EPI_ISL_938069, EPI_ISL_938070, EPI_ISL_938071, EPI_ISL_938072, EPI_ISL_938073, EPI_ISL_938074, EPI_ISL_938075, EPI_ISL_938076, EPI_ISL_938077, EPI_ISL_938078, EPI_ISL_938079, EPI_ISL_938080, EPI_ISL_938081, EPI_ISL_938082, EPI_ISL_938083, EPI_ISL_938084, EPI_ISL_938085, EPI_ISL_938086, EPI_ISL_938087, EPI_ISL_938088, EPI_ISL_938089, EPI_ISL_938090, EPI_ISL_938091, EPI_ISL_938092, EPI_ISL_938093, EPI_ISL_938094, EPI_ISL_938095, EPI_ISL_938096, EPI_ISL_938097, EPI_ISL_938098, EPI_ISL_938099, EPI_ISL_938100, EPI_ISL_938101, EPI_ISL_938102, EPI_ISL_938103, EPI_ISL_938104, EPI_ISL_938105, EPI_ISL_938106, EPI_ISL_938107, EPI_ISL_938108, EPI_ISL_938109, EPI_ISL_938110, EPI_ISL_938111, EPI_ISL_938112, EPI_ISL_938113, EPI_ISL_938114, EPI_ISL_938115, EPI_ISL_938116, EPI_ISL_938117, EPI_ISL_938118, EPI_ISL_938119, EPI_ISL_938121, EPI_ISL_938122, EPI_ISL_938123, EPI_ISL_938124, EPI_ISL_938125, EPI_ISL_938126, EPI_ISL_938127, EPI_ISL_938128, EPI_ISL_938129, EPI_ISL_938130, EPI_ISL_938131, EPI_ISL_938132, EPI_ISL_938133, EPI_ISL_938134, EPI_ISL_938135, EPI_ISL_938136, EPI_ISL_938137, EPI_ISL_938138, EPI_ISL_938139, EPI_ISL_938140, EPI_ISL_938141, EPI_ISL_938142, EPI_ISL_938143, EPI_ISL_938144, EPI_ISL_938145, EPI_ISL_938146, EPI_ISL_938147, EPI_ISL_938148, EPI_ISL_938149, EPI_ISL_938150, EPI_ISL_938152, EPI_ISL_938153, EPI_ISL_938154, EPI_ISL_938155, EPI_ISL_938156, EPI_ISL_938157, EPI_ISL_938158, EPI_ISL_938159, EPI_ISL_938160, EPI_ISL_938161, EPI_ISL_938162, EPI_ISL_938163, EPI_ISL_938164, EPI_ISL_938165, EPI_ISL_938166, EPI_ISL_938167, EPI_ISL_938168, EPI_ISL_938169, EPI_ISL_938170, EPI_ISL_938171, EPI_ISL_938172, EPI_ISL_938173, EPI_ISL_938174, EPI_ISL_938175, EPI_ISL_938176, EPI_ISL_938177, EPI_ISL_938178, EPI_ISL_938179, EPI_ISL_938180, EPI_ISL_938181, EPI_ISL_938182, EPI_ISL_938183, EPI_ISL_938184, EPI_ISL_938185, EPI_ISL_938186, EPI_ISL_938187, EPI_ISL_938188, EPI_ISL_938189, EPI_ISL_938190, EPI_ISL_938191, EPI_ISL_938192, EPI_ISL_938193, EPI_ISL_938194, EPI_ISL_938195, EPI_ISL_938196, EPI_ISL_938197, EPI_ISL_938198, EPI_ISL_938199, EPI_ISL_938200, EPI_ISL_938201, EPI_ISL_938202, EPI_ISL_938203, EPI_ISL_938204, EPI_ISL_938205, EPI_ISL_938206, EPI_ISL_938207, EPI_ISL_938208, EPI_ISL_938209, EPI_ISL_938210, EPI_ISL_938211, EPI_ISL_938212, EPI_ISL_938213, EPI_ISL_938214, EPI_ISL_938215, EPI_ISL_938216, EPI_ISL_938217, EPI_ISL_938218, EPI_ISL_938219, EPI_ISL_938220, EPI_ISL_938221, EPI_ISL_938222, EPI_ISL_938223, EPI_ISL_938224, EPI_ISL_938225, EPI_ISL_938226, EPI_ISL_938227, EPI_ISL_938228, EPI_ISL_938229, EPI_ISL_938230, EPI_ISL_938231, EPI_ISL_938232, EPI_ISL_938233, EPI_ISL_938581, EPI_ISL_938587, EPI_ISL_938592, EPI_ISL_938596, EPI_ISL_938599, EPI_ISL_938602, EPI_ISL_938603, EPI_ISL_938607, EPI_ISL_938610, EPI_ISL_938612, EPI_ISL_938613, EPI_ISL_938614, EPI_ISL_938619, EPI_ISL_938630, EPI_ISL_938631, EPI_ISL_938632, EPI_ISL_938633, EPI_ISL_938636, EPI_ISL_938637, EPI_ISL_938643, EPI_ISL_938645, EPI_ISL_938648, EPI_ISL_938650, EPI_ISL_938652, EPI_ISL_938657, EPI_ISL_938666, EPI_ISL_938667, EPI_ISL_938676, EPI_ISL_938684, EPI_ISL_938692, EPI_ISL_938694, EPI_ISL_938700, EPI_ISL_938701, EPI_ISL_938707, EPI_ISL_938721, EPI_ISL_938729, EPI_ISL_938731, EPI_ISL_938732, EPI_ISL_938733, EPI_ISL_938737, EPI_ISL_938741, EPI_ISL_938743, EPI_ISL_938744, EPI_ISL_938746, EPI_ISL_938747, EPI_ISL_938750, EPI_ISL_938751, EPI_ISL_938752, EPI_ISL_938753, EPI_ISL_938756, EPI_ISL_938757, EPI_ISL_938758, EPI_ISL_938762, EPI_ISL_938764, EPI_ISL_938769, EPI_ISL_938771, EPI_ISL_938777, EPI_ISL_938783, EPI_ISL_938785, EPI_ISL_938787, EPI_ISL_938794, EPI_ISL_938796, EPI_ISL_938797, EPI_ISL_938800, EPI_ISL_938801, EPI_ISL_938803, EPI_ISL_938804, EPI_ISL_938806, EPI_ISL_938807, EPI_ISL_938810, EPI_ISL_938811, EPI_ISL_938815, EPI_ISL_938829, EPI_ISL_938837, EPI_ISL_938840, EPI_ISL_938845, EPI_ISL_938846, EPI_ISL_938854, EPI_ISL_938865, EPI_ISL_938875, EPI_ISL_938876, EPI_ISL_938882, EPI_ISL_938885, EPI_ISL_938886, EPI_ISL_938900, EPI_ISL_938901, EPI_ISL_938903, EPI_ISL_938908, EPI_ISL_938911, EPI_ISL_938913, EPI_ISL_938916, EPI_ISL_938921, EPI_ISL_938931, EPI_ISL_938933, EPI_ISL_938934, EPI_ISL_938944, EPI_ISL_938945, EPI_ISL_938946, EPI_ISL_938947, EPI_ISL_938948, EPI_ISL_938949, EPI_ISL_938950, EPI_ISL_938951, EPI_ISL_938952, EPI_ISL_938953, EPI_ISL_938954, EPI_ISL_938955, EPI_ISL_938956, EPI_ISL_938957, EPI_ISL_938958, EPI_ISL_938959, EPI_ISL_938960, EPI_ISL_938961, EPI_ISL_938962, EPI_ISL_938963, EPI_ISL_938964, EPI_ISL_938965, EPI_ISL_938966, EPI_ISL_938967, EPI_ISL_938968, EPI_ISL_938969, EPI_ISL_938970, EPI_ISL_938971, EPI_ISL_938972, EPI_ISL_938973, EPI_ISL_938974, EPI_ISL_938975, EPI_ISL_938976, EPI_ISL_938977, EPI_ISL_938978, EPI_ISL_938979, EPI_ISL_938980, EPI_ISL_938981, EPI_ISL_938982, EPI_ISL_938983, EPI_ISL_938984, EPI_ISL_938985, EPI_ISL_938986, EPI_ISL_938987, EPI_ISL_938988, EPI_ISL_938989, EPI_ISL_938990, EPI_ISL_938991, EPI_ISL_938992, EPI_ISL_938993, EPI_ISL_938994, EPI_ISL_938995, EPI_ISL_938996, EPI_ISL_938997, EPI_ISL_938998, EPI_ISL_938999, EPI_ISL_939000, EPI_ISL_939001, EPI_ISL_939002, EPI_ISL_939003, EPI_ISL_939004, EPI_ISL_939005, EPI_ISL_939006, EPI_ISL_939007, EPI_ISL_939008, EPI_ISL_939009, EPI_ISL_939010, EPI_ISL_939011, EPI_ISL_939012, EPI_ISL_939013, EPI_ISL_939014, EPI_ISL_939015, EPI_ISL_939016, EPI_ISL_939017, EPI_ISL_939018, EPI_ISL_939019, EPI_ISL_939020, EPI_ISL_939021, EPI_ISL_939022, EPI_ISL_939023, EPI_ISL_939024, EPI_ISL_939025, EPI_ISL_939026, EPI_ISL_939027, EPI_ISL_939028, EPI_ISL_939029, EPI_ISL_939030, EPI_ISL_939031, EPI_ISL_939032, EPI_ISL_939033, EPI_ISL_939034, EPI_ISL_939035, EPI_ISL_939036, EPI_ISL_939037, EPI_ISL_939038, EPI_ISL_939039, EPI_ISL_939040, EPI_ISL_939041, EPI_ISL_939042, EPI_ISL_939043, EPI_ISL_939044, EPI_ISL_939045, EPI_ISL_939046, EPI_ISL_939047, EPI_ISL_939048, EPI_ISL_939049, EPI_ISL_939050, EPI_ISL_939051, EPI_ISL_939052, EPI_ISL_939053, EPI_ISL_939054, EPI_ISL_939055, EPI_ISL_939056, EPI_ISL_939057, EPI_ISL_939058, EPI_ISL_939059, EPI_ISL_939060, EPI_ISL_939061, EPI_ISL_939062, EPI_ISL_939063, EPI_ISL_939064, EPI_ISL_939065, EPI_ISL_939066, EPI_ISL_939067, EPI_ISL_939068, EPI_ISL_939069, EPI_ISL_939070, EPI_ISL_939071, EPI_ISL_939072, EPI_ISL_939073, EPI_ISL_939074, EPI_ISL_939075, EPI_ISL_939076, EPI_ISL_939077, EPI_ISL_939078, EPI_ISL_939079, EPI_ISL_939080, EPI_ISL_939081, EPI_ISL_939082, EPI_ISL_939083, EPI_ISL_939084, EPI_ISL_939085, EPI_ISL_939086, EPI_ISL_939087, EPI_ISL_939088, EPI_ISL_939089, EPI_ISL_939090, EPI_ISL_939091, EPI_ISL_939092, EPI_ISL_939093, EPI_ISL_939094, EPI_ISL_939095, EPI_ISL_939096, EPI_ISL_939097, EPI_ISL_939098, EPI_ISL_939099, EPI_ISL_939100, EPI_ISL_939101, EPI_ISL_939102, EPI_ISL_939103, EPI_ISL_939104, EPI_ISL_939105, EPI_ISL_939106, EPI_ISL_939107, EPI_ISL_939108, EPI_ISL_939109, EPI_ISL_939110, EPI_ISL_939111, EPI_ISL_939112, EPI_ISL_939113, EPI_ISL_939114, EPI_ISL_939115, EPI_ISL_939116, EPI_ISL_939117, EPI_ISL_939118, EPI_ISL_939119, EPI_ISL_939120, EPI_ISL_939121, EPI_ISL_939122, EPI_ISL_939123, EPI_ISL_939124, EPI_ISL_939125, EPI_ISL_939126, EPI_ISL_939127, EPI_ISL_939128, EPI_ISL_939129, EPI_ISL_939130, EPI_ISL_939131, EPI_ISL_939132, EPI_ISL_939133, EPI_ISL_939134, EPI_ISL_939135, EPI_ISL_939136, EPI_ISL_939137, EPI_ISL_939138, EPI_ISL_939139, EPI_ISL_939140, EPI_ISL_939141, EPI_ISL_939142, EPI_ISL_939143, EPI_ISL_939144, EPI_ISL_939145, EPI_ISL_939146, EPI_ISL_939147, EPI_ISL_939148, EPI_ISL_939149, EPI_ISL_939150, EPI_ISL_939151, EPI_ISL_939152, EPI_ISL_939153, EPI_ISL_939154, EPI_ISL_939155, EPI_ISL_939156, EPI_ISL_939157, EPI_ISL_939158, EPI_ISL_939159, EPI_ISL_939160, EPI_ISL_939161, EPI_ISL_939162, EPI_ISL_939163, EPI_ISL_939164, EPI_ISL_939165, EPI_ISL_939166, EPI_ISL_939167, EPI_ISL_939168, EPI_ISL_939169, EPI_ISL_939170, EPI_ISL_939171, EPI_ISL_939172, EPI_ISL_939173, EPI_ISL_939174, EPI_ISL_939175, EPI_ISL_939176, EPI_ISL_939177, EPI_ISL_939178, EPI_ISL_939179, EPI_ISL_939180, EPI_ISL_939181, EPI_ISL_939182, EPI_ISL_939183, EPI_ISL_939184, EPI_ISL_939185, EPI_ISL_939186, EPI_ISL_939187, EPI_ISL_939188, EPI_ISL_939189, EPI_ISL_939190, EPI_ISL_939191, EPI_ISL_939192, EPI_ISL_939193, EPI_ISL_939194, EPI_ISL_939195, EPI_ISL_939196, EPI_ISL_939197, EPI_ISL_939198, EPI_ISL_939199, EPI_ISL_939200, EPI_ISL_939201, EPI_ISL_939202, EPI_ISL_939203, EPI_ISL_939204, EPI_ISL_939205, EPI_ISL_939206, EPI_ISL_939207, EPI_ISL_939208, EPI_ISL_939209, EPI_ISL_939210, EPI_ISL_939211, EPI_ISL_939212, EPI_ISL_939213, EPI_ISL_939214, EPI_ISL_939215, EPI_ISL_939216, EPI_ISL_939217, EPI_ISL_939218, EPI_ISL_939219, EPI_ISL_939220, EPI_ISL_939221, EPI_ISL_939222, EPI_ISL_939223, EPI_ISL_939224, EPI_ISL_939225, EPI_ISL_939226, EPI_ISL_939227, EPI_ISL_939228, EPI_ISL_939229, EPI_ISL_939230, EPI_ISL_939231, EPI_ISL_939232, EPI_ISL_939233, EPI_ISL_939234, EPI_ISL_939235, EPI_ISL_939236, EPI_ISL_939237, EPI_ISL_939238, EPI_ISL_939239, EPI_ISL_939240, EPI_ISL_939241, EPI_ISL_939242, EPI_ISL_939243, EPI_ISL_939244, EPI_ISL_939245, EPI_ISL_939246, EPI_ISL_939247, EPI_ISL_939248, EPI_ISL_939249, EPI_ISL_939250, EPI_ISL_939251, EPI_ISL_939252, EPI_ISL_939253, EPI_ISL_939254, EPI_ISL_939255, EPI_ISL_939256, EPI_ISL_939257, EPI_ISL_939258, EPI_ISL_939259, EPI_ISL_939260, EPI_ISL_939261, EPI_ISL_939262, EPI_ISL_939263, EPI_ISL_939264, EPI_ISL_939265, EPI_ISL_939266, EPI_ISL_939267, EPI_ISL_939268, EPI_ISL_939269, EPI_ISL_939270, EPI_ISL_939271, EPI_ISL_939272, EPI_ISL_939273, EPI_ISL_939274, EPI_ISL_939275, EPI_ISL_939276, EPI_ISL_939277, EPI_ISL_939278, EPI_ISL_939279, EPI_ISL_939280, EPI_ISL_939281, EPI_ISL_939282, EPI_ISL_939286, EPI_ISL_939288, EPI_ISL_939290, EPI_ISL_939291, EPI_ISL_939292, EPI_ISL_939293, EPI_ISL_939294, EPI_ISL_939296, EPI_ISL_939300, EPI_ISL_939301, EPI_ISL_939302, EPI_ISL_939306, EPI_ISL_939307, EPI_ISL_939313, EPI_ISL_939314, EPI_ISL_939315, EPI_ISL_939318, EPI_ISL_939319, EPI_ISL_939320, EPI_ISL_939321, EPI_ISL_939327, EPI_ISL_939329, EPI_ISL_939332, EPI_ISL_939334, EPI_ISL_939337, EPI_ISL_939339, EPI_ISL_939340, EPI_ISL_939343, EPI_ISL_939346, EPI_ISL_939348, EPI_ISL_939350, EPI_ISL_939351, EPI_ISL_939352, EPI_ISL_939354, EPI_ISL_939355, EPI_ISL_939356, EPI_ISL_939357, EPI_ISL_939358, EPI_ISL_939359, EPI_ISL_939361, EPI_ISL_939364, EPI_ISL_939365, EPI_ISL_939368, EPI_ISL_939369, EPI_ISL_939370, EPI_ISL_939372, EPI_ISL_939373, EPI_ISL_939378, EPI_ISL_939380, EPI_ISL_939388, EPI_ISL_939389, EPI_ISL_939390, EPI_ISL_939391, EPI_ISL_939393, EPI_ISL_939395, EPI_ISL_939396, EPI_ISL_939397, EPI_ISL_939398, EPI_ISL_939399, EPI_ISL_939401, EPI_ISL_939402, EPI_ISL_939403, EPI_ISL_939404, EPI_ISL_939405, EPI_ISL_939406, EPI_ISL_939407, EPI_ISL_939413, EPI_ISL_939414, EPI_ISL_939415, EPI_ISL_939416, EPI_ISL_939418, EPI_ISL_939420, EPI_ISL_939422, EPI_ISL_939425, EPI_ISL_939428, EPI_ISL_939429, EPI_ISL_939431, EPI_ISL_939432, EPI_ISL_939438, EPI_ISL_939439, EPI_ISL_939443, EPI_ISL_939450, EPI_ISL_939454, EPI_ISL_939456, EPI_ISL_939458, EPI_ISL_939459, EPI_ISL_939460, EPI_ISL_939461, EPI_ISL_939462, EPI_ISL_939466, EPI_ISL_939468, EPI_ISL_939469, EPI_ISL_939472, EPI_ISL_939473, EPI_ISL_939477, EPI_ISL_939478, EPI_ISL_939481, EPI_ISL_939483, EPI_ISL_939485, EPI_ISL_939487, EPI_ISL_939492, EPI_ISL_939493, EPI_ISL_939494, EPI_ISL_939497, EPI_ISL_939498, EPI_ISL_939500, EPI_ISL_939504, EPI_ISL_939505, EPI_ISL_939506, EPI_ISL_939507, EPI_ISL_939508, EPI_ISL_939509, EPI_ISL_939510, EPI_ISL_939512, EPI_ISL_939513, EPI_ISL_939514, EPI_ISL_939517, EPI_ISL_939519, EPI_ISL_939520, EPI_ISL_939521, EPI_ISL_939522, EPI_ISL_939523, EPI_ISL_939524, EPI_ISL_939526, EPI_ISL_939528, EPI_ISL_939532, EPI_ISL_939533, EPI_ISL_939535, EPI_ISL_939536, EPI_ISL_939537, EPI_ISL_939541, EPI_ISL_939542, EPI_ISL_939547, EPI_ISL_939548, EPI_ISL_939550, EPI_ISL_939551, EPI_ISL_939552, EPI_ISL_939554, EPI_ISL_939555, EPI_ISL_939556, EPI_ISL_939559, EPI_ISL_939561, EPI_ISL_939562, EPI_ISL_939563, EPI_ISL_939565, EPI_ISL_939566, EPI_ISL_939567, EPI_ISL_939571, EPI_ISL_939572, EPI_ISL_939573, EPI_ISL_939574, EPI_ISL_939576, EPI_ISL_939577, EPI_ISL_939578, EPI_ISL_939579, EPI_ISL_939581, EPI_ISL_939582, EPI_ISL_939585, EPI_ISL_939588, EPI_ISL_939594, EPI_ISL_939595, EPI_ISL_939596, EPI_ISL_939598, EPI_ISL_939599, EPI_ISL_939600, EPI_ISL_939601, EPI_ISL_939603, EPI_ISL_939605, EPI_ISL_939606, EPI_ISL_945752, EPI_ISL_945837, EPI_ISL_946094, EPI_ISL_946153, EPI_ISL_946269, EPI_ISL_946323, EPI_ISL_946363, EPI_ISL_946381, EPI_ISL_946403, EPI_ISL_947110, EPI_ISL_947113, EPI_ISL_947120, EPI_ISL_947140, EPI_ISL_947149, EPI_ISL_947172, EPI_ISL_947184, EPI_ISL_947189, EPI_ISL_947201, EPI_ISL_947240, EPI_ISL_947256, EPI_ISL_947261, EPI_ISL_947274, EPI_ISL_947318, EPI_ISL_947337, EPI_ISL_947354, EPI_ISL_947361, EPI_ISL_947368, EPI_ISL_947374, EPI_ISL_947400, EPI_ISL_958276, EPI_ISL_958398, EPI_ISL_963428 | | | |
| see above | Lighthouse Lab in Milton Keynes | Wellcome Sanger Institute for the COVID-19 Genomics UK (COG-UK) Consortium | Cordelia Langford; David K. Jackson; Dominic Kwiatkowski; Ewan Harrison; Ian Johnston; John Sillitoe on behalf of the Wellcome Sanger Institute COVID-19 Surveillance Team; Roberto Amato; Sonia Goncalves; The Lighthouse Lab in Milton Keynes and Alex Alderton |
| EPI_ISL_922169, EPI_ISL_922170, EPI_ISL_922171, EPI_ISL_950729, EPI_ISL_950730, EPI_ISL_950731, EPI_ISL_950732, EPI_ISL_950733, EPI_ISL_950734, EPI_ISL_950735, EPI_ISL_950736, EPI_ISL_950737, EPI_ISL_950738, EPI_ISL_950739, EPI_ISL_950740, EPI_ISL_950741, EPI_ISL_950742, EPI_ISL_950743, EPI_ISL_950744, EPI_ISL_950745, EPI_ISL_950746, EPI_ISL_950747, EPI_ISL_950748, EPI_ISL_950749, EPI_ISL_950750, EPI_ISL_950751, EPI_ISL_950752, EPI_ISL_950753, EPI_ISL_950754, EPI_ISL_998972, EPI_ISL_998973, EPI_ISL_998974, EPI_ISL_998975, EPI_ISL_998976, EPI_ISL_998977, EPI_ISL_998978, EPI_ISL_998979, EPI_ISL_998980, EPI_ISL_998981, EPI_ISL_998982, EPI_ISL_998983, EPI_ISL_998984, EPI_ISL_998985, EPI_ISL_1105847, EPI_ISL_1105848, EPI_ISL_1105849, EPI_ISL_1105850, EPI_ISL_1178688 | | | |
| see above | Lincolnshire Hospitals and DeepSeq Nottingham | COVID-19 Genomics UK (COG-UK) Consortium | Christopher Moore; Fei Sang; Joeseph Chappell; Johnny Debebe; Jonathan Ball; Matthew Carlisle; Matthew Loose; Nadine Holmes; Nichola Duckworth; Patrick McClure; Sarah Walsh; Tim Sloan; Victoria Wright |
| EPI_ISL_919519, EPI_ISL_919520, EPI_ISL_919521, EPI_ISL_919522, EPI_ISL_919523, EPI_ISL_919610, EPI_ISL_919620, EPI_ISL_919621, EPI_ISL_919622, EPI_ISL_919624, EPI_ISL_919625, EPI_ISL_919626, EPI_ISL_919627, EPI_ISL_919633, EPI_ISL_919636, EPI_ISL_919640, EPI_ISL_919664, EPI_ISL_919665, EPI_ISL_919666, EPI_ISL_919667, EPI_ISL_919668, EPI_ISL_919669, EPI_ISL_919670, EPI_ISL_919671, EPI_ISL_919672, EPI_ISL_919673, EPI_ISL_919674, EPI_ISL_919675, EPI_ISL_919676, EPI_ISL_919677, EPI_ISL_919678, EPI_ISL_919679, EPI_ISL_919680, EPI_ISL_919681, EPI_ISL_919682, EPI_ISL_919683, EPI_ISL_919684, EPI_ISL_919685, EPI_ISL_919686, EPI_ISL_919687, EPI_ISL_919688, EPI_ISL_919689, EPI_ISL_919692, EPI_ISL_919693, EPI_ISL_919694, EPI_ISL_919696, EPI_ISL_919697, EPI_ISL_919698, EPI_ISL_919699, EPI_ISL_919700, EPI_ISL_919701, EPI_ISL_919702, EPI_ISL_919703, EPI_ISL_919704, EPI_ISL_919705, EPI_ISL_949671, EPI_ISL_949672, EPI_ISL_949684, EPI_ISL_949685, EPI_ISL_949686, EPI_ISL_949687, EPI_ISL_997381, EPI_ISL_997406, EPI_ISL_1104339, EPI_ISL_1104341, EPI_ISL_1104343, EPI_ISL_1104344, EPI_ISL_1104354, EPI_ISL_1104356, EPI_ISL_1104357, EPI_ISL_1104363, EPI_ISL_1104369, EPI_ISL_1104370, EPI_ISL_1104371, EPI_ISL_1104372, EPI_ISL_1104374, EPI_ISL_1104375, EPI_ISL_1104376, EPI_ISL_1104377, EPI_ISL_1104379, EPI_ISL_1177888, EPI_ISL_1177889, EPI_ISL_1177904, EPI_ISL_1177908, EPI_ISL_1247769, EPI_ISL_1247770, EPI_ISL_1247771, EPI_ISL_1247773, EPI_ISL_1247774, EPI_ISL_1247777, EPI_ISL_1247778, EPI_ISL_1247780, EPI_ISL_1247782, EPI_ISL_1247783, EPI_ISL_1247785, EPI_ISL_1247786, EPI_ISL_1247787, EPI_ISL_1247791, EPI_ISL_1247793, EPI_ISL_1247794, EPI_ISL_1247795, EPI_ISL_1247796, EPI_ISL_1247797, EPI_ISL_1247798, EPI_ISL_1247799, EPI_ISL_1247800, EPI_ISL_1247802, EPI_ISL_1247803, EPI_ISL_1247804, EPI_ISL_1247805, EPI_ISL_1247806, EPI_ISL_1247807, EPI_ISL_1247808, EPI_ISL_1247810, EPI_ISL_1247811, EPI_ISL_1247812, EPI_ISL_1247815, EPI_ISL_1247816, EPI_ISL_1247817, EPI_ISL_1247818, EPI_ISL_1546008, EPI_ISL_1546009, EPI_ISL_1546010, EPI_ISL_1546011, EPI_ISL_1546013, EPI_ISL_1546014, EPI_ISL_1546015, EPI_ISL_1546016, EPI_ISL_1546017, EPI_ISL_1546018, EPI_ISL_1546019, EPI_ISL_1546020, EPI_ISL_1546021, EPI_ISL_1546022, EPI_ISL_1546023, EPI_ISL_1832399 | | | |
| see above | Liverpool Clinical Laboratories | COVID-19 Genomics UK (COG-UK) Consortium | A Alrezaihi; Alessandro Gerada; Alistair Darby; Angela Cowell; Anita Lucaci; Anu Chawla; Cassie Olateju; Catherine Hartley; Charlotte Nelson; Ecaterina Vamos; Elaine O'Toole; Eleanor G Bentley; Ghada T Shawli; Hermione Webster; Isabel García-Dorival; James Johnson; James P Stewart; Jenifer Manson; Joanne Watts; Jones Benjamin; Jordan J Clark; Julian Hiscox; L Luu; Lucille Rainbow; M Almsaud; Margaret Hughes; Mark Whitehead; Matthew Gemmell; Miren Iturriza-Gomara; Muhannad Alruwaili; N.P Randle; Neil Swainston; PKF Gilmore; Parul Sharma; Rebekah Penrice-Randal; Richard Eccles; Richard Gregory; Sam Haldenby; Steve Paterson; Stuart D Armstrong; Trevor Ian Robinson; Ximeng Han |
| EPI_ISL_921631, EPI_ISL_921632, EPI_ISL_921633, EPI_ISL_921634, EPI_ISL_921635, EPI_ISL_921636, EPI_ISL_921637, EPI_ISL_921638, EPI_ISL_921639, EPI_ISL_921640, EPI_ISL_921641, EPI_ISL_921642, EPI_ISL_921643, EPI_ISL_921644, EPI_ISL_921645, EPI_ISL_921646, EPI_ISL_921647, EPI_ISL_921648, EPI_ISL_921649, EPI_ISL_921650, EPI_ISL_921651, EPI_ISL_921652, EPI_ISL_921653, EPI_ISL_921654, EPI_ISL_921655, EPI_ISL_921656, EPI_ISL_921657, EPI_ISL_921658, EPI_ISL_950337, EPI_ISL_950338, EPI_ISL_950342, EPI_ISL_950347, EPI_ISL_950348, EPI_ISL_950349, EPI_ISL_950350, EPI_ISL_950494, EPI_ISL_950495, EPI_ISL_950496, EPI_ISL_950497, EPI_ISL_950498, EPI_ISL_950499, EPI_ISL_950500, EPI_ISL_950501, EPI_ISL_950502, EPI_ISL_950503, EPI_ISL_950504, EPI_ISL_950505, EPI_ISL_950506, EPI_ISL_950507, EPI_ISL_950508, EPI_ISL_950509, EPI_ISL_950510, EPI_ISL_950511, EPI_ISL_950512, EPI_ISL_950513, EPI_ISL_950514, EPI_ISL_950515, EPI_ISL_950516, EPI_ISL_950517, EPI_ISL_950518, EPI_ISL_950519, EPI_ISL_950520, EPI_ISL_950521, EPI_ISL_950522, EPI_ISL_998414, EPI_ISL_998415, EPI_ISL_998416, EPI_ISL_998417, EPI_ISL_998418, EPI_ISL_998419, EPI_ISL_998420, EPI_ISL_998421, EPI_ISL_998422, EPI_ISL_998423, EPI_ISL_998424, EPI_ISL_998425, EPI_ISL_998426, EPI_ISL_998427, EPI_ISL_998428, EPI_ISL_998429, EPI_ISL_998430, EPI_ISL_998431, EPI_ISL_998432, EPI_ISL_998433, EPI_ISL_998434, EPI_ISL_998435, EPI_ISL_998436, EPI_ISL_998437, EPI_ISL_998438, EPI_ISL_998439, EPI_ISL_998440, EPI_ISL_998445, EPI_ISL_998446, EPI_ISL_998448, EPI_ISL_998449, EPI_ISL_998450, EPI_ISL_998451, EPI_ISL_998452, EPI_ISL_998453, EPI_ISL_998454, EPI_ISL_998455, EPI_ISL_998456, EPI_ISL_998457, EPI_ISL_998458, EPI_ISL_998459, EPI_ISL_998460, EPI_ISL_998464, EPI_ISL_998465, EPI_ISL_998466, EPI_ISL_998467, EPI_ISL_998468, EPI_ISL_998469, EPI_ISL_998470, EPI_ISL_998471, EPI_ISL_998472, EPI_ISL_998473, EPI_ISL_998474, EPI_ISL_998476, EPI_ISL_998477, EPI_ISL_998478, EPI_ISL_998479, EPI_ISL_998480, EPI_ISL_998481, EPI_ISL_998482, EPI_ISL_998483, EPI_ISL_998485, EPI_ISL_998496, EPI_ISL_998499, EPI_ISL_1178171, EPI_ISL_1178172, EPI_ISL_1178173, EPI_ISL_1178174, EPI_ISL_1178175, EPI_ISL_1178176, EPI_ISL_1178177, EPI_ISL_1178178, EPI_ISL_1178179, EPI_ISL_1178180, EPI_ISL_1178181, EPI_ISL_1178182, EPI_ISL_1178183, EPI_ISL_1178184, EPI_ISL_1178185, EPI_ISL_1178186, EPI_ISL_1178187, EPI_ISL_1178188, EPI_ISL_1178189, EPI_ISL_1178190, EPI_ISL_1178191, EPI_ISL_1178192, EPI_ISL_1178232, EPI_ISL_1178237, EPI_ISL_1178240, EPI_ISL_1178244, EPI_ISL_1178247, EPI_ISL_1178253, EPI_ISL_1178258, EPI_ISL_1178267, EPI_ISL_1178269, EPI_ISL_1178285, EPI_ISL_1178287, EPI_ISL_1178288, EPI_ISL_1178289, EPI_ISL_1178290, EPI_ISL_1178314, EPI_ISL_1178324, EPI_ISL_1178325, EPI_ISL_1178399, EPI_ISL_1248877, EPI_ISL_1248902, EPI_ISL_1248903, EPI_ISL_1248904, EPI_ISL_1248905, EPI_ISL_1248906, EPI_ISL_1248907, EPI_ISL_1248908, EPI_ISL_1248909, EPI_ISL_1248910, EPI_ISL_1248911, EPI_ISL_1248912, EPI_ISL_1248913, EPI_ISL_1248914 | | | |
| see above | Northumbria University / South Tees Hospitals NHS Foundation Trust / North Cumbria Integrated Care NHS Foundation Trust / North Tees and Hartlepool NHS Foundation Trust / Newcastle Hospitals NHS Foundation Trust | COVID-19 Genomics UK (COG-UK) Consortium | Andrew Nelson; Brendan Payne; Clive Graham; Darren L Smith; Debra Padgett; Edward Barton; Emma Swindells; Garren Scott; Gary Black; Gary Eltringham; Giles S Holt; Greg R Young; Jane Greenaway; Jennifer Collins; John Allan; Joshua Loh; Lynn Dover; Matthew Bashton; Mohammad A Tariq; Paul Baker; Sarah Essex; Steve Liggett; Wen C Yew; Yusri Taha |
| EPI_ISL_951500, EPI_ISL_951510, EPI_ISL_951511, EPI_ISL_951563, EPI_ISL_951589, EPI_ISL_951594, EPI_ISL_951595, EPI_ISL_951596, EPI_ISL_951597, EPI_ISL_951598, EPI_ISL_951601, EPI_ISL_951602, EPI_ISL_951603, EPI_ISL_951604, EPI_ISL_951605, EPI_ISL_951606, EPI_ISL_951607, EPI_ISL_951608, EPI_ISL_951609, EPI_ISL_951610, EPI_ISL_951611, EPI_ISL_951612, EPI_ISL_951613, EPI_ISL_951614, EPI_ISL_951615, EPI_ISL_951616, EPI_ISL_951617, EPI_ISL_951618, EPI_ISL_951619, EPI_ISL_951620, EPI_ISL_951624, EPI_ISL_951625, EPI_ISL_951639, EPI_ISL_951640, EPI_ISL_951643, EPI_ISL_951644, EPI_ISL_951645, EPI_ISL_951646, EPI_ISL_951649, EPI_ISL_951650, EPI_ISL_951651, EPI_ISL_951652, EPI_ISL_951653, EPI_ISL_951654, EPI_ISL_951655, EPI_ISL_951658, EPI_ISL_951660, EPI_ISL_951661, EPI_ISL_951664, EPI_ISL_951667, EPI_ISL_951668, EPI_ISL_951669, EPI_ISL_951670, EPI_ISL_951672, EPI_ISL_951673, EPI_ISL_951674, EPI_ISL_951675, EPI_ISL_951676, EPI_ISL_951677, EPI_ISL_951679, EPI_ISL_951680, EPI_ISL_951681, EPI_ISL_951684, EPI_ISL_951685, EPI_ISL_951686, EPI_ISL_951687, EPI_ISL_951688, EPI_ISL_951689, EPI_ISL_951690, EPI_ISL_951691, EPI_ISL_951693, EPI_ISL_951694, EPI_ISL_951695, EPI_ISL_951696, EPI_ISL_951698, EPI_ISL_951700, EPI_ISL_951701, EPI_ISL_951702, EPI_ISL_951703, EPI_ISL_951704, EPI_ISL_951705, EPI_ISL_951706, EPI_ISL_951708, EPI_ISL_951709, EPI_ISL_951711, EPI_ISL_951712, EPI_ISL_951713, EPI_ISL_951714, EPI_ISL_951715, EPI_ISL_951717, EPI_ISL_951718, EPI_ISL_951719, EPI_ISL_951720, EPI_ISL_951721, EPI_ISL_951725, EPI_ISL_951728, EPI_ISL_951730, EPI_ISL_951731, EPI_ISL_951732, EPI_ISL_951733, EPI_ISL_951735, EPI_ISL_951736, EPI_ISL_951737, EPI_ISL_951741, EPI_ISL_951743, EPI_ISL_951746, EPI_ISL_951747, EPI_ISL_951748, EPI_ISL_951749, EPI_ISL_951752, EPI_ISL_951755, EPI_ISL_951757, EPI_ISL_951762, EPI_ISL_951763, EPI_ISL_951764, EPI_ISL_951768, EPI_ISL_951770, EPI_ISL_951771, EPI_ISL_951772, EPI_ISL_951773, EPI_ISL_951774, EPI_ISL_951775, EPI_ISL_951776, EPI_ISL_951777, EPI_ISL_951784, EPI_ISL_951823, EPI_ISL_951825, EPI_ISL_951826, EPI_ISL_951827, EPI_ISL_951829, EPI_ISL_951831, EPI_ISL_951832, EPI_ISL_951833, EPI_ISL_951834, EPI_ISL_951842, EPI_ISL_951845, EPI_ISL_951846, EPI_ISL_951847, EPI_ISL_951848, EPI_ISL_951849, EPI_ISL_951850, EPI_ISL_951987, EPI_ISL_951992, EPI_ISL_951998, EPI_ISL_951999, EPI_ISL_952000, EPI_ISL_952002, EPI_ISL_952168, EPI_ISL_952170, EPI_ISL_952171, EPI_ISL_952172, EPI_ISL_952173, EPI_ISL_952174, EPI_ISL_952175, EPI_ISL_952176, EPI_ISL_952177, EPI_ISL_952178, EPI_ISL_952179, EPI_ISL_952180, EPI_ISL_952192, EPI_ISL_952194, EPI_ISL_952195, EPI_ISL_952196, EPI_ISL_952199, EPI_ISL_952201, EPI_ISL_952202, EPI_ISL_952203, EPI_ISL_952204, EPI_ISL_952205, EPI_ISL_952207, EPI_ISL_952213, EPI_ISL_952214, EPI_ISL_952215, EPI_ISL_952216, EPI_ISL_952217, EPI_ISL_952223, EPI_ISL_952224, EPI_ISL_952226, EPI_ISL_952231, EPI_ISL_952234, EPI_ISL_952235, EPI_ISL_952238, EPI_ISL_952239, EPI_ISL_952241, EPI_ISL_952244, EPI_ISL_952246, EPI_ISL_952248, EPI_ISL_952250, EPI_ISL_1052463, EPI_ISL_1052464, EPI_ISL_1052466, EPI_ISL_1052467, EPI_ISL_1052468, EPI_ISL_1052489, EPI_ISL_1052490, EPI_ISL_1052491, EPI_ISL_1052496, EPI_ISL_1052499, EPI_ISL_1052500, EPI_ISL_1052501, EPI_ISL_1052502, EPI_ISL_1052504, EPI_ISL_1052505, EPI_ISL_1052506, EPI_ISL_1052507, EPI_ISL_1052508, EPI_ISL_1052509, EPI_ISL_1052510, EPI_ISL_1053525, EPI_ISL_1053547, EPI_ISL_1178890, EPI_ISL_1178891, EPI_ISL_1476451, EPI_ISL_1476452, EPI_ISL_1476453, EPI_ISL_1476454, EPI_ISL_1476456, EPI_ISL_1476457, EPI_ISL_1476458, EPI_ISL_1476460, EPI_ISL_1476464, EPI_ISL_1476465, EPI_ISL_1476466, EPI_ISL_1476467, EPI_ISL_1476471, EPI_ISL_1476472, EPI_ISL_1476474, EPI_ISL_1476476, EPI_ISL_1476477, EPI_ISL_1476504, EPI_ISL_1476514 | | | |
| see above | Originating lab: Wales Specialist Virology Centre Sequencing lab: Pathogen Genomics Unit | Public Health Wales Microbiology Cardiff Wales Specialist Virology Centre | Alec Birchley; Alexander Adams; Amy Gaskin; Angela Marchbank; Bree Gatica-Wilcox; Catherine Moore; Jason Coombes; Joanne Watkins; Joel Southgate; Johnathan Evans; Laura Gifford; Lauren Gilbert; Lee Graham; Malorie Perry; Matthew Bull; Nicole Pacchiarini; Sally Corden; Sara Kumziene-Summerhayes; Sara Rey; Sarah Taylor; Simon Cottrell; Sophie Jones; Tom Connor |
| EPI_ISL_951486, EPI_ISL_951487, EPI_ISL_951488, EPI_ISL_998995, EPI_ISL_999185, EPI_ISL_999186, EPI_ISL_999187, EPI_ISL_999188, EPI_ISL_999189, EPI_ISL_999190, EPI_ISL_999191, EPI_ISL_999192, EPI_ISL_999193, EPI_ISL_999194, EPI_ISL_999195, EPI_ISL_999197, EPI_ISL_999217, EPI_ISL_999224, EPI_ISL_999227, EPI_ISL_999228, EPI_ISL_999229, EPI_ISL_999230, EPI_ISL_999232, EPI_ISL_999233, EPI_ISL_999234, EPI_ISL_999235, EPI_ISL_999236, EPI_ISL_999238, EPI_ISL_999239, EPI_ISL_999241, EPI_ISL_999242, EPI_ISL_999243, EPI_ISL_999244, EPI_ISL_999245, EPI_ISL_999246, EPI_ISL_999248, EPI_ISL_999249, EPI_ISL_999250, EPI_ISL_999251, EPI_ISL_999252, EPI_ISL_999253, EPI_ISL_999254, EPI_ISL_999256, EPI_ISL_999257, EPI_ISL_999258, EPI_ISL_999259, EPI_ISL_999260, EPI_ISL_999261, EPI_ISL_999262, EPI_ISL_999263, EPI_ISL_999265, EPI_ISL_999267, EPI_ISL_999268, EPI_ISL_999269, EPI_ISL_999273, EPI_ISL_999274, EPI_ISL_999275, EPI_ISL_999276, EPI_ISL_999278, EPI_ISL_999279, EPI_ISL_999281, EPI_ISL_999282, EPI_ISL_999283, EPI_ISL_999284, EPI_ISL_999287, EPI_ISL_999288, EPI_ISL_999289, EPI_ISL_999293, EPI_ISL_999296, EPI_ISL_999297, EPI_ISL_999298, EPI_ISL_999299, EPI_ISL_999300, EPI_ISL_999301, EPI_ISL_999314, EPI_ISL_999483, EPI_ISL_999505, EPI_ISL_999510, EPI_ISL_999511, EPI_ISL_1051594, EPI_ISL_1051601, EPI_ISL_1051860, EPI_ISL_1051861, EPI_ISL_1051862, EPI_ISL_1051870, EPI_ISL_1051871, EPI_ISL_1051885, EPI_ISL_1051886, EPI_ISL_1051887, EPI_ISL_1051891, EPI_ISL_1178732, EPI_ISL_1178748, EPI_ISL_1249219, EPI_ISL_1309741, EPI_ISL_1309742, EPI_ISL_1309743, EPI_ISL_1309744, EPI_ISL_1309754, EPI_ISL_1309755, EPI_ISL_1387221, EPI_ISL_1833073, EPI_ISL_1833077, EPI_ISL_1833086, EPI_ISL_1833098 | | | |
| see above | Oxford Viromics, NDM, University of Oxford; Oxford University Hospitals; Basingstoke and North Hampshire Hospital | COVID-19 Genomics UK (COG-UK) Consortium | Alex Mobbs; Amy Trebes; Anita Justice; Catrin Moore; Christophe Fraser; David Bonsall; David Buck; Emma Wise; George Macintyre; Jessica Lynch; John Todd; Mariateresa de Cesare; Matilde Mori; Monique Andersson; Nathan Moore; Nick Cortes; Robert Shaw; Stephen Kidd; Tanya Golubchik; Timothy Peto |
| EPI_ISL_921851, EPI_ISL_921852, EPI_ISL_921853, EPI_ISL_921857, EPI_ISL_921858, EPI_ISL_921859, EPI_ISL_921861, EPI_ISL_921863, EPI_ISL_921864, EPI_ISL_921865, EPI_ISL_921866, EPI_ISL_921867, EPI_ISL_921868, EPI_ISL_921870, EPI_ISL_921872, EPI_ISL_921874, EPI_ISL_921875, EPI_ISL_921876, EPI_ISL_921877, EPI_ISL_921904, EPI_ISL_921909, EPI_ISL_921910, EPI_ISL_921912, EPI_ISL_921913, EPI_ISL_921914, EPI_ISL_921917, EPI_ISL_921923, EPI_ISL_921931, EPI_ISL_921933, EPI_ISL_921934, EPI_ISL_921935, EPI_ISL_921938, EPI_ISL_921940, EPI_ISL_921942, EPI_ISL_921947, EPI_ISL_921952, EPI_ISL_921953, EPI_ISL_921966, EPI_ISL_921968, EPI_ISL_921969, EPI_ISL_921978, EPI_ISL_998824, EPI_ISL_998826, EPI_ISL_998831, EPI_ISL_998833, EPI_ISL_1051425, EPI_ISL_1051426, EPI_ISL_1051431, EPI_ISL_1051433, EPI_ISL_1051435, EPI_ISL_1051436, EPI_ISL_1051441, EPI_ISL_1051452, EPI_ISL_1051459, EPI_ISL_1051462, EPI_ISL_1051465, EPI_ISL_1051468, EPI_ISL_1051473, EPI_ISL_1051475, EPI_ISL_1051476, EPI_ISL_1051479, EPI_ISL_1051481, EPI_ISL_1051484, EPI_ISL_1051485, EPI_ISL_1051489, EPI_ISL_1051490, EPI_ISL_1051501, EPI_ISL_1051503, EPI_ISL_1051504, EPI_ISL_1051506, EPI_ISL_1051507, EPI_ISL_1051510, EPI_ISL_1051514, EPI_ISL_1051516, EPI_ISL_1051519, EPI_ISL_1051520, EPI_ISL_1051521, EPI_ISL_1051522, EPI_ISL_1178440, EPI_ISL_1178451, EPI_ISL_1178461, EPI_ISL_1178466, EPI_ISL_1178473, EPI_ISL_1178475, EPI_ISL_1178479, EPI_ISL_1178481, EPI_ISL_1178485, EPI_ISL_1178486, EPI_ISL_1178489, EPI_ISL_1178490, EPI_ISL_1178491, EPI_ISL_1178492, EPI_ISL_1178494, EPI_ISL_1248963, EPI_ISL_1248978, EPI_ISL_1248980, EPI_ISL_1248983, EPI_ISL_1248985, EPI_ISL_1248986, EPI_ISL_1248989, EPI_ISL_1248991, EPI_ISL_1248992, EPI_ISL_1248995, EPI_ISL_1249000, EPI_ISL_1249005, EPI_ISL_1249009, EPI_ISL_1249010, EPI_ISL_1249015, EPI_ISL_1249016, EPI_ISL_1249017, EPI_ISL_1249021, EPI_ISL_1249022, EPI_ISL_1249023, EPI_ISL_1249030, EPI_ISL_1249031, EPI_ISL_1249032, EPI_ISL_1249033, EPI_ISL_1249037, EPI_ISL_1249041, EPI_ISL_1249044, EPI_ISL_1249045, EPI_ISL_1249047, EPI_ISL_1249048, EPI_ISL_1249050, EPI_ISL_1249051, EPI_ISL_1249052, EPI_ISL_1249053, EPI_ISL_1249054, EPI_ISL_1309667, EPI_ISL_1832948 | | | |
| see above | Quadram Institute Bioscience | COVID-19 Genomics UK (COG-UK) Consortium | Alexander J Trotter; Alison E. Mather; Alp Aydin; Ana P. Tedim; Anastasia Kolyva; Andrew Bell; Andrew J. Page; Claire Stuart; Dave J. Baker; Gemma L. Kay; John Wain; Justin O'Grady; Leonardo de Oliveira Martins; Lizzie Meadows; Maria Diaz; Mark Webber; Muhammed Yasir; Nabil-Fareed Alikhan; Ngozi Elumogo; Nicholas M. Thomson; Rachael Stanley; Rachel Gilroy; Reenesh Prakash; Samir Dervisevic; Samuel Bloomfield; Steven Rudder; Thanh Le-Viet |
| EPI_ISL_921999, EPI_ISL_922000, EPI_ISL_922001, EPI_ISL_922002, EPI_ISL_922003, EPI_ISL_922004, EPI_ISL_922005, EPI_ISL_922006, EPI_ISL_922007, EPI_ISL_922008, EPI_ISL_922009, EPI_ISL_922010, EPI_ISL_922011, EPI_ISL_922012, EPI_ISL_922013, EPI_ISL_922014, EPI_ISL_922015, EPI_ISL_922016, EPI_ISL_922017, EPI_ISL_922018, EPI_ISL_922019, EPI_ISL_922020, EPI_ISL_922021, EPI_ISL_922022, EPI_ISL_922023, EPI_ISL_922024, EPI_ISL_922025, EPI_ISL_922026, EPI_ISL_922027, EPI_ISL_922028, EPI_ISL_922029, EPI_ISL_922030, EPI_ISL_922031, EPI_ISL_922032, EPI_ISL_922033, EPI_ISL_922034, EPI_ISL_922035, EPI_ISL_922036, EPI_ISL_950638 | | | |
| see above | Queens Medical Centre, Clinical Microbiology Department / DeepSeq Nottingham | COVID-19 Genomics UK (COG-UK) Consortium | Christopher Moore; Fei Sang; Gemma Clark; Hannah Howson-Wells; Johnny Debebe; Jonathan Ball; Joseph Chappell; Manjinder Khakh; Matthew Carlisle; Matthew Loose; Michelle M Lister; Nadine Holmes; Patrick McClure; Theocharis Tsoleridis; Vicki M Fleming; Victoria Wright; Wendy Smith |
| EPI_ISL_1046238, EPI_ISL_1046310, EPI_ISL_1046322 | Randox Laboratories | Wellcome Sanger Institute for the COVID-19 Genomics | Cordelia Langford; David K. Jackson; Dominic Kwiatkowski; Ewan Harrison; Ian Johnston; Jeffrey Barrett; John Sillitoe on behalf of the Wellcome Sanger Institute COVID-19 Surveillance Team; Randox Laboratories and Alex Alderton; Roberto Amato; Sonia Goncalves |

UK (COG-UK) Consortium

EPI_ISL_998176, EPI_ISL_998177, EPI_ISL_998181, EPI_ISL_998182, EPI_ISL_998183, EPI_ISL_998184, EPI_ISL_998185, EPI_ISL_998186, EPI_ISL_998187, EPI_ISL_998188, EPI_ISL_998225, EPI_ISL_998226, EPI_ISL_998227, EPI_ISL_998228, EPI_ISL_998268, EPI_ISL_998270, EPI_ISL_998271, EPI_ISL_998272, EPI_ISL_998273, EPI_ISL_1475031, EPI_ISL_1475032, EPI_ISL_1475033, EPI_ISL_1475035, EPI_ISL_1475048, EPI_ISL_1475051, EPI_ISL_1475052, EPI_ISL_1475054, EPI_ISL_1475055, EPI_ISL_1475056, EPI_ISL_1475061, EPI_ISL_1475084, EPI_ISL_1475086, EPI_ISL_1475087, EPI_ISL_1475088, EPI_ISL_1475089, EPI_ISL_1475090, EPI_ISL_1475122, EPI_ISL_1475123, EPI_ISL_1475128, EPI_ISL_1475129, EPI_ISL_1475130

| see above | Regional Virus Laboratory, Belfast Health and Social Care Trust | COVID-19 Genomics UK (COG-UK) Consortium | Alison Watt; Ciara Cox; Conall McCaughey; David Simpson; Derek Fairley; James McKenna; Mairead Connor; Susan Feeney; Tanya Curran; Zoltan Molnar |
|---|---|---|---|

EPI_ISL_1296563, EPI_ISL_1296564, EPI_ISL_1296565, EPI_ISL_1296566, EPI_ISL_1296567, EPI_ISL_1296568, EPI_ISL_1296569, EPI_ISL_1296570, EPI_ISL_1296572, EPI_ISL_1296573, EPI_ISL_1296574, EPI_ISL_1296575, EPI_ISL_1296576, EPI_ISL_1296577, EPI_ISL_1296579, EPI_ISL_1296580, EPI_ISL_1296583, EPI_ISL_1296584, EPI_ISL_1296599

| see above | Respiratory Virus Unit, National Infection Service, Public Health England | COVID-19 Genomics UK (COG-UK) Consortium | PHE Covid Sequencing Team |
|---|---|---|---|

EPI_ISL_920410, EPI_ISL_920424, EPI_ISL_920427, EPI_ISL_920437, EPI_ISL_920440, EPI_ISL_920447, EPI_ISL_920454, EPI_ISL_920455, EPI_ISL_920463, EPI_ISL_920472, EPI_ISL_920480, EPI_ISL_920488, EPI_ISL_920522, EPI_ISL_920523, EPI_ISL_920524, EPI_ISL_920525, EPI_ISL_920531, EPI_ISL_920532, EPI_ISL_920533, EPI_ISL_920534, EPI_ISL_920535, EPI_ISL_920536, EPI_ISL_920541, EPI_ISL_920542, EPI_ISL_920543, EPI_ISL_920544, EPI_ISL_920545, EPI_ISL_920546, EPI_ISL_920552, EPI_ISL_920553, EPI_ISL_920554, EPI_ISL_920555, EPI_ISL_920556, EPI_ISL_920557, EPI_ISL_920561, EPI_ISL_920562, EPI_ISL_920563, EPI_ISL_920564, EPI_ISL_920565, EPI_ISL_920566, EPI_ISL_920570, EPI_ISL_920571, EPI_ISL_920572, EPI_ISL_920573, EPI_ISL_920578, EPI_ISL_920579, EPI_ISL_920580, EPI_ISL_920581, EPI_ISL_920587, EPI_ISL_920588, EPI_ISL_920589, EPI_ISL_920590, EPI_ISL_920591, EPI_ISL_920592, EPI_ISL_920593, EPI_ISL_920597, EPI_ISL_920598, EPI_ISL_920599, EPI_ISL_920600, EPI_ISL_920601, EPI_ISL_920602, EPI_ISL_920604, EPI_ISL_920605, EPI_ISL_920606, EPI_ISL_920607, EPI_ISL_920608, EPI_ISL_920609, EPI_ISL_920610, EPI_ISL_920611, EPI_ISL_920614, EPI_ISL_920615, EPI_ISL_920616, EPI_ISL_920617, EPI_ISL_920618, EPI_ISL_920619, EPI_ISL_920622, EPI_ISL_920623, EPI_ISL_920624, EPI_ISL_920625, EPI_ISL_920626, EPI_ISL_920628, EPI_ISL_920632, EPI_ISL_920633, EPI_ISL_920634, EPI_ISL_920635, EPI_ISL_920638, EPI_ISL_920639, EPI_ISL_920640, EPI_ISL_920641, EPI_ISL_920642, EPI_ISL_920643, EPI_ISL_920644, EPI_ISL_920645, EPI_ISL_920648, EPI_ISL_920649, EPI_ISL_920650, EPI_ISL_920651, EPI_ISL_920652, EPI_ISL_920654, EPI_ISL_920655, EPI_ISL_920656, EPI_ISL_920657, EPI_ISL_920658, EPI_ISL_920662, EPI_ISL_920663, EPI_ISL_920664, EPI_ISL_920665, EPI_ISL_920668, EPI_ISL_920669, EPI_ISL_920672, EPI_ISL_920673, EPI_ISL_920675, EPI_ISL_920677, EPI_ISL_920678, EPI_ISL_920679, EPI_ISL_920681, EPI_ISL_920682, EPI_ISL_920686, EPI_ISL_920687, EPI_ISL_920690, EPI_ISL_920692, EPI_ISL_920694, EPI_ISL_920695, EPI_ISL_920698, EPI_ISL_920699, EPI_ISL_920703, EPI_ISL_920704, EPI_ISL_920707, EPI_ISL_920708, EPI_ISL_920712, EPI_ISL_920713, EPI_ISL_920715, EPI_ISL_920716, EPI_ISL_920718, EPI_ISL_920726, EPI_ISL_920727, EPI_ISL_920740, EPI_ISL_920741, EPI_ISL_920742, EPI_ISL_920743, EPI_ISL_920749, EPI_ISL_920750, EPI_ISL_920751, EPI_ISL_920759, EPI_ISL_920760, EPI_ISL_920764, EPI_ISL_920765, EPI_ISL_920766, EPI_ISL_920769, EPI_ISL_920772, EPI_ISL_920781, EPI_ISL_1104772, EPI_ISL_1104774, EPI_ISL_1104778, EPI_ISL_1104784, EPI_ISL_1104785, EPI_ISL_1104788, EPI_ISL_1104790, EPI_ISL_1104791, EPI_ISL_1104792, EPI_ISL_1104795, EPI_ISL_1104797, EPI_ISL_1104798, EPI_ISL_1104799, EPI_ISL_1104801, EPI_ISL_1104803, EPI_ISL_1104804, EPI_ISL_1104806, EPI_ISL_1104809, EPI_ISL_1104811, EPI_ISL_1104812, EPI_ISL_1104814, EPI_ISL_1104820, EPI_ISL_1104822, EPI_ISL_1104827, EPI_ISL_1104828, EPI_ISL_1104831, EPI_ISL_1105043, EPI_ISL_1105079, EPI_ISL_1105084, EPI_ISL_1105091, EPI_ISL_1105115, EPI_ISL_1105159, EPI_ISL_1105168, EPI_ISL_1105170, EPI_ISL_1105176, EPI_ISL_1105185, EPI_ISL_1105219, EPI_ISL_1105250, EPI_ISL_1105259, EPI_ISL_1105292, EPI_ISL_1105324, EPI_ISL_1105329, EPI_ISL_1105341, EPI_ISL_1248451, EPI_ISL_1248452, EPI_ISL_1248488, EPI_ISL_1637056, EPI_ISL_1637067

| see above | University College London Hospital | COVID-19 Genomics UK (COG-UK) Consortium | Catherine Houlihan; Dan Frampton; Dr Catherine Houlihan; Dr Daniel Frampton; Dr Judith Heaney; Dr Moira Spyer; Dr Paul Grant and Dr Eleni Nastouli; Judith Heaney; Matthew Byott; Moira Spyer and Eleni Nastouli; Stuart Kirk |
|---|---|---|---|

EPI_ISL_865614, EPI_ISL_865615, EPI_ISL_865616, EPI_ISL_865617, EPI_ISL_865618, EPI_ISL_865623, EPI_ISL_865630, EPI_ISL_865639, EPI_ISL_866021, EPI_ISL_866024, EPI_ISL_866027, EPI_ISL_866028, EPI_ISL_866030, EPI_ISL_866031, EPI_ISL_866032, EPI_ISL_866033, EPI_ISL_866034, EPI_ISL_866035, EPI_ISL_866036, EPI_ISL_866037, EPI_ISL_866039, EPI_ISL_866041, EPI_ISL_866042, EPI_ISL_866045, EPI_ISL_866046, EPI_ISL_866047, EPI_ISL_919776, EPI_ISL_919777, EPI_ISL_919778, EPI_ISL_919856, EPI_ISL_919857, EPI_ISL_919858, EPI_ISL_919859, EPI_ISL_919860, EPI_ISL_919861, EPI_ISL_919862, EPI_ISL_919863, EPI_ISL_919864, EPI_ISL_919865, EPI_ISL_919866, EPI_ISL_919867, EPI_ISL_919868, EPI_ISL_919869, EPI_ISL_919870, EPI_ISL_919874, EPI_ISL_919875, EPI_ISL_919876, EPI_ISL_919877, EPI_ISL_919878, EPI_ISL_919879, EPI_ISL_919880, EPI_ISL_919882, EPI_ISL_919883, EPI_ISL_919884, EPI_ISL_919885, EPI_ISL_919886, EPI_ISL_919887, EPI_ISL_919888, EPI_ISL_919889, EPI_ISL_919890, EPI_ISL_919891, EPI_ISL_919892, EPI_ISL_919893, EPI_ISL_919894, EPI_ISL_919895, EPI_ISL_919896, EPI_ISL_919897, EPI_ISL_919898, EPI_ISL_919899, EPI_ISL_919900, EPI_ISL_919901, EPI_ISL_919902, EPI_ISL_919920, EPI_ISL_919921, EPI_ISL_919922, EPI_ISL_919923, EPI_ISL_919924, EPI_ISL_919925, EPI_ISL_919926, EPI_ISL_919933, EPI_ISL_919945, EPI_ISL_919952, EPI_ISL_919955, EPI_ISL_919957, EPI_ISL_920067, EPI_ISL_920069, EPI_ISL_920070, EPI_ISL_920073, EPI_ISL_920074, EPI_ISL_920075, EPI_ISL_920076, EPI_ISL_920077, EPI_ISL_920078, EPI_ISL_920081, EPI_ISL_920082, EPI_ISL_920083, EPI_ISL_920084, EPI_ISL_920085, EPI_ISL_920086, EPI_ISL_920087, EPI_ISL_920088, EPI_ISL_920089, EPI_ISL_920090, EPI_ISL_920091, EPI_ISL_920117, EPI_ISL_920118, EPI_ISL_920119, EPI_ISL_920120, EPI_ISL_920121, EPI_ISL_920122, EPI_ISL_920123, EPI_ISL_920124, EPI_ISL_920125, EPI_ISL_920126, EPI_ISL_920127, EPI_ISL_920128, EPI_ISL_920129, EPI_ISL_920130, EPI_ISL_920131, EPI_ISL_920132, EPI_ISL_920133, EPI_ISL_920134, EPI_ISL_920135, EPI_ISL_920136, EPI_ISL_920137, EPI_ISL_920138, EPI_ISL_920139, EPI_ISL_920140, EPI_ISL_920141, EPI_ISL_920142, EPI_ISL_920143, EPI_ISL_920144, EPI_ISL_920145, EPI_ISL_920146, EPI_ISL_920147, EPI_ISL_920148, EPI_ISL_920149, EPI_ISL_920150, EPI_ISL_920151, EPI_ISL_920153, EPI_ISL_920154, EPI_ISL_920155, EPI_ISL_920156, EPI_ISL_920157, EPI_ISL_920158, EPI_ISL_920159, EPI_ISL_920160, EPI_ISL_920161, EPI_ISL_920162, EPI_ISL_920163, EPI_ISL_920164, EPI_ISL_920165, EPI_ISL_920166, EPI_ISL_920167, EPI_ISL_920168, EPI_ISL_920169, EPI_ISL_920170, EPI_ISL_920171, EPI_ISL_920172, EPI_ISL_920173, EPI_ISL_920810, EPI_ISL_920818, EPI_ISL_920819, EPI_ISL_920820, EPI_ISL_920821, EPI_ISL_920822, EPI_ISL_920823, EPI_ISL_920824, EPI_ISL_920825, EPI_ISL_920826, EPI_ISL_920827, EPI_ISL_920828, EPI_ISL_920829, EPI_ISL_920830, EPI_ISL_920833, EPI_ISL_920834, EPI_ISL_920835, EPI_ISL_920836, EPI_ISL_920837, EPI_ISL_920838, EPI_ISL_949792, EPI_ISL_949906, EPI_ISL_949907, EPI_ISL_949908, EPI_ISL_949963, EPI_ISL_949975, EPI_ISL_949976, EPI_ISL_949977, EPI_ISL_949978, EPI_ISL_949980, EPI_ISL_949981, EPI_ISL_949982, EPI_ISL_949983, EPI_ISL_949984, EPI_ISL_949985, EPI_ISL_949986, EPI_ISL_949987, EPI_ISL_949988, EPI_ISL_949989, EPI_ISL_949990, EPI_ISL_949991, EPI_ISL_949992, EPI_ISL_949993, EPI_ISL_949994, EPI_ISL_949995, EPI_ISL_949996, EPI_ISL_949997, EPI_ISL_949998, EPI_ISL_949999, EPI_ISL_950000, EPI_ISL_950001, EPI_ISL_950002, EPI_ISL_950003, EPI_ISL_950004, EPI_ISL_950005, EPI_ISL_950006, EPI_ISL_950007, EPI_ISL_950008, EPI_ISL_950009, EPI_ISL_950010, EPI_ISL_950011, EPI_ISL_950012, EPI_ISL_950013, EPI_ISL_950014, EPI_ISL_950015, EPI_ISL_950016, EPI_ISL_950017, EPI_ISL_950018, EPI_ISL_950020, EPI_ISL_950021, EPI_ISL_950022, EPI_ISL_950023, EPI_ISL_950024, EPI_ISL_950025, EPI_ISL_950026, EPI_ISL_950027, EPI_ISL_950029, EPI_ISL_950030, EPI_ISL_950031, EPI_ISL_950032, EPI_ISL_950033, EPI_ISL_950035, EPI_ISL_950036, EPI_ISL_950037, EPI_ISL_950038, EPI_ISL_950170, EPI_ISL_997679, EPI_ISL_997683, EPI_ISL_997684, EPI_ISL_997685, EPI_ISL_997686, EPI_ISL_997688, EPI_ISL_997689, EPI_ISL_997690, EPI_ISL_997693, EPI_ISL_1105470, EPI_ISL_1248214, EPI_ISL_1248215, EPI_ISL_1248216, EPI_ISL_1248217, EPI_ISL_1248218, EPI_ISL_1248219, EPI_ISL_1248220, EPI_ISL_1248221, EPI_ISL_1248222, EPI_ISL_1248223, EPI_ISL_1248224, EPI_ISL_1248225, EPI_ISL_1248226, EPI_ISL_1248227, EPI_ISL_1248228, EPI_ISL_1248229, EPI_ISL_1248232, EPI_ISL_1248233, EPI_ISL_1248234, EPI_ISL_1248239, EPI_ISL_1248242, EPI_ISL_1248246, EPI_ISL_1248249, EPI_ISL_1248251, EPI_ISL_1248253, EPI_ISL_1248255, EPI_ISL_1248257, EPI_ISL_1309026, EPI_ISL_1309027, EPI_ISL_1309038, EPI_ISL_1309052, EPI_ISL_1309076, EPI_ISL_1309101, EPI_ISL_1309104, EPI_ISL_1474859, EPI_ISL_1474867, EPI_ISL_1637113, EPI_ISL_1637115, EPI_ISL_1637141, EPI_ISL_1730398, EPI_ISL_1832572

| see above | University College London, Great Ormond Street Hospital for Children NHS Foundation Trust, Imperial College Healthcare NHS Trust | COVID-19 Genomics UK (COG-UK) Consortium | Alison Holmes; Charlotte Williams; Helena Tutill; Jacqueline Findlay; James Price; Judith Breuer; Julianne Brown; Kathryn Harris; Leysa Forrest; Mark Kristiansen; Paola Niola; Paola Resende Silva; Patricia Dyal; Paul Randell; Rachel Williams; Samuel Weeks; Sergi Castellano; Sunando Roy; Tony Brooks; Yasmin Panchbhaya |
|---|---|---|---|
| EPI_ISL_997370, EPI_ISL_1047959, EPI_ISL_2125913, EPI_ISL_2125916, EPI_ISL_2125931 | University Hospitals Of Leicester NHS Trust and DeepSeq Nottingham | COVID-19 Genomics UK (COG-UK) Consortium | Christopher Holmes; Christopher Moore; Fei Sang; Joeseph Chappell; Johnny Debebe; Jonathan Ball; Julian Tang; Karlie Fallon; Matthew Carlisle; Matthew Loose; Nadine Holmes; Patrick McClure; Paul Bird; Thomas Helmer; Victoria Wright |

EPI_ISL_949409, EPI_ISL_949412, EPI_ISL_949413, EPI_ISL_949414, EPI_ISL_949415, EPI_ISL_949416, EPI_ISL_996395, EPI_ISL_996402, EPI_ISL_996403, EPI_ISL_996404, EPI_ISL_996405, EPI_ISL_996406, EPI_ISL_996409, EPI_ISL_996410, EPI_ISL_996411, EPI_ISL_996448, EPI_ISL_996460, EPI_ISL_996461, EPI_ISL_1046988, EPI_ISL_1046989, EPI_ISL_1046991, EPI_ISL_1047001, EPI_ISL_1047004, EPI_ISL_1047005, EPI_ISL_1047007, EPI_ISL_1047008, EPI_ISL_1047009, EPI_ISL_1047022, EPI_ISL_1047061, EPI_ISL_1047062, EPI_ISL_1386509, EPI_ISL_1474519

| see above | University of Birmingham | COVID-19 Genomics UK (COG-UK) Consortium | Alex Richter; Andrew Bosworth; Andrew Bosworth. Queen Elizabeth Hospital: Anna Casey; Andrew D Beggs PHE Heartlands Lab: Husam Osman; Charlotte Poxon; Institute of Microbiology; Joanne Stockton; Josh Quick; Kasun Wanigasooriya; Mike Kidd; Nicholas Loman. University of Birmingham Testing Laboratory: Celina M Whalley; Oliver Pickles; Radoslaw Poplawski; Samuel Nicholls; University of Birmingham: Claire McMurray; Will Rowe |
|---|---|---|---|

EPI_ISL_996577, EPI_ISL_996578, EPI_ISL_997251, EPI_ISL_997252, EPI_ISL_997253, EPI_ISL_997254, EPI_ISL_997255, EPI_ISL_997256, EPI_ISL_997257, EPI_ISL_997259, EPI_ISL_997261, EPI_ISL_997262, EPI_ISL_997263, EPI_ISL_997264, EPI_ISL_997265, EPI_ISL_997269, EPI_ISL_997270, EPI_ISL_997271, EPI_ISL_997272, EPI_ISL_997273, EPI_ISL_997275, EPI_ISL_997276, EPI_ISL_997277, EPI_ISL_997278, EPI_ISL_997279, EPI_ISL_997280, EPI_ISL_997281, EPI_ISL_997333, EPI_ISL_997334, EPI_ISL_997335, EPI_ISL_1308536, EPI_ISL_1308548, EPI_ISL_1308549, EPI_ISL_1308550, EPI_ISL_1308551, EPI_ISL_1308552, EPI_ISL_1308553, EPI_ISL_1308554, EPI_ISL_1308555, EPI_ISL_1308556, EPI_ISL_1308561, EPI_ISL_1308562, EPI_ISL_1308566, EPI_ISL_1308569, EPI_ISL_1308572, EPI_ISL_1308573, EPI_ISL_1308574, EPI_ISL_1308582, EPI_ISL_1545817, EPI_ISL_1545820

| see above | University of Exeter | COVID-19 Genomics UK (COG-UK) Consortium | Aaron Jeffries; Audrey Farbos; Ben Temperton; Jane Masoli; Joanna Warwick-Dugdale; Michelle Michelsen; Robyn Manley; Stephen Michell |
|---|---|---|---|

EPI_ISL_997080, EPI_ISL_997081, EPI_ISL_997082, EPI_ISL_997083, EPI_ISL_997084, EPI_ISL_997085, EPI_ISL_997086, EPI_ISL_997087, EPI_ISL_997088, EPI_ISL_997089, EPI_ISL_997090, EPI_ISL_997091, EPI_ISL_997092, EPI_ISL_997093, EPI_ISL_997106, EPI_ISL_997112, EPI_ISL_997140, EPI_ISL_997141, EPI_ISL_997142, EPI_ISL_997143, EPI_ISL_997144, EPI_ISL_997145, EPI_ISL_997146, EPI_ISL_997147, EPI_ISL_997148, EPI_ISL_997149, EPI_ISL_997150, EPI_ISL_1047893, EPI_ISL_1104219, EPI_ISL_1104228, EPI_ISL_1104229, EPI_ISL_1104262, EPI_ISL_1104263, EPI_ISL_1177691, EPI_ISL_1177727, EPI_ISL_1177728, EPI_ISL_1177730, EPI_ISL_1177732, EPI_ISL_1247719, EPI_ISL_1247721, EPI_ISL_1247722, EPI_ISL_1247732, EPI_ISL_1247733, EPI_ISL_1308818, EPI_ISL_1308821, EPI_ISL_1308853, EPI_ISL_1730102, EPI_ISL_1730232

| see above | Virology Department, Royal Infirmary of Edinburgh, NHS Lothian / School of Biological Sciences, University of Edinburgh | COVID-19 Genomics UK (COG-UK) Consortium | Colquhoun R; Cotton S; Dewar R; Gallagher A; Hill V; Jackson B; Maloney D; McCrone JT; McHugh M; O'Toole Á; Rambaut A; Rooke S; Scher E; Templeton K; Yu X |
|---|---|---|---|
| EPI_ISL_919404, EPI_ISL_949642 | Virology Department, Royal Infirmary of Edinburgh, NHS Lothian / School of Biological Sciences, University of Edinburgh / Institute of Genetics and Molecular Medicine, University of Edinburgh | COVID-19 Genomics UK (COG-UK) Consortium | Balcaza C; Colquhoun R; Dewar R; Gallagher M; Hill V; Jackson B; McCrone JT; McHugh M; O'Toole Á; Rambaut A; Rooke S; Scher E; Templeton K; Williams TC; Yu X |

EPI_ISL_868359, EPI_ISL_868360, EPI_ISL_868377, EPI_ISL_868393, EPI_ISL_868403, EPI_ISL_868418, EPI_ISL_868433, EPI_ISL_868443, EPI_ISL_868449, EPI_ISL_868466, EPI_ISL_868472, EPI_ISL_868505, EPI_ISL_868511, EPI_ISL_868514, EPI_ISL_868520, EPI_ISL_868534, EPI_ISL_868538, EPI_ISL_868539, EPI_ISL_868540, EPI_ISL_868549, EPI_ISL_868558, EPI_ISL_868567, EPI_ISL_868575, EPI_ISL_868586, EPI_ISL_868606, EPI_ISL_868615, EPI_ISL_868647, EPI_ISL_868653, EPI_ISL_868658, EPI_ISL_868663, EPI_ISL_868670, EPI_ISL_868671, EPI_ISL_868686, EPI_ISL_868702, EPI_ISL_868710, EPI_ISL_868713, EPI_ISL_924080, EPI_ISL_924081, EPI_ISL_924086, EPI_ISL_924088, EPI_ISL_924090, EPI_ISL_924093, EPI_ISL_924095, EPI_ISL_924100, EPI_ISL_924110, EPI_ISL_924113, EPI_ISL_924115, EPI_ISL_924116, EPI_ISL_924120, EPI_ISL_924124, EPI_ISL_924127, EPI_ISL_924130, EPI_ISL_924133, EPI_ISL_924134, EPI_ISL_924135, EPI_ISL_924137, EPI_ISL_924138, EPI_ISL_924139, EPI_ISL_924140, EPI_ISL_924143, EPI_ISL_924147, EPI_ISL_924149, EPI_ISL_924151, EPI_ISL_924152, EPI_ISL_924154, EPI_ISL_924155, EPI_ISL_924156, EPI_ISL_924158, EPI_ISL_924161, EPI_ISL_924162, EPI_ISL_924167, EPI_ISL_924171, EPI_ISL_924172, EPI_ISL_924175, EPI_ISL_924179, EPI_ISL_924180, EPI_ISL_924187, EPI_ISL_924188, EPI_ISL_924190, EPI_ISL_924192, EPI_ISL_924194, EPI_ISL_924195, EPI_ISL_924202, EPI_ISL_924205, EPI_ISL_924210, EPI_ISL_924211, EPI_ISL_924214, EPI_ISL_924219, EPI_ISL_924221, EPI_ISL_924222, EPI_ISL_924226, EPI_ISL_924232, EPI_ISL_924234, EPI_ISL_924236, EPI_ISL_924241, EPI_ISL_924246, EPI_ISL_924248, EPI_ISL_924254, EPI_ISL_924257, EPI_ISL_924259, EPI_ISL_924260, EPI_ISL_924261, EPI_ISL_924262, EPI_ISL_924264, EPI_ISL_924267, EPI_ISL_924271, EPI_ISL_924272, EPI_ISL_924275, EPI_ISL_924286, EPI_ISL_924288, EPI_ISL_924289, EPI_ISL_924294, EPI_ISL_924300, EPI_ISL_924301, EPI_ISL_924309, EPI_ISL_924317, EPI_ISL_924318, EPI_ISL_924320, EPI_ISL_924321, EPI_ISL_924322, EPI_ISL_924327, EPI_ISL_924329, EPI_ISL_924331, EPI_ISL_924336, EPI_ISL_924338, EPI_ISL_924344, EPI_ISL_924346, EPI_ISL_924347, EPI_ISL_924349, EPI_ISL_924353, EPI_ISL_924355, EPI_ISL_924362, EPI_ISL_924365, EPI_ISL_924367, EPI_ISL_924370, EPI_ISL_924371, EPI_ISL_924372, EPI_ISL_924373, EPI_ISL_924375, EPI_ISL_924378, EPI_ISL_924381, EPI_ISL_924391, EPI_ISL_924393, EPI_ISL_924395, EPI_ISL_924396, EPI_ISL_924398, EPI_ISL_924401, EPI_ISL_924403, EPI_ISL_924404, EPI_ISL_924405, EPI_ISL_924407, EPI_ISL_924411, EPI_ISL_924415, EPI_ISL_924416, EPI_ISL_924417, EPI_ISL_1000641, EPI_ISL_1000703, EPI_ISL_1000706, EPI_ISL_1000725, EPI_ISL_1047647, EPI_ISL_1047648, EPI_ISL_1047649, EPI_ISL_1047650, EPI_ISL_1047651, EPI_ISL_1047652, EPI_ISL_1047653, EPI_ISL_1047654, EPI_ISL_1047655, EPI_ISL_1047656, EPI_ISL_1047657, EPI_ISL_1047658, EPI_ISL_1047659, EPI_ISL_1047660, EPI_ISL_1047661, EPI_ISL_1047662, EPI_ISL_1047663, EPI_ISL_1047664, EPI_ISL_1047665, EPI_ISL_1047666, EPI_ISL_1047667, EPI_ISL_1047668, EPI_ISL_1047669, EPI_ISL_1047670, EPI_ISL_1047671, EPI_ISL_1047672, EPI_ISL_1047673, EPI_ISL_1047674, EPI_ISL_1047675, EPI_ISL_1047676, EPI_ISL_1047677, EPI_ISL_1047678, EPI_ISL_1054664, EPI_ISL_1054701, EPI_ISL_1180072, EPI_ISL_1180075, EPI_ISL_1180079, EPI_ISL_1180082

| see above | Virology Department, Sheffield Teaching Hospitals NHS Foundation Trust/Department of Infection, Immunity and Cardiovascular Disease, The Medical School, University of Sheffield | COVID-19 Genomics UK (COG-UK) Consortium | Adri Angyal; Alex Keeley; Benjamin Lindsey; Cariad Evans; Danielle Groves; Dave Partridge; Katie Johnson; Laura Carrilero; Luke Green; Matthew Parker; Matthew Wyles; Mehmet Yavuz; Mohammad Raza; Nikki Smith; Paul Parsons; Rachel Tucker; Rebecca Brown; Thushan de Silva |
|---|---|---|---|

EPI_ISL_922884, EPI_ISL_922887, EPI_ISL_922891, EPI_ISL_922938, EPI_ISL_922941, EPI_ISL_922942, EPI_ISL_922943, EPI_ISL_922944, EPI_ISL_922946, EPI_ISL_922947, EPI_ISL_922948, EPI_ISL_922949, EPI_ISL_922950, EPI_ISL_922953, EPI_ISL_922954, EPI_ISL_922955, EPI_ISL_922957, EPI_ISL_922958, EPI_ISL_922959, EPI_ISL_922960, EPI_ISL_922961, EPI_ISL_922964, EPI_ISL_923051, EPI_ISL_923153, EPI_ISL_923158, EPI_ISL_923159, EPI_ISL_923169, EPI_ISL_923171, EPI_ISL_923173, EPI_ISL_923174, EPI_ISL_923175, EPI_ISL_923176, EPI_ISL_923177, EPI_ISL_923178, EPI_ISL_923179, EPI_ISL_923180, EPI_ISL_923181, EPI_ISL_923182, EPI_ISL_923183, EPI_ISL_923184, EPI_ISL_923185, EPI_ISL_923186, EPI_ISL_923187, EPI_ISL_923188, EPI_ISL_923189, EPI_ISL_923190, EPI_ISL_923191, EPI_ISL_923192, EPI_ISL_923193, EPI_ISL_923194, EPI_ISL_923195, EPI_ISL_923196, EPI_ISL_923197, EPI_ISL_923198, EPI_ISL_923199, EPI_ISL_923200, EPI_ISL_923201, EPI_ISL_923203, EPI_ISL_923205, EPI_ISL_923206, EPI_ISL_923208, EPI_ISL_923209, EPI_ISL_923210, EPI_ISL_923211, EPI_ISL_999986, EPI_ISL_1052503, EPI_ISL_1476450, EPI_ISL_1833132, EPI_ISL_1833160, EPI_ISL_1833162, EPI_ISL_1833168, EPI_ISL_1833177

| see above | Wales Specialist Virology Centre Sequencing lab: Pathogen Genomics Unit | Public Health Wales Microbiology Cardiff Wales Specialist Virology Centre | Alec Birchley; Alexander Adams; Amy Gaskin; Angela Marchbank; Bree Gatica-Wilcox; Catherine Moore; Jason Coombes; Joanne Watkins; Joel Southgate; Johnathan Evans; Laura Gifford; Lauren Gilbert; Lee Graham; Malorie Perry; Matthew Bull; Nicole Pacchiarini; Sally Corden; Sara Kumziene-Summerhayes; Sara Rey; Sarah Taylor; Simon Cottrell; Sophie Jones; Tom Connor |
|---|---|---|---|

EPI_ISL_919281, EPI_ISL_919282, EPI_ISL_919283, EPI_ISL_919284, EPI_ISL_919285, EPI_ISL_919286, EPI_ISL_919287, EPI_ISL_919288, EPI_ISL_919289, EPI_ISL_919290, EPI_ISL_919291, EPI_ISL_919292, EPI_ISL_919293, EPI_ISL_919294, EPI_ISL_919295, EPI_ISL_919296, EPI_ISL_919297, EPI_ISL_919298, EPI_ISL_919299, EPI_ISL_919300, EPI_ISL_919302, EPI_ISL_919303, EPI_ISL_919304, EPI_ISL_919305, EPI_ISL_919306, EPI_ISL_919307, EPI_ISL_949608, EPI_ISL_949609, EPI_ISL_949610, EPI_ISL_949614, EPI_ISL_997050, EPI_ISL_1047836, EPI_ISL_1047837, EPI_ISL_1050241, EPI_ISL_2125497

| see above | West of Scotland Specialist Virology Centre, NHSGGC / MRC-University of Glasgow Centre for Virus Research | COVID-19 Genomics UK (COG-UK) Consortium | Alasdair MacLean; Alice Broos; Ana da Silva Filipe; Antonia Ho; Daniel Mair; David L Robertson; Emma Thomson; Guy Mollett; Ioulia Tsatsani; James Shepherd; Jenna Nichols; Joseph Hughes; Kathy Li; Kathy Smollett; Kyriaki Nomikou; Lily Tong; Matthew Holden; Natasha Johnson; Rachel Blacow; Richard Orton; Rory Gunson; Sarah McDonald; Sharif Shaaban; Sreenu Vattipally; Stephen Carmichael |
|---|---|---|---|

We gratefully acknowledge the following Authors from the Originating laboratories responsible for obtaining the specimens, as well as the Submitting laboratories where the genome data were generated and shared via GISAID, on which this research is based.

All Submitters of data may be contacted directly via www.gisaid.org

Authors are sorted alphabetically.

| Accession ID | Originating Laboratory | Submitting Laboratory | Authors |
| --- | --- | --- | --- |
| EPI_ISL_1391414 | Barts Health NHS Trust | Barts Health NHS Trust | BROAD; Beatrix; CUTINO-MOGUEL; Claire; David; Dola; Esin; HARRINGTON; KARAA; KELE; KULASEGARAN-SHYLINI; Maria-Teresa; OWOYEMI; Raghavendran |

EPI_ISL_997615, EPI_ISL_997652, EPI_ISL_997653, EPI_ISL_997654, EPI_ISL_997655, EPI_ISL_997656, EPI_ISL_997662, EPI_ISL_997663, EPI_ISL_997664, EPI_ISL_997666, EPI_ISL_997667, EPI_ISL_997669, EPI_ISL_997670, EPI_ISL_997671, EPI_ISL_997673, EPI_ISL_997674, EPI_ISL_1048054, EPI_ISL_1048055, EPI_ISL_1048057, EPI_ISL_1048058, EPI_ISL_1048059, EPI_ISL_1048062, EPI_ISL_1048066, EPI_ISL_1048069, EPI_ISL_1048076, EPI_ISL_1048089, EPI_ISL_1048097, EPI_ISL_1050356

| | | | |
| --- | --- | --- | --- |
| see above | Barts Health NHS Trust | COVID-19 Genomics UK (COG-UK) Consortium | BROAD; Beatrix; CUTINO-MOGUEL; Claire; David; Dola; HARRINGTON; KELE; KULASEGARAN-SHYLINI; Maria-Teresa; OWOYEMI; Raghavendran |

EPI_ISL_953241, EPI_ISL_953243, EPI_ISL_953244, EPI_ISL_953246, EPI_ISL_953247, EPI_ISL_953251, EPI_ISL_953254, EPI_ISL_953259, EPI_ISL_953261, EPI_ISL_953262, EPI_ISL_953263, EPI_ISL_953267, EPI_ISL_953272, EPI_ISL_953281, EPI_ISL_1000806, EPI_ISL_1000808, EPI_ISL_1000816, EPI_ISL_1000819, EPI_ISL_1000822, EPI_ISL_1000826, EPI_ISL_1000832, EPI_ISL_1000833, EPI_ISL_1000834, EPI_ISL_1000835, EPI_ISL_1000836, EPI_ISL_1000837, EPI_ISL_1000845, EPI_ISL_1000964, EPI_ISL_1054815, EPI_ISL_1054830, EPI_ISL_1054834

| | | | |
| --- | --- | --- | --- |
| see above | Bioinformatics and Biostatistics Lab, Advanced Sequencing Facility | COVID-19 Genomics UK (COG-UK) Consortium | Aengus Stewart; Chelsea Sawyer; Harshil Patel; Jerome Nicod; Laura Cubitt; Margaret Crawford |

EPI_ISL_923349, EPI_ISL_952453, EPI_ISL_952454, EPI_ISL_952456, EPI_ISL_952459, EPI_ISL_952460, EPI_ISL_952462, EPI_ISL_952464, EPI_ISL_952465, EPI_ISL_952466, EPI_ISL_952467, EPI_ISL_952469, EPI_ISL_952470, EPI_ISL_952477, EPI_ISL_952478, EPI_ISL_952479, EPI_ISL_952480, EPI_ISL_952481, EPI_ISL_952482, EPI_ISL_952483, EPI_ISL_952484, EPI_ISL_952485, EPI_ISL_952558, EPI_ISL_1000240, EPI_ISL_1000241, EPI_ISL_1000242, EPI_ISL_1000243, EPI_ISL_1000257, EPI_ISL_1000260, EPI_ISL_1000262, EPI_ISL_1000267, EPI_ISL_1000269, EPI_ISL_1000271, EPI_ISL_1000273, EPI_ISL_1000281, EPI_ISL_1000282, EPI_ISL_1000292, EPI_ISL_1000302, EPI_ISL_1000308, EPI_ISL_1000314, EPI_ISL_1000456, EPI_ISL_1000457, EPI_ISL_1000458, EPI_ISL_1000459, EPI_ISL_1000460, EPI_ISL_1000482, EPI_ISL_1000483, EPI_ISL_1000601, EPI_ISL_1000605, EPI_ISL_1000606, EPI_ISL_1000607, EPI_ISL_1000608, EPI_ISL_1000609, EPI_ISL_1000610, EPI_ISL_1000618, EPI_ISL_1000619, EPI_ISL_1000620, EPI_ISL_1000621, EPI_ISL_1000622, EPI_ISL_1000623, EPI_ISL_1000624, EPI_ISL_1000625, EPI_ISL_1000627, EPI_ISL_1053844, EPI_ISL_1053845, EPI_ISL_1053846, EPI_ISL_1053847, EPI_ISL_1053848, EPI_ISL_1053849, EPI_ISL_1053850, EPI_ISL_1053853, EPI_ISL_1053854, EPI_ISL_1053855, EPI_ISL_1053892, EPI_ISL_1053898, EPI_ISL_1053899, EPI_ISL_1053900, EPI_ISL_1053901, EPI_ISL_1053902, EPI_ISL_1053933, EPI_ISL_1107634, EPI_ISL_1107635, EPI_ISL_1107636, EPI_ISL_1107650, EPI_ISL_1107651, EPI_ISL_1107652, EPI_ISL_1107653, EPI_ISL_1107654, EPI_ISL_1179831, EPI_ISL_1179833, EPI_ISL_1310349, EPI_ISL_1730877, EPI_ISL_1979864, EPI_ISL_1979865, EPI_ISL_1979973, EPI_ISL_1979975

| | | | |
| --- | --- | --- | --- |
| see above | Centre for Enzyme Innovation, University of Portsmouth / Translational Research Laboratory, Portsmouth Hospitals NHS Trust | COVID-19 Genomics UK (COG-UK) Consortium | Angela Beckett; Christopher Fearn; Kate Cook; Katie Loveson; Salman Goudarzi; Samuel Robson; Scott Elliott; Sharon Glaysher |

EPI_ISL_919149, EPI_ISL_919150, EPI_ISL_919151, EPI_ISL_919152, EPI_ISL_919153, EPI_ISL_919154, EPI_ISL_919155, EPI_ISL_919156, EPI_ISL_919157, EPI_ISL_919158, EPI_ISL_919159, EPI_ISL_919160, EPI_ISL_919161, EPI_ISL_919162, EPI_ISL_919163, EPI_ISL_919164, EPI_ISL_919165, EPI_ISL_919166, EPI_ISL_919167, EPI_ISL_919168, EPI_ISL_919169, EPI_ISL_919170, EPI_ISL_919171, EPI_ISL_919172, EPI_ISL_919173, EPI_ISL_919174, EPI_ISL_919175, EPI_ISL_919176, EPI_ISL_919177, EPI_ISL_919179, EPI_ISL_919180, EPI_ISL_919181, EPI_ISL_919182, EPI_ISL_919183, EPI_ISL_919184, EPI_ISL_919185, EPI_ISL_919186, EPI_ISL_919187, EPI_ISL_919188, EPI_ISL_919189, EPI_ISL_949462, EPI_ISL_949463, EPI_ISL_949464, EPI_ISL_949465, EPI_ISL_949467, EPI_ISL_949468, EPI_ISL_949469, EPI_ISL_949473, EPI_ISL_949475, EPI_ISL_949476, EPI_ISL_949477, EPI_ISL_949524, EPI_ISL_949525, EPI_ISL_949526, EPI_ISL_949527, EPI_ISL_949528, EPI_ISL_949529, EPI_ISL_949530, EPI_ISL_949531, EPI_ISL_949532, EPI_ISL_949533, EPI_ISL_949534, EPI_ISL_949535, EPI_ISL_949536, EPI_ISL_949537, EPI_ISL_949538, EPI_ISL_949539, EPI_ISL_949540, EPI_ISL_949542, EPI_ISL_949544, EPI_ISL_949546, EPI_ISL_949548, EPI_ISL_949549, EPI_ISL_949550, EPI_ISL_949553, EPI_ISL_949554, EPI_ISL_949555, EPI_ISL_949556, EPI_ISL_996673, EPI_ISL_996675, EPI_ISL_996677, EPI_ISL_996679, EPI_ISL_996687, EPI_ISL_996691, EPI_ISL_996693, EPI_ISL_996697, EPI_ISL_996699, EPI_ISL_996703, EPI_ISL_996705, EPI_ISL_996709, EPI_ISL_996713, EPI_ISL_996715, EPI_ISL_996717, EPI_ISL_996720, EPI_ISL_996722, EPI_ISL_996732, EPI_ISL_996734, EPI_ISL_996736, EPI_ISL_996865, EPI_ISL_996866, EPI_ISL_996867, EPI_ISL_996868, EPI_ISL_996869, EPI_ISL_996870, EPI_ISL_996871, EPI_ISL_996872, EPI_ISL_996873, EPI_ISL_996874, EPI_ISL_996875, EPI_ISL_996876, EPI_ISL_996877, EPI_ISL_996878, EPI_ISL_996879, EPI_ISL_996880, EPI_ISL_996881, EPI_ISL_996882, EPI_ISL_996883, EPI_ISL_996884, EPI_ISL_996885, EPI_ISL_996886, EPI_ISL_996887, EPI_ISL_996888, EPI_ISL_996889, EPI_ISL_996890, EPI_ISL_996891, EPI_ISL_996892, EPI_ISL_996893, EPI_ISL_996894, EPI_ISL_996895, EPI_ISL_996896, EPI_ISL_996897, EPI_ISL_996898, EPI_ISL_996899, EPI_ISL_996900, EPI_ISL_996901, EPI_ISL_996902, EPI_ISL_996903, EPI_ISL_996904, EPI_ISL_996905, EPI_ISL_996906, EPI_ISL_996907, EPI_ISL_996908, EPI_ISL_996909, EPI_ISL_996910, EPI_ISL_996911, EPI_ISL_996912, EPI_ISL_996913, EPI_ISL_996914, EPI_ISL_996915, EPI_ISL_996916, EPI_ISL_996917, EPI_ISL_996918, EPI_ISL_996919, EPI_ISL_996920, EPI_ISL_996921, EPI_ISL_996922, EPI_ISL_996923, EPI_ISL_996924, EPI_ISL_996925, EPI_ISL_996926, EPI_ISL_996927, EPI_ISL_996928, EPI_ISL_996929, EPI_ISL_996930, EPI_ISL_996931, EPI_ISL_996932, EPI_ISL_996933, EPI_ISL_996934, EPI_ISL_996935, EPI_ISL_996936, EPI_ISL_996937, EPI_ISL_996938, EPI_ISL_996939, EPI_ISL_996940, EPI_ISL_996941, EPI_ISL_996942, EPI_ISL_996943, EPI_ISL_996944, EPI_ISL_996945, EPI_ISL_996946, EPI_ISL_996947, EPI_ISL_996948, EPI_ISL_996949, EPI_ISL_996950, EPI_ISL_996951, EPI_ISL_996952, EPI_ISL_996953, EPI_ISL_996954, EPI_ISL_996955, EPI_ISL_996956, EPI_ISL_996957, EPI_ISL_996958, EPI_ISL_996959, EPI_ISL_996960, EPI_ISL_996961, EPI_ISL_996962, EPI_ISL_996963, EPI_ISL_996964, EPI_ISL_996965, EPI_ISL_996966, EPI_ISL_996967, EPI_ISL_996968, EPI_ISL_996969, EPI_ISL_996970, EPI_ISL_996971, EPI_ISL_996972, EPI_ISL_996973, EPI_ISL_996974, EPI_ISL_996975, EPI_ISL_996976, EPI_ISL_996977, EPI_ISL_996978, EPI_ISL_996979, EPI_ISL_996980, EPI_ISL_996981, EPI_ISL_996982, EPI_ISL_996983, EPI_ISL_996984, EPI_ISL_996985, EPI_ISL_996986, EPI_ISL_996987, EPI_ISL_996988, EPI_ISL_996989, EPI_ISL_996990, EPI_ISL_996991, EPI_ISL_996992, EPI_ISL_996993, EPI_ISL_996994, EPI_ISL_996995, EPI_ISL_996996, EPI_ISL_996997, EPI_ISL_996998, EPI_ISL_996999, EPI_ISL_997000, EPI_ISL_997001, EPI_ISL_997002, EPI_ISL_997003, EPI_ISL_997004, EPI_ISL_997005, EPI_ISL_997006, EPI_ISL_997007, EPI_ISL_997008, EPI_ISL_997009, EPI_ISL_997010, EPI_ISL_997011, EPI_ISL_997012, EPI_ISL_997013, EPI_ISL_997014, EPI_ISL_997015, EPI_ISL_997016, EPI_ISL_997017, EPI_ISL_997018, EPI_ISL_997019, EPI_ISL_997020, EPI_ISL_997021, EPI_ISL_997022, EPI_ISL_997023, EPI_ISL_997024, EPI_ISL_997025, EPI_ISL_997026, EPI_ISL_997027, EPI_ISL_997028, EPI_ISL_997029, EPI_ISL_997030, EPI_ISL_997031, EPI_ISL_997032, EPI_ISL_997033, EPI_ISL_997034, EPI_ISL_997035, EPI_ISL_997036, EPI_ISL_997037, EPI_ISL_997038, EPI_ISL_997039, EPI_ISL_997040, EPI_ISL_997041, EPI_ISL_997042, EPI_ISL_997043, EPI_ISL_997044, EPI_ISL_997045, EPI_ISL_997046, EPI_ISL_997047, EPI_ISL_997048, EPI_ISL_997049, EPI_ISL_1047148, EPI_ISL_1047149, EPI_ISL_1047150, EPI_ISL_1047151, EPI_ISL_1047152, EPI_ISL_1047153, EPI_ISL_1047154, EPI_ISL_1047155, EPI_ISL_1047156, EPI_ISL_1047157, EPI_ISL_1047158, EPI_ISL_1047159, EPI_ISL_1047160, EPI_ISL_1047161, EPI_ISL_1047162, EPI_ISL_1047163, EPI_ISL_1047164, EPI_ISL_1047165, EPI_ISL_1047166, EPI_ISL_1047167, EPI_ISL_1047168, EPI_ISL_1047169, EPI_ISL_1047170, EPI_ISL_1047171, EPI_ISL_1047172, EPI_ISL_1047173, EPI_ISL_1047174, EPI_ISL_1047175, EPI_ISL_1047176, EPI_ISL_1047177, EPI_ISL_1047178, EPI_ISL_1047179, EPI_ISL_1047180, EPI_ISL_1047181, EPI_ISL_1047182, EPI_ISL_1047183, EPI_ISL_1047185, EPI_ISL_1047186, EPI_ISL_1047190, EPI_ISL_1047194, EPI_ISL_1047195, EPI_ISL_1047196, EPI_ISL_1047197, EPI_ISL_1047198, EPI_ISL_1047199, EPI_ISL_1047200, EPI_ISL_1047201, EPI_ISL_1047202, EPI_ISL_1047203, EPI_ISL_1047204, EPI_ISL_1047205, EPI_ISL_1047206, EPI_ISL_1047207, EPI_ISL_1047208, EPI_ISL_1047209, EPI_ISL_1047210, EPI_ISL_1047211, EPI_ISL_1047212, EPI_ISL_1047213, EPI_ISL_1047214, EPI_ISL_1047215, EPI_ISL_1047216, EPI_ISL_1047217, EPI_ISL_1047218, EPI_ISL_1047219, EPI_ISL_1047220, EPI_ISL_1047221, EPI_ISL_1047222, EPI_ISL_1047223, EPI_ISL_1047224, EPI_ISL_1047225, EPI_ISL_1047226, EPI_ISL_1047227, EPI_ISL_1047228, EPI_ISL_1047229, EPI_ISL_1047230, EPI_ISL_1047231, EPI_ISL_1047232, EPI_ISL_1047233, EPI_ISL_1047234, EPI_ISL_1047235, EPI_ISL_1047236, EPI_ISL_1047237, EPI_ISL_1047238, EPI_ISL_1047239, EPI_ISL_1047240, EPI_ISL_1047241, EPI_ISL_1047242, EPI_ISL_1047243, EPI_ISL_1047244, EPI_ISL_1047245, EPI_ISL_1047246, EPI_ISL_1047247, EPI_ISL_1047248, EPI_ISL_1047249, EPI_ISL_1047250, EPI_ISL_1047251, EPI_ISL_1047252, EPI_ISL_1047253, EPI_ISL_1047254, EPI_ISL_1047255, EPI_ISL_1047256, EPI_ISL_1047257, EPI_ISL_1047258, EPI_ISL_1047259, EPI_ISL_1047260, EPI_ISL_1047261, EPI_ISL_1047262, EPI_ISL_1047263, EPI_ISL_1047264, EPI_ISL_1047265, EPI_ISL_1047266, EPI_ISL_1047267, EPI_ISL_1047268, EPI_ISL_1047269, EPI_ISL_1047270, EPI_ISL_1047271, EPI_ISL_1047272, EPI_ISL_1047273, EPI_ISL_1047274, EPI_ISL_1047275, EPI_ISL_1047276, EPI_ISL_1047277, EPI_ISL_1047278, EPI_ISL_1047279, EPI_ISL_1047280, EPI_ISL_1047281, EPI_ISL_1047282, EPI_ISL_1047283, EPI_ISL_1047284, EPI_ISL_1047285, EPI_ISL_1047286, EPI_ISL_1047287, EPI_ISL_1047288, EPI_ISL_1047289, EPI_ISL_1047290, EPI_ISL_1047291, EPI_ISL_1047292, EPI_ISL_1047293, EPI_ISL_1047294, EPI_ISL_1047295, EPI_ISL_1047296, EPI_ISL_1047297, EPI_ISL_1047298, EPI_ISL_1047299, EPI_ISL_1047300, EPI_ISL_1047301, EPI_ISL_1047302, EPI_ISL_1047303, EPI_ISL_1047304, EPI_ISL_1047305, EPI_ISL_1047306, EPI_ISL_1047307, EPI_ISL_1047308, EPI_ISL_1047309, EPI_ISL_1047310, EPI_ISL_1047311, EPI_ISL_1047312, EPI_ISL_1047313, EPI_ISL_1047314, EPI_ISL_1047315, EPI_ISL_1047316, EPI_ISL_1047317, EPI_ISL_1047318, EPI_ISL_1047319, EPI_ISL_1047320, EPI_ISL_1047321, EPI_ISL_1047322, EPI_ISL_1047323, EPI_ISL_1047324, EPI_ISL_1047325, EPI_ISL_1047326, EPI_ISL_1047327, EPI_ISL_1047328, EPI_ISL_1047329, EPI_ISL_1047330, EPI_ISL_1047332, EPI_ISL_1047333, EPI_ISL_1047334, EPI_ISL_1047335, EPI_ISL_1047336, EPI_ISL_1047337, EPI_ISL_1047338, EPI_ISL_1047339, EPI_ISL_1047340, EPI_ISL_1047341, EPI_ISL_1047342, EPI_ISL_1047343, EPI_ISL_1047344, EPI_ISL_1047345, EPI_ISL_1047346, EPI_ISL_1047347, EPI_ISL_1047348, EPI_ISL_1047352, EPI_ISL_1047355, EPI_ISL_1047356, EPI_ISL_1047357, EPI_ISL_1047358, EPI_ISL_1047359, EPI_ISL_1047360, EPI_ISL_1047361, EPI_ISL_1047362, EPI_ISL_1047363, EPI_ISL_1047364, EPI_ISL_1047365, EPI_ISL_1047366, EPI_ISL_1047367, EPI_ISL_1047368, EPI_ISL_1047369, EPI_ISL_1047370, EPI_ISL_1047371, EPI_ISL_1047372, EPI_ISL_1047373, EPI_ISL_1047374, EPI_ISL_1047375, EPI_ISL_1047376, EPI_ISL_1047377, EPI_ISL_1047378, EPI_ISL_1047379, EPI_ISL_1047380, EPI_ISL_1047381, EPI_ISL_1047382, EPI_ISL_1047383, EPI_ISL_1047384, EPI_ISL_1047385, EPI_ISL_1047386, EPI_ISL_1047387, EPI_ISL_1047388, EPI_ISL_1047389, EPI_ISL_1047390, EPI_ISL_1047391, EPI_ISL_1047392, EPI_ISL_1047393, EPI_ISL_1047394, EPI_ISL_1047395, EPI_ISL_1047396, EPI_ISL_1047397, EPI_ISL_1047398, EPI_ISL_1047399, EPI_ISL_1047400, EPI_ISL_1047401, EPI_ISL_1047402, EPI_ISL_1047403, EPI_ISL_1047404, EPI_ISL_1047405, EPI_ISL_1047406, EPI_ISL_1047407, EPI_ISL_1047408, EPI_ISL_1047409, EPI_ISL_1047410, EPI_ISL_1047411, EPI_ISL_1047412, EPI_ISL_1047413, EPI_ISL_1047414, EPI_ISL_1047415, EPI_ISL_1047416, EPI_ISL_1047417, EPI_ISL_1047418, EPI_ISL_1047419, EPI_ISL_1047420, EPI_ISL_1047421, EPI_ISL_1047422, EPI_ISL_1047423, EPI_ISL_1047424, EPI_ISL_1047425, EPI_ISL_1047426, EPI_ISL_1047427, EPI_ISL_1047428, EPI_ISL_1047429, EPI_ISL_1047430, EPI_ISL_1047431, EPI_ISL_1047432, EPI_ISL_1047433, EPI_ISL_1047434, EPI_ISL_1047435, EPI_ISL_1047436, EPI_ISL_1047437, EPI_ISL_1047438, EPI_ISL_1047439, EPI_ISL_1047440, EPI_ISL_1047441, EPI_ISL_1047442, EPI_ISL_1047443, EPI_ISL_1047444, EPI_ISL_1047445, EPI_ISL_1047446, EPI_ISL_1047447, EPI_ISL_1047448, EPI_ISL_1047449, EPI_ISL_1047450, EPI_ISL_1047451, EPI_ISL_1047452, EPI_ISL_1047453, EPI_ISL_1047454, EPI_ISL_1047455, EPI_ISL_1047456, EPI_ISL_1047457, EPI_ISL_1047458, EPI_ISL_1047459, EPI_ISL_1047460, EPI_ISL_1047461, EPI_ISL_1047462, EPI_ISL_1047463, EPI_ISL_1047464, EPI_ISL_1047465, EPI_ISL_1047466, EPI_ISL_1047467, EPI_ISL_1047468, EPI_ISL_1047469, EPI_ISL_1047470, EPI_ISL_1047471, EPI_ISL_1047472, EPI_ISL_1047473, EPI_ISL_1047474, EPI_ISL_1047475, EPI_ISL_1047476, EPI_ISL_1047477, EPI_ISL_1047478, EPI_ISL_1047479, EPI_ISL_1047480, EPI_ISL_1047481, EPI_ISL_1047482, EPI_ISL_1047483, EPI_ISL_1047484, EPI_ISL_1047485, EPI_ISL_1047486, EPI_ISL_1047487, EPI_ISL_1047488, EPI_ISL_1047489, EPI_ISL_1047490, EPI_ISL_1047491, EPI_ISL_1047492, EPI_ISL_1047493, EPI_ISL_1047494, EPI_ISL_1047495, EPI_ISL_1047496, EPI_ISL_1047497, EPI_ISL_1047498, EPI_ISL_1047499, EPI_ISL_1047500, EPI_ISL_1047501, EPI_ISL_1047502, EPI_ISL_1047503, EPI_ISL_1047504, EPI_ISL_1047505, EPI_ISL_1047506, EPI_ISL_1047507, EPI_ISL_1047508, EPI_ISL_1047509, EPI_ISL_1047510, EPI_ISL_1047512, EPI_ISL_1047513, EPI_ISL_1047514, EPI_ISL_1047515, EPI_ISL_1047516, EPI_ISL_1047517, EPI_ISL_1047518, EPI_ISL_1047519, EPI_ISL_1047520, EPI_ISL_1047521, EPI_ISL_1047522, EPI_ISL_1047523, EPI_ISL_1047524, EPI_ISL_1047525, EPI_ISL_1047526, EPI_ISL_1047527, EPI_ISL_1047528, EPI_ISL_1047529, EPI_ISL_1047530, EPI_ISL_1047531, EPI_ISL_1047532, EPI_ISL_1047533, EPI_ISL_1047534, EPI_ISL_1047535, EPI_ISL_1047536, EPI_ISL_1047537, EPI_ISL_1047538, EPI_ISL_1047539, EPI_ISL_1047540, EPI_ISL_1047541, EPI_ISL_1047542, EPI_ISL_1047543, EPI_ISL_1047544, EPI_ISL_1047545, EPI_ISL_1047546, EPI_ISL_1047547, EPI_ISL_1047548, EPI_ISL_1047549, EPI_ISL_1047550, EPI_ISL_1047551, EPI_ISL_1047552, EPI_ISL_1047553, EPI_ISL_1047554, EPI_ISL_1047555, EPI_ISL_1047556, EPI_ISL_1047557, EPI_ISL_1047558, EPI_ISL_1047559, EPI_ISL_1047560, EPI_ISL_1047561, EPI_ISL_1047562, EPI_ISL_1047563, EPI_ISL_1047564, EPI_ISL_1047565, EPI_ISL_1047566, EPI_ISL_1047567, EPI_ISL_1047568, EPI_ISL_1047569, EPI_ISL_1047570, EPI_ISL_1047571, EPI_ISL_1047572, EPI_ISL_1047573, EPI_ISL_1047574, EPI_ISL_1047575, EPI_ISL_1047576, EPI_ISL_1047577, EPI_ISL_1047578, EPI_ISL_1047579, EPI_ISL_1047580, EPI_ISL_1047581, EPI_ISL_1103936, EPI_ISL_1103937, EPI_ISL_1103938, EPI_ISL_1103939, EPI_ISL_1103940, EPI_ISL_1103941, EPI_ISL_1103942, EPI_ISL_1103943, EPI_ISL_1103944, EPI_ISL_1103946, EPI_ISL_1103947, EPI_ISL_1103948, EPI_ISL_1179911, EPI_ISL_1179933, EPI_ISL_1179935, EPI_ISL_1179936, EPI_ISL_1179951, EPI_ISL_1179952, EPI_ISL_1179953, EPI_ISL_1179954, EPI_ISL_1179955, EPI_ISL_1179956, EPI_ISL_1179957, EPI_ISL_1179958, EPI_ISL_1179959, EPI_ISL_1179960, EPI_ISL_1179961, EPI_ISL_1179962, EPI_ISL_1179963, EPI_ISL_1179975, EPI_ISL_1179976, EPI_ISL_1179977, EPI_ISL_1179978, EPI_ISL_1179983, EPI_ISL_1179990, EPI_ISL_1179991, EPI_ISL_1179992, EPI_ISL_1179993, EPI_ISL_1179994, EPI_ISL_1179995, EPI_ISL_1179996, EPI_ISL_1180017, EPI_ISL_1180023, EPI_ISL_1180024, EPI_ISL_1180025, EPI_ISL_1180026, EPI_ISL_1180027, EPI_ISL_1180028

| | | | |
| --- | --- | --- | --- |
| see above | Department of Pathology, University of Cambridge | COVID-19 Genomics UK (COG-UK) Consortium | Aminu S. Jahun; Ian Goodfellow; Iliana Georgana; Martin D. Curran; Myra Hosmillo; Rhys Izu; Rhys Izuagbe; Surendra Parmar; William L. Hamilton; Yasmin Chaudhry |

EPI_ISL_931586, EPI_ISL_931587, EPI_ISL_931588, EPI_ISL_931590, EPI_ISL_931591, EPI_ISL_931592, EPI_ISL_931593, EPI_ISL_931594, EPI_ISL_931596, EPI_ISL_931597, EPI_ISL_931598, EPI_ISL_931599, EPI_ISL_931600, EPI_ISL_931601, EPI_ISL_931602, EPI_ISL_931603, EPI_ISL_931604, EPI_ISL_931605, EPI_ISL_931606, EPI_ISL_931607, EPI_ISL_931608, EPI_ISL_931609, EPI_ISL_931610, EPI_ISL_931611, EPI_ISL_931612, EPI_ISL_931613, EPI_ISL_931614, EPI_ISL_931615, EPI_ISL_931616, EPI_ISL_931617, EPI_ISL_931618, EPI_ISL_931619, EPI_ISL_931620, EPI_ISL_931621, EPI_ISL_931622, EPI_ISL_931623, EPI_ISL_931624, EPI_ISL_931625, EPI_ISL_931626, EPI_ISL_931627, EPI_ISL_931628, EPI_ISL_931629, EPI_ISL_931630, EPI_ISL_931631, EPI_ISL_931632, EPI_ISL_931633, EPI_ISL_931634, EPI_ISL_931635, EPI_ISL_931636, EPI_ISL_931637, EPI_ISL_931638, EPI_ISL_931639, EPI_ISL_931640, EPI_ISL_931641, EPI_ISL_931642, EPI_ISL_931643, EPI_ISL_931644, EPI_ISL_931645, EPI_ISL_931646, EPI_ISL_931647, EPI_ISL_931648, EPI_ISL_931649, EPI_ISL_931650, EPI_ISL_931651, EPI_ISL_931652, EPI_ISL_931653, EPI_ISL_931654, EPI_ISL_931655, EPI_ISL_931656, EPI_ISL_931657, EPI_ISL_931658, EPI_ISL_931659, EPI_ISL_931660, EPI_ISL_931661, EPI_ISL_931662, EPI_ISL_931663, EPI_ISL_931664, EPI_ISL_931665, EPI_ISL_931666, EPI_ISL_931667, EPI_ISL_931668, EPI_ISL_931669, EPI_ISL_931670, EPI_ISL_931671, EPI_ISL_931672, EPI_ISL_931673, EPI_ISL_931674, EPI_ISL_931675, EPI_ISL_931676, EPI_ISL_931677, EPI_ISL_931678, EPI_ISL_931679, EPI_ISL_931680, EPI_ISL_931681, EPI_ISL_931682, EPI_ISL_931683, EPI_ISL_931684, EPI_ISL_931685, EPI_ISL_931686, EPI_ISL_931687, EPI_ISL_931688, EPI_ISL_931689, EPI_ISL_931691, EPI_ISL_931692, EPI_ISL_931693, EPI_ISL_931694, EPI_ISL_931695, EPI_ISL_931696, EPI_ISL_931697, EPI_ISL_931698, EPI_ISL_931699, EPI_ISL_931700, EPI_ISL_931701, EPI_ISL_931702, EPI_ISL_931703, EPI_ISL_931704, EPI_ISL_931705, EPI_ISL_931706, EPI_ISL_931707, EPI_ISL_931708, EPI_ISL_931709, EPI_ISL_931710, EPI_ISL_931711, EPI_ISL_931712, EPI_ISL_931713, EPI_ISL_931714, EPI_ISL_931715, EPI_ISL_931716, EPI_ISL_931717, EPI_ISL_931718, EPI_ISL_931719, EPI_ISL_931720, EPI_ISL_931721, EPI_ISL_931722, EPI_ISL_931723, EPI_ISL_931724, EPI_ISL_931725, EPI_ISL_931727, EPI_ISL_931728, EPI_ISL_931729, EPI_ISL_931730, EPI_ISL_931731, EPI_ISL_931732, EPI_ISL_931733, EPI_ISL_931734, EPI_ISL_931735, EPI_ISL_931736, EPI_ISL_931737, EPI_ISL_931738, EPI_ISL_931739, EPI_ISL_931740, EPI_ISL_931741, EPI_ISL_931742, EPI_ISL_931743, EPI_ISL_931744, EPI_ISL_931745, EPI_ISL_931746, EPI_ISL_931747, EPI_ISL_931748, EPI_ISL_931749, EPI_ISL_931750, EPI_ISL_931751, EPI_ISL_931752, EPI_ISL_931753, EPI_ISL_931754, EPI_ISL_931755, EPI_ISL_931756, EPI_ISL_931757, EPI_ISL_931758, EPI_ISL_931759, EPI_ISL_931760, EPI_ISL_931761, EPI_ISL_931762, EPI_ISL_931763, EPI_ISL_931764, EPI_ISL_931765, EPI_ISL_931766, EPI_ISL_931767, EPI_ISL_931768, EPI_ISL_931769, EPI_ISL_931770, EPI_ISL_931771, EPI_ISL_931772, EPI_ISL_931773, EPI_ISL_931774, EPI_ISL_931775, EPI_ISL_931776, EPI_ISL_931777, EPI_ISL_931778, EPI_ISL_931779, EPI_ISL_931780, EPI_ISL_931781, EPI_ISL_931782, EPI_ISL_931783, EPI_ISL_931784, EPI_ISL_931785, EPI_ISL_931786, EPI_ISL_931787, EPI_ISL_931788, EPI_ISL_931789, EPI_ISL_931790, EPI_ISL_931791, EPI_ISL_931792, EPI_ISL_931793, EPI_ISL_931794, EPI_ISL_931795, EPI_ISL_931796, EPI_ISL_931797, EPI_ISL_931798, EPI_ISL_931799, EPI_ISL_931800, EPI_ISL_931801, EPI_ISL_931802, EPI_ISL_931803, EPI_ISL_931804, EPI_ISL_931805, EPI_ISL_931806, EPI_ISL_931807, EPI_ISL_931808, EPI_ISL_931809, EPI_ISL_931810, EPI_ISL_931811, EPI_ISL_931812, EPI_ISL_931813, EPI_ISL_931814, EPI_ISL_931815, EPI_ISL_931816, EPI_ISL_931817, EPI_ISL_931819, EPI_ISL_931820, EPI_ISL_931821, EPI_ISL_931823, EPI_ISL_931824, EPI_ISL_931825, EPI_ISL_931826, EPI_ISL_931828, EPI_ISL_931829, EPI_ISL_931830, EPI_ISL_931831, EPI_ISL_931832, EPI_ISL_931833, EPI_ISL_931834, EPI_ISL_931835, EPI_ISL_931836, EPI_ISL_931837, EPI_ISL_931838, EPI_ISL_931839, EPI_ISL_931840, EPI_ISL_931841, EPI_ISL_931842, EPI_ISL_931843, EPI_ISL_931844, EPI_ISL_931845, EPI_ISL_931846, EPI_ISL_931847, EPI_ISL_931848, EPI_ISL_931849, EPI_ISL_931850, EPI_ISL_931851, EPI_ISL_931852, EPI_ISL_931853, EPI_ISL_931854, EPI_ISL_931855, EPI_ISL_931856, EPI_ISL_931857, EPI_ISL_931858, EPI_ISL_931859, EPI_ISL_931860, EPI_ISL_931861, EPI_ISL_931862, EPI_ISL_931863, EPI_ISL_931864, EPI_ISL_931865, EPI_ISL_931866, EPI_ISL_931867, EPI_ISL_931868, EPI_ISL_931869, EPI_ISL_931870, EPI_ISL_931871, EPI_ISL_931872, EPI_ISL_931873, EPI_ISL_931875, EPI_ISL_931876, EPI_ISL_931877, EPI_ISL_931878, EPI_ISL_931879, EPI_ISL_931880, EPI_ISL_931881, EPI_ISL_931882, EPI_ISL_931883, EPI_ISL_931884, EPI_ISL_931885, EPI_ISL_931886, EPI_ISL_931887, EPI_ISL_931888, EPI_ISL_931889, EPI_ISL_931890, EPI_ISL_931891, EPI_ISL_931892, EPI_ISL_931893, EPI_ISL_931894, EPI_ISL_931895, EPI_ISL_931896, EPI_ISL_931897, EPI_ISL_931898, EPI_ISL_931899, EPI_ISL_931900, EPI_ISL_931901, EPI_ISL_931902, EPI_ISL_931903, EPI_ISL_931904, EPI_ISL_931905, EPI_ISL_931906, EPI_ISL_931907, EPI_ISL_931908, EPI_ISL_931909, EPI_ISL_931910, EPI_ISL_931911, EPI_ISL_931912, EPI_ISL_931913, EPI_ISL_931914, EPI_ISL_931915, EPI_ISL_931916, EPI_ISL_931917, EPI_ISL_931919, EPI_ISL_931920, EPI_ISL_931922, EPI_ISL_931923, EPI_ISL_931924, EPI_ISL_931925, EPI_ISL_931926, EPI_ISL_931927, EPI_ISL_931928, EPI_ISL_931929, EPI_ISL_931930, EPI_ISL_931931, EPI_ISL_931932, EPI_ISL_931933, EPI_ISL_931934, EPI_ISL_931935, EPI_ISL_931936, EPI_ISL_931938, EPI_ISL_931940, EPI_ISL_931941, EPI_ISL_931942, EPI_ISL_931944, EPI_ISL_931945, EPI_ISL_931946, EPI_ISL_931947, EPI_ISL_931948, EPI_ISL_931949, EPI_ISL_931950, EPI_ISL_931951, EPI_ISL_931952, EPI_ISL_931954, EPI_ISL_931955, EPI_ISL_931956, EPI_ISL_931958, EPI_ISL_931960, EPI_ISL_931961, EPI_ISL_931962, EPI_ISL_931963, EPI_ISL_931964, EPI_ISL_931965, EPI_ISL_931966, EPI_ISL_931967, EPI_ISL_931968, EPI_ISL_931969, EPI_ISL_931970, EPI_ISL_931971, EPI_ISL_931972, EPI_ISL_931973, EPI_ISL_931974, EPI_ISL_931975, EPI_ISL_931976, EPI_ISL_931977, EPI_ISL_931978, EPI_ISL_931980, EPI_ISL_931981, EPI_ISL_931982, EPI_ISL_931983, EPI_ISL_931984, EPI_ISL_931985, EPI_ISL_931986, EPI_ISL_931987, EPI_ISL_931988, EPI_ISL_931989, EPI_ISL_931990, EPI_ISL_931991, EPI_ISL_931992, EPI_ISL_931993, EPI_ISL_931994, EPI_ISL_931995, EPI_ISL_931996, EPI_ISL_931997, EPI_ISL_931998, EPI_ISL_931999, EPI_ISL_932000, EPI_ISL_932001, EPI_ISL_932002, EPI_ISL_932003, EPI_ISL_932004, EPI_ISL_932005, EPI_ISL_932006, EPI_ISL_932007, EPI_ISL_932008, EPI_ISL_932009, EPI_ISL_932010, EPI_ISL_932011, EPI_ISL_932012, EPI_ISL_932013, EPI_ISL_932014, EPI_ISL_932015, EPI_ISL_932016, EPI_ISL_932018, EPI_ISL_932019, EPI_ISL_932020, EPI_ISL_932021, EPI_ISL_932022, EPI_ISL_932023, EPI_ISL_932024, EPI_ISL_932025, EPI_ISL_932026, EPI_ISL_932027, EPI_ISL_932028, EPI_ISL_932029, EPI_ISL_932030, EPI_ISL_932031, EPI_ISL_932032, EPI_ISL_932033, EPI_ISL_932034, EPI_ISL_932035, EPI_ISL_932036, EPI_ISL_932037, EPI_ISL_932038, EPI_ISL_932040, EPI_ISL_932041, EPI_ISL_932042, EPI_ISL_932043, EPI_ISL_932044, EPI_ISL_932045, EPI_ISL_932047, EPI_ISL_932048, EPI_ISL_932049, EPI_ISL_932050, EPI_ISL_932051, EPI_ISL_932052, EPI_ISL_932053, EPI_ISL_932054, EPI_ISL_932055, EPI_ISL_932056, EPI_ISL_932058, EPI_ISL_932059, EPI_ISL_932060, EPI_ISL_932062, EPI_ISL_932063, EPI_ISL_932064, EPI_ISL_932065, EPI_ISL_932066, EPI_ISL_932067, EPI_ISL_932069, EPI_ISL_932070, EPI_ISL_932071, EPI_ISL_932072, EPI_ISL_932073, EPI_ISL_932074, EPI_ISL_932075, EPI_ISL_932076, EPI_ISL_932077, EPI_ISL_932078, EPI_ISL_932079, EPI_ISL_932080, EPI_ISL_932081, EPI_ISL_932082, EPI_ISL_932083, EPI_ISL_932084, EPI_ISL_932085, EPI_ISL_932087, EPI_ISL_932088, EPI_ISL_932089, EPI_ISL_932090, EPI_ISL_932091, EPI_ISL_932092, EPI_ISL_932093, EPI_ISL_932094, EPI_ISL_932095, EPI_ISL_932096, EPI_ISL_932097, EPI_ISL_932098, EPI_ISL_932099, EPI_ISL_932100, EPI_ISL_932101, EPI_ISL_932102, EPI_ISL_932103, EPI_ISL_932104, EPI_ISL_932105, EPI_ISL_932106, EPI_ISL_932107, EPI_ISL_932109, EPI_ISL_932110, EPI_ISL_932111, EPI_ISL_932112, EPI_ISL_932113, EPI_ISL_932115, EPI_ISL_932116, EPI_ISL_932117, EPI_ISL_932118, EPI_ISL_932119, EPI_ISL_932120, EPI_ISL_932121, EPI_ISL_932122, EPI_ISL_932123, EPI_ISL_932124, EPI_ISL_932125, EPI_ISL_932126, EPI_ISL_932127, EPI_ISL_932128, EPI_ISL_932129, EPI_ISL_932130, EPI_ISL_932131, EPI_ISL_932132, EPI_ISL_932133, EPI_ISL_932134, EPI_ISL_932135, EPI_ISL_932136, EPI_ISL_932137, EPI_ISL_932138, EPI_ISL_932139, EPI_ISL_932140, EPI_ISL_932141, EPI_ISL_932142, EPI_ISL_932143, EPI_ISL_932144, EPI_ISL_932145, EPI_ISL_932146, EPI_ISL_932147, EPI_ISL_932148, EPI_ISL_932149, EPI_ISL_932150, EPI_ISL_932151, EPI_ISL_932152, EPI_ISL_932153, EPI_ISL_932154, EPI_ISL_932155, EPI_ISL_932156, EPI_ISL_932158, EPI_ISL_932159, EPI_ISL_932160, EPI_ISL_932161, EPI_ISL_932162, EPI_ISL_932163, EPI_ISL_932164, EPI_ISL_932165, EPI_ISL_932166, EPI_ISL_932167, EPI_ISL_932168, EPI_ISL_932169, EPI_ISL_932170, EPI_ISL_932171, EPI_ISL_932172, EPI_ISL_932173, EPI_ISL_932174, EPI_ISL_932175, EPI_ISL_932176, EPI_ISL_932177, EPI_ISL_932178, EPI_ISL_932179, EPI_ISL_932180, EPI_ISL_932181, EPI_ISL_932182, EPI_ISL_932183, EPI_ISL_932184, EPI_ISL_932185, EPI_ISL_932186, EPI_ISL_932187, EPI_ISL_932188, EPI_ISL_932189, EPI_ISL_932190, EPI_ISL_932191, EPI_ISL_932192, EPI_ISL_932193, EPI_ISL_932194, EPI_ISL_932195, EPI_ISL_932196, EPI_ISL_932197, EPI_ISL_932198, EPI_ISL_932199, EPI_ISL_932200, EPI_ISL_932202, EPI_ISL_932203, EPI_ISL_932204, EPI_ISL_932205, EPI_ISL_932206, EPI_ISL_932207, EPI_ISL_932208, EPI_ISL_932209, EPI_ISL_932210, EPI_ISL_932211, EPI_ISL_932212, EPI_ISL_932213, EPI_ISL_932214, EPI_ISL_932215, EPI_ISL_932216, EPI_ISL_932217, EPI_ISL_932218, EPI_ISL_932219, EPI_ISL_932220, EPI_ISL_932221, EPI_ISL_932223, EPI_ISL_932224, EPI_ISL_932225, EPI_ISL_932226, EPI_ISL_932227, EPI_ISL_932228, EPI_ISL_932229, EPI_ISL_932230, EPI_ISL_932232, EPI_ISL_932233, EPI_ISL_932234, EPI_ISL_932235, EPI_ISL_932236, EPI_ISL_932237, EPI_ISL_932238, EPI_ISL_932239, EPI_ISL_932242, EPI_ISL_932243, EPI_ISL_932244, EPI_ISL_932245, EPI_ISL_932246, EPI_ISL_932247, EPI_ISL_932248, EPI_ISL_932249, EPI_ISL_932250, EPI_ISL_937545, EPI_ISL_937546, EPI_ISL_937547, EPI_ISL_937548, EPI_ISL_937552, EPI_ISL_937555, EPI_ISL_937556, EPI_ISL_937560, EPI_ISL_937561, EPI_ISL_937563, EPI_ISL_937565, EPI_ISL_937566, EPI_ISL_937567, EPI_ISL_937572, EPI_ISL_937574, EPI_ISL_937575, EPI_ISL_937577, EPI_ISL_937582, EPI_ISL_937584, EPI_ISL_937585, EPI_ISL_937589, EPI_ISL_937591, EPI_ISL_937593, EPI_ISL_937597, EPI_ISL_937598, EPI_ISL_937600, EPI_ISL_937602, EPI_ISL_937603, EPI_ISL_937606, EPI_ISL_937607, EPI_ISL_937608, EPI_ISL_937611, EPI_ISL_937613, EPI_ISL_937615, EPI_ISL_937617, EPI_ISL_937619, EPI_ISL_937622, EPI_ISL_937624, EPI_ISL_937625, EPI_ISL_937626, EPI_ISL_937628, EPI_ISL_937629, EPI_ISL_937631, EPI_ISL_937632, EPI_ISL_937633, EPI_ISL_937635, EPI_ISL_937637, EPI_ISL_937642, EPI_ISL_937644, EPI_ISL_937645, EPI_ISL_937646, EPI_ISL_937647, EPI_ISL_937652, EPI_ISL_937654, EPI_ISL_937655, EPI_ISL_937656, EPI_ISL_937657, EPI_ISL_937659, EPI_ISL_937660, EPI_ISL_937661, EPI_ISL_937662, EPI_ISL_937667, EPI_ISL_937671, EPI_ISL_937673, EPI_ISL_937676, EPI_ISL_937677, EPI_ISL_937678, EPI_ISL_937679, EPI_ISL_937681, EPI_ISL_937683, EPI_ISL_937684, EPI_ISL_937686, EPI_ISL_937690, EPI_ISL_937691, EPI_ISL_937692, EPI_ISL_937694, EPI_ISL_937695, EPI_ISL_937696, EPI_ISL_937698, EPI_ISL_937701, EPI_ISL_937702, EPI_ISL_937705, EPI_ISL_937706, EPI_ISL_937708, EPI_ISL_937713, EPI_ISL_937718, EPI_ISL_937719, EPI_ISL_937722, EPI_ISL_937723, EPI_ISL_937725, EPI_ISL_937727, EPI_ISL_937728, EPI_ISL_937729, EPI_ISL_937731, EPI_ISL_937732, EPI_ISL_937733, EPI_ISL_937735, EPI_ISL_937736, EPI_ISL_937739, EPI_ISL_937740, EPI_ISL_937743, EPI_ISL_937747, EPI_ISL_937750, EPI_ISL_937754, EPI_ISL_937755, EPI_ISL_937756, EPI_ISL_937757, EPI_ISL_937758, EPI_ISL_937759, EPI_ISL_937760, EPI_ISL_937764, EPI_ISL_937771, EPI_ISL_937772, EPI_ISL_937773, EPI_ISL_937774, EPI_ISL_937775, EPI_ISL_937776, EPI_ISL_937778, EPI_ISL_937779, EPI_ISL_937782, EPI_ISL_937783, EPI_ISL_937785, EPI_ISL_937786, EPI_ISL_937788, EPI_ISL_937789, EPI_ISL_937791, EPI_ISL_937793, EPI_ISL_937795, EPI_ISL_937796, EPI_ISL_937797, EPI_ISL_937804, EPI_ISL_937807, EPI_ISL_937809, EPI_ISL_937810, EPI_ISL_937812, EPI_ISL_937813, EPI_ISL_937814, EPI_ISL_937816, EPI_ISL_937819, EPI_ISL_937821, EPI_ISL_937823, EPI_ISL_937825, EPI_ISL_937827, EPI_ISL_937828, EPI_ISL_937832, EPI_ISL_937833, EPI_ISL_937834, EPI_ISL_937836, EPI_ISL_937838, EPI_ISL_937841, EPI_ISL_937842, EPI_ISL_937844, EPI_ISL_937845, EPI_ISL_937846, EPI_ISL_937847, EPI_ISL_937848, EPI_ISL_937851, EPI_ISL_937855, EPI_ISL_937856, EPI_ISL_937857, EPI_ISL_937858, EPI_ISL_937859, EPI_ISL_937861, EPI_ISL_937866, EPI_ISL_937868, EPI_ISL_937869, EPI_ISL_937870, EPI_ISL_937871, EPI_ISL_937876, EPI_ISL_937878, EPI_ISL_937879, EPI_ISL_937880, EPI_ISL_937882, EPI_ISL_937884, EPI_ISL_937885, EPI_ISL_938935, EPI_ISL_938936, EPI_ISL_938937, EPI_ISL_938938, EPI_ISL_938939, EPI_ISL_938941, EPI_ISL_938942, EPI_ISL_938943, EPI_ISL_945377, EPI_ISL_945378, EPI_ISL_945380, EPI_ISL_945381, EPI_ISL_945382, EPI_ISL_945383, EPI_ISL_945384, EPI_ISL_945385, EPI_ISL_945388, EPI_ISL_945390, EPI_ISL_945391, EPI_ISL_945392, EPI_ISL_945393, EPI_ISL_945394, EPI_ISL_945395, EPI_ISL_945396, EPI_ISL_945397, EPI_ISL_945398, EPI_ISL_945399, EPI_ISL_945400, EPI_ISL_945401, EPI_ISL_945403, EPI_ISL_945404, EPI_ISL_945405, EPI_ISL_945406, EPI_ISL_945407, EPI_ISL_945410, EPI_ISL_945411, EPI_ISL_945412, EPI_ISL_945413, EPI_ISL_945414, EPI_ISL_945415, EPI_ISL_945416, EPI_ISL_945417, EPI_ISL_945418, EPI_ISL_945421, EPI_ISL_945424, EPI_ISL_945425, EPI_ISL_945426, EPI_ISL_945427, EPI_ISL_945428, EPI_ISL_945429, EPI_ISL_945430, EPI_ISL_945431, EPI_ISL_945432, EPI_ISL_945434, EPI_ISL_945435, EPI_ISL_945436, EPI_ISL_945437, EPI_ISL_945438, EPI_ISL_945439, EPI_ISL_945440, EPI_ISL_945441, EPI_ISL_945442, EPI_ISL_945443, EPI_ISL_945445, EPI_ISL_945447, EPI_ISL_945448, EPI_ISL_945450, EPI_ISL_945451, EPI_ISL_945452, EPI_ISL_945453, EPI_ISL_945454, EPI_ISL_945455, EPI_ISL_945456, EPI_ISL_945457, EPI_ISL_945460, EPI_ISL_945461, EPI_ISL_945462, EPI_ISL_945463, EPI_ISL_945464, EPI_ISL_945465, EPI_ISL_945470, EPI_ISL_945471, EPI_ISL_945472, EPI_ISL_945473, EPI_ISL_945474, EPI_ISL_945476, EPI_ISL_945477, EPI_ISL_945478, EPI_ISL_945479, EPI_ISL_945481, EPI_ISL_945482, EPI_ISL_945483, EPI_ISL_945484, EPI_ISL_945486, EPI_ISL_945487, EPI_ISL_945488, EPI_ISL_945489, EPI_ISL_945491, EPI_ISL_945492, EPI_ISL_945493, EPI_ISL_945496, EPI_ISL_945497, EPI_ISL_945499, EPI_ISL_945500, EPI_ISL_945501, EPI_ISL_945503, EPI_ISL_945504, EPI_ISL_945505, EPI_ISL_945507, EPI_ISL_945509, EPI_ISL_945510, EPI_ISL_945511, EPI_ISL_945512, EPI_ISL_945513, EPI_ISL_945515, EPI_ISL_945516, EPI_ISL_945517, EPI_ISL_945519, EPI_ISL_945520, EPI_ISL_945521, EPI_ISL_945523, EPI_ISL_945524, EPI_ISL_945526, EPI_ISL_945527, EPI_ISL_945528, EPI_ISL_945530, EPI_ISL_945531, EPI_ISL_945532, EPI_ISL_945533, EPI_ISL_945534, EPI_ISL_945535, EPI_ISL_945536, EPI_ISL_945537, EPI_ISL_945540, EPI_ISL_945541, EPI_ISL_945542, EPI_ISL_945544, EPI_ISL_945550, EPI_ISL_945551, EPI_ISL_945552, EPI_ISL_945553, EPI_ISL_945554, EPI_ISL_945556, EPI_ISL_945557, EPI_ISL_945558, EPI_ISL_945559, EPI_ISL_945560, EPI_ISL_945562, EPI_ISL_945564, EPI_ISL_945565, EPI_ISL_945566, EPI_ISL_945567, EPI_ISL_945568, EPI_ISL_945570, EPI_ISL_945571, EPI_ISL_945572, EPI_ISL_945573, EPI_ISL_945574, EPI_ISL_945575, EPI_ISL_945576, EPI_ISL_945577, EPI_ISL_945578, EPI_ISL_945580, EPI_ISL_945581, EPI_ISL_945583, EPI_ISL_945584, EPI_ISL_945585, EPI_ISL_945586, EPI_ISL_945588, EPI_ISL_945590, EPI_ISL_945592, EPI_ISL_945593, EPI_ISL_945594, EPI_ISL_945595, EPI_ISL_945596, EPI_ISL_945598, EPI_ISL_945599, EPI_ISL_945600, EPI_ISL_945601, EPI_ISL_945602, EPI_ISL_945603, EPI_ISL_945604, EPI_ISL_945605, EPI_ISL_945606, EPI_ISL_945607, EPI_ISL_945608, EPI_ISL_945609, EPI_ISL_945610, EPI_ISL_945611, EPI_ISL_945612, EPI_ISL_945613, EPI_ISL_945614, EPI_ISL_945615, EPI_ISL_945616, EPI_ISL_945617, EPI_ISL_945618, EPI_ISL_945620, EPI_ISL_945622, EPI_ISL_945623, EPI_ISL_945624, EPI_ISL_945625, EPI_ISL_945626, EPI_ISL_945628, EPI_ISL_945629, EPI_ISL_945630, EPI_ISL_945631, EPI_ISL_945632, EPI_ISL_945633, EPI_ISL_945634, EPI_ISL_945637, EPI_ISL_945638, EPI_ISL_945639, EPI_ISL_945640, EPI_ISL_945641, EPI_ISL_945642, EPI_ISL_945643, EPI_ISL_945644, EPI_ISL_945645, EPI_ISL_945646, EPI_ISL_945647, EPI_ISL_945649, EPI_ISL_945651, EPI_ISL_945652, EPI_ISL_945653,

EPI_ISL_945654, EPI_ISL_945655, EPI_ISL_945657, EPI_ISL_945658, EPI_ISL_945659, EPI_ISL_945660, EPI_ISL_945661, EPI_ISL_945662, EPI_ISL_945663, EPI_ISL_945665, EPI_ISL_945666, EPI_ISL_945667, EPI_ISL_945669, EPI_ISL_945670, EPI_ISL_945671, EPI_ISL_945673, EPI_ISL_945675, EPI_ISL_945676, EPI_ISL_945679, EPI_ISL_945680, EPI_ISL_945683, EPI_ISL_945684, EPI_ISL_945685, EPI_ISL_945686,
EPI_ISL_945687, EPI_ISL_945688, EPI_ISL_945689, EPI_ISL_945690, EPI_ISL_945691, EPI_ISL_945693, EPI_ISL_945694, EPI_ISL_945695, EPI_ISL_945699, EPI_ISL_945700, EPI_ISL_945702, EPI_ISL_945703, EPI_ISL_945705, EPI_ISL_945706, EPI_ISL_945707, EPI_ISL_945708, EPI_ISL_945709, EPI_ISL_945710, EPI_ISL_945711, EPI_ISL_945712, EPI_ISL_945713, EPI_ISL_945714, EPI_ISL_946422, EPI_ISL_946423,
EPI_ISL_946424, EPI_ISL_946425, EPI_ISL_946426, EPI_ISL_946428, EPI_ISL_946429, EPI_ISL_946430, EPI_ISL_946431, EPI_ISL_946432, EPI_ISL_946433, EPI_ISL_946434, EPI_ISL_946435, EPI_ISL_946436, EPI_ISL_946437, EPI_ISL_946438, EPI_ISL_946439, EPI_ISL_946440, EPI_ISL_946441, EPI_ISL_946442, EPI_ISL_946443, EPI_ISL_946444, EPI_ISL_946446, EPI_ISL_946447, EPI_ISL_946448, EPI_ISL_946449,
EPI_ISL_946450, EPI_ISL_946451, EPI_ISL_946452, EPI_ISL_946453, EPI_ISL_946455, EPI_ISL_946456, EPI_ISL_946457, EPI_ISL_946458, EPI_ISL_946459, EPI_ISL_946460, EPI_ISL_946461, EPI_ISL_946462, EPI_ISL_946463, EPI_ISL_946464, EPI_ISL_946465, EPI_ISL_946466, EPI_ISL_946467, EPI_ISL_946468, EPI_ISL_946469, EPI_ISL_946470, EPI_ISL_946472, EPI_ISL_946473, EPI_ISL_946474, EPI_ISL_946475,
EPI_ISL_946476, EPI_ISL_946477, EPI_ISL_946478, EPI_ISL_946479, EPI_ISL_946480, EPI_ISL_946481, EPI_ISL_946482, EPI_ISL_946483, EPI_ISL_946484, EPI_ISL_946485, EPI_ISL_946486, EPI_ISL_946487, EPI_ISL_946488, EPI_ISL_946489, EPI_ISL_946490, EPI_ISL_946491, EPI_ISL_946492, EPI_ISL_946494, EPI_ISL_946495, EPI_ISL_946496, EPI_ISL_946497, EPI_ISL_946498, EPI_ISL_946499, EPI_ISL_946500,
EPI_ISL_946501, EPI_ISL_946502, EPI_ISL_946503, EPI_ISL_946504, EPI_ISL_946505, EPI_ISL_946506, EPI_ISL_946507, EPI_ISL_946508, EPI_ISL_946509, EPI_ISL_946510, EPI_ISL_946511, EPI_ISL_946512, EPI_ISL_946513, EPI_ISL_946514, EPI_ISL_946515, EPI_ISL_946516, EPI_ISL_946517, EPI_ISL_946518, EPI_ISL_946519, EPI_ISL_946520, EPI_ISL_946521, EPI_ISL_946522, EPI_ISL_946523, EPI_ISL_946524,
EPI_ISL_946525, EPI_ISL_946526, EPI_ISL_946527, EPI_ISL_946528, EPI_ISL_946529, EPI_ISL_946530, EPI_ISL_946531, EPI_ISL_946532, EPI_ISL_946533, EPI_ISL_946534, EPI_ISL_946535, EPI_ISL_946536, EPI_ISL_946537, EPI_ISL_946538, EPI_ISL_946539, EPI_ISL_946540, EPI_ISL_946541, EPI_ISL_946542, EPI_ISL_946543, EPI_ISL_946544, EPI_ISL_946545, EPI_ISL_946546, EPI_ISL_946548, EPI_ISL_946549,
EPI_ISL_946550, EPI_ISL_946551, EPI_ISL_946552, EPI_ISL_946553, EPI_ISL_946554, EPI_ISL_946555, EPI_ISL_946556, EPI_ISL_946557, EPI_ISL_946558, EPI_ISL_946559, EPI_ISL_946560, EPI_ISL_946561, EPI_ISL_946562, EPI_ISL_946563, EPI_ISL_946564, EPI_ISL_946565, EPI_ISL_946566, EPI_ISL_946567, EPI_ISL_946568, EPI_ISL_946569, EPI_ISL_946570, EPI_ISL_946571, EPI_ISL_946572, EPI_ISL_946573,
EPI_ISL_946574, EPI_ISL_946575, EPI_ISL_946576, EPI_ISL_946577, EPI_ISL_946578, EPI_ISL_946579, EPI_ISL_946581, EPI_ISL_946582, EPI_ISL_946583, EPI_ISL_946584, EPI_ISL_946586, EPI_ISL_946587, EPI_ISL_946588, EPI_ISL_946589, EPI_ISL_946590, EPI_ISL_946591, EPI_ISL_946592, EPI_ISL_946593, EPI_ISL_946594, EPI_ISL_946595, EPI_ISL_946596, EPI_ISL_946597, EPI_ISL_946598, EPI_ISL_946599,
EPI_ISL_946600, EPI_ISL_946601, EPI_ISL_946602, EPI_ISL_946603, EPI_ISL_946604, EPI_ISL_946605, EPI_ISL_946606, EPI_ISL_946607, EPI_ISL_946608, EPI_ISL_946609, EPI_ISL_946610, EPI_ISL_946611, EPI_ISL_946612, EPI_ISL_946613, EPI_ISL_946614, EPI_ISL_946615, EPI_ISL_946616, EPI_ISL_946617, EPI_ISL_946618, EPI_ISL_946619, EPI_ISL_946620, EPI_ISL_946621, EPI_ISL_946622, EPI_ISL_946623,
EPI_ISL_946624, EPI_ISL_946625, EPI_ISL_946626, EPI_ISL_946627, EPI_ISL_946628, EPI_ISL_946629, EPI_ISL_946630, EPI_ISL_946631, EPI_ISL_946632, EPI_ISL_946633, EPI_ISL_946634, EPI_ISL_946635, EPI_ISL_946636, EPI_ISL_946637, EPI_ISL_946638, EPI_ISL_946639, EPI_ISL_946640, EPI_ISL_946641, EPI_ISL_946642, EPI_ISL_946643, EPI_ISL_946644, EPI_ISL_946645, EPI_ISL_946647, EPI_ISL_946648,
EPI_ISL_946649, EPI_ISL_946650, EPI_ISL_946651, EPI_ISL_946652, EPI_ISL_946653, EPI_ISL_946654, EPI_ISL_946655, EPI_ISL_946656, EPI_ISL_946657, EPI_ISL_946658, EPI_ISL_946659, EPI_ISL_946660, EPI_ISL_946661, EPI_ISL_946662, EPI_ISL_946663, EPI_ISL_946664, EPI_ISL_946665, EPI_ISL_946666, EPI_ISL_946667, EPI_ISL_946668, EPI_ISL_946669, EPI_ISL_946670, EPI_ISL_946671, EPI_ISL_946673,
EPI_ISL_946674, EPI_ISL_946675, EPI_ISL_946676, EPI_ISL_946677, EPI_ISL_946678, EPI_ISL_946679, EPI_ISL_946680, EPI_ISL_946681, EPI_ISL_946682, EPI_ISL_946683, EPI_ISL_946684, EPI_ISL_946685, EPI_ISL_946686, EPI_ISL_946687, EPI_ISL_946688, EPI_ISL_946689, EPI_ISL_946690, EPI_ISL_946691, EPI_ISL_946692, EPI_ISL_946693, EPI_ISL_946694, EPI_ISL_946695, EPI_ISL_946696, EPI_ISL_946697,
EPI_ISL_946698, EPI_ISL_946699, EPI_ISL_946700, EPI_ISL_946702, EPI_ISL_946703, EPI_ISL_946704, EPI_ISL_946705, EPI_ISL_946706, EPI_ISL_946707, EPI_ISL_946708, EPI_ISL_946709, EPI_ISL_946710, EPI_ISL_946711, EPI_ISL_946712, EPI_ISL_946713, EPI_ISL_946714, EPI_ISL_946715, EPI_ISL_946716, EPI_ISL_946717, EPI_ISL_946718, EPI_ISL_946719, EPI_ISL_946720, EPI_ISL_946721, EPI_ISL_946722,
EPI_ISL_946723, EPI_ISL_946724, EPI_ISL_946725, EPI_ISL_946726, EPI_ISL_946727, EPI_ISL_946728, EPI_ISL_946729, EPI_ISL_946730, EPI_ISL_946731, EPI_ISL_946732, EPI_ISL_946733, EPI_ISL_946734, EPI_ISL_946735, EPI_ISL_946736, EPI_ISL_946737, EPI_ISL_946738, EPI_ISL_946739, EPI_ISL_946740, EPI_ISL_946741, EPI_ISL_946742, EPI_ISL_946743, EPI_ISL_946744, EPI_ISL_946745, EPI_ISL_946747,
EPI_ISL_946748, EPI_ISL_946749, EPI_ISL_946750, EPI_ISL_946751, EPI_ISL_946752, EPI_ISL_946753, EPI_ISL_946754, EPI_ISL_946755, EPI_ISL_946756, EPI_ISL_946757, EPI_ISL_946758, EPI_ISL_946759, EPI_ISL_946760, EPI_ISL_946761, EPI_ISL_946762, EPI_ISL_946763, EPI_ISL_946764, EPI_ISL_946765, EPI_ISL_946766, EPI_ISL_946767, EPI_ISL_946768, EPI_ISL_946769, EPI_ISL_946770, EPI_ISL_946771,
EPI_ISL_946772, EPI_ISL_946773, EPI_ISL_946774, EPI_ISL_946775, EPI_ISL_946776, EPI_ISL_946777, EPI_ISL_946778, EPI_ISL_946779, EPI_ISL_946781, EPI_ISL_946782, EPI_ISL_946783, EPI_ISL_946784, EPI_ISL_946785, EPI_ISL_946786, EPI_ISL_946787, EPI_ISL_946788, EPI_ISL_946789, EPI_ISL_946790, EPI_ISL_946791, EPI_ISL_946792, EPI_ISL_946793, EPI_ISL_946794, EPI_ISL_946795, EPI_ISL_946796,
EPI_ISL_946797, EPI_ISL_946798, EPI_ISL_946799, EPI_ISL_946800, EPI_ISL_946801, EPI_ISL_946802, EPI_ISL_946803, EPI_ISL_946804, EPI_ISL_946805, EPI_ISL_946806, EPI_ISL_946807, EPI_ISL_946808, EPI_ISL_946809, EPI_ISL_946810, EPI_ISL_946811, EPI_ISL_946812, EPI_ISL_946813, EPI_ISL_946814, EPI_ISL_946815, EPI_ISL_946816, EPI_ISL_946817, EPI_ISL_946818, EPI_ISL_946819, EPI_ISL_946820,
EPI_ISL_946821, EPI_ISL_946822, EPI_ISL_946823, EPI_ISL_946824, EPI_ISL_946825, EPI_ISL_946826, EPI_ISL_946827, EPI_ISL_946828, EPI_ISL_946829, EPI_ISL_946830, EPI_ISL_946831, EPI_ISL_946832, EPI_ISL_946833, EPI_ISL_946834, EPI_ISL_946835, EPI_ISL_946836, EPI_ISL_946837, EPI_ISL_946838, EPI_ISL_946839, EPI_ISL_946840, EPI_ISL_946841, EPI_ISL_946842, EPI_ISL_946843, EPI_ISL_946844,
EPI_ISL_946845, EPI_ISL_946846, EPI_ISL_946847, EPI_ISL_946848, EPI_ISL_946849, EPI_ISL_946850, EPI_ISL_946851, EPI_ISL_946852, EPI_ISL_946853, EPI_ISL_946854, EPI_ISL_946855, EPI_ISL_946856, EPI_ISL_946857, EPI_ISL_946858, EPI_ISL_946859, EPI_ISL_946860, EPI_ISL_946861, EPI_ISL_946862, EPI_ISL_946863, EPI_ISL_946864, EPI_ISL_946865, EPI_ISL_946866, EPI_ISL_946867, EPI_ISL_946868,
EPI_ISL_946869, EPI_ISL_946870, EPI_ISL_946871, EPI_ISL_946872, EPI_ISL_946873, EPI_ISL_946874, EPI_ISL_946875, EPI_ISL_946876, EPI_ISL_946877, EPI_ISL_946878, EPI_ISL_946879, EPI_ISL_946880, EPI_ISL_946881, EPI_ISL_946882, EPI_ISL_946883, EPI_ISL_946884, EPI_ISL_946885, EPI_ISL_946886, EPI_ISL_946887, EPI_ISL_946888, EPI_ISL_946889, EPI_ISL_946890, EPI_ISL_946891, EPI_ISL_946892,
EPI_ISL_946893, EPI_ISL_946894, EPI_ISL_946895, EPI_ISL_946896, EPI_ISL_946897, EPI_ISL_946898, EPI_ISL_946899, EPI_ISL_946900, EPI_ISL_946901, EPI_ISL_946902, EPI_ISL_946903, EPI_ISL_946904, EPI_ISL_946905, EPI_ISL_946906, EPI_ISL_946907, EPI_ISL_946908, EPI_ISL_946909, EPI_ISL_946910, EPI_ISL_946911, EPI_ISL_946912, EPI_ISL_946914, EPI_ISL_946915, EPI_ISL_946916, EPI_ISL_946917,
EPI_ISL_946918, EPI_ISL_946919, EPI_ISL_946920, EPI_ISL_946921, EPI_ISL_946922, EPI_ISL_946923, EPI_ISL_946924, EPI_ISL_946925, EPI_ISL_946926, EPI_ISL_946927, EPI_ISL_946928, EPI_ISL_946929, EPI_ISL_946930, EPI_ISL_946931, EPI_ISL_946932, EPI_ISL_946933, EPI_ISL_946934, EPI_ISL_946935, EPI_ISL_946936, EPI_ISL_946937, EPI_ISL_946938, EPI_ISL_946939, EPI_ISL_946940, EPI_ISL_946941,
EPI_ISL_946942, EPI_ISL_946943, EPI_ISL_946944, EPI_ISL_946945, EPI_ISL_946946, EPI_ISL_946947, EPI_ISL_946948, EPI_ISL_946949, EPI_ISL_946950, EPI_ISL_946951, EPI_ISL_946952, EPI_ISL_946953, EPI_ISL_946954, EPI_ISL_946955, EPI_ISL_946957, EPI_ISL_946958, EPI_ISL_946959, EPI_ISL_946960, EPI_ISL_946961, EPI_ISL_946962, EPI_ISL_946964, EPI_ISL_946965, EPI_ISL_946966, EPI_ISL_946967,
EPI_ISL_946968, EPI_ISL_946969, EPI_ISL_946970, EPI_ISL_946971, EPI_ISL_946972, EPI_ISL_946973, EPI_ISL_946974, EPI_ISL_946975, EPI_ISL_946976, EPI_ISL_946977, EPI_ISL_946978, EPI_ISL_946979, EPI_ISL_946980, EPI_ISL_946981, EPI_ISL_946982, EPI_ISL_946983, EPI_ISL_946984, EPI_ISL_946985, EPI_ISL_946986, EPI_ISL_946987, EPI_ISL_946988, EPI_ISL_946989, EPI_ISL_946990, EPI_ISL_946991,
EPI_ISL_946993, EPI_ISL_946994, EPI_ISL_946995, EPI_ISL_946996, EPI_ISL_946997, EPI_ISL_946998, EPI_ISL_946999, EPI_ISL_947000, EPI_ISL_947001, EPI_ISL_947002, EPI_ISL_947003, EPI_ISL_947004, EPI_ISL_947005, EPI_ISL_947006, EPI_ISL_947007, EPI_ISL_947008, EPI_ISL_947009, EPI_ISL_947010, EPI_ISL_947011, EPI_ISL_947012, EPI_ISL_947013, EPI_ISL_947014, EPI_ISL_947015, EPI_ISL_947016,
EPI_ISL_947017, EPI_ISL_947018, EPI_ISL_947019, EPI_ISL_947020, EPI_ISL_947021, EPI_ISL_947022, EPI_ISL_947023, EPI_ISL_947024, EPI_ISL_947025, EPI_ISL_947026, EPI_ISL_947027, EPI_ISL_947028, EPI_ISL_947029, EPI_ISL_947030, EPI_ISL_947031, EPI_ISL_947032, EPI_ISL_947033, EPI_ISL_947034, EPI_ISL_947035, EPI_ISL_947036, EPI_ISL_947037, EPI_ISL_947038, EPI_ISL_947039, EPI_ISL_947040,
EPI_ISL_947041, EPI_ISL_947042, EPI_ISL_947043, EPI_ISL_947044, EPI_ISL_947045, EPI_ISL_947046, EPI_ISL_947047, EPI_ISL_947048, EPI_ISL_947049, EPI_ISL_947050, EPI_ISL_947051, EPI_ISL_947052, EPI_ISL_947053, EPI_ISL_947054, EPI_ISL_947055, EPI_ISL_947056, EPI_ISL_947057, EPI_ISL_947058, EPI_ISL_947059, EPI_ISL_947060, EPI_ISL_947061, EPI_ISL_947062, EPI_ISL_947063, EPI_ISL_947064,
EPI_ISL_947065, EPI_ISL_947066, EPI_ISL_947067, EPI_ISL_947068, EPI_ISL_947069, EPI_ISL_947070, EPI_ISL_947071, EPI_ISL_947072, EPI_ISL_947073, EPI_ISL_947074, EPI_ISL_947075, EPI_ISL_947076, EPI_ISL_947077, EPI_ISL_947078, EPI_ISL_947079, EPI_ISL_947080, EPI_ISL_947081, EPI_ISL_947082, EPI_ISL_947083, EPI_ISL_947084, EPI_ISL_947085, EPI_ISL_947086, EPI_ISL_947087, EPI_ISL_947088,
EPI_ISL_947089, EPI_ISL_947090, EPI_ISL_947091, EPI_ISL_947092, EPI_ISL_947093, EPI_ISL_947094, EPI_ISL_947095, EPI_ISL_947096, EPI_ISL_947097, EPI_ISL_947098, EPI_ISL_947099, EPI_ISL_947100, EPI_ISL_947101, EPI_ISL_947102, EPI_ISL_947103, EPI_ISL_947104, EPI_ISL_947105, EPI_ISL_947106, EPI_ISL_956446, EPI_ISL_956487, EPI_ISL_956508, EPI_ISL_956562, EPI_ISL_956576, EPI_ISL_956585,
EPI_ISL_956587, EPI_ISL_956592, EPI_ISL_956601, EPI_ISL_956602, EPI_ISL_956606, EPI_ISL_956618, EPI_ISL_956625, EPI_ISL_956626, EPI_ISL_956631, EPI_ISL_956636, EPI_ISL_956651, EPI_ISL_956690, EPI_ISL_956691, EPI_ISL_956694, EPI_ISL_956699, EPI_ISL_956708, EPI_ISL_956729, EPI_ISL_958566, EPI_ISL_958917, EPI_ISL_958918, EPI_ISL_958919, EPI_ISL_958920, EPI_ISL_958921, EPI_ISL_958922,
EPI_ISL_958923, EPI_ISL_958924, EPI_ISL_958925, EPI_ISL_958926, EPI_ISL_958927, EPI_ISL_958928, EPI_ISL_958929, EPI_ISL_958930, EPI_ISL_958931, EPI_ISL_958932, EPI_ISL_958933, EPI_ISL_958934, EPI_ISL_958935, EPI_ISL_958936, EPI_ISL_958937, EPI_ISL_958938, EPI_ISL_958939, EPI_ISL_958940, EPI_ISL_958941, EPI_ISL_958942, EPI_ISL_958943, EPI_ISL_958944, EPI_ISL_958945, EPI_ISL_958946,
EPI_ISL_958947, EPI_ISL_958948, EPI_ISL_958949, EPI_ISL_958950, EPI_ISL_958951, EPI_ISL_958952, EPI_ISL_958953, EPI_ISL_958954, EPI_ISL_958955, EPI_ISL_958956, EPI_ISL_958957, EPI_ISL_958958, EPI_ISL_958959, EPI_ISL_958960, EPI_ISL_958961, EPI_ISL_958962, EPI_ISL_958963, EPI_ISL_958964, EPI_ISL_958965, EPI_ISL_958966, EPI_ISL_958967, EPI_ISL_958968, EPI_ISL_958969, EPI_ISL_958970,
EPI_ISL_958971, EPI_ISL_958972, EPI_ISL_958973, EPI_ISL_958974, EPI_ISL_958975, EPI_ISL_958976, EPI_ISL_958977, EPI_ISL_958978, EPI_ISL_958979, EPI_ISL_958980, EPI_ISL_958981, EPI_ISL_958982, EPI_ISL_958983, EPI_ISL_958984, EPI_ISL_958985, EPI_ISL_958986, EPI_ISL_958987, EPI_ISL_958988, EPI_ISL_958989, EPI_ISL_958990, EPI_ISL_958991, EPI_ISL_958992, EPI_ISL_958993, EPI_ISL_958994,
EPI_ISL_958995, EPI_ISL_958996, EPI_ISL_958997, EPI_ISL_958998, EPI_ISL_958999, EPI_ISL_959000, EPI_ISL_959001, EPI_ISL_959002, EPI_ISL_959003, EPI_ISL_959004, EPI_ISL_959005, EPI_ISL_959006, EPI_ISL_959007, EPI_ISL_959008, EPI_ISL_959009, EPI_ISL_959010, EPI_ISL_959011, EPI_ISL_959012, EPI_ISL_959013, EPI_ISL_959014, EPI_ISL_959015, EPI_ISL_959016, EPI_ISL_959017, EPI_ISL_959018,
EPI_ISL_959019, EPI_ISL_959020, EPI_ISL_959021, EPI_ISL_959022, EPI_ISL_959023, EPI_ISL_959024, EPI_ISL_959025, EPI_ISL_959026, EPI_ISL_959027, EPI_ISL_959028, EPI_ISL_959029, EPI_ISL_959030, EPI_ISL_959031, EPI_ISL_959032, EPI_ISL_959033, EPI_ISL_959034, EPI_ISL_959035, EPI_ISL_959036, EPI_ISL_959037, EPI_ISL_959038, EPI_ISL_959039, EPI_ISL_959040, EPI_ISL_959041, EPI_ISL_959042,
EPI_ISL_959043, EPI_ISL_959044, EPI_ISL_959045, EPI_ISL_959046, EPI_ISL_959047, EPI_ISL_959048, EPI_ISL_959049, EPI_ISL_959050, EPI_ISL_959051, EPI_ISL_959052, EPI_ISL_959053, EPI_ISL_959054, EPI_ISL_959055, EPI_ISL_959056, EPI_ISL_959057, EPI_ISL_959058, EPI_ISL_959059, EPI_ISL_959060, EPI_ISL_959061, EPI_ISL_959062, EPI_ISL_959063, EPI_ISL_959064, EPI_ISL_959065, EPI_ISL_959066,
EPI_ISL_959067, EPI_ISL_959068, EPI_ISL_959069, EPI_ISL_959070, EPI_ISL_959071, EPI_ISL_959072, EPI_ISL_959073, EPI_ISL_959074, EPI_ISL_959075, EPI_ISL_959076, EPI_ISL_959077, EPI_ISL_959078, EPI_ISL_959079, EPI_ISL_959080, EPI_ISL_959081, EPI_ISL_959082, EPI_ISL_959083, EPI_ISL_959084, EPI_ISL_959085, EPI_ISL_959086, EPI_ISL_959087, EPI_ISL_959088, EPI_ISL_959089, EPI_ISL_959090,
EPI_ISL_959091, EPI_ISL_959092, EPI_ISL_959093, EPI_ISL_959094, EPI_ISL_959095, EPI_ISL_959096, EPI_ISL_959097, EPI_ISL_959098, EPI_ISL_959099, EPI_ISL_959100, EPI_ISL_959101, EPI_ISL_959102, EPI_ISL_959103, EPI_ISL_959104, EPI_ISL_959105, EPI_ISL_959106, EPI_ISL_959107, EPI_ISL_959108, EPI_ISL_959109, EPI_ISL_959110, EPI_ISL_959111, EPI_ISL_959112, EPI_ISL_959113, EPI_ISL_959114,
EPI_ISL_959115, EPI_ISL_959116, EPI_ISL_959117, EPI_ISL_959118, EPI_ISL_959119, EPI_ISL_959120, EPI_ISL_959121, EPI_ISL_959122, EPI_ISL_959123, EPI_ISL_959124, EPI_ISL_959125, EPI_ISL_959126, EPI_ISL_959127, EPI_ISL_959128, EPI_ISL_959129, EPI_ISL_959130, EPI_ISL_959131, EPI_ISL_959132, EPI_ISL_959133, EPI_ISL_959134, EPI_ISL_959135, EPI_ISL_959136, EPI_ISL_959137, EPI_ISL_959138,
EPI_ISL_959139, EPI_ISL_959140, EPI_ISL_959141, EPI_ISL_959142, EPI_ISL_959144, EPI_ISL_959145, EPI_ISL_959146, EPI_ISL_959147, EPI_ISL_959148, EPI_ISL_959149, EPI_ISL_959150, EPI_ISL_959151, EPI_ISL_959152, EPI_ISL_959153, EPI_ISL_959154, EPI_ISL_959155, EPI_ISL_959156, EPI_ISL_959157, EPI_ISL_959158, EPI_ISL_959159, EPI_ISL_959160, EPI_ISL_959161, EPI_ISL_959162,
EPI_ISL_959163, EPI_ISL_959164, EPI_ISL_959165, EPI_ISL_959166, EPI_ISL_959167, EPI_ISL_959168, EPI_ISL_959169, EPI_ISL_959170, EPI_ISL_959171, EPI_ISL_959172, EPI_ISL_959173, EPI_ISL_959174, EPI_ISL_959175, EPI_ISL_959176, EPI_ISL_959177, EPI_ISL_959178, EPI_ISL_959179, EPI_ISL_959180, EPI_ISL_959181, EPI_ISL_959182, EPI_ISL_959183, EPI_ISL_959184, EPI_ISL_959185, EPI_ISL_959186,
EPI_ISL_959187, EPI_ISL_959188, EPI_ISL_959189, EPI_ISL_959190, EPI_ISL_959191, EPI_ISL_959192, EPI_ISL_959193, EPI_ISL_959194, EPI_ISL_959195, EPI_ISL_959196, EPI_ISL_959197, EPI_ISL_959198, EPI_ISL_959199, EPI_ISL_959200, EPI_ISL_959201, EPI_ISL_959202, EPI_ISL_959203, EPI_ISL_959204, EPI_ISL_959205, EPI_ISL_959206, EPI_ISL_959207, EPI_ISL_959208, EPI_ISL_959209, EPI_ISL_959210,
EPI_ISL_959211, EPI_ISL_959212, EPI_ISL_959213, EPI_ISL_959214, EPI_ISL_959215, EPI_ISL_959216, EPI_ISL_959217, EPI_ISL_959218, EPI_ISL_959219, EPI_ISL_959220, EPI_ISL_959221, EPI_ISL_959222, EPI_ISL_959223, EPI_ISL_959224, EPI_ISL_959225, EPI_ISL_959226, EPI_ISL_959227, EPI_ISL_959228, EPI_ISL_959229, EPI_ISL_959230, EPI_ISL_959231, EPI_ISL_959232, EPI_ISL_959233, EPI_ISL_959234,
EPI_ISL_959235, EPI_ISL_959236, EPI_ISL_959237, EPI_ISL_959238, EPI_ISL_959239, EPI_ISL_959240, EPI_ISL_959241, EPI_ISL_959242, EPI_ISL_959243, EPI_ISL_959244, EPI_ISL_959245, EPI_ISL_959246, EPI_ISL_959247, EPI_ISL_959248, EPI_ISL_959249, EPI_ISL_959250, EPI_ISL_959251, EPI_ISL_959252, EPI_ISL_959253, EPI_ISL_959254, EPI_ISL_959255, EPI_ISL_959256, EPI_ISL_959257, EPI_ISL_959258,
EPI_ISL_959259, EPI_ISL_959260, EPI_ISL_959261, EPI_ISL_959262, EPI_ISL_959263, EPI_ISL_959264, EPI_ISL_959265, EPI_ISL_959266, EPI_ISL_959268, EPI_ISL_959269, EPI_ISL_959270, EPI_ISL_963651, EPI_ISL_963654, EPI_ISL_963658, EPI_ISL_963659, EPI_ISL_963665, EPI_ISL_963670, EPI_ISL_963671, EPI_ISL_963672, EPI_ISL_963673, EPI_ISL_963676, EPI_ISL_963677, EPI_ISL_963679, EPI_ISL_963681,
EPI_ISL_963685, EPI_ISL_963686, EPI_ISL_963687, EPI_ISL_963689, EPI_ISL_963690, EPI_ISL_963694, EPI_ISL_963695, EPI_ISL_963700, EPI_ISL_963701, EPI_ISL_963703, EPI_ISL_963704, EPI_ISL_963706, EPI_ISL_963707, EPI_ISL_963709, EPI_ISL_963718, EPI_ISL_963720, EPI_ISL_963721, EPI_ISL_963724, EPI_ISL_963726, EPI_ISL_963727, EPI_ISL_963731, EPI_ISL_963732, EPI_ISL_963733, EPI_ISL_963734,
EPI_ISL_963735, EPI_ISL_963736, EPI_ISL_963737, EPI_ISL_963738, EPI_ISL_963740, EPI_ISL_963741, EPI_ISL_963747, EPI_ISL_963750, EPI_ISL_963753, EPI_ISL_963754, EPI_ISL_963758, EPI_ISL_963759, EPI_ISL_963761, EPI_ISL_963764, EPI_ISL_963765, EPI_ISL_963766, EPI_ISL_963768, EPI_ISL_963769, EPI_ISL_963772, EPI_ISL_963773, EPI_ISL_963774, EPI_ISL_963775, EPI_ISL_963779, EPI_ISL_963780,
EPI_ISL_963781, EPI_ISL_963783, EPI_ISL_963786, EPI_ISL_963789, EPI_ISL_963790, EPI_ISL_963794, EPI_ISL_963798, EPI_ISL_963800, EPI_ISL_963803, EPI_ISL_963805, EPI_ISL_963806, EPI_ISL_963807, EPI_ISL_963808, EPI_ISL_963810, EPI_ISL_963813, EPI_ISL_963814, EPI_ISL_963815, EPI_ISL_963816, EPI_ISL_963819, EPI_ISL_963821, EPI_ISL_963822, EPI_ISL_963823, EPI_ISL_963824, EPI_ISL_963825,
EPI_ISL_963826, EPI_ISL_963831, EPI_ISL_963832, EPI_ISL_963833, EPI_ISL_963835, EPI_ISL_963838, EPI_ISL_963839, EPI_ISL_963841, EPI_ISL_963844, EPI_ISL_963845, EPI_ISL_963846, EPI_ISL_963849, EPI_ISL_963850, EPI_ISL_963851, EPI_ISL_963852, EPI_ISL_963853, EPI_ISL_963855, EPI_ISL_963857, EPI_ISL_963859, EPI_ISL_963861, EPI_ISL_963863, EPI_ISL_963864, EPI_ISL_963865, EPI_ISL_963866,
EPI_ISL_963867, EPI_ISL_963868, EPI_ISL_963869, EPI_ISL_963870, EPI_ISL_963872, EPI_ISL_963873, EPI_ISL_963874, EPI_ISL_963875, EPI_ISL_963877, EPI_ISL_963878, EPI_ISL_963879, EPI_ISL_963880, EPI_ISL_963881, EPI_ISL_963882, EPI_ISL_963883, EPI_ISL_963884, EPI_ISL_963889, EPI_ISL_963890, EPI_ISL_963891, EPI_ISL_963893, EPI_ISL_963895, EPI_ISL_963896, EPI_ISL_963900, EPI_ISL_963901,
EPI_ISL_963902, EPI_ISL_963903, EPI_ISL_963904, EPI_ISL_963906, EPI_ISL_963908, EPI_ISL_963909, EPI_ISL_963911, EPI_ISL_963914, EPI_ISL_963915, EPI_ISL_963916, EPI_ISL_963921, EPI_ISL_963922, EPI_ISL_963923, EPI_ISL_963925, EPI_ISL_963927, EPI_ISL_963932, EPI_ISL_963934, EPI_ISL_963936, EPI_ISL_963938, EPI_ISL_963941, EPI_ISL_963942, EPI_ISL_963945, EPI_ISL_963947, EPI_ISL_963949,
EPI_ISL_963952, EPI_ISL_963953, EPI_ISL_963958, EPI_ISL_963960, EPI_ISL_963962, EPI_ISL_963964, EPI_ISL_963968, EPI_ISL_963970, EPI_ISL_963971, EPI_ISL_963973, EPI_ISL_963976, EPI_ISL_963977, EPI_ISL_963978, EPI_ISL_963979, EPI_ISL_963981, EPI_ISL_963984, EPI_ISL_964067, EPI_ISL_964087, EPI_ISL_964236, EPI_ISL_964710, EPI_ISL_964749, EPI_ISL_964759, EPI_ISL_964817,
EPI_ISL_964877, EPI_ISL_969454, EPI_ISL_970032, EPI_ISL_970055, EPI_ISL_970142, EPI_ISL_989482, EPI_ISL_989483, EPI_ISL_989485, EPI_ISL_989489, EPI_ISL_989490, EPI_ISL_989491, EPI_ISL_989494, EPI_ISL_989495, EPI_ISL_989496, EPI_ISL_989504, EPI_ISL_989505, EPI_ISL_989506, EPI_ISL_989507, EPI_ISL_989522, EPI_ISL_989742, EPI_ISL_989751, EPI_ISL_989769, EPI_ISL_989856, EPI_ISL_989857,
EPI_ISL_989858, EPI_ISL_989859, EPI_ISL_989861, EPI_ISL_989862, EPI_ISL_989865, EPI_ISL_989866, EPI_ISL_989867, EPI_ISL_989868, EPI_ISL_989869, EPI_ISL_989870, EPI_ISL_989871, EPI_ISL_989873, EPI_ISL_989874, EPI_ISL_989876, EPI_ISL_989877, EPI_ISL_989879, EPI_ISL_989881, EPI_ISL_989998, EPI_ISL_989999, EPI_ISL_990000, EPI_ISL_990001, EPI_ISL_990002, EPI_ISL_990004, EPI_ISL_990005,
EPI_ISL_990006, EPI_ISL_990007, EPI_ISL_990008, EPI_ISL_990009, EPI_ISL_990010, EPI_ISL_990011, EPI_ISL_990012, EPI_ISL_990013, EPI_ISL_990014, EPI_ISL_990015, EPI_ISL_990016, EPI_ISL_990017, EPI_ISL_990018, EPI_ISL_990019, EPI_ISL_990020, EPI_ISL_990021, EPI_ISL_990022, EPI_ISL_990023, EPI_ISL_990024, EPI_ISL_990025, EPI_ISL_990026, EPI_ISL_990027, EPI_ISL_990028, EPI_ISL_990029,
EPI_ISL_990030, EPI_ISL_990031, EPI_ISL_990032, EPI_ISL_990033, EPI_ISL_990034, EPI_ISL_990035, EPI_ISL_990036, EPI_ISL_990037, EPI_ISL_990038, EPI_ISL_990039, EPI_ISL_990040, EPI_ISL_990041, EPI_ISL_990042, EPI_ISL_990141, EPI_ISL_990142, EPI_ISL_990143, EPI_ISL_990144, EPI_ISL_990145, EPI_ISL_990146, EPI_ISL_990147, EPI_ISL_990148, EPI_ISL_990149, EPI_ISL_990150, EPI_ISL_990151,
EPI_ISL_990152, EPI_ISL_990153, EPI_ISL_990154, EPI_ISL_990155, EPI_ISL_990156, EPI_ISL_990157, EPI_ISL_990158, EPI_ISL_990159, EPI_ISL_990160, EPI_ISL_990161, EPI_ISL_990162, EPI_ISL_990163, EPI_ISL_990164, EPI_ISL_990165, EPI_ISL_990166, EPI_ISL_990167, EPI_ISL_990168, EPI_ISL_990169, EPI_ISL_990170, EPI_ISL_990171, EPI_ISL_990172, EPI_ISL_990173, EPI_ISL_990174, EPI_ISL_990175,
EPI_ISL_990176, EPI_ISL_990177, EPI_ISL_990178, EPI_ISL_990179, EPI_ISL_990180, EPI_ISL_990181, EPI_ISL_990182, EPI_ISL_990183, EPI_ISL_990184, EPI_ISL_990185, EPI_ISL_990186, EPI_ISL_990187, EPI_ISL_990188, EPI_ISL_990189, EPI_ISL_990190, EPI_ISL_990191, EPI_ISL_990192, EPI_ISL_990193, EPI_ISL_990194, EPI_ISL_990195, EPI_ISL_990196, EPI_ISL_990197, EPI_ISL_990199, EPI_ISL_990200,
EPI_ISL_990201, EPI_ISL_990202, EPI_ISL_990203, EPI_ISL_990204, EPI_ISL_990205, EPI_ISL_990206, EPI_ISL_990207, EPI_ISL_990208, EPI_ISL_990209, EPI_ISL_990210, EPI_ISL_990211, EPI_ISL_990212, EPI_ISL_990213, EPI_ISL_990214, EPI_ISL_990215, EPI_ISL_990216, EPI_ISL_990217, EPI_ISL_990218, EPI_ISL_990219, EPI_ISL_990220, EPI_ISL_990221, EPI_ISL_990222, EPI_ISL_990223, EPI_ISL_990224,
EPI_ISL_990225, EPI_ISL_990226, EPI_ISL_990227, EPI_ISL_990228, EPI_ISL_990229, EPI_ISL_990230, EPI_ISL_990231, EPI_ISL_990232, EPI_ISL_990233, EPI_ISL_990234, EPI_ISL_990235, EPI_ISL_990236, EPI_ISL_990237, EPI_ISL_990238, EPI_ISL_990239, EPI_ISL_990240, EPI_ISL_990241, EPI_ISL_990242, EPI_ISL_990243, EPI_ISL_990244, EPI_ISL_990245, EPI_ISL_990246, EPI_ISL_990247, EPI_ISL_990248,
EPI_ISL_990249, EPI_ISL_990250, EPI_ISL_990251, EPI_ISL_990252, EPI_ISL_990253, EPI_ISL_990254, EPI_ISL_990255, EPI_ISL_990256, EPI_ISL_990257, EPI_ISL_990258, EPI_ISL_990259, EPI_ISL_990260, EPI_ISL_990261, EPI_ISL_990262, EPI_ISL_990263, EPI_ISL_990264, EPI_ISL_990265, EPI_ISL_990266, EPI_ISL_990267, EPI_ISL_990268, EPI_ISL_990269, EPI_ISL_990270, EPI_ISL_990271, EPI_ISL_990272,
EPI_ISL_990273, EPI_ISL_990274, EPI_ISL_990275, EPI_ISL_990276, EPI_ISL_990277, EPI_ISL_990278, EPI_ISL_990279, EPI_ISL_990280, EPI_ISL_990281, EPI_ISL_990282, EPI_ISL_990283, EPI_ISL_990284, EPI_ISL_990285, EPI_ISL_990287, EPI_ISL_990288, EPI_ISL_990289, EPI_ISL_990290, EPI_ISL_990291, EPI_ISL_990292, EPI_ISL_990293, EPI_ISL_990294, EPI_ISL_990295, EPI_ISL_990296, EPI_ISL_990297,
EPI_ISL_990298, EPI_ISL_990299, EPI_ISL_990300, EPI_ISL_990301, EPI_ISL_990302, EPI_ISL_990303, EPI_ISL_990304, EPI_ISL_990305, EPI_ISL_990306, EPI_ISL_990307, EPI_ISL_990308, EPI_ISL_990309, EPI_ISL_990310, EPI_ISL_990311, EPI_ISL_990312, EPI_ISL_990313, EPI_ISL_990314, EPI_ISL_990315, EPI_ISL_990316, EPI_ISL_990317, EPI_ISL_990318, EPI_ISL_990319, EPI_ISL_990320, EPI_ISL_990321,
EPI_ISL_990322, EPI_ISL_990323, EPI_ISL_990324, EPI_ISL_990325, EPI_ISL_990326, EPI_ISL_990327, EPI_ISL_990328, EPI_ISL_990329, EPI_ISL_990330, EPI_ISL_990331, EPI_ISL_990332, EPI_ISL_990333, EPI_ISL_990334, EPI_ISL_990335, EPI_ISL_990336, EPI_ISL_990337, EPI_ISL_990338, EPI_ISL_990339, EPI_ISL_990340, EPI_ISL_990341, EPI_ISL_990342, EPI_ISL_990343, EPI_ISL_990344, EPI_ISL_990345,
EPI_ISL_990346, EPI_ISL_990347, EPI_ISL_990348, EPI_ISL_990349, EPI_ISL_990350, EPI_ISL_990351, EPI_ISL_990352, EPI_ISL_990353, EPI_ISL_990354, EPI_ISL_990355, EPI_ISL_990356, EPI_ISL_990357, EPI_ISL_990358, EPI_ISL_990359, EPI_ISL_990360, EPI_ISL_990361, EPI_ISL_990362, EPI_ISL_990363, EPI_ISL_990364, EPI_ISL_990365, EPI_ISL_990366, EPI_ISL_990367, EPI_ISL_990368, EPI_ISL_990369,
EPI_ISL_990370, EPI_ISL_990371, EPI_ISL_990372, EPI_ISL_990373, EPI_ISL_990374, EPI_ISL_990375, EPI_ISL_990376, EPI_ISL_990377, EPI_ISL_990378, EPI_ISL_990379, EPI_ISL_990380, EPI_ISL_990381, EPI_ISL_990382, EPI_ISL_990383, EPI_ISL_990384, EPI_ISL_990385, EPI_ISL_990386, EPI_ISL_990387, EPI_ISL_990388, EPI_ISL_990389, EPI_ISL_990390, EPI_ISL_990391, EPI_ISL_990392, EPI_ISL_990393,
EPI_ISL_990394, EPI_ISL_990395, EPI_ISL_990396, EPI_ISL_990397, EPI_ISL_990398, EPI_ISL_990399, EPI_ISL_990400, EPI_ISL_990401, EPI_ISL_990402, EPI_ISL_990403, EPI_ISL_990404, EPI_ISL_990405, EPI_ISL_990406, EPI_ISL_990407, EPI_ISL_990408, EPI_ISL_990409, EPI_ISL_990410, EPI_ISL_990411, EPI_ISL_990412, EPI_ISL_990413, EPI_ISL_990414, EPI_ISL_990415, EPI_ISL_990416, EPI_ISL_990417,
EPI_ISL_990418, EPI_ISL_990419, EPI_ISL_990420, EPI_ISL_990421, EPI_ISL_990422, EPI_ISL_990423, EPI_ISL_990424, EPI_ISL_990425, EPI_ISL_990426, EPI_ISL_990427, EPI_ISL_990428, EPI_ISL_990429, EPI_ISL_990430, EPI_ISL_990431, EPI_ISL_990432, EPI_ISL_990433, EPI_ISL_990434, EPI_ISL_990436, EPI_ISL_990437, EPI_ISL_990438, EPI_ISL_990439, EPI_ISL_990440, EPI_ISL_990441, EPI_ISL_990442,
EPI_ISL_990443, EPI_ISL_990444, EPI_ISL_990445, EPI_ISL_990446, EPI_ISL_990447, EPI_ISL_990448, EPI_ISL_990449, EPI_ISL_990450, EPI_ISL_990451, EPI_ISL_990452, EPI_ISL_990453, EPI_ISL_990454, EPI_ISL_990455, EPI_ISL_990456, EPI_ISL_990457, EPI_ISL_990458, EPI_ISL_990459, EPI_ISL_990460, EPI_ISL_990461, EPI_ISL_990462, EPI_ISL_990463, EPI_ISL_990464, EPI_ISL_990465, EPI_ISL_990466,
EPI_ISL_990467, EPI_ISL_990468, EPI_ISL_990469, EPI_ISL_990470, EPI_ISL_990471, EPI_ISL_990472, EPI_ISL_990473, EPI_ISL_990474, EPI_ISL_990475, EPI_ISL_990476, EPI_ISL_990477, EPI_ISL_990478, EPI_ISL_990479, EPI_ISL_990480, EPI_ISL_990481, EPI_ISL_990482, EPI_ISL_990483, EPI_ISL_990484, EPI_ISL_990485, EPI_ISL_990486, EPI_ISL_990487, EPI_ISL_990488, EPI_ISL_990489, EPI_ISL_990490,
EPI_ISL_990491, EPI_ISL_990492, EPI_ISL_990493, EPI_ISL_990494, EPI_ISL_990495, EPI_ISL_990496, EPI_ISL_990497, EPI_ISL_990498, EPI_ISL_990501, EPI_ISL_990503, EPI_ISL_990505, EPI_ISL_990506, EPI_ISL_990507, EPI_ISL_990508, EPI_ISL_990509, EPI_ISL_990511, EPI_ISL_990512, EPI_ISL_990513, EPI_ISL_990514, EPI_ISL_990515, EPI_ISL_990517,
EPI_ISL_990518, EPI_ISL_990519, EPI_ISL_990520, EPI_ISL_990521, EPI_ISL_990522, EPI_ISL_990523, EPI_ISL_990524, EPI_ISL_990525, EPI_ISL_990526, EPI_ISL_990527, EPI_ISL_990528, EPI_ISL_990529, EPI_ISL_990530, EPI_ISL_990531, EPI_ISL_990532, EPI_ISL_990533, EPI_ISL_990534, EPI_ISL_990535, EPI_ISL_990536, EPI_ISL_990537, EPI_ISL_990538, EPI_ISL_990539, EPI_ISL_990540, EPI_ISL_990541,
EPI_ISL_990542, EPI_ISL_990543, EPI_ISL_990544, EPI_ISL_990546, EPI_ISL_990547, EPI_ISL_990548, EPI_ISL_990549, EPI_ISL_990550, EPI_ISL_990551, EPI_ISL_990552, EPI_ISL_990553, EPI_ISL_990554, EPI_ISL_990555, EPI_ISL_990556, EPI_ISL_990558, EPI_ISL_990559, EPI_ISL_990560, EPI_ISL_990561, EPI_ISL_990562, EPI_ISL_990563, EPI_ISL_990564, EPI_ISL_990565, EPI_ISL_990566, EPI_ISL_990567,
EPI_ISL_990568, EPI_ISL_990569, EPI_ISL_990570, EPI_ISL_990572, EPI_ISL_990573, EPI_ISL_990574, EPI_ISL_990575, EPI_ISL_990577, EPI_ISL_990578, EPI_ISL_990580, EPI_ISL_990581, EPI_ISL_990582, EPI_ISL_990583, EPI_ISL_990584, EPI_ISL_990585, EPI_ISL_990586, EPI_ISL_990587, EPI_ISL_990588, EPI_ISL_990589, EPI_ISL_990590, EPI_ISL_990591, EPI_ISL_990592, EPI_ISL_990593, EPI_ISL_990594,
EPI_ISL_990595, EPI_ISL_990596, EPI_ISL_990597, EPI_ISL_990598, EPI_ISL_990599, EPI_ISL_990600, EPI_ISL_990601, EPI_ISL_990602, EPI_ISL_990603, EPI_ISL_990604, EPI_ISL_990605, EPI_ISL_990606, EPI_ISL_990607, EPI_ISL_990608, EPI_ISL_990609, EPI_ISL_990610, EPI_ISL_990611, EPI_ISL_990612, EPI_ISL_990613, EPI_ISL_990614, EPI_ISL_990615, EPI_ISL_990617, EPI_ISL_990619, EPI_ISL_990620,
EPI_ISL_990621, EPI_ISL_990622, EPI_ISL_990623, EPI_ISL_990624, EPI_ISL_990625, EPI_ISL_990626, EPI_ISL_990627, EPI_ISL_990628, EPI_ISL_990629, EPI_ISL_990630, EPI_ISL_990631, EPI_ISL_990632, EPI_ISL_990633, EPI_ISL_990634, EPI_ISL_990635, EPI_ISL_990636, EPI_ISL_990637, EPI_ISL_990638, EPI_ISL_990639, EPI_ISL_990640, EPI_ISL_990641, EPI_ISL_990642, EPI_ISL_990643, EPI_ISL_990644,
EPI_ISL_990645, EPI_ISL_990646, EPI_ISL_990647, EPI_ISL_990648, EPI_ISL_990649, EPI_ISL_990650, EPI_ISL_990651, EPI_ISL_990653, EPI_ISL_990654, EPI_ISL_990655, EPI_ISL_990656, EPI_ISL_990657, EPI_ISL_990659, EPI_ISL_990660, EPI_ISL_990661, EPI_ISL_990663, EPI_ISL_990665, EPI_ISL_990666, EPI_ISL_990667, EPI_ISL_990668, EPI_ISL_990669, EPI_ISL_990670, EPI_ISL_990671, EPI_ISL_990672,
EPI_ISL_990673, EPI_ISL_990674, EPI_ISL_990675, EPI_ISL_990676, EPI_ISL_990677, EPI_ISL_990678, EPI_ISL_990679, EPI_ISL_990681, EPI_ISL_990682, EPI_ISL_990683, EPI_ISL_990684, EPI_ISL_990685, EPI_ISL_990686, EPI_ISL_990687, EPI_ISL_990688, EPI_ISL_990689, EPI_ISL_990690, EPI_ISL_990691, EPI_ISL_990692, EPI_ISL_990693, EPI_ISL_990694, EPI_ISL_990695, EPI_ISL_990696, EPI_ISL_990697,
EPI_ISL_990698, EPI_ISL_990699, EPI_ISL_990700, EPI_ISL_990701, EPI_ISL_990702, EPI_ISL_990703, EPI_ISL_990704, EPI_ISL_990705, EPI_ISL_990706, EPI_ISL_990707, EPI_ISL_990708, EPI_ISL_990709, EPI_ISL_990710, EPI_ISL_990711, EPI_ISL_990713, EPI_ISL_990714, EPI_ISL_990715, EPI_ISL_990716, EPI_ISL_990717, EPI_ISL_990718, EPI_ISL_990719, EPI_ISL_990720, EPI_ISL_990721, EPI_ISL_990722,
EPI_ISL_990723, EPI_ISL_990724, EPI_ISL_990725, EPI_ISL_990727, EPI_ISL_990728, EPI_ISL_990729, EPI_ISL_990730, EPI_ISL_990731, EPI_ISL_990732, EPI_ISL_990733, EPI_ISL_990734, EPI_ISL_990735, EPI_ISL_990736, EPI_ISL_990737, EPI_ISL_990738, EPI_ISL_990739, EPI_ISL_990740, EPI_ISL_990741, EPI_ISL_990742, EPI_ISL_990743, EPI_ISL_990744, EPI_ISL_990746, EPI_ISL_990747, EPI_ISL_990748,
EPI_ISL_990749, EPI_ISL_990750, EPI_ISL_990751, EPI_ISL_990752, EPI_ISL_990753, EPI_ISL_990754, EPI_ISL_990755, EPI_ISL_990756, EPI_ISL_990757, EPI_ISL_990758, EPI_ISL_990759, EPI_ISL_990760, EPI_ISL_990761, EPI_ISL_990762, EPI_ISL_990763, EPI_ISL_990764, EPI_ISL_990765, EPI_ISL_990766, EPI_ISL_990767, EPI_ISL_990768, EPI_ISL_990769, EPI_ISL_990770, EPI_ISL_990771, EPI_ISL_990772,
EPI_ISL_990773, EPI_ISL_990774, EPI_ISL_990775, EPI_ISL_990776, EPI_ISL_990777, EPI_ISL_990778, EPI_ISL_990780, EPI_ISL_990781, EPI_ISL_990782, EPI_ISL_990783, EPI_ISL_990784, EPI_ISL_990785, EPI_ISL_990786, EPI_ISL_990787, EPI_ISL_990788, EPI_ISL_990789, EPI_ISL_990790, EPI_ISL_990791, EPI_ISL_990792, EPI_ISL_990793, EPI_ISL_990794, EPI_ISL_990795, EPI_ISL_990796, EPI_ISL_990797,
EPI_ISL_990798, EPI_ISL_990799, EPI_ISL_990800, EPI_ISL_990801, EPI_ISL_990802, EPI_ISL_990803, EPI_ISL_990804, EPI_ISL_990805, EPI_ISL_990806, EPI_ISL_990807, EPI_ISL_990808, EPI_ISL_990809, EPI_ISL_990810, EPI_ISL_990811, EPI_ISL_990812, EPI_ISL_990813, EPI_ISL_990814, EPI_ISL_990815, EPI_ISL_990816, EPI_ISL_990817, EPI_ISL_990818, EPI_ISL_990819, EPI_ISL_990820, EPI_ISL_990821,
EPI_ISL_990822, EPI_ISL_990823, EPI_ISL_990824, EPI_ISL_990825, EPI_ISL_990826, EPI_ISL_990827, EPI_ISL_990828, EPI_ISL_990829, EPI_ISL_990830, EPI_ISL_990831, EPI_ISL_990832, EPI_ISL_990833, EPI_ISL_990834, EPI_ISL_990835, EPI_ISL_990836, EPI_ISL_990837, EPI_ISL_990838, EPI_ISL_990839, EPI_ISL_990840, EPI_ISL_990841, EPI_ISL_990842, EPI_ISL_990843, EPI_ISL_990844, EPI_ISL_990845,
EPI_ISL_990846, EPI_ISL_990847, EPI_ISL_990848, EPI_ISL_990849, EPI_ISL_990850, EPI_ISL_990851, EPI_ISL_990852, EPI_ISL_990853, EPI_ISL_990854, EPI_ISL_990855, EPI_ISL_990856, EPI_ISL_990857, EPI_ISL_990858, EPI_ISL_990859, EPI_ISL_990860, EPI_ISL_990861, EPI_ISL_990862, EPI_ISL_990863, EPI_ISL_990864, EPI_ISL_990865, EPI_ISL_990866, EPI_ISL_990867, EPI_ISL_990868, EPI_ISL_990869,
EPI_ISL_990870, EPI_ISL_990871, EPI_ISL_990872, EPI_ISL_990873, EPI_ISL_990874, EPI_ISL_990875, EPI_ISL_990876, EPI_ISL_990877, EPI_ISL_990878, EPI_ISL_990879, EPI_ISL_990880, EPI_ISL_990882, EPI_ISL_990883, EPI_ISL_990884, EPI_ISL_990885, EPI_ISL_990886, EPI_ISL_990887, EPI_ISL_990888, EPI_ISL_990889, EPI_ISL_990890, EPI_ISL_990891, EPI_ISL_990892, EPI_ISL_990893, EPI_ISL_990894,
EPI_ISL_990895, EPI_ISL_990896, EPI_ISL_990897, EPI_ISL_990898, EPI_ISL_990899, EPI_ISL_990900, EPI_ISL_990901, EPI_ISL_990902, EPI_ISL_990903, EPI_ISL_990904, EPI_ISL_990905, EPI_ISL_990906, EPI_ISL_990907, EPI_ISL_990908, EPI_ISL_990909, EPI_ISL_990910, EPI_ISL_990911, EPI_ISL_990912, EPI_ISL_990913, EPI_ISL_990914, EPI_ISL_990915, EPI_ISL_990916, EPI_ISL_990917, EPI_ISL_990918,
EPI_ISL_990919, EPI_ISL_990920, EPI_ISL_990921, EPI_ISL_990922, EPI_ISL_990923, EPI_ISL_990924, EPI_ISL_990925, EPI_ISL_990926, EPI_ISL_990927, EPI_ISL_990928, EPI_ISL_990929, EPI_ISL_990930, EPI_ISL_990931, EPI_ISL_990932, EPI_ISL_990933, EPI_ISL_990934, EPI_ISL_990935, EPI_ISL_990936, EPI_ISL_990937, EPI_ISL_990938, EPI_ISL_990939, EPI_ISL_990940, EPI_ISL_990941, EPI_ISL_990942,
EPI_ISL_990943, EPI_ISL_990944, EPI_ISL_990945, EPI_ISL_990946, EPI_ISL_990947, EPI_ISL_990948, EPI_ISL_990949, EPI_ISL_990950, EPI_ISL_990951, EPI_ISL_990952, EPI_ISL_990953, EPI_ISL_990954, EPI_ISL_990955, EPI_ISL_990956, EPI_ISL_990957, EPI_ISL_990958, EPI_ISL_990959, EPI_ISL_990960, EPI_ISL_990961, EPI_ISL_990962, EPI_ISL_990963, EPI_ISL_990964, EPI_ISL_990965, EPI_ISL_990966,
EPI_ISL_990967, EPI_ISL_990968, EPI_ISL_990969, EPI_ISL_990970, EPI_ISL_990971, EPI_ISL_990972, EPI_ISL_990973, EPI_ISL_990974, EPI_ISL_990975, EPI_ISL_990976, EPI_ISL_990977, EPI_ISL_990978, EPI_ISL_990979, EPI_ISL_990980, EPI_ISL_990981, EPI_ISL_990982, EPI_ISL_990983, EPI_ISL_990984, EPI_ISL_990985, EPI_ISL_990986, EPI_ISL_990987, EPI_ISL_990988, EPI_ISL_990989, EPI_ISL_990990,
EPI_ISL_990991, EPI_ISL_990992, EPI_ISL_990993, EPI_ISL_990994, EPI_ISL_990995, EPI_ISL_990996, EPI_ISL_990997, EPI_ISL_990998, EPI_ISL_990999, EPI_ISL_991000, EPI_ISL_991001, EPI_ISL_991002, EPI_ISL_991003, EPI_ISL_991004, EPI_ISL_991005, EPI_ISL_991006, EPI_ISL_991007, EPI_ISL_991008, EPI_ISL_991009, EPI_ISL_991010, EPI_ISL_991011, EPI_ISL_991012, EPI_ISL_991013, EPI_ISL_991014,
EPI_ISL_991015, EPI_ISL_991016, EPI_ISL_991017, EPI_ISL_991018, EPI_ISL_991019, EPI_ISL_991020, EPI_ISL_991021, EPI_ISL_991022, EPI_ISL_991023, EPI_ISL_991024, EPI_ISL_991025, EPI_ISL_991026, EPI_ISL_991027, EPI_ISL_991028, EPI_ISL_991029, EPI_ISL_991030, EPI_ISL_991031, EPI_ISL_991032, EPI_ISL_991033, EPI_ISL_991034, EPI_ISL_991035, EPI_ISL_991036, EPI_ISL_991037, EPI_ISL_991038,
EPI_ISL_991039, EPI_ISL_991040, EPI_ISL_991041, EPI_ISL_991042, EPI_ISL_991043, EPI_ISL_991044, EPI_ISL_991045, EPI_ISL_991046, EPI_ISL_991047, EPI_ISL_991048, EPI_ISL_991049, EPI_ISL_991050, EPI_ISL_991051, EPI_ISL_991052, EPI_ISL_991053, EPI_ISL_991054, EPI_ISL_991055, EPI_ISL_991056, EPI_ISL_991057, EPI_ISL_991058, EPI_ISL_991059, EPI_ISL_991060, EPI_ISL_991061, EPI_ISL_991062,
EPI_ISL_991063, EPI_ISL_991064, EPI_ISL_991065, EPI_ISL_991066, EPI_ISL_991067, EPI_ISL_991068, EPI_ISL_991069, EPI_ISL_991070, EPI_ISL_991071, EPI_ISL_991072, EPI_ISL_991073, EPI_ISL_991074, EPI_ISL_991075, EPI_ISL_991076, EPI_ISL_991077, EPI_ISL_991078, EPI_ISL_991079, EPI_ISL_991080, EPI_ISL_991081, EPI_ISL_991082, EPI_ISL_991083, EPI_ISL_991084, EPI_ISL_991085, EPI_ISL_991086,
EPI_ISL_991087, EPI_ISL_991088, EPI_ISL_991089, EPI_ISL_991090, EPI_ISL_991091, EPI_ISL_991092, EPI_ISL_991093, EPI_ISL_991094, EPI_ISL_991095, EPI_ISL_991096, EPI_ISL_991097, EPI_ISL_991098, EPI_ISL_991099, EPI_ISL_991100, EPI_ISL_991101, EPI_ISL_991102, EPI_ISL_991103, EPI_ISL_991104, EPI_ISL_991105, EPI_ISL_991106, EPI_ISL_991107, EPI_ISL_991108, EPI_ISL_991109, EPI_ISL_991110,
EPI_ISL_991111, EPI_ISL_991112, EPI_ISL_991113, EPI_ISL_991114, EPI_ISL_991115, EPI_ISL_991116, EPI_ISL_991117, EPI_ISL_991118, EPI_ISL_991119, EPI_ISL_991120, EPI_ISL_991121, EPI_ISL_991122, EPI_ISL_991123, EPI_ISL_991124, EPI_ISL_991125, EPI_ISL_991126, EPI_ISL_991127, EPI_ISL_991128, EPI_ISL_991129, EPI_ISL_991130, EPI_ISL_991131, EPI_ISL_991132, EPI_ISL_991133, EPI_ISL_991134,
EPI_ISL_991135, EPI_ISL_991136, EPI_ISL_991137, EPI_ISL_991138, EPI_ISL_991139, EPI_ISL_991140, EPI_ISL_991141, EPI_ISL_991142, EPI_ISL_991143, EPI_ISL_991144, EPI_ISL_991145, EPI_ISL_991146, EPI_ISL_991147, EPI_ISL_991148, EPI_ISL_991149, EPI_ISL_991150, EPI_ISL_991151, EPI_ISL_991152, EPI_ISL_991153, EPI_ISL_991154, EPI_ISL_991155, EPI_ISL_991156, EPI_ISL_991157, EPI_ISL_991158,
EPI_ISL_991159, EPI_ISL_991160, EPI_ISL_991161, EPI_ISL_991162, EPI_ISL_991163, EPI_ISL_991164, EPI_ISL_991165, EPI_ISL_991166, EPI_ISL_991167, EPI_ISL_991168, EPI_ISL_991169, EPI_ISL_991170, EPI_ISL_991171, EPI_ISL_991172, EPI_ISL_991222, EPI_ISL_991223, EPI_ISL_991224, EPI_ISL_991225, EPI_ISL_991226, EPI_ISL_991227, EPI_ISL_991228, EPI_ISL_991229, EPI_ISL_991230, EPI_ISL_991231,
EPI_ISL_991232, EPI_ISL_991233, EPI_ISL_991234, EPI_ISL_991235, EPI_ISL_991236, EPI_ISL_991237, EPI_ISL_991238, EPI_ISL_991239, EPI_ISL_991240, EPI_ISL_991241, EPI_ISL_991242, EPI_ISL_991243, EPI_ISL_991244, EPI_ISL_991245, EPI_ISL_991247, EPI_ISL_991248, EPI_ISL_991249, EPI_ISL_991250, EPI_ISL_991251, EPI_ISL_991252, EPI_ISL_991253, EPI_ISL_991254, EPI_ISL_991255,
EPI_ISL_991256, EPI_ISL_991257, EPI_ISL_991258, EPI_ISL_991259, EPI_ISL_991260, EPI_ISL_991261, EPI_ISL_991262, EPI_ISL_991263, EPI_ISL_991264, EPI_ISL_991265, EPI_ISL_991266, EPI_ISL_991267, EPI_ISL_991268, EPI_ISL_991270, EPI_ISL_991271, EPI_ISL_991273, EPI_ISL_991275, EPI_ISL_991276, EPI_ISL_991277, EPI_ISL_991278, EPI_ISL_991279, EPI_ISL_991280, EPI_ISL_991281, EPI_ISL_991282,
EPI_ISL_991283, EPI_ISL_991284, EPI_ISL_991285, EPI_ISL_991286, EPI_ISL_991287, EPI_ISL_991288, EPI_ISL_991289, EPI_ISL_991290, EPI_ISL_991296, EPI_ISL_991297, EPI_ISL_991298, EPI_ISL_991299, EPI_ISL_991301, EPI_ISL_991303, EPI_ISL_991304, EPI_ISL_991307, EPI_ISL_991308, EPI_ISL_991565, EPI_ISL_991595, EPI_ISL_991629, EPI_ISL_991637, EPI_ISL_991653, EPI_ISL_991680,
EPI_ISL_991692, EPI_ISL_991694, EPI_ISL_991703, EPI_ISL_991715, EPI_ISL_991755, EPI_ISL_991781, EPI_ISL_991793, EPI_ISL_991817, EPI_ISL_991826, EPI_ISL_991834, EPI_ISL_991846, EPI_ISL_991848, EPI_ISL_991860, EPI_ISL_991873, EPI_ISL_991900, EPI_ISL_993004, EPI_ISL_1007549, EPI_ISL_1007550, EPI_ISL_1007551, EPI_ISL_1007552, EPI_ISL_1007553, EPI_ISL_1007554, EPI_ISL_1007555,
EPI_ISL_1007556, EPI_ISL_1007557, EPI_ISL_1007559, EPI_ISL_1007560, EPI_ISL_1007563, EPI_ISL_1012713, EPI_ISL_1012714, EPI_ISL_1012715, EPI_ISL_1012716, EPI_ISL_1012717, EPI_ISL_1012718, EPI_ISL_1012719, EPI_ISL_1012720, EPI_ISL_1012721, EPI_ISL_1012722, EPI_ISL_1012724, EPI_ISL_1012725, EPI_ISL_1012726, EPI_ISL_1012727, EPI_ISL_1012728,
EPI_ISL_1012729, EPI_ISL_1012730, EPI_ISL_1012731, EPI_ISL_1012732, EPI_ISL_1012733, EPI_ISL_1012734, EPI_ISL_1012735, EPI_ISL_1012736, EPI_ISL_1012737, EPI_ISL_1012738, EPI_ISL_1012739, EPI_ISL_1012740, EPI_ISL_1012741, EPI_ISL_1012742, EPI_ISL_1012743, EPI_ISL_1012744, EPI_ISL_1012745, EPI_ISL_1012746, EPI_ISL_1012747, EPI_ISL_1012748, EPI_ISL_1012749, EPI_ISL_1012750,
EPI_ISL_1012751, EPI_ISL_1012752, EPI_ISL_1012753, EPI_ISL_1012755, EPI_ISL_1012756, EPI_ISL_1012757, EPI_ISL_1012758, EPI_ISL_1012759, EPI_ISL_1012760, EPI_ISL_1012761, EPI_ISL_1012763, EPI_ISL_1012764, EPI_ISL_1012765, EPI_ISL_1012766, EPI_ISL_1012767, EPI_ISL_1012768, EPI_ISL_1012769, EPI_ISL_1012770, EPI_ISL_1012771, EPI_ISL_1012772, EPI_ISL_1012773, EPI_ISL_1012774,
EPI_ISL_1012775, EPI_ISL_1012776, EPI_ISL_1012777, EPI_ISL_1012778, EPI_ISL_1012779, EPI_ISL_1012780, EPI_ISL_1012781, EPI_ISL_1012782, EPI_ISL_1012783, EPI_ISL_1012784, EPI_ISL_1012785, EPI_ISL_1012786, EPI_ISL_1043899, EPI_ISL_1043900, EPI_ISL_1043902, EPI_ISL_1043903, EPI_ISL_1043904, EPI_ISL_1043905, EPI_ISL_1043906, EPI_ISL_1043907, EPI_ISL_1043908, EPI_ISL_1043909,
EPI_ISL_1043910, EPI_ISL_1043911, EPI_ISL_1043912, EPI_ISL_1043913, EPI_ISL_1043914, EPI_ISL_1043915, EPI_ISL_1043916, EPI_ISL_1043917, EPI_ISL_1043918, EPI_ISL_1043919, EPI_ISL_1043920, EPI_ISL_1043921, EPI_ISL_1043922, EPI_ISL_1043923, EPI_ISL_1043924, EPI_ISL_1043925, EPI_ISL_1043926, EPI_ISL_1043927, EPI_ISL_1043928, EPI_ISL_1043929, EPI_ISL_1043930, EPI_ISL_1043931,
EPI_ISL_1043932, EPI_ISL_1043933, EPI_ISL_1043934, EPI_ISL_1043935, EPI_ISL_1043936, EPI_ISL_1043937, EPI_ISL_1043938, EPI_ISL_1043939, EPI_ISL_1043940, EPI_ISL_1043941, EPI_ISL_1043942, EPI_ISL_1043943, EPI_ISL_1043944, EPI_ISL_1043945, EPI_ISL_1043946, EPI_ISL_1043947, EPI_ISL_1043948, EPI_ISL_1043949, EPI_ISL_1043950, EPI_ISL_1043951, EPI_ISL_1043952, EPI_ISL_1043953,
EPI_ISL_1043954, EPI_ISL_1043955, EPI_ISL_1043956, EPI_ISL_1043957, EPI_ISL_1043958, EPI_ISL_1043959, EPI_ISL_1043960, EPI_ISL_1043961, EPI_ISL_1043962, EPI_ISL_1043963, EPI_ISL_1043964, EPI_ISL_1043965, EPI_ISL_1043966, EPI_ISL_1043967, EPI_ISL_1043968, EPI_ISL_1043969, EPI_ISL_1043970, EPI_ISL_1043971, EPI_ISL_1043972, EPI_ISL_1043973, EPI_ISL_1043974, EPI_ISL_1043975,

EPI_ISL_1043976, EPI_ISL_1043977, EPI_ISL_1043978, EPI_ISL_1043979, EPI_ISL_1043980, EPI_ISL_1043981, EPI_ISL_1043982, EPI_ISL_1043983, EPI_ISL_1043984, EPI_ISL_1043985, EPI_ISL_1043986, EPI_ISL_1043987, EPI_ISL_1043988, EPI_ISL_1043989, EPI_ISL_1043990, EPI_ISL_1043991, EPI_ISL_1043992, EPI_ISL_1043993, EPI_ISL_1043994, EPI_ISL_1043995, EPI_ISL_1043996, EPI_ISL_1043997,
EPI_ISL_1043998, EPI_ISL_1043999, EPI_ISL_1044000, EPI_ISL_1044001, EPI_ISL_1044002, EPI_ISL_1044003, EPI_ISL_1044004, EPI_ISL_1044005, EPI_ISL_1044006, EPI_ISL_1044007, EPI_ISL_1044008, EPI_ISL_1044009, EPI_ISL_1044010, EPI_ISL_1044011, EPI_ISL_1044012, EPI_ISL_1044013, EPI_ISL_1044014, EPI_ISL_1044015, EPI_ISL_1044016, EPI_ISL_1044017, EPI_ISL_1044018, EPI_ISL_1044019,
EPI_ISL_1044020, EPI_ISL_1044021, EPI_ISL_1044022, EPI_ISL_1044023, EPI_ISL_1044024, EPI_ISL_1044025, EPI_ISL_1044026, EPI_ISL_1044027, EPI_ISL_1044028, EPI_ISL_1044029, EPI_ISL_1044030, EPI_ISL_1044031, EPI_ISL_1044032, EPI_ISL_1044033, EPI_ISL_1044034, EPI_ISL_1044035, EPI_ISL_1044036, EPI_ISL_1044037, EPI_ISL_1044038, EPI_ISL_1044039, EPI_ISL_1044040, EPI_ISL_1044041,
EPI_ISL_1044042, EPI_ISL_1044043, EPI_ISL_1044044, EPI_ISL_1044045, EPI_ISL_1044046, EPI_ISL_1044047, EPI_ISL_1044048, EPI_ISL_1044049, EPI_ISL_1044050, EPI_ISL_1044051, EPI_ISL_1044052, EPI_ISL_1044053, EPI_ISL_1044054, EPI_ISL_1044055, EPI_ISL_1044056, EPI_ISL_1044057, EPI_ISL_1044058, EPI_ISL_1044059, EPI_ISL_1044060, EPI_ISL_1044061, EPI_ISL_1044062, EPI_ISL_1044063,
EPI_ISL_1044064, EPI_ISL_1044065, EPI_ISL_1044066, EPI_ISL_1044067, EPI_ISL_1044068, EPI_ISL_1044069, EPI_ISL_1044070, EPI_ISL_1044071, EPI_ISL_1044072, EPI_ISL_1044073, EPI_ISL_1044074, EPI_ISL_1044075, EPI_ISL_1044076, EPI_ISL_1044077, EPI_ISL_1044078, EPI_ISL_1044079, EPI_ISL_1044080, EPI_ISL_1044081, EPI_ISL_1044082, EPI_ISL_1044083, EPI_ISL_1044084, EPI_ISL_1044085,
EPI_ISL_1044086, EPI_ISL_1044087, EPI_ISL_1044088, EPI_ISL_1044089, EPI_ISL_1044090, EPI_ISL_1044091, EPI_ISL_1044092, EPI_ISL_1044093, EPI_ISL_1044094, EPI_ISL_1044095, EPI_ISL_1044096, EPI_ISL_1044097, EPI_ISL_1044098, EPI_ISL_1044099, EPI_ISL_1044100, EPI_ISL_1044101, EPI_ISL_1044102, EPI_ISL_1044103, EPI_ISL_1044104, EPI_ISL_1044105, EPI_ISL_1044106, EPI_ISL_1044107,
EPI_ISL_1044108, EPI_ISL_1044109, EPI_ISL_1044110, EPI_ISL_1044111, EPI_ISL_1044112, EPI_ISL_1044113, EPI_ISL_1044114, EPI_ISL_1044115, EPI_ISL_1044116, EPI_ISL_1044117, EPI_ISL_1044118, EPI_ISL_1044119, EPI_ISL_1044120, EPI_ISL_1044121, EPI_ISL_1044122, EPI_ISL_1044123, EPI_ISL_1044124, EPI_ISL_1044125, EPI_ISL_1044126, EPI_ISL_1044127, EPI_ISL_1044128, EPI_ISL_1044129,
EPI_ISL_1044130, EPI_ISL_1044131, EPI_ISL_1044132, EPI_ISL_1044133, EPI_ISL_1044134, EPI_ISL_1044135, EPI_ISL_1044136, EPI_ISL_1044137, EPI_ISL_1044138, EPI_ISL_1044139, EPI_ISL_1044140, EPI_ISL_1044141, EPI_ISL_1044142, EPI_ISL_1044143, EPI_ISL_1044144, EPI_ISL_1044145, EPI_ISL_1044146, EPI_ISL_1044147, EPI_ISL_1044148, EPI_ISL_1044149, EPI_ISL_1044150, EPI_ISL_1044151,
EPI_ISL_1044152, EPI_ISL_1044153, EPI_ISL_1044154, EPI_ISL_1044155, EPI_ISL_1044156, EPI_ISL_1044157, EPI_ISL_1044158, EPI_ISL_1044159, EPI_ISL_1044160, EPI_ISL_1044161, EPI_ISL_1044162, EPI_ISL_1044163, EPI_ISL_1044164, EPI_ISL_1044165, EPI_ISL_1044166, EPI_ISL_1044167, EPI_ISL_1044168, EPI_ISL_1044169, EPI_ISL_1044170, EPI_ISL_1044171, EPI_ISL_1044172, EPI_ISL_1044173,
EPI_ISL_1044174, EPI_ISL_1044175, EPI_ISL_1044176, EPI_ISL_1044177, EPI_ISL_1044178, EPI_ISL_1044179, EPI_ISL_1044180, EPI_ISL_1044181, EPI_ISL_1044182, EPI_ISL_1044183, EPI_ISL_1044184, EPI_ISL_1044185, EPI_ISL_1044186, EPI_ISL_1044187, EPI_ISL_1044188, EPI_ISL_1044189, EPI_ISL_1044190, EPI_ISL_1044191, EPI_ISL_1044192, EPI_ISL_1044193, EPI_ISL_1044194, EPI_ISL_1044195,
EPI_ISL_1044196, EPI_ISL_1044197, EPI_ISL_1044198, EPI_ISL_1044199, EPI_ISL_1044200, EPI_ISL_1044201, EPI_ISL_1044202, EPI_ISL_1044203, EPI_ISL_1044204, EPI_ISL_1044205, EPI_ISL_1044206, EPI_ISL_1044207, EPI_ISL_1044208, EPI_ISL_1044209, EPI_ISL_1044210, EPI_ISL_1044211, EPI_ISL_1044212, EPI_ISL_1044213, EPI_ISL_1044214, EPI_ISL_1044215, EPI_ISL_1044216, EPI_ISL_1044217,
EPI_ISL_1044218, EPI_ISL_1044219, EPI_ISL_1044220, EPI_ISL_1044221, EPI_ISL_1044222, EPI_ISL_1044223, EPI_ISL_1044224, EPI_ISL_1057726, EPI_ISL_1057727, EPI_ISL_1057728, EPI_ISL_1057729, EPI_ISL_1057730, EPI_ISL_1057731, EPI_ISL_1057733, EPI_ISL_1057734, EPI_ISL_1057735, EPI_ISL_1057736, EPI_ISL_1057737, EPI_ISL_1057738, EPI_ISL_1057739, EPI_ISL_1057740, EPI_ISL_1057741,
EPI_ISL_1057742, EPI_ISL_1057743, EPI_ISL_1057744, EPI_ISL_1057746, EPI_ISL_1057747, EPI_ISL_1057748, EPI_ISL_1057749, EPI_ISL_1057750, EPI_ISL_1057751, EPI_ISL_1057752, EPI_ISL_1057753, EPI_ISL_1057754, EPI_ISL_1057755, EPI_ISL_1057756, EPI_ISL_1057757, EPI_ISL_1057758, EPI_ISL_1057759, EPI_ISL_1057760, EPI_ISL_1057761, EPI_ISL_1057762, EPI_ISL_1057763

| | | | |
|---|---|---|---|
| see above | Lighthouse Lab in Alderley Park | Wellcome Sanger Institute for the COVID-19 Genomics UK (COG-UK) Consortium | Cordelia Langford; David K. Jackson; Dominic Kwiatkowski; Ewan Harrison; Ian Johnston; Jacquelyn Wynn; Jeffrey Barrett; John Sillitoe on behalf of the Wellcome Sanger Institute COVID-19 Surveillance Team; John Sillitoe on behalf of the Wellcome Sanger Institute COVID-19 Surveillance Team (http://www.sanger.ac.uk/covid-team); Mairead Hyland; Roberto Amato; Sonia Goncalves; The Lighthouse Lab in Alderley Park and Alex Alderton |

EPI_ISL_948102, EPI_ISL_948104, EPI_ISL_948107, EPI_ISL_948110, EPI_ISL_948114, EPI_ISL_948117, EPI_ISL_948118, EPI_ISL_948120, EPI_ISL_948122, EPI_ISL_948123, EPI_ISL_948124, EPI_ISL_948131, EPI_ISL_948134, EPI_ISL_948136, EPI_ISL_948137, EPI_ISL_948139, EPI_ISL_948145, EPI_ISL_948147, EPI_ISL_948149, EPI_ISL_948151, EPI_ISL_948153, EPI_ISL_948163, EPI_ISL_948164, EPI_ISL_948165,
EPI_ISL_948166, EPI_ISL_948172, EPI_ISL_948174, EPI_ISL_948176, EPI_ISL_948180, EPI_ISL_948181, EPI_ISL_948183, EPI_ISL_948187, EPI_ISL_948188, EPI_ISL_948189, EPI_ISL_948191, EPI_ISL_948192, EPI_ISL_948193, EPI_ISL_948197, EPI_ISL_948198, EPI_ISL_948199, EPI_ISL_948200, EPI_ISL_948201, EPI_ISL_948203, EPI_ISL_948205, EPI_ISL_948207, EPI_ISL_948208, EPI_ISL_948209, EPI_ISL_948211,
EPI_ISL_948212, EPI_ISL_948213, EPI_ISL_948214, EPI_ISL_948216, EPI_ISL_948219, EPI_ISL_948220, EPI_ISL_948223, EPI_ISL_948224, EPI_ISL_948227, EPI_ISL_948228, EPI_ISL_948230, EPI_ISL_948233, EPI_ISL_948234, EPI_ISL_948235, EPI_ISL_948237, EPI_ISL_948239, EPI_ISL_948240, EPI_ISL_948241, EPI_ISL_948244, EPI_ISL_948247, EPI_ISL_948249, EPI_ISL_948250, EPI_ISL_948251, EPI_ISL_948253,
EPI_ISL_948255, EPI_ISL_948257, EPI_ISL_948258, EPI_ISL_948259, EPI_ISL_948261, EPI_ISL_948264, EPI_ISL_948265, EPI_ISL_948269, EPI_ISL_948274, EPI_ISL_948277, EPI_ISL_948278, EPI_ISL_948279, EPI_ISL_948280, EPI_ISL_948281, EPI_ISL_948282, EPI_ISL_948283, EPI_ISL_948284, EPI_ISL_948286, EPI_ISL_948288, EPI_ISL_948296, EPI_ISL_948298, EPI_ISL_948300, EPI_ISL_948301, EPI_ISL_948302,
EPI_ISL_948305, EPI_ISL_948306, EPI_ISL_948308, EPI_ISL_948310, EPI_ISL_948311, EPI_ISL_948312, EPI_ISL_948313, EPI_ISL_948314, EPI_ISL_948316, EPI_ISL_948321, EPI_ISL_948322, EPI_ISL_948323, EPI_ISL_948326, EPI_ISL_948328, EPI_ISL_948331, EPI_ISL_948332, EPI_ISL_948337, EPI_ISL_948338, EPI_ISL_948340, EPI_ISL_948341, EPI_ISL_948342, EPI_ISL_948344, EPI_ISL_948345, EPI_ISL_948346,
EPI_ISL_948350, EPI_ISL_948355, EPI_ISL_948359, EPI_ISL_948361, EPI_ISL_948364, EPI_ISL_948366, EPI_ISL_948372, EPI_ISL_948378, EPI_ISL_948380, EPI_ISL_948381, EPI_ISL_948382, EPI_ISL_948383, EPI_ISL_948391, EPI_ISL_948392, EPI_ISL_948393, EPI_ISL_948395, EPI_ISL_948396, EPI_ISL_948397, EPI_ISL_948402, EPI_ISL_948408, EPI_ISL_948412, EPI_ISL_948413, EPI_ISL_948414, EPI_ISL_948420,
EPI_ISL_948421, EPI_ISL_948422, EPI_ISL_948423, EPI_ISL_948424, EPI_ISL_948431, EPI_ISL_948433, EPI_ISL_948434, EPI_ISL_948435, EPI_ISL_948437, EPI_ISL_948441, EPI_ISL_948442, EPI_ISL_948445, EPI_ISL_959267, EPI_ISL_971425, EPI_ISL_987288, EPI_ISL_987289, EPI_ISL_987292, EPI_ISL_987293, EPI_ISL_987295, EPI_ISL_987296, EPI_ISL_987301, EPI_ISL_987302, EPI_ISL_987304, EPI_ISL_987306,
EPI_ISL_987307, EPI_ISL_987309, EPI_ISL_987313, EPI_ISL_987315, EPI_ISL_987318, EPI_ISL_987321, EPI_ISL_987325, EPI_ISL_987344, EPI_ISL_987348, EPI_ISL_987361, EPI_ISL_987365, EPI_ISL_987367, EPI_ISL_987368, EPI_ISL_987372, EPI_ISL_987379, EPI_ISL_987383, EPI_ISL_987387, EPI_ISL_987392, EPI_ISL_987397, EPI_ISL_987401, EPI_ISL_987407, EPI_ISL_987408, EPI_ISL_987409, EPI_ISL_987412,
EPI_ISL_987413, EPI_ISL_987416, EPI_ISL_987417, EPI_ISL_987421, EPI_ISL_987425, EPI_ISL_987430, EPI_ISL_987432, EPI_ISL_987449, EPI_ISL_987451, EPI_ISL_987456, EPI_ISL_987458, EPI_ISL_987462, EPI_ISL_987467, EPI_ISL_987472, EPI_ISL_987476, EPI_ISL_987478, EPI_ISL_987480, EPI_ISL_987482, EPI_ISL_987489, EPI_ISL_987490, EPI_ISL_987493, EPI_ISL_987505, EPI_ISL_987508, EPI_ISL_987513,
EPI_ISL_987518, EPI_ISL_987519, EPI_ISL_987520, EPI_ISL_987521, EPI_ISL_987523, EPI_ISL_987525, EPI_ISL_987526, EPI_ISL_987527, EPI_ISL_987540, EPI_ISL_987541, EPI_ISL_987542, EPI_ISL_987545, EPI_ISL_987546, EPI_ISL_987553, EPI_ISL_987554, EPI_ISL_987559, EPI_ISL_987561, EPI_ISL_987562, EPI_ISL_987566, EPI_ISL_987578, EPI_ISL_987579, EPI_ISL_987581, EPI_ISL_987583, EPI_ISL_987595,
EPI_ISL_987596, EPI_ISL_987599, EPI_ISL_987603, EPI_ISL_987604, EPI_ISL_987605, EPI_ISL_987607, EPI_ISL_987611, EPI_ISL_987613, EPI_ISL_987615, EPI_ISL_987617, EPI_ISL_987623, EPI_ISL_1187053, EPI_ISL_1187064, EPI_ISL_1187068, EPI_ISL_1187069, EPI_ISL_1187073, EPI_ISL_1187103, EPI_ISL_1187113, EPI_ISL_1187114, EPI_ISL_1187130, EPI_ISL_1187136, EPI_ISL_1187140, EPI_ISL_1187146,
EPI_ISL_1187165, EPI_ISL_1187171, EPI_ISL_1187176, EPI_ISL_1187177, EPI_ISL_1187192, EPI_ISL_1187193, EPI_ISL_1187204, EPI_ISL_1187218, EPI_ISL_1187219, EPI_ISL_1187221, EPI_ISL_1187225, EPI_ISL_1187226, EPI_ISL_1187237, EPI_ISL_1187240, EPI_ISL_1187245, EPI_ISL_1187253, EPI_ISL_1206041, EPI_ISL_1206042, EPI_ISL_1206043, EPI_ISL_1206044, EPI_ISL_1206046, EPI_ISL_1206048,
EPI_ISL_1206049, EPI_ISL_1206052, EPI_ISL_1206053, EPI_ISL_1206055, EPI_ISL_1206057, EPI_ISL_1206059, EPI_ISL_1206060, EPI_ISL_1206061, EPI_ISL_1206062, EPI_ISL_1206063, EPI_ISL_1206064, EPI_ISL_1206069, EPI_ISL_1206070, EPI_ISL_1206071, EPI_ISL_1206072, EPI_ISL_1206074, EPI_ISL_1206083, EPI_ISL_1206085, EPI_ISL_1206086, EPI_ISL_1206088, EPI_ISL_1206089, EPI_ISL_1206094,
EPI_ISL_1206095, EPI_ISL_1206098, EPI_ISL_1206099, EPI_ISL_1206101, EPI_ISL_1206102, EPI_ISL_1206103, EPI_ISL_1206105, EPI_ISL_1206106, EPI_ISL_1206107, EPI_ISL_1206116, EPI_ISL_1206119, EPI_ISL_1206120, EPI_ISL_1206121, EPI_ISL_1206122, EPI_ISL_1206126, EPI_ISL_1206129, EPI_ISL_1206131, EPI_ISL_1206135, EPI_ISL_1206138, EPI_ISL_1206140, EPI_ISL_1206141, EPI_ISL_1206142,
EPI_ISL_1206144, EPI_ISL_1206146, EPI_ISL_1206147, EPI_ISL_1206149, EPI_ISL_1206154, EPI_ISL_1206155, EPI_ISL_1206158, EPI_ISL_1206160, EPI_ISL_1206162, EPI_ISL_1206163, EPI_ISL_1206165, EPI_ISL_1206168, EPI_ISL_1206172, EPI_ISL_1206173, EPI_ISL_1206178, EPI_ISL_1206180, EPI_ISL_1206182, EPI_ISL_1206183, EPI_ISL_1206186, EPI_ISL_1206187, EPI_ISL_1206191, EPI_ISL_1206192,
EPI_ISL_1206196, EPI_ISL_1206198, EPI_ISL_1206200, EPI_ISL_1206203, EPI_ISL_1206207, EPI_ISL_1206213, EPI_ISL_1206217, EPI_ISL_1206221, EPI_ISL_1206222, EPI_ISL_1206226, EPI_ISL_1206230, EPI_ISL_1206233, EPI_ISL_1206237, EPI_ISL_1206244, EPI_ISL_1206245, EPI_ISL_1206248, EPI_ISL_1206251, EPI_ISL_1206258, EPI_ISL_1206262, EPI_ISL_1206263, EPI_ISL_1206266, EPI_ISL_1206268,
EPI_ISL_1206272, EPI_ISL_1206275, EPI_ISL_1206279, EPI_ISL_1206280, EPI_ISL_1206284, EPI_ISL_1206291, EPI_ISL_1223161, EPI_ISL_1223164, EPI_ISL_1223174, EPI_ISL_1223224, EPI_ISL_1223240, EPI_ISL_1223245, EPI_ISL_1223288, EPI_ISL_1223291, EPI_ISL_1223383, EPI_ISL_1223391, EPI_ISL_1223392, EPI_ISL_1223401, EPI_ISL_1223404, EPI_ISL_1223411, EPI_ISL_1223424, EPI_ISL_1223426,
EPI_ISL_1223435, EPI_ISL_1242052, EPI_ISL_1242057, EPI_ISL_1242066, EPI_ISL_1242096, EPI_ISL_1242124, EPI_ISL_1242132, EPI_ISL_1242136, EPI_ISL_1242145, EPI_ISL_1242149, EPI_ISL_1242162, EPI_ISL_1242179, EPI_ISL_1242187, EPI_ISL_1242188, EPI_ISL_1242211, EPI_ISL_1242225, EPI_ISL_1242227, EPI_ISL_1242280, EPI_ISL_1333481, EPI_ISL_1333488, EPI_ISL_1333490, EPI_ISL_1333491,
EPI_ISL_1333492, EPI_ISL_1333500, EPI_ISL_1333502, EPI_ISL_1333504, EPI_ISL_1333505, EPI_ISL_1333507, EPI_ISL_1333508, EPI_ISL_1333834, EPI_ISL_1333835, EPI_ISL_1333836, EPI_ISL_1333839, EPI_ISL_1333935, EPI_ISL_1333941

| | | | |
|---|---|---|---|
| see above | Lighthouse Lab in Cambridge | Wellcome Sanger Institute for the COVID-19 Genomics UK (COG-UK) Consortium | Cordelia Langford; David K. Jackson; Dominic Kwiatkowski; Ewan Harrison; Ian Johnston; Jeffrey Barrett; John Sillitoe on behalf of the Wellcome Sanger Institute COVID-19 Surveillance Team; John Sillitoe on behalf of the Wellcome Sanger Institute COVID-19 Surveillance Team (http://www.sanger.ac.uk/covid-team); Rob Howes; Roberto Amato; Sonia Goncalves; The Lighthouse Lab in Cambridge and Alex Alderton |

EPI_ISL_932850, EPI_ISL_932853, EPI_ISL_932855, EPI_ISL_932856, EPI_ISL_932857, EPI_ISL_932858, EPI_ISL_932859, EPI_ISL_932860, EPI_ISL_932863, EPI_ISL_932864, EPI_ISL_932865, EPI_ISL_932870, EPI_ISL_932871, EPI_ISL_932879, EPI_ISL_932884, EPI_ISL_932885, EPI_ISL_932894, EPI_ISL_932896, EPI_ISL_932897, EPI_ISL_932899, EPI_ISL_932903, EPI_ISL_932905, EPI_ISL_932908, EPI_ISL_932914,
EPI_ISL_932915, EPI_ISL_932917, EPI_ISL_932918, EPI_ISL_932919, EPI_ISL_932920, EPI_ISL_932921, EPI_ISL_932925, EPI_ISL_932929, EPI_ISL_932930, EPI_ISL_932931, EPI_ISL_932932, EPI_ISL_932936, EPI_ISL_932939, EPI_ISL_932944, EPI_ISL_932948, EPI_ISL_932949, EPI_ISL_932955, EPI_ISL_932957, EPI_ISL_932958, EPI_ISL_932965, EPI_ISL_932966, EPI_ISL_932967, EPI_ISL_932970, EPI_ISL_932980,
EPI_ISL_932986, EPI_ISL_932987, EPI_ISL_932991, EPI_ISL_932994, EPI_ISL_932995, EPI_ISL_932996, EPI_ISL_933000, EPI_ISL_933002, EPI_ISL_933004, EPI_ISL_933006, EPI_ISL_933007, EPI_ISL_933009, EPI_ISL_933011, EPI_ISL_933018, EPI_ISL_933020, EPI_ISL_933021, EPI_ISL_933028, EPI_ISL_933033, EPI_ISL_933035, EPI_ISL_933036, EPI_ISL_933037, EPI_ISL_933038, EPI_ISL_933040, EPI_ISL_933042,
EPI_ISL_933044, EPI_ISL_933050, EPI_ISL_933051, EPI_ISL_933053, EPI_ISL_933054, EPI_ISL_933056, EPI_ISL_933058, EPI_ISL_933065, EPI_ISL_933067, EPI_ISL_933069, EPI_ISL_933070, EPI_ISL_933072, EPI_ISL_933073, EPI_ISL_933074, EPI_ISL_933079, EPI_ISL_933080, EPI_ISL_933096, EPI_ISL_933099, EPI_ISL_933100, EPI_ISL_933109, EPI_ISL_933110, EPI_ISL_939283, EPI_ISL_939284, EPI_ISL_939285,
EPI_ISL_939287, EPI_ISL_939289, EPI_ISL_939295, EPI_ISL_939297, EPI_ISL_939298, EPI_ISL_939299, EPI_ISL_939303, EPI_ISL_939304, EPI_ISL_939305, EPI_ISL_939308, EPI_ISL_939309, EPI_ISL_939310, EPI_ISL_939311, EPI_ISL_939312, EPI_ISL_939316, EPI_ISL_939317, EPI_ISL_939322, EPI_ISL_939323, EPI_ISL_939324, EPI_ISL_939325, EPI_ISL_939326, EPI_ISL_939328, EPI_ISL_939330, EPI_ISL_939331,
EPI_ISL_939333, EPI_ISL_939335, EPI_ISL_939336, EPI_ISL_939338, EPI_ISL_939341, EPI_ISL_939342, EPI_ISL_939344, EPI_ISL_939345, EPI_ISL_939347, EPI_ISL_939349, EPI_ISL_939353, EPI_ISL_939360, EPI_ISL_939362, EPI_ISL_939363, EPI_ISL_939366, EPI_ISL_939367, EPI_ISL_939371, EPI_ISL_939374, EPI_ISL_939375, EPI_ISL_939376, EPI_ISL_939377, EPI_ISL_939379, EPI_ISL_939381, EPI_ISL_939382,
EPI_ISL_939383, EPI_ISL_939384, EPI_ISL_939385, EPI_ISL_939386, EPI_ISL_939387, EPI_ISL_939392, EPI_ISL_939394, EPI_ISL_939400, EPI_ISL_939408, EPI_ISL_939409, EPI_ISL_939410, EPI_ISL_939411, EPI_ISL_939412, EPI_ISL_939417, EPI_ISL_939419, EPI_ISL_939421, EPI_ISL_939423, EPI_ISL_939424, EPI_ISL_939426, EPI_ISL_939427, EPI_ISL_939430, EPI_ISL_939433, EPI_ISL_939434, EPI_ISL_939435,
EPI_ISL_939436, EPI_ISL_939437, EPI_ISL_939440, EPI_ISL_939441, EPI_ISL_939442, EPI_ISL_939444, EPI_ISL_939445, EPI_ISL_939446, EPI_ISL_939447, EPI_ISL_939448, EPI_ISL_939449, EPI_ISL_939451, EPI_ISL_939452, EPI_ISL_939453, EPI_ISL_939455, EPI_ISL_939457, EPI_ISL_939463, EPI_ISL_939464, EPI_ISL_939465, EPI_ISL_939467, EPI_ISL_939470, EPI_ISL_939471, EPI_ISL_939474, EPI_ISL_939475,
EPI_ISL_939476, EPI_ISL_939479, EPI_ISL_939480, EPI_ISL_939482, EPI_ISL_939484, EPI_ISL_939486, EPI_ISL_939488, EPI_ISL_939489, EPI_ISL_939490, EPI_ISL_939491, EPI_ISL_939495, EPI_ISL_939496, EPI_ISL_939499, EPI_ISL_939501, EPI_ISL_939502, EPI_ISL_939503, EPI_ISL_939511, EPI_ISL_939515, EPI_ISL_939516, EPI_ISL_939518, EPI_ISL_939525, EPI_ISL_939527, EPI_ISL_939529, EPI_ISL_939530,
EPI_ISL_939531, EPI_ISL_939534, EPI_ISL_939538, EPI_ISL_939539, EPI_ISL_939540, EPI_ISL_939543, EPI_ISL_939544, EPI_ISL_939545, EPI_ISL_939546, EPI_ISL_939549, EPI_ISL_939553, EPI_ISL_939557, EPI_ISL_939558, EPI_ISL_939560, EPI_ISL_939564, EPI_ISL_939568, EPI_ISL_939569, EPI_ISL_939570, EPI_ISL_939575, EPI_ISL_939580, EPI_ISL_939583, EPI_ISL_939584, EPI_ISL_939586, EPI_ISL_939587,
EPI_ISL_939589, EPI_ISL_939590, EPI_ISL_939591, EPI_ISL_939592, EPI_ISL_939593, EPI_ISL_939597, EPI_ISL_939602, EPI_ISL_944799, EPI_ISL_944805, EPI_ISL_944824, EPI_ISL_944863, EPI_ISL_944883, EPI_ISL_944908, EPI_ISL_944914, EPI_ISL_944923, EPI_ISL_944953, EPI_ISL_944976, EPI_ISL_944978, EPI_ISL_945023, EPI_ISL_945032, EPI_ISL_945034, EPI_ISL_945761, EPI_ISL_945858, EPI_ISL_945875,
EPI_ISL_945897, EPI_ISL_945923, EPI_ISL_945977, EPI_ISL_946003, EPI_ISL_946032, EPI_ISL_946047, EPI_ISL_946117, EPI_ISL_946219, EPI_ISL_946259, EPI_ISL_946289, EPI_ISL_946290, EPI_ISL_946303, EPI_ISL_946331, EPI_ISL_946373, EPI_ISL_946387, EPI_ISL_946395, EPI_ISL_946398, EPI_ISL_946408, EPI_ISL_946419, EPI_ISL_947107, EPI_ISL_947108, EPI_ISL_947118, EPI_ISL_947119, EPI_ISL_947121,
EPI_ISL_947126, EPI_ISL_947127, EPI_ISL_947131, EPI_ISL_947134, EPI_ISL_947137, EPI_ISL_947139, EPI_ISL_947141, EPI_ISL_947147, EPI_ISL_947150, EPI_ISL_947153, EPI_ISL_947163, EPI_ISL_947165, EPI_ISL_947169, EPI_ISL_947179, EPI_ISL_947182, EPI_ISL_947183, EPI_ISL_947186, EPI_ISL_947187, EPI_ISL_947191, EPI_ISL_947197, EPI_ISL_947202, EPI_ISL_947204, EPI_ISL_947211, EPI_ISL_947212,
EPI_ISL_947213, EPI_ISL_947214, EPI_ISL_947228, EPI_ISL_947229, EPI_ISL_947230, EPI_ISL_947233, EPI_ISL_947236, EPI_ISL_947238, EPI_ISL_947239, EPI_ISL_947245, EPI_ISL_947250, EPI_ISL_947253, EPI_ISL_947263, EPI_ISL_947267, EPI_ISL_947269, EPI_ISL_947271, EPI_ISL_947272, EPI_ISL_947280, EPI_ISL_947283, EPI_ISL_947294, EPI_ISL_947295, EPI_ISL_947305, EPI_ISL_947308, EPI_ISL_947314,
EPI_ISL_947316, EPI_ISL_947331, EPI_ISL_947332, EPI_ISL_947335, EPI_ISL_947338, EPI_ISL_947341, EPI_ISL_947343, EPI_ISL_947344, EPI_ISL_947347, EPI_ISL_947353, EPI_ISL_947356, EPI_ISL_947357, EPI_ISL_947360, EPI_ISL_947362, EPI_ISL_947363, EPI_ISL_947364, EPI_ISL_947365, EPI_ISL_947370, EPI_ISL_947377, EPI_ISL_947380, EPI_ISL_947383, EPI_ISL_947386, EPI_ISL_947389, EPI_ISL_947391,
EPI_ISL_947394, EPI_ISL_947396, EPI_ISL_947398, EPI_ISL_947399, EPI_ISL_947402, EPI_ISL_947404, EPI_ISL_947405, EPI_ISL_948100, EPI_ISL_948103, EPI_ISL_948105, EPI_ISL_948106, EPI_ISL_948108, EPI_ISL_948109, EPI_ISL_948111, EPI_ISL_948112, EPI_ISL_948113, EPI_ISL_948115, EPI_ISL_948119, EPI_ISL_948121, EPI_ISL_948126, EPI_ISL_948127, EPI_ISL_948129, EPI_ISL_948130, EPI_ISL_948132,
EPI_ISL_948138, EPI_ISL_948140, EPI_ISL_948141, EPI_ISL_948142, EPI_ISL_948143, EPI_ISL_948144, EPI_ISL_948146, EPI_ISL_948148, EPI_ISL_948152, EPI_ISL_948154, EPI_ISL_948156, EPI_ISL_948157, EPI_ISL_948158, EPI_ISL_948161, EPI_ISL_948162, EPI_ISL_948167, EPI_ISL_948168, EPI_ISL_948169, EPI_ISL_948170, EPI_ISL_948171, EPI_ISL_948173, EPI_ISL_948175, EPI_ISL_948179, EPI_ISL_948182,
EPI_ISL_948184, EPI_ISL_948190, EPI_ISL_948195, EPI_ISL_948196, EPI_ISL_948202, EPI_ISL_948206, EPI_ISL_948217, EPI_ISL_948218, EPI_ISL_948222, EPI_ISL_948225, EPI_ISL_948226, EPI_ISL_948229, EPI_ISL_948232, EPI_ISL_948236, EPI_ISL_948238, EPI_ISL_948242, EPI_ISL_948243, EPI_ISL_948245, EPI_ISL_948246, EPI_ISL_948248, EPI_ISL_948254, EPI_ISL_948256, EPI_ISL_948260, EPI_ISL_948262,
EPI_ISL_948263, EPI_ISL_948267, EPI_ISL_948268, EPI_ISL_948270, EPI_ISL_948271, EPI_ISL_948272, EPI_ISL_948273, EPI_ISL_948275, EPI_ISL_948276, EPI_ISL_948285, EPI_ISL_948287, EPI_ISL_948290, EPI_ISL_948291, EPI_ISL_948293, EPI_ISL_948295, EPI_ISL_948297, EPI_ISL_948299, EPI_ISL_948303, EPI_ISL_948304, EPI_ISL_948307, EPI_ISL_948309, EPI_ISL_948315, EPI_ISL_948317, EPI_ISL_948318,
EPI_ISL_948319, EPI_ISL_948320, EPI_ISL_948324, EPI_ISL_948325, EPI_ISL_948329, EPI_ISL_948333, EPI_ISL_948334, EPI_ISL_948335, EPI_ISL_948336, EPI_ISL_948339, EPI_ISL_948343, EPI_ISL_948347, EPI_ISL_948348, EPI_ISL_948349, EPI_ISL_948351, EPI_ISL_948352, EPI_ISL_948353, EPI_ISL_948354, EPI_ISL_948356, EPI_ISL_948357, EPI_ISL_948358, EPI_ISL_948360, EPI_ISL_948362, EPI_ISL_948363,
EPI_ISL_948365, EPI_ISL_948367, EPI_ISL_948369, EPI_ISL_948370, EPI_ISL_948371, EPI_ISL_948373, EPI_ISL_948375, EPI_ISL_948379, EPI_ISL_948384, EPI_ISL_948385, EPI_ISL_948386, EPI_ISL_948387, EPI_ISL_948389, EPI_ISL_948390, EPI_ISL_948394, EPI_ISL_948398, EPI_ISL_948399, EPI_ISL_948400, EPI_ISL_948401, EPI_ISL_948403, EPI_ISL_948404, EPI_ISL_948405, EPI_ISL_948406, EPI_ISL_948409,
EPI_ISL_948410, EPI_ISL_948411, EPI_ISL_948415, EPI_ISL_948416, EPI_ISL_948417, EPI_ISL_948418, EPI_ISL_948419, EPI_ISL_948425, EPI_ISL_948426, EPI_ISL_948427, EPI_ISL_948428, EPI_ISL_948429, EPI_ISL_948430, EPI_ISL_948436, EPI_ISL_948438, EPI_ISL_948439, EPI_ISL_948440, EPI_ISL_948443, EPI_ISL_948444, EPI_ISL_948446, EPI_ISL_948447, EPI_ISL_948448, EPI_ISL_948787, EPI_ISL_948788,
EPI_ISL_948789, EPI_ISL_948790, EPI_ISL_948791, EPI_ISL_948792, EPI_ISL_948793, EPI_ISL_948794, EPI_ISL_948795, EPI_ISL_948796, EPI_ISL_948797, EPI_ISL_948798, EPI_ISL_948799, EPI_ISL_948800, EPI_ISL_948801, EPI_ISL_948802, EPI_ISL_948803, EPI_ISL_948804, EPI_ISL_948805, EPI_ISL_948806, EPI_ISL_948807, EPI_ISL_948808, EPI_ISL_948809, EPI_ISL_948810, EPI_ISL_948811, EPI_ISL_948812,
EPI_ISL_948813, EPI_ISL_948814, EPI_ISL_948815, EPI_ISL_948816, EPI_ISL_948817, EPI_ISL_948818, EPI_ISL_948819, EPI_ISL_948820, EPI_ISL_948821, EPI_ISL_948822, EPI_ISL_948823, EPI_ISL_948824, EPI_ISL_948825, EPI_ISL_948826, EPI_ISL_948827, EPI_ISL_948828, EPI_ISL_948831, EPI_ISL_948832, EPI_ISL_948833, EPI_ISL_948835, EPI_ISL_948836, EPI_ISL_948837, EPI_ISL_948838, EPI_ISL_948839,
EPI_ISL_948840, EPI_ISL_948841, EPI_ISL_948842, EPI_ISL_948843, EPI_ISL_948844, EPI_ISL_948846, EPI_ISL_948847, EPI_ISL_948849, EPI_ISL_948850, EPI_ISL_948851, EPI_ISL_948852, EPI_ISL_948853, EPI_ISL_948854, EPI_ISL_948855, EPI_ISL_948856, EPI_ISL_948857, EPI_ISL_948858, EPI_ISL_948859, EPI_ISL_948860, EPI_ISL_948861, EPI_ISL_948862, EPI_ISL_948863, EPI_ISL_948864, EPI_ISL_948865,
EPI_ISL_948866, EPI_ISL_948867, EPI_ISL_948868, EPI_ISL_948869, EPI_ISL_948870, EPI_ISL_948871, EPI_ISL_948872, EPI_ISL_948873, EPI_ISL_948874, EPI_ISL_948875, EPI_ISL_948876, EPI_ISL_948877, EPI_ISL_948878, EPI_ISL_948879, EPI_ISL_948880, EPI_ISL_948881, EPI_ISL_948882, EPI_ISL_948883, EPI_ISL_948884, EPI_ISL_948885, EPI_ISL_948886, EPI_ISL_948887, EPI_ISL_948888, EPI_ISL_948889,
EPI_ISL_948890, EPI_ISL_948891, EPI_ISL_948892, EPI_ISL_948893, EPI_ISL_948894, EPI_ISL_948895, EPI_ISL_948896, EPI_ISL_948897, EPI_ISL_948898, EPI_ISL_948899, EPI_ISL_948900, EPI_ISL_948901, EPI_ISL_948902, EPI_ISL_948903, EPI_ISL_948904, EPI_ISL_948905, EPI_ISL_948906, EPI_ISL_948907, EPI_ISL_948908, EPI_ISL_948909, EPI_ISL_948910, EPI_ISL_948911, EPI_ISL_948912, EPI_ISL_948913,
EPI_ISL_948914, EPI_ISL_948915, EPI_ISL_948916, EPI_ISL_948917, EPI_ISL_948918, EPI_ISL_948919, EPI_ISL_948920, EPI_ISL_948921, EPI_ISL_948922, EPI_ISL_948923, EPI_ISL_948924, EPI_ISL_948925, EPI_ISL_948926, EPI_ISL_948927, EPI_ISL_948928, EPI_ISL_948929, EPI_ISL_948930, EPI_ISL_948931, EPI_ISL_948932, EPI_ISL_948933, EPI_ISL_948934, EPI_ISL_948935, EPI_ISL_948936, EPI_ISL_948937,
EPI_ISL_948938, EPI_ISL_948939, EPI_ISL_948940, EPI_ISL_948941, EPI_ISL_948942, EPI_ISL_948943, EPI_ISL_948944, EPI_ISL_990058, EPI_ISL_990059, EPI_ISL_990060, EPI_ISL_990061, EPI_ISL_990062, EPI_ISL_990063, EPI_ISL_990064, EPI_ISL_990065, EPI_ISL_990066, EPI_ISL_990067, EPI_ISL_990068, EPI_ISL_990069, EPI_ISL_990070, EPI_ISL_990071, EPI_ISL_990072, EPI_ISL_990073, EPI_ISL_990074,
EPI_ISL_990075, EPI_ISL_990076, EPI_ISL_990077, EPI_ISL_990078, EPI_ISL_990079, EPI_ISL_990080, EPI_ISL_990081, EPI_ISL_990082, EPI_ISL_990083, EPI_ISL_990084, EPI_ISL_990085, EPI_ISL_990086, EPI_ISL_990087, EPI_ISL_990088, EPI_ISL_990089, EPI_ISL_990090, EPI_ISL_990091, EPI_ISL_990092, EPI_ISL_990093, EPI_ISL_990094, EPI_ISL_990095, EPI_ISL_990096, EPI_ISL_990097, EPI_ISL_990098,
EPI_ISL_990099, EPI_ISL_990100, EPI_ISL_990101, EPI_ISL_990102, EPI_ISL_990103, EPI_ISL_990104, EPI_ISL_990105, EPI_ISL_990106, EPI_ISL_990107, EPI_ISL_990108, EPI_ISL_990109, EPI_ISL_990110, EPI_ISL_990111, EPI_ISL_990112, EPI_ISL_990113, EPI_ISL_990114, EPI_ISL_990115, EPI_ISL_990116, EPI_ISL_990117, EPI_ISL_990118, EPI_ISL_990119, EPI_ISL_990120, EPI_ISL_990121, EPI_ISL_990122,
EPI_ISL_990123, EPI_ISL_990124, EPI_ISL_990125, EPI_ISL_990126, EPI_ISL_990127, EPI_ISL_990128, EPI_ISL_990129, EPI_ISL_990130, EPI_ISL_990131, EPI_ISL_990132, EPI_ISL_990133, EPI_ISL_990134, EPI_ISL_990135, EPI_ISL_990136, EPI_ISL_990137, EPI_ISL_990138, EPI_ISL_990139, EPI_ISL_990140, EPI_ISL_991352, EPI_ISL_991353, EPI_ISL_991356, EPI_ISL_991357, EPI_ISL_991360, EPI_ISL_991362,
EPI_ISL_991363, EPI_ISL_991364, EPI_ISL_991365, EPI_ISL_991369, EPI_ISL_991372, EPI_ISL_991373, EPI_ISL_991374, EPI_ISL_991375, EPI_ISL_991376, EPI_ISL_991378, EPI_ISL_991384, EPI_ISL_991388, EPI_ISL_991390, EPI_ISL_991391, EPI_ISL_991393, EPI_ISL_991396, EPI_ISL_991401, EPI_ISL_991402, EPI_ISL_991404, EPI_ISL_991406, EPI_ISL_991407, EPI_ISL_991408, EPI_ISL_991410, EPI_ISL_991411,
EPI_ISL_991413, EPI_ISL_991414, EPI_ISL_991416, EPI_ISL_991420, EPI_ISL_991421, EPI_ISL_991424, EPI_ISL_991426, EPI_ISL_991427, EPI_ISL_991429, EPI_ISL_991430, EPI_ISL_991431, EPI_ISL_991433, EPI_ISL_991436, EPI_ISL_991438, EPI_ISL_991439, EPI_ISL_991441, EPI_ISL_991442, EPI_ISL_991444, EPI_ISL_991445, EPI_ISL_991449, EPI_ISL_991452, EPI_ISL_991456, EPI_ISL_991457, EPI_ISL_991459,
EPI_ISL_991461, EPI_ISL_991462, EPI_ISL_991466, EPI_ISL_991468, EPI_ISL_991469, EPI_ISL_991471, EPI_ISL_991472, EPI_ISL_991473, EPI_ISL_991474, EPI_ISL_991480, EPI_ISL_991484, EPI_ISL_991485, EPI_ISL_991487, EPI_ISL_991488, EPI_ISL_991490, EPI_ISL_991492, EPI_ISL_991495, EPI_ISL_991497, EPI_ISL_991504, EPI_ISL_991505, EPI_ISL_991507, EPI_ISL_991508, EPI_ISL_991509, EPI_ISL_991512,
EPI_ISL_991513, EPI_ISL_991514, EPI_ISL_991516, EPI_ISL_991518, EPI_ISL_991519, EPI_ISL_991520, EPI_ISL_991521, EPI_ISL_991522, EPI_ISL_991524, EPI_ISL_991526, EPI_ISL_991529, EPI_ISL_991535, EPI_ISL_991538, EPI_ISL_991539, EPI_ISL_991542, EPI_ISL_991545, EPI_ISL_991550, EPI_ISL_991552, EPI_ISL_991554, EPI_ISL_991555, EPI_ISL_991557, EPI_ISL_991559, EPI_ISL_991560, EPI_ISL_991562,
EPI_ISL_991563, EPI_ISL_991564, EPI_ISL_1007561, EPI_ISL_1007562, EPI_ISL_1019587, EPI_ISL_1019588, EPI_ISL_1019589, EPI_ISL_1019590, EPI_ISL_1019591, EPI_ISL_1019592, EPI_ISL_1019593, EPI_ISL_1019594, EPI_ISL_1019595, EPI_ISL_1019596, EPI_ISL_1019597, EPI_ISL_1019598, EPI_ISL_1019599, EPI_ISL_1019600, EPI_ISL_1019601, EPI_ISL_1019602, EPI_ISL_1019603, EPI_ISL_1019604,
EPI_ISL_1019605, EPI_ISL_1019606, EPI_ISL_1019607, EPI_ISL_1019609, EPI_ISL_1019610, EPI_ISL_1019611, EPI_ISL_1019612, EPI_ISL_1019613, EPI_ISL_1019614, EPI_ISL_1019616, EPI_ISL_1019617, EPI_ISL_1019618, EPI_ISL_1019619, EPI_ISL_1019620, EPI_ISL_1019621, EPI_ISL_1019622, EPI_ISL_1019623, EPI_ISL_1019624, EPI_ISL_1019625, EPI_ISL_1019626, EPI_ISL_1019627, EPI_ISL_1019629,
EPI_ISL_1019630, EPI_ISL_1019631, EPI_ISL_1019632, EPI_ISL_1019633, EPI_ISL_1019634, EPI_ISL_1019635, EPI_ISL_1019636, EPI_ISL_1019638, EPI_ISL_1019639, EPI_ISL_1019643, EPI_ISL_1019644, EPI_ISL_1019646, EPI_ISL_1019648, EPI_ISL_1019649, EPI_ISL_1019651, EPI_ISL_1019652, EPI_ISL_1019654, EPI_ISL_1019656, EPI_ISL_1019657, EPI_ISL_1019659, EPI_ISL_1019661, EPI_ISL_1019662,
EPI_ISL_1019664, EPI_ISL_1019669, EPI_ISL_1019670, EPI_ISL_1019672, EPI_ISL_1019673, EPI_ISL_1019675, EPI_ISL_1019677, EPI_ISL_1019679, EPI_ISL_1019680, EPI_ISL_1019682, EPI_ISL_1019684, EPI_ISL_1019685, EPI_ISL_1019687, EPI_ISL_1019689, EPI_ISL_1019692, EPI_ISL_1019694, EPI_ISL_1019695, EPI_ISL_1019697, EPI_ISL_1019698, EPI_ISL_1019700, EPI_ISL_1019701, EPI_ISL_1019703,
EPI_ISL_1019704, EPI_ISL_1019708, EPI_ISL_1019709, EPI_ISL_1019712, EPI_ISL_1019714, EPI_ISL_1019716, EPI_ISL_1019717, EPI_ISL_1019719, EPI_ISL_1019721, EPI_ISL_1019723, EPI_ISL_1019725, EPI_ISL_1019727, EPI_ISL_1019728, EPI_ISL_1019733, EPI_ISL_1019735, EPI_ISL_1019736, EPI_ISL_1019738, EPI_ISL_1019740, EPI_ISL_1019743, EPI_ISL_1019746, EPI_ISL_1019749, EPI_ISL_1019751,
EPI_ISL_1019752, EPI_ISL_1019755, EPI_ISL_1019757, EPI_ISL_1019758, EPI_ISL_1019760, EPI_ISL_1019761, EPI_ISL_1019763, EPI_ISL_1019765, EPI_ISL_1019766, EPI_ISL_1019768, EPI_ISL_1019769, EPI_ISL_1019771, EPI_ISL_1019773, EPI_ISL_1019774, EPI_ISL_1019776, EPI_ISL_1019777, EPI_ISL_1019779

| | | | |
|---|---|---|---|
| see above | Lighthouse Lab in Glasgow | Wellcome Sanger Institute for the COVID-19 Genomics UK (COG-UK) Consortium | Anna Dominiczak and Alex Alderton; Carol Clugston; Cordelia Langford; David Gray; David K. Jackson; Dominic Kwiatkowski; Ewan Harrison; Harper VanSteenhouse; Ian Johnston; Jeffrey Barrett; John Sillitoe on behalf of the Wellcome Sanger Institute COVID-19 Surveillance Team; Roberto Amato; Sonia Goncalves; Yumi Kasai |
| EPI_ISL_1050225, EPI_ISL_1050227 | Lighthouse Lab in Glasgow / MRC-University of Glasgow Centre for Virus Research | COVID-19 Genomics UK (COG-UK) Consortium | Alasdair MacLean; Alice Broos; Ana da Silva Filipe; Anna Dominiczak; Antonia Ho; Carol Clugston; Daniel Mair; David Gray; David L Robertson; Emma Thomson; Harper VanSteenhouse; James Shepherd; Jenna Nichols; Joseph Hughes; Kathy Li; Kathy Smollett; Kyriaki Nomikou; Lily Tong; Matthew Holden; Natasha Johnson; Richard Orton; Rory Gunson; Sarah McDonald; Sharif Shaaban; Sreenu Vattipally; Stephen Carmichael; Yumi Kasai |

EPI_ISL_938579, EPI_ISL_938580, EPI_ISL_938582, EPI_ISL_938583, EPI_ISL_938584, EPI_ISL_938585, EPI_ISL_938586, EPI_ISL_938588, EPI_ISL_938589, EPI_ISL_938590, EPI_ISL_938591, EPI_ISL_938593, EPI_ISL_938594, EPI_ISL_938595, EPI_ISL_938597, EPI_ISL_938598, EPI_ISL_938600, EPI_ISL_938601, EPI_ISL_938604, EPI_ISL_938605, EPI_ISL_938606, EPI_ISL_938608, EPI_ISL_938609, EPI_ISL_938611,
EPI_ISL_938615, EPI_ISL_938616, EPI_ISL_938617, EPI_ISL_938618, EPI_ISL_938620, EPI_ISL_938621, EPI_ISL_938622, EPI_ISL_938623, EPI_ISL_938624, EPI_ISL_938625, EPI_ISL_938626, EPI_ISL_938627, EPI_ISL_938628, EPI_ISL_938629, EPI_ISL_938634, EPI_ISL_938635, EPI_ISL_938638, EPI_ISL_938639, EPI_ISL_938640, EPI_ISL_938641, EPI_ISL_938642, EPI_ISL_938644, EPI_ISL_938646, EPI_ISL_938647,
EPI_ISL_938649, EPI_ISL_938651, EPI_ISL_938653, EPI_ISL_938654, EPI_ISL_938655, EPI_ISL_938656, EPI_ISL_938658, EPI_ISL_938659, EPI_ISL_938660, EPI_ISL_938661, EPI_ISL_938662, EPI_ISL_938663, EPI_ISL_938664, EPI_ISL_938665, EPI_ISL_938668, EPI_ISL_938669, EPI_ISL_938670, EPI_ISL_938671, EPI_ISL_938672, EPI_ISL_938673, EPI_ISL_938674, EPI_ISL_938675, EPI_ISL_938677, EPI_ISL_938678,
EPI_ISL_938679, EPI_ISL_938680, EPI_ISL_938681, EPI_ISL_938682, EPI_ISL_938683, EPI_ISL_938685, EPI_ISL_938686, EPI_ISL_938687, EPI_ISL_938688, EPI_ISL_938689, EPI_ISL_938690, EPI_ISL_938691, EPI_ISL_938693, EPI_ISL_938695, EPI_ISL_938696, EPI_ISL_938697, EPI_ISL_938698, EPI_ISL_938699, EPI_ISL_938702, EPI_ISL_938703, EPI_ISL_938704, EPI_ISL_938705, EPI_ISL_938706, EPI_ISL_938708,
EPI_ISL_938709, EPI_ISL_938710, EPI_ISL_938711, EPI_ISL_938712, EPI_ISL_938713, EPI_ISL_938714, EPI_ISL_938715, EPI_ISL_938716, EPI_ISL_938717, EPI_ISL_938718, EPI_ISL_938719, EPI_ISL_938720, EPI_ISL_938722, EPI_ISL_938723, EPI_ISL_938724, EPI_ISL_938725, EPI_ISL_938726, EPI_ISL_938727, EPI_ISL_938728, EPI_ISL_938730, EPI_ISL_938734, EPI_ISL_938735, EPI_ISL_938736, EPI_ISL_938738,
EPI_ISL_938739, EPI_ISL_938740, EPI_ISL_938742, EPI_ISL_938745, EPI_ISL_938748, EPI_ISL_938749, EPI_ISL_938754, EPI_ISL_938755, EPI_ISL_938759, EPI_ISL_938760, EPI_ISL_938761, EPI_ISL_938763, EPI_ISL_938765, EPI_ISL_938766, EPI_ISL_938767, EPI_ISL_938768, EPI_ISL_938770, EPI_ISL_938772, EPI_ISL_938773, EPI_ISL_938774, EPI_ISL_938775, EPI_ISL_938776, EPI_ISL_938778, EPI_ISL_938779,
EPI_ISL_938780, EPI_ISL_938781, EPI_ISL_938782, EPI_ISL_938784, EPI_ISL_938786, EPI_ISL_938788, EPI_ISL_938789, EPI_ISL_938790, EPI_ISL_938791, EPI_ISL_938792, EPI_ISL_938793, EPI_ISL_938795, EPI_ISL_938798, EPI_ISL_938799, EPI_ISL_938802, EPI_ISL_938805, EPI_ISL_938808, EPI_ISL_938809, EPI_ISL_938812, EPI_ISL_938813, EPI_ISL_938814, EPI_ISL_938816, EPI_ISL_938817, EPI_ISL_938818,
EPI_ISL_938819, EPI_ISL_938820, EPI_ISL_938821, EPI_ISL_938822, EPI_ISL_938823, EPI_ISL_938824, EPI_ISL_938825, EPI_ISL_938826, EPI_ISL_938827, EPI_ISL_938828, EPI_ISL_938830, EPI_ISL_938831, EPI_ISL_938832, EPI_ISL_938833, EPI_ISL_938834, EPI_ISL_938835, EPI_ISL_938836, EPI_ISL_938838, EPI_ISL_938839, EPI_ISL_938841, EPI_ISL_938842, EPI_ISL_938843, EPI_ISL_938844, EPI_ISL_938847,
EPI_ISL_938848, EPI_ISL_938849, EPI_ISL_938850, EPI_ISL_938851, EPI_ISL_938852, EPI_ISL_938853, EPI_ISL_938855, EPI_ISL_938856, EPI_ISL_938857, EPI_ISL_938858, EPI_ISL_938859, EPI_ISL_938860, EPI_ISL_938861, EPI_ISL_938862, EPI_ISL_938863, EPI_ISL_938864, EPI_ISL_938866, EPI_ISL_938867, EPI_ISL_938868, EPI_ISL_938869, EPI_ISL_938870, EPI_ISL_938871, EPI_ISL_938872, EPI_ISL_938873,
EPI_ISL_938874, EPI_ISL_938877, EPI_ISL_938878, EPI_ISL_938879, EPI_ISL_938880, EPI_ISL_938881, EPI_ISL_938883, EPI_ISL_938884, EPI_ISL_938887, EPI_ISL_938888, EPI_ISL_938889, EPI_ISL_938890, EPI_ISL_938891, EPI_ISL_938892, EPI_ISL_938893, EPI_ISL_938894, EPI_ISL_938895, EPI_ISL_938896, EPI_ISL_938897, EPI_ISL_938898, EPI_ISL_938899, EPI_ISL_938902, EPI_ISL_938904, EPI_ISL_938905,
EPI_ISL_938906, EPI_ISL_938907, EPI_ISL_938909, EPI_ISL_938910, EPI_ISL_938912, EPI_ISL_938914, EPI_ISL_938915, EPI_ISL_938917, EPI_ISL_938918, EPI_ISL_938919, EPI_ISL_938920, EPI_ISL_938922, EPI_ISL_938923, EPI_ISL_938924, EPI_ISL_938925, EPI_ISL_938926, EPI_ISL_938927, EPI_ISL_938928, EPI_ISL_938929, EPI_ISL_938930, EPI_ISL_938932, EPI_ISL_945715, EPI_ISL_945717, EPI_ISL_945719,
EPI_ISL_945720, EPI_ISL_945721, EPI_ISL_945723, EPI_ISL_945724, EPI_ISL_945727, EPI_ISL_945731, EPI_ISL_945734, EPI_ISL_945739, EPI_ISL_945740, EPI_ISL_945745, EPI_ISL_945746, EPI_ISL_945748, EPI_ISL_945749, EPI_ISL_945750, EPI_ISL_945751, EPI_ISL_945754, EPI_ISL_945756, EPI_ISL_945760, EPI_ISL_945764, EPI_ISL_945765, EPI_ISL_945766, EPI_ISL_945767, EPI_ISL_945769, EPI_ISL_945770,
EPI_ISL_945773, EPI_ISL_945774, EPI_ISL_945775, EPI_ISL_945778, EPI_ISL_945779, EPI_ISL_945780, EPI_ISL_945783, EPI_ISL_945784, EPI_ISL_945785, EPI_ISL_945786, EPI_ISL_945787, EPI_ISL_945789, EPI_ISL_945790, EPI_ISL_945791, EPI_ISL_945793, EPI_ISL_945794, EPI_ISL_945797, EPI_ISL_945798, EPI_ISL_945801, EPI_ISL_945805, EPI_ISL_945810, EPI_ISL_945811, EPI_ISL_945814, EPI_ISL_945819,
EPI_ISL_945820, EPI_ISL_945823, EPI_ISL_945825, EPI_ISL_945827, EPI_ISL_945829, EPI_ISL_945830, EPI_ISL_945832, EPI_ISL_945833, EPI_ISL_945834, EPI_ISL_945838, EPI_ISL_945840, EPI_ISL_945842, EPI_ISL_945845, EPI_ISL_945847, EPI_ISL_945852, EPI_ISL_945854, EPI_ISL_945855, EPI_ISL_945856, EPI_ISL_945857, EPI_ISL_945859, EPI_ISL_945861, EPI_ISL_945864, EPI_ISL_945866, EPI_ISL_945876,
EPI_ISL_945880, EPI_ISL_945881, EPI_ISL_945883, EPI_ISL_945886, EPI_ISL_945887, EPI_ISL_945888, EPI_ISL_945890, EPI_ISL_945891, EPI_ISL_945892, EPI_ISL_945893, EPI_ISL_945894, EPI_ISL_945896, EPI_ISL_945902, EPI_ISL_945903, EPI_ISL_945905, EPI_ISL_945912, EPI_ISL_945913, EPI_ISL_945915, EPI_ISL_945918, EPI_ISL_945919, EPI_ISL_945922, EPI_ISL_945924, EPI_ISL_945925, EPI_ISL_945927,
EPI_ISL_945928, EPI_ISL_945929, EPI_ISL_945930, EPI_ISL_945931, EPI_ISL_945932, EPI_ISL_945934, EPI_ISL_945936, EPI_ISL_945937, EPI_ISL_945941, EPI_ISL_945942, EPI_ISL_945943, EPI_ISL_945946, EPI_ISL_945947, EPI_ISL_945948, EPI_ISL_945949, EPI_ISL_945952, EPI_ISL_945953, EPI_ISL_945955, EPI_ISL_945957, EPI_ISL_945959, EPI_ISL_945964, EPI_ISL_945966, EPI_ISL_945968, EPI_ISL_945973,
EPI_ISL_945974, EPI_ISL_945975, EPI_ISL_945978, EPI_ISL_945981, EPI_ISL_945982, EPI_ISL_945985, EPI_ISL_945986, EPI_ISL_945987, EPI_ISL_945989, EPI_ISL_945991, EPI_ISL_945992, EPI_ISL_945995, EPI_ISL_945996, EPI_ISL_946001, EPI_ISL_946004, EPI_ISL_946012, EPI_ISL_946013, EPI_ISL_946015, EPI_ISL_946016, EPI_ISL_946017, EPI_ISL_946018, EPI_ISL_946019, EPI_ISL_946020, EPI_ISL_946022,
EPI_ISL_946024, EPI_ISL_946025, EPI_ISL_946027, EPI_ISL_946029, EPI_ISL_946031, EPI_ISL_946033, EPI_ISL_946034, EPI_ISL_946036, EPI_ISL_946038, EPI_ISL_946039, EPI_ISL_946040, EPI_ISL_946042, EPI_ISL_946043, EPI_ISL_946044, EPI_ISL_946045, EPI_ISL_946046, EPI_ISL_946050, EPI_ISL_946051, EPI_ISL_946053, EPI_ISL_946054, EPI_ISL_946056, EPI_ISL_946060, EPI_ISL_946061, EPI_ISL_946063,
EPI_ISL_946064, EPI_ISL_946065, EPI_ISL_946074, EPI_ISL_946075, EPI_ISL_946077, EPI_ISL_946078, EPI_ISL_946079, EPI_ISL_946081, EPI_ISL_946082, EPI_ISL_946083, EPI_ISL_946084, EPI_ISL_946085, EPI_ISL_946089, EPI_ISL_946091, EPI_ISL_946092, EPI_ISL_946095, EPI_ISL_946097, EPI_ISL_946098, EPI_ISL_946099, EPI_ISL_946100, EPI_ISL_946101, EPI_ISL_946102, EPI_ISL_946103, EPI_ISL_946104,
EPI_ISL_946105, EPI_ISL_946107, EPI_ISL_946108, EPI_ISL_946111, EPI_ISL_946114, EPI_ISL_946116, EPI_ISL_946118, EPI_ISL_946119, EPI_ISL_946120, EPI_ISL_946121, EPI_ISL_946122, EPI_ISL_946125, EPI_ISL_946128, EPI_ISL_946130, EPI_ISL_946131, EPI_ISL_946132, EPI_ISL_946133, EPI_ISL_946134, EPI_ISL_946136, EPI_ISL_946138, EPI_ISL_946139, EPI_ISL_946140, EPI_ISL_946141, EPI_ISL_946142,
EPI_ISL_946143, EPI_ISL_946144, EPI_ISL_946146, EPI_ISL_946148, EPI_ISL_946151, EPI_ISL_946152, EPI_ISL_946154, EPI_ISL_946155, EPI_ISL_946156, EPI_ISL_946158, EPI_ISL_946159, EPI_ISL_946161, EPI_ISL_946163, EPI_ISL_946164, EPI_ISL_946165, EPI_ISL_946166, EPI_ISL_946167, EPI_ISL_946168, EPI_ISL_946170, EPI_ISL_946171, EPI_ISL_946172, EPI_ISL_946175, EPI_ISL_946178, EPI_ISL_946179,

EPI_ISL_946183, EPI_ISL_946184, EPI_ISL_946185, EPI_ISL_946186, EPI_ISL_946187, EPI_ISL_946189, EPI_ISL_946191, EPI_ISL_946193, EPI_ISL_946194, EPI_ISL_946195, EPI_ISL_946197, EPI_ISL_946198, EPI_ISL_946200, EPI_ISL_946201, EPI_ISL_946203, EPI_ISL_946204, EPI_ISL_946205, EPI_ISL_946207, EPI_ISL_946209, EPI_ISL_946212, EPI_ISL_946214, EPI_ISL_946216, EPI_ISL_946217, EPI_ISL_946222, EPI_ISL_946223, EPI_ISL_946226, EPI_ISL_946228, EPI_ISL_946230, EPI_ISL_946235, EPI_ISL_946236, EPI_ISL_946237, EPI_ISL_946238, EPI_ISL_946239, EPI_ISL_946240, EPI_ISL_946241, EPI_ISL_946242, EPI_ISL_946244, EPI_ISL_946245, EPI_ISL_946246, EPI_ISL_946247, EPI_ISL_946248, EPI_ISL_946249, EPI_ISL_946251, EPI_ISL_946252, EPI_ISL_946255, EPI_ISL_946256, EPI_ISL_946257, EPI_ISL_946258, EPI_ISL_946261, EPI_ISL_946262, EPI_ISL_946263, EPI_ISL_946265, EPI_ISL_946266, EPI_ISL_946267, EPI_ISL_946268, EPI_ISL_946272, EPI_ISL_946273, EPI_ISL_946275, EPI_ISL_946276, EPI_ISL_946277, EPI_ISL_946278, EPI_ISL_946279, EPI_ISL_946280, EPI_ISL_946282, EPI_ISL_946283, EPI_ISL_946287, EPI_ISL_946288, EPI_ISL_946291, EPI_ISL_946293, EPI_ISL_946294, EPI_ISL_946295, EPI_ISL_946296, EPI_ISL_946297, EPI_ISL_946299, EPI_ISL_946300, EPI_ISL_946305, EPI_ISL_946306, EPI_ISL_946307, EPI_ISL_946308, EPI_ISL_946309, EPI_ISL_946310, EPI_ISL_946311, EPI_ISL_946312, EPI_ISL_946313, EPI_ISL_946314, EPI_ISL_946315, EPI_ISL_946316, EPI_ISL_946317, EPI_ISL_946320, EPI_ISL_946321, EPI_ISL_946322, EPI_ISL_946324, EPI_ISL_946325, EPI_ISL_946327, EPI_ISL_946329, EPI_ISL_946330, EPI_ISL_946332, EPI_ISL_946334, EPI_ISL_946336, EPI_ISL_946337, EPI_ISL_946340, EPI_ISL_946341, EPI_ISL_946342, EPI_ISL_946343, EPI_ISL_946344, EPI_ISL_946345, EPI_ISL_946346, EPI_ISL_946347, EPI_ISL_946348, EPI_ISL_946349, EPI_ISL_946350, EPI_ISL_946352, EPI_ISL_946353, EPI_ISL_946354, EPI_ISL_946355, EPI_ISL_946358, EPI_ISL_946359, EPI_ISL_946362, EPI_ISL_946364, EPI_ISL_946368, EPI_ISL_946369, EPI_ISL_946370, EPI_ISL_946372, EPI_ISL_946376, EPI_ISL_946377, EPI_ISL_946382, EPI_ISL_946383, EPI_ISL_946384, EPI_ISL_946386, EPI_ISL_946388, EPI_ISL_946389, EPI_ISL_946390, EPI_ISL_946392, EPI_ISL_946393, EPI_ISL_946394, EPI_ISL_946396, EPI_ISL_946399, EPI_ISL_946401, EPI_ISL_946402, EPI_ISL_946404, EPI_ISL_946406, EPI_ISL_946409, EPI_ISL_946412, EPI_ISL_946413, EPI_ISL_946414, EPI_ISL_946415, EPI_ISL_946416, EPI_ISL_946417, EPI_ISL_946418, EPI_ISL_946420, EPI_ISL_947109, EPI_ISL_947111, EPI_ISL_947112, EPI_ISL_947114, EPI_ISL_947115, EPI_ISL_947116, EPI_ISL_947117, EPI_ISL_947122, EPI_ISL_947123, EPI_ISL_947124, EPI_ISL_947125, EPI_ISL_947128, EPI_ISL_947129, EPI_ISL_947130, EPI_ISL_947132, EPI_ISL_947133, EPI_ISL_947135, EPI_ISL_947136, EPI_ISL_947138, EPI_ISL_947142, EPI_ISL_947143, EPI_ISL_947144, EPI_ISL_947145, EPI_ISL_947146, EPI_ISL_947148, EPI_ISL_947151, EPI_ISL_947152, EPI_ISL_947154, EPI_ISL_947155, EPI_ISL_947156, EPI_ISL_947157, EPI_ISL_947158, EPI_ISL_947159, EPI_ISL_947160, EPI_ISL_947161, EPI_ISL_947162, EPI_ISL_947164, EPI_ISL_947166, EPI_ISL_947167, EPI_ISL_947168, EPI_ISL_947170, EPI_ISL_947171, EPI_ISL_947173, EPI_ISL_947174, EPI_ISL_947175, EPI_ISL_947176, EPI_ISL_947177, EPI_ISL_947178, EPI_ISL_947180, EPI_ISL_947181, EPI_ISL_947185, EPI_ISL_947188, EPI_ISL_947190, EPI_ISL_947192, EPI_ISL_947193, EPI_ISL_947194, EPI_ISL_947195, EPI_ISL_947196, EPI_ISL_947198, EPI_ISL_947199, EPI_ISL_947200, EPI_ISL_947203, EPI_ISL_947205, EPI_ISL_947206, EPI_ISL_947207, EPI_ISL_947209, EPI_ISL_947210, EPI_ISL_947215, EPI_ISL_947216, EPI_ISL_947217, EPI_ISL_947218, EPI_ISL_947219, EPI_ISL_947220, EPI_ISL_947221, EPI_ISL_947222, EPI_ISL_947223, EPI_ISL_947224, EPI_ISL_947225, EPI_ISL_947226, EPI_ISL_947227, EPI_ISL_947231, EPI_ISL_947232, EPI_ISL_947234, EPI_ISL_947235, EPI_ISL_947237, EPI_ISL_947241, EPI_ISL_947242, EPI_ISL_947243, EPI_ISL_947244, EPI_ISL_947246, EPI_ISL_947248, EPI_ISL_947251, EPI_ISL_947254, EPI_ISL_947258, EPI_ISL_947259, EPI_ISL_947264, EPI_ISL_947266, EPI_ISL_947276, EPI_ISL_947277, EPI_ISL_947279, EPI_ISL_947282, EPI_ISL_947285, EPI_ISL_947286, EPI_ISL_947288, EPI_ISL_947289, EPI_ISL_947291, EPI_ISL_947292, EPI_ISL_947297, EPI_ISL_947299, EPI_ISL_947300, EPI_ISL_947302, EPI_ISL_947303, EPI_ISL_947306, EPI_ISL_947310, EPI_ISL_947311, EPI_ISL_947313, EPI_ISL_947319, EPI_ISL_947321, EPI_ISL_947333, EPI_ISL_947334, EPI_ISL_947336, EPI_ISL_947339, EPI_ISL_947340, EPI_ISL_947342, EPI_ISL_947345, EPI_ISL_947346, EPI_ISL_947348, EPI_ISL_947349, EPI_ISL_947351, EPI_ISL_947355, EPI_ISL_947358, EPI_ISL_947359, EPI_ISL_947366, EPI_ISL_947367, EPI_ISL_947369, EPI_ISL_947371, EPI_ISL_947372, EPI_ISL_947373, EPI_ISL_947375, EPI_ISL_947376, EPI_ISL_947378, EPI_ISL_947379, EPI_ISL_947381, EPI_ISL_947382, EPI_ISL_947384, EPI_ISL_947385, EPI_ISL_947387, EPI_ISL_947388, EPI_ISL_947390, EPI_ISL_947392, EPI_ISL_947393, EPI_ISL_947395, EPI_ISL_947397, EPI_ISL_947401, EPI_ISL_947403, EPI_ISL_948135, EPI_ISL_948252, EPI_ISL_948327, EPI_ISL_957613, EPI_ISL_957616, EPI_ISL_957620, EPI_ISL_957621, EPI_ISL_957624, EPI_ISL_957627, EPI_ISL_957630, EPI_ISL_957635, EPI_ISL_957641, EPI_ISL_957643, EPI_ISL_957645, EPI_ISL_957650, EPI_ISL_957652, EPI_ISL_957654, EPI_ISL_957655, EPI_ISL_957656, EPI_ISL_957660, EPI_ISL_957670, EPI_ISL_957672, EPI_ISL_957675, EPI_ISL_957677, EPI_ISL_957680, EPI_ISL_957683, EPI_ISL_957686, EPI_ISL_957687, EPI_ISL_957691, EPI_ISL_957698, EPI_ISL_957700, EPI_ISL_957701, EPI_ISL_957707, EPI_ISL_957721, EPI_ISL_957722, EPI_ISL_957723, EPI_ISL_957726, EPI_ISL_957735, EPI_ISL_957740, EPI_ISL_957741, EPI_ISL_957745, EPI_ISL_957747, EPI_ISL_957748, EPI_ISL_957753, EPI_ISL_957755, EPI_ISL_957757, EPI_ISL_957759, EPI_ISL_957766, EPI_ISL_957769, EPI_ISL_957771, EPI_ISL_957772, EPI_ISL_957780, EPI_ISL_957783, EPI_ISL_957787, EPI_ISL_957797, EPI_ISL_957799, EPI_ISL_957813, EPI_ISL_957817, EPI_ISL_957822, EPI_ISL_957826, EPI_ISL_957828, EPI_ISL_957829, EPI_ISL_957830, EPI_ISL_957832, EPI_ISL_957838, EPI_ISL_957841, EPI_ISL_957844, EPI_ISL_957856, EPI_ISL_957859, EPI_ISL_957862, EPI_ISL_957863, EPI_ISL_957866, EPI_ISL_957867, EPI_ISL_957868, EPI_ISL_957872, EPI_ISL_957877, EPI_ISL_957878, EPI_ISL_957886, EPI_ISL_957892, EPI_ISL_957895, EPI_ISL_957897, EPI_ISL_957904, EPI_ISL_957907, EPI_ISL_957921, EPI_ISL_957961, EPI_ISL_957962, EPI_ISL_957971, EPI_ISL_958044, EPI_ISL_958194, EPI_ISL_958206, EPI_ISL_958215, EPI_ISL_958216, EPI_ISL_958217, EPI_ISL_958218, EPI_ISL_958219, EPI_ISL_958220, EPI_ISL_958221, EPI_ISL_958222, EPI_ISL_958223, EPI_ISL_958224, EPI_ISL_958225, EPI_ISL_958226, EPI_ISL_958227, EPI_ISL_958228, EPI_ISL_958229, EPI_ISL_958230, EPI_ISL_958231, EPI_ISL_958232, EPI_ISL_958233, EPI_ISL_958234, EPI_ISL_958235, EPI_ISL_958236, EPI_ISL_958237, EPI_ISL_958238, EPI_ISL_958239, EPI_ISL_958240, EPI_ISL_958241, EPI_ISL_958242, EPI_ISL_958243, EPI_ISL_958244, EPI_ISL_958245, EPI_ISL_958246, EPI_ISL_958247, EPI_ISL_958248, EPI_ISL_958249, EPI_ISL_958250, EPI_ISL_958251, EPI_ISL_958252, EPI_ISL_958253, EPI_ISL_958254, EPI_ISL_958255, EPI_ISL_958256, EPI_ISL_958257, EPI_ISL_958258, EPI_ISL_958259, EPI_ISL_958260, EPI_ISL_958261, EPI_ISL_958262, EPI_ISL_958263, EPI_ISL_958264, EPI_ISL_958265, EPI_ISL_958266, EPI_ISL_958267, EPI_ISL_958268, EPI_ISL_958269, EPI_ISL_958270, EPI_ISL_958271, EPI_ISL_958272, EPI_ISL_958273, EPI_ISL_958274, EPI_ISL_958275, EPI_ISL_958277, EPI_ISL_958278, EPI_ISL_958279, EPI_ISL_958280, EPI_ISL_958281, EPI_ISL_958282, EPI_ISL_958283, EPI_ISL_958284, EPI_ISL_958285, EPI_ISL_958286, EPI_ISL_958287, EPI_ISL_958288, EPI_ISL_958289, EPI_ISL_958290, EPI_ISL_958291, EPI_ISL_958292, EPI_ISL_958293, EPI_ISL_958294, EPI_ISL_958295, EPI_ISL_958296, EPI_ISL_958297, EPI_ISL_958298, EPI_ISL_958299, EPI_ISL_958300, EPI_ISL_958301, EPI_ISL_958302, EPI_ISL_958303, EPI_ISL_958304, EPI_ISL_958305, EPI_ISL_958306, EPI_ISL_958307, EPI_ISL_958308, EPI_ISL_958309, EPI_ISL_958310, EPI_ISL_958311, EPI_ISL_958312, EPI_ISL_958313, EPI_ISL_958314, EPI_ISL_958315, EPI_ISL_958316, EPI_ISL_958317, EPI_ISL_958318, EPI_ISL_958319, EPI_ISL_958320, EPI_ISL_958321, EPI_ISL_958322, EPI_ISL_958323, EPI_ISL_958324, EPI_ISL_958325, EPI_ISL_958326, EPI_ISL_958327, EPI_ISL_958328, EPI_ISL_958329, EPI_ISL_958330, EPI_ISL_958331, EPI_ISL_958332, EPI_ISL_958333, EPI_ISL_958334, EPI_ISL_958335, EPI_ISL_958336, EPI_ISL_958337, EPI_ISL_958338, EPI_ISL_958339, EPI_ISL_958340, EPI_ISL_958341, EPI_ISL_958342, EPI_ISL_958343, EPI_ISL_958344, EPI_ISL_958345, EPI_ISL_958346, EPI_ISL_958347, EPI_ISL_958348, EPI_ISL_958349, EPI_ISL_958350, EPI_ISL_958351, EPI_ISL_958352, EPI_ISL_958353, EPI_ISL_958354, EPI_ISL_958355, EPI_ISL_958356, EPI_ISL_958357, EPI_ISL_958358, EPI_ISL_958359, EPI_ISL_958360, EPI_ISL_958361, EPI_ISL_958362, EPI_ISL_958363, EPI_ISL_958364, EPI_ISL_958365, EPI_ISL_958366, EPI_ISL_958367, EPI_ISL_958368, EPI_ISL_958369, EPI_ISL_958370, EPI_ISL_958371, EPI_ISL_958372, EPI_ISL_958373, EPI_ISL_958374, EPI_ISL_958375, EPI_ISL_958376, EPI_ISL_958377, EPI_ISL_958378, EPI_ISL_958379, EPI_ISL_958380, EPI_ISL_958381, EPI_ISL_958382, EPI_ISL_958383, EPI_ISL_958384, EPI_ISL_958385, EPI_ISL_958386, EPI_ISL_958387, EPI_ISL_958388, EPI_ISL_958389, EPI_ISL_958390, EPI_ISL_958391, EPI_ISL_958392, EPI_ISL_958393, EPI_ISL_958394, EPI_ISL_958395, EPI_ISL_958396, EPI_ISL_958397, EPI_ISL_958399, EPI_ISL_958400, EPI_ISL_958401, EPI_ISL_958402, EPI_ISL_958403, EPI_ISL_958404, EPI_ISL_958405, EPI_ISL_958406, EPI_ISL_958407, EPI_ISL_958408, EPI_ISL_958409, EPI_ISL_958410, EPI_ISL_958411, EPI_ISL_958412, EPI_ISL_958413, EPI_ISL_958414, EPI_ISL_958415, EPI_ISL_958416, EPI_ISL_958417, EPI_ISL_958418, EPI_ISL_958419, EPI_ISL_958420, EPI_ISL_958421, EPI_ISL_958422, EPI_ISL_958423, EPI_ISL_958424, EPI_ISL_958425, EPI_ISL_958426, EPI_ISL_958427, EPI_ISL_958428, EPI_ISL_958429, EPI_ISL_958430, EPI_ISL_958431, EPI_ISL_958432, EPI_ISL_958433, EPI_ISL_958434, EPI_ISL_958435, EPI_ISL_958436, EPI_ISL_958437, EPI_ISL_958438, EPI_ISL_958439, EPI_ISL_958440, EPI_ISL_958441, EPI_ISL_958442, EPI_ISL_958443, EPI_ISL_958444, EPI_ISL_958445, EPI_ISL_958446, EPI_ISL_958447, EPI_ISL_958448, EPI_ISL_958449, EPI_ISL_958450, EPI_ISL_958451, EPI_ISL_958452, EPI_ISL_958453, EPI_ISL_958454, EPI_ISL_958455, EPI_ISL_958456, EPI_ISL_958457, EPI_ISL_958458, EPI_ISL_958459, EPI_ISL_958460, EPI_ISL_958461, EPI_ISL_958462, EPI_ISL_958463, EPI_ISL_958464, EPI_ISL_958465, EPI_ISL_958466, EPI_ISL_958467, EPI_ISL_958468, EPI_ISL_958469, EPI_ISL_958470, EPI_ISL_958471, EPI_ISL_958472, EPI_ISL_958473, EPI_ISL_958474, EPI_ISL_958475, EPI_ISL_958476, EPI_ISL_958477, EPI_ISL_958478, EPI_ISL_958479, EPI_ISL_958480, EPI_ISL_958481, EPI_ISL_958482, EPI_ISL_958483, EPI_ISL_958484, EPI_ISL_958485, EPI_ISL_958486, EPI_ISL_958487, EPI_ISL_958488, EPI_ISL_958489, EPI_ISL_958490, EPI_ISL_958491, EPI_ISL_958492, EPI_ISL_958493, EPI_ISL_958494, EPI_ISL_958495, EPI_ISL_958496, EPI_ISL_958497, EPI_ISL_958498, EPI_ISL_958499, EPI_ISL_958500, EPI_ISL_958501, EPI_ISL_958502, EPI_ISL_958503, EPI_ISL_958504, EPI_ISL_958505, EPI_ISL_958506, EPI_ISL_958507, EPI_ISL_958508, EPI_ISL_958509, EPI_ISL_958510, EPI_ISL_958511, EPI_ISL_958512, EPI_ISL_958513, EPI_ISL_958514, EPI_ISL_958515, EPI_ISL_958516, EPI_ISL_958517, EPI_ISL_958518, EPI_ISL_958519, EPI_ISL_958520, EPI_ISL_958521, EPI_ISL_958522, EPI_ISL_958523, EPI_ISL_958524, EPI_ISL_958525, EPI_ISL_958526, EPI_ISL_958527, EPI_ISL_958528, EPI_ISL_958529, EPI_ISL_958530, EPI_ISL_958531, EPI_ISL_958532, EPI_ISL_958533, EPI_ISL_958534, EPI_ISL_958535, EPI_ISL_958536, EPI_ISL_958537, EPI_ISL_958538, EPI_ISL_958539, EPI_ISL_958540, EPI_ISL_958541, EPI_ISL_958542, EPI_ISL_958543, EPI_ISL_958544, EPI_ISL_958545, EPI_ISL_958546, EPI_ISL_958547, EPI_ISL_958548, EPI_ISL_958549, EPI_ISL_958550, EPI_ISL_958551, EPI_ISL_958552, EPI_ISL_958553, EPI_ISL_958554, EPI_ISL_958555, EPI_ISL_958556, EPI_ISL_958557, EPI_ISL_958558, EPI_ISL_958559, EPI_ISL_958560, EPI_ISL_958561, EPI_ISL_958562, EPI_ISL_958563, EPI_ISL_958569, EPI_ISL_958572, EPI_ISL_958578, EPI_ISL_958579, EPI_ISL_958586, EPI_ISL_958587, EPI_ISL_958590, EPI_ISL_958591, EPI_ISL_958595, EPI_ISL_958600, EPI_ISL_958602, EPI_ISL_958610, EPI_ISL_958624, EPI_ISL_958632, EPI_ISL_958633, EPI_ISL_958634, EPI_ISL_958636, EPI_ISL_958638, EPI_ISL_958639, EPI_ISL_958641, EPI_ISL_958646, EPI_ISL_958650, EPI_ISL_958659, EPI_ISL_958662, EPI_ISL_958663, EPI_ISL_958670, EPI_ISL_958671, EPI_ISL_958673, EPI_ISL_958680, EPI_ISL_958684, EPI_ISL_958690, EPI_ISL_958691, EPI_ISL_958696, EPI_ISL_958705, EPI_ISL_958706, EPI_ISL_958713, EPI_ISL_958715, EPI_ISL_958716, EPI_ISL_958723, EPI_ISL_958725, EPI_ISL_958726, EPI_ISL_958731, EPI_ISL_958733, EPI_ISL_958735, EPI_ISL_958745, EPI_ISL_958747, EPI_ISL_958748, EPI_ISL_958754, EPI_ISL_958756, EPI_ISL_958760, EPI_ISL_958763, EPI_ISL_958767, EPI_ISL_958769, EPI_ISL_958775, EPI_ISL_958779, EPI_ISL_958784, EPI_ISL_958787, EPI_ISL_958789, EPI_ISL_958791, EPI_ISL_958792, EPI_ISL_958793, EPI_ISL_958803, EPI_ISL_958807, EPI_ISL_958808, EPI_ISL_958811, EPI_ISL_958815, EPI_ISL_958823, EPI_ISL_958826, EPI_ISL_958828, EPI_ISL_958833, EPI_ISL_958836, EPI_ISL_958840, EPI_ISL_958842, EPI_ISL_958849, EPI_ISL_958851, EPI_ISL_958863, EPI_ISL_958866, EPI_ISL_958873, EPI_ISL_958878, EPI_ISL_958884, EPI_ISL_958885, EPI_ISL_958887, EPI_ISL_958889, EPI_ISL_958891, EPI_ISL_958893, EPI_ISL_958895, EPI_ISL_958898, EPI_ISL_958902, EPI_ISL_958903, EPI_ISL_958904, EPI_ISL_958908, EPI_ISL_958911, EPI_ISL_958912, EPI_ISL_958915, EPI_ISL_958916, EPI_ISL_959271, EPI_ISL_959272, EPI_ISL_963325, EPI_ISL_963408, EPI_ISL_963471, EPI_ISL_963524, EPI_ISL_963555, EPI_ISL_970589, EPI_ISL_970591, EPI_ISL_970593, EPI_ISL_970595, EPI_ISL_970599, EPI_ISL_970601, EPI_ISL_970602, EPI_ISL_970605, EPI_ISL_970607, EPI_ISL_970609, EPI_ISL_970612, EPI_ISL_970614, EPI_ISL_970618, EPI_ISL_970619, EPI_ISL_970623, EPI_ISL_970625, EPI_ISL_970630, EPI_ISL_970632, EPI_ISL_970633, EPI_ISL_970637, EPI_ISL_970641, EPI_ISL_970643, EPI_ISL_970648, EPI_ISL_970650, EPI_ISL_970652, EPI_ISL_970654, EPI_ISL_970656, EPI_ISL_970658, EPI_ISL_970660, EPI_ISL_970661, EPI_ISL_970666, EPI_ISL_970668, EPI_ISL_970670, EPI_ISL_970675, EPI_ISL_970677, EPI_ISL_970683, EPI_ISL_970685, EPI_ISL_970689, EPI_ISL_970690, EPI_ISL_970692, EPI_ISL_970694, EPI_ISL_970697, EPI_ISL_970699, EPI_ISL_970703, EPI_ISL_970704, EPI_ISL_970706, EPI_ISL_970711, EPI_ISL_970713, EPI_ISL_970717, EPI_ISL_970720, EPI_ISL_970722, EPI_ISL_970726, EPI_ISL_970727, EPI_ISL_970729, EPI_ISL_970734, EPI_ISL_970736, EPI_ISL_970738, EPI_ISL_970739, EPI_ISL_970741, EPI_ISL_970746, EPI_ISL_970749, EPI_ISL_970755, EPI_ISL_970757, EPI_ISL_970758, EPI_ISL_970760, EPI_ISL_970762, EPI_ISL_970764, EPI_ISL_970766, EPI_ISL_970770, EPI_ISL_970772, EPI_ISL_970774, EPI_ISL_970777, EPI_ISL_970779, EPI_ISL_970781, EPI_ISL_970782, EPI_ISL_970784, EPI_ISL_970786, EPI_ISL_970788, EPI_ISL_970792, EPI_ISL_970794, EPI_ISL_970796, EPI_ISL_970797, EPI_ISL_970800, EPI_ISL_970802, EPI_ISL_970803, EPI_ISL_970805, EPI_ISL_970807, EPI_ISL_970808, EPI_ISL_970845, EPI_ISL_970899, EPI_ISL_970925, EPI_ISL_970944, EPI_ISL_970968, EPI_ISL_971008, EPI_ISL_971023, EPI_ISL_971039, EPI_ISL_971050, EPI_ISL_971055, EPI_ISL_971059, EPI_ISL_971063, EPI_ISL_971066, EPI_ISL_971069, EPI_ISL_971074, EPI_ISL_971079, EPI_ISL_971083, EPI_ISL_971087, EPI_ISL_971090, EPI_ISL_971099, EPI_ISL_971102, EPI_ISL_971108, EPI_ISL_971109, EPI_ISL_971113, EPI_ISL_971117, EPI_ISL_971119, EPI_ISL_971123, EPI_ISL_971128, EPI_ISL_971130, EPI_ISL_971134, EPI_ISL_971138, EPI_ISL_971146, EPI_ISL_971149, EPI_ISL_971153, EPI_ISL_971156, EPI_ISL_971161, EPI_ISL_971164, EPI_ISL_971165, EPI_ISL_971180, EPI_ISL_971185, EPI_ISL_971190, EPI_ISL_971197, EPI_ISL_971200, EPI_ISL_971204, EPI_ISL_971209, EPI_ISL_971211, EPI_ISL_971215, EPI_ISL_971218, EPI_ISL_971221, EPI_ISL_971224, EPI_ISL_971228, EPI_ISL_971234, EPI_ISL_971236, EPI_ISL_971246, EPI_ISL_971248, EPI_ISL_971251, EPI_ISL_971254, EPI_ISL_971258, EPI_ISL_971264, EPI_ISL_971267, EPI_ISL_971273, EPI_ISL_971277, EPI_ISL_971284, EPI_ISL_971286, EPI_ISL_971289, EPI_ISL_971295, EPI_ISL_971301, EPI_ISL_971304, EPI_ISL_971307, EPI_ISL_971310, EPI_ISL_971313, EPI_ISL_971316, EPI_ISL_971319, EPI_ISL_971320, EPI_ISL_971324, EPI_ISL_971330, EPI_ISL_971336, EPI_ISL_971343, EPI_ISL_971350, EPI_ISL_971352, EPI_ISL_971357, EPI_ISL_971373, EPI_ISL_971376, EPI_ISL_971378, EPI_ISL_971382, EPI_ISL_971388, EPI_ISL_971392, EPI_ISL_971397, EPI_ISL_971400, EPI_ISL_971402, EPI_ISL_971405, EPI_ISL_971407, EPI_ISL_971410, EPI_ISL_971413, EPI_ISL_971417, EPI_ISL_971420, EPI_ISL_971423, EPI_ISL_971428, EPI_ISL_988483, EPI_ISL_988485, EPI_ISL_988488, EPI_ISL_988490, EPI_ISL_988493, EPI_ISL_988513, EPI_ISL_988520, EPI_ISL_988522, EPI_ISL_988526, EPI_ISL_988534, EPI_ISL_988539, EPI_ISL_988545, EPI_ISL_988551, EPI_ISL_988568, EPI_ISL_988577, EPI_ISL_988585, EPI_ISL_988586, EPI_ISL_988601, EPI_ISL_988605, EPI_ISL_988611, EPI_ISL_988615, EPI_ISL_988619, EPI_ISL_988620, EPI_ISL_988624, EPI_ISL_988632, EPI_ISL_988647, EPI_ISL_988648, EPI_ISL_988649, EPI_ISL_988652, EPI_ISL_988655, EPI_ISL_988661, EPI_ISL_988663, EPI_ISL_988665, EPI_ISL_988671, EPI_ISL_988681, EPI_ISL_988683, EPI_ISL_1006341, EPI_ISL_1006344, EPI_ISL_1006345, EPI_ISL_1006348, EPI_ISL_1006354, EPI_ISL_1006357, EPI_ISL_1006358, EPI_ISL_1006365, EPI_ISL_1006366, EPI_ISL_1006367, EPI_ISL_1006373, EPI_ISL_1006374, EPI_ISL_1006378, EPI_ISL_1006385, EPI_ISL_1006387, EPI_ISL_1006390, EPI_ISL_1006391, EPI_ISL_1006395, EPI_ISL_1006399, EPI_ISL_1006409, EPI_ISL_1006412, EPI_ISL_1006422, EPI_ISL_1006423, EPI_ISL_1006426, EPI_ISL_1006428, EPI_ISL_1006435, EPI_ISL_1006438, EPI_ISL_1006439, EPI_ISL_1006445, EPI_ISL_1006453, EPI_ISL_1006456, EPI_ISL_1006457, EPI_ISL_1006458, EPI_ISL_1006467, EPI_ISL_1006471, EPI_ISL_1006474, EPI_ISL_1006475, EPI_ISL_1006476, EPI_ISL_1006477, EPI_ISL_1006479, EPI_ISL_1006482, EPI_ISL_1006488, EPI_ISL_1006489, EPI_ISL_1006490, EPI_ISL_1006501, EPI_ISL_1006511, EPI_ISL_1006516, EPI_ISL_1006521, EPI_ISL_1006529, EPI_ISL_1006530, EPI_ISL_1006533, EPI_ISL_1006535, EPI_ISL_1006538, EPI_ISL_1830701

| | | | |
|---|---|---|---|
| see above | Lighthouse Lab in Milton Keynes | Wellcome Sanger Institute for the COVID-19 Genomics UK (COG-UK) Consortium | Cordelia Langford; David K. Jackson; Dominic Kwiatkowski; Ewan Harrison; Ian Johnston; Jeffrey Barrett; John Sillitoe on behalf of the Wellcome Sanger Institute COVID-19 Surveillance Team; John Sillitoe on behalf of the Wellcome Sanger Institute COVID-19 Surveillance Team (http://www.sanger.ac.uk/covid-team); Roberto Amato; Sonia Goncalves; The Lighthouse Lab in Milton Keynes and Alex Alderton |

EPI_ISL_950755, EPI_ISL_950756, EPI_ISL_950757, EPI_ISL_950758, EPI_ISL_950759, EPI_ISL_950760, EPI_ISL_950761, EPI_ISL_1105851, EPI_ISL_1105852, EPI_ISL_1105853, EPI_ISL_1105854, EPI_ISL_1105855, EPI_ISL_1105856, EPI_ISL_1105857, EPI_ISL_1105858, EPI_ISL_1105859, EPI_ISL_1105860

| | | | |
|---|---|---|---|
| see above | Lincolnshire Hospitals and DeepSeq Nottingham | COVID-19 Genomics UK (COG-UK) Consortium | Christopher Moore; Fei Sang; Joeseph Chappell; Johnny Debebe; Jonathan Ball; Matthew Carlisle; Matthew Loose; Nadine Holmes; Nichola Duckworth; Patrick McClure; Sarah Walsh; Tim Sloan; Victoria Wright |

EPI_ISL_919629, EPI_ISL_919630, EPI_ISL_919663, EPI_ISL_919706, EPI_ISL_919707, EPI_ISL_919708, EPI_ISL_919709, EPI_ISL_919710, EPI_ISL_919712, EPI_ISL_919713, EPI_ISL_919714, EPI_ISL_919715, EPI_ISL_919716, EPI_ISL_919717, EPI_ISL_919718, EPI_ISL_919719, EPI_ISL_919720, EPI_ISL_919721, EPI_ISL_919722, EPI_ISL_919723, EPI_ISL_919724, EPI_ISL_949655, EPI_ISL_949656, EPI_ISL_949657, EPI_ISL_949658, EPI_ISL_949659, EPI_ISL_949660, EPI_ISL_949661, EPI_ISL_949665, EPI_ISL_949667, EPI_ISL_949673, EPI_ISL_949674, EPI_ISL_949675, EPI_ISL_949676, EPI_ISL_949677, EPI_ISL_949678, EPI_ISL_949679, EPI_ISL_949680, EPI_ISL_949681, EPI_ISL_949682, EPI_ISL_949683, EPI_ISL_949688, EPI_ISL_949689, EPI_ISL_949690, EPI_ISL_949691, EPI_ISL_949692, EPI_ISL_949693, EPI_ISL_949694, EPI_ISL_949695, EPI_ISL_949696, EPI_ISL_949697, EPI_ISL_949698, EPI_ISL_949699, EPI_ISL_949700, EPI_ISL_949701, EPI_ISL_949702, EPI_ISL_949703, EPI_ISL_949704, EPI_ISL_949705, EPI_ISL_949706, EPI_ISL_949707, EPI_ISL_997382, EPI_ISL_997393, EPI_ISL_997394, EPI_ISL_997395, EPI_ISL_997396, EPI_ISL_997397, EPI_ISL_997398, EPI_ISL_997399, EPI_ISL_997400, EPI_ISL_997401, EPI_ISL_997402, EPI_ISL_997403, EPI_ISL_997404, EPI_ISL_997407, EPI_ISL_997408, EPI_ISL_997409, EPI_ISL_997410, EPI_ISL_997411, EPI_ISL_997439, EPI_ISL_997442, EPI_ISL_997467, EPI_ISL_997468, EPI_ISL_997469, EPI_ISL_997470, EPI_ISL_997471, EPI_ISL_997472, EPI_ISL_997474, EPI_ISL_997475, EPI_ISL_997476, EPI_ISL_997477, EPI_ISL_997478, EPI_ISL_997479, EPI_ISL_997480, EPI_ISL_997481, EPI_ISL_997482, EPI_ISL_1048017, EPI_ISL_1048018, EPI_ISL_1048019, EPI_ISL_1048020, EPI_ISL_1048021, EPI_ISL_1048022, EPI_ISL_1104322, EPI_ISL_1104323, EPI_ISL_1104387, EPI_ISL_1104498, EPI_ISL_1104504, EPI_ISL_1177890, EPI_ISL_1247819, EPI_ISL_1247820, EPI_ISL_1247821, EPI_ISL_1247822, EPI_ISL_1474666, EPI_ISL_1546024, EPI_ISL_1546025, EPI_ISL_1546026, EPI_ISL_1546027, EPI_ISL_1546028, EPI_ISL_1546029, EPI_ISL_1546030, EPI_ISL_1546031, EPI_ISL_1546032, EPI_ISL_1546033

| | | | |
|---|---|---|---|
| see above | Liverpool Clinical Laboratories | COVID-19 Genomics UK (COG-UK) Consortium | A Alrezaihi; Alessandro Gerada; Alistair Darby; Angela Cowell; Anita Lucaci; Anu Chawla; Cassie Olateju; Catherine Hartley; Charlotte Nelson; Ecaterina Vamos; Elaine O'Toole; Eleanor G Bentley; Ghada T Shawli; Hermione Webster; Isabel García-Dorival; James Johnson; James P Stewart; Jenifer Manson; Joanne Watts; Jones Benjamin; Jordan J Clark; Julian Hiscox; L Luu; Lucille Rainbow; M Almsaud; Margaret Hughes; Mark Whitehead; Matthew Gemmell; Miren Iturriza-Gomara; Muhannad Alruwaili; N.P Randle; Neil Swainston; PKF Gilmore; Parul Sharma; Rebekah Penrice-Randal; Richard Eccles; Richard Gregory; Sam Haldenby; Steve Paterson; Stuart D Armstrong; Trevor Ian Robinson; Ximeng Han |

EPI_ISL_921659, EPI_ISL_921660, EPI_ISL_921661, EPI_ISL_921662, EPI_ISL_950373, EPI_ISL_950376, EPI_ISL_950379, EPI_ISL_950382, EPI_ISL_950391, EPI_ISL_950392, EPI_ISL_950393, EPI_ISL_950394, EPI_ISL_950395, EPI_ISL_950396, EPI_ISL_950397, EPI_ISL_950398, EPI_ISL_950399, EPI_ISL_950400, EPI_ISL_950401, EPI_ISL_950523, EPI_ISL_950524, EPI_ISL_950525, EPI_ISL_950526, EPI_ISL_950527, EPI_ISL_950528, EPI_ISL_950529, EPI_ISL_950530, EPI_ISL_950531, EPI_ISL_950532, EPI_ISL_950533, EPI_ISL_950534, EPI_ISL_950535, EPI_ISL_950536, EPI_ISL_950537, EPI_ISL_950538, EPI_ISL_950539, EPI_ISL_950540, EPI_ISL_950541, EPI_ISL_950542, EPI_ISL_950543, EPI_ISL_998484, EPI_ISL_998486, EPI_ISL_998487, EPI_ISL_998488, EPI_ISL_998489, EPI_ISL_998490, EPI_ISL_998491, EPI_ISL_998493, EPI_ISL_998494, EPI_ISL_998495, EPI_ISL_998497, EPI_ISL_998498, EPI_ISL_998500, EPI_ISL_998501, EPI_ISL_998502, EPI_ISL_998503, EPI_ISL_998559, EPI_ISL_998560, EPI_ISL_998561, EPI_ISL_998562, EPI_ISL_998563, EPI_ISL_998564, EPI_ISL_998565, EPI_ISL_1051222, EPI_ISL_1051223, EPI_ISL_1051224, EPI_ISL_1051225, EPI_ISL_1051226, EPI_ISL_1051227, EPI_ISL_1051228, EPI_ISL_1051229, EPI_ISL_1051230, EPI_ISL_1051231, EPI_ISL_1051232, EPI_ISL_1051233, EPI_ISL_1051234, EPI_ISL_1051235, EPI_ISL_1051236, EPI_ISL_1051237, EPI_ISL_1051238, EPI_ISL_1051239, EPI_ISL_1051240, EPI_ISL_1051241, EPI_ISL_1051242, EPI_ISL_1051243, EPI_ISL_1178143, EPI_ISL_1178144, EPI_ISL_1178145, EPI_ISL_1178146, EPI_ISL_1178147, EPI_ISL_1178193, EPI_ISL_1178194, EPI_ISL_1178195, EPI_ISL_1178196, EPI_ISL_1178197, EPI_ISL_1178198, EPI_ISL_1178199, EPI_ISL_1178200, EPI_ISL_1178201, EPI_ISL_1178202, EPI_ISL_1178203, EPI_ISL_1178204, EPI_ISL_1178205, EPI_ISL_1178207, EPI_ISL_1178208, EPI_ISL_1178209, EPI_ISL_1178210, EPI_ISL_1178211, EPI_ISL_1178212, EPI_ISL_1178213, EPI_ISL_1178214, EPI_ISL_1178215, EPI_ISL_1178216, EPI_ISL_1178217, EPI_ISL_1178218, EPI_ISL_1178219, EPI_ISL_1178220, EPI_ISL_1178221, EPI_ISL_1178222

| | | | |
|---|---|---|---|
| see above | Northumbria University / South Tees Hospitals NHS Foundation Trust / North Cumbria Integrated Care NHS Foundation Trust / North Tees and Hartlepool NHS Foundation Trust / Newcastle Hospitals NHS Foundation Trust | COVID-19 Genomics UK (COG-UK) Consortium | Andrew Nelson; Brendan Payne; Clive Graham; Darren L Smith; Debra Padgett; Edward Barton; Emma Swindells; Garren Scott; Gary Black; Gary Eltringham; Giles S Holt; Greg R Young; Jane Greenaway; Jennifer Collins; John Allan; Joshua Loh; Lynn Dover; Matthew Bashton; Mohammad A Tariq; Paul Baker; Sarah Essex; Steve Liggett; Wen C Yew; Yusri Taha |

EPI_ISL_951492, EPI_ISL_951493, EPI_ISL_951494, EPI_ISL_951495, EPI_ISL_951496, EPI_ISL_951497, EPI_ISL_951498, EPI_ISL_951499, EPI_ISL_951501, EPI_ISL_951502, EPI_ISL_951503, EPI_ISL_951504, EPI_ISL_951505, EPI_ISL_951506, EPI_ISL_951508, EPI_ISL_951512, EPI_ISL_951513, EPI_ISL_951514, EPI_ISL_951515, EPI_ISL_951517, EPI_ISL_951518, EPI_ISL_951519, EPI_ISL_951521, EPI_ISL_951522, EPI_ISL_951523, EPI_ISL_951524, EPI_ISL_951525, EPI_ISL_951527, EPI_ISL_951528, EPI_ISL_951529, EPI_ISL_951530, EPI_ISL_951531, EPI_ISL_951532, EPI_ISL_951533, EPI_ISL_951534, EPI_ISL_951535, EPI_ISL_951536, EPI_ISL_951537, EPI_ISL_951538, EPI_ISL_951541, EPI_ISL_951542, EPI_ISL_951543, EPI_ISL_951544, EPI_ISL_951545, EPI_ISL_951546, EPI_ISL_951547, EPI_ISL_951548, EPI_ISL_951549, EPI_ISL_951550, EPI_ISL_951552, EPI_ISL_951553, EPI_ISL_951554, EPI_ISL_951556, EPI_ISL_951557, EPI_ISL_951558, EPI_ISL_951559, EPI_ISL_951560, EPI_ISL_951561, EPI_ISL_951562, EPI_ISL_951564, EPI_ISL_951567, EPI_ISL_951569, EPI_ISL_951570, EPI_ISL_951571, EPI_ISL_951572, EPI_ISL_951573, EPI_ISL_951574, EPI_ISL_951575, EPI_ISL_951576, EPI_ISL_951577, EPI_ISL_951578, EPI_ISL_951579, EPI_ISL_951580, EPI_ISL_951581, EPI_ISL_951582, EPI_ISL_951583, EPI_ISL_951585, EPI_ISL_951738, EPI_ISL_951739, EPI_ISL_951742, EPI_ISL_951744, EPI_ISL_951745, EPI_ISL_951753, EPI_ISL_951758, EPI_ISL_951759, EPI_ISL_951760, EPI_ISL_951761, EPI_ISL_951765, EPI_ISL_951766, EPI_ISL_951767, EPI_ISL_951769, EPI_ISL_951778, EPI_ISL_951779, EPI_ISL_951780, EPI_ISL_951781, EPI_ISL_951782, EPI_ISL_951785, EPI_ISL_951786, EPI_ISL_951787, EPI_ISL_951820, EPI_ISL_951821, EPI_ISL_951843, EPI_ISL_951844, EPI_ISL_951851, EPI_ISL_951852, EPI_ISL_951853, EPI_ISL_951855, EPI_ISL_951856, EPI_ISL_951857, EPI_ISL_951858, EPI_ISL_951859, EPI_ISL_951861, EPI_ISL_951862, EPI_ISL_951863, EPI_ISL_951866, EPI_ISL_951867, EPI_ISL_951869, EPI_ISL_951875, EPI_ISL_951876, EPI_ISL_951877, EPI_ISL_951878, EPI_ISL_951881, EPI_ISL_951882, EPI_ISL_951884, EPI_ISL_951886, EPI_ISL_951887, EPI_ISL_951888, EPI_ISL_951890, EPI_ISL_951893, EPI_ISL_951894, EPI_ISL_951895, EPI_ISL_951896, EPI_ISL_951897, EPI_ISL_951898, EPI_ISL_951900, EPI_ISL_951901, EPI_ISL_951902, EPI_ISL_951903, EPI_ISL_951904, EPI_ISL_951905, EPI_ISL_951906, EPI_ISL_951907, EPI_ISL_951908, EPI_ISL_951909, EPI_ISL_951910, EPI_ISL_951911, EPI_ISL_951912, EPI_ISL_951913, EPI_ISL_951916, EPI_ISL_951917, EPI_ISL_951918, EPI_ISL_951919, EPI_ISL_951920, EPI_ISL_951921, EPI_ISL_951922, EPI_ISL_951923, EPI_ISL_951924, EPI_ISL_951925, EPI_ISL_951926, EPI_ISL_951927, EPI_ISL_951928, EPI_ISL_951929, EPI_ISL_951930, EPI_ISL_951931, EPI_ISL_951932, EPI_ISL_951933, EPI_ISL_951934, EPI_ISL_951935, EPI_ISL_951936, EPI_ISL_951937, EPI_ISL_951938, EPI_ISL_951939, EPI_ISL_951940, EPI_ISL_951941, EPI_ISL_951942, EPI_ISL_951943, EPI_ISL_951944, EPI_ISL_951945, EPI_ISL_951946, EPI_ISL_951947, EPI_ISL_951948, EPI_ISL_951949, EPI_ISL_951950, EPI_ISL_951951, EPI_ISL_951952, EPI_ISL_951953, EPI_ISL_951954, EPI_ISL_951955, EPI_ISL_951956, EPI_ISL_951957, EPI_ISL_951958, EPI_ISL_951959, EPI_ISL_951960, EPI_ISL_951961, EPI_ISL_951962, EPI_ISL_951963, EPI_ISL_951964, EPI_ISL_951965, EPI_ISL_951966, EPI_ISL_951968, EPI_ISL_951969, EPI_ISL_951972, EPI_ISL_951973, EPI_ISL_952006, EPI_ISL_952007, EPI_ISL_952008, EPI_ISL_952009, EPI_ISL_952010, EPI_ISL_952011, EPI_ISL_952012, EPI_ISL_952013, EPI_ISL_952014, EPI_ISL_952015, EPI_ISL_952018, EPI_ISL_952019, EPI_ISL_952020, EPI_ISL_952022, EPI_ISL_952025, EPI_ISL_952027, EPI_ISL_952029, EPI_ISL_952030, EPI_ISL_952031, EPI_ISL_952032, EPI_ISL_952033, EPI_ISL_952035, EPI_ISL_952036, EPI_ISL_952037, EPI_ISL_952038, EPI_ISL_952039, EPI_ISL_952040, EPI_ISL_952041, EPI_ISL_952061, EPI_ISL_952062, EPI_ISL_952063, EPI_ISL_952064, EPI_ISL_952065, EPI_ISL_952066, EPI_ISL_952067, EPI_ISL_952069, EPI_ISL_952070, EPI_ISL_952071, EPI_ISL_952072, EPI_ISL_952073, EPI_ISL_952074, EPI_ISL_952075, EPI_ISL_952076, EPI_ISL_952077, EPI_ISL_952078, EPI_ISL_952079, EPI_ISL_952082, EPI_ISL_952083, EPI_ISL_952084, EPI_ISL_952085, EPI_ISL_952086, EPI_ISL_952087, EPI_ISL_952088, EPI_ISL_952089, EPI_ISL_952090, EPI_ISL_952091, EPI_ISL_952092, EPI_ISL_952093, EPI_ISL_952095, EPI_ISL_952096, EPI_ISL_952097, EPI_ISL_952100, EPI_ISL_952102, EPI_ISL_952104, EPI_ISL_952108, EPI_ISL_952109, EPI_ISL_952110, EPI_ISL_952111, EPI_ISL_952112, EPI_ISL_952113, EPI_ISL_952114, EPI_ISL_952119, EPI_ISL_952120, EPI_ISL_952122, EPI_ISL_952123, EPI_ISL_952124, EPI_ISL_952126, EPI_ISL_952128, EPI_ISL_952129, EPI_ISL_952130, EPI_ISL_952131, EPI_ISL_952132, EPI_ISL_952133, EPI_ISL_952134, EPI_ISL_952136, EPI_ISL_952137, EPI_ISL_952138, EPI_ISL_952139, EPI_ISL_952140, EPI_ISL_952141, EPI_ISL_952143, EPI_ISL_952144, EPI_ISL_952145, EPI_ISL_952146, EPI_ISL_952147, EPI_ISL_952148, EPI_ISL_952149, EPI_ISL_952150, EPI_ISL_952151, EPI_ISL_952152, EPI_ISL_952153, EPI_ISL_952154, EPI_ISL_952156, EPI_ISL_952157, EPI_ISL_952158, EPI_ISL_952159, EPI_ISL_952161, EPI_ISL_952162, EPI_ISL_952163, EPI_ISL_952164, EPI_ISL_952165, EPI_ISL_952166, EPI_ISL_952167, EPI_ISL_999646, EPI_ISL_999647, EPI_ISL_999652, EPI_ISL_999659, EPI_ISL_999664, EPI_ISL_999669, EPI_ISL_999683, EPI_ISL_999684, EPI_ISL_999685, EPI_ISL_999686, EPI_ISL_999687, EPI_ISL_999694, EPI_ISL_999695, EPI_ISL_999808, EPI_ISL_1051920, EPI_ISL_1051922, EPI_ISL_1051923, EPI_ISL_1051924, EPI_ISL_1051925, EPI_ISL_1051926, EPI_ISL_1051927, EPI_ISL_1051928, EPI_ISL_1051929, EPI_ISL_1051930, EPI_ISL_1051932, EPI_ISL_1051933, EPI_ISL_1051934, EPI_ISL_1051935, EPI_ISL_1051936, EPI_ISL_1051937, EPI_ISL_1051938, EPI_ISL_1051939, EPI_ISL_1051940, EPI_ISL_1051941, EPI_ISL_1051942, EPI_ISL_1051943, EPI_ISL_1051944, EPI_ISL_1051946, EPI_ISL_1051947, EPI_ISL_1051948, EPI_ISL_1051949, EPI_ISL_1051950, EPI_ISL_1051952, EPI_ISL_1051953, EPI_ISL_1051954, EPI_ISL_1051955, EPI_ISL_1051956, EPI_ISL_1051957, EPI_ISL_1051958, EPI_ISL_1051959, EPI_ISL_1051961, EPI_ISL_1051962, EPI_ISL_1051963, EPI_ISL_1051964, EPI_ISL_1051966, EPI_ISL_1051967, EPI_ISL_1051968, EPI_ISL_1051969, EPI_ISL_1051970, EPI_ISL_1051971, EPI_ISL_1051972, EPI_ISL_1051973, EPI_ISL_1051974, EPI_ISL_1051975, EPI_ISL_1051976, EPI_ISL_1051978, EPI_ISL_1051979, EPI_ISL_1051981, EPI_ISL_1051982, EPI_ISL_1051983, EPI_ISL_1051984, EPI_ISL_1051985, EPI_ISL_1051986, EPI_ISL_1051987, EPI_ISL_1051988, EPI_ISL_1051989, EPI_ISL_1051990, EPI_ISL_1051991, EPI_ISL_1051992, EPI_ISL_1051993, EPI_ISL_1051994, EPI_ISL_1051995, EPI_ISL_1051997, EPI_ISL_1051998, EPI_ISL_1051999, EPI_ISL_1052000, EPI_ISL_1052001, EPI_ISL_1052002, EPI_ISL_1052003, EPI_ISL_1052004, EPI_ISL_1052005, EPI_ISL_1052006, EPI_ISL_1052007, EPI_ISL_1052008, EPI_ISL_1052009, EPI_ISL_1052010, EPI_ISL_1052011, EPI_ISL_1052012, EPI_ISL_1052013, EPI_ISL_1052014, EPI_ISL_1052015, EPI_ISL_1052016, EPI_ISL_1052017, EPI_ISL_1052018, EPI_ISL_1052021, EPI_ISL_1052022, EPI_ISL_1052024, EPI_ISL_1052025, EPI_ISL_1052026, EPI_ISL_1052027, EPI_ISL_1052029, EPI_ISL_1052031, EPI_ISL_1052033, EPI_ISL_1052034, EPI_ISL_1052035, EPI_ISL_1052036, EPI_ISL_1052038, EPI_ISL_1052039, EPI_ISL_1052040, EPI_ISL_1052041, EPI_ISL_1052042, EPI_ISL_1052043, EPI_ISL_1052044, EPI_ISL_1052045, EPI_ISL_1052047, EPI_ISL_1052048, EPI_ISL_1052050, EPI_ISL_1052051, EPI_ISL_1052052, EPI_ISL_1052053, EPI_ISL_1052055, EPI_ISL_1052056, EPI_ISL_1052058, EPI_ISL_1052060, EPI_ISL_1052061, EPI_ISL_1052062, EPI_ISL_1052063, EPI_ISL_1052064, EPI_ISL_1052065, EPI_ISL_1052066, EPI_ISL_1052067, EPI_ISL_1052068, EPI_ISL_1052070, EPI_ISL_1052071, EPI_ISL_1052072, EPI_ISL_1052073, EPI_ISL_1052074, EPI_ISL_1052075, EPI_ISL_1052076, EPI_ISL_1052077, EPI_ISL_1052078, EPI_ISL_1052079, EPI_ISL_1052080, EPI_ISL_1052081, EPI_ISL_1052082, EPI_ISL_1052083, EPI_ISL_1052084, EPI_ISL_1052086, EPI_ISL_1052087, EPI_ISL_1052088, EPI_ISL_1052089, EPI_ISL_1052090, EPI_ISL_1052091, EPI_ISL_1052092, EPI_ISL_1052094, EPI_ISL_1052095, EPI_ISL_1052098, EPI_ISL_1052099, EPI_ISL_1052100, EPI_ISL_1052101, EPI_ISL_1052102, EPI_ISL_1052103, EPI_ISL_1052104, EPI_ISL_1052105, EPI_ISL_1052106, EPI_ISL_1052107, EPI_ISL_1052109, EPI_ISL_1052110, EPI_ISL_1052111, EPI_ISL_1052112, EPI_ISL_1052116, EPI_ISL_1052117, EPI_ISL_1052118, EPI_ISL_1052119, EPI_ISL_1052120, EPI_ISL_1052121, EPI_ISL_1052122, EPI_ISL_1052123, EPI_ISL_1052124, EPI_ISL_1052125, EPI_ISL_1052126, EPI_ISL_1052127, EPI_ISL_1052128, EPI_ISL_1052129, EPI_ISL_1052132, EPI_ISL_1052133, EPI_ISL_1052134, EPI_ISL_1052136, EPI_ISL_1052137, EPI_ISL_1052203, EPI_ISL_1052204, EPI_ISL_1052462, EPI_ISL_1052465, EPI_ISL_1052473, EPI_ISL_1052474, EPI_ISL_1052480, EPI_ISL_1052511, EPI_ISL_1052512, EPI_ISL_1052513, EPI_ISL_1052514, EPI_ISL_1052515, EPI_ISL_1052516, EPI_ISL_1052521, EPI_ISL_1052523, EPI_ISL_1052524, EPI_ISL_1052525, EPI_ISL_1053502, EPI_ISL_1053521, EPI_ISL_1053522, EPI_ISL_1053548, EPI_ISL_1053549, EPI_ISL_1053550, EPI_ISL_1053551, EPI_ISL_1053553, EPI_ISL_1053555, EPI_ISL_1053556, EPI_ISL_1053557, EPI_ISL_1053559, EPI_ISL_1053560, EPI_ISL_1053561, EPI_ISL_1053563, EPI_ISL_1053564, EPI_ISL_1106645, EPI_ISL_1179155, EPI_ISL_1476455, EPI_ISL_1476484, EPI_ISL_1476486, EPI_ISL_1476488, EPI_ISL_1476499

| | | | |
|---|---|---|---|
| see above | Originating lab: Wales Specialist Virology Centre Sequencing lab: Pathogen Genomics Unit | Public Health Wales Microbiology Cardiff Wales Specialist Virology Centre | Alec Birchley; Alexander Adams; Amy Gaskin; Angela Marchbank; Bree Gatica-Wilcox; Catherine Moore; Jason Coombes; Joanne Watkins; Joel Southgate; Johnathan Evans; Laura Gifford; Lauren Gilbert; Lee Graham; Malorie Perry; Matthew Bull; Nicole Pacchiarini; Sally Corden; Sara Kumziene-Summerhayes; Sara Rey; Sarah Taylor; Simon Cottrell; Sophie Jones; Tom Connor |

EPI_ISL_998994, EPI_ISL_999302, EPI_ISL_999303, EPI_ISL_999307, EPI_ISL_999310, EPI_ISL_999311, EPI_ISL_999317, EPI_ISL_999318, EPI_ISL_999320, EPI_ISL_999322, EPI_ISL_999323, EPI_ISL_999324, EPI_ISL_999325, EPI_ISL_999326, EPI_ISL_999327, EPI_ISL_999329, EPI_ISL_999330, EPI_ISL_999331, EPI_ISL_999332, EPI_ISL_999335, EPI_ISL_999337, EPI_ISL_999339, EPI_ISL_999340, EPI_ISL_999342, EPI_ISL_999343, EPI_ISL_999344, EPI_ISL_999345, EPI_ISL_999346, EPI_ISL_999347, EPI_ISL_999484, EPI_ISL_999485, EPI_ISL_999486, EPI_ISL_999487, EPI_ISL_999488, EPI_ISL_999489, EPI_ISL_999490, EPI_ISL_999491, EPI_ISL_999492, EPI_ISL_999493, EPI_ISL_999494, EPI_ISL_1051597, EPI_ISL_1051609, EPI_ISL_1051610, EPI_ISL_1051611, EPI_ISL_1051612, EPI_ISL_1051613, EPI_ISL_1051615, EPI_ISL_1051616, EPI_ISL_1051617, EPI_ISL_1051619, EPI_ISL_1051621, EPI_ISL_1051622, EPI_ISL_1051623, EPI_ISL_1051624, EPI_ISL_1051625, EPI_ISL_1051628, EPI_ISL_1051632, EPI_ISL_1051633, EPI_ISL_1051636, EPI_ISL_1051660, EPI_ISL_1051670, EPI_ISL_1051675, EPI_ISL_1051676, EPI_ISL_1051681, EPI_ISL_1051682, EPI_ISL_1051690, EPI_ISL_1051893, EPI_ISL_1051894, EPI_ISL_1051895, EPI_ISL_1051896, EPI_ISL_1051897, EPI_ISL_1051904, EPI_ISL_1051905, EPI_ISL_1178826, EPI_ISL_1249220, EPI_ISL_1249221, EPI_ISL_1249222, EPI_ISL_1249223, EPI_ISL_1387219, EPI_ISL_1833065, EPI_ISL_1833075, EPI_ISL_1833104

| | | | |
|---|---|---|---|
| see above | Oxford Viromics, NDM, University of Oxford; Oxford University Hospitals; Basingstoke and North Hampshire Hospital | COVID-19 Genomics UK (COG-UK) Consortium | Alex Mobbs; Amy Trebes; Anita Justice; Catrin Moore; Christophe Fraser; David Bonsall; David Buck; Emma Wise; George Macintyre; Jessica Lynch; John Todd; Mariateresa de Cesare; Matilde Mori; Monique Andersson; Nathan Moore; Nick Cortes; Robert Shaw; Stephen Kidd; Tanya Golubchik; Timothy Peto |

EPI_ISL_998772, EPI_ISL_998773, EPI_ISL_998776, EPI_ISL_998777, EPI_ISL_998780, EPI_ISL_998781, EPI_ISL_998785, EPI_ISL_998786, EPI_ISL_998788, EPI_ISL_998795, EPI_ISL_998798, EPI_ISL_998801, EPI_ISL_998802, EPI_ISL_998804, EPI_ISL_998806, EPI_ISL_998816, EPI_ISL_998817, EPI_ISL_998838, EPI_ISL_998839, EPI_ISL_998841, EPI_ISL_998842, EPI_ISL_998843, EPI_ISL_998844, EPI_ISL_998846,

| | | | |
|---|---|---|---|
| EPI_ISL_998847, EPI_ISL_998849, EPI_ISL_998853, EPI_ISL_998857, EPI_ISL_998858, EPI_ISL_998859, EPI_ISL_998860, EPI_ISL_998861, EPI_ISL_998864, EPI_ISL_998865, EPI_ISL_998866, EPI_ISL_998867, EPI_ISL_998869, EPI_ISL_998870, EPI_ISL_998871, EPI_ISL_998872, EPI_ISL_998874, EPI_ISL_998875, EPI_ISL_998880, EPI_ISL_998881, EPI_ISL_998882, EPI_ISL_998884, EPI_ISL_998885, EPI_ISL_998888, EPI_ISL_1178435, EPI_ISL_1178438, EPI_ISL_1178439, EPI_ISL_1178441, EPI_ISL_1178445, EPI_ISL_1178447, EPI_ISL_1178449, EPI_ISL_1178452, EPI_ISL_1178453, EPI_ISL_1178455, EPI_ISL_1178457, EPI_ISL_1178459, EPI_ISL_1178463, EPI_ISL_1178472, EPI_ISL_1178474, EPI_ISL_1178477, EPI_ISL_1178478, EPI_ISL_1178480, EPI_ISL_1178482, EPI_ISL_1178483, EPI_ISL_1178487, EPI_ISL_1178493, EPI_ISL_1178495, EPI_ISL_1178507, EPI_ISL_1248939, EPI_ISL_1248940, EPI_ISL_1248941, EPI_ISL_1248942, EPI_ISL_1248943, EPI_ISL_1248944, EPI_ISL_1248946, EPI_ISL_1248948, EPI_ISL_1248952, EPI_ISL_1248954, EPI_ISL_1248957, EPI_ISL_1248960, EPI_ISL_1248974, EPI_ISL_1249038, EPI_ISL_1249039, EPI_ISL_1249042, EPI_ISL_1309668, EPI_ISL_1309669, EPI_ISL_1309673 | | | |
| see above | Quadram Institute Bioscience | COVID-19 Genomics UK (COG-UK) Consortium | Alexander J Trotter; Alison E. Mather; Alp Aydin; Ana P. Tedim; Anastasia Kolyva; Andrew Bell; Andrew J. Page; Claire Stuart; Dave J. Baker; Gemma L. Kay; John Wain; Justin O'Grady; Leonardo de Oliveira Martins; Lizzie Meadows; Maria Diaz; Mark Webber; Muhammed Yasir; Nabil-Fareed Alikhan; Ngozi Elumogo; Nicholas M. Thomson; Rachael Stanley; Rachel Gilroy; Reenesh Prakash; Samir Dervisevic; Samuel Bloomfield; Steven Rudder; Thanh Le-Viet |
| EPI_ISL_922037, EPI_ISL_922038, EPI_ISL_922039, EPI_ISL_922040, EPI_ISL_950639, EPI_ISL_950640, EPI_ISL_950641, EPI_ISL_950642, EPI_ISL_950643, EPI_ISL_950644, EPI_ISL_950645, EPI_ISL_1054396, EPI_ISL_1108970, EPI_ISL_1108983, EPI_ISL_1109013, EPI_ISL_1109024, EPI_ISL_1109025, EPI_ISL_1109027, EPI_ISL_1109029, EPI_ISL_1109031, EPI_ISL_1109033, EPI_ISL_1109034, EPI_ISL_1109038, EPI_ISL_1109039, EPI_ISL_1109040, EPI_ISL_1109041, EPI_ISL_1387166, EPI_ISL_2127626 | | | |
| see above | Queens Medical Centre, Clinical Microbiology Department / DeepSeq Nottingham | COVID-19 Genomics UK (COG-UK) Consortium | Christopher Moore; Emily Park; Fei Sang; Gemma Clark; Hannah Howson-Wells; Jack Hill; Jayasree Dey; Johnny Debebe; Jonathan Ball; Joseph Chappell; Manjinder Khakh; Matthew Carlisle; Matthew Loose; Michelle M Lister; Nadine Holmes; Patrick McClure; Theocharis Tsoleridis; Timothy Byaruhanga; Vicki M Fleming; Victoria Wright; Wendy Smith |
| EPI_ISL_1045980, EPI_ISL_1045981, EPI_ISL_1045982, EPI_ISL_1045984, EPI_ISL_1045985, EPI_ISL_1045986, EPI_ISL_1045987, EPI_ISL_1045988, EPI_ISL_1045989, EPI_ISL_1045990, EPI_ISL_1045991, EPI_ISL_1045992, EPI_ISL_1045993, EPI_ISL_1045994, EPI_ISL_1045995, EPI_ISL_1045996, EPI_ISL_1045997, EPI_ISL_1045998, EPI_ISL_1045999, EPI_ISL_1046000, EPI_ISL_1046001, EPI_ISL_1046002, EPI_ISL_1046003, EPI_ISL_1046004, EPI_ISL_1046005, EPI_ISL_1046006, EPI_ISL_1046007, EPI_ISL_1046008, EPI_ISL_1046009, EPI_ISL_1046010, EPI_ISL_1046011, EPI_ISL_1046012, EPI_ISL_1046013, EPI_ISL_1046014, EPI_ISL_1046015, EPI_ISL_1046016, EPI_ISL_1046017, EPI_ISL_1046018, EPI_ISL_1046019, EPI_ISL_1046020, EPI_ISL_1046021, EPI_ISL_1046022, EPI_ISL_1046023, EPI_ISL_1046024, EPI_ISL_1046025, EPI_ISL_1046026, EPI_ISL_1046027, EPI_ISL_1046028, EPI_ISL_1046029, EPI_ISL_1046030, EPI_ISL_1046031, EPI_ISL_1046032, EPI_ISL_1046033, EPI_ISL_1046034, EPI_ISL_1046035, EPI_ISL_1046036, EPI_ISL_1046037, EPI_ISL_1046038, EPI_ISL_1046039, EPI_ISL_1046040, EPI_ISL_1046041, EPI_ISL_1046042, EPI_ISL_1046043, EPI_ISL_1046044, EPI_ISL_1046045, EPI_ISL_1046046, EPI_ISL_1046047, EPI_ISL_1046048, EPI_ISL_1046049, EPI_ISL_1046050, EPI_ISL_1046051, EPI_ISL_1046052, EPI_ISL_1046053, EPI_ISL_1046054, EPI_ISL_1046055, EPI_ISL_1046056, EPI_ISL_1046057, EPI_ISL_1046058, EPI_ISL_1046059, EPI_ISL_1046060, EPI_ISL_1046061, EPI_ISL_1046062, EPI_ISL_1046063, EPI_ISL_1046064, EPI_ISL_1046065, EPI_ISL_1046066, EPI_ISL_1046067, EPI_ISL_1046068, EPI_ISL_1046069, EPI_ISL_1046070, EPI_ISL_1046071, EPI_ISL_1046072, EPI_ISL_1046073, EPI_ISL_1046074, EPI_ISL_1046075, EPI_ISL_1046076, EPI_ISL_1046077, EPI_ISL_1046078, EPI_ISL_1046079, EPI_ISL_1046080, EPI_ISL_1046081, EPI_ISL_1046082, EPI_ISL_1046083, EPI_ISL_1046084, EPI_ISL_1046085, EPI_ISL_1046086, EPI_ISL_1046087, EPI_ISL_1046088, EPI_ISL_1046089, EPI_ISL_1046090, EPI_ISL_1046091, EPI_ISL_1046092, EPI_ISL_1046093, EPI_ISL_1046094, EPI_ISL_1046095, EPI_ISL_1046096, EPI_ISL_1046097, EPI_ISL_1046098, EPI_ISL_1046099, EPI_ISL_1046100, EPI_ISL_1046101, EPI_ISL_1046102, EPI_ISL_1046103, EPI_ISL_1046104, EPI_ISL_1046105, EPI_ISL_1046106, EPI_ISL_1046107, EPI_ISL_1046108, EPI_ISL_1046109, EPI_ISL_1046110, EPI_ISL_1046111, EPI_ISL_1046112, EPI_ISL_1046113, EPI_ISL_1046114, EPI_ISL_1046115, EPI_ISL_1046117, EPI_ISL_1046118, EPI_ISL_1046119, EPI_ISL_1046120, EPI_ISL_1046121, EPI_ISL_1046122, EPI_ISL_1046123, EPI_ISL_1046124, EPI_ISL_1046125, EPI_ISL_1046126, EPI_ISL_1046127, EPI_ISL_1046128, EPI_ISL_1046129, EPI_ISL_1046130, EPI_ISL_1046131, EPI_ISL_1046132, EPI_ISL_1046133, EPI_ISL_1046134, EPI_ISL_1046135, EPI_ISL_1046136, EPI_ISL_1046137, EPI_ISL_1046138, EPI_ISL_1046139, EPI_ISL_1046140, EPI_ISL_1046141, EPI_ISL_1046142, EPI_ISL_1046143, EPI_ISL_1046144, EPI_ISL_1046145, EPI_ISL_1046146, EPI_ISL_1046147, EPI_ISL_1046148, EPI_ISL_1046149, EPI_ISL_1046150, EPI_ISL_1046151, EPI_ISL_1046152, EPI_ISL_1046153, EPI_ISL_1046154, EPI_ISL_1046155, EPI_ISL_1046156, EPI_ISL_1046157, EPI_ISL_1046158, EPI_ISL_1046159, EPI_ISL_1046160, EPI_ISL_1046161, EPI_ISL_1046162, EPI_ISL_1046163, EPI_ISL_1046164, EPI_ISL_1046165, EPI_ISL_1046167, EPI_ISL_1046168, EPI_ISL_1046169, EPI_ISL_1046170, EPI_ISL_1046171, EPI_ISL_1046172, EPI_ISL_1046173, EPI_ISL_1046174, EPI_ISL_1046175, EPI_ISL_1046176, EPI_ISL_1046177, EPI_ISL_1046178, EPI_ISL_1046179, EPI_ISL_1046180, EPI_ISL_1046181, EPI_ISL_1046182, EPI_ISL_1046183, EPI_ISL_1046184, EPI_ISL_1046185, EPI_ISL_1046186, EPI_ISL_1046187, EPI_ISL_1046188, EPI_ISL_1046189, EPI_ISL_1046190, EPI_ISL_1046191, EPI_ISL_1046192, EPI_ISL_1046193, EPI_ISL_1046194, EPI_ISL_1046195, EPI_ISL_1046196, EPI_ISL_1046197, EPI_ISL_1046198, EPI_ISL_1046199, EPI_ISL_1046200, EPI_ISL_1046201, EPI_ISL_1046202, EPI_ISL_1046203, EPI_ISL_1046204, EPI_ISL_1046205, EPI_ISL_1046206, EPI_ISL_1046207, EPI_ISL_1046208, EPI_ISL_1046209, EPI_ISL_1046210, EPI_ISL_1046211, EPI_ISL_1046212, EPI_ISL_1046213, EPI_ISL_1046214, EPI_ISL_1046215, EPI_ISL_1046216, EPI_ISL_1046217, EPI_ISL_1046218, EPI_ISL_1046219, EPI_ISL_1046220, EPI_ISL_1046221, EPI_ISL_1046222, EPI_ISL_1046223, EPI_ISL_1046224, EPI_ISL_1046225, EPI_ISL_1046226, EPI_ISL_1046227, EPI_ISL_1046228, EPI_ISL_1046229, EPI_ISL_1046230, EPI_ISL_1046231, EPI_ISL_1046232, EPI_ISL_1046233, EPI_ISL_1046234, EPI_ISL_1046235, EPI_ISL_1046236, EPI_ISL_1046237, EPI_ISL_1046239, EPI_ISL_1046240, EPI_ISL_1046241, EPI_ISL_1046242, EPI_ISL_1046243, EPI_ISL_1046244, EPI_ISL_1046245, EPI_ISL_1046246, EPI_ISL_1046247, EPI_ISL_1046248, EPI_ISL_1046249, EPI_ISL_1046250, EPI_ISL_1046251, EPI_ISL_1046252, EPI_ISL_1046253, EPI_ISL_1046254, EPI_ISL_1046255, EPI_ISL_1046256, EPI_ISL_1046257, EPI_ISL_1046258, EPI_ISL_1046259, EPI_ISL_1046260, EPI_ISL_1046261, EPI_ISL_1046262, EPI_ISL_1046263, EPI_ISL_1046264, EPI_ISL_1046265, EPI_ISL_1046266, EPI_ISL_1046267, EPI_ISL_1046268, EPI_ISL_1046269, EPI_ISL_1046270, EPI_ISL_1046271, EPI_ISL_1046272, EPI_ISL_1046273, EPI_ISL_1046274, EPI_ISL_1046275, EPI_ISL_1046276, EPI_ISL_1046277, EPI_ISL_1046278, EPI_ISL_1046279, EPI_ISL_1046280, EPI_ISL_1046281, EPI_ISL_1046282, EPI_ISL_1046283, EPI_ISL_1046284, EPI_ISL_1046285, EPI_ISL_1046286, EPI_ISL_1046287, EPI_ISL_1046288, EPI_ISL_1046289, EPI_ISL_1046290, EPI_ISL_1046291, EPI_ISL_1046292, EPI_ISL_1046293, EPI_ISL_1046294, EPI_ISL_1046295, EPI_ISL_1046296, EPI_ISL_1046297, EPI_ISL_1046298, EPI_ISL_1046299, EPI_ISL_1046300, EPI_ISL_1046301, EPI_ISL_1046302, EPI_ISL_1046303, EPI_ISL_1046304, EPI_ISL_1046305, EPI_ISL_1046306, EPI_ISL_1046307, EPI_ISL_1046308, EPI_ISL_1046309, EPI_ISL_1046311, EPI_ISL_1046312, EPI_ISL_1046313, EPI_ISL_1046314, EPI_ISL_1046315, EPI_ISL_1046316, EPI_ISL_1046317, EPI_ISL_1046318, EPI_ISL_1046319, EPI_ISL_1046320, EPI_ISL_1046321, EPI_ISL_1046323, EPI_ISL_1046324, EPI_ISL_1046325, EPI_ISL_1046326, EPI_ISL_1046327, EPI_ISL_1046328, EPI_ISL_1046329, EPI_ISL_1046330, EPI_ISL_1046331, EPI_ISL_1046332, EPI_ISL_1046333, EPI_ISL_1046334, EPI_ISL_1046335 | | | |
| see above | Randox Laboratories | Wellcome Sanger Institute for the COVID-19 Genomics UK (COG-UK) Consortium | Cordelia Langford; David K. Jackson; Dominic Kwiatkowski; Ewan Harrison; Ian Johnston; Jeffrey Barrett; John Sillitoe on behalf of the Wellcome Sanger Institute COVID-19 Surveillance Team; Randox Laboratories and Alex Alderton; Roberto Amato; Sonia Goncalves |
| EPI_ISL_998178, EPI_ISL_998179, EPI_ISL_998211, EPI_ISL_998229, EPI_ISL_998230, EPI_ISL_998280, EPI_ISL_998281, EPI_ISL_998283, EPI_ISL_998284, EPI_ISL_998285, EPI_ISL_998286, EPI_ISL_998288, EPI_ISL_998289, EPI_ISL_998290, EPI_ISL_998291, EPI_ISL_998292, EPI_ISL_998293, EPI_ISL_998294, EPI_ISL_998295, EPI_ISL_998296, EPI_ISL_998297, EPI_ISL_998298, EPI_ISL_1475010, EPI_ISL_1475011, EPI_ISL_1475012, EPI_ISL_1475013, EPI_ISL_1475014, EPI_ISL_1475015, EPI_ISL_1475016, EPI_ISL_1475040, EPI_ISL_1475042, EPI_ISL_1475045, EPI_ISL_1475046, EPI_ISL_1475049, EPI_ISL_1475050, EPI_ISL_1475057, EPI_ISL_1475058, EPI_ISL_1475059, EPI_ISL_1475060, EPI_ISL_1475119, EPI_ISL_1475120, EPI_ISL_1475124, EPI_ISL_1475125, EPI_ISL_1475127, EPI_ISL_1475131 | | | |
| see above | Regional Virus Laboratory, Belfast Health and Social Care Trust | COVID-19 Genomics UK (COG-UK) Consortium | Alison Watt; Ciara Cox; Conall McCaughey; David Simpson; Derek Fairley; James McKenna; Mairead Connor; Susan Feeney; Tanya Curran; Zoltan Molnar |
| EPI_ISL_1296578, EPI_ISL_1296581, EPI_ISL_1296582, EPI_ISL_1296585, EPI_ISL_1296586, EPI_ISL_1296588 | Respiratory Virus Unit, National Infection Service, Public Health England | COVID-19 Genomics UK (COG-UK) Consortium | PHE Covid Sequencing Team |
| EPI_ISL_1833616 | St George's University Hospitals NHS Foundation Trust | COVID-19 Genomics UK (COG-UK) Consortium | Adam Witney; Cassie Pope; Irene Monahan; Joshua Taylor; Ken Laing; NgeeKeong Tan |
| EPI_ISL_920671, EPI_ISL_920685, EPI_ISL_920693, EPI_ISL_920701, EPI_ISL_920702, EPI_ISL_920723, EPI_ISL_920724, EPI_ISL_920725, EPI_ISL_920728, EPI_ISL_920731, EPI_ISL_920732, EPI_ISL_920733, EPI_ISL_920734, EPI_ISL_920752, EPI_ISL_920756, EPI_ISL_920757, EPI_ISL_920758, EPI_ISL_920761, EPI_ISL_920763, EPI_ISL_920771, EPI_ISL_920776, EPI_ISL_920777, EPI_ISL_920778, EPI_ISL_920779, EPI_ISL_920780, EPI_ISL_1104771, EPI_ISL_1104775, EPI_ISL_1104776, EPI_ISL_1104777, EPI_ISL_1104779, EPI_ISL_1104780, EPI_ISL_1104786, EPI_ISL_1104793, EPI_ISL_1104805, EPI_ISL_1104807, EPI_ISL_1104808, EPI_ISL_1104813, EPI_ISL_1104815, EPI_ISL_1104816, EPI_ISL_1104817, EPI_ISL_1104821, EPI_ISL_1104823, EPI_ISL_1104829, EPI_ISL_1104833, EPI_ISL_1104835, EPI_ISL_1104837, EPI_ISL_1104839, EPI_ISL_1104840, EPI_ISL_1104845, EPI_ISL_1104850, EPI_ISL_1104851, EPI_ISL_1104852, EPI_ISL_1104853, EPI_ISL_1104858, EPI_ISL_1104860, EPI_ISL_1104861, EPI_ISL_1104862, EPI_ISL_1104865, EPI_ISL_1104866, EPI_ISL_1104870, EPI_ISL_1104871, EPI_ISL_1105163, EPI_ISL_1105201, EPI_ISL_1105220, EPI_ISL_1105332, EPI_ISL_1105342, EPI_ISL_1105346, EPI_ISL_1105347 | | | |
| see above | University College London Hospital | COVID-19 Genomics UK (COG-UK) Consortium | Catherine Houlihan; Dan Frampton; Judith Heaney; Matthew Byott; Moira Spyer and Eleni Nastouli; Stuart Kirk |
| EPI_ISL_919871, EPI_ISL_919872, EPI_ISL_919873, EPI_ISL_919881, EPI_ISL_919903, EPI_ISL_919904, EPI_ISL_919905, EPI_ISL_919906, EPI_ISL_919907, EPI_ISL_919908, EPI_ISL_919909, EPI_ISL_919910, EPI_ISL_919911, EPI_ISL_919912, EPI_ISL_919913, EPI_ISL_919914, EPI_ISL_919915, EPI_ISL_919916, EPI_ISL_919917, EPI_ISL_919918, EPI_ISL_919919, EPI_ISL_919927, EPI_ISL_919928, EPI_ISL_919929, EPI_ISL_919930, EPI_ISL_919931, EPI_ISL_919932, EPI_ISL_919934, EPI_ISL_919935, EPI_ISL_919936, EPI_ISL_919937, EPI_ISL_919938, EPI_ISL_919939, EPI_ISL_919940, EPI_ISL_919941, EPI_ISL_919942, EPI_ISL_919943, EPI_ISL_919944, EPI_ISL_919946, EPI_ISL_919947, EPI_ISL_919948, EPI_ISL_919949, EPI_ISL_919950, EPI_ISL_919951, EPI_ISL_919953, EPI_ISL_919954, EPI_ISL_919956, EPI_ISL_919958, EPI_ISL_919959, EPI_ISL_919960, EPI_ISL_920092, EPI_ISL_920093, EPI_ISL_920094, EPI_ISL_920095, EPI_ISL_920096, EPI_ISL_920097, EPI_ISL_920098, EPI_ISL_920099, EPI_ISL_920100, EPI_ISL_920101, EPI_ISL_920102, EPI_ISL_920103, EPI_ISL_920104, EPI_ISL_920105, EPI_ISL_920106, EPI_ISL_920107, EPI_ISL_920108, EPI_ISL_920109, EPI_ISL_920110, EPI_ISL_920111, EPI_ISL_920112, EPI_ISL_920113, EPI_ISL_920114, EPI_ISL_920115, EPI_ISL_920116, EPI_ISL_920152, EPI_ISL_949793, EPI_ISL_949794, EPI_ISL_949802, EPI_ISL_949909, EPI_ISL_949910, EPI_ISL_949911, EPI_ISL_949912, EPI_ISL_949913, EPI_ISL_949914, EPI_ISL_949917, EPI_ISL_949919, EPI_ISL_949948, EPI_ISL_950019, EPI_ISL_950028, EPI_ISL_950034, EPI_ISL_950039, EPI_ISL_950040, EPI_ISL_950041, EPI_ISL_950042, EPI_ISL_950043, EPI_ISL_950044, EPI_ISL_950045, EPI_ISL_950046, EPI_ISL_950047, EPI_ISL_950049, EPI_ISL_950050, EPI_ISL_950051, EPI_ISL_950052, EPI_ISL_950053, EPI_ISL_950054, EPI_ISL_950055, EPI_ISL_950056, EPI_ISL_950057, EPI_ISL_950058, EPI_ISL_950059, EPI_ISL_950060, EPI_ISL_950061, EPI_ISL_950062, EPI_ISL_950063, EPI_ISL_950064, EPI_ISL_950065, EPI_ISL_950066, EPI_ISL_950067, EPI_ISL_950171, EPI_ISL_950172, EPI_ISL_950174, EPI_ISL_950175, EPI_ISL_950176, EPI_ISL_950178, EPI_ISL_950179, EPI_ISL_950180, EPI_ISL_950181, EPI_ISL_950182, EPI_ISL_950183, EPI_ISL_950184, EPI_ISL_950185, EPI_ISL_950186, EPI_ISL_950187, EPI_ISL_950188, EPI_ISL_950189, EPI_ISL_950190, EPI_ISL_950191, EPI_ISL_950192, EPI_ISL_950193, EPI_ISL_950194, EPI_ISL_950196, EPI_ISL_950197, EPI_ISL_950199, EPI_ISL_997687, EPI_ISL_997691, EPI_ISL_997692, EPI_ISL_997753, EPI_ISL_997879, EPI_ISL_997880, EPI_ISL_997881, EPI_ISL_997887, EPI_ISL_997901, EPI_ISL_997904, EPI_ISL_997908, EPI_ISL_997909, EPI_ISL_997916, EPI_ISL_997922, EPI_ISL_997923, EPI_ISL_1048139, EPI_ISL_1048140, EPI_ISL_1048141, EPI_ISL_1048142, EPI_ISL_1048143, EPI_ISL_1048144, EPI_ISL_1048145, EPI_ISL_1048146, EPI_ISL_1048147, EPI_ISL_1048148, EPI_ISL_1048149, EPI_ISL_1048150, EPI_ISL_1048151, EPI_ISL_1048152, EPI_ISL_1048153, EPI_ISL_1048154, EPI_ISL_1048155, EPI_ISL_1048156, EPI_ISL_1048157, EPI_ISL_1048158, EPI_ISL_1048159, EPI_ISL_1048160, EPI_ISL_1048161, EPI_ISL_1048162, EPI_ISL_1048163, EPI_ISL_1048164, EPI_ISL_1048165, EPI_ISL_1048166, EPI_ISL_1048167, EPI_ISL_1048168, EPI_ISL_1048169, EPI_ISL_1048170, EPI_ISL_1048171, EPI_ISL_1048172, EPI_ISL_1048173, EPI_ISL_1048174, EPI_ISL_1048175, EPI_ISL_1048176, EPI_ISL_1048177, EPI_ISL_1048178, EPI_ISL_1048179, EPI_ISL_1048180, EPI_ISL_1048181, EPI_ISL_1048182, EPI_ISL_1048183, EPI_ISL_1048184, EPI_ISL_1048185, EPI_ISL_1048186, EPI_ISL_1048187, EPI_ISL_1048188, EPI_ISL_1048189, EPI_ISL_1048190, EPI_ISL_1048191, EPI_ISL_1048192, EPI_ISL_1048193, EPI_ISL_1048194, EPI_ISL_1048195, EPI_ISL_1048196, EPI_ISL_1048197, EPI_ISL_1048198, EPI_ISL_1048199, EPI_ISL_1048200, EPI_ISL_1048201, EPI_ISL_1048202, EPI_ISL_1048203, EPI_ISL_1048204, EPI_ISL_1048205, EPI_ISL_1048206, EPI_ISL_1048207, EPI_ISL_1048208, EPI_ISL_1048209, EPI_ISL_1048210, EPI_ISL_1048211, EPI_ISL_1048212, EPI_ISL_1048213, EPI_ISL_1048214, EPI_ISL_1048215, EPI_ISL_1048233, EPI_ISL_1048234, EPI_ISL_1048235, EPI_ISL_1048236, EPI_ISL_1048237, EPI_ISL_1050369, EPI_ISL_1050392, EPI_ISL_1104611, EPI_ISL_1104612, EPI_ISL_1104613, EPI_ISL_1104614, EPI_ISL_1104615, EPI_ISL_1104616, EPI_ISL_1104617, EPI_ISL_1104618, EPI_ISL_1104619, EPI_ISL_1105463, EPI_ISL_1105465, EPI_ISL_1248167, EPI_ISL_1248188, EPI_ISL_1248207, EPI_ISL_1248208, EPI_ISL_1248209, EPI_ISL_1248210, EPI_ISL_1248211, EPI_ISL_1248212, EPI_ISL_1248213, EPI_ISL_1309055, EPI_ISL_1309063, EPI_ISL_1309066, EPI_ISL_1309110, EPI_ISL_1386883, EPI_ISL_1474838, EPI_ISL_1637110, EPI_ISL_1637126, EPI_ISL_1637128, EPI_ISL_1637157, EPI_ISL_1637158 | | | |
| see above | University College London, Great Ormond Street Hospital for Children NHS Foundation Trust, Imperial College Healthcare NHS Trust | COVID-19 Genomics UK (COG-UK) Consortium | Alison Holmes; Charlotte Williams; Helena Tutill; Jacqueline Findlay; James Price; Judith Breuer; Julianne Brown; Kathryn Harris; Leysa Forrest; Mark Kristiansen; Paola Niola; Paola Resende Silva; Patricia Dyal; Paul Randell; Rachel Williams; Samuel Weeks; Sergi Castellano; Sunando Roy; Tony Brooks; Yasmin Panchbhaya |
| EPI_ISL_949410, EPI_ISL_996401, EPI_ISL_996408, EPI_ISL_996412, EPI_ISL_996456, EPI_ISL_996458, EPI_ISL_996462, EPI_ISL_996463, EPI_ISL_996469, EPI_ISL_1046990, EPI_ISL_1046995, EPI_ISL_1047011, EPI_ISL_1047019, EPI_ISL_1047034, EPI_ISL_1047036, EPI_ISL_1047045, EPI_ISL_1047049 | | | |
| see above | University of Birmingham | COVID-19 Genomics UK (COG-UK) Consortium | Alex Richter; Andrew Bosworth; Andrew Bosworth. Queen Elizabeth Hospital: Anna Casey; Andrew D Beggs PHE Heartlands Lab: Husam Osman; Charlotte Poxon; Institute of Microbiology; Joanne Stockton; Josh Quick; Kasun Wanigasooriya; Mike Kidd; Nicholas Loman. University of Birmingham Testing Laboratory: Celina M Whalley; Oliver Pickles; Radoslaw Poplawski; Samuel Nicholls; University of Birmingham: Claire McMurray; Will Rowe |
| EPI_ISL_996543, EPI_ISL_996544, EPI_ISL_996545, EPI_ISL_996546, EPI_ISL_996551, EPI_ISL_996552, EPI_ISL_996557, EPI_ISL_996595, EPI_ISL_996597, EPI_ISL_997165, EPI_ISL_997166, EPI_ISL_997167, EPI_ISL_997168, EPI_ISL_997169, EPI_ISL_997170, EPI_ISL_997171, EPI_ISL_997172, EPI_ISL_997174, EPI_ISL_997175, EPI_ISL_997176, EPI_ISL_997177, EPI_ISL_997178, EPI_ISL_997179, EPI_ISL_997186, EPI_ISL_997187, EPI_ISL_997266, EPI_ISL_997267, EPI_ISL_997268, EPI_ISL_997274, EPI_ISL_997282, EPI_ISL_997283, EPI_ISL_997284, EPI_ISL_997336, EPI_ISL_997338, EPI_ISL_1054531, EPI_ISL_1103759, EPI_ISL_1308546, EPI_ISL_1308557, EPI_ISL_1308558, EPI_ISL_1308560, EPI_ISL_1308571, EPI_ISL_1308575, EPI_ISL_1308576, EPI_ISL_1308577, EPI_ISL_1308639, EPI_ISL_1545818 | | | |
| see above | University of Exeter | COVID-19 Genomics UK (COG-UK) Consortium | Aaron Jeffries; Audrey Farbos; Ben Temperton; Jane Masoli; Joanna Warwick-Dugdale; Michelle Michelsen; Robyn Manley; Stephen Michell |
| EPI_ISL_997094, EPI_ISL_997095, EPI_ISL_997096, EPI_ISL_997097, EPI_ISL_997098, EPI_ISL_997099, EPI_ISL_997100, EPI_ISL_997101, EPI_ISL_997102, EPI_ISL_997103, EPI_ISL_997132, EPI_ISL_997133, EPI_ISL_997134, EPI_ISL_997135, EPI_ISL_997136, EPI_ISL_997137, EPI_ISL_997138, EPI_ISL_997139, EPI_ISL_1047860, EPI_ISL_1104222, EPI_ISL_1104225, EPI_ISL_1104226, EPI_ISL_1104227, EPI_ISL_1177707, EPI_ISL_1177708, EPI_ISL_1177729, EPI_ISL_1177731, EPI_ISL_1177733, EPI_ISL_1247689, EPI_ISL_1247690, EPI_ISL_1247693, EPI_ISL_1247729, EPI_ISL_1308819, EPI_ISL_1308820, EPI_ISL_1308823, EPI_ISL_1308824, EPI_ISL_1308850, EPI_ISL_1308854, EPI_ISL_1308855, EPI_ISL_1730104 | | | |
| see above | Virology Department, Royal Infirmary of Edinburgh, NHS Lothian / School of Biological Sciences, University of Edinburgh | COVID-19 Genomics UK (COG-UK) Consortium | Colquhoun R; Cotton S; Dewar R; Gallagher A; Hill V; Jackson B; Maloney D; McCrone JT; McHugh M; O'Toole Á; Rambaut A; Rooke S; Scher E; Templeton K; Yu X |
| EPI_ISL_919405, EPI_ISL_949645, EPI_ISL_949646 | Virology Department, Royal Infirmary of Edinburgh, NHS Lothian / School of Biological Sciences, University of Edinburgh / Institute of Genetics and Molecular Medicine, University of Edinburgh | COVID-19 Genomics UK (COG-UK) Consortium | Balcaza C; Colquhoun R; Dewar R; Gallagher M; Hill V; Jackson B; McCrone JT; McHugh M; O'Toole Á; Rambaut A; Rooke S; Scher E; Templeton K; Williams TC; Yu X |
| EPI_ISL_924082, EPI_ISL_924085, EPI_ISL_924087, EPI_ISL_924089, EPI_ISL_924092, EPI_ISL_924097, EPI_ISL_924099, EPI_ISL_924103, EPI_ISL_924105, EPI_ISL_924109, EPI_ISL_924111, EPI_ISL_924112, EPI_ISL_924117, EPI_ISL_924118, EPI_ISL_924125, EPI_ISL_924126, EPI_ISL_924129, EPI_ISL_924131, EPI_ISL_924136, EPI_ISL_924142, EPI_ISL_924144, EPI_ISL_924146, EPI_ISL_924153, EPI_ISL_924160, EPI_ISL_924163, EPI_ISL_924165, EPI_ISL_924173, EPI_ISL_924183, EPI_ISL_924184, EPI_ISL_924185, EPI_ISL_924186, EPI_ISL_924191, EPI_ISL_924193, EPI_ISL_924198, EPI_ISL_924200, EPI_ISL_924201, EPI_ISL_924209, EPI_ISL_924212, EPI_ISL_924216, EPI_ISL_924217, EPI_ISL_924223, EPI_ISL_924227, EPI_ISL_924228, EPI_ISL_924229, EPI_ISL_924235, EPI_ISL_924238, EPI_ISL_924243, EPI_ISL_924249, EPI_ISL_924251, EPI_ISL_924258, EPI_ISL_924263, EPI_ISL_924269, EPI_ISL_924273, EPI_ISL_924276, EPI_ISL_924277, EPI_ISL_924282, EPI_ISL_924287, EPI_ISL_924293, EPI_ISL_924297, EPI_ISL_924298, EPI_ISL_924304, EPI_ISL_924305, EPI_ISL_924306, EPI_ISL_924308, EPI_ISL_924313, EPI_ISL_924330, EPI_ISL_924335, EPI_ISL_924339, EPI_ISL_924340, EPI_ISL_924350, EPI_ISL_924352, EPI_ISL_924356, EPI_ISL_924359, EPI_ISL_924361, EPI_ISL_924366, EPI_ISL_924368, EPI_ISL_924374, EPI_ISL_924377, EPI_ISL_924379, EPI_ISL_924382, EPI_ISL_924390, EPI_ISL_924410, EPI_ISL_924414, EPI_ISL_952953, EPI_ISL_952978, EPI_ISL_1000763, EPI_ISL_1000787, EPI_ISL_1047679, EPI_ISL_1047680, EPI_ISL_1047681, EPI_ISL_1047682, EPI_ISL_1047683, EPI_ISL_1047684, EPI_ISL_1047686, EPI_ISL_1047687, EPI_ISL_1047688, EPI_ISL_1047689, EPI_ISL_1047690, EPI_ISL_1047691, EPI_ISL_1047692, EPI_ISL_1047693, EPI_ISL_1047694, EPI_ISL_1047695, EPI_ISL_1047696, EPI_ISL_1047697, EPI_ISL_1047698, EPI_ISL_1047699, EPI_ISL_1047700, EPI_ISL_1047701, EPI_ISL_1047702, EPI_ISL_1047703, EPI_ISL_1047705, EPI_ISL_1047706, EPI_ISL_1047707, EPI_ISL_1047708, EPI_ISL_1047709, EPI_ISL_1047710, EPI_ISL_1047711, EPI_ISL_1050065, EPI_ISL_1180083, EPI_ISL_1180087 | | | |
| see above | Virology Department, Sheffield Teaching Hospitals NHS Foundation Trust/Department of Infection, Immunity and Cardiovascular Disease, The Medical School, University of Sheffield | COVID-19 Genomics UK (COG-UK) Consortium | Adri Angyal; Alex Keeley; Benjamin Lindsey; Cariad Evans; Danielle Groves; Dave Partridge; Katie Johnson; Laura Carrilero; Luke Green; Matthew Parker; Matthew Wyles; Mehmet Yavuz; Mohammad Raza; Nikki Smith; Paul Parsons; Rachel Tucker; Rebecca Brown; Thushan de Silva |
| EPI_ISL_999987, EPI_ISL_1051921, EPI_ISL_1051931, EPI_ISL_1051945, EPI_ISL_1051951, EPI_ISL_1051960, EPI_ISL_1051965, EPI_ISL_1051977, EPI_ISL_1051980, EPI_ISL_1051996, EPI_ISL_1052019, EPI_ISL_1052020, EPI_ISL_1052023, EPI_ISL_1052028, EPI_ISL_1052030, EPI_ISL_1052032, EPI_ISL_1052037, EPI_ISL_1052046, EPI_ISL_1052049, EPI_ISL_1052054, EPI_ISL_1052057, EPI_ISL_1052059, EPI_ISL_1052069, EPI_ISL_1052085, EPI_ISL_1052093, EPI_ISL_1052096, EPI_ISL_1052097, EPI_ISL_1052108, EPI_ISL_1052113, EPI_ISL_1052114, EPI_ISL_1052115, EPI_ISL_1052130, EPI_ISL_1052131, EPI_ISL_1052135, EPI_ISL_1052517, EPI_ISL_1052518, EPI_ISL_1052519, EPI_ISL_1052520, EPI_ISL_1053552, EPI_ISL_1053554, EPI_ISL_1053558, EPI_ISL_1178885, EPI_ISL_1178892, EPI_ISL_1476495, EPI_ISL_1833135, EPI_ISL_1833144, EPI_ISL_1833161 | | | |
| see above | Wales Specialist Virology Centre Sequencing lab: Pathogen Genomics Unit | Public Health Wales Microbiology Cardiff Wales Specialist Virology Centre | Alec Birchley; Alexander Adams; Amy Gaskin; Angela Marchbank; Bree Gatica-Wilcox; Catherine Moore; Jason Coombes; Joanne Watkins; Joel Southgate; Johnathan Evans; Laura Gifford; Lauren Gilbert; Lee Graham; Malorie Perry; Matthew Bull; Nicole Pacchiarini; Sally Corden; Sara Kumziene-Summerhayes; Sara Rey; Sarah Taylor; Simon Cottrell; Sophie Jones; Tom Connor |
| EPI_ISL_949611, EPI_ISL_949615, EPI_ISL_997052, EPI_ISL_997053, EPI_ISL_997054, EPI_ISL_997055, EPI_ISL_997056, EPI_ISL_997057, EPI_ISL_997058, EPI_ISL_997059, EPI_ISL_997060, EPI_ISL_1047846 | | | |
| see above | West of Scotland Specialist Virology Centre, NHSGGC / MRC-University of Glasgow Centre for Virus Research | COVID-19 Genomics UK (COG-UK) Consortium | Alasdair MacLean; Alice Broos; Ana da Silva Filipe; Antonia Ho; Daniel Mair; David L Robertson; Emma Thomson; Guy Mollett; James Shepherd; Jenna Nichols; Joseph Hughes; Kathy Li; Kathy Smollett; Kyriaki Nomikou; Lily Tong; Matthew Holden; Natasha Johnson; Rachel Blacow; Richard Orton; Rory Gunson; Sarah McDonald; Sharif Shaaban; Sreenu Vattipally; Stephen Carmichael |

We gratefully acknowledge the following Authors from the Originating laboratories responsible for obtaining the specimens, as well as the Submitting laboratories where the genome data were generated and shared via GISAID, on which this research is based.

All Submitters of data may be contacted directly via www.gisaid.org

Authors are sorted alphabetically.

| Accession ID | Originating Laboratory | Submitting Laboratory | Authors |
|---|---|---|---|
| EPI_ISL_997657, EPI_ISL_997658, EPI_ISL_997659, EPI_ISL_997660, EPI_ISL_997665, EPI_ISL_997672, EPI_ISL_1048060, EPI_ISL_1048063, EPI_ISL_1048064, EPI_ISL_1048070, EPI_ISL_1048079, EPI_ISL_1048080, EPI_ISL_1048082, EPI_ISL_1048084, EPI_ISL_1048104, EPI_ISL_1048135 | | | |
| see above | Barts Health NHS Trust | COVID-19 Genomics UK (COG-UK) Consortium | BROAD; Beatrix; CUTINO-MOGUEL; Claire; David; Dola; HARRINGTON; KELE; KULASEGARAN-SHYLINI; Maria-Teresa; OWOYEMI; Raghavendran |
| EPI_ISL_953240, EPI_ISL_953245, EPI_ISL_953248, EPI_ISL_953249, EPI_ISL_953250, EPI_ISL_953252, EPI_ISL_953253, EPI_ISL_953255, EPI_ISL_953256, EPI_ISL_953257, EPI_ISL_953258, EPI_ISL_953264, EPI_ISL_953265, EPI_ISL_953266, EPI_ISL_953268, EPI_ISL_953269, EPI_ISL_953270, EPI_ISL_953271, EPI_ISL_953275, EPI_ISL_953276, EPI_ISL_953277, EPI_ISL_953279, EPI_ISL_953280, EPI_ISL_953282, EPI_ISL_953283, EPI_ISL_953284, EPI_ISL_953285, EPI_ISL_1000864, EPI_ISL_1000923, EPI_ISL_1000933, EPI_ISL_1054825, EPI_ISL_1054826, EPI_ISL_1054827, EPI_ISL_1054828, EPI_ISL_1054829, EPI_ISL_1054831 | | | |
| see above | Bioinformatics and Biostatistics Lab, Advanced Sequencing Facility | COVID-19 Genomics UK (COG-UK) Consortium | Aengus Stewart; Chelsea Sawyer; Harshil Patel; Jerome Nicod; Laura Cubitt; Margaret Crawford |
| EPI_ISL_952488, EPI_ISL_1000191, EPI_ISL_1000288, EPI_ISL_1000289, EPI_ISL_1000290, EPI_ISL_1000293, EPI_ISL_1000294, EPI_ISL_1000295, EPI_ISL_1000296, EPI_ISL_1000299, EPI_ISL_1000300, EPI_ISL_1000301, EPI_ISL_1000303, EPI_ISL_1000304, EPI_ISL_1000305, EPI_ISL_1000306, EPI_ISL_1000313, EPI_ISL_1000315, EPI_ISL_1000316, EPI_ISL_1000323, EPI_ISL_1000461, EPI_ISL_1000462, EPI_ISL_1000463, EPI_ISL_1000464, EPI_ISL_1000465, EPI_ISL_1000525, EPI_ISL_1000542, EPI_ISL_1000546, EPI_ISL_1000595, EPI_ISL_1000596, EPI_ISL_1000597, EPI_ISL_1000598, EPI_ISL_1000599, EPI_ISL_1000600, EPI_ISL_1000602, EPI_ISL_1000603, EPI_ISL_1000604, EPI_ISL_1000612, EPI_ISL_1000613, EPI_ISL_1000614, EPI_ISL_1000615, EPI_ISL_1000616, EPI_ISL_1000617, EPI_ISL_1050793, EPI_ISL_1050799, EPI_ISL_1050816, EPI_ISL_1050817, EPI_ISL_1050818, EPI_ISL_1050854, EPI_ISL_1050874, EPI_ISL_1050879, EPI_ISL_1050880, EPI_ISL_1050911, EPI_ISL_1053851, EPI_ISL_1053852, EPI_ISL_1053856, EPI_ISL_1053857, EPI_ISL_1053893, EPI_ISL_1053932, EPI_ISL_1053955, EPI_ISL_1053956, EPI_ISL_1107632, EPI_ISL_1107655, EPI_ISL_1107656, EPI_ISL_1107657, EPI_ISL_1107658, EPI_ISL_1107659, EPI_ISL_1107660, EPI_ISL_1107661, EPI_ISL_1107662, EPI_ISL_1107663, EPI_ISL_1107785, EPI_ISL_1179828, EPI_ISL_1179829, EPI_ISL_1250029, EPI_ISL_1979867, EPI_ISL_1979868, EPI_ISL_1979870, EPI_ISL_1979871, EPI_ISL_1979873, EPI_ISL_1979874, EPI_ISL_1979876, EPI_ISL_1979877, EPI_ISL_1979879 | | | |
| see above | Centre for Enzyme Innovation, University of Portsmouth / Translational Research Laboratory, Portsmouth Hospitals NHS Trust | COVID-19 Genomics UK (COG-UK) Consortium | Angela Beckett; Christopher Fearn; Kate Cook; Katie Loveson; Salman Goudarzi; Samuel Robson; Scott Elliott; Sharon Glaysher |
| EPI_ISL_949543, EPI_ISL_949545, EPI_ISL_949547, EPI_ISL_949551, EPI_ISL_949552, EPI_ISL_949557, EPI_ISL_949558, EPI_ISL_949559, EPI_ISL_949560, EPI_ISL_949561, EPI_ISL_949562, EPI_ISL_949563, EPI_ISL_949564, EPI_ISL_949565, EPI_ISL_949566, EPI_ISL_949567, EPI_ISL_949568, EPI_ISL_949569, EPI_ISL_949570, EPI_ISL_949571, EPI_ISL_949572, EPI_ISL_949573, EPI_ISL_949574, EPI_ISL_949575, EPI_ISL_949576, EPI_ISL_949577, EPI_ISL_949578, EPI_ISL_949579, EPI_ISL_949580, EPI_ISL_949581, EPI_ISL_949582, EPI_ISL_949583, EPI_ISL_949584, EPI_ISL_949585, EPI_ISL_949586, EPI_ISL_949587, EPI_ISL_949588, EPI_ISL_949589, EPI_ISL_949590, EPI_ISL_949591, EPI_ISL_949592, EPI_ISL_949593, EPI_ISL_949594, EPI_ISL_949595, EPI_ISL_949596, EPI_ISL_949597, EPI_ISL_949598, EPI_ISL_949599, EPI_ISL_949600, EPI_ISL_949601, EPI_ISL_949602, EPI_ISL_949603, EPI_ISL_949604, EPI_ISL_949605, EPI_ISL_949606, EPI_ISL_949607, EPI_ISL_996613, EPI_ISL_996615, EPI_ISL_996616, EPI_ISL_996617, EPI_ISL_996618, EPI_ISL_996619, EPI_ISL_996620, EPI_ISL_996621, EPI_ISL_996622, EPI_ISL_996623, EPI_ISL_996624, EPI_ISL_996625, EPI_ISL_996626, EPI_ISL_996628, EPI_ISL_996629, EPI_ISL_996631, EPI_ISL_996632, EPI_ISL_996633, EPI_ISL_996634, EPI_ISL_996635, EPI_ISL_996636, EPI_ISL_996644, EPI_ISL_996701, EPI_ISL_996711, EPI_ISL_1047188, EPI_ISL_1047189, EPI_ISL_1047192, EPI_ISL_1047193, EPI_ISL_1047349, EPI_ISL_1047350, EPI_ISL_1047351, EPI_ISL_1047353, EPI_ISL_1047354, EPI_ISL_1050605, EPI_ISL_1050606, EPI_ISL_1050610, EPI_ISL_1050614, EPI_ISL_1050619, EPI_ISL_1050632, EPI_ISL_1050633, EPI_ISL_1050634, EPI_ISL_1050635, EPI_ISL_1050636, EPI_ISL_1050637, EPI_ISL_1050638, EPI_ISL_1050639, EPI_ISL_1050640, EPI_ISL_1050641, EPI_ISL_1050642, EPI_ISL_1050643, EPI_ISL_1050644, EPI_ISL_1050645, EPI_ISL_1050646, EPI_ISL_1050647, EPI_ISL_1050648, EPI_ISL_1050649, EPI_ISL_1050650, EPI_ISL_1050651, EPI_ISL_1050652, EPI_ISL_1050653, EPI_ISL_1050654, EPI_ISL_1050655, EPI_ISL_1050656, EPI_ISL_1050657, EPI_ISL_1050658, EPI_ISL_1050659, EPI_ISL_1050660, EPI_ISL_1050661, EPI_ISL_1050662, EPI_ISL_1050663, EPI_ISL_1050664, EPI_ISL_1050665, EPI_ISL_1050666, EPI_ISL_1050667, EPI_ISL_1050668, EPI_ISL_1050669, EPI_ISL_1050670, EPI_ISL_1050671, EPI_ISL_1050672, EPI_ISL_1050673, EPI_ISL_1050674, EPI_ISL_1050675, EPI_ISL_1050685, EPI_ISL_1050686, EPI_ISL_1050687, EPI_ISL_1050688, EPI_ISL_1050689, EPI_ISL_1050690, EPI_ISL_1050691, EPI_ISL_1050692, EPI_ISL_1050693, EPI_ISL_1050694, EPI_ISL_1050695, EPI_ISL_1050696, EPI_ISL_1050697, EPI_ISL_1050698, EPI_ISL_1050699, EPI_ISL_1050700, EPI_ISL_1050701, EPI_ISL_1050702, EPI_ISL_1050704, EPI_ISL_1050705, EPI_ISL_1050706, EPI_ISL_1050707, EPI_ISL_1050708, EPI_ISL_1050709, EPI_ISL_1050710, EPI_ISL_1050711, EPI_ISL_1050712, EPI_ISL_1050713, EPI_ISL_1050714, EPI_ISL_1050715, EPI_ISL_1050716, EPI_ISL_1050717, EPI_ISL_1050718, EPI_ISL_1050719, EPI_ISL_1050720, EPI_ISL_1050721, EPI_ISL_1050722, EPI_ISL_1103884, EPI_ISL_1103888, EPI_ISL_1103890, EPI_ISL_1103945, EPI_ISL_1179891, EPI_ISL_1179892, EPI_ISL_1179893, EPI_ISL_1179894, EPI_ISL_1179895, EPI_ISL_1179896, EPI_ISL_1179897, EPI_ISL_1179898, EPI_ISL_1179899, EPI_ISL_1179900, EPI_ISL_1179901, EPI_ISL_1179902, EPI_ISL_1179903, EPI_ISL_1179904, EPI_ISL_1179905, EPI_ISL_1179906, EPI_ISL_1179907, EPI_ISL_1179908, EPI_ISL_1179909, EPI_ISL_1179910, EPI_ISL_1179912, EPI_ISL_1179913, EPI_ISL_1179914, EPI_ISL_1179915, EPI_ISL_1179916, EPI_ISL_1179917, EPI_ISL_1179918, EPI_ISL_1179919, EPI_ISL_1179920, EPI_ISL_1179921, EPI_ISL_1179922, EPI_ISL_1179923, EPI_ISL_1179924, EPI_ISL_1179925, EPI_ISL_1179926, EPI_ISL_1179927, EPI_ISL_1179928, EPI_ISL_1179929, EPI_ISL_1179930, EPI_ISL_1179931, EPI_ISL_1179932, EPI_ISL_1179934, EPI_ISL_1179937, EPI_ISL_1179938, EPI_ISL_1179939, EPI_ISL_1179940, EPI_ISL_1179941, EPI_ISL_1179942, EPI_ISL_1179943, EPI_ISL_1179944, EPI_ISL_1179945, EPI_ISL_1179946, EPI_ISL_1179947, EPI_ISL_1179948, EPI_ISL_1179949, EPI_ISL_1179950, EPI_ISL_1179964, EPI_ISL_1179965, EPI_ISL_1179966, EPI_ISL_1179967, EPI_ISL_1179968, EPI_ISL_1179969, EPI_ISL_1179970, EPI_ISL_1179971, EPI_ISL_1179972, EPI_ISL_1179973, EPI_ISL_1179974, EPI_ISL_1179979, EPI_ISL_1179980, EPI_ISL_1179981, EPI_ISL_1179982, EPI_ISL_1179984, EPI_ISL_1179985, EPI_ISL_1179986, EPI_ISL_1179987, EPI_ISL_1179988, EPI_ISL_1179989, EPI_ISL_1179997, EPI_ISL_1179998, EPI_ISL_1179999, EPI_ISL_1180000, EPI_ISL_1180001, EPI_ISL_1180002, EPI_ISL_1180003, EPI_ISL_1180004, EPI_ISL_1180005, EPI_ISL_1180006, EPI_ISL_1180007, EPI_ISL_1180008, EPI_ISL_1180009, EPI_ISL_1180010, EPI_ISL_1180011, EPI_ISL_1180012, EPI_ISL_1180013, EPI_ISL_1180014, EPI_ISL_1180015, EPI_ISL_1180016, EPI_ISL_1180018, EPI_ISL_1180019, EPI_ISL_1180020, EPI_ISL_1180021, EPI_ISL_1180022, EPI_ISL_1180029, EPI_ISL_1180030, EPI_ISL_1180031, EPI_ISL_1180032, EPI_ISL_1180033, EPI_ISL_1180034, EPI_ISL_1180035, EPI_ISL_1180036, EPI_ISL_1180037, EPI_ISL_1180038, EPI_ISL_1180039, EPI_ISL_1180040, EPI_ISL_1180041, EPI_ISL_1180042, EPI_ISL_1180043, EPI_ISL_1180044, EPI_ISL_1180045, EPI_ISL_1180046, EPI_ISL_1180047, EPI_ISL_1180048, EPI_ISL_1180049, EPI_ISL_1180050, EPI_ISL_1180051, EPI_ISL_1180052 | | | |
| see above | Department of Pathology, University of Cambridge | COVID-19 Genomics UK (COG-UK) Consortium | Aminu S. Jahun; Ian Goodfellow; Iliana Georgana; Martin D. Curran; Myra Hosmillo; Rhys Izu; Rhys Izuagbe; Surendra Parmar; William L. Hamilton; Yasmin Chaudhry |
| EPI_ISL_945374, EPI_ISL_945375, EPI_ISL_945376, EPI_ISL_945379, EPI_ISL_945386, EPI_ISL_945387, EPI_ISL_945389, EPI_ISL_945402, EPI_ISL_945408, EPI_ISL_945409, EPI_ISL_945419, EPI_ISL_945420, EPI_ISL_945422, EPI_ISL_945423, EPI_ISL_945433, EPI_ISL_945444, EPI_ISL_945446, EPI_ISL_945449, EPI_ISL_945458, EPI_ISL_945459, EPI_ISL_945466, EPI_ISL_945467, EPI_ISL_945468, EPI_ISL_945469, EPI_ISL_945475, EPI_ISL_945480, EPI_ISL_945485, EPI_ISL_945490, EPI_ISL_945494, EPI_ISL_945495, EPI_ISL_945498, EPI_ISL_945502, EPI_ISL_945506, EPI_ISL_945508, EPI_ISL_945514, EPI_ISL_945518, EPI_ISL_945522, EPI_ISL_945525, EPI_ISL_945529, EPI_ISL_945538, EPI_ISL_945539, EPI_ISL_945543, EPI_ISL_945545, EPI_ISL_945546, EPI_ISL_945548, EPI_ISL_945549, EPI_ISL_945555, EPI_ISL_945561, EPI_ISL_945563, EPI_ISL_945569, EPI_ISL_945579, EPI_ISL_945582, EPI_ISL_945587, EPI_ISL_945589, EPI_ISL_945591, EPI_ISL_945597, EPI_ISL_945619, EPI_ISL_945621, EPI_ISL_945627, EPI_ISL_945635, EPI_ISL_945636, EPI_ISL_945648, EPI_ISL_945650, EPI_ISL_945656, EPI_ISL_945664, EPI_ISL_945668, EPI_ISL_945672, EPI_ISL_945674, EPI_ISL_945677, EPI_ISL_945678, EPI_ISL_945681, EPI_ISL_945682, EPI_ISL_945692, EPI_ISL_945696, EPI_ISL_945697, EPI_ISL_945698, EPI_ISL_945701, EPI_ISL_945704, EPI_ISL_956422, EPI_ISL_956423, EPI_ISL_956424, EPI_ISL_956425, EPI_ISL_956426, EPI_ISL_956427, EPI_ISL_956428, EPI_ISL_956429, EPI_ISL_956430, EPI_ISL_956431, EPI_ISL_956432, EPI_ISL_956433, EPI_ISL_956434, EPI_ISL_956435, EPI_ISL_956436, EPI_ISL_956437, EPI_ISL_956438, EPI_ISL_956439, EPI_ISL_956440, EPI_ISL_956441, EPI_ISL_956442, EPI_ISL_956443, EPI_ISL_956444, EPI_ISL_956445, EPI_ISL_956447, EPI_ISL_956448, EPI_ISL_956449, EPI_ISL_956450, EPI_ISL_956451, EPI_ISL_956452, EPI_ISL_956453, EPI_ISL_956454, EPI_ISL_956455, EPI_ISL_956456, EPI_ISL_956457, EPI_ISL_956458, EPI_ISL_956459, EPI_ISL_956460, EPI_ISL_956461, EPI_ISL_956462, EPI_ISL_956463, EPI_ISL_956464, EPI_ISL_956465, EPI_ISL_956466, EPI_ISL_956467, EPI_ISL_956468, EPI_ISL_956469, EPI_ISL_956470, EPI_ISL_956471, EPI_ISL_956472, EPI_ISL_956473, EPI_ISL_956474, EPI_ISL_956475, EPI_ISL_956476, EPI_ISL_956477, EPI_ISL_956478, EPI_ISL_956479, EPI_ISL_956480, EPI_ISL_956481, EPI_ISL_956482, EPI_ISL_956483, EPI_ISL_956484, EPI_ISL_956485, EPI_ISL_956486, EPI_ISL_956488, EPI_ISL_956489, EPI_ISL_956490, EPI_ISL_956491, EPI_ISL_956492, EPI_ISL_956493, EPI_ISL_956494, EPI_ISL_956495, EPI_ISL_956496, EPI_ISL_956497, EPI_ISL_956498, EPI_ISL_956499, EPI_ISL_956500, EPI_ISL_956501, EPI_ISL_956502, EPI_ISL_956503, EPI_ISL_956504, EPI_ISL_956505, EPI_ISL_956506, EPI_ISL_956507, EPI_ISL_956509, EPI_ISL_956510, EPI_ISL_956511, EPI_ISL_956512, EPI_ISL_956513, EPI_ISL_956514, EPI_ISL_956515, EPI_ISL_956516, EPI_ISL_956517, EPI_ISL_956518, EPI_ISL_956519, EPI_ISL_956520, EPI_ISL_956521, EPI_ISL_956522, EPI_ISL_956523, EPI_ISL_956524, EPI_ISL_956525, EPI_ISL_956526, EPI_ISL_956527, EPI_ISL_956528, EPI_ISL_956529, EPI_ISL_956530, EPI_ISL_956531, EPI_ISL_956532, EPI_ISL_956533, EPI_ISL_956534, EPI_ISL_956535, EPI_ISL_956536, EPI_ISL_956537, EPI_ISL_956538, EPI_ISL_956539, EPI_ISL_956540, EPI_ISL_956541, EPI_ISL_956542, EPI_ISL_956543, EPI_ISL_956544, EPI_ISL_956545, EPI_ISL_956546, EPI_ISL_956547, EPI_ISL_956548, EPI_ISL_956549, EPI_ISL_956550, EPI_ISL_956551, EPI_ISL_956552, EPI_ISL_956553, EPI_ISL_956554, EPI_ISL_956555, EPI_ISL_956556, EPI_ISL_956557, EPI_ISL_956558, EPI_ISL_956559, EPI_ISL_956560, EPI_ISL_956561, EPI_ISL_956563, EPI_ISL_956564, EPI_ISL_956565, EPI_ISL_956566, EPI_ISL_956567, EPI_ISL_956568, EPI_ISL_956569, EPI_ISL_956570, EPI_ISL_956571, EPI_ISL_956572, EPI_ISL_956573, EPI_ISL_956574, EPI_ISL_956575, EPI_ISL_956577, EPI_ISL_956578, EPI_ISL_956579, EPI_ISL_956580, EPI_ISL_956581, EPI_ISL_956582, EPI_ISL_956583, EPI_ISL_956584, EPI_ISL_956586, EPI_ISL_956588, EPI_ISL_956589, EPI_ISL_956590, EPI_ISL_956591, EPI_ISL_956593, EPI_ISL_956594, EPI_ISL_956595, EPI_ISL_956596, EPI_ISL_956597, EPI_ISL_956598, EPI_ISL_956599, EPI_ISL_956600, EPI_ISL_956603, EPI_ISL_956604, EPI_ISL_956605, EPI_ISL_956607, EPI_ISL_956608, EPI_ISL_956609, EPI_ISL_956610, EPI_ISL_956611, EPI_ISL_956612, EPI_ISL_956613, EPI_ISL_956614, EPI_ISL_956615, EPI_ISL_956616, EPI_ISL_956617, EPI_ISL_956619, EPI_ISL_956620, EPI_ISL_956621, EPI_ISL_956622, EPI_ISL_956623, EPI_ISL_956624, EPI_ISL_956627, EPI_ISL_956628, EPI_ISL_956629, EPI_ISL_956630, EPI_ISL_956632, EPI_ISL_956633, EPI_ISL_956634, EPI_ISL_956635, EPI_ISL_956637, EPI_ISL_956638, EPI_ISL_956639, EPI_ISL_956640, EPI_ISL_956641, EPI_ISL_956642, EPI_ISL_956643, EPI_ISL_956644, EPI_ISL_956645, EPI_ISL_956646, EPI_ISL_956647, EPI_ISL_956648, EPI_ISL_956649, EPI_ISL_956650, EPI_ISL_956652, EPI_ISL_956653, EPI_ISL_956654, EPI_ISL_956655, EPI_ISL_956656, EPI_ISL_956657, EPI_ISL_956658, EPI_ISL_956659, EPI_ISL_956660, EPI_ISL_956661, EPI_ISL_956662, EPI_ISL_956663, EPI_ISL_956664, EPI_ISL_956665, EPI_ISL_956666, EPI_ISL_956667, EPI_ISL_956669, EPI_ISL_956670, EPI_ISL_956671, EPI_ISL_956672, EPI_ISL_956673, EPI_ISL_956674, EPI_ISL_956675, EPI_ISL_956676, EPI_ISL_956677, EPI_ISL_956678, EPI_ISL_956679, EPI_ISL_956680, EPI_ISL_956681, EPI_ISL_956682, EPI_ISL_956683, EPI_ISL_956684, EPI_ISL_956685, EPI_ISL_956686, EPI_ISL_956687, EPI_ISL_956688, EPI_ISL_956689, EPI_ISL_956692, EPI_ISL_956693, EPI_ISL_956695, EPI_ISL_956696, EPI_ISL_956697, EPI_ISL_956698, EPI_ISL_956700, EPI_ISL_956701, EPI_ISL_956702, EPI_ISL_956703, EPI_ISL_956704, EPI_ISL_956705, EPI_ISL_956706, EPI_ISL_956707, EPI_ISL_956709, EPI_ISL_956710, EPI_ISL_956711, EPI_ISL_956712, EPI_ISL_956713, EPI_ISL_956714, EPI_ISL_956715, EPI_ISL_956716, EPI_ISL_956717, EPI_ISL_956718, EPI_ISL_956719, EPI_ISL_956720, EPI_ISL_956721, EPI_ISL_956722, EPI_ISL_956723, EPI_ISL_956724, EPI_ISL_956725, EPI_ISL_956726, EPI_ISL_956727, EPI_ISL_956728, EPI_ISL_956730, EPI_ISL_956731, EPI_ISL_956732, EPI_ISL_956733, EPI_ISL_956734, EPI_ISL_956735, EPI_ISL_956736, EPI_ISL_956737, EPI_ISL_956738, EPI_ISL_956739, EPI_ISL_956740, EPI_ISL_956741, EPI_ISL_956742, EPI_ISL_956743, EPI_ISL_956744, EPI_ISL_956745, EPI_ISL_956746, EPI_ISL_956747, EPI_ISL_956748, EPI_ISL_956749, EPI_ISL_956750, EPI_ISL_956751, EPI_ISL_956752, EPI_ISL_956753, EPI_ISL_956754, EPI_ISL_956755, EPI_ISL_956756, EPI_ISL_956757, EPI_ISL_956758, EPI_ISL_956759, EPI_ISL_956760, EPI_ISL_956761, EPI_ISL_956762, EPI_ISL_956763, EPI_ISL_956764, EPI_ISL_962975, EPI_ISL_962976, EPI_ISL_962977, EPI_ISL_962980, EPI_ISL_962981, EPI_ISL_962982, EPI_ISL_962983, EPI_ISL_962984, EPI_ISL_962985, EPI_ISL_962986, EPI_ISL_962988, EPI_ISL_962990, EPI_ISL_962991, EPI_ISL_962992, EPI_ISL_962993, EPI_ISL_962994, EPI_ISL_962996, EPI_ISL_962998, EPI_ISL_963000, EPI_ISL_963001, EPI_ISL_963002, EPI_ISL_963003, EPI_ISL_963005, EPI_ISL_963009, EPI_ISL_963010, EPI_ISL_963014, EPI_ISL_963015, EPI_ISL_963017, EPI_ISL_963018, EPI_ISL_963020, EPI_ISL_963021, EPI_ISL_963023, EPI_ISL_963024, EPI_ISL_963025, EPI_ISL_963028, EPI_ISL_963029, EPI_ISL_963031, EPI_ISL_963033, EPI_ISL_963034, EPI_ISL_963035, EPI_ISL_963036, EPI_ISL_963037, EPI_ISL_963039, EPI_ISL_963041, EPI_ISL_963043, EPI_ISL_963045, EPI_ISL_963046, EPI_ISL_963047, EPI_ISL_963050, EPI_ISL_963051, EPI_ISL_963052, EPI_ISL_963053, EPI_ISL_963054, EPI_ISL_963055, EPI_ISL_963057, EPI_ISL_963062, EPI_ISL_963063, EPI_ISL_963064, EPI_ISL_963065, EPI_ISL_963066, EPI_ISL_963067, EPI_ISL_963069, EPI_ISL_963073, EPI_ISL_963074, EPI_ISL_963075, EPI_ISL_963076, EPI_ISL_963077, EPI_ISL_963078, EPI_ISL_963080, EPI_ISL_963082, EPI_ISL_963083, EPI_ISL_963084, EPI_ISL_963086, EPI_ISL_963087, EPI_ISL_963089, EPI_ISL_963091, EPI_ISL_963093, EPI_ISL_963095, EPI_ISL_963096, EPI_ISL_963097, EPI_ISL_963099, EPI_ISL_963100, EPI_ISL_963101, EPI_ISL_963103, EPI_ISL_963104, EPI_ISL_963107, EPI_ISL_963108, EPI_ISL_963109, EPI_ISL_963113, EPI_ISL_963114, EPI_ISL_963116, EPI_ISL_963117, EPI_ISL_963118, EPI_ISL_963120, EPI_ISL_963121, EPI_ISL_963122, EPI_ISL_963125, EPI_ISL_963126, EPI_ISL_963128, EPI_ISL_963131, EPI_ISL_963132, EPI_ISL_963135, EPI_ISL_963136, EPI_ISL_963138, EPI_ISL_963139, EPI_ISL_963143, EPI_ISL_963145, EPI_ISL_963147, EPI_ISL_963148, EPI_ISL_963149, EPI_ISL_963151, EPI_ISL_963153, EPI_ISL_963154, EPI_ISL_963157, EPI_ISL_963158, EPI_ISL_963159, EPI_ISL_963160, EPI_ISL_963162, EPI_ISL_963165, EPI_ISL_963167, EPI_ISL_963168, EPI_ISL_963169, EPI_ISL_963170, EPI_ISL_963172, EPI_ISL_963174, EPI_ISL_963177, EPI_ISL_963179, EPI_ISL_963180, EPI_ISL_963181, EPI_ISL_963185, EPI_ISL_963186, EPI_ISL_963187, EPI_ISL_963189, EPI_ISL_963190, EPI_ISL_963191, EPI_ISL_963192, EPI_ISL_963193, EPI_ISL_963196, EPI_ISL_963197, EPI_ISL_963198, EPI_ISL_963200, EPI_ISL_963201, EPI_ISL_963202, EPI_ISL_963203, EPI_ISL_963206, EPI_ISL_963207, EPI_ISL_963208, EPI_ISL_963209, EPI_ISL_963210, EPI_ISL_963211, EPI_ISL_963212, EPI_ISL_963213, EPI_ISL_963214, EPI_ISL_963215, EPI_ISL_963217, EPI_ISL_963218, EPI_ISL_963220, EPI_ISL_963221, EPI_ISL_963224, EPI_ISL_963225, EPI_ISL_963227, EPI_ISL_963228, EPI_ISL_963230, EPI_ISL_963231, EPI_ISL_963232, EPI_ISL_963233, EPI_ISL_963235, EPI_ISL_963236, EPI_ISL_963237, EPI_ISL_963239, EPI_ISL_963240, EPI_ISL_963241, EPI_ISL_963242, EPI_ISL_963245, EPI_ISL_963247, EPI_ISL_963248, EPI_ISL_963249, EPI_ISL_963250, EPI_ISL_963252, EPI_ISL_963253, EPI_ISL_963254, EPI_ISL_963255, EPI_ISL_963256, EPI_ISL_963259, EPI_ISL_963260, EPI_ISL_963261, EPI_ISL_963263, EPI_ISL_963264, EPI_ISL_963266, EPI_ISL_963267, EPI_ISL_963268, EPI_ISL_963269, EPI_ISL_963270, EPI_ISL_963271, EPI_ISL_963272, EPI_ISL_963273, EPI_ISL_963274, EPI_ISL_963276, EPI_ISL_963277, EPI_ISL_963278, EPI_ISL_963280, EPI_ISL_963281, EPI_ISL_963282, EPI_ISL_963283, EPI_ISL_963285, EPI_ISL_963286, EPI_ISL_963292, EPI_ISL_963295, EPI_ISL_963299, EPI_ISL_963301, EPI_ISL_963306, EPI_ISL_963307, EPI_ISL_963308, EPI_ISL_963309, EPI_ISL_963310, EPI_ISL_963312, EPI_ISL_963313, EPI_ISL_963314, EPI_ISL_963316, EPI_ISL_963317, EPI_ISL_963318, EPI_ISL_963322, EPI_ISL_963323, EPI_ISL_963652, EPI_ISL_963653, EPI_ISL_963655, EPI_ISL_963656, EPI_ISL_963657, EPI_ISL_963660, EPI_ISL_963661, EPI_ISL_963662, EPI_ISL_963663, EPI_ISL_963664, EPI_ISL_963666, EPI_ISL_963667, EPI_ISL_963668, EPI_ISL_963669, EPI_ISL_963674, EPI_ISL_963675, EPI_ISL_963678, EPI_ISL_963680, EPI_ISL_963682, EPI_ISL_963683, EPI_ISL_963684, EPI_ISL_963688, EPI_ISL_963691, EPI_ISL_963692, EPI_ISL_963693, EPI_ISL_963696, EPI_ISL_963697, EPI_ISL_963698, EPI_ISL_963699, EPI_ISL_963702, EPI_ISL_963705, EPI_ISL_963708, EPI_ISL_963710, EPI_ISL_963711, EPI_ISL_963712, EPI_ISL_963713, EPI_ISL_963714, EPI_ISL_963715, EPI_ISL_963716, EPI_ISL_963717, EPI_ISL_963719, EPI_ISL_963722, EPI_ISL_963723, EPI_ISL_963725, EPI_ISL_963728, EPI_ISL_963729, EPI_ISL_963730, EPI_ISL_963739, EPI_ISL_963742, EPI_ISL_963743, EPI_ISL_963744, EPI_ISL_963745, EPI_ISL_963746, EPI_ISL_963748, EPI_ISL_963749, EPI_ISL_963751, EPI_ISL_963752, EPI_ISL_963755, EPI_ISL_963756, EPI_ISL_963757, EPI_ISL_963760, EPI_ISL_963762, EPI_ISL_963763, EPI_ISL_963767, EPI_ISL_963770, EPI_ISL_963771, EPI_ISL_963776, EPI_ISL_963777, EPI_ISL_963778, EPI_ISL_963782, EPI_ISL_963784, EPI_ISL_963785, EPI_ISL_963787, EPI_ISL_963788, EPI_ISL_963791, EPI_ISL_963792, EPI_ISL_963793, EPI_ISL_963795, EPI_ISL_963796, EPI_ISL_963797, EPI_ISL_963799, EPI_ISL_963801, EPI_ISL_963802, EPI_ISL_963804, EPI_ISL_963809, EPI_ISL_963811, EPI_ISL_963812, EPI_ISL_963817, EPI_ISL_963818, EPI_ISL_963820, EPI_ISL_963827, EPI_ISL_963828, EPI_ISL_963829, EPI_ISL_963830, EPI_ISL_963834, EPI_ISL_963836, EPI_ISL_963837, EPI_ISL_963840, EPI_ISL_963842, EPI_ISL_963843, EPI_ISL_963847, EPI_ISL_963848, EPI_ISL_963854, EPI_ISL_963856, EPI_ISL_963858, EPI_ISL_963860, EPI_ISL_963862, EPI_ISL_963871, EPI_ISL_963876, EPI_ISL_963885, EPI_ISL_963886, EPI_ISL_963887, EPI_ISL_963888, EPI_ISL_963892, EPI_ISL_963894, EPI_ISL_963897, EPI_ISL_963898, EPI_ISL_963899, EPI_ISL_963905, EPI_ISL_963907, EPI_ISL_963910, EPI_ISL_963912, EPI_ISL_963913, EPI_ISL_963917, EPI_ISL_963918, EPI_ISL_963919, EPI_ISL_963920, EPI_ISL_963924, EPI_ISL_963926, EPI_ISL_963928, EPI_ISL_963929, EPI_ISL_963930, EPI_ISL_963931, EPI_ISL_963933, EPI_ISL_963935, EPI_ISL_963937, EPI_ISL_963939, EPI_ISL_963940, EPI_ISL_963943, EPI_ISL_963944, EPI_ISL_963946, EPI_ISL_963948, EPI_ISL_963950, EPI_ISL_963951, EPI_ISL_963954, EPI_ISL_963956, EPI_ISL_963957, EPI_ISL_963959, EPI_ISL_963961, EPI_ISL_963963, EPI_ISL_963965, EPI_ISL_963966, EPI_ISL_963967, EPI_ISL_963969, EPI_ISL_963972, EPI_ISL_963974, EPI_ISL_963975, EPI_ISL_963982, EPI_ISL_963983, EPI_ISL_963985, EPI_ISL_963986, EPI_ISL_963987, EPI_ISL_963988, EPI_ISL_963989, EPI_ISL_963990, EPI_ISL_963991, EPI_ISL_963992, EPI_ISL_963993, EPI_ISL_963994, EPI_ISL_963995, EPI_ISL_963996, EPI_ISL_963997, EPI_ISL_963998, EPI_ISL_963999, EPI_ISL_964000, EPI_ISL_964001, EPI_ISL_964002, EPI_ISL_964003, EPI_ISL_964004, EPI_ISL_964005, EPI_ISL_964006, EPI_ISL_964007, EPI_ISL_964008, EPI_ISL_964009, EPI_ISL_964010, EPI_ISL_964011, EPI_ISL_964012, EPI_ISL_964013, EPI_ISL_964014, EPI_ISL_964015, EPI_ISL_964016, EPI_ISL_964017, EPI_ISL_964018, EPI_ISL_964019, EPI_ISL_964020, EPI_ISL_964021, EPI_ISL_964022, EPI_ISL_964023, EPI_ISL_964024, EPI_ISL_964025, EPI_ISL_964026, EPI_ISL_964027, EPI_ISL_964028, EPI_ISL_964029, EPI_ISL_964030, EPI_ISL_964031, EPI_ISL_964032, EPI_ISL_964033, EPI_ISL_964034, EPI_ISL_964035, EPI_ISL_964036, EPI_ISL_964037, EPI_ISL_964038, EPI_ISL_964039, EPI_ISL_964040, EPI_ISL_964041, EPI_ISL_964042, EPI_ISL_964043, EPI_ISL_964044, EPI_ISL_964045, EPI_ISL_964046, EPI_ISL_964047, EPI_ISL_964048, EPI_ISL_964049, EPI_ISL_964050, EPI_ISL_964051, EPI_ISL_964052, EPI_ISL_964053, EPI_ISL_964054, EPI_ISL_964055, EPI_ISL_964056, EPI_ISL_964057, EPI_ISL_964058, EPI_ISL_964059, EPI_ISL_964060, EPI_ISL_964061, EPI_ISL_964062, EPI_ISL_964063, EPI_ISL_964064, EPI_ISL_964065, EPI_ISL_964066, EPI_ISL_964068, EPI_ISL_964069, EPI_ISL_964070, EPI_ISL_964071, EPI_ISL_964072, EPI_ISL_964073, EPI_ISL_964074, EPI_ISL_964075, EPI_ISL_964076, EPI_ISL_964077, EPI_ISL_964078, EPI_ISL_964079, EPI_ISL_964080, EPI_ISL_964081, EPI_ISL_964082, EPI_ISL_964083, EPI_ISL_964084, EPI_ISL_964085, EPI_ISL_964086, EPI_ISL_964088, EPI_ISL_964089, EPI_ISL_964090, EPI_ISL_964091, EPI_ISL_964092, EPI_ISL_964093, EPI_ISL_964094, EPI_ISL_964095, EPI_ISL_964096, EPI_ISL_964097, EPI_ISL_964098, EPI_ISL_964099, EPI_ISL_964100, EPI_ISL_964101, EPI_ISL_964102, EPI_ISL_964103, EPI_ISL_964104, EPI_ISL_964105, EPI_ISL_964106, EPI_ISL_964108, EPI_ISL_964109, EPI_ISL_964110, EPI_ISL_964111, EPI_ISL_964112, EPI_ISL_964113, EPI_ISL_964114, EPI_ISL_964115, EPI_ISL_964116, EPI_ISL_964117, EPI_ISL_964118, EPI_ISL_964119, EPI_ISL_964120, EPI_ISL_964121, EPI_ISL_964122, EPI_ISL_964123, EPI_ISL_964124, EPI_ISL_964125, EPI_ISL_964126, EPI_ISL_964127, EPI_ISL_964128, EPI_ISL_964129, EPI_ISL_964130, EPI_ISL_964131, EPI_ISL_964132, EPI_ISL_964133, EPI_ISL_964134, EPI_ISL_964135, EPI_ISL_964136, EPI_ISL_964137, EPI_ISL_964138, EPI_ISL_964139, EPI_ISL_964140, EPI_ISL_964141, EPI_ISL_964142, EPI_ISL_964143, EPI_ISL_964144, EPI_ISL_964145, EPI_ISL_964146, EPI_ISL_964147, EPI_ISL_964148, EPI_ISL_964149, EPI_ISL_964150, EPI_ISL_964151, EPI_ISL_964152, EPI_ISL_964153, EPI_ISL_964154, EPI_ISL_964155, EPI_ISL_964156, EPI_ISL_964157, EPI_ISL_964158, EPI_ISL_964159, EPI_ISL_964160, EPI_ISL_964161, EPI_ISL_964162, EPI_ISL_964163, EPI_ISL_964164, EPI_ISL_964165, EPI_ISL_964166, EPI_ISL_964167, EPI_ISL_964168, EPI_ISL_964169, EPI_ISL_964170, EPI_ISL_964171, EPI_ISL_964172, EPI_ISL_964173, EPI_ISL_964174, EPI_ISL_964175, EPI_ISL_964176, EPI_ISL_964177, EPI_ISL_964178, EPI_ISL_964179, EPI_ISL_964180, EPI_ISL_964181, EPI_ISL_964182, EPI_ISL_964183, EPI_ISL_964184, EPI_ISL_964185, EPI_ISL_964186, EPI_ISL_964187, EPI_ISL_964188, EPI_ISL_964189, EPI_ISL_964190, EPI_ISL_964191, EPI_ISL_964192, EPI_ISL_964193, EPI_ISL_964194, EPI_ISL_964195, EPI_ISL_964196, EPI_ISL_964197, EPI_ISL_964198, EPI_ISL_964199, EPI_ISL_964200, EPI_ISL_964201, EPI_ISL_964202, EPI_ISL_964203, EPI_ISL_964204, EPI_ISL_964205, EPI_ISL_964206, EPI_ISL_964207, EPI_ISL_964208, EPI_ISL_964209, EPI_ISL_964210, EPI_ISL_964211, EPI_ISL_964212, EPI_ISL_964213, EPI_ISL_964214, EPI_ISL_964215, EPI_ISL_964216, EPI_ISL_964217, EPI_ISL_964218, EPI_ISL_964220, EPI_ISL_964221, EPI_ISL_964222, EPI_ISL_964223, EPI_ISL_964224, EPI_ISL_964225, EPI_ISL_964226, EPI_ISL_964227, EPI_ISL_964228, EPI_ISL_964229, EPI_ISL_964230, EPI_ISL_964231, EPI_ISL_964232, EPI_ISL_964233, EPI_ISL_964235, EPI_ISL_964237, EPI_ISL_964238, EPI_ISL_964239, EPI_ISL_964240, EPI_ISL_964241, EPI_ISL_964242, EPI_ISL_964243, EPI_ISL_964244, EPI_ISL_964245, EPI_ISL_964246, EPI_ISL_964247, EPI_ISL_964248, EPI_ISL_964249, EPI_ISL_964250, EPI_ISL_964251, EPI_ISL_964252, EPI_ISL_964253, EPI_ISL_964255, EPI_ISL_964256, EPI_ISL_964258, EPI_ISL_964260, EPI_ISL_964261, EPI_ISL_964263, EPI_ISL_964264, EPI_ISL_964266, EPI_ISL_964268, EPI_ISL_964269, EPI_ISL_964271, EPI_ISL_964273, EPI_ISL_964274, EPI_ISL_964276, EPI_ISL_964277, EPI_ISL_964279, EPI_ISL_964280, EPI_ISL_964282, EPI_ISL_964283, EPI_ISL_964285, EPI_ISL_964286, EPI_ISL_964288, EPI_ISL_964290, EPI_ISL_964291, EPI_ISL_964293, EPI_ISL_964294, EPI_ISL_964296, EPI_ISL_964298, EPI_ISL_964299, EPI_ISL_964301, EPI_ISL_964302, EPI_ISL_964304, EPI_ISL_964306, EPI_ISL_964307, EPI_ISL_964309, EPI_ISL_964310, EPI_ISL_964311, EPI_ISL_964312, EPI_ISL_964313, EPI_ISL_964314, EPI_ISL_964315, EPI_ISL_964316, EPI_ISL_964317, EPI_ISL_964318, EPI_ISL_964319, EPI_ISL_964320, EPI_ISL_964321, EPI_ISL_964322, EPI_ISL_964323, EPI_ISL_964324, EPI_ISL_964325, EPI_ISL_964326, EPI_ISL_964327, EPI_ISL_964328, EPI_ISL_964329, EPI_ISL_964330, EPI_ISL_964331, EPI_ISL_964332, EPI_ISL_964333, EPI_ISL_964334, EPI_ISL_964335, EPI_ISL_964336, EPI_ISL_964337, EPI_ISL_964338, EPI_ISL_964339, EPI_ISL_964340, EPI_ISL_964341, EPI_ISL_964342, EPI_ISL_964343, EPI_ISL_964344, EPI_ISL_964345, EPI_ISL_964543, EPI_ISL_964544, EPI_ISL_964545, EPI_ISL_964546, EPI_ISL_964547, EPI_ISL_964548, EPI_ISL_964549, EPI_ISL_964550, EPI_ISL_964551, EPI_ISL_964552, EPI_ISL_964553, EPI_ISL_964554, EPI_ISL_964555, EPI_ISL_964556, EPI_ISL_964557, EPI_ISL_964558, EPI_ISL_964560, EPI_ISL_964561, EPI_ISL_964562, EPI_ISL_964563, EPI_ISL_964564, EPI_ISL_964565, EPI_ISL_964566, EPI_ISL_964567, EPI_ISL_964568, EPI_ISL_964569, EPI_ISL_964570, EPI_ISL_964571, EPI_ISL_964572, EPI_ISL_964573, EPI_ISL_964574, EPI_ISL_964575, EPI_ISL_964576, EPI_ISL_964577, EPI_ISL_964578, EPI_ISL_964579, EPI_ISL_964580, EPI_ISL_964581, EPI_ISL_964582, EPI_ISL_964583, EPI_ISL_964584, EPI_ISL_964585, EPI_ISL_964586, EPI_ISL_964587, EPI_ISL_964588, EPI_ISL_964589, EPI_ISL_964590, EPI_ISL_964591, EPI_ISL_964592, EPI_ISL_964593, EPI_ISL_964594, EPI_ISL_964595, EPI_ISL_964596, EPI_ISL_964597, EPI_ISL_964598, EPI_ISL_964599, EPI_ISL_964600, EPI_ISL_964601, EPI_ISL_964602, EPI_ISL_964603, EPI_ISL_964604, EPI_ISL_964605, EPI_ISL_964606, EPI_ISL_964607, EPI_ISL_964608, EPI_ISL_964609, EPI_ISL_964610, EPI_ISL_964611, EPI_ISL_964612, EPI_ISL_964613, EPI_ISL_964614, EPI_ISL_964615, EPI_ISL_964616, EPI_ISL_964617, EPI_ISL_964618, EPI_ISL_964619, EPI_ISL_964620, EPI_ISL_964621, EPI_ISL_964622, EPI_ISL_964623, EPI_ISL_964624, EPI_ISL_964625, EPI_ISL_964626, EPI_ISL_964627, EPI_ISL_964628, EPI_ISL_964629, EPI_ISL_964630, EPI_ISL_964631, EPI_ISL_964632, EPI_ISL_964633, EPI_ISL_964634, EPI_ISL_964635, EPI_ISL_964636, EPI_ISL_964637, EPI_ISL_964638, EPI_ISL_964639, EPI_ISL_964640, EPI_ISL_964641, EPI_ISL_964642, EPI_ISL_964643, EPI_ISL_964644, EPI_ISL_964645, EPI_ISL_964646, EPI_ISL_964647, EPI_ISL_964648, EPI_ISL_964649, EPI_ISL_964650, EPI_ISL_964651, EPI_ISL_964652, EPI_ISL_964653, EPI_ISL_964654, EPI_ISL_964655, EPI_ISL_964656, EPI_ISL_964657, EPI_ISL_964658, EPI_ISL_964659, EPI_ISL_964660, EPI_ISL_964661, EPI_ISL_964662, EPI_ISL_964663, EPI_ISL_964664, EPI_ISL_964665, EPI_ISL_964666, EPI_ISL_964667, EPI_ISL_964668, EPI_ISL_964669, EPI_ISL_964670, EPI_ISL_964671, EPI_ISL_964672, EPI_ISL_964673, EPI_ISL_964674, EPI_ISL_964675, EPI_ISL_964676, EPI_ISL_964677, EPI_ISL_964678, EPI_ISL_964679, EPI_ISL_964680, EPI_ISL_964681, EPI_ISL_964682, EPI_ISL_964683, EPI_ISL_964684, EPI_ISL_964685, EPI_ISL_964686, EPI_ISL_964687, EPI_ISL_964688, EPI_ISL_964689, EPI_ISL_964690, EPI_ISL_964691, EPI_ISL_964692, EPI_ISL_964693, EPI_ISL_964694, EPI_ISL_964695, EPI_ISL_964696, EPI_ISL_964697, EPI_ISL_964698, EPI_ISL_964699, EPI_ISL_964700, EPI_ISL_964701, EPI_ISL_964702, EPI_ISL_964703, EPI_ISL_964704, EPI_ISL_964705, EPI_ISL_964706, EPI_ISL_964707, EPI_ISL_964708, EPI_ISL_964709, EPI_ISL_964711, EPI_ISL_964712, EPI_ISL_964713, EPI_ISL_964714, EPI_ISL_964715, EPI_ISL_964716, EPI_ISL_964717, EPI_ISL_964718, EPI_ISL_964719, EPI_ISL_964720, EPI_ISL_964721, EPI_ISL_964722, EPI_ISL_964723, EPI_ISL_964724, EPI_ISL_964725, EPI_ISL_964727, EPI_ISL_964728, EPI_ISL_964729, EPI_ISL_964730, EPI_ISL_964731, EPI_ISL_964732, EPI_ISL_964733, EPI_ISL_964734, EPI_ISL_964735, EPI_ISL_964736, EPI_ISL_964737, EPI_ISL_964738, EPI_ISL_964739, EPI_ISL_964740, EPI_ISL_964741, EPI_ISL_964742, EPI_ISL_964743, EPI_ISL_964744, EPI_ISL_964745, EPI_ISL_964746, EPI_ISL_964747, EPI_ISL_964748, EPI_ISL_964750, EPI_ISL_964751, EPI_ISL_964752, EPI_ISL_964753, EPI_ISL_964754, EPI_ISL_964755, EPI_ISL_964756, EPI_ISL_964757, EPI_ISL_964758, EPI_ISL_964760, EPI_ISL_964761, EPI_ISL_964762, EPI_ISL_964763, EPI_ISL_964764, EPI_ISL_964765, EPI_ISL_964766, EPI_ISL_964767, EPI_ISL_964768, EPI_ISL_964769, EPI_ISL_964770, EPI_ISL_964771, EPI_ISL_964772, EPI_ISL_964773, EPI_ISL_964774, EPI_ISL_964775, EPI_ISL_964776, EPI_ISL_964777, EPI_ISL_964778, EPI_ISL_964779, EPI_ISL_964780, EPI_ISL_964781, EPI_ISL_964782, EPI_ISL_964783, EPI_ISL_964784, EPI_ISL_964785, EPI_ISL_964786, EPI_ISL_964787, EPI_ISL_964788, EPI_ISL_964789, EPI_ISL_964790, EPI_ISL_964791, EPI_ISL_964792, EPI_ISL_964793, EPI_ISL_964794, EPI_ISL_964795, EPI_ISL_964796, EPI_ISL_964797, EPI_ISL_964798, EPI_ISL_964799, EPI_ISL_964800, EPI_ISL_964801, EPI_ISL_964802, EPI_ISL_964803, EPI_ISL_964804, EPI_ISL_964805, EPI_ISL_964806, EPI_ISL_964807, EPI_ISL_964808, EPI_ISL_964809, EPI_ISL_964810, EPI_ISL_964811, EPI_ISL_964812, EPI_ISL_964813, EPI_ISL_964814, EPI_ISL_964815, EPI_ISL_964816, EPI_ISL_964818, EPI_ISL_964819, EPI_ISL_964820, EPI_ISL_964821, EPI_ISL_964822, EPI_ISL_964823, EPI_ISL_964824, EPI_ISL_964825, EPI_ISL_964826, EPI_ISL_964827, EPI_ISL_964828, EPI_ISL_964829, EPI_ISL_964830, EPI_ISL_964831, EPI_ISL_964832, EPI_ISL_964833, EPI_ISL_964834, EPI_ISL_964835, EPI_ISL_964836, EPI_ISL_964838, EPI_ISL_964839, EPI_ISL_964840, EPI_ISL_964841, EPI_ISL_964842, EPI_ISL_964843, EPI_ISL_964844, EPI_ISL_964845, EPI_ISL_964846, EPI_ISL_964847, EPI_ISL_964848, EPI_ISL_964849, EPI_ISL_964850, EPI_ISL_964851, EPI_ISL_964852, EPI_ISL_964853, EPI_ISL_964854, EPI_ISL_964855, EPI_ISL_964856, EPI_ISL_964857, EPI_ISL_964858, EPI_ISL_964859, EPI_ISL_964860, EPI_ISL_964861, EPI_ISL_964862, EPI_ISL_964863, EPI_ISL_964864, EPI_ISL_964865, EPI_ISL_964866, EPI_ISL_964867, EPI_ISL_964868, EPI_ISL_964869, EPI_ISL_964870, EPI_ISL_964871, EPI_ISL_964872, EPI_ISL_964873, EPI_ISL_964874, EPI_ISL_964875, EPI_ISL_969457, EPI_ISL_969458, EPI_ISL_969459, EPI_ISL_969460, EPI_ISL_969461, EPI_ISL_969462, EPI_ISL_969463, EPI_ISL_969464, EPI_ISL_969465, EPI_ISL_969466, EPI_ISL_969467, EPI_ISL_969468, EPI_ISL_969469, EPI_ISL_969471, EPI_ISL_969472, EPI_ISL_969473, EPI_ISL_969474, EPI_ISL_969475, EPI_ISL_969476, EPI_ISL_969477, EPI_ISL_969478, EPI_ISL_969479, EPI_ISL_969480, EPI_ISL_969481, EPI_ISL_969482, EPI_ISL_969483, EPI_ISL_969484, EPI_ISL_969485, EPI_ISL_969486, EPI_ISL_969487, EPI_ISL_969488, EPI_ISL_969489, EPI_ISL_969490, EPI_ISL_969491, EPI_ISL_969492, EPI_ISL_969493, EPI_ISL_969494, EPI_ISL_969495, EPI_ISL_969496, EPI_ISL_969497, EPI_ISL_969498, EPI_ISL_969500, EPI_ISL_969501, EPI_ISL_969502, EPI_ISL_969503, EPI_ISL_969504, EPI_ISL_969505, EPI_ISL_969506, EPI_ISL_969507, EPI_ISL_969508, EPI_ISL_969510, EPI_ISL_969511, EPI_ISL_969512, EPI_ISL_969513, EPI_ISL_969514, EPI_ISL_969515, EPI_ISL_969516, EPI_ISL_969517, EPI_ISL_969518, EPI_ISL_969519, EPI_ISL_969520, EPI_ISL_969521, EPI_ISL_969522, EPI_ISL_969523, EPI_ISL_969524, EPI_ISL_969525, EPI_ISL_969526, EPI_ISL_969527, EPI_ISL_969528, EPI_ISL_969529, EPI_ISL_969530, EPI_ISL_969531, EPI_ISL_969532, EPI_ISL_969533, EPI_ISL_969534, EPI_ISL_969535, EPI_ISL_969536, EPI_ISL_969537, EPI_ISL_969538, EPI_ISL_969539, EPI_ISL_969540, EPI_ISL_969541, EPI_ISL_969542, EPI_ISL_969543, EPI_ISL_969544, EPI_ISL_969545, EPI_ISL_969546, EPI_ISL_969548, EPI_ISL_969549, EPI_ISL_969550, EPI_ISL_969551, EPI_ISL_969552, EPI_ISL_969553, EPI_ISL_969554, EPI_ISL_969555, EPI_ISL_969556, EPI_ISL_969557, EPI_ISL_969558, EPI_ISL_969559, EPI_ISL_969560, EPI_ISL_969561, EPI_ISL_969562, EPI_ISL_969563, EPI_ISL_969564, EPI_ISL_969565, EPI_ISL_969566, EPI_ISL_969567, EPI_ISL_969568, EPI_ISL_969569, EPI_ISL_969570, EPI_ISL_969571, EPI_ISL_969572, EPI_ISL_969573, EPI_ISL_969574, EPI_ISL_969575, EPI_ISL_969576, EPI_ISL_969577, EPI_ISL_969578, EPI_ISL_969579, EPI_ISL_969580, EPI_ISL_969581, EPI_ISL_969582, EPI_ISL_969583, EPI_ISL_969584, EPI_ISL_969585, EPI_ISL_969586, EPI_ISL_969587, EPI_ISL_969588, EPI_ISL_969589, EPI_ISL_969591, EPI_ISL_969592, EPI_ISL_969593, EPI_ISL_969594, EPI_ISL_969595, EPI_ISL_969596, EPI_ISL_969597, EPI_ISL_969598, EPI_ISL_969599, EPI_ISL_969600, EPI_ISL_969602, EPI_ISL_969603, EPI_ISL_969604, EPI_ISL_969605, EPI_ISL_969606, EPI_ISL_969607, | | | |

EPI_ISL_969608, EPI_ISL_969609, EPI_ISL_969610, EPI_ISL_969611, EPI_ISL_969612, EPI_ISL_969613, EPI_ISL_969614, EPI_ISL_969615, EPI_ISL_969616, EPI_ISL_969617, EPI_ISL_969618, EPI_ISL_969619, EPI_ISL_969620, EPI_ISL_969621, EPI_ISL_969622, EPI_ISL_969623, EPI_ISL_969624, EPI_ISL_969625, EPI_ISL_969626, EPI_ISL_969627, EPI_ISL_969628, EPI_ISL_969629, EPI_ISL_969630, EPI_ISL_969631, EPI_ISL_969632, EPI_ISL_969633, EPI_ISL_969634, EPI_ISL_969635, EPI_ISL_969636, EPI_ISL_969637, EPI_ISL_969638, EPI_ISL_969639, EPI_ISL_969640, EPI_ISL_969641, EPI_ISL_969642, EPI_ISL_969643, EPI_ISL_969644, EPI_ISL_969645, EPI_ISL_969646, EPI_ISL_969647, EPI_ISL_969649, EPI_ISL_969650, EPI_ISL_969651, EPI_ISL_969652, EPI_ISL_969653, EPI_ISL_969654, EPI_ISL_969655, EPI_ISL_969656, EPI_ISL_969657, EPI_ISL_969658, EPI_ISL_969659, EPI_ISL_969660, EPI_ISL_969661, EPI_ISL_969662, EPI_ISL_969663, EPI_ISL_969664, EPI_ISL_969665, EPI_ISL_969666, EPI_ISL_969667, EPI_ISL_969668, EPI_ISL_969669, EPI_ISL_969670, EPI_ISL_969671, EPI_ISL_969672, EPI_ISL_969673, EPI_ISL_969674, EPI_ISL_969675, EPI_ISL_969677, EPI_ISL_969679, EPI_ISL_969682, EPI_ISL_969683, EPI_ISL_969685, EPI_ISL_969688, EPI_ISL_969690, EPI_ISL_969691, EPI_ISL_969693, EPI_ISL_969695, EPI_ISL_969697, EPI_ISL_969699, EPI_ISL_969701, EPI_ISL_969702, EPI_ISL_969704, EPI_ISL_969706, EPI_ISL_969708, EPI_ISL_969710, EPI_ISL_969712, EPI_ISL_969714, EPI_ISL_969716, EPI_ISL_969718, EPI_ISL_969720, EPI_ISL_969722, EPI_ISL_969724, EPI_ISL_969726, EPI_ISL_969728, EPI_ISL_969730, EPI_ISL_969732, EPI_ISL_969734, EPI_ISL_969736, EPI_ISL_969739, EPI_ISL_969742, EPI_ISL_969744, EPI_ISL_969746, EPI_ISL_969748, EPI_ISL_969750, EPI_ISL_969752, EPI_ISL_969754, EPI_ISL_969756, EPI_ISL_969759, EPI_ISL_969760, EPI_ISL_969762, EPI_ISL_969764, EPI_ISL_969766, EPI_ISL_969768, EPI_ISL_969769, EPI_ISL_969771, EPI_ISL_969774, EPI_ISL_969776, EPI_ISL_969779, EPI_ISL_969780, EPI_ISL_969782, EPI_ISL_969785, EPI_ISL_969786, EPI_ISL_969789, EPI_ISL_969791, EPI_ISL_969793, EPI_ISL_969795, EPI_ISL_969797, EPI_ISL_969799, EPI_ISL_969801, EPI_ISL_969802, EPI_ISL_969804, EPI_ISL_969806, EPI_ISL_969808, EPI_ISL_969809, EPI_ISL_969812, EPI_ISL_969813, EPI_ISL_969815, EPI_ISL_969817, EPI_ISL_969819, EPI_ISL_969821, EPI_ISL_969823, EPI_ISL_969824, EPI_ISL_969826, EPI_ISL_969828, EPI_ISL_969831, EPI_ISL_969833, EPI_ISL_969834, EPI_ISL_969836, EPI_ISL_969838, EPI_ISL_969840, EPI_ISL_969841, EPI_ISL_969843, EPI_ISL_969846, EPI_ISL_969848, EPI_ISL_969849, EPI_ISL_969852, EPI_ISL_969853, EPI_ISL_969856, EPI_ISL_969858, EPI_ISL_969859, EPI_ISL_969861, EPI_ISL_969863, EPI_ISL_969865, EPI_ISL_969867, EPI_ISL_969869, EPI_ISL_969871, EPI_ISL_969873, EPI_ISL_969875, EPI_ISL_969877, EPI_ISL_969879, EPI_ISL_969881, EPI_ISL_969883, EPI_ISL_969886, EPI_ISL_969887, EPI_ISL_969889, EPI_ISL_969891, EPI_ISL_969893, EPI_ISL_969895, EPI_ISL_969897, EPI_ISL_969899, EPI_ISL_969901, EPI_ISL_969903, EPI_ISL_969906, EPI_ISL_969907, EPI_ISL_969909, EPI_ISL_969911, EPI_ISL_969914, EPI_ISL_969916, EPI_ISL_969917, EPI_ISL_969919, EPI_ISL_969921, EPI_ISL_969923, EPI_ISL_969925, EPI_ISL_969928, EPI_ISL_969930, EPI_ISL_969932, EPI_ISL_969934, EPI_ISL_969936, EPI_ISL_969937, EPI_ISL_969939, EPI_ISL_969940, EPI_ISL_969943, EPI_ISL_969945, EPI_ISL_969947, EPI_ISL_969948, EPI_ISL_969951, EPI_ISL_969952, EPI_ISL_969955, EPI_ISL_969956, EPI_ISL_969958, EPI_ISL_969961, EPI_ISL_969963, EPI_ISL_969965, EPI_ISL_969967, EPI_ISL_969969, EPI_ISL_969971, EPI_ISL_969974, EPI_ISL_969975, EPI_ISL_969977, EPI_ISL_969979, EPI_ISL_969982, EPI_ISL_969983, EPI_ISL_969986, EPI_ISL_969988, EPI_ISL_969990, EPI_ISL_969992, EPI_ISL_969994, EPI_ISL_969996, EPI_ISL_969998, EPI_ISL_970000, EPI_ISL_970002, EPI_ISL_970004, EPI_ISL_970005, EPI_ISL_970007, EPI_ISL_970009, EPI_ISL_970011, EPI_ISL_970013, EPI_ISL_970015, EPI_ISL_970017, EPI_ISL_970019, EPI_ISL_970022, EPI_ISL_970024, EPI_ISL_970026, EPI_ISL_970028, EPI_ISL_970030, EPI_ISL_970033, EPI_ISL_970035, EPI_ISL_970037, EPI_ISL_970039, EPI_ISL_970041, EPI_ISL_970043, EPI_ISL_970045, EPI_ISL_970047, EPI_ISL_970049, EPI_ISL_970050, EPI_ISL_970052, EPI_ISL_970057, EPI_ISL_970059, EPI_ISL_970061, EPI_ISL_970064, EPI_ISL_970066, EPI_ISL_970068, EPI_ISL_970070, EPI_ISL_970073, EPI_ISL_970075, EPI_ISL_970076, EPI_ISL_970079, EPI_ISL_970080, EPI_ISL_970082, EPI_ISL_970084, EPI_ISL_970086, EPI_ISL_970087, EPI_ISL_970089, EPI_ISL_970091, EPI_ISL_970094, EPI_ISL_970096, EPI_ISL_970098, EPI_ISL_970099, EPI_ISL_970101, EPI_ISL_970103, EPI_ISL_970105, EPI_ISL_970107, EPI_ISL_970109, EPI_ISL_970111, EPI_ISL_970113, EPI_ISL_970115, EPI_ISL_970117, EPI_ISL_970119, EPI_ISL_970120, EPI_ISL_970122, EPI_ISL_970124, EPI_ISL_970126, EPI_ISL_970128, EPI_ISL_970130, EPI_ISL_970132, EPI_ISL_970134, EPI_ISL_970136, EPI_ISL_970138, EPI_ISL_970140, EPI_ISL_970145, EPI_ISL_970147, EPI_ISL_970148, EPI_ISL_970150, EPI_ISL_970152, EPI_ISL_970153, EPI_ISL_970155, EPI_ISL_970157, EPI_ISL_970160, EPI_ISL_970161, EPI_ISL_970163, EPI_ISL_970165, EPI_ISL_970166, EPI_ISL_970168, EPI_ISL_970170, EPI_ISL_970172, EPI_ISL_970173, EPI_ISL_970175, EPI_ISL_970177, EPI_ISL_970181, EPI_ISL_970183, EPI_ISL_970184, EPI_ISL_970186, EPI_ISL_970188, EPI_ISL_970190, EPI_ISL_970192, EPI_ISL_970195, EPI_ISL_970197, EPI_ISL_970198, EPI_ISL_970200, EPI_ISL_970202, EPI_ISL_970205, EPI_ISL_970206, EPI_ISL_970209, EPI_ISL_970211, EPI_ISL_970213, EPI_ISL_970215, EPI_ISL_970217, EPI_ISL_970219, EPI_ISL_970221, EPI_ISL_970224, EPI_ISL_970225, EPI_ISL_970227, EPI_ISL_970229, EPI_ISL_970231, EPI_ISL_970233, EPI_ISL_970235, EPI_ISL_970237, EPI_ISL_970239, EPI_ISL_970242, EPI_ISL_970247, EPI_ISL_970249, EPI_ISL_970251, EPI_ISL_970252, EPI_ISL_970254, EPI_ISL_970256, EPI_ISL_970258, EPI_ISL_970260, EPI_ISL_970262, EPI_ISL_970264, EPI_ISL_970266, EPI_ISL_970268, EPI_ISL_970270, EPI_ISL_970272, EPI_ISL_970274, EPI_ISL_970275, EPI_ISL_970278, EPI_ISL_970279, EPI_ISL_970281, EPI_ISL_970283, EPI_ISL_970285, EPI_ISL_970287, EPI_ISL_970288, EPI_ISL_970291, EPI_ISL_970292, EPI_ISL_970295, EPI_ISL_970297, EPI_ISL_970299, EPI_ISL_970301, EPI_ISL_970303, EPI_ISL_970306, EPI_ISL_970307, EPI_ISL_970309, EPI_ISL_970312, EPI_ISL_970314, EPI_ISL_970316, EPI_ISL_970319, EPI_ISL_970321, EPI_ISL_970323, EPI_ISL_970324, EPI_ISL_970326, EPI_ISL_970328, EPI_ISL_970330, EPI_ISL_970332, EPI_ISL_970334, EPI_ISL_970337, EPI_ISL_970338, EPI_ISL_970340, EPI_ISL_970342, EPI_ISL_970344, EPI_ISL_970346, EPI_ISL_970348, EPI_ISL_970350, EPI_ISL_970352, EPI_ISL_970354, EPI_ISL_970356, EPI_ISL_970358, EPI_ISL_970360, EPI_ISL_970362, EPI_ISL_970364, EPI_ISL_970365, EPI_ISL_970368, EPI_ISL_970370, EPI_ISL_970371, EPI_ISL_970373, EPI_ISL_970376, EPI_ISL_970377, EPI_ISL_970380, EPI_ISL_970382, EPI_ISL_970383, EPI_ISL_970386, EPI_ISL_970387, EPI_ISL_970390, EPI_ISL_970392, EPI_ISL_970394, EPI_ISL_970396, EPI_ISL_970398, EPI_ISL_970400, EPI_ISL_970402, EPI_ISL_970404, EPI_ISL_970406, EPI_ISL_970409, EPI_ISL_970411, EPI_ISL_970412, EPI_ISL_970415, EPI_ISL_970416, EPI_ISL_970419, EPI_ISL_970421, EPI_ISL_970422, EPI_ISL_970424, EPI_ISL_970426, EPI_ISL_970428, EPI_ISL_970430, EPI_ISL_970432, EPI_ISL_970435, EPI_ISL_970436, EPI_ISL_970439, EPI_ISL_970441, EPI_ISL_970443, EPI_ISL_970444, EPI_ISL_970446, EPI_ISL_970448, EPI_ISL_970450, EPI_ISL_970452, EPI_ISL_970454, EPI_ISL_970457, EPI_ISL_970460, EPI_ISL_970462, EPI_ISL_970464, EPI_ISL_970466, EPI_ISL_970468, EPI_ISL_970470, EPI_ISL_970473, EPI_ISL_970475, EPI_ISL_970477, EPI_ISL_970478, EPI_ISL_970481, EPI_ISL_970483, EPI_ISL_970486, EPI_ISL_970488, EPI_ISL_970490, EPI_ISL_970492, EPI_ISL_970493, EPI_ISL_970495, EPI_ISL_970497, EPI_ISL_970499, EPI_ISL_970500, EPI_ISL_970502, EPI_ISL_970504, EPI_ISL_970506, EPI_ISL_970508, EPI_ISL_970510, EPI_ISL_970512, EPI_ISL_970513, EPI_ISL_970515, EPI_ISL_970517, EPI_ISL_970519, EPI_ISL_970521, EPI_ISL_970523, EPI_ISL_970525, EPI_ISL_970527, EPI_ISL_970528, EPI_ISL_970530, EPI_ISL_970533, EPI_ISL_970535, EPI_ISL_970537, EPI_ISL_970540, EPI_ISL_970541, EPI_ISL_970543, EPI_ISL_970546, EPI_ISL_970548, EPI_ISL_970551, EPI_ISL_970553, EPI_ISL_970555, EPI_ISL_970558, EPI_ISL_970560, EPI_ISL_970562, EPI_ISL_970564, EPI_ISL_970566, EPI_ISL_970568, EPI_ISL_970569, EPI_ISL_970571, EPI_ISL_970573, EPI_ISL_970575, EPI_ISL_970577, EPI_ISL_970579, EPI_ISL_970581, EPI_ISL_970583, EPI_ISL_970585, EPI_ISL_987005, EPI_ISL_987104, EPI_ISL_988811, EPI_ISL_989021, EPI_ISL_989030, EPI_ISL_989032, EPI_ISL_989033, EPI_ISL_989042, EPI_ISL_989043, EPI_ISL_989044, EPI_ISL_989045, EPI_ISL_989052, EPI_ISL_989053, EPI_ISL_989056, EPI_ISL_989057, EPI_ISL_989059, EPI_ISL_989061, EPI_ISL_989063, EPI_ISL_989064, EPI_ISL_989066, EPI_ISL_989068, EPI_ISL_989069, EPI_ISL_989070, EPI_ISL_989071, EPI_ISL_989072, EPI_ISL_989073, EPI_ISL_989075, EPI_ISL_989077, EPI_ISL_989080, EPI_ISL_989081, EPI_ISL_989082, EPI_ISL_989084, EPI_ISL_989085, EPI_ISL_989086, EPI_ISL_989088, EPI_ISL_989089, EPI_ISL_989092, EPI_ISL_989093, EPI_ISL_989094, EPI_ISL_989095, EPI_ISL_989096, EPI_ISL_989099, EPI_ISL_989100, EPI_ISL_989102, EPI_ISL_989103, EPI_ISL_989104, EPI_ISL_989105, EPI_ISL_989107, EPI_ISL_989108, EPI_ISL_989109, EPI_ISL_989110, EPI_ISL_989111, EPI_ISL_989114, EPI_ISL_989116, EPI_ISL_989117, EPI_ISL_989118, EPI_ISL_989120, EPI_ISL_989121, EPI_ISL_989123, EPI_ISL_989125, EPI_ISL_989126, EPI_ISL_989127, EPI_ISL_989129, EPI_ISL_989130, EPI_ISL_989131, EPI_ISL_989132, EPI_ISL_989136, EPI_ISL_989138, EPI_ISL_989139, EPI_ISL_989141, EPI_ISL_989142, EPI_ISL_989143, EPI_ISL_989145, EPI_ISL_989146, EPI_ISL_989147, EPI_ISL_989148, EPI_ISL_989149, EPI_ISL_989150, EPI_ISL_989152, EPI_ISL_989153, EPI_ISL_989156, EPI_ISL_989159, EPI_ISL_989162, EPI_ISL_989164, EPI_ISL_989165, EPI_ISL_989166, EPI_ISL_989170, EPI_ISL_989171, EPI_ISL_989172, EPI_ISL_989173, EPI_ISL_989174, EPI_ISL_989175, EPI_ISL_989176, EPI_ISL_989178, EPI_ISL_989179, EPI_ISL_989180, EPI_ISL_989181, EPI_ISL_989182, EPI_ISL_989183, EPI_ISL_989184, EPI_ISL_989185, EPI_ISL_989186, EPI_ISL_989187, EPI_ISL_989188, EPI_ISL_989190, EPI_ISL_989191, EPI_ISL_989192, EPI_ISL_989194, EPI_ISL_989195, EPI_ISL_989199, EPI_ISL_989200, EPI_ISL_989201, EPI_ISL_989203, EPI_ISL_989204, EPI_ISL_989206, EPI_ISL_989207, EPI_ISL_989209, EPI_ISL_989212, EPI_ISL_989213, EPI_ISL_989214, EPI_ISL_989215, EPI_ISL_989216, EPI_ISL_989217, EPI_ISL_989218, EPI_ISL_989221, EPI_ISL_989222, EPI_ISL_989223, EPI_ISL_989225, EPI_ISL_989226, EPI_ISL_989227, EPI_ISL_989228, EPI_ISL_989229, EPI_ISL_989230, EPI_ISL_989231, EPI_ISL_989233, EPI_ISL_989234, EPI_ISL_989236, EPI_ISL_989238, EPI_ISL_989239, EPI_ISL_989240, EPI_ISL_989242, EPI_ISL_989246, EPI_ISL_989247, EPI_ISL_989248, EPI_ISL_989249, EPI_ISL_989250, EPI_ISL_989251, EPI_ISL_989252, EPI_ISL_989253, EPI_ISL_989254, EPI_ISL_989255, EPI_ISL_989256, EPI_ISL_989257, EPI_ISL_989258, EPI_ISL_989259, EPI_ISL_989260, EPI_ISL_989263, EPI_ISL_989264, EPI_ISL_989265, EPI_ISL_989266, EPI_ISL_989267, EPI_ISL_989268, EPI_ISL_989270, EPI_ISL_989271, EPI_ISL_989272, EPI_ISL_989276, EPI_ISL_989278, EPI_ISL_989279, EPI_ISL_989281, EPI_ISL_989282, EPI_ISL_989283, EPI_ISL_989291, EPI_ISL_989292, EPI_ISL_989295, EPI_ISL_989296, EPI_ISL_989297, EPI_ISL_989298, EPI_ISL_989299, EPI_ISL_989300, EPI_ISL_989301, EPI_ISL_989302, EPI_ISL_989303, EPI_ISL_989305, EPI_ISL_989306, EPI_ISL_989308, EPI_ISL_989309, EPI_ISL_989310, EPI_ISL_989311, EPI_ISL_989314, EPI_ISL_989316, EPI_ISL_989317, EPI_ISL_989318, EPI_ISL_989319, EPI_ISL_989320, EPI_ISL_989321, EPI_ISL_989323, EPI_ISL_989325, EPI_ISL_989326, EPI_ISL_989327, EPI_ISL_989328, EPI_ISL_989330, EPI_ISL_989333, EPI_ISL_989335, EPI_ISL_989336, EPI_ISL_989337, EPI_ISL_989338, EPI_ISL_989339, EPI_ISL_989341, EPI_ISL_989343, EPI_ISL_989344, EPI_ISL_989345, EPI_ISL_989346, EPI_ISL_989347, EPI_ISL_989355, EPI_ISL_989356, EPI_ISL_989357, EPI_ISL_989358, EPI_ISL_989360, EPI_ISL_989444, EPI_ISL_989445, EPI_ISL_989447, EPI_ISL_989448, EPI_ISL_989484, EPI_ISL_989486, EPI_ISL_989487, EPI_ISL_989488, EPI_ISL_989492, EPI_ISL_989493, EPI_ISL_989497, EPI_ISL_989498, EPI_ISL_989499, EPI_ISL_989500, EPI_ISL_989501, EPI_ISL_989502, EPI_ISL_989503, EPI_ISL_989508, EPI_ISL_989509, EPI_ISL_989510, EPI_ISL_989511, EPI_ISL_989512, EPI_ISL_989513, EPI_ISL_989514, EPI_ISL_989515, EPI_ISL_989516, EPI_ISL_989517, EPI_ISL_989518, EPI_ISL_989519, EPI_ISL_989520, EPI_ISL_989521, EPI_ISL_989523, EPI_ISL_989524, EPI_ISL_989525, EPI_ISL_989526, EPI_ISL_989527, EPI_ISL_989528, EPI_ISL_989529, EPI_ISL_989696, EPI_ISL_989697, EPI_ISL_989698, EPI_ISL_989699, EPI_ISL_989700, EPI_ISL_989701, EPI_ISL_989702, EPI_ISL_989703, EPI_ISL_989704, EPI_ISL_989705, EPI_ISL_989706, EPI_ISL_989707, EPI_ISL_989708, EPI_ISL_989709, EPI_ISL_989710, EPI_ISL_989711, EPI_ISL_989712, EPI_ISL_989713, EPI_ISL_989714, EPI_ISL_989715, EPI_ISL_989716, EPI_ISL_989717, EPI_ISL_989718, EPI_ISL_989719, EPI_ISL_989720, EPI_ISL_989721, EPI_ISL_989722, EPI_ISL_989723, EPI_ISL_989724, EPI_ISL_989725, EPI_ISL_989726, EPI_ISL_989727, EPI_ISL_989728, EPI_ISL_989729, EPI_ISL_989730, EPI_ISL_989731, EPI_ISL_989732, EPI_ISL_989733, EPI_ISL_989734, EPI_ISL_989735, EPI_ISL_989736, EPI_ISL_989737, EPI_ISL_989738, EPI_ISL_989739, EPI_ISL_989740, EPI_ISL_989741, EPI_ISL_989743, EPI_ISL_989744, EPI_ISL_989745, EPI_ISL_989746, EPI_ISL_989747, EPI_ISL_989748, EPI_ISL_989749, EPI_ISL_989750, EPI_ISL_989752, EPI_ISL_989753, EPI_ISL_989754, EPI_ISL_989755, EPI_ISL_989756, EPI_ISL_989757, EPI_ISL_989758, EPI_ISL_989759, EPI_ISL_989760, EPI_ISL_989761, EPI_ISL_989762, EPI_ISL_989763, EPI_ISL_989764, EPI_ISL_989765, EPI_ISL_989766, EPI_ISL_989767, EPI_ISL_989768, EPI_ISL_989770, EPI_ISL_989771, EPI_ISL_989772, EPI_ISL_989773, EPI_ISL_989774, EPI_ISL_989775, EPI_ISL_989776, EPI_ISL_989777, EPI_ISL_989778, EPI_ISL_989779, EPI_ISL_989780, EPI_ISL_989781, EPI_ISL_989782, EPI_ISL_989783, EPI_ISL_989784, EPI_ISL_989785, EPI_ISL_989786, EPI_ISL_989787, EPI_ISL_989788, EPI_ISL_989789, EPI_ISL_989790, EPI_ISL_989791, EPI_ISL_989792, EPI_ISL_989793, EPI_ISL_989794, EPI_ISL_989860, EPI_ISL_989863, EPI_ISL_989864, EPI_ISL_989872, EPI_ISL_989875, EPI_ISL_989878, EPI_ISL_989880, EPI_ISL_1007440, EPI_ISL_1007441, EPI_ISL_1007442, EPI_ISL_1007443, EPI_ISL_1007444, EPI_ISL_1012695, EPI_ISL_1012697, EPI_ISL_1012698, EPI_ISL_1012699, EPI_ISL_1012700, EPI_ISL_1012701, EPI_ISL_1012703, EPI_ISL_1012705, EPI_ISL_1012706, EPI_ISL_1012707, EPI_ISL_1012708, EPI_ISL_1012709, EPI_ISL_1012712, EPI_ISL_1449684

| | | | |
|---|---|---|---|
| see above | Lighthouse Lab in Alderley Park | Wellcome Sanger Institute for the COVID-19 Genomics UK (COG-UK) Consortium | Cordelia Langford; David K. Jackson; Dominic Kwiatkowski; Ewan Harrison; Ian Johnston; Jacquelyn Wynn; Jeffrey Barrett; John Sillitoe on behalf of the Wellcome Sanger Institute COVID-19 Surveillance Team; John Sillitoe on behalf of the Wellcome Sanger Institute COVID-19 Surveillance Team (http://www.sanger.ac.uk/covid-team); Mairead Hyland; Roberto Amato; Sonia Goncalves; The Lighthouse Lab in Alderley Park and Alex Alderton |

EPI_ISL_957612, EPI_ISL_957615, EPI_ISL_957617, EPI_ISL_957618, EPI_ISL_957622, EPI_ISL_957625, EPI_ISL_957626, EPI_ISL_957634, EPI_ISL_957636, EPI_ISL_957638, EPI_ISL_957639, EPI_ISL_957640, EPI_ISL_957642, EPI_ISL_957644, EPI_ISL_957646, EPI_ISL_957647, EPI_ISL_957648, EPI_ISL_957657, EPI_ISL_957658, EPI_ISL_957661, EPI_ISL_957665, EPI_ISL_957676, EPI_ISL_957688, EPI_ISL_957689, EPI_ISL_957696, EPI_ISL_957702, EPI_ISL_957703, EPI_ISL_957704, EPI_ISL_957706, EPI_ISL_957709, EPI_ISL_957714, EPI_ISL_957716, EPI_ISL_957719, EPI_ISL_957728, EPI_ISL_957731, EPI_ISL_957733, EPI_ISL_957734, EPI_ISL_957737, EPI_ISL_957738, EPI_ISL_957739, EPI_ISL_957742, EPI_ISL_957746, EPI_ISL_957751, EPI_ISL_957752, EPI_ISL_957754, EPI_ISL_957763, EPI_ISL_957764, EPI_ISL_957776, EPI_ISL_957779, EPI_ISL_957781, EPI_ISL_957782, EPI_ISL_957784, EPI_ISL_957788, EPI_ISL_957789, EPI_ISL_957798, EPI_ISL_957805, EPI_ISL_957807, EPI_ISL_957808, EPI_ISL_957809, EPI_ISL_957811, EPI_ISL_957812, EPI_ISL_957815, EPI_ISL_957818, EPI_ISL_957820, EPI_ISL_957821, EPI_ISL_957825, EPI_ISL_957831, EPI_ISL_957833, EPI_ISL_957834, EPI_ISL_957840, EPI_ISL_957846, EPI_ISL_957852, EPI_ISL_957855, EPI_ISL_957857, EPI_ISL_957861, EPI_ISL_957869, EPI_ISL_957870, EPI_ISL_957873, EPI_ISL_957874, EPI_ISL_957876, EPI_ISL_957880, EPI_ISL_957881, EPI_ISL_957882, EPI_ISL_957883, EPI_ISL_957887, EPI_ISL_957890, EPI_ISL_957891, EPI_ISL_957896, EPI_ISL_957898, EPI_ISL_957902, EPI_ISL_957903, EPI_ISL_957905, EPI_ISL_957906, EPI_ISL_957908, EPI_ISL_987638, EPI_ISL_987639, EPI_ISL_987640, EPI_ISL_987641, EPI_ISL_987642, EPI_ISL_987643, EPI_ISL_987644, EPI_ISL_987645, EPI_ISL_987646, EPI_ISL_987647, EPI_ISL_987648, EPI_ISL_987649, EPI_ISL_987650, EPI_ISL_987651, EPI_ISL_987652, EPI_ISL_987653, EPI_ISL_987654, EPI_ISL_987655, EPI_ISL_987656, EPI_ISL_987657, EPI_ISL_987658, EPI_ISL_987659, EPI_ISL_987660, EPI_ISL_987661, EPI_ISL_987662, EPI_ISL_987663, EPI_ISL_987664, EPI_ISL_987665, EPI_ISL_987666, EPI_ISL_987667, EPI_ISL_987668, EPI_ISL_987669, EPI_ISL_987670, EPI_ISL_987671, EPI_ISL_987672, EPI_ISL_987673, EPI_ISL_987674, EPI_ISL_987675, EPI_ISL_987676, EPI_ISL_987677, EPI_ISL_987678, EPI_ISL_987679, EPI_ISL_987680, EPI_ISL_987681, EPI_ISL_987682, EPI_ISL_987683, EPI_ISL_987684, EPI_ISL_987685, EPI_ISL_987686, EPI_ISL_987687, EPI_ISL_987688, EPI_ISL_987689, EPI_ISL_987690, EPI_ISL_987691, EPI_ISL_987692, EPI_ISL_987693, EPI_ISL_987694, EPI_ISL_987695, EPI_ISL_987696, EPI_ISL_987697, EPI_ISL_987698, EPI_ISL_987699, EPI_ISL_987700, EPI_ISL_987701, EPI_ISL_987702, EPI_ISL_987703, EPI_ISL_987704, EPI_ISL_987705, EPI_ISL_987706, EPI_ISL_987707, EPI_ISL_987708, EPI_ISL_987709, EPI_ISL_987710, EPI_ISL_987711, EPI_ISL_987712, EPI_ISL_987713, EPI_ISL_987714, EPI_ISL_987715, EPI_ISL_987716, EPI_ISL_987717, EPI_ISL_987718, EPI_ISL_987719, EPI_ISL_987720, EPI_ISL_987721, EPI_ISL_987722, EPI_ISL_987723, EPI_ISL_987724, EPI_ISL_987725, EPI_ISL_987726, EPI_ISL_987727, EPI_ISL_987728, EPI_ISL_987729, EPI_ISL_987730, EPI_ISL_987731, EPI_ISL_987732, EPI_ISL_987733, EPI_ISL_987734, EPI_ISL_987735, EPI_ISL_987736, EPI_ISL_987737, EPI_ISL_987738, EPI_ISL_987739, EPI_ISL_987740, EPI_ISL_987741, EPI_ISL_987742, EPI_ISL_987743, EPI_ISL_987744, EPI_ISL_987745, EPI_ISL_987746, EPI_ISL_987747, EPI_ISL_987748, EPI_ISL_987749, EPI_ISL_987750, EPI_ISL_987751, EPI_ISL_987752, EPI_ISL_987753, EPI_ISL_987754, EPI_ISL_987755, EPI_ISL_987756, EPI_ISL_987757, EPI_ISL_987758, EPI_ISL_987759, EPI_ISL_987760, EPI_ISL_987761, EPI_ISL_987762, EPI_ISL_987763, EPI_ISL_987764, EPI_ISL_987765, EPI_ISL_987766, EPI_ISL_987767, EPI_ISL_987768, EPI_ISL_987769, EPI_ISL_987770, EPI_ISL_987771, EPI_ISL_987772, EPI_ISL_987773, EPI_ISL_987774, EPI_ISL_987775, EPI_ISL_987776, EPI_ISL_987777, EPI_ISL_987778, EPI_ISL_987779, EPI_ISL_987780, EPI_ISL_987781, EPI_ISL_987782, EPI_ISL_987783, EPI_ISL_987784, EPI_ISL_987785, EPI_ISL_987786, EPI_ISL_987787, EPI_ISL_987788, EPI_ISL_987789, EPI_ISL_987790, EPI_ISL_987791, EPI_ISL_987792, EPI_ISL_987793, EPI_ISL_987794, EPI_ISL_987795, EPI_ISL_987796, EPI_ISL_987797, EPI_ISL_987798, EPI_ISL_987799, EPI_ISL_987800, EPI_ISL_987801, EPI_ISL_987802, EPI_ISL_987803, EPI_ISL_987804, EPI_ISL_987805, EPI_ISL_987806, EPI_ISL_987807, EPI_ISL_987808, EPI_ISL_987809, EPI_ISL_987810, EPI_ISL_987811, EPI_ISL_987812, EPI_ISL_987813, EPI_ISL_987814, EPI_ISL_987815, EPI_ISL_987816, EPI_ISL_987817, EPI_ISL_987818, EPI_ISL_987819, EPI_ISL_987820, EPI_ISL_987821, EPI_ISL_987822, EPI_ISL_987823, EPI_ISL_987824, EPI_ISL_987825, EPI_ISL_987826, EPI_ISL_987827, EPI_ISL_987828, EPI_ISL_987829, EPI_ISL_987830, EPI_ISL_987831, EPI_ISL_987832, EPI_ISL_987833, EPI_ISL_987834, EPI_ISL_987835, EPI_ISL_987836, EPI_ISL_987837, EPI_ISL_987838, EPI_ISL_987839, EPI_ISL_987840, EPI_ISL_987841, EPI_ISL_987842, EPI_ISL_987843, EPI_ISL_987844, EPI_ISL_987845, EPI_ISL_987846, EPI_ISL_987847, EPI_ISL_987848, EPI_ISL_987849, EPI_ISL_987850, EPI_ISL_987851, EPI_ISL_987852, EPI_ISL_987853, EPI_ISL_987854, EPI_ISL_987855, EPI_ISL_987856, EPI_ISL_987857, EPI_ISL_987858, EPI_ISL_987859, EPI_ISL_987860, EPI_ISL_987861, EPI_ISL_987862, EPI_ISL_987863, EPI_ISL_987864, EPI_ISL_987865, EPI_ISL_987866, EPI_ISL_987867, EPI_ISL_987868, EPI_ISL_987869, EPI_ISL_987870, EPI_ISL_987871, EPI_ISL_987872, EPI_ISL_987873, EPI_ISL_987874, EPI_ISL_987875, EPI_ISL_987876, EPI_ISL_987877, EPI_ISL_987878, EPI_ISL_987879, EPI_ISL_987880, EPI_ISL_987881, EPI_ISL_987882, EPI_ISL_987883, EPI_ISL_987884, EPI_ISL_987885, EPI_ISL_987886, EPI_ISL_987887, EPI_ISL_987888, EPI_ISL_987889, EPI_ISL_987890, EPI_ISL_987891, EPI_ISL_987892, EPI_ISL_987893, EPI_ISL_987894, EPI_ISL_987895, EPI_ISL_987896, EPI_ISL_987897, EPI_ISL_987898, EPI_ISL_987899, EPI_ISL_987900, EPI_ISL_987901, EPI_ISL_987902, EPI_ISL_987903, EPI_ISL_987904, EPI_ISL_987905, EPI_ISL_987906, EPI_ISL_987907, EPI_ISL_987908, EPI_ISL_987909, EPI_ISL_987910, EPI_ISL_987911, EPI_ISL_987912, EPI_ISL_987913, EPI_ISL_987914, EPI_ISL_987915, EPI_ISL_987916, EPI_ISL_987917, EPI_ISL_987918, EPI_ISL_987919, EPI_ISL_987920, EPI_ISL_987921, EPI_ISL_987922, EPI_ISL_987923, EPI_ISL_987924, EPI_ISL_987925, EPI_ISL_987926, EPI_ISL_987927, EPI_ISL_987928, EPI_ISL_987929, EPI_ISL_987930, EPI_ISL_987931, EPI_ISL_987932, EPI_ISL_987933, EPI_ISL_987934, EPI_ISL_987935, EPI_ISL_987936, EPI_ISL_987937, EPI_ISL_987938, EPI_ISL_987939, EPI_ISL_987940, EPI_ISL_987941, EPI_ISL_987942, EPI_ISL_987943, EPI_ISL_987944, EPI_ISL_987945, EPI_ISL_987946, EPI_ISL_987947, EPI_ISL_987948, EPI_ISL_987949, EPI_ISL_987950, EPI_ISL_987951, EPI_ISL_987952, EPI_ISL_987953, EPI_ISL_987954, EPI_ISL_987955, EPI_ISL_987956, EPI_ISL_987957, EPI_ISL_987958, EPI_ISL_987959, EPI_ISL_987960, EPI_ISL_987961, EPI_ISL_987962, EPI_ISL_987963, EPI_ISL_987964, EPI_ISL_987965, EPI_ISL_987966, EPI_ISL_987967, EPI_ISL_987968, EPI_ISL_987969, EPI_ISL_987970, EPI_ISL_987971, EPI_ISL_987972, EPI_ISL_987973, EPI_ISL_987974, EPI_ISL_987975, EPI_ISL_987976, EPI_ISL_987977, EPI_ISL_987978, EPI_ISL_987979, EPI_ISL_987980, EPI_ISL_987981, EPI_ISL_987982, EPI_ISL_987983, EPI_ISL_987984, EPI_ISL_987985, EPI_ISL_987986, EPI_ISL_987987, EPI_ISL_987988, EPI_ISL_987989, EPI_ISL_987990, EPI_ISL_987991, EPI_ISL_987992, EPI_ISL_987993, EPI_ISL_987994, EPI_ISL_987995, EPI_ISL_987996, EPI_ISL_987997, EPI_ISL_987998, EPI_ISL_987999, EPI_ISL_988000, EPI_ISL_988001, EPI_ISL_988002, EPI_ISL_988003, EPI_ISL_988004, EPI_ISL_988005, EPI_ISL_988006, EPI_ISL_988007, EPI_ISL_988008, EPI_ISL_988009, EPI_ISL_988010, EPI_ISL_988257, EPI_ISL_988348, EPI_ISL_1007435, EPI_ISL_1018766, EPI_ISL_1223167, EPI_ISL_1223193, EPI_ISL_1223236, EPI_ISL_1223246, EPI_ISL_1223258, EPI_ISL_1223265, EPI_ISL_1223270, EPI_ISL_1223283, EPI_ISL_1223285, EPI_ISL_1223357, EPI_ISL_1224517, EPI_ISL_1224537, EPI_ISL_1224567, EPI_ISL_1224585, EPI_ISL_1224594, EPI_ISL_1224604, EPI_ISL_1224608, EPI_ISL_1224614, EPI_ISL_1224626, EPI_ISL_1224639, EPI_ISL_1224642, EPI_ISL_1224670, EPI_ISL_1224700, EPI_ISL_1224705, EPI_ISL_1242051, EPI_ISL_1242056, EPI_ISL_1242082, EPI_ISL_1242087, EPI_ISL_1242088, EPI_ISL_1242091, EPI_ISL_1242117, EPI_ISL_1242150, EPI_ISL_1242169, EPI_ISL_1242170, EPI_ISL_1242174, EPI_ISL_1242177, EPI_ISL_1242178, EPI_ISL_1242182, EPI_ISL_1242197, EPI_ISL_1242221, EPI_ISL_1242236, EPI_ISL_1242252, EPI_ISL_1333480, EPI_ISL_1489495, EPI_ISL_1489496, EPI_ISL_1489497, EPI_ISL_1489498, EPI_ISL_1489500, EPI_ISL_1489501, EPI_ISL_1489502, EPI_ISL_1489504, EPI_ISL_1489506, EPI_ISL_1489508, EPI_ISL_1489512, EPI_ISL_1489513, EPI_ISL_1489514, EPI_ISL_1489516, EPI_ISL_1489519, EPI_ISL_1489521, EPI_ISL_1489522, EPI_ISL_1489523, EPI_ISL_1489524, EPI_ISL_1489525

| | | | |
|---|---|---|---|
| see above | Lighthouse Lab in Cambridge | Wellcome Sanger Institute for the COVID-19 Genomics UK (COG-UK) Consortium | Cordelia Langford; David K. Jackson; Dominic Kwiatkowski; Ewan Harrison; Ian Johnston; Jeffrey Barrett; John Sillitoe on behalf of the Wellcome Sanger Institute COVID-19 Surveillance Team; John Sillitoe on behalf of the Wellcome Sanger Institute COVID-19 Surveillance Team (http://www.sanger.ac.uk/covid-team); Rob Howes; Roberto Amato; Sonia Goncalves; The Lighthouse Lab in Cambridge and Alex Alderton |

EPI_ISL_907237, EPI_ISL_944792, EPI_ISL_944793, EPI_ISL_944794, EPI_ISL_944795, EPI_ISL_944796, EPI_ISL_944797, EPI_ISL_944798, EPI_ISL_944800, EPI_ISL_944801, EPI_ISL_944802, EPI_ISL_944803, EPI_ISL_944804, EPI_ISL_944806, EPI_ISL_944807, EPI_ISL_944808, EPI_ISL_944809, EPI_ISL_944810, EPI_ISL_944811, EPI_ISL_944812, EPI_ISL_944813, EPI_ISL_944814, EPI_ISL_944815, EPI_ISL_944816, EPI_ISL_944817, EPI_ISL_944818, EPI_ISL_944819, EPI_ISL_944820, EPI_ISL_944821, EPI_ISL_944822, EPI_ISL_944823, EPI_ISL_944825, EPI_ISL_944826, EPI_ISL_944827, EPI_ISL_944828, EPI_ISL_944829, EPI_ISL_944830, EPI_ISL_944831, EPI_ISL_944832, EPI_ISL_944833, EPI_ISL_944834, EPI_ISL_944835, EPI_ISL_944836, EPI_ISL_944837, EPI_ISL_944838, EPI_ISL_944839, EPI_ISL_944840, EPI_ISL_944841, EPI_ISL_944842, EPI_ISL_944843, EPI_ISL_944845, EPI_ISL_944846, EPI_ISL_944847, EPI_ISL_944848, EPI_ISL_944849, EPI_ISL_944850, EPI_ISL_944851, EPI_ISL_944852, EPI_ISL_944853, EPI_ISL_944854, EPI_ISL_944855, EPI_ISL_944856, EPI_ISL_944857, EPI_ISL_944858, EPI_ISL_944859, EPI_ISL_944860, EPI_ISL_944861, EPI_ISL_944862, EPI_ISL_944864, EPI_ISL_944865, EPI_ISL_944866, EPI_ISL_944867, EPI_ISL_944868, EPI_ISL_944869, EPI_ISL_944870, EPI_ISL_944871, EPI_ISL_944872, EPI_ISL_944873, EPI_ISL_944874, EPI_ISL_944875, EPI_ISL_944876, EPI_ISL_944877, EPI_ISL_944878, EPI_ISL_944879, EPI_ISL_944880, EPI_ISL_944881, EPI_ISL_944882, EPI_ISL_944884, EPI_ISL_944885, EPI_ISL_944886, EPI_ISL_944887, EPI_ISL_944888, EPI_ISL_944889, EPI_ISL_944890, EPI_ISL_944891, EPI_ISL_944892, EPI_ISL_944893, EPI_ISL_944894, EPI_ISL_944895, EPI_ISL_944896, EPI_ISL_944897, EPI_ISL_944898, EPI_ISL_944899, EPI_ISL_944900, EPI_ISL_944901, EPI_ISL_944902, EPI_ISL_944903, EPI_ISL_944904, EPI_ISL_944905, EPI_ISL_944906, EPI_ISL_944907, EPI_ISL_944909, EPI_ISL_944910, EPI_ISL_944911, EPI_ISL_944912, EPI_ISL_944913, EPI_ISL_944915, EPI_ISL_944916, EPI_ISL_944917, EPI_ISL_944918, EPI_ISL_944919, EPI_ISL_944920, EPI_ISL_944921, EPI_ISL_944922, EPI_ISL_944924, EPI_ISL_944925, EPI_ISL_944926, EPI_ISL_944927, EPI_ISL_944928, EPI_ISL_944929, EPI_ISL_944930, EPI_ISL_944931, EPI_ISL_944932, EPI_ISL_944933, EPI_ISL_944934, EPI_ISL_944935, EPI_ISL_944936, EPI_ISL_944937, EPI_ISL_944938, EPI_ISL_944939, EPI_ISL_944940, EPI_ISL_944941, EPI_ISL_944942, EPI_ISL_944943, EPI_ISL_944944, EPI_ISL_944945, EPI_ISL_944946, EPI_ISL_944947, EPI_ISL_944948, EPI_ISL_944949, EPI_ISL_944950, EPI_ISL_944951, EPI_ISL_944952, EPI_ISL_944954, EPI_ISL_944955, EPI_ISL_944956, EPI_ISL_944957, EPI_ISL_944958, EPI_ISL_944959, EPI_ISL_944960, EPI_ISL_944961, EPI_ISL_944962, EPI_ISL_944963, EPI_ISL_944964, EPI_ISL_944965, EPI_ISL_944966, EPI_ISL_944967, EPI_ISL_944968, EPI_ISL_944969, EPI_ISL_944970, EPI_ISL_944971, EPI_ISL_944972, EPI_ISL_944973, EPI_ISL_944974, EPI_ISL_944975, EPI_ISL_944977, EPI_ISL_944979, EPI_ISL_944980, EPI_ISL_944981, EPI_ISL_944982, EPI_ISL_944983, EPI_ISL_944984, EPI_ISL_944985, EPI_ISL_944986, EPI_ISL_944987, EPI_ISL_944988, EPI_ISL_944989, EPI_ISL_944990, EPI_ISL_944991, EPI_ISL_944992, EPI_ISL_944993, EPI_ISL_944994, EPI_ISL_944995, EPI_ISL_944996, EPI_ISL_944997, EPI_ISL_944998, EPI_ISL_944999, EPI_ISL_945000, EPI_ISL_945001, EPI_ISL_945002, EPI_ISL_945003, EPI_ISL_945004, EPI_ISL_945005, EPI_ISL_945006, EPI_ISL_945007, EPI_ISL_945008, EPI_ISL_945009, EPI_ISL_945010, EPI_ISL_945011, EPI_ISL_945012, EPI_ISL_945013, EPI_ISL_945014, EPI_ISL_945015, EPI_ISL_945016, EPI_ISL_945017, EPI_ISL_945018, EPI_ISL_945019, EPI_ISL_945020, EPI_ISL_945021, EPI_ISL_945022, EPI_ISL_945024, EPI_ISL_945025, EPI_ISL_945026, EPI_ISL_945027, EPI_ISL_945028, EPI_ISL_945029, EPI_ISL_945030, EPI_ISL_945031, EPI_ISL_945033, EPI_ISL_945035, EPI_ISL_945036, EPI_ISL_945037, EPI_ISL_945038, EPI_ISL_945039, EPI_ISL_945040, EPI_ISL_945041, EPI_ISL_945042, EPI_ISL_945043, EPI_ISL_945044, EPI_ISL_945045, EPI_ISL_945046, EPI_ISL_945047, EPI_ISL_945048, EPI_ISL_945716, EPI_ISL_945718, EPI_ISL_945722, EPI_ISL_945725, EPI_ISL_945726, EPI_ISL_945728, EPI_ISL_945729, EPI_ISL_945730, EPI_ISL_945732, EPI_ISL_945733, EPI_ISL_945735, EPI_ISL_945736, EPI_ISL_945737, EPI_ISL_945738, EPI_ISL_945741, EPI_ISL_945742, EPI_ISL_945743, EPI_ISL_945744, EPI_ISL_945747, EPI_ISL_945753, EPI_ISL_945755, EPI_ISL_945757, EPI_ISL_945758, EPI_ISL_945759, EPI_ISL_945762, EPI_ISL_945763, EPI_ISL_945768, EPI_ISL_945771, EPI_ISL_945772, EPI_ISL_945776, EPI_ISL_945777, EPI_ISL_945781, EPI_ISL_945782, EPI_ISL_945788, EPI_ISL_945792, EPI_ISL_945795, EPI_ISL_945796, EPI_ISL_945799, EPI_ISL_945800, EPI_ISL_945802, EPI_ISL_945803, EPI_ISL_945804, EPI_ISL_945806, EPI_ISL_945807, EPI_ISL_945808, EPI_ISL_945809, EPI_ISL_945812, EPI_ISL_945813, EPI_ISL_945815, EPI_ISL_945816, EPI_ISL_945817, EPI_ISL_945818, EPI_ISL_945821, EPI_ISL_945822, EPI_ISL_945824, EPI_ISL_945826, EPI_ISL_945828, EPI_ISL_945831, EPI_ISL_945835, EPI_ISL_945836, EPI_ISL_945839, EPI_ISL_945841, EPI_ISL_945843, EPI_ISL_945844, EPI_ISL_945846, EPI_ISL_945848, EPI_ISL_945849, EPI_ISL_945850, EPI_ISL_945851, EPI_ISL_945853, EPI_ISL_945860, EPI_ISL_945862, EPI_ISL_945863, EPI_ISL_945865, EPI_ISL_945867, EPI_ISL_945868, EPI_ISL_945869, EPI_ISL_945870, EPI_ISL_945871, EPI_ISL_945872, EPI_ISL_945873, EPI_ISL_945874, EPI_ISL_945877, EPI_ISL_945878, EPI_ISL_945879, EPI_ISL_945882, EPI_ISL_945884, EPI_ISL_945885, EPI_ISL_945889, EPI_ISL_945895, EPI_ISL_945898, EPI_ISL_945899, EPI_ISL_945900, EPI_ISL_945901, EPI_ISL_945904, EPI_ISL_945906, EPI_ISL_945907, EPI_ISL_945908, EPI_ISL_945909, EPI_ISL_945910, EPI_ISL_945911, EPI_ISL_945914, EPI_ISL_945916, EPI_ISL_945917, EPI_ISL_945920, EPI_ISL_945921, EPI_ISL_945926, EPI_ISL_945933, EPI_ISL_945935, EPI_ISL_945938, EPI_ISL_945939, EPI_ISL_945940, EPI_ISL_945944, EPI_ISL_945945, EPI_ISL_945950, EPI_ISL_945951, EPI_ISL_945954, EPI_ISL_945956, EPI_ISL_945958, EPI_ISL_945960, EPI_ISL_945961, EPI_ISL_945962, EPI_ISL_945963, EPI_ISL_945965, EPI_ISL_945967, EPI_ISL_945969, EPI_ISL_945970, EPI_ISL_945971, EPI_ISL_945972, EPI_ISL_945976, EPI_ISL_945979, EPI_ISL_945980, EPI_ISL_945983, EPI_ISL_945984, EPI_ISL_945988, EPI_ISL_945990, EPI_ISL_945993, EPI_ISL_945994, EPI_ISL_945997, EPI_ISL_945998, EPI_ISL_945999, EPI_ISL_946000, EPI_ISL_946002, EPI_ISL_946005, EPI_ISL_946006, EPI_ISL_946007, EPI_ISL_946008, EPI_ISL_946009, EPI_ISL_946010, EPI_ISL_946011, EPI_ISL_946014, EPI_ISL_946021, EPI_ISL_946023, EPI_ISL_946026, EPI_ISL_946028, EPI_ISL_946030, EPI_ISL_946035, EPI_ISL_946037, EPI_ISL_946041, EPI_ISL_946048, EPI_ISL_946049, EPI_ISL_946052, EPI_ISL_946055, EPI_ISL_946057, EPI_ISL_946058, EPI_ISL_946059, EPI_ISL_946062, EPI_ISL_946066, EPI_ISL_946067, EPI_ISL_946068, EPI_ISL_946069, EPI_ISL_946070, EPI_ISL_946071, EPI_ISL_946072, EPI_ISL_946073, EPI_ISL_946076, EPI_ISL_946080, EPI_ISL_946086, EPI_ISL_946087, EPI_ISL_946088, EPI_ISL_946090, EPI_ISL_946093, EPI_ISL_946096, EPI_ISL_946106, EPI_ISL_946109, EPI_ISL_946110, EPI_ISL_946112, EPI_ISL_946113, EPI_ISL_946115, EPI_ISL_946123, EPI_ISL_946124, EPI_ISL_946126, EPI_ISL_946127, EPI_ISL_946129, EPI_ISL_946135, EPI_ISL_946137, EPI_ISL_946145, EPI_ISL_946147, EPI_ISL_946149, EPI_ISL_946150, EPI_ISL_946157, EPI_ISL_946160, EPI_ISL_946162, EPI_ISL_946169, EPI_ISL_946173, EPI_ISL_946174, EPI_ISL_946176, EPI_ISL_946177, EPI_ISL_946180, EPI_ISL_946181, EPI_ISL_946182, EPI_ISL_946188, EPI_ISL_946190, EPI_ISL_946192, EPI_ISL_946196, EPI_ISL_946199, EPI_ISL_946202, EPI_ISL_946206, EPI_ISL_946208, EPI_ISL_946210, EPI_ISL_946211, EPI_ISL_946213, EPI_ISL_946215, EPI_ISL_946218, EPI_ISL_946220, EPI_ISL_946221, EPI_ISL_946224, EPI_ISL_946225, EPI_ISL_946227, EPI_ISL_946229, EPI_ISL_946231, EPI_ISL_946232, EPI_ISL_946233, EPI_ISL_946234, EPI_ISL_946243, EPI_ISL_946250, EPI_ISL_946253, EPI_ISL_946254, EPI_ISL_946260, EPI_ISL_946264, EPI_ISL_946270, EPI_ISL_946271, EPI_ISL_946274, EPI_ISL_946281, EPI_ISL_946284, EPI_ISL_946285, EPI_ISL_946286, EPI_ISL_946292, EPI_ISL_946298, EPI_ISL_946301, EPI_ISL_946302, EPI_ISL_946304, EPI_ISL_946318, EPI_ISL_946319, EPI_ISL_946326, EPI_ISL_946328, EPI_ISL_946333, EPI_ISL_946335, EPI_ISL_946338, EPI_ISL_946339, EPI_ISL_946351, EPI_ISL_946356, EPI_ISL_946357, EPI_ISL_946360, EPI_ISL_946361, EPI_ISL_946365, EPI_ISL_946366, EPI_ISL_946367, EPI_ISL_946371, EPI_ISL_946374, EPI_ISL_946375, EPI_ISL_946378, EPI_ISL_946379, EPI_ISL_946380, EPI_ISL_946385, EPI_ISL_946391, EPI_ISL_946397, EPI_ISL_946400, EPI_ISL_946405, EPI_ISL_946407, EPI_ISL_946410, EPI_ISL_946411, EPI_ISL_957057, EPI_ISL_957080, EPI_ISL_957084, EPI_ISL_957090, EPI_ISL_957093, EPI_ISL_957094, EPI_ISL_957095, EPI_ISL_957130, EPI_ISL_957149, EPI_ISL_957154, EPI_ISL_957162, EPI_ISL_957171, EPI_ISL_957177, EPI_ISL_957183, EPI_ISL_957188, EPI_ISL_957201, EPI_ISL_957227, EPI_ISL_957248, EPI_ISL_957256, EPI_ISL_957270, EPI_ISL_957275, EPI_ISL_957294, EPI_ISL_957312, EPI_ISL_957674, EPI_ISL_963326, EPI_ISL_963330, EPI_ISL_963341, EPI_ISL_963344, EPI_ISL_963345, EPI_ISL_963346, EPI_ISL_963348, EPI_ISL_963355, EPI_ISL_963356, EPI_ISL_963357, EPI_ISL_963358, EPI_ISL_963370, EPI_ISL_963371, EPI_ISL_963376, EPI_ISL_963380, EPI_ISL_963386, EPI_ISL_963397, EPI_ISL_963419, EPI_ISL_963421, EPI_ISL_963425, EPI_ISL_963426, EPI_ISL_963431, EPI_ISL_963433, EPI_ISL_963434, EPI_ISL_963449, EPI_ISL_963452, EPI_ISL_963458, EPI_ISL_963475, EPI_ISL_963481, EPI_ISL_963489, EPI_ISL_963490, EPI_ISL_963495, EPI_ISL_963509, EPI_ISL_963511, EPI_ISL_963520, EPI_ISL_963521, EPI_ISL_963529, EPI_ISL_963530, EPI_ISL_963538, EPI_ISL_963554, EPI_ISL_963559, EPI_ISL_963560, EPI_ISL_963562, EPI_ISL_963570, EPI_ISL_963573, EPI_ISL_963576, EPI_ISL_963581, EPI_ISL_963584, EPI_ISL_963591, EPI_ISL_963594, EPI_ISL_963601, EPI_ISL_963603, EPI_ISL_963607, EPI_ISL_963618, EPI_ISL_963626, EPI_ISL_963633, EPI_ISL_963639, EPI_ISL_963641, EPI_ISL_963649, EPI_ISL_964364, EPI_ISL_964390, EPI_ISL_964393, EPI_ISL_964421, EPI_ISL_964439, EPI_ISL_964451, EPI_ISL_964465, EPI_ISL_964487, EPI_ISL_964512, EPI_ISL_989449, EPI_ISL_989450, EPI_ISL_989451, EPI_ISL_989452, EPI_ISL_989453, EPI_ISL_989454, EPI_ISL_989455, EPI_ISL_989456, EPI_ISL_989457, EPI_ISL_989458, EPI_ISL_989459, EPI_ISL_989460, EPI_ISL_989461, EPI_ISL_989462, EPI_ISL_989463, EPI_ISL_989464, EPI_ISL_989465, EPI_ISL_989466, EPI_ISL_989467, EPI_ISL_989468, EPI_ISL_989469, EPI_ISL_989470, EPI_ISL_989471, EPI_ISL_989472, EPI_ISL_989473, EPI_ISL_989474, EPI_ISL_989475, EPI_ISL_989476, EPI_ISL_989477, EPI_ISL_989478, EPI_ISL_989479, EPI_ISL_989480, EPI_ISL_989481, EPI_ISL_989808, EPI_ISL_989834, EPI_ISL_989836, EPI_ISL_989844, EPI_ISL_989852, EPI_ISL_989854, EPI_ISL_989882, EPI_ISL_989883, EPI_ISL_989884, EPI_ISL_989885, EPI_ISL_989886, EPI_ISL_989887, EPI_ISL_989888, EPI_ISL_989889, EPI_ISL_989890, EPI_ISL_989891, EPI_ISL_989892, EPI_ISL_989893, EPI_ISL_989894, EPI_ISL_989895, EPI_ISL_989896, EPI_ISL_989897, EPI_ISL_989898, EPI_ISL_989899, EPI_ISL_989900, EPI_ISL_989901, EPI_ISL_989902, EPI_ISL_989903, EPI_ISL_989904, EPI_ISL_989905, EPI_ISL_989906, EPI_ISL_989907, EPI_ISL_989908, EPI_ISL_989909, EPI_ISL_989910, EPI_ISL_989911, EPI_ISL_989912, EPI_ISL_989913, EPI_ISL_989914, EPI_ISL_989915, EPI_ISL_989916, EPI_ISL_989917, EPI_ISL_989918, EPI_ISL_989919, EPI_ISL_989920, EPI_ISL_989921, EPI_ISL_989922, EPI_ISL_989923, EPI_ISL_989924, EPI_ISL_989925, EPI_ISL_989926, EPI_ISL_989927, EPI_ISL_989928, EPI_ISL_989929, EPI_ISL_989930, EPI_ISL_989931, EPI_ISL_989932, EPI_ISL_989933, EPI_ISL_989934, EPI_ISL_989935, EPI_ISL_989936, EPI_ISL_989937, EPI_ISL_989938, EPI_ISL_989939, EPI_ISL_989940, EPI_ISL_989941, EPI_ISL_989942, EPI_ISL_989943, EPI_ISL_989944, EPI_ISL_989945, EPI_ISL_989946, EPI_ISL_989947, EPI_ISL_989948, EPI_ISL_989949, EPI_ISL_989950, EPI_ISL_989951, EPI_ISL_989952, EPI_ISL_989953, EPI_ISL_989954, EPI_ISL_989955, EPI_ISL_989956, EPI_ISL_989957, EPI_ISL_989958, EPI_ISL_989959, EPI_ISL_989960, EPI_ISL_989961, EPI_ISL_989962, EPI_ISL_989963, EPI_ISL_989964, EPI_ISL_989965, EPI_ISL_989966, EPI_ISL_989967, EPI_ISL_989968, EPI_ISL_989969, EPI_ISL_989970, EPI_ISL_989971, EPI_ISL_989972, EPI_ISL_989973, EPI_ISL_989974, EPI_ISL_989975, EPI_ISL_989976, EPI_ISL_989977, EPI_ISL_989978, EPI_ISL_989979, EPI_ISL_989980, EPI_ISL_989981, EPI_ISL_989982

| | | | |
|---|---|---|---|
| see above | Lighthouse Lab in Glasgow | Wellcome Sanger Institute for the COVID-19 Genomics UK (COG-UK) Consortium | Anna Dominiczak and Alex Alderton; Carol Clugston; Cordelia Langford; David Gray; David K. Jackson; Dominic Kwiatkowski; Ewan Harrison; Harper VanSteenhouse; Ian Johnston; John Sillitoe on behalf of the Wellcome Sanger Institute COVID-19 Surveillance Team; John Sillitoe on behalf of the Wellcome Sanger Institute COVID-19 Surveillance Team (http://www.sanger.ac.uk/covid-team); Roberto Amato; Sonia Goncalves; Yumi Kasai |

EPI_ISL_956765, EPI_ISL_956766, EPI_ISL_956767, EPI_ISL_956768, EPI_ISL_956769, EPI_ISL_956770, EPI_ISL_956771, EPI_ISL_956772, EPI_ISL_956773, EPI_ISL_956774, EPI_ISL_956775, EPI_ISL_956776, EPI_ISL_956777, EPI_ISL_956778, EPI_ISL_956779, EPI_ISL_956780, EPI_ISL_956781, EPI_ISL_956782, EPI_ISL_956783, EPI_ISL_956784, EPI_ISL_956785, EPI_ISL_956786, EPI_ISL_956787, EPI_ISL_956788, EPI_ISL_956789, EPI_ISL_956790, EPI_ISL_956791, EPI_ISL_956792, EPI_ISL_956793, EPI_ISL_956794, EPI_ISL_956795, EPI_ISL_956796, EPI_ISL_956797, EPI_ISL_956798, EPI_ISL_956799, EPI_ISL_956800, EPI_ISL_956801, EPI_ISL_956802, EPI_ISL_956803, EPI_ISL_956804, EPI_ISL_956805, EPI_ISL_956806, EPI_ISL_956807, EPI_ISL_956808, EPI_ISL_956809, EPI_ISL_956810, EPI_ISL_956811, EPI_ISL_956812, EPI_ISL_956813, EPI_ISL_956814, EPI_ISL_956815, EPI_ISL_956816, EPI_ISL_956817, EPI_ISL_956818, EPI_ISL_956819, EPI_ISL_956820, EPI_ISL_956821, EPI_ISL_956822, EPI_ISL_956823, EPI_ISL_956824, EPI_ISL_956825, EPI_ISL_956826, EPI_ISL_956827, EPI_ISL_956828, EPI_ISL_956829, EPI_ISL_956830, EPI_ISL_956831, EPI_ISL_956832, EPI_ISL_956833, EPI_ISL_956834, EPI_ISL_956835, EPI_ISL_956836, EPI_ISL_956837, EPI_ISL_956838, EPI_ISL_956839, EPI_ISL_956840, EPI_ISL_956841, EPI_ISL_956842, EPI_ISL_956843, EPI_ISL_956844, EPI_ISL_956845, EPI_ISL_956846, EPI_ISL_956847, EPI_ISL_956848, EPI_ISL_956849, EPI_ISL_956850, EPI_ISL_956851, EPI_ISL_956852, EPI_ISL_956853, EPI_ISL_956854, EPI_ISL_956855, EPI_ISL_956856, EPI_ISL_956857, EPI_ISL_956858, EPI_ISL_956859, EPI_ISL_956860, EPI_ISL_956861, EPI_ISL_956862, EPI_ISL_956863, EPI_ISL_956864, EPI_ISL_956865, EPI_ISL_956866, EPI_ISL_956867, EPI_ISL_956868, EPI_ISL_956869, EPI_ISL_956870, EPI_ISL_956871, EPI_ISL_956872, EPI_ISL_956873, EPI_ISL_956874, EPI_ISL_956875, EPI_ISL_956876, EPI_ISL_956877, EPI_ISL_956878, EPI_ISL_956879, EPI_ISL_956880, EPI_ISL_956881, EPI_ISL_956882, EPI_ISL_956883, EPI_ISL_956884, EPI_ISL_956885, EPI_ISL_956886, EPI_ISL_956887, EPI_ISL_956888, EPI_ISL_956889, EPI_ISL_956890, EPI_ISL_956891, EPI_ISL_956892, EPI_ISL_956893, EPI_ISL_956894, EPI_ISL_956895, EPI_ISL_956896, EPI_ISL_956897, EPI_ISL_956898, EPI_ISL_956899, EPI_ISL_956900, EPI_ISL_956901, EPI_ISL_956902, EPI_ISL_956903, EPI_ISL_956904, EPI_ISL_956905, EPI_ISL_956906, EPI_ISL_956907, EPI_ISL_956908,

EPI_ISL_956909, EPI_ISL_956910, EPI_ISL_956911, EPI_ISL_956912, EPI_ISL_956913, EPI_ISL_956914, EPI_ISL_956915, EPI_ISL_956916, EPI_ISL_956917, EPI_ISL_956918, EPI_ISL_956919, EPI_ISL_956920, EPI_ISL_956921, EPI_ISL_956922, EPI_ISL_956923, EPI_ISL_956924, EPI_ISL_956925, EPI_ISL_956926, EPI_ISL_956927, EPI_ISL_956928, EPI_ISL_956929, EPI_ISL_956930, EPI_ISL_956931, EPI_ISL_956932,
EPI_ISL_956933, EPI_ISL_956934, EPI_ISL_956935, EPI_ISL_956936, EPI_ISL_956937, EPI_ISL_956938, EPI_ISL_956939, EPI_ISL_956940, EPI_ISL_956941, EPI_ISL_956942, EPI_ISL_956943, EPI_ISL_956944, EPI_ISL_956945, EPI_ISL_956946, EPI_ISL_956947, EPI_ISL_956948, EPI_ISL_956949, EPI_ISL_956950, EPI_ISL_956951, EPI_ISL_956952, EPI_ISL_956953, EPI_ISL_956954, EPI_ISL_956955, EPI_ISL_956956,
EPI_ISL_956957, EPI_ISL_956958, EPI_ISL_956959, EPI_ISL_956960, EPI_ISL_956961, EPI_ISL_956962, EPI_ISL_956963, EPI_ISL_956964, EPI_ISL_956965, EPI_ISL_956966, EPI_ISL_956967, EPI_ISL_956968, EPI_ISL_956969, EPI_ISL_956970, EPI_ISL_956971, EPI_ISL_956972, EPI_ISL_956973, EPI_ISL_956974, EPI_ISL_956975, EPI_ISL_956976, EPI_ISL_956977, EPI_ISL_956978, EPI_ISL_956979, EPI_ISL_956980,
EPI_ISL_956981, EPI_ISL_956982, EPI_ISL_956983, EPI_ISL_956984, EPI_ISL_956985, EPI_ISL_956986, EPI_ISL_956987, EPI_ISL_956988, EPI_ISL_956989, EPI_ISL_956990, EPI_ISL_956991, EPI_ISL_956992, EPI_ISL_956993, EPI_ISL_956994, EPI_ISL_956995, EPI_ISL_956996, EPI_ISL_956997, EPI_ISL_956998, EPI_ISL_956999, EPI_ISL_957000, EPI_ISL_957001, EPI_ISL_957002, EPI_ISL_957003, EPI_ISL_957004,
EPI_ISL_957005, EPI_ISL_957006, EPI_ISL_957007, EPI_ISL_957008, EPI_ISL_957009, EPI_ISL_957010, EPI_ISL_957011, EPI_ISL_957012, EPI_ISL_957013, EPI_ISL_957014, EPI_ISL_957015, EPI_ISL_957016, EPI_ISL_957017, EPI_ISL_957018, EPI_ISL_957019, EPI_ISL_957020, EPI_ISL_957021, EPI_ISL_957022, EPI_ISL_957023, EPI_ISL_957024, EPI_ISL_957025, EPI_ISL_957026, EPI_ISL_957027, EPI_ISL_957028,
EPI_ISL_957029, EPI_ISL_957030, EPI_ISL_957031, EPI_ISL_957032, EPI_ISL_957033, EPI_ISL_957034, EPI_ISL_957035, EPI_ISL_957036, EPI_ISL_957037, EPI_ISL_957038, EPI_ISL_957039, EPI_ISL_957040, EPI_ISL_957041, EPI_ISL_957042, EPI_ISL_957043, EPI_ISL_957044, EPI_ISL_957045, EPI_ISL_957046, EPI_ISL_957047, EPI_ISL_957048, EPI_ISL_957049, EPI_ISL_957050, EPI_ISL_957051, EPI_ISL_957052,
EPI_ISL_957053, EPI_ISL_957056, EPI_ISL_957058, EPI_ISL_957059, EPI_ISL_957062, EPI_ISL_957063, EPI_ISL_957064, EPI_ISL_957066, EPI_ISL_957069, EPI_ISL_957070, EPI_ISL_957074, EPI_ISL_957076, EPI_ISL_957085, EPI_ISL_957087, EPI_ISL_957088, EPI_ISL_957096, EPI_ISL_957098, EPI_ISL_957100, EPI_ISL_957106, EPI_ISL_957108, EPI_ISL_957109, EPI_ISL_957111, EPI_ISL_957112, EPI_ISL_957113,
EPI_ISL_957116, EPI_ISL_957119, EPI_ISL_957120, EPI_ISL_957123, EPI_ISL_957125, EPI_ISL_957126, EPI_ISL_957127, EPI_ISL_957128, EPI_ISL_957129, EPI_ISL_957131, EPI_ISL_957132, EPI_ISL_957134, EPI_ISL_957135, EPI_ISL_957136, EPI_ISL_957137, EPI_ISL_957138, EPI_ISL_957139, EPI_ISL_957142, EPI_ISL_957143, EPI_ISL_957144, EPI_ISL_957145, EPI_ISL_957146, EPI_ISL_957147, EPI_ISL_957148,
EPI_ISL_957152, EPI_ISL_957153, EPI_ISL_957156, EPI_ISL_957159, EPI_ISL_957161, EPI_ISL_957163, EPI_ISL_957164, EPI_ISL_957167, EPI_ISL_957169, EPI_ISL_957173, EPI_ISL_957174, EPI_ISL_957175, EPI_ISL_957178, EPI_ISL_957179, EPI_ISL_957181, EPI_ISL_957182, EPI_ISL_957184, EPI_ISL_957185, EPI_ISL_957186, EPI_ISL_957189, EPI_ISL_957192, EPI_ISL_957193, EPI_ISL_957195, EPI_ISL_957196,
EPI_ISL_957197, EPI_ISL_957199, EPI_ISL_957202, EPI_ISL_957203, EPI_ISL_957204, EPI_ISL_957207, EPI_ISL_957209, EPI_ISL_957213, EPI_ISL_957214, EPI_ISL_957217, EPI_ISL_957218, EPI_ISL_957220, EPI_ISL_957221, EPI_ISL_957222, EPI_ISL_957226, EPI_ISL_957228, EPI_ISL_957230, EPI_ISL_957232, EPI_ISL_957233, EPI_ISL_957234, EPI_ISL_957235, EPI_ISL_957236, EPI_ISL_957238, EPI_ISL_957240,
EPI_ISL_957241, EPI_ISL_957244, EPI_ISL_957247, EPI_ISL_957249, EPI_ISL_957251, EPI_ISL_957252, EPI_ISL_957258, EPI_ISL_957261, EPI_ISL_957264, EPI_ISL_957265, EPI_ISL_957267, EPI_ISL_957269, EPI_ISL_957273, EPI_ISL_957277, EPI_ISL_957278, EPI_ISL_957280, EPI_ISL_957281, EPI_ISL_957282, EPI_ISL_957284, EPI_ISL_957288, EPI_ISL_957290, EPI_ISL_957293, EPI_ISL_957295, EPI_ISL_957296,
EPI_ISL_957298, EPI_ISL_957300, EPI_ISL_957301, EPI_ISL_957303, EPI_ISL_957305, EPI_ISL_957307, EPI_ISL_957309, EPI_ISL_957313, EPI_ISL_957316, EPI_ISL_957317, EPI_ISL_957318, EPI_ISL_957321, EPI_ISL_957324, EPI_ISL_957325, EPI_ISL_957326, EPI_ISL_957329, EPI_ISL_957332, EPI_ISL_957333, EPI_ISL_957335, EPI_ISL_957338, EPI_ISL_957339, EPI_ISL_957340, EPI_ISL_957341, EPI_ISL_957342,
EPI_ISL_957343, EPI_ISL_957344, EPI_ISL_957345, EPI_ISL_957346, EPI_ISL_957347, EPI_ISL_957348, EPI_ISL_957349, EPI_ISL_957350, EPI_ISL_957351, EPI_ISL_957352, EPI_ISL_957353, EPI_ISL_957354, EPI_ISL_957355, EPI_ISL_957356, EPI_ISL_957357, EPI_ISL_957358, EPI_ISL_957359, EPI_ISL_957360, EPI_ISL_957361, EPI_ISL_957362, EPI_ISL_957363, EPI_ISL_957364, EPI_ISL_957365, EPI_ISL_957366,
EPI_ISL_957367, EPI_ISL_957368, EPI_ISL_957369, EPI_ISL_957370, EPI_ISL_957371, EPI_ISL_957372, EPI_ISL_957373, EPI_ISL_957374, EPI_ISL_957375, EPI_ISL_957376, EPI_ISL_957377, EPI_ISL_957378, EPI_ISL_957379, EPI_ISL_957380, EPI_ISL_957381, EPI_ISL_957382, EPI_ISL_957383, EPI_ISL_957384, EPI_ISL_957385, EPI_ISL_957386, EPI_ISL_957387, EPI_ISL_957388, EPI_ISL_957389, EPI_ISL_957390,
EPI_ISL_957391, EPI_ISL_957392, EPI_ISL_957393, EPI_ISL_957394, EPI_ISL_957395, EPI_ISL_957396, EPI_ISL_957397, EPI_ISL_957398, EPI_ISL_957399, EPI_ISL_957400, EPI_ISL_957401, EPI_ISL_957402, EPI_ISL_957403, EPI_ISL_957404, EPI_ISL_957405, EPI_ISL_957406, EPI_ISL_957407, EPI_ISL_957408, EPI_ISL_957409, EPI_ISL_957410, EPI_ISL_957411, EPI_ISL_957412, EPI_ISL_957413, EPI_ISL_957414,
EPI_ISL_957415, EPI_ISL_957416, EPI_ISL_957417, EPI_ISL_957418, EPI_ISL_957419, EPI_ISL_957420, EPI_ISL_957421, EPI_ISL_957422, EPI_ISL_957424, EPI_ISL_957425, EPI_ISL_957426, EPI_ISL_957427, EPI_ISL_957428, EPI_ISL_957429, EPI_ISL_957430, EPI_ISL_957431, EPI_ISL_957432, EPI_ISL_957433, EPI_ISL_957434, EPI_ISL_957435, EPI_ISL_957436, EPI_ISL_957437, EPI_ISL_957438, EPI_ISL_957439,
EPI_ISL_957440, EPI_ISL_957441, EPI_ISL_957442, EPI_ISL_957443, EPI_ISL_957444, EPI_ISL_957445, EPI_ISL_957446, EPI_ISL_957447, EPI_ISL_957448, EPI_ISL_957449, EPI_ISL_957450, EPI_ISL_957451, EPI_ISL_957452, EPI_ISL_957453, EPI_ISL_957454, EPI_ISL_957455, EPI_ISL_957456, EPI_ISL_957457, EPI_ISL_957458, EPI_ISL_957459, EPI_ISL_957460, EPI_ISL_957461, EPI_ISL_957462, EPI_ISL_957463,
EPI_ISL_957464, EPI_ISL_957465, EPI_ISL_957466, EPI_ISL_957467, EPI_ISL_957468, EPI_ISL_957469, EPI_ISL_957470, EPI_ISL_957471, EPI_ISL_957472, EPI_ISL_957473, EPI_ISL_957474, EPI_ISL_957475, EPI_ISL_957476, EPI_ISL_957477, EPI_ISL_957478, EPI_ISL_957479, EPI_ISL_957480, EPI_ISL_957481, EPI_ISL_957482, EPI_ISL_957483, EPI_ISL_957484, EPI_ISL_957485, EPI_ISL_957486, EPI_ISL_957487,
EPI_ISL_957488, EPI_ISL_957489, EPI_ISL_957490, EPI_ISL_957491, EPI_ISL_957492, EPI_ISL_957493, EPI_ISL_957494, EPI_ISL_957495, EPI_ISL_957496, EPI_ISL_957497, EPI_ISL_957498, EPI_ISL_957499, EPI_ISL_957500, EPI_ISL_957501, EPI_ISL_957502, EPI_ISL_957503, EPI_ISL_957504, EPI_ISL_957505, EPI_ISL_957506, EPI_ISL_957507, EPI_ISL_957508, EPI_ISL_957509, EPI_ISL_957510, EPI_ISL_957511,
EPI_ISL_957512, EPI_ISL_957513, EPI_ISL_957514, EPI_ISL_957515, EPI_ISL_957516, EPI_ISL_957517, EPI_ISL_957518, EPI_ISL_957519, EPI_ISL_957520, EPI_ISL_957521, EPI_ISL_957522, EPI_ISL_957523, EPI_ISL_957524, EPI_ISL_957525, EPI_ISL_957526, EPI_ISL_957527, EPI_ISL_957528, EPI_ISL_957529, EPI_ISL_957530, EPI_ISL_957531, EPI_ISL_957532, EPI_ISL_957533, EPI_ISL_957534, EPI_ISL_957535,
EPI_ISL_957536, EPI_ISL_957537, EPI_ISL_957538, EPI_ISL_957539, EPI_ISL_957540, EPI_ISL_957541, EPI_ISL_957542, EPI_ISL_957543, EPI_ISL_957544, EPI_ISL_957545, EPI_ISL_957546, EPI_ISL_957547, EPI_ISL_957548, EPI_ISL_957549, EPI_ISL_957550, EPI_ISL_957551, EPI_ISL_957552, EPI_ISL_957553, EPI_ISL_957554, EPI_ISL_957555, EPI_ISL_957556, EPI_ISL_957557, EPI_ISL_957558, EPI_ISL_957559,
EPI_ISL_957560, EPI_ISL_957561, EPI_ISL_957562, EPI_ISL_957563, EPI_ISL_957564, EPI_ISL_957565, EPI_ISL_957566, EPI_ISL_957567, EPI_ISL_957568, EPI_ISL_957569, EPI_ISL_957570, EPI_ISL_957571, EPI_ISL_957572, EPI_ISL_957573, EPI_ISL_957574, EPI_ISL_957575, EPI_ISL_957576, EPI_ISL_957577, EPI_ISL_957578, EPI_ISL_957579, EPI_ISL_957580, EPI_ISL_957581, EPI_ISL_957582, EPI_ISL_957583,
EPI_ISL_957584, EPI_ISL_957585, EPI_ISL_957586, EPI_ISL_957587, EPI_ISL_957588, EPI_ISL_957589, EPI_ISL_957590, EPI_ISL_957591, EPI_ISL_957592, EPI_ISL_957593, EPI_ISL_957594, EPI_ISL_957595, EPI_ISL_957596, EPI_ISL_957597, EPI_ISL_957598, EPI_ISL_957599, EPI_ISL_957600, EPI_ISL_957601, EPI_ISL_957602, EPI_ISL_957603, EPI_ISL_957604, EPI_ISL_957605, EPI_ISL_957606, EPI_ISL_957607,
EPI_ISL_957608, EPI_ISL_957609, EPI_ISL_957610, EPI_ISL_957611, EPI_ISL_957614, EPI_ISL_957619, EPI_ISL_957623, EPI_ISL_957628, EPI_ISL_957629, EPI_ISL_957631, EPI_ISL_957632, EPI_ISL_957659, EPI_ISL_957662, EPI_ISL_957667, EPI_ISL_957668, EPI_ISL_957671, EPI_ISL_957684, EPI_ISL_957685, EPI_ISL_957690, EPI_ISL_957692, EPI_ISL_957695, EPI_ISL_957699, EPI_ISL_957705, EPI_ISL_957710,
EPI_ISL_957711, EPI_ISL_957712, EPI_ISL_957724, EPI_ISL_957727, EPI_ISL_957744, EPI_ISL_957760, EPI_ISL_957761, EPI_ISL_957762, EPI_ISL_957767, EPI_ISL_957775, EPI_ISL_957777, EPI_ISL_957786, EPI_ISL_957790, EPI_ISL_957791, EPI_ISL_957795, EPI_ISL_957800, EPI_ISL_957801, EPI_ISL_957803, EPI_ISL_957804, EPI_ISL_957810, EPI_ISL_957816, EPI_ISL_957823, EPI_ISL_957827, EPI_ISL_957835,
EPI_ISL_957842, EPI_ISL_957843, EPI_ISL_957849, EPI_ISL_957850, EPI_ISL_957858, EPI_ISL_957885, EPI_ISL_957888, EPI_ISL_957893, EPI_ISL_957899, EPI_ISL_957900, EPI_ISL_957901, EPI_ISL_957909, EPI_ISL_957910, EPI_ISL_957911, EPI_ISL_957912, EPI_ISL_957913, EPI_ISL_957914, EPI_ISL_957915, EPI_ISL_957916, EPI_ISL_957917, EPI_ISL_957918, EPI_ISL_957919, EPI_ISL_957920, EPI_ISL_957922,
EPI_ISL_957923, EPI_ISL_957924, EPI_ISL_957925, EPI_ISL_957926, EPI_ISL_957927, EPI_ISL_957928, EPI_ISL_957929, EPI_ISL_957930, EPI_ISL_957931, EPI_ISL_957932, EPI_ISL_957933, EPI_ISL_957934, EPI_ISL_957935, EPI_ISL_957936, EPI_ISL_957937, EPI_ISL_957938, EPI_ISL_957939, EPI_ISL_957940, EPI_ISL_957941, EPI_ISL_957942, EPI_ISL_957943, EPI_ISL_957944, EPI_ISL_957945, EPI_ISL_957946,
EPI_ISL_957947, EPI_ISL_957948, EPI_ISL_957949, EPI_ISL_957950, EPI_ISL_957951, EPI_ISL_957952, EPI_ISL_957953, EPI_ISL_957954, EPI_ISL_957955, EPI_ISL_957956, EPI_ISL_957957, EPI_ISL_957958, EPI_ISL_957959, EPI_ISL_957960, EPI_ISL_957963, EPI_ISL_957964, EPI_ISL_957965, EPI_ISL_957967, EPI_ISL_957968, EPI_ISL_957969, EPI_ISL_957970, EPI_ISL_957972, EPI_ISL_957973, EPI_ISL_957974,
EPI_ISL_957975, EPI_ISL_957976, EPI_ISL_957977, EPI_ISL_957978, EPI_ISL_957979, EPI_ISL_957980, EPI_ISL_957981, EPI_ISL_957982, EPI_ISL_957983, EPI_ISL_957984, EPI_ISL_957985, EPI_ISL_957986, EPI_ISL_957987, EPI_ISL_957988, EPI_ISL_957989, EPI_ISL_957990, EPI_ISL_957991, EPI_ISL_957992, EPI_ISL_957993, EPI_ISL_957994, EPI_ISL_957995, EPI_ISL_957996, EPI_ISL_957997, EPI_ISL_957998,
EPI_ISL_957999, EPI_ISL_958000, EPI_ISL_958001, EPI_ISL_958002, EPI_ISL_958003, EPI_ISL_958004, EPI_ISL_958005, EPI_ISL_958006, EPI_ISL_958007, EPI_ISL_958008, EPI_ISL_958009, EPI_ISL_958010, EPI_ISL_958011, EPI_ISL_958012, EPI_ISL_958013, EPI_ISL_958014, EPI_ISL_958015, EPI_ISL_958016, EPI_ISL_958017, EPI_ISL_958018, EPI_ISL_958019, EPI_ISL_958020, EPI_ISL_958021, EPI_ISL_958022,
EPI_ISL_958023, EPI_ISL_958024, EPI_ISL_958025, EPI_ISL_958026, EPI_ISL_958027, EPI_ISL_958028, EPI_ISL_958029, EPI_ISL_958030, EPI_ISL_958031, EPI_ISL_958032, EPI_ISL_958033, EPI_ISL_958034, EPI_ISL_958035, EPI_ISL_958036, EPI_ISL_958037, EPI_ISL_958038, EPI_ISL_958039, EPI_ISL_958040, EPI_ISL_958041, EPI_ISL_958042, EPI_ISL_958043, EPI_ISL_958045, EPI_ISL_958046, EPI_ISL_958047,
EPI_ISL_958048, EPI_ISL_958049, EPI_ISL_958050, EPI_ISL_958051, EPI_ISL_958052, EPI_ISL_958053, EPI_ISL_958054, EPI_ISL_958055, EPI_ISL_958056, EPI_ISL_958057, EPI_ISL_958058, EPI_ISL_958059, EPI_ISL_958060, EPI_ISL_958061, EPI_ISL_958062, EPI_ISL_958063, EPI_ISL_958064, EPI_ISL_958065, EPI_ISL_958066, EPI_ISL_958067, EPI_ISL_958068, EPI_ISL_958069, EPI_ISL_958070, EPI_ISL_958071,
EPI_ISL_958072, EPI_ISL_958073, EPI_ISL_958074, EPI_ISL_958075, EPI_ISL_958076, EPI_ISL_958077, EPI_ISL_958078, EPI_ISL_958079, EPI_ISL_958080, EPI_ISL_958081, EPI_ISL_958082, EPI_ISL_958083, EPI_ISL_958084, EPI_ISL_958085, EPI_ISL_958086, EPI_ISL_958087, EPI_ISL_958088, EPI_ISL_958089, EPI_ISL_958090, EPI_ISL_958091, EPI_ISL_958092, EPI_ISL_958093, EPI_ISL_958094, EPI_ISL_958095,
EPI_ISL_958096, EPI_ISL_958097, EPI_ISL_958098, EPI_ISL_958099, EPI_ISL_958100, EPI_ISL_958101, EPI_ISL_958102, EPI_ISL_958103, EPI_ISL_958104, EPI_ISL_958105, EPI_ISL_958106, EPI_ISL_958107, EPI_ISL_958108, EPI_ISL_958109, EPI_ISL_958110, EPI_ISL_958111, EPI_ISL_958112, EPI_ISL_958113, EPI_ISL_958114, EPI_ISL_958115, EPI_ISL_958116, EPI_ISL_958117, EPI_ISL_958118, EPI_ISL_958119,
EPI_ISL_958120, EPI_ISL_958121, EPI_ISL_958122, EPI_ISL_958123, EPI_ISL_958124, EPI_ISL_958125, EPI_ISL_958126, EPI_ISL_958127, EPI_ISL_958128, EPI_ISL_958129, EPI_ISL_958130, EPI_ISL_958131, EPI_ISL_958132, EPI_ISL_958133, EPI_ISL_958134, EPI_ISL_958135, EPI_ISL_958136, EPI_ISL_958137, EPI_ISL_958138, EPI_ISL_958139, EPI_ISL_958140, EPI_ISL_958141, EPI_ISL_958142, EPI_ISL_958143,
EPI_ISL_958144, EPI_ISL_958145, EPI_ISL_958146, EPI_ISL_958147, EPI_ISL_958148, EPI_ISL_958149, EPI_ISL_958150, EPI_ISL_958151, EPI_ISL_958152, EPI_ISL_958153, EPI_ISL_958154, EPI_ISL_958155, EPI_ISL_958156, EPI_ISL_958157, EPI_ISL_958158, EPI_ISL_958159, EPI_ISL_958160, EPI_ISL_958161, EPI_ISL_958162, EPI_ISL_958163, EPI_ISL_958164, EPI_ISL_958165, EPI_ISL_958166, EPI_ISL_958167,
EPI_ISL_958168, EPI_ISL_958169, EPI_ISL_958170, EPI_ISL_958171, EPI_ISL_958172, EPI_ISL_958173, EPI_ISL_958174, EPI_ISL_958175, EPI_ISL_958176, EPI_ISL_958177, EPI_ISL_958178, EPI_ISL_958179, EPI_ISL_958180, EPI_ISL_958181, EPI_ISL_958182, EPI_ISL_958183, EPI_ISL_958184, EPI_ISL_958185, EPI_ISL_958186, EPI_ISL_958187, EPI_ISL_958188, EPI_ISL_958189, EPI_ISL_958190, EPI_ISL_958191,
EPI_ISL_958192, EPI_ISL_958193, EPI_ISL_958195, EPI_ISL_958196, EPI_ISL_958197, EPI_ISL_958198, EPI_ISL_958199, EPI_ISL_958200, EPI_ISL_958201, EPI_ISL_958202, EPI_ISL_958203, EPI_ISL_958204, EPI_ISL_958205, EPI_ISL_958207, EPI_ISL_958208, EPI_ISL_958209, EPI_ISL_958210, EPI_ISL_958211, EPI_ISL_958212, EPI_ISL_958213, EPI_ISL_958214, EPI_ISL_958567, EPI_ISL_958568, EPI_ISL_958570,
EPI_ISL_958571, EPI_ISL_958573, EPI_ISL_958574, EPI_ISL_958575, EPI_ISL_958576, EPI_ISL_958577, EPI_ISL_958580, EPI_ISL_958581, EPI_ISL_958582, EPI_ISL_958583, EPI_ISL_958584, EPI_ISL_958585, EPI_ISL_958588, EPI_ISL_958589, EPI_ISL_958592, EPI_ISL_958593, EPI_ISL_958594, EPI_ISL_958596, EPI_ISL_958597, EPI_ISL_958598, EPI_ISL_958599, EPI_ISL_958601, EPI_ISL_958603, EPI_ISL_958604,
EPI_ISL_958605, EPI_ISL_958606, EPI_ISL_958607, EPI_ISL_958608, EPI_ISL_958609, EPI_ISL_958611, EPI_ISL_958612, EPI_ISL_958613, EPI_ISL_958614, EPI_ISL_958615, EPI_ISL_958616, EPI_ISL_958617, EPI_ISL_958618, EPI_ISL_958619, EPI_ISL_958620, EPI_ISL_958621, EPI_ISL_958622, EPI_ISL_958623, EPI_ISL_958625, EPI_ISL_958626, EPI_ISL_958627, EPI_ISL_958628, EPI_ISL_958629, EPI_ISL_958630,
EPI_ISL_958631, EPI_ISL_958635, EPI_ISL_958637, EPI_ISL_958640, EPI_ISL_958642, EPI_ISL_958643, EPI_ISL_958644, EPI_ISL_958645, EPI_ISL_958647, EPI_ISL_958648, EPI_ISL_958649, EPI_ISL_958651, EPI_ISL_958652, EPI_ISL_958653, EPI_ISL_958654, EPI_ISL_958655, EPI_ISL_958656, EPI_ISL_958657, EPI_ISL_958658, EPI_ISL_958660, EPI_ISL_958661, EPI_ISL_958664, EPI_ISL_958665, EPI_ISL_958666,
EPI_ISL_958667, EPI_ISL_958668, EPI_ISL_958669, EPI_ISL_958672, EPI_ISL_958674, EPI_ISL_958675, EPI_ISL_958676, EPI_ISL_958677, EPI_ISL_958678, EPI_ISL_958679, EPI_ISL_958681, EPI_ISL_958682, EPI_ISL_958683, EPI_ISL_958685, EPI_ISL_958686, EPI_ISL_958687, EPI_ISL_958688, EPI_ISL_958689, EPI_ISL_958692, EPI_ISL_958693, EPI_ISL_958694, EPI_ISL_958695, EPI_ISL_958697, EPI_ISL_958698,
EPI_ISL_958699, EPI_ISL_958700, EPI_ISL_958701, EPI_ISL_958702, EPI_ISL_958703, EPI_ISL_958704, EPI_ISL_958707, EPI_ISL_958708, EPI_ISL_958709, EPI_ISL_958710, EPI_ISL_958711, EPI_ISL_958712, EPI_ISL_958714, EPI_ISL_958717, EPI_ISL_958718, EPI_ISL_958719, EPI_ISL_958720, EPI_ISL_958721, EPI_ISL_958722, EPI_ISL_958724, EPI_ISL_958727, EPI_ISL_958728, EPI_ISL_958729, EPI_ISL_958730,
EPI_ISL_958732, EPI_ISL_958734, EPI_ISL_958736, EPI_ISL_958737, EPI_ISL_958738, EPI_ISL_958739, EPI_ISL_958740, EPI_ISL_958741, EPI_ISL_958742, EPI_ISL_958743, EPI_ISL_958744, EPI_ISL_958746, EPI_ISL_958749, EPI_ISL_958750, EPI_ISL_958752, EPI_ISL_958753, EPI_ISL_958755, EPI_ISL_958757, EPI_ISL_958758, EPI_ISL_958759, EPI_ISL_958761, EPI_ISL_958762, EPI_ISL_958764, EPI_ISL_958765,
EPI_ISL_958766, EPI_ISL_958768, EPI_ISL_958770, EPI_ISL_958771, EPI_ISL_958772, EPI_ISL_958773, EPI_ISL_958774, EPI_ISL_958776, EPI_ISL_958777, EPI_ISL_958778, EPI_ISL_958780, EPI_ISL_958781, EPI_ISL_958782, EPI_ISL_958783, EPI_ISL_958785, EPI_ISL_958786, EPI_ISL_958788, EPI_ISL_958790, EPI_ISL_958794, EPI_ISL_958795, EPI_ISL_958796, EPI_ISL_958797, EPI_ISL_958798, EPI_ISL_958799,
EPI_ISL_958800, EPI_ISL_958801, EPI_ISL_958802, EPI_ISL_958804, EPI_ISL_958805, EPI_ISL_958806, EPI_ISL_958809, EPI_ISL_958810, EPI_ISL_958812, EPI_ISL_958813, EPI_ISL_958814, EPI_ISL_958816, EPI_ISL_958817, EPI_ISL_958818, EPI_ISL_958819, EPI_ISL_958820, EPI_ISL_958821, EPI_ISL_958822, EPI_ISL_958824, EPI_ISL_958825, EPI_ISL_958827, EPI_ISL_958829, EPI_ISL_958830, EPI_ISL_958831,
EPI_ISL_958832, EPI_ISL_958834, EPI_ISL_958835, EPI_ISL_958837, EPI_ISL_958838, EPI_ISL_958839, EPI_ISL_958841, EPI_ISL_958843, EPI_ISL_958844, EPI_ISL_958845, EPI_ISL_958846, EPI_ISL_958847, EPI_ISL_958848, EPI_ISL_958850, EPI_ISL_958852, EPI_ISL_958853, EPI_ISL_958854, EPI_ISL_958855, EPI_ISL_958856, EPI_ISL_958857, EPI_ISL_958858, EPI_ISL_958859, EPI_ISL_958860, EPI_ISL_958861,
EPI_ISL_958862, EPI_ISL_958864, EPI_ISL_958865, EPI_ISL_958867, EPI_ISL_958868, EPI_ISL_958869, EPI_ISL_958870, EPI_ISL_958871, EPI_ISL_958872, EPI_ISL_958874, EPI_ISL_958875, EPI_ISL_958876, EPI_ISL_958877, EPI_ISL_958879, EPI_ISL_958880, EPI_ISL_958881, EPI_ISL_958882, EPI_ISL_958883, EPI_ISL_958886, EPI_ISL_958888, EPI_ISL_958890, EPI_ISL_958892, EPI_ISL_958894, EPI_ISL_958896,
EPI_ISL_958897, EPI_ISL_958899, EPI_ISL_958900, EPI_ISL_958901, EPI_ISL_958905, EPI_ISL_958906, EPI_ISL_958907, EPI_ISL_958909, EPI_ISL_958910, EPI_ISL_958913, EPI_ISL_958914, EPI_ISL_963324, EPI_ISL_963327, EPI_ISL_963328, EPI_ISL_963329, EPI_ISL_963331, EPI_ISL_963332, EPI_ISL_963333, EPI_ISL_963334, EPI_ISL_963335, EPI_ISL_963336, EPI_ISL_963337, EPI_ISL_963338, EPI_ISL_963339,
EPI_ISL_963340, EPI_ISL_963342, EPI_ISL_963343, EPI_ISL_963347, EPI_ISL_963349, EPI_ISL_963350, EPI_ISL_963351, EPI_ISL_963352, EPI_ISL_963353, EPI_ISL_963354, EPI_ISL_963359, EPI_ISL_963360, EPI_ISL_963361, EPI_ISL_963362, EPI_ISL_963363, EPI_ISL_963364, EPI_ISL_963365, EPI_ISL_963366, EPI_ISL_963367, EPI_ISL_963368, EPI_ISL_963369, EPI_ISL_963372, EPI_ISL_963373, EPI_ISL_963374,
EPI_ISL_963375, EPI_ISL_963377, EPI_ISL_963378, EPI_ISL_963379, EPI_ISL_963381, EPI_ISL_963382, EPI_ISL_963383, EPI_ISL_963384, EPI_ISL_963385, EPI_ISL_963387, EPI_ISL_963388, EPI_ISL_963389, EPI_ISL_963390, EPI_ISL_963391, EPI_ISL_963392, EPI_ISL_963393, EPI_ISL_963394, EPI_ISL_963395, EPI_ISL_963396, EPI_ISL_963398, EPI_ISL_963399, EPI_ISL_963400, EPI_ISL_963401, EPI_ISL_963402,
EPI_ISL_963403, EPI_ISL_963404, EPI_ISL_963405, EPI_ISL_963406, EPI_ISL_963407, EPI_ISL_963409, EPI_ISL_963410, EPI_ISL_963411, EPI_ISL_963412, EPI_ISL_963413, EPI_ISL_963414, EPI_ISL_963415, EPI_ISL_963416, EPI_ISL_963417, EPI_ISL_963418, EPI_ISL_963420, EPI_ISL_963422, EPI_ISL_963423, EPI_ISL_963424, EPI_ISL_963427, EPI_ISL_963429, EPI_ISL_963430, EPI_ISL_963432, EPI_ISL_963435,
EPI_ISL_963436, EPI_ISL_963437, EPI_ISL_963438, EPI_ISL_963439, EPI_ISL_963440, EPI_ISL_963441, EPI_ISL_963442, EPI_ISL_963443, EPI_ISL_963444, EPI_ISL_963445, EPI_ISL_963446, EPI_ISL_963447, EPI_ISL_963448, EPI_ISL_963450, EPI_ISL_963451, EPI_ISL_963453, EPI_ISL_963454, EPI_ISL_963455, EPI_ISL_963456, EPI_ISL_963457, EPI_ISL_963459, EPI_ISL_963460, EPI_ISL_963461, EPI_ISL_963462,
EPI_ISL_963463, EPI_ISL_963464, EPI_ISL_963465, EPI_ISL_963466, EPI_ISL_963467, EPI_ISL_963468, EPI_ISL_963469, EPI_ISL_963470, EPI_ISL_963472, EPI_ISL_963473, EPI_ISL_963474, EPI_ISL_963476, EPI_ISL_963477, EPI_ISL_963478, EPI_ISL_963479, EPI_ISL_963480, EPI_ISL_963482, EPI_ISL_963483, EPI_ISL_963484, EPI_ISL_963485, EPI_ISL_963486, EPI_ISL_963487, EPI_ISL_963488, EPI_ISL_963491,
EPI_ISL_963492, EPI_ISL_963493, EPI_ISL_963494, EPI_ISL_963496, EPI_ISL_963497, EPI_ISL_963498, EPI_ISL_963499, EPI_ISL_963500, EPI_ISL_963501, EPI_ISL_963502, EPI_ISL_963503, EPI_ISL_963504, EPI_ISL_963505, EPI_ISL_963506, EPI_ISL_963507, EPI_ISL_963508, EPI_ISL_963510, EPI_ISL_963512, EPI_ISL_963513, EPI_ISL_963514, EPI_ISL_963515, EPI_ISL_963516, EPI_ISL_963517, EPI_ISL_963518,
EPI_ISL_963519, EPI_ISL_963522, EPI_ISL_963523, EPI_ISL_963525, EPI_ISL_963526, EPI_ISL_963527, EPI_ISL_963528, EPI_ISL_963531, EPI_ISL_963532, EPI_ISL_963533, EPI_ISL_963534, EPI_ISL_963535, EPI_ISL_963536, EPI_ISL_963537, EPI_ISL_963539, EPI_ISL_963540, EPI_ISL_963541, EPI_ISL_963542, EPI_ISL_963543, EPI_ISL_963544, EPI_ISL_963545, EPI_ISL_963546, EPI_ISL_963547, EPI_ISL_963548,
EPI_ISL_963549, EPI_ISL_963550, EPI_ISL_963551, EPI_ISL_963552, EPI_ISL_963553, EPI_ISL_963556, EPI_ISL_963557, EPI_ISL_963558, EPI_ISL_963561, EPI_ISL_963563, EPI_ISL_963564, EPI_ISL_963565, EPI_ISL_963566, EPI_ISL_963567, EPI_ISL_963568, EPI_ISL_963569, EPI_ISL_963571, EPI_ISL_963572, EPI_ISL_963574, EPI_ISL_963575, EPI_ISL_963577, EPI_ISL_963578, EPI_ISL_963579, EPI_ISL_963580,
EPI_ISL_963582, EPI_ISL_963583, EPI_ISL_963585, EPI_ISL_963586, EPI_ISL_963587, EPI_ISL_963588, EPI_ISL_963589, EPI_ISL_963590, EPI_ISL_963592, EPI_ISL_963593, EPI_ISL_963595, EPI_ISL_963596, EPI_ISL_963597, EPI_ISL_963598, EPI_ISL_963599, EPI_ISL_963600, EPI_ISL_963602, EPI_ISL_963604, EPI_ISL_963605, EPI_ISL_963606, EPI_ISL_963608, EPI_ISL_963609, EPI_ISL_963610, EPI_ISL_963611,
EPI_ISL_963612, EPI_ISL_963613, EPI_ISL_963614, EPI_ISL_963615, EPI_ISL_963616, EPI_ISL_963617, EPI_ISL_963619, EPI_ISL_963620, EPI_ISL_963621, EPI_ISL_963622, EPI_ISL_963623, EPI_ISL_963624, EPI_ISL_963625, EPI_ISL_963627, EPI_ISL_963628, EPI_ISL_963629, EPI_ISL_963630, EPI_ISL_963631, EPI_ISL_963632, EPI_ISL_963634, EPI_ISL_963635, EPI_ISL_963636, EPI_ISL_963637, EPI_ISL_963638,
EPI_ISL_963640, EPI_ISL_963642, EPI_ISL_963643, EPI_ISL_963644, EPI_ISL_963645, EPI_ISL_963646, EPI_ISL_963647, EPI_ISL_963648, EPI_ISL_963650, EPI_ISL_964370, EPI_ISL_964376, EPI_ISL_964497, EPI_ISL_964498, EPI_ISL_964506, EPI_ISL_964522, EPI_ISL_964876, EPI_ISL_970587, EPI_ISL_970597, EPI_ISL_970616, EPI_ISL_970621, EPI_ISL_970628, EPI_ISL_970635, EPI_ISL_970639, EPI_ISL_970645,
EPI_ISL_970664, EPI_ISL_970673, EPI_ISL_970680, EPI_ISL_970681, EPI_ISL_970687, EPI_ISL_970701, EPI_ISL_970708, EPI_ISL_970709, EPI_ISL_970715, EPI_ISL_970724, EPI_ISL_970732, EPI_ISL_970744, EPI_ISL_970747, EPI_ISL_970751, EPI_ISL_970753, EPI_ISL_970768, EPI_ISL_970775, EPI_ISL_970790, EPI_ISL_970826, EPI_ISL_970860, EPI_ISL_970875, EPI_ISL_970986, EPI_ISL_971037, EPI_ISL_971042,
EPI_ISL_971044, EPI_ISL_971047, EPI_ISL_971053, EPI_ISL_971060, EPI_ISL_971071, EPI_ISL_971076, EPI_ISL_971093, EPI_ISL_971096, EPI_ISL_971105, EPI_ISL_971125, EPI_ISL_971135, EPI_ISL_971142, EPI_ISL_971168, EPI_ISL_971171, EPI_ISL_971174, EPI_ISL_971176, EPI_ISL_971183, EPI_ISL_971191, EPI_ISL_971194, EPI_ISL_971206, EPI_ISL_971231, EPI_ISL_971239, EPI_ISL_971242, EPI_ISL_971255,
EPI_ISL_971261, EPI_ISL_971269, EPI_ISL_971281, EPI_ISL_971292, EPI_ISL_971297, EPI_ISL_971327, EPI_ISL_971333, EPI_ISL_971339, EPI_ISL_971347, EPI_ISL_971360, EPI_ISL_971364, EPI_ISL_971365, EPI_ISL_971370, EPI_ISL_971385, EPI_ISL_971394, EPI_ISL_979974, EPI_ISL_979975, EPI_ISL_979976, EPI_ISL_979977, EPI_ISL_979978, EPI_ISL_979979, EPI_ISL_979980, EPI_ISL_979981, EPI_ISL_979982,
EPI_ISL_979983, EPI_ISL_979984, EPI_ISL_979985, EPI_ISL_979986, EPI_ISL_979987, EPI_ISL_979988, EPI_ISL_979989, EPI_ISL_979990, EPI_ISL_979991, EPI_ISL_979992, EPI_ISL_979993, EPI_ISL_979994, EPI_ISL_979995, EPI_ISL_979996, EPI_ISL_979997, EPI_ISL_980000, EPI_ISL_980001, EPI_ISL_980002, EPI_ISL_980003, EPI_ISL_980004, EPI_ISL_980005, EPI_ISL_980007, EPI_ISL_980008, EPI_ISL_980009,
EPI_ISL_980010, EPI_ISL_980011, EPI_ISL_980013, EPI_ISL_980014, EPI_ISL_980015, EPI_ISL_980016, EPI_ISL_980018, EPI_ISL_980020, EPI_ISL_980021, EPI_ISL_980022, EPI_ISL_980023, EPI_ISL_980024, EPI_ISL_980025, EPI_ISL_980026, EPI_ISL_980027, EPI_ISL_980029, EPI_ISL_980031, EPI_ISL_980032, EPI_ISL_980033, EPI_ISL_980034, EPI_ISL_980037, EPI_ISL_980038, EPI_ISL_980039, EPI_ISL_980040,
EPI_ISL_980041, EPI_ISL_980042, EPI_ISL_980043, EPI_ISL_980044, EPI_ISL_980045, EPI_ISL_980047, EPI_ISL_980050, EPI_ISL_980053, EPI_ISL_980054, EPI_ISL_980055, EPI_ISL_980056, EPI_ISL_980057, EPI_ISL_980058, EPI_ISL_980059, EPI_ISL_980060, EPI_ISL_980061, EPI_ISL_980063, EPI_ISL_980064, EPI_ISL_980065, EPI_ISL_980066, EPI_ISL_980067, EPI_ISL_980068, EPI_ISL_980069, EPI_ISL_980070,
EPI_ISL_980071, EPI_ISL_980072, EPI_ISL_980073, EPI_ISL_980074, EPI_ISL_980075, EPI_ISL_980076, EPI_ISL_980077, EPI_ISL_980078, EPI_ISL_980079, EPI_ISL_980080, EPI_ISL_980081, EPI_ISL_980082, EPI_ISL_980083, EPI_ISL_980084, EPI_ISL_980085, EPI_ISL_980087, EPI_ISL_980088, EPI_ISL_980089, EPI_ISL_980090, EPI_ISL_980091, EPI_ISL_980092, EPI_ISL_980093, EPI_ISL_980094, EPI_ISL_980095,
EPI_ISL_980097, EPI_ISL_980098, EPI_ISL_980099, EPI_ISL_980100, EPI_ISL_980101, EPI_ISL_980102, EPI_ISL_980103, EPI_ISL_980104, EPI_ISL_980105, EPI_ISL_980107, EPI_ISL_980108, EPI_ISL_980109, EPI_ISL_980110, EPI_ISL_980111, EPI_ISL_980113, EPI_ISL_980114, EPI_ISL_980115, EPI_ISL_980116, EPI_ISL_980117, EPI_ISL_980118, EPI_ISL_980119, EPI_ISL_980120, EPI_ISL_980121, EPI_ISL_980122,
EPI_ISL_980123, EPI_ISL_980124, EPI_ISL_980125, EPI_ISL_980126, EPI_ISL_980127, EPI_ISL_980128, EPI_ISL_980129, EPI_ISL_980130, EPI_ISL_980131, EPI_ISL_980132, EPI_ISL_980133, EPI_ISL_980134, EPI_ISL_980135, EPI_ISL_980136, EPI_ISL_980137, EPI_ISL_980138, EPI_ISL_980139, EPI_ISL_980140, EPI_ISL_980141, EPI_ISL_980142, EPI_ISL_980143, EPI_ISL_980144, EPI_ISL_980145, EPI_ISL_980147,
EPI_ISL_980148, EPI_ISL_980149, EPI_ISL_980150, EPI_ISL_980151, EPI_ISL_980152, EPI_ISL_980153, EPI_ISL_980154, EPI_ISL_980155, EPI_ISL_980156, EPI_ISL_980157, EPI_ISL_980158, EPI_ISL_980159, EPI_ISL_980160, EPI_ISL_980161, EPI_ISL_980162, EPI_ISL_980163, EPI_ISL_980164, EPI_ISL_980165, EPI_ISL_980167, EPI_ISL_980168, EPI_ISL_980169, EPI_ISL_980170, EPI_ISL_980171, EPI_ISL_980172,
EPI_ISL_980173, EPI_ISL_980174, EPI_ISL_980175, EPI_ISL_980176, EPI_ISL_980177, EPI_ISL_980178, EPI_ISL_980179, EPI_ISL_980180, EPI_ISL_980181, EPI_ISL_980182, EPI_ISL_980183, EPI_ISL_980184, EPI_ISL_980185, EPI_ISL_980186, EPI_ISL_980187, EPI_ISL_980188, EPI_ISL_980189, EPI_ISL_980190, EPI_ISL_980192, EPI_ISL_980193, EPI_ISL_980194, EPI_ISL_980195, EPI_ISL_980196, EPI_ISL_980198,
EPI_ISL_980199, EPI_ISL_980200, EPI_ISL_980201, EPI_ISL_980202, EPI_ISL_980203, EPI_ISL_980204, EPI_ISL_980205, EPI_ISL_980206, EPI_ISL_980207, EPI_ISL_980208, EPI_ISL_980210, EPI_ISL_980211, EPI_ISL_980212, EPI_ISL_980213, EPI_ISL_980215, EPI_ISL_980217, EPI_ISL_980218, EPI_ISL_980219, EPI_ISL_980220, EPI_ISL_980221, EPI_ISL_980222, EPI_ISL_980224, EPI_ISL_980225, EPI_ISL_980226,
EPI_ISL_980227, EPI_ISL_980228, EPI_ISL_980229, EPI_ISL_980230, EPI_ISL_980231, EPI_ISL_980232, EPI_ISL_980233, EPI_ISL_980234, EPI_ISL_980551, EPI_ISL_980552, EPI_ISL_980553, EPI_ISL_980554, EPI_ISL_980555, EPI_ISL_980556, EPI_ISL_980557, EPI_ISL_980558, EPI_ISL_980559, EPI_ISL_980560, EPI_ISL_980561, EPI_ISL_980562, EPI_ISL_980563, EPI_ISL_980564, EPI_ISL_980565, EPI_ISL_980567,
EPI_ISL_980568, EPI_ISL_980569, EPI_ISL_980570, EPI_ISL_980571, EPI_ISL_980572, EPI_ISL_980573, EPI_ISL_980574, EPI_ISL_980575, EPI_ISL_980577, EPI_ISL_980579, EPI_ISL_980580, EPI_ISL_980581, EPI_ISL_980582, EPI_ISL_980583, EPI_ISL_980584, EPI_ISL_980585, EPI_ISL_980586, EPI_ISL_980588, EPI_ISL_980589, EPI_ISL_980590, EPI_ISL_980591, EPI_ISL_980593, EPI_ISL_980594, EPI_ISL_980595,
EPI_ISL_980596, EPI_ISL_980597, EPI_ISL_980598, EPI_ISL_980599, EPI_ISL_980600, EPI_ISL_980601, EPI_ISL_980602, EPI_ISL_980603, EPI_ISL_980604, EPI_ISL_980606, EPI_ISL_980607, EPI_ISL_980608, EPI_ISL_980609, EPI_ISL_980610, EPI_ISL_980611, EPI_ISL_980612, EPI_ISL_980613, EPI_ISL_980614, EPI_ISL_980615, EPI_ISL_980616, EPI_ISL_980617, EPI_ISL_980618, EPI_ISL_980619, EPI_ISL_980621,
EPI_ISL_980622, EPI_ISL_980623, EPI_ISL_980624, EPI_ISL_980625, EPI_ISL_980626, EPI_ISL_980627, EPI_ISL_980628, EPI_ISL_980629, EPI_ISL_980630, EPI_ISL_980631, EPI_ISL_980632, EPI_ISL_980633, EPI_ISL_980634, EPI_ISL_980635, EPI_ISL_980636, EPI_ISL_980637, EPI_ISL_980638, EPI_ISL_980639, EPI_ISL_980640, EPI_ISL_980641, EPI_ISL_980642, EPI_ISL_980643, EPI_ISL_980644, EPI_ISL_980645,
EPI_ISL_980646, EPI_ISL_980647, EPI_ISL_980648, EPI_ISL_980649, EPI_ISL_980650, EPI_ISL_980651, EPI_ISL_980652, EPI_ISL_980653, EPI_ISL_980654, EPI_ISL_980655, EPI_ISL_980656, EPI_ISL_980657, EPI_ISL_980658, EPI_ISL_980659, EPI_ISL_980660, EPI_ISL_980661, EPI_ISL_980662, EPI_ISL_980663, EPI_ISL_980664, EPI_ISL_980665, EPI_ISL_980666, EPI_ISL_980667, EPI_ISL_980668, EPI_ISL_980669,
EPI_ISL_980670, EPI_ISL_980671, EPI_ISL_980672, EPI_ISL_980673, EPI_ISL_980674, EPI_ISL_980675, EPI_ISL_980676, EPI_ISL_980677, EPI_ISL_980679, EPI_ISL_980680, EPI_ISL_980681, EPI_ISL_980683, EPI_ISL_980684, EPI_ISL_980685, EPI_ISL_980686, EPI_ISL_980687, EPI_ISL_980688, EPI_ISL_980689, EPI_ISL_980690, EPI_ISL_980691, EPI_ISL_980692, EPI_ISL_980693, EPI_ISL_980694, EPI_ISL_980695,
EPI_ISL_980696, EPI_ISL_980697, EPI_ISL_980699, EPI_ISL_980700, EPI_ISL_980701, EPI_ISL_980702, EPI_ISL_980703, EPI_ISL_980704, EPI_ISL_980705, EPI_ISL_980706, EPI_ISL_980707, EPI_ISL_980708, EPI_ISL_980709, EPI_ISL_980710, EPI_ISL_980711, EPI_ISL_980712, EPI_ISL_980713, EPI_ISL_980714, EPI_ISL_980715, EPI_ISL_980716, EPI_ISL_980717, EPI_ISL_980718, EPI_ISL_980719, EPI_ISL_980720,
EPI_ISL_980722, EPI_ISL_980723, EPI_ISL_980724, EPI_ISL_980725, EPI_ISL_980726, EPI_ISL_980727, EPI_ISL_980728, EPI_ISL_980729, EPI_ISL_980730, EPI_ISL_980731, EPI_ISL_980732, EPI_ISL_980733, EPI_ISL_980734, EPI_ISL_980735, EPI_ISL_980736, EPI_ISL_980737, EPI_ISL_980738, EPI_ISL_980739, EPI_ISL_980740, EPI_ISL_980741, EPI_ISL_980742, EPI_ISL_980743, EPI_ISL_980744, EPI_ISL_980745,
EPI_ISL_980746, EPI_ISL_980747, EPI_ISL_980748, EPI_ISL_980749, EPI_ISL_980750, EPI_ISL_980751, EPI_ISL_980754, EPI_ISL_980755, EPI_ISL_980756, EPI_ISL_980757, EPI_ISL_980758, EPI_ISL_980759, EPI_ISL_980760, EPI_ISL_980761, EPI_ISL_980763, EPI_ISL_980764, EPI_ISL_980765, EPI_ISL_980766, EPI_ISL_980767, EPI_ISL_980768, EPI_ISL_980769, EPI_ISL_980770, EPI_ISL_980771, EPI_ISL_980772,
EPI_ISL_980773, EPI_ISL_980774, EPI_ISL_980775, EPI_ISL_980776, EPI_ISL_980777, EPI_ISL_980778, EPI_ISL_980779, EPI_ISL_980780, EPI_ISL_980781, EPI_ISL_980782, EPI_ISL_980783, EPI_ISL_980785, EPI_ISL_980786, EPI_ISL_980787, EPI_ISL_980788, EPI_ISL_980789, EPI_ISL_980790, EPI_ISL_980791, EPI_ISL_980792, EPI_ISL_980793, EPI_ISL_980794, EPI_ISL_980795, EPI_ISL_980796, EPI_ISL_980797,
EPI_ISL_980798, EPI_ISL_980800, EPI_ISL_980801, EPI_ISL_980802, EPI_ISL_980804, EPI_ISL_980805, EPI_ISL_980806, EPI_ISL_985985, EPI_ISL_986002, EPI_ISL_986039, EPI_ISL_986049, EPI_ISL_986061, EPI_ISL_986075, EPI_ISL_986090, EPI_ISL_986096, EPI_ISL_988241, EPI_ISL_988314, EPI_ISL_988486, EPI_ISL_988495, EPI_ISL_988504, EPI_ISL_988515, EPI_ISL_988516, EPI_ISL_988517, EPI_ISL_988537,
EPI_ISL_988538, EPI_ISL_988552, EPI_ISL_988554, EPI_ISL_988555, EPI_ISL_988558, EPI_ISL_988564, EPI_ISL_988576, EPI_ISL_988589, EPI_ISL_988592, EPI_ISL_988600, EPI_ISL_988604, EPI_ISL_988623, EPI_ISL_988631, EPI_ISL_988640, EPI_ISL_988646, EPI_ISL_988651, EPI_ISL_988685, EPI_ISL_989362, EPI_ISL_989831, EPI_ISL_1006342, EPI_ISL_1006343, EPI_ISL_1006346, EPI_ISL_1006347,
EPI_ISL_1006349, EPI_ISL_1006350, EPI_ISL_1006351, EPI_ISL_1006352, EPI_ISL_1006353, EPI_ISL_1006355, EPI_ISL_1006356, EPI_ISL_1006359, EPI_ISL_1006360, EPI_ISL_1006361, EPI_ISL_1006362, EPI_ISL_1006363, EPI_ISL_1006364, EPI_ISL_1006368, EPI_ISL_1006369, EPI_ISL_1006370, EPI_ISL_1006371, EPI_ISL_1006372, EPI_ISL_1006375, EPI_ISL_1006376, EPI_ISL_1006377, EPI_ISL_1006379,
EPI_ISL_1006380, EPI_ISL_1006381, EPI_ISL_1006382, EPI_ISL_1006383, EPI_ISL_1006384, EPI_ISL_1006386, EPI_ISL_1006388, EPI_ISL_1006389, EPI_ISL_1006392, EPI_ISL_1006393, EPI_ISL_1006394, EPI_ISL_1006396, EPI_ISL_1006397, EPI_ISL_1006398, EPI_ISL_1006400, EPI_ISL_1006401, EPI_ISL_1006402, EPI_ISL_1006403, EPI_ISL_1006404, EPI_ISL_1006405, EPI_ISL_1006406, EPI_ISL_1006407,
EPI_ISL_1006408, EPI_ISL_1006410, EPI_ISL_1006411, EPI_ISL_1006413, EPI_ISL_1006414, EPI_ISL_1006415, EPI_ISL_1006416, EPI_ISL_1006417, EPI_ISL_1006418, EPI_ISL_1006419, EPI_ISL_1006420, EPI_ISL_1006421, EPI_ISL_1006424, EPI_ISL_1006425, EPI_ISL_1006427, EPI_ISL_1006429, EPI_ISL_1006430, EPI_ISL_1006431, EPI_ISL_1006432, EPI_ISL_1006433, EPI_ISL_1006434, EPI_ISL_1006436,
EPI_ISL_1006437, EPI_ISL_1006440, EPI_ISL_1006441, EPI_ISL_1006442, EPI_ISL_1006443, EPI_ISL_1006444, EPI_ISL_1006446, EPI_ISL_1006447, EPI_ISL_1006448, EPI_ISL_1006449, EPI_ISL_1006450, EPI_ISL_1006451, EPI_ISL_1006452, EPI_ISL_1006454, EPI_ISL_1006455, EPI_ISL_1006459, EPI_ISL_1006460, EPI_ISL_1006461, EPI_ISL_1006462, EPI_ISL_1006463, EPI_ISL_1006464, EPI_ISL_1006465,
EPI_ISL_1006466, EPI_ISL_1006468, EPI_ISL_1006469, EPI_ISL_1006470, EPI_ISL_1006472, EPI_ISL_1006473, EPI_ISL_1006478, EPI_ISL_1006480, EPI_ISL_1006481, EPI_ISL_1006483, EPI_ISL_1006484, EPI_ISL_1006485, EPI_ISL_1006486, EPI_ISL_1006487, EPI_ISL_1006491, EPI_ISL_1006492, EPI_ISL_1006493, EPI_ISL_1006494, EPI_ISL_1006495, EPI_ISL_1006496, EPI_ISL_1006497, EPI_ISL_1006498,
EPI_ISL_1006499, EPI_ISL_1006500, EPI_ISL_1006502, EPI_ISL_1006503, EPI_ISL_1006504, EPI_ISL_1006505, EPI_ISL_1006506, EPI_ISL_1006507, EPI_ISL_1006508, EPI_ISL_1006509, EPI_ISL_1006510, EPI_ISL_1006512, EPI_ISL_1006513, EPI_ISL_1006514, EPI_ISL_1006515, EPI_ISL_1006517, EPI_ISL_1006518, EPI_ISL_1006519, EPI_ISL_1006520, EPI_ISL_1006522, EPI_ISL_1006523, EPI_ISL_1006524,
EPI_ISL_1006525, EPI_ISL_1006526, EPI_ISL_1006527, EPI_ISL_1006528, EPI_ISL_1006531, EPI_ISL_1006532, EPI_ISL_1006534, EPI_ISL_1006536, EPI_ISL_1006537, EPI_ISL_1006539, EPI_ISL_1006546, EPI_ISL_1012403, EPI_ISL_1012608, EPI_ISL_1019586

| | | | |
|---|---|---|---|
| see above | Lighthouse Lab in Milton Keynes | Wellcome Sanger Institute for the COVID-19 Genomics UK (COG-UK) Consortium | Cordelia Langford; David K. Jackson; Dominic Kwiatkowski; Ewan Harrison; Ian Johnston; Jeffrey Barrett; John Sillitoe on behalf of the Wellcome Sanger Institute COVID-19 Surveillance Team; John Sillitoe on behalf of the Wellcome Sanger Institute COVID-19 Surveillance Team (http://www.sanger.ac.uk/covid-team); Roberto Amato; Sonia Goncalves; The Lighthouse Lab in Milton Keynes and Alex Alderton |

EPI_ISL_1105861, EPI_ISL_1105862, EPI_ISL_1105863, EPI_ISL_1105864, EPI_ISL_1105865, EPI_ISL_1105866, EPI_ISL_1105867, EPI_ISL_1105868, EPI_ISL_1105869, EPI_ISL_1105870, EPI_ISL_1105871, EPI_ISL_1105883, EPI_ISL_1105884, EPI_ISL_1105885, EPI_ISL_1105886, EPI_ISL_1105887, EPI_ISL_1105888, EPI_ISL_1105889, EPI_ISL_1105890

| | | | |
|---|---|---|---|
| see above | Lincolnshire Hospitals and DeepSeq Nottingham | COVID-19 Genomics UK (COG-UK) Consortium | Christopher Moore; Fei Sang; Joeseph Chappell; Johnny Debebe; Jonathan Ball; Matthew Carlisle; Matthew Loose; Nadine Holmes; Nichola Duckworth; Patrick McClure; Sarah Walsh; Tim Sloan; Victoria Wright |

EPI_ISL_949662, EPI_ISL_949663, EPI_ISL_949664, EPI_ISL_949666, EPI_ISL_949668, EPI_ISL_949669, EPI_ISL_949670, EPI_ISL_949708, EPI_ISL_949709, EPI_ISL_949710, EPI_ISL_949711, EPI_ISL_949712, EPI_ISL_949713, EPI_ISL_949714, EPI_ISL_949717, EPI_ISL_949718, EPI_ISL_949719, EPI_ISL_949720, EPI_ISL_949721, EPI_ISL_949722, EPI_ISL_949723, EPI_ISL_949724, EPI_ISL_949725, EPI_ISL_949726,
EPI_ISL_949727, EPI_ISL_949728, EPI_ISL_949729, EPI_ISL_949730, EPI_ISL_949731, EPI_ISL_949732, EPI_ISL_949733, EPI_ISL_949734, EPI_ISL_949735, EPI_ISL_949736, EPI_ISL_949737, EPI_ISL_949738, EPI_ISL_949739, EPI_ISL_949740, EPI_ISL_949741, EPI_ISL_949742, EPI_ISL_949743, EPI_ISL_949744, EPI_ISL_949745, EPI_ISL_949746, EPI_ISL_997423, EPI_ISL_997424, EPI_ISL_997425, EPI_ISL_997426,
EPI_ISL_997427, EPI_ISL_997428, EPI_ISL_997429, EPI_ISL_997430, EPI_ISL_997431, EPI_ISL_997432, EPI_ISL_997433, EPI_ISL_997434, EPI_ISL_997435, EPI_ISL_997436, EPI_ISL_997437, EPI_ISL_997438, EPI_ISL_997440, EPI_ISL_997441, EPI_ISL_997444, EPI_ISL_997446, EPI_ISL_997447, EPI_ISL_997448, EPI_ISL_997449, EPI_ISL_997450, EPI_ISL_997463, EPI_ISL_997465, EPI_ISL_997486, EPI_ISL_997487,
EPI_ISL_997489, EPI_ISL_997492, EPI_ISL_997493, EPI_ISL_997494, EPI_ISL_997495, EPI_ISL_997496, EPI_ISL_997497, EPI_ISL_997498, EPI_ISL_997499, EPI_ISL_997500, EPI_ISL_997501, EPI_ISL_1047968, EPI_ISL_1047969, EPI_ISL_1047970, EPI_ISL_1047972, EPI_ISL_1048026, EPI_ISL_1048027, EPI_ISL_1048029, EPI_ISL_1104500, EPI_ISL_1104501, EPI_ISL_1104502, EPI_ISL_1104503, EPI_ISL_1104505,
EPI_ISL_1104506, EPI_ISL_1104507, EPI_ISL_1104568, EPI_ISL_1177891, EPI_ISL_1177892, EPI_ISL_1247823, EPI_ISL_1247824, EPI_ISL_1247825, EPI_ISL_1247827, EPI_ISL_1249257, EPI_ISL_1546034, EPI_ISL_1546035, EPI_ISL_1546036, EPI_ISL_1546037, EPI_ISL_1546038, EPI_ISL_1546039, EPI_ISL_1546040

| | | | |
|---|---|---|---|
| see above | Liverpool Clinical Laboratories | COVID-19 Genomics UK (COG-UK) Consortium | A Alrezaihi; Alessandro Gerada; Alistair Darby; Angela Cowell; Anita Lucaci; Anu Chawla; Cassie Olateju; Catherine Hartley; Charlotte Nelson; Ecaterina Vamos; Elaine O'Toole; Eleanor G Bentley; Ghada T Shawli; Hermione Webster; Isabel García-Dorival; James Johnson; James P Stewart; Jenifer Manson; Joanne Watts; Jones Benjamin; Jordan J Clark; Julian Hiscox; L Luu; Lucille Rainbow; M Almsaud; Margaret Hughes; Mark Whitehead; Matthew Gemmell; Miren Iturriza-Gomara; Muhannad Alruwaili; N.P Randle; Neil Swainston; PKF Gilmore; Parul Sharma; Rebekah Penrice-Randal; Richard Eccles; Richard Gregory; Sam Haldenby; Steve Paterson; Stuart D Armstrong; Trevor Ian Robinson; Ximeng Han |

EPI_ISL_950402, EPI_ISL_950403, EPI_ISL_950404, EPI_ISL_950405, EPI_ISL_950406, EPI_ISL_950407, EPI_ISL_950408, EPI_ISL_950409, EPI_ISL_950410, EPI_ISL_950411, EPI_ISL_950412, EPI_ISL_950413, EPI_ISL_950414, EPI_ISL_950415, EPI_ISL_950424, EPI_ISL_950425, EPI_ISL_950426, EPI_ISL_950427, EPI_ISL_950428, EPI_ISL_950429, EPI_ISL_950430, EPI_ISL_950431, EPI_ISL_950433, EPI_ISL_998566,
EPI_ISL_998567, EPI_ISL_998568, EPI_ISL_998569, EPI_ISL_998570, EPI_ISL_998571, EPI_ISL_1051244, EPI_ISL_1051245, EPI_ISL_1051246, EPI_ISL_1051247, EPI_ISL_1051248, EPI_ISL_1051249, EPI_ISL_1051250, EPI_ISL_1051251, EPI_ISL_1051252, EPI_ISL_1051253, EPI_ISL_1051254, EPI_ISL_1051255, EPI_ISL_1051256, EPI_ISL_1051257, EPI_ISL_1051258, EPI_ISL_1051259, EPI_ISL_1051260,

| | | | |
|---|---|---|---|
| EPI_ISL_1051261, EPI_ISL_1051262, EPI_ISL_1051263, EPI_ISL_1051264, EPI_ISL_1051265, EPI_ISL_1051266, EPI_ISL_1051267, EPI_ISL_1054141, EPI_ISL_1054192, EPI_ISL_1054194, EPI_ISL_1054233, EPI_ISL_1054239, EPI_ISL_1054241, EPI_ISL_1054243, EPI_ISL_1054248, EPI_ISL_1054250, EPI_ISL_1054252, EPI_ISL_1054254, EPI_ISL_1103949, EPI_ISL_1103950, EPI_ISL_1103951, EPI_ISL_1103952, EPI_ISL_1103953, EPI_ISL_1103954, EPI_ISL_1103955, EPI_ISL_1103956, EPI_ISL_1103957, EPI_ISL_1103958, EPI_ISL_1103959, EPI_ISL_1103960, EPI_ISL_1103961, EPI_ISL_1103962, EPI_ISL_1103963, EPI_ISL_1103964, EPI_ISL_1103965, EPI_ISL_1103966, EPI_ISL_1103967, EPI_ISL_1103968, EPI_ISL_1103969, EPI_ISL_1103970, EPI_ISL_1103971, EPI_ISL_1103972, EPI_ISL_1103973, EPI_ISL_1103974, EPI_ISL_1103975, EPI_ISL_1103976, EPI_ISL_1103977, EPI_ISL_1103978, EPI_ISL_1103979, EPI_ISL_1103980, EPI_ISL_1103981, EPI_ISL_1103982, EPI_ISL_1103983, EPI_ISL_1103984, EPI_ISL_1103985, EPI_ISL_1103986, EPI_ISL_1103987, EPI_ISL_1103988, EPI_ISL_1103989, EPI_ISL_1103990, EPI_ISL_1103991, EPI_ISL_1103993, EPI_ISL_1103994, EPI_ISL_1103995, EPI_ISL_1103996, EPI_ISL_1103997, EPI_ISL_1103998, EPI_ISL_1103999, EPI_ISL_1104000, EPI_ISL_1104001, EPI_ISL_1104002, EPI_ISL_1104003, EPI_ISL_1104004, EPI_ISL_1104005, EPI_ISL_1104006, EPI_ISL_1104007, EPI_ISL_1104008, EPI_ISL_1104009, EPI_ISL_1104010, EPI_ISL_1104011, EPI_ISL_1104012, EPI_ISL_1104013, EPI_ISL_1104014, EPI_ISL_1104015, EPI_ISL_1104016, EPI_ISL_1104017, EPI_ISL_1104018, EPI_ISL_1104019, EPI_ISL_1104020, EPI_ISL_1104021, EPI_ISL_1104022, EPI_ISL_1104023, EPI_ISL_1104024, EPI_ISL_1105486, EPI_ISL_1105488, EPI_ISL_1105519, EPI_ISL_1105520, EPI_ISL_1178148, EPI_ISL_1178149, EPI_ISL_1178150, EPI_ISL_1178151, EPI_ISL_1178152, EPI_ISL_1178153, EPI_ISL_1178154, EPI_ISL_1178155, EPI_ISL_1178156, EPI_ISL_1178157 | | | |
| see above | Northumbria University / South Tees Hospitals NHS Foundation Trust / North Cumbria Integrated Care NHS Foundation Trust / North Tees and Hartlepool NHS Foundation Trust / Newcastle Hospitals NHS Foundation Trust | COVID-19 Genomics UK (COG-UK) Consortium | Andrew Nelson; Brendan Payne; Clive Graham; Darren L Smith; Debra Padgett; Edward Barton; Emma Swindells; Garren Scott; Gary Black; Gary Eltringham; Giles S Holt; Greg R Young; Jane Greenaway; Jennifer Collins; John Allan; Joshua Loh; Lynn Dover; Matthew Bashton; Mohammad A Tariq; Paul Baker; Sarah Essex; Steve Liggett; Wen C Yew; Yusri Taha |
| EPI_ISL_951783, EPI_ISL_951971, EPI_ISL_951975, EPI_ISL_951976, EPI_ISL_951977, EPI_ISL_951978, EPI_ISL_951979, EPI_ISL_951980, EPI_ISL_951981, EPI_ISL_951982, EPI_ISL_951983, EPI_ISL_951984, EPI_ISL_951985, EPI_ISL_951986, EPI_ISL_952021, EPI_ISL_952023, EPI_ISL_952028, EPI_ISL_952044, EPI_ISL_952046, EPI_ISL_952047, EPI_ISL_952048, EPI_ISL_952049, EPI_ISL_952050, EPI_ISL_952052, EPI_ISL_952053, EPI_ISL_952054, EPI_ISL_952055, EPI_ISL_952056, EPI_ISL_952057, EPI_ISL_952058, EPI_ISL_952060, EPI_ISL_952252, EPI_ISL_952253, EPI_ISL_952255, EPI_ISL_952256, EPI_ISL_952257, EPI_ISL_952258, EPI_ISL_952259, EPI_ISL_952262, EPI_ISL_952266, EPI_ISL_952267, EPI_ISL_952268, EPI_ISL_952269, EPI_ISL_952270, EPI_ISL_952271, EPI_ISL_952272, EPI_ISL_952273, EPI_ISL_952274, EPI_ISL_952277, EPI_ISL_952278, EPI_ISL_952279, EPI_ISL_952280, EPI_ISL_952281, EPI_ISL_952282, EPI_ISL_952283, EPI_ISL_952284, EPI_ISL_952285, EPI_ISL_952286, EPI_ISL_952289, EPI_ISL_952290, EPI_ISL_952291, EPI_ISL_952299, EPI_ISL_952300, EPI_ISL_952301, EPI_ISL_952302, EPI_ISL_952303, EPI_ISL_952304, EPI_ISL_952305, EPI_ISL_952306, EPI_ISL_952307, EPI_ISL_952308, EPI_ISL_952309, EPI_ISL_952310, EPI_ISL_952311, EPI_ISL_952312, EPI_ISL_952313, EPI_ISL_952314, EPI_ISL_952315, EPI_ISL_952316, EPI_ISL_952317, EPI_ISL_952318, EPI_ISL_952319, EPI_ISL_952320, EPI_ISL_952321, EPI_ISL_952322, EPI_ISL_952323, EPI_ISL_952327, EPI_ISL_952328, EPI_ISL_952331, EPI_ISL_952332, EPI_ISL_952333, EPI_ISL_952334, EPI_ISL_952335, EPI_ISL_952337, EPI_ISL_952338, EPI_ISL_952340, EPI_ISL_952341, EPI_ISL_952351, EPI_ISL_952352, EPI_ISL_952360, EPI_ISL_952361, EPI_ISL_952366, EPI_ISL_952367, EPI_ISL_952368, EPI_ISL_952370, EPI_ISL_952371, EPI_ISL_952372, EPI_ISL_952373, EPI_ISL_952374, EPI_ISL_952375, EPI_ISL_952376, EPI_ISL_999516, EPI_ISL_999638, EPI_ISL_999642, EPI_ISL_999643, EPI_ISL_999644, EPI_ISL_999701, EPI_ISL_999702, EPI_ISL_999704, EPI_ISL_999713, EPI_ISL_999714, EPI_ISL_999717, EPI_ISL_999720, EPI_ISL_999724, EPI_ISL_999725, EPI_ISL_999729, EPI_ISL_999735, EPI_ISL_999741, EPI_ISL_999742, EPI_ISL_999747, EPI_ISL_999750, EPI_ISL_999755, EPI_ISL_999756, EPI_ISL_999757, EPI_ISL_999758, EPI_ISL_999762, EPI_ISL_999774, EPI_ISL_999824, EPI_ISL_999825, EPI_ISL_999826, EPI_ISL_999830, EPI_ISL_999832, EPI_ISL_1052138, EPI_ISL_1052139, EPI_ISL_1052140, EPI_ISL_1052143, EPI_ISL_1052145, EPI_ISL_1052146, EPI_ISL_1052149, EPI_ISL_1052150, EPI_ISL_1052151, EPI_ISL_1052152, EPI_ISL_1052153, EPI_ISL_1052154, EPI_ISL_1052155, EPI_ISL_1052156, EPI_ISL_1052157, EPI_ISL_1052159, EPI_ISL_1052160, EPI_ISL_1052161, EPI_ISL_1052162, EPI_ISL_1052163, EPI_ISL_1052165, EPI_ISL_1052166, EPI_ISL_1052167, EPI_ISL_1052168, EPI_ISL_1052169, EPI_ISL_1052170, EPI_ISL_1052171, EPI_ISL_1052172, EPI_ISL_1052173, EPI_ISL_1052174, EPI_ISL_1052175, EPI_ISL_1052176, EPI_ISL_1052177, EPI_ISL_1052183, EPI_ISL_1052188, EPI_ISL_1052190, EPI_ISL_1052191, EPI_ISL_1052192, EPI_ISL_1052193, EPI_ISL_1052195, EPI_ISL_1052261, EPI_ISL_1052262, EPI_ISL_1052263, EPI_ISL_1052268, EPI_ISL_1052270, EPI_ISL_1052271, EPI_ISL_1052470, EPI_ISL_1052472, EPI_ISL_1052484, EPI_ISL_1052485, EPI_ISL_1052486, EPI_ISL_1052487, EPI_ISL_1052488, EPI_ISL_1052492, EPI_ISL_1052493, EPI_ISL_1052494, EPI_ISL_1052495, EPI_ISL_1052522, EPI_ISL_1052526, EPI_ISL_1052527, EPI_ISL_1052528, EPI_ISL_1052530, EPI_ISL_1052531, EPI_ISL_1052532, EPI_ISL_1052533, EPI_ISL_1052534, EPI_ISL_1052535, EPI_ISL_1052536, EPI_ISL_1052537, EPI_ISL_1053513, EPI_ISL_1053515, EPI_ISL_1053516, EPI_ISL_1053517, EPI_ISL_1053518, EPI_ISL_1053530, EPI_ISL_1053531, EPI_ISL_1053532, EPI_ISL_1053533, EPI_ISL_1053534, EPI_ISL_1053535, EPI_ISL_1053539, EPI_ISL_1053540, EPI_ISL_1053562, EPI_ISL_1053565, EPI_ISL_1053567, EPI_ISL_1053568, EPI_ISL_1053569, EPI_ISL_1053571, EPI_ISL_1053572, EPI_ISL_1053573, EPI_ISL_1053574, EPI_ISL_1053575, EPI_ISL_1053579, EPI_ISL_1053581, EPI_ISL_1053588, EPI_ISL_1053589, EPI_ISL_1106646, EPI_ISL_1106647 | | | |
| see above | Originating lab: Wales Specialist Virology Centre Sequencing lab: Pathogen Genomics Unit | Public Health Wales Microbiology Cardiff Wales Specialist Virology Centre | Alec Birchley; Alexander Adams; Amy Gaskin; Angela Marchbank; Bree Gatica-Wilcox; Catherine Moore; Jason Coombes; Joanne Watkins; Joel Southgate; Johnathan Evans; Laura Gifford; Lauren Gilbert; Lee Graham; Malorie Perry; Matthew Bull; Nicole Pacchiarini; Sally Corden; Sara Kumziene-Summerhayes; Sara Rey; Sarah Taylor; Simon Cottrell; Sophie Jones; Tom Connor |
| EPI_ISL_998997, EPI_ISL_998999, EPI_ISL_999000, EPI_ISL_999328, EPI_ISL_1051596, EPI_ISL_1051627, EPI_ISL_1051630, EPI_ISL_1051631, EPI_ISL_1051634, EPI_ISL_1051635, EPI_ISL_1051639, EPI_ISL_1051640, EPI_ISL_1051642, EPI_ISL_1051643, EPI_ISL_1051645, EPI_ISL_1051646, EPI_ISL_1051647, EPI_ISL_1051649, EPI_ISL_1051651, EPI_ISL_1051652, EPI_ISL_1051653, EPI_ISL_1051654, EPI_ISL_1051655, EPI_ISL_1051658, EPI_ISL_1051659, EPI_ISL_1051661, EPI_ISL_1051662, EPI_ISL_1051663, EPI_ISL_1051664, EPI_ISL_1051667, EPI_ISL_1051668, EPI_ISL_1051669, EPI_ISL_1051672, EPI_ISL_1051674, EPI_ISL_1051677, EPI_ISL_1051678, EPI_ISL_1051679, EPI_ISL_1051680, EPI_ISL_1051683, EPI_ISL_1051684, EPI_ISL_1051686, EPI_ISL_1051687, EPI_ISL_1051689, EPI_ISL_1051691, EPI_ISL_1051693, EPI_ISL_1051715, EPI_ISL_1051716, EPI_ISL_1051717, EPI_ISL_1051718, EPI_ISL_1051719, EPI_ISL_1051721, EPI_ISL_1051722, EPI_ISL_1051726, EPI_ISL_1051728, EPI_ISL_1051738, EPI_ISL_1051877, EPI_ISL_1051878, EPI_ISL_1051880, EPI_ISL_1051881, EPI_ISL_1051882, EPI_ISL_1051883, EPI_ISL_1051884, EPI_ISL_1051899, EPI_ISL_1051900, EPI_ISL_1051901, EPI_ISL_1051903, EPI_ISL_1051908, EPI_ISL_1051909, EPI_ISL_1051910, EPI_ISL_1051911, EPI_ISL_1051912, EPI_ISL_1051913, EPI_ISL_1051914, EPI_ISL_1051916, EPI_ISL_1051917, EPI_ISL_1051918, EPI_ISL_1178854, EPI_ISL_1249224, EPI_ISL_1249241, EPI_ISL_1387213, EPI_ISL_1387216, EPI_ISL_1387217, EPI_ISL_1387225, EPI_ISL_1833063, EPI_ISL_1833093 | | | |
| see above | Oxford Viromics, NDM, University of Oxford; Oxford University Hospitals; Basingstoke and North Hampshire Hospital | COVID-19 Genomics UK (COG-UK) Consortium | Alex Mobbs; Amy Trebes; Anita Justice; Catrin Moore; Christophe Fraser; David Bonsall; David Buck; Emma Wise; George Macintyre; Jessica Lynch; John Todd; Mariateresa de Cesare; Matilde Mori; Monique Andersson; Nathan Moore; Nick Cortes; Robert Shaw; Stephen Kidd; Tanya Golubchik; Timothy Peto |
| EPI_ISL_998808, EPI_ISL_998819, EPI_ISL_998845, EPI_ISL_998848, EPI_ISL_998862, EPI_ISL_998863, EPI_ISL_998886, EPI_ISL_1178433, EPI_ISL_1178434, EPI_ISL_1178436, EPI_ISL_1178437, EPI_ISL_1178444, EPI_ISL_1178448, EPI_ISL_1178450, EPI_ISL_1178456, EPI_ISL_1178458, EPI_ISL_1178460, EPI_ISL_1178462, EPI_ISL_1178464, EPI_ISL_1178467, EPI_ISL_1178468, EPI_ISL_1178469, EPI_ISL_1178470, EPI_ISL_1178484, EPI_ISL_1178488, EPI_ISL_1178501, EPI_ISL_1248945, EPI_ISL_1248947, EPI_ISL_1248950, EPI_ISL_1248951, EPI_ISL_1248953, EPI_ISL_1248955, EPI_ISL_1248956, EPI_ISL_1248958, EPI_ISL_1248959, EPI_ISL_1248961, EPI_ISL_1248962, EPI_ISL_1248979, EPI_ISL_1248984, EPI_ISL_1248987, EPI_ISL_1248994, EPI_ISL_1248999, EPI_ISL_1249004, EPI_ISL_1249008, EPI_ISL_1249013, EPI_ISL_1249014, EPI_ISL_1249019, EPI_ISL_1249025, EPI_ISL_1249027, EPI_ISL_1249028, EPI_ISL_1832999 | | | |
| see above | Quadram Institute Bioscience | COVID-19 Genomics UK (COG-UK) Consortium | Alexander J Trotter; Alison E. Mather; Alp Aydin; Ana P. Tedim; Anastasia Kolyva; Andrew Bell; Andrew J. Page; Claire Stuart; Dave J. Baker; Gemma L. Kay; John Wain; Justin O'Grady; Leonardo de Oliveira Martins; Lizzie Meadows; Maria Diaz; Mark Webber; Muhammed Yasir; Nabil-Fareed Alikhan; Ngozi Elumogo; Nicholas M. Thomson; Rachael Stanley; Rachel Gilroy; Reenesh Prakash; Samir Dervisevic; Samuel Bloomfield; Steven Rudder; Thanh Le-Viet |
| EPI_ISL_950646, EPI_ISL_950647, EPI_ISL_950648, EPI_ISL_950649, EPI_ISL_950650, EPI_ISL_950651, EPI_ISL_950652, EPI_ISL_950653, EPI_ISL_950654, EPI_ISL_950655, EPI_ISL_950656, EPI_ISL_950657, EPI_ISL_950658, EPI_ISL_950659, EPI_ISL_950660, EPI_ISL_950661, EPI_ISL_950662, EPI_ISL_950663, EPI_ISL_950664, EPI_ISL_950665, EPI_ISL_950666, EPI_ISL_950667, EPI_ISL_950668, EPI_ISL_950669, EPI_ISL_950670, EPI_ISL_950762, EPI_ISL_1051590, EPI_ISL_1054372, EPI_ISL_1054373, EPI_ISL_1054374, EPI_ISL_1054375, EPI_ISL_1054376, EPI_ISL_1054377, EPI_ISL_1054378, EPI_ISL_1054379, EPI_ISL_1054380, EPI_ISL_1054381, EPI_ISL_1054382, EPI_ISL_1054383, EPI_ISL_1054384, EPI_ISL_1054385, EPI_ISL_1054386, EPI_ISL_1054387, EPI_ISL_1054388, EPI_ISL_1054389, EPI_ISL_1054390, EPI_ISL_1054391, EPI_ISL_1054392, EPI_ISL_1054393, EPI_ISL_1054394, EPI_ISL_1054395, EPI_ISL_1054397, EPI_ISL_1054398, EPI_ISL_1054399, EPI_ISL_1054400, EPI_ISL_1054401, EPI_ISL_1054402, EPI_ISL_1054403, EPI_ISL_1054404, EPI_ISL_1054405, EPI_ISL_1054406, EPI_ISL_1054407, EPI_ISL_1054408, EPI_ISL_1054409, EPI_ISL_1054410, EPI_ISL_1054411, EPI_ISL_1054412, EPI_ISL_1054413, EPI_ISL_1054414, EPI_ISL_1054415, EPI_ISL_1054416, EPI_ISL_1054417, EPI_ISL_1054418, EPI_ISL_1054419, EPI_ISL_1054420, EPI_ISL_1054421, EPI_ISL_1054422, EPI_ISL_1054423, EPI_ISL_1054424, EPI_ISL_1054425, EPI_ISL_1054426, EPI_ISL_1054427, EPI_ISL_1054428, EPI_ISL_1054429, EPI_ISL_1054430, EPI_ISL_1054431, EPI_ISL_1054432, EPI_ISL_1054433, EPI_ISL_1054434, EPI_ISL_1054435, EPI_ISL_1054436, EPI_ISL_1054437, EPI_ISL_1054438, EPI_ISL_1054439, EPI_ISL_1054440, EPI_ISL_1054441, EPI_ISL_1054442, EPI_ISL_1054443, EPI_ISL_1054444, EPI_ISL_1054445, EPI_ISL_1054446, EPI_ISL_1054447, EPI_ISL_1054448, EPI_ISL_1054449, EPI_ISL_1054450, EPI_ISL_1054451, EPI_ISL_1054452, EPI_ISL_1054453, EPI_ISL_1054454, EPI_ISL_1054455, EPI_ISL_1054456, EPI_ISL_1054457, EPI_ISL_1054458, EPI_ISL_1054459, EPI_ISL_1054460, EPI_ISL_1054461, EPI_ISL_1108957, EPI_ISL_1108960, EPI_ISL_1108961, EPI_ISL_1108962, EPI_ISL_1108963, EPI_ISL_1108964, EPI_ISL_1108965, EPI_ISL_1108966, EPI_ISL_1108967, EPI_ISL_1108968, EPI_ISL_1108969, EPI_ISL_1108971, EPI_ISL_1108973, EPI_ISL_1108974, EPI_ISL_1108975, EPI_ISL_1108976, EPI_ISL_1108977, EPI_ISL_1108978, EPI_ISL_1108979, EPI_ISL_1108980, EPI_ISL_1108981, EPI_ISL_1108982, EPI_ISL_1108984, EPI_ISL_1108985, EPI_ISL_1108986, EPI_ISL_1108987, EPI_ISL_1108988, EPI_ISL_1108989, EPI_ISL_1108990, EPI_ISL_1108991, EPI_ISL_1108992, EPI_ISL_1108993, EPI_ISL_1108994, EPI_ISL_1108995, EPI_ISL_1108996, EPI_ISL_1108997, EPI_ISL_1108998, EPI_ISL_1108999, EPI_ISL_1109000, EPI_ISL_1109001, EPI_ISL_1109002, EPI_ISL_1109003, EPI_ISL_1109004, EPI_ISL_1109005, EPI_ISL_1109006, EPI_ISL_1109007, EPI_ISL_1109008, EPI_ISL_1109009, EPI_ISL_1109010, EPI_ISL_1109011, EPI_ISL_1109012, EPI_ISL_1109014, EPI_ISL_1109015, EPI_ISL_1109016, EPI_ISL_1109017, EPI_ISL_1109018, EPI_ISL_1109019, EPI_ISL_1109020, EPI_ISL_1109021, EPI_ISL_1109022, EPI_ISL_1109023, EPI_ISL_1109026, EPI_ISL_1109028, EPI_ISL_1109030, EPI_ISL_1109032, EPI_ISL_1109035, EPI_ISL_1109037, EPI_ISL_1109042, EPI_ISL_1109043, EPI_ISL_1109044 | | | |
| see above | Queens Medical Centre, Clinical Microbiology Department / DeepSeq Nottingham | COVID-19 Genomics UK (COG-UK) Consortium | Christopher Moore; Fei Sang; Gemma Clark; Hannah Howson-Wells; Johnny Debebe; Jonathan Ball; Joseph Chappell; Manjinder Khakh; Matthew Carlisle; Matthew Loose; Michelle M Lister; Nadine Holmes; Patrick McClure; Theocharis Tsoleridis; Vicki M Fleming; Victoria Wright; Wendy Smith |
| EPI_ISL_1045983, EPI_ISL_1046337, EPI_ISL_1046338, EPI_ISL_1046339, EPI_ISL_1046340, EPI_ISL_1046341, EPI_ISL_1046342, EPI_ISL_1046343, EPI_ISL_1046344, EPI_ISL_1046345, EPI_ISL_1046346, EPI_ISL_1046347, EPI_ISL_1046348, EPI_ISL_1046349, EPI_ISL_1046350, EPI_ISL_1046351, EPI_ISL_1046352, EPI_ISL_1046353, EPI_ISL_1046354, EPI_ISL_1046355, EPI_ISL_1046356, EPI_ISL_1046357, EPI_ISL_1046358, EPI_ISL_1046359, EPI_ISL_1046360, EPI_ISL_1046361, EPI_ISL_1046362, EPI_ISL_1046363, EPI_ISL_1046364, EPI_ISL_1046365, EPI_ISL_1046366, EPI_ISL_1046367, EPI_ISL_1046368, EPI_ISL_1046369, EPI_ISL_1046370, EPI_ISL_1046371, EPI_ISL_1046372, EPI_ISL_1046373, EPI_ISL_1046374, EPI_ISL_1046375, EPI_ISL_1046376, EPI_ISL_1046377, EPI_ISL_1046378, EPI_ISL_1046379, EPI_ISL_1046380, EPI_ISL_1046381, EPI_ISL_1046382, EPI_ISL_1046383, EPI_ISL_1046384, EPI_ISL_1046385, EPI_ISL_1046386, EPI_ISL_1046387, EPI_ISL_1046388, EPI_ISL_1046389, EPI_ISL_1046390, EPI_ISL_1046391, EPI_ISL_1046392, EPI_ISL_1046393, EPI_ISL_1046394, EPI_ISL_1046395, EPI_ISL_1046396, EPI_ISL_1046397, EPI_ISL_1046398, EPI_ISL_1046399, EPI_ISL_1046400, EPI_ISL_1046401, EPI_ISL_1046402, EPI_ISL_1046403, EPI_ISL_1046404, EPI_ISL_1046405, EPI_ISL_1046406, EPI_ISL_1046407, EPI_ISL_1046408, EPI_ISL_1046409, EPI_ISL_1046410, EPI_ISL_1046411, EPI_ISL_1046412, EPI_ISL_1046413, EPI_ISL_1046414, EPI_ISL_1046415, EPI_ISL_1046416, EPI_ISL_1046417, EPI_ISL_1046418, EPI_ISL_1046419, EPI_ISL_1046420, EPI_ISL_1046421, EPI_ISL_1046422, EPI_ISL_1046423, EPI_ISL_1046424, EPI_ISL_1046425, EPI_ISL_1046426, EPI_ISL_1046427, EPI_ISL_1046428, EPI_ISL_1046429, EPI_ISL_1046430, EPI_ISL_1046431, EPI_ISL_1046432, EPI_ISL_1046433, EPI_ISL_1046434, EPI_ISL_1046435, EPI_ISL_1046436, EPI_ISL_1046437, EPI_ISL_1046438, EPI_ISL_1046439, EPI_ISL_1046440, EPI_ISL_1046441, EPI_ISL_1046442, EPI_ISL_1046443, EPI_ISL_1046444, EPI_ISL_1046445, EPI_ISL_1046446, EPI_ISL_1046447, EPI_ISL_1046448, EPI_ISL_1046449, EPI_ISL_1046450, EPI_ISL_1046451, EPI_ISL_1046452, EPI_ISL_1046453, EPI_ISL_1046454, EPI_ISL_1046455, EPI_ISL_1046456, EPI_ISL_1046457, EPI_ISL_1046458, EPI_ISL_1046459, EPI_ISL_1046460, EPI_ISL_1046461, EPI_ISL_1046462, EPI_ISL_1046463, EPI_ISL_1046464, EPI_ISL_1046465, EPI_ISL_1046466, EPI_ISL_1046467, EPI_ISL_1046468, EPI_ISL_1046469, EPI_ISL_1046470, EPI_ISL_1046471, EPI_ISL_1046472, EPI_ISL_1046473, EPI_ISL_1046474, EPI_ISL_1046475, EPI_ISL_1046476, EPI_ISL_1046477, EPI_ISL_1046478, EPI_ISL_1046479, EPI_ISL_1046480, EPI_ISL_1046481, EPI_ISL_1046482, EPI_ISL_1046483, EPI_ISL_1046484, EPI_ISL_1046485, EPI_ISL_1046486, EPI_ISL_1046487, EPI_ISL_1046488, EPI_ISL_1046489, EPI_ISL_1046490, EPI_ISL_1046491, EPI_ISL_1046492, EPI_ISL_1046493, EPI_ISL_1046494, EPI_ISL_1046495, EPI_ISL_1046496, EPI_ISL_1046497, EPI_ISL_1046498, EPI_ISL_1046499, EPI_ISL_1046500, EPI_ISL_1046501, EPI_ISL_1046502, EPI_ISL_1046503, EPI_ISL_1046504, EPI_ISL_1046505, EPI_ISL_1046506, EPI_ISL_1046507, EPI_ISL_1046508, EPI_ISL_1046509, EPI_ISL_1046510, EPI_ISL_1046511, EPI_ISL_1046512, EPI_ISL_1046513, EPI_ISL_1046514, EPI_ISL_1046515, EPI_ISL_1046516, EPI_ISL_1046517, EPI_ISL_1046518, EPI_ISL_1046519, EPI_ISL_1046520, EPI_ISL_1046521, EPI_ISL_1046522, EPI_ISL_1046523, EPI_ISL_1046524, EPI_ISL_1046525, EPI_ISL_1046526, EPI_ISL_1046527, EPI_ISL_1046528, EPI_ISL_1046529, EPI_ISL_1046530, EPI_ISL_1046531, EPI_ISL_1046532, EPI_ISL_1046533, EPI_ISL_1046534, EPI_ISL_1046535, EPI_ISL_1046536, EPI_ISL_1046537, EPI_ISL_1046538, EPI_ISL_1046539, EPI_ISL_1046540, EPI_ISL_1046541, EPI_ISL_1046542, EPI_ISL_1046543, EPI_ISL_1046544, EPI_ISL_1046545, EPI_ISL_1046546, EPI_ISL_1046547, EPI_ISL_1046548, EPI_ISL_1046549, EPI_ISL_1046550, EPI_ISL_1046551, EPI_ISL_1046552, EPI_ISL_1046553, EPI_ISL_1046554, EPI_ISL_1046555, EPI_ISL_1046556, EPI_ISL_1046558, EPI_ISL_1046559, EPI_ISL_1046560, EPI_ISL_1046561, EPI_ISL_1046562, EPI_ISL_1046563, EPI_ISL_1046564, EPI_ISL_1046565, EPI_ISL_1046566, EPI_ISL_1046567, EPI_ISL_1046568, EPI_ISL_1046570, EPI_ISL_1046571, EPI_ISL_1046572, EPI_ISL_1046573, EPI_ISL_1046574, EPI_ISL_1046575, EPI_ISL_1046576, EPI_ISL_1046577, EPI_ISL_1046578, EPI_ISL_1046579, EPI_ISL_1046580, EPI_ISL_1046581, EPI_ISL_1046582, EPI_ISL_1046583, EPI_ISL_1046584, EPI_ISL_1046585, EPI_ISL_1046586, EPI_ISL_1046587, EPI_ISL_1046588, EPI_ISL_1046589, EPI_ISL_1046590, EPI_ISL_1046591, EPI_ISL_1046592, EPI_ISL_1046593, EPI_ISL_1046594, EPI_ISL_1046595, EPI_ISL_1046596, EPI_ISL_1046597, EPI_ISL_1046598, EPI_ISL_1046599, EPI_ISL_1046600, EPI_ISL_1046601, EPI_ISL_1046602, EPI_ISL_1046603, EPI_ISL_1046604, EPI_ISL_1046605, EPI_ISL_1046606, EPI_ISL_1046607, EPI_ISL_1046608, EPI_ISL_1046609, EPI_ISL_1046610, EPI_ISL_1046611, EPI_ISL_1046612, EPI_ISL_1046613, EPI_ISL_1046614, EPI_ISL_1046615, EPI_ISL_1046616, EPI_ISL_1046617, EPI_ISL_1046618, EPI_ISL_1046619, EPI_ISL_1046620, EPI_ISL_1046621, EPI_ISL_1046622, EPI_ISL_1046623, EPI_ISL_1046624, EPI_ISL_1046625, EPI_ISL_1046626, EPI_ISL_1046627, EPI_ISL_1046628, EPI_ISL_1046629, EPI_ISL_1046630, EPI_ISL_1046631, EPI_ISL_1046632, EPI_ISL_1046633, EPI_ISL_1046634, EPI_ISL_1046635, EPI_ISL_1046636, EPI_ISL_1046637, EPI_ISL_1046638, EPI_ISL_1046639, EPI_ISL_1046640, EPI_ISL_1046641, EPI_ISL_1046642, EPI_ISL_1046643, EPI_ISL_1046644, EPI_ISL_1046645, EPI_ISL_1046646, EPI_ISL_1046647, EPI_ISL_1046648, EPI_ISL_1046649, EPI_ISL_1046650, EPI_ISL_1046651, EPI_ISL_1046652, EPI_ISL_1046653, EPI_ISL_1046654, EPI_ISL_1046655, EPI_ISL_1046656, EPI_ISL_1046657, EPI_ISL_1046659, EPI_ISL_1046660, EPI_ISL_1046661, EPI_ISL_1046662, EPI_ISL_1046663, EPI_ISL_1046664, EPI_ISL_1046665, EPI_ISL_1046667, EPI_ISL_1046668, EPI_ISL_1046669, EPI_ISL_1046670, EPI_ISL_1046671, EPI_ISL_1046672, EPI_ISL_1046673, EPI_ISL_1046674, EPI_ISL_1046675, EPI_ISL_1046677, EPI_ISL_1046678, EPI_ISL_1046679, EPI_ISL_1046680, EPI_ISL_1046681, EPI_ISL_1046683, EPI_ISL_1046684, EPI_ISL_1046685, EPI_ISL_1046686, EPI_ISL_1046687, EPI_ISL_1046688, EPI_ISL_1046689, EPI_ISL_1046690, EPI_ISL_1046691, EPI_ISL_1046692, EPI_ISL_1046693, EPI_ISL_1046694, EPI_ISL_1046695, EPI_ISL_1046696, EPI_ISL_1046697, EPI_ISL_1046699, EPI_ISL_1046700, EPI_ISL_1046701, EPI_ISL_1046702, EPI_ISL_1046703, EPI_ISL_1046704, EPI_ISL_1046705, EPI_ISL_1046706, EPI_ISL_1046708, EPI_ISL_1046709, EPI_ISL_1046710, EPI_ISL_1046711, EPI_ISL_1046712, EPI_ISL_1046713, EPI_ISL_1046714, EPI_ISL_1046715, EPI_ISL_1046716, EPI_ISL_1046717, EPI_ISL_1046718, EPI_ISL_1046719, EPI_ISL_1046720, EPI_ISL_1046721, EPI_ISL_1046722, EPI_ISL_1046723, EPI_ISL_1046724, EPI_ISL_1046725, EPI_ISL_1046726, EPI_ISL_1046727, EPI_ISL_1046729, EPI_ISL_1046730, EPI_ISL_1046731, EPI_ISL_1046732, EPI_ISL_1046733, EPI_ISL_1046734, EPI_ISL_1046735, EPI_ISL_1046737, EPI_ISL_1046738, EPI_ISL_1046739, EPI_ISL_1046740, EPI_ISL_1046741, EPI_ISL_1046742, EPI_ISL_1046743, EPI_ISL_1046744, EPI_ISL_1046745, EPI_ISL_1046746, EPI_ISL_1046747, EPI_ISL_1046748, EPI_ISL_1046749, EPI_ISL_1046750, EPI_ISL_1046751, EPI_ISL_1046752, EPI_ISL_1046753, EPI_ISL_1046754, EPI_ISL_1046755, EPI_ISL_1046756, EPI_ISL_1046757, EPI_ISL_1046759, EPI_ISL_1046760 | | | |
| see above | Randox Laboratories | Wellcome Sanger Institute for the COVID-19 Genomics UK (COG-UK) Consortium | Cordelia Langford; David K. Jackson; Dominic Kwiatkowski; Ewan Harrison; Ian Johnston; Jeffrey Barrett; John Sillitoe on behalf of the Wellcome Sanger Institute COVID-19 Surveillance Team; Randox Laboratories and Alex Alderton; Roberto Amato; Sonia Goncalves |
| EPI_ISL_998232, EPI_ISL_998299, EPI_ISL_998300, EPI_ISL_998301, EPI_ISL_998303, EPI_ISL_1474996, EPI_ISL_1474997, EPI_ISL_1474998, EPI_ISL_1475000, EPI_ISL_1475017, EPI_ISL_1475036, EPI_ISL_1475037, EPI_ISL_1475038, EPI_ISL_1475039, EPI_ISL_1475043, EPI_ISL_1475044, EPI_ISL_1475132, EPI_ISL_1475134, EPI_ISL_1475138, EPI_ISL_1475140, EPI_ISL_1475141, EPI_ISL_1475142 | | | |
| see above | Regional Virus Laboratory, Belfast Health and Social Care Trust | COVID-19 Genomics UK (COG-UK) Consortium | Alison Watt; Ciara Cox; Conall McCaughey; David Simpson; Derek Fairley; James McKenna; Mairead Connor; Susan Feeney; Tanya Curran; Zoltan Molnar |
| EPI_ISL_1296587, EPI_ISL_1296589, EPI_ISL_1296590, EPI_ISL_1296591, EPI_ISL_1296592, EPI_ISL_1296593, EPI_ISL_1296594, EPI_ISL_1296595, EPI_ISL_1296596, EPI_ISL_1296597, EPI_ISL_1296598 | | | |
| see above | Respiratory Virus Unit, National Infection Service, Public Health England | COVID-19 Genomics UK (COG-UK) Consortium | PHE Covid Sequencing Team |
| EPI_ISL_1833617, EPI_ISL_1833618, EPI_ISL_1833621, EPI_ISL_1833622, EPI_ISL_1833625, EPI_ISL_1833626, EPI_ISL_1833627, EPI_ISL_1833628, EPI_ISL_1833629, EPI_ISL_1833631, EPI_ISL_1833632, EPI_ISL_1833633, EPI_ISL_1833634, EPI_ISL_1833635, EPI_ISL_1833637, EPI_ISL_1833638, EPI_ISL_1833639, EPI_ISL_1833640, EPI_ISL_1833642, EPI_ISL_1833645, EPI_ISL_1833646, EPI_ISL_1833648, EPI_ISL_1833680, EPI_ISL_1833694 | | | |
| see above | St George's University Hospitals NHS Foundation Trust | COVID-19 Genomics UK (COG-UK) Consortium | Adam Witney; Cassie Pope; Irene Monahan; Joshua Taylor; Ken Laing; NgeeKeong Tan |
| EPI_ISL_1104832, EPI_ISL_1104836, EPI_ISL_1104838, EPI_ISL_1104843, EPI_ISL_1104844, EPI_ISL_1104847, EPI_ISL_1104848, EPI_ISL_1104849, EPI_ISL_1104855, EPI_ISL_1104856, EPI_ISL_1104857, EPI_ISL_1104859, EPI_ISL_1104863, EPI_ISL_1104864, EPI_ISL_1104867, EPI_ISL_1104872, EPI_ISL_1104874, EPI_ISL_1104879, EPI_ISL_1104880, EPI_ISL_1104881, EPI_ISL_1104886, EPI_ISL_1104887, EPI_ISL_1104899, EPI_ISL_1104900, EPI_ISL_1104901, EPI_ISL_1104902, EPI_ISL_1104908, EPI_ISL_1104909, EPI_ISL_1104910, EPI_ISL_1104911, EPI_ISL_1104916, EPI_ISL_1104917, EPI_ISL_1104918, EPI_ISL_1104919, EPI_ISL_1104920, EPI_ISL_1104925, EPI_ISL_1104933, EPI_ISL_1104934, EPI_ISL_1104939, EPI_ISL_1104940, EPI_ISL_1104943, EPI_ISL_1104944, EPI_ISL_1104945, EPI_ISL_1104946, EPI_ISL_1104950, EPI_ISL_1104952, EPI_ISL_1104953, EPI_ISL_1104955, EPI_ISL_1104956, EPI_ISL_1104959, EPI_ISL_1104960, EPI_ISL_1104961, EPI_ISL_1104965, EPI_ISL_1104966, EPI_ISL_1104967, EPI_ISL_1105182, EPI_ISL_1105191, EPI_ISL_1105192, EPI_ISL_1105194, EPI_ISL_1105198, EPI_ISL_1105225, EPI_ISL_1105227 | | | |
| see above | University College London Hospital | COVID-19 Genomics UK (COG-UK) Consortium | Catherine Houlihan; Dan Frampton; Judith Heaney; Matthew Byott; Moira Spyer and Eleni Nastouli; Stuart Kirk |
| EPI_ISL_949795, EPI_ISL_949796, EPI_ISL_949797, EPI_ISL_949798, EPI_ISL_949799, EPI_ISL_949800, EPI_ISL_949801, EPI_ISL_949803, EPI_ISL_949804, EPI_ISL_949805, EPI_ISL_949806, EPI_ISL_949807, EPI_ISL_949808, EPI_ISL_949809, EPI_ISL_949810, EPI_ISL_949812, EPI_ISL_949813, EPI_ISL_949845, EPI_ISL_949847, EPI_ISL_949848, EPI_ISL_949855, EPI_ISL_949866, EPI_ISL_949915, EPI_ISL_949916, EPI_ISL_949918, EPI_ISL_949921, EPI_ISL_949922, EPI_ISL_949923, EPI_ISL_949928, EPI_ISL_949929, EPI_ISL_949930, EPI_ISL_949931, EPI_ISL_949932, EPI_ISL_949933, EPI_ISL_949935, EPI_ISL_949936, EPI_ISL_949940, EPI_ISL_949941, EPI_ISL_949942, EPI_ISL_949943, EPI_ISL_949944, EPI_ISL_949947, EPI_ISL_949949, EPI_ISL_949950, EPI_ISL_949951, EPI_ISL_949953, EPI_ISL_949954, EPI_ISL_949957, EPI_ISL_949961, EPI_ISL_950173, EPI_ISL_950177, EPI_ISL_950195, EPI_ISL_950198, EPI_ISL_950200, EPI_ISL_950201, EPI_ISL_950202, EPI_ISL_950203, EPI_ISL_950204, EPI_ISL_950205, EPI_ISL_950206, EPI_ISL_950207, EPI_ISL_950208, EPI_ISL_950209, EPI_ISL_950210, EPI_ISL_997726, EPI_ISL_997833, EPI_ISL_997834, EPI_ISL_997882, EPI_ISL_997883, EPI_ISL_997884, EPI_ISL_997885, EPI_ISL_997886, EPI_ISL_997888, EPI_ISL_997889, EPI_ISL_997893, EPI_ISL_997895, EPI_ISL_997896, EPI_ISL_997897, EPI_ISL_997898, EPI_ISL_997899, EPI_ISL_997900, EPI_ISL_997902, EPI_ISL_997903, EPI_ISL_997910, EPI_ISL_997911, EPI_ISL_997912, EPI_ISL_997913, EPI_ISL_997914, EPI_ISL_997915, EPI_ISL_997917, EPI_ISL_997918, EPI_ISL_997919, EPI_ISL_997920, EPI_ISL_997921, EPI_ISL_997924, EPI_ISL_997925, EPI_ISL_997926, EPI_ISL_997931, EPI_ISL_997951, EPI_ISL_998012, EPI_ISL_998013, EPI_ISL_998014, EPI_ISL_998015, EPI_ISL_998016, EPI_ISL_998017, EPI_ISL_998018, EPI_ISL_998019, EPI_ISL_998020, EPI_ISL_998021, EPI_ISL_998022, EPI_ISL_998023, EPI_ISL_998024, EPI_ISL_998025, EPI_ISL_998026, EPI_ISL_998027, EPI_ISL_998028, EPI_ISL_998029, EPI_ISL_998030, EPI_ISL_998031, EPI_ISL_998032, EPI_ISL_998033, EPI_ISL_998034, EPI_ISL_998035, EPI_ISL_998036, EPI_ISL_998037, EPI_ISL_998038, EPI_ISL_998039, EPI_ISL_998040, EPI_ISL_998041, EPI_ISL_998042, EPI_ISL_998043, EPI_ISL_998045, EPI_ISL_998046, EPI_ISL_998047, EPI_ISL_998048, EPI_ISL_998049, EPI_ISL_998050, EPI_ISL_998051, EPI_ISL_998052, EPI_ISL_998053, EPI_ISL_998054, EPI_ISL_998055, EPI_ISL_998056, EPI_ISL_998057, EPI_ISL_998058, EPI_ISL_998059, EPI_ISL_998060, EPI_ISL_998061, EPI_ISL_998062, EPI_ISL_998063, EPI_ISL_998064, EPI_ISL_998065, EPI_ISL_998066, EPI_ISL_998067, EPI_ISL_998068, EPI_ISL_998069, EPI_ISL_998070, EPI_ISL_998071, EPI_ISL_998072, EPI_ISL_998073, EPI_ISL_998074, EPI_ISL_998075, EPI_ISL_998076, EPI_ISL_998077, EPI_ISL_998078, EPI_ISL_998079, EPI_ISL_998080, EPI_ISL_998081, EPI_ISL_998082, EPI_ISL_998083, EPI_ISL_998084, EPI_ISL_998085, EPI_ISL_998086, EPI_ISL_998087, EPI_ISL_998088, EPI_ISL_998089, EPI_ISL_998090, EPI_ISL_998091, EPI_ISL_998092, EPI_ISL_998093, EPI_ISL_998094, EPI_ISL_998095, EPI_ISL_998096, EPI_ISL_998097, EPI_ISL_998098, EPI_ISL_1050582, EPI_ISL_1050583, EPI_ISL_1050584, EPI_ISL_1050585, EPI_ISL_1050586, EPI_ISL_1050587, EPI_ISL_1050588, EPI_ISL_1050589, EPI_ISL_1050590, EPI_ISL_1050591, EPI_ISL_1050592, EPI_ISL_1050593, EPI_ISL_1050594, EPI_ISL_1050595, EPI_ISL_1050596, EPI_ISL_1050597, EPI_ISL_1050598, EPI_ISL_1050599, EPI_ISL_1050600, EPI_ISL_1050601, EPI_ISL_1050602, EPI_ISL_1050603, EPI_ISL_1050604, EPI_ISL_1050607, EPI_ISL_1050608, EPI_ISL_1050609, EPI_ISL_1050611, EPI_ISL_1050612, EPI_ISL_1050613, EPI_ISL_1050615, EPI_ISL_1050616, EPI_ISL_1050617, EPI_ISL_1050618, EPI_ISL_1050620, EPI_ISL_1050621, EPI_ISL_1050622, EPI_ISL_1050623, EPI_ISL_1050624, EPI_ISL_1050625, EPI_ISL_1050626, EPI_ISL_1050627, EPI_ISL_1050628, EPI_ISL_1050629, EPI_ISL_1050630, EPI_ISL_1050631, EPI_ISL_1050676, EPI_ISL_1050677, EPI_ISL_1050678, EPI_ISL_1050679, EPI_ISL_1050680, EPI_ISL_1050681, EPI_ISL_1050682, EPI_ISL_1050683, EPI_ISL_1050684, EPI_ISL_1050685, EPI_ISL_1050723, EPI_ISL_1050724, EPI_ISL_1050725, EPI_ISL_1050726, EPI_ISL_1050727, EPI_ISL_1050728, EPI_ISL_1050729, EPI_ISL_1050730, EPI_ISL_1050731, EPI_ISL_1050732, EPI_ISL_1050733, EPI_ISL_1050734, EPI_ISL_1050735, EPI_ISL_1050736, EPI_ISL_1050737, EPI_ISL_1050738, EPI_ISL_1050739, EPI_ISL_1050740, EPI_ISL_1050741, EPI_ISL_1050742, EPI_ISL_1050743, EPI_ISL_1050744, EPI_ISL_1050745, EPI_ISL_1050746, EPI_ISL_1050747, EPI_ISL_1105447, EPI_ISL_1105448, EPI_ISL_1105467, EPI_ISL_1248160, EPI_ISL_1248161, EPI_ISL_1248162, EPI_ISL_1248169, EPI_ISL_1248170, EPI_ISL_1248171, EPI_ISL_1248177, EPI_ISL_1248180, EPI_ISL_1248189, EPI_ISL_1248197, EPI_ISL_1248199, EPI_ISL_1248200, EPI_ISL_1248262, EPI_ISL_1248531, EPI_ISL_1309060, EPI_ISL_1309122 | | | |
| see above | University College London, Great Ormond Street Hospital for Children NHS Foundation Trust, Imperial College Healthcare NHS Trust | COVID-19 Genomics UK (COG-UK) Consortium | Alison Holmes; Charlotte Williams; Helena Tutill; Jacqueline Findlay; James Price; Judith Breuer; Julianne Brown; Kathryn Harris; Leysa Forrest; Mark Kristiansen; Paola Niola; Paola Resende Silva; Patricia Dyal; Paul Randell; Rachel Williams; Samuel Weeks; Sergi Castellano; Sunando Roy; Tony Brooks; Yasmin Panchbhaya |
| EPI_ISL_949405, EPI_ISL_996407, EPI_ISL_996442, EPI_ISL_996443, EPI_ISL_996444, EPI_ISL_996445, EPI_ISL_996464, EPI_ISL_996465, EPI_ISL_996466, EPI_ISL_996467, EPI_ISL_996468, EPI_ISL_996471, EPI_ISL_996472, EPI_ISL_996473, EPI_ISL_996474, EPI_ISL_1047000, EPI_ISL_1047002, EPI_ISL_1047003, EPI_ISL_1047006, EPI_ISL_1047010, EPI_ISL_1047013, EPI_ISL_1047014, EPI_ISL_1047021, EPI_ISL_1047031, EPI_ISL_1047037, EPI_ISL_1047038, EPI_ISL_1047041, EPI_ISL_1047055, EPI_ISL_1047056, EPI_ISL_1636594 | | | |
| see above | University of Birmingham | COVID-19 Genomics UK (COG-UK) Consortium | Alex Richter; Andrew Bosworth; Andrew Bosworth. Queen Elizabeth Hospital: Anna Casey; Andrew D Beggs PHE Heartlands Lab: Husam Osman; Charlotte Poxon; Institute of Microbiology; Joanne Stockton; Josh Quick; Kasun Wanigasooriya; Mike Kidd; Nicholas Loman. University of Birmingham Testing Laboratory: Celina M Whalley; Oliver Pickles; Radoslaw Poplawski; Samuel Nicholls; University of Birmingham: Claire McMurray; Will Rowe |

EPI_ISL_996480, EPI_ISL_996547, EPI_ISL_996548, EPI_ISL_996549, EPI_ISL_996555, EPI_ISL_996556, EPI_ISL_996558, EPI_ISL_996559, EPI_ISL_996561, EPI_ISL_996562, EPI_ISL_996563, EPI_ISL_996565, EPI_ISL_996567, EPI_ISL_996568, EPI_ISL_997180, EPI_ISL_997181, EPI_ISL_997182, EPI_ISL_997183, EPI_ISL_997185, EPI_ISL_997188, EPI_ISL_997189, EPI_ISL_997190, EPI_ISL_997191, EPI_ISL_997192, EPI_ISL_997193, EPI_ISL_997194, EPI_ISL_997195, EPI_ISL_997196, EPI_ISL_997197, EPI_ISL_997198, EPI_ISL_997199, EPI_ISL_997200, EPI_ISL_997202, EPI_ISL_997203, EPI_ISL_997204, EPI_ISL_997205, EPI_ISL_997207, EPI_ISL_997208, EPI_ISL_997286, EPI_ISL_997287, EPI_ISL_997339, EPI_ISL_1050293, EPI_ISL_1050294, EPI_ISL_1050295, EPI_ISL_1050296, EPI_ISL_1050299, EPI_ISL_1050300, EPI_ISL_1050301, EPI_ISL_1050302, EPI_ISL_1050303, EPI_ISL_1053977, EPI_ISL_1053978, EPI_ISL_1053979, EPI_ISL_1053980, EPI_ISL_1053982, EPI_ISL_1053983, EPI_ISL_1053984, EPI_ISL_1053985, EPI_ISL_1053986, EPI_ISL_1053987, EPI_ISL_1053989, EPI_ISL_1053990, EPI_ISL_1053991, EPI_ISL_1053992, EPI_ISL_1053993, EPI_ISL_1053994, EPI_ISL_1053995, EPI_ISL_1053996, EPI_ISL_1053998, EPI_ISL_1053999, EPI_ISL_1054001, EPI_ISL_1054002, EPI_ISL_1054004, EPI_ISL_1054005, EPI_ISL_1054006, EPI_ISL_1054007, EPI_ISL_1054009, EPI_ISL_1054010, EPI_ISL_1054012, EPI_ISL_1054013, EPI_ISL_1054014, EPI_ISL_1054015, EPI_ISL_1054016, EPI_ISL_1054018, EPI_ISL_1054019, EPI_ISL_1054020, EPI_ISL_1054022, EPI_ISL_1054023, EPI_ISL_1054024, EPI_ISL_1054025, EPI_ISL_1054026, EPI_ISL_1054027, EPI_ISL_1054028, EPI_ISL_1054029, EPI_ISL_1054030, EPI_ISL_1054031, EPI_ISL_1054032, EPI_ISL_1054033, EPI_ISL_1054034, EPI_ISL_1054035, EPI_ISL_1054036, EPI_ISL_1054037, EPI_ISL_1054040, EPI_ISL_1054042, EPI_ISL_1054043, EPI_ISL_1054044, EPI_ISL_1054045, EPI_ISL_1054046, EPI_ISL_1054047, EPI_ISL_1054048, EPI_ISL_1054049, EPI_ISL_1054050, EPI_ISL_1054051, EPI_ISL_1054053, EPI_ISL_1054054, EPI_ISL_1054055, EPI_ISL_1054056, EPI_ISL_1054057, EPI_ISL_1054058, EPI_ISL_1054059, EPI_ISL_1054060, EPI_ISL_1054065, EPI_ISL_1054066, EPI_ISL_1054067, EPI_ISL_1054069, EPI_ISL_1054070, EPI_ISL_1054071, EPI_ISL_1054072, EPI_ISL_1054073, EPI_ISL_1054074, EPI_ISL_1054075, EPI_ISL_1054076, EPI_ISL_1054077, EPI_ISL_1054078, EPI_ISL_1054079, EPI_ISL_1054081, EPI_ISL_1054082, EPI_ISL_1054083, EPI_ISL_1054084, EPI_ISL_1054085, EPI_ISL_1054086, EPI_ISL_1054087, EPI_ISL_1054088, EPI_ISL_1054089, EPI_ISL_1054090, EPI_ISL_1054091, EPI_ISL_1054092, EPI_ISL_1054093, EPI_ISL_1054094, EPI_ISL_1054096, EPI_ISL_1054097, EPI_ISL_1054098, EPI_ISL_1054099, EPI_ISL_1054100, EPI_ISL_1054101, EPI_ISL_1054102, EPI_ISL_1054103, EPI_ISL_1054104, EPI_ISL_1054105, EPI_ISL_1054106, EPI_ISL_1054107, EPI_ISL_1054110, EPI_ISL_1054111, EPI_ISL_1054112, EPI_ISL_1054114, EPI_ISL_1054115, EPI_ISL_1054116, EPI_ISL_1054117, EPI_ISL_1054118, EPI_ISL_1054120, EPI_ISL_1054121, EPI_ISL_1054122, EPI_ISL_1054123, EPI_ISL_1054125, EPI_ISL_1054126, EPI_ISL_1054127, EPI_ISL_1054128, EPI_ISL_1054130, EPI_ISL_1054462, EPI_ISL_1054463, EPI_ISL_1054464, EPI_ISL_1054465, EPI_ISL_1054466, EPI_ISL_1054467, EPI_ISL_1054469, EPI_ISL_1054470, EPI_ISL_1054472, EPI_ISL_1054473, EPI_ISL_1054474, EPI_ISL_1054475, EPI_ISL_1054476, EPI_ISL_1054478, EPI_ISL_1054479, EPI_ISL_1054480, EPI_ISL_1054481, EPI_ISL_1054482, EPI_ISL_1054483, EPI_ISL_1054484, EPI_ISL_1054485, EPI_ISL_1054486, EPI_ISL_1054487, EPI_ISL_1054488, EPI_ISL_1054489, EPI_ISL_1054490, EPI_ISL_1054491, EPI_ISL_1054492, EPI_ISL_1054493, EPI_ISL_1054494, EPI_ISL_1054495, EPI_ISL_1054496, EPI_ISL_1054497, EPI_ISL_1054498, EPI_ISL_1054499, EPI_ISL_1054500, EPI_ISL_1054501, EPI_ISL_1054502, EPI_ISL_1054503, EPI_ISL_1054504, EPI_ISL_1054505, EPI_ISL_1054506, EPI_ISL_1054507, EPI_ISL_1054508, EPI_ISL_1054510, EPI_ISL_1054511, EPI_ISL_1054512, EPI_ISL_1054513, EPI_ISL_1054514, EPI_ISL_1054515, EPI_ISL_1054516, EPI_ISL_1054517, EPI_ISL_1054518, EPI_ISL_1054519, EPI_ISL_1054520, EPI_ISL_1054521, EPI_ISL_1054522, EPI_ISL_1054523, EPI_ISL_1054524, EPI_ISL_1054525, EPI_ISL_1054526, EPI_ISL_1054527, EPI_ISL_1054528, EPI_ISL_1054529, EPI_ISL_1054530, EPI_ISL_1054532, EPI_ISL_1054533, EPI_ISL_1054534, EPI_ISL_1054535, EPI_ISL_1054536, EPI_ISL_1054537, EPI_ISL_1054538, EPI_ISL_1054539, EPI_ISL_1054540, EPI_ISL_1054541, EPI_ISL_1103757, EPI_ISL_1103792, EPI_ISL_1103797, EPI_ISL_1247526, EPI_ISL_1250123, EPI_ISL_1308567, EPI_ISL_1308568, EPI_ISL_1308579, EPI_ISL_1308642

| | | | |
|---|---|---|---|
| see above | University of Exeter | COVID-19 Genomics UK (COG-UK) Consortium | Aaron Jeffries; Audrey Farbos; Ben Temperton; Jane Masoli; Joanna Warwick-Dugdale; Michelle Michelsen; Robyn Manley; Stephen Michell |

EPI_ISL_997104, EPI_ISL_997105, EPI_ISL_997107, EPI_ISL_997108, EPI_ISL_997109, EPI_ISL_997110, EPI_ISL_997111, EPI_ISL_997126, EPI_ISL_997127, EPI_ISL_997128, EPI_ISL_997129, EPI_ISL_997130, EPI_ISL_997131, EPI_ISL_997164, EPI_ISL_1047861, EPI_ISL_1047862, EPI_ISL_1047863, EPI_ISL_1047864, EPI_ISL_1047865, EPI_ISL_1047892, EPI_ISL_1104223, EPI_ISL_1177709, EPI_ISL_1177710, EPI_ISL_1177720, EPI_ISL_1177721, EPI_ISL_1177722, EPI_ISL_1247691, EPI_ISL_1247692, EPI_ISL_1247728, EPI_ISL_1247730, EPI_ISL_1308706, EPI_ISL_1308822, EPI_ISL_1308841, EPI_ISL_1974838

| | | | |
|---|---|---|---|
| see above | Virology Department, Royal Infirmary of Edinburgh, NHS Lothian / School of Biological Sciences, University of Edinburgh | COVID-19 Genomics UK (COG-UK) Consortium | Colquhoun R; Cotton S; Dewar R; Gallagher A; Hill V; Jackson B; Maloney D; McCrone JT; McHugh M; O'Toole Á; Rambaut A; Rooke S; Scher E; Templeton K; Yu X |

EPI_ISL_924084, EPI_ISL_924101, EPI_ISL_924104, EPI_ISL_924108, EPI_ISL_924119, EPI_ISL_924122, EPI_ISL_924128, EPI_ISL_924148, EPI_ISL_924164, EPI_ISL_924176, EPI_ISL_924196, EPI_ISL_924197, EPI_ISL_924203, EPI_ISL_924215, EPI_ISL_924225, EPI_ISL_924240, EPI_ISL_924245, EPI_ISL_924252, EPI_ISL_924265, EPI_ISL_924274, EPI_ISL_924281, EPI_ISL_924296, EPI_ISL_924302, EPI_ISL_924328, EPI_ISL_924332, EPI_ISL_924334, EPI_ISL_924376, EPI_ISL_924384, EPI_ISL_924389, EPI_ISL_924392, EPI_ISL_924397, EPI_ISL_924406, EPI_ISL_952957, EPI_ISL_952959, EPI_ISL_952964, EPI_ISL_952972, EPI_ISL_952986, EPI_ISL_952988, EPI_ISL_952991, EPI_ISL_952994, EPI_ISL_953003, EPI_ISL_953006, EPI_ISL_953012, EPI_ISL_953013, EPI_ISL_953017, EPI_ISL_998109, EPI_ISL_998120, EPI_ISL_998121, EPI_ISL_998129, EPI_ISL_998134, EPI_ISL_1000653, EPI_ISL_1000687, EPI_ISL_1000695, EPI_ISL_1047582, EPI_ISL_1047583, EPI_ISL_1047584, EPI_ISL_1047585, EPI_ISL_1047586, EPI_ISL_1047587, EPI_ISL_1047588, EPI_ISL_1047589, EPI_ISL_1047590, EPI_ISL_1047591, EPI_ISL_1047592, EPI_ISL_1047593, EPI_ISL_1047595, EPI_ISL_1047597, EPI_ISL_1047598, EPI_ISL_1047599, EPI_ISL_1047600, EPI_ISL_1047601, EPI_ISL_1047602, EPI_ISL_1047603, EPI_ISL_1047604, EPI_ISL_1047605, EPI_ISL_1047606, EPI_ISL_1047607, EPI_ISL_1047608, EPI_ISL_1047609, EPI_ISL_1047610, EPI_ISL_1047611, EPI_ISL_1047612, EPI_ISL_1047613, EPI_ISL_1047614, EPI_ISL_1047615, EPI_ISL_1047616, EPI_ISL_1047617, EPI_ISL_1047618, EPI_ISL_1047619, EPI_ISL_1047620, EPI_ISL_1047621, EPI_ISL_1047622, EPI_ISL_1047623, EPI_ISL_1047624, EPI_ISL_1047625, EPI_ISL_1047626, EPI_ISL_1047627, EPI_ISL_1047628, EPI_ISL_1047629, EPI_ISL_1047630, EPI_ISL_1047631, EPI_ISL_1047632, EPI_ISL_1047634, EPI_ISL_1047635, EPI_ISL_1047636, EPI_ISL_1047637, EPI_ISL_1047638, EPI_ISL_1047639, EPI_ISL_1047640, EPI_ISL_1047641, EPI_ISL_1047642, EPI_ISL_1047643, EPI_ISL_1047644, EPI_ISL_1047645, EPI_ISL_1047646, EPI_ISL_1047719, EPI_ISL_1047720, EPI_ISL_1047721, EPI_ISL_1047743, EPI_ISL_1047754, EPI_ISL_1047755, EPI_ISL_1047756, EPI_ISL_1047757, EPI_ISL_1047758, EPI_ISL_1047759, EPI_ISL_1047781, EPI_ISL_1047782, EPI_ISL_1047783, EPI_ISL_1047784, EPI_ISL_1047797, EPI_ISL_1047798, EPI_ISL_1047799, EPI_ISL_1047800, EPI_ISL_1047801, EPI_ISL_1047802, EPI_ISL_1047803, EPI_ISL_1050066, EPI_ISL_1050067, EPI_ISL_1050887, EPI_ISL_1050892, EPI_ISL_1050894, EPI_ISL_1050905, EPI_ISL_1104025, EPI_ISL_1104026, EPI_ISL_1104027, EPI_ISL_1104028, EPI_ISL_1104029, EPI_ISL_1104030, EPI_ISL_1104031, EPI_ISL_1104032, EPI_ISL_1104033, EPI_ISL_1104034, EPI_ISL_1104035, EPI_ISL_1104036, EPI_ISL_1104037, EPI_ISL_1104038, EPI_ISL_1104039, EPI_ISL_1104040, EPI_ISL_1104048, EPI_ISL_1104049, EPI_ISL_1104050, EPI_ISL_1104051, EPI_ISL_1104056, EPI_ISL_1104070, EPI_ISL_1104071, EPI_ISL_1180059, EPI_ISL_1180089

| | | | |
|---|---|---|---|
| see above | Virology Department, Sheffield Teaching Hospitals NHS Foundation Trust/Department of Infection, Immunity and Cardiovascular Disease, The Medical School, University of Sheffield | COVID-19 Genomics UK (COG-UK) Consortium | Adri Angyal; Alex Keeley; Benjamin Lindsey; Cariad Evans; Danielle Groves; Dave Partridge; Katie Johnson; Laura Carrilero; Luke Green; Matthew Parker; Matthew Wyles; Mehmet Yavuz; Mohammad Raza; Nikki Smith; Paul Parsons; Rachel Tucker; Rebecca Brown; Thushan de Silva |

EPI_ISL_1052142, EPI_ISL_1052144, EPI_ISL_1052185, EPI_ISL_1052194, EPI_ISL_1052264, EPI_ISL_1052265, EPI_ISL_1052266, EPI_ISL_1052267, EPI_ISL_1052269, EPI_ISL_1052272, EPI_ISL_1052469, EPI_ISL_1052471, EPI_ISL_1053490, EPI_ISL_1053503, EPI_ISL_1053504, EPI_ISL_1053514, EPI_ISL_1053546, EPI_ISL_1053566, EPI_ISL_1053576, EPI_ISL_1053577, EPI_ISL_1053580, EPI_ISL_1053582, EPI_ISL_1053583, EPI_ISL_1053584, EPI_ISL_1053585, EPI_ISL_1833157, EPI_ISL_1833165, EPI_ISL_1833166, EPI_ISL_1833167, EPI_ISL_1833170, EPI_ISL_1833171, EPI_ISL_2127726

| | | | |
|---|---|---|---|
| see above | Wales Specialist Virology Centre Sequencing lab: Pathogen Genomics Unit | Public Health Wales Microbiology Cardiff Wales Specialist Virology Centre | Alec Birchley; Alexander Adams; Amy Gaskin; Angela Marchbank; Bree Gatica-Wilcox; Catherine Moore; Jason Coombes; Joanne Watkins; Joel Southgate; Johnathan Evans; Laura Gifford; Lauren Gilbert; Lee Graham; Malorie Perry; Matthew Bull; Nicole Pacchiarini; Sally Corden; Sara Kumziene-Summerhayes; Sara Rey; Sarah Taylor; Simon Cottrell; Sophie Jones; Tom Connor |

EPI_ISL_949613, EPI_ISL_949616, EPI_ISL_949617, EPI_ISL_949619, EPI_ISL_949621, EPI_ISL_949623, EPI_ISL_949625, EPI_ISL_949627, EPI_ISL_949629, EPI_ISL_997063, EPI_ISL_997068, EPI_ISL_997069, EPI_ISL_1047804, EPI_ISL_1047805, EPI_ISL_1047806, EPI_ISL_1047807, EPI_ISL_1047808, EPI_ISL_1047809, EPI_ISL_1047810, EPI_ISL_1047811, EPI_ISL_1047812, EPI_ISL_1047813, EPI_ISL_1047814, EPI_ISL_1047815, EPI_ISL_1047816, EPI_ISL_1047817, EPI_ISL_1047818, EPI_ISL_1047819, EPI_ISL_1047820, EPI_ISL_1047821, EPI_ISL_1047822, EPI_ISL_1047823, EPI_ISL_1047824, EPI_ISL_1047825, EPI_ISL_1047826, EPI_ISL_1047827, EPI_ISL_1047828, EPI_ISL_1047829, EPI_ISL_1047830, EPI_ISL_1047831, EPI_ISL_1047832, EPI_ISL_1047833, EPI_ISL_1050174, EPI_ISL_1050219, EPI_ISL_1050220, EPI_ISL_1050222, EPI_ISL_2125500

| | | | |
|---|---|---|---|
| see above | West of Scotland Specialist Virology Centre, NHSGGC / MRC-University of Glasgow Centre for Virus Research | COVID-19 Genomics UK (COG-UK) Consortium | Alasdair MacLean; Alice Broos; Ana da Silva Filipe; Antonia Ho; Daniel Mair; David L Robertson; Emma Thomson; Guy Mollett; Ioulia Tsatsani; James Shepherd; Jenna Nichols; Joseph Hughes; Kathy Li; Kathy Smollett; Kyriaki Nomikou; Lily Tong; Matthew Holden; Natasha Johnson; Rachel Blacow; Richard Orton; Rory Gunson; Sarah McDonald; Sharif Shaaban; Sreenu Vattipally; Stephen Carmichael |

We gratefully acknowledge the following Authors from the Originating laboratories responsible for obtaining the specimens, as well as the Submitting laboratories where the genome data were generated and shared via GISAID, on which this research is based.

All Submitters of data may be contacted directly via www.gisaid.org

Authors are sorted alphabetically.

| Accession ID | Originating Laboratory | Submitting Laboratory | Authors |
|---|---|---|---|
| EPI_ISL_997661, EPI_ISL_997668, EPI_ISL_1048053, EPI_ISL_1048061, EPI_ISL_1048072, EPI_ISL_1048074, EPI_ISL_1048077, EPI_ISL_1048081, EPI_ISL_1048085, EPI_ISL_1048091, EPI_ISL_1048092, EPI_ISL_1048093, EPI_ISL_1048098, EPI_ISL_1048099, EPI_ISL_1048100, EPI_ISL_1048101, EPI_ISL_1048129, EPI_ISL_1177990 | | | |
| see above | Barts Health NHS Trust | COVID-19 Genomics UK (COG-UK) Consortium | BROAD; Beatrix; CUTINO-MOGUEL; Claire; David; Dola; HARRINGTON; KELE; KULASEGARAN-SHYLINI; Maria-Teresa; OWOYEMI; Raghavendran |
| EPI_ISL_1000838, EPI_ISL_1000839, EPI_ISL_1000840, EPI_ISL_1000841, EPI_ISL_1000842, EPI_ISL_1000843, EPI_ISL_1000844, EPI_ISL_1000846, EPI_ISL_1000847, EPI_ISL_1000848, EPI_ISL_1000849, EPI_ISL_1000850, EPI_ISL_1000851, EPI_ISL_1000852, EPI_ISL_1000853, EPI_ISL_1000854, EPI_ISL_1000855, EPI_ISL_1000856, EPI_ISL_1000857, EPI_ISL_1000858, EPI_ISL_1000859, EPI_ISL_1000860, EPI_ISL_1000861, EPI_ISL_1000862, EPI_ISL_1000863, EPI_ISL_1000865, EPI_ISL_1000866, EPI_ISL_1000867, EPI_ISL_1000868, EPI_ISL_1000869, EPI_ISL_1000870, EPI_ISL_1000871, EPI_ISL_1000872, EPI_ISL_1000873, EPI_ISL_1000874, EPI_ISL_1000875, EPI_ISL_1000876, EPI_ISL_1000877, EPI_ISL_1000878, EPI_ISL_1000879, EPI_ISL_1000880, EPI_ISL_1000917, EPI_ISL_1000918, EPI_ISL_1000921, EPI_ISL_1000924, EPI_ISL_1000925, EPI_ISL_1000927, EPI_ISL_1000928, EPI_ISL_1000929, EPI_ISL_1000930, EPI_ISL_1000931, EPI_ISL_1000932, EPI_ISL_1000935, EPI_ISL_1054757, EPI_ISL_1054759, EPI_ISL_1054761, EPI_ISL_1054762, EPI_ISL_1054765, EPI_ISL_1054766, EPI_ISL_1054767, EPI_ISL_1054772, EPI_ISL_1054773, EPI_ISL_1054774, EPI_ISL_1054787, EPI_ISL_1054832 | | | |
| see above | Bioinformatics and Biostatistics Lab, Advanced Sequencing Facility | COVID-19 Genomics UK (COG-UK) Consortium | Aengus Stewart; Chelsea Sawyer; Harshil Patel; Jerome Nicod; Laura Cubitt; Margaret Crawford |
| EPI_ISL_1832335 | Centre for Clinical Infection and Diagnostics Research and Genomics Innovation Unit, Guy's and St. Thomas' NHS Trust | COVID-19 Genomics UK (COG-UK) Consortium | Ali Raza Awan; Chloe Fisher; Jonathan Edgeworth; Luke Snell; Penny Cliff; Rahul Batra |
| EPI_ISL_952486, EPI_ISL_952487, EPI_ISL_1000187, EPI_ISL_1000188, EPI_ISL_1000189, EPI_ISL_1000190, EPI_ISL_1000192, EPI_ISL_1000193, EPI_ISL_1000194, EPI_ISL_1000195, EPI_ISL_1000196, EPI_ISL_1000197, EPI_ISL_1000198, EPI_ISL_1000199, EPI_ISL_1000200, EPI_ISL_1000201, EPI_ISL_1000202, EPI_ISL_1000203, EPI_ISL_1000213, EPI_ISL_1000218, EPI_ISL_1000219, EPI_ISL_1000291, EPI_ISL_1000297, EPI_ISL_1000298, EPI_ISL_1000307, EPI_ISL_1000309, EPI_ISL_1000310, EPI_ISL_1000311, EPI_ISL_1000312, EPI_ISL_1000317, EPI_ISL_1000318, EPI_ISL_1000319, EPI_ISL_1000320, EPI_ISL_1000321, EPI_ISL_1000322, EPI_ISL_1000324, EPI_ISL_1000466, EPI_ISL_1000467, EPI_ISL_1000468, EPI_ISL_1000469, EPI_ISL_1000470, EPI_ISL_1000477, EPI_ISL_1000573, EPI_ISL_1000576, EPI_ISL_1000611, EPI_ISL_1000628, EPI_ISL_1000629, EPI_ISL_1000630, EPI_ISL_1050748, EPI_ISL_1050749, EPI_ISL_1050750, EPI_ISL_1050751, EPI_ISL_1050752, EPI_ISL_1050753, EPI_ISL_1050754, EPI_ISL_1050755, EPI_ISL_1050756, EPI_ISL_1050757, EPI_ISL_1050758, EPI_ISL_1050759, EPI_ISL_1050760, EPI_ISL_1050761, EPI_ISL_1050762, EPI_ISL_1050763, EPI_ISL_1050764, EPI_ISL_1050765, EPI_ISL_1050766, EPI_ISL_1050767, EPI_ISL_1050768, EPI_ISL_1050769, EPI_ISL_1050770, EPI_ISL_1050771, EPI_ISL_1050772, EPI_ISL_1050773, EPI_ISL_1050774, EPI_ISL_1050775, EPI_ISL_1050776, EPI_ISL_1050777, EPI_ISL_1050778, EPI_ISL_1050779, EPI_ISL_1050780, EPI_ISL_1050781, EPI_ISL_1050782, EPI_ISL_1050783, EPI_ISL_1050784, EPI_ISL_1050785, EPI_ISL_1050786, EPI_ISL_1050787, EPI_ISL_1050788, EPI_ISL_1050789, EPI_ISL_1050790, EPI_ISL_1050791, EPI_ISL_1050792, EPI_ISL_1050794, EPI_ISL_1050795, EPI_ISL_1050796, EPI_ISL_1050797, EPI_ISL_1050798, EPI_ISL_1050800, EPI_ISL_1050801, EPI_ISL_1050802, EPI_ISL_1050803, EPI_ISL_1050804, EPI_ISL_1050805, EPI_ISL_1050806, EPI_ISL_1050807, EPI_ISL_1050808, EPI_ISL_1050809, EPI_ISL_1050810, EPI_ISL_1050811, EPI_ISL_1050812, EPI_ISL_1050813, EPI_ISL_1050814, EPI_ISL_1050815, EPI_ISL_1050819, EPI_ISL_1050820, EPI_ISL_1050821, EPI_ISL_1050822, EPI_ISL_1050823, EPI_ISL_1050824, EPI_ISL_1050825, EPI_ISL_1050826, EPI_ISL_1050827, EPI_ISL_1050835, EPI_ISL_1050836, EPI_ISL_1050837, EPI_ISL_1050838, EPI_ISL_1050839, EPI_ISL_1050842, EPI_ISL_1050843, EPI_ISL_1050844, EPI_ISL_1050845, EPI_ISL_1050846, EPI_ISL_1050847, EPI_ISL_1050848, EPI_ISL_1050849, EPI_ISL_1050850, EPI_ISL_1050851, EPI_ISL_1050852, EPI_ISL_1050853, EPI_ISL_1050855, EPI_ISL_1050856, EPI_ISL_1050857, EPI_ISL_1050858, EPI_ISL_1050859, EPI_ISL_1050860, EPI_ISL_1050861, EPI_ISL_1050871, EPI_ISL_1050872, EPI_ISL_1050873, EPI_ISL_1050875, EPI_ISL_1050876, EPI_ISL_1050877, EPI_ISL_1050878, EPI_ISL_1050881, EPI_ISL_1050882, EPI_ISL_1050909, EPI_ISL_1050910, EPI_ISL_1050912, EPI_ISL_1050913, EPI_ISL_1050914, EPI_ISL_1053749, EPI_ISL_1053750, EPI_ISL_1053821, EPI_ISL_1053823, EPI_ISL_1053858, EPI_ISL_1053859, EPI_ISL_1053860, EPI_ISL_1053938, EPI_ISL_1053939, EPI_ISL_1053940, EPI_ISL_1053941, EPI_ISL_1053942, EPI_ISL_1053943, EPI_ISL_1053944, EPI_ISL_1053945, EPI_ISL_1053946, EPI_ISL_1053947, EPI_ISL_1053950, EPI_ISL_1053953, EPI_ISL_1053954, EPI_ISL_1053957, EPI_ISL_1053958, EPI_ISL_1053959, EPI_ISL_1053960, EPI_ISL_1053965, EPI_ISL_1053966, EPI_ISL_1053967, EPI_ISL_1053968, EPI_ISL_1053969, EPI_ISL_1053970, EPI_ISL_1053971, EPI_ISL_1054572, EPI_ISL_1105494, EPI_ISL_1105495, EPI_ISL_1105511, EPI_ISL_1105512, EPI_ISL_1107586, EPI_ISL_1107638, EPI_ISL_1107639, EPI_ISL_1107640, EPI_ISL_1107641, EPI_ISL_1107664, EPI_ISL_1107665, EPI_ISL_1107666, EPI_ISL_1107667, EPI_ISL_1107668, EPI_ISL_1107789, EPI_ISL_1109230, EPI_ISL_1179827, EPI_ISL_1388736, EPI_ISL_1388738, EPI_ISL_1388739, EPI_ISL_1730879, EPI_ISL_1979789 | | | |
| see above | Centre for Enzyme Innovation, University of Portsmouth / Translational Research Laboratory, Portsmouth Hospitals NHS Trust | COVID-19 Genomics UK (COG-UK) Consortium | Angela Beckett; Christopher Fearn; Kate Cook; Katie Loveson; Salman Goudarzi; Samuel Robson; Scott Elliott; Sharon Glaysher |
| EPI_ISL_996604, EPI_ISL_996605, EPI_ISL_996606, EPI_ISL_996607, EPI_ISL_996608, EPI_ISL_996609, EPI_ISL_996610, EPI_ISL_996611, EPI_ISL_996612, EPI_ISL_996614, EPI_ISL_996637, EPI_ISL_996638, EPI_ISL_996639, EPI_ISL_996640, EPI_ISL_996641, EPI_ISL_996642, EPI_ISL_996643, EPI_ISL_996645, EPI_ISL_996646, EPI_ISL_996647, EPI_ISL_996648, EPI_ISL_996649, EPI_ISL_996650, EPI_ISL_996651, EPI_ISL_996652, EPI_ISL_996654, EPI_ISL_996655, EPI_ISL_996656, EPI_ISL_996657, EPI_ISL_996658, EPI_ISL_996659, EPI_ISL_996660, EPI_ISL_996661, EPI_ISL_996662, EPI_ISL_996663, EPI_ISL_996664, EPI_ISL_996665, EPI_ISL_996666, EPI_ISL_996667, EPI_ISL_996668, EPI_ISL_996669, EPI_ISL_996670, EPI_ISL_996671, EPI_ISL_996743, EPI_ISL_996744, EPI_ISL_996745, EPI_ISL_996746, EPI_ISL_996747, EPI_ISL_996748, EPI_ISL_996749, EPI_ISL_996750, EPI_ISL_996751, EPI_ISL_996752, EPI_ISL_996753, EPI_ISL_996754, EPI_ISL_996755, EPI_ISL_996756, EPI_ISL_996757, EPI_ISL_996758, EPI_ISL_996759, EPI_ISL_996760, EPI_ISL_996761, EPI_ISL_996762, EPI_ISL_996763, EPI_ISL_996764, EPI_ISL_996765, EPI_ISL_996766, EPI_ISL_996767, EPI_ISL_996768, EPI_ISL_996769, EPI_ISL_996770, EPI_ISL_996771, EPI_ISL_996772, EPI_ISL_996773, EPI_ISL_996774, EPI_ISL_996775, EPI_ISL_996776, EPI_ISL_996777, EPI_ISL_996779, EPI_ISL_996780, EPI_ISL_996781, EPI_ISL_996782, EPI_ISL_996783, EPI_ISL_996784, EPI_ISL_996785, EPI_ISL_996786, EPI_ISL_996794, EPI_ISL_996801, EPI_ISL_996813, EPI_ISL_996815, EPI_ISL_996817, EPI_ISL_996819, EPI_ISL_996820, EPI_ISL_996822, EPI_ISL_996825, EPI_ISL_996828, EPI_ISL_996832, EPI_ISL_996833, EPI_ISL_996835, EPI_ISL_996836, EPI_ISL_1047070, EPI_ISL_1047072, EPI_ISL_1047073, EPI_ISL_1047074, EPI_ISL_1047075, EPI_ISL_1047076, EPI_ISL_1047077, EPI_ISL_1047080, EPI_ISL_1047082, EPI_ISL_1047083, EPI_ISL_1047084, EPI_ISL_1047086, EPI_ISL_1047087, EPI_ISL_1047088, EPI_ISL_1047090, EPI_ISL_1047091, EPI_ISL_1054259, EPI_ISL_1054265, EPI_ISL_1054292, EPI_ISL_1054326, EPI_ISL_1054327, EPI_ISL_1054352, EPI_ISL_1103893, EPI_ISL_1107804 | | | |
| see above | Department of Pathology, University of Cambridge | COVID-19 Genomics UK (COG-UK) Consortium | Aminu S. Jahun; Ian Goodfellow; Iliana Georgana; Martin D. Curran; Myra Hosmillo; Rhys Izuagbe; Surendra Parmar; William L. Hamilton; Yasmin Chaudhry |
| EPI_ISL_962978, EPI_ISL_962979, EPI_ISL_962987, EPI_ISL_962989, EPI_ISL_962995, EPI_ISL_962997, EPI_ISL_962999, EPI_ISL_963004, EPI_ISL_963006, EPI_ISL_963007, EPI_ISL_963008, EPI_ISL_963011, EPI_ISL_963012, EPI_ISL_963013, EPI_ISL_963016, EPI_ISL_963019, EPI_ISL_963022, EPI_ISL_963026, EPI_ISL_963027, EPI_ISL_963030, EPI_ISL_963032, EPI_ISL_963038, EPI_ISL_963040, EPI_ISL_963042, EPI_ISL_963044, EPI_ISL_963048, EPI_ISL_963049, EPI_ISL_963056, EPI_ISL_963058, EPI_ISL_963059, EPI_ISL_963060, EPI_ISL_963068, EPI_ISL_963070, EPI_ISL_963071, EPI_ISL_963072, EPI_ISL_963079, EPI_ISL_963081, EPI_ISL_963085, EPI_ISL_963088, EPI_ISL_963090, EPI_ISL_963092, EPI_ISL_963094, EPI_ISL_963098, EPI_ISL_963102, EPI_ISL_963105, EPI_ISL_963110, EPI_ISL_963111, EPI_ISL_963112, EPI_ISL_963115, EPI_ISL_963119, EPI_ISL_963123, EPI_ISL_963124, EPI_ISL_963127, EPI_ISL_963129, EPI_ISL_963130, EPI_ISL_963133, EPI_ISL_963134, EPI_ISL_963137, EPI_ISL_963140, EPI_ISL_963141, EPI_ISL_963142, EPI_ISL_963144, EPI_ISL_963146, EPI_ISL_963150, EPI_ISL_963152, EPI_ISL_963155, EPI_ISL_963156, EPI_ISL_963161, EPI_ISL_963163, EPI_ISL_963164, EPI_ISL_963166, EPI_ISL_963171, EPI_ISL_963173, EPI_ISL_963175, EPI_ISL_963176, EPI_ISL_963178, EPI_ISL_963182, EPI_ISL_963183, EPI_ISL_963184, EPI_ISL_963188, EPI_ISL_963194, EPI_ISL_963195, EPI_ISL_963199, EPI_ISL_963204, EPI_ISL_963205, EPI_ISL_963216, EPI_ISL_963219, EPI_ISL_963222, EPI_ISL_963223, EPI_ISL_963226, EPI_ISL_963229, EPI_ISL_963234, EPI_ISL_963238, EPI_ISL_963243, EPI_ISL_963244, EPI_ISL_963246, EPI_ISL_963251, EPI_ISL_963257, EPI_ISL_963258, EPI_ISL_963262, EPI_ISL_963265, EPI_ISL_963275, EPI_ISL_963279, EPI_ISL_963284, EPI_ISL_963287, EPI_ISL_963288, EPI_ISL_963289, EPI_ISL_963290, EPI_ISL_963291, EPI_ISL_963293, EPI_ISL_963294, EPI_ISL_963297, EPI_ISL_963298, EPI_ISL_963300, EPI_ISL_963302, EPI_ISL_963303, EPI_ISL_963304, EPI_ISL_963305, EPI_ISL_963311, EPI_ISL_963315, EPI_ISL_963319, EPI_ISL_963320, EPI_ISL_964559, EPI_ISL_964837, EPI_ISL_985613, EPI_ISL_985614, EPI_ISL_985616, EPI_ISL_985618, EPI_ISL_985619, EPI_ISL_985620, EPI_ISL_985622, EPI_ISL_985623, EPI_ISL_985624, EPI_ISL_985626, EPI_ISL_985627, EPI_ISL_985628, EPI_ISL_985631, EPI_ISL_985632, EPI_ISL_985634, EPI_ISL_985635, EPI_ISL_985637, EPI_ISL_985638, EPI_ISL_985639, EPI_ISL_985640, EPI_ISL_985643, EPI_ISL_985644, EPI_ISL_985645, EPI_ISL_985646, EPI_ISL_985647, EPI_ISL_985648, EPI_ISL_985649, EPI_ISL_985650, EPI_ISL_985651, EPI_ISL_985652, EPI_ISL_985653, EPI_ISL_985654, EPI_ISL_985655, EPI_ISL_985656, EPI_ISL_985657, EPI_ISL_985658, EPI_ISL_985659, EPI_ISL_985660, EPI_ISL_985661, EPI_ISL_985662, EPI_ISL_985663, EPI_ISL_985664, EPI_ISL_985665, EPI_ISL_985666, EPI_ISL_985667, EPI_ISL_985668, EPI_ISL_985669, EPI_ISL_985670, EPI_ISL_985671, EPI_ISL_985672, EPI_ISL_985673, EPI_ISL_985674, EPI_ISL_985676, EPI_ISL_985677, EPI_ISL_985680, EPI_ISL_985681, EPI_ISL_985682, EPI_ISL_985683, EPI_ISL_985684, EPI_ISL_985685, EPI_ISL_985686, EPI_ISL_985687, EPI_ISL_985689, EPI_ISL_985691, EPI_ISL_985692, EPI_ISL_985693, EPI_ISL_985694, EPI_ISL_985695, EPI_ISL_985697, EPI_ISL_985698, EPI_ISL_985699, EPI_ISL_985700, EPI_ISL_985702, EPI_ISL_985703, EPI_ISL_985704, EPI_ISL_985705, EPI_ISL_985706, EPI_ISL_985708, EPI_ISL_985709, EPI_ISL_985710, EPI_ISL_985712, EPI_ISL_985713, EPI_ISL_985714, EPI_ISL_985717, EPI_ISL_985718, EPI_ISL_985719, EPI_ISL_985721, EPI_ISL_985722, EPI_ISL_985723, EPI_ISL_985724, EPI_ISL_985725, EPI_ISL_985726, EPI_ISL_985727, EPI_ISL_985729, EPI_ISL_985730, EPI_ISL_985731, EPI_ISL_985732, EPI_ISL_985734, EPI_ISL_985735, EPI_ISL_985736, EPI_ISL_985739, EPI_ISL_985740, EPI_ISL_985741, EPI_ISL_985742, EPI_ISL_985743, EPI_ISL_985744, EPI_ISL_985745, EPI_ISL_985747, EPI_ISL_985748, EPI_ISL_985749, EPI_ISL_985750, EPI_ISL_985751, EPI_ISL_985752, EPI_ISL_985753, EPI_ISL_985754, EPI_ISL_985755, EPI_ISL_985756, EPI_ISL_985757, EPI_ISL_985758, EPI_ISL_985760, EPI_ISL_985761, EPI_ISL_985762, EPI_ISL_985763, EPI_ISL_985764, EPI_ISL_985765, EPI_ISL_985766, EPI_ISL_985767, EPI_ISL_985768, EPI_ISL_985769, EPI_ISL_985770, EPI_ISL_985771, EPI_ISL_985772, EPI_ISL_985775, EPI_ISL_985776, EPI_ISL_985777, EPI_ISL_985778, EPI_ISL_985779, EPI_ISL_985780, EPI_ISL_985781, EPI_ISL_985783, EPI_ISL_985784, EPI_ISL_985787, EPI_ISL_985789, EPI_ISL_985790, EPI_ISL_985791, EPI_ISL_985792, EPI_ISL_985794, EPI_ISL_985796, EPI_ISL_985797, EPI_ISL_985798, EPI_ISL_985799, EPI_ISL_985802, EPI_ISL_985804, EPI_ISL_985808, EPI_ISL_985809, EPI_ISL_985811, EPI_ISL_985812, EPI_ISL_985813, EPI_ISL_985814, EPI_ISL_985815, EPI_ISL_985817, EPI_ISL_985818, EPI_ISL_985820, EPI_ISL_985821, EPI_ISL_985822, EPI_ISL_985823, EPI_ISL_985824, EPI_ISL_985825, EPI_ISL_985826, EPI_ISL_985827, EPI_ISL_985828, EPI_ISL_985829, EPI_ISL_985830, EPI_ISL_985831, EPI_ISL_985832, EPI_ISL_985833, EPI_ISL_985834, EPI_ISL_985835, EPI_ISL_985836, EPI_ISL_985837, EPI_ISL_985838, EPI_ISL_985839, EPI_ISL_985840, EPI_ISL_985841, EPI_ISL_985842, EPI_ISL_985844, EPI_ISL_985845, EPI_ISL_985847, EPI_ISL_985848, EPI_ISL_985849, EPI_ISL_985850, EPI_ISL_985851, EPI_ISL_985852, EPI_ISL_985853, EPI_ISL_985854, EPI_ISL_985855, EPI_ISL_985856, EPI_ISL_985858, EPI_ISL_985859, EPI_ISL_985861, EPI_ISL_985862, EPI_ISL_985864, EPI_ISL_985865, EPI_ISL_985869, EPI_ISL_985871, EPI_ISL_985873, EPI_ISL_985875, EPI_ISL_985876, EPI_ISL_985877, EPI_ISL_985878, EPI_ISL_985879, EPI_ISL_985881, EPI_ISL_985882, EPI_ISL_985883, EPI_ISL_985884, EPI_ISL_985885, EPI_ISL_985886, EPI_ISL_985887, EPI_ISL_985889, EPI_ISL_985890, EPI_ISL_985892, EPI_ISL_985893, EPI_ISL_985894, EPI_ISL_985895, EPI_ISL_985896, EPI_ISL_985897, EPI_ISL_985898, EPI_ISL_985899, EPI_ISL_985900, EPI_ISL_985901, EPI_ISL_985902, EPI_ISL_985904, EPI_ISL_985905, EPI_ISL_985906, EPI_ISL_985907, EPI_ISL_985909, EPI_ISL_985910, EPI_ISL_985911, EPI_ISL_985912, EPI_ISL_985913, EPI_ISL_985915, EPI_ISL_985916, EPI_ISL_985917, EPI_ISL_985918, EPI_ISL_985919, EPI_ISL_985920, EPI_ISL_985921, EPI_ISL_985922, EPI_ISL_985923, EPI_ISL_985924, EPI_ISL_985925, EPI_ISL_985926, EPI_ISL_985927, EPI_ISL_985928, EPI_ISL_985929, EPI_ISL_985930, EPI_ISL_985931, EPI_ISL_985932, EPI_ISL_985933, EPI_ISL_985934, EPI_ISL_985935, EPI_ISL_985936, EPI_ISL_985937, EPI_ISL_985938, EPI_ISL_985940, EPI_ISL_985941, EPI_ISL_985942, EPI_ISL_985945, EPI_ISL_986824, EPI_ISL_986825, EPI_ISL_986826, EPI_ISL_986827, EPI_ISL_986828, EPI_ISL_986829, EPI_ISL_986830, EPI_ISL_986831, EPI_ISL_986832, EPI_ISL_986833, EPI_ISL_986834, EPI_ISL_986835, EPI_ISL_986836, EPI_ISL_986837, EPI_ISL_986838, EPI_ISL_986839, EPI_ISL_986840, EPI_ISL_986841, EPI_ISL_986842, EPI_ISL_986843, EPI_ISL_986844, EPI_ISL_986845, EPI_ISL_986846, EPI_ISL_986847, EPI_ISL_986848, EPI_ISL_986849, EPI_ISL_986850, EPI_ISL_986851, EPI_ISL_986852, EPI_ISL_986853, EPI_ISL_986855, EPI_ISL_986856, EPI_ISL_986857, EPI_ISL_986858, EPI_ISL_986859, EPI_ISL_986860, EPI_ISL_986861, EPI_ISL_986862, EPI_ISL_986863, EPI_ISL_986864, EPI_ISL_986865, EPI_ISL_986866, EPI_ISL_986867, EPI_ISL_986868, EPI_ISL_986869, EPI_ISL_986870, EPI_ISL_986871, EPI_ISL_986872, EPI_ISL_986873, EPI_ISL_986874, EPI_ISL_986875, EPI_ISL_986876, EPI_ISL_986877, EPI_ISL_986878, EPI_ISL_986879, EPI_ISL_986880, EPI_ISL_986881, EPI_ISL_986882, EPI_ISL_986883, EPI_ISL_986884, EPI_ISL_986885, EPI_ISL_986886, EPI_ISL_986887, EPI_ISL_986888, EPI_ISL_986889, EPI_ISL_986890, EPI_ISL_986891, EPI_ISL_986892, EPI_ISL_986893, EPI_ISL_986894, EPI_ISL_986896, EPI_ISL_986897, EPI_ISL_986898, EPI_ISL_986899, EPI_ISL_986901, EPI_ISL_986902, EPI_ISL_986903, EPI_ISL_986905, EPI_ISL_986906, EPI_ISL_986907, EPI_ISL_986908, EPI_ISL_986909, EPI_ISL_986910, EPI_ISL_986911, EPI_ISL_986913, EPI_ISL_986915, EPI_ISL_986916, EPI_ISL_986917, EPI_ISL_986919, EPI_ISL_986920, EPI_ISL_986921, EPI_ISL_986922, EPI_ISL_986923, EPI_ISL_986924, EPI_ISL_986925, EPI_ISL_986926, EPI_ISL_986927, EPI_ISL_986928, EPI_ISL_986929, EPI_ISL_986930, EPI_ISL_986931, EPI_ISL_986932, EPI_ISL_986933, EPI_ISL_986934, EPI_ISL_986935, EPI_ISL_986936, EPI_ISL_986938, EPI_ISL_986939, EPI_ISL_986940, EPI_ISL_986941, EPI_ISL_986942, EPI_ISL_986943, EPI_ISL_986944, EPI_ISL_986945, EPI_ISL_986946, EPI_ISL_986947, EPI_ISL_986948, EPI_ISL_986949, EPI_ISL_986950, EPI_ISL_986951, EPI_ISL_986952, EPI_ISL_986953, EPI_ISL_986954, EPI_ISL_986955, EPI_ISL_986956, EPI_ISL_986957, EPI_ISL_986958, EPI_ISL_986959, EPI_ISL_986960, EPI_ISL_986961, EPI_ISL_986962, EPI_ISL_986963, EPI_ISL_986964, EPI_ISL_986965, EPI_ISL_986966, EPI_ISL_986967, EPI_ISL_986968, EPI_ISL_986969, EPI_ISL_986970, EPI_ISL_986971, EPI_ISL_986972, EPI_ISL_986973, EPI_ISL_986974, EPI_ISL_986975, EPI_ISL_986976, EPI_ISL_986977, EPI_ISL_986978, EPI_ISL_986979, EPI_ISL_986980, EPI_ISL_986981, EPI_ISL_986982, EPI_ISL_986983, EPI_ISL_986984, EPI_ISL_986985, EPI_ISL_986986, EPI_ISL_986987, EPI_ISL_986988, EPI_ISL_986989, EPI_ISL_986990, EPI_ISL_986991, EPI_ISL_986992, EPI_ISL_986993, EPI_ISL_986994, EPI_ISL_986995, EPI_ISL_986996, EPI_ISL_986997, EPI_ISL_986998, EPI_ISL_986999, EPI_ISL_987000, EPI_ISL_987001, EPI_ISL_987003, EPI_ISL_987004, EPI_ISL_987006, EPI_ISL_987007, EPI_ISL_987008, EPI_ISL_987009, EPI_ISL_987010, EPI_ISL_987011, EPI_ISL_987012, EPI_ISL_987013, EPI_ISL_987014, EPI_ISL_987015, EPI_ISL_987016, EPI_ISL_987017, EPI_ISL_987018, EPI_ISL_987019, EPI_ISL_987020, EPI_ISL_987021, EPI_ISL_987022, EPI_ISL_987023, EPI_ISL_987024, EPI_ISL_987025, EPI_ISL_987026, EPI_ISL_987027, EPI_ISL_987028, EPI_ISL_987029, EPI_ISL_987030, EPI_ISL_987031, EPI_ISL_987032, EPI_ISL_987033, EPI_ISL_987034, EPI_ISL_987035, EPI_ISL_987036, EPI_ISL_987037, EPI_ISL_987038, EPI_ISL_987039, EPI_ISL_987040, EPI_ISL_987041, EPI_ISL_987042, EPI_ISL_987043, EPI_ISL_987044, EPI_ISL_987046, EPI_ISL_987047, EPI_ISL_987048, EPI_ISL_987049, EPI_ISL_987050, EPI_ISL_987051, EPI_ISL_987052, EPI_ISL_987055, EPI_ISL_987056, EPI_ISL_987057, EPI_ISL_987058, EPI_ISL_987059, EPI_ISL_987060, EPI_ISL_987061, EPI_ISL_987062, EPI_ISL_987063, EPI_ISL_987064, EPI_ISL_987065, EPI_ISL_987066, EPI_ISL_987067, EPI_ISL_987068, EPI_ISL_987069, EPI_ISL_987070, EPI_ISL_987071, EPI_ISL_987072, EPI_ISL_987073, EPI_ISL_987074, EPI_ISL_987075, EPI_ISL_987076, EPI_ISL_987077, EPI_ISL_987078, EPI_ISL_987079, EPI_ISL_987080, EPI_ISL_987081, EPI_ISL_987082, EPI_ISL_987083, EPI_ISL_987084, EPI_ISL_987085, EPI_ISL_987086, EPI_ISL_987087, EPI_ISL_987088, EPI_ISL_987089, EPI_ISL_987090, EPI_ISL_987091, EPI_ISL_987092, EPI_ISL_987093, EPI_ISL_987094, EPI_ISL_987095, EPI_ISL_987096, EPI_ISL_987098, EPI_ISL_987101, EPI_ISL_987102, EPI_ISL_987103, EPI_ISL_987106, EPI_ISL_987108, EPI_ISL_987109, EPI_ISL_987110, EPI_ISL_987111, EPI_ISL_987113, EPI_ISL_987114, EPI_ISL_987115, EPI_ISL_987116, EPI_ISL_987117, EPI_ISL_987118, EPI_ISL_987119, EPI_ISL_987120, EPI_ISL_987121, EPI_ISL_987122, EPI_ISL_987123, EPI_ISL_987125, EPI_ISL_987126, EPI_ISL_987127, EPI_ISL_987128, EPI_ISL_987129, EPI_ISL_987130, EPI_ISL_987131, EPI_ISL_987132, EPI_ISL_987133, EPI_ISL_987135, EPI_ISL_987136, EPI_ISL_987137, EPI_ISL_987138, EPI_ISL_987139, EPI_ISL_987140, EPI_ISL_987141, EPI_ISL_987142, EPI_ISL_987143, EPI_ISL_987145, EPI_ISL_987146, EPI_ISL_987147, EPI_ISL_987148, EPI_ISL_987149, EPI_ISL_987150, EPI_ISL_987151, EPI_ISL_987152, EPI_ISL_987153, EPI_ISL_987154, EPI_ISL_987155, EPI_ISL_987156, EPI_ISL_987157, EPI_ISL_987158, EPI_ISL_987159, EPI_ISL_987160, EPI_ISL_988686, EPI_ISL_988687, EPI_ISL_988688, EPI_ISL_988689, EPI_ISL_988690, EPI_ISL_988691, EPI_ISL_988692, EPI_ISL_988693, EPI_ISL_988694, EPI_ISL_988695, EPI_ISL_988696, EPI_ISL_988697, EPI_ISL_988698, EPI_ISL_988699, EPI_ISL_988700, EPI_ISL_988701, EPI_ISL_988702, EPI_ISL_988703, EPI_ISL_988704, EPI_ISL_988705, EPI_ISL_988706, EPI_ISL_988707, EPI_ISL_988708, EPI_ISL_988709, EPI_ISL_988710, EPI_ISL_988711, EPI_ISL_988712, EPI_ISL_988713, EPI_ISL_988714, EPI_ISL_988715, EPI_ISL_988716, EPI_ISL_988717, EPI_ISL_988718, EPI_ISL_988719, EPI_ISL_988720, EPI_ISL_988721, EPI_ISL_988722, EPI_ISL_988723, EPI_ISL_988724, EPI_ISL_988725, EPI_ISL_988726, EPI_ISL_988727, EPI_ISL_988728, EPI_ISL_988729, EPI_ISL_988730, EPI_ISL_988731, EPI_ISL_988732, EPI_ISL_988733, EPI_ISL_988734, EPI_ISL_988735, EPI_ISL_988736, EPI_ISL_988737, EPI_ISL_988738, EPI_ISL_988739, EPI_ISL_988740, EPI_ISL_988741, EPI_ISL_988742, EPI_ISL_988743, EPI_ISL_988744, EPI_ISL_988745, EPI_ISL_988746, EPI_ISL_988747, EPI_ISL_988748, EPI_ISL_988749, EPI_ISL_988750, EPI_ISL_988751, EPI_ISL_988752, EPI_ISL_988753, EPI_ISL_988754, EPI_ISL_988755, EPI_ISL_988756, EPI_ISL_988757, EPI_ISL_988758, EPI_ISL_988759, EPI_ISL_988760, EPI_ISL_988761, EPI_ISL_988762, EPI_ISL_988763, EPI_ISL_988764, EPI_ISL_988765, EPI_ISL_988766, EPI_ISL_988767, EPI_ISL_988768, EPI_ISL_988769, EPI_ISL_988770, EPI_ISL_988771, EPI_ISL_988772, EPI_ISL_988773, EPI_ISL_988774, EPI_ISL_988775, EPI_ISL_988776, EPI_ISL_988777, EPI_ISL_988778, EPI_ISL_988779, EPI_ISL_988780, EPI_ISL_988781, EPI_ISL_988782, EPI_ISL_988783, EPI_ISL_988784, EPI_ISL_988785, EPI_ISL_988786, EPI_ISL_988787, EPI_ISL_988788, EPI_ISL_988789, EPI_ISL_988790, EPI_ISL_988791, EPI_ISL_988792, EPI_ISL_988793, EPI_ISL_988794, EPI_ISL_988795, EPI_ISL_988796, EPI_ISL_988797, EPI_ISL_988798, EPI_ISL_988799, EPI_ISL_988800, EPI_ISL_988801, EPI_ISL_988802, EPI_ISL_988803, EPI_ISL_988804, EPI_ISL_988805, EPI_ISL_988806, EPI_ISL_988807, EPI_ISL_988808, EPI_ISL_988809, EPI_ISL_988810, EPI_ISL_988812, EPI_ISL_988813, EPI_ISL_988814, EPI_ISL_988815, EPI_ISL_988816, EPI_ISL_988817, EPI_ISL_988818, EPI_ISL_988819, EPI_ISL_988820, EPI_ISL_988821, EPI_ISL_988822, EPI_ISL_988823, EPI_ISL_988824, EPI_ISL_988825, EPI_ISL_988826, EPI_ISL_988827, EPI_ISL_988828, EPI_ISL_988829, EPI_ISL_988830, EPI_ISL_988831, EPI_ISL_988832, EPI_ISL_988833, EPI_ISL_988834, EPI_ISL_988835, EPI_ISL_988836, EPI_ISL_988837, EPI_ISL_988838, EPI_ISL_988839, EPI_ISL_988840, EPI_ISL_988841, EPI_ISL_988842, EPI_ISL_988843, EPI_ISL_988844, EPI_ISL_988845, EPI_ISL_988846, EPI_ISL_988847, EPI_ISL_988848, EPI_ISL_988849, EPI_ISL_988850, EPI_ISL_988851, EPI_ISL_988852, EPI_ISL_988853, EPI_ISL_988854, EPI_ISL_988855, EPI_ISL_988856, EPI_ISL_988857, EPI_ISL_988858, EPI_ISL_988859, EPI_ISL_988860, EPI_ISL_988861, EPI_ISL_988862, EPI_ISL_988863, EPI_ISL_988864, EPI_ISL_988865, EPI_ISL_988866, EPI_ISL_988867, EPI_ISL_988868, EPI_ISL_988869, EPI_ISL_988870, EPI_ISL_988871, EPI_ISL_988872, EPI_ISL_988873, EPI_ISL_988874, EPI_ISL_988875, EPI_ISL_988876, EPI_ISL_988877, EPI_ISL_988878, EPI_ISL_988879, EPI_ISL_988880, EPI_ISL_988881, EPI_ISL_988882, EPI_ISL_988883, EPI_ISL_988884, EPI_ISL_988885, EPI_ISL_988886, EPI_ISL_988887, EPI_ISL_988888, EPI_ISL_988889, EPI_ISL_988890, EPI_ISL_988891, EPI_ISL_988892, EPI_ISL_988893, EPI_ISL_988894, EPI_ISL_988895, EPI_ISL_988896, EPI_ISL_988897, EPI_ISL_988898, EPI_ISL_988899, EPI_ISL_988900, EPI_ISL_988901, EPI_ISL_988902, EPI_ISL_988903, EPI_ISL_988904, EPI_ISL_988905, EPI_ISL_988906, EPI_ISL_988907, EPI_ISL_988908, EPI_ISL_988909, EPI_ISL_988910, EPI_ISL_988911, EPI_ISL_988912, EPI_ISL_988913, EPI_ISL_988914, EPI_ISL_988915, EPI_ISL_988916, EPI_ISL_988917, EPI_ISL_988918, EPI_ISL_988919, EPI_ISL_988920, EPI_ISL_988921, EPI_ISL_988922, EPI_ISL_988923, EPI_ISL_988924, EPI_ISL_988925, EPI_ISL_988926, EPI_ISL_988927, EPI_ISL_988928, EPI_ISL_988929, EPI_ISL_988930, EPI_ISL_988931, EPI_ISL_988932, EPI_ISL_988933, EPI_ISL_988934, EPI_ISL_988935, EPI_ISL_988936, EPI_ISL_988937, EPI_ISL_988938, EPI_ISL_988939, EPI_ISL_988940, EPI_ISL_988941, EPI_ISL_988942, EPI_ISL_988943, EPI_ISL_988944, EPI_ISL_988945, EPI_ISL_988946, EPI_ISL_988947, EPI_ISL_988948, EPI_ISL_988949, EPI_ISL_988950, EPI_ISL_988951, EPI_ISL_988952, EPI_ISL_988953, EPI_ISL_988954, EPI_ISL_988955, EPI_ISL_988956, EPI_ISL_988957, EPI_ISL_988958, EPI_ISL_988959, EPI_ISL_988960, EPI_ISL_988961, EPI_ISL_988962, EPI_ISL_988963, EPI_ISL_988964, EPI_ISL_988965, EPI_ISL_988966, EPI_ISL_988967, EPI_ISL_988968, EPI_ISL_988969, EPI_ISL_988970, EPI_ISL_988971, EPI_ISL_988972, EPI_ISL_988973, EPI_ISL_988974, EPI_ISL_988975, EPI_ISL_988976, EPI_ISL_988977, EPI_ISL_988978, EPI_ISL_988979, EPI_ISL_988980, EPI_ISL_988981, EPI_ISL_988982, EPI_ISL_988983, EPI_ISL_988984, EPI_ISL_988985, EPI_ISL_988986, EPI_ISL_988987, EPI_ISL_988988, EPI_ISL_988989, EPI_ISL_988990, EPI_ISL_988991, EPI_ISL_988992, EPI_ISL_988993, EPI_ISL_988994, EPI_ISL_988995, EPI_ISL_988996, EPI_ISL_988997, EPI_ISL_988998, EPI_ISL_988999, EPI_ISL_989000, EPI_ISL_989001, EPI_ISL_989002, EPI_ISL_989003, EPI_ISL_989004, EPI_ISL_989005, EPI_ISL_989006, EPI_ISL_989007, EPI_ISL_989008, EPI_ISL_989009, EPI_ISL_989010, EPI_ISL_989011, EPI_ISL_989012, EPI_ISL_989013, EPI_ISL_989014, EPI_ISL_989015, EPI_ISL_989016, EPI_ISL_989017, EPI_ISL_989018, EPI_ISL_989019, EPI_ISL_989020, EPI_ISL_989022, EPI_ISL_989023, EPI_ISL_989024, EPI_ISL_989025, EPI_ISL_989026, EPI_ISL_989027, EPI_ISL_989028, EPI_ISL_989029, EPI_ISL_989031, EPI_ISL_989034, EPI_ISL_989035, EPI_ISL_989036, EPI_ISL_989037, EPI_ISL_989038, EPI_ISL_989039, EPI_ISL_989040, EPI_ISL_989041, EPI_ISL_989046, EPI_ISL_989047, EPI_ISL_989048, EPI_ISL_989049, EPI_ISL_989050, EPI_ISL_989051, EPI_ISL_989054, EPI_ISL_989055, EPI_ISL_989058, EPI_ISL_989060, EPI_ISL_989062, EPI_ISL_989065, EPI_ISL_989067, EPI_ISL_989074, EPI_ISL_989076, EPI_ISL_989078, EPI_ISL_989079, EPI_ISL_989083, EPI_ISL_989087, EPI_ISL_989090, EPI_ISL_989091, EPI_ISL_989097, EPI_ISL_989098, EPI_ISL_989101, EPI_ISL_989106, EPI_ISL_989112, EPI_ISL_989113, EPI_ISL_989115, EPI_ISL_989119, EPI_ISL_989122, EPI_ISL_989124, EPI_ISL_989128, EPI_ISL_989133, EPI_ISL_989134, EPI_ISL_989135, EPI_ISL_989137, EPI_ISL_989140, EPI_ISL_989144, EPI_ISL_989151, EPI_ISL_989154, EPI_ISL_989155, EPI_ISL_989157, EPI_ISL_989158, EPI_ISL_989160, EPI_ISL_989161, EPI_ISL_989163, EPI_ISL_989167, EPI_ISL_989168, EPI_ISL_989169, EPI_ISL_989177, EPI_ISL_989189, EPI_ISL_989193, EPI_ISL_989196, EPI_ISL_989197, EPI_ISL_989198, EPI_ISL_989202, EPI_ISL_989205, EPI_ISL_989208, EPI_ISL_989210, EPI_ISL_989211, EPI_ISL_989219, EPI_ISL_989220, EPI_ISL_989224, EPI_ISL_989232, EPI_ISL_989235, EPI_ISL_989237, EPI_ISL_989243, EPI_ISL_989244, EPI_ISL_989245, EPI_ISL_989261, EPI_ISL_989262, EPI_ISL_989269, EPI_ISL_989273, EPI_ISL_989274, EPI_ISL_989275, EPI_ISL_989277, EPI_ISL_989280, EPI_ISL_989284, EPI_ISL_989285, EPI_ISL_989286, EPI_ISL_989287, EPI_ISL_989288, EPI_ISL_989289, EPI_ISL_989290, EPI_ISL_989293, EPI_ISL_989294, EPI_ISL_989304, EPI_ISL_989307, EPI_ISL_989312, EPI_ISL_989313, EPI_ISL_989315, EPI_ISL_989322, EPI_ISL_989324, EPI_ISL_989329, EPI_ISL_989331, EPI_ISL_989332, EPI_ISL_989334, EPI_ISL_989340, EPI_ISL_989342, EPI_ISL_989348, EPI_ISL_989349, EPI_ISL_989350, EPI_ISL_989351, EPI_ISL_989352, EPI_ISL_989353, EPI_ISL_989354, EPI_ISL_989359, EPI_ISL_989361, EPI_ISL_989446, EPI_ISL_1006540, EPI_ISL_1006541, EPI_ISL_1006542, EPI_ISL_1006543, EPI_ISL_1006937, EPI_ISL_1006938, EPI_ISL_1006939, EPI_ISL_1006940, EPI_ISL_1007436, EPI_ISL_1007437, EPI_ISL_1007438, EPI_ISL_1007439, EPI_ISL_1012550, EPI_ISL_1012551, EPI_ISL_1012552, EPI_ISL_1012553, EPI_ISL_1012554, EPI_ISL_1012555, EPI_ISL_1012556, EPI_ISL_1012557, EPI_ISL_1012558, EPI_ISL_1012559, EPI_ISL_1012560, EPI_ISL_1012561, EPI_ISL_1012562, EPI_ISL_1012563, EPI_ISL_1012564, EPI_ISL_1012565, EPI_ISL_1012566, EPI_ISL_1012567, EPI_ISL_1012568, EPI_ISL_1012569, EPI_ISL_1012570, EPI_ISL_1012571, EPI_ISL_1012572, EPI_ISL_1012573, EPI_ISL_1012574, EPI_ISL_1012575, EPI_ISL_1012576, EPI_ISL_1012577, EPI_ISL_1012578, EPI_ISL_1012579, EPI_ISL_1012580, EPI_ISL_1012581, EPI_ISL_1012582, EPI_ISL_1012583, EPI_ISL_1012584, EPI_ISL_1012585, EPI_ISL_1012586, EPI_ISL_1012587, EPI_ISL_1012588, EPI_ISL_1012589, EPI_ISL_1012590, EPI_ISL_1012591, EPI_ISL_1012592, EPI_ISL_1012593, EPI_ISL_1012594, EPI_ISL_1012595, EPI_ISL_1012597, EPI_ISL_1012598, EPI_ISL_1012599, EPI_ISL_1012600, EPI_ISL_1012601, EPI_ISL_1012602, EPI_ISL_1012603, EPI_ISL_1012604, EPI_ISL_1012605, EPI_ISL_1012606, EPI_ISL_1012607, EPI_ISL_1012672, EPI_ISL_1012673, EPI_ISL_1012674, EPI_ISL_1012675, EPI_ISL_1012676, EPI_ISL_1012677, EPI_ISL_1012678, EPI_ISL_1012679, EPI_ISL_1012680, EPI_ISL_1012681, EPI_ISL_1012682, EPI_ISL_1012683, EPI_ISL_1012684, EPI_ISL_1012685, EPI_ISL_1012686, EPI_ISL_1012687, EPI_ISL_1012688, EPI_ISL_1012689, EPI_ISL_1012690, EPI_ISL_1012691, EPI_ISL_1012692, EPI_ISL_1012693, EPI_ISL_1012694, EPI_ISL_1012696, EPI_ISL_1012702, EPI_ISL_1012704, EPI_ISL_1012710, EPI_ISL_1012711, EPI_ISL_1245907 | | | |
| see above | Lighthouse Lab in Alderley Park | Wellcome Sanger Institute for the COVID-19 Genomics UK (COG-UK) Consortium | Cordelia Langford; David K. Jackson; Dominic Kwiatkowski; Ewan Harrison; Ian Johnston; Jacquelyn Wynn; Jeffrey Barrett; John Sillitoe on behalf of the Wellcome Sanger Institute COVID-19 Surveillance Team; John Sillitoe on behalf of the Wellcome Sanger Institute COVID-19 Surveillance Team (http://www.sanger.ac.uk/covid-team); Mairead Hyland; Roberto Amato; Sonia Goncalves; The Lighthouse Lab in Alderley Park and Alex Alderton |
| EPI_ISL_980235, EPI_ISL_980236, EPI_ISL_980237, EPI_ISL_980238, EPI_ISL_980239, EPI_ISL_980240, EPI_ISL_980241, EPI_ISL_980242, EPI_ISL_980243, EPI_ISL_980244, EPI_ISL_980245, EPI_ISL_980246, EPI_ISL_980247, EPI_ISL_980248, EPI_ISL_980249, EPI_ISL_980250, EPI_ISL_980251, EPI_ISL_980252, EPI_ISL_980253, EPI_ISL_980254, EPI_ISL_980255, EPI_ISL_980256, EPI_ISL_980257, EPI_ISL_980258, EPI_ISL_980259, EPI_ISL_980260, EPI_ISL_980261, EPI_ISL_980262, EPI_ISL_980263, EPI_ISL_980264, EPI_ISL_980265, EPI_ISL_980266, EPI_ISL_980267, EPI_ISL_980268, EPI_ISL_980269, EPI_ISL_980270, EPI_ISL_980271, EPI_ISL_980272, EPI_ISL_980273, EPI_ISL_980274, EPI_ISL_980275, EPI_ISL_980276, EPI_ISL_980277, EPI_ISL_980278, EPI_ISL_980279, EPI_ISL_980280, EPI_ISL_980281, EPI_ISL_980282, EPI_ISL_980283, EPI_ISL_980284, EPI_ISL_980285, EPI_ISL_980286, EPI_ISL_980287, EPI_ISL_980288, EPI_ISL_980289, EPI_ISL_980290, EPI_ISL_980291, EPI_ISL_980292, EPI_ISL_980293, EPI_ISL_980294, EPI_ISL_980295, EPI_ISL_980296, EPI_ISL_980297, EPI_ISL_980298, EPI_ISL_980299, EPI_ISL_980300, EPI_ISL_980301, EPI_ISL_980302, EPI_ISL_980303, EPI_ISL_980304, EPI_ISL_980305, EPI_ISL_980306, EPI_ISL_980307, EPI_ISL_980308, EPI_ISL_980309, EPI_ISL_980310, EPI_ISL_980311, EPI_ISL_980312, EPI_ISL_980313, EPI_ISL_980314, EPI_ISL_980315, EPI_ISL_980316, EPI_ISL_980317, EPI_ISL_980318, EPI_ISL_980319, EPI_ISL_980320, EPI_ISL_980321, EPI_ISL_980322, EPI_ISL_980323, EPI_ISL_980324, EPI_ISL_980325, EPI_ISL_980326, EPI_ISL_980327, EPI_ISL_980328, EPI_ISL_980329, EPI_ISL_980330, EPI_ISL_980331, EPI_ISL_980332, EPI_ISL_980333, EPI_ISL_980334, EPI_ISL_980335, EPI_ISL_980336, EPI_ISL_980337, EPI_ISL_980338, EPI_ISL_980339, EPI_ISL_980340, EPI_ISL_980341, EPI_ISL_980342, EPI_ISL_980343, EPI_ISL_980344, EPI_ISL_980345, EPI_ISL_980346, EPI_ISL_980347, EPI_ISL_980348, EPI_ISL_980349, EPI_ISL_980350, EPI_ISL_980351, EPI_ISL_980352, EPI_ISL_980353, EPI_ISL_980354, EPI_ISL_980355, EPI_ISL_980356, EPI_ISL_980357, EPI_ISL_980358, EPI_ISL_980359, EPI_ISL_980360, EPI_ISL_980361, EPI_ISL_980362, EPI_ISL_980363, EPI_ISL_980364, EPI_ISL_980365, EPI_ISL_980366, EPI_ISL_980367, EPI_ISL_980368, EPI_ISL_980369, EPI_ISL_980370, EPI_ISL_980371, EPI_ISL_980372, EPI_ISL_980373, EPI_ISL_980374, EPI_ISL_980375, EPI_ISL_980376, EPI_ISL_980377, EPI_ISL_980378, EPI_ISL_980379, EPI_ISL_980380, EPI_ISL_980381, EPI_ISL_980382, EPI_ISL_980383, EPI_ISL_980384, EPI_ISL_980385, EPI_ISL_980386, EPI_ISL_980387, EPI_ISL_980388, EPI_ISL_980389, EPI_ISL_980390, EPI_ISL_980391, EPI_ISL_980392, EPI_ISL_980393, EPI_ISL_980394, EPI_ISL_980395, EPI_ISL_980396, EPI_ISL_980397, EPI_ISL_980398, EPI_ISL_980399, EPI_ISL_980400, EPI_ISL_980401, EPI_ISL_980402, | | | |

| | | | |
|---|---|---|---|
| EPI_ISL_980403, EPI_ISL_980404, EPI_ISL_980405, EPI_ISL_980406, EPI_ISL_980407, EPI_ISL_980408, EPI_ISL_980409, EPI_ISL_980410, EPI_ISL_980411, EPI_ISL_980412, EPI_ISL_980413, EPI_ISL_980414, EPI_ISL_980415, EPI_ISL_980416, EPI_ISL_980417, EPI_ISL_980418, EPI_ISL_980419, EPI_ISL_980420, EPI_ISL_980421, EPI_ISL_980422, EPI_ISL_980423, EPI_ISL_980424, EPI_ISL_980425, EPI_ISL_980426, EPI_ISL_980427, EPI_ISL_980428, EPI_ISL_980429, EPI_ISL_980430, EPI_ISL_980431, EPI_ISL_980432, EPI_ISL_980433, EPI_ISL_980434, EPI_ISL_980435, EPI_ISL_980436, EPI_ISL_980437, EPI_ISL_980438, EPI_ISL_980439, EPI_ISL_980440, EPI_ISL_980441, EPI_ISL_980442, EPI_ISL_980443, EPI_ISL_980444, EPI_ISL_980445, EPI_ISL_980446, EPI_ISL_980447, EPI_ISL_980448, EPI_ISL_980449, EPI_ISL_980450, EPI_ISL_980451, EPI_ISL_980452, EPI_ISL_980453, EPI_ISL_980454, EPI_ISL_980455, EPI_ISL_980456, EPI_ISL_980457, EPI_ISL_980458, EPI_ISL_980459, EPI_ISL_980460, EPI_ISL_980461, EPI_ISL_980462, EPI_ISL_980463, EPI_ISL_980464, EPI_ISL_980465, EPI_ISL_980466, EPI_ISL_980467, EPI_ISL_980468, EPI_ISL_980469, EPI_ISL_980470, EPI_ISL_980471, EPI_ISL_980472, EPI_ISL_980473, EPI_ISL_980474, EPI_ISL_980475, EPI_ISL_980476, EPI_ISL_980477, EPI_ISL_980478, EPI_ISL_980479, EPI_ISL_980480, EPI_ISL_980481, EPI_ISL_980482, EPI_ISL_980483, EPI_ISL_980484, EPI_ISL_980485, EPI_ISL_980486, EPI_ISL_980487, EPI_ISL_980488, EPI_ISL_980489, EPI_ISL_980490, EPI_ISL_980491, EPI_ISL_980492, EPI_ISL_980493, EPI_ISL_980494, EPI_ISL_980495, EPI_ISL_980496, EPI_ISL_980497, EPI_ISL_980498, EPI_ISL_980499, EPI_ISL_980500, EPI_ISL_980501, EPI_ISL_980502, EPI_ISL_980503, EPI_ISL_980504, EPI_ISL_980505, EPI_ISL_980506, EPI_ISL_980507, EPI_ISL_980508, EPI_ISL_980509, EPI_ISL_980510, EPI_ISL_980511, EPI_ISL_980512, EPI_ISL_980513, EPI_ISL_980514, EPI_ISL_980515, EPI_ISL_980516, EPI_ISL_980517, EPI_ISL_980518, EPI_ISL_980519, EPI_ISL_980520, EPI_ISL_980521, EPI_ISL_980522, EPI_ISL_980523, EPI_ISL_980524, EPI_ISL_980525, EPI_ISL_980526, EPI_ISL_980527, EPI_ISL_980528, EPI_ISL_980529, EPI_ISL_980530, EPI_ISL_980531, EPI_ISL_980532, EPI_ISL_980533, EPI_ISL_980534, EPI_ISL_980535, EPI_ISL_980536, EPI_ISL_980537, EPI_ISL_980538, EPI_ISL_980539, EPI_ISL_980540, EPI_ISL_980541, EPI_ISL_980542, EPI_ISL_980543, EPI_ISL_980544, EPI_ISL_980545, EPI_ISL_980546, EPI_ISL_980547, EPI_ISL_980548, EPI_ISL_980549, EPI_ISL_980550, EPI_ISL_985417, EPI_ISL_985418, EPI_ISL_985420, EPI_ISL_985422, EPI_ISL_985423, EPI_ISL_985426, EPI_ISL_985430, EPI_ISL_985436, EPI_ISL_985440, EPI_ISL_985445, EPI_ISL_985451, EPI_ISL_985453, EPI_ISL_985478, EPI_ISL_985480, EPI_ISL_985489, EPI_ISL_985495, EPI_ISL_985512, EPI_ISL_985517, EPI_ISL_985518, EPI_ISL_985533, EPI_ISL_985534, EPI_ISL_985537, EPI_ISL_985541, EPI_ISL_985542, EPI_ISL_985548, EPI_ISL_985550, EPI_ISL_985551, EPI_ISL_985556, EPI_ISL_985563, EPI_ISL_985564, EPI_ISL_985574, EPI_ISL_985581, EPI_ISL_985585, EPI_ISL_985588, EPI_ISL_985593, EPI_ISL_985594, EPI_ISL_985597, EPI_ISL_985599, EPI_ISL_985602, EPI_ISL_985607, EPI_ISL_985609, EPI_ISL_988011, EPI_ISL_988012, EPI_ISL_988013, EPI_ISL_988014, EPI_ISL_988015, EPI_ISL_988016, EPI_ISL_988017, EPI_ISL_988018, EPI_ISL_988019, EPI_ISL_988021, EPI_ISL_988025, EPI_ISL_988026, EPI_ISL_988027, EPI_ISL_988028, EPI_ISL_988029, EPI_ISL_988031, EPI_ISL_988032, EPI_ISL_988033, EPI_ISL_988034, EPI_ISL_988035, EPI_ISL_988036, EPI_ISL_988037, EPI_ISL_988038, EPI_ISL_988040, EPI_ISL_988041, EPI_ISL_988045, EPI_ISL_988046, EPI_ISL_988047, EPI_ISL_988049, EPI_ISL_988050, EPI_ISL_988051, EPI_ISL_988052, EPI_ISL_988053, EPI_ISL_988054, EPI_ISL_988055, EPI_ISL_988056, EPI_ISL_988057, EPI_ISL_988058, EPI_ISL_988059, EPI_ISL_988063, EPI_ISL_988064, EPI_ISL_988065, EPI_ISL_988066, EPI_ISL_988068, EPI_ISL_988069, EPI_ISL_988070, EPI_ISL_988071, EPI_ISL_988072, EPI_ISL_988073, EPI_ISL_988074, EPI_ISL_988075, EPI_ISL_988077, EPI_ISL_988078, EPI_ISL_988080, EPI_ISL_988081, EPI_ISL_988082, EPI_ISL_988083, EPI_ISL_988084, EPI_ISL_988085, EPI_ISL_988087, EPI_ISL_988088, EPI_ISL_988089, EPI_ISL_988090, EPI_ISL_988091, EPI_ISL_988094, EPI_ISL_988095, EPI_ISL_988096, EPI_ISL_988098, EPI_ISL_988099, EPI_ISL_988100, EPI_ISL_988101, EPI_ISL_988103, EPI_ISL_988104, EPI_ISL_988105, EPI_ISL_988106, EPI_ISL_988108, EPI_ISL_988109, EPI_ISL_988110, EPI_ISL_988111, EPI_ISL_988112, EPI_ISL_988114, EPI_ISL_988115, EPI_ISL_988116, EPI_ISL_988119, EPI_ISL_988120, EPI_ISL_988121, EPI_ISL_988122, EPI_ISL_988123, EPI_ISL_988124, EPI_ISL_988125, EPI_ISL_988126, EPI_ISL_988127, EPI_ISL_988129, EPI_ISL_988130, EPI_ISL_988131, EPI_ISL_988132, EPI_ISL_988133, EPI_ISL_988134, EPI_ISL_988135, EPI_ISL_988136, EPI_ISL_988138, EPI_ISL_988139, EPI_ISL_988140, EPI_ISL_988141, EPI_ISL_988142, EPI_ISL_988144, EPI_ISL_988145, EPI_ISL_988146, EPI_ISL_988148, EPI_ISL_988149, EPI_ISL_988150, EPI_ISL_988151, EPI_ISL_988152, EPI_ISL_988153, EPI_ISL_988155, EPI_ISL_988157, EPI_ISL_988158, EPI_ISL_988159, EPI_ISL_988161, EPI_ISL_988165, EPI_ISL_988166, EPI_ISL_988167, EPI_ISL_988169, EPI_ISL_988170, EPI_ISL_988172, EPI_ISL_988173, EPI_ISL_988174, EPI_ISL_988175, EPI_ISL_988176, EPI_ISL_988177, EPI_ISL_988178, EPI_ISL_988179, EPI_ISL_988180, EPI_ISL_988181, EPI_ISL_988182, EPI_ISL_988183, EPI_ISL_988184, EPI_ISL_988185, EPI_ISL_988186, EPI_ISL_988187, EPI_ISL_988188, EPI_ISL_988190, EPI_ISL_988191, EPI_ISL_988197, EPI_ISL_988198, EPI_ISL_988201, EPI_ISL_988202, EPI_ISL_988204, EPI_ISL_988206, EPI_ISL_988208, EPI_ISL_988211, EPI_ISL_988212, EPI_ISL_988215, EPI_ISL_988216, EPI_ISL_988219, EPI_ISL_988220, EPI_ISL_988227, EPI_ISL_988230, EPI_ISL_988231, EPI_ISL_988233, EPI_ISL_988234, EPI_ISL_988237, EPI_ISL_988240, EPI_ISL_988244, EPI_ISL_988245, EPI_ISL_988246, EPI_ISL_988251, EPI_ISL_988252, EPI_ISL_988258, EPI_ISL_988260, EPI_ISL_988265, EPI_ISL_988266, EPI_ISL_988267, EPI_ISL_988269, EPI_ISL_988272, EPI_ISL_988273, EPI_ISL_988275, EPI_ISL_988277, EPI_ISL_988278, EPI_ISL_988279, EPI_ISL_988281, EPI_ISL_988282, EPI_ISL_988283, EPI_ISL_988284, EPI_ISL_988287, EPI_ISL_988288, EPI_ISL_988289, EPI_ISL_988291, EPI_ISL_988292, EPI_ISL_988294, EPI_ISL_988295, EPI_ISL_988297, EPI_ISL_988301, EPI_ISL_988303, EPI_ISL_988311, EPI_ISL_988316, EPI_ISL_988318, EPI_ISL_988319, EPI_ISL_988320, EPI_ISL_988322, EPI_ISL_988326, EPI_ISL_988327, EPI_ISL_988328, EPI_ISL_988330, EPI_ISL_988331, EPI_ISL_988334, EPI_ISL_988336, EPI_ISL_988338, EPI_ISL_988341, EPI_ISL_988342, EPI_ISL_988344, EPI_ISL_988345, EPI_ISL_988346, EPI_ISL_988349, EPI_ISL_988350, EPI_ISL_988353, EPI_ISL_988356, EPI_ISL_988360, EPI_ISL_988361, EPI_ISL_988365, EPI_ISL_988366, EPI_ISL_988367, EPI_ISL_988368, EPI_ISL_988369, EPI_ISL_988371, EPI_ISL_988372, EPI_ISL_988373, EPI_ISL_988374, EPI_ISL_988375, EPI_ISL_988376, EPI_ISL_988379, EPI_ISL_988380, EPI_ISL_988382, EPI_ISL_988386, EPI_ISL_988387, EPI_ISL_988388, EPI_ISL_988392, EPI_ISL_988393, EPI_ISL_988394, EPI_ISL_988396, EPI_ISL_988400, EPI_ISL_988401, EPI_ISL_988402, EPI_ISL_988405, EPI_ISL_988406, EPI_ISL_988407, EPI_ISL_988408, EPI_ISL_988410, EPI_ISL_988412, EPI_ISL_988413, EPI_ISL_988414, EPI_ISL_988415, EPI_ISL_988416, EPI_ISL_988419, EPI_ISL_988421, EPI_ISL_988422, EPI_ISL_988424, EPI_ISL_988425, EPI_ISL_988431, EPI_ISL_988432, EPI_ISL_988438, EPI_ISL_988439, EPI_ISL_988440, EPI_ISL_988441, EPI_ISL_988443, EPI_ISL_988444, EPI_ISL_988445, EPI_ISL_988447, EPI_ISL_988450, EPI_ISL_988454, EPI_ISL_988455, EPI_ISL_988458, EPI_ISL_988461, EPI_ISL_988462, EPI_ISL_988464, EPI_ISL_988466, EPI_ISL_988469, EPI_ISL_988470, EPI_ISL_988471, EPI_ISL_988472, EPI_ISL_988473, EPI_ISL_988474, EPI_ISL_988476, EPI_ISL_988479, EPI_ISL_989363, EPI_ISL_989364, EPI_ISL_989365, EPI_ISL_989366, EPI_ISL_989367, EPI_ISL_989368, EPI_ISL_989369, EPI_ISL_989370, EPI_ISL_989371, EPI_ISL_989372, EPI_ISL_989373, EPI_ISL_989374, EPI_ISL_989375, EPI_ISL_989376, EPI_ISL_989377, EPI_ISL_989378, EPI_ISL_989379, EPI_ISL_989380, EPI_ISL_989381, EPI_ISL_989382, EPI_ISL_989383, EPI_ISL_989384, EPI_ISL_989385, EPI_ISL_989386, EPI_ISL_989387, EPI_ISL_989388, EPI_ISL_989389, EPI_ISL_989390, EPI_ISL_989391, EPI_ISL_989392, EPI_ISL_989393, EPI_ISL_989394, EPI_ISL_989395, EPI_ISL_989396, EPI_ISL_989397, EPI_ISL_989398, EPI_ISL_989399, EPI_ISL_989400, EPI_ISL_989401, EPI_ISL_989402, EPI_ISL_989403, EPI_ISL_989404, EPI_ISL_989405, EPI_ISL_989406, EPI_ISL_989407, EPI_ISL_989408, EPI_ISL_989409, EPI_ISL_989410, EPI_ISL_989411, EPI_ISL_989412, EPI_ISL_989413, EPI_ISL_989414, EPI_ISL_989415, EPI_ISL_989416, EPI_ISL_989417, EPI_ISL_989418, EPI_ISL_989419, EPI_ISL_989420, EPI_ISL_989421, EPI_ISL_989422, EPI_ISL_989423, EPI_ISL_989424, EPI_ISL_989425, EPI_ISL_989426, EPI_ISL_989427, EPI_ISL_989428, EPI_ISL_989429, EPI_ISL_989430, EPI_ISL_989431, EPI_ISL_989432, EPI_ISL_989433, EPI_ISL_989434, EPI_ISL_989435, EPI_ISL_989436, EPI_ISL_989437, EPI_ISL_989438, EPI_ISL_989439, EPI_ISL_989440, EPI_ISL_989441, EPI_ISL_989442, EPI_ISL_989443, EPI_ISL_1006249, EPI_ISL_1006262, EPI_ISL_1006267, EPI_ISL_1006269, EPI_ISL_1006272, EPI_ISL_1006275, EPI_ISL_1006280, EPI_ISL_1006283, EPI_ISL_1006291, EPI_ISL_1006301, EPI_ISL_1006308, EPI_ISL_1006311, EPI_ISL_1006312, EPI_ISL_1006313, EPI_ISL_1006317, EPI_ISL_1006322, EPI_ISL_1006323, EPI_ISL_1006324, EPI_ISL_1006331, EPI_ISL_1006873, EPI_ISL_1007192, EPI_ISL_1007200, EPI_ISL_1007207, EPI_ISL_1007242, EPI_ISL_1007245, EPI_ISL_1007262, EPI_ISL_1007269, EPI_ISL_1007292, EPI_ISL_1007296, EPI_ISL_1007313, EPI_ISL_1007337, EPI_ISL_1007346, EPI_ISL_1007349, EPI_ISL_1007357, EPI_ISL_1007367, EPI_ISL_1007369, EPI_ISL_1007381, EPI_ISL_1007404, EPI_ISL_1007405, EPI_ISL_1007407, EPI_ISL_1007415, EPI_ISL_1007416, EPI_ISL_1007417, EPI_ISL_1007428, EPI_ISL_1007430, EPI_ISL_1007445, EPI_ISL_1007446, EPI_ISL_1007447, EPI_ISL_1007448, EPI_ISL_1007449, EPI_ISL_1007450, EPI_ISL_1007451, EPI_ISL_1007452, EPI_ISL_1007453, EPI_ISL_1007454, EPI_ISL_1007456, EPI_ISL_1007457, EPI_ISL_1007458, EPI_ISL_1007459, EPI_ISL_1007461, EPI_ISL_1007463, EPI_ISL_1007464, EPI_ISL_1007467, EPI_ISL_1007469, EPI_ISL_1007471, EPI_ISL_1007472, EPI_ISL_1007473, EPI_ISL_1007474, EPI_ISL_1007476, EPI_ISL_1007477, EPI_ISL_1007478, EPI_ISL_1007479, EPI_ISL_1007480, EPI_ISL_1007481, EPI_ISL_1007483, EPI_ISL_1007485, EPI_ISL_1007486, EPI_ISL_1007488, EPI_ISL_1007491, EPI_ISL_1007492, EPI_ISL_1007494, EPI_ISL_1007495, EPI_ISL_1007496, EPI_ISL_1007497, EPI_ISL_1007498, EPI_ISL_1007500, EPI_ISL_1007502, EPI_ISL_1007504, EPI_ISL_1007505, EPI_ISL_1007506, EPI_ISL_1007508, EPI_ISL_1007509, EPI_ISL_1007510, EPI_ISL_1007511, EPI_ISL_1007512, EPI_ISL_1007513, EPI_ISL_1007514, EPI_ISL_1007515, EPI_ISL_1007516, EPI_ISL_1007517, EPI_ISL_1007519, EPI_ISL_1007521, EPI_ISL_1007522, EPI_ISL_1007523, EPI_ISL_1007524, EPI_ISL_1007528, EPI_ISL_1007529, EPI_ISL_1007530, EPI_ISL_1007531, EPI_ISL_1007532, EPI_ISL_1007533, EPI_ISL_1007534, EPI_ISL_1007535, EPI_ISL_1007536, EPI_ISL_1007537, EPI_ISL_1007538, EPI_ISL_1007539, EPI_ISL_1007540, EPI_ISL_1007541, EPI_ISL_1007542, EPI_ISL_1007543, EPI_ISL_1007544, EPI_ISL_1007545, EPI_ISL_1007546, EPI_ISL_1007547, EPI_ISL_1007548, EPI_ISL_1012616, EPI_ISL_1012621, EPI_ISL_1012627, EPI_ISL_1012637, EPI_ISL_1012643, EPI_ISL_1012650, EPI_ISL_1012655, EPI_ISL_1012661, EPI_ISL_1012663, EPI_ISL_1012665, EPI_ISL_1012666, EPI_ISL_1012668, EPI_ISL_1034143, EPI_ISL_1187062, EPI_ISL_1187091, EPI_ISL_1187129, EPI_ISL_1187169, EPI_ISL_1206045, EPI_ISL_1206067, EPI_ISL_1206073, EPI_ISL_1206079, EPI_ISL_1206080, EPI_ISL_1206090, EPI_ISL_1206097, EPI_ISL_1206100, EPI_ISL_1206109, EPI_ISL_1206113, EPI_ISL_1206118, EPI_ISL_1206132, EPI_ISL_1206139, EPI_ISL_1206148, EPI_ISL_1206152, EPI_ISL_1206157, EPI_ISL_1206159, EPI_ISL_1206176, EPI_ISL_1206179, EPI_ISL_1206181, EPI_ISL_1206193, EPI_ISL_1206215, EPI_ISL_1206219, EPI_ISL_1206231, EPI_ISL_1206250, EPI_ISL_1206256, EPI_ISL_1206265, EPI_ISL_1206270, EPI_ISL_1206273, EPI_ISL_1223169, EPI_ISL_1223212, EPI_ISL_1223261, EPI_ISL_1223330, EPI_ISL_1223337, EPI_ISL_1223345, EPI_ISL_1223410, EPI_ISL_1242049, EPI_ISL_1242050, EPI_ISL_1242070, EPI_ISL_1242073, EPI_ISL_1242093, EPI_ISL_1242094, EPI_ISL_1242110, EPI_ISL_1242115, EPI_ISL_1242116, EPI_ISL_1242131, EPI_ISL_1242141, EPI_ISL_1242147, EPI_ISL_1242158, EPI_ISL_1242159, EPI_ISL_1242166, EPI_ISL_1242168, EPI_ISL_1242198, EPI_ISL_1242201, EPI_ISL_1242217, EPI_ISL_1242223, EPI_ISL_1242228, EPI_ISL_1242229, EPI_ISL_1242237, EPI_ISL_1242250, EPI_ISL_1242254, EPI_ISL_1242264, EPI_ISL_1242266, EPI_ISL_1242275, EPI_ISL_1244871, EPI_ISL_1244872, EPI_ISL_1244875, EPI_ISL_1244877, EPI_ISL_1244880, EPI_ISL_1244882, EPI_ISL_1244905, EPI_ISL_1244913, EPI_ISL_1244915, EPI_ISL_1244920, EPI_ISL_1244922, EPI_ISL_1244932, EPI_ISL_1244936, EPI_ISL_1244939, EPI_ISL_1244941, EPI_ISL_1244949, EPI_ISL_1244979, EPI_ISL_1245002, EPI_ISL_1245004, EPI_ISL_1467594, EPI_ISL_1467596, EPI_ISL_1467623, EPI_ISL_1467694, EPI_ISL_1467702, EPI_ISL_1467703, EPI_ISL_1467731, EPI_ISL_1467774, EPI_ISL_1467777 | | | |
| see above | Lighthouse Lab in Cambridge | Wellcome Sanger Institute for the COVID-19 Genomics UK (COG-UK) Consortium | Cordelia Langford; David K. Jackson; Dominic Kwiatkowski; Ewan Harrison; Ian Johnston; Jeffrey Barrett; John Sillitoe on behalf of the Wellcome Sanger Institute COVID-19 Surveillance Team; John Sillitoe on behalf of the Wellcome Sanger Institute COVID-19 Surveillance Team (http://www.sanger.ac.uk/covid-team); Rob Howes; Roberto Amato; Sonia Goncalves; The Lighthouse Lab in Cambridge and Alex Alderton |
| EPI_ISL_957054, EPI_ISL_957055, EPI_ISL_957060, EPI_ISL_957061, EPI_ISL_957065, EPI_ISL_957067, EPI_ISL_957068, EPI_ISL_957071, EPI_ISL_957072, EPI_ISL_957073, EPI_ISL_957075, EPI_ISL_957077, EPI_ISL_957078, EPI_ISL_957079, EPI_ISL_957081, EPI_ISL_957083, EPI_ISL_957086, EPI_ISL_957089, EPI_ISL_957091, EPI_ISL_957092, EPI_ISL_957097, EPI_ISL_957099, EPI_ISL_957102, EPI_ISL_957103, EPI_ISL_957104, EPI_ISL_957105, EPI_ISL_957107, EPI_ISL_957110, EPI_ISL_957114, EPI_ISL_957115, EPI_ISL_957117, EPI_ISL_957118, EPI_ISL_957121, EPI_ISL_957122, EPI_ISL_957124, EPI_ISL_957133, EPI_ISL_957140, EPI_ISL_957141, EPI_ISL_957150, EPI_ISL_957151, EPI_ISL_957155, EPI_ISL_957157, EPI_ISL_957158, EPI_ISL_957160, EPI_ISL_957165, EPI_ISL_957166, EPI_ISL_957168, EPI_ISL_957170, EPI_ISL_957172, EPI_ISL_957176, EPI_ISL_957180, EPI_ISL_957187, EPI_ISL_957190, EPI_ISL_957191, EPI_ISL_957194, EPI_ISL_957198, EPI_ISL_957200, EPI_ISL_957205, EPI_ISL_957206, EPI_ISL_957208, EPI_ISL_957210, EPI_ISL_957211, EPI_ISL_957212, EPI_ISL_957215, EPI_ISL_957216, EPI_ISL_957219, EPI_ISL_957223, EPI_ISL_957224, EPI_ISL_957225, EPI_ISL_957229, EPI_ISL_957231, EPI_ISL_957237, EPI_ISL_957239, EPI_ISL_957242, EPI_ISL_957243, EPI_ISL_957245, EPI_ISL_957246, EPI_ISL_957250, EPI_ISL_957253, EPI_ISL_957254, EPI_ISL_957255, EPI_ISL_957257, EPI_ISL_957259, EPI_ISL_957260, EPI_ISL_957262, EPI_ISL_957263, EPI_ISL_957266, EPI_ISL_957268, EPI_ISL_957271, EPI_ISL_957272, EPI_ISL_957274, EPI_ISL_957276, EPI_ISL_957279, EPI_ISL_957283, EPI_ISL_957285, EPI_ISL_957286, EPI_ISL_957287, EPI_ISL_957289, EPI_ISL_957291, EPI_ISL_957295, EPI_ISL_957297, EPI_ISL_957299, EPI_ISL_957302, EPI_ISL_957304, EPI_ISL_957306, EPI_ISL_957308, EPI_ISL_957310, EPI_ISL_957311, EPI_ISL_957314, EPI_ISL_957315, EPI_ISL_957319, EPI_ISL_957320, EPI_ISL_957322, EPI_ISL_957323, EPI_ISL_957327, EPI_ISL_957328, EPI_ISL_957330, EPI_ISL_957331, EPI_ISL_957334, EPI_ISL_957336, EPI_ISL_957337, EPI_ISL_957633, EPI_ISL_957637, EPI_ISL_957649, EPI_ISL_957651, EPI_ISL_957653, EPI_ISL_957663, EPI_ISL_957664, EPI_ISL_957666, EPI_ISL_957669, EPI_ISL_957673, EPI_ISL_957678, EPI_ISL_957679, EPI_ISL_957681, EPI_ISL_957682, EPI_ISL_957693, EPI_ISL_957694, EPI_ISL_957697, EPI_ISL_957708, EPI_ISL_957713, EPI_ISL_957715, EPI_ISL_957717, EPI_ISL_957718, EPI_ISL_957720, EPI_ISL_957725, EPI_ISL_957729, EPI_ISL_957730, EPI_ISL_957732, EPI_ISL_957736, EPI_ISL_957743, EPI_ISL_957749, EPI_ISL_957750, EPI_ISL_957756, EPI_ISL_957758, EPI_ISL_957765, EPI_ISL_957768, EPI_ISL_957770, EPI_ISL_957773, EPI_ISL_957774, EPI_ISL_957778, EPI_ISL_957785, EPI_ISL_957792, EPI_ISL_957793, EPI_ISL_957796, EPI_ISL_957802, EPI_ISL_957806, EPI_ISL_957814, EPI_ISL_957819, EPI_ISL_957824, EPI_ISL_957836, EPI_ISL_957837, EPI_ISL_957839, EPI_ISL_957845, EPI_ISL_957847, EPI_ISL_957848, EPI_ISL_957851, EPI_ISL_957853, EPI_ISL_957854, EPI_ISL_957860, EPI_ISL_957864, EPI_ISL_957865, EPI_ISL_957871, EPI_ISL_957875, EPI_ISL_957879, EPI_ISL_957884, EPI_ISL_957889, EPI_ISL_957894, EPI_ISL_964348, EPI_ISL_964349, EPI_ISL_964351, EPI_ISL_964352, EPI_ISL_964355, EPI_ISL_964357, EPI_ISL_964358, EPI_ISL_964359, EPI_ISL_964360, EPI_ISL_964362, EPI_ISL_964366, EPI_ISL_964367, EPI_ISL_964368, EPI_ISL_964369, EPI_ISL_964371, EPI_ISL_964372, EPI_ISL_964375, EPI_ISL_964377, EPI_ISL_964378, EPI_ISL_964380, EPI_ISL_964382, EPI_ISL_964384, EPI_ISL_964387, EPI_ISL_964388, EPI_ISL_964391, EPI_ISL_964396, EPI_ISL_964397, EPI_ISL_964399, EPI_ISL_964401, EPI_ISL_964404, EPI_ISL_964407, EPI_ISL_964408, EPI_ISL_964409, EPI_ISL_964410, EPI_ISL_964411, EPI_ISL_964413, EPI_ISL_964415, EPI_ISL_964416, EPI_ISL_964417, EPI_ISL_964422, EPI_ISL_964424, EPI_ISL_964425, EPI_ISL_964427, EPI_ISL_964430, EPI_ISL_964433, EPI_ISL_964434, EPI_ISL_964436, EPI_ISL_964437, EPI_ISL_964440, EPI_ISL_964441, EPI_ISL_964442, EPI_ISL_964446, EPI_ISL_964449, EPI_ISL_964452, EPI_ISL_964456, EPI_ISL_964457, EPI_ISL_964458, EPI_ISL_964460, EPI_ISL_964468, EPI_ISL_964469, EPI_ISL_964470, EPI_ISL_964471, EPI_ISL_964472, EPI_ISL_964473, EPI_ISL_964474, EPI_ISL_964476, EPI_ISL_964477, EPI_ISL_964480, EPI_ISL_964482, EPI_ISL_964483, EPI_ISL_964484, EPI_ISL_964488, EPI_ISL_964489, EPI_ISL_964490, EPI_ISL_964493, EPI_ISL_964494, EPI_ISL_964500, EPI_ISL_964501, EPI_ISL_964503, EPI_ISL_964504, EPI_ISL_964508, EPI_ISL_964509, EPI_ISL_964511, EPI_ISL_964514, EPI_ISL_964516, EPI_ISL_964518, EPI_ISL_964520, EPI_ISL_964521, EPI_ISL_964523, EPI_ISL_964525, EPI_ISL_964526, EPI_ISL_964527, EPI_ISL_964528, EPI_ISL_964532, EPI_ISL_964533, EPI_ISL_964537, EPI_ISL_964539, EPI_ISL_964540, EPI_ISL_964541, EPI_ISL_964542, EPI_ISL_969455, EPI_ISL_986104, EPI_ISL_986120, EPI_ISL_986126, EPI_ISL_986128, EPI_ISL_986132, EPI_ISL_986134, EPI_ISL_986139, EPI_ISL_986152, EPI_ISL_986154, EPI_ISL_986159, EPI_ISL_986163, EPI_ISL_986167, EPI_ISL_986171, EPI_ISL_986172, EPI_ISL_986178, EPI_ISL_986184, EPI_ISL_986190, EPI_ISL_986197, EPI_ISL_986199, EPI_ISL_986209, EPI_ISL_986212, EPI_ISL_986229, EPI_ISL_986230, EPI_ISL_986238, EPI_ISL_986239, EPI_ISL_986255, EPI_ISL_986260, EPI_ISL_986262, EPI_ISL_986263, EPI_ISL_986265, EPI_ISL_986266, EPI_ISL_986268, EPI_ISL_986271, EPI_ISL_986273, EPI_ISL_986274, EPI_ISL_986275, EPI_ISL_986276, EPI_ISL_986277, EPI_ISL_986278, EPI_ISL_986279, EPI_ISL_986280, EPI_ISL_986281, EPI_ISL_986282, EPI_ISL_986283, EPI_ISL_986285, EPI_ISL_986286, EPI_ISL_986287, EPI_ISL_986288, EPI_ISL_986289, EPI_ISL_986292, EPI_ISL_986293, EPI_ISL_986295, EPI_ISL_986296, EPI_ISL_986298, EPI_ISL_986301, EPI_ISL_986303, EPI_ISL_986304, EPI_ISL_986305, EPI_ISL_986307, EPI_ISL_986310, EPI_ISL_986311, EPI_ISL_986312, EPI_ISL_986313, EPI_ISL_986314, EPI_ISL_986315, EPI_ISL_986317, EPI_ISL_986319, EPI_ISL_986321, EPI_ISL_986322, EPI_ISL_986323, EPI_ISL_986324, EPI_ISL_986325, EPI_ISL_986327, EPI_ISL_986329, EPI_ISL_986332, EPI_ISL_986333, EPI_ISL_986336, EPI_ISL_986338, EPI_ISL_986339, EPI_ISL_986340, EPI_ISL_986341, EPI_ISL_986343, EPI_ISL_986344, EPI_ISL_986345, EPI_ISL_986347, EPI_ISL_986349, EPI_ISL_986351, EPI_ISL_986357, EPI_ISL_986358, EPI_ISL_986359, EPI_ISL_986360, EPI_ISL_986361, EPI_ISL_986362, EPI_ISL_986363, EPI_ISL_986364, EPI_ISL_986366, EPI_ISL_986373, EPI_ISL_986376, EPI_ISL_986377, EPI_ISL_986378, EPI_ISL_986379, EPI_ISL_986380, EPI_ISL_986383, EPI_ISL_986385, EPI_ISL_986386, EPI_ISL_986387, EPI_ISL_986390, EPI_ISL_986391, EPI_ISL_986392, EPI_ISL_986393, EPI_ISL_986394, EPI_ISL_986398, EPI_ISL_986399, EPI_ISL_986400, EPI_ISL_986401, EPI_ISL_986402, EPI_ISL_986403, EPI_ISL_986404, EPI_ISL_986405, EPI_ISL_986406, EPI_ISL_986408, EPI_ISL_986412, EPI_ISL_986413, EPI_ISL_986414, EPI_ISL_986416, EPI_ISL_986417, EPI_ISL_986418, EPI_ISL_986421, EPI_ISL_986422, EPI_ISL_986423, EPI_ISL_986424, EPI_ISL_986425, EPI_ISL_986428, EPI_ISL_986429, EPI_ISL_986431, EPI_ISL_986432, EPI_ISL_986433, EPI_ISL_986434, EPI_ISL_986435, EPI_ISL_986436, EPI_ISL_986437, EPI_ISL_986438, EPI_ISL_989530, EPI_ISL_989531, EPI_ISL_989532, EPI_ISL_989533, EPI_ISL_989534, EPI_ISL_989535, EPI_ISL_989536, EPI_ISL_989537, EPI_ISL_989538, EPI_ISL_989539, EPI_ISL_989540, EPI_ISL_989541, EPI_ISL_989542, EPI_ISL_989544, EPI_ISL_989545, EPI_ISL_989546, EPI_ISL_989547, EPI_ISL_989548, EPI_ISL_989549, EPI_ISL_989550, EPI_ISL_989551, EPI_ISL_989552, EPI_ISL_989553, EPI_ISL_989554, EPI_ISL_989555, EPI_ISL_989556, EPI_ISL_989557, EPI_ISL_989558, EPI_ISL_989559, EPI_ISL_989560, EPI_ISL_989561, EPI_ISL_989562, EPI_ISL_989563, EPI_ISL_989564, EPI_ISL_989565, EPI_ISL_989566, EPI_ISL_989567, EPI_ISL_989568, EPI_ISL_989569, EPI_ISL_989570, EPI_ISL_989571, EPI_ISL_989572, EPI_ISL_989573, EPI_ISL_989574, EPI_ISL_989575, EPI_ISL_989576, EPI_ISL_989577, EPI_ISL_989578, EPI_ISL_989579, EPI_ISL_989580, EPI_ISL_989581, EPI_ISL_989583, EPI_ISL_989584, EPI_ISL_989585, EPI_ISL_989586, EPI_ISL_989587, EPI_ISL_989588, EPI_ISL_989589, EPI_ISL_989590, EPI_ISL_989592, EPI_ISL_989593, EPI_ISL_989595, EPI_ISL_989596, EPI_ISL_989597, EPI_ISL_989598, EPI_ISL_989599, EPI_ISL_989600, EPI_ISL_989601, EPI_ISL_989602, EPI_ISL_989603, EPI_ISL_989604, EPI_ISL_989605, EPI_ISL_989606, EPI_ISL_989607, EPI_ISL_989608, EPI_ISL_989609, EPI_ISL_989610, EPI_ISL_989611, EPI_ISL_989612, EPI_ISL_989613, EPI_ISL_989614, EPI_ISL_989615, EPI_ISL_989616, EPI_ISL_989617, EPI_ISL_989618, EPI_ISL_989619, EPI_ISL_989620, EPI_ISL_989621, EPI_ISL_989622, EPI_ISL_989623, EPI_ISL_989624, EPI_ISL_989625, EPI_ISL_989626, EPI_ISL_989627, EPI_ISL_989628, EPI_ISL_989629, EPI_ISL_989630, EPI_ISL_989631, EPI_ISL_989632, EPI_ISL_989633, EPI_ISL_989634, EPI_ISL_989635, EPI_ISL_989636, EPI_ISL_989637, EPI_ISL_989638, EPI_ISL_989639, EPI_ISL_989640, EPI_ISL_989641, EPI_ISL_989642, EPI_ISL_989643, EPI_ISL_989644, EPI_ISL_989645, EPI_ISL_989646, EPI_ISL_989647, EPI_ISL_989648, EPI_ISL_989649, EPI_ISL_989650, EPI_ISL_989651, EPI_ISL_989652, EPI_ISL_989653, EPI_ISL_989654, EPI_ISL_989655, EPI_ISL_989656, EPI_ISL_989657, EPI_ISL_989658, EPI_ISL_989659, EPI_ISL_989660, EPI_ISL_989661, EPI_ISL_989662, EPI_ISL_989663, EPI_ISL_989664, EPI_ISL_989665, EPI_ISL_989666, EPI_ISL_989667, EPI_ISL_989668, EPI_ISL_989669, EPI_ISL_989670, EPI_ISL_989671, EPI_ISL_989672, EPI_ISL_989673, EPI_ISL_989674, EPI_ISL_989675, EPI_ISL_989676, EPI_ISL_989677, EPI_ISL_989678, EPI_ISL_989679, EPI_ISL_989680, EPI_ISL_989681, EPI_ISL_989682, EPI_ISL_989683, EPI_ISL_989684, EPI_ISL_989685, EPI_ISL_989686, EPI_ISL_989687, EPI_ISL_989688, EPI_ISL_989689, EPI_ISL_989690, EPI_ISL_989691, EPI_ISL_989692, EPI_ISL_989693, EPI_ISL_989694, EPI_ISL_989695, EPI_ISL_989795, EPI_ISL_989796, EPI_ISL_989797, EPI_ISL_989798, EPI_ISL_989799, EPI_ISL_989800, EPI_ISL_989801, EPI_ISL_989802, EPI_ISL_989803, EPI_ISL_989804, EPI_ISL_989805, EPI_ISL_989806, EPI_ISL_989807, EPI_ISL_989809, EPI_ISL_989810, EPI_ISL_989811, EPI_ISL_989812, EPI_ISL_989813, EPI_ISL_989814, EPI_ISL_989815, EPI_ISL_989816, EPI_ISL_989817, EPI_ISL_989818, EPI_ISL_989819, EPI_ISL_989820, EPI_ISL_989821, EPI_ISL_989822, EPI_ISL_989823, EPI_ISL_989824, EPI_ISL_989825, EPI_ISL_989826, EPI_ISL_989827, EPI_ISL_989828, EPI_ISL_989829, EPI_ISL_989830, EPI_ISL_989832, EPI_ISL_989833, EPI_ISL_989835, EPI_ISL_989837, EPI_ISL_989838, EPI_ISL_989839, EPI_ISL_989840, EPI_ISL_989841, EPI_ISL_989842, EPI_ISL_989843, EPI_ISL_989845, EPI_ISL_989846, EPI_ISL_989847, EPI_ISL_989848, EPI_ISL_989849, EPI_ISL_989850, EPI_ISL_989851, EPI_ISL_989853, EPI_ISL_989855, EPI_ISL_1006139, EPI_ISL_1190727, EPI_ISL_1698751 | | | |
| see above | Lighthouse Lab in Glasgow | Wellcome Sanger Institute for the COVID-19 Genomics UK (COG-UK) Consortium | Anna Dominiczak and Alex Alderton; Carol Clugston; Cordelia Langford; David Gray; David K. Jackson; Dominic Kwiatkowski; Ewan Harrison; Harper VanSteenhouse; Ian Johnston; Jeffrey Barrett; John Sillitoe on behalf of the Wellcome Sanger Institute COVID-19 Surveillance Team; John Sillitoe on behalf of the Wellcome Sanger Institute COVID-19 Surveillance Team (http://www.sanger.ac.uk/covid-team); Roberto Amato; Sonia Goncalves; Yumi Kasai |
| EPI_ISL_1050223 | Lighthouse Lab in Glasgow / MRC-University of Glasgow Centre for Virus Research | COVID-19 Genomics UK (COG-UK) Consortium | Alasdair MacLean; Alice Broos; Ana da Silva Filipe; Anna Dominiczak; Antonia Ho; Carol Clugston; Daniel Mair; David Gray; David L Robertson; Emma Thomson; Harper VanSteenhouse; James Shepherd; Jenna Nichols; Joseph Hughes; Kathy Li; Kathy Smollett; Kyriaki Nomikou; Lily Tong; Matthew Holden; Natasha Johnson; Richard Orton; Rory Gunson; Sarah McDonald; Sharif Shaaban; Sreenu Vattipally; Stephen Carmichael; Yumi Kasai |
| EPI_ISL_964346, EPI_ISL_964347, EPI_ISL_964350, EPI_ISL_964353, EPI_ISL_964354, EPI_ISL_964356, EPI_ISL_964361, EPI_ISL_964363, EPI_ISL_964365, EPI_ISL_964373, EPI_ISL_964374, EPI_ISL_964379, EPI_ISL_964381, EPI_ISL_964383, EPI_ISL_964385, EPI_ISL_964386, EPI_ISL_964389, EPI_ISL_964392, EPI_ISL_964394, EPI_ISL_964395, EPI_ISL_964398, EPI_ISL_964400, EPI_ISL_964402, EPI_ISL_964403, EPI_ISL_964405, EPI_ISL_964406, EPI_ISL_964412, EPI_ISL_964414, EPI_ISL_964418, EPI_ISL_964419, EPI_ISL_964420, EPI_ISL_964423, EPI_ISL_964426, EPI_ISL_964428, EPI_ISL_964431, EPI_ISL_964432, EPI_ISL_964435, EPI_ISL_964438, EPI_ISL_964443, EPI_ISL_964444, EPI_ISL_964445, EPI_ISL_964447, EPI_ISL_964448, EPI_ISL_964450, EPI_ISL_964453, EPI_ISL_964454, EPI_ISL_964455, EPI_ISL_964459, EPI_ISL_964461, EPI_ISL_964462, EPI_ISL_964463, EPI_ISL_964464, EPI_ISL_964466, EPI_ISL_964467, EPI_ISL_964478, EPI_ISL_964479, EPI_ISL_964481, EPI_ISL_964485, EPI_ISL_964486, EPI_ISL_964491, EPI_ISL_964492, EPI_ISL_964495, EPI_ISL_964496, EPI_ISL_964499, EPI_ISL_964502, EPI_ISL_964505, EPI_ISL_964507, EPI_ISL_964510, EPI_ISL_964513, EPI_ISL_964515, EPI_ISL_964517, EPI_ISL_964519, EPI_ISL_964524, EPI_ISL_964529, EPI_ISL_964530, EPI_ISL_964531, EPI_ISL_964534, EPI_ISL_964535, EPI_ISL_964536, EPI_ISL_964538, EPI_ISL_979998, EPI_ISL_979999, EPI_ISL_980006, EPI_ISL_980012, EPI_ISL_980017, EPI_ISL_980028, EPI_ISL_980030, EPI_ISL_980035, EPI_ISL_980036, EPI_ISL_980046, EPI_ISL_980048, EPI_ISL_980049, EPI_ISL_980051, EPI_ISL_980052, EPI_ISL_980062, EPI_ISL_980086, EPI_ISL_980096, EPI_ISL_980106, EPI_ISL_980112, EPI_ISL_980146, EPI_ISL_980166, EPI_ISL_980191, EPI_ISL_980197, EPI_ISL_980209, EPI_ISL_980214, EPI_ISL_980216, EPI_ISL_980223, EPI_ISL_980566, EPI_ISL_980576, EPI_ISL_980578, EPI_ISL_980587, EPI_ISL_980592, EPI_ISL_980605, EPI_ISL_980620, EPI_ISL_980678, EPI_ISL_980682, EPI_ISL_980698, EPI_ISL_980721, EPI_ISL_980752, EPI_ISL_980753, EPI_ISL_980762, EPI_ISL_980784, EPI_ISL_980799, EPI_ISL_980803, EPI_ISL_985946, EPI_ISL_985947, EPI_ISL_985948, EPI_ISL_985949, EPI_ISL_985950, EPI_ISL_985951, EPI_ISL_985952, EPI_ISL_985953, EPI_ISL_985954, EPI_ISL_985955, EPI_ISL_985956, EPI_ISL_985957, EPI_ISL_985958, EPI_ISL_985959, EPI_ISL_985960, EPI_ISL_985961, EPI_ISL_985962, EPI_ISL_985963, EPI_ISL_985964, EPI_ISL_985965, EPI_ISL_985966, EPI_ISL_985967, EPI_ISL_985968, EPI_ISL_985969, EPI_ISL_985970, EPI_ISL_985971, EPI_ISL_985972, EPI_ISL_985973, EPI_ISL_985974, EPI_ISL_985975, EPI_ISL_985976, EPI_ISL_985977, EPI_ISL_985978, EPI_ISL_985979, EPI_ISL_985980, EPI_ISL_985981, EPI_ISL_985982, EPI_ISL_985983, EPI_ISL_985984, EPI_ISL_985986, EPI_ISL_985987, EPI_ISL_985988, EPI_ISL_985989, EPI_ISL_985990, EPI_ISL_985991, EPI_ISL_985992, EPI_ISL_985993, EPI_ISL_985994, EPI_ISL_985995, EPI_ISL_985996, EPI_ISL_985997, EPI_ISL_985998, EPI_ISL_985999, EPI_ISL_986000, EPI_ISL_986001, EPI_ISL_986003, EPI_ISL_986004, EPI_ISL_986005, EPI_ISL_986006, EPI_ISL_986007, EPI_ISL_986008, EPI_ISL_986009, EPI_ISL_986010, EPI_ISL_986011, EPI_ISL_986012, EPI_ISL_986013, EPI_ISL_986014, EPI_ISL_986015, EPI_ISL_986016, EPI_ISL_986017, EPI_ISL_986018, EPI_ISL_986019, EPI_ISL_986020, EPI_ISL_986021, EPI_ISL_986022, EPI_ISL_986023, EPI_ISL_986024, EPI_ISL_986025, EPI_ISL_986026, EPI_ISL_986027, EPI_ISL_986028, EPI_ISL_986029, EPI_ISL_986030, EPI_ISL_986031, EPI_ISL_986032, EPI_ISL_986033, EPI_ISL_986034, EPI_ISL_986035, EPI_ISL_986036, EPI_ISL_986037, EPI_ISL_986038, EPI_ISL_986040, EPI_ISL_986041, EPI_ISL_986042, EPI_ISL_986043, EPI_ISL_986044, EPI_ISL_986045, EPI_ISL_986046, EPI_ISL_986047, EPI_ISL_986048, EPI_ISL_986050, EPI_ISL_986051, EPI_ISL_986052, EPI_ISL_986053, EPI_ISL_986054, EPI_ISL_986055, EPI_ISL_986056, EPI_ISL_986057, EPI_ISL_986058, EPI_ISL_986059, EPI_ISL_986060, EPI_ISL_986062, EPI_ISL_986063, EPI_ISL_986064, EPI_ISL_986065, EPI_ISL_986066, EPI_ISL_986067, EPI_ISL_986068, EPI_ISL_986069, EPI_ISL_986070, EPI_ISL_986071, EPI_ISL_986072, EPI_ISL_986073, EPI_ISL_986074, EPI_ISL_986076, EPI_ISL_986077, EPI_ISL_986078, EPI_ISL_986079, EPI_ISL_986080, EPI_ISL_986081, EPI_ISL_986082, EPI_ISL_986083, EPI_ISL_986084, EPI_ISL_986085, EPI_ISL_986086, EPI_ISL_986087, EPI_ISL_986088, EPI_ISL_986089, EPI_ISL_986091, EPI_ISL_986092, EPI_ISL_986093, EPI_ISL_986094, EPI_ISL_986095, EPI_ISL_986097, EPI_ISL_986098, EPI_ISL_986099, EPI_ISL_986100, EPI_ISL_986101, EPI_ISL_986102, EPI_ISL_986103, EPI_ISL_986105, EPI_ISL_986107, EPI_ISL_986108, EPI_ISL_986110, EPI_ISL_986111, EPI_ISL_986113, EPI_ISL_986121, EPI_ISL_986122, EPI_ISL_986124, EPI_ISL_986127, EPI_ISL_986129, EPI_ISL_986130, EPI_ISL_986131, EPI_ISL_986135, EPI_ISL_986136, EPI_ISL_986137, EPI_ISL_986138, EPI_ISL_986140, EPI_ISL_986141, EPI_ISL_986142, EPI_ISL_986143, EPI_ISL_986145, EPI_ISL_986146, EPI_ISL_986147, EPI_ISL_986148, EPI_ISL_986149, EPI_ISL_986150, EPI_ISL_986151, EPI_ISL_986153, EPI_ISL_986155, EPI_ISL_986158, EPI_ISL_986161, EPI_ISL_986164, EPI_ISL_986166, EPI_ISL_986168, EPI_ISL_986170, EPI_ISL_986173, EPI_ISL_986175, EPI_ISL_986176, EPI_ISL_986177, EPI_ISL_986179, EPI_ISL_986183, EPI_ISL_986186, EPI_ISL_986187, EPI_ISL_986188, EPI_ISL_986189, EPI_ISL_986191, EPI_ISL_986192, EPI_ISL_986193, EPI_ISL_986194, EPI_ISL_986195, EPI_ISL_986196, EPI_ISL_986198, EPI_ISL_986202, EPI_ISL_986203, EPI_ISL_986206, EPI_ISL_986207, EPI_ISL_986210, EPI_ISL_986214, EPI_ISL_986216, EPI_ISL_986219, EPI_ISL_986220, EPI_ISL_986221, EPI_ISL_986223, EPI_ISL_986224, EPI_ISL_986225, EPI_ISL_986226, EPI_ISL_986227, EPI_ISL_986228, EPI_ISL_986231, EPI_ISL_986232, EPI_ISL_986233, EPI_ISL_986235, EPI_ISL_986237, EPI_ISL_986240, EPI_ISL_986242, EPI_ISL_986245, EPI_ISL_986246, EPI_ISL_986247, EPI_ISL_986251, EPI_ISL_986252, EPI_ISL_986253, EPI_ISL_986256, EPI_ISL_986258, EPI_ISL_986259, EPI_ISL_986261, EPI_ISL_986264, EPI_ISL_986269, EPI_ISL_988205, EPI_ISL_988207, EPI_ISL_988209, EPI_ISL_988210, EPI_ISL_988213, EPI_ISL_988217, EPI_ISL_988218, EPI_ISL_988221, EPI_ISL_988222, EPI_ISL_988223, EPI_ISL_988224, EPI_ISL_988225, EPI_ISL_988229, EPI_ISL_988235, EPI_ISL_988236, EPI_ISL_988238, EPI_ISL_988239, EPI_ISL_988242, EPI_ISL_988243, EPI_ISL_988248, EPI_ISL_988249, EPI_ISL_988250, EPI_ISL_988254, EPI_ISL_988256, EPI_ISL_988261, EPI_ISL_988262, EPI_ISL_988263, EPI_ISL_988270, EPI_ISL_988271, EPI_ISL_988276, EPI_ISL_988286, EPI_ISL_988290, EPI_ISL_988296, EPI_ISL_988299, EPI_ISL_988305, EPI_ISL_988306, EPI_ISL_988307, EPI_ISL_988308, EPI_ISL_988309, EPI_ISL_988310, EPI_ISL_988312, EPI_ISL_988313, EPI_ISL_988317, EPI_ISL_988321, EPI_ISL_988325, EPI_ISL_988329, EPI_ISL_988332, EPI_ISL_988335, EPI_ISL_988337, EPI_ISL_988347, EPI_ISL_988351, EPI_ISL_988354, EPI_ISL_988357, EPI_ISL_988358, EPI_ISL_988362, EPI_ISL_988363, EPI_ISL_988364, EPI_ISL_988370, EPI_ISL_988377, EPI_ISL_988378, EPI_ISL_988383, EPI_ISL_988384, EPI_ISL_988385, EPI_ISL_988389, EPI_ISL_988391, EPI_ISL_988395, EPI_ISL_988398, EPI_ISL_988399, EPI_ISL_988403, EPI_ISL_988404, EPI_ISL_988417, EPI_ISL_988418, EPI_ISL_988423, EPI_ISL_988426, EPI_ISL_988427, EPI_ISL_988428, EPI_ISL_988430, EPI_ISL_988434, EPI_ISL_988436, EPI_ISL_988446, EPI_ISL_988448, EPI_ISL_988449, EPI_ISL_988452, EPI_ISL_988453, EPI_ISL_988459, EPI_ISL_988460, EPI_ISL_988463, EPI_ISL_988468, EPI_ISL_988480, EPI_ISL_988482, EPI_ISL_988484, EPI_ISL_988487, EPI_ISL_988489, EPI_ISL_988491, EPI_ISL_988492, EPI_ISL_988494, EPI_ISL_988496, EPI_ISL_988497, EPI_ISL_988498, EPI_ISL_988499, EPI_ISL_988500, EPI_ISL_988501, EPI_ISL_988502, EPI_ISL_988503, EPI_ISL_988505, EPI_ISL_988506, EPI_ISL_988507, EPI_ISL_988508, EPI_ISL_988509, EPI_ISL_988510, EPI_ISL_988511, EPI_ISL_988512, EPI_ISL_988514, EPI_ISL_988518, EPI_ISL_988519, EPI_ISL_988521, EPI_ISL_988523, EPI_ISL_988524, EPI_ISL_988525, EPI_ISL_988527, EPI_ISL_988528, EPI_ISL_988529, EPI_ISL_988530, EPI_ISL_988531, EPI_ISL_988532, EPI_ISL_988533, EPI_ISL_988535, EPI_ISL_988536, EPI_ISL_988540, EPI_ISL_988541, EPI_ISL_988542, EPI_ISL_988543, EPI_ISL_988544, EPI_ISL_988546, EPI_ISL_988547, EPI_ISL_988548, EPI_ISL_988549, EPI_ISL_988550, EPI_ISL_988553, EPI_ISL_988556, EPI_ISL_988557, EPI_ISL_988559, EPI_ISL_988560, EPI_ISL_988561, EPI_ISL_988562, EPI_ISL_988563, EPI_ISL_988565, EPI_ISL_988566, EPI_ISL_988567, EPI_ISL_988569, EPI_ISL_988570, EPI_ISL_988571, EPI_ISL_988572, EPI_ISL_988573, EPI_ISL_988574, EPI_ISL_988575, EPI_ISL_988578, EPI_ISL_988579, EPI_ISL_988580, EPI_ISL_988581, EPI_ISL_988582, EPI_ISL_988583, EPI_ISL_988584, EPI_ISL_988587, EPI_ISL_988588, EPI_ISL_988590, EPI_ISL_988591, EPI_ISL_988593, EPI_ISL_988594, EPI_ISL_988595, EPI_ISL_988596, EPI_ISL_988597, EPI_ISL_988598, EPI_ISL_988599, EPI_ISL_988602, EPI_ISL_988603, EPI_ISL_988606, EPI_ISL_988607, EPI_ISL_988608, EPI_ISL_988609, EPI_ISL_988610, EPI_ISL_988612, EPI_ISL_988613, EPI_ISL_988614, EPI_ISL_988616, EPI_ISL_988617, EPI_ISL_988618, EPI_ISL_988621, EPI_ISL_988622, EPI_ISL_988625, EPI_ISL_988626, EPI_ISL_988627, EPI_ISL_988628, EPI_ISL_988629, EPI_ISL_988630, EPI_ISL_988633, EPI_ISL_988634, EPI_ISL_988635, EPI_ISL_988636, EPI_ISL_988637, EPI_ISL_988638, EPI_ISL_988639, EPI_ISL_988641, EPI_ISL_988642, EPI_ISL_988643, EPI_ISL_988644, EPI_ISL_988645, EPI_ISL_988650, EPI_ISL_988653, EPI_ISL_988654, EPI_ISL_988656, EPI_ISL_988657, EPI_ISL_988658, EPI_ISL_988659, EPI_ISL_988660, EPI_ISL_988662, EPI_ISL_988664, EPI_ISL_988666, EPI_ISL_988667, EPI_ISL_988668, EPI_ISL_988669, EPI_ISL_988670, EPI_ISL_988672, EPI_ISL_988673, EPI_ISL_988674, EPI_ISL_988675, EPI_ISL_988676, EPI_ISL_988677, EPI_ISL_988678, EPI_ISL_988679, EPI_ISL_988680, EPI_ISL_988682, EPI_ISL_988684, EPI_ISL_1005961, EPI_ISL_1005963, EPI_ISL_1005965, EPI_ISL_1005969, EPI_ISL_1005970, EPI_ISL_1005972, EPI_ISL_1005973, EPI_ISL_1005975, EPI_ISL_1005977, EPI_ISL_1005978, EPI_ISL_1005980, EPI_ISL_1005982, EPI_ISL_1005983, EPI_ISL_1005984, EPI_ISL_1005985, EPI_ISL_1005987, EPI_ISL_1005988, EPI_ISL_1005990, EPI_ISL_1005991, EPI_ISL_1005992, EPI_ISL_1005994, EPI_ISL_1005996, EPI_ISL_1005999, EPI_ISL_1006000, EPI_ISL_1006002, EPI_ISL_1006003, EPI_ISL_1006005, EPI_ISL_1006007, EPI_ISL_1006009, EPI_ISL_1006010, EPI_ISL_1006011, EPI_ISL_1006012, EPI_ISL_1006019, EPI_ISL_1006021, EPI_ISL_1006022, EPI_ISL_1006023, EPI_ISL_1006024, EPI_ISL_1006026, EPI_ISL_1006027, EPI_ISL_1006028, EPI_ISL_1006029, EPI_ISL_1006031, EPI_ISL_1006032, EPI_ISL_1006034, EPI_ISL_1006036, EPI_ISL_1006037, EPI_ISL_1006039, EPI_ISL_1006040, EPI_ISL_1006043, EPI_ISL_1006044, EPI_ISL_1006045, EPI_ISL_1006048, EPI_ISL_1006049, EPI_ISL_1006051, EPI_ISL_1006053, EPI_ISL_1006058, EPI_ISL_1006060, EPI_ISL_1006067, EPI_ISL_1006069, EPI_ISL_1006071, EPI_ISL_1006072, EPI_ISL_1006076, EPI_ISL_1006078, EPI_ISL_1006079, EPI_ISL_1006081, EPI_ISL_1006082, EPI_ISL_1006084, EPI_ISL_1006085, EPI_ISL_1006086, EPI_ISL_1006088, EPI_ISL_1006089, EPI_ISL_1006092, EPI_ISL_1006095, EPI_ISL_1006099, EPI_ISL_1006100, EPI_ISL_1006101, EPI_ISL_1006102, EPI_ISL_1006103, EPI_ISL_1006104, EPI_ISL_1006105, EPI_ISL_1006107, EPI_ISL_1006110, EPI_ISL_1006111, EPI_ISL_1006113, EPI_ISL_1006115, EPI_ISL_1006116, EPI_ISL_1006118, EPI_ISL_1006120, EPI_ISL_1006121, EPI_ISL_1006123, EPI_ISL_1006124, EPI_ISL_1006125, EPI_ISL_1006126, EPI_ISL_1006127, EPI_ISL_1006130, EPI_ISL_1006131, EPI_ISL_1006132, EPI_ISL_1006133, EPI_ISL_1006135, EPI_ISL_1006137, EPI_ISL_1006142, EPI_ISL_1006143, EPI_ISL_1006144, EPI_ISL_1006146, EPI_ISL_1006147, EPI_ISL_1006148, EPI_ISL_1006149, EPI_ISL_1006153, EPI_ISL_1006154, EPI_ISL_1006155, EPI_ISL_1006157, EPI_ISL_1006158, EPI_ISL_1006159, EPI_ISL_1006162, EPI_ISL_1006163, EPI_ISL_1006166, EPI_ISL_1006171, EPI_ISL_1006172, EPI_ISL_1006173, EPI_ISL_1006175, EPI_ISL_1006176, EPI_ISL_1006177, EPI_ISL_1006179, EPI_ISL_1006180, EPI_ISL_1006181, EPI_ISL_1006183, EPI_ISL_1006184, EPI_ISL_1006185, EPI_ISL_1006544, EPI_ISL_1006545, EPI_ISL_1006547, EPI_ISL_1006550, EPI_ISL_1006551, EPI_ISL_1006553, EPI_ISL_1006555, EPI_ISL_1006556, EPI_ISL_1006559, EPI_ISL_1006560, EPI_ISL_1006565, EPI_ISL_1006566, EPI_ISL_1006568, EPI_ISL_1006570, EPI_ISL_1006573, EPI_ISL_1006574, EPI_ISL_1006576, EPI_ISL_1006579, EPI_ISL_1006580, EPI_ISL_1006582, EPI_ISL_1006583, EPI_ISL_1006584, EPI_ISL_1006586, EPI_ISL_1006589, EPI_ISL_1006590, EPI_ISL_1006591, EPI_ISL_1006594, EPI_ISL_1006601, EPI_ISL_1006602, EPI_ISL_1006604, EPI_ISL_1006605, EPI_ISL_1006606, EPI_ISL_1006607, EPI_ISL_1006609, EPI_ISL_1006613, EPI_ISL_1006614, EPI_ISL_1006616, EPI_ISL_1006617, EPI_ISL_1006620, EPI_ISL_1006621, EPI_ISL_1006624, EPI_ISL_1006626, EPI_ISL_1006630, EPI_ISL_1006632, EPI_ISL_1006634, EPI_ISL_1006636, EPI_ISL_1006637, EPI_ISL_1006639, EPI_ISL_1006642, EPI_ISL_1006645, EPI_ISL_1006646, EPI_ISL_1006649, EPI_ISL_1006650, EPI_ISL_1006651, EPI_ISL_1006656, EPI_ISL_1006657, EPI_ISL_1006658, EPI_ISL_1006659, EPI_ISL_1006660, EPI_ISL_1006661, EPI_ISL_1006662, EPI_ISL_1006663, EPI_ISL_1006664, EPI_ISL_1006665, EPI_ISL_1006666, EPI_ISL_1006667, EPI_ISL_1006668, EPI_ISL_1006669, EPI_ISL_1006670, EPI_ISL_1006671, EPI_ISL_1006672, EPI_ISL_1006673, EPI_ISL_1006674, EPI_ISL_1006675, EPI_ISL_1006676, EPI_ISL_1006677, EPI_ISL_1006678, EPI_ISL_1006679, EPI_ISL_1006680, EPI_ISL_1006681, EPI_ISL_1006682, EPI_ISL_1006683, EPI_ISL_1006684, EPI_ISL_1006685, EPI_ISL_1006686, EPI_ISL_1006687, EPI_ISL_1006688, EPI_ISL_1006689, EPI_ISL_1006690, EPI_ISL_1006691, EPI_ISL_1006692, EPI_ISL_1006693, EPI_ISL_1006694, EPI_ISL_1006695, EPI_ISL_1006696, EPI_ISL_1006697, EPI_ISL_1006698, EPI_ISL_1006699, EPI_ISL_1006700, EPI_ISL_1006701, EPI_ISL_1006702, EPI_ISL_1006703, EPI_ISL_1006704, EPI_ISL_1006705, EPI_ISL_1006706, EPI_ISL_1006707, EPI_ISL_1006708, EPI_ISL_1006709, EPI_ISL_1006710, EPI_ISL_1006711, EPI_ISL_1006712, EPI_ISL_1006713, EPI_ISL_1006714, EPI_ISL_1006715, EPI_ISL_1006716, EPI_ISL_1006717, EPI_ISL_1006718, EPI_ISL_1006719, EPI_ISL_1006720, EPI_ISL_1006721, EPI_ISL_1006722, EPI_ISL_1006723, EPI_ISL_1006724, EPI_ISL_1006725, EPI_ISL_1006726, EPI_ISL_1006727, EPI_ISL_1006728, EPI_ISL_1006729, EPI_ISL_1006730, EPI_ISL_1006731, EPI_ISL_1006732, EPI_ISL_1006733, EPI_ISL_1006734, EPI_ISL_1006735, EPI_ISL_1006736, EPI_ISL_1006737, EPI_ISL_1006738, EPI_ISL_1006739, EPI_ISL_1006740, EPI_ISL_1006741, EPI_ISL_1006742, EPI_ISL_1006743, EPI_ISL_1006744, EPI_ISL_1006745, EPI_ISL_1006746, EPI_ISL_1006747, EPI_ISL_1006748, EPI_ISL_1006749, EPI_ISL_1006750, EPI_ISL_1006751, EPI_ISL_1006752, EPI_ISL_1006753, EPI_ISL_1006754, EPI_ISL_1006755, EPI_ISL_1006756, EPI_ISL_1006757, | | | |

EPI_ISL_1006758, EPI_ISL_1006759, EPI_ISL_1006760, EPI_ISL_1006761, EPI_ISL_1006762, EPI_ISL_1006763, EPI_ISL_1006764, EPI_ISL_1006765, EPI_ISL_1006766, EPI_ISL_1006767, EPI_ISL_1006768, EPI_ISL_1006769, EPI_ISL_1006770, EPI_ISL_1006771, EPI_ISL_1006772, EPI_ISL_1006773, EPI_ISL_1006774, EPI_ISL_1006775, EPI_ISL_1006776, EPI_ISL_1006777, EPI_ISL_1006778, EPI_ISL_1006779, EPI_ISL_1006780, EPI_ISL_1006781, EPI_ISL_1006782, EPI_ISL_1006783, EPI_ISL_1006784, EPI_ISL_1006785, EPI_ISL_1006786, EPI_ISL_1006787, EPI_ISL_1006788, EPI_ISL_1006789, EPI_ISL_1006790, EPI_ISL_1006791, EPI_ISL_1006792, EPI_ISL_1006793, EPI_ISL_1006794, EPI_ISL_1006795, EPI_ISL_1006796, EPI_ISL_1006797, EPI_ISL_1006798, EPI_ISL_1006799, EPI_ISL_1006800, EPI_ISL_1006801, EPI_ISL_1006802, EPI_ISL_1006803, EPI_ISL_1006804, EPI_ISL_1006805, EPI_ISL_1006806, EPI_ISL_1006807, EPI_ISL_1006808, EPI_ISL_1006809, EPI_ISL_1006810, EPI_ISL_1006811, EPI_ISL_1006812, EPI_ISL_1006813, EPI_ISL_1006814, EPI_ISL_1006815, EPI_ISL_1006816, EPI_ISL_1006817, EPI_ISL_1006818, EPI_ISL_1006819, EPI_ISL_1006820, EPI_ISL_1006821, EPI_ISL_1006822, EPI_ISL_1006823, EPI_ISL_1006824, EPI_ISL_1006825, EPI_ISL_1006826, EPI_ISL_1006827, EPI_ISL_1006828, EPI_ISL_1006829, EPI_ISL_1006830, EPI_ISL_1006831, EPI_ISL_1006832, EPI_ISL_1006833, EPI_ISL_1006834, EPI_ISL_1006835, EPI_ISL_1006836, EPI_ISL_1006837, EPI_ISL_1006838, EPI_ISL_1006839, EPI_ISL_1006840, EPI_ISL_1006841, EPI_ISL_1006842, EPI_ISL_1006843, EPI_ISL_1006844, EPI_ISL_1006845, EPI_ISL_1006846, EPI_ISL_1006847, EPI_ISL_1006848, EPI_ISL_1006849, EPI_ISL_1006850, EPI_ISL_1006851, EPI_ISL_1006852, EPI_ISL_1006853, EPI_ISL_1006854, EPI_ISL_1006855, EPI_ISL_1006856, EPI_ISL_1006857, EPI_ISL_1006858, EPI_ISL_1006859, EPI_ISL_1006860, EPI_ISL_1006861, EPI_ISL_1006862, EPI_ISL_1006863, EPI_ISL_1006941, EPI_ISL_1006942, EPI_ISL_1006943, EPI_ISL_1006944, EPI_ISL_1006945, EPI_ISL_1006946, EPI_ISL_1006947, EPI_ISL_1006948, EPI_ISL_1006949, EPI_ISL_1006950, EPI_ISL_1006951, EPI_ISL_1006952, EPI_ISL_1006953, EPI_ISL_1006954, EPI_ISL_1006955, EPI_ISL_1006956, EPI_ISL_1006957, EPI_ISL_1006958, EPI_ISL_1006959, EPI_ISL_1006960, EPI_ISL_1006961, EPI_ISL_1006962, EPI_ISL_1006963, EPI_ISL_1006964, EPI_ISL_1006965, EPI_ISL_1006966, EPI_ISL_1006967, EPI_ISL_1006968, EPI_ISL_1006969, EPI_ISL_1006970, EPI_ISL_1006971, EPI_ISL_1006972, EPI_ISL_1006973, EPI_ISL_1006974, EPI_ISL_1006975, EPI_ISL_1006976, EPI_ISL_1006977, EPI_ISL_1006978, EPI_ISL_1006979, EPI_ISL_1006980, EPI_ISL_1006981, EPI_ISL_1006982, EPI_ISL_1006983, EPI_ISL_1006984, EPI_ISL_1006985, EPI_ISL_1006986, EPI_ISL_1006987, EPI_ISL_1006988, EPI_ISL_1006989, EPI_ISL_1006990, EPI_ISL_1006991, EPI_ISL_1006992, EPI_ISL_1006993, EPI_ISL_1006994, EPI_ISL_1006995, EPI_ISL_1006996, EPI_ISL_1006997, EPI_ISL_1006998, EPI_ISL_1006999, EPI_ISL_1007000, EPI_ISL_1007001, EPI_ISL_1007002, EPI_ISL_1007003, EPI_ISL_1007004, EPI_ISL_1007005, EPI_ISL_1007006, EPI_ISL_1007007, EPI_ISL_1007008, EPI_ISL_1007009, EPI_ISL_1007010, EPI_ISL_1007011, EPI_ISL_1007012, EPI_ISL_1007013, EPI_ISL_1007014, EPI_ISL_1007015, EPI_ISL_1007016, EPI_ISL_1007017, EPI_ISL_1007018, EPI_ISL_1007019, EPI_ISL_1007020, EPI_ISL_1007021, EPI_ISL_1007022, EPI_ISL_1007023, EPI_ISL_1007024, EPI_ISL_1007025, EPI_ISL_1007026, EPI_ISL_1007027, EPI_ISL_1007028, EPI_ISL_1007029, EPI_ISL_1007030, EPI_ISL_1007031, EPI_ISL_1007032, EPI_ISL_1007033, EPI_ISL_1007034, EPI_ISL_1007035, EPI_ISL_1007036, EPI_ISL_1007037, EPI_ISL_1007038, EPI_ISL_1007039, EPI_ISL_1007040, EPI_ISL_1007041, EPI_ISL_1007042, EPI_ISL_1007043, EPI_ISL_1007044, EPI_ISL_1007045, EPI_ISL_1007046, EPI_ISL_1007047, EPI_ISL_1007048, EPI_ISL_1007049, EPI_ISL_1007050, EPI_ISL_1007051, EPI_ISL_1007052, EPI_ISL_1007053, EPI_ISL_1007054, EPI_ISL_1007055, EPI_ISL_1007056, EPI_ISL_1007057, EPI_ISL_1007058, EPI_ISL_1007059, EPI_ISL_1007060, EPI_ISL_1007061, EPI_ISL_1007062, EPI_ISL_1007063, EPI_ISL_1007064, EPI_ISL_1007065, EPI_ISL_1007066, EPI_ISL_1007067, EPI_ISL_1007068, EPI_ISL_1007069, EPI_ISL_1007070, EPI_ISL_1007071, EPI_ISL_1007072, EPI_ISL_1007073, EPI_ISL_1007074, EPI_ISL_1007075, EPI_ISL_1007076, EPI_ISL_1007077, EPI_ISL_1007078, EPI_ISL_1007079, EPI_ISL_1007080, EPI_ISL_1007081, EPI_ISL_1007082, EPI_ISL_1007083, EPI_ISL_1007084, EPI_ISL_1007085, EPI_ISL_1007086, EPI_ISL_1007087, EPI_ISL_1007088, EPI_ISL_1007089, EPI_ISL_1007090, EPI_ISL_1007091, EPI_ISL_1007092, EPI_ISL_1007093, EPI_ISL_1007094, EPI_ISL_1007095, EPI_ISL_1007096, EPI_ISL_1007097, EPI_ISL_1007098, EPI_ISL_1007099, EPI_ISL_1007100, EPI_ISL_1007101, EPI_ISL_1007102, EPI_ISL_1007103, EPI_ISL_1007104, EPI_ISL_1007105, EPI_ISL_1007106, EPI_ISL_1007107, EPI_ISL_1007108, EPI_ISL_1007109, EPI_ISL_1007110, EPI_ISL_1007111, EPI_ISL_1007112, EPI_ISL_1007113, EPI_ISL_1007114, EPI_ISL_1007115, EPI_ISL_1007116, EPI_ISL_1007117, EPI_ISL_1007118, EPI_ISL_1007119, EPI_ISL_1007120, EPI_ISL_1007121, EPI_ISL_1007122, EPI_ISL_1007123, EPI_ISL_1007124, EPI_ISL_1007125, EPI_ISL_1007126, EPI_ISL_1007127, EPI_ISL_1007128, EPI_ISL_1007129, EPI_ISL_1007130, EPI_ISL_1007131, EPI_ISL_1007132, EPI_ISL_1007133, EPI_ISL_1007134, EPI_ISL_1007135, EPI_ISL_1007136, EPI_ISL_1007137, EPI_ISL_1007138, EPI_ISL_1007139, EPI_ISL_1007140, EPI_ISL_1007141, EPI_ISL_1007142, EPI_ISL_1007143, EPI_ISL_1007144, EPI_ISL_1007145, EPI_ISL_1007146, EPI_ISL_1007147, EPI_ISL_1007148, EPI_ISL_1007149, EPI_ISL_1007150, EPI_ISL_1007151, EPI_ISL_1007152, EPI_ISL_1007153, EPI_ISL_1007154, EPI_ISL_1007155, EPI_ISL_1007156, EPI_ISL_1007157, EPI_ISL_1007158, EPI_ISL_1007159, EPI_ISL_1007160, EPI_ISL_1007161, EPI_ISL_1007162, EPI_ISL_1007163, EPI_ISL_1007164, EPI_ISL_1007165, EPI_ISL_1007166, EPI_ISL_1007167, EPI_ISL_1007168, EPI_ISL_1007169, EPI_ISL_1007170, EPI_ISL_1007171, EPI_ISL_1007172, EPI_ISL_1007173, EPI_ISL_1007174, EPI_ISL_1007175, EPI_ISL_1007176, EPI_ISL_1007177, EPI_ISL_1007178, EPI_ISL_1007179, EPI_ISL_1007180, EPI_ISL_1007526, EPI_ISL_1007527, EPI_ISL_1012404, EPI_ISL_1012405, EPI_ISL_1012406, EPI_ISL_1012408, EPI_ISL_1012410, EPI_ISL_1012411, EPI_ISL_1012412, EPI_ISL_1012413, EPI_ISL_1012415, EPI_ISL_1012417, EPI_ISL_1012418, EPI_ISL_1012419, EPI_ISL_1012420, EPI_ISL_1012422, EPI_ISL_1012424, EPI_ISL_1012425, EPI_ISL_1012426, EPI_ISL_1012427, EPI_ISL_1012429, EPI_ISL_1012430, EPI_ISL_1012432, EPI_ISL_1012434, EPI_ISL_1012435, EPI_ISL_1012436, EPI_ISL_1012437, EPI_ISL_1012439, EPI_ISL_1012440, EPI_ISL_1012441, EPI_ISL_1012442, EPI_ISL_1012443, EPI_ISL_1012444, EPI_ISL_1012445, EPI_ISL_1012446, EPI_ISL_1012447, EPI_ISL_1012448, EPI_ISL_1012451, EPI_ISL_1012452, EPI_ISL_1012454, EPI_ISL_1012455, EPI_ISL_1012456, EPI_ISL_1012459, EPI_ISL_1012460, EPI_ISL_1012461, EPI_ISL_1012462, EPI_ISL_1012463, EPI_ISL_1012464, EPI_ISL_1012465, EPI_ISL_1012466, EPI_ISL_1012467, EPI_ISL_1012468, EPI_ISL_1012469, EPI_ISL_1012471, EPI_ISL_1012472, EPI_ISL_1012473, EPI_ISL_1012474, EPI_ISL_1012476, EPI_ISL_1012477, EPI_ISL_1012478, EPI_ISL_1012479, EPI_ISL_1012480, EPI_ISL_1012481, EPI_ISL_1012483, EPI_ISL_1012485, EPI_ISL_1012486, EPI_ISL_1012487, EPI_ISL_1012488, EPI_ISL_1012489, EPI_ISL_1012490, EPI_ISL_1012491, EPI_ISL_1012492, EPI_ISL_1012494, EPI_ISL_1012495, EPI_ISL_1012496, EPI_ISL_1012497, EPI_ISL_1012501, EPI_ISL_1012502, EPI_ISL_1012503, EPI_ISL_1012504, EPI_ISL_1012505, EPI_ISL_1012506, EPI_ISL_1012507, EPI_ISL_1012508, EPI_ISL_1012509, EPI_ISL_1012510, EPI_ISL_1012511, EPI_ISL_1012512, EPI_ISL_1012513, EPI_ISL_1012515, EPI_ISL_1012516, EPI_ISL_1012517, EPI_ISL_1012518, EPI_ISL_1012519, EPI_ISL_1012520, EPI_ISL_1012522, EPI_ISL_1012523, EPI_ISL_1012524, EPI_ISL_1012525, EPI_ISL_1012526, EPI_ISL_1012527, EPI_ISL_1012528, EPI_ISL_1012529, EPI_ISL_1012530, EPI_ISL_1012533, EPI_ISL_1012534, EPI_ISL_1012536, EPI_ISL_1012537, EPI_ISL_1012538, EPI_ISL_1012539, EPI_ISL_1012540, EPI_ISL_1012542, EPI_ISL_1012543, EPI_ISL_1012544, EPI_ISL_1012545, EPI_ISL_1012546, EPI_ISL_1012547, EPI_ISL_1012548, EPI_ISL_1012669, EPI_ISL_1012670, EPI_ISL_1012671, EPI_ISL_1019481, EPI_ISL_1033566, EPI_ISL_1033574, EPI_ISL_1033579, EPI_ISL_1033583, EPI_ISL_1033584, EPI_ISL_1033589, EPI_ISL_1033590, EPI_ISL_1033592, EPI_ISL_1033595, EPI_ISL_1033598, EPI_ISL_1033600, EPI_ISL_1033602, EPI_ISL_1033603, EPI_ISL_1033607, EPI_ISL_1033611, EPI_ISL_1033612, EPI_ISL_1033614, EPI_ISL_1033619, EPI_ISL_1033622, EPI_ISL_1033625, EPI_ISL_1033626, EPI_ISL_1033627, EPI_ISL_1033628, EPI_ISL_1033631, EPI_ISL_1033632, EPI_ISL_1033636, EPI_ISL_1033638, EPI_ISL_1033641, EPI_ISL_1033645, EPI_ISL_1033648, EPI_ISL_1033649, EPI_ISL_1033650, EPI_ISL_1033651, EPI_ISL_1033655, EPI_ISL_1033656, EPI_ISL_1033658, EPI_ISL_1033661, EPI_ISL_1033664, EPI_ISL_1033668, EPI_ISL_1033671, EPI_ISL_1033676, EPI_ISL_1033679, EPI_ISL_1033680, EPI_ISL_1033683, EPI_ISL_1033686, EPI_ISL_1033688, EPI_ISL_1033691, EPI_ISL_1033692, EPI_ISL_1033698, EPI_ISL_1033703, EPI_ISL_1033707, EPI_ISL_1033710, EPI_ISL_1033714, EPI_ISL_1033715, EPI_ISL_1033722, EPI_ISL_1033723, EPI_ISL_1033725, EPI_ISL_1033727, EPI_ISL_1033728, EPI_ISL_1033729, EPI_ISL_1033730, EPI_ISL_1033733, EPI_ISL_1033740, EPI_ISL_1033742, EPI_ISL_1033746, EPI_ISL_1033747, EPI_ISL_1033748, EPI_ISL_1033751, EPI_ISL_1033753, EPI_ISL_1033754, EPI_ISL_1033756, EPI_ISL_1033757, EPI_ISL_1033763, EPI_ISL_1033770, EPI_ISL_1033777, EPI_ISL_1033779, EPI_ISL_1033787, EPI_ISL_1033794, EPI_ISL_1033795, EPI_ISL_1033798, EPI_ISL_1033799, EPI_ISL_1033803, EPI_ISL_1033813, EPI_ISL_1033814, EPI_ISL_1033821, EPI_ISL_1033822, EPI_ISL_1033825, EPI_ISL_1033830, EPI_ISL_1033835, EPI_ISL_1033840, EPI_ISL_1033843, EPI_ISL_1033844, EPI_ISL_1033846, EPI_ISL_1033854, EPI_ISL_1033856, EPI_ISL_1033864, EPI_ISL_1033865, EPI_ISL_1033867, EPI_ISL_1033869, EPI_ISL_1033872, EPI_ISL_1033874, EPI_ISL_1033875, EPI_ISL_1033881, EPI_ISL_1033885, EPI_ISL_1033886, EPI_ISL_1033887, EPI_ISL_1033891, EPI_ISL_1033896, EPI_ISL_1033897, EPI_ISL_1033900, EPI_ISL_1033901, EPI_ISL_1033903, EPI_ISL_1033905, EPI_ISL_1033906, EPI_ISL_1033914, EPI_ISL_1033916, EPI_ISL_1033917, EPI_ISL_1033948, EPI_ISL_1033954, EPI_ISL_1033974, EPI_ISL_1034006, EPI_ISL_1034019, EPI_ISL_1034021, EPI_ISL_1034030, EPI_ISL_1034044, EPI_ISL_1034045, EPI_ISL_1034056, EPI_ISL_1034067, EPI_ISL_1034078, EPI_ISL_1034080, EPI_ISL_1034082, EPI_ISL_1034090, EPI_ISL_1034097, EPI_ISL_1034099, EPI_ISL_1034111, EPI_ISL_1034112, EPI_ISL_1034115, EPI_ISL_1034134, EPI_ISL_1042632, EPI_ISL_1042796, EPI_ISL_1043308, EPI_ISL_1045520, EPI_ISL_1045888, EPI_ISL_1045905, EPI_ISL_1045906, EPI_ISL_1045909, EPI_ISL_1045910, EPI_ISL_1045915, EPI_ISL_1045916, EPI_ISL_1045919, EPI_ISL_1045922, EPI_ISL_1045928, EPI_ISL_1045929, EPI_ISL_1045930, EPI_ISL_1045935, EPI_ISL_1045941, EPI_ISL_1045946, EPI_ISL_1045950, EPI_ISL_1045951, EPI_ISL_1045953, EPI_ISL_1045963, EPI_ISL_1045964, EPI_ISL_1045966, EPI_ISL_1072959, EPI_ISL_1245398, EPI_ISL_1264168

| | | | |
|---|---|---|---|
| see above | Lighthouse Lab in Milton Keynes | Wellcome Sanger Institute for the COVID-19 Genomics UK (COG-UK) Consortium | Cordelia Langford; David K. Jackson; Dominic Kwiatkowski; Ewan Harrison; Ian Johnston; Jeffrey Barrett; John Sillitoe on behalf of the Wellcome Sanger Institute COVID-19 Surveillance Team; John Sillitoe on behalf of the Wellcome Sanger Institute COVID-19 Surveillance Team (http://www.sanger.ac.uk/covid-team); Roberto Amato; Sonia Goncalves; The Lighthouse Lab in Milton Keynes and Alex Alderton |

EPI_ISL_1105872, EPI_ISL_1105873, EPI_ISL_1105874, EPI_ISL_1105875, EPI_ISL_1105876, EPI_ISL_1105877, EPI_ISL_1105878, EPI_ISL_1105879, EPI_ISL_1105880, EPI_ISL_1105881, EPI_ISL_1105882, EPI_ISL_1105891, EPI_ISL_1105892, EPI_ISL_1105893, EPI_ISL_1105894, EPI_ISL_1105895, EPI_ISL_1105896, EPI_ISL_1105897, EPI_ISL_1105898, EPI_ISL_1105899, EPI_ISL_1105900, EPI_ISL_1105901, EPI_ISL_1105902, EPI_ISL_1105903, EPI_ISL_1105904, EPI_ISL_1105905, EPI_ISL_1105906, EPI_ISL_1105907, EPI_ISL_1105908, EPI_ISL_1105909, EPI_ISL_1178591, EPI_ISL_1178592

| | | | |
|---|---|---|---|
| see above | Lincolnshire Hospitals and DeepSeq Nottingham | COVID-19 Genomics UK (COG-UK) Consortium | Christopher Moore; Fei Sang; Joeseph Chappell; Johnny Debebe; Jonathan Ball; Matthew Carlisle; Matthew Loose; Nadine Holmes; Nichola Duckworth; Patrick McClure; Sarah Walsh; Tim Sloan; Victoria Wright |

EPI_ISL_949715, EPI_ISL_949716, EPI_ISL_997418, EPI_ISL_997419, EPI_ISL_997420, EPI_ISL_997421, EPI_ISL_997422, EPI_ISL_997443, EPI_ISL_997445, EPI_ISL_997451, EPI_ISL_997452, EPI_ISL_997453, EPI_ISL_997454, EPI_ISL_997455, EPI_ISL_997457, EPI_ISL_997458, EPI_ISL_997459, EPI_ISL_997460, EPI_ISL_997461, EPI_ISL_997464, EPI_ISL_997466, EPI_ISL_997483, EPI_ISL_997484, EPI_ISL_997485, EPI_ISL_997488, EPI_ISL_997490, EPI_ISL_997503, EPI_ISL_997504, EPI_ISL_997505, EPI_ISL_997506, EPI_ISL_997507, EPI_ISL_997508, EPI_ISL_997509, EPI_ISL_997510, EPI_ISL_997511, EPI_ISL_997512, EPI_ISL_997513, EPI_ISL_997514, EPI_ISL_997517, EPI_ISL_997518, EPI_ISL_997521, EPI_ISL_997522, EPI_ISL_997523, EPI_ISL_997524, EPI_ISL_997525, EPI_ISL_997526, EPI_ISL_997527, EPI_ISL_997528, EPI_ISL_997529, EPI_ISL_997532, EPI_ISL_997533, EPI_ISL_997540, EPI_ISL_997541, EPI_ISL_997542, EPI_ISL_997543, EPI_ISL_997544, EPI_ISL_997545, EPI_ISL_997546, EPI_ISL_997547, EPI_ISL_997548, EPI_ISL_997549, EPI_ISL_997550, EPI_ISL_997551, EPI_ISL_997552, EPI_ISL_997553, EPI_ISL_997554, EPI_ISL_997555, EPI_ISL_997556, EPI_ISL_997557, EPI_ISL_997558, EPI_ISL_997559, EPI_ISL_997560, EPI_ISL_997561, EPI_ISL_997562, EPI_ISL_997563, EPI_ISL_997564, EPI_ISL_997565, EPI_ISL_997566, EPI_ISL_997567, EPI_ISL_997568, EPI_ISL_997569, EPI_ISL_997570, EPI_ISL_1047960, EPI_ISL_1047961, EPI_ISL_1047962, EPI_ISL_1047963, EPI_ISL_1047964, EPI_ISL_1047965, EPI_ISL_1047966, EPI_ISL_1047967, EPI_ISL_1047971, EPI_ISL_1047973, EPI_ISL_1048028, EPI_ISL_1048030, EPI_ISL_1048031, EPI_ISL_1048032, EPI_ISL_1048033, EPI_ISL_1048034, EPI_ISL_1048035, EPI_ISL_1048036, EPI_ISL_1048041, EPI_ISL_1048042, EPI_ISL_1048044, EPI_ISL_1048045, EPI_ISL_1048046, EPI_ISL_1050355, EPI_ISL_1104324, EPI_ISL_1104325, EPI_ISL_1104326, EPI_ISL_1104327, EPI_ISL_1104328, EPI_ISL_1104329, EPI_ISL_1104330, EPI_ISL_1104331, EPI_ISL_1104332, EPI_ISL_1104385, EPI_ISL_1104386, EPI_ISL_1104389, EPI_ISL_1104395, EPI_ISL_1104433, EPI_ISL_1104435, EPI_ISL_1104437, EPI_ISL_1104438, EPI_ISL_1104439, EPI_ISL_1104441, EPI_ISL_1104442, EPI_ISL_1104443, EPI_ISL_1104444, EPI_ISL_1104445, EPI_ISL_1104446, EPI_ISL_1104447, EPI_ISL_1104448, EPI_ISL_1104449, EPI_ISL_1104450, EPI_ISL_1104452, EPI_ISL_1104453, EPI_ISL_1104454, EPI_ISL_1104455, EPI_ISL_1104508, EPI_ISL_1104509, EPI_ISL_1104510, EPI_ISL_1104511, EPI_ISL_1104512, EPI_ISL_1104513, EPI_ISL_1104515, EPI_ISL_1104516, EPI_ISL_1104517, EPI_ISL_1104518, EPI_ISL_1104519, EPI_ISL_1104520, EPI_ISL_1104535, EPI_ISL_1104536, EPI_ISL_1104537, EPI_ISL_1104538, EPI_ISL_1104539, EPI_ISL_1104540, EPI_ISL_1104541, EPI_ISL_1104569, EPI_ISL_1105524, EPI_ISL_1105525, EPI_ISL_1105526, EPI_ISL_1105528, EPI_ISL_1105529, EPI_ISL_1105530, EPI_ISL_1105531, EPI_ISL_1105532, EPI_ISL_1105533, EPI_ISL_1105534, EPI_ISL_1105535, EPI_ISL_1105536, EPI_ISL_1105537, EPI_ISL_1107793, EPI_ISL_1107794, EPI_ISL_1107795, EPI_ISL_1107796, EPI_ISL_1107797, EPI_ISL_1107798, EPI_ISL_1107799, EPI_ISL_1107800, EPI_ISL_1107802, EPI_ISL_1107803, EPI_ISL_1177894, EPI_ISL_1177897, EPI_ISL_1177898, EPI_ISL_1177907, EPI_ISL_1177914, EPI_ISL_1177957, EPI_ISL_1247826, EPI_ISL_1247864, EPI_ISL_1247865, EPI_ISL_1247866, EPI_ISL_1247867, EPI_ISL_1247869, EPI_ISL_1249253, EPI_ISL_1249254, EPI_ISL_1249255, EPI_ISL_1249256, EPI_ISL_1249258, EPI_ISL_1249259, EPI_ISL_1249261, EPI_ISL_1546041, EPI_ISL_1546042, EPI_ISL_1546043, EPI_ISL_1546045, EPI_ISL_1546047, EPI_ISL_1546049, EPI_ISL_1546050, EPI_ISL_1546052, EPI_ISL_1546054, EPI_ISL_1546056, EPI_ISL_1546058, EPI_ISL_1546059, EPI_ISL_1546061, EPI_ISL_1546063, EPI_ISL_1546065, EPI_ISL_1546067, EPI_ISL_1546069, EPI_ISL_1546080, EPI_ISL_1546085, EPI_ISL_1832342, EPI_ISL_1832343, EPI_ISL_1832344, EPI_ISL_1832345, EPI_ISL_1832346, EPI_ISL_1979131, EPI_ISL_1979132, EPI_ISL_1979134, EPI_ISL_1979135, EPI_ISL_1979137, EPI_ISL_1979142, EPI_ISL_1979143, EPI_ISL_1979145, EPI_ISL_1979154

| | | | |
|---|---|---|---|
| see above | Liverpool Clinical Laboratories | COVID-19 Genomics UK (COG-UK) Consortium | A Alrezaihi; Alessandro Gerada; Alistair Darby; Angela Cowell; Anita Lucaci; Anu Chawla; Cassie Olateju; Catherine Hartley; Charlotte Nelson; Ecaterina Vamos; Elaine O'Toole; Eleanor G Bentley; Ghada T Shawli; Hermione Webster; Isabel García-Dorival; James Johnson; James P Stewart; Jenifer Manson; Joanne Watts; Jones Benjamin; Jordan J Clark; Julian Hiscox; L Luu; Lucille Rainbow; M Almsaud; Margaret Hughes; Mark Whitehead; Matthew Gemmell; Miren Iturriza-Gomara; Muhannad Alruwaili; N.P Randle; Neil Swainston; PKF Gilmore; Parul Sharma; Rebekah Penrice-Randal; Richard Eccles; Richard Gregory; Sam Haldenby; Steve Paterson; Stuart D Armstrong; Trevor Ian Robinson; Ximeng Han |

EPI_ISL_950432, EPI_ISL_950434, EPI_ISL_950435, EPI_ISL_998504, EPI_ISL_998507, EPI_ISL_998508, EPI_ISL_998509, EPI_ISL_998510, EPI_ISL_998511, EPI_ISL_998512, EPI_ISL_998513, EPI_ISL_998514, EPI_ISL_998515, EPI_ISL_998516, EPI_ISL_998518, EPI_ISL_998519, EPI_ISL_998521, EPI_ISL_998522, EPI_ISL_998523, EPI_ISL_998524, EPI_ISL_998527, EPI_ISL_998528, EPI_ISL_998529, EPI_ISL_998530, EPI_ISL_998531, EPI_ISL_998532, EPI_ISL_998533, EPI_ISL_998534, EPI_ISL_998556, EPI_ISL_998572, EPI_ISL_998573, EPI_ISL_998574, EPI_ISL_998575, EPI_ISL_998576, EPI_ISL_998577, EPI_ISL_998578, EPI_ISL_998579, EPI_ISL_998580, EPI_ISL_998581, EPI_ISL_998582, EPI_ISL_998583, EPI_ISL_998584, EPI_ISL_998585, EPI_ISL_998586, EPI_ISL_1050993, EPI_ISL_1051268, EPI_ISL_1051269, EPI_ISL_1051270, EPI_ISL_1051271, EPI_ISL_1051272, EPI_ISL_1051273, EPI_ISL_1051274, EPI_ISL_1051275, EPI_ISL_1051276, EPI_ISL_1051277, EPI_ISL_1051278, EPI_ISL_1051279, EPI_ISL_1051280, EPI_ISL_1051281, EPI_ISL_1051282, EPI_ISL_1051283, EPI_ISL_1051284, EPI_ISL_1051285, EPI_ISL_1051286, EPI_ISL_1051287, EPI_ISL_1051288, EPI_ISL_1051289, EPI_ISL_1051290, EPI_ISL_1051291, EPI_ISL_1051292, EPI_ISL_1051293, EPI_ISL_1051294, EPI_ISL_1051295, EPI_ISL_1051296, EPI_ISL_1051297, EPI_ISL_1051298, EPI_ISL_1051299, EPI_ISL_1051300, EPI_ISL_1051301, EPI_ISL_1051302, EPI_ISL_1051303, EPI_ISL_1051304, EPI_ISL_1051305, EPI_ISL_1051306, EPI_ISL_1051307, EPI_ISL_1051308, EPI_ISL_1051309, EPI_ISL_1051310, EPI_ISL_1051311, EPI_ISL_1051312, EPI_ISL_1051313, EPI_ISL_1051314, EPI_ISL_1051315, EPI_ISL_1051316, EPI_ISL_1051317, EPI_ISL_1051318, EPI_ISL_1051319, EPI_ISL_1051320, EPI_ISL_1051321, EPI_ISL_1051322, EPI_ISL_1051323, EPI_ISL_1051324, EPI_ISL_1051325, EPI_ISL_1051326, EPI_ISL_1051327, EPI_ISL_1051328, EPI_ISL_1051329, EPI_ISL_1051330, EPI_ISL_1051331, EPI_ISL_1051332, EPI_ISL_1051333, EPI_ISL_1051334, EPI_ISL_1051335, EPI_ISL_1051336, EPI_ISL_1051337, EPI_ISL_1051338, EPI_ISL_1051339, EPI_ISL_1051340, EPI_ISL_1051403, EPI_ISL_1051404, EPI_ISL_1051405, EPI_ISL_1051406, EPI_ISL_1051407, EPI_ISL_1051408, EPI_ISL_1051409, EPI_ISL_1051410, EPI_ISL_1054131, EPI_ISL_1054132, EPI_ISL_1054133, EPI_ISL_1054134, EPI_ISL_1054135, EPI_ISL_1054136, EPI_ISL_1054137, EPI_ISL_1054138, EPI_ISL_1054139, EPI_ISL_1054140, EPI_ISL_1054142, EPI_ISL_1054143, EPI_ISL_1054144, EPI_ISL_1054145, EPI_ISL_1054146, EPI_ISL_1054147, EPI_ISL_1054148, EPI_ISL_1054149, EPI_ISL_1054150, EPI_ISL_1054151, EPI_ISL_1054152, EPI_ISL_1054153, EPI_ISL_1054154, EPI_ISL_1054155, EPI_ISL_1054156, EPI_ISL_1054157, EPI_ISL_1054158, EPI_ISL_1054159, EPI_ISL_1054160, EPI_ISL_1054161, EPI_ISL_1054162, EPI_ISL_1054163, EPI_ISL_1054164, EPI_ISL_1054165, EPI_ISL_1054166, EPI_ISL_1054167, EPI_ISL_1054168, EPI_ISL_1054169, EPI_ISL_1054170, EPI_ISL_1054171, EPI_ISL_1054172, EPI_ISL_1054173, EPI_ISL_1054174, EPI_ISL_1054175, EPI_ISL_1054176, EPI_ISL_1054177, EPI_ISL_1054178, EPI_ISL_1054179, EPI_ISL_1054180, EPI_ISL_1054181, EPI_ISL_1054182, EPI_ISL_1054183, EPI_ISL_1054184, EPI_ISL_1054185, EPI_ISL_1054186, EPI_ISL_1054187, EPI_ISL_1054188, EPI_ISL_1054189, EPI_ISL_1054190, EPI_ISL_1054191, EPI_ISL_1054193, EPI_ISL_1054195, EPI_ISL_1054196, EPI_ISL_1054197, EPI_ISL_1054198, EPI_ISL_1054199, EPI_ISL_1054200, EPI_ISL_1054201, EPI_ISL_1054202, EPI_ISL_1054203, EPI_ISL_1054204, EPI_ISL_1054205, EPI_ISL_1054206, EPI_ISL_1054207, EPI_ISL_1054208, EPI_ISL_1054209, EPI_ISL_1054210, EPI_ISL_1054211, EPI_ISL_1054212, EPI_ISL_1054213, EPI_ISL_1054214, EPI_ISL_1054215, EPI_ISL_1054216, EPI_ISL_1054217, EPI_ISL_1054218, EPI_ISL_1054219, EPI_ISL_1054220, EPI_ISL_1054221, EPI_ISL_1054222, EPI_ISL_1054223, EPI_ISL_1054224, EPI_ISL_1054225, EPI_ISL_1054226, EPI_ISL_1054227, EPI_ISL_1054228, EPI_ISL_1054229, EPI_ISL_1054230, EPI_ISL_1054231, EPI_ISL_1054232, EPI_ISL_1054234, EPI_ISL_1054235, EPI_ISL_1054236, EPI_ISL_1054237, EPI_ISL_1054238, EPI_ISL_1054240, EPI_ISL_1054242, EPI_ISL_1054244, EPI_ISL_1054245, EPI_ISL_1054246, EPI_ISL_1054247, EPI_ISL_1054249, EPI_ISL_1054251, EPI_ISL_1054253, EPI_ISL_1054255, EPI_ISL_1054256, EPI_ISL_1054257, EPI_ISL_1105473, EPI_ISL_1105474, EPI_ISL_1105475, EPI_ISL_1105476, EPI_ISL_1105477, EPI_ISL_1105478, EPI_ISL_1105479, EPI_ISL_1105480, EPI_ISL_1105481, EPI_ISL_1105482, EPI_ISL_1105483, EPI_ISL_1105484, EPI_ISL_1105485, EPI_ISL_1105487, EPI_ISL_1105489, EPI_ISL_1105490, EPI_ISL_1105491, EPI_ISL_1105492, EPI_ISL_1105493, EPI_ISL_1105496, EPI_ISL_1105497, EPI_ISL_1105498, EPI_ISL_1105499, EPI_ISL_1105500, EPI_ISL_1105501, EPI_ISL_1105502, EPI_ISL_1105503, EPI_ISL_1105504, EPI_ISL_1105505, EPI_ISL_1105506, EPI_ISL_1105507, EPI_ISL_1105508, EPI_ISL_1105509, EPI_ISL_1105510, EPI_ISL_1105513, EPI_ISL_1105514, EPI_ISL_1105515, EPI_ISL_1105516, EPI_ISL_1105517, EPI_ISL_1105518, EPI_ISL_1105521, EPI_ISL_1105522, EPI_ISL_1105523, EPI_ISL_1105572, EPI_ISL_1105731, EPI_ISL_1105732, EPI_ISL_1105733, EPI_ISL_1109063, EPI_ISL_1109078, EPI_ISL_1109089, EPI_ISL_1109099, EPI_ISL_1109107, EPI_ISL_1109132, EPI_ISL_1109163, EPI_ISL_1109166, EPI_ISL_1109174, EPI_ISL_1109179, EPI_ISL_1109180, EPI_ISL_1109181, EPI_ISL_1109182, EPI_ISL_1109187, EPI_ISL_1109188, EPI_ISL_1109190, EPI_ISL_1109192, EPI_ISL_1178121, EPI_ISL_1178122, EPI_ISL_1178123, EPI_ISL_1178158, EPI_ISL_1178159, EPI_ISL_1178160, EPI_ISL_1178161, EPI_ISL_1178162, EPI_ISL_1178163, EPI_ISL_1178164, EPI_ISL_1178165, EPI_ISL_1178166, EPI_ISL_1178167, EPI_ISL_1178168, EPI_ISL_1178206, EPI_ISL_1248822, EPI_ISL_1248823, EPI_ISL_1248824, EPI_ISL_1248825, EPI_ISL_1248830, EPI_ISL_1248831, EPI_ISL_1248832, EPI_ISL_1248901, EPI_ISL_1309647, EPI_ISL_1309648, EPI_ISL_1309649, EPI_ISL_1309650, EPI_ISL_1309651, EPI_ISL_1309654

| | | | |
|---|---|---|---|
| see above | Northumbria University / South Tees Hospitals NHS Foundation Trust / North Cumbria Integrated Care NHS Foundation Trust / North Tees and Hartlepool NHS Foundation Trust / Newcastle Hospitals NHS Foundation Trust | COVID-19 Genomics UK (COG-UK) Consortium | Andrew Nelson; Brendan Payne; Clive Graham; Darren L Smith; Debra Padgett; Edward Barton; Emma Swindells; Garren Scott; Gary Black; Gary Eltringham; Giles S Holt; Greg R Young; Jane Greenaway; Jennifer Collins; John Allan; Joshua Loh; Lynn Dover; Matthew Bashton; Mohammad A Tariq; Paul Baker; Sarah Essex; Steve Liggett; Wen C Yew; Yusri Taha |

EPI_ISL_952251, EPI_ISL_952261, EPI_ISL_952263, EPI_ISL_952264, EPI_ISL_952275, EPI_ISL_952276, EPI_ISL_952287, EPI_ISL_952288, EPI_ISL_952292, EPI_ISL_952293, EPI_ISL_952294, EPI_ISL_952295, EPI_ISL_952296, EPI_ISL_952297, EPI_ISL_952324, EPI_ISL_952325, EPI_ISL_952326, EPI_ISL_952329, EPI_ISL_952330, EPI_ISL_952339, EPI_ISL_952342, EPI_ISL_952343, EPI_ISL_952344, EPI_ISL_952345, EPI_ISL_952346, EPI_ISL_952347, EPI_ISL_952348, EPI_ISL_952349, EPI_ISL_952350, EPI_ISL_952353, EPI_ISL_952354, EPI_ISL_952355, EPI_ISL_952356, EPI_ISL_952357, EPI_ISL_952358, EPI_ISL_952359, EPI_ISL_952362, EPI_ISL_952363, EPI_ISL_952364, EPI_ISL_999517, EPI_ISL_999518, EPI_ISL_999519, EPI_ISL_999520, EPI_ISL_999521, EPI_ISL_999522, EPI_ISL_999523, EPI_ISL_999524, EPI_ISL_999525, EPI_ISL_999526, EPI_ISL_999528, EPI_ISL_999529, EPI_ISL_999530, EPI_ISL_999531, EPI_ISL_999532, EPI_ISL_999533, EPI_ISL_999534, EPI_ISL_999537, EPI_ISL_999538, EPI_ISL_999540, EPI_ISL_999541, EPI_ISL_999542, EPI_ISL_999543, EPI_ISL_999544, EPI_ISL_999545, EPI_ISL_999546, EPI_ISL_999547, EPI_ISL_999548, EPI_ISL_999549, EPI_ISL_999550, EPI_ISL_999552, EPI_ISL_999553, EPI_ISL_999554, EPI_ISL_999556, EPI_ISL_999557, EPI_ISL_999559, EPI_ISL_999560, EPI_ISL_999561, EPI_ISL_999562, EPI_ISL_999563, EPI_ISL_999565, EPI_ISL_999566, EPI_ISL_999567, EPI_ISL_999569, EPI_ISL_999571, EPI_ISL_999572, EPI_ISL_999574, EPI_ISL_999575, EPI_ISL_999576, EPI_ISL_999577, EPI_ISL_999578, EPI_ISL_999579, EPI_ISL_999580, EPI_ISL_999581, EPI_ISL_999582, EPI_ISL_999583, EPI_ISL_999585, EPI_ISL_999586, EPI_ISL_999588, EPI_ISL_999589, EPI_ISL_999590, EPI_ISL_999591, EPI_ISL_999592, EPI_ISL_999593, EPI_ISL_999595, EPI_ISL_999596, EPI_ISL_999597, EPI_ISL_999598, EPI_ISL_999599, EPI_ISL_999601, EPI_ISL_999602, EPI_ISL_999603, EPI_ISL_999605, EPI_ISL_999606, EPI_ISL_999607, EPI_ISL_999608, EPI_ISL_999609, EPI_ISL_999611, EPI_ISL_999612, EPI_ISL_999613, EPI_ISL_999614, EPI_ISL_999615, EPI_ISL_999616, EPI_ISL_999618, EPI_ISL_999620, EPI_ISL_999621, EPI_ISL_999622, EPI_ISL_999623, EPI_ISL_999625, EPI_ISL_999626, EPI_ISL_999627, EPI_ISL_999628, EPI_ISL_999630, EPI_ISL_999633, EPI_ISL_999634, EPI_ISL_999648, EPI_ISL_999650, EPI_ISL_999651, EPI_ISL_999655, EPI_ISL_999656, EPI_ISL_999657, EPI_ISL_999658, EPI_ISL_999752, EPI_ISL_999753, EPI_ISL_999764, EPI_ISL_999765, EPI_ISL_999767, EPI_ISL_999770, EPI_ISL_999771, EPI_ISL_999772, EPI_ISL_999773, EPI_ISL_999775, EPI_ISL_999776, EPI_ISL_999777, EPI_ISL_999778, EPI_ISL_999779, EPI_ISL_999781, EPI_ISL_999782, EPI_ISL_999783, EPI_ISL_999784, EPI_ISL_999785, EPI_ISL_999786, EPI_ISL_999787, EPI_ISL_999788, EPI_ISL_999789, EPI_ISL_999790, EPI_ISL_999791, EPI_ISL_999792, EPI_ISL_999793, EPI_ISL_999794, EPI_ISL_999795, EPI_ISL_999796, EPI_ISL_999797, EPI_ISL_999798, EPI_ISL_999799, EPI_ISL_999800, EPI_ISL_999801, EPI_ISL_999802, EPI_ISL_999803, EPI_ISL_999804, EPI_ISL_999805, EPI_ISL_999806, EPI_ISL_999807, EPI_ISL_999809, EPI_ISL_999810, EPI_ISL_999811, EPI_ISL_999818, EPI_ISL_999819, EPI_ISL_999829, EPI_ISL_999833, EPI_ISL_999834, EPI_ISL_999847, EPI_ISL_999848, EPI_ISL_999852, EPI_ISL_999853, EPI_ISL_999858, EPI_ISL_999859, EPI_ISL_999870, EPI_ISL_999871, EPI_ISL_999878, EPI_ISL_999879, EPI_ISL_999881, EPI_ISL_999886, EPI_ISL_999891, EPI_ISL_999892, EPI_ISL_999894, EPI_ISL_999900, EPI_ISL_999905, EPI_ISL_999906, EPI_ISL_999907, EPI_ISL_999908, EPI_ISL_999910, EPI_ISL_999911, EPI_ISL_999912, EPI_ISL_999915, EPI_ISL_999916, EPI_ISL_999917, EPI_ISL_999918, EPI_ISL_999919, EPI_ISL_999920, EPI_ISL_999921, EPI_ISL_999922, EPI_ISL_999923, EPI_ISL_999924, EPI_ISL_999925, EPI_ISL_999926, EPI_ISL_999927, EPI_ISL_999929, EPI_ISL_999930, EPI_ISL_999931, EPI_ISL_999933, EPI_ISL_999935, EPI_ISL_999936, EPI_ISL_999937, EPI_ISL_999938, EPI_ISL_999943, EPI_ISL_999944, EPI_ISL_1052141, EPI_ISL_1052148, EPI_ISL_1052158, EPI_ISL_1052164, EPI_ISL_1052178, EPI_ISL_1052179, EPI_ISL_1052180, EPI_ISL_1052181, EPI_ISL_1052182, EPI_ISL_1052184, EPI_ISL_1052186, EPI_ISL_1052187, EPI_ISL_1052189, EPI_ISL_1052196, EPI_ISL_1052197, EPI_ISL_1052198, EPI_ISL_1052199, EPI_ISL_1052200, EPI_ISL_1052201, EPI_ISL_1052202, EPI_ISL_1052205, EPI_ISL_1052206, EPI_ISL_1052208, EPI_ISL_1052209, EPI_ISL_1052210, EPI_ISL_1052211, EPI_ISL_1052212, EPI_ISL_1052214, EPI_ISL_1052217, EPI_ISL_1052218, EPI_ISL_1052219, EPI_ISL_1052220, EPI_ISL_1052221, EPI_ISL_1052223, EPI_ISL_1052224, EPI_ISL_1052226, EPI_ISL_1052227, EPI_ISL_1052228, EPI_ISL_1052229, EPI_ISL_1052230, EPI_ISL_1052231, EPI_ISL_1052232, EPI_ISL_1052233, EPI_ISL_1052235, EPI_ISL_1052236, EPI_ISL_1052237, EPI_ISL_1052238, EPI_ISL_1052239, EPI_ISL_1052240, EPI_ISL_1052241, EPI_ISL_1052242, EPI_ISL_1052243, EPI_ISL_1052244, EPI_ISL_1052245, EPI_ISL_1052246, EPI_ISL_1052247, EPI_ISL_1052248, EPI_ISL_1052249, EPI_ISL_1052254, EPI_ISL_1052255, EPI_ISL_1052256, EPI_ISL_1052257, EPI_ISL_1052258, EPI_ISL_1052260, EPI_ISL_1052285, EPI_ISL_1052286, EPI_ISL_1052288, EPI_ISL_1052292, EPI_ISL_1052293, EPI_ISL_1052296, EPI_ISL_1052298, EPI_ISL_1052299, EPI_ISL_1052304, EPI_ISL_1052312, EPI_ISL_1052320, EPI_ISL_1052322, EPI_ISL_1052323, EPI_ISL_1052324, EPI_ISL_1052327, EPI_ISL_1052328, EPI_ISL_1052329, EPI_ISL_1052330, EPI_ISL_1052332, EPI_ISL_1052333, EPI_ISL_1052334, EPI_ISL_1052335, EPI_ISL_1052336, EPI_ISL_1052337, EPI_ISL_1052339, EPI_ISL_1052340, EPI_ISL_1052341, EPI_ISL_1052342, EPI_ISL_1052343, EPI_ISL_1052345, EPI_ISL_1052350, EPI_ISL_1052352, EPI_ISL_1052355, EPI_ISL_1052357, EPI_ISL_1052358, EPI_ISL_1052359, EPI_ISL_1052360, EPI_ISL_1052361, EPI_ISL_1052362, EPI_ISL_1052365, EPI_ISL_1052366, EPI_ISL_1052367, EPI_ISL_1052371, EPI_ISL_1052373, EPI_ISL_1052375, EPI_ISL_1052377, EPI_ISL_1052379, EPI_ISL_1052380, EPI_ISL_1052381, EPI_ISL_1052382, EPI_ISL_1052383, EPI_ISL_1052384, EPI_ISL_1052385, EPI_ISL_1052386, EPI_ISL_1052390, EPI_ISL_1052392, EPI_ISL_1052393, EPI_ISL_1052395, EPI_ISL_1052397, EPI_ISL_1052399, EPI_ISL_1052401, EPI_ISL_1052404, EPI_ISL_1052405, EPI_ISL_1052407, EPI_ISL_1052410, EPI_ISL_1052411, EPI_ISL_1052414, EPI_ISL_1052418, EPI_ISL_1052419, EPI_ISL_1052420, EPI_ISL_1052421, EPI_ISL_1052426, EPI_ISL_1052427, EPI_ISL_1052429, EPI_ISL_1052430, EPI_ISL_1052431, EPI_ISL_1052432, EPI_ISL_1052433, EPI_ISL_1052435, EPI_ISL_1052437, EPI_ISL_1052438, EPI_ISL_1052440, EPI_ISL_1052443, EPI_ISL_1052444, EPI_ISL_1052446, EPI_ISL_1052447, EPI_ISL_1052448, EPI_ISL_1052449, EPI_ISL_1052451, EPI_ISL_1052452, EPI_ISL_1052454, EPI_ISL_1052456, EPI_ISL_1052457, EPI_ISL_1052458, EPI_ISL_1052460, EPI_ISL_1052476, EPI_ISL_1052481, EPI_ISL_1052498, EPI_ISL_1052529, EPI_ISL_1052538, EPI_ISL_1052539, EPI_ISL_1052540, EPI_ISL_1052541, EPI_ISL_1052542, EPI_ISL_1052544, EPI_ISL_1052545, EPI_ISL_1052547, EPI_ISL_1052548, EPI_ISL_1052549, EPI_ISL_1052553, EPI_ISL_1052554, EPI_ISL_1052555, EPI_ISL_1052557, EPI_ISL_1052558, EPI_ISL_1052559, EPI_ISL_1052560, EPI_ISL_1052597, EPI_ISL_1052599, EPI_ISL_1052601, EPI_ISL_1052606, EPI_ISL_1052607, EPI_ISL_1052612, EPI_ISL_1052615, EPI_ISL_1052619, EPI_ISL_1052620, EPI_ISL_1052621, EPI_ISL_1052629, EPI_ISL_1052646, EPI_ISL_1052654, EPI_ISL_1052666, EPI_ISL_1052669, EPI_ISL_1052670, EPI_ISL_1052671, EPI_ISL_1052675, EPI_ISL_1052676, EPI_ISL_1052681, EPI_ISL_1052715, EPI_ISL_1052753, EPI_ISL_1052762, EPI_ISL_1053105, EPI_ISL_1053106, EPI_ISL_1053107, EPI_ISL_1053108, EPI_ISL_1053492, EPI_ISL_1053494, EPI_ISL_1053519, EPI_ISL_1053520, EPI_ISL_1053527, EPI_ISL_1053528, EPI_ISL_1053529, EPI_ISL_1053537, EPI_ISL_1053590, EPI_ISL_1053591, EPI_ISL_1053592, EPI_ISL_1053593, EPI_ISL_1053595, EPI_ISL_1053597, EPI_ISL_1053598, EPI_ISL_1053599, EPI_ISL_1053600, EPI_ISL_1053601, EPI_ISL_1053602, EPI_ISL_1053603, EPI_ISL_1053604, EPI_ISL_1053605, EPI_ISL_1053606, EPI_ISL_1053607, EPI_ISL_1053625, EPI_ISL_1053629, EPI_ISL_1053630, EPI_ISL_1053631, EPI_ISL_1106649, EPI_ISL_1178893, EPI_ISL_1178894, EPI_ISL_1178895, EPI_ISL_1179144, EPI_ISL_1179145, EPI_ISL_1179146, EPI_ISL_1179156, EPI_ISL_1179157, EPI_ISL_1179158, EPI_ISL_1179159, EPI_ISL_1179160, EPI_ISL_1179161, EPI_ISL_1179162, EPI_ISL_1179163, EPI_ISL_1179164, EPI_ISL_1179165, EPI_ISL_1179166, EPI_ISL_1179167, EPI_ISL_1179168, EPI_ISL_1179169, EPI_ISL_1179170, EPI_ISL_1179171, EPI_ISL_1179172, EPI_ISL_1179173, EPI_ISL_1179174, EPI_ISL_1179175, EPI_ISL_1179176, EPI_ISL_1179177, EPI_ISL_1179178, EPI_ISL_1179179, EPI_ISL_1179180, EPI_ISL_1179181, EPI_ISL_1179182, EPI_ISL_1179183, EPI_ISL_1179184, EPI_ISL_1179185, EPI_ISL_1179186, EPI_ISL_1179187, EPI_ISL_1387502

| | | | |
|---|---|---|---|
| see above | Originating lab: Wales Specialist Virology Centre Sequencing lab: Pathogen Genomics Unit | Public Health Wales Microbiology Cardiff Wales Specialist Virology Centre | Alec Birchley; Alexander Adams; Amy Gaskin; Angela Marchbank; Bree Gatica-Wilcox; Catherine Moore; Jason Coombes; Joanne Watkins; Joel Southgate; Johnathan Evans; Laura Gifford; Lauren Gilbert; Lee Graham; Malorie Perry; Matthew Bull; Nicole Pacchiarini; Sally Corden; Sara Kumziene-Summerhayes; Sara Rey; Sarah Taylor; Simon Cottrell; Sophie Jones; Tom Connor |

EPI_ISL_1051694, EPI_ISL_1051695, EPI_ISL_1051696, EPI_ISL_1051698, EPI_ISL_1051700, EPI_ISL_1051701, EPI_ISL_1051702, EPI_ISL_1051703, EPI_ISL_1051704, EPI_ISL_1051705, EPI_ISL_1051706, EPI_ISL_1051707, EPI_ISL_1051708, EPI_ISL_1051709, EPI_ISL_1051711, EPI_ISL_1051713, EPI_ISL_1051714, EPI_ISL_1051723, EPI_ISL_1051724, EPI_ISL_1051729, EPI_ISL_1051730, EPI_ISL_1051731, EPI_ISL_1051732, EPI_ISL_1051733, EPI_ISL_1051734, EPI_ISL_1051735, EPI_ISL_1051736, EPI_ISL_1051737, EPI_ISL_1051741, EPI_ISL_1051742, EPI_ISL_1051743, EPI_ISL_1051744, EPI_ISL_1051745, EPI_ISL_1051747, EPI_ISL_1051749, EPI_ISL_1051750, EPI_ISL_1051751, EPI_ISL_1051752, EPI_ISL_1051753, EPI_ISL_1051754, EPI_ISL_1051755, EPI_ISL_1051756, EPI_ISL_1051757, EPI_ISL_1051759, EPI_ISL_1051760, EPI_ISL_1051761, EPI_ISL_1051763, EPI_ISL_1051764, EPI_ISL_1051765, EPI_ISL_1051766, EPI_ISL_1051767, EPI_ISL_1051769, EPI_ISL_1051770, EPI_ISL_1051771, EPI_ISL_1051772, EPI_ISL_1051773, EPI_ISL_1051775, EPI_ISL_1051776, EPI_ISL_1051778, EPI_ISL_1051780, EPI_ISL_1051781, EPI_ISL_1051782, EPI_ISL_1051783, EPI_ISL_1051784, EPI_ISL_1178717, EPI_ISL_1178723, EPI_ISL_1178727, EPI_ISL_1178735, EPI_ISL_1178740, EPI_ISL_1178742, EPI_ISL_1178744, EPI_ISL_1178747, EPI_ISL_1178751, EPI_ISL_1178759, EPI_ISL_1178765, EPI_ISL_1178766, EPI_ISL_1178770, EPI_ISL_1178772, EPI_ISL_1178773, EPI_ISL_1178779, EPI_ISL_1178781, EPI_ISL_1178792, EPI_ISL_1178797, EPI_ISL_1178805, EPI_ISL_1178816, EPI_ISL_1178819, EPI_ISL_1178827, EPI_ISL_1178829, EPI_ISL_1178848, EPI_ISL_1178853, EPI_ISL_1178856, EPI_ISL_1178857, EPI_ISL_1178867, EPI_ISL_1178870, EPI_ISL_1249210, EPI_ISL_1249212, EPI_ISL_1249213, EPI_ISL_1249214, EPI_ISL_1249229, EPI_ISL_1249230, EPI_ISL_1249231, EPI_ISL_1249232, EPI_ISL_1249233, EPI_ISL_1249236, EPI_ISL_1249238, EPI_ISL_1249240, EPI_ISL_1309751, EPI_ISL_1309752, EPI_ISL_1309782, EPI_ISL_1387226, EPI_ISL_1387227, EPI_ISL_1387229, EPI_ISL_1387230, EPI_ISL_1387231, EPI_ISL_1387232, EPI_ISL_1387233, EPI_ISL_1387234, EPI_ISL_1730499, EPI_ISL_1730500, EPI_ISL_1730501, EPI_ISL_1730522, EPI_ISL_1833094

| | | | |
|---|---|---|---|
| see above | Oxford Viromics, NDM, University of Oxford; Oxford University Hospitals; Basingstoke and North Hampshire Hospital | COVID-19 Genomics UK (COG-UK) Consortium | Alex Mobbs; Amy Trebes; Anita Justice; Catrin Moore; Christophe Fraser; David Bonsall; David Buck; Emma Wise; George Macintyre; Jessica Lynch; John Todd; Mariateresa de Cesare; Matilde Mori; Monique Andersson; Nathan Moore; Nick Cortes; Robert Shaw; Stephen Kidd; Tanya Golubchik; Timothy Peto |

EPI_ISL_998774, EPI_ISL_998775, EPI_ISL_998778, EPI_ISL_998779, EPI_ISL_998782, EPI_ISL_998783, EPI_ISL_998784, EPI_ISL_998787, EPI_ISL_998789, EPI_ISL_998790, EPI_ISL_998791, EPI_ISL_998792, EPI_ISL_998793, EPI_ISL_998794, EPI_ISL_998796, EPI_ISL_998797, EPI_ISL_998799, EPI_ISL_998800, EPI_ISL_998803, EPI_ISL_998805, EPI_ISL_998807, EPI_ISL_998809, EPI_ISL_998810, EPI_ISL_998811,

EPI_ISL_998812, EPI_ISL_998813, EPI_ISL_998814, EPI_ISL_998815, EPI_ISL_998818, EPI_ISL_998820, EPI_ISL_998837, EPI_ISL_998840, EPI_ISL_998850, EPI_ISL_998851, EPI_ISL_998852, EPI_ISL_998854, EPI_ISL_998855, EPI_ISL_998856, EPI_ISL_998868, EPI_ISL_998873, EPI_ISL_998876, EPI_ISL_998877, EPI_ISL_998879, EPI_ISL_998883, EPI_ISL_998887, EPI_ISL_1105538, EPI_ISL_1105539, EPI_ISL_1105540, EPI_ISL_1105541, EPI_ISL_1105542, EPI_ISL_1105543, EPI_ISL_1105544, EPI_ISL_1105545, EPI_ISL_1105546, EPI_ISL_1105547, EPI_ISL_1105548, EPI_ISL_1105549, EPI_ISL_1105550, EPI_ISL_1105551, EPI_ISL_1105552, EPI_ISL_1105553, EPI_ISL_1105554, EPI_ISL_1105555, EPI_ISL_1105556, EPI_ISL_1105557, EPI_ISL_1105558, EPI_ISL_1105559, EPI_ISL_1105560, EPI_ISL_1105561, EPI_ISL_1105562, EPI_ISL_1105563, EPI_ISL_1105564, EPI_ISL_1105565, EPI_ISL_1105566, EPI_ISL_1105567, EPI_ISL_1105568, EPI_ISL_1105569, EPI_ISL_1105570, EPI_ISL_1105571, EPI_ISL_1107835, EPI_ISL_1107998, EPI_ISL_1108075, EPI_ISL_1108387, EPI_ISL_1178496, EPI_ISL_1178497, EPI_ISL_1178498, EPI_ISL_1178499, EPI_ISL_1178500, EPI_ISL_1178502, EPI_ISL_1178503, EPI_ISL_1178504, EPI_ISL_1178505, EPI_ISL_1178506, EPI_ISL_1178508, EPI_ISL_1178509, EPI_ISL_1178510, EPI_ISL_1178511, EPI_ISL_1178512, EPI_ISL_1178513, EPI_ISL_1178514, EPI_ISL_1178515, EPI_ISL_1178516, EPI_ISL_1178517, EPI_ISL_1248949, EPI_ISL_1248964, EPI_ISL_1248965, EPI_ISL_1248966, EPI_ISL_1248967, EPI_ISL_1248968, EPI_ISL_1248969, EPI_ISL_1248970, EPI_ISL_1248971, EPI_ISL_1248972, EPI_ISL_1248973, EPI_ISL_1248975, EPI_ISL_1248976, EPI_ISL_1248977, EPI_ISL_1248997, EPI_ISL_1248998, EPI_ISL_1249011, EPI_ISL_1249012, EPI_ISL_1249029, EPI_ISL_1249034, EPI_ISL_1249035, EPI_ISL_1249036, EPI_ISL_1249040, EPI_ISL_1249043, EPI_ISL_1249046, EPI_ISL_1249049, EPI_ISL_1249056, EPI_ISL_1249061, EPI_ISL_1309670, EPI_ISL_1309671, EPI_ISL_1309672, EPI_ISL_1309674, EPI_ISL_1309675, EPI_ISL_1309676, EPI_ISL_1309677, EPI_ISL_1309678, EPI_ISL_1832844, EPI_ISL_1832896, EPI_ISL_1832920, EPI_ISL_1832921, EPI_ISL_1832926, EPI_ISL_1978904, EPI_ISL_1978908, EPI_ISL_1978918, EPI_ISL_1978925, EPI_ISL_1978928, EPI_ISL_1978931, EPI_ISL_1978933, EPI_ISL_1978935, EPI_ISL_1978940, EPI_ISL_1978943

see above | Quadram Institute Bioscience | COVID-19 Genomics UK (COG-UK) Consortium | Alexander J Trotter; Alison E. Mather; Alp Aydin; Ana P. Tedim; Anastasia Kolyva; Andrew Bell; Andrew J. Page; Claire Stuart; Dave J. Baker; Gemma L. Kay; John Wain; Justin O'Grady; Leonardo de Oliveira Martins; Lizzie Meadows; Maria Diaz; Mark Webber; Muhammed Yasir; Nabil-Fareed Alikhan; Ngozi Elumogo; Nicholas M. Thomson; Rachael Stanley; Rachel Gilroy; Reenesh Prakash; Samir Dervisevic; Samuel Bloomfield; Steven Rudder; Thanh Le-Viet

EPI_ISL_950671, EPI_ISL_950672, EPI_ISL_950673, EPI_ISL_950674, EPI_ISL_950675, EPI_ISL_950676, EPI_ISL_950677, EPI_ISL_950678, EPI_ISL_950679, EPI_ISL_950680, EPI_ISL_950681, EPI_ISL_950682, EPI_ISL_950683, EPI_ISL_950684, EPI_ISL_950685, EPI_ISL_950686, EPI_ISL_950687, EPI_ISL_950688, EPI_ISL_950689, EPI_ISL_950690, EPI_ISL_950691, EPI_ISL_950692, EPI_ISL_950693, EPI_ISL_950694, EPI_ISL_950695, EPI_ISL_950696, EPI_ISL_950697, EPI_ISL_950717, EPI_ISL_950718, EPI_ISL_950719, EPI_ISL_950720, EPI_ISL_950721, EPI_ISL_950722, EPI_ISL_950723, EPI_ISL_950724, EPI_ISL_950725, EPI_ISL_950726, EPI_ISL_950727, EPI_ISL_950728, EPI_ISL_998889, EPI_ISL_998890, EPI_ISL_998891, EPI_ISL_998893, EPI_ISL_998894, EPI_ISL_998895, EPI_ISL_998896, EPI_ISL_998897, EPI_ISL_998898, EPI_ISL_998899, EPI_ISL_998900, EPI_ISL_998901, EPI_ISL_998902, EPI_ISL_998903, EPI_ISL_998904, EPI_ISL_998905, EPI_ISL_998906, EPI_ISL_998907, EPI_ISL_998908, EPI_ISL_998910, EPI_ISL_1051556, EPI_ISL_1108972

see above | Queens Medical Centre, Clinical Microbiology Department / DeepSeq Nottingham | COVID-19 Genomics UK (COG-UK) Consortium | Christopher Moore; Fei Sang; Gemma Clark; Hannah Howson-Wells; Johnny Debebe; Jonathan Ball; Joseph Chappell; Manjinder Khakh; Matthew Carlisle; Matthew Loose; Michelle M Lister; Nadine Holmes; Patrick McClure; Theocharis Tsoleridis; Vicki M Fleming; Victoria Wright; Wendy Smith

EPI_ISL_1042002, EPI_ISL_1042004, EPI_ISL_1042005, EPI_ISL_1042006, EPI_ISL_1042007, EPI_ISL_1042009, EPI_ISL_1042010, EPI_ISL_1042012, EPI_ISL_1042013, EPI_ISL_1042015, EPI_ISL_1042021, EPI_ISL_1042022, EPI_ISL_1042025, EPI_ISL_1042026, EPI_ISL_1042027, EPI_ISL_1042029, EPI_ISL_1042032, EPI_ISL_1042034, EPI_ISL_1042038, EPI_ISL_1042039, EPI_ISL_1042041, EPI_ISL_1042044, EPI_ISL_1042052, EPI_ISL_1042054, EPI_ISL_1042055, EPI_ISL_1042056, EPI_ISL_1042058, EPI_ISL_1042061, EPI_ISL_1042063, EPI_ISL_1042068, EPI_ISL_1042070, EPI_ISL_1042072, EPI_ISL_1042078, EPI_ISL_1042079, EPI_ISL_1042083, EPI_ISL_1042086, EPI_ISL_1042087, EPI_ISL_1042088, EPI_ISL_1042089, EPI_ISL_1042090, EPI_ISL_1042093, EPI_ISL_1042095, EPI_ISL_1042099, EPI_ISL_1042104, EPI_ISL_1042105, EPI_ISL_1042106, EPI_ISL_1042107, EPI_ISL_1042110, EPI_ISL_1042111, EPI_ISL_1042114, EPI_ISL_1042116, EPI_ISL_1042118, EPI_ISL_1042119, EPI_ISL_1042121, EPI_ISL_1042124, EPI_ISL_1042125, EPI_ISL_1042127, EPI_ISL_1042128, EPI_ISL_1042129, EPI_ISL_1042131, EPI_ISL_1042132, EPI_ISL_1042133, EPI_ISL_1042136, EPI_ISL_1042137, EPI_ISL_1042138, EPI_ISL_1042139, EPI_ISL_1042142, EPI_ISL_1042144, EPI_ISL_1042145, EPI_ISL_1042146, EPI_ISL_1042147, EPI_ISL_1042148, EPI_ISL_1042151, EPI_ISL_1042154, EPI_ISL_1042155, EPI_ISL_1042156, EPI_ISL_1042159, EPI_ISL_1042161, EPI_ISL_1042166, EPI_ISL_1042167, EPI_ISL_1042169, EPI_ISL_1042170, EPI_ISL_1042171, EPI_ISL_1042173, EPI_ISL_1042178, EPI_ISL_1042180, EPI_ISL_1042183, EPI_ISL_1042185, EPI_ISL_1042186, EPI_ISL_1042188, EPI_ISL_1042192, EPI_ISL_1042193, EPI_ISL_1042195, EPI_ISL_1042197, EPI_ISL_1042198, EPI_ISL_1042199, EPI_ISL_1042200, EPI_ISL_1042201, EPI_ISL_1042203, EPI_ISL_1042204, EPI_ISL_1042205, EPI_ISL_1042207, EPI_ISL_1042209, EPI_ISL_1042210, EPI_ISL_1042211, EPI_ISL_1042212, EPI_ISL_1042217, EPI_ISL_1042219, EPI_ISL_1042220, EPI_ISL_1042221, EPI_ISL_1042224, EPI_ISL_1042229, EPI_ISL_1042230, EPI_ISL_1042235, EPI_ISL_1042236, EPI_ISL_1042237, EPI_ISL_1042239, EPI_ISL_1042244, EPI_ISL_1042246, EPI_ISL_1042247, EPI_ISL_1042251, EPI_ISL_1042254, EPI_ISL_1042255, EPI_ISL_1042256, EPI_ISL_1042257, EPI_ISL_1042258, EPI_ISL_1042261, EPI_ISL_1042262, EPI_ISL_1042263, EPI_ISL_1042265, EPI_ISL_1042268, EPI_ISL_1042270, EPI_ISL_1042273, EPI_ISL_1042275, EPI_ISL_1042276, EPI_ISL_1042277, EPI_ISL_1042278, EPI_ISL_1042283, EPI_ISL_1042285, EPI_ISL_1042287, EPI_ISL_1042290, EPI_ISL_1042292, EPI_ISL_1042294, EPI_ISL_1042296, EPI_ISL_1042301, EPI_ISL_1042302, EPI_ISL_1042303, EPI_ISL_1042304, EPI_ISL_1042305, EPI_ISL_1042309, EPI_ISL_1042311, EPI_ISL_1042313, EPI_ISL_1042316, EPI_ISL_1042318, EPI_ISL_1042319, EPI_ISL_1042323, EPI_ISL_1042324, EPI_ISL_1042327, EPI_ISL_1042328, EPI_ISL_1042330, EPI_ISL_1042331, EPI_ISL_1042332, EPI_ISL_1042336, EPI_ISL_1042337, EPI_ISL_1042338, EPI_ISL_1042339, EPI_ISL_1042340, EPI_ISL_1042341, EPI_ISL_1042343, EPI_ISL_1042346, EPI_ISL_1042347, EPI_ISL_1042348, EPI_ISL_1042349, EPI_ISL_1042351, EPI_ISL_1042352, EPI_ISL_1042353, EPI_ISL_1042354, EPI_ISL_1042355, EPI_ISL_1042567, EPI_ISL_1042581, EPI_ISL_1042586, EPI_ISL_1042592, EPI_ISL_1042596, EPI_ISL_1042598, EPI_ISL_1042599, EPI_ISL_1042601, EPI_ISL_1042603, EPI_ISL_1042604, EPI_ISL_1042606, EPI_ISL_1042609, EPI_ISL_1042614, EPI_ISL_1042617, EPI_ISL_1042619, EPI_ISL_1042621, EPI_ISL_1042626, EPI_ISL_1042627, EPI_ISL_1042631, EPI_ISL_1042637, EPI_ISL_1042640, EPI_ISL_1042643, EPI_ISL_1042647, EPI_ISL_1042659, EPI_ISL_1042661, EPI_ISL_1042666, EPI_ISL_1042673, EPI_ISL_1042674, EPI_ISL_1042677, EPI_ISL_1042684, EPI_ISL_1042691, EPI_ISL_1042696, EPI_ISL_1042720, EPI_ISL_1042721, EPI_ISL_1042730, EPI_ISL_1042731, EPI_ISL_1042734, EPI_ISL_1042742, EPI_ISL_1042744, EPI_ISL_1042756, EPI_ISL_1042765, EPI_ISL_1042769, EPI_ISL_1042774, EPI_ISL_1042777, EPI_ISL_1042779, EPI_ISL_1042782, EPI_ISL_1042783, EPI_ISL_1042784, EPI_ISL_1042786, EPI_ISL_1042789, EPI_ISL_1042793, EPI_ISL_1042800, EPI_ISL_1042801, EPI_ISL_1042803, EPI_ISL_1042807, EPI_ISL_1042809, EPI_ISL_1042813, EPI_ISL_1042815, EPI_ISL_1042819, EPI_ISL_1042821, EPI_ISL_1042824, EPI_ISL_1042828, EPI_ISL_1043177, EPI_ISL_1043178, EPI_ISL_1043187, EPI_ISL_1043191, EPI_ISL_1043198, EPI_ISL_1043200, EPI_ISL_1043202, EPI_ISL_1043212, EPI_ISL_1043220, EPI_ISL_1043227, EPI_ISL_1043239, EPI_ISL_1043245, EPI_ISL_1043266, EPI_ISL_1043268, EPI_ISL_1043277, EPI_ISL_1043280, EPI_ISL_1043282, EPI_ISL_1043291, EPI_ISL_1043301, EPI_ISL_1043307, EPI_ISL_1043310, EPI_ISL_1043315, EPI_ISL_1043317, EPI_ISL_1043319, EPI_ISL_1043327, EPI_ISL_1043342, EPI_ISL_1043346, EPI_ISL_1043349, EPI_ISL_1043351, EPI_ISL_1043352, EPI_ISL_1043354, EPI_ISL_1043357, EPI_ISL_1043359, EPI_ISL_1043361, EPI_ISL_1043362, EPI_ISL_1043364, EPI_ISL_1043368, EPI_ISL_1043369, EPI_ISL_1043370, EPI_ISL_1043371, EPI_ISL_1043376, EPI_ISL_1043377, EPI_ISL_1043378, EPI_ISL_1043379, EPI_ISL_1043381, EPI_ISL_1043383, EPI_ISL_1043387, EPI_ISL_1043390, EPI_ISL_1043391, EPI_ISL_1043392, EPI_ISL_1043393, EPI_ISL_1043395, EPI_ISL_1043397, EPI_ISL_1043401, EPI_ISL_1043404, EPI_ISL_1043406, EPI_ISL_1043408, EPI_ISL_1043409, EPI_ISL_1043410, EPI_ISL_1043412, EPI_ISL_1043415, EPI_ISL_1043418, EPI_ISL_1043420, EPI_ISL_1043423, EPI_ISL_1043424, EPI_ISL_1043425, EPI_ISL_1043428, EPI_ISL_1043431, EPI_ISL_1043432, EPI_ISL_1043433, EPI_ISL_1043434, EPI_ISL_1043436, EPI_ISL_1043437, EPI_ISL_1043438, EPI_ISL_1043442, EPI_ISL_1043446, EPI_ISL_1043447, EPI_ISL_1043448, EPI_ISL_1043449, EPI_ISL_1043450, EPI_ISL_1043451, EPI_ISL_1043454, EPI_ISL_1043456, EPI_ISL_1043457, EPI_ISL_1043458, EPI_ISL_1043462, EPI_ISL_1043463, EPI_ISL_1043466, EPI_ISL_1043467, EPI_ISL_1043469, EPI_ISL_1043470, EPI_ISL_1043473, EPI_ISL_1043475, EPI_ISL_1043478, EPI_ISL_1043479, EPI_ISL_1043480, EPI_ISL_1043484, EPI_ISL_1043486, EPI_ISL_1043487, EPI_ISL_1043489, EPI_ISL_1043490, EPI_ISL_1043492, EPI_ISL_1043498, EPI_ISL_1043499, EPI_ISL_1043502, EPI_ISL_1043503, EPI_ISL_1043505, EPI_ISL_1043508, EPI_ISL_1043510, EPI_ISL_1043511, EPI_ISL_1043513, EPI_ISL_1043514, EPI_ISL_1043516, EPI_ISL_1043517, EPI_ISL_1043518, EPI_ISL_1043519, EPI_ISL_1043520, EPI_ISL_1043523, EPI_ISL_1043525, EPI_ISL_1043528, EPI_ISL_1043530, EPI_ISL_1043531, EPI_ISL_1043533, EPI_ISL_1043534, EPI_ISL_1043535, EPI_ISL_1043537, EPI_ISL_1043540, EPI_ISL_1043541, EPI_ISL_1043542, EPI_ISL_1043543, EPI_ISL_1043546, EPI_ISL_1043547, EPI_ISL_1043552, EPI_ISL_1043553, EPI_ISL_1043555, EPI_ISL_1043559, EPI_ISL_1043566, EPI_ISL_1043568, EPI_ISL_1045199, EPI_ISL_1045201, EPI_ISL_1045207, EPI_ISL_1045210, EPI_ISL_1045213, EPI_ISL_1045218, EPI_ISL_1045221, EPI_ISL_1045225, EPI_ISL_1045226, EPI_ISL_1045229, EPI_ISL_1045231, EPI_ISL_1045237, EPI_ISL_1045240, EPI_ISL_1045242, EPI_ISL_1045243, EPI_ISL_1045247, EPI_ISL_1045251, EPI_ISL_1045253, EPI_ISL_1045254, EPI_ISL_1045256, EPI_ISL_1045259, EPI_ISL_1045261, EPI_ISL_1045268, EPI_ISL_1045274, EPI_ISL_1045276, EPI_ISL_1045277, EPI_ISL_1045280, EPI_ISL_1045286, EPI_ISL_1045287, EPI_ISL_1045289, EPI_ISL_1045290, EPI_ISL_1045293, EPI_ISL_1045298, EPI_ISL_1045299, EPI_ISL_1045300, EPI_ISL_1045305, EPI_ISL_1045306, EPI_ISL_1045319, EPI_ISL_1045329, EPI_ISL_1045333, EPI_ISL_1045334, EPI_ISL_1045335, EPI_ISL_1045336, EPI_ISL_1045339, EPI_ISL_1045348, EPI_ISL_1045349, EPI_ISL_1045350, EPI_ISL_1045351, EPI_ISL_1045353, EPI_ISL_1045355, EPI_ISL_1045361, EPI_ISL_1045364, EPI_ISL_1045365, EPI_ISL_1045366, EPI_ISL_1045367, EPI_ISL_1045371, EPI_ISL_1045372, EPI_ISL_1045374, EPI_ISL_1045379, EPI_ISL_1045381, EPI_ISL_1045382, EPI_ISL_1045384, EPI_ISL_1045386, EPI_ISL_1045389, EPI_ISL_1045393, EPI_ISL_1045397, EPI_ISL_1045398, EPI_ISL_1045400, EPI_ISL_1045405, EPI_ISL_1045406, EPI_ISL_1045407, EPI_ISL_1045412, EPI_ISL_1045418, EPI_ISL_1045420, EPI_ISL_1045422, EPI_ISL_1045435, EPI_ISL_1045439, EPI_ISL_1045443, EPI_ISL_1045444, EPI_ISL_1045446, EPI_ISL_1045447, EPI_ISL_1045451, EPI_ISL_1045452, EPI_ISL_1045458, EPI_ISL_1045459, EPI_ISL_1045462, EPI_ISL_1045471, EPI_ISL_1045709, EPI_ISL_1045710, EPI_ISL_1045712, EPI_ISL_1045715, EPI_ISL_1045716, EPI_ISL_1045717, EPI_ISL_1045719, EPI_ISL_1045720, EPI_ISL_1045721, EPI_ISL_1045722, EPI_ISL_1045723, EPI_ISL_1045724, EPI_ISL_1045725, EPI_ISL_1045726, EPI_ISL_1045729, EPI_ISL_1045732, EPI_ISL_1045735, EPI_ISL_1045736, EPI_ISL_1045737, EPI_ISL_1045739, EPI_ISL_1045741, EPI_ISL_1045744, EPI_ISL_1045746, EPI_ISL_1045749, EPI_ISL_1045752, EPI_ISL_1045754, EPI_ISL_1045758, EPI_ISL_1045759, EPI_ISL_1045760, EPI_ISL_1045764, EPI_ISL_1045765, EPI_ISL_1045766, EPI_ISL_1045769, EPI_ISL_1045770, EPI_ISL_1045771, EPI_ISL_1045772, EPI_ISL_1045774, EPI_ISL_1045776, EPI_ISL_1045780, EPI_ISL_1045785, EPI_ISL_1045787, EPI_ISL_1045790, EPI_ISL_1045792, EPI_ISL_1045793, EPI_ISL_1045795, EPI_ISL_1045797, EPI_ISL_1045798, EPI_ISL_1045802, EPI_ISL_1045804, EPI_ISL_1045806, EPI_ISL_1045807, EPI_ISL_1045808, EPI_ISL_1045811, EPI_ISL_1045813, EPI_ISL_1045814, EPI_ISL_1045816, EPI_ISL_1045818, EPI_ISL_1045821, EPI_ISL_1045823, EPI_ISL_1045824, EPI_ISL_1045828, EPI_ISL_1045830, EPI_ISL_1045832, EPI_ISL_1045833, EPI_ISL_1045834, EPI_ISL_1045835, EPI_ISL_1045837, EPI_ISL_1045839, EPI_ISL_1045840, EPI_ISL_1045843, EPI_ISL_1045847, EPI_ISL_1045849, EPI_ISL_1045851, EPI_ISL_1045852, EPI_ISL_1045856, EPI_ISL_1045858, EPI_ISL_1045860, EPI_ISL_1045861, EPI_ISL_1045863, EPI_ISL_1045864, EPI_ISL_1045867, EPI_ISL_1045868, EPI_ISL_1045871, EPI_ISL_1045875, EPI_ISL_1046116, EPI_ISL_1046166, EPI_ISL_1046557, EPI_ISL_1046658, EPI_ISL_1046666, EPI_ISL_1046676, EPI_ISL_1046682, EPI_ISL_1046698, EPI_ISL_1046707, EPI_ISL_1046728, EPI_ISL_1046736, EPI_ISL_1046758, EPI_ISL_1057665, EPI_ISL_1057666, EPI_ISL_1057667, EPI_ISL_1057669, EPI_ISL_1057670, EPI_ISL_1057671, EPI_ISL_1057672, EPI_ISL_1057673, EPI_ISL_1057674, EPI_ISL_1057677, EPI_ISL_1057678, EPI_ISL_1057679, EPI_ISL_1057680, EPI_ISL_1057681, EPI_ISL_1057682, EPI_ISL_1057683, EPI_ISL_1057764, EPI_ISL_1057765, EPI_ISL_1057766, EPI_ISL_1057767, EPI_ISL_1057768, EPI_ISL_1057769, EPI_ISL_1057770, EPI_ISL_1057771, EPI_ISL_1057772, EPI_ISL_1057773, EPI_ISL_1057774, EPI_ISL_1057775, EPI_ISL_1057776, EPI_ISL_1057777, EPI_ISL_1057778, EPI_ISL_1057779, EPI_ISL_1057780, EPI_ISL_1057781, EPI_ISL_1057782, EPI_ISL_1057783, EPI_ISL_1057784, EPI_ISL_1057785, EPI_ISL_1057786, EPI_ISL_1057787, EPI_ISL_1057788, EPI_ISL_1057789, EPI_ISL_1057790, EPI_ISL_1057792, EPI_ISL_1057793, EPI_ISL_1057794, EPI_ISL_1057795, EPI_ISL_1057796, EPI_ISL_1057797, EPI_ISL_1057798, EPI_ISL_1057799, EPI_ISL_1057800, EPI_ISL_1057801, EPI_ISL_1057802, EPI_ISL_1057803, EPI_ISL_1057804, EPI_ISL_1057805, EPI_ISL_1057806, EPI_ISL_1057807, EPI_ISL_1057808, EPI_ISL_1057809, EPI_ISL_1057810, EPI_ISL_1057811, EPI_ISL_1057812, EPI_ISL_1057813, EPI_ISL_1057814, EPI_ISL_1057815, EPI_ISL_1057816, EPI_ISL_1057817, EPI_ISL_1063190, EPI_ISL_1063192, EPI_ISL_1063194, EPI_ISL_1063214, EPI_ISL_1063218, EPI_ISL_1063230, EPI_ISL_1063231, EPI_ISL_1063233, EPI_ISL_1063237, EPI_ISL_1063238, EPI_ISL_1063242, EPI_ISL_1063251, EPI_ISL_1063252, EPI_ISL_1063253, EPI_ISL_1063254, EPI_ISL_1063255, EPI_ISL_1063256, EPI_ISL_1063257, EPI_ISL_1063258, EPI_ISL_1063259, EPI_ISL_1063260, EPI_ISL_1063261, EPI_ISL_1063262, EPI_ISL_1063263, EPI_ISL_1063264, EPI_ISL_1063265, EPI_ISL_1063266, EPI_ISL_1063267, EPI_ISL_1063268, EPI_ISL_1063269, EPI_ISL_1063270, EPI_ISL_1063271, EPI_ISL_1063272, EPI_ISL_1063273, EPI_ISL_1063274, EPI_ISL_1063275, EPI_ISL_1063276, EPI_ISL_1063277, EPI_ISL_1063278, EPI_ISL_1063279, EPI_ISL_1063280, EPI_ISL_1063281, EPI_ISL_1063282, EPI_ISL_1063283, EPI_ISL_1063284, EPI_ISL_1063285, EPI_ISL_1063286, EPI_ISL_1063287, EPI_ISL_1063292, EPI_ISL_1063293, EPI_ISL_1063295, EPI_ISL_1063296, EPI_ISL_1063297, EPI_ISL_1063298, EPI_ISL_1063307, EPI_ISL_1063310, EPI_ISL_1063311, EPI_ISL_1063312, EPI_ISL_1063313, EPI_ISL_1063322, EPI_ISL_1063323, EPI_ISL_1063324, EPI_ISL_1063326, EPI_ISL_1063327, EPI_ISL_1063331, EPI_ISL_1063336, EPI_ISL_1063338, EPI_ISL_1063340, EPI_ISL_1063342, EPI_ISL_1063345, EPI_ISL_1063347, EPI_ISL_1063348, EPI_ISL_1063350, EPI_ISL_1063351, EPI_ISL_1063352, EPI_ISL_1063353, EPI_ISL_1063354, EPI_ISL_1063355, EPI_ISL_1063356, EPI_ISL_1063370, EPI_ISL_1063386, EPI_ISL_1063388, EPI_ISL_1063390, EPI_ISL_1063395, EPI_ISL_1063397, EPI_ISL_1069991, EPI_ISL_1069992, EPI_ISL_1069995, EPI_ISL_1069998, EPI_ISL_1070001, EPI_ISL_1070005, EPI_ISL_1070006, EPI_ISL_1070008, EPI_ISL_1070010, EPI_ISL_1070019, EPI_ISL_1070022, EPI_ISL_1070037, EPI_ISL_1070038, EPI_ISL_1070042, EPI_ISL_1070045, EPI_ISL_1070047, EPI_ISL_1070048, EPI_ISL_1070050, EPI_ISL_1070053, EPI_ISL_1070058, EPI_ISL_1070061, EPI_ISL_1070064, EPI_ISL_1070071, EPI_ISL_1070073, EPI_ISL_1070082, EPI_ISL_1070083, EPI_ISL_1070084, EPI_ISL_1070085, EPI_ISL_1070090, EPI_ISL_1070094, EPI_ISL_1070095, EPI_ISL_1070100, EPI_ISL_1070106, EPI_ISL_1070107, EPI_ISL_1070109, EPI_ISL_1070113, EPI_ISL_1070115, EPI_ISL_1070118, EPI_ISL_1070121, EPI_ISL_1070122, EPI_ISL_1070124, EPI_ISL_1070125, EPI_ISL_1070129, EPI_ISL_1070733, EPI_ISL_1070735, EPI_ISL_1070736, EPI_ISL_1070743, EPI_ISL_1070744, EPI_ISL_1070745, EPI_ISL_1070749, EPI_ISL_1070756, EPI_ISL_1070759, EPI_ISL_1070762, EPI_ISL_1070765, EPI_ISL_1070766, EPI_ISL_1070767, EPI_ISL_1070769, EPI_ISL_1070770, EPI_ISL_1070771, EPI_ISL_1070773, EPI_ISL_1070777, EPI_ISL_1070779, EPI_ISL_1070781, EPI_ISL_1070782, EPI_ISL_1070786, EPI_ISL_1070791, EPI_ISL_1070794, EPI_ISL_1070795, EPI_ISL_1070804, EPI_ISL_1070807, EPI_ISL_1070809, EPI_ISL_1070810, EPI_ISL_1070814, EPI_ISL_1070815, EPI_ISL_1070818, EPI_ISL_1070819, EPI_ISL_1070822, EPI_ISL_1070824, EPI_ISL_1070832, EPI_ISL_1070834, EPI_ISL_1070835, EPI_ISL_1070839, EPI_ISL_1070842, EPI_ISL_1070843, EPI_ISL_1070845, EPI_ISL_1070849, EPI_ISL_1070859, EPI_ISL_1070860, EPI_ISL_1070861, EPI_ISL_1070863, EPI_ISL_1070865, EPI_ISL_1070867, EPI_ISL_1070873, EPI_ISL_1070877, EPI_ISL_1070879, EPI_ISL_1070883, EPI_ISL_1070887, EPI_ISL_1070888, EPI_ISL_1070898, EPI_ISL_1070899, EPI_ISL_1070901, EPI_ISL_1070903, EPI_ISL_1070907, EPI_ISL_1070908, EPI_ISL_1070912, EPI_ISL_1070913, EPI_ISL_1070914, EPI_ISL_1070917, EPI_ISL_1070919, EPI_ISL_1070924, EPI_ISL_1070927, EPI_ISL_1070930, EPI_ISL_1070936, EPI_ISL_1070944, EPI_ISL_1070945, EPI_ISL_1070949, EPI_ISL_1070956, EPI_ISL_1070957, EPI_ISL_1070969, EPI_ISL_1070989, EPI_ISL_1071004, EPI_ISL_1071007, EPI_ISL_1071010, EPI_ISL_1071014, EPI_ISL_1071017, EPI_ISL_1071021, EPI_ISL_1071022, EPI_ISL_1071025, EPI_ISL_1071026, EPI_ISL_1071029, EPI_ISL_1071032, EPI_ISL_1071035, EPI_ISL_1071040, EPI_ISL_1116311, EPI_ISL_1116312, EPI_ISL_1116313, EPI_ISL_1116314, EPI_ISL_1116315, EPI_ISL_1116316, EPI_ISL_1116317, EPI_ISL_1116318, EPI_ISL_1116319, EPI_ISL_1116320, EPI_ISL_1116321, EPI_ISL_1116322, EPI_ISL_1116323, EPI_ISL_1116324, EPI_ISL_1116325, EPI_ISL_1116326, EPI_ISL_1116327, EPI_ISL_1116328, EPI_ISL_1116329, EPI_ISL_1116330, EPI_ISL_1116331, EPI_ISL_1116332, EPI_ISL_1116333, EPI_ISL_1116334, EPI_ISL_1116335, EPI_ISL_1116336, EPI_ISL_1116337, EPI_ISL_1116338, EPI_ISL_1116339, EPI_ISL_1116340, EPI_ISL_1116341, EPI_ISL_1116342, EPI_ISL_1116343, EPI_ISL_1116344, EPI_ISL_1538404

see above | Randox Laboratories | Wellcome Sanger Institute for the COVID-19 Genomics UK (COG-UK) Consortium | Cordelia Langford; David K. Jackson; Dominic Kwiatkowski; Ewan Harrison; Ian Johnston; Jeffrey Barrett; John Sillitoe on behalf of the Wellcome Sanger Institute COVID-19 Surveillance Team; Randox Laboratories and Alex Alderton; Roberto Amato; Sonia Goncalves

EPI_ISL_998234, EPI_ISL_998235, EPI_ISL_998236, EPI_ISL_1475001, EPI_ISL_1475002, EPI_ISL_1475022, EPI_ISL_1475023, EPI_ISL_1475024, EPI_ISL_1475025, EPI_ISL_1475026, EPI_ISL_1475027, EPI_ISL_1475028, EPI_ISL_1475029, EPI_ISL_1475030, EPI_ISL_1475135, EPI_ISL_1475145, EPI_ISL_1475146, EPI_ISL_1475147, EPI_ISL_1475148, EPI_ISL_1475149, EPI_ISL_1475151, EPI_ISL_1475153, EPI_ISL_1475154, EPI_ISL_1475155, EPI_ISL_1475156, EPI_ISL_1475157, EPI_ISL_1475158, EPI_ISL_1475159, EPI_ISL_1475162, EPI_ISL_1475164, EPI_ISL_1475165, EPI_ISL_1475166, EPI_ISL_1475169, EPI_ISL_1475184, EPI_ISL_1475185, EPI_ISL_1475186, EPI_ISL_1475187

see above | Regional Virus Laboratory, Belfast Health and Social Care Trust | COVID-19 Genomics UK (COG-UK) Consortium | Alison Watt; Ciara Cox; Conall McCaughey; David Simpson; Derek Fairley; James McKenna; Mairead Connor; Susan Feeney; Tanya Curran; Zoltan Molnar

EPI_ISL_1296600, EPI_ISL_1296601, EPI_ISL_1296602, EPI_ISL_1296603, EPI_ISL_1296604, EPI_ISL_1296605, EPI_ISL_1296606, EPI_ISL_1296608, EPI_ISL_1296609, EPI_ISL_1296610, EPI_ISL_1296612, EPI_ISL_1296613, EPI_ISL_1296615, EPI_ISL_1296621, EPI_ISL_1296627

see above | Respiratory Virus Unit, National Infection Service, Public Health England | COVID-19 Genomics UK (COG-UK) Consortium | PHE Covid Sequencing Team

EPI_ISL_1833619, EPI_ISL_1833620, EPI_ISL_1833650, EPI_ISL_1833651, EPI_ISL_1833654, EPI_ISL_1833655, EPI_ISL_1833657, EPI_ISL_1833659, EPI_ISL_1833660, EPI_ISL_1833662, EPI_ISL_1833664, EPI_ISL_1833665, EPI_ISL_1833666, EPI_ISL_1833667, EPI_ISL_1833669, EPI_ISL_1833670, EPI_ISL_1833671, EPI_ISL_1833672, EPI_ISL_1833674, EPI_ISL_1833675, EPI_ISL_1833677, EPI_ISL_1833678, EPI_ISL_1833681, EPI_ISL_1833685, EPI_ISL_1833686, EPI_ISL_1833688, EPI_ISL_1833689, EPI_ISL_1833691, EPI_ISL_1833696, EPI_ISL_1833697, EPI_ISL_1833699, EPI_ISL_1833712, EPI_ISL_1833713, EPI_ISL_1833715, EPI_ISL_1833717, EPI_ISL_1833719, EPI_ISL_1833720, EPI_ISL_1833722, EPI_ISL_1833723, EPI_ISL_1833725, EPI_ISL_1833727, EPI_ISL_1833728, EPI_ISL_1833730, EPI_ISL_1833732, EPI_ISL_1833733, EPI_ISL_1833735, EPI_ISL_1833736, EPI_ISL_1833737, EPI_ISL_1833738, EPI_ISL_1833740, EPI_ISL_1833742, EPI_ISL_1833743, EPI_ISL_1833744, EPI_ISL_1833747, EPI_ISL_1833749, EPI_ISL_1833756, EPI_ISL_1833767, EPI_ISL_1833768, EPI_ISL_1833769, EPI_ISL_1833770, EPI_ISL_1833792, EPI_ISL_1833793

see above | St George's University Hospitals NHS Foundation Trust | COVID-19 Genomics UK (COG-UK) Consortium | Adam Witney; Cassie Pope; Irene Monahan; Joshua Taylor; Ken Laing; NgeeKeong Tan

EPI_ISL_2125504, EPI_ISL_2125506, EPI_ISL_2125510, EPI_ISL_2125512, EPI_ISL_2125514, EPI_ISL_2125516, EPI_ISL_2125518, EPI_ISL_2125520, EPI_ISL_2125522

see above | The Roslin Institute and R(D)SVS, University of Edinburgh / Virology Department, Royal Infirmary of Edinburgh, NHS Lothian / School of Biological Sciences, University of Edinburgh | COVID-19 Genomics UK (COG-UK) Consortium | C; Colquhoun R; Cotton S; Dewar R; Gallagher A; Hill V; Jackson B; Maloney D; McCrone JT; McHugh M; Newman; O'Toole Á; Rambaut A; Rooke S; Scher E; Tait-Burkhard C; Templeton K; Warr A; Yu X

EPI_ISL_1104930, EPI_ISL_1104936, EPI_ISL_1104942, EPI_ISL_1104957, EPI_ISL_1104962, EPI_ISL_1104964, EPI_ISL_1104970, EPI_ISL_1104971, EPI_ISL_1104985, EPI_ISL_1104994, EPI_ISL_1104995, EPI_ISL_1105004, EPI_ISL_1105013, EPI_ISL_1105014, EPI_ISL_1105015, EPI_ISL_1105022, EPI_ISL_1105023, EPI_ISL_1105028, EPI_ISL_1105029, EPI_ISL_1105030, EPI_ISL_1105038, EPI_ISL_1105039, EPI_ISL_1105040, EPI_ISL_1105041, EPI_ISL_1105044, EPI_ISL_1105046, EPI_ISL_1105047, EPI_ISL_1105048, EPI_ISL_1105049, EPI_ISL_1105050, EPI_ISL_1105052, EPI_ISL_1105053, EPI_ISL_1105054, EPI_ISL_1105055, EPI_ISL_1105057, EPI_ISL_1105059, EPI_ISL_1105061, EPI_ISL_1105062, EPI_ISL_1105063, EPI_ISL_1105064, EPI_ISL_1105066, EPI_ISL_1105067, EPI_ISL_1105068, EPI_ISL_1105070, EPI_ISL_1105071, EPI_ISL_1105073, EPI_ISL_1105074, EPI_ISL_1105075, EPI_ISL_1105076, EPI_ISL_1105077, EPI_ISL_1105078, EPI_ISL_1105080, EPI_ISL_1105081, EPI_ISL_1105082, EPI_ISL_1105086, EPI_ISL_1105087, EPI_ISL_1105088, EPI_ISL_1105089, EPI_ISL_1105090, EPI_ISL_1105093, EPI_ISL_1105094, EPI_ISL_1105095, EPI_ISL_1105096, EPI_ISL_1105097, EPI_ISL_1105098, EPI_ISL_1105100, EPI_ISL_1105101, EPI_ISL_1105102, EPI_ISL_1105105, EPI_ISL_1105106, EPI_ISL_1105107, EPI_ISL_1105108, EPI_ISL_1105110, EPI_ISL_1105111, EPI_ISL_1105112, EPI_ISL_1105113, EPI_ISL_1105114, EPI_ISL_1105118, EPI_ISL_1105119, EPI_ISL_1105120, EPI_ISL_1105125, EPI_ISL_1105129, EPI_ISL_1105130, EPI_ISL_1105131, EPI_ISL_1105134, EPI_ISL_1105135, EPI_ISL_1105136, EPI_ISL_1105137, EPI_ISL_1105138, EPI_ISL_1105139, EPI_ISL_1105142, EPI_ISL_1105146, EPI_ISL_1105150, EPI_ISL_1105151, EPI_ISL_1105155, EPI_ISL_1105156, EPI_ISL_1105158, EPI_ISL_1105162, EPI_ISL_1105173, EPI_ISL_1105174, EPI_ISL_1105180, EPI_ISL_1105189, EPI_ISL_1105197, EPI_ISL_1248496

see above | University College London Hospital | COVID-19 Genomics UK (COG-UK) Consortium | Catherine Houlihan; Dan Frampton; Dr Catherine Houlihan; Dr Daniel Frampton; Dr Judith Heaney; Dr Moira Spyer; Dr Paul Grant and Dr Eleni Nastouli; Judith Heaney; Matthew Byott; Moira Spyer and Eleni Nastouli; Stuart Kirk

EPI_ISL_949811, EPI_ISL_949814, EPI_ISL_949815, EPI_ISL_949816, EPI_ISL_949817, EPI_ISL_949818, EPI_ISL_949819, EPI_ISL_949820, EPI_ISL_949821, EPI_ISL_949822, EPI_ISL_949823, EPI_ISL_949824, EPI_ISL_949825, EPI_ISL_949826, EPI_ISL_949827, EPI_ISL_949828, EPI_ISL_949829, EPI_ISL_949830, EPI_ISL_949831, EPI_ISL_949832, EPI_ISL_949833, EPI_ISL_949834, EPI_ISL_949835, EPI_ISL_949836, EPI_ISL_949837, EPI_ISL_949838, EPI_ISL_949839, EPI_ISL_949840, EPI_ISL_949841, EPI_ISL_949842, EPI_ISL_949843, EPI_ISL_949844, EPI_ISL_949846, EPI_ISL_949849, EPI_ISL_949850, EPI_ISL_949851, EPI_ISL_949852, EPI_ISL_949853, EPI_ISL_949854, EPI_ISL_949856, EPI_ISL_949857, EPI_ISL_949858, EPI_ISL_949859, EPI_ISL_949860, EPI_ISL_949861, EPI_ISL_949862, EPI_ISL_949863, EPI_ISL_949864, EPI_ISL_949865, EPI_ISL_949867, EPI_ISL_949868, EPI_ISL_949869, EPI_ISL_949870, EPI_ISL_949871, EPI_ISL_949872, EPI_ISL_949873, EPI_ISL_949874, EPI_ISL_949875, EPI_ISL_949876, EPI_ISL_949877, EPI_ISL_949878, EPI_ISL_949879, EPI_ISL_949880, EPI_ISL_949881, EPI_ISL_949882, EPI_ISL_949883, EPI_ISL_949884, EPI_ISL_949885, EPI_ISL_949886, EPI_ISL_949887, EPI_ISL_949888, EPI_ISL_949889, EPI_ISL_949890, EPI_ISL_949891, EPI_ISL_949892, EPI_ISL_949893, EPI_ISL_949894, EPI_ISL_949895, EPI_ISL_949896, EPI_ISL_949897, EPI_ISL_949898, EPI_ISL_949899, EPI_ISL_949900, EPI_ISL_949901, EPI_ISL_949902, EPI_ISL_949903, EPI_ISL_949904, EPI_ISL_949920, EPI_ISL_949926, EPI_ISL_949934, EPI_ISL_949937, EPI_ISL_949938, EPI_ISL_949939, EPI_ISL_949945, EPI_ISL_949946, EPI_ISL_949952, EPI_ISL_949955, EPI_ISL_949956, EPI_ISL_949958, EPI_ISL_949960, EPI_ISL_949962, EPI_ISL_949964, EPI_ISL_949965, EPI_ISL_949966, EPI_ISL_949967, EPI_ISL_949968, EPI_ISL_949969, EPI_ISL_949970, EPI_ISL_949971, EPI_ISL_949972, EPI_ISL_949973, EPI_ISL_949974, EPI_ISL_997694, EPI_ISL_997701, EPI_ISL_997703, EPI_ISL_997704, EPI_ISL_997705, EPI_ISL_997735, EPI_ISL_997736, EPI_ISL_997737, EPI_ISL_997738, EPI_ISL_997739, EPI_ISL_997740, EPI_ISL_997741, EPI_ISL_997742, EPI_ISL_997743, EPI_ISL_997744, EPI_ISL_997745, EPI_ISL_997746, EPI_ISL_997747, EPI_ISL_997748, EPI_ISL_997749, EPI_ISL_997750, EPI_ISL_997835, EPI_ISL_997836, EPI_ISL_997837, EPI_ISL_997838, EPI_ISL_997843, EPI_ISL_997844, EPI_ISL_997845, EPI_ISL_997847, EPI_ISL_997866, EPI_ISL_997867, EPI_ISL_997868, EPI_ISL_997869, EPI_ISL_997870, EPI_ISL_997871, EPI_ISL_997878, EPI_ISL_997890, EPI_ISL_997891, EPI_ISL_997892, EPI_ISL_997894, EPI_ISL_997905, EPI_ISL_997906, EPI_ISL_997907, EPI_ISL_997927, EPI_ISL_997928, EPI_ISL_997929, EPI_ISL_997930, EPI_ISL_997932, EPI_ISL_997933, EPI_ISL_997934, EPI_ISL_997935, EPI_ISL_997936, EPI_ISL_997937, EPI_ISL_997938, EPI_ISL_997939, EPI_ISL_997940, EPI_ISL_997941, EPI_ISL_997942, EPI_ISL_997943, EPI_ISL_997944, EPI_ISL_997945, EPI_ISL_997946, EPI_ISL_997947, EPI_ISL_997948, EPI_ISL_997949, EPI_ISL_997950, EPI_ISL_997952, EPI_ISL_997953, EPI_ISL_997954, EPI_ISL_997955, EPI_ISL_997956, EPI_ISL_997957, EPI_ISL_997958, EPI_ISL_997959, EPI_ISL_997960, EPI_ISL_997961, EPI_ISL_997962, EPI_ISL_997963, EPI_ISL_997964, EPI_ISL_997965, EPI_ISL_997966, EPI_ISL_997967, EPI_ISL_997968, EPI_ISL_997969, EPI_ISL_997970, EPI_ISL_997971, EPI_ISL_997972, EPI_ISL_997973, EPI_ISL_997974, EPI_ISL_997976, EPI_ISL_997977, EPI_ISL_997979, EPI_ISL_997981, EPI_ISL_997984, EPI_ISL_997986, EPI_ISL_997988, EPI_ISL_1050568, EPI_ISL_1105449, EPI_ISL_1105466, EPI_ISL_1105468, EPI_ISL_1248190, EPI_ISL_1248198, EPI_ISL_1248258, EPI_ISL_1248260, EPI_ISL_1248264, EPI_ISL_1248266, EPI_ISL_1248268, EPI_ISL_1248269, EPI_ISL_1248271, EPI_ISL_1248273, EPI_ISL_1248275, EPI_ISL_1248276, EPI_ISL_1248278, EPI_ISL_1248280, EPI_ISL_1248521, EPI_ISL_1309039, EPI_ISL_1309043, EPI_ISL_1309056, EPI_ISL_1309067, EPI_ISL_1309072, EPI_ISL_1309073, EPI_ISL_1474862, EPI_ISL_1474863, EPI_ISL_1637162

see above | University College London, Great Ormond Street Hospital for Children NHS Foundation Trust, Imperial College Healthcare NHS Trust | COVID-19 Genomics UK (COG-UK) Consortium | Alison Holmes; Charlotte Williams; Helena Tutill; Jacqueline Findlay; James Price; Judith Breuer; Julianne Brown; Kathryn Harris; Leysa Forrest; Mark Kristiansen; Paola Niola; Paola Resende Silva; Patricia Dyal; Paul Randell; Rachel Williams; Samuel Weeks; Sergi Castellano; Sunando Roy; Tony Brooks; Yasmin Panchbhaya

EPI_ISL_997373, EPI_ISL_997375, EPI_ISL_1104297, EPI_ISL_1104300 | University Hospitals Of Leicester NHS Trust and DeepSeq Nottingham | COVID-19 Genomics UK (COG-UK) Consortium | Christopher Holmes; Christopher Moore; Fei Sang; Joeseph Chappell; Johnny Debebe; Jonathan Ball; Julian Tang; Karlie Fallon; Matthew Carlisle; Matthew Loose; Nadine Holmes; Patrick McClure; Paul Bird; Thomas Helmer; Victoria Wright

EPI_ISL_996413, EPI_ISL_996414, EPI_ISL_996416, EPI_ISL_996418, EPI_ISL_996420, EPI_ISL_996421, EPI_ISL_996440, EPI_ISL_996446, EPI_ISL_996452, EPI_ISL_996457, EPI_ISL_996470, EPI_ISL_996476, EPI_ISL_996478, EPI_ISL_996479, EPI_ISL_1047012, EPI_ISL_1047026, EPI_ISL_1047027, EPI_ISL_1047028, EPI_ISL_1047029, EPI_ISL_1047032, EPI_ISL_1047033, EPI_ISL_1047035, EPI_ISL_1047040, EPI_ISL_1047043, EPI_ISL_1047046, EPI_ISL_1047047, EPI_ISL_1047048, EPI_ISL_1047050, EPI_ISL_1103752, EPI_ISL_1729987, EPI_ISL_1729988, EPI_ISL_1832075, EPI_ISL_1832077, EPI_ISL_1832079, EPI_ISL_1832083, EPI_ISL_1832087, EPI_ISL_1832089

see above | University of Birmingham | COVID-19 Genomics UK (COG-UK) Consortium | Alex Richter; Andrew Bosworth; Andrew Bosworth. Queen Elizabeth Hospital: Anna Casey; Andrew D Beggs PHE Heartlands Lab: Husam Osman; Charlotte Poxon; Institute of Microbiology: Joanne Stockton; Josh Quick; Kasun Wanigasooriya; Mike Kidd; Nicholas Loman. University of Birmingham Testing Laboratory: Celina M Whalley; Oliver Pickles; Radoslaw Poplawski; Samuel Nicholls; University of Birmingham: Claire McMurray; Will Rowe

EPI_ISL_996481, EPI_ISL_996538, EPI_ISL_996539, EPI_ISL_996540, EPI_ISL_996541, EPI_ISL_996542, EPI_ISL_996550, EPI_ISL_1050297, EPI_ISL_1050298, EPI_ISL_1050304, EPI_ISL_1050305, EPI_ISL_1050306, EPI_ISL_1050326, EPI_ISL_1050330, EPI_ISL_1050331, EPI_ISL_1050332, EPI_ISL_1050333, EPI_ISL_1050335, EPI_ISL_1050336, EPI_ISL_1050337, EPI_ISL_1050338, EPI_ISL_1050339,

| | | | |
|---|---|---|---|
| EPI_ISL_1050340, EPI_ISL_1050341, EPI_ISL_1050342, EPI_ISL_1050343, EPI_ISL_1050344, EPI_ISL_1050345, EPI_ISL_1103758, EPI_ISL_1103760, EPI_ISL_1103761, EPI_ISL_1103763, EPI_ISL_1103764, EPI_ISL_1103766, EPI_ISL_1103768, EPI_ISL_1103771, EPI_ISL_1103772, EPI_ISL_1103773, EPI_ISL_1103775, EPI_ISL_1103776, EPI_ISL_1103777, EPI_ISL_1103781, EPI_ISL_1103782, EPI_ISL_1103783, EPI_ISL_1103785, EPI_ISL_1103787, EPI_ISL_1103788, EPI_ISL_1103789, EPI_ISL_1103790, EPI_ISL_1103791, EPI_ISL_1103793, EPI_ISL_1103794, EPI_ISL_1103795, EPI_ISL_1103796, EPI_ISL_1103798, EPI_ISL_1103801, EPI_ISL_1103802, EPI_ISL_1308537, EPI_ISL_1308538, EPI_ISL_1308540, EPI_ISL_1308541, EPI_ISL_1308542, EPI_ISL_1308544, EPI_ISL_1308545, EPI_ISL_1308565, EPI_ISL_1308570, EPI_ISL_1308581, EPI_ISL_1545771 | | | |
| see above | University of Exeter | COVID-19 Genomics UK (COG-UK) Consortium | Aaron Jeffries; Audrey Farbos; Ben Temperton; Jane Masoli; Joanna Warwick-Dugdale; Michelle Michelsen; Robyn Manley; Stephen Michell |
| EPI_ISL_997125, EPI_ISL_1047866, EPI_ISL_1047867, EPI_ISL_1047868, EPI_ISL_1047869, EPI_ISL_1047870, EPI_ISL_1047871, EPI_ISL_1047872, EPI_ISL_1047873, EPI_ISL_1047874, EPI_ISL_1047875, EPI_ISL_1047876, EPI_ISL_1047877, EPI_ISL_1047878, EPI_ISL_1047879, EPI_ISL_1047880, EPI_ISL_1047881, EPI_ISL_1047882, EPI_ISL_1047883, EPI_ISL_1104198, EPI_ISL_1104199, EPI_ISL_1104200, EPI_ISL_1104201, EPI_ISL_1104202, EPI_ISL_1104205, EPI_ISL_1104209, EPI_ISL_1104210, EPI_ISL_1104211, EPI_ISL_1104214, EPI_ISL_1104216, EPI_ISL_1104224, EPI_ISL_1177711, EPI_ISL_1177712, EPI_ISL_1177713, EPI_ISL_1177717, EPI_ISL_1177723, EPI_ISL_1177724, EPI_ISL_1177749, EPI_ISL_1247686, EPI_ISL_1247687, EPI_ISL_1247688, EPI_ISL_1247694, EPI_ISL_1247695, EPI_ISL_1247723, EPI_ISL_1247742, EPI_ISL_1247743, EPI_ISL_1247745, EPI_ISL_1308707, EPI_ISL_1308708, EPI_ISL_1308721, EPI_ISL_1308722, EPI_ISL_1308723, EPI_ISL_1308831, EPI_ISL_1308837, EPI_ISL_1308840, EPI_ISL_1308844, EPI_ISL_1308859, EPI_ISL_1386687, EPI_ISL_1474592, EPI_ISL_1474593, EPI_ISL_1730224, EPI_ISL_1730225 | | | |
| see above | Virology Department, Royal Infirmary of Edinburgh, NHS Lothian / School of Biological Sciences, University of Edinburgh | COVID-19 Genomics UK (COG-UK) Consortium | Colquhoun R; Cotton S; Dewar R; Gallagher A; Hill V; Jackson B; Maloney D; McCrone JT; McHugh M; O'Toole Á; Rambaut A; Rooke S; Scher E; Templeton K; Yu X |
| EPI_ISL_924166, EPI_ISL_924231, EPI_ISL_924290, EPI_ISL_924351, EPI_ISL_952954, EPI_ISL_952955, EPI_ISL_952956, EPI_ISL_952958, EPI_ISL_952960, EPI_ISL_952961, EPI_ISL_952962, EPI_ISL_952963, EPI_ISL_952965, EPI_ISL_952966, EPI_ISL_952967, EPI_ISL_952968, EPI_ISL_952970, EPI_ISL_952971, EPI_ISL_952973, EPI_ISL_952974, EPI_ISL_952975, EPI_ISL_952976, EPI_ISL_952977, EPI_ISL_952979, EPI_ISL_952980, EPI_ISL_952981, EPI_ISL_952982, EPI_ISL_952984, EPI_ISL_952985, EPI_ISL_952987, EPI_ISL_952989, EPI_ISL_952990, EPI_ISL_952992, EPI_ISL_952993, EPI_ISL_952995, EPI_ISL_952996, EPI_ISL_952997, EPI_ISL_952998, EPI_ISL_952999, EPI_ISL_953000, EPI_ISL_953001, EPI_ISL_953002, EPI_ISL_953004, EPI_ISL_953005, EPI_ISL_953007, EPI_ISL_953008, EPI_ISL_953009, EPI_ISL_953010, EPI_ISL_953011, EPI_ISL_953015, EPI_ISL_953016, EPI_ISL_953018, EPI_ISL_953019, EPI_ISL_953020, EPI_ISL_953021, EPI_ISL_953022, EPI_ISL_953023, EPI_ISL_998099, EPI_ISL_998100, EPI_ISL_998101, EPI_ISL_998102, EPI_ISL_998103, EPI_ISL_998104, EPI_ISL_998105, EPI_ISL_998106, EPI_ISL_998107, EPI_ISL_998108, EPI_ISL_998110, EPI_ISL_998111, EPI_ISL_998112, EPI_ISL_998113, EPI_ISL_998114, EPI_ISL_998115, EPI_ISL_998116, EPI_ISL_998117, EPI_ISL_998118, EPI_ISL_998119, EPI_ISL_998122, EPI_ISL_998123, EPI_ISL_998124, EPI_ISL_998125, EPI_ISL_998126, EPI_ISL_998127, EPI_ISL_998128, EPI_ISL_998130, EPI_ISL_998131, EPI_ISL_998132, EPI_ISL_998133, EPI_ISL_998135, EPI_ISL_1000642, EPI_ISL_1000646, EPI_ISL_1000647, EPI_ISL_1000666, EPI_ISL_1000676, EPI_ISL_1000679, EPI_ISL_1000689, EPI_ISL_1000698, EPI_ISL_1000710, EPI_ISL_1000713, EPI_ISL_1000722, EPI_ISL_1000728, EPI_ISL_1000732, EPI_ISL_1000735, EPI_ISL_1000743, EPI_ISL_1000744, EPI_ISL_1000750, EPI_ISL_1000757, EPI_ISL_1000758, EPI_ISL_1000774, EPI_ISL_1000786, EPI_ISL_1000789, EPI_ISL_1000790, EPI_ISL_1047594, EPI_ISL_1047596, EPI_ISL_1047712, EPI_ISL_1047713, EPI_ISL_1047714, EPI_ISL_1047715, EPI_ISL_1047716, EPI_ISL_1047717, EPI_ISL_1047718, EPI_ISL_1047722, EPI_ISL_1047723, EPI_ISL_1047724, EPI_ISL_1047725, EPI_ISL_1047726, EPI_ISL_1047727, EPI_ISL_1047728, EPI_ISL_1047729, EPI_ISL_1047730, EPI_ISL_1047731, EPI_ISL_1047732, EPI_ISL_1047733, EPI_ISL_1047734, EPI_ISL_1047735, EPI_ISL_1047736, EPI_ISL_1047737, EPI_ISL_1047738, EPI_ISL_1047739, EPI_ISL_1047740, EPI_ISL_1047741, EPI_ISL_1047742, EPI_ISL_1047744, EPI_ISL_1047745, EPI_ISL_1047746, EPI_ISL_1047747, EPI_ISL_1047748, EPI_ISL_1047749, EPI_ISL_1047750, EPI_ISL_1047751, EPI_ISL_1047752, EPI_ISL_1047753, EPI_ISL_1047760, EPI_ISL_1047761, EPI_ISL_1047762, EPI_ISL_1047763, EPI_ISL_1047764, EPI_ISL_1047765, EPI_ISL_1047766, EPI_ISL_1047767, EPI_ISL_1047768, EPI_ISL_1047769, EPI_ISL_1047770, EPI_ISL_1047771, EPI_ISL_1047773, EPI_ISL_1047774, EPI_ISL_1047775, EPI_ISL_1047776, EPI_ISL_1047777, EPI_ISL_1047778, EPI_ISL_1047779, EPI_ISL_1047780, EPI_ISL_1047785, EPI_ISL_1047786, EPI_ISL_1047787, EPI_ISL_1047788, EPI_ISL_1047789, EPI_ISL_1047790, EPI_ISL_1047791, EPI_ISL_1047792, EPI_ISL_1047793, EPI_ISL_1047794, EPI_ISL_1047795, EPI_ISL_1047796, EPI_ISL_1050828, EPI_ISL_1050829, EPI_ISL_1050830, EPI_ISL_1050831, EPI_ISL_1050832, EPI_ISL_1050833, EPI_ISL_1050834, EPI_ISL_1050840, EPI_ISL_1050841, EPI_ISL_1050863, EPI_ISL_1050864, EPI_ISL_1050865, EPI_ISL_1050866, EPI_ISL_1050867, EPI_ISL_1050868, EPI_ISL_1050870, EPI_ISL_1050883, EPI_ISL_1050884, EPI_ISL_1050885, EPI_ISL_1050886, EPI_ISL_1050888, EPI_ISL_1050889, EPI_ISL_1050890, EPI_ISL_1050891, EPI_ISL_1050893, EPI_ISL_1050895, EPI_ISL_1050896, EPI_ISL_1050897, EPI_ISL_1050898, EPI_ISL_1050899, EPI_ISL_1050900, EPI_ISL_1050901, EPI_ISL_1050903, EPI_ISL_1050904, EPI_ISL_1050906, EPI_ISL_1050907, EPI_ISL_1050908, EPI_ISL_1104041, EPI_ISL_1104042, EPI_ISL_1104043, EPI_ISL_1104044, EPI_ISL_1104045, EPI_ISL_1104046, EPI_ISL_1104052, EPI_ISL_1104053, EPI_ISL_1104054, EPI_ISL_1104055, EPI_ISL_1104057, EPI_ISL_1104058, EPI_ISL_1104059, EPI_ISL_1104060, EPI_ISL_1104062, EPI_ISL_1104063, EPI_ISL_1104065, EPI_ISL_1104066, EPI_ISL_1104067, EPI_ISL_1104068, EPI_ISL_1104069, EPI_ISL_1105573, EPI_ISL_1105574, EPI_ISL_1105575, EPI_ISL_1109243, EPI_ISL_1109244, EPI_ISL_1109245, EPI_ISL_1109246, EPI_ISL_1109247, EPI_ISL_1109248, EPI_ISL_1109249, EPI_ISL_1109250, EPI_ISL_1109251, EPI_ISL_1109252, EPI_ISL_1109253, EPI_ISL_1109254, EPI_ISL_1109255, EPI_ISL_1109256, EPI_ISL_1109257, EPI_ISL_1109258, EPI_ISL_1109259, EPI_ISL_1109260, EPI_ISL_1109261, EPI_ISL_1109262, EPI_ISL_1109263, EPI_ISL_1109264, EPI_ISL_1109265, EPI_ISL_1109266, EPI_ISL_1109267, EPI_ISL_1109268, EPI_ISL_1109269, EPI_ISL_1109270, EPI_ISL_1109271, EPI_ISL_1109272, EPI_ISL_1109273, EPI_ISL_1109274, EPI_ISL_1109275, EPI_ISL_1109276, EPI_ISL_1109277, EPI_ISL_1109278, EPI_ISL_1109279, EPI_ISL_1109280, EPI_ISL_1109281, EPI_ISL_1109282, EPI_ISL_1109284, EPI_ISL_1109285, EPI_ISL_1109286, EPI_ISL_1109287, EPI_ISL_1109288, EPI_ISL_1109289, EPI_ISL_1109290, EPI_ISL_1109291, EPI_ISL_1109292, EPI_ISL_1109293, EPI_ISL_1109294, EPI_ISL_1109295, EPI_ISL_1109296, EPI_ISL_1109297, EPI_ISL_1109298, EPI_ISL_1109299, EPI_ISL_1109300, EPI_ISL_1109301, EPI_ISL_1109302, EPI_ISL_1109303, EPI_ISL_1109304, EPI_ISL_1109305, EPI_ISL_1109306, EPI_ISL_1109307, EPI_ISL_1109308, EPI_ISL_1109309, EPI_ISL_1109310, EPI_ISL_1109311, EPI_ISL_1109312, EPI_ISL_1109313, EPI_ISL_1109314, EPI_ISL_1109315, EPI_ISL_1109316, EPI_ISL_1109317, EPI_ISL_1180058, EPI_ISL_1250296 | | | |
| see above | Virology Department, Sheffield Teaching Hospitals NHS Foundation Trust/Department of Infection, Immunity and Cardiovascular Disease, The Medical School, University of Sheffield | COVID-19 Genomics UK (COG-UK) Consortium | Adri Angyal; Alex Keeley; Benjamin Lindsey; Cariad Evans; Danielle Groves; Dave Partridge; Katie Johnson; Laura Carrilero; Luke Green; Matthew Parker; Matthew Wyles; Mehmet Yavuz; Mohammad Raza; Nikki Smith; Paul Parsons; Rachel Tucker; Rebecca Brown; Thushan de Silva |
| EPI_ISL_999945, EPI_ISL_999947, EPI_ISL_999948, EPI_ISL_999949, EPI_ISL_999952, EPI_ISL_999957, EPI_ISL_999958, EPI_ISL_999959, EPI_ISL_999961, EPI_ISL_999963, EPI_ISL_999964, EPI_ISL_999965, EPI_ISL_999969, EPI_ISL_999970, EPI_ISL_999973, EPI_ISL_999974, EPI_ISL_999975, EPI_ISL_999976, EPI_ISL_999977, EPI_ISL_999978, EPI_ISL_999979, EPI_ISL_999980, EPI_ISL_999982, EPI_ISL_999983, EPI_ISL_999984, EPI_ISL_999988, EPI_ISL_999989, EPI_ISL_999990, EPI_ISL_999991, EPI_ISL_999993, EPI_ISL_999994, EPI_ISL_999995, EPI_ISL_999999, EPI_ISL_1000000, EPI_ISL_1000001, EPI_ISL_1000007, EPI_ISL_1000010, EPI_ISL_1000011, EPI_ISL_1000017, EPI_ISL_1000025, EPI_ISL_1000028, EPI_ISL_1000035, EPI_ISL_1000039, EPI_ISL_1000043, EPI_ISL_1000049, EPI_ISL_1000051, EPI_ISL_1000057, EPI_ISL_1000064, EPI_ISL_1000066, EPI_ISL_1000068, EPI_ISL_1000070, EPI_ISL_1000078, EPI_ISL_1000079, EPI_ISL_1000080, EPI_ISL_1000081, EPI_ISL_1000082, EPI_ISL_1000083, EPI_ISL_1000084, EPI_ISL_1000085, EPI_ISL_1000087, EPI_ISL_1000088, EPI_ISL_1000092, EPI_ISL_1000093, EPI_ISL_1000094, EPI_ISL_1000095, EPI_ISL_1000096, EPI_ISL_1000097, EPI_ISL_1000098, EPI_ISL_1000102, EPI_ISL_1000103, EPI_ISL_1000104, EPI_ISL_1000105, EPI_ISL_1000109, EPI_ISL_1000111, EPI_ISL_1000112, EPI_ISL_1000113, EPI_ISL_1000115, EPI_ISL_1000117, EPI_ISL_1000118, EPI_ISL_1000122, EPI_ISL_1000123, EPI_ISL_1000124, EPI_ISL_1000125, EPI_ISL_1000126, EPI_ISL_1000127, EPI_ISL_1000129, EPI_ISL_1000130, EPI_ISL_1000131, EPI_ISL_1000133, EPI_ISL_1000134, EPI_ISL_1000135, EPI_ISL_1000137, EPI_ISL_1000138, EPI_ISL_1000140, EPI_ISL_1000142, EPI_ISL_1000143, EPI_ISL_1000144, EPI_ISL_1000146, EPI_ISL_1000151, EPI_ISL_1000152, EPI_ISL_1000153, EPI_ISL_1000154, EPI_ISL_1000155, EPI_ISL_1000156, EPI_ISL_1000157, EPI_ISL_1000158, EPI_ISL_1000159, EPI_ISL_1000160, EPI_ISL_1000161, EPI_ISL_1000162, EPI_ISL_1000163, EPI_ISL_1000164, EPI_ISL_1000165, EPI_ISL_1000166, EPI_ISL_1000167, EPI_ISL_1000168, EPI_ISL_1000171, EPI_ISL_1000173, EPI_ISL_1000174, EPI_ISL_1000179, EPI_ISL_1000181, EPI_ISL_1000182, EPI_ISL_1000183, EPI_ISL_1000184, EPI_ISL_1000185, EPI_ISL_1000186, EPI_ISL_1052147, EPI_ISL_1052207, EPI_ISL_1052213, EPI_ISL_1052215, EPI_ISL_1052216, EPI_ISL_1052222, EPI_ISL_1052225, EPI_ISL_1052234, EPI_ISL_1052250, EPI_ISL_1052251, EPI_ISL_1052252, EPI_ISL_1052253, EPI_ISL_1052259, EPI_ISL_1052273, EPI_ISL_1052274, EPI_ISL_1052275, EPI_ISL_1052276, EPI_ISL_1052277, EPI_ISL_1052278, EPI_ISL_1052279, EPI_ISL_1052280, EPI_ISL_1052281, EPI_ISL_1052282, EPI_ISL_1052283, EPI_ISL_1052284, EPI_ISL_1052287, EPI_ISL_1052289, EPI_ISL_1052290, EPI_ISL_1052291, EPI_ISL_1052294, EPI_ISL_1052295, EPI_ISL_1052297, EPI_ISL_1052300, EPI_ISL_1052301, EPI_ISL_1052302, EPI_ISL_1052303, EPI_ISL_1052305, EPI_ISL_1052306, EPI_ISL_1052307, EPI_ISL_1052308, EPI_ISL_1052309, EPI_ISL_1052310, EPI_ISL_1052311, EPI_ISL_1052313, EPI_ISL_1052314, EPI_ISL_1052315, EPI_ISL_1052316, EPI_ISL_1052317, EPI_ISL_1052318, EPI_ISL_1052319, EPI_ISL_1052321, EPI_ISL_1052325, EPI_ISL_1052326, EPI_ISL_1052331, EPI_ISL_1052338, EPI_ISL_1052344, EPI_ISL_1052346, EPI_ISL_1052347, EPI_ISL_1052348, EPI_ISL_1052349, EPI_ISL_1052351, EPI_ISL_1052353, EPI_ISL_1052354, EPI_ISL_1052356, EPI_ISL_1052363, EPI_ISL_1052364, EPI_ISL_1052368, EPI_ISL_1052369, EPI_ISL_1052370, EPI_ISL_1052372, EPI_ISL_1052374, EPI_ISL_1052376, EPI_ISL_1052378, EPI_ISL_1052387, EPI_ISL_1052388, EPI_ISL_1052389, EPI_ISL_1052391, EPI_ISL_1052394, EPI_ISL_1052396, EPI_ISL_1052398, EPI_ISL_1052400, EPI_ISL_1052402, EPI_ISL_1052403, EPI_ISL_1052406, EPI_ISL_1052408, EPI_ISL_1052409, EPI_ISL_1052412, EPI_ISL_1052413, EPI_ISL_1052415, EPI_ISL_1052416, EPI_ISL_1052417, EPI_ISL_1052422, EPI_ISL_1052423, EPI_ISL_1052424, EPI_ISL_1052425, EPI_ISL_1052428, EPI_ISL_1052434, EPI_ISL_1052436, EPI_ISL_1052439, EPI_ISL_1052441, EPI_ISL_1052442, EPI_ISL_1052445, EPI_ISL_1052450, EPI_ISL_1052453, EPI_ISL_1052455, EPI_ISL_1052459, EPI_ISL_1052461, EPI_ISL_1052543, EPI_ISL_1052546, EPI_ISL_1052551, EPI_ISL_1052552, EPI_ISL_1052600, EPI_ISL_1052608, EPI_ISL_1052618, EPI_ISL_1052622, EPI_ISL_1052628, EPI_ISL_1052630, EPI_ISL_1052631, EPI_ISL_1052636, EPI_ISL_1052637, EPI_ISL_1052638, EPI_ISL_1052639, EPI_ISL_1052640, EPI_ISL_1052641, EPI_ISL_1052642, EPI_ISL_1052643, EPI_ISL_1052644, EPI_ISL_1052645, EPI_ISL_1052647, EPI_ISL_1052655, EPI_ISL_1052656, EPI_ISL_1052664, EPI_ISL_1052667, EPI_ISL_1052668, EPI_ISL_1052672, EPI_ISL_1052674, EPI_ISL_1052682, EPI_ISL_1052683, EPI_ISL_1052706, EPI_ISL_1052708, EPI_ISL_1052710, EPI_ISL_1052711, EPI_ISL_1052712, EPI_ISL_1052730, EPI_ISL_1052731, EPI_ISL_1052736, EPI_ISL_1052741, EPI_ISL_1052750, EPI_ISL_1052761, EPI_ISL_1053491, EPI_ISL_1053493, EPI_ISL_1053495, EPI_ISL_1053496, EPI_ISL_1053536, EPI_ISL_1053570, EPI_ISL_1053578, EPI_ISL_1053586, EPI_ISL_1053587, EPI_ISL_1106255, EPI_ISL_1106648, EPI_ISL_1106650, EPI_ISL_1106651, EPI_ISL_1106652, EPI_ISL_1106653, EPI_ISL_1106654, EPI_ISL_1106655, EPI_ISL_1106656, EPI_ISL_1106657, EPI_ISL_1106658, EPI_ISL_1106660, EPI_ISL_1106661, EPI_ISL_1106662 | | | |
| see above | Wales Specialist Virology Centre Sequencing lab: Pathogen Genomics Unit | Public Health Wales Microbiology Cardiff Wales Specialist Virology Centre | Alec Birchley; Alexander Adams; Amy Gaskin; Angela Marchbank; Bree Gatica-Wilcox; Catherine Moore; Jason Coombes; Joanne Watkins; Joel Southgate; Johnathan Evans; Laura Gifford; Lauren Gilbert; Lee Graham; Malorie Perry; Matthew Bull; Nicole Pacchiarini; Sally Corden; Sara Kumziene-Summerhayes; Sara Rey; Sarah Taylor; Simon Cottrell; Sophie Jones; Tom Connor |
| EPI_ISL_997062, EPI_ISL_997064, EPI_ISL_997065, EPI_ISL_997066, EPI_ISL_997070, EPI_ISL_997071, EPI_ISL_1047834, EPI_ISL_1047835, EPI_ISL_1047847, EPI_ISL_1047848, EPI_ISL_1047849, EPI_ISL_1047850, EPI_ISL_1050175, EPI_ISL_1050176, EPI_ISL_1050177, EPI_ISL_1050178, EPI_ISL_1050179, EPI_ISL_1050180, EPI_ISL_1050181, EPI_ISL_1050182, EPI_ISL_1050183, EPI_ISL_1050184, EPI_ISL_1050185, EPI_ISL_1050186, EPI_ISL_1050187, EPI_ISL_1050188, EPI_ISL_1050189, EPI_ISL_1050190, EPI_ISL_1050191, EPI_ISL_1050192, EPI_ISL_1050193, EPI_ISL_1050194, EPI_ISL_1104192 | | | |
| see above | West of Scotland Specialist Virology Centre, NHSGGC / MRC-University of Glasgow Centre for Virus Research | COVID-19 Genomics UK (COG-UK) Consortium | Alasdair MacLean; Alice Broos; Ana da Silva Filipe; Antonia Ho; Daniel Mair; David L Robertson; Emma Thomson; Guy Mollett; James Shepherd; Jenna Nichols; Joseph Hughes; Kathy Li; Kathy Smollett; Kyriaki Nomikou; Lily Tong; Matthew Holden; Natasha Johnson; Rachel Blacow; Richard Orton; Rory Gunson; Sarah McDonald; Sharif Shaaban; Sreenu Vattipally; Stephen Carmichael |

We gratefully acknowledge the following Authors from the Originating laboratories responsible for obtaining the specimens, as well as the Submitting laboratories where the genome data were generated and shared via GISAID, on which this research is based.

All Submitters of data may be contacted directly via www.gisaid.org

Authors are sorted alphabetically.

| Accession ID | Originating Laboratory | Submitting Laboratory | Authors |
|---|---|---|---|
| EPI_ISL_1048065, EPI_ISL_1048068, EPI_ISL_1048073, EPI_ISL_1048075, EPI_ISL_1048086, EPI_ISL_1048087, EPI_ISL_1048103, EPI_ISL_1048107, EPI_ISL_1048108, EPI_ISL_1048109, EPI_ISL_1048110, EPI_ISL_1048119, EPI_ISL_1048120, EPI_ISL_1048123, EPI_ISL_1048126, EPI_ISL_1048127, EPI_ISL_1048128, EPI_ISL_1048130, EPI_ISL_1048131, EPI_ISL_1048133, EPI_ISL_1048134, EPI_ISL_1177989, EPI_ISL_1177991, EPI_ISL_1177992, EPI_ISL_1177993, EPI_ISL_1730308, EPI_ISL_1975109 | | | |
| see above | Barts Health NHS Trust | COVID-19 Genomics UK (COG-UK) Consortium | BROAD; Beatrix; CUTINO-MOGUEL; Claire; David; Dola; HARRINGTON; KELE; KULASEGARAN-SHYLINI; Maria-Teresa; OWOYEMI; Raghavendran |
| EPI_ISL_1000914, EPI_ISL_1000915, EPI_ISL_1000916, EPI_ISL_1000919, EPI_ISL_1000920, EPI_ISL_1000922, EPI_ISL_1054763, EPI_ISL_1054771, EPI_ISL_1054776, EPI_ISL_1054778, EPI_ISL_1054781, EPI_ISL_1054783, EPI_ISL_1054784, EPI_ISL_1054785, EPI_ISL_1054786, EPI_ISL_1054788, EPI_ISL_1054789, EPI_ISL_1054790, EPI_ISL_1054791 | | | |
| see above | Bioinformatics and Biostatistics Lab, Advanced Sequencing Facility | COVID-19 Genomics UK (COG-UK) Consortium | Aengus Stewart; Chelsea Sawyer; Harshil Patel; Jerome Nicod; Laura Cubitt; Margaret Crawford |
| EPI_ISL_1000204, EPI_ISL_1000205, EPI_ISL_1000206, EPI_ISL_1000207, EPI_ISL_1000208, EPI_ISL_1000209, EPI_ISL_1000210, EPI_ISL_1000211, EPI_ISL_1000212, EPI_ISL_1000214, EPI_ISL_1000215, EPI_ISL_1000216, EPI_ISL_1000217, EPI_ISL_1000220, EPI_ISL_1000221, EPI_ISL_1000222, EPI_ISL_1000223, EPI_ISL_1000224, EPI_ISL_1000225, EPI_ISL_1000226, EPI_ISL_1000227, EPI_ISL_1000228, EPI_ISL_1000229, EPI_ISL_1000230, EPI_ISL_1000231, EPI_ISL_1000233, EPI_ISL_1000416, EPI_ISL_1000471, EPI_ISL_1050075, EPI_ISL_1050076, EPI_ISL_1050077, EPI_ISL_1050078, EPI_ISL_1050079, EPI_ISL_1050080, EPI_ISL_1050081, EPI_ISL_1050082, EPI_ISL_1050083, EPI_ISL_1050084, EPI_ISL_1050085, EPI_ISL_1050086, EPI_ISL_1050087, EPI_ISL_1050088, EPI_ISL_1050089, EPI_ISL_1050090, EPI_ISL_1050091, EPI_ISL_1050092, EPI_ISL_1050093, EPI_ISL_1050094, EPI_ISL_1050095, EPI_ISL_1050096, EPI_ISL_1050097, EPI_ISL_1050098, EPI_ISL_1050099, EPI_ISL_1050100, EPI_ISL_1050101, EPI_ISL_1050102, EPI_ISL_1050103, EPI_ISL_1050104, EPI_ISL_1050105, EPI_ISL_1050106, EPI_ISL_1050107, EPI_ISL_1050108, EPI_ISL_1050109, EPI_ISL_1050110, EPI_ISL_1050111, EPI_ISL_1050112, EPI_ISL_1050113, EPI_ISL_1050114, EPI_ISL_1050115, EPI_ISL_1050116, EPI_ISL_1050117, EPI_ISL_1050118, EPI_ISL_1050119, EPI_ISL_1050120, EPI_ISL_1050121, EPI_ISL_1050122, EPI_ISL_1050123, EPI_ISL_1050124, EPI_ISL_1050125, EPI_ISL_1050126, EPI_ISL_1050127, EPI_ISL_1050128, EPI_ISL_1050129, EPI_ISL_1050130, EPI_ISL_1050131, EPI_ISL_1050132, EPI_ISL_1050133, EPI_ISL_1050134, EPI_ISL_1050135, EPI_ISL_1050136, EPI_ISL_1050137, EPI_ISL_1050138, EPI_ISL_1050139, EPI_ISL_1050140, EPI_ISL_1050141, EPI_ISL_1050142, EPI_ISL_1050143, EPI_ISL_1050144, EPI_ISL_1050145, EPI_ISL_1050146, EPI_ISL_1050147, EPI_ISL_1050148, EPI_ISL_1050149, EPI_ISL_1050150, EPI_ISL_1050151, EPI_ISL_1050152, EPI_ISL_1050153, EPI_ISL_1050154, EPI_ISL_1050155, EPI_ISL_1050156, EPI_ISL_1050157, EPI_ISL_1050158, EPI_ISL_1050159, EPI_ISL_1050160, EPI_ISL_1050161, EPI_ISL_1050162, EPI_ISL_1050163, EPI_ISL_1050164, EPI_ISL_1050165, EPI_ISL_1050166, EPI_ISL_1050167, EPI_ISL_1050168, EPI_ISL_1050169, EPI_ISL_1050170, EPI_ISL_1050171, EPI_ISL_1050172, EPI_ISL_1050173, EPI_ISL_1053906, EPI_ISL_1053907, EPI_ISL_1053948, EPI_ISL_1053949, EPI_ISL_1053951, EPI_ISL_1053952, EPI_ISL_1053961, EPI_ISL_1053962, EPI_ISL_1053963, EPI_ISL_1053964, EPI_ISL_1053972, EPI_ISL_1053973, EPI_ISL_1053974, EPI_ISL_1053975, EPI_ISL_1054542, EPI_ISL_1054543, EPI_ISL_1054544, EPI_ISL_1054545, EPI_ISL_1054546, EPI_ISL_1054547, EPI_ISL_1054548, EPI_ISL_1054549, EPI_ISL_1054550, EPI_ISL_1054551, EPI_ISL_1054552, EPI_ISL_1054553, EPI_ISL_1054554, EPI_ISL_1054555, EPI_ISL_1054556, EPI_ISL_1054557, EPI_ISL_1054558, EPI_ISL_1054559, EPI_ISL_1054560, EPI_ISL_1054561, EPI_ISL_1054562, EPI_ISL_1054563, EPI_ISL_1054564, EPI_ISL_1054565, EPI_ISL_1054566, EPI_ISL_1054567, EPI_ISL_1054568, EPI_ISL_1054569, EPI_ISL_1054570, EPI_ISL_1054571, EPI_ISL_1054573, EPI_ISL_1054574, EPI_ISL_1054575, EPI_ISL_1054576, EPI_ISL_1054577, EPI_ISL_1054578, EPI_ISL_1054579, EPI_ISL_1054580, EPI_ISL_1054581, EPI_ISL_1054582, EPI_ISL_1054583, EPI_ISL_1054584, EPI_ISL_1054585, EPI_ISL_1054586, EPI_ISL_1054587, EPI_ISL_1054588, EPI_ISL_1054589, EPI_ISL_1054590, EPI_ISL_1054591, EPI_ISL_1054592, EPI_ISL_1054593, EPI_ISL_1054594, EPI_ISL_1054595, EPI_ISL_1054596, EPI_ISL_1054597, EPI_ISL_1054598, EPI_ISL_1054599, EPI_ISL_1054600, EPI_ISL_1054601, EPI_ISL_1054602, EPI_ISL_1054603, EPI_ISL_1054604, EPI_ISL_1054605, EPI_ISL_1054606, EPI_ISL_1054607, EPI_ISL_1054608, EPI_ISL_1054609, EPI_ISL_1054610, EPI_ISL_1054611, EPI_ISL_1054612, EPI_ISL_1054613, EPI_ISL_1054614, EPI_ISL_1054615, EPI_ISL_1054616, EPI_ISL_1054617, EPI_ISL_1054618, EPI_ISL_1054619, EPI_ISL_1104085, EPI_ISL_1104088, EPI_ISL_1107642, EPI_ISL_1107643, EPI_ISL_1107644, EPI_ISL_1107645, EPI_ISL_1107669, EPI_ISL_1107670, EPI_ISL_1107671, EPI_ISL_1107672, EPI_ISL_1107673, EPI_ISL_1107674, EPI_ISL_1107675, EPI_ISL_1107676, EPI_ISL_1107677, EPI_ISL_1107716, EPI_ISL_1107717, EPI_ISL_1107718, EPI_ISL_1107719, EPI_ISL_1107720, EPI_ISL_1107721, EPI_ISL_1107722, EPI_ISL_1107734, EPI_ISL_1107735, EPI_ISL_1107736, EPI_ISL_1107737, EPI_ISL_1107738, EPI_ISL_1107739, EPI_ISL_1107740, EPI_ISL_1107741, EPI_ISL_1107742, EPI_ISL_1107743, EPI_ISL_1107744, EPI_ISL_1107786, EPI_ISL_1107787, EPI_ISL_1107788, EPI_ISL_1109229, EPI_ISL_1109237, EPI_ISL_1109238, EPI_ISL_1109239, EPI_ISL_1109240, EPI_ISL_1109241, EPI_ISL_1109242, EPI_ISL_1179824, EPI_ISL_1388710, EPI_ISL_1388712, EPI_ISL_1388714, EPI_ISL_1388715, EPI_ISL_1388717, EPI_ISL_1388719, EPI_ISL_1388721, EPI_ISL_1388724, EPI_ISL_1388725, EPI_ISL_1388728, EPI_ISL_1388730, EPI_ISL_1388756, EPI_ISL_1388758, EPI_ISL_1388760, EPI_ISL_1388764, EPI_ISL_1730913 | | | |
| see above | Centre for Enzyme Innovation, University of Portsmouth / Translational Research Laboratory, Portsmouth Hospitals NHS Trust | COVID-19 Genomics UK (COG-UK) Consortium | Angela Beckett; Christopher Fearn; Kate Cook; Katie Loveson; Salman Goudarzi; Samuel Robson; Scott Elliott; Sharon Glaysher |
| EPI_ISL_996653, EPI_ISL_996787, EPI_ISL_996788, EPI_ISL_996789, EPI_ISL_996790, EPI_ISL_996792, EPI_ISL_996793, EPI_ISL_996795, EPI_ISL_996796, EPI_ISL_996797, EPI_ISL_996798, EPI_ISL_996799, EPI_ISL_996800, EPI_ISL_996802, EPI_ISL_996803, EPI_ISL_996804, EPI_ISL_996805, EPI_ISL_996806, EPI_ISL_996808, EPI_ISL_996809, EPI_ISL_996810, EPI_ISL_996811, EPI_ISL_996812, EPI_ISL_996814, EPI_ISL_996816, EPI_ISL_996818, EPI_ISL_996821, EPI_ISL_996823, EPI_ISL_996824, EPI_ISL_996826, EPI_ISL_996827, EPI_ISL_996829, EPI_ISL_996830, EPI_ISL_996831, EPI_ISL_996837, EPI_ISL_996838, EPI_ISL_996839, EPI_ISL_996840, EPI_ISL_996841, EPI_ISL_996842, EPI_ISL_996843, EPI_ISL_996844, EPI_ISL_996845, EPI_ISL_996846, EPI_ISL_996847, EPI_ISL_996848, EPI_ISL_996849, EPI_ISL_996850, EPI_ISL_996851, EPI_ISL_996854, EPI_ISL_996855, EPI_ISL_996856, EPI_ISL_996857, EPI_ISL_996858, EPI_ISL_996859, EPI_ISL_996860, EPI_ISL_996861, EPI_ISL_996862, EPI_ISL_996863, EPI_ISL_996864, EPI_ISL_1047081, EPI_ISL_1054258, EPI_ISL_1054260, EPI_ISL_1054261, EPI_ISL_1054262, EPI_ISL_1054263, EPI_ISL_1054264, EPI_ISL_1054266, EPI_ISL_1054267, EPI_ISL_1054268, EPI_ISL_1054269, EPI_ISL_1054270, EPI_ISL_1054271, EPI_ISL_1054272, EPI_ISL_1054273, EPI_ISL_1054274, EPI_ISL_1054275, EPI_ISL_1054276, EPI_ISL_1054277, EPI_ISL_1054278, EPI_ISL_1054279, EPI_ISL_1054280, EPI_ISL_1054281, EPI_ISL_1054282, EPI_ISL_1054283, EPI_ISL_1054284, EPI_ISL_1054285, EPI_ISL_1054286, EPI_ISL_1054287, EPI_ISL_1054288, EPI_ISL_1054289, EPI_ISL_1054290, EPI_ISL_1054291, EPI_ISL_1054293, EPI_ISL_1054294, EPI_ISL_1054295, EPI_ISL_1054296, EPI_ISL_1054297, EPI_ISL_1054298, EPI_ISL_1054299, EPI_ISL_1054300, EPI_ISL_1054301, EPI_ISL_1054302, EPI_ISL_1054303, EPI_ISL_1054304, EPI_ISL_1054305, EPI_ISL_1054306, EPI_ISL_1054307, EPI_ISL_1054308, EPI_ISL_1054309, EPI_ISL_1054310, EPI_ISL_1054311, EPI_ISL_1054312, EPI_ISL_1054313, EPI_ISL_1054314, EPI_ISL_1054315, EPI_ISL_1054316, EPI_ISL_1054317, EPI_ISL_1054318, EPI_ISL_1054319, EPI_ISL_1054320, EPI_ISL_1054321, EPI_ISL_1054322, EPI_ISL_1054323, EPI_ISL_1054324, EPI_ISL_1054325, EPI_ISL_1054328, EPI_ISL_1054329, EPI_ISL_1054330, EPI_ISL_1054331, EPI_ISL_1054332, EPI_ISL_1054333, EPI_ISL_1054334, EPI_ISL_1054335, EPI_ISL_1054336, EPI_ISL_1054337, EPI_ISL_1054338, EPI_ISL_1054339, EPI_ISL_1054340, EPI_ISL_1054341, EPI_ISL_1054342, EPI_ISL_1054343, EPI_ISL_1054344, EPI_ISL_1054345, EPI_ISL_1054346, EPI_ISL_1054347, EPI_ISL_1054348, EPI_ISL_1054349, EPI_ISL_1054350, EPI_ISL_1054351, EPI_ISL_1054353, EPI_ISL_1054354, EPI_ISL_1054355, EPI_ISL_1054356, EPI_ISL_1054357, EPI_ISL_1054358, EPI_ISL_1054359, EPI_ISL_1054360, EPI_ISL_1054361, EPI_ISL_1054362, EPI_ISL_1054363, EPI_ISL_1054364, EPI_ISL_1054365, EPI_ISL_1054366, EPI_ISL_1054367, EPI_ISL_1054368, EPI_ISL_1054369, EPI_ISL_1054370, EPI_ISL_1054371, EPI_ISL_1103885, EPI_ISL_1103891, EPI_ISL_1107805, EPI_ISL_1107806, EPI_ISL_1107807, EPI_ISL_1107808, EPI_ISL_1107809, EPI_ISL_1107810, EPI_ISL_1107811, EPI_ISL_1107812, EPI_ISL_1107813, EPI_ISL_1107814, EPI_ISL_1107815, EPI_ISL_1107816, EPI_ISL_1107817, EPI_ISL_1107818, EPI_ISL_1107819, EPI_ISL_1107820, EPI_ISL_1107821, EPI_ISL_1107822, EPI_ISL_1107823, EPI_ISL_1107824, EPI_ISL_1107825, EPI_ISL_1107830, EPI_ISL_1107831, EPI_ISL_1107839, EPI_ISL_1176408, EPI_ISL_1176418, EPI_ISL_1176478, EPI_ISL_1176491, EPI_ISL_1176500, EPI_ISL_1176509, EPI_ISL_1176515, EPI_ISL_1176523, EPI_ISL_1176533, EPI_ISL_1176581, EPI_ISL_1176660, EPI_ISL_1176677, EPI_ISL_1176698, EPI_ISL_1247026, EPI_ISL_1247031 | | | |
| see above | Department of Pathology, University of Cambridge | COVID-19 Genomics UK (COG-UK) Consortium | Aminu S. Jahun; Ian Goodfellow; Iliana Georgana; Martin D. Curran; Myra Hosmillo; Rhys Izuagbe; Surendra Parmar; William L. Hamilton; Yasmin Chaudhry |
| EPI_ISL_985612, EPI_ISL_985615, EPI_ISL_985617, EPI_ISL_985621, EPI_ISL_985625, EPI_ISL_985629, EPI_ISL_985630, EPI_ISL_985633, EPI_ISL_985636, EPI_ISL_985641, EPI_ISL_985642, EPI_ISL_985675, EPI_ISL_985678, EPI_ISL_985679, EPI_ISL_985688, EPI_ISL_985690, EPI_ISL_985696, EPI_ISL_985701, EPI_ISL_985707, EPI_ISL_985711, EPI_ISL_985715, EPI_ISL_985716, EPI_ISL_985720, EPI_ISL_985728, EPI_ISL_985733, EPI_ISL_985737, EPI_ISL_985738, EPI_ISL_985746, EPI_ISL_985759, EPI_ISL_985773, EPI_ISL_985774, EPI_ISL_985782, EPI_ISL_985785, EPI_ISL_985786, EPI_ISL_985793, EPI_ISL_985795, EPI_ISL_985800, EPI_ISL_985801, EPI_ISL_985803, EPI_ISL_985805, EPI_ISL_985806, EPI_ISL_985807, EPI_ISL_985810, EPI_ISL_985816, EPI_ISL_985819, EPI_ISL_985843, EPI_ISL_985846, EPI_ISL_985857, EPI_ISL_985860, EPI_ISL_985863, EPI_ISL_985866, EPI_ISL_985867, EPI_ISL_985868, EPI_ISL_985870, EPI_ISL_985872, EPI_ISL_985874, EPI_ISL_985880, EPI_ISL_985888, EPI_ISL_985891, EPI_ISL_985903, EPI_ISL_985908, EPI_ISL_985914, EPI_ISL_985939, EPI_ISL_985943, EPI_ISL_985944, EPI_ISL_986854, EPI_ISL_986895, EPI_ISL_986900, EPI_ISL_986904, EPI_ISL_986912, EPI_ISL_986914, EPI_ISL_986918, EPI_ISL_986937, EPI_ISL_987002, EPI_ISL_987045, EPI_ISL_987053, EPI_ISL_987054, EPI_ISL_987097, EPI_ISL_987099, EPI_ISL_987100, EPI_ISL_987105, EPI_ISL_987107, EPI_ISL_987112, EPI_ISL_987124, EPI_ISL_987134, EPI_ISL_987144, EPI_ISL_1011757, EPI_ISL_1011758, EPI_ISL_1011759, EPI_ISL_1011760, EPI_ISL_1011761, EPI_ISL_1011762, EPI_ISL_1011763, EPI_ISL_1011764, EPI_ISL_1011765, EPI_ISL_1011766, EPI_ISL_1011767, EPI_ISL_1011768, EPI_ISL_1011769, EPI_ISL_1011770, EPI_ISL_1011771, EPI_ISL_1011772, EPI_ISL_1011773, EPI_ISL_1011774, EPI_ISL_1011775, EPI_ISL_1011776, EPI_ISL_1011777, EPI_ISL_1011778, EPI_ISL_1011779, EPI_ISL_1011780, EPI_ISL_1011781, EPI_ISL_1011782, EPI_ISL_1011783, EPI_ISL_1011784, EPI_ISL_1011785, EPI_ISL_1011786, EPI_ISL_1011787, EPI_ISL_1011788, EPI_ISL_1011789, EPI_ISL_1011790, EPI_ISL_1011791, EPI_ISL_1011792, EPI_ISL_1011793, EPI_ISL_1011794, EPI_ISL_1011795, EPI_ISL_1011796, EPI_ISL_1011797, EPI_ISL_1011798, EPI_ISL_1011799, EPI_ISL_1011800, EPI_ISL_1011801, EPI_ISL_1011802, EPI_ISL_1011803, EPI_ISL_1011804, EPI_ISL_1011805, EPI_ISL_1011806, EPI_ISL_1011807, EPI_ISL_1011808, EPI_ISL_1011809, EPI_ISL_1011810, EPI_ISL_1011811, EPI_ISL_1011812, EPI_ISL_1011813, EPI_ISL_1011814, EPI_ISL_1011815, EPI_ISL_1011816, EPI_ISL_1011817, EPI_ISL_1011818, EPI_ISL_1011819, EPI_ISL_1011820, EPI_ISL_1011821, EPI_ISL_1011822, EPI_ISL_1011823, EPI_ISL_1011824, EPI_ISL_1011825, EPI_ISL_1011826, EPI_ISL_1011827, EPI_ISL_1011828, EPI_ISL_1011829, EPI_ISL_1011830, EPI_ISL_1011831, EPI_ISL_1011832, EPI_ISL_1011833, EPI_ISL_1011834, EPI_ISL_1011835, EPI_ISL_1011836, EPI_ISL_1011837, EPI_ISL_1011838, EPI_ISL_1011839, EPI_ISL_1011840, EPI_ISL_1011841, EPI_ISL_1011842, EPI_ISL_1011843, EPI_ISL_1011844, EPI_ISL_1011845, EPI_ISL_1011846, EPI_ISL_1011847, EPI_ISL_1011848, EPI_ISL_1011849, EPI_ISL_1011850, EPI_ISL_1011851, EPI_ISL_1011852, EPI_ISL_1011853, EPI_ISL_1011854, EPI_ISL_1011855, EPI_ISL_1011856, EPI_ISL_1011857, EPI_ISL_1011858, EPI_ISL_1011859, EPI_ISL_1011860, EPI_ISL_1011861, EPI_ISL_1011862, EPI_ISL_1011863, EPI_ISL_1011864, EPI_ISL_1011865, EPI_ISL_1011866, EPI_ISL_1011867, EPI_ISL_1011868, EPI_ISL_1011869, EPI_ISL_1011870, EPI_ISL_1011871, EPI_ISL_1011872, EPI_ISL_1011873, EPI_ISL_1011874, EPI_ISL_1011875, EPI_ISL_1011876, EPI_ISL_1011877, EPI_ISL_1011878, EPI_ISL_1011879, EPI_ISL_1011880, EPI_ISL_1011881, EPI_ISL_1011882, EPI_ISL_1011883, EPI_ISL_1011884, EPI_ISL_1011885, EPI_ISL_1011886, EPI_ISL_1011887, EPI_ISL_1011888, EPI_ISL_1011889, EPI_ISL_1011890, EPI_ISL_1011891, EPI_ISL_1011892, EPI_ISL_1011893, EPI_ISL_1011894, EPI_ISL_1011895, EPI_ISL_1011896, EPI_ISL_1011897, EPI_ISL_1011898, EPI_ISL_1011899, EPI_ISL_1011900, EPI_ISL_1011901, EPI_ISL_1011902, EPI_ISL_1011903, EPI_ISL_1011904, EPI_ISL_1011905, EPI_ISL_1011906, EPI_ISL_1011907, EPI_ISL_1011908, EPI_ISL_1011909, EPI_ISL_1011910, EPI_ISL_1011911, EPI_ISL_1011912, EPI_ISL_1011913, EPI_ISL_1011914, EPI_ISL_1011915, EPI_ISL_1011916, EPI_ISL_1011917, EPI_ISL_1011918, EPI_ISL_1011919, EPI_ISL_1011920, EPI_ISL_1011921, EPI_ISL_1011922, EPI_ISL_1011923, EPI_ISL_1011924, EPI_ISL_1011925, EPI_ISL_1011926, EPI_ISL_1011927, EPI_ISL_1011928, EPI_ISL_1011929, EPI_ISL_1011930, EPI_ISL_1011931, EPI_ISL_1011932, EPI_ISL_1011933, EPI_ISL_1011934, EPI_ISL_1011935, EPI_ISL_1011936, EPI_ISL_1011937, EPI_ISL_1011938, EPI_ISL_1011939, EPI_ISL_1011940, EPI_ISL_1011941, EPI_ISL_1011942, EPI_ISL_1011943, EPI_ISL_1011944, EPI_ISL_1011945, EPI_ISL_1011946, EPI_ISL_1011947, EPI_ISL_1011948, EPI_ISL_1011949, EPI_ISL_1011950, EPI_ISL_1011951, EPI_ISL_1011952, EPI_ISL_1011953, EPI_ISL_1011954, EPI_ISL_1011955, EPI_ISL_1011956, EPI_ISL_1011957, EPI_ISL_1011958, EPI_ISL_1011959, EPI_ISL_1011960, EPI_ISL_1011961, EPI_ISL_1011962, EPI_ISL_1011963, EPI_ISL_1011964, EPI_ISL_1011965, EPI_ISL_1011966, EPI_ISL_1011967, EPI_ISL_1011968, EPI_ISL_1011969, EPI_ISL_1011970, EPI_ISL_1011971, EPI_ISL_1011972, EPI_ISL_1011973, EPI_ISL_1011974, EPI_ISL_1011975, EPI_ISL_1011976, EPI_ISL_1011977, EPI_ISL_1011978, EPI_ISL_1011979, EPI_ISL_1011980, EPI_ISL_1011981, EPI_ISL_1011982, EPI_ISL_1011983, EPI_ISL_1011984, EPI_ISL_1011985, EPI_ISL_1011986, EPI_ISL_1011987, EPI_ISL_1011988, EPI_ISL_1011989, EPI_ISL_1011990, EPI_ISL_1011991, EPI_ISL_1011992, EPI_ISL_1011993, EPI_ISL_1011994, EPI_ISL_1011995, EPI_ISL_1011996, EPI_ISL_1011997, EPI_ISL_1011998, EPI_ISL_1011999, EPI_ISL_1012000, EPI_ISL_1012001, EPI_ISL_1012002, EPI_ISL_1012003, EPI_ISL_1012004, EPI_ISL_1012005, EPI_ISL_1012006, EPI_ISL_1012007, EPI_ISL_1012008, EPI_ISL_1012009, EPI_ISL_1012010, EPI_ISL_1012011, EPI_ISL_1012012, EPI_ISL_1012013, EPI_ISL_1012014, EPI_ISL_1012015, EPI_ISL_1012016, EPI_ISL_1012017, EPI_ISL_1012018, EPI_ISL_1012019, EPI_ISL_1012020, EPI_ISL_1012021, EPI_ISL_1012022, EPI_ISL_1012023, EPI_ISL_1012024, EPI_ISL_1012025, EPI_ISL_1012026, EPI_ISL_1012027, EPI_ISL_1012028, EPI_ISL_1012029, EPI_ISL_1012030, EPI_ISL_1012031, EPI_ISL_1012032, EPI_ISL_1012033, EPI_ISL_1012034, EPI_ISL_1012035, EPI_ISL_1012036, EPI_ISL_1012037, EPI_ISL_1012038, EPI_ISL_1012039, EPI_ISL_1012040, EPI_ISL_1012041, EPI_ISL_1012042, EPI_ISL_1012043, EPI_ISL_1012044, EPI_ISL_1012045, EPI_ISL_1012046, EPI_ISL_1012047, EPI_ISL_1012048, EPI_ISL_1012049, EPI_ISL_1012050, EPI_ISL_1012051, EPI_ISL_1012052, EPI_ISL_1012053, EPI_ISL_1012054, EPI_ISL_1012055, EPI_ISL_1012056, EPI_ISL_1012057, EPI_ISL_1012058, EPI_ISL_1012059, EPI_ISL_1012060, EPI_ISL_1012061, EPI_ISL_1012062, EPI_ISL_1012063, EPI_ISL_1012064, EPI_ISL_1012065, EPI_ISL_1012066, EPI_ISL_1012067, EPI_ISL_1012068, EPI_ISL_1012069, EPI_ISL_1012070, EPI_ISL_1012071, EPI_ISL_1012072, EPI_ISL_1012073, EPI_ISL_1012074, EPI_ISL_1012075, EPI_ISL_1012076, EPI_ISL_1012077, EPI_ISL_1012078, EPI_ISL_1012079, EPI_ISL_1012080, EPI_ISL_1012081, EPI_ISL_1012082, EPI_ISL_1012083, EPI_ISL_1012084, EPI_ISL_1012085, EPI_ISL_1012086, EPI_ISL_1012087, EPI_ISL_1012088, EPI_ISL_1012089, EPI_ISL_1012090, EPI_ISL_1012091, EPI_ISL_1012092, EPI_ISL_1012093, EPI_ISL_1012094, EPI_ISL_1012095, EPI_ISL_1012096, EPI_ISL_1012097, EPI_ISL_1012098, EPI_ISL_1012596, EPI_ISL_1018329, EPI_ISL_1018330, EPI_ISL_1018331, EPI_ISL_1018332, EPI_ISL_1018333, EPI_ISL_1018334, EPI_ISL_1018335, EPI_ISL_1018336, EPI_ISL_1018337, EPI_ISL_1018338, EPI_ISL_1018339, EPI_ISL_1018340, EPI_ISL_1018341, EPI_ISL_1018342, EPI_ISL_1018343, EPI_ISL_1018344, EPI_ISL_1018345, EPI_ISL_1018346, EPI_ISL_1018347, EPI_ISL_1018348, EPI_ISL_1018349, EPI_ISL_1018350, EPI_ISL_1018351, EPI_ISL_1018352, EPI_ISL_1018353, EPI_ISL_1018354, EPI_ISL_1018355, EPI_ISL_1018356, EPI_ISL_1018357, EPI_ISL_1018358, EPI_ISL_1018359, EPI_ISL_1018360, EPI_ISL_1018361, EPI_ISL_1018362, EPI_ISL_1018363, EPI_ISL_1018364, EPI_ISL_1018366, EPI_ISL_1018367, EPI_ISL_1018368, EPI_ISL_1018369, EPI_ISL_1018370, EPI_ISL_1018371, EPI_ISL_1018372, EPI_ISL_1018373, EPI_ISL_1018374, EPI_ISL_1018375, EPI_ISL_1018376, EPI_ISL_1018377, EPI_ISL_1018378, EPI_ISL_1018379, EPI_ISL_1018380, EPI_ISL_1018381, EPI_ISL_1018382, EPI_ISL_1018383, EPI_ISL_1018384, EPI_ISL_1018385, EPI_ISL_1018386, EPI_ISL_1018387, EPI_ISL_1018388, EPI_ISL_1018389, EPI_ISL_1018390, EPI_ISL_1018391, EPI_ISL_1018392, EPI_ISL_1018393, EPI_ISL_1018394, EPI_ISL_1018395, EPI_ISL_1018396, EPI_ISL_1018397, EPI_ISL_1018398, EPI_ISL_1018399, EPI_ISL_1018400, EPI_ISL_1018401, EPI_ISL_1018402, EPI_ISL_1018403, EPI_ISL_1018404, EPI_ISL_1018405, EPI_ISL_1018406, EPI_ISL_1018407, EPI_ISL_1018408, EPI_ISL_1018409, EPI_ISL_1018410, EPI_ISL_1018411, EPI_ISL_1018412, EPI_ISL_1018413, EPI_ISL_1018414, EPI_ISL_1018415, EPI_ISL_1018416, EPI_ISL_1018417, EPI_ISL_1018418, EPI_ISL_1018419, EPI_ISL_1018420, EPI_ISL_1018421, EPI_ISL_1018422, EPI_ISL_1018423, EPI_ISL_1018424, EPI_ISL_1018425, EPI_ISL_1018426, EPI_ISL_1018427, EPI_ISL_1018428, EPI_ISL_1018429, EPI_ISL_1018430, EPI_ISL_1018431, EPI_ISL_1018432, EPI_ISL_1018433, EPI_ISL_1018434, EPI_ISL_1018435, EPI_ISL_1018436, EPI_ISL_1018437, EPI_ISL_1018438, EPI_ISL_1018439, EPI_ISL_1018440, EPI_ISL_1018441, EPI_ISL_1018442, EPI_ISL_1018443, EPI_ISL_1018444, EPI_ISL_1018445, EPI_ISL_1018446, EPI_ISL_1018447, EPI_ISL_1018448, EPI_ISL_1018449, EPI_ISL_1018450, EPI_ISL_1018451, EPI_ISL_1018452, EPI_ISL_1018453, EPI_ISL_1018454, EPI_ISL_1018455, EPI_ISL_1018456, EPI_ISL_1018457, EPI_ISL_1018458, EPI_ISL_1018459, EPI_ISL_1018460, EPI_ISL_1018461, EPI_ISL_1018462, EPI_ISL_1018463, EPI_ISL_1018465, EPI_ISL_1018466, EPI_ISL_1018467, EPI_ISL_1018468, EPI_ISL_1018469, EPI_ISL_1018470, EPI_ISL_1018471, EPI_ISL_1018472, EPI_ISL_1018473, EPI_ISL_1018474, EPI_ISL_1018475, EPI_ISL_1018476, EPI_ISL_1018477, EPI_ISL_1018478, EPI_ISL_1018479, EPI_ISL_1018480, EPI_ISL_1018481, EPI_ISL_1018482, EPI_ISL_1018483, EPI_ISL_1018484, EPI_ISL_1018485, EPI_ISL_1018486, EPI_ISL_1018487, EPI_ISL_1018488, EPI_ISL_1018489, EPI_ISL_1018490, EPI_ISL_1018491, EPI_ISL_1018492, EPI_ISL_1018493, EPI_ISL_1018494, EPI_ISL_1018495, EPI_ISL_1018496, EPI_ISL_1018497, EPI_ISL_1018498, EPI_ISL_1018499, EPI_ISL_1018500, EPI_ISL_1018501, EPI_ISL_1018502, EPI_ISL_1018503, EPI_ISL_1018504, EPI_ISL_1018505, EPI_ISL_1018506, EPI_ISL_1018507, EPI_ISL_1018508, EPI_ISL_1018509, EPI_ISL_1018510, EPI_ISL_1018511, EPI_ISL_1018512, EPI_ISL_1018513, EPI_ISL_1018514, EPI_ISL_1018515, EPI_ISL_1018516, EPI_ISL_1018517, EPI_ISL_1018518, EPI_ISL_1018519, EPI_ISL_1018521, EPI_ISL_1018522, EPI_ISL_1018523, EPI_ISL_1018524, EPI_ISL_1018525, EPI_ISL_1018526, EPI_ISL_1018527, EPI_ISL_1018528, EPI_ISL_1018529, EPI_ISL_1018530, EPI_ISL_1018531, EPI_ISL_1018532, EPI_ISL_1018533, EPI_ISL_1018534, EPI_ISL_1018535, EPI_ISL_1018536, EPI_ISL_1018537, EPI_ISL_1018538, EPI_ISL_1018539, EPI_ISL_1018540, EPI_ISL_1018541, EPI_ISL_1018542, EPI_ISL_1018543, EPI_ISL_1018544, EPI_ISL_1018545, EPI_ISL_1018546, EPI_ISL_1018547, EPI_ISL_1018548, EPI_ISL_1018549, EPI_ISL_1018550, EPI_ISL_1018551, EPI_ISL_1018552, EPI_ISL_1018553, EPI_ISL_1018554, EPI_ISL_1018555, EPI_ISL_1018556, EPI_ISL_1018557, EPI_ISL_1018558, EPI_ISL_1018559, EPI_ISL_1018560, EPI_ISL_1018561, EPI_ISL_1018562, EPI_ISL_1018563, EPI_ISL_1018564, EPI_ISL_1018565, EPI_ISL_1018566, EPI_ISL_1018567, EPI_ISL_1018568, EPI_ISL_1018569, EPI_ISL_1018570, EPI_ISL_1018571, EPI_ISL_1018572, EPI_ISL_1018573, EPI_ISL_1018574, EPI_ISL_1018575, EPI_ISL_1018576, EPI_ISL_1018577, EPI_ISL_1018578, EPI_ISL_1018579, EPI_ISL_1018580, EPI_ISL_1018581, EPI_ISL_1018582, EPI_ISL_1018583, EPI_ISL_1018584, EPI_ISL_1018585, EPI_ISL_1018586, EPI_ISL_1018587, EPI_ISL_1018588, EPI_ISL_1018589, EPI_ISL_1018590, EPI_ISL_1018591, EPI_ISL_1018592, EPI_ISL_1018593, EPI_ISL_1018594, EPI_ISL_1018595, EPI_ISL_1018596, EPI_ISL_1018597, EPI_ISL_1018598, EPI_ISL_1018599, EPI_ISL_1018600, EPI_ISL_1018601, EPI_ISL_1018602, EPI_ISL_1018603, EPI_ISL_1018604, EPI_ISL_1018605, EPI_ISL_1018607, EPI_ISL_1018608, EPI_ISL_1018609, EPI_ISL_1018610, EPI_ISL_1018611, EPI_ISL_1018612, EPI_ISL_1018613, EPI_ISL_1018614, EPI_ISL_1018615, EPI_ISL_1018616, EPI_ISL_1018617, EPI_ISL_1018618, EPI_ISL_1018619, EPI_ISL_1018620, EPI_ISL_1018621, EPI_ISL_1018622, EPI_ISL_1018623, EPI_ISL_1018624, EPI_ISL_1018625, EPI_ISL_1018626, EPI_ISL_1018627, EPI_ISL_1018628, EPI_ISL_1018629, EPI_ISL_1018630, EPI_ISL_1018631, EPI_ISL_1018632, EPI_ISL_1018633, EPI_ISL_1018634, EPI_ISL_1018635, EPI_ISL_1018636, EPI_ISL_1018637, EPI_ISL_1018638, EPI_ISL_1018639, EPI_ISL_1018640, EPI_ISL_1018641, EPI_ISL_1018642, EPI_ISL_1018643, EPI_ISL_1018644, EPI_ISL_1018645, EPI_ISL_1018646, EPI_ISL_1018647, EPI_ISL_1018648, EPI_ISL_1018649, EPI_ISL_1018650, EPI_ISL_1018651, EPI_ISL_1018652, EPI_ISL_1018653, EPI_ISL_1018654, EPI_ISL_1018655, EPI_ISL_1018656, EPI_ISL_1018657, EPI_ISL_1018658, EPI_ISL_1018659, EPI_ISL_1018660, EPI_ISL_1018661, EPI_ISL_1018662, EPI_ISL_1018663, EPI_ISL_1018664, EPI_ISL_1018665, EPI_ISL_1018666, EPI_ISL_1018667, EPI_ISL_1018668, EPI_ISL_1018669, EPI_ISL_1018670, EPI_ISL_1018671, EPI_ISL_1018672, EPI_ISL_1018673, EPI_ISL_1018674, EPI_ISL_1018675, EPI_ISL_1018676, EPI_ISL_1018677, EPI_ISL_1018678, EPI_ISL_1018679, EPI_ISL_1018680, EPI_ISL_1018681, EPI_ISL_1018682, EPI_ISL_1018683, EPI_ISL_1018684, EPI_ISL_1018685, EPI_ISL_1018686, EPI_ISL_1018687, EPI_ISL_1018688, EPI_ISL_1018950, EPI_ISL_1018951, EPI_ISL_1018952, EPI_ISL_1018953, EPI_ISL_1018954, EPI_ISL_1018955, EPI_ISL_1018956, EPI_ISL_1018957, EPI_ISL_1018958, EPI_ISL_1018959, EPI_ISL_1018960, EPI_ISL_1018961, EPI_ISL_1018962, EPI_ISL_1018963, EPI_ISL_1018964, EPI_ISL_1018965, EPI_ISL_1018966, EPI_ISL_1018967, EPI_ISL_1018968, EPI_ISL_1018969, EPI_ISL_1018970, EPI_ISL_1018971, EPI_ISL_1018972, EPI_ISL_1018973, EPI_ISL_1018974, EPI_ISL_1018975, EPI_ISL_1018976, EPI_ISL_1018977, EPI_ISL_1018978, EPI_ISL_1018979, EPI_ISL_1018980, EPI_ISL_1018981, EPI_ISL_1018982, EPI_ISL_1018983, EPI_ISL_1018984, EPI_ISL_1018985, EPI_ISL_1018986, EPI_ISL_1018987, EPI_ISL_1018988, EPI_ISL_1018989, EPI_ISL_1018990, EPI_ISL_1018991, EPI_ISL_1018992, EPI_ISL_1018993, EPI_ISL_1018994, EPI_ISL_1018995, EPI_ISL_1018996, EPI_ISL_1018997, EPI_ISL_1018998, EPI_ISL_1018999, EPI_ISL_1019000, EPI_ISL_1019001, EPI_ISL_1019002, EPI_ISL_1019003, EPI_ISL_1019004, EPI_ISL_1019005, EPI_ISL_1019006, EPI_ISL_1019007, EPI_ISL_1019008, EPI_ISL_1019009, EPI_ISL_1019010, EPI_ISL_1019011, EPI_ISL_1019012, EPI_ISL_1019013, EPI_ISL_1019014, EPI_ISL_1019015, EPI_ISL_1019016, EPI_ISL_1019017, EPI_ISL_1019018, EPI_ISL_1019019, EPI_ISL_1019020, EPI_ISL_1019021, EPI_ISL_1019022, EPI_ISL_1019023, EPI_ISL_1019024, EPI_ISL_1019025, EPI_ISL_1019026, EPI_ISL_1019027, EPI_ISL_1019028, EPI_ISL_1019029, EPI_ISL_1019030, EPI_ISL_1019031, EPI_ISL_1019032, EPI_ISL_1019033, EPI_ISL_1019034, EPI_ISL_1019035, EPI_ISL_1019036, EPI_ISL_1019037, EPI_ISL_1019038, EPI_ISL_1019039, EPI_ISL_1019040, EPI_ISL_1019041, EPI_ISL_1019042, EPI_ISL_1019043, EPI_ISL_1019044, EPI_ISL_1019045, EPI_ISL_1019046, EPI_ISL_1019047, EPI_ISL_1019048, EPI_ISL_1019049, EPI_ISL_1019050, EPI_ISL_1019051, EPI_ISL_1019052, EPI_ISL_1019053, EPI_ISL_1019054, EPI_ISL_1019055, EPI_ISL_1019056, EPI_ISL_1019057, EPI_ISL_1019058, EPI_ISL_1019059, EPI_ISL_1019060, EPI_ISL_1019061, EPI_ISL_1019062, EPI_ISL_1019063, EPI_ISL_1019064, EPI_ISL_1019065, EPI_ISL_1019066, EPI_ISL_1019067, EPI_ISL_1019068, EPI_ISL_1019069, EPI_ISL_1019070, EPI_ISL_1019071, EPI_ISL_1019072, EPI_ISL_1019073, EPI_ISL_1019074, EPI_ISL_1019075, EPI_ISL_1019076, EPI_ISL_1019077, EPI_ISL_1019078, EPI_ISL_1019079, EPI_ISL_1019080, EPI_ISL_1019081, EPI_ISL_1019082, EPI_ISL_1019083, EPI_ISL_1019084, EPI_ISL_1019085, EPI_ISL_1019086, EPI_ISL_1019087, EPI_ISL_1019088, EPI_ISL_1019089, EPI_ISL_1019090, EPI_ISL_1019091, EPI_ISL_1019092, EPI_ISL_1019093, EPI_ISL_1019094, EPI_ISL_1019095, EPI_ISL_1019096, EPI_ISL_1019097, EPI_ISL_1019098, EPI_ISL_1019099, EPI_ISL_1019100, EPI_ISL_1019101, EPI_ISL_1019102, EPI_ISL_1019103, EPI_ISL_1019104, EPI_ISL_1019105, EPI_ISL_1019106, EPI_ISL_1019107, EPI_ISL_1019108, EPI_ISL_1019109, EPI_ISL_1019110, EPI_ISL_1019111, EPI_ISL_1019112, EPI_ISL_1019113, EPI_ISL_1019114, EPI_ISL_1019115, EPI_ISL_1019116, EPI_ISL_1019117, EPI_ISL_1019118, EPI_ISL_1019119, EPI_ISL_1019120, EPI_ISL_1019121, EPI_ISL_1019122, EPI_ISL_1019123, EPI_ISL_1019124, EPI_ISL_1019125, EPI_ISL_1019126, EPI_ISL_1019127, EPI_ISL_1019128, EPI_ISL_1019129, EPI_ISL_1019130, EPI_ISL_1019131, EPI_ISL_1019132, EPI_ISL_1019133, EPI_ISL_1019134, EPI_ISL_1019135, EPI_ISL_1019136, EPI_ISL_1019137, EPI_ISL_1019138, EPI_ISL_1019139, EPI_ISL_1019140, EPI_ISL_1019141, EPI_ISL_1019142, EPI_ISL_1019143, EPI_ISL_1019144, EPI_ISL_1019145, EPI_ISL_1019146, EPI_ISL_1019147, EPI_ISL_1019148, EPI_ISL_1019149, EPI_ISL_1019150, EPI_ISL_1019151, EPI_ISL_1019152, EPI_ISL_1019153, EPI_ISL_1019154, EPI_ISL_1019155, EPI_ISL_1019156, EPI_ISL_1019157, EPI_ISL_1019158, EPI_ISL_1019159, EPI_ISL_1019160, EPI_ISL_1019161, EPI_ISL_1019162, EPI_ISL_1019163, EPI_ISL_1019164, EPI_ISL_1019165, EPI_ISL_1019166, EPI_ISL_1019167, EPI_ISL_1019168, EPI_ISL_1019169, EPI_ISL_1019170, EPI_ISL_1019171, EPI_ISL_1019172, EPI_ISL_1019173, EPI_ISL_1019174, EPI_ISL_1019175, EPI_ISL_1019176, EPI_ISL_1019177, EPI_ISL_1019178, EPI_ISL_1019179, EPI_ISL_1019180, EPI_ISL_1019181, EPI_ISL_1019182, EPI_ISL_1019183, EPI_ISL_1019184, EPI_ISL_1019185, EPI_ISL_1019186, EPI_ISL_1019187, EPI_ISL_1019188, EPI_ISL_1019189, EPI_ISL_1019190, EPI_ISL_1019191, EPI_ISL_1019192, EPI_ISL_1019193, EPI_ISL_1019194, EPI_ISL_1019195, EPI_ISL_1019196, EPI_ISL_1019197, EPI_ISL_1019198, EPI_ISL_1019199, EPI_ISL_1019200, EPI_ISL_1019201, EPI_ISL_1019202, EPI_ISL_1019203, EPI_ISL_1019204, EPI_ISL_1019205, EPI_ISL_1019206, EPI_ISL_1019207, EPI_ISL_1019208, EPI_ISL_1019209, EPI_ISL_1019210, EPI_ISL_1019211, EPI_ISL_1019212, EPI_ISL_1019213, EPI_ISL_1019214, EPI_ISL_1019215, EPI_ISL_1019216, EPI_ISL_1019217, EPI_ISL_1019218, EPI_ISL_1019219, EPI_ISL_1019220, EPI_ISL_1019221, EPI_ISL_1019222, EPI_ISL_1019223, EPI_ISL_1019224, EPI_ISL_1019225, EPI_ISL_1019226, EPI_ISL_1019227, EPI_ISL_1019228, EPI_ISL_1019229, EPI_ISL_1019230, EPI_ISL_1019231, EPI_ISL_1019232, EPI_ISL_1019233, EPI_ISL_1019234, EPI_ISL_1019235, EPI_ISL_1019236, EPI_ISL_1019237, EPI_ISL_1019238, EPI_ISL_1019239, EPI_ISL_1019240, EPI_ISL_1019241, EPI_ISL_1019242, EPI_ISL_1019243, EPI_ISL_1019244, EPI_ISL_1019245, EPI_ISL_1019246, EPI_ISL_1019247, EPI_ISL_1019248, EPI_ISL_1019249, EPI_ISL_1019250, EPI_ISL_1019251, EPI_ISL_1019252, EPI_ISL_1019253, EPI_ISL_1019254, EPI_ISL_1019255, EPI_ISL_1019256, EPI_ISL_1019257, EPI_ISL_1019258, EPI_ISL_1019259, EPI_ISL_1019260, EPI_ISL_1019261, EPI_ISL_1019262, EPI_ISL_1019263, EPI_ISL_1019264, EPI_ISL_1019265, EPI_ISL_1019266, EPI_ISL_1019267, EPI_ISL_1019268, EPI_ISL_1019269, EPI_ISL_1019270, EPI_ISL_1019271, EPI_ISL_1019272, EPI_ISL_1019273, EPI_ISL_1019274, EPI_ISL_1019275, EPI_ISL_1019276, EPI_ISL_1019277, EPI_ISL_1019278, EPI_ISL_1019279, EPI_ISL_1019280, EPI_ISL_1019281, EPI_ISL_1019282, EPI_ISL_1019283, EPI_ISL_1019284, EPI_ISL_1019285, EPI_ISL_1019482, EPI_ISL_1019483, EPI_ISL_1019484, EPI_ISL_1019485, EPI_ISL_1019486, EPI_ISL_1019487, EPI_ISL_1019488, EPI_ISL_1019489, EPI_ISL_1019490, EPI_ISL_1019491, EPI_ISL_1019492, EPI_ISL_1019493, EPI_ISL_1019494, EPI_ISL_1019495, EPI_ISL_1019496, EPI_ISL_1019497, EPI_ISL_1019498, EPI_ISL_1019499, EPI_ISL_1019500, EPI_ISL_1019501, EPI_ISL_1019502, EPI_ISL_1019503, EPI_ISL_1033537, EPI_ISL_1033538, EPI_ISL_1033539, EPI_ISL_1033540, EPI_ISL_1033541, EPI_ISL_1033542, EPI_ISL_1033543, EPI_ISL_1033544, EPI_ISL_1033545, EPI_ISL_1033546, EPI_ISL_1033547, EPI_ISL_1033548, EPI_ISL_1033549, EPI_ISL_1033550, EPI_ISL_1033551, EPI_ISL_1033552, EPI_ISL_1033553, EPI_ISL_1033554, EPI_ISL_1033555, EPI_ISL_1033556, EPI_ISL_1033557, EPI_ISL_1033558, EPI_ISL_1033559, EPI_ISL_1033560, EPI_ISL_1033561, EPI_ISL_1033562, EPI_ISL_1033563, EPI_ISL_1033564, EPI_ISL_1033565, EPI_ISL_1034141, EPI_ISL_1034142, EPI_ISL_1043724, EPI_ISL_1044399, EPI_ISL_1044448, EPI_ISL_1044482, EPI_ISL_1044492, EPI_ISL_1044516, EPI_ISL_1044521, EPI_ISL_1044542, EPI_ISL_1044632, EPI_ISL_1044647, EPI_ISL_1044657, EPI_ISL_1044702, EPI_ISL_1044908, EPI_ISL_1044988, EPI_ISL_1045125, EPI_ISL_1045970, EPI_ISL_1045971, EPI_ISL_1045972, EPI_ISL_1045973, EPI_ISL_1045974, EPI_ISL_1045975, EPI_ISL_1045976, EPI_ISL_1045977, EPI_ISL_1045978, EPI_ISL_1045979, EPI_ISL_1057723, EPI_ISL_1057859, EPI_ISL_1057860, EPI_ISL_1057869, EPI_ISL_1057989, EPI_ISL_1057994, EPI_ISL_1057995, EPI_ISL_1057997, EPI_ISL_1058000, EPI_ISL_1058017 | | | |
| see above | Lighthouse Lab in Alderley Park | Wellcome Sanger Institute for the COVID-19 Genomics UK (COG-UK) Consortium | Cordelia Langford; David K. Jackson; Dominic Kwiatkowski; Ewan Harrison; Ian Johnston; Jacquelyn Wynn; Jeffrey Barrett; John Sillitoe on behalf of the Wellcome Sanger Institute COVID-19 Surveillance Team; John Sillitoe on behalf of the Wellcome Sanger Institute COVID-19 Surveillance Team (http://www.sanger.ac.uk/covid-team); Mairead Hyland; Roberto Amato; Sonia Goncalves; The Lighthouse Lab in Alderley Park and Alex Alderton |
| EPI_ISL_985411, EPI_ISL_985412, EPI_ISL_985413, EPI_ISL_985414, EPI_ISL_985415, EPI_ISL_985416, EPI_ISL_985419, EPI_ISL_985421, EPI_ISL_985424, EPI_ISL_985425, EPI_ISL_985427, EPI_ISL_985428, EPI_ISL_985429, EPI_ISL_985431, EPI_ISL_985432, EPI_ISL_985434, EPI_ISL_985435, EPI_ISL_985437, EPI_ISL_985438, EPI_ISL_985439, EPI_ISL_985441, EPI_ISL_985442, EPI_ISL_985443, EPI_ISL_985444, EPI_ISL_985446, EPI_ISL_985447, EPI_ISL_985448, EPI_ISL_985449, EPI_ISL_985450, EPI_ISL_985452, EPI_ISL_985453, EPI_ISL_985454, EPI_ISL_985455, EPI_ISL_985456, EPI_ISL_985457, EPI_ISL_985458, EPI_ISL_985459, EPI_ISL_985460, EPI_ISL_985461, EPI_ISL_985462, EPI_ISL_985463, EPI_ISL_985464, EPI_ISL_985465, EPI_ISL_985466, EPI_ISL_985467, EPI_ISL_985468, EPI_ISL_985469, EPI_ISL_985470, | | | |

EPI_ISL_985471, EPI_ISL_985472, EPI_ISL_985473, EPI_ISL_985474, EPI_ISL_985475, EPI_ISL_985476, EPI_ISL_985477, EPI_ISL_985479, EPI_ISL_985481, EPI_ISL_985482, EPI_ISL_985483, EPI_ISL_985484, EPI_ISL_985485, EPI_ISL_985486, EPI_ISL_985487, EPI_ISL_985488, EPI_ISL_985490, EPI_ISL_985491, EPI_ISL_985492, EPI_ISL_985493, EPI_ISL_985494, EPI_ISL_985496, EPI_ISL_985497, EPI_ISL_985498, EPI_ISL_985499, EPI_ISL_985500, EPI_ISL_985501, EPI_ISL_985502, EPI_ISL_985503, EPI_ISL_985504, EPI_ISL_985505, EPI_ISL_985506, EPI_ISL_985507, EPI_ISL_985508, EPI_ISL_985509, EPI_ISL_985510, EPI_ISL_985511, EPI_ISL_985513, EPI_ISL_985514, EPI_ISL_985515, EPI_ISL_985516, EPI_ISL_985519, EPI_ISL_985520, EPI_ISL_985521, EPI_ISL_985522, EPI_ISL_985523, EPI_ISL_985524, EPI_ISL_985525, EPI_ISL_985526, EPI_ISL_985527, EPI_ISL_985528, EPI_ISL_985529, EPI_ISL_985530, EPI_ISL_985531, EPI_ISL_985532, EPI_ISL_985535, EPI_ISL_985536, EPI_ISL_985538, EPI_ISL_985539, EPI_ISL_985540, EPI_ISL_985543, EPI_ISL_985544, EPI_ISL_985545, EPI_ISL_985546, EPI_ISL_985547, EPI_ISL_985549, EPI_ISL_985552, EPI_ISL_985553, EPI_ISL_985554, EPI_ISL_985555, EPI_ISL_985557, EPI_ISL_985558, EPI_ISL_985559, EPI_ISL_985560, EPI_ISL_985561, EPI_ISL_985562, EPI_ISL_985565, EPI_ISL_985566, EPI_ISL_985567, EPI_ISL_985568, EPI_ISL_985569, EPI_ISL_985570, EPI_ISL_985571, EPI_ISL_985572, EPI_ISL_985573, EPI_ISL_985575, EPI_ISL_985576, EPI_ISL_985577, EPI_ISL_985578, EPI_ISL_985579, EPI_ISL_985580, EPI_ISL_985582, EPI_ISL_985583, EPI_ISL_985584, EPI_ISL_985586, EPI_ISL_985587, EPI_ISL_985589, EPI_ISL_985590, EPI_ISL_985591, EPI_ISL_985592, EPI_ISL_985595, EPI_ISL_985596, EPI_ISL_985598, EPI_ISL_985600, EPI_ISL_985601, EPI_ISL_985603, EPI_ISL_985604, EPI_ISL_985605, EPI_ISL_985606, EPI_ISL_985608, EPI_ISL_985610, EPI_ISL_985611, EPI_ISL_986439, EPI_ISL_986440, EPI_ISL_986441, EPI_ISL_986442, EPI_ISL_986443, EPI_ISL_986444, EPI_ISL_986445, EPI_ISL_986446, EPI_ISL_986447, EPI_ISL_986448, EPI_ISL_986449, EPI_ISL_986450, EPI_ISL_986451, EPI_ISL_986452, EPI_ISL_986453, EPI_ISL_986454, EPI_ISL_986455, EPI_ISL_986456, EPI_ISL_986457, EPI_ISL_986458, EPI_ISL_986459, EPI_ISL_986460, EPI_ISL_986461, EPI_ISL_986462, EPI_ISL_986463, EPI_ISL_986464, EPI_ISL_986465, EPI_ISL_986466, EPI_ISL_986467, EPI_ISL_986468, EPI_ISL_986469, EPI_ISL_986470, EPI_ISL_986471, EPI_ISL_986472, EPI_ISL_986473, EPI_ISL_986474, EPI_ISL_986475, EPI_ISL_986476, EPI_ISL_986477, EPI_ISL_986478, EPI_ISL_986479, EPI_ISL_986480, EPI_ISL_986481, EPI_ISL_986482, EPI_ISL_986483, EPI_ISL_986484, EPI_ISL_986485, EPI_ISL_986486, EPI_ISL_986487, EPI_ISL_986488, EPI_ISL_986489, EPI_ISL_986490, EPI_ISL_986491, EPI_ISL_986492, EPI_ISL_986493, EPI_ISL_986494, EPI_ISL_986495, EPI_ISL_986496, EPI_ISL_986497, EPI_ISL_986498, EPI_ISL_986499, EPI_ISL_986500, EPI_ISL_986501, EPI_ISL_986502, EPI_ISL_986503, EPI_ISL_986504, EPI_ISL_986505, EPI_ISL_986506, EPI_ISL_986507, EPI_ISL_986508, EPI_ISL_986509, EPI_ISL_986510, EPI_ISL_986511, EPI_ISL_986512, EPI_ISL_986513, EPI_ISL_986514, EPI_ISL_986515, EPI_ISL_986516, EPI_ISL_986517, EPI_ISL_986518, EPI_ISL_986519, EPI_ISL_986520, EPI_ISL_986521, EPI_ISL_986522, EPI_ISL_986523, EPI_ISL_986524, EPI_ISL_986525, EPI_ISL_986526, EPI_ISL_986527, EPI_ISL_986528, EPI_ISL_986529, EPI_ISL_986530, EPI_ISL_986531, EPI_ISL_986532, EPI_ISL_986533, EPI_ISL_986534, EPI_ISL_986535, EPI_ISL_986536, EPI_ISL_986537, EPI_ISL_986538, EPI_ISL_986539, EPI_ISL_986540, EPI_ISL_986541, EPI_ISL_986542, EPI_ISL_986543, EPI_ISL_986544, EPI_ISL_986545, EPI_ISL_986546, EPI_ISL_986547, EPI_ISL_986548, EPI_ISL_986549, EPI_ISL_986550, EPI_ISL_986551, EPI_ISL_986552, EPI_ISL_986553, EPI_ISL_986554, EPI_ISL_986555, EPI_ISL_986556, EPI_ISL_986557, EPI_ISL_986558, EPI_ISL_986559, EPI_ISL_986560, EPI_ISL_986561, EPI_ISL_986562, EPI_ISL_986564, EPI_ISL_986565, EPI_ISL_986566, EPI_ISL_986567, EPI_ISL_986568, EPI_ISL_986569, EPI_ISL_986570, EPI_ISL_986571, EPI_ISL_986572, EPI_ISL_986573, EPI_ISL_986574, EPI_ISL_986575, EPI_ISL_986576, EPI_ISL_986577, EPI_ISL_986578, EPI_ISL_986579, EPI_ISL_986580, EPI_ISL_986581, EPI_ISL_986582, EPI_ISL_986583, EPI_ISL_986584, EPI_ISL_986585, EPI_ISL_986586, EPI_ISL_986587, EPI_ISL_986588, EPI_ISL_986589, EPI_ISL_986590, EPI_ISL_986591, EPI_ISL_986592, EPI_ISL_986593, EPI_ISL_986594, EPI_ISL_986595, EPI_ISL_986596, EPI_ISL_986597, EPI_ISL_986598, EPI_ISL_986599, EPI_ISL_986600, EPI_ISL_986601, EPI_ISL_986602, EPI_ISL_986603, EPI_ISL_986604, EPI_ISL_986605, EPI_ISL_986606, EPI_ISL_986607, EPI_ISL_986608, EPI_ISL_986609, EPI_ISL_986610, EPI_ISL_986611, EPI_ISL_986612, EPI_ISL_986613, EPI_ISL_986614, EPI_ISL_986615, EPI_ISL_986616, EPI_ISL_986617, EPI_ISL_986618, EPI_ISL_986619, EPI_ISL_986620, EPI_ISL_986621, EPI_ISL_986622, EPI_ISL_986623, EPI_ISL_986624, EPI_ISL_986625, EPI_ISL_986626, EPI_ISL_986627, EPI_ISL_986628, EPI_ISL_986629, EPI_ISL_986630, EPI_ISL_986631, EPI_ISL_986632, EPI_ISL_986633, EPI_ISL_986634, EPI_ISL_986635, EPI_ISL_986636, EPI_ISL_986637, EPI_ISL_986638, EPI_ISL_986639, EPI_ISL_986640, EPI_ISL_986641, EPI_ISL_986642, EPI_ISL_986643, EPI_ISL_986644, EPI_ISL_986645, EPI_ISL_986646, EPI_ISL_986647, EPI_ISL_986648, EPI_ISL_986649, EPI_ISL_986650, EPI_ISL_986651, EPI_ISL_986652, EPI_ISL_986653, EPI_ISL_986654, EPI_ISL_986655, EPI_ISL_986656, EPI_ISL_986657, EPI_ISL_986658, EPI_ISL_986659, EPI_ISL_986660, EPI_ISL_986661, EPI_ISL_986662, EPI_ISL_986663, EPI_ISL_986664, EPI_ISL_986665, EPI_ISL_986666, EPI_ISL_986667, EPI_ISL_986668, EPI_ISL_986669, EPI_ISL_986670, EPI_ISL_986671, EPI_ISL_986672, EPI_ISL_986673, EPI_ISL_986674, EPI_ISL_986675, EPI_ISL_986676, EPI_ISL_986677, EPI_ISL_986678, EPI_ISL_986679, EPI_ISL_986680, EPI_ISL_986681, EPI_ISL_986682, EPI_ISL_986683, EPI_ISL_986684, EPI_ISL_986685, EPI_ISL_986686, EPI_ISL_986687, EPI_ISL_986688, EPI_ISL_986689, EPI_ISL_986690, EPI_ISL_986691, EPI_ISL_986692, EPI_ISL_986693, EPI_ISL_986694, EPI_ISL_986695, EPI_ISL_986696, EPI_ISL_986697, EPI_ISL_986698, EPI_ISL_986699, EPI_ISL_986700, EPI_ISL_986701, EPI_ISL_986702, EPI_ISL_986703, EPI_ISL_986704, EPI_ISL_986705, EPI_ISL_986706, EPI_ISL_986707, EPI_ISL_986708, EPI_ISL_986709, EPI_ISL_986710, EPI_ISL_986711, EPI_ISL_986712, EPI_ISL_986713, EPI_ISL_986714, EPI_ISL_986715, EPI_ISL_986716, EPI_ISL_986717, EPI_ISL_986718, EPI_ISL_986719, EPI_ISL_986720, EPI_ISL_986721, EPI_ISL_986722, EPI_ISL_986723, EPI_ISL_986724, EPI_ISL_986725, EPI_ISL_986726, EPI_ISL_986727, EPI_ISL_986728, EPI_ISL_986729, EPI_ISL_986730, EPI_ISL_986731, EPI_ISL_986732, EPI_ISL_986733, EPI_ISL_986734, EPI_ISL_986735, EPI_ISL_986736, EPI_ISL_986737, EPI_ISL_986738, EPI_ISL_986739, EPI_ISL_986740, EPI_ISL_986741, EPI_ISL_986742, EPI_ISL_986743, EPI_ISL_986744, EPI_ISL_986745, EPI_ISL_986746, EPI_ISL_986747, EPI_ISL_986748, EPI_ISL_986749, EPI_ISL_986750, EPI_ISL_986751, EPI_ISL_986752, EPI_ISL_986753, EPI_ISL_986754, EPI_ISL_986755, EPI_ISL_986756, EPI_ISL_986757, EPI_ISL_986758, EPI_ISL_986759, EPI_ISL_986760, EPI_ISL_986761, EPI_ISL_986762, EPI_ISL_986763, EPI_ISL_986764, EPI_ISL_986765, EPI_ISL_986766, EPI_ISL_986767, EPI_ISL_986768, EPI_ISL_986769, EPI_ISL_986770, EPI_ISL_986771, EPI_ISL_986772, EPI_ISL_986773, EPI_ISL_986774, EPI_ISL_986775, EPI_ISL_986776, EPI_ISL_986777, EPI_ISL_986778, EPI_ISL_986779, EPI_ISL_986780, EPI_ISL_986781, EPI_ISL_986782, EPI_ISL_986783, EPI_ISL_986784, EPI_ISL_986785, EPI_ISL_986786, EPI_ISL_986787, EPI_ISL_986788, EPI_ISL_986789, EPI_ISL_986790, EPI_ISL_986791, EPI_ISL_986792, EPI_ISL_986793, EPI_ISL_986794, EPI_ISL_986795, EPI_ISL_986796, EPI_ISL_986797, EPI_ISL_986798, EPI_ISL_986799, EPI_ISL_986800, EPI_ISL_986801, EPI_ISL_986802, EPI_ISL_986803, EPI_ISL_986804, EPI_ISL_986805, EPI_ISL_986806, EPI_ISL_986807, EPI_ISL_986808, EPI_ISL_986809, EPI_ISL_986810, EPI_ISL_986811, EPI_ISL_986812, EPI_ISL_986813, EPI_ISL_986814, EPI_ISL_986815, EPI_ISL_986816, EPI_ISL_986817, EPI_ISL_986818, EPI_ISL_986819, EPI_ISL_986820, EPI_ISL_986821, EPI_ISL_986822, EPI_ISL_986823, EPI_ISL_987161, EPI_ISL_987162, EPI_ISL_987163, EPI_ISL_987164, EPI_ISL_987165, EPI_ISL_987166, EPI_ISL_987167, EPI_ISL_987168, EPI_ISL_987169, EPI_ISL_987170, EPI_ISL_987171, EPI_ISL_987172, EPI_ISL_987173, EPI_ISL_987174, EPI_ISL_987175, EPI_ISL_987176, EPI_ISL_987177, EPI_ISL_987178, EPI_ISL_987179, EPI_ISL_987180, EPI_ISL_987181, EPI_ISL_987182, EPI_ISL_987183, EPI_ISL_987184, EPI_ISL_987185, EPI_ISL_987186, EPI_ISL_987187, EPI_ISL_987188, EPI_ISL_987189, EPI_ISL_987190, EPI_ISL_987191, EPI_ISL_987192, EPI_ISL_987193, EPI_ISL_987194, EPI_ISL_987195, EPI_ISL_987196, EPI_ISL_987197, EPI_ISL_987198, EPI_ISL_987199, EPI_ISL_987200, EPI_ISL_987201, EPI_ISL_987202, EPI_ISL_987203, EPI_ISL_987204, EPI_ISL_987205, EPI_ISL_987206, EPI_ISL_987207, EPI_ISL_987208, EPI_ISL_987209, EPI_ISL_987210, EPI_ISL_987211, EPI_ISL_987212, EPI_ISL_987213, EPI_ISL_987214, EPI_ISL_987215, EPI_ISL_987216, EPI_ISL_987217, EPI_ISL_987218, EPI_ISL_987219, EPI_ISL_987220, EPI_ISL_987221, EPI_ISL_987222, EPI_ISL_987223, EPI_ISL_987224, EPI_ISL_987225, EPI_ISL_987226, EPI_ISL_987227, EPI_ISL_987228, EPI_ISL_987229, EPI_ISL_987230, EPI_ISL_987231, EPI_ISL_987232, EPI_ISL_987233, EPI_ISL_987234, EPI_ISL_987235, EPI_ISL_987236, EPI_ISL_987237, EPI_ISL_987238, EPI_ISL_987239, EPI_ISL_987240, EPI_ISL_987241, EPI_ISL_987242, EPI_ISL_987243, EPI_ISL_987244, EPI_ISL_987245, EPI_ISL_987246, EPI_ISL_987247, EPI_ISL_987248, EPI_ISL_987249, EPI_ISL_987250, EPI_ISL_987251, EPI_ISL_987252, EPI_ISL_987253, EPI_ISL_987254, EPI_ISL_987255, EPI_ISL_987256, EPI_ISL_987257, EPI_ISL_987258, EPI_ISL_987259, EPI_ISL_987260, EPI_ISL_987261, EPI_ISL_987262, EPI_ISL_987263, EPI_ISL_987264, EPI_ISL_987265, EPI_ISL_987266, EPI_ISL_987267, EPI_ISL_987268, EPI_ISL_987269, EPI_ISL_987270, EPI_ISL_987271, EPI_ISL_987272, EPI_ISL_987273, EPI_ISL_987274, EPI_ISL_987275, EPI_ISL_987276, EPI_ISL_987277, EPI_ISL_987278, EPI_ISL_987279, EPI_ISL_987280, EPI_ISL_987281, EPI_ISL_988020, EPI_ISL_988022, EPI_ISL_988023, EPI_ISL_988024, EPI_ISL_988030, EPI_ISL_988039, EPI_ISL_988042, EPI_ISL_988043, EPI_ISL_988044, EPI_ISL_988048, EPI_ISL_988060, EPI_ISL_988061, EPI_ISL_988062, EPI_ISL_988067, EPI_ISL_988076, EPI_ISL_988079, EPI_ISL_988086, EPI_ISL_988092, EPI_ISL_988093, EPI_ISL_988097, EPI_ISL_988102, EPI_ISL_988107, EPI_ISL_988113, EPI_ISL_988117, EPI_ISL_988118, EPI_ISL_988128, EPI_ISL_988137, EPI_ISL_988143, EPI_ISL_988147, EPI_ISL_988154, EPI_ISL_988156, EPI_ISL_988160, EPI_ISL_988162, EPI_ISL_988163, EPI_ISL_988164, EPI_ISL_988168, EPI_ISL_988171, EPI_ISL_988189, EPI_ISL_988192, EPI_ISL_988193, EPI_ISL_988194, EPI_ISL_988195, EPI_ISL_988196, EPI_ISL_988199, EPI_ISL_988200, EPI_ISL_988203, EPI_ISL_1005700, EPI_ISL_1005701, EPI_ISL_1005702, EPI_ISL_1005703, EPI_ISL_1005704, EPI_ISL_1005705, EPI_ISL_1005706, EPI_ISL_1005707, EPI_ISL_1005708, EPI_ISL_1005709, EPI_ISL_1005710, EPI_ISL_1005711, EPI_ISL_1005712, EPI_ISL_1005713, EPI_ISL_1005714, EPI_ISL_1005715, EPI_ISL_1005716, EPI_ISL_1005719, EPI_ISL_1005720, EPI_ISL_1005721, EPI_ISL_1005722, EPI_ISL_1005723, EPI_ISL_1005724, EPI_ISL_1005725, EPI_ISL_1005726, EPI_ISL_1005728, EPI_ISL_1005729, EPI_ISL_1005730, EPI_ISL_1005732, EPI_ISL_1005733, EPI_ISL_1005734, EPI_ISL_1005735, EPI_ISL_1005736, EPI_ISL_1005738, EPI_ISL_1005739, EPI_ISL_1005740, EPI_ISL_1005744, EPI_ISL_1005745, EPI_ISL_1005747, EPI_ISL_1005748, EPI_ISL_1005749, EPI_ISL_1005750, EPI_ISL_1005751, EPI_ISL_1005752, EPI_ISL_1005754, EPI_ISL_1005755, EPI_ISL_1005756, EPI_ISL_1005757, EPI_ISL_1005758, EPI_ISL_1005759, EPI_ISL_1005761, EPI_ISL_1005762, EPI_ISL_1005763, EPI_ISL_1005764, EPI_ISL_1005766, EPI_ISL_1005767, EPI_ISL_1005768, EPI_ISL_1005769, EPI_ISL_1005770, EPI_ISL_1005771, EPI_ISL_1005772, EPI_ISL_1005775, EPI_ISL_1005776, EPI_ISL_1005779, EPI_ISL_1005780, EPI_ISL_1005781, EPI_ISL_1005783, EPI_ISL_1005784, EPI_ISL_1005786, EPI_ISL_1005788, EPI_ISL_1005789, EPI_ISL_1005790, EPI_ISL_1005791, EPI_ISL_1005792, EPI_ISL_1005793, EPI_ISL_1005794, EPI_ISL_1005795, EPI_ISL_1005796, EPI_ISL_1005797, EPI_ISL_1005798, EPI_ISL_1005800, EPI_ISL_1005801, EPI_ISL_1005802, EPI_ISL_1005803, EPI_ISL_1005805, EPI_ISL_1005806, EPI_ISL_1005808, EPI_ISL_1005810, EPI_ISL_1005811, EPI_ISL_1005812, EPI_ISL_1005813, EPI_ISL_1005815, EPI_ISL_1005818, EPI_ISL_1005819, EPI_ISL_1005820, EPI_ISL_1005822, EPI_ISL_1005823, EPI_ISL_1005824, EPI_ISL_1005826, EPI_ISL_1005827, EPI_ISL_1005828, EPI_ISL_1005829, EPI_ISL_1005830, EPI_ISL_1005832, EPI_ISL_1005833, EPI_ISL_1005834, EPI_ISL_1005835, EPI_ISL_1005837, EPI_ISL_1005839, EPI_ISL_1005840, EPI_ISL_1005842, EPI_ISL_1005844, EPI_ISL_1005845, EPI_ISL_1005846, EPI_ISL_1005847, EPI_ISL_1005848, EPI_ISL_1005849, EPI_ISL_1005850, EPI_ISL_1005851, EPI_ISL_1005852, EPI_ISL_1005853, EPI_ISL_1005854, EPI_ISL_1005855, EPI_ISL_1005856, EPI_ISL_1005858, EPI_ISL_1005859, EPI_ISL_1005860, EPI_ISL_1005861, EPI_ISL_1005863, EPI_ISL_1005864, EPI_ISL_1005865, EPI_ISL_1005866, EPI_ISL_1005867, EPI_ISL_1005868, EPI_ISL_1005869, EPI_ISL_1005870, EPI_ISL_1005871, EPI_ISL_1005872, EPI_ISL_1005875, EPI_ISL_1005876, EPI_ISL_1005877, EPI_ISL_1005878, EPI_ISL_1005879, EPI_ISL_1005880, EPI_ISL_1005881, EPI_ISL_1005882, EPI_ISL_1005884, EPI_ISL_1005886, EPI_ISL_1005887, EPI_ISL_1005888, EPI_ISL_1005890, EPI_ISL_1005893, EPI_ISL_1005896, EPI_ISL_1005897, EPI_ISL_1005898, EPI_ISL_1005899, EPI_ISL_1005904, EPI_ISL_1005905, EPI_ISL_1005906, EPI_ISL_1005907, EPI_ISL_1005908, EPI_ISL_1005910, EPI_ISL_1005913, EPI_ISL_1005915, EPI_ISL_1005916, EPI_ISL_1005917, EPI_ISL_1005918, EPI_ISL_1005919, EPI_ISL_1005920, EPI_ISL_1005921, EPI_ISL_1005923, EPI_ISL_1005924, EPI_ISL_1005925, EPI_ISL_1005926, EPI_ISL_1005928, EPI_ISL_1005929, EPI_ISL_1005930, EPI_ISL_1005931, EPI_ISL_1005932, EPI_ISL_1005933, EPI_ISL_1005934, EPI_ISL_1005936, EPI_ISL_1005937, EPI_ISL_1005938, EPI_ISL_1005941, EPI_ISL_1005942, EPI_ISL_1005943, EPI_ISL_1005944, EPI_ISL_1005945, EPI_ISL_1005946, EPI_ISL_1005948, EPI_ISL_1005949, EPI_ISL_1005950, EPI_ISL_1005953, EPI_ISL_1005954, EPI_ISL_1005955, EPI_ISL_1005956, EPI_ISL_1005959, EPI_ISL_1005960, EPI_ISL_1006186, EPI_ISL_1006187, EPI_ISL_1006188, EPI_ISL_1006189, EPI_ISL_1006190, EPI_ISL_1006191, EPI_ISL_1006192, EPI_ISL_1006193, EPI_ISL_1006194, EPI_ISL_1006195, EPI_ISL_1006196, EPI_ISL_1006197, EPI_ISL_1006198, EPI_ISL_1006199, EPI_ISL_1006200, EPI_ISL_1006201, EPI_ISL_1006202, EPI_ISL_1006203, EPI_ISL_1006204, EPI_ISL_1006205, EPI_ISL_1006206, EPI_ISL_1006207, EPI_ISL_1006208, EPI_ISL_1006209, EPI_ISL_1006210, EPI_ISL_1006211, EPI_ISL_1006212, EPI_ISL_1006213, EPI_ISL_1006214, EPI_ISL_1006215, EPI_ISL_1006216, EPI_ISL_1006217, EPI_ISL_1006218, EPI_ISL_1006219, EPI_ISL_1006220, EPI_ISL_1006221, EPI_ISL_1006222, EPI_ISL_1006223, EPI_ISL_1006224, EPI_ISL_1006225, EPI_ISL_1006226, EPI_ISL_1006227, EPI_ISL_1006228, EPI_ISL_1006229, EPI_ISL_1006230, EPI_ISL_1006231, EPI_ISL_1006232, EPI_ISL_1006233, EPI_ISL_1006234, EPI_ISL_1006235, EPI_ISL_1006236, EPI_ISL_1006237, EPI_ISL_1006238, EPI_ISL_1006239, EPI_ISL_1006240, EPI_ISL_1006241, EPI_ISL_1006242, EPI_ISL_1006243, EPI_ISL_1006244, EPI_ISL_1006245, EPI_ISL_1006246, EPI_ISL_1006247, EPI_ISL_1006248, EPI_ISL_1006250, EPI_ISL_1006251, EPI_ISL_1006252, EPI_ISL_1006253, EPI_ISL_1006254, EPI_ISL_1006255, EPI_ISL_1006256, EPI_ISL_1006258, EPI_ISL_1006259, EPI_ISL_1006260, EPI_ISL_1006261, EPI_ISL_1006263, EPI_ISL_1006264, EPI_ISL_1006265, EPI_ISL_1006266, EPI_ISL_1006268, EPI_ISL_1006270, EPI_ISL_1006271, EPI_ISL_1006273, EPI_ISL_1006274, EPI_ISL_1006276, EPI_ISL_1006277, EPI_ISL_1006278, EPI_ISL_1006279, EPI_ISL_1006281, EPI_ISL_1006282, EPI_ISL_1006284, EPI_ISL_1006285, EPI_ISL_1006286, EPI_ISL_1006287, EPI_ISL_1006288, EPI_ISL_1006289, EPI_ISL_1006290, EPI_ISL_1006292, EPI_ISL_1006293, EPI_ISL_1006294, EPI_ISL_1006295, EPI_ISL_1006296, EPI_ISL_1006297, EPI_ISL_1006298, EPI_ISL_1006299, EPI_ISL_1006300, EPI_ISL_1006302, EPI_ISL_1006303, EPI_ISL_1006304, EPI_ISL_1006305, EPI_ISL_1006306, EPI_ISL_1006307, EPI_ISL_1006309, EPI_ISL_1006310, EPI_ISL_1006314, EPI_ISL_1006315, EPI_ISL_1006316, EPI_ISL_1006318, EPI_ISL_1006319, EPI_ISL_1006320, EPI_ISL_1006321, EPI_ISL_1006325, EPI_ISL_1006326, EPI_ISL_1006327, EPI_ISL_1006328, EPI_ISL_1006329, EPI_ISL_1006330, EPI_ISL_1006332, EPI_ISL_1006333, EPI_ISL_1006334, EPI_ISL_1006335, EPI_ISL_1006336, EPI_ISL_1006337, EPI_ISL_1006338, EPI_ISL_1006339, EPI_ISL_1006548, EPI_ISL_1006549, EPI_ISL_1006552, EPI_ISL_1006554, EPI_ISL_1006557, EPI_ISL_1006558, EPI_ISL_1006561, EPI_ISL_1006562, EPI_ISL_1006563, EPI_ISL_1006564, EPI_ISL_1006567, EPI_ISL_1006569, EPI_ISL_1006571, EPI_ISL_1006572, EPI_ISL_1006575, EPI_ISL_1006577, EPI_ISL_1006578, EPI_ISL_1006581, EPI_ISL_1006585, EPI_ISL_1006587, EPI_ISL_1006588, EPI_ISL_1006592, EPI_ISL_1006593, EPI_ISL_1006595, EPI_ISL_1006596, EPI_ISL_1006597, EPI_ISL_1006598, EPI_ISL_1006599, EPI_ISL_1006600, EPI_ISL_1006603, EPI_ISL_1006608, EPI_ISL_1006610, EPI_ISL_1006611, EPI_ISL_1006612, EPI_ISL_1006615, EPI_ISL_1006618, EPI_ISL_1006619, EPI_ISL_1006622, EPI_ISL_1006623, EPI_ISL_1006625, EPI_ISL_1006627, EPI_ISL_1006628, EPI_ISL_1006629, EPI_ISL_1006631, EPI_ISL_1006633, EPI_ISL_1006635, EPI_ISL_1006638, EPI_ISL_1006640, EPI_ISL_1006641, EPI_ISL_1006643, EPI_ISL_1006644, EPI_ISL_1006647, EPI_ISL_1006652, EPI_ISL_1006653, EPI_ISL_1006654, EPI_ISL_1006655, EPI_ISL_1006864, EPI_ISL_1006865, EPI_ISL_1006866, EPI_ISL_1006867, EPI_ISL_1006868, EPI_ISL_1006869, EPI_ISL_1006870, EPI_ISL_1006871, EPI_ISL_1006872, EPI_ISL_1006874, EPI_ISL_1006875, EPI_ISL_1006876, EPI_ISL_1006877, EPI_ISL_1006878, EPI_ISL_1006879, EPI_ISL_1006880, EPI_ISL_1006881, EPI_ISL_1006882, EPI_ISL_1006883, EPI_ISL_1006884, EPI_ISL_1006885, EPI_ISL_1006886, EPI_ISL_1006887, EPI_ISL_1006888, EPI_ISL_1006889, EPI_ISL_1006890, EPI_ISL_1006891, EPI_ISL_1006892, EPI_ISL_1006893, EPI_ISL_1006894, EPI_ISL_1006895, EPI_ISL_1006896, EPI_ISL_1006897, EPI_ISL_1006898, EPI_ISL_1006899, EPI_ISL_1006900, EPI_ISL_1006901, EPI_ISL_1006902, EPI_ISL_1006903, EPI_ISL_1006904, EPI_ISL_1006905, EPI_ISL_1006906, EPI_ISL_1006907, EPI_ISL_1006908, EPI_ISL_1006909, EPI_ISL_1006910, EPI_ISL_1006911, EPI_ISL_1006912, EPI_ISL_1006913, EPI_ISL_1006914, EPI_ISL_1006915, EPI_ISL_1006916, EPI_ISL_1006917, EPI_ISL_1006918, EPI_ISL_1006919, EPI_ISL_1006920, EPI_ISL_1006921, EPI_ISL_1006922, EPI_ISL_1006923, EPI_ISL_1006924, EPI_ISL_1006925, EPI_ISL_1006926, EPI_ISL_1006927, EPI_ISL_1006928, EPI_ISL_1006929, EPI_ISL_1006930, EPI_ISL_1006931, EPI_ISL_1006932, EPI_ISL_1006933, EPI_ISL_1006934, EPI_ISL_1006935, EPI_ISL_1006936, EPI_ISL_1007181, EPI_ISL_1007182, EPI_ISL_1007183, EPI_ISL_1007185, EPI_ISL_1007186, EPI_ISL_1007187, EPI_ISL_1007188, EPI_ISL_1007189, EPI_ISL_1007190, EPI_ISL_1007191, EPI_ISL_1007193, EPI_ISL_1007194, EPI_ISL_1007195, EPI_ISL_1007196, EPI_ISL_1007197, EPI_ISL_1007198, EPI_ISL_1007199, EPI_ISL_1007201, EPI_ISL_1007202, EPI_ISL_1007203, EPI_ISL_1007204, EPI_ISL_1007205, EPI_ISL_1007206, EPI_ISL_1007207, EPI_ISL_1007208, EPI_ISL_1007209, EPI_ISL_1007210, EPI_ISL_1007211, EPI_ISL_1007212, EPI_ISL_1007213, EPI_ISL_1007214, EPI_ISL_1007215, EPI_ISL_1007216, EPI_ISL_1007217, EPI_ISL_1007218, EPI_ISL_1007219, EPI_ISL_1007220, EPI_ISL_1007221, EPI_ISL_1007222, EPI_ISL_1007223, EPI_ISL_1007224, EPI_ISL_1007225, EPI_ISL_1007226, EPI_ISL_1007227, EPI_ISL_1007228, EPI_ISL_1007229, EPI_ISL_1007230, EPI_ISL_1007231, EPI_ISL_1007232, EPI_ISL_1007233, EPI_ISL_1007234, EPI_ISL_1007235, EPI_ISL_1007236, EPI_ISL_1007237, EPI_ISL_1007238, EPI_ISL_1007239, EPI_ISL_1007240, EPI_ISL_1007241, EPI_ISL_1007243, EPI_ISL_1007244, EPI_ISL_1007246, EPI_ISL_1007247, EPI_ISL_1007248, EPI_ISL_1007249, EPI_ISL_1007250, EPI_ISL_1007251, EPI_ISL_1007252, EPI_ISL_1007253, EPI_ISL_1007254, EPI_ISL_1007255, EPI_ISL_1007256, EPI_ISL_1007257, EPI_ISL_1007258, EPI_ISL_1007259, EPI_ISL_1007260, EPI_ISL_1007261, EPI_ISL_1007263, EPI_ISL_1007264, EPI_ISL_1007265, EPI_ISL_1007266, EPI_ISL_1007267, EPI_ISL_1007268, EPI_ISL_1007270, EPI_ISL_1007271, EPI_ISL_1007272, EPI_ISL_1007273, EPI_ISL_1007274, EPI_ISL_1007275, EPI_ISL_1007276, EPI_ISL_1007277, EPI_ISL_1007278, EPI_ISL_1007279, EPI_ISL_1007280, EPI_ISL_1007281, EPI_ISL_1007282, EPI_ISL_1007283, EPI_ISL_1007284, EPI_ISL_1007285, EPI_ISL_1007286, EPI_ISL_1007287, EPI_ISL_1007288, EPI_ISL_1007289, EPI_ISL_1007290, EPI_ISL_1007291, EPI_ISL_1007293, EPI_ISL_1007294, EPI_ISL_1007295, EPI_ISL_1007297, EPI_ISL_1007298, EPI_ISL_1007299, EPI_ISL_1007300, EPI_ISL_1007301, EPI_ISL_1007302, EPI_ISL_1007303, EPI_ISL_1007304, EPI_ISL_1007305, EPI_ISL_1007306, EPI_ISL_1007307, EPI_ISL_1007308, EPI_ISL_1007309, EPI_ISL_1007310, EPI_ISL_1007311, EPI_ISL_1007312, EPI_ISL_1007314, EPI_ISL_1007315, EPI_ISL_1007316, EPI_ISL_1007317, EPI_ISL_1007318, EPI_ISL_1007319, EPI_ISL_1007320, EPI_ISL_1007321, EPI_ISL_1007322, EPI_ISL_1007323, EPI_ISL_1007324, EPI_ISL_1007325, EPI_ISL_1007326, EPI_ISL_1007327, EPI_ISL_1007328, EPI_ISL_1007329, EPI_ISL_1007330, EPI_ISL_1007331, EPI_ISL_1007332, EPI_ISL_1007333, EPI_ISL_1007334, EPI_ISL_1007335, EPI_ISL_1007336, EPI_ISL_1007338, EPI_ISL_1007339, EPI_ISL_1007340, EPI_ISL_1007341, EPI_ISL_1007342, EPI_ISL_1007343, EPI_ISL_1007344, EPI_ISL_1007345, EPI_ISL_1007347, EPI_ISL_1007348, EPI_ISL_1007350, EPI_ISL_1007351, EPI_ISL_1007352, EPI_ISL_1007353, EPI_ISL_1007354, EPI_ISL_1007355, EPI_ISL_1007356, EPI_ISL_1007358, EPI_ISL_1007359, EPI_ISL_1007360, EPI_ISL_1007361, EPI_ISL_1007362, EPI_ISL_1007363, EPI_ISL_1007364, EPI_ISL_1007365, EPI_ISL_1007366, EPI_ISL_1007368, EPI_ISL_1007370, EPI_ISL_1007371, EPI_ISL_1007372, EPI_ISL_1007373, EPI_ISL_1007374, EPI_ISL_1007375, EPI_ISL_1007376, EPI_ISL_1007377, EPI_ISL_1007378, EPI_ISL_1007379, EPI_ISL_1007380, EPI_ISL_1007382, EPI_ISL_1007383, EPI_ISL_1007384, EPI_ISL_1007385, EPI_ISL_1007387, EPI_ISL_1007388, EPI_ISL_1007389, EPI_ISL_1007390, EPI_ISL_1007391, EPI_ISL_1007393, EPI_ISL_1007394, EPI_ISL_1007395, EPI_ISL_1007396, EPI_ISL_1007397, EPI_ISL_1007398, EPI_ISL_1007399, EPI_ISL_1007400, EPI_ISL_1007401, EPI_ISL_1007402, EPI_ISL_1007403, EPI_ISL_1007406, EPI_ISL_1007408, EPI_ISL_1007409, EPI_ISL_1007410, EPI_ISL_1007411, EPI_ISL_1007412, EPI_ISL_1007413, EPI_ISL_1007414, EPI_ISL_1007418, EPI_ISL_1007419, EPI_ISL_1007420, EPI_ISL_1007421, EPI_ISL_1007422, EPI_ISL_1007423, EPI_ISL_1007424, EPI_ISL_1007425, EPI_ISL_1007426, EPI_ISL_1007427, EPI_ISL_1007429, EPI_ISL_1007431, EPI_ISL_1007432, EPI_ISL_1007455, EPI_ISL_1007460, EPI_ISL_1007462, EPI_ISL_1007465, EPI_ISL_1007466, EPI_ISL_1007468, EPI_ISL_1007470, EPI_ISL_1007475, EPI_ISL_1007482, EPI_ISL_1007484, EPI_ISL_1007487, EPI_ISL_1007489, EPI_ISL_1007490, EPI_ISL_1007493, EPI_ISL_1007499, EPI_ISL_1007501, EPI_ISL_1007503, EPI_ISL_1007507, EPI_ISL_1007518, EPI_ISL_1007520, EPI_ISL_1012099, EPI_ISL_1012100, EPI_ISL_1012101, EPI_ISL_1012102, EPI_ISL_1012103, EPI_ISL_1012104, EPI_ISL_1012105, EPI_ISL_1012106, EPI_ISL_1012107, EPI_ISL_1012108, EPI_ISL_1012109, EPI_ISL_1012110, EPI_ISL_1012111, EPI_ISL_1012112, EPI_ISL_1012113, EPI_ISL_1012114, EPI_ISL_1012115, EPI_ISL_1012116, EPI_ISL_1012117, EPI_ISL_1012118, EPI_ISL_1012119, EPI_ISL_1012120, EPI_ISL_1012121, EPI_ISL_1012122, EPI_ISL_1012123, EPI_ISL_1012124, EPI_ISL_1012125, EPI_ISL_1012126, EPI_ISL_1012127, EPI_ISL_1012128, EPI_ISL_1012129, EPI_ISL_1012130, EPI_ISL_1012131, EPI_ISL_1012132, EPI_ISL_1012133, EPI_ISL_1012134, EPI_ISL_1012135, EPI_ISL_1012136, EPI_ISL_1012137, EPI_ISL_1012138, EPI_ISL_1012139, EPI_ISL_1012140, EPI_ISL_1012141, EPI_ISL_1012142, EPI_ISL_1012143, EPI_ISL_1012144, EPI_ISL_1012145, EPI_ISL_1012146, EPI_ISL_1012147, EPI_ISL_1012148, EPI_ISL_1012149, EPI_ISL_1012150, EPI_ISL_1012151, EPI_ISL_1012152, EPI_ISL_1012153, EPI_ISL_1012154, EPI_ISL_1012155, EPI_ISL_1012156, EPI_ISL_1012157, EPI_ISL_1012158, EPI_ISL_1012159, EPI_ISL_1012160, EPI_ISL_1012161, EPI_ISL_1012162, EPI_ISL_1012163, EPI_ISL_1012164, EPI_ISL_1012165, EPI_ISL_1012166, EPI_ISL_1012167, EPI_ISL_1012168, EPI_ISL_1012169, EPI_ISL_1012170, EPI_ISL_1012171, EPI_ISL_1012172, EPI_ISL_1012173, EPI_ISL_1012174, EPI_ISL_1012175, EPI_ISL_1012176, EPI_ISL_1012177, EPI_ISL_1012178, EPI_ISL_1012179, EPI_ISL_1012180, EPI_ISL_1012181, EPI_ISL_1012182, EPI_ISL_1012183, EPI_ISL_1012184, EPI_ISL_1012185, EPI_ISL_1012186, EPI_ISL_1012187, EPI_ISL_1012188, EPI_ISL_1012189, EPI_ISL_1012190, EPI_ISL_1012191, EPI_ISL_1012192, EPI_ISL_1012193, EPI_ISL_1012194, EPI_ISL_1012195, EPI_ISL_1012196, EPI_ISL_1012197, EPI_ISL_1012198, EPI_ISL_1012199, EPI_ISL_1012200, EPI_ISL_1012201, EPI_ISL_1012202, EPI_ISL_1012203, EPI_ISL_1012204, EPI_ISL_1012205, EPI_ISL_1012206, EPI_ISL_1012207, EPI_ISL_1012208, EPI_ISL_1012209, EPI_ISL_1012210, EPI_ISL_1012211, EPI_ISL_1012212, EPI_ISL_1012213, EPI_ISL_1012214, EPI_ISL_1012215, EPI_ISL_1012216, EPI_ISL_1012217, EPI_ISL_1012218, EPI_ISL_1012219, EPI_ISL_1012220, EPI_ISL_1012221, EPI_ISL_1012222, EPI_ISL_1012223, EPI_ISL_1012224, EPI_ISL_1012225, EPI_ISL_1012226, EPI_ISL_1012227, EPI_ISL_1012228, EPI_ISL_1012229, EPI_ISL_1012230, EPI_ISL_1012231, EPI_ISL_1012232, EPI_ISL_1012233, EPI_ISL_1012234, EPI_ISL_1012235, EPI_ISL_1012236, EPI_ISL_1012237, EPI_ISL_1012238, EPI_ISL_1012239, EPI_ISL_1012240, EPI_ISL_1012241, EPI_ISL_1012242, EPI_ISL_1012243, EPI_ISL_1012244, EPI_ISL_1012245, EPI_ISL_1012246, EPI_ISL_1012247, EPI_ISL_1012248, EPI_ISL_1012249, EPI_ISL_1012250, EPI_ISL_1012251, EPI_ISL_1012252, EPI_ISL_1012253, EPI_ISL_1012254, EPI_ISL_1012255, EPI_ISL_1012256, EPI_ISL_1012257, EPI_ISL_1012258, EPI_ISL_1012259, EPI_ISL_1012260, EPI_ISL_1012261, EPI_ISL_1012262, EPI_ISL_1012263, EPI_ISL_1012264, EPI_ISL_1012265, EPI_ISL_1012266, EPI_ISL_1012267, EPI_ISL_1012268, EPI_ISL_1012269, EPI_ISL_1012270, EPI_ISL_1012271, EPI_ISL_1012272, EPI_ISL_1012273, EPI_ISL_1012274, EPI_ISL_1012275, EPI_ISL_1012276, EPI_ISL_1012277, EPI_ISL_1012278, EPI_ISL_1012279, EPI_ISL_1012280, EPI_ISL_1012281, EPI_ISL_1012282, EPI_ISL_1012283, EPI_ISL_1012284, EPI_ISL_1012285, EPI_ISL_1012286, EPI_ISL_1012287, EPI_ISL_1012288, EPI_ISL_1012289, EPI_ISL_1012290, EPI_ISL_1012291, EPI_ISL_1012292, EPI_ISL_1012293, EPI_ISL_1012294, EPI_ISL_1012295, EPI_ISL_1012296, EPI_ISL_1012297, EPI_ISL_1012298, EPI_ISL_1012299, EPI_ISL_1012300, EPI_ISL_1012301, EPI_ISL_1012302, EPI_ISL_1012303, EPI_ISL_1012304, EPI_ISL_1012305, EPI_ISL_1012306, EPI_ISL_1012307, EPI_ISL_1012308, EPI_ISL_1012309, EPI_ISL_1012310, EPI_ISL_1012311, EPI_ISL_1012312, EPI_ISL_1012313, EPI_ISL_1012314, EPI_ISL_1012315, EPI_ISL_1012316, EPI_ISL_1012317, EPI_ISL_1012318, EPI_ISL_1012319, EPI_ISL_1012320, EPI_ISL_1012321, EPI_ISL_1012322, EPI_ISL_1012323, EPI_ISL_1012324, EPI_ISL_1012325, EPI_ISL_1012326, EPI_ISL_1012327, EPI_ISL_1012328, EPI_ISL_1012329, EPI_ISL_1012330, EPI_ISL_1012331, EPI_ISL_1012332, EPI_ISL_1012333, EPI_ISL_1012334, EPI_ISL_1012335, EPI_ISL_1012336, EPI_ISL_1012337, EPI_ISL_1012338, EPI_ISL_1012339, EPI_ISL_1012340, EPI_ISL_1012341, EPI_ISL_1012342, EPI_ISL_1012343, EPI_ISL_1012344, EPI_ISL_1012345, EPI_ISL_1012346, EPI_ISL_1012347, EPI_ISL_1012348, EPI_ISL_1012349, EPI_ISL_1012350, EPI_ISL_1012351, EPI_ISL_1012352, EPI_ISL_1012353, EPI_ISL_1012354, EPI_ISL_1012355, EPI_ISL_1012356, EPI_ISL_1012357, EPI_ISL_1012358, EPI_ISL_1012359, EPI_ISL_1012360, EPI_ISL_1012361, EPI_ISL_1012362, EPI_ISL_1012363, EPI_ISL_1012364, EPI_ISL_1012365, EPI_ISL_1012366, EPI_ISL_1012367, EPI_ISL_1012368, EPI_ISL_1012369, EPI_ISL_1012370, EPI_ISL_1012371, EPI_ISL_1012372, EPI_ISL_1012373, EPI_ISL_1012374, EPI_ISL_1012375, EPI_ISL_1012377, EPI_ISL_1012378, EPI_ISL_1012379, EPI_ISL_1012380, EPI_ISL_1012381, EPI_ISL_1012382, EPI_ISL_1012383, EPI_ISL_1012384, EPI_ISL_1012385, EPI_ISL_1012386, EPI_ISL_1012387, EPI_ISL_1012389, EPI_ISL_1012390, EPI_ISL_1012391, EPI_ISL_1012392, EPI_ISL_1012393, EPI_ISL_1012394, EPI_ISL_1012395, EPI_ISL_1012397, EPI_ISL_1012398, EPI_ISL_1012399, EPI_ISL_1012400, EPI_ISL_1012401, EPI_ISL_1012402, EPI_ISL_1012407, EPI_ISL_1012409, EPI_ISL_1012414, EPI_ISL_1012416, EPI_ISL_1012428, EPI_ISL_1012431, EPI_ISL_1012438, EPI_ISL_1012449, EPI_ISL_1012450, EPI_ISL_1012458, EPI_ISL_1012482, EPI_ISL_1012484, EPI_ISL_1012493, EPI_ISL_1012498, EPI_ISL_1012499, EPI_ISL_1012514, EPI_ISL_1012521, EPI_ISL_1012531, EPI_ISL_1012541, EPI_ISL_1012609, EPI_ISL_1012610, EPI_ISL_1012611, EPI_ISL_1012612, EPI_ISL_1012613, EPI_ISL_1012614, EPI_ISL_1012615, EPI_ISL_1012617, EPI_ISL_1012618, EPI_ISL_1012619, EPI_ISL_1012620, EPI_ISL_1012622, EPI_ISL_1012623, EPI_ISL_1012624, EPI_ISL_1012625, EPI_ISL_1012626, EPI_ISL_1012628, EPI_ISL_1012630, EPI_ISL_1012631, EPI_ISL_1012632, EPI_ISL_1012633, EPI_ISL_1012634, EPI_ISL_1012635, EPI_ISL_1012636, EPI_ISL_1012638, EPI_ISL_1012639, EPI_ISL_1012640, EPI_ISL_1012641, EPI_ISL_1012642, EPI_ISL_1012644, EPI_ISL_1012645, EPI_ISL_1012646, EPI_ISL_1012647, EPI_ISL_1012648, EPI_ISL_1012649, EPI_ISL_1012651, EPI_ISL_1012652, EPI_ISL_1012653, EPI_ISL_1012654, EPI_ISL_1012656, EPI_ISL_1012657, EPI_ISL_1012658, EPI_ISL_1012659, EPI_ISL_1012660, EPI_ISL_1012662, EPI_ISL_1012664, EPI_ISL_1018693, EPI_ISL_1018694, EPI_ISL_1018701, EPI_ISL_1018713, EPI_ISL_1018725, EPI_ISL_1018730, EPI_ISL_1018755, EPI_ISL_1018756, EPI_ISL_1018762, EPI_ISL_1018776, EPI_ISL_1018800, EPI_ISL_1018803, EPI_ISL_1018807, EPI_ISL_1018808, EPI_ISL_1018814, EPI_ISL_1018838, EPI_ISL_1018844, EPI_ISL_1018858, EPI_ISL_1018865, EPI_ISL_1018872, EPI_ISL_1018874, EPI_ISL_1018876, EPI_ISL_1018880, EPI_ISL_1018882, EPI_ISL_1018890, EPI_ISL_1018907, EPI_ISL_1018912, EPI_ISL_1018914, EPI_ISL_1018915, EPI_ISL_1018917, EPI_ISL_1018920, EPI_ISL_1018921, EPI_ISL_1018929, EPI_ISL_1018935, EPI_ISL_1018941, EPI_ISL_1019300, EPI_ISL_1019305, EPI_ISL_1019311, EPI_ISL_1019331, EPI_ISL_1019349, EPI_ISL_1019352, EPI_ISL_1019353, EPI_ISL_1019354, EPI_ISL_1019355, EPI_ISL_1019356, EPI_ISL_1019357, EPI_ISL_1019358, EPI_ISL_1019359, EPI_ISL_1019360, EPI_ISL_1019361, EPI_ISL_1019362, EPI_ISL_1019363, EPI_ISL_1019364, EPI_ISL_1019365, EPI_ISL_1019367, EPI_ISL_1019368, EPI_ISL_1019369, EPI_ISL_1019370, EPI_ISL_1019371, EPI_ISL_1019372, EPI_ISL_1019373, EPI_ISL_1019374, EPI_ISL_1019375, EPI_ISL_1019376, EPI_ISL_1019377, EPI_ISL_1019378, EPI_ISL_1019379, EPI_ISL_1019380, EPI_ISL_1019381, EPI_ISL_1019383, EPI_ISL_1019384, EPI_ISL_1019385, EPI_ISL_1019386, EPI_ISL_1019387, EPI_ISL_1019388, EPI_ISL_1019389, EPI_ISL_1019390, EPI_ISL_1019391, EPI_ISL_1019392, EPI_ISL_1019393, EPI_ISL_1019394, EPI_ISL_1019395, EPI_ISL_1019396, EPI_ISL_1019397, EPI_ISL_1019398, EPI_ISL_1019399, EPI_ISL_1019400, EPI_ISL_1019401, EPI_ISL_1019402, EPI_ISL_1019403, EPI_ISL_1019404, EPI_ISL_1019405, EPI_ISL_1019406, EPI_ISL_1019407, EPI_ISL_1019408, EPI_ISL_1019504, EPI_ISL_1019505, EPI_ISL_1019506, EPI_ISL_1033166, EPI_ISL_1033175, EPI_ISL_1033216, EPI_ISL_1033244, EPI_ISL_1033270, EPI_ISL_1033299, EPI_ISL_1033428, EPI_ISL_1033484, EPI_ISL_1033492, EPI_ISL_1033616, EPI_ISL_1033670, EPI_ISL_1033699, EPI_ISL_1033704, EPI_ISL_1033755, EPI_ISL_1033766, EPI_ISL_1033767, EPI_ISL_1033772, EPI_ISL_1033773, EPI_ISL_1033780, EPI_ISL_1033791, EPI_ISL_1033793, EPI_ISL_1033797, EPI_ISL_1033802, EPI_ISL_1033806, EPI_ISL_1033807, EPI_ISL_1033823, EPI_ISL_1033836, EPI_ISL_1033845, EPI_ISL_1033850, EPI_ISL_1033853, EPI_ISL_1033855, EPI_ISL_1033862, EPI_ISL_1033868, EPI_ISL_1033880, EPI_ISL_1033884, EPI_ISL_1033889, EPI_ISL_1033893, EPI_ISL_1033895, EPI_ISL_1033898, EPI_ISL_1033904, EPI_ISL_1033909, EPI_ISL_1042322, EPI_ISL_1045844, EPI_ISL_1045898, EPI_ISL_1045917, EPI_ISL_1045926, EPI_ISL_1045958, EPI_ISL_1045965, EPI_ISL_1187054, EPI_ISL_1187055, EPI_ISL_1187058, EPI_ISL_1187060, EPI_ISL_1187065, EPI_ISL_1187070, EPI_ISL_1187071, EPI_ISL_1187072, EPI_ISL_1187083, EPI_ISL_1187084, EPI_ISL_1187085, EPI_ISL_1187090, EPI_ISL_1187094, EPI_ISL_1187106, EPI_ISL_1187107, EPI_ISL_1187108, EPI_ISL_1187109, EPI_ISL_1187111, EPI_ISL_1187112, EPI_ISL_1187121, EPI_ISL_1187122, EPI_ISL_1187126, EPI_ISL_1187128, EPI_ISL_1187132, EPI_ISL_1187138, EPI_ISL_1187142, EPI_ISL_1187152, EPI_ISL_1187153, EPI_ISL_1187154, EPI_ISL_1187157, EPI_ISL_1187158, EPI_ISL_1187159, EPI_ISL_1187163, EPI_ISL_1187164, EPI_ISL_1187167, EPI_ISL_1187173, EPI_ISL_1187175, EPI_ISL_1187180, EPI_ISL_1187182, EPI_ISL_1187185, EPI_ISL_1187187, EPI_ISL_1187200, EPI_ISL_1187202, EPI_ISL_1187205, EPI_ISL_1187206, EPI_ISL_1187208, EPI_ISL_1187209, EPI_ISL_1187211, EPI_ISL_1187216, EPI_ISL_1187222, EPI_ISL_1187223, EPI_ISL_1187224, EPI_ISL_1187235, EPI_ISL_1187236, EPI_ISL_1187241, EPI_ISL_1187243, EPI_ISL_1187246, EPI_ISL_1187248, EPI_ISL_1187249, EPI_ISL_1206058, EPI_ISL_1206065, EPI_ISL_1206081, EPI_ISL_1206115, EPI_ISL_1206136, EPI_ISL_1206150, EPI_ISL_1206167, EPI_ISL_1206169, EPI_ISL_1206174, EPI_ISL_1206197, EPI_ISL_1206209, EPI_ISL_1206210, EPI_ISL_1206243, EPI_ISL_1206249, EPI_ISL_1206267, EPI_ISL_1206314, EPI_ISL_1206317, EPI_ISL_1206320, EPI_ISL_1206342, EPI_ISL_1206348, EPI_ISL_1206386, EPI_ISL_1206389, EPI_ISL_1206391, EPI_ISL_1206452, EPI_ISL_1206459, EPI_ISL_1206462, EPI_ISL_1206463, EPI_ISL_1206468, EPI_ISL_1206484, EPI_ISL_1206497, EPI_ISL_1223162, EPI_ISL_1223171, EPI_ISL_1223185, EPI_ISL_1223186, EPI_ISL_1223190, EPI_ISL_1223198, EPI_ISL_1223199, EPI_ISL_1223200, EPI_ISL_1223201, EPI_ISL_1223210, EPI_ISL_1223211, EPI_ISL_1223216, EPI_ISL_1223231, EPI_ISL_1223234, EPI_ISL_1223237, EPI_ISL_1223255, EPI_ISL_1223266, EPI_ISL_1223271, EPI_ISL_1223274, EPI_ISL_1223282, EPI_ISL_1223287, EPI_ISL_1223314, EPI_ISL_1223316, EPI_ISL_1223326, EPI_ISL_1223348, EPI_ISL_1223350, EPI_ISL_1223379, EPI_ISL_1223403, EPI_ISL_1223409, EPI_ISL_1223421, EPI_ISL_1223434, EPI_ISL_1242249, EPI_ISL_1256428, EPI_ISL_1333801, EPI_ISL_1333803, EPI_ISL_1333805, EPI_ISL_1333806, EPI_ISL_1333807, EPI_ISL_1333808, EPI_ISL_1333809, EPI_ISL_1333811, EPI_ISL_1333812, EPI_ISL_1333813, EPI_ISL_1333814, EPI_ISL_1333817, EPI_ISL_1333818, EPI_ISL_1333819, EPI_ISL_1333820, EPI_ISL_1333824, EPI_ISL_1333825, EPI_ISL_1333826, EPI_ISL_1333837, EPI_ISL_1333890, EPI_ISL_1333906, EPI_ISL_1333908, EPI_ISL_1333914, EPI_ISL_1333924, EPI_ISL_1333930, EPI_ISL_1333939, EPI_ISL_1333953, EPI_ISL_1333954, EPI_ISL_1467633, EPI_ISL_1467644, EPI_ISL_1467654, EPI_ISL_1467680, EPI_ISL_1467686, EPI_ISL_1467691, EPI_ISL_1467706, EPI_ISL_1467727, EPI_ISL_1467739, EPI_ISL_1467752, EPI_ISL_1467759

| | | | |
|---|---|---|---|
| see above | Lighthouse Lab in Cambridge | Wellcome Sanger Institute for the COVID-19 Genomics UK (COG-UK) Consortium | Cordelia Langford; David K. Jackson; Dominic Kwiatkowski; Ewan Harrison; Ian Johnston; Jeffrey Barrett; John Sillitoe on behalf of the Wellcome Sanger Institute COVID-19 Surveillance Team; John Sillitoe on behalf of the Wellcome Sanger Institute COVID-19 Surveillance Team (http://www.sanger.ac.uk/covid-team); Rob Howes; Roberto Amato; Sonia Goncalves; The Lighthouse Lab in Cambridge and Alex Alderton |

EPI_ISL_986106, EPI_ISL_986109, EPI_ISL_986112, EPI_ISL_986114, EPI_ISL_986115, EPI_ISL_986116, EPI_ISL_986117, EPI_ISL_986118, EPI_ISL_986119, EPI_ISL_986125, EPI_ISL_986133, EPI_ISL_986144, EPI_ISL_986156, EPI_ISL_986157, EPI_ISL_986160, EPI_ISL_986162, EPI_ISL_986165, EPI_ISL_986169, EPI_ISL_986174, EPI_ISL_986180, EPI_ISL_986181, EPI_ISL_986182, EPI_ISL_986185, EPI_ISL_986191, EPI_ISL_986200, EPI_ISL_986201, EPI_ISL_986204, EPI_ISL_986205, EPI_ISL_986208, EPI_ISL_986211, EPI_ISL_986213, EPI_ISL_986215, EPI_ISL_986217, EPI_ISL_986218, EPI_ISL_986222, EPI_ISL_986234, EPI_ISL_986236, EPI_ISL_986241, EPI_ISL_986243, EPI_ISL_986244, EPI_ISL_986248, EPI_ISL_986249, EPI_ISL_986250, EPI_ISL_986254, EPI_ISL_986257, EPI_ISL_986267, EPI_ISL_986270, EPI_ISL_986272, EPI_ISL_986284, EPI_ISL_986290, EPI_ISL_986291, EPI_ISL_986294, EPI_ISL_986297, EPI_ISL_986299, EPI_ISL_986300, EPI_ISL_986302, EPI_ISL_986306, EPI_ISL_986308, EPI_ISL_986309, EPI_ISL_986316, EPI_ISL_986318, EPI_ISL_986320, EPI_ISL_986326, EPI_ISL_986328, EPI_ISL_986330, EPI_ISL_986331, EPI_ISL_986334, EPI_ISL_986335, EPI_ISL_986337, EPI_ISL_986342, EPI_ISL_986346, EPI_ISL_986348, EPI_ISL_986350, EPI_ISL_986352, EPI_ISL_986353, EPI_ISL_986355, EPI_ISL_986356, EPI_ISL_986365, EPI_ISL_986367, EPI_ISL_986368, EPI_ISL_986369, EPI_ISL_986370, EPI_ISL_986371, EPI_ISL_986372, EPI_ISL_986374, EPI_ISL_986375, EPI_ISL_986381, EPI_ISL_986382, EPI_ISL_986384, EPI_ISL_986388, EPI_ISL_986389, EPI_ISL_986395, EPI_ISL_986396, EPI_ISL_986397, EPI_ISL_986407, EPI_ISL_986409, EPI_ISL_986410, EPI_ISL_986411, EPI_ISL_986415, EPI_ISL_986419, EPI_ISL_986420, EPI_ISL_986426, EPI_ISL_986427, EPI_ISL_986430, EPI_ISL_1005966, EPI_ISL_1005968, EPI_ISL_1005981, EPI_ISL_1005989, EPI_ISL_1005995, EPI_ISL_1005998, EPI_ISL_1006006, EPI_ISL_1006015, EPI_ISL_1006018, EPI_ISL_1006020, EPI_ISL_1006025, EPI_ISL_1006030, EPI_ISL_1006035, EPI_ISL_1006046, EPI_ISL_1006047, EPI_ISL_1006052, EPI_ISL_1006057, EPI_ISL_1006059, EPI_ISL_1006064, EPI_ISL_1006066, EPI_ISL_1006070, EPI_ISL_1006073, EPI_ISL_1006077, EPI_ISL_1006080, EPI_ISL_1006083, EPI_ISL_1006087, EPI_ISL_1006090, EPI_ISL_1006094, EPI_ISL_1006096, EPI_ISL_1006106, EPI_ISL_1006108, EPI_ISL_1006109, EPI_ISL_1006114, EPI_ISL_1006117, EPI_ISL_1006136, EPI_ISL_1006140, EPI_ISL_1006150, EPI_ISL_1006156, EPI_ISL_1006164, EPI_ISL_1006168, EPI_ISL_1006169, EPI_ISL_1019507, EPI_ISL_1019509, EPI_ISL_1019510, EPI_ISL_1019511, EPI_ISL_1019512, EPI_ISL_1019513, EPI_ISL_1019514, EPI_ISL_1019515, EPI_ISL_1019517, EPI_ISL_1019518, EPI_ISL_1019519, EPI_ISL_1019520, EPI_ISL_1019521, EPI_ISL_1019522, EPI_ISL_1019523, EPI_ISL_1019524, EPI_ISL_1019525, EPI_ISL_1019526, EPI_ISL_1019527, EPI_ISL_1019528, EPI_ISL_1019529, EPI_ISL_1019530, EPI_ISL_1019531, EPI_ISL_1019532, EPI_ISL_1019533, EPI_ISL_1019534, EPI_ISL_1019535, EPI_ISL_1019536, EPI_ISL_1019537, EPI_ISL_1019538, EPI_ISL_1019539, EPI_ISL_1019540, EPI_ISL_1019541, EPI_ISL_1019542, EPI_ISL_1019543, EPI_ISL_1019544, EPI_ISL_1019545, EPI_ISL_1019546, EPI_ISL_1019547, EPI_ISL_1019548, EPI_ISL_1019549, EPI_ISL_1019550, EPI_ISL_1019551, EPI_ISL_1019552, EPI_ISL_1019553, EPI_ISL_1019554, EPI_ISL_1019555, EPI_ISL_1019556, EPI_ISL_1019557, EPI_ISL_1019558, EPI_ISL_1019559, EPI_ISL_1019560, EPI_ISL_1019561, EPI_ISL_1019562, EPI_ISL_1019563, EPI_ISL_1019564, EPI_ISL_1019565, EPI_ISL_1019566, EPI_ISL_1019567, EPI_ISL_1019568, EPI_ISL_1019569, EPI_ISL_1019570, EPI_ISL_1019571, EPI_ISL_1019572, EPI_ISL_1019573, EPI_ISL_1019574, EPI_ISL_1019575, EPI_ISL_1019576, EPI_ISL_1019577, EPI_ISL_1019578, EPI_ISL_1019579, EPI_ISL_1019580, EPI_ISL_1019581, EPI_ISL_1019582, EPI_ISL_1042575, EPI_ISL_1042583, EPI_ISL_1042658, EPI_ISL_1042672, EPI_ISL_1042722, EPI_ISL_1042759, EPI_ISL_1042766, EPI_ISL_1042772, EPI_ISL_1042781, EPI_ISL_1062875, EPI_ISL_1063112, EPI_ISL_1063120, EPI_ISL_1063127, EPI_ISL_1063162, EPI_ISL_1063188, EPI_ISL_1063253, EPI_ISL_1070372, EPI_ISL_1070459, EPI_ISL_1070626, EPI_ISL_1103391, EPI_ISL_1103394, EPI_ISL_1245337, EPI_ISL_1758778, EPI_ISL_1758781, EPI_ISL_1758784, EPI_ISL_1758788, EPI_ISL_1758790, EPI_ISL_1758792, EPI_ISL_1758795, EPI_ISL_1758799, EPI_ISL_1758824, EPI_ISL_1758831, EPI_ISL_1758843, EPI_ISL_1758848, EPI_ISL_1758869, EPI_ISL_1758878, EPI_ISL_1807099, EPI_ISL_1807126

| | | | |
|---|---|---|---|
| see above | Lighthouse Lab in Glasgow | Wellcome Sanger Institute for the COVID-19 Genomics UK (COG-UK) Consortium | Anna Dominiczak and Alex Alderton; Carol Clugston; Cordelia Langford; David Gray; David K. Jackson; Dominic Kwiatkowski; Ewan Harrison; Harper VanSteenhouse; Ian Johnston; Jeffrey Barrett; John Sillitoe on behalf of the Wellcome Sanger Institute COVID-19 Surveillance Team; John Sillitoe on behalf of the Wellcome Sanger Institute COVID-19 Surveillance Team (http://www.sanger.ac.uk/covid-team); Roberto Amato; Sonia Goncalves; Yumi Kasai |

EPI_ISL_969456, EPI_ISL_1005718, EPI_ISL_1005727, EPI_ISL_1005731, EPI_ISL_1005741, EPI_ISL_1005742, EPI_ISL_1005743, EPI_ISL_1005746, EPI_ISL_1005753, EPI_ISL_1005760, EPI_ISL_1005765, EPI_ISL_1005773, EPI_ISL_1005774, EPI_ISL_1005778, EPI_ISL_1005782, EPI_ISL_1005785, EPI_ISL_1005799, EPI_ISL_1005804, EPI_ISL_1005807, EPI_ISL_1005809, EPI_ISL_1005816, EPI_ISL_1005817,
EPI_ISL_1005831, EPI_ISL_1005836, EPI_ISL_1005838, EPI_ISL_1005841, EPI_ISL_1005843, EPI_ISL_1005857, EPI_ISL_1005862, EPI_ISL_1005873, EPI_ISL_1005874, EPI_ISL_1005883, EPI_ISL_1005885, EPI_ISL_1005889, EPI_ISL_1005892, EPI_ISL_1005895, EPI_ISL_1005901, EPI_ISL_1005902, EPI_ISL_1005903, EPI_ISL_1005909, EPI_ISL_1005911, EPI_ISL_1005912, EPI_ISL_1005914, EPI_ISL_1005922,
EPI_ISL_1005927, EPI_ISL_1005935, EPI_ISL_1005939, EPI_ISL_1005940, EPI_ISL_1005951, EPI_ISL_1005952, EPI_ISL_1005957, EPI_ISL_1005958, EPI_ISL_1005962, EPI_ISL_1005964, EPI_ISL_1005967, EPI_ISL_1005971, EPI_ISL_1005974, EPI_ISL_1005986, EPI_ISL_1005993, EPI_ISL_1005997, EPI_ISL_1006001, EPI_ISL_1006004, EPI_ISL_1006008, EPI_ISL_1006013, EPI_ISL_1006014, EPI_ISL_1006016,
EPI_ISL_1006017, EPI_ISL_1006033, EPI_ISL_1006041, EPI_ISL_1006042, EPI_ISL_1006050, EPI_ISL_1006054, EPI_ISL_1006055, EPI_ISL_1006056, EPI_ISL_1006061, EPI_ISL_1006062, EPI_ISL_1006063, EPI_ISL_1006065, EPI_ISL_1006068, EPI_ISL_1006074, EPI_ISL_1006075, EPI_ISL_1006091, EPI_ISL_1006093, EPI_ISL_1006097, EPI_ISL_1006098, EPI_ISL_1006112, EPI_ISL_1006119, EPI_ISL_1006122,
EPI_ISL_1006128, EPI_ISL_1006129, EPI_ISL_1006134, EPI_ISL_1006138, EPI_ISL_1006141, EPI_ISL_1006145, EPI_ISL_1006151, EPI_ISL_1006152, EPI_ISL_1006160, EPI_ISL_1006161, EPI_ISL_1006165, EPI_ISL_1006167, EPI_ISL_1006170, EPI_ISL_1006174, EPI_ISL_1006178, EPI_ISL_1006182, EPI_ISL_1012421, EPI_ISL_1012423, EPI_ISL_1012433, EPI_ISL_1012453, EPI_ISL_1012457, EPI_ISL_1012470,
EPI_ISL_1012475, EPI_ISL_1012500, EPI_ISL_1012532, EPI_ISL_1012535, EPI_ISL_1019409, EPI_ISL_1019410, EPI_ISL_1019411, EPI_ISL_1019412, EPI_ISL_1019413, EPI_ISL_1019414, EPI_ISL_1019415, EPI_ISL_1019416, EPI_ISL_1019417, EPI_ISL_1019418, EPI_ISL_1019419, EPI_ISL_1019420, EPI_ISL_1019421, EPI_ISL_1019422, EPI_ISL_1019423, EPI_ISL_1019424, EPI_ISL_1019425, EPI_ISL_1019426,
EPI_ISL_1019427, EPI_ISL_1019428, EPI_ISL_1019429, EPI_ISL_1019430, EPI_ISL_1019431, EPI_ISL_1019432, EPI_ISL_1019433, EPI_ISL_1019434, EPI_ISL_1019435, EPI_ISL_1019436, EPI_ISL_1019437, EPI_ISL_1019438, EPI_ISL_1019439, EPI_ISL_1019440, EPI_ISL_1019441, EPI_ISL_1019442, EPI_ISL_1019443, EPI_ISL_1019444, EPI_ISL_1019445, EPI_ISL_1019446, EPI_ISL_1019447, EPI_ISL_1019448,
EPI_ISL_1019449, EPI_ISL_1019450, EPI_ISL_1019451, EPI_ISL_1019452, EPI_ISL_1019453, EPI_ISL_1019454, EPI_ISL_1019455, EPI_ISL_1019456, EPI_ISL_1019457, EPI_ISL_1019458, EPI_ISL_1019459, EPI_ISL_1019460, EPI_ISL_1019461, EPI_ISL_1019462, EPI_ISL_1019463, EPI_ISL_1019464, EPI_ISL_1019465, EPI_ISL_1019466, EPI_ISL_1019467, EPI_ISL_1019468, EPI_ISL_1019469, EPI_ISL_1019470,
EPI_ISL_1019471, EPI_ISL_1019472, EPI_ISL_1019473, EPI_ISL_1019474, EPI_ISL_1019475, EPI_ISL_1019476, EPI_ISL_1019477, EPI_ISL_1019478, EPI_ISL_1019479, EPI_ISL_1019480, EPI_ISL_1033567, EPI_ISL_1033568, EPI_ISL_1033569, EPI_ISL_1033570, EPI_ISL_1033571, EPI_ISL_1033572, EPI_ISL_1033573, EPI_ISL_1033575, EPI_ISL_1033576, EPI_ISL_1033577, EPI_ISL_1033578, EPI_ISL_1033580,
EPI_ISL_1033581, EPI_ISL_1033582, EPI_ISL_1033585, EPI_ISL_1033586, EPI_ISL_1033587, EPI_ISL_1033588, EPI_ISL_1033591, EPI_ISL_1033593, EPI_ISL_1033594, EPI_ISL_1033596, EPI_ISL_1033597, EPI_ISL_1033599, EPI_ISL_1033604, EPI_ISL_1033605, EPI_ISL_1033606, EPI_ISL_1033608, EPI_ISL_1033609, EPI_ISL_1033613, EPI_ISL_1033615, EPI_ISL_1033617, EPI_ISL_1033618, EPI_ISL_1033621,
EPI_ISL_1033623, EPI_ISL_1033624, EPI_ISL_1033629, EPI_ISL_1033630, EPI_ISL_1033633, EPI_ISL_1033634, EPI_ISL_1033635, EPI_ISL_1033637, EPI_ISL_1033639, EPI_ISL_1033640, EPI_ISL_1033642, EPI_ISL_1033643, EPI_ISL_1033646, EPI_ISL_1033647, EPI_ISL_1033652, EPI_ISL_1033653, EPI_ISL_1033654, EPI_ISL_1033657, EPI_ISL_1033659, EPI_ISL_1033662, EPI_ISL_1033663, EPI_ISL_1033665,
EPI_ISL_1033666, EPI_ISL_1033667, EPI_ISL_1033672, EPI_ISL_1033674, EPI_ISL_1033675, EPI_ISL_1033677, EPI_ISL_1033678, EPI_ISL_1033681, EPI_ISL_1033682, EPI_ISL_1033684, EPI_ISL_1033685, EPI_ISL_1033687, EPI_ISL_1033689, EPI_ISL_1033690, EPI_ISL_1033693, EPI_ISL_1033694, EPI_ISL_1033695, EPI_ISL_1033696, EPI_ISL_1033697, EPI_ISL_1033700, EPI_ISL_1033702, EPI_ISL_1033705,
EPI_ISL_1033706, EPI_ISL_1033708, EPI_ISL_1033709, EPI_ISL_1033711, EPI_ISL_1033712, EPI_ISL_1033713, EPI_ISL_1033716, EPI_ISL_1033717, EPI_ISL_1033718, EPI_ISL_1033719, EPI_ISL_1033720, EPI_ISL_1033721, EPI_ISL_1033724, EPI_ISL_1033726, EPI_ISL_1033731, EPI_ISL_1033732, EPI_ISL_1033734, EPI_ISL_1033735, EPI_ISL_1033736, EPI_ISL_1033737, EPI_ISL_1033738, EPI_ISL_1033739,
EPI_ISL_1033741, EPI_ISL_1033743, EPI_ISL_1033744, EPI_ISL_1033745, EPI_ISL_1033749, EPI_ISL_1033750, EPI_ISL_1033752, EPI_ISL_1033758, EPI_ISL_1033759, EPI_ISL_1033760, EPI_ISL_1033761, EPI_ISL_1033762, EPI_ISL_1033765, EPI_ISL_1033768, EPI_ISL_1033769, EPI_ISL_1033771, EPI_ISL_1033774, EPI_ISL_1033775, EPI_ISL_1033776, EPI_ISL_1033778, EPI_ISL_1033781, EPI_ISL_1033782,
EPI_ISL_1033783, EPI_ISL_1033784, EPI_ISL_1033785, EPI_ISL_1033786, EPI_ISL_1033788, EPI_ISL_1033789, EPI_ISL_1033790, EPI_ISL_1033792, EPI_ISL_1033796, EPI_ISL_1033800, EPI_ISL_1033801, EPI_ISL_1033804, EPI_ISL_1033805, EPI_ISL_1033808, EPI_ISL_1033809, EPI_ISL_1033810, EPI_ISL_1033811, EPI_ISL_1033812, EPI_ISL_1033815, EPI_ISL_1033816, EPI_ISL_1033817, EPI_ISL_1033818,
EPI_ISL_1033819, EPI_ISL_1033820, EPI_ISL_1033824, EPI_ISL_1033826, EPI_ISL_1033827, EPI_ISL_1033828, EPI_ISL_1033829, EPI_ISL_1033831, EPI_ISL_1033832, EPI_ISL_1033833, EPI_ISL_1033834, EPI_ISL_1033837, EPI_ISL_1033838, EPI_ISL_1033839, EPI_ISL_1033841, EPI_ISL_1033842, EPI_ISL_1033847, EPI_ISL_1033848, EPI_ISL_1033849, EPI_ISL_1033851, EPI_ISL_1033852, EPI_ISL_1033857,
EPI_ISL_1033858, EPI_ISL_1033859, EPI_ISL_1033860, EPI_ISL_1033861, EPI_ISL_1033863, EPI_ISL_1033866, EPI_ISL_1033870, EPI_ISL_1033871, EPI_ISL_1033873, EPI_ISL_1033876, EPI_ISL_1033877, EPI_ISL_1033878, EPI_ISL_1033879, EPI_ISL_1033882, EPI_ISL_1033883, EPI_ISL_1033888, EPI_ISL_1033890, EPI_ISL_1033892, EPI_ISL_1033894, EPI_ISL_1033899, EPI_ISL_1033902, EPI_ISL_1033907,
EPI_ISL_1033908, EPI_ISL_1033910, EPI_ISL_1033911, EPI_ISL_1033912, EPI_ISL_1033913, EPI_ISL_1033915, EPI_ISL_1033918, EPI_ISL_1033919, EPI_ISL_1033920, EPI_ISL_1033921, EPI_ISL_1033922, EPI_ISL_1033923, EPI_ISL_1033924, EPI_ISL_1033925, EPI_ISL_1033926, EPI_ISL_1033927, EPI_ISL_1033928, EPI_ISL_1033929, EPI_ISL_1033930, EPI_ISL_1033931, EPI_ISL_1033932, EPI_ISL_1033933,
EPI_ISL_1033934, EPI_ISL_1033935, EPI_ISL_1033936, EPI_ISL_1033937, EPI_ISL_1033938, EPI_ISL_1033939, EPI_ISL_1033940, EPI_ISL_1033941, EPI_ISL_1033942, EPI_ISL_1033943, EPI_ISL_1033944, EPI_ISL_1033945, EPI_ISL_1033946, EPI_ISL_1033947, EPI_ISL_1033949, EPI_ISL_1033950, EPI_ISL_1033951, EPI_ISL_1033952, EPI_ISL_1033953, EPI_ISL_1033955, EPI_ISL_1033956, EPI_ISL_1033957,
EPI_ISL_1033958, EPI_ISL_1033959, EPI_ISL_1033960, EPI_ISL_1033961, EPI_ISL_1033962, EPI_ISL_1033963, EPI_ISL_1033964, EPI_ISL_1033965, EPI_ISL_1033966, EPI_ISL_1033967, EPI_ISL_1033968, EPI_ISL_1033969, EPI_ISL_1033970, EPI_ISL_1033971, EPI_ISL_1033972, EPI_ISL_1033973, EPI_ISL_1033975, EPI_ISL_1033976, EPI_ISL_1033977, EPI_ISL_1033978, EPI_ISL_1033979, EPI_ISL_1033980,
EPI_ISL_1033981, EPI_ISL_1033982, EPI_ISL_1033983, EPI_ISL_1033984, EPI_ISL_1033985, EPI_ISL_1033986, EPI_ISL_1033987, EPI_ISL_1033988, EPI_ISL_1033989, EPI_ISL_1033990, EPI_ISL_1033991, EPI_ISL_1033992, EPI_ISL_1033993, EPI_ISL_1033994, EPI_ISL_1033995, EPI_ISL_1033996, EPI_ISL_1033997, EPI_ISL_1033998, EPI_ISL_1033999, EPI_ISL_1034000, EPI_ISL_1034001, EPI_ISL_1034002,
EPI_ISL_1034003, EPI_ISL_1034004, EPI_ISL_1034005, EPI_ISL_1034007, EPI_ISL_1034008, EPI_ISL_1034009, EPI_ISL_1034010, EPI_ISL_1034011, EPI_ISL_1034012, EPI_ISL_1034013, EPI_ISL_1034014, EPI_ISL_1034015, EPI_ISL_1034016, EPI_ISL_1034017, EPI_ISL_1034018, EPI_ISL_1034020, EPI_ISL_1034022, EPI_ISL_1034023, EPI_ISL_1034024, EPI_ISL_1034025, EPI_ISL_1034026, EPI_ISL_1034027,
EPI_ISL_1034028, EPI_ISL_1034029, EPI_ISL_1034031, EPI_ISL_1034032, EPI_ISL_1034033, EPI_ISL_1034034, EPI_ISL_1034035, EPI_ISL_1034036, EPI_ISL_1034037, EPI_ISL_1034038, EPI_ISL_1034039, EPI_ISL_1034040, EPI_ISL_1034041, EPI_ISL_1034042, EPI_ISL_1034043, EPI_ISL_1034046, EPI_ISL_1034047, EPI_ISL_1034048, EPI_ISL_1034049, EPI_ISL_1034050, EPI_ISL_1034051, EPI_ISL_1034052,
EPI_ISL_1034053, EPI_ISL_1034054, EPI_ISL_1034055, EPI_ISL_1034057, EPI_ISL_1034058, EPI_ISL_1034059, EPI_ISL_1034060, EPI_ISL_1034061, EPI_ISL_1034062, EPI_ISL_1034063, EPI_ISL_1034064, EPI_ISL_1034065, EPI_ISL_1034066, EPI_ISL_1034068, EPI_ISL_1034069, EPI_ISL_1034070, EPI_ISL_1034071, EPI_ISL_1034072, EPI_ISL_1034073, EPI_ISL_1034074, EPI_ISL_1034075, EPI_ISL_1034076,
EPI_ISL_1034077, EPI_ISL_1034079, EPI_ISL_1034081, EPI_ISL_1034083, EPI_ISL_1034084, EPI_ISL_1034085, EPI_ISL_1034086, EPI_ISL_1034087, EPI_ISL_1034088, EPI_ISL_1034089, EPI_ISL_1034091, EPI_ISL_1034092, EPI_ISL_1034093, EPI_ISL_1034094, EPI_ISL_1034095, EPI_ISL_1034096, EPI_ISL_1034098, EPI_ISL_1034100, EPI_ISL_1034101, EPI_ISL_1034102, EPI_ISL_1034103, EPI_ISL_1034104,
EPI_ISL_1034105, EPI_ISL_1034106, EPI_ISL_1034107, EPI_ISL_1034108, EPI_ISL_1034109, EPI_ISL_1034110, EPI_ISL_1034113, EPI_ISL_1034114, EPI_ISL_1034116, EPI_ISL_1034117, EPI_ISL_1034118, EPI_ISL_1034119, EPI_ISL_1034120, EPI_ISL_1034121, EPI_ISL_1034122, EPI_ISL_1034123, EPI_ISL_1034124, EPI_ISL_1034125, EPI_ISL_1034126, EPI_ISL_1034127, EPI_ISL_1034128, EPI_ISL_1034129,
EPI_ISL_1034130, EPI_ISL_1034131, EPI_ISL_1034132, EPI_ISL_1034133, EPI_ISL_1034135, EPI_ISL_1034136, EPI_ISL_1034137, EPI_ISL_1034138, EPI_ISL_1034139, EPI_ISL_1034140, EPI_ISL_1042568, EPI_ISL_1042569, EPI_ISL_1042573, EPI_ISL_1042577, EPI_ISL_1042578, EPI_ISL_1042579, EPI_ISL_1042582, EPI_ISL_1042585, EPI_ISL_1042588, EPI_ISL_1042589, EPI_ISL_1042590, EPI_ISL_1042593,
EPI_ISL_1042594, EPI_ISL_1042602, EPI_ISL_1042605, EPI_ISL_1042607, EPI_ISL_1042608, EPI_ISL_1042610, EPI_ISL_1042618, EPI_ISL_1042622, EPI_ISL_1042625, EPI_ISL_1042628, EPI_ISL_1042629, EPI_ISL_1042634, EPI_ISL_1042635, EPI_ISL_1042636, EPI_ISL_1042638, EPI_ISL_1042639, EPI_ISL_1042641, EPI_ISL_1042642, EPI_ISL_1042646, EPI_ISL_1042648, EPI_ISL_1042651,
EPI_ISL_1042652, EPI_ISL_1042653, EPI_ISL_1042655, EPI_ISL_1042656, EPI_ISL_1042657, EPI_ISL_1042660, EPI_ISL_1042662, EPI_ISL_1042664, EPI_ISL_1042665, EPI_ISL_1042667, EPI_ISL_1042668, EPI_ISL_1042670, EPI_ISL_1042671, EPI_ISL_1042675, EPI_ISL_1042678, EPI_ISL_1042679, EPI_ISL_1042683, EPI_ISL_1042685, EPI_ISL_1042688, EPI_ISL_1042689, EPI_ISL_1042692, EPI_ISL_1042693,
EPI_ISL_1042694, EPI_ISL_1042695, EPI_ISL_1042699, EPI_ISL_1042700, EPI_ISL_1042701, EPI_ISL_1042702, EPI_ISL_1042704, EPI_ISL_1042705, EPI_ISL_1042707, EPI_ISL_1042708, EPI_ISL_1042709, EPI_ISL_1042710, EPI_ISL_1042711, EPI_ISL_1042714, EPI_ISL_1042715, EPI_ISL_1042724, EPI_ISL_1042725, EPI_ISL_1042733, EPI_ISL_1042735, EPI_ISL_1042739, EPI_ISL_1042740, EPI_ISL_1042743,
EPI_ISL_1042748, EPI_ISL_1042749, EPI_ISL_1042750, EPI_ISL_1042754, EPI_ISL_1042755, EPI_ISL_1042757, EPI_ISL_1042758, EPI_ISL_1042762, EPI_ISL_1042767, EPI_ISL_1042768, EPI_ISL_1042770, EPI_ISL_1042771, EPI_ISL_1042773, EPI_ISL_1042775, EPI_ISL_1042778, EPI_ISL_1042787, EPI_ISL_1042790, EPI_ISL_1042791, EPI_ISL_1042792, EPI_ISL_1042794, EPI_ISL_1042795, EPI_ISL_1042797,
EPI_ISL_1042798, EPI_ISL_1042802, EPI_ISL_1042805, EPI_ISL_1042806, EPI_ISL_1042808, EPI_ISL_1042811, EPI_ISL_1042814, EPI_ISL_1042816, EPI_ISL_1042820, EPI_ISL_1042822, EPI_ISL_1042823, EPI_ISL_1042827, EPI_ISL_1042829, EPI_ISL_1042831, EPI_ISL_1042832, EPI_ISL_1043165, EPI_ISL_1043166, EPI_ISL_1043167, EPI_ISL_1043168, EPI_ISL_1043169, EPI_ISL_1043170, EPI_ISL_1043171,
EPI_ISL_1043172, EPI_ISL_1043173, EPI_ISL_1043174, EPI_ISL_1043175, EPI_ISL_1043176, EPI_ISL_1043179, EPI_ISL_1043180, EPI_ISL_1043181, EPI_ISL_1043182, EPI_ISL_1043183, EPI_ISL_1043184, EPI_ISL_1043185, EPI_ISL_1043186, EPI_ISL_1043188, EPI_ISL_1043189, EPI_ISL_1043190, EPI_ISL_1043192, EPI_ISL_1043193, EPI_ISL_1043194, EPI_ISL_1043196, EPI_ISL_1043197, EPI_ISL_1043199,
EPI_ISL_1043201, EPI_ISL_1043203, EPI_ISL_1043204, EPI_ISL_1043205, EPI_ISL_1043206, EPI_ISL_1043208, EPI_ISL_1043209, EPI_ISL_1043210, EPI_ISL_1043211, EPI_ISL_1043213, EPI_ISL_1043214, EPI_ISL_1043215, EPI_ISL_1043216, EPI_ISL_1043217, EPI_ISL_1043218, EPI_ISL_1043219, EPI_ISL_1043221, EPI_ISL_1043222, EPI_ISL_1043223, EPI_ISL_1043224, EPI_ISL_1043225, EPI_ISL_1043226,
EPI_ISL_1043228, EPI_ISL_1043229, EPI_ISL_1043230, EPI_ISL_1043231, EPI_ISL_1043232, EPI_ISL_1043233, EPI_ISL_1043234, EPI_ISL_1043235, EPI_ISL_1043236, EPI_ISL_1043237, EPI_ISL_1043238, EPI_ISL_1043240, EPI_ISL_1043241, EPI_ISL_1043242, EPI_ISL_1043243, EPI_ISL_1043244, EPI_ISL_1043246, EPI_ISL_1043247, EPI_ISL_1043248, EPI_ISL_1043249, EPI_ISL_1043250, EPI_ISL_1043251,
EPI_ISL_1043252, EPI_ISL_1043253, EPI_ISL_1043254, EPI_ISL_1043255, EPI_ISL_1043256, EPI_ISL_1043257, EPI_ISL_1043258, EPI_ISL_1043259, EPI_ISL_1043260, EPI_ISL_1043261, EPI_ISL_1043262, EPI_ISL_1043263, EPI_ISL_1043264, EPI_ISL_1043265, EPI_ISL_1043267, EPI_ISL_1043269, EPI_ISL_1043270, EPI_ISL_1043271, EPI_ISL_1043272, EPI_ISL_1043273, EPI_ISL_1043274, EPI_ISL_1043275,
EPI_ISL_1043276, EPI_ISL_1043278, EPI_ISL_1043279, EPI_ISL_1043281, EPI_ISL_1043283, EPI_ISL_1043284, EPI_ISL_1043285, EPI_ISL_1043286, EPI_ISL_1043287, EPI_ISL_1043288, EPI_ISL_1043289, EPI_ISL_1043290, EPI_ISL_1043292, EPI_ISL_1043293, EPI_ISL_1043294, EPI_ISL_1043295, EPI_ISL_1043296, EPI_ISL_1043297, EPI_ISL_1043298, EPI_ISL_1043299, EPI_ISL_1043300, EPI_ISL_1043302,
EPI_ISL_1043303, EPI_ISL_1043304, EPI_ISL_1043305, EPI_ISL_1043306, EPI_ISL_1043309, EPI_ISL_1043311, EPI_ISL_1043312, EPI_ISL_1043313, EPI_ISL_1043314, EPI_ISL_1043316, EPI_ISL_1043318, EPI_ISL_1043320, EPI_ISL_1043321, EPI_ISL_1043322, EPI_ISL_1043323, EPI_ISL_1043324, EPI_ISL_1043325, EPI_ISL_1043326, EPI_ISL_1043328, EPI_ISL_1043330, EPI_ISL_1043331, EPI_ISL_1043332,
EPI_ISL_1043333, EPI_ISL_1043334, EPI_ISL_1043335, EPI_ISL_1043336, EPI_ISL_1043337, EPI_ISL_1043338, EPI_ISL_1043339, EPI_ISL_1043340, EPI_ISL_1043341, EPI_ISL_1043343, EPI_ISL_1043348, EPI_ISL_1043353, EPI_ISL_1043380, EPI_ISL_1043400, EPI_ISL_1043407, EPI_ISL_1043413, EPI_ISL_1043416, EPI_ISL_1043417, EPI_ISL_1043427, EPI_ISL_1043459, EPI_ISL_1043460, EPI_ISL_1043464,
EPI_ISL_1043483, EPI_ISL_1043485, EPI_ISL_1043494, EPI_ISL_1043495, EPI_ISL_1043515, EPI_ISL_1043526, EPI_ISL_1043527, EPI_ISL_1043536, EPI_ISL_1045198, EPI_ISL_1045200, EPI_ISL_1045202, EPI_ISL_1045203, EPI_ISL_1045204, EPI_ISL_1045205, EPI_ISL_1045206, EPI_ISL_1045208, EPI_ISL_1045209, EPI_ISL_1045211, EPI_ISL_1045212, EPI_ISL_1045214, EPI_ISL_1045215, EPI_ISL_1045216,
EPI_ISL_1045217, EPI_ISL_1045219, EPI_ISL_1045220, EPI_ISL_1045222, EPI_ISL_1045223, EPI_ISL_1045224, EPI_ISL_1045227, EPI_ISL_1045228, EPI_ISL_1045230, EPI_ISL_1045232, EPI_ISL_1045233, EPI_ISL_1045234, EPI_ISL_1045235, EPI_ISL_1045236, EPI_ISL_1045238, EPI_ISL_1045239, EPI_ISL_1045241, EPI_ISL_1045244, EPI_ISL_1045245, EPI_ISL_1045246, EPI_ISL_1045248, EPI_ISL_1045249,
EPI_ISL_1045250, EPI_ISL_1045252, EPI_ISL_1045255, EPI_ISL_1045257, EPI_ISL_1045258, EPI_ISL_1045260, EPI_ISL_1045262, EPI_ISL_1045263, EPI_ISL_1045264, EPI_ISL_1045265, EPI_ISL_1045266, EPI_ISL_1045267, EPI_ISL_1045269, EPI_ISL_1045270, EPI_ISL_1045271, EPI_ISL_1045272, EPI_ISL_1045273, EPI_ISL_1045275, EPI_ISL_1045278, EPI_ISL_1045279, EPI_ISL_1045281, EPI_ISL_1045282,
EPI_ISL_1045283, EPI_ISL_1045284, EPI_ISL_1045285, EPI_ISL_1045288, EPI_ISL_1045291, EPI_ISL_1045292, EPI_ISL_1045294, EPI_ISL_1045295, EPI_ISL_1045296, EPI_ISL_1045297, EPI_ISL_1045301, EPI_ISL_1045302, EPI_ISL_1045303, EPI_ISL_1045304, EPI_ISL_1045307, EPI_ISL_1045308, EPI_ISL_1045309, EPI_ISL_1045310, EPI_ISL_1045311, EPI_ISL_1045312, EPI_ISL_1045313, EPI_ISL_1045314,
EPI_ISL_1045315, EPI_ISL_1045316, EPI_ISL_1045317, EPI_ISL_1045318, EPI_ISL_1045320, EPI_ISL_1045321, EPI_ISL_1045322, EPI_ISL_1045323, EPI_ISL_1045324, EPI_ISL_1045325, EPI_ISL_1045326, EPI_ISL_1045327, EPI_ISL_1045328, EPI_ISL_1045330, EPI_ISL_1045331, EPI_ISL_1045332, EPI_ISL_1045337, EPI_ISL_1045338, EPI_ISL_1045340, EPI_ISL_1045341, EPI_ISL_1045342, EPI_ISL_1045343,
EPI_ISL_1045344, EPI_ISL_1045345, EPI_ISL_1045346, EPI_ISL_1045347, EPI_ISL_1045352, EPI_ISL_1045354, EPI_ISL_1045356, EPI_ISL_1045357, EPI_ISL_1045358, EPI_ISL_1045359, EPI_ISL_1045360, EPI_ISL_1045362, EPI_ISL_1045363, EPI_ISL_1045368, EPI_ISL_1045369, EPI_ISL_1045370, EPI_ISL_1045373, EPI_ISL_1045375, EPI_ISL_1045376, EPI_ISL_1045377, EPI_ISL_1045378, EPI_ISL_1045380,
EPI_ISL_1045383, EPI_ISL_1045385, EPI_ISL_1045387, EPI_ISL_1045388, EPI_ISL_1045390, EPI_ISL_1045391, EPI_ISL_1045392, EPI_ISL_1045394, EPI_ISL_1045395, EPI_ISL_1045396, EPI_ISL_1045399, EPI_ISL_1045401, EPI_ISL_1045402, EPI_ISL_1045403, EPI_ISL_1045404, EPI_ISL_1045408, EPI_ISL_1045409, EPI_ISL_1045410, EPI_ISL_1045411, EPI_ISL_1045413, EPI_ISL_1045414, EPI_ISL_1045415,
EPI_ISL_1045416, EPI_ISL_1045417, EPI_ISL_1045419, EPI_ISL_1045421, EPI_ISL_1045423, EPI_ISL_1045424, EPI_ISL_1045425, EPI_ISL_1045426, EPI_ISL_1045427, EPI_ISL_1045428, EPI_ISL_1045429, EPI_ISL_1045430, EPI_ISL_1045431, EPI_ISL_1045432, EPI_ISL_1045433, EPI_ISL_1045434, EPI_ISL_1045436, EPI_ISL_1045437, EPI_ISL_1045438, EPI_ISL_1045440, EPI_ISL_1045441, EPI_ISL_1045442,
EPI_ISL_1045445, EPI_ISL_1045448, EPI_ISL_1045449, EPI_ISL_1045450, EPI_ISL_1045453, EPI_ISL_1045454, EPI_ISL_1045455, EPI_ISL_1045456, EPI_ISL_1045457, EPI_ISL_1045460, EPI_ISL_1045461, EPI_ISL_1045463, EPI_ISL_1045464, EPI_ISL_1045465, EPI_ISL_1045466, EPI_ISL_1045467, EPI_ISL_1045468, EPI_ISL_1045469, EPI_ISL_1045470, EPI_ISL_1045472, EPI_ISL_1045474, EPI_ISL_1045475,
EPI_ISL_1045476, EPI_ISL_1045477, EPI_ISL_1045479, EPI_ISL_1045480, EPI_ISL_1045481, EPI_ISL_1045482, EPI_ISL_1045484, EPI_ISL_1045486, EPI_ISL_1045487, EPI_ISL_1045488, EPI_ISL_1045489, EPI_ISL_1045490, EPI_ISL_1045494, EPI_ISL_1045497, EPI_ISL_1045500, EPI_ISL_1045502, EPI_ISL_1045503, EPI_ISL_1045505, EPI_ISL_1045508, EPI_ISL_1045509, EPI_ISL_1045511, EPI_ISL_1045513,
EPI_ISL_1045514, EPI_ISL_1045515, EPI_ISL_1045516, EPI_ISL_1045517, EPI_ISL_1045519, EPI_ISL_1045521, EPI_ISL_1045522, EPI_ISL_1045523, EPI_ISL_1045524, EPI_ISL_1045525, EPI_ISL_1045526, EPI_ISL_1045527, EPI_ISL_1045530, EPI_ISL_1045531, EPI_ISL_1045532, EPI_ISL_1045534, EPI_ISL_1045537, EPI_ISL_1045538, EPI_ISL_1045540, EPI_ISL_1045541, EPI_ISL_1045542, EPI_ISL_1045543,
EPI_ISL_1045544, EPI_ISL_1045545, EPI_ISL_1045548, EPI_ISL_1045549, EPI_ISL_1045551, EPI_ISL_1045552, EPI_ISL_1045553, EPI_ISL_1045558, EPI_ISL_1045559, EPI_ISL_1045560, EPI_ISL_1045562, EPI_ISL_1045563, EPI_ISL_1045564, EPI_ISL_1045565, EPI_ISL_1045566, EPI_ISL_1045567, EPI_ISL_1045568, EPI_ISL_1045569, EPI_ISL_1045570, EPI_ISL_1045571, EPI_ISL_1045572, EPI_ISL_1045573,
EPI_ISL_1045576, EPI_ISL_1045578, EPI_ISL_1045579, EPI_ISL_1045580, EPI_ISL_1045581, EPI_ISL_1045582, EPI_ISL_1045583, EPI_ISL_1045584, EPI_ISL_1045585, EPI_ISL_1045588, EPI_ISL_1045590, EPI_ISL_1045591, EPI_ISL_1045592, EPI_ISL_1045593, EPI_ISL_1045594, EPI_ISL_1045595, EPI_ISL_1045598, EPI_ISL_1045600, EPI_ISL_1045601, EPI_ISL_1045603, EPI_ISL_1045604, EPI_ISL_1045606,
EPI_ISL_1045608, EPI_ISL_1045609, EPI_ISL_1045610, EPI_ISL_1045611, EPI_ISL_1045614, EPI_ISL_1045615, EPI_ISL_1045617, EPI_ISL_1045620, EPI_ISL_1045621, EPI_ISL_1045622, EPI_ISL_1045624, EPI_ISL_1045625, EPI_ISL_1045627, EPI_ISL_1045628, EPI_ISL_1045629, EPI_ISL_1045630, EPI_ISL_1045634, EPI_ISL_1045635, EPI_ISL_1045636, EPI_ISL_1045637, EPI_ISL_1045639, EPI_ISL_1045640,
EPI_ISL_1045641, EPI_ISL_1045644, EPI_ISL_1045646, EPI_ISL_1045647, EPI_ISL_1045648, EPI_ISL_1045649, EPI_ISL_1045650, EPI_ISL_1045651, EPI_ISL_1045652, EPI_ISL_1045653, EPI_ISL_1045654, EPI_ISL_1045657, EPI_ISL_1045658, EPI_ISL_1045660, EPI_ISL_1045663, EPI_ISL_1045665, EPI_ISL_1045666, EPI_ISL_1045669, EPI_ISL_1045670, EPI_ISL_1045672, EPI_ISL_1045674, EPI_ISL_1045675,
EPI_ISL_1045676, EPI_ISL_1045677, EPI_ISL_1045678, EPI_ISL_1045679, EPI_ISL_1045680, EPI_ISL_1045681, EPI_ISL_1045705, EPI_ISL_1045707, EPI_ISL_1045711, EPI_ISL_1045713, EPI_ISL_1045714, EPI_ISL_1045718, EPI_ISL_1045731, EPI_ISL_1045734, EPI_ISL_1045738, EPI_ISL_1045740, EPI_ISL_1045742, EPI_ISL_1045743, EPI_ISL_1045745, EPI_ISL_1045748, EPI_ISL_1045750, EPI_ISL_1045753,
EPI_ISL_1045756, EPI_ISL_1045763, EPI_ISL_1045767, EPI_ISL_1045775, EPI_ISL_1045779, EPI_ISL_1045781, EPI_ISL_1045782, EPI_ISL_1045783, EPI_ISL_1045784, EPI_ISL_1045786, EPI_ISL_1045789, EPI_ISL_1045791, EPI_ISL_1045794, EPI_ISL_1045796, EPI_ISL_1045799, EPI_ISL_1045800, EPI_ISL_1045801, EPI_ISL_1045803, EPI_ISL_1045805, EPI_ISL_1045810, EPI_ISL_1045817, EPI_ISL_1045825,
EPI_ISL_1045826, EPI_ISL_1045827, EPI_ISL_1045829, EPI_ISL_1045836, EPI_ISL_1045838, EPI_ISL_1045841, EPI_ISL_1045855, EPI_ISL_1045857, EPI_ISL_1045859, EPI_ISL_1045862, EPI_ISL_1045865, EPI_ISL_1045866, EPI_ISL_1045869, EPI_ISL_1045873, EPI_ISL_1045874, EPI_ISL_1045879, EPI_ISL_1045883, EPI_ISL_1045885, EPI_ISL_1045886, EPI_ISL_1045889, EPI_ISL_1045895, EPI_ISL_1045900,
EPI_ISL_1045902, EPI_ISL_1045911, EPI_ISL_1045913, EPI_ISL_1045921, EPI_ISL_1045923, EPI_ISL_1045925, EPI_ISL_1045934, EPI_ISL_1045936, EPI_ISL_1045938, EPI_ISL_1045940, EPI_ISL_1045942, EPI_ISL_1045944, EPI_ISL_1045952, EPI_ISL_1045954, EPI_ISL_1045957, EPI_ISL_1045959, EPI_ISL_1045960, EPI_ISL_1045961, EPI_ISL_1045968, EPI_ISL_1045969, EPI_ISL_1057430, EPI_ISL_1057431,
EPI_ISL_1057432, EPI_ISL_1057434, EPI_ISL_1057436, EPI_ISL_1057437, EPI_ISL_1057438, EPI_ISL_1057440, EPI_ISL_1057442, EPI_ISL_1057444, EPI_ISL_1057445, EPI_ISL_1057446, EPI_ISL_1057447, EPI_ISL_1057448, EPI_ISL_1057449, EPI_ISL_1057450, EPI_ISL_1057451, EPI_ISL_1057452, EPI_ISL_1057453, EPI_ISL_1057456, EPI_ISL_1057457, EPI_ISL_1057458, EPI_ISL_1057459, EPI_ISL_1057461,
EPI_ISL_1057463, EPI_ISL_1057465, EPI_ISL_1057466, EPI_ISL_1057468, EPI_ISL_1057469, EPI_ISL_1057470, EPI_ISL_1057471, EPI_ISL_1057473, EPI_ISL_1057474, EPI_ISL_1057475, EPI_ISL_1057476, EPI_ISL_1057477, EPI_ISL_1057478, EPI_ISL_1057479, EPI_ISL_1057480, EPI_ISL_1057481, EPI_ISL_1057482, EPI_ISL_1057483, EPI_ISL_1057484, EPI_ISL_1057485, EPI_ISL_1057488, EPI_ISL_1057489,
EPI_ISL_1057491, EPI_ISL_1057492, EPI_ISL_1057494, EPI_ISL_1057495, EPI_ISL_1057497, EPI_ISL_1057498, EPI_ISL_1057499, EPI_ISL_1057500, EPI_ISL_1057501, EPI_ISL_1057503, EPI_ISL_1057504, EPI_ISL_1057506, EPI_ISL_1057507, EPI_ISL_1057508, EPI_ISL_1057511, EPI_ISL_1057512, EPI_ISL_1057513, EPI_ISL_1057515, EPI_ISL_1057516, EPI_ISL_1057517, EPI_ISL_1057519, EPI_ISL_1057521,
EPI_ISL_1057523, EPI_ISL_1057524, EPI_ISL_1057526, EPI_ISL_1057527, EPI_ISL_1057528, EPI_ISL_1057529, EPI_ISL_1057530, EPI_ISL_1057531, EPI_ISL_1057532, EPI_ISL_1057533, EPI_ISL_1057534, EPI_ISL_1057535, EPI_ISL_1057537, EPI_ISL_1057538, EPI_ISL_1057540, EPI_ISL_1057541, EPI_ISL_1057542, EPI_ISL_1057543, EPI_ISL_1057544, EPI_ISL_1057545, EPI_ISL_1057550, EPI_ISL_1057551,
EPI_ISL_1057552, EPI_ISL_1057553, EPI_ISL_1057554, EPI_ISL_1057556, EPI_ISL_1057558, EPI_ISL_1057560, EPI_ISL_1057562, EPI_ISL_1057563, EPI_ISL_1057564, EPI_ISL_1057566, EPI_ISL_1057567, EPI_ISL_1057568, EPI_ISL_1057569, EPI_ISL_1057570, EPI_ISL_1057571, EPI_ISL_1057572, EPI_ISL_1057573, EPI_ISL_1057574, EPI_ISL_1057576, EPI_ISL_1057577, EPI_ISL_1057578, EPI_ISL_1057580,
EPI_ISL_1057581, EPI_ISL_1057583, EPI_ISL_1057584, EPI_ISL_1057586, EPI_ISL_1057587, EPI_ISL_1057588, EPI_ISL_1057590, EPI_ISL_1057591, EPI_ISL_1057593, EPI_ISL_1057594, EPI_ISL_1057595, EPI_ISL_1057596, EPI_ISL_1057597, EPI_ISL_1057598, EPI_ISL_1057599, EPI_ISL_1057600, EPI_ISL_1057601, EPI_ISL_1057603, EPI_ISL_1057604, EPI_ISL_1057606, EPI_ISL_1057608, EPI_ISL_1057609,
EPI_ISL_1057610, EPI_ISL_1057611, EPI_ISL_1057612, EPI_ISL_1057614, EPI_ISL_1057615, EPI_ISL_1057616, EPI_ISL_1057617, EPI_ISL_1057619, EPI_ISL_1057620, EPI_ISL_1057621, EPI_ISL_1057622, EPI_ISL_1057625, EPI_ISL_1057626, EPI_ISL_1057627, EPI_ISL_1057628, EPI_ISL_1057630, EPI_ISL_1057631, EPI_ISL_1057632, EPI_ISL_1057633, EPI_ISL_1057634, EPI_ISL_1057635, EPI_ISL_1057636,
EPI_ISL_1057637, EPI_ISL_1057638, EPI_ISL_1057639, EPI_ISL_1057640, EPI_ISL_1057641, EPI_ISL_1057642, EPI_ISL_1057643, EPI_ISL_1057644, EPI_ISL_1057645, EPI_ISL_1057647, EPI_ISL_1057648, EPI_ISL_1057651, EPI_ISL_1057652, EPI_ISL_1057653, EPI_ISL_1057655, EPI_ISL_1057656, EPI_ISL_1057657, EPI_ISL_1057658, EPI_ISL_1057659, EPI_ISL_1057660, EPI_ISL_1057662, EPI_ISL_1057668,
EPI_ISL_1057675, EPI_ISL_1057676, EPI_ISL_1057877, EPI_ISL_1057878, EPI_ISL_1057880, EPI_ISL_1057882, EPI_ISL_1057883, EPI_ISL_1057884, EPI_ISL_1057885, EPI_ISL_1057886, EPI_ISL_1057887, EPI_ISL_1057889, EPI_ISL_1057890, EPI_ISL_1057891, EPI_ISL_1057892, EPI_ISL_1057894, EPI_ISL_1057895, EPI_ISL_1057896, EPI_ISL_1057897, EPI_ISL_1057898, EPI_ISL_1057900, EPI_ISL_1057901,
EPI_ISL_1057903, EPI_ISL_1057905, EPI_ISL_1057906, EPI_ISL_1057908, EPI_ISL_1057909, EPI_ISL_1057910, EPI_ISL_1057911, EPI_ISL_1057912, EPI_ISL_1057913, EPI_ISL_1057915, EPI_ISL_1057916, EPI_ISL_1057919, EPI_ISL_1057920, EPI_ISL_1057921, EPI_ISL_1057923, EPI_ISL_1057925, EPI_ISL_1057926, EPI_ISL_1057927, EPI_ISL_1057928, EPI_ISL_1057929, EPI_ISL_1057930, EPI_ISL_1057933,
EPI_ISL_1057935, EPI_ISL_1057937, EPI_ISL_1057938, EPI_ISL_1057940, EPI_ISL_1057941, EPI_ISL_1057943, EPI_ISL_1057947, EPI_ISL_1057948, EPI_ISL_1057950, EPI_ISL_1057952, EPI_ISL_1057953, EPI_ISL_1057954, EPI_ISL_1057955, EPI_ISL_1057959, EPI_ISL_1057962, EPI_ISL_1057963, EPI_ISL_1057964, EPI_ISL_1057967, EPI_ISL_1057968, EPI_ISL_1057969, EPI_ISL_1057970, EPI_ISL_1057971,
EPI_ISL_1057973, EPI_ISL_1057975, EPI_ISL_1063046, EPI_ISL_1070343, EPI_ISL_1070346, EPI_ISL_1070351, EPI_ISL_1070358, EPI_ISL_1070367, EPI_ISL_1070374, EPI_ISL_1070381, EPI_ISL_1070383, EPI_ISL_1070384, EPI_ISL_1070392, EPI_ISL_1070402, EPI_ISL_1070411, EPI_ISL_1070412, EPI_ISL_1070415, EPI_ISL_1070419, EPI_ISL_1070431, EPI_ISL_1070434, EPI_ISL_1070442, EPI_ISL_1070444,
EPI_ISL_1070448, EPI_ISL_1070449, EPI_ISL_1070450, EPI_ISL_1070458, EPI_ISL_1070463, EPI_ISL_1070465, EPI_ISL_1070469, EPI_ISL_1070475, EPI_ISL_1070476, EPI_ISL_1070481, EPI_ISL_1070482, EPI_ISL_1070491, EPI_ISL_1070497, EPI_ISL_1070514, EPI_ISL_1070530, EPI_ISL_1070543, EPI_ISL_1070546, EPI_ISL_1070548, EPI_ISL_1070550, EPI_ISL_1070551, EPI_ISL_1070556, EPI_ISL_1070562,
EPI_ISL_1070566, EPI_ISL_1070568, EPI_ISL_1070575, EPI_ISL_1070584, EPI_ISL_1070590, EPI_ISL_1070607, EPI_ISL_1070608, EPI_ISL_1070611, EPI_ISL_1070615, EPI_ISL_1070727, EPI_ISL_1070748, EPI_ISL_1070780, EPI_ISL_1070870, EPI_ISL_1070894, EPI_ISL_1070910, EPI_ISL_1070940, EPI_ISL_1070991, EPI_ISL_1071034, EPI_ISL_1071039, EPI_ISL_1071056, EPI_ISL_1071075, EPI_ISL_1071081,
EPI_ISL_1071115, EPI_ISL_1071130, EPI_ISL_1071154, EPI_ISL_1071180, EPI_ISL_1071256, EPI_ISL_1071281, EPI_ISL_1071294, EPI_ISL_1071302, EPI_ISL_1071306, EPI_ISL_1071322, EPI_ISL_1071337, EPI_ISL_1071350, EPI_ISL_1072394, EPI_ISL_1072461, EPI_ISL_1099549, EPI_ISL_1099579, EPI_ISL_1099677, EPI_ISL_1099690, EPI_ISL_1099703, EPI_ISL_1099726, EPI_ISL_1334131, EPI_ISL_1334132,
EPI_ISL_1334133, EPI_ISL_1334134, EPI_ISL_1334135, EPI_ISL_1334136, EPI_ISL_1334137, EPI_ISL_1334138, EPI_ISL_1334139, EPI_ISL_1334140, EPI_ISL_1334141, EPI_ISL_1334142, EPI_ISL_1334143, EPI_ISL_1334144, EPI_ISL_1334145, EPI_ISL_1334146, EPI_ISL_1334147, EPI_ISL_1334148, EPI_ISL_1334149, EPI_ISL_1334150, EPI_ISL_1334151, EPI_ISL_1334152, EPI_ISL_1334153, EPI_ISL_1334154,
EPI_ISL_1334155, EPI_ISL_1334156, EPI_ISL_1334157, EPI_ISL_1334158, EPI_ISL_1334159, EPI_ISL_1334160, EPI_ISL_1334161, EPI_ISL_1334162, EPI_ISL_1334163, EPI_ISL_1334164, EPI_ISL_1334165, EPI_ISL_1334166, EPI_ISL_1334167, EPI_ISL_1334168, EPI_ISL_1334169, EPI_ISL_1334170, EPI_ISL_1334171, EPI_ISL_1334172, EPI_ISL_1334173, EPI_ISL_1334174, EPI_ISL_1334175, EPI_ISL_1334176,
EPI_ISL_1334177, EPI_ISL_1334178, EPI_ISL_1334179, EPI_ISL_1334180, EPI_ISL_1334181, EPI_ISL_1334182, EPI_ISL_1334183, EPI_ISL_1334184, EPI_ISL_1334185, EPI_ISL_1334186, EPI_ISL_1334187, EPI_ISL_1334188, EPI_ISL_1334189, EPI_ISL_1334190, EPI_ISL_1334191, EPI_ISL_1334192, EPI_ISL_1334193, EPI_ISL_1334194, EPI_ISL_1334195, EPI_ISL_1334196, EPI_ISL_1334197, EPI_ISL_1334198,
EPI_ISL_1334199, EPI_ISL_1334200, EPI_ISL_1334201, EPI_ISL_1334202, EPI_ISL_1334203, EPI_ISL_1334204, EPI_ISL_1334205, EPI_ISL_1334206, EPI_ISL_1334207, EPI_ISL_1334208, EPI_ISL_1334209, EPI_ISL_1334210, EPI_ISL_1334211, EPI_ISL_1334212, EPI_ISL_1334213, EPI_ISL_1334214, EPI_ISL_1334215, EPI_ISL_1334216, EPI_ISL_1334217, EPI_ISL_1334218, EPI_ISL_1334219, EPI_ISL_1334220,
EPI_ISL_1334221, EPI_ISL_1334222, EPI_ISL_1334223, EPI_ISL_1334224, EPI_ISL_1334225, EPI_ISL_1334226, EPI_ISL_1334227, EPI_ISL_1334228, EPI_ISL_1334229, EPI_ISL_1334230, EPI_ISL_1334231, EPI_ISL_1334232, EPI_ISL_1334233, EPI_ISL_1334234, EPI_ISL_1334235, EPI_ISL_1334236, EPI_ISL_1334237, EPI_ISL_1334238, EPI_ISL_1334239, EPI_ISL_1334240, EPI_ISL_1334241, EPI_ISL_1334242,
EPI_ISL_1334243, EPI_ISL_1334244, EPI_ISL_1334245, EPI_ISL_1334246, EPI_ISL_1334247, EPI_ISL_1334248, EPI_ISL_1334249, EPI_ISL_1334250, EPI_ISL_1334251, EPI_ISL_1334252, EPI_ISL_1334253, EPI_ISL_1334254, EPI_ISL_1334255, EPI_ISL_1334256, EPI_ISL_1334257, EPI_ISL_1334258, EPI_ISL_1334259, EPI_ISL_1334260, EPI_ISL_1334261, EPI_ISL_1334262, EPI_ISL_1334263, EPI_ISL_1334264,
EPI_ISL_1334265, EPI_ISL_1334266, EPI_ISL_1334267, EPI_ISL_1334268, EPI_ISL_1334269, EPI_ISL_1334270, EPI_ISL_1334271, EPI_ISL_1334272, EPI_ISL_1334273, EPI_ISL_1334274, EPI_ISL_1334275, EPI_ISL_1334276, EPI_ISL_1334277, EPI_ISL_1334278, EPI_ISL_1334279, EPI_ISL_1334280, EPI_ISL_1334281, EPI_ISL_1334282, EPI_ISL_1334283, EPI_ISL_1334284, EPI_ISL_1334285, EPI_ISL_1334286,
EPI_ISL_1376469, EPI_ISL_1790248

| | | | |
|---|---|---|---|
| see above | Lighthouse Lab in Milton Keynes | Wellcome Sanger Institute for the COVID-19 Genomics UK (COG-UK) Consortium | Cordelia Langford; David K. Jackson; Dominic Kwiatkowski; Ewan Harrison; Ian Johnston; Jeffrey Barrett; John Sillitoe on behalf of the Wellcome Sanger Institute COVID-19 Surveillance Team; Roberto Amato; Sonia Goncalves; The Lighthouse Lab in Milton Keynes and Alex Alderton |

EPI_ISL_1105910, EPI_ISL_1105911, EPI_ISL_1105912, EPI_ISL_1105913, EPI_ISL_1105914, EPI_ISL_1105915, EPI_ISL_1105916, EPI_ISL_1105917, EPI_ISL_1105918, EPI_ISL_1105919, EPI_ISL_1105920, EPI_ISL_1105921, EPI_ISL_1105922, EPI_ISL_1105923, EPI_ISL_1105924, EPI_ISL_1105925, EPI_ISL_1105926, EPI_ISL_1105927, EPI_ISL_1105928, EPI_ISL_1105929, EPI_ISL_1105930, EPI_ISL_1105931,
EPI_ISL_1178593, EPI_ISL_1178594, EPI_ISL_1178595, EPI_ISL_1178596, EPI_ISL_1178597, EPI_ISL_1178598, EPI_ISL_1178599, EPI_ISL_1178600, EPI_ISL_1178601, EPI_ISL_1178602, EPI_ISL_1178603

| | | | |
|---|---|---|---|
| see above | Lincolnshire Hospitals and DeepSeq Nottingham | COVID-19 Genomics UK (COG-UK) Consortium | Christopher Moore; Fei Sang; Joeseph Chappell; Johnny Debebe; Jonathan Ball; Matthew Carlisle; Matthew Loose; Nadine Holmes; Nichola Duckworth; Patrick McClure; Sarah Walsh; Tim Sloan; Victoria Wright |

EPI_ISL_997530, EPI_ISL_997531, EPI_ISL_997534, EPI_ISL_997535, EPI_ISL_997536, EPI_ISL_997537, EPI_ISL_997538, EPI_ISL_997539, EPI_ISL_1047975, EPI_ISL_1047976, EPI_ISL_1047977, EPI_ISL_1047979, EPI_ISL_1047980, EPI_ISL_1047981, EPI_ISL_1047982, EPI_ISL_1047983, EPI_ISL_1047985, EPI_ISL_1047986, EPI_ISL_1047987, EPI_ISL_1047990, EPI_ISL_1047991, EPI_ISL_1047992,
EPI_ISL_1047993, EPI_ISL_1047994, EPI_ISL_1047995, EPI_ISL_1047996, EPI_ISL_1047998, EPI_ISL_1047999, EPI_ISL_1048000, EPI_ISL_1048001, EPI_ISL_1048002, EPI_ISL_1048003, EPI_ISL_1048004, EPI_ISL_1048007, EPI_ISL_1048009, EPI_ISL_1048010, EPI_ISL_1048011, EPI_ISL_1048012, EPI_ISL_1048014, EPI_ISL_1048015, EPI_ISL_1048037, EPI_ISL_1048038, EPI_ISL_1048039, EPI_ISL_1048040,
EPI_ISL_1048043, EPI_ISL_1048047, EPI_ISL_1103590, EPI_ISL_1103591, EPI_ISL_1103592, EPI_ISL_1104390, EPI_ISL_1104391, EPI_ISL_1104392, EPI_ISL_1104393, EPI_ISL_1104394, EPI_ISL_1104400, EPI_ISL_1104401, EPI_ISL_1104408, EPI_ISL_1104410, EPI_ISL_1104419, EPI_ISL_1104422, EPI_ISL_1104423, EPI_ISL_1104424, EPI_ISL_1104425, EPI_ISL_1104426, EPI_ISL_1104427, EPI_ISL_1104429,
EPI_ISL_1104430, EPI_ISL_1104431, EPI_ISL_1104432, EPI_ISL_1104440, EPI_ISL_1104499, EPI_ISL_1104514, EPI_ISL_1104521, EPI_ISL_1104522, EPI_ISL_1104523, EPI_ISL_1104525, EPI_ISL_1104526, EPI_ISL_1104527, EPI_ISL_1104528, EPI_ISL_1104529, EPI_ISL_1104530, EPI_ISL_1104531, EPI_ISL_1104532, EPI_ISL_1104533, EPI_ISL_1104534, EPI_ISL_1104542, EPI_ISL_1104571, EPI_ISL_1105577,
EPI_ISL_1105579, EPI_ISL_1177893, EPI_ISL_1177895, EPI_ISL_1177896, EPI_ISL_1177917, EPI_ISL_1177918, EPI_ISL_1177919, EPI_ISL_1177920, EPI_ISL_1177921, EPI_ISL_1177922, EPI_ISL_1177924, EPI_ISL_1177925, EPI_ISL_1247832, EPI_ISL_1247833, EPI_ISL_1247834, EPI_ISL_1247876, EPI_ISL_1247877, EPI_ISL_1247878, EPI_ISL_1247879, EPI_ISL_1247880, EPI_ISL_1247881, EPI_ISL_1474665,
EPI_ISL_1979139, EPI_ISL_1979140, EPI_ISL_1979147, EPI_ISL_1979149, EPI_ISL_1979150, EPI_ISL_1979152, EPI_ISL_1979155, EPI_ISL_1979157, EPI_ISL_1979159, EPI_ISL_1979161, EPI_ISL_1979162

| | | | |
|---|---|---|---|
| see above | Liverpool Clinical Laboratories | COVID-19 Genomics UK (COG-UK) Consortium | A Alrezaihi; Alessandro Gerada; Alistair Darby; Angela Cowell; Anita Lucaci; Anu Chawla; Cassie Olateju; Catherine Hartley; Charlotte Nelson; Ecaterina Vamos; Elaine O'Toole; Eleanor G Bentley; Ghada T Shawli; Hermione Webster; Isabel García-Dorival; James Johnson; James P Stewart; Jenifer Manson; Joanne Watts; Jones Benjamin; Jordan J Clark; Julian Hiscox; L Luu; Lucille Rainbow; M Almsaud; Margaret Hughes; Mark Whitehead; Matthew Gemmell; Miren Iturriza-Gomara; Muhannad Alruwaili; N.P Randle; Neil Swainston; PKF Gilmore; Parul Sharma; Rebekah Penrice-Randal; Richard Eccles; Richard Gregory; Sam Haldenby; Steve Paterson; Stuart D Armstrong; Trevor Ian Robinson; Ximeng Han |

EPI_ISL_998535, EPI_ISL_998542, EPI_ISL_998543, EPI_ISL_998544, EPI_ISL_998546, EPI_ISL_998552, EPI_ISL_998553, EPI_ISL_998554, EPI_ISL_998555, EPI_ISL_998557, EPI_ISL_998558, EPI_ISL_1050994, EPI_ISL_1050995, EPI_ISL_1051001, EPI_ISL_1051002, EPI_ISL_1051003, EPI_ISL_1051004, EPI_ISL_1051005, EPI_ISL_1051007, EPI_ISL_1051008, EPI_ISL_1051009, EPI_ISL_1051010, EPI_ISL_1051011,
EPI_ISL_1051012, EPI_ISL_1051013, EPI_ISL_1051014, EPI_ISL_1051015, EPI_ISL_1051016, EPI_ISL_1051019, EPI_ISL_1051022, EPI_ISL_1051402, EPI_ISL_1105734, EPI_ISL_1105735, EPI_ISL_1105736, EPI_ISL_1105737, EPI_ISL_1105738, EPI_ISL_1105739, EPI_ISL_1105740, EPI_ISL_1105741, EPI_ISL_1105742, EPI_ISL_1105743, EPI_ISL_1105812, EPI_ISL_1105813, EPI_ISL_1108441, EPI_ISL_1108442,
EPI_ISL_1108443, EPI_ISL_1108444, EPI_ISL_1108445, EPI_ISL_1108446, EPI_ISL_1108447, EPI_ISL_1108448, EPI_ISL_1108453, EPI_ISL_1108458, EPI_ISL_1108459, EPI_ISL_1108461, EPI_ISL_1108462, EPI_ISL_1108463, EPI_ISL_1108465, EPI_ISL_1108468, EPI_ISL_1108469, EPI_ISL_1108472, EPI_ISL_1108473, EPI_ISL_1108474, EPI_ISL_1108475, EPI_ISL_1108476, EPI_ISL_1108477, EPI_ISL_1108478,
EPI_ISL_1108488, EPI_ISL_1108489, EPI_ISL_1108490, EPI_ISL_1108491, EPI_ISL_1108492, EPI_ISL_1108493, EPI_ISL_1108494, EPI_ISL_1108495, EPI_ISL_1108496, EPI_ISL_1108497, EPI_ISL_1108498, EPI_ISL_1108569, EPI_ISL_1108570, EPI_ISL_1108571, EPI_ISL_1108572, EPI_ISL_1108573, EPI_ISL_1108574, EPI_ISL_1108575, EPI_ISL_1108576, EPI_ISL_1108577, EPI_ISL_1108578, EPI_ISL_1108579,
EPI_ISL_1108580, EPI_ISL_1108615, EPI_ISL_1108616, EPI_ISL_1108617, EPI_ISL_1108618, EPI_ISL_1108619, EPI_ISL_1108620, EPI_ISL_1108621, EPI_ISL_1108622, EPI_ISL_1108623, EPI_ISL_1108624, EPI_ISL_1108625, EPI_ISL_1108626, EPI_ISL_1108627, EPI_ISL_1108628, EPI_ISL_1108629, EPI_ISL_1109045, EPI_ISL_1109046, EPI_ISL_1109047, EPI_ISL_1109048, EPI_ISL_1109049, EPI_ISL_1109050,
EPI_ISL_1109051, EPI_ISL_1109052, EPI_ISL_1109053, EPI_ISL_1109054, EPI_ISL_1109055, EPI_ISL_1109056, EPI_ISL_1109057, EPI_ISL_1109058, EPI_ISL_1109059, EPI_ISL_1109060, EPI_ISL_1109061, EPI_ISL_1109062, EPI_ISL_1109064, EPI_ISL_1109065, EPI_ISL_1109066, EPI_ISL_1109067, EPI_ISL_1109068, EPI_ISL_1109069, EPI_ISL_1109070, EPI_ISL_1109071, EPI_ISL_1109072, EPI_ISL_1109073,
EPI_ISL_1109074, EPI_ISL_1109075, EPI_ISL_1109076, EPI_ISL_1109077, EPI_ISL_1109079, EPI_ISL_1109080, EPI_ISL_1109081, EPI_ISL_1109082, EPI_ISL_1109083, EPI_ISL_1109084, EPI_ISL_1109085, EPI_ISL_1109086, EPI_ISL_1109087, EPI_ISL_1109088, EPI_ISL_1109090, EPI_ISL_1109091, EPI_ISL_1109092, EPI_ISL_1109093, EPI_ISL_1109094, EPI_ISL_1109095, EPI_ISL_1109096, EPI_ISL_1109097,
EPI_ISL_1109098, EPI_ISL_1109100, EPI_ISL_1109101, EPI_ISL_1109102, EPI_ISL_1109103, EPI_ISL_1109104, EPI_ISL_1109105, EPI_ISL_1109106, EPI_ISL_1109108, EPI_ISL_1109109, EPI_ISL_1109110, EPI_ISL_1109111, EPI_ISL_1109112, EPI_ISL_1109113, EPI_ISL_1109114, EPI_ISL_1109115, EPI_ISL_1109116, EPI_ISL_1109117, EPI_ISL_1109118, EPI_ISL_1109119, EPI_ISL_1109120, EPI_ISL_1109121,
EPI_ISL_1109122, EPI_ISL_1109123, EPI_ISL_1109124, EPI_ISL_1109125, EPI_ISL_1109126, EPI_ISL_1109127, EPI_ISL_1109128, EPI_ISL_1109129, EPI_ISL_1109130, EPI_ISL_1109131, EPI_ISL_1109133, EPI_ISL_1109134, EPI_ISL_1109135, EPI_ISL_1109136, EPI_ISL_1109137, EPI_ISL_1109138, EPI_ISL_1109139, EPI_ISL_1109140, EPI_ISL_1109141, EPI_ISL_1109142, EPI_ISL_1109143, EPI_ISL_1109144,
EPI_ISL_1109145, EPI_ISL_1109146, EPI_ISL_1109147, EPI_ISL_1109148, EPI_ISL_1109149, EPI_ISL_1109150, EPI_ISL_1109151, EPI_ISL_1109152, EPI_ISL_1109153, EPI_ISL_1109154, EPI_ISL_1109155, EPI_ISL_1109156, EPI_ISL_1109157, EPI_ISL_1109158, EPI_ISL_1109159, EPI_ISL_1109160, EPI_ISL_1109161, EPI_ISL_1109162, EPI_ISL_1109164, EPI_ISL_1109165, EPI_ISL_1109167, EPI_ISL_1109168,

EPI_ISL_1109169, EPI_ISL_1109170, EPI_ISL_1109171, EPI_ISL_1109172, EPI_ISL_1109173, EPI_ISL_1109175, EPI_ISL_1109176, EPI_ISL_1109177, EPI_ISL_1109178, EPI_ISL_1109183, EPI_ISL_1109184, EPI_ISL_1109185, EPI_ISL_1109186, EPI_ISL_1109189, EPI_ISL_1109191, EPI_ISL_1109193, EPI_ISL_1109194, EPI_ISL_1109195, EPI_ISL_1109196, EPI_ISL_1109197, EPI_ISL_1109198, EPI_ISL_1109199, EPI_ISL_1109200, EPI_ISL_1109201, EPI_ISL_1109202, EPI_ISL_1109203, EPI_ISL_1109204, EPI_ISL_1109205, EPI_ISL_1109206, EPI_ISL_1109207, EPI_ISL_1109208, EPI_ISL_1109209, EPI_ISL_1109210, EPI_ISL_1109211, EPI_ISL_1109212, EPI_ISL_1109213, EPI_ISL_1109214, EPI_ISL_1109215, EPI_ISL_1109216, EPI_ISL_1109217, EPI_ISL_1109218, EPI_ISL_1109219, EPI_ISL_1109220, EPI_ISL_1109221, EPI_ISL_1109222, EPI_ISL_1109223, EPI_ISL_1109224, EPI_ISL_1109225, EPI_ISL_1109226, EPI_ISL_1109227, EPI_ISL_1109228, EPI_ISL_1109231, EPI_ISL_1109232, EPI_ISL_1109233, EPI_ISL_1109234, EPI_ISL_1109235, EPI_ISL_1109236, EPI_ISL_1178169, EPI_ISL_1178170, EPI_ISL_1248826, EPI_ISL_1248827, EPI_ISL_1248828, EPI_ISL_1248829, EPI_ISL_1248833, EPI_ISL_1248834, EPI_ISL_1248835, EPI_ISL_1248836, EPI_ISL_1248837, EPI_ISL_1248838, EPI_ISL_1248839, EPI_ISL_1248840, EPI_ISL_1248841, EPI_ISL_1248842, EPI_ISL_1248843, EPI_ISL_1248844, EPI_ISL_1248845, EPI_ISL_1248846, EPI_ISL_1248847, EPI_ISL_1309491, EPI_ISL_1309492, EPI_ISL_1309493, EPI_ISL_1309652, EPI_ISL_1309653, EPI_ISL_1309655, EPI_ISL_1309656, EPI_ISL_1309657, EPI_ISL_1309658, EPI_ISL_1309659, EPI_ISL_1309660, EPI_ISL_1387113, EPI_ISL_1387124, EPI_ISL_1387125, EPI_ISL_1387126, EPI_ISL_1387127, EPI_ISL_1475408, EPI_ISL_1475409, EPI_ISL_1475411, EPI_ISL_1475425, EPI_ISL_1475426, EPI_ISL_1475427, EPI_ISL_1475428, EPI_ISL_1475429, EPI_ISL_1475430, EPI_ISL_1475431

| | | | |
|---|---|---|---|
| see above | Northumbria University / South Tees Hospitals NHS Foundation Trust / North Cumbria Integrated Care NHS Foundation Trust / North Tees and Hartlepool NHS Foundation Trust / Newcastle Hospitals NHS Foundation Trust | COVID-19 Genomics UK (COG-UK) Consortium | Andrew Nelson; Brendan Payne; Clive Graham; Darren L Smith; Debra Padgett; Edward Barton; Emma Swindells; Garren Scott; Gary Black; Gary Eltringham; Giles S Holt; Greg R Young; Jane Greenaway; Jennifer Collins; John Allan; Joshua Loh; Lynn Dover; Matthew Bashton; Mohammad A Tariq; Paul Baker; Sarah Essex; Steve Liggett; Wen C Yew; Yusri Taha |

EPI_ISL_1052475, EPI_ISL_1052477, EPI_ISL_1052478, EPI_ISL_1052479, EPI_ISL_1052482, EPI_ISL_1052483, EPI_ISL_1052556, EPI_ISL_1052561, EPI_ISL_1052562, EPI_ISL_1052563, EPI_ISL_1052564, EPI_ISL_1052565, EPI_ISL_1052566, EPI_ISL_1052568, EPI_ISL_1052569, EPI_ISL_1052570, EPI_ISL_1052571, EPI_ISL_1052572, EPI_ISL_1052573, EPI_ISL_1052574, EPI_ISL_1052575, EPI_ISL_1052578, EPI_ISL_1052579, EPI_ISL_1052580, EPI_ISL_1052583, EPI_ISL_1052584, EPI_ISL_1052585, EPI_ISL_1052586, EPI_ISL_1052587, EPI_ISL_1052588, EPI_ISL_1052589, EPI_ISL_1052590, EPI_ISL_1052593, EPI_ISL_1052594, EPI_ISL_1052595, EPI_ISL_1052596, EPI_ISL_1052598, EPI_ISL_1052605, EPI_ISL_1052609, EPI_ISL_1052610, EPI_ISL_1052611, EPI_ISL_1052613, EPI_ISL_1052614, EPI_ISL_1052617, EPI_ISL_1052623, EPI_ISL_1052624, EPI_ISL_1052625, EPI_ISL_1052626, EPI_ISL_1052627, EPI_ISL_1052632, EPI_ISL_1052633, EPI_ISL_1052635, EPI_ISL_1052648, EPI_ISL_1052649, EPI_ISL_1052650, EPI_ISL_1052652, EPI_ISL_1052653, EPI_ISL_1052657, EPI_ISL_1052658, EPI_ISL_1052659, EPI_ISL_1052660, EPI_ISL_1052661, EPI_ISL_1052662, EPI_ISL_1052663, EPI_ISL_1052678, EPI_ISL_1052679, EPI_ISL_1052680, EPI_ISL_1052686, EPI_ISL_1052687, EPI_ISL_1052688, EPI_ISL_1052689, EPI_ISL_1052690, EPI_ISL_1052691, EPI_ISL_1052692, EPI_ISL_1052693, EPI_ISL_1052695, EPI_ISL_1052696, EPI_ISL_1052698, EPI_ISL_1052699, EPI_ISL_1052700, EPI_ISL_1052701, EPI_ISL_1052702, EPI_ISL_1052703, EPI_ISL_1052704, EPI_ISL_1052705, EPI_ISL_1052707, EPI_ISL_1052713, EPI_ISL_1052714, EPI_ISL_1052716, EPI_ISL_1052717, EPI_ISL_1052718, EPI_ISL_1052719, EPI_ISL_1052720, EPI_ISL_1052721, EPI_ISL_1052722, EPI_ISL_1052723, EPI_ISL_1052725, EPI_ISL_1052733, EPI_ISL_1052734, EPI_ISL_1052735, EPI_ISL_1052737, EPI_ISL_1052738, EPI_ISL_1052739, EPI_ISL_1052740, EPI_ISL_1052742, EPI_ISL_1052743, EPI_ISL_1052744, EPI_ISL_1052747, EPI_ISL_1052748, EPI_ISL_1052749, EPI_ISL_1052751, EPI_ISL_1052752, EPI_ISL_1052754, EPI_ISL_1052756, EPI_ISL_1052757, EPI_ISL_1052764, EPI_ISL_1052765, EPI_ISL_1052766, EPI_ISL_1052771, EPI_ISL_1052772, EPI_ISL_1052773, EPI_ISL_1052776, EPI_ISL_1052779, EPI_ISL_1052780, EPI_ISL_1052781, EPI_ISL_1052782, EPI_ISL_1052784, EPI_ISL_1052785, EPI_ISL_1052787, EPI_ISL_1052789, EPI_ISL_1052791, EPI_ISL_1052792, EPI_ISL_1052793, EPI_ISL_1052794, EPI_ISL_1052795, EPI_ISL_1052796, EPI_ISL_1052797, EPI_ISL_1052798, EPI_ISL_1052799, EPI_ISL_1052800, EPI_ISL_1052801, EPI_ISL_1052803, EPI_ISL_1052804, EPI_ISL_1052805, EPI_ISL_1052807, EPI_ISL_1052814, EPI_ISL_1052815, EPI_ISL_1052816, EPI_ISL_1052817, EPI_ISL_1052818, EPI_ISL_1052819, EPI_ISL_1052821, EPI_ISL_1052823, EPI_ISL_1052824, EPI_ISL_1052825, EPI_ISL_1052827, EPI_ISL_1052829, EPI_ISL_1052830, EPI_ISL_1052831, EPI_ISL_1052832, EPI_ISL_1052833, EPI_ISL_1052834, EPI_ISL_1052835, EPI_ISL_1052836, EPI_ISL_1052837, EPI_ISL_1052839, EPI_ISL_1052840, EPI_ISL_1052842, EPI_ISL_1052843, EPI_ISL_1052845, EPI_ISL_1052846, EPI_ISL_1052850, EPI_ISL_1052851, EPI_ISL_1052852, EPI_ISL_1052853, EPI_ISL_1052854, EPI_ISL_1052855, EPI_ISL_1052856, EPI_ISL_1052857, EPI_ISL_1052858, EPI_ISL_1052859, EPI_ISL_1052860, EPI_ISL_1052861, EPI_ISL_1052865, EPI_ISL_1052866, EPI_ISL_1052868, EPI_ISL_1052870, EPI_ISL_1052871, EPI_ISL_1052872, EPI_ISL_1052873, EPI_ISL_1052875, EPI_ISL_1052877, EPI_ISL_1052879, EPI_ISL_1052880, EPI_ISL_1052881, EPI_ISL_1052882, EPI_ISL_1052884, EPI_ISL_1052885, EPI_ISL_1052886, EPI_ISL_1052888, EPI_ISL_1052889, EPI_ISL_1052890, EPI_ISL_1052891, EPI_ISL_1052892, EPI_ISL_1052893, EPI_ISL_1052894, EPI_ISL_1052895, EPI_ISL_1052896, EPI_ISL_1052897, EPI_ISL_1052900, EPI_ISL_1052901, EPI_ISL_1052902, EPI_ISL_1052903, EPI_ISL_1052904, EPI_ISL_1052906, EPI_ISL_1052907, EPI_ISL_1052908, EPI_ISL_1052909, EPI_ISL_1052912, EPI_ISL_1052913, EPI_ISL_1052914, EPI_ISL_1052915, EPI_ISL_1052918, EPI_ISL_1052920, EPI_ISL_1052921, EPI_ISL_1052922, EPI_ISL_1052924, EPI_ISL_1052926, EPI_ISL_1052927, EPI_ISL_1052928, EPI_ISL_1052929, EPI_ISL_1052930, EPI_ISL_1052931, EPI_ISL_1052932, EPI_ISL_1052933, EPI_ISL_1052934, EPI_ISL_1052935, EPI_ISL_1052936, EPI_ISL_1052937, EPI_ISL_1052941, EPI_ISL_1052942, EPI_ISL_1052943, EPI_ISL_1052945, EPI_ISL_1052946, EPI_ISL_1052947, EPI_ISL_1052948, EPI_ISL_1052950, EPI_ISL_1052951, EPI_ISL_1052952, EPI_ISL_1052953, EPI_ISL_1052954, EPI_ISL_1052956, EPI_ISL_1052957, EPI_ISL_1052958, EPI_ISL_1052959, EPI_ISL_1052960, EPI_ISL_1052961, EPI_ISL_1052975, EPI_ISL_1052976, EPI_ISL_1052977, EPI_ISL_1052978, EPI_ISL_1052979, EPI_ISL_1052982, EPI_ISL_1052983, EPI_ISL_1052984, EPI_ISL_1052985, EPI_ISL_1052986, EPI_ISL_1052987, EPI_ISL_1052988, EPI_ISL_1052989, EPI_ISL_1052990, EPI_ISL_1052991, EPI_ISL_1052992, EPI_ISL_1052993, EPI_ISL_1052994, EPI_ISL_1052995, EPI_ISL_1052996, EPI_ISL_1052997, EPI_ISL_1052998, EPI_ISL_1052999, EPI_ISL_1053000, EPI_ISL_1053001, EPI_ISL_1053002, EPI_ISL_1053003, EPI_ISL_1053004, EPI_ISL_1053005, EPI_ISL_1053006, EPI_ISL_1053007, EPI_ISL_1053008, EPI_ISL_1053009, EPI_ISL_1053010, EPI_ISL_1053011, EPI_ISL_1053012, EPI_ISL_1053013, EPI_ISL_1053014, EPI_ISL_1053015, EPI_ISL_1053016, EPI_ISL_1053017, EPI_ISL_1053018, EPI_ISL_1053019, EPI_ISL_1053020, EPI_ISL_1053021, EPI_ISL_1053023, EPI_ISL_1053024, EPI_ISL_1053025, EPI_ISL_1053026, EPI_ISL_1053027, EPI_ISL_1053029, EPI_ISL_1053030, EPI_ISL_1053031, EPI_ISL_1053032, EPI_ISL_1053033, EPI_ISL_1053034, EPI_ISL_1053035, EPI_ISL_1053036, EPI_ISL_1053037, EPI_ISL_1053038, EPI_ISL_1053040, EPI_ISL_1053041, EPI_ISL_1053042, EPI_ISL_1053043, EPI_ISL_1053045, EPI_ISL_1053046, EPI_ISL_1053047, EPI_ISL_1053049, EPI_ISL_1053050, EPI_ISL_1053051, EPI_ISL_1053052, EPI_ISL_1053053, EPI_ISL_1053055, EPI_ISL_1053056, EPI_ISL_1053057, EPI_ISL_1053060, EPI_ISL_1053062, EPI_ISL_1053063, EPI_ISL_1053064, EPI_ISL_1053065, EPI_ISL_1053066, EPI_ISL_1053067, EPI_ISL_1053068, EPI_ISL_1053069, EPI_ISL_1053070, EPI_ISL_1053071, EPI_ISL_1053072, EPI_ISL_1053073, EPI_ISL_1053074, EPI_ISL_1053075, EPI_ISL_1053076, EPI_ISL_1053078, EPI_ISL_1053079, EPI_ISL_1053085, EPI_ISL_1053086, EPI_ISL_1053087, EPI_ISL_1053088, EPI_ISL_1053089, EPI_ISL_1053090, EPI_ISL_1053109, EPI_ISL_1053110, EPI_ISL_1053112, EPI_ISL_1053113, EPI_ISL_1053114, EPI_ISL_1053115, EPI_ISL_1053116, EPI_ISL_1053118, EPI_ISL_1053119, EPI_ISL_1053120, EPI_ISL_1053121, EPI_ISL_1053122, EPI_ISL_1053123, EPI_ISL_1053124, EPI_ISL_1053125, EPI_ISL_1053126, EPI_ISL_1053127, EPI_ISL_1053130, EPI_ISL_1053131, EPI_ISL_1053132, EPI_ISL_1053133, EPI_ISL_1053134, EPI_ISL_1053135, EPI_ISL_1053136, EPI_ISL_1053137, EPI_ISL_1053138, EPI_ISL_1053141, EPI_ISL_1053143, EPI_ISL_1053144, EPI_ISL_1053236, EPI_ISL_1053238, EPI_ISL_1053239, EPI_ISL_1053260, EPI_ISL_1053261, EPI_ISL_1053262, EPI_ISL_1053263, EPI_ISL_1053264, EPI_ISL_1053265, EPI_ISL_1053266, EPI_ISL_1053267, EPI_ISL_1053268, EPI_ISL_1053269, EPI_ISL_1053270, EPI_ISL_1053273, EPI_ISL_1053275, EPI_ISL_1053277, EPI_ISL_1053279, EPI_ISL_1053280, EPI_ISL_1053281, EPI_ISL_1053282, EPI_ISL_1053286, EPI_ISL_1053316, EPI_ISL_1053497, EPI_ISL_1053523, EPI_ISL_1053541, EPI_ISL_1053542, EPI_ISL_1053543, EPI_ISL_1053544, EPI_ISL_1053545, EPI_ISL_1053638, EPI_ISL_1053642, EPI_ISL_1053653, EPI_ISL_1053654, EPI_ISL_1053655, EPI_ISL_1053656, EPI_ISL_1053657, EPI_ISL_1053658, EPI_ISL_1053697, EPI_ISL_1053699, EPI_ISL_1053700, EPI_ISL_1053701, EPI_ISL_1053707, EPI_ISL_1053708, EPI_ISL_1053710, EPI_ISL_1053711, EPI_ISL_1053712, EPI_ISL_1053713, EPI_ISL_1053717, EPI_ISL_1053718, EPI_ISL_1053722, EPI_ISL_1053723, EPI_ISL_1053724, EPI_ISL_1053725, EPI_ISL_1053726, EPI_ISL_1053728, EPI_ISL_1053729, EPI_ISL_1053730, EPI_ISL_1053731, EPI_ISL_1053732, EPI_ISL_1053734, EPI_ISL_1053735, EPI_ISL_1053736, EPI_ISL_1053737, EPI_ISL_1053738, EPI_ISL_1053739, EPI_ISL_1053740, EPI_ISL_1053742, EPI_ISL_1053744, EPI_ISL_1053745, EPI_ISL_1053746, EPI_ISL_1053747, EPI_ISL_1053748, EPI_ISL_1106267, EPI_ISL_1106300, EPI_ISL_1106603, EPI_ISL_1106604, EPI_ISL_1106606, EPI_ISL_1106609, EPI_ISL_1106610, EPI_ISL_1106611, EPI_ISL_1106612, EPI_ISL_1106613, EPI_ISL_1106614, EPI_ISL_1106615, EPI_ISL_1106616, EPI_ISL_1106617, EPI_ISL_1106618, EPI_ISL_1106619, EPI_ISL_1106620, EPI_ISL_1106621, EPI_ISL_1106626, EPI_ISL_1106628, EPI_ISL_1106629, EPI_ISL_1106630, EPI_ISL_1106631, EPI_ISL_1106632, EPI_ISL_1106633, EPI_ISL_1106634, EPI_ISL_1106635, EPI_ISL_1106638, EPI_ISL_1106663, EPI_ISL_1106664, EPI_ISL_1106668, EPI_ISL_1106670, EPI_ISL_1106672, EPI_ISL_1106674, EPI_ISL_1106675, EPI_ISL_1106676, EPI_ISL_1106678, EPI_ISL_1106679, EPI_ISL_1106681, EPI_ISL_1106682, EPI_ISL_1106683, EPI_ISL_1106684, EPI_ISL_1106690, EPI_ISL_1106692, EPI_ISL_1106693, EPI_ISL_1106694, EPI_ISL_1106695, EPI_ISL_1106696, EPI_ISL_1106698, EPI_ISL_1106699, EPI_ISL_1106701, EPI_ISL_1106702, EPI_ISL_1106703, EPI_ISL_1106704, EPI_ISL_1178896, EPI_ISL_1178928, EPI_ISL_1179147, EPI_ISL_1179148, EPI_ISL_1179149, EPI_ISL_1179150, EPI_ISL_1179151, EPI_ISL_1179152, EPI_ISL_1179153, EPI_ISL_1179154, EPI_ISL_1179188, EPI_ISL_1179189, EPI_ISL_1179190, EPI_ISL_1179191, EPI_ISL_1179192, EPI_ISL_1179193, EPI_ISL_1179194, EPI_ISL_1179195, EPI_ISL_1179196, EPI_ISL_1179197, EPI_ISL_1179198, EPI_ISL_1179199, EPI_ISL_1179200, EPI_ISL_1179201, EPI_ISL_1179202, EPI_ISL_1179203, EPI_ISL_1179204, EPI_ISL_1179205, EPI_ISL_1179206, EPI_ISL_1179207, EPI_ISL_1179208, EPI_ISL_1179209, EPI_ISL_1179210, EPI_ISL_1179211, EPI_ISL_1179212, EPI_ISL_1179213, EPI_ISL_1179214, EPI_ISL_1179215, EPI_ISL_1179216, EPI_ISL_1179217, EPI_ISL_1179218, EPI_ISL_1179219, EPI_ISL_1179220, EPI_ISL_1179221, EPI_ISL_1179222, EPI_ISL_1179223, EPI_ISL_1179224, EPI_ISL_1179225, EPI_ISL_1179226, EPI_ISL_1179227, EPI_ISL_1179239, EPI_ISL_1179242, EPI_ISL_1179247, EPI_ISL_1179248, EPI_ISL_1179252, EPI_ISL_1179253, EPI_ISL_1179254, EPI_ISL_1179259, EPI_ISL_1179260, EPI_ISL_1179269, EPI_ISL_1179300, EPI_ISL_1179321, EPI_ISL_1179323, EPI_ISL_1179325, EPI_ISL_1179326, EPI_ISL_1179327, EPI_ISL_1179333, EPI_ISL_1179334, EPI_ISL_1179373, EPI_ISL_1387237, EPI_ISL_1387238, EPI_ISL_1387239, EPI_ISL_1387240, EPI_ISL_1387241, EPI_ISL_1387243, EPI_ISL_1387244, EPI_ISL_1387245, EPI_ISL_1387246, EPI_ISL_1387247, EPI_ISL_1387248, EPI_ISL_1387249, EPI_ISL_1387250, EPI_ISL_1387251, EPI_ISL_1387252, EPI_ISL_1387253, EPI_ISL_1387254, EPI_ISL_1387255, EPI_ISL_1387256, EPI_ISL_1387257, EPI_ISL_1387258, EPI_ISL_1387259, EPI_ISL_1387261, EPI_ISL_1387262, EPI_ISL_1387263, EPI_ISL_1387264, EPI_ISL_1387265, EPI_ISL_1387266, EPI_ISL_1387267, EPI_ISL_1387268, EPI_ISL_1387269, EPI_ISL_1387270, EPI_ISL_1387271, EPI_ISL_1387272, EPI_ISL_1387273, EPI_ISL_1387274, EPI_ISL_1387275, EPI_ISL_1387277, EPI_ISL_1387279, EPI_ISL_1387280, EPI_ISL_1387281, EPI_ISL_1387282, EPI_ISL_1387284, EPI_ISL_1387285, EPI_ISL_1387286, EPI_ISL_1387287, EPI_ISL_1387288, EPI_ISL_1387289, EPI_ISL_1387290, EPI_ISL_1387291, EPI_ISL_1387292, EPI_ISL_1387293, EPI_ISL_1387294, EPI_ISL_1387295, EPI_ISL_1387296, EPI_ISL_1387298, EPI_ISL_1387299, EPI_ISL_1387301, EPI_ISL_1387302, EPI_ISL_1387303, EPI_ISL_1387306, EPI_ISL_1387310, EPI_ISL_1387311, EPI_ISL_1387313, EPI_ISL_1387314, EPI_ISL_1387316, EPI_ISL_1387327, EPI_ISL_1387500, EPI_ISL_1387501, EPI_ISL_1387503, EPI_ISL_1387504, EPI_ISL_1387505, EPI_ISL_1387506, EPI_ISL_1387507, EPI_ISL_1387508, EPI_ISL_1387509, EPI_ISL_1387510, EPI_ISL_1387513, EPI_ISL_1387515, EPI_ISL_1387516, EPI_ISL_1387517, EPI_ISL_1387518, EPI_ISL_1387519, EPI_ISL_1387520, EPI_ISL_1387521, EPI_ISL_1387522, EPI_ISL_1387523, EPI_ISL_1387525, EPI_ISL_1387526, EPI_ISL_1387527, EPI_ISL_1387528, EPI_ISL_1387535, EPI_ISL_1387539, EPI_ISL_1387540, EPI_ISL_1387541, EPI_ISL_1387543, EPI_ISL_1387544, EPI_ISL_1387545, EPI_ISL_1387548, EPI_ISL_1387549, EPI_ISL_1387550, EPI_ISL_1387566, EPI_ISL_1387567, EPI_ISL_1387568, EPI_ISL_1387569, EPI_ISL_1387570, EPI_ISL_1387578, EPI_ISL_1387582, EPI_ISL_1387586, EPI_ISL_1387589, EPI_ISL_1387591

| | | | |
|---|---|---|---|
| see above | Originating lab: Wales Specialist Virology Centre Sequencing lab: Pathogen Genomics Unit | Public Health Wales Microbiology Cardiff Wales Specialist Virology Centre | Alec Birchley; Alexander Adams; Amy Gaskin; Angela Marchbank; Bree Gatica-Wilcox; Catherine Moore; Jason Coombes; Joanne Watkins; Joel Southgate; Johnathan Evans; Laura Gifford; Lauren Gilbert; Lee Graham; Malorie Perry; Matthew Bull; Nicole Pacchiarini; Sally Corden; Sara Kumziene-Summerhayes; Sara Rey; Sarah Taylor; Simon Cottrell; Sophie Jones; Tom Connor |

EPI_ISL_1178712, EPI_ISL_1178719, EPI_ISL_1178722, EPI_ISL_1178729, EPI_ISL_1178731, EPI_ISL_1178734, EPI_ISL_1178738, EPI_ISL_1178753, EPI_ISL_1178756, EPI_ISL_1178769, EPI_ISL_1178784, EPI_ISL_1178796, EPI_ISL_1178809, EPI_ISL_1178817, EPI_ISL_1178820, EPI_ISL_1178823, EPI_ISL_1178824, EPI_ISL_1178825, EPI_ISL_1178831, EPI_ISL_1178832, EPI_ISL_1178833, EPI_ISL_1178834, EPI_ISL_1178836, EPI_ISL_1178843, EPI_ISL_1178845, EPI_ISL_1178851, EPI_ISL_1178861, EPI_ISL_1249088, EPI_ISL_1249091, EPI_ISL_1249092, EPI_ISL_1249093, EPI_ISL_1249096, EPI_ISL_1249101, EPI_ISL_1249102, EPI_ISL_1249103, EPI_ISL_1249106, EPI_ISL_1249108, EPI_ISL_1249109, EPI_ISL_1249110, EPI_ISL_1249111, EPI_ISL_1249112, EPI_ISL_1249114, EPI_ISL_1249115, EPI_ISL_1249116, EPI_ISL_1249117, EPI_ISL_1249118, EPI_ISL_1249129, EPI_ISL_1249215, EPI_ISL_1249216, EPI_ISL_1249218, EPI_ISL_1249245, EPI_ISL_1249246, EPI_ISL_1249249, EPI_ISL_1249250, EPI_ISL_1249252, EPI_ISL_1309749, EPI_ISL_1309750, EPI_ISL_1309778, EPI_ISL_1309779, EPI_ISL_1309780, EPI_ISL_1309781, EPI_ISL_1730523, EPI_ISL_1730524, EPI_ISL_1833074, EPI_ISL_2127679

| | | | |
|---|---|---|---|
| see above | Oxford Viromics, NDM, University of Oxford; Oxford University Hospitals; Basingstoke and North Hampshire Hospital | COVID-19 Genomics UK (COG-UK) Consortium | Alex Mobbs; Amy Trebes; Anita Justice; Catrin Moore; Christophe Fraser; David Bonsall; David Buck; Emma Wise; George Macintyre; Jessica Lynch; John Todd; Mariateresa de Cesare; Matilde Mori; Monique Andersson; Nathan Moore; Nick Cortes; Robert Shaw; Stephen Kidd; Tanya Golubchik; Timothy Peto |

EPI_ISL_998878, EPI_ISL_1051419, EPI_ISL_1051420, EPI_ISL_1051421, EPI_ISL_1051422, EPI_ISL_1051423, EPI_ISL_1051424, EPI_ISL_1051530, EPI_ISL_1051534, EPI_ISL_1051536, EPI_ISL_1051537, EPI_ISL_1051539, EPI_ISL_1051541, EPI_ISL_1051543, EPI_ISL_1051545, EPI_ISL_1051548, EPI_ISL_1051549, EPI_ISL_1051550, EPI_ISL_1051551, EPI_ISL_1051552, EPI_ISL_1051555, EPI_ISL_1103649, EPI_ISL_1103650, EPI_ISL_1103651, EPI_ISL_1103652, EPI_ISL_1103653, EPI_ISL_1103654, EPI_ISL_1103655, EPI_ISL_1103656, EPI_ISL_1103657, EPI_ISL_1103659, EPI_ISL_1103660, EPI_ISL_1103661, EPI_ISL_1103662, EPI_ISL_1103663, EPI_ISL_1103664, EPI_ISL_1103665, EPI_ISL_1103666, EPI_ISL_1103667, EPI_ISL_1103668, EPI_ISL_1103669, EPI_ISL_1103670, EPI_ISL_1103671, EPI_ISL_1103672, EPI_ISL_1103673, EPI_ISL_1103674, EPI_ISL_1103675, EPI_ISL_1103676, EPI_ISL_1103677, EPI_ISL_1103678, EPI_ISL_1103679, EPI_ISL_1103680, EPI_ISL_1103688, EPI_ISL_1103697, EPI_ISL_1103700, EPI_ISL_1103703, EPI_ISL_1107826, EPI_ISL_1107827, EPI_ISL_1107828, EPI_ISL_1107829, EPI_ISL_1107832, EPI_ISL_1107833, EPI_ISL_1107834, EPI_ISL_1107836, EPI_ISL_1107837, EPI_ISL_1107838, EPI_ISL_1107840, EPI_ISL_1107841, EPI_ISL_1107842, EPI_ISL_1107843, EPI_ISL_1107844, EPI_ISL_1107845, EPI_ISL_1107846, EPI_ISL_1107847, EPI_ISL_1107848, EPI_ISL_1107849, EPI_ISL_1107850, EPI_ISL_1107851, EPI_ISL_1107852, EPI_ISL_1107853, EPI_ISL_1107854, EPI_ISL_1107855, EPI_ISL_1107856, EPI_ISL_1107857, EPI_ISL_1107858, EPI_ISL_1107859, EPI_ISL_1107860, EPI_ISL_1107861, EPI_ISL_1107862, EPI_ISL_1107863, EPI_ISL_1107864, EPI_ISL_1107865, EPI_ISL_1107866, EPI_ISL_1107867, EPI_ISL_1107868, EPI_ISL_1107869, EPI_ISL_1107870, EPI_ISL_1107871, EPI_ISL_1107872, EPI_ISL_1107873, EPI_ISL_1107874, EPI_ISL_1107875, EPI_ISL_1107876, EPI_ISL_1107877, EPI_ISL_1107878, EPI_ISL_1107879, EPI_ISL_1107880, EPI_ISL_1107881, EPI_ISL_1107882, EPI_ISL_1107883, EPI_ISL_1107884, EPI_ISL_1107885, EPI_ISL_1107886, EPI_ISL_1107887, EPI_ISL_1107888, EPI_ISL_1107889, EPI_ISL_1107890, EPI_ISL_1107891, EPI_ISL_1107892, EPI_ISL_1107893, EPI_ISL_1107894, EPI_ISL_1107895, EPI_ISL_1107896, EPI_ISL_1107897, EPI_ISL_1107898, EPI_ISL_1107899, EPI_ISL_1107900, EPI_ISL_1107901, EPI_ISL_1107902, EPI_ISL_1107903, EPI_ISL_1107904, EPI_ISL_1107905, EPI_ISL_1107906, EPI_ISL_1107907, EPI_ISL_1107908, EPI_ISL_1107909, EPI_ISL_1107910, EPI_ISL_1107911, EPI_ISL_1107912, EPI_ISL_1107913, EPI_ISL_1107914, EPI_ISL_1107915, EPI_ISL_1107916, EPI_ISL_1107917, EPI_ISL_1107918, EPI_ISL_1107919, EPI_ISL_1107920, EPI_ISL_1107921, EPI_ISL_1107922, EPI_ISL_1107923, EPI_ISL_1107924, EPI_ISL_1107925, EPI_ISL_1107926, EPI_ISL_1107927, EPI_ISL_1107928, EPI_ISL_1107929, EPI_ISL_1107930, EPI_ISL_1107931, EPI_ISL_1107932, EPI_ISL_1107933, EPI_ISL_1107934, EPI_ISL_1107935, EPI_ISL_1107936, EPI_ISL_1107937, EPI_ISL_1107938, EPI_ISL_1107939, EPI_ISL_1107940, EPI_ISL_1107941, EPI_ISL_1107942, EPI_ISL_1107943, EPI_ISL_1107944, EPI_ISL_1107945, EPI_ISL_1107946, EPI_ISL_1107947, EPI_ISL_1107948, EPI_ISL_1107949, EPI_ISL_1107950, EPI_ISL_1107951, EPI_ISL_1107952, EPI_ISL_1107953, EPI_ISL_1107954, EPI_ISL_1107955, EPI_ISL_1107956, EPI_ISL_1107957, EPI_ISL_1107958, EPI_ISL_1107959, EPI_ISL_1107960, EPI_ISL_1107961, EPI_ISL_1107962, EPI_ISL_1107963, EPI_ISL_1107964, EPI_ISL_1107965, EPI_ISL_1107966, EPI_ISL_1107967, EPI_ISL_1107968, EPI_ISL_1107969, EPI_ISL_1107970, EPI_ISL_1107971, EPI_ISL_1107972, EPI_ISL_1107973, EPI_ISL_1107974, EPI_ISL_1107975, EPI_ISL_1107976, EPI_ISL_1107977, EPI_ISL_1107978, EPI_ISL_1107979, EPI_ISL_1107980, EPI_ISL_1107981, EPI_ISL_1107982, EPI_ISL_1107983, EPI_ISL_1107984, EPI_ISL_1107985, EPI_ISL_1107986, EPI_ISL_1107987, EPI_ISL_1107988, EPI_ISL_1107989, EPI_ISL_1107990, EPI_ISL_1107991, EPI_ISL_1107992, EPI_ISL_1107993, EPI_ISL_1107994, EPI_ISL_1107995, EPI_ISL_1107996, EPI_ISL_1107997, EPI_ISL_1107999, EPI_ISL_1108000, EPI_ISL_1108001, EPI_ISL_1108002, EPI_ISL_1108003, EPI_ISL_1108004, EPI_ISL_1108005, EPI_ISL_1108006, EPI_ISL_1108007, EPI_ISL_1108008, EPI_ISL_1108009, EPI_ISL_1108010, EPI_ISL_1108011, EPI_ISL_1108012, EPI_ISL_1108013, EPI_ISL_1108014, EPI_ISL_1108015, EPI_ISL_1108016, EPI_ISL_1108018, EPI_ISL_1108019, EPI_ISL_1108020, EPI_ISL_1108021, EPI_ISL_1108022, EPI_ISL_1108023, EPI_ISL_1108024, EPI_ISL_1108025, EPI_ISL_1108026, EPI_ISL_1108027, EPI_ISL_1108028, EPI_ISL_1108029, EPI_ISL_1108030, EPI_ISL_1108031, EPI_ISL_1108032, EPI_ISL_1108033, EPI_ISL_1108034, EPI_ISL_1108035, EPI_ISL_1108036, EPI_ISL_1108037, EPI_ISL_1108038, EPI_ISL_1108039, EPI_ISL_1108040, EPI_ISL_1108041, EPI_ISL_1108042, EPI_ISL_1108043, EPI_ISL_1108044, EPI_ISL_1108045, EPI_ISL_1108046, EPI_ISL_1108047, EPI_ISL_1108048, EPI_ISL_1108049, EPI_ISL_1108050, EPI_ISL_1108051, EPI_ISL_1108052, EPI_ISL_1108053, EPI_ISL_1108054, EPI_ISL_1108055, EPI_ISL_1108056, EPI_ISL_1108057, EPI_ISL_1108058, EPI_ISL_1108059, EPI_ISL_1108060, EPI_ISL_1108061, EPI_ISL_1108062, EPI_ISL_1108063, EPI_ISL_1108064, EPI_ISL_1108065, EPI_ISL_1108066, EPI_ISL_1108067, EPI_ISL_1108068, EPI_ISL_1108069, EPI_ISL_1108070, EPI_ISL_1108071, EPI_ISL_1108072, EPI_ISL_1108073, EPI_ISL_1108074, EPI_ISL_1108076, EPI_ISL_1108077, EPI_ISL_1108078, EPI_ISL_1108079, EPI_ISL_1108080, EPI_ISL_1108081, EPI_ISL_1108082, EPI_ISL_1108083, EPI_ISL_1108084, EPI_ISL_1108085, EPI_ISL_1108086, EPI_ISL_1108087, EPI_ISL_1108088, EPI_ISL_1108089, EPI_ISL_1108090, EPI_ISL_1108091, EPI_ISL_1108092, EPI_ISL_1108093, EPI_ISL_1108094, EPI_ISL_1108095, EPI_ISL_1108096, EPI_ISL_1108097, EPI_ISL_1108098, EPI_ISL_1108099, EPI_ISL_1108100, EPI_ISL_1108101, EPI_ISL_1108102, EPI_ISL_1108103, EPI_ISL_1108104, EPI_ISL_1108105, EPI_ISL_1108106, EPI_ISL_1108107, EPI_ISL_1108108, EPI_ISL_1108109, EPI_ISL_1108110, EPI_ISL_1108111, EPI_ISL_1108112, EPI_ISL_1108113, EPI_ISL_1108114, EPI_ISL_1108115, EPI_ISL_1108116, EPI_ISL_1108117, EPI_ISL_1108118, EPI_ISL_1108119, EPI_ISL_1108120, EPI_ISL_1108121, EPI_ISL_1108122, EPI_ISL_1108123, EPI_ISL_1108124, EPI_ISL_1108125, EPI_ISL_1108126, EPI_ISL_1108127, EPI_ISL_1108128, EPI_ISL_1108129, EPI_ISL_1108130, EPI_ISL_1108131, EPI_ISL_1108132, EPI_ISL_1108133, EPI_ISL_1108134, EPI_ISL_1108135, EPI_ISL_1108136, EPI_ISL_1108137, EPI_ISL_1108138, EPI_ISL_1108139, EPI_ISL_1108140, EPI_ISL_1108141, EPI_ISL_1108142, EPI_ISL_1108143, EPI_ISL_1108144, EPI_ISL_1108145, EPI_ISL_1108146, EPI_ISL_1108147, EPI_ISL_1108148, EPI_ISL_1108149, EPI_ISL_1108150, EPI_ISL_1108151, EPI_ISL_1108152, EPI_ISL_1108153, EPI_ISL_1108154, EPI_ISL_1108155, EPI_ISL_1108156, EPI_ISL_1108157, EPI_ISL_1108158, EPI_ISL_1108159, EPI_ISL_1108160, EPI_ISL_1108161, EPI_ISL_1108162, EPI_ISL_1108163, EPI_ISL_1108164, EPI_ISL_1108165, EPI_ISL_1108166, EPI_ISL_1108167, EPI_ISL_1108168, EPI_ISL_1108169, EPI_ISL_1108170, EPI_ISL_1108171, EPI_ISL_1108172, EPI_ISL_1108173, EPI_ISL_1108174, EPI_ISL_1108175, EPI_ISL_1108176, EPI_ISL_1108177, EPI_ISL_1108178, EPI_ISL_1108179, EPI_ISL_1108180, EPI_ISL_1108181, EPI_ISL_1108182, EPI_ISL_1108183, EPI_ISL_1108184, EPI_ISL_1108185, EPI_ISL_1108186, EPI_ISL_1108187, EPI_ISL_1108188, EPI_ISL_1108189, EPI_ISL_1108190, EPI_ISL_1108191, EPI_ISL_1108192, EPI_ISL_1108193, EPI_ISL_1108194, EPI_ISL_1108195, EPI_ISL_1108196, EPI_ISL_1108197, EPI_ISL_1108198, EPI_ISL_1108199, EPI_ISL_1108200, EPI_ISL_1108201, EPI_ISL_1108202, EPI_ISL_1108203, EPI_ISL_1108204, EPI_ISL_1108205, EPI_ISL_1108206, EPI_ISL_1108207, EPI_ISL_1108208, EPI_ISL_1108209, EPI_ISL_1108210, EPI_ISL_1108211, EPI_ISL_1108212, EPI_ISL_1108213, EPI_ISL_1108214, EPI_ISL_1108215, EPI_ISL_1108216, EPI_ISL_1108217, EPI_ISL_1108218, EPI_ISL_1108219, EPI_ISL_1108220, EPI_ISL_1108221, EPI_ISL_1108222, EPI_ISL_1108223, EPI_ISL_1108224, EPI_ISL_1108225, EPI_ISL_1108226, EPI_ISL_1108227, EPI_ISL_1108228, EPI_ISL_1108229, EPI_ISL_1108230, EPI_ISL_1108231, EPI_ISL_1108232, EPI_ISL_1108233, EPI_ISL_1108234, EPI_ISL_1108235, EPI_ISL_1108236, EPI_ISL_1108237, EPI_ISL_1108238, EPI_ISL_1108239, EPI_ISL_1108240, EPI_ISL_1108241, EPI_ISL_1108242, EPI_ISL_1108243, EPI_ISL_1108244, EPI_ISL_1108245, EPI_ISL_1108246, EPI_ISL_1108247, EPI_ISL_1108248, EPI_ISL_1108249, EPI_ISL_1108250, EPI_ISL_1108251, EPI_ISL_1108252, EPI_ISL_1108253, EPI_ISL_1108254, EPI_ISL_1108256, EPI_ISL_1108257, EPI_ISL_1108258, EPI_ISL_1108259, EPI_ISL_1108260, EPI_ISL_1108261, EPI_ISL_1108262, EPI_ISL_1108263, EPI_ISL_1108264, EPI_ISL_1108265, EPI_ISL_1108266, EPI_ISL_1108267, EPI_ISL_1108268, EPI_ISL_1108269, EPI_ISL_1108270, EPI_ISL_1108271, EPI_ISL_1108272, EPI_ISL_1108273, EPI_ISL_1108274, EPI_ISL_1108275, EPI_ISL_1108276, EPI_ISL_1108277, EPI_ISL_1108278, EPI_ISL_1108279, EPI_ISL_1108280, EPI_ISL_1108281, EPI_ISL_1108282, EPI_ISL_1108283, EPI_ISL_1108284, EPI_ISL_1108285, EPI_ISL_1108286, EPI_ISL_1108287, EPI_ISL_1108288, EPI_ISL_1108289, EPI_ISL_1108290, EPI_ISL_1108291, EPI_ISL_1108292, EPI_ISL_1108293, EPI_ISL_1108294, EPI_ISL_1108295, EPI_ISL_1108296, EPI_ISL_1108297, EPI_ISL_1108298, EPI_ISL_1108299, EPI_ISL_1108300, EPI_ISL_1108301, EPI_ISL_1108302, EPI_ISL_1108303, EPI_ISL_1108304, EPI_ISL_1108305, EPI_ISL_1108306, EPI_ISL_1108307, EPI_ISL_1108308, EPI_ISL_1108309, EPI_ISL_1108310, EPI_ISL_1108311, EPI_ISL_1108312, EPI_ISL_1108313, EPI_ISL_1108314, EPI_ISL_1108315, EPI_ISL_1108316, EPI_ISL_1108317, EPI_ISL_1108318, EPI_ISL_1108319, EPI_ISL_1108320, EPI_ISL_1108321, EPI_ISL_1108322, EPI_ISL_1108323, EPI_ISL_1108324, EPI_ISL_1108325, EPI_ISL_1108326, EPI_ISL_1108327, EPI_ISL_1108328, EPI_ISL_1108329, EPI_ISL_1108330, EPI_ISL_1108331, EPI_ISL_1108332, EPI_ISL_1108333, EPI_ISL_1108334, EPI_ISL_1108335, EPI_ISL_1108336, EPI_ISL_1108337, EPI_ISL_1108338, EPI_ISL_1108339, EPI_ISL_1108340, EPI_ISL_1108341, EPI_ISL_1108342, EPI_ISL_1108343, EPI_ISL_1108344, EPI_ISL_1108345, EPI_ISL_1108346, EPI_ISL_1108347, EPI_ISL_1108348, EPI_ISL_1108349, EPI_ISL_1108350, EPI_ISL_1108351, EPI_ISL_1108352, EPI_ISL_1108353, EPI_ISL_1108354, EPI_ISL_1108355, EPI_ISL_1108356, EPI_ISL_1108357, EPI_ISL_1108358, EPI_ISL_1108359, EPI_ISL_1108360, EPI_ISL_1108361, EPI_ISL_1108362, EPI_ISL_1108363, EPI_ISL_1108364, EPI_ISL_1108365, EPI_ISL_1108366, EPI_ISL_1108367, EPI_ISL_1108368, EPI_ISL_1108369, EPI_ISL_1108370, EPI_ISL_1108371, EPI_ISL_1108372, EPI_ISL_1108373, EPI_ISL_1108374, EPI_ISL_1108375, EPI_ISL_1108376, EPI_ISL_1108377, EPI_ISL_1108378, EPI_ISL_1108379, EPI_ISL_1108380, EPI_ISL_1108381, EPI_ISL_1108382, EPI_ISL_1108383, EPI_ISL_1108384, EPI_ISL_1108385, EPI_ISL_1108386, EPI_ISL_1108388, EPI_ISL_1108389, EPI_ISL_1108390, EPI_ISL_1108391, EPI_ISL_1108392, EPI_ISL_1108393, EPI_ISL_1108394, EPI_ISL_1108395, EPI_ISL_1108396, EPI_ISL_1108397, EPI_ISL_1108398, EPI_ISL_1108399, EPI_ISL_1108400, EPI_ISL_1108401, EPI_ISL_1108402, EPI_ISL_1108403, EPI_ISL_1108404, EPI_ISL_1108405, EPI_ISL_1108406, EPI_ISL_1108407, EPI_ISL_1108408, EPI_ISL_1108409, EPI_ISL_1108410, EPI_ISL_1108411, EPI_ISL_1108412, EPI_ISL_1108413, EPI_ISL_1108414, EPI_ISL_1108415, EPI_ISL_1108416, EPI_ISL_1108417, EPI_ISL_1108418, EPI_ISL_1108419, EPI_ISL_1108420, EPI_ISL_1108421, EPI_ISL_1108422, EPI_ISL_1108423, EPI_ISL_1108424, EPI_ISL_1108425, EPI_ISL_1108426, EPI_ISL_1108427, EPI_ISL_1108428, EPI_ISL_1108429, EPI_ISL_1108430, EPI_ISL_1108431, EPI_ISL_1108432, EPI_ISL_1108433, EPI_ISL_1108434, EPI_ISL_1108435, EPI_ISL_1108436, EPI_ISL_1108437, EPI_ISL_1108438, EPI_ISL_1108439, EPI_ISL_1108440, EPI_ISL_1108449, EPI_ISL_1108450, EPI_ISL_1108451, EPI_ISL_1108452, EPI_ISL_1108454, EPI_ISL_1108455, EPI_ISL_1108456, EPI_ISL_1108457, EPI_ISL_1108460, EPI_ISL_1108464, EPI_ISL_1108466, EPI_ISL_1108467, EPI_ISL_1108470, EPI_ISL_1108471, EPI_ISL_1108479, EPI_ISL_1108480, EPI_ISL_1108481, EPI_ISL_1108482, EPI_ISL_1108483, EPI_ISL_1108484, EPI_ISL_1108485, EPI_ISL_1108486, EPI_ISL_1108487, EPI_ISL_1108499, EPI_ISL_1108500, EPI_ISL_1108501, EPI_ISL_1108502, EPI_ISL_1108504, EPI_ISL_1108505, EPI_ISL_1108506, EPI_ISL_1108507, EPI_ISL_1108508, EPI_ISL_1108509, EPI_ISL_1108510, EPI_ISL_1108511, EPI_ISL_1108512, EPI_ISL_1108513, EPI_ISL_1108514, EPI_ISL_1108515, EPI_ISL_1108516, EPI_ISL_1108517, EPI_ISL_1108518, EPI_ISL_1108519, EPI_ISL_1108520, EPI_ISL_1108521, EPI_ISL_1108522, EPI_ISL_1108523, EPI_ISL_1108524, EPI_ISL_1108525, EPI_ISL_1108526, EPI_ISL_1108527, EPI_ISL_1108528, EPI_ISL_1108529, EPI_ISL_1108530, EPI_ISL_1108531, EPI_ISL_1108532, EPI_ISL_1108533, EPI_ISL_1108534, EPI_ISL_1108535, EPI_ISL_1108536, EPI_ISL_1108537, EPI_ISL_1108538, EPI_ISL_1108539, EPI_ISL_1108540, EPI_ISL_1108541, EPI_ISL_1108542, EPI_ISL_1108543, EPI_ISL_1108544, EPI_ISL_1108545, EPI_ISL_1108546, EPI_ISL_1108547, EPI_ISL_1108548, EPI_ISL_1108549, EPI_ISL_1108550, EPI_ISL_1108551, EPI_ISL_1108552, EPI_ISL_1108553, EPI_ISL_1108554, EPI_ISL_1108555, EPI_ISL_1108556, EPI_ISL_1108557, EPI_ISL_1108558, EPI_ISL_1108559, EPI_ISL_1108560, EPI_ISL_1108561, EPI_ISL_1108562, EPI_ISL_1108563, EPI_ISL_1108564, EPI_ISL_1108565, EPI_ISL_1108566, EPI_ISL_1108567, EPI_ISL_1108568, EPI_ISL_1108581, EPI_ISL_1108582, EPI_ISL_1108583, EPI_ISL_1108584, EPI_ISL_1108585, EPI_ISL_1108586, EPI_ISL_1108587, EPI_ISL_1108588, EPI_ISL_1108589, EPI_ISL_1108590, EPI_ISL_1108591, EPI_ISL_1108592, EPI_ISL_1108593, EPI_ISL_1108594, EPI_ISL_1108595, EPI_ISL_1108596, EPI_ISL_1108597, EPI_ISL_1108598, EPI_ISL_1108599, EPI_ISL_1108600, EPI_ISL_1108601, EPI_ISL_1108602, EPI_ISL_1108603, EPI_ISL_1108604, EPI_ISL_1108605, EPI_ISL_1108606, EPI_ISL_1108607, EPI_ISL_1108608, EPI_ISL_1108609, EPI_ISL_1108610, EPI_ISL_1108611, EPI_ISL_1108612, EPI_ISL_1108613, EPI_ISL_1108614, EPI_ISL_1178520, EPI_ISL_1178534, EPI_ISL_1178535, EPI_ISL_1178541, EPI_ISL_1247395, EPI_ISL_1248981, EPI_ISL_1249001, EPI_ISL_1249003, EPI_ISL_1249024, EPI_ISL_1249058, EPI_ISL_1249060, EPI_ISL_1249062, EPI_ISL_1386410, EPI_ISL_1386411, EPI_ISL_1386416, EPI_ISL_1386426, EPI_ISL_1386433, EPI_ISL_1386434, EPI_ISL_1386439, EPI_ISL_1386451, EPI_ISL_1387150, EPI_ISL_1476686, EPI_ISL_1476687, EPI_ISL_1832892, EPI_ISL_1832893, EPI_ISL_1832894, EPI_ISL_1832895, EPI_ISL_1832897, EPI_ISL_1832924, EPI_ISL_1978906, EPI_ISL_1978910, EPI_ISL_1978911, EPI_ISL_1978920, EPI_ISL_1978926, EPI_ISL_1978929

| | | | |
|---|---|---|---|
| see above | Quadram Institute Bioscience | COVID-19 Genomics UK (COG-UK) Consortium | Alexander J Trotter; Alison E. Mather; Alp Aydin; Ana P. Tedim; Anastasia Kolyva; Andrew Bell; Andrew J. Page; Claire Stuart; Dave J. Baker; Gemma L. Kay; John Wain; Justin O'Grady; Leonardo de Oliveira Martins; Lizzie Meadows; Maria Diaz; Mark Webber; Muhammed Yasir; Nabil-Fareed Alikhan; Ngozi Elumogo; Nicholas M. Thomson; Rachael Stanley; Rachel Gilroy; Reenesh Prakash; Samir Dervisevic; Samuel Bloomfield; Steven Rudder; Thanh Le-Viet |

EPI_ISL_998909, EPI_ISL_998911, EPI_ISL_998912, EPI_ISL_998913, EPI_ISL_998914, EPI_ISL_998915, EPI_ISL_998916, EPI_ISL_998917, EPI_ISL_998918, EPI_ISL_998919, EPI_ISL_998920, EPI_ISL_1051557

| | | | |
|---|---|---|---|
| see above | Queens Medical Centre, Clinical Microbiology Department / DeepSeq Nottingham | COVID-19 Genomics UK (COG-UK) Consortium | Christopher Moore; Fei Sang; Gemma Clark; Hannah Howson-Wells; Johnny Debebe; Jonathan Ball; Joseph Chappell; Manjinder Khakh; Matthew Carlisle; Matthew Loose; Michelle M Lister; Nadine Holmes; Patrick McClure; Theocharis Tsoleridis; Vicki M Fleming; Victoria Wright; Wendy Smith |

EPI_ISL_1042615, EPI_ISL_1043548, EPI_ISL_1046569, EPI_ISL_1070146, EPI_ISL_1070163, EPI_ISL_1070178, EPI_ISL_1070216, EPI_ISL_1070269, EPI_ISL_1070307, EPI_ISL_1072613, EPI_ISL_1072618, EPI_ISL_1072626, EPI_ISL_1072635, EPI_ISL_1072692, EPI_ISL_1072705, EPI_ISL_1072715, EPI_ISL_1072739, EPI_ISL_1072744, EPI_ISL_1072750, EPI_ISL_1072754, EPI_ISL_1072782, EPI_ISL_1072795, EPI_ISL_1072817, EPI_ISL_1072818, EPI_ISL_1072824, EPI_ISL_1072834, EPI_ISL_1072852, EPI_ISL_1072888, EPI_ISL_1072902, EPI_ISL_1072921, EPI_ISL_1072934, EPI_ISL_1072935, EPI_ISL_1072943, EPI_ISL_1099757, EPI_ISL_1099772, EPI_ISL_1099784, EPI_ISL_1099806, EPI_ISL_1099808, EPI_ISL_1099813, EPI_ISL_1099814, EPI_ISL_1099823, EPI_ISL_1099853, EPI_ISL_1099855, EPI_ISL_1099863, EPI_ISL_1099866, EPI_ISL_1099870, EPI_ISL_1099872, EPI_ISL_1099877, EPI_ISL_1099882, EPI_ISL_1099885, EPI_ISL_1099889, EPI_ISL_1099894, EPI_ISL_1099910, EPI_ISL_1099916, EPI_ISL_1099918, EPI_ISL_1099921, EPI_ISL_1099952, EPI_ISL_1099973, EPI_ISL_1099974, EPI_ISL_1099992, EPI_ISL_1100008, EPI_ISL_1100026, EPI_ISL_1100027, EPI_ISL_1100031, EPI_ISL_1100032, EPI_ISL_1100033, EPI_ISL_1100038, EPI_ISL_1100040, EPI_ISL_1100042, EPI_ISL_1100068, EPI_ISL_1100069, EPI_ISL_1101682, EPI_ISL_1101691, EPI_ISL_1101695, EPI_ISL_1101722, EPI_ISL_1101725, EPI_ISL_1101759, EPI_ISL_1101767, EPI_ISL_1101819, EPI_ISL_1101851, EPI_ISL_1101880, EPI_ISL_1101901, EPI_ISL_1101933, EPI_ISL_1102040, EPI_ISL_1102117, EPI_ISL_1102143, EPI_ISL_1102177, EPI_ISL_1102206, EPI_ISL_1102246, EPI_ISL_1102290, EPI_ISL_1102381, EPI_ISL_1102382, EPI_ISL_1102402, EPI_ISL_1102486, EPI_ISL_1102532, EPI_ISL_1102544, EPI_ISL_1102561, EPI_ISL_1102596, EPI_ISL_1102722, EPI_ISL_1102733, EPI_ISL_1102742, EPI_ISL_1102756, EPI_ISL_1102759, EPI_ISL_1102800, EPI_ISL_1102804, EPI_ISL_1102819, EPI_ISL_1102820, EPI_ISL_1102876, EPI_ISL_1102882, EPI_ISL_1102894, EPI_ISL_1102907, EPI_ISL_1102936, EPI_ISL_1102940, EPI_ISL_1102961, EPI_ISL_1102968, EPI_ISL_1102979, EPI_ISL_1102984, EPI_ISL_1102997, EPI_ISL_1103016, EPI_ISL_1103023, EPI_ISL_1103027, EPI_ISL_1103037, EPI_ISL_1103041, EPI_ISL_1116227, EPI_ISL_1116357, EPI_ISL_1116385, EPI_ISL_1116402, EPI_ISL_1116410

| | | | |
|---|---|---|---|
| see above | Randox Laboratories | Wellcome Sanger Institute for the COVID-19 Genomics UK (COG-UK) Consortium | Cordelia Langford; David K. Jackson; Dominic Kwiatkowski; Ewan Harrison; Ian Johnston; Jeffrey Barrett; John Sillitoe on behalf of the Wellcome Sanger Institute COVID-19 Surveillance Team; Randox Laboratories and Alex Alderton; Roberto Amato; Sonia Goncalves |

EPI_ISL_1475137, EPI_ISL_1475171, EPI_ISL_1475172, EPI_ISL_1475174, EPI_ISL_1475176, EPI_ISL_1475177, EPI_ISL_1475179, EPI_ISL_1475180, EPI_ISL_1475181, EPI_ISL_1475183

| | | | |
|---|---|---|---|
| see above | Regional Virus Laboratory, Belfast Health and Social Care Trust | COVID-19 Genomics UK (COG-UK) Consortium | Alison Watt; Ciara Cox; Conall McCaughey; David Simpson; Derek Fairley; James McKenna; Mairead Connor; Susan Feeney; Tanya Curran; Zoltan Molnar |

EPI_ISL_1296607, EPI_ISL_1296611, EPI_ISL_1296614, EPI_ISL_1296616, EPI_ISL_1296617, EPI_ISL_1296619, EPI_ISL_1296620

| | | | |
|---|---|---|---|
| see above | Respiratory Virus Unit, National Infection Service, Public Health England | COVID-19 Genomics UK (COG-UK) Consortium | PHE Covid Sequencing Team |

EPI_ISL_1833700, EPI_ISL_1833702, EPI_ISL_1833703, EPI_ISL_1833705, EPI_ISL_1833706, EPI_ISL_1833708, EPI_ISL_1833710, EPI_ISL_1833750, EPI_ISL_1833753, EPI_ISL_1833754, EPI_ISL_1833755, EPI_ISL_1833757, EPI_ISL_1833758, EPI_ISL_1833759, EPI_ISL_1833761, EPI_ISL_1833762, EPI_ISL_1833763, EPI_ISL_1833764, EPI_ISL_1833765, EPI_ISL_1833766, EPI_ISL_1833771, EPI_ISL_1833772, EPI_ISL_1833773, EPI_ISL_1833774, EPI_ISL_1833776, EPI_ISL_1833777, EPI_ISL_1833779, EPI_ISL_1833780, EPI_ISL_1833781, EPI_ISL_1833785, EPI_ISL_1833786, EPI_ISL_1833788, EPI_ISL_1833794

| | | | |
|---|---|---|---|
| see above | St George's University Hospitals NHS Foundation Trust | COVID-19 Genomics UK (COG-UK) Consortium | Adam Witney; Cassie Pope; Irene Monahan; Joshua Taylor; Ken Laing; NgeeKeong Tan |

EPI_ISL_2125523, EPI_ISL_2125525, EPI_ISL_2125527, EPI_ISL_2125529, EPI_ISL_2125531, EPI_ISL_2125533, EPI_ISL_2125535, EPI_ISL_2125537, EPI_ISL_2125538, EPI_ISL_2125540, EPI_ISL_2125543

| | | | |
|---|---|---|---|
| see above | The Roslin Institute and R(D)SVS, University of Edinburgh / Virology Department, Royal Infirmary of Edinburgh, NHS Lothian / School of Biological Sciences, University of Edinburgh | COVID-19 Genomics UK (COG-UK) Consortium | C; Colquhoun R; Cotton S; Dewar R; Gallagher A; Hill V; Jackson B; Maloney D; McCrone JT; McHugh M; Newman; O'Toole Á; Rambaut A; Rooke S; Scher E; Tait-Burkhard C; Templeton K; Warr A; Yu X |

EPI_ISL_1105124, EPI_ISL_1105144, EPI_ISL_1105145, EPI_ISL_1105147, EPI_ISL_1105152, EPI_ISL_1105153, EPI_ISL_1105154, EPI_ISL_1105160, EPI_ISL_1105161, EPI_ISL_1105166, EPI_ISL_1105167, EPI_ISL_1105171, EPI_ISL_1105172, EPI_ISL_1105181, EPI_ISL_1105183, EPI_ISL_1105187, EPI_ISL_1105188, EPI_ISL_1105195, EPI_ISL_1105196, EPI_ISL_1105202, EPI_ISL_1105203, EPI_ISL_1105204, EPI_ISL_1105205, EPI_ISL_1105206, EPI_ISL_1105207, EPI_ISL_1105209, EPI_ISL_1105210, EPI_ISL_1105216, EPI_ISL_1105217, EPI_ISL_1105218, EPI_ISL_1105222, EPI_ISL_1105223, EPI_ISL_1105224, EPI_ISL_1105228, EPI_ISL_1105229, EPI_ISL_1105230, EPI_ISL_1105233, EPI_ISL_1105236, EPI_ISL_1105237, EPI_ISL_1105238, EPI_ISL_1105239, EPI_ISL_1105242, EPI_ISL_1105243, EPI_ISL_1105247, EPI_ISL_1105248, EPI_ISL_1105249, EPI_ISL_1105253, EPI_ISL_1105254, EPI_ISL_1105304, EPI_ISL_1105323, EPI_ISL_1105325, EPI_ISL_1248454, EPI_ISL_1248477

| | | | |
|---|---|---|---|
| see above | University College London Hospital | COVID-19 Genomics UK (COG-UK) Consortium | Catherine Houlihan; Dan Frampton; Dr Catherine Houlihan; Dr Daniel Frampton; Dr Judith Heaney; Dr Moira Spyer; Dr Paul Grant and Dr Eleni Nastouli; Judith Heaney; Matthew Byott; Moira Spyer and Eleni Nastouli; Stuart Kirk |

EPI_ISL_997695, EPI_ISL_997696, EPI_ISL_997697, EPI_ISL_997698, EPI_ISL_997699, EPI_ISL_997700, EPI_ISL_997702, EPI_ISL_997706, EPI_ISL_997707, EPI_ISL_997708, EPI_ISL_997709, EPI_ISL_997710, EPI_ISL_997711, EPI_ISL_997712, EPI_ISL_997713, EPI_ISL_997714, EPI_ISL_997715, EPI_ISL_997716, EPI_ISL_997717, EPI_ISL_997718, EPI_ISL_997719, EPI_ISL_997720, EPI_ISL_997721, EPI_ISL_997722, EPI_ISL_997723, EPI_ISL_997724, EPI_ISL_997725, EPI_ISL_997727, EPI_ISL_997728, EPI_ISL_997729, EPI_ISL_997730, EPI_ISL_997731, EPI_ISL_997732, EPI_ISL_997733, EPI_ISL_997734, EPI_ISL_997754, EPI_ISL_997755, EPI_ISL_997772, EPI_ISL_997773, EPI_ISL_997774, EPI_ISL_997775, EPI_ISL_997776, EPI_ISL_997777, EPI_ISL_997778, EPI_ISL_997779, EPI_ISL_997783, EPI_ISL_997784, EPI_ISL_997785, EPI_ISL_997786, EPI_ISL_997787, EPI_ISL_997788, EPI_ISL_997789, EPI_ISL_997790, EPI_ISL_997791, EPI_ISL_997792, EPI_ISL_997793, EPI_ISL_997794, EPI_ISL_997795, EPI_ISL_997796, EPI_ISL_997797, EPI_ISL_997806, EPI_ISL_997807, EPI_ISL_997808, EPI_ISL_997809, EPI_ISL_997810, EPI_ISL_997811, EPI_ISL_997812, EPI_ISL_997813, EPI_ISL_997814, EPI_ISL_997815, EPI_ISL_997816, EPI_ISL_997817, EPI_ISL_997818, EPI_ISL_997819, EPI_ISL_997820, EPI_ISL_997821, EPI_ISL_997822, EPI_ISL_997823, EPI_ISL_997824, EPI_ISL_997825, EPI_ISL_997826, EPI_ISL_997827, EPI_ISL_997828, EPI_ISL_997829, EPI_ISL_997830, EPI_ISL_997831, EPI_ISL_997832, EPI_ISL_997846, EPI_ISL_997848, EPI_ISL_997849, EPI_ISL_997850, EPI_ISL_997851, EPI_ISL_997852, EPI_ISL_997853, EPI_ISL_997854, EPI_ISL_997855, EPI_ISL_997858, EPI_ISL_997860, EPI_ISL_997861, EPI_ISL_997862, EPI_ISL_997863, EPI_ISL_997864, EPI_ISL_997865, EPI_ISL_997872, EPI_ISL_997873, EPI_ISL_997874, EPI_ISL_997875, EPI_ISL_997876, EPI_ISL_997975, EPI_ISL_997978, EPI_ISL_997980, EPI_ISL_997982, EPI_ISL_997983, EPI_ISL_997985, EPI_ISL_997987, EPI_ISL_997989, EPI_ISL_997990, EPI_ISL_997991, EPI_ISL_997992, EPI_ISL_997993, EPI_ISL_997994, EPI_ISL_1050426, EPI_ISL_1050427, EPI_ISL_1050428, EPI_ISL_1050429, EPI_ISL_1050565, EPI_ISL_1050566, EPI_ISL_1050567, EPI_ISL_1050569, EPI_ISL_1050570, EPI_ISL_1050571, EPI_ISL_1050572, EPI_ISL_1050573, EPI_ISL_1050574, EPI_ISL_1050575, EPI_ISL_1050576, EPI_ISL_1050577, EPI_ISL_1050578, EPI_ISL_1050579, EPI_ISL_1050580, EPI_ISL_1050581, EPI_ISL_1104620, EPI_ISL_1104621, EPI_ISL_1104622, EPI_ISL_1104623, EPI_ISL_1104624, EPI_ISL_1104625, EPI_ISL_1104626, EPI_ISL_1104627, EPI_ISL_1104628, EPI_ISL_1104629, EPI_ISL_1105348, EPI_ISL_1105349, EPI_ISL_1105350, EPI_ISL_1105351, EPI_ISL_1105352, EPI_ISL_1105353, EPI_ISL_1105354, EPI_ISL_1105355, EPI_ISL_1105356, EPI_ISL_1105357, EPI_ISL_1105358, EPI_ISL_1105359, EPI_ISL_1105360, EPI_ISL_1105361, EPI_ISL_1105362, EPI_ISL_1105363, EPI_ISL_1105364, EPI_ISL_1105365, EPI_ISL_1105366, EPI_ISL_1105367, EPI_ISL_1105368, EPI_ISL_1105369, EPI_ISL_1105370, EPI_ISL_1105371, EPI_ISL_1105373, EPI_ISL_1105374, EPI_ISL_1105375, EPI_ISL_1105376, EPI_ISL_1105377, EPI_ISL_1105378, EPI_ISL_1105384, EPI_ISL_1105452, EPI_ISL_1105464, EPI_ISL_1105469, EPI_ISL_1105471, EPI_ISL_1247948, EPI_ISL_1247949, EPI_ISL_1247983, EPI_ISL_1250223, EPI_ISL_1250235, EPI_ISL_1250238, EPI_ISL_1308960, EPI_ISL_1308961, EPI_ISL_1308962, EPI_ISL_1308963, EPI_ISL_1308964, EPI_ISL_1308965, EPI_ISL_1308966, EPI_ISL_1309040, EPI_ISL_1309062, EPI_ISL_1474864, EPI_ISL_1637159, EPI_ISL_1637161, EPI_ISL_1637163

| | | | |
|---|---|---|---|
| see above | University College London, Great Ormond Street Hospital for Children NHS Foundation Trust, Imperial College Healthcare NHS Trust | COVID-19 Genomics UK (COG-UK) Consortium | Alison Holmes; Charlotte Williams; Helena Tutill; Jacqueline Findlay; James Price; Judith Breuer; Julianne Brown; Kathryn Harris; Leysa Forrest; Mark Kristiansen; Paola Niola; Paola Resende Silva; Patricia Dyal; Paul Randell; Rachel Williams; Samuel Weeks; Sergi Castellano; Sunando Roy; Tony Brooks; Yasmin Panchbhaya |

EPI_ISL_997367, EPI_ISL_997368, EPI_ISL_997369, EPI_ISL_997371, EPI_ISL_997372, EPI_ISL_997374, EPI_ISL_1104294, EPI_ISL_1177792

| | | | |
|---|---|---|---|
| see above | University Hospitals Of Leicester NHS Trust and DeepSeq Nottingham | COVID-19 Genomics UK (COG-UK) Consortium | Christopher Holmes; Christopher Moore; Fei Sang; Joeseph Chappell; Johnny Debebe; Jonathan Ball; Julian Tang; Karlie Fallon; Matthew Carlisle; Matthew Loose; Nadine Holmes; Patrick McClure; Paul Bird; Thomas Helmer; Victoria Wright |

EPI_ISL_996439, EPI_ISL_996475, EPI_ISL_996477, EPI_ISL_1047016, EPI_ISL_1047017, EPI_ISL_1047018, EPI_ISL_1047020, EPI_ISL_1047023, EPI_ISL_1047025, EPI_ISL_1047030, EPI_ISL_1047039, EPI_ISL_1047051, EPI_ISL_1047052, EPI_ISL_1047053, EPI_ISL_1047054, EPI_ISL_1047057, EPI_ISL_1047058, EPI_ISL_1047059, EPI_ISL_1103747, EPI_ISL_1103748, EPI_ISL_1103749, EPI_ISL_1103750, EPI_ISL_1103753, EPI_ISL_1103755, EPI_ISL_1103756, EPI_ISL_1177616, EPI_ISL_1177617, EPI_ISL_1177618, EPI_ISL_1177619, EPI_ISL_1177620, EPI_ISL_1474475, EPI_ISL_1832081, EPI_ISL_1832085

| | | | |
|---|---|---|---|
| see above | University of Birmingham | COVID-19 Genomics UK (COG-UK) Consortium | Alex Richter; Andrew Bosworth; Andrew Bosworth. Queen Elizabeth Hospital: Anna Casey; Andrew D Beggs PHE Heartlands Lab: Husam Osman; Charlotte Poxon; Institute of Microbiology; Joanne Stockton; Josh Quick; Kasun Wanigasooriya; Mike Kidd; Nicholas Loman. University of Birmingham Testing Laboratory: Celina M Whalley; Oliver Pickles; Radoslaw Poplawski; Samuel Nicholls; University of Birmingham: Claire McMurray; Will Rowe |

EPI_ISL_1050068, EPI_ISL_1050069, EPI_ISL_1050070, EPI_ISL_1050071, EPI_ISL_1050072, EPI_ISL_1050073, EPI_ISL_1050074, EPI_ISL_1050323, EPI_ISL_1050324, EPI_ISL_1050327, EPI_ISL_1050328, EPI_ISL_1050329, EPI_ISL_1103769, EPI_ISL_1103770, EPI_ISL_1103778, EPI_ISL_1103780, EPI_ISL_1103784, EPI_ISL_1103786, EPI_ISL_1103799, EPI_ISL_1103800, EPI_ISL_1103807, EPI_ISL_1103830, EPI_ISL_1103840, EPI_ISL_1103864, EPI_ISL_1103866, EPI_ISL_1308535, EPI_ISL_1308539, EPI_ISL_1308543, EPI_ISL_1308643

| | | | |
|---|---|---|---|
| see above | University of Exeter | COVID-19 Genomics UK (COG-UK) Consortium | Aaron Jeffries; Audrey Farbos; Ben Temperton; Jane Masoli; Joanna Warwick-Dugdale; Michelle Michelsen; Robyn Manley; Stephen Michell |

EPI_ISL_1047884, EPI_ISL_1047885, EPI_ISL_1047886, EPI_ISL_1047887, EPI_ISL_1047888, EPI_ISL_1047889, EPI_ISL_1047890, EPI_ISL_1047891, EPI_ISL_1047894, EPI_ISL_1047895, EPI_ISL_1047896, EPI_ISL_1047897, EPI_ISL_1047898, EPI_ISL_1047899, EPI_ISL_1047900, EPI_ISL_1047901, EPI_ISL_1047902, EPI_ISL_1050243, EPI_ISL_1104203, EPI_ISL_1104204, EPI_ISL_1104206, EPI_ISL_1104207, EPI_ISL_1104208, EPI_ISL_1104212, EPI_ISL_1104213, EPI_ISL_1104215, EPI_ISL_1177703, EPI_ISL_1177714, EPI_ISL_1177715, EPI_ISL_1177718, EPI_ISL_1177748, EPI_ISL_1177750, EPI_ISL_1247731, EPI_ISL_1247744, EPI_ISL_1308829, EPI_ISL_1308830, EPI_ISL_1308851, EPI_ISL_1386676, EPI_ISL_1386677, EPI_ISL_1386678, EPI_ISL_1386679, EPI_ISL_1386680, EPI_ISL_1386681, EPI_ISL_1386682, EPI_ISL_1386688, EPI_ISL_1636701, EPI_ISL_1730182, EPI_ISL_1730186, EPI_ISL_1730187, EPI_ISL_1730194, EPI_ISL_1730195, EPI_ISL_1730196, EPI_ISL_1730197, EPI_ISL_1730198, EPI_ISL_1730199, EPI_ISL_1730200, EPI_ISL_1730207, EPI_ISL_1730208, EPI_ISL_1730209, EPI_ISL_1730210, EPI_ISL_1730211, EPI_ISL_1730212, EPI_ISL_1730213, EPI_ISL_1730214, EPI_ISL_1730216, EPI_ISL_1730217, EPI_ISL_1730218, EPI_ISL_1730219, EPI_ISL_1730220, EPI_ISL_1730221, EPI_ISL_1730233

| | | | |
|---|---|---|---|
| see above | Virology Department, Royal Infirmary of Edinburgh, NHS Lothian / School of Biological Sciences, University of Edinburgh | COVID-19 Genomics UK (COG-UK) Consortium | Colquhoun R; Cotton S; Dewar R; Gallagher A; Hill V; Jackson B; Maloney D; McCrone JT; McHugh M; O'Toole Á; Rambaut A; Rooke S; Scher E; Templeton K; Yu X |

EPI_ISL_1000644, EPI_ISL_1000645, EPI_ISL_1000648, EPI_ISL_1000649, EPI_ISL_1000652, EPI_ISL_1000654, EPI_ISL_1000655, EPI_ISL_1000656, EPI_ISL_1000657, EPI_ISL_1000658, EPI_ISL_1000659, EPI_ISL_1000661, EPI_ISL_1000662, EPI_ISL_1000663, EPI_ISL_1000664, EPI_ISL_1000669, EPI_ISL_1000672, EPI_ISL_1000674, EPI_ISL_1000678, EPI_ISL_1000680, EPI_ISL_1000682, EPI_ISL_1000683, EPI_ISL_1000684, EPI_ISL_1000685, EPI_ISL_1000686, EPI_ISL_1000690, EPI_ISL_1000691, EPI_ISL_1000692, EPI_ISL_1000693, EPI_ISL_1000696, EPI_ISL_1000697, EPI_ISL_1000699, EPI_ISL_1000700, EPI_ISL_1000701, EPI_ISL_1000704, EPI_ISL_1000708, EPI_ISL_1000709, EPI_ISL_1000711, EPI_ISL_1000714, EPI_ISL_1000715, EPI_ISL_1000716, EPI_ISL_1000717, EPI_ISL_1000718, EPI_ISL_1000719, EPI_ISL_1000720, EPI_ISL_1000721, EPI_ISL_1000724, EPI_ISL_1000726, EPI_ISL_1000727, EPI_ISL_1000729, EPI_ISL_1000730, EPI_ISL_1000736, EPI_ISL_1000737, EPI_ISL_1000739, EPI_ISL_1000740, EPI_ISL_1000741, EPI_ISL_1000742, EPI_ISL_1000746, EPI_ISL_1000748, EPI_ISL_1000749, EPI_ISL_1000751, EPI_ISL_1000752, EPI_ISL_1000753, EPI_ISL_1000755, EPI_ISL_1000759, EPI_ISL_1000760, EPI_ISL_1000761, EPI_ISL_1000762, EPI_ISL_1000764, EPI_ISL_1000765, EPI_ISL_1000766, EPI_ISL_1000767, EPI_ISL_1000768, EPI_ISL_1000770, EPI_ISL_1000771, EPI_ISL_1000772, EPI_ISL_1000773, EPI_ISL_1000777, EPI_ISL_1000778, EPI_ISL_1000779, EPI_ISL_1000780, EPI_ISL_1000781, EPI_ISL_1000783, EPI_ISL_1000784, EPI_ISL_1000791, EPI_ISL_1054640, EPI_ISL_1104072, EPI_ISL_1104073, EPI_ISL_1104074, EPI_ISL_1104075, EPI_ISL_1104076, EPI_ISL_1104077, EPI_ISL_1104078, EPI_ISL_1104079, EPI_ISL_1104080, EPI_ISL_1104081, EPI_ISL_1104082, EPI_ISL_1104083, EPI_ISL_1104084, EPI_ISL_1104086, EPI_ISL_1104087, EPI_ISL_1104089, EPI_ISL_1104090, EPI_ISL_1104091, EPI_ISL_1104092, EPI_ISL_1104093, EPI_ISL_1104094, EPI_ISL_1104095, EPI_ISL_1104096, EPI_ISL_1104097, EPI_ISL_1104098, EPI_ISL_1104100, EPI_ISL_1104101, EPI_ISL_1104102, EPI_ISL_1108630, EPI_ISL_1108631, EPI_ISL_1108632, EPI_ISL_1108633, EPI_ISL_1108634, EPI_ISL_1108635, EPI_ISL_1108637, EPI_ISL_1108638, EPI_ISL_1108639, EPI_ISL_1108640, EPI_ISL_1108641, EPI_ISL_1108642, EPI_ISL_1108643, EPI_ISL_1108644, EPI_ISL_1108645, EPI_ISL_1108646, EPI_ISL_1108647, EPI_ISL_1108648, EPI_ISL_1108649, EPI_ISL_1108650, EPI_ISL_1108653, EPI_ISL_1108655, EPI_ISL_1108656, EPI_ISL_1108658, EPI_ISL_1108659, EPI_ISL_1108660, EPI_ISL_1108661, EPI_ISL_1108663, EPI_ISL_1108664, EPI_ISL_1108666, EPI_ISL_1108667, EPI_ISL_1108668, EPI_ISL_1108669, EPI_ISL_1108670, EPI_ISL_1108671, EPI_ISL_1108672, EPI_ISL_1108673, EPI_ISL_1108674, EPI_ISL_1108675, EPI_ISL_1108676, EPI_ISL_1108677, EPI_ISL_1108678, EPI_ISL_1108679, EPI_ISL_1108680, EPI_ISL_1108681, EPI_ISL_1108682, EPI_ISL_1108683, EPI_ISL_1108684, EPI_ISL_1108686, EPI_ISL_1108687, EPI_ISL_1108688, EPI_ISL_1108689, EPI_ISL_1108690, EPI_ISL_1108691, EPI_ISL_1108692, EPI_ISL_1108693, EPI_ISL_1108694, EPI_ISL_1108695, EPI_ISL_1108696, EPI_ISL_1108697, EPI_ISL_1108698, EPI_ISL_1108699, EPI_ISL_1108700, EPI_ISL_1108701, EPI_ISL_1108702, EPI_ISL_1108703, EPI_ISL_1108705, EPI_ISL_1108706, EPI_ISL_1108707, EPI_ISL_1108709, EPI_ISL_1108710, EPI_ISL_1108712, EPI_ISL_1108713, EPI_ISL_1108714, EPI_ISL_1108715, EPI_ISL_1108717, EPI_ISL_1108718, EPI_ISL_1108719, EPI_ISL_1108720, EPI_ISL_1108721, EPI_ISL_1108722, EPI_ISL_1108723, EPI_ISL_1108724, EPI_ISL_1108726, EPI_ISL_1108728, EPI_ISL_1108730, EPI_ISL_1108731, EPI_ISL_1108732, EPI_ISL_1108733, EPI_ISL_1108734, EPI_ISL_1108735, EPI_ISL_1108736, EPI_ISL_1108737, EPI_ISL_1108739, EPI_ISL_1108740, EPI_ISL_1108741, EPI_ISL_1108742, EPI_ISL_1108743, EPI_ISL_1108744, EPI_ISL_1108745, EPI_ISL_1108746, EPI_ISL_1108747, EPI_ISL_1108748, EPI_ISL_1108749, EPI_ISL_1108750, EPI_ISL_1108751, EPI_ISL_1108752, EPI_ISL_1108753, EPI_ISL_1108801, EPI_ISL_1180073, EPI_ISL_1180090, EPI_ISL_1180098, EPI_ISL_1638033

| | | | |
|---|---|---|---|
| see above | Virology Department, Sheffield Teaching Hospitals NHS Foundation Trust/Department of Infection, Immunity and Cardiovascular Disease, The Medical School, University of Sheffield | COVID-19 Genomics UK (COG-UK) Consortium | Adri Angyal; Alex Keeley; Benjamin Lindsey; Cariad Evans; Danielle Groves; Dave Partridge; Katie Johnson; Laura Carrilero; Luke Green; Matthew Parker; Matthew Wyles; Mehmet Yavuz; Mohammad Raza; Nikki Smith; Paul Parsons; Rachel Tucker; Rebecca Brown; Thushan de Silva |

EPI_ISL_1052550, EPI_ISL_1052567, EPI_ISL_1052576, EPI_ISL_1052577, EPI_ISL_1052581, EPI_ISL_1052582, EPI_ISL_1052591, EPI_ISL_1052592, EPI_ISL_1052602, EPI_ISL_1052603, EPI_ISL_1052604, EPI_ISL_1052616, EPI_ISL_1052634, EPI_ISL_1052651, EPI_ISL_1052665, EPI_ISL_1052673, EPI_ISL_1052677, EPI_ISL_1052684, EPI_ISL_1052685, EPI_ISL_1052694, EPI_ISL_1052697, EPI_ISL_1052709, EPI_ISL_1052724, EPI_ISL_1052726, EPI_ISL_1052727, EPI_ISL_1052728, EPI_ISL_1052729, EPI_ISL_1052732, EPI_ISL_1052745, EPI_ISL_1052746, EPI_ISL_1052755, EPI_ISL_1052758, EPI_ISL_1052759, EPI_ISL_1052760, EPI_ISL_1052763, EPI_ISL_1052767, EPI_ISL_1052768, EPI_ISL_1052769, EPI_ISL_1052770, EPI_ISL_1052774, EPI_ISL_1052775, EPI_ISL_1052777, EPI_ISL_1052778, EPI_ISL_1052783, EPI_ISL_1052786, EPI_ISL_1052788, EPI_ISL_1052790, EPI_ISL_1052802, EPI_ISL_1052806, EPI_ISL_1052808, EPI_ISL_1052809, EPI_ISL_1052810, EPI_ISL_1052811, EPI_ISL_1052812, EPI_ISL_1052813, EPI_ISL_1052820, EPI_ISL_1052822, EPI_ISL_1052826, EPI_ISL_1052828, EPI_ISL_1052838, EPI_ISL_1052841, EPI_ISL_1052844, EPI_ISL_1052847, EPI_ISL_1052848, EPI_ISL_1052849, EPI_ISL_1052862, EPI_ISL_1052863, EPI_ISL_1052864, EPI_ISL_1052867, EPI_ISL_1052869, EPI_ISL_1052874, EPI_ISL_1052876, EPI_ISL_1052878, EPI_ISL_1052883, EPI_ISL_1052887, EPI_ISL_1052898, EPI_ISL_1052899, EPI_ISL_1052905, EPI_ISL_1052910, EPI_ISL_1052911, EPI_ISL_1052916, EPI_ISL_1052917, EPI_ISL_1052919, EPI_ISL_1052923, EPI_ISL_1052925, EPI_ISL_1052938, EPI_ISL_1052939, EPI_ISL_1052940, EPI_ISL_1052944, EPI_ISL_1052949, EPI_ISL_1052955, EPI_ISL_1052980, EPI_ISL_1052981, EPI_ISL_1053022, EPI_ISL_1053028, EPI_ISL_1053039, EPI_ISL_1053044, EPI_ISL_1053048, EPI_ISL_1053054, EPI_ISL_1053058, EPI_ISL_1053059, EPI_ISL_1053061, EPI_ISL_1053077, EPI_ISL_1053084, EPI_ISL_1053117, EPI_ISL_1053128, EPI_ISL_1053139, EPI_ISL_1053237, EPI_ISL_1053271, EPI_ISL_1053272, EPI_ISL_1053274, EPI_ISL_1053278, EPI_ISL_1053283, EPI_ISL_1053284, EPI_ISL_1053287, EPI_ISL_1053288, EPI_ISL_1053498, EPI_ISL_1106015, EPI_ISL_1106017, EPI_ISL_1106019, EPI_ISL_1106020, EPI_ISL_1106025, EPI_ISL_1106027, EPI_ISL_1106028, EPI_ISL_1106029, EPI_ISL_1106030, EPI_ISL_1106032, EPI_ISL_1106033, EPI_ISL_1106034, EPI_ISL_1106036, EPI_ISL_1106037, EPI_ISL_1106038, EPI_ISL_1106041, EPI_ISL_1106042, EPI_ISL_1106044, EPI_ISL_1106045, EPI_ISL_1106046, EPI_ISL_1106047, EPI_ISL_1106048, EPI_ISL_1106049, EPI_ISL_1106050, EPI_ISL_1106051, EPI_ISL_1106052, EPI_ISL_1106053, EPI_ISL_1106054, EPI_ISL_1106055, EPI_ISL_1106056, EPI_ISL_1106057, EPI_ISL_1106058, EPI_ISL_1106059, EPI_ISL_1106060, EPI_ISL_1106061, EPI_ISL_1106062, EPI_ISL_1106063, EPI_ISL_1106065, EPI_ISL_1106066, EPI_ISL_1106073, EPI_ISL_1106074, EPI_ISL_1106075, EPI_ISL_1106076, EPI_ISL_1106254, EPI_ISL_1106282, EPI_ISL_1106286, EPI_ISL_1106287, EPI_ISL_1106288, EPI_ISL_1106289, EPI_ISL_1106290, EPI_ISL_1106291, EPI_ISL_1106292, EPI_ISL_1106293, EPI_ISL_1106294, EPI_ISL_1106296, EPI_ISL_1106297, EPI_ISL_1106605, EPI_ISL_1106627, EPI_ISL_1106659, EPI_ISL_1106665, EPI_ISL_1106666, EPI_ISL_1106667, EPI_ISL_1106669, EPI_ISL_1106671, EPI_ISL_1106673, EPI_ISL_1106680, EPI_ISL_1106685, EPI_ISL_1106686, EPI_ISL_1106687, EPI_ISL_1106688, EPI_ISL_1106689, EPI_ISL_1106691, EPI_ISL_1106697, EPI_ISL_1106700, EPI_ISL_1106771, EPI_ISL_1106772, EPI_ISL_1106773, EPI_ISL_1178927, EPI_ISL_1387242, EPI_ISL_1387260, EPI_ISL_1387276, EPI_ISL_1387278, EPI_ISL_1387283, EPI_ISL_1387297, EPI_ISL_1387300, EPI_ISL_1387305, EPI_ISL_1387315, EPI_ISL_1387514, EPI_ISL_1387524, EPI_ISL_1387542, EPI_ISL_1387546, EPI_ISL_1387547, EPI_ISL_1387587

| | | | |
|---|---|---|---|
| see above | Wales Specialist Virology Centre Sequencing lab: Pathogen Genomics Unit | Public Health Wales Microbiology Cardiff Wales Specialist Virology Centre | Alec Birchley; Alexander Adams; Amy Gaskin; Angela Marchbank; Bree Gatica-Wilcox; Catherine Moore; Jason Coombes; Joanne Watkins; Joel Southgate; Johnathan Evans; Laura Gifford; Lauren Gilbert; Lee Graham; Malorie Perry; Matthew Bull; Nicole Pacchiarini; Sally Corden; Sara Kumziene-Summerhayes; Sara Rey; Sarah Taylor; Simon Cottrell; Sophie Jones; Tom Connor |

EPI_ISL_1047841, EPI_ISL_1047842, EPI_ISL_1047843, EPI_ISL_1047844, EPI_ISL_1047845, EPI_ISL_1050195, EPI_ISL_1050196, EPI_ISL_1050197, EPI_ISL_1050198, EPI_ISL_1050199, EPI_ISL_1050200, EPI_ISL_1050201, EPI_ISL_1050202, EPI_ISL_1050203, EPI_ISL_1050204, EPI_ISL_1050205, EPI_ISL_1050206, EPI_ISL_1050207, EPI_ISL_1050208, EPI_ISL_1050209, EPI_ISL_1050210, EPI_ISL_1050211, EPI_ISL_1050212, EPI_ISL_1050213, EPI_ISL_1050214, EPI_ISL_1050215, EPI_ISL_1050217, EPI_ISL_1050240, EPI_ISL_1104123, EPI_ISL_1104124, EPI_ISL_1104125, EPI_ISL_1104126, EPI_ISL_1104127, EPI_ISL_1104128, EPI_ISL_1104129, EPI_ISL_1104130, EPI_ISL_1104131, EPI_ISL_1104132, EPI_ISL_1104133, EPI_ISL_1104134, EPI_ISL_1104135, EPI_ISL_1104136, EPI_ISL_1104137, EPI_ISL_1104138, EPI_ISL_1104139, EPI_ISL_1104140, EPI_ISL_1104141, EPI_ISL_1104142, EPI_ISL_1104143, EPI_ISL_1104144, EPI_ISL_1104145, EPI_ISL_1104146, EPI_ISL_1104147, EPI_ISL_1104149, EPI_ISL_1386595

| | | | |
|---|---|---|---|
| see above | West of Scotland Specialist Virology Centre, NHSGGC / MRC-University of Glasgow Centre for Virus Research | COVID-19 Genomics UK (COG-UK) Consortium | Alasdair MacLean; Alice Broos; Ana da Silva Filipe; Antonia Ho; Daniel Mair; David L Robertson; Emma Thomson; Guy Mollett; James Shepherd; Jenna Nichols; Joseph Hughes; Kathy Li; Kathy Smollett; Kyriaki Nomikou; Lily Tong; Matthew Holden; Natasha Johnson; Rachel Blacow; Richard Orton; Rory Gunson; Sarah McDonald; Sharif Shaaban; Sreenu Vattipally; Stephen Carmichael |

We gratefully acknowledge the following Authors from the Originating laboratories responsible for obtaining the specimens, as well as the Submitting laboratories where the genome data were generated and shared via GISAID, on which this research is based.

All Submitters of data may be contacted directly via www.gisaid.org

Authors are sorted alphabetically.

| Accession ID | Originating Laboratory | Submitting Laboratory | Authors |
|---|---|---|---|
| EPI_ISL_1048111, EPI_ISL_1048112, EPI_ISL_1048113, EPI_ISL_1048114, EPI_ISL_1048115, EPI_ISL_1048116, EPI_ISL_1048117, EPI_ISL_1048118, EPI_ISL_1048121, EPI_ISL_1048122, EPI_ISL_1048124, EPI_ISL_1048125, EPI_ISL_1048132, EPI_ISL_1474803, EPI_ISL_1730309 | | | |
| see above | Barts Health NHS Trust | COVID-19 Genomics UK (COG-UK) Consortium | BROAD; Beatrix; CUTINO-MOGUEL; Claire; David; Dola; HARRINGTON; KELE; KULASEGARAN-SHYLINI; Maria-Teresa; OWOYEMI; Raghavendran |
| EPI_ISL_1054758, EPI_ISL_1054764, EPI_ISL_1054768, EPI_ISL_1054769, EPI_ISL_1054777, EPI_ISL_1054780, EPI_ISL_1054782, EPI_ISL_1054797, EPI_ISL_1054800, EPI_ISL_1054802, EPI_ISL_1054808 | | | |
| see above | Bioinformatics and Biostatistics Lab, Advanced Sequencing Facility | COVID-19 Genomics UK (COG-UK) Consortium | Aengus Stewart; Chelsea Sawyer; Harshil Patel; Jerome Nicod; Laura Cubitt; Margaret Crawford |
| EPI_ISL_1000232, EPI_ISL_1000234, EPI_ISL_1050915, EPI_ISL_1053751, EPI_ISL_1053752, EPI_ISL_1053753, EPI_ISL_1053894, EPI_ISL_1053895, EPI_ISL_1053896, EPI_ISL_1053897, EPI_ISL_1053915, EPI_ISL_1053916, EPI_ISL_1053922, EPI_ISL_1053923, EPI_ISL_1053926, EPI_ISL_1103600, EPI_ISL_1103615, EPI_ISL_1107516, EPI_ISL_1107517, EPI_ISL_1107518, EPI_ISL_1107519, EPI_ISL_1107520, EPI_ISL_1107521, EPI_ISL_1107522, EPI_ISL_1107523, EPI_ISL_1107524, EPI_ISL_1107525, EPI_ISL_1107526, EPI_ISL_1107646, EPI_ISL_1107647, EPI_ISL_1107648, EPI_ISL_1107678, EPI_ISL_1107679, EPI_ISL_1107680, EPI_ISL_1107713, EPI_ISL_1107714, EPI_ISL_1107715, EPI_ISL_1107723, EPI_ISL_1107724, EPI_ISL_1107725, EPI_ISL_1107726, EPI_ISL_1107727, EPI_ISL_1107728, EPI_ISL_1107729, EPI_ISL_1107730, EPI_ISL_1107731, EPI_ISL_1107732, EPI_ISL_1107733, EPI_ISL_1107745, EPI_ISL_1107746, EPI_ISL_1107747, EPI_ISL_1107748, EPI_ISL_1177245, EPI_ISL_1177345, EPI_ISL_1177395, EPI_ISL_1179652, EPI_ISL_1179653, EPI_ISL_1179654, EPI_ISL_1179655, EPI_ISL_1179656, EPI_ISL_1179657, EPI_ISL_1179658, EPI_ISL_1179659, EPI_ISL_1179660, EPI_ISL_1179661, EPI_ISL_1179662, EPI_ISL_1179663, EPI_ISL_1179664, EPI_ISL_1179665, EPI_ISL_1179666, EPI_ISL_1179667, EPI_ISL_1179668, EPI_ISL_1179669, EPI_ISL_1179674, EPI_ISL_1179685, EPI_ISL_1179699, EPI_ISL_1179704, EPI_ISL_1179705, EPI_ISL_1179707, EPI_ISL_1179709, EPI_ISL_1179711, EPI_ISL_1179712, EPI_ISL_1179714, EPI_ISL_1179716, EPI_ISL_1179717, EPI_ISL_1179719, EPI_ISL_1179721, EPI_ISL_1179722, EPI_ISL_1179724, EPI_ISL_1179726, EPI_ISL_1179728, EPI_ISL_1179729, EPI_ISL_1179731, EPI_ISL_1179738, EPI_ISL_1388706, EPI_ISL_1388708, EPI_ISL_1388732, EPI_ISL_1388734, EPI_ISL_1388743, EPI_ISL_1388745, EPI_ISL_1388747, EPI_ISL_1388749, EPI_ISL_1388752, EPI_ISL_1730841, EPI_ISL_1730842, EPI_ISL_1979791, EPI_ISL_1979793 | | | |
| see above | Centre for Enzyme Innovation, University of Portsmouth / Translational Research Laboratory, Portsmouth Hospitals NHS Trust | COVID-19 Genomics UK (COG-UK) Consortium | Angela Beckett; Christopher Fearn; Kate Cook; Katie Loveson; Salman Goudarzi; Samuel Robson; Scott Elliott; Sharon Glaysher |
| EPI_ISL_1047094, EPI_ISL_1047097, EPI_ISL_1047098, EPI_ISL_1047100, EPI_ISL_1047101, EPI_ISL_1047102, EPI_ISL_1047103, EPI_ISL_1047104, EPI_ISL_1047105, EPI_ISL_1047106, EPI_ISL_1047107, EPI_ISL_1047108, EPI_ISL_1047109, EPI_ISL_1047110, EPI_ISL_1047111, EPI_ISL_1047112, EPI_ISL_1047113, EPI_ISL_1103883, EPI_ISL_1103892, EPI_ISL_1103894, EPI_ISL_1176402, EPI_ISL_1176403, EPI_ISL_1176404, EPI_ISL_1176405, EPI_ISL_1176406, EPI_ISL_1176407, EPI_ISL_1176409, EPI_ISL_1176410, EPI_ISL_1176411, EPI_ISL_1176412, EPI_ISL_1176413, EPI_ISL_1176414, EPI_ISL_1176415, EPI_ISL_1176416, EPI_ISL_1176417, EPI_ISL_1176419, EPI_ISL_1176420, EPI_ISL_1176421, EPI_ISL_1176422, EPI_ISL_1176423, EPI_ISL_1176424, EPI_ISL_1176425, EPI_ISL_1176426, EPI_ISL_1176427, EPI_ISL_1176428, EPI_ISL_1176429, EPI_ISL_1176430, EPI_ISL_1176431, EPI_ISL_1176432, EPI_ISL_1176433, EPI_ISL_1176434, EPI_ISL_1176435, EPI_ISL_1176436, EPI_ISL_1176437, EPI_ISL_1176438, EPI_ISL_1176439, EPI_ISL_1176440, EPI_ISL_1176441, EPI_ISL_1176442, EPI_ISL_1176443, EPI_ISL_1176444, EPI_ISL_1176445, EPI_ISL_1176446, EPI_ISL_1176447, EPI_ISL_1176448, EPI_ISL_1176449, EPI_ISL_1176450, EPI_ISL_1176451, EPI_ISL_1176452, EPI_ISL_1176453, EPI_ISL_1176454, EPI_ISL_1176455, EPI_ISL_1176456, EPI_ISL_1176457, EPI_ISL_1176458, EPI_ISL_1176459, EPI_ISL_1176460, EPI_ISL_1176461, EPI_ISL_1176462, EPI_ISL_1176463, EPI_ISL_1176464, EPI_ISL_1176465, EPI_ISL_1176466, EPI_ISL_1176467, EPI_ISL_1176468, EPI_ISL_1176469, EPI_ISL_1176470, EPI_ISL_1176471, EPI_ISL_1176472, EPI_ISL_1176473, EPI_ISL_1176474, EPI_ISL_1176475, EPI_ISL_1176476, EPI_ISL_1176477, EPI_ISL_1176479, EPI_ISL_1176480, EPI_ISL_1176481, EPI_ISL_1176482, EPI_ISL_1176483, EPI_ISL_1176484, EPI_ISL_1176485, EPI_ISL_1176486, EPI_ISL_1176487, EPI_ISL_1176488, EPI_ISL_1176489, EPI_ISL_1176490, EPI_ISL_1176492, EPI_ISL_1176493, EPI_ISL_1176494, EPI_ISL_1176495, EPI_ISL_1176496, EPI_ISL_1176497, EPI_ISL_1176498, EPI_ISL_1176499, EPI_ISL_1176501, EPI_ISL_1176502, EPI_ISL_1176503, EPI_ISL_1176504, EPI_ISL_1176505, EPI_ISL_1176506, EPI_ISL_1176507, EPI_ISL_1176508, EPI_ISL_1176510, EPI_ISL_1176511, EPI_ISL_1176512, EPI_ISL_1176513, EPI_ISL_1176514, EPI_ISL_1176516, EPI_ISL_1176517, EPI_ISL_1176518, EPI_ISL_1176519, EPI_ISL_1176520, EPI_ISL_1176521, EPI_ISL_1176522, EPI_ISL_1176524, EPI_ISL_1176525, EPI_ISL_1176526, EPI_ISL_1176527, EPI_ISL_1176528, EPI_ISL_1176529, EPI_ISL_1176530, EPI_ISL_1176531, EPI_ISL_1176532, EPI_ISL_1176534, EPI_ISL_1176535, EPI_ISL_1176536, EPI_ISL_1176537, EPI_ISL_1176538, EPI_ISL_1176539, EPI_ISL_1176540, EPI_ISL_1176541, EPI_ISL_1176542, EPI_ISL_1176543, EPI_ISL_1176544, EPI_ISL_1176545, EPI_ISL_1176546, EPI_ISL_1176547, EPI_ISL_1176548, EPI_ISL_1176549, EPI_ISL_1176550, EPI_ISL_1176551, EPI_ISL_1176552, EPI_ISL_1176553, EPI_ISL_1176554, EPI_ISL_1176555, EPI_ISL_1176556, EPI_ISL_1176557, EPI_ISL_1176558, EPI_ISL_1176559, EPI_ISL_1176560, EPI_ISL_1176561, EPI_ISL_1176562, EPI_ISL_1176563, EPI_ISL_1176564, EPI_ISL_1176565, EPI_ISL_1176566, EPI_ISL_1176567, EPI_ISL_1176568, EPI_ISL_1176569, EPI_ISL_1176570, EPI_ISL_1176571, EPI_ISL_1176572, EPI_ISL_1176573, EPI_ISL_1176574, EPI_ISL_1176575, EPI_ISL_1176576, EPI_ISL_1176577, EPI_ISL_1176578, EPI_ISL_1176579, EPI_ISL_1176580, EPI_ISL_1176582, EPI_ISL_1176583, EPI_ISL_1176584, EPI_ISL_1176585, EPI_ISL_1176586, EPI_ISL_1176587, EPI_ISL_1176588, EPI_ISL_1176589, EPI_ISL_1176590, EPI_ISL_1176591, EPI_ISL_1176592, EPI_ISL_1176593, EPI_ISL_1176594, EPI_ISL_1176595, EPI_ISL_1176596, EPI_ISL_1176597, EPI_ISL_1176598, EPI_ISL_1176599, EPI_ISL_1176600, EPI_ISL_1176601, EPI_ISL_1176602, EPI_ISL_1176603, EPI_ISL_1176604, EPI_ISL_1176605, EPI_ISL_1176606, EPI_ISL_1176607, EPI_ISL_1176608, EPI_ISL_1176609, EPI_ISL_1176610, EPI_ISL_1176611, EPI_ISL_1176612, EPI_ISL_1176613, EPI_ISL_1176614, EPI_ISL_1176615, EPI_ISL_1176616, EPI_ISL_1176617, EPI_ISL_1176618, EPI_ISL_1176619, EPI_ISL_1176620, EPI_ISL_1176621, EPI_ISL_1176622, EPI_ISL_1176623, EPI_ISL_1176624, EPI_ISL_1176625, EPI_ISL_1176626, EPI_ISL_1176627, EPI_ISL_1176628, EPI_ISL_1176629, EPI_ISL_1176630, EPI_ISL_1176631, EPI_ISL_1176632, EPI_ISL_1176633, EPI_ISL_1176634, EPI_ISL_1176635, EPI_ISL_1176636, EPI_ISL_1176637, EPI_ISL_1176638, EPI_ISL_1176639, EPI_ISL_1176640, EPI_ISL_1176641, EPI_ISL_1176642, EPI_ISL_1176643, EPI_ISL_1176644, EPI_ISL_1176645, EPI_ISL_1176646, EPI_ISL_1176647, EPI_ISL_1176648, EPI_ISL_1176649, EPI_ISL_1176650, EPI_ISL_1176651, EPI_ISL_1176652, EPI_ISL_1176653, EPI_ISL_1176654, EPI_ISL_1176655, EPI_ISL_1176656, EPI_ISL_1176657, EPI_ISL_1176658, EPI_ISL_1176659, EPI_ISL_1176661, EPI_ISL_1176662, EPI_ISL_1176663, EPI_ISL_1176664, EPI_ISL_1176665, EPI_ISL_1176666, EPI_ISL_1176667, EPI_ISL_1176668, EPI_ISL_1176669, EPI_ISL_1176670, EPI_ISL_1176671, EPI_ISL_1176672, EPI_ISL_1176673, EPI_ISL_1176674, EPI_ISL_1176675, EPI_ISL_1176676, EPI_ISL_1176678, EPI_ISL_1176679, EPI_ISL_1176680, EPI_ISL_1176681, EPI_ISL_1176682, EPI_ISL_1176683, EPI_ISL_1176684, EPI_ISL_1176685, EPI_ISL_1176686, EPI_ISL_1176687, EPI_ISL_1176688, EPI_ISL_1176689, EPI_ISL_1176690, EPI_ISL_1176691, EPI_ISL_1176692, EPI_ISL_1176693, EPI_ISL_1176694, EPI_ISL_1176695, EPI_ISL_1176696, EPI_ISL_1176697, EPI_ISL_1176699, EPI_ISL_1176700, EPI_ISL_1176701, EPI_ISL_1176702, EPI_ISL_1176703, EPI_ISL_1176704, EPI_ISL_1176705, EPI_ISL_1176706, EPI_ISL_1176707, EPI_ISL_1176708, EPI_ISL_1176709, EPI_ISL_1176710, EPI_ISL_1176711, EPI_ISL_1176712, EPI_ISL_1176713, EPI_ISL_1176714, EPI_ISL_1176715, EPI_ISL_1176716, EPI_ISL_1176717, EPI_ISL_1176718, EPI_ISL_1176719, EPI_ISL_1176720, EPI_ISL_1176721, EPI_ISL_1176722, EPI_ISL_1176723, EPI_ISL_1176729, EPI_ISL_1176730, EPI_ISL_1176735, EPI_ISL_1176741, EPI_ISL_1176742, EPI_ISL_1176753, EPI_ISL_1176755, EPI_ISL_1176764, EPI_ISL_1176765, EPI_ISL_1176766, EPI_ISL_1176767, EPI_ISL_1176776, EPI_ISL_1176785, EPI_ISL_1176786, EPI_ISL_1176796, EPI_ISL_1176797, EPI_ISL_1176807, EPI_ISL_1247021, EPI_ISL_1247022, EPI_ISL_1247023, EPI_ISL_1247024, EPI_ISL_1247025, EPI_ISL_1247027, EPI_ISL_1247028, EPI_ISL_1247029, EPI_ISL_1247030, EPI_ISL_1247032 | | | |
| see above | Department of Pathology, University of Cambridge | COVID-19 Genomics UK (COG-UK) Consortium | Aminu S. Jahun; Ian Goodfellow; Iliana Georgana; Martin D. Curran; Myra Hosmillo; Rhys Izuagbe; Surendra Parmar; William L. Hamilton; Yasmin Chaudhry |
| EPI_ISL_1018365, EPI_ISL_1018464, EPI_ISL_1018520, EPI_ISL_1018606, EPI_ISL_1042834, EPI_ISL_1042835, EPI_ISL_1042836, EPI_ISL_1042837, EPI_ISL_1042838, EPI_ISL_1042839, EPI_ISL_1042840, EPI_ISL_1042841, EPI_ISL_1042842, EPI_ISL_1042843, EPI_ISL_1042844, EPI_ISL_1042845, EPI_ISL_1042846, EPI_ISL_1042847, EPI_ISL_1042848, EPI_ISL_1042849, EPI_ISL_1042850, EPI_ISL_1042851, EPI_ISL_1042852, EPI_ISL_1042853, EPI_ISL_1042854, EPI_ISL_1042855, EPI_ISL_1042856, EPI_ISL_1042857, EPI_ISL_1042858, EPI_ISL_1042859, EPI_ISL_1042860, EPI_ISL_1042861, EPI_ISL_1042862, EPI_ISL_1042863, EPI_ISL_1042864, EPI_ISL_1042865, EPI_ISL_1042866, EPI_ISL_1042867, EPI_ISL_1042868, EPI_ISL_1042869, EPI_ISL_1042870, EPI_ISL_1042871, EPI_ISL_1042872, EPI_ISL_1042873, EPI_ISL_1042874, EPI_ISL_1042875, EPI_ISL_1042876, EPI_ISL_1042877, EPI_ISL_1042878, EPI_ISL_1042879, EPI_ISL_1042880, EPI_ISL_1042881, EPI_ISL_1042882, EPI_ISL_1042883, EPI_ISL_1042884, EPI_ISL_1042885, EPI_ISL_1042886, EPI_ISL_1042887, EPI_ISL_1042888, EPI_ISL_1042889, EPI_ISL_1042890, EPI_ISL_1042891, EPI_ISL_1042892, EPI_ISL_1042893, EPI_ISL_1042894, EPI_ISL_1042895, EPI_ISL_1042896, EPI_ISL_1042897, EPI_ISL_1042898, EPI_ISL_1042899, EPI_ISL_1042900, EPI_ISL_1042901, EPI_ISL_1042902, EPI_ISL_1042903, EPI_ISL_1042904, EPI_ISL_1042905, EPI_ISL_1042906, EPI_ISL_1042907, EPI_ISL_1042908, EPI_ISL_1042909, EPI_ISL_1042910, EPI_ISL_1042911, EPI_ISL_1042912, EPI_ISL_1042913, EPI_ISL_1042914, EPI_ISL_1042915, EPI_ISL_1042916, EPI_ISL_1042917, EPI_ISL_1042918, EPI_ISL_1042919, EPI_ISL_1042920, EPI_ISL_1042921, EPI_ISL_1042922, EPI_ISL_1042923, EPI_ISL_1042924, EPI_ISL_1042925, EPI_ISL_1042926, EPI_ISL_1042927, EPI_ISL_1042928, EPI_ISL_1042929, EPI_ISL_1042930, EPI_ISL_1042931, EPI_ISL_1042932, EPI_ISL_1042933, EPI_ISL_1042934, EPI_ISL_1042935, EPI_ISL_1042936, EPI_ISL_1042937, EPI_ISL_1042938, EPI_ISL_1042939, EPI_ISL_1042940, EPI_ISL_1042941, EPI_ISL_1042942, EPI_ISL_1042943, EPI_ISL_1042944, EPI_ISL_1042945, EPI_ISL_1042946, EPI_ISL_1042947, EPI_ISL_1042948, EPI_ISL_1042949, EPI_ISL_1042950, EPI_ISL_1042951, EPI_ISL_1042952, EPI_ISL_1042953, EPI_ISL_1042954, EPI_ISL_1042955, EPI_ISL_1042956, EPI_ISL_1042957, EPI_ISL_1042958, EPI_ISL_1042959, EPI_ISL_1042960, EPI_ISL_1042961, EPI_ISL_1042962, EPI_ISL_1042963, EPI_ISL_1042964, EPI_ISL_1042965, EPI_ISL_1042966, EPI_ISL_1042967, EPI_ISL_1042968, EPI_ISL_1042969, EPI_ISL_1042970, EPI_ISL_1042971, EPI_ISL_1042972, EPI_ISL_1042973, EPI_ISL_1042974, EPI_ISL_1042975, EPI_ISL_1042976, EPI_ISL_1042977, EPI_ISL_1042978, EPI_ISL_1042979, EPI_ISL_1042980, EPI_ISL_1042981, EPI_ISL_1042982, EPI_ISL_1042983, EPI_ISL_1042984, EPI_ISL_1042985, EPI_ISL_1042986, EPI_ISL_1042987, EPI_ISL_1042988, EPI_ISL_1042989, EPI_ISL_1042990, EPI_ISL_1042991, EPI_ISL_1042992, EPI_ISL_1042993, EPI_ISL_1042994, EPI_ISL_1042995, EPI_ISL_1042996, EPI_ISL_1042997, EPI_ISL_1042998, EPI_ISL_1042999, EPI_ISL_1043000, EPI_ISL_1043001, EPI_ISL_1043002, EPI_ISL_1043003, EPI_ISL_1043004, EPI_ISL_1043005, EPI_ISL_1043006, EPI_ISL_1043007, EPI_ISL_1043008, EPI_ISL_1043009, EPI_ISL_1043010, EPI_ISL_1043011, EPI_ISL_1043012, EPI_ISL_1043013, EPI_ISL_1043014, EPI_ISL_1043015, EPI_ISL_1043016, EPI_ISL_1043017, EPI_ISL_1043018, EPI_ISL_1043019, EPI_ISL_1043020, EPI_ISL_1043021, EPI_ISL_1043022, EPI_ISL_1043023, EPI_ISL_1043024, EPI_ISL_1043025, EPI_ISL_1043026, EPI_ISL_1043027, EPI_ISL_1043028, EPI_ISL_1043029, EPI_ISL_1043030, EPI_ISL_1043031, EPI_ISL_1043032, EPI_ISL_1043033, EPI_ISL_1043034, EPI_ISL_1043035, EPI_ISL_1043036, EPI_ISL_1043037, EPI_ISL_1043038, EPI_ISL_1043039, EPI_ISL_1043040, EPI_ISL_1043041, EPI_ISL_1043042, EPI_ISL_1043043, EPI_ISL_1043044, EPI_ISL_1043045, EPI_ISL_1043046, EPI_ISL_1043047, EPI_ISL_1043048, EPI_ISL_1043049, EPI_ISL_1043050, EPI_ISL_1043051, EPI_ISL_1043052, EPI_ISL_1043053, EPI_ISL_1043054, EPI_ISL_1043055, EPI_ISL_1043056, EPI_ISL_1043057, EPI_ISL_1043058, EPI_ISL_1043059, EPI_ISL_1043060, EPI_ISL_1043061, EPI_ISL_1043062, EPI_ISL_1043063, EPI_ISL_1043064, EPI_ISL_1043065, EPI_ISL_1043066, EPI_ISL_1043067, EPI_ISL_1043068, EPI_ISL_1043069, EPI_ISL_1043070, EPI_ISL_1043071, EPI_ISL_1043072, EPI_ISL_1043073, EPI_ISL_1043074, EPI_ISL_1043075, EPI_ISL_1043076, EPI_ISL_1043077, EPI_ISL_1043078, EPI_ISL_1043079, EPI_ISL_1043080, EPI_ISL_1043081, EPI_ISL_1043082, EPI_ISL_1043083, EPI_ISL_1043084, EPI_ISL_1043085, EPI_ISL_1043086, EPI_ISL_1043087, EPI_ISL_1043088, EPI_ISL_1043089, EPI_ISL_1043090, EPI_ISL_1043091, EPI_ISL_1043092, EPI_ISL_1043093, EPI_ISL_1043094, EPI_ISL_1043095, EPI_ISL_1043096, EPI_ISL_1043097, EPI_ISL_1043098, EPI_ISL_1043099, EPI_ISL_1043100, EPI_ISL_1043101, EPI_ISL_1043102, EPI_ISL_1043103, EPI_ISL_1043104, EPI_ISL_1043105, EPI_ISL_1043106, EPI_ISL_1043107, EPI_ISL_1043108, EPI_ISL_1043109, EPI_ISL_1043110, EPI_ISL_1043111, EPI_ISL_1043112, EPI_ISL_1043113, EPI_ISL_1043114, EPI_ISL_1043115, EPI_ISL_1043116, EPI_ISL_1043117, EPI_ISL_1043118, EPI_ISL_1043119, EPI_ISL_1043120, EPI_ISL_1043121, EPI_ISL_1043122, EPI_ISL_1043123, EPI_ISL_1043124, EPI_ISL_1043125, EPI_ISL_1043126, EPI_ISL_1043127, EPI_ISL_1043128, EPI_ISL_1043129, EPI_ISL_1043130, EPI_ISL_1043131, EPI_ISL_1043132, EPI_ISL_1043133, EPI_ISL_1043134, EPI_ISL_1043135, EPI_ISL_1043136, EPI_ISL_1043137, EPI_ISL_1043138, EPI_ISL_1043139, EPI_ISL_1043140, EPI_ISL_1043141, EPI_ISL_1043142, EPI_ISL_1043143, EPI_ISL_1043144, EPI_ISL_1043145, EPI_ISL_1043146, EPI_ISL_1043147, EPI_ISL_1043148, EPI_ISL_1043149, EPI_ISL_1043150, EPI_ISL_1043151, EPI_ISL_1043152, EPI_ISL_1043153, EPI_ISL_1043154, EPI_ISL_1043155, EPI_ISL_1043156, EPI_ISL_1043157, EPI_ISL_1043158, EPI_ISL_1043159, EPI_ISL_1043160, EPI_ISL_1043161, EPI_ISL_1043162, EPI_ISL_1043163, EPI_ISL_1043164, EPI_ISL_1043569, EPI_ISL_1043570, EPI_ISL_1043571, EPI_ISL_1043572, EPI_ISL_1043573, EPI_ISL_1043574, EPI_ISL_1043575, EPI_ISL_1043576, EPI_ISL_1043577, EPI_ISL_1043578, EPI_ISL_1043579, EPI_ISL_1043580, EPI_ISL_1043581, EPI_ISL_1043582, EPI_ISL_1043583, EPI_ISL_1043584, EPI_ISL_1043585, EPI_ISL_1043586, EPI_ISL_1043587, EPI_ISL_1043588, EPI_ISL_1043589, EPI_ISL_1043590, EPI_ISL_1043591, EPI_ISL_1043592, EPI_ISL_1043593, EPI_ISL_1043594, EPI_ISL_1043595, EPI_ISL_1043596, EPI_ISL_1043597, EPI_ISL_1043598, EPI_ISL_1043599, EPI_ISL_1043600, EPI_ISL_1043601, EPI_ISL_1043602, EPI_ISL_1043603, EPI_ISL_1043604, EPI_ISL_1043605, EPI_ISL_1043606, EPI_ISL_1043607, EPI_ISL_1043608, EPI_ISL_1043609, EPI_ISL_1043610, EPI_ISL_1043611, EPI_ISL_1043612, EPI_ISL_1043613, EPI_ISL_1043614, EPI_ISL_1043615, EPI_ISL_1043616, EPI_ISL_1043617, EPI_ISL_1043618, EPI_ISL_1043619, EPI_ISL_1043620, EPI_ISL_1043621, EPI_ISL_1043622, EPI_ISL_1043623, EPI_ISL_1043624, EPI_ISL_1043625, EPI_ISL_1043626, EPI_ISL_1043627, EPI_ISL_1043628, EPI_ISL_1043629, EPI_ISL_1043630, EPI_ISL_1043631, EPI_ISL_1043632, EPI_ISL_1043633, EPI_ISL_1043634, EPI_ISL_1043635, EPI_ISL_1043636, EPI_ISL_1043637, EPI_ISL_1043638, EPI_ISL_1043639, EPI_ISL_1043640, EPI_ISL_1043641, EPI_ISL_1043642, EPI_ISL_1043643, EPI_ISL_1043644, EPI_ISL_1043645, EPI_ISL_1043646, EPI_ISL_1043647, EPI_ISL_1043648, EPI_ISL_1043649, EPI_ISL_1043650, EPI_ISL_1043651, EPI_ISL_1043652, EPI_ISL_1043653, EPI_ISL_1043654, EPI_ISL_1043655, EPI_ISL_1043656, EPI_ISL_1043657, EPI_ISL_1043658, EPI_ISL_1043659, EPI_ISL_1043660, EPI_ISL_1043661, EPI_ISL_1043662, EPI_ISL_1043663, EPI_ISL_1043664, EPI_ISL_1043665, EPI_ISL_1043666, EPI_ISL_1043667, EPI_ISL_1043668, EPI_ISL_1043669, EPI_ISL_1043670, EPI_ISL_1043671, EPI_ISL_1043672, EPI_ISL_1043673, EPI_ISL_1043674, EPI_ISL_1043675, EPI_ISL_1043676, EPI_ISL_1043677, EPI_ISL_1043678, EPI_ISL_1043679, EPI_ISL_1043680, EPI_ISL_1043681, EPI_ISL_1043682, EPI_ISL_1043683, EPI_ISL_1043684, EPI_ISL_1043685, EPI_ISL_1043686, EPI_ISL_1043687, EPI_ISL_1043688, EPI_ISL_1043689, EPI_ISL_1043690, EPI_ISL_1043691, EPI_ISL_1043692, EPI_ISL_1043693, EPI_ISL_1043694, EPI_ISL_1043695, EPI_ISL_1043696, EPI_ISL_1043697, EPI_ISL_1043698, EPI_ISL_1043699, EPI_ISL_1043700, EPI_ISL_1043701, EPI_ISL_1043702, EPI_ISL_1043703, EPI_ISL_1043704, EPI_ISL_1043705, EPI_ISL_1043706, EPI_ISL_1043707, EPI_ISL_1043708, EPI_ISL_1043709, EPI_ISL_1043710, EPI_ISL_1043711, EPI_ISL_1043712, EPI_ISL_1043713, EPI_ISL_1043714, EPI_ISL_1043715, EPI_ISL_1043716, EPI_ISL_1043717, EPI_ISL_1043718, EPI_ISL_1043719, EPI_ISL_1043720, EPI_ISL_1043721, EPI_ISL_1043722, EPI_ISL_1043723, EPI_ISL_1043725, EPI_ISL_1043726, EPI_ISL_1043727, EPI_ISL_1043728, EPI_ISL_1043729, EPI_ISL_1043730, EPI_ISL_1043731, EPI_ISL_1043732, EPI_ISL_1043733, EPI_ISL_1043734, EPI_ISL_1043735, EPI_ISL_1043736, EPI_ISL_1043737, EPI_ISL_1043738, EPI_ISL_1043739, EPI_ISL_1043740, EPI_ISL_1043741, EPI_ISL_1043742, EPI_ISL_1043743, EPI_ISL_1043744, EPI_ISL_1043745, EPI_ISL_1043746, EPI_ISL_1043747, EPI_ISL_1043748, EPI_ISL_1043749, EPI_ISL_1043750, EPI_ISL_1043751, EPI_ISL_1043752, EPI_ISL_1043753, EPI_ISL_1043754, EPI_ISL_1043755, EPI_ISL_1043756, EPI_ISL_1043757, EPI_ISL_1043758, EPI_ISL_1043759, EPI_ISL_1043760, EPI_ISL_1043761, EPI_ISL_1043762, EPI_ISL_1043763, EPI_ISL_1043764, EPI_ISL_1043765, EPI_ISL_1043766, EPI_ISL_1043767, EPI_ISL_1043768, EPI_ISL_1043769, EPI_ISL_1043770, EPI_ISL_1043771, EPI_ISL_1043772, EPI_ISL_1043773, EPI_ISL_1043774, EPI_ISL_1043775, EPI_ISL_1043776, EPI_ISL_1043777, EPI_ISL_1043778, EPI_ISL_1043779, EPI_ISL_1043780, EPI_ISL_1043781, EPI_ISL_1043782, EPI_ISL_1043783, EPI_ISL_1043784, EPI_ISL_1043785, EPI_ISL_1043786, EPI_ISL_1043787, EPI_ISL_1043788, EPI_ISL_1043789, EPI_ISL_1043790, EPI_ISL_1043791, EPI_ISL_1043792, EPI_ISL_1043793, EPI_ISL_1043794, EPI_ISL_1043795, EPI_ISL_1043796, EPI_ISL_1043797, EPI_ISL_1043798, EPI_ISL_1043799, EPI_ISL_1043800, EPI_ISL_1043801, EPI_ISL_1043802, EPI_ISL_1043803, EPI_ISL_1043804, EPI_ISL_1043805, EPI_ISL_1043806, EPI_ISL_1043807, EPI_ISL_1043808, EPI_ISL_1043809, EPI_ISL_1043810, EPI_ISL_1043811, EPI_ISL_1043812, EPI_ISL_1043813, EPI_ISL_1043814, EPI_ISL_1043815, EPI_ISL_1043816, EPI_ISL_1043817, EPI_ISL_1043818, EPI_ISL_1043819, EPI_ISL_1043820, EPI_ISL_1043821, EPI_ISL_1043822, EPI_ISL_1043823, EPI_ISL_1043824, EPI_ISL_1043825, EPI_ISL_1043826, EPI_ISL_1043827, EPI_ISL_1043828, EPI_ISL_1043829, EPI_ISL_1043830, EPI_ISL_1043831, EPI_ISL_1043832, EPI_ISL_1043833, EPI_ISL_1043834, EPI_ISL_1043835, EPI_ISL_1043836, EPI_ISL_1043837, EPI_ISL_1043838, EPI_ISL_1043839, EPI_ISL_1043840, EPI_ISL_1043841, EPI_ISL_1043842, EPI_ISL_1043843, EPI_ISL_1043844, EPI_ISL_1043845, EPI_ISL_1043846, EPI_ISL_1043847, EPI_ISL_1043848, EPI_ISL_1043849, EPI_ISL_1043850, EPI_ISL_1043851, EPI_ISL_1043852, EPI_ISL_1043853, EPI_ISL_1043854, EPI_ISL_1043855, EPI_ISL_1043856, EPI_ISL_1043857, EPI_ISL_1043858, EPI_ISL_1043859, EPI_ISL_1043860, EPI_ISL_1043861, EPI_ISL_1043862, EPI_ISL_1043863, EPI_ISL_1043864, EPI_ISL_1043865, EPI_ISL_1043866, EPI_ISL_1043867, EPI_ISL_1043868, EPI_ISL_1043869, EPI_ISL_1043870, EPI_ISL_1043871, EPI_ISL_1043872, EPI_ISL_1043873, EPI_ISL_1043874, EPI_ISL_1043875, EPI_ISL_1043876, EPI_ISL_1043877, EPI_ISL_1043878, EPI_ISL_1043879, EPI_ISL_1043880, EPI_ISL_1043881, EPI_ISL_1043882, EPI_ISL_1043883, EPI_ISL_1043884, EPI_ISL_1043885, EPI_ISL_1043886, EPI_ISL_1043887, EPI_ISL_1043888, EPI_ISL_1043889, EPI_ISL_1043890, EPI_ISL_1043891, EPI_ISL_1043892, EPI_ISL_1043893, EPI_ISL_1043894, EPI_ISL_1043895, EPI_ISL_1043896, EPI_ISL_1043897, EPI_ISL_1043898, EPI_ISL_1044386, EPI_ISL_1044387, EPI_ISL_1044388, EPI_ISL_1044389, EPI_ISL_1044390, EPI_ISL_1044391, EPI_ISL_1044392, EPI_ISL_1044393, EPI_ISL_1044394, EPI_ISL_1044395, EPI_ISL_1044396, EPI_ISL_1044397, EPI_ISL_1044398, EPI_ISL_1044400, EPI_ISL_1044401, EPI_ISL_1044402, EPI_ISL_1044403, EPI_ISL_1044404, EPI_ISL_1044405, EPI_ISL_1044406, EPI_ISL_1044407, EPI_ISL_1044408, EPI_ISL_1044409, EPI_ISL_1044410, EPI_ISL_1044411, EPI_ISL_1044412, EPI_ISL_1044413, EPI_ISL_1044414, EPI_ISL_1044415, EPI_ISL_1044416, EPI_ISL_1044417, EPI_ISL_1044418, EPI_ISL_1044419, EPI_ISL_1044420, EPI_ISL_1044421, EPI_ISL_1044422, EPI_ISL_1044423, EPI_ISL_1044424, EPI_ISL_1044425, EPI_ISL_1044426, EPI_ISL_1044427, EPI_ISL_1044428, EPI_ISL_1044429, EPI_ISL_1044430, EPI_ISL_1044431, EPI_ISL_1044432, EPI_ISL_1044433, EPI_ISL_1044434, EPI_ISL_1044435, EPI_ISL_1044436, EPI_ISL_1044437, EPI_ISL_1044438, EPI_ISL_1044439, EPI_ISL_1044440, EPI_ISL_1044441, EPI_ISL_1044442, EPI_ISL_1044443, EPI_ISL_1044444, EPI_ISL_1044445, EPI_ISL_1044446, EPI_ISL_1044447, EPI_ISL_1044449, EPI_ISL_1044450, EPI_ISL_1044451, EPI_ISL_1044452, EPI_ISL_1044453, EPI_ISL_1044454, EPI_ISL_1044455, EPI_ISL_1044456, EPI_ISL_1044457, EPI_ISL_1044458, EPI_ISL_1044459, EPI_ISL_1044460, EPI_ISL_1044461, EPI_ISL_1044462, EPI_ISL_1044463, EPI_ISL_1044464, EPI_ISL_1044465, EPI_ISL_1044466, EPI_ISL_1044467, EPI_ISL_1044468, EPI_ISL_1044469, EPI_ISL_1044470, EPI_ISL_1044471, EPI_ISL_1044472, EPI_ISL_1044473, EPI_ISL_1044474, EPI_ISL_1044475, EPI_ISL_1044476, EPI_ISL_1044477, EPI_ISL_1044478, EPI_ISL_1044479, EPI_ISL_1044480, EPI_ISL_1044481, EPI_ISL_1044483, EPI_ISL_1044484, EPI_ISL_1044485, EPI_ISL_1044486, EPI_ISL_1044487, EPI_ISL_1044488, EPI_ISL_1044489, EPI_ISL_1044490, EPI_ISL_1044491, EPI_ISL_1044493, EPI_ISL_1044494, EPI_ISL_1044495, EPI_ISL_1044496, EPI_ISL_1044497, EPI_ISL_1044498, EPI_ISL_1044499, EPI_ISL_1044500, EPI_ISL_1044501, EPI_ISL_1044502, EPI_ISL_1044503, EPI_ISL_1044504, EPI_ISL_1044505, EPI_ISL_1044506, EPI_ISL_1044507, EPI_ISL_1044508, EPI_ISL_1044509, EPI_ISL_1044510, EPI_ISL_1044511, EPI_ISL_1044512, EPI_ISL_1044513, EPI_ISL_1044514, EPI_ISL_1044515, EPI_ISL_1044517, EPI_ISL_1044518, EPI_ISL_1044519, EPI_ISL_1044520, EPI_ISL_1044522, EPI_ISL_1044523, EPI_ISL_1044524, EPI_ISL_1044525, EPI_ISL_1044526, EPI_ISL_1044527, EPI_ISL_1044528, EPI_ISL_1044529, EPI_ISL_1044530, EPI_ISL_1044531, EPI_ISL_1044532, EPI_ISL_1044533, EPI_ISL_1044534, EPI_ISL_1044535, EPI_ISL_1044536, EPI_ISL_1044537, EPI_ISL_1044538, EPI_ISL_1044539, EPI_ISL_1044540, EPI_ISL_1044541, EPI_ISL_1044543, EPI_ISL_1044544, EPI_ISL_1044545, EPI_ISL_1044546, EPI_ISL_1044547, EPI_ISL_1044548, EPI_ISL_1044549, EPI_ISL_1044550, EPI_ISL_1044551, EPI_ISL_1044552, EPI_ISL_1044553, EPI_ISL_1044554, EPI_ISL_1044555, EPI_ISL_1044556, EPI_ISL_1044557, EPI_ISL_1044558, EPI_ISL_1044559, EPI_ISL_1044560, EPI_ISL_1044561, EPI_ISL_1044562, EPI_ISL_1044563, EPI_ISL_1044564, EPI_ISL_1044565, EPI_ISL_1044566, EPI_ISL_1044567, EPI_ISL_1044568, EPI_ISL_1044569, EPI_ISL_1044570, EPI_ISL_1044571, EPI_ISL_1044572, EPI_ISL_1044573, EPI_ISL_1044574, EPI_ISL_1044575, EPI_ISL_1044576, EPI_ISL_1044577, EPI_ISL_1044578, EPI_ISL_1044579, EPI_ISL_1044580, EPI_ISL_1044581, EPI_ISL_1044582, EPI_ISL_1044583, EPI_ISL_1044584, EPI_ISL_1044585, EPI_ISL_1044586, EPI_ISL_1044587, EPI_ISL_1044588, EPI_ISL_1044589, EPI_ISL_1044590, EPI_ISL_1044591, EPI_ISL_1044592, EPI_ISL_1044593, EPI_ISL_1044594, EPI_ISL_1044595, EPI_ISL_1044596, EPI_ISL_1044597, EPI_ISL_1044598, EPI_ISL_1044599, EPI_ISL_1044600, EPI_ISL_1044601, EPI_ISL_1044602, EPI_ISL_1044603, EPI_ISL_1044604, EPI_ISL_1044605, EPI_ISL_1044606, EPI_ISL_1044607, EPI_ISL_1044608, EPI_ISL_1044609, EPI_ISL_1044610, EPI_ISL_1044611, EPI_ISL_1044612, EPI_ISL_1044613, EPI_ISL_1044614, EPI_ISL_1044615, EPI_ISL_1044616, EPI_ISL_1044617, EPI_ISL_1044618, EPI_ISL_1044619, EPI_ISL_1044620, EPI_ISL_1044621, EPI_ISL_1044622, EPI_ISL_1044623, EPI_ISL_1044624, EPI_ISL_1044625, EPI_ISL_1044626, EPI_ISL_1044627, EPI_ISL_1044628, EPI_ISL_1044629, EPI_ISL_1044630, EPI_ISL_1044631, EPI_ISL_1044633, EPI_ISL_1044634, EPI_ISL_1044635, EPI_ISL_1044636, EPI_ISL_1044637, EPI_ISL_1044638, EPI_ISL_1044639, EPI_ISL_1044640, EPI_ISL_1044641, EPI_ISL_1044642, EPI_ISL_1044643, EPI_ISL_1044644, EPI_ISL_1044645, EPI_ISL_1044646, EPI_ISL_1044648, EPI_ISL_1044649, EPI_ISL_1044650, EPI_ISL_1044651, EPI_ISL_1044652, EPI_ISL_1044653, EPI_ISL_1044654, EPI_ISL_1044655, EPI_ISL_1044656, EPI_ISL_1044658, EPI_ISL_1044659, EPI_ISL_1044660, EPI_ISL_1044661, EPI_ISL_1044662, EPI_ISL_1044663, EPI_ISL_1044664, EPI_ISL_1044665, EPI_ISL_1044666, EPI_ISL_1044667, EPI_ISL_1044668, EPI_ISL_1044669, EPI_ISL_1044670, EPI_ISL_1044671, EPI_ISL_1044672, EPI_ISL_1044673, EPI_ISL_1044674, EPI_ISL_1044675, EPI_ISL_1044676, EPI_ISL_1044677, EPI_ISL_1044678, EPI_ISL_1044679, EPI_ISL_1044680, EPI_ISL_1044681, EPI_ISL_1044682, EPI_ISL_1044683, EPI_ISL_1044684, EPI_ISL_1044685, EPI_ISL_1044686, EPI_ISL_1044687, EPI_ISL_1044688, EPI_ISL_1044689, EPI_ISL_1044690, EPI_ISL_1044691, EPI_ISL_1044692, EPI_ISL_1044693, EPI_ISL_1044694, EPI_ISL_1044695, EPI_ISL_1044696, EPI_ISL_1044697, EPI_ISL_1044698, EPI_ISL_1044699, EPI_ISL_1044700, EPI_ISL_1044701, EPI_ISL_1044703, EPI_ISL_1044704, EPI_ISL_1044705, EPI_ISL_1044706, EPI_ISL_1044707, EPI_ISL_1044708, EPI_ISL_1044709, EPI_ISL_1044710, EPI_ISL_1044711, EPI_ISL_1044712, EPI_ISL_1044713, EPI_ISL_1044714, EPI_ISL_1044715, EPI_ISL_1044716, EPI_ISL_1044717, EPI_ISL_1044718, EPI_ISL_1044719, EPI_ISL_1044720, EPI_ISL_1044721, EPI_ISL_1044722, EPI_ISL_1044723, EPI_ISL_1044724, EPI_ISL_1044865, EPI_ISL_1044866, EPI_ISL_1044867, EPI_ISL_1044868, EPI_ISL_1044869, EPI_ISL_1044870, EPI_ISL_1044871, EPI_ISL_1044872, EPI_ISL_1044873, EPI_ISL_1044874, EPI_ISL_1044875, EPI_ISL_1044876, EPI_ISL_1044877, EPI_ISL_1044878, EPI_ISL_1044879, EPI_ISL_1044880, EPI_ISL_1044881, EPI_ISL_1044882, EPI_ISL_1044883, EPI_ISL_1044884, EPI_ISL_1044885, EPI_ISL_1044886, EPI_ISL_1044887, EPI_ISL_1044888, EPI_ISL_1044889, EPI_ISL_1044890, EPI_ISL_1044891, EPI_ISL_1044892, EPI_ISL_1044893, EPI_ISL_1044894, EPI_ISL_1044895, EPI_ISL_1044896, EPI_ISL_1044897, EPI_ISL_1044898, EPI_ISL_1044899, EPI_ISL_1044900, EPI_ISL_1044901, EPI_ISL_1044902, EPI_ISL_1044903, EPI_ISL_1044904, EPI_ISL_1044905, EPI_ISL_1044906, EPI_ISL_1044907, EPI_ISL_1044909, EPI_ISL_1044910, EPI_ISL_1044911, EPI_ISL_1044912, EPI_ISL_1044913, EPI_ISL_1044914, EPI_ISL_1044915, EPI_ISL_1044916, EPI_ISL_1044917, EPI_ISL_1044918, EPI_ISL_1044919, EPI_ISL_1044920, EPI_ISL_1044921, EPI_ISL_1044922, EPI_ISL_1044923, EPI_ISL_1044924, EPI_ISL_1044925, EPI_ISL_1044926, EPI_ISL_1044927, EPI_ISL_1044928, EPI_ISL_1044929, EPI_ISL_1044930, EPI_ISL_1044931, EPI_ISL_1044932, EPI_ISL_1044933, EPI_ISL_1044934, EPI_ISL_1044935, EPI_ISL_1044936, EPI_ISL_1044937, EPI_ISL_1044938, EPI_ISL_1044939, EPI_ISL_1044940, EPI_ISL_1044941, EPI_ISL_1044942, EPI_ISL_1044943, EPI_ISL_1044944, EPI_ISL_1044945, EPI_ISL_1044946, EPI_ISL_1044947, EPI_ISL_1044948, EPI_ISL_1044949, EPI_ISL_1044950, EPI_ISL_1044951, EPI_ISL_1044952, EPI_ISL_1044953, EPI_ISL_1044954, EPI_ISL_1044955, EPI_ISL_1044956, EPI_ISL_1044957, EPI_ISL_1044958, EPI_ISL_1044959, EPI_ISL_1044960, EPI_ISL_1044961, EPI_ISL_1044962, EPI_ISL_1044963, EPI_ISL_1044964, EPI_ISL_1044965, EPI_ISL_1044966, EPI_ISL_1044967, EPI_ISL_1044968, EPI_ISL_1044969, EPI_ISL_1044970, EPI_ISL_1044971, EPI_ISL_1044972, EPI_ISL_1044973, EPI_ISL_1044974, EPI_ISL_1044975, EPI_ISL_1044976, EPI_ISL_1044977, EPI_ISL_1044978, EPI_ISL_1044979, EPI_ISL_1044980, EPI_ISL_1044981, EPI_ISL_1044982, EPI_ISL_1044983, EPI_ISL_1044984, EPI_ISL_1044985, EPI_ISL_1044986, EPI_ISL_1044987, EPI_ISL_1044989, EPI_ISL_1044990, EPI_ISL_1044991, EPI_ISL_1044992, EPI_ISL_1044993, EPI_ISL_1044994, EPI_ISL_1044995, EPI_ISL_1044996, EPI_ISL_1044997, EPI_ISL_1044998, EPI_ISL_1044999, EPI_ISL_1045000, EPI_ISL_1045001, EPI_ISL_1045002, EPI_ISL_1045003, EPI_ISL_1045004, EPI_ISL_1045005, EPI_ISL_1045006, EPI_ISL_1045007, EPI_ISL_1045008, EPI_ISL_1045009, EPI_ISL_1045010, EPI_ISL_1045011, EPI_ISL_1045012, EPI_ISL_1045013, EPI_ISL_1045014, EPI_ISL_1045015, EPI_ISL_1045016, EPI_ISL_1045017, EPI_ISL_1045018, EPI_ISL_1045019, EPI_ISL_1045020, EPI_ISL_1045021, EPI_ISL_1045022, EPI_ISL_1045023, EPI_ISL_1045024, EPI_ISL_1045025, EPI_ISL_1045026, EPI_ISL_1045027, EPI_ISL_1045028, EPI_ISL_1045029, EPI_ISL_1045030, EPI_ISL_1045031, EPI_ISL_1045032, EPI_ISL_1045033, EPI_ISL_1045034, EPI_ISL_1045035, EPI_ISL_1045036, EPI_ISL_1045037, EPI_ISL_1045038, EPI_ISL_1045039, EPI_ISL_1045040, EPI_ISL_1045041, EPI_ISL_1045042, EPI_ISL_1045043, EPI_ISL_1045044, EPI_ISL_1045045, EPI_ISL_1045046, EPI_ISL_1045047, EPI_ISL_1045048, EPI_ISL_1045049, EPI_ISL_1045050, EPI_ISL_1045051, EPI_ISL_1045052, EPI_ISL_1045053, EPI_ISL_1045054, EPI_ISL_1045055, EPI_ISL_1045056, EPI_ISL_1045057, EPI_ISL_1045058, EPI_ISL_1045059, EPI_ISL_1045060, EPI_ISL_1045061, EPI_ISL_1045062, EPI_ISL_1045063, EPI_ISL_1045064, EPI_ISL_1045065, EPI_ISL_1045066, EPI_ISL_1045067, EPI_ISL_1045068, EPI_ISL_1045069, EPI_ISL_1045070, EPI_ISL_1045071, EPI_ISL_1045072, EPI_ISL_1045073, EPI_ISL_1045074, EPI_ISL_1045075, EPI_ISL_1045076, EPI_ISL_1045077, EPI_ISL_1045078, EPI_ISL_1045079, EPI_ISL_1045080, EPI_ISL_1045081, EPI_ISL_1045082, EPI_ISL_1045083, EPI_ISL_1045084, EPI_ISL_1045085, EPI_ISL_1045086, EPI_ISL_1045087, EPI_ISL_1045088, EPI_ISL_1045089, EPI_ISL_1045090, EPI_ISL_1045091, EPI_ISL_1045092, EPI_ISL_1045093, EPI_ISL_1045094, EPI_ISL_1045095, EPI_ISL_1045096, EPI_ISL_1045097, EPI_ISL_1045098, EPI_ISL_1045099, EPI_ISL_1045100, EPI_ISL_1045101, EPI_ISL_1045102, EPI_ISL_1045103, EPI_ISL_1045104, EPI_ISL_1045105, EPI_ISL_1045106, EPI_ISL_1045107, EPI_ISL_1045108, EPI_ISL_1045109, EPI_ISL_1045110, EPI_ISL_1045111, EPI_ISL_1045112, EPI_ISL_1045113, EPI_ISL_1045114, EPI_ISL_1045115, EPI_ISL_1045116, EPI_ISL_1045117, EPI_ISL_1045118, EPI_ISL_1045119, EPI_ISL_1045120, EPI_ISL_1045121, EPI_ISL_1045122, EPI_ISL_1045123, EPI_ISL_1045124, EPI_ISL_1045126, EPI_ISL_1045127, EPI_ISL_1045128, EPI_ISL_1045129, EPI_ISL_1045130, EPI_ISL_1045131, EPI_ISL_1045132, EPI_ISL_1045133, EPI_ISL_1045134, EPI_ISL_1045135, EPI_ISL_1045136, EPI_ISL_1045137, EPI_ISL_1045138, EPI_ISL_1045139, EPI_ISL_1045140, EPI_ISL_1045141, EPI_ISL_1045142, EPI_ISL_1045143, EPI_ISL_1045144, EPI_ISL_1045145, EPI_ISL_1045146, EPI_ISL_1045147, EPI_ISL_1045148, EPI_ISL_1045149, EPI_ISL_1045150, EPI_ISL_1045151, EPI_ISL_1045152, EPI_ISL_1045153, EPI_ISL_1045154, EPI_ISL_1045155, EPI_ISL_1045156, EPI_ISL_1045157, EPI_ISL_1045158, EPI_ISL_1045159, EPI_ISL_1045160, EPI_ISL_1045161, EPI_ISL_1045162, EPI_ISL_1045163, EPI_ISL_1045164, EPI_ISL_1045165, EPI_ISL_1045166, EPI_ISL_1045167, EPI_ISL_1045168, EPI_ISL_1045169, EPI_ISL_1045170, EPI_ISL_1045171, EPI_ISL_1045172, EPI_ISL_1045173, EPI_ISL_1045174, EPI_ISL_1045175, EPI_ISL_1045176, EPI_ISL_1045177, EPI_ISL_1045178, EPI_ISL_1045179, EPI_ISL_1045180, EPI_ISL_1045181, EPI_ISL_1045182, EPI_ISL_1045183, EPI_ISL_1045184, EPI_ISL_1045185, EPI_ISL_1045186, EPI_ISL_1045187, EPI_ISL_1045188, EPI_ISL_1045189, EPI_ISL_1045190, EPI_ISL_1045191, EPI_ISL_1045192, EPI_ISL_1045193, EPI_ISL_1045194, EPI_ISL_1045195, EPI_ISL_1045196, EPI_ISL_1045197, EPI_ISL_1057684, EPI_ISL_1057685, EPI_ISL_1057686, EPI_ISL_1057687, EPI_ISL_1057688, EPI_ISL_1057689, EPI_ISL_1057690, EPI_ISL_1057691, EPI_ISL_1057692, EPI_ISL_1057693, EPI_ISL_1057694, EPI_ISL_1057695, EPI_ISL_1057696, EPI_ISL_1057697, EPI_ISL_1057698, EPI_ISL_1057699, EPI_ISL_1057700, EPI_ISL_1057701, EPI_ISL_1057702, EPI_ISL_1057703, EPI_ISL_1057704, EPI_ISL_1057705, EPI_ISL_1057706, EPI_ISL_1057707, EPI_ISL_1057708, EPI_ISL_1057709, EPI_ISL_1057710, EPI_ISL_1057711, EPI_ISL_1057712, EPI_ISL_1057713, EPI_ISL_1057714, EPI_ISL_1057715, EPI_ISL_1057716, EPI_ISL_1057717, EPI_ISL_1057718, EPI_ISL_1057719, EPI_ISL_1057720, EPI_ISL_1057721, EPI_ISL_1057722, EPI_ISL_1057724, EPI_ISL_1057725, EPI_ISL_1057857, EPI_ISL_1057858, EPI_ISL_1057861, EPI_ISL_1057862, EPI_ISL_1057863, EPI_ISL_1057864, EPI_ISL_1057865, EPI_ISL_1057866, EPI_ISL_1057867, EPI_ISL_1057868, EPI_ISL_1057870, EPI_ISL_1057871, EPI_ISL_1057872, EPI_ISL_1057873, EPI_ISL_1057874, EPI_ISL_1057875, EPI_ISL_1057876, EPI_ISL_1057979, EPI_ISL_1057980, EPI_ISL_1057981, EPI_ISL_1057982, EPI_ISL_1057983, EPI_ISL_1057984, | | | |

EPI_ISL_1057985, EPI_ISL_1057986, EPI_ISL_1057987, EPI_ISL_1057988, EPI_ISL_1057990, EPI_ISL_1057991, EPI_ISL_1057992, EPI_ISL_1057993, EPI_ISL_1057996, EPI_ISL_1057998, EPI_ISL_1057999, EPI_ISL_1058001, EPI_ISL_1058002, EPI_ISL_1058003, EPI_ISL_1058004, EPI_ISL_1058005, EPI_ISL_1058006, EPI_ISL_1058007, EPI_ISL_1058008, EPI_ISL_1058009, EPI_ISL_1058010, EPI_ISL_1058011, EPI_ISL_1058012, EPI_ISL_1058013, EPI_ISL_1058014, EPI_ISL_1058015, EPI_ISL_1058016, EPI_ISL_1058018, EPI_ISL_1061697, EPI_ISL_1061698, EPI_ISL_1061699, EPI_ISL_1061703, EPI_ISL_1061707, EPI_ISL_1061708, EPI_ISL_1061710, EPI_ISL_1061711, EPI_ISL_1061716, EPI_ISL_1061719, EPI_ISL_1061723, EPI_ISL_1061728, EPI_ISL_1061730, EPI_ISL_1061731, EPI_ISL_1061733, EPI_ISL_1061740, EPI_ISL_1061747, EPI_ISL_1061753, EPI_ISL_1061756, EPI_ISL_1061757, EPI_ISL_1061758, EPI_ISL_1061760, EPI_ISL_1061762, EPI_ISL_1061763, EPI_ISL_1061764, EPI_ISL_1061765, EPI_ISL_1061767, EPI_ISL_1061770, EPI_ISL_1061775, EPI_ISL_1061776, EPI_ISL_1061780, EPI_ISL_1061781, EPI_ISL_1061782, EPI_ISL_1061787, EPI_ISL_1061788, EPI_ISL_1061790, EPI_ISL_1061793, EPI_ISL_1061796, EPI_ISL_1061799, EPI_ISL_1061801, EPI_ISL_1061803, EPI_ISL_1061807, EPI_ISL_1061812, EPI_ISL_1061814, EPI_ISL_1061816, EPI_ISL_1061820, EPI_ISL_1061822, EPI_ISL_1061823, EPI_ISL_1061827, EPI_ISL_1061831, EPI_ISL_1061832, EPI_ISL_1061833, EPI_ISL_1061838, EPI_ISL_1061840, EPI_ISL_1061847, EPI_ISL_1061848, EPI_ISL_1061849, EPI_ISL_1061853, EPI_ISL_1061854, EPI_ISL_1061856, EPI_ISL_1061859, EPI_ISL_1061862, EPI_ISL_1061865, EPI_ISL_1061871, EPI_ISL_1061872, EPI_ISL_1061874, EPI_ISL_1061875, EPI_ISL_1061876, EPI_ISL_1061880, EPI_ISL_1061882, EPI_ISL_1061883, EPI_ISL_1061884, EPI_ISL_1061885, EPI_ISL_1061889, EPI_ISL_1061892, EPI_ISL_1061897, EPI_ISL_1061905, EPI_ISL_1061909, EPI_ISL_1061911, EPI_ISL_1061919, EPI_ISL_1061920, EPI_ISL_1061927, EPI_ISL_1061928, EPI_ISL_1061930, EPI_ISL_1061932, EPI_ISL_1061933, EPI_ISL_1061937, EPI_ISL_1061938, EPI_ISL_1061943, EPI_ISL_1061945, EPI_ISL_1061947, EPI_ISL_1061949, EPI_ISL_1061954, EPI_ISL_1061956, EPI_ISL_1061959, EPI_ISL_1061964, EPI_ISL_1061967, EPI_ISL_1061975, EPI_ISL_1061979, EPI_ISL_1061984, EPI_ISL_1061986, EPI_ISL_1061988, EPI_ISL_1061992, EPI_ISL_1061993, EPI_ISL_1061994, EPI_ISL_1061995, EPI_ISL_1062000, EPI_ISL_1062003, EPI_ISL_1062004, EPI_ISL_1062011, EPI_ISL_1062017, EPI_ISL_1062019, EPI_ISL_1062023, EPI_ISL_1062029, EPI_ISL_1062039, EPI_ISL_1062040, EPI_ISL_1062043, EPI_ISL_1062048, EPI_ISL_1062050, EPI_ISL_1062052, EPI_ISL_1062053, EPI_ISL_1062054, EPI_ISL_1062055, EPI_ISL_1062056, EPI_ISL_1062057, EPI_ISL_1062058, EPI_ISL_1063016, EPI_ISL_1063017, EPI_ISL_1063018, EPI_ISL_1063019, EPI_ISL_1063020, EPI_ISL_1063021, EPI_ISL_1063022, EPI_ISL_1063023, EPI_ISL_1063024, EPI_ISL_1063025, EPI_ISL_1063026, EPI_ISL_1063027, EPI_ISL_1063028, EPI_ISL_1063029, EPI_ISL_1063030, EPI_ISL_1063031, EPI_ISL_1063032, EPI_ISL_1063033, EPI_ISL_1063034, EPI_ISL_1063035, EPI_ISL_1063036, EPI_ISL_1063037, EPI_ISL_1063038, EPI_ISL_1063039, EPI_ISL_1063040, EPI_ISL_1063041, EPI_ISL_1063042, EPI_ISL_1063043, EPI_ISL_1063044, EPI_ISL_1063045, EPI_ISL_1071640, EPI_ISL_1071651, EPI_ISL_1071670, EPI_ISL_1071870, EPI_ISL_1071911, EPI_ISL_1071921, EPI_ISL_1072034, EPI_ISL_1072041, EPI_ISL_1072051, EPI_ISL_1072072, EPI_ISL_1072080, EPI_ISL_1072093, EPI_ISL_1072116, EPI_ISL_1072125, EPI_ISL_1072132, EPI_ISL_1072160, EPI_ISL_1072189, EPI_ISL_1072247, EPI_ISL_1072249, EPI_ISL_1072269, EPI_ISL_1072288, EPI_ISL_1072313, EPI_ISL_1072332, EPI_ISL_1083317, EPI_ISL_1084701, EPI_ISL_1084709, EPI_ISL_1101619, EPI_ISL_1374602

| | | | |
|---|---|---|---|
| see above | Lighthouse Lab in Alderley Park | Wellcome Sanger Institute for the COVID-19 Genomics UK (COG-UK) Consortium | Cordelia Langford; David K. Jackson; Dominic Kwiatkowski; Ewan Harrison; Ian Johnston; Jacquelyn Wynn; Jeffrey Barrett; John Sillitoe on behalf of the Wellcome Sanger Institute COVID-19 Surveillance Team; Mairead Hyland; Roberto Amato; Sonia Goncalves; The Lighthouse Lab in Alderley Park and Alex Alderton |

EPI_ISL_986563, EPI_ISL_1005737, EPI_ISL_1005777, EPI_ISL_1005787, EPI_ISL_1005814, EPI_ISL_1005821, EPI_ISL_1005825, EPI_ISL_1005891, EPI_ISL_1005894, EPI_ISL_1005900, EPI_ISL_1006257, EPI_ISL_1012133, EPI_ISL_1012549, EPI_ISL_1018689, EPI_ISL_1018690, EPI_ISL_1018691, EPI_ISL_1018692, EPI_ISL_1018695, EPI_ISL_1018696, EPI_ISL_1018697, EPI_ISL_1018698, EPI_ISL_1018699, EPI_ISL_1018700, EPI_ISL_1018702, EPI_ISL_1018703, EPI_ISL_1018704, EPI_ISL_1018705, EPI_ISL_1018706, EPI_ISL_1018707, EPI_ISL_1018708, EPI_ISL_1018709, EPI_ISL_1018710, EPI_ISL_1018711, EPI_ISL_1018712, EPI_ISL_1018714, EPI_ISL_1018715, EPI_ISL_1018716, EPI_ISL_1018717, EPI_ISL_1018718, EPI_ISL_1018719, EPI_ISL_1018720, EPI_ISL_1018721, EPI_ISL_1018722, EPI_ISL_1018723, EPI_ISL_1018724, EPI_ISL_1018726, EPI_ISL_1018727, EPI_ISL_1018728, EPI_ISL_1018729, EPI_ISL_1018731, EPI_ISL_1018732, EPI_ISL_1018733, EPI_ISL_1018734, EPI_ISL_1018735, EPI_ISL_1018736, EPI_ISL_1018737, EPI_ISL_1018738, EPI_ISL_1018739, EPI_ISL_1018740, EPI_ISL_1018741, EPI_ISL_1018742, EPI_ISL_1018743, EPI_ISL_1018744, EPI_ISL_1018745, EPI_ISL_1018746, EPI_ISL_1018747, EPI_ISL_1018748, EPI_ISL_1018749, EPI_ISL_1018750, EPI_ISL_1018751, EPI_ISL_1018752, EPI_ISL_1018753, EPI_ISL_1018754, EPI_ISL_1018757, EPI_ISL_1018758, EPI_ISL_1018759, EPI_ISL_1018760, EPI_ISL_1018761, EPI_ISL_1018763, EPI_ISL_1018764, EPI_ISL_1018765, EPI_ISL_1018767, EPI_ISL_1018768, EPI_ISL_1018769, EPI_ISL_1018770, EPI_ISL_1018771, EPI_ISL_1018772, EPI_ISL_1018773, EPI_ISL_1018774, EPI_ISL_1018775, EPI_ISL_1018777, EPI_ISL_1018778, EPI_ISL_1018779, EPI_ISL_1018780, EPI_ISL_1018781, EPI_ISL_1018782, EPI_ISL_1018783, EPI_ISL_1018784, EPI_ISL_1018785, EPI_ISL_1018786, EPI_ISL_1018787, EPI_ISL_1018788, EPI_ISL_1018789, EPI_ISL_1018790, EPI_ISL_1018791, EPI_ISL_1018792, EPI_ISL_1018793, EPI_ISL_1018794, EPI_ISL_1018795, EPI_ISL_1018796, EPI_ISL_1018797, EPI_ISL_1018798, EPI_ISL_1018799, EPI_ISL_1018801, EPI_ISL_1018802, EPI_ISL_1018804, EPI_ISL_1018805, EPI_ISL_1018806, EPI_ISL_1018809, EPI_ISL_1018810, EPI_ISL_1018811, EPI_ISL_1018812, EPI_ISL_1018813, EPI_ISL_1018815, EPI_ISL_1018817, EPI_ISL_1018818, EPI_ISL_1018819, EPI_ISL_1018820, EPI_ISL_1018821, EPI_ISL_1018822, EPI_ISL_1018823, EPI_ISL_1018824, EPI_ISL_1018825, EPI_ISL_1018826, EPI_ISL_1018827, EPI_ISL_1018828, EPI_ISL_1018829, EPI_ISL_1018830, EPI_ISL_1018831, EPI_ISL_1018832, EPI_ISL_1018833, EPI_ISL_1018834, EPI_ISL_1018835, EPI_ISL_1018836, EPI_ISL_1018837, EPI_ISL_1018839, EPI_ISL_1018840, EPI_ISL_1018841, EPI_ISL_1018842, EPI_ISL_1018843, EPI_ISL_1018845, EPI_ISL_1018846, EPI_ISL_1018847, EPI_ISL_1018848, EPI_ISL_1018849, EPI_ISL_1018850, EPI_ISL_1018851, EPI_ISL_1018852, EPI_ISL_1018853, EPI_ISL_1018854, EPI_ISL_1018855, EPI_ISL_1018856, EPI_ISL_1018857, EPI_ISL_1018859, EPI_ISL_1018860, EPI_ISL_1018861, EPI_ISL_1018862, EPI_ISL_1018863, EPI_ISL_1018864, EPI_ISL_1018866, EPI_ISL_1018867, EPI_ISL_1018868, EPI_ISL_1018869, EPI_ISL_1018870, EPI_ISL_1018871, EPI_ISL_1018873, EPI_ISL_1018875, EPI_ISL_1018877, EPI_ISL_1018878, EPI_ISL_1018879, EPI_ISL_1018881, EPI_ISL_1018883, EPI_ISL_1018884, EPI_ISL_1018885, EPI_ISL_1018886, EPI_ISL_1018887, EPI_ISL_1018888, EPI_ISL_1018889, EPI_ISL_1018891, EPI_ISL_1018892, EPI_ISL_1018893, EPI_ISL_1018894, EPI_ISL_1018895, EPI_ISL_1018896, EPI_ISL_1018897, EPI_ISL_1018898, EPI_ISL_1018899, EPI_ISL_1018900, EPI_ISL_1018901, EPI_ISL_1018902, EPI_ISL_1018903, EPI_ISL_1018904, EPI_ISL_1018905, EPI_ISL_1018906, EPI_ISL_1018908, EPI_ISL_1018910, EPI_ISL_1018911, EPI_ISL_1018913, EPI_ISL_1018916, EPI_ISL_1018918, EPI_ISL_1018919, EPI_ISL_1018922, EPI_ISL_1018923, EPI_ISL_1018925, EPI_ISL_1018926, EPI_ISL_1018927, EPI_ISL_1018928, EPI_ISL_1018930, EPI_ISL_1018931, EPI_ISL_1018932, EPI_ISL_1018933, EPI_ISL_1018934, EPI_ISL_1018936, EPI_ISL_1018937, EPI_ISL_1018938, EPI_ISL_1018939, EPI_ISL_1018940, EPI_ISL_1018942, EPI_ISL_1018943, EPI_ISL_1018944, EPI_ISL_1018945, EPI_ISL_1018946, EPI_ISL_1018947, EPI_ISL_1018948, EPI_ISL_1018949, EPI_ISL_1019286, EPI_ISL_1019287, EPI_ISL_1019288, EPI_ISL_1019289, EPI_ISL_1019290, EPI_ISL_1019291, EPI_ISL_1019292, EPI_ISL_1019293, EPI_ISL_1019294, EPI_ISL_1019295, EPI_ISL_1019296, EPI_ISL_1019297, EPI_ISL_1019298, EPI_ISL_1019299, EPI_ISL_1019301, EPI_ISL_1019302, EPI_ISL_1019303, EPI_ISL_1019304, EPI_ISL_1019306, EPI_ISL_1019307, EPI_ISL_1019308, EPI_ISL_1019309, EPI_ISL_1019310, EPI_ISL_1019312, EPI_ISL_1019313, EPI_ISL_1019314, EPI_ISL_1019315, EPI_ISL_1019316, EPI_ISL_1019317, EPI_ISL_1019318, EPI_ISL_1019319, EPI_ISL_1019320, EPI_ISL_1019321, EPI_ISL_1019322, EPI_ISL_1019323, EPI_ISL_1019324, EPI_ISL_1019325, EPI_ISL_1019326, EPI_ISL_1019327, EPI_ISL_1019328, EPI_ISL_1019329, EPI_ISL_1019330, EPI_ISL_1019332, EPI_ISL_1019333, EPI_ISL_1019334, EPI_ISL_1019335, EPI_ISL_1019336, EPI_ISL_1019337, EPI_ISL_1019338, EPI_ISL_1019339, EPI_ISL_1019340, EPI_ISL_1019341, EPI_ISL_1019342, EPI_ISL_1019343, EPI_ISL_1019344, EPI_ISL_1019345, EPI_ISL_1019346, EPI_ISL_1019347, EPI_ISL_1019348, EPI_ISL_1019350, EPI_ISL_1019351, EPI_ISL_1033163, EPI_ISL_1033164, EPI_ISL_1033165, EPI_ISL_1033167, EPI_ISL_1033168, EPI_ISL_1033169, EPI_ISL_1033170, EPI_ISL_1033171, EPI_ISL_1033172, EPI_ISL_1033173, EPI_ISL_1033174, EPI_ISL_1033176, EPI_ISL_1033177, EPI_ISL_1033178, EPI_ISL_1033179, EPI_ISL_1033180, EPI_ISL_1033181, EPI_ISL_1033182, EPI_ISL_1033183, EPI_ISL_1033184, EPI_ISL_1033185, EPI_ISL_1033186, EPI_ISL_1033187, EPI_ISL_1033188, EPI_ISL_1033189, EPI_ISL_1033190, EPI_ISL_1033191, EPI_ISL_1033192, EPI_ISL_1033193, EPI_ISL_1033194, EPI_ISL_1033195, EPI_ISL_1033196, EPI_ISL_1033197, EPI_ISL_1033198, EPI_ISL_1033199, EPI_ISL_1033200, EPI_ISL_1033201, EPI_ISL_1033202, EPI_ISL_1033203, EPI_ISL_1033204, EPI_ISL_1033205, EPI_ISL_1033206, EPI_ISL_1033207, EPI_ISL_1033208, EPI_ISL_1033209, EPI_ISL_1033210, EPI_ISL_1033211, EPI_ISL_1033212, EPI_ISL_1033213, EPI_ISL_1033214, EPI_ISL_1033215, EPI_ISL_1033217, EPI_ISL_1033218, EPI_ISL_1033219, EPI_ISL_1033220, EPI_ISL_1033221, EPI_ISL_1033222, EPI_ISL_1033223, EPI_ISL_1033224, EPI_ISL_1033225, EPI_ISL_1033226, EPI_ISL_1033227, EPI_ISL_1033228, EPI_ISL_1033229, EPI_ISL_1033230, EPI_ISL_1033231, EPI_ISL_1033232, EPI_ISL_1033233, EPI_ISL_1033234, EPI_ISL_1033235, EPI_ISL_1033236, EPI_ISL_1033237, EPI_ISL_1033238, EPI_ISL_1033239, EPI_ISL_1033240, EPI_ISL_1033241, EPI_ISL_1033242, EPI_ISL_1033243, EPI_ISL_1033245, EPI_ISL_1033246, EPI_ISL_1033247, EPI_ISL_1033248, EPI_ISL_1033249, EPI_ISL_1033250, EPI_ISL_1033251, EPI_ISL_1033252, EPI_ISL_1033253, EPI_ISL_1033254, EPI_ISL_1033255, EPI_ISL_1033256, EPI_ISL_1033257, EPI_ISL_1033258, EPI_ISL_1033259, EPI_ISL_1033260, EPI_ISL_1033261, EPI_ISL_1033262, EPI_ISL_1033263, EPI_ISL_1033264, EPI_ISL_1033265, EPI_ISL_1033266, EPI_ISL_1033267, EPI_ISL_1033268, EPI_ISL_1033269, EPI_ISL_1033271, EPI_ISL_1033272, EPI_ISL_1033273, EPI_ISL_1033274, EPI_ISL_1033275, EPI_ISL_1033276, EPI_ISL_1033277, EPI_ISL_1033278, EPI_ISL_1033279, EPI_ISL_1033280, EPI_ISL_1033281, EPI_ISL_1033282, EPI_ISL_1033283, EPI_ISL_1033284, EPI_ISL_1033285, EPI_ISL_1033286, EPI_ISL_1033287, EPI_ISL_1033288, EPI_ISL_1033290, EPI_ISL_1033291, EPI_ISL_1033292, EPI_ISL_1033293, EPI_ISL_1033294, EPI_ISL_1033295, EPI_ISL_1033296, EPI_ISL_1033297, EPI_ISL_1033298, EPI_ISL_1033300, EPI_ISL_1033301, EPI_ISL_1033302, EPI_ISL_1033303, EPI_ISL_1033304, EPI_ISL_1033305, EPI_ISL_1033306, EPI_ISL_1033307, EPI_ISL_1033308, EPI_ISL_1033309, EPI_ISL_1033310, EPI_ISL_1033311, EPI_ISL_1033312, EPI_ISL_1033313, EPI_ISL_1033314, EPI_ISL_1033315, EPI_ISL_1033316, EPI_ISL_1033317, EPI_ISL_1033318, EPI_ISL_1033319, EPI_ISL_1033320, EPI_ISL_1033321, EPI_ISL_1033322, EPI_ISL_1033323, EPI_ISL_1033324, EPI_ISL_1033325, EPI_ISL_1033326, EPI_ISL_1033327, EPI_ISL_1033328, EPI_ISL_1033329, EPI_ISL_1033330, EPI_ISL_1033331, EPI_ISL_1033332, EPI_ISL_1033333, EPI_ISL_1033334, EPI_ISL_1033335, EPI_ISL_1033336, EPI_ISL_1033337, EPI_ISL_1033338, EPI_ISL_1033339, EPI_ISL_1033340, EPI_ISL_1033341, EPI_ISL_1033342, EPI_ISL_1033343, EPI_ISL_1033344, EPI_ISL_1033345, EPI_ISL_1033346, EPI_ISL_1033347, EPI_ISL_1033348, EPI_ISL_1033349, EPI_ISL_1033350, EPI_ISL_1033351, EPI_ISL_1033352, EPI_ISL_1033353, EPI_ISL_1033354, EPI_ISL_1033355, EPI_ISL_1033356, EPI_ISL_1033357, EPI_ISL_1033358, EPI_ISL_1033359, EPI_ISL_1033360, EPI_ISL_1033361, EPI_ISL_1033362, EPI_ISL_1033363, EPI_ISL_1033364, EPI_ISL_1033365, EPI_ISL_1033366, EPI_ISL_1033367, EPI_ISL_1033368, EPI_ISL_1033369, EPI_ISL_1033370, EPI_ISL_1033371, EPI_ISL_1033372, EPI_ISL_1033373, EPI_ISL_1033374, EPI_ISL_1033375, EPI_ISL_1033376, EPI_ISL_1033377, EPI_ISL_1033378, EPI_ISL_1033379, EPI_ISL_1033380, EPI_ISL_1033381, EPI_ISL_1033382, EPI_ISL_1033383, EPI_ISL_1033384, EPI_ISL_1033385, EPI_ISL_1033386, EPI_ISL_1033387, EPI_ISL_1033388, EPI_ISL_1033389, EPI_ISL_1033390, EPI_ISL_1033391, EPI_ISL_1033392, EPI_ISL_1033393, EPI_ISL_1033394, EPI_ISL_1033395, EPI_ISL_1033396, EPI_ISL_1033397, EPI_ISL_1033398, EPI_ISL_1033399, EPI_ISL_1033400, EPI_ISL_1033401, EPI_ISL_1033402, EPI_ISL_1033403, EPI_ISL_1033404, EPI_ISL_1033405, EPI_ISL_1033406, EPI_ISL_1033407, EPI_ISL_1033408, EPI_ISL_1033409, EPI_ISL_1033410, EPI_ISL_1033411, EPI_ISL_1033412, EPI_ISL_1033413, EPI_ISL_1033414, EPI_ISL_1033415, EPI_ISL_1033416, EPI_ISL_1033417, EPI_ISL_1033418, EPI_ISL_1033419, EPI_ISL_1033420, EPI_ISL_1033421, EPI_ISL_1033422, EPI_ISL_1033423, EPI_ISL_1033424, EPI_ISL_1033425, EPI_ISL_1033426, EPI_ISL_1033427, EPI_ISL_1033429, EPI_ISL_1033430, EPI_ISL_1033431, EPI_ISL_1033432, EPI_ISL_1033433, EPI_ISL_1033434, EPI_ISL_1033435, EPI_ISL_1033436, EPI_ISL_1033437, EPI_ISL_1033438, EPI_ISL_1033439, EPI_ISL_1033440, EPI_ISL_1033441, EPI_ISL_1033442, EPI_ISL_1033443, EPI_ISL_1033444, EPI_ISL_1033445, EPI_ISL_1033446, EPI_ISL_1033447, EPI_ISL_1033448, EPI_ISL_1033449, EPI_ISL_1033450, EPI_ISL_1033451, EPI_ISL_1033452, EPI_ISL_1033453, EPI_ISL_1033454, EPI_ISL_1033455, EPI_ISL_1033456, EPI_ISL_1033457, EPI_ISL_1033458, EPI_ISL_1033459, EPI_ISL_1033460, EPI_ISL_1033461, EPI_ISL_1033462, EPI_ISL_1033463, EPI_ISL_1033464, EPI_ISL_1033465, EPI_ISL_1033466, EPI_ISL_1033467, EPI_ISL_1033468, EPI_ISL_1033469, EPI_ISL_1033470, EPI_ISL_1033471, EPI_ISL_1033472, EPI_ISL_1033473, EPI_ISL_1033474, EPI_ISL_1033475, EPI_ISL_1033476, EPI_ISL_1033477, EPI_ISL_1033478, EPI_ISL_1033479, EPI_ISL_1033480, EPI_ISL_1033481, EPI_ISL_1033482, EPI_ISL_1033483, EPI_ISL_1033485, EPI_ISL_1033486, EPI_ISL_1033487, EPI_ISL_1033488, EPI_ISL_1033489, EPI_ISL_1033490, EPI_ISL_1033491, EPI_ISL_1033493, EPI_ISL_1033494, EPI_ISL_1033495, EPI_ISL_1033496, EPI_ISL_1033497, EPI_ISL_1033498, EPI_ISL_1033499, EPI_ISL_1033500, EPI_ISL_1033501, EPI_ISL_1033502, EPI_ISL_1033503, EPI_ISL_1033504, EPI_ISL_1033505, EPI_ISL_1033506, EPI_ISL_1033507, EPI_ISL_1033508, EPI_ISL_1033509, EPI_ISL_1033510, EPI_ISL_1033511, EPI_ISL_1033512, EPI_ISL_1033513, EPI_ISL_1033514, EPI_ISL_1033515, EPI_ISL_1033516, EPI_ISL_1033517, EPI_ISL_1033518, EPI_ISL_1033519, EPI_ISL_1033520, EPI_ISL_1033521, EPI_ISL_1033522, EPI_ISL_1033523, EPI_ISL_1033524, EPI_ISL_1033525, EPI_ISL_1033526, EPI_ISL_1033527, EPI_ISL_1033528, EPI_ISL_1033529, EPI_ISL_1033530, EPI_ISL_1033531, EPI_ISL_1033532, EPI_ISL_1033533, EPI_ISL_1033534, EPI_ISL_1033535, EPI_ISL_1033536, EPI_ISL_1033601, EPI_ISL_1033610, EPI_ISL_1033620, EPI_ISL_1033644, EPI_ISL_1033660, EPI_ISL_1033669, EPI_ISL_1033673, EPI_ISL_1033701, EPI_ISL_1042014, EPI_ISL_1042017, EPI_ISL_1042020, EPI_ISL_1042028, EPI_ISL_1042031, EPI_ISL_1042033, EPI_ISL_1042036, EPI_ISL_1042037, EPI_ISL_1042040, EPI_ISL_1042043, EPI_ISL_1042045, EPI_ISL_1042046, EPI_ISL_1042047, EPI_ISL_1042048, EPI_ISL_1042050, EPI_ISL_1042066, EPI_ISL_1042067, EPI_ISL_1042074, EPI_ISL_1042077, EPI_ISL_1042081, EPI_ISL_1042084, EPI_ISL_1042085, EPI_ISL_1042091, EPI_ISL_1042094, EPI_ISL_1042096, EPI_ISL_1042100, EPI_ISL_1042109, EPI_ISL_1042113, EPI_ISL_1042115, EPI_ISL_1042120, EPI_ISL_1042122, EPI_ISL_1042130, EPI_ISL_1042143, EPI_ISL_1042149, EPI_ISL_1042150, EPI_ISL_1042157, EPI_ISL_1042160, EPI_ISL_1042164, EPI_ISL_1042168, EPI_ISL_1042176, EPI_ISL_1042184, EPI_ISL_1042189, EPI_ISL_1042196, EPI_ISL_1042208, EPI_ISL_1042223, EPI_ISL_1042228, EPI_ISL_1042231, EPI_ISL_1042238, EPI_ISL_1042243, EPI_ISL_1042245, EPI_ISL_1042249, EPI_ISL_1042252, EPI_ISL_1042266, EPI_ISL_1042271, EPI_ISL_1042272, EPI_ISL_1042279, EPI_ISL_1042280, EPI_ISL_1042282, EPI_ISL_1042298, EPI_ISL_1042299, EPI_ISL_1042300, EPI_ISL_1042306, EPI_ISL_1042315, EPI_ISL_1042333, EPI_ISL_1042335, EPI_ISL_1042342, EPI_ISL_1042344, EPI_ISL_1042345, EPI_ISL_1044225, EPI_ISL_1044226, EPI_ISL_1044227, EPI_ISL_1044228, EPI_ISL_1044229, EPI_ISL_1044230, EPI_ISL_1044231, EPI_ISL_1044232, EPI_ISL_1044233, EPI_ISL_1044234, EPI_ISL_1044235, EPI_ISL_1044236, EPI_ISL_1044237, EPI_ISL_1044238, EPI_ISL_1044239, EPI_ISL_1044240, EPI_ISL_1044241, EPI_ISL_1044243, EPI_ISL_1044244, EPI_ISL_1044245, EPI_ISL_1044246, EPI_ISL_1044247, EPI_ISL_1044248, EPI_ISL_1044249, EPI_ISL_1044250, EPI_ISL_1044251, EPI_ISL_1044252, EPI_ISL_1044253, EPI_ISL_1044254, EPI_ISL_1044255, EPI_ISL_1044256, EPI_ISL_1044257, EPI_ISL_1044258, EPI_ISL_1044259, EPI_ISL_1044260, EPI_ISL_1044261, EPI_ISL_1044262, EPI_ISL_1044263, EPI_ISL_1044264, EPI_ISL_1044265, EPI_ISL_1044266, EPI_ISL_1044267, EPI_ISL_1044268, EPI_ISL_1044269, EPI_ISL_1044270, EPI_ISL_1044271, EPI_ISL_1044272, EPI_ISL_1044273, EPI_ISL_1044274, EPI_ISL_1044275, EPI_ISL_1044276, EPI_ISL_1044277, EPI_ISL_1044278, EPI_ISL_1044279, EPI_ISL_1044280, EPI_ISL_1044281, EPI_ISL_1044282, EPI_ISL_1044283, EPI_ISL_1044284, EPI_ISL_1044285, EPI_ISL_1044286, EPI_ISL_1044287, EPI_ISL_1044289, EPI_ISL_1044290, EPI_ISL_1044291, EPI_ISL_1044292, EPI_ISL_1044293, EPI_ISL_1044294, EPI_ISL_1044295, EPI_ISL_1044296, EPI_ISL_1044297, EPI_ISL_1044298, EPI_ISL_1044299, EPI_ISL_1044300, EPI_ISL_1044301, EPI_ISL_1044302, EPI_ISL_1044303, EPI_ISL_1044304, EPI_ISL_1044305, EPI_ISL_1044306, EPI_ISL_1044307, EPI_ISL_1044308, EPI_ISL_1044309, EPI_ISL_1044310, EPI_ISL_1044311, EPI_ISL_1044312, EPI_ISL_1044313, EPI_ISL_1044314, EPI_ISL_1044315, EPI_ISL_1044316, EPI_ISL_1044317, EPI_ISL_1044318, EPI_ISL_1044319, EPI_ISL_1044320, EPI_ISL_1044321, EPI_ISL_1044322, EPI_ISL_1044323, EPI_ISL_1044324, EPI_ISL_1044325, EPI_ISL_1044326, EPI_ISL_1044327, EPI_ISL_1044328, EPI_ISL_1044329, EPI_ISL_1044330, EPI_ISL_1044331, EPI_ISL_1044332, EPI_ISL_1044333, EPI_ISL_1044334, EPI_ISL_1044335, EPI_ISL_1044336, EPI_ISL_1044337, EPI_ISL_1044338, EPI_ISL_1044339, EPI_ISL_1044340, EPI_ISL_1044341, EPI_ISL_1044342, EPI_ISL_1044343, EPI_ISL_1044344, EPI_ISL_1044345, EPI_ISL_1044346, EPI_ISL_1044347, EPI_ISL_1044348, EPI_ISL_1044349, EPI_ISL_1044350, EPI_ISL_1044351, EPI_ISL_1044352, EPI_ISL_1044353, EPI_ISL_1044354, EPI_ISL_1044355, EPI_ISL_1044356, EPI_ISL_1044357, EPI_ISL_1044358, EPI_ISL_1044359, EPI_ISL_1044360, EPI_ISL_1044361, EPI_ISL_1044362, EPI_ISL_1044363, EPI_ISL_1044364, EPI_ISL_1044365, EPI_ISL_1044366, EPI_ISL_1044367, EPI_ISL_1044368, EPI_ISL_1044369, EPI_ISL_1044370, EPI_ISL_1044371, EPI_ISL_1044372, EPI_ISL_1044373, EPI_ISL_1044374, EPI_ISL_1044375, EPI_ISL_1044376, EPI_ISL_1044377, EPI_ISL_1044378, EPI_ISL_1044379, EPI_ISL_1044380, EPI_ISL_1044381, EPI_ISL_1044382, EPI_ISL_1044383, EPI_ISL_1044384, EPI_ISL_1044385, EPI_ISL_1044725, EPI_ISL_1044726, EPI_ISL_1044727, EPI_ISL_1044728, EPI_ISL_1044729, EPI_ISL_1044730, EPI_ISL_1044731, EPI_ISL_1044732, EPI_ISL_1044733, EPI_ISL_1044734, EPI_ISL_1044735, EPI_ISL_1044736, EPI_ISL_1044737, EPI_ISL_1044738, EPI_ISL_1044739, EPI_ISL_1044740, EPI_ISL_1044741, EPI_ISL_1044742, EPI_ISL_1044743, EPI_ISL_1044744, EPI_ISL_1044745, EPI_ISL_1044746, EPI_ISL_1044747, EPI_ISL_1044748, EPI_ISL_1044749, EPI_ISL_1044750, EPI_ISL_1044751, EPI_ISL_1044752, EPI_ISL_1044753, EPI_ISL_1044754, EPI_ISL_1044755, EPI_ISL_1044756, EPI_ISL_1044757, EPI_ISL_1044758, EPI_ISL_1044759, EPI_ISL_1044760, EPI_ISL_1044761, EPI_ISL_1044762, EPI_ISL_1044763, EPI_ISL_1044764, EPI_ISL_1044765, EPI_ISL_1044766, EPI_ISL_1044767, EPI_ISL_1044768, EPI_ISL_1044769, EPI_ISL_1044770, EPI_ISL_1044771, EPI_ISL_1044772, EPI_ISL_1044773, EPI_ISL_1044774, EPI_ISL_1044775, EPI_ISL_1044776, EPI_ISL_1044777, EPI_ISL_1044778, EPI_ISL_1044779, EPI_ISL_1044780, EPI_ISL_1044781, EPI_ISL_1044782, EPI_ISL_1044783, EPI_ISL_1044784, EPI_ISL_1044785, EPI_ISL_1044786, EPI_ISL_1044787, EPI_ISL_1044788, EPI_ISL_1044789, EPI_ISL_1044790, EPI_ISL_1044791, EPI_ISL_1044792, EPI_ISL_1044793, EPI_ISL_1044794, EPI_ISL_1044795, EPI_ISL_1044796, EPI_ISL_1044797, EPI_ISL_1044798, EPI_ISL_1044799, EPI_ISL_1044800, EPI_ISL_1044801, EPI_ISL_1044802, EPI_ISL_1044803, EPI_ISL_1044804, EPI_ISL_1044805, EPI_ISL_1044806, EPI_ISL_1044807, EPI_ISL_1044808, EPI_ISL_1044809, EPI_ISL_1044810, EPI_ISL_1044811, EPI_ISL_1044812, EPI_ISL_1044813, EPI_ISL_1044814, EPI_ISL_1044815, EPI_ISL_1044816, EPI_ISL_1044817, EPI_ISL_1044818, EPI_ISL_1044819, EPI_ISL_1044820, EPI_ISL_1044821, EPI_ISL_1044822, EPI_ISL_1044823, EPI_ISL_1044824, EPI_ISL_1044825, EPI_ISL_1044826, EPI_ISL_1044827, EPI_ISL_1044828, EPI_ISL_1044829, EPI_ISL_1044830, EPI_ISL_1044831, EPI_ISL_1044832, EPI_ISL_1044833, EPI_ISL_1044834, EPI_ISL_1044835, EPI_ISL_1044836, EPI_ISL_1044837, EPI_ISL_1044838, EPI_ISL_1044839, EPI_ISL_1044840, EPI_ISL_1044841, EPI_ISL_1044842, EPI_ISL_1044843, EPI_ISL_1044844, EPI_ISL_1044845, EPI_ISL_1044846, EPI_ISL_1044847, EPI_ISL_1044848, EPI_ISL_1044849, EPI_ISL_1044850, EPI_ISL_1044851, EPI_ISL_1044852, EPI_ISL_1044853, EPI_ISL_1044854, EPI_ISL_1044855, EPI_ISL_1044856, EPI_ISL_1044857, EPI_ISL_1044858, EPI_ISL_1044859, EPI_ISL_1044860, EPI_ISL_1044861, EPI_ISL_1044862, EPI_ISL_1044863, EPI_ISL_1044864, EPI_ISL_1045682, EPI_ISL_1045683, EPI_ISL_1045684, EPI_ISL_1045685, EPI_ISL_1045686, EPI_ISL_1045687, EPI_ISL_1045688, EPI_ISL_1045689, EPI_ISL_1045690, EPI_ISL_1045691, EPI_ISL_1045692, EPI_ISL_1045693, EPI_ISL_1045694, EPI_ISL_1045695, EPI_ISL_1045696, EPI_ISL_1045697, EPI_ISL_1045698, EPI_ISL_1045699, EPI_ISL_1045700, EPI_ISL_1045701, EPI_ISL_1045702, EPI_ISL_1045703, EPI_ISL_1045704, EPI_ISL_1045706, EPI_ISL_1045708, EPI_ISL_1045727, EPI_ISL_1045728, EPI_ISL_1045730, EPI_ISL_1045733, EPI_ISL_1045747, EPI_ISL_1045751, EPI_ISL_1045755, EPI_ISL_1045757, EPI_ISL_1045761, EPI_ISL_1045762, EPI_ISL_1045768, EPI_ISL_1045773, EPI_ISL_1045777, EPI_ISL_1045778, EPI_ISL_1045788, EPI_ISL_1045809, EPI_ISL_1045812, EPI_ISL_1045815, EPI_ISL_1045819, EPI_ISL_1045820, EPI_ISL_1045822, EPI_ISL_1045831, EPI_ISL_1045842, EPI_ISL_1045845, EPI_ISL_1045846, EPI_ISL_1045848, EPI_ISL_1045850, EPI_ISL_1045853, EPI_ISL_1045854, EPI_ISL_1045870, EPI_ISL_1045872, EPI_ISL_1045876, EPI_ISL_1045877, EPI_ISL_1045878, EPI_ISL_1045881, EPI_ISL_1045882, EPI_ISL_1045884, EPI_ISL_1045887, EPI_ISL_1045890, EPI_ISL_1045891, EPI_ISL_1045892, EPI_ISL_1045893, EPI_ISL_1045894, EPI_ISL_1045896, EPI_ISL_1045897, EPI_ISL_1045899, EPI_ISL_1045901, EPI_ISL_1045903, EPI_ISL_1045904, EPI_ISL_1045907, EPI_ISL_1045908, EPI_ISL_1045912, EPI_ISL_1045918, EPI_ISL_1045920, EPI_ISL_1045924, EPI_ISL_1045927, EPI_ISL_1045931, EPI_ISL_1045932, EPI_ISL_1045933, EPI_ISL_1045937, EPI_ISL_1045939, EPI_ISL_1045943, EPI_ISL_1045945, EPI_ISL_1045947, EPI_ISL_1045948, EPI_ISL_1045949, EPI_ISL_1045955, EPI_ISL_1045956, EPI_ISL_1045962, EPI_ISL_1045967, EPI_ISL_1057818, EPI_ISL_1057819, EPI_ISL_1057820, EPI_ISL_1057821, EPI_ISL_1057822, EPI_ISL_1057823, EPI_ISL_1057824, EPI_ISL_1057825, EPI_ISL_1057826, EPI_ISL_1057827, EPI_ISL_1057828, EPI_ISL_1057829, EPI_ISL_1057830, EPI_ISL_1057831, EPI_ISL_1057832, EPI_ISL_1057833, EPI_ISL_1057834, EPI_ISL_1057835, EPI_ISL_1057836, EPI_ISL_1057837, EPI_ISL_1057838, EPI_ISL_1057839, EPI_ISL_1057840, EPI_ISL_1057841, EPI_ISL_1057842, EPI_ISL_1057843, EPI_ISL_1057844, EPI_ISL_1057845, EPI_ISL_1057846, EPI_ISL_1057847, EPI_ISL_1057848, EPI_ISL_1057849, EPI_ISL_1057850, EPI_ISL_1057851, EPI_ISL_1057852, EPI_ISL_1057853, EPI_ISL_1057854, EPI_ISL_1057855, EPI_ISL_1057856, EPI_ISL_1057879, EPI_ISL_1057881, EPI_ISL_1057888, EPI_ISL_1057893, EPI_ISL_1057899, EPI_ISL_1057902, EPI_ISL_1057904, EPI_ISL_1057907, EPI_ISL_1057914, EPI_ISL_1057917, EPI_ISL_1057918, EPI_ISL_1057922, EPI_ISL_1057924, EPI_ISL_1057931, EPI_ISL_1057932, EPI_ISL_1057934, EPI_ISL_1057936, EPI_ISL_1057939, EPI_ISL_1057942, EPI_ISL_1057944, EPI_ISL_1057945, EPI_ISL_1057946, EPI_ISL_1057949, EPI_ISL_1057951, EPI_ISL_1057956, EPI_ISL_1057957, EPI_ISL_1057958, EPI_ISL_1057960, EPI_ISL_1057961, EPI_ISL_1057965, EPI_ISL_1057966, EPI_ISL_1057972, EPI_ISL_1057974, EPI_ISL_1057976, EPI_ISL_1057977, EPI_ISL_1057978, EPI_ISL_1061661, EPI_ISL_1061674, EPI_ISL_1061687, EPI_ISL_1063396, EPI_ISL_1069994, EPI_ISL_1070011, EPI_ISL_1070016, EPI_ISL_1070023, EPI_ISL_1070029, EPI_ISL_1070039, EPI_ISL_1070051, EPI_ISL_1070052, EPI_ISL_1070055, EPI_ISL_1070068, EPI_ISL_1070074, EPI_ISL_1070077, EPI_ISL_1070088, EPI_ISL_1070341, EPI_ISL_1070348, EPI_ISL_1070350, EPI_ISL_1070352, EPI_ISL_1070353, EPI_ISL_1070355, EPI_ISL_1070357, EPI_ISL_1070359, EPI_ISL_1070361, EPI_ISL_1070366, EPI_ISL_1070369, EPI_ISL_1070371, EPI_ISL_1070376, EPI_ISL_1070385, EPI_ISL_1070386, EPI_ISL_1070390, EPI_ISL_1070391, EPI_ISL_1070399, EPI_ISL_1070403, EPI_ISL_1070405, EPI_ISL_1070413, EPI_ISL_1070420, EPI_ISL_1070421, EPI_ISL_1070424, EPI_ISL_1070426, EPI_ISL_1070428, EPI_ISL_1070430, EPI_ISL_1070435, EPI_ISL_1070436, EPI_ISL_1070440, EPI_ISL_1070441, EPI_ISL_1070451, EPI_ISL_1070455, EPI_ISL_1070456, EPI_ISL_1070460, EPI_ISL_1070464, EPI_ISL_1070466, EPI_ISL_1070467, EPI_ISL_1070472, EPI_ISL_1070474, EPI_ISL_1070480, EPI_ISL_1070484, EPI_ISL_1070487, EPI_ISL_1070489, EPI_ISL_1070490, EPI_ISL_1070493, EPI_ISL_1070494, EPI_ISL_1070498, EPI_ISL_1070499, EPI_ISL_1070500, EPI_ISL_1070501, EPI_ISL_1070503, EPI_ISL_1070504, EPI_ISL_1070507, EPI_ISL_1070508, EPI_ISL_1070510, EPI_ISL_1070516, EPI_ISL_1070520, EPI_ISL_1070525, EPI_ISL_1070527, EPI_ISL_1070531, EPI_ISL_1070532, EPI_ISL_1070540, EPI_ISL_1070544, EPI_ISL_1070545, EPI_ISL_1070547, EPI_ISL_1070549, EPI_ISL_1070552, EPI_ISL_1070555, EPI_ISL_1070558, EPI_ISL_1070561, EPI_ISL_1070567, EPI_ISL_1070570, EPI_ISL_1070574, EPI_ISL_1070577, EPI_ISL_1070578, EPI_ISL_1070579, EPI_ISL_1070580, EPI_ISL_1070581, EPI_ISL_1070591, EPI_ISL_1070594, EPI_ISL_1070597, EPI_ISL_1070598, EPI_ISL_1070601, EPI_ISL_1070612, EPI_ISL_1070616, EPI_ISL_1070617, EPI_ISL_1070619, EPI_ISL_1070621, EPI_ISL_1070622, EPI_ISL_1070625, EPI_ISL_1082783, EPI_ISL_1102750, EPI_ISL_1102864, EPI_ISL_1102913, EPI_ISL_1102963, EPI_ISL_1116310, EPI_ISL_1187078, EPI_ISL_1187151, EPI_ISL_1187168, EPI_ISL_1187181, EPI_ISL_1187212, EPI_ISL_1190726, EPI_ISL_1206082, EPI_ISL_1206123, EPI_ISL_1206125, EPI_ISL_1206127, EPI_ISL_1206128, EPI_ISL_1206133, EPI_ISL_1206134, EPI_ISL_1206143, EPI_ISL_1206153, EPI_ISL_1206161, EPI_ISL_1206194, EPI_ISL_1206211, EPI_ISL_1206212, EPI_ISL_1206214, EPI_ISL_1206227, EPI_ISL_1206229, EPI_ISL_1206235, EPI_ISL_1206236, EPI_ISL_1206242, EPI_ISL_1206246, EPI_ISL_1206247, EPI_ISL_1206255, EPI_ISL_1206259, EPI_ISL_1206276, EPI_ISL_1206277, EPI_ISL_1206281, EPI_ISL_1206283, EPI_ISL_1206300, EPI_ISL_1206437, EPI_ISL_1223170, EPI_ISL_1223173, EPI_ISL_1223181, EPI_ISL_1223188, EPI_ISL_1223191, EPI_ISL_1223197, EPI_ISL_1223202, EPI_ISL_1223207, EPI_ISL_1223213, EPI_ISL_1223220, EPI_ISL_1223226, EPI_ISL_1223229, EPI_ISL_1223233, EPI_ISL_1223241, EPI_ISL_1223251, EPI_ISL_1223268, EPI_ISL_1223305, EPI_ISL_1223320, EPI_ISL_1223328, EPI_ISL_1223339, EPI_ISL_1223342, EPI_ISL_1223354, EPI_ISL_1223359, EPI_ISL_1223372, EPI_ISL_1223386, EPI_ISL_1223395, EPI_ISL_1223397, EPI_ISL_1223416, EPI_ISL_1223431, EPI_ISL_1224536, EPI_ISL_1224578, EPI_ISL_1224582, EPI_ISL_1224611, EPI_ISL_1224676, EPI_ISL_1224677, EPI_ISL_1242053, EPI_ISL_1242059, EPI_ISL_1242072, EPI_ISL_1242109, EPI_ISL_1242144, EPI_ISL_1242152, EPI_ISL_1242161, EPI_ISL_1242209, EPI_ISL_1242234, EPI_ISL_1244870, EPI_ISL_1244940, EPI_ISL_1244983, EPI_ISL_1256378, EPI_ISL_1256410, EPI_ISL_1256416, EPI_ISL_1256431, EPI_ISL_1256433, EPI_ISL_1256439, EPI_ISL_1256440, EPI_ISL_1256465, EPI_ISL_1332943, EPI_ISL_1333185, EPI_ISL_1333187, EPI_ISL_1333195, EPI_ISL_1333202, EPI_ISL_1333223, EPI_ISL_1333230, EPI_ISL_1333246, EPI_ISL_1333278, EPI_ISL_1333279, EPI_ISL_1333283, EPI_ISL_1333284, EPI_ISL_1333292, EPI_ISL_1333318, EPI_ISL_1333329, EPI_ISL_1333496, EPI_ISL_1333521, EPI_ISL_1333556, EPI_ISL_1333572, EPI_ISL_1333608, EPI_ISL_1333802, EPI_ISL_1333804, EPI_ISL_1333810, EPI_ISL_1333816, EPI_ISL_1333821, EPI_ISL_1333822, EPI_ISL_1333823, EPI_ISL_1467582, EPI_ISL_1467586, EPI_ISL_1467599, EPI_ISL_1467608, EPI_ISL_1467612, EPI_ISL_1467616, EPI_ISL_1467622, EPI_ISL_1467626, EPI_ISL_1467629, EPI_ISL_1467646, EPI_ISL_1467653, EPI_ISL_1467655, EPI_ISL_1467665, EPI_ISL_1467675, EPI_ISL_1467677, EPI_ISL_1467692, EPI_ISL_1467697, EPI_ISL_1467708, EPI_ISL_1467726, EPI_ISL_1467728, EPI_ISL_1467735, EPI_ISL_1467741, EPI_ISL_1467743, EPI_ISL_1467756, EPI_ISL_1467767, EPI_ISL_1467768, EPI_ISL_1467769, EPI_ISL_1467770, EPI_ISL_1467783, EPI_ISL_1467785, EPI_ISL_1467794, EPI_ISL_1467801, EPI_ISL_1474163

| | | | |
|---|---|---|---|
| see above | Lighthouse Lab in Cambridge | Wellcome Sanger Institute for the COVID-19 Genomics UK (COG-UK) Consortium | Cordelia Langford; David K. Jackson; Dominic Kwiatkowski; Ewan Harrison; Ian Johnston; Jeffrey Barrett; John Sillitoe on behalf of the Wellcome Sanger Institute COVID-19 Surveillance Team; John Sillitoe on behalf of the Wellcome Sanger Institute COVID-19 Surveillance Team (http://www.sanger.ac.uk/covid-team); Rob Howes; Roberto Amato; Sonia Goncalves; The Lighthouse Lab in Cambridge and Alex Alderton |

EPI_ISL_1042018, EPI_ISL_1042060, EPI_ISL_1042065, EPI_ISL_1042102, EPI_ISL_1042141, EPI_ISL_1042174, EPI_ISL_1042182, EPI_ISL_1042206, EPI_ISL_1042214, EPI_ISL_1042215, EPI_ISL_1042216, EPI_ISL_1042218, EPI_ISL_1042225, EPI_ISL_1042227, EPI_ISL_1042234, EPI_ISL_1042260, EPI_ISL_1042264, EPI_ISL_1042267, EPI_ISL_1042281, EPI_ISL_1042310, EPI_ISL_1042321, EPI_ISL_1042460, EPI_ISL_1042473, EPI_ISL_1042498, EPI_ISL_1042534, EPI_ISL_1042543, EPI_ISL_1042570, EPI_ISL_1042571, EPI_ISL_1042572, EPI_ISL_1042574, EPI_ISL_1042576, EPI_ISL_1042580, EPI_ISL_1042584, EPI_ISL_1042587, EPI_ISL_1042591, EPI_ISL_1042595, EPI_ISL_1042597, EPI_ISL_1042600, EPI_ISL_1042611, EPI_ISL_1042612, EPI_ISL_1042613, EPI_ISL_1042616, EPI_ISL_1042620, EPI_ISL_1042623, EPI_ISL_1042624, EPI_ISL_1042630, EPI_ISL_1042633, EPI_ISL_1042645, EPI_ISL_1042649, EPI_ISL_1042650, EPI_ISL_1042654, EPI_ISL_1042663, EPI_ISL_1042669, EPI_ISL_1042676, EPI_ISL_1042680, EPI_ISL_1042681, EPI_ISL_1042682, EPI_ISL_1042686, EPI_ISL_1042687, EPI_ISL_1042690, EPI_ISL_1042697, EPI_ISL_1042698, EPI_ISL_1042703, EPI_ISL_1042706, EPI_ISL_1042712, EPI_ISL_1042713, EPI_ISL_1042716, EPI_ISL_1042717, EPI_ISL_1042718, EPI_ISL_1042719, EPI_ISL_1042723, EPI_ISL_1042726, EPI_ISL_1042727, EPI_ISL_1042728, EPI_ISL_1042729, EPI_ISL_1042732, EPI_ISL_1042736, EPI_ISL_1042737, EPI_ISL_1042738, EPI_ISL_1042741, EPI_ISL_1042745, EPI_ISL_1042746, EPI_ISL_1042747, EPI_ISL_1042751, EPI_ISL_1042752, EPI_ISL_1042753, EPI_ISL_1042760, EPI_ISL_1042761, EPI_ISL_1042763, EPI_ISL_1042764, EPI_ISL_1042776, EPI_ISL_1042780, EPI_ISL_1042785, EPI_ISL_1042788, EPI_ISL_1042799, EPI_ISL_1042804, EPI_ISL_1042810, EPI_ISL_1042812, EPI_ISL_1042817, EPI_ISL_1042818, EPI_ISL_1042825, EPI_ISL_1042826, EPI_ISL_1042830, EPI_ISL_1042833, EPI_ISL_1043344, EPI_ISL_1043374, EPI_ISL_1043375, EPI_ISL_1043385, EPI_ISL_1043386, EPI_ISL_1043388, EPI_ISL_1043389, EPI_ISL_1043398, EPI_ISL_1043411, EPI_ISL_1043414, EPI_ISL_1043419, EPI_ISL_1043429, EPI_ISL_1043430, EPI_ISL_1043444, EPI_ISL_1043445, EPI_ISL_1043452, EPI_ISL_1043461, EPI_ISL_1043481, EPI_ISL_1043497, EPI_ISL_1043550, EPI_ISL_1043562, EPI_ISL_1043563, EPI_ISL_1045473, EPI_ISL_1045478, EPI_ISL_1045483, EPI_ISL_1045485, EPI_ISL_1045491, EPI_ISL_1045492, EPI_ISL_1045493, EPI_ISL_1045495, EPI_ISL_1045496, EPI_ISL_1045498, EPI_ISL_1045499, EPI_ISL_1045501, EPI_ISL_1045506, EPI_ISL_1045507, EPI_ISL_1045510, EPI_ISL_1045512, EPI_ISL_1045518, EPI_ISL_1045528, EPI_ISL_1045529, EPI_ISL_1045533, EPI_ISL_1045535, EPI_ISL_1045536, EPI_ISL_1045539, EPI_ISL_1045546, EPI_ISL_1045547, EPI_ISL_1045550, EPI_ISL_1045554, EPI_ISL_1045555, EPI_ISL_1045556, EPI_ISL_1045557, EPI_ISL_1045561, EPI_ISL_1045574, EPI_ISL_1045575, EPI_ISL_1045577, EPI_ISL_1045586, EPI_ISL_1045587, EPI_ISL_1045589, EPI_ISL_1045596, EPI_ISL_1045597, EPI_ISL_1045599, EPI_ISL_1045602, EPI_ISL_1045605, EPI_ISL_1045607, EPI_ISL_1045612, EPI_ISL_1045613, EPI_ISL_1045616, EPI_ISL_1045618, EPI_ISL_1045619, EPI_ISL_1045623, EPI_ISL_1045626, EPI_ISL_1045631, EPI_ISL_1045638, EPI_ISL_1045642, EPI_ISL_1045643, EPI_ISL_1045645, EPI_ISL_1045655, EPI_ISL_1045656, EPI_ISL_1045659, EPI_ISL_1045661, EPI_ISL_1045662, EPI_ISL_1045664, EPI_ISL_1045667, EPI_ISL_1045668, EPI_ISL_1045671, EPI_ISL_1045673, EPI_ISL_1057254, EPI_ISL_1057337, EPI_ISL_1057426, EPI_ISL_1057429, EPI_ISL_1057433, EPI_ISL_1057435, EPI_ISL_1057439, EPI_ISL_1057441, EPI_ISL_1057443, EPI_ISL_1057454, EPI_ISL_1057455, EPI_ISL_1057460, EPI_ISL_1057462, EPI_ISL_1057464, EPI_ISL_1057467, EPI_ISL_1057472, EPI_ISL_1057486, EPI_ISL_1057487, EPI_ISL_1057490, EPI_ISL_1057493, EPI_ISL_1057496, EPI_ISL_1057502, EPI_ISL_1057505, EPI_ISL_1057509, EPI_ISL_1057510, EPI_ISL_1057514, EPI_ISL_1057518, EPI_ISL_1057520, EPI_ISL_1057522, EPI_ISL_1057525, EPI_ISL_1057536, EPI_ISL_1057539, EPI_ISL_1057547, EPI_ISL_1057548, EPI_ISL_1057549, EPI_ISL_1057555, EPI_ISL_1057557, EPI_ISL_1057559, EPI_ISL_1057561, EPI_ISL_1057565, EPI_ISL_1057575, EPI_ISL_1057579, EPI_ISL_1057582, EPI_ISL_1057589, EPI_ISL_1057592, EPI_ISL_1057602, EPI_ISL_1057605, EPI_ISL_1057607, EPI_ISL_1057613, EPI_ISL_1057618, EPI_ISL_1057623, EPI_ISL_1057624, EPI_ISL_1057629, EPI_ISL_1057646, EPI_ISL_1057649, EPI_ISL_1057650, EPI_ISL_1057654, EPI_ISL_1057661, EPI_ISL_1063047, EPI_ISL_1063048, EPI_ISL_1063049, EPI_ISL_1063050, EPI_ISL_1063051, EPI_ISL_1063052, EPI_ISL_1063058, EPI_ISL_1063063, EPI_ISL_1063082, EPI_ISL_1063087, EPI_ISL_1063101, EPI_ISL_1063189, EPI_ISL_1063191, EPI_ISL_1063193, EPI_ISL_1063195, EPI_ISL_1063196, EPI_ISL_1063197, EPI_ISL_1063198, EPI_ISL_1063199, EPI_ISL_1063200, EPI_ISL_1063201, EPI_ISL_1063202, EPI_ISL_1063203, EPI_ISL_1063204, EPI_ISL_1063206, EPI_ISL_1063207, EPI_ISL_1063208, EPI_ISL_1063209, EPI_ISL_1063210, EPI_ISL_1063211, EPI_ISL_1063212, EPI_ISL_1063213, EPI_ISL_1063215, EPI_ISL_1063216, EPI_ISL_1063217, EPI_ISL_1063219, EPI_ISL_1063220, EPI_ISL_1063221, EPI_ISL_1063222, EPI_ISL_1063223, EPI_ISL_1063224, EPI_ISL_1063225, EPI_ISL_1063226, EPI_ISL_1063227, EPI_ISL_1063228, EPI_ISL_1063229, EPI_ISL_1063232, EPI_ISL_1063234, EPI_ISL_1063235, EPI_ISL_1063236, EPI_ISL_1063239, EPI_ISL_1063240, EPI_ISL_1063241, EPI_ISL_1063243, EPI_ISL_1063244, EPI_ISL_1063245, EPI_ISL_1063247, EPI_ISL_1063248, EPI_ISL_1063249, EPI_ISL_1063250, EPI_ISL_1063288, EPI_ISL_1063289, EPI_ISL_1063290, EPI_ISL_1063291, EPI_ISL_1063294, EPI_ISL_1063299, EPI_ISL_1063300, EPI_ISL_1063301, EPI_ISL_1063302, EPI_ISL_1063303, EPI_ISL_1063304, EPI_ISL_1063305, EPI_ISL_1063306, EPI_ISL_1063308, EPI_ISL_1063309, EPI_ISL_1063314, EPI_ISL_1063315, EPI_ISL_1063316, EPI_ISL_1063317, EPI_ISL_1063318, EPI_ISL_1063319, EPI_ISL_1063320, EPI_ISL_1063321, EPI_ISL_1063325, EPI_ISL_1063328, EPI_ISL_1063329, EPI_ISL_1063330, EPI_ISL_1063332, EPI_ISL_1063333, EPI_ISL_1063334, EPI_ISL_1063335, EPI_ISL_1063337, EPI_ISL_1063339, EPI_ISL_1063341, EPI_ISL_1063343, EPI_ISL_1063344, EPI_ISL_1063346, EPI_ISL_1063349, EPI_ISL_1063357, EPI_ISL_1063358, EPI_ISL_1063359, EPI_ISL_1063360, EPI_ISL_1063361, EPI_ISL_1063362, EPI_ISL_1063363, EPI_ISL_1063364, EPI_ISL_1063365, EPI_ISL_1063366, EPI_ISL_1063367, EPI_ISL_1063368, EPI_ISL_1063369, EPI_ISL_1063371, EPI_ISL_1063372, EPI_ISL_1063373, EPI_ISL_1063374, EPI_ISL_1063376, EPI_ISL_1063377, EPI_ISL_1063378, EPI_ISL_1063379, EPI_ISL_1063380, EPI_ISL_1063381, EPI_ISL_1063382, EPI_ISL_1063383, EPI_ISL_1063384, EPI_ISL_1063385, EPI_ISL_1063387, EPI_ISL_1063389, EPI_ISL_1063391, EPI_ISL_1063392, EPI_ISL_1063393, EPI_ISL_1063394, EPI_ISL_1069661, EPI_ISL_1070336, EPI_ISL_1070342, EPI_ISL_1070344, EPI_ISL_1070345, EPI_ISL_1070347, EPI_ISL_1070349, EPI_ISL_1070354, EPI_ISL_1070356, EPI_ISL_1070360, EPI_ISL_1070362, EPI_ISL_1070363, EPI_ISL_1070364, EPI_ISL_1070368, EPI_ISL_1070373, EPI_ISL_1070375, EPI_ISL_1070377, EPI_ISL_1070378, EPI_ISL_1070379, EPI_ISL_1070380, EPI_ISL_1070382, EPI_ISL_1070387, EPI_ISL_1070388, EPI_ISL_1070389, EPI_ISL_1070393, EPI_ISL_1070394, EPI_ISL_1070395, EPI_ISL_1070396, EPI_ISL_1070397, EPI_ISL_1070398, EPI_ISL_1070404, EPI_ISL_1070406, EPI_ISL_1070407, EPI_ISL_1070408, EPI_ISL_1070409, EPI_ISL_1070410, EPI_ISL_1070414, EPI_ISL_1070416, EPI_ISL_1070417, EPI_ISL_1070418, EPI_ISL_1070422, EPI_ISL_1070423, EPI_ISL_1070425, EPI_ISL_1070427, EPI_ISL_1070429, EPI_ISL_1070432, EPI_ISL_1070433, EPI_ISL_1070438, EPI_ISL_1070439, EPI_ISL_1070443, EPI_ISL_1070445, EPI_ISL_1070446, EPI_ISL_1070447, EPI_ISL_1070452, EPI_ISL_1070453, EPI_ISL_1070457, EPI_ISL_1070462, EPI_ISL_1070468, EPI_ISL_1070470, EPI_ISL_1070471, EPI_ISL_1070473, EPI_ISL_1070477, EPI_ISL_1070478, EPI_ISL_1070479, EPI_ISL_1070483, EPI_ISL_1070485, EPI_ISL_1070486, EPI_ISL_1070488, EPI_ISL_1070495, EPI_ISL_1070496, EPI_ISL_1070502, EPI_ISL_1070505, EPI_ISL_1070506, EPI_ISL_1070509, EPI_ISL_1070511, EPI_ISL_1070513, EPI_ISL_1070517, EPI_ISL_1070518, EPI_ISL_1070521, EPI_ISL_1070522, EPI_ISL_1070523, EPI_ISL_1070524, EPI_ISL_1070528, EPI_ISL_1070529, EPI_ISL_1070533, EPI_ISL_1070534, EPI_ISL_1070535, EPI_ISL_1070536, EPI_ISL_1070537, EPI_ISL_1070538, EPI_ISL_1070539, EPI_ISL_1070541, EPI_ISL_1070542, EPI_ISL_1070553, EPI_ISL_1070554, EPI_ISL_1070557, EPI_ISL_1070559, EPI_ISL_1070560, EPI_ISL_1070563, EPI_ISL_1070564, EPI_ISL_1070565, EPI_ISL_1070569, EPI_ISL_1070571, EPI_ISL_1070572, EPI_ISL_1070573, EPI_ISL_1070576, EPI_ISL_1070583, EPI_ISL_1070585, EPI_ISL_1070586, EPI_ISL_1070587, EPI_ISL_1070588, EPI_ISL_1070589, EPI_ISL_1070592, EPI_ISL_1070593, EPI_ISL_1070595, EPI_ISL_1070596, EPI_ISL_1070599, EPI_ISL_1070600, EPI_ISL_1070602, EPI_ISL_1070603, EPI_ISL_1070604, EPI_ISL_1070605, EPI_ISL_1070606, EPI_ISL_1070609, EPI_ISL_1070610, EPI_ISL_1070613, EPI_ISL_1070614, EPI_ISL_1070618, EPI_ISL_1070620, EPI_ISL_1070623, EPI_ISL_1070624, EPI_ISL_1070627, EPI_ISL_1070729, EPI_ISL_1070734, EPI_ISL_1070737, EPI_ISL_1070739, EPI_ISL_1070741, EPI_ISL_1070750, EPI_ISL_1070751, EPI_ISL_1070753, EPI_ISL_1070761, EPI_ISL_1070763, EPI_ISL_1070768, EPI_ISL_1070774, EPI_ISL_1070775, EPI_ISL_1070778, EPI_ISL_1070797, EPI_ISL_1070798, EPI_ISL_1070799, EPI_ISL_1070800, EPI_ISL_1070812, EPI_ISL_1070817, EPI_ISL_1070823, EPI_ISL_1070828, EPI_ISL_1070838, EPI_ISL_1070846, EPI_ISL_1070847, EPI_ISL_1070852, EPI_ISL_1070853, EPI_ISL_1070875, EPI_ISL_1070878, EPI_ISL_1070880, EPI_ISL_1070881, EPI_ISL_1070884, EPI_ISL_1070891, EPI_ISL_1070893, EPI_ISL_1070895, EPI_ISL_1070897, EPI_ISL_1070915, EPI_ISL_1070918, EPI_ISL_1070920, EPI_ISL_1070921, EPI_ISL_1070922, EPI_ISL_1070929, EPI_ISL_1070932, EPI_ISL_1070935, EPI_ISL_1070942, EPI_ISL_1070948, EPI_ISL_1070950, EPI_ISL_1070951, EPI_ISL_1070955, EPI_ISL_1070959,

EPI_ISL_1070964, EPI_ISL_1070966, EPI_ISL_1070968, EPI_ISL_1070971, EPI_ISL_1070972, EPI_ISL_1070973, EPI_ISL_1070976, EPI_ISL_1070979, EPI_ISL_1070980, EPI_ISL_1070981, EPI_ISL_1070982, EPI_ISL_1070983, EPI_ISL_1070984, EPI_ISL_1070985, EPI_ISL_1070986, EPI_ISL_1070990, EPI_ISL_1070996, EPI_ISL_1070997, EPI_ISL_1070998, EPI_ISL_1070999, EPI_ISL_1071006, EPI_ISL_1071008, EPI_ISL_1071011, EPI_ISL_1071013, EPI_ISL_1071015, EPI_ISL_1071023, EPI_ISL_1071027, EPI_ISL_1071033, EPI_ISL_1071041, EPI_ISL_1071043, EPI_ISL_1071044, EPI_ISL_1083050, EPI_ISL_1084216, EPI_ISL_1084478, EPI_ISL_1099506, EPI_ISL_1099507, EPI_ISL_1099511, EPI_ISL_1099512, EPI_ISL_1099518, EPI_ISL_1099519, EPI_ISL_1099522, EPI_ISL_1099523, EPI_ISL_1099530, EPI_ISL_1099540, EPI_ISL_1099552, EPI_ISL_1099553, EPI_ISL_1099555, EPI_ISL_1099559, EPI_ISL_1099562, EPI_ISL_1099576, EPI_ISL_1099583, EPI_ISL_1099585, EPI_ISL_1099589, EPI_ISL_1099592, EPI_ISL_1099593, EPI_ISL_1099594, EPI_ISL_1099595, EPI_ISL_1099596, EPI_ISL_1099600, EPI_ISL_1099609, EPI_ISL_1099616, EPI_ISL_1099617, EPI_ISL_1099629, EPI_ISL_1099632, EPI_ISL_1099633, EPI_ISL_1099635, EPI_ISL_1099639, EPI_ISL_1099643, EPI_ISL_1099645, EPI_ISL_1099647, EPI_ISL_1099652, EPI_ISL_1099654, EPI_ISL_1099656, EPI_ISL_1099659, EPI_ISL_1099663, EPI_ISL_1099666, EPI_ISL_1099667, EPI_ISL_1099669, EPI_ISL_1099670, EPI_ISL_1099674, EPI_ISL_1099689, EPI_ISL_1099699, EPI_ISL_1099706, EPI_ISL_1099714, EPI_ISL_1099718, EPI_ISL_1099729, EPI_ISL_1099741, EPI_ISL_1101977, EPI_ISL_1101980, EPI_ISL_1103054, EPI_ISL_1103055, EPI_ISL_1103056, EPI_ISL_1103057, EPI_ISL_1103058, EPI_ISL_1103059, EPI_ISL_1103060, EPI_ISL_1103061, EPI_ISL_1103062, EPI_ISL_1103063, EPI_ISL_1103064, EPI_ISL_1103065, EPI_ISL_1103066, EPI_ISL_1103067, EPI_ISL_1103068, EPI_ISL_1103069, EPI_ISL_1103070, EPI_ISL_1103071, EPI_ISL_1103072, EPI_ISL_1103073, EPI_ISL_1103074, EPI_ISL_1103075, EPI_ISL_1103076, EPI_ISL_1103077, EPI_ISL_1103078, EPI_ISL_1103079, EPI_ISL_1103080, EPI_ISL_1103081, EPI_ISL_1103082, EPI_ISL_1103083, EPI_ISL_1103084, EPI_ISL_1103085, EPI_ISL_1103086, EPI_ISL_1103087, EPI_ISL_1103088, EPI_ISL_1103089, EPI_ISL_1103090, EPI_ISL_1103091, EPI_ISL_1103092, EPI_ISL_1103093, EPI_ISL_1103094, EPI_ISL_1103095, EPI_ISL_1103096, EPI_ISL_1103097, EPI_ISL_1103098, EPI_ISL_1103099, EPI_ISL_1103100, EPI_ISL_1103101, EPI_ISL_1103102, EPI_ISL_1103103, EPI_ISL_1123111, EPI_ISL_1123143, EPI_ISL_1123153, EPI_ISL_1123157, EPI_ISL_1123164, EPI_ISL_1123168, EPI_ISL_1123188, EPI_ISL_1123192, EPI_ISL_1257203

| | | | |
|---|---|---|---|
| see above | Lighthouse Lab in Glasgow | Wellcome Sanger Institute for the COVID-19 Genomics UK (COG-UK) Consortium | Anna Dominiczak and Alex Alderton; Carol Clugston; Cordelia Langford; David Gray; David K. Jackson; Dominic Kwiatkowski; Ewan Harrison; Harper VanSteenhouse; Ian Johnston; Jeffrey Barrett; John Sillitoe on behalf of the Wellcome Sanger Institute COVID-19 Surveillance Team; Roberto Amato; Sonia Goncalves; Yumi Kasai |

EPI_ISL_1043195, EPI_ISL_1043329, EPI_ISL_1043345, EPI_ISL_1043347, EPI_ISL_1043350, EPI_ISL_1043355, EPI_ISL_1043356, EPI_ISL_1043358, EPI_ISL_1043360, EPI_ISL_1043363, EPI_ISL_1043365, EPI_ISL_1043366, EPI_ISL_1043367, EPI_ISL_1043372, EPI_ISL_1043373, EPI_ISL_1043382, EPI_ISL_1043384, EPI_ISL_1043394, EPI_ISL_1043396, EPI_ISL_1043399, EPI_ISL_1043402, EPI_ISL_1043403, EPI_ISL_1043405, EPI_ISL_1043421, EPI_ISL_1043422, EPI_ISL_1043426, EPI_ISL_1043435, EPI_ISL_1043439, EPI_ISL_1043440, EPI_ISL_1043441, EPI_ISL_1043443, EPI_ISL_1043453, EPI_ISL_1043455, EPI_ISL_1043465, EPI_ISL_1043468, EPI_ISL_1043471, EPI_ISL_1043472, EPI_ISL_1043476, EPI_ISL_1043477, EPI_ISL_1043482, EPI_ISL_1043488, EPI_ISL_1043491, EPI_ISL_1043493, EPI_ISL_1043496, EPI_ISL_1043500, EPI_ISL_1043501, EPI_ISL_1043504, EPI_ISL_1043509, EPI_ISL_1043512, EPI_ISL_1043521, EPI_ISL_1043522, EPI_ISL_1043524, EPI_ISL_1043529, EPI_ISL_1043532, EPI_ISL_1043538, EPI_ISL_1043539, EPI_ISL_1043544, EPI_ISL_1043545, EPI_ISL_1043549, EPI_ISL_1043551, EPI_ISL_1043554, EPI_ISL_1043556, EPI_ISL_1043557, EPI_ISL_1043558, EPI_ISL_1043560, EPI_ISL_1043561, EPI_ISL_1043564, EPI_ISL_1043565, EPI_ISL_1043567, EPI_ISL_1057107, EPI_ISL_1057108, EPI_ISL_1057109, EPI_ISL_1057110, EPI_ISL_1057111, EPI_ISL_1057112, EPI_ISL_1057113, EPI_ISL_1057114, EPI_ISL_1057115, EPI_ISL_1057116, EPI_ISL_1057117, EPI_ISL_1057118, EPI_ISL_1057119, EPI_ISL_1057120, EPI_ISL_1057123, EPI_ISL_1057124, EPI_ISL_1057126, EPI_ISL_1057129, EPI_ISL_1057130, EPI_ISL_1057131, EPI_ISL_1057132, EPI_ISL_1057133, EPI_ISL_1057134, EPI_ISL_1057135, EPI_ISL_1057137, EPI_ISL_1057138, EPI_ISL_1057139, EPI_ISL_1057143, EPI_ISL_1057144, EPI_ISL_1057145, EPI_ISL_1057146, EPI_ISL_1057147, EPI_ISL_1057148, EPI_ISL_1057149, EPI_ISL_1057151, EPI_ISL_1057153, EPI_ISL_1057155, EPI_ISL_1057156, EPI_ISL_1057157, EPI_ISL_1057159, EPI_ISL_1057161, EPI_ISL_1057162, EPI_ISL_1057164, EPI_ISL_1057165, EPI_ISL_1057166, EPI_ISL_1057167, EPI_ISL_1057168, EPI_ISL_1057169, EPI_ISL_1057170, EPI_ISL_1057175, EPI_ISL_1057176, EPI_ISL_1057177, EPI_ISL_1057178, EPI_ISL_1057179, EPI_ISL_1057180, EPI_ISL_1057181, EPI_ISL_1057182, EPI_ISL_1057183, EPI_ISL_1057184, EPI_ISL_1057185, EPI_ISL_1057186, EPI_ISL_1057188, EPI_ISL_1057190, EPI_ISL_1057192, EPI_ISL_1057193, EPI_ISL_1057195, EPI_ISL_1057197, EPI_ISL_1057198, EPI_ISL_1057199, EPI_ISL_1057201, EPI_ISL_1057203, EPI_ISL_1057204, EPI_ISL_1057205, EPI_ISL_1057206, EPI_ISL_1057207, EPI_ISL_1057208, EPI_ISL_1057209, EPI_ISL_1057212, EPI_ISL_1057214, EPI_ISL_1057216, EPI_ISL_1057218, EPI_ISL_1057219, EPI_ISL_1057220, EPI_ISL_1057221, EPI_ISL_1057222, EPI_ISL_1057224, EPI_ISL_1057226, EPI_ISL_1057227, EPI_ISL_1057228, EPI_ISL_1057229, EPI_ISL_1057230, EPI_ISL_1057232, EPI_ISL_1057233, EPI_ISL_1057234, EPI_ISL_1057235, EPI_ISL_1057236, EPI_ISL_1057237, EPI_ISL_1057239, EPI_ISL_1057240, EPI_ISL_1057241, EPI_ISL_1057242, EPI_ISL_1057243, EPI_ISL_1057245, EPI_ISL_1057246, EPI_ISL_1057247, EPI_ISL_1057250, EPI_ISL_1057251, EPI_ISL_1057252, EPI_ISL_1057255, EPI_ISL_1057256, EPI_ISL_1057258, EPI_ISL_1057259, EPI_ISL_1057260, EPI_ISL_1057261, EPI_ISL_1057262, EPI_ISL_1057264, EPI_ISL_1057265, EPI_ISL_1057267, EPI_ISL_1057268, EPI_ISL_1057269, EPI_ISL_1057270, EPI_ISL_1057272, EPI_ISL_1057273, EPI_ISL_1057274, EPI_ISL_1057278, EPI_ISL_1057279, EPI_ISL_1057280, EPI_ISL_1057281, EPI_ISL_1057282, EPI_ISL_1057283, EPI_ISL_1057284, EPI_ISL_1057285, EPI_ISL_1057287, EPI_ISL_1057288, EPI_ISL_1057289, EPI_ISL_1057290, EPI_ISL_1057291, EPI_ISL_1057292, EPI_ISL_1057293, EPI_ISL_1057294, EPI_ISL_1057295, EPI_ISL_1057296, EPI_ISL_1057297, EPI_ISL_1057300, EPI_ISL_1057304, EPI_ISL_1057305, EPI_ISL_1057307, EPI_ISL_1057308, EPI_ISL_1057310, EPI_ISL_1057311, EPI_ISL_1057312, EPI_ISL_1057313, EPI_ISL_1057314, EPI_ISL_1057315, EPI_ISL_1057316, EPI_ISL_1057317, EPI_ISL_1057318, EPI_ISL_1057320, EPI_ISL_1057322, EPI_ISL_1057323, EPI_ISL_1057324, EPI_ISL_1057326, EPI_ISL_1057327, EPI_ISL_1057328, EPI_ISL_1057329, EPI_ISL_1057330, EPI_ISL_1057331, EPI_ISL_1057333, EPI_ISL_1057334, EPI_ISL_1057335, EPI_ISL_1057336, EPI_ISL_1057338, EPI_ISL_1057339, EPI_ISL_1057340, EPI_ISL_1057342, EPI_ISL_1057343, EPI_ISL_1057344, EPI_ISL_1057346, EPI_ISL_1057347, EPI_ISL_1057348, EPI_ISL_1057349, EPI_ISL_1057350, EPI_ISL_1057351, EPI_ISL_1057352, EPI_ISL_1057353, EPI_ISL_1057355, EPI_ISL_1057356, EPI_ISL_1057357, EPI_ISL_1057358, EPI_ISL_1057360, EPI_ISL_1057361, EPI_ISL_1057362, EPI_ISL_1057364, EPI_ISL_1057365, EPI_ISL_1057367, EPI_ISL_1057368, EPI_ISL_1057369, EPI_ISL_1057370, EPI_ISL_1057371, EPI_ISL_1057372, EPI_ISL_1057373, EPI_ISL_1057374, EPI_ISL_1057375, EPI_ISL_1057376, EPI_ISL_1057377, EPI_ISL_1057378, EPI_ISL_1057379, EPI_ISL_1057380, EPI_ISL_1057381, EPI_ISL_1057382, EPI_ISL_1057384, EPI_ISL_1057385, EPI_ISL_1057387, EPI_ISL_1057388, EPI_ISL_1057389, EPI_ISL_1057391, EPI_ISL_1057392, EPI_ISL_1057393, EPI_ISL_1057395, EPI_ISL_1057397, EPI_ISL_1057401, EPI_ISL_1057402, EPI_ISL_1057403, EPI_ISL_1057404, EPI_ISL_1057405, EPI_ISL_1057406, EPI_ISL_1057407, EPI_ISL_1057408, EPI_ISL_1057409, EPI_ISL_1057410, EPI_ISL_1057411, EPI_ISL_1057412, EPI_ISL_1057413, EPI_ISL_1057415, EPI_ISL_1057416, EPI_ISL_1057417, EPI_ISL_1057418, EPI_ISL_1057420, EPI_ISL_1057422, EPI_ISL_1057423, EPI_ISL_1057424, EPI_ISL_1057425, EPI_ISL_1057428, EPI_ISL_1062059, EPI_ISL_1062061, EPI_ISL_1062063, EPI_ISL_1062067, EPI_ISL_1062073, EPI_ISL_1062080, EPI_ISL_1062082, EPI_ISL_1062084, EPI_ISL_1062086, EPI_ISL_1062087, EPI_ISL_1062095, EPI_ISL_1062101, EPI_ISL_1062103, EPI_ISL_1062106, EPI_ISL_1062108, EPI_ISL_1062113, EPI_ISL_1062115, EPI_ISL_1062118, EPI_ISL_1062119, EPI_ISL_1062120, EPI_ISL_1062122, EPI_ISL_1062125, EPI_ISL_1062127, EPI_ISL_1062130, EPI_ISL_1062131, EPI_ISL_1062132, EPI_ISL_1062135, EPI_ISL_1062138, EPI_ISL_1062140, EPI_ISL_1062147, EPI_ISL_1062159, EPI_ISL_1062162, EPI_ISL_1062163, EPI_ISL_1062165, EPI_ISL_1062170, EPI_ISL_1062171, EPI_ISL_1062172, EPI_ISL_1062173, EPI_ISL_1062185, EPI_ISL_1062200, EPI_ISL_1062202, EPI_ISL_1062203, EPI_ISL_1062205, EPI_ISL_1062216, EPI_ISL_1062219, EPI_ISL_1062222, EPI_ISL_1062225, EPI_ISL_1062234, EPI_ISL_1062239, EPI_ISL_1062241, EPI_ISL_1062248, EPI_ISL_1062254, EPI_ISL_1062255, EPI_ISL_1062258, EPI_ISL_1062262, EPI_ISL_1062269, EPI_ISL_1062273, EPI_ISL_1062277, EPI_ISL_1062280, EPI_ISL_1062285, EPI_ISL_1062288, EPI_ISL_1062289, EPI_ISL_1062290, EPI_ISL_1062291, EPI_ISL_1062294, EPI_ISL_1062300, EPI_ISL_1062301, EPI_ISL_1062302, EPI_ISL_1062305, EPI_ISL_1062307, EPI_ISL_1062308, EPI_ISL_1062313, EPI_ISL_1062314, EPI_ISL_1062315, EPI_ISL_1062316, EPI_ISL_1062323, EPI_ISL_1062324, EPI_ISL_1062340, EPI_ISL_1062346, EPI_ISL_1062349, EPI_ISL_1062350, EPI_ISL_1062355, EPI_ISL_1062363, EPI_ISL_1062364, EPI_ISL_1062367, EPI_ISL_1062370, EPI_ISL_1062373, EPI_ISL_1062383, EPI_ISL_1062391, EPI_ISL_1062396, EPI_ISL_1062404, EPI_ISL_1062408, EPI_ISL_1062410, EPI_ISL_1062411, EPI_ISL_1062414, EPI_ISL_1062415, EPI_ISL_1062417, EPI_ISL_1062418, EPI_ISL_1062420, EPI_ISL_1062439, EPI_ISL_1062450, EPI_ISL_1062456, EPI_ISL_1062457, EPI_ISL_1062460, EPI_ISL_1062464, EPI_ISL_1062473, EPI_ISL_1062477, EPI_ISL_1062495, EPI_ISL_1062496, EPI_ISL_1062498, EPI_ISL_1062503, EPI_ISL_1062504, EPI_ISL_1062508, EPI_ISL_1062509, EPI_ISL_1062522, EPI_ISL_1062529, EPI_ISL_1062535, EPI_ISL_1062540, EPI_ISL_1062541, EPI_ISL_1062548, EPI_ISL_1062552, EPI_ISL_1062554, EPI_ISL_1062569, EPI_ISL_1062572, EPI_ISL_1062573, EPI_ISL_1062584, EPI_ISL_1062593, EPI_ISL_1062606, EPI_ISL_1062613, EPI_ISL_1062617, EPI_ISL_1062624, EPI_ISL_1062625, EPI_ISL_1062632, EPI_ISL_1062638, EPI_ISL_1062640, EPI_ISL_1062643, EPI_ISL_1062647, EPI_ISL_1062651, EPI_ISL_1062667, EPI_ISL_1062675, EPI_ISL_1062692, EPI_ISL_1062694, EPI_ISL_1062707, EPI_ISL_1062734, EPI_ISL_1062740, EPI_ISL_1062741, EPI_ISL_1062743, EPI_ISL_1062747, EPI_ISL_1062753, EPI_ISL_1062755, EPI_ISL_1062769, EPI_ISL_1062791, EPI_ISL_1062795, EPI_ISL_1062805, EPI_ISL_1062812, EPI_ISL_1062826, EPI_ISL_1062848, EPI_ISL_1062850, EPI_ISL_1062857, EPI_ISL_1062858, EPI_ISL_1062863, EPI_ISL_1062868, EPI_ISL_1062872, EPI_ISL_1062897, EPI_ISL_1062914, EPI_ISL_1062915, EPI_ISL_1062919, EPI_ISL_1062928, EPI_ISL_1062931, EPI_ISL_1062954, EPI_ISL_1062957, EPI_ISL_1062975, EPI_ISL_1062979, EPI_ISL_1063009, EPI_ISL_1069405, EPI_ISL_1069407, EPI_ISL_1069422, EPI_ISL_1069437, EPI_ISL_1069457, EPI_ISL_1069459, EPI_ISL_1069461, EPI_ISL_1069468, EPI_ISL_1069498, EPI_ISL_1069502, EPI_ISL_1069505, EPI_ISL_1069506, EPI_ISL_1069513, EPI_ISL_1069531, EPI_ISL_1069537, EPI_ISL_1069555, EPI_ISL_1069558, EPI_ISL_1069562, EPI_ISL_1069570, EPI_ISL_1069576, EPI_ISL_1069589, EPI_ISL_1069590, EPI_ISL_1069596, EPI_ISL_1069609, EPI_ISL_1069636, EPI_ISL_1069642, EPI_ISL_1069643, EPI_ISL_1069644, EPI_ISL_1069650, EPI_ISL_1069658, EPI_ISL_1069664, EPI_ISL_1069673, EPI_ISL_1069692, EPI_ISL_1069709, EPI_ISL_1069720, EPI_ISL_1069725, EPI_ISL_1069738, EPI_ISL_1069818, EPI_ISL_1069820, EPI_ISL_1069834, EPI_ISL_1069854, EPI_ISL_1069872, EPI_ISL_1069881, EPI_ISL_1069906, EPI_ISL_1069974, EPI_ISL_1069978, EPI_ISL_1070365, EPI_ISL_1070370, EPI_ISL_1070437, EPI_ISL_1070461, EPI_ISL_1070492, EPI_ISL_1070512, EPI_ISL_1070515, EPI_ISL_1070519, EPI_ISL_1070526, EPI_ISL_1070582, EPI_ISL_1070726, EPI_ISL_1070728, EPI_ISL_1070730, EPI_ISL_1070731, EPI_ISL_1070732, EPI_ISL_1070738, EPI_ISL_1070740, EPI_ISL_1070742, EPI_ISL_1070746, EPI_ISL_1070747, EPI_ISL_1070752, EPI_ISL_1070754, EPI_ISL_1070755, EPI_ISL_1070757, EPI_ISL_1070758, EPI_ISL_1070760, EPI_ISL_1070764, EPI_ISL_1070772, EPI_ISL_1070776, EPI_ISL_1070783, EPI_ISL_1070784, EPI_ISL_1070785, EPI_ISL_1070787, EPI_ISL_1070788, EPI_ISL_1070789, EPI_ISL_1070790, EPI_ISL_1070792, EPI_ISL_1070793, EPI_ISL_1070796, EPI_ISL_1070801, EPI_ISL_1070802, EPI_ISL_1070803, EPI_ISL_1070806, EPI_ISL_1070808, EPI_ISL_1070811, EPI_ISL_1070813, EPI_ISL_1070816, EPI_ISL_1070820, EPI_ISL_1070821, EPI_ISL_1070825, EPI_ISL_1070826, EPI_ISL_1070827, EPI_ISL_1070829, EPI_ISL_1070830, EPI_ISL_1070831, EPI_ISL_1070833, EPI_ISL_1070836, EPI_ISL_1070837, EPI_ISL_1070840, EPI_ISL_1070841, EPI_ISL_1070844, EPI_ISL_1070851, EPI_ISL_1070854, EPI_ISL_1070855, EPI_ISL_1070856, EPI_ISL_1070857, EPI_ISL_1070858, EPI_ISL_1070862, EPI_ISL_1070864, EPI_ISL_1070866, EPI_ISL_1070868, EPI_ISL_1070869, EPI_ISL_1070871, EPI_ISL_1070872, EPI_ISL_1070874, EPI_ISL_1070876, EPI_ISL_1070882, EPI_ISL_1070885, EPI_ISL_1070886, EPI_ISL_1070889, EPI_ISL_1070890, EPI_ISL_1070892, EPI_ISL_1070896, EPI_ISL_1070900, EPI_ISL_1070902, EPI_ISL_1070904, EPI_ISL_1070905, EPI_ISL_1070906, EPI_ISL_1070909, EPI_ISL_1070911, EPI_ISL_1070916, EPI_ISL_1070923, EPI_ISL_1070925, EPI_ISL_1070926, EPI_ISL_1070928, EPI_ISL_1070931, EPI_ISL_1070933, EPI_ISL_1070934, EPI_ISL_1070937, EPI_ISL_1070938, EPI_ISL_1070939, EPI_ISL_1070941, EPI_ISL_1070943, EPI_ISL_1070946, EPI_ISL_1070947, EPI_ISL_1070952, EPI_ISL_1070953, EPI_ISL_1070954, EPI_ISL_1070958, EPI_ISL_1070960, EPI_ISL_1070961, EPI_ISL_1070962, EPI_ISL_1070963, EPI_ISL_1070965, EPI_ISL_1070967, EPI_ISL_1070970, EPI_ISL_1070974, EPI_ISL_1070975, EPI_ISL_1070977, EPI_ISL_1070978, EPI_ISL_1070987, EPI_ISL_1070988, EPI_ISL_1070992, EPI_ISL_1070993, EPI_ISL_1070994, EPI_ISL_1070995, EPI_ISL_1071000, EPI_ISL_1071001, EPI_ISL_1071002, EPI_ISL_1071003, EPI_ISL_1071005, EPI_ISL_1071009, EPI_ISL_1071012, EPI_ISL_1071016, EPI_ISL_1071018, EPI_ISL_1071019, EPI_ISL_1071020, EPI_ISL_1071024, EPI_ISL_1071028, EPI_ISL_1071030, EPI_ISL_1071031, EPI_ISL_1071036, EPI_ISL_1071037, EPI_ISL_1071038, EPI_ISL_1071042, EPI_ISL_1071045, EPI_ISL_1071046, EPI_ISL_1071047, EPI_ISL_1071048, EPI_ISL_1071049, EPI_ISL_1071050, EPI_ISL_1071051, EPI_ISL_1071052, EPI_ISL_1071053, EPI_ISL_1071054, EPI_ISL_1071055, EPI_ISL_1071057, EPI_ISL_1071058, EPI_ISL_1071059, EPI_ISL_1071060, EPI_ISL_1071061, EPI_ISL_1071062, EPI_ISL_1071063, EPI_ISL_1071064, EPI_ISL_1071065, EPI_ISL_1071066, EPI_ISL_1071067, EPI_ISL_1071068, EPI_ISL_1071069, EPI_ISL_1071070, EPI_ISL_1071071, EPI_ISL_1071072, EPI_ISL_1071073, EPI_ISL_1071074, EPI_ISL_1071076, EPI_ISL_1071077, EPI_ISL_1071078, EPI_ISL_1071079, EPI_ISL_1071080, EPI_ISL_1071082, EPI_ISL_1071083, EPI_ISL_1071084, EPI_ISL_1071085, EPI_ISL_1071086, EPI_ISL_1071087, EPI_ISL_1071088, EPI_ISL_1071089, EPI_ISL_1071090, EPI_ISL_1071091, EPI_ISL_1071092, EPI_ISL_1071093, EPI_ISL_1071094, EPI_ISL_1071095, EPI_ISL_1071096, EPI_ISL_1071097, EPI_ISL_1071098, EPI_ISL_1071099, EPI_ISL_1071100, EPI_ISL_1071101, EPI_ISL_1071102, EPI_ISL_1071103, EPI_ISL_1071104, EPI_ISL_1071105, EPI_ISL_1071106, EPI_ISL_1071107, EPI_ISL_1071108, EPI_ISL_1071109, EPI_ISL_1071110, EPI_ISL_1071112, EPI_ISL_1071113, EPI_ISL_1071114, EPI_ISL_1071116, EPI_ISL_1071117, EPI_ISL_1071118, EPI_ISL_1071119, EPI_ISL_1071120, EPI_ISL_1071121, EPI_ISL_1071122, EPI_ISL_1071123, EPI_ISL_1071124, EPI_ISL_1071125, EPI_ISL_1071126, EPI_ISL_1071127, EPI_ISL_1071128, EPI_ISL_1071129, EPI_ISL_1071131, EPI_ISL_1071132, EPI_ISL_1071133, EPI_ISL_1071134, EPI_ISL_1071135, EPI_ISL_1071136, EPI_ISL_1071137, EPI_ISL_1071138, EPI_ISL_1071139, EPI_ISL_1071140, EPI_ISL_1071141, EPI_ISL_1071142, EPI_ISL_1071143, EPI_ISL_1071144, EPI_ISL_1071145, EPI_ISL_1071146, EPI_ISL_1071147, EPI_ISL_1071148, EPI_ISL_1071149, EPI_ISL_1071150, EPI_ISL_1071151, EPI_ISL_1071152, EPI_ISL_1071153, EPI_ISL_1071155, EPI_ISL_1071156, EPI_ISL_1071157, EPI_ISL_1071158, EPI_ISL_1071159, EPI_ISL_1071160, EPI_ISL_1071161, EPI_ISL_1071162, EPI_ISL_1071163, EPI_ISL_1071164, EPI_ISL_1071165, EPI_ISL_1071166, EPI_ISL_1071167, EPI_ISL_1071168, EPI_ISL_1071169, EPI_ISL_1071170, EPI_ISL_1071171, EPI_ISL_1071172, EPI_ISL_1071173, EPI_ISL_1071174, EPI_ISL_1071175, EPI_ISL_1071176, EPI_ISL_1071177, EPI_ISL_1071178, EPI_ISL_1071179, EPI_ISL_1071181, EPI_ISL_1071182, EPI_ISL_1071183, EPI_ISL_1071184, EPI_ISL_1071185, EPI_ISL_1071186, EPI_ISL_1071187, EPI_ISL_1071188, EPI_ISL_1071189, EPI_ISL_1071190, EPI_ISL_1071191, EPI_ISL_1071192, EPI_ISL_1071193, EPI_ISL_1071194, EPI_ISL_1071195, EPI_ISL_1071196, EPI_ISL_1071197, EPI_ISL_1071198, EPI_ISL_1071199, EPI_ISL_1071200, EPI_ISL_1071201, EPI_ISL_1071202, EPI_ISL_1071203, EPI_ISL_1071204, EPI_ISL_1071205, EPI_ISL_1071206, EPI_ISL_1071207, EPI_ISL_1071208, EPI_ISL_1071209, EPI_ISL_1071210, EPI_ISL_1071211, EPI_ISL_1071212, EPI_ISL_1071213, EPI_ISL_1071214, EPI_ISL_1071215, EPI_ISL_1071216, EPI_ISL_1071217, EPI_ISL_1071218, EPI_ISL_1071219, EPI_ISL_1071220, EPI_ISL_1071221, EPI_ISL_1071222, EPI_ISL_1071224, EPI_ISL_1071225, EPI_ISL_1071226, EPI_ISL_1071227, EPI_ISL_1071228, EPI_ISL_1071229, EPI_ISL_1071230, EPI_ISL_1071231, EPI_ISL_1071232, EPI_ISL_1071233, EPI_ISL_1071234, EPI_ISL_1071235, EPI_ISL_1071236, EPI_ISL_1071237, EPI_ISL_1071238, EPI_ISL_1071239, EPI_ISL_1071240, EPI_ISL_1071241, EPI_ISL_1071242, EPI_ISL_1071243, EPI_ISL_1071244, EPI_ISL_1071245, EPI_ISL_1071246, EPI_ISL_1071247, EPI_ISL_1071248, EPI_ISL_1071249, EPI_ISL_1071250, EPI_ISL_1071251, EPI_ISL_1071252, EPI_ISL_1071253, EPI_ISL_1071254, EPI_ISL_1071255, EPI_ISL_1071257, EPI_ISL_1071258, EPI_ISL_1071259, EPI_ISL_1071260, EPI_ISL_1071261, EPI_ISL_1071262, EPI_ISL_1071263, EPI_ISL_1071264, EPI_ISL_1071265, EPI_ISL_1071266, EPI_ISL_1071267, EPI_ISL_1071268, EPI_ISL_1071269, EPI_ISL_1071270, EPI_ISL_1071271, EPI_ISL_1071272, EPI_ISL_1071273, EPI_ISL_1071274, EPI_ISL_1071275, EPI_ISL_1071276, EPI_ISL_1071277, EPI_ISL_1071278, EPI_ISL_1071279, EPI_ISL_1071280, EPI_ISL_1071282, EPI_ISL_1071283, EPI_ISL_1071284, EPI_ISL_1071285, EPI_ISL_1071286, EPI_ISL_1071287, EPI_ISL_1071288, EPI_ISL_1071289, EPI_ISL_1071290, EPI_ISL_1071291, EPI_ISL_1071292, EPI_ISL_1071293, EPI_ISL_1071295, EPI_ISL_1071296, EPI_ISL_1071297, EPI_ISL_1071298, EPI_ISL_1071299, EPI_ISL_1071300, EPI_ISL_1071301, EPI_ISL_1071303, EPI_ISL_1071304, EPI_ISL_1071305, EPI_ISL_1071307, EPI_ISL_1071308, EPI_ISL_1071309, EPI_ISL_1071310, EPI_ISL_1071311, EPI_ISL_1071312, EPI_ISL_1071313, EPI_ISL_1071314, EPI_ISL_1071315, EPI_ISL_1071316, EPI_ISL_1071317, EPI_ISL_1071318, EPI_ISL_1071319, EPI_ISL_1071320, EPI_ISL_1071321, EPI_ISL_1071323, EPI_ISL_1071324, EPI_ISL_1071325, EPI_ISL_1071326, EPI_ISL_1071327, EPI_ISL_1071328, EPI_ISL_1071329, EPI_ISL_1071330, EPI_ISL_1071331, EPI_ISL_1071332, EPI_ISL_1071333, EPI_ISL_1071334, EPI_ISL_1071335, EPI_ISL_1071336, EPI_ISL_1071338, EPI_ISL_1071339, EPI_ISL_1071340, EPI_ISL_1071341, EPI_ISL_1071342, EPI_ISL_1071343, EPI_ISL_1071344, EPI_ISL_1071345, EPI_ISL_1071346, EPI_ISL_1071347, EPI_ISL_1071348, EPI_ISL_1071349, EPI_ISL_1071351, EPI_ISL_1071352, EPI_ISL_1071353, EPI_ISL_1071354, EPI_ISL_1071355, EPI_ISL_1071356, EPI_ISL_1071357, EPI_ISL_1071358, EPI_ISL_1071359, EPI_ISL_1071360, EPI_ISL_1071361, EPI_ISL_1071362, EPI_ISL_1071363, EPI_ISL_1071364, EPI_ISL_1071365, EPI_ISL_1071366, EPI_ISL_1071367, EPI_ISL_1071368, EPI_ISL_1071957, EPI_ISL_1072343, EPI_ISL_1072344, EPI_ISL_1072346, EPI_ISL_1072347, EPI_ISL_1072348, EPI_ISL_1072349, EPI_ISL_1072350, EPI_ISL_1072351, EPI_ISL_1072352, EPI_ISL_1072353, EPI_ISL_1072355, EPI_ISL_1072356, EPI_ISL_1072357, EPI_ISL_1072358, EPI_ISL_1072359, EPI_ISL_1072360, EPI_ISL_1072361, EPI_ISL_1072362, EPI_ISL_1072363, EPI_ISL_1072364, EPI_ISL_1072365, EPI_ISL_1072366, EPI_ISL_1072367, EPI_ISL_1072370, EPI_ISL_1072371, EPI_ISL_1072372, EPI_ISL_1072373, EPI_ISL_1072374, EPI_ISL_1072375, EPI_ISL_1072376, EPI_ISL_1072377, EPI_ISL_1072378, EPI_ISL_1072379, EPI_ISL_1072381, EPI_ISL_1072382, EPI_ISL_1072383, EPI_ISL_1072384, EPI_ISL_1072385, EPI_ISL_1072386, EPI_ISL_1072387, EPI_ISL_1072388, EPI_ISL_1072389, EPI_ISL_1072390, EPI_ISL_1072391, EPI_ISL_1072392, EPI_ISL_1072393, EPI_ISL_1072395, EPI_ISL_1072396, EPI_ISL_1072397, EPI_ISL_1072398, EPI_ISL_1072400, EPI_ISL_1072401, EPI_ISL_1072402, EPI_ISL_1072403, EPI_ISL_1072404, EPI_ISL_1072405, EPI_ISL_1072406, EPI_ISL_1072407, EPI_ISL_1072409, EPI_ISL_1072410, EPI_ISL_1072411, EPI_ISL_1072414, EPI_ISL_1072415, EPI_ISL_1072416, EPI_ISL_1072417, EPI_ISL_1072418, EPI_ISL_1072419, EPI_ISL_1072420, EPI_ISL_1072421, EPI_ISL_1072423, EPI_ISL_1072424, EPI_ISL_1072425, EPI_ISL_1072426, EPI_ISL_1072427, EPI_ISL_1072428, EPI_ISL_1072429, EPI_ISL_1072430, EPI_ISL_1072431, EPI_ISL_1072433, EPI_ISL_1072434, EPI_ISL_1072435, EPI_ISL_1072436, EPI_ISL_1072437, EPI_ISL_1072438, EPI_ISL_1072440, EPI_ISL_1072441, EPI_ISL_1072442, EPI_ISL_1072443, EPI_ISL_1072444, EPI_ISL_1072445, EPI_ISL_1072446, EPI_ISL_1072447, EPI_ISL_1072448, EPI_ISL_1072449, EPI_ISL_1072450, EPI_ISL_1072451, EPI_ISL_1072452, EPI_ISL_1072453, EPI_ISL_1072454, EPI_ISL_1072455, EPI_ISL_1072457, EPI_ISL_1072458, EPI_ISL_1072460, EPI_ISL_1072462, EPI_ISL_1072463, EPI_ISL_1072464, EPI_ISL_1072465, EPI_ISL_1072467, EPI_ISL_1072469, EPI_ISL_1072470, EPI_ISL_1072472, EPI_ISL_1072473, EPI_ISL_1072474, EPI_ISL_1072475, EPI_ISL_1072476, EPI_ISL_1072477, EPI_ISL_1072479, EPI_ISL_1072480, EPI_ISL_1072481, EPI_ISL_1072483, EPI_ISL_1072484, EPI_ISL_1072485, EPI_ISL_1072486, EPI_ISL_1072487, EPI_ISL_1072488, EPI_ISL_1072489, EPI_ISL_1072490, EPI_ISL_1072491, EPI_ISL_1072492, EPI_ISL_1072493, EPI_ISL_1072494, EPI_ISL_1072495, EPI_ISL_1072496, EPI_ISL_1072498, EPI_ISL_1072499, EPI_ISL_1072500, EPI_ISL_1072501, EPI_ISL_1072502, EPI_ISL_1072503, EPI_ISL_1072504, EPI_ISL_1072505, EPI_ISL_1072506, EPI_ISL_1072507, EPI_ISL_1072508, EPI_ISL_1072509, EPI_ISL_1072510, EPI_ISL_1072513, EPI_ISL_1072514, EPI_ISL_1072516, EPI_ISL_1072517, EPI_ISL_1072518, EPI_ISL_1072519, EPI_ISL_1072520, EPI_ISL_1072521, EPI_ISL_1072522, EPI_ISL_1072523, EPI_ISL_1072524, EPI_ISL_1072525, EPI_ISL_1072526, EPI_ISL_1072527, EPI_ISL_1072528, EPI_ISL_1072529, EPI_ISL_1072531, EPI_ISL_1072532, EPI_ISL_1072533, EPI_ISL_1072534, EPI_ISL_1072535, EPI_ISL_1072536, EPI_ISL_1072537, EPI_ISL_1072538, EPI_ISL_1072539, EPI_ISL_1072540, EPI_ISL_1072541, EPI_ISL_1072542, EPI_ISL_1072543, EPI_ISL_1072544, EPI_ISL_1072545, EPI_ISL_1072546, EPI_ISL_1072547, EPI_ISL_1072548, EPI_ISL_1072549, EPI_ISL_1072550, EPI_ISL_1072551, EPI_ISL_1072552, EPI_ISL_1072554, EPI_ISL_1072555, EPI_ISL_1072556, EPI_ISL_1072557, EPI_ISL_1072558, EPI_ISL_1072559, EPI_ISL_1072560, EPI_ISL_1072562, EPI_ISL_1072563, EPI_ISL_1072564, EPI_ISL_1072565, EPI_ISL_1072566, EPI_ISL_1072567, EPI_ISL_1072568, EPI_ISL_1072569, EPI_ISL_1072570, EPI_ISL_1072573, EPI_ISL_1072574, EPI_ISL_1072575, EPI_ISL_1072576, EPI_ISL_1072577, EPI_ISL_1072578, EPI_ISL_1072579, EPI_ISL_1072580, EPI_ISL_1072581, EPI_ISL_1072582, EPI_ISL_1072583, EPI_ISL_1072585, EPI_ISL_1072586, EPI_ISL_1072587, EPI_ISL_1072588, EPI_ISL_1072589, EPI_ISL_1072590, EPI_ISL_1072592, EPI_ISL_1072593, EPI_ISL_1072595, EPI_ISL_1072596, EPI_ISL_1072598, EPI_ISL_1072599, EPI_ISL_1072600, EPI_ISL_1072602, EPI_ISL_1082533, EPI_ISL_1083823, EPI_ISL_1083837, EPI_ISL_1083848, EPI_ISL_1083877, EPI_ISL_1083878, EPI_ISL_1083881, EPI_ISL_1083882, EPI_ISL_1083888, EPI_ISL_1083903, EPI_ISL_1083905, EPI_ISL_1083924, EPI_ISL_1083928, EPI_ISL_1083931, EPI_ISL_1083986, EPI_ISL_1084009, EPI_ISL_1084021, EPI_ISL_1084044, EPI_ISL_1084045, EPI_ISL_1084079, EPI_ISL_1084091, EPI_ISL_1084125, EPI_ISL_1084151, EPI_ISL_1084245, EPI_ISL_1084255, EPI_ISL_1084306, EPI_ISL_1084316, EPI_ISL_1084333, EPI_ISL_1084344, EPI_ISL_1084376, EPI_ISL_1084380, EPI_ISL_1084382, EPI_ISL_1084400, EPI_ISL_1084406, EPI_ISL_1084430, EPI_ISL_1084719, EPI_ISL_1084720, EPI_ISL_1099504, EPI_ISL_1099505, EPI_ISL_1099508, EPI_ISL_1099510, EPI_ISL_1099534, EPI_ISL_1099535, EPI_ISL_1099547, EPI_ISL_1099565, EPI_ISL_1099571, EPI_ISL_1099581, EPI_ISL_1099603, EPI_ISL_1099604, EPI_ISL_1099613, EPI_ISL_1099626, EPI_ISL_1099681, EPI_ISL_1099687, EPI_ISL_1099688, EPI_ISL_1099691, EPI_ISL_1099705, EPI_ISL_1099713, EPI_ISL_1099721, EPI_ISL_1099737, EPI_ISL_1099739, EPI_ISL_1101101, EPI_ISL_1101224, EPI_ISL_1101250, EPI_ISL_1101265, EPI_ISL_1101286, EPI_ISL_1122644

| | | | |
|---|---|---|---|
| see above | Lighthouse Lab in Milton Keynes | Wellcome Sanger Institute for the COVID-19 Genomics UK (COG-UK) Consortium | Cordelia Langford; David K. Jackson; Dominic Kwiatkowski; Ewan Harrison; Ian Johnston; Jeffrey Barrett; John Sillitoe on behalf of the Wellcome Sanger Institute COVID-19 Surveillance Team; Roberto Amato; Sonia Goncalves; The Lighthouse Lab in Milton Keynes and Alex Alderton |

EPI_ISL_1178604, EPI_ISL_1178605, EPI_ISL_1178606, EPI_ISL_1178607, EPI_ISL_1178608, EPI_ISL_1178609, EPI_ISL_1178610, EPI_ISL_1178611, EPI_ISL_1178612, EPI_ISL_1178613, EPI_ISL_1178614, EPI_ISL_1178615, EPI_ISL_1178616, EPI_ISL_1178617, EPI_ISL_1178618, EPI_ISL_1178619, EPI_ISL_1178622, EPI_ISL_1178654, EPI_ISL_1178655, EPI_ISL_1178656, EPI_ISL_1178657, EPI_ISL_1178658, EPI_ISL_1178659, EPI_ISL_1178660, EPI_ISL_1178661, EPI_ISL_1178662, EPI_ISL_1178663, EPI_ISL_1309697

| | | | |
|---|---|---|---|
| see above | Lincolnshire Hospitals and DeepSeq Nottingham | COVID-19 Genomics UK (COG-UK) Consortium | Christopher Moore; Fei Sang; Joeseph Chappell; Johnny Debebe; Jonathan Ball; Matthew Carlisle; Matthew Loose; Nadine Holmes; Nichola Duckworth; Patrick McClure; Sarah Walsh; Tim Sloan; Victoria Wright |

EPI_ISL_1104396, EPI_ISL_1104397, EPI_ISL_1104398, EPI_ISL_1104399, EPI_ISL_1104402, EPI_ISL_1104403, EPI_ISL_1104404, EPI_ISL_1104405, EPI_ISL_1104406, EPI_ISL_1104407, EPI_ISL_1104409, EPI_ISL_1104411, EPI_ISL_1104412, EPI_ISL_1104413, EPI_ISL_1104414, EPI_ISL_1104416, EPI_ISL_1104417, EPI_ISL_1104418, EPI_ISL_1104420, EPI_ISL_1104421, EPI_ISL_1104456, EPI_ISL_1104457, EPI_ISL_1104458, EPI_ISL_1104459, EPI_ISL_1104463, EPI_ISL_1104465, EPI_ISL_1104469, EPI_ISL_1104470, EPI_ISL_1104473, EPI_ISL_1104474, EPI_ISL_1104475, EPI_ISL_1104476, EPI_ISL_1104477, EPI_ISL_1104486, EPI_ISL_1104487, EPI_ISL_1104488, EPI_ISL_1104490, EPI_ISL_1104492, EPI_ISL_1104493, EPI_ISL_1104494, EPI_ISL_1104495, EPI_ISL_1104496, EPI_ISL_1104497, EPI_ISL_1104524, EPI_ISL_1104562, EPI_ISL_1105576, EPI_ISL_1108834, EPI_ISL_1177900, EPI_ISL_1177901, EPI_ISL_1177902, EPI_ISL_1177903, EPI_ISL_1247835, EPI_ISL_1247836, EPI_ISL_1247837, EPI_ISL_1247845, EPI_ISL_1247846, EPI_ISL_1247847, EPI_ISL_1247848, EPI_ISL_1247849, EPI_ISL_1247850, EPI_ISL_1247851, EPI_ISL_1247852, EPI_ISL_1247854, EPI_ISL_1247855, EPI_ISL_1247856, EPI_ISL_1247857, EPI_ISL_1247858, EPI_ISL_1247859, EPI_ISL_1247860, EPI_ISL_1247861, EPI_ISL_1247862, EPI_ISL_1247863, EPI_ISL_1247882, EPI_ISL_1249263, EPI_ISL_1249265, EPI_ISL_1249266, EPI_ISL_1249267, EPI_ISL_1249268, EPI_ISL_1249269, EPI_ISL_1249270, EPI_ISL_1249271, EPI_ISL_1249272, EPI_ISL_1249274, EPI_ISL_1249275, EPI_ISL_1308939, EPI_ISL_1546071, EPI_ISL_1546072, EPI_ISL_1546076, EPI_ISL_1546078, EPI_ISL_1546092, EPI_ISL_1546093, EPI_ISL_1546095, EPI_ISL_1546097, EPI_ISL_1546099, EPI_ISL_1546101, EPI_ISL_1546103, EPI_ISL_1546105, EPI_ISL_1546107, EPI_ISL_1546108, EPI_ISL_1546110, EPI_ISL_1546112, EPI_ISL_1546115, EPI_ISL_1546118, EPI_ISL_1979164

| | | | |
|---|---|---|---|
| see above | Liverpool Clinical Laboratories | COVID-19 Genomics UK (COG-UK) Consortium | A Alrezaihi; Alessandro Gerada; Alistair Darby; Angela Cowell; Anita Lucaci; Anu Chawla; Cassie Olateju; Catherine Hartley; Charlotte Nelson; Ecaterina Vamos; Elaine O'Toole; Eleanor G Bentley; Ghada T Shawli; Hermione Webster; Isabel García-Dorival; James Johnson; James P Stewart; Jenifer Manson; Joanne Watts; Jones Benjamin; Jordan J Clark; Julian Hiscox; L Luu; Lucille Rainbow; M Almsaud; Margaret Hughes; Mark Whitehead; Matthew Gemmell; Miren Iturriza-Gomara; Muhannad Alruwaili; N.P Randle; Neil Swainston; PKF Gilmore; Parul Sharma; Rebekah Penrice-Randal; Richard Eccles; Richard Gregory; Sam Haldenby; Steve Paterson; Stuart D Armstrong; Trevor Ian Robinson; Ximeng Han |

EPI_ISL_1051017, EPI_ISL_1051023, EPI_ISL_1051024, EPI_ISL_1051025, EPI_ISL_1051026, EPI_ISL_1051027, EPI_ISL_1051031, EPI_ISL_1105710, EPI_ISL_1105713, EPI_ISL_1105714, EPI_ISL_1105744, EPI_ISL_1105745, EPI_ISL_1105746, EPI_ISL_1105747, EPI_ISL_1105748, EPI_ISL_1105749, EPI_ISL_1105750, EPI_ISL_1105751, EPI_ISL_1105752, EPI_ISL_1105753, EPI_ISL_1176913, EPI_ISL_1176918, EPI_ISL_1176944, EPI_ISL_1176945, EPI_ISL_1176997, EPI_ISL_1177008, EPI_ISL_1177068, EPI_ISL_1177077, EPI_ISL_1177176, EPI_ISL_1177185, EPI_ISL_1177200, EPI_ISL_1177211, EPI_ISL_1177224, EPI_ISL_1248848, EPI_ISL_1248849, EPI_ISL_1248850, EPI_ISL_1248851, EPI_ISL_1248852, EPI_ISL_1248853, EPI_ISL_1248854, EPI_ISL_1248855, EPI_ISL_1248856, EPI_ISL_1248857, EPI_ISL_1248858, EPI_ISL_1248859, EPI_ISL_1248860, EPI_ISL_1248861, EPI_ISL_1248862, EPI_ISL_1309468, EPI_ISL_1309469, EPI_ISL_1309470, EPI_ISL_1309471, EPI_ISL_1309472, EPI_ISL_1309473, EPI_ISL_1309474, EPI_ISL_1309475, EPI_ISL_1309612, EPI_ISL_1309613, EPI_ISL_1309614, EPI_ISL_1309615, EPI_ISL_1309616, EPI_ISL_1309617, EPI_ISL_1309618, EPI_ISL_1309620, EPI_ISL_1309622, EPI_ISL_1309640, EPI_ISL_1309642, EPI_ISL_1309643, EPI_ISL_1309644, EPI_ISL_1309645, EPI_ISL_1309646, EPI_ISL_1387114, EPI_ISL_1387115, EPI_ISL_1387116, EPI_ISL_1387117, EPI_ISL_1387118, EPI_ISL_1387119, EPI_ISL_1387121, EPI_ISL_1387122, EPI_ISL_1387123, EPI_ISL_1475339, EPI_ISL_1475340, EPI_ISL_1475410, EPI_ISL_1475412, EPI_ISL_1475413, EPI_ISL_1475414, EPI_ISL_1475419, EPI_ISL_1475420, EPI_ISL_1475421, EPI_ISL_1475422, EPI_ISL_1475423, EPI_ISL_1475424

| | | | |
|---|---|---|---|
| see above | Northumbria University / South Tees Hospitals NHS Foundation Trust / North Cumbria Integrated Care NHS Foundation Trust / North Tees and Hartlepool NHS Foundation Trust / Newcastle Hospitals NHS Foundation Trust | COVID-19 Genomics UK (COG-UK) Consortium | Andrew Nelson; Brendan Payne; Clive Graham; Darren L Smith; Debra Padgett; Edward Barton; Emma Swindells; Garren Scott; Gary Black; Gary Eltringham; Giles S Holt; Greg R Young; Jane Greenaway; Jennifer Collins; John Allan; Joshua Loh; Lynn Dover; Matthew Bashton; Mohammad A Tariq; Paul Baker; Sarah Essex; Steve Liggett; Wen C Yew; Yusri Taha |

EPI_ISL_1052962, EPI_ISL_1052963, EPI_ISL_1052964, EPI_ISL_1052965, EPI_ISL_1052966, EPI_ISL_1052967, EPI_ISL_1052968, EPI_ISL_1052969, EPI_ISL_1052970, EPI_ISL_1052972, EPI_ISL_1053080, EPI_ISL_1053081, EPI_ISL_1053083, EPI_ISL_1053091, EPI_ISL_1053092, EPI_ISL_1053093, EPI_ISL_1053094, EPI_ISL_1053095, EPI_ISL_1053096, EPI_ISL_1053097, EPI_ISL_1053098, EPI_ISL_1053099, EPI_ISL_1053100, EPI_ISL_1053101, EPI_ISL_1053102, EPI_ISL_1053103, EPI_ISL_1053104, EPI_ISL_1053111, EPI_ISL_1053140, EPI_ISL_1053142, EPI_ISL_1053145, EPI_ISL_1053146, EPI_ISL_1053147, EPI_ISL_1053148, EPI_ISL_1053149, EPI_ISL_1053150, EPI_ISL_1053151, EPI_ISL_1053152, EPI_ISL_1053153, EPI_ISL_1053154, EPI_ISL_1053155, EPI_ISL_1053156, EPI_ISL_1053160, EPI_ISL_1053162, EPI_ISL_1053163, EPI_ISL_1053164, EPI_ISL_1053166, EPI_ISL_1053169, EPI_ISL_1053172, EPI_ISL_1053173, EPI_ISL_1053174, EPI_ISL_1053175, EPI_ISL_1053178, EPI_ISL_1053179, EPI_ISL_1053180, EPI_ISL_1053181, EPI_ISL_1053183, EPI_ISL_1053184, EPI_ISL_1053185, EPI_ISL_1053187, EPI_ISL_1053188, EPI_ISL_1053189, EPI_ISL_1053191, EPI_ISL_1053192, EPI_ISL_1053193, EPI_ISL_1053195, EPI_ISL_1053196, EPI_ISL_1053197, EPI_ISL_1053198, EPI_ISL_1053199, EPI_ISL_1053200, EPI_ISL_1053203, EPI_ISL_1053204, EPI_ISL_1053205, EPI_ISL_1053206, EPI_ISL_1053208, EPI_ISL_1053209, EPI_ISL_1053210, EPI_ISL_1053211, EPI_ISL_1053212, EPI_ISL_1053213, EPI_ISL_1053214, EPI_ISL_1053215, EPI_ISL_1053216, EPI_ISL_1053217, EPI_ISL_1053218, EPI_ISL_1053219, EPI_ISL_1053220, EPI_ISL_1053221, EPI_ISL_1053222, EPI_ISL_1053223, EPI_ISL_1053224, EPI_ISL_1053225, EPI_ISL_1053226, EPI_ISL_1053227, EPI_ISL_1053228, EPI_ISL_1053231, EPI_ISL_1053232, EPI_ISL_1053233, EPI_ISL_1053234, EPI_ISL_1053235, EPI_ISL_1053240, EPI_ISL_1053242, EPI_ISL_1053243, EPI_ISL_1053247, EPI_ISL_1053248, EPI_ISL_1053249, EPI_ISL_1053251, EPI_ISL_1053252, EPI_ISL_1053253, EPI_ISL_1053254, EPI_ISL_1053255, EPI_ISL_1053285, EPI_ISL_1053290, EPI_ISL_1053291, EPI_ISL_1053292, EPI_ISL_1053293, EPI_ISL_1053294, EPI_ISL_1053295, EPI_ISL_1053296, EPI_ISL_1053298, EPI_ISL_1053299, EPI_ISL_1053300, EPI_ISL_1053301, EPI_ISL_1053302, EPI_ISL_1053303, EPI_ISL_1053304, EPI_ISL_1053305, EPI_ISL_1053306, EPI_ISL_1053307, EPI_ISL_1053308, EPI_ISL_1053309, EPI_ISL_1053310, EPI_ISL_1053311, EPI_ISL_1053312, EPI_ISL_1053313, EPI_ISL_1053315, EPI_ISL_1053320, EPI_ISL_1053321, EPI_ISL_1053329, EPI_ISL_1053330, EPI_ISL_1053332, EPI_ISL_1053334, EPI_ISL_1053335, EPI_ISL_1053337, EPI_ISL_1053338, EPI_ISL_1053339, EPI_ISL_1053340, EPI_ISL_1053341, EPI_ISL_1053343, EPI_ISL_1053345, EPI_ISL_1053346, EPI_ISL_1053347, EPI_ISL_1053349, EPI_ISL_1053353, EPI_ISL_1053355, EPI_ISL_1053356, EPI_ISL_1053358, EPI_ISL_1053359, EPI_ISL_1053361, EPI_ISL_1053362, EPI_ISL_1053363, EPI_ISL_1053365, EPI_ISL_1053366, EPI_ISL_1053367, EPI_ISL_1053368, EPI_ISL_1053369, EPI_ISL_1053371, EPI_ISL_1053372, EPI_ISL_1053373, EPI_ISL_1053374, EPI_ISL_1053375, EPI_ISL_1053376, EPI_ISL_1053377, EPI_ISL_1053378, EPI_ISL_1053379, EPI_ISL_1053381, EPI_ISL_1053382, EPI_ISL_1053383, EPI_ISL_1053384, EPI_ISL_1053385, EPI_ISL_1053386, EPI_ISL_1053389, EPI_ISL_1053390, EPI_ISL_1053391, EPI_ISL_1053392, EPI_ISL_1053393, EPI_ISL_1053395, EPI_ISL_1053396, EPI_ISL_1053397, EPI_ISL_1053398, EPI_ISL_1053399, EPI_ISL_1053400, EPI_ISL_1053403, EPI_ISL_1053405, EPI_ISL_1053406, EPI_ISL_1053407, EPI_ISL_1053408, EPI_ISL_1053416, EPI_ISL_1053420, EPI_ISL_1053422, EPI_ISL_1053425, EPI_ISL_1053427, EPI_ISL_1053429, EPI_ISL_1053430, EPI_ISL_1053441, EPI_ISL_1053443, EPI_ISL_1053445, EPI_ISL_1053447, EPI_ISL_1053450, EPI_ISL_1053451, EPI_ISL_1053452, EPI_ISL_1053454, EPI_ISL_1053455, EPI_ISL_1053457, EPI_ISL_1053462, EPI_ISL_1053463, EPI_ISL_1053464, EPI_ISL_1053465, EPI_ISL_1053466, EPI_ISL_1053467, EPI_ISL_1053468, EPI_ISL_1053469, EPI_ISL_1053471, EPI_ISL_1053472, EPI_ISL_1053475, EPI_ISL_1053476, EPI_ISL_1053478, EPI_ISL_1053479, EPI_ISL_1053480, EPI_ISL_1053483, EPI_ISL_1053484, EPI_ISL_1053485, EPI_ISL_1053486, EPI_ISL_1053487, EPI_ISL_1053488, EPI_ISL_1053505, EPI_ISL_1053506, EPI_ISL_1053507, EPI_ISL_1053508, EPI_ISL_1053509, EPI_ISL_1053659, EPI_ISL_1053664, EPI_ISL_1053669, EPI_ISL_1053675, EPI_ISL_1053679, EPI_ISL_1106212, EPI_ISL_1106213, EPI_ISL_1106214, EPI_ISL_1106215, EPI_ISL_1106216, EPI_ISL_1106217, EPI_ISL_1106218, EPI_ISL_1106219, EPI_ISL_1106220, EPI_ISL_1106221, EPI_ISL_1106222, EPI_ISL_1106223, EPI_ISL_1106224, EPI_ISL_1106225, EPI_ISL_1106226, EPI_ISL_1106227, EPI_ISL_1106228, EPI_ISL_1106229, EPI_ISL_1106230, EPI_ISL_1106231, EPI_ISL_1106232, EPI_ISL_1106233, EPI_ISL_1106234, EPI_ISL_1106235, EPI_ISL_1106236, EPI_ISL_1106237, EPI_ISL_1106239, EPI_ISL_1106240, EPI_ISL_1106241, EPI_ISL_1106242, EPI_ISL_1106243, EPI_ISL_1106245, EPI_ISL_1106246, EPI_ISL_1106248, EPI_ISL_1106249, EPI_ISL_1106250, EPI_ISL_1106251, EPI_ISL_1106252, EPI_ISL_1106256, EPI_ISL_1106257, EPI_ISL_1106258, EPI_ISL_1106259, EPI_ISL_1106268, EPI_ISL_1106269, EPI_ISL_1106270, EPI_ISL_1106271, EPI_ISL_1106275, EPI_ISL_1106276, EPI_ISL_1106277, EPI_ISL_1106278, EPI_ISL_1106281, EPI_ISL_1106295, EPI_ISL_1106299, EPI_ISL_1106301, EPI_ISL_1106303, EPI_ISL_1106304, EPI_ISL_1106306, EPI_ISL_1106309, EPI_ISL_1106310,

| | | | |
|---|---|---|---|
| EPI_ISL_1106311, EPI_ISL_1106313, EPI_ISL_1106318, EPI_ISL_1106319, EPI_ISL_1106320, EPI_ISL_1106321, EPI_ISL_1106324, EPI_ISL_1106326, EPI_ISL_1106327, EPI_ISL_1106328, EPI_ISL_1106330, EPI_ISL_1106331, EPI_ISL_1106332, EPI_ISL_1106333, EPI_ISL_1106339, EPI_ISL_1106341, EPI_ISL_1106342, EPI_ISL_1106343, EPI_ISL_1106344, EPI_ISL_1106345, EPI_ISL_1106346, EPI_ISL_1106347, EPI_ISL_1106348, EPI_ISL_1106349, EPI_ISL_1106350, EPI_ISL_1106351, EPI_ISL_1106354, EPI_ISL_1106705, EPI_ISL_1106706, EPI_ISL_1106708, EPI_ISL_1106709, EPI_ISL_1178886, EPI_ISL_1178897, EPI_ISL_1387304, EPI_ISL_1387307, EPI_ISL_1387308, EPI_ISL_1387309, EPI_ISL_1387317, EPI_ISL_1387318, EPI_ISL_1387320, EPI_ISL_1387322, EPI_ISL_1387323, EPI_ISL_1387324, EPI_ISL_1387325, EPI_ISL_1387328, EPI_ISL_1387329, EPI_ISL_1387332, EPI_ISL_1387333, EPI_ISL_1387334, EPI_ISL_1387335, EPI_ISL_1387336, EPI_ISL_1387337, EPI_ISL_1387338, EPI_ISL_1387339, EPI_ISL_1387340, EPI_ISL_1387342, EPI_ISL_1387343, EPI_ISL_1387344, EPI_ISL_1387345, EPI_ISL_1387346, EPI_ISL_1387347, EPI_ISL_1387348, EPI_ISL_1387349, EPI_ISL_1387351, EPI_ISL_1387352, EPI_ISL_1387354, EPI_ISL_1387355, EPI_ISL_1387356, EPI_ISL_1387357, EPI_ISL_1387358, EPI_ISL_1387359, EPI_ISL_1387360, EPI_ISL_1387361, EPI_ISL_1387363, EPI_ISL_1387364, EPI_ISL_1387367, EPI_ISL_1387368, EPI_ISL_1387369, EPI_ISL_1387370, EPI_ISL_1546999, EPI_ISL_1547008, EPI_ISL_1547010, EPI_ISL_1547015, EPI_ISL_1547016, EPI_ISL_1547017, EPI_ISL_1547019, EPI_ISL_1547020, EPI_ISL_1547021, EPI_ISL_1547024, EPI_ISL_1547027 | | | |
| see above | Originating lab: Wales Specialist Virology Centre Sequencing lab: Pathogen Genomics Unit | Public Health Wales Microbiology Cardiff Wales Specialist Virology Centre | Alec Birchley; Alexander Adams; Amy Gaskin; Angela Marchbank; Bree Gatica-Wilcox; Catherine Moore; Jason Coombes; Joanne Watkins; Joel Southgate; Johnathan Evans; Laura Gifford; Lauren Gilbert; Lee Graham; Malorie Perry; Matthew Bull; Nicole Pacchiarini; Sally Corden; Sara Kumziene-Summerhayes; Sara Rey; Sarah Taylor; Simon Cottrell; Sophie Jones; Tom Connor |
| EPI_ISL_1178704, EPI_ISL_1178709, EPI_ISL_1178760, EPI_ISL_1178775, EPI_ISL_1178810, EPI_ISL_1178818, EPI_ISL_1249072, EPI_ISL_1249073, EPI_ISL_1249074, EPI_ISL_1249075, EPI_ISL_1249076, EPI_ISL_1249077, EPI_ISL_1249078, EPI_ISL_1249079, EPI_ISL_1249080, EPI_ISL_1249081, EPI_ISL_1249082, EPI_ISL_1249083, EPI_ISL_1249084, EPI_ISL_1249086, EPI_ISL_1249087, EPI_ISL_1249090, EPI_ISL_1249094, EPI_ISL_1249097, EPI_ISL_1249098, EPI_ISL_1249099, EPI_ISL_1249100, EPI_ISL_1249119, EPI_ISL_1249120, EPI_ISL_1249122, EPI_ISL_1249123, EPI_ISL_1249124, EPI_ISL_1249126, EPI_ISL_1249127, EPI_ISL_1249128, EPI_ISL_1249130, EPI_ISL_1249131, EPI_ISL_1249132, EPI_ISL_1249133, EPI_ISL_1249134, EPI_ISL_1249135, EPI_ISL_1249136, EPI_ISL_1249137, EPI_ISL_1249138, EPI_ISL_1249141, EPI_ISL_1249144, EPI_ISL_1249145, EPI_ISL_1249146, EPI_ISL_1249149, EPI_ISL_1249150, EPI_ISL_1309756, EPI_ISL_1309758, EPI_ISL_1309761, EPI_ISL_1309762, EPI_ISL_1309775, EPI_ISL_1387218, EPI_ISL_1833064 | | | |
| see above | Oxford Viromics, NDM, University of Oxford; Oxford University Hospitals; Basingstoke and North Hampshire Hospital | COVID-19 Genomics UK (COG-UK) Consortium | Alex Mobbs; Amy Trebes; Anita Justice; Catrin Moore; Christophe Fraser; David Bonsall; David Buck; Emma Wise; George Macintyre; Jessica Lynch; John Todd; Mariateresa de Cesare; Matilde Mori; Monique Andersson; Nathan Moore; Nick Cortes; Robert Shaw; Stephen Kidd; Tanya Golubchik; Timothy Peto |
| EPI_ISL_1051413, EPI_ISL_1051414, EPI_ISL_1051415, EPI_ISL_1051416, EPI_ISL_1051417, EPI_ISL_1051418, EPI_ISL_1051531, EPI_ISL_1051532, EPI_ISL_1051533, EPI_ISL_1051535, EPI_ISL_1051538, EPI_ISL_1051540, EPI_ISL_1051542, EPI_ISL_1051544, EPI_ISL_1051546, EPI_ISL_1051547, EPI_ISL_1051553, EPI_ISL_1051554, EPI_ISL_1103658, EPI_ISL_1103681, EPI_ISL_1103682, EPI_ISL_1103683, EPI_ISL_1103684, EPI_ISL_1103685, EPI_ISL_1103686, EPI_ISL_1103687, EPI_ISL_1103689, EPI_ISL_1103690, EPI_ISL_1103691, EPI_ISL_1103692, EPI_ISL_1103693, EPI_ISL_1103694, EPI_ISL_1103695, EPI_ISL_1103696, EPI_ISL_1103698, EPI_ISL_1103699, EPI_ISL_1103701, EPI_ISL_1103702, EPI_ISL_1103704, EPI_ISL_1103705, EPI_ISL_1103706, EPI_ISL_1103707, EPI_ISL_1103708, EPI_ISL_1103709, EPI_ISL_1103710, EPI_ISL_1103711, EPI_ISL_1103712, EPI_ISL_1103713, EPI_ISL_1103714, EPI_ISL_1103715, EPI_ISL_1103716, EPI_ISL_1103717, EPI_ISL_1103718, EPI_ISL_1103719, EPI_ISL_1103720, EPI_ISL_1103721, EPI_ISL_1103722, EPI_ISL_1103723, EPI_ISL_1103724, EPI_ISL_1103725, EPI_ISL_1103726, EPI_ISL_1103727, EPI_ISL_1103728, EPI_ISL_1178518, EPI_ISL_1178519, EPI_ISL_1178521, EPI_ISL_1178524, EPI_ISL_1178525, EPI_ISL_1178527, EPI_ISL_1178528, EPI_ISL_1178529, EPI_ISL_1178531, EPI_ISL_1178532, EPI_ISL_1178533, EPI_ISL_1178537, EPI_ISL_1178538, EPI_ISL_1178539, EPI_ISL_1178540, EPI_ISL_1178542, EPI_ISL_1178543, EPI_ISL_1178544, EPI_ISL_1178545, EPI_ISL_1178546, EPI_ISL_1178547, EPI_ISL_1178548, EPI_ISL_1247373, EPI_ISL_1308023, EPI_ISL_1308024, EPI_ISL_1308025, EPI_ISL_1308033, EPI_ISL_1308034, EPI_ISL_1308035, EPI_ISL_1308045, EPI_ISL_1308046, EPI_ISL_1308053, EPI_ISL_1308054, EPI_ISL_1308055, EPI_ISL_1308065, EPI_ISL_1308066, EPI_ISL_1308076, EPI_ISL_1308077, EPI_ISL_1308078, EPI_ISL_1308086, EPI_ISL_1308087, EPI_ISL_1308088, EPI_ISL_1308098, EPI_ISL_1308099, EPI_ISL_1386427, EPI_ISL_1386453, EPI_ISL_1386460, EPI_ISL_1386464, EPI_ISL_1474166, EPI_ISL_1474205, EPI_ISL_1474225, EPI_ISL_1474226, EPI_ISL_1474893, EPI_ISL_1474946, EPI_ISL_1474955, EPI_ISL_1476688, EPI_ISL_1476689, EPI_ISL_1476690, EPI_ISL_1832908, EPI_ISL_1832911, EPI_ISL_1832913, EPI_ISL_1832914, EPI_ISL_1832916, EPI_ISL_1832918, EPI_ISL_1832919, EPI_ISL_1832922, EPI_ISL_1832925, EPI_ISL_1832927, EPI_ISL_1832937, EPI_ISL_1832938, EPI_ISL_1832939, EPI_ISL_1832977, EPI_ISL_1832981, EPI_ISL_1832986 | | | |
| see above | Quadram Institute Bioscience | COVID-19 Genomics UK (COG-UK) Consortium | Alexander J Trotter; Alison E. Mather; Alp Aydin; Ana P. Tedim; Anastasia Kolyva; Andrew Bell; Andrew J. Page; Claire Stuart; Dave J. Baker; Gemma L. Kay; John Wain; Justin O'Grady; Leonardo de Oliveira Martins; Lizzie Meadows; Maria Diaz; Mark Webber; Muhammed Yasir; Nabil-Fareed Alikhan; Ngozi Elumogo; Nicholas M. Thomson; Rachael Stanley; Rachel Gilroy; Reenesh Prakash; Samir Dervisevic; Samuel Bloomfield; Steven Rudder; Thanh Le-Viet |
| EPI_ISL_1051558, EPI_ISL_1051559, EPI_ISL_1051560, EPI_ISL_1051561, EPI_ISL_1051562, EPI_ISL_1051563, EPI_ISL_1051564, EPI_ISL_1051565, EPI_ISL_1051566, EPI_ISL_1051567, EPI_ISL_1051568, EPI_ISL_1051569, EPI_ISL_1051570, EPI_ISL_1051571, EPI_ISL_1051572, EPI_ISL_1051573, EPI_ISL_1051574, EPI_ISL_1051575, EPI_ISL_1051576, EPI_ISL_1051577, EPI_ISL_1051578, EPI_ISL_1051579, EPI_ISL_1051580, EPI_ISL_1051581, EPI_ISL_1051582, EPI_ISL_1051583, EPI_ISL_1051586, EPI_ISL_1051587 | | | |
| see above | Queens Medical Centre, Clinical Microbiology Department / DeepSeq Nottingham | COVID-19 Genomics UK (COG-UK) Consortium | Christopher Moore; Fei Sang; Gemma Clark; Hannah Howson-Wells; Johnny Debebe; Jonathan Ball; Joseph Chappell; Manjinder Khakh; Matthew Carlisle; Matthew Loose; Michelle M Lister; Nadine Holmes; Patrick McClure; Theocharis Tsoleridis; Vicki M Fleming; Victoria Wright; Wendy Smith |
| EPI_ISL_1042126, EPI_ISL_1043506, EPI_ISL_1070152, EPI_ISL_1070154, EPI_ISL_1070155, EPI_ISL_1070156, EPI_ISL_1070157, EPI_ISL_1070159, EPI_ISL_1070161, EPI_ISL_1070164, EPI_ISL_1070165, EPI_ISL_1070167, EPI_ISL_1070168, EPI_ISL_1070169, EPI_ISL_1070175, EPI_ISL_1070182, EPI_ISL_1070188, EPI_ISL_1070201, EPI_ISL_1070202, EPI_ISL_1070212, EPI_ISL_1070217, EPI_ISL_1070219, EPI_ISL_1070220, EPI_ISL_1070222, EPI_ISL_1070228, EPI_ISL_1070230, EPI_ISL_1070231, EPI_ISL_1070241, EPI_ISL_1070245, EPI_ISL_1070249, EPI_ISL_1070258, EPI_ISL_1070261, EPI_ISL_1070263, EPI_ISL_1070265, EPI_ISL_1070270, EPI_ISL_1070273, EPI_ISL_1070274, EPI_ISL_1070276, EPI_ISL_1070279, EPI_ISL_1070289, EPI_ISL_1070318, EPI_ISL_1070329, EPI_ISL_1070335, EPI_ISL_1070339, EPI_ISL_1072606, EPI_ISL_1072609, EPI_ISL_1072616, EPI_ISL_1072617, EPI_ISL_1072620, EPI_ISL_1072627, EPI_ISL_1072638, EPI_ISL_1072641, EPI_ISL_1072656, EPI_ISL_1072672, EPI_ISL_1072678, EPI_ISL_1072685, EPI_ISL_1072690, EPI_ISL_1072691, EPI_ISL_1072693, EPI_ISL_1072694, EPI_ISL_1072696, EPI_ISL_1072701, EPI_ISL_1072707, EPI_ISL_1072708, EPI_ISL_1072709, EPI_ISL_1072710, EPI_ISL_1072713, EPI_ISL_1072717, EPI_ISL_1072721, EPI_ISL_1072723, EPI_ISL_1072727, EPI_ISL_1072733, EPI_ISL_1072740, EPI_ISL_1072742, EPI_ISL_1072745, EPI_ISL_1072751, EPI_ISL_1072778, EPI_ISL_1072784, EPI_ISL_1072792, EPI_ISL_1072800, EPI_ISL_1072802, EPI_ISL_1072804, EPI_ISL_1072812, EPI_ISL_1072814, EPI_ISL_1072820, EPI_ISL_1072828, EPI_ISL_1072836, EPI_ISL_1072841, EPI_ISL_1072842, EPI_ISL_1072843, EPI_ISL_1072849, EPI_ISL_1072850, EPI_ISL_1072867, EPI_ISL_1072871, EPI_ISL_1072876, EPI_ISL_1072877, EPI_ISL_1072881, EPI_ISL_1072882, EPI_ISL_1072894, EPI_ISL_1072896, EPI_ISL_1072907, EPI_ISL_1072909, EPI_ISL_1072912, EPI_ISL_1072933, EPI_ISL_1072944, EPI_ISL_1072947, EPI_ISL_1072950, EPI_ISL_1072951, EPI_ISL_1072952, EPI_ISL_1082527, EPI_ISL_1082528, EPI_ISL_1082540, EPI_ISL_1082545, EPI_ISL_1082546, EPI_ISL_1082556, EPI_ISL_1082557, EPI_ISL_1082564, EPI_ISL_1082566, EPI_ISL_1082568, EPI_ISL_1082570, EPI_ISL_1082581, EPI_ISL_1082585, EPI_ISL_1082586, EPI_ISL_1082587, EPI_ISL_1082589, EPI_ISL_1082595, EPI_ISL_1082597, EPI_ISL_1082602, EPI_ISL_1082603, EPI_ISL_1082607, EPI_ISL_1082608, EPI_ISL_1082615, EPI_ISL_1082616, EPI_ISL_1082622, EPI_ISL_1082626, EPI_ISL_1082628, EPI_ISL_1082631, EPI_ISL_1082637, EPI_ISL_1082639, EPI_ISL_1082642, EPI_ISL_1082643, EPI_ISL_1082644, EPI_ISL_1082647, EPI_ISL_1082648, EPI_ISL_1082655, EPI_ISL_1082660, EPI_ISL_1082661, EPI_ISL_1082662, EPI_ISL_1082664, EPI_ISL_1082666, EPI_ISL_1082667, EPI_ISL_1082668, EPI_ISL_1082672, EPI_ISL_1082673, EPI_ISL_1082675, EPI_ISL_1082678, EPI_ISL_1082682, EPI_ISL_1082683, EPI_ISL_1082684, EPI_ISL_1082686, EPI_ISL_1082691, EPI_ISL_1082696, EPI_ISL_1082697, EPI_ISL_1082702, EPI_ISL_1082703, EPI_ISL_1082707, EPI_ISL_1082713, EPI_ISL_1082722, EPI_ISL_1082728, EPI_ISL_1082735, EPI_ISL_1082739, EPI_ISL_1082742, EPI_ISL_1082743, EPI_ISL_1082750, EPI_ISL_1082757, EPI_ISL_1082760, EPI_ISL_1082769, EPI_ISL_1082770, EPI_ISL_1082771, EPI_ISL_1082786, EPI_ISL_1082789, EPI_ISL_1082794, EPI_ISL_1082795, EPI_ISL_1082798, EPI_ISL_1082799, EPI_ISL_1082801, EPI_ISL_1082803, EPI_ISL_1082806, EPI_ISL_1082810, EPI_ISL_1082818, EPI_ISL_1082824, EPI_ISL_1082825, EPI_ISL_1082829, EPI_ISL_1082830, EPI_ISL_1099742, EPI_ISL_1099743, EPI_ISL_1099745, EPI_ISL_1099749, EPI_ISL_1099750, EPI_ISL_1099752, EPI_ISL_1099753, EPI_ISL_1099754, EPI_ISL_1099756, EPI_ISL_1099759, EPI_ISL_1099760, EPI_ISL_1099767, EPI_ISL_1099768, EPI_ISL_1099769, EPI_ISL_1099773, EPI_ISL_1099776, EPI_ISL_1099783, EPI_ISL_1099793, EPI_ISL_1099799, EPI_ISL_1099802, EPI_ISL_1099803, EPI_ISL_1099804, EPI_ISL_1099805, EPI_ISL_1099807, EPI_ISL_1099809, EPI_ISL_1099810, EPI_ISL_1099811, EPI_ISL_1099812, EPI_ISL_1099815, EPI_ISL_1099820, EPI_ISL_1099822, EPI_ISL_1099827, EPI_ISL_1099829, EPI_ISL_1099832, EPI_ISL_1099834, EPI_ISL_1099836, EPI_ISL_1099839, EPI_ISL_1099840, EPI_ISL_1099843, EPI_ISL_1099844, EPI_ISL_1099847, EPI_ISL_1099849, EPI_ISL_1099851, EPI_ISL_1099856, EPI_ISL_1099859, EPI_ISL_1099860, EPI_ISL_1099861, EPI_ISL_1099862, EPI_ISL_1099864, EPI_ISL_1099865, EPI_ISL_1099867, EPI_ISL_1099869, EPI_ISL_1099871, EPI_ISL_1099874, EPI_ISL_1099875, EPI_ISL_1099881, EPI_ISL_1099883, EPI_ISL_1099886, EPI_ISL_1099891, EPI_ISL_1099892, EPI_ISL_1099893, EPI_ISL_1099897, EPI_ISL_1099898, EPI_ISL_1099906, EPI_ISL_1099907, EPI_ISL_1099911, EPI_ISL_1099915, EPI_ISL_1099923, EPI_ISL_1099924, EPI_ISL_1099926, EPI_ISL_1099933, EPI_ISL_1099937, EPI_ISL_1099939, EPI_ISL_1099942, EPI_ISL_1099944, EPI_ISL_1099946, EPI_ISL_1099947, EPI_ISL_1099948, EPI_ISL_1099953, EPI_ISL_1099958, EPI_ISL_1099961, EPI_ISL_1099963, EPI_ISL_1099964, EPI_ISL_1099966, EPI_ISL_1099968, EPI_ISL_1099969, EPI_ISL_1099970, EPI_ISL_1099971, EPI_ISL_1099975, EPI_ISL_1099976, EPI_ISL_1099979, EPI_ISL_1099980, EPI_ISL_1099982, EPI_ISL_1099989, EPI_ISL_1099994, EPI_ISL_1099998, EPI_ISL_1099999, EPI_ISL_1100000, EPI_ISL_1100001, EPI_ISL_1100003, EPI_ISL_1100004, EPI_ISL_1100012, EPI_ISL_1100018, EPI_ISL_1100019, EPI_ISL_1100028, EPI_ISL_1100034, EPI_ISL_1100035, EPI_ISL_1100039, EPI_ISL_1100043, EPI_ISL_1100044, EPI_ISL_1100046, EPI_ISL_1100048, EPI_ISL_1100049, EPI_ISL_1100053, EPI_ISL_1100054, EPI_ISL_1100058, EPI_ISL_1100059, EPI_ISL_1100066, EPI_ISL_1100070, EPI_ISL_1100072, EPI_ISL_1100759, EPI_ISL_1100761, EPI_ISL_1100763, EPI_ISL_1100764, EPI_ISL_1100768, EPI_ISL_1100770, EPI_ISL_1100771, EPI_ISL_1100772, EPI_ISL_1100773, EPI_ISL_1100775, EPI_ISL_1100781, EPI_ISL_1100793, EPI_ISL_1100800, EPI_ISL_1100801, EPI_ISL_1100803, EPI_ISL_1100805, EPI_ISL_1100812, EPI_ISL_1100813, EPI_ISL_1100820, EPI_ISL_1100824, EPI_ISL_1100826, EPI_ISL_1100827, EPI_ISL_1100832, EPI_ISL_1100837, EPI_ISL_1100838, EPI_ISL_1100840, EPI_ISL_1100841, EPI_ISL_1100842, EPI_ISL_1100844, EPI_ISL_1100845, EPI_ISL_1100847, EPI_ISL_1100853, EPI_ISL_1100855, EPI_ISL_1100857, EPI_ISL_1100867, EPI_ISL_1100868, EPI_ISL_1100869, EPI_ISL_1100872, EPI_ISL_1100875, EPI_ISL_1100881, EPI_ISL_1100883, EPI_ISL_1100889, EPI_ISL_1100892, EPI_ISL_1100894, EPI_ISL_1100896, EPI_ISL_1100897, EPI_ISL_1100902, EPI_ISL_1100908, EPI_ISL_1100917, EPI_ISL_1100921, EPI_ISL_1100922, EPI_ISL_1100926, EPI_ISL_1100937, EPI_ISL_1100939, EPI_ISL_1100940, EPI_ISL_1100941, EPI_ISL_1100942, EPI_ISL_1100944, EPI_ISL_1100946, EPI_ISL_1100948, EPI_ISL_1100960, EPI_ISL_1100962, EPI_ISL_1100972, EPI_ISL_1100980, EPI_ISL_1100982, EPI_ISL_1100983, EPI_ISL_1100984, EPI_ISL_1100986, EPI_ISL_1101666, EPI_ISL_1101667, EPI_ISL_1101668, EPI_ISL_1101670, EPI_ISL_1101674, EPI_ISL_1101676, EPI_ISL_1101677, EPI_ISL_1101683, EPI_ISL_1101684, EPI_ISL_1101689, EPI_ISL_1101692, EPI_ISL_1101701, EPI_ISL_1101707, EPI_ISL_1101708, EPI_ISL_1101711, EPI_ISL_1101712, EPI_ISL_1101718, EPI_ISL_1101726, EPI_ISL_1101727, EPI_ISL_1101731, EPI_ISL_1101732, EPI_ISL_1101734, EPI_ISL_1101735, EPI_ISL_1101736, EPI_ISL_1101739, EPI_ISL_1101740, EPI_ISL_1101741, EPI_ISL_1101748, EPI_ISL_1101750, EPI_ISL_1101752, EPI_ISL_1101753, EPI_ISL_1101754, EPI_ISL_1101755, EPI_ISL_1101760, EPI_ISL_1101762, EPI_ISL_1101766, EPI_ISL_1101768, EPI_ISL_1101770, EPI_ISL_1101771, EPI_ISL_1101774, EPI_ISL_1101775, EPI_ISL_1101777, EPI_ISL_1101780, EPI_ISL_1101782, EPI_ISL_1101786, EPI_ISL_1101787, EPI_ISL_1101794, EPI_ISL_1101799, EPI_ISL_1101800, EPI_ISL_1101802, EPI_ISL_1101805, EPI_ISL_1101806, EPI_ISL_1101808, EPI_ISL_1101809, EPI_ISL_1101814, EPI_ISL_1101816, EPI_ISL_1101823, EPI_ISL_1101824, EPI_ISL_1101830, EPI_ISL_1101831, EPI_ISL_1101836, EPI_ISL_1101838, EPI_ISL_1101844, EPI_ISL_1101845, EPI_ISL_1101848, EPI_ISL_1101859, EPI_ISL_1101861, EPI_ISL_1101863, EPI_ISL_1101865, EPI_ISL_1101871, EPI_ISL_1101873, EPI_ISL_1101879, EPI_ISL_1101884, EPI_ISL_1101885, EPI_ISL_1101888, EPI_ISL_1101889, EPI_ISL_1101892, EPI_ISL_1101893, EPI_ISL_1101895, EPI_ISL_1101896, EPI_ISL_1101897, EPI_ISL_1101898, EPI_ISL_1101900, EPI_ISL_1101903, EPI_ISL_1101905, EPI_ISL_1101909, EPI_ISL_1101914, EPI_ISL_1101920, EPI_ISL_1101922, EPI_ISL_1101923, EPI_ISL_1101924, EPI_ISL_1101938, EPI_ISL_1101941, EPI_ISL_1101943, EPI_ISL_1101947, EPI_ISL_1101949, EPI_ISL_1101954, EPI_ISL_1101956, EPI_ISL_1101958, EPI_ISL_1101961, EPI_ISL_1101963, EPI_ISL_1101965, EPI_ISL_1102026, EPI_ISL_1102029, EPI_ISL_1102030, EPI_ISL_1102033, EPI_ISL_1102037, EPI_ISL_1102039, EPI_ISL_1102044, EPI_ISL_1102047, EPI_ISL_1102051, EPI_ISL_1102053, EPI_ISL_1102058, EPI_ISL_1102059, EPI_ISL_1102060, EPI_ISL_1102066, EPI_ISL_1102067, EPI_ISL_1102070, EPI_ISL_1102071, EPI_ISL_1102074, EPI_ISL_1102075, EPI_ISL_1102076, EPI_ISL_1102077, EPI_ISL_1102083, EPI_ISL_1102086, EPI_ISL_1102095, EPI_ISL_1102097, EPI_ISL_1102105, EPI_ISL_1102110, EPI_ISL_1102113, EPI_ISL_1102115, EPI_ISL_1102119, EPI_ISL_1102121, EPI_ISL_1102127, EPI_ISL_1102128, EPI_ISL_1102137, EPI_ISL_1102140, EPI_ISL_1102144, EPI_ISL_1102146, EPI_ISL_1102147, EPI_ISL_1102150, EPI_ISL_1102151, EPI_ISL_1102155, EPI_ISL_1102157, EPI_ISL_1102158, EPI_ISL_1102161, EPI_ISL_1102162, EPI_ISL_1102166, EPI_ISL_1102173, EPI_ISL_1102178, EPI_ISL_1102179, EPI_ISL_1102180, EPI_ISL_1102181, EPI_ISL_1102187, EPI_ISL_1102190, EPI_ISL_1102192, EPI_ISL_1102203, EPI_ISL_1102204, EPI_ISL_1102207, EPI_ISL_1102209, EPI_ISL_1102214, EPI_ISL_1102217, EPI_ISL_1102218, EPI_ISL_1102220, EPI_ISL_1102227, EPI_ISL_1102229, EPI_ISL_1102233, EPI_ISL_1102236, EPI_ISL_1102238, EPI_ISL_1102239, EPI_ISL_1102242, EPI_ISL_1102247, EPI_ISL_1102251, EPI_ISL_1102256, EPI_ISL_1102258, EPI_ISL_1102261, EPI_ISL_1102262, EPI_ISL_1102271, EPI_ISL_1102274, EPI_ISL_1102275, EPI_ISL_1102277, EPI_ISL_1102281, EPI_ISL_1102282, EPI_ISL_1102283, EPI_ISL_1102284, EPI_ISL_1102287, EPI_ISL_1102288, EPI_ISL_1102291, EPI_ISL_1102292, EPI_ISL_1102293, EPI_ISL_1102295, EPI_ISL_1102301, EPI_ISL_1102306, EPI_ISL_1102308, EPI_ISL_1102311, EPI_ISL_1102317, EPI_ISL_1102321, EPI_ISL_1102327, EPI_ISL_1102332, EPI_ISL_1102334, EPI_ISL_1102340, EPI_ISL_1102357, EPI_ISL_1102359, EPI_ISL_1102378, EPI_ISL_1102379, EPI_ISL_1102383, EPI_ISL_1102385, EPI_ISL_1102390, EPI_ISL_1102391, EPI_ISL_1102398, EPI_ISL_1102406, EPI_ISL_1102407, EPI_ISL_1102411, EPI_ISL_1102421, EPI_ISL_1102424, EPI_ISL_1102427, EPI_ISL_1102433, EPI_ISL_1102434, EPI_ISL_1102435, EPI_ISL_1102439, EPI_ISL_1102449, EPI_ISL_1102452, EPI_ISL_1102455, EPI_ISL_1102457, EPI_ISL_1102458, EPI_ISL_1102459, EPI_ISL_1102475, EPI_ISL_1102480, EPI_ISL_1102483, EPI_ISL_1102495, EPI_ISL_1102496, EPI_ISL_1102514, EPI_ISL_1102515, EPI_ISL_1102517, EPI_ISL_1102519, EPI_ISL_1102523, EPI_ISL_1102524, EPI_ISL_1102528, EPI_ISL_1102529, EPI_ISL_1102538, EPI_ISL_1102542, EPI_ISL_1102549, EPI_ISL_1102552, EPI_ISL_1102563, EPI_ISL_1102571, EPI_ISL_1102574, EPI_ISL_1102575, EPI_ISL_1102583, EPI_ISL_1102591, EPI_ISL_1102593, EPI_ISL_1102609, EPI_ISL_1102612, EPI_ISL_1102613, EPI_ISL_1102624, EPI_ISL_1102715, EPI_ISL_1102716, EPI_ISL_1102717, EPI_ISL_1102720, EPI_ISL_1102724, EPI_ISL_1102725, EPI_ISL_1102726, EPI_ISL_1102728, EPI_ISL_1102729, EPI_ISL_1102732, EPI_ISL_1102736, EPI_ISL_1102737, EPI_ISL_1102741, EPI_ISL_1102744, EPI_ISL_1102748, EPI_ISL_1102752, EPI_ISL_1102762, EPI_ISL_1102764, EPI_ISL_1102768, EPI_ISL_1102772, EPI_ISL_1102775, EPI_ISL_1102776, EPI_ISL_1102777, EPI_ISL_1102780, EPI_ISL_1102781, EPI_ISL_1102782, EPI_ISL_1102783, EPI_ISL_1102786, EPI_ISL_1102787, EPI_ISL_1102793, EPI_ISL_1102795, EPI_ISL_1102797, EPI_ISL_1102798, EPI_ISL_1102801, EPI_ISL_1102811, EPI_ISL_1102813, EPI_ISL_1102815, EPI_ISL_1102818, EPI_ISL_1102821, EPI_ISL_1102822, EPI_ISL_1102823, EPI_ISL_1102827, EPI_ISL_1102830, EPI_ISL_1102832, EPI_ISL_1102837, EPI_ISL_1102840, EPI_ISL_1102844, EPI_ISL_1102850, EPI_ISL_1102851, EPI_ISL_1102852, EPI_ISL_1102853, EPI_ISL_1102855, EPI_ISL_1102856, EPI_ISL_1102859, EPI_ISL_1102860, EPI_ISL_1102861, EPI_ISL_1102862, EPI_ISL_1102868, EPI_ISL_1102870, EPI_ISL_1102872, EPI_ISL_1102874, EPI_ISL_1102875, EPI_ISL_1102880, EPI_ISL_1102881, EPI_ISL_1102885, EPI_ISL_1102888, EPI_ISL_1102889, EPI_ISL_1102890, EPI_ISL_1102891, EPI_ISL_1102892, EPI_ISL_1102893, EPI_ISL_1102895, EPI_ISL_1102896, EPI_ISL_1102898, EPI_ISL_1102900, EPI_ISL_1102902, EPI_ISL_1102903, EPI_ISL_1102904, EPI_ISL_1102909, EPI_ISL_1102910, EPI_ISL_1102911, EPI_ISL_1102912, EPI_ISL_1102914, EPI_ISL_1102915, EPI_ISL_1102917, EPI_ISL_1102918, EPI_ISL_1102920, EPI_ISL_1102925, EPI_ISL_1102926, EPI_ISL_1102932, EPI_ISL_1102937, EPI_ISL_1102939, EPI_ISL_1102942, EPI_ISL_1102946, EPI_ISL_1102957, EPI_ISL_1102958, EPI_ISL_1102965, EPI_ISL_1102966, EPI_ISL_1102969, EPI_ISL_1102970, EPI_ISL_1102971, EPI_ISL_1102973, EPI_ISL_1102974, EPI_ISL_1102975, EPI_ISL_1102976, EPI_ISL_1102977, EPI_ISL_1102978, EPI_ISL_1102986, EPI_ISL_1102987, EPI_ISL_1102991, EPI_ISL_1102995, EPI_ISL_1103001, EPI_ISL_1103004, EPI_ISL_1103005, EPI_ISL_1103006, EPI_ISL_1103022, EPI_ISL_1103024, EPI_ISL_1103026, EPI_ISL_1103029, EPI_ISL_1103031, EPI_ISL_1103032, EPI_ISL_1103033, EPI_ISL_1103034, EPI_ISL_1103036, EPI_ISL_1103038, EPI_ISL_1103040, EPI_ISL_1103042, EPI_ISL_1103043, EPI_ISL_1103047, EPI_ISL_1103049, EPI_ISL_1115673, EPI_ISL_1115674, EPI_ISL_1115675, EPI_ISL_1115676, EPI_ISL_1115677, EPI_ISL_1115678, EPI_ISL_1115679, EPI_ISL_1115680, EPI_ISL_1115681, EPI_ISL_1115682, EPI_ISL_1115683, EPI_ISL_1115684, EPI_ISL_1115718, EPI_ISL_1115723, EPI_ISL_1115724, EPI_ISL_1115725, EPI_ISL_1115726, EPI_ISL_1115733, EPI_ISL_1115743, EPI_ISL_1115744, EPI_ISL_1115748, EPI_ISL_1115751, EPI_ISL_1115754, EPI_ISL_1115759, EPI_ISL_1115762, EPI_ISL_1115765, EPI_ISL_1115771, EPI_ISL_1115772, EPI_ISL_1115773, EPI_ISL_1115778, EPI_ISL_1115781, EPI_ISL_1115784, EPI_ISL_1115794, EPI_ISL_1115825, EPI_ISL_1115826, EPI_ISL_1115827, EPI_ISL_1115828, EPI_ISL_1115829, EPI_ISL_1115830, EPI_ISL_1115831, EPI_ISL_1115832, EPI_ISL_1115833, EPI_ISL_1115834, EPI_ISL_1115835, EPI_ISL_1115836, EPI_ISL_1115837, EPI_ISL_1115838, EPI_ISL_1115839, EPI_ISL_1115840, EPI_ISL_1115841, EPI_ISL_1115842, EPI_ISL_1115843, EPI_ISL_1115844, EPI_ISL_1115845, EPI_ISL_1115846, EPI_ISL_1115847, EPI_ISL_1115848, EPI_ISL_1115849, EPI_ISL_1115850, EPI_ISL_1115851, EPI_ISL_1115852, EPI_ISL_1115853, EPI_ISL_1115854, EPI_ISL_1115855, EPI_ISL_1115856, EPI_ISL_1115857, EPI_ISL_1115858, EPI_ISL_1115859, EPI_ISL_1115860, EPI_ISL_1115861, EPI_ISL_1115862, EPI_ISL_1115863, EPI_ISL_1115864, EPI_ISL_1115865, EPI_ISL_1115866, EPI_ISL_1115867, EPI_ISL_1115868, EPI_ISL_1115869, EPI_ISL_1115870, EPI_ISL_1115871, EPI_ISL_1115872, EPI_ISL_1115873, EPI_ISL_1115874, EPI_ISL_1115875, EPI_ISL_1115876, EPI_ISL_1115877, EPI_ISL_1115878, EPI_ISL_1115879, EPI_ISL_1115880, EPI_ISL_1115881, EPI_ISL_1115882, EPI_ISL_1115883, EPI_ISL_1115884, EPI_ISL_1115885, EPI_ISL_1115886, EPI_ISL_1115887, EPI_ISL_1115888, EPI_ISL_1115889, EPI_ISL_1115890, EPI_ISL_1115891, EPI_ISL_1116143, EPI_ISL_1116145, EPI_ISL_1116146, EPI_ISL_1116150, EPI_ISL_1116151, EPI_ISL_1116153, EPI_ISL_1116159, EPI_ISL_1116160, EPI_ISL_1116161, EPI_ISL_1116166, EPI_ISL_1116169, EPI_ISL_1116171, EPI_ISL_1116174, EPI_ISL_1116182, EPI_ISL_1116186, EPI_ISL_1116189, EPI_ISL_1116193, EPI_ISL_1116198, EPI_ISL_1116209, EPI_ISL_1116211, EPI_ISL_1116213, EPI_ISL_1116214, EPI_ISL_1116221, EPI_ISL_1116222, EPI_ISL_1116223, EPI_ISL_1116225, EPI_ISL_1116226, EPI_ISL_1116228, EPI_ISL_1116229, EPI_ISL_1116230, EPI_ISL_1116231, EPI_ISL_1116232, EPI_ISL_1116233, EPI_ISL_1116234, EPI_ISL_1116235, EPI_ISL_1116236, EPI_ISL_1116237, EPI_ISL_1116238, EPI_ISL_1116239, EPI_ISL_1116240, EPI_ISL_1116242, EPI_ISL_1116243, EPI_ISL_1116245, EPI_ISL_1116246, EPI_ISL_1116247, EPI_ISL_1116248, EPI_ISL_1116249, EPI_ISL_1116250, EPI_ISL_1116251, EPI_ISL_1116252, EPI_ISL_1116253, EPI_ISL_1116254, EPI_ISL_1116255, EPI_ISL_1116256, EPI_ISL_1116257, EPI_ISL_1116258, EPI_ISL_1116259, EPI_ISL_1116260, EPI_ISL_1116261, EPI_ISL_1116262, EPI_ISL_1116263, EPI_ISL_1116345, EPI_ISL_1116346, EPI_ISL_1116347, EPI_ISL_1116348, EPI_ISL_1116349, EPI_ISL_1116350, EPI_ISL_1116351, EPI_ISL_1116352, EPI_ISL_1116353, EPI_ISL_1116354, EPI_ISL_1116355, EPI_ISL_1116356, EPI_ISL_1116358, EPI_ISL_1116359, EPI_ISL_1116360, EPI_ISL_1116361, EPI_ISL_1116362, EPI_ISL_1116363, EPI_ISL_1116364, EPI_ISL_1116365, EPI_ISL_1116366, EPI_ISL_1116367, EPI_ISL_1116368, EPI_ISL_1116369, EPI_ISL_1116370, EPI_ISL_1116371, EPI_ISL_1116372, EPI_ISL_1116373, EPI_ISL_1116374, EPI_ISL_1116375, EPI_ISL_1116376, EPI_ISL_1116377, EPI_ISL_1116378, EPI_ISL_1116379, EPI_ISL_1116380, EPI_ISL_1116381, EPI_ISL_1116382, EPI_ISL_1116383, EPI_ISL_1116384, EPI_ISL_1116386, EPI_ISL_1116387, EPI_ISL_1116388, EPI_ISL_1116389, EPI_ISL_1116390, EPI_ISL_1116391, EPI_ISL_1116392, EPI_ISL_1116393, EPI_ISL_1116394, EPI_ISL_1116395, EPI_ISL_1116396, EPI_ISL_1116397, EPI_ISL_1116398, EPI_ISL_1116399, EPI_ISL_1116400, EPI_ISL_1116401, EPI_ISL_1116403, EPI_ISL_1116404, EPI_ISL_1116405, EPI_ISL_1116406, EPI_ISL_1116407, EPI_ISL_1116408, EPI_ISL_1116409, EPI_ISL_1116411, EPI_ISL_1116412, EPI_ISL_1116413, EPI_ISL_1116414, EPI_ISL_1116415, EPI_ISL_1116416, EPI_ISL_1116417, EPI_ISL_1116418, EPI_ISL_1116419, EPI_ISL_1116420, EPI_ISL_1116421, EPI_ISL_1116422, EPI_ISL_1116423, EPI_ISL_1116424, EPI_ISL_1116425, EPI_ISL_1116426, EPI_ISL_1116427, EPI_ISL_1116428, EPI_ISL_1174101, EPI_ISL_1174220, EPI_ISL_1189843, EPI_ISL_1189864, EPI_ISL_1189884, EPI_ISL_1189891, EPI_ISL_1189962, EPI_ISL_1189967, EPI_ISL_1189970, EPI_ISL_1189971, EPI_ISL_1189985, EPI_ISL_1189995, EPI_ISL_1190003, EPI_ISL_1190004, EPI_ISL_1190011, EPI_ISL_1190046, EPI_ISL_1190066, EPI_ISL_1242343, EPI_ISL_1243460, EPI_ISL_1243540, EPI_ISL_1243552, EPI_ISL_1243568, EPI_ISL_1243596, EPI_ISL_1257064, EPI_ISL_1257074, EPI_ISL_1257075, EPI_ISL_1257077, EPI_ISL_1257083, EPI_ISL_1257090, EPI_ISL_1257094, EPI_ISL_1257095, EPI_ISL_1257099, EPI_ISL_1257100, EPI_ISL_1257101, EPI_ISL_1257111, EPI_ISL_1257122, EPI_ISL_1257124, EPI_ISL_1257126, EPI_ISL_1257128, EPI_ISL_1257137, EPI_ISL_1257140, EPI_ISL_1257141, EPI_ISL_1257142, EPI_ISL_1257143, EPI_ISL_1257149, EPI_ISL_1257155, EPI_ISL_1257157, EPI_ISL_1257170, EPI_ISL_1257177, EPI_ISL_1257179, EPI_ISL_1257187, EPI_ISL_1257195, EPI_ISL_1257201, EPI_ISL_1257210, EPI_ISL_1257215, EPI_ISL_1257217, EPI_ISL_1257234, EPI_ISL_1257238, EPI_ISL_1257239, EPI_ISL_1257244, EPI_ISL_1257250, EPI_ISL_1257270, EPI_ISL_1257271, EPI_ISL_1257277, EPI_ISL_1264906, EPI_ISL_1264907, EPI_ISL_1264908, EPI_ISL_1264909, EPI_ISL_1264910, EPI_ISL_1264911, EPI_ISL_1264912, EPI_ISL_1264913, EPI_ISL_1264914, EPI_ISL_1264915, EPI_ISL_1264916, EPI_ISL_1264917, EPI_ISL_1264918, EPI_ISL_1264919, EPI_ISL_1264920, EPI_ISL_1264921, EPI_ISL_1264922, EPI_ISL_1264923, EPI_ISL_1264924, EPI_ISL_1264925, EPI_ISL_1264926, EPI_ISL_1264927, EPI_ISL_1264928, EPI_ISL_1264929, EPI_ISL_1264930, EPI_ISL_1264931, EPI_ISL_1264932, EPI_ISL_1264933, EPI_ISL_1264934, EPI_ISL_1264935, EPI_ISL_1264936, EPI_ISL_1264937, EPI_ISL_1264938, EPI_ISL_1264939, EPI_ISL_1264940, EPI_ISL_1264941, EPI_ISL_1264942, EPI_ISL_1264943, EPI_ISL_1264944, EPI_ISL_1264945, EPI_ISL_1264946, EPI_ISL_1264947, EPI_ISL_1264948, EPI_ISL_1264949, EPI_ISL_1264950, EPI_ISL_1264951, EPI_ISL_1264952, EPI_ISL_1264953, EPI_ISL_1264954, EPI_ISL_1265034 | | | |
| see above | Randox Laboratories | Wellcome Sanger Institute for the COVID-19 Genomics UK (COG-UK) Consortium | Cordelia Langford; David K. Jackson; Dominic Kwiatkowski; Ewan Harrison; Ian Johnston; Jeffrey Barrett; John Sillitoe on behalf of the Wellcome Sanger Institute COVID-19 Surveillance Team; Randox Laboratories and Alex Alderton; Roberto Amato; Sonia Goncalves |
| EPI_ISL_1296618, EPI_ISL_1296622, EPI_ISL_1296623, EPI_ISL_1296624 | Respiratory Virus Unit, National Infection Service, Public Health England | COVID-19 Genomics UK (COG-UK) Consortium | PHE Covid Sequencing Team |
| EPI_ISL_1833783, EPI_ISL_1833784, EPI_ISL_1833787, EPI_ISL_1833795, EPI_ISL_1833796, EPI_ISL_1833797, EPI_ISL_1833798 | | | |
| see above | St George's University Hospitals NHS Foundation Trust | COVID-19 Genomics UK (COG-UK) Consortium | Adam Witney; Cassie Pope; Irene Monahan; Joshua Taylor; Ken Laing; NgeeKeong Tan |
| EPI_ISL_2125541, EPI_ISL_2125545, EPI_ISL_2125547, EPI_ISL_2125548 | The Roslin Institute and R(D)SVS, University of Edinburgh / Virology Department, Royal Infirmary of Edinburgh, NHS Lothian / School of Biological Sciences, University of Edinburgh | COVID-19 Genomics UK (COG-UK) Consortium | C; Colquhoun R; Cotton S; Dewar R; Gallagher A; Hill V; Jackson B; Maloney D; McCrone JT; McHugh M; Newman; O'Toole Á; Rambaut A; Rooke S; Scher E; Tait-Burkhard C; Templeton K; Warr A; Yu X |
| EPI_ISL_1105211, EPI_ISL_1105212, EPI_ISL_1105213, EPI_ISL_1105214, EPI_ISL_1105215, EPI_ISL_1105221, EPI_ISL_1105226, EPI_ISL_1105231, EPI_ISL_1105232, EPI_ISL_1105234, EPI_ISL_1105235, EPI_ISL_1105240, EPI_ISL_1105241, EPI_ISL_1105244, EPI_ISL_1105245, EPI_ISL_1105246, EPI_ISL_1105251, EPI_ISL_1105252, EPI_ISL_1105256, EPI_ISL_1105262, EPI_ISL_1105263, EPI_ISL_1105269, EPI_ISL_1105270, EPI_ISL_1105271, EPI_ISL_1105277, EPI_ISL_1105278, EPI_ISL_1105279, EPI_ISL_1105285, EPI_ISL_1105286, EPI_ISL_1105290, EPI_ISL_1105291, EPI_ISL_1105295, EPI_ISL_1105298, EPI_ISL_1105299, EPI_ISL_1105300, EPI_ISL_1105308, EPI_ISL_1105309, EPI_ISL_1105314, EPI_ISL_1105316, EPI_ISL_1105330, EPI_ISL_1105339, EPI_ISL_1105340, EPI_ISL_1105343 | | | |
| see above | University College London Hospital | COVID-19 Genomics UK (COG-UK) Consortium | Catherine Houlihan; Dan Frampton; Judith Heaney; Matthew Byott; Moira Spyer and Eleni Nastouli; Stuart Kirk |
| EPI_ISL_997751, EPI_ISL_997752, EPI_ISL_997756, EPI_ISL_997757, EPI_ISL_997758, EPI_ISL_997759, EPI_ISL_997760, EPI_ISL_997761, EPI_ISL_997762, EPI_ISL_997763, EPI_ISL_997764, EPI_ISL_997765, EPI_ISL_997766, EPI_ISL_997767, EPI_ISL_997768, EPI_ISL_997769, EPI_ISL_997770, EPI_ISL_997771, EPI_ISL_997780, EPI_ISL_997781, EPI_ISL_997782, EPI_ISL_997798, EPI_ISL_997799, EPI_ISL_997800, EPI_ISL_997801, EPI_ISL_997802, EPI_ISL_997803, EPI_ISL_997804, EPI_ISL_997805, EPI_ISL_997856, EPI_ISL_997857, EPI_ISL_997859, EPI_ISL_997877, EPI_ISL_1048216, EPI_ISL_1048217, EPI_ISL_1048218, EPI_ISL_1048219, EPI_ISL_1048220, EPI_ISL_1048221, EPI_ISL_1048222, EPI_ISL_1048223, EPI_ISL_1048224, EPI_ISL_1048225, EPI_ISL_1048226, EPI_ISL_1048227, EPI_ISL_1048228, EPI_ISL_1048229, EPI_ISL_1048230, EPI_ISL_1048231, EPI_ISL_1048232, EPI_ISL_1050417, EPI_ISL_1050418, EPI_ISL_1050419, EPI_ISL_1050420, EPI_ISL_1050421, EPI_ISL_1050422, EPI_ISL_1050423, EPI_ISL_1050430, EPI_ISL_1050526, EPI_ISL_1050527, EPI_ISL_1050528, EPI_ISL_1050557, EPI_ISL_1050558, EPI_ISL_1050559, EPI_ISL_1050560, EPI_ISL_1104682, EPI_ISL_1104683, EPI_ISL_1104684, EPI_ISL_1104685, EPI_ISL_1104686, EPI_ISL_1104687, EPI_ISL_1104688, EPI_ISL_1104689, EPI_ISL_1104691, EPI_ISL_1104697, EPI_ISL_1104714, EPI_ISL_1104721, EPI_ISL_1104736, EPI_ISL_1104740, EPI_ISL_1104749, EPI_ISL_1104750, EPI_ISL_1104751, EPI_ISL_1104753, EPI_ISL_1104754, EPI_ISL_1104756, EPI_ISL_1104759, EPI_ISL_1104760, EPI_ISL_1104761, EPI_ISL_1104762, EPI_ISL_1105372, EPI_ISL_1105379, EPI_ISL_1105380, EPI_ISL_1105381, EPI_ISL_1105382, EPI_ISL_1105383, EPI_ISL_1105385, EPI_ISL_1105386, EPI_ISL_1105387, EPI_ISL_1105388, EPI_ISL_1105389, EPI_ISL_1105390, EPI_ISL_1105391, EPI_ISL_1105392, EPI_ISL_1105393, EPI_ISL_1105394, EPI_ISL_1105395, EPI_ISL_1105396, EPI_ISL_1105397, EPI_ISL_1105398, EPI_ISL_1105399, EPI_ISL_1105400, EPI_ISL_1105401, EPI_ISL_1105402, EPI_ISL_1105403, EPI_ISL_1105404, EPI_ISL_1105406, EPI_ISL_1105407, EPI_ISL_1105408, EPI_ISL_1105409, EPI_ISL_1105410, EPI_ISL_1105411, EPI_ISL_1105412, EPI_ISL_1105413, EPI_ISL_1105414, EPI_ISL_1105415, EPI_ISL_1105416, EPI_ISL_1105417, EPI_ISL_1105446, EPI_ISL_1105453, EPI_ISL_1105454, EPI_ISL_1105455, EPI_ISL_1105456, EPI_ISL_1105457, EPI_ISL_1105458, EPI_ISL_1105459, EPI_ISL_1105460, EPI_ISL_1105461, EPI_ISL_1105472, EPI_ISL_1247955, EPI_ISL_1247989, EPI_ISL_1248332, EPI_ISL_1250224, EPI_ISL_1250225, EPI_ISL_1250226, EPI_ISL_1250227, EPI_ISL_1250228, EPI_ISL_1250229, EPI_ISL_1250239, EPI_ISL_1250240, EPI_ISL_1309059, EPI_ISL_1309089, EPI_ISL_1386918, EPI_ISL_1386919, EPI_ISL_1386920, EPI_ISL_1386921, EPI_ISL_1386922, | | | |

| | | | |
|---|---|---|---|
| EPI_ISL_1386923, EPI_ISL_1386924, EPI_ISL_1386925, EPI_ISL_1637171, EPI_ISL_1975111 | | | |
| see above | University College London, Great Ormond Street Hospital for Children NHS Foundation Trust, Imperial College Healthcare NHS Trust | COVID-19 Genomics UK (COG-UK) Consortium | Alison Holmes; Charlotte Williams; Helena Tutill; Jacqueline Findlay; James Price; Judith Breuer; Julianne Brown; Kathryn Harris; Leysa Forrest; Mark Kristiansen; Paola Niola; Paola Resende Silva; Patricia Dyal; Paul Randell; Rachel Williams; Samuel Weeks; Sergi Castellano; Sunando Roy; Tony Brooks; Yasmin Panchbhaya |
| EPI_ISL_1104298, EPI_ISL_1104299, EPI_ISL_1104301, EPI_ISL_1104302, EPI_ISL_1104303, EPI_ISL_1104304, EPI_ISL_1104307 | | | |
| see above | University Hospitals Of Leicester NHS Trust and DeepSeq Nottingham | COVID-19 Genomics UK (COG-UK) Consortium | Christopher Holmes; Christopher Moore; Fei Sang; Joeseph Chappell; Johnny Debebe; Jonathan Ball; Julian Tang; Karlie Fallon; Matthew Carlisle; Matthew Loose; Nadine Holmes; Patrick McClure; Paul Bird; Thomas Helmer; Victoria Wright |
| EPI_ISL_1046953, EPI_ISL_1046954, EPI_ISL_1046955, EPI_ISL_1047024, EPI_ISL_1103744, EPI_ISL_1103745, EPI_ISL_1103751, EPI_ISL_1308508, EPI_ISL_1474472 | | | |
| see above | University of Birmingham | COVID-19 Genomics UK (COG-UK) Consortium | Alex Richter; Andrew Bosworth; Andrew Bosworth. Queen Elizabeth Hospital: Anna Casey; Andrew D Beggs PHE Heartlands Lab: Husam Osman; Charlotte Poxon; Institute of Microbiology; Joanne Stockton; Josh Quick; Kasun Wanigasooriya; Mike Kidd; Nicholas Loman. University of Birmingham Testing Laboratory: Celina M Whalley; Oliver Pickles; Radoslaw Poplawski; Samuel Nicholls; University of Birmingham: Claire McMurray; Will Rowe |
| EPI_ISL_1103808, EPI_ISL_1103809, EPI_ISL_1103810, EPI_ISL_1103811, EPI_ISL_1103812, EPI_ISL_1103813, EPI_ISL_1103814, EPI_ISL_1103815, EPI_ISL_1103816, EPI_ISL_1103817, EPI_ISL_1103818, EPI_ISL_1103819, EPI_ISL_1103820, EPI_ISL_1103821, EPI_ISL_1103822, EPI_ISL_1103823, EPI_ISL_1103824, EPI_ISL_1103825, EPI_ISL_1103826, EPI_ISL_1103827, EPI_ISL_1103828, EPI_ISL_1103829, EPI_ISL_1103831, EPI_ISL_1103833, EPI_ISL_1103834, EPI_ISL_1103835, EPI_ISL_1103836, EPI_ISL_1103837, EPI_ISL_1103838, EPI_ISL_1103839, EPI_ISL_1103841, EPI_ISL_1103842, EPI_ISL_1103844, EPI_ISL_1103846, EPI_ISL_1103848, EPI_ISL_1103849, EPI_ISL_1103850, EPI_ISL_1103851, EPI_ISL_1103852, EPI_ISL_1103853, EPI_ISL_1103854, EPI_ISL_1103855, EPI_ISL_1103856, EPI_ISL_1103857, EPI_ISL_1103858, EPI_ISL_1103860, EPI_ISL_1103862, EPI_ISL_1103863, EPI_ISL_1103867, EPI_ISL_1103868, EPI_ISL_1103869, EPI_ISL_1103870, EPI_ISL_1103872, EPI_ISL_1103874, EPI_ISL_1103876, EPI_ISL_1177751, EPI_ISL_1177752, EPI_ISL_1177753, EPI_ISL_1177754, EPI_ISL_1177755, EPI_ISL_1177756, EPI_ISL_1177757, EPI_ISL_1177761, EPI_ISL_1177762, EPI_ISL_1177763, EPI_ISL_1247443, EPI_ISL_1247444, EPI_ISL_1247447, EPI_ISL_1247448, EPI_ISL_1247449, EPI_ISL_1247450, EPI_ISL_1247451, EPI_ISL_1247453, EPI_ISL_1247454, EPI_ISL_1247455, EPI_ISL_1247456, EPI_ISL_1247457, EPI_ISL_1247458, EPI_ISL_1247459, EPI_ISL_1247460, EPI_ISL_1247486, EPI_ISL_1247487, EPI_ISL_1247488, EPI_ISL_1247491, EPI_ISL_1308600, EPI_ISL_1308601 | | | |
| see above | University of Exeter | COVID-19 Genomics UK (COG-UK) Consortium | Aaron Jeffries; Audrey Farbos; Ben Temperton; Jane Masoli; Joanna Warwick-Dugdale; Michelle Michelsen; Robyn Manley; Stephen Michell |
| EPI_ISL_1047903, EPI_ISL_1047904, EPI_ISL_1047905, EPI_ISL_1047906, EPI_ISL_1047907, EPI_ISL_1047908, EPI_ISL_1047909, EPI_ISL_1047910, EPI_ISL_1047911, EPI_ISL_1047912, EPI_ISL_1047913, EPI_ISL_1050244, EPI_ISL_1104264, EPI_ISL_1104265, EPI_ISL_1104266, EPI_ISL_1104267, EPI_ISL_1104270, EPI_ISL_1104271, EPI_ISL_1177716, EPI_ISL_1177719, EPI_ISL_1177725, EPI_ISL_1177745, EPI_ISL_1247699, EPI_ISL_1247726, EPI_ISL_1247734, EPI_ISL_1247735, EPI_ISL_1247736, EPI_ISL_1247737, EPI_ISL_1247747, EPI_ISL_1308760, EPI_ISL_1308761, EPI_ISL_1308762, EPI_ISL_1308763, EPI_ISL_1308764, EPI_ISL_1308765, EPI_ISL_1308766, EPI_ISL_1308767, EPI_ISL_1308768, EPI_ISL_1308825, EPI_ISL_1308827, EPI_ISL_1308828, EPI_ISL_1308836, EPI_ISL_1308852, EPI_ISL_1386650, EPI_ISL_1386684, EPI_ISL_1386689, EPI_ISL_1386690, EPI_ISL_1730073, EPI_ISL_1730177, EPI_ISL_1730178, EPI_ISL_1730179, EPI_ISL_1730180, EPI_ISL_1730181, EPI_ISL_1730183, EPI_ISL_1730184, EPI_ISL_1730185, EPI_ISL_1730188, EPI_ISL_1730189, EPI_ISL_1730190, EPI_ISL_1730191, EPI_ISL_1730192, EPI_ISL_1730193, EPI_ISL_1730201, EPI_ISL_1730203, EPI_ISL_1730204, EPI_ISL_1730205, EPI_ISL_1730206, EPI_ISL_1730215, EPI_ISL_1730222, EPI_ISL_1730226, EPI_ISL_1730227, EPI_ISL_1730228, EPI_ISL_1730229, EPI_ISL_1730230, EPI_ISL_2125731, EPI_ISL_2125867 | | | |
| see above | Virology Department, Royal Infirmary of Edinburgh, NHS Lothian / School of Biological Sciences, University of Edinburgh | COVID-19 Genomics UK (COG-UK) Consortium | Colquhoun R; Cotton S; Dewar R; Gallagher A; Hill V; Jackson B; Maloney D; McCrone JT; McHugh M; O'Toole Á; Rambaut A; Rooke S; Scher E; Templeton K; Yu X |
| EPI_ISL_1000643, EPI_ISL_1000660, EPI_ISL_1000667, EPI_ISL_1000681, EPI_ISL_1000688, EPI_ISL_1000707, EPI_ISL_1000712, EPI_ISL_1000723, EPI_ISL_1000731, EPI_ISL_1000733, EPI_ISL_1000745, EPI_ISL_1000754, EPI_ISL_1000756, EPI_ISL_1000769, EPI_ISL_1000775, EPI_ISL_1000776, EPI_ISL_1000782, EPI_ISL_1000785, EPI_ISL_1000792, EPI_ISL_1054626, EPI_ISL_1054635, EPI_ISL_1054645, EPI_ISL_1054646, EPI_ISL_1054648, EPI_ISL_1054651, EPI_ISL_1054652, EPI_ISL_1054653, EPI_ISL_1054657, EPI_ISL_1054661, EPI_ISL_1054662, EPI_ISL_1054663, EPI_ISL_1054671, EPI_ISL_1054672, EPI_ISL_1054683, EPI_ISL_1054684, EPI_ISL_1054689, EPI_ISL_1054690, EPI_ISL_1054691, EPI_ISL_1054694, EPI_ISL_1054696, EPI_ISL_1054698, EPI_ISL_1054702, EPI_ISL_1054705, EPI_ISL_1054707, EPI_ISL_1054708, EPI_ISL_1054709, EPI_ISL_1105580, EPI_ISL_1105587, EPI_ISL_1105598, EPI_ISL_1105624, EPI_ISL_1108754, EPI_ISL_1108755, EPI_ISL_1108756, EPI_ISL_1108757, EPI_ISL_1108758, EPI_ISL_1108759, EPI_ISL_1108760, EPI_ISL_1108761, EPI_ISL_1108762, EPI_ISL_1108763, EPI_ISL_1108764, EPI_ISL_1108765, EPI_ISL_1108766, EPI_ISL_1108767, EPI_ISL_1108768, EPI_ISL_1108769, EPI_ISL_1108771, EPI_ISL_1108772, EPI_ISL_1108773, EPI_ISL_1108774, EPI_ISL_1108775, EPI_ISL_1108776, EPI_ISL_1108777, EPI_ISL_1108778, EPI_ISL_1108779, EPI_ISL_1108780, EPI_ISL_1108781, EPI_ISL_1108782, EPI_ISL_1108783, EPI_ISL_1108784, EPI_ISL_1108785, EPI_ISL_1108786, EPI_ISL_1108787, EPI_ISL_1108788, EPI_ISL_1108789, EPI_ISL_1108791, EPI_ISL_1108792, EPI_ISL_1108793, EPI_ISL_1108795, EPI_ISL_1108796, EPI_ISL_1108797, EPI_ISL_1108798, EPI_ISL_1108799, EPI_ISL_1108800, EPI_ISL_1108802, EPI_ISL_1108803, EPI_ISL_1108804, EPI_ISL_1108805, EPI_ISL_1108806, EPI_ISL_1108807, EPI_ISL_1108808, EPI_ISL_1108809, EPI_ISL_1108810, EPI_ISL_1108811, EPI_ISL_1108812, EPI_ISL_1108813, EPI_ISL_1108814, EPI_ISL_1108816, EPI_ISL_1108817, EPI_ISL_1108818, EPI_ISL_1108819, EPI_ISL_1108820, EPI_ISL_1108821, EPI_ISL_1108822, EPI_ISL_1108823, EPI_ISL_1108824, EPI_ISL_1108825, EPI_ISL_1108826, EPI_ISL_1108827, EPI_ISL_1108828, EPI_ISL_1108829, EPI_ISL_1108830, EPI_ISL_1108831, EPI_ISL_1108832, EPI_ISL_1108833, EPI_ISL_1176815, EPI_ISL_1176827 | | | |
| see above | Virology Department, Sheffield Teaching Hospitals NHS Foundation Trust/Department of Infection, Immunity and Cardiovascular Disease, The Medical School, University of Sheffield | COVID-19 Genomics UK (COG-UK) Consortium | Adri Angyal; Alex Keeley; Benjamin Lindsey; Cariad Evans; Danielle Groves; Dave Partridge; Katie Johnson; Laura Carrilero; Luke Green; Matthew Parker; Matthew Wyles; Mehmet Yavuz; Mohammad Raza; Nikki Smith; Paul Parsons; Rachel Tucker; Rebecca Brown; Thushan de Silva |
| EPI_ISL_1052971, EPI_ISL_1052973, EPI_ISL_1052974, EPI_ISL_1053082, EPI_ISL_1053157, EPI_ISL_1053158, EPI_ISL_1053159, EPI_ISL_1053161, EPI_ISL_1053165, EPI_ISL_1053167, EPI_ISL_1053168, EPI_ISL_1053170, EPI_ISL_1053171, EPI_ISL_1053176, EPI_ISL_1053177, EPI_ISL_1053182, EPI_ISL_1053186, EPI_ISL_1053190, EPI_ISL_1053194, EPI_ISL_1053201, EPI_ISL_1053202, EPI_ISL_1053207, EPI_ISL_1053229, EPI_ISL_1053230, EPI_ISL_1053241, EPI_ISL_1053244, EPI_ISL_1053245, EPI_ISL_1053246, EPI_ISL_1053276, EPI_ISL_1053289, EPI_ISL_1053297, EPI_ISL_1053317, EPI_ISL_1053318, EPI_ISL_1053319, EPI_ISL_1053323, EPI_ISL_1053324, EPI_ISL_1053325, EPI_ISL_1053326, EPI_ISL_1053333, EPI_ISL_1053336, EPI_ISL_1053342, EPI_ISL_1053344, EPI_ISL_1053350, EPI_ISL_1053352, EPI_ISL_1053357, EPI_ISL_1053360, EPI_ISL_1053370, EPI_ISL_1053380, EPI_ISL_1053394, EPI_ISL_1053401, EPI_ISL_1053404, EPI_ISL_1053409, EPI_ISL_1053410, EPI_ISL_1053412, EPI_ISL_1053453, EPI_ISL_1053456, EPI_ISL_1053458, EPI_ISL_1053470, EPI_ISL_1053473, EPI_ISL_1053474, EPI_ISL_1053481, EPI_ISL_1053489, EPI_ISL_1105932, EPI_ISL_1105933, EPI_ISL_1105934, EPI_ISL_1105935, EPI_ISL_1105936, EPI_ISL_1105938, EPI_ISL_1105939, EPI_ISL_1105940, EPI_ISL_1105941, EPI_ISL_1105942, EPI_ISL_1105943, EPI_ISL_1105944, EPI_ISL_1105945, EPI_ISL_1105947, EPI_ISL_1105948, EPI_ISL_1105949, EPI_ISL_1105950, EPI_ISL_1105951, EPI_ISL_1105952, EPI_ISL_1105953, EPI_ISL_1105954, EPI_ISL_1105955, EPI_ISL_1105956, EPI_ISL_1105958, EPI_ISL_1105959, EPI_ISL_1105960, EPI_ISL_1105961, EPI_ISL_1105962, EPI_ISL_1105963, EPI_ISL_1105965, EPI_ISL_1105968, EPI_ISL_1105969, EPI_ISL_1105970, EPI_ISL_1105971, EPI_ISL_1105972, EPI_ISL_1105974, EPI_ISL_1105975, EPI_ISL_1105976, EPI_ISL_1105977, EPI_ISL_1105978, EPI_ISL_1105979, EPI_ISL_1105980, EPI_ISL_1105981, EPI_ISL_1105982, EPI_ISL_1105983, EPI_ISL_1105984, EPI_ISL_1105985, EPI_ISL_1105986, EPI_ISL_1105987, EPI_ISL_1105988, EPI_ISL_1105989, EPI_ISL_1105990, EPI_ISL_1105991, EPI_ISL_1105992, EPI_ISL_1105993, EPI_ISL_1105995, EPI_ISL_1105996, EPI_ISL_1105997, EPI_ISL_1105998, EPI_ISL_1105999, EPI_ISL_1106000, EPI_ISL_1106001, EPI_ISL_1106002, EPI_ISL_1106003, EPI_ISL_1106004, EPI_ISL_1106005, EPI_ISL_1106006, EPI_ISL_1106007, EPI_ISL_1106008, EPI_ISL_1106009, EPI_ISL_1106010, EPI_ISL_1106011, EPI_ISL_1106012, EPI_ISL_1106013, EPI_ISL_1106077, EPI_ISL_1106078, EPI_ISL_1106079, EPI_ISL_1106080, EPI_ISL_1106081, EPI_ISL_1106082, EPI_ISL_1106083, EPI_ISL_1106084, EPI_ISL_1106085, EPI_ISL_1106086, EPI_ISL_1106087, EPI_ISL_1106088, EPI_ISL_1106089, EPI_ISL_1106090, EPI_ISL_1106091, EPI_ISL_1106092, EPI_ISL_1106093, EPI_ISL_1106094, EPI_ISL_1106095, EPI_ISL_1106096, EPI_ISL_1106097, EPI_ISL_1106098, EPI_ISL_1106099, EPI_ISL_1106100, EPI_ISL_1106101, EPI_ISL_1106102, EPI_ISL_1106103, EPI_ISL_1106104, EPI_ISL_1106105, EPI_ISL_1106106, EPI_ISL_1106107, EPI_ISL_1106108, EPI_ISL_1106109, EPI_ISL_1106110, EPI_ISL_1106111, EPI_ISL_1106112, EPI_ISL_1106113, EPI_ISL_1106114, EPI_ISL_1106115, EPI_ISL_1106116, EPI_ISL_1106117, EPI_ISL_1106118, EPI_ISL_1106119, EPI_ISL_1106120, EPI_ISL_1106121, EPI_ISL_1106122, EPI_ISL_1106123, EPI_ISL_1106124, EPI_ISL_1106125, EPI_ISL_1106126, EPI_ISL_1106128, EPI_ISL_1106129, EPI_ISL_1106130, EPI_ISL_1106131, EPI_ISL_1106132, EPI_ISL_1106133, EPI_ISL_1106134, EPI_ISL_1106137, EPI_ISL_1106138, EPI_ISL_1106139, EPI_ISL_1106140, EPI_ISL_1106141, EPI_ISL_1106142, EPI_ISL_1106143, EPI_ISL_1106144, EPI_ISL_1106145, EPI_ISL_1106146, EPI_ISL_1106147, EPI_ISL_1106148, EPI_ISL_1106149, EPI_ISL_1106150, EPI_ISL_1106151, EPI_ISL_1106152, EPI_ISL_1106153, EPI_ISL_1106154, EPI_ISL_1106155, EPI_ISL_1106156, EPI_ISL_1106157, EPI_ISL_1106158, EPI_ISL_1106159, EPI_ISL_1106160, EPI_ISL_1106161, EPI_ISL_1106163, EPI_ISL_1106164, EPI_ISL_1106166, EPI_ISL_1106167, EPI_ISL_1106168, EPI_ISL_1106169, EPI_ISL_1106170, EPI_ISL_1106171, EPI_ISL_1106173, EPI_ISL_1106174, EPI_ISL_1106175, EPI_ISL_1106176, EPI_ISL_1106177, EPI_ISL_1106178, EPI_ISL_1106180, EPI_ISL_1106181, EPI_ISL_1106182, EPI_ISL_1106183, EPI_ISL_1106184, EPI_ISL_1106189, EPI_ISL_1106190, EPI_ISL_1106191, EPI_ISL_1106192, EPI_ISL_1106193, EPI_ISL_1106194, EPI_ISL_1106195, EPI_ISL_1106196, EPI_ISL_1106199, EPI_ISL_1106200, EPI_ISL_1106201, EPI_ISL_1106202, EPI_ISL_1106207, EPI_ISL_1106208, EPI_ISL_1106209, EPI_ISL_1106210, EPI_ISL_1106211, EPI_ISL_1106238, EPI_ISL_1106283, EPI_ISL_1106284, EPI_ISL_1106285, EPI_ISL_1106302, EPI_ISL_1106305, EPI_ISL_1106307, EPI_ISL_1106308, EPI_ISL_1106312, EPI_ISL_1106314, EPI_ISL_1106315, EPI_ISL_1106316, EPI_ISL_1106317, EPI_ISL_1106322, EPI_ISL_1106323, EPI_ISL_1106325, EPI_ISL_1106329, EPI_ISL_1106334, EPI_ISL_1106335, EPI_ISL_1106336, EPI_ISL_1106337, EPI_ISL_1106338, EPI_ISL_1106340, EPI_ISL_1106438, EPI_ISL_1106607, EPI_ISL_1106707, EPI_ISL_1106710, EPI_ISL_1106711, EPI_ISL_1106712, EPI_ISL_1106713, EPI_ISL_1106714, EPI_ISL_1106921, EPI_ISL_1387312, EPI_ISL_1387319, EPI_ISL_1387321, EPI_ISL_1387326, EPI_ISL_1387330, EPI_ISL_1387331, EPI_ISL_1387341, EPI_ISL_1387350, EPI_ISL_1387362, EPI_ISL_1387365, EPI_ISL_1387366, EPI_ISL_1387371 | | | |
| see above | Wales Specialist Virology Centre Sequencing lab: Pathogen Genomics Unit | Public Health Wales Microbiology Cardiff Wales Specialist Virology Centre | Alec Birchley; Alexander Adams; Amy Gaskin; Angela Marchbank; Bree Gatica-Wilcox; Catherine Moore; Jason Coombes; Joanne Watkins; Joel Southgate; Johnathan Evans; Laura Gifford; Lauren Gilbert; Lee Graham; Malorie Perry; Matthew Bull; Nicole Pacchiarini; Sally Corden; Sara Kumziene-Summerhayes; Sara Rey; Sarah Taylor; Simon Cottrell; Sophie Jones; Tom Connor |
| EPI_ISL_1047851, EPI_ISL_1047852, EPI_ISL_1047853, EPI_ISL_1047854, EPI_ISL_1050216, EPI_ISL_1050218, EPI_ISL_1050242, EPI_ISL_1104103, EPI_ISL_1104148, EPI_ISL_1104150, EPI_ISL_1104151, EPI_ISL_1104152, EPI_ISL_1104153, EPI_ISL_1104154, EPI_ISL_1104155, EPI_ISL_1104156, EPI_ISL_1104157, EPI_ISL_1104158, EPI_ISL_1104159, EPI_ISL_1104160, EPI_ISL_1104161, EPI_ISL_1104162, EPI_ISL_1104163, EPI_ISL_1104164, EPI_ISL_1104165, EPI_ISL_1104166, EPI_ISL_1104167, EPI_ISL_1104168, EPI_ISL_1104169, EPI_ISL_1104170, EPI_ISL_1104171, EPI_ISL_1104172, EPI_ISL_1104173, EPI_ISL_1104174, EPI_ISL_1104175, EPI_ISL_1104176, EPI_ISL_1104177, EPI_ISL_1104178, EPI_ISL_1104186, EPI_ISL_1247664 | | | |
| see above | West of Scotland Specialist Virology Centre, NHSGGC / MRC-University of Glasgow Centre for Virus Research | COVID-19 Genomics UK (COG-UK) Consortium | Alasdair MacLean; Alice Broos; Ana da Silva Filipe; Antonia Ho; Daniel Mair; David L Robertson; Emma Thomson; Guy Mollett; James Shepherd; Jenna Nichols; Joseph Hughes; Kathy Li; Kathy Smollett; Kyriaki Nomikou; Lily Tong; Matthew Holden; Natasha Johnson; Rachel Blacow; Richard Orton; Rory Gunson; Sarah McDonald; Sharif Shaaban; Sreenu Vattipally; Stephen Carmichael |

We gratefully acknowledge the following Authors from the Originating laboratories responsible for obtaining the specimens, as well as the Submitting laboratories where the genome data were generated and shared via GISAID, on which this research is based.

All Submitters of data may be contacted directly via www.gisaid.org

Authors are sorted alphabetically.

| Accession ID | Originating Laboratory | Submitting Laboratory | Authors |
|---|---|---|---|
| EPI_ISL_1048106, EPI_ISL_1177995, EPI_ISL_1177996, EPI_ISL_1177997 | Barts Health NHS Trust | COVID-19 Genomics UK (COG-UK) Consortium | BROAD; Beatrix; CUTINO-MOGUEL; Claire; David; Dola; HARRINGTON; KELE; KULASEGARAN-SHYLINI; Maria-Teresa; OWOYEMI; Raghavendran |
| EPI_ISL_1054799, EPI_ISL_1054820 | Bioinformatics and Biostatistics Lab, Advanced Sequencing Facility | COVID-19 Genomics UK (COG-UK) Consortium | Aengus Stewart; Chelsea Sawyer; Harshil Patel; Jerome Nicod; Laura Cubitt; Margaret Crawford |
| EPI_ISL_1053927, EPI_ISL_1103595, EPI_ISL_1103596, EPI_ISL_1103598, EPI_ISL_1103605, EPI_ISL_1103607, EPI_ISL_1103608, EPI_ISL_1103610, EPI_ISL_1103611, EPI_ISL_1103613, EPI_ISL_1103620, EPI_ISL_1103625, EPI_ISL_1103628, EPI_ISL_1103629, EPI_ISL_1103630, EPI_ISL_1103631, EPI_ISL_1103633, EPI_ISL_1103634, EPI_ISL_1103636, EPI_ISL_1103638, EPI_ISL_1103639, EPI_ISL_1103644, EPI_ISL_1103645, EPI_ISL_1103646, EPI_ISL_1103648, EPI_ISL_1107527, EPI_ISL_1107528, EPI_ISL_1107529, EPI_ISL_1107530, EPI_ISL_1107681, EPI_ISL_1107682, EPI_ISL_1107683, EPI_ISL_1107684, EPI_ISL_1107685, EPI_ISL_1107758, EPI_ISL_1107778, EPI_ISL_1107779, EPI_ISL_1107780, EPI_ISL_1107781, EPI_ISL_1107782, EPI_ISL_1107783, EPI_ISL_1177231, EPI_ISL_1177232, EPI_ISL_1177235, EPI_ISL_1177236, EPI_ISL_1177237, EPI_ISL_1177238, EPI_ISL_1177239, EPI_ISL_1177240, EPI_ISL_1177241, EPI_ISL_1177243, EPI_ISL_1177246, EPI_ISL_1177247, EPI_ISL_1177251, EPI_ISL_1177252, EPI_ISL_1177253, EPI_ISL_1177262, EPI_ISL_1177263, EPI_ISL_1177270, EPI_ISL_1177273, EPI_ISL_1177281, EPI_ISL_1177300, EPI_ISL_1177311, EPI_ISL_1177323, EPI_ISL_1177332, EPI_ISL_1177344, EPI_ISL_1177349, EPI_ISL_1177359, EPI_ISL_1177367, EPI_ISL_1177387, EPI_ISL_1177411, EPI_ISL_1177420, EPI_ISL_1177421, EPI_ISL_1177429, EPI_ISL_1177438, EPI_ISL_1177441, EPI_ISL_1177444, EPI_ISL_1177446, EPI_ISL_1177450, EPI_ISL_1179646, EPI_ISL_1179647, EPI_ISL_1179648, EPI_ISL_1179649, EPI_ISL_1179650, EPI_ISL_1179651, EPI_ISL_1179670, EPI_ISL_1179671, EPI_ISL_1179672, EPI_ISL_1179673, EPI_ISL_1179675, EPI_ISL_1179676, EPI_ISL_1179677, EPI_ISL_1179678, EPI_ISL_1179679, EPI_ISL_1179681, EPI_ISL_1179683, EPI_ISL_1179687, EPI_ISL_1179689, EPI_ISL_1179691, EPI_ISL_1179692, EPI_ISL_1179694, EPI_ISL_1179695, EPI_ISL_1179697, EPI_ISL_1179702, EPI_ISL_1310303 | | | |
| see above | Centre for Enzyme Innovation, University of Portsmouth / Translational Research Laboratory, Portsmouth Hospitals NHS Trust | COVID-19 Genomics UK (COG-UK) Consortium | Angela Beckett; Christopher Fearn; Kate Cook; Katie Loveson; Salman Goudarzi; Samuel Robson; Scott Elliott; Sharon Glaysher |
| EPI_ISL_1047092, EPI_ISL_1047093, EPI_ISL_1047095, EPI_ISL_1047096, EPI_ISL_1047114, EPI_ISL_1047115, EPI_ISL_1047116, EPI_ISL_1047117, EPI_ISL_1047118, EPI_ISL_1047119, EPI_ISL_1047120, EPI_ISL_1047121, EPI_ISL_1047122, EPI_ISL_1047123, EPI_ISL_1047124, EPI_ISL_1047125, EPI_ISL_1047126, EPI_ISL_1047127, EPI_ISL_1047128, EPI_ISL_1047129, EPI_ISL_1047130, EPI_ISL_1047131, EPI_ISL_1047132, EPI_ISL_1047133, EPI_ISL_1047134, EPI_ISL_1047136, EPI_ISL_1047138, EPI_ISL_1047140, EPI_ISL_1047144, EPI_ISL_1047145, EPI_ISL_1176724, EPI_ISL_1176725, EPI_ISL_1176726, EPI_ISL_1176727, EPI_ISL_1176728, EPI_ISL_1176731, EPI_ISL_1176732, EPI_ISL_1176733, EPI_ISL_1176734, EPI_ISL_1176736, EPI_ISL_1176737, EPI_ISL_1176738, EPI_ISL_1176739, EPI_ISL_1176740, EPI_ISL_1176743, EPI_ISL_1176744, EPI_ISL_1176745, EPI_ISL_1176746, EPI_ISL_1176747, EPI_ISL_1176748, EPI_ISL_1176749, EPI_ISL_1176750, EPI_ISL_1176751, EPI_ISL_1176752, EPI_ISL_1176754, EPI_ISL_1176756, EPI_ISL_1176757, EPI_ISL_1176758, EPI_ISL_1176759, EPI_ISL_1176760, EPI_ISL_1176761, EPI_ISL_1176762, EPI_ISL_1176763, EPI_ISL_1176768, EPI_ISL_1176769, EPI_ISL_1176770, EPI_ISL_1176771, EPI_ISL_1176772, EPI_ISL_1176773, EPI_ISL_1176774, EPI_ISL_1176775, EPI_ISL_1176777, EPI_ISL_1176778, EPI_ISL_1176779, EPI_ISL_1176780, EPI_ISL_1176781, EPI_ISL_1176782, EPI_ISL_1176783, EPI_ISL_1176784, EPI_ISL_1176787, EPI_ISL_1176788, EPI_ISL_1176789, EPI_ISL_1176790, EPI_ISL_1176791, EPI_ISL_1176792, EPI_ISL_1176793, EPI_ISL_1176794, EPI_ISL_1176795, EPI_ISL_1176798, EPI_ISL_1176799, EPI_ISL_1176800, EPI_ISL_1176801, EPI_ISL_1176802, EPI_ISL_1176803, EPI_ISL_1176804, EPI_ISL_1176805, EPI_ISL_1176806, EPI_ISL_1176808, EPI_ISL_1176809, EPI_ISL_1176810, EPI_ISL_1176811, EPI_ISL_1176812, EPI_ISL_1177640, EPI_ISL_1177651, EPI_ISL_1730025 | | | |
| see above | Department of Pathology, University of Cambridge | COVID-19 Genomics UK (COG-UK) Consortium | Aminu S. Jahun; Ian Goodfellow; Iliana Georgana; Martin D. Curran; Myra Hosmillo; Rhys Izuagbe; Surendra Parmar; William L. Hamilton; Yasmin Chaudhry |
| EPI_ISL_1061700, EPI_ISL_1061701, EPI_ISL_1061702, EPI_ISL_1061704, EPI_ISL_1061705, EPI_ISL_1061706, EPI_ISL_1061709, EPI_ISL_1061712, EPI_ISL_1061713, EPI_ISL_1061714, EPI_ISL_1061715, EPI_ISL_1061717, EPI_ISL_1061718, EPI_ISL_1061720, EPI_ISL_1061721, EPI_ISL_1061722, EPI_ISL_1061724, EPI_ISL_1061725, EPI_ISL_1061726, EPI_ISL_1061727, EPI_ISL_1061729, EPI_ISL_1061732, EPI_ISL_1061734, EPI_ISL_1061735, EPI_ISL_1061736, EPI_ISL_1061737, EPI_ISL_1061738, EPI_ISL_1061739, EPI_ISL_1061741, EPI_ISL_1061742, EPI_ISL_1061743, EPI_ISL_1061744, EPI_ISL_1061745, EPI_ISL_1061746, EPI_ISL_1061748, EPI_ISL_1061749, EPI_ISL_1061750, EPI_ISL_1061751, EPI_ISL_1061752, EPI_ISL_1061754, EPI_ISL_1061755, EPI_ISL_1061759, EPI_ISL_1061761, EPI_ISL_1061766, EPI_ISL_1061768, EPI_ISL_1061769, EPI_ISL_1061771, EPI_ISL_1061772, EPI_ISL_1061773, EPI_ISL_1061774, EPI_ISL_1061777, EPI_ISL_1061778, EPI_ISL_1061779, EPI_ISL_1061783, EPI_ISL_1061784, EPI_ISL_1061785, EPI_ISL_1061786, EPI_ISL_1061789, EPI_ISL_1061791, EPI_ISL_1061792, EPI_ISL_1061794, EPI_ISL_1061795, EPI_ISL_1061797, EPI_ISL_1061798, EPI_ISL_1061800, EPI_ISL_1061802, EPI_ISL_1061804, EPI_ISL_1061805, EPI_ISL_1061806, EPI_ISL_1061808, EPI_ISL_1061809, EPI_ISL_1061810, EPI_ISL_1061811, EPI_ISL_1061813, EPI_ISL_1061815, EPI_ISL_1061817, EPI_ISL_1061818, EPI_ISL_1061819, EPI_ISL_1061821, EPI_ISL_1061824, EPI_ISL_1061825, EPI_ISL_1061826, EPI_ISL_1061828, EPI_ISL_1061829, EPI_ISL_1061830, EPI_ISL_1061834, EPI_ISL_1061835, EPI_ISL_1061836, EPI_ISL_1061837, EPI_ISL_1061839, EPI_ISL_1061841, EPI_ISL_1061842, EPI_ISL_1061843, EPI_ISL_1061844, EPI_ISL_1061845, EPI_ISL_1061846, EPI_ISL_1061850, EPI_ISL_1061851, EPI_ISL_1061852, EPI_ISL_1061855, EPI_ISL_1061857, EPI_ISL_1061858, EPI_ISL_1061860, EPI_ISL_1061861, EPI_ISL_1061863, EPI_ISL_1061864, EPI_ISL_1061866, EPI_ISL_1061867, EPI_ISL_1061868, EPI_ISL_1061869, EPI_ISL_1061870, EPI_ISL_1061873, EPI_ISL_1061877, EPI_ISL_1061878, EPI_ISL_1061879, EPI_ISL_1061881, EPI_ISL_1061886, EPI_ISL_1061887, EPI_ISL_1061888, EPI_ISL_1061890, EPI_ISL_1061891, EPI_ISL_1061893, EPI_ISL_1061894, EPI_ISL_1061895, EPI_ISL_1061896, EPI_ISL_1061898, EPI_ISL_1061899, EPI_ISL_1061900, EPI_ISL_1061901, EPI_ISL_1061902, EPI_ISL_1061903, EPI_ISL_1061904, EPI_ISL_1061906, EPI_ISL_1061907, EPI_ISL_1061908, EPI_ISL_1061910, EPI_ISL_1061912, EPI_ISL_1061913, EPI_ISL_1061914, EPI_ISL_1061915, EPI_ISL_1061916, EPI_ISL_1061917, EPI_ISL_1061918, EPI_ISL_1061921, EPI_ISL_1061922, EPI_ISL_1061923, EPI_ISL_1061924, EPI_ISL_1061925, EPI_ISL_1061926, EPI_ISL_1061929, EPI_ISL_1061931, EPI_ISL_1061934, EPI_ISL_1061935, EPI_ISL_1061936, EPI_ISL_1061939, EPI_ISL_1061940, EPI_ISL_1061941, EPI_ISL_1061942, EPI_ISL_1061944, EPI_ISL_1061946, EPI_ISL_1061948, EPI_ISL_1061950, EPI_ISL_1061951, EPI_ISL_1061952, EPI_ISL_1061953, EPI_ISL_1061955, EPI_ISL_1061957, EPI_ISL_1061958, EPI_ISL_1061960, EPI_ISL_1061961, EPI_ISL_1061962, EPI_ISL_1061963, EPI_ISL_1061965, EPI_ISL_1061966, EPI_ISL_1061968, EPI_ISL_1061969, EPI_ISL_1061970, EPI_ISL_1061971, EPI_ISL_1061972, EPI_ISL_1061973, EPI_ISL_1061974, EPI_ISL_1061976, EPI_ISL_1061977, EPI_ISL_1061978, EPI_ISL_1061980, EPI_ISL_1061981, EPI_ISL_1061982, EPI_ISL_1061983, EPI_ISL_1061985, EPI_ISL_1061987, EPI_ISL_1061989, EPI_ISL_1061990, EPI_ISL_1061991, EPI_ISL_1061996, EPI_ISL_1061997, EPI_ISL_1061998, EPI_ISL_1061999, EPI_ISL_1062001, EPI_ISL_1062002, EPI_ISL_1062005, EPI_ISL_1062006, EPI_ISL_1062007, EPI_ISL_1062008, EPI_ISL_1062009, EPI_ISL_1062010, EPI_ISL_1062012, EPI_ISL_1062013, EPI_ISL_1062014, EPI_ISL_1062015, EPI_ISL_1062016, EPI_ISL_1062018, EPI_ISL_1062020, EPI_ISL_1062021, EPI_ISL_1062022, EPI_ISL_1062024, EPI_ISL_1062025, EPI_ISL_1062026, EPI_ISL_1062027, EPI_ISL_1062028, EPI_ISL_1062030, EPI_ISL_1062031, EPI_ISL_1062032, EPI_ISL_1062033, EPI_ISL_1062034, EPI_ISL_1062035, EPI_ISL_1062036, EPI_ISL_1062037, EPI_ISL_1062038, EPI_ISL_1062041, EPI_ISL_1062042, EPI_ISL_1062044, EPI_ISL_1062045, EPI_ISL_1062047, EPI_ISL_1062049, EPI_ISL_1062051, EPI_ISL_1071638, EPI_ISL_1071639, EPI_ISL_1071644, EPI_ISL_1071645, EPI_ISL_1071646, EPI_ISL_1071647, EPI_ISL_1071653, EPI_ISL_1071655, EPI_ISL_1071657, EPI_ISL_1071658, EPI_ISL_1071660, EPI_ISL_1071661, EPI_ISL_1071663, EPI_ISL_1071664, EPI_ISL_1071665, EPI_ISL_1071667, EPI_ISL_1071668, EPI_ISL_1071673, EPI_ISL_1071674, EPI_ISL_1071675, EPI_ISL_1071678, EPI_ISL_1071684, EPI_ISL_1071685, EPI_ISL_1071687, EPI_ISL_1071688, EPI_ISL_1071689, EPI_ISL_1071690, EPI_ISL_1071691, EPI_ISL_1071696, EPI_ISL_1071698, EPI_ISL_1071699, EPI_ISL_1071700, EPI_ISL_1071701, EPI_ISL_1071702, EPI_ISL_1071703, EPI_ISL_1071704, EPI_ISL_1071705, EPI_ISL_1071707, EPI_ISL_1071708, EPI_ISL_1071711, EPI_ISL_1071712, EPI_ISL_1071714, EPI_ISL_1071716, EPI_ISL_1071717, EPI_ISL_1071718, EPI_ISL_1071721, EPI_ISL_1071722, EPI_ISL_1071723, EPI_ISL_1071724, EPI_ISL_1071725, EPI_ISL_1071726, EPI_ISL_1071729, EPI_ISL_1071732, EPI_ISL_1071733, EPI_ISL_1071734, EPI_ISL_1071735, EPI_ISL_1071736, EPI_ISL_1071740, EPI_ISL_1071743, EPI_ISL_1071744, EPI_ISL_1071747, EPI_ISL_1071748, EPI_ISL_1071751, EPI_ISL_1071752, EPI_ISL_1071753, EPI_ISL_1071754, EPI_ISL_1071756, EPI_ISL_1071760, EPI_ISL_1071762, EPI_ISL_1071763, EPI_ISL_1071767, EPI_ISL_1071768, EPI_ISL_1071769, EPI_ISL_1071770, EPI_ISL_1071771, EPI_ISL_1071775, EPI_ISL_1071776, EPI_ISL_1071779, EPI_ISL_1071780, EPI_ISL_1071782, EPI_ISL_1071787, EPI_ISL_1071788, EPI_ISL_1071789, EPI_ISL_1071790, EPI_ISL_1071791, EPI_ISL_1071792, EPI_ISL_1071796, EPI_ISL_1071798, EPI_ISL_1071808, EPI_ISL_1071809, EPI_ISL_1071814, EPI_ISL_1071817, EPI_ISL_1071818, EPI_ISL_1071820, EPI_ISL_1071821, EPI_ISL_1071822, EPI_ISL_1071826, EPI_ISL_1071827, EPI_ISL_1071829, EPI_ISL_1071834, EPI_ISL_1071835, EPI_ISL_1071836, EPI_ISL_1071840, EPI_ISL_1071842, EPI_ISL_1071843, EPI_ISL_1071844, EPI_ISL_1071845, EPI_ISL_1071847, EPI_ISL_1071850, EPI_ISL_1071851, EPI_ISL_1071852, EPI_ISL_1071853, EPI_ISL_1071854, EPI_ISL_1071855, EPI_ISL_1071856, EPI_ISL_1071857, EPI_ISL_1071860, EPI_ISL_1071861, EPI_ISL_1071863, EPI_ISL_1071864, EPI_ISL_1071866, EPI_ISL_1071871, EPI_ISL_1071876, EPI_ISL_1071877, EPI_ISL_1071879, EPI_ISL_1071880, EPI_ISL_1071884, EPI_ISL_1071885, EPI_ISL_1071886, EPI_ISL_1071888, EPI_ISL_1071889, EPI_ISL_1071890, EPI_ISL_1071891, EPI_ISL_1071893, EPI_ISL_1071899, EPI_ISL_1071901, EPI_ISL_1071902, EPI_ISL_1071903, EPI_ISL_1071905, EPI_ISL_1071906, EPI_ISL_1071907, EPI_ISL_1071908, EPI_ISL_1071909, EPI_ISL_1071917, EPI_ISL_1071918, EPI_ISL_1071919, EPI_ISL_1071920, EPI_ISL_1071922, EPI_ISL_1071925, EPI_ISL_1071927, EPI_ISL_1071935, EPI_ISL_1071936, EPI_ISL_1071941, EPI_ISL_1071943, EPI_ISL_1071948, EPI_ISL_1071949, EPI_ISL_1071952, EPI_ISL_1071953, EPI_ISL_1071955, EPI_ISL_1071956, EPI_ISL_1071964, EPI_ISL_1071965, EPI_ISL_1071966, EPI_ISL_1071967, EPI_ISL_1071969, EPI_ISL_1071972, EPI_ISL_1071973, EPI_ISL_1071974, EPI_ISL_1071975, EPI_ISL_1071976, EPI_ISL_1071977, EPI_ISL_1071980, EPI_ISL_1071981, EPI_ISL_1071986, EPI_ISL_1071990, EPI_ISL_1071993, EPI_ISL_1071994, EPI_ISL_1071997, EPI_ISL_1071998, EPI_ISL_1071999, EPI_ISL_1072000, EPI_ISL_1072001, EPI_ISL_1072002, EPI_ISL_1072003, EPI_ISL_1072004, EPI_ISL_1072005, EPI_ISL_1072006, EPI_ISL_1072007, EPI_ISL_1072008, EPI_ISL_1072009, EPI_ISL_1072010, EPI_ISL_1072011, EPI_ISL_1072012, EPI_ISL_1072013, EPI_ISL_1072014, EPI_ISL_1072015, EPI_ISL_1072016, EPI_ISL_1072017, EPI_ISL_1072018, EPI_ISL_1072019, EPI_ISL_1072021, EPI_ISL_1072022, EPI_ISL_1072023, EPI_ISL_1072024, EPI_ISL_1072025, EPI_ISL_1072026, EPI_ISL_1072027, EPI_ISL_1072028, EPI_ISL_1072029, EPI_ISL_1072030, EPI_ISL_1072031, EPI_ISL_1072032, EPI_ISL_1072033, EPI_ISL_1072035, EPI_ISL_1072036, EPI_ISL_1072037, EPI_ISL_1072038, EPI_ISL_1072039, EPI_ISL_1072040, EPI_ISL_1072042, EPI_ISL_1072043, EPI_ISL_1072044, EPI_ISL_1072045, EPI_ISL_1072046, EPI_ISL_1072047, EPI_ISL_1072048, EPI_ISL_1072049, EPI_ISL_1072050, EPI_ISL_1072052, EPI_ISL_1072053, EPI_ISL_1072054, EPI_ISL_1072055, EPI_ISL_1072056, EPI_ISL_1072057, EPI_ISL_1072058, EPI_ISL_1072059, EPI_ISL_1072060, EPI_ISL_1072061, EPI_ISL_1072062, EPI_ISL_1072063, EPI_ISL_1072064, EPI_ISL_1072065, EPI_ISL_1072066, EPI_ISL_1072067, EPI_ISL_1072068, EPI_ISL_1072069, EPI_ISL_1072070, EPI_ISL_1072071, EPI_ISL_1072073, EPI_ISL_1072074, EPI_ISL_1072075, EPI_ISL_1072076, EPI_ISL_1072077, EPI_ISL_1072078, EPI_ISL_1072079, EPI_ISL_1072081, EPI_ISL_1072082, EPI_ISL_1072083, EPI_ISL_1072084, EPI_ISL_1072085, EPI_ISL_1072086, EPI_ISL_1072087, EPI_ISL_1072088, EPI_ISL_1072089, EPI_ISL_1072090, EPI_ISL_1072091, EPI_ISL_1072092, EPI_ISL_1072094, EPI_ISL_1072095, EPI_ISL_1072096, EPI_ISL_1072097, EPI_ISL_1072098, EPI_ISL_1072099, EPI_ISL_1072100, EPI_ISL_1072101, EPI_ISL_1072102, EPI_ISL_1072103, EPI_ISL_1072104, EPI_ISL_1072105, EPI_ISL_1072106, EPI_ISL_1072107, EPI_ISL_1072108, EPI_ISL_1072109, EPI_ISL_1072110, EPI_ISL_1072111, EPI_ISL_1072112, EPI_ISL_1072113, EPI_ISL_1072114, EPI_ISL_1072115, EPI_ISL_1072117, EPI_ISL_1072118, EPI_ISL_1072119, EPI_ISL_1072120, EPI_ISL_1072121, EPI_ISL_1072122, EPI_ISL_1072123, EPI_ISL_1072124, EPI_ISL_1072126, EPI_ISL_1072127, EPI_ISL_1072128, EPI_ISL_1072129, EPI_ISL_1072130, EPI_ISL_1072131, EPI_ISL_1072133, EPI_ISL_1072134, EPI_ISL_1072135, EPI_ISL_1072136, EPI_ISL_1072137, EPI_ISL_1072138, EPI_ISL_1072139, EPI_ISL_1072140, EPI_ISL_1072141, EPI_ISL_1072142, EPI_ISL_1072143, EPI_ISL_1072144, EPI_ISL_1072145, EPI_ISL_1072146, EPI_ISL_1072147, EPI_ISL_1072148, EPI_ISL_1072149, EPI_ISL_1072150, EPI_ISL_1072151, EPI_ISL_1072152, EPI_ISL_1072153, EPI_ISL_1072154, EPI_ISL_1072155, EPI_ISL_1072156, EPI_ISL_1072157, EPI_ISL_1072158, EPI_ISL_1072159, EPI_ISL_1072161, EPI_ISL_1072162, EPI_ISL_1072163, EPI_ISL_1072164, EPI_ISL_1072165, EPI_ISL_1072166, EPI_ISL_1072167, EPI_ISL_1072168, EPI_ISL_1072169, EPI_ISL_1072170, EPI_ISL_1072171, EPI_ISL_1072172, EPI_ISL_1072173, EPI_ISL_1072174, EPI_ISL_1072175, EPI_ISL_1072176, EPI_ISL_1072177, EPI_ISL_1072178, EPI_ISL_1072179, EPI_ISL_1072180, EPI_ISL_1072181, EPI_ISL_1072182, EPI_ISL_1072183, EPI_ISL_1072184, EPI_ISL_1072185, EPI_ISL_1072186, EPI_ISL_1072187, EPI_ISL_1072188, EPI_ISL_1072190, EPI_ISL_1072191, EPI_ISL_1072192, EPI_ISL_1072193, EPI_ISL_1072194, EPI_ISL_1072195, EPI_ISL_1072196, EPI_ISL_1072197, EPI_ISL_1072198, EPI_ISL_1072199, EPI_ISL_1072200, EPI_ISL_1072201, EPI_ISL_1072202, EPI_ISL_1072203, EPI_ISL_1072204, EPI_ISL_1072205, EPI_ISL_1072206, EPI_ISL_1072207, EPI_ISL_1072208, EPI_ISL_1072209, EPI_ISL_1072210, EPI_ISL_1072211, EPI_ISL_1072212, EPI_ISL_1072213, EPI_ISL_1072214, EPI_ISL_1072215, EPI_ISL_1072216, EPI_ISL_1072217, EPI_ISL_1072218, EPI_ISL_1072219, EPI_ISL_1072220, EPI_ISL_1072221, EPI_ISL_1072222, EPI_ISL_1072223, EPI_ISL_1072224, EPI_ISL_1072225, EPI_ISL_1072226, EPI_ISL_1072227, EPI_ISL_1072228, EPI_ISL_1072229, EPI_ISL_1072230, EPI_ISL_1072231, EPI_ISL_1072232, EPI_ISL_1072233, EPI_ISL_1072234, EPI_ISL_1072235, EPI_ISL_1072236, EPI_ISL_1072237, EPI_ISL_1072238, EPI_ISL_1072239, EPI_ISL_1072240, EPI_ISL_1072241, EPI_ISL_1072242, EPI_ISL_1072243, EPI_ISL_1072244, EPI_ISL_1072245, EPI_ISL_1072246, EPI_ISL_1072248, EPI_ISL_1072250, EPI_ISL_1072251, EPI_ISL_1072252, EPI_ISL_1072253, EPI_ISL_1072254, EPI_ISL_1072255, EPI_ISL_1072256, EPI_ISL_1072257, EPI_ISL_1072258, EPI_ISL_1072259, EPI_ISL_1072260, EPI_ISL_1072261, EPI_ISL_1072262, EPI_ISL_1072263, EPI_ISL_1072264, EPI_ISL_1072265, EPI_ISL_1072266, EPI_ISL_1072267, EPI_ISL_1072268, EPI_ISL_1072270, EPI_ISL_1072271, EPI_ISL_1072272, EPI_ISL_1072273, EPI_ISL_1072274, EPI_ISL_1072275, EPI_ISL_1072276, EPI_ISL_1072277, EPI_ISL_1072278, EPI_ISL_1072279, EPI_ISL_1072280, EPI_ISL_1072281, EPI_ISL_1072282, EPI_ISL_1072283, EPI_ISL_1072284, EPI_ISL_1072285, EPI_ISL_1072286, EPI_ISL_1072287, EPI_ISL_1072289, EPI_ISL_1072290, EPI_ISL_1072291, EPI_ISL_1072292, EPI_ISL_1072293, EPI_ISL_1072294, EPI_ISL_1072295, EPI_ISL_1072296, EPI_ISL_1072297, EPI_ISL_1072298, EPI_ISL_1072299, EPI_ISL_1072300, EPI_ISL_1072301, EPI_ISL_1072302, EPI_ISL_1072303, EPI_ISL_1072304, EPI_ISL_1072305, EPI_ISL_1072306, EPI_ISL_1072307, EPI_ISL_1072308, EPI_ISL_1072309, EPI_ISL_1072310, EPI_ISL_1072311, EPI_ISL_1072312, EPI_ISL_1072314, EPI_ISL_1072315, EPI_ISL_1072316, EPI_ISL_1072317, EPI_ISL_1072318, EPI_ISL_1072319, EPI_ISL_1072320, EPI_ISL_1072321, EPI_ISL_1072322, EPI_ISL_1072323, EPI_ISL_1072324, EPI_ISL_1072325, EPI_ISL_1072326, EPI_ISL_1072327, EPI_ISL_1072328, EPI_ISL_1072329, EPI_ISL_1072330, EPI_ISL_1072331, EPI_ISL_1072333, EPI_ISL_1072334, EPI_ISL_1072335, EPI_ISL_1072336, EPI_ISL_1072337, EPI_ISL_1072338, EPI_ISL_1072339, EPI_ISL_1072340, EPI_ISL_1072341, EPI_ISL_1072342, EPI_ISL_1072603, EPI_ISL_1072604, EPI_ISL_1072605, EPI_ISL_1072606, EPI_ISL_1072607, EPI_ISL_1072608, EPI_ISL_1072610, EPI_ISL_1072611, EPI_ISL_1072612, EPI_ISL_1072614, EPI_ISL_1072615, EPI_ISL_1072619, EPI_ISL_1072621, EPI_ISL_1072622, EPI_ISL_1072623, EPI_ISL_1072624, EPI_ISL_1072625, EPI_ISL_1072628, EPI_ISL_1072629, EPI_ISL_1072630, EPI_ISL_1072631, EPI_ISL_1072632, EPI_ISL_1072633, EPI_ISL_1072634, EPI_ISL_1072636, EPI_ISL_1072637, EPI_ISL_1072639, EPI_ISL_1072640, EPI_ISL_1072642, EPI_ISL_1072643, EPI_ISL_1072644, EPI_ISL_1072645, EPI_ISL_1072646, EPI_ISL_1072647, EPI_ISL_1072648, EPI_ISL_1072649, EPI_ISL_1072650, EPI_ISL_1072651, EPI_ISL_1072652, EPI_ISL_1072653, EPI_ISL_1072654, EPI_ISL_1072655, EPI_ISL_1072657, EPI_ISL_1072658, EPI_ISL_1072659, EPI_ISL_1072660, EPI_ISL_1072661, EPI_ISL_1072662, EPI_ISL_1072663, EPI_ISL_1072664, EPI_ISL_1072665, EPI_ISL_1072666, EPI_ISL_1072667, EPI_ISL_1072668, EPI_ISL_1072669, EPI_ISL_1072670, EPI_ISL_1072671, EPI_ISL_1072673, EPI_ISL_1072674, EPI_ISL_1072675, EPI_ISL_1072676, EPI_ISL_1072677, EPI_ISL_1072679, EPI_ISL_1072680, EPI_ISL_1072681, EPI_ISL_1072682, EPI_ISL_1072683, EPI_ISL_1072684, EPI_ISL_1072686, EPI_ISL_1072687, EPI_ISL_1072688, EPI_ISL_1072689, EPI_ISL_1072695, EPI_ISL_1072697, EPI_ISL_1072698, EPI_ISL_1072699, EPI_ISL_1072700, EPI_ISL_1072702, EPI_ISL_1072703, EPI_ISL_1072704, EPI_ISL_1072706, EPI_ISL_1072711, EPI_ISL_1072712, EPI_ISL_1072714, EPI_ISL_1072716, EPI_ISL_1072718, EPI_ISL_1072719, EPI_ISL_1072720, EPI_ISL_1072722, EPI_ISL_1072724, EPI_ISL_1072725, EPI_ISL_1072726, EPI_ISL_1072728, EPI_ISL_1072729, EPI_ISL_1072730, EPI_ISL_1072731, EPI_ISL_1072732, EPI_ISL_1072734, EPI_ISL_1072735, EPI_ISL_1072736, EPI_ISL_1072737, EPI_ISL_1072738, EPI_ISL_1072741, EPI_ISL_1072743, EPI_ISL_1072746, EPI_ISL_1072747, EPI_ISL_1072748, EPI_ISL_1072749, EPI_ISL_1072752, EPI_ISL_1072753, EPI_ISL_1072755, EPI_ISL_1072756, EPI_ISL_1072757, EPI_ISL_1072758, EPI_ISL_1072759, EPI_ISL_1072760, EPI_ISL_1072761, EPI_ISL_1072762, EPI_ISL_1072763, EPI_ISL_1072764, EPI_ISL_1072765, EPI_ISL_1072766, EPI_ISL_1072767, EPI_ISL_1072768, EPI_ISL_1072769, EPI_ISL_1072770, EPI_ISL_1072771, EPI_ISL_1072772, EPI_ISL_1072773, EPI_ISL_1072774, EPI_ISL_1072775, EPI_ISL_1072776, EPI_ISL_1072777, EPI_ISL_1072779, EPI_ISL_1072780, EPI_ISL_1072781, EPI_ISL_1072783, EPI_ISL_1072785, EPI_ISL_1072786, EPI_ISL_1072787, EPI_ISL_1072788, EPI_ISL_1072789, EPI_ISL_1072790, EPI_ISL_1072791, EPI_ISL_1072793, EPI_ISL_1072794, EPI_ISL_1072796, EPI_ISL_1072797, EPI_ISL_1072798, EPI_ISL_1072799, EPI_ISL_1072801, EPI_ISL_1072803, EPI_ISL_1072805, EPI_ISL_1072806, EPI_ISL_1072807, EPI_ISL_1072808, EPI_ISL_1072809, EPI_ISL_1072810, EPI_ISL_1072811, EPI_ISL_1072813, EPI_ISL_1072815, EPI_ISL_1072816, EPI_ISL_1072819, EPI_ISL_1072821, EPI_ISL_1072822, EPI_ISL_1072823, EPI_ISL_1072825, EPI_ISL_1072826, EPI_ISL_1072827, EPI_ISL_1072829, EPI_ISL_1072830, EPI_ISL_1072831, EPI_ISL_1072832, EPI_ISL_1072833, EPI_ISL_1072835, EPI_ISL_1072837, EPI_ISL_1072838, EPI_ISL_1072839, EPI_ISL_1072840, EPI_ISL_1072844, EPI_ISL_1072846, EPI_ISL_1072847, EPI_ISL_1072848, EPI_ISL_1072851, EPI_ISL_1072853, EPI_ISL_1072854, EPI_ISL_1072855, EPI_ISL_1072856, EPI_ISL_1072857, EPI_ISL_1072858, EPI_ISL_1072859, EPI_ISL_1072860, EPI_ISL_1072861, EPI_ISL_1072862, EPI_ISL_1072863, EPI_ISL_1072864, EPI_ISL_1072865, EPI_ISL_1072866, EPI_ISL_1072868, EPI_ISL_1072869, EPI_ISL_1072870, EPI_ISL_1072872, EPI_ISL_1072873, EPI_ISL_1072874, EPI_ISL_1072875, EPI_ISL_1072878, EPI_ISL_1072879, EPI_ISL_1072880, EPI_ISL_1072883, EPI_ISL_1072884, EPI_ISL_1072885, EPI_ISL_1072886, EPI_ISL_1072887, EPI_ISL_1072889, EPI_ISL_1072890, EPI_ISL_1072891, EPI_ISL_1072892, EPI_ISL_1072893, EPI_ISL_1072895, EPI_ISL_1072897, EPI_ISL_1072898, EPI_ISL_1072899, EPI_ISL_1072900, EPI_ISL_1072901, EPI_ISL_1072903, EPI_ISL_1072904, EPI_ISL_1072905, EPI_ISL_1072906, EPI_ISL_1072908, EPI_ISL_1072910, EPI_ISL_1072911, EPI_ISL_1072913, EPI_ISL_1072914, EPI_ISL_1072915, EPI_ISL_1072916, EPI_ISL_1072917, EPI_ISL_1072918, EPI_ISL_1072919, EPI_ISL_1072920, EPI_ISL_1072922, EPI_ISL_1072923, EPI_ISL_1072924, EPI_ISL_1072925, EPI_ISL_1072926, EPI_ISL_1072927, EPI_ISL_1072928, EPI_ISL_1072929, EPI_ISL_1072930, EPI_ISL_1072931, EPI_ISL_1072932, EPI_ISL_1072936, EPI_ISL_1072937, EPI_ISL_1072938, EPI_ISL_1072939, EPI_ISL_1072940, EPI_ISL_1072941, EPI_ISL_1072942, EPI_ISL_1072945, EPI_ISL_1072946, EPI_ISL_1072948, EPI_ISL_1072949, EPI_ISL_1072953, EPI_ISL_1072954, EPI_ISL_1072955, EPI_ISL_1072956, EPI_ISL_1072957, EPI_ISL_1072958, EPI_ISL_1083131, EPI_ISL_1083147, EPI_ISL_1083190, EPI_ISL_1083313, EPI_ISL_1083326, EPI_ISL_1083353, EPI_ISL_1083408, EPI_ISL_1083423, EPI_ISL_1083431, EPI_ISL_1083452, EPI_ISL_1083611, EPI_ISL_1084677, EPI_ISL_1084678, EPI_ISL_1084679, EPI_ISL_1084680, EPI_ISL_1084681, EPI_ISL_1084682, EPI_ISL_1084683, EPI_ISL_1084684, EPI_ISL_1084685, EPI_ISL_1084686, EPI_ISL_1084687, EPI_ISL_1084688, EPI_ISL_1084689, EPI_ISL_1084690, EPI_ISL_1084691, EPI_ISL_1084692, EPI_ISL_1084693, EPI_ISL_1084694, EPI_ISL_1084695, EPI_ISL_1084696, EPI_ISL_1084697, EPI_ISL_1084698, EPI_ISL_1084699, EPI_ISL_1084700, EPI_ISL_1084702, EPI_ISL_1084703, EPI_ISL_1084704, EPI_ISL_1084705, EPI_ISL_1084706, EPI_ISL_1084707, EPI_ISL_1084708, EPI_ISL_1084710, EPI_ISL_1084711, EPI_ISL_1084712, EPI_ISL_1084713, EPI_ISL_1084714, EPI_ISL_1084715, EPI_ISL_1084716, EPI_ISL_1084717, EPI_ISL_1084718, EPI_ISL_1101338, EPI_ISL_1116284, EPI_ISL_1116287, EPI_ISL_1175747, EPI_ISL_1190725 | | | |
| see above | Lighthouse Lab in Alderley Park | Wellcome Sanger Institute for the COVID-19 Genomics UK (COG-UK) Consortium | Cordelia Langford; David K. Jackson; Dominic Kwiatkowski; Ewan Harrison; Ian Johnston; Jacquelyn Wynn; Jeffrey Barrett; John Sillitoe on behalf of the Wellcome Sanger Institute COVID-19 Surveillance Team; Mairead Hyland; Roberto Amato; Sonia Goncalves; The Lighthouse Lab in Alderley Park and Alex Alderton |
| EPI_ISL_1042003, EPI_ISL_1042008, EPI_ISL_1042011, EPI_ISL_1042016, EPI_ISL_1042019, EPI_ISL_1042023, EPI_ISL_1042024, EPI_ISL_1042030, EPI_ISL_1042035, EPI_ISL_1042042, EPI_ISL_1042049, EPI_ISL_1042051, EPI_ISL_1042053, EPI_ISL_1042057, EPI_ISL_1042059, EPI_ISL_1042062, EPI_ISL_1042064, EPI_ISL_1042069, EPI_ISL_1042071, EPI_ISL_1042073, EPI_ISL_1042075, EPI_ISL_1042076, EPI_ISL_1042080, EPI_ISL_1042082, EPI_ISL_1042092, EPI_ISL_1042097, EPI_ISL_1042098, EPI_ISL_1042101, EPI_ISL_1042103, EPI_ISL_1042108, EPI_ISL_1042112, EPI_ISL_1042117, EPI_ISL_1042123, EPI_ISL_1042134, EPI_ISL_1042135, EPI_ISL_1042140, EPI_ISL_1042152, EPI_ISL_1042153, EPI_ISL_1042158, EPI_ISL_1042162, EPI_ISL_1042163, EPI_ISL_1042165, EPI_ISL_1042172, EPI_ISL_1042175, EPI_ISL_1042177, EPI_ISL_1042179, EPI_ISL_1042181, EPI_ISL_1042187, EPI_ISL_1042190, EPI_ISL_1042191, EPI_ISL_1042194, EPI_ISL_1042202, EPI_ISL_1042213, EPI_ISL_1042222, EPI_ISL_1042226, EPI_ISL_1042232, EPI_ISL_1042233, EPI_ISL_1042240, EPI_ISL_1042241, EPI_ISL_1042242, EPI_ISL_1042248, EPI_ISL_1042250, EPI_ISL_1042253, EPI_ISL_1042259, EPI_ISL_1042269, EPI_ISL_1042274, EPI_ISL_1042284, EPI_ISL_1042286, EPI_ISL_1042288, EPI_ISL_1042289, EPI_ISL_1042291, EPI_ISL_1042293, EPI_ISL_1042295, EPI_ISL_1042297, EPI_ISL_1042307, EPI_ISL_1042308, EPI_ISL_1042312, EPI_ISL_1042314, EPI_ISL_1042317, EPI_ISL_1042320, EPI_ISL_1042325, EPI_ISL_1042326, EPI_ISL_1042329, EPI_ISL_1042334, EPI_ISL_1042350, EPI_ISL_1042356, EPI_ISL_1045880, EPI_ISL_1046336, EPI_ISL_1061435, EPI_ISL_1061436, EPI_ISL_1061437, EPI_ISL_1061438, EPI_ISL_1061439, EPI_ISL_1061440, EPI_ISL_1061441, EPI_ISL_1061442, EPI_ISL_1061443, EPI_ISL_1061444, EPI_ISL_1061445, EPI_ISL_1061446, EPI_ISL_1061447, EPI_ISL_1061448, EPI_ISL_1061449, EPI_ISL_1061450, EPI_ISL_1061451, EPI_ISL_1061452, EPI_ISL_1061453, EPI_ISL_1061454, EPI_ISL_1061455, EPI_ISL_1061456, EPI_ISL_1061457, EPI_ISL_1061458, EPI_ISL_1061459, EPI_ISL_1061460, EPI_ISL_1061461, EPI_ISL_1061462, EPI_ISL_1061463, EPI_ISL_1061464, EPI_ISL_1061465, EPI_ISL_1061466, EPI_ISL_1061467, EPI_ISL_1061468, EPI_ISL_1061469, EPI_ISL_1061470, EPI_ISL_1061471, EPI_ISL_1061472, EPI_ISL_1061473, EPI_ISL_1061474, EPI_ISL_1061475, EPI_ISL_1061476, EPI_ISL_1061477, EPI_ISL_1061478, EPI_ISL_1061479, EPI_ISL_1061480, EPI_ISL_1061481, EPI_ISL_1061482, EPI_ISL_1061483, EPI_ISL_1061484, EPI_ISL_1061485, EPI_ISL_1061486, EPI_ISL_1061487, EPI_ISL_1061488, EPI_ISL_1061489, EPI_ISL_1061490, EPI_ISL_1061491, EPI_ISL_1061492, EPI_ISL_1061493, EPI_ISL_1061494, EPI_ISL_1061495, EPI_ISL_1061496, EPI_ISL_1061497, EPI_ISL_1061498, EPI_ISL_1061499, EPI_ISL_1061500, EPI_ISL_1061501, EPI_ISL_1061502, EPI_ISL_1061503, EPI_ISL_1061504, EPI_ISL_1061505, EPI_ISL_1061506, EPI_ISL_1061507, EPI_ISL_1061508, EPI_ISL_1061509, EPI_ISL_1061510, EPI_ISL_1061511, EPI_ISL_1061512, EPI_ISL_1061513, EPI_ISL_1061514, EPI_ISL_1061515, EPI_ISL_1061516, EPI_ISL_1061517, EPI_ISL_1061518, EPI_ISL_1061519, EPI_ISL_1061520, EPI_ISL_1061521, EPI_ISL_1061522, EPI_ISL_1061523, EPI_ISL_1061524, EPI_ISL_1061525, EPI_ISL_1061526, EPI_ISL_1061527, EPI_ISL_1061528, EPI_ISL_1061529, EPI_ISL_1061530, EPI_ISL_1061531, EPI_ISL_1061532, EPI_ISL_1061533, EPI_ISL_1061534, EPI_ISL_1061535, EPI_ISL_1061536, EPI_ISL_1061537, EPI_ISL_1061538, EPI_ISL_1061539, EPI_ISL_1061540, EPI_ISL_1061541, EPI_ISL_1061542, EPI_ISL_1061543, EPI_ISL_1061544, EPI_ISL_1061545, EPI_ISL_1061546, EPI_ISL_1061547, EPI_ISL_1061548, EPI_ISL_1061549, EPI_ISL_1061550, EPI_ISL_1061551, EPI_ISL_1061552, EPI_ISL_1061553, EPI_ISL_1061554, EPI_ISL_1061555, EPI_ISL_1061556, EPI_ISL_1061557, EPI_ISL_1061558, EPI_ISL_1061559, EPI_ISL_1061560, EPI_ISL_1061561, EPI_ISL_1061562, EPI_ISL_1061563, EPI_ISL_1061564, EPI_ISL_1061565, EPI_ISL_1061566, EPI_ISL_1061567, EPI_ISL_1061568, EPI_ISL_1061569, EPI_ISL_1061570, EPI_ISL_1061571, EPI_ISL_1061572, EPI_ISL_1061573, EPI_ISL_1061574, EPI_ISL_1061575, EPI_ISL_1061576, EPI_ISL_1061577, EPI_ISL_1061578, EPI_ISL_1061579, EPI_ISL_1061580, EPI_ISL_1061581, EPI_ISL_1061582, EPI_ISL_1061583, EPI_ISL_1061584, EPI_ISL_1061586, EPI_ISL_1061587, EPI_ISL_1061588, EPI_ISL_1061589, EPI_ISL_1061590, EPI_ISL_1061591, EPI_ISL_1061592, EPI_ISL_1061593, EPI_ISL_1061594, EPI_ISL_1061595, EPI_ISL_1061596, EPI_ISL_1061597, EPI_ISL_1061598, EPI_ISL_1061599, EPI_ISL_1061600, EPI_ISL_1061601, EPI_ISL_1061602, EPI_ISL_1061603, EPI_ISL_1061604, EPI_ISL_1061605, EPI_ISL_1061606, EPI_ISL_1061607, EPI_ISL_1061608, EPI_ISL_1061609, EPI_ISL_1061610, EPI_ISL_1061611, EPI_ISL_1061612, EPI_ISL_1061613, EPI_ISL_1061614, EPI_ISL_1061615, EPI_ISL_1061616, EPI_ISL_1061617, EPI_ISL_1061618, EPI_ISL_1061619, EPI_ISL_1061620, EPI_ISL_1061621, EPI_ISL_1061622, EPI_ISL_1061623, EPI_ISL_1061624, EPI_ISL_1061625, EPI_ISL_1061626, EPI_ISL_1061627, EPI_ISL_1061628, EPI_ISL_1061629, EPI_ISL_1061630, EPI_ISL_1061631, EPI_ISL_1061632, EPI_ISL_1061633, EPI_ISL_1061634, EPI_ISL_1061635, EPI_ISL_1061636, EPI_ISL_1061637, EPI_ISL_1061638, EPI_ISL_1061639, EPI_ISL_1061640, EPI_ISL_1061641, EPI_ISL_1061642, EPI_ISL_1061643, EPI_ISL_1061644, EPI_ISL_1061645, EPI_ISL_1061646, EPI_ISL_1061647, EPI_ISL_1061648, EPI_ISL_1061649, EPI_ISL_1061650, EPI_ISL_1061651, EPI_ISL_1061652, EPI_ISL_1061653, EPI_ISL_1061654, EPI_ISL_1061655, EPI_ISL_1061656, EPI_ISL_1061657, EPI_ISL_1061658, EPI_ISL_1061659, EPI_ISL_1061660, EPI_ISL_1061662, EPI_ISL_1061663, EPI_ISL_1061664, EPI_ISL_1061665, EPI_ISL_1061666, EPI_ISL_1061667, EPI_ISL_1061668, EPI_ISL_1061669, EPI_ISL_1061670, EPI_ISL_1061671, EPI_ISL_1061672, EPI_ISL_1061673, EPI_ISL_1061674, EPI_ISL_1061676, EPI_ISL_1061677, EPI_ISL_1061678, EPI_ISL_1061679, EPI_ISL_1061680, EPI_ISL_1061681, EPI_ISL_1061682, EPI_ISL_1061683, EPI_ISL_1061684, EPI_ISL_1061685, EPI_ISL_1061686, EPI_ISL_1061688, EPI_ISL_1061689, EPI_ISL_1061690, EPI_ISL_1061691, EPI_ISL_1061692, EPI_ISL_1061693, EPI_ISL_1061694, EPI_ISL_1061695, EPI_ISL_1061696, EPI_ISL_1062412, EPI_ISL_1062416, EPI_ISL_1062419, EPI_ISL_1062429, EPI_ISL_1062435, EPI_ISL_1062438, EPI_ISL_1062442, EPI_ISL_1062443, EPI_ISL_1062452, EPI_ISL_1062453, EPI_ISL_1062462, EPI_ISL_1062463, EPI_ISL_1062467, EPI_ISL_1062468, EPI_ISL_1062476, EPI_ISL_1062482, EPI_ISL_1062483, EPI_ISL_1062485, EPI_ISL_1062491, EPI_ISL_1062500, EPI_ISL_1062507, EPI_ISL_1062519, EPI_ISL_1062520, EPI_ISL_1062528, EPI_ISL_1062532, EPI_ISL_1062536, EPI_ISL_1062537, EPI_ISL_1062547, EPI_ISL_1062550, EPI_ISL_1062553, EPI_ISL_1062556, EPI_ISL_1062557, EPI_ISL_1062561, EPI_ISL_1062565, EPI_ISL_1062574, EPI_ISL_1062578, EPI_ISL_1062586, EPI_ISL_1062592, EPI_ISL_1062597, EPI_ISL_1062602, EPI_ISL_1062607, EPI_ISL_1062608, EPI_ISL_1062609, EPI_ISL_1062611, EPI_ISL_1062615, EPI_ISL_1062618, EPI_ISL_1062622, EPI_ISL_1062623, EPI_ISL_1062627, EPI_ISL_1062629, EPI_ISL_1062642, EPI_ISL_1062648, EPI_ISL_1062649, EPI_ISL_1062657, EPI_ISL_1062658, EPI_ISL_1062662, EPI_ISL_1062664, EPI_ISL_1062665, EPI_ISL_1062672, EPI_ISL_1062677, EPI_ISL_1062683, EPI_ISL_1062690, EPI_ISL_1062698, EPI_ISL_1062699, EPI_ISL_1062705, EPI_ISL_1062708, EPI_ISL_1062725, EPI_ISL_1062730, EPI_ISL_1062748, EPI_ISL_1062763, EPI_ISL_1062765, EPI_ISL_1062767, EPI_ISL_1069993, EPI_ISL_1069996, EPI_ISL_1069999, EPI_ISL_1070000, EPI_ISL_1070007, EPI_ISL_1070009, EPI_ISL_1070013, EPI_ISL_1070014, EPI_ISL_1070015, EPI_ISL_1070021, EPI_ISL_1070024, EPI_ISL_1070026, EPI_ISL_1070028, EPI_ISL_1070030, EPI_ISL_1070032, EPI_ISL_1070033, EPI_ISL_1070040, EPI_ISL_1070041, EPI_ISL_1070043, EPI_ISL_1070044, EPI_ISL_1070046, EPI_ISL_1070049, EPI_ISL_1070056, EPI_ISL_1070057, EPI_ISL_1070059, EPI_ISL_1070062, EPI_ISL_1070063, EPI_ISL_1070065, EPI_ISL_1070066, EPI_ISL_1070067, EPI_ISL_1070076, EPI_ISL_1070079, EPI_ISL_1070080, EPI_ISL_1070086, EPI_ISL_1070089, EPI_ISL_1070091, EPI_ISL_1070092, EPI_ISL_1070096, EPI_ISL_1070101, EPI_ISL_1070104, EPI_ISL_1070105, EPI_ISL_1070110, EPI_ISL_1070112, EPI_ISL_1070114, EPI_ISL_1070117, EPI_ISL_1070119, EPI_ISL_1070120, EPI_ISL_1070123, EPI_ISL_1070126, EPI_ISL_1070127, EPI_ISL_1070128, EPI_ISL_1070130, EPI_ISL_1070211, EPI_ISL_1071372, EPI_ISL_1071373, EPI_ISL_1071388, EPI_ISL_1071393, EPI_ISL_1071396, EPI_ISL_1071399, EPI_ISL_1071419, EPI_ISL_1071438, EPI_ISL_1071441, EPI_ISL_1071444, EPI_ISL_1071445, EPI_ISL_1071450, EPI_ISL_1071463, EPI_ISL_1071466, EPI_ISL_1071499, EPI_ISL_1071506, EPI_ISL_1071536, EPI_ISL_1071538, EPI_ISL_1071555, EPI_ISL_1071565, EPI_ISL_1071571, EPI_ISL_1071582, EPI_ISL_1071601, EPI_ISL_1071625, EPI_ISL_1084474, EPI_ISL_1084477, EPI_ISL_1084483, EPI_ISL_1084486, EPI_ISL_1084489, EPI_ISL_1084494, EPI_ISL_1084497, EPI_ISL_1084499, EPI_ISL_1084512, EPI_ISL_1084513, EPI_ISL_1084514, EPI_ISL_1084516, EPI_ISL_1084518, EPI_ISL_1084522, EPI_ISL_1102774, EPI_ISL_1102814, EPI_ISL_1102829, EPI_ISL_1102871, EPI_ISL_1102886, EPI_ISL_1102944, EPI_ISL_1102948, EPI_ISL_1102955, EPI_ISL_1102960, EPI_ISL_1103007, EPI_ISL_1187089, EPI_ISL_1187102, EPI_ISL_1187125, EPI_ISL_1187135, EPI_ISL_1187144, EPI_ISL_1187179, EPI_ISL_1187194, EPI_ISL_1187250, EPI_ISL_1206056, EPI_ISL_1206078, EPI_ISL_1206084, EPI_ISL_1206096, EPI_ISL_1206108, EPI_ISL_1206111, EPI_ISL_1206114, EPI_ISL_1206185, EPI_ISL_1206199, EPI_ISL_1206201, EPI_ISL_1206206, EPI_ISL_1206218, EPI_ISL_1206228, EPI_ISL_1206252, EPI_ISL_1206399, EPI_ISL_1206486, EPI_ISL_1223165, EPI_ISL_1223175, EPI_ISL_1223177, EPI_ISL_1223180, EPI_ISL_1223196, | | | |

EPI_ISL_1223203, EPI_ISL_1223205, EPI_ISL_1223208, EPI_ISL_1223243, EPI_ISL_1223244, EPI_ISL_1223250, EPI_ISL_1223252, EPI_ISL_1223303, EPI_ISL_1223304, EPI_ISL_1223307, EPI_ISL_1223309, EPI_ISL_1223310, EPI_ISL_1223315, EPI_ISL_1223324, EPI_ISL_1223331, EPI_ISL_1223353, EPI_ISL_1223374, EPI_ISL_1223375, EPI_ISL_1223388, EPI_ISL_1223389, EPI_ISL_1223414, EPI_ISL_1223418, EPI_ISL_1224542, EPI_ISL_1224663, EPI_ISL_1244867, EPI_ISL_1244886, EPI_ISL_1244894, EPI_ISL_1244898, EPI_ISL_1244906, EPI_ISL_1244912, EPI_ISL_1244921, EPI_ISL_1244933, EPI_ISL_1244954, EPI_ISL_1244961, EPI_ISL_1244995, EPI_ISL_1256395, EPI_ISL_1256417, EPI_ISL_1256442, EPI_ISL_1256447, EPI_ISL_1275572, EPI_ISL_1275622, EPI_ISL_1275689, EPI_ISL_1275699, EPI_ISL_1275718, EPI_ISL_1275868, EPI_ISL_1275935, EPI_ISL_1275949, EPI_ISL_1276010, EPI_ISL_1276928, EPI_ISL_1276943, EPI_ISL_1276950, EPI_ISL_1276964, EPI_ISL_1276967, EPI_ISL_1276986, EPI_ISL_1276991, EPI_ISL_1277014, EPI_ISL_1277018, EPI_ISL_1277065, EPI_ISL_1277067, EPI_ISL_1277072, EPI_ISL_1277094, EPI_ISL_1277102, EPI_ISL_1277134, EPI_ISL_1277174, EPI_ISL_1333872, EPI_ISL_1467587, EPI_ISL_1467613, EPI_ISL_1467620, EPI_ISL_1467651, EPI_ISL_1467658, EPI_ISL_1467668, EPI_ISL_1467674, EPI_ISL_1467695, EPI_ISL_1467712, EPI_ISL_1467738, EPI_ISL_1467740, EPI_ISL_1467750, EPI_ISL_1467762, EPI_ISL_1467778, EPI_ISL_1467798, EPI_ISL_1467802

| | | | |
|---|---|---|---|
| see above | Lighthouse Lab in Cambridge | Wellcome Sanger Institute for the COVID-19 Genomics UK (COG-UK) Consortium | Cordelia Langford; David K. Jackson; Dominic Kwiatkowski; Ewan Harrison; Ian Johnston; Jeffrey Barrett; John Sillitoe on behalf of the Wellcome Sanger Institute COVID-19 Surveillance Team; Rob Howes; Roberto Amato; Sonia Goncalves; The Lighthouse Lab in Cambridge and Alex Alderton |

EPI_ISL_1042357, EPI_ISL_1042358, EPI_ISL_1042359, EPI_ISL_1042360, EPI_ISL_1042361, EPI_ISL_1042362, EPI_ISL_1042363, EPI_ISL_1042364, EPI_ISL_1042365, EPI_ISL_1042366, EPI_ISL_1042367, EPI_ISL_1042368, EPI_ISL_1042369, EPI_ISL_1042370, EPI_ISL_1042371, EPI_ISL_1042372, EPI_ISL_1042373, EPI_ISL_1042374, EPI_ISL_1042375, EPI_ISL_1042376, EPI_ISL_1042377, EPI_ISL_1042378, EPI_ISL_1042379, EPI_ISL_1042380, EPI_ISL_1042381, EPI_ISL_1042382, EPI_ISL_1042383, EPI_ISL_1042384, EPI_ISL_1042385, EPI_ISL_1042386, EPI_ISL_1042387, EPI_ISL_1042388, EPI_ISL_1042389, EPI_ISL_1042390, EPI_ISL_1042391, EPI_ISL_1042392, EPI_ISL_1042393, EPI_ISL_1042394, EPI_ISL_1042395, EPI_ISL_1042396, EPI_ISL_1042397, EPI_ISL_1042398, EPI_ISL_1042399, EPI_ISL_1042400, EPI_ISL_1042401, EPI_ISL_1042402, EPI_ISL_1042403, EPI_ISL_1042404, EPI_ISL_1042405, EPI_ISL_1042406, EPI_ISL_1042407, EPI_ISL_1042408, EPI_ISL_1042409, EPI_ISL_1042410, EPI_ISL_1042411, EPI_ISL_1042412, EPI_ISL_1042413, EPI_ISL_1042414, EPI_ISL_1042415, EPI_ISL_1042416, EPI_ISL_1042417, EPI_ISL_1042418, EPI_ISL_1042419, EPI_ISL_1042420, EPI_ISL_1042421, EPI_ISL_1042422, EPI_ISL_1042423, EPI_ISL_1042424, EPI_ISL_1042425, EPI_ISL_1042426, EPI_ISL_1042427, EPI_ISL_1042428, EPI_ISL_1042429, EPI_ISL_1042430, EPI_ISL_1042431, EPI_ISL_1042432, EPI_ISL_1042433, EPI_ISL_1042434, EPI_ISL_1042435, EPI_ISL_1042436, EPI_ISL_1042437, EPI_ISL_1042438, EPI_ISL_1042439, EPI_ISL_1042440, EPI_ISL_1042441, EPI_ISL_1042442, EPI_ISL_1042443, EPI_ISL_1042444, EPI_ISL_1042445, EPI_ISL_1042446, EPI_ISL_1042447, EPI_ISL_1042448, EPI_ISL_1042449, EPI_ISL_1042450, EPI_ISL_1042451, EPI_ISL_1042452, EPI_ISL_1042453, EPI_ISL_1042454, EPI_ISL_1042455, EPI_ISL_1042456, EPI_ISL_1042457, EPI_ISL_1042458, EPI_ISL_1042459, EPI_ISL_1042461, EPI_ISL_1042462, EPI_ISL_1042463, EPI_ISL_1042464, EPI_ISL_1042465, EPI_ISL_1042466, EPI_ISL_1042467, EPI_ISL_1042468, EPI_ISL_1042469, EPI_ISL_1042470, EPI_ISL_1042471, EPI_ISL_1042472, EPI_ISL_1042474, EPI_ISL_1042475, EPI_ISL_1042476, EPI_ISL_1042477, EPI_ISL_1042478, EPI_ISL_1042479, EPI_ISL_1042480, EPI_ISL_1042481, EPI_ISL_1042482, EPI_ISL_1042483, EPI_ISL_1042484, EPI_ISL_1042485, EPI_ISL_1042486, EPI_ISL_1042487, EPI_ISL_1042488, EPI_ISL_1042489, EPI_ISL_1042490, EPI_ISL_1042491, EPI_ISL_1042492, EPI_ISL_1042493, EPI_ISL_1042494, EPI_ISL_1042495, EPI_ISL_1042496, EPI_ISL_1042497, EPI_ISL_1042499, EPI_ISL_1042500, EPI_ISL_1042501, EPI_ISL_1042502, EPI_ISL_1042503, EPI_ISL_1042504, EPI_ISL_1042505, EPI_ISL_1042506, EPI_ISL_1042507, EPI_ISL_1042508, EPI_ISL_1042509, EPI_ISL_1042510, EPI_ISL_1042511, EPI_ISL_1042512, EPI_ISL_1042513, EPI_ISL_1042514, EPI_ISL_1042515, EPI_ISL_1042516, EPI_ISL_1042517, EPI_ISL_1042518, EPI_ISL_1042519, EPI_ISL_1042520, EPI_ISL_1042521, EPI_ISL_1042522, EPI_ISL_1042523, EPI_ISL_1042524, EPI_ISL_1042525, EPI_ISL_1042526, EPI_ISL_1042527, EPI_ISL_1042528, EPI_ISL_1042529, EPI_ISL_1042530, EPI_ISL_1042531, EPI_ISL_1042532, EPI_ISL_1042533, EPI_ISL_1042535, EPI_ISL_1042536, EPI_ISL_1042537, EPI_ISL_1042538, EPI_ISL_1042539, EPI_ISL_1042540, EPI_ISL_1042541, EPI_ISL_1042542, EPI_ISL_1042544, EPI_ISL_1042545, EPI_ISL_1042546, EPI_ISL_1042547, EPI_ISL_1042548, EPI_ISL_1042549, EPI_ISL_1042550, EPI_ISL_1042551, EPI_ISL_1042552, EPI_ISL_1042553, EPI_ISL_1042554, EPI_ISL_1042555, EPI_ISL_1042556, EPI_ISL_1042557, EPI_ISL_1042558, EPI_ISL_1042559, EPI_ISL_1042560, EPI_ISL_1042561, EPI_ISL_1042562, EPI_ISL_1042563, EPI_ISL_1042564, EPI_ISL_1042565, EPI_ISL_1042566, EPI_ISL_1057121, EPI_ISL_1057122, EPI_ISL_1057125, EPI_ISL_1057127, EPI_ISL_1057128, EPI_ISL_1057136, EPI_ISL_1057140, EPI_ISL_1057141, EPI_ISL_1057142, EPI_ISL_1057150, EPI_ISL_1057152, EPI_ISL_1057154, EPI_ISL_1057158, EPI_ISL_1057160, EPI_ISL_1057163, EPI_ISL_1057171, EPI_ISL_1057172, EPI_ISL_1057173, EPI_ISL_1057174, EPI_ISL_1057187, EPI_ISL_1057189, EPI_ISL_1057191, EPI_ISL_1057194, EPI_ISL_1057196, EPI_ISL_1057200, EPI_ISL_1057202, EPI_ISL_1057210, EPI_ISL_1057211, EPI_ISL_1057213, EPI_ISL_1057215, EPI_ISL_1057217, EPI_ISL_1057223, EPI_ISL_1057225, EPI_ISL_1057231, EPI_ISL_1057238, EPI_ISL_1057244, EPI_ISL_1057248, EPI_ISL_1057249, EPI_ISL_1057253, EPI_ISL_1057257, EPI_ISL_1057263, EPI_ISL_1057266, EPI_ISL_1057271, EPI_ISL_1057275, EPI_ISL_1057276, EPI_ISL_1057277, EPI_ISL_1057286, EPI_ISL_1057298, EPI_ISL_1057299, EPI_ISL_1057301, EPI_ISL_1057302, EPI_ISL_1057303, EPI_ISL_1057306, EPI_ISL_1057309, EPI_ISL_1057319, EPI_ISL_1057321, EPI_ISL_1057325, EPI_ISL_1057332, EPI_ISL_1057341, EPI_ISL_1057345, EPI_ISL_1057354, EPI_ISL_1057359, EPI_ISL_1057363, EPI_ISL_1057366, EPI_ISL_1057383, EPI_ISL_1057386, EPI_ISL_1057390, EPI_ISL_1057394, EPI_ISL_1057396, EPI_ISL_1057398, EPI_ISL_1057399, EPI_ISL_1057400, EPI_ISL_1057414, EPI_ISL_1057419, EPI_ISL_1057421, EPI_ISL_1057427, EPI_ISL_1057663, EPI_ISL_1062064, EPI_ISL_1062071, EPI_ISL_1062078, EPI_ISL_1062079, EPI_ISL_1062085, EPI_ISL_1062088, EPI_ISL_1062089, EPI_ISL_1062092, EPI_ISL_1062094, EPI_ISL_1062096, EPI_ISL_1062097, EPI_ISL_1062098, EPI_ISL_1062099, EPI_ISL_1062100, EPI_ISL_1062105, EPI_ISL_1062110, EPI_ISL_1062112, EPI_ISL_1062114, EPI_ISL_1062123, EPI_ISL_1062126, EPI_ISL_1062136, EPI_ISL_1062139, EPI_ISL_1062142, EPI_ISL_1062144, EPI_ISL_1062150, EPI_ISL_1062151, EPI_ISL_1062152, EPI_ISL_1062153, EPI_ISL_1062156, EPI_ISL_1062158, EPI_ISL_1062161, EPI_ISL_1062164, EPI_ISL_1062168, EPI_ISL_1062174, EPI_ISL_1062177, EPI_ISL_1062178, EPI_ISL_1062190, EPI_ISL_1062192, EPI_ISL_1062193, EPI_ISL_1062196, EPI_ISL_1062201, EPI_ISL_1062211, EPI_ISL_1062212, EPI_ISL_1062213, EPI_ISL_1062215, EPI_ISL_1062218, EPI_ISL_1062226, EPI_ISL_1062228, EPI_ISL_1062232, EPI_ISL_1062244, EPI_ISL_1062245, EPI_ISL_1062251, EPI_ISL_1062252, EPI_ISL_1062253, EPI_ISL_1062272, EPI_ISL_1062276, EPI_ISL_1062278, EPI_ISL_1062279, EPI_ISL_1062281, EPI_ISL_1062283, EPI_ISL_1062284, EPI_ISL_1062287, EPI_ISL_1062297, EPI_ISL_1062298, EPI_ISL_1062304, EPI_ISL_1062306, EPI_ISL_1062310, EPI_ISL_1062319, EPI_ISL_1062328, EPI_ISL_1062330, EPI_ISL_1062331, EPI_ISL_1062333, EPI_ISL_1062334, EPI_ISL_1062337, EPI_ISL_1062338, EPI_ISL_1062341, EPI_ISL_1062343, EPI_ISL_1062348, EPI_ISL_1062352, EPI_ISL_1062356, EPI_ISL_1062357, EPI_ISL_1062360, EPI_ISL_1062368, EPI_ISL_1062369, EPI_ISL_1062375, EPI_ISL_1062380, EPI_ISL_1062390, EPI_ISL_1062393, EPI_ISL_1062398, EPI_ISL_1062399, EPI_ISL_1062405, EPI_ISL_1062406, EPI_ISL_1062407, EPI_ISL_1062409, EPI_ISL_1062413, EPI_ISL_1062421, EPI_ISL_1062422, EPI_ISL_1062424, EPI_ISL_1062425, EPI_ISL_1062426, EPI_ISL_1062427, EPI_ISL_1062428, EPI_ISL_1062430, EPI_ISL_1062431, EPI_ISL_1062432, EPI_ISL_1062433, EPI_ISL_1062434, EPI_ISL_1062437, EPI_ISL_1062440, EPI_ISL_1062441, EPI_ISL_1062444, EPI_ISL_1062445, EPI_ISL_1062446, EPI_ISL_1062447, EPI_ISL_1062448, EPI_ISL_1062449, EPI_ISL_1062451, EPI_ISL_1062454, EPI_ISL_1062455, EPI_ISL_1062458, EPI_ISL_1062459, EPI_ISL_1062461, EPI_ISL_1062466, EPI_ISL_1062469, EPI_ISL_1062470, EPI_ISL_1062471, EPI_ISL_1062472, EPI_ISL_1062474, EPI_ISL_1062479, EPI_ISL_1062480, EPI_ISL_1062481, EPI_ISL_1062484, EPI_ISL_1062486, EPI_ISL_1062487, EPI_ISL_1062488, EPI_ISL_1062489, EPI_ISL_1062490, EPI_ISL_1062492, EPI_ISL_1062493, EPI_ISL_1062494, EPI_ISL_1062497, EPI_ISL_1062499, EPI_ISL_1062501, EPI_ISL_1062502, EPI_ISL_1062505, EPI_ISL_1062506, EPI_ISL_1062510, EPI_ISL_1062511, EPI_ISL_1062512, EPI_ISL_1062513, EPI_ISL_1062514, EPI_ISL_1062515, EPI_ISL_1062516, EPI_ISL_1062517, EPI_ISL_1062518, EPI_ISL_1062521, EPI_ISL_1062523, EPI_ISL_1062524, EPI_ISL_1062525, EPI_ISL_1062526, EPI_ISL_1062527, EPI_ISL_1062530, EPI_ISL_1062531, EPI_ISL_1062533, EPI_ISL_1062534, EPI_ISL_1062538, EPI_ISL_1062543, EPI_ISL_1062544, EPI_ISL_1062545, EPI_ISL_1062546, EPI_ISL_1062549, EPI_ISL_1062551, EPI_ISL_1062555, EPI_ISL_1062558, EPI_ISL_1062559, EPI_ISL_1062560, EPI_ISL_1062563, EPI_ISL_1062564, EPI_ISL_1062566, EPI_ISL_1062567, EPI_ISL_1062568, EPI_ISL_1062570, EPI_ISL_1062571, EPI_ISL_1062575, EPI_ISL_1062576, EPI_ISL_1062577, EPI_ISL_1062579, EPI_ISL_1062580, EPI_ISL_1062581, EPI_ISL_1062582, EPI_ISL_1062583, EPI_ISL_1062585, EPI_ISL_1062587, EPI_ISL_1062588, EPI_ISL_1062590, EPI_ISL_1062591, EPI_ISL_1062594, EPI_ISL_1062595, EPI_ISL_1062596, EPI_ISL_1062598, EPI_ISL_1062601, EPI_ISL_1062603, EPI_ISL_1062604, EPI_ISL_1062605, EPI_ISL_1062610, EPI_ISL_1062612, EPI_ISL_1062616, EPI_ISL_1062619, EPI_ISL_1062620, EPI_ISL_1062621, EPI_ISL_1062628, EPI_ISL_1062630, EPI_ISL_1062631, EPI_ISL_1062633, EPI_ISL_1062634, EPI_ISL_1062635, EPI_ISL_1062636, EPI_ISL_1062637, EPI_ISL_1062639, EPI_ISL_1062646, EPI_ISL_1062650, EPI_ISL_1062652, EPI_ISL_1062653, EPI_ISL_1062654, EPI_ISL_1062655, EPI_ISL_1062656, EPI_ISL_1062659, EPI_ISL_1062660, EPI_ISL_1062661, EPI_ISL_1062663, EPI_ISL_1062666, EPI_ISL_1062668, EPI_ISL_1062669, EPI_ISL_1062670, EPI_ISL_1062671, EPI_ISL_1062673, EPI_ISL_1062674, EPI_ISL_1062676, EPI_ISL_1062678, EPI_ISL_1062680, EPI_ISL_1062681, EPI_ISL_1062682, EPI_ISL_1062684, EPI_ISL_1062685, EPI_ISL_1062687, EPI_ISL_1062688, EPI_ISL_1062689, EPI_ISL_1062691, EPI_ISL_1062693, EPI_ISL_1062696, EPI_ISL_1062697, EPI_ISL_1062700, EPI_ISL_1062701, EPI_ISL_1062702, EPI_ISL_1062703, EPI_ISL_1062704, EPI_ISL_1062706, EPI_ISL_1062709, EPI_ISL_1062710, EPI_ISL_1062711, EPI_ISL_1062712, EPI_ISL_1062713, EPI_ISL_1062714, EPI_ISL_1062715, EPI_ISL_1062716, EPI_ISL_1062717, EPI_ISL_1062718, EPI_ISL_1062719, EPI_ISL_1062720, EPI_ISL_1062721, EPI_ISL_1062722, EPI_ISL_1062723, EPI_ISL_1062724, EPI_ISL_1062726, EPI_ISL_1062727, EPI_ISL_1062728, EPI_ISL_1062729, EPI_ISL_1062731, EPI_ISL_1062732, EPI_ISL_1062733, EPI_ISL_1062735, EPI_ISL_1062736, EPI_ISL_1062737, EPI_ISL_1062738, EPI_ISL_1062739, EPI_ISL_1062744, EPI_ISL_1062745, EPI_ISL_1062746, EPI_ISL_1062749, EPI_ISL_1062750, EPI_ISL_1062751, EPI_ISL_1062752, EPI_ISL_1062754, EPI_ISL_1062756, EPI_ISL_1062757, EPI_ISL_1062758, EPI_ISL_1062759, EPI_ISL_1062760, EPI_ISL_1062761, EPI_ISL_1062762, EPI_ISL_1062764, EPI_ISL_1062766, EPI_ISL_1062768, EPI_ISL_1062770, EPI_ISL_1062771, EPI_ISL_1062772, EPI_ISL_1062773, EPI_ISL_1062774, EPI_ISL_1062775, EPI_ISL_1062776, EPI_ISL_1062777, EPI_ISL_1062778, EPI_ISL_1062779, EPI_ISL_1062780, EPI_ISL_1062781, EPI_ISL_1062782, EPI_ISL_1062783, EPI_ISL_1062784, EPI_ISL_1062785, EPI_ISL_1062786, EPI_ISL_1062787, EPI_ISL_1062788, EPI_ISL_1062789, EPI_ISL_1062790, EPI_ISL_1062792, EPI_ISL_1062793, EPI_ISL_1062794, EPI_ISL_1062796, EPI_ISL_1062797, EPI_ISL_1062798, EPI_ISL_1062799, EPI_ISL_1062800, EPI_ISL_1062801, EPI_ISL_1062802, EPI_ISL_1062803, EPI_ISL_1062804, EPI_ISL_1062806, EPI_ISL_1062807, EPI_ISL_1062808, EPI_ISL_1062809, EPI_ISL_1062810, EPI_ISL_1062811, EPI_ISL_1062813, EPI_ISL_1062814, EPI_ISL_1062815, EPI_ISL_1062816, EPI_ISL_1062817, EPI_ISL_1062818, EPI_ISL_1062819, EPI_ISL_1062820, EPI_ISL_1062821, EPI_ISL_1062822, EPI_ISL_1062823, EPI_ISL_1062824, EPI_ISL_1062825, EPI_ISL_1062827, EPI_ISL_1062828, EPI_ISL_1062829, EPI_ISL_1062830, EPI_ISL_1062831, EPI_ISL_1062832, EPI_ISL_1062833, EPI_ISL_1062834, EPI_ISL_1062835, EPI_ISL_1062836, EPI_ISL_1062837, EPI_ISL_1062838, EPI_ISL_1062839, EPI_ISL_1062840, EPI_ISL_1062841, EPI_ISL_1062842, EPI_ISL_1062843, EPI_ISL_1062844, EPI_ISL_1062845, EPI_ISL_1062846, EPI_ISL_1062847, EPI_ISL_1062849, EPI_ISL_1062851, EPI_ISL_1062852, EPI_ISL_1062853, EPI_ISL_1062854, EPI_ISL_1062855, EPI_ISL_1062856, EPI_ISL_1062859, EPI_ISL_1062860, EPI_ISL_1062861, EPI_ISL_1062862, EPI_ISL_1062864, EPI_ISL_1062865, EPI_ISL_1062866, EPI_ISL_1062867, EPI_ISL_1062869, EPI_ISL_1062870, EPI_ISL_1062871, EPI_ISL_1062873, EPI_ISL_1062874, EPI_ISL_1062876, EPI_ISL_1062877, EPI_ISL_1062878, EPI_ISL_1062879, EPI_ISL_1062880, EPI_ISL_1062881, EPI_ISL_1062882, EPI_ISL_1062883, EPI_ISL_1062884, EPI_ISL_1062885, EPI_ISL_1062886, EPI_ISL_1062887, EPI_ISL_1062888, EPI_ISL_1062889, EPI_ISL_1062890, EPI_ISL_1062891, EPI_ISL_1062892, EPI_ISL_1062893, EPI_ISL_1062894, EPI_ISL_1062895, EPI_ISL_1062896, EPI_ISL_1062898, EPI_ISL_1062899, EPI_ISL_1062900, EPI_ISL_1062901, EPI_ISL_1062902, EPI_ISL_1062903, EPI_ISL_1062904, EPI_ISL_1062905, EPI_ISL_1062906, EPI_ISL_1062907, EPI_ISL_1062908, EPI_ISL_1062909, EPI_ISL_1062910, EPI_ISL_1062911, EPI_ISL_1062912, EPI_ISL_1062913, EPI_ISL_1062916, EPI_ISL_1062917, EPI_ISL_1062918, EPI_ISL_1062920, EPI_ISL_1062921, EPI_ISL_1062922, EPI_ISL_1062923, EPI_ISL_1062924, EPI_ISL_1062925, EPI_ISL_1062926, EPI_ISL_1062927, EPI_ISL_1062929, EPI_ISL_1062930, EPI_ISL_1062932, EPI_ISL_1062933, EPI_ISL_1062934, EPI_ISL_1062937, EPI_ISL_1062938, EPI_ISL_1062939, EPI_ISL_1062940, EPI_ISL_1062941, EPI_ISL_1062942, EPI_ISL_1062943, EPI_ISL_1062944, EPI_ISL_1062945, EPI_ISL_1062946, EPI_ISL_1062947, EPI_ISL_1062948, EPI_ISL_1062949, EPI_ISL_1062950, EPI_ISL_1062951, EPI_ISL_1062952, EPI_ISL_1062953, EPI_ISL_1062955, EPI_ISL_1062956, EPI_ISL_1062958, EPI_ISL_1062959, EPI_ISL_1062960, EPI_ISL_1062961, EPI_ISL_1062962, EPI_ISL_1062963, EPI_ISL_1062964, EPI_ISL_1062965, EPI_ISL_1062966, EPI_ISL_1062967, EPI_ISL_1062968, EPI_ISL_1062969, EPI_ISL_1062970, EPI_ISL_1062971, EPI_ISL_1062972, EPI_ISL_1062973, EPI_ISL_1062974, EPI_ISL_1062976, EPI_ISL_1062977, EPI_ISL_1062978, EPI_ISL_1062980, EPI_ISL_1062981, EPI_ISL_1062982, EPI_ISL_1062983, EPI_ISL_1062984, EPI_ISL_1062985, EPI_ISL_1062986, EPI_ISL_1062987, EPI_ISL_1062988, EPI_ISL_1062989, EPI_ISL_1062990, EPI_ISL_1062991, EPI_ISL_1062992, EPI_ISL_1062993, EPI_ISL_1062994, EPI_ISL_1062995, EPI_ISL_1062996, EPI_ISL_1062997, EPI_ISL_1062998, EPI_ISL_1062999, EPI_ISL_1063000, EPI_ISL_1063001, EPI_ISL_1063002, EPI_ISL_1063003, EPI_ISL_1063004, EPI_ISL_1063005, EPI_ISL_1063006, EPI_ISL_1063007, EPI_ISL_1063008, EPI_ISL_1063010, EPI_ISL_1063011, EPI_ISL_1063012, EPI_ISL_1063013, EPI_ISL_1063014, EPI_ISL_1063015, EPI_ISL_1063053, EPI_ISL_1063054, EPI_ISL_1063055, EPI_ISL_1063056, EPI_ISL_1063057, EPI_ISL_1063059, EPI_ISL_1063060, EPI_ISL_1063061, EPI_ISL_1063062, EPI_ISL_1063064, EPI_ISL_1063065, EPI_ISL_1063066, EPI_ISL_1063067, EPI_ISL_1063068, EPI_ISL_1063069, EPI_ISL_1063070, EPI_ISL_1063071, EPI_ISL_1063072, EPI_ISL_1063073, EPI_ISL_1063074, EPI_ISL_1063075, EPI_ISL_1063076, EPI_ISL_1063077, EPI_ISL_1063078, EPI_ISL_1063079, EPI_ISL_1063080, EPI_ISL_1063081, EPI_ISL_1063083, EPI_ISL_1063084, EPI_ISL_1063085, EPI_ISL_1063086, EPI_ISL_1063088, EPI_ISL_1063089, EPI_ISL_1063090, EPI_ISL_1063091, EPI_ISL_1063092, EPI_ISL_1063093, EPI_ISL_1063094, EPI_ISL_1063095, EPI_ISL_1063096, EPI_ISL_1063097, EPI_ISL_1063098, EPI_ISL_1063099, EPI_ISL_1063100, EPI_ISL_1063102, EPI_ISL_1063103, EPI_ISL_1063104, EPI_ISL_1063105, EPI_ISL_1063106, EPI_ISL_1063107, EPI_ISL_1063108, EPI_ISL_1063109, EPI_ISL_1063110, EPI_ISL_1063111, EPI_ISL_1063113, EPI_ISL_1063114, EPI_ISL_1063115, EPI_ISL_1063116, EPI_ISL_1063117, EPI_ISL_1063118, EPI_ISL_1063119, EPI_ISL_1063121, EPI_ISL_1063122, EPI_ISL_1063123, EPI_ISL_1063124, EPI_ISL_1063125, EPI_ISL_1063126, EPI_ISL_1063128, EPI_ISL_1063129, EPI_ISL_1063130, EPI_ISL_1063131, EPI_ISL_1063132, EPI_ISL_1063133, EPI_ISL_1063134, EPI_ISL_1063135, EPI_ISL_1063136, EPI_ISL_1063137, EPI_ISL_1063138, EPI_ISL_1063139, EPI_ISL_1063140, EPI_ISL_1063141, EPI_ISL_1063142, EPI_ISL_1063143, EPI_ISL_1063144, EPI_ISL_1063145, EPI_ISL_1063146, EPI_ISL_1063147, EPI_ISL_1063148, EPI_ISL_1063149, EPI_ISL_1063150, EPI_ISL_1063151, EPI_ISL_1063152, EPI_ISL_1063153, EPI_ISL_1063154, EPI_ISL_1063155, EPI_ISL_1063156, EPI_ISL_1063157, EPI_ISL_1063158, EPI_ISL_1063159, EPI_ISL_1063160, EPI_ISL_1063161, EPI_ISL_1063163, EPI_ISL_1063164, EPI_ISL_1063165, EPI_ISL_1063166, EPI_ISL_1063167, EPI_ISL_1063168, EPI_ISL_1063169, EPI_ISL_1063170, EPI_ISL_1063171, EPI_ISL_1063172, EPI_ISL_1063173, EPI_ISL_1063174, EPI_ISL_1063175, EPI_ISL_1063176, EPI_ISL_1063177, EPI_ISL_1063178, EPI_ISL_1063179, EPI_ISL_1063180, EPI_ISL_1063181, EPI_ISL_1063182, EPI_ISL_1063183, EPI_ISL_1063184, EPI_ISL_1063185, EPI_ISL_1063186, EPI_ISL_1063187, EPI_ISL_1069541, EPI_ISL_1069561, EPI_ISL_1069581, EPI_ISL_1070192, EPI_ISL_1070204, EPI_ISL_1070206, EPI_ISL_1070297, EPI_ISL_1070330, EPI_ISL_1071428, EPI_ISL_1072345, EPI_ISL_1072354, EPI_ISL_1072368, EPI_ISL_1072369, EPI_ISL_1072380, EPI_ISL_1072399, EPI_ISL_1072408, EPI_ISL_1072412, EPI_ISL_1072413, EPI_ISL_1072422, EPI_ISL_1072432, EPI_ISL_1072439, EPI_ISL_1072456, EPI_ISL_1072459, EPI_ISL_1072466, EPI_ISL_1072468, EPI_ISL_1072471, EPI_ISL_1072478, EPI_ISL_1072482, EPI_ISL_1072497, EPI_ISL_1072511, EPI_ISL_1072512, EPI_ISL_1072515, EPI_ISL_1072530, EPI_ISL_1072553, EPI_ISL_1072571, EPI_ISL_1072572, EPI_ISL_1072584, EPI_ISL_1072591, EPI_ISL_1072594, EPI_ISL_1072597, EPI_ISL_1072601, EPI_ISL_1084203, EPI_ISL_1084272, EPI_ISL_1084273, EPI_ISL_1084296, EPI_ISL_1084298, EPI_ISL_1084321, EPI_ISL_1084332, EPI_ISL_1084353, EPI_ISL_1084377, EPI_ISL_1084415, EPI_ISL_1084416, EPI_ISL_1084448, EPI_ISL_1084470, EPI_ISL_1084471, EPI_ISL_1084472, EPI_ISL_1084473, EPI_ISL_1084475, EPI_ISL_1084476, EPI_ISL_1084480, EPI_ISL_1084481, EPI_ISL_1084482, EPI_ISL_1084484, EPI_ISL_1084485, EPI_ISL_1084487, EPI_ISL_1084488, EPI_ISL_1084491, EPI_ISL_1084492, EPI_ISL_1084493, EPI_ISL_1084495, EPI_ISL_1084496, EPI_ISL_1084500, EPI_ISL_1084502, EPI_ISL_1084503, EPI_ISL_1084504, EPI_ISL_1084505, EPI_ISL_1084506, EPI_ISL_1084507, EPI_ISL_1084508, EPI_ISL_1084509, EPI_ISL_1084510, EPI_ISL_1084511, EPI_ISL_1084515, EPI_ISL_1084517, EPI_ISL_1084519, EPI_ISL_1084520, EPI_ISL_1084521, EPI_ISL_1084524, EPI_ISL_1102165, EPI_ISL_1103206, EPI_ISL_1103209, EPI_ISL_1103210, EPI_ISL_1103250, EPI_ISL_1103253, EPI_ISL_1103258, EPI_ISL_1103292, EPI_ISL_1103365, EPI_ISL_1103366, EPI_ISL_1103367, EPI_ISL_1103368, EPI_ISL_1103369, EPI_ISL_1103370, EPI_ISL_1103371, EPI_ISL_1103372, EPI_ISL_1103373, EPI_ISL_1103374, EPI_ISL_1103375, EPI_ISL_1103376, EPI_ISL_1103377, EPI_ISL_1103378, EPI_ISL_1103379, EPI_ISL_1103380, EPI_ISL_1103381, EPI_ISL_1103382, EPI_ISL_1103383, EPI_ISL_1103384, EPI_ISL_1103385, EPI_ISL_1103386, EPI_ISL_1103387, EPI_ISL_1103388, EPI_ISL_1103389, EPI_ISL_1103390, EPI_ISL_1103392, EPI_ISL_1103393, EPI_ISL_1103395, EPI_ISL_1103396, EPI_ISL_1103397, EPI_ISL_1103398, EPI_ISL_1103399, EPI_ISL_1103400, EPI_ISL_1103401, EPI_ISL_1103402, EPI_ISL_1103403, EPI_ISL_1103404, EPI_ISL_1103405, EPI_ISL_1103406, EPI_ISL_1103407, EPI_ISL_1103408, EPI_ISL_1103409, EPI_ISL_1103410, EPI_ISL_1103411, EPI_ISL_1103412, EPI_ISL_1103413, EPI_ISL_1103414, EPI_ISL_1103415, EPI_ISL_1103416, EPI_ISL_1103417, EPI_ISL_1103418, EPI_ISL_1103419, EPI_ISL_1103420, EPI_ISL_1103421, EPI_ISL_1103422, EPI_ISL_1103423, EPI_ISL_1103424, EPI_ISL_1103425, EPI_ISL_1103426, EPI_ISL_1103427, EPI_ISL_1103428, EPI_ISL_1103429, EPI_ISL_1103430, EPI_ISL_1103431, EPI_ISL_1103432, EPI_ISL_1103433, EPI_ISL_1103434, EPI_ISL_1103435, EPI_ISL_1103436, EPI_ISL_1103437, EPI_ISL_1103438, EPI_ISL_1103439, EPI_ISL_1103440, EPI_ISL_1103441, EPI_ISL_1103443, EPI_ISL_1103444, EPI_ISL_1103445, EPI_ISL_1103446, EPI_ISL_1103447, EPI_ISL_1103448, EPI_ISL_1103449, EPI_ISL_1103450, EPI_ISL_1103451, EPI_ISL_1103452, EPI_ISL_1103453, EPI_ISL_1103454, EPI_ISL_1103455, EPI_ISL_1103456, EPI_ISL_1103457, EPI_ISL_1103458, EPI_ISL_1103459, EPI_ISL_1103460, EPI_ISL_1103461, EPI_ISL_1103462, EPI_ISL_1103463, EPI_ISL_1103464, EPI_ISL_1103465, EPI_ISL_1103466, EPI_ISL_1103467, EPI_ISL_1103468, EPI_ISL_1103469, EPI_ISL_1103470, EPI_ISL_1103471, EPI_ISL_1103472, EPI_ISL_1103473, EPI_ISL_1103474, EPI_ISL_1103475, EPI_ISL_1103476, EPI_ISL_1103477, EPI_ISL_1103478, EPI_ISL_1103479, EPI_ISL_1103480, EPI_ISL_1103481, EPI_ISL_1103482, EPI_ISL_1103483, EPI_ISL_1103484, EPI_ISL_1103485, EPI_ISL_1103486, EPI_ISL_1103487, EPI_ISL_1103488, EPI_ISL_1103489, EPI_ISL_1103490, EPI_ISL_1103491, EPI_ISL_1103492, EPI_ISL_1103493, EPI_ISL_1103494, EPI_ISL_1103495, EPI_ISL_1103496, EPI_ISL_1103497, EPI_ISL_1103498, EPI_ISL_1103499, EPI_ISL_1103500, EPI_ISL_1103501, EPI_ISL_1103502, EPI_ISL_1103503, EPI_ISL_1103504, EPI_ISL_1103505, EPI_ISL_1103506, EPI_ISL_1103507, EPI_ISL_1103508, EPI_ISL_1103509, EPI_ISL_1103510, EPI_ISL_1103511, EPI_ISL_1103512, EPI_ISL_1103513, EPI_ISL_1103514, EPI_ISL_1103515, EPI_ISL_1103516, EPI_ISL_1103517, EPI_ISL_1103518, EPI_ISL_1103519, EPI_ISL_1103520, EPI_ISL_1103521, EPI_ISL_1103522, EPI_ISL_1103523, EPI_ISL_1103524, EPI_ISL_1103525, EPI_ISL_1103526, EPI_ISL_1103527, EPI_ISL_1103528, EPI_ISL_1103529, EPI_ISL_1103530, EPI_ISL_1103531, EPI_ISL_1103532, EPI_ISL_1103533, EPI_ISL_1103534, EPI_ISL_1103535, EPI_ISL_1103536, EPI_ISL_1103537, EPI_ISL_1103538, EPI_ISL_1103539, EPI_ISL_1103540, EPI_ISL_1103541, EPI_ISL_1103542, EPI_ISL_1190134, EPI_ISL_1208144, EPI_ISL_1246692, EPI_ISL_2122934

| | | | |
|---|---|---|---|
| see above | Lighthouse Lab in Glasgow | Wellcome Sanger Institute for the COVID-19 Genomics UK (COG-UK) Consortium | Anna Dominiczak and Alex Alderton; Carol Clugston; Cordelia Langford; David Gray; David K. Jackson; Dominic Kwiatkowski; Ewan Harrison; Harper VanSteenhouse; Ian Johnston; Jeffrey Barrett; John Sillitoe on behalf of the Wellcome Sanger Institute COVID-19 Surveillance Team; Roberto Amato; Sonia Goncalves; Yumi Kasai |

EPI_ISL_1062060, EPI_ISL_1062062, EPI_ISL_1062065, EPI_ISL_1062066, EPI_ISL_1062068, EPI_ISL_1062069, EPI_ISL_1062070, EPI_ISL_1062072, EPI_ISL_1062074, EPI_ISL_1062075, EPI_ISL_1062076, EPI_ISL_1062077, EPI_ISL_1062081, EPI_ISL_1062083, EPI_ISL_1062090, EPI_ISL_1062091, EPI_ISL_1062093, EPI_ISL_1062102, EPI_ISL_1062104, EPI_ISL_1062107, EPI_ISL_1062109, EPI_ISL_1062111, EPI_ISL_1062116, EPI_ISL_1062117, EPI_ISL_1062121, EPI_ISL_1062124, EPI_ISL_1062128, EPI_ISL_1062129, EPI_ISL_1062133, EPI_ISL_1062134, EPI_ISL_1062137, EPI_ISL_1062141, EPI_ISL_1062143, EPI_ISL_1062145, EPI_ISL_1062146, EPI_ISL_1062148, EPI_ISL_1062149, EPI_ISL_1062154, EPI_ISL_1062155, EPI_ISL_1062157, EPI_ISL_1062160, EPI_ISL_1062166, EPI_ISL_1062167, EPI_ISL_1062169, EPI_ISL_1062175, EPI_ISL_1062176, EPI_ISL_1062179, EPI_ISL_1062180, EPI_ISL_1062181, EPI_ISL_1062182, EPI_ISL_1062183, EPI_ISL_1062184, EPI_ISL_1062186, EPI_ISL_1062187, EPI_ISL_1062188, EPI_ISL_1062189, EPI_ISL_1062191, EPI_ISL_1062194, EPI_ISL_1062195, EPI_ISL_1062197, EPI_ISL_1062198, EPI_ISL_1062199, EPI_ISL_1062204, EPI_ISL_1062206, EPI_ISL_1062207, EPI_ISL_1062208, EPI_ISL_1062209, EPI_ISL_1062210, EPI_ISL_1062214, EPI_ISL_1062217, EPI_ISL_1062220, EPI_ISL_1062221, EPI_ISL_1062223, EPI_ISL_1062224, EPI_ISL_1062227, EPI_ISL_1062229, EPI_ISL_1062230, EPI_ISL_1062231, EPI_ISL_1062233, EPI_ISL_1062235, EPI_ISL_1062236, EPI_ISL_1062237, EPI_ISL_1062238, EPI_ISL_1062240, EPI_ISL_1062242, EPI_ISL_1062243, EPI_ISL_1062246, EPI_ISL_1062247, EPI_ISL_1062249, EPI_ISL_1062250, EPI_ISL_1062256, EPI_ISL_1062257, EPI_ISL_1062259, EPI_ISL_1062260, EPI_ISL_1062261, EPI_ISL_1062263, EPI_ISL_1062264, EPI_ISL_1062265, EPI_ISL_1062266, EPI_ISL_1062267, EPI_ISL_1062268, EPI_ISL_1062270, EPI_ISL_1062271, EPI_ISL_1062274, EPI_ISL_1062275, EPI_ISL_1062282, EPI_ISL_1062286, EPI_ISL_1062292, EPI_ISL_1062293, EPI_ISL_1062295, EPI_ISL_1062296, EPI_ISL_1062299, EPI_ISL_1062303, EPI_ISL_1062309, EPI_ISL_1062311, EPI_ISL_1062312, EPI_ISL_1062317, EPI_ISL_1062318, EPI_ISL_1062320, EPI_ISL_1062321, EPI_ISL_1062322, EPI_ISL_1062325, EPI_ISL_1062326, EPI_ISL_1062327, EPI_ISL_1062329, EPI_ISL_1062332, EPI_ISL_1062335, EPI_ISL_1062336, EPI_ISL_1062339, EPI_ISL_1062342, EPI_ISL_1062344, EPI_ISL_1062345, EPI_ISL_1062347, EPI_ISL_1062351, EPI_ISL_1062353, EPI_ISL_1062354, EPI_ISL_1062358, EPI_ISL_1062359, EPI_ISL_1062361, EPI_ISL_1062362, EPI_ISL_1062365, EPI_ISL_1062366, EPI_ISL_1062371, EPI_ISL_1062372, EPI_ISL_1062374, EPI_ISL_1062376, EPI_ISL_1062377, EPI_ISL_1062378, EPI_ISL_1062379, EPI_ISL_1062381, EPI_ISL_1062382, EPI_ISL_1062384, EPI_ISL_1062385, EPI_ISL_1062386, EPI_ISL_1062387, EPI_ISL_1062388, EPI_ISL_1062389, EPI_ISL_1062392, EPI_ISL_1062394, EPI_ISL_1062395, EPI_ISL_1062397, EPI_ISL_1062400, EPI_ISL_1062401, EPI_ISL_1062402, EPI_ISL_1062403, EPI_ISL_1062423, EPI_ISL_1062436, EPI_ISL_1062465, EPI_ISL_1062475, EPI_ISL_1062478, EPI_ISL_1062539, EPI_ISL_1062542, EPI_ISL_1062562, EPI_ISL_1062589, EPI_ISL_1062599, EPI_ISL_1062600, EPI_ISL_1062614, EPI_ISL_1062626, EPI_ISL_1062641, EPI_ISL_1062644, EPI_ISL_1062645, EPI_ISL_1062679, EPI_ISL_1062686, EPI_ISL_1062695, EPI_ISL_1062742, EPI_ISL_1069400, EPI_ISL_1069406, EPI_ISL_1069408, EPI_ISL_1069411, EPI_ISL_1069412, EPI_ISL_1069413, EPI_ISL_1069414, EPI_ISL_1069416, EPI_ISL_1069417, EPI_ISL_1069418, EPI_ISL_1069419, EPI_ISL_1069421, EPI_ISL_1069423, EPI_ISL_1069426, EPI_ISL_1069427, EPI_ISL_1069428, EPI_ISL_1069429, EPI_ISL_1069430, EPI_ISL_1069431, EPI_ISL_1069432, EPI_ISL_1069433, EPI_ISL_1069434, EPI_ISL_1069435, EPI_ISL_1069436, EPI_ISL_1069438, EPI_ISL_1069439, EPI_ISL_1069440, EPI_ISL_1069443, EPI_ISL_1069444, EPI_ISL_1069445, EPI_ISL_1069446, EPI_ISL_1069447, EPI_ISL_1069448, EPI_ISL_1069451, EPI_ISL_1069452, EPI_ISL_1069454, EPI_ISL_1069455, EPI_ISL_1069458, EPI_ISL_1069462, EPI_ISL_1069463, EPI_ISL_1069465, EPI_ISL_1069466, EPI_ISL_1069467, EPI_ISL_1069469, EPI_ISL_1069470, EPI_ISL_1069472, EPI_ISL_1069473, EPI_ISL_1069474, EPI_ISL_1069475, EPI_ISL_1069476, EPI_ISL_1069477, EPI_ISL_1069478, EPI_ISL_1069479, EPI_ISL_1069480, EPI_ISL_1069481, EPI_ISL_1069482, EPI_ISL_1069485, EPI_ISL_1069486, EPI_ISL_1069488, EPI_ISL_1069489, EPI_ISL_1069490, EPI_ISL_1069492, EPI_ISL_1069493, EPI_ISL_1069494, EPI_ISL_1069497, EPI_ISL_1069499, EPI_ISL_1069500, EPI_ISL_1069503, EPI_ISL_1069504, EPI_ISL_1069508, EPI_ISL_1069509, EPI_ISL_1069510, EPI_ISL_1069511, EPI_ISL_1069512, EPI_ISL_1069519, EPI_ISL_1069520, EPI_ISL_1069522, EPI_ISL_1069524, EPI_ISL_1069525, EPI_ISL_1069526, EPI_ISL_1069527, EPI_ISL_1069528, EPI_ISL_1069530, EPI_ISL_1069532, EPI_ISL_1069533, EPI_ISL_1069535, EPI_ISL_1069539, EPI_ISL_1069540, EPI_ISL_1069542, EPI_ISL_1069543, EPI_ISL_1069544, EPI_ISL_1069546, EPI_ISL_1069547, EPI_ISL_1069549, EPI_ISL_1069550, EPI_ISL_1069551, EPI_ISL_1069552, EPI_ISL_1069553, EPI_ISL_1069554, EPI_ISL_1069556, EPI_ISL_1069559, EPI_ISL_1069560, EPI_ISL_1069563, EPI_ISL_1069565, EPI_ISL_1069566, EPI_ISL_1069567, EPI_ISL_1069568, EPI_ISL_1069569, EPI_ISL_1069572, EPI_ISL_1069574, EPI_ISL_1069578, EPI_ISL_1069580, EPI_ISL_1069582, EPI_ISL_1069584, EPI_ISL_1069585, EPI_ISL_1069586, EPI_ISL_1069588, EPI_ISL_1069591, EPI_ISL_1069592, EPI_ISL_1069593, EPI_ISL_1069594, EPI_ISL_1069595, EPI_ISL_1069597, EPI_ISL_1069598, EPI_ISL_1069599, EPI_ISL_1069601, EPI_ISL_1069602, EPI_ISL_1069605, EPI_ISL_1069610, EPI_ISL_1069611, EPI_ISL_1069612, EPI_ISL_1069613, EPI_ISL_1069614, EPI_ISL_1069616, EPI_ISL_1069617, EPI_ISL_1069618, EPI_ISL_1069619, EPI_ISL_1069620, EPI_ISL_1069622, EPI_ISL_1069623, EPI_ISL_1069624, EPI_ISL_1069625, EPI_ISL_1069626, EPI_ISL_1069627, EPI_ISL_1069630, EPI_ISL_1069631, EPI_ISL_1069632, EPI_ISL_1069633, EPI_ISL_1069634, EPI_ISL_1069635, EPI_ISL_1069639, EPI_ISL_1069641, EPI_ISL_1069645, EPI_ISL_1069647, EPI_ISL_1069648, EPI_ISL_1069651, EPI_ISL_1069652, EPI_ISL_1069653, EPI_ISL_1069654, EPI_ISL_1069655, EPI_ISL_1069657, EPI_ISL_1069660, EPI_ISL_1069665, EPI_ISL_1069666, EPI_ISL_1069667, EPI_ISL_1069668, EPI_ISL_1069669, EPI_ISL_1069670, EPI_ISL_1069671, EPI_ISL_1069672, EPI_ISL_1069675, EPI_ISL_1069677, EPI_ISL_1069678, EPI_ISL_1069679, EPI_ISL_1069682, EPI_ISL_1069684, EPI_ISL_1069685, EPI_ISL_1069687, EPI_ISL_1069688, EPI_ISL_1069690, EPI_ISL_1069691, EPI_ISL_1069694, EPI_ISL_1069695, EPI_ISL_1069697, EPI_ISL_1069699, EPI_ISL_1069700, EPI_ISL_1069702, EPI_ISL_1069703, EPI_ISL_1069704, EPI_ISL_1069706, EPI_ISL_1069707, EPI_ISL_1069710, EPI_ISL_1069711, EPI_ISL_1069713, EPI_ISL_1069714, EPI_ISL_1069716, EPI_ISL_1069718, EPI_ISL_1069719, EPI_ISL_1069721, EPI_ISL_1069722, EPI_ISL_1069723, EPI_ISL_1069724, EPI_ISL_1069726, EPI_ISL_1069728, EPI_ISL_1069729, EPI_ISL_1069732, EPI_ISL_1069735, EPI_ISL_1069737, EPI_ISL_1069741, EPI_ISL_1069743, EPI_ISL_1069746, EPI_ISL_1069747, EPI_ISL_1069748, EPI_ISL_1069753, EPI_ISL_1069755, EPI_ISL_1069756, EPI_ISL_1069757, EPI_ISL_1069759, EPI_ISL_1069762, EPI_ISL_1069765, EPI_ISL_1069770, EPI_ISL_1069771, EPI_ISL_1069776, EPI_ISL_1069777, EPI_ISL_1069778, EPI_ISL_1069783, EPI_ISL_1069784, EPI_ISL_1069788, EPI_ISL_1069789, EPI_ISL_1069793, EPI_ISL_1069794, EPI_ISL_1069798, EPI_ISL_1069800, EPI_ISL_1069802, EPI_ISL_1069803, EPI_ISL_1069804, EPI_ISL_1069806, EPI_ISL_1069808, EPI_ISL_1069809, EPI_ISL_1069811, EPI_ISL_1069817, EPI_ISL_1069819, EPI_ISL_1069823, EPI_ISL_1069825, EPI_ISL_1069826, EPI_ISL_1069828, EPI_ISL_1069829, EPI_ISL_1069830, EPI_ISL_1069848, EPI_ISL_1069858, EPI_ISL_1069859, EPI_ISL_1069862, EPI_ISL_1069863, EPI_ISL_1069867, EPI_ISL_1069869, EPI_ISL_1069870, EPI_ISL_1069871, EPI_ISL_1069874, EPI_ISL_1069875, EPI_ISL_1069878, EPI_ISL_1069892, EPI_ISL_1069894, EPI_ISL_1069895, EPI_ISL_1069897, EPI_ISL_1069898, EPI_ISL_1069900, EPI_ISL_1069901, EPI_ISL_1069904, EPI_ISL_1069905, EPI_ISL_1069907, EPI_ISL_1069912, EPI_ISL_1069919, EPI_ISL_1069920, EPI_ISL_1069924, EPI_ISL_1069925, EPI_ISL_1069929, EPI_ISL_1069930, EPI_ISL_1069933, EPI_ISL_1069938, EPI_ISL_1069939, EPI_ISL_1069940, EPI_ISL_1069946, EPI_ISL_1069949, EPI_ISL_1069952, EPI_ISL_1069954, EPI_ISL_1069955, EPI_ISL_1069958, EPI_ISL_1069960, EPI_ISL_1069962, EPI_ISL_1069965, EPI_ISL_1069967, EPI_ISL_1069972, EPI_ISL_1069973, EPI_ISL_1069980, EPI_ISL_1069981, EPI_ISL_1069985, EPI_ISL_1069987, EPI_ISL_1069990, EPI_ISL_1071643, EPI_ISL_1071650, EPI_ISL_1071693, EPI_ISL_1071731, EPI_ISL_1071750, EPI_ISL_1071819, EPI_ISL_1071823, EPI_ISL_1071883, EPI_ISL_1071900, EPI_ISL_1071924, EPI_ISL_1071944, EPI_ISL_1071971, EPI_ISL_1071984, EPI_ISL_1071995, EPI_ISL_1082534, EPI_ISL_1082580, EPI_ISL_1082582, EPI_ISL_1082620, EPI_ISL_1082625, EPI_ISL_1082650, EPI_ISL_1082659, EPI_ISL_1082680, EPI_ISL_1082681, EPI_ISL_1082721, EPI_ISL_1082734, EPI_ISL_1082740, EPI_ISL_1082777, EPI_ISL_1082814, EPI_ISL_1082973, EPI_ISL_1083029, EPI_ISL_1083078, EPI_ISL_1083818, EPI_ISL_1083819, EPI_ISL_1083820, EPI_ISL_1083821, EPI_ISL_1083822, EPI_ISL_1083824, EPI_ISL_1083825, EPI_ISL_1083827, EPI_ISL_1083828, EPI_ISL_1083830, EPI_ISL_1083831, EPI_ISL_1083832, EPI_ISL_1083833, EPI_ISL_1083835, EPI_ISL_1083836, EPI_ISL_1083838, EPI_ISL_1083840, EPI_ISL_1083849, EPI_ISL_1083851, EPI_ISL_1083852, EPI_ISL_1083853, EPI_ISL_1083854, EPI_ISL_1083858, EPI_ISL_1083859, EPI_ISL_1083860, EPI_ISL_1083866, EPI_ISL_1083868, EPI_ISL_1083870, EPI_ISL_1083871, EPI_ISL_1083891, EPI_ISL_1083892, EPI_ISL_1083893, EPI_ISL_1083894, EPI_ISL_1083895, EPI_ISL_1083897, EPI_ISL_1083899, EPI_ISL_1083901, EPI_ISL_1083902, EPI_ISL_1083907, EPI_ISL_1083908, EPI_ISL_1083911, EPI_ISL_1083914, EPI_ISL_1083918, EPI_ISL_1083920, EPI_ISL_1083921, EPI_ISL_1083922, EPI_ISL_1083923, EPI_ISL_1083927, EPI_ISL_1083936, EPI_ISL_1083939, EPI_ISL_1083942, EPI_ISL_1083947, EPI_ISL_1083949, EPI_ISL_1083951, EPI_ISL_1083952, EPI_ISL_1083955, EPI_ISL_1083957, EPI_ISL_1083959, EPI_ISL_1083960, EPI_ISL_1083962, EPI_ISL_1083964, EPI_ISL_1083966, EPI_ISL_1083969, EPI_ISL_1083971, EPI_ISL_1083973, EPI_ISL_1083976, EPI_ISL_1083977, EPI_ISL_1083978, EPI_ISL_1083979, EPI_ISL_1083984, EPI_ISL_1083988, EPI_ISL_1083989, EPI_ISL_1083993, EPI_ISL_1083995, EPI_ISL_1083997, EPI_ISL_1083998, EPI_ISL_1084001, EPI_ISL_1084004, EPI_ISL_1084007, EPI_ISL_1084008, EPI_ISL_1084010, EPI_ISL_1084011, EPI_ISL_1084012, EPI_ISL_1084013, EPI_ISL_1084020, EPI_ISL_1084022, EPI_ISL_1084029, EPI_ISL_1084031, EPI_ISL_1084037, EPI_ISL_1084040, EPI_ISL_1084047, EPI_ISL_1084048, EPI_ISL_1084052, EPI_ISL_1084055, EPI_ISL_1084059, EPI_ISL_1084062, EPI_ISL_1084065, EPI_ISL_1084066, EPI_ISL_1084069, EPI_ISL_1084071, EPI_ISL_1084074, EPI_ISL_1084076, EPI_ISL_1084080, EPI_ISL_1084088, EPI_ISL_1084093, EPI_ISL_1084095, EPI_ISL_1084099, EPI_ISL_1084102, EPI_ISL_1084104, EPI_ISL_1084106, EPI_ISL_1084107, EPI_ISL_1084108, EPI_ISL_1084110, EPI_ISL_1084115, EPI_ISL_1084118, EPI_ISL_1084121, EPI_ISL_1084122, EPI_ISL_1084124, EPI_ISL_1084126, EPI_ISL_1084129, EPI_ISL_1084130, EPI_ISL_1084133, EPI_ISL_1084135, EPI_ISL_1084137, EPI_ISL_1084141, EPI_ISL_1084143, EPI_ISL_1084145, EPI_ISL_1084146, EPI_ISL_1084148, EPI_ISL_1084149, EPI_ISL_1084160, EPI_ISL_1084162, EPI_ISL_1084164, EPI_ISL_1084165, EPI_ISL_1084167, EPI_ISL_1084169, EPI_ISL_1084170, EPI_ISL_1084171, EPI_ISL_1084172, EPI_ISL_1084173, EPI_ISL_1084174, EPI_ISL_1084175, EPI_ISL_1084178, EPI_ISL_1084180, EPI_ISL_1084182, EPI_ISL_1084184, EPI_ISL_1084185, EPI_ISL_1084186, EPI_ISL_1084187, EPI_ISL_1084188, EPI_ISL_1084189, EPI_ISL_1084192, EPI_ISL_1084201, EPI_ISL_1084202, EPI_ISL_1084204, EPI_ISL_1084210, EPI_ISL_1084211, EPI_ISL_1084213, EPI_ISL_1084214, EPI_ISL_1084215, EPI_ISL_1084220, EPI_ISL_1084222, EPI_ISL_1084223, EPI_ISL_1084226, EPI_ISL_1084227, EPI_ISL_1084228, EPI_ISL_1084230, EPI_ISL_1084234, EPI_ISL_1084238, EPI_ISL_1084239, EPI_ISL_1084242, EPI_ISL_1084243, EPI_ISL_1084246, EPI_ISL_1084247, EPI_ISL_1084248, EPI_ISL_1084251, EPI_ISL_1084252, EPI_ISL_1084253, EPI_ISL_1084259, EPI_ISL_1084260, EPI_ISL_1084261, EPI_ISL_1084262, EPI_ISL_1084263, EPI_ISL_1084265, EPI_ISL_1084266, EPI_ISL_1084268, EPI_ISL_1084269, EPI_ISL_1084277, EPI_ISL_1084278, EPI_ISL_1084283, EPI_ISL_1084284, EPI_ISL_1084285, EPI_ISL_1084287, EPI_ISL_1084288, EPI_ISL_1084290, EPI_ISL_1084292, EPI_ISL_1084293, EPI_ISL_1084295, EPI_ISL_1084297, EPI_ISL_1084300, EPI_ISL_1084304, EPI_ISL_1084308, EPI_ISL_1084309, EPI_ISL_1084310, EPI_ISL_1084311, EPI_ISL_1084313, EPI_ISL_1084317, EPI_ISL_1084320, EPI_ISL_1084322, EPI_ISL_1084324, EPI_ISL_1084325, EPI_ISL_1084335, EPI_ISL_1084339, EPI_ISL_1084341, EPI_ISL_1084342, EPI_ISL_1084347, EPI_ISL_1084349, EPI_ISL_1084350, EPI_ISL_1084351, EPI_ISL_1084354, EPI_ISL_1084356, EPI_ISL_1084357, EPI_ISL_1084359, EPI_ISL_1084364, EPI_ISL_1084365, EPI_ISL_1084366, EPI_ISL_1084370, EPI_ISL_1084371, EPI_ISL_1084372, EPI_ISL_1084374, EPI_ISL_1084383, EPI_ISL_1084386, EPI_ISL_1084387, EPI_ISL_1084388, EPI_ISL_1084389, EPI_ISL_1084392, EPI_ISL_1084395, EPI_ISL_1084396, EPI_ISL_1084397, EPI_ISL_1084398, EPI_ISL_1084399, EPI_ISL_1084403, EPI_ISL_1084404, EPI_ISL_1084405, EPI_ISL_1084408, EPI_ISL_1084409, EPI_ISL_1084410, EPI_ISL_1084411, EPI_ISL_1084414, EPI_ISL_1084417, EPI_ISL_1084418, EPI_ISL_1084419, EPI_ISL_1084420, EPI_ISL_1084421, EPI_ISL_1084422, EPI_ISL_1084423, EPI_ISL_1084424, EPI_ISL_1084426, EPI_ISL_1084427, EPI_ISL_1084429, EPI_ISL_1084432, EPI_ISL_1084435, EPI_ISL_1084439, EPI_ISL_1084442, EPI_ISL_1084445, EPI_ISL_1084446, EPI_ISL_1084452, EPI_ISL_1084453, EPI_ISL_1084456, EPI_ISL_1084457, EPI_ISL_1084458, EPI_ISL_1084459, EPI_ISL_1084462, EPI_ISL_1084463, EPI_ISL_1084464, EPI_ISL_1084465, EPI_ISL_1084466, EPI_ISL_1084467, EPI_ISL_1098980, EPI_ISL_1100432, EPI_ISL_1100851, EPI_ISL_1101003, EPI_ISL_1101043, EPI_ISL_1101056, EPI_ISL_1101089, EPI_ISL_1101090, EPI_ISL_1101102, EPI_ISL_1101156, EPI_ISL_1101164, EPI_ISL_1101165, EPI_ISL_1101166, EPI_ISL_1101168, EPI_ISL_1101183, EPI_ISL_1101194, EPI_ISL_1101197, EPI_ISL_1101198, EPI_ISL_1101199, EPI_ISL_1101216, EPI_ISL_1101217, EPI_ISL_1101218, EPI_ISL_1101235, EPI_ISL_1101244, EPI_ISL_1101261, EPI_ISL_1101262, EPI_ISL_1101263, EPI_ISL_1101274, EPI_ISL_1101285, EPI_ISL_1101291, EPI_ISL_1101301, EPI_ISL_1102035, EPI_ISL_1102126, EPI_ISL_1102142, EPI_ISL_1102149, EPI_ISL_1102200, EPI_ISL_1102210, EPI_ISL_1102230, EPI_ISL_1102252, EPI_ISL_1102265, EPI_ISL_1102278, EPI_ISL_1102280, EPI_ISL_1102285, EPI_ISL_1102431, EPI_ISL_1122533, EPI_ISL_1122621, EPI_ISL_1122722, EPI_ISL_1122726, EPI_ISL_1175435, EPI_ISL_1175470, EPI_ISL_1175719, EPI_ISL_1206525, EPI_ISL_1206542, EPI_ISL_1206602, EPI_ISL_1206707, EPI_ISL_1206738, EPI_ISL_1206757, EPI_ISL_1206764, EPI_ISL_1207025, EPI_ISL_1207587, EPI_ISL_1207704, EPI_ISL_1257200, EPI_ISL_1806485

| | | | |
|---|---|---|---|
| see above | Lighthouse Lab in Milton Keynes | Wellcome Sanger Institute for the COVID-19 Genomics UK (COG-UK) Consortium | Cordelia Langford; David K. Jackson; Dominic Kwiatkowski; Ewan Harrison; Ian Johnston; Jeffrey Barrett; John Sillitoe on behalf of the Wellcome Sanger Institute COVID-19 Surveillance Team; Roberto Amato; Sonia Goncalves; The Lighthouse Lab in Milton Keynes and Alex Alderton |

| | | | |
|---|---|---|---|
| EPI_ISL_1178620, EPI_ISL_1178621, EPI_ISL_1178625, EPI_ISL_1178626, EPI_ISL_1178664, EPI_ISL_1178665, EPI_ISL_1178666, EPI_ISL_1178667, EPI_ISL_1178668 | | | |
| see above | Lincolnshire Hospitals and DeepSeq Nottingham | COVID-19 Genomics UK (COG-UK) Consortium | Christopher Moore; Fei Sang; Joeseph Chappell; Johnny Debebe; Jonathan Ball; Matthew Carlisle; Matthew Loose; Nadine Holmes; Nichola Duckworth; Patrick McClure; Sarah Walsh; Tim Sloan; Victoria Wright |
| EPI_ISL_1104460, EPI_ISL_1104461, EPI_ISL_1104462, EPI_ISL_1104464, EPI_ISL_1104466, EPI_ISL_1104467, EPI_ISL_1104468, EPI_ISL_1104471, EPI_ISL_1104472, EPI_ISL_1104478, EPI_ISL_1104479, EPI_ISL_1104480, EPI_ISL_1104481, EPI_ISL_1104482, EPI_ISL_1104483, EPI_ISL_1104484, EPI_ISL_1104485, EPI_ISL_1104551, EPI_ISL_1104552, EPI_ISL_1104553, EPI_ISL_1104557, EPI_ISL_1104559, EPI_ISL_1104563, EPI_ISL_1104564, EPI_ISL_1104565, EPI_ISL_1104566, EPI_ISL_1104567, EPI_ISL_1177899, EPI_ISL_1177916, EPI_ISL_1177923, EPI_ISL_1247844, EPI_ISL_1247853, EPI_ISL_1247870, EPI_ISL_1249273, EPI_ISL_1249276, EPI_ISL_1249277, EPI_ISL_1249278, EPI_ISL_1249279, EPI_ISL_1249280, EPI_ISL_1249281, EPI_ISL_1249283, EPI_ISL_1308902, EPI_ISL_1308903, EPI_ISL_1308906, EPI_ISL_1308907, EPI_ISL_1308916, EPI_ISL_1546081, EPI_ISL_1546083 | | | |
| see above | Liverpool Clinical Laboratories | COVID-19 Genomics UK (COG-UK) Consortium | A Alrezaihi; Alessandro Gerada; Alistair Darby; Angela Cowell; Anita Lucaci; Anu Chawla; Cassie Olateju; Catherine Hartley; Charlotte Nelson; Ecaterina Vamos; Elaine O'Toole; Eleanor G Bentley; Ghada T Shawli; Hermione Webster; Isabel García-Dorival; James Johnson; James P Stewart; Jenifer Manson; Joanne Watts; Jones Benjamin; Jordan J Clark; Julian Hiscox; L Luu; Lucille Rainbow; M Almsaud; Margaret Hughes; Mark Whitehead; Matthew Gemmell; Miren Iturriza-Gomara; Muhannad Alruwaili; N P Randle; Neil Swainston; PKF Gilmore; Parul Sharma; Rebekah Penrice-Randal; Richard Eccles; Richard Gregory; Sam Haldenby; Steve Paterson; Stuart D Armstrong; Trevor Ian Robinson; Ximeng Han |
| EPI_ISL_1051032, EPI_ISL_1051033, EPI_ISL_1051034, EPI_ISL_1051035, EPI_ISL_1051036, EPI_ISL_1051037, EPI_ISL_1105754, EPI_ISL_1105755, EPI_ISL_1105756, EPI_ISL_1105758, EPI_ISL_1176904, EPI_ISL_1176906, EPI_ISL_1176908, EPI_ISL_1176910, EPI_ISL_1176911, EPI_ISL_1176912, EPI_ISL_1176916, EPI_ISL_1176919, EPI_ISL_1176920, EPI_ISL_1176921, EPI_ISL_1176922, EPI_ISL_1176923, EPI_ISL_1176928, EPI_ISL_1176930, EPI_ISL_1176931, EPI_ISL_1176932, EPI_ISL_1176933, EPI_ISL_1176941, EPI_ISL_1176942, EPI_ISL_1176943, EPI_ISL_1176947, EPI_ISL_1176950, EPI_ISL_1176951, EPI_ISL_1176957, EPI_ISL_1176961, EPI_ISL_1176962, EPI_ISL_1176963, EPI_ISL_1176965, EPI_ISL_1176970, EPI_ISL_1176971, EPI_ISL_1176972, EPI_ISL_1176973, EPI_ISL_1176974, EPI_ISL_1176976, EPI_ISL_1176980, EPI_ISL_1176982, EPI_ISL_1176983, EPI_ISL_1176984, EPI_ISL_1176985, EPI_ISL_1176988, EPI_ISL_1176989, EPI_ISL_1176990, EPI_ISL_1176992, EPI_ISL_1176993, EPI_ISL_1176994, EPI_ISL_1176995, EPI_ISL_1176996, EPI_ISL_1176998, EPI_ISL_1176999, EPI_ISL_1177002, EPI_ISL_1177003, EPI_ISL_1177004, EPI_ISL_1177007, EPI_ISL_1177011, EPI_ISL_1177012, EPI_ISL_1177013, EPI_ISL_1177014, EPI_ISL_1177015, EPI_ISL_1177018, EPI_ISL_1177019, EPI_ISL_1177020, EPI_ISL_1177024, EPI_ISL_1177025, EPI_ISL_1177026, EPI_ISL_1177029, EPI_ISL_1177030, EPI_ISL_1177031, EPI_ISL_1177032, EPI_ISL_1177034, EPI_ISL_1177035, EPI_ISL_1177036, EPI_ISL_1177037, EPI_ISL_1177038, EPI_ISL_1177039, EPI_ISL_1177040, EPI_ISL_1177041, EPI_ISL_1177043, EPI_ISL_1177045, EPI_ISL_1177046, EPI_ISL_1177047, EPI_ISL_1177048, EPI_ISL_1177049, EPI_ISL_1177050, EPI_ISL_1177051, EPI_ISL_1177052, EPI_ISL_1177053, EPI_ISL_1177055, EPI_ISL_1177057, EPI_ISL_1177058, EPI_ISL_1177059, EPI_ISL_1177060, EPI_ISL_1177061, EPI_ISL_1177063, EPI_ISL_1177065, EPI_ISL_1177067, EPI_ISL_1177069, EPI_ISL_1177070, EPI_ISL_1177071, EPI_ISL_1177078, EPI_ISL_1177079, EPI_ISL_1177080, EPI_ISL_1177081, EPI_ISL_1177086, EPI_ISL_1177087, EPI_ISL_1177088, EPI_ISL_1177089, EPI_ISL_1177094, EPI_ISL_1177095, EPI_ISL_1177096, EPI_ISL_1177097, EPI_ISL_1177098, EPI_ISL_1177099, EPI_ISL_1177100, EPI_ISL_1177101, EPI_ISL_1177105, EPI_ISL_1177106, EPI_ISL_1177107, EPI_ISL_1177108, EPI_ISL_1177109, EPI_ISL_1177110, EPI_ISL_1177115, EPI_ISL_1177116, EPI_ISL_1177117, EPI_ISL_1177118, EPI_ISL_1177119, EPI_ISL_1177123, EPI_ISL_1177124, EPI_ISL_1177125, EPI_ISL_1177126, EPI_ISL_1177131, EPI_ISL_1177132, EPI_ISL_1177133, EPI_ISL_1177134, EPI_ISL_1177135, EPI_ISL_1177136, EPI_ISL_1177137, EPI_ISL_1177140, EPI_ISL_1177141, EPI_ISL_1177142, EPI_ISL_1177143, EPI_ISL_1177144, EPI_ISL_1177145, EPI_ISL_1177146, EPI_ISL_1177147, EPI_ISL_1177148, EPI_ISL_1177149, EPI_ISL_1177150, EPI_ISL_1177151, EPI_ISL_1177152, EPI_ISL_1177153, EPI_ISL_1177154, EPI_ISL_1177155, EPI_ISL_1177156, EPI_ISL_1177157, EPI_ISL_1177158, EPI_ISL_1177159, EPI_ISL_1177160, EPI_ISL_1177161, EPI_ISL_1177162, EPI_ISL_1177163, EPI_ISL_1177164, EPI_ISL_1177165, EPI_ISL_1177166, EPI_ISL_1177167, EPI_ISL_1177168, EPI_ISL_1177169, EPI_ISL_1177170, EPI_ISL_1177171, EPI_ISL_1177172, EPI_ISL_1177173, EPI_ISL_1177174, EPI_ISL_1177175, EPI_ISL_1177177, EPI_ISL_1177178, EPI_ISL_1177179, EPI_ISL_1177180, EPI_ISL_1177181, EPI_ISL_1177182, EPI_ISL_1177183, EPI_ISL_1177184, EPI_ISL_1177186, EPI_ISL_1177187, EPI_ISL_1177188, EPI_ISL_1177189, EPI_ISL_1177190, EPI_ISL_1177191, EPI_ISL_1177192, EPI_ISL_1177193, EPI_ISL_1177194, EPI_ISL_1177195, EPI_ISL_1177196, EPI_ISL_1177197, EPI_ISL_1177198, EPI_ISL_1177199, EPI_ISL_1177201, EPI_ISL_1177202, EPI_ISL_1177203, EPI_ISL_1177204, EPI_ISL_1177205, EPI_ISL_1177206, EPI_ISL_1177207, EPI_ISL_1177208, EPI_ISL_1177209, EPI_ISL_1177210, EPI_ISL_1177212, EPI_ISL_1177213, EPI_ISL_1177214, EPI_ISL_1177215, EPI_ISL_1177216, EPI_ISL_1177217, EPI_ISL_1177218, EPI_ISL_1177219, EPI_ISL_1177220, EPI_ISL_1177221, EPI_ISL_1177222, EPI_ISL_1177223, EPI_ISL_1177225, EPI_ISL_1177226, EPI_ISL_1177227, EPI_ISL_1177228, EPI_ISL_1177229, EPI_ISL_1177230, EPI_ISL_1248863, EPI_ISL_1248864, EPI_ISL_1248865, EPI_ISL_1309494, EPI_ISL_1309495, EPI_ISL_1309619, EPI_ISL_1309621, EPI_ISL_1309623, EPI_ISL_1309624, EPI_ISL_1309625, EPI_ISL_1309626, EPI_ISL_1309627, EPI_ISL_1309628, EPI_ISL_1309629, EPI_ISL_1309630, EPI_ISL_1309631, EPI_ISL_1309632, EPI_ISL_1309633, EPI_ISL_1309637, EPI_ISL_1309638, EPI_ISL_1309639, EPI_ISL_1309641, EPI_ISL_1387120, EPI_ISL_1475415, EPI_ISL_1475416, EPI_ISL_1475417, EPI_ISL_1475418, EPI_ISL_1475432, EPI_ISL_1475435 | | | |
| see above | Northumbria University / South Tees Hospitals NHS Foundation Trust / North Cumbria Integrated Care NHS Foundation Trust / North Tees and Hartlepool NHS Foundation Trust / Newcastle Hospitals NHS Foundation Trust | COVID-19 Genomics UK (COG-UK) Consortium | Andrew Nelson; Brendan Payne; Clive Graham; Darren L Smith; Debra Padgett; Edward Barton; Emma Swindells; Garren Scott; Gary Black; Gary Eltringham; Giles S Holt; Greg R Young; Jane Greenaway; Jennifer Collins; John Allan; Joshua Loh; Lynn Dover; Matthew Bashton; Mohammad A Tariq; Paul Baker; Sarah Essex; Steve Liggett; Wen C Yew; Yusri Taha |
| EPI_ISL_1053250, EPI_ISL_1053256, EPI_ISL_1053257, EPI_ISL_1053258, EPI_ISL_1053314, EPI_ISL_1053331, EPI_ISL_1053348, EPI_ISL_1053351, EPI_ISL_1053387, EPI_ISL_1053388, EPI_ISL_1053402, EPI_ISL_1053414, EPI_ISL_1053418, EPI_ISL_1053432, EPI_ISL_1053435, EPI_ISL_1053437, EPI_ISL_1053439, EPI_ISL_1053459, EPI_ISL_1053460, EPI_ISL_1053461, EPI_ISL_1053477, EPI_ISL_1053482, EPI_ISL_1053501, EPI_ISL_1053510, EPI_ISL_1053511, EPI_ISL_1053512, EPI_ISL_1053524, EPI_ISL_1053538, EPI_ISL_1053680, EPI_ISL_1053684, EPI_ISL_1053689, EPI_ISL_1053692, EPI_ISL_1053694, EPI_ISL_1053695, EPI_ISL_1053698, EPI_ISL_1053709, EPI_ISL_1053715, EPI_ISL_1053716, EPI_ISL_1053720, EPI_ISL_1053727, EPI_ISL_1053743, EPI_ISL_1106253, EPI_ISL_1106260, EPI_ISL_1106261, EPI_ISL_1106262, EPI_ISL_1106263, EPI_ISL_1106264, EPI_ISL_1106265, EPI_ISL_1106272, EPI_ISL_1106279, EPI_ISL_1106280, EPI_ISL_1106353, EPI_ISL_1106357, EPI_ISL_1106358, EPI_ISL_1106360, EPI_ISL_1106361, EPI_ISL_1106362, EPI_ISL_1106363, EPI_ISL_1106364, EPI_ISL_1106365, EPI_ISL_1106366, EPI_ISL_1106371, EPI_ISL_1106372, EPI_ISL_1106375, EPI_ISL_1106376, EPI_ISL_1106383, EPI_ISL_1106387, EPI_ISL_1106392, EPI_ISL_1106397, EPI_ISL_1106400, EPI_ISL_1106403, EPI_ISL_1106404, EPI_ISL_1106405, EPI_ISL_1106411, EPI_ISL_1106412, EPI_ISL_1106413, EPI_ISL_1106416, EPI_ISL_1106418, EPI_ISL_1106419, EPI_ISL_1106420, EPI_ISL_1106421, EPI_ISL_1106422, EPI_ISL_1106423, EPI_ISL_1106425, EPI_ISL_1106426, EPI_ISL_1106428, EPI_ISL_1106429, EPI_ISL_1106430, EPI_ISL_1106431, EPI_ISL_1106440, EPI_ISL_1106442, EPI_ISL_1106443, EPI_ISL_1106444, EPI_ISL_1106446, EPI_ISL_1106447, EPI_ISL_1106449, EPI_ISL_1106450, EPI_ISL_1106451, EPI_ISL_1106452, EPI_ISL_1106453, EPI_ISL_1106454, EPI_ISL_1106455, EPI_ISL_1106457, EPI_ISL_1106460, EPI_ISL_1106461, EPI_ISL_1106462, EPI_ISL_1106463, EPI_ISL_1106464, EPI_ISL_1106465, EPI_ISL_1106466, EPI_ISL_1106467, EPI_ISL_1106468, EPI_ISL_1106469, EPI_ISL_1106470, EPI_ISL_1106475, EPI_ISL_1106477, EPI_ISL_1106478, EPI_ISL_1106479, EPI_ISL_1106480, EPI_ISL_1106481, EPI_ISL_1106482, EPI_ISL_1106483, EPI_ISL_1106484, EPI_ISL_1106485, EPI_ISL_1106487, EPI_ISL_1106489, EPI_ISL_1106491, EPI_ISL_1106493, EPI_ISL_1106494, EPI_ISL_1106495, EPI_ISL_1106496, EPI_ISL_1106497, EPI_ISL_1106498, EPI_ISL_1106501, EPI_ISL_1106502, EPI_ISL_1106503, EPI_ISL_1106504, EPI_ISL_1106506, EPI_ISL_1106507, EPI_ISL_1106508, EPI_ISL_1106510, EPI_ISL_1106511, EPI_ISL_1106513, EPI_ISL_1106514, EPI_ISL_1106515, EPI_ISL_1106527, EPI_ISL_1106528, EPI_ISL_1106531, EPI_ISL_1106532, EPI_ISL_1106534, EPI_ISL_1106536, EPI_ISL_1106538, EPI_ISL_1106539, EPI_ISL_1106540, EPI_ISL_1106541, EPI_ISL_1106542, EPI_ISL_1106543, EPI_ISL_1106544, EPI_ISL_1106545, EPI_ISL_1106546, EPI_ISL_1106547, EPI_ISL_1106548, EPI_ISL_1106550, EPI_ISL_1106552, EPI_ISL_1106555, EPI_ISL_1106557, EPI_ISL_1106558, EPI_ISL_1106559, EPI_ISL_1106561, EPI_ISL_1106562, EPI_ISL_1106563, EPI_ISL_1106564, EPI_ISL_1106566, EPI_ISL_1106568, EPI_ISL_1106570, EPI_ISL_1106573, EPI_ISL_1106574, EPI_ISL_1106575, EPI_ISL_1106576, EPI_ISL_1106579, EPI_ISL_1106580, EPI_ISL_1106581, EPI_ISL_1106582, EPI_ISL_1106583, EPI_ISL_1106584, EPI_ISL_1106585, EPI_ISL_1106588, EPI_ISL_1106589, EPI_ISL_1106590, EPI_ISL_1106591, EPI_ISL_1106595, EPI_ISL_1106597, EPI_ISL_1106598, EPI_ISL_1106600, EPI_ISL_1106601, EPI_ISL_1106640, EPI_ISL_1106641, EPI_ISL_1107264, EPI_ISL_1178898, EPI_ISL_1178899, EPI_ISL_1178930, EPI_ISL_1547025, EPI_ISL_1547026 | | | |
| see above | Originating lab: Wales Specialist Virology Centre Sequencing lab: Pathogen Genomics Unit | Public Health Wales Microbiology Cardiff Wales Specialist Virology Centre | Alec Birchley; Alexander Adams; Amy Gaskin; Angela Marchbank; Bree Gatica-Wilcox; Catherine Moore; Jason Coombes; Joanne Watkins; Joel Southgate; Johnathan Evans; Laura Gifford; Lauren Gilbert; Lee Graham; Malorie Perry; Matthew Bull; Nicole Pacchiarini; Sally Corden; Sara Kumziene-Summerhayes; Sara Rey; Sarah Taylor; Simon Cottrell; Sophie Jones; Tom Connor |
| EPI_ISL_1249147, EPI_ISL_1249154, EPI_ISL_1249156, EPI_ISL_1249159, EPI_ISL_1249160, EPI_ISL_1249162, EPI_ISL_1309757, EPI_ISL_1309765, EPI_ISL_1309766, EPI_ISL_1309768, EPI_ISL_1309769, EPI_ISL_1309770, EPI_ISL_1309772, EPI_ISL_1309773, EPI_ISL_1309774, EPI_ISL_1309776, EPI_ISL_1309777, EPI_ISL_1387235, EPI_ISL_1475543 | | | |
| see above | Oxford Viromics, NDM, University of Oxford; Oxford University Hospitals; Basingstoke and North Hampshire Hospital | COVID-19 Genomics UK (COG-UK) Consortium | Alex Mobbs; Amy Trebes; Anita Justice; Catrin Moore; Christophe Fraser; David Bonsall; David Buck; Emma Wise; George Macintyre; Jessica Lynch; John Todd; Mariateresa de Cesare; Matilde Mori; Monique Andersson; Nathan Moore; Nick Cortes; Robert Shaw; Stephen Kidd; Tanya Golubchik; Timothy Peto |
| EPI_ISL_1178425, EPI_ISL_1178427, EPI_ISL_1178428, EPI_ISL_1178429, EPI_ISL_1178522, EPI_ISL_1178523, EPI_ISL_1178526, EPI_ISL_1178530, EPI_ISL_1178549, EPI_ISL_1247386, EPI_ISL_1247391, EPI_ISL_1247394, EPI_ISL_1249007, EPI_ISL_1249018, EPI_ISL_1249020, EPI_ISL_1308014, EPI_ISL_1308015, EPI_ISL_1308016, EPI_ISL_1308017, EPI_ISL_1308018, EPI_ISL_1308019, EPI_ISL_1308020, EPI_ISL_1308021, EPI_ISL_1308022, EPI_ISL_1308026, EPI_ISL_1308027, EPI_ISL_1308028, EPI_ISL_1308029, EPI_ISL_1308030, EPI_ISL_1308031, EPI_ISL_1308032, EPI_ISL_1308036, EPI_ISL_1308037, EPI_ISL_1308038, EPI_ISL_1308039, EPI_ISL_1308040, EPI_ISL_1308041, EPI_ISL_1308042, EPI_ISL_1308043, EPI_ISL_1308044, EPI_ISL_1308048, EPI_ISL_1308049, EPI_ISL_1308050, EPI_ISL_1308051, EPI_ISL_1308052, EPI_ISL_1308056, EPI_ISL_1308057, EPI_ISL_1308058, EPI_ISL_1308059, EPI_ISL_1308060, EPI_ISL_1308061, EPI_ISL_1308062, EPI_ISL_1308063, EPI_ISL_1308064, EPI_ISL_1308067, EPI_ISL_1308068, EPI_ISL_1308069, EPI_ISL_1308070, EPI_ISL_1308071, EPI_ISL_1308072, EPI_ISL_1308073, EPI_ISL_1308074, EPI_ISL_1308075, EPI_ISL_1308079, EPI_ISL_1308080, EPI_ISL_1308081, EPI_ISL_1308082, EPI_ISL_1308083, EPI_ISL_1308084, EPI_ISL_1308085, EPI_ISL_1308089, EPI_ISL_1308090, EPI_ISL_1308091, EPI_ISL_1308092, EPI_ISL_1308093, EPI_ISL_1308094, EPI_ISL_1308095, EPI_ISL_1308096, EPI_ISL_1308097, EPI_ISL_1387145, EPI_ISL_1387149, EPI_ISL_1387152, EPI_ISL_1387154, EPI_ISL_1387155, EPI_ISL_1387156, EPI_ISL_1474167, EPI_ISL_1474168, EPI_ISL_1474169, EPI_ISL_1474170, EPI_ISL_1474171, EPI_ISL_1474172, EPI_ISL_1474173, EPI_ISL_1474174, EPI_ISL_1474175, EPI_ISL_1474176, EPI_ISL_1474177, EPI_ISL_1474178, EPI_ISL_1474179, EPI_ISL_1474180, EPI_ISL_1474181, EPI_ISL_1474182, EPI_ISL_1474183, EPI_ISL_1474184, EPI_ISL_1474185, EPI_ISL_1474186, EPI_ISL_1474187, EPI_ISL_1474188, EPI_ISL_1474189, EPI_ISL_1474190, EPI_ISL_1474191, EPI_ISL_1474192, EPI_ISL_1474193, EPI_ISL_1474194, EPI_ISL_1474195, EPI_ISL_1474196, EPI_ISL_1474197, EPI_ISL_1474198, EPI_ISL_1474199, EPI_ISL_1474200, EPI_ISL_1474201, EPI_ISL_1474202, EPI_ISL_1474203, EPI_ISL_1474204, EPI_ISL_1474206, EPI_ISL_1474207, EPI_ISL_1474208, EPI_ISL_1474209, EPI_ISL_1474210, EPI_ISL_1474211, EPI_ISL_1474212, EPI_ISL_1474213, EPI_ISL_1474214, EPI_ISL_1474215, EPI_ISL_1474216, EPI_ISL_1474217, EPI_ISL_1474218, EPI_ISL_1474219, EPI_ISL_1474220, EPI_ISL_1474221, EPI_ISL_1474222, EPI_ISL_1474223, EPI_ISL_1474224, EPI_ISL_1474227, EPI_ISL_1474228, EPI_ISL_1474229, EPI_ISL_1474230, EPI_ISL_1474231, EPI_ISL_1474232, EPI_ISL_1474233, EPI_ISL_1474234, EPI_ISL_1474235, EPI_ISL_1474900, EPI_ISL_1474903, EPI_ISL_1474910, EPI_ISL_1474924, EPI_ISL_1474925, EPI_ISL_1474930, EPI_ISL_1474931, EPI_ISL_1474933, EPI_ISL_1474937, EPI_ISL_1474938, EPI_ISL_1474940, EPI_ISL_1474945, EPI_ISL_1474949, EPI_ISL_1474956, EPI_ISL_1474957, EPI_ISL_1832769 | | | |
| see above | Quadram Institute Bioscience | COVID-19 Genomics UK (COG-UK) Consortium | Alexander J Trotter; Alison E. Mather; Alp Aydin; Ana P. Tedim; Anastasia Kolyva; Andrew Bell; Andrew J. Page; Claire Stuart; Dave J. Baker; Gemma L. Kay; John Wain; Justin O'Grady; Leonardo de Oliveira Martins; Lizzie Meadows; Maria Diaz; Mark Webber; Muhammed Yasir; Nabil-Fareed Alikhan; Ngozi Elumogo; Nicholas M. Thomson; Rachael Stanley; Rachel Gilroy; Reenesh Prakash; Samir Dervisevic; Samuel Bloomfield; Steven Rudder; Thanh Le-Viet |
| EPI_ISL_1051584, EPI_ISL_1051585, EPI_ISL_1051588, EPI_ISL_1051589, EPI_ISL_1051591 | Queens Medical Centre, Clinical Microbiology Department / DeepSeq Nottingham | COVID-19 Genomics UK (COG-UK) Consortium | Christopher Moore; Fei Sang; Gemma Clark; Hannah Howson-Wells; Johnny Debebe; Jonathan Ball; Joseph Chappell; Manjinder Khakh; Matthew Carlisle; Matthew Loose; Michelle M Lister; Nadine Holmes; Patrick McClure; Theocharis Tsoleridis; Vicki M Fleming; Victoria Wright; Wendy Smith |
| EPI_ISL_1101916, EPI_ISL_1115719, EPI_ISL_1115720, EPI_ISL_1115721, EPI_ISL_1115722, EPI_ISL_1115727, EPI_ISL_1115728, EPI_ISL_1115729, EPI_ISL_1115730, EPI_ISL_1115731, EPI_ISL_1115732, EPI_ISL_1115734, EPI_ISL_1115735, EPI_ISL_1115736, EPI_ISL_1115737, EPI_ISL_1115738, EPI_ISL_1115739, EPI_ISL_1115740, EPI_ISL_1115741, EPI_ISL_1115742, EPI_ISL_1115745, EPI_ISL_1115746, EPI_ISL_1115747, EPI_ISL_1115749, EPI_ISL_1115750, EPI_ISL_1115752, EPI_ISL_1115753, EPI_ISL_1115755, EPI_ISL_1115756, EPI_ISL_1115757, EPI_ISL_1115758, EPI_ISL_1115760, EPI_ISL_1115761, EPI_ISL_1115763, EPI_ISL_1115764, EPI_ISL_1115766, EPI_ISL_1115767, EPI_ISL_1115768, EPI_ISL_1115769, EPI_ISL_1115770, EPI_ISL_1115774, EPI_ISL_1115775, EPI_ISL_1115776, EPI_ISL_1115777, EPI_ISL_1115779, EPI_ISL_1115780, EPI_ISL_1115782, EPI_ISL_1115783, EPI_ISL_1115785, EPI_ISL_1115786, EPI_ISL_1115787, EPI_ISL_1115788, EPI_ISL_1115789, EPI_ISL_1115790, EPI_ISL_1115791, EPI_ISL_1115792, EPI_ISL_1115793, EPI_ISL_1115795, EPI_ISL_1115796, EPI_ISL_1116139, EPI_ISL_1116140, EPI_ISL_1116141, EPI_ISL_1116142, EPI_ISL_1116144, EPI_ISL_1116147, EPI_ISL_1116148, EPI_ISL_1116149, EPI_ISL_1116152, EPI_ISL_1116155, EPI_ISL_1116156, EPI_ISL_1116157, EPI_ISL_1116158, EPI_ISL_1116162, EPI_ISL_1116163, EPI_ISL_1116164, EPI_ISL_1116165, EPI_ISL_1116167, EPI_ISL_1116168, EPI_ISL_1116170, EPI_ISL_1116172, EPI_ISL_1116173, EPI_ISL_1116175, EPI_ISL_1116176, EPI_ISL_1116177, EPI_ISL_1116178, EPI_ISL_1116179, EPI_ISL_1116180, EPI_ISL_1116181, EPI_ISL_1116183, EPI_ISL_1116184, EPI_ISL_1116185, EPI_ISL_1116187, EPI_ISL_1116188, EPI_ISL_1116190, EPI_ISL_1116191, EPI_ISL_1116192, EPI_ISL_1116194, EPI_ISL_1116195, EPI_ISL_1116196, EPI_ISL_1116197, EPI_ISL_1116199, EPI_ISL_1116200, EPI_ISL_1116201, EPI_ISL_1116202, EPI_ISL_1116203, EPI_ISL_1116204, EPI_ISL_1116205, EPI_ISL_1116206, EPI_ISL_1116207, EPI_ISL_1116208, EPI_ISL_1116210, EPI_ISL_1116212, EPI_ISL_1116215, EPI_ISL_1116216, EPI_ISL_1116217, EPI_ISL_1116218, EPI_ISL_1116219, EPI_ISL_1116220, EPI_ISL_1116244, EPI_ISL_1174050, EPI_ISL_1174059, EPI_ISL_1174060, EPI_ISL_1174068, EPI_ISL_1174082, EPI_ISL_1174092, EPI_ISL_1174109, EPI_ISL_1174111, EPI_ISL_1174113, EPI_ISL_1174122, EPI_ISL_1174182, EPI_ISL_1174190, EPI_ISL_1174205, EPI_ISL_1174212, EPI_ISL_1174228, EPI_ISL_1174231, EPI_ISL_1174236, EPI_ISL_1174240, EPI_ISL_1174244, EPI_ISL_1174255, EPI_ISL_1174259, EPI_ISL_1174297, EPI_ISL_1174306, EPI_ISL_1174308, EPI_ISL_1174310, EPI_ISL_1174930, EPI_ISL_1174958, EPI_ISL_1174990, EPI_ISL_1175021, EPI_ISL_1175105, EPI_ISL_1186858, EPI_ISL_1186869, EPI_ISL_1186889, EPI_ISL_1186903, EPI_ISL_1186916, EPI_ISL_1186925, EPI_ISL_1186935, EPI_ISL_1186937, EPI_ISL_1186959, EPI_ISL_1186971, EPI_ISL_1186973, EPI_ISL_1186975, EPI_ISL_1187002, EPI_ISL_1187010, EPI_ISL_1189844, EPI_ISL_1189845, EPI_ISL_1189846, EPI_ISL_1189847, EPI_ISL_1189849, EPI_ISL_1189852, EPI_ISL_1189857, EPI_ISL_1189858, EPI_ISL_1189859, EPI_ISL_1189863, EPI_ISL_1189868, EPI_ISL_1189874, EPI_ISL_1189876, EPI_ISL_1189878, EPI_ISL_1189880, EPI_ISL_1189882, EPI_ISL_1189885, EPI_ISL_1189892, EPI_ISL_1189894, EPI_ISL_1189896, EPI_ISL_1189897, EPI_ISL_1189898, EPI_ISL_1189905, EPI_ISL_1189909, EPI_ISL_1189915, EPI_ISL_1189916, EPI_ISL_1189928, EPI_ISL_1189932, EPI_ISL_1189933, EPI_ISL_1189934, EPI_ISL_1189940, EPI_ISL_1189946, EPI_ISL_1189948, EPI_ISL_1189957, EPI_ISL_1189963, EPI_ISL_1189964, EPI_ISL_1189965, EPI_ISL_1189968, EPI_ISL_1189973, EPI_ISL_1189976, EPI_ISL_1189978, EPI_ISL_1189981, EPI_ISL_1189984, EPI_ISL_1189988, EPI_ISL_1189990, EPI_ISL_1189992, EPI_ISL_1189994, EPI_ISL_1189996, EPI_ISL_1189998, EPI_ISL_1190000, EPI_ISL_1190005, EPI_ISL_1190009, EPI_ISL_1190010, EPI_ISL_1190014, EPI_ISL_1190017, EPI_ISL_1190019, EPI_ISL_1190020, EPI_ISL_1190021, EPI_ISL_1190024, EPI_ISL_1190026, EPI_ISL_1190027, EPI_ISL_1190028, EPI_ISL_1190034, EPI_ISL_1190035, EPI_ISL_1190037, EPI_ISL_1190038, EPI_ISL_1190039, EPI_ISL_1190042, EPI_ISL_1190044, EPI_ISL_1190047, EPI_ISL_1190048, EPI_ISL_1190049, EPI_ISL_1190050, EPI_ISL_1190055, EPI_ISL_1190057, EPI_ISL_1190064, EPI_ISL_1190067, EPI_ISL_1190068, EPI_ISL_1190076, EPI_ISL_1190078, EPI_ISL_1190079, EPI_ISL_1190081, EPI_ISL_1190083, EPI_ISL_1190084, EPI_ISL_1190086, EPI_ISL_1190087, EPI_ISL_1190088, EPI_ISL_1190090, EPI_ISL_1190093, EPI_ISL_1190094, EPI_ISL_1190095, EPI_ISL_1190096, EPI_ISL_1190100, EPI_ISL_1190103, EPI_ISL_1190110, EPI_ISL_1190113, EPI_ISL_1190115, EPI_ISL_1242284, EPI_ISL_1242336, EPI_ISL_1242374, EPI_ISL_1242387, EPI_ISL_1242392, EPI_ISL_1242399, EPI_ISL_1242434, EPI_ISL_1242435, EPI_ISL_1242439, EPI_ISL_1242444, EPI_ISL_1242446, EPI_ISL_1242449, EPI_ISL_1242452, EPI_ISL_1242460, EPI_ISL_1242491, EPI_ISL_1242532, EPI_ISL_1242540, EPI_ISL_1242542, EPI_ISL_1242544, EPI_ISL_1243435, EPI_ISL_1243437, EPI_ISL_1243453, EPI_ISL_1243461, EPI_ISL_1243468, EPI_ISL_1243469, EPI_ISL_1243473, EPI_ISL_1243475, EPI_ISL_1243481, EPI_ISL_1243485, EPI_ISL_1243489, EPI_ISL_1243492, EPI_ISL_1243501, EPI_ISL_1243503, EPI_ISL_1243505, EPI_ISL_1243506, EPI_ISL_1243515, EPI_ISL_1243528, EPI_ISL_1243529, EPI_ISL_1243533, EPI_ISL_1243551, EPI_ISL_1243555, EPI_ISL_1243556, EPI_ISL_1243562, EPI_ISL_1243577, EPI_ISL_1243579, EPI_ISL_1243581, EPI_ISL_1243604, EPI_ISL_1265017, EPI_ISL_1265018, EPI_ISL_1265019, EPI_ISL_1265021, EPI_ISL_1265022, EPI_ISL_1265023, EPI_ISL_1265024, EPI_ISL_1265027, EPI_ISL_1265030, EPI_ISL_1265032, EPI_ISL_1265036, EPI_ISL_1265039, EPI_ISL_1265043, EPI_ISL_1265044, EPI_ISL_1265045, EPI_ISL_1265046, EPI_ISL_1265047, EPI_ISL_1265048, EPI_ISL_1265051, EPI_ISL_1265053, EPI_ISL_1265054, EPI_ISL_1265056, EPI_ISL_1265057, EPI_ISL_1265061, EPI_ISL_1265065, EPI_ISL_1265073, EPI_ISL_1265076, EPI_ISL_1265077, EPI_ISL_1265080, EPI_ISL_1265082, EPI_ISL_1265083, EPI_ISL_1265086, EPI_ISL_1265089, EPI_ISL_1265090, EPI_ISL_1265093, EPI_ISL_1265094, EPI_ISL_1265095, EPI_ISL_1265097, EPI_ISL_1265098, EPI_ISL_1265100, EPI_ISL_1265101, EPI_ISL_1265104, EPI_ISL_1265105, EPI_ISL_1265106, EPI_ISL_1265108, EPI_ISL_1265110, EPI_ISL_1265113, EPI_ISL_1265115, EPI_ISL_1265116, EPI_ISL_1265118, EPI_ISL_1265119, EPI_ISL_1265120, EPI_ISL_1265121, EPI_ISL_1265123, EPI_ISL_1265126, EPI_ISL_1265129, EPI_ISL_1265130, EPI_ISL_1265131, EPI_ISL_1265136, EPI_ISL_1265139, EPI_ISL_1265140 | | | |
| see above | Randox Laboratories | Wellcome Sanger Institute for the COVID-19 Genomics UK (COG-UK) Consortium | Cordelia Langford; David K. Jackson; Dominic Kwiatkowski; Ewan Harrison; Ian Johnston; Jeffrey Barrett; John Sillitoe on behalf of the Wellcome Sanger Institute COVID-19 Surveillance Team; Randox Laboratories and Alex Alderton; Roberto Amato; Sonia Goncalves |
| EPI_ISL_1105307, EPI_ISL_1105311, EPI_ISL_1105315, EPI_ISL_1105321, EPI_ISL_1105322, EPI_ISL_1105327, EPI_ISL_1105328, EPI_ISL_1105334, EPI_ISL_1105337 | | | |
| see above | University College London Hospital | COVID-19 Genomics UK (COG-UK) Consortium | Catherine Houlihan; Dan Frampton; Judith Heaney; Matthew Byott; Moira Spyer and Eleni Nastouli; Stuart Kirk |
| EPI_ISL_1050529, EPI_ISL_1050549, EPI_ISL_1050556, EPI_ISL_1104690, EPI_ISL_1104692, EPI_ISL_1104693, EPI_ISL_1104706, EPI_ISL_1104707, EPI_ISL_1104708, EPI_ISL_1104710, EPI_ISL_1104711, EPI_ISL_1104712, EPI_ISL_1104713, EPI_ISL_1104715, EPI_ISL_1104716, EPI_ISL_1104717, EPI_ISL_1104718, EPI_ISL_1104719, EPI_ISL_1104720, EPI_ISL_1104722, EPI_ISL_1104723, EPI_ISL_1104724, EPI_ISL_1104725, EPI_ISL_1104726, EPI_ISL_1104727, EPI_ISL_1104728, EPI_ISL_1104729, EPI_ISL_1104730, EPI_ISL_1104731, EPI_ISL_1104732, EPI_ISL_1104733, EPI_ISL_1104734, EPI_ISL_1104735, EPI_ISL_1104737, EPI_ISL_1104738, EPI_ISL_1104739, EPI_ISL_1104743, EPI_ISL_1104744, EPI_ISL_1104745, EPI_ISL_1104748, EPI_ISL_1104752, EPI_ISL_1104755, EPI_ISL_1104757, EPI_ISL_1104758, EPI_ISL_1105405, EPI_ISL_1105418, EPI_ISL_1105419, EPI_ISL_1105420, EPI_ISL_1105421, EPI_ISL_1105422, EPI_ISL_1105423, EPI_ISL_1105424, EPI_ISL_1105425, EPI_ISL_1105426, EPI_ISL_1105427, EPI_ISL_1105428, EPI_ISL_1105429, EPI_ISL_1105430, EPI_ISL_1105431, EPI_ISL_1105432, EPI_ISL_1105433, EPI_ISL_1105434, EPI_ISL_1105435, EPI_ISL_1105436, EPI_ISL_1105437, EPI_ISL_1105438, EPI_ISL_1105440, EPI_ISL_1105462, EPI_ISL_1247990, EPI_ISL_1250230, EPI_ISL_1250231, EPI_ISL_1250232, EPI_ISL_1250233, EPI_ISL_1250234, EPI_ISL_1250236, EPI_ISL_1250241, EPI_ISL_1386927, EPI_ISL_1637164 | | | |
| see above | University College London, Great Ormond Street Hospital for Children NHS Foundation Trust, Imperial College Healthcare NHS Trust | COVID-19 Genomics UK (COG-UK) Consortium | Alison Holmes; Charlotte Williams; Helena Tutill; Jacqueline Findlay; James Price; Judith Breuer; Julianne Brown; Kathryn Harris; Leysa Forrest; Mark Kristiansen; Paola Niola; Paola Resende Silva; Patricia Dyal; Paul Randell; Rachel Williams; Samuel Weeks; Sergi Castellano; Sunando Roy; Tony Brooks; Yasmin Panchbhaya |
| EPI_ISL_1177787, EPI_ISL_1177790, EPI_ISL_1177794, EPI_ISL_1177795, EPI_ISL_1177798, EPI_ISL_1177800 | University Hospitals Of Leicester NHS Trust and DeepSeq Nottingham | COVID-19 Genomics UK (COG-UK) Consortium | Christopher Holmes; Christopher Moore; Fei Sang; Joeseph Chappell; Johnny Debebe; Jonathan Ball; Julian Tang; Karlie Fallon; Matthew Carlisle; Matthew Loose; Nadine Holmes; Patrick McClure; Paul Bird; Thomas Helmer; Victoria Wright |
| EPI_ISL_1046951, EPI_ISL_1046952, EPI_ISL_1046957, EPI_ISL_1046959, EPI_ISL_1046960, EPI_ISL_1046961, EPI_ISL_1046962, EPI_ISL_1046963, EPI_ISL_1046964, EPI_ISL_1046965, EPI_ISL_1046966, EPI_ISL_1046967, EPI_ISL_1046968, EPI_ISL_1046969, EPI_ISL_1046971, EPI_ISL_1046972, EPI_ISL_1046973, EPI_ISL_1046974, EPI_ISL_1046980, EPI_ISL_1046983, EPI_ISL_1103746, EPI_ISL_1474473 | | | |
| see above | University of Birmingham | COVID-19 Genomics UK (COG-UK) Consortium | Alex Richter; Andrew Bosworth; Andrew Bosworth. Queen Elizabeth Hospital: Anna Casey; Andrew D Beggs PHE Heartlands Lab: Husam Osman; Charlotte Poxon; Institute of Microbiology; Joanne Stockton; Josh Quick; Kasun Wanigasooriya; Mike Kidd; Nicholas Loman. University of Birmingham Testing Laboratory: Celina M Whalley; Oliver Pickles; Radoslaw Poplawski; Samuel Nicholls; University of Birmingham: Claire McMurray; Will Rowe |
| EPI_ISL_1103843, EPI_ISL_1103845, EPI_ISL_1177759, EPI_ISL_1177760, EPI_ISL_1177770, EPI_ISL_1177771, EPI_ISL_1247445, EPI_ISL_1247446, EPI_ISL_1247452, EPI_ISL_1247490, EPI_ISL_1247527, EPI_ISL_1247528, EPI_ISL_1308602, EPI_ISL_1308603, EPI_ISL_1308629, EPI_ISL_1308631, EPI_ISL_1308635 | | | |
| see above | University of Exeter | COVID-19 Genomics UK (COG-UK) Consortium | Aaron Jeffries; Audrey Farbos; Ben Temperton; Jane Masoli; Joanna Warwick-Dugdale; Michelle Michelsen; Robyn Manley; Stephen Michell |
| EPI_ISL_1104268, EPI_ISL_1104269, EPI_ISL_1104272, EPI_ISL_1104273, EPI_ISL_1104274, EPI_ISL_1177726, EPI_ISL_1177735, EPI_ISL_1247698, EPI_ISL_1247700, EPI_ISL_1247727, EPI_ISL_1247738, EPI_ISL_1247739, EPI_ISL_1308709, EPI_ISL_1308710, EPI_ISL_1308711, EPI_ISL_1308712, EPI_ISL_1308727, EPI_ISL_1308752, EPI_ISL_1308753, EPI_ISL_1308754, EPI_ISL_1308755, EPI_ISL_1308756, EPI_ISL_1308757, EPI_ISL_1308758, EPI_ISL_1308759, EPI_ISL_1308826, EPI_ISL_1386686, EPI_ISL_1386691, EPI_ISL_1474574, EPI_ISL_1474591, EPI_ISL_1474594, EPI_ISL_1730074, EPI_ISL_1730075 | | | |
| see above | Virology Department, Royal Infirmary of Edinburgh, NHS Lothian / School of Biological Sciences, University of Edinburgh | COVID-19 Genomics UK (COG-UK) Consortium | Colquhoun R; Cotton S; Dewar R; Gallagher A; Hill V; Jackson B; Maloney D; McCrone JT; McHugh M; O'Toole Á; Rambaut A; Rooke S; Scher E; Templeton K; Yu X |
| EPI_ISL_1054621, EPI_ISL_1054622, EPI_ISL_1054623, EPI_ISL_1054632, EPI_ISL_1054636, EPI_ISL_1054638, EPI_ISL_1054644, EPI_ISL_1054650, EPI_ISL_1054660, EPI_ISL_1054666, EPI_ISL_1054667, EPI_ISL_1054669, EPI_ISL_1054670, EPI_ISL_1054673, EPI_ISL_1054675, EPI_ISL_1054676, EPI_ISL_1054685, EPI_ISL_1054688, EPI_ISL_1054693, EPI_ISL_1054695, EPI_ISL_1054697, EPI_ISL_1054699, EPI_ISL_1054703, EPI_ISL_1054706, EPI_ISL_1105581, EPI_ISL_1105584, EPI_ISL_1105590, EPI_ISL_1105591, EPI_ISL_1105600, EPI_ISL_1105601, EPI_ISL_1105603, EPI_ISL_1105604, EPI_ISL_1105607, EPI_ISL_1105608, EPI_ISL_1105609, EPI_ISL_1105611, EPI_ISL_1105616, EPI_ISL_1105622, EPI_ISL_1105623, EPI_ISL_1105626, EPI_ISL_1105627, EPI_ISL_1105634, EPI_ISL_1105635, EPI_ISL_1105636, EPI_ISL_1105637, EPI_ISL_1105642, EPI_ISL_1105643, EPI_ISL_1105645, EPI_ISL_1105646, EPI_ISL_1105653, EPI_ISL_1105655, EPI_ISL_1176816, EPI_ISL_1176818, EPI_ISL_1176820, EPI_ISL_1176821, EPI_ISL_1176822, EPI_ISL_1176826, EPI_ISL_1176828, EPI_ISL_1176829, EPI_ISL_1176830, EPI_ISL_1176831, EPI_ISL_1176832, EPI_ISL_1176833, EPI_ISL_1176836, EPI_ISL_1176837, EPI_ISL_1176838, EPI_ISL_1176839, EPI_ISL_1176840, EPI_ISL_1176841, EPI_ISL_1176843, EPI_ISL_1176844, EPI_ISL_1176849, EPI_ISL_1176850, EPI_ISL_1176851, EPI_ISL_1176852, EPI_ISL_1176853, EPI_ISL_1176855, EPI_ISL_1176860, EPI_ISL_1176861, EPI_ISL_1176864, EPI_ISL_1176866, EPI_ISL_1176867, EPI_ISL_1176871, EPI_ISL_1176872, EPI_ISL_1176873, EPI_ISL_1176874, EPI_ISL_1176875, EPI_ISL_1176877, | | | |

EPI_ISL_1176878, EPI_ISL_1176881, EPI_ISL_1176882, EPI_ISL_1176883, EPI_ISL_1176884, EPI_ISL_1176885, EPI_ISL_1176886, EPI_ISL_1176888, EPI_ISL_1176889, EPI_ISL_1176893, EPI_ISL_1176894, EPI_ISL_1176895, EPI_ISL_1176896, EPI_ISL_1176897, EPI_ISL_1176898, EPI_ISL_1176899, EPI_ISL_1178068, EPI_ISL_1178069, EPI_ISL_1178070, EPI_ISL_1178073, EPI_ISL_1178074, EPI_ISL_1178076, EPI_ISL_1178078, EPI_ISL_1178079, EPI_ISL_1178081, EPI_ISL_1178082, EPI_ISL_1178084, EPI_ISL_1178085, EPI_ISL_1178089, EPI_ISL_1178093, EPI_ISL_1180053, EPI_ISL_1180068, EPI_ISL_1180088, EPI_ISL_1180097, EPI_ISL_1248578

| | | | |
|---|---|---|---|
| see above | Virology Department, Sheffield Teaching Hospitals NHS Foundation Trust/Department of Infection, Immunity and Cardiovascular Disease, The Medical School, University of Sheffield | COVID-19 Genomics UK (COG-UK) Consortium | Adri Angyal; Alex Keeley; Benjamin Lindsey; Cariad Evans; Danielle Groves; Dave Partridge; Katie Johnson; Laura Carrilero; Luke Green; Matthew Parker; Matthew Wyles; Mehmet Yavuz; Mohammad Raza; Nikki Smith; Paul Parsons; Rachel Tucker; Rebecca Brown; Thushan de Silva |

EPI_ISL_1053322, EPI_ISL_1053327, EPI_ISL_1053328, EPI_ISL_1053364, EPI_ISL_1053499, EPI_ISL_1053500, EPI_ISL_1105937, EPI_ISL_1105957, EPI_ISL_1105964, EPI_ISL_1105966, EPI_ISL_1105967, EPI_ISL_1105973, EPI_ISL_1105994, EPI_ISL_1106014, EPI_ISL_1106016, EPI_ISL_1106018, EPI_ISL_1106021, EPI_ISL_1106022, EPI_ISL_1106023, EPI_ISL_1106024, EPI_ISL_1106026, EPI_ISL_1106031, EPI_ISL_1106035, EPI_ISL_1106039, EPI_ISL_1106040, EPI_ISL_1106043, EPI_ISL_1106064, EPI_ISL_1106067, EPI_ISL_1106068, EPI_ISL_1106069, EPI_ISL_1106070, EPI_ISL_1106071, EPI_ISL_1106072, EPI_ISL_1106127, EPI_ISL_1106135, EPI_ISL_1106136, EPI_ISL_1106162, EPI_ISL_1106165, EPI_ISL_1106172, EPI_ISL_1106179, EPI_ISL_1106185, EPI_ISL_1106186, EPI_ISL_1106187, EPI_ISL_1106188, EPI_ISL_1106197, EPI_ISL_1106198, EPI_ISL_1106203, EPI_ISL_1106204, EPI_ISL_1106205, EPI_ISL_1106206, EPI_ISL_1106247, EPI_ISL_1106273, EPI_ISL_1106274, EPI_ISL_1106298, EPI_ISL_1106352, EPI_ISL_1106355, EPI_ISL_1106356, EPI_ISL_1106359, EPI_ISL_1106367, EPI_ISL_1106368, EPI_ISL_1106369, EPI_ISL_1106370, EPI_ISL_1106373, EPI_ISL_1106374, EPI_ISL_1106378, EPI_ISL_1106379, EPI_ISL_1106382, EPI_ISL_1106384, EPI_ISL_1106385, EPI_ISL_1106386, EPI_ISL_1106388, EPI_ISL_1106389, EPI_ISL_1106390, EPI_ISL_1106391, EPI_ISL_1106393, EPI_ISL_1106394, EPI_ISL_1106396, EPI_ISL_1106398, EPI_ISL_1106399, EPI_ISL_1106402, EPI_ISL_1106406, EPI_ISL_1106410, EPI_ISL_1106414, EPI_ISL_1106415, EPI_ISL_1106417, EPI_ISL_1106424, EPI_ISL_1106427, EPI_ISL_1106432, EPI_ISL_1106433, EPI_ISL_1106434, EPI_ISL_1106435, EPI_ISL_1106436, EPI_ISL_1106437, EPI_ISL_1106439, EPI_ISL_1106441, EPI_ISL_1106445, EPI_ISL_1106448, EPI_ISL_1106456, EPI_ISL_1106458, EPI_ISL_1106459, EPI_ISL_1106471, EPI_ISL_1106472, EPI_ISL_1106473, EPI_ISL_1106474, EPI_ISL_1106476, EPI_ISL_1106486, EPI_ISL_1106488, EPI_ISL_1106490, EPI_ISL_1106492, EPI_ISL_1106499, EPI_ISL_1106500, EPI_ISL_1106505, EPI_ISL_1106509, EPI_ISL_1106512, EPI_ISL_1106516, EPI_ISL_1106517, EPI_ISL_1106518, EPI_ISL_1106519, EPI_ISL_1106520, EPI_ISL_1106521, EPI_ISL_1106522, EPI_ISL_1106523, EPI_ISL_1106524, EPI_ISL_1106525, EPI_ISL_1106526, EPI_ISL_1106529, EPI_ISL_1106530, EPI_ISL_1106533, EPI_ISL_1106535, EPI_ISL_1106537, EPI_ISL_1106549, EPI_ISL_1106551, EPI_ISL_1106553, EPI_ISL_1106554, EPI_ISL_1106556, EPI_ISL_1106560, EPI_ISL_1106565, EPI_ISL_1106567, EPI_ISL_1106569, EPI_ISL_1106571, EPI_ISL_1106572, EPI_ISL_1106577, EPI_ISL_1106578, EPI_ISL_1106586, EPI_ISL_1106587, EPI_ISL_1106592, EPI_ISL_1106593, EPI_ISL_1106594, EPI_ISL_1106596, EPI_ISL_1106599, EPI_ISL_1106602, EPI_ISL_1106608, EPI_ISL_1106715, EPI_ISL_1106716, EPI_ISL_1106717, EPI_ISL_1106718, EPI_ISL_1106719, EPI_ISL_1106720, EPI_ISL_1106721, EPI_ISL_1106726, EPI_ISL_1106727, EPI_ISL_1106728, EPI_ISL_1106729, EPI_ISL_1106730, EPI_ISL_1106731, EPI_ISL_1106732, EPI_ISL_1106733, EPI_ISL_1106734, EPI_ISL_1106735, EPI_ISL_1106736, EPI_ISL_1106737, EPI_ISL_1106746, EPI_ISL_1106748, EPI_ISL_1106749, EPI_ISL_1106750, EPI_ISL_1106751, EPI_ISL_1106754, EPI_ISL_1106755, EPI_ISL_1106770, EPI_ISL_1106779, EPI_ISL_1106781, EPI_ISL_1106782, EPI_ISL_1106783, EPI_ISL_1106791, EPI_ISL_1106922, EPI_ISL_1106923, EPI_ISL_1106924, EPI_ISL_1106937, EPI_ISL_1106938, EPI_ISL_1106939, EPI_ISL_1106940, EPI_ISL_1106941, EPI_ISL_1106943, EPI_ISL_1106944, EPI_ISL_1106953, EPI_ISL_1106954, EPI_ISL_1106955, EPI_ISL_1106956, EPI_ISL_1106957, EPI_ISL_1107001, EPI_ISL_1107263, EPI_ISL_1178887, EPI_ISL_1178929

| | | | |
|---|---|---|---|
| see above | Wales Specialist Virology Centre Sequencing lab: Pathogen Genomics Unit | Public Health Wales Microbiology Cardiff Wales Specialist Virology Centre | Alec Birchley; Alexander Adams; Amy Gaskin; Angela Marchbank; Bree Gatica-Wilcox; Catherine Moore; Jason Coombes; Joanne Watkins; Joel Southgate; Johnathan Evans; Laura Gifford; Lauren Gilbert; Lee Graham; Malorie Perry; Matthew Bull; Nicole Pacchiarini; Sally Corden; Sara Kumziene-Summerhayes; Sara Rey; Sarah Taylor; Simon Cottrell; Sophie Jones; Tom Connor |

EPI_ISL_1104179, EPI_ISL_1104180, EPI_ISL_1104181, EPI_ISL_1104182, EPI_ISL_1104183, EPI_ISL_1104184, EPI_ISL_1104185, EPI_ISL_1104187, EPI_ISL_1104188, EPI_ISL_1104189, EPI_ISL_1104190, EPI_ISL_1247662

| | | | |
|---|---|---|---|
| see above | West of Scotland Specialist Virology Centre, NHSGGC / MRC-University of Glasgow Centre for Virus Research | COVID-19 Genomics UK (COG-UK) Consortium | Alasdair MacLean; Alice Broos; Ana da Silva Filipe; Antonia Ho; Daniel Mair; David L Robertson; Emma Thomson; Guy Mollett; James Shepherd; Jenna Nichols; Joseph Hughes; Kathy Li; Kathy Smollett; Kyriaki Nomikou; Lily Tong; Matthew Holden; Natasha Johnson; Rachel Blacow; Richard Orton; Rory Gunson; Sarah McDonald; Sharif Shaaban; Sreenu Vattipally; Stephen Carmichael |

We gratefully acknowledge the following Authors from the Originating laboratories responsible for obtaining the specimens, as well as the Submitting laboratories where the genome data were generated and shared via GISAID, on which this research is based.

All Submitters of data may be contacted directly via www.gisaid.org

Authors are sorted alphabetically.

| Accession ID | Originating Laboratory | Submitting Laboratory | Authors |
|---|---|---|---|
| EPI_ISL_1730317, EPI_ISL_1730320, EPI_ISL_1730322 | Barts Health NHS Trust | COVID-19 Genomics UK (COG-UK) Consortium | BROAD; Beatrix; CUTINO-MOGUEL; Claire; David; Dola; HARRINGTON; KELE; KULASEGARAN-SHYLINI; Maria-Teresa; OWOYEMI; Raghavendran |
| EPI_ISL_1638255, EPI_ISL_1638257, EPI_ISL_1638258, EPI_ISL_1833927 | Bioinformatics and Biostatistics Lab, Advanced Sequencing Facility | COVID-19 Genomics UK (COG-UK) Consortium | Aengus Stewart; Chelsea Sawyer; Harshil Patel; Jerome Nicod; Laura Cubitt; Margaret Crawford |
| EPI_ISL_1637980, EPI_ISL_1730822, EPI_ISL_1730823, EPI_ISL_1730824, EPI_ISL_1730825, EPI_ISL_1730826, EPI_ISL_1730827, EPI_ISL_1730828, EPI_ISL_1730829, EPI_ISL_1730830, EPI_ISL_1730889, EPI_ISL_1730890, EPI_ISL_1730891, EPI_ISL_1730892, EPI_ISL_1730893, EPI_ISL_1730894, EPI_ISL_1730895, EPI_ISL_1730897, EPI_ISL_1730900, EPI_ISL_1730905, EPI_ISL_1730906, EPI_ISL_1730907, EPI_ISL_1730914, EPI_ISL_1730915, EPI_ISL_1730916, EPI_ISL_1730925 | | | |
| see above | Centre for Enzyme Innovation, University of Portsmouth / Translational Research Laboratory, Portsmouth Hospitals NHS Trust | COVID-19 Genomics UK (COG-UK) Consortium | Angela Beckett; Christopher Fearn; Kate Cook; Katie Loveson; Salman Goudarzi; Samuel Robson; Scott Elliott; Sharon Glaysher |
| EPI_ISL_1545849, EPI_ISL_1636622, EPI_ISL_1636623, EPI_ISL_1636624, EPI_ISL_1730043, EPI_ISL_1730045, EPI_ISL_1730046, EPI_ISL_1730048, EPI_ISL_1730049, EPI_ISL_1730050, EPI_ISL_1730051, EPI_ISL_1730052, EPI_ISL_1730053, EPI_ISL_1730054, EPI_ISL_1730056, EPI_ISL_1730057, EPI_ISL_1730058, EPI_ISL_1730059, EPI_ISL_1730060, EPI_ISL_1730061, EPI_ISL_1730062, EPI_ISL_1730063, EPI_ISL_1730066 | | | |
| see above | Department of Pathology, University of Cambridge | COVID-19 Genomics UK (COG-UK) Consortium | Aminu S. Jahun; Ian Goodfellow; Iliana Georgana; Martin D. Curran; Myra Hosmillo; Rhys Izuagbe; Surendra Parmar; William L. Hamilton; Yasmin Chaudhry |
| EPI_ISL_1831313, EPI_ISL_1831315, EPI_ISL_1831317, EPI_ISL_1831322, EPI_ISL_1831323, EPI_ISL_1831329, EPI_ISL_1831330, EPI_ISL_1831334, EPI_ISL_1831342, EPI_ISL_1831344, EPI_ISL_1831345, EPI_ISL_1831348, EPI_ISL_1831349, EPI_ISL_1831350, EPI_ISL_1831354, EPI_ISL_1831357, EPI_ISL_1831362, EPI_ISL_1831364, EPI_ISL_1831376, EPI_ISL_1831381, EPI_ISL_1831382, EPI_ISL_1831383, EPI_ISL_1831385 | | | |
| see above | Health Services Laboratories | Wellcome Sanger Institute for the COVID-19 Genomics UK (COG-UK) Consortium | Cordelia Langford; David K. Jackson; Dominic Kwiatkowski; Ewan Harrison; Health Services Laboratories and Alex Alderton; Ian Johnston; Jeffrey Barrett; John Sillitoe on behalf of the Wellcome Sanger Institute COVID-19 Surveillance Team; Roberto Amato; Sonia Goncalves |
| EPI_ISL_1564333, EPI_ISL_1564334, EPI_ISL_1564336, EPI_ISL_1564337, EPI_ISL_1564338, EPI_ISL_1564339, EPI_ISL_1564340, EPI_ISL_1564342, EPI_ISL_1564343, EPI_ISL_1564346, EPI_ISL_1564347, EPI_ISL_1564348, EPI_ISL_1564350, EPI_ISL_1564351, EPI_ISL_1564352, EPI_ISL_1564353, EPI_ISL_1564358, EPI_ISL_1564359, EPI_ISL_1564360, EPI_ISL_1564361, EPI_ISL_1564363, EPI_ISL_1564365, EPI_ISL_1564366, EPI_ISL_1564367, EPI_ISL_1564368, EPI_ISL_1564369, EPI_ISL_1564370, EPI_ISL_1564372, EPI_ISL_1564374, EPI_ISL_1564376, EPI_ISL_1564379, EPI_ISL_1564380, EPI_ISL_1564381, EPI_ISL_1564382, EPI_ISL_1564383, EPI_ISL_1564384, EPI_ISL_1564386, EPI_ISL_1564387, EPI_ISL_1564388, EPI_ISL_1564390, EPI_ISL_1564391, EPI_ISL_1564392, EPI_ISL_1564393, EPI_ISL_1564394, EPI_ISL_1564395, EPI_ISL_1564396, EPI_ISL_1564398, EPI_ISL_1564399, EPI_ISL_1564400, EPI_ISL_1564402, EPI_ISL_1564403, EPI_ISL_1564406, EPI_ISL_1564408, EPI_ISL_1564409, EPI_ISL_1564410, EPI_ISL_1564413, EPI_ISL_1564414, EPI_ISL_1564415, EPI_ISL_1564416, EPI_ISL_1564417, EPI_ISL_1564418, EPI_ISL_1564421, EPI_ISL_1564424, EPI_ISL_1564425, EPI_ISL_1564426, EPI_ISL_1564427, EPI_ISL_1564428, EPI_ISL_1564429, EPI_ISL_1564431, EPI_ISL_1564432, EPI_ISL_1564433, EPI_ISL_1564434, EPI_ISL_1564435, EPI_ISL_1564439, EPI_ISL_1564440, EPI_ISL_1564442, EPI_ISL_1564443, EPI_ISL_1564444, EPI_ISL_1564445, EPI_ISL_1564446, EPI_ISL_1564447, EPI_ISL_1564448, EPI_ISL_1564451, EPI_ISL_1564452, EPI_ISL_1564453, EPI_ISL_1564455, EPI_ISL_1564456, EPI_ISL_1564457, EPI_ISL_1564458, EPI_ISL_1564459, EPI_ISL_1564461, EPI_ISL_1564462, EPI_ISL_1564463, EPI_ISL_1564464, EPI_ISL_1564466, EPI_ISL_1564467, EPI_ISL_1564468, EPI_ISL_1564469, EPI_ISL_1564471, EPI_ISL_1564472, EPI_ISL_1564473, EPI_ISL_1564474, EPI_ISL_1564476, EPI_ISL_1564477, EPI_ISL_1564478, EPI_ISL_1564482, EPI_ISL_1564483, EPI_ISL_1564484, EPI_ISL_1564485, EPI_ISL_1564486, EPI_ISL_1564488, EPI_ISL_1564490, EPI_ISL_1564491, EPI_ISL_1564492, EPI_ISL_1564496, EPI_ISL_1564497, EPI_ISL_1564499, EPI_ISL_1564500, EPI_ISL_1564501, EPI_ISL_1564502, EPI_ISL_1564504, EPI_ISL_1564505, EPI_ISL_1564506, EPI_ISL_1564507, EPI_ISL_1564508, EPI_ISL_1564509, EPI_ISL_1564511, EPI_ISL_1564512, EPI_ISL_1564513, EPI_ISL_1564515, EPI_ISL_1564517, EPI_ISL_1564519, EPI_ISL_1564520, EPI_ISL_1564521, EPI_ISL_1564523, EPI_ISL_1564524, EPI_ISL_1564525, EPI_ISL_1564526, EPI_ISL_1564527, EPI_ISL_1564528, EPI_ISL_1564531, EPI_ISL_1564533, EPI_ISL_1564534, EPI_ISL_1564535, EPI_ISL_1564538, EPI_ISL_1564539, EPI_ISL_1564540, EPI_ISL_1564541, EPI_ISL_1564542, EPI_ISL_1564544, EPI_ISL_1564545, EPI_ISL_1564546, EPI_ISL_1564547, EPI_ISL_1564548, EPI_ISL_1564549, EPI_ISL_1564551, EPI_ISL_1564552, EPI_ISL_1564553, EPI_ISL_1564554, EPI_ISL_1564555, EPI_ISL_1564556, EPI_ISL_1564560, EPI_ISL_1564563, EPI_ISL_1564565, EPI_ISL_1564566, EPI_ISL_1564568, EPI_ISL_1564569, EPI_ISL_1564570, EPI_ISL_1564571, EPI_ISL_1564572, EPI_ISL_1564573, EPI_ISL_1564574, EPI_ISL_1564576, EPI_ISL_1564577, EPI_ISL_1564578, EPI_ISL_1564579, EPI_ISL_1564580, EPI_ISL_1564581, EPI_ISL_1564583, EPI_ISL_1564585, EPI_ISL_1564587, EPI_ISL_1564588, EPI_ISL_1564589, EPI_ISL_1564591, EPI_ISL_1564592, EPI_ISL_1564593, EPI_ISL_1564594, EPI_ISL_1564596, EPI_ISL_1564597, EPI_ISL_1564598, EPI_ISL_1564601, EPI_ISL_1564602, EPI_ISL_1564603, EPI_ISL_1564604, EPI_ISL_1564605, EPI_ISL_1564608, EPI_ISL_1564609, EPI_ISL_1564627, EPI_ISL_1564628, EPI_ISL_1564629, EPI_ISL_1564630, EPI_ISL_1564631, EPI_ISL_1564634, EPI_ISL_1564635, EPI_ISL_1564636, EPI_ISL_1564637, EPI_ISL_1564638, EPI_ISL_1564639, EPI_ISL_1564640, EPI_ISL_1564642, EPI_ISL_1564643, EPI_ISL_1564644, EPI_ISL_1564646, EPI_ISL_1564647, EPI_ISL_1564648, EPI_ISL_1564650, EPI_ISL_1564652, EPI_ISL_1564653, EPI_ISL_1564654, EPI_ISL_1564655, EPI_ISL_1564656, EPI_ISL_1564657, EPI_ISL_1564659, EPI_ISL_1564660, EPI_ISL_1564661, EPI_ISL_1564662, EPI_ISL_1564663, EPI_ISL_1564664, EPI_ISL_1564665, EPI_ISL_1564666, EPI_ISL_1564668, EPI_ISL_1564670, EPI_ISL_1564671, EPI_ISL_1564672, EPI_ISL_1564673, EPI_ISL_1564675, EPI_ISL_1564676, EPI_ISL_1564677, EPI_ISL_1564678, EPI_ISL_1564679, EPI_ISL_1564680, EPI_ISL_1564681, EPI_ISL_1564682, EPI_ISL_1564685, EPI_ISL_1564686, EPI_ISL_1564687, EPI_ISL_1564688, EPI_ISL_1564689, EPI_ISL_1564690, EPI_ISL_1564691, EPI_ISL_1564692, EPI_ISL_1564695, EPI_ISL_1564696, EPI_ISL_1564697, EPI_ISL_1564698, EPI_ISL_1564700, EPI_ISL_1564702, EPI_ISL_1564703, EPI_ISL_1564705, EPI_ISL_1564708, EPI_ISL_1564709, EPI_ISL_1564710, EPI_ISL_1564711, EPI_ISL_1564712, EPI_ISL_1564713, EPI_ISL_1564714, EPI_ISL_1564716, EPI_ISL_1564717, EPI_ISL_1564718, EPI_ISL_1564719, EPI_ISL_1564720, EPI_ISL_1564722, EPI_ISL_1564723, EPI_ISL_1564724, EPI_ISL_1564726, EPI_ISL_1564727, EPI_ISL_1564729, EPI_ISL_1564730, EPI_ISL_1564733, EPI_ISL_1564735, EPI_ISL_1564736, EPI_ISL_1564737, EPI_ISL_1564738, EPI_ISL_1564740, EPI_ISL_1564741, EPI_ISL_1564742, EPI_ISL_1564743, EPI_ISL_1564745, EPI_ISL_1564747, EPI_ISL_1564748, EPI_ISL_1564749, EPI_ISL_1564750, EPI_ISL_1564751, EPI_ISL_1564752, EPI_ISL_1564753, EPI_ISL_1564755, EPI_ISL_1564756, EPI_ISL_1564757, EPI_ISL_1564758, EPI_ISL_1564760, EPI_ISL_1564761, EPI_ISL_1564762, EPI_ISL_1564763, EPI_ISL_1565338, EPI_ISL_1583685, EPI_ISL_1583693, EPI_ISL_1583696, EPI_ISL_1583698, EPI_ISL_1583699, EPI_ISL_1583709, EPI_ISL_1583712, EPI_ISL_1583720, EPI_ISL_1583723, EPI_ISL_1583728, EPI_ISL_1583742, EPI_ISL_1583743, EPI_ISL_1583744, EPI_ISL_1583748, EPI_ISL_1583750, EPI_ISL_1583752, EPI_ISL_1583753, EPI_ISL_1583755, EPI_ISL_1583756, EPI_ISL_1583757, EPI_ISL_1583758, EPI_ISL_1583759, EPI_ISL_1583763, EPI_ISL_1583764, EPI_ISL_1583769, EPI_ISL_1583770, EPI_ISL_1583771, EPI_ISL_1583773, EPI_ISL_1583774, EPI_ISL_1583776, EPI_ISL_1583777, EPI_ISL_1583780, EPI_ISL_1583781, EPI_ISL_1583783, EPI_ISL_1583785, EPI_ISL_1583787, EPI_ISL_1583790, EPI_ISL_1583792, EPI_ISL_1583797, EPI_ISL_1583799, EPI_ISL_1583807, EPI_ISL_1583808, EPI_ISL_1583811, EPI_ISL_1583813, EPI_ISL_1583816, EPI_ISL_1583817, EPI_ISL_1583819, EPI_ISL_1583824, EPI_ISL_1583828, EPI_ISL_1583829, EPI_ISL_1583832, EPI_ISL_1583844, EPI_ISL_1583845, EPI_ISL_1583852, EPI_ISL_1583853, EPI_ISL_1583857, EPI_ISL_1583858, EPI_ISL_1583859, EPI_ISL_1583861, EPI_ISL_1583864, EPI_ISL_1583865, EPI_ISL_1583869, EPI_ISL_1583870, EPI_ISL_1583872, EPI_ISL_1583875, EPI_ISL_1583878, EPI_ISL_1583879, EPI_ISL_1583880, EPI_ISL_1583884, EPI_ISL_1583886, EPI_ISL_1583889, EPI_ISL_1583891, EPI_ISL_1583892, EPI_ISL_1583897, EPI_ISL_1583898, EPI_ISL_1583900, EPI_ISL_1583902, EPI_ISL_1583905, EPI_ISL_1583906, EPI_ISL_1583910, EPI_ISL_1583912, EPI_ISL_1583914, EPI_ISL_1583916, EPI_ISL_1583917, EPI_ISL_1583919, EPI_ISL_1584167, EPI_ISL_1584169, EPI_ISL_1584170, EPI_ISL_1584174, EPI_ISL_1584184, EPI_ISL_1584194, EPI_ISL_1584196, EPI_ISL_1584198, EPI_ISL_1584202, EPI_ISL_1584214, EPI_ISL_1584220, EPI_ISL_1584225, EPI_ISL_1584230, EPI_ISL_1584231, EPI_ISL_1584236, EPI_ISL_1584237, EPI_ISL_1584243, EPI_ISL_1584247, EPI_ISL_1584249, EPI_ISL_1584251, EPI_ISL_1584261, EPI_ISL_1584262, EPI_ISL_1584265, EPI_ISL_1584267, EPI_ISL_1584270, EPI_ISL_1584273, EPI_ISL_1584276, EPI_ISL_1584278, EPI_ISL_1584281, EPI_ISL_1584285, EPI_ISL_1584291, EPI_ISL_1584296, EPI_ISL_1584303, EPI_ISL_1584304, EPI_ISL_1584312, EPI_ISL_1584319, EPI_ISL_1584321, EPI_ISL_1584323, EPI_ISL_1584324, EPI_ISL_1584332, EPI_ISL_1584339, EPI_ISL_1584343, EPI_ISL_1584344, EPI_ISL_1584347, EPI_ISL_1584349, EPI_ISL_1584351, EPI_ISL_1584352, EPI_ISL_1584355, EPI_ISL_1584359, EPI_ISL_1584361, EPI_ISL_1584362, EPI_ISL_1584366, EPI_ISL_1584372, EPI_ISL_1584373, EPI_ISL_1584376, EPI_ISL_1584377, EPI_ISL_1584381, EPI_ISL_1584384, EPI_ISL_1584386, EPI_ISL_1584393, EPI_ISL_1584397, EPI_ISL_1584400, EPI_ISL_1584401, EPI_ISL_1584405, EPI_ISL_1584411, EPI_ISL_1584412, EPI_ISL_1584413, EPI_ISL_1584423, EPI_ISL_1584427, EPI_ISL_1584433, EPI_ISL_1584435, EPI_ISL_1584438, EPI_ISL_1584439, EPI_ISL_1584528, EPI_ISL_1594714, EPI_ISL_1594715, EPI_ISL_1594716, EPI_ISL_1594717, EPI_ISL_1594722, EPI_ISL_1594724, EPI_ISL_1594725, EPI_ISL_1594732, EPI_ISL_1594733, EPI_ISL_1594734, EPI_ISL_1594737, EPI_ISL_1594738, EPI_ISL_1594739, EPI_ISL_1594740, EPI_ISL_1594741, EPI_ISL_1594742, EPI_ISL_1594745, EPI_ISL_1594747, EPI_ISL_1594750, EPI_ISL_1594762, EPI_ISL_1594763, EPI_ISL_1594764, EPI_ISL_1594766, EPI_ISL_1594767, EPI_ISL_1594768, EPI_ISL_1594773, EPI_ISL_1594774, EPI_ISL_1594775, EPI_ISL_1594776, EPI_ISL_1594778, EPI_ISL_1594780, EPI_ISL_1594781, EPI_ISL_1594782, EPI_ISL_1594784, EPI_ISL_1594785, EPI_ISL_1594786, EPI_ISL_1594789, EPI_ISL_1594791, EPI_ISL_1594792, EPI_ISL_1594793, EPI_ISL_1594800, EPI_ISL_1594802, EPI_ISL_1594804, EPI_ISL_1594811, EPI_ISL_1594812, EPI_ISL_1594813, EPI_ISL_1594815, EPI_ISL_1594817, EPI_ISL_1594818, EPI_ISL_1594821, EPI_ISL_1594822, EPI_ISL_1594824, EPI_ISL_1594825, EPI_ISL_1594827, EPI_ISL_1594828, EPI_ISL_1594830, EPI_ISL_1594834, EPI_ISL_1594835, EPI_ISL_1594836, EPI_ISL_1594838, EPI_ISL_1594841, EPI_ISL_1594846, EPI_ISL_1594849, EPI_ISL_1594850, EPI_ISL_1594853, EPI_ISL_1594855, EPI_ISL_1594857, EPI_ISL_1594860, EPI_ISL_1594863, EPI_ISL_1594864, EPI_ISL_1594865, EPI_ISL_1594867, EPI_ISL_1594868, EPI_ISL_1594869, EPI_ISL_1594870, EPI_ISL_1594872, EPI_ISL_1594873, EPI_ISL_1594875, EPI_ISL_1594879, EPI_ISL_1594881, EPI_ISL_1594882, EPI_ISL_1594883, EPI_ISL_1594884, EPI_ISL_1594885, EPI_ISL_1594886, EPI_ISL_1594887, EPI_ISL_1594889, EPI_ISL_1594890, EPI_ISL_1594891, EPI_ISL_1594893, EPI_ISL_1594894, EPI_ISL_1594897, EPI_ISL_1594899, EPI_ISL_1594902, EPI_ISL_1594905, EPI_ISL_1594906, EPI_ISL_1594907, EPI_ISL_1594908, EPI_ISL_1594914, EPI_ISL_1594915, EPI_ISL_1594917, EPI_ISL_1594919, EPI_ISL_1594921, EPI_ISL_1594926, EPI_ISL_1594931, EPI_ISL_1594935, EPI_ISL_1594937, EPI_ISL_1594941, EPI_ISL_1594944, EPI_ISL_1594946, EPI_ISL_1594948, EPI_ISL_1594949, EPI_ISL_1594950, EPI_ISL_1594951, EPI_ISL_1594954, EPI_ISL_1594955, EPI_ISL_1594958, EPI_ISL_1594961, EPI_ISL_1594963, EPI_ISL_1594964, EPI_ISL_1594965, EPI_ISL_1594967, EPI_ISL_1594969, EPI_ISL_1594970, EPI_ISL_1594973, EPI_ISL_1594974, EPI_ISL_1594976, EPI_ISL_1594977, EPI_ISL_1594982, EPI_ISL_1594985, EPI_ISL_1594987, EPI_ISL_1594989, EPI_ISL_1594990, EPI_ISL_1594992, EPI_ISL_1594993, EPI_ISL_1595470, EPI_ISL_1615688, EPI_ISL_1615697, EPI_ISL_1615698, EPI_ISL_1615706, EPI_ISL_1615707, EPI_ISL_1615716, EPI_ISL_1615729, EPI_ISL_1615733, EPI_ISL_1615735, EPI_ISL_1615736, EPI_ISL_1615747, EPI_ISL_1615749, EPI_ISL_1615757, EPI_ISL_1615759, EPI_ISL_1615763, EPI_ISL_1615778, EPI_ISL_1615783, EPI_ISL_1615790, EPI_ISL_1615792, EPI_ISL_1615794, EPI_ISL_1615796, EPI_ISL_1615805, EPI_ISL_1615815, EPI_ISL_1615817, EPI_ISL_1615837, EPI_ISL_1615843, EPI_ISL_1615847, EPI_ISL_1615855, EPI_ISL_1615858, EPI_ISL_1615864, EPI_ISL_1615872, EPI_ISL_1615881, EPI_ISL_1615882, EPI_ISL_1615885, EPI_ISL_1615899, EPI_ISL_1615901, EPI_ISL_1615905, EPI_ISL_1615918, EPI_ISL_1616267, EPI_ISL_1616283, EPI_ISL_1616593, EPI_ISL_1630822, EPI_ISL_1630823, EPI_ISL_1630824, EPI_ISL_1630825, EPI_ISL_1630826, EPI_ISL_1630827, EPI_ISL_1630828, EPI_ISL_1630829, EPI_ISL_1630831, EPI_ISL_1630832, EPI_ISL_1630833, EPI_ISL_1630835, EPI_ISL_1630836, EPI_ISL_1630837, EPI_ISL_1630838, EPI_ISL_1630839, EPI_ISL_1630840, EPI_ISL_1630841, EPI_ISL_1630842, EPI_ISL_1630843, EPI_ISL_1630844, EPI_ISL_1630845, EPI_ISL_1630849, EPI_ISL_1630851, EPI_ISL_1630852, EPI_ISL_1630854, EPI_ISL_1630856, EPI_ISL_1630857, EPI_ISL_1630858, EPI_ISL_1630859, EPI_ISL_1630860, EPI_ISL_1630861, EPI_ISL_1630862, EPI_ISL_1630863, EPI_ISL_1630864, EPI_ISL_1630865, EPI_ISL_1630866, EPI_ISL_1630867, EPI_ISL_1630868, EPI_ISL_1630869, EPI_ISL_1630870, EPI_ISL_1630873, EPI_ISL_1630874, EPI_ISL_1630875, EPI_ISL_1630876, EPI_ISL_1630877, EPI_ISL_1630878, EPI_ISL_1630880, EPI_ISL_1630881, EPI_ISL_1630882, EPI_ISL_1630883, EPI_ISL_1630884, EPI_ISL_1630885, EPI_ISL_1630886, EPI_ISL_1630888, EPI_ISL_1630890, EPI_ISL_1630891, EPI_ISL_1630892, EPI_ISL_1630893, EPI_ISL_1630894, EPI_ISL_1630895, EPI_ISL_1630897, EPI_ISL_1630898, EPI_ISL_1630899, EPI_ISL_1630900, EPI_ISL_1630901, EPI_ISL_1630902, EPI_ISL_1630903, EPI_ISL_1630904, EPI_ISL_1630905, EPI_ISL_1630906, EPI_ISL_1630907, EPI_ISL_1630909, EPI_ISL_1630911, EPI_ISL_1630912, EPI_ISL_1630913, EPI_ISL_1630914, EPI_ISL_1630915, EPI_ISL_1630916, EPI_ISL_1630917, EPI_ISL_1630918, EPI_ISL_1630919, EPI_ISL_1630920, EPI_ISL_1630921, EPI_ISL_1630922, EPI_ISL_1630923, EPI_ISL_1630924, EPI_ISL_1630925, EPI_ISL_1630926, EPI_ISL_1630927, EPI_ISL_1630928, EPI_ISL_1630929, EPI_ISL_1630930, EPI_ISL_1630931, EPI_ISL_1630932, EPI_ISL_1630934, EPI_ISL_1630935, EPI_ISL_1630936, EPI_ISL_1630937, EPI_ISL_1630938, EPI_ISL_1630939, EPI_ISL_1630940, EPI_ISL_1630941, EPI_ISL_1630942, EPI_ISL_1630943, EPI_ISL_1630945, EPI_ISL_1630946, EPI_ISL_1630947, EPI_ISL_1630950, EPI_ISL_1630951, EPI_ISL_1630952, EPI_ISL_1630953, EPI_ISL_1630954, EPI_ISL_1630955, EPI_ISL_1630956, EPI_ISL_1630957, EPI_ISL_1630959, EPI_ISL_1630960, EPI_ISL_1630961, EPI_ISL_1630962, EPI_ISL_1630963, EPI_ISL_1630964, EPI_ISL_1630965, EPI_ISL_1630966, EPI_ISL_1630967, EPI_ISL_1630969, EPI_ISL_1630970, EPI_ISL_1630971, EPI_ISL_1630972, EPI_ISL_1630973, EPI_ISL_1630974, EPI_ISL_1630975, EPI_ISL_1630976, EPI_ISL_1630977, EPI_ISL_1630979, EPI_ISL_1630980, EPI_ISL_1630981, EPI_ISL_1630982, EPI_ISL_1630983, EPI_ISL_1630985, EPI_ISL_1630986, EPI_ISL_1630987, EPI_ISL_1630990, EPI_ISL_1630991, EPI_ISL_1630992, EPI_ISL_1630993, EPI_ISL_1630994, EPI_ISL_1630995, EPI_ISL_1630996, EPI_ISL_1630997, EPI_ISL_1630998, EPI_ISL_1630999, EPI_ISL_1631001, EPI_ISL_1631002, EPI_ISL_1631003, EPI_ISL_1631004, EPI_ISL_1631005, EPI_ISL_1631006, EPI_ISL_1631007, EPI_ISL_1631010, EPI_ISL_1631011, EPI_ISL_1631012, EPI_ISL_1631013, EPI_ISL_1631014, EPI_ISL_1631015, EPI_ISL_1631016, EPI_ISL_1631019, EPI_ISL_1631020, EPI_ISL_1631021, EPI_ISL_1631022, EPI_ISL_1631023, EPI_ISL_1631025, EPI_ISL_1631026, EPI_ISL_1631027, EPI_ISL_1631029, EPI_ISL_1631031, EPI_ISL_1631032, EPI_ISL_1631033, EPI_ISL_1631034, EPI_ISL_1631035, EPI_ISL_1631036, EPI_ISL_1631037, EPI_ISL_1631038, EPI_ISL_1631040, EPI_ISL_1631041, EPI_ISL_1631043, EPI_ISL_1631044, EPI_ISL_1631045, EPI_ISL_1631046, EPI_ISL_1631048, EPI_ISL_1631049, EPI_ISL_1631050, EPI_ISL_1631052, EPI_ISL_1631053, EPI_ISL_1631054, EPI_ISL_1631055, EPI_ISL_1631057, EPI_ISL_1631058, EPI_ISL_1631059, EPI_ISL_1631060, EPI_ISL_1631061, EPI_ISL_1631062, EPI_ISL_1631063, EPI_ISL_1631064, EPI_ISL_1631065, EPI_ISL_1631066, EPI_ISL_1631068, EPI_ISL_1631069, EPI_ISL_1631070, EPI_ISL_1631071, EPI_ISL_1631072, EPI_ISL_1631074, EPI_ISL_1631075, EPI_ISL_1631076, EPI_ISL_1631077, EPI_ISL_1631078, EPI_ISL_1631079, EPI_ISL_1631080, EPI_ISL_1631081, EPI_ISL_1631082, EPI_ISL_1631084, EPI_ISL_1631085, EPI_ISL_1631086, EPI_ISL_1631087, EPI_ISL_1631088, EPI_ISL_1631096, EPI_ISL_1631098, EPI_ISL_1631099, EPI_ISL_1631103, EPI_ISL_1631115, EPI_ISL_1631120, EPI_ISL_1631121, EPI_ISL_1631127, EPI_ISL_1631130, EPI_ISL_1631132, EPI_ISL_1631135, EPI_ISL_1631137, EPI_ISL_1631139, EPI_ISL_1631143, EPI_ISL_1631148, EPI_ISL_1631152, EPI_ISL_1631154, EPI_ISL_1631163, EPI_ISL_1631166, EPI_ISL_1631171, EPI_ISL_1631182, EPI_ISL_1631184, EPI_ISL_1631186, EPI_ISL_1631191, EPI_ISL_1631198, EPI_ISL_1631207, EPI_ISL_1631208, EPI_ISL_1631221, EPI_ISL_1631223, EPI_ISL_1631228, EPI_ISL_1631231, EPI_ISL_1631236, EPI_ISL_1631239, EPI_ISL_1631240, EPI_ISL_1631241, EPI_ISL_1631251, EPI_ISL_1631255, EPI_ISL_1631260, EPI_ISL_1631267, EPI_ISL_1631273, EPI_ISL_1631280, EPI_ISL_1631281, EPI_ISL_1631283, EPI_ISL_1631289, EPI_ISL_1631293, EPI_ISL_1631299, EPI_ISL_1631300, EPI_ISL_1631313, EPI_ISL_1631316, EPI_ISL_1631321, EPI_ISL_1631326, EPI_ISL_1631333, EPI_ISL_1631758, EPI_ISL_1631778, EPI_ISL_1631780, EPI_ISL_1631783, EPI_ISL_1631784, EPI_ISL_1631804, EPI_ISL_1631813, EPI_ISL_1631819, EPI_ISL_1631852, EPI_ISL_1631854, EPI_ISL_1631864, EPI_ISL_1631873, EPI_ISL_1631895, EPI_ISL_1631903, EPI_ISL_1631930, EPI_ISL_1631942, EPI_ISL_1631956, EPI_ISL_1631963, EPI_ISL_1631965, EPI_ISL_1631966, EPI_ISL_1631972, EPI_ISL_1631978, EPI_ISL_1631985, EPI_ISL_1631990, EPI_ISL_1631995, EPI_ISL_1631999, EPI_ISL_1632000, EPI_ISL_1632002, EPI_ISL_1632003, EPI_ISL_1632010, EPI_ISL_1632017, EPI_ISL_1632019, EPI_ISL_1632020, EPI_ISL_1632029, EPI_ISL_1632038, EPI_ISL_1632040, EPI_ISL_1632044, EPI_ISL_1632047, EPI_ISL_1632054, EPI_ISL_1632055, EPI_ISL_1632060, EPI_ISL_1632066, EPI_ISL_1632073, EPI_ISL_1632075, EPI_ISL_1632077, EPI_ISL_1632080, EPI_ISL_1632082, EPI_ISL_1632086, EPI_ISL_1632087, EPI_ISL_1632088, EPI_ISL_1632093, EPI_ISL_1632098, EPI_ISL_1632100, EPI_ISL_1632101, EPI_ISL_1632102, EPI_ISL_1632103, EPI_ISL_1632105, EPI_ISL_1632107, EPI_ISL_1632108, EPI_ISL_1632109, EPI_ISL_1632110, EPI_ISL_1632113, EPI_ISL_1632116, EPI_ISL_1632120, EPI_ISL_1632121, EPI_ISL_1632124, EPI_ISL_1632125, EPI_ISL_1632129, EPI_ISL_1632132, EPI_ISL_1632133, EPI_ISL_1632134, EPI_ISL_1632137, EPI_ISL_1632139, EPI_ISL_1632147, EPI_ISL_1632148, EPI_ISL_1632149, EPI_ISL_1632153, EPI_ISL_1632165, EPI_ISL_1632169, EPI_ISL_1632176, EPI_ISL_1632180, EPI_ISL_1632184, EPI_ISL_1632186, EPI_ISL_1632188, EPI_ISL_1632189, EPI_ISL_1632190, EPI_ISL_1632192, EPI_ISL_1632194, EPI_ISL_1632197, EPI_ISL_1632199, EPI_ISL_1632202, EPI_ISL_1632204, EPI_ISL_1632208, EPI_ISL_1632209, EPI_ISL_1632211, EPI_ISL_1632214, EPI_ISL_1632221, EPI_ISL_1632227, EPI_ISL_1632236, EPI_ISL_1632241, EPI_ISL_1632259, EPI_ISL_1632267, EPI_ISL_1632273, EPI_ISL_1632280, EPI_ISL_1632281, EPI_ISL_1632284, EPI_ISL_1632285, EPI_ISL_1632292, EPI_ISL_1632296, EPI_ISL_1632297, EPI_ISL_1634714, EPI_ISL_1634716, EPI_ISL_1635122, EPI_ISL_1635149, EPI_ISL_1635155, EPI_ISL_1635321, EPI_ISL_1652694, EPI_ISL_1652697, EPI_ISL_1652701, EPI_ISL_1652703, EPI_ISL_1652708, EPI_ISL_1652713, EPI_ISL_1652719, EPI_ISL_1652720, EPI_ISL_1652722, EPI_ISL_1652727, EPI_ISL_1652728, EPI_ISL_1652729, EPI_ISL_1652733, EPI_ISL_1652734, EPI_ISL_1652735, EPI_ISL_1652736, EPI_ISL_1652738, EPI_ISL_1652739, EPI_ISL_1652743, EPI_ISL_1652745, EPI_ISL_1652746, EPI_ISL_1652747, EPI_ISL_1652748, EPI_ISL_1652750, EPI_ISL_1652753, EPI_ISL_1652756, EPI_ISL_1652760, EPI_ISL_1652761, EPI_ISL_1652763, EPI_ISL_1652764, EPI_ISL_1652766, EPI_ISL_1652769, EPI_ISL_1652770, EPI_ISL_1652771, EPI_ISL_1652773, EPI_ISL_1652774, EPI_ISL_1652775, EPI_ISL_1652777, EPI_ISL_1652779, EPI_ISL_1652781, EPI_ISL_1652788, EPI_ISL_1652789, EPI_ISL_1652791, EPI_ISL_1652793, EPI_ISL_1652794, EPI_ISL_1652796, EPI_ISL_1652797, EPI_ISL_1652802, EPI_ISL_1652808, EPI_ISL_1652809, EPI_ISL_1652811, EPI_ISL_1652813, EPI_ISL_1652814, EPI_ISL_1652818, EPI_ISL_1652819, EPI_ISL_1652821, EPI_ISL_1652825, EPI_ISL_1652826, EPI_ISL_1652831, EPI_ISL_1652834, EPI_ISL_1652836, EPI_ISL_1652837, EPI_ISL_1652839, EPI_ISL_1652840, EPI_ISL_1652841, EPI_ISL_1652843, EPI_ISL_1652845, EPI_ISL_1652846, EPI_ISL_1652848, EPI_ISL_1652849, EPI_ISL_1652850, EPI_ISL_1652853, EPI_ISL_1652854, EPI_ISL_1652856, EPI_ISL_1652857, EPI_ISL_1652861, EPI_ISL_1652862, EPI_ISL_1652864, EPI_ISL_1652865, EPI_ISL_1652867, EPI_ISL_1652869, EPI_ISL_1652871, EPI_ISL_1652873, EPI_ISL_1652874, EPI_ISL_1652875, EPI_ISL_1652877, EPI_ISL_1652878, EPI_ISL_1652879, EPI_ISL_1652888, EPI_ISL_1652890, EPI_ISL_1652898, EPI_ISL_1652900, EPI_ISL_1652901, EPI_ISL_1652903, EPI_ISL_1652904, EPI_ISL_1652907, EPI_ISL_1652910, EPI_ISL_1652912, EPI_ISL_1652913, EPI_ISL_1652915, EPI_ISL_1652916, EPI_ISL_1652917, EPI_ISL_1652920, EPI_ISL_1652924, EPI_ISL_1652929, EPI_ISL_1652930, EPI_ISL_1652933, EPI_ISL_1652937, EPI_ISL_1652938, EPI_ISL_1652939, EPI_ISL_1652952, EPI_ISL_1652979, EPI_ISL_1652986, EPI_ISL_1653009, EPI_ISL_1653011, EPI_ISL_1653024, EPI_ISL_1653025, EPI_ISL_1653040, EPI_ISL_1653058, EPI_ISL_1653108, EPI_ISL_1653125, EPI_ISL_1653175, EPI_ISL_1653179, EPI_ISL_1653180, EPI_ISL_1653187, EPI_ISL_1653195, EPI_ISL_1653225, EPI_ISL_1653227, EPI_ISL_1653229, EPI_ISL_1653230, EPI_ISL_1653232, EPI_ISL_1653234, EPI_ISL_1653235, EPI_ISL_1653236, EPI_ISL_1653242, EPI_ISL_1653243, EPI_ISL_1653245, EPI_ISL_1653247, EPI_ISL_1653250, EPI_ISL_1653259, EPI_ISL_1653260, EPI_ISL_1653269, EPI_ISL_1653270, EPI_ISL_1653271, EPI_ISL_1653277, EPI_ISL_1653279, EPI_ISL_1653280, EPI_ISL_1653284, EPI_ISL_1653288, EPI_ISL_1653290, EPI_ISL_1653297, EPI_ISL_1653298, EPI_ISL_1653301, EPI_ISL_1653317, EPI_ISL_1653327, EPI_ISL_1653330, EPI_ISL_1653332, EPI_ISL_1653334, EPI_ISL_1653338, EPI_ISL_1653342, EPI_ISL_1653346, EPI_ISL_1653347, EPI_ISL_1653348, EPI_ISL_1653349, EPI_ISL_1653351, EPI_ISL_1653352, EPI_ISL_1653358, EPI_ISL_1653361, EPI_ISL_1653364, EPI_ISL_1653370, EPI_ISL_1653377, EPI_ISL_1653379, EPI_ISL_1653380, EPI_ISL_1653387, EPI_ISL_1653391, EPI_ISL_1653392, EPI_ISL_1653394, EPI_ISL_1653396, EPI_ISL_1653400, EPI_ISL_1653401, EPI_ISL_1653405, EPI_ISL_1653413, EPI_ISL_1653415, EPI_ISL_1653421, EPI_ISL_1653433, EPI_ISL_1653435, EPI_ISL_1653438, EPI_ISL_1653439, EPI_ISL_1653440, EPI_ISL_1653441, EPI_ISL_1653446, EPI_ISL_1653447, EPI_ISL_1653453, EPI_ISL_1653458, EPI_ISL_1653460, EPI_ISL_1653461, EPI_ISL_1653468, EPI_ISL_1653474, EPI_ISL_1653479, EPI_ISL_1653481, EPI_ISL_1653483, EPI_ISL_1653484, EPI_ISL_1653487, EPI_ISL_1653489, EPI_ISL_1653490, EPI_ISL_1653492, EPI_ISL_1653494, EPI_ISL_1653495, EPI_ISL_1653496, EPI_ISL_1653497, EPI_ISL_1653499, EPI_ISL_1653500, EPI_ISL_1653501, EPI_ISL_1653503, EPI_ISL_1653504, EPI_ISL_1653508, EPI_ISL_1653510, EPI_ISL_1653511, EPI_ISL_1653512, EPI_ISL_1653513, EPI_ISL_1653514, EPI_ISL_1653515, EPI_ISL_1653517, EPI_ISL_1653518, EPI_ISL_1653519, EPI_ISL_1653520, EPI_ISL_1653521, EPI_ISL_1653522, EPI_ISL_1653525, EPI_ISL_1653531, EPI_ISL_1653532, EPI_ISL_1653534, EPI_ISL_1653535, EPI_ISL_1653537, EPI_ISL_1653538, EPI_ISL_1653541, EPI_ISL_1653542, EPI_ISL_1653543, EPI_ISL_1653544, EPI_ISL_1653545, EPI_ISL_1653546, EPI_ISL_1653547, EPI_ISL_1653548, EPI_ISL_1653549, EPI_ISL_1653551, EPI_ISL_1653554, EPI_ISL_1653556, EPI_ISL_1653557, EPI_ISL_1653558, EPI_ISL_1653559, EPI_ISL_1653560, EPI_ISL_1653561, EPI_ISL_1653564, EPI_ISL_1653565, EPI_ISL_1653567, EPI_ISL_1653569, EPI_ISL_1653570, EPI_ISL_1653571, EPI_ISL_1653572, EPI_ISL_1653573, EPI_ISL_1653574, EPI_ISL_1653575, EPI_ISL_1653577, EPI_ISL_1653578, EPI_ISL_1653579, EPI_ISL_1653580, EPI_ISL_1653581, EPI_ISL_1653582, EPI_ISL_1653583, EPI_ISL_1653584, EPI_ISL_1653585, EPI_ISL_1653590, EPI_ISL_1653591, EPI_ISL_1653592, EPI_ISL_1653593, EPI_ISL_1653595, EPI_ISL_1653596, EPI_ISL_1653599, EPI_ISL_1653600, EPI_ISL_1653603, EPI_ISL_1653604, EPI_ISL_1653605, EPI_ISL_1653606, EPI_ISL_1653607, EPI_ISL_1653611, EPI_ISL_1653613, EPI_ISL_1653614, EPI_ISL_1653616, EPI_ISL_1653618, EPI_ISL_1653619, EPI_ISL_1653620, EPI_ISL_1653623, EPI_ISL_1653624, EPI_ISL_1653625, EPI_ISL_1653626, EPI_ISL_1653627, EPI_ISL_1653629, EPI_ISL_1653630, EPI_ISL_1653631, EPI_ISL_1653632, EPI_ISL_1653633, EPI_ISL_1653634, EPI_ISL_1653635, EPI_ISL_1653636, EPI_ISL_1653637, EPI_ISL_1653640, EPI_ISL_1653641, EPI_ISL_1653642, EPI_ISL_1653643, EPI_ISL_1653644, EPI_ISL_1653645, EPI_ISL_1653646, EPI_ISL_1653647, EPI_ISL_1653649, EPI_ISL_1653652, EPI_ISL_1653655, EPI_ISL_1653656, EPI_ISL_1653657, EPI_ISL_1653658, EPI_ISL_1653659, EPI_ISL_1653660, EPI_ISL_1653661, EPI_ISL_1653662, EPI_ISL_1653663, EPI_ISL_1653668, EPI_ISL_1653669, EPI_ISL_1653670, EPI_ISL_1653672, EPI_ISL_1653673, EPI_ISL_1653676, EPI_ISL_1653677, EPI_ISL_1653679, EPI_ISL_1653682, EPI_ISL_1653683, EPI_ISL_1653686, EPI_ISL_1653687, EPI_ISL_1653688, EPI_ISL_1653690, EPI_ISL_1653697, EPI_ISL_1653698, EPI_ISL_1653700, EPI_ISL_1653702, EPI_ISL_1653703, EPI_ISL_1653704, EPI_ISL_1653705, EPI_ISL_1653706, EPI_ISL_1653707, EPI_ISL_1653708, EPI_ISL_1653709, EPI_ISL_1653710, EPI_ISL_1653711, EPI_ISL_1653712, EPI_ISL_1653713, EPI_ISL_1653714, EPI_ISL_1653715, EPI_ISL_1653716, EPI_ISL_1653728, EPI_ISL_1653758, EPI_ISL_1672977, EPI_ISL_1673041, EPI_ISL_1697446, EPI_ISL_1697475, EPI_ISL_1697487, EPI_ISL_1697498, EPI_ISL_1697511, EPI_ISL_1697560, EPI_ISL_1697602, EPI_ISL_1697619, EPI_ISL_1697643, EPI_ISL_1697707, EPI_ISL_1697718, EPI_ISL_1697733, EPI_ISL_1698277, EPI_ISL_1698321, EPI_ISL_1698341, EPI_ISL_1698472, EPI_ISL_1698512, EPI_ISL_1698771, EPI_ISL_1698775, EPI_ISL_1698778, EPI_ISL_1698783, EPI_ISL_1698790, EPI_ISL_1698795, EPI_ISL_1698798, EPI_ISL_1698802, EPI_ISL_1698816, EPI_ISL_1698818, EPI_ISL_1698828, EPI_ISL_1698830, EPI_ISL_1698835, EPI_ISL_1698844, EPI_ISL_1698850, EPI_ISL_1698852, EPI_ISL_1698858, EPI_ISL_1698859, EPI_ISL_1698868, EPI_ISL_1698875, EPI_ISL_1698894, EPI_ISL_1698905, EPI_ISL_1698909, EPI_ISL_1698923, EPI_ISL_1698924, EPI_ISL_1698942, EPI_ISL_1698949, EPI_ISL_1698963, EPI_ISL_1698968, EPI_ISL_1698979, EPI_ISL_1698985, EPI_ISL_1698987, EPI_ISL_1698993, EPI_ISL_1698995, EPI_ISL_1698998, EPI_ISL_1698999, EPI_ISL_1699013, EPI_ISL_1699017, EPI_ISL_1699022, EPI_ISL_1699027, EPI_ISL_1699045, EPI_ISL_1699047, EPI_ISL_1699050, EPI_ISL_1699053, EPI_ISL_1699056, EPI_ISL_1699061, EPI_ISL_1699063, EPI_ISL_1699081, EPI_ISL_1699084, EPI_ISL_1699088, EPI_ISL_1699089, EPI_ISL_1699094, EPI_ISL_1699099, EPI_ISL_1699101, EPI_ISL_1699104, EPI_ISL_1699105, EPI_ISL_1699107, EPI_ISL_1699110, EPI_ISL_1699112, EPI_ISL_1699118, EPI_ISL_1699120, EPI_ISL_1699122, EPI_ISL_1699127, EPI_ISL_1699133, EPI_ISL_1699142, EPI_ISL_1699157, EPI_ISL_1699172, EPI_ISL_1699180, EPI_ISL_1699192, EPI_ISL_1699203, EPI_ISL_1699207, EPI_ISL_1699209, EPI_ISL_1699210, EPI_ISL_1699211, EPI_ISL_1699214, EPI_ISL_1699215, EPI_ISL_1699228, EPI_ISL_1699229, EPI_ISL_1699231, EPI_ISL_1699232, EPI_ISL_1699238, EPI_ISL_1699250, EPI_ISL_1699254, EPI_ISL_1699290, EPI_ISL_1699292, EPI_ISL_1699296, EPI_ISL_1699334, EPI_ISL_1699341, EPI_ISL_1699362, EPI_ISL_1699372, EPI_ISL_1699380, EPI_ISL_1699399, EPI_ISL_1699411, EPI_ISL_1699425, EPI_ISL_1699441, EPI_ISL_1699494, EPI_ISL_1699513, EPI_ISL_1699535, EPI_ISL_1699536, EPI_ISL_1699540, EPI_ISL_1699542, EPI_ISL_1699554, EPI_ISL_1699569, EPI_ISL_1699589, EPI_ISL_1699709, EPI_ISL_1699721, EPI_ISL_1699842, EPI_ISL_1699844, EPI_ISL_1699845, EPI_ISL_1699849, EPI_ISL_1699851, EPI_ISL_1699853, EPI_ISL_1699854, EPI_ISL_1699856, EPI_ISL_1699857, EPI_ISL_1699859, EPI_ISL_1699863, EPI_ISL_1699864, EPI_ISL_1699865, EPI_ISL_1699866, EPI_ISL_1699870, EPI_ISL_1699871, EPI_ISL_1699873, EPI_ISL_1699875, EPI_ISL_1699877, EPI_ISL_1699879, EPI_ISL_1699881, EPI_ISL_1699882, EPI_ISL_1699884, EPI_ISL_1699886, EPI_ISL_1699887, EPI_ISL_1699888, EPI_ISL_1699891, EPI_ISL_1699892, EPI_ISL_1699893, EPI_ISL_1699896, EPI_ISL_1699900, EPI_ISL_1699901, EPI_ISL_1699903, EPI_ISL_1699904, EPI_ISL_1699905, EPI_ISL_1699906, EPI_ISL_1699908, EPI_ISL_1699909, EPI_ISL_1699911, EPI_ISL_1699912, EPI_ISL_1699918, EPI_ISL_1699919, EPI_ISL_1699920, EPI_ISL_1699923, EPI_ISL_1699924, EPI_ISL_1699925, EPI_ISL_1699926, EPI_ISL_1699935, EPI_ISL_1699936, EPI_ISL_1699938, EPI_ISL_1699939, EPI_ISL_1699940, EPI_ISL_1699944, EPI_ISL_1699945, EPI_ISL_1699946, EPI_ISL_1699947, EPI_ISL_1699948, EPI_ISL_1699949, EPI_ISL_1699951, EPI_ISL_1699952, EPI_ISL_1699953, EPI_ISL_1699954, EPI_ISL_1699956, EPI_ISL_1699957, EPI_ISL_1699958, EPI_ISL_1699959, EPI_ISL_1699961, EPI_ISL_1699962, EPI_ISL_1699963, EPI_ISL_1699965, EPI_ISL_1699966, EPI_ISL_1699968, EPI_ISL_1699969, EPI_ISL_1699970, EPI_ISL_1699971, EPI_ISL_1699972, EPI_ISL_1699973, EPI_ISL_1699975, EPI_ISL_1699976, EPI_ISL_1699978, EPI_ISL_1699981, EPI_ISL_1699982, EPI_ISL_1699983, EPI_ISL_1699985, EPI_ISL_1699987, EPI_ISL_1699988, EPI_ISL_1699989, EPI_ISL_1699990, EPI_ISL_1699993, EPI_ISL_1699996, EPI_ISL_1699999, EPI_ISL_1700000, EPI_ISL_1700001, EPI_ISL_1700002, EPI_ISL_1700003, EPI_ISL_1700004, EPI_ISL_1700005, EPI_ISL_1700008, EPI_ISL_1700009, EPI_ISL_1700011, EPI_ISL_1700012, EPI_ISL_1700014, EPI_ISL_1700016, EPI_ISL_1700017, EPI_ISL_1700018, EPI_ISL_1700019, EPI_ISL_1700023, EPI_ISL_1700024, EPI_ISL_1700026, EPI_ISL_1700032, EPI_ISL_1700033, EPI_ISL_1700035, EPI_ISL_1700038, EPI_ISL_1700039, EPI_ISL_1700040, EPI_ISL_1700043, EPI_ISL_1700045, EPI_ISL_1700046, EPI_ISL_1700049, EPI_ISL_1700051, EPI_ISL_1700054, EPI_ISL_1700055, EPI_ISL_1700056, EPI_ISL_1700058, EPI_ISL_1700061, EPI_ISL_1700062, EPI_ISL_1700064, EPI_ISL_1700065, EPI_ISL_1700066, EPI_ISL_1700072, EPI_ISL_1700073, EPI_ISL_1700075, EPI_ISL_1700077, EPI_ISL_1700082, EPI_ISL_1700083, EPI_ISL_1700084, EPI_ISL_1700086, EPI_ISL_1700087, EPI_ISL_1700088, EPI_ISL_1700089, EPI_ISL_1700091, EPI_ISL_1700092, EPI_ISL_1700093, EPI_ISL_1700094, EPI_ISL_1700095, EPI_ISL_1700097, EPI_ISL_1700099, EPI_ISL_1700101, EPI_ISL_1700104, EPI_ISL_1700105, EPI_ISL_1700106, EPI_ISL_1700107, EPI_ISL_1700110, EPI_ISL_1700111, EPI_ISL_1700114, EPI_ISL_1700115, EPI_ISL_1700117, EPI_ISL_1700119, EPI_ISL_1700120, EPI_ISL_1700121, EPI_ISL_1700122, EPI_ISL_1700123, EPI_ISL_1700124, EPI_ISL_1700125, EPI_ISL_1700126, EPI_ISL_1700128, EPI_ISL_1700129, EPI_ISL_1700131, EPI_ISL_1700132, EPI_ISL_1700133, EPI_ISL_1700134, EPI_ISL_1700137, EPI_ISL_1700142, EPI_ISL_1700146, EPI_ISL_1700147, EPI_ISL_1700150, EPI_ISL_1700155, EPI_ISL_1700156, EPI_ISL_1700158, EPI_ISL_1700160, EPI_ISL_1700161, EPI_ISL_1700162, EPI_ISL_1700163, EPI_ISL_1700168, EPI_ISL_1700171, EPI_ISL_1700172, | | | |

EPI_ISL_1700175, EPI_ISL_1700176, EPI_ISL_1700178, EPI_ISL_1700180, EPI_ISL_1700181, EPI_ISL_1700186, EPI_ISL_1700194, EPI_ISL_1700195, EPI_ISL_1700197, EPI_ISL_1700199, EPI_ISL_1700206, EPI_ISL_1700210, EPI_ISL_1700211, EPI_ISL_1700215, EPI_ISL_1700218, EPI_ISL_1700219, EPI_ISL_1700225, EPI_ISL_1700230, EPI_ISL_1700231, EPI_ISL_1700232, EPI_ISL_1700234, EPI_ISL_1700235, EPI_ISL_1700236, EPI_ISL_1700239, EPI_ISL_1700242, EPI_ISL_1700247, EPI_ISL_1700249, EPI_ISL_1700252, EPI_ISL_1700259, EPI_ISL_1700260, EPI_ISL_1700261, EPI_ISL_1700264, EPI_ISL_1700266, EPI_ISL_1700267, EPI_ISL_1700271, EPI_ISL_1700273, EPI_ISL_1700276, EPI_ISL_1700279, EPI_ISL_1700281, EPI_ISL_1700282, EPI_ISL_1700284, EPI_ISL_1700287, EPI_ISL_1700289, EPI_ISL_1700290, EPI_ISL_1700291, EPI_ISL_1700298, EPI_ISL_1700304, EPI_ISL_1700305, EPI_ISL_1700306, EPI_ISL_1700309, EPI_ISL_1700311, EPI_ISL_1700314, EPI_ISL_1700315, EPI_ISL_1700316, EPI_ISL_1700321, EPI_ISL_1700322, EPI_ISL_1700323, EPI_ISL_1700324, EPI_ISL_1700327, EPI_ISL_1700328, EPI_ISL_1700330, EPI_ISL_1700334, EPI_ISL_1700335, EPI_ISL_1700337, EPI_ISL_1700339, EPI_ISL_1700341, EPI_ISL_1700344, EPI_ISL_1700345, EPI_ISL_1700346, EPI_ISL_1700349, EPI_ISL_1700350, EPI_ISL_1700351, EPI_ISL_1700352, EPI_ISL_1700354, EPI_ISL_1700358, EPI_ISL_1700362, EPI_ISL_1700368, EPI_ISL_1700372, EPI_ISL_1700375, EPI_ISL_1700376, EPI_ISL_1700377, EPI_ISL_1700379, EPI_ISL_1700381, EPI_ISL_1700383, EPI_ISL_1700384, EPI_ISL_1700388, EPI_ISL_1700390, EPI_ISL_1700391, EPI_ISL_1700395, EPI_ISL_1700398, EPI_ISL_1700400, EPI_ISL_1700402, EPI_ISL_1700403, EPI_ISL_1700404, EPI_ISL_1700405, EPI_ISL_1700407, EPI_ISL_1700412, EPI_ISL_1700416, EPI_ISL_1700418, EPI_ISL_1700419, EPI_ISL_1700420, EPI_ISL_1700422, EPI_ISL_1700424, EPI_ISL_1700427, EPI_ISL_1700428, EPI_ISL_1700429, EPI_ISL_1700432, EPI_ISL_1700434, EPI_ISL_1700437, EPI_ISL_1700438, EPI_ISL_1700439, EPI_ISL_1700440, EPI_ISL_1700445, EPI_ISL_1700446, EPI_ISL_1700447, EPI_ISL_1700450, EPI_ISL_1700451, EPI_ISL_1700452, EPI_ISL_1700453, EPI_ISL_1700455, EPI_ISL_1700462, EPI_ISL_1700463, EPI_ISL_1700464, EPI_ISL_1700465, EPI_ISL_1700467, EPI_ISL_1700472, EPI_ISL_1700474, EPI_ISL_1700477, EPI_ISL_1700478, EPI_ISL_1700481, EPI_ISL_1700482, EPI_ISL_1700483, EPI_ISL_1700489, EPI_ISL_1700491, EPI_ISL_1700495, EPI_ISL_1700496, EPI_ISL_1700497, EPI_ISL_1700498, EPI_ISL_1700499, EPI_ISL_1700500, EPI_ISL_1700501, EPI_ISL_1700502, EPI_ISL_1700503, EPI_ISL_1700504, EPI_ISL_1700505, EPI_ISL_1700506, EPI_ISL_1700507, EPI_ISL_1700508, EPI_ISL_1700509, EPI_ISL_1700512, EPI_ISL_1700513, EPI_ISL_1718330, EPI_ISL_1719743

| | | | |
|---|---|---|---|
| see above | Lighthouse Lab in Alderley Park | Wellcome Sanger Institute for the COVID-19 Genomics UK (COG-UK) Consortium | Cordelia Langford; David K. Jackson; Dominic Kwiatkowski; Ewan Harrison; Ian Johnston; Jacquelyn Wynn; Jeffrey Barrett; John Sillitoe on behalf of the Wellcome Sanger Institute COVID-19 Surveillance Team; Mairead Hyland; Roberto Amato; Sonia Goncalves; The Lighthouse Lab in Alderley Park and Alex Alderton |

EPI_ISL_1564038, EPI_ISL_1564047, EPI_ISL_1564053, EPI_ISL_1564062, EPI_ISL_1564065, EPI_ISL_1564066, EPI_ISL_1564073, EPI_ISL_1564078, EPI_ISL_1564086, EPI_ISL_1564088, EPI_ISL_1564097, EPI_ISL_1564102, EPI_ISL_1564112, EPI_ISL_1564115, EPI_ISL_1564124, EPI_ISL_1564125, EPI_ISL_1564130, EPI_ISL_1564132, EPI_ISL_1564139, EPI_ISL_1564172, EPI_ISL_1564178, EPI_ISL_1564179, EPI_ISL_1564181, EPI_ISL_1564185, EPI_ISL_1564187, EPI_ISL_1564200, EPI_ISL_1564203, EPI_ISL_1564215, EPI_ISL_1564221, EPI_ISL_1564223, EPI_ISL_1564224, EPI_ISL_1564258, EPI_ISL_1564270, EPI_ISL_1564271, EPI_ISL_1564276, EPI_ISL_1564281, EPI_ISL_1564284, EPI_ISL_1564286, EPI_ISL_1564287, EPI_ISL_1564300, EPI_ISL_1564304, EPI_ISL_1564314, EPI_ISL_1564319, EPI_ISL_1564326, EPI_ISL_1564328, EPI_ISL_1583924, EPI_ISL_1583925, EPI_ISL_1583931, EPI_ISL_1583938, EPI_ISL_1583940, EPI_ISL_1583942, EPI_ISL_1583944, EPI_ISL_1583947, EPI_ISL_1583948, EPI_ISL_1583950, EPI_ISL_1583955, EPI_ISL_1583962, EPI_ISL_1583964, EPI_ISL_1583966, EPI_ISL_1583968, EPI_ISL_1583970, EPI_ISL_1583972, EPI_ISL_1583974, EPI_ISL_1583981, EPI_ISL_1583984, EPI_ISL_1583985, EPI_ISL_1583986, EPI_ISL_1583987, EPI_ISL_1583996, EPI_ISL_1583997, EPI_ISL_1583998, EPI_ISL_1584000, EPI_ISL_1584008, EPI_ISL_1584010, EPI_ISL_1584011, EPI_ISL_1584017, EPI_ISL_1584032, EPI_ISL_1584036, EPI_ISL_1584043, EPI_ISL_1584047, EPI_ISL_1584051, EPI_ISL_1584055, EPI_ISL_1584067, EPI_ISL_1584068, EPI_ISL_1584069, EPI_ISL_1584070, EPI_ISL_1584071, EPI_ISL_1584072, EPI_ISL_1584076, EPI_ISL_1584077, EPI_ISL_1584082, EPI_ISL_1584089, EPI_ISL_1584090, EPI_ISL_1584091, EPI_ISL_1584092, EPI_ISL_1584096, EPI_ISL_1584099, EPI_ISL_1584100, EPI_ISL_1584101, EPI_ISL_1584106, EPI_ISL_1584111, EPI_ISL_1584114, EPI_ISL_1584115, EPI_ISL_1584118, EPI_ISL_1584121, EPI_ISL_1584123, EPI_ISL_1584125, EPI_ISL_1584131, EPI_ISL_1584135, EPI_ISL_1584440, EPI_ISL_1584444, EPI_ISL_1584470, EPI_ISL_1584476, EPI_ISL_1594464, EPI_ISL_1594466, EPI_ISL_1594471, EPI_ISL_1594482, EPI_ISL_1594500, EPI_ISL_1594512, EPI_ISL_1594520, EPI_ISL_1594522, EPI_ISL_1594528, EPI_ISL_1594540, EPI_ISL_1594569, EPI_ISL_1594590, EPI_ISL_1594596, EPI_ISL_1594613, EPI_ISL_1594635, EPI_ISL_1594641, EPI_ISL_1594650, EPI_ISL_1594659, EPI_ISL_1594660, EPI_ISL_1594703, EPI_ISL_1594719, EPI_ISL_1594726, EPI_ISL_1594728, EPI_ISL_1594730, EPI_ISL_1594731, EPI_ISL_1594736, EPI_ISL_1594756, EPI_ISL_1594758, EPI_ISL_1594759, EPI_ISL_1594760, EPI_ISL_1594765, EPI_ISL_1594769, EPI_ISL_1594770, EPI_ISL_1594787, EPI_ISL_1594788, EPI_ISL_1594794, EPI_ISL_1594798, EPI_ISL_1594801, EPI_ISL_1594803, EPI_ISL_1594810, EPI_ISL_1594819, EPI_ISL_1594820, EPI_ISL_1594823, EPI_ISL_1594826, EPI_ISL_1594832, EPI_ISL_1594837, EPI_ISL_1594842, EPI_ISL_1594843, EPI_ISL_1594847, EPI_ISL_1594848, EPI_ISL_1594854, EPI_ISL_1594862, EPI_ISL_1594866, EPI_ISL_1594871, EPI_ISL_1594874, EPI_ISL_1594876, EPI_ISL_1594892, EPI_ISL_1594895, EPI_ISL_1594898, EPI_ISL_1594900, EPI_ISL_1594901, EPI_ISL_1594904, EPI_ISL_1594911, EPI_ISL_1594918, EPI_ISL_1594924, EPI_ISL_1594927, EPI_ISL_1594933, EPI_ISL_1594936, EPI_ISL_1594956, EPI_ISL_1594966, EPI_ISL_1594983, EPI_ISL_1594988, EPI_ISL_1595450, EPI_ISL_1595472, EPI_ISL_1615689, EPI_ISL_1615692, EPI_ISL_1615693, EPI_ISL_1615694, EPI_ISL_1615695, EPI_ISL_1615699, EPI_ISL_1615700, EPI_ISL_1615702, EPI_ISL_1615703, EPI_ISL_1615705, EPI_ISL_1615709, EPI_ISL_1615714, EPI_ISL_1615717, EPI_ISL_1615719, EPI_ISL_1615723, EPI_ISL_1615726, EPI_ISL_1615728, EPI_ISL_1615731, EPI_ISL_1615732, EPI_ISL_1615734, EPI_ISL_1615738, EPI_ISL_1615740, EPI_ISL_1615742, EPI_ISL_1615744, EPI_ISL_1615745, EPI_ISL_1615751, EPI_ISL_1615753, EPI_ISL_1615754, EPI_ISL_1615755, EPI_ISL_1615756, EPI_ISL_1615761, EPI_ISL_1615762, EPI_ISL_1615764, EPI_ISL_1615765, EPI_ISL_1615768, EPI_ISL_1615769, EPI_ISL_1615771, EPI_ISL_1615775, EPI_ISL_1615776, EPI_ISL_1615780, EPI_ISL_1615781, EPI_ISL_1615784, EPI_ISL_1615787, EPI_ISL_1615789, EPI_ISL_1615793, EPI_ISL_1615797, EPI_ISL_1615798, EPI_ISL_1615800, EPI_ISL_1615807, EPI_ISL_1615808, EPI_ISL_1615812, EPI_ISL_1615816, EPI_ISL_1615818, EPI_ISL_1615820, EPI_ISL_1615822, EPI_ISL_1615823, EPI_ISL_1615827, EPI_ISL_1615828, EPI_ISL_1615829, EPI_ISL_1615830, EPI_ISL_1615831, EPI_ISL_1615832, EPI_ISL_1615836, EPI_ISL_1615838, EPI_ISL_1615839, EPI_ISL_1615842, EPI_ISL_1615848, EPI_ISL_1615854, EPI_ISL_1615856, EPI_ISL_1615857, EPI_ISL_1615861, EPI_ISL_1615862, EPI_ISL_1615863, EPI_ISL_1615865, EPI_ISL_1615866, EPI_ISL_1615870, EPI_ISL_1615875, EPI_ISL_1615877, EPI_ISL_1615878, EPI_ISL_1615879, EPI_ISL_1615880, EPI_ISL_1615883, EPI_ISL_1615884, EPI_ISL_1615886, EPI_ISL_1615888, EPI_ISL_1615889, EPI_ISL_1615890, EPI_ISL_1615891, EPI_ISL_1615893, EPI_ISL_1615894, EPI_ISL_1615896, EPI_ISL_1615897, EPI_ISL_1615900, EPI_ISL_1615904, EPI_ISL_1615906, EPI_ISL_1615907, EPI_ISL_1615909, EPI_ISL_1615910, EPI_ISL_1615911, EPI_ISL_1615912, EPI_ISL_1615914, EPI_ISL_1615915, EPI_ISL_1615916, EPI_ISL_1615921, EPI_ISL_1615922, EPI_ISL_1615923, EPI_ISL_1615926, EPI_ISL_1615928, EPI_ISL_1615929, EPI_ISL_1615932, EPI_ISL_1616188, EPI_ISL_1616211, EPI_ISL_1616261, EPI_ISL_1616262, EPI_ISL_1616265, EPI_ISL_1616272, EPI_ISL_1616273, EPI_ISL_1616281, EPI_ISL_1616285, EPI_ISL_1616286, EPI_ISL_1616287, EPI_ISL_1630292, EPI_ISL_1630314, EPI_ISL_1630333, EPI_ISL_1630366, EPI_ISL_1630372, EPI_ISL_1630387, EPI_ISL_1630416, EPI_ISL_1630432, EPI_ISL_1630435, EPI_ISL_1631097, EPI_ISL_1631100, EPI_ISL_1631102, EPI_ISL_1631106, EPI_ISL_1631124, EPI_ISL_1631134, EPI_ISL_1631155, EPI_ISL_1631158, EPI_ISL_1631162, EPI_ISL_1631179, EPI_ISL_1631190, EPI_ISL_1631201, EPI_ISL_1631215, EPI_ISL_1631219, EPI_ISL_1631227, EPI_ISL_1631246, EPI_ISL_1631263, EPI_ISL_1631279, EPI_ISL_1631284, EPI_ISL_1631291, EPI_ISL_1631305, EPI_ISL_1631330, EPI_ISL_1631332, EPI_ISL_1631424, EPI_ISL_1631997, EPI_ISL_1631998, EPI_ISL_1632001, EPI_ISL_1632004, EPI_ISL_1632008, EPI_ISL_1632023, EPI_ISL_1632069, EPI_ISL_1632152, EPI_ISL_1632155, EPI_ISL_1632185, EPI_ISL_1632237, EPI_ISL_1632299, EPI_ISL_1632305, EPI_ISL_1632306, EPI_ISL_1632311, EPI_ISL_1632314, EPI_ISL_1632316, EPI_ISL_1632318, EPI_ISL_1632320, EPI_ISL_1632325, EPI_ISL_1632326, EPI_ISL_1632332, EPI_ISL_1632333, EPI_ISL_1632335, EPI_ISL_1632343, EPI_ISL_1632359, EPI_ISL_1632360, EPI_ISL_1632361, EPI_ISL_1632365, EPI_ISL_1632368, EPI_ISL_1632371, EPI_ISL_1632372, EPI_ISL_1632379, EPI_ISL_1632381, EPI_ISL_1632383, EPI_ISL_1632384, EPI_ISL_1632386, EPI_ISL_1632390, EPI_ISL_1632394, EPI_ISL_1632395, EPI_ISL_1632401, EPI_ISL_1632405, EPI_ISL_1632407, EPI_ISL_1632410, EPI_ISL_1632412, EPI_ISL_1632419, EPI_ISL_1632426, EPI_ISL_1632433, EPI_ISL_1632434, EPI_ISL_1632437, EPI_ISL_1632448, EPI_ISL_1632449, EPI_ISL_1632450, EPI_ISL_1632461, EPI_ISL_1632463, EPI_ISL_1632467, EPI_ISL_1632469, EPI_ISL_1632471, EPI_ISL_1632475, EPI_ISL_1632476, EPI_ISL_1632481, EPI_ISL_1634718, EPI_ISL_1634743, EPI_ISL_1635163, EPI_ISL_1635165, EPI_ISL_1635195, EPI_ISL_1635207, EPI_ISL_1635223, EPI_ISL_1635344, EPI_ISL_1635351, EPI_ISL_1635356, EPI_ISL_1635359, EPI_ISL_1635361, EPI_ISL_1635371, EPI_ISL_1635379, EPI_ISL_1635392, EPI_ISL_1635400, EPI_ISL_1635403, EPI_ISL_1635404, EPI_ISL_1652954, EPI_ISL_1652967, EPI_ISL_1652968, EPI_ISL_1652970, EPI_ISL_1652971, EPI_ISL_1652976, EPI_ISL_1652978, EPI_ISL_1652980, EPI_ISL_1652981, EPI_ISL_1652982, EPI_ISL_1652983, EPI_ISL_1652984, EPI_ISL_1652989, EPI_ISL_1652994, EPI_ISL_1652995, EPI_ISL_1652996, EPI_ISL_1653005, EPI_ISL_1653010, EPI_ISL_1653015, EPI_ISL_1653020, EPI_ISL_1653021, EPI_ISL_1653027, EPI_ISL_1653028, EPI_ISL_1653033, EPI_ISL_1653042, EPI_ISL_1653043, EPI_ISL_1653045, EPI_ISL_1653051, EPI_ISL_1653069, EPI_ISL_1653073, EPI_ISL_1653074, EPI_ISL_1653081, EPI_ISL_1653083, EPI_ISL_1653090, EPI_ISL_1653093, EPI_ISL_1653096, EPI_ISL_1653097, EPI_ISL_1653099, EPI_ISL_1653100, EPI_ISL_1653102, EPI_ISL_1653104, EPI_ISL_1653109, EPI_ISL_1653110, EPI_ISL_1653111, EPI_ISL_1653114, EPI_ISL_1653116, EPI_ISL_1653119, EPI_ISL_1653120, EPI_ISL_1653121, EPI_ISL_1653130, EPI_ISL_1653132, EPI_ISL_1653141, EPI_ISL_1653146, EPI_ISL_1653147, EPI_ISL_1653156, EPI_ISL_1653159, EPI_ISL_1653167, EPI_ISL_1653174, EPI_ISL_1653178, EPI_ISL_1653181, EPI_ISL_1653197, EPI_ISL_1653201, EPI_ISL_1653233, EPI_ISL_1653244, EPI_ISL_1653251, EPI_ISL_1653257, EPI_ISL_1653268, EPI_ISL_1653272, EPI_ISL_1653276, EPI_ISL_1653278, EPI_ISL_1653281, EPI_ISL_1653282, EPI_ISL_1653286, EPI_ISL_1653287, EPI_ISL_1653289, EPI_ISL_1653300, EPI_ISL_1653305, EPI_ISL_1653310, EPI_ISL_1653315, EPI_ISL_1653316, EPI_ISL_1653320, EPI_ISL_1653326, EPI_ISL_1653345, EPI_ISL_1653350, EPI_ISL_1653359, EPI_ISL_1653362, EPI_ISL_1653368, EPI_ISL_1653371, EPI_ISL_1653374, EPI_ISL_1653375, EPI_ISL_1653378, EPI_ISL_1653382, EPI_ISL_1653386, EPI_ISL_1653389, EPI_ISL_1653390, EPI_ISL_1653403, EPI_ISL_1653406, EPI_ISL_1653417, EPI_ISL_1653419, EPI_ISL_1653422, EPI_ISL_1653423, EPI_ISL_1653424, EPI_ISL_1653426, EPI_ISL_1653428, EPI_ISL_1653429, EPI_ISL_1653431, EPI_ISL_1653449, EPI_ISL_1653455, EPI_ISL_1653457, EPI_ISL_1653463, EPI_ISL_1653469, EPI_ISL_1653472, EPI_ISL_1653473, EPI_ISL_1653480, EPI_ISL_1653498, EPI_ISL_1653553, EPI_ISL_1653598, EPI_ISL_1653601, EPI_ISL_1653639, EPI_ISL_1653650, EPI_ISL_1653666, EPI_ISL_1653675, EPI_ISL_1653680, EPI_ISL_1653681, EPI_ISL_1653719, EPI_ISL_1653755, EPI_ISL_1653757, EPI_ISL_1653759, EPI_ISL_1653769, EPI_ISL_1653770, EPI_ISL_1673053, EPI_ISL_1673056, EPI_ISL_1673058, EPI_ISL_1673061, EPI_ISL_1673087, EPI_ISL_1673094, EPI_ISL_1673103, EPI_ISL_1673115, EPI_ISL_1673132, EPI_ISL_1673140, EPI_ISL_1673141, EPI_ISL_1673142, EPI_ISL_1673147, EPI_ISL_1697443, EPI_ISL_1697453, EPI_ISL_1697468, EPI_ISL_1697469, EPI_ISL_1697472, EPI_ISL_1697482, EPI_ISL_1697484, EPI_ISL_1697486, EPI_ISL_1697491, EPI_ISL_1697495, EPI_ISL_1697496, EPI_ISL_1697508, EPI_ISL_1697514, EPI_ISL_1697517, EPI_ISL_1697519, EPI_ISL_1697523, EPI_ISL_1697525, EPI_ISL_1697529, EPI_ISL_1697532, EPI_ISL_1697534, EPI_ISL_1697546, EPI_ISL_1697564, EPI_ISL_1697573, EPI_ISL_1697576, EPI_ISL_1697588, EPI_ISL_1697595, EPI_ISL_1697607, EPI_ISL_1697620, EPI_ISL_1697623, EPI_ISL_1697625, EPI_ISL_1697630, EPI_ISL_1697636, EPI_ISL_1697639, EPI_ISL_1697648, EPI_ISL_1697652, EPI_ISL_1697655, EPI_ISL_1697657, EPI_ISL_1697660, EPI_ISL_1697664, EPI_ISL_1697677, EPI_ISL_1697695, EPI_ISL_1697698, EPI_ISL_1697704, EPI_ISL_1697709, EPI_ISL_1697710, EPI_ISL_1697717, EPI_ISL_1697722, EPI_ISL_1697723, EPI_ISL_1697735, EPI_ISL_1697736, EPI_ISL_1697749, EPI_ISL_1697761, EPI_ISL_1697772, EPI_ISL_1697779, EPI_ISL_1697787, EPI_ISL_1697802, EPI_ISL_1697813, EPI_ISL_1697823, EPI_ISL_1697829, EPI_ISL_1697837, EPI_ISL_1697851, EPI_ISL_1697852, EPI_ISL_1697864, EPI_ISL_1697867, EPI_ISL_1697886, EPI_ISL_1697887, EPI_ISL_1697898, EPI_ISL_1697907, EPI_ISL_1697908, EPI_ISL_1697912, EPI_ISL_1697914, EPI_ISL_1697918, EPI_ISL_1697919, EPI_ISL_1697923, EPI_ISL_1697925, EPI_ISL_1697937, EPI_ISL_1697942, EPI_ISL_1697946, EPI_ISL_1697960, EPI_ISL_1697962, EPI_ISL_1697963, EPI_ISL_1697965, EPI_ISL_1698261, EPI_ISL_1698270, EPI_ISL_1698324, EPI_ISL_1698392, EPI_ISL_1698394, EPI_ISL_1698431, EPI_ISL_1700177, EPI_ISL_1700182, EPI_ISL_1700183, EPI_ISL_1700187, EPI_ISL_1700217, EPI_ISL_1700220, EPI_ISL_1700237, EPI_ISL_1700238, EPI_ISL_1700246, EPI_ISL_1700248, EPI_ISL_1700256, EPI_ISL_1700262, EPI_ISL_1700268, EPI_ISL_1700283, EPI_ISL_1700288, EPI_ISL_1700292, EPI_ISL_1700293, EPI_ISL_1700297, EPI_ISL_1700318, EPI_ISL_1700325, EPI_ISL_1700326, EPI_ISL_1700336, EPI_ISL_1700342, EPI_ISL_1700360, EPI_ISL_1700365, EPI_ISL_1700366, EPI_ISL_1700367, EPI_ISL_1700393, EPI_ISL_1700399, EPI_ISL_1700408, EPI_ISL_1700411, EPI_ISL_1700414, EPI_ISL_1700423, EPI_ISL_1700426, EPI_ISL_1700442, EPI_ISL_1700443, EPI_ISL_1700444, EPI_ISL_1700449, EPI_ISL_1700458, EPI_ISL_1700485, EPI_ISL_1700510, EPI_ISL_1700511, EPI_ISL_1700514, EPI_ISL_1700515, EPI_ISL_1719144, EPI_ISL_1719439, EPI_ISL_1719487, EPI_ISL_1719697, EPI_ISL_1719732, EPI_ISL_1719734, EPI_ISL_1719736, EPI_ISL_1719737, EPI_ISL_1719738, EPI_ISL_1719744, EPI_ISL_1742176, EPI_ISL_1742178, EPI_ISL_1742179, EPI_ISL_1742203, EPI_ISL_1742229, EPI_ISL_1742242, EPI_ISL_1987383

| | | | |
|---|---|---|---|
| see above | Lighthouse Lab in Cambridge | Wellcome Sanger Institute for the COVID-19 Genomics UK (COG-UK) Consortium | Cordelia Langford; David K. Jackson; Dominic Kwiatkowski; Ewan Harrison; Ian Johnston; Jeffrey Barrett; John Sillitoe on behalf of the Wellcome Sanger Institute COVID-19 Surveillance Team; Rob Howes; Roberto Amato; Sonia Goncalves; The Lighthouse Lab in Cambridge and Alex Alderton |

EPI_ISL_1544499, EPI_ISL_1544502, EPI_ISL_1544506, EPI_ISL_1544517, EPI_ISL_1544519, EPI_ISL_1544534, EPI_ISL_1544543, EPI_ISL_1544545, EPI_ISL_1544558, EPI_ISL_1544560, EPI_ISL_1544581, EPI_ISL_1544599, EPI_ISL_1544604, EPI_ISL_1544606, EPI_ISL_1544621, EPI_ISL_1544629, EPI_ISL_1544639, EPI_ISL_1544646, EPI_ISL_1544648, EPI_ISL_1544649, EPI_ISL_1544656, EPI_ISL_1544658, EPI_ISL_1544666, EPI_ISL_1544670, EPI_ISL_1544673, EPI_ISL_1544678, EPI_ISL_1544682, EPI_ISL_1544685, EPI_ISL_1544696, EPI_ISL_1544715, EPI_ISL_1544725, EPI_ISL_1564026, EPI_ISL_1564027, EPI_ISL_1564028, EPI_ISL_1564029, EPI_ISL_1564031, EPI_ISL_1564032, EPI_ISL_1564033, EPI_ISL_1564034, EPI_ISL_1564036, EPI_ISL_1564037, EPI_ISL_1564039, EPI_ISL_1564041, EPI_ISL_1564042, EPI_ISL_1564044, EPI_ISL_1564048, EPI_ISL_1564049, EPI_ISL_1564054, EPI_ISL_1564055, EPI_ISL_1564056, EPI_ISL_1564058, EPI_ISL_1564059, EPI_ISL_1564069, EPI_ISL_1564074, EPI_ISL_1564075, EPI_ISL_1564079, EPI_ISL_1564082, EPI_ISL_1564083, EPI_ISL_1564085, EPI_ISL_1564090, EPI_ISL_1564091, EPI_ISL_1564096, EPI_ISL_1564104, EPI_ISL_1564105, EPI_ISL_1564109, EPI_ISL_1564111, EPI_ISL_1564114, EPI_ISL_1564117, EPI_ISL_1564120, EPI_ISL_1564122, EPI_ISL_1564128, EPI_ISL_1564133, EPI_ISL_1564134, EPI_ISL_1564135, EPI_ISL_1564137, EPI_ISL_1564138, EPI_ISL_1564141, EPI_ISL_1564142, EPI_ISL_1564143, EPI_ISL_1564145, EPI_ISL_1564146, EPI_ISL_1564148, EPI_ISL_1564149, EPI_ISL_1564151, EPI_ISL_1564154, EPI_ISL_1564155, EPI_ISL_1564156, EPI_ISL_1564157, EPI_ISL_1564158, EPI_ISL_1564161, EPI_ISL_1564162, EPI_ISL_1564163, EPI_ISL_1564164, EPI_ISL_1564165, EPI_ISL_1564167, EPI_ISL_1564168, EPI_ISL_1564169, EPI_ISL_1564170, EPI_ISL_1564174, EPI_ISL_1564177, EPI_ISL_1564180, EPI_ISL_1564184, EPI_ISL_1564186, EPI_ISL_1564189, EPI_ISL_1564191, EPI_ISL_1564192, EPI_ISL_1564195, EPI_ISL_1564196, EPI_ISL_1564198, EPI_ISL_1564199, EPI_ISL_1564201, EPI_ISL_1564207, EPI_ISL_1564208, EPI_ISL_1564209, EPI_ISL_1564212, EPI_ISL_1564213, EPI_ISL_1564214, EPI_ISL_1564216, EPI_ISL_1564217, EPI_ISL_1564218, EPI_ISL_1564220, EPI_ISL_1564225, EPI_ISL_1564226, EPI_ISL_1564227, EPI_ISL_1564230, EPI_ISL_1564232, EPI_ISL_1564233, EPI_ISL_1564235, EPI_ISL_1564238, EPI_ISL_1564242, EPI_ISL_1564243, EPI_ISL_1564245, EPI_ISL_1564248, EPI_ISL_1564249, EPI_ISL_1564250, EPI_ISL_1564252, EPI_ISL_1564255, EPI_ISL_1564257, EPI_ISL_1564260, EPI_ISL_1564261, EPI_ISL_1564263, EPI_ISL_1564264, EPI_ISL_1564266, EPI_ISL_1564267, EPI_ISL_1564268, EPI_ISL_1564269, EPI_ISL_1564273, EPI_ISL_1564274, EPI_ISL_1564275, EPI_ISL_1564278, EPI_ISL_1564279, EPI_ISL_1564282, EPI_ISL_1564283, EPI_ISL_1564288, EPI_ISL_1564290, EPI_ISL_1564292, EPI_ISL_1564293, EPI_ISL_1564305, EPI_ISL_1564306, EPI_ISL_1564308, EPI_ISL_1564309, EPI_ISL_1564311, EPI_ISL_1564312, EPI_ISL_1564313, EPI_ISL_1564317, EPI_ISL_1564318, EPI_ISL_1564320, EPI_ISL_1564321, EPI_ISL_1564322, EPI_ISL_1564325, EPI_ISL_1564327, EPI_ISL_1564329, EPI_ISL_1564330, EPI_ISL_1564331, EPI_ISL_1583688, EPI_ISL_1583692, EPI_ISL_1583695, EPI_ISL_1583701, EPI_ISL_1583702, EPI_ISL_1583704, EPI_ISL_1583706, EPI_ISL_1583707, EPI_ISL_1583711, EPI_ISL_1583718, EPI_ISL_1583721, EPI_ISL_1583724, EPI_ISL_1583726, EPI_ISL_1583729, EPI_ISL_1583735, EPI_ISL_1583737, EPI_ISL_1583738, EPI_ISL_1583740, EPI_ISL_1583741, EPI_ISL_1583745, EPI_ISL_1583747, EPI_ISL_1583749, EPI_ISL_1583751, EPI_ISL_1583754, EPI_ISL_1583761, EPI_ISL_1583762, EPI_ISL_1583765, EPI_ISL_1583766, EPI_ISL_1583768, EPI_ISL_1583772, EPI_ISL_1583779, EPI_ISL_1583788, EPI_ISL_1583794, EPI_ISL_1583795, EPI_ISL_1583800, EPI_ISL_1583802, EPI_ISL_1583805, EPI_ISL_1583806, EPI_ISL_1583810, EPI_ISL_1583812, EPI_ISL_1583814, EPI_ISL_1583818, EPI_ISL_1583820, EPI_ISL_1583821, EPI_ISL_1583827, EPI_ISL_1583831, EPI_ISL_1583838, EPI_ISL_1583842, EPI_ISL_1583846, EPI_ISL_1583847, EPI_ISL_1583849, EPI_ISL_1583850, EPI_ISL_1583851, EPI_ISL_1583856, EPI_ISL_1583860, EPI_ISL_1583866, EPI_ISL_1583868, EPI_ISL_1583871, EPI_ISL_1583876, EPI_ISL_1583877, EPI_ISL_1583881, EPI_ISL_1583882, EPI_ISL_1583883, EPI_ISL_1583885, EPI_ISL_1583887, EPI_ISL_1583888, EPI_ISL_1583894, EPI_ISL_1583899, EPI_ISL_1583903, EPI_ISL_1583907, EPI_ISL_1583913, EPI_ISL_1583921, EPI_ISL_1583926, EPI_ISL_1583945, EPI_ISL_1583946, EPI_ISL_1583951, EPI_ISL_1583954, EPI_ISL_1583960, EPI_ISL_1583963, EPI_ISL_1583967, EPI_ISL_1583983, EPI_ISL_1583999, EPI_ISL_1584002, EPI_ISL_1584007, EPI_ISL_1584012, EPI_ISL_1584015, EPI_ISL_1584016, EPI_ISL_1584022, EPI_ISL_1584034, EPI_ISL_1584039, EPI_ISL_1584040, EPI_ISL_1584046, EPI_ISL_1584050, EPI_ISL_1584052, EPI_ISL_1584053, EPI_ISL_1584056, EPI_ISL_1584062, EPI_ISL_1584066, EPI_ISL_1584073, EPI_ISL_1584084, EPI_ISL_1584087, EPI_ISL_1584094, EPI_ISL_1584102, EPI_ISL_1584103, EPI_ISL_1584108, EPI_ISL_1584110, EPI_ISL_1584112, EPI_ISL_1584116, EPI_ISL_1584119, EPI_ISL_1584122, EPI_ISL_1584127, EPI_ISL_1584128, EPI_ISL_1584130, EPI_ISL_1584132, EPI_ISL_1584168, EPI_ISL_1584177, EPI_ISL_1584182, EPI_ISL_1584183, EPI_ISL_1584185, EPI_ISL_1584187, EPI_ISL_1584188, EPI_ISL_1584189, EPI_ISL_1584191, EPI_ISL_1584192, EPI_ISL_1584195, EPI_ISL_1584200, EPI_ISL_1584204, EPI_ISL_1584206, EPI_ISL_1584208, EPI_ISL_1584213, EPI_ISL_1584215, EPI_ISL_1584216, EPI_ISL_1584217, EPI_ISL_1584228, EPI_ISL_1584232, EPI_ISL_1584233, EPI_ISL_1584235, EPI_ISL_1584239, EPI_ISL_1584240, EPI_ISL_1584241, EPI_ISL_1584245, EPI_ISL_1584248, EPI_ISL_1584250, EPI_ISL_1584253, EPI_ISL_1584255, EPI_ISL_1584256, EPI_ISL_1584258, EPI_ISL_1584259, EPI_ISL_1584260, EPI_ISL_1584277, EPI_ISL_1584279, EPI_ISL_1584288, EPI_ISL_1584290, EPI_ISL_1584293, EPI_ISL_1584298, EPI_ISL_1584309, EPI_ISL_1584310, EPI_ISL_1584327, EPI_ISL_1584329, EPI_ISL_1584330, EPI_ISL_1584334, EPI_ISL_1584337, EPI_ISL_1584345, EPI_ISL_1584346, EPI_ISL_1584348, EPI_ISL_1584353, EPI_ISL_1584356, EPI_ISL_1584357, EPI_ISL_1584368, EPI_ISL_1584369, EPI_ISL_1584371, EPI_ISL_1584375, EPI_ISL_1584380, EPI_ISL_1584383, EPI_ISL_1584391, EPI_ISL_1584398, EPI_ISL_1584402, EPI_ISL_1584408, EPI_ISL_1584414, EPI_ISL_1584415, EPI_ISL_1584418, EPI_ISL_1584419, EPI_ISL_1584422, EPI_ISL_1584436, EPI_ISL_1584437, EPI_ISL_1584441, EPI_ISL_1584442, EPI_ISL_1584443, EPI_ISL_1584445, EPI_ISL_1584446, EPI_ISL_1584447, EPI_ISL_1584448, EPI_ISL_1584449, EPI_ISL_1584450, EPI_ISL_1584451, EPI_ISL_1584452, EPI_ISL_1584453, EPI_ISL_1584454, EPI_ISL_1584455, EPI_ISL_1584456, EPI_ISL_1584457, EPI_ISL_1584458, EPI_ISL_1584459, EPI_ISL_1584461, EPI_ISL_1584462, EPI_ISL_1584463, EPI_ISL_1584464, EPI_ISL_1584465, EPI_ISL_1584467, EPI_ISL_1584468, EPI_ISL_1584471, EPI_ISL_1584473, EPI_ISL_1584474, EPI_ISL_1584477, EPI_ISL_1584479, EPI_ISL_1584480, EPI_ISL_1584482, EPI_ISL_1584485, EPI_ISL_1584486, EPI_ISL_1584488, EPI_ISL_1584491, EPI_ISL_1584492, EPI_ISL_1584494, EPI_ISL_1584495, EPI_ISL_1584496, EPI_ISL_1584499, EPI_ISL_1584500, EPI_ISL_1584501, EPI_ISL_1584502, EPI_ISL_1584503, EPI_ISL_1584504, EPI_ISL_1584505, EPI_ISL_1584506, EPI_ISL_1584507, EPI_ISL_1584508, EPI_ISL_1584509, EPI_ISL_1584510, EPI_ISL_1584512, EPI_ISL_1584514, EPI_ISL_1584516, EPI_ISL_1584517, EPI_ISL_1584518, EPI_ISL_1584519, EPI_ISL_1584520, EPI_ISL_1584521, EPI_ISL_1584522, EPI_ISL_1584523, EPI_ISL_1584524, EPI_ISL_1584525, EPI_ISL_1584526, EPI_ISL_1584527, EPI_ISL_1584577, EPI_ISL_1584589, EPI_ISL_1584590, EPI_ISL_1584597, EPI_ISL_1584600, EPI_ISL_1584601, EPI_ISL_1584607, EPI_ISL_1584608, EPI_ISL_1584612, EPI_ISL_1584614, EPI_ISL_1584625, EPI_ISL_1584626, EPI_ISL_1584627, EPI_ISL_1584628, EPI_ISL_1584634, EPI_ISL_1584636, EPI_ISL_1584637, EPI_ISL_1584639, EPI_ISL_1584642, EPI_ISL_1584648, EPI_ISL_1584678, EPI_ISL_1584685, EPI_ISL_1584693, EPI_ISL_1584705, EPI_ISL_1594453, EPI_ISL_1594454, EPI_ISL_1594455, EPI_ISL_1594456, EPI_ISL_1594458, EPI_ISL_1594459, EPI_ISL_1594460, EPI_ISL_1594461, EPI_ISL_1594462, EPI_ISL_1594467, EPI_ISL_1594468, EPI_ISL_1594469, EPI_ISL_1594470, EPI_ISL_1594472, EPI_ISL_1594473, EPI_ISL_1594474, EPI_ISL_1594475, EPI_ISL_1594476, EPI_ISL_1594477, EPI_ISL_1594479, EPI_ISL_1594480, EPI_ISL_1594481, EPI_ISL_1594483, EPI_ISL_1594485, EPI_ISL_1594486, EPI_ISL_1594487, EPI_ISL_1594488, EPI_ISL_1594489, EPI_ISL_1594490, EPI_ISL_1594491, EPI_ISL_1594493, EPI_ISL_1594495, EPI_ISL_1594496, EPI_ISL_1594497, EPI_ISL_1594498, EPI_ISL_1594499, EPI_ISL_1594501, EPI_ISL_1594502, EPI_ISL_1594503, EPI_ISL_1594504, EPI_ISL_1594505, EPI_ISL_1594506, EPI_ISL_1594508, EPI_ISL_1594509, EPI_ISL_1594510, EPI_ISL_1594511, EPI_ISL_1594513, EPI_ISL_1594514, EPI_ISL_1594515, EPI_ISL_1594517, EPI_ISL_1594518, EPI_ISL_1594519, EPI_ISL_1594521, EPI_ISL_1594523, EPI_ISL_1594524, EPI_ISL_1594525, EPI_ISL_1594526, EPI_ISL_1594527, EPI_ISL_1594529, EPI_ISL_1594531, EPI_ISL_1594532, EPI_ISL_1594533, EPI_ISL_1594534, EPI_ISL_1594535, EPI_ISL_1594537, EPI_ISL_1594538, EPI_ISL_1594539, EPI_ISL_1594542, EPI_ISL_1594543, EPI_ISL_1594544, EPI_ISL_1594545, EPI_ISL_1594546, EPI_ISL_1594548, EPI_ISL_1594550, EPI_ISL_1594552, EPI_ISL_1594553, EPI_ISL_1594554, EPI_ISL_1594555, EPI_ISL_1594556, EPI_ISL_1594558, EPI_ISL_1594559, EPI_ISL_1594560, EPI_ISL_1594561, EPI_ISL_1594562, EPI_ISL_1594564, EPI_ISL_1594565, EPI_ISL_1594566, EPI_ISL_1594568, EPI_ISL_1594570, EPI_ISL_1594572, EPI_ISL_1594573, EPI_ISL_1594575, EPI_ISL_1594576, EPI_ISL_1594577, EPI_ISL_1594578, EPI_ISL_1594579, EPI_ISL_1594580, EPI_ISL_1594581, EPI_ISL_1594583, EPI_ISL_1594584, EPI_ISL_1594585, EPI_ISL_1594586, EPI_ISL_1594587, EPI_ISL_1594589, EPI_ISL_1594591, EPI_ISL_1594592, EPI_ISL_1594593, EPI_ISL_1594594, EPI_ISL_1594595, EPI_ISL_1594597, EPI_ISL_1594598, EPI_ISL_1594599, EPI_ISL_1594600, EPI_ISL_1594601, EPI_ISL_1594602, EPI_ISL_1594603, EPI_ISL_1594604, EPI_ISL_1594605, EPI_ISL_1594606, EPI_ISL_1594607, EPI_ISL_1594608, EPI_ISL_1594609, EPI_ISL_1594610, EPI_ISL_1594611, EPI_ISL_1594614, EPI_ISL_1594616, EPI_ISL_1594617, EPI_ISL_1594618, EPI_ISL_1594619, EPI_ISL_1594620, EPI_ISL_1594621, EPI_ISL_1594623, EPI_ISL_1594626, EPI_ISL_1594627, EPI_ISL_1594628, EPI_ISL_1594629, EPI_ISL_1594630, EPI_ISL_1594631, EPI_ISL_1594632, EPI_ISL_1594633, EPI_ISL_1594634, EPI_ISL_1594636, EPI_ISL_1594637, EPI_ISL_1594638, EPI_ISL_1594640, EPI_ISL_1594642, EPI_ISL_1594644, EPI_ISL_1594645, EPI_ISL_1594646, EPI_ISL_1594647, EPI_ISL_1594648, EPI_ISL_1594651, EPI_ISL_1594652, EPI_ISL_1594653, EPI_ISL_1594654, EPI_ISL_1594656, EPI_ISL_1594657, EPI_ISL_1594658, EPI_ISL_1594663, EPI_ISL_1594664, EPI_ISL_1594665, EPI_ISL_1594666, EPI_ISL_1594670, EPI_ISL_1594671, EPI_ISL_1594672, EPI_ISL_1594673, EPI_ISL_1594674, EPI_ISL_1594676, EPI_ISL_1594677, EPI_ISL_1594683, EPI_ISL_1594684, EPI_ISL_1594685, EPI_ISL_1594686, EPI_ISL_1594688, EPI_ISL_1594690, EPI_ISL_1594691, EPI_ISL_1594692, EPI_ISL_1594693, EPI_ISL_1594695, EPI_ISL_1594696, EPI_ISL_1594697, EPI_ISL_1594698, EPI_ISL_1594699, EPI_ISL_1594701, EPI_ISL_1594702, EPI_ISL_1594704, EPI_ISL_1594705, EPI_ISL_1594706, EPI_ISL_1594707, EPI_ISL_1594708, EPI_ISL_1594709, EPI_ISL_1594710, EPI_ISL_1594711, EPI_ISL_1594713, EPI_ISL_1594718, EPI_ISL_1594720, EPI_ISL_1594721, EPI_ISL_1594723, EPI_ISL_1594727, EPI_ISL_1594729, EPI_ISL_1594735, EPI_ISL_1594743, EPI_ISL_1594744, EPI_ISL_1594746, EPI_ISL_1594752, EPI_ISL_1594754, EPI_ISL_1594755, EPI_ISL_1594757, EPI_ISL_1594761, EPI_ISL_1594771, EPI_ISL_1594772, EPI_ISL_1594777, EPI_ISL_1594779, EPI_ISL_1594783, EPI_ISL_1594790, EPI_ISL_1594795, EPI_ISL_1594796, EPI_ISL_1594805, EPI_ISL_1594806, EPI_ISL_1594808, EPI_ISL_1594809, EPI_ISL_1594816, EPI_ISL_1594829, EPI_ISL_1594839, EPI_ISL_1594840, EPI_ISL_1594845, EPI_ISL_1594851, EPI_ISL_1594852, EPI_ISL_1594856, EPI_ISL_1594858, EPI_ISL_1594859, EPI_ISL_1594878, EPI_ISL_1594880, EPI_ISL_1594896, EPI_ISL_1594903, EPI_ISL_1594909, EPI_ISL_1594912, EPI_ISL_1594913, EPI_ISL_1594916, EPI_ISL_1594920, EPI_ISL_1594922, EPI_ISL_1594923, EPI_ISL_1594925, EPI_ISL_1594928, EPI_ISL_1594930, EPI_ISL_1594932, EPI_ISL_1594939, EPI_ISL_1594940, EPI_ISL_1594942, EPI_ISL_1594943, EPI_ISL_1594952, EPI_ISL_1594959, EPI_ISL_1594960, EPI_ISL_1594968, EPI_ISL_1594971, EPI_ISL_1594972, EPI_ISL_1594978, EPI_ISL_1594979, EPI_ISL_1594984, EPI_ISL_1594986, EPI_ISL_1594991, EPI_ISL_1594995, EPI_ISL_1594996, EPI_ISL_1594999, EPI_ISL_1595002, EPI_ISL_1595004, EPI_ISL_1595005, EPI_ISL_1595008, EPI_ISL_1595010, EPI_ISL_1595011, EPI_ISL_1595012, EPI_ISL_1595014, EPI_ISL_1595015, EPI_ISL_1595017, EPI_ISL_1595018, EPI_ISL_1595020, EPI_ISL_1595021, EPI_ISL_1595022, EPI_ISL_1595023, EPI_ISL_1595025, EPI_ISL_1595026, EPI_ISL_1595027, EPI_ISL_1595028, EPI_ISL_1595030, EPI_ISL_1595031, EPI_ISL_1595032, EPI_ISL_1595034, EPI_ISL_1595035, EPI_ISL_1595036, EPI_ISL_1595037, EPI_ISL_1595038, EPI_ISL_1595039, EPI_ISL_1595040, EPI_ISL_1595041, EPI_ISL_1595045, EPI_ISL_1595046, EPI_ISL_1595047, EPI_ISL_1595049, EPI_ISL_1595050, EPI_ISL_1595051, EPI_ISL_1595052, EPI_ISL_1595053, EPI_ISL_1595055, EPI_ISL_1595056, EPI_ISL_1595057, EPI_ISL_1595060, EPI_ISL_1595061, EPI_ISL_1595062, EPI_ISL_1595063, EPI_ISL_1595064, EPI_ISL_1595065, EPI_ISL_1595067, EPI_ISL_1595068, EPI_ISL_1595069, EPI_ISL_1595071, EPI_ISL_1595072, EPI_ISL_1595074, EPI_ISL_1595076, EPI_ISL_1595077, EPI_ISL_1595078, EPI_ISL_1595079, EPI_ISL_1595080, EPI_ISL_1595081, EPI_ISL_1595082, EPI_ISL_1595085, EPI_ISL_1595086, EPI_ISL_1595087, EPI_ISL_1595088, EPI_ISL_1595089, EPI_ISL_1595091, EPI_ISL_1595093, EPI_ISL_1595094, EPI_ISL_1595095, EPI_ISL_1595096, EPI_ISL_1595097, EPI_ISL_1595099, EPI_ISL_1595102, EPI_ISL_1595103, EPI_ISL_1595104, EPI_ISL_1595105, EPI_ISL_1595108, EPI_ISL_1595109, EPI_ISL_1595110, EPI_ISL_1595112, EPI_ISL_1595113, EPI_ISL_1595114, EPI_ISL_1595115, EPI_ISL_1595116, EPI_ISL_1595117, EPI_ISL_1595118, EPI_ISL_1595119, EPI_ISL_1595121, EPI_ISL_1595124, EPI_ISL_1595125, EPI_ISL_1595126, EPI_ISL_1595127, EPI_ISL_1595129, EPI_ISL_1595130, EPI_ISL_1595131, EPI_ISL_1595133, EPI_ISL_1595134, EPI_ISL_1595136, EPI_ISL_1595137, EPI_ISL_1595138, EPI_ISL_1595139, EPI_ISL_1595143, EPI_ISL_1595145, EPI_ISL_1595148, EPI_ISL_1595149, EPI_ISL_1595150, EPI_ISL_1595152, EPI_ISL_1595153, EPI_ISL_1595155, EPI_ISL_1595156, EPI_ISL_1595157, EPI_ISL_1595158, EPI_ISL_1595159, EPI_ISL_1595162, EPI_ISL_1595163, EPI_ISL_1595166, EPI_ISL_1595170, EPI_ISL_1595171, EPI_ISL_1595174, EPI_ISL_1595175, EPI_ISL_1595176, EPI_ISL_1595177, EPI_ISL_1595178, EPI_ISL_1595179, EPI_ISL_1595181, EPI_ISL_1595182, EPI_ISL_1595183, EPI_ISL_1595184, EPI_ISL_1595186, EPI_ISL_1595187, EPI_ISL_1595188, EPI_ISL_1595190, EPI_ISL_1595191, EPI_ISL_1595192, EPI_ISL_1595194, EPI_ISL_1595200, EPI_ISL_1595473, EPI_ISL_1615691, EPI_ISL_1615721, EPI_ISL_1615727, EPI_ISL_1615739, EPI_ISL_1615741, EPI_ISL_1615746, EPI_ISL_1615750, EPI_ISL_1615801, EPI_ISL_1615803, EPI_ISL_1615814, EPI_ISL_1615833, EPI_ISL_1615844, EPI_ISL_1615852, EPI_ISL_1615868, EPI_ISL_1615873, EPI_ISL_1615887, EPI_ISL_1615895, EPI_ISL_1615898, EPI_ISL_1615902, EPI_ISL_1615908, EPI_ISL_1615917, EPI_ISL_1615924, EPI_ISL_1615931, EPI_ISL_1615933, EPI_ISL_1615935, EPI_ISL_1615936, EPI_ISL_1615938, EPI_ISL_1615940, EPI_ISL_1615941, EPI_ISL_1615947, EPI_ISL_1615948, EPI_ISL_1615953, EPI_ISL_1615956, EPI_ISL_1615977, EPI_ISL_1615978, EPI_ISL_1615982, EPI_ISL_1615986, EPI_ISL_1615988, EPI_ISL_1615989, EPI_ISL_1615990, EPI_ISL_1615993, EPI_ISL_1615994, EPI_ISL_1615996, EPI_ISL_1615998, EPI_ISL_1615999, EPI_ISL_1616001, EPI_ISL_1616004, EPI_ISL_1616007, EPI_ISL_1616020, EPI_ISL_1616022, EPI_ISL_1616023, EPI_ISL_1616024, EPI_ISL_1616025, EPI_ISL_1616027, EPI_ISL_1616029, EPI_ISL_1616031, EPI_ISL_1616032, EPI_ISL_1616039, EPI_ISL_1616044, EPI_ISL_1616048, EPI_ISL_1616049, EPI_ISL_1616051, EPI_ISL_1616055, EPI_ISL_1616057, EPI_ISL_1616059, EPI_ISL_1616060, EPI_ISL_1616061, EPI_ISL_1616063, EPI_ISL_1616064, EPI_ISL_1616069, EPI_ISL_1616073, EPI_ISL_1616075, EPI_ISL_1616076, EPI_ISL_1616077, EPI_ISL_1616079, EPI_ISL_1616082, EPI_ISL_1616087, EPI_ISL_1616091, EPI_ISL_1616092, EPI_ISL_1616097, EPI_ISL_1616099, EPI_ISL_1616100, EPI_ISL_1616101, EPI_ISL_1616102, EPI_ISL_1616103, EPI_ISL_1616104, EPI_ISL_1616106, EPI_ISL_1616107, EPI_ISL_1616109, EPI_ISL_1616110, EPI_ISL_1616111, EPI_ISL_1616112, EPI_ISL_1616113, EPI_ISL_1616114, EPI_ISL_1616117, EPI_ISL_1616120, EPI_ISL_1616121, EPI_ISL_1616123, EPI_ISL_1616127, EPI_ISL_1616128, EPI_ISL_1616130, EPI_ISL_1616134, EPI_ISL_1616137, EPI_ISL_1616139, EPI_ISL_1616140, EPI_ISL_1616143, EPI_ISL_1616146, EPI_ISL_1616150, EPI_ISL_1616152, EPI_ISL_1616153, EPI_ISL_1616155, EPI_ISL_1616157, EPI_ISL_1616159, EPI_ISL_1616160, EPI_ISL_1616162, EPI_ISL_1616163, EPI_ISL_1616165, EPI_ISL_1616167, EPI_ISL_1616170, EPI_ISL_1616171, EPI_ISL_1616177, EPI_ISL_1616179, EPI_ISL_1616181, EPI_ISL_1616186, EPI_ISL_1616187, EPI_ISL_1616189, EPI_ISL_1616190, EPI_ISL_1616191, EPI_ISL_1616192, EPI_ISL_1616193, EPI_ISL_1616195, EPI_ISL_1616196, EPI_ISL_1616197, EPI_ISL_1616198, EPI_ISL_1616199, EPI_ISL_1616200, EPI_ISL_1616201, EPI_ISL_1616202, EPI_ISL_1616203, EPI_ISL_1616204, EPI_ISL_1616205, EPI_ISL_1616206, EPI_ISL_1616207, EPI_ISL_1616208, EPI_ISL_1616209, EPI_ISL_1616210, EPI_ISL_1616212, EPI_ISL_1616213, EPI_ISL_1616214, EPI_ISL_1616215, EPI_ISL_1616216, EPI_ISL_1616217, EPI_ISL_1616218, EPI_ISL_1616219, EPI_ISL_1616220, EPI_ISL_1616221, EPI_ISL_1616222, EPI_ISL_1616223, EPI_ISL_1616224, EPI_ISL_1616225, EPI_ISL_1616226, EPI_ISL_1616227, EPI_ISL_1616228, EPI_ISL_1616229, EPI_ISL_1616230, EPI_ISL_1616231, EPI_ISL_1616232, EPI_ISL_1616233, EPI_ISL_1616234, EPI_ISL_1616235, EPI_ISL_1616236, EPI_ISL_1616237, EPI_ISL_1616238, EPI_ISL_1616239, EPI_ISL_1616240, EPI_ISL_1616241, EPI_ISL_1616242, EPI_ISL_1616243, EPI_ISL_1616244, EPI_ISL_1616245, EPI_ISL_1616246, EPI_ISL_1616247, EPI_ISL_1616248, EPI_ISL_1616249, EPI_ISL_1616250, EPI_ISL_1616251, EPI_ISL_1616252, EPI_ISL_1616253, EPI_ISL_1616254, EPI_ISL_1616255, EPI_ISL_1616256, EPI_ISL_1616257, EPI_ISL_1616258, EPI_ISL_1616259, EPI_ISL_1616260, EPI_ISL_1616263, EPI_ISL_1616264, EPI_ISL_1616266, EPI_ISL_1616268, EPI_ISL_1616269, EPI_ISL_1616270, EPI_ISL_1616271, EPI_ISL_1616274, EPI_ISL_1616275, EPI_ISL_1616276, EPI_ISL_1616277, EPI_ISL_1616278, EPI_ISL_1616280, EPI_ISL_1616282, EPI_ISL_1616284, EPI_ISL_1616288, EPI_ISL_1616289, EPI_ISL_1616290, EPI_ISL_1616291, EPI_ISL_1616292, EPI_ISL_1616293, EPI_ISL_1616294, EPI_ISL_1616296, EPI_ISL_1616297, EPI_ISL_1616298, EPI_ISL_1616299, EPI_ISL_1616301, EPI_ISL_1616302, EPI_ISL_1616304, EPI_ISL_1616305, EPI_ISL_1616306, EPI_ISL_1616307, EPI_ISL_1616308, EPI_ISL_1616309, EPI_ISL_1616310, EPI_ISL_1616311, EPI_ISL_1616312, EPI_ISL_1616313, EPI_ISL_1616314, EPI_ISL_1616315, EPI_ISL_1616316, EPI_ISL_1616317, EPI_ISL_1616318, EPI_ISL_1616319, EPI_ISL_1616320, EPI_ISL_1616321, EPI_ISL_1616322, EPI_ISL_1616323, EPI_ISL_1616324, EPI_ISL_1616325, EPI_ISL_1616326, EPI_ISL_1616327, EPI_ISL_1616328, EPI_ISL_1616329, EPI_ISL_1616330, EPI_ISL_1616331, EPI_ISL_1616332, EPI_ISL_1616333, EPI_ISL_1616334, EPI_ISL_1616335, EPI_ISL_1616336, EPI_ISL_1616337, EPI_ISL_1616338, EPI_ISL_1616339, EPI_ISL_1616340, EPI_ISL_1616341, EPI_ISL_1616342, EPI_ISL_1616343, EPI_ISL_1616344, EPI_ISL_1616345, EPI_ISL_1616346, EPI_ISL_1616347, EPI_ISL_1616348, EPI_ISL_1616349, EPI_ISL_1616350, EPI_ISL_1616351, EPI_ISL_1616352, EPI_ISL_1616353, EPI_ISL_1616354, EPI_ISL_1616355, EPI_ISL_1616356, EPI_ISL_1616357, EPI_ISL_1616358, EPI_ISL_1616359, EPI_ISL_1616360, EPI_ISL_1616361, EPI_ISL_1616362, EPI_ISL_1616363, EPI_ISL_1616364, EPI_ISL_1616365, EPI_ISL_1616366, EPI_ISL_1616367, EPI_ISL_1616368, EPI_ISL_1616369, EPI_ISL_1616370, EPI_ISL_1616371, EPI_ISL_1616372, EPI_ISL_1616373, EPI_ISL_1616374, EPI_ISL_1616375, EPI_ISL_1616376, EPI_ISL_1616377, EPI_ISL_1616378, EPI_ISL_1616379, EPI_ISL_1616380, EPI_ISL_1616381, EPI_ISL_1616382, EPI_ISL_1616383, EPI_ISL_1616384, EPI_ISL_1616385, EPI_ISL_1616386, EPI_ISL_1616387, EPI_ISL_1616388, EPI_ISL_1616389, EPI_ISL_1616390, EPI_ISL_1616391, EPI_ISL_1616392, EPI_ISL_1616393, EPI_ISL_1616394, EPI_ISL_1616395, EPI_ISL_1616396, EPI_ISL_1616397, EPI_ISL_1616398, EPI_ISL_1616399, EPI_ISL_1616400, EPI_ISL_1616401, EPI_ISL_1616402, EPI_ISL_1616404, EPI_ISL_1616405, EPI_ISL_1616406, EPI_ISL_1616407, EPI_ISL_1616408, EPI_ISL_1616409, EPI_ISL_1616410, EPI_ISL_1616411, EPI_ISL_1616412, EPI_ISL_1616413, EPI_ISL_1616414, EPI_ISL_1616416, EPI_ISL_1616417, EPI_ISL_1616418, EPI_ISL_1616419, EPI_ISL_1616420, EPI_ISL_1616421, EPI_ISL_1616422, EPI_ISL_1616423, EPI_ISL_1616424, EPI_ISL_1616425, EPI_ISL_1616426, EPI_ISL_1616427, EPI_ISL_1616428, EPI_ISL_1616429, EPI_ISL_1616430, EPI_ISL_1616431, EPI_ISL_1616432, EPI_ISL_1616433, EPI_ISL_1616436, EPI_ISL_1616437, EPI_ISL_1616440, EPI_ISL_1616441, EPI_ISL_1616442, EPI_ISL_1616443, EPI_ISL_1616444, EPI_ISL_1616445, EPI_ISL_1616446, EPI_ISL_1616447, EPI_ISL_1616448, EPI_ISL_1616449, EPI_ISL_1616450, EPI_ISL_1616452, EPI_ISL_1616453, EPI_ISL_1616455, EPI_ISL_1616457, EPI_ISL_1616459, EPI_ISL_1616460, EPI_ISL_1616462, EPI_ISL_1616463, EPI_ISL_1616465, EPI_ISL_1616466, EPI_ISL_1616467, EPI_ISL_1616468, EPI_ISL_1616469, EPI_ISL_1616470, EPI_ISL_1616471, EPI_ISL_1616474, EPI_ISL_1616477, EPI_ISL_1616478, EPI_ISL_1616479, EPI_ISL_1616480, EPI_ISL_1616483, EPI_ISL_1616484, EPI_ISL_1616486, EPI_ISL_1616487, EPI_ISL_1616488, EPI_ISL_1616489, EPI_ISL_1616491, EPI_ISL_1616492, EPI_ISL_1616493, EPI_ISL_1616494, EPI_ISL_1616495, EPI_ISL_1616496, EPI_ISL_1616497, EPI_ISL_1616498, EPI_ISL_1616499, EPI_ISL_1616500, EPI_ISL_1616501, EPI_ISL_1616502, EPI_ISL_1616503, EPI_ISL_1616504, EPI_ISL_1616505, EPI_ISL_1616508, EPI_ISL_1616510, EPI_ISL_1616511, EPI_ISL_1616512, EPI_ISL_1616513, EPI_ISL_1616516, EPI_ISL_1616517, EPI_ISL_1616518, EPI_ISL_1616519, EPI_ISL_1616520, EPI_ISL_1616521, EPI_ISL_1616522, EPI_ISL_1616524, EPI_ISL_1616525, EPI_ISL_1616527, EPI_ISL_1616528, EPI_ISL_1616529, EPI_ISL_1616532, EPI_ISL_1616533, EPI_ISL_1616535, EPI_ISL_1616536, EPI_ISL_1616537, EPI_ISL_1616539, EPI_ISL_1616541, EPI_ISL_1616542, EPI_ISL_1616543, EPI_ISL_1616544, EPI_ISL_1616545, EPI_ISL_1616547, EPI_ISL_1616548, EPI_ISL_1616549, EPI_ISL_1616550, EPI_ISL_1616553, EPI_ISL_1616554, EPI_ISL_1616555, EPI_ISL_1616556, EPI_ISL_1616557, EPI_ISL_1616558, EPI_ISL_1616559, EPI_ISL_1616560, EPI_ISL_1616561, EPI_ISL_1616562, EPI_ISL_1616563, EPI_ISL_1616565, EPI_ISL_1616566, EPI_ISL_1616575, EPI_ISL_1616576, EPI_ISL_1616577, EPI_ISL_1616578, EPI_ISL_1616579, EPI_ISL_1616580, EPI_ISL_1616581, EPI_ISL_1616582, EPI_ISL_1616583, EPI_ISL_1616586, EPI_ISL_1616587, EPI_ISL_1616589, EPI_ISL_1616590, EPI_ISL_1616591, EPI_ISL_1616592, EPI_ISL_1616594, EPI_ISL_1616595, EPI_ISL_1616596, EPI_ISL_1616597, EPI_ISL_1616598, EPI_ISL_1616599, EPI_ISL_1616600, EPI_ISL_1616601, EPI_ISL_1616602, EPI_ISL_1616603, EPI_ISL_1630282, EPI_ISL_1630283, EPI_ISL_1630285, EPI_ISL_1630286, EPI_ISL_1630287, EPI_ISL_1630288, EPI_ISL_1630290, EPI_ISL_1630296,

EPI_ISL_1630297, EPI_ISL_1630300, EPI_ISL_1630301, EPI_ISL_1630302, EPI_ISL_1630303, EPI_ISL_1630306, EPI_ISL_1630307, EPI_ISL_1630308, EPI_ISL_1630309, EPI_ISL_1630310, EPI_ISL_1630315, EPI_ISL_1630316, EPI_ISL_1630317, EPI_ISL_1630318, EPI_ISL_1630321, EPI_ISL_1630322, EPI_ISL_1630323, EPI_ISL_1630325, EPI_ISL_1630327, EPI_ISL_1630330, EPI_ISL_1630331, EPI_ISL_1630332, EPI_ISL_1630334, EPI_ISL_1630335, EPI_ISL_1630337, EPI_ISL_1630340, EPI_ISL_1630341, EPI_ISL_1630343, EPI_ISL_1630344, EPI_ISL_1630345, EPI_ISL_1630346, EPI_ISL_1630347, EPI_ISL_1630350, EPI_ISL_1630351, EPI_ISL_1630354, EPI_ISL_1630355, EPI_ISL_1630356, EPI_ISL_1630359, EPI_ISL_1630361, EPI_ISL_1630362, EPI_ISL_1630364, EPI_ISL_1630365, EPI_ISL_1630369, EPI_ISL_1630370, EPI_ISL_1630371, EPI_ISL_1630373, EPI_ISL_1630376, EPI_ISL_1630379, EPI_ISL_1630381, EPI_ISL_1630382, EPI_ISL_1630385, EPI_ISL_1630389, EPI_ISL_1630392, EPI_ISL_1630393, EPI_ISL_1630394, EPI_ISL_1630398, EPI_ISL_1630399, EPI_ISL_1630400, EPI_ISL_1630405, EPI_ISL_1630407, EPI_ISL_1630408, EPI_ISL_1630409, EPI_ISL_1630411, EPI_ISL_1630413, EPI_ISL_1630415, EPI_ISL_1630417, EPI_ISL_1630418, EPI_ISL_1630419, EPI_ISL_1630421, EPI_ISL_1630422, EPI_ISL_1630423, EPI_ISL_1630424, EPI_ISL_1630425, EPI_ISL_1630426, EPI_ISL_1630428, EPI_ISL_1630429, EPI_ISL_1630430, EPI_ISL_1630433, EPI_ISL_1630436, EPI_ISL_1630437, EPI_ISL_1630440, EPI_ISL_1630441, EPI_ISL_1630442, EPI_ISL_1630443, EPI_ISL_1630445, EPI_ISL_1630446, EPI_ISL_1630447, EPI_ISL_1630448, EPI_ISL_1630449, EPI_ISL_1630452, EPI_ISL_1630453, EPI_ISL_1630454, EPI_ISL_1630456, EPI_ISL_1630457, EPI_ISL_1630458, EPI_ISL_1630459, EPI_ISL_1630460, EPI_ISL_1630462, EPI_ISL_1630466, EPI_ISL_1630467, EPI_ISL_1630468, EPI_ISL_1630470, EPI_ISL_1630471, EPI_ISL_1630472, EPI_ISL_1630473, EPI_ISL_1630477, EPI_ISL_1630481, EPI_ISL_1630482, EPI_ISL_1630483, EPI_ISL_1630484, EPI_ISL_1630485, EPI_ISL_1630487, EPI_ISL_1630495, EPI_ISL_1630496, EPI_ISL_1630498, EPI_ISL_1630499, EPI_ISL_1630501, EPI_ISL_1630502, EPI_ISL_1630504, EPI_ISL_1630505, EPI_ISL_1630507, EPI_ISL_1630509, EPI_ISL_1630511, EPI_ISL_1630512, EPI_ISL_1630514, EPI_ISL_1630515, EPI_ISL_1630830, EPI_ISL_1630834, EPI_ISL_1630846, EPI_ISL_1630847, EPI_ISL_1630848, EPI_ISL_1630850, EPI_ISL_1630853, EPI_ISL_1630855, EPI_ISL_1630871, EPI_ISL_1630872, EPI_ISL_1630879, EPI_ISL_1630889, EPI_ISL_1630896, EPI_ISL_1630908, EPI_ISL_1630910, EPI_ISL_1630933, EPI_ISL_1630944, EPI_ISL_1630948, EPI_ISL_1630949, EPI_ISL_1630958, EPI_ISL_1630968, EPI_ISL_1630978, EPI_ISL_1630984, EPI_ISL_1630988, EPI_ISL_1630989, EPI_ISL_1631000, EPI_ISL_1631008, EPI_ISL_1631009, EPI_ISL_1631017, EPI_ISL_1631018, EPI_ISL_1631024, EPI_ISL_1631028, EPI_ISL_1631030, EPI_ISL_1631039, EPI_ISL_1631047, EPI_ISL_1631051, EPI_ISL_1631056, EPI_ISL_1631067, EPI_ISL_1631073, EPI_ISL_1631083, EPI_ISL_1631089, EPI_ISL_1631090, EPI_ISL_1631091, EPI_ISL_1631095, EPI_ISL_1631104, EPI_ISL_1631107, EPI_ISL_1631108, EPI_ISL_1631109, EPI_ISL_1631110, EPI_ISL_1631111, EPI_ISL_1631112, EPI_ISL_1631113, EPI_ISL_1631114, EPI_ISL_1631119, EPI_ISL_1631125, EPI_ISL_1631128, EPI_ISL_1631131, EPI_ISL_1631133, EPI_ISL_1631136, EPI_ISL_1631138, EPI_ISL_1631140, EPI_ISL_1631145, EPI_ISL_1631147, EPI_ISL_1631150, EPI_ISL_1631151, EPI_ISL_1631153, EPI_ISL_1631156, EPI_ISL_1631157, EPI_ISL_1631165, EPI_ISL_1631167, EPI_ISL_1631168, EPI_ISL_1631169, EPI_ISL_1631172, EPI_ISL_1631173, EPI_ISL_1631174, EPI_ISL_1631175, EPI_ISL_1631176, EPI_ISL_1631178, EPI_ISL_1631180, EPI_ISL_1631187, EPI_ISL_1631192, EPI_ISL_1631194, EPI_ISL_1631195, EPI_ISL_1631204, EPI_ISL_1631205, EPI_ISL_1631206, EPI_ISL_1631209, EPI_ISL_1631211, EPI_ISL_1631214, EPI_ISL_1631216, EPI_ISL_1631220, EPI_ISL_1631222, EPI_ISL_1631224, EPI_ISL_1631226, EPI_ISL_1631229, EPI_ISL_1631232, EPI_ISL_1631235, EPI_ISL_1631237, EPI_ISL_1631242, EPI_ISL_1631243, EPI_ISL_1631244, EPI_ISL_1631245, EPI_ISL_1631247, EPI_ISL_1631248, EPI_ISL_1631253, EPI_ISL_1631254, EPI_ISL_1631256, EPI_ISL_1631258, EPI_ISL_1631259, EPI_ISL_1631261, EPI_ISL_1631262, EPI_ISL_1631264, EPI_ISL_1631265, EPI_ISL_1631266, EPI_ISL_1631268, EPI_ISL_1631269, EPI_ISL_1631270, EPI_ISL_1631272, EPI_ISL_1631275, EPI_ISL_1631278, EPI_ISL_1631282, EPI_ISL_1631285, EPI_ISL_1631288, EPI_ISL_1631294, EPI_ISL_1631295, EPI_ISL_1631296, EPI_ISL_1631297, EPI_ISL_1631298, EPI_ISL_1631301, EPI_ISL_1631302, EPI_ISL_1631304, EPI_ISL_1631306, EPI_ISL_1631307, EPI_ISL_1631308, EPI_ISL_1631309, EPI_ISL_1631310, EPI_ISL_1631311, EPI_ISL_1631312, EPI_ISL_1631317, EPI_ISL_1631318, EPI_ISL_1631320, EPI_ISL_1631322, EPI_ISL_1631323, EPI_ISL_1631324, EPI_ISL_1631325, EPI_ISL_1631327, EPI_ISL_1631328, EPI_ISL_1631329, EPI_ISL_1631335, EPI_ISL_1631337, EPI_ISL_1631338, EPI_ISL_1631339, EPI_ISL_1631340, EPI_ISL_1631341, EPI_ISL_1631342, EPI_ISL_1631343, EPI_ISL_1631344, EPI_ISL_1631345, EPI_ISL_1631346, EPI_ISL_1631347, EPI_ISL_1631348, EPI_ISL_1631349, EPI_ISL_1631350, EPI_ISL_1631351, EPI_ISL_1631352, EPI_ISL_1631353, EPI_ISL_1631354, EPI_ISL_1631355, EPI_ISL_1631356, EPI_ISL_1631357, EPI_ISL_1631358, EPI_ISL_1631359, EPI_ISL_1631360, EPI_ISL_1631361, EPI_ISL_1631362, EPI_ISL_1631363, EPI_ISL_1631364, EPI_ISL_1631365, EPI_ISL_1631366, EPI_ISL_1631367, EPI_ISL_1631368, EPI_ISL_1631369, EPI_ISL_1631370, EPI_ISL_1631371, EPI_ISL_1631372, EPI_ISL_1631373, EPI_ISL_1631374, EPI_ISL_1631375, EPI_ISL_1631376, EPI_ISL_1631377, EPI_ISL_1631378, EPI_ISL_1631379, EPI_ISL_1631380, EPI_ISL_1631381, EPI_ISL_1631382, EPI_ISL_1631383, EPI_ISL_1631384, EPI_ISL_1631385, EPI_ISL_1631386, EPI_ISL_1631387, EPI_ISL_1631389, EPI_ISL_1631390, EPI_ISL_1631391, EPI_ISL_1631392, EPI_ISL_1631393, EPI_ISL_1631394, EPI_ISL_1631395, EPI_ISL_1631396, EPI_ISL_1631397, EPI_ISL_1631398, EPI_ISL_1631399, EPI_ISL_1631400, EPI_ISL_1631401, EPI_ISL_1631402, EPI_ISL_1631403, EPI_ISL_1631404, EPI_ISL_1631405, EPI_ISL_1631406, EPI_ISL_1631407, EPI_ISL_1631408, EPI_ISL_1631409, EPI_ISL_1631410, EPI_ISL_1631411, EPI_ISL_1631412, EPI_ISL_1631413, EPI_ISL_1631414, EPI_ISL_1631415, EPI_ISL_1631416, EPI_ISL_1631417, EPI_ISL_1631418, EPI_ISL_1631419, EPI_ISL_1631420, EPI_ISL_1631421, EPI_ISL_1631422, EPI_ISL_1631423, EPI_ISL_1631426, EPI_ISL_1631764, EPI_ISL_1631771, EPI_ISL_1631774, EPI_ISL_1631777, EPI_ISL_1631787, EPI_ISL_1631794, EPI_ISL_1631816, EPI_ISL_1631825, EPI_ISL_1631828, EPI_ISL_1631831, EPI_ISL_1631848, EPI_ISL_1631849, EPI_ISL_1631853, EPI_ISL_1631855, EPI_ISL_1631857, EPI_ISL_1631861, EPI_ISL_1631862, EPI_ISL_1631874, EPI_ISL_1631881, EPI_ISL_1631884, EPI_ISL_1631889, EPI_ISL_1631890, EPI_ISL_1631893, EPI_ISL_1631898, EPI_ISL_1631899, EPI_ISL_1631905, EPI_ISL_1631907, EPI_ISL_1631908, EPI_ISL_1631915, EPI_ISL_1631916, EPI_ISL_1631919, EPI_ISL_1631925, EPI_ISL_1631926, EPI_ISL_1631927, EPI_ISL_1631931, EPI_ISL_1631935, EPI_ISL_1631937, EPI_ISL_1631938, EPI_ISL_1631939, EPI_ISL_1631940, EPI_ISL_1631947, EPI_ISL_1631952, EPI_ISL_1631954, EPI_ISL_1631955, EPI_ISL_1631957, EPI_ISL_1631960, EPI_ISL_1631968, EPI_ISL_1631974, EPI_ISL_1631975, EPI_ISL_1631979, EPI_ISL_1631980, EPI_ISL_1631984, EPI_ISL_1631987, EPI_ISL_1631988, EPI_ISL_1631989, EPI_ISL_1631993, EPI_ISL_1632007, EPI_ISL_1632013, EPI_ISL_1632014, EPI_ISL_1632015, EPI_ISL_1632022, EPI_ISL_1632024, EPI_ISL_1632025, EPI_ISL_1632026, EPI_ISL_1632027, EPI_ISL_1632028, EPI_ISL_1632036, EPI_ISL_1632037, EPI_ISL_1632039, EPI_ISL_1632041, EPI_ISL_1632042, EPI_ISL_1632043, EPI_ISL_1632046, EPI_ISL_1632048, EPI_ISL_1632049, EPI_ISL_1632051, EPI_ISL_1632052, EPI_ISL_1632058, EPI_ISL_1632059, EPI_ISL_1632065, EPI_ISL_1632068, EPI_ISL_1632070, EPI_ISL_1632071, EPI_ISL_1632072, EPI_ISL_1632074, EPI_ISL_1632085, EPI_ISL_1632089, EPI_ISL_1632092, EPI_ISL_1632094, EPI_ISL_1632095, EPI_ISL_1632097, EPI_ISL_1632112, EPI_ISL_1632114, EPI_ISL_1632115, EPI_ISL_1632135, EPI_ISL_1632138, EPI_ISL_1632140, EPI_ISL_1632143, EPI_ISL_1632150, EPI_ISL_1632151, EPI_ISL_1632154, EPI_ISL_1632157, EPI_ISL_1632163, EPI_ISL_1632166, EPI_ISL_1632167, EPI_ISL_1632171, EPI_ISL_1632172, EPI_ISL_1632173, EPI_ISL_1632179, EPI_ISL_1632182, EPI_ISL_1632195, EPI_ISL_1632196, EPI_ISL_1632201, EPI_ISL_1632205, EPI_ISL_1632207, EPI_ISL_1632212, EPI_ISL_1632213, EPI_ISL_1632216, EPI_ISL_1632218, EPI_ISL_1632220, EPI_ISL_1632222, EPI_ISL_1632224, EPI_ISL_1632226, EPI_ISL_1632233, EPI_ISL_1632234, EPI_ISL_1632238, EPI_ISL_1632240, EPI_ISL_1632242, EPI_ISL_1632244, EPI_ISL_1632246, EPI_ISL_1632248, EPI_ISL_1632249, EPI_ISL_1632250, EPI_ISL_1632255, EPI_ISL_1632258, EPI_ISL_1632260, EPI_ISL_1632265, EPI_ISL_1632266, EPI_ISL_1632269, EPI_ISL_1632274, EPI_ISL_1632276, EPI_ISL_1632283, EPI_ISL_1632290, EPI_ISL_1632291, EPI_ISL_1632294, EPI_ISL_1632298, EPI_ISL_1632300, EPI_ISL_1632301, EPI_ISL_1632302, EPI_ISL_1632303, EPI_ISL_1632304, EPI_ISL_1632307, EPI_ISL_1632308, EPI_ISL_1632309, EPI_ISL_1632312, EPI_ISL_1632313, EPI_ISL_1632317, EPI_ISL_1632319, EPI_ISL_1632321, EPI_ISL_1632322, EPI_ISL_1632323, EPI_ISL_1632324, EPI_ISL_1632327, EPI_ISL_1632329, EPI_ISL_1632330, EPI_ISL_1632334, EPI_ISL_1632336, EPI_ISL_1632337, EPI_ISL_1632338, EPI_ISL_1632339, EPI_ISL_1632340, EPI_ISL_1632341, EPI_ISL_1632344, EPI_ISL_1632345, EPI_ISL_1632346, EPI_ISL_1632347, EPI_ISL_1632349, EPI_ISL_1632350, EPI_ISL_1632351, EPI_ISL_1632352, EPI_ISL_1632356, EPI_ISL_1632357, EPI_ISL_1632358, EPI_ISL_1632362, EPI_ISL_1632363, EPI_ISL_1632364, EPI_ISL_1632366, EPI_ISL_1632367, EPI_ISL_1632369, EPI_ISL_1632370, EPI_ISL_1632373, EPI_ISL_1632374, EPI_ISL_1632375, EPI_ISL_1632376, EPI_ISL_1632378, EPI_ISL_1632380, EPI_ISL_1632382, EPI_ISL_1632385, EPI_ISL_1632387, EPI_ISL_1632388, EPI_ISL_1632392, EPI_ISL_1632393, EPI_ISL_1632396, EPI_ISL_1632397, EPI_ISL_1632398, EPI_ISL_1632399, EPI_ISL_1632400, EPI_ISL_1632402, EPI_ISL_1632403, EPI_ISL_1632404, EPI_ISL_1632406, EPI_ISL_1632408, EPI_ISL_1632409, EPI_ISL_1632411, EPI_ISL_1632413, EPI_ISL_1632414, EPI_ISL_1632415, EPI_ISL_1632416, EPI_ISL_1632417, EPI_ISL_1632418, EPI_ISL_1632420, EPI_ISL_1632421, EPI_ISL_1632422, EPI_ISL_1632423, EPI_ISL_1632424, EPI_ISL_1632427, EPI_ISL_1632428, EPI_ISL_1632429, EPI_ISL_1632430, EPI_ISL_1632431, EPI_ISL_1632432, EPI_ISL_1632435, EPI_ISL_1632438, EPI_ISL_1632440, EPI_ISL_1632441, EPI_ISL_1632442, EPI_ISL_1632444, EPI_ISL_1632445, EPI_ISL_1632446, EPI_ISL_1632447, EPI_ISL_1632453, EPI_ISL_1632454, EPI_ISL_1632455, EPI_ISL_1632456, EPI_ISL_1632457, EPI_ISL_1632458, EPI_ISL_1632460, EPI_ISL_1632464, EPI_ISL_1632465, EPI_ISL_1632468, EPI_ISL_1632470, EPI_ISL_1632473, EPI_ISL_1632474, EPI_ISL_1632477, EPI_ISL_1632478, EPI_ISL_1632479, EPI_ISL_1634489, EPI_ISL_1634490, EPI_ISL_1634492, EPI_ISL_1634493, EPI_ISL_1634495, EPI_ISL_1634496, EPI_ISL_1634497, EPI_ISL_1634498, EPI_ISL_1634501, EPI_ISL_1634502, EPI_ISL_1634503, EPI_ISL_1634504, EPI_ISL_1634508, EPI_ISL_1634512, EPI_ISL_1634513, EPI_ISL_1634518, EPI_ISL_1634522, EPI_ISL_1634527, EPI_ISL_1634528, EPI_ISL_1634532, EPI_ISL_1634537, EPI_ISL_1634539, EPI_ISL_1634540, EPI_ISL_1634543, EPI_ISL_1634544, EPI_ISL_1634545, EPI_ISL_1634547, EPI_ISL_1634548, EPI_ISL_1634550, EPI_ISL_1634554, EPI_ISL_1634555, EPI_ISL_1634558, EPI_ISL_1634559, EPI_ISL_1634566, EPI_ISL_1634568, EPI_ISL_1634571, EPI_ISL_1634572, EPI_ISL_1634574, EPI_ISL_1634575, EPI_ISL_1634577, EPI_ISL_1634578, EPI_ISL_1634582, EPI_ISL_1634583, EPI_ISL_1634584, EPI_ISL_1634586, EPI_ISL_1634587, EPI_ISL_1634589, EPI_ISL_1634595, EPI_ISL_1634600, EPI_ISL_1634601, EPI_ISL_1634602, EPI_ISL_1634603, EPI_ISL_1634604, EPI_ISL_1634605, EPI_ISL_1634607, EPI_ISL_1634610, EPI_ISL_1634613, EPI_ISL_1634615, EPI_ISL_1634617, EPI_ISL_1634620, EPI_ISL_1634622, EPI_ISL_1634623, EPI_ISL_1634624, EPI_ISL_1634626, EPI_ISL_1634628, EPI_ISL_1634631, EPI_ISL_1634632, EPI_ISL_1634633, EPI_ISL_1634634, EPI_ISL_1634635, EPI_ISL_1634643, EPI_ISL_1634649, EPI_ISL_1634650, EPI_ISL_1634651, EPI_ISL_1634653, EPI_ISL_1634656, EPI_ISL_1634661, EPI_ISL_1634664, EPI_ISL_1634665, EPI_ISL_1634667, EPI_ISL_1634669, EPI_ISL_1634674, EPI_ISL_1634676, EPI_ISL_1634677, EPI_ISL_1634680, EPI_ISL_1634681, EPI_ISL_1634683, EPI_ISL_1634685, EPI_ISL_1634690, EPI_ISL_1634691, EPI_ISL_1634695, EPI_ISL_1634696, EPI_ISL_1634697, EPI_ISL_1634700, EPI_ISL_1634701, EPI_ISL_1634702, EPI_ISL_1634703, EPI_ISL_1634704, EPI_ISL_1634705, EPI_ISL_1634706, EPI_ISL_1634707, EPI_ISL_1634708, EPI_ISL_1634709, EPI_ISL_1634710, EPI_ISL_1634711, EPI_ISL_1634712, EPI_ISL_1634713, EPI_ISL_1634715, EPI_ISL_1634717, EPI_ISL_1634719, EPI_ISL_1634720, EPI_ISL_1634721, EPI_ISL_1634722, EPI_ISL_1634723, EPI_ISL_1634724, EPI_ISL_1634725, EPI_ISL_1634726, EPI_ISL_1634727, EPI_ISL_1634728, EPI_ISL_1634729, EPI_ISL_1634730, EPI_ISL_1634731, EPI_ISL_1634732, EPI_ISL_1634733, EPI_ISL_1634734, EPI_ISL_1634735, EPI_ISL_1634736, EPI_ISL_1634738, EPI_ISL_1634739, EPI_ISL_1634740, EPI_ISL_1634741, EPI_ISL_1634742, EPI_ISL_1634744, EPI_ISL_1634745, EPI_ISL_1634746, EPI_ISL_1634747, EPI_ISL_1634748, EPI_ISL_1634749, EPI_ISL_1634750, EPI_ISL_1634751, EPI_ISL_1634752, EPI_ISL_1634753, EPI_ISL_1634754, EPI_ISL_1634755, EPI_ISL_1634756, EPI_ISL_1634757, EPI_ISL_1634758, EPI_ISL_1634759, EPI_ISL_1634760, EPI_ISL_1634761, EPI_ISL_1634762, EPI_ISL_1634763, EPI_ISL_1634764, EPI_ISL_1634765, EPI_ISL_1634766, EPI_ISL_1634767, EPI_ISL_1634768, EPI_ISL_1634769, EPI_ISL_1634770, EPI_ISL_1634771, EPI_ISL_1634772, EPI_ISL_1634773, EPI_ISL_1634774, EPI_ISL_1634775, EPI_ISL_1634776, EPI_ISL_1634777, EPI_ISL_1634779, EPI_ISL_1634780, EPI_ISL_1634781, EPI_ISL_1634782, EPI_ISL_1634785, EPI_ISL_1634787, EPI_ISL_1634788, EPI_ISL_1634789, EPI_ISL_1634790, EPI_ISL_1634791, EPI_ISL_1634792, EPI_ISL_1634793, EPI_ISL_1634795, EPI_ISL_1634797, EPI_ISL_1634798, EPI_ISL_1634799, EPI_ISL_1634800, EPI_ISL_1634801, EPI_ISL_1634802, EPI_ISL_1634803, EPI_ISL_1634804, EPI_ISL_1634805, EPI_ISL_1634806, EPI_ISL_1634809, EPI_ISL_1634810, EPI_ISL_1634811, EPI_ISL_1634812, EPI_ISL_1634815, EPI_ISL_1634817, EPI_ISL_1634819, EPI_ISL_1634820, EPI_ISL_1634822, EPI_ISL_1634823, EPI_ISL_1634824, EPI_ISL_1634825, EPI_ISL_1634826, EPI_ISL_1634827, EPI_ISL_1634828, EPI_ISL_1634829, EPI_ISL_1634833, EPI_ISL_1634834, EPI_ISL_1634835, EPI_ISL_1634837, EPI_ISL_1634838, EPI_ISL_1634839, EPI_ISL_1634840, EPI_ISL_1634841, EPI_ISL_1634842, EPI_ISL_1634843, EPI_ISL_1634844, EPI_ISL_1634845, EPI_ISL_1634847, EPI_ISL_1634849, EPI_ISL_1634851, EPI_ISL_1634853, EPI_ISL_1634854, EPI_ISL_1634856, EPI_ISL_1634857, EPI_ISL_1634858, EPI_ISL_1634859, EPI_ISL_1634860, EPI_ISL_1634861, EPI_ISL_1634862, EPI_ISL_1634863, EPI_ISL_1634864, EPI_ISL_1634865, EPI_ISL_1634866, EPI_ISL_1634867, EPI_ISL_1634868, EPI_ISL_1634869, EPI_ISL_1634870, EPI_ISL_1634871, EPI_ISL_1634872, EPI_ISL_1634873, EPI_ISL_1634874, EPI_ISL_1634875, EPI_ISL_1634876, EPI_ISL_1634877, EPI_ISL_1634878, EPI_ISL_1634879, EPI_ISL_1634880, EPI_ISL_1634881, EPI_ISL_1634883, EPI_ISL_1634884, EPI_ISL_1634885, EPI_ISL_1634886, EPI_ISL_1634887, EPI_ISL_1634888, EPI_ISL_1634890, EPI_ISL_1634891, EPI_ISL_1634893, EPI_ISL_1634894, EPI_ISL_1634895, EPI_ISL_1634896, EPI_ISL_1634899, EPI_ISL_1634901, EPI_ISL_1634902, EPI_ISL_1634904, EPI_ISL_1634905, EPI_ISL_1634906, EPI_ISL_1634907, EPI_ISL_1634908, EPI_ISL_1634909, EPI_ISL_1634910, EPI_ISL_1634911, EPI_ISL_1634912, EPI_ISL_1634914, EPI_ISL_1634915, EPI_ISL_1634916, EPI_ISL_1634917, EPI_ISL_1634918, EPI_ISL_1634919, EPI_ISL_1634920, EPI_ISL_1634922, EPI_ISL_1634923, EPI_ISL_1634924, EPI_ISL_1634925, EPI_ISL_1634927, EPI_ISL_1634928, EPI_ISL_1634929, EPI_ISL_1634930, EPI_ISL_1634931, EPI_ISL_1634933, EPI_ISL_1634934, EPI_ISL_1634935, EPI_ISL_1634936, EPI_ISL_1634937, EPI_ISL_1634939, EPI_ISL_1634941, EPI_ISL_1634942, EPI_ISL_1634943, EPI_ISL_1634944, EPI_ISL_1634945, EPI_ISL_1634946, EPI_ISL_1634948, EPI_ISL_1634949, EPI_ISL_1634951, EPI_ISL_1634952, EPI_ISL_1634953, EPI_ISL_1634954, EPI_ISL_1634955, EPI_ISL_1634958, EPI_ISL_1634959, EPI_ISL_1634960, EPI_ISL_1634961, EPI_ISL_1634963, EPI_ISL_1634964, EPI_ISL_1634965, EPI_ISL_1634966, EPI_ISL_1634968, EPI_ISL_1634970, EPI_ISL_1634971, EPI_ISL_1634972, EPI_ISL_1634973, EPI_ISL_1634974, EPI_ISL_1634975, EPI_ISL_1634976, EPI_ISL_1634977, EPI_ISL_1634978, EPI_ISL_1634979, EPI_ISL_1634980, EPI_ISL_1634981, EPI_ISL_1634982, EPI_ISL_1634983, EPI_ISL_1634984, EPI_ISL_1634985, EPI_ISL_1634986, EPI_ISL_1634988, EPI_ISL_1634989, EPI_ISL_1634990, EPI_ISL_1634991, EPI_ISL_1634992, EPI_ISL_1634993, EPI_ISL_1634994, EPI_ISL_1634995, EPI_ISL_1634996, EPI_ISL_1634997, EPI_ISL_1634998, EPI_ISL_1634999, EPI_ISL_1635000, EPI_ISL_1635001, EPI_ISL_1635002, EPI_ISL_1635003, EPI_ISL_1635004, EPI_ISL_1635005, EPI_ISL_1635006, EPI_ISL_1635007, EPI_ISL_1635008, EPI_ISL_1635009, EPI_ISL_1635010, EPI_ISL_1635011, EPI_ISL_1635012, EPI_ISL_1635013, EPI_ISL_1635014, EPI_ISL_1635015, EPI_ISL_1635016, EPI_ISL_1635017, EPI_ISL_1635018, EPI_ISL_1635019, EPI_ISL_1635020, EPI_ISL_1635021, EPI_ISL_1635022, EPI_ISL_1635023, EPI_ISL_1635024, EPI_ISL_1635025, EPI_ISL_1635026, EPI_ISL_1635027, EPI_ISL_1635028, EPI_ISL_1635029, EPI_ISL_1635030, EPI_ISL_1635031, EPI_ISL_1635032, EPI_ISL_1635033, EPI_ISL_1635034, EPI_ISL_1635035, EPI_ISL_1635036, EPI_ISL_1635037, EPI_ISL_1635038, EPI_ISL_1635039, EPI_ISL_1635040, EPI_ISL_1635041, EPI_ISL_1635042, EPI_ISL_1635043, EPI_ISL_1635044, EPI_ISL_1635045, EPI_ISL_1635046, EPI_ISL_1635047, EPI_ISL_1635048, EPI_ISL_1635049, EPI_ISL_1635050, EPI_ISL_1635051, EPI_ISL_1635052, EPI_ISL_1635053, EPI_ISL_1635054, EPI_ISL_1635055, EPI_ISL_1635056, EPI_ISL_1635057, EPI_ISL_1635058, EPI_ISL_1635059, EPI_ISL_1635060, EPI_ISL_1635061, EPI_ISL_1635062, EPI_ISL_1635063, EPI_ISL_1635064, EPI_ISL_1635065, EPI_ISL_1635066, EPI_ISL_1635067, EPI_ISL_1635068, EPI_ISL_1635069, EPI_ISL_1635120, EPI_ISL_1635121, EPI_ISL_1635123, EPI_ISL_1635125, EPI_ISL_1635126, EPI_ISL_1635127, EPI_ISL_1635128, EPI_ISL_1635129, EPI_ISL_1635130, EPI_ISL_1635131, EPI_ISL_1635132, EPI_ISL_1635133, EPI_ISL_1635134, EPI_ISL_1635135, EPI_ISL_1635136, EPI_ISL_1635137, EPI_ISL_1635138, EPI_ISL_1635139, EPI_ISL_1635140, EPI_ISL_1635141, EPI_ISL_1635142, EPI_ISL_1635143, EPI_ISL_1635144, EPI_ISL_1635145, EPI_ISL_1635146, EPI_ISL_1635147, EPI_ISL_1635148, EPI_ISL_1635150, EPI_ISL_1635151, EPI_ISL_1635152, EPI_ISL_1635153, EPI_ISL_1635154, EPI_ISL_1635156, EPI_ISL_1635157, EPI_ISL_1635158, EPI_ISL_1635159, EPI_ISL_1635160, EPI_ISL_1635161, EPI_ISL_1635162, EPI_ISL_1635164, EPI_ISL_1635166, EPI_ISL_1635167, EPI_ISL_1635168, EPI_ISL_1635169, EPI_ISL_1635170, EPI_ISL_1635171, EPI_ISL_1635172, EPI_ISL_1635173, EPI_ISL_1635174, EPI_ISL_1635175, EPI_ISL_1635176, EPI_ISL_1635177, EPI_ISL_1635178, EPI_ISL_1635179, EPI_ISL_1635180, EPI_ISL_1635181, EPI_ISL_1635182, EPI_ISL_1635183, EPI_ISL_1635184, EPI_ISL_1635185, EPI_ISL_1635186, EPI_ISL_1635187, EPI_ISL_1635188, EPI_ISL_1635189, EPI_ISL_1635190, EPI_ISL_1635191, EPI_ISL_1635192, EPI_ISL_1635193, EPI_ISL_1635194, EPI_ISL_1635195, EPI_ISL_1635197, EPI_ISL_1635198, EPI_ISL_1635199, EPI_ISL_1635200, EPI_ISL_1635201, EPI_ISL_1635202, EPI_ISL_1635203, EPI_ISL_1635204, EPI_ISL_1635205, EPI_ISL_1635206, EPI_ISL_1635208, EPI_ISL_1635209, EPI_ISL_1635210, EPI_ISL_1635211, EPI_ISL_1635212, EPI_ISL_1635213, EPI_ISL_1635214, EPI_ISL_1635215, EPI_ISL_1635216, EPI_ISL_1635217, EPI_ISL_1635218, EPI_ISL_1635219, EPI_ISL_1635220, EPI_ISL_1635221, EPI_ISL_1635222, EPI_ISL_1635224, EPI_ISL_1635225, EPI_ISL_1635226, EPI_ISL_1635227, EPI_ISL_1635228, EPI_ISL_1635229, EPI_ISL_1635230, EPI_ISL_1635231, EPI_ISL_1635232, EPI_ISL_1635233, EPI_ISL_1635234, EPI_ISL_1635235, EPI_ISL_1635236, EPI_ISL_1635237, EPI_ISL_1635238, EPI_ISL_1635239, EPI_ISL_1635240, EPI_ISL_1635241, EPI_ISL_1635242, EPI_ISL_1635243, EPI_ISL_1635244, EPI_ISL_1635246, EPI_ISL_1635247, EPI_ISL_1635248, EPI_ISL_1635250, EPI_ISL_1635251, EPI_ISL_1635252, EPI_ISL_1635253, EPI_ISL_1635254, EPI_ISL_1635255, EPI_ISL_1635256, EPI_ISL_1635257, EPI_ISL_1635258, EPI_ISL_1635259, EPI_ISL_1635260, EPI_ISL_1635261, EPI_ISL_1635262, EPI_ISL_1635263, EPI_ISL_1635264, EPI_ISL_1635265, EPI_ISL_1635266, EPI_ISL_1635267, EPI_ISL_1635268, EPI_ISL_1635269, EPI_ISL_1635270, EPI_ISL_1635271, EPI_ISL_1635272, EPI_ISL_1635273, EPI_ISL_1635274, EPI_ISL_1635275, EPI_ISL_1635276, EPI_ISL_1635277, EPI_ISL_1635278, EPI_ISL_1635279, EPI_ISL_1635280, EPI_ISL_1635281, EPI_ISL_1635282, EPI_ISL_1635283, EPI_ISL_1635284, EPI_ISL_1635285, EPI_ISL_1635286, EPI_ISL_1635287, EPI_ISL_1635288, EPI_ISL_1635289, EPI_ISL_1635290, EPI_ISL_1635291, EPI_ISL_1635292, EPI_ISL_1635293, EPI_ISL_1635294, EPI_ISL_1635296, EPI_ISL_1635297, EPI_ISL_1635298, EPI_ISL_1635300, EPI_ISL_1635301, EPI_ISL_1635302, EPI_ISL_1635303, EPI_ISL_1635304, EPI_ISL_1635305, EPI_ISL_1635306, EPI_ISL_1635307, EPI_ISL_1635308, EPI_ISL_1635309, EPI_ISL_1635310, EPI_ISL_1635311, EPI_ISL_1635312, EPI_ISL_1635314, EPI_ISL_1635315, EPI_ISL_1635316, EPI_ISL_1635317, EPI_ISL_1635318, EPI_ISL_1635319, EPI_ISL_1635320, EPI_ISL_1635322, EPI_ISL_1635323, EPI_ISL_1635324, EPI_ISL_1635326, EPI_ISL_1635327, EPI_ISL_1635328, EPI_ISL_1635329, EPI_ISL_1635330, EPI_ISL_1635331, EPI_ISL_1635332, EPI_ISL_1635333, EPI_ISL_1635334, EPI_ISL_1635335, EPI_ISL_1635336, EPI_ISL_1635337, EPI_ISL_1635338, EPI_ISL_1635339, EPI_ISL_1635340, EPI_ISL_1635341, EPI_ISL_1635342, EPI_ISL_1635343, EPI_ISL_1635345, EPI_ISL_1635346, EPI_ISL_1635347, EPI_ISL_1635348, EPI_ISL_1635349, EPI_ISL_1635350, EPI_ISL_1635352, EPI_ISL_1635353, EPI_ISL_1635354, EPI_ISL_1635355, EPI_ISL_1635357, EPI_ISL_1635358, EPI_ISL_1635360, EPI_ISL_1635362, EPI_ISL_1635363, EPI_ISL_1635364, EPI_ISL_1635365, EPI_ISL_1635366, EPI_ISL_1635367, EPI_ISL_1635368, EPI_ISL_1635369, EPI_ISL_1635370, EPI_ISL_1635372, EPI_ISL_1635373, EPI_ISL_1635374, EPI_ISL_1635375, EPI_ISL_1635376, EPI_ISL_1635377, EPI_ISL_1635378, EPI_ISL_1635380, EPI_ISL_1635381, EPI_ISL_1635382, EPI_ISL_1635383, EPI_ISL_1635384, EPI_ISL_1635385, EPI_ISL_1635386, EPI_ISL_1635387, EPI_ISL_1635388, EPI_ISL_1635389, EPI_ISL_1635390, EPI_ISL_1635391, EPI_ISL_1635393, EPI_ISL_1635394, EPI_ISL_1635395, EPI_ISL_1635396, EPI_ISL_1635397, EPI_ISL_1635398, EPI_ISL_1635399, EPI_ISL_1635401, EPI_ISL_1635402, EPI_ISL_1635405, EPI_ISL_1635406, EPI_ISL_1635407, EPI_ISL_1635408, EPI_ISL_1635409, EPI_ISL_1635410, EPI_ISL_1635411, EPI_ISL_1635412, EPI_ISL_1635413, EPI_ISL_1635414, EPI_ISL_1635415, EPI_ISL_1635416, EPI_ISL_1635417, EPI_ISL_1635418, EPI_ISL_1635419, EPI_ISL_1635420, EPI_ISL_1635421, EPI_ISL_1635422, EPI_ISL_1652691, EPI_ISL_1652702, EPI_ISL_1652704, EPI_ISL_1652721, EPI_ISL_1652723, EPI_ISL_1652726, EPI_ISL_1652731, EPI_ISL_1652737, EPI_ISL_1652740, EPI_ISL_1652741, EPI_ISL_1652752, EPI_ISL_1652776, EPI_ISL_1652778, EPI_ISL_1652787, EPI_ISL_1652792, EPI_ISL_1652798, EPI_ISL_1652801, EPI_ISL_1652815, EPI_ISL_1652820, EPI_ISL_1652832, EPI_ISL_1652844, EPI_ISL_1652851, EPI_ISL_1652858, EPI_ISL_1652866, EPI_ISL_1652872, EPI_ISL_1652876, EPI_ISL_1652894, EPI_ISL_1652895, EPI_ISL_1652896, EPI_ISL_1652899, EPI_ISL_1652918, EPI_ISL_1652919, EPI_ISL_1652922, EPI_ISL_1652926, EPI_ISL_1652928, EPI_ISL_1652935, EPI_ISL_1652936, EPI_ISL_1652943, EPI_ISL_1652944, EPI_ISL_1652945, EPI_ISL_1652946, EPI_ISL_1652947, EPI_ISL_1652948, EPI_ISL_1652949, EPI_ISL_1652950, EPI_ISL_1652953, EPI_ISL_1652955, EPI_ISL_1652956, EPI_ISL_1652957, EPI_ISL_1652959, EPI_ISL_1652960, EPI_ISL_1652961, EPI_ISL_1652962, EPI_ISL_1652963, EPI_ISL_1652965, EPI_ISL_1652966, EPI_ISL_1652969, EPI_ISL_1652972, EPI_ISL_1652974, EPI_ISL_1652975, EPI_ISL_1652977, EPI_ISL_1652985, EPI_ISL_1652987, EPI_ISL_1652988, EPI_ISL_1652990, EPI_ISL_1652992, EPI_ISL_1652997, EPI_ISL_1652999, EPI_ISL_1653000, EPI_ISL_1653002, EPI_ISL_1653003, EPI_ISL_1653004, EPI_ISL_1653007, EPI_ISL_1653008, EPI_ISL_1653013, EPI_ISL_1653014, EPI_ISL_1653016, EPI_ISL_1653017, EPI_ISL_1653019, EPI_ISL_1653023, EPI_ISL_1653026, EPI_ISL_1653029, EPI_ISL_1653030, EPI_ISL_1653031, EPI_ISL_1653032, EPI_ISL_1653034, EPI_ISL_1653035, EPI_ISL_1653036, EPI_ISL_1653037, EPI_ISL_1653038, EPI_ISL_1653039, EPI_ISL_1653041, EPI_ISL_1653044, EPI_ISL_1653046, EPI_ISL_1653047, EPI_ISL_1653048, EPI_ISL_1653049, EPI_ISL_1653054, EPI_ISL_1653055, EPI_ISL_1653056, EPI_ISL_1653057, EPI_ISL_1653060, EPI_ISL_1653063, EPI_ISL_1653064, EPI_ISL_1653065, EPI_ISL_1653066, EPI_ISL_1653067, EPI_ISL_1653068, EPI_ISL_1653070, EPI_ISL_1653071, EPI_ISL_1653072, EPI_ISL_1653075, EPI_ISL_1653076, EPI_ISL_1653077, EPI_ISL_1653079, EPI_ISL_1653082, EPI_ISL_1653084, EPI_ISL_1653085, EPI_ISL_1653086, EPI_ISL_1653087, EPI_ISL_1653088, EPI_ISL_1653089, EPI_ISL_1653091, EPI_ISL_1653092, EPI_ISL_1653094, EPI_ISL_1653095, EPI_ISL_1653098, EPI_ISL_1653101, EPI_ISL_1653103, EPI_ISL_1653105, EPI_ISL_1653106, EPI_ISL_1653107, EPI_ISL_1653112, EPI_ISL_1653113, EPI_ISL_1653115, EPI_ISL_1653117, EPI_ISL_1653118, EPI_ISL_1653122, EPI_ISL_1653123, EPI_ISL_1653124, EPI_ISL_1653127, EPI_ISL_1653128, EPI_ISL_1653129, EPI_ISL_1653131, EPI_ISL_1653134, EPI_ISL_1653135, EPI_ISL_1653137, EPI_ISL_1653139, EPI_ISL_1653140, EPI_ISL_1653142, EPI_ISL_1653143, EPI_ISL_1653144, EPI_ISL_1653145, EPI_ISL_1653148, EPI_ISL_1653150, EPI_ISL_1653151, EPI_ISL_1653152, EPI_ISL_1653153, EPI_ISL_1653154, EPI_ISL_1653155, EPI_ISL_1653158, EPI_ISL_1653160, EPI_ISL_1653161, EPI_ISL_1653162, EPI_ISL_1653163, EPI_ISL_1653164, EPI_ISL_1653165, EPI_ISL_1653166, EPI_ISL_1653168, EPI_ISL_1653169, EPI_ISL_1653170, EPI_ISL_1653171, EPI_ISL_1653172, EPI_ISL_1653173, EPI_ISL_1653177, EPI_ISL_1653182, EPI_ISL_1653184, EPI_ISL_1653186, EPI_ISL_1653188, EPI_ISL_1653189, EPI_ISL_1653190, EPI_ISL_1653192, EPI_ISL_1653193, EPI_ISL_1653195, EPI_ISL_1653196, EPI_ISL_1653198, EPI_ISL_1653199, EPI_ISL_1653202, EPI_ISL_1653203, EPI_ISL_1653204, EPI_ISL_1653205, EPI_ISL_1653206, EPI_ISL_1653207, EPI_ISL_1653208, EPI_ISL_1653211, EPI_ISL_1653212, EPI_ISL_1653215, EPI_ISL_1653217, EPI_ISL_1653220, EPI_ISL_1653221, EPI_ISL_1653224, EPI_ISL_1653226, EPI_ISL_1653231, EPI_ISL_1653237, EPI_ISL_1653239, EPI_ISL_1653241, EPI_ISL_1653249, EPI_ISL_1653252, EPI_ISL_1653253, EPI_ISL_1653255, EPI_ISL_1653256, EPI_ISL_1653262, EPI_ISL_1653264, EPI_ISL_1653273, EPI_ISL_1653293, EPI_ISL_1653294, EPI_ISL_1653296, EPI_ISL_1653299, EPI_ISL_1653302, EPI_ISL_1653303, EPI_ISL_1653304, EPI_ISL_1653307, EPI_ISL_1653308, EPI_ISL_1653309, EPI_ISL_1653312, EPI_ISL_1653313, EPI_ISL_1653314, EPI_ISL_1653319, EPI_ISL_1653321, EPI_ISL_1653325, EPI_ISL_1653329, EPI_ISL_1653331, EPI_ISL_1653333, EPI_ISL_1653335, EPI_ISL_1653337, EPI_ISL_1653339, EPI_ISL_1653340, EPI_ISL_1653341, EPI_ISL_1653343, EPI_ISL_1653353, EPI_ISL_1653354, EPI_ISL_1653356, EPI_ISL_1653357, EPI_ISL_1653360, EPI_ISL_1653365, EPI_ISL_1653369, EPI_ISL_1653381, EPI_ISL_1653385, EPI_ISL_1653388, EPI_ISL_1653395, EPI_ISL_1653402, EPI_ISL_1653404, EPI_ISL_1653409, EPI_ISL_1653412, EPI_ISL_1653414, EPI_ISL_1653418, EPI_ISL_1653427, EPI_ISL_1653432, EPI_ISL_1653436, EPI_ISL_1653445, EPI_ISL_1653456, EPI_ISL_1653464, EPI_ISL_1653465, EPI_ISL_1653471, EPI_ISL_1653475, EPI_ISL_1653476, EPI_ISL_1653477, EPI_ISL_1653478, EPI_ISL_1653482, EPI_ISL_1653488, EPI_ISL_1653491, EPI_ISL_1653493, EPI_ISL_1653502, EPI_ISL_1653505, EPI_ISL_1653506, EPI_ISL_1653507, EPI_ISL_1653516, EPI_ISL_1653523, EPI_ISL_1653524, EPI_ISL_1653526, EPI_ISL_1653527, EPI_ISL_1653528, EPI_ISL_1653529, EPI_ISL_1653530, EPI_ISL_1653536, EPI_ISL_1653539, EPI_ISL_1653540, EPI_ISL_1653550, EPI_ISL_1653552, EPI_ISL_1653562, EPI_ISL_1653566, EPI_ISL_1653568, EPI_ISL_1653576, EPI_ISL_1653586, EPI_ISL_1653587, EPI_ISL_1653588, EPI_ISL_1653602, EPI_ISL_1653608, EPI_ISL_1653609, EPI_ISL_1653610, EPI_ISL_1653612, EPI_ISL_1653617, EPI_ISL_1653621, EPI_ISL_1653622, EPI_ISL_1653648, EPI_ISL_1653651, EPI_ISL_1653653, EPI_ISL_1653664, EPI_ISL_1653665, EPI_ISL_1653671, EPI_ISL_1653674, EPI_ISL_1653678, EPI_ISL_1653684, EPI_ISL_1653685, EPI_ISL_1653689, EPI_ISL_1653699, EPI_ISL_1653701, EPI_ISL_1653729, EPI_ISL_1672972, EPI_ISL_1672984, EPI_ISL_1672986, EPI_ISL_1672988, EPI_ISL_1672990, EPI_ISL_1672997, EPI_ISL_1673002, EPI_ISL_1673005, EPI_ISL_1673006, EPI_ISL_1673008, EPI_ISL_1673010, EPI_ISL_1673017, EPI_ISL_1673018, EPI_ISL_1673019, EPI_ISL_1673023, EPI_ISL_1673031, EPI_ISL_1673035, EPI_ISL_1673042, EPI_ISL_1673044, EPI_ISL_1673045, EPI_ISL_1673046, EPI_ISL_1673047, EPI_ISL_1673048, EPI_ISL_1673049, EPI_ISL_1673050, EPI_ISL_1673051, EPI_ISL_1673052, EPI_ISL_1673054, EPI_ISL_1673057, EPI_ISL_1673059, EPI_ISL_1673060, EPI_ISL_1673062, EPI_ISL_1673063, EPI_ISL_1673064, EPI_ISL_1673065, EPI_ISL_1673066, EPI_ISL_1673067, EPI_ISL_1673069, EPI_ISL_1673070, EPI_ISL_1673071, EPI_ISL_1673072, EPI_ISL_1673073, EPI_ISL_1673074, EPI_ISL_1673075, EPI_ISL_1673076, EPI_ISL_1673077, EPI_ISL_1673078, EPI_ISL_1673079, EPI_ISL_1673080, EPI_ISL_1673081, EPI_ISL_1673082, EPI_ISL_1673083, EPI_ISL_1673084, EPI_ISL_1673085, EPI_ISL_1673086, EPI_ISL_1673088, EPI_ISL_1673089, EPI_ISL_1673090, EPI_ISL_1673091, EPI_ISL_1673092, EPI_ISL_1673093, EPI_ISL_1673095, EPI_ISL_1673096, EPI_ISL_1673097, EPI_ISL_1673098, EPI_ISL_1673099, EPI_ISL_1673101, EPI_ISL_1673102, EPI_ISL_1673104, EPI_ISL_1673105, EPI_ISL_1673106, EPI_ISL_1673107, EPI_ISL_1673108, EPI_ISL_1673109, EPI_ISL_1673110, EPI_ISL_1673111, EPI_ISL_1673112, EPI_ISL_1673113, EPI_ISL_1673116, EPI_ISL_1673117, EPI_ISL_1673119, EPI_ISL_1673120, EPI_ISL_1673122, EPI_ISL_1673123, EPI_ISL_1673124, EPI_ISL_1673125, EPI_ISL_1673126, EPI_ISL_1673127, EPI_ISL_1673128, EPI_ISL_1673129, EPI_ISL_1673130, EPI_ISL_1673131, EPI_ISL_1673133, EPI_ISL_1673134, EPI_ISL_1673135, EPI_ISL_1673136, EPI_ISL_1673137, EPI_ISL_1673138, EPI_ISL_1673139, EPI_ISL_1673143, EPI_ISL_1673144, EPI_ISL_1673145, EPI_ISL_1673146, EPI_ISL_1673148, EPI_ISL_1673149, EPI_ISL_1673150, EPI_ISL_1673151, EPI_ISL_1673152, EPI_ISL_1673153, EPI_ISL_1673154, EPI_ISL_1673155, EPI_ISL_1673156, EPI_ISL_1673157, EPI_ISL_1673158, EPI_ISL_1673159, EPI_ISL_1673160, EPI_ISL_1673161, EPI_ISL_1673162, EPI_ISL_1673163, EPI_ISL_1673164, EPI_ISL_1673165, EPI_ISL_1673166, EPI_ISL_1673167, EPI_ISL_1673169, EPI_ISL_1673170, EPI_ISL_1673171, EPI_ISL_1673172, EPI_ISL_1673173, EPI_ISL_1673174, EPI_ISL_1673175, EPI_ISL_1673176, EPI_ISL_1673177, EPI_ISL_1673178, EPI_ISL_1673179, EPI_ISL_1673180, EPI_ISL_1673181, EPI_ISL_1673182, EPI_ISL_1673183, EPI_ISL_1673184, EPI_ISL_1673185, EPI_ISL_1673186, EPI_ISL_1673187, EPI_ISL_1673188, EPI_ISL_1673189, EPI_ISL_1673190, EPI_ISL_1673191, EPI_ISL_1673192, EPI_ISL_1673193, EPI_ISL_1673194, EPI_ISL_1673195, EPI_ISL_1673196, EPI_ISL_1673197, EPI_ISL_1673198, EPI_ISL_1673199, EPI_ISL_1673200, EPI_ISL_1673201, EPI_ISL_1673202, EPI_ISL_1673203, EPI_ISL_1673204, EPI_ISL_1673205, EPI_ISL_1673206, EPI_ISL_1673207, EPI_ISL_1673208, EPI_ISL_1673209, EPI_ISL_1673210, EPI_ISL_1673211, EPI_ISL_1673212, EPI_ISL_1673213, EPI_ISL_1673214, EPI_ISL_1673215, EPI_ISL_1673216, EPI_ISL_1673217, EPI_ISL_1673218, EPI_ISL_1673219, EPI_ISL_1673220, EPI_ISL_1673221, EPI_ISL_1673222, EPI_ISL_1673223, EPI_ISL_1673224, EPI_ISL_1673225, EPI_ISL_1673226, EPI_ISL_1673227, EPI_ISL_1673228, EPI_ISL_1673229, EPI_ISL_1673230, EPI_ISL_1673231, EPI_ISL_1673232, EPI_ISL_1673233, EPI_ISL_1673234, EPI_ISL_1673235, EPI_ISL_1673236, EPI_ISL_1673237, EPI_ISL_1673238, EPI_ISL_1673239, EPI_ISL_1673240, EPI_ISL_1673241, EPI_ISL_1673242, EPI_ISL_1673243, EPI_ISL_1673244, EPI_ISL_1673245, EPI_ISL_1673246, EPI_ISL_1673247, EPI_ISL_1673248, EPI_ISL_1673249, EPI_ISL_1673250, EPI_ISL_1673251, EPI_ISL_1673252, EPI_ISL_1673253, EPI_ISL_1673254, EPI_ISL_1697746, EPI_ISL_1697784, EPI_ISL_1697785, EPI_ISL_1697798, EPI_ISL_1697809, EPI_ISL_1697815, EPI_ISL_1697825, EPI_ISL_1697834, EPI_ISL_1697836, EPI_ISL_1697842, EPI_ISL_1697845, EPI_ISL_1697860, EPI_ISL_1697889, EPI_ISL_1697890, EPI_ISL_1697930, EPI_ISL_1697935, EPI_ISL_1697966, EPI_ISL_1697968, EPI_ISL_1698113, EPI_ISL_1698615, EPI_ISL_1698621, EPI_ISL_1698650, EPI_ISL_1698662, EPI_ISL_1698663, EPI_ISL_1698683, EPI_ISL_1698937, EPI_ISL_1698980, EPI_ISL_1699014, EPI_ISL_1699019, EPI_ISL_1699023, EPI_ISL_1699024, EPI_ISL_1699025, EPI_ISL_1699026, EPI_ISL_1699029, EPI_ISL_1699031, EPI_ISL_1699032, EPI_ISL_1699034, EPI_ISL_1699038, EPI_ISL_1699041, EPI_ISL_1699043, EPI_ISL_1699046, EPI_ISL_1699049, EPI_ISL_1699051, EPI_ISL_1699055, EPI_ISL_1699057, EPI_ISL_1699064, EPI_ISL_1699066, EPI_ISL_1699073, EPI_ISL_1699074, EPI_ISL_1699076, EPI_ISL_1699077, EPI_ISL_1699078, EPI_ISL_1699085, EPI_ISL_1699096, EPI_ISL_1699100, EPI_ISL_1699102, EPI_ISL_1699109, EPI_ISL_1699111, EPI_ISL_1699113, EPI_ISL_1699114, EPI_ISL_1699115, EPI_ISL_1699117, EPI_ISL_1699123, EPI_ISL_1699124, EPI_ISL_1699125, EPI_ISL_1699129, EPI_ISL_1699132, EPI_ISL_1699136, EPI_ISL_1699140, EPI_ISL_1699145, EPI_ISL_1699146, EPI_ISL_1699147, EPI_ISL_1699149, EPI_ISL_1699151, EPI_ISL_1699152, EPI_ISL_1699153, EPI_ISL_1699158, EPI_ISL_1699159, EPI_ISL_1699161, EPI_ISL_1699162, EPI_ISL_1699164, EPI_ISL_1699166, EPI_ISL_1699167, EPI_ISL_1699169, EPI_ISL_1699171, EPI_ISL_1699173, EPI_ISL_1699175, EPI_ISL_1699177, EPI_ISL_1699184, EPI_ISL_1699188, EPI_ISL_1699189, EPI_ISL_1699194, EPI_ISL_1699198, EPI_ISL_1699200, EPI_ISL_1699201, EPI_ISL_1699202, EPI_ISL_1699212, EPI_ISL_1699213, EPI_ISL_1699217, EPI_ISL_1699219, EPI_ISL_1699222, EPI_ISL_1699224, EPI_ISL_1699225, EPI_ISL_1699226, EPI_ISL_1699233, EPI_ISL_1699235, EPI_ISL_1699236, EPI_ISL_1699243, EPI_ISL_1699244, EPI_ISL_1699245, EPI_ISL_1699247, EPI_ISL_1699249, EPI_ISL_1699251, EPI_ISL_1699253, EPI_ISL_1699256, EPI_ISL_1699257, EPI_ISL_1699258, EPI_ISL_1699259, EPI_ISL_1699260, EPI_ISL_1699261, EPI_ISL_1699262, EPI_ISL_1699265, EPI_ISL_1699268, EPI_ISL_1699274, EPI_ISL_1699275, EPI_ISL_1699277, EPI_ISL_1699280, EPI_ISL_1699283, EPI_ISL_1699285, EPI_ISL_1699286, EPI_ISL_1699307, EPI_ISL_1699365, EPI_ISL_1699451, EPI_ISL_1699452, EPI_ISL_1699456, EPI_ISL_1699478, EPI_ISL_1699481, EPI_ISL_1699496, EPI_ISL_1699519, EPI_ISL_1699530, EPI_ISL_1699531, EPI_ISL_1699575, EPI_ISL_1699595, EPI_ISL_1699611, EPI_ISL_1699618, EPI_ISL_1699668, EPI_ISL_1699683, EPI_ISL_1699700, EPI_ISL_1699708, EPI_ISL_1699763, EPI_ISL_1699846, EPI_ISL_1699848, EPI_ISL_1699850, EPI_ISL_1699852, EPI_ISL_1699860, EPI_ISL_1699861, EPI_ISL_1699862, EPI_ISL_1699868, EPI_ISL_1699874, EPI_ISL_1699876, EPI_ISL_1699878, EPI_ISL_1699889, EPI_ISL_1699890, EPI_ISL_1699894, EPI_ISL_1699897, EPI_ISL_1699899, EPI_ISL_1699907, EPI_ISL_1699910, EPI_ISL_1699915, EPI_ISL_1699916, EPI_ISL_1699917, EPI_ISL_1699921, EPI_ISL_1699922, EPI_ISL_1699928, EPI_ISL_1699929, EPI_ISL_1699930, EPI_ISL_1699932, EPI_ISL_1699933, EPI_ISL_1699934, EPI_ISL_1699941, EPI_ISL_1699942, EPI_ISL_1699943, EPI_ISL_1699950, EPI_ISL_1699955, EPI_ISL_1699964, EPI_ISL_1699974, EPI_ISL_1699986, EPI_ISL_1699991, EPI_ISL_1699994, EPI_ISL_1699995, EPI_ISL_1699998, EPI_ISL_1700006, EPI_ISL_1700010, EPI_ISL_1700015, EPI_ISL_1700027, EPI_ISL_1700028, EPI_ISL_1700030, EPI_ISL_1700031, EPI_ISL_1700034, EPI_ISL_1700036, EPI_ISL_1700037, EPI_ISL_1700041, EPI_ISL_1700042, EPI_ISL_1700047, EPI_ISL_1700048, EPI_ISL_1700052, EPI_ISL_1700053, EPI_ISL_1700057, EPI_ISL_1700059, EPI_ISL_1700060, EPI_ISL_1700063, EPI_ISL_1700067, EPI_ISL_1700068, EPI_ISL_1700069, EPI_ISL_1700070, EPI_ISL_1700079, EPI_ISL_1700081, EPI_ISL_1700085, EPI_ISL_1700090, EPI_ISL_1700096, EPI_ISL_1700100, EPI_ISL_1700103, EPI_ISL_1700109, EPI_ISL_1700112, EPI_ISL_1700113, EPI_ISL_1700118, EPI_ISL_1700127, EPI_ISL_1700135, EPI_ISL_1700136, EPI_ISL_1700139, EPI_ISL_1700141, EPI_ISL_1700143, EPI_ISL_1700144, EPI_ISL_1700145, EPI_ISL_1700151, EPI_ISL_1700154, EPI_ISL_1700159, EPI_ISL_1700164, EPI_ISL_1700166, EPI_ISL_1700170, EPI_ISL_1700173, EPI_ISL_1700174, EPI_ISL_1700179, EPI_ISL_1700184, EPI_ISL_1700190, EPI_ISL_1700191, EPI_ISL_1700192, EPI_ISL_1700193, EPI_ISL_1700196, EPI_ISL_1700200, EPI_ISL_1700201, EPI_ISL_1700204, EPI_ISL_1700207, EPI_ISL_1700208, EPI_ISL_1700209, EPI_ISL_1700212, EPI_ISL_1700213, EPI_ISL_1700214, EPI_ISL_1700216, EPI_ISL_1700221, EPI_ISL_1700223, EPI_ISL_1700227, EPI_ISL_1700228, EPI_ISL_1700240, EPI_ISL_1700241, EPI_ISL_1700243, EPI_ISL_1700244, EPI_ISL_1700250, EPI_ISL_1700251, EPI_ISL_1700253, EPI_ISL_1700254, EPI_ISL_1700255, EPI_ISL_1700257, EPI_ISL_1700258, EPI_ISL_1700265, EPI_ISL_1700269, EPI_ISL_1700270, EPI_ISL_1700274, EPI_ISL_1700275, EPI_ISL_1700277, EPI_ISL_1700278, EPI_ISL_1700286, EPI_ISL_1700294, EPI_ISL_1700300, EPI_ISL_1700302, EPI_ISL_1700307, EPI_ISL_1700308, EPI_ISL_1700312, EPI_ISL_1700319, EPI_ISL_1700320, EPI_ISL_1700331, EPI_ISL_1700332, EPI_ISL_1700333, EPI_ISL_1700338, EPI_ISL_1700343, EPI_ISL_1700347, EPI_ISL_1700355, EPI_ISL_1700356, EPI_ISL_1700357, EPI_ISL_1700361, EPI_ISL_1700363, EPI_ISL_1700364, EPI_ISL_1700369, EPI_ISL_1700370, EPI_ISL_1700373, EPI_ISL_1700378, EPI_ISL_1700380, EPI_ISL_1700385, EPI_ISL_1700386, EPI_ISL_1700389, EPI_ISL_1700392, EPI_ISL_1700396, EPI_ISL_1700410, EPI_ISL_1700413, EPI_ISL_1700415, EPI_ISL_1700433, EPI_ISL_1700436, EPI_ISL_1700441, EPI_ISL_1700456, EPI_ISL_1700457, EPI_ISL_1700459, EPI_ISL_1700461, EPI_ISL_1700469, EPI_ISL_1700470, EPI_ISL_1700473, EPI_ISL_1700476, EPI_ISL_1700479, EPI_ISL_1700484, EPI_ISL_1700487, EPI_ISL_1700490, EPI_ISL_1700492, EPI_ISL_1700493, EPI_ISL_1741139, EPI_ISL_1741787, EPI_ISL_1741794, EPI_ISL_1741799, EPI_ISL_1741806, EPI_ISL_1741812, EPI_ISL_1741818, EPI_ISL_1741825, EPI_ISL_1741826, EPI_ISL_1741832, EPI_ISL_1741840, EPI_ISL_1741846, EPI_ISL_1741850, EPI_ISL_1741853, EPI_ISL_1741854, EPI_ISL_1741855, EPI_ISL_1741861, EPI_ISL_1741869, EPI_ISL_1741870, EPI_ISL_1741881, EPI_ISL_1741902, EPI_ISL_1741905, EPI_ISL_1741911, EPI_ISL_1741913, EPI_ISL_1741921, EPI_ISL_1741923, EPI_ISL_1741929, EPI_ISL_1741937, EPI_ISL_1741965, EPI_ISL_1741976, EPI_ISL_1741983, EPI_ISL_1741995, EPI_ISL_1742013, EPI_ISL_1742014, EPI_ISL_1742015, EPI_ISL_1742016, EPI_ISL_1742017, EPI_ISL_1742018, EPI_ISL_1742020, EPI_ISL_1742021, EPI_ISL_1742022, EPI_ISL_1742023, EPI_ISL_1742024, EPI_ISL_1742025, EPI_ISL_1742026, EPI_ISL_1742027, EPI_ISL_1742029, EPI_ISL_1742030, EPI_ISL_1742031, EPI_ISL_1742032, EPI_ISL_1742033, EPI_ISL_1742035, EPI_ISL_1742036, EPI_ISL_1742037, EPI_ISL_1742038, EPI_ISL_1742039, EPI_ISL_1742040, EPI_ISL_1742041, EPI_ISL_1742042, EPI_ISL_1742043, EPI_ISL_1742044, EPI_ISL_1742045, EPI_ISL_1742046, EPI_ISL_1742047, EPI_ISL_1742048, EPI_ISL_1742049, EPI_ISL_1742050, EPI_ISL_1742051, EPI_ISL_1742052, EPI_ISL_1742053, EPI_ISL_1742054, EPI_ISL_1742056, EPI_ISL_1742057, EPI_ISL_1742058, EPI_ISL_1742059, EPI_ISL_1742061, EPI_ISL_1742063, EPI_ISL_1742064, EPI_ISL_1742065, EPI_ISL_1742066, EPI_ISL_1742067, EPI_ISL_1742068, EPI_ISL_1742069, EPI_ISL_1742070, EPI_ISL_1742071, EPI_ISL_1742072, EPI_ISL_1742073, EPI_ISL_1742074, EPI_ISL_1742075, EPI_ISL_1742076, EPI_ISL_1742077, EPI_ISL_1742078, EPI_ISL_1742079, EPI_ISL_1742080, EPI_ISL_1742081, EPI_ISL_1742082, EPI_ISL_1742083, EPI_ISL_1742084, EPI_ISL_1742085, EPI_ISL_1742086, EPI_ISL_1742091, EPI_ISL_1742093, EPI_ISL_1742101, EPI_ISL_1742102, EPI_ISL_1742104, EPI_ISL_1742122, EPI_ISL_1742125, EPI_ISL_1742132, EPI_ISL_1742133, EPI_ISL_1742146, EPI_ISL_1742147, EPI_ISL_1742152, EPI_ISL_1742153, EPI_ISL_1742156, EPI_ISL_1742157, EPI_ISL_1742164, EPI_ISL_1742182, EPI_ISL_1742191, EPI_ISL_1742197, EPI_ISL_1742204, EPI_ISL_1742210, EPI_ISL_1742211, EPI_ISL_1742221, EPI_ISL_1742226, EPI_ISL_1742231, EPI_ISL_1742232, EPI_ISL_1742233, EPI_ISL_1742239, EPI_ISL_1742243, EPI_ISL_1742249, EPI_ISL_1742250, EPI_ISL_1742254, EPI_ISL_1742258, EPI_ISL_1742259, EPI_ISL_1742260, EPI_ISL_1742261, EPI_ISL_1758948

| | | | |
|---|---|---|---|
| see above | Lighthouse Lab in Glasgow | Wellcome Sanger Institute for the COVID-19 Genomics UK (COG-UK) Consortium | Anna Dominiczak and Alex Alderton; Carol Clugston; Cordelia Langford; David Gray; David K. Jackson; Dominic Kwiatkowski; Ewan Harrison; Harper VanSteenhouse; Ian Johnston; Jeffrey Barrett; John Sillitoe on behalf of the Wellcome Sanger Institute COVID-19 Surveillance Team; Roberto Amato; Sonia Goncalves; Yumi Kasai |

EPI_ISL_1538402, EPI_ISL_1544220, EPI_ISL_1564040, EPI_ISL_1564046, EPI_ISL_1564063, EPI_ISL_1564087, EPI_ISL_1564100, EPI_ISL_1564121, EPI_ISL_1564159, EPI_ISL_1564190, EPI_ISL_1564193, EPI_ISL_1564222, EPI_ISL_1564228, EPI_ISL_1564229, EPI_ISL_1564236, EPI_ISL_1564237, EPI_ISL_1564240, EPI_ISL_1564244, EPI_ISL_1564246, EPI_ISL_1564254, EPI_ISL_1564285, EPI_ISL_1564294, EPI_ISL_1564297, EPI_ISL_1564298, EPI_ISL_1564299, EPI_ISL_1564302, EPI_ISL_1564310, EPI_ISL_1564316, EPI_ISL_1584165, EPI_ISL_1584166, EPI_ISL_1584179, EPI_ISL_1584180, EPI_ISL_1584181, EPI_ISL_1584186, EPI_ISL_1584190, EPI_ISL_1584199, EPI_ISL_1584203, EPI_ISL_1584205, EPI_ISL_1584207, EPI_ISL_1584209, EPI_ISL_1584210, EPI_ISL_1584211, EPI_ISL_1584212, EPI_ISL_1584218, EPI_ISL_1584221, EPI_ISL_1584224, EPI_ISL_1584226, EPI_ISL_1584227, EPI_ISL_1584228, EPI_ISL_1584229, EPI_ISL_1584234, EPI_ISL_1584238, EPI_ISL_1584246, EPI_ISL_1584252, EPI_ISL_1584263, EPI_ISL_1584266, EPI_ISL_1584269, EPI_ISL_1584271, EPI_ISL_1584272, EPI_ISL_1584274, EPI_ISL_1584283, EPI_ISL_1584284, EPI_ISL_1584286, EPI_ISL_1584287, EPI_ISL_1584289, EPI_ISL_1584292, EPI_ISL_1584294, EPI_ISL_1584295, EPI_ISL_1584300, EPI_ISL_1584301, EPI_ISL_1584302, EPI_ISL_1584306, EPI_ISL_1584307, EPI_ISL_1584313, EPI_ISL_1584315, EPI_ISL_1584317, EPI_ISL_1584318, EPI_ISL_1584322, EPI_ISL_1584331, EPI_ISL_1584335, EPI_ISL_1584336, EPI_ISL_1584340, EPI_ISL_1584341, EPI_ISL_1584342, EPI_ISL_1584350, EPI_ISL_1584354, EPI_ISL_1584358, EPI_ISL_1584360, EPI_ISL_1584364, EPI_ISL_1584370, EPI_ISL_1584374, EPI_ISL_1584378, EPI_ISL_1584385, EPI_ISL_1584387, EPI_ISL_1584388, EPI_ISL_1584390, EPI_ISL_1584392, EPI_ISL_1584395, EPI_ISL_1584396, EPI_ISL_1584399, EPI_ISL_1584404, EPI_ISL_1584406, EPI_ISL_1584407, EPI_ISL_1584409, EPI_ISL_1584416, EPI_ISL_1584420, EPI_ISL_1584424, EPI_ISL_1584425, EPI_ISL_1584428, EPI_ISL_1584429, EPI_ISL_1584430, EPI_ISL_1584432, EPI_ISL_1584434, EPI_ISL_1594457, EPI_ISL_1594463, EPI_ISL_1594465, EPI_ISL_1594478, EPI_ISL_1594492, EPI_ISL_1594494, EPI_ISL_1594507, EPI_ISL_1594530, EPI_ISL_1594536, EPI_ISL_1594541, EPI_ISL_1594547, EPI_ISL_1594549, EPI_ISL_1594551, EPI_ISL_1594557, EPI_ISL_1594563, EPI_ISL_1594567, EPI_ISL_1594571, EPI_ISL_1594574, EPI_ISL_1594582, EPI_ISL_1594588, EPI_ISL_1594612, EPI_ISL_1594615, EPI_ISL_1594622, EPI_ISL_1594624, EPI_ISL_1594625, EPI_ISL_1594639, EPI_ISL_1594643, EPI_ISL_1594649, EPI_ISL_1594655, EPI_ISL_1594661, EPI_ISL_1594662, EPI_ISL_1594667, EPI_ISL_1594668, EPI_ISL_1594669, EPI_ISL_1594675, EPI_ISL_1594678, EPI_ISL_1594679, EPI_ISL_1594680, EPI_ISL_1594681, EPI_ISL_1594682, EPI_ISL_1594687, EPI_ISL_1594689, EPI_ISL_1594694, EPI_ISL_1594700, EPI_ISL_1594712, EPI_ISL_1594753, EPI_ISL_1594799, EPI_ISL_1594831, EPI_ISL_1594833, EPI_ISL_1594888, EPI_ISL_1594910, EPI_ISL_1594938, EPI_ISL_1594945, EPI_ISL_1594947, EPI_ISL_1594953, EPI_ISL_1594975, EPI_ISL_1595009, EPI_ISL_1595054, EPI_ISL_1595058, EPI_ISL_1595090, EPI_ISL_1595100, EPI_ISL_1595154, EPI_ISL_1595164, EPI_ISL_1595169, EPI_ISL_1595189, EPI_ISL_1595471, EPI_ISL_1615690, EPI_ISL_1615696, EPI_ISL_1615701, EPI_ISL_1615704, EPI_ISL_1615708, EPI_ISL_1615710, EPI_ISL_1615711, EPI_ISL_1615712, EPI_ISL_1615715, EPI_ISL_1615718, EPI_ISL_1615720, EPI_ISL_1615722, EPI_ISL_1615724, EPI_ISL_1615725, EPI_ISL_1615730, EPI_ISL_1615737, EPI_ISL_1615743, EPI_ISL_1615748, EPI_ISL_1615758, EPI_ISL_1615760, EPI_ISL_1615767, EPI_ISL_1615770, EPI_ISL_1615772, EPI_ISL_1615773, EPI_ISL_1615774, EPI_ISL_1615777, EPI_ISL_1615779, EPI_ISL_1615782, EPI_ISL_1615785, EPI_ISL_1615786, EPI_ISL_1615788, EPI_ISL_1615791, EPI_ISL_1615795, EPI_ISL_1615799, EPI_ISL_1615802, EPI_ISL_1615804, EPI_ISL_1615806, EPI_ISL_1615809, EPI_ISL_1615810, EPI_ISL_1615811, EPI_ISL_1615813, EPI_ISL_1615819, EPI_ISL_1615821, EPI_ISL_1615824, EPI_ISL_1615825, EPI_ISL_1615826, EPI_ISL_1615834, EPI_ISL_1615835, EPI_ISL_1615840, EPI_ISL_1615841, EPI_ISL_1615845, EPI_ISL_1615846, EPI_ISL_1615849, EPI_ISL_1615850, EPI_ISL_1615851, EPI_ISL_1615853, EPI_ISL_1615859, EPI_ISL_1615860, EPI_ISL_1615867, EPI_ISL_1615871, EPI_ISL_1615874, EPI_ISL_1615876, EPI_ISL_1615892, EPI_ISL_1615903, EPI_ISL_1615913, EPI_ISL_1615920, EPI_ISL_1615925, EPI_ISL_1615927, EPI_ISL_1615930, EPI_ISL_1615934, EPI_ISL_1615937, EPI_ISL_1615942, EPI_ISL_1615943, EPI_ISL_1615944, EPI_ISL_1615945, EPI_ISL_1615946, EPI_ISL_1615949, EPI_ISL_1615950, EPI_ISL_1615951, EPI_ISL_1615952, EPI_ISL_1615954, EPI_ISL_1615955, EPI_ISL_1615957, EPI_ISL_1615958, EPI_ISL_1615959, EPI_ISL_1615960, EPI_ISL_1615961, EPI_ISL_1615962, EPI_ISL_1615963, EPI_ISL_1615964, EPI_ISL_1615966, EPI_ISL_1615967, EPI_ISL_1615968, EPI_ISL_1615969, EPI_ISL_1615970, EPI_ISL_1615971, EPI_ISL_1615972, EPI_ISL_1615973, EPI_ISL_1615974, EPI_ISL_1615975, EPI_ISL_1615976, EPI_ISL_1615979, EPI_ISL_1615980, EPI_ISL_1615981, EPI_ISL_1615983, EPI_ISL_1615984, EPI_ISL_1615985, EPI_ISL_1615991, EPI_ISL_1615992, EPI_ISL_1615995, EPI_ISL_1615997, EPI_ISL_1616002, EPI_ISL_1616003, EPI_ISL_1616005, EPI_ISL_1616006, EPI_ISL_1616009, EPI_ISL_1616010, EPI_ISL_1616011, EPI_ISL_1616012, EPI_ISL_1616013, EPI_ISL_1616014, EPI_ISL_1616015, EPI_ISL_1616016, EPI_ISL_1616018, EPI_ISL_1616019, EPI_ISL_1616021, EPI_ISL_1616026, EPI_ISL_1616028, EPI_ISL_1616030, EPI_ISL_1616033, EPI_ISL_1616034, EPI_ISL_1616035, EPI_ISL_1616036, EPI_ISL_1616037, EPI_ISL_1616038, EPI_ISL_1616040, EPI_ISL_1616041, EPI_ISL_1616042, EPI_ISL_1616043, EPI_ISL_1616045, EPI_ISL_1616046, EPI_ISL_1616047, EPI_ISL_1616050, EPI_ISL_1616052, EPI_ISL_1616053, EPI_ISL_1616054, EPI_ISL_1616056, EPI_ISL_1616058, EPI_ISL_1616062, EPI_ISL_1616065, EPI_ISL_1616066, EPI_ISL_1616067, EPI_ISL_1616068, EPI_ISL_1616070, EPI_ISL_1616071, EPI_ISL_1616072, EPI_ISL_1616074, EPI_ISL_1616078, EPI_ISL_1616080, EPI_ISL_1616081, EPI_ISL_1616083, EPI_ISL_1616084, EPI_ISL_1616085, EPI_ISL_1616086, EPI_ISL_1616089, EPI_ISL_1616090, EPI_ISL_1616093, EPI_ISL_1616094, EPI_ISL_1616095, EPI_ISL_1616096, EPI_ISL_1616098, EPI_ISL_1616105, EPI_ISL_1616108, EPI_ISL_1616115, EPI_ISL_1616116, EPI_ISL_1616118, EPI_ISL_1616119, EPI_ISL_1616122, EPI_ISL_1616124, EPI_ISL_1616125, EPI_ISL_1616126, EPI_ISL_1616129, EPI_ISL_1616131, EPI_ISL_1616132, EPI_ISL_1616133, EPI_ISL_1616135, EPI_ISL_1616136, EPI_ISL_1616138, EPI_ISL_1616141, EPI_ISL_1616142, EPI_ISL_1616144, EPI_ISL_1616145, EPI_ISL_1616147, EPI_ISL_1616148, EPI_ISL_1616149, EPI_ISL_1616154, EPI_ISL_1616158, EPI_ISL_1616161, EPI_ISL_1616164, EPI_ISL_1616166, EPI_ISL_1616168, EPI_ISL_1616169, EPI_ISL_1616172, EPI_ISL_1616173, EPI_ISL_1616174, EPI_ISL_1616175, EPI_ISL_1616176, EPI_ISL_1616178, EPI_ISL_1616180, EPI_ISL_1616182, EPI_ISL_1616183, EPI_ISL_1616184, EPI_ISL_1616185, EPI_ISL_1630284, EPI_ISL_1630289, EPI_ISL_1630291, EPI_ISL_1630298, EPI_ISL_1630305, EPI_ISL_1630320, EPI_ISL_1630326, EPI_ISL_1630336, EPI_ISL_1630339, EPI_ISL_1630349, EPI_ISL_1630357, EPI_ISL_1630358, EPI_ISL_1630360, EPI_ISL_1630363, EPI_ISL_1630374, EPI_ISL_1630375, EPI_ISL_1630377, EPI_ISL_1630378, EPI_ISL_1630380, EPI_ISL_1630383, EPI_ISL_1630388, EPI_ISL_1630390, EPI_ISL_1630395, EPI_ISL_1630402, EPI_ISL_1630406, EPI_ISL_1630412, EPI_ISL_1630444, EPI_ISL_1630455, EPI_ISL_1630461, EPI_ISL_1630463, EPI_ISL_1630464, EPI_ISL_1630469, EPI_ISL_1630474, EPI_ISL_1630475, EPI_ISL_1630478, EPI_ISL_1630479, EPI_ISL_1630488, EPI_ISL_1630493, EPI_ISL_1630494, EPI_ISL_1630508, EPI_ISL_1631101, EPI_ISL_1631118, EPI_ISL_1631122, EPI_ISL_1631126, EPI_ISL_1631129, EPI_ISL_1631141, EPI_ISL_1631142, EPI_ISL_1631159, EPI_ISL_1631161, EPI_ISL_1631181, EPI_ISL_1631188, EPI_ISL_1631193, EPI_ISL_1631197, EPI_ISL_1631199, EPI_ISL_1631218, EPI_ISL_1631233, EPI_ISL_1631234, EPI_ISL_1631252, EPI_ISL_1631277, EPI_ISL_1631290, EPI_ISL_1631292, EPI_ISL_1631315, EPI_ISL_1631425, EPI_ISL_1631754, EPI_ISL_1631755, EPI_ISL_1631756, EPI_ISL_1631757, EPI_ISL_1631759, EPI_ISL_1631760, EPI_ISL_1631767, EPI_ISL_1631775, EPI_ISL_1631776, EPI_ISL_1631781, EPI_ISL_1631792, EPI_ISL_1631793, EPI_ISL_1631797, EPI_ISL_1631798, EPI_ISL_1631799, EPI_ISL_1631801, EPI_ISL_1631802, EPI_ISL_1631803, EPI_ISL_1631806, EPI_ISL_1631811, EPI_ISL_1631812, EPI_ISL_1631814, EPI_ISL_1631815, EPI_ISL_1631818, EPI_ISL_1631820, EPI_ISL_1631821, EPI_ISL_1631824, EPI_ISL_1631826, EPI_ISL_1631827, EPI_ISL_1631832, EPI_ISL_1631837, EPI_ISL_1631838, EPI_ISL_1631856, EPI_ISL_1631858, EPI_ISL_1631859, EPI_ISL_1631860, EPI_ISL_1631865, EPI_ISL_1631866, EPI_ISL_1631869, EPI_ISL_1631876, EPI_ISL_1631877, EPI_ISL_1631880, EPI_ISL_1631882, EPI_ISL_1631887, EPI_ISL_1631891, EPI_ISL_1631892, EPI_ISL_1631900, EPI_ISL_1631902, EPI_ISL_1631912, EPI_ISL_1631913, EPI_ISL_1631918, EPI_ISL_1631922, EPI_ISL_1631923, EPI_ISL_1631924, EPI_ISL_1631929, EPI_ISL_1631941, EPI_ISL_1631943, EPI_ISL_1631945, EPI_ISL_1631949, EPI_ISL_1631953, EPI_ISL_1631961, EPI_ISL_1631970, EPI_ISL_1631981, EPI_ISL_1631991, EPI_ISL_1631994, EPI_ISL_1631996, EPI_ISL_1632011, EPI_ISL_1632021, EPI_ISL_1632031, EPI_ISL_1632032, EPI_ISL_1632034, EPI_ISL_1632035, EPI_ISL_1632081, EPI_ISL_1632106, EPI_ISL_1632111, EPI_ISL_1632117, EPI_ISL_1632118, EPI_ISL_1632126, EPI_ISL_1632127, EPI_ISL_1632136, EPI_ISL_1632142, EPI_ISL_1632161, EPI_ISL_1632174, EPI_ISL_1632175, EPI_ISL_1632181, EPI_ISL_1632191, EPI_ISL_1632198, EPI_ISL_1632200, EPI_ISL_1632215, EPI_ISL_1632217, EPI_ISL_1632223, EPI_ISL_1632229, EPI_ISL_1632231, EPI_ISL_1632232, EPI_ISL_1632235, EPI_ISL_1632245, EPI_ISL_1632252, EPI_ISL_1632254, EPI_ISL_1632257, EPI_ISL_1632264, EPI_ISL_1632268, EPI_ISL_1632271, EPI_ISL_1632279, EPI_ISL_1632286, EPI_ISL_1632288, EPI_ISL_1632293, EPI_ISL_1632295, EPI_ISL_1632328, EPI_ISL_1632348, EPI_ISL_1632377, EPI_ISL_1632391, EPI_ISL_1632436, EPI_ISL_1632439, EPI_ISL_1632459, EPI_ISL_1632472, EPI_ISL_1632480, EPI_ISL_1634487, EPI_ISL_1634488, EPI_ISL_1634491, EPI_ISL_1634494, EPI_ISL_1634499, EPI_ISL_1634500, EPI_ISL_1634505, EPI_ISL_1634506, EPI_ISL_1634507, EPI_ISL_1634510, EPI_ISL_1634511, EPI_ISL_1634514, EPI_ISL_1634515, EPI_ISL_1634516, EPI_ISL_1634517, EPI_ISL_1634519, EPI_ISL_1634520, EPI_ISL_1634521, EPI_ISL_1634523, EPI_ISL_1634524, EPI_ISL_1634525, EPI_ISL_1634526, EPI_ISL_1634529, EPI_ISL_1634530, EPI_ISL_1634531, EPI_ISL_1634533, EPI_ISL_1634534, EPI_ISL_1634535, EPI_ISL_1634536, EPI_ISL_1634538, EPI_ISL_1634541, EPI_ISL_1634542, EPI_ISL_1634546, EPI_ISL_1634549, EPI_ISL_1634551, EPI_ISL_1634552, EPI_ISL_1634553, EPI_ISL_1634556, EPI_ISL_1634557, EPI_ISL_1634560, EPI_ISL_1634561, EPI_ISL_1634562, EPI_ISL_1634563, EPI_ISL_1634564, EPI_ISL_1634565, EPI_ISL_1634567, EPI_ISL_1634569, EPI_ISL_1634570, EPI_ISL_1634573, EPI_ISL_1634576, EPI_ISL_1634579, EPI_ISL_1634580, EPI_ISL_1634581, EPI_ISL_1634585, EPI_ISL_1634588, EPI_ISL_1634590, EPI_ISL_1634591, EPI_ISL_1634592, EPI_ISL_1634593, EPI_ISL_1634594, EPI_ISL_1634596, EPI_ISL_1634597, EPI_ISL_1634598, EPI_ISL_1634599, EPI_ISL_1634606, EPI_ISL_1634608, EPI_ISL_1634609, EPI_ISL_1634611, EPI_ISL_1634612, EPI_ISL_1634614, EPI_ISL_1634616, EPI_ISL_1634618, EPI_ISL_1634619, EPI_ISL_1634621, EPI_ISL_1634625, EPI_ISL_1634627, EPI_ISL_1634629, EPI_ISL_1634630, EPI_ISL_1634636, EPI_ISL_1634637, EPI_ISL_1634638, EPI_ISL_1634639, EPI_ISL_1634640, EPI_ISL_1634641, EPI_ISL_1634642, EPI_ISL_1634644, EPI_ISL_1634645, EPI_ISL_1634646, EPI_ISL_1634647, EPI_ISL_1634648, EPI_ISL_1634652, EPI_ISL_1634654, EPI_ISL_1634655, EPI_ISL_1634657, EPI_ISL_1634658, EPI_ISL_1634659, EPI_ISL_1634660, EPI_ISL_1634662, EPI_ISL_1634663, EPI_ISL_1634666, EPI_ISL_1634668, EPI_ISL_1634670, EPI_ISL_1634671, EPI_ISL_1634672, EPI_ISL_1634673, EPI_ISL_1634675, EPI_ISL_1634678, EPI_ISL_1634679, EPI_ISL_1634682, EPI_ISL_1634684, EPI_ISL_1634686, EPI_ISL_1634687, EPI_ISL_1634688, EPI_ISL_1634689, EPI_ISL_1634692, EPI_ISL_1634693, EPI_ISL_1634694, EPI_ISL_1634698, EPI_ISL_1634699, EPI_ISL_1635313, EPI_ISL_1652998, EPI_ISL_1653176, EPI_ISL_1653183, EPI_ISL_1653185, EPI_ISL_1653209, EPI_ISL_1653210, EPI_ISL_1653213, EPI_ISL_1653214, EPI_ISL_1653216, EPI_ISL_1653218, EPI_ISL_1653219, EPI_ISL_1653222, EPI_ISL_1653223, EPI_ISL_1653228, EPI_ISL_1653238, EPI_ISL_1653240, EPI_ISL_1653246, EPI_ISL_1653248, EPI_ISL_1653254, EPI_ISL_1653258, EPI_ISL_1653261, EPI_ISL_1653263, EPI_ISL_1653265, EPI_ISL_1653266, EPI_ISL_1653267, EPI_ISL_1653274, EPI_ISL_1653275, EPI_ISL_1653283, EPI_ISL_1653285, EPI_ISL_1653291, EPI_ISL_1653292, EPI_ISL_1653295, EPI_ISL_1653306, EPI_ISL_1653311, EPI_ISL_1653318, EPI_ISL_1653322, EPI_ISL_1653323, EPI_ISL_1653324, EPI_ISL_1653328, EPI_ISL_1653336, EPI_ISL_1653344, EPI_ISL_1653355, EPI_ISL_1653366, EPI_ISL_1653367, EPI_ISL_1653373, EPI_ISL_1653376, EPI_ISL_1653383, EPI_ISL_1653384, EPI_ISL_1653393, EPI_ISL_1653397, EPI_ISL_1653398, EPI_ISL_1653399, EPI_ISL_1653407, EPI_ISL_1653410, EPI_ISL_1653411, EPI_ISL_1653416, EPI_ISL_1653420, EPI_ISL_1653425, EPI_ISL_1653430, EPI_ISL_1653434, EPI_ISL_1653437, EPI_ISL_1653442, EPI_ISL_1653443, EPI_ISL_1653444, EPI_ISL_1653448, EPI_ISL_1653450, EPI_ISL_1653451, EPI_ISL_1653452, EPI_ISL_1653454, EPI_ISL_1653459, EPI_ISL_1653462, EPI_ISL_1653466, EPI_ISL_1653467, EPI_ISL_1653470, EPI_ISL_1653485, EPI_ISL_1673121, EPI_ISL_1697456, EPI_ISL_1697497, EPI_ISL_1697516, EPI_ISL_1697518, EPI_ISL_1697522, EPI_ISL_1697526, EPI_ISL_1697552, EPI_ISL_1697565, EPI_ISL_1697571, EPI_ISL_1697579, EPI_ISL_1697592, EPI_ISL_1697596, EPI_ISL_1697617, EPI_ISL_1697645, EPI_ISL_1697650, EPI_ISL_1697684, EPI_ISL_1697748, EPI_ISL_1697756, EPI_ISL_1697759, EPI_ISL_1697762, EPI_ISL_1697768, EPI_ISL_1697769, EPI_ISL_1697775, EPI_ISL_1697776, EPI_ISL_1697777, EPI_ISL_1697783, EPI_ISL_1697788, EPI_ISL_1697791, EPI_ISL_1697792, EPI_ISL_1697795, EPI_ISL_1697799, EPI_ISL_1697800, EPI_ISL_1697805, EPI_ISL_1697817, EPI_ISL_1697819, EPI_ISL_1697821, EPI_ISL_1697828, EPI_ISL_1697835, EPI_ISL_1697843, EPI_ISL_1697846, EPI_ISL_1697853, EPI_ISL_1697854, EPI_ISL_1697856, EPI_ISL_1697857, EPI_ISL_1697859, EPI_ISL_1697863, EPI_ISL_1697871, EPI_ISL_1697874, EPI_ISL_1697875, EPI_ISL_1697883, EPI_ISL_1697885, EPI_ISL_1697896, EPI_ISL_1697899, EPI_ISL_1697900, EPI_ISL_1697904, EPI_ISL_1697917, EPI_ISL_1697921, EPI_ISL_1697924, EPI_ISL_1697928, EPI_ISL_1697934, EPI_ISL_1697948, EPI_ISL_1697949, EPI_ISL_1697959, EPI_ISL_1697961, EPI_ISL_1697969, EPI_ISL_1698054, EPI_ISL_1698784, EPI_ISL_1698786, EPI_ISL_1698793, EPI_ISL_1698794, EPI_ISL_1698799, EPI_ISL_1698804, EPI_ISL_1698805, EPI_ISL_1698806, EPI_ISL_1698809, EPI_ISL_1698815, EPI_ISL_1698821, EPI_ISL_1698824, EPI_ISL_1698832, EPI_ISL_1698840, EPI_ISL_1698846, EPI_ISL_1698857, EPI_ISL_1698860, EPI_ISL_1698861, EPI_ISL_1698865, EPI_ISL_1698870, EPI_ISL_1698873, EPI_ISL_1698876, EPI_ISL_1698879, EPI_ISL_1698883, EPI_ISL_1698886, EPI_ISL_1698889, EPI_ISL_1698890, EPI_ISL_1698891, EPI_ISL_1698902, EPI_ISL_1698906, EPI_ISL_1698911, EPI_ISL_1698912, EPI_ISL_1698916, EPI_ISL_1698921, EPI_ISL_1698945, EPI_ISL_1698956, EPI_ISL_1698959, EPI_ISL_1698966, EPI_ISL_1698967, EPI_ISL_1698974, EPI_ISL_1698977, EPI_ISL_1698983, EPI_ISL_1698989, EPI_ISL_1698994, EPI_ISL_1699000, EPI_ISL_1699007, EPI_ISL_1699012, EPI_ISL_1699015, EPI_ISL_1699016, EPI_ISL_1699018, EPI_ISL_1699020, EPI_ISL_1699021, EPI_ISL_1699028, EPI_ISL_1699030, EPI_ISL_1699042, EPI_ISL_1699054, EPI_ISL_1699059, EPI_ISL_1699062, EPI_ISL_1699065, EPI_ISL_1699075, EPI_ISL_1699083, EPI_ISL_1699086, EPI_ISL_1699087, EPI_ISL_1699090, EPI_ISL_1699092, EPI_ISL_1699093, EPI_ISL_1699095, EPI_ISL_1699097, EPI_ISL_1699098, EPI_ISL_1699103, EPI_ISL_1699116, EPI_ISL_1699121, EPI_ISL_1699130, EPI_ISL_1699135, EPI_ISL_1699137, EPI_ISL_1699139, EPI_ISL_1699144, EPI_ISL_1699154, EPI_ISL_1699160, EPI_ISL_1699163, EPI_ISL_1699165, EPI_ISL_1699168, EPI_ISL_1699170, EPI_ISL_1699174, EPI_ISL_1699176, EPI_ISL_1699178, EPI_ISL_1699179, EPI_ISL_1699181, EPI_ISL_1699183, EPI_ISL_1699185, EPI_ISL_1699186, EPI_ISL_1699187, EPI_ISL_1699190, EPI_ISL_1699191, EPI_ISL_1699193, EPI_ISL_1699205, EPI_ISL_1699206, EPI_ISL_1699208, EPI_ISL_1699216, EPI_ISL_1699221, EPI_ISL_1699223, EPI_ISL_1699227, EPI_ISL_1699230, EPI_ISL_1699234, EPI_ISL_1699241, EPI_ISL_1699255, EPI_ISL_1699264, EPI_ISL_1699267, EPI_ISL_1699272, EPI_ISL_1699273, EPI_ISL_1699276, EPI_ISL_1699278, EPI_ISL_1699279, EPI_ISL_1699319, EPI_ISL_1699348, EPI_ISL_1699414, EPI_ISL_1699417, EPI_ISL_1699450, EPI_ISL_1699574, EPI_ISL_1699590, EPI_ISL_1699601, EPI_ISL_1699651, EPI_ISL_1699657, EPI_ISL_1699665, EPI_ISL_1699669, EPI_ISL_1699674, EPI_ISL_1699695, EPI_ISL_1699705, EPI_ISL_1699739, EPI_ISL_1699766, EPI_ISL_1699801, EPI_ISL_1699825, EPI_ISL_1699841, EPI_ISL_1699843, EPI_ISL_1699847, EPI_ISL_1699855, EPI_ISL_1699858, EPI_ISL_1699867, EPI_ISL_1699869, EPI_ISL_1699872, EPI_ISL_1699880, EPI_ISL_1699883, EPI_ISL_1699885, EPI_ISL_1699895, EPI_ISL_1699898, EPI_ISL_1699902, EPI_ISL_1699913, EPI_ISL_1699914, EPI_ISL_1699927, EPI_ISL_1699931, EPI_ISL_1699937, EPI_ISL_1699960, EPI_ISL_1699967, EPI_ISL_1699977, EPI_ISL_1699979, EPI_ISL_1699980, EPI_ISL_1699984, EPI_ISL_1699990, EPI_ISL_1699997, EPI_ISL_1700013, EPI_ISL_1700020, EPI_ISL_1700021, EPI_ISL_1700022, EPI_ISL_1700025, EPI_ISL_1700029, EPI_ISL_1700044, EPI_ISL_1700050, EPI_ISL_1700071, EPI_ISL_1700074, EPI_ISL_1700076, EPI_ISL_1700078, EPI_ISL_1700080, EPI_ISL_1700102, EPI_ISL_1700108, EPI_ISL_1700116, EPI_ISL_1700130, EPI_ISL_1700138, EPI_ISL_1700140, EPI_ISL_1700148, EPI_ISL_1700149, EPI_ISL_1700152, EPI_ISL_1700153, EPI_ISL_1700157, EPI_ISL_1700165, EPI_ISL_1700167, EPI_ISL_1700169, EPI_ISL_1719197, EPI_ISL_1758205, EPI_ISL_1758208, EPI_ISL_1758209, EPI_ISL_1758210, EPI_ISL_1758213, EPI_ISL_1758215, EPI_ISL_1758221, EPI_ISL_1758222, EPI_ISL_1758224, EPI_ISL_1758226, EPI_ISL_1758228, EPI_ISL_1758231, EPI_ISL_1758232, EPI_ISL_1758253, EPI_ISL_1758258, EPI_ISL_1758260, EPI_ISL_1758262, EPI_ISL_1758263, EPI_ISL_1758265, EPI_ISL_1758269, EPI_ISL_1758274, EPI_ISL_1758276, EPI_ISL_1758277, EPI_ISL_1758278, EPI_ISL_1758281, EPI_ISL_1758282, EPI_ISL_1758283, EPI_ISL_1758288, EPI_ISL_1758300, EPI_ISL_1758302, EPI_ISL_1758309, EPI_ISL_1758310, EPI_ISL_1758323, EPI_ISL_1758327, EPI_ISL_1758329, EPI_ISL_1758331, EPI_ISL_1758335, EPI_ISL_1758341, EPI_ISL_1758347, EPI_ISL_1758350, EPI_ISL_1758351, EPI_ISL_1758354, EPI_ISL_1758362, EPI_ISL_1758363, EPI_ISL_1758367, EPI_ISL_1758375, EPI_ISL_1758388, EPI_ISL_1758389, EPI_ISL_1758390, EPI_ISL_1758398, EPI_ISL_1758402, EPI_ISL_1758405, EPI_ISL_1758417, EPI_ISL_1758419, EPI_ISL_1758422, EPI_ISL_1758425, EPI_ISL_1758426, EPI_ISL_1758427, EPI_ISL_1758428, EPI_ISL_1758430, EPI_ISL_1758453, EPI_ISL_1758454, EPI_ISL_1758457, EPI_ISL_1758458, EPI_ISL_1758459, EPI_ISL_1758464, EPI_ISL_1758467, EPI_ISL_1758468, EPI_ISL_1758471, EPI_ISL_1758473, EPI_ISL_1758474, EPI_ISL_1758475, EPI_ISL_1758477, EPI_ISL_1758478, EPI_ISL_1758480, EPI_ISL_1758481, EPI_ISL_1758487, EPI_ISL_1758488, EPI_ISL_1758489, EPI_ISL_1758490, EPI_ISL_1758491, EPI_ISL_1758494, EPI_ISL_1758498, EPI_ISL_1758499, EPI_ISL_1758500, EPI_ISL_1758501, EPI_ISL_1758502, EPI_ISL_1758504, EPI_ISL_1758506, EPI_ISL_1758508, EPI_ISL_1758512, EPI_ISL_1758513, EPI_ISL_1758514, EPI_ISL_1758515, EPI_ISL_1758516, EPI_ISL_1758517, EPI_ISL_1758519, EPI_ISL_1758524, EPI_ISL_1758528, EPI_ISL_1758534, EPI_ISL_1758535, EPI_ISL_1758537, EPI_ISL_1758539, EPI_ISL_1758542, EPI_ISL_1758543, EPI_ISL_1758544, EPI_ISL_1758546, EPI_ISL_1758548, EPI_ISL_1758551, EPI_ISL_1758552, EPI_ISL_1758553, EPI_ISL_1758554, EPI_ISL_1758557, EPI_ISL_1758560, EPI_ISL_1758561, EPI_ISL_1758564, EPI_ISL_1758565, EPI_ISL_1758566, EPI_ISL_1758571, EPI_ISL_1758574, EPI_ISL_1758575, EPI_ISL_1758576, EPI_ISL_1758577, EPI_ISL_1758579, EPI_ISL_1758580, EPI_ISL_1758585, EPI_ISL_1758586, EPI_ISL_1758588, EPI_ISL_1758589, EPI_ISL_1758591, EPI_ISL_1758592, EPI_ISL_1758593, EPI_ISL_1758594, EPI_ISL_1758595, EPI_ISL_1758596, EPI_ISL_1758599, EPI_ISL_1758600, EPI_ISL_1758601, EPI_ISL_1758602, EPI_ISL_1758607, EPI_ISL_1758610, EPI_ISL_1758611, EPI_ISL_1758613, EPI_ISL_1758615, EPI_ISL_1758617, EPI_ISL_1758618, EPI_ISL_1758620, EPI_ISL_1758621, EPI_ISL_1758622, EPI_ISL_1758623, EPI_ISL_1758626, EPI_ISL_1758627, EPI_ISL_1758629, EPI_ISL_1758631, EPI_ISL_1758633, EPI_ISL_1758634, EPI_ISL_1758637, EPI_ISL_1758638, EPI_ISL_1758639, EPI_ISL_1758640, EPI_ISL_1758641, EPI_ISL_1758642, EPI_ISL_1758643, EPI_ISL_1758644, EPI_ISL_1758646, EPI_ISL_1758647, EPI_ISL_1758648, EPI_ISL_1758650, EPI_ISL_1758651, EPI_ISL_1758652, EPI_ISL_1758653, EPI_ISL_1758654, EPI_ISL_1758655, EPI_ISL_1758657, EPI_ISL_1758659, EPI_ISL_1758661, EPI_ISL_1758663, EPI_ISL_1758665, EPI_ISL_1759194, EPI_ISL_1759199, EPI_ISL_1759204, EPI_ISL_1759216, EPI_ISL_1759218, EPI_ISL_1759229, EPI_ISL_1759231, EPI_ISL_1759234, EPI_ISL_1759243, EPI_ISL_1759245, EPI_ISL_1759261, EPI_ISL_1759275, EPI_ISL_1759277, EPI_ISL_1759280, EPI_ISL_1759302, EPI_ISL_1759315, EPI_ISL_1759322, EPI_ISL_1759327, EPI_ISL_1759331, EPI_ISL_1759333, EPI_ISL_1759337, EPI_ISL_1759341, EPI_ISL_1759813, EPI_ISL_1759831, EPI_ISL_1759848, EPI_ISL_1759854, EPI_ISL_1759865, EPI_ISL_1759866, EPI_ISL_1759869, EPI_ISL_1759878, EPI_ISL_1759883, EPI_ISL_1759891, EPI_ISL_1759892, EPI_ISL_1790214, EPI_ISL_1790218, EPI_ISL_1790219, EPI_ISL_1790220, EPI_ISL_1790221, EPI_ISL_1790223, EPI_ISL_1790224, EPI_ISL_1790225, EPI_ISL_1790226, EPI_ISL_1790227, EPI_ISL_1790233, EPI_ISL_1790234, EPI_ISL_1790236, EPI_ISL_1790237, EPI_ISL_1790239, EPI_ISL_1790240, EPI_ISL_1790241, EPI_ISL_1790242, EPI_ISL_1790243, EPI_ISL_1790244, EPI_ISL_1790245, EPI_ISL_1790246, EPI_ISL_1790247, EPI_ISL_1790250, EPI_ISL_1790251, EPI_ISL_1790254, EPI_ISL_1790257, EPI_ISL_1790259, EPI_ISL_1790261, EPI_ISL_1790262, EPI_ISL_1790264, EPI_ISL_1790265, EPI_ISL_1790267, EPI_ISL_1790268, EPI_ISL_1790270, EPI_ISL_1790271, EPI_ISL_1790273, EPI_ISL_1790276, EPI_ISL_1790277, EPI_ISL_1790278, EPI_ISL_1790279, EPI_ISL_1790280, EPI_ISL_1790281, EPI_ISL_1790282, EPI_ISL_1790283, EPI_ISL_1790284, EPI_ISL_1790288, EPI_ISL_1790291, EPI_ISL_1790293, EPI_ISL_1790294, EPI_ISL_1790296, EPI_ISL_1790297, EPI_ISL_1790298, EPI_ISL_1790299, EPI_ISL_1790301, EPI_ISL_1790302, EPI_ISL_1790304, EPI_ISL_1790305, EPI_ISL_1790309, EPI_ISL_1790310, EPI_ISL_1790311, EPI_ISL_1790312, EPI_ISL_1790313, EPI_ISL_1790317, EPI_ISL_1790318, EPI_ISL_1790319, EPI_ISL_1790320, EPI_ISL_1790326, EPI_ISL_1790329, EPI_ISL_1790330, EPI_ISL_1790334, EPI_ISL_1790335, EPI_ISL_1790338, EPI_ISL_1790339, EPI_ISL_1790341, EPI_ISL_1790343, EPI_ISL_1790345, EPI_ISL_1790346, EPI_ISL_1790348, EPI_ISL_1790350, EPI_ISL_1790352, EPI_ISL_1790353, EPI_ISL_1790354, EPI_ISL_1790355, EPI_ISL_1790356, EPI_ISL_1790358, EPI_ISL_1790360, EPI_ISL_1790363, EPI_ISL_1790365, EPI_ISL_1790366, EPI_ISL_1790368, EPI_ISL_1790374, EPI_ISL_1790376, EPI_ISL_1790377, EPI_ISL_1790379, EPI_ISL_1790383, EPI_ISL_1790384, EPI_ISL_1790385, EPI_ISL_1790386, EPI_ISL_1790389, EPI_ISL_1790390, EPI_ISL_1790391, EPI_ISL_1790392, EPI_ISL_1790396, EPI_ISL_1790400, EPI_ISL_1790401, EPI_ISL_1790403, EPI_ISL_1790404, EPI_ISL_1790405, EPI_ISL_1790406, EPI_ISL_1790407, EPI_ISL_1790410, EPI_ISL_1790412, EPI_ISL_1790416, EPI_ISL_1790417, EPI_ISL_1790418, EPI_ISL_1790420, EPI_ISL_1790421, EPI_ISL_1790422, EPI_ISL_1790424, EPI_ISL_1790425, EPI_ISL_1790429, EPI_ISL_1790430, EPI_ISL_1791019, EPI_ISL_1806217, EPI_ISL_1806224, EPI_ISL_1806225, EPI_ISL_1806228, EPI_ISL_1806233, EPI_ISL_1806239, EPI_ISL_1806242, EPI_ISL_1806245, EPI_ISL_1806248, EPI_ISL_1806249, EPI_ISL_1806252, EPI_ISL_1806254, EPI_ISL_1806273, EPI_ISL_1806282, EPI_ISL_1806284, EPI_ISL_1806285, EPI_ISL_1806297, EPI_ISL_1806299, EPI_ISL_1806304, EPI_ISL_1806309, EPI_ISL_1806312, EPI_ISL_1806316, EPI_ISL_1806319, EPI_ISL_1806321, EPI_ISL_1806328, EPI_ISL_1806330, EPI_ISL_1806336, EPI_ISL_1806337, EPI_ISL_1806339, EPI_ISL_1806346, EPI_ISL_1806348, EPI_ISL_1806354, EPI_ISL_1806355, EPI_ISL_1806357, EPI_ISL_1806358, EPI_ISL_1806359, EPI_ISL_1806365, EPI_ISL_1806369, EPI_ISL_1806370, EPI_ISL_1806383, EPI_ISL_1806386, EPI_ISL_1806387, EPI_ISL_1806389, EPI_ISL_1806396, EPI_ISL_1806401, EPI_ISL_1806415, EPI_ISL_1806416, EPI_ISL_1806418, EPI_ISL_1806424, EPI_ISL_1806425, EPI_ISL_1806431, EPI_ISL_1806434, EPI_ISL_1806448, EPI_ISL_1806449, EPI_ISL_1806451, EPI_ISL_1806456, EPI_ISL_1806458, EPI_ISL_1806459, EPI_ISL_1806462, EPI_ISL_1806463, EPI_ISL_1806464, EPI_ISL_1806466, EPI_ISL_1806467, EPI_ISL_1806472, EPI_ISL_1806478, EPI_ISL_1806488, EPI_ISL_1806496, EPI_ISL_1806500, EPI_ISL_1806501, EPI_ISL_1829215, EPI_ISL_1829269, EPI_ISL_1829327, EPI_ISL_1829363, EPI_ISL_1829432, EPI_ISL_1829442, EPI_ISL_1829461, EPI_ISL_1829500, EPI_ISL_1829525, EPI_ISL_1829532, EPI_ISL_1829540, EPI_ISL_1829561, EPI_ISL_1829570, EPI_ISL_1829593, EPI_ISL_1829601, EPI_ISL_1829643, EPI_ISL_1829666, EPI_ISL_1829673, EPI_ISL_1829700, EPI_ISL_1829703, EPI_ISL_1829723, EPI_ISL_1829801, EPI_ISL_1830235, EPI_ISL_1830405, EPI_ISL_1830409, EPI_ISL_1830414, EPI_ISL_1830416, EPI_ISL_1830429, EPI_ISL_1830440, EPI_ISL_1830441, EPI_ISL_1830448, EPI_ISL_1830461, EPI_ISL_1830470, EPI_ISL_1830476, EPI_ISL_1830488, EPI_ISL_1830517, EPI_ISL_1830556, EPI_ISL_1830557, EPI_ISL_1830580, EPI_ISL_1830581, EPI_ISL_1830583, EPI_ISL_1830584, EPI_ISL_1830596, EPI_ISL_1830605, EPI_ISL_1830621, EPI_ISL_1830632, EPI_ISL_1830634, EPI_ISL_1830644, EPI_ISL_1830664, EPI_ISL_1830669, EPI_ISL_1830687, EPI_ISL_1830694, EPI_ISL_1830703, EPI_ISL_1830709, EPI_ISL_1830763, EPI_ISL_1830998, EPI_ISL_1831194, EPI_ISL_1831261, EPI_ISL_1831262, EPI_ISL_1912023, EPI_ISL_1912056, EPI_ISL_1912057, EPI_ISL_1912068, EPI_ISL_1912099, EPI_ISL_1912102, EPI_ISL_1912107, EPI_ISL_1912142, EPI_ISL_1912167, EPI_ISL_1912200, EPI_ISL_1912220, EPI_ISL_1912273, EPI_ISL_1912292, EPI_ISL_1912303, EPI_ISL_1912317, EPI_ISL_1986586, EPI_ISL_1986588, EPI_ISL_1986596, EPI_ISL_1986600, EPI_ISL_1986602, EPI_ISL_1986608, EPI_ISL_1986615, EPI_ISL_1986621, EPI_ISL_1986622, EPI_ISL_1986624, EPI_ISL_1986636, EPI_ISL_1986640, EPI_ISL_1986641, EPI_ISL_1986642, EPI_ISL_1986643, EPI_ISL_1986645, EPI_ISL_1986646, EPI_ISL_1986647, EPI_ISL_1986650, EPI_ISL_1986657, EPI_ISL_1986670, EPI_ISL_1986672, EPI_ISL_1986683, EPI_ISL_1986685, EPI_ISL_1986694, EPI_ISL_1986696, EPI_ISL_1986700, EPI_ISL_1986709, EPI_ISL_1986713, EPI_ISL_1986719, EPI_ISL_1986723, EPI_ISL_1986728, EPI_ISL_1986734, EPI_ISL_1986738, EPI_ISL_1986745, EPI_ISL_1986765, EPI_ISL_1986766, EPI_ISL_1986767, EPI_ISL_1986776, EPI_ISL_1986777, EPI_ISL_1986784, EPI_ISL_1986788, EPI_ISL_1986789, EPI_ISL_1986796, EPI_ISL_1986799, EPI_ISL_1986800, EPI_ISL_1986808, EPI_ISL_1986815, EPI_ISL_1986818, EPI_ISL_1986826, EPI_ISL_1986828, EPI_ISL_1986835, EPI_ISL_1986836, EPI_ISL_1986838, EPI_ISL_1986839, EPI_ISL_1986843, EPI_ISL_1986844, EPI_ISL_1986849, EPI_ISL_1986852, EPI_ISL_1986872, EPI_ISL_1986883, EPI_ISL_1986884, EPI_ISL_1986892, EPI_ISL_1986898, EPI_ISL_1986905, EPI_ISL_1986910, EPI_ISL_1986911, EPI_ISL_1986913, EPI_ISL_1986915, EPI_ISL_1986917, EPI_ISL_1986918, EPI_ISL_1986919, EPI_ISL_1986926, EPI_ISL_1986934, EPI_ISL_1986938, EPI_ISL_1986940, EPI_ISL_1986942, EPI_ISL_2005346, EPI_ISL_2005348, EPI_ISL_2005351, EPI_ISL_2005352, EPI_ISL_2005371, EPI_ISL_2005374, EPI_ISL_2005390, EPI_ISL_2005397, EPI_ISL_2005398, EPI_ISL_2005401, EPI_ISL_2005405, EPI_ISL_2005408, EPI_ISL_2005410, EPI_ISL_2005411, EPI_ISL_2005427, EPI_ISL_2005431, EPI_ISL_2005433, EPI_ISL_2005436, EPI_ISL_2005454, EPI_ISL_2005456, EPI_ISL_2005474, EPI_ISL_2005482, EPI_ISL_2005485, EPI_ISL_2005488, EPI_ISL_2005491, EPI_ISL_2005492, EPI_ISL_2005500, EPI_ISL_2005505, EPI_ISL_2005508, EPI_ISL_2005526, EPI_ISL_2005528, EPI_ISL_2005530, EPI_ISL_2005541, EPI_ISL_2005543, EPI_ISL_2005547, EPI_ISL_2005549, EPI_ISL_2005565, EPI_ISL_2005570, EPI_ISL_2005574, EPI_ISL_2005576, EPI_ISL_2005580, EPI_ISL_2005586, EPI_ISL_2005595, EPI_ISL_2005596, EPI_ISL_2005602, EPI_ISL_2005615, EPI_ISL_2005621, EPI_ISL_2005623, EPI_ISL_2005626, EPI_ISL_2005627, EPI_ISL_2005628, EPI_ISL_2005634, EPI_ISL_2005636, EPI_ISL_2005644, EPI_ISL_2005645, EPI_ISL_2005648, EPI_ISL_2005653, EPI_ISL_2005654, EPI_ISL_2005658, EPI_ISL_2005662, EPI_ISL_2005665, EPI_ISL_2005667, EPI_ISL_2005669, EPI_ISL_2005670, EPI_ISL_2005673, EPI_ISL_2005674, EPI_ISL_2005677, EPI_ISL_2005678, EPI_ISL_2005680, EPI_ISL_2005681, EPI_ISL_2005686, EPI_ISL_2005687, EPI_ISL_2005688, EPI_ISL_2005689, EPI_ISL_2005693, EPI_ISL_2005696, EPI_ISL_2005699, EPI_ISL_2005701, EPI_ISL_2005704, EPI_ISL_2005708, EPI_ISL_2005713, EPI_ISL_2005717, EPI_ISL_2005718, EPI_ISL_2005719, EPI_ISL_2005720, EPI_ISL_2005723, EPI_ISL_2005724, EPI_ISL_2005726, EPI_ISL_2005727, EPI_ISL_2005731, EPI_ISL_2005735, EPI_ISL_2005737, EPI_ISL_2005738, EPI_ISL_2005742, EPI_ISL_2005744, EPI_ISL_2005745, EPI_ISL_2005750, EPI_ISL_2005752, EPI_ISL_2005753, EPI_ISL_2005755, EPI_ISL_2005759, EPI_ISL_2005762, EPI_ISL_2005763, EPI_ISL_2005765, EPI_ISL_2005766, EPI_ISL_2005767, EPI_ISL_2005768, EPI_ISL_2005770, EPI_ISL_2005772, EPI_ISL_2005773, EPI_ISL_2005774, EPI_ISL_2005775, EPI_ISL_2005776, EPI_ISL_2005777, EPI_ISL_2005778, EPI_ISL_2005779, EPI_ISL_2005780, EPI_ISL_2005781, EPI_ISL_2005782, EPI_ISL_2005783, EPI_ISL_2005787, EPI_ISL_2005788, EPI_ISL_2005789, EPI_ISL_2005790, EPI_ISL_2005791, EPI_ISL_2005793, EPI_ISL_2005794, EPI_ISL_2005798, EPI_ISL_2005800, EPI_ISL_2005801, EPI_ISL_2005805, EPI_ISL_2005806, EPI_ISL_2005807, EPI_ISL_2005809, EPI_ISL_2006302, EPI_ISL_2006312, EPI_ISL_2006328, EPI_ISL_2006333, EPI_ISL_2006334, EPI_ISL_2006345, EPI_ISL_2006358, EPI_ISL_2006369, EPI_ISL_2006392, EPI_ISL_2006405, EPI_ISL_2006438, EPI_ISL_2006442, EPI_ISL_2006452, EPI_ISL_2006462, EPI_ISL_2006471, EPI_ISL_2006485, EPI_ISL_2006487, EPI_ISL_2006493, EPI_ISL_2006508, EPI_ISL_2022179, EPI_ISL_2022278, EPI_ISL_2022346, EPI_ISL_2022432

| | | | |
|---|---|---|---|
| see above | Lighthouse Lab in Milton Keynes | Wellcome Sanger Institute for the COVID-19 Genomics UK (COG-UK) Consortium | Cordelia Langford; David K. Jackson; Dominic Kwiatkowski; Ewan Harrison; Ian Johnston; Jeffrey Barrett; John Sillitoe on behalf of the Wellcome Sanger Institute COVID-19 Surveillance Team; Roberto Amato; Sonia Goncalves; The Lighthouse Lab in Milton Keynes and Alex Alderton |
| EPI_ISL_1833011, EPI_ISL_1833012, EPI_ISL_1833017, EPI_ISL_1833018, EPI_ISL_1833019, EPI_ISL_1833020 | Lincolnshire Hospitals and DeepSeq Nottingham | COVID-19 Genomics UK (COG-UK) Consortium | Christopher Moore; Fei Sang; Joeseph Chappell; Johnny Debebe; Jonathan Ball; Matthew Carlisle; Matthew Loose; Nadine Holmes; Nichola Duckworth; Patrick McClure; Sarah Walsh; Tim Sloan; Victoria Wright |

EPI_ISL_1636866, EPI_ISL_1636867, EPI_ISL_1636868, EPI_ISL_1636869, EPI_ISL_1636870, EPI_ISL_1636871, EPI_ISL_1636872, EPI_ISL_1636876, EPI_ISL_1636877, EPI_ISL_1636883, EPI_ISL_1832353, EPI_ISL_1832355, EPI_ISL_1832363, EPI_ISL_1832366, EPI_ISL_1832370, EPI_ISL_1832372, EPI_ISL_1832374, EPI_ISL_1832380, EPI_ISL_1832383, EPI_ISL_1832393, EPI_ISL_1832395, EPI_ISL_1832397, EPI_ISL_1832422, EPI_ISL_1832424, EPI_ISL_1832426, EPI_ISL_1832429, EPI_ISL_1832431, EPI_ISL_1832435, EPI_ISL_1832466, EPI_ISL_1832468, EPI_ISL_1832470, EPI_ISL_1832478, EPI_ISL_1832480, EPI_ISL_1832482, EPI_ISL_1832486, EPI_ISL_1832488, EPI_ISL_1832490, EPI_ISL_1832494, EPI_ISL_1832496, EPI_ISL_1832498, EPI_ISL_1832500, EPI_ISL_1832502, EPI_ISL_1832507, EPI_ISL_1832510, EPI_ISL_1832512, EPI_ISL_1974929, EPI_ISL_1974931, EPI_ISL_1974938, EPI_ISL_1974949, EPI_ISL_1974960, EPI_ISL_1974962, EPI_ISL_1974963, EPI_ISL_1974965, EPI_ISL_1974976

| | | | |
|---|---|---|---|
| see above | Liverpool Clinical Laboratories | COVID-19 Genomics UK (COG-UK) Consortium | A Alrezaihi; Alessandro Gerada; Alistair Darby; Angela Cowell; Anita Lucaci; Anu Chawla; Cassie Olateju; Catherine Hartley; Charlotte Nelson; Ecaterina Vamos; Elaine O'Toole; Eleanor G Bentley; Ghada T Shawli; Hermione Webster; Isabel García-Dorival; James Johnson; James P Stewart; Jenifer Manson; Joanne Watts; Jones Benjamin; Jordan J Clark; Julian Hiscox; L Luu; Lucille Rainbow; M Almsaud; Margaret Hughes; Mark Whitehead; Matthew Gemmell; Miren Iturriza-Gomara; Muhannad Alruwaili; N.P Randle; Neil Swainston; PKF Gilmore; Parul Sharma; Rebekah Penrice-Randal; Richard Eccles; Richard Gregory; Sam Haldenby; Steve Paterson; Stuart D Armstrong; Trevor Ian Robinson; Ximeng Han |

EPI_ISL_1637182, EPI_ISL_1637185, EPI_ISL_1637186, EPI_ISL_1637187, EPI_ISL_1637189, EPI_ISL_1637190, EPI_ISL_1637191, EPI_ISL_1637192, EPI_ISL_1637193, EPI_ISL_1637225, EPI_ISL_1637232, EPI_ISL_1637233, EPI_ISL_1637234, EPI_ISL_1730399, EPI_ISL_1730400, EPI_ISL_1730405, EPI_ISL_1978536, EPI_ISL_1978538, EPI_ISL_1978540, EPI_ISL_1978542, EPI_ISL_1978543, EPI_ISL_1978545, EPI_ISL_1978546, EPI_ISL_1978547, EPI_ISL_1978548, EPI_ISL_1978551, EPI_ISL_1978552, EPI_ISL_1978553, EPI_ISL_1978554, EPI_ISL_1978555, EPI_ISL_1978557, EPI_ISL_1978559, EPI_ISL_1978560, EPI_ISL_1978562, EPI_ISL_1978624, EPI_ISL_1978630, EPI_ISL_1978633, EPI_ISL_1978635, EPI_ISL_1978637, EPI_ISL_1978639, EPI_ISL_1978640, EPI_ISL_1978643, EPI_ISL_1978645, EPI_ISL_1978647, EPI_ISL_1978648, EPI_ISL_1978650, EPI_ISL_1978652, EPI_ISL_1978655, EPI_ISL_1978657, EPI_ISL_1978737, EPI_ISL_1978739, EPI_ISL_1978744, EPI_ISL_1978746, EPI_ISL_1978893

| | | | |
|---|---|---|---|
| see above | Northumbria University / South Tees Hospitals NHS Foundation Trust / North Cumbria Integrated Care NHS Foundation Trust / North Tees and Hartlepool NHS Foundation Trust / Newcastle Hospitals NHS Foundation Trust | COVID-19 Genomics UK (COG-UK) Consortium | Andrew Nelson; Brendan Payne; Clive Graham; Darren L Smith; Debra Padgett; Edward Barton; Emma Swindells; Garren Scott; Gary Black; Gary Eltringham; Giles S Holt; Greg R Young; Jane Greenaway; Jennifer Collins; John Allan; Joshua Loh; Lynn Dover; Matthew Bashton; Mohammad A Tariq; Paul Baker; Sarah Essex; Steve Liggett; Wen C Yew; Yusri Taha |

EPI_ISL_1546251, EPI_ISL_1546253, EPI_ISL_1546254, EPI_ISL_1546255, EPI_ISL_1546257, EPI_ISL_1546258, EPI_ISL_1546259, EPI_ISL_1546260, EPI_ISL_1546261, EPI_ISL_1546262, EPI_ISL_1546263, EPI_ISL_1546264, EPI_ISL_1546265, EPI_ISL_1546266, EPI_ISL_1546267, EPI_ISL_1546268, EPI_ISL_1546304, EPI_ISL_1546305, EPI_ISL_1546309, EPI_ISL_1546428, EPI_ISL_1546536, EPI_ISL_1546541, EPI_ISL_1546547, EPI_ISL_1546549, EPI_ISL_1546553, EPI_ISL_1546557, EPI_ISL_1546558, EPI_ISL_1546559, EPI_ISL_1546560, EPI_ISL_1546561, EPI_ISL_1546562, EPI_ISL_1546564, EPI_ISL_1546565, EPI_ISL_1546566, EPI_ISL_1546567, EPI_ISL_1546568, EPI_ISL_1546569, EPI_ISL_1546570, EPI_ISL_1546571, EPI_ISL_1546572, EPI_ISL_1546573, EPI_ISL_1546574, EPI_ISL_1546590, EPI_ISL_1546591, EPI_ISL_1546593, EPI_ISL_1546594, EPI_ISL_1546595, EPI_ISL_1546596, EPI_ISL_1546597, EPI_ISL_1546599, EPI_ISL_1546600, EPI_ISL_1546601, EPI_ISL_1546602, EPI_ISL_1546603, EPI_ISL_1546604, EPI_ISL_1546605, EPI_ISL_1546606, EPI_ISL_1546607, EPI_ISL_1546609, EPI_ISL_1546610, EPI_ISL_1546611, EPI_ISL_1546612, EPI_ISL_1546613, EPI_ISL_1546614, EPI_ISL_1546615, EPI_ISL_1546616, EPI_ISL_1546617, EPI_ISL_1546619, EPI_ISL_1546621, EPI_ISL_1546622, EPI_ISL_1546623, EPI_ISL_1546625, EPI_ISL_1546626, EPI_ISL_1546627, EPI_ISL_1546628, EPI_ISL_1546631, EPI_ISL_1546632, EPI_ISL_1546633, EPI_ISL_1546635, EPI_ISL_1546637, EPI_ISL_1546638, EPI_ISL_1730596, EPI_ISL_1730598, EPI_ISL_1730600, EPI_ISL_1730601, EPI_ISL_1730602, EPI_ISL_1730603, EPI_ISL_1730604, EPI_ISL_1730605, EPI_ISL_1730639, EPI_ISL_1730667, EPI_ISL_1730669, EPI_ISL_1730670, EPI_ISL_1730671, EPI_ISL_1730672, EPI_ISL_1730679, EPI_ISL_1730680, EPI_ISL_1730681, EPI_ISL_1730682, EPI_ISL_1730684, EPI_ISL_1730685, EPI_ISL_1730686, EPI_ISL_1730687, EPI_ISL_1730688, EPI_ISL_1730689, EPI_ISL_1730765, EPI_ISL_1730773, EPI_ISL_1730784, EPI_ISL_1730786, EPI_ISL_1730789

| | | | |
|---|---|---|---|
| see above | Originating lab: Wales Specialist Virology Centre Sequencing lab: Pathogen Genomics Unit | Public Health Wales Microbiology Cardiff Wales Specialist Virology Centre | Alec Birchley; Alexander Adams; Amy Gaskin; Angela Marchbank; Bree Gatica-Wilcox; Catherine Moore; Jason Coombes; Joanne Watkins; Joel Southgate; Johnathan Evans; Laura Gifford; Lauren Gilbert; Lee Graham; Malorie Perry; Matthew Bull; Nicole Pacchiarini; Sally Corden; Sara Kumziene-Summerhayes; Sara Rey; Sarah Taylor; Simon Cottrell; Sophie Jones; Tom Connor |

EPI_ISL_1730561, EPI_ISL_1730562, EPI_ISL_1730567, EPI_ISL_1730568, EPI_ISL_1730569, EPI_ISL_1730573, EPI_ISL_1730574, EPI_ISL_1730575, EPI_ISL_1730580, EPI_ISL_1730581, EPI_ISL_1730582, EPI_ISL_1730588, EPI_ISL_1730589, EPI_ISL_1730591, EPI_ISL_1730592, EPI_ISL_1730593, EPI_ISL_1979029, EPI_ISL_1979032, EPI_ISL_1979033, EPI_ISL_1979040, EPI_ISL_1979041, EPI_ISL_1979043,

| | | | |
|---|---|---|---|
| EPI_ISL_1979048, EPI_ISL_1979053, EPI_ISL_1979054, EPI_ISL_1979072, EPI_ISL_1979074, EPI_ISL_1979079, EPI_ISL_1979080, EPI_ISL_1979087, EPI_ISL_1979088, EPI_ISL_1979094, EPI_ISL_2127687 | | | |
| see above | Oxford Viromics, NDM, University of Oxford; Oxford University Hospitals; Basingstoke and North Hampshire Hospital | COVID-19 Genomics UK (COG-UK) Consortium | Alex Mobbs; Amy Trebes; Anita Justice; Catrin Moore; Christophe Fraser; David Bonsall; David Buck; Emma Wise; George Macintyre; Jessica Lynch; John Todd; Mariateresa de Cesare; Matilde Mori; Monique Andersson; Nathan Moore; Nick Cortes; Robert Shaw; Stephen Kidd; Tanya Golubchik; Timothy Peto |
| EPI_ISL_2080492 | POOLE MICROBIOLOGY LABORATORY | COVID-19 Genomics UK (COG-UK) Consortium | PHE Covid Sequencing Team |
| EPI_ISL_1832746, EPI_ISL_1832755, EPI_ISL_1832764, EPI_ISL_1832842, EPI_ISL_1832846, EPI_ISL_1832852, EPI_ISL_1832854, EPI_ISL_1832856, EPI_ISL_1832858, EPI_ISL_1832860, EPI_ISL_1832862, EPI_ISL_1832863, EPI_ISL_1832864, EPI_ISL_1832865, EPI_ISL_1832866, EPI_ISL_1832868, EPI_ISL_1832869, EPI_ISL_1832870, EPI_ISL_1832875, EPI_ISL_1832891, EPI_ISL_1832898, EPI_ISL_1832899, EPI_ISL_1832900, EPI_ISL_1832901, EPI_ISL_1832902, EPI_ISL_1832903, EPI_ISL_1832906, EPI_ISL_1832907, EPI_ISL_1832909, EPI_ISL_1832910, EPI_ISL_1832912, EPI_ISL_1832915 | | | |
| see above | Quadram Institute Bioscience | COVID-19 Genomics UK (COG-UK) Consortium | Alexander J Trotter; Alison E. Mather; Alp Aydin; Ana P. Tedim; Anastasia Kolyva; Andrew Bell; Andrew J. Page; Claire Stuart; Dave J. Baker; Gemma L. Kay; John Wain; Justin O'Grady; Leonardo de Oliveira Martins; Lizzie Meadows; Maria Diaz; Mark Webber; Muhammed Yasir; Nabil-Fareed Alikhan; Ngozi Elumogo; Nicholas M. Thomson; Rachael Stanley; Rachel Gilroy; Reenesh Prakash; Samir Dervisevic; Samuel Bloomfield; Steven Rudder; Thanh Le-Viet |
| EPI_ISL_1637237, EPI_ISL_1637238, EPI_ISL_1637239, EPI_ISL_1637240, EPI_ISL_1730411, EPI_ISL_1730412, EPI_ISL_1730413, EPI_ISL_1730414, EPI_ISL_1730415, EPI_ISL_1730416, EPI_ISL_1730417, EPI_ISL_1730418, EPI_ISL_1833026, EPI_ISL_1833027, EPI_ISL_1833028 | | | |
| see above | Queens Medical Centre, Clinical Microbiology Department / DeepSeq Nottingham | COVID-19 Genomics UK (COG-UK) Consortium | Christopher Moore; Emily Park; Fei Sang; Gemma Clark; Hannah Howson-Wells; Jack Hill; Jayasree Dey; Johnny Debebe; Jonathan Ball; Joseph Chappell; Manjinder Khakh; Matthew Carlisle; Matthew Loose; Michelle M Lister; Nadine Holmes; Patrick McClure; Theocharis Tsoleridis; Timothy Byaruhanga; Vicki M Fleming; Victoria Wright; Wendy Smith |
| EPI_ISL_1538049, EPI_ISL_1564356, EPI_ISL_1564385, EPI_ISL_1564419, EPI_ISL_1564438, EPI_ISL_1564530, EPI_ISL_1564536, EPI_ISL_1564584, EPI_ISL_1564599, EPI_ISL_1564684, EPI_ISL_1564706, EPI_ISL_1564764, EPI_ISL_1584559, EPI_ISL_1630293, EPI_ISL_1630294, EPI_ISL_1630295, EPI_ISL_1630299, EPI_ISL_1630304, EPI_ISL_1630311, EPI_ISL_1630312, EPI_ISL_1630313, EPI_ISL_1630319, EPI_ISL_1630324, EPI_ISL_1630328, EPI_ISL_1630329, EPI_ISL_1630338, EPI_ISL_1630342, EPI_ISL_1630348, EPI_ISL_1630352, EPI_ISL_1630353, EPI_ISL_1630367, EPI_ISL_1630368, EPI_ISL_1630384, EPI_ISL_1630386, EPI_ISL_1630391, EPI_ISL_1630397, EPI_ISL_1630401, EPI_ISL_1630403, EPI_ISL_1630404, EPI_ISL_1630410, EPI_ISL_1630414, EPI_ISL_1630420, EPI_ISL_1630427, EPI_ISL_1630431, EPI_ISL_1630434, EPI_ISL_1630438, EPI_ISL_1630439, EPI_ISL_1630450, EPI_ISL_1630451, EPI_ISL_1630465, EPI_ISL_1630476, EPI_ISL_1630480, EPI_ISL_1630486, EPI_ISL_1630489, EPI_ISL_1630490, EPI_ISL_1630491, EPI_ISL_1630492, EPI_ISL_1630497, EPI_ISL_1630500, EPI_ISL_1630503, EPI_ISL_1630506, EPI_ISL_1630510, EPI_ISL_1630513, EPI_ISL_1631092, EPI_ISL_1631093, EPI_ISL_1631105, EPI_ISL_1631144, EPI_ISL_1631146, EPI_ISL_1631160, EPI_ISL_1631164, EPI_ISL_1631170, EPI_ISL_1631202, EPI_ISL_1631203, EPI_ISL_1631210, EPI_ISL_1631212, EPI_ISL_1631213, EPI_ISL_1631217, EPI_ISL_1631225, EPI_ISL_1631230, EPI_ISL_1631238, EPI_ISL_1631249, EPI_ISL_1631257, EPI_ISL_1631271, EPI_ISL_1631276, EPI_ISL_1631314, EPI_ISL_1631319, EPI_ISL_1631331, EPI_ISL_1631334, EPI_ISL_1631753, EPI_ISL_1631761, EPI_ISL_1631762, EPI_ISL_1631763, EPI_ISL_1631765, EPI_ISL_1631766, EPI_ISL_1631768, EPI_ISL_1631769, EPI_ISL_1631770, EPI_ISL_1631773, EPI_ISL_1631779, EPI_ISL_1631782, EPI_ISL_1631785, EPI_ISL_1631786, EPI_ISL_1631788, EPI_ISL_1631789, EPI_ISL_1631790, EPI_ISL_1631791, EPI_ISL_1631795, EPI_ISL_1631796, EPI_ISL_1631800, EPI_ISL_1631805, EPI_ISL_1631807, EPI_ISL_1631809, EPI_ISL_1631817, EPI_ISL_1631822, EPI_ISL_1631823, EPI_ISL_1631829, EPI_ISL_1631830, EPI_ISL_1631833, EPI_ISL_1631835, EPI_ISL_1631836, EPI_ISL_1631839, EPI_ISL_1631840, EPI_ISL_1631841, EPI_ISL_1631842, EPI_ISL_1631843, EPI_ISL_1631845, EPI_ISL_1631846, EPI_ISL_1631847, EPI_ISL_1631850, EPI_ISL_1631851, EPI_ISL_1631863, EPI_ISL_1631867, EPI_ISL_1631868, EPI_ISL_1631871, EPI_ISL_1631872, EPI_ISL_1631875, EPI_ISL_1631879, EPI_ISL_1631883, EPI_ISL_1631885, EPI_ISL_1631886, EPI_ISL_1631888, EPI_ISL_1631894, EPI_ISL_1631897, EPI_ISL_1631901, EPI_ISL_1631906, EPI_ISL_1631909, EPI_ISL_1631910, EPI_ISL_1631911, EPI_ISL_1631914, EPI_ISL_1631917, EPI_ISL_1631920, EPI_ISL_1631932, EPI_ISL_1631933, EPI_ISL_1631936, EPI_ISL_1631944, EPI_ISL_1631946, EPI_ISL_1631950, EPI_ISL_1631951, EPI_ISL_1631958, EPI_ISL_1631962, EPI_ISL_1631964, EPI_ISL_1631967, EPI_ISL_1631969, EPI_ISL_1631971, EPI_ISL_1631973, EPI_ISL_1631976, EPI_ISL_1631977, EPI_ISL_1631982, EPI_ISL_1631983, EPI_ISL_1631992, EPI_ISL_1632005, EPI_ISL_1632006, EPI_ISL_1632009, EPI_ISL_1632012, EPI_ISL_1632016, EPI_ISL_1632018, EPI_ISL_1632030, EPI_ISL_1632045, EPI_ISL_1632050, EPI_ISL_1632053, EPI_ISL_1632056, EPI_ISL_1632057, EPI_ISL_1632061, EPI_ISL_1632062, EPI_ISL_1632063, EPI_ISL_1632064, EPI_ISL_1632067, EPI_ISL_1632076, EPI_ISL_1632078, EPI_ISL_1632079, EPI_ISL_1632083, EPI_ISL_1632084, EPI_ISL_1632091, EPI_ISL_1632096, EPI_ISL_1632099, EPI_ISL_1632104, EPI_ISL_1632119, EPI_ISL_1632122, EPI_ISL_1632123, EPI_ISL_1632128, EPI_ISL_1632130, EPI_ISL_1632131, EPI_ISL_1632141, EPI_ISL_1632144, EPI_ISL_1632145, EPI_ISL_1632156, EPI_ISL_1632158, EPI_ISL_1632159, EPI_ISL_1632160, EPI_ISL_1632162, EPI_ISL_1632164, EPI_ISL_1632177, EPI_ISL_1632178, EPI_ISL_1632183, EPI_ISL_1632193, EPI_ISL_1632203, EPI_ISL_1632206, EPI_ISL_1632210, EPI_ISL_1632219, EPI_ISL_1632225, EPI_ISL_1632230, EPI_ISL_1632239, EPI_ISL_1632243, EPI_ISL_1632247, EPI_ISL_1632251, EPI_ISL_1632253, EPI_ISL_1632256, EPI_ISL_1632261, EPI_ISL_1632262, EPI_ISL_1632263, EPI_ISL_1632270, EPI_ISL_1632272, EPI_ISL_1632275, EPI_ISL_1632277, EPI_ISL_1632278, EPI_ISL_1632282, EPI_ISL_1632287, EPI_ISL_1632331, EPI_ISL_1632342, EPI_ISL_1632353, EPI_ISL_1632425, EPI_ISL_1632462, EPI_ISL_1634737, EPI_ISL_1635325, EPI_ISL_1652692, EPI_ISL_1652693, EPI_ISL_1652698, EPI_ISL_1652700, EPI_ISL_1652705, EPI_ISL_1652711, EPI_ISL_1652712, EPI_ISL_1652714, EPI_ISL_1652716, EPI_ISL_1652717, EPI_ISL_1652718, EPI_ISL_1652732, EPI_ISL_1652742, EPI_ISL_1652744, EPI_ISL_1652754, EPI_ISL_1652755, EPI_ISL_1652759, EPI_ISL_1652762, EPI_ISL_1652765, EPI_ISL_1652767, EPI_ISL_1652768, EPI_ISL_1652772, EPI_ISL_1652782, EPI_ISL_1652790, EPI_ISL_1652804, EPI_ISL_1652807, EPI_ISL_1652812, EPI_ISL_1652822, EPI_ISL_1652824, EPI_ISL_1652827, EPI_ISL_1652828, EPI_ISL_1652830, EPI_ISL_1652833, EPI_ISL_1652835, EPI_ISL_1652838, EPI_ISL_1652847, EPI_ISL_1652852, EPI_ISL_1652855, EPI_ISL_1652859, EPI_ISL_1652860, EPI_ISL_1652863, EPI_ISL_1652870, EPI_ISL_1652880, EPI_ISL_1652881, EPI_ISL_1652882, EPI_ISL_1652883, EPI_ISL_1652884, EPI_ISL_1652887, EPI_ISL_1652891, EPI_ISL_1652892, EPI_ISL_1652893, EPI_ISL_1652897, EPI_ISL_1652902, EPI_ISL_1652905, EPI_ISL_1652906, EPI_ISL_1652921, EPI_ISL_1652923, EPI_ISL_1652927, EPI_ISL_1652931, EPI_ISL_1652941, EPI_ISL_1652942, EPI_ISL_1653718, EPI_ISL_1653720, EPI_ISL_1653722, EPI_ISL_1653723, EPI_ISL_1653724, EPI_ISL_1653725, EPI_ISL_1653726, EPI_ISL_1653727, EPI_ISL_1653730, EPI_ISL_1653731, EPI_ISL_1653732, EPI_ISL_1653733, EPI_ISL_1653734, EPI_ISL_1653735, EPI_ISL_1653736, EPI_ISL_1653737, EPI_ISL_1653740, EPI_ISL_1653741, EPI_ISL_1653742, EPI_ISL_1653743, EPI_ISL_1653744, EPI_ISL_1653745, EPI_ISL_1653746, EPI_ISL_1653747, EPI_ISL_1653748, EPI_ISL_1653749, EPI_ISL_1653750, EPI_ISL_1653751, EPI_ISL_1653752, EPI_ISL_1653753, EPI_ISL_1653756, EPI_ISL_1653760, EPI_ISL_1653761, EPI_ISL_1653762, EPI_ISL_1653763, EPI_ISL_1653764, EPI_ISL_1653765, EPI_ISL_1653766, EPI_ISL_1653767, EPI_ISL_1653768, EPI_ISL_1673255, EPI_ISL_1699044, EPI_ISL_1699058, EPI_ISL_1699080, EPI_ISL_1699082, EPI_ISL_1699091, EPI_ISL_1699128, EPI_ISL_1699131, EPI_ISL_1699138, EPI_ISL_1699204, EPI_ISL_1699237, EPI_ISL_1699248, EPI_ISL_1699263, EPI_ISL_1699271, EPI_ISL_1699281, EPI_ISL_1699284, EPI_ISL_1700185, EPI_ISL_1700188, EPI_ISL_1700189, EPI_ISL_1700198, EPI_ISL_1700202, EPI_ISL_1700205, EPI_ISL_1700222, EPI_ISL_1700224, EPI_ISL_1700226, EPI_ISL_1700229, EPI_ISL_1700233, EPI_ISL_1700272, EPI_ISL_1700280, EPI_ISL_1700285, EPI_ISL_1700295, EPI_ISL_1700296, EPI_ISL_1700299, EPI_ISL_1700303, EPI_ISL_1700310, EPI_ISL_1700313, EPI_ISL_1700317, EPI_ISL_1700329, EPI_ISL_1700340, EPI_ISL_1700348, EPI_ISL_1700353, EPI_ISL_1700359, EPI_ISL_1700374, EPI_ISL_1700382, EPI_ISL_1700387, EPI_ISL_1700394, EPI_ISL_1700397, EPI_ISL_1700401, EPI_ISL_1700406, EPI_ISL_1700409, EPI_ISL_1700417, EPI_ISL_1700425, EPI_ISL_1700430, EPI_ISL_1700431, EPI_ISL_1700435, EPI_ISL_1700448, EPI_ISL_1700454, EPI_ISL_1700468, EPI_ISL_1700471, EPI_ISL_1700480, EPI_ISL_1700486, EPI_ISL_1700488, EPI_ISL_1700494, EPI_ISL_1742294, EPI_ISL_1742295, EPI_ISL_1742296, EPI_ISL_1742297, EPI_ISL_1742298, EPI_ISL_1742299, EPI_ISL_1742300, EPI_ISL_1742301, EPI_ISL_1742302, EPI_ISL_1742303, EPI_ISL_1742304, EPI_ISL_1742305, EPI_ISL_1742306, EPI_ISL_1742307, EPI_ISL_1830165, EPI_ISL_1830286 | | | |
| see above | Randox Laboratories | Wellcome Sanger Institute for the COVID-19 Genomics UK (COG-UK) Consortium | Cordelia Langford; David K. Jackson; Dominic Kwiatkowski; Ewan Harrison; Ian Johnston; Jeffrey Barrett; John Sillitoe on behalf of the Wellcome Sanger Institute COVID-19 Surveillance Team; Randox Laboratories and Alex Alderton; Roberto Amato; Sonia Goncalves |
| EPI_ISL_1976555, EPI_ISL_1976557, EPI_ISL_1976559, EPI_ISL_1976563, EPI_ISL_1976568, EPI_ISL_1976570, EPI_ISL_1976572, EPI_ISL_1976573, EPI_ISL_1976575, EPI_ISL_1976577, EPI_ISL_1976579, EPI_ISL_1976580, EPI_ISL_1976582, EPI_ISL_1976584, EPI_ISL_1976586, EPI_ISL_1976589, EPI_ISL_1976590, EPI_ISL_1976594, EPI_ISL_1976596, EPI_ISL_1976599, EPI_ISL_1976601, EPI_ISL_1976602, EPI_ISL_1976604, EPI_ISL_1976606, EPI_ISL_1976609, EPI_ISL_1976611, EPI_ISL_1976612, EPI_ISL_1976616, EPI_ISL_1976619, EPI_ISL_1976621, EPI_ISL_1976623, EPI_ISL_1976626, EPI_ISL_1976628, EPI_ISL_1976631, EPI_ISL_1976633, EPI_ISL_1976635, EPI_ISL_1976636, EPI_ISL_1976638, EPI_ISL_1976640, EPI_ISL_1976642, EPI_ISL_1976644, EPI_ISL_1976645, EPI_ISL_1976647, EPI_ISL_1976650, EPI_ISL_1976652, EPI_ISL_1976655, EPI_ISL_1976657 | | | |
| see above | Regional Virus Laboratory, Belfast Health and Social Care Trust | COVID-19 Genomics UK (COG-UK) Consortium | Alison Watt; Ciara Cox; Conall McCaughey; David Simpson; Derek Fairley; James McKenna; Mairead Connor; Susan Feeney; Tanya Curran; Zoltan Molnar |
| EPI_ISL_1833822, EPI_ISL_1833824, EPI_ISL_1833825, EPI_ISL_1833826 | St George's University Hospitals NHS Foundation Trust | COVID-19 Genomics UK (COG-UK) Consortium | Adam Witney; Cassie Pope; Irene Monahan; Joshua Taylor; Ken Laing; NgeeKeong Tan |
| EPI_ISL_1546237, EPI_ISL_1637001, EPI_ISL_1637002, EPI_ISL_1637003, EPI_ISL_1637004, EPI_ISL_1637006, EPI_ISL_1637007, EPI_ISL_1637008, EPI_ISL_1637009, EPI_ISL_1637010, EPI_ISL_1637011, EPI_ISL_1637012, EPI_ISL_1637013, EPI_ISL_1637014, EPI_ISL_1637015, EPI_ISL_1637016, EPI_ISL_1637017, EPI_ISL_1637018, EPI_ISL_1637019, EPI_ISL_1637091, EPI_ISL_1637092, EPI_ISL_1637093, EPI_ISL_1637094, EPI_ISL_1637095, EPI_ISL_1637096, EPI_ISL_1637097, EPI_ISL_1637098, EPI_ISL_1637099, EPI_ISL_1637177, EPI_ISL_1637181, EPI_ISL_1730326, EPI_ISL_1730328, EPI_ISL_1730329, EPI_ISL_1730342, EPI_ISL_1730343, EPI_ISL_1730344, EPI_ISL_1730345, EPI_ISL_1730369, EPI_ISL_1730370, EPI_ISL_1730371, EPI_ISL_1730372, EPI_ISL_1730373, EPI_ISL_1730377, EPI_ISL_1730379, EPI_ISL_1730380, EPI_ISL_1730382, EPI_ISL_1730384, EPI_ISL_1730386, EPI_ISL_1730387, EPI_ISL_1730388, EPI_ISL_1730389, EPI_ISL_1730390 | | | |
| see above | University College London, Great Ormond Street Hospital for Children NHS Foundation Trust, Imperial College Healthcare NHS Trust | COVID-19 Genomics UK (COG-UK) Consortium | Alison Holmes; Charlotte Williams; Helena Tutill; Jacqueline Findlay; James Price; Judith Breuer; Julianne Brown; Kathryn Harris; Leysa Forrest; Mark Kristiansen; Paola Niola; Paola Resende Silva; Patricia Dyal; Paul Randell; Rachel Williams; Samuel Weeks; Sergi Castellano; Sunando Roy; Tony Brooks; Yasmin Panchbhaya |
| EPI_ISL_1636721, EPI_ISL_1636723, EPI_ISL_1730304 | University Hospitals Of Leicester NHS Trust and DeepSeq Nottingham | COVID-19 Genomics UK (COG-UK) Consortium | Christopher Holmes; Christopher Moore; Fei Sang; Joeseph Chappell; Johnny Debebe; Jonathan Ball; Julian Tang; Karlie Fallon; Matthew Carlisle; Matthew Loose; Nadine Holmes; Patrick McClure; Paul Bird; Thomas Helmer; Victoria Wright |
| EPI_ISL_1636529, EPI_ISL_1636530, EPI_ISL_1636531, EPI_ISL_1636538, EPI_ISL_1636539, EPI_ISL_1636546, EPI_ISL_1636548, EPI_ISL_1636549, EPI_ISL_1636574, EPI_ISL_1636577, EPI_ISL_1636592, EPI_ISL_1636597, EPI_ISL_1636598, EPI_ISL_1636600, EPI_ISL_1729990, EPI_ISL_1729991, EPI_ISL_1729992, EPI_ISL_1729994, EPI_ISL_1729996, EPI_ISL_1729998, EPI_ISL_1730000, EPI_ISL_1730009, EPI_ISL_1730016 | | | |
| see above | University of Birmingham | COVID-19 Genomics UK (COG-UK) Consortium | Alex Richter; Andrew Bosworth; Andrew Bosworth. Queen Elizabeth Hospital: Anna Casey; Andrew D Beggs PHE Heartlands Lab: Husam Osman; Charlotte Poxon; Institute of Microbiology; Joanne Stockton; Josh Quick; Kasun Wanigasooriya; Mike Kidd; Nicholas Loman. University of Birmingham Testing Laboratory: Celina M Whalley; Oliver Pickles; Radoslaw Poplawski; Samuel Nicholls; University of Birmingham: Claire McMurray; Will Rowe |
| EPI_ISL_1730149, EPI_ISL_1730150, EPI_ISL_1730151, EPI_ISL_1730152, EPI_ISL_1730153 | University of Exeter | COVID-19 Genomics UK (COG-UK) Consortium | Aaron Jeffries; Audrey Farbos; Ben Temperton; Jane Masoli; Joanna Warwick-Dugdale; Michelle Michelsen; Robyn Manley; Stephen Michell |
| EPI_ISL_1636697, EPI_ISL_1636700, EPI_ISL_1636705, EPI_ISL_1636706, EPI_ISL_1636707, EPI_ISL_1636708, EPI_ISL_1730106, EPI_ISL_1730107, EPI_ISL_1730108, EPI_ISL_1730109, EPI_ISL_1730110, EPI_ISL_1730111, EPI_ISL_1730112, EPI_ISL_1730113, EPI_ISL_1730114, EPI_ISL_1730115, EPI_ISL_1730116, EPI_ISL_1730117, EPI_ISL_1730118, EPI_ISL_1730119, EPI_ISL_1730124, EPI_ISL_1730125, EPI_ISL_1730126, EPI_ISL_1730130, EPI_ISL_1832248, EPI_ISL_1832272, EPI_ISL_1974814, EPI_ISL_1974890, EPI_ISL_1974898, EPI_ISL_2125694, EPI_ISL_2125696, EPI_ISL_2125701, EPI_ISL_2125878, EPI_ISL_2125879, EPI_ISL_2125881 | | | |
| see above | Virology Department, Royal Infirmary of Edinburgh, NHS Lothian / School of Biological Sciences, University of Edinburgh | COVID-19 Genomics UK (COG-UK) Consortium | Colquhoun R; Cotton S; Dewar R; Gallagher A; Hill V; Jackson B; Maloney D; McCrone JT; McHugh M; O'Toole Á; Rambaut A; Rooke S; Scher E; Templeton K; Yu X |
| EPI_ISL_1547319, EPI_ISL_1547321, EPI_ISL_1547324, EPI_ISL_1547336, EPI_ISL_1638036, EPI_ISL_1638037, EPI_ISL_1638039, EPI_ISL_1638040, EPI_ISL_1638042, EPI_ISL_1638043, EPI_ISL_1638044, EPI_ISL_1638045, EPI_ISL_1638046, EPI_ISL_1638047, EPI_ISL_1638048, EPI_ISL_1638049, EPI_ISL_1638050, EPI_ISL_1638053, EPI_ISL_1638054, EPI_ISL_1638055, EPI_ISL_1638056, EPI_ISL_1638057, EPI_ISL_1638059, EPI_ISL_1638061, EPI_ISL_1638065, EPI_ISL_1638066, EPI_ISL_1638067, EPI_ISL_1638071, EPI_ISL_1638072, EPI_ISL_1638073, EPI_ISL_1638077, EPI_ISL_1638078, EPI_ISL_1638080, EPI_ISL_1730939, EPI_ISL_1730940, EPI_ISL_1730946, EPI_ISL_1730950, EPI_ISL_1730953, EPI_ISL_1730954, EPI_ISL_1730957, EPI_ISL_1730959, EPI_ISL_1730961, EPI_ISL_1730962, EPI_ISL_1730963, EPI_ISL_1730972, EPI_ISL_1730975, EPI_ISL_1730981, EPI_ISL_1730984, EPI_ISL_1730988, EPI_ISL_1730989, EPI_ISL_1730990, EPI_ISL_1730992, EPI_ISL_1730993, EPI_ISL_1730994, EPI_ISL_1730996, EPI_ISL_1730998, EPI_ISL_1730999, EPI_ISL_1731005, EPI_ISL_1731009, EPI_ISL_1731010, EPI_ISL_1731011, EPI_ISL_1731016, EPI_ISL_1731018, EPI_ISL_1731019, EPI_ISL_1731021, EPI_ISL_1731022, EPI_ISL_1731025, EPI_ISL_1731028, EPI_ISL_1731030, EPI_ISL_1731031, EPI_ISL_1731032, EPI_ISL_1731033, EPI_ISL_1731039, EPI_ISL_1833835, EPI_ISL_1833856, EPI_ISL_1833878 | | | |
| see above | Virology Department, Sheffield Teaching Hospitals NHS Foundation Trust/Department of Infection, Immunity and Cardiovascular Disease, The Medical School, University of Sheffield | COVID-19 Genomics UK (COG-UK) Consortium | Adri Angyal; Alex Keeley; Benjamin Lindsey; Cariad Evans; Danielle Groves; Dave Partridge; Katie Johnson; Laura Carrilero; Luke Green; Matthew Parker; Matthew Wyles; Mehmet Yavuz; Mohammad Raza; Nikki Smith; Paul Parsons; Rachel Tucker; Rebecca Brown; Thushan de Silva |
| EPI_ISL_1546252, EPI_ISL_1546256, EPI_ISL_1546427, EPI_ISL_1546471, EPI_ISL_1546548, EPI_ISL_1546550, EPI_ISL_1546551, EPI_ISL_1546552, EPI_ISL_1546555, EPI_ISL_1546592, EPI_ISL_1546624, EPI_ISL_1546634, EPI_ISL_1546636, EPI_ISL_1637458, EPI_ISL_1637459, EPI_ISL_1637460, EPI_ISL_1637461, EPI_ISL_1637462, EPI_ISL_1637463, EPI_ISL_1637464, EPI_ISL_1637465, EPI_ISL_1637466, EPI_ISL_1637467, EPI_ISL_1637468, EPI_ISL_1637469, EPI_ISL_1637470, EPI_ISL_1637471, EPI_ISL_1637472, EPI_ISL_1637473, EPI_ISL_1637474, EPI_ISL_1637475, EPI_ISL_1637476, EPI_ISL_1637477, EPI_ISL_1637478, EPI_ISL_1637479, EPI_ISL_1637480, EPI_ISL_1637481, EPI_ISL_1637482, EPI_ISL_1637483, EPI_ISL_1637484, EPI_ISL_1637485, EPI_ISL_1637486, EPI_ISL_1637488, EPI_ISL_1637489, EPI_ISL_1637491, EPI_ISL_1637492, EPI_ISL_1637494, EPI_ISL_1637495, EPI_ISL_1637496, EPI_ISL_1637497, EPI_ISL_1637498, EPI_ISL_1637499, EPI_ISL_1637501, EPI_ISL_1637505, EPI_ISL_1637506, EPI_ISL_1637507, EPI_ISL_1637508, EPI_ISL_1637509, EPI_ISL_1637511, EPI_ISL_1637513, EPI_ISL_1637515, EPI_ISL_1637516, EPI_ISL_1637517, EPI_ISL_1637518, EPI_ISL_1637519, EPI_ISL_1637520, EPI_ISL_1637521, EPI_ISL_1637522, EPI_ISL_1637524, EPI_ISL_1637525, EPI_ISL_1637526, EPI_ISL_1637527, EPI_ISL_1637528, EPI_ISL_1637530, EPI_ISL_1637531, EPI_ISL_1637532, EPI_ISL_1637533, EPI_ISL_1637534, EPI_ISL_1637535, EPI_ISL_1637536, EPI_ISL_1637537, EPI_ISL_1637540, EPI_ISL_1637541, EPI_ISL_1637543, EPI_ISL_1637544, EPI_ISL_1637547, EPI_ISL_1637552, EPI_ISL_1637553, EPI_ISL_1637554, EPI_ISL_1637555, EPI_ISL_1637556, EPI_ISL_1637557, EPI_ISL_1637558, EPI_ISL_1637559, EPI_ISL_1637560, EPI_ISL_1637561, EPI_ISL_1637562, EPI_ISL_1637563, EPI_ISL_1637564, EPI_ISL_1637565, EPI_ISL_1637566, EPI_ISL_1637567, EPI_ISL_1637568, EPI_ISL_1637569, EPI_ISL_1637570, EPI_ISL_1637571, EPI_ISL_1637572, EPI_ISL_1637573, EPI_ISL_1637574, EPI_ISL_1637575, EPI_ISL_1637576, EPI_ISL_1637577, EPI_ISL_1637578, EPI_ISL_1637579, EPI_ISL_1637580, EPI_ISL_1637581, EPI_ISL_1637603, EPI_ISL_1637604, EPI_ISL_1637605, EPI_ISL_1637606, EPI_ISL_1637610, EPI_ISL_1637611, EPI_ISL_1637614, EPI_ISL_1637615, EPI_ISL_1637616, EPI_ISL_1637619, EPI_ISL_1637621, EPI_ISL_1637622, EPI_ISL_1637627, EPI_ISL_1637628, EPI_ISL_1637629, EPI_ISL_1637630, EPI_ISL_1637631, EPI_ISL_1637632, EPI_ISL_1637633, EPI_ISL_1637634, EPI_ISL_1637635, EPI_ISL_1637636, EPI_ISL_1637637, EPI_ISL_1637638, EPI_ISL_1637639, EPI_ISL_1637640, EPI_ISL_1637641, EPI_ISL_1637642, EPI_ISL_1637643, EPI_ISL_1637644, EPI_ISL_1637645, EPI_ISL_1637646, EPI_ISL_1637647, EPI_ISL_1637648, EPI_ISL_1637649, EPI_ISL_1637650, EPI_ISL_1637651, EPI_ISL_1637652, EPI_ISL_1637653, EPI_ISL_1637668, EPI_ISL_1637669, EPI_ISL_1637670, EPI_ISL_1637671, EPI_ISL_1637672, EPI_ISL_1637673, EPI_ISL_1637674, EPI_ISL_1637675, EPI_ISL_1637676, EPI_ISL_1637677, EPI_ISL_1637678, EPI_ISL_1637679, EPI_ISL_1637680, EPI_ISL_1637681, EPI_ISL_1637682, EPI_ISL_1637683, EPI_ISL_1637684, EPI_ISL_1637685, EPI_ISL_1637686, EPI_ISL_1637687, EPI_ISL_1637688, EPI_ISL_1637689, EPI_ISL_1637690, EPI_ISL_1637691, EPI_ISL_1637692, EPI_ISL_1637693, EPI_ISL_1637694, EPI_ISL_1637695, EPI_ISL_1637696, EPI_ISL_1637698, EPI_ISL_1637699, EPI_ISL_1637700, EPI_ISL_1637701, EPI_ISL_1637702, EPI_ISL_1637703, EPI_ISL_1637704, EPI_ISL_1637705, EPI_ISL_1637706, EPI_ISL_1637707, EPI_ISL_1637708, EPI_ISL_1637709, EPI_ISL_1637710, EPI_ISL_1637711, EPI_ISL_1637712, EPI_ISL_1637713, EPI_ISL_1637714, EPI_ISL_1637715, EPI_ISL_1637716, EPI_ISL_1637717, EPI_ISL_1637718, EPI_ISL_1637719, EPI_ISL_1637720, EPI_ISL_1637721, EPI_ISL_1637722, EPI_ISL_1637723, EPI_ISL_1637724, EPI_ISL_1637725, EPI_ISL_1637726, EPI_ISL_1637727, EPI_ISL_1637728, EPI_ISL_1637729, EPI_ISL_1637730, EPI_ISL_1637731, EPI_ISL_1637732, EPI_ISL_1637733, EPI_ISL_1637734, EPI_ISL_1637735, EPI_ISL_1637736, EPI_ISL_1637737, EPI_ISL_1637738, EPI_ISL_1637739, EPI_ISL_1637740, EPI_ISL_1637741, EPI_ISL_1637743, EPI_ISL_1637744, EPI_ISL_1637745, EPI_ISL_1637746, EPI_ISL_1637747, EPI_ISL_1637748, EPI_ISL_1637749, EPI_ISL_1637750, EPI_ISL_1637751, EPI_ISL_1637752, EPI_ISL_1637753, EPI_ISL_1637754, EPI_ISL_1637755, EPI_ISL_1637756, EPI_ISL_1637757, EPI_ISL_1637758, EPI_ISL_1637759, EPI_ISL_1637760, EPI_ISL_1637761, EPI_ISL_1637762, EPI_ISL_1637763, EPI_ISL_1637764, EPI_ISL_1637765, EPI_ISL_1637766, EPI_ISL_1637767, EPI_ISL_1637768, EPI_ISL_1833186, EPI_ISL_1833190, EPI_ISL_1833200 | | | |
| see above | Wales Specialist Virology Centre Sequencing lab: Pathogen Genomics Unit | Public Health Wales Microbiology Cardiff Wales Specialist Virology Centre | Alec Birchley; Alexander Adams; Amy Gaskin; Angela Marchbank; Bree Gatica-Wilcox; Catherine Moore; Jason Coombes; Joanne Watkins; Joel Southgate; Johnathan Evans; Laura Gifford; Lauren Gilbert; Lee Graham; Malorie Perry; Matthew Bull; Nicole Pacchiarini; Sally Corden; Sara Kumziene-Summerhayes; Sara Rey; Sarah Taylor; Simon Cottrell; Sophie Jones; Tom Connor |
| EPI_ISL_1832281, EPI_ISL_1832282, EPI_ISL_1832283, EPI_ISL_1832284, EPI_ISL_1832285, EPI_ISL_1832286, EPI_ISL_1832288, EPI_ISL_1832289, EPI_ISL_1832290, EPI_ISL_1832291, EPI_ISL_1832292, EPI_ISL_1832293, EPI_ISL_1832294, EPI_ISL_1832295, EPI_ISL_1832296, EPI_ISL_1832298, EPI_ISL_1832299 | | | |
| see above | West of Scotland Specialist Virology Centre, NHSGGC / MRC-University of Glasgow Centre for Virus Research | COVID-19 Genomics UK (COG-UK) Consortium | Alasdair MacLean; Alice Broos; Ana da Silva Filipe; Antonia Ho; Daniel Mair; David L Robertson; Emma Thomson; Guy Mollett; James Shepherd; Jenna Nichols; Joseph Hughes; Kathy Li; Kathy Smollett; Kyriaki Nomikou; Lily Tong; Matthew Holden; Natasha Johnson; Rachel Blacow; Richard Orton; Rory Gunson; Sarah McDonald; Sharif Shaaban; Sreenu Vattipally; Stephen Carmichael |

We gratefully acknowledge the following Authors from the Originating laboratories responsible for obtaining the specimens, as well as the Submitting laboratories where the genome data were generated and shared via GISAID, on which this research is based.

All Submitters of data may be contacted directly via www.gisaid.org

Authors are sorted alphabetically.

| Accession ID | Originating Laboratory | Submitting Laboratory | Authors |
|---|---|---|---|
| EPI_ISL_1730321, EPI_ISL_1730323, EPI_ISL_1832539, EPI_ISL_1832543, EPI_ISL_1832545, EPI_ISL_1832547, EPI_ISL_1832549, EPI_ISL_1832551, EPI_ISL_1832553, EPI_ISL_1832555, EPI_ISL_1832557, EPI_ISL_1832559, EPI_ISL_1832562, EPI_ISL_1832564, EPI_ISL_1832566, EPI_ISL_1832568, EPI_ISL_1832570, EPI_ISL_1975069, EPI_ISL_1975070, EPI_ISL_1975072, EPI_ISL_1975074, EPI_ISL_1975075, EPI_ISL_1975101 | | | |
| see above | Barts Health NHS Trust | COVID-19 Genomics UK (COG-UK) Consortium | BROAD; Beatrix; CUTINO-MOGUEL; Claire; David; Dola; HARRINGTON; KELE; KULASEGARAN-SHYLINI; Maria-Teresa; OWOYEMI; Raghavendran |
| EPI_ISL_1833923, EPI_ISL_1833931, EPI_ISL_1833933, EPI_ISL_1833947 | Bioinformatics and Biostatistics Lab, Advanced Sequencing Facility | COVID-19 Genomics UK (COG-UK) Consortium | Aengus Stewart; Chelsea Sawyer; Harshil Patel; Jerome Nicod; Laura Cubitt; Margaret Crawford |
| EPI_ISL_1730896, EPI_ISL_1730898, EPI_ISL_1730899, EPI_ISL_1730901, EPI_ISL_1730902, EPI_ISL_1730903, EPI_ISL_1730904, EPI_ISL_1730908, EPI_ISL_1730909, EPI_ISL_1730910, EPI_ISL_1730912, EPI_ISL_1730922, EPI_ISL_1730923, EPI_ISL_1730924, EPI_ISL_1833279, EPI_ISL_1833287, EPI_ISL_1833340, EPI_ISL_1833347, EPI_ISL_1833349, EPI_ISL_1833350, EPI_ISL_2127895 | | | |
| see above | Centre for Enzyme Innovation, University of Portsmouth / Translational Research Laboratory, Portsmouth Hospitals NHS Trust | COVID-19 Genomics UK (COG-UK) Consortium | Angela Beckett; Christopher Fearn; Kate Cook; Katie Loveson; Salman Goudarzi; Samuel Robson; Scott Elliott; Sharon Glaysher |
| EPI_ISL_1636625, EPI_ISL_1730047, EPI_ISL_1730064, EPI_ISL_1730065, EPI_ISL_1730067, EPI_ISL_1730068, EPI_ISL_1730070, EPI_ISL_1730071, EPI_ISL_1730072, EPI_ISL_1832176, EPI_ISL_1832178, EPI_ISL_1832180, EPI_ISL_1832182, EPI_ISL_1832184, EPI_ISL_1832187, EPI_ISL_1832189, EPI_ISL_1832191, EPI_ISL_1832193, EPI_ISL_1832194, EPI_ISL_1832195, EPI_ISL_1832196, EPI_ISL_1832197, EPI_ISL_1832198, EPI_ISL_1832199, EPI_ISL_1832200, EPI_ISL_1832201, EPI_ISL_1832202, EPI_ISL_1974789 | | | |
| see above | Department of Pathology, University of Cambridge | COVID-19 Genomics UK (COG-UK) Consortium | Aminu S. Jahun; Ian Goodfellow; Iliana Georgana; Martin D. Curran; Myra Hosmillo; Rhys Izuagbe; Surendra Parmar; William L. Hamilton; Yasmin Chaudhry |
| EPI_ISL_1831284, EPI_ISL_1831285, EPI_ISL_1831286, EPI_ISL_1831287, EPI_ISL_1831288, EPI_ISL_1831289, EPI_ISL_1831290, EPI_ISL_1831291, EPI_ISL_1831292, EPI_ISL_1831293, EPI_ISL_1831294, EPI_ISL_1831295, EPI_ISL_1831296, EPI_ISL_1831297, EPI_ISL_1831298, EPI_ISL_1831299, EPI_ISL_1831300, EPI_ISL_1831301, EPI_ISL_1831302, EPI_ISL_1831303, EPI_ISL_1831304, EPI_ISL_1831305, EPI_ISL_1831306, EPI_ISL_1831307, EPI_ISL_1831308, EPI_ISL_1831309, EPI_ISL_1831310, EPI_ISL_1831311, EPI_ISL_1831312, EPI_ISL_1831314, EPI_ISL_1831316, EPI_ISL_1831318, EPI_ISL_1831319, EPI_ISL_1831320, EPI_ISL_1831321, EPI_ISL_1831324, EPI_ISL_1831325, EPI_ISL_1831326, EPI_ISL_1831327, EPI_ISL_1831328, EPI_ISL_1831331, EPI_ISL_1831332, EPI_ISL_1831333, EPI_ISL_1831335, EPI_ISL_1831336, EPI_ISL_1831337, EPI_ISL_1831338, EPI_ISL_1831339, EPI_ISL_1831340, EPI_ISL_1831341, EPI_ISL_1831343, EPI_ISL_1831346, EPI_ISL_1831347, EPI_ISL_1831351, EPI_ISL_1831352, EPI_ISL_1831353, EPI_ISL_1831355, EPI_ISL_1831356, EPI_ISL_1831358, EPI_ISL_1831359, EPI_ISL_1831360, EPI_ISL_1831361, EPI_ISL_1831363, EPI_ISL_1831365, EPI_ISL_1831366, EPI_ISL_1831367, EPI_ISL_1831368, EPI_ISL_1831369, EPI_ISL_1831370, EPI_ISL_1831371, EPI_ISL_1831372, EPI_ISL_1831373, EPI_ISL_1831374, EPI_ISL_1831375, EPI_ISL_1831377, EPI_ISL_1831378, EPI_ISL_1831379, EPI_ISL_1831380, EPI_ISL_1831384, EPI_ISL_1831386, EPI_ISL_2006541, EPI_ISL_2006545, EPI_ISL_2006546, EPI_ISL_2006547, EPI_ISL_2006549, EPI_ISL_2006553, EPI_ISL_2006554, EPI_ISL_2006555, EPI_ISL_2006557, EPI_ISL_2006558, EPI_ISL_2006559, EPI_ISL_2006560, EPI_ISL_2006561, EPI_ISL_2006562, EPI_ISL_2006564, EPI_ISL_2006568, EPI_ISL_2006570, EPI_ISL_2006571, EPI_ISL_2006572, EPI_ISL_2006573, EPI_ISL_2006574, EPI_ISL_2006575, EPI_ISL_2006576, EPI_ISL_2006579, EPI_ISL_2006581, EPI_ISL_2006583, EPI_ISL_2006585, EPI_ISL_2006587, EPI_ISL_2006588, EPI_ISL_2006590, EPI_ISL_2006593, EPI_ISL_2006596, EPI_ISL_2006598, EPI_ISL_2006601, EPI_ISL_2006604, EPI_ISL_2006605, EPI_ISL_2006606, EPI_ISL_2006611, EPI_ISL_2006615, EPI_ISL_2006617, EPI_ISL_2006622, EPI_ISL_2006624, EPI_ISL_2006625, EPI_ISL_2006627, EPI_ISL_2006628, EPI_ISL_2006629, EPI_ISL_2006630, EPI_ISL_2006633, EPI_ISL_2006634, EPI_ISL_2006635, EPI_ISL_2006638, EPI_ISL_2006639, EPI_ISL_2006641, EPI_ISL_2006643, EPI_ISL_2006647, EPI_ISL_2006650, EPI_ISL_2006651, EPI_ISL_2006654, EPI_ISL_2006655, EPI_ISL_2006657, EPI_ISL_2006658, EPI_ISL_2006660, EPI_ISL_2006663, EPI_ISL_2006665 | | | |
| see above | Health Services Laboratories | Wellcome Sanger Institute for the COVID-19 Genomics UK (COG-UK) Consortium | Cordelia Langford; David K. Jackson; Dominic Kwiatkowski; Ewan Harrison; Health Services Laboratories and Alex Alderton; Ian Johnston; Jeffrey Barrett; John Sillitoe on behalf of the Wellcome Sanger Institute COVID-19 Surveillance Team; Roberto Amato; Sonia Goncalves |
| EPI_ISL_1652715, EPI_ISL_1652725, EPI_ISL_1652757, EPI_ISL_1652886, EPI_ISL_1652908, EPI_ISL_1697444, EPI_ISL_1697445, EPI_ISL_1697447, EPI_ISL_1697449, EPI_ISL_1697450, EPI_ISL_1697451, EPI_ISL_1697452, EPI_ISL_1697454, EPI_ISL_1697457, EPI_ISL_1697458, EPI_ISL_1697459, EPI_ISL_1697460, EPI_ISL_1697461, EPI_ISL_1697462, EPI_ISL_1697463, EPI_ISL_1697464, EPI_ISL_1697467, EPI_ISL_1697470, EPI_ISL_1697471, EPI_ISL_1697473, EPI_ISL_1697474, EPI_ISL_1697476, EPI_ISL_1697477, EPI_ISL_1697478, EPI_ISL_1697479, EPI_ISL_1697481, EPI_ISL_1697483, EPI_ISL_1697488, EPI_ISL_1697489, EPI_ISL_1697490, EPI_ISL_1697492, EPI_ISL_1697499, EPI_ISL_1697500, EPI_ISL_1697501, EPI_ISL_1697502, EPI_ISL_1697504, EPI_ISL_1697505, EPI_ISL_1697507, EPI_ISL_1697509, EPI_ISL_1697510, EPI_ISL_1697512, EPI_ISL_1697513, EPI_ISL_1697515, EPI_ISL_1697520, EPI_ISL_1697521, EPI_ISL_1697524, EPI_ISL_1697528, EPI_ISL_1697533, EPI_ISL_1697535, EPI_ISL_1697537, EPI_ISL_1697540, EPI_ISL_1697541, EPI_ISL_1697542, EPI_ISL_1697545, EPI_ISL_1697548, EPI_ISL_1697550, EPI_ISL_1697551, EPI_ISL_1697553, EPI_ISL_1697555, EPI_ISL_1697556, EPI_ISL_1697557, EPI_ISL_1697558, EPI_ISL_1697559, EPI_ISL_1697561, EPI_ISL_1697562, EPI_ISL_1697563, EPI_ISL_1697567, EPI_ISL_1697569, EPI_ISL_1697570, EPI_ISL_1697572, EPI_ISL_1697574, EPI_ISL_1697575, EPI_ISL_1697581, EPI_ISL_1697582, EPI_ISL_1697583, EPI_ISL_1697584, EPI_ISL_1697585, EPI_ISL_1697586, EPI_ISL_1697589, EPI_ISL_1697590, EPI_ISL_1697593, EPI_ISL_1697597, EPI_ISL_1697598, EPI_ISL_1697600, EPI_ISL_1697604, EPI_ISL_1697605, EPI_ISL_1697608, EPI_ISL_1697609, EPI_ISL_1697610, EPI_ISL_1697611, EPI_ISL_1697612, EPI_ISL_1697614, EPI_ISL_1697615, EPI_ISL_1697616, EPI_ISL_1697618, EPI_ISL_1697621, EPI_ISL_1697624, EPI_ISL_1697626, EPI_ISL_1697628, EPI_ISL_1697629, EPI_ISL_1697631, EPI_ISL_1697633, EPI_ISL_1697634, EPI_ISL_1697637, EPI_ISL_1697638, EPI_ISL_1697641, EPI_ISL_1697642, EPI_ISL_1697646, EPI_ISL_1697647, EPI_ISL_1697649, EPI_ISL_1697651, EPI_ISL_1697653, EPI_ISL_1697658, EPI_ISL_1697659, EPI_ISL_1697661, EPI_ISL_1697662, EPI_ISL_1697663, EPI_ISL_1697665, EPI_ISL_1697668, EPI_ISL_1697669, EPI_ISL_1697670, EPI_ISL_1697671, EPI_ISL_1697672, EPI_ISL_1697675, EPI_ISL_1697676, EPI_ISL_1697679, EPI_ISL_1697680, EPI_ISL_1697681, EPI_ISL_1697682, EPI_ISL_1697683, EPI_ISL_1697686, EPI_ISL_1697687, EPI_ISL_1697688, EPI_ISL_1697692, EPI_ISL_1697694, EPI_ISL_1697697, EPI_ISL_1697699, EPI_ISL_1697700, EPI_ISL_1697701, EPI_ISL_1697703, EPI_ISL_1697705, EPI_ISL_1697706, EPI_ISL_1697708, EPI_ISL_1697712, EPI_ISL_1697713, EPI_ISL_1697714, EPI_ISL_1697715, EPI_ISL_1697716, EPI_ISL_1697720, EPI_ISL_1697721, EPI_ISL_1697724, EPI_ISL_1697727, EPI_ISL_1697730, EPI_ISL_1697731, EPI_ISL_1697732, EPI_ISL_1697737, EPI_ISL_1697738, EPI_ISL_1697739, EPI_ISL_1697740, EPI_ISL_1697741, EPI_ISL_1697742, EPI_ISL_1697971, EPI_ISL_1697980, EPI_ISL_1697998, EPI_ISL_1698019, EPI_ISL_1698026, EPI_ISL_1698030, EPI_ISL_1698045, EPI_ISL_1698049, EPI_ISL_1698056, EPI_ISL_1698071, EPI_ISL_1698075, EPI_ISL_1698080, EPI_ISL_1698091, EPI_ISL_1698100, EPI_ISL_1698117, EPI_ISL_1698130, EPI_ISL_1698146, EPI_ISL_1698157, EPI_ISL_1698165, EPI_ISL_1698169, EPI_ISL_1698173, EPI_ISL_1698186, EPI_ISL_1698188, EPI_ISL_1698190, EPI_ISL_1698203, EPI_ISL_1698204, EPI_ISL_1698205, EPI_ISL_1698209, EPI_ISL_1698213, EPI_ISL_1698219, EPI_ISL_1698227, EPI_ISL_1698229, EPI_ISL_1698231, EPI_ISL_1698247, EPI_ISL_1698252, EPI_ISL_1698254, EPI_ISL_1698256, EPI_ISL_1698257, EPI_ISL_1698259, EPI_ISL_1698263, EPI_ISL_1698265, EPI_ISL_1698266, EPI_ISL_1698269, EPI_ISL_1698271, EPI_ISL_1698275, EPI_ISL_1698276, EPI_ISL_1698278, EPI_ISL_1698281, EPI_ISL_1698282, EPI_ISL_1698286, EPI_ISL_1698287, EPI_ISL_1698288, EPI_ISL_1698289, EPI_ISL_1698290, EPI_ISL_1698291, EPI_ISL_1698292, EPI_ISL_1698293, EPI_ISL_1698294, EPI_ISL_1698296, EPI_ISL_1698297, EPI_ISL_1698298, EPI_ISL_1698299, EPI_ISL_1698300, EPI_ISL_1698301, EPI_ISL_1698302, EPI_ISL_1698303, EPI_ISL_1698304, EPI_ISL_1698306, EPI_ISL_1698308, EPI_ISL_1698309, EPI_ISL_1698310, EPI_ISL_1698311, EPI_ISL_1698312, EPI_ISL_1698313, EPI_ISL_1698316, EPI_ISL_1698318, EPI_ISL_1698319, EPI_ISL_1698320, EPI_ISL_1698323, EPI_ISL_1698325, EPI_ISL_1698326, EPI_ISL_1698329, EPI_ISL_1698330, EPI_ISL_1698331, EPI_ISL_1698333, EPI_ISL_1698334, EPI_ISL_1698335, EPI_ISL_1698336, EPI_ISL_1698337, EPI_ISL_1698338, EPI_ISL_1698340, EPI_ISL_1698343, EPI_ISL_1698344, EPI_ISL_1698345, EPI_ISL_1698348, EPI_ISL_1698350, EPI_ISL_1698351, EPI_ISL_1698352, EPI_ISL_1698353, EPI_ISL_1698354, EPI_ISL_1698355, EPI_ISL_1698356, EPI_ISL_1698358, EPI_ISL_1698360, EPI_ISL_1698362, EPI_ISL_1698363, EPI_ISL_1698365, EPI_ISL_1698367, EPI_ISL_1698368, EPI_ISL_1698369, EPI_ISL_1698371, EPI_ISL_1698372, EPI_ISL_1698374, EPI_ISL_1698375, EPI_ISL_1698376, EPI_ISL_1698377, EPI_ISL_1698380, EPI_ISL_1698381, EPI_ISL_1698382, EPI_ISL_1698385, EPI_ISL_1698386, EPI_ISL_1698388, EPI_ISL_1698389, EPI_ISL_1698390, EPI_ISL_1698395, EPI_ISL_1698396, EPI_ISL_1698397, EPI_ISL_1698399, EPI_ISL_1698400, EPI_ISL_1698401, EPI_ISL_1698402, EPI_ISL_1698404, EPI_ISL_1698408, EPI_ISL_1698411, EPI_ISL_1698413, EPI_ISL_1698414, EPI_ISL_1698415, EPI_ISL_1698416, EPI_ISL_1698419, EPI_ISL_1698420, EPI_ISL_1698421, EPI_ISL_1698422, EPI_ISL_1698423, EPI_ISL_1698424, EPI_ISL_1698425, EPI_ISL_1698427, EPI_ISL_1698433, EPI_ISL_1698434, EPI_ISL_1698437, EPI_ISL_1698438, EPI_ISL_1698440, EPI_ISL_1698441, EPI_ISL_1698443, EPI_ISL_1698445, EPI_ISL_1698446, EPI_ISL_1698448, EPI_ISL_1698449, EPI_ISL_1698451, EPI_ISL_1698453, EPI_ISL_1698454, EPI_ISL_1698455, EPI_ISL_1698456, EPI_ISL_1698457, EPI_ISL_1698460, EPI_ISL_1698462, EPI_ISL_1698464, EPI_ISL_1698466, EPI_ISL_1698467, EPI_ISL_1698468, EPI_ISL_1698470, EPI_ISL_1698471, EPI_ISL_1698473, EPI_ISL_1698474, EPI_ISL_1698477, EPI_ISL_1698478, EPI_ISL_1698480, EPI_ISL_1698481, EPI_ISL_1698483, EPI_ISL_1698484, EPI_ISL_1698485, EPI_ISL_1698488, EPI_ISL_1698489, EPI_ISL_1698491, EPI_ISL_1698494, EPI_ISL_1698495, EPI_ISL_1698496, EPI_ISL_1698497, EPI_ISL_1698499, EPI_ISL_1698500, EPI_ISL_1698503, EPI_ISL_1698507, EPI_ISL_1698509, EPI_ISL_1698513, EPI_ISL_1698514, EPI_ISL_1698515, EPI_ISL_1698516, EPI_ISL_1698517, EPI_ISL_1698518, EPI_ISL_1698519, EPI_ISL_1698522, EPI_ISL_1698523, EPI_ISL_1698525, EPI_ISL_1698528, EPI_ISL_1698529, EPI_ISL_1698530, EPI_ISL_1698531, EPI_ISL_1698535, EPI_ISL_1698536, EPI_ISL_1698538, EPI_ISL_1698540, EPI_ISL_1698541, EPI_ISL_1698542, EPI_ISL_1698543, EPI_ISL_1698544, EPI_ISL_1698545, EPI_ISL_1698546, EPI_ISL_1698764, EPI_ISL_1698792, EPI_ISL_1698817, EPI_ISL_1698825, EPI_ISL_1698826, EPI_ISL_1698831, EPI_ISL_1698834, EPI_ISL_1698843, EPI_ISL_1698849, EPI_ISL_1698867, EPI_ISL_1698869, EPI_ISL_1698871, EPI_ISL_1698878, EPI_ISL_1698884, EPI_ISL_1698896, EPI_ISL_1698898, EPI_ISL_1698910, EPI_ISL_1698913, EPI_ISL_1698918, EPI_ISL_1698931, EPI_ISL_1698946, EPI_ISL_1698955, EPI_ISL_1698978, EPI_ISL_1698986, EPI_ISL_1698988, EPI_ISL_1698996, EPI_ISL_1699001, EPI_ISL_1699033, EPI_ISL_1699035, EPI_ISL_1699036, EPI_ISL_1699037, EPI_ISL_1699039, EPI_ISL_1699040, EPI_ISL_1699048, EPI_ISL_1699052, EPI_ISL_1699060, EPI_ISL_1699067, EPI_ISL_1699068, EPI_ISL_1699069, EPI_ISL_1699070, EPI_ISL_1699071, EPI_ISL_1699072, EPI_ISL_1699079, EPI_ISL_1699106, EPI_ISL_1699108, EPI_ISL_1699119, EPI_ISL_1699126, EPI_ISL_1699134, EPI_ISL_1699141, EPI_ISL_1699148, EPI_ISL_1699150, EPI_ISL_1699155, EPI_ISL_1699156, EPI_ISL_1699182, EPI_ISL_1699195, EPI_ISL_1699196, EPI_ISL_1699197, EPI_ISL_1699199, EPI_ISL_1699218, EPI_ISL_1699220, EPI_ISL_1699239, EPI_ISL_1699240, EPI_ISL_1699242, EPI_ISL_1699246, EPI_ISL_1699252, EPI_ISL_1699266, EPI_ISL_1699269, EPI_ISL_1699270, EPI_ISL_1699282, EPI_ISL_1699291, EPI_ISL_1699294, EPI_ISL_1699295, EPI_ISL_1699298, EPI_ISL_1699299, EPI_ISL_1699300, EPI_ISL_1699301, EPI_ISL_1699303, EPI_ISL_1699304, EPI_ISL_1699305, EPI_ISL_1699306, EPI_ISL_1699308, EPI_ISL_1699310, EPI_ISL_1699311, EPI_ISL_1699312, EPI_ISL_1699313, EPI_ISL_1699314, EPI_ISL_1699315, EPI_ISL_1699316, EPI_ISL_1699317, EPI_ISL_1699320, EPI_ISL_1699323, EPI_ISL_1699324, EPI_ISL_1699325, EPI_ISL_1699329, EPI_ISL_1699330, EPI_ISL_1699331, EPI_ISL_1699332, EPI_ISL_1699337, EPI_ISL_1699338, EPI_ISL_1699340, EPI_ISL_1699344, EPI_ISL_1699345, EPI_ISL_1699346, EPI_ISL_1699347, EPI_ISL_1699350, EPI_ISL_1699351, EPI_ISL_1699352, EPI_ISL_1699357, EPI_ISL_1699359, EPI_ISL_1699361, EPI_ISL_1699364, EPI_ISL_1699366, EPI_ISL_1699368, EPI_ISL_1699369, EPI_ISL_1699370, EPI_ISL_1699371, EPI_ISL_1699375, EPI_ISL_1699376, EPI_ISL_1699377, EPI_ISL_1699378, EPI_ISL_1699381, EPI_ISL_1699384, EPI_ISL_1699386, EPI_ISL_1699387, EPI_ISL_1699388, EPI_ISL_1699389, EPI_ISL_1699391, EPI_ISL_1699392, EPI_ISL_1699393, EPI_ISL_1699394, EPI_ISL_1699395, EPI_ISL_1699396, EPI_ISL_1699398, EPI_ISL_1699400, EPI_ISL_1699401, EPI_ISL_1699402, EPI_ISL_1699403, EPI_ISL_1699405, EPI_ISL_1699406, EPI_ISL_1699407, EPI_ISL_1699409, EPI_ISL_1699410, EPI_ISL_1699412, EPI_ISL_1699415, EPI_ISL_1699416, EPI_ISL_1699418, EPI_ISL_1699419, EPI_ISL_1699421, EPI_ISL_1699422, EPI_ISL_1699424, EPI_ISL_1699426, EPI_ISL_1699428, EPI_ISL_1699429, EPI_ISL_1699431, EPI_ISL_1699432, EPI_ISL_1699433, EPI_ISL_1699435, EPI_ISL_1699436, EPI_ISL_1699437, EPI_ISL_1699438, EPI_ISL_1699439, EPI_ISL_1699440, EPI_ISL_1699442, EPI_ISL_1699453, EPI_ISL_1699455, EPI_ISL_1699457, EPI_ISL_1699458, EPI_ISL_1699461, EPI_ISL_1699462, EPI_ISL_1699465, EPI_ISL_1699466, EPI_ISL_1699467, EPI_ISL_1699470, EPI_ISL_1699474, EPI_ISL_1699477, EPI_ISL_1699482, EPI_ISL_1699483, EPI_ISL_1699485, EPI_ISL_1699488, EPI_ISL_1699489, EPI_ISL_1699491, EPI_ISL_1699493, EPI_ISL_1699495, EPI_ISL_1699497, EPI_ISL_1699500, EPI_ISL_1699501, EPI_ISL_1699502, EPI_ISL_1699503, EPI_ISL_1699506, EPI_ISL_1699508, EPI_ISL_1699509, EPI_ISL_1699510, EPI_ISL_1699511, EPI_ISL_1699512, EPI_ISL_1699514, EPI_ISL_1699515, EPI_ISL_1699516, EPI_ISL_1699518, EPI_ISL_1699521, EPI_ISL_1699523, EPI_ISL_1699525, EPI_ISL_1699526, EPI_ISL_1699528, EPI_ISL_1699529, EPI_ISL_1699532, EPI_ISL_1699534, EPI_ISL_1699537, EPI_ISL_1699539, EPI_ISL_1699543, EPI_ISL_1699544, EPI_ISL_1699545, EPI_ISL_1699546, EPI_ISL_1699547, EPI_ISL_1699550, EPI_ISL_1699551, EPI_ISL_1699552, EPI_ISL_1699553, EPI_ISL_1699556, EPI_ISL_1699557, EPI_ISL_1699558, EPI_ISL_1699561, EPI_ISL_1699562, EPI_ISL_1699563, EPI_ISL_1699564, EPI_ISL_1699565, EPI_ISL_1699568, EPI_ISL_1699570, EPI_ISL_1699586, EPI_ISL_1699603, EPI_ISL_1699616, EPI_ISL_1699623, EPI_ISL_1699629, EPI_ISL_1699646, EPI_ISL_1699650, EPI_ISL_1699660, EPI_ISL_1699713, EPI_ISL_1699717, EPI_ISL_1699723, EPI_ISL_1699738, EPI_ISL_1699751, EPI_ISL_1699752, EPI_ISL_1699774, EPI_ISL_1699775, EPI_ISL_1699817, EPI_ISL_1699820, EPI_ISL_1699824, EPI_ISL_1699836, EPI_ISL_1699838, EPI_ISL_1718322, EPI_ISL_1718324, EPI_ISL_1718325, EPI_ISL_1718326, EPI_ISL_1718327, EPI_ISL_1718329, EPI_ISL_1718331, EPI_ISL_1718335, EPI_ISL_1718339, EPI_ISL_1718340, EPI_ISL_1718341, EPI_ISL_1718342, EPI_ISL_1718345, EPI_ISL_1718346, EPI_ISL_1718348, EPI_ISL_1718352, EPI_ISL_1718354, EPI_ISL_1718356, EPI_ISL_1718357, EPI_ISL_1718363, EPI_ISL_1718367, EPI_ISL_1718368, EPI_ISL_1718369, EPI_ISL_1718370, EPI_ISL_1718372, EPI_ISL_1718373, EPI_ISL_1718375, EPI_ISL_1718376, EPI_ISL_1718380, EPI_ISL_1718381, EPI_ISL_1718383, EPI_ISL_1718389, EPI_ISL_1718390, EPI_ISL_1718392, EPI_ISL_1718393, EPI_ISL_1718394, EPI_ISL_1718396, EPI_ISL_1718398, EPI_ISL_1718399, EPI_ISL_1718400, EPI_ISL_1718406, EPI_ISL_1718409, EPI_ISL_1718411, EPI_ISL_1718413, EPI_ISL_1718414, EPI_ISL_1718419, EPI_ISL_1718421, EPI_ISL_1718422, EPI_ISL_1718425, EPI_ISL_1718426, EPI_ISL_1718428, EPI_ISL_1718429, EPI_ISL_1718430, EPI_ISL_1718431, EPI_ISL_1718433, EPI_ISL_1718434, EPI_ISL_1718436, EPI_ISL_1718437, EPI_ISL_1718438, EPI_ISL_1718442, EPI_ISL_1718444, EPI_ISL_1718446, EPI_ISL_1718450, EPI_ISL_1718452, EPI_ISL_1718454, EPI_ISL_1718455, EPI_ISL_1718458, EPI_ISL_1718459, EPI_ISL_1718460, EPI_ISL_1718465, EPI_ISL_1718469, EPI_ISL_1718470, EPI_ISL_1718476, EPI_ISL_1718479, EPI_ISL_1718480, EPI_ISL_1718482, EPI_ISL_1718483, EPI_ISL_1718484, EPI_ISL_1718486, EPI_ISL_1718490, EPI_ISL_1718496, EPI_ISL_1718497, EPI_ISL_1718500, EPI_ISL_1718501, EPI_ISL_1718504, EPI_ISL_1718507, EPI_ISL_1718510, EPI_ISL_1718511, EPI_ISL_1718517, EPI_ISL_1718518, EPI_ISL_1718519, EPI_ISL_1718527, EPI_ISL_1718528, EPI_ISL_1718531, EPI_ISL_1718532, EPI_ISL_1718540, EPI_ISL_1718541, EPI_ISL_1718542, EPI_ISL_1718543, EPI_ISL_1718544, EPI_ISL_1718545, EPI_ISL_1718548, EPI_ISL_1718549, EPI_ISL_1718555, EPI_ISL_1718560, EPI_ISL_1718561, EPI_ISL_1718565, EPI_ISL_1718569, EPI_ISL_1718576, EPI_ISL_1718578, EPI_ISL_1718580, EPI_ISL_1718582, EPI_ISL_1718583, EPI_ISL_1718584, EPI_ISL_1718585, EPI_ISL_1718588, EPI_ISL_1718589, EPI_ISL_1718592, EPI_ISL_1718593, EPI_ISL_1718595, EPI_ISL_1718597, EPI_ISL_1718598, EPI_ISL_1718599, EPI_ISL_1718601, EPI_ISL_1718603, EPI_ISL_1718604, EPI_ISL_1718605, EPI_ISL_1718607, EPI_ISL_1718609, EPI_ISL_1718614, EPI_ISL_1718615, EPI_ISL_1718616, EPI_ISL_1718619, EPI_ISL_1718620, EPI_ISL_1718621, EPI_ISL_1718623, EPI_ISL_1718624, EPI_ISL_1718625, EPI_ISL_1718626, EPI_ISL_1718627, EPI_ISL_1718628, EPI_ISL_1718629, EPI_ISL_1718630, EPI_ISL_1718631, EPI_ISL_1718632, EPI_ISL_1718633, EPI_ISL_1718634, EPI_ISL_1718635, EPI_ISL_1718636, EPI_ISL_1718637, EPI_ISL_1718638, EPI_ISL_1718639, EPI_ISL_1718641, EPI_ISL_1718643, EPI_ISL_1718646, EPI_ISL_1718647, EPI_ISL_1718648, EPI_ISL_1718650, EPI_ISL_1718651, EPI_ISL_1718652, EPI_ISL_1718653, EPI_ISL_1718654, EPI_ISL_1718655, EPI_ISL_1718656, EPI_ISL_1718657, EPI_ISL_1718658, EPI_ISL_1718659, EPI_ISL_1718660, EPI_ISL_1718661, EPI_ISL_1718662, EPI_ISL_1718664, EPI_ISL_1718665, EPI_ISL_1718666, EPI_ISL_1718667, EPI_ISL_1718669, EPI_ISL_1718670, EPI_ISL_1718671, EPI_ISL_1718673, EPI_ISL_1718674, EPI_ISL_1718675, EPI_ISL_1718676, EPI_ISL_1718677, EPI_ISL_1718678, EPI_ISL_1718680, EPI_ISL_1718681, EPI_ISL_1718682, EPI_ISL_1718683, EPI_ISL_1718684, EPI_ISL_1718687, EPI_ISL_1718689, EPI_ISL_1718690, EPI_ISL_1718693, EPI_ISL_1718694, EPI_ISL_1718696, EPI_ISL_1718698, EPI_ISL_1718699, EPI_ISL_1718702, EPI_ISL_1718705, EPI_ISL_1718706, EPI_ISL_1718708, EPI_ISL_1718709, EPI_ISL_1718710, EPI_ISL_1718711, EPI_ISL_1718712, EPI_ISL_1718713, EPI_ISL_1718714, EPI_ISL_1718715, EPI_ISL_1718716, EPI_ISL_1718717, EPI_ISL_1718718, EPI_ISL_1718719, EPI_ISL_1718720, EPI_ISL_1718721, EPI_ISL_1718722, EPI_ISL_1718723, EPI_ISL_1718724, EPI_ISL_1718725, EPI_ISL_1718726, EPI_ISL_1718728, EPI_ISL_1718729, EPI_ISL_1718730, EPI_ISL_1718731, EPI_ISL_1718732, EPI_ISL_1718733, EPI_ISL_1718734, EPI_ISL_1718735, EPI_ISL_1718736, EPI_ISL_1718737, EPI_ISL_1718738, EPI_ISL_1718739, EPI_ISL_1718740, EPI_ISL_1718741, EPI_ISL_1718743, EPI_ISL_1718745, EPI_ISL_1718747, EPI_ISL_1718748, EPI_ISL_1718749, EPI_ISL_1718752, EPI_ISL_1718753, EPI_ISL_1718754, EPI_ISL_1718755, EPI_ISL_1718756, EPI_ISL_1718757, EPI_ISL_1718758, EPI_ISL_1718759, EPI_ISL_1718761, EPI_ISL_1718762, EPI_ISL_1718763, EPI_ISL_1718764, EPI_ISL_1718765, EPI_ISL_1718766, EPI_ISL_1718767, EPI_ISL_1718768, EPI_ISL_1718769, EPI_ISL_1718770, EPI_ISL_1718771, EPI_ISL_1718772, EPI_ISL_1718773, EPI_ISL_1718774, EPI_ISL_1718775, EPI_ISL_1718776, EPI_ISL_1718777, EPI_ISL_1718778, EPI_ISL_1718779, EPI_ISL_1718780, EPI_ISL_1718781, EPI_ISL_1718782, EPI_ISL_1718785, EPI_ISL_1718786, EPI_ISL_1718787, EPI_ISL_1718788, EPI_ISL_1718790, EPI_ISL_1718792, EPI_ISL_1718793, EPI_ISL_1718794, EPI_ISL_1718795, EPI_ISL_1718796, EPI_ISL_1718797, EPI_ISL_1718798, EPI_ISL_1718799, EPI_ISL_1718800, EPI_ISL_1718802, EPI_ISL_1718803, EPI_ISL_1718804, EPI_ISL_1718805, EPI_ISL_1718807, EPI_ISL_1718808, EPI_ISL_1718809, EPI_ISL_1718811, EPI_ISL_1718812, EPI_ISL_1718813, EPI_ISL_1718814, EPI_ISL_1718815, EPI_ISL_1718816, EPI_ISL_1718817, EPI_ISL_1718819, EPI_ISL_1718820, EPI_ISL_1718821, EPI_ISL_1718823, EPI_ISL_1718824, EPI_ISL_1718825, EPI_ISL_1718826, EPI_ISL_1718827, EPI_ISL_1718828, EPI_ISL_1718829, EPI_ISL_1718830, EPI_ISL_1718831, EPI_ISL_1718832, EPI_ISL_1718833, EPI_ISL_1718834, EPI_ISL_1718835, EPI_ISL_1718836, EPI_ISL_1718837, EPI_ISL_1718839, EPI_ISL_1718840, EPI_ISL_1718841, EPI_ISL_1718842, EPI_ISL_1718843, EPI_ISL_1718844, EPI_ISL_1718845, EPI_ISL_1718847, EPI_ISL_1718848, EPI_ISL_1718849, EPI_ISL_1718850, EPI_ISL_1718851, EPI_ISL_1718852, EPI_ISL_1718853, EPI_ISL_1718854, EPI_ISL_1718855, EPI_ISL_1718856, EPI_ISL_1718859, EPI_ISL_1718861, EPI_ISL_1718862, EPI_ISL_1718863, EPI_ISL_1718864, EPI_ISL_1718865, EPI_ISL_1718866, EPI_ISL_1718867, EPI_ISL_1718868, EPI_ISL_1718870, EPI_ISL_1718871, EPI_ISL_1718873, EPI_ISL_1718874, EPI_ISL_1718877, EPI_ISL_1718878, EPI_ISL_1718879, EPI_ISL_1718880, EPI_ISL_1718881, EPI_ISL_1718882, EPI_ISL_1718883, EPI_ISL_1718884, EPI_ISL_1719433, EPI_ISL_1719443, EPI_ISL_1719446, EPI_ISL_1719449, EPI_ISL_1719450, EPI_ISL_1719451, EPI_ISL_1719454, EPI_ISL_1719455, EPI_ISL_1719459, EPI_ISL_1719461, EPI_ISL_1719465, EPI_ISL_1719469, EPI_ISL_1719470, EPI_ISL_1719480, EPI_ISL_1719481, EPI_ISL_1719488, EPI_ISL_1719489, EPI_ISL_1719496, EPI_ISL_1719498, EPI_ISL_1719500, EPI_ISL_1719501, EPI_ISL_1719505, EPI_ISL_1719506, EPI_ISL_1719507, EPI_ISL_1719512, EPI_ISL_1719513, EPI_ISL_1719514, EPI_ISL_1719519, EPI_ISL_1719526, EPI_ISL_1719533, EPI_ISL_1719542, EPI_ISL_1719544, EPI_ISL_1719547, EPI_ISL_1719548, EPI_ISL_1719549, EPI_ISL_1719558, EPI_ISL_1719560, EPI_ISL_1719571, EPI_ISL_1719576, EPI_ISL_1719582, EPI_ISL_1719588, EPI_ISL_1719591, EPI_ISL_1719593, EPI_ISL_1719595, EPI_ISL_1719598, EPI_ISL_1719599, EPI_ISL_1719600, EPI_ISL_1719603, EPI_ISL_1719604, EPI_ISL_1719605, EPI_ISL_1719606, EPI_ISL_1719609, EPI_ISL_1719613, EPI_ISL_1719616, EPI_ISL_1719624, EPI_ISL_1719625, EPI_ISL_1719639, EPI_ISL_1719643, EPI_ISL_1719647, EPI_ISL_1719650, EPI_ISL_1719656, EPI_ISL_1719659, EPI_ISL_1719665, EPI_ISL_1719666, EPI_ISL_1719671, EPI_ISL_1719679, EPI_ISL_1719685, EPI_ISL_1719688, EPI_ISL_1719691, EPI_ISL_1719699, EPI_ISL_1719710, EPI_ISL_1719711, EPI_ISL_1719712, EPI_ISL_1719713, EPI_ISL_1719717, EPI_ISL_1719721, EPI_ISL_1719726, EPI_ISL_1719733, EPI_ISL_1719741, EPI_ISL_1719742, EPI_ISL_1740532, EPI_ISL_1740538, EPI_ISL_1740552, EPI_ISL_1740553, EPI_ISL_1740555, EPI_ISL_1740556, EPI_ISL_1740571, EPI_ISL_1740572, EPI_ISL_1740574, EPI_ISL_1740576, EPI_ISL_1740585, EPI_ISL_1740590, EPI_ISL_1740592, EPI_ISL_1740595, EPI_ISL_1740599, EPI_ISL_1740600, EPI_ISL_1740604, EPI_ISL_1740605, EPI_ISL_1740607, EPI_ISL_1740609, EPI_ISL_1740616, EPI_ISL_1740629, EPI_ISL_1740630, EPI_ISL_1740632, EPI_ISL_1740637, EPI_ISL_1740641, EPI_ISL_1740642, EPI_ISL_1740651, EPI_ISL_1740652, EPI_ISL_1740654, EPI_ISL_1740655, EPI_ISL_1740657, EPI_ISL_1740660, EPI_ISL_1740662, EPI_ISL_1740664, EPI_ISL_1740666, EPI_ISL_1740669, EPI_ISL_1740671, EPI_ISL_1740674, EPI_ISL_1740675, EPI_ISL_1740681, EPI_ISL_1740682, EPI_ISL_1740688, EPI_ISL_1740697, EPI_ISL_1740703, EPI_ISL_1740704, EPI_ISL_1740706, EPI_ISL_1740707, EPI_ISL_1740714, EPI_ISL_1740715, EPI_ISL_1740722, EPI_ISL_1740725, EPI_ISL_1740726, EPI_ISL_1740731, EPI_ISL_1740732, EPI_ISL_1740736, EPI_ISL_1740746, EPI_ISL_1740748, EPI_ISL_1740750, EPI_ISL_1740751, EPI_ISL_1740753, EPI_ISL_1740760, EPI_ISL_1740761, EPI_ISL_1740763, EPI_ISL_1740764, EPI_ISL_1740769, EPI_ISL_1740778, EPI_ISL_1740783, EPI_ISL_1740786, EPI_ISL_1740787, EPI_ISL_1740788, EPI_ISL_1740790, EPI_ISL_1740792, EPI_ISL_1740794, EPI_ISL_1740796, EPI_ISL_1740797, EPI_ISL_1740798, EPI_ISL_1740800, EPI_ISL_1740801, EPI_ISL_1740803, EPI_ISL_1740804, EPI_ISL_1740805, EPI_ISL_1740808, EPI_ISL_1740810, EPI_ISL_1740811, EPI_ISL_1740812, EPI_ISL_1740815, EPI_ISL_1740816, EPI_ISL_1740818, EPI_ISL_1740819, EPI_ISL_1740820, EPI_ISL_1740823, EPI_ISL_1740826, EPI_ISL_1740827, EPI_ISL_1740829, EPI_ISL_1740830, EPI_ISL_1740831, EPI_ISL_1740832, EPI_ISL_1740833, EPI_ISL_1740834, EPI_ISL_1740835, EPI_ISL_1740836, EPI_ISL_1740837, EPI_ISL_1740838, EPI_ISL_1740839, EPI_ISL_1740840, EPI_ISL_1740841, EPI_ISL_1740845, EPI_ISL_1740846, EPI_ISL_1740847, EPI_ISL_1740848, EPI_ISL_1740850, EPI_ISL_1740852, EPI_ISL_1740853, EPI_ISL_1740857, EPI_ISL_1740859, EPI_ISL_1740863, EPI_ISL_1740866, EPI_ISL_1740867, EPI_ISL_1740868, EPI_ISL_1740869, EPI_ISL_1740870, EPI_ISL_1740871, EPI_ISL_1740872, EPI_ISL_1740873, EPI_ISL_1740876, EPI_ISL_1740877, EPI_ISL_1740878, EPI_ISL_1740879, EPI_ISL_1740880, EPI_ISL_1740881, EPI_ISL_1740883, EPI_ISL_1740884, EPI_ISL_1740885, EPI_ISL_1740886, EPI_ISL_1740889, EPI_ISL_1740890, EPI_ISL_1740894, EPI_ISL_1740895, EPI_ISL_1740896, EPI_ISL_1740899, EPI_ISL_1740902, EPI_ISL_1740903, EPI_ISL_1740904, EPI_ISL_1740906, EPI_ISL_1740908, EPI_ISL_1740910, EPI_ISL_1740911, EPI_ISL_1740912, EPI_ISL_1740913, EPI_ISL_1740914, EPI_ISL_1740916, EPI_ISL_1740917, EPI_ISL_1740918, EPI_ISL_1740919, EPI_ISL_1740921, EPI_ISL_1740923, EPI_ISL_1740925, EPI_ISL_1740926, EPI_ISL_1740927, EPI_ISL_1740928, EPI_ISL_1740930, EPI_ISL_1740932, EPI_ISL_1740933, EPI_ISL_1740938, EPI_ISL_1740940, EPI_ISL_1740942, EPI_ISL_1740946, EPI_ISL_1740949, EPI_ISL_1740950, EPI_ISL_1740952, EPI_ISL_1740955, EPI_ISL_1740959, EPI_ISL_1740960, EPI_ISL_1740961, EPI_ISL_1740966, EPI_ISL_1740967, EPI_ISL_1740968, EPI_ISL_1740973, EPI_ISL_1740977, EPI_ISL_1740978, EPI_ISL_1740980, EPI_ISL_1740983, EPI_ISL_1740984, EPI_ISL_1740985, EPI_ISL_1740986, EPI_ISL_1740987, EPI_ISL_1740988, EPI_ISL_1740990, EPI_ISL_1740991, EPI_ISL_1740992, EPI_ISL_1740995, EPI_ISL_1740996, EPI_ISL_1740997, EPI_ISL_1741000, EPI_ISL_1741001, EPI_ISL_1741002, EPI_ISL_1741003, EPI_ISL_1741005, EPI_ISL_1741006, EPI_ISL_1741010, EPI_ISL_1741011, EPI_ISL_1741012, EPI_ISL_1741013, EPI_ISL_1741015, EPI_ISL_1741017, EPI_ISL_1741018, EPI_ISL_1741019, EPI_ISL_1741022, EPI_ISL_1741023, EPI_ISL_1741025, EPI_ISL_1741026, EPI_ISL_1741027, EPI_ISL_1741028, EPI_ISL_1741029, EPI_ISL_1741030, EPI_ISL_1741033, EPI_ISL_1741034, EPI_ISL_1741037, EPI_ISL_1741038, EPI_ISL_1741039, EPI_ISL_1741041, EPI_ISL_1741043, EPI_ISL_1741046, EPI_ISL_1741047, EPI_ISL_1741049, EPI_ISL_1741050, EPI_ISL_1741051, EPI_ISL_1741052, EPI_ISL_1741053, EPI_ISL_1741056, EPI_ISL_1741059, EPI_ISL_1741062, EPI_ISL_1741063, EPI_ISL_1741066, EPI_ISL_1741074, EPI_ISL_1741076, EPI_ISL_1741083, EPI_ISL_1741085, EPI_ISL_1741087, EPI_ISL_1741088, EPI_ISL_1741090, EPI_ISL_1741093, EPI_ISL_1741094, EPI_ISL_1741101, EPI_ISL_1741102, EPI_ISL_1741104, EPI_ISL_1741110, EPI_ISL_1741111, EPI_ISL_1741114, EPI_ISL_1741115, EPI_ISL_1741116, EPI_ISL_1741117, EPI_ISL_1741120, EPI_ISL_1741122, EPI_ISL_1741125, EPI_ISL_1741127, EPI_ISL_1741130, EPI_ISL_1741131, EPI_ISL_1741133, EPI_ISL_1741134, EPI_ISL_1741135, EPI_ISL_1741140, EPI_ISL_1741148, EPI_ISL_1741150, EPI_ISL_1741151, EPI_ISL_1741154, EPI_ISL_1741156, EPI_ISL_1741159, EPI_ISL_1741160, EPI_ISL_1741161, EPI_ISL_1741163, EPI_ISL_1741164, EPI_ISL_1741165, EPI_ISL_1741166, EPI_ISL_1741167, EPI_ISL_1741168, EPI_ISL_1741169, EPI_ISL_1741175, EPI_ISL_1741177, EPI_ISL_1741180, EPI_ISL_1741186, EPI_ISL_1741187, EPI_ISL_1741189, EPI_ISL_1741195, EPI_ISL_1741197, EPI_ISL_1741198, EPI_ISL_1741202, EPI_ISL_1741207, EPI_ISL_1741208, EPI_ISL_1741210, EPI_ISL_1741218, EPI_ISL_1741226, EPI_ISL_1741227, EPI_ISL_1741229, EPI_ISL_1741233, EPI_ISL_1741234, EPI_ISL_1741236, EPI_ISL_1741238, EPI_ISL_1741239, EPI_ISL_1741241, EPI_ISL_1741245, EPI_ISL_1741246, EPI_ISL_1741247, EPI_ISL_1741248, EPI_ISL_1741249, EPI_ISL_1741251, EPI_ISL_1741252, EPI_ISL_1741254, EPI_ISL_1741255, EPI_ISL_1741256, EPI_ISL_1741260, EPI_ISL_1741261, EPI_ISL_1741263, EPI_ISL_1741268, EPI_ISL_1741272, EPI_ISL_1741514, EPI_ISL_1741524, EPI_ISL_1741542, EPI_ISL_1741545, EPI_ISL_1741547, EPI_ISL_1741548, EPI_ISL_1741725, EPI_ISL_1741737, EPI_ISL_1741765, EPI_ISL_1742171, EPI_ISL_1742285, EPI_ISL_1742290, EPI_ISL_1758188, EPI_ISL_1758190, EPI_ISL_1758195, EPI_ISL_1758196, EPI_ISL_1758198, EPI_ISL_1758202, EPI_ISL_1758203, EPI_ISL_1758206, EPI_ISL_1758212, EPI_ISL_1758214, EPI_ISL_1758217, EPI_ISL_1758218, EPI_ISL_1758233, EPI_ISL_1758234, EPI_ISL_1758237, EPI_ISL_1758238, EPI_ISL_1758241, EPI_ISL_1758243, EPI_ISL_1758245, EPI_ISL_1758247, EPI_ISL_1758252, EPI_ISL_1758254, EPI_ISL_1758256, EPI_ISL_1758259, EPI_ISL_1758261, EPI_ISL_1758266, EPI_ISL_1758268, EPI_ISL_1758270, EPI_ISL_1758273, EPI_ISL_1758284, EPI_ISL_1758289, EPI_ISL_1758290, EPI_ISL_1758292, EPI_ISL_1758294, EPI_ISL_1758296, EPI_ISL_1758297, EPI_ISL_1758301, EPI_ISL_1758303, EPI_ISL_1758306, EPI_ISL_1758308, EPI_ISL_1758313, EPI_ISL_1758314, EPI_ISL_1758316, EPI_ISL_1758317, EPI_ISL_1758319, EPI_ISL_1758322, EPI_ISL_1758324, EPI_ISL_1758325, EPI_ISL_1758328, EPI_ISL_1758332, EPI_ISL_1758342, EPI_ISL_1758343, EPI_ISL_1758348, EPI_ISL_1758349, EPI_ISL_1758352, EPI_ISL_1758353, EPI_ISL_1758355, EPI_ISL_1758356, EPI_ISL_1758360, EPI_ISL_1758361, EPI_ISL_1758364, EPI_ISL_1758368, EPI_ISL_1758372, EPI_ISL_1758373, EPI_ISL_1758376, EPI_ISL_1758378, EPI_ISL_1758379, EPI_ISL_1758387, EPI_ISL_1758397, EPI_ISL_1758400, EPI_ISL_1758401, EPI_ISL_1758404, EPI_ISL_1758406, EPI_ISL_1758412, EPI_ISL_1758414, EPI_ISL_1758415, EPI_ISL_1758418, EPI_ISL_1758420, EPI_ISL_1758423, EPI_ISL_1758424, EPI_ISL_1758433, EPI_ISL_1758437, EPI_ISL_1758440, EPI_ISL_1758442, EPI_ISL_1758443, EPI_ISL_1758444, EPI_ISL_1758446, EPI_ISL_1758448, EPI_ISL_1758449, EPI_ISL_1758450, EPI_ISL_1758451, EPI_ISL_1758879, EPI_ISL_1758880, EPI_ISL_1758881, EPI_ISL_1758884, EPI_ISL_1758885, EPI_ISL_1758888, EPI_ISL_1758890, EPI_ISL_1758891, EPI_ISL_1758892, EPI_ISL_1758895, EPI_ISL_1758896, EPI_ISL_1758898, EPI_ISL_1758899, EPI_ISL_1758900, EPI_ISL_1758903, EPI_ISL_1758905, EPI_ISL_1758906, EPI_ISL_1758907, EPI_ISL_1758908, EPI_ISL_1758909, EPI_ISL_1758912, EPI_ISL_1758914, EPI_ISL_1758915, EPI_ISL_1758917, EPI_ISL_1758918, EPI_ISL_1758922, EPI_ISL_1758923, EPI_ISL_1758925, EPI_ISL_1758926, EPI_ISL_1758927, EPI_ISL_1758929, EPI_ISL_1758930, EPI_ISL_1758931, EPI_ISL_1758934, EPI_ISL_1758935, EPI_ISL_1758936, EPI_ISL_1758937, EPI_ISL_1758941, EPI_ISL_1758944, EPI_ISL_1758950, EPI_ISL_1758955, EPI_ISL_1758956, EPI_ISL_1758958, EPI_ISL_1758959, EPI_ISL_1758960, EPI_ISL_1758962, EPI_ISL_1758963, EPI_ISL_1758965, EPI_ISL_1758967, EPI_ISL_1758968, EPI_ISL_1758970, EPI_ISL_1758971, EPI_ISL_1758972, EPI_ISL_1758973, EPI_ISL_1758974, EPI_ISL_1758975, EPI_ISL_1758977, EPI_ISL_1758978, EPI_ISL_1758983, EPI_ISL_1758984, | | | |

EPI_ISL_1758985, EPI_ISL_1758986, EPI_ISL_1758988, EPI_ISL_1758989, EPI_ISL_1758991, EPI_ISL_1758992, EPI_ISL_1758995, EPI_ISL_1758997, EPI_ISL_1758998, EPI_ISL_1759001, EPI_ISL_1759002, EPI_ISL_1759004, EPI_ISL_1759006, EPI_ISL_1759007, EPI_ISL_1759008, EPI_ISL_1759009, EPI_ISL_1759010, EPI_ISL_1759013, EPI_ISL_1759014, EPI_ISL_1759015, EPI_ISL_1759016, EPI_ISL_1759017,
EPI_ISL_1759018, EPI_ISL_1759019, EPI_ISL_1759020, EPI_ISL_1759023, EPI_ISL_1759024, EPI_ISL_1759025, EPI_ISL_1759027, EPI_ISL_1759028, EPI_ISL_1759029, EPI_ISL_1759031, EPI_ISL_1759032, EPI_ISL_1759033, EPI_ISL_1759034, EPI_ISL_1759035, EPI_ISL_1759038, EPI_ISL_1759039, EPI_ISL_1759042, EPI_ISL_1759043, EPI_ISL_1759045, EPI_ISL_1759048, EPI_ISL_1759049, EPI_ISL_1759050,
EPI_ISL_1759053, EPI_ISL_1759054, EPI_ISL_1759056, EPI_ISL_1759057, EPI_ISL_1759058, EPI_ISL_1759060, EPI_ISL_1759062, EPI_ISL_1759063, EPI_ISL_1759064, EPI_ISL_1759069, EPI_ISL_1759070, EPI_ISL_1759071, EPI_ISL_1759073, EPI_ISL_1759075, EPI_ISL_1759077, EPI_ISL_1759079, EPI_ISL_1759080, EPI_ISL_1759081, EPI_ISL_1759083, EPI_ISL_1759086, EPI_ISL_1759087, EPI_ISL_1759088,
EPI_ISL_1759089, EPI_ISL_1759090, EPI_ISL_1759091, EPI_ISL_1759092, EPI_ISL_1759093, EPI_ISL_1759094, EPI_ISL_1759097, EPI_ISL_1759098, EPI_ISL_1759101, EPI_ISL_1759102, EPI_ISL_1759104, EPI_ISL_1759105, EPI_ISL_1759107, EPI_ISL_1759109, EPI_ISL_1759110, EPI_ISL_1759111, EPI_ISL_1759112, EPI_ISL_1759113, EPI_ISL_1759115, EPI_ISL_1759118, EPI_ISL_1759122, EPI_ISL_1759124,
EPI_ISL_1759127, EPI_ISL_1759129, EPI_ISL_1759130, EPI_ISL_1759131, EPI_ISL_1759133, EPI_ISL_1759134, EPI_ISL_1759135, EPI_ISL_1759136, EPI_ISL_1759137, EPI_ISL_1759139, EPI_ISL_1759140, EPI_ISL_1759142, EPI_ISL_1759143, EPI_ISL_1759146, EPI_ISL_1759148, EPI_ISL_1759149, EPI_ISL_1759150, EPI_ISL_1759153, EPI_ISL_1759155, EPI_ISL_1759156, EPI_ISL_1759157, EPI_ISL_1759161,
EPI_ISL_1759165, EPI_ISL_1759167, EPI_ISL_1759168, EPI_ISL_1759171, EPI_ISL_1759173, EPI_ISL_1759174, EPI_ISL_1759187, EPI_ISL_1759190, EPI_ISL_1759266, EPI_ISL_1759274, EPI_ISL_1759286, EPI_ISL_1759293, EPI_ISL_1759301, EPI_ISL_1759320, EPI_ISL_1759325, EPI_ISL_1759812, EPI_ISL_1759815, EPI_ISL_1759828, EPI_ISL_1759842, EPI_ISL_1759879, EPI_ISL_1790432, EPI_ISL_1790438,
EPI_ISL_1790439, EPI_ISL_1790442, EPI_ISL_1790445, EPI_ISL_1790451, EPI_ISL_1790457, EPI_ISL_1790461, EPI_ISL_1790462, EPI_ISL_1790463, EPI_ISL_1790465, EPI_ISL_1790472, EPI_ISL_1790474, EPI_ISL_1790482, EPI_ISL_1790483, EPI_ISL_1790484, EPI_ISL_1790497, EPI_ISL_1790500, EPI_ISL_1790504, EPI_ISL_1790506, EPI_ISL_1790525, EPI_ISL_1790534, EPI_ISL_1790538, EPI_ISL_1790542,
EPI_ISL_1790543, EPI_ISL_1790546, EPI_ISL_1790547, EPI_ISL_1790548, EPI_ISL_1790549, EPI_ISL_1790554, EPI_ISL_1790563, EPI_ISL_1790571, EPI_ISL_1790576, EPI_ISL_1790577, EPI_ISL_1790579, EPI_ISL_1790584, EPI_ISL_1790585, EPI_ISL_1790589, EPI_ISL_1790591, EPI_ISL_1790596, EPI_ISL_1790603, EPI_ISL_1790616, EPI_ISL_1790617, EPI_ISL_1790618, EPI_ISL_1790621, EPI_ISL_1790626,
EPI_ISL_1790628, EPI_ISL_1790630, EPI_ISL_1790632, EPI_ISL_1790639, EPI_ISL_1790644, EPI_ISL_1790651, EPI_ISL_1790653, EPI_ISL_1790660, EPI_ISL_1790662, EPI_ISL_1790663, EPI_ISL_1790665, EPI_ISL_1790666, EPI_ISL_1790670, EPI_ISL_1790682, EPI_ISL_1790685, EPI_ISL_1790692, EPI_ISL_1790693, EPI_ISL_1790704, EPI_ISL_1790718, EPI_ISL_1790723, EPI_ISL_1790726, EPI_ISL_1790731,
EPI_ISL_1790736, EPI_ISL_1790745, EPI_ISL_1790747, EPI_ISL_1790748, EPI_ISL_1790750, EPI_ISL_1790759, EPI_ISL_1790760, EPI_ISL_1790762, EPI_ISL_1790765, EPI_ISL_1790767, EPI_ISL_1790771, EPI_ISL_1790775, EPI_ISL_1790776, EPI_ISL_1790777, EPI_ISL_1790787, EPI_ISL_1790788, EPI_ISL_1790790, EPI_ISL_1790791, EPI_ISL_1790793, EPI_ISL_1790794, EPI_ISL_1790797, EPI_ISL_1790805,
EPI_ISL_1790806, EPI_ISL_1790807, EPI_ISL_1790808, EPI_ISL_1790809, EPI_ISL_1790813, EPI_ISL_1790823, EPI_ISL_1790828, EPI_ISL_1790830, EPI_ISL_1790831, EPI_ISL_1790835, EPI_ISL_1790842, EPI_ISL_1790843, EPI_ISL_1790844, EPI_ISL_1790846, EPI_ISL_1790851, EPI_ISL_1790854, EPI_ISL_1790855, EPI_ISL_1790858, EPI_ISL_1790860, EPI_ISL_1790863, EPI_ISL_1790872, EPI_ISL_1790874,
EPI_ISL_1790875, EPI_ISL_1790876, EPI_ISL_1790880, EPI_ISL_1790882, EPI_ISL_1790884, EPI_ISL_1790887, EPI_ISL_1790889, EPI_ISL_1790890, EPI_ISL_1790891, EPI_ISL_1790892, EPI_ISL_1790897, EPI_ISL_1790906, EPI_ISL_1790912, EPI_ISL_1790913, EPI_ISL_1790916, EPI_ISL_1790918, EPI_ISL_1790919, EPI_ISL_1790921, EPI_ISL_1790922, EPI_ISL_1790926, EPI_ISL_1790929, EPI_ISL_1790930,
EPI_ISL_1790934, EPI_ISL_1790936, EPI_ISL_1790937, EPI_ISL_1790938, EPI_ISL_1790939, EPI_ISL_1790941, EPI_ISL_1790943, EPI_ISL_1790945, EPI_ISL_1790946, EPI_ISL_1790947, EPI_ISL_1790951, EPI_ISL_1790953, EPI_ISL_1790955, EPI_ISL_1790965, EPI_ISL_1790966, EPI_ISL_1790968, EPI_ISL_1790969, EPI_ISL_1790970, EPI_ISL_1790971, EPI_ISL_1790973, EPI_ISL_1790975, EPI_ISL_1790978,
EPI_ISL_1790982, EPI_ISL_1790986, EPI_ISL_1790987, EPI_ISL_1790988, EPI_ISL_1790990, EPI_ISL_1790992, EPI_ISL_1790995, EPI_ISL_1791001, EPI_ISL_1791008, EPI_ISL_1791010, EPI_ISL_1791011, EPI_ISL_1791014, EPI_ISL_1791016, EPI_ISL_1791017, EPI_ISL_1791021, EPI_ISL_1791022, EPI_ISL_1805964, EPI_ISL_1805965, EPI_ISL_1805969, EPI_ISL_1805970, EPI_ISL_1805974, EPI_ISL_1805979,
EPI_ISL_1805982, EPI_ISL_1805986, EPI_ISL_1805988, EPI_ISL_1805993, EPI_ISL_1805994, EPI_ISL_1805995, EPI_ISL_1805997, EPI_ISL_1805999, EPI_ISL_1806001, EPI_ISL_1806004, EPI_ISL_1806007, EPI_ISL_1806008, EPI_ISL_1806010, EPI_ISL_1806013, EPI_ISL_1806017, EPI_ISL_1806019, EPI_ISL_1806021, EPI_ISL_1806027, EPI_ISL_1806033, EPI_ISL_1806034, EPI_ISL_1806038, EPI_ISL_1806043,
EPI_ISL_1806045, EPI_ISL_1806046, EPI_ISL_1806048, EPI_ISL_1806051, EPI_ISL_1806052, EPI_ISL_1806053, EPI_ISL_1806063, EPI_ISL_1806064, EPI_ISL_1806069, EPI_ISL_1806073, EPI_ISL_1806075, EPI_ISL_1806076, EPI_ISL_1806080, EPI_ISL_1806082, EPI_ISL_1806083, EPI_ISL_1806087, EPI_ISL_1806089, EPI_ISL_1806093, EPI_ISL_1806095, EPI_ISL_1806096, EPI_ISL_1806100, EPI_ISL_1806102,
EPI_ISL_1806104, EPI_ISL_1806106, EPI_ISL_1806108, EPI_ISL_1806111, EPI_ISL_1806112, EPI_ISL_1806119, EPI_ISL_1806121, EPI_ISL_1806122, EPI_ISL_1806123, EPI_ISL_1806124, EPI_ISL_1806125, EPI_ISL_1806131, EPI_ISL_1806142, EPI_ISL_1806143, EPI_ISL_1806144, EPI_ISL_1806147, EPI_ISL_1806149, EPI_ISL_1806152, EPI_ISL_1806153, EPI_ISL_1806156, EPI_ISL_1806157, EPI_ISL_1806158,
EPI_ISL_1806161, EPI_ISL_1806167, EPI_ISL_1806169, EPI_ISL_1806170, EPI_ISL_1806171, EPI_ISL_1806172, EPI_ISL_1806174, EPI_ISL_1806175, EPI_ISL_1806176, EPI_ISL_1806189, EPI_ISL_1806195, EPI_ISL_1806202, EPI_ISL_1806205, EPI_ISL_1806504, EPI_ISL_1806505, EPI_ISL_1806507, EPI_ISL_1806510, EPI_ISL_1806512, EPI_ISL_1806513, EPI_ISL_1806514, EPI_ISL_1806518, EPI_ISL_1806519,
EPI_ISL_1806520, EPI_ISL_1806521, EPI_ISL_1806522, EPI_ISL_1806525, EPI_ISL_1806527, EPI_ISL_1806528, EPI_ISL_1806529, EPI_ISL_1806534, EPI_ISL_1806535, EPI_ISL_1806537, EPI_ISL_1806538, EPI_ISL_1806539, EPI_ISL_1806540, EPI_ISL_1806541, EPI_ISL_1806542, EPI_ISL_1806543, EPI_ISL_1806545, EPI_ISL_1806547, EPI_ISL_1806548, EPI_ISL_1806549, EPI_ISL_1806550, EPI_ISL_1806551,
EPI_ISL_1806552, EPI_ISL_1806553, EPI_ISL_1806554, EPI_ISL_1806556, EPI_ISL_1806560, EPI_ISL_1806562, EPI_ISL_1806564, EPI_ISL_1806565, EPI_ISL_1806567, EPI_ISL_1806568, EPI_ISL_1806570, EPI_ISL_1806571, EPI_ISL_1806573, EPI_ISL_1806574, EPI_ISL_1806575, EPI_ISL_1806576, EPI_ISL_1806577, EPI_ISL_1806578, EPI_ISL_1806579, EPI_ISL_1806580, EPI_ISL_1806581, EPI_ISL_1806582,
EPI_ISL_1806583, EPI_ISL_1806584, EPI_ISL_1806585, EPI_ISL_1806586, EPI_ISL_1806588, EPI_ISL_1806589, EPI_ISL_1806590, EPI_ISL_1806591, EPI_ISL_1806594, EPI_ISL_1806595, EPI_ISL_1806597, EPI_ISL_1806598, EPI_ISL_1806600, EPI_ISL_1806602, EPI_ISL_1806603, EPI_ISL_1806604, EPI_ISL_1806605, EPI_ISL_1806607, EPI_ISL_1806608, EPI_ISL_1806609, EPI_ISL_1806610, EPI_ISL_1806613,
EPI_ISL_1806614, EPI_ISL_1806615, EPI_ISL_1806617, EPI_ISL_1806618, EPI_ISL_1806619, EPI_ISL_1806620, EPI_ISL_1806622, EPI_ISL_1806623, EPI_ISL_1806624, EPI_ISL_1806625, EPI_ISL_1806626, EPI_ISL_1806627, EPI_ISL_1806628, EPI_ISL_1806630, EPI_ISL_1806633, EPI_ISL_1806634, EPI_ISL_1806637, EPI_ISL_1806640, EPI_ISL_1806644, EPI_ISL_1806645, EPI_ISL_1806647, EPI_ISL_1806648,
EPI_ISL_1806649, EPI_ISL_1806650, EPI_ISL_1806654, EPI_ISL_1806655, EPI_ISL_1806658, EPI_ISL_1806659, EPI_ISL_1806660, EPI_ISL_1806661, EPI_ISL_1806663, EPI_ISL_1806664, EPI_ISL_1806665, EPI_ISL_1806668, EPI_ISL_1806669, EPI_ISL_1806670, EPI_ISL_1806671, EPI_ISL_1806672, EPI_ISL_1806676, EPI_ISL_1806677, EPI_ISL_1806680, EPI_ISL_1806682, EPI_ISL_1806683, EPI_ISL_1806685,
EPI_ISL_1806686, EPI_ISL_1806689, EPI_ISL_1806690, EPI_ISL_1806691, EPI_ISL_1806692, EPI_ISL_1806693, EPI_ISL_1806694, EPI_ISL_1806695, EPI_ISL_1806696, EPI_ISL_1806697, EPI_ISL_1806698, EPI_ISL_1806699, EPI_ISL_1806700, EPI_ISL_1806701, EPI_ISL_1806702, EPI_ISL_1806704, EPI_ISL_1806705, EPI_ISL_1806706, EPI_ISL_1806707, EPI_ISL_1806708, EPI_ISL_1806710, EPI_ISL_1806711,
EPI_ISL_1806712, EPI_ISL_1806713, EPI_ISL_1806714, EPI_ISL_1806715, EPI_ISL_1806717, EPI_ISL_1806718, EPI_ISL_1806722, EPI_ISL_1806724, EPI_ISL_1806726, EPI_ISL_1806727, EPI_ISL_1806730, EPI_ISL_1806731, EPI_ISL_1806732, EPI_ISL_1806733, EPI_ISL_1806734, EPI_ISL_1806736, EPI_ISL_1806739, EPI_ISL_1806740, EPI_ISL_1806742, EPI_ISL_1806743, EPI_ISL_1806744, EPI_ISL_1806745,
EPI_ISL_1806746, EPI_ISL_1806747, EPI_ISL_1806749, EPI_ISL_1806750, EPI_ISL_1806751, EPI_ISL_1806752, EPI_ISL_1806754, EPI_ISL_1806755, EPI_ISL_1806757, EPI_ISL_1806759, EPI_ISL_1806760, EPI_ISL_1806762, EPI_ISL_1806763, EPI_ISL_1806764, EPI_ISL_1806765, EPI_ISL_1806766, EPI_ISL_1806768, EPI_ISL_1806769, EPI_ISL_1806770, EPI_ISL_1806771, EPI_ISL_1806774, EPI_ISL_1806776,
EPI_ISL_1806779, EPI_ISL_1806780, EPI_ISL_1806781, EPI_ISL_1806782, EPI_ISL_1806783, EPI_ISL_1806786, EPI_ISL_1806787, EPI_ISL_1806789, EPI_ISL_1806790, EPI_ISL_1806791, EPI_ISL_1806793, EPI_ISL_1806794, EPI_ISL_1806796, EPI_ISL_1806797, EPI_ISL_1806799, EPI_ISL_1806800, EPI_ISL_1806804, EPI_ISL_1806805, EPI_ISL_1806807, EPI_ISL_1806809, EPI_ISL_1806810, EPI_ISL_1806811,
EPI_ISL_1806812, EPI_ISL_1806814, EPI_ISL_1806815, EPI_ISL_1806816, EPI_ISL_1806817, EPI_ISL_1806818, EPI_ISL_1806819, EPI_ISL_1806820, EPI_ISL_1806822, EPI_ISL_1806823, EPI_ISL_1806824, EPI_ISL_1806825, EPI_ISL_1806827, EPI_ISL_1806829, EPI_ISL_1806831, EPI_ISL_1806834, EPI_ISL_1806836, EPI_ISL_1806838, EPI_ISL_1806839, EPI_ISL_1806841, EPI_ISL_1806842, EPI_ISL_1806843,
EPI_ISL_1806845, EPI_ISL_1806846, EPI_ISL_1806849, EPI_ISL_1806850, EPI_ISL_1806851, EPI_ISL_1806853, EPI_ISL_1806898, EPI_ISL_1807081, EPI_ISL_1829916, EPI_ISL_1829934, EPI_ISL_1830110, EPI_ISL_1830121, EPI_ISL_1830173, EPI_ISL_1830183, EPI_ISL_1830300, EPI_ISL_1830325, EPI_ISL_1830339, EPI_ISL_1830360, EPI_ISL_1830613, EPI_ISL_1830805, EPI_ISL_1830911, EPI_ISL_1830959,
EPI_ISL_1830979, EPI_ISL_1831048, EPI_ISL_1831070, EPI_ISL_1831263, EPI_ISL_1831264, EPI_ISL_1831265, EPI_ISL_1831266, EPI_ISL_1831267, EPI_ISL_1831268, EPI_ISL_1912785, EPI_ISL_1912786, EPI_ISL_1912787, EPI_ISL_1912788, EPI_ISL_1912789, EPI_ISL_1985195, EPI_ISL_1985199, EPI_ISL_1985208, EPI_ISL_1985209, EPI_ISL_1985217, EPI_ISL_1985219, EPI_ISL_1985221, EPI_ISL_1985222,
EPI_ISL_1985229, EPI_ISL_1985236, EPI_ISL_1985237, EPI_ISL_1985240, EPI_ISL_1985246, EPI_ISL_1985249, EPI_ISL_1985250, EPI_ISL_1985255, EPI_ISL_1985257, EPI_ISL_1985258, EPI_ISL_1985259, EPI_ISL_1985263, EPI_ISL_1985269, EPI_ISL_1985272, EPI_ISL_1985280, EPI_ISL_1985291, EPI_ISL_1985293, EPI_ISL_1985301, EPI_ISL_1985306, EPI_ISL_1985307, EPI_ISL_1985309, EPI_ISL_1985311,
EPI_ISL_1985314, EPI_ISL_1985318, EPI_ISL_1985320, EPI_ISL_1985323, EPI_ISL_1985324, EPI_ISL_1985336, EPI_ISL_1985337, EPI_ISL_1985338, EPI_ISL_1985340, EPI_ISL_1985342, EPI_ISL_1985347, EPI_ISL_1985348, EPI_ISL_1985349, EPI_ISL_1985354, EPI_ISL_1985357, EPI_ISL_1985359, EPI_ISL_1985360, EPI_ISL_1985362, EPI_ISL_1985364, EPI_ISL_1985370, EPI_ISL_1985373, EPI_ISL_1985377,
EPI_ISL_1985383, EPI_ISL_1985384, EPI_ISL_1985387, EPI_ISL_1985389, EPI_ISL_1985390, EPI_ISL_1985394, EPI_ISL_1985396, EPI_ISL_1985398, EPI_ISL_1985401, EPI_ISL_1985402, EPI_ISL_1985411, EPI_ISL_1985413, EPI_ISL_1985414, EPI_ISL_1985419, EPI_ISL_1985420, EPI_ISL_1985421, EPI_ISL_1985427, EPI_ISL_1985429, EPI_ISL_1985431, EPI_ISL_1985436, EPI_ISL_1985437, EPI_ISL_1985454,
EPI_ISL_1985461, EPI_ISL_1985462, EPI_ISL_1985470, EPI_ISL_1985473, EPI_ISL_1985474, EPI_ISL_1985477, EPI_ISL_1985479, EPI_ISL_1985488, EPI_ISL_1985491, EPI_ISL_1985494, EPI_ISL_1985495, EPI_ISL_1985499, EPI_ISL_1985503, EPI_ISL_1985504, EPI_ISL_2005614, EPI_ISL_2005620, EPI_ISL_2005622, EPI_ISL_2005629, EPI_ISL_2005630, EPI_ISL_2005632, EPI_ISL_2005633, EPI_ISL_2005635,
EPI_ISL_2005637, EPI_ISL_2005639, EPI_ISL_2005640, EPI_ISL_2005641, EPI_ISL_2005643, EPI_ISL_2005655, EPI_ISL_2005657, EPI_ISL_2005661, EPI_ISL_2005668, EPI_ISL_2005682, EPI_ISL_2005684, EPI_ISL_2005691, EPI_ISL_2005692, EPI_ISL_2005694, EPI_ISL_2005697, EPI_ISL_2005698, EPI_ISL_2005702, EPI_ISL_2005707, EPI_ISL_2005709, EPI_ISL_2005710, EPI_ISL_2005711, EPI_ISL_2005722,
EPI_ISL_2005728, EPI_ISL_2005730, EPI_ISL_2005741, EPI_ISL_2005743, EPI_ISL_2005746, EPI_ISL_2005747, EPI_ISL_2005748, EPI_ISL_2005749, EPI_ISL_2005756, EPI_ISL_2005760, EPI_ISL_2005771, EPI_ISL_2005785, EPI_ISL_2005786, EPI_ISL_2005797, EPI_ISL_2006090, EPI_ISL_2006105, EPI_ISL_2006128, EPI_ISL_2006169, EPI_ISL_2006216, EPI_ISL_2006242, EPI_ISL_2006278, EPI_ISL_2006536,
EPI_ISL_2080485

| | | | |
|---|---|---|---|
| see above | Lighthouse Lab in Alderley Park | Wellcome Sanger Institute for the COVID-19 Genomics UK (COG-UK) Consortium | Cordelia Langford; David K. Jackson; Dominic Kwiatkowski; Ewan Harrison; Ian Johnston; Jacquelyn Wynn; Jeffrey Barrett; John Sillitoe on behalf of the Wellcome Sanger Institute COVID-19 Surveillance Team; Mairead Hyland; Roberto Amato; Sonia Goncalves; The Lighthouse Lab in Alderley Park and Alex Alderton |

EPI_ISL_1652951, EPI_ISL_1652958, EPI_ISL_1652973, EPI_ISL_1652991, EPI_ISL_1652993, EPI_ISL_1653001, EPI_ISL_1653012, EPI_ISL_1653022, EPI_ISL_1653050, EPI_ISL_1653052, EPI_ISL_1653053, EPI_ISL_1653061, EPI_ISL_1653062, EPI_ISL_1653080, EPI_ISL_1653133, EPI_ISL_1653138, EPI_ISL_1653149, EPI_ISL_1653157, EPI_ISL_1653191, EPI_ISL_1653200, EPI_ISL_1673055, EPI_ISL_1697503,
EPI_ISL_1697531, EPI_ISL_1697536, EPI_ISL_1697554, EPI_ISL_1697599, EPI_ISL_1697606, EPI_ISL_1697613, EPI_ISL_1697635, EPI_ISL_1697640, EPI_ISL_1697666, EPI_ISL_1697673, EPI_ISL_1697678, EPI_ISL_1697693, EPI_ISL_1697719, EPI_ISL_1697725, EPI_ISL_1697773, EPI_ISL_1697803, EPI_ISL_1697807, EPI_ISL_1697818, EPI_ISL_1697824, EPI_ISL_1697826, EPI_ISL_1697848, EPI_ISL_1697855,
EPI_ISL_1697858, EPI_ISL_1697868, EPI_ISL_1697870, EPI_ISL_1697911, EPI_ISL_1697922, EPI_ISL_1697932, EPI_ISL_1697956, EPI_ISL_1697964, EPI_ISL_1697967, EPI_ISL_1698255, EPI_ISL_1698258, EPI_ISL_1698260, EPI_ISL_1698262, EPI_ISL_1698264, EPI_ISL_1698267, EPI_ISL_1698268, EPI_ISL_1698272, EPI_ISL_1698273, EPI_ISL_1698274, EPI_ISL_1698279, EPI_ISL_1698280, EPI_ISL_1698283,
EPI_ISL_1698284, EPI_ISL_1698285, EPI_ISL_1698295, EPI_ISL_1698305, EPI_ISL_1698307, EPI_ISL_1698314, EPI_ISL_1698315, EPI_ISL_1698317, EPI_ISL_1698327, EPI_ISL_1698328, EPI_ISL_1698332, EPI_ISL_1698339, EPI_ISL_1698342, EPI_ISL_1698346, EPI_ISL_1698347, EPI_ISL_1698349, EPI_ISL_1698357, EPI_ISL_1698359, EPI_ISL_1698361, EPI_ISL_1698364, EPI_ISL_1698366, EPI_ISL_1698370,
EPI_ISL_1698373, EPI_ISL_1698378, EPI_ISL_1698379, EPI_ISL_1698383, EPI_ISL_1698384, EPI_ISL_1698387, EPI_ISL_1698391, EPI_ISL_1698393, EPI_ISL_1698398, EPI_ISL_1698403, EPI_ISL_1698405, EPI_ISL_1698406, EPI_ISL_1698407, EPI_ISL_1698409, EPI_ISL_1698410, EPI_ISL_1698412, EPI_ISL_1698417, EPI_ISL_1698418, EPI_ISL_1698426, EPI_ISL_1698428, EPI_ISL_1698429, EPI_ISL_1698430,
EPI_ISL_1698432, EPI_ISL_1698435, EPI_ISL_1698436, EPI_ISL_1698439, EPI_ISL_1698442, EPI_ISL_1698444, EPI_ISL_1698447, EPI_ISL_1698450, EPI_ISL_1698452, EPI_ISL_1698458, EPI_ISL_1698459, EPI_ISL_1698461, EPI_ISL_1698463, EPI_ISL_1698465, EPI_ISL_1698469, EPI_ISL_1698475, EPI_ISL_1698476, EPI_ISL_1698479, EPI_ISL_1698482, EPI_ISL_1698486, EPI_ISL_1698487, EPI_ISL_1698490,
EPI_ISL_1698492, EPI_ISL_1698493, EPI_ISL_1698498, EPI_ISL_1698501, EPI_ISL_1698502, EPI_ISL_1698504, EPI_ISL_1698505, EPI_ISL_1698506, EPI_ISL_1698508, EPI_ISL_1698510, EPI_ISL_1698511, EPI_ISL_1698520, EPI_ISL_1698521, EPI_ISL_1698524, EPI_ISL_1698526, EPI_ISL_1698527, EPI_ISL_1698532, EPI_ISL_1698533, EPI_ISL_1698534, EPI_ISL_1698537, EPI_ISL_1698539, EPI_ISL_1700203,
EPI_ISL_1700245, EPI_ISL_1700263, EPI_ISL_1700301, EPI_ISL_1700371, EPI_ISL_1700421, EPI_ISL_1700460, EPI_ISL_1700466, EPI_ISL_1700475, EPI_ISL_1718897, EPI_ISL_1718904, EPI_ISL_1718912, EPI_ISL_1718913, EPI_ISL_1718921, EPI_ISL_1718927, EPI_ISL_1718928, EPI_ISL_1718929, EPI_ISL_1718930, EPI_ISL_1718931, EPI_ISL_1718932, EPI_ISL_1718942, EPI_ISL_1718943, EPI_ISL_1718947,
EPI_ISL_1718959, EPI_ISL_1718968, EPI_ISL_1718971, EPI_ISL_1718973, EPI_ISL_1718979, EPI_ISL_1718980, EPI_ISL_1718981, EPI_ISL_1718989, EPI_ISL_1718991, EPI_ISL_1718992, EPI_ISL_1718997, EPI_ISL_1719004, EPI_ISL_1719007, EPI_ISL_1719010, EPI_ISL_1719011, EPI_ISL_1719013, EPI_ISL_1719017, EPI_ISL_1719022, EPI_ISL_1719026, EPI_ISL_1719027, EPI_ISL_1719033, EPI_ISL_1719036,
EPI_ISL_1719042, EPI_ISL_1719045, EPI_ISL_1719048, EPI_ISL_1719051, EPI_ISL_1719062, EPI_ISL_1719065, EPI_ISL_1719066, EPI_ISL_1719071, EPI_ISL_1719084, EPI_ISL_1719088, EPI_ISL_1719089, EPI_ISL_1719099, EPI_ISL_1719107, EPI_ISL_1719115, EPI_ISL_1719118, EPI_ISL_1719120, EPI_ISL_1719127, EPI_ISL_1719128, EPI_ISL_1719134, EPI_ISL_1719142, EPI_ISL_1719145, EPI_ISL_1719150,
EPI_ISL_1719157, EPI_ISL_1719159, EPI_ISL_1719167, EPI_ISL_1719168, EPI_ISL_1719175, EPI_ISL_1719177, EPI_ISL_1719191, EPI_ISL_1719195, EPI_ISL_1719196, EPI_ISL_1719214, EPI_ISL_1719216, EPI_ISL_1719434, EPI_ISL_1719435, EPI_ISL_1719438, EPI_ISL_1719441, EPI_ISL_1719444, EPI_ISL_1719445, EPI_ISL_1719447, EPI_ISL_1719452, EPI_ISL_1719458, EPI_ISL_1719460, EPI_ISL_1719466,
EPI_ISL_1719467, EPI_ISL_1719468, EPI_ISL_1719479, EPI_ISL_1719482, EPI_ISL_1719484, EPI_ISL_1719485, EPI_ISL_1719486, EPI_ISL_1719493, EPI_ISL_1719499, EPI_ISL_1719503, EPI_ISL_1719504, EPI_ISL_1719508, EPI_ISL_1719515, EPI_ISL_1719517, EPI_ISL_1719518, EPI_ISL_1719521, EPI_ISL_1719522, EPI_ISL_1719524, EPI_ISL_1719527, EPI_ISL_1719528, EPI_ISL_1719529, EPI_ISL_1719534,
EPI_ISL_1719536, EPI_ISL_1719539, EPI_ISL_1719543, EPI_ISL_1719546, EPI_ISL_1719551, EPI_ISL_1719561, EPI_ISL_1719569, EPI_ISL_1719570, EPI_ISL_1719574, EPI_ISL_1719575, EPI_ISL_1719583, EPI_ISL_1719584, EPI_ISL_1719585, EPI_ISL_1719587, EPI_ISL_1719592, EPI_ISL_1719594, EPI_ISL_1719596, EPI_ISL_1719607, EPI_ISL_1719618, EPI_ISL_1719623, EPI_ISL_1719629, EPI_ISL_1719630,
EPI_ISL_1719635, EPI_ISL_1719636, EPI_ISL_1719640, EPI_ISL_1719642, EPI_ISL_1719645, EPI_ISL_1719648, EPI_ISL_1719651, EPI_ISL_1719655, EPI_ISL_1719658, EPI_ISL_1719660, EPI_ISL_1719668, EPI_ISL_1719670, EPI_ISL_1719676, EPI_ISL_1719680, EPI_ISL_1719684, EPI_ISL_1719686, EPI_ISL_1719687, EPI_ISL_1719690, EPI_ISL_1719692, EPI_ISL_1719696, EPI_ISL_1719703, EPI_ISL_1719709,
EPI_ISL_1719719, EPI_ISL_1719725, EPI_ISL_1719727, EPI_ISL_1719729, EPI_ISL_1719735, EPI_ISL_1719739, EPI_ISL_1719740, EPI_ISL_1740534, EPI_ISL_1740565, EPI_ISL_1740618, EPI_ISL_1740672, EPI_ISL_1740689, EPI_ISL_1740729, EPI_ISL_1740734, EPI_ISL_1740758, EPI_ISL_1740779, EPI_ISL_1740780, EPI_ISL_1741274, EPI_ISL_1741283, EPI_ISL_1741336, EPI_ISL_1741358, EPI_ISL_1741363,
EPI_ISL_1741390, EPI_ISL_1741405, EPI_ISL_1741412, EPI_ISL_1741427, EPI_ISL_1741433, EPI_ISL_1741440, EPI_ISL_1741446, EPI_ISL_1741550, EPI_ISL_1741551, EPI_ISL_1741553, EPI_ISL_1741755, EPI_ISL_1742168, EPI_ISL_1742183, EPI_ISL_1742215, EPI_ISL_1742284, EPI_ISL_1742286, EPI_ISL_1742287, EPI_ISL_1742288, EPI_ISL_1742289, EPI_ISL_1742291, EPI_ISL_1742292, EPI_ISL_1742293,
EPI_ISL_1758455, EPI_ISL_1758463, EPI_ISL_1758509, EPI_ISL_1758530, EPI_ISL_1758532, EPI_ISL_1758536, EPI_ISL_1758547, EPI_ISL_1758558, EPI_ISL_1758625, EPI_ISL_1758645, EPI_ISL_1759180, EPI_ISL_1759202, EPI_ISL_1759215, EPI_ISL_1759219, EPI_ISL_1759242, EPI_ISL_1759257, EPI_ISL_1759265, EPI_ISL_1759267, EPI_ISL_1759268, EPI_ISL_1759290, EPI_ISL_1759295, EPI_ISL_1759306,
EPI_ISL_1759307, EPI_ISL_1759308, EPI_ISL_1759340, EPI_ISL_1759346, EPI_ISL_1759350, EPI_ISL_1759823, EPI_ISL_1759824, EPI_ISL_1759827, EPI_ISL_1759832, EPI_ISL_1759844, EPI_ISL_1759850, EPI_ISL_1759863, EPI_ISL_1759864, EPI_ISL_1759884, EPI_ISL_1759888, EPI_ISL_1759894, EPI_ISL_1791020, EPI_ISL_1807055, EPI_ISL_1807068, EPI_ISL_1807155, EPI_ISL_1807184, EPI_ISL_1912016,
EPI_ISL_1912019, EPI_ISL_1912026, EPI_ISL_1912027, EPI_ISL_1912028, EPI_ISL_1912029, EPI_ISL_1912036, EPI_ISL_1912038, EPI_ISL_1912041, EPI_ISL_1912049, EPI_ISL_1912064, EPI_ISL_1912066, EPI_ISL_1912074, EPI_ISL_1912076, EPI_ISL_1912078, EPI_ISL_1912085, EPI_ISL_1912086, EPI_ISL_1912088, EPI_ISL_1912091, EPI_ISL_1912093, EPI_ISL_1912111, EPI_ISL_1912114, EPI_ISL_1912119,
EPI_ISL_1912124, EPI_ISL_1912157, EPI_ISL_1912173, EPI_ISL_1912174, EPI_ISL_1912175, EPI_ISL_1912203, EPI_ISL_1912205, EPI_ISL_1912218, EPI_ISL_1912237, EPI_ISL_1912251, EPI_ISL_1912253, EPI_ISL_1912267, EPI_ISL_1912300, EPI_ISL_1912306, EPI_ISL_1912322, EPI_ISL_1912337, EPI_ISL_1912338, EPI_ISL_1939720

| | | | |
|---|---|---|---|
| see above | Lighthouse Lab in Cambridge | Wellcome Sanger Institute for the COVID-19 Genomics UK (COG-UK) Consortium | Cordelia Langford; David K. Jackson; Dominic Kwiatkowski; Ewan Harrison; Ian Johnston; Jeffrey Barrett; John Sillitoe on behalf of the Wellcome Sanger Institute COVID-19 Surveillance Team; Rob Howes; Roberto Amato; Sonia Goncalves; The Lighthouse Lab in Cambridge and Alex Alderton |

EPI_ISL_1652695, EPI_ISL_1652696, EPI_ISL_1652699, EPI_ISL_1652706, EPI_ISL_1652707, EPI_ISL_1652709, EPI_ISL_1652710, EPI_ISL_1652730, EPI_ISL_1652749, EPI_ISL_1652751, EPI_ISL_1652758, EPI_ISL_1652780, EPI_ISL_1652783, EPI_ISL_1652784, EPI_ISL_1652785, EPI_ISL_1652786, EPI_ISL_1652795, EPI_ISL_1652800, EPI_ISL_1652803, EPI_ISL_1652806, EPI_ISL_1652810, EPI_ISL_1652816,
EPI_ISL_1652817, EPI_ISL_1652823, EPI_ISL_1652829, EPI_ISL_1652842, EPI_ISL_1652868, EPI_ISL_1652885, EPI_ISL_1652889, EPI_ISL_1652909, EPI_ISL_1652911, EPI_ISL_1652914, EPI_ISL_1652925, EPI_ISL_1652932, EPI_ISL_1652934, EPI_ISL_1652940, EPI_ISL_1672971, EPI_ISL_1672973, EPI_ISL_1672974, EPI_ISL_1672975, EPI_ISL_1672976, EPI_ISL_1672978, EPI_ISL_1672979, EPI_ISL_1672980,
EPI_ISL_1672981, EPI_ISL_1672982, EPI_ISL_1672983, EPI_ISL_1672985, EPI_ISL_1672987, EPI_ISL_1672989, EPI_ISL_1672991, EPI_ISL_1672992, EPI_ISL_1672993, EPI_ISL_1672994, EPI_ISL_1672995, EPI_ISL_1672996, EPI_ISL_1672998, EPI_ISL_1672999, EPI_ISL_1673000, EPI_ISL_1673001, EPI_ISL_1673003, EPI_ISL_1673004, EPI_ISL_1673007, EPI_ISL_1673009, EPI_ISL_1673011, EPI_ISL_1673012,
EPI_ISL_1673013, EPI_ISL_1673014, EPI_ISL_1673015, EPI_ISL_1673016, EPI_ISL_1673020, EPI_ISL_1673021, EPI_ISL_1673022, EPI_ISL_1673024, EPI_ISL_1673025, EPI_ISL_1673026, EPI_ISL_1673027, EPI_ISL_1673028, EPI_ISL_1673029, EPI_ISL_1673030, EPI_ISL_1673032, EPI_ISL_1673033, EPI_ISL_1673034, EPI_ISL_1673036, EPI_ISL_1673037, EPI_ISL_1673038, EPI_ISL_1673039, EPI_ISL_1673040,
EPI_ISL_1673043, EPI_ISL_1697448, EPI_ISL_1697455, EPI_ISL_1697465, EPI_ISL_1697466, EPI_ISL_1697485, EPI_ISL_1697493, EPI_ISL_1697494, EPI_ISL_1697527, EPI_ISL_1697538, EPI_ISL_1697543, EPI_ISL_1697544, EPI_ISL_1697566, EPI_ISL_1697578, EPI_ISL_1697580, EPI_ISL_1697587, EPI_ISL_1697632, EPI_ISL_1697644, EPI_ISL_1697667, EPI_ISL_1697674, EPI_ISL_1697689, EPI_ISL_1697691,
EPI_ISL_1697726, EPI_ISL_1697728, EPI_ISL_1697729, EPI_ISL_1697734, EPI_ISL_1697744, EPI_ISL_1697745, EPI_ISL_1697750, EPI_ISL_1697751, EPI_ISL_1697753, EPI_ISL_1697765, EPI_ISL_1697766, EPI_ISL_1697767, EPI_ISL_1697771, EPI_ISL_1697774, EPI_ISL_1697778, EPI_ISL_1697780, EPI_ISL_1697781, EPI_ISL_1697786, EPI_ISL_1697790, EPI_ISL_1697793, EPI_ISL_1697794, EPI_ISL_1697796,
EPI_ISL_1697797, EPI_ISL_1697808, EPI_ISL_1697810, EPI_ISL_1697811, EPI_ISL_1697816, EPI_ISL_1697822, EPI_ISL_1697827, EPI_ISL_1697830, EPI_ISL_1697832, EPI_ISL_1697833, EPI_ISL_1697838, EPI_ISL_1697840, EPI_ISL_1697841, EPI_ISL_1697850, EPI_ISL_1697862, EPI_ISL_1697865, EPI_ISL_1697869, EPI_ISL_1697872, EPI_ISL_1697873, EPI_ISL_1697879, EPI_ISL_1697880, EPI_ISL_1697888,
EPI_ISL_1697891, EPI_ISL_1697892, EPI_ISL_1697893, EPI_ISL_1697894, EPI_ISL_1697901, EPI_ISL_1697905, EPI_ISL_1697906, EPI_ISL_1697910, EPI_ISL_1697913, EPI_ISL_1697916, EPI_ISL_1697929, EPI_ISL_1697931, EPI_ISL_1697933, EPI_ISL_1697936, EPI_ISL_1697937, EPI_ISL_1697938, EPI_ISL_1697939, EPI_ISL_1697940, EPI_ISL_1697944, EPI_ISL_1697950, EPI_ISL_1697951, EPI_ISL_1697952,
EPI_ISL_1697955, EPI_ISL_1697957, EPI_ISL_1697958, EPI_ISL_1698011, EPI_ISL_1698046, EPI_ISL_1698061, EPI_ISL_1698078, EPI_ISL_1698115, EPI_ISL_1698116, EPI_ISL_1698132, EPI_ISL_1698182, EPI_ISL_1698185, EPI_ISL_1698194, EPI_ISL_1698195, EPI_ISL_1698199, EPI_ISL_1698207, EPI_ISL_1698225, EPI_ISL_1698242, EPI_ISL_1698547, EPI_ISL_1698548, EPI_ISL_1698549, EPI_ISL_1698550,
EPI_ISL_1698551, EPI_ISL_1698552, EPI_ISL_1698553, EPI_ISL_1698554, EPI_ISL_1698555, EPI_ISL_1698556, EPI_ISL_1698557, EPI_ISL_1698559, EPI_ISL_1698560, EPI_ISL_1698561, EPI_ISL_1698562, EPI_ISL_1698563, EPI_ISL_1698564, EPI_ISL_1698565, EPI_ISL_1698567, EPI_ISL_1698568, EPI_ISL_1698569, EPI_ISL_1698570, EPI_ISL_1698571, EPI_ISL_1698572, EPI_ISL_1698573, EPI_ISL_1698574,
EPI_ISL_1698575, EPI_ISL_1698576, EPI_ISL_1698577, EPI_ISL_1698578, EPI_ISL_1698579, EPI_ISL_1698580, EPI_ISL_1698581, EPI_ISL_1698582, EPI_ISL_1698583, EPI_ISL_1698584, EPI_ISL_1698585, EPI_ISL_1698586, EPI_ISL_1698587, EPI_ISL_1698588, EPI_ISL_1698589, EPI_ISL_1698591, EPI_ISL_1698592, EPI_ISL_1698593, EPI_ISL_1698594, EPI_ISL_1698595, EPI_ISL_1698596, EPI_ISL_1698597,
EPI_ISL_1698598, EPI_ISL_1698599, EPI_ISL_1698600, EPI_ISL_1698601, EPI_ISL_1698602, EPI_ISL_1698603, EPI_ISL_1698604, EPI_ISL_1698605, EPI_ISL_1698606, EPI_ISL_1698607, EPI_ISL_1698608, EPI_ISL_1698609, EPI_ISL_1698611, EPI_ISL_1698612, EPI_ISL_1698613, EPI_ISL_1698614, EPI_ISL_1698616, EPI_ISL_1698617, EPI_ISL_1698618, EPI_ISL_1698619, EPI_ISL_1698620, EPI_ISL_1698622,
EPI_ISL_1698623, EPI_ISL_1698625, EPI_ISL_1698627, EPI_ISL_1698628, EPI_ISL_1698629, EPI_ISL_1698630, EPI_ISL_1698631, EPI_ISL_1698632, EPI_ISL_1698633, EPI_ISL_1698634, EPI_ISL_1698635, EPI_ISL_1698637, EPI_ISL_1698638, EPI_ISL_1698639, EPI_ISL_1698640, EPI_ISL_1698641, EPI_ISL_1698642, EPI_ISL_1698643, EPI_ISL_1698644, EPI_ISL_1698645, EPI_ISL_1698646, EPI_ISL_1698647,
EPI_ISL_1698648, EPI_ISL_1698649, EPI_ISL_1698651, EPI_ISL_1698652, EPI_ISL_1698654, EPI_ISL_1698655, EPI_ISL_1698656, EPI_ISL_1698657, EPI_ISL_1698658, EPI_ISL_1698659, EPI_ISL_1698660, EPI_ISL_1698661, EPI_ISL_1698664, EPI_ISL_1698665, EPI_ISL_1698666, EPI_ISL_1698667, EPI_ISL_1698668, EPI_ISL_1698669, EPI_ISL_1698670, EPI_ISL_1698671, EPI_ISL_1698672, EPI_ISL_1698673,
EPI_ISL_1698674, EPI_ISL_1698675, EPI_ISL_1698676, EPI_ISL_1698677, EPI_ISL_1698678, EPI_ISL_1698679, EPI_ISL_1698680, EPI_ISL_1698681, EPI_ISL_1698682, EPI_ISL_1698684, EPI_ISL_1698685, EPI_ISL_1698686, EPI_ISL_1698687, EPI_ISL_1698688, EPI_ISL_1698689, EPI_ISL_1698690, EPI_ISL_1698691, EPI_ISL_1698692, EPI_ISL_1698693, EPI_ISL_1698694, EPI_ISL_1698695, EPI_ISL_1698696,
EPI_ISL_1698697, EPI_ISL_1698698, EPI_ISL_1698699, EPI_ISL_1698700, EPI_ISL_1698701, EPI_ISL_1698702, EPI_ISL_1698703, EPI_ISL_1698704, EPI_ISL_1698705, EPI_ISL_1698706, EPI_ISL_1698707, EPI_ISL_1698708, EPI_ISL_1698709, EPI_ISL_1698710, EPI_ISL_1698711, EPI_ISL_1698712, EPI_ISL_1698713, EPI_ISL_1698714, EPI_ISL_1698715, EPI_ISL_1698716, EPI_ISL_1698717, EPI_ISL_1698718,
EPI_ISL_1698719, EPI_ISL_1698720, EPI_ISL_1698721, EPI_ISL_1698722, EPI_ISL_1698723, EPI_ISL_1698724, EPI_ISL_1698725, EPI_ISL_1698726, EPI_ISL_1698727, EPI_ISL_1698728, EPI_ISL_1698729, EPI_ISL_1698730, EPI_ISL_1698731, EPI_ISL_1698732, EPI_ISL_1698733, EPI_ISL_1698734, EPI_ISL_1698735, EPI_ISL_1698736, EPI_ISL_1698737, EPI_ISL_1698738, EPI_ISL_1698739, EPI_ISL_1698740,
EPI_ISL_1698741, EPI_ISL_1698742, EPI_ISL_1698743, EPI_ISL_1698744, EPI_ISL_1698745, EPI_ISL_1698746, EPI_ISL_1698747, EPI_ISL_1698748, EPI_ISL_1698749, EPI_ISL_1698750, EPI_ISL_1698752, EPI_ISL_1698753, EPI_ISL_1698754, EPI_ISL_1698755, EPI_ISL_1698756, EPI_ISL_1698757, EPI_ISL_1698758, EPI_ISL_1698760, EPI_ISL_1698761, EPI_ISL_1698762, EPI_ISL_1698763, EPI_ISL_1698765,
EPI_ISL_1698766, EPI_ISL_1698768, EPI_ISL_1698769, EPI_ISL_1698772, EPI_ISL_1698773, EPI_ISL_1698774, EPI_ISL_1698776, EPI_ISL_1698779, EPI_ISL_1698781, EPI_ISL_1698782, EPI_ISL_1698785, EPI_ISL_1698787, EPI_ISL_1698788, EPI_ISL_1698800, EPI_ISL_1698801, EPI_ISL_1698803, EPI_ISL_1698807, EPI_ISL_1698810, EPI_ISL_1698811, EPI_ISL_1698812, EPI_ISL_1698813, EPI_ISL_1698819,
EPI_ISL_1698820, EPI_ISL_1698822, EPI_ISL_1698823, EPI_ISL_1698827, EPI_ISL_1698836, EPI_ISL_1698837, EPI_ISL_1698839, EPI_ISL_1698841, EPI_ISL_1698842, EPI_ISL_1698845, EPI_ISL_1698847, EPI_ISL_1698848, EPI_ISL_1698851, EPI_ISL_1698853, EPI_ISL_1698854, EPI_ISL_1698856, EPI_ISL_1698863, EPI_ISL_1698872, EPI_ISL_1698874, EPI_ISL_1698881, EPI_ISL_1698882, EPI_ISL_1698885,
EPI_ISL_1698887, EPI_ISL_1698892, EPI_ISL_1698893, EPI_ISL_1698897, EPI_ISL_1698900, EPI_ISL_1698903, EPI_ISL_1698904, EPI_ISL_1698915, EPI_ISL_1698920, EPI_ISL_1698922, EPI_ISL_1698925, EPI_ISL_1698926, EPI_ISL_1698928, EPI_ISL_1698930, EPI_ISL_1698932, EPI_ISL_1698933, EPI_ISL_1698935, EPI_ISL_1698936, EPI_ISL_1698938, EPI_ISL_1698939, EPI_ISL_1698940, EPI_ISL_1698941,
EPI_ISL_1698947, EPI_ISL_1698948, EPI_ISL_1698951, EPI_ISL_1698952, EPI_ISL_1698953, EPI_ISL_1698954, EPI_ISL_1698957, EPI_ISL_1698958, EPI_ISL_1698960, EPI_ISL_1698961, EPI_ISL_1698962, EPI_ISL_1698964, EPI_ISL_1698969, EPI_ISL_1698970, EPI_ISL_1698973, EPI_ISL_1698976, EPI_ISL_1698981, EPI_ISL_1698982, EPI_ISL_1698984, EPI_ISL_1698991, EPI_ISL_1698992, EPI_ISL_1698997,
EPI_ISL_1699002, EPI_ISL_1699003, EPI_ISL_1699004, EPI_ISL_1699005, EPI_ISL_1699006, EPI_ISL_1699010, EPI_ISL_1699011, EPI_ISL_1699287, EPI_ISL_1699288, EPI_ISL_1699293, EPI_ISL_1699309, EPI_ISL_1699318, EPI_ISL_1699326, EPI_ISL_1699327, EPI_ISL_1699333, EPI_ISL_1699342, EPI_ISL_1699343, EPI_ISL_1699349, EPI_ISL_1699353, EPI_ISL_1699354, EPI_ISL_1699356, EPI_ISL_1699358,
EPI_ISL_1699360, EPI_ISL_1699367, EPI_ISL_1699373, EPI_ISL_1699374, EPI_ISL_1699379, EPI_ISL_1699382, EPI_ISL_1699383, EPI_ISL_1699390, EPI_ISL_1699397, EPI_ISL_1699408, EPI_ISL_1699423, EPI_ISL_1699427, EPI_ISL_1699430, EPI_ISL_1699434, EPI_ISL_1699454, EPI_ISL_1699459, EPI_ISL_1699460, EPI_ISL_1699463, EPI_ISL_1699464, EPI_ISL_1699468, EPI_ISL_1699469, EPI_ISL_1699471,
EPI_ISL_1699472, EPI_ISL_1699473, EPI_ISL_1699480, EPI_ISL_1699484, EPI_ISL_1699486, EPI_ISL_1699487, EPI_ISL_1699490, EPI_ISL_1699492, EPI_ISL_1699498, EPI_ISL_1699499, EPI_ISL_1699504, EPI_ISL_1699505, EPI_ISL_1699507, EPI_ISL_1699517, EPI_ISL_1699520, EPI_ISL_1699522, EPI_ISL_1699527, EPI_ISL_1699533, EPI_ISL_1699538, EPI_ISL_1699548, EPI_ISL_1699549, EPI_ISL_1699567,
EPI_ISL_1699621, EPI_ISL_1699633, EPI_ISL_1699649, EPI_ISL_1699670, EPI_ISL_1699677, EPI_ISL_1699688, EPI_ISL_1699697, EPI_ISL_1699711, EPI_ISL_1699714, EPI_ISL_1699718, EPI_ISL_1699719, EPI_ISL_1699732, EPI_ISL_1699741, EPI_ISL_1699748, EPI_ISL_1699749, EPI_ISL_1699762, EPI_ISL_1699764, EPI_ISL_1699778, EPI_ISL_1699783, EPI_ISL_1699790, EPI_ISL_1699808, EPI_ISL_1699813,
EPI_ISL_1699816, EPI_ISL_1718332, EPI_ISL_1718336, EPI_ISL_1718338, EPI_ISL_1718343, EPI_ISL_1718351, EPI_ISL_1718360, EPI_ISL_1718361, EPI_ISL_1718364, EPI_ISL_1718366, EPI_ISL_1718379, EPI_ISL_1718384, EPI_ISL_1718385, EPI_ISL_1718387, EPI_ISL_1718388, EPI_ISL_1718401, EPI_ISL_1718416, EPI_ISL_1718417, EPI_ISL_1718423, EPI_ISL_1718439, EPI_ISL_1718453, EPI_ISL_1718464,
EPI_ISL_1718466, EPI_ISL_1718471, EPI_ISL_1718474, EPI_ISL_1718492, EPI_ISL_1718493, EPI_ISL_1718498, EPI_ISL_1718499, EPI_ISL_1718505, EPI_ISL_1718512, EPI_ISL_1718521, EPI_ISL_1718522, EPI_ISL_1718535, EPI_ISL_1718536, EPI_ISL_1718538, EPI_ISL_1718552, EPI_ISL_1718557, EPI_ISL_1718563, EPI_ISL_1718567, EPI_ISL_1718570, EPI_ISL_1718579, EPI_ISL_1718591, EPI_ISL_1718608,
EPI_ISL_1718679, EPI_ISL_1718727, EPI_ISL_1718784, EPI_ISL_1718806, EPI_ISL_1719218, EPI_ISL_1719219, EPI_ISL_1719220, EPI_ISL_1719221, EPI_ISL_1719222, EPI_ISL_1719223, EPI_ISL_1719224, EPI_ISL_1719225, EPI_ISL_1719226, EPI_ISL_1719227, EPI_ISL_1719228, EPI_ISL_1719229, EPI_ISL_1719230, EPI_ISL_1719231, EPI_ISL_1719232, EPI_ISL_1719233, EPI_ISL_1719234, EPI_ISL_1719235,
EPI_ISL_1719236, EPI_ISL_1719237, EPI_ISL_1719238, EPI_ISL_1719239, EPI_ISL_1719240, EPI_ISL_1719241, EPI_ISL_1719242, EPI_ISL_1719243, EPI_ISL_1719244, EPI_ISL_1719245, EPI_ISL_1719246, EPI_ISL_1719247, EPI_ISL_1719248, EPI_ISL_1719249, EPI_ISL_1719250, EPI_ISL_1719251, EPI_ISL_1719252, EPI_ISL_1719253, EPI_ISL_1719254, EPI_ISL_1719255, EPI_ISL_1719256, EPI_ISL_1719257,
EPI_ISL_1719258, EPI_ISL_1719259, EPI_ISL_1719260, EPI_ISL_1719261, EPI_ISL_1719262, EPI_ISL_1719263, EPI_ISL_1719264, EPI_ISL_1719265, EPI_ISL_1719266, EPI_ISL_1719267, EPI_ISL_1719268, EPI_ISL_1719269, EPI_ISL_1719270, EPI_ISL_1719271, EPI_ISL_1719272, EPI_ISL_1719273, EPI_ISL_1719274, EPI_ISL_1719275, EPI_ISL_1719276, EPI_ISL_1719277, EPI_ISL_1719278, EPI_ISL_1719279,
EPI_ISL_1719280, EPI_ISL_1719281, EPI_ISL_1719282, EPI_ISL_1719283, EPI_ISL_1719284, EPI_ISL_1719285, EPI_ISL_1719286, EPI_ISL_1719287, EPI_ISL_1719288, EPI_ISL_1719289, EPI_ISL_1719290, EPI_ISL_1719291, EPI_ISL_1719292, EPI_ISL_1719293, EPI_ISL_1719294, EPI_ISL_1719295, EPI_ISL_1719296, EPI_ISL_1719297, EPI_ISL_1719298, EPI_ISL_1719299, EPI_ISL_1719300, EPI_ISL_1719301,
EPI_ISL_1719302, EPI_ISL_1719303, EPI_ISL_1719304, EPI_ISL_1719305, EPI_ISL_1719306, EPI_ISL_1719307, EPI_ISL_1719308, EPI_ISL_1719309, EPI_ISL_1719310, EPI_ISL_1719311, EPI_ISL_1719312, EPI_ISL_1719313, EPI_ISL_1719314, EPI_ISL_1719315, EPI_ISL_1719316, EPI_ISL_1719317, EPI_ISL_1719318, EPI_ISL_1719319, EPI_ISL_1719320, EPI_ISL_1719321, EPI_ISL_1719322, EPI_ISL_1719323,
EPI_ISL_1719324, EPI_ISL_1719325, EPI_ISL_1719326, EPI_ISL_1719327, EPI_ISL_1719328, EPI_ISL_1719329, EPI_ISL_1719330, EPI_ISL_1719331, EPI_ISL_1719332, EPI_ISL_1719333, EPI_ISL_1719334, EPI_ISL_1719335, EPI_ISL_1719336, EPI_ISL_1719337, EPI_ISL_1719338, EPI_ISL_1719339, EPI_ISL_1719340, EPI_ISL_1719341, EPI_ISL_1719342, EPI_ISL_1719343, EPI_ISL_1719344, EPI_ISL_1719345,
EPI_ISL_1719346, EPI_ISL_1719347, EPI_ISL_1719348, EPI_ISL_1719349, EPI_ISL_1719350, EPI_ISL_1719351, EPI_ISL_1719352, EPI_ISL_1719353, EPI_ISL_1719354, EPI_ISL_1719355, EPI_ISL_1719356, EPI_ISL_1719357, EPI_ISL_1719358, EPI_ISL_1719359, EPI_ISL_1719360, EPI_ISL_1719361, EPI_ISL_1719362, EPI_ISL_1719363, EPI_ISL_1719364, EPI_ISL_1719365, EPI_ISL_1719366, EPI_ISL_1719367,
EPI_ISL_1719368, EPI_ISL_1719369, EPI_ISL_1719370, EPI_ISL_1719371, EPI_ISL_1719372, EPI_ISL_1719373, EPI_ISL_1719374, EPI_ISL_1719375, EPI_ISL_1719376, EPI_ISL_1719377, EPI_ISL_1719378, EPI_ISL_1719379, EPI_ISL_1719380, EPI_ISL_1719381, EPI_ISL_1719382, EPI_ISL_1719383, EPI_ISL_1719384, EPI_ISL_1719385, EPI_ISL_1719386, EPI_ISL_1719387, EPI_ISL_1719388, EPI_ISL_1719389,
EPI_ISL_1719390, EPI_ISL_1719391, EPI_ISL_1719392, EPI_ISL_1719393, EPI_ISL_1719394, EPI_ISL_1719395, EPI_ISL_1719396, EPI_ISL_1719397, EPI_ISL_1719398, EPI_ISL_1719399, EPI_ISL_1719400, EPI_ISL_1719401, EPI_ISL_1719402, EPI_ISL_1719403, EPI_ISL_1719404, EPI_ISL_1719405, EPI_ISL_1719406, EPI_ISL_1719407, EPI_ISL_1719408, EPI_ISL_1719409, EPI_ISL_1719410, EPI_ISL_1719411,
EPI_ISL_1719412, EPI_ISL_1719413, EPI_ISL_1719414, EPI_ISL_1719415, EPI_ISL_1719416, EPI_ISL_1719417, EPI_ISL_1719418, EPI_ISL_1719419, EPI_ISL_1719420, EPI_ISL_1719421, EPI_ISL_1719422, EPI_ISL_1719423, EPI_ISL_1719424, EPI_ISL_1719425, EPI_ISL_1719426, EPI_ISL_1719427, EPI_ISL_1719428, EPI_ISL_1719429, EPI_ISL_1719430, EPI_ISL_1719431, EPI_ISL_1719432, EPI_ISL_1719436,
EPI_ISL_1719440, EPI_ISL_1719442, EPI_ISL_1719448, EPI_ISL_1719453, EPI_ISL_1719456, EPI_ISL_1719457, EPI_ISL_1719462, EPI_ISL_1719463, EPI_ISL_1719464, EPI_ISL_1719471, EPI_ISL_1719472, EPI_ISL_1719473, EPI_ISL_1719474, EPI_ISL_1719475, EPI_ISL_1719476, EPI_ISL_1719477, EPI_ISL_1719478, EPI_ISL_1719483, EPI_ISL_1719490, EPI_ISL_1719491, EPI_ISL_1719492, EPI_ISL_1719494,
EPI_ISL_1719495, EPI_ISL_1719497, EPI_ISL_1719502, EPI_ISL_1719509, EPI_ISL_1719510, EPI_ISL_1719511, EPI_ISL_1719516, EPI_ISL_1719520, EPI_ISL_1719523, EPI_ISL_1719525, EPI_ISL_1719530, EPI_ISL_1719531, EPI_ISL_1719532, EPI_ISL_1719535, EPI_ISL_1719537, EPI_ISL_1719538, EPI_ISL_1719540, EPI_ISL_1719541, EPI_ISL_1719545, EPI_ISL_1719550, EPI_ISL_1719552, EPI_ISL_1719553,
EPI_ISL_1719554, EPI_ISL_1719555, EPI_ISL_1719556, EPI_ISL_1719557, EPI_ISL_1719559, EPI_ISL_1719562, EPI_ISL_1719563, EPI_ISL_1719564, EPI_ISL_1719565, EPI_ISL_1719566, EPI_ISL_1719567, EPI_ISL_1719568, EPI_ISL_1719572, EPI_ISL_1719573, EPI_ISL_1719577, EPI_ISL_1719578, EPI_ISL_1719579, EPI_ISL_1719580, EPI_ISL_1719581, EPI_ISL_1719586, EPI_ISL_1719589, EPI_ISL_1719590,
EPI_ISL_1719597, EPI_ISL_1719601, EPI_ISL_1719602, EPI_ISL_1719608, EPI_ISL_1719610, EPI_ISL_1719611, EPI_ISL_1719612, EPI_ISL_1719614, EPI_ISL_1719615, EPI_ISL_1719617, EPI_ISL_1719619, EPI_ISL_1719620, EPI_ISL_1719621, EPI_ISL_1719622, EPI_ISL_1719626, EPI_ISL_1719627, EPI_ISL_1719628, EPI_ISL_1719631, EPI_ISL_1719632, EPI_ISL_1719633, EPI_ISL_1719634, EPI_ISL_1719637,
EPI_ISL_1719638, EPI_ISL_1719641, EPI_ISL_1719644, EPI_ISL_1719646, EPI_ISL_1719649, EPI_ISL_1719652, EPI_ISL_1719653, EPI_ISL_1719654, EPI_ISL_1719657, EPI_ISL_1719661, EPI_ISL_1719662, EPI_ISL_1719663, EPI_ISL_1719664, EPI_ISL_1719667, EPI_ISL_1719669, EPI_ISL_1719672, EPI_ISL_1719673, EPI_ISL_1719674, EPI_ISL_1719675, EPI_ISL_1719677, EPI_ISL_1719678, EPI_ISL_1719681,
EPI_ISL_1719682, EPI_ISL_1719683, EPI_ISL_1719689, EPI_ISL_1719693, EPI_ISL_1719694, EPI_ISL_1719695, EPI_ISL_1719698, EPI_ISL_1719700, EPI_ISL_1719701, EPI_ISL_1719702, EPI_ISL_1719704, EPI_ISL_1719705, EPI_ISL_1719706, EPI_ISL_1719707, EPI_ISL_1719708, EPI_ISL_1719714, EPI_ISL_1719715, EPI_ISL_1719716, EPI_ISL_1719718, EPI_ISL_1719720, EPI_ISL_1719722, EPI_ISL_1719723,
EPI_ISL_1719724, EPI_ISL_1719728, EPI_ISL_1740536, EPI_ISL_1740537, EPI_ISL_1740539, EPI_ISL_1740540, EPI_ISL_1740542, EPI_ISL_1740543, EPI_ISL_1740544, EPI_ISL_1740545, EPI_ISL_1740546, EPI_ISL_1740547, EPI_ISL_1740548, EPI_ISL_1740549, EPI_ISL_1740554, EPI_ISL_1740557, EPI_ISL_1740559, EPI_ISL_1740560, EPI_ISL_1740561, EPI_ISL_1740562, EPI_ISL_1740563, EPI_ISL_1740566,
EPI_ISL_1740568, EPI_ISL_1740569, EPI_ISL_1740570, EPI_ISL_1740573, EPI_ISL_1740575, EPI_ISL_1740577, EPI_ISL_1740581, EPI_ISL_1740582, EPI_ISL_1740583, EPI_ISL_1740584, EPI_ISL_1740586, EPI_ISL_1740587, EPI_ISL_1740591, EPI_ISL_1740593, EPI_ISL_1740594, EPI_ISL_1740596, EPI_ISL_1740598, EPI_ISL_1740602, EPI_ISL_1740603, EPI_ISL_1740606, EPI_ISL_1740608, EPI_ISL_1740612,

EPI_ISL_1740613, EPI_ISL_1740615, EPI_ISL_1740617, EPI_ISL_1740620, EPI_ISL_1740622, EPI_ISL_1740623, EPI_ISL_1740625, EPI_ISL_1740626, EPI_ISL_1740627, EPI_ISL_1740628, EPI_ISL_1740631, EPI_ISL_1740634, EPI_ISL_1740635, EPI_ISL_1740638, EPI_ISL_1740639, EPI_ISL_1740640, EPI_ISL_1740644, EPI_ISL_1740645, EPI_ISL_1740647, EPI_ISL_1740648, EPI_ISL_1740649, EPI_ISL_1740650, EPI_ISL_1740653, EPI_ISL_1740659, EPI_ISL_1740665, EPI_ISL_1740667, EPI_ISL_1740670, EPI_ISL_1740676, EPI_ISL_1740677, EPI_ISL_1740679, EPI_ISL_1740680, EPI_ISL_1740683, EPI_ISL_1740684, EPI_ISL_1740685, EPI_ISL_1740686, EPI_ISL_1740692, EPI_ISL_1740693, EPI_ISL_1740694, EPI_ISL_1740695, EPI_ISL_1740696, EPI_ISL_1740700, EPI_ISL_1740702, EPI_ISL_1740705, EPI_ISL_1740709, EPI_ISL_1740711, EPI_ISL_1740717, EPI_ISL_1740718, EPI_ISL_1740720, EPI_ISL_1740721, EPI_ISL_1740723, EPI_ISL_1740724, EPI_ISL_1740727, EPI_ISL_1740728, EPI_ISL_1740733, EPI_ISL_1740735, EPI_ISL_1740738, EPI_ISL_1740739, EPI_ISL_1740742, EPI_ISL_1740743, EPI_ISL_1740747, EPI_ISL_1740749, EPI_ISL_1740752, EPI_ISL_1740756, EPI_ISL_1740757, EPI_ISL_1740765, EPI_ISL_1740767, EPI_ISL_1740770, EPI_ISL_1740771, EPI_ISL_1740775, EPI_ISL_1740776, EPI_ISL_1740777, EPI_ISL_1740789, EPI_ISL_1740791, EPI_ISL_1740795, EPI_ISL_1740806, EPI_ISL_1740809, EPI_ISL_1740817, EPI_ISL_1740825, EPI_ISL_1740828, EPI_ISL_1740849, EPI_ISL_1740854, EPI_ISL_1740855, EPI_ISL_1740861, EPI_ISL_1740865, EPI_ISL_1740892, EPI_ISL_1740897, EPI_ISL_1740901, EPI_ISL_1740905, EPI_ISL_1740907, EPI_ISL_1740920, EPI_ISL_1740924, EPI_ISL_1740929, EPI_ISL_1740934, EPI_ISL_1740937, EPI_ISL_1740939, EPI_ISL_1740941, EPI_ISL_1740943, EPI_ISL_1740944, EPI_ISL_1740948, EPI_ISL_1740951, EPI_ISL_1740956, EPI_ISL_1740957, EPI_ISL_1740958, EPI_ISL_1740962, EPI_ISL_1740969, EPI_ISL_1740974, EPI_ISL_1740975, EPI_ISL_1740979, EPI_ISL_1740981, EPI_ISL_1740982, EPI_ISL_1740993, EPI_ISL_1740994, EPI_ISL_1740998, EPI_ISL_1741004, EPI_ISL_1741007, EPI_ISL_1741009, EPI_ISL_1741014, EPI_ISL_1741031, EPI_ISL_1741032, EPI_ISL_1741035, EPI_ISL_1741045, EPI_ISL_1741055, EPI_ISL_1741058, EPI_ISL_1741060, EPI_ISL_1741061, EPI_ISL_1741064, EPI_ISL_1741065, EPI_ISL_1741067, EPI_ISL_1741068, EPI_ISL_1741069, EPI_ISL_1741070, EPI_ISL_1741071, EPI_ISL_1741072, EPI_ISL_1741073, EPI_ISL_1741075, EPI_ISL_1741077, EPI_ISL_1741078, EPI_ISL_1741079, EPI_ISL_1741080, EPI_ISL_1741081, EPI_ISL_1741082, EPI_ISL_1741084, EPI_ISL_1741086, EPI_ISL_1741089, EPI_ISL_1741091, EPI_ISL_1741092, EPI_ISL_1741095, EPI_ISL_1741096, EPI_ISL_1741097, EPI_ISL_1741098, EPI_ISL_1741099, EPI_ISL_1741100, EPI_ISL_1741103, EPI_ISL_1741105, EPI_ISL_1741106, EPI_ISL_1741107, EPI_ISL_1741108, EPI_ISL_1741109, EPI_ISL_1741112, EPI_ISL_1741113, EPI_ISL_1741118, EPI_ISL_1741119, EPI_ISL_1741121, EPI_ISL_1741123, EPI_ISL_1741124, EPI_ISL_1741126, EPI_ISL_1741128, EPI_ISL_1741129, EPI_ISL_1741132, EPI_ISL_1741136, EPI_ISL_1741137, EPI_ISL_1741138, EPI_ISL_1741141, EPI_ISL_1741142, EPI_ISL_1741143, EPI_ISL_1741144, EPI_ISL_1741145, EPI_ISL_1741146, EPI_ISL_1741147, EPI_ISL_1741149, EPI_ISL_1741152, EPI_ISL_1741153, EPI_ISL_1741155, EPI_ISL_1741157, EPI_ISL_1741158, EPI_ISL_1741162, EPI_ISL_1741170, EPI_ISL_1741171, EPI_ISL_1741172, EPI_ISL_1741173, EPI_ISL_1741174, EPI_ISL_1741176, EPI_ISL_1741178, EPI_ISL_1741179, EPI_ISL_1741181, EPI_ISL_1741182, EPI_ISL_1741183, EPI_ISL_1741184, EPI_ISL_1741185, EPI_ISL_1741188, EPI_ISL_1741190, EPI_ISL_1741191, EPI_ISL_1741192, EPI_ISL_1741193, EPI_ISL_1741194, EPI_ISL_1741196, EPI_ISL_1741199, EPI_ISL_1741200, EPI_ISL_1741201, EPI_ISL_1741203, EPI_ISL_1741204, EPI_ISL_1741205, EPI_ISL_1741206, EPI_ISL_1741209, EPI_ISL_1741211, EPI_ISL_1741212, EPI_ISL_1741213, EPI_ISL_1741214, EPI_ISL_1741215, EPI_ISL_1741216, EPI_ISL_1741217, EPI_ISL_1741219, EPI_ISL_1741220, EPI_ISL_1741221, EPI_ISL_1741222, EPI_ISL_1741223, EPI_ISL_1741224, EPI_ISL_1741225, EPI_ISL_1741228, EPI_ISL_1741230, EPI_ISL_1741231, EPI_ISL_1741232, EPI_ISL_1741235, EPI_ISL_1741237, EPI_ISL_1741240, EPI_ISL_1741242, EPI_ISL_1741243, EPI_ISL_1741244, EPI_ISL_1741250, EPI_ISL_1741253, EPI_ISL_1741257, EPI_ISL_1741258, EPI_ISL_1741259, EPI_ISL_1741262, EPI_ISL_1741264, EPI_ISL_1741265, EPI_ISL_1741266, EPI_ISL_1741267, EPI_ISL_1741269, EPI_ISL_1741270, EPI_ISL_1741271, EPI_ISL_1741287, EPI_ISL_1741299, EPI_ISL_1741303, EPI_ISL_1741304, EPI_ISL_1741305, EPI_ISL_1741308, EPI_ISL_1741309, EPI_ISL_1741311, EPI_ISL_1741312, EPI_ISL_1741314, EPI_ISL_1741323, EPI_ISL_1741326, EPI_ISL_1741331, EPI_ISL_1741338, EPI_ISL_1741352, EPI_ISL_1741368, EPI_ISL_1741370, EPI_ISL_1741371, EPI_ISL_1741372, EPI_ISL_1741373, EPI_ISL_1741374, EPI_ISL_1741375, EPI_ISL_1741382, EPI_ISL_1741392, EPI_ISL_1741393, EPI_ISL_1741394, EPI_ISL_1741404, EPI_ISL_1741406, EPI_ISL_1741407, EPI_ISL_1741410, EPI_ISL_1741417, EPI_ISL_1741420, EPI_ISL_1741421, EPI_ISL_1741438, EPI_ISL_1741439, EPI_ISL_1741451, EPI_ISL_1741452, EPI_ISL_1741453, EPI_ISL_1741463, EPI_ISL_1741469, EPI_ISL_1741482, EPI_ISL_1741486, EPI_ISL_1741487, EPI_ISL_1741495, EPI_ISL_1741496, EPI_ISL_1741497, EPI_ISL_1741498, EPI_ISL_1741499, EPI_ISL_1741500, EPI_ISL_1741501, EPI_ISL_1741502, EPI_ISL_1741503, EPI_ISL_1741504, EPI_ISL_1741505, EPI_ISL_1741506, EPI_ISL_1741507, EPI_ISL_1741508, EPI_ISL_1741509, EPI_ISL_1741510, EPI_ISL_1741511, EPI_ISL_1741512, EPI_ISL_1741513, EPI_ISL_1741515, EPI_ISL_1741516, EPI_ISL_1741517, EPI_ISL_1741518, EPI_ISL_1741519, EPI_ISL_1741520, EPI_ISL_1741521, EPI_ISL_1741522, EPI_ISL_1741523, EPI_ISL_1741525, EPI_ISL_1741526, EPI_ISL_1741527, EPI_ISL_1741528, EPI_ISL_1741529, EPI_ISL_1741530, EPI_ISL_1741531, EPI_ISL_1741532, EPI_ISL_1741534, EPI_ISL_1741535, EPI_ISL_1741536, EPI_ISL_1741537, EPI_ISL_1741538, EPI_ISL_1741539, EPI_ISL_1741540, EPI_ISL_1741541, EPI_ISL_1741554, EPI_ISL_1741556, EPI_ISL_1741557, EPI_ISL_1741559, EPI_ISL_1741561, EPI_ISL_1741562, EPI_ISL_1741564, EPI_ISL_1741565, EPI_ISL_1741567, EPI_ISL_1741568, EPI_ISL_1741570, EPI_ISL_1741571, EPI_ISL_1741573, EPI_ISL_1741574, EPI_ISL_1741576, EPI_ISL_1741577, EPI_ISL_1741579, EPI_ISL_1741580, EPI_ISL_1741582, EPI_ISL_1741584, EPI_ISL_1741585, EPI_ISL_1741587, EPI_ISL_1741588, EPI_ISL_1741590, EPI_ISL_1741591, EPI_ISL_1741593, EPI_ISL_1741594, EPI_ISL_1741596, EPI_ISL_1741597, EPI_ISL_1741599, EPI_ISL_1741600, EPI_ISL_1741602, EPI_ISL_1741603, EPI_ISL_1741605, EPI_ISL_1741606, EPI_ISL_1741608, EPI_ISL_1741609, EPI_ISL_1741611, EPI_ISL_1741612, EPI_ISL_1741613, EPI_ISL_1741614, EPI_ISL_1741615, EPI_ISL_1741616, EPI_ISL_1741617, EPI_ISL_1741618, EPI_ISL_1741619, EPI_ISL_1741620, EPI_ISL_1741621, EPI_ISL_1741622, EPI_ISL_1741623, EPI_ISL_1741624, EPI_ISL_1741625, EPI_ISL_1741626, EPI_ISL_1741627, EPI_ISL_1741628, EPI_ISL_1741629, EPI_ISL_1741630, EPI_ISL_1741631, EPI_ISL_1741632, EPI_ISL_1741633, EPI_ISL_1741634, EPI_ISL_1741635, EPI_ISL_1741636, EPI_ISL_1741637, EPI_ISL_1741638, EPI_ISL_1741639, EPI_ISL_1741640, EPI_ISL_1741641, EPI_ISL_1741642, EPI_ISL_1741643, EPI_ISL_1741644, EPI_ISL_1741645, EPI_ISL_1741646, EPI_ISL_1741647, EPI_ISL_1741648, EPI_ISL_1741649, EPI_ISL_1741650, EPI_ISL_1741651, EPI_ISL_1741652, EPI_ISL_1741653, EPI_ISL_1741654, EPI_ISL_1741655, EPI_ISL_1741656, EPI_ISL_1741657, EPI_ISL_1741658, EPI_ISL_1741659, EPI_ISL_1741660, EPI_ISL_1741661, EPI_ISL_1741662, EPI_ISL_1741663, EPI_ISL_1741664, EPI_ISL_1741665, EPI_ISL_1741666, EPI_ISL_1741667, EPI_ISL_1741668, EPI_ISL_1741669, EPI_ISL_1741670, EPI_ISL_1741671, EPI_ISL_1741672, EPI_ISL_1741673, EPI_ISL_1741674, EPI_ISL_1741675, EPI_ISL_1741676, EPI_ISL_1741677, EPI_ISL_1741678, EPI_ISL_1741679, EPI_ISL_1741680, EPI_ISL_1741681, EPI_ISL_1741682, EPI_ISL_1741683, EPI_ISL_1741684, EPI_ISL_1741685, EPI_ISL_1741686, EPI_ISL_1741687, EPI_ISL_1741688, EPI_ISL_1741689, EPI_ISL_1741690, EPI_ISL_1741691, EPI_ISL_1741692, EPI_ISL_1741693, EPI_ISL_1741694, EPI_ISL_1741695, EPI_ISL_1741696, EPI_ISL_1741697, EPI_ISL_1741698, EPI_ISL_1741699, EPI_ISL_1741700, EPI_ISL_1741701, EPI_ISL_1741702, EPI_ISL_1741703, EPI_ISL_1741704, EPI_ISL_1741705, EPI_ISL_1741706, EPI_ISL_1741707, EPI_ISL_1741708, EPI_ISL_1741709, EPI_ISL_1741710, EPI_ISL_1741711, EPI_ISL_1741712, EPI_ISL_1741713, EPI_ISL_1741714, EPI_ISL_1741715, EPI_ISL_1741716, EPI_ISL_1741717, EPI_ISL_1741718, EPI_ISL_1741719, EPI_ISL_1741720, EPI_ISL_1741721, EPI_ISL_1741722, EPI_ISL_1741723, EPI_ISL_1741724, EPI_ISL_1741726, EPI_ISL_1741727, EPI_ISL_1741728, EPI_ISL_1741729, EPI_ISL_1741730, EPI_ISL_1741731, EPI_ISL_1741732, EPI_ISL_1741733, EPI_ISL_1741734, EPI_ISL_1741735, EPI_ISL_1741736, EPI_ISL_1741738, EPI_ISL_1741739, EPI_ISL_1741740, EPI_ISL_1741741, EPI_ISL_1741742, EPI_ISL_1741743, EPI_ISL_1741744, EPI_ISL_1741745, EPI_ISL_1741746, EPI_ISL_1741747, EPI_ISL_1741748, EPI_ISL_1741749, EPI_ISL_1741750, EPI_ISL_1741751, EPI_ISL_1741752, EPI_ISL_1741753, EPI_ISL_1741754, EPI_ISL_1741756, EPI_ISL_1741757, EPI_ISL_1741758, EPI_ISL_1741759, EPI_ISL_1741760, EPI_ISL_1741761, EPI_ISL_1741762, EPI_ISL_1741763, EPI_ISL_1741764, EPI_ISL_1741766, EPI_ISL_1741767, EPI_ISL_1741768, EPI_ISL_1741769, EPI_ISL_1741770, EPI_ISL_1741771, EPI_ISL_1741772, EPI_ISL_1741773, EPI_ISL_1741774, EPI_ISL_1741775, EPI_ISL_1741776, EPI_ISL_1741777, EPI_ISL_1741778, EPI_ISL_1741779, EPI_ISL_1741780, EPI_ISL_1741781, EPI_ISL_1741782, EPI_ISL_1741783, EPI_ISL_1741785, EPI_ISL_1741786, EPI_ISL_1741788, EPI_ISL_1741789, EPI_ISL_1741790, EPI_ISL_1741791, EPI_ISL_1741792, EPI_ISL_1741793, EPI_ISL_1741795, EPI_ISL_1741797, EPI_ISL_1741798, EPI_ISL_1741800, EPI_ISL_1741801, EPI_ISL_1741802, EPI_ISL_1741803, EPI_ISL_1741804, EPI_ISL_1741805, EPI_ISL_1741807, EPI_ISL_1741808, EPI_ISL_1741809, EPI_ISL_1741810, EPI_ISL_1741811, EPI_ISL_1741813, EPI_ISL_1741815, EPI_ISL_1741816, EPI_ISL_1741817, EPI_ISL_1741819, EPI_ISL_1741820, EPI_ISL_1741821, EPI_ISL_1741822, EPI_ISL_1741823, EPI_ISL_1741827, EPI_ISL_1741828, EPI_ISL_1741829, EPI_ISL_1741830, EPI_ISL_1741831, EPI_ISL_1741833, EPI_ISL_1741834, EPI_ISL_1741835, EPI_ISL_1741836, EPI_ISL_1741837, EPI_ISL_1741838, EPI_ISL_1741839, EPI_ISL_1741841, EPI_ISL_1741842, EPI_ISL_1741843, EPI_ISL_1741845, EPI_ISL_1741847, EPI_ISL_1741848, EPI_ISL_1741849, EPI_ISL_1741851, EPI_ISL_1741852, EPI_ISL_1741856, EPI_ISL_1741857, EPI_ISL_1741858, EPI_ISL_1741859, EPI_ISL_1741860, EPI_ISL_1741862, EPI_ISL_1741863, EPI_ISL_1741864, EPI_ISL_1741865, EPI_ISL_1741867, EPI_ISL_1741868, EPI_ISL_1741871, EPI_ISL_1741872, EPI_ISL_1741873, EPI_ISL_1741874, EPI_ISL_1741875, EPI_ISL_1741876, EPI_ISL_1741877, EPI_ISL_1741878, EPI_ISL_1741879, EPI_ISL_1741880, EPI_ISL_1741882, EPI_ISL_1741883, EPI_ISL_1741884, EPI_ISL_1741885, EPI_ISL_1741886, EPI_ISL_1741887, EPI_ISL_1741889, EPI_ISL_1741890, EPI_ISL_1741891, EPI_ISL_1741892, EPI_ISL_1741893, EPI_ISL_1741894, EPI_ISL_1741895, EPI_ISL_1741896, EPI_ISL_1741897, EPI_ISL_1741898, EPI_ISL_1741899, EPI_ISL_1741900, EPI_ISL_1741901, EPI_ISL_1741903, EPI_ISL_1741904, EPI_ISL_1741906, EPI_ISL_1741907, EPI_ISL_1741908, EPI_ISL_1741909, EPI_ISL_1741910, EPI_ISL_1741912, EPI_ISL_1741914, EPI_ISL_1741915, EPI_ISL_1741916, EPI_ISL_1741917, EPI_ISL_1741918, EPI_ISL_1741919, EPI_ISL_1741920, EPI_ISL_1741922, EPI_ISL_1741924, EPI_ISL_1741925, EPI_ISL_1741926, EPI_ISL_1741927, EPI_ISL_1741928, EPI_ISL_1741930, EPI_ISL_1741931, EPI_ISL_1741932, EPI_ISL_1741933, EPI_ISL_1741934, EPI_ISL_1741935, EPI_ISL_1741936, EPI_ISL_1741938, EPI_ISL_1741939, EPI_ISL_1741940, EPI_ISL_1741941, EPI_ISL_1741942, EPI_ISL_1741943, EPI_ISL_1741944, EPI_ISL_1741945, EPI_ISL_1741946, EPI_ISL_1741947, EPI_ISL_1741948, EPI_ISL_1741949, EPI_ISL_1741950, EPI_ISL_1741951, EPI_ISL_1741952, EPI_ISL_1741953, EPI_ISL_1741954, EPI_ISL_1741955, EPI_ISL_1741956, EPI_ISL_1741957, EPI_ISL_1741958, EPI_ISL_1741959, EPI_ISL_1741960, EPI_ISL_1741961, EPI_ISL_1741962, EPI_ISL_1741963, EPI_ISL_1741964, EPI_ISL_1741966, EPI_ISL_1741967, EPI_ISL_1741968, EPI_ISL_1741969, EPI_ISL_1741970, EPI_ISL_1741971, EPI_ISL_1741972, EPI_ISL_1741973, EPI_ISL_1741974, EPI_ISL_1741975, EPI_ISL_1741977, EPI_ISL_1741978, EPI_ISL_1741979, EPI_ISL_1741980, EPI_ISL_1741981, EPI_ISL_1741982, EPI_ISL_1741984, EPI_ISL_1741985, EPI_ISL_1741986, EPI_ISL_1741987, EPI_ISL_1741989, EPI_ISL_1741990, EPI_ISL_1741992, EPI_ISL_1741993, EPI_ISL_1741994, EPI_ISL_1741996, EPI_ISL_1741997, EPI_ISL_1741998, EPI_ISL_1741999, EPI_ISL_1742000, EPI_ISL_1742001, EPI_ISL_1742002, EPI_ISL_1742003, EPI_ISL_1742004, EPI_ISL_1742005, EPI_ISL_1742006, EPI_ISL_1742007, EPI_ISL_1742008, EPI_ISL_1742009, EPI_ISL_1742010, EPI_ISL_1742011, EPI_ISL_1742028, EPI_ISL_1742087, EPI_ISL_1742088, EPI_ISL_1742090, EPI_ISL_1742092, EPI_ISL_1742094, EPI_ISL_1742095, EPI_ISL_1742096, EPI_ISL_1742097, EPI_ISL_1742098, EPI_ISL_1742099, EPI_ISL_1742100, EPI_ISL_1742103, EPI_ISL_1742105, EPI_ISL_1742106, EPI_ISL_1742107, EPI_ISL_1742108, EPI_ISL_1742109, EPI_ISL_1742110, EPI_ISL_1742111, EPI_ISL_1742112, EPI_ISL_1742113, EPI_ISL_1742114, EPI_ISL_1742115, EPI_ISL_1742116, EPI_ISL_1742117, EPI_ISL_1742118, EPI_ISL_1742119, EPI_ISL_1742120, EPI_ISL_1742123, EPI_ISL_1742124, EPI_ISL_1742126, EPI_ISL_1742127, EPI_ISL_1742128, EPI_ISL_1742129, EPI_ISL_1742130, EPI_ISL_1742131, EPI_ISL_1742134, EPI_ISL_1742135, EPI_ISL_1742137, EPI_ISL_1742138, EPI_ISL_1742139, EPI_ISL_1742140, EPI_ISL_1742141, EPI_ISL_1742142, EPI_ISL_1742143, EPI_ISL_1742144, EPI_ISL_1742145, EPI_ISL_1742148, EPI_ISL_1742149, EPI_ISL_1742150, EPI_ISL_1742151, EPI_ISL_1742154, EPI_ISL_1742158, EPI_ISL_1742159, EPI_ISL_1742160, EPI_ISL_1742161, EPI_ISL_1742163, EPI_ISL_1742165, EPI_ISL_1742166, EPI_ISL_1742167, EPI_ISL_1742169, EPI_ISL_1742170, EPI_ISL_1742172, EPI_ISL_1742173, EPI_ISL_1742174, EPI_ISL_1742175, EPI_ISL_1742177, EPI_ISL_1742180, EPI_ISL_1742181, EPI_ISL_1742184, EPI_ISL_1742185, EPI_ISL_1742186, EPI_ISL_1742187, EPI_ISL_1742188, EPI_ISL_1742189, EPI_ISL_1742190, EPI_ISL_1742192, EPI_ISL_1742193, EPI_ISL_1742194, EPI_ISL_1742195, EPI_ISL_1742196, EPI_ISL_1742198, EPI_ISL_1742199, EPI_ISL_1742200, EPI_ISL_1742201, EPI_ISL_1742202, EPI_ISL_1742206, EPI_ISL_1742207, EPI_ISL_1742208, EPI_ISL_1742209, EPI_ISL_1742212, EPI_ISL_1742213, EPI_ISL_1742214, EPI_ISL_1742216, EPI_ISL_1742217, EPI_ISL_1742218, EPI_ISL_1742219, EPI_ISL_1742220, EPI_ISL_1742223, EPI_ISL_1742224, EPI_ISL_1742225, EPI_ISL_1742227, EPI_ISL_1742228, EPI_ISL_1742230, EPI_ISL_1742234, EPI_ISL_1742235, EPI_ISL_1742236, EPI_ISL_1742237, EPI_ISL_1742238, EPI_ISL_1742240, EPI_ISL_1742241, EPI_ISL_1742244, EPI_ISL_1742245, EPI_ISL_1742246, EPI_ISL_1742247, EPI_ISL_1742248, EPI_ISL_1742251, EPI_ISL_1742252, EPI_ISL_1742253, EPI_ISL_1742255, EPI_ISL_1742256, EPI_ISL_1742257, EPI_ISL_1742262, EPI_ISL_1742263, EPI_ISL_1742264, EPI_ISL_1742265, EPI_ISL_1742266, EPI_ISL_1742267, EPI_ISL_1742268, EPI_ISL_1742269, EPI_ISL_1742270, EPI_ISL_1742271, EPI_ISL_1742272, EPI_ISL_1742273, EPI_ISL_1742275, EPI_ISL_1742276, EPI_ISL_1742277, EPI_ISL_1742278, EPI_ISL_1742279, EPI_ISL_1742280, EPI_ISL_1742281, EPI_ISL_1742282, EPI_ISL_1742283, EPI_ISL_1758187, EPI_ISL_1758189, EPI_ISL_1758192, EPI_ISL_1758193, EPI_ISL_1758194, EPI_ISL_1758197, EPI_ISL_1758199, EPI_ISL_1758200, EPI_ISL_1758201, EPI_ISL_1758204, EPI_ISL_1758207, EPI_ISL_1758211, EPI_ISL_1758216, EPI_ISL_1758219, EPI_ISL_1758220, EPI_ISL_1758223, EPI_ISL_1758225, EPI_ISL_1758227, EPI_ISL_1758229, EPI_ISL_1758230, EPI_ISL_1758235, EPI_ISL_1758236, EPI_ISL_1758239, EPI_ISL_1758240, EPI_ISL_1758242, EPI_ISL_1758244, EPI_ISL_1758246, EPI_ISL_1758248, EPI_ISL_1758249, EPI_ISL_1758250, EPI_ISL_1758251, EPI_ISL_1758255, EPI_ISL_1758257, EPI_ISL_1758264, EPI_ISL_1758267, EPI_ISL_1758271, EPI_ISL_1758272, EPI_ISL_1758275, EPI_ISL_1758279, EPI_ISL_1758280, EPI_ISL_1758285, EPI_ISL_1758286, EPI_ISL_1758287, EPI_ISL_1758291, EPI_ISL_1758295, EPI_ISL_1758298, EPI_ISL_1758299, EPI_ISL_1758304, EPI_ISL_1758305, EPI_ISL_1758307, EPI_ISL_1758312, EPI_ISL_1758315, EPI_ISL_1758318, EPI_ISL_1758321, EPI_ISL_1758326, EPI_ISL_1758330, EPI_ISL_1758333, EPI_ISL_1758334, EPI_ISL_1758336, EPI_ISL_1758337, EPI_ISL_1758338, EPI_ISL_1758339, EPI_ISL_1758340, EPI_ISL_1758345, EPI_ISL_1758346, EPI_ISL_1758357, EPI_ISL_1758358, EPI_ISL_1758359, EPI_ISL_1758365, EPI_ISL_1758366, EPI_ISL_1758369, EPI_ISL_1758370, EPI_ISL_1758371, EPI_ISL_1758374, EPI_ISL_1758377, EPI_ISL_1758380, EPI_ISL_1758381, EPI_ISL_1758382, EPI_ISL_1758384, EPI_ISL_1758385, EPI_ISL_1758386, EPI_ISL_1758391, EPI_ISL_1758392, EPI_ISL_1758393, EPI_ISL_1758394, EPI_ISL_1758395, EPI_ISL_1758396, EPI_ISL_1758399, EPI_ISL_1758403, EPI_ISL_1758407, EPI_ISL_1758408, EPI_ISL_1758409, EPI_ISL_1758410, EPI_ISL_1758413, EPI_ISL_1758421, EPI_ISL_1758431, EPI_ISL_1758432, EPI_ISL_1758434, EPI_ISL_1758435, EPI_ISL_1758436, EPI_ISL_1758438, EPI_ISL_1758439, EPI_ISL_1758441, EPI_ISL_1758445, EPI_ISL_1758447, EPI_ISL_1758452, EPI_ISL_1758460, EPI_ISL_1758462, EPI_ISL_1758465, EPI_ISL_1758469, EPI_ISL_1758470, EPI_ISL_1758479, EPI_ISL_1758482, EPI_ISL_1758483, EPI_ISL_1758486, EPI_ISL_1758493, EPI_ISL_1758496, EPI_ISL_1758505, EPI_ISL_1758507, EPI_ISL_1758510, EPI_ISL_1758511, EPI_ISL_1758518, EPI_ISL_1758522, EPI_ISL_1758525, EPI_ISL_1758527, EPI_ISL_1758529, EPI_ISL_1758531, EPI_ISL_1758538, EPI_ISL_1758541, EPI_ISL_1758555, EPI_ISL_1758563, EPI_ISL_1758567, EPI_ISL_1758568, EPI_ISL_1758572, EPI_ISL_1758573, EPI_ISL_1758578, EPI_ISL_1758582, EPI_ISL_1758597, EPI_ISL_1758606, EPI_ISL_1758609, EPI_ISL_1758612, EPI_ISL_1758614, EPI_ISL_1758624, EPI_ISL_1758630, EPI_ISL_1758635, EPI_ISL_1758656, EPI_ISL_1758658, EPI_ISL_1758660, EPI_ISL_1758666, EPI_ISL_1758667, EPI_ISL_1758668, EPI_ISL_1758669, EPI_ISL_1758670, EPI_ISL_1758671, EPI_ISL_1758672, EPI_ISL_1758673, EPI_ISL_1758674, EPI_ISL_1758675, EPI_ISL_1758676, EPI_ISL_1758677, EPI_ISL_1758678, EPI_ISL_1758679, EPI_ISL_1758680, EPI_ISL_1758681, EPI_ISL_1758682, EPI_ISL_1758683, EPI_ISL_1758684, EPI_ISL_1758685, EPI_ISL_1758686, EPI_ISL_1758687, EPI_ISL_1758688, EPI_ISL_1758689, EPI_ISL_1758690, EPI_ISL_1758691, EPI_ISL_1758692, EPI_ISL_1758693, EPI_ISL_1758694, EPI_ISL_1758695, EPI_ISL_1758696, EPI_ISL_1758697, EPI_ISL_1758698, EPI_ISL_1758699, EPI_ISL_1758700, EPI_ISL_1758701, EPI_ISL_1758702, EPI_ISL_1758703, EPI_ISL_1758704, EPI_ISL_1758705, EPI_ISL_1758706, EPI_ISL_1758707, EPI_ISL_1758708, EPI_ISL_1758709, EPI_ISL_1758710, EPI_ISL_1758711, EPI_ISL_1758712, EPI_ISL_1758713, EPI_ISL_1758714, EPI_ISL_1758715, EPI_ISL_1758716, EPI_ISL_1758717, EPI_ISL_1758718, EPI_ISL_1758719, EPI_ISL_1758720, EPI_ISL_1758721, EPI_ISL_1758722, EPI_ISL_1758723, EPI_ISL_1758724, EPI_ISL_1758725, EPI_ISL_1758726, EPI_ISL_1758727, EPI_ISL_1758728, EPI_ISL_1758729, EPI_ISL_1758730, EPI_ISL_1758731, EPI_ISL_1758732, EPI_ISL_1758733, EPI_ISL_1758734, EPI_ISL_1758735, EPI_ISL_1758736, EPI_ISL_1758737, EPI_ISL_1758738, EPI_ISL_1758739, EPI_ISL_1758740, EPI_ISL_1758741, EPI_ISL_1758742, EPI_ISL_1758744, EPI_ISL_1758745, EPI_ISL_1758746, EPI_ISL_1758747, EPI_ISL_1758748, EPI_ISL_1758749, EPI_ISL_1758750, EPI_ISL_1758751, EPI_ISL_1758752, EPI_ISL_1758753, EPI_ISL_1758754, EPI_ISL_1758755, EPI_ISL_1758756, EPI_ISL_1758757, EPI_ISL_1758758, EPI_ISL_1758759, EPI_ISL_1758760, EPI_ISL_1758761, EPI_ISL_1758762, EPI_ISL_1758763, EPI_ISL_1758764, EPI_ISL_1758765, EPI_ISL_1758766, EPI_ISL_1758767, EPI_ISL_1758768, EPI_ISL_1758769, EPI_ISL_1758770, EPI_ISL_1758771, EPI_ISL_1758772, EPI_ISL_1758773, EPI_ISL_1758774, EPI_ISL_1758775, EPI_ISL_1758776, EPI_ISL_1758777, EPI_ISL_1758779, EPI_ISL_1758780, EPI_ISL_1758782, EPI_ISL_1758783, EPI_ISL_1758785, EPI_ISL_1758786, EPI_ISL_1758787, EPI_ISL_1758789, EPI_ISL_1758791, EPI_ISL_1758793, EPI_ISL_1758794, EPI_ISL_1758796, EPI_ISL_1758797, EPI_ISL_1758798, EPI_ISL_1758800, EPI_ISL_1758801, EPI_ISL_1758802, EPI_ISL_1758803, EPI_ISL_1758804, EPI_ISL_1758805, EPI_ISL_1758806, EPI_ISL_1758807, EPI_ISL_1758808, EPI_ISL_1758809, EPI_ISL_1758810, EPI_ISL_1758811, EPI_ISL_1758812, EPI_ISL_1758813, EPI_ISL_1758814, EPI_ISL_1758815, EPI_ISL_1758816, EPI_ISL_1758817, EPI_ISL_1758818, EPI_ISL_1758819, EPI_ISL_1758820, EPI_ISL_1758821, EPI_ISL_1758822, EPI_ISL_1758823, EPI_ISL_1758825, EPI_ISL_1758826, EPI_ISL_1758827, EPI_ISL_1758828, EPI_ISL_1758829, EPI_ISL_1758830, EPI_ISL_1758832, EPI_ISL_1758833, EPI_ISL_1758834, EPI_ISL_1758835, EPI_ISL_1758836, EPI_ISL_1758837, EPI_ISL_1758838, EPI_ISL_1758839, EPI_ISL_1758840, EPI_ISL_1758841, EPI_ISL_1758842, EPI_ISL_1758844, EPI_ISL_1758845, EPI_ISL_1758846, EPI_ISL_1758847, EPI_ISL_1758849, EPI_ISL_1758850, EPI_ISL_1758851, EPI_ISL_1758852, EPI_ISL_1758853, EPI_ISL_1758854, EPI_ISL_1758855, EPI_ISL_1758856, EPI_ISL_1758857, EPI_ISL_1758858, EPI_ISL_1758859, EPI_ISL_1758860, EPI_ISL_1758861, EPI_ISL_1758862, EPI_ISL_1758863, EPI_ISL_1758864, EPI_ISL_1758865, EPI_ISL_1758866, EPI_ISL_1758867, EPI_ISL_1758868, EPI_ISL_1758870, EPI_ISL_1758871, EPI_ISL_1758872, EPI_ISL_1758873, EPI_ISL_1758874, EPI_ISL_1758875, EPI_ISL_1758876, EPI_ISL_1758877, EPI_ISL_1758883, EPI_ISL_1758887, EPI_ISL_1758889, EPI_ISL_1758893, EPI_ISL_1758894, EPI_ISL_1758897, EPI_ISL_1758901, EPI_ISL_1758913, EPI_ISL_1758916, EPI_ISL_1758920, EPI_ISL_1758924, EPI_ISL_1758928, EPI_ISL_1758933, EPI_ISL_1758938, EPI_ISL_1758939, EPI_ISL_1758940, EPI_ISL_1758942, EPI_ISL_1758943, EPI_ISL_1758945, EPI_ISL_1758947, EPI_ISL_1758949, EPI_ISL_1758951, EPI_ISL_1758952, EPI_ISL_1758953, EPI_ISL_1758954, EPI_ISL_1758957, EPI_ISL_1758961, EPI_ISL_1758966, EPI_ISL_1758969, EPI_ISL_1758976, EPI_ISL_1758979, EPI_ISL_1758981, EPI_ISL_1758987, EPI_ISL_1758990, EPI_ISL_1758993, EPI_ISL_1758994, EPI_ISL_1758996, EPI_ISL_1758999, EPI_ISL_1759003, EPI_ISL_1759011, EPI_ISL_1759012, EPI_ISL_1759021, EPI_ISL_1759022, EPI_ISL_1759030, EPI_ISL_1759036, EPI_ISL_1759037, EPI_ISL_1759040, EPI_ISL_1759044, EPI_ISL_1759047, EPI_ISL_1759051, EPI_ISL_1759052, EPI_ISL_1759061, EPI_ISL_1759066, EPI_ISL_1759067, EPI_ISL_1759068, EPI_ISL_1759072, EPI_ISL_1759074, EPI_ISL_1759076, EPI_ISL_1759078, EPI_ISL_1759082, EPI_ISL_1759085, EPI_ISL_1759096, EPI_ISL_1759099, EPI_ISL_1759116, EPI_ISL_1759119, EPI_ISL_1759120, EPI_ISL_1759125, EPI_ISL_1759126, EPI_ISL_1759128, EPI_ISL_1759138, EPI_ISL_1759141, EPI_ISL_1759144, EPI_ISL_1759145, EPI_ISL_1759147, EPI_ISL_1759152, EPI_ISL_1759159, EPI_ISL_1759160, EPI_ISL_1759162, EPI_ISL_1759163, EPI_ISL_1759166, EPI_ISL_1759169, EPI_ISL_1759172, EPI_ISL_1759184, EPI_ISL_1759186, EPI_ISL_1759208, EPI_ISL_1759209, EPI_ISL_1759221, EPI_ISL_1759222, EPI_ISL_1759227, EPI_ISL_1759237, EPI_ISL_1759248, EPI_ISL_1759251, EPI_ISL_1759269, EPI_ISL_1759270, EPI_ISL_1759282, EPI_ISL_1759287, EPI_ISL_1759288, EPI_ISL_1759289, EPI_ISL_1759294, EPI_ISL_1759297, EPI_ISL_1759300, EPI_ISL_1759305, EPI_ISL_1759309, EPI_ISL_1759314, EPI_ISL_1759317, EPI_ISL_1759329, EPI_ISL_1759339, EPI_ISL_1759347, EPI_ISL_1759814, EPI_ISL_1759817, EPI_ISL_1759829, EPI_ISL_1759833, EPI_ISL_1759840, EPI_ISL_1759859, EPI_ISL_1759861, EPI_ISL_1759867, EPI_ISL_1759873, EPI_ISL_1759877, EPI_ISL_1759881, EPI_ISL_1759890, EPI_ISL_1759896, EPI_ISL_1759897, EPI_ISL_1759898, EPI_ISL_1759899, EPI_ISL_1759900, EPI_ISL_1759901, EPI_ISL_1759902, EPI_ISL_1759903, EPI_ISL_1759904, EPI_ISL_1759905, EPI_ISL_1759906, EPI_ISL_1759907, EPI_ISL_1759908, EPI_ISL_1759909, EPI_ISL_1759910, EPI_ISL_1759911, EPI_ISL_1759912, EPI_ISL_1759913, EPI_ISL_1759914, EPI_ISL_1759915, EPI_ISL_1759916, EPI_ISL_1759917, EPI_ISL_1759918, EPI_ISL_1759919, EPI_ISL_1759920, EPI_ISL_1759921, EPI_ISL_1759922, EPI_ISL_1759923, EPI_ISL_1759924, EPI_ISL_1759925, EPI_ISL_1759926, EPI_ISL_1759927, EPI_ISL_1759928, EPI_ISL_1759929, EPI_ISL_1759930, EPI_ISL_1759931, EPI_ISL_1759932, EPI_ISL_1759933, EPI_ISL_1759934, EPI_ISL_1759935, EPI_ISL_1759936, EPI_ISL_1759937, EPI_ISL_1759938, EPI_ISL_1759939, EPI_ISL_1759940, EPI_ISL_1759941, EPI_ISL_1759942, EPI_ISL_1790216, EPI_ISL_1790222, EPI_ISL_1790228, EPI_ISL_1790229, EPI_ISL_1790230, EPI_ISL_1790231, EPI_ISL_1790235, EPI_ISL_1790238, EPI_ISL_1790249, EPI_ISL_1790252, EPI_ISL_1790253, EPI_ISL_1790255, EPI_ISL_1790256, EPI_ISL_1790263, EPI_ISL_1790266, EPI_ISL_1790274, EPI_ISL_1790275, EPI_ISL_1790285, EPI_ISL_1790290, EPI_ISL_1790292, EPI_ISL_1790295, EPI_ISL_1790306, EPI_ISL_1790314, EPI_ISL_1790315, EPI_ISL_1790316, EPI_ISL_1790321, EPI_ISL_1790327, EPI_ISL_1790331, EPI_ISL_1790332, EPI_ISL_1790336, EPI_ISL_1790337, EPI_ISL_1790342, EPI_ISL_1790344, EPI_ISL_1790351, EPI_ISL_1790357, EPI_ISL_1790359, EPI_ISL_1790362, EPI_ISL_1790371, EPI_ISL_1790373, EPI_ISL_1790375, EPI_ISL_1790380, EPI_ISL_1790387, EPI_ISL_1790394, EPI_ISL_1790395, EPI_ISL_1790397, EPI_ISL_1790398, EPI_ISL_1790399, EPI_ISL_1790402, EPI_ISL_1790408, EPI_ISL_1790409, EPI_ISL_1790411, EPI_ISL_1790413, EPI_ISL_1790419, EPI_ISL_1790426, EPI_ISL_1790447, EPI_ISL_1790448, EPI_ISL_1790456, EPI_ISL_1790459, EPI_ISL_1790467, EPI_ISL_1790480, EPI_ISL_1790491, EPI_ISL_1790493, EPI_ISL_1790502, EPI_ISL_1790505, EPI_ISL_1790515, EPI_ISL_1790524, EPI_ISL_1790531, EPI_ISL_1790533, EPI_ISL_1790536, EPI_ISL_1790544, EPI_ISL_1790552, EPI_ISL_1790553, EPI_ISL_1790555, EPI_ISL_1790557, EPI_ISL_1790558, EPI_ISL_1790567, EPI_ISL_1790569, EPI_ISL_1790574, EPI_ISL_1790581, EPI_ISL_1790601, EPI_ISL_1790606, EPI_ISL_1790615, EPI_ISL_1790627, EPI_ISL_1790634, EPI_ISL_1790635, EPI_ISL_1790636, EPI_ISL_1790647, EPI_ISL_1790650, EPI_ISL_1790652, EPI_ISL_1790659, EPI_ISL_1790667, EPI_ISL_1790676, EPI_ISL_1790684, EPI_ISL_1790691, EPI_ISL_1790694, EPI_ISL_1790705, EPI_ISL_1790710, EPI_ISL_1790717, EPI_ISL_1790721, EPI_ISL_1790722, EPI_ISL_1790739, EPI_ISL_1790755, EPI_ISL_1790756, EPI_ISL_1790772, EPI_ISL_1790773, EPI_ISL_1790795, EPI_ISL_1790811, EPI_ISL_1790838, EPI_ISL_1790849, EPI_ISL_1790859, EPI_ISL_1790871, EPI_ISL_1790873, EPI_ISL_1790895, EPI_ISL_1790920, EPI_ISL_1790932, EPI_ISL_1790933, EPI_ISL_1790950, EPI_ISL_1790952, EPI_ISL_1790956, EPI_ISL_1790958, EPI_ISL_1790961, EPI_ISL_1790963, EPI_ISL_1790981, EPI_ISL_1790983, EPI_ISL_1790994, EPI_ISL_1791004, EPI_ISL_1791015, EPI_ISL_1805968, EPI_ISL_1805973, EPI_ISL_1805978, EPI_ISL_1805981, EPI_ISL_1805983, EPI_ISL_1805985, EPI_ISL_1805987, EPI_ISL_1805991, EPI_ISL_1805992, EPI_ISL_1806002, EPI_ISL_1806005, EPI_ISL_1806015, EPI_ISL_1806016, EPI_ISL_1806020, EPI_ISL_1806025, EPI_ISL_1806026, EPI_ISL_1806029, EPI_ISL_1806030, EPI_ISL_1806035, EPI_ISL_1806039, EPI_ISL_1806044, EPI_ISL_1806047, EPI_ISL_1806049, EPI_ISL_1806050, EPI_ISL_1806055, EPI_ISL_1806056, EPI_ISL_1806058, EPI_ISL_1806061, EPI_ISL_1806062, EPI_ISL_1806065, EPI_ISL_1806067, EPI_ISL_1806071, EPI_ISL_1806074, EPI_ISL_1806077, EPI_ISL_1806078, EPI_ISL_1806085, EPI_ISL_1806086, EPI_ISL_1806088, EPI_ISL_1806090, EPI_ISL_1806091, EPI_ISL_1806092, EPI_ISL_1806094, EPI_ISL_1806099, EPI_ISL_1806126, EPI_ISL_1806128, EPI_ISL_1806134, EPI_ISL_1806135, EPI_ISL_1806136, EPI_ISL_1806139, EPI_ISL_1806141, EPI_ISL_1806145, EPI_ISL_1806146, EPI_ISL_1806150, EPI_ISL_1806154, EPI_ISL_1806155, EPI_ISL_1806159, EPI_ISL_1806162, EPI_ISL_1806163, EPI_ISL_1806166, EPI_ISL_1806168, EPI_ISL_1806178, EPI_ISL_1806180, EPI_ISL_1806181, EPI_ISL_1806182, EPI_ISL_1806185, EPI_ISL_1806199, EPI_ISL_1806201, EPI_ISL_1806204, EPI_ISL_1806207, EPI_ISL_1806208, EPI_ISL_1806210, EPI_ISL_1806211, EPI_ISL_1806213, EPI_ISL_1806214, EPI_ISL_1806219, EPI_ISL_1806227, EPI_ISL_1806231, EPI_ISL_1806234, EPI_ISL_1806236, EPI_ISL_1806237, EPI_ISL_1806246, EPI_ISL_1806257, EPI_ISL_1806258, EPI_ISL_1806260, EPI_ISL_1806261, EPI_ISL_1806263, EPI_ISL_1806267, EPI_ISL_1806269, EPI_ISL_1806275, EPI_ISL_1806278, EPI_ISL_1806279, EPI_ISL_1806281, EPI_ISL_1806287, EPI_ISL_1806288, EPI_ISL_1806293, EPI_ISL_1806295, EPI_ISL_1806300, EPI_ISL_1806302, EPI_ISL_1806303, EPI_ISL_1806308, EPI_ISL_1806311, EPI_ISL_1806318, EPI_ISL_1806322, EPI_ISL_1806324, EPI_ISL_1806325, EPI_ISL_1806331, EPI_ISL_1806333, EPI_ISL_1806334, EPI_ISL_1806340, EPI_ISL_1806349, EPI_ISL_1806350, EPI_ISL_1806351, EPI_ISL_1806353, EPI_ISL_1806356, EPI_ISL_1806360, EPI_ISL_1806361, EPI_ISL_1806362, EPI_ISL_1806364, EPI_ISL_1806368, EPI_ISL_1806371, EPI_ISL_1806372, EPI_ISL_1806373, EPI_ISL_1806374, EPI_ISL_1806375, EPI_ISL_1806376, EPI_ISL_1806378, EPI_ISL_1806381, EPI_ISL_1806382, EPI_ISL_1806384, EPI_ISL_1806385, EPI_ISL_1806388, EPI_ISL_1806391, EPI_ISL_1806395, EPI_ISL_1806398, EPI_ISL_1806399, EPI_ISL_1806400, EPI_ISL_1806403, EPI_ISL_1806404, EPI_ISL_1806408, EPI_ISL_1806409, EPI_ISL_1806411, EPI_ISL_1806413, EPI_ISL_1806414, EPI_ISL_1806421, EPI_ISL_1806422, EPI_ISL_1806423, EPI_ISL_1806427, EPI_ISL_1806428, EPI_ISL_1806429, EPI_ISL_1806432, EPI_ISL_1806435, EPI_ISL_1806436, EPI_ISL_1806437, EPI_ISL_1806438, EPI_ISL_1806439, EPI_ISL_1806440, EPI_ISL_1806441, EPI_ISL_1806442, EPI_ISL_1806444, EPI_ISL_1806445, EPI_ISL_1806452, EPI_ISL_1806453, EPI_ISL_1806457, EPI_ISL_1806460, EPI_ISL_1806461, EPI_ISL_1806465, EPI_ISL_1806471, EPI_ISL_1806474, EPI_ISL_1806477, EPI_ISL_1806479, EPI_ISL_1806482, EPI_ISL_1806483, EPI_ISL_1806484, EPI_ISL_1806486, EPI_ISL_1806487, EPI_ISL_1806489, EPI_ISL_1806490, EPI_ISL_1806492, EPI_ISL_1806493, EPI_ISL_1806495, EPI_ISL_1806497, EPI_ISL_1806498, EPI_ISL_1806499, EPI_ISL_1806502, EPI_ISL_1806854, EPI_ISL_1806855, EPI_ISL_1806856, EPI_ISL_1806857, EPI_ISL_1806858, EPI_ISL_1806859, EPI_ISL_1806860, EPI_ISL_1806861, EPI_ISL_1806862, EPI_ISL_1806863, EPI_ISL_1806864, EPI_ISL_1806865, EPI_ISL_1806866, EPI_ISL_1806867, EPI_ISL_1806868, EPI_ISL_1806869, EPI_ISL_1806870, EPI_ISL_1806871, EPI_ISL_1806872, EPI_ISL_1806873, EPI_ISL_1806874, EPI_ISL_1806875, EPI_ISL_1806876, EPI_ISL_1806877, EPI_ISL_1806878, EPI_ISL_1806879, EPI_ISL_1806880, EPI_ISL_1806881, EPI_ISL_1806882, EPI_ISL_1806883, EPI_ISL_1806884, EPI_ISL_1806885, EPI_ISL_1806886, EPI_ISL_1806887, EPI_ISL_1806888, EPI_ISL_1806889, EPI_ISL_1806890, EPI_ISL_1806891, EPI_ISL_1806892, EPI_ISL_1806893, EPI_ISL_1806894, EPI_ISL_1806895, EPI_ISL_1806896, EPI_ISL_1806906, EPI_ISL_1806907, EPI_ISL_1806908, EPI_ISL_1806909, EPI_ISL_1806911, EPI_ISL_1806912, EPI_ISL_1806913, EPI_ISL_1806914, EPI_ISL_1806915, EPI_ISL_1806916, EPI_ISL_1806917, EPI_ISL_1806919, EPI_ISL_1806920, EPI_ISL_1806921, EPI_ISL_1806922, EPI_ISL_1806924, EPI_ISL_1806925, EPI_ISL_1806926, EPI_ISL_1806929, EPI_ISL_1806931, EPI_ISL_1806932, EPI_ISL_1806933, EPI_ISL_1806934, EPI_ISL_1806936, EPI_ISL_1806938, EPI_ISL_1806939, EPI_ISL_1806940, EPI_ISL_1806942, EPI_ISL_1806944, EPI_ISL_1806946, EPI_ISL_1806947, EPI_ISL_1806948, EPI_ISL_1806951, EPI_ISL_1806952, EPI_ISL_1806973, EPI_ISL_1806974, EPI_ISL_1806975, EPI_ISL_1806976, EPI_ISL_1806977, EPI_ISL_1806978, EPI_ISL_1806979, EPI_ISL_1806980, EPI_ISL_1806981, EPI_ISL_1806982, EPI_ISL_1806983, EPI_ISL_1806984, EPI_ISL_1806985, EPI_ISL_1806986, EPI_ISL_1806987, EPI_ISL_1806988, EPI_ISL_1806989, EPI_ISL_1806990, EPI_ISL_1806991, EPI_ISL_1806992, EPI_ISL_1806993, EPI_ISL_1806994, EPI_ISL_1806995, EPI_ISL_1806996, EPI_ISL_1806997, EPI_ISL_1806998, EPI_ISL_1806999, EPI_ISL_1807000, EPI_ISL_1807001, EPI_ISL_1807002, EPI_ISL_1807003, EPI_ISL_1807004, EPI_ISL_1807005, EPI_ISL_1807006, EPI_ISL_1807007, EPI_ISL_1807008, EPI_ISL_1807009, EPI_ISL_1807010, EPI_ISL_1807011, EPI_ISL_1807012, EPI_ISL_1807013, EPI_ISL_1807014, EPI_ISL_1807015, EPI_ISL_1807016, EPI_ISL_1807017, EPI_ISL_1807018, EPI_ISL_1807019, EPI_ISL_1807020, EPI_ISL_1807021, EPI_ISL_1807022, EPI_ISL_1807023, EPI_ISL_1807024, EPI_ISL_1807025, EPI_ISL_1807026, EPI_ISL_1807027, EPI_ISL_1807028, EPI_ISL_1807029, EPI_ISL_1807030, EPI_ISL_1807031, EPI_ISL_1807032, EPI_ISL_1807033, EPI_ISL_1807034, EPI_ISL_1807035, EPI_ISL_1807036, EPI_ISL_1807037, EPI_ISL_1807038, EPI_ISL_1807039, EPI_ISL_1807040, EPI_ISL_1807041, EPI_ISL_1807042, EPI_ISL_1807043, EPI_ISL_1807044, EPI_ISL_1807045, EPI_ISL_1807046, EPI_ISL_1807047, EPI_ISL_1807048, EPI_ISL_1807049, EPI_ISL_1807050, EPI_ISL_1807051, EPI_ISL_1807052, EPI_ISL_1807053, EPI_ISL_1807054, EPI_ISL_1807056, EPI_ISL_1807057, EPI_ISL_1807058, EPI_ISL_1807059, EPI_ISL_1807060, EPI_ISL_1807061, EPI_ISL_1807062, EPI_ISL_1807063, EPI_ISL_1807064, EPI_ISL_1807065, EPI_ISL_1807066, EPI_ISL_1807067, EPI_ISL_1807069, EPI_ISL_1807070, EPI_ISL_1807071, EPI_ISL_1807072, EPI_ISL_1807073, EPI_ISL_1807074, EPI_ISL_1807075, EPI_ISL_1807076, EPI_ISL_1807077, EPI_ISL_1807078, EPI_ISL_1807079, EPI_ISL_1807080, EPI_ISL_1807082, EPI_ISL_1807083, EPI_ISL_1807084, EPI_ISL_1807085, EPI_ISL_1807086, EPI_ISL_1807087, EPI_ISL_1807088, EPI_ISL_1807089, EPI_ISL_1807090, EPI_ISL_1807091, EPI_ISL_1807092, EPI_ISL_1807093, EPI_ISL_1807094, EPI_ISL_1807095, EPI_ISL_1807096, EPI_ISL_1807097, EPI_ISL_1807098, EPI_ISL_1807100, EPI_ISL_1807101, EPI_ISL_1807102, EPI_ISL_1807103, EPI_ISL_1807104, EPI_ISL_1807105, EPI_ISL_1807106, EPI_ISL_1807107, EPI_ISL_1807108, EPI_ISL_1807109, EPI_ISL_1807110, EPI_ISL_1807111, EPI_ISL_1807112, EPI_ISL_1807114, EPI_ISL_1807115, EPI_ISL_1807116, EPI_ISL_1807117, EPI_ISL_1807118, EPI_ISL_1807119, EPI_ISL_1807120, EPI_ISL_1807121, EPI_ISL_1807122, EPI_ISL_1807123, EPI_ISL_1807124, EPI_ISL_1807125, EPI_ISL_1807127, EPI_ISL_1807128, EPI_ISL_1807129, EPI_ISL_1807130, EPI_ISL_1807131, EPI_ISL_1807132, EPI_ISL_1807133, EPI_ISL_1807134, EPI_ISL_1807135, EPI_ISL_1807136, EPI_ISL_1807137, EPI_ISL_1807138, EPI_ISL_1807139, EPI_ISL_1807140, EPI_ISL_1807141, EPI_ISL_1807142, EPI_ISL_1807143, EPI_ISL_1807144, EPI_ISL_1807145, EPI_ISL_1807146, EPI_ISL_1807147, EPI_ISL_1807148, EPI_ISL_1807149, EPI_ISL_1807150, EPI_ISL_1807151, EPI_ISL_1807152, EPI_ISL_1807153, EPI_ISL_1807154, EPI_ISL_1807156, EPI_ISL_1807157, EPI_ISL_1807158, EPI_ISL_1807159, EPI_ISL_1807160, EPI_ISL_1807161, EPI_ISL_1807162, EPI_ISL_1807163, EPI_ISL_1807164, EPI_ISL_1807165, EPI_ISL_1807166, EPI_ISL_1807167, EPI_ISL_1807168, EPI_ISL_1807169, EPI_ISL_1807170, EPI_ISL_1807171, EPI_ISL_1807172, EPI_ISL_1807173, EPI_ISL_1807174, EPI_ISL_1807175, EPI_ISL_1807176, EPI_ISL_1807177, EPI_ISL_1807178, EPI_ISL_1829485, EPI_ISL_1829490, EPI_ISL_1829502, EPI_ISL_1829503, EPI_ISL_1829505, EPI_ISL_1829521, EPI_ISL_1829543, EPI_ISL_1829553, EPI_ISL_1829604, EPI_ISL_1829606, EPI_ISL_1829624, EPI_ISL_1829642, EPI_ISL_1829648, EPI_ISL_1829649, EPI_ISL_1829661, EPI_ISL_1829699, EPI_ISL_1829710, EPI_ISL_1829737, EPI_ISL_1829747, EPI_ISL_1829769, EPI_ISL_1829780, EPI_ISL_1829796, EPI_ISL_1830119, EPI_ISL_1830187, EPI_ISL_1830253, EPI_ISL_1830256, EPI_ISL_1830274, EPI_ISL_1830299, EPI_ISL_1830436, EPI_ISL_1830546, EPI_ISL_1830599, EPI_ISL_1830876, EPI_ISL_1830917, EPI_ISL_1830997, EPI_ISL_1831068, EPI_ISL_1831088, EPI_ISL_1831089, EPI_ISL_1831090, EPI_ISL_1831091, EPI_ISL_1831092, EPI_ISL_1831093, EPI_ISL_1831094, EPI_ISL_1831095, EPI_ISL_1831096, EPI_ISL_1831097, EPI_ISL_1831098, EPI_ISL_1831099, EPI_ISL_1831100, EPI_ISL_1831101, EPI_ISL_1831102, EPI_ISL_1831103, EPI_ISL_1831104, EPI_ISL_1831105, EPI_ISL_1831106, EPI_ISL_1831107, EPI_ISL_1831108, EPI_ISL_1831109, EPI_ISL_1831110, EPI_ISL_1831111, EPI_ISL_1831112, EPI_ISL_1831113, EPI_ISL_1831114, EPI_ISL_1831115, EPI_ISL_1831116, EPI_ISL_1831117, EPI_ISL_1831118, EPI_ISL_1831119, EPI_ISL_1831120, EPI_ISL_1831121, EPI_ISL_1831122, EPI_ISL_1831123, EPI_ISL_1831124, EPI_ISL_1831125, EPI_ISL_1831126, EPI_ISL_1831127, EPI_ISL_1831128, EPI_ISL_1831129, EPI_ISL_1831130, EPI_ISL_1831131, EPI_ISL_1831132, EPI_ISL_1831133, EPI_ISL_1831134, EPI_ISL_1831135, EPI_ISL_1831136, EPI_ISL_1831137, EPI_ISL_1831138, EPI_ISL_1831139, EPI_ISL_1831140, EPI_ISL_1831141, EPI_ISL_1831142, EPI_ISL_1831143, EPI_ISL_1831144, EPI_ISL_1831145, EPI_ISL_1831146, EPI_ISL_1831147, EPI_ISL_1831148, EPI_ISL_1831149, EPI_ISL_1831150, EPI_ISL_1831151, EPI_ISL_1831152, EPI_ISL_1831153, EPI_ISL_1831154, EPI_ISL_1831155, EPI_ISL_1831156, EPI_ISL_1831157, EPI_ISL_1831158, EPI_ISL_1831159, EPI_ISL_1831160, EPI_ISL_1831161, EPI_ISL_1831162, EPI_ISL_1831163, EPI_ISL_1831164, EPI_ISL_1831165, EPI_ISL_1831166, EPI_ISL_1831167, EPI_ISL_1831168, EPI_ISL_1831169, EPI_ISL_1831170, EPI_ISL_1831171, EPI_ISL_1831172, EPI_ISL_1831173, EPI_ISL_1831174, EPI_ISL_1831175, EPI_ISL_1831176, EPI_ISL_1831177, EPI_ISL_1831178, EPI_ISL_1831179, EPI_ISL_1831180, EPI_ISL_1831181, EPI_ISL_1831182, EPI_ISL_1831183, EPI_ISL_1831184, EPI_ISL_1831185, EPI_ISL_1831186, EPI_ISL_1831187, EPI_ISL_1831188, EPI_ISL_1831189, EPI_ISL_1831190, EPI_ISL_1831191, EPI_ISL_1831195, EPI_ISL_1831196, EPI_ISL_1831197, EPI_ISL_1831198, EPI_ISL_1831199, EPI_ISL_1831200, EPI_ISL_1831201, EPI_ISL_1831202, EPI_ISL_1831203, EPI_ISL_1831204, EPI_ISL_1831205, EPI_ISL_1831206, EPI_ISL_1831207, EPI_ISL_1831208, EPI_ISL_1831209, EPI_ISL_1831210, EPI_ISL_1831211, EPI_ISL_1831212, EPI_ISL_1831213, EPI_ISL_1831214, EPI_ISL_1831215, EPI_ISL_1831216, EPI_ISL_1831217, EPI_ISL_1831218, EPI_ISL_1831219, EPI_ISL_1831220, EPI_ISL_1831221, EPI_ISL_1831222, EPI_ISL_1831223, EPI_ISL_1831224, EPI_ISL_1831225, EPI_ISL_1831226, EPI_ISL_1831227, EPI_ISL_1831228, EPI_ISL_1831229, EPI_ISL_1831230, EPI_ISL_1831231, EPI_ISL_1831232, EPI_ISL_1831233, EPI_ISL_1831234, EPI_ISL_1831235, EPI_ISL_1831236, EPI_ISL_1831237, EPI_ISL_1831238, EPI_ISL_1831239, EPI_ISL_1831240, EPI_ISL_1831241, EPI_ISL_1831242, EPI_ISL_1831243, EPI_ISL_1831244, EPI_ISL_1831245, EPI_ISL_1831246, EPI_ISL_1831247, EPI_ISL_1831248, EPI_ISL_1831249, EPI_ISL_1831250, EPI_ISL_1831251, EPI_ISL_1831252, EPI_ISL_1831253, EPI_ISL_1831254, EPI_ISL_1831255, EPI_ISL_1831257, EPI_ISL_1831258, EPI_ISL_1831259, EPI_ISL_1831260, EPI_ISL_1912008, EPI_ISL_1912015, EPI_ISL_1912024, EPI_ISL_1912046, EPI_ISL_1912060, EPI_ISL_1912063, EPI_ISL_1912065, EPI_ISL_1912069, EPI_ISL_1912083, EPI_ISL_1912087, EPI_ISL_1912104, EPI_ISL_1912127, EPI_ISL_1912137, EPI_ISL_1912147, EPI_ISL_1912155, EPI_ISL_1912181, EPI_ISL_1912187, EPI_ISL_1912190, EPI_ISL_1912209, EPI_ISL_1912228, EPI_ISL_1912229, EPI_ISL_1912231, EPI_ISL_1912235, EPI_ISL_1912238, EPI_ISL_1912246, EPI_ISL_1912254, EPI_ISL_1912259, EPI_ISL_1912275, EPI_ISL_1912278, EPI_ISL_1912280, EPI_ISL_1912289, EPI_ISL_1912312, EPI_ISL_1912320, EPI_ISL_1912324, EPI_ISL_1912326, EPI_ISL_1912330, EPI_ISL_1985444, EPI_ISL_2005631, EPI_ISL_2005679, EPI_ISL_2005700, EPI_ISL_2005761, EPI_ISL_2006118, EPI_ISL_2006133, EPI_ISL_2006195, EPI_ISL_2006205, EPI_ISL_2006241, EPI_ISL_2006399

| | | | |
|---|---|---|---|
| see above | Lighthouse Lab in Glasgow | Wellcome Sanger Institute for the COVID-19 Genomics UK (COG-UK) Consortium | Anna Dominiczak and Alex Alderton; Carol Clugston; Cordelia Langford; David Gray; David K. Jackson; Dominic Kwiatkowski; Ewan Harrison; Harper VanSteenhouse; Ian Johnston; Jeffrey Barrett; John Sillitoe on behalf of the Wellcome Sanger Institute COVID-19 Surveillance Team; Roberto Amato; Sonia Goncalves; Yumi Kasai |

EPI_ISL_1697480, EPI_ISL_1697506, EPI_ISL_1697530, EPI_ISL_1697539, EPI_ISL_1697547, EPI_ISL_1697549, EPI_ISL_1697568, EPI_ISL_1697577, EPI_ISL_1697591, EPI_ISL_1697594, EPI_ISL_1697601, EPI_ISL_1697603, EPI_ISL_1697622, EPI_ISL_1697627, EPI_ISL_1697654, EPI_ISL_1697656, EPI_ISL_1697685, EPI_ISL_1697690, EPI_ISL_1697696, EPI_ISL_1697702, EPI_ISL_1697711, EPI_ISL_1697754, EPI_ISL_1697755, EPI_ISL_1697757, EPI_ISL_1697758, EPI_ISL_1697763, EPI_ISL_1697839, EPI_ISL_1697844, EPI_ISL_1697847, EPI_ISL_1697849, EPI_ISL_1697866, EPI_ISL_1697882, EPI_ISL_1697884, EPI_ISL_1697902, EPI_ISL_1697903, EPI_ISL_1697941, EPI_ISL_1697947, EPI_ISL_1697970, EPI_ISL_1697972, EPI_ISL_1697973, EPI_ISL_1697974, EPI_ISL_1697977, EPI_ISL_1697979, EPI_ISL_1697981, EPI_ISL_1697983, EPI_ISL_1697984, EPI_ISL_1697985, EPI_ISL_1697987, EPI_ISL_1697988, EPI_ISL_1697990, EPI_ISL_1697991, EPI_ISL_1697992, EPI_ISL_1697993, EPI_ISL_1697994, EPI_ISL_1697995, EPI_ISL_1697997, EPI_ISL_1697999, EPI_ISL_1698000, EPI_ISL_1698001, EPI_ISL_1698002, EPI_ISL_1698003, EPI_ISL_1698004, EPI_ISL_1698005, EPI_ISL_1698006, EPI_ISL_1698008, EPI_ISL_1698009,

| | | | |
|---|---|---|---|
| EPI_ISL_1698010, EPI_ISL_1698013, EPI_ISL_1698014, EPI_ISL_1698017, EPI_ISL_1698018, EPI_ISL_1698020, EPI_ISL_1698021, EPI_ISL_1698022, EPI_ISL_1698023, EPI_ISL_1698024, EPI_ISL_1698025, EPI_ISL_1698027, EPI_ISL_1698029, EPI_ISL_1698031, EPI_ISL_1698032, EPI_ISL_1698033, EPI_ISL_1698035, EPI_ISL_1698038, EPI_ISL_1698039, EPI_ISL_1698040, EPI_ISL_1698042, EPI_ISL_1698043, EPI_ISL_1698047, EPI_ISL_1698048, EPI_ISL_1698050, EPI_ISL_1698051, EPI_ISL_1698052, EPI_ISL_1698053, EPI_ISL_1698055, EPI_ISL_1698059, EPI_ISL_1698060, EPI_ISL_1698064, EPI_ISL_1698065, EPI_ISL_1698068, EPI_ISL_1698070, EPI_ISL_1698072, EPI_ISL_1698073, EPI_ISL_1698074, EPI_ISL_1698076, EPI_ISL_1698079, EPI_ISL_1698081, EPI_ISL_1698083, EPI_ISL_1698084, EPI_ISL_1698085, EPI_ISL_1698086, EPI_ISL_1698088, EPI_ISL_1698089, EPI_ISL_1698092, EPI_ISL_1698093, EPI_ISL_1698094, EPI_ISL_1698095, EPI_ISL_1698096, EPI_ISL_1698097, EPI_ISL_1698098, EPI_ISL_1698101, EPI_ISL_1698102, EPI_ISL_1698104, EPI_ISL_1698105, EPI_ISL_1698106, EPI_ISL_1698108, EPI_ISL_1698110, EPI_ISL_1698111, EPI_ISL_1698112, EPI_ISL_1698119, EPI_ISL_1698120, EPI_ISL_1698121, EPI_ISL_1698122, EPI_ISL_1698123, EPI_ISL_1698124, EPI_ISL_1698125, EPI_ISL_1698126, EPI_ISL_1698127, EPI_ISL_1698128, EPI_ISL_1698133, EPI_ISL_1698134, EPI_ISL_1698135, EPI_ISL_1698137, EPI_ISL_1698138, EPI_ISL_1698139, EPI_ISL_1698140, EPI_ISL_1698141, EPI_ISL_1698142, EPI_ISL_1698143, EPI_ISL_1698144, EPI_ISL_1698145, EPI_ISL_1698147, EPI_ISL_1698148, EPI_ISL_1698149, EPI_ISL_1698151, EPI_ISL_1698152, EPI_ISL_1698154, EPI_ISL_1698155, EPI_ISL_1698158, EPI_ISL_1698159, EPI_ISL_1698160, EPI_ISL_1698161, EPI_ISL_1698162, EPI_ISL_1698163, EPI_ISL_1698167, EPI_ISL_1698168, EPI_ISL_1698170, EPI_ISL_1698171, EPI_ISL_1698172, EPI_ISL_1698174, EPI_ISL_1698175, EPI_ISL_1698177, EPI_ISL_1698179, EPI_ISL_1698181, EPI_ISL_1698184, EPI_ISL_1698187, EPI_ISL_1698189, EPI_ISL_1698191, EPI_ISL_1698192, EPI_ISL_1698193, EPI_ISL_1698196, EPI_ISL_1698198, EPI_ISL_1698200, EPI_ISL_1698201, EPI_ISL_1698206, EPI_ISL_1698208, EPI_ISL_1698210, EPI_ISL_1698211, EPI_ISL_1698212, EPI_ISL_1698215, EPI_ISL_1698216, EPI_ISL_1698217, EPI_ISL_1698218, EPI_ISL_1698220, EPI_ISL_1698222, EPI_ISL_1698224, EPI_ISL_1698230, EPI_ISL_1698232, EPI_ISL_1698233, EPI_ISL_1698235, EPI_ISL_1698236, EPI_ISL_1698237, EPI_ISL_1698238, EPI_ISL_1698239, EPI_ISL_1698240, EPI_ISL_1698243, EPI_ISL_1698244, EPI_ISL_1698245, EPI_ISL_1698246, EPI_ISL_1698248, EPI_ISL_1698249, EPI_ISL_1698250, EPI_ISL_1698251, EPI_ISL_1698253, EPI_ISL_1698322, EPI_ISL_1698767, EPI_ISL_1698770, EPI_ISL_1698777, EPI_ISL_1698780, EPI_ISL_1698789, EPI_ISL_1698791, EPI_ISL_1698796, EPI_ISL_1698797, EPI_ISL_1698808, EPI_ISL_1698814, EPI_ISL_1698829, EPI_ISL_1698833, EPI_ISL_1698838, EPI_ISL_1698855, EPI_ISL_1698862, EPI_ISL_1698864, EPI_ISL_1698866, EPI_ISL_1698877, EPI_ISL_1698880, EPI_ISL_1698888, EPI_ISL_1698895, EPI_ISL_1698899, EPI_ISL_1698901, EPI_ISL_1698907, EPI_ISL_1698908, EPI_ISL_1698914, EPI_ISL_1698917, EPI_ISL_1698919, EPI_ISL_1698927, EPI_ISL_1698929, EPI_ISL_1698934, EPI_ISL_1698943, EPI_ISL_1698944, EPI_ISL_1698950, EPI_ISL_1698965, EPI_ISL_1698971, EPI_ISL_1698972, EPI_ISL_1698975, EPI_ISL_1698990, EPI_ISL_1699008, EPI_ISL_1699009, EPI_ISL_1699289, EPI_ISL_1699297, EPI_ISL_1699302, EPI_ISL_1699321, EPI_ISL_1699322, EPI_ISL_1699328, EPI_ISL_1699335, EPI_ISL_1699336, EPI_ISL_1699339, EPI_ISL_1699355, EPI_ISL_1699363, EPI_ISL_1699385, EPI_ISL_1699413, EPI_ISL_1699420, EPI_ISL_1699475, EPI_ISL_1699555, EPI_ISL_1699559, EPI_ISL_1699566, EPI_ISL_1699571, EPI_ISL_1699572, EPI_ISL_1699573, EPI_ISL_1699576, EPI_ISL_1699577, EPI_ISL_1699578, EPI_ISL_1699579, EPI_ISL_1699580, EPI_ISL_1699582, EPI_ISL_1699583, EPI_ISL_1699584, EPI_ISL_1699588, EPI_ISL_1699591, EPI_ISL_1699592, EPI_ISL_1699593, EPI_ISL_1699594, EPI_ISL_1699596, EPI_ISL_1699598, EPI_ISL_1699599, EPI_ISL_1699600, EPI_ISL_1699602, EPI_ISL_1699604, EPI_ISL_1699605, EPI_ISL_1699607, EPI_ISL_1699608, EPI_ISL_1699609, EPI_ISL_1699610, EPI_ISL_1699613, EPI_ISL_1699614, EPI_ISL_1699615, EPI_ISL_1699617, EPI_ISL_1699619, EPI_ISL_1699620, EPI_ISL_1699622, EPI_ISL_1699624, EPI_ISL_1699625, EPI_ISL_1699626, EPI_ISL_1699627, EPI_ISL_1699628, EPI_ISL_1699632, EPI_ISL_1699634, EPI_ISL_1699635, EPI_ISL_1699636, EPI_ISL_1699638, EPI_ISL_1699639, EPI_ISL_1699640, EPI_ISL_1699641, EPI_ISL_1699642, EPI_ISL_1699643, EPI_ISL_1699644, EPI_ISL_1699645, EPI_ISL_1699647, EPI_ISL_1699648, EPI_ISL_1699652, EPI_ISL_1699653, EPI_ISL_1699654, EPI_ISL_1699656, EPI_ISL_1699658, EPI_ISL_1699659, EPI_ISL_1699661, EPI_ISL_1699662, EPI_ISL_1699663, EPI_ISL_1699664, EPI_ISL_1699666, EPI_ISL_1699667, EPI_ISL_1699672, EPI_ISL_1699673, EPI_ISL_1699675, EPI_ISL_1699676, EPI_ISL_1699678, EPI_ISL_1699680, EPI_ISL_1699681, EPI_ISL_1699682, EPI_ISL_1699684, EPI_ISL_1699686, EPI_ISL_1699689, EPI_ISL_1699690, EPI_ISL_1699691, EPI_ISL_1699692, EPI_ISL_1699693, EPI_ISL_1699698, EPI_ISL_1699699, EPI_ISL_1699702, EPI_ISL_1699703, EPI_ISL_1699706, EPI_ISL_1699707, EPI_ISL_1699710, EPI_ISL_1699715, EPI_ISL_1699716, EPI_ISL_1699720, EPI_ISL_1699722, EPI_ISL_1699724, EPI_ISL_1699725, EPI_ISL_1699726, EPI_ISL_1699728, EPI_ISL_1699729, EPI_ISL_1699730, EPI_ISL_1699731, EPI_ISL_1699733, EPI_ISL_1699735, EPI_ISL_1699736, EPI_ISL_1699737, EPI_ISL_1699740, EPI_ISL_1699742, EPI_ISL_1699743, EPI_ISL_1699744, EPI_ISL_1699746, EPI_ISL_1699747, EPI_ISL_1699750, EPI_ISL_1699753, EPI_ISL_1699754, EPI_ISL_1699756, EPI_ISL_1699757, EPI_ISL_1699758, EPI_ISL_1699759, EPI_ISL_1699760, EPI_ISL_1699761, EPI_ISL_1699767, EPI_ISL_1699768, EPI_ISL_1699770, EPI_ISL_1699771, EPI_ISL_1699772, EPI_ISL_1699773, EPI_ISL_1699776, EPI_ISL_1699777, EPI_ISL_1699779, EPI_ISL_1699780, EPI_ISL_1699781, EPI_ISL_1699782, EPI_ISL_1699784, EPI_ISL_1699785, EPI_ISL_1699786, EPI_ISL_1699787, EPI_ISL_1699788, EPI_ISL_1699791, EPI_ISL_1699794, EPI_ISL_1699796, EPI_ISL_1699797, EPI_ISL_1699800, EPI_ISL_1699802, EPI_ISL_1699803, EPI_ISL_1699804, EPI_ISL_1699805, EPI_ISL_1699806, EPI_ISL_1699807, EPI_ISL_1699809, EPI_ISL_1699810, EPI_ISL_1699811, EPI_ISL_1699812, EPI_ISL_1699814, EPI_ISL_1699815, EPI_ISL_1699818, EPI_ISL_1699819, EPI_ISL_1699821, EPI_ISL_1699822, EPI_ISL_1699823, EPI_ISL_1699826, EPI_ISL_1699827, EPI_ISL_1699828, EPI_ISL_1699829, EPI_ISL_1699830, EPI_ISL_1699831, EPI_ISL_1699833, EPI_ISL_1699834, EPI_ISL_1699835, EPI_ISL_1699837, EPI_ISL_1699839, EPI_ISL_1699840, EPI_ISL_1718323, EPI_ISL_1718328, EPI_ISL_1718333, EPI_ISL_1718334, EPI_ISL_1718337, EPI_ISL_1718344, EPI_ISL_1718347, EPI_ISL_1718349, EPI_ISL_1718350, EPI_ISL_1718353, EPI_ISL_1718355, EPI_ISL_1718358, EPI_ISL_1718359, EPI_ISL_1718365, EPI_ISL_1718371, EPI_ISL_1718374, EPI_ISL_1718377, EPI_ISL_1718378, EPI_ISL_1718382, EPI_ISL_1718386, EPI_ISL_1718391, EPI_ISL_1718395, EPI_ISL_1718397, EPI_ISL_1718402, EPI_ISL_1718403, EPI_ISL_1718404, EPI_ISL_1718405, EPI_ISL_1718407, EPI_ISL_1718410, EPI_ISL_1718412, EPI_ISL_1718415, EPI_ISL_1718418, EPI_ISL_1718420, EPI_ISL_1718424, EPI_ISL_1718427, EPI_ISL_1718432, EPI_ISL_1718435, EPI_ISL_1718440, EPI_ISL_1718441, EPI_ISL_1718443, EPI_ISL_1718445, EPI_ISL_1718447, EPI_ISL_1718448, EPI_ISL_1718449, EPI_ISL_1718451, EPI_ISL_1718456, EPI_ISL_1718457, EPI_ISL_1718461, EPI_ISL_1718462, EPI_ISL_1718463, EPI_ISL_1718467, EPI_ISL_1718468, EPI_ISL_1718472, EPI_ISL_1718473, EPI_ISL_1718475, EPI_ISL_1718478, EPI_ISL_1718481, EPI_ISL_1718485, EPI_ISL_1718487, EPI_ISL_1718488, EPI_ISL_1718489, EPI_ISL_1718491, EPI_ISL_1718494, EPI_ISL_1718495, EPI_ISL_1718502, EPI_ISL_1718503, EPI_ISL_1718506, EPI_ISL_1718508, EPI_ISL_1718509, EPI_ISL_1718513, EPI_ISL_1718514, EPI_ISL_1718515, EPI_ISL_1718516, EPI_ISL_1718520, EPI_ISL_1718523, EPI_ISL_1718524, EPI_ISL_1718525, EPI_ISL_1718526, EPI_ISL_1718529, EPI_ISL_1718530, EPI_ISL_1718533, EPI_ISL_1718534, EPI_ISL_1718537, EPI_ISL_1718539, EPI_ISL_1718546, EPI_ISL_1718547, EPI_ISL_1718550, EPI_ISL_1718551, EPI_ISL_1718553, EPI_ISL_1718554, EPI_ISL_1718556, EPI_ISL_1718558, EPI_ISL_1718559, EPI_ISL_1718562, EPI_ISL_1718564, EPI_ISL_1718566, EPI_ISL_1718568, EPI_ISL_1718572, EPI_ISL_1718573, EPI_ISL_1718574, EPI_ISL_1718575, EPI_ISL_1718577, EPI_ISL_1718581, EPI_ISL_1718586, EPI_ISL_1718587, EPI_ISL_1718590, EPI_ISL_1718594, EPI_ISL_1718596, EPI_ISL_1718600, EPI_ISL_1718602, EPI_ISL_1718606, EPI_ISL_1718610, EPI_ISL_1718611, EPI_ISL_1718612, EPI_ISL_1718613, EPI_ISL_1718617, EPI_ISL_1718618, EPI_ISL_1718622, EPI_ISL_1718640, EPI_ISL_1718642, EPI_ISL_1718644, EPI_ISL_1718645, EPI_ISL_1718663, EPI_ISL_1718668, EPI_ISL_1718685, EPI_ISL_1718686, EPI_ISL_1718688, EPI_ISL_1718691, EPI_ISL_1718692, EPI_ISL_1718695, EPI_ISL_1718697, EPI_ISL_1718701, EPI_ISL_1718707, EPI_ISL_1718742, EPI_ISL_1718744, EPI_ISL_1718746, EPI_ISL_1718750, EPI_ISL_1718760, EPI_ISL_1718783, EPI_ISL_1718789, EPI_ISL_1718791, EPI_ISL_1718810, EPI_ISL_1718818, EPI_ISL_1718822, EPI_ISL_1718838, EPI_ISL_1718846, EPI_ISL_1718857, EPI_ISL_1718858, EPI_ISL_1718860, EPI_ISL_1718869, EPI_ISL_1718875, EPI_ISL_1718885, EPI_ISL_1718886, EPI_ISL_1718887, EPI_ISL_1718888, EPI_ISL_1718889, EPI_ISL_1718890, EPI_ISL_1718891, EPI_ISL_1718892, EPI_ISL_1718893, EPI_ISL_1718894, EPI_ISL_1718895, EPI_ISL_1718898, EPI_ISL_1718899, EPI_ISL_1718900, EPI_ISL_1718901, EPI_ISL_1718902, EPI_ISL_1718903, EPI_ISL_1718905, EPI_ISL_1718906, EPI_ISL_1718907, EPI_ISL_1718908, EPI_ISL_1718909, EPI_ISL_1718910, EPI_ISL_1718911, EPI_ISL_1718914, EPI_ISL_1718915, EPI_ISL_1718916, EPI_ISL_1718917, EPI_ISL_1718918, EPI_ISL_1718919, EPI_ISL_1718920, EPI_ISL_1718922, EPI_ISL_1718923, EPI_ISL_1718924, EPI_ISL_1718925, EPI_ISL_1718926, EPI_ISL_1718933, EPI_ISL_1718934, EPI_ISL_1718935, EPI_ISL_1718936, EPI_ISL_1718937, EPI_ISL_1718938, EPI_ISL_1718939, EPI_ISL_1718940, EPI_ISL_1718941, EPI_ISL_1718944, EPI_ISL_1718945, EPI_ISL_1718946, EPI_ISL_1718948, EPI_ISL_1718949, EPI_ISL_1718950, EPI_ISL_1718951, EPI_ISL_1718952, EPI_ISL_1718953, EPI_ISL_1718954, EPI_ISL_1718955, EPI_ISL_1718956, EPI_ISL_1718957, EPI_ISL_1718958, EPI_ISL_1718960, EPI_ISL_1718961, EPI_ISL_1718962, EPI_ISL_1718963, EPI_ISL_1718964, EPI_ISL_1718965, EPI_ISL_1718966, EPI_ISL_1718967, EPI_ISL_1718969, EPI_ISL_1718970, EPI_ISL_1718972, EPI_ISL_1718974, EPI_ISL_1718975, EPI_ISL_1718976, EPI_ISL_1718977, EPI_ISL_1718978, EPI_ISL_1718982, EPI_ISL_1718983, EPI_ISL_1718984, EPI_ISL_1718985, EPI_ISL_1718986, EPI_ISL_1718987, EPI_ISL_1718988, EPI_ISL_1718990, EPI_ISL_1718993, EPI_ISL_1718994, EPI_ISL_1718995, EPI_ISL_1718996, EPI_ISL_1718998, EPI_ISL_1718999, EPI_ISL_1719000, EPI_ISL_1719001, EPI_ISL_1719002, EPI_ISL_1719003, EPI_ISL_1719005, EPI_ISL_1719006, EPI_ISL_1719008, EPI_ISL_1719009, EPI_ISL_1719012, EPI_ISL_1719014, EPI_ISL_1719015, EPI_ISL_1719016, EPI_ISL_1719018, EPI_ISL_1719019, EPI_ISL_1719020, EPI_ISL_1719021, EPI_ISL_1719023, EPI_ISL_1719024, EPI_ISL_1719025, EPI_ISL_1719028, EPI_ISL_1719029, EPI_ISL_1719030, EPI_ISL_1719031, EPI_ISL_1719032, EPI_ISL_1719034, EPI_ISL_1719035, EPI_ISL_1719037, EPI_ISL_1719038, EPI_ISL_1719039, EPI_ISL_1719040, EPI_ISL_1719041, EPI_ISL_1719043, EPI_ISL_1719044, EPI_ISL_1719046, EPI_ISL_1719047, EPI_ISL_1719049, EPI_ISL_1719050, EPI_ISL_1719052, EPI_ISL_1719053, EPI_ISL_1719054, EPI_ISL_1719055, EPI_ISL_1719056, EPI_ISL_1719057, EPI_ISL_1719058, EPI_ISL_1719059, EPI_ISL_1719060, EPI_ISL_1719061, EPI_ISL_1719063, EPI_ISL_1719064, EPI_ISL_1719067, EPI_ISL_1719068, EPI_ISL_1719069, EPI_ISL_1719070, EPI_ISL_1719072, EPI_ISL_1719073, EPI_ISL_1719074, EPI_ISL_1719075, EPI_ISL_1719076, EPI_ISL_1719077, EPI_ISL_1719078, EPI_ISL_1719079, EPI_ISL_1719080, EPI_ISL_1719081, EPI_ISL_1719082, EPI_ISL_1719083, EPI_ISL_1719085, EPI_ISL_1719086, EPI_ISL_1719087, EPI_ISL_1719090, EPI_ISL_1719091, EPI_ISL_1719092, EPI_ISL_1719093, EPI_ISL_1719094, EPI_ISL_1719095, EPI_ISL_1719096, EPI_ISL_1719097, EPI_ISL_1719098, EPI_ISL_1719100, EPI_ISL_1719101, EPI_ISL_1719102, EPI_ISL_1719103, EPI_ISL_1719104, EPI_ISL_1719105, EPI_ISL_1719106, EPI_ISL_1719108, EPI_ISL_1719109, EPI_ISL_1719110, EPI_ISL_1719111, EPI_ISL_1719112, EPI_ISL_1719113, EPI_ISL_1719114, EPI_ISL_1719116, EPI_ISL_1719119, EPI_ISL_1719121, EPI_ISL_1719122, EPI_ISL_1719123, EPI_ISL_1719124, EPI_ISL_1719125, EPI_ISL_1719126, EPI_ISL_1719129, EPI_ISL_1719130, EPI_ISL_1719131, EPI_ISL_1719132, EPI_ISL_1719133, EPI_ISL_1719135, EPI_ISL_1719136, EPI_ISL_1719137, EPI_ISL_1719138, EPI_ISL_1719139, EPI_ISL_1719140, EPI_ISL_1719141, EPI_ISL_1719143, EPI_ISL_1719146, EPI_ISL_1719147, EPI_ISL_1719148, EPI_ISL_1719149, EPI_ISL_1719151, EPI_ISL_1719152, EPI_ISL_1719153, EPI_ISL_1719154, EPI_ISL_1719155, EPI_ISL_1719156, EPI_ISL_1719158, EPI_ISL_1719160, EPI_ISL_1719161, EPI_ISL_1719162, EPI_ISL_1719163, EPI_ISL_1719164, EPI_ISL_1719165, EPI_ISL_1719166, EPI_ISL_1719169, EPI_ISL_1719170, EPI_ISL_1719171, EPI_ISL_1719172, EPI_ISL_1719173, EPI_ISL_1719174, EPI_ISL_1719176, EPI_ISL_1719178, EPI_ISL_1719179, EPI_ISL_1719180, EPI_ISL_1719181, EPI_ISL_1719182, EPI_ISL_1719183, EPI_ISL_1719184, EPI_ISL_1719185, EPI_ISL_1719186, EPI_ISL_1719187, EPI_ISL_1719188, EPI_ISL_1719189, EPI_ISL_1719190, EPI_ISL_1719192, EPI_ISL_1719193, EPI_ISL_1719194, EPI_ISL_1719198, EPI_ISL_1719199, EPI_ISL_1719200, EPI_ISL_1719201, EPI_ISL_1719202, EPI_ISL_1719203, EPI_ISL_1719204, EPI_ISL_1719205, EPI_ISL_1719206, EPI_ISL_1719207, EPI_ISL_1719208, EPI_ISL_1719209, EPI_ISL_1719210, EPI_ISL_1719211, EPI_ISL_1719212, EPI_ISL_1719213, EPI_ISL_1719215, EPI_ISL_1719217, EPI_ISL_1719730, EPI_ISL_1719731, EPI_ISL_1740785, EPI_ISL_1740793, EPI_ISL_1740799, EPI_ISL_1740802, EPI_ISL_1740807, EPI_ISL_1740813, EPI_ISL_1740814, EPI_ISL_1740821, EPI_ISL_1740822, EPI_ISL_1740824, EPI_ISL_1740842, EPI_ISL_1740843, EPI_ISL_1740844, EPI_ISL_1740851, EPI_ISL_1740856, EPI_ISL_1740858, EPI_ISL_1740860, EPI_ISL_1740862, EPI_ISL_1740864, EPI_ISL_1740874, EPI_ISL_1740875, EPI_ISL_1740882, EPI_ISL_1740887, EPI_ISL_1740888, EPI_ISL_1740891, EPI_ISL_1740893, EPI_ISL_1740898, EPI_ISL_1740900, EPI_ISL_1740909, EPI_ISL_1740915, EPI_ISL_1740922, EPI_ISL_1740931, EPI_ISL_1740935, EPI_ISL_1740936, EPI_ISL_1740945, EPI_ISL_1740947, EPI_ISL_1740953, EPI_ISL_1740954, EPI_ISL_1740963, EPI_ISL_1740964, EPI_ISL_1740965, EPI_ISL_1740970, EPI_ISL_1740971, EPI_ISL_1740972, EPI_ISL_1740976, EPI_ISL_1740989, EPI_ISL_1740999, EPI_ISL_1741008, EPI_ISL_1741016, EPI_ISL_1741020, EPI_ISL_1741021, EPI_ISL_1741024, EPI_ISL_1741036, EPI_ISL_1741040, EPI_ISL_1741042, EPI_ISL_1741044, EPI_ISL_1741048, EPI_ISL_1741054, EPI_ISL_1741057, EPI_ISL_1741276, EPI_ISL_1741277, EPI_ISL_1741278, EPI_ISL_1741279, EPI_ISL_1741280, EPI_ISL_1741281, EPI_ISL_1741282, EPI_ISL_1741284, EPI_ISL_1741285, EPI_ISL_1741286, EPI_ISL_1741288, EPI_ISL_1741289, EPI_ISL_1741290, EPI_ISL_1741291, EPI_ISL_1741292, EPI_ISL_1741293, EPI_ISL_1741295, EPI_ISL_1741296, EPI_ISL_1741297, EPI_ISL_1741300, EPI_ISL_1741302, EPI_ISL_1741307, EPI_ISL_1741313, EPI_ISL_1741315, EPI_ISL_1741316, EPI_ISL_1741317, EPI_ISL_1741318, EPI_ISL_1741319, EPI_ISL_1741320, EPI_ISL_1741321, EPI_ISL_1741322, EPI_ISL_1741324, EPI_ISL_1741327, EPI_ISL_1741328, EPI_ISL_1741329, EPI_ISL_1741330, EPI_ISL_1741332, EPI_ISL_1741333, EPI_ISL_1741335, EPI_ISL_1741337, EPI_ISL_1741339, EPI_ISL_1741340, EPI_ISL_1741341, EPI_ISL_1741342, EPI_ISL_1741343, EPI_ISL_1741344, EPI_ISL_1741345, EPI_ISL_1741346, EPI_ISL_1741347, EPI_ISL_1741348, EPI_ISL_1741349, EPI_ISL_1741350, EPI_ISL_1741353, EPI_ISL_1741354, EPI_ISL_1741355, EPI_ISL_1741356, EPI_ISL_1741357, EPI_ISL_1741359, EPI_ISL_1741360, EPI_ISL_1741361, EPI_ISL_1741364, EPI_ISL_1741365, EPI_ISL_1741367, EPI_ISL_1741369, EPI_ISL_1741377, EPI_ISL_1741379, EPI_ISL_1741380, EPI_ISL_1741381, EPI_ISL_1741383, EPI_ISL_1741385, EPI_ISL_1741386, EPI_ISL_1741387, EPI_ISL_1741388, EPI_ISL_1741389, EPI_ISL_1741395, EPI_ISL_1741397, EPI_ISL_1741398, EPI_ISL_1741400, EPI_ISL_1741401, EPI_ISL_1741402, EPI_ISL_1741403, EPI_ISL_1741408, EPI_ISL_1741409, EPI_ISL_1741411, EPI_ISL_1741413, EPI_ISL_1741415, EPI_ISL_1741416, EPI_ISL_1741418, EPI_ISL_1741419, EPI_ISL_1741422, EPI_ISL_1741423, EPI_ISL_1741424, EPI_ISL_1741425, EPI_ISL_1741426, EPI_ISL_1741428, EPI_ISL_1741430, EPI_ISL_1741431, EPI_ISL_1741434, EPI_ISL_1741436, EPI_ISL_1741437, EPI_ISL_1741441, EPI_ISL_1741442, EPI_ISL_1741443, EPI_ISL_1741444, EPI_ISL_1741445, EPI_ISL_1741447, EPI_ISL_1741448, EPI_ISL_1741449, EPI_ISL_1741450, EPI_ISL_1741454, EPI_ISL_1741455, EPI_ISL_1741456, EPI_ISL_1741457, EPI_ISL_1741458, EPI_ISL_1741459, EPI_ISL_1741460, EPI_ISL_1741461, EPI_ISL_1741462, EPI_ISL_1741465, EPI_ISL_1741467, EPI_ISL_1741470, EPI_ISL_1741471, EPI_ISL_1741472, EPI_ISL_1741473, EPI_ISL_1741474, EPI_ISL_1741476, EPI_ISL_1741477, EPI_ISL_1741478, EPI_ISL_1741479, EPI_ISL_1741480, EPI_ISL_1741483, EPI_ISL_1741484, EPI_ISL_1741485, EPI_ISL_1741489, EPI_ISL_1741490, EPI_ISL_1741491, EPI_ISL_1741492, EPI_ISL_1741493, EPI_ISL_1741533, EPI_ISL_1741991, EPI_ISL_1742274, EPI_ISL_1758456, EPI_ISL_1758466, EPI_ISL_1758472, EPI_ISL_1758476, EPI_ISL_1758484, EPI_ISL_1758485, EPI_ISL_1758492, EPI_ISL_1758495, EPI_ISL_1758497, EPI_ISL_1758503, EPI_ISL_1758523, EPI_ISL_1758526, EPI_ISL_1758533, EPI_ISL_1758540, EPI_ISL_1758549, EPI_ISL_1758550, EPI_ISL_1758559, EPI_ISL_1758569, EPI_ISL_1758570, EPI_ISL_1758587, EPI_ISL_1758598, EPI_ISL_1758608, EPI_ISL_1758616, EPI_ISL_1758619, EPI_ISL_1758628, EPI_ISL_1758632, EPI_ISL_1758636, EPI_ISL_1758662, EPI_ISL_1758664, EPI_ISL_1758886, EPI_ISL_1758904, EPI_ISL_1758910, EPI_ISL_1758911, EPI_ISL_1758919, EPI_ISL_1758932, EPI_ISL_1758946, EPI_ISL_1758964, EPI_ISL_1758980, EPI_ISL_1759000, EPI_ISL_1759041, EPI_ISL_1759046, EPI_ISL_1759055, EPI_ISL_1759059, EPI_ISL_1759065, EPI_ISL_1759084, EPI_ISL_1759100, EPI_ISL_1759103, EPI_ISL_1759106, EPI_ISL_1759108, EPI_ISL_1759114, EPI_ISL_1759117, EPI_ISL_1759121, EPI_ISL_1759123, EPI_ISL_1759151, EPI_ISL_1759158, EPI_ISL_1759164, EPI_ISL_1759170, EPI_ISL_1759175, EPI_ISL_1759176, EPI_ISL_1759177, EPI_ISL_1759178, EPI_ISL_1759179, EPI_ISL_1759181, EPI_ISL_1759182, EPI_ISL_1759183, EPI_ISL_1759185, EPI_ISL_1759188, EPI_ISL_1759189, EPI_ISL_1759191, EPI_ISL_1759192, EPI_ISL_1759193, EPI_ISL_1759195, EPI_ISL_1759196, EPI_ISL_1759197, EPI_ISL_1759198, EPI_ISL_1759200, EPI_ISL_1759201, EPI_ISL_1759203, EPI_ISL_1759205, EPI_ISL_1759206, EPI_ISL_1759207, EPI_ISL_1759210, EPI_ISL_1759211, EPI_ISL_1759212, EPI_ISL_1759213, EPI_ISL_1759214, EPI_ISL_1759217, EPI_ISL_1759220, EPI_ISL_1759223, EPI_ISL_1759224, EPI_ISL_1759225, EPI_ISL_1759226, EPI_ISL_1759228, EPI_ISL_1759230, EPI_ISL_1759232, EPI_ISL_1759233, EPI_ISL_1759235, EPI_ISL_1759236, EPI_ISL_1759238, EPI_ISL_1759239, EPI_ISL_1759240, EPI_ISL_1759241, EPI_ISL_1759244, EPI_ISL_1759247, EPI_ISL_1759249, EPI_ISL_1759250, EPI_ISL_1759252, EPI_ISL_1759253, EPI_ISL_1759254, EPI_ISL_1759255, EPI_ISL_1759256, EPI_ISL_1759258, EPI_ISL_1759259, EPI_ISL_1759260, EPI_ISL_1759262, EPI_ISL_1759263, EPI_ISL_1759264, EPI_ISL_1759271, EPI_ISL_1759272, EPI_ISL_1759273, EPI_ISL_1759276, EPI_ISL_1759278, EPI_ISL_1759279, EPI_ISL_1759281, EPI_ISL_1759283, EPI_ISL_1759284, EPI_ISL_1759285, EPI_ISL_1759291, EPI_ISL_1759292, EPI_ISL_1759296, EPI_ISL_1759298, EPI_ISL_1759299, EPI_ISL_1759303, EPI_ISL_1759304, EPI_ISL_1759310, EPI_ISL_1759311, EPI_ISL_1759312, EPI_ISL_1759313, EPI_ISL_1759316, EPI_ISL_1759318, EPI_ISL_1759319, EPI_ISL_1759321, EPI_ISL_1759323, EPI_ISL_1759324, EPI_ISL_1759326, EPI_ISL_1759328, EPI_ISL_1759330, EPI_ISL_1759332, EPI_ISL_1759334, EPI_ISL_1759335, EPI_ISL_1759336, EPI_ISL_1759338, EPI_ISL_1759342, EPI_ISL_1759343, EPI_ISL_1759344, EPI_ISL_1759345, EPI_ISL_1759348, EPI_ISL_1759349, EPI_ISL_1759816, EPI_ISL_1759818, EPI_ISL_1759819, EPI_ISL_1759820, EPI_ISL_1759821, EPI_ISL_1759822, EPI_ISL_1759825, EPI_ISL_1759826, EPI_ISL_1759830, EPI_ISL_1759834, EPI_ISL_1759836, EPI_ISL_1759837, EPI_ISL_1759838, EPI_ISL_1759839, EPI_ISL_1759841, EPI_ISL_1759843, EPI_ISL_1759845, EPI_ISL_1759846, EPI_ISL_1759847, EPI_ISL_1759849, EPI_ISL_1759851, EPI_ISL_1759852, EPI_ISL_1759853, EPI_ISL_1759855, EPI_ISL_1759856, EPI_ISL_1759857, EPI_ISL_1759858, EPI_ISL_1759860, EPI_ISL_1759862, EPI_ISL_1759868, EPI_ISL_1759870, EPI_ISL_1759871, EPI_ISL_1759872, EPI_ISL_1759874, EPI_ISL_1759875, EPI_ISL_1759876, EPI_ISL_1759880, EPI_ISL_1759885, EPI_ISL_1759887, EPI_ISL_1759889, EPI_ISL_1759893, EPI_ISL_1759895, EPI_ISL_1790431, EPI_ISL_1790433, EPI_ISL_1790434, EPI_ISL_1790435, EPI_ISL_1790436, EPI_ISL_1790437, EPI_ISL_1790440, EPI_ISL_1790441, EPI_ISL_1790443, EPI_ISL_1790444, EPI_ISL_1790449, EPI_ISL_1790450, EPI_ISL_1790452, EPI_ISL_1790453, EPI_ISL_1790454, EPI_ISL_1790455, EPI_ISL_1790458, EPI_ISL_1790460, EPI_ISL_1790466, EPI_ISL_1790468, EPI_ISL_1790469, EPI_ISL_1790470, EPI_ISL_1790471, EPI_ISL_1790473, EPI_ISL_1790475, EPI_ISL_1790476, EPI_ISL_1790477, EPI_ISL_1790478, EPI_ISL_1790479, EPI_ISL_1790481, EPI_ISL_1790485, EPI_ISL_1790486, EPI_ISL_1790487, EPI_ISL_1790488, EPI_ISL_1790489, EPI_ISL_1790490, EPI_ISL_1790492, EPI_ISL_1790494, EPI_ISL_1790495, EPI_ISL_1790496, EPI_ISL_1790498, EPI_ISL_1790499, EPI_ISL_1790501, EPI_ISL_1790503, EPI_ISL_1790507, EPI_ISL_1790508, EPI_ISL_1790509, EPI_ISL_1790510, EPI_ISL_1790511, EPI_ISL_1790512, EPI_ISL_1790513, EPI_ISL_1790514, EPI_ISL_1790516, EPI_ISL_1790517, EPI_ISL_1790518, EPI_ISL_1790519, EPI_ISL_1790520, EPI_ISL_1790521, EPI_ISL_1790522, EPI_ISL_1790523, EPI_ISL_1790526, EPI_ISL_1790527, EPI_ISL_1790528, EPI_ISL_1790529, EPI_ISL_1790530, EPI_ISL_1790532, EPI_ISL_1790535, EPI_ISL_1790537, EPI_ISL_1790539, EPI_ISL_1790540, EPI_ISL_1790541, EPI_ISL_1790545, EPI_ISL_1790550, EPI_ISL_1790551, EPI_ISL_1790556, EPI_ISL_1790559, EPI_ISL_1790560, EPI_ISL_1790561, EPI_ISL_1790562, EPI_ISL_1790564, EPI_ISL_1790565, EPI_ISL_1790566, EPI_ISL_1790568, EPI_ISL_1790570, EPI_ISL_1790572, EPI_ISL_1790573, EPI_ISL_1790575, EPI_ISL_1790578, EPI_ISL_1790580, EPI_ISL_1790582, EPI_ISL_1790583, EPI_ISL_1790586, EPI_ISL_1790587, EPI_ISL_1790588, EPI_ISL_1790590, EPI_ISL_1790592, EPI_ISL_1790593, EPI_ISL_1790594, EPI_ISL_1790595, EPI_ISL_1790597, EPI_ISL_1790598, EPI_ISL_1790599, EPI_ISL_1790600, EPI_ISL_1790602, EPI_ISL_1790604, EPI_ISL_1790605, EPI_ISL_1790607, EPI_ISL_1790608, EPI_ISL_1790609, EPI_ISL_1790610, EPI_ISL_1790611, EPI_ISL_1790612, EPI_ISL_1790613, EPI_ISL_1790614, EPI_ISL_1790619, EPI_ISL_1790620, EPI_ISL_1790622, EPI_ISL_1790623, EPI_ISL_1790624, EPI_ISL_1790625, EPI_ISL_1790629, EPI_ISL_1790631, EPI_ISL_1790633, EPI_ISL_1790637, EPI_ISL_1790638, EPI_ISL_1790640, EPI_ISL_1790641, EPI_ISL_1790642, EPI_ISL_1790643, EPI_ISL_1790645, EPI_ISL_1790646, EPI_ISL_1790648, EPI_ISL_1790649, EPI_ISL_1790654, EPI_ISL_1790655, EPI_ISL_1790656, EPI_ISL_1790657, EPI_ISL_1790658, EPI_ISL_1790661, EPI_ISL_1790664, EPI_ISL_1790668, EPI_ISL_1790669, EPI_ISL_1790671, EPI_ISL_1790672, EPI_ISL_1790673, EPI_ISL_1790674, EPI_ISL_1790675, EPI_ISL_1790677, EPI_ISL_1790678, EPI_ISL_1790679, EPI_ISL_1790680, EPI_ISL_1790681, EPI_ISL_1790683, EPI_ISL_1790686, EPI_ISL_1790687, EPI_ISL_1790688, EPI_ISL_1790689, EPI_ISL_1790690, EPI_ISL_1790695, EPI_ISL_1790696, EPI_ISL_1790697, EPI_ISL_1790698, EPI_ISL_1790699, EPI_ISL_1790700, EPI_ISL_1790701, EPI_ISL_1790702, EPI_ISL_1790703, EPI_ISL_1790706, EPI_ISL_1790707, EPI_ISL_1790708, EPI_ISL_1790709, EPI_ISL_1790711, EPI_ISL_1790712, EPI_ISL_1790713, EPI_ISL_1790714, EPI_ISL_1790715, EPI_ISL_1790716, EPI_ISL_1790719, EPI_ISL_1790720, EPI_ISL_1790725, EPI_ISL_1790727, EPI_ISL_1790728, EPI_ISL_1790729, EPI_ISL_1790730, EPI_ISL_1790732, EPI_ISL_1790733, EPI_ISL_1790734, EPI_ISL_1790735, EPI_ISL_1790737, EPI_ISL_1790738, EPI_ISL_1790740, EPI_ISL_1790741, EPI_ISL_1790743, EPI_ISL_1790744, EPI_ISL_1790746, EPI_ISL_1790749, EPI_ISL_1790751, EPI_ISL_1790752, EPI_ISL_1790753, EPI_ISL_1790754, EPI_ISL_1790757, EPI_ISL_1790758, EPI_ISL_1790761, EPI_ISL_1790763, EPI_ISL_1790764, EPI_ISL_1790766, EPI_ISL_1790768, EPI_ISL_1790769, EPI_ISL_1790770, EPI_ISL_1790774, EPI_ISL_1790778, EPI_ISL_1790779, EPI_ISL_1790780, EPI_ISL_1790781, EPI_ISL_1790782, EPI_ISL_1790783, EPI_ISL_1790784, EPI_ISL_1790785, EPI_ISL_1790786, EPI_ISL_1790789, EPI_ISL_1790792, EPI_ISL_1790796, EPI_ISL_1790798, EPI_ISL_1790799, EPI_ISL_1790800, EPI_ISL_1790801, EPI_ISL_1790802, EPI_ISL_1790803, EPI_ISL_1790804, EPI_ISL_1790810, EPI_ISL_1790812, EPI_ISL_1790814, EPI_ISL_1790815, EPI_ISL_1790816, EPI_ISL_1790817, EPI_ISL_1790818, EPI_ISL_1790819, EPI_ISL_1790820, EPI_ISL_1790821, EPI_ISL_1790822, EPI_ISL_1790824, EPI_ISL_1790825, EPI_ISL_1790826, EPI_ISL_1790827, EPI_ISL_1790829, EPI_ISL_1790832, EPI_ISL_1790834, EPI_ISL_1790837, EPI_ISL_1790839, EPI_ISL_1790840, EPI_ISL_1790841, EPI_ISL_1790845, EPI_ISL_1790847, EPI_ISL_1790848, EPI_ISL_1790850, EPI_ISL_1790852, EPI_ISL_1790853, EPI_ISL_1790856, EPI_ISL_1790857, EPI_ISL_1790861, EPI_ISL_1790862, EPI_ISL_1790864, EPI_ISL_1790865, EPI_ISL_1790866, EPI_ISL_1790867, EPI_ISL_1790868, EPI_ISL_1790869, EPI_ISL_1790870, EPI_ISL_1790877, EPI_ISL_1790878, EPI_ISL_1790879, EPI_ISL_1790881, EPI_ISL_1790883, EPI_ISL_1790885, EPI_ISL_1790886, EPI_ISL_1790888, EPI_ISL_1790893, EPI_ISL_1790894, EPI_ISL_1790896, EPI_ISL_1790898, EPI_ISL_1790899, EPI_ISL_1790900, EPI_ISL_1790901, EPI_ISL_1790902, EPI_ISL_1790904, EPI_ISL_1790905, EPI_ISL_1790908, EPI_ISL_1790909, EPI_ISL_1790910, EPI_ISL_1790911, EPI_ISL_1790914, EPI_ISL_1790915, EPI_ISL_1790917, EPI_ISL_1790923, EPI_ISL_1790924, EPI_ISL_1790925, EPI_ISL_1790927, EPI_ISL_1790928, EPI_ISL_1790931, EPI_ISL_1790935, EPI_ISL_1790940, EPI_ISL_1790942, EPI_ISL_1790944, EPI_ISL_1790948, EPI_ISL_1790949, EPI_ISL_1790954, EPI_ISL_1790957, EPI_ISL_1790959, EPI_ISL_1790960, EPI_ISL_1790962, EPI_ISL_1790964, EPI_ISL_1790967, EPI_ISL_1790972, EPI_ISL_1790974, EPI_ISL_1790976, EPI_ISL_1790979, EPI_ISL_1790980, EPI_ISL_1790984, EPI_ISL_1790985, EPI_ISL_1790989, EPI_ISL_1790991, EPI_ISL_1790993, EPI_ISL_1790996, EPI_ISL_1790997, EPI_ISL_1790998, EPI_ISL_1790999, EPI_ISL_1791000, EPI_ISL_1791002, EPI_ISL_1791003, EPI_ISL_1791005, EPI_ISL_1791006, EPI_ISL_1791009, EPI_ISL_1791012, EPI_ISL_1791013, EPI_ISL_1791018, EPI_ISL_1805963, EPI_ISL_1805966, EPI_ISL_1805967, EPI_ISL_1805971, EPI_ISL_1805972, EPI_ISL_1805975, EPI_ISL_1805976, EPI_ISL_1805977, EPI_ISL_1805980, EPI_ISL_1805984, EPI_ISL_1805989, EPI_ISL_1805990, EPI_ISL_1805996, EPI_ISL_1805998, EPI_ISL_1806000, EPI_ISL_1806003, EPI_ISL_1806006, EPI_ISL_1806009, EPI_ISL_1806011, EPI_ISL_1806012, EPI_ISL_1806014, EPI_ISL_1806018, EPI_ISL_1806022, EPI_ISL_1806024, EPI_ISL_1806028, EPI_ISL_1806031, EPI_ISL_1806032, EPI_ISL_1806036, EPI_ISL_1806037, EPI_ISL_1806040, EPI_ISL_1806041, EPI_ISL_1806042, EPI_ISL_1806054, EPI_ISL_1806057, EPI_ISL_1806059, EPI_ISL_1806060, EPI_ISL_1806066, EPI_ISL_1806068, EPI_ISL_1806070, EPI_ISL_1806072, EPI_ISL_1806079, EPI_ISL_1806081, EPI_ISL_1806084, EPI_ISL_1806097, EPI_ISL_1806098, EPI_ISL_1806101, EPI_ISL_1806105, EPI_ISL_1806107, EPI_ISL_1806110, EPI_ISL_1806113, EPI_ISL_1806114, EPI_ISL_1806115, EPI_ISL_1806116, EPI_ISL_1806117, EPI_ISL_1806118, EPI_ISL_1806120, EPI_ISL_1806127, EPI_ISL_1806129, EPI_ISL_1806130, EPI_ISL_1806132, EPI_ISL_1806133, EPI_ISL_1806137, EPI_ISL_1806138, EPI_ISL_1806148, EPI_ISL_1806151, EPI_ISL_1806160, EPI_ISL_1806164, EPI_ISL_1806165, EPI_ISL_1806173, EPI_ISL_1806177, EPI_ISL_1806179, EPI_ISL_1806187, EPI_ISL_1806190, EPI_ISL_1806192, EPI_ISL_1806193, EPI_ISL_1806196, EPI_ISL_1806198, EPI_ISL_1806216, EPI_ISL_1806221, EPI_ISL_1806230, EPI_ISL_1806240, EPI_ISL_1806243, EPI_ISL_1806251, EPI_ISL_1806255, EPI_ISL_1806264, EPI_ISL_1806266, EPI_ISL_1806271, EPI_ISL_1806276, EPI_ISL_1806290, EPI_ISL_1806291, EPI_ISL_1806306, EPI_ISL_1806314, EPI_ISL_1806343, EPI_ISL_1806344, EPI_ISL_1806352, EPI_ISL_1806366, EPI_ISL_1806377, EPI_ISL_1806390, EPI_ISL_1806392, EPI_ISL_1806394, EPI_ISL_1806397, EPI_ISL_1806402, EPI_ISL_1806405, EPI_ISL_1806410, EPI_ISL_1806412, EPI_ISL_1806417, EPI_ISL_1806420, EPI_ISL_1806426, EPI_ISL_1806430, EPI_ISL_1806433, EPI_ISL_1806443, EPI_ISL_1806446, EPI_ISL_1806447, EPI_ISL_1806454, EPI_ISL_1806455, EPI_ISL_1806468, EPI_ISL_1806469, EPI_ISL_1806470, EPI_ISL_1806473, EPI_ISL_1806475, EPI_ISL_1806476, EPI_ISL_1806480, EPI_ISL_1806481, EPI_ISL_1806491, EPI_ISL_1806494, EPI_ISL_1806503, EPI_ISL_1806508, EPI_ISL_1806526, EPI_ISL_1806536, EPI_ISL_1806558, EPI_ISL_1806566, EPI_ISL_1806601, EPI_ISL_1806621, EPI_ISL_1806639, EPI_ISL_1806642, EPI_ISL_1806643, EPI_ISL_1806653, EPI_ISL_1806667, EPI_ISL_1806674, EPI_ISL_1806675, EPI_ISL_1806720, EPI_ISL_1806723, EPI_ISL_1806741, EPI_ISL_1806761, EPI_ISL_1806772, EPI_ISL_1806777, EPI_ISL_1806798, EPI_ISL_1806802, EPI_ISL_1806806, EPI_ISL_1806808, EPI_ISL_1806832, EPI_ISL_1806835, EPI_ISL_1806848, EPI_ISL_1806897, EPI_ISL_1806899, EPI_ISL_1806900, EPI_ISL_1806901, EPI_ISL_1806902, EPI_ISL_1806903, EPI_ISL_1806904, EPI_ISL_1806905, EPI_ISL_1829206, EPI_ISL_1829208, EPI_ISL_1829239, EPI_ISL_1829264, EPI_ISL_1829287, EPI_ISL_1829315, EPI_ISL_1829316, EPI_ISL_1829372, EPI_ISL_1829421, EPI_ISL_1829534, EPI_ISL_1829651, EPI_ISL_1829773, EPI_ISL_1830111, EPI_ISL_1830112, EPI_ISL_1830116, EPI_ISL_1830117, EPI_ISL_1830123, EPI_ISL_1830132, EPI_ISL_1830136, EPI_ISL_1830142, EPI_ISL_1830152, EPI_ISL_1830156, EPI_ISL_1830168, EPI_ISL_1830172, EPI_ISL_1830177, EPI_ISL_1830178, EPI_ISL_1830179, EPI_ISL_1830186, EPI_ISL_1830196, EPI_ISL_1830197, EPI_ISL_1830200, EPI_ISL_1830202, EPI_ISL_1830210, EPI_ISL_1830213, EPI_ISL_1830222, EPI_ISL_1830223, EPI_ISL_1830226, EPI_ISL_1830229, EPI_ISL_1830230, EPI_ISL_1830231, EPI_ISL_1830239, EPI_ISL_1830249, EPI_ISL_1830260, EPI_ISL_1830262, EPI_ISL_1830264, EPI_ISL_1830269, EPI_ISL_1830272, EPI_ISL_1830278, EPI_ISL_1830281, EPI_ISL_1830282, EPI_ISL_1830284, EPI_ISL_1830288, EPI_ISL_1830305, EPI_ISL_1830310, EPI_ISL_1830317, EPI_ISL_1830324, EPI_ISL_1830327, EPI_ISL_1830328, EPI_ISL_1830335, EPI_ISL_1830337, EPI_ISL_1830341, EPI_ISL_1830344, EPI_ISL_1830355, EPI_ISL_1830359, EPI_ISL_1830370, EPI_ISL_1830372, EPI_ISL_1830374, EPI_ISL_1830376, EPI_ISL_1830462, EPI_ISL_1830525, EPI_ISL_1830529, EPI_ISL_1830565, EPI_ISL_1830569, EPI_ISL_1830603, EPI_ISL_1830708, EPI_ISL_1831269, EPI_ISL_1831270, EPI_ISL_1831271, EPI_ISL_1831272, EPI_ISL_1831273, EPI_ISL_1831274, EPI_ISL_1831275, EPI_ISL_1831276, EPI_ISL_1831277, EPI_ISL_1831278, EPI_ISL_1831279, EPI_ISL_1831280, EPI_ISL_1831281, EPI_ISL_1831282, EPI_ISL_1831283, EPI_ISL_1857956, EPI_ISL_1912017, EPI_ISL_1912070, EPI_ISL_1912128, EPI_ISL_1912130, EPI_ISL_1912201, EPI_ISL_1912240, EPI_ISL_1912773, EPI_ISL_1912778, EPI_ISL_1912779, EPI_ISL_1912780, EPI_ISL_1921520, EPI_ISL_1985193, EPI_ISL_1985194, EPI_ISL_1985196, EPI_ISL_1985198, EPI_ISL_1985200, EPI_ISL_1985201, EPI_ISL_1985202, EPI_ISL_1985203, EPI_ISL_1985204, EPI_ISL_1985206, EPI_ISL_1985207, EPI_ISL_1985210, EPI_ISL_1985211, EPI_ISL_1985213, EPI_ISL_1985215, EPI_ISL_1985216, EPI_ISL_1985220, EPI_ISL_1985223, EPI_ISL_1985225, EPI_ISL_1985226, EPI_ISL_1985227, EPI_ISL_1985228, EPI_ISL_1985230, EPI_ISL_1985231, EPI_ISL_1985232, EPI_ISL_1985234, EPI_ISL_1985235, EPI_ISL_1985238, EPI_ISL_1985239, EPI_ISL_1985241, EPI_ISL_1985242, EPI_ISL_1985243, EPI_ISL_1985245, EPI_ISL_1985247, EPI_ISL_1985248, EPI_ISL_1985251, EPI_ISL_1985254, EPI_ISL_1985256, EPI_ISL_1985260, EPI_ISL_1985261, EPI_ISL_1985264, EPI_ISL_1985265, EPI_ISL_1985267, EPI_ISL_1985268, EPI_ISL_1985270, EPI_ISL_1985273, EPI_ISL_1985274, EPI_ISL_1985275, EPI_ISL_1985277, EPI_ISL_1985278, EPI_ISL_1985281, EPI_ISL_1985282, EPI_ISL_1985283, EPI_ISL_1985284, EPI_ISL_1985285, EPI_ISL_1985287, EPI_ISL_1985288, EPI_ISL_1985289, EPI_ISL_1985290, EPI_ISL_1985292, EPI_ISL_1985294, EPI_ISL_1985295, EPI_ISL_1985296, EPI_ISL_1985298, EPI_ISL_1985299, EPI_ISL_1985300, EPI_ISL_1985302, EPI_ISL_1985303, EPI_ISL_1985305, EPI_ISL_1985308, EPI_ISL_1985312, EPI_ISL_1985313, EPI_ISL_1985315, EPI_ISL_1985316, EPI_ISL_1985317, EPI_ISL_1985321, EPI_ISL_1985322, EPI_ISL_1985325, EPI_ISL_1985327, EPI_ISL_1985328, EPI_ISL_1985329, EPI_ISL_1985330, EPI_ISL_1985331, EPI_ISL_1985332, EPI_ISL_1985334, EPI_ISL_1985335, EPI_ISL_1985339, EPI_ISL_1985344, EPI_ISL_1985345, EPI_ISL_1985350, EPI_ISL_1985351, EPI_ISL_1985352, EPI_ISL_1985353, EPI_ISL_1985356, EPI_ISL_1985358, EPI_ISL_1985361, EPI_ISL_1985363, EPI_ISL_1985365, EPI_ISL_1985367, EPI_ISL_1985368, EPI_ISL_1985369, EPI_ISL_1985371, EPI_ISL_1985372, EPI_ISL_1985375, EPI_ISL_1985376, EPI_ISL_1985378, EPI_ISL_1985379, EPI_ISL_1985380, EPI_ISL_1985386, EPI_ISL_1985388, EPI_ISL_1985392, EPI_ISL_1985393, EPI_ISL_1985395, EPI_ISL_1985397, EPI_ISL_1985399, EPI_ISL_1985403, EPI_ISL_1985404, EPI_ISL_1985405, EPI_ISL_1985406, EPI_ISL_1985408, EPI_ISL_1985409, EPI_ISL_1985410, EPI_ISL_1985412, EPI_ISL_1985415, EPI_ISL_1985416, EPI_ISL_1985418, EPI_ISL_1985422, EPI_ISL_1985423, EPI_ISL_1985425, EPI_ISL_1985428, EPI_ISL_1985430, EPI_ISL_1985433, EPI_ISL_1985434, EPI_ISL_1985435, EPI_ISL_1985438, EPI_ISL_1985439, EPI_ISL_1985441, EPI_ISL_1985442, EPI_ISL_1985443, EPI_ISL_1985446, EPI_ISL_1985447, EPI_ISL_1985449, EPI_ISL_1985450, EPI_ISL_1985452, EPI_ISL_1985453, EPI_ISL_1985456, EPI_ISL_1985457, EPI_ISL_1985459, EPI_ISL_1985463, EPI_ISL_1985464, EPI_ISL_1985465, EPI_ISL_1985467, EPI_ISL_1985468, EPI_ISL_1985469, EPI_ISL_1985471, EPI_ISL_1985475, EPI_ISL_1985476, EPI_ISL_1985478, EPI_ISL_1985481, EPI_ISL_1985483, EPI_ISL_1985484, EPI_ISL_1985486, EPI_ISL_1985487, EPI_ISL_1985490, EPI_ISL_1985492, EPI_ISL_1985498, EPI_ISL_1985500, EPI_ISL_1985502, EPI_ISL_1985506, EPI_ISL_1985507, EPI_ISL_1985508, EPI_ISL_1985510, EPI_ISL_1985512, EPI_ISL_1986801, EPI_ISL_1986869, EPI_ISL_1986877, EPI_ISL_1986941, EPI_ISL_2004625, EPI_ISL_2004639, EPI_ISL_2004757, EPI_ISL_2004804, EPI_ISL_2004834, EPI_ISL_2004840, EPI_ISL_2004841, EPI_ISL_2004869, EPI_ISL_2005650, EPI_ISL_2005739, EPI_ISL_2005795, EPI_ISL_2006523, EPI_ISL_2006528, EPI_ISL_2006530, EPI_ISL_2006531, EPI_ISL_2006535, EPI_ISL_2006537, EPI_ISL_2006538, EPI_ISL_2022501, EPI_ISL_2022504, EPI_ISL_2022513, EPI_ISL_2022514, EPI_ISL_2022523, EPI_ISL_2022526, EPI_ISL_2023027, EPI_ISL_2023029, EPI_ISL_2023030, EPI_ISL_2023031, EPI_ISL_2119908 | | | |
| see above | Lighthouse Lab in Milton Keynes | Wellcome Sanger Institute for the COVID-19 Genomics UK (COG-UK) Consortium | Cordelia Langford; David K. Jackson; Dominic Kwiatkowski; Ewan Harrison; Ian Johnston; Jeffrey Barrett; John Sillitoe on behalf of the Wellcome Sanger Institute COVID-19 Surveillance Team; Roberto Amato; Sonia Goncalves; The Lighthouse Lab in Milton Keynes and Alex Alderton |
| EPI_ISL_1833013, EPI_ISL_1833021, EPI_ISL_1833022 | Lincolnshire Hospitals and DeepSeq Nottingham | COVID-19 Genomics UK (COG-UK) Consortium | Christopher Moore; Fei Sang; Joeseph Chappell; Johnny Debebe; Jonathan Ball; Matthew Carlisle; Matthew Loose; Nadine Holmes; Nichola Duckworth; Patrick McClure; Sarah Walsh; Tim Sloan; Victoria Wright |
| EPI_ISL_1832410, EPI_ISL_1832412, EPI_ISL_1832414, EPI_ISL_1832416, EPI_ISL_1832418, EPI_ISL_1832420, EPI_ISL_1832433, EPI_ISL_1832436, EPI_ISL_1832438, EPI_ISL_1832441, EPI_ISL_1832443, EPI_ISL_1832445, EPI_ISL_1832447, EPI_ISL_1832449, EPI_ISL_1832451, EPI_ISL_1832453, EPI_ISL_1832455, EPI_ISL_1832457, EPI_ISL_1832461, EPI_ISL_1832463, EPI_ISL_1832471, EPI_ISL_1832474, EPI_ISL_1832476, EPI_ISL_1832514, EPI_ISL_1832516, EPI_ISL_1832518, EPI_ISL_1832520, EPI_ISL_1974928, EPI_ISL_1974932, EPI_ISL_1974944, EPI_ISL_1974945, EPI_ISL_1974947, EPI_ISL_1974951, EPI_ISL_1974952, EPI_ISL_1974954, EPI_ISL_1974955, EPI_ISL_1974957, EPI_ISL_1974958, EPI_ISL_1974966, EPI_ISL_1974967, EPI_ISL_1974969, EPI_ISL_1974971, EPI_ISL_1974973, EPI_ISL_1974974, EPI_ISL_1974978, EPI_ISL_1974980, EPI_ISL_1974982, EPI_ISL_1974986, EPI_ISL_1974988, EPI_ISL_1974990, EPI_ISL_1974991, EPI_ISL_1974993, EPI_ISL_1975011, EPI_ISL_1975012, EPI_ISL_1975014, EPI_ISL_1975017, EPI_ISL_2125954, EPI_ISL_2125971, EPI_ISL_2125975, EPI_ISL_2125980 | | | |
| see above | Liverpool Clinical Laboratories | COVID-19 Genomics UK (COG-UK) Consortium | A Alrezaihi; Alessandro Gerada; Alistair Darby; Angela Cowell; Anita Lucaci; Anu Chawla; Cassie Olateju; Catherine Hartley; Charlotte Nelson; Ecaterina Vamos; Elaine O'Toole; Eleanor G Bentley; Ghada T Shawli; Hermione Webster; Isabel García-Dorival; James Johnson; James P Stewart; Jenifer Manson; Joanne Watts; Jones Benjamin; Jordan J Clark; Julian Hiscox; L Luu; Lucille Rainbow; M Almsaud; Margaret Hughes; Mark Whitehead; Matthew Gemmell; Miren Iturriza-Gomara; Muhannad Alruwaili; N.P Randle; Neil Swainston; PKF Gilmore; Parul Sharma; Rebekah Penrice-Randal; Richard Eccles; Richard Gregory; Sam Haldenby; Steve Paterson; Stuart D Armstrong; Trevor Ian Robinson; Ximeng Han |
| EPI_ISL_1730402, EPI_ISL_1730403, EPI_ISL_1730404, EPI_ISL_1730406, EPI_ISL_1730407, EPI_ISL_1832707, EPI_ISL_1832710, EPI_ISL_1832714, EPI_ISL_1978358, EPI_ISL_1978364, EPI_ISL_1978366, EPI_ISL_1978367, EPI_ISL_1978369, EPI_ISL_1978371, EPI_ISL_1978372, EPI_ISL_1978374, EPI_ISL_1978375, EPI_ISL_1978376, EPI_ISL_1978377, EPI_ISL_1978378, EPI_ISL_1978379, EPI_ISL_1978380, EPI_ISL_1978381, EPI_ISL_1978382, EPI_ISL_1978383, EPI_ISL_1978384, EPI_ISL_1978385, EPI_ISL_1978387, EPI_ISL_1978388, EPI_ISL_1978389, EPI_ISL_1978390, EPI_ISL_1978391, EPI_ISL_1978392, EPI_ISL_1978393, EPI_ISL_1978394, EPI_ISL_1978395, EPI_ISL_1978397, EPI_ISL_1978412, EPI_ISL_1978434, EPI_ISL_1978435, EPI_ISL_1978437, EPI_ISL_1978438, EPI_ISL_1978439, EPI_ISL_1978440, EPI_ISL_1978441, EPI_ISL_1978442, EPI_ISL_1978444, EPI_ISL_1978445, EPI_ISL_1978446, EPI_ISL_1978447, EPI_ISL_1978448, EPI_ISL_1978449, EPI_ISL_1978450, EPI_ISL_1978451, EPI_ISL_1978452, EPI_ISL_1978455, EPI_ISL_1978456, EPI_ISL_1978457, EPI_ISL_1978459, EPI_ISL_1978461, EPI_ISL_1978462, EPI_ISL_1978466, EPI_ISL_1978470, EPI_ISL_1978471, EPI_ISL_1978473, EPI_ISL_1978475, EPI_ISL_1978476, EPI_ISL_1978478, EPI_ISL_1978480, EPI_ISL_1978481, EPI_ISL_1978483, EPI_ISL_1978485, EPI_ISL_1978487, EPI_ISL_1978488, EPI_ISL_1978489, EPI_ISL_1978491, EPI_ISL_1978493, EPI_ISL_1978494, EPI_ISL_1978496, EPI_ISL_1978497, EPI_ISL_1978499, EPI_ISL_1978500, EPI_ISL_1978502, EPI_ISL_1978503, EPI_ISL_1978505, EPI_ISL_1978506, EPI_ISL_1978507, EPI_ISL_1978509, | | | |

| | | | |
|---|---|---|---|
| EPI_ISL_1978510, EPI_ISL_1978512, EPI_ISL_1978514, EPI_ISL_1978515, EPI_ISL_1978517, EPI_ISL_1978518, EPI_ISL_1978520, EPI_ISL_1978525, EPI_ISL_1978526, EPI_ISL_1978528, EPI_ISL_1978529, EPI_ISL_1978531, EPI_ISL_1978533, EPI_ISL_1978556, EPI_ISL_1978561, EPI_ISL_1978563, EPI_ISL_1978564, EPI_ISL_1978565, EPI_ISL_1978566, EPI_ISL_1978567, EPI_ISL_1978570, EPI_ISL_1978572, EPI_ISL_1978574, EPI_ISL_1978577, EPI_ISL_1978579, EPI_ISL_1978581, EPI_ISL_1978583, EPI_ISL_1978585, EPI_ISL_1978586, EPI_ISL_1978588, EPI_ISL_1978590, EPI_ISL_1978591, EPI_ISL_1978593, EPI_ISL_1978595, EPI_ISL_1978597, EPI_ISL_1978598, EPI_ISL_1978600, EPI_ISL_1978602, EPI_ISL_1978604, EPI_ISL_1978606, EPI_ISL_1978607, EPI_ISL_1978609, EPI_ISL_1978611, EPI_ISL_1978613, EPI_ISL_1978614, EPI_ISL_1978616, EPI_ISL_1978617, EPI_ISL_1978619, EPI_ISL_1978621, EPI_ISL_1978623, EPI_ISL_1978626, EPI_ISL_1978660, EPI_ISL_1978662, EPI_ISL_1978663, EPI_ISL_1978665, EPI_ISL_1978669, EPI_ISL_1978670, EPI_ISL_1978682, EPI_ISL_1978686, EPI_ISL_1978736, EPI_ISL_1978741, EPI_ISL_1978743, EPI_ISL_1978891 | | | |
| see above | Northumbria University / South Tees Hospitals NHS Foundation Trust / North Cumbria Integrated Care NHS Foundation Trust / North Tees and Hartlepool NHS Foundation Trust / Newcastle Hospitals NHS Foundation Trust | COVID-19 Genomics UK (COG-UK) Consortium | Andrew Nelson; Brendan Payne; Clive Graham; Darren L Smith; Debra Padgett; Edward Barton; Emma Swindells; Garren Scott; Gary Black; Gary Eltringham; Giles S Holt; Greg R Young; Jane Greenaway; Jennifer Collins; John Allan; Joshua Loh; Lynn Dover; Matthew Bashton; Mohammad A Tariq; Paul Baker; Sarah Essex; Steve Liggett; Wen C Yew; Yusri Taha |
| EPI_ISL_1730597, EPI_ISL_1730599, EPI_ISL_1730606, EPI_ISL_1730607, EPI_ISL_1730608, EPI_ISL_1730609, EPI_ISL_1730610, EPI_ISL_1730611, EPI_ISL_1730612, EPI_ISL_1730613, EPI_ISL_1730614, EPI_ISL_1730615, EPI_ISL_1730616, EPI_ISL_1730617, EPI_ISL_1730618, EPI_ISL_1730619, EPI_ISL_1730620, EPI_ISL_1730621, EPI_ISL_1730622, EPI_ISL_1730623, EPI_ISL_1730624, EPI_ISL_1730625, EPI_ISL_1730626, EPI_ISL_1730627, EPI_ISL_1730628, EPI_ISL_1730629, EPI_ISL_1730630, EPI_ISL_1730631, EPI_ISL_1730632, EPI_ISL_1730633, EPI_ISL_1730634, EPI_ISL_1730635, EPI_ISL_1730636, EPI_ISL_1730637, EPI_ISL_1730638, EPI_ISL_1730640, EPI_ISL_1730641, EPI_ISL_1730642, EPI_ISL_1730643, EPI_ISL_1730644, EPI_ISL_1730645, EPI_ISL_1730646, EPI_ISL_1730647, EPI_ISL_1730648, EPI_ISL_1730649, EPI_ISL_1730650, EPI_ISL_1730651, EPI_ISL_1730652, EPI_ISL_1730653, EPI_ISL_1730654, EPI_ISL_1730655, EPI_ISL_1730656, EPI_ISL_1730657, EPI_ISL_1730658, EPI_ISL_1730659, EPI_ISL_1730660, EPI_ISL_1730661, EPI_ISL_1730662, EPI_ISL_1730663, EPI_ISL_1730664, EPI_ISL_1730665, EPI_ISL_1730666, EPI_ISL_1730690, EPI_ISL_1730691, EPI_ISL_1730692, EPI_ISL_1730693, EPI_ISL_1730694, EPI_ISL_1730695, EPI_ISL_1730696, EPI_ISL_1730697, EPI_ISL_1730698, EPI_ISL_1730699, EPI_ISL_1730700, EPI_ISL_1730701, EPI_ISL_1730702, EPI_ISL_1730703, EPI_ISL_1730704, EPI_ISL_1730705, EPI_ISL_1730706, EPI_ISL_1730707, EPI_ISL_1730708, EPI_ISL_1730709, EPI_ISL_1730710, EPI_ISL_1730711, EPI_ISL_1730712, EPI_ISL_1730713, EPI_ISL_1730714, EPI_ISL_1730715, EPI_ISL_1730716, EPI_ISL_1730717, EPI_ISL_1730718, EPI_ISL_1730719, EPI_ISL_1730720, EPI_ISL_1730721, EPI_ISL_1730722, EPI_ISL_1730723, EPI_ISL_1730724, EPI_ISL_1730725, EPI_ISL_1730726, EPI_ISL_1730727, EPI_ISL_1730728, EPI_ISL_1730729, EPI_ISL_1730730, EPI_ISL_1730731, EPI_ISL_1730732, EPI_ISL_1730733, EPI_ISL_1730734, EPI_ISL_1730735, EPI_ISL_1730736, EPI_ISL_1730737, EPI_ISL_1730738, EPI_ISL_1730739, EPI_ISL_1730740, EPI_ISL_1730741, EPI_ISL_1730742, EPI_ISL_1730743, EPI_ISL_1730744, EPI_ISL_1730745, EPI_ISL_1730746, EPI_ISL_1730747, EPI_ISL_1730748, EPI_ISL_1730749, EPI_ISL_1730750, EPI_ISL_1730751, EPI_ISL_1730752, EPI_ISL_1730753, EPI_ISL_1730754, EPI_ISL_1730755, EPI_ISL_1730756, EPI_ISL_1730757, EPI_ISL_1730758, EPI_ISL_1730759, EPI_ISL_1730760, EPI_ISL_1730761, EPI_ISL_1730762, EPI_ISL_1730763, EPI_ISL_1730764, EPI_ISL_1730766, EPI_ISL_1730767, EPI_ISL_1730768, EPI_ISL_1730769, EPI_ISL_1730770, EPI_ISL_1730771, EPI_ISL_1730772, EPI_ISL_1730774, EPI_ISL_1730775, EPI_ISL_1730776, EPI_ISL_1730777, EPI_ISL_1730778, EPI_ISL_1730779, EPI_ISL_1730780, EPI_ISL_1730781, EPI_ISL_1730782, EPI_ISL_1730783, EPI_ISL_1730785, EPI_ISL_1730787, EPI_ISL_1730788, EPI_ISL_1730790, EPI_ISL_1730791, EPI_ISL_1730792, EPI_ISL_1730793, EPI_ISL_1730794, EPI_ISL_1730795, EPI_ISL_1730796, EPI_ISL_1730797, EPI_ISL_1730798, EPI_ISL_1730799, EPI_ISL_1730800, EPI_ISL_1730801, EPI_ISL_1730802, EPI_ISL_1730803, EPI_ISL_1730804, EPI_ISL_1730805, EPI_ISL_1730806, EPI_ISL_1730807, EPI_ISL_1730808, EPI_ISL_1730809, EPI_ISL_1730810, EPI_ISL_1730811, EPI_ISL_1730812, EPI_ISL_1730813, EPI_ISL_1730814, EPI_ISL_1730815, EPI_ISL_1730816, EPI_ISL_1730817, EPI_ISL_1730818, EPI_ISL_1730819, EPI_ISL_1979341, EPI_ISL_1979343, EPI_ISL_1979344, EPI_ISL_1979346, EPI_ISL_1979347, EPI_ISL_1979349, EPI_ISL_1979351, EPI_ISL_1979364, EPI_ISL_1979470, EPI_ISL_1979472, EPI_ISL_1979672, EPI_ISL_1979693 | | | |
| see above | Originating lab: Wales Specialist Virology Centre Sequencing lab: Pathogen Genomics Unit | Public Health Wales Microbiology Cardiff Wales Specialist Virology Centre | Alec Birchley; Alexander Adams; Amy Gaskin; Angela Marchbank; Bree Gatica-Wilcox; Catherine Moore; Jason Coombes; Joanne Watkins; Joel Southgate; Johnathan Evans; Laura Gifford; Lauren Gilbert; Lee Graham; Malorie Perry; Matthew Bull; Nicole Pacchiarini; Sally Corden; Sara Kumziene-Summerhayes; Sara Rey; Sarah Taylor; Simon Cottrell; Sophie Jones; Tom Connor |
| EPI_ISL_1979038, EPI_ISL_1979050, EPI_ISL_1979051, EPI_ISL_1979056, EPI_ISL_1979058, EPI_ISL_1979060, EPI_ISL_1979063, EPI_ISL_1979064, EPI_ISL_1979066, EPI_ISL_1979068, EPI_ISL_1979069, EPI_ISL_1979082, EPI_ISL_1979090, EPI_ISL_1979095, EPI_ISL_1979097 | | | |
| see above | Oxford Viromics, NDM, University of Oxford; Oxford University Hospitals; Basingstoke and North Hampshire Hospital | COVID-19 Genomics UK (COG-UK) Consortium | Alex Mobbs; Amy Trebes; Anita Justice; Catrin Moore; Christophe Fraser; David Bonsall; David Buck; Emma Wise; George Macintyre; Jessica Lynch; John Todd; Mariateresa de Cesare; Matilde Mori; Monique Andersson; Nathan Moore; Nick Cortes; Robert Shaw; Stephen Kidd; Tanya Golubchik; Timothy Peto |
| EPI_ISL_1832723, EPI_ISL_1832742, EPI_ISL_1832763, EPI_ISL_1832771, EPI_ISL_1832773, EPI_ISL_1832775, EPI_ISL_1832778, EPI_ISL_1832780, EPI_ISL_1832782, EPI_ISL_1832784, EPI_ISL_1832786, EPI_ISL_1832788, EPI_ISL_1832791, EPI_ISL_1832793, EPI_ISL_1832794, EPI_ISL_1832796, EPI_ISL_1832797, EPI_ISL_1832798, EPI_ISL_1832799, EPI_ISL_1832801, EPI_ISL_1832804, EPI_ISL_1832805, EPI_ISL_1832812, EPI_ISL_1832814, EPI_ISL_1832816, EPI_ISL_1832818, EPI_ISL_1832820, EPI_ISL_1832822, EPI_ISL_1832824, EPI_ISL_1832826, EPI_ISL_1832828, EPI_ISL_1832830, EPI_ISL_1832832, EPI_ISL_1832834, EPI_ISL_1832836, EPI_ISL_1832839, EPI_ISL_1832840, EPI_ISL_1832848, EPI_ISL_1832850, EPI_ISL_1832867, EPI_ISL_1832871, EPI_ISL_1832872, EPI_ISL_1832873, EPI_ISL_1832874, EPI_ISL_1832876, EPI_ISL_1832877, EPI_ISL_1832878, EPI_ISL_1832879, EPI_ISL_1832880, EPI_ISL_1832881, EPI_ISL_1832882, EPI_ISL_1832883, EPI_ISL_1832884, EPI_ISL_1832885, EPI_ISL_1832886, EPI_ISL_1832887, EPI_ISL_1832888, EPI_ISL_1832889, EPI_ISL_1832890, EPI_ISL_1978900, EPI_ISL_1978944, EPI_ISL_1978948, EPI_ISL_1978949 | | | |
| see above | Quadram Institute Bioscience | COVID-19 Genomics UK (COG-UK) Consortium | Alexander J Trotter; Alison E. Mather; Alp Aydin; Ana P. Tedim; Anastasia Kolyva; Andrew Bell; Andrew J. Page; Claire Stuart; Dave J. Baker; Gemma L. Kay; John Wain; Justin O'Grady; Leonardo de Oliveira Martins; Lizzie Meadows; Maria Diaz; Mark Webber; Muhammed Yasir; Nabil-Fareed Alikhan; Ngozi Elumogo; Nicholas M. Thomson; Rachael Stanley; Rachel Gilroy; Reenesh Prakash; Samir Dervisevic; Samuel Bloomfield; Steven Rudder; Thanh Le-Viet |
| EPI_ISL_1730408, EPI_ISL_1730409, EPI_ISL_1730410, EPI_ISL_1730423, EPI_ISL_1730424, EPI_ISL_1730425, EPI_ISL_1833003, EPI_ISL_1833029, EPI_ISL_1833030, EPI_ISL_1833031, EPI_ISL_1833032 | | | |
| see above | Queens Medical Centre, Clinical Microbiology Department / DeepSeq Nottingham | COVID-19 Genomics UK (COG-UK) Consortium | Christopher Moore; Emily Park; Fei Sang; Gemma Clark; Hannah Howson-Wells; Jack Hill; Jayasree Dey; Johnny Debebe; Jonathan Ball; Joseph Chappell; Manjinder Khakh; Matthew Carlisle; Matthew Loose; Michelle M Lister; Nadine Holmes; Patrick McClure; Theocharis Tsoleridis; Timothy Byaruhanga; Vicki M Fleming; Victoria Wright; Wendy Smith |
| EPI_ISL_1652799, EPI_ISL_1652805, EPI_ISL_1697975, EPI_ISL_1697976, EPI_ISL_1697978, EPI_ISL_1697982, EPI_ISL_1697986, EPI_ISL_1697989, EPI_ISL_1697996, EPI_ISL_1698007, EPI_ISL_1698012, EPI_ISL_1698015, EPI_ISL_1698016, EPI_ISL_1698028, EPI_ISL_1698034, EPI_ISL_1698036, EPI_ISL_1698037, EPI_ISL_1698041, EPI_ISL_1698044, EPI_ISL_1698057, EPI_ISL_1698058, EPI_ISL_1698062, EPI_ISL_1698063, EPI_ISL_1698066, EPI_ISL_1698067, EPI_ISL_1698069, EPI_ISL_1698077, EPI_ISL_1698082, EPI_ISL_1698087, EPI_ISL_1698090, EPI_ISL_1698099, EPI_ISL_1698103, EPI_ISL_1698107, EPI_ISL_1698109, EPI_ISL_1698114, EPI_ISL_1698118, EPI_ISL_1698129, EPI_ISL_1698131, EPI_ISL_1698136, EPI_ISL_1698150, EPI_ISL_1698153, EPI_ISL_1698156, EPI_ISL_1698164, EPI_ISL_1698166, EPI_ISL_1698176, EPI_ISL_1698180, EPI_ISL_1698183, EPI_ISL_1698197, EPI_ISL_1698202, EPI_ISL_1698214, EPI_ISL_1698221, EPI_ISL_1698223, EPI_ISL_1698226, EPI_ISL_1698228, EPI_ISL_1698234, EPI_ISL_1698241, EPI_ISL_1718649, EPI_ISL_1718672, EPI_ISL_1718700, EPI_ISL_1718703, EPI_ISL_1718704, EPI_ISL_1718751, EPI_ISL_1718801, EPI_ISL_1718872, EPI_ISL_1718876, EPI_ISL_1740533, EPI_ISL_1740535, EPI_ISL_1740551, EPI_ISL_1740558, EPI_ISL_1740564, EPI_ISL_1740567, EPI_ISL_1740578, EPI_ISL_1740579, EPI_ISL_1740588, EPI_ISL_1740589, EPI_ISL_1740597, EPI_ISL_1740601, EPI_ISL_1740610, EPI_ISL_1740611, EPI_ISL_1740614, EPI_ISL_1740619, EPI_ISL_1740621, EPI_ISL_1740624, EPI_ISL_1740633, EPI_ISL_1740636, EPI_ISL_1740646, EPI_ISL_1740656, EPI_ISL_1740661, EPI_ISL_1740663, EPI_ISL_1740668, EPI_ISL_1740673, EPI_ISL_1740678, EPI_ISL_1740690, EPI_ISL_1740691, EPI_ISL_1740701, EPI_ISL_1740708, EPI_ISL_1740710, EPI_ISL_1740712, EPI_ISL_1740716, EPI_ISL_1740719, EPI_ISL_1740730, EPI_ISL_1740740, EPI_ISL_1740744, EPI_ISL_1740745, EPI_ISL_1740754, EPI_ISL_1740759, EPI_ISL_1740762, EPI_ISL_1740766, EPI_ISL_1740772, EPI_ISL_1740773, EPI_ISL_1740774, EPI_ISL_1740782, EPI_ISL_1740784, EPI_ISL_1741273, EPI_ISL_1741275, EPI_ISL_1741294, EPI_ISL_1741298, EPI_ISL_1741301, EPI_ISL_1741306, EPI_ISL_1741310, EPI_ISL_1741325, EPI_ISL_1741334, EPI_ISL_1741351, EPI_ISL_1741362, EPI_ISL_1741366, EPI_ISL_1741376, EPI_ISL_1741378, EPI_ISL_1741384, EPI_ISL_1741391, EPI_ISL_1741396, EPI_ISL_1741399, EPI_ISL_1741414, EPI_ISL_1741429, EPI_ISL_1741432, EPI_ISL_1741435, EPI_ISL_1741466, EPI_ISL_1741468, EPI_ISL_1741475, EPI_ISL_1741481, EPI_ISL_1741488, EPI_ISL_1741543, EPI_ISL_1741544, EPI_ISL_1742012, EPI_ISL_1742019, EPI_ISL_1742034, EPI_ISL_1742055, EPI_ISL_1742060, EPI_ISL_1742062, EPI_ISL_1758882, EPI_ISL_1758902, EPI_ISL_1758921, EPI_ISL_1758982, EPI_ISL_1759005, EPI_ISL_1759026, EPI_ISL_1759095, EPI_ISL_1759132, EPI_ISL_1790215, EPI_ISL_1790217, EPI_ISL_1790232, EPI_ISL_1790258, EPI_ISL_1790260, EPI_ISL_1790269, EPI_ISL_1790272, EPI_ISL_1790286, EPI_ISL_1790287, EPI_ISL_1790289, EPI_ISL_1790300, EPI_ISL_1790303, EPI_ISL_1790307, EPI_ISL_1790308, EPI_ISL_1790322, EPI_ISL_1790323, EPI_ISL_1790324, EPI_ISL_1790325, EPI_ISL_1790328, EPI_ISL_1790333, EPI_ISL_1790340, EPI_ISL_1790347, EPI_ISL_1790349, EPI_ISL_1790361, EPI_ISL_1790364, EPI_ISL_1790367, EPI_ISL_1790369, EPI_ISL_1790370, EPI_ISL_1790372, EPI_ISL_1790378, EPI_ISL_1790381, EPI_ISL_1790382, EPI_ISL_1790388, EPI_ISL_1790393, EPI_ISL_1790414, EPI_ISL_1790415, EPI_ISL_1790423, EPI_ISL_1790427, EPI_ISL_1790428, EPI_ISL_1806910, EPI_ISL_1806918, EPI_ISL_1806923, EPI_ISL_1806927, EPI_ISL_1806928, EPI_ISL_1806930, EPI_ISL_1806935, EPI_ISL_1806937, EPI_ISL_1806941, EPI_ISL_1806943, EPI_ISL_1806945, EPI_ISL_1806949, EPI_ISL_1806950, EPI_ISL_1806953, EPI_ISL_1806954, EPI_ISL_1806955, EPI_ISL_1806956, EPI_ISL_1806957, EPI_ISL_1806958, EPI_ISL_1806959, EPI_ISL_1806960, EPI_ISL_1806961, EPI_ISL_1806962, EPI_ISL_1806963, EPI_ISL_1806964, EPI_ISL_1806965, EPI_ISL_1806966, EPI_ISL_1806967, EPI_ISL_1806968, EPI_ISL_1806969, EPI_ISL_1806970, EPI_ISL_1806971, EPI_ISL_1806972, EPI_ISL_1807179, EPI_ISL_1807180, EPI_ISL_1807181, EPI_ISL_1807182, EPI_ISL_1807183, EPI_ISL_1829807, EPI_ISL_1829837, EPI_ISL_1829864, EPI_ISL_1829892, EPI_ISL_1829924, EPI_ISL_1829933, EPI_ISL_1829936, EPI_ISL_1829942, EPI_ISL_1829960, EPI_ISL_1829977, EPI_ISL_1830001, EPI_ISL_1830010, EPI_ISL_1830032, EPI_ISL_1830033, EPI_ISL_1830067, EPI_ISL_1830108, EPI_ISL_1830115, EPI_ISL_1830118, EPI_ISL_1830120, EPI_ISL_1830127, EPI_ISL_1830131, EPI_ISL_1830138, EPI_ISL_1830141, EPI_ISL_1830146, EPI_ISL_1830157, EPI_ISL_1830164, EPI_ISL_1830189, EPI_ISL_1830194, EPI_ISL_1830206, EPI_ISL_1830207, EPI_ISL_1830209, EPI_ISL_1830216, EPI_ISL_1830218, EPI_ISL_1830221, EPI_ISL_1830224, EPI_ISL_1830227, EPI_ISL_1830233, EPI_ISL_1830238, EPI_ISL_1830240, EPI_ISL_1830242, EPI_ISL_1830245, EPI_ISL_1830247, EPI_ISL_1830248, EPI_ISL_1830267, EPI_ISL_1830270, EPI_ISL_1830275, EPI_ISL_1830280, EPI_ISL_1830289, EPI_ISL_1830290, EPI_ISL_1830291, EPI_ISL_1830292, EPI_ISL_1830296, EPI_ISL_1830304, EPI_ISL_1830313, EPI_ISL_1830331, EPI_ISL_1830343, EPI_ISL_1830351, EPI_ISL_1830366, EPI_ISL_1830375, EPI_ISL_1830378, EPI_ISL_1830382, EPI_ISL_1830386, EPI_ISL_1912772, EPI_ISL_1912774, EPI_ISL_1912775, EPI_ISL_1912776, EPI_ISL_1912777, EPI_ISL_1912781, EPI_ISL_1912783, EPI_ISL_1921440, EPI_ISL_1921441, EPI_ISL_1921443, EPI_ISL_1921449, EPI_ISL_1921463, EPI_ISL_1921467, EPI_ISL_1921485, EPI_ISL_1921488, EPI_ISL_1921489, EPI_ISL_1921490, EPI_ISL_1921502, EPI_ISL_1921513, EPI_ISL_1921522, EPI_ISL_1921523, EPI_ISL_1921538, EPI_ISL_1921545, EPI_ISL_1921557, EPI_ISL_1921558, EPI_ISL_1921559, EPI_ISL_1921561, EPI_ISL_1921567, EPI_ISL_1921568, EPI_ISL_1921575, EPI_ISL_1921582, EPI_ISL_1921590, EPI_ISL_1921596, EPI_ISL_1921597, EPI_ISL_1921600, EPI_ISL_1921604, EPI_ISL_1921626, EPI_ISL_1921627, EPI_ISL_1921631, EPI_ISL_1921648, EPI_ISL_1921660, EPI_ISL_1921673, EPI_ISL_1921679, EPI_ISL_1921684, EPI_ISL_1921691, EPI_ISL_1921699, EPI_ISL_1921702, EPI_ISL_1921709, EPI_ISL_1921711, EPI_ISL_1921713, EPI_ISL_1921717, EPI_ISL_1921722, EPI_ISL_1921731, EPI_ISL_1921732, EPI_ISL_1985214, EPI_ISL_1985253, EPI_ISL_1985266, EPI_ISL_1985276, EPI_ISL_1985385, EPI_ISL_1985448, EPI_ISL_1985480, EPI_ISL_1985493, EPI_ISL_1985505, EPI_ISL_1985511, EPI_ISL_1987380, EPI_ISL_2005613, EPI_ISL_2005616, EPI_ISL_2005617, EPI_ISL_2005618, EPI_ISL_2005619, EPI_ISL_2005624, EPI_ISL_2005625, EPI_ISL_2005638, EPI_ISL_2005642, EPI_ISL_2005646, EPI_ISL_2005647, EPI_ISL_2005649, EPI_ISL_2005651, EPI_ISL_2005652, EPI_ISL_2005656, EPI_ISL_2005659, EPI_ISL_2005660, EPI_ISL_2005663, EPI_ISL_2005664, EPI_ISL_2005666, EPI_ISL_2005671, EPI_ISL_2005672, EPI_ISL_2005675, EPI_ISL_2005683, EPI_ISL_2005690, EPI_ISL_2005695, EPI_ISL_2005703, EPI_ISL_2005705, EPI_ISL_2005706, EPI_ISL_2005712, EPI_ISL_2005714, EPI_ISL_2005715, EPI_ISL_2005716, EPI_ISL_2005721, EPI_ISL_2005729, EPI_ISL_2005732, EPI_ISL_2005733, EPI_ISL_2005734, EPI_ISL_2005736, EPI_ISL_2005740, EPI_ISL_2005764, EPI_ISL_2005769, EPI_ISL_2005784, EPI_ISL_2005792, EPI_ISL_2005802, EPI_ISL_2005803, EPI_ISL_2005804, EPI_ISL_2005808, EPI_ISL_2022496, EPI_ISL_2022497, EPI_ISL_2022498, EPI_ISL_2022499, EPI_ISL_2022500, EPI_ISL_2022502, EPI_ISL_2022503, EPI_ISL_2022505, EPI_ISL_2022506, EPI_ISL_2022508, EPI_ISL_2022509, EPI_ISL_2022510, EPI_ISL_2022511, EPI_ISL_2022512, EPI_ISL_2022515, EPI_ISL_2022516, EPI_ISL_2022517, EPI_ISL_2022518, EPI_ISL_2022519, EPI_ISL_2022520, EPI_ISL_2022521, EPI_ISL_2022522, EPI_ISL_2022524, EPI_ISL_2022525, EPI_ISL_2022527, EPI_ISL_2022528, EPI_ISL_2022529, EPI_ISL_2023028 | | | |
| see above | Randox Laboratories | Wellcome Sanger Institute for the COVID-19 Genomics UK (COG-UK) Consortium | Cordelia Langford; David K. Jackson; Dominic Kwiatkowski; Ewan Harrison; Ian Johnston; Jeffrey Barrett; John Sillitoe on behalf of the Wellcome Sanger Institute COVID-19 Surveillance Team; Randox Laboratories and Alex Alderton; Roberto Amato; Sonia Goncalves |
| EPI_ISL_1978258, EPI_ISL_1978259, EPI_ISL_1978261, EPI_ISL_1978263, EPI_ISL_1978265, EPI_ISL_1978266, EPI_ISL_1978268, EPI_ISL_1978270, EPI_ISL_1978271, EPI_ISL_1978273, EPI_ISL_1978275, EPI_ISL_1978276, EPI_ISL_1978278, EPI_ISL_1978280, EPI_ISL_1978282, EPI_ISL_1978283, EPI_ISL_1978285, EPI_ISL_1978287, EPI_ISL_1978289, EPI_ISL_1978290, EPI_ISL_1978292, EPI_ISL_1978293, EPI_ISL_1978295, EPI_ISL_1978297, EPI_ISL_1978299, EPI_ISL_1978300, EPI_ISL_1978302, EPI_ISL_1978304, EPI_ISL_1978306, EPI_ISL_1978307, EPI_ISL_1978309, EPI_ISL_1978310, EPI_ISL_1978312, EPI_ISL_1978313, EPI_ISL_1978315, EPI_ISL_1978316, EPI_ISL_1978318, EPI_ISL_1978319, EPI_ISL_1978321, EPI_ISL_1978322, EPI_ISL_1978324, EPI_ISL_1978326, EPI_ISL_1978327, EPI_ISL_1978329, EPI_ISL_1978330, EPI_ISL_1978332, EPI_ISL_1978334, EPI_ISL_1978335, EPI_ISL_1978337, EPI_ISL_1978339, EPI_ISL_1978341, EPI_ISL_1978342, EPI_ISL_1978344, EPI_ISL_1978345, EPI_ISL_1978347, EPI_ISL_1978349, EPI_ISL_1978351, EPI_ISL_1978352, EPI_ISL_1978354, EPI_ISL_1978356 | | | |
| see above | Regional Virus Laboratory, Belfast Health and Social Care Trust | COVID-19 Genomics UK (COG-UK) Consortium | Alison Watt; Ciara Cox; Conall McCaughey; David Simpson; Derek Fairley; James McKenna; Mairead Connor; Susan Feeney; Tanya Curran; Zoltan Molnar |
| EPI_ISL_1833828, EPI_ISL_1833829 | St George's University Hospitals NHS Foundation Trust | COVID-19 Genomics UK (COG-UK) Consortium | Adam Witney; Cassie Pope; Irene Monahan; Joshua Taylor; Ken Laing; NgeeKeong Tan |
| EPI_ISL_1637005, EPI_ISL_1637100, EPI_ISL_1637101, EPI_ISL_1637102, EPI_ISL_1637103, EPI_ISL_1637104, EPI_ISL_1730324, EPI_ISL_1730325, EPI_ISL_1730327, EPI_ISL_1730330, EPI_ISL_1730331, EPI_ISL_1730332, EPI_ISL_1730333, EPI_ISL_1730335, EPI_ISL_1730340, EPI_ISL_1730341, EPI_ISL_1730346, EPI_ISL_1730347, EPI_ISL_1730348, EPI_ISL_1730349, EPI_ISL_1730350, EPI_ISL_1730351, EPI_ISL_1730352, EPI_ISL_1730353, EPI_ISL_1730354, EPI_ISL_1730363, EPI_ISL_1730364, EPI_ISL_1730365, EPI_ISL_1730366, EPI_ISL_1730374, EPI_ISL_1730376, EPI_ISL_1730378, EPI_ISL_1730385, EPI_ISL_1832585, EPI_ISL_1832590, EPI_ISL_1832591, EPI_ISL_1832598, EPI_ISL_1832602, EPI_ISL_1832604, EPI_ISL_1832606, EPI_ISL_1832611, EPI_ISL_1832646, EPI_ISL_1832649, EPI_ISL_1832651, EPI_ISL_1832657, EPI_ISL_1832659, EPI_ISL_1832661, EPI_ISL_1832663, EPI_ISL_1832665, EPI_ISL_1832667, EPI_ISL_1832669, EPI_ISL_1832671, EPI_ISL_1832672, EPI_ISL_1832674, EPI_ISL_1832676, EPI_ISL_1832701 | | | |
| see above | University College London, Great Ormond Street Hospital for Children NHS Foundation Trust, Imperial College Healthcare NHS Trust | COVID-19 Genomics UK (COG-UK) Consortium | Alison Holmes; Charlotte Williams; Helena Tutill; Jacqueline Findlay; James Price; Judith Breuer; Julianne Brown; Kathryn Harris; Leysa Forrest; Mark Kristiansen; Paola Niola; Paola Resende Silva; Patricia Dyal; Paul Randell; Rachel Williams; Samuel Weeks; Sergi Castellano; Sunando Roy; Tony Brooks; Yasmin Panchbhaya |
| EPI_ISL_1832336, EPI_ISL_1832337, EPI_ISL_1832338, EPI_ISL_1832339, EPI_ISL_1832340, EPI_ISL_2125909, EPI_ISL_2125911 | | | |
| see above | University Hospitals Of Leicester NHS Trust and DeepSeq Nottingham | COVID-19 Genomics UK (COG-UK) Consortium | Christopher Holmes; Christopher Moore; Fei Sang; Joeseph Chappell; Johnny Debebe; Jonathan Ball; Julian Tang; Karlie Fallon; Matthew Carlisle; Matthew Loose; Nadine Holmes; Patrick McClure; Paul Bird; Thomas Helmer; Victoria Wright |
| EPI_ISL_1729993, EPI_ISL_1729995, EPI_ISL_1729997, EPI_ISL_1729999, EPI_ISL_1730001, EPI_ISL_1730002, EPI_ISL_1730004, EPI_ISL_1730005, EPI_ISL_1730006, EPI_ISL_1730007, EPI_ISL_1730008, EPI_ISL_1730010, EPI_ISL_1730011, EPI_ISL_1730012, EPI_ISL_1730013, EPI_ISL_1730014, EPI_ISL_1730015, EPI_ISL_1832137, EPI_ISL_1832139, EPI_ISL_1832141, EPI_ISL_1832143, EPI_ISL_1832147, EPI_ISL_1832149 | | | |
| see above | University of Birmingham | COVID-19 Genomics UK (COG-UK) Consortium | Alex Richter; Andrew Bosworth; Andrew Bosworth. Queen Elizabeth Hospital: Anna Casey; Andrew D Beggs PHE Heartlands Lab: Husam Osman; Charlotte Poxon; Institute of Microbiology; Joanne Stockton; Josh Quick; Kasun Wanigasooriya; Mike Kidd; Nicholas Loman. University of Birmingham Testing Laboratory: Celina M Whalley; Oliver Pickles; Radoslaw Poplawski; Samuel Nicholls; University of Birmingham: Claire McMurray; Will Rowe |
| EPI_ISL_1730127, EPI_ISL_1730128, EPI_ISL_1730129, EPI_ISL_1730131, EPI_ISL_1730132, EPI_ISL_1730133, EPI_ISL_1730134, EPI_ISL_1730135, EPI_ISL_1832247, EPI_ISL_1832252, EPI_ISL_1832253, EPI_ISL_1832254, EPI_ISL_1832255, EPI_ISL_1832256, EPI_ISL_1832257, EPI_ISL_1832258, EPI_ISL_1832259, EPI_ISL_1832261, EPI_ISL_1832262, EPI_ISL_1832263, EPI_ISL_1832273, EPI_ISL_1974829, EPI_ISL_1974831, EPI_ISL_1974832, EPI_ISL_1974862, EPI_ISL_1974863, EPI_ISL_1974864, EPI_ISL_1974866, EPI_ISL_1974867, EPI_ISL_1974869, EPI_ISL_1974871, EPI_ISL_1974872, EPI_ISL_1974874, EPI_ISL_1974876, EPI_ISL_1974878, EPI_ISL_1974879, EPI_ISL_1974900, EPI_ISL_1974901, EPI_ISL_1974913, EPI_ISL_2125698, EPI_ISL_2125700, EPI_ISL_2125703, EPI_ISL_2125704, EPI_ISL_2125706, EPI_ISL_2125870, EPI_ISL_2125893 | | | |
| see above | Virology Department, Royal Infirmary of Edinburgh, NHS Lothian / School of Biological Sciences, University of Edinburgh | COVID-19 Genomics UK (COG-UK) Consortium | Colquhoun R; Cotton S; Dewar R; Gallagher A; Hill V; Jackson B; Maloney D; McCrone JT; McHugh M; O'Toole Á; Rambaut A; Rooke S; Scher E; Templeton K; Yu X |
| EPI_ISL_1730932, EPI_ISL_1730933, EPI_ISL_1730934, EPI_ISL_1730937, EPI_ISL_1730938, EPI_ISL_1730941, EPI_ISL_1730942, EPI_ISL_1730943, EPI_ISL_1730944, EPI_ISL_1730945, EPI_ISL_1730947, EPI_ISL_1730949, EPI_ISL_1730952, EPI_ISL_1730956, EPI_ISL_1730958, EPI_ISL_1730960, EPI_ISL_1730964, EPI_ISL_1730966, EPI_ISL_1730967, EPI_ISL_1730968, EPI_ISL_1730970, EPI_ISL_1730971, EPI_ISL_1730973, EPI_ISL_1730977, EPI_ISL_1730980, EPI_ISL_1730983, EPI_ISL_1730986, EPI_ISL_1730991, EPI_ISL_1730997, EPI_ISL_1731000, EPI_ISL_1731001, EPI_ISL_1731003, EPI_ISL_1731004, EPI_ISL_1731006, EPI_ISL_1731007, EPI_ISL_1731008, EPI_ISL_1731013, EPI_ISL_1731014, EPI_ISL_1731015, EPI_ISL_1731017, EPI_ISL_1731023, EPI_ISL_1731035, EPI_ISL_1731037, EPI_ISL_1731038, EPI_ISL_1731040, EPI_ISL_1731041, EPI_ISL_1833831, EPI_ISL_1833832, EPI_ISL_1833834, EPI_ISL_1833837, EPI_ISL_1833839, EPI_ISL_1833843, EPI_ISL_1833848, EPI_ISL_1833850, EPI_ISL_1833851, EPI_ISL_1833852, EPI_ISL_1833853, EPI_ISL_1833855, EPI_ISL_1833857, EPI_ISL_1833859, EPI_ISL_1833860, EPI_ISL_1833862, EPI_ISL_1833863, EPI_ISL_1833864, EPI_ISL_1833867, EPI_ISL_1833870, EPI_ISL_1833873, EPI_ISL_1833874, EPI_ISL_1833875, EPI_ISL_1833879, EPI_ISL_1833882, EPI_ISL_1833884, EPI_ISL_1833886, EPI_ISL_1833887, EPI_ISL_1833890, EPI_ISL_1833893, EPI_ISL_1833895, EPI_ISL_1833897, EPI_ISL_1833898, EPI_ISL_1833900, EPI_ISL_1833901, EPI_ISL_1833902, EPI_ISL_1833905, EPI_ISL_1833907, EPI_ISL_1833909, EPI_ISL_1833910, EPI_ISL_1833911, EPI_ISL_1833912, EPI_ISL_1833914, EPI_ISL_1833915, EPI_ISL_1833916, EPI_ISL_1980360, EPI_ISL_1980366, EPI_ISL_1980396, EPI_ISL_1980411, EPI_ISL_1980415, EPI_ISL_1980431, EPI_ISL_1980454 | | | |
| see above | Virology Department, Sheffield Teaching Hospitals NHS Foundation Trust/Department of Infection, Immunity and Cardiovascular Disease, The Medical School, University of Sheffield | COVID-19 Genomics UK (COG-UK) Consortium | Adri Angyal; Alex Keeley; Benjamin Lindsey; Cariad Evans; Danielle Groves; Dave Partridge; Katie Johnson; Laura Carrilero; Luke Green; Matthew Parker; Matthew Wyles; Mehmet Yavuz; Mohammad Raza; Nikki Smith; Paul Parsons; Rachel Tucker; Rebecca Brown; Thushan de Silva |
| EPI_ISL_1637487, EPI_ISL_1637490, EPI_ISL_1637493, EPI_ISL_1637500, EPI_ISL_1637502, EPI_ISL_1637503, EPI_ISL_1637504, EPI_ISL_1637510, EPI_ISL_1637512, EPI_ISL_1637514, EPI_ISL_1637523, EPI_ISL_1637529, EPI_ISL_1637538, EPI_ISL_1637539, EPI_ISL_1637542, EPI_ISL_1637545, EPI_ISL_1637546, EPI_ISL_1637548, EPI_ISL_1637549, EPI_ISL_1637601, EPI_ISL_1637602, EPI_ISL_1637607, EPI_ISL_1637608, EPI_ISL_1637609, EPI_ISL_1637612, EPI_ISL_1637613, EPI_ISL_1637617, EPI_ISL_1637618, EPI_ISL_1637620, EPI_ISL_1637623, EPI_ISL_1637624, EPI_ISL_1637625, EPI_ISL_1637626, EPI_ISL_1833137, EPI_ISL_1833148, EPI_ISL_1833179, EPI_ISL_1833180, EPI_ISL_1833182, EPI_ISL_1833183, EPI_ISL_1833184, EPI_ISL_1833185, EPI_ISL_1833187, EPI_ISL_1833188, EPI_ISL_1833191, EPI_ISL_1833192, EPI_ISL_1833193, EPI_ISL_1833194, EPI_ISL_1833195, EPI_ISL_1833197, EPI_ISL_1833198, EPI_ISL_1833199, EPI_ISL_1833201, EPI_ISL_1833202, EPI_ISL_1833204, EPI_ISL_1833206, EPI_ISL_1833207, EPI_ISL_1833208, EPI_ISL_1833209, EPI_ISL_1833210, EPI_ISL_1833211, EPI_ISL_1833212, EPI_ISL_1833213, EPI_ISL_1833214, EPI_ISL_1833216, EPI_ISL_1833217, EPI_ISL_1833218, EPI_ISL_1833219, EPI_ISL_1833220, EPI_ISL_1833221, EPI_ISL_1833222, EPI_ISL_1833223, EPI_ISL_1833224, EPI_ISL_1833225, EPI_ISL_1833226, EPI_ISL_1833228, EPI_ISL_1833229, EPI_ISL_1833230, EPI_ISL_1833234, EPI_ISL_1833236, EPI_ISL_1833237, EPI_ISL_1833238, EPI_ISL_1833239, EPI_ISL_1833240, EPI_ISL_1833241, EPI_ISL_1833242, EPI_ISL_1833243, EPI_ISL_1833244, EPI_ISL_1833245, EPI_ISL_1833246, EPI_ISL_1833247, EPI_ISL_1833248, EPI_ISL_1833249, EPI_ISL_1833250, EPI_ISL_1833251, EPI_ISL_1833252, EPI_ISL_1833253, EPI_ISL_1833254, EPI_ISL_1833255, EPI_ISL_1833256, EPI_ISL_1833257, EPI_ISL_1833258, EPI_ISL_1833259 | | | |
| see above | Wales Specialist Virology Centre Sequencing lab: Pathogen Genomics Unit | Public Health Wales Microbiology Cardiff Wales Specialist Virology Centre | Alec Birchley; Alexander Adams; Amy Gaskin; Angela Marchbank; Bree Gatica-Wilcox; Catherine Moore; Jason Coombes; Joanne Watkins; Joel Southgate; Johnathan Evans; Laura Gifford; Lauren Gilbert; Lee Graham; Malorie Perry; Matthew Bull; Nicole Pacchiarini; Sally Corden; Sara Kumziene-Summerhayes; Sara Rey; Sarah Taylor; Simon Cottrell; Sophie Jones; Tom Connor |
| EPI_ISL_1832301, EPI_ISL_1832302, EPI_ISL_1832303, EPI_ISL_1832304, EPI_ISL_1832306, EPI_ISL_1832307, EPI_ISL_1832308, EPI_ISL_1832309, EPI_ISL_1832310, EPI_ISL_1832311, EPI_ISL_1832312, EPI_ISL_2125498 | | | |
| see above | West of Scotland Specialist Virology Centre, NHSGGC / MRC-University of Glasgow Centre for Virus Research | COVID-19 Genomics UK (COG-UK) Consortium | Alasdair MacLean; Alice Broos; Ana da Silva Filipe; Antonia Ho; Daniel Mair; David L Robertson; Emma Thomson; Guy Mollett; Ioulia Tsatsani; James Shepherd; Jenna Nichols; Joseph Hughes; Kathy Li; Kathy Smollett; Kyriaki Nomikou; Lily Tong; Matthew Holden; Natasha Johnson; Rachel Blacow; Richard Orton; Rory Gunson; Sarah McDonald; Sharif Shaaban; Sreenu Vattipally; Stephen Carmichael |

We gratefully acknowledge the following Authors from the Originating laboratories responsible for obtaining the specimens, as well as the Submitting laboratories where the genome data were generated and shared via GISAID, on which this research is based.

All Submitters of data may be contacted directly via www.gisaid.org

Authors are sorted alphabetically.

| Accession ID | Originating Laboratory | Submitting Laboratory | Authors |
|---|---|---|---|
| EPI_ISL_1975057, EPI_ISL_1975059, EPI_ISL_1975060, EPI_ISL_1975062, EPI_ISL_1975064, EPI_ISL_1975066, EPI_ISL_1975067, EPI_ISL_1975077, EPI_ISL_1975102, EPI_ISL_1975104, EPI_ISL_1975106, EPI_ISL_1975107, EPI_ISL_2126006, EPI_ISL_2126007, EPI_ISL_2126010, EPI_ISL_2126018, EPI_ISL_2126019, EPI_ISL_2126023 | | | |
| see above | Barts Health NHS Trust | COVID-19 Genomics UK (COG-UK) Consortium | BROAD; Beatrix; CUTINO-MOGUEL; Claire; David; Dola; HARRINGTON; KELE; KULASEGARAN-SHYLINI; Maria-Teresa; OWOYEMI; Raghavendran |
| EPI_ISL_1833946, EPI_ISL_2128517 | Bioinformatics and Biostatistics Lab, Advanced Sequencing Facility | COVID-19 Genomics UK (COG-UK) Consortium | Aengus Stewart; Chelsea Sawyer; Harshil Patel; Jerome Nicod; Laura Cubitt; Margaret Crawford |
| EPI_ISL_1833278, EPI_ISL_1833280, EPI_ISL_1833281, EPI_ISL_1833282, EPI_ISL_1833283, EPI_ISL_1833284, EPI_ISL_1833285, EPI_ISL_1833286, EPI_ISL_1833288, EPI_ISL_1833341, EPI_ISL_1833345, EPI_ISL_1833346, EPI_ISL_1833348, EPI_ISL_1833351, EPI_ISL_1979798, EPI_ISL_1980109, EPI_ISL_1980111, EPI_ISL_1980112, EPI_ISL_1980210, EPI_ISL_1980212, EPI_ISL_1980213, EPI_ISL_2127897, EPI_ISL_2127899, EPI_ISL_2128176 | | | |
| see above | Centre for Enzyme Innovation, University of Portsmouth / Translational Research Laboratory, Portsmouth Hospitals NHS Trust | COVID-19 Genomics UK (COG-UK) Consortium | Angela Beckett; Christopher Fearn; Kate Cook; Katie Loveson; Salman Goudarzi; Samuel Robson; Scott Elliott; Sharon Glaysher |
| EPI_ISL_1832205, EPI_ISL_1832207, EPI_ISL_1832210, EPI_ISL_1832216, EPI_ISL_1832218, EPI_ISL_1832219, EPI_ISL_1832220, EPI_ISL_1832221, EPI_ISL_1832222, EPI_ISL_1832223, EPI_ISL_1832224, EPI_ISL_1832225, EPI_ISL_1832226, EPI_ISL_1832227, EPI_ISL_1832228, EPI_ISL_1832229, EPI_ISL_1832230, EPI_ISL_1832231, EPI_ISL_1832232, EPI_ISL_1832233, EPI_ISL_1974765, EPI_ISL_1974766, EPI_ISL_1974768, EPI_ISL_1974769, EPI_ISL_1974771, EPI_ISL_1974773, EPI_ISL_1974775, EPI_ISL_1974777, EPI_ISL_1974778, EPI_ISL_1974779, EPI_ISL_1974781, EPI_ISL_1974783, EPI_ISL_1974786 | | | |
| see above | Department of Pathology, University of Cambridge | COVID-19 Genomics UK (COG-UK) Consortium | Aminu S. Jahun; Ian Goodfellow; Iliana Georgana; Martin D. Curran; Myra Hosmillo; Rhys Izuagbe; Surendra Parmar; William L. Hamilton; Yasmin Chaudhry |
| EPI_ISL_2005342, EPI_ISL_2005344, EPI_ISL_2005345, EPI_ISL_2005350, EPI_ISL_2005354, EPI_ISL_2005356, EPI_ISL_2005368, EPI_ISL_2005376, EPI_ISL_2005383, EPI_ISL_2005399, EPI_ISL_2005402, EPI_ISL_2005403, EPI_ISL_2005407, EPI_ISL_2005409, EPI_ISL_2005415, EPI_ISL_2005416, EPI_ISL_2005417, EPI_ISL_2005419, EPI_ISL_2005420, EPI_ISL_2005424, EPI_ISL_2005428, EPI_ISL_2005432, EPI_ISL_2005440, EPI_ISL_2005453, EPI_ISL_2005455, EPI_ISL_2005465, EPI_ISL_2005470, EPI_ISL_2005471, EPI_ISL_2005473, EPI_ISL_2005475, EPI_ISL_2005480, EPI_ISL_2005483, EPI_ISL_2005487, EPI_ISL_2005490, EPI_ISL_2005494, EPI_ISL_2005497, EPI_ISL_2005498, EPI_ISL_2005501, EPI_ISL_2005524, EPI_ISL_2005527, EPI_ISL_2005534, EPI_ISL_2005535, EPI_ISL_2005536, EPI_ISL_2005542, EPI_ISL_2005544, EPI_ISL_2005545, EPI_ISL_2005546, EPI_ISL_2005554, EPI_ISL_2005559, EPI_ISL_2005562, EPI_ISL_2005563, EPI_ISL_2005566, EPI_ISL_2005569, EPI_ISL_2005572, EPI_ISL_2005575, EPI_ISL_2005583, EPI_ISL_2005587, EPI_ISL_2005591, EPI_ISL_2005592, EPI_ISL_2005594, EPI_ISL_2005604, EPI_ISL_2005607, EPI_ISL_2006542, EPI_ISL_2006543, EPI_ISL_2006544, EPI_ISL_2006548, EPI_ISL_2006550, EPI_ISL_2006551, EPI_ISL_2006552, EPI_ISL_2006556, EPI_ISL_2006563, EPI_ISL_2006565, EPI_ISL_2006566, EPI_ISL_2006567, EPI_ISL_2006569, EPI_ISL_2006577, EPI_ISL_2006578, EPI_ISL_2006580, EPI_ISL_2006582, EPI_ISL_2006584, EPI_ISL_2006586, EPI_ISL_2006589, EPI_ISL_2006591, EPI_ISL_2006592, EPI_ISL_2006594, EPI_ISL_2006595, EPI_ISL_2006597, EPI_ISL_2006599, EPI_ISL_2006600, EPI_ISL_2006602, EPI_ISL_2006603, EPI_ISL_2006607, EPI_ISL_2006608, EPI_ISL_2006609, EPI_ISL_2006610, EPI_ISL_2006612, EPI_ISL_2006613, EPI_ISL_2006614, EPI_ISL_2006616, EPI_ISL_2006618, EPI_ISL_2006619, EPI_ISL_2006620, EPI_ISL_2006621, EPI_ISL_2006623, EPI_ISL_2006626, EPI_ISL_2006631, EPI_ISL_2006632, EPI_ISL_2006636, EPI_ISL_2006637, EPI_ISL_2006640, EPI_ISL_2006642, EPI_ISL_2006644, EPI_ISL_2006645, EPI_ISL_2006646, EPI_ISL_2006648, EPI_ISL_2006649, EPI_ISL_2006652, EPI_ISL_2006653, EPI_ISL_2006656, EPI_ISL_2006659, EPI_ISL_2006661, EPI_ISL_2006662, EPI_ISL_2006664, EPI_ISL_2006666, EPI_ISL_2022532, EPI_ISL_2022533, EPI_ISL_2022534, EPI_ISL_2022535, EPI_ISL_2022536, EPI_ISL_2022537, EPI_ISL_2022538, EPI_ISL_2022539, EPI_ISL_2022541, EPI_ISL_2022545, EPI_ISL_2022547, EPI_ISL_2022548, EPI_ISL_2022549, EPI_ISL_2022550, EPI_ISL_2022552, EPI_ISL_2022554, EPI_ISL_2022556, EPI_ISL_2022557, EPI_ISL_2022558, EPI_ISL_2022559, EPI_ISL_2022560, EPI_ISL_2022561, EPI_ISL_2022562, EPI_ISL_2022564, EPI_ISL_2022565, EPI_ISL_2022566, EPI_ISL_2022567, EPI_ISL_2022568, EPI_ISL_2022569, EPI_ISL_2022570, EPI_ISL_2022571, EPI_ISL_2022572, EPI_ISL_2022573, EPI_ISL_2022574, EPI_ISL_2022575, EPI_ISL_2022576, EPI_ISL_2022580, EPI_ISL_2022581, EPI_ISL_2022583, EPI_ISL_2022587, EPI_ISL_2022588, EPI_ISL_2022589, EPI_ISL_2022590, EPI_ISL_2080361, EPI_ISL_2080364, EPI_ISL_2080365, EPI_ISL_2080366, EPI_ISL_2080368, EPI_ISL_2080369, EPI_ISL_2080370, EPI_ISL_2080372, EPI_ISL_2080373, EPI_ISL_2080374, EPI_ISL_2080379, EPI_ISL_2080385, EPI_ISL_2080387, EPI_ISL_2080388, EPI_ISL_2080391, EPI_ISL_2080393, EPI_ISL_2080394, EPI_ISL_2080396, EPI_ISL_2080398, EPI_ISL_2080399, EPI_ISL_2080400, EPI_ISL_2080402, EPI_ISL_2080403, EPI_ISL_2080404, EPI_ISL_2080407, EPI_ISL_2080409, EPI_ISL_2080412, EPI_ISL_2080413, EPI_ISL_2080417, EPI_ISL_2080418, EPI_ISL_2080419, EPI_ISL_2080420, EPI_ISL_2080424, EPI_ISL_2080425, EPI_ISL_2080429, EPI_ISL_2080431, EPI_ISL_2080432, EPI_ISL_2080433, EPI_ISL_2080434, EPI_ISL_2080438, EPI_ISL_2080440, EPI_ISL_2080441, EPI_ISL_2080442, EPI_ISL_2080446, EPI_ISL_2080451, EPI_ISL_2080452, EPI_ISL_2080453, EPI_ISL_2080454, EPI_ISL_2080455, EPI_ISL_2080457, EPI_ISL_2080458, EPI_ISL_2080459, EPI_ISL_2080461, EPI_ISL_2080462, EPI_ISL_2080464, EPI_ISL_2080465, EPI_ISL_2080468, EPI_ISL_2080470, EPI_ISL_2080474 | | | |
| see above | Health Services Laboratories | Wellcome Sanger Institute for the COVID-19 Genomics UK (COG-UK) Consortium | Cordelia Langford; David K. Jackson; Dominic Kwiatkowski; Ewan Harrison; Health Services Laboratories and Alex Alderton; Ian Johnston; Jeffrey Barrett; John Sillitoe on behalf of the Wellcome Sanger Institute COVID-19 Surveillance Team; Roberto Amato; Sonia Goncalves |
| EPI_ISL_1790464, EPI_ISL_1806509, EPI_ISL_1806511, EPI_ISL_1806515, EPI_ISL_1806516, EPI_ISL_1806517, EPI_ISL_1806523, EPI_ISL_1806530, EPI_ISL_1806531, EPI_ISL_1806532, EPI_ISL_1806533, EPI_ISL_1806544, EPI_ISL_1806546, EPI_ISL_1806555, EPI_ISL_1806557, EPI_ISL_1806559, EPI_ISL_1806561, EPI_ISL_1806563, EPI_ISL_1806572, EPI_ISL_1806593, EPI_ISL_1806599, EPI_ISL_1806612, EPI_ISL_1806616, EPI_ISL_1806629, EPI_ISL_1806635, EPI_ISL_1806638, EPI_ISL_1806641, EPI_ISL_1806651, EPI_ISL_1806656, EPI_ISL_1806657, EPI_ISL_1806662, EPI_ISL_1806666, EPI_ISL_1806678, EPI_ISL_1806679, EPI_ISL_1806684, EPI_ISL_1806688, EPI_ISL_1806703, EPI_ISL_1806716, EPI_ISL_1806719, EPI_ISL_1806721, EPI_ISL_1806728, EPI_ISL_1806729, EPI_ISL_1806737, EPI_ISL_1806738, EPI_ISL_1806756, EPI_ISL_1806758, EPI_ISL_1806773, EPI_ISL_1806775, EPI_ISL_1806778, EPI_ISL_1806784, EPI_ISL_1806785, EPI_ISL_1806788, EPI_ISL_1806795, EPI_ISL_1806801, EPI_ISL_1806803, EPI_ISL_1806813, EPI_ISL_1806826, EPI_ISL_1806833, EPI_ISL_1806837, EPI_ISL_1806844, EPI_ISL_1806852, EPI_ISL_1829209, EPI_ISL_1829211, EPI_ISL_1829212, EPI_ISL_1829213, EPI_ISL_1829214, EPI_ISL_1829216, EPI_ISL_1829220, EPI_ISL_1829222, EPI_ISL_1829223, EPI_ISL_1829227, EPI_ISL_1829229, EPI_ISL_1829232, EPI_ISL_1829233, EPI_ISL_1829237, EPI_ISL_1829242, EPI_ISL_1829243, EPI_ISL_1829245, EPI_ISL_1829252, EPI_ISL_1829254, EPI_ISL_1829255, EPI_ISL_1829258, EPI_ISL_1829260, EPI_ISL_1829262, EPI_ISL_1829271, EPI_ISL_1829272, EPI_ISL_1829273, EPI_ISL_1829274, EPI_ISL_1829276, EPI_ISL_1829278, EPI_ISL_1829279, EPI_ISL_1829281, EPI_ISL_1829282, EPI_ISL_1829285, EPI_ISL_1829289, EPI_ISL_1829291, EPI_ISL_1829292, EPI_ISL_1829296, EPI_ISL_1829297, EPI_ISL_1829299, EPI_ISL_1829300, EPI_ISL_1829302, EPI_ISL_1829304, EPI_ISL_1829307, EPI_ISL_1829310, EPI_ISL_1829311, EPI_ISL_1829318, EPI_ISL_1829319, EPI_ISL_1829320, EPI_ISL_1829325, EPI_ISL_1829333, EPI_ISL_1829337, EPI_ISL_1829342, EPI_ISL_1829344, EPI_ISL_1829345, EPI_ISL_1829352, EPI_ISL_1829353, EPI_ISL_1829356, EPI_ISL_1829357, EPI_ISL_1829359, EPI_ISL_1829360, EPI_ISL_1829362, EPI_ISL_1829373, EPI_ISL_1829374, EPI_ISL_1829375, EPI_ISL_1829376, EPI_ISL_1829382, EPI_ISL_1829400, EPI_ISL_1829403, EPI_ISL_1829406, EPI_ISL_1829408, EPI_ISL_1829411, EPI_ISL_1829412, EPI_ISL_1829413, EPI_ISL_1829419, EPI_ISL_1829420, EPI_ISL_1829427, EPI_ISL_1829430, EPI_ISL_1829433, EPI_ISL_1829434, EPI_ISL_1829435, EPI_ISL_1829437, EPI_ISL_1829438, EPI_ISL_1829440, EPI_ISL_1829444, EPI_ISL_1829452, EPI_ISL_1829456, EPI_ISL_1829457, EPI_ISL_1829458, EPI_ISL_1829460, EPI_ISL_1829462, EPI_ISL_1829474, EPI_ISL_1829804, EPI_ISL_1829806, EPI_ISL_1829808, EPI_ISL_1829809, EPI_ISL_1829811, EPI_ISL_1829813, EPI_ISL_1829815, EPI_ISL_1829816, EPI_ISL_1829818, EPI_ISL_1829819, EPI_ISL_1829821, EPI_ISL_1829822, EPI_ISL_1829823, EPI_ISL_1829825, EPI_ISL_1829828, EPI_ISL_1829830, EPI_ISL_1829835, EPI_ISL_1829838, EPI_ISL_1829839, EPI_ISL_1829840, EPI_ISL_1829842, EPI_ISL_1829843, EPI_ISL_1829848, EPI_ISL_1829850, EPI_ISL_1829851, EPI_ISL_1829853, EPI_ISL_1829854, EPI_ISL_1829855, EPI_ISL_1829859, EPI_ISL_1829860, EPI_ISL_1829862, EPI_ISL_1829863, EPI_ISL_1829865, EPI_ISL_1829866, EPI_ISL_1829867, EPI_ISL_1829869, EPI_ISL_1829871, EPI_ISL_1829872, EPI_ISL_1829874, EPI_ISL_1829875, EPI_ISL_1829878, EPI_ISL_1829879, EPI_ISL_1829880, EPI_ISL_1829881, EPI_ISL_1829882, EPI_ISL_1829883, EPI_ISL_1829884, EPI_ISL_1829885, EPI_ISL_1829886, EPI_ISL_1829887, EPI_ISL_1829889, EPI_ISL_1829896, EPI_ISL_1829898, EPI_ISL_1829899, EPI_ISL_1829900, EPI_ISL_1829902, EPI_ISL_1829904, EPI_ISL_1829905, EPI_ISL_1829909, EPI_ISL_1829913, EPI_ISL_1829914, EPI_ISL_1829915, EPI_ISL_1829917, EPI_ISL_1829920, EPI_ISL_1829921, EPI_ISL_1829922, EPI_ISL_1829925, EPI_ISL_1829929, EPI_ISL_1829930, EPI_ISL_1829935, EPI_ISL_1829937, EPI_ISL_1829938, EPI_ISL_1829939, EPI_ISL_1829940, EPI_ISL_1829943, EPI_ISL_1829944, EPI_ISL_1829946, EPI_ISL_1829950, EPI_ISL_1829952, EPI_ISL_1829953, EPI_ISL_1829962, EPI_ISL_1829963, EPI_ISL_1829964, EPI_ISL_1829965, EPI_ISL_1829966, EPI_ISL_1829969, EPI_ISL_1829970, EPI_ISL_1829973, EPI_ISL_1829974, EPI_ISL_1829976, EPI_ISL_1829979, EPI_ISL_1829982, EPI_ISL_1829983, EPI_ISL_1829984, EPI_ISL_1829985, EPI_ISL_1829986, EPI_ISL_1829987, EPI_ISL_1829988, EPI_ISL_1829994, EPI_ISL_1829995, EPI_ISL_1829996, EPI_ISL_1830003, EPI_ISL_1830004, EPI_ISL_1830006, EPI_ISL_1830007, EPI_ISL_1830011, EPI_ISL_1830012, EPI_ISL_1830013, EPI_ISL_1830019, EPI_ISL_1830020, EPI_ISL_1830021, EPI_ISL_1830023, EPI_ISL_1830024, EPI_ISL_1830025, EPI_ISL_1830026, EPI_ISL_1830031, EPI_ISL_1830035, EPI_ISL_1830036, EPI_ISL_1830039, EPI_ISL_1830040, EPI_ISL_1830042, EPI_ISL_1830044, EPI_ISL_1830046, EPI_ISL_1830048, EPI_ISL_1830049, EPI_ISL_1830052, EPI_ISL_1830053, EPI_ISL_1830054, EPI_ISL_1830055, EPI_ISL_1830058, EPI_ISL_1830061, EPI_ISL_1830062, EPI_ISL_1830064, EPI_ISL_1830065, EPI_ISL_1830066, EPI_ISL_1830069, EPI_ISL_1830071, EPI_ISL_1830072, EPI_ISL_1830074, EPI_ISL_1830075, EPI_ISL_1830077, EPI_ISL_1830078, EPI_ISL_1830080, EPI_ISL_1830082, EPI_ISL_1830083, EPI_ISL_1830085, EPI_ISL_1830089, EPI_ISL_1830090, EPI_ISL_1830092, EPI_ISL_1830093, EPI_ISL_1830094, EPI_ISL_1830095, EPI_ISL_1830098, EPI_ISL_1830101, EPI_ISL_1830105, EPI_ISL_1830109, EPI_ISL_1830114, EPI_ISL_1830122, EPI_ISL_1830125, EPI_ISL_1830126, EPI_ISL_1830128, EPI_ISL_1830133, EPI_ISL_1830137, EPI_ISL_1830140, EPI_ISL_1830150, EPI_ISL_1830154, EPI_ISL_1830155, EPI_ISL_1830158, EPI_ISL_1830159, EPI_ISL_1830166, EPI_ISL_1830167, EPI_ISL_1830170, EPI_ISL_1830171, EPI_ISL_1830174, EPI_ISL_1830175, EPI_ISL_1830176, EPI_ISL_1830181, EPI_ISL_1830182, EPI_ISL_1830184, EPI_ISL_1830185, EPI_ISL_1830188, EPI_ISL_1830192, EPI_ISL_1830195, EPI_ISL_1830201, EPI_ISL_1830203, EPI_ISL_1830208, EPI_ISL_1830211, EPI_ISL_1830214, EPI_ISL_1830217, EPI_ISL_1830219, EPI_ISL_1830220, EPI_ISL_1830225, EPI_ISL_1830234, EPI_ISL_1830237, EPI_ISL_1830241, EPI_ISL_1830251, EPI_ISL_1830252, EPI_ISL_1830255, EPI_ISL_1830259, EPI_ISL_1830271, EPI_ISL_1830298, EPI_ISL_1830303, EPI_ISL_1830306, EPI_ISL_1830308, EPI_ISL_1830309, EPI_ISL_1830312, EPI_ISL_1830314, EPI_ISL_1830315, EPI_ISL_1830316, EPI_ISL_1830319, EPI_ISL_1830321, EPI_ISL_1830326, EPI_ISL_1830330, EPI_ISL_1830332, EPI_ISL_1830338, EPI_ISL_1830340, EPI_ISL_1830347, EPI_ISL_1830348, EPI_ISL_1830350, EPI_ISL_1830353, EPI_ISL_1830356, EPI_ISL_1830357, EPI_ISL_1830361, EPI_ISL_1830363, EPI_ISL_1830364, EPI_ISL_1830365, EPI_ISL_1830368, EPI_ISL_1830371, EPI_ISL_1830377, EPI_ISL_1830383, EPI_ISL_1830388, EPI_ISL_1830391, EPI_ISL_1830393, EPI_ISL_1830395, EPI_ISL_1830396, EPI_ISL_1830397, EPI_ISL_1830398, EPI_ISL_1830402, EPI_ISL_1830407, EPI_ISL_1830415, EPI_ISL_1830420, EPI_ISL_1830422, EPI_ISL_1830425, EPI_ISL_1830426, EPI_ISL_1830427, EPI_ISL_1830433, EPI_ISL_1830434, EPI_ISL_1830439, EPI_ISL_1830443, EPI_ISL_1830444, EPI_ISL_1830445, EPI_ISL_1830455, EPI_ISL_1830458, EPI_ISL_1830460, EPI_ISL_1830463, EPI_ISL_1830464, EPI_ISL_1830466, EPI_ISL_1830467, EPI_ISL_1830468, EPI_ISL_1830471, EPI_ISL_1830478, EPI_ISL_1830479, EPI_ISL_1830480, EPI_ISL_1830484, EPI_ISL_1830486, EPI_ISL_1830487, EPI_ISL_1830490, EPI_ISL_1830491, EPI_ISL_1830500, EPI_ISL_1830504, EPI_ISL_1830505, EPI_ISL_1830507, EPI_ISL_1830513, EPI_ISL_1830516, EPI_ISL_1830519, EPI_ISL_1830520, EPI_ISL_1830521, EPI_ISL_1830530, EPI_ISL_1830533, EPI_ISL_1830534, EPI_ISL_1830535, EPI_ISL_1830536, EPI_ISL_1830538, EPI_ISL_1830539, EPI_ISL_1830541, EPI_ISL_1830543, EPI_ISL_1830547, EPI_ISL_1830548, EPI_ISL_1830550, EPI_ISL_1830552, EPI_ISL_1830553, EPI_ISL_1830558, EPI_ISL_1830561, EPI_ISL_1830566, EPI_ISL_1830567, EPI_ISL_1830571, EPI_ISL_1830573, EPI_ISL_1830575, EPI_ISL_1830577, EPI_ISL_1830582, EPI_ISL_1830586, EPI_ISL_1830587, EPI_ISL_1830588, EPI_ISL_1830590, EPI_ISL_1830591, EPI_ISL_1830598, EPI_ISL_1830602, EPI_ISL_1830610, EPI_ISL_1830611, EPI_ISL_1830616, EPI_ISL_1830617, EPI_ISL_1830618, EPI_ISL_1830620, EPI_ISL_1830622, EPI_ISL_1830625, EPI_ISL_1830627, EPI_ISL_1830628, EPI_ISL_1830636, EPI_ISL_1830639, EPI_ISL_1830640, EPI_ISL_1830641, EPI_ISL_1830642, EPI_ISL_1830645, EPI_ISL_1830646, EPI_ISL_1830647, EPI_ISL_1830653, EPI_ISL_1830655, EPI_ISL_1830658, EPI_ISL_1830662, EPI_ISL_1830663, EPI_ISL_1830666, EPI_ISL_1830678, EPI_ISL_1830680, EPI_ISL_1830683, EPI_ISL_1830684, EPI_ISL_1830686, EPI_ISL_1830690, EPI_ISL_1830691, EPI_ISL_1830696, EPI_ISL_1830697, EPI_ISL_1830698, EPI_ISL_1830699, EPI_ISL_1830705, EPI_ISL_1830711, EPI_ISL_1830721, EPI_ISL_1830723, EPI_ISL_1830724, EPI_ISL_1830728, EPI_ISL_1830730, EPI_ISL_1830731, EPI_ISL_1830735, EPI_ISL_1830740, EPI_ISL_1830741, EPI_ISL_1830746, EPI_ISL_1830748, EPI_ISL_1830752, EPI_ISL_1830758, EPI_ISL_1830761, EPI_ISL_1830762, EPI_ISL_1830764, EPI_ISL_1830769, EPI_ISL_1830772, EPI_ISL_1830775, EPI_ISL_1830778, EPI_ISL_1830780, EPI_ISL_1830789, EPI_ISL_1830791, EPI_ISL_1830793, EPI_ISL_1830799, EPI_ISL_1830803, EPI_ISL_1830806, EPI_ISL_1830807, EPI_ISL_1830808, EPI_ISL_1830811, EPI_ISL_1830812, EPI_ISL_1830814, EPI_ISL_1830815, EPI_ISL_1830817, EPI_ISL_1830818, EPI_ISL_1830819, EPI_ISL_1830823, EPI_ISL_1830828, EPI_ISL_1830832, EPI_ISL_1830833, EPI_ISL_1830836, EPI_ISL_1830840, EPI_ISL_1830844, EPI_ISL_1830846, EPI_ISL_1830848, EPI_ISL_1830850, EPI_ISL_1830856, EPI_ISL_1830858, EPI_ISL_1830859, EPI_ISL_1830863, EPI_ISL_1830868, EPI_ISL_1830869, EPI_ISL_1830870, EPI_ISL_1830878, EPI_ISL_1830880, EPI_ISL_1830881, EPI_ISL_1830883, EPI_ISL_1830888, EPI_ISL_1830895, EPI_ISL_1830896, EPI_ISL_1830897, EPI_ISL_1830898, EPI_ISL_1830914, EPI_ISL_1830921, EPI_ISL_1830925, EPI_ISL_1830927, EPI_ISL_1830929, EPI_ISL_1830935, EPI_ISL_1830943, EPI_ISL_1830944, EPI_ISL_1830947, EPI_ISL_1830949, EPI_ISL_1830956, EPI_ISL_1830961, EPI_ISL_1830966, EPI_ISL_1830968, EPI_ISL_1830970, EPI_ISL_1830981, EPI_ISL_1830983, EPI_ISL_1830985, EPI_ISL_1831001, EPI_ISL_1831002, EPI_ISL_1831003, EPI_ISL_1831005, EPI_ISL_1831006, EPI_ISL_1831008, EPI_ISL_1831011, EPI_ISL_1831015, EPI_ISL_1831018, EPI_ISL_1831019, EPI_ISL_1831023, EPI_ISL_1831027, EPI_ISL_1831029, EPI_ISL_1831034, EPI_ISL_1831039, EPI_ISL_1831043, EPI_ISL_1831045, EPI_ISL_1831049, EPI_ISL_1831050, EPI_ISL_1831052, EPI_ISL_1831054, EPI_ISL_1831055, EPI_ISL_1831057, EPI_ISL_1831058, EPI_ISL_1831060, EPI_ISL_1831061, EPI_ISL_1831071, EPI_ISL_1831073, EPI_ISL_1831074, EPI_ISL_1831079, EPI_ISL_1831080, EPI_ISL_1831082, EPI_ISL_1831084, EPI_ISL_1857806, EPI_ISL_1857807, EPI_ISL_1857809, EPI_ISL_1857811, EPI_ISL_1857812, EPI_ISL_1857815, EPI_ISL_1857816, EPI_ISL_1857817, EPI_ISL_1857819, EPI_ISL_1857821, EPI_ISL_1857822, EPI_ISL_1857824, EPI_ISL_1857825, EPI_ISL_1857828, EPI_ISL_1857829, EPI_ISL_1857830, EPI_ISL_1857832, EPI_ISL_1857833, EPI_ISL_1857834, EPI_ISL_1857835, EPI_ISL_1857836, EPI_ISL_1857837, EPI_ISL_1857838, EPI_ISL_1857839, EPI_ISL_1857840, EPI_ISL_1857841, EPI_ISL_1857843, EPI_ISL_1857850, EPI_ISL_1857854, EPI_ISL_1857856, EPI_ISL_1857857, EPI_ISL_1857859, EPI_ISL_1857860, EPI_ISL_1857863, EPI_ISL_1857864, EPI_ISL_1857868, EPI_ISL_1857869, EPI_ISL_1857872, EPI_ISL_1857875, EPI_ISL_1857876, EPI_ISL_1857877, EPI_ISL_1857878, EPI_ISL_1857879, EPI_ISL_1857880, EPI_ISL_1857883, EPI_ISL_1857886, EPI_ISL_1857887, EPI_ISL_1857890, EPI_ISL_1857891, EPI_ISL_1857892, EPI_ISL_1857893, EPI_ISL_1857896, EPI_ISL_1857904, EPI_ISL_1857908, EPI_ISL_1857912, EPI_ISL_1857913, EPI_ISL_1857914, EPI_ISL_1857917, EPI_ISL_1857920, EPI_ISL_1857921, EPI_ISL_1857922, EPI_ISL_1857923, EPI_ISL_1857926, EPI_ISL_1857927, EPI_ISL_1857928, EPI_ISL_1857932, EPI_ISL_1857933, EPI_ISL_1857934, EPI_ISL_1857937, EPI_ISL_1857941, EPI_ISL_1857942, EPI_ISL_1857943, EPI_ISL_1857948, EPI_ISL_1857952, EPI_ISL_1857954, EPI_ISL_1857955, EPI_ISL_1857957, EPI_ISL_1857960, EPI_ISL_1857961, EPI_ISL_1857963, EPI_ISL_1857964, EPI_ISL_1857967, EPI_ISL_1857969, EPI_ISL_1857970, EPI_ISL_1857972, EPI_ISL_1857974, EPI_ISL_1857977, EPI_ISL_1857980, EPI_ISL_1857982, EPI_ISL_1857986, EPI_ISL_1857988, EPI_ISL_1857989, EPI_ISL_1857993, EPI_ISL_1857994, EPI_ISL_1858001, EPI_ISL_1858006, EPI_ISL_1858009, EPI_ISL_1858010, EPI_ISL_1858012, EPI_ISL_1858015, EPI_ISL_1858018, EPI_ISL_1858020, EPI_ISL_1858021, EPI_ISL_1858023, EPI_ISL_1858026, EPI_ISL_1858031, EPI_ISL_1858032, EPI_ISL_1858034, EPI_ISL_1858035, EPI_ISL_1858037, EPI_ISL_1858041, EPI_ISL_1858048, EPI_ISL_1858049, EPI_ISL_1858051, EPI_ISL_1858054, EPI_ISL_1858057, EPI_ISL_1858060, EPI_ISL_1858065, EPI_ISL_1858066, EPI_ISL_1858070, EPI_ISL_1858071, EPI_ISL_1858073, EPI_ISL_1858075, EPI_ISL_1858076, EPI_ISL_1858079, EPI_ISL_1858081, EPI_ISL_1858084, EPI_ISL_1858086, EPI_ISL_1858087, EPI_ISL_1858089, EPI_ISL_1858092, EPI_ISL_1858094, EPI_ISL_1858097, EPI_ISL_1858099, EPI_ISL_1858101, EPI_ISL_1858104, EPI_ISL_1858107, EPI_ISL_1858108, EPI_ISL_1858110, EPI_ISL_1858113, EPI_ISL_1858114, EPI_ISL_1858119, EPI_ISL_1858121, EPI_ISL_1858124, EPI_ISL_1858127, EPI_ISL_1858132, EPI_ISL_1858134, EPI_ISL_1858136, EPI_ISL_1858138, EPI_ISL_1858139, EPI_ISL_1858143, EPI_ISL_1858147, EPI_ISL_1858152, EPI_ISL_1858155, EPI_ISL_1858156, EPI_ISL_1858158, EPI_ISL_1858207, EPI_ISL_1858208, EPI_ISL_1858211, EPI_ISL_1858213, EPI_ISL_1858217, EPI_ISL_1858221, EPI_ISL_1858228, EPI_ISL_1858237, EPI_ISL_1912007, EPI_ISL_1912014, EPI_ISL_1912025, EPI_ISL_1912040, EPI_ISL_1912042, EPI_ISL_1912043, EPI_ISL_1912054, EPI_ISL_1912062, EPI_ISL_1912067, EPI_ISL_1912077, EPI_ISL_1912079, EPI_ISL_1912080, EPI_ISL_1912089, EPI_ISL_1912090, EPI_ISL_1912092, EPI_ISL_1912095, EPI_ISL_1912097, EPI_ISL_1912098, EPI_ISL_1912100, EPI_ISL_1912101, EPI_ISL_1912106, EPI_ISL_1912112, EPI_ISL_1912115, EPI_ISL_1912120, EPI_ISL_1912121, EPI_ISL_1912122, EPI_ISL_1912123, EPI_ISL_1912133, EPI_ISL_1912134, EPI_ISL_1912135, EPI_ISL_1912136, EPI_ISL_1912138, EPI_ISL_1912139, EPI_ISL_1912146, EPI_ISL_1912148, EPI_ISL_1912156, EPI_ISL_1912160, EPI_ISL_1912161, EPI_ISL_1912162, EPI_ISL_1912163, EPI_ISL_1912166, EPI_ISL_1912169, EPI_ISL_1912170, EPI_ISL_1912176, EPI_ISL_1912178, EPI_ISL_1912180, EPI_ISL_1912182, EPI_ISL_1912183, EPI_ISL_1912184, EPI_ISL_1912185, EPI_ISL_1912188, EPI_ISL_1912191, EPI_ISL_1912193, EPI_ISL_1912194, EPI_ISL_1912197, EPI_ISL_1912202, EPI_ISL_1912204, EPI_ISL_1912206, EPI_ISL_1912214, EPI_ISL_1912223, EPI_ISL_1912225, EPI_ISL_1912226, EPI_ISL_1912248, EPI_ISL_1912261, EPI_ISL_1912262, EPI_ISL_1912268, EPI_ISL_1912283, EPI_ISL_1912286, EPI_ISL_1912288, EPI_ISL_1912294, EPI_ISL_1912297, EPI_ISL_1912307, EPI_ISL_1912309, EPI_ISL_1912314, EPI_ISL_1912318, EPI_ISL_1912321, EPI_ISL_1912325, EPI_ISL_1912328, EPI_ISL_1912329, EPI_ISL_1912332, EPI_ISL_1912333, EPI_ISL_1912342, EPI_ISL_1912344, EPI_ISL_1912347, EPI_ISL_1912348, EPI_ISL_1912351, EPI_ISL_1912352, EPI_ISL_1912353, EPI_ISL_1912354, EPI_ISL_1912355, EPI_ISL_1912358, EPI_ISL_1912359, EPI_ISL_1912360, EPI_ISL_1912361, EPI_ISL_1912362, EPI_ISL_1912364, EPI_ISL_1912365, EPI_ISL_1912369, EPI_ISL_1912371, EPI_ISL_1912373, EPI_ISL_1912374, EPI_ISL_1912375, EPI_ISL_1912376, EPI_ISL_1912377, EPI_ISL_1912379, EPI_ISL_1912381, EPI_ISL_1912382, EPI_ISL_1912383, EPI_ISL_1912384, EPI_ISL_1912385, EPI_ISL_1912387, EPI_ISL_1912389, EPI_ISL_1912390, EPI_ISL_1912393, EPI_ISL_1912394, EPI_ISL_1912396, EPI_ISL_1912397, EPI_ISL_1912398, EPI_ISL_1912399, EPI_ISL_1912401, EPI_ISL_1912402, EPI_ISL_1912403, EPI_ISL_1912405, EPI_ISL_1912407, EPI_ISL_1912408, EPI_ISL_1912409, EPI_ISL_1912413, EPI_ISL_1912414, EPI_ISL_1912415, EPI_ISL_1912416, EPI_ISL_1912418, EPI_ISL_1912422, EPI_ISL_1912425, EPI_ISL_1912426, EPI_ISL_1912427, EPI_ISL_1912428, EPI_ISL_1912429, EPI_ISL_1912430, EPI_ISL_1912431, EPI_ISL_1912432, EPI_ISL_1912434, EPI_ISL_1912435, EPI_ISL_1912436, EPI_ISL_1912437, EPI_ISL_1912438, EPI_ISL_1912439, EPI_ISL_1912442, EPI_ISL_1912443, EPI_ISL_1912444, EPI_ISL_1912446, EPI_ISL_1912447, EPI_ISL_1912448, EPI_ISL_1912449, EPI_ISL_1912452, EPI_ISL_1912453, EPI_ISL_1912454, EPI_ISL_1912455, EPI_ISL_1912456, EPI_ISL_1912458, EPI_ISL_1912459, EPI_ISL_1912460, EPI_ISL_1912461, EPI_ISL_1912463, EPI_ISL_1912464, EPI_ISL_1912465, EPI_ISL_1912466, EPI_ISL_1912467, EPI_ISL_1912468, EPI_ISL_1912469, EPI_ISL_1912471, EPI_ISL_1912474, EPI_ISL_1912475, EPI_ISL_1912479, EPI_ISL_1912480, EPI_ISL_1912481, EPI_ISL_1912482, EPI_ISL_1912483, EPI_ISL_1912484, EPI_ISL_1912486, EPI_ISL_1912487, EPI_ISL_1912488, EPI_ISL_1912490, EPI_ISL_1912491, EPI_ISL_1912492, EPI_ISL_1912493, EPI_ISL_1912494, EPI_ISL_1912495, EPI_ISL_1912496, EPI_ISL_1912498, EPI_ISL_1912499, EPI_ISL_1912503, EPI_ISL_1912504, EPI_ISL_1912505, EPI_ISL_1912506, EPI_ISL_1912507, EPI_ISL_1912508, EPI_ISL_1912509, EPI_ISL_1912510, EPI_ISL_1912514, EPI_ISL_1912515, EPI_ISL_1912516, EPI_ISL_1912517, EPI_ISL_1912521, EPI_ISL_1912522, EPI_ISL_1912523, EPI_ISL_1912524, EPI_ISL_1912526, EPI_ISL_1912528, EPI_ISL_1912529, EPI_ISL_1912530, EPI_ISL_1912531, EPI_ISL_1912532, EPI_ISL_1912533, EPI_ISL_1912534, EPI_ISL_1912535, EPI_ISL_1912536, EPI_ISL_1912537, EPI_ISL_1912538, EPI_ISL_1912539, EPI_ISL_1912540, EPI_ISL_1912541, EPI_ISL_1912542, EPI_ISL_1912543, EPI_ISL_1912544, EPI_ISL_1912545, EPI_ISL_1912548, EPI_ISL_1912551, EPI_ISL_1912552, EPI_ISL_1912553, EPI_ISL_1912554, EPI_ISL_1912555, EPI_ISL_1912556, EPI_ISL_1912557, EPI_ISL_1912559, EPI_ISL_1912560, EPI_ISL_1912561, EPI_ISL_1912562, EPI_ISL_1912563, EPI_ISL_1912564, EPI_ISL_1912566, EPI_ISL_1912567, EPI_ISL_1912568, EPI_ISL_1912570, EPI_ISL_1912571, EPI_ISL_1912572, EPI_ISL_1912574, EPI_ISL_1912575, EPI_ISL_1912577, EPI_ISL_1912578, EPI_ISL_1912579, EPI_ISL_1912580, EPI_ISL_1912582, EPI_ISL_1912583, EPI_ISL_1912584, EPI_ISL_1912585, EPI_ISL_1912586, EPI_ISL_1912587, EPI_ISL_1912590, EPI_ISL_1912592, EPI_ISL_1912593, EPI_ISL_1912729, EPI_ISL_1912737, EPI_ISL_1912740, EPI_ISL_1912751, EPI_ISL_1912767, EPI_ISL_1912784, EPI_ISL_1921166, EPI_ISL_1921167, EPI_ISL_1921168, EPI_ISL_1921169, EPI_ISL_1921170, EPI_ISL_1921171, EPI_ISL_1921172, EPI_ISL_1921173, EPI_ISL_1921174, EPI_ISL_1921175, EPI_ISL_1921176, EPI_ISL_1921177, EPI_ISL_1921178, EPI_ISL_1921179, EPI_ISL_1921180, EPI_ISL_1921181, EPI_ISL_1921182, EPI_ISL_1921183, EPI_ISL_1921184, EPI_ISL_1921185, EPI_ISL_1921186, EPI_ISL_1921187, EPI_ISL_1921188, EPI_ISL_1921189, EPI_ISL_1921190, EPI_ISL_1921191, EPI_ISL_1921192, EPI_ISL_1921194, EPI_ISL_1921195, EPI_ISL_1921196, EPI_ISL_1921197, EPI_ISL_1921198, EPI_ISL_1921199, EPI_ISL_1921200, EPI_ISL_1921201, EPI_ISL_1921202, EPI_ISL_1921203, EPI_ISL_1921204, EPI_ISL_1921205, EPI_ISL_1921206, EPI_ISL_1921207, EPI_ISL_1921208, EPI_ISL_1921209, EPI_ISL_1921210, EPI_ISL_1921211, EPI_ISL_1921212, EPI_ISL_1921213, EPI_ISL_1921214, EPI_ISL_1921215, EPI_ISL_1921216, EPI_ISL_1921217, EPI_ISL_1921218, EPI_ISL_1921219, EPI_ISL_1921220, EPI_ISL_1921221, EPI_ISL_1921222, EPI_ISL_1921223, EPI_ISL_1921224, EPI_ISL_1921225, EPI_ISL_1921226, EPI_ISL_1921227, EPI_ISL_1921228, EPI_ISL_1921229, EPI_ISL_1921230, EPI_ISL_1921231, EPI_ISL_1921232, EPI_ISL_1921233, EPI_ISL_1921234, EPI_ISL_1921235, EPI_ISL_1921236, EPI_ISL_1921237, EPI_ISL_1921238, EPI_ISL_1921239, EPI_ISL_1921240, EPI_ISL_1921241, EPI_ISL_1921242, EPI_ISL_1921243, EPI_ISL_1921244, EPI_ISL_1921245, EPI_ISL_1921246, EPI_ISL_1921247, EPI_ISL_1921248, EPI_ISL_1921249, EPI_ISL_1921250, EPI_ISL_1921251, EPI_ISL_1921252, EPI_ISL_1921253, EPI_ISL_1921254, EPI_ISL_1921255, EPI_ISL_1921256, EPI_ISL_1921257, EPI_ISL_1921258, EPI_ISL_1921259, EPI_ISL_1921261, EPI_ISL_1921262, EPI_ISL_1921263, EPI_ISL_1921265, EPI_ISL_1921266, EPI_ISL_1921267, EPI_ISL_1921268, EPI_ISL_1921269, EPI_ISL_1921270, EPI_ISL_1921271, EPI_ISL_1921272, EPI_ISL_1921273, EPI_ISL_1921274, EPI_ISL_1921275, EPI_ISL_1921276, EPI_ISL_1921277, EPI_ISL_1921278, EPI_ISL_1921279, EPI_ISL_1921280, EPI_ISL_1921281, EPI_ISL_1921282, EPI_ISL_1921283, EPI_ISL_1921284, EPI_ISL_1921285, EPI_ISL_1921286, EPI_ISL_1921287, EPI_ISL_1921289, EPI_ISL_1921290, EPI_ISL_1921291, EPI_ISL_1921292, EPI_ISL_1921293, EPI_ISL_1921294, EPI_ISL_1921295, EPI_ISL_1921296, EPI_ISL_1921297, EPI_ISL_1921298, EPI_ISL_1921299, EPI_ISL_1921300, EPI_ISL_1921301, EPI_ISL_1921302, EPI_ISL_1921303, EPI_ISL_1921304, EPI_ISL_1921305, EPI_ISL_1921306, EPI_ISL_1921307, EPI_ISL_1921308, EPI_ISL_1921309, EPI_ISL_1921310, EPI_ISL_1921311, EPI_ISL_1921312, EPI_ISL_1921313, EPI_ISL_1921314, EPI_ISL_1921315, EPI_ISL_1921316, EPI_ISL_1921317, EPI_ISL_1921318, EPI_ISL_1921319, EPI_ISL_1921320, EPI_ISL_1921321, EPI_ISL_1921322, EPI_ISL_1921323, EPI_ISL_1921324, EPI_ISL_1921325, EPI_ISL_1921326, EPI_ISL_1921327, EPI_ISL_1921328, EPI_ISL_1921329, EPI_ISL_1921330, EPI_ISL_1921331, EPI_ISL_1921332, EPI_ISL_1921333, EPI_ISL_1921334, EPI_ISL_1921335, EPI_ISL_1921336, EPI_ISL_1921337, EPI_ISL_1921338, EPI_ISL_1921339, EPI_ISL_1921340, EPI_ISL_1921341, EPI_ISL_1921342, EPI_ISL_1921343, EPI_ISL_1921344, EPI_ISL_1921345, EPI_ISL_1921346, EPI_ISL_1921347, EPI_ISL_1921348, EPI_ISL_1921349, EPI_ISL_1921350, EPI_ISL_1921351, EPI_ISL_1921352, EPI_ISL_1921353, EPI_ISL_1921354, EPI_ISL_1921355, EPI_ISL_1921356, EPI_ISL_1921357, EPI_ISL_1921358, EPI_ISL_1921359, EPI_ISL_1921360, EPI_ISL_1921361, EPI_ISL_1921362, EPI_ISL_1921363, EPI_ISL_1921364, EPI_ISL_1921365, EPI_ISL_1921366, EPI_ISL_1921367, EPI_ISL_1921368, EPI_ISL_1921369, EPI_ISL_1921370, EPI_ISL_1921371, EPI_ISL_1921372, EPI_ISL_1921373, EPI_ISL_1921374, EPI_ISL_1921375, EPI_ISL_1921376, EPI_ISL_1921377, EPI_ISL_1921378, EPI_ISL_1921379, EPI_ISL_1921380, EPI_ISL_1921381, EPI_ISL_1921382, EPI_ISL_1921383, EPI_ISL_1921384, EPI_ISL_1921385, EPI_ISL_1921386, EPI_ISL_1921387, EPI_ISL_1921388, EPI_ISL_1921389, EPI_ISL_1921390, EPI_ISL_1921391, EPI_ISL_1921392, EPI_ISL_1921393, EPI_ISL_1921394, EPI_ISL_1921395, EPI_ISL_1921396, EPI_ISL_1921397, EPI_ISL_1921398, EPI_ISL_1921399, EPI_ISL_1921400, EPI_ISL_1921401, EPI_ISL_1921402, EPI_ISL_1921403, EPI_ISL_1921404, EPI_ISL_1921405, EPI_ISL_1921406, EPI_ISL_1921407, EPI_ISL_1921408, EPI_ISL_1921409, EPI_ISL_1921410, EPI_ISL_1921411, EPI_ISL_1921412, EPI_ISL_1921413, EPI_ISL_1921414, EPI_ISL_1921415, EPI_ISL_1921416, EPI_ISL_1921417, EPI_ISL_1921418, EPI_ISL_1921419, EPI_ISL_1921420, EPI_ISL_1921421, EPI_ISL_1921422, EPI_ISL_1921423, EPI_ISL_1921424, EPI_ISL_1921426, EPI_ISL_1921427, EPI_ISL_1921428, EPI_ISL_1921429, EPI_ISL_1921430, EPI_ISL_1921431, EPI_ISL_1921432, EPI_ISL_1921433, EPI_ISL_1921434, EPI_ISL_1921435, EPI_ISL_1921436, EPI_ISL_1921437, EPI_ISL_1921438, EPI_ISL_1921444, EPI_ISL_1921446, EPI_ISL_1921459, EPI_ISL_1921465, EPI_ISL_1921486, EPI_ISL_1921494, EPI_ISL_1921495, EPI_ISL_1921496, EPI_ISL_1921506, EPI_ISL_1921507, EPI_ISL_1921509, EPI_ISL_1921515, EPI_ISL_1921525, EPI_ISL_1921533, EPI_ISL_1921552, EPI_ISL_1921553, EPI_ISL_1921555, EPI_ISL_1921572, EPI_ISL_1921588, EPI_ISL_1921598, EPI_ISL_1921613, EPI_ISL_1921618, EPI_ISL_1921619, EPI_ISL_1921634, EPI_ISL_1921637, EPI_ISL_1921645, EPI_ISL_1921656, EPI_ISL_1921662, EPI_ISL_1921666, EPI_ISL_1921667, EPI_ISL_1921669, EPI_ISL_1921676, EPI_ISL_1921685, EPI_ISL_1921686, EPI_ISL_1921698, EPI_ISL_1921700, EPI_ISL_1921703, EPI_ISL_1921725, EPI_ISL_1921735, EPI_ISL_1921738, EPI_ISL_1939677, EPI_ISL_1939690, EPI_ISL_1939719, EPI_ISL_1985497, EPI_ISL_1985685, EPI_ISL_1985688, EPI_ISL_1985689, EPI_ISL_1985692, EPI_ISL_1985694, EPI_ISL_1985695, EPI_ISL_1985696, EPI_ISL_1985709, EPI_ISL_1985711, EPI_ISL_1985712, EPI_ISL_1985716, EPI_ISL_1985719, EPI_ISL_1985734, EPI_ISL_1985744, EPI_ISL_1985748, EPI_ISL_1985755, EPI_ISL_1985757, EPI_ISL_1985762, EPI_ISL_1985765, EPI_ISL_1985767, EPI_ISL_1985803, EPI_ISL_1985811, EPI_ISL_1985820, EPI_ISL_1985822, EPI_ISL_1985825, EPI_ISL_1985826, EPI_ISL_1985828, EPI_ISL_1985835, EPI_ISL_1985841, EPI_ISL_1985848, EPI_ISL_1985861, EPI_ISL_1985862, EPI_ISL_1985864, EPI_ISL_1985871, EPI_ISL_1985872, EPI_ISL_1985873, EPI_ISL_1985876, EPI_ISL_1985886, EPI_ISL_1985890, EPI_ISL_1985892, EPI_ISL_1985896, EPI_ISL_1985901, EPI_ISL_1985902, EPI_ISL_1985909, EPI_ISL_1985910, EPI_ISL_1985914, EPI_ISL_1985920, EPI_ISL_1985921, EPI_ISL_1985925, EPI_ISL_1985929, EPI_ISL_1985948, EPI_ISL_1985953, EPI_ISL_1985954, EPI_ISL_1985957, EPI_ISL_1985965, EPI_ISL_1985967, EPI_ISL_1985971, EPI_ISL_1985976, EPI_ISL_1985979, EPI_ISL_1985980, EPI_ISL_1985983, EPI_ISL_1985984, EPI_ISL_1985989, EPI_ISL_1985990, EPI_ISL_1986006, EPI_ISL_1986009, EPI_ISL_1986013, EPI_ISL_1986018, EPI_ISL_1986021, EPI_ISL_1986028, EPI_ISL_1986030, EPI_ISL_1986032, EPI_ISL_1986036, EPI_ISL_1986041, EPI_ISL_1986045, EPI_ISL_1986046, EPI_ISL_1986058, EPI_ISL_1986062, EPI_ISL_1986064, EPI_ISL_1986068, EPI_ISL_1986069, EPI_ISL_1986071, EPI_ISL_1986074, EPI_ISL_1986075, EPI_ISL_1986076, EPI_ISL_1986082, EPI_ISL_1986092, EPI_ISL_1986096, EPI_ISL_1986097, EPI_ISL_1986099, EPI_ISL_1986105, EPI_ISL_1986107, EPI_ISL_1986113, EPI_ISL_1986114, EPI_ISL_1986115, EPI_ISL_1986118, EPI_ISL_1986124, EPI_ISL_1986125, EPI_ISL_1986128, EPI_ISL_1986129, EPI_ISL_1986130, EPI_ISL_1986136, EPI_ISL_1986139, EPI_ISL_1986144, EPI_ISL_1986155, EPI_ISL_1986159, EPI_ISL_1986163, EPI_ISL_1986166, EPI_ISL_1986167, EPI_ISL_1986168, EPI_ISL_1986171, EPI_ISL_1986172, EPI_ISL_1986180, EPI_ISL_1986184, EPI_ISL_1986185, EPI_ISL_1986186, EPI_ISL_1986203, EPI_ISL_1986206, EPI_ISL_1986208, EPI_ISL_1986210, EPI_ISL_1986213, EPI_ISL_1986218, EPI_ISL_1986220, EPI_ISL_1986224, EPI_ISL_1986227, EPI_ISL_1986233, EPI_ISL_1986237, EPI_ISL_1986252, EPI_ISL_1986255, EPI_ISL_1986267, EPI_ISL_1986275, EPI_ISL_1986278, EPI_ISL_1986281, EPI_ISL_1986283, EPI_ISL_1986284, EPI_ISL_1986290, EPI_ISL_1986297, | | | |

EPI_ISL_1986304, EPI_ISL_1986305, EPI_ISL_1986310, EPI_ISL_1986311, EPI_ISL_1986314, EPI_ISL_1986315, EPI_ISL_1986317, EPI_ISL_1986318, EPI_ISL_1986323, EPI_ISL_1986331, EPI_ISL_1986334, EPI_ISL_1986336, EPI_ISL_1986338, EPI_ISL_1986339, EPI_ISL_1986360, EPI_ISL_1986362, EPI_ISL_1986365, EPI_ISL_1986372, EPI_ISL_1986376, EPI_ISL_1986390, EPI_ISL_1986494, EPI_ISL_1986502, EPI_ISL_1986511, EPI_ISL_1986516, EPI_ISL_1986519, EPI_ISL_1986523, EPI_ISL_1986528, EPI_ISL_1986539, EPI_ISL_1986542, EPI_ISL_1986550, EPI_ISL_1986563, EPI_ISL_1986567, EPI_ISL_1986572, EPI_ISL_1986573, EPI_ISL_1986574, EPI_ISL_1986576, EPI_ISL_1986577, EPI_ISL_1986579, EPI_ISL_1986580, EPI_ISL_1986582, EPI_ISL_1986585, EPI_ISL_1986589, EPI_ISL_1986593, EPI_ISL_1986595, EPI_ISL_1986601, EPI_ISL_1986607, EPI_ISL_1986611, EPI_ISL_1986612, EPI_ISL_1986617, EPI_ISL_1986618, EPI_ISL_1986626, EPI_ISL_1986628, EPI_ISL_1986632, EPI_ISL_1986634, EPI_ISL_1986637, EPI_ISL_1986639, EPI_ISL_1986649, EPI_ISL_1986658, EPI_ISL_1986660, EPI_ISL_1986663, EPI_ISL_1986666, EPI_ISL_1986668, EPI_ISL_1986669, EPI_ISL_1986671, EPI_ISL_1986677, EPI_ISL_1986678, EPI_ISL_1986679, EPI_ISL_1986682, EPI_ISL_1986684, EPI_ISL_1986689, EPI_ISL_1986690, EPI_ISL_1986692, EPI_ISL_1986693, EPI_ISL_1986699, EPI_ISL_1986701, EPI_ISL_1986702, EPI_ISL_1986703, EPI_ISL_1986707, EPI_ISL_1986712, EPI_ISL_1986716, EPI_ISL_1986718, EPI_ISL_1986720, EPI_ISL_1986721, EPI_ISL_1986722, EPI_ISL_1986730, EPI_ISL_1986733, EPI_ISL_1986739, EPI_ISL_1986740, EPI_ISL_1986741, EPI_ISL_1986743, EPI_ISL_1986746, EPI_ISL_1986748, EPI_ISL_1986749, EPI_ISL_1986756, EPI_ISL_1986761, EPI_ISL_1986763, EPI_ISL_1986768, EPI_ISL_1986771, EPI_ISL_1986773, EPI_ISL_1986775, EPI_ISL_1986780, EPI_ISL_1986781, EPI_ISL_1986783, EPI_ISL_1986790, EPI_ISL_1986792, EPI_ISL_1986793, EPI_ISL_1986803, EPI_ISL_1986810, EPI_ISL_1986812, EPI_ISL_1986813, EPI_ISL_1986820, EPI_ISL_1986822, EPI_ISL_1986829, EPI_ISL_1986840, EPI_ISL_1986845, EPI_ISL_1986853, EPI_ISL_1986854, EPI_ISL_1986858, EPI_ISL_1986860, EPI_ISL_1986861, EPI_ISL_1986864, EPI_ISL_1986865, EPI_ISL_1986866, EPI_ISL_1986871, EPI_ISL_1986873, EPI_ISL_1986876, EPI_ISL_1986878, EPI_ISL_1986879, EPI_ISL_1986886, EPI_ISL_1986887, EPI_ISL_1986891, EPI_ISL_1986893, EPI_ISL_1986895, EPI_ISL_1986896, EPI_ISL_1986900, EPI_ISL_1986904, EPI_ISL_1986921, EPI_ISL_1986930, EPI_ISL_1986933, EPI_ISL_1986944, EPI_ISL_1986957, EPI_ISL_1986958, EPI_ISL_1986961, EPI_ISL_1986962, EPI_ISL_1986963, EPI_ISL_1986964, EPI_ISL_1986970, EPI_ISL_1986972, EPI_ISL_1986974, EPI_ISL_1986976, EPI_ISL_1986980, EPI_ISL_1986986, EPI_ISL_1986992, EPI_ISL_1986993, EPI_ISL_1986995, EPI_ISL_1986996, EPI_ISL_1987000, EPI_ISL_1987001, EPI_ISL_1987006, EPI_ISL_1987007, EPI_ISL_1987009, EPI_ISL_1987011, EPI_ISL_1987012, EPI_ISL_1987016, EPI_ISL_1987018, EPI_ISL_1987020, EPI_ISL_1987024, EPI_ISL_1987025, EPI_ISL_1987028, EPI_ISL_1987030, EPI_ISL_1987032, EPI_ISL_1987034, EPI_ISL_1987040, EPI_ISL_1987041, EPI_ISL_1987046, EPI_ISL_1987048, EPI_ISL_1987049, EPI_ISL_1987050, EPI_ISL_1987054, EPI_ISL_1987055, EPI_ISL_1987056, EPI_ISL_1987061, EPI_ISL_1987062, EPI_ISL_1987063, EPI_ISL_1987068, EPI_ISL_1987070, EPI_ISL_1987073, EPI_ISL_1987074, EPI_ISL_1987123, EPI_ISL_1987126, EPI_ISL_1987128, EPI_ISL_1987129, EPI_ISL_1987130, EPI_ISL_1987136, EPI_ISL_1987140, EPI_ISL_1987146, EPI_ISL_1987149, EPI_ISL_1987151, EPI_ISL_1987152, EPI_ISL_1987153, EPI_ISL_1987154, EPI_ISL_1987156, EPI_ISL_1987157, EPI_ISL_1987161, EPI_ISL_1987162, EPI_ISL_1987165, EPI_ISL_1987167, EPI_ISL_1987168, EPI_ISL_1987172, EPI_ISL_1987173, EPI_ISL_1987175, EPI_ISL_1987179, EPI_ISL_1987180, EPI_ISL_1987188, EPI_ISL_1987189, EPI_ISL_1987192, EPI_ISL_1987196, EPI_ISL_1987197, EPI_ISL_1987198, EPI_ISL_1987203, EPI_ISL_1987204, EPI_ISL_1987206, EPI_ISL_1987207, EPI_ISL_1987211, EPI_ISL_1987212, EPI_ISL_1987213, EPI_ISL_1987214, EPI_ISL_1987218, EPI_ISL_1987221, EPI_ISL_1987223, EPI_ISL_1987224, EPI_ISL_1987225, EPI_ISL_1987227, EPI_ISL_1987230, EPI_ISL_1987233, EPI_ISL_1987234, EPI_ISL_1987235, EPI_ISL_1987236, EPI_ISL_1987239, EPI_ISL_1987241, EPI_ISL_1987250, EPI_ISL_1987253, EPI_ISL_1987255, EPI_ISL_1987258, EPI_ISL_1987264, EPI_ISL_1987265, EPI_ISL_1987267, EPI_ISL_1987269, EPI_ISL_1987275, EPI_ISL_1987276, EPI_ISL_1987278, EPI_ISL_1987287, EPI_ISL_1987289, EPI_ISL_1987294, EPI_ISL_1987295, EPI_ISL_1987297, EPI_ISL_1987298, EPI_ISL_1987299, EPI_ISL_1987303, EPI_ISL_1987311, EPI_ISL_1987321, EPI_ISL_1987325, EPI_ISL_1987326, EPI_ISL_1987333, EPI_ISL_1987334, EPI_ISL_1987337, EPI_ISL_1987341, EPI_ISL_1987348, EPI_ISL_1987354, EPI_ISL_1987364, EPI_ISL_1987366, EPI_ISL_1987369, EPI_ISL_1987375, EPI_ISL_1987376, EPI_ISL_1987377, EPI_ISL_2004346, EPI_ISL_2004347, EPI_ISL_2004352, EPI_ISL_2004354, EPI_ISL_2004357, EPI_ISL_2004358, EPI_ISL_2004359, EPI_ISL_2004363, EPI_ISL_2004364, EPI_ISL_2004366, EPI_ISL_2004367, EPI_ISL_2004369, EPI_ISL_2004370, EPI_ISL_2004371, EPI_ISL_2004375, EPI_ISL_2004376, EPI_ISL_2004382, EPI_ISL_2004383, EPI_ISL_2004385, EPI_ISL_2004386, EPI_ISL_2004388, EPI_ISL_2004391, EPI_ISL_2004392, EPI_ISL_2004393, EPI_ISL_2004400, EPI_ISL_2004404, EPI_ISL_2004406, EPI_ISL_2004407, EPI_ISL_2004410, EPI_ISL_2004414, EPI_ISL_2004416, EPI_ISL_2004417, EPI_ISL_2004421, EPI_ISL_2004424, EPI_ISL_2004425, EPI_ISL_2004426, EPI_ISL_2004428, EPI_ISL_2004429, EPI_ISL_2004431, EPI_ISL_2004432, EPI_ISL_2004433, EPI_ISL_2004437, EPI_ISL_2004443, EPI_ISL_2004445, EPI_ISL_2004449, EPI_ISL_2004450, EPI_ISL_2004452, EPI_ISL_2004456, EPI_ISL_2004457, EPI_ISL_2004461, EPI_ISL_2004463, EPI_ISL_2004467, EPI_ISL_2004468, EPI_ISL_2004470, EPI_ISL_2004471, EPI_ISL_2004482, EPI_ISL_2004483, EPI_ISL_2004486, EPI_ISL_2004488, EPI_ISL_2004490, EPI_ISL_2004493, EPI_ISL_2004497, EPI_ISL_2004499, EPI_ISL_2004500, EPI_ISL_2004504, EPI_ISL_2004509, EPI_ISL_2004513, EPI_ISL_2004517, EPI_ISL_2004518, EPI_ISL_2004523, EPI_ISL_2004524, EPI_ISL_2004526, EPI_ISL_2004528, EPI_ISL_2004531, EPI_ISL_2004532, EPI_ISL_2004533, EPI_ISL_2004537, EPI_ISL_2004539, EPI_ISL_2004540, EPI_ISL_2004544, EPI_ISL_2004552, EPI_ISL_2004554, EPI_ISL_2004560, EPI_ISL_2004561, EPI_ISL_2004562, EPI_ISL_2004563, EPI_ISL_2004570, EPI_ISL_2004578, EPI_ISL_2004579, EPI_ISL_2004581, EPI_ISL_2004582, EPI_ISL_2004583, EPI_ISL_2004586, EPI_ISL_2004589, EPI_ISL_2004590, EPI_ISL_2004593, EPI_ISL_2004594, EPI_ISL_2004597, EPI_ISL_2004599, EPI_ISL_2004600, EPI_ISL_2004603, EPI_ISL_2004606, EPI_ISL_2004615, EPI_ISL_2004616, EPI_ISL_2004704, EPI_ISL_2004774, EPI_ISL_2004884, EPI_ISL_2004887, EPI_ISL_2004888, EPI_ISL_2004889, EPI_ISL_2004891, EPI_ISL_2004892, EPI_ISL_2004904, EPI_ISL_2004905, EPI_ISL_2004906, EPI_ISL_2004912, EPI_ISL_2004915, EPI_ISL_2004918, EPI_ISL_2004923, EPI_ISL_2004924, EPI_ISL_2004932, EPI_ISL_2004934, EPI_ISL_2004938, EPI_ISL_2004939, EPI_ISL_2004941, EPI_ISL_2004945, EPI_ISL_2004952, EPI_ISL_2004953, EPI_ISL_2004955, EPI_ISL_2004958, EPI_ISL_2004962, EPI_ISL_2004963, EPI_ISL_2004967, EPI_ISL_2004969, EPI_ISL_2004970, EPI_ISL_2004974, EPI_ISL_2004980, EPI_ISL_2004987, EPI_ISL_2004995, EPI_ISL_2004999, EPI_ISL_2005004, EPI_ISL_2005007, EPI_ISL_2005008, EPI_ISL_2005009, EPI_ISL_2005010, EPI_ISL_2005012, EPI_ISL_2005013, EPI_ISL_2005014, EPI_ISL_2005017, EPI_ISL_2005020, EPI_ISL_2005022, EPI_ISL_2005023, EPI_ISL_2005025, EPI_ISL_2005031, EPI_ISL_2005034, EPI_ISL_2005040, EPI_ISL_2005043, EPI_ISL_2005048, EPI_ISL_2005049, EPI_ISL_2005050, EPI_ISL_2005060, EPI_ISL_2005063, EPI_ISL_2005065, EPI_ISL_2005066, EPI_ISL_2005068, EPI_ISL_2005072, EPI_ISL_2005073, EPI_ISL_2005075, EPI_ISL_2005079, EPI_ISL_2005081, EPI_ISL_2005087, EPI_ISL_2005088, EPI_ISL_2005093, EPI_ISL_2005096, EPI_ISL_2005105, EPI_ISL_2005107, EPI_ISL_2005110, EPI_ISL_2005111, EPI_ISL_2005112, EPI_ISL_2005114, EPI_ISL_2005116, EPI_ISL_2005117, EPI_ISL_2005119, EPI_ISL_2005120, EPI_ISL_2005122, EPI_ISL_2005123, EPI_ISL_2005126, EPI_ISL_2005129, EPI_ISL_2005136, EPI_ISL_2005139, EPI_ISL_2005140, EPI_ISL_2005141, EPI_ISL_2005145, EPI_ISL_2005146, EPI_ISL_2005151, EPI_ISL_2005152, EPI_ISL_2005160, EPI_ISL_2005163, EPI_ISL_2005166, EPI_ISL_2005167, EPI_ISL_2005172, EPI_ISL_2005173, EPI_ISL_2005174, EPI_ISL_2005176, EPI_ISL_2005178, EPI_ISL_2005180, EPI_ISL_2005181, EPI_ISL_2005184, EPI_ISL_2005185, EPI_ISL_2005188, EPI_ISL_2005189, EPI_ISL_2005197, EPI_ISL_2005199, EPI_ISL_2005200, EPI_ISL_2005201, EPI_ISL_2005202, EPI_ISL_2005204, EPI_ISL_2005207, EPI_ISL_2005209, EPI_ISL_2005212, EPI_ISL_2005213, EPI_ISL_2005214, EPI_ISL_2005218, EPI_ISL_2005220, EPI_ISL_2005227, EPI_ISL_2005228, EPI_ISL_2005229, EPI_ISL_2005230, EPI_ISL_2005232, EPI_ISL_2005237, EPI_ISL_2005238, EPI_ISL_2005239, EPI_ISL_2005241, EPI_ISL_2005242, EPI_ISL_2005243, EPI_ISL_2005250, EPI_ISL_2005251, EPI_ISL_2005252, EPI_ISL_2005253, EPI_ISL_2005254, EPI_ISL_2005258, EPI_ISL_2005259, EPI_ISL_2005260, EPI_ISL_2005261, EPI_ISL_2005268, EPI_ISL_2005271, EPI_ISL_2005272, EPI_ISL_2005273, EPI_ISL_2005274, EPI_ISL_2005278, EPI_ISL_2005280, EPI_ISL_2005285, EPI_ISL_2005288, EPI_ISL_2005289, EPI_ISL_2005291, EPI_ISL_2005292, EPI_ISL_2005295, EPI_ISL_2005299, EPI_ISL_2005301, EPI_ISL_2005302, EPI_ISL_2005309, EPI_ISL_2005310, EPI_ISL_2005312, EPI_ISL_2005318, EPI_ISL_2005321, EPI_ISL_2005324, EPI_ISL_2005325, EPI_ISL_2005327, EPI_ISL_2005329, EPI_ISL_2005330, EPI_ISL_2005331, EPI_ISL_2005333, EPI_ISL_2005335, EPI_ISL_2005337, EPI_ISL_2005347, EPI_ISL_2005349, EPI_ISL_2005353, EPI_ISL_2005357, EPI_ISL_2005358, EPI_ISL_2005360, EPI_ISL_2005362, EPI_ISL_2005363, EPI_ISL_2005364, EPI_ISL_2005369, EPI_ISL_2005378, EPI_ISL_2005379, EPI_ISL_2005382, EPI_ISL_2005384, EPI_ISL_2005385, EPI_ISL_2005391, EPI_ISL_2005392, EPI_ISL_2005393, EPI_ISL_2005395, EPI_ISL_2005396, EPI_ISL_2005404, EPI_ISL_2005413, EPI_ISL_2005418, EPI_ISL_2005422, EPI_ISL_2005425, EPI_ISL_2005426, EPI_ISL_2005430, EPI_ISL_2005435, EPI_ISL_2005437, EPI_ISL_2005438, EPI_ISL_2005442, EPI_ISL_2005443, EPI_ISL_2005448, EPI_ISL_2005450, EPI_ISL_2005452, EPI_ISL_2005462, EPI_ISL_2005464, EPI_ISL_2005468, EPI_ISL_2005469, EPI_ISL_2005476, EPI_ISL_2005478, EPI_ISL_2005486, EPI_ISL_2005493, EPI_ISL_2005496, EPI_ISL_2005499, EPI_ISL_2005504, EPI_ISL_2005513, EPI_ISL_2005515, EPI_ISL_2005516, EPI_ISL_2005519, EPI_ISL_2005520, EPI_ISL_2005521, EPI_ISL_2005522, EPI_ISL_2005531, EPI_ISL_2005532, EPI_ISL_2005533, EPI_ISL_2005538, EPI_ISL_2005539, EPI_ISL_2005540, EPI_ISL_2005548, EPI_ISL_2005550, EPI_ISL_2005551, EPI_ISL_2005552, EPI_ISL_2005556, EPI_ISL_2005557, EPI_ISL_2005558, EPI_ISL_2005560, EPI_ISL_2005561, EPI_ISL_2005564, EPI_ISL_2005567, EPI_ISL_2005571, EPI_ISL_2005573, EPI_ISL_2005577, EPI_ISL_2005582, EPI_ISL_2005584, EPI_ISL_2005585, EPI_ISL_2005588, EPI_ISL_2005589, EPI_ISL_2005590, EPI_ISL_2005593, EPI_ISL_2005598, EPI_ISL_2005599, EPI_ISL_2005603, EPI_ISL_2005608, EPI_ISL_2005610, EPI_ISL_2005611, EPI_ISL_2005851, EPI_ISL_2005931, EPI_ISL_2006072, EPI_ISL_2006074, EPI_ISL_2006075, EPI_ISL_2006078, EPI_ISL_2006080, EPI_ISL_2006096, EPI_ISL_2006102, EPI_ISL_2006112, EPI_ISL_2006113, EPI_ISL_2006114, EPI_ISL_2006120, EPI_ISL_2006121, EPI_ISL_2006124, EPI_ISL_2006126, EPI_ISL_2006132, EPI_ISL_2006134, EPI_ISL_2006135, EPI_ISL_2006138, EPI_ISL_2006139, EPI_ISL_2006149, EPI_ISL_2006150, EPI_ISL_2006153, EPI_ISL_2006159, EPI_ISL_2006163, EPI_ISL_2006166, EPI_ISL_2006167, EPI_ISL_2006176, EPI_ISL_2006185, EPI_ISL_2006187, EPI_ISL_2006188, EPI_ISL_2006191, EPI_ISL_2006192, EPI_ISL_2006197, EPI_ISL_2006200, EPI_ISL_2006203, EPI_ISL_2006206, EPI_ISL_2006208, EPI_ISL_2006209, EPI_ISL_2006210, EPI_ISL_2006214, EPI_ISL_2006220, EPI_ISL_2006221, EPI_ISL_2006228, EPI_ISL_2006234, EPI_ISL_2006236, EPI_ISL_2006247, EPI_ISL_2006248, EPI_ISL_2006251, EPI_ISL_2006252, EPI_ISL_2006255, EPI_ISL_2006257, EPI_ISL_2006259, EPI_ISL_2006260, EPI_ISL_2006261, EPI_ISL_2006263, EPI_ISL_2006268, EPI_ISL_2006269, EPI_ISL_2006270, EPI_ISL_2006272, EPI_ISL_2006274, EPI_ISL_2006282, EPI_ISL_2006289, EPI_ISL_2006291, EPI_ISL_2006294, EPI_ISL_2006669, EPI_ISL_2006670, EPI_ISL_2006671, EPI_ISL_2006672, EPI_ISL_2021319, EPI_ISL_2021463, EPI_ISL_2021649, EPI_ISL_2021689, EPI_ISL_2021697, EPI_ISL_2021701, EPI_ISL_2021703, EPI_ISL_2021704, EPI_ISL_2021707, EPI_ISL_2021710, EPI_ISL_2021711, EPI_ISL_2021712, EPI_ISL_2021713, EPI_ISL_2021717, EPI_ISL_2021719, EPI_ISL_2021728, EPI_ISL_2021733, EPI_ISL_2021735, EPI_ISL_2021736, EPI_ISL_2021739, EPI_ISL_2021742, EPI_ISL_2021743, EPI_ISL_2021746, EPI_ISL_2021747, EPI_ISL_2021748, EPI_ISL_2021749, EPI_ISL_2021752, EPI_ISL_2021754, EPI_ISL_2021757, EPI_ISL_2021764, EPI_ISL_2021767, EPI_ISL_2021769, EPI_ISL_2021771, EPI_ISL_2021773, EPI_ISL_2021774, EPI_ISL_2021775, EPI_ISL_2021778, EPI_ISL_2021780, EPI_ISL_2021782, EPI_ISL_2021784, EPI_ISL_2021791, EPI_ISL_2021793, EPI_ISL_2021794, EPI_ISL_2021797, EPI_ISL_2021805, EPI_ISL_2021807, EPI_ISL_2021809, EPI_ISL_2021813, EPI_ISL_2021814, EPI_ISL_2021820, EPI_ISL_2021822, EPI_ISL_2021826, EPI_ISL_2021828, EPI_ISL_2021829, EPI_ISL_2021835, EPI_ISL_2021843, EPI_ISL_2021847, EPI_ISL_2021853, EPI_ISL_2021857, EPI_ISL_2021865, EPI_ISL_2021867, EPI_ISL_2021891, EPI_ISL_2021900, EPI_ISL_2021901, EPI_ISL_2021904, EPI_ISL_2021908, EPI_ISL_2021912, EPI_ISL_2021923, EPI_ISL_2021934, EPI_ISL_2021935, EPI_ISL_2021952, EPI_ISL_2021956, EPI_ISL_2021961, EPI_ISL_2021988, EPI_ISL_2021993, EPI_ISL_2021994, EPI_ISL_2022000, EPI_ISL_2022002, EPI_ISL_2022018, EPI_ISL_2022023, EPI_ISL_2022027, EPI_ISL_2022041, EPI_ISL_2022055, EPI_ISL_2022066, EPI_ISL_2022072, EPI_ISL_2022080, EPI_ISL_2022091, EPI_ISL_2022115, EPI_ISL_2022120, EPI_ISL_2022127, EPI_ISL_2022129, EPI_ISL_2022149, EPI_ISL_2022154, EPI_ISL_2022261, EPI_ISL_2022495, EPI_ISL_2022645, EPI_ISL_2022667, EPI_ISL_2022805, EPI_ISL_2022840, EPI_ISL_2022979, EPI_ISL_2080360, EPI_ISL_2080421, EPI_ISL_2080483, EPI_ISL_2080484, EPI_ISL_2091463, EPI_ISL_2091468, EPI_ISL_2091473, EPI_ISL_2091480, EPI_ISL_2091482, EPI_ISL_2091484, EPI_ISL_2091486, EPI_ISL_2091493, EPI_ISL_2091496, EPI_ISL_2091500, EPI_ISL_2091511, EPI_ISL_2091515, EPI_ISL_2091525, EPI_ISL_2091536, EPI_ISL_2091539, EPI_ISL_2091543, EPI_ISL_2091547, EPI_ISL_2091555, EPI_ISL_2091562, EPI_ISL_2091564, EPI_ISL_2091565, EPI_ISL_2091566, EPI_ISL_2091569, EPI_ISL_2091571, EPI_ISL_2091581, EPI_ISL_2091585, EPI_ISL_2091587, EPI_ISL_2091592, EPI_ISL_2091596, EPI_ISL_2091602, EPI_ISL_2091604, EPI_ISL_2091613, EPI_ISL_2091624, EPI_ISL_2091632, EPI_ISL_2091638, EPI_ISL_2091640, EPI_ISL_2091649, EPI_ISL_2091650, EPI_ISL_2091652, EPI_ISL_2091656, EPI_ISL_2091664, EPI_ISL_2091669, EPI_ISL_2091670, EPI_ISL_2091672, EPI_ISL_2091679, EPI_ISL_2091685, EPI_ISL_2091687, EPI_ISL_2091693, EPI_ISL_2091694, EPI_ISL_2091695, EPI_ISL_2091698, EPI_ISL_2091702, EPI_ISL_2091704, EPI_ISL_2091710, EPI_ISL_2091711, EPI_ISL_2091724, EPI_ISL_2091729, EPI_ISL_2091730, EPI_ISL_2091733, EPI_ISL_2091735, EPI_ISL_2091741, EPI_ISL_2091742, EPI_ISL_2091744, EPI_ISL_2091749, EPI_ISL_2091751, EPI_ISL_2091754, EPI_ISL_2091756, EPI_ISL_2091759, EPI_ISL_2091760, EPI_ISL_2091763, EPI_ISL_2091765, EPI_ISL_2091766, EPI_ISL_2091769, EPI_ISL_2091772, EPI_ISL_2091774, EPI_ISL_2091779, EPI_ISL_2091781, EPI_ISL_2091783, EPI_ISL_2091784, EPI_ISL_2091786, EPI_ISL_2091788, EPI_ISL_2091789, EPI_ISL_2091791, EPI_ISL_2092467, EPI_ISL_2092468, EPI_ISL_2092469, EPI_ISL_2092470, EPI_ISL_2092471, EPI_ISL_2092472, EPI_ISL_2092473, EPI_ISL_2092474, EPI_ISL_2092475, EPI_ISL_2092476, EPI_ISL_2092477, EPI_ISL_2092478, EPI_ISL_2092479, EPI_ISL_2092480, EPI_ISL_2092482, EPI_ISL_2092483, EPI_ISL_2092484, EPI_ISL_2092485, EPI_ISL_2092486, EPI_ISL_2092487, EPI_ISL_2092488, EPI_ISL_2092489, EPI_ISL_2092490, EPI_ISL_2092491, EPI_ISL_2092492, EPI_ISL_2092493, EPI_ISL_2092494, EPI_ISL_2092496, EPI_ISL_2092498, EPI_ISL_2092499, EPI_ISL_2092500, EPI_ISL_2092501, EPI_ISL_2092502, EPI_ISL_2092503, EPI_ISL_2092504, EPI_ISL_2092505, EPI_ISL_2092506, EPI_ISL_2092507, EPI_ISL_2092509, EPI_ISL_2092511, EPI_ISL_2092512, EPI_ISL_2092514, EPI_ISL_2092515, EPI_ISL_2092516, EPI_ISL_2092518, EPI_ISL_2092519, EPI_ISL_2092520, EPI_ISL_2092521, EPI_ISL_2092522, EPI_ISL_2092523, EPI_ISL_2092524, EPI_ISL_2092526, EPI_ISL_2092527, EPI_ISL_2092528, EPI_ISL_2092531, EPI_ISL_2092533, EPI_ISL_2092534, EPI_ISL_2092535, EPI_ISL_2092537, EPI_ISL_2092539, EPI_ISL_2092540, EPI_ISL_2092541, EPI_ISL_2092543, EPI_ISL_2092544, EPI_ISL_2092545, EPI_ISL_2092546, EPI_ISL_2092547, EPI_ISL_2092548, EPI_ISL_2092550, EPI_ISL_2092551, EPI_ISL_2092552, EPI_ISL_2092553, EPI_ISL_2092554, EPI_ISL_2092555, EPI_ISL_2092556, EPI_ISL_2092557, EPI_ISL_2092558, EPI_ISL_2092559, EPI_ISL_2092560, EPI_ISL_2092561, EPI_ISL_2092562, EPI_ISL_2092563, EPI_ISL_2092564, EPI_ISL_2092565, EPI_ISL_2092566, EPI_ISL_2092568, EPI_ISL_2092569, EPI_ISL_2092571, EPI_ISL_2092575, EPI_ISL_2092577, EPI_ISL_2092578, EPI_ISL_2092580, EPI_ISL_2092581, EPI_ISL_2092582, EPI_ISL_2092584, EPI_ISL_2092585, EPI_ISL_2092586, EPI_ISL_2092587, EPI_ISL_2092588, EPI_ISL_2092589, EPI_ISL_2092590, EPI_ISL_2092591, EPI_ISL_2092592, EPI_ISL_2092593, EPI_ISL_2092595, EPI_ISL_2092596, EPI_ISL_2092597, EPI_ISL_2092598, EPI_ISL_2092599, EPI_ISL_2092600, EPI_ISL_2092601, EPI_ISL_2092602, EPI_ISL_2092604, EPI_ISL_2092605, EPI_ISL_2092606, EPI_ISL_2092607, EPI_ISL_2092609, EPI_ISL_2092611, EPI_ISL_2092612, EPI_ISL_2092613, EPI_ISL_2092614, EPI_ISL_2092615, EPI_ISL_2092618, EPI_ISL_2092619, EPI_ISL_2092620, EPI_ISL_2092621, EPI_ISL_2092622, EPI_ISL_2092624, EPI_ISL_2092625, EPI_ISL_2092626, EPI_ISL_2092628, EPI_ISL_2092629, EPI_ISL_2092630, EPI_ISL_2092631, EPI_ISL_2092635, EPI_ISL_2092636, EPI_ISL_2092637, EPI_ISL_2092638, EPI_ISL_2092639, EPI_ISL_2092640, EPI_ISL_2092641, EPI_ISL_2092642, EPI_ISL_2092643, EPI_ISL_2092644, EPI_ISL_2092645, EPI_ISL_2092646, EPI_ISL_2119082, EPI_ISL_2120233, EPI_ISL_2120347, EPI_ISL_2138718

| | | | |
|---|---|---|---|
| see above | Lighthouse Lab in Alderley Park | Wellcome Sanger Institute for the COVID-19 Genomics UK (COG-UK) Consortium | Cordelia Langford; David K. Jackson; Dominic Kwiatkowski; Ewan Harrison; Ian Johnston; Jacquelyn Wynn; Jeffrey Barrett; John Sillitoe on behalf of the Wellcome Sanger Institute COVID-19 Surveillance Team; Mairead Hyland; Roberto Amato; Sonia Goncalves; The Lighthouse Lab in Alderley Park and Alex Alderton |

EPI_ISL_1719745, EPI_ISL_1719746, EPI_ISL_1740541, EPI_ISL_1740550, EPI_ISL_1740580, EPI_ISL_1740643, EPI_ISL_1740658, EPI_ISL_1740687, EPI_ISL_1740698, EPI_ISL_1740699, EPI_ISL_1740713, EPI_ISL_1740737, EPI_ISL_1740741, EPI_ISL_1740768, EPI_ISL_1740781, EPI_ISL_1806592, EPI_ISL_1806792, EPI_ISL_1831086

| | | | |
|---|---|---|---|
| see above | Lighthouse Lab in Cambridge | Wellcome Sanger Institute for the COVID-19 Genomics UK (COG-UK) Consortium | Cordelia Langford; David K. Jackson; Dominic Kwiatkowski; Ewan Harrison; Ian Johnston; Jeffrey Barrett; John Sillitoe on behalf of the Wellcome Sanger Institute COVID-19 Surveillance Team; Rob Howes; Roberto Amato; Sonia Goncalves; The Lighthouse Lab in Cambridge and Alex Alderton |

EPI_ISL_1829484, EPI_ISL_1829486, EPI_ISL_1829487, EPI_ISL_1829489, EPI_ISL_1829491, EPI_ISL_1829492, EPI_ISL_1829493, EPI_ISL_1829494, EPI_ISL_1829495, EPI_ISL_1829496, EPI_ISL_1829497, EPI_ISL_1829498, EPI_ISL_1829499, EPI_ISL_1829501, EPI_ISL_1829504, EPI_ISL_1829506, EPI_ISL_1829507, EPI_ISL_1829508, EPI_ISL_1829509, EPI_ISL_1829510, EPI_ISL_1829512, EPI_ISL_1829514, EPI_ISL_1829515, EPI_ISL_1829516, EPI_ISL_1829517, EPI_ISL_1829518, EPI_ISL_1829520, EPI_ISL_1829523, EPI_ISL_1829524, EPI_ISL_1829526, EPI_ISL_1829529, EPI_ISL_1829531, EPI_ISL_1829533, EPI_ISL_1829535, EPI_ISL_1829537, EPI_ISL_1829538, EPI_ISL_1829539, EPI_ISL_1829541, EPI_ISL_1829542, EPI_ISL_1829544, EPI_ISL_1829545, EPI_ISL_1829547, EPI_ISL_1829548, EPI_ISL_1829549, EPI_ISL_1829550, EPI_ISL_1829551, EPI_ISL_1829552, EPI_ISL_1829554, EPI_ISL_1829555, EPI_ISL_1829556, EPI_ISL_1829558, EPI_ISL_1829559, EPI_ISL_1829560, EPI_ISL_1829562, EPI_ISL_1829563, EPI_ISL_1829564, EPI_ISL_1829565, EPI_ISL_1829567, EPI_ISL_1829568, EPI_ISL_1829569, EPI_ISL_1829571, EPI_ISL_1829573, EPI_ISL_1829575, EPI_ISL_1829576, EPI_ISL_1829577, EPI_ISL_1829578, EPI_ISL_1829580, EPI_ISL_1829581, EPI_ISL_1829582, EPI_ISL_1829583, EPI_ISL_1829584, EPI_ISL_1829585, EPI_ISL_1829588, EPI_ISL_1829589, EPI_ISL_1829590, EPI_ISL_1829591, EPI_ISL_1829592, EPI_ISL_1829594, EPI_ISL_1829595, EPI_ISL_1829597, EPI_ISL_1829598, EPI_ISL_1829599, EPI_ISL_1829600, EPI_ISL_1829602, EPI_ISL_1829603, EPI_ISL_1829605, EPI_ISL_1829607, EPI_ISL_1829608, EPI_ISL_1829609, EPI_ISL_1829610, EPI_ISL_1829611, EPI_ISL_1829612, EPI_ISL_1829613, EPI_ISL_1829614, EPI_ISL_1829615, EPI_ISL_1829616, EPI_ISL_1829617, EPI_ISL_1829618, EPI_ISL_1829619, EPI_ISL_1829620, EPI_ISL_1829621, EPI_ISL_1829622, EPI_ISL_1829623, EPI_ISL_1829625, EPI_ISL_1829626, EPI_ISL_1829627, EPI_ISL_1829628, EPI_ISL_1829629, EPI_ISL_1829630, EPI_ISL_1829631, EPI_ISL_1829634, EPI_ISL_1829636, EPI_ISL_1829637, EPI_ISL_1829638, EPI_ISL_1829639, EPI_ISL_1829640, EPI_ISL_1829641, EPI_ISL_1829644, EPI_ISL_1829645, EPI_ISL_1829646, EPI_ISL_1829647, EPI_ISL_1829650, EPI_ISL_1829652, EPI_ISL_1829653, EPI_ISL_1829654, EPI_ISL_1829655, EPI_ISL_1829657, EPI_ISL_1829658, EPI_ISL_1829659, EPI_ISL_1829660, EPI_ISL_1829662, EPI_ISL_1829663, EPI_ISL_1829664, EPI_ISL_1829665, EPI_ISL_1829667, EPI_ISL_1829669, EPI_ISL_1829670, EPI_ISL_1829671, EPI_ISL_1829672, EPI_ISL_1829674, EPI_ISL_1829675, EPI_ISL_1829676, EPI_ISL_1829677, EPI_ISL_1829678, EPI_ISL_1829679, EPI_ISL_1829681, EPI_ISL_1829682, EPI_ISL_1829684, EPI_ISL_1829685, EPI_ISL_1829686, EPI_ISL_1829687, EPI_ISL_1829688, EPI_ISL_1829690, EPI_ISL_1829691, EPI_ISL_1829692, EPI_ISL_1829693, EPI_ISL_1829694, EPI_ISL_1829695, EPI_ISL_1829696, EPI_ISL_1829697, EPI_ISL_1829698, EPI_ISL_1829701, EPI_ISL_1829702, EPI_ISL_1829704, EPI_ISL_1829706, EPI_ISL_1829707, EPI_ISL_1829708, EPI_ISL_1829711, EPI_ISL_1829712, EPI_ISL_1829714, EPI_ISL_1829715, EPI_ISL_1829716, EPI_ISL_1829717, EPI_ISL_1829718, EPI_ISL_1829719, EPI_ISL_1829720, EPI_ISL_1829721, EPI_ISL_1829722, EPI_ISL_1829724, EPI_ISL_1829725, EPI_ISL_1829726, EPI_ISL_1829727, EPI_ISL_1829728, EPI_ISL_1829730, EPI_ISL_1829731, EPI_ISL_1829732, EPI_ISL_1829733, EPI_ISL_1829734, EPI_ISL_1829735, EPI_ISL_1829736, EPI_ISL_1829738, EPI_ISL_1829739, EPI_ISL_1829740, EPI_ISL_1829741, EPI_ISL_1829743, EPI_ISL_1829745, EPI_ISL_1829746, EPI_ISL_1829748, EPI_ISL_1829749, EPI_ISL_1829751, EPI_ISL_1829753, EPI_ISL_1829754, EPI_ISL_1829755, EPI_ISL_1829756, EPI_ISL_1829757, EPI_ISL_1829758, EPI_ISL_1829759, EPI_ISL_1829760, EPI_ISL_1829761, EPI_ISL_1829762, EPI_ISL_1829764, EPI_ISL_1829765, EPI_ISL_1829767, EPI_ISL_1829768, EPI_ISL_1829772, EPI_ISL_1829775, EPI_ISL_1829778, EPI_ISL_1829779, EPI_ISL_1829781, EPI_ISL_1829782, EPI_ISL_1829783, EPI_ISL_1829784, EPI_ISL_1829785, EPI_ISL_1829786, EPI_ISL_1829787, EPI_ISL_1829789, EPI_ISL_1829790, EPI_ISL_1829791, EPI_ISL_1829792, EPI_ISL_1829793, EPI_ISL_1829794, EPI_ISL_1829795, EPI_ISL_1829797, EPI_ISL_1829798, EPI_ISL_1829799, EPI_ISL_1829802, EPI_ISL_1829812, EPI_ISL_1829824, EPI_ISL_1829833, EPI_ISL_1829836, EPI_ISL_1829844, EPI_ISL_1829856, EPI_ISL_1829857, EPI_ISL_1829870, EPI_ISL_1829873, EPI_ISL_1829888, EPI_ISL_1829890, EPI_ISL_1829901, EPI_ISL_1829903, EPI_ISL_1829906, EPI_ISL_1829907, EPI_ISL_1829908, EPI_ISL_1829919, EPI_ISL_1829923, EPI_ISL_1829927, EPI_ISL_1829928, EPI_ISL_1829932, EPI_ISL_1829941, EPI_ISL_1829948, EPI_ISL_1829949, EPI_ISL_1829957, EPI_ISL_1829959, EPI_ISL_1829972, EPI_ISL_1829989, EPI_ISL_1829997, EPI_ISL_1829998, EPI_ISL_1830000, EPI_ISL_1830005, EPI_ISL_1830009, EPI_ISL_1830015, EPI_ISL_1830016, EPI_ISL_1830018, EPI_ISL_1830027, EPI_ISL_1830028, EPI_ISL_1830043, EPI_ISL_1830050, EPI_ISL_1830056, EPI_ISL_1830063, EPI_ISL_1830068, EPI_ISL_1830070, EPI_ISL_1830079, EPI_ISL_1830081, EPI_ISL_1830088, EPI_ISL_1830096, EPI_ISL_1830097, EPI_ISL_1830099, EPI_ISL_1830102, EPI_ISL_1830103, EPI_ISL_1830106, EPI_ISL_1830107, EPI_ISL_1830113, EPI_ISL_1830124, EPI_ISL_1830129, EPI_ISL_1830130, EPI_ISL_1830134, EPI_ISL_1830135, EPI_ISL_1830139, EPI_ISL_1830143, EPI_ISL_1830144, EPI_ISL_1830145, EPI_ISL_1830147, EPI_ISL_1830148, EPI_ISL_1830149, EPI_ISL_1830151, EPI_ISL_1830153, EPI_ISL_1830160, EPI_ISL_1830161, EPI_ISL_1830163, EPI_ISL_1830169, EPI_ISL_1830190, EPI_ISL_1830191, EPI_ISL_1830193, EPI_ISL_1830198, EPI_ISL_1830199, EPI_ISL_1830204, EPI_ISL_1830205, EPI_ISL_1830212, EPI_ISL_1830215, EPI_ISL_1830228, EPI_ISL_1830232, EPI_ISL_1830236, EPI_ISL_1830243, EPI_ISL_1830244, EPI_ISL_1830246, EPI_ISL_1830250, EPI_ISL_1830254, EPI_ISL_1830258, EPI_ISL_1830261, EPI_ISL_1830263, EPI_ISL_1830265, EPI_ISL_1830266, EPI_ISL_1830268, EPI_ISL_1830273, EPI_ISL_1830276, EPI_ISL_1830277, EPI_ISL_1830279, EPI_ISL_1830283, EPI_ISL_1830285, EPI_ISL_1830287, EPI_ISL_1830293, EPI_ISL_1830294, EPI_ISL_1830295, EPI_ISL_1830297, EPI_ISL_1830301, EPI_ISL_1830302, EPI_ISL_1830307, EPI_ISL_1830311, EPI_ISL_1830318, EPI_ISL_1830320, EPI_ISL_1830322, EPI_ISL_1830329, EPI_ISL_1830334, EPI_ISL_1830336, EPI_ISL_1830342, EPI_ISL_1830345, EPI_ISL_1830346, EPI_ISL_1830349, EPI_ISL_1830352, EPI_ISL_1830354, EPI_ISL_1830358, EPI_ISL_1830362, EPI_ISL_1830367, EPI_ISL_1830369, EPI_ISL_1830373, EPI_ISL_1830379, EPI_ISL_1830380, EPI_ISL_1830381, EPI_ISL_1830384, EPI_ISL_1830385, EPI_ISL_1830387, EPI_ISL_1830392, EPI_ISL_1830399, EPI_ISL_1830418, EPI_ISL_1830421, EPI_ISL_1830428, EPI_ISL_1830430, EPI_ISL_1830435, EPI_ISL_1830442, EPI_ISL_1830451, EPI_ISL_1830452, EPI_ISL_1830453, EPI_ISL_1830456, EPI_ISL_1830465, EPI_ISL_1830469, EPI_ISL_1830474, EPI_ISL_1830475, EPI_ISL_1830477, EPI_ISL_1830493, EPI_ISL_1830494, EPI_ISL_1830497, EPI_ISL_1830502, EPI_ISL_1830508, EPI_ISL_1830509, EPI_ISL_1830522, EPI_ISL_1830523, EPI_ISL_1830527, EPI_ISL_1830528, EPI_ISL_1830532, EPI_ISL_1830540, EPI_ISL_1830544, EPI_ISL_1830549, EPI_ISL_1830551, EPI_ISL_1830554, EPI_ISL_1830570, EPI_ISL_1830592, EPI_ISL_1830593, EPI_ISL_1830595, EPI_ISL_1830600, EPI_ISL_1830608, EPI_ISL_1830609, EPI_ISL_1830612, EPI_ISL_1830615, EPI_ISL_1830629, EPI_ISL_1830638, EPI_ISL_1830643, EPI_ISL_1830657, EPI_ISL_1830660, EPI_ISL_1830668, EPI_ISL_1830670, EPI_ISL_1830672, EPI_ISL_1830674, EPI_ISL_1830685, EPI_ISL_1830700, EPI_ISL_1830702, EPI_ISL_1830707, EPI_ISL_1830717, EPI_ISL_1830729, EPI_ISL_1830732, EPI_ISL_1830734, EPI_ISL_1830736, EPI_ISL_1830737, EPI_ISL_1830738, EPI_ISL_1830739, EPI_ISL_1830742, EPI_ISL_1830743, EPI_ISL_1830744, EPI_ISL_1830745, EPI_ISL_1830747, EPI_ISL_1830749, EPI_ISL_1830750, EPI_ISL_1830751, EPI_ISL_1830753, EPI_ISL_1830754, EPI_ISL_1830755, EPI_ISL_1830756, EPI_ISL_1830757, EPI_ISL_1830759, EPI_ISL_1830760, EPI_ISL_1830765, EPI_ISL_1830766, EPI_ISL_1830767, EPI_ISL_1830768, EPI_ISL_1830770, EPI_ISL_1830771, EPI_ISL_1830773, EPI_ISL_1830774, EPI_ISL_1830776, EPI_ISL_1830777, EPI_ISL_1830779, EPI_ISL_1830781, EPI_ISL_1830782, EPI_ISL_1830783, EPI_ISL_1830784, EPI_ISL_1830785, EPI_ISL_1830786, EPI_ISL_1830787, EPI_ISL_1830788, EPI_ISL_1830790, EPI_ISL_1830792, EPI_ISL_1830794, EPI_ISL_1830795, EPI_ISL_1830796, EPI_ISL_1830797, EPI_ISL_1830798, EPI_ISL_1830800, EPI_ISL_1830801, EPI_ISL_1830804, EPI_ISL_1830809, EPI_ISL_1830810, EPI_ISL_1830816, EPI_ISL_1830820, EPI_ISL_1830821, EPI_ISL_1830822, EPI_ISL_1830824, EPI_ISL_1830825, EPI_ISL_1830826, EPI_ISL_1830827, EPI_ISL_1830829, EPI_ISL_1830830, EPI_ISL_1830831, EPI_ISL_1830834, EPI_ISL_1830835, EPI_ISL_1830837, EPI_ISL_1830838, EPI_ISL_1830839, EPI_ISL_1830841, EPI_ISL_1830842, EPI_ISL_1830843, EPI_ISL_1830845, EPI_ISL_1830847, EPI_ISL_1830849, EPI_ISL_1830851, EPI_ISL_1830852, EPI_ISL_1830853, EPI_ISL_1830854, EPI_ISL_1830855, EPI_ISL_1830857, EPI_ISL_1830860, EPI_ISL_1830861, EPI_ISL_1830862, EPI_ISL_1830864, EPI_ISL_1830865, EPI_ISL_1830866, EPI_ISL_1830867, EPI_ISL_1830871, EPI_ISL_1830872, EPI_ISL_1830873, EPI_ISL_1830874, EPI_ISL_1830875, EPI_ISL_1830877, EPI_ISL_1830879, EPI_ISL_1830882, EPI_ISL_1830884, EPI_ISL_1830885, EPI_ISL_1830886, EPI_ISL_1830887, EPI_ISL_1830889, EPI_ISL_1830890, EPI_ISL_1830891, EPI_ISL_1830892, EPI_ISL_1830893, EPI_ISL_1830894, EPI_ISL_1830899, EPI_ISL_1830900, EPI_ISL_1830901, EPI_ISL_1830902, EPI_ISL_1830903, EPI_ISL_1830904, EPI_ISL_1830905, EPI_ISL_1830906, EPI_ISL_1830907, EPI_ISL_1830908, EPI_ISL_1830909, EPI_ISL_1830910, EPI_ISL_1830912, EPI_ISL_1830913, EPI_ISL_1830915, EPI_ISL_1830916, EPI_ISL_1830918, EPI_ISL_1830919, EPI_ISL_1830920, EPI_ISL_1830922, EPI_ISL_1830923, EPI_ISL_1830924, EPI_ISL_1830926, EPI_ISL_1830928, EPI_ISL_1830930, EPI_ISL_1830931, EPI_ISL_1830932, EPI_ISL_1830933, EPI_ISL_1830934, EPI_ISL_1830936, EPI_ISL_1830937, EPI_ISL_1830938, EPI_ISL_1830939, EPI_ISL_1830940, EPI_ISL_1830941, EPI_ISL_1830945, EPI_ISL_1830946, EPI_ISL_1830948, EPI_ISL_1830950, EPI_ISL_1830951, EPI_ISL_1830952, EPI_ISL_1830953, EPI_ISL_1830954, EPI_ISL_1830955, EPI_ISL_1830957, EPI_ISL_1830958, EPI_ISL_1830960, EPI_ISL_1830962, EPI_ISL_1830963, EPI_ISL_1830964, EPI_ISL_1830965, EPI_ISL_1830967, EPI_ISL_1830969, EPI_ISL_1830971, EPI_ISL_1830972, EPI_ISL_1830973, EPI_ISL_1830974, EPI_ISL_1830975, EPI_ISL_1830976, EPI_ISL_1830977, EPI_ISL_1830978, EPI_ISL_1830980, EPI_ISL_1830982, EPI_ISL_1830984, EPI_ISL_1830986, EPI_ISL_1830987, EPI_ISL_1830988, EPI_ISL_1830989, EPI_ISL_1830990, EPI_ISL_1830991, EPI_ISL_1830992, EPI_ISL_1830993, EPI_ISL_1830994, EPI_ISL_1830995, EPI_ISL_1830996, EPI_ISL_1830999, EPI_ISL_1831000, EPI_ISL_1831004, EPI_ISL_1831007, EPI_ISL_1831009, EPI_ISL_1831010, EPI_ISL_1831012, EPI_ISL_1831013, EPI_ISL_1831014, EPI_ISL_1831016, EPI_ISL_1831017, EPI_ISL_1831020, EPI_ISL_1831021, EPI_ISL_1831022, EPI_ISL_1831024, EPI_ISL_1831025, EPI_ISL_1831026, EPI_ISL_1831028, EPI_ISL_1831030, EPI_ISL_1831031, EPI_ISL_1831032, EPI_ISL_1831033, EPI_ISL_1831035, EPI_ISL_1831036, EPI_ISL_1831037, EPI_ISL_1831038, EPI_ISL_1831040, EPI_ISL_1831041, EPI_ISL_1831042, EPI_ISL_1831044, EPI_ISL_1831046, EPI_ISL_1831047, EPI_ISL_1831051, EPI_ISL_1831053, EPI_ISL_1831056, EPI_ISL_1831059, EPI_ISL_1831062, EPI_ISL_1831063, EPI_ISL_1831064, EPI_ISL_1831065, EPI_ISL_1831066, EPI_ISL_1831067, EPI_ISL_1831069, EPI_ISL_1831072, EPI_ISL_1831075, EPI_ISL_1831076, EPI_ISL_1831077, EPI_ISL_1831078, EPI_ISL_1831081, EPI_ISL_1831083, EPI_ISL_1831085, EPI_ISL_1831193, EPI_ISL_1857588, EPI_ISL_1857590, EPI_ISL_1857591, EPI_ISL_1857592, EPI_ISL_1857594, EPI_ISL_1857596, EPI_ISL_1857598, EPI_ISL_1857599, EPI_ISL_1857600, EPI_ISL_1857601, EPI_ISL_1857602, EPI_ISL_1857604, EPI_ISL_1857605, EPI_ISL_1857606, EPI_ISL_1857607, EPI_ISL_1857608, EPI_ISL_1857609, EPI_ISL_1857610, EPI_ISL_1857611, EPI_ISL_1857612, EPI_ISL_1857613, EPI_ISL_1857614, EPI_ISL_1857617, EPI_ISL_1857618, EPI_ISL_1857619, EPI_ISL_1857620, EPI_ISL_1857621, EPI_ISL_1857622, EPI_ISL_1857623, EPI_ISL_1857624, EPI_ISL_1857625, EPI_ISL_1857626, EPI_ISL_1857627, EPI_ISL_1857628, EPI_ISL_1857629, EPI_ISL_1857632, EPI_ISL_1857637, EPI_ISL_1857638, EPI_ISL_1857642, EPI_ISL_1857644, EPI_ISL_1857645, EPI_ISL_1857646, EPI_ISL_1857647, EPI_ISL_1857648, EPI_ISL_1857651, EPI_ISL_1857652, EPI_ISL_1857653, EPI_ISL_1857654, EPI_ISL_1857656, EPI_ISL_1857657, EPI_ISL_1857658, EPI_ISL_1857659, EPI_ISL_1857660, EPI_ISL_1857662, EPI_ISL_1857663, EPI_ISL_1857666, EPI_ISL_1857667, EPI_ISL_1857668, EPI_ISL_1857669, EPI_ISL_1857670, EPI_ISL_1857671, EPI_ISL_1857672, EPI_ISL_1857675, EPI_ISL_1857676, EPI_ISL_1857677, EPI_ISL_1857678, EPI_ISL_1857679, EPI_ISL_1857681, EPI_ISL_1857682, EPI_ISL_1857683, EPI_ISL_1857684, EPI_ISL_1857685, EPI_ISL_1857686, EPI_ISL_1857687, EPI_ISL_1857688, EPI_ISL_1857689, EPI_ISL_1857690, EPI_ISL_1857691, EPI_ISL_1857692, EPI_ISL_1857693, EPI_ISL_1857694, EPI_ISL_1857696, EPI_ISL_1857697, EPI_ISL_1857698, EPI_ISL_1857701, EPI_ISL_1857702, EPI_ISL_1857704, EPI_ISL_1857705, EPI_ISL_1857706, EPI_ISL_1857707, EPI_ISL_1857708, EPI_ISL_1857709, EPI_ISL_1857710, EPI_ISL_1857711, EPI_ISL_1857712, EPI_ISL_1857713, EPI_ISL_1857714, EPI_ISL_1857715, EPI_ISL_1857716, EPI_ISL_1857718, EPI_ISL_1857719, EPI_ISL_1857720, EPI_ISL_1857723, EPI_ISL_1857724, EPI_ISL_1857725, EPI_ISL_1857726, EPI_ISL_1857727, EPI_ISL_1857730, EPI_ISL_1857731, EPI_ISL_1857732, EPI_ISL_1857733, EPI_ISL_1857734, EPI_ISL_1857735, EPI_ISL_1857736, EPI_ISL_1857737, EPI_ISL_1857738, EPI_ISL_1857739, EPI_ISL_1857740, EPI_ISL_1857741, EPI_ISL_1857742, EPI_ISL_1857743, EPI_ISL_1857744, EPI_ISL_1857746, EPI_ISL_1857747, EPI_ISL_1857748, EPI_ISL_1857749, EPI_ISL_1857750, EPI_ISL_1857751, EPI_ISL_1857752, EPI_ISL_1857753, EPI_ISL_1857754, EPI_ISL_1857755, EPI_ISL_1857759, EPI_ISL_1857761, EPI_ISL_1857762, EPI_ISL_1857764, EPI_ISL_1857765, EPI_ISL_1857768, EPI_ISL_1857769, EPI_ISL_1857771, EPI_ISL_1857772, EPI_ISL_1857773, EPI_ISL_1857775, EPI_ISL_1857776, EPI_ISL_1857777, EPI_ISL_1857778, EPI_ISL_1857780, EPI_ISL_1857781, EPI_ISL_1857782, EPI_ISL_1857783, EPI_ISL_1857784, EPI_ISL_1857785, EPI_ISL_1857787, EPI_ISL_1857788, EPI_ISL_1857789, EPI_ISL_1857791, EPI_ISL_1857792, EPI_ISL_1857793, EPI_ISL_1857795, EPI_ISL_1857796, EPI_ISL_1857797, EPI_ISL_1857798, EPI_ISL_1857799, EPI_ISL_1857800, EPI_ISL_1857801, EPI_ISL_1857802, EPI_ISL_1857803, EPI_ISL_1857804, EPI_ISL_1857805, EPI_ISL_1857810, EPI_ISL_1857813, EPI_ISL_1857814, EPI_ISL_1857818, EPI_ISL_1857820, EPI_ISL_1857823, EPI_ISL_1857826, EPI_ISL_1857844, EPI_ISL_1857845, EPI_ISL_1857853, EPI_ISL_1857865, EPI_ISL_1857867, EPI_ISL_1857881, EPI_ISL_1857882, EPI_ISL_1857888, EPI_ISL_1857889, EPI_ISL_1857894, EPI_ISL_1857898, EPI_ISL_1857900, EPI_ISL_1857901, EPI_ISL_1857902, EPI_ISL_1857907, EPI_ISL_1857909, EPI_ISL_1857910, EPI_ISL_1857911, EPI_ISL_1857916, EPI_ISL_1857925, EPI_ISL_1857929, EPI_ISL_1857930, EPI_ISL_1857936, EPI_ISL_1857939, EPI_ISL_1857947, EPI_ISL_1857949, EPI_ISL_1857950, EPI_ISL_1857951, EPI_ISL_1857962, EPI_ISL_1857975, EPI_ISL_1857983, EPI_ISL_1857985, EPI_ISL_1857991, EPI_ISL_1857996, EPI_ISL_1857997, EPI_ISL_1858003, EPI_ISL_1858028, EPI_ISL_1858038, EPI_ISL_1858040, EPI_ISL_1858044, EPI_ISL_1858046, EPI_ISL_1858052, EPI_ISL_1858055, EPI_ISL_1858063, EPI_ISL_1858068, EPI_ISL_1858078, EPI_ISL_1858083, EPI_ISL_1858096, EPI_ISL_1858102, EPI_ISL_1858105, EPI_ISL_1858116, EPI_ISL_1858123, EPI_ISL_1858126, EPI_ISL_1858146, EPI_ISL_1858149, EPI_ISL_1858151, EPI_ISL_1858199, EPI_ISL_1858200, EPI_ISL_1858203, EPI_ISL_1858205, EPI_ISL_1858210, EPI_ISL_1858218, EPI_ISL_1858220, EPI_ISL_1858223, EPI_ISL_1858225, EPI_ISL_1858226, EPI_ISL_1858229, EPI_ISL_1858231, EPI_ISL_1858233, EPI_ISL_1858234, EPI_ISL_1912350, EPI_ISL_1912356, EPI_ISL_1912357, EPI_ISL_1912363, EPI_ISL_1912366, EPI_ISL_1912367, EPI_ISL_1912368, EPI_ISL_1912370, EPI_ISL_1912372, EPI_ISL_1912378, EPI_ISL_1912380, EPI_ISL_1912386, EPI_ISL_1912388, EPI_ISL_1912391, EPI_ISL_1912392, EPI_ISL_1912395, EPI_ISL_1912400, EPI_ISL_1912404, EPI_ISL_1912406, EPI_ISL_1912411, EPI_ISL_1912412, EPI_ISL_1912417, EPI_ISL_1912419, EPI_ISL_1912420, EPI_ISL_1912421, EPI_ISL_1912423, EPI_ISL_1912424, EPI_ISL_1912433, EPI_ISL_1912440, EPI_ISL_1912441, EPI_ISL_1912445, EPI_ISL_1912450, EPI_ISL_1912451, EPI_ISL_1912457, EPI_ISL_1912462, EPI_ISL_1912470, EPI_ISL_1912472, EPI_ISL_1912473, EPI_ISL_1912476, EPI_ISL_1912477, EPI_ISL_1912478, EPI_ISL_1912485, EPI_ISL_1912489, EPI_ISL_1912497, EPI_ISL_1912500, EPI_ISL_1912501, EPI_ISL_1912502, EPI_ISL_1912511, EPI_ISL_1912512, EPI_ISL_1912513, EPI_ISL_1912518, EPI_ISL_1912519, EPI_ISL_1912520, EPI_ISL_1912525, EPI_ISL_1912527, EPI_ISL_1912546, EPI_ISL_1912547, EPI_ISL_1912549, EPI_ISL_1912550, EPI_ISL_1912558, EPI_ISL_1912565, EPI_ISL_1912569, EPI_ISL_1912573, EPI_ISL_1912576, EPI_ISL_1912581, EPI_ISL_1912588, EPI_ISL_1912589, EPI_ISL_1912591, EPI_ISL_1912594, EPI_ISL_1912595, EPI_ISL_1912596, EPI_ISL_1912597, EPI_ISL_1912598, EPI_ISL_1912599, EPI_ISL_1912600, EPI_ISL_1912601, EPI_ISL_1912602, EPI_ISL_1912603, EPI_ISL_1912604, EPI_ISL_1912605, EPI_ISL_1912606, EPI_ISL_1912607, EPI_ISL_1912608, EPI_ISL_1912609, EPI_ISL_1912610, EPI_ISL_1912611, EPI_ISL_1912612, EPI_ISL_1912613, EPI_ISL_1912614, EPI_ISL_1912615, EPI_ISL_1912616, EPI_ISL_1912617, EPI_ISL_1912618, EPI_ISL_1912619, EPI_ISL_1912620, EPI_ISL_1912621, EPI_ISL_1912622, EPI_ISL_1912623, EPI_ISL_1912624, EPI_ISL_1912625, EPI_ISL_1912627, EPI_ISL_1912628, EPI_ISL_1912629, EPI_ISL_1912630, EPI_ISL_1912631, EPI_ISL_1912632, EPI_ISL_1912633, EPI_ISL_1912634, EPI_ISL_1912635, EPI_ISL_1912636, EPI_ISL_1912637, EPI_ISL_1912638, EPI_ISL_1912640, EPI_ISL_1912641, EPI_ISL_1912642, EPI_ISL_1912643, EPI_ISL_1912644, EPI_ISL_1912645, EPI_ISL_1912646, EPI_ISL_1912647, EPI_ISL_1912648, EPI_ISL_1912649, EPI_ISL_1912650, EPI_ISL_1912651, EPI_ISL_1912652, EPI_ISL_1912653, EPI_ISL_1912654, EPI_ISL_1912655, EPI_ISL_1912656, EPI_ISL_1912657, EPI_ISL_1912658, EPI_ISL_1912659, EPI_ISL_1912660, EPI_ISL_1912661, EPI_ISL_1912662, EPI_ISL_1912663, EPI_ISL_1912664, EPI_ISL_1912666, EPI_ISL_1912667, EPI_ISL_1912668, EPI_ISL_1912669, EPI_ISL_1912670, EPI_ISL_1912671, EPI_ISL_1912672, EPI_ISL_1912673, EPI_ISL_1912674, EPI_ISL_1912675, EPI_ISL_1912676, EPI_ISL_1912677, EPI_ISL_1912679, EPI_ISL_1912680, EPI_ISL_1912681, EPI_ISL_1912682, EPI_ISL_1912683, EPI_ISL_1912684, EPI_ISL_1912685, EPI_ISL_1912686, EPI_ISL_1912687, EPI_ISL_1912688, EPI_ISL_1912689, EPI_ISL_1912691, EPI_ISL_1912692, EPI_ISL_1912693, EPI_ISL_1912694, EPI_ISL_1912695, EPI_ISL_1912696, EPI_ISL_1912697, EPI_ISL_1912698, EPI_ISL_1912699, EPI_ISL_1912700, EPI_ISL_1912701, EPI_ISL_1912702, EPI_ISL_1912703, EPI_ISL_1912704, EPI_ISL_1912705, EPI_ISL_1912706, EPI_ISL_1912707, EPI_ISL_1912708, EPI_ISL_1912709, EPI_ISL_1912710, EPI_ISL_1912711, EPI_ISL_1912712, EPI_ISL_1912713, EPI_ISL_1912714, EPI_ISL_1912715, EPI_ISL_1912716, EPI_ISL_1912717, EPI_ISL_1912718, EPI_ISL_1912719, EPI_ISL_1912720, EPI_ISL_1912721, EPI_ISL_1912722, EPI_ISL_1912723, EPI_ISL_1912724, EPI_ISL_1912725, EPI_ISL_1912726, EPI_ISL_1912727, EPI_ISL_1912728, EPI_ISL_1912730, EPI_ISL_1912731, EPI_ISL_1912732, EPI_ISL_1912733, EPI_ISL_1912734, EPI_ISL_1912735, EPI_ISL_1912736, EPI_ISL_1912738, EPI_ISL_1912739, EPI_ISL_1912741, EPI_ISL_1912742, EPI_ISL_1912743, EPI_ISL_1912744, EPI_ISL_1912745, EPI_ISL_1912746, EPI_ISL_1912747, EPI_ISL_1912748, EPI_ISL_1912749, EPI_ISL_1912750, EPI_ISL_1921260, EPI_ISL_1921425, EPI_ISL_1921737, EPI_ISL_1939618, EPI_ISL_1939619, EPI_ISL_1939620, EPI_ISL_1939621, EPI_ISL_1939623, EPI_ISL_1939624, EPI_ISL_1939625, EPI_ISL_1939626, EPI_ISL_1939627, EPI_ISL_1939628, EPI_ISL_1939629, EPI_ISL_1939630, EPI_ISL_1939631, EPI_ISL_1939632, EPI_ISL_1939633, EPI_ISL_1939634, EPI_ISL_1939635, EPI_ISL_1939636, EPI_ISL_1939637, EPI_ISL_1939638, EPI_ISL_1939639, EPI_ISL_1939640, EPI_ISL_1939641, EPI_ISL_1939642, EPI_ISL_1939643, EPI_ISL_1939644, EPI_ISL_1939645, EPI_ISL_1939646, EPI_ISL_1939647, EPI_ISL_1939648, EPI_ISL_1939649, EPI_ISL_1939650, EPI_ISL_1939651, EPI_ISL_1939652, EPI_ISL_1939653, EPI_ISL_1939654, EPI_ISL_1939655, EPI_ISL_1939656, EPI_ISL_1939657, EPI_ISL_1939658,

EPI_ISL_1939659, EPI_ISL_1939660, EPI_ISL_1939661, EPI_ISL_1939662, EPI_ISL_1939663, EPI_ISL_1939664, EPI_ISL_1939665, EPI_ISL_1939666, EPI_ISL_1939667, EPI_ISL_1939668, EPI_ISL_1939669, EPI_ISL_1939670, EPI_ISL_1939671, EPI_ISL_1939672, EPI_ISL_1939673, EPI_ISL_1939674, EPI_ISL_1939675, EPI_ISL_1939676, EPI_ISL_1939678, EPI_ISL_1939679, EPI_ISL_1939680, EPI_ISL_1939681,
EPI_ISL_1939682, EPI_ISL_1939683, EPI_ISL_1939684, EPI_ISL_1939685, EPI_ISL_1939686, EPI_ISL_1939687, EPI_ISL_1939688, EPI_ISL_1939689, EPI_ISL_1939691, EPI_ISL_1939692, EPI_ISL_1939693, EPI_ISL_1939694, EPI_ISL_1939695, EPI_ISL_1939696, EPI_ISL_1939697, EPI_ISL_1939698, EPI_ISL_1939699, EPI_ISL_1939700, EPI_ISL_1939701, EPI_ISL_1939702, EPI_ISL_1939703, EPI_ISL_1939704,
EPI_ISL_1939705, EPI_ISL_1939706, EPI_ISL_1939707, EPI_ISL_1939721, EPI_ISL_1939722, EPI_ISL_1939723, EPI_ISL_1939724, EPI_ISL_1939725, EPI_ISL_1939726, EPI_ISL_1939727, EPI_ISL_1939728, EPI_ISL_1939729, EPI_ISL_1939730, EPI_ISL_1939731, EPI_ISL_1939732, EPI_ISL_1939733, EPI_ISL_1939734, EPI_ISL_1939735, EPI_ISL_1939736, EPI_ISL_1939737, EPI_ISL_1939738, EPI_ISL_1939739,
EPI_ISL_1939740, EPI_ISL_1939741, EPI_ISL_1939742, EPI_ISL_1939743, EPI_ISL_1939744, EPI_ISL_1939745, EPI_ISL_1939746, EPI_ISL_1939747, EPI_ISL_1939748, EPI_ISL_1939749, EPI_ISL_1939750, EPI_ISL_1939751, EPI_ISL_1939752, EPI_ISL_1939753, EPI_ISL_1939754, EPI_ISL_1939755, EPI_ISL_1939756, EPI_ISL_1939757, EPI_ISL_1939758, EPI_ISL_1939759, EPI_ISL_1939760, EPI_ISL_1939761,
EPI_ISL_1939762, EPI_ISL_1939763, EPI_ISL_1939764, EPI_ISL_1939765, EPI_ISL_1939766, EPI_ISL_1939767, EPI_ISL_1939768, EPI_ISL_1939769, EPI_ISL_1939770, EPI_ISL_1939771, EPI_ISL_1939772, EPI_ISL_1939773, EPI_ISL_1939774, EPI_ISL_1939775, EPI_ISL_1939776, EPI_ISL_1939777, EPI_ISL_1939778, EPI_ISL_1939779, EPI_ISL_1939780, EPI_ISL_1939781, EPI_ISL_1939782, EPI_ISL_1939783,
EPI_ISL_1939784, EPI_ISL_1939785, EPI_ISL_1939786, EPI_ISL_1939787, EPI_ISL_1939788, EPI_ISL_1939789, EPI_ISL_1939790, EPI_ISL_1939791, EPI_ISL_1939792, EPI_ISL_1939793, EPI_ISL_1939794, EPI_ISL_1939795, EPI_ISL_1939796, EPI_ISL_1939797, EPI_ISL_1939798, EPI_ISL_1939799, EPI_ISL_1939800, EPI_ISL_1939801, EPI_ISL_1939802, EPI_ISL_1939803, EPI_ISL_1939804, EPI_ISL_1939805,
EPI_ISL_1939806, EPI_ISL_1939807, EPI_ISL_1939808, EPI_ISL_1939809, EPI_ISL_1939810, EPI_ISL_1939811, EPI_ISL_1939812, EPI_ISL_1939813, EPI_ISL_1939814, EPI_ISL_1939815, EPI_ISL_1939816, EPI_ISL_1939817, EPI_ISL_1939818, EPI_ISL_1939819, EPI_ISL_1939820, EPI_ISL_1939821, EPI_ISL_1939822, EPI_ISL_1939823, EPI_ISL_1939824, EPI_ISL_1939825, EPI_ISL_1939826, EPI_ISL_1986111,
EPI_ISL_1986112, EPI_ISL_1986116, EPI_ISL_1986126, EPI_ISL_1986131, EPI_ISL_1986132, EPI_ISL_1986134, EPI_ISL_1986138, EPI_ISL_1986148, EPI_ISL_1986149, EPI_ISL_1986151, EPI_ISL_1986152, EPI_ISL_1986156, EPI_ISL_1986157, EPI_ISL_1986160, EPI_ISL_1986175, EPI_ISL_1986176, EPI_ISL_1986179, EPI_ISL_1986182, EPI_ISL_1986189, EPI_ISL_1986192, EPI_ISL_1986194, EPI_ISL_1986200,
EPI_ISL_1986201, EPI_ISL_1986204, EPI_ISL_1986209, EPI_ISL_1986215, EPI_ISL_1986216, EPI_ISL_1986219, EPI_ISL_1986228, EPI_ISL_1986231, EPI_ISL_1986238, EPI_ISL_1986241, EPI_ISL_1986251, EPI_ISL_1986254, EPI_ISL_1986256, EPI_ISL_1986264, EPI_ISL_1986265, EPI_ISL_1986266, EPI_ISL_1986268, EPI_ISL_1986270, EPI_ISL_1986272, EPI_ISL_1986273, EPI_ISL_1986276, EPI_ISL_1986279,
EPI_ISL_1986282, EPI_ISL_1986294, EPI_ISL_1986296, EPI_ISL_1986298, EPI_ISL_1986324, EPI_ISL_1986325, EPI_ISL_1986326, EPI_ISL_1986332, EPI_ISL_1986349, EPI_ISL_1986353, EPI_ISL_1986354, EPI_ISL_1986355, EPI_ISL_1986366, EPI_ISL_1986370, EPI_ISL_1986373, EPI_ISL_1986374, EPI_ISL_1986378, EPI_ISL_1986381, EPI_ISL_1986386, EPI_ISL_1986389, EPI_ISL_1986493, EPI_ISL_1986497,
EPI_ISL_1986505, EPI_ISL_1986508, EPI_ISL_1986509, EPI_ISL_1986510, EPI_ISL_1986512, EPI_ISL_1986517, EPI_ISL_1986518, EPI_ISL_1986520, EPI_ISL_1986522, EPI_ISL_1986524, EPI_ISL_1986526, EPI_ISL_1986533, EPI_ISL_1986534, EPI_ISL_1986536, EPI_ISL_1986538, EPI_ISL_1986552, EPI_ISL_1986553, EPI_ISL_1986558, EPI_ISL_1986559, EPI_ISL_1986560, EPI_ISL_1986569, EPI_ISL_1986571,
EPI_ISL_1986594, EPI_ISL_1986605, EPI_ISL_1986609, EPI_ISL_1986619, EPI_ISL_1986623, EPI_ISL_1986627, EPI_ISL_1986648, EPI_ISL_1986664, EPI_ISL_1986665, EPI_ISL_1986686, EPI_ISL_1986710, EPI_ISL_1986726, EPI_ISL_1986736, EPI_ISL_1986753, EPI_ISL_1986762, EPI_ISL_1986770, EPI_ISL_1986778, EPI_ISL_1986785, EPI_ISL_1986797, EPI_ISL_1986804, EPI_ISL_1986805, EPI_ISL_1986806,
EPI_ISL_1986807, EPI_ISL_1986816, EPI_ISL_1986823, EPI_ISL_1986831, EPI_ISL_1986842, EPI_ISL_1986859, EPI_ISL_1986874, EPI_ISL_1986880, EPI_ISL_1986882, EPI_ISL_1986889, EPI_ISL_1986890, EPI_ISL_1986899, EPI_ISL_1986907, EPI_ISL_1986912, EPI_ISL_1986923, EPI_ISL_1986924, EPI_ISL_1986925, EPI_ISL_1986927, EPI_ISL_1986928, EPI_ISL_1986935, EPI_ISL_1986939, EPI_ISL_1986946,
EPI_ISL_1986952, EPI_ISL_1986954, EPI_ISL_1986967, EPI_ISL_1986971, EPI_ISL_1986977, EPI_ISL_1986981, EPI_ISL_1986987, EPI_ISL_1986988, EPI_ISL_1986989, EPI_ISL_1987002, EPI_ISL_1987004, EPI_ISL_1987005, EPI_ISL_1987013, EPI_ISL_1987023, EPI_ISL_1987031, EPI_ISL_1987033, EPI_ISL_1987036, EPI_ISL_1987042, EPI_ISL_1987051, EPI_ISL_1987058, EPI_ISL_1987059, EPI_ISL_1987069,
EPI_ISL_1987072, EPI_ISL_1987132, EPI_ISL_1987135, EPI_ISL_1987138, EPI_ISL_1987139, EPI_ISL_1987142, EPI_ISL_1987145, EPI_ISL_1987166, EPI_ISL_1987169, EPI_ISL_1987191, EPI_ISL_1987194, EPI_ISL_1987205, EPI_ISL_1987209, EPI_ISL_1987219, EPI_ISL_1987231, EPI_ISL_1987232, EPI_ISL_1987244, EPI_ISL_1987246, EPI_ISL_1987251, EPI_ISL_1987262, EPI_ISL_1987268, EPI_ISL_1987271,
EPI_ISL_1987273, EPI_ISL_1987274, EPI_ISL_1987279, EPI_ISL_1987280, EPI_ISL_1987281, EPI_ISL_1987285, EPI_ISL_1987292, EPI_ISL_1987300, EPI_ISL_1987302, EPI_ISL_1987307, EPI_ISL_1987309, EPI_ISL_1987312, EPI_ISL_1987314, EPI_ISL_1987315, EPI_ISL_1987323, EPI_ISL_1987330, EPI_ISL_1987339, EPI_ISL_1987340, EPI_ISL_1987345, EPI_ISL_1987355, EPI_ISL_1987356, EPI_ISL_1987362,
EPI_ISL_1987363, EPI_ISL_1987373, EPI_ISL_2004641, EPI_ISL_2004880, EPI_ISL_2004881, EPI_ISL_2004882, EPI_ISL_2004883, EPI_ISL_2004885, EPI_ISL_2004886, EPI_ISL_2004890, EPI_ISL_2004893, EPI_ISL_2004894, EPI_ISL_2004895, EPI_ISL_2004896, EPI_ISL_2004897, EPI_ISL_2004898, EPI_ISL_2004899, EPI_ISL_2004900, EPI_ISL_2004901, EPI_ISL_2004902, EPI_ISL_2004903, EPI_ISL_2004907,
EPI_ISL_2004908, EPI_ISL_2004909, EPI_ISL_2004910, EPI_ISL_2004911, EPI_ISL_2004913, EPI_ISL_2004914, EPI_ISL_2004916, EPI_ISL_2004917, EPI_ISL_2004919, EPI_ISL_2004920, EPI_ISL_2004921, EPI_ISL_2004922, EPI_ISL_2004925, EPI_ISL_2004926, EPI_ISL_2004927, EPI_ISL_2004928, EPI_ISL_2004929, EPI_ISL_2004930, EPI_ISL_2004931, EPI_ISL_2004933, EPI_ISL_2004935, EPI_ISL_2004936,
EPI_ISL_2004937, EPI_ISL_2004940, EPI_ISL_2004942, EPI_ISL_2004943, EPI_ISL_2004944, EPI_ISL_2004946, EPI_ISL_2004947, EPI_ISL_2004948, EPI_ISL_2004949, EPI_ISL_2004950, EPI_ISL_2004951, EPI_ISL_2004954, EPI_ISL_2004956, EPI_ISL_2004957, EPI_ISL_2004959, EPI_ISL_2004960, EPI_ISL_2004961, EPI_ISL_2004964, EPI_ISL_2004965, EPI_ISL_2004966, EPI_ISL_2004968, EPI_ISL_2004971,
EPI_ISL_2004972, EPI_ISL_2004973, EPI_ISL_2004975, EPI_ISL_2004976, EPI_ISL_2004977, EPI_ISL_2004978, EPI_ISL_2004979, EPI_ISL_2004981, EPI_ISL_2004982, EPI_ISL_2004983, EPI_ISL_2004984, EPI_ISL_2004985, EPI_ISL_2004986, EPI_ISL_2004988, EPI_ISL_2004989, EPI_ISL_2004990, EPI_ISL_2004991, EPI_ISL_2004992, EPI_ISL_2004993, EPI_ISL_2004994, EPI_ISL_2004996, EPI_ISL_2004997,
EPI_ISL_2004998, EPI_ISL_2005000, EPI_ISL_2005001, EPI_ISL_2005002, EPI_ISL_2005003, EPI_ISL_2005005, EPI_ISL_2005006, EPI_ISL_2005011, EPI_ISL_2005015, EPI_ISL_2005016, EPI_ISL_2005018, EPI_ISL_2005019, EPI_ISL_2005021, EPI_ISL_2005024, EPI_ISL_2005026, EPI_ISL_2005027, EPI_ISL_2005028, EPI_ISL_2005029, EPI_ISL_2005030, EPI_ISL_2005032, EPI_ISL_2005033, EPI_ISL_2005035,
EPI_ISL_2005036, EPI_ISL_2005037, EPI_ISL_2005038, EPI_ISL_2005039, EPI_ISL_2005041, EPI_ISL_2005042, EPI_ISL_2005044, EPI_ISL_2005045, EPI_ISL_2005046, EPI_ISL_2005047, EPI_ISL_2005051, EPI_ISL_2005052, EPI_ISL_2005053, EPI_ISL_2005054, EPI_ISL_2005055, EPI_ISL_2005056, EPI_ISL_2005057, EPI_ISL_2005058, EPI_ISL_2005059, EPI_ISL_2005061, EPI_ISL_2005062, EPI_ISL_2005064,
EPI_ISL_2005067, EPI_ISL_2005069, EPI_ISL_2005070, EPI_ISL_2005071, EPI_ISL_2005074, EPI_ISL_2005076, EPI_ISL_2005077, EPI_ISL_2005078, EPI_ISL_2005080, EPI_ISL_2005082, EPI_ISL_2005083, EPI_ISL_2005084, EPI_ISL_2005085, EPI_ISL_2005086, EPI_ISL_2005089, EPI_ISL_2005090, EPI_ISL_2005091, EPI_ISL_2005092, EPI_ISL_2005094, EPI_ISL_2005095, EPI_ISL_2005097, EPI_ISL_2005098,
EPI_ISL_2005099, EPI_ISL_2005100, EPI_ISL_2005101, EPI_ISL_2005102, EPI_ISL_2005103, EPI_ISL_2005104, EPI_ISL_2005106, EPI_ISL_2005108, EPI_ISL_2005109, EPI_ISL_2005113, EPI_ISL_2005115, EPI_ISL_2005118, EPI_ISL_2005121, EPI_ISL_2005124, EPI_ISL_2005125, EPI_ISL_2005127, EPI_ISL_2005128, EPI_ISL_2005130, EPI_ISL_2005131, EPI_ISL_2005132, EPI_ISL_2005133, EPI_ISL_2005134,
EPI_ISL_2005135, EPI_ISL_2005137, EPI_ISL_2005138, EPI_ISL_2005142, EPI_ISL_2005143, EPI_ISL_2005144, EPI_ISL_2005147, EPI_ISL_2005148, EPI_ISL_2005149, EPI_ISL_2005150, EPI_ISL_2005153, EPI_ISL_2005154, EPI_ISL_2005155, EPI_ISL_2005156, EPI_ISL_2005157, EPI_ISL_2005158, EPI_ISL_2005159, EPI_ISL_2005161, EPI_ISL_2005164, EPI_ISL_2005165, EPI_ISL_2005168, EPI_ISL_2005169,
EPI_ISL_2005170, EPI_ISL_2005171, EPI_ISL_2005175, EPI_ISL_2005177, EPI_ISL_2005179, EPI_ISL_2005182, EPI_ISL_2005183, EPI_ISL_2005186, EPI_ISL_2005190, EPI_ISL_2005191, EPI_ISL_2005193, EPI_ISL_2005194, EPI_ISL_2005195, EPI_ISL_2005196, EPI_ISL_2005198, EPI_ISL_2005203, EPI_ISL_2005205, EPI_ISL_2005206, EPI_ISL_2005208, EPI_ISL_2005210, EPI_ISL_2005211, EPI_ISL_2005215,
EPI_ISL_2005216, EPI_ISL_2005217, EPI_ISL_2005219, EPI_ISL_2005221, EPI_ISL_2005222, EPI_ISL_2005223, EPI_ISL_2005224, EPI_ISL_2005225, EPI_ISL_2005226, EPI_ISL_2005231, EPI_ISL_2005233, EPI_ISL_2005234, EPI_ISL_2005235, EPI_ISL_2005236, EPI_ISL_2005240, EPI_ISL_2005244, EPI_ISL_2005245, EPI_ISL_2005246, EPI_ISL_2005247, EPI_ISL_2005248, EPI_ISL_2005249, EPI_ISL_2005255,
EPI_ISL_2005256, EPI_ISL_2005257, EPI_ISL_2005262, EPI_ISL_2005263, EPI_ISL_2005264, EPI_ISL_2005265, EPI_ISL_2005266, EPI_ISL_2005267, EPI_ISL_2005270, EPI_ISL_2005275, EPI_ISL_2005276, EPI_ISL_2005277, EPI_ISL_2005279, EPI_ISL_2005281, EPI_ISL_2005282, EPI_ISL_2005283, EPI_ISL_2005284, EPI_ISL_2005286, EPI_ISL_2005287, EPI_ISL_2005290, EPI_ISL_2005293, EPI_ISL_2005294,
EPI_ISL_2005296, EPI_ISL_2005297, EPI_ISL_2005298, EPI_ISL_2005300, EPI_ISL_2005303, EPI_ISL_2005304, EPI_ISL_2005305, EPI_ISL_2005306, EPI_ISL_2005307, EPI_ISL_2005308, EPI_ISL_2005311, EPI_ISL_2005313, EPI_ISL_2005314, EPI_ISL_2005315, EPI_ISL_2005316, EPI_ISL_2005317, EPI_ISL_2005319, EPI_ISL_2005320, EPI_ISL_2005322, EPI_ISL_2005323, EPI_ISL_2005326, EPI_ISL_2005328,
EPI_ISL_2005332, EPI_ISL_2005334, EPI_ISL_2005336, EPI_ISL_2005338, EPI_ISL_2005339, EPI_ISL_2005340, EPI_ISL_2005814, EPI_ISL_2005818, EPI_ISL_2005820, EPI_ISL_2005834, EPI_ISL_2005841, EPI_ISL_2005867, EPI_ISL_2005884, EPI_ISL_2005893, EPI_ISL_2005895, EPI_ISL_2005900, EPI_ISL_2005914, EPI_ISL_2005918, EPI_ISL_2005923, EPI_ISL_2005927, EPI_ISL_2005930, EPI_ISL_2005932,
EPI_ISL_2005940, EPI_ISL_2005945, EPI_ISL_2005957, EPI_ISL_2005965, EPI_ISL_2005967, EPI_ISL_2005968, EPI_ISL_2005976, EPI_ISL_2005979, EPI_ISL_2005982, EPI_ISL_2005998, EPI_ISL_2006000, EPI_ISL_2006006, EPI_ISL_2006013, EPI_ISL_2006043, EPI_ISL_2006047, EPI_ISL_2006050, EPI_ISL_2006054, EPI_ISL_2006058, EPI_ISL_2006059, EPI_ISL_2006068, EPI_ISL_2006071, EPI_ISL_2006073,
EPI_ISL_2006076, EPI_ISL_2006077, EPI_ISL_2006079, EPI_ISL_2006081, EPI_ISL_2006082, EPI_ISL_2006083, EPI_ISL_2006084, EPI_ISL_2006085, EPI_ISL_2006086, EPI_ISL_2006087, EPI_ISL_2006088, EPI_ISL_2006089, EPI_ISL_2006091, EPI_ISL_2006092, EPI_ISL_2006093, EPI_ISL_2006094, EPI_ISL_2006095, EPI_ISL_2006097, EPI_ISL_2006098, EPI_ISL_2006099, EPI_ISL_2006100, EPI_ISL_2006101,
EPI_ISL_2006103, EPI_ISL_2006104, EPI_ISL_2006106, EPI_ISL_2006107, EPI_ISL_2006108, EPI_ISL_2006109, EPI_ISL_2006110, EPI_ISL_2006111, EPI_ISL_2006115, EPI_ISL_2006116, EPI_ISL_2006117, EPI_ISL_2006119, EPI_ISL_2006122, EPI_ISL_2006123, EPI_ISL_2006125, EPI_ISL_2006127, EPI_ISL_2006129, EPI_ISL_2006130, EPI_ISL_2006131, EPI_ISL_2006136, EPI_ISL_2006137, EPI_ISL_2006140,
EPI_ISL_2006141, EPI_ISL_2006142, EPI_ISL_2006143, EPI_ISL_2006144, EPI_ISL_2006145, EPI_ISL_2006146, EPI_ISL_2006147, EPI_ISL_2006148, EPI_ISL_2006151, EPI_ISL_2006152, EPI_ISL_2006154, EPI_ISL_2006155, EPI_ISL_2006156, EPI_ISL_2006157, EPI_ISL_2006158, EPI_ISL_2006160, EPI_ISL_2006161, EPI_ISL_2006162, EPI_ISL_2006164, EPI_ISL_2006165, EPI_ISL_2006168, EPI_ISL_2006170,
EPI_ISL_2006171, EPI_ISL_2006172, EPI_ISL_2006173, EPI_ISL_2006174, EPI_ISL_2006175, EPI_ISL_2006177, EPI_ISL_2006178, EPI_ISL_2006179, EPI_ISL_2006180, EPI_ISL_2006181, EPI_ISL_2006182, EPI_ISL_2006183, EPI_ISL_2006184, EPI_ISL_2006186, EPI_ISL_2006189, EPI_ISL_2006190, EPI_ISL_2006193, EPI_ISL_2006194, EPI_ISL_2006196, EPI_ISL_2006198, EPI_ISL_2006199, EPI_ISL_2006201,
EPI_ISL_2006202, EPI_ISL_2006204, EPI_ISL_2006207, EPI_ISL_2006211, EPI_ISL_2006212, EPI_ISL_2006213, EPI_ISL_2006215, EPI_ISL_2006217, EPI_ISL_2006218, EPI_ISL_2006219, EPI_ISL_2006222, EPI_ISL_2006223, EPI_ISL_2006224, EPI_ISL_2006225, EPI_ISL_2006226, EPI_ISL_2006227, EPI_ISL_2006229, EPI_ISL_2006230, EPI_ISL_2006231, EPI_ISL_2006232, EPI_ISL_2006233, EPI_ISL_2006235,
EPI_ISL_2006237, EPI_ISL_2006238, EPI_ISL_2006239, EPI_ISL_2006240, EPI_ISL_2006243, EPI_ISL_2006244, EPI_ISL_2006245, EPI_ISL_2006246, EPI_ISL_2006249, EPI_ISL_2006250, EPI_ISL_2006253, EPI_ISL_2006254, EPI_ISL_2006256, EPI_ISL_2006258, EPI_ISL_2006262, EPI_ISL_2006264, EPI_ISL_2006265, EPI_ISL_2006266, EPI_ISL_2006267, EPI_ISL_2006271, EPI_ISL_2006273, EPI_ISL_2006275,
EPI_ISL_2006276, EPI_ISL_2006277, EPI_ISL_2006279, EPI_ISL_2006280, EPI_ISL_2006281, EPI_ISL_2006283, EPI_ISL_2006284, EPI_ISL_2006285, EPI_ISL_2006286, EPI_ISL_2006287, EPI_ISL_2006288, EPI_ISL_2006290, EPI_ISL_2006292, EPI_ISL_2006293, EPI_ISL_2006295, EPI_ISL_2006296, EPI_ISL_2006297, EPI_ISL_2006298, EPI_ISL_2006299, EPI_ISL_2006300, EPI_ISL_2006301, EPI_ISL_2006303,
EPI_ISL_2006304, EPI_ISL_2006305, EPI_ISL_2006306, EPI_ISL_2006307, EPI_ISL_2006308, EPI_ISL_2006309, EPI_ISL_2006310, EPI_ISL_2006311, EPI_ISL_2006313, EPI_ISL_2006314, EPI_ISL_2006315, EPI_ISL_2006316, EPI_ISL_2006317, EPI_ISL_2006318, EPI_ISL_2006319, EPI_ISL_2006320, EPI_ISL_2006321, EPI_ISL_2006322, EPI_ISL_2006323, EPI_ISL_2006324, EPI_ISL_2006325, EPI_ISL_2006326,
EPI_ISL_2006327, EPI_ISL_2006329, EPI_ISL_2006330, EPI_ISL_2006331, EPI_ISL_2006332, EPI_ISL_2006335, EPI_ISL_2006336, EPI_ISL_2006337, EPI_ISL_2006338, EPI_ISL_2006339, EPI_ISL_2006340, EPI_ISL_2006341, EPI_ISL_2006342, EPI_ISL_2006343, EPI_ISL_2006346, EPI_ISL_2006347, EPI_ISL_2006348, EPI_ISL_2006349, EPI_ISL_2006350, EPI_ISL_2006351, EPI_ISL_2006352, EPI_ISL_2006353,
EPI_ISL_2006354, EPI_ISL_2006355, EPI_ISL_2006356, EPI_ISL_2006357, EPI_ISL_2006359, EPI_ISL_2006360, EPI_ISL_2006361, EPI_ISL_2006362, EPI_ISL_2006363, EPI_ISL_2006364, EPI_ISL_2006365, EPI_ISL_2006366, EPI_ISL_2006367, EPI_ISL_2006368, EPI_ISL_2006370, EPI_ISL_2006371, EPI_ISL_2006372, EPI_ISL_2006373, EPI_ISL_2006374, EPI_ISL_2006375, EPI_ISL_2006376, EPI_ISL_2006377,
EPI_ISL_2006378, EPI_ISL_2006379, EPI_ISL_2006380, EPI_ISL_2006381, EPI_ISL_2006382, EPI_ISL_2006383, EPI_ISL_2006384, EPI_ISL_2006385, EPI_ISL_2006386, EPI_ISL_2006387, EPI_ISL_2006388, EPI_ISL_2006389, EPI_ISL_2006390, EPI_ISL_2006391, EPI_ISL_2006393, EPI_ISL_2006394, EPI_ISL_2006395, EPI_ISL_2006396, EPI_ISL_2006397, EPI_ISL_2006398, EPI_ISL_2006400, EPI_ISL_2006401,
EPI_ISL_2006402, EPI_ISL_2006403, EPI_ISL_2006404, EPI_ISL_2006406, EPI_ISL_2006407, EPI_ISL_2006408, EPI_ISL_2006409, EPI_ISL_2006410, EPI_ISL_2006411, EPI_ISL_2006412, EPI_ISL_2006413, EPI_ISL_2006414, EPI_ISL_2006415, EPI_ISL_2006416, EPI_ISL_2006417, EPI_ISL_2006418, EPI_ISL_2006419, EPI_ISL_2006420, EPI_ISL_2006421, EPI_ISL_2006422, EPI_ISL_2006423, EPI_ISL_2006424,
EPI_ISL_2006425, EPI_ISL_2006426, EPI_ISL_2006427, EPI_ISL_2006428, EPI_ISL_2006429, EPI_ISL_2006430, EPI_ISL_2006431, EPI_ISL_2006432, EPI_ISL_2006433, EPI_ISL_2006434, EPI_ISL_2006435, EPI_ISL_2006436, EPI_ISL_2006437, EPI_ISL_2006439, EPI_ISL_2006440, EPI_ISL_2006441, EPI_ISL_2006443, EPI_ISL_2006444, EPI_ISL_2006445, EPI_ISL_2006446, EPI_ISL_2006447, EPI_ISL_2006448,
EPI_ISL_2006449, EPI_ISL_2006450, EPI_ISL_2006451, EPI_ISL_2006453, EPI_ISL_2006454, EPI_ISL_2006455, EPI_ISL_2006456, EPI_ISL_2006457, EPI_ISL_2006459, EPI_ISL_2006460, EPI_ISL_2006461, EPI_ISL_2006464, EPI_ISL_2006465, EPI_ISL_2006466, EPI_ISL_2006467, EPI_ISL_2006468, EPI_ISL_2006469, EPI_ISL_2006470, EPI_ISL_2006472, EPI_ISL_2006473, EPI_ISL_2006474, EPI_ISL_2006475,
EPI_ISL_2006476, EPI_ISL_2006477, EPI_ISL_2006478, EPI_ISL_2006479, EPI_ISL_2006480, EPI_ISL_2006481, EPI_ISL_2006482, EPI_ISL_2006483, EPI_ISL_2006484, EPI_ISL_2006486, EPI_ISL_2006488, EPI_ISL_2006489, EPI_ISL_2006490, EPI_ISL_2006491, EPI_ISL_2006492, EPI_ISL_2006494, EPI_ISL_2006495, EPI_ISL_2006496, EPI_ISL_2006497, EPI_ISL_2006498, EPI_ISL_2006499, EPI_ISL_2006500,
EPI_ISL_2006501, EPI_ISL_2006502, EPI_ISL_2006503, EPI_ISL_2006504, EPI_ISL_2006505, EPI_ISL_2006506, EPI_ISL_2006507, EPI_ISL_2006509, EPI_ISL_2006510, EPI_ISL_2006511, EPI_ISL_2006512, EPI_ISL_2006513, EPI_ISL_2006514, EPI_ISL_2006515, EPI_ISL_2006516, EPI_ISL_2006517, EPI_ISL_2006518, EPI_ISL_2006519, EPI_ISL_2006520, EPI_ISL_2006521, EPI_ISL_2021091, EPI_ISL_2021098,
EPI_ISL_2021113, EPI_ISL_2021117, EPI_ISL_2021166, EPI_ISL_2021168, EPI_ISL_2021250, EPI_ISL_2021312, EPI_ISL_2021328, EPI_ISL_2021698, EPI_ISL_2021699, EPI_ISL_2021725, EPI_ISL_2021726, EPI_ISL_2021727, EPI_ISL_2021729, EPI_ISL_2021734, EPI_ISL_2021737, EPI_ISL_2021756, EPI_ISL_2021758, EPI_ISL_2021760, EPI_ISL_2021765, EPI_ISL_2021770, EPI_ISL_2021777, EPI_ISL_2021779,
EPI_ISL_2021785, EPI_ISL_2021789, EPI_ISL_2021792, EPI_ISL_2021798, EPI_ISL_2021806, EPI_ISL_2021808, EPI_ISL_2021811, EPI_ISL_2021812, EPI_ISL_2021816, EPI_ISL_2021818, EPI_ISL_2021819, EPI_ISL_2021831, EPI_ISL_2021837, EPI_ISL_2021842, EPI_ISL_2022439, EPI_ISL_2022440, EPI_ISL_2022441, EPI_ISL_2022442, EPI_ISL_2022443, EPI_ISL_2022444, EPI_ISL_2022446, EPI_ISL_2022447,
EPI_ISL_2022448, EPI_ISL_2022449, EPI_ISL_2022450, EPI_ISL_2022451, EPI_ISL_2022453, EPI_ISL_2022454, EPI_ISL_2022455, EPI_ISL_2022456, EPI_ISL_2022457, EPI_ISL_2022458, EPI_ISL_2022460, EPI_ISL_2022461, EPI_ISL_2022462, EPI_ISL_2022463, EPI_ISL_2022465, EPI_ISL_2022466, EPI_ISL_2022467, EPI_ISL_2022468, EPI_ISL_2022469, EPI_ISL_2022470, EPI_ISL_2022471, EPI_ISL_2022472,
EPI_ISL_2022473, EPI_ISL_2022474, EPI_ISL_2022475, EPI_ISL_2022476, EPI_ISL_2022477, EPI_ISL_2022478, EPI_ISL_2022479, EPI_ISL_2022480, EPI_ISL_2022481, EPI_ISL_2022482, EPI_ISL_2022483, EPI_ISL_2022484, EPI_ISL_2022485, EPI_ISL_2022486, EPI_ISL_2022487, EPI_ISL_2022488, EPI_ISL_2022489, EPI_ISL_2022490, EPI_ISL_2022491, EPI_ISL_2022492, EPI_ISL_2022493, EPI_ISL_2022540,
EPI_ISL_2022544, EPI_ISL_2022551, EPI_ISL_2022553, EPI_ISL_2022555, EPI_ISL_2022582, EPI_ISL_2022586, EPI_ISL_2022591, EPI_ISL_2022592, EPI_ISL_2022593, EPI_ISL_2022594, EPI_ISL_2022595, EPI_ISL_2022596, EPI_ISL_2022597, EPI_ISL_2022598, EPI_ISL_2022599, EPI_ISL_2022600, EPI_ISL_2022601, EPI_ISL_2022602, EPI_ISL_2022603, EPI_ISL_2022604, EPI_ISL_2022605, EPI_ISL_2022606,
EPI_ISL_2022607, EPI_ISL_2022608, EPI_ISL_2022609, EPI_ISL_2022610, EPI_ISL_2022611, EPI_ISL_2022612, EPI_ISL_2022613, EPI_ISL_2022614, EPI_ISL_2022615, EPI_ISL_2022617, EPI_ISL_2022618, EPI_ISL_2022619, EPI_ISL_2022620, EPI_ISL_2022621, EPI_ISL_2022622, EPI_ISL_2022623, EPI_ISL_2022624, EPI_ISL_2022625, EPI_ISL_2022626, EPI_ISL_2022627, EPI_ISL_2022628,
EPI_ISL_2022629, EPI_ISL_2022630, EPI_ISL_2022631, EPI_ISL_2022632, EPI_ISL_2022633, EPI_ISL_2022634, EPI_ISL_2022635, EPI_ISL_2022636, EPI_ISL_2022637, EPI_ISL_2022638, EPI_ISL_2022639, EPI_ISL_2022640, EPI_ISL_2022641, EPI_ISL_2022642, EPI_ISL_2022643, EPI_ISL_2022644, EPI_ISL_2022646, EPI_ISL_2022647, EPI_ISL_2022648, EPI_ISL_2022649, EPI_ISL_2022650, EPI_ISL_2022651,
EPI_ISL_2022652, EPI_ISL_2022653, EPI_ISL_2022654, EPI_ISL_2022655, EPI_ISL_2022656, EPI_ISL_2022657, EPI_ISL_2022658, EPI_ISL_2022659, EPI_ISL_2022661, EPI_ISL_2022662, EPI_ISL_2022663, EPI_ISL_2022664, EPI_ISL_2022665, EPI_ISL_2022666, EPI_ISL_2022669, EPI_ISL_2022670, EPI_ISL_2022671, EPI_ISL_2022672, EPI_ISL_2022673, EPI_ISL_2022674, EPI_ISL_2022675,
EPI_ISL_2022676, EPI_ISL_2022677, EPI_ISL_2022678, EPI_ISL_2022679, EPI_ISL_2022680, EPI_ISL_2022681, EPI_ISL_2022683, EPI_ISL_2022684, EPI_ISL_2022685, EPI_ISL_2022686, EPI_ISL_2022687, EPI_ISL_2022688, EPI_ISL_2022689, EPI_ISL_2022690, EPI_ISL_2022691, EPI_ISL_2022692, EPI_ISL_2022693, EPI_ISL_2022694, EPI_ISL_2022695, EPI_ISL_2022696, EPI_ISL_2022697,
EPI_ISL_2022698, EPI_ISL_2022699, EPI_ISL_2022700, EPI_ISL_2022701, EPI_ISL_2022702, EPI_ISL_2022703, EPI_ISL_2022704, EPI_ISL_2022705, EPI_ISL_2022706, EPI_ISL_2022707, EPI_ISL_2022708, EPI_ISL_2022709, EPI_ISL_2022710, EPI_ISL_2022711, EPI_ISL_2022712, EPI_ISL_2022713, EPI_ISL_2022714, EPI_ISL_2022715, EPI_ISL_2022716, EPI_ISL_2022717, EPI_ISL_2022718, EPI_ISL_2022719,
EPI_ISL_2022720, EPI_ISL_2022721, EPI_ISL_2022722, EPI_ISL_2022723, EPI_ISL_2022724, EPI_ISL_2022725, EPI_ISL_2022726, EPI_ISL_2022727, EPI_ISL_2022728, EPI_ISL_2022729, EPI_ISL_2022730, EPI_ISL_2022731, EPI_ISL_2022732, EPI_ISL_2022733, EPI_ISL_2022734, EPI_ISL_2022735, EPI_ISL_2022736, EPI_ISL_2022737, EPI_ISL_2022738, EPI_ISL_2022739, EPI_ISL_2022740, EPI_ISL_2022741,
EPI_ISL_2022742, EPI_ISL_2022743, EPI_ISL_2022744, EPI_ISL_2022745, EPI_ISL_2022746, EPI_ISL_2022747, EPI_ISL_2022748, EPI_ISL_2022749, EPI_ISL_2022750, EPI_ISL_2022751, EPI_ISL_2022752, EPI_ISL_2022753, EPI_ISL_2022754, EPI_ISL_2022755, EPI_ISL_2022756, EPI_ISL_2022757, EPI_ISL_2022758, EPI_ISL_2022759, EPI_ISL_2022760, EPI_ISL_2022761, EPI_ISL_2022762, EPI_ISL_2022763,
EPI_ISL_2022764, EPI_ISL_2022765, EPI_ISL_2022766, EPI_ISL_2022767, EPI_ISL_2022768, EPI_ISL_2022769, EPI_ISL_2022770, EPI_ISL_2022771, EPI_ISL_2022772, EPI_ISL_2022773, EPI_ISL_2022774, EPI_ISL_2022775, EPI_ISL_2022776, EPI_ISL_2022777, EPI_ISL_2022778, EPI_ISL_2022779, EPI_ISL_2022780, EPI_ISL_2022799, EPI_ISL_2022810, EPI_ISL_2022817, EPI_ISL_2022821, EPI_ISL_2022832,
EPI_ISL_2022834, EPI_ISL_2022842, EPI_ISL_2022850, EPI_ISL_2022948, EPI_ISL_2022949, EPI_ISL_2022950, EPI_ISL_2022951, EPI_ISL_2022952, EPI_ISL_2022953, EPI_ISL_2022954, EPI_ISL_2022955, EPI_ISL_2022956, EPI_ISL_2022957, EPI_ISL_2022958, EPI_ISL_2022959, EPI_ISL_2022960, EPI_ISL_2022961, EPI_ISL_2022962, EPI_ISL_2022963, EPI_ISL_2022964, EPI_ISL_2022965, EPI_ISL_2022966,
EPI_ISL_2022967, EPI_ISL_2022968, EPI_ISL_2022969, EPI_ISL_2022970, EPI_ISL_2022971, EPI_ISL_2022972, EPI_ISL_2022973, EPI_ISL_2022974, EPI_ISL_2022975, EPI_ISL_2022976, EPI_ISL_2022977, EPI_ISL_2022978, EPI_ISL_2022980, EPI_ISL_2022981, EPI_ISL_2022982, EPI_ISL_2022983, EPI_ISL_2022984, EPI_ISL_2022985, EPI_ISL_2022986, EPI_ISL_2022987, EPI_ISL_2022988, EPI_ISL_2022989,
EPI_ISL_2022990, EPI_ISL_2022991, EPI_ISL_2022992, EPI_ISL_2022993, EPI_ISL_2022994, EPI_ISL_2022995, EPI_ISL_2022996, EPI_ISL_2022997, EPI_ISL_2022998, EPI_ISL_2022999, EPI_ISL_2023000, EPI_ISL_2023001, EPI_ISL_2023002, EPI_ISL_2023003, EPI_ISL_2023004, EPI_ISL_2023005, EPI_ISL_2023006, EPI_ISL_2023007, EPI_ISL_2023008, EPI_ISL_2023009, EPI_ISL_2023010, EPI_ISL_2023011,
EPI_ISL_2023012, EPI_ISL_2023013, EPI_ISL_2023014, EPI_ISL_2023015, EPI_ISL_2023016, EPI_ISL_2023017, EPI_ISL_2023018, EPI_ISL_2023019, EPI_ISL_2023020, EPI_ISL_2023021, EPI_ISL_2023022, EPI_ISL_2023023, EPI_ISL_2023024, EPI_ISL_2023025, EPI_ISL_2023026, EPI_ISL_2092481, EPI_ISL_2092495, EPI_ISL_2092497, EPI_ISL_2092508, EPI_ISL_2092510, EPI_ISL_2092517, EPI_ISL_2092525,
EPI_ISL_2092529, EPI_ISL_2092530, EPI_ISL_2092532, EPI_ISL_2092536, EPI_ISL_2092538, EPI_ISL_2092542, EPI_ISL_2092549, EPI_ISL_2092567, EPI_ISL_2092570, EPI_ISL_2092572, EPI_ISL_2092573, EPI_ISL_2092574, EPI_ISL_2092576, EPI_ISL_2092579, EPI_ISL_2092583, EPI_ISL_2092594, EPI_ISL_2092603, EPI_ISL_2092608, EPI_ISL_2092610, EPI_ISL_2092616, EPI_ISL_2092617, EPI_ISL_2092627,
EPI_ISL_2092632, EPI_ISL_2092633, EPI_ISL_2092634, EPI_ISL_2092647, EPI_ISL_2092648, EPI_ISL_2092649, EPI_ISL_2092650, EPI_ISL_2092651, EPI_ISL_2092652, EPI_ISL_2092653, EPI_ISL_2092654, EPI_ISL_2092655, EPI_ISL_2092656, EPI_ISL_2092658, EPI_ISL_2092659, EPI_ISL_2092660, EPI_ISL_2092661, EPI_ISL_2092662, EPI_ISL_2092663, EPI_ISL_2092664, EPI_ISL_2092665,
EPI_ISL_2092666, EPI_ISL_2092667, EPI_ISL_2092668, EPI_ISL_2092669, EPI_ISL_2092670, EPI_ISL_2092671, EPI_ISL_2092672, EPI_ISL_2092673, EPI_ISL_2092674, EPI_ISL_2120019, EPI_ISL_2120332, EPI_ISL_2122707, EPI_ISL_2122708, EPI_ISL_2122710, EPI_ISL_2122712, EPI_ISL_2122713, EPI_ISL_2122714, EPI_ISL_2122716, EPI_ISL_2122718, EPI_ISL_2122719, EPI_ISL_2122721, EPI_ISL_2122722,
EPI_ISL_2122724, EPI_ISL_2122725, EPI_ISL_2122727, EPI_ISL_2122728, EPI_ISL_2122730, EPI_ISL_2122731, EPI_ISL_2122733, EPI_ISL_2122735, EPI_ISL_2122736, EPI_ISL_2122738, EPI_ISL_2122739, EPI_ISL_2122741, EPI_ISL_2122742, EPI_ISL_2122744, EPI_ISL_2122745, EPI_ISL_2122747, EPI_ISL_2122748, EPI_ISL_2122750, EPI_ISL_2122751, EPI_ISL_2122752, EPI_ISL_2122754, EPI_ISL_2122756,
EPI_ISL_2122758, EPI_ISL_2122760, EPI_ISL_2122761, EPI_ISL_2122763, EPI_ISL_2122764, EPI_ISL_2122766, EPI_ISL_2122767, EPI_ISL_2122769, EPI_ISL_2122771, EPI_ISL_2122772, EPI_ISL_2122773, EPI_ISL_2122774, EPI_ISL_2122775, EPI_ISL_2122776, EPI_ISL_2122779, EPI_ISL_2122781, EPI_ISL_2122782, EPI_ISL_2122784, EPI_ISL_2122789, EPI_ISL_2122792, EPI_ISL_2122794, EPI_ISL_2122796,
EPI_ISL_2122797, EPI_ISL_2122798, EPI_ISL_2122800, EPI_ISL_2122801, EPI_ISL_2122803, EPI_ISL_2122804, EPI_ISL_2122806, EPI_ISL_2122807, EPI_ISL_2122809, EPI_ISL_2122810, EPI_ISL_2122811, EPI_ISL_2122813, EPI_ISL_2122814, EPI_ISL_2122817, EPI_ISL_2122820, EPI_ISL_2122822, EPI_ISL_2122823, EPI_ISL_2122825, EPI_ISL_2122826, EPI_ISL_2122827, EPI_ISL_2122829,
EPI_ISL_2122830, EPI_ISL_2122833, EPI_ISL_2122835, EPI_ISL_2122836, EPI_ISL_2122837, EPI_ISL_2122839, EPI_ISL_2122840, EPI_ISL_2122841, EPI_ISL_2122842, EPI_ISL_2122844, EPI_ISL_2122846, EPI_ISL_2122847, EPI_ISL_2122849, EPI_ISL_2122850, EPI_ISL_2122851, EPI_ISL_2122853, EPI_ISL_2122854, EPI_ISL_2122856, EPI_ISL_2122857, EPI_ISL_2122859, EPI_ISL_2122860, EPI_ISL_2122862,
EPI_ISL_2122864, EPI_ISL_2122865, EPI_ISL_2122866, EPI_ISL_2122868, EPI_ISL_2122869, EPI_ISL_2122872, EPI_ISL_2122874, EPI_ISL_2122876, EPI_ISL_2122878, EPI_ISL_2122879, EPI_ISL_2122881, EPI_ISL_2122882, EPI_ISL_2122884, EPI_ISL_2122885, EPI_ISL_2122887, EPI_ISL_2122888, EPI_ISL_2122891, EPI_ISL_2122893, EPI_ISL_2122896, EPI_ISL_2122897,
EPI_ISL_2122899, EPI_ISL_2122900, EPI_ISL_2122902, EPI_ISL_2122903, EPI_ISL_2122905, EPI_ISL_2122906, EPI_ISL_2122908, EPI_ISL_2122909, EPI_ISL_2122910, EPI_ISL_2122912, EPI_ISL_2122913, EPI_ISL_2122915, EPI_ISL_2122916, EPI_ISL_2122917, EPI_ISL_2122919, EPI_ISL_2122920, EPI_ISL_2122922, EPI_ISL_2122923, EPI_ISL_2122925, EPI_ISL_2122926, EPI_ISL_2122928, EPI_ISL_2122930,
EPI_ISL_2122931, EPI_ISL_2122933

| | | | |
|---|---|---|---|
| see above | Lighthouse Lab in Glasgow | Wellcome Sanger Institute for the COVID-19 Genomics UK (COG-UK) Consortium | Anna Dominiczak and Alex Alderton; Carol Clugston; Cordelia Langford; David Gray; David K. Jackson; Dominic Kwiatkowski; Ewan Harrison; Harper VanSteenhouse; Ian Johnston; Jeffrey Barrett; John Sillitoe on behalf of the Wellcome Sanger Institute COVID-19 Surveillance Team; Roberto Amato; Sonia Goncalves; Yumi Kasai |

EPI_ISL_1790724, EPI_ISL_1790833, EPI_ISL_1790836, EPI_ISL_1790903, EPI_ISL_1790907, EPI_ISL_1790977, EPI_ISL_1791007, EPI_ISL_1806184, EPI_ISL_1806222, EPI_ISL_1806272, EPI_ISL_1806294, EPI_ISL_1806315, EPI_ISL_1806327, EPI_ISL_1806342, EPI_ISL_1806363, EPI_ISL_1806367, EPI_ISL_1806379, EPI_ISL_1806380, EPI_ISL_1806393, EPI_ISL_1806406, EPI_ISL_1806407, EPI_ISL_1806450,
EPI_ISL_1806506, EPI_ISL_1806524, EPI_ISL_1806569, EPI_ISL_1806587, EPI_ISL_1806596, EPI_ISL_1806606, EPI_ISL_1806611, EPI_ISL_1806632, EPI_ISL_1806636, EPI_ISL_1806646, EPI_ISL_1806652, EPI_ISL_1806673, EPI_ISL_1806681, EPI_ISL_1806687, EPI_ISL_1806709, EPI_ISL_1806725, EPI_ISL_1806735, EPI_ISL_1806753, EPI_ISL_1806767, EPI_ISL_1806821,
EPI_ISL_1806828, EPI_ISL_1806830, EPI_ISL_1806840, EPI_ISL_1806847, EPI_ISL_1829207, EPI_ISL_1829210, EPI_ISL_1829218, EPI_ISL_1829219, EPI_ISL_1829221, EPI_ISL_1829224, EPI_ISL_1829225, EPI_ISL_1829226, EPI_ISL_1829228, EPI_ISL_1829230, EPI_ISL_1829231, EPI_ISL_1829234, EPI_ISL_1829236, EPI_ISL_1829238, EPI_ISL_1829240, EPI_ISL_1829241,
EPI_ISL_1829244, EPI_ISL_1829246, EPI_ISL_1829247, EPI_ISL_1829248, EPI_ISL_1829249, EPI_ISL_1829250, EPI_ISL_1829251, EPI_ISL_1829253, EPI_ISL_1829256, EPI_ISL_1829257, EPI_ISL_1829259, EPI_ISL_1829261, EPI_ISL_1829263, EPI_ISL_1829265, EPI_ISL_1829266, EPI_ISL_1829267, EPI_ISL_1829268, EPI_ISL_1829270, EPI_ISL_1829275, EPI_ISL_1829277, EPI_ISL_1829280, EPI_ISL_1829283,
EPI_ISL_1829284, EPI_ISL_1829286, EPI_ISL_1829288, EPI_ISL_1829290, EPI_ISL_1829293, EPI_ISL_1829294, EPI_ISL_1829295, EPI_ISL_1829298, EPI_ISL_1829301, EPI_ISL_1829303, EPI_ISL_1829305, EPI_ISL_1829306, EPI_ISL_1829308, EPI_ISL_1829309, EPI_ISL_1829312, EPI_ISL_1829313, EPI_ISL_1829314, EPI_ISL_1829317, EPI_ISL_1829321, EPI_ISL_1829322, EPI_ISL_1829323, EPI_ISL_1829324,
EPI_ISL_1829326, EPI_ISL_1829328, EPI_ISL_1829329, EPI_ISL_1829330, EPI_ISL_1829331, EPI_ISL_1829332, EPI_ISL_1829334, EPI_ISL_1829335, EPI_ISL_1829336, EPI_ISL_1829338, EPI_ISL_1829339, EPI_ISL_1829340, EPI_ISL_1829341, EPI_ISL_1829343, EPI_ISL_1829346, EPI_ISL_1829347, EPI_ISL_1829348, EPI_ISL_1829349, EPI_ISL_1829350, EPI_ISL_1829351, EPI_ISL_1829354, EPI_ISL_1829355,
EPI_ISL_1829358, EPI_ISL_1829361, EPI_ISL_1829364, EPI_ISL_1829365, EPI_ISL_1829366, EPI_ISL_1829367, EPI_ISL_1829368, EPI_ISL_1829369, EPI_ISL_1829370, EPI_ISL_1829371, EPI_ISL_1829377, EPI_ISL_1829378, EPI_ISL_1829379, EPI_ISL_1829380, EPI_ISL_1829381, EPI_ISL_1829383, EPI_ISL_1829384, EPI_ISL_1829385, EPI_ISL_1829386, EPI_ISL_1829387, EPI_ISL_1829388, EPI_ISL_1829389,
EPI_ISL_1829390, EPI_ISL_1829391, EPI_ISL_1829392, EPI_ISL_1829393, EPI_ISL_1829394, EPI_ISL_1829395, EPI_ISL_1829396, EPI_ISL_1829397, EPI_ISL_1829398, EPI_ISL_1829399, EPI_ISL_1829401, EPI_ISL_1829402, EPI_ISL_1829404, EPI_ISL_1829405, EPI_ISL_1829407, EPI_ISL_1829409, EPI_ISL_1829410, EPI_ISL_1829414, EPI_ISL_1829415, EPI_ISL_1829416, EPI_ISL_1829417, EPI_ISL_1829418,
EPI_ISL_1829422, EPI_ISL_1829423, EPI_ISL_1829424, EPI_ISL_1829425, EPI_ISL_1829426, EPI_ISL_1829428, EPI_ISL_1829429, EPI_ISL_1829431, EPI_ISL_1829436, EPI_ISL_1829439, EPI_ISL_1829441, EPI_ISL_1829443, EPI_ISL_1829445, EPI_ISL_1829446, EPI_ISL_1829447, EPI_ISL_1829448, EPI_ISL_1829449, EPI_ISL_1829450, EPI_ISL_1829451, EPI_ISL_1829453, EPI_ISL_1829454, EPI_ISL_1829455,
EPI_ISL_1829459, EPI_ISL_1829463, EPI_ISL_1829464, EPI_ISL_1829465, EPI_ISL_1829466, EPI_ISL_1829467, EPI_ISL_1829468, EPI_ISL_1829469, EPI_ISL_1829470, EPI_ISL_1829471, EPI_ISL_1829472, EPI_ISL_1829473, EPI_ISL_1829476, EPI_ISL_1829477, EPI_ISL_1829478, EPI_ISL_1829479, EPI_ISL_1829480, EPI_ISL_1829481, EPI_ISL_1829482, EPI_ISL_1829483, EPI_ISL_1829488,
EPI_ISL_1829511, EPI_ISL_1829513, EPI_ISL_1829519, EPI_ISL_1829522, EPI_ISL_1829527, EPI_ISL_1829528, EPI_ISL_1829530, EPI_ISL_1829536, EPI_ISL_1829557, EPI_ISL_1829566, EPI_ISL_1829572, EPI_ISL_1829574, EPI_ISL_1829579, EPI_ISL_1829586, EPI_ISL_1829587, EPI_ISL_1829632, EPI_ISL_1829635, EPI_ISL_1829656, EPI_ISL_1829668, EPI_ISL_1829680, EPI_ISL_1829683, EPI_ISL_1829689,
EPI_ISL_1829705, EPI_ISL_1829710, EPI_ISL_1829713, EPI_ISL_1829729, EPI_ISL_1829742, EPI_ISL_1829744, EPI_ISL_1829750, EPI_ISL_1829763, EPI_ISL_1829770, EPI_ISL_1829771, EPI_ISL_1829774, EPI_ISL_1829777, EPI_ISL_1829788, EPI_ISL_1829800, EPI_ISL_1830104, EPI_ISL_1830162, EPI_ISL_1830180, EPI_ISL_1830257, EPI_ISL_1830323, EPI_ISL_1830333, EPI_ISL_1830389, EPI_ISL_1830390,
EPI_ISL_1830394, EPI_ISL_1830400, EPI_ISL_1830401, EPI_ISL_1830403, EPI_ISL_1830404, EPI_ISL_1830406, EPI_ISL_1830408, EPI_ISL_1830410, EPI_ISL_1830411, EPI_ISL_1830412, EPI_ISL_1830413, EPI_ISL_1830417, EPI_ISL_1830419, EPI_ISL_1830423, EPI_ISL_1830424, EPI_ISL_1830431, EPI_ISL_1830432, EPI_ISL_1830437, EPI_ISL_1830438, EPI_ISL_1830447, EPI_ISL_1830449, EPI_ISL_1830450,
EPI_ISL_1830454, EPI_ISL_1830457, EPI_ISL_1830459, EPI_ISL_1830472, EPI_ISL_1830473, EPI_ISL_1830481, EPI_ISL_1830482, EPI_ISL_1830483, EPI_ISL_1830485, EPI_ISL_1830489, EPI_ISL_1830492, EPI_ISL_1830495, EPI_ISL_1830498, EPI_ISL_1830499, EPI_ISL_1830501, EPI_ISL_1830503, EPI_ISL_1830506, EPI_ISL_1830508, EPI_ISL_1830510, EPI_ISL_1830511, EPI_ISL_1830512, EPI_ISL_1830514,
EPI_ISL_1830515, EPI_ISL_1830518, EPI_ISL_1830524, EPI_ISL_1830526, EPI_ISL_1830531, EPI_ISL_1830537, EPI_ISL_1830542, EPI_ISL_1830545, EPI_ISL_1830555, EPI_ISL_1830559, EPI_ISL_1830562, EPI_ISL_1830563, EPI_ISL_1830564, EPI_ISL_1830568, EPI_ISL_1830572, EPI_ISL_1830574, EPI_ISL_1830576, EPI_ISL_1830578, EPI_ISL_1830579, EPI_ISL_1830585, EPI_ISL_1830589, EPI_ISL_1830594,
EPI_ISL_1830597, EPI_ISL_1830601, EPI_ISL_1830604, EPI_ISL_1830606, EPI_ISL_1830607, EPI_ISL_1830614, EPI_ISL_1830619, EPI_ISL_1830623, EPI_ISL_1830624, EPI_ISL_1830626, EPI_ISL_1830630, EPI_ISL_1830631, EPI_ISL_1830633, EPI_ISL_1830635, EPI_ISL_1830637, EPI_ISL_1830648, EPI_ISL_1830649, EPI_ISL_1830650, EPI_ISL_1830651, EPI_ISL_1830652, EPI_ISL_1830654, EPI_ISL_1830656,
EPI_ISL_1830659, EPI_ISL_1830661, EPI_ISL_1830665, EPI_ISL_1830667, EPI_ISL_1830671, EPI_ISL_1830673, EPI_ISL_1830675, EPI_ISL_1830676, EPI_ISL_1830677, EPI_ISL_1830679, EPI_ISL_1830681, EPI_ISL_1830682, EPI_ISL_1830688, EPI_ISL_1830689, EPI_ISL_1830692, EPI_ISL_1830693, EPI_ISL_1830695, EPI_ISL_1830704, EPI_ISL_1830706, EPI_ISL_1830710, EPI_ISL_1830712, EPI_ISL_1830713,
EPI_ISL_1830714, EPI_ISL_1830715, EPI_ISL_1830716, EPI_ISL_1830718, EPI_ISL_1830719, EPI_ISL_1830720, EPI_ISL_1830722, EPI_ISL_1830725, EPI_ISL_1830726, EPI_ISL_1830727, EPI_ISL_1830813, EPI_ISL_1831087, EPI_ISL_1857827, EPI_ISL_1857831, EPI_ISL_1857842, EPI_ISL_1857846, EPI_ISL_1857847, EPI_ISL_1857848, EPI_ISL_1857849, EPI_ISL_1857851, EPI_ISL_1857852, EPI_ISL_1857855,
EPI_ISL_1857858, EPI_ISL_1857861, EPI_ISL_1857862, EPI_ISL_1857866, EPI_ISL_1857870, EPI_ISL_1857871, EPI_ISL_1857873, EPI_ISL_1857874, EPI_ISL_1857884, EPI_ISL_1857895, EPI_ISL_1857897, EPI_ISL_1857899, EPI_ISL_1857903, EPI_ISL_1857905, EPI_ISL_1857906, EPI_ISL_1857915, EPI_ISL_1857918, EPI_ISL_1857919, EPI_ISL_1857924, EPI_ISL_1857931, EPI_ISL_1857935, EPI_ISL_1857938,
EPI_ISL_1857940, EPI_ISL_1857944, EPI_ISL_1857946, EPI_ISL_1857953, EPI_ISL_1857958, EPI_ISL_1857959, EPI_ISL_1857965, EPI_ISL_1857966, EPI_ISL_1857979, EPI_ISL_1857999, EPI_ISL_1858004, EPI_ISL_1858008, EPI_ISL_1858017, EPI_ISL_1858024, EPI_ISL_1858043, EPI_ISL_1858059, EPI_ISL_1858062, EPI_ISL_1858091, EPI_ISL_1858111, EPI_ISL_1858118, EPI_ISL_1858129,
EPI_ISL_1858131, EPI_ISL_1858141, EPI_ISL_1858144, EPI_ISL_1858154, EPI_ISL_1858202, EPI_ISL_1858215, EPI_ISL_1858236, EPI_ISL_1858239, EPI_ISL_1912003, EPI_ISL_1912004, EPI_ISL_1912005, EPI_ISL_1912006, EPI_ISL_1912009, EPI_ISL_1912010, EPI_ISL_1912011, EPI_ISL_1912012, EPI_ISL_1912013, EPI_ISL_1912018, EPI_ISL_1912020, EPI_ISL_1912021, EPI_ISL_1912022, EPI_ISL_1912030,
EPI_ISL_1912031, EPI_ISL_1912032, EPI_ISL_1912033, EPI_ISL_1912034, EPI_ISL_1912035, EPI_ISL_1912037, EPI_ISL_1912039, EPI_ISL_1912044, EPI_ISL_1912045, EPI_ISL_1912047, EPI_ISL_1912048, EPI_ISL_1912050, EPI_ISL_1912051, EPI_ISL_1912052, EPI_ISL_1912053, EPI_ISL_1912055, EPI_ISL_1912058, EPI_ISL_1912059, EPI_ISL_1912061, EPI_ISL_1912071, EPI_ISL_1912072, EPI_ISL_1912073,
EPI_ISL_1912075, EPI_ISL_1912081, EPI_ISL_1912082, EPI_ISL_1912084, EPI_ISL_1912094, EPI_ISL_1912096, EPI_ISL_1912103, EPI_ISL_1912105, EPI_ISL_1912108, EPI_ISL_1912109, EPI_ISL_1912110, EPI_ISL_1912113, EPI_ISL_1912116, EPI_ISL_1912117, EPI_ISL_1912118, EPI_ISL_1912125, EPI_ISL_1912126, EPI_ISL_1912129, EPI_ISL_1912131, EPI_ISL_1912132, EPI_ISL_1912140, EPI_ISL_1912141,
EPI_ISL_1912143, EPI_ISL_1912144, EPI_ISL_1912145, EPI_ISL_1912149, EPI_ISL_1912150, EPI_ISL_1912151, EPI_ISL_1912152, EPI_ISL_1912153, EPI_ISL_1912154, EPI_ISL_1912158, EPI_ISL_1912159, EPI_ISL_1912164, EPI_ISL_1912165, EPI_ISL_1912168, EPI_ISL_1912171, EPI_ISL_1912172, EPI_ISL_1912177, EPI_ISL_1912179, EPI_ISL_1912186, EPI_ISL_1912189, EPI_ISL_1912192, EPI_ISL_1912195,
EPI_ISL_1912196, EPI_ISL_1912199, EPI_ISL_1912207, EPI_ISL_1912211, EPI_ISL_1912212, EPI_ISL_1912215, EPI_ISL_1912217, EPI_ISL_1912221, EPI_ISL_1912232, EPI_ISL_1912234, EPI_ISL_1912242, EPI_ISL_1912243, EPI_ISL_1912245, EPI_ISL_1912249, EPI_ISL_1912256, EPI_ISL_1912257, EPI_ISL_1912264, EPI_ISL_1912265, EPI_ISL_1912270, EPI_ISL_1912272, EPI_ISL_1912276, EPI_ISL_1912281,
EPI_ISL_1912284, EPI_ISL_1912291, EPI_ISL_1912295, EPI_ISL_1912298, EPI_ISL_1912301, EPI_ISL_1912304, EPI_ISL_1912311, EPI_ISL_1912316, EPI_ISL_1912319, EPI_ISL_1912323, EPI_ISL_1912327, EPI_ISL_1912331, EPI_ISL_1912335, EPI_ISL_1912336, EPI_ISL_1912339, EPI_ISL_1912341, EPI_ISL_1912343, EPI_ISL_1912345, EPI_ISL_1912349, EPI_ISL_1912752, EPI_ISL_1912753,
EPI_ISL_1912754, EPI_ISL_1912755, EPI_ISL_1912756, EPI_ISL_1912757, EPI_ISL_1912758, EPI_ISL_1921439, EPI_ISL_1921442, EPI_ISL_1921445, EPI_ISL_1921447, EPI_ISL_1921448, EPI_ISL_1921450, EPI_ISL_1921451, EPI_ISL_1921452, EPI_ISL_1921453, EPI_ISL_1921454, EPI_ISL_1921455, EPI_ISL_1921456, EPI_ISL_1921457, EPI_ISL_1921458, EPI_ISL_1921460, EPI_ISL_1921461, EPI_ISL_1921462,
EPI_ISL_1921464, EPI_ISL_1921466, EPI_ISL_1921469, EPI_ISL_1921470, EPI_ISL_1921471, EPI_ISL_1921472, EPI_ISL_1921473, EPI_ISL_1921474, EPI_ISL_1921475, EPI_ISL_1921476, EPI_ISL_1921477, EPI_ISL_1921478, EPI_ISL_1921479, EPI_ISL_1921480, EPI_ISL_1921481, EPI_ISL_1921482, EPI_ISL_1921483, EPI_ISL_1921487, EPI_ISL_1921491, EPI_ISL_1921492,
EPI_ISL_1921493, EPI_ISL_1921497, EPI_ISL_1921498, EPI_ISL_1921499, EPI_ISL_1921500, EPI_ISL_1921501, EPI_ISL_1921503, EPI_ISL_1921504, EPI_ISL_1921505, EPI_ISL_1921508, EPI_ISL_1921510, EPI_ISL_1921511, EPI_ISL_1921512, EPI_ISL_1921514, EPI_ISL_1921516, EPI_ISL_1921517, EPI_ISL_1921518, EPI_ISL_1921519, EPI_ISL_1921521, EPI_ISL_1921524, EPI_ISL_1921526, EPI_ISL_1921527,
EPI_ISL_1921528, EPI_ISL_1921529, EPI_ISL_1921530, EPI_ISL_1921531, EPI_ISL_1921532, EPI_ISL_1921534, EPI_ISL_1921535, EPI_ISL_1921537, EPI_ISL_1921539, EPI_ISL_1921540, EPI_ISL_1921541, EPI_ISL_1921542, EPI_ISL_1921543, EPI_ISL_1921544, EPI_ISL_1921546, EPI_ISL_1921547, EPI_ISL_1921548, EPI_ISL_1921549, EPI_ISL_1921550, EPI_ISL_1921551, EPI_ISL_1921554,
EPI_ISL_1921556, EPI_ISL_1921560, EPI_ISL_1921562, EPI_ISL_1921563, EPI_ISL_1921565, EPI_ISL_1921566, EPI_ISL_1921569, EPI_ISL_1921570, EPI_ISL_1921571, EPI_ISL_1921573, EPI_ISL_1921574, EPI_ISL_1921576, EPI_ISL_1921577, EPI_ISL_1921578, EPI_ISL_1921579, EPI_ISL_1921581, EPI_ISL_1921584, EPI_ISL_1921585, EPI_ISL_1921586,
EPI_ISL_1921587, EPI_ISL_1921589, EPI_ISL_1921591, EPI_ISL_1921592, EPI_ISL_1921593, EPI_ISL_1921594, EPI_ISL_1921595, EPI_ISL_1921599, EPI_ISL_1921601, EPI_ISL_1921602, EPI_ISL_1921603, EPI_ISL_1921605, EPI_ISL_1921606, EPI_ISL_1921607, EPI_ISL_1921608, EPI_ISL_1921609, EPI_ISL_1921610, EPI_ISL_1921611, EPI_ISL_1921612, EPI_ISL_1921614, EPI_ISL_1921615, EPI_ISL_1921616,

EPI_ISL_1921617, EPI_ISL_1921620, EPI_ISL_1921621, EPI_ISL_1921622, EPI_ISL_1921623, EPI_ISL_1921624, EPI_ISL_1921625, EPI_ISL_1921628, EPI_ISL_1921629, EPI_ISL_1921630, EPI_ISL_1921632, EPI_ISL_1921633, EPI_ISL_1921635, EPI_ISL_1921636, EPI_ISL_1921638, EPI_ISL_1921639, EPI_ISL_1921640, EPI_ISL_1921641, EPI_ISL_1921642, EPI_ISL_1921643, EPI_ISL_1921644, EPI_ISL_1921646, EPI_ISL_1921647, EPI_ISL_1921649, EPI_ISL_1921650, EPI_ISL_1921651, EPI_ISL_1921652, EPI_ISL_1921653, EPI_ISL_1921654, EPI_ISL_1921655, EPI_ISL_1921657, EPI_ISL_1921658, EPI_ISL_1921659, EPI_ISL_1921661, EPI_ISL_1921663, EPI_ISL_1921664, EPI_ISL_1921665, EPI_ISL_1921668, EPI_ISL_1921670, EPI_ISL_1921671, EPI_ISL_1921672, EPI_ISL_1921674, EPI_ISL_1921675, EPI_ISL_1921677, EPI_ISL_1921678, EPI_ISL_1921680, EPI_ISL_1921681, EPI_ISL_1921682, EPI_ISL_1921683, EPI_ISL_1921687, EPI_ISL_1921688, EPI_ISL_1921689, EPI_ISL_1921690, EPI_ISL_1921692, EPI_ISL_1921693, EPI_ISL_1921694, EPI_ISL_1921695, EPI_ISL_1921696, EPI_ISL_1921697, EPI_ISL_1921701, EPI_ISL_1921704, EPI_ISL_1921705, EPI_ISL_1921706, EPI_ISL_1921707, EPI_ISL_1921708, EPI_ISL_1921710, EPI_ISL_1921712, EPI_ISL_1921714, EPI_ISL_1921715, EPI_ISL_1921716, EPI_ISL_1921718, EPI_ISL_1921719, EPI_ISL_1921720, EPI_ISL_1921721, EPI_ISL_1921723, EPI_ISL_1921724, EPI_ISL_1921726, EPI_ISL_1921727, EPI_ISL_1921728, EPI_ISL_1921729, EPI_ISL_1921730, EPI_ISL_1921733, EPI_ISL_1921734, EPI_ISL_1921736, EPI_ISL_1939708, EPI_ISL_1939709, EPI_ISL_1939710, EPI_ISL_1939711, EPI_ISL_1939712, EPI_ISL_1939713, EPI_ISL_1939714, EPI_ISL_1939715, EPI_ISL_1939716, EPI_ISL_1939717, EPI_ISL_1939718, EPI_ISL_1985343, EPI_ISL_1985455, EPI_ISL_1985460, EPI_ISL_1985485, EPI_ISL_1985513, EPI_ISL_1985514, EPI_ISL_1985515, EPI_ISL_1985516, EPI_ISL_1985518, EPI_ISL_1985519, EPI_ISL_1985520, EPI_ISL_1985521, EPI_ISL_1985522, EPI_ISL_1985523, EPI_ISL_1985525, EPI_ISL_1985526, EPI_ISL_1985527, EPI_ISL_1985528, EPI_ISL_1985529, EPI_ISL_1985531, EPI_ISL_1985532, EPI_ISL_1985533, EPI_ISL_1985534, EPI_ISL_1985535, EPI_ISL_1985536, EPI_ISL_1985538, EPI_ISL_1985539, EPI_ISL_1985540, EPI_ISL_1985541, EPI_ISL_1985543, EPI_ISL_1985544, EPI_ISL_1985545, EPI_ISL_1985546, EPI_ISL_1985547, EPI_ISL_1985548, EPI_ISL_1985549, EPI_ISL_1985551, EPI_ISL_1985552, EPI_ISL_1985553, EPI_ISL_1985554, EPI_ISL_1985555, EPI_ISL_1985556, EPI_ISL_1985558, EPI_ISL_1985559, EPI_ISL_1985560, EPI_ISL_1985561, EPI_ISL_1985562, EPI_ISL_1985563, EPI_ISL_1985565, EPI_ISL_1985566, EPI_ISL_1985567, EPI_ISL_1985568, EPI_ISL_1985570, EPI_ISL_1985571, EPI_ISL_1985572, EPI_ISL_1985573, EPI_ISL_1985574, EPI_ISL_1985575, EPI_ISL_1985576, EPI_ISL_1985578, EPI_ISL_1985579, EPI_ISL_1985580, EPI_ISL_1985581, EPI_ISL_1985582, EPI_ISL_1985583, EPI_ISL_1985584, EPI_ISL_1985586, EPI_ISL_1985587, EPI_ISL_1985588, EPI_ISL_1985589, EPI_ISL_1985590, EPI_ISL_1985592, EPI_ISL_1985593, EPI_ISL_1985594, EPI_ISL_1985595, EPI_ISL_1985596, EPI_ISL_1985597, EPI_ISL_1985599, EPI_ISL_1985600, EPI_ISL_1985601, EPI_ISL_1985602, EPI_ISL_1985603, EPI_ISL_1985604, EPI_ISL_1985605, EPI_ISL_1985607, EPI_ISL_1985608, EPI_ISL_1985609, EPI_ISL_1985610, EPI_ISL_1985611, EPI_ISL_1985612, EPI_ISL_1985613, EPI_ISL_1985614, EPI_ISL_1985616, EPI_ISL_1985617, EPI_ISL_1985618, EPI_ISL_1985619, EPI_ISL_1985620, EPI_ISL_1985622, EPI_ISL_1985623, EPI_ISL_1985624, EPI_ISL_1985625, EPI_ISL_1985626, EPI_ISL_1985627, EPI_ISL_1985629, EPI_ISL_1985630, EPI_ISL_1985631, EPI_ISL_1985632, EPI_ISL_1985633, EPI_ISL_1985635, EPI_ISL_1985636, EPI_ISL_1985637, EPI_ISL_1985638, EPI_ISL_1985639, EPI_ISL_1985640, EPI_ISL_1985642, EPI_ISL_1985643, EPI_ISL_1985644, EPI_ISL_1985645, EPI_ISL_1985646, EPI_ISL_1985647, EPI_ISL_1985648, EPI_ISL_1985650, EPI_ISL_1985651, EPI_ISL_1985652, EPI_ISL_1985653, EPI_ISL_1985654, EPI_ISL_1985656, EPI_ISL_1985657, EPI_ISL_1985658, EPI_ISL_1985659, EPI_ISL_1985660, EPI_ISL_1985661, EPI_ISL_1985663, EPI_ISL_1985664, EPI_ISL_1985665, EPI_ISL_1985666, EPI_ISL_1985667, EPI_ISL_1985668, EPI_ISL_1985669, EPI_ISL_1985670, EPI_ISL_1985671, EPI_ISL_1985673, EPI_ISL_1985674, EPI_ISL_1985675, EPI_ISL_1985676, EPI_ISL_1985677, EPI_ISL_1985678, EPI_ISL_1985680, EPI_ISL_1985681, EPI_ISL_1985682, EPI_ISL_1985683, EPI_ISL_1985684, EPI_ISL_1985687, EPI_ISL_1985690, EPI_ISL_1985691, EPI_ISL_1985697, EPI_ISL_1985698, EPI_ISL_1985699, EPI_ISL_1985701, EPI_ISL_1985702, EPI_ISL_1985703, EPI_ISL_1985704, EPI_ISL_1985705, EPI_ISL_1985706, EPI_ISL_1985708, EPI_ISL_1985710, EPI_ISL_1985713, EPI_ISL_1985715, EPI_ISL_1985717, EPI_ISL_1985718, EPI_ISL_1985720, EPI_ISL_1985722, EPI_ISL_1985723, EPI_ISL_1985724, EPI_ISL_1985725, EPI_ISL_1985726, EPI_ISL_1985727, EPI_ISL_1985728, EPI_ISL_1985730, EPI_ISL_1985731, EPI_ISL_1985732, EPI_ISL_1985733, EPI_ISL_1985735, EPI_ISL_1985736, EPI_ISL_1985738, EPI_ISL_1985739, EPI_ISL_1985740, EPI_ISL_1985741, EPI_ISL_1985742, EPI_ISL_1985743, EPI_ISL_1985746, EPI_ISL_1985747, EPI_ISL_1985749, EPI_ISL_1985750, EPI_ISL_1985752, EPI_ISL_1985753, EPI_ISL_1985754, EPI_ISL_1985756, EPI_ISL_1985758, EPI_ISL_1985760, EPI_ISL_1985761, EPI_ISL_1985763, EPI_ISL_1985764, EPI_ISL_1985766, EPI_ISL_1985769, EPI_ISL_1985804, EPI_ISL_1985806, EPI_ISL_1985807, EPI_ISL_1985808, EPI_ISL_1985809, EPI_ISL_1985812, EPI_ISL_1985813, EPI_ISL_1985814, EPI_ISL_1985815, EPI_ISL_1985816, EPI_ISL_1985817, EPI_ISL_1985819, EPI_ISL_1985821, EPI_ISL_1985823, EPI_ISL_1985827, EPI_ISL_1985829, EPI_ISL_1985831, EPI_ISL_1985832, EPI_ISL_1985833, EPI_ISL_1985834, EPI_ISL_1985836, EPI_ISL_1985837, EPI_ISL_1985839, EPI_ISL_1985840, EPI_ISL_1985842, EPI_ISL_1985843, EPI_ISL_1985844, EPI_ISL_1985846, EPI_ISL_1985847, EPI_ISL_1985849, EPI_ISL_1985850, EPI_ISL_1985851, EPI_ISL_1985853, EPI_ISL_1985854, EPI_ISL_1985855, EPI_ISL_1985856, EPI_ISL_1985857, EPI_ISL_1985858, EPI_ISL_1985860, EPI_ISL_1985863, EPI_ISL_1985865, EPI_ISL_1985867, EPI_ISL_1985868, EPI_ISL_1985869, EPI_ISL_1985870, EPI_ISL_1985875, EPI_ISL_1985877, EPI_ISL_1985878, EPI_ISL_1985879, EPI_ISL_1985880, EPI_ISL_1985882, EPI_ISL_1985883, EPI_ISL_1985884, EPI_ISL_1985885, EPI_ISL_1985887, EPI_ISL_1985888, EPI_ISL_1985891, EPI_ISL_1985893, EPI_ISL_1985894, EPI_ISL_1985895, EPI_ISL_1985898, EPI_ISL_1985899, EPI_ISL_1985900, EPI_ISL_1985903, EPI_ISL_1985905, EPI_ISL_1985906, EPI_ISL_1985907, EPI_ISL_1985908, EPI_ISL_1985911, EPI_ISL_1985913, EPI_ISL_1985915, EPI_ISL_1985917, EPI_ISL_1985918, EPI_ISL_1985922, EPI_ISL_1985923, EPI_ISL_1985924, EPI_ISL_1985926, EPI_ISL_1985928, EPI_ISL_1985930, EPI_ISL_1985931, EPI_ISL_1985932, EPI_ISL_1985933, EPI_ISL_1985935, EPI_ISL_1985936, EPI_ISL_1985937, EPI_ISL_1985938, EPI_ISL_1985939, EPI_ISL_1985940, EPI_ISL_1985942, EPI_ISL_1985943, EPI_ISL_1985944, EPI_ISL_1985945, EPI_ISL_1985946, EPI_ISL_1985947, EPI_ISL_1985949, EPI_ISL_1985951, EPI_ISL_1985952, EPI_ISL_1985955, EPI_ISL_1985956, EPI_ISL_1985958, EPI_ISL_1985960, EPI_ISL_1985961, EPI_ISL_1985962, EPI_ISL_1985963, EPI_ISL_1985964, EPI_ISL_1985968, EPI_ISL_1985969, EPI_ISL_1985970, EPI_ISL_1985973, EPI_ISL_1985974, EPI_ISL_1985975, EPI_ISL_1985978, EPI_ISL_1985981, EPI_ISL_1985982, EPI_ISL_1985985, EPI_ISL_1985987, EPI_ISL_1985988, EPI_ISL_1985991, EPI_ISL_1985993, EPI_ISL_1985994, EPI_ISL_1985995, EPI_ISL_1985996, EPI_ISL_1985997, EPI_ISL_1985998, EPI_ISL_1986000, EPI_ISL_1986001, EPI_ISL_1986002, EPI_ISL_1986003, EPI_ISL_1986004, EPI_ISL_1986005, EPI_ISL_1986007, EPI_ISL_1986008, EPI_ISL_1986011, EPI_ISL_1986012, EPI_ISL_1986014, EPI_ISL_1986015, EPI_ISL_1986016, EPI_ISL_1986017, EPI_ISL_1986019, EPI_ISL_1986022, EPI_ISL_1986023, EPI_ISL_1986024, EPI_ISL_1986025, EPI_ISL_1986026, EPI_ISL_1986029, EPI_ISL_1986031, EPI_ISL_1986033, EPI_ISL_1986035, EPI_ISL_1986037, EPI_ISL_1986038, EPI_ISL_1986039, EPI_ISL_1986040, EPI_ISL_1986042, EPI_ISL_1986043, EPI_ISL_1986044, EPI_ISL_1986048, EPI_ISL_1986049, EPI_ISL_1986050, EPI_ISL_1986052, EPI_ISL_1986053, EPI_ISL_1986054, EPI_ISL_1986055, EPI_ISL_1986056, EPI_ISL_1986057, EPI_ISL_1986059, EPI_ISL_1986061, EPI_ISL_1986063, EPI_ISL_1986065, EPI_ISL_1986066, EPI_ISL_1986067, EPI_ISL_1986072, EPI_ISL_1986073, EPI_ISL_1986077, EPI_ISL_1986078, EPI_ISL_1986080, EPI_ISL_1986081, EPI_ISL_1986083, EPI_ISL_1986084, EPI_ISL_1986085, EPI_ISL_1986086, EPI_ISL_1986088, EPI_ISL_1986089, EPI_ISL_1986090, EPI_ISL_1986091, EPI_ISL_1986093, EPI_ISL_1986095, EPI_ISL_1986098, EPI_ISL_1986100, EPI_ISL_1986101, EPI_ISL_1986103, EPI_ISL_1986104, EPI_ISL_1986106, EPI_ISL_1986108, EPI_ISL_1986109, EPI_ISL_1986119, EPI_ISL_1986120, EPI_ISL_1986121, EPI_ISL_1986122, EPI_ISL_1986127, EPI_ISL_1986135, EPI_ISL_1986137, EPI_ISL_1986140, EPI_ISL_1986142, EPI_ISL_1986143, EPI_ISL_1986145, EPI_ISL_1986146, EPI_ISL_1986150, EPI_ISL_1986153, EPI_ISL_1986158, EPI_ISL_1986161, EPI_ISL_1986164, EPI_ISL_1986165, EPI_ISL_1986170, EPI_ISL_1986173, EPI_ISL_1986177, EPI_ISL_1986178, EPI_ISL_1986183, EPI_ISL_1986187, EPI_ISL_1986188, EPI_ISL_1986190, EPI_ISL_1986193, EPI_ISL_1986195, EPI_ISL_1986196, EPI_ISL_1986198, EPI_ISL_1986199, EPI_ISL_1986202, EPI_ISL_1986207, EPI_ISL_1986211, EPI_ISL_1986214, EPI_ISL_1986217, EPI_ISL_1986222, EPI_ISL_1986223, EPI_ISL_1986225, EPI_ISL_1986226, EPI_ISL_1986229, EPI_ISL_1986232, EPI_ISL_1986234, EPI_ISL_1986236, EPI_ISL_1986239, EPI_ISL_1986240, EPI_ISL_1986243, EPI_ISL_1986244, EPI_ISL_1986245, EPI_ISL_1986246, EPI_ISL_1986247, EPI_ISL_1986248, EPI_ISL_1986249, EPI_ISL_1986253, EPI_ISL_1986258, EPI_ISL_1986259, EPI_ISL_1986260, EPI_ISL_1986261, EPI_ISL_1986262, EPI_ISL_1986271, EPI_ISL_1986274, EPI_ISL_1986280, EPI_ISL_1986286, EPI_ISL_1986287, EPI_ISL_1986288, EPI_ISL_1986289, EPI_ISL_1986291, EPI_ISL_1986293, EPI_ISL_1986295, EPI_ISL_1986300, EPI_ISL_1986301, EPI_ISL_1986303, EPI_ISL_1986306, EPI_ISL_1986308, EPI_ISL_1986309, EPI_ISL_1986313, EPI_ISL_1986316, EPI_ISL_1986321, EPI_ISL_1986322, EPI_ISL_1986329, EPI_ISL_1986330, EPI_ISL_1986333, EPI_ISL_1986340, EPI_ISL_1986341, EPI_ISL_1986344, EPI_ISL_1986345, EPI_ISL_1986346, EPI_ISL_1986347, EPI_ISL_1986348, EPI_ISL_1986352, EPI_ISL_1986356, EPI_ISL_1986358, EPI_ISL_1986359, EPI_ISL_1986363, EPI_ISL_1986367, EPI_ISL_1986368, EPI_ISL_1986375, EPI_ISL_1986380, EPI_ISL_1986382, EPI_ISL_1986383, EPI_ISL_1986384, EPI_ISL_1986388, EPI_ISL_1986391, EPI_ISL_1986496, EPI_ISL_1986498, EPI_ISL_1986499, EPI_ISL_1986500, EPI_ISL_1986503, EPI_ISL_1986504, EPI_ISL_1986506, EPI_ISL_1986513, EPI_ISL_1986514, EPI_ISL_1986525, EPI_ISL_1986527, EPI_ISL_1986530, EPI_ISL_1986531, EPI_ISL_1986532, EPI_ISL_1986537, EPI_ISL_1986540, EPI_ISL_1986543, EPI_ISL_1986544, EPI_ISL_1986545, EPI_ISL_1986546, EPI_ISL_1986547, EPI_ISL_1986549, EPI_ISL_1986551, EPI_ISL_1986554, EPI_ISL_1986555, EPI_ISL_1986557, EPI_ISL_1986561, EPI_ISL_1986564, EPI_ISL_1986565, EPI_ISL_1986566, EPI_ISL_1986568, EPI_ISL_1986578, EPI_ISL_1986581, EPI_ISL_1986584, EPI_ISL_1986587, EPI_ISL_1986591, EPI_ISL_1986598, EPI_ISL_1986599, EPI_ISL_1986603, EPI_ISL_1986606, EPI_ISL_1986614, EPI_ISL_1986620, EPI_ISL_1986629, EPI_ISL_1986630, EPI_ISL_1986633, EPI_ISL_1986651, EPI_ISL_1986652, EPI_ISL_1986653, EPI_ISL_1986656, EPI_ISL_1986662, EPI_ISL_1986674, EPI_ISL_1986676, EPI_ISL_1986680, EPI_ISL_1986697, EPI_ISL_1986704, EPI_ISL_1986705, EPI_ISL_1986714, EPI_ISL_1986715, EPI_ISL_1986725, EPI_ISL_1986727, EPI_ISL_1986729, EPI_ISL_1986732, EPI_ISL_1986735, EPI_ISL_1986744, EPI_ISL_1986747, EPI_ISL_1986752, EPI_ISL_1986754, EPI_ISL_1986755, EPI_ISL_1986760, EPI_ISL_1986769, EPI_ISL_1986774, EPI_ISL_1986782, EPI_ISL_1986787, EPI_ISL_1986794, EPI_ISL_1986798, EPI_ISL_1986809, EPI_ISL_1986814, EPI_ISL_1986819, EPI_ISL_1986821, EPI_ISL_1986825, EPI_ISL_1986827, EPI_ISL_1986830, EPI_ISL_1986834, EPI_ISL_1986837, EPI_ISL_1986846, EPI_ISL_1986848, EPI_ISL_1986850, EPI_ISL_1986851, EPI_ISL_1986856, EPI_ISL_1986857, EPI_ISL_1986885, EPI_ISL_1986897, EPI_ISL_1986901, EPI_ISL_1986903, EPI_ISL_1986906, EPI_ISL_1986909, EPI_ISL_1986914, EPI_ISL_1986920, EPI_ISL_1986931, EPI_ISL_1986932, EPI_ISL_1986937, EPI_ISL_1986948, EPI_ISL_1986950, EPI_ISL_1986951, EPI_ISL_1986955, EPI_ISL_1986965, EPI_ISL_1986966, EPI_ISL_1986969, EPI_ISL_1986973, EPI_ISL_1986978, EPI_ISL_1986982, EPI_ISL_1986985, EPI_ISL_1986991, EPI_ISL_1986994, EPI_ISL_1986997, EPI_ISL_1986999, EPI_ISL_1987010, EPI_ISL_1987014, EPI_ISL_1987017, EPI_ISL_1987019, EPI_ISL_1987027, EPI_ISL_1987038, EPI_ISL_1987039, EPI_ISL_1987043, EPI_ISL_1987052, EPI_ISL_1987064, EPI_ISL_1987066, EPI_ISL_1987067, EPI_ISL_1987075, EPI_ISL_1987076, EPI_ISL_1987122, EPI_ISL_1987124, EPI_ISL_1987141, EPI_ISL_1987144, EPI_ISL_1987158, EPI_ISL_1987159, EPI_ISL_1987164, EPI_ISL_1987176, EPI_ISL_1987181, EPI_ISL_1987184, EPI_ISL_1987186, EPI_ISL_1987187, EPI_ISL_1987199, EPI_ISL_1987201, EPI_ISL_1987202, EPI_ISL_1987217, EPI_ISL_1987226, EPI_ISL_1987228, EPI_ISL_1987237, EPI_ISL_1987240, EPI_ISL_1987247, EPI_ISL_1987248, EPI_ISL_1987252, EPI_ISL_1987257, EPI_ISL_1987263, EPI_ISL_1987266, EPI_ISL_1987272, EPI_ISL_1987288, EPI_ISL_1987290, EPI_ISL_1987304, EPI_ISL_1987305, EPI_ISL_1987306, EPI_ISL_1987310, EPI_ISL_1987313, EPI_ISL_1987316, EPI_ISL_1987318, EPI_ISL_1987319, EPI_ISL_1987320, EPI_ISL_1987327, EPI_ISL_1987328, EPI_ISL_1987332, EPI_ISL_1987342, EPI_ISL_1987346, EPI_ISL_1987347, EPI_ISL_1987349, EPI_ISL_1987350, EPI_ISL_1987357, EPI_ISL_1987360, EPI_ISL_1987361, EPI_ISL_1987368, EPI_ISL_1987370, EPI_ISL_1987372, EPI_ISL_1987374, EPI_ISL_1987379, EPI_ISL_1987381, EPI_ISL_1987382, EPI_ISL_2004372, EPI_ISL_2004398, EPI_ISL_2004627, EPI_ISL_2004632, EPI_ISL_2004633, EPI_ISL_2004635, EPI_ISL_2004637, EPI_ISL_2004643, EPI_ISL_2004646, EPI_ISL_2004648, EPI_ISL_2004651, EPI_ISL_2004652, EPI_ISL_2004653, EPI_ISL_2004657, EPI_ISL_2004660, EPI_ISL_2004662, EPI_ISL_2004667, EPI_ISL_2004668, EPI_ISL_2004670, EPI_ISL_2004674, EPI_ISL_2004675, EPI_ISL_2004676, EPI_ISL_2004677, EPI_ISL_2004679, EPI_ISL_2004680, EPI_ISL_2004682, EPI_ISL_2004685, EPI_ISL_2004687, EPI_ISL_2004691, EPI_ISL_2004693, EPI_ISL_2004694, EPI_ISL_2004695, EPI_ISL_2004697, EPI_ISL_2004701, EPI_ISL_2004702, EPI_ISL_2004703, EPI_ISL_2004709, EPI_ISL_2004713, EPI_ISL_2004715, EPI_ISL_2004717, EPI_ISL_2004724, EPI_ISL_2004726, EPI_ISL_2004728, EPI_ISL_2004729, EPI_ISL_2004734, EPI_ISL_2004735, EPI_ISL_2004738, EPI_ISL_2004743, EPI_ISL_2004744, EPI_ISL_2004745, EPI_ISL_2004748, EPI_ISL_2004749, EPI_ISL_2004751, EPI_ISL_2004752, EPI_ISL_2004753, EPI_ISL_2004755, EPI_ISL_2004758, EPI_ISL_2004759, EPI_ISL_2004762, EPI_ISL_2004764, EPI_ISL_2004767, EPI_ISL_2004768, EPI_ISL_2004769, EPI_ISL_2004773, EPI_ISL_2004775, EPI_ISL_2004783, EPI_ISL_2004787, EPI_ISL_2004788, EPI_ISL_2004789, EPI_ISL_2004790, EPI_ISL_2004792, EPI_ISL_2004794, EPI_ISL_2004795, EPI_ISL_2004796, EPI_ISL_2004798, EPI_ISL_2004799, EPI_ISL_2004805, EPI_ISL_2004806, EPI_ISL_2004807, EPI_ISL_2004810, EPI_ISL_2004812, EPI_ISL_2004813, EPI_ISL_2004815, EPI_ISL_2004816, EPI_ISL_2004817, EPI_ISL_2004818, EPI_ISL_2004823, EPI_ISL_2004824, EPI_ISL_2004830, EPI_ISL_2004833, EPI_ISL_2004836, EPI_ISL_2004837, EPI_ISL_2004842, EPI_ISL_2004846, EPI_ISL_2004847, EPI_ISL_2004849, EPI_ISL_2004850, EPI_ISL_2004852, EPI_ISL_2004854, EPI_ISL_2004860, EPI_ISL_2004862, EPI_ISL_2004864, EPI_ISL_2004865, EPI_ISL_2004868, EPI_ISL_2004871, EPI_ISL_2004876, EPI_ISL_2004878, EPI_ISL_2005269, EPI_ISL_2005341, EPI_ISL_2005343, EPI_ISL_2005355, EPI_ISL_2005359, EPI_ISL_2005361, EPI_ISL_2005365, EPI_ISL_2005366, EPI_ISL_2005367, EPI_ISL_2005370, EPI_ISL_2005372, EPI_ISL_2005373, EPI_ISL_2005375, EPI_ISL_2005377, EPI_ISL_2005380, EPI_ISL_2005381, EPI_ISL_2005386, EPI_ISL_2005387, EPI_ISL_2005388, EPI_ISL_2005389, EPI_ISL_2005394, EPI_ISL_2005400, EPI_ISL_2005406, EPI_ISL_2005412, EPI_ISL_2005414, EPI_ISL_2005421, EPI_ISL_2005423, EPI_ISL_2005429, EPI_ISL_2005434, EPI_ISL_2005439, EPI_ISL_2005441, EPI_ISL_2005444, EPI_ISL_2005445, EPI_ISL_2005446, EPI_ISL_2005447, EPI_ISL_2005449, EPI_ISL_2005451, EPI_ISL_2005457, EPI_ISL_2005458, EPI_ISL_2005459, EPI_ISL_2005460, EPI_ISL_2005461, EPI_ISL_2005463, EPI_ISL_2005466, EPI_ISL_2005467, EPI_ISL_2005472, EPI_ISL_2005477, EPI_ISL_2005479, EPI_ISL_2005481, EPI_ISL_2005484, EPI_ISL_2005489, EPI_ISL_2005495, EPI_ISL_2005502, EPI_ISL_2005503, EPI_ISL_2005506, EPI_ISL_2005507, EPI_ISL_2005509, EPI_ISL_2005510, EPI_ISL_2005511, EPI_ISL_2005512, EPI_ISL_2005514, EPI_ISL_2005517, EPI_ISL_2005518, EPI_ISL_2005523, EPI_ISL_2005525, EPI_ISL_2005529, EPI_ISL_2005537, EPI_ISL_2005553, EPI_ISL_2005555, EPI_ISL_2005568, EPI_ISL_2005578, EPI_ISL_2005579, EPI_ISL_2005581, EPI_ISL_2005597, EPI_ISL_2005600, EPI_ISL_2005601, EPI_ISL_2005605, EPI_ISL_2005606, EPI_ISL_2005609, EPI_ISL_2005612, EPI_ISL_2005676, EPI_ISL_2005685, EPI_ISL_2005725, EPI_ISL_2005754, EPI_ISL_2005757, EPI_ISL_2005796, EPI_ISL_2005799, EPI_ISL_2005810, EPI_ISL_2005821, EPI_ISL_2005824, EPI_ISL_2005833, EPI_ISL_2005835, EPI_ISL_2005842, EPI_ISL_2005843, EPI_ISL_2005844, EPI_ISL_2005847, EPI_ISL_2005856, EPI_ISL_2005862, EPI_ISL_2005869, EPI_ISL_2005871, EPI_ISL_2005875, EPI_ISL_2005876, EPI_ISL_2005878, EPI_ISL_2005879, EPI_ISL_2005881, EPI_ISL_2005882, EPI_ISL_2005887, EPI_ISL_2005889, EPI_ISL_2005891, EPI_ISL_2005894, EPI_ISL_2005896, EPI_ISL_2005904, EPI_ISL_2005910, EPI_ISL_2005912, EPI_ISL_2006055, EPI_ISL_2005925, EPI_ISL_2005936, EPI_ISL_2005938, EPI_ISL_2005954, EPI_ISL_2005959, EPI_ISL_2005960, EPI_ISL_2005961, EPI_ISL_2005969, EPI_ISL_2005970, EPI_ISL_2005986, EPI_ISL_2005994, EPI_ISL_2006003, EPI_ISL_2006015, EPI_ISL_2006016, EPI_ISL_2006027, EPI_ISL_2006029, EPI_ISL_2006031, EPI_ISL_2006032, EPI_ISL_2006036, EPI_ISL_2006045, EPI_ISL_2006051, EPI_ISL_2006052, EPI_ISL_2006055, EPI_ISL_2006057, EPI_ISL_2006061, EPI_ISL_2006067, EPI_ISL_2006344, EPI_ISL_2006458, EPI_ISL_2006463, EPI_ISL_2006522, EPI_ISL_2006524, EPI_ISL_2006525, EPI_ISL_2006526, EPI_ISL_2006527, EPI_ISL_2006529, EPI_ISL_2006532, EPI_ISL_2006533, EPI_ISL_2006534, EPI_ISL_2006539, EPI_ISL_2006540, EPI_ISL_2006667, EPI_ISL_2006668, EPI_ISL_2021041, EPI_ISL_2021046, EPI_ISL_2021047, EPI_ISL_2021055, EPI_ISL_2021062, EPI_ISL_2021063, EPI_ISL_2021065, EPI_ISL_2021067, EPI_ISL_2021068, EPI_ISL_2021071, EPI_ISL_2021073, EPI_ISL_2021074, EPI_ISL_2021081, EPI_ISL_2021086, EPI_ISL_2021087, EPI_ISL_2021097, EPI_ISL_2021099, EPI_ISL_2021104, EPI_ISL_2021107, EPI_ISL_2021109, EPI_ISL_2021115, EPI_ISL_2021120, EPI_ISL_2021123, EPI_ISL_2021128, EPI_ISL_2021130, EPI_ISL_2021132, EPI_ISL_2021133, EPI_ISL_2021136, EPI_ISL_2021139, EPI_ISL_2021141, EPI_ISL_2021149, EPI_ISL_2021157, EPI_ISL_2021167, EPI_ISL_2021171, EPI_ISL_2021182, EPI_ISL_2021185, EPI_ISL_2021191, EPI_ISL_2021219, EPI_ISL_2021221, EPI_ISL_2021224, EPI_ISL_2021227, EPI_ISL_2021263, EPI_ISL_2021267, EPI_ISL_2021268, EPI_ISL_2021270, EPI_ISL_2021278, EPI_ISL_2021285, EPI_ISL_2021293, EPI_ISL_2021294, EPI_ISL_2021301, EPI_ISL_2021309, EPI_ISL_2021334, EPI_ISL_2021351, EPI_ISL_2021352, EPI_ISL_2021356, EPI_ISL_2021371, EPI_ISL_2021381, EPI_ISL_2021384, EPI_ISL_2021391, EPI_ISL_2021404, EPI_ISL_2021406, EPI_ISL_2021407, EPI_ISL_2021409, EPI_ISL_2021412, EPI_ISL_2021416, EPI_ISL_2021417, EPI_ISL_2021424, EPI_ISL_2021429, EPI_ISL_2021433, EPI_ISL_2021435, EPI_ISL_2021436, EPI_ISL_2021438, EPI_ISL_2021440, EPI_ISL_2021441, EPI_ISL_2021442, EPI_ISL_2021445, EPI_ISL_2021447, EPI_ISL_2021452, EPI_ISL_2021454, EPI_ISL_2021458, EPI_ISL_2021471, EPI_ISL_2021479, EPI_ISL_2021480, EPI_ISL_2021481, EPI_ISL_2021495, EPI_ISL_2021501, EPI_ISL_2021502, EPI_ISL_2021504, EPI_ISL_2021506, EPI_ISL_2021507, EPI_ISL_2021510, EPI_ISL_2021517, EPI_ISL_2021518, EPI_ISL_2021522, EPI_ISL_2021534, EPI_ISL_2021535, EPI_ISL_2021538, EPI_ISL_2021539, EPI_ISL_2021545, EPI_ISL_2021551, EPI_ISL_2021555, EPI_ISL_2021556, EPI_ISL_2021557, EPI_ISL_2021566, EPI_ISL_2021567, EPI_ISL_2021571, EPI_ISL_2021577, EPI_ISL_2021578, EPI_ISL_2021580, EPI_ISL_2021584, EPI_ISL_2021586, EPI_ISL_2021587, EPI_ISL_2021588, EPI_ISL_2021590, EPI_ISL_2021595, EPI_ISL_2021597, EPI_ISL_2021611, EPI_ISL_2021626, EPI_ISL_2021630, EPI_ISL_2021632, EPI_ISL_2021642, EPI_ISL_2021645, EPI_ISL_2021646, EPI_ISL_2021655, EPI_ISL_2021666, EPI_ISL_2021668, EPI_ISL_2021673, EPI_ISL_2021676, EPI_ISL_2021677, EPI_ISL_2021678, EPI_ISL_2021681, EPI_ISL_2021685, EPI_ISL_2021686, EPI_ISL_2021687, EPI_ISL_2021688, EPI_ISL_2021691, EPI_ISL_2021695, EPI_ISL_2021696, EPI_ISL_2021700, EPI_ISL_2021702, EPI_ISL_2021705, EPI_ISL_2021706, EPI_ISL_2021708, EPI_ISL_2021709, EPI_ISL_2021716, EPI_ISL_2021718, EPI_ISL_2021720, EPI_ISL_2021723, EPI_ISL_2021724, EPI_ISL_2021730, EPI_ISL_2021731, EPI_ISL_2021732, EPI_ISL_2021738, EPI_ISL_2021741, EPI_ISL_2021744, EPI_ISL_2021753, EPI_ISL_2021761, EPI_ISL_2021762, EPI_ISL_2021766, EPI_ISL_2021772, EPI_ISL_2021781, EPI_ISL_2021783, EPI_ISL_2021787, EPI_ISL_2021790, EPI_ISL_2021795, EPI_ISL_2021796, EPI_ISL_2021799, EPI_ISL_2021800, EPI_ISL_2021804, EPI_ISL_2021810, EPI_ISL_2021817, EPI_ISL_2021821, EPI_ISL_2021824, EPI_ISL_2021827, EPI_ISL_2021830, EPI_ISL_2021832, EPI_ISL_2021833, EPI_ISL_2021834, EPI_ISL_2021836, EPI_ISL_2021838, EPI_ISL_2021839, EPI_ISL_2021840, EPI_ISL_2021841, EPI_ISL_2021844, EPI_ISL_2021845, EPI_ISL_2021846, EPI_ISL_2021848, EPI_ISL_2021855, EPI_ISL_2021858, EPI_ISL_2021859, EPI_ISL_2021860, EPI_ISL_2021863, EPI_ISL_2021866, EPI_ISL_2021871, EPI_ISL_2021875, EPI_ISL_2021880, EPI_ISL_2021885, EPI_ISL_2021886, EPI_ISL_2021888, EPI_ISL_2021889, EPI_ISL_2021899, EPI_ISL_2021905, EPI_ISL_2021906, EPI_ISL_2021907, EPI_ISL_2021909, EPI_ISL_2021915, EPI_ISL_2021919, EPI_ISL_2021920, EPI_ISL_2021921, EPI_ISL_2021925, EPI_ISL_2021927, EPI_ISL_2021928, EPI_ISL_2021929, EPI_ISL_2021936, EPI_ISL_2021937, EPI_ISL_2021942, EPI_ISL_2021945, EPI_ISL_2021946, EPI_ISL_2021950, EPI_ISL_2021953, EPI_ISL_2021959, EPI_ISL_2021960, EPI_ISL_2021964, EPI_ISL_2021966, EPI_ISL_2021967, EPI_ISL_2021968, EPI_ISL_2021970, EPI_ISL_2021975, EPI_ISL_2021976, EPI_ISL_2021979, EPI_ISL_2021982, EPI_ISL_2021985, EPI_ISL_2021987, EPI_ISL_2021992, EPI_ISL_2021997, EPI_ISL_2021999, EPI_ISL_2022001, EPI_ISL_2022004, EPI_ISL_2022007, EPI_ISL_2022010, EPI_ISL_2022012, EPI_ISL_2022017, EPI_ISL_2022024, EPI_ISL_2022029, EPI_ISL_2022030, EPI_ISL_2022034, EPI_ISL_2022037, EPI_ISL_2022040, EPI_ISL_2022042, EPI_ISL_2022047, EPI_ISL_2022051, EPI_ISL_2022052, EPI_ISL_2022056, EPI_ISL_2022057, EPI_ISL_2022058, EPI_ISL_2022063, EPI_ISL_2022064, EPI_ISL_2022067, EPI_ISL_2022068, EPI_ISL_2022069, EPI_ISL_2022070, EPI_ISL_2022073, EPI_ISL_2022076, EPI_ISL_2022078, EPI_ISL_2022079, EPI_ISL_2022081, EPI_ISL_2022083, EPI_ISL_2022085, EPI_ISL_2022088, EPI_ISL_2022089, EPI_ISL_2022093, EPI_ISL_2022098, EPI_ISL_2022099, EPI_ISL_2022100, EPI_ISL_2022111, EPI_ISL_2022114, EPI_ISL_2022117, EPI_ISL_2022118, EPI_ISL_2022119, EPI_ISL_2022121, EPI_ISL_2022133, EPI_ISL_2022136, EPI_ISL_2022139, EPI_ISL_2022140, EPI_ISL_2022143, EPI_ISL_2022144, EPI_ISL_2022145, EPI_ISL_2022147, EPI_ISL_2022150, EPI_ISL_2022152, EPI_ISL_2022153, EPI_ISL_2022156, EPI_ISL_2022204, EPI_ISL_2022245, EPI_ISL_2022445, EPI_ISL_2022452, EPI_ISL_2022494, EPI_ISL_2022531, EPI_ISL_2080477, EPI_ISL_2091469, EPI_ISL_2091472, EPI_ISL_2091477, EPI_ISL_2091479, EPI_ISL_2091481, EPI_ISL_2091487, EPI_ISL_2091489, EPI_ISL_2091492, EPI_ISL_2091494, EPI_ISL_2091495, EPI_ISL_2091497, EPI_ISL_2091502, EPI_ISL_2091510, EPI_ISL_2091514, EPI_ISL_2091518, EPI_ISL_2091521, EPI_ISL_2091522, EPI_ISL_2091524, EPI_ISL_2091526, EPI_ISL_2091528, EPI_ISL_2091532, EPI_ISL_2091533, EPI_ISL_2091534, EPI_ISL_2091540, EPI_ISL_2091542, EPI_ISL_2091549, EPI_ISL_2091553, EPI_ISL_2091554, EPI_ISL_2091557, EPI_ISL_2091558, EPI_ISL_2091582, EPI_ISL_2091583, EPI_ISL_2091586, EPI_ISL_2091590, EPI_ISL_2091603, EPI_ISL_2091605, EPI_ISL_2091614, EPI_ISL_2091622, EPI_ISL_2091625, EPI_ISL_2091627, EPI_ISL_2091630, EPI_ISL_2091633, EPI_ISL_2091636, EPI_ISL_2091639, EPI_ISL_2091642, EPI_ISL_2091643, EPI_ISL_2091651, EPI_ISL_2091660, EPI_ISL_2091665, EPI_ISL_2091666, EPI_ISL_2091671, EPI_ISL_2091678, EPI_ISL_2091681, EPI_ISL_2091691, EPI_ISL_2091699, EPI_ISL_2091703, EPI_ISL_2091713, EPI_ISL_2091719, EPI_ISL_2091728, EPI_ISL_2091736, EPI_ISL_2091738, EPI_ISL_2091740, EPI_ISL_2091743, EPI_ISL_2091745, EPI_ISL_2091746, EPI_ISL_2091750, EPI_ISL_2091757, EPI_ISL_2091761, EPI_ISL_2091764, EPI_ISL_2091785, EPI_ISL_2091793, EPI_ISL_2091983, EPI_ISL_2092072, EPI_ISL_2092084, EPI_ISL_2118877, EPI_ISL_2118944, EPI_ISL_2119009, EPI_ISL_2119293, EPI_ISL_2120084, EPI_ISL_2120106, EPI_ISL_2120271, EPI_ISL_2120581, EPI_ISL_2120648, EPI_ISL_2120830, EPI_ISL_2121029, EPI_ISL_2121052, EPI_ISL_2121153, EPI_ISL_2121516, EPI_ISL_2138175, EPI_ISL_2138187, EPI_ISL_2138192, EPI_ISL_2138280, EPI_ISL_2138301, EPI_ISL_2138497

see above | Lighthouse Lab in Milton Keynes | Wellcome Sanger Institute for the COVID-19 Genomics UK (COG-UK) Consortium | Cordelia Langford; David K. Jackson; Dominic Kwiatkowski; Ewan Harrison; Ian Johnston; Jeffrey Barrett; John Sillitoe on behalf of the Wellcome Sanger Institute COVID-19 Surveillance Team; Roberto Amato; Sonia Goncalves; The Lighthouse Lab in Milton Keynes and Alex Alderton

EPI_ISL_1832522, EPI_ISL_1832524, EPI_ISL_1832526, EPI_ISL_1832528, EPI_ISL_1974983, EPI_ISL_1974984, EPI_ISL_1974994, EPI_ISL_1974996, EPI_ISL_1974998, EPI_ISL_1974999, EPI_ISL_1975001, EPI_ISL_1975003, EPI_ISL_1975004, EPI_ISL_1975006, EPI_ISL_1975008, EPI_ISL_1975009, EPI_ISL_1975015, EPI_ISL_1975018, EPI_ISL_1975020, EPI_ISL_1975022, EPI_ISL_1975023, EPI_ISL_1975025, EPI_ISL_1975026, EPI_ISL_1975028, EPI_ISL_1975030, EPI_ISL_1975031, EPI_ISL_1975033, EPI_ISL_1975034, EPI_ISL_1975036, EPI_ISL_1975038, EPI_ISL_1975039, EPI_ISL_2125946, EPI_ISL_2125948, EPI_ISL_2125950, EPI_ISL_2125952, EPI_ISL_2125957, EPI_ISL_2125958, EPI_ISL_2125960, EPI_ISL_2125962, EPI_ISL_2125964, EPI_ISL_2125966, EPI_ISL_2125968, EPI_ISL_2125969, EPI_ISL_2125973, EPI_ISL_2125977, EPI_ISL_2125978, EPI_ISL_2125982, EPI_ISL_2125984, EPI_ISL_2125986

see above | Liverpool Clinical Laboratories | COVID-19 Genomics UK (COG-UK) Consortium | A Alrezaihi; Alessandro Gerada; Alistair Darby; Angela Cowell; Anita Lucaci; Anu Chawla; Cassie Olateju; Catherine Hartley; Charlotte Nelson; Ecaterina Vamos; Elaine O'Toole; Eleanor G Bentley; Ghada T Shawli; Hermione Webster; Isabel García-Dorival; James Johnson; James P Stewart; Jenifer Manson; Joanne Watts; Jones Benjamin; Jordan J Clark; Julian Hiscox; L Luu; Lucille Rainbow; M Almsaud; Margaret Hughes; Mark Whitehead; Matthew Gemmell; Miren Iturriza-Gomara; Muhannad Alruwaili; N.P Randle; Neil Swainston; PKF Gilmore; Parul Sharma; Rebekah Penrice-Randal; Richard Eccles; Richard Gregory; Sam Haldenby; Steve Paterson; Stuart D Armstrong; Trevor Ian Robinson; Ximeng Han

EPI_ISL_1832716, EPI_ISL_1832718, EPI_ISL_1832720, EPI_ISL_1832722, EPI_ISL_1978360, EPI_ISL_1978363, EPI_ISL_1978402, EPI_ISL_1978403, EPI_ISL_1978405, EPI_ISL_1978406, EPI_ISL_1978408, EPI_ISL_1978413, EPI_ISL_1978415, EPI_ISL_1978417, EPI_ISL_1978419, EPI_ISL_1978420, EPI_ISL_1978424, EPI_ISL_1978425, EPI_ISL_1978431, EPI_ISL_1978432, EPI_ISL_1978628, EPI_ISL_1978631, EPI_ISL_1978672, EPI_ISL_1978674, EPI_ISL_1978675, EPI_ISL_1978677, EPI_ISL_1978679, EPI_ISL_1978680, EPI_ISL_1978684, EPI_ISL_1978687, EPI_ISL_1978688, EPI_ISL_1978690, EPI_ISL_1978692, EPI_ISL_1978693, EPI_ISL_1978695, EPI_ISL_1978697, EPI_ISL_1978698, EPI_ISL_1978700, EPI_ISL_1978702, EPI_ISL_1978703, EPI_ISL_1978705, EPI_ISL_1978708, EPI_ISL_1978710, EPI_ISL_1978711, EPI_ISL_1978713, EPI_ISL_1978715, EPI_ISL_1978717, EPI_ISL_1978718, EPI_ISL_1978720, EPI_ISL_1978721, EPI_ISL_1978723, EPI_ISL_1978724, EPI_ISL_1978726, EPI_ISL_1978728, EPI_ISL_1978729, EPI_ISL_1978733, EPI_ISL_1978734, EPI_ISL_1978889, EPI_ISL_2126188, EPI_ISL_2126190, EPI_ISL_2126191, EPI_ISL_2126195, EPI_ISL_2126197, EPI_ISL_2126199, EPI_ISL_2126201, EPI_ISL_2126203, EPI_ISL_2126205, EPI_ISL_2126207, EPI_ISL_2126209, EPI_ISL_2126211, EPI_ISL_2126212, EPI_ISL_2126214, EPI_ISL_2126216, EPI_ISL_2126218, EPI_ISL_2126220, EPI_ISL_2126222, EPI_ISL_2126224, EPI_ISL_2126226, EPI_ISL_2126228, EPI_ISL_2126230, EPI_ISL_2126232, EPI_ISL_2126233, EPI_ISL_2126235, EPI_ISL_2126237, EPI_ISL_2126240, EPI_ISL_2126242, EPI_ISL_2126244, EPI_ISL_2126246, EPI_ISL_2126248, EPI_ISL_2126250, EPI_ISL_2126251, EPI_ISL_2126255, EPI_ISL_2126258, EPI_ISL_2126260, EPI_ISL_2126262, EPI_ISL_2126263, EPI_ISL_2126265, EPI_ISL_2126267, EPI_ISL_2126269, EPI_ISL_2126270, EPI_ISL_2126272, EPI_ISL_2126274, EPI_ISL_2126276, EPI_ISL_2126278, EPI_ISL_2126280, EPI_ISL_2126282, EPI_ISL_2126283, EPI_ISL_2126285, EPI_ISL_2126287, EPI_ISL_2126289, EPI_ISL_2126291, EPI_ISL_2126292, EPI_ISL_2126294, EPI_ISL_2126296, EPI_ISL_2126298, EPI_ISL_2126300, EPI_ISL_2126302, EPI_ISL_2126303, EPI_ISL_2126305, EPI_ISL_2126307, EPI_ISL_2126309, EPI_ISL_2126311, EPI_ISL_2126314, EPI_ISL_2126316, EPI_ISL_2126317, EPI_ISL_2126319, EPI_ISL_2126321, EPI_ISL_2126323, EPI_ISL_2126325, EPI_ISL_2126326, EPI_ISL_2126327, EPI_ISL_2126329, EPI_ISL_2126331, EPI_ISL_2126333, EPI_ISL_2126334, EPI_ISL_2126336, EPI_ISL_2126338, EPI_ISL_2126340, EPI_ISL_2126342, EPI_ISL_2126344, EPI_ISL_2126345, EPI_ISL_2126347, EPI_ISL_2126349, EPI_ISL_2126350, EPI_ISL_2126351, EPI_ISL_2126353, EPI_ISL_2126355, EPI_ISL_2126356, EPI_ISL_2126358, EPI_ISL_2126360, EPI_ISL_2126361, EPI_ISL_2126363, EPI_ISL_2126365, EPI_ISL_2126367, EPI_ISL_2126369, EPI_ISL_2126370, EPI_ISL_2126372, EPI_ISL_2126374, EPI_ISL_2126376, EPI_ISL_2126380, EPI_ISL_2126381, EPI_ISL_2126383, EPI_ISL_2126384, EPI_ISL_2126389, EPI_ISL_2126391, EPI_ISL_2126393, EPI_ISL_2126395, EPI_ISL_2126397, EPI_ISL_2126398, EPI_ISL_2126400, EPI_ISL_2126402, EPI_ISL_2126403, EPI_ISL_2126405, EPI_ISL_2126407, EPI_ISL_2126409, EPI_ISL_2126411, EPI_ISL_2126412, EPI_ISL_2126414, EPI_ISL_2126416, EPI_ISL_2126418, EPI_ISL_2126419, EPI_ISL_2126421, EPI_ISL_2126423, EPI_ISL_2126425, EPI_ISL_2126427, EPI_ISL_2126429, EPI_ISL_2126431, EPI_ISL_2126433, EPI_ISL_2126435, EPI_ISL_2126437, EPI_ISL_2126439, EPI_ISL_2126440, EPI_ISL_2126442, EPI_ISL_2126444, EPI_ISL_2126446, EPI_ISL_2126448, EPI_ISL_2126450, EPI_ISL_2126452, EPI_ISL_2126454, EPI_ISL_2126456, EPI_ISL_2126458, EPI_ISL_2126460, EPI_ISL_2126462, EPI_ISL_2126464, EPI_ISL_2126466, EPI_ISL_2126467, EPI_ISL_2126469, EPI_ISL_2126471, EPI_ISL_2126473, EPI_ISL_2126474, EPI_ISL_2126477, EPI_ISL_2126478, EPI_ISL_2126480, EPI_ISL_2126482, EPI_ISL_2126484, EPI_ISL_2126486, EPI_ISL_2126488, EPI_ISL_2126490, EPI_ISL_2126491, EPI_ISL_2126493, EPI_ISL_2126495, EPI_ISL_2126497, EPI_ISL_2126499, EPI_ISL_2126501, EPI_ISL_2126502, EPI_ISL_2126504, EPI_ISL_2126508, EPI_ISL_2126510, EPI_ISL_2126511, EPI_ISL_2126513, EPI_ISL_2126515, EPI_ISL_2126517, EPI_ISL_2126519, EPI_ISL_2126520, EPI_ISL_2126522, EPI_ISL_2126524, EPI_ISL_2126525, EPI_ISL_2126527, EPI_ISL_2126530, EPI_ISL_2126532, EPI_ISL_2126534, EPI_ISL_2126535, EPI_ISL_2126537, EPI_ISL_2126539, EPI_ISL_2126541, EPI_ISL_2126544, EPI_ISL_2126546, EPI_ISL_2126548, EPI_ISL_2126550, EPI_ISL_2126551, EPI_ISL_2126553, EPI_ISL_2126555, EPI_ISL_2126557, EPI_ISL_2126559, EPI_ISL_2126560, EPI_ISL_2126562, EPI_ISL_2126564, EPI_ISL_2126567, EPI_ISL_2126569, EPI_ISL_2126570, EPI_ISL_2126572, EPI_ISL_2126574, EPI_ISL_2126576, EPI_ISL_2126578, EPI_ISL_2126580, EPI_ISL_2126582, EPI_ISL_2126584, EPI_ISL_2126586, EPI_ISL_2126588, EPI_ISL_2126590, EPI_ISL_2126591, EPI_ISL_2126592, EPI_ISL_2126594, EPI_ISL_2126596, EPI_ISL_2126598, EPI_ISL_2126600, EPI_ISL_2126602, EPI_ISL_2126603, EPI_ISL_2126605, EPI_ISL_2126607, EPI_ISL_2126609, EPI_ISL_2126610, EPI_ISL_2126611, EPI_ISL_2126613, EPI_ISL_2126615, EPI_ISL_2126617, EPI_ISL_2126620, EPI_ISL_2126622, EPI_ISL_2126624, EPI_ISL_2126626, EPI_ISL_2126628, EPI_ISL_2126630, EPI_ISL_2126631, EPI_ISL_2126634, EPI_ISL_2126635, EPI_ISL_2126637, EPI_ISL_2126641, EPI_ISL_2126643, EPI_ISL_2126646, EPI_ISL_2126648, EPI_ISL_2126650, EPI_ISL_2126652, EPI_ISL_2126654, EPI_ISL_2126656, EPI_ISL_2126657, EPI_ISL_2126659, EPI_ISL_2126660, EPI_ISL_2126662, EPI_ISL_2126664, EPI_ISL_2126666, EPI_ISL_2126668, EPI_ISL_2126670, EPI_ISL_2126672, EPI_ISL_2126673, EPI_ISL_2126677, EPI_ISL_2126679, EPI_ISL_2126681, EPI_ISL_2126683, EPI_ISL_2126685, EPI_ISL_2126689, EPI_ISL_2126691, EPI_ISL_2126692, EPI_ISL_2126696, EPI_ISL_2126698, EPI_ISL_2126699, EPI_ISL_2126702, EPI_ISL_2126704, EPI_ISL_2126706, EPI_ISL_2126707, EPI_ISL_2126709, EPI_ISL_2126712, EPI_ISL_2126714, EPI_ISL_2126715, EPI_ISL_2126720, EPI_ISL_2126722, EPI_ISL_2126724, EPI_ISL_2126732, EPI_ISL_2126733, EPI_ISL_2126735, EPI_ISL_2126737, EPI_ISL_2126739, EPI_ISL_2126742, EPI_ISL_2126744, EPI_ISL_2126745, EPI_ISL_2126747, EPI_ISL_2126749, EPI_ISL_2126751, EPI_ISL_2126753, EPI_ISL_2126757, EPI_ISL_2126759, EPI_ISL_2126760, EPI_ISL_2126762, EPI_ISL_2126764, EPI_ISL_2126766, EPI_ISL_2126768, EPI_ISL_2126769, EPI_ISL_2126771, EPI_ISL_2126773, EPI_ISL_2126775, EPI_ISL_2126777, EPI_ISL_2126779, EPI_ISL_2126781, EPI_ISL_2126783, EPI_ISL_2126784, EPI_ISL_2126786, EPI_ISL_2126788, EPI_ISL_2126790, EPI_ISL_2126792, EPI_ISL_2126794, EPI_ISL_2126796, EPI_ISL_2126798, EPI_ISL_2127594, EPI_ISL_2127595, EPI_ISL_2127601, EPI_ISL_2127602

see above | Northumbria University / South Tees Hospitals NHS Foundation Trust / North Cumbria Integrated Care NHS Foundation Trust / North Tees and Hartlepool NHS Foundation Trust / Newcastle Hospitals NHS Foundation Trust | COVID-19 Genomics UK (COG-UK) Consortium | Andrew Nelson; Brendan Payne; Clive Graham; Darren L Smith; Debra Padgett; Edward Barton; Emma Swindells; Garren Scott; Gary Black; Gary Eltringham; Giles S Holt; Greg R Young; Jane Greenaway; Jennifer Collins; John Allan; Joshua Loh; Lynn Dover; Matthew Bashton; Mohammad A Tariq; Paul Baker; Sarah Essex; Steve Liggett; Wen C Yew; Yusri Taha

EPI_ISL_1979228, EPI_ISL_1979230, EPI_ISL_1979231, EPI_ISL_1979233, EPI_ISL_1979234, EPI_ISL_1979236, EPI_ISL_1979237, EPI_ISL_1979239, EPI_ISL_1979241, EPI_ISL_1979242, EPI_ISL_1979244, EPI_ISL_1979246, EPI_ISL_1979247, EPI_ISL_1979249, EPI_ISL_1979251, EPI_ISL_1979252, EPI_ISL_1979254, EPI_ISL_1979255, EPI_ISL_1979257, EPI_ISL_1979258, EPI_ISL_1979259, EPI_ISL_1979261, EPI_ISL_1979263, EPI_ISL_1979264, EPI_ISL_1979266, EPI_ISL_1979267, EPI_ISL_1979269, EPI_ISL_1979270, EPI_ISL_1979274, EPI_ISL_1979275, EPI_ISL_1979276, EPI_ISL_1979278, EPI_ISL_1979280, EPI_ISL_1979281, EPI_ISL_1979283, EPI_ISL_1979284, EPI_ISL_1979285, EPI_ISL_1979287, EPI_ISL_1979289, EPI_ISL_1979290, EPI_ISL_1979292, EPI_ISL_1979293, EPI_ISL_1979295, EPI_ISL_1979296, EPI_ISL_1979298, EPI_ISL_1979300, EPI_ISL_1979303, EPI_ISL_1979305, EPI_ISL_1979306, EPI_ISL_1979308, EPI_ISL_1979309, EPI_ISL_1979312, EPI_ISL_1979318, EPI_ISL_1979320, EPI_ISL_1979322, EPI_ISL_1979323, EPI_ISL_1979325, EPI_ISL_1979328, EPI_ISL_1979329, EPI_ISL_1979331, EPI_ISL_1979333, EPI_ISL_1979334, EPI_ISL_1979336, EPI_ISL_1979339, EPI_ISL_1979353, EPI_ISL_1979354, EPI_ISL_1979356, EPI_ISL_1979357, EPI_ISL_1979359, EPI_ISL_1979361, EPI_ISL_1979362, EPI_ISL_1979366, EPI_ISL_1979368, EPI_ISL_1979369, EPI_ISL_1979371, EPI_ISL_1979372, EPI_ISL_1979374, EPI_ISL_1979375, EPI_ISL_1979377, EPI_ISL_1979378, EPI_ISL_1979380, EPI_ISL_1979381, EPI_ISL_1979383, EPI_ISL_1979384, EPI_ISL_1979386, EPI_ISL_1979387, EPI_ISL_1979389, EPI_ISL_1979390, EPI_ISL_1979391, EPI_ISL_1979392, EPI_ISL_1979393, EPI_ISL_1979394, EPI_ISL_1979395, EPI_ISL_1979396, EPI_ISL_1979398, EPI_ISL_1979400, EPI_ISL_1979401, EPI_ISL_1979403, EPI_ISL_1979405, EPI_ISL_1979406, EPI_ISL_1979408, EPI_ISL_1979410, EPI_ISL_1979412, EPI_ISL_1979413, EPI_ISL_1979415, EPI_ISL_1979416, EPI_ISL_1979418, EPI_ISL_1979420, EPI_ISL_1979421, EPI_ISL_1979423, EPI_ISL_1979424, EPI_ISL_1979426, EPI_ISL_1979428, EPI_ISL_1979430, EPI_ISL_1979431, EPI_ISL_1979433, EPI_ISL_1979434, EPI_ISL_1979436, EPI_ISL_1979437, EPI_ISL_1979439, EPI_ISL_1979441, EPI_ISL_1979442, EPI_ISL_1979444, EPI_ISL_1979445, EPI_ISL_1979447, EPI_ISL_1979448, EPI_ISL_1979449, EPI_ISL_1979450, EPI_ISL_1979452, EPI_ISL_1979453, EPI_ISL_1979455, EPI_ISL_1979456, EPI_ISL_1979458, EPI_ISL_1979460, EPI_ISL_1979461, EPI_ISL_1979463, EPI_ISL_1979464, EPI_ISL_1979466, EPI_ISL_1979467, EPI_ISL_1979469, EPI_ISL_1979473, EPI_ISL_1979475, EPI_ISL_1979477, EPI_ISL_1979478, EPI_ISL_1979480, EPI_ISL_1979481, EPI_ISL_1979483, EPI_ISL_1979484, EPI_ISL_1979485, EPI_ISL_1979487, EPI_ISL_1979488, EPI_ISL_1979490, EPI_ISL_1979492, EPI_ISL_1979495, EPI_ISL_1979501, EPI_ISL_1979506, EPI_ISL_1979507, EPI_ISL_1979509, EPI_ISL_1979510, EPI_ISL_1979514, EPI_ISL_1979522, EPI_ISL_1979523, EPI_ISL_1979525, EPI_ISL_1979526, EPI_ISL_1979528, EPI_ISL_1979529, EPI_ISL_1979531, EPI_ISL_1979532, EPI_ISL_1979534, EPI_ISL_1979536, EPI_ISL_1979537, EPI_ISL_1979539, EPI_ISL_1979541, EPI_ISL_1979542, EPI_ISL_1979544, EPI_ISL_1979545, EPI_ISL_1979547, EPI_ISL_1979548, EPI_ISL_1979550, EPI_ISL_1979552, EPI_ISL_1979553, EPI_ISL_1979555, EPI_ISL_1979556, EPI_ISL_1979558, EPI_ISL_1979559, EPI_ISL_1979561, EPI_ISL_1979562, EPI_ISL_1979564, EPI_ISL_1979565, EPI_ISL_1979567, EPI_ISL_1979568, EPI_ISL_1979570, EPI_ISL_1979573, EPI_ISL_1979575, EPI_ISL_1979577, EPI_ISL_1979578, EPI_ISL_1979580, EPI_ISL_1979581, EPI_ISL_1979583, EPI_ISL_1979585, EPI_ISL_1979586, EPI_ISL_1979588, EPI_ISL_1979590, EPI_ISL_1979591, EPI_ISL_1979593, EPI_ISL_1979594, EPI_ISL_1979596, EPI_ISL_1979597, EPI_ISL_1979599, EPI_ISL_1979601, EPI_ISL_1979604, EPI_ISL_1979607, EPI_ISL_1979609, EPI_ISL_1979613, EPI_ISL_1979615, EPI_ISL_1979621, EPI_ISL_1979627, EPI_ISL_1979629, EPI_ISL_1979630, EPI_ISL_1979638, EPI_ISL_1979642, EPI_ISL_1979645, EPI_ISL_1979650, EPI_ISL_1979651, EPI_ISL_1979654, EPI_ISL_1979656, EPI_ISL_1979657, EPI_ISL_1979659, EPI_ISL_1979661, EPI_ISL_1979662, EPI_ISL_1979664, EPI_ISL_1979665, EPI_ISL_1979667, EPI_ISL_1979669, EPI_ISL_1979671, EPI_ISL_1979674, EPI_ISL_1979675, EPI_ISL_1979677, EPI_ISL_1979678, EPI_ISL_1979680, EPI_ISL_1979682,

| | | | |
|---|---|---|---|
| EPI_ISL_1979683, EPI_ISL_1979685, EPI_ISL_1979686, EPI_ISL_1979688, EPI_ISL_1979690, EPI_ISL_1979691, EPI_ISL_1979695, EPI_ISL_1979696, EPI_ISL_1979698, EPI_ISL_1979703, EPI_ISL_1979704, EPI_ISL_1979708, EPI_ISL_1979711, EPI_ISL_1979717, EPI_ISL_1979718, EPI_ISL_1979720, EPI_ISL_1979722, EPI_ISL_1979730, EPI_ISL_1979731, EPI_ISL_1979733, EPI_ISL_1979735, EPI_ISL_1979738, EPI_ISL_1979739 | | | |
| see above | Originating lab: Wales Specialist Virology Centre Sequencing lab: Pathogen Genomics Unit | Public Health Wales Microbiology Cardiff Wales Specialist Virology Centre | Alec Birchley; Alexander Adams; Amy Gaskin; Angela Marchbank; Bree Gatica-Wilcox; Catherine Moore; Jason Coombes; Joanne Watkins; Joel Southgate; Johnathan Evans; Laura Gifford; Lauren Gilbert; Lee Graham; Malorie Perry; Matthew Bull; Nicole Pacchiarini; Sally Corden; Sara Kumziene-Summerhayes; Sara Rey; Sarah Taylor; Simon Cottrell; Sophie Jones; Tom Connor |
| EPI_ISL_1832726, EPI_ISL_1974718, EPI_ISL_1978901, EPI_ISL_1978903, EPI_ISL_1978923, EPI_ISL_1978946, EPI_ISL_1978947, EPI_ISL_1978950, EPI_ISL_1978951 | | | |
| see above | Quadram Institute Bioscience | COVID-19 Genomics UK (COG-UK) Consortium | Alexander J Trotter; Alison E. Mather; Alp Aydin; Ana P. Tedim; Anastasia Kolyva; Andrew Bell; Andrew J. Page; Claire Stuart; Dave J. Baker; Gemma L. Kay; John Wain; Justin O'Grady; Leonardo de Oliveira Martins; Lizzie Meadows; Maria Diaz; Mark Webber; Muhammed Yasir; Nabil-Fareed Alikhan; Ngozi Elumogo; Nicholas M. Thomson; Rachael Stanley; Rachel Gilroy; Reenesh Prakash; Samir Dervisevic; Samuel Bloomfield; Steven Rudder; Thanh Le-Viet |
| EPI_ISL_1833002, EPI_ISL_1833023, EPI_ISL_1833024, EPI_ISL_1833033, EPI_ISL_1833034, EPI_ISL_1978958, EPI_ISL_1978961, EPI_ISL_1978962, EPI_ISL_1978963 | | | |
| see above | Queens Medical Centre, Clinical Microbiology Department / DeepSeq Nottingham | COVID-19 Genomics UK (COG-UK) Consortium | Christopher Moore; Emily Park; Fei Sang; Gemma Clark; Hannah Howson-Wells; Jack Hill; Jayasree Dey; Johnny Debebe; Jonathan Ball; Joseph Chappell; Manjinder Khakh; Matthew Carlisle; Matthew Loose; Michelle M Lister; Nadine Holmes; Patrick McClure; Theocharis Tsoleridis; Timothy Byaruhanga; Vicki M Fleming; Victoria Wright; Wendy Smith |
| EPI_ISL_1829776, EPI_ISL_1829803, EPI_ISL_1829805, EPI_ISL_1829810, EPI_ISL_1829814, EPI_ISL_1829817, EPI_ISL_1829820, EPI_ISL_1829826, EPI_ISL_1829827, EPI_ISL_1829829, EPI_ISL_1829831, EPI_ISL_1829832, EPI_ISL_1829841, EPI_ISL_1829845, EPI_ISL_1829846, EPI_ISL_1829847, EPI_ISL_1829852, EPI_ISL_1829858, EPI_ISL_1829861, EPI_ISL_1829868, EPI_ISL_1829876, EPI_ISL_1829877, EPI_ISL_1829891, EPI_ISL_1829893, EPI_ISL_1829894, EPI_ISL_1829897, EPI_ISL_1829910, EPI_ISL_1829911, EPI_ISL_1829912, EPI_ISL_1829918, EPI_ISL_1829926, EPI_ISL_1829931, EPI_ISL_1829945, EPI_ISL_1829947, EPI_ISL_1829951, EPI_ISL_1829954, EPI_ISL_1829955, EPI_ISL_1829956, EPI_ISL_1829958, EPI_ISL_1829961, EPI_ISL_1829967, EPI_ISL_1829968, EPI_ISL_1829971, EPI_ISL_1829975, EPI_ISL_1829978, EPI_ISL_1829980, EPI_ISL_1829981, EPI_ISL_1829990, EPI_ISL_1829991, EPI_ISL_1829992, EPI_ISL_1829993, EPI_ISL_1829999, EPI_ISL_1830002, EPI_ISL_1830008, EPI_ISL_1830017, EPI_ISL_1830022, EPI_ISL_1830029, EPI_ISL_1830030, EPI_ISL_1830034, EPI_ISL_1830037, EPI_ISL_1830038, EPI_ISL_1830041, EPI_ISL_1830045, EPI_ISL_1830047, EPI_ISL_1830051, EPI_ISL_1830057, EPI_ISL_1830059, EPI_ISL_1830060, EPI_ISL_1830073, EPI_ISL_1830076, EPI_ISL_1830084, EPI_ISL_1830086, EPI_ISL_1830087, EPI_ISL_1830091, EPI_ISL_1830100, EPI_ISL_1857593, EPI_ISL_1857595, EPI_ISL_1857603, EPI_ISL_1857615, EPI_ISL_1857616, EPI_ISL_1857630, EPI_ISL_1857631, EPI_ISL_1857634, EPI_ISL_1857635, EPI_ISL_1857636, EPI_ISL_1857639, EPI_ISL_1857640, EPI_ISL_1857641, EPI_ISL_1857643, EPI_ISL_1857649, EPI_ISL_1857650, EPI_ISL_1857655, EPI_ISL_1857661, EPI_ISL_1857664, EPI_ISL_1857665, EPI_ISL_1857673, EPI_ISL_1857674, EPI_ISL_1857680, EPI_ISL_1857695, EPI_ISL_1857699, EPI_ISL_1857700, EPI_ISL_1857703, EPI_ISL_1857717, EPI_ISL_1857721, EPI_ISL_1857722, EPI_ISL_1857728, EPI_ISL_1857729, EPI_ISL_1857756, EPI_ISL_1857757, EPI_ISL_1857758, EPI_ISL_1857760, EPI_ISL_1857766, EPI_ISL_1857767, EPI_ISL_1857770, EPI_ISL_1857774, EPI_ISL_1857779, EPI_ISL_1857786, EPI_ISL_1857790, EPI_ISL_1857794, EPI_ISL_1912639, EPI_ISL_1912690, EPI_ISL_1912759, EPI_ISL_1912760, EPI_ISL_1912761, EPI_ISL_1912762, EPI_ISL_1912763, EPI_ISL_1912764, EPI_ISL_1912765, EPI_ISL_1912766, EPI_ISL_1912768, EPI_ISL_1912769, EPI_ISL_1912770, EPI_ISL_1912771, EPI_ISL_1912782, EPI_ISL_1986592, EPI_ISL_1986613, EPI_ISL_1986655, EPI_ISL_1986661, EPI_ISL_1986673, EPI_ISL_1986688, EPI_ISL_1986695, EPI_ISL_1986751, EPI_ISL_1986757, EPI_ISL_1986759, EPI_ISL_1986832, EPI_ISL_1986863, EPI_ISL_1986867, EPI_ISL_1986870, EPI_ISL_1986943, EPI_ISL_1986947, EPI_ISL_1986949, EPI_ISL_1986956, EPI_ISL_1986959, EPI_ISL_1986979, EPI_ISL_1986983, EPI_ISL_1987003, EPI_ISL_1987022, EPI_ISL_1987026, EPI_ISL_1987035, EPI_ISL_1987044, EPI_ISL_1987047, EPI_ISL_1987057, EPI_ISL_1987127, EPI_ISL_1987133, EPI_ISL_1987134, EPI_ISL_1987143, EPI_ISL_1987147, EPI_ISL_1987150, EPI_ISL_1987160, EPI_ISL_1987170, EPI_ISL_1987174, EPI_ISL_1987178, EPI_ISL_1987182, EPI_ISL_1987183, EPI_ISL_1987190, EPI_ISL_1987195, EPI_ISL_1987210, EPI_ISL_1987216, EPI_ISL_1987220, EPI_ISL_1987242, EPI_ISL_1987243, EPI_ISL_1987249, EPI_ISL_1987256, EPI_ISL_1987259, EPI_ISL_1987260, EPI_ISL_1987282, EPI_ISL_1987283, EPI_ISL_1987284, EPI_ISL_1987291, EPI_ISL_1987296, EPI_ISL_1987322, EPI_ISL_1987329, EPI_ISL_1987335, EPI_ISL_1987336, EPI_ISL_1987343, EPI_ISL_1987351, EPI_ISL_1987353, EPI_ISL_1987358, EPI_ISL_1987365, EPI_ISL_1987367, EPI_ISL_2004344, EPI_ISL_2004348, EPI_ISL_2004350, EPI_ISL_2004355, EPI_ISL_2004396, EPI_ISL_2004409, EPI_ISL_2004413, EPI_ISL_2004423, EPI_ISL_2004434, EPI_ISL_2004447, EPI_ISL_2004448, EPI_ISL_2004455, EPI_ISL_2004472, EPI_ISL_2004477, EPI_ISL_2004478, EPI_ISL_2004491, EPI_ISL_2004527, EPI_ISL_2004546, EPI_ISL_2004548, EPI_ISL_2004574, EPI_ISL_2004595, EPI_ISL_2004604, EPI_ISL_2004611, EPI_ISL_2005162, EPI_ISL_2005187, EPI_ISL_2005192, EPI_ISL_2005816, EPI_ISL_2005823, EPI_ISL_2005828, EPI_ISL_2005837, EPI_ISL_2005838, EPI_ISL_2005839, EPI_ISL_2005845, EPI_ISL_2005846, EPI_ISL_2005850, EPI_ISL_2005852, EPI_ISL_2005858, EPI_ISL_2005863, EPI_ISL_2005865, EPI_ISL_2005866, EPI_ISL_2005868, EPI_ISL_2005877, EPI_ISL_2005886, EPI_ISL_2005890, EPI_ISL_2005898, EPI_ISL_2005901, EPI_ISL_2005902, EPI_ISL_2005905, EPI_ISL_2005906, EPI_ISL_2005911, EPI_ISL_2005913, EPI_ISL_2005919, EPI_ISL_2005924, EPI_ISL_2005926, EPI_ISL_2005928, EPI_ISL_2005939, EPI_ISL_2005941, EPI_ISL_2005946, EPI_ISL_2005947, EPI_ISL_2005953, EPI_ISL_2005955, EPI_ISL_2005958, EPI_ISL_2005962, EPI_ISL_2005964, EPI_ISL_2005984, EPI_ISL_2005985, EPI_ISL_2005988, EPI_ISL_2005989, EPI_ISL_2005992, EPI_ISL_2005993, EPI_ISL_2005996, EPI_ISL_2006002, EPI_ISL_2006007, EPI_ISL_2006008, EPI_ISL_2006010, EPI_ISL_2006012, EPI_ISL_2006018, EPI_ISL_2006021, EPI_ISL_2006024, EPI_ISL_2006026, EPI_ISL_2006034, EPI_ISL_2006035, EPI_ISL_2006039, EPI_ISL_2006040, EPI_ISL_2006044, EPI_ISL_2006053, EPI_ISL_2006062, EPI_ISL_2021715, EPI_ISL_2021786, EPI_ISL_2021802, EPI_ISL_2021825, EPI_ISL_2022459, EPI_ISL_2022464, EPI_ISL_2022507, EPI_ISL_2022543, EPI_ISL_2022546, EPI_ISL_2022578, EPI_ISL_2022585, EPI_ISL_2091889, EPI_ISL_2091907, EPI_ISL_2091998, EPI_ISL_2092038 | | | |
| see above | Randox Laboratories | Wellcome Sanger Institute for the COVID-19 Genomics UK (COG-UK) Consortium | Cordelia Langford; David K. Jackson; Dominic Kwiatkowski; Ewan Harrison; Ian Johnston; Jeffrey Barrett; John Sillitoe on behalf of the Wellcome Sanger Institute COVID-19 Surveillance Team; Randox Laboratories and Alex Alderton; Roberto Amato; Sonia Goncalves |
| EPI_ISL_1832580, EPI_ISL_1832582, EPI_ISL_1832584, EPI_ISL_1832588, EPI_ISL_1832596, EPI_ISL_1832599, EPI_ISL_1832608, EPI_ISL_1832610, EPI_ISL_1832614, EPI_ISL_1832615, EPI_ISL_1832617, EPI_ISL_1832619, EPI_ISL_1832623, EPI_ISL_1832625, EPI_ISL_1832627, EPI_ISL_1832629, EPI_ISL_1832631, EPI_ISL_1832633, EPI_ISL_1832636, EPI_ISL_1832638, EPI_ISL_1832640, EPI_ISL_1832641, EPI_ISL_1832643, EPI_ISL_1832653, EPI_ISL_1832655, EPI_ISL_1832679, EPI_ISL_1832681, EPI_ISL_1832682, EPI_ISL_1832684, EPI_ISL_1832686, EPI_ISL_1832688, EPI_ISL_1832690, EPI_ISL_1832692, EPI_ISL_1832694, EPI_ISL_1832696, EPI_ISL_1975116, EPI_ISL_1975120, EPI_ISL_1975121, EPI_ISL_1975124, EPI_ISL_1975126, EPI_ISL_1975127, EPI_ISL_1975128, EPI_ISL_1975131, EPI_ISL_1975134, EPI_ISL_1975136, EPI_ISL_1975138, EPI_ISL_1975139, EPI_ISL_1975141, EPI_ISL_1975142, EPI_ISL_1975144, EPI_ISL_1975145, EPI_ISL_1975147, EPI_ISL_1975149, EPI_ISL_1975151, EPI_ISL_1975152, EPI_ISL_1975153, EPI_ISL_1975155, EPI_ISL_1975157, EPI_ISL_1975158, EPI_ISL_1975160, EPI_ISL_1975162, EPI_ISL_1975163, EPI_ISL_1975165, EPI_ISL_1975166, EPI_ISL_1975168, EPI_ISL_1975170, EPI_ISL_1975172, EPI_ISL_1975174, EPI_ISL_1975175, EPI_ISL_1975177, EPI_ISL_2126134, EPI_ISL_2126144 | | | |
| see above | University College London, Great Ormond Street Hospital for Children NHS Foundation Trust, Imperial College Healthcare NHS Trust | COVID-19 Genomics UK (COG-UK) Consortium | Alison Holmes; Charlotte Williams; Helena Tutill; Jacqueline Findlay; James Price; Judith Breuer; Julianne Brown; Kathryn Harris; Leysa Forrest; Mark Kristiansen; Paola Niola; Paola Resende Silva; Patricia Dyal; Paul Randell; Rachel Williams; Samuel Weeks; Sergi Castellano; Sunando Roy; Tony Brooks; Yasmin Panchbhaya |
| EPI_ISL_2125896, EPI_ISL_2125898, EPI_ISL_2125899, EPI_ISL_2125901, EPI_ISL_2125903, EPI_ISL_2125905, EPI_ISL_2125921, EPI_ISL_2125927 | | | |
| see above | University Hospitals Of Leicester NHS Trust and DeepSeq Nottingham | COVID-19 Genomics UK (COG-UK) Consortium | Christopher Holmes; Christopher Moore; Fei Sang; Joeseph Chappell; Johnny Debebe; Jonathan Ball; Julian Tang; Karlie Fallon; Matthew Carlisle; Matthew Loose; Nadine Holmes; Patrick McClure; Paul Bird; Thomas Helmer; Victoria Wright |
| EPI_ISL_1832145, EPI_ISL_1832151, EPI_ISL_1832153, EPI_ISL_1832155, EPI_ISL_1832157, EPI_ISL_1832159, EPI_ISL_1974723, EPI_ISL_1974724, EPI_ISL_1974726, EPI_ISL_1974728, EPI_ISL_1974729, EPI_ISL_1974731, EPI_ISL_1974733, EPI_ISL_1974734, EPI_ISL_1974736, EPI_ISL_1974737, EPI_ISL_1974739, EPI_ISL_1974741, EPI_ISL_1974742, EPI_ISL_1974744, EPI_ISL_1974745, EPI_ISL_1974747, EPI_ISL_1974749, EPI_ISL_1974750, EPI_ISL_1974752, EPI_ISL_1974753, EPI_ISL_1974755, EPI_ISL_1974757, EPI_ISL_2125412, EPI_ISL_2125414, EPI_ISL_2125416, EPI_ISL_2125417, EPI_ISL_2125420, EPI_ISL_2125421, EPI_ISL_2125423, EPI_ISL_2125425 | | | |
| see above | University of Birmingham | COVID-19 Genomics UK (COG-UK) Consortium | Alex Richter; Andrew Bosworth; Andrew Bosworth. Queen Elizabeth Hospital: Anna Casey; Andrew D Beggs PHE Heartlands Lab: Husam Osman; Charlotte Poxon; Institute of Microbiology; Joanne Stockton; Josh Quick; Kasun Wanigasooriya; Mike Kidd; Nicholas Loman. University of Birmingham Testing Laboratory: Celina M Whalley; Oliver Pickles; Radoslaw Poplawski; Samuel Nicholls; University of Birmingham: Claire McMurray; Will Rowe |
| EPI_ISL_1832274, EPI_ISL_1832275, EPI_ISL_1832276, EPI_ISL_1974834, EPI_ISL_1974836, EPI_ISL_1974837, EPI_ISL_1974840, EPI_ISL_1974841, EPI_ISL_1974842, EPI_ISL_1974844, EPI_ISL_1974846, EPI_ISL_1974847, EPI_ISL_1974849, EPI_ISL_1974850, EPI_ISL_1974852, EPI_ISL_1974853, EPI_ISL_1974855, EPI_ISL_1974856, EPI_ISL_1974857, EPI_ISL_1974859, EPI_ISL_1974860, EPI_ISL_1974881, EPI_ISL_1974882, EPI_ISL_1974884, EPI_ISL_1974885, EPI_ISL_1974887, EPI_ISL_1974889, EPI_ISL_1974903, EPI_ISL_1974905, EPI_ISL_2125635, EPI_ISL_2125637, EPI_ISL_2125638, EPI_ISL_2125640, EPI_ISL_2125642, EPI_ISL_2125644, EPI_ISL_2125646, EPI_ISL_2125647, EPI_ISL_2125649, EPI_ISL_2125651, EPI_ISL_2125652, EPI_ISL_2125654, EPI_ISL_2125655, EPI_ISL_2125657, EPI_ISL_2125659, EPI_ISL_2125661, EPI_ISL_2125662, EPI_ISL_2125664, EPI_ISL_2125666, EPI_ISL_2125668, EPI_ISL_2125689, EPI_ISL_2125708, EPI_ISL_2125710, EPI_ISL_2125711, EPI_ISL_2125713, EPI_ISL_2125715, EPI_ISL_2125717, EPI_ISL_2125751, EPI_ISL_2125865, EPI_ISL_2125866, EPI_ISL_2125874, EPI_ISL_2125875, EPI_ISL_2125876, EPI_ISL_2125883, EPI_ISL_2125894 | | | |
| see above | Virology Department, Royal Infirmary of Edinburgh, NHS Lothian / School of Biological Sciences, University of Edinburgh | COVID-19 Genomics UK (COG-UK) Consortium | Colquhoun R; Cotton S; Dewar R; Gallagher A; Hill V; Jackson B; Maloney D; McCrone JT; McHugh M; O'Toole Á; Rambaut A; Rooke S; Scher E; Templeton K; Yu X |
| EPI_ISL_1833833, EPI_ISL_1833836, EPI_ISL_1833838, EPI_ISL_1833845, EPI_ISL_1833861, EPI_ISL_1833868, EPI_ISL_1833876, EPI_ISL_1833883, EPI_ISL_1833891, EPI_ISL_1833899, EPI_ISL_1833906, EPI_ISL_1980340, EPI_ISL_1980342, EPI_ISL_1980344, EPI_ISL_1980345, EPI_ISL_1980347, EPI_ISL_1980348, EPI_ISL_1980349, EPI_ISL_1980351, EPI_ISL_1980358, EPI_ISL_1980359, EPI_ISL_1980362, EPI_ISL_1980363, EPI_ISL_1980372, EPI_ISL_1980374, EPI_ISL_1980377, EPI_ISL_1980379, EPI_ISL_1980382, EPI_ISL_1980384, EPI_ISL_1980385, EPI_ISL_1980387, EPI_ISL_1980389, EPI_ISL_1980390, EPI_ISL_1980393, EPI_ISL_1980395, EPI_ISL_1980398, EPI_ISL_1980399, EPI_ISL_1980401, EPI_ISL_1980403, EPI_ISL_1980406, EPI_ISL_1980407, EPI_ISL_1980409, EPI_ISL_1980418, EPI_ISL_1980420, EPI_ISL_1980425, EPI_ISL_1980426, EPI_ISL_1980428, EPI_ISL_1980430, EPI_ISL_1980435, EPI_ISL_1980437, EPI_ISL_1980438, EPI_ISL_1980440, EPI_ISL_1980443, EPI_ISL_1980445, EPI_ISL_1980448, EPI_ISL_1980449, EPI_ISL_1980451, EPI_ISL_1980456, EPI_ISL_1980457, EPI_ISL_2128465, EPI_ISL_2128473, EPI_ISL_2128476, EPI_ISL_2128478, EPI_ISL_2128479, EPI_ISL_2128483, EPI_ISL_2128485, EPI_ISL_2128487, EPI_ISL_2128492, EPI_ISL_2128494, EPI_ISL_2128497, EPI_ISL_2128499, EPI_ISL_2128503 | | | |
| see above | Virology Department, Sheffield Teaching Hospitals NHS Foundation Trust/Department of Infection, Immunity and Cardiovascular Disease, The Medical School, University of Sheffield | COVID-19 Genomics UK (COG-UK) Consortium | Adri Angyal; Alex Keeley; Benjamin Lindsey; Cariad Evans; Danielle Groves; Dave Partridge; Katie Johnson; Laura Carrilero; Luke Green; Matthew Parker; Matthew Wyles; Mehmet Yavuz; Mohammad Raza; Nikki Smith; Paul Parsons; Rachel Tucker; Rebecca Brown; Thushan de Silva |
| EPI_ISL_1833181, EPI_ISL_1833203, EPI_ISL_1833215, EPI_ISL_1833227, EPI_ISL_1833231, EPI_ISL_1833232, EPI_ISL_1833233, EPI_ISL_1833235, EPI_ISL_2127876, EPI_ISL_2127878, EPI_ISL_2127882, EPI_ISL_2127885, EPI_ISL_2127888, EPI_ISL_2127889, EPI_ISL_2127892, EPI_ISL_2127893 | | | |
| see above | Wales Specialist Virology Centre Sequencing lab: Pathogen Genomics Unit | Public Health Wales Microbiology Cardiff Wales Specialist Virology Centre | Alec Birchley; Alexander Adams; Amy Gaskin; Angela Marchbank; Bree Gatica-Wilcox; Catherine Moore; Jason Coombes; Joanne Watkins; Joel Southgate; Johnathan Evans; Laura Gifford; Lauren Gilbert; Lee Graham; Malorie Perry; Matthew Bull; Nicole Pacchiarini; Sally Corden; Sara Kumziene-Summerhayes; Sara Rey; Sarah Taylor; Simon Cottrell; Sophie Jones; Tom Connor |

We gratefully acknowledge the following Authors from the Originating laboratories responsible for obtaining the specimens, as well as the Submitting laboratories where the genome data were generated and shared via GISAID, on which this research is based.

All Submitters of data may be contacted directly via www.gisaid.org

Authors are sorted alphabetically.

| Accession ID | Originating Laboratory | Submitting Laboratory | Authors |
|---|---|---|---|
| EPI_ISL_2126011, EPI_ISL_2126013, EPI_ISL_2126024, EPI_ISL_2126026, EPI_ISL_2126029, EPI_ISL_2126031, EPI_ISL_2126033, EPI_ISL_2126035, EPI_ISL_2126062, EPI_ISL_2126115, EPI_ISL_2126117, EPI_ISL_2126119, EPI_ISL_2126121, EPI_ISL_2126123, EPI_ISL_2126125 | | | |
| see above | Barts Health NHS Trust | COVID-19 Genomics UK (COG-UK) Consortium | BROAD; Beatrix; CUTINO-MOGUEL; Claire; David; Dola; HARRINGTON; KELE; KULASEGARAN-SHYLINI; Maria-Teresa; OWOYEMI; Raghavendran |
| EPI_ISL_2128554, EPI_ISL_2128558, EPI_ISL_2128565, EPI_ISL_2128582, EPI_ISL_2128595, EPI_ISL_2128599 | Bioinformatics and Biostatistics Lab, Advanced Sequencing Facility | COVID-19 Genomics UK (COG-UK) Consortium | Aengus Stewart; Chelsea Sawyer; Harshil Patel; Jerome Nicod; Laura Cubitt; Margaret Crawford |
| EPI_ISL_1980114, EPI_ISL_1980116, EPI_ISL_1980118, EPI_ISL_1980119, EPI_ISL_1980121, EPI_ISL_1980122, EPI_ISL_1980215, EPI_ISL_2128178, EPI_ISL_2128180, EPI_ISL_2128182, EPI_ISL_2128225, EPI_ISL_2128227, EPI_ISL_2128229, EPI_ISL_2128231, EPI_ISL_2128233, EPI_ISL_2128234, EPI_ISL_2128236, EPI_ISL_2128238, EPI_ISL_2128370, EPI_ISL_2128372 | | | |
| see above | Centre for Enzyme Innovation, University of Portsmouth / Translational Research Laboratory, Portsmouth Hospitals NHS Trust | COVID-19 Genomics UK (COG-UK) Consortium | Angela Beckett; Christopher Fearn; Kate Cook; Katie Loveson; Salman Goudarzi; Samuel Robson; Scott Elliott; Sharon Glaysher |
| EPI_ISL_1974763, EPI_ISL_1974784, EPI_ISL_1974796, EPI_ISL_2125456, EPI_ISL_2125458, EPI_ISL_2125459, EPI_ISL_2125461, EPI_ISL_2125463, EPI_ISL_2125465, EPI_ISL_2125467, EPI_ISL_2125469, EPI_ISL_2125471, EPI_ISL_2125472, EPI_ISL_2125474, EPI_ISL_2125476, EPI_ISL_2125479, EPI_ISL_2125480, EPI_ISL_2125482, EPI_ISL_2125484, EPI_ISL_2125486 | | | |
| see above | Department of Pathology, University of Cambridge | COVID-19 Genomics UK (COG-UK) Consortium | Aminu S. Jahun; Ian Goodfellow; Iliana Georgana; Martin D. Curran; Myra Hosmillo; Rhys Izuagbe; Surendra Parmar; William L. Hamilton; Yasmin Chaudhry |
| EPI_ISL_2080362, EPI_ISL_2080363, EPI_ISL_2080367, EPI_ISL_2080371, EPI_ISL_2080375, EPI_ISL_2080376, EPI_ISL_2080377, EPI_ISL_2080378, EPI_ISL_2080380, EPI_ISL_2080381, EPI_ISL_2080382, EPI_ISL_2080383, EPI_ISL_2080384, EPI_ISL_2080386, EPI_ISL_2080389, EPI_ISL_2080390, EPI_ISL_2080392, EPI_ISL_2080395, EPI_ISL_2080397, EPI_ISL_2080401, EPI_ISL_2080405, EPI_ISL_2080406, EPI_ISL_2080408, EPI_ISL_2080410, EPI_ISL_2080411, EPI_ISL_2080414, EPI_ISL_2080415, EPI_ISL_2080416, EPI_ISL_2080422, EPI_ISL_2080423, EPI_ISL_2080426, EPI_ISL_2080427, EPI_ISL_2080428, EPI_ISL_2080430, EPI_ISL_2080435, EPI_ISL_2080436, EPI_ISL_2080437, EPI_ISL_2080439, EPI_ISL_2080443, EPI_ISL_2080444, EPI_ISL_2080445, EPI_ISL_2080447, EPI_ISL_2080448, EPI_ISL_2080449, EPI_ISL_2080450, EPI_ISL_2080456, EPI_ISL_2080460, EPI_ISL_2080463, EPI_ISL_2080466, EPI_ISL_2080467, EPI_ISL_2080469, EPI_ISL_2080471, EPI_ISL_2080472, EPI_ISL_2080473, EPI_ISL_2138370, EPI_ISL_2138382, EPI_ISL_2138385, EPI_ISL_2138388, EPI_ISL_2138408, EPI_ISL_2138416, EPI_ISL_2138418, EPI_ISL_2138425, EPI_ISL_2138429, EPI_ISL_2138434, EPI_ISL_2138436, EPI_ISL_2138442, EPI_ISL_2138455, EPI_ISL_2138459, EPI_ISL_2138469, EPI_ISL_2138471, EPI_ISL_2138475, EPI_ISL_2138485, EPI_ISL_2138503, EPI_ISL_2138516, EPI_ISL_2138517, EPI_ISL_2138521, EPI_ISL_2138523, EPI_ISL_2138542, EPI_ISL_2138543, EPI_ISL_2138549, EPI_ISL_2138561, EPI_ISL_2138572, EPI_ISL_2138580, EPI_ISL_2138582, EPI_ISL_2138597, EPI_ISL_2138600, EPI_ISL_2138617, EPI_ISL_2138622, EPI_ISL_2138649, EPI_ISL_2138678, EPI_ISL_2138681, EPI_ISL_2138684, EPI_ISL_2138688, EPI_ISL_2138691, EPI_ISL_2138694 | | | |
| see above | Health Services Laboratories | Wellcome Sanger Institute for the COVID-19 Genomics UK (COG-UK) Consortium | Cordelia Langford; David K. Jackson; Dominic Kwiatkowski; Ewan Harrison; Health Services Laboratories and Alex Alderton; Ian Johnston; Jeffrey Barrett; John Sillitoe on behalf of the Wellcome Sanger Institute COVID-19 Surveillance Team; Roberto Amato; Sonia Goncalves |
| EPI_ISL_2004345, EPI_ISL_2004351, EPI_ISL_2004353, EPI_ISL_2004356, EPI_ISL_2004360, EPI_ISL_2004361, EPI_ISL_2004362, EPI_ISL_2004365, EPI_ISL_2004368, EPI_ISL_2004373, EPI_ISL_2004377, EPI_ISL_2004378, EPI_ISL_2004379, EPI_ISL_2004380, EPI_ISL_2004381, EPI_ISL_2004384, EPI_ISL_2004387, EPI_ISL_2004389, EPI_ISL_2004390, EPI_ISL_2004394, EPI_ISL_2004395, EPI_ISL_2004397, EPI_ISL_2004399, EPI_ISL_2004401, EPI_ISL_2004403, EPI_ISL_2004405, EPI_ISL_2004411, EPI_ISL_2004412, EPI_ISL_2004415, EPI_ISL_2004419, EPI_ISL_2004420, EPI_ISL_2004422, EPI_ISL_2004427, EPI_ISL_2004430, EPI_ISL_2004435, EPI_ISL_2004439, EPI_ISL_2004440, EPI_ISL_2004441, EPI_ISL_2004442, EPI_ISL_2004444, EPI_ISL_2004446, EPI_ISL_2004451, EPI_ISL_2004454, EPI_ISL_2004458, EPI_ISL_2004459, EPI_ISL_2004460, EPI_ISL_2004462, EPI_ISL_2004464, EPI_ISL_2004465, EPI_ISL_2004466, EPI_ISL_2004469, EPI_ISL_2004474, EPI_ISL_2004475, EPI_ISL_2004479, EPI_ISL_2004480, EPI_ISL_2004481, EPI_ISL_2004487, EPI_ISL_2004489, EPI_ISL_2004492, EPI_ISL_2004495, EPI_ISL_2004496, EPI_ISL_2004498, EPI_ISL_2004501, EPI_ISL_2004502, EPI_ISL_2004503, EPI_ISL_2004505, EPI_ISL_2004506, EPI_ISL_2004507, EPI_ISL_2004510, EPI_ISL_2004511, EPI_ISL_2004512, EPI_ISL_2004514, EPI_ISL_2004515, EPI_ISL_2004516, EPI_ISL_2004519, EPI_ISL_2004520, EPI_ISL_2004521, EPI_ISL_2004522, EPI_ISL_2004525, EPI_ISL_2004529, EPI_ISL_2004530, EPI_ISL_2004534, EPI_ISL_2004535, EPI_ISL_2004536, EPI_ISL_2004538, EPI_ISL_2004541, EPI_ISL_2004542, EPI_ISL_2004543, EPI_ISL_2004547, EPI_ISL_2004550, EPI_ISL_2004551, EPI_ISL_2004553, EPI_ISL_2004555, EPI_ISL_2004556, EPI_ISL_2004557, EPI_ISL_2004558, EPI_ISL_2004564, EPI_ISL_2004565, EPI_ISL_2004566, EPI_ISL_2004567, EPI_ISL_2004568, EPI_ISL_2004569, EPI_ISL_2004571, EPI_ISL_2004572, EPI_ISL_2004573, EPI_ISL_2004575, EPI_ISL_2004576, EPI_ISL_2004577, EPI_ISL_2004580, EPI_ISL_2004585, EPI_ISL_2004587, EPI_ISL_2004588, EPI_ISL_2004591, EPI_ISL_2004596, EPI_ISL_2004598, EPI_ISL_2004601, EPI_ISL_2004602, EPI_ISL_2004607, EPI_ISL_2004608, EPI_ISL_2004609, EPI_ISL_2004610, EPI_ISL_2004612, EPI_ISL_2004613, EPI_ISL_2004614, EPI_ISL_2004617, EPI_ISL_2004618, EPI_ISL_2004619, EPI_ISL_2004658, EPI_ISL_2004671, EPI_ISL_2004673, EPI_ISL_2004688, EPI_ISL_2004725, EPI_ISL_2004732, EPI_ISL_2004747, EPI_ISL_2004754, EPI_ISL_2004779, EPI_ISL_2004811, EPI_ISL_2004821, EPI_ISL_2004845, EPI_ISL_2021043, EPI_ISL_2021044, EPI_ISL_2021052, EPI_ISL_2021053, EPI_ISL_2021054, EPI_ISL_2021056, EPI_ISL_2021059, EPI_ISL_2021061, EPI_ISL_2021064, EPI_ISL_2021077, EPI_ISL_2021082, EPI_ISL_2021083, EPI_ISL_2021090, EPI_ISL_2021092, EPI_ISL_2021110, EPI_ISL_2021111, EPI_ISL_2021119, EPI_ISL_2021124, EPI_ISL_2021127, EPI_ISL_2021131, EPI_ISL_2021144, EPI_ISL_2021145, EPI_ISL_2021147, EPI_ISL_2021151, EPI_ISL_2021152, EPI_ISL_2021154, EPI_ISL_2021158, EPI_ISL_2021159, EPI_ISL_2021170, EPI_ISL_2021181, EPI_ISL_2021183, EPI_ISL_2021186, EPI_ISL_2021190, EPI_ISL_2021193, EPI_ISL_2021194, EPI_ISL_2021195, EPI_ISL_2021197, EPI_ISL_2021198, EPI_ISL_2021200, EPI_ISL_2021205, EPI_ISL_2021209, EPI_ISL_2021215, EPI_ISL_2021218, EPI_ISL_2021222, EPI_ISL_2021225, EPI_ISL_2021228, EPI_ISL_2021231, EPI_ISL_2021235, EPI_ISL_2021236, EPI_ISL_2021241, EPI_ISL_2021242, EPI_ISL_2021248, EPI_ISL_2021249, EPI_ISL_2021252, EPI_ISL_2021254, EPI_ISL_2021255, EPI_ISL_2021258, EPI_ISL_2021262, EPI_ISL_2021271, EPI_ISL_2021274, EPI_ISL_2021275, EPI_ISL_2021277, EPI_ISL_2021280, EPI_ISL_2021281, EPI_ISL_2021282, EPI_ISL_2021287, EPI_ISL_2021291, EPI_ISL_2021292, EPI_ISL_2021296, EPI_ISL_2021298, EPI_ISL_2021310, EPI_ISL_2021311, EPI_ISL_2021315, EPI_ISL_2021316, EPI_ISL_2021317, EPI_ISL_2021318, EPI_ISL_2021326, EPI_ISL_2021327, EPI_ISL_2021329, EPI_ISL_2021331, EPI_ISL_2021332, EPI_ISL_2021339, EPI_ISL_2021343, EPI_ISL_2021348, EPI_ISL_2021349, EPI_ISL_2021358, EPI_ISL_2021362, EPI_ISL_2021363, EPI_ISL_2021364, EPI_ISL_2021365, EPI_ISL_2021366, EPI_ISL_2021367, EPI_ISL_2021369, EPI_ISL_2021370, EPI_ISL_2021373, EPI_ISL_2021374, EPI_ISL_2021375, EPI_ISL_2021376, EPI_ISL_2021377, EPI_ISL_2021379, EPI_ISL_2021380, EPI_ISL_2021385, EPI_ISL_2021386, EPI_ISL_2021388, EPI_ISL_2021389, EPI_ISL_2021390, EPI_ISL_2021393, EPI_ISL_2021394, EPI_ISL_2021395, EPI_ISL_2021396, EPI_ISL_2021397, EPI_ISL_2021400, EPI_ISL_2021402, EPI_ISL_2021403, EPI_ISL_2021408, EPI_ISL_2021410, EPI_ISL_2021411, EPI_ISL_2021413, EPI_ISL_2021414, EPI_ISL_2021418, EPI_ISL_2021420, EPI_ISL_2021423, EPI_ISL_2021426, EPI_ISL_2021428, EPI_ISL_2021431, EPI_ISL_2021432, EPI_ISL_2021437, EPI_ISL_2021443, EPI_ISL_2021446, EPI_ISL_2021449, EPI_ISL_2021450, EPI_ISL_2021451, EPI_ISL_2021455, EPI_ISL_2021456, EPI_ISL_2021457, EPI_ISL_2021459, EPI_ISL_2021460, EPI_ISL_2021464, EPI_ISL_2021466, EPI_ISL_2021468, EPI_ISL_2021469, EPI_ISL_2021470, EPI_ISL_2021472, EPI_ISL_2021473, EPI_ISL_2021474, EPI_ISL_2021475, EPI_ISL_2021476, EPI_ISL_2021482, EPI_ISL_2021484, EPI_ISL_2021485, EPI_ISL_2021486, EPI_ISL_2021487, EPI_ISL_2021488, EPI_ISL_2021489, EPI_ISL_2021490, EPI_ISL_2021491, EPI_ISL_2021493, EPI_ISL_2021497, EPI_ISL_2021498, EPI_ISL_2021499, EPI_ISL_2021500, EPI_ISL_2021503, EPI_ISL_2021505, EPI_ISL_2021512, EPI_ISL_2021513, EPI_ISL_2021514, EPI_ISL_2021515, EPI_ISL_2021516, EPI_ISL_2021519, EPI_ISL_2021520, EPI_ISL_2021521, EPI_ISL_2021524, EPI_ISL_2021525, EPI_ISL_2021526, EPI_ISL_2021527, EPI_ISL_2021528, EPI_ISL_2021529, EPI_ISL_2021530, EPI_ISL_2021531, EPI_ISL_2021532, EPI_ISL_2021533, EPI_ISL_2021537, EPI_ISL_2021540, EPI_ISL_2021542, EPI_ISL_2021543, EPI_ISL_2021547, EPI_ISL_2021550, EPI_ISL_2021551, EPI_ISL_2021552, EPI_ISL_2021553, EPI_ISL_2021559, EPI_ISL_2021560, EPI_ISL_2021561, EPI_ISL_2021562, EPI_ISL_2021563, EPI_ISL_2021564, EPI_ISL_2021565, EPI_ISL_2021568, EPI_ISL_2021569, EPI_ISL_2021570, EPI_ISL_2021579, EPI_ISL_2021581, EPI_ISL_2021582, EPI_ISL_2021583, EPI_ISL_2021585, EPI_ISL_2021589, EPI_ISL_2021591, EPI_ISL_2021592, EPI_ISL_2021594, EPI_ISL_2021596, EPI_ISL_2021598, EPI_ISL_2021599, EPI_ISL_2021600, EPI_ISL_2021602, EPI_ISL_2021604, EPI_ISL_2021605, EPI_ISL_2021606, EPI_ISL_2021607, EPI_ISL_2021608, EPI_ISL_2021609, EPI_ISL_2021615, EPI_ISL_2021616, EPI_ISL_2021617, EPI_ISL_2021618, EPI_ISL_2021619, EPI_ISL_2021621, EPI_ISL_2021623, EPI_ISL_2021624, EPI_ISL_2021627, EPI_ISL_2021628, EPI_ISL_2021629, EPI_ISL_2021631, EPI_ISL_2021633, EPI_ISL_2021634, EPI_ISL_2021637, EPI_ISL_2021640, EPI_ISL_2021641, EPI_ISL_2021643, EPI_ISL_2021644, EPI_ISL_2021647, EPI_ISL_2021648, EPI_ISL_2021650, EPI_ISL_2021651, EPI_ISL_2021652, EPI_ISL_2021653, EPI_ISL_2021656, EPI_ISL_2021657, EPI_ISL_2021660, EPI_ISL_2021662, EPI_ISL_2021664, EPI_ISL_2021667, EPI_ISL_2021669, EPI_ISL_2021670, EPI_ISL_2021672, EPI_ISL_2021674, EPI_ISL_2021680, EPI_ISL_2021682, EPI_ISL_2021684, EPI_ISL_2021692, EPI_ISL_2021693, EPI_ISL_2021694, EPI_ISL_2021852, EPI_ISL_2021861, EPI_ISL_2021864, EPI_ISL_2021869, EPI_ISL_2021870, EPI_ISL_2021873, EPI_ISL_2021876, EPI_ISL_2021882, EPI_ISL_2021883, EPI_ISL_2021890, EPI_ISL_2021892, EPI_ISL_2021895, EPI_ISL_2021903, EPI_ISL_2021911, EPI_ISL_2021913, EPI_ISL_2021916, EPI_ISL_2021918, EPI_ISL_2021933, EPI_ISL_2021938, EPI_ISL_2021941, EPI_ISL_2021943, EPI_ISL_2021949, EPI_ISL_2021954, EPI_ISL_2021963, EPI_ISL_2021971, EPI_ISL_2021977, EPI_ISL_2021980, EPI_ISL_2021984, EPI_ISL_2021986, EPI_ISL_2021989, EPI_ISL_2022003, EPI_ISL_2022005, EPI_ISL_2022009, EPI_ISL_2022011, EPI_ISL_2022020, EPI_ISL_2022021, EPI_ISL_2022033, EPI_ISL_2022035, EPI_ISL_2022038, EPI_ISL_2022053, EPI_ISL_2022061, EPI_ISL_2022062, EPI_ISL_2022074, EPI_ISL_2022075, EPI_ISL_2022077, EPI_ISL_2022086, EPI_ISL_2022087, EPI_ISL_2022092, EPI_ISL_2022097, EPI_ISL_2022101, EPI_ISL_2022109, EPI_ISL_2022112, EPI_ISL_2022124, EPI_ISL_2022132, EPI_ISL_2022135, EPI_ISL_2022138, EPI_ISL_2022142, EPI_ISL_2022160, EPI_ISL_2022162, EPI_ISL_2022163, EPI_ISL_2022164, EPI_ISL_2022165, EPI_ISL_2022166, EPI_ISL_2022168, EPI_ISL_2022171, EPI_ISL_2022172, EPI_ISL_2022178, EPI_ISL_2022181, EPI_ISL_2022182, EPI_ISL_2022183, EPI_ISL_2022184, EPI_ISL_2022185, EPI_ISL_2022188, EPI_ISL_2022189, EPI_ISL_2022190, EPI_ISL_2022192, EPI_ISL_2022193, EPI_ISL_2022195, EPI_ISL_2022196, EPI_ISL_2022197, EPI_ISL_2022198, EPI_ISL_2022205, EPI_ISL_2022206, EPI_ISL_2022208, EPI_ISL_2022209, EPI_ISL_2022210, EPI_ISL_2022211, EPI_ISL_2022212, EPI_ISL_2022214, EPI_ISL_2022215, EPI_ISL_2022216, EPI_ISL_2022217, EPI_ISL_2022218, EPI_ISL_2022219, EPI_ISL_2022220, EPI_ISL_2022221, EPI_ISL_2022222, EPI_ISL_2022223, EPI_ISL_2022225, EPI_ISL_2022226, EPI_ISL_2022227, EPI_ISL_2022228, EPI_ISL_2022229, EPI_ISL_2022230, EPI_ISL_2022232, EPI_ISL_2022233, EPI_ISL_2022234, EPI_ISL_2022235, EPI_ISL_2022237, EPI_ISL_2022243, EPI_ISL_2022246, EPI_ISL_2022247, EPI_ISL_2022248, EPI_ISL_2022249, EPI_ISL_2022252, EPI_ISL_2022253, EPI_ISL_2022254, EPI_ISL_2022255, EPI_ISL_2022257, EPI_ISL_2022259, EPI_ISL_2022260, EPI_ISL_2022266, EPI_ISL_2022267, EPI_ISL_2022268, EPI_ISL_2022269, EPI_ISL_2022270, EPI_ISL_2022271, EPI_ISL_2022273, EPI_ISL_2022274, EPI_ISL_2022275, EPI_ISL_2022276, EPI_ISL_2022282, EPI_ISL_2022283, EPI_ISL_2022284, EPI_ISL_2022287, EPI_ISL_2022289, EPI_ISL_2022291, EPI_ISL_2022292, EPI_ISL_2022293, EPI_ISL_2022294, EPI_ISL_2022297, EPI_ISL_2022298, EPI_ISL_2022300, EPI_ISL_2022301, EPI_ISL_2022302, EPI_ISL_2022303, EPI_ISL_2022307, EPI_ISL_2022309, EPI_ISL_2022310, EPI_ISL_2022311, EPI_ISL_2022312, EPI_ISL_2022313, EPI_ISL_2022314, EPI_ISL_2022315, EPI_ISL_2022316, EPI_ISL_2022318, EPI_ISL_2022320, EPI_ISL_2022321, EPI_ISL_2022322, EPI_ISL_2022323, EPI_ISL_2022324, EPI_ISL_2022325, EPI_ISL_2022326, EPI_ISL_2022328, EPI_ISL_2022329, EPI_ISL_2022331, EPI_ISL_2022333, EPI_ISL_2022334, EPI_ISL_2022335, EPI_ISL_2022336, EPI_ISL_2022338, EPI_ISL_2022339, EPI_ISL_2022341, EPI_ISL_2022344, EPI_ISL_2022345, EPI_ISL_2022347, EPI_ISL_2022349, EPI_ISL_2022350, EPI_ISL_2022353, EPI_ISL_2022354, EPI_ISL_2022355, EPI_ISL_2022357, EPI_ISL_2022360, EPI_ISL_2022361, EPI_ISL_2022362, EPI_ISL_2022363, EPI_ISL_2022365, EPI_ISL_2022366, EPI_ISL_2022368, EPI_ISL_2022370, EPI_ISL_2022371, EPI_ISL_2022372, EPI_ISL_2022377, EPI_ISL_2022378, EPI_ISL_2022379, EPI_ISL_2022380, EPI_ISL_2022382, EPI_ISL_2022384, EPI_ISL_2022385, EPI_ISL_2022386, EPI_ISL_2022389, EPI_ISL_2022390, EPI_ISL_2022391, EPI_ISL_2022392, EPI_ISL_2022393, EPI_ISL_2022394, EPI_ISL_2022395, EPI_ISL_2022396, EPI_ISL_2022401, EPI_ISL_2022402, EPI_ISL_2022403, EPI_ISL_2022404, EPI_ISL_2022405, EPI_ISL_2022406, EPI_ISL_2022407, EPI_ISL_2022409, EPI_ISL_2022411, EPI_ISL_2022412, EPI_ISL_2022413, EPI_ISL_2022414, EPI_ISL_2022416, EPI_ISL_2022417, EPI_ISL_2022418, EPI_ISL_2022421, EPI_ISL_2022422, EPI_ISL_2022424, EPI_ISL_2022425, EPI_ISL_2022426, EPI_ISL_2022427, EPI_ISL_2022430, EPI_ISL_2022431, EPI_ISL_2022433, EPI_ISL_2022434, EPI_ISL_2022435, EPI_ISL_2022436, EPI_ISL_2022438, EPI_ISL_2022784, EPI_ISL_2022845, EPI_ISL_2080475, EPI_ISL_2080481, EPI_ISL_2080482, EPI_ISL_2091462, EPI_ISL_2091464, EPI_ISL_2091465, EPI_ISL_2091466, EPI_ISL_2091467, EPI_ISL_2091470, EPI_ISL_2091471, EPI_ISL_2091474, EPI_ISL_2091475, EPI_ISL_2091476, EPI_ISL_2091478, EPI_ISL_2091483, EPI_ISL_2091485, EPI_ISL_2091488, EPI_ISL_2091490, EPI_ISL_2091491, EPI_ISL_2091498, EPI_ISL_2091499, EPI_ISL_2091501, EPI_ISL_2091503, EPI_ISL_2091504, EPI_ISL_2091505, EPI_ISL_2091506, EPI_ISL_2091507, EPI_ISL_2091508, EPI_ISL_2091509, EPI_ISL_2091512, EPI_ISL_2091513, EPI_ISL_2091516, EPI_ISL_2091517, EPI_ISL_2091519, EPI_ISL_2091520, EPI_ISL_2091523, EPI_ISL_2091527, EPI_ISL_2091529, EPI_ISL_2091530, EPI_ISL_2091531, EPI_ISL_2091535, EPI_ISL_2091537, EPI_ISL_2091538, EPI_ISL_2091541, EPI_ISL_2091544, EPI_ISL_2091545, EPI_ISL_2091546, EPI_ISL_2091548, EPI_ISL_2091550, EPI_ISL_2091551, EPI_ISL_2091552, EPI_ISL_2091556, EPI_ISL_2091559, EPI_ISL_2091560, EPI_ISL_2091561, EPI_ISL_2091563, EPI_ISL_2091567, EPI_ISL_2091568, EPI_ISL_2091570, EPI_ISL_2091572, EPI_ISL_2091573, EPI_ISL_2091574, EPI_ISL_2091575, EPI_ISL_2091576, EPI_ISL_2091577, EPI_ISL_2091578, EPI_ISL_2091579, EPI_ISL_2091580, EPI_ISL_2091584, EPI_ISL_2091588, EPI_ISL_2091589, EPI_ISL_2091591, EPI_ISL_2091593, EPI_ISL_2091594, EPI_ISL_2091595, EPI_ISL_2091597, EPI_ISL_2091598, EPI_ISL_2091599, EPI_ISL_2091600, EPI_ISL_2091601, EPI_ISL_2091606, EPI_ISL_2091607, EPI_ISL_2091608, EPI_ISL_2091609, EPI_ISL_2091610, EPI_ISL_2091611, EPI_ISL_2091612, EPI_ISL_2091615, EPI_ISL_2091616, EPI_ISL_2091617, EPI_ISL_2091618, EPI_ISL_2091619, EPI_ISL_2091620, EPI_ISL_2091621, EPI_ISL_2091623, EPI_ISL_2091626, EPI_ISL_2091628, EPI_ISL_2091629, EPI_ISL_2091631, EPI_ISL_2091634, EPI_ISL_2091635, EPI_ISL_2091637, EPI_ISL_2091641, EPI_ISL_2091644, EPI_ISL_2091645, EPI_ISL_2091646, EPI_ISL_2091647, EPI_ISL_2091648, EPI_ISL_2091653, EPI_ISL_2091654, EPI_ISL_2091655, EPI_ISL_2091657, EPI_ISL_2091658, EPI_ISL_2091661, EPI_ISL_2091662, EPI_ISL_2091663, EPI_ISL_2091667, EPI_ISL_2091668, EPI_ISL_2091673, EPI_ISL_2091674, EPI_ISL_2091675, EPI_ISL_2091676, EPI_ISL_2091677, EPI_ISL_2091680, EPI_ISL_2091682, EPI_ISL_2091683, EPI_ISL_2091684, EPI_ISL_2091686, EPI_ISL_2091688, EPI_ISL_2091689, EPI_ISL_2091690, EPI_ISL_2091692, EPI_ISL_2091696, EPI_ISL_2091697, EPI_ISL_2091700, EPI_ISL_2091701, EPI_ISL_2091705, EPI_ISL_2091706, EPI_ISL_2091707, EPI_ISL_2091708, EPI_ISL_2091709, EPI_ISL_2091712, EPI_ISL_2091714, EPI_ISL_2091715, EPI_ISL_2091716, EPI_ISL_2091717, EPI_ISL_2091718, EPI_ISL_2091720, EPI_ISL_2091721, EPI_ISL_2091722, EPI_ISL_2091723, EPI_ISL_2091725, EPI_ISL_2091726, EPI_ISL_2091727, EPI_ISL_2091731, EPI_ISL_2091732, EPI_ISL_2091734, EPI_ISL_2091737, EPI_ISL_2091739, EPI_ISL_2091747, EPI_ISL_2091748, EPI_ISL_2091752, EPI_ISL_2091753, EPI_ISL_2091755, EPI_ISL_2091758, EPI_ISL_2091762, EPI_ISL_2091767, EPI_ISL_2091768, EPI_ISL_2091770, EPI_ISL_2091771, EPI_ISL_2091773, EPI_ISL_2091775, EPI_ISL_2091776, EPI_ISL_2091777, EPI_ISL_2091778, EPI_ISL_2091780, EPI_ISL_2091782, EPI_ISL_2091787, EPI_ISL_2091790, EPI_ISL_2091792, EPI_ISL_2091800, EPI_ISL_2091801, EPI_ISL_2091804, EPI_ISL_2091808, EPI_ISL_2091810, EPI_ISL_2091812, EPI_ISL_2091814, EPI_ISL_2091816, EPI_ISL_2091819, EPI_ISL_2091823, EPI_ISL_2091825, EPI_ISL_2091827, EPI_ISL_2091828, EPI_ISL_2091829, EPI_ISL_2091830, EPI_ISL_2091831, EPI_ISL_2091833, EPI_ISL_2091836, EPI_ISL_2091841, EPI_ISL_2091842, EPI_ISL_2091844, EPI_ISL_2091845, EPI_ISL_2091847, EPI_ISL_2091851, EPI_ISL_2091852, EPI_ISL_2091854, EPI_ISL_2091856, EPI_ISL_2091858, EPI_ISL_2091859, EPI_ISL_2091863, EPI_ISL_2091864, EPI_ISL_2091865, EPI_ISL_2091866, EPI_ISL_2091868, EPI_ISL_2091869, EPI_ISL_2091870, EPI_ISL_2091874, EPI_ISL_2091877, EPI_ISL_2091878, EPI_ISL_2091880, EPI_ISL_2091881, EPI_ISL_2091884, EPI_ISL_2091885, EPI_ISL_2091888, EPI_ISL_2091891, EPI_ISL_2091892, EPI_ISL_2091893, EPI_ISL_2091894, EPI_ISL_2091896, EPI_ISL_2091897, EPI_ISL_2091898, EPI_ISL_2091904, EPI_ISL_2091905, EPI_ISL_2091910, EPI_ISL_2091911, EPI_ISL_2091915, EPI_ISL_2091917, EPI_ISL_2091919, EPI_ISL_2091921, EPI_ISL_2091922, EPI_ISL_2091924, EPI_ISL_2091929, EPI_ISL_2091930, EPI_ISL_2091931, EPI_ISL_2091934, EPI_ISL_2091936, EPI_ISL_2091937, EPI_ISL_2091940, EPI_ISL_2091943, EPI_ISL_2091945, EPI_ISL_2091947, EPI_ISL_2091951, EPI_ISL_2091954, EPI_ISL_2091955, EPI_ISL_2091958, EPI_ISL_2091959, EPI_ISL_2091961, EPI_ISL_2091962, EPI_ISL_2091964, EPI_ISL_2091965, EPI_ISL_2091966, EPI_ISL_2091967, EPI_ISL_2091968, EPI_ISL_2091970, EPI_ISL_2091974, EPI_ISL_2091977, EPI_ISL_2091978, EPI_ISL_2091979, EPI_ISL_2091980, EPI_ISL_2091982, EPI_ISL_2091987, EPI_ISL_2091995, EPI_ISL_2091996, EPI_ISL_2091999, EPI_ISL_2092001, EPI_ISL_2092002, EPI_ISL_2092003, EPI_ISL_2092005, EPI_ISL_2092006, EPI_ISL_2092008, EPI_ISL_2092010, EPI_ISL_2092011, EPI_ISL_2092013, EPI_ISL_2092020, EPI_ISL_2092022, EPI_ISL_2092023, EPI_ISL_2092026, EPI_ISL_2092034, EPI_ISL_2092039, EPI_ISL_2092040, EPI_ISL_2092046, EPI_ISL_2092047, EPI_ISL_2092048, EPI_ISL_2092049, EPI_ISL_2092050, EPI_ISL_2092052, EPI_ISL_2092054, EPI_ISL_2092055, EPI_ISL_2092057, EPI_ISL_2092058, EPI_ISL_2092059, EPI_ISL_2092060, EPI_ISL_2092061, EPI_ISL_2092075, EPI_ISL_2092076, EPI_ISL_2092082, EPI_ISL_2092085, EPI_ISL_2092091, EPI_ISL_2092092, EPI_ISL_2092096, EPI_ISL_2092099, EPI_ISL_2092100, EPI_ISL_2092102, EPI_ISL_2092103, EPI_ISL_2092104, EPI_ISL_2092105, EPI_ISL_2092106, EPI_ISL_2092107, EPI_ISL_2092108, EPI_ISL_2092109, EPI_ISL_2092110, EPI_ISL_2092111, EPI_ISL_2092113, EPI_ISL_2092115, EPI_ISL_2092116, EPI_ISL_2092117, EPI_ISL_2092119, EPI_ISL_2092121, EPI_ISL_2092122, EPI_ISL_2092123, EPI_ISL_2092124, EPI_ISL_2092125, EPI_ISL_2092126, EPI_ISL_2092127, EPI_ISL_2092128, EPI_ISL_2092129, EPI_ISL_2092130, EPI_ISL_2092131, EPI_ISL_2092132, EPI_ISL_2092133, EPI_ISL_2092134, EPI_ISL_2092135, EPI_ISL_2092136, EPI_ISL_2092137, EPI_ISL_2092138, EPI_ISL_2092139, EPI_ISL_2092142, EPI_ISL_2092144, EPI_ISL_2092145, EPI_ISL_2092146, EPI_ISL_2092147, EPI_ISL_2092148, EPI_ISL_2092149, EPI_ISL_2092150, EPI_ISL_2092151, EPI_ISL_2092153, EPI_ISL_2092154, EPI_ISL_2092155, EPI_ISL_2092157, EPI_ISL_2092158, EPI_ISL_2092159, EPI_ISL_2092160, EPI_ISL_2092161, EPI_ISL_2092164, EPI_ISL_2092165, EPI_ISL_2092166, EPI_ISL_2092168, EPI_ISL_2092169, EPI_ISL_2092170, EPI_ISL_2092172, EPI_ISL_2092173, EPI_ISL_2092174, EPI_ISL_2092175, EPI_ISL_2092176, EPI_ISL_2092177, EPI_ISL_2092178, EPI_ISL_2092179, EPI_ISL_2092180, EPI_ISL_2092182, EPI_ISL_2092184, EPI_ISL_2092185, EPI_ISL_2092186, EPI_ISL_2092187, EPI_ISL_2092189, EPI_ISL_2092190, EPI_ISL_2092191, EPI_ISL_2092192, EPI_ISL_2092194, EPI_ISL_2092196, EPI_ISL_2092197, EPI_ISL_2092198, EPI_ISL_2092199, EPI_ISL_2092201, EPI_ISL_2092204, EPI_ISL_2092205, EPI_ISL_2092206, EPI_ISL_2092207, EPI_ISL_2092208, EPI_ISL_2092209, EPI_ISL_2092210, EPI_ISL_2092211, EPI_ISL_2092212, EPI_ISL_2092213, EPI_ISL_2092215, EPI_ISL_2092216, EPI_ISL_2092217, EPI_ISL_2092218, EPI_ISL_2092219, EPI_ISL_2092220, EPI_ISL_2092221, EPI_ISL_2092224, EPI_ISL_2092225, EPI_ISL_2092226, EPI_ISL_2092227, EPI_ISL_2092228, EPI_ISL_2092229, EPI_ISL_2092230, EPI_ISL_2092231, EPI_ISL_2092232, EPI_ISL_2092234, EPI_ISL_2092235, EPI_ISL_2092236, EPI_ISL_2092237, EPI_ISL_2092238, EPI_ISL_2092241, EPI_ISL_2092242, EPI_ISL_2092244, EPI_ISL_2092246, EPI_ISL_2092247, EPI_ISL_2092248, EPI_ISL_2092249, EPI_ISL_2092250, EPI_ISL_2092251, EPI_ISL_2092252, EPI_ISL_2092253, EPI_ISL_2092254, EPI_ISL_2092255, EPI_ISL_2092257, EPI_ISL_2092258, EPI_ISL_2092260, EPI_ISL_2092261, EPI_ISL_2092262, EPI_ISL_2092263, EPI_ISL_2092267, EPI_ISL_2092268, EPI_ISL_2092269, EPI_ISL_2092271, EPI_ISL_2092274, EPI_ISL_2092275, EPI_ISL_2092276, EPI_ISL_2092277, EPI_ISL_2092279, EPI_ISL_2092281, EPI_ISL_2092282, EPI_ISL_2092283, EPI_ISL_2092284, EPI_ISL_2092285, EPI_ISL_2092286, EPI_ISL_2092287, EPI_ISL_2092288, EPI_ISL_2092289, EPI_ISL_2092290, EPI_ISL_2092291, EPI_ISL_2092292, EPI_ISL_2092293, EPI_ISL_2092294, EPI_ISL_2092295, EPI_ISL_2092297, EPI_ISL_2092298, EPI_ISL_2092299, EPI_ISL_2092300, EPI_ISL_2092301, EPI_ISL_2092302, EPI_ISL_2092303, EPI_ISL_2092304, EPI_ISL_2092305, EPI_ISL_2092306, EPI_ISL_2092307, EPI_ISL_2092308, EPI_ISL_2092309, EPI_ISL_2092310, EPI_ISL_2092311, EPI_ISL_2092313, EPI_ISL_2092314, EPI_ISL_2092315, EPI_ISL_2092316, EPI_ISL_2092317, EPI_ISL_2092318, EPI_ISL_2092319, EPI_ISL_2092320, EPI_ISL_2092321, EPI_ISL_2092322, EPI_ISL_2092323, EPI_ISL_2092324, EPI_ISL_2092325, EPI_ISL_2092326, EPI_ISL_2092327, EPI_ISL_2092328, EPI_ISL_2092329, EPI_ISL_2092330, EPI_ISL_2092331, EPI_ISL_2092332, EPI_ISL_2092333, EPI_ISL_2092334, EPI_ISL_2092335, EPI_ISL_2092336, EPI_ISL_2092337, EPI_ISL_2092340, EPI_ISL_2092343, EPI_ISL_2092344, EPI_ISL_2092345, EPI_ISL_2092346, EPI_ISL_2092348, EPI_ISL_2092349, EPI_ISL_2092350, EPI_ISL_2092351, EPI_ISL_2092352, EPI_ISL_2092353, EPI_ISL_2092354, EPI_ISL_2092355, EPI_ISL_2092356, EPI_ISL_2092357, EPI_ISL_2092358, EPI_ISL_2092359, EPI_ISL_2092360, EPI_ISL_2092361, EPI_ISL_2092363, EPI_ISL_2092364, EPI_ISL_2092366, EPI_ISL_2092367, EPI_ISL_2092368, EPI_ISL_2092369, EPI_ISL_2092370, EPI_ISL_2092371, EPI_ISL_2092372, EPI_ISL_2092374, EPI_ISL_2092375, EPI_ISL_2092376, EPI_ISL_2092377, EPI_ISL_2092378, EPI_ISL_2092379, EPI_ISL_2092381, EPI_ISL_2092382, EPI_ISL_2092383, EPI_ISL_2092384, EPI_ISL_2092385, EPI_ISL_2092386, EPI_ISL_2092387, EPI_ISL_2092388, EPI_ISL_2092389, EPI_ISL_2092390, EPI_ISL_2092391, EPI_ISL_2092392, EPI_ISL_2092393, EPI_ISL_2092394, EPI_ISL_2092395, EPI_ISL_2092396, EPI_ISL_2092397, EPI_ISL_2092398, EPI_ISL_2092399, EPI_ISL_2092400, EPI_ISL_2092402, EPI_ISL_2092403, EPI_ISL_2092404, EPI_ISL_2092405, EPI_ISL_2092407, EPI_ISL_2092408, EPI_ISL_2092409, EPI_ISL_2092410, EPI_ISL_2092412, EPI_ISL_2092413, EPI_ISL_2092415, EPI_ISL_2092418, EPI_ISL_2092420, EPI_ISL_2092421, EPI_ISL_2092422, EPI_ISL_2092423, EPI_ISL_2092424, EPI_ISL_2092426, EPI_ISL_2092427, EPI_ISL_2092429, EPI_ISL_2092430, EPI_ISL_2092431, EPI_ISL_2092432, EPI_ISL_2092433, EPI_ISL_2092434, EPI_ISL_2092435, EPI_ISL_2092436, EPI_ISL_2092437, EPI_ISL_2092439, EPI_ISL_2092443, EPI_ISL_2092444, EPI_ISL_2092445, EPI_ISL_2092446, EPI_ISL_2092447, EPI_ISL_2092448, EPI_ISL_2092449, EPI_ISL_2092450, EPI_ISL_2092452, EPI_ISL_2092453, EPI_ISL_2092454, EPI_ISL_2092455, EPI_ISL_2092456, EPI_ISL_2092457, EPI_ISL_2092458, EPI_ISL_2092460, EPI_ISL_2092461, EPI_ISL_2092462, EPI_ISL_2092463, EPI_ISL_2092464, EPI_ISL_2092465, EPI_ISL_2092466, EPI_ISL_2092741, EPI_ISL_2092742, EPI_ISL_2117512, EPI_ISL_2117525, EPI_ISL_2117528, EPI_ISL_2117533, EPI_ISL_2117539, EPI_ISL_2117546, EPI_ISL_2117552, EPI_ISL_2117553, EPI_ISL_2117557, EPI_ISL_2117560, EPI_ISL_2117565, EPI_ISL_2117568, EPI_ISL_2117569, EPI_ISL_2117573, EPI_ISL_2117577, EPI_ISL_2117580, EPI_ISL_2117588, EPI_ISL_2117590, EPI_ISL_2117594, EPI_ISL_2117595, EPI_ISL_2117603, EPI_ISL_2117604, EPI_ISL_2117610, EPI_ISL_2117614, EPI_ISL_2117619, EPI_ISL_2117620, EPI_ISL_2117622, EPI_ISL_2117623, EPI_ISL_2117624, EPI_ISL_2117626, EPI_ISL_2117633, EPI_ISL_2117647, EPI_ISL_2117650, EPI_ISL_2117652, EPI_ISL_2117654, EPI_ISL_2117658, EPI_ISL_2117661, EPI_ISL_2117663, EPI_ISL_2117665, EPI_ISL_2117670, EPI_ISL_2117672, EPI_ISL_2117673, EPI_ISL_2117674, EPI_ISL_2117684, EPI_ISL_2117685, EPI_ISL_2117687, EPI_ISL_2117688, EPI_ISL_2117690, EPI_ISL_2117691, EPI_ISL_2117693, EPI_ISL_2117694, EPI_ISL_2117696, EPI_ISL_2117699, EPI_ISL_2117702, EPI_ISL_2117708, EPI_ISL_2117717, EPI_ISL_2117719, EPI_ISL_2117720, EPI_ISL_2117723, EPI_ISL_2117729, EPI_ISL_2117742, EPI_ISL_2117748, EPI_ISL_2117754, EPI_ISL_2117756, EPI_ISL_2117758, EPI_ISL_2117762, EPI_ISL_2117764, EPI_ISL_2117769, EPI_ISL_2117770, EPI_ISL_2117772, EPI_ISL_2117784, EPI_ISL_2117787, EPI_ISL_2117794, EPI_ISL_2117796, EPI_ISL_2117809, EPI_ISL_2117819, EPI_ISL_2117829, EPI_ISL_2117831, EPI_ISL_2117832, EPI_ISL_2117834, EPI_ISL_2117838, EPI_ISL_2117843, EPI_ISL_2117845, EPI_ISL_2117851, EPI_ISL_2117860, EPI_ISL_2117865, EPI_ISL_2117871, EPI_ISL_2117877, EPI_ISL_2117879, EPI_ISL_2117880, EPI_ISL_2117895, EPI_ISL_2117903, EPI_ISL_2117908, EPI_ISL_2117919, EPI_ISL_2117931, EPI_ISL_2117933, EPI_ISL_2117934, EPI_ISL_2117935, EPI_ISL_2117938, EPI_ISL_2117939, EPI_ISL_2117941, EPI_ISL_2117949, EPI_ISL_2117954, EPI_ISL_2117957, EPI_ISL_2117966, EPI_ISL_2117967, EPI_ISL_2117969, EPI_ISL_2117974, EPI_ISL_2117980, EPI_ISL_2117981, EPI_ISL_2117984, EPI_ISL_2117993, EPI_ISL_2117994, EPI_ISL_2117996, EPI_ISL_2118001, EPI_ISL_2118002, EPI_ISL_2118005, EPI_ISL_2118010, EPI_ISL_2118011, EPI_ISL_2118016, EPI_ISL_2118027, EPI_ISL_2118029, EPI_ISL_2118041, EPI_ISL_2118052, EPI_ISL_2118059, EPI_ISL_2118060, EPI_ISL_2118061, EPI_ISL_2118064, EPI_ISL_2118065, EPI_ISL_2118068, EPI_ISL_2118070, EPI_ISL_2118072, EPI_ISL_2118076, EPI_ISL_2118083, EPI_ISL_2118099, EPI_ISL_2118100, EPI_ISL_2118102, EPI_ISL_2118106, EPI_ISL_2118108, EPI_ISL_2118111, EPI_ISL_2118114, EPI_ISL_2118120, EPI_ISL_2118129, EPI_ISL_2118132, EPI_ISL_2118142, EPI_ISL_2118148, EPI_ISL_2118158, EPI_ISL_2118162, EPI_ISL_2118170, EPI_ISL_2118171, EPI_ISL_2118173, EPI_ISL_2118174, EPI_ISL_2118185, EPI_ISL_2118191, EPI_ISL_2118192, EPI_ISL_2118195, EPI_ISL_2118211, EPI_ISL_2118219, EPI_ISL_2118220, EPI_ISL_2118233, EPI_ISL_2118235, EPI_ISL_2118237, EPI_ISL_2118238, EPI_ISL_2118244, EPI_ISL_2118249, EPI_ISL_2118250, EPI_ISL_2118252, EPI_ISL_2118254, EPI_ISL_2118255, EPI_ISL_2118256, EPI_ISL_2118261, EPI_ISL_2118262, EPI_ISL_2118264, EPI_ISL_2118265, EPI_ISL_2118267, EPI_ISL_2118268, EPI_ISL_2118270, EPI_ISL_2118271, EPI_ISL_2118272, EPI_ISL_2118274, EPI_ISL_2118310, EPI_ISL_2118312, EPI_ISL_2118314, EPI_ISL_2118317, EPI_ISL_2118318, EPI_ISL_2118320, EPI_ISL_2118321, EPI_ISL_2118322, EPI_ISL_2118327, EPI_ISL_2118329, EPI_ISL_2118330, EPI_ISL_2118331, EPI_ISL_2118333, EPI_ISL_2118334, EPI_ISL_2118336, EPI_ISL_2118341, EPI_ISL_2118342, EPI_ISL_2118346, EPI_ISL_2118348, EPI_ISL_2118350, EPI_ISL_2118351, EPI_ISL_2118353, EPI_ISL_2118354, EPI_ISL_2118357, EPI_ISL_2118360, EPI_ISL_2118362, EPI_ISL_2118365, EPI_ISL_2118366, EPI_ISL_2118369, EPI_ISL_2118372, EPI_ISL_2118374, EPI_ISL_2118375, EPI_ISL_2118377, EPI_ISL_2118378, EPI_ISL_2118380, EPI_ISL_2118381, EPI_ISL_2118384, EPI_ISL_2118385, EPI_ISL_2118387, EPI_ISL_2118388, EPI_ISL_2118391, EPI_ISL_2118393, EPI_ISL_2118394, EPI_ISL_2118396, EPI_ISL_2118397, EPI_ISL_2118404, EPI_ISL_2118408, EPI_ISL_2118411, EPI_ISL_2118414, EPI_ISL_2118416, EPI_ISL_2118419, EPI_ISL_2118421, EPI_ISL_2118422, EPI_ISL_2118424, EPI_ISL_2118427, EPI_ISL_2118428, EPI_ISL_2118430, EPI_ISL_2118431, EPI_ISL_2118433, EPI_ISL_2118434, EPI_ISL_2118437, EPI_ISL_2118441, EPI_ISL_2118443, EPI_ISL_2118444, EPI_ISL_2118446, EPI_ISL_2118447, EPI_ISL_2118448, EPI_ISL_2118456, EPI_ISL_2118457, EPI_ISL_2118460, EPI_ISL_2118462, EPI_ISL_2118465, EPI_ISL_2118469, EPI_ISL_2118471, EPI_ISL_2118472, EPI_ISL_2118473, EPI_ISL_2118475, EPI_ISL_2118476, EPI_ISL_2118478, EPI_ISL_2118479, EPI_ISL_2118481, EPI_ISL_2118482, EPI_ISL_2118484, EPI_ISL_2118485, EPI_ISL_2118487, EPI_ISL_2118488, EPI_ISL_2118489, EPI_ISL_2118491, EPI_ISL_2118492, EPI_ISL_2118494, EPI_ISL_2118496, EPI_ISL_2118497, EPI_ISL_2118500, EPI_ISL_2118502, EPI_ISL_2118504, EPI_ISL_2118506, EPI_ISL_2118509, EPI_ISL_2118510, EPI_ISL_2118512, EPI_ISL_2118513, EPI_ISL_2118515, EPI_ISL_2118516, EPI_ISL_2118518, EPI_ISL_2118521, EPI_ISL_2118522, EPI_ISL_2118523, EPI_ISL_2118525, EPI_ISL_2118528, EPI_ISL_2118529, EPI_ISL_2118531, EPI_ISL_2118532, EPI_ISL_2118534, EPI_ISL_2118535, EPI_ISL_2118537, EPI_ISL_2118538, EPI_ISL_2118540, EPI_ISL_2118541, EPI_ISL_2118543, EPI_ISL_2118544, EPI_ISL_2118546, EPI_ISL_2118551, EPI_ISL_2118552, EPI_ISL_2118554, EPI_ISL_2118556, EPI_ISL_2118558, EPI_ISL_2118560, EPI_ISL_2118561, EPI_ISL_2118564, EPI_ISL_2118566, EPI_ISL_2118567, EPI_ISL_2118569, EPI_ISL_2118573, EPI_ISL_2118575, EPI_ISL_2118579, EPI_ISL_2118581, EPI_ISL_2118582, EPI_ISL_2118585, EPI_ISL_2118586, EPI_ISL_2118589, EPI_ISL_2118590, EPI_ISL_2118593, EPI_ISL_2118595, EPI_ISL_2118596, EPI_ISL_2118598, EPI_ISL_2118599, EPI_ISL_2118601, EPI_ISL_2118604, EPI_ISL_2118609, EPI_ISL_2118610, EPI_ISL_2118612, EPI_ISL_2118613, EPI_ISL_2118615, EPI_ISL_2118618, EPI_ISL_2118622, EPI_ISL_2118624, EPI_ISL_2118625, EPI_ISL_2118628, EPI_ISL_2118630, EPI_ISL_2118631, EPI_ISL_2118634, EPI_ISL_2118636, EPI_ISL_2118638, EPI_ISL_2118639, EPI_ISL_2118642, EPI_ISL_2118643, EPI_ISL_2118645, EPI_ISL_2118648, EPI_ISL_2118649, EPI_ISL_2118652, EPI_ISL_2118654, EPI_ISL_2118655, EPI_ISL_2118657, EPI_ISL_2118658, EPI_ISL_2118659, EPI_ISL_2118661, EPI_ISL_2118662, EPI_ISL_2118667, EPI_ISL_2118668, EPI_ISL_2118670, EPI_ISL_2118671, EPI_ISL_2118673, EPI_ISL_2118675, EPI_ISL_2118676, EPI_ISL_2118678, EPI_ISL_2118679, EPI_ISL_2118682, EPI_ISL_2118684, EPI_ISL_2118685, EPI_ISL_2118687, EPI_ISL_2118689, EPI_ISL_2118690, EPI_ISL_2118692, EPI_ISL_2118694, EPI_ISL_2118696, EPI_ISL_2118698, EPI_ISL_2118700, EPI_ISL_2118701, EPI_ISL_2118704, EPI_ISL_2118705, EPI_ISL_2118708, EPI_ISL_2118709, EPI_ISL_2118711, EPI_ISL_2118712, EPI_ISL_2118713, EPI_ISL_2118716, EPI_ISL_2118718, EPI_ISL_2118721, EPI_ISL_2118722, EPI_ISL_2118724, EPI_ISL_2118725, EPI_ISL_2118728, EPI_ISL_2118730, EPI_ISL_2118732, EPI_ISL_2118733, EPI_ISL_2118735, EPI_ISL_2118738, EPI_ISL_2118739, EPI_ISL_2118740, | | | |

EPI_ISL_2118742, EPI_ISL_2118744, EPI_ISL_2118745, EPI_ISL_2118748, EPI_ISL_2118750, EPI_ISL_2118751, EPI_ISL_2118756, EPI_ISL_2118758, EPI_ISL_2118762, EPI_ISL_2118763, EPI_ISL_2118765, EPI_ISL_2118766, EPI_ISL_2118769, EPI_ISL_2118770, EPI_ISL_2118773, EPI_ISL_2118775, EPI_ISL_2118776, EPI_ISL_2118778, EPI_ISL_2118779, EPI_ISL_2118781, EPI_ISL_2118785, EPI_ISL_2118786, EPI_ISL_2118788, EPI_ISL_2118791, EPI_ISL_2118793, EPI_ISL_2118796, EPI_ISL_2118797, EPI_ISL_2118803, EPI_ISL_2118807, EPI_ISL_2118811, EPI_ISL_2118813, EPI_ISL_2118814, EPI_ISL_2118816, EPI_ISL_2118817, EPI_ISL_2118819, EPI_ISL_2118822, EPI_ISL_2118828, EPI_ISL_2118829, EPI_ISL_2118830, EPI_ISL_2118831, EPI_ISL_2118832, EPI_ISL_2118835, EPI_ISL_2118843, EPI_ISL_2118844, EPI_ISL_2118849, EPI_ISL_2118851, EPI_ISL_2118852, EPI_ISL_2118855, EPI_ISL_2118864, EPI_ISL_2118865, EPI_ISL_2118868, EPI_ISL_2118876, EPI_ISL_2118882, EPI_ISL_2118885, EPI_ISL_2118890, EPI_ISL_2118891, EPI_ISL_2118897, EPI_ISL_2118903, EPI_ISL_2118908, EPI_ISL_2118912, EPI_ISL_2118915, EPI_ISL_2118918, EPI_ISL_2118922, EPI_ISL_2118923, EPI_ISL_2118924, EPI_ISL_2118926, EPI_ISL_2118928, EPI_ISL_2118930, EPI_ISL_2118933, EPI_ISL_2118935, EPI_ISL_2118939, EPI_ISL_2118945, EPI_ISL_2118955, EPI_ISL_2118958, EPI_ISL_2118960, EPI_ISL_2118965, EPI_ISL_2118967, EPI_ISL_2118970, EPI_ISL_2118973, EPI_ISL_2118979, EPI_ISL_2118983, EPI_ISL_2118985, EPI_ISL_2118988, EPI_ISL_2118989, EPI_ISL_2118991, EPI_ISL_2118992, EPI_ISL_2118998, EPI_ISL_2119006, EPI_ISL_2119013, EPI_ISL_2119015, EPI_ISL_2119017, EPI_ISL_2119020, EPI_ISL_2119022, EPI_ISL_2119024, EPI_ISL_2119027, EPI_ISL_2119039, EPI_ISL_2119040, EPI_ISL_2119041, EPI_ISL_2119043, EPI_ISL_2119045, EPI_ISL_2119083, EPI_ISL_2119086, EPI_ISL_2119088, EPI_ISL_2119089, EPI_ISL_2119091, EPI_ISL_2119097, EPI_ISL_2119099, EPI_ISL_2119100, EPI_ISL_2119102, EPI_ISL_2119113, EPI_ISL_2119121, EPI_ISL_2119122, EPI_ISL_2119124, EPI_ISL_2119127, EPI_ISL_2119128, EPI_ISL_2119130, EPI_ISL_2119133, EPI_ISL_2119134, EPI_ISL_2119138, EPI_ISL_2119141, EPI_ISL_2119142, EPI_ISL_2119146, EPI_ISL_2119149, EPI_ISL_2119150, EPI_ISL_2119152, EPI_ISL_2119155, EPI_ISL_2119158, EPI_ISL_2119160, EPI_ISL_2119163, EPI_ISL_2119166, EPI_ISL_2119169, EPI_ISL_2119172, EPI_ISL_2119183, EPI_ISL_2119185, EPI_ISL_2119192, EPI_ISL_2119196, EPI_ISL_2119199, EPI_ISL_2119204, EPI_ISL_2119206, EPI_ISL_2119210, EPI_ISL_2119211, EPI_ISL_2119216, EPI_ISL_2119218, EPI_ISL_2119220, EPI_ISL_2119222, EPI_ISL_2119226, EPI_ISL_2119230, EPI_ISL_2119234, EPI_ISL_2119237, EPI_ISL_2119243, EPI_ISL_2119244, EPI_ISL_2119247, EPI_ISL_2119250, EPI_ISL_2119256, EPI_ISL_2119260, EPI_ISL_2119265, EPI_ISL_2119266, EPI_ISL_2119268, EPI_ISL_2119271, EPI_ISL_2119274, EPI_ISL_2119275, EPI_ISL_2119277, EPI_ISL_2119278, EPI_ISL_2119280, EPI_ISL_2119285, EPI_ISL_2119289, EPI_ISL_2119294, EPI_ISL_2119301, EPI_ISL_2119309, EPI_ISL_2119311, EPI_ISL_2119320, EPI_ISL_2119324, EPI_ISL_2119327, EPI_ISL_2119330, EPI_ISL_2119336, EPI_ISL_2119343, EPI_ISL_2119346, EPI_ISL_2119349, EPI_ISL_2119370, EPI_ISL_2119395, EPI_ISL_2119406, EPI_ISL_2119410, EPI_ISL_2119416, EPI_ISL_2119428, EPI_ISL_2119432, EPI_ISL_2119439, EPI_ISL_2119442, EPI_ISL_2119445, EPI_ISL_2119448, EPI_ISL_2119452, EPI_ISL_2119471, EPI_ISL_2119473, EPI_ISL_2119476, EPI_ISL_2119480, EPI_ISL_2119484, EPI_ISL_2119487, EPI_ISL_2119504, EPI_ISL_2119518, EPI_ISL_2119534, EPI_ISL_2119537, EPI_ISL_2119539, EPI_ISL_2119541, EPI_ISL_2119543, EPI_ISL_2119551, EPI_ISL_2119554, EPI_ISL_2119559, EPI_ISL_2119561, EPI_ISL_2119569, EPI_ISL_2119571, EPI_ISL_2119574, EPI_ISL_2119582, EPI_ISL_2119584, EPI_ISL_2119587, EPI_ISL_2119591, EPI_ISL_2119595, EPI_ISL_2119599, EPI_ISL_2119609, EPI_ISL_2119620, EPI_ISL_2119622, EPI_ISL_2119623, EPI_ISL_2119626, EPI_ISL_2119628, EPI_ISL_2119631, EPI_ISL_2119633, EPI_ISL_2119638, EPI_ISL_2119639, EPI_ISL_2119646, EPI_ISL_2119651, EPI_ISL_2119679, EPI_ISL_2119691, EPI_ISL_2119709, EPI_ISL_2119710, EPI_ISL_2119713, EPI_ISL_2119715, EPI_ISL_2119726, EPI_ISL_2119729, EPI_ISL_2119734, EPI_ISL_2119736, EPI_ISL_2119738, EPI_ISL_2119762, EPI_ISL_2119765, EPI_ISL_2119769, EPI_ISL_2119772, EPI_ISL_2119775, EPI_ISL_2119780, EPI_ISL_2119781, EPI_ISL_2119794, EPI_ISL_2119822, EPI_ISL_2119835, EPI_ISL_2119838, EPI_ISL_2119849, EPI_ISL_2119852, EPI_ISL_2119855, EPI_ISL_2119859, EPI_ISL_2119867, EPI_ISL_2119872, EPI_ISL_2119873, EPI_ISL_2119902, EPI_ISL_2119915, EPI_ISL_2119919, EPI_ISL_2119943, EPI_ISL_2119951, EPI_ISL_2119958, EPI_ISL_2119961, EPI_ISL_2119988, EPI_ISL_2119994, EPI_ISL_2119999, EPI_ISL_2120012, EPI_ISL_2120015, EPI_ISL_2120017, EPI_ISL_2120022, EPI_ISL_2120031, EPI_ISL_2120043, EPI_ISL_2120051, EPI_ISL_2120069, EPI_ISL_2120072, EPI_ISL_2120087, EPI_ISL_2120092, EPI_ISL_2120121, EPI_ISL_2120124, EPI_ISL_2120127, EPI_ISL_2120129, EPI_ISL_2120130, EPI_ISL_2120136, EPI_ISL_2120149, EPI_ISL_2120161, EPI_ISL_2120169, EPI_ISL_2120175, EPI_ISL_2120178, EPI_ISL_2120186, EPI_ISL_2120199, EPI_ISL_2120201, EPI_ISL_2120204, EPI_ISL_2120208, EPI_ISL_2120248, EPI_ISL_2120261, EPI_ISL_2120265, EPI_ISL_2120304, EPI_ISL_2120336, EPI_ISL_2120361, EPI_ISL_2120364, EPI_ISL_2120370, EPI_ISL_2120374, EPI_ISL_2120378, EPI_ISL_2120382, EPI_ISL_2120385, EPI_ISL_2120387, EPI_ISL_2120415, EPI_ISL_2120425, EPI_ISL_2120434, EPI_ISL_2120445, EPI_ISL_2120453, EPI_ISL_2120455, EPI_ISL_2120458, EPI_ISL_2120460, EPI_ISL_2120461, EPI_ISL_2120463, EPI_ISL_2120464, EPI_ISL_2120468, EPI_ISL_2120469, EPI_ISL_2120479, EPI_ISL_2120480, EPI_ISL_2120482, EPI_ISL_2120483, EPI_ISL_2120486, EPI_ISL_2120489, EPI_ISL_2120492, EPI_ISL_2120494, EPI_ISL_2120495, EPI_ISL_2120498, EPI_ISL_2120500, EPI_ISL_2120505, EPI_ISL_2120506, EPI_ISL_2120509, EPI_ISL_2120511, EPI_ISL_2120512, EPI_ISL_2120517, EPI_ISL_2120518, EPI_ISL_2120523, EPI_ISL_2120525, EPI_ISL_2120529, EPI_ISL_2120530, EPI_ISL_2120533, EPI_ISL_2120536, EPI_ISL_2120539, EPI_ISL_2120542, EPI_ISL_2120544, EPI_ISL_2120547, EPI_ISL_2120551, EPI_ISL_2120552, EPI_ISL_2120554, EPI_ISL_2120556, EPI_ISL_2120559, EPI_ISL_2120562, EPI_ISL_2120563, EPI_ISL_2120567, EPI_ISL_2120569, EPI_ISL_2120572, EPI_ISL_2120579, EPI_ISL_2120582, EPI_ISL_2120587, EPI_ISL_2120588, EPI_ISL_2120591, EPI_ISL_2120592, EPI_ISL_2120595, EPI_ISL_2120600, EPI_ISL_2120602, EPI_ISL_2120604, EPI_ISL_2120611, EPI_ISL_2120617, EPI_ISL_2120619, EPI_ISL_2120620, EPI_ISL_2120624, EPI_ISL_2120625, EPI_ISL_2120627, EPI_ISL_2120633, EPI_ISL_2120637, EPI_ISL_2120640, EPI_ISL_2120645, EPI_ISL_2120651, EPI_ISL_2120691, EPI_ISL_2120692, EPI_ISL_2120694, EPI_ISL_2120702, EPI_ISL_2120705, EPI_ISL_2120706, EPI_ISL_2120709, EPI_ISL_2120710, EPI_ISL_2120712, EPI_ISL_2120715, EPI_ISL_2120721, EPI_ISL_2120727, EPI_ISL_2120728, EPI_ISL_2120730, EPI_ISL_2120732, EPI_ISL_2120737, EPI_ISL_2120742, EPI_ISL_2120743, EPI_ISL_2120748, EPI_ISL_2120750, EPI_ISL_2120757, EPI_ISL_2120759, EPI_ISL_2120763, EPI_ISL_2120764, EPI_ISL_2120766, EPI_ISL_2120767, EPI_ISL_2120768, EPI_ISL_2120769, EPI_ISL_2120771, EPI_ISL_2120772, EPI_ISL_2120778, EPI_ISL_2120780, EPI_ISL_2120784, EPI_ISL_2120785, EPI_ISL_2120789, EPI_ISL_2120796, EPI_ISL_2120799, EPI_ISL_2120803, EPI_ISL_2120805, EPI_ISL_2120809, EPI_ISL_2120811, EPI_ISL_2120815, EPI_ISL_2120818, EPI_ISL_2120819, EPI_ISL_2120821, EPI_ISL_2120822, EPI_ISL_2120824, EPI_ISL_2120825, EPI_ISL_2120831, EPI_ISL_2120834, EPI_ISL_2120843, EPI_ISL_2120845, EPI_ISL_2120851, EPI_ISL_2120854, EPI_ISL_2120856, EPI_ISL_2120857, EPI_ISL_2120860, EPI_ISL_2120865, EPI_ISL_2120869, EPI_ISL_2120872, EPI_ISL_2120879, EPI_ISL_2120885, EPI_ISL_2120888, EPI_ISL_2120889, EPI_ISL_2120894, EPI_ISL_2120895, EPI_ISL_2120897, EPI_ISL_2120901, EPI_ISL_2120909, EPI_ISL_2120911, EPI_ISL_2120912, EPI_ISL_2120913, EPI_ISL_2120916, EPI_ISL_2120919, EPI_ISL_2120923, EPI_ISL_2120925, EPI_ISL_2120930, EPI_ISL_2120933, EPI_ISL_2120935, EPI_ISL_2120936, EPI_ISL_2120938, EPI_ISL_2120939, EPI_ISL_2120941, EPI_ISL_2120942, EPI_ISL_2120944, EPI_ISL_2120945, EPI_ISL_2120953, EPI_ISL_2120957, EPI_ISL_2120960, EPI_ISL_2120962, EPI_ISL_2120963, EPI_ISL_2120972, EPI_ISL_2120976, EPI_ISL_2120977, EPI_ISL_2120981, EPI_ISL_2120984, EPI_ISL_2120989, EPI_ISL_2120990, EPI_ISL_2120993, EPI_ISL_2120995, EPI_ISL_2120998, EPI_ISL_2120999, EPI_ISL_2121001, EPI_ISL_2121008, EPI_ISL_2121017, EPI_ISL_2121019, EPI_ISL_2121095, EPI_ISL_2121188, EPI_ISL_2121370, EPI_ISL_2121546, EPI_ISL_2122128, EPI_ISL_2122130, EPI_ISL_2122131, EPI_ISL_2122133, EPI_ISL_2122135, EPI_ISL_2122136, EPI_ISL_2122139, EPI_ISL_2122140, EPI_ISL_2122149, EPI_ISL_2122151, EPI_ISL_2122158, EPI_ISL_2122161, EPI_ISL_2122165, EPI_ISL_2122181, EPI_ISL_2122183, EPI_ISL_2122189, EPI_ISL_2122191, EPI_ISL_2122194, EPI_ISL_2122195, EPI_ISL_2122206, EPI_ISL_2122208, EPI_ISL_2122210, EPI_ISL_2122215, EPI_ISL_2122217, EPI_ISL_2122218, EPI_ISL_2122221, EPI_ISL_2122229, EPI_ISL_2122235, EPI_ISL_2122240, EPI_ISL_2122241, EPI_ISL_2122245, EPI_ISL_2122246, EPI_ISL_2122248, EPI_ISL_2122251, EPI_ISL_2122254, EPI_ISL_2122275, EPI_ISL_2122276, EPI_ISL_2122282, EPI_ISL_2122286, EPI_ISL_2122289, EPI_ISL_2122298, EPI_ISL_2122301, EPI_ISL_2122310, EPI_ISL_2122322, EPI_ISL_2122325, EPI_ISL_2122328, EPI_ISL_2122334, EPI_ISL_2122335, EPI_ISL_2122341, EPI_ISL_2122343, EPI_ISL_2122345, EPI_ISL_2122348, EPI_ISL_2122349, EPI_ISL_2122352, EPI_ISL_2122354, EPI_ISL_2122355, EPI_ISL_2122358, EPI_ISL_2122360, EPI_ISL_2122363, EPI_ISL_2122365, EPI_ISL_2122375, EPI_ISL_2122378, EPI_ISL_2122381, EPI_ISL_2122383, EPI_ISL_2122384, EPI_ISL_2122386, EPI_ISL_2122390, EPI_ISL_2122397, EPI_ISL_2122398, EPI_ISL_2122400, EPI_ISL_2122403, EPI_ISL_2122408, EPI_ISL_2122409, EPI_ISL_2122412, EPI_ISL_2122414, EPI_ISL_2122420, EPI_ISL_2122421, EPI_ISL_2122424, EPI_ISL_2122429, EPI_ISL_2122430, EPI_ISL_2122432, EPI_ISL_2122433, EPI_ISL_2122435, EPI_ISL_2122438, EPI_ISL_2122439, EPI_ISL_2122441, EPI_ISL_2122442, EPI_ISL_2122444, EPI_ISL_2122445, EPI_ISL_2122448, EPI_ISL_2122455, EPI_ISL_2122456, EPI_ISL_2122461, EPI_ISL_2122462, EPI_ISL_2122463, EPI_ISL_2122478, EPI_ISL_2122490, EPI_ISL_2122499, EPI_ISL_2122500, EPI_ISL_2122504, EPI_ISL_2122508, EPI_ISL_2122510, EPI_ISL_2122516, EPI_ISL_2122523, EPI_ISL_2122524, EPI_ISL_2122529, EPI_ISL_2122530, EPI_ISL_2122540, EPI_ISL_2122541, EPI_ISL_2122546, EPI_ISL_2122547, EPI_ISL_2122548, EPI_ISL_2122553, EPI_ISL_2122557, EPI_ISL_2122560, EPI_ISL_2122563, EPI_ISL_2122565, EPI_ISL_2122583, EPI_ISL_2122587, EPI_ISL_2122593, EPI_ISL_2122597, EPI_ISL_2122602, EPI_ISL_2122604, EPI_ISL_2122606, EPI_ISL_2122609, EPI_ISL_2122623, EPI_ISL_2122629, EPI_ISL_2122630, EPI_ISL_2122638, EPI_ISL_2122644, EPI_ISL_2122646, EPI_ISL_2122647, EPI_ISL_2122654, EPI_ISL_2122655, EPI_ISL_2122662, EPI_ISL_2122664, EPI_ISL_2122668, EPI_ISL_2122671, EPI_ISL_2122672, EPI_ISL_2122676, EPI_ISL_2122677, EPI_ISL_2122679, EPI_ISL_2122680, EPI_ISL_2122683, EPI_ISL_2122685, EPI_ISL_2122687, EPI_ISL_2122689, EPI_ISL_2122698, EPI_ISL_2122699, EPI_ISL_2122705, EPI_ISL_2137844, EPI_ISL_2137845, EPI_ISL_2137846, EPI_ISL_2137848, EPI_ISL_2137850, EPI_ISL_2137852, EPI_ISL_2137854, EPI_ISL_2137875, EPI_ISL_2137876, EPI_ISL_2137877, EPI_ISL_2137879, EPI_ISL_2137882, EPI_ISL_2137885, EPI_ISL_2137887, EPI_ISL_2137888, EPI_ISL_2137890, EPI_ISL_2137891, EPI_ISL_2137893, EPI_ISL_2137894, EPI_ISL_2137896, EPI_ISL_2137898, EPI_ISL_2137899, EPI_ISL_2137900, EPI_ISL_2137903, EPI_ISL_2137904, EPI_ISL_2137907, EPI_ISL_2137910, EPI_ISL_2137911, EPI_ISL_2137919, EPI_ISL_2137920, EPI_ISL_2137925, EPI_ISL_2137928, EPI_ISL_2137929, EPI_ISL_2137933, EPI_ISL_2137947, EPI_ISL_2137950, EPI_ISL_2137952, EPI_ISL_2137954, EPI_ISL_2137956, EPI_ISL_2137963, EPI_ISL_2137967, EPI_ISL_2137968, EPI_ISL_2137972, EPI_ISL_2137976, EPI_ISL_2137978, EPI_ISL_2137979, EPI_ISL_2137983, EPI_ISL_2137990, EPI_ISL_2137993, EPI_ISL_2137997, EPI_ISL_2137998, EPI_ISL_2138000, EPI_ISL_2138001, EPI_ISL_2138002, EPI_ISL_2138003, EPI_ISL_2138004, EPI_ISL_2138006, EPI_ISL_2138012, EPI_ISL_2138019, EPI_ISL_2138020, EPI_ISL_2138022, EPI_ISL_2138023, EPI_ISL_2138024, EPI_ISL_2138028, EPI_ISL_2138031, EPI_ISL_2138032, EPI_ISL_2138034, EPI_ISL_2138041, EPI_ISL_2138043, EPI_ISL_2138044, EPI_ISL_2138046, EPI_ISL_2138047, EPI_ISL_2138048, EPI_ISL_2138049, EPI_ISL_2138054, EPI_ISL_2138062, EPI_ISL_2138063, EPI_ISL_2138066, EPI_ISL_2138071, EPI_ISL_2138073, EPI_ISL_2138075, EPI_ISL_2138076, EPI_ISL_2138078, EPI_ISL_2138082, EPI_ISL_2138087, EPI_ISL_2138092, EPI_ISL_2138093, EPI_ISL_2138094, EPI_ISL_2138096, EPI_ISL_2138098, EPI_ISL_2138106, EPI_ISL_2138107, EPI_ISL_2138109, EPI_ISL_2138111, EPI_ISL_2138112, EPI_ISL_2138120, EPI_ISL_2138122, EPI_ISL_2138128, EPI_ISL_2138129, EPI_ISL_2138136, EPI_ISL_2138137, EPI_ISL_2138139, EPI_ISL_2138142, EPI_ISL_2138143, EPI_ISL_2138144, EPI_ISL_2138147, EPI_ISL_2138155, EPI_ISL_2138158, EPI_ISL_2138159, EPI_ISL_2138163, EPI_ISL_2138164, EPI_ISL_2138167, EPI_ISL_2138171, EPI_ISL_2138173, EPI_ISL_2138176, EPI_ISL_2138178, EPI_ISL_2138181, EPI_ISL_2138183, EPI_ISL_2138184, EPI_ISL_2138193, EPI_ISL_2138198, EPI_ISL_2138205, EPI_ISL_2138206, EPI_ISL_2138208, EPI_ISL_2138210, EPI_ISL_2138214, EPI_ISL_2138215, EPI_ISL_2138219, EPI_ISL_2138224, EPI_ISL_2138226, EPI_ISL_2138232, EPI_ISL_2138237, EPI_ISL_2138238, EPI_ISL_2138242, EPI_ISL_2138246, EPI_ISL_2138247, EPI_ISL_2138248, EPI_ISL_2138255, EPI_ISL_2138260, EPI_ISL_2138262, EPI_ISL_2138264, EPI_ISL_2138268, EPI_ISL_2138272, EPI_ISL_2138273, EPI_ISL_2138274, EPI_ISL_2138276, EPI_ISL_2138283, EPI_ISL_2138285, EPI_ISL_2138287, EPI_ISL_2138288, EPI_ISL_2138289, EPI_ISL_2138291, EPI_ISL_2138293, EPI_ISL_2138294, EPI_ISL_2138296, EPI_ISL_2138300, EPI_ISL_2138306, EPI_ISL_2138307, EPI_ISL_2138311, EPI_ISL_2138315, EPI_ISL_2138318, EPI_ISL_2138319, EPI_ISL_2138322, EPI_ISL_2138325, EPI_ISL_2138328, EPI_ISL_2138331, EPI_ISL_2138337, EPI_ISL_2138339, EPI_ISL_2138344, EPI_ISL_2138353, EPI_ISL_2138354, EPI_ISL_2138355, EPI_ISL_2138356, EPI_ISL_2138359, EPI_ISL_2138360, EPI_ISL_2138363, EPI_ISL_2138372, EPI_ISL_2138379, EPI_ISL_2138386, EPI_ISL_2138390, EPI_ISL_2138391, EPI_ISL_2138398, EPI_ISL_2138406, EPI_ISL_2138407, EPI_ISL_2138409, EPI_ISL_2138414, EPI_ISL_2138415, EPI_ISL_2138420, EPI_ISL_2138428, EPI_ISL_2138430, EPI_ISL_2138431, EPI_ISL_2138440, EPI_ISL_2138441, EPI_ISL_2138446, EPI_ISL_2138448, EPI_ISL_2138451, EPI_ISL_2138456, EPI_ISL_2138458, EPI_ISL_2138460, EPI_ISL_2138461, EPI_ISL_2138467, EPI_ISL_2138470, EPI_ISL_2138472, EPI_ISL_2138476, EPI_ISL_2138477, EPI_ISL_2138479, EPI_ISL_2138480, EPI_ISL_2138481, EPI_ISL_2138483, EPI_ISL_2138484, EPI_ISL_2138487, EPI_ISL_2138488, EPI_ISL_2138489, EPI_ISL_2138491, EPI_ISL_2138492, EPI_ISL_2138493, EPI_ISL_2138494, EPI_ISL_2138501, EPI_ISL_2138505, EPI_ISL_2138508, EPI_ISL_2138511, EPI_ISL_2138512, EPI_ISL_2138520, EPI_ISL_2138522, EPI_ISL_2138524, EPI_ISL_2138528, EPI_ISL_2138532, EPI_ISL_2138533, EPI_ISL_2138538, EPI_ISL_2138544, EPI_ISL_2138553, EPI_ISL_2138558, EPI_ISL_2138559, EPI_ISL_2138560, EPI_ISL_2138562, EPI_ISL_2138563, EPI_ISL_2138565, EPI_ISL_2138568, EPI_ISL_2138569, EPI_ISL_2138571, EPI_ISL_2138577, EPI_ISL_2138578, EPI_ISL_2138579, EPI_ISL_2138583, EPI_ISL_2138584, EPI_ISL_2138586, EPI_ISL_2138587, EPI_ISL_2138589, EPI_ISL_2138594, EPI_ISL_2138596, EPI_ISL_2138601, EPI_ISL_2138602, EPI_ISL_2138603, EPI_ISL_2138604, EPI_ISL_2138606, EPI_ISL_2138607, EPI_ISL_2138608, EPI_ISL_2138609, EPI_ISL_2138610, EPI_ISL_2138612, EPI_ISL_2138616, EPI_ISL_2138619, EPI_ISL_2138620, EPI_ISL_2138624, EPI_ISL_2138633, EPI_ISL_2138635, EPI_ISL_2138636, EPI_ISL_2138641, EPI_ISL_2138644, EPI_ISL_2138647, EPI_ISL_2138650, EPI_ISL_2138653, EPI_ISL_2138663, EPI_ISL_2138665, EPI_ISL_2138670, EPI_ISL_2138674, EPI_ISL_2138675, EPI_ISL_2138683, EPI_ISL_2138686, EPI_ISL_2138689, EPI_ISL_2138692, EPI_ISL_2138693, EPI_ISL_2138697, EPI_ISL_2138698, EPI_ISL_2138705, EPI_ISL_2138710, EPI_ISL_2138713, EPI_ISL_2138715, EPI_ISL_2138717

see above | Lighthouse Lab in Alderley Park | Wellcome Sanger Institute for the COVID-19 Genomics UK (COG-UK) Consortium | Cordelia Langford; David K. Jackson; Dominic Kwiatkowski; Ewan Harrison; Ian Johnston; Jacquelyn Wynn; Jeffrey Barrett; John Sillitoe on behalf of the Wellcome Sanger Institute COVID-19 Surveillance Team; Mairead Hyland; Roberto Amato; Sonia Goncalves; The Lighthouse Lab in Alderley Park and Alex Alderton

EPI_ISL_2004349, EPI_ISL_2004402, EPI_ISL_2004408, EPI_ISL_2004418, EPI_ISL_2004438, EPI_ISL_2004484, EPI_ISL_2004508, EPI_ISL_2004549, EPI_ISL_2004559, EPI_ISL_2004605, EPI_ISL_2004644, EPI_ISL_2004711, EPI_ISL_2004716, EPI_ISL_2004721, EPI_ISL_2004814, EPI_ISL_2004844, EPI_ISL_2004866, EPI_ISL_2004873, EPI_ISL_2004877, EPI_ISL_2021040, EPI_ISL_2021042, EPI_ISL_2021045, EPI_ISL_2021048, EPI_ISL_2021049, EPI_ISL_2021050, EPI_ISL_2021051, EPI_ISL_2021057, EPI_ISL_2021058, EPI_ISL_2021060, EPI_ISL_2021066, EPI_ISL_2021070, EPI_ISL_2021072, EPI_ISL_2021075, EPI_ISL_2021076, EPI_ISL_2021078, EPI_ISL_2021079, EPI_ISL_2021080, EPI_ISL_2021084, EPI_ISL_2021085, EPI_ISL_2021088, EPI_ISL_2021089, EPI_ISL_2021093, EPI_ISL_2021094, EPI_ISL_2021095, EPI_ISL_2021096, EPI_ISL_2021100, EPI_ISL_2021101, EPI_ISL_2021102, EPI_ISL_2021103, EPI_ISL_2021105, EPI_ISL_2021106, EPI_ISL_2021108, EPI_ISL_2021112, EPI_ISL_2021116, EPI_ISL_2021118, EPI_ISL_2021121, EPI_ISL_2021122, EPI_ISL_2021125, EPI_ISL_2021126, EPI_ISL_2021129, EPI_ISL_2021134, EPI_ISL_2021135, EPI_ISL_2021138, EPI_ISL_2021140, EPI_ISL_2021142, EPI_ISL_2021146, EPI_ISL_2021148, EPI_ISL_2021150, EPI_ISL_2021153, EPI_ISL_2021155, EPI_ISL_2021156, EPI_ISL_2021160, EPI_ISL_2021162, EPI_ISL_2021163, EPI_ISL_2021164, EPI_ISL_2021165, EPI_ISL_2021169, EPI_ISL_2021172, EPI_ISL_2021173, EPI_ISL_2021174, EPI_ISL_2021175, EPI_ISL_2021176, EPI_ISL_2021177, EPI_ISL_2021178, EPI_ISL_2021179, EPI_ISL_2021180, EPI_ISL_2021184, EPI_ISL_2021187, EPI_ISL_2021188, EPI_ISL_2021192, EPI_ISL_2021196, EPI_ISL_2021199, EPI_ISL_2021201, EPI_ISL_2021202, EPI_ISL_2021203, EPI_ISL_2021204, EPI_ISL_2021206, EPI_ISL_2021207, EPI_ISL_2021208, EPI_ISL_2021210, EPI_ISL_2021211, EPI_ISL_2021212, EPI_ISL_2021213, EPI_ISL_2021214, EPI_ISL_2021216, EPI_ISL_2021217, EPI_ISL_2021220, EPI_ISL_2021223, EPI_ISL_2021226, EPI_ISL_2021230, EPI_ISL_2021232, EPI_ISL_2021233, EPI_ISL_2021234, EPI_ISL_2021237, EPI_ISL_2021238, EPI_ISL_2021239, EPI_ISL_2021240, EPI_ISL_2021243, EPI_ISL_2021244, EPI_ISL_2021245, EPI_ISL_2021246, EPI_ISL_2021247, EPI_ISL_2021251, EPI_ISL_2021256, EPI_ISL_2021257, EPI_ISL_2021259, EPI_ISL_2021260, EPI_ISL_2021261, EPI_ISL_2021264, EPI_ISL_2021265, EPI_ISL_2021266, EPI_ISL_2021269, EPI_ISL_2021272, EPI_ISL_2021273, EPI_ISL_2021276, EPI_ISL_2021279, EPI_ISL_2021283, EPI_ISL_2021284, EPI_ISL_2021286, EPI_ISL_2021288, EPI_ISL_2021289, EPI_ISL_2021290, EPI_ISL_2021295, EPI_ISL_2021297, EPI_ISL_2021299, EPI_ISL_2021300, EPI_ISL_2021302, EPI_ISL_2021303, EPI_ISL_2021304, EPI_ISL_2021305, EPI_ISL_2021306, EPI_ISL_2021307, EPI_ISL_2021308, EPI_ISL_2021313, EPI_ISL_2021314, EPI_ISL_2021320, EPI_ISL_2021321, EPI_ISL_2021322, EPI_ISL_2021323, EPI_ISL_2021324, EPI_ISL_2021325, EPI_ISL_2021330, EPI_ISL_2021333, EPI_ISL_2021335, EPI_ISL_2021336, EPI_ISL_2021337, EPI_ISL_2021338, EPI_ISL_2021340, EPI_ISL_2021341, EPI_ISL_2021342, EPI_ISL_2021344, EPI_ISL_2021345, EPI_ISL_2021346, EPI_ISL_2021347, EPI_ISL_2021350, EPI_ISL_2021353, EPI_ISL_2021354, EPI_ISL_2021355, EPI_ISL_2021357, EPI_ISL_2021359, EPI_ISL_2021360, EPI_ISL_2021361, EPI_ISL_2022340, EPI_ISL_2022781, EPI_ISL_2022782, EPI_ISL_2022783, EPI_ISL_2022785, EPI_ISL_2022786, EPI_ISL_2022787, EPI_ISL_2022788, EPI_ISL_2022789, EPI_ISL_2022790, EPI_ISL_2022791, EPI_ISL_2022792, EPI_ISL_2022793, EPI_ISL_2022794, EPI_ISL_2022795, EPI_ISL_2022796, EPI_ISL_2022797, EPI_ISL_2022798, EPI_ISL_2022800, EPI_ISL_2022801, EPI_ISL_2022802, EPI_ISL_2022803, EPI_ISL_2022804, EPI_ISL_2022806, EPI_ISL_2022807, EPI_ISL_2022808, EPI_ISL_2022809, EPI_ISL_2022811, EPI_ISL_2022812, EPI_ISL_2022813, EPI_ISL_2022814, EPI_ISL_2022815, EPI_ISL_2022816, EPI_ISL_2022818, EPI_ISL_2022819, EPI_ISL_2022820, EPI_ISL_2022822, EPI_ISL_2022823, EPI_ISL_2022824, EPI_ISL_2022825, EPI_ISL_2022827, EPI_ISL_2022828, EPI_ISL_2022829, EPI_ISL_2022830, EPI_ISL_2022831, EPI_ISL_2022833, EPI_ISL_2022835, EPI_ISL_2022836, EPI_ISL_2022837, EPI_ISL_2022838, EPI_ISL_2022839, EPI_ISL_2022841, EPI_ISL_2022843, EPI_ISL_2022844, EPI_ISL_2022846, EPI_ISL_2022847, EPI_ISL_2022848, EPI_ISL_2022849, EPI_ISL_2022851, EPI_ISL_2022852, EPI_ISL_2022853, EPI_ISL_2022854, EPI_ISL_2022855, EPI_ISL_2022856, EPI_ISL_2022857, EPI_ISL_2022858, EPI_ISL_2022859, EPI_ISL_2022860, EPI_ISL_2022861, EPI_ISL_2022862, EPI_ISL_2022863, EPI_ISL_2022864, EPI_ISL_2022865, EPI_ISL_2022866, EPI_ISL_2022867, EPI_ISL_2022868, EPI_ISL_2022869, EPI_ISL_2022870, EPI_ISL_2022871, EPI_ISL_2022872, EPI_ISL_2022873, EPI_ISL_2022874, EPI_ISL_2022875, EPI_ISL_2022876, EPI_ISL_2022877, EPI_ISL_2022878, EPI_ISL_2022879, EPI_ISL_2022880, EPI_ISL_2022881, EPI_ISL_2022882, EPI_ISL_2022883, EPI_ISL_2022884, EPI_ISL_2022885, EPI_ISL_2022886, EPI_ISL_2022887, EPI_ISL_2022888, EPI_ISL_2022889, EPI_ISL_2022890, EPI_ISL_2022891, EPI_ISL_2022892, EPI_ISL_2022893, EPI_ISL_2022894, EPI_ISL_2022895, EPI_ISL_2022896, EPI_ISL_2022897, EPI_ISL_2022898, EPI_ISL_2022899, EPI_ISL_2022900, EPI_ISL_2022901, EPI_ISL_2022903, EPI_ISL_2022904, EPI_ISL_2022905, EPI_ISL_2022906, EPI_ISL_2022907, EPI_ISL_2022908, EPI_ISL_2022909, EPI_ISL_2022910, EPI_ISL_2022911, EPI_ISL_2022914, EPI_ISL_2022915, EPI_ISL_2022916, EPI_ISL_2022917, EPI_ISL_2022918, EPI_ISL_2022919, EPI_ISL_2022920, EPI_ISL_2022921, EPI_ISL_2022923, EPI_ISL_2022924, EPI_ISL_2022925, EPI_ISL_2022926, EPI_ISL_2022927, EPI_ISL_2022928, EPI_ISL_2022929, EPI_ISL_2022930, EPI_ISL_2022931, EPI_ISL_2022932, EPI_ISL_2022933, EPI_ISL_2022934, EPI_ISL_2022935, EPI_ISL_2022936, EPI_ISL_2022937, EPI_ISL_2022938, EPI_ISL_2022939, EPI_ISL_2022940, EPI_ISL_2022941, EPI_ISL_2022942, EPI_ISL_2022943, EPI_ISL_2022944, EPI_ISL_2022945, EPI_ISL_2022946, EPI_ISL_2022947, EPI_ISL_2092676, EPI_ISL_2092677, EPI_ISL_2092678, EPI_ISL_2092679, EPI_ISL_2092680, EPI_ISL_2092681, EPI_ISL_2092682, EPI_ISL_2092683, EPI_ISL_2092684, EPI_ISL_2092685, EPI_ISL_2092686, EPI_ISL_2092687, EPI_ISL_2092688, EPI_ISL_2092689, EPI_ISL_2092690, EPI_ISL_2092691, EPI_ISL_2092692, EPI_ISL_2092693, EPI_ISL_2092694, EPI_ISL_2092695, EPI_ISL_2092696, EPI_ISL_2092697, EPI_ISL_2092698, EPI_ISL_2092699, EPI_ISL_2092700, EPI_ISL_2092701, EPI_ISL_2092702, EPI_ISL_2092703, EPI_ISL_2092704, EPI_ISL_2092705, EPI_ISL_2092706, EPI_ISL_2092707, EPI_ISL_2092708, EPI_ISL_2092709, EPI_ISL_2092710, EPI_ISL_2092711, EPI_ISL_2092712, EPI_ISL_2092713, EPI_ISL_2092714, EPI_ISL_2092715, EPI_ISL_2092716, EPI_ISL_2092717, EPI_ISL_2092718, EPI_ISL_2092719, EPI_ISL_2092720, EPI_ISL_2092721, EPI_ISL_2092722, EPI_ISL_2092723, EPI_ISL_2092724, EPI_ISL_2092725, EPI_ISL_2092726, EPI_ISL_2092727, EPI_ISL_2092728, EPI_ISL_2092729, EPI_ISL_2092730, EPI_ISL_2092731, EPI_ISL_2092732, EPI_ISL_2092733, EPI_ISL_2092734, EPI_ISL_2092735, EPI_ISL_2092736, EPI_ISL_2092737, EPI_ISL_2092738, EPI_ISL_2092739, EPI_ISL_2092740, EPI_ISL_2092743, EPI_ISL_2092744, EPI_ISL_2092745, EPI_ISL_2092746, EPI_ISL_2092747, EPI_ISL_2092748, EPI_ISL_2092749, EPI_ISL_2092750, EPI_ISL_2092751, EPI_ISL_2092752, EPI_ISL_2092753, EPI_ISL_2092754, EPI_ISL_2092755, EPI_ISL_2092756, EPI_ISL_2117519, EPI_ISL_2117521, EPI_ISL_2117532, EPI_ISL_2117536, EPI_ISL_2117554, EPI_ISL_2117563, EPI_ISL_2117571, EPI_ISL_2117578, EPI_ISL_2117579, EPI_ISL_2117581, EPI_ISL_2117597, EPI_ISL_2117600, EPI_ISL_2117621, EPI_ISL_2117627, EPI_ISL_2117632, EPI_ISL_2117639, EPI_ISL_2117643, EPI_ISL_2117644, EPI_ISL_2117645, EPI_ISL_2117648, EPI_ISL_2117649, EPI_ISL_2117651, EPI_ISL_2117657, EPI_ISL_2117660, EPI_ISL_2117667, EPI_ISL_2117668, EPI_ISL_2117679, EPI_ISL_2117681, EPI_ISL_2117682, EPI_ISL_2117697, EPI_ISL_2117703, EPI_ISL_2117713, EPI_ISL_2117714, EPI_ISL_2117716, EPI_ISL_2117726, EPI_ISL_2117727, EPI_ISL_2117731, EPI_ISL_2117734, EPI_ISL_2117736, EPI_ISL_2117745, EPI_ISL_2117749, EPI_ISL_2117751, EPI_ISL_2117752, EPI_ISL_2117760, EPI_ISL_2117761, EPI_ISL_2117765, EPI_ISL_2117767, EPI_ISL_2117773, EPI_ISL_2117779, EPI_ISL_2117781, EPI_ISL_2117799, EPI_ISL_2117804, EPI_ISL_2117806, EPI_ISL_2117808, EPI_ISL_2117811, EPI_ISL_2117814, EPI_ISL_2117817, EPI_ISL_2117820, EPI_ISL_2117823, EPI_ISL_2117828, EPI_ISL_2117837, EPI_ISL_2117842, EPI_ISL_2117854, EPI_ISL_2117855, EPI_ISL_2117857, EPI_ISL_2117864, EPI_ISL_2117870, EPI_ISL_2117873, EPI_ISL_2117883, EPI_ISL_2117885, EPI_ISL_2117893, EPI_ISL_2117900, EPI_ISL_2117904, EPI_ISL_2117906, EPI_ISL_2117907, EPI_ISL_2117909, EPI_ISL_2117912, EPI_ISL_2117915, EPI_ISL_2117921, EPI_ISL_2117922, EPI_ISL_2117925, EPI_ISL_2117927, EPI_ISL_2117930, EPI_ISL_2117945, EPI_ISL_2117946, EPI_ISL_2117948, EPI_ISL_2117951, EPI_ISL_2117958, EPI_ISL_2117963, EPI_ISL_2117977, EPI_ISL_2117978, EPI_ISL_2117985, EPI_ISL_2117988, EPI_ISL_2117990, EPI_ISL_2117997, EPI_ISL_2117998, EPI_ISL_2118004, EPI_ISL_2118008, EPI_ISL_2118013, EPI_ISL_2118017, EPI_ISL_2118020, EPI_ISL_2118022, EPI_ISL_2118025, EPI_ISL_2118030, EPI_ISL_2118033, EPI_ISL_2118038, EPI_ISL_2118042, EPI_ISL_2118046, EPI_ISL_2118048, EPI_ISL_2118053, EPI_ISL_2118054, EPI_ISL_2118056, EPI_ISL_2118062, EPI_ISL_2118067, EPI_ISL_2118073, EPI_ISL_2118078, EPI_ISL_2118081, EPI_ISL_2118086, EPI_ISL_2118087, EPI_ISL_2118088, EPI_ISL_2118090, EPI_ISL_2118091, EPI_ISL_2118094, EPI_ISL_2118103, EPI_ISL_2118105, EPI_ISL_2118109, EPI_ISL_2118125, EPI_ISL_2118126, EPI_ISL_2118127, EPI_ISL_2118130, EPI_ISL_2118133, EPI_ISL_2118135, EPI_ISL_2118137, EPI_ISL_2118139, EPI_ISL_2118143, EPI_ISL_2118145, EPI_ISL_2118146, EPI_ISL_2118149, EPI_ISL_2118150, EPI_ISL_2118153, EPI_ISL_2118159, EPI_ISL_2118167, EPI_ISL_2118176, EPI_ISL_2118177, EPI_ISL_2118180, EPI_ISL_2118193, EPI_ISL_2118199, EPI_ISL_2118200, EPI_ISL_2118212, EPI_ISL_2118222, EPI_ISL_2118232, EPI_ISL_2118240, EPI_ISL_2118241, EPI_ISL_2118243, EPI_ISL_2118246, EPI_ISL_2118247, EPI_ISL_2118806, EPI_ISL_2119351, EPI_ISL_2119354, EPI_ISL_2119359, EPI_ISL_2119364, EPI_ISL_2119375, EPI_ISL_2119378, EPI_ISL_2119386, EPI_ISL_2119387, EPI_ISL_2119389, EPI_ISL_2119392, EPI_ISL_2119398, EPI_ISL_2119403, EPI_ISL_2119420, EPI_ISL_2119424, EPI_ISL_2119425, EPI_ISL_2119430, EPI_ISL_2119431, EPI_ISL_2119435, EPI_ISL_2119444, EPI_ISL_2119449, EPI_ISL_2119460, EPI_ISL_2119462, EPI_ISL_2119463, EPI_ISL_2119470, EPI_ISL_2119474, EPI_ISL_2119482, EPI_ISL_2119485, EPI_ISL_2119491, EPI_ISL_2119498, EPI_ISL_2119502, EPI_ISL_2119512, EPI_ISL_2119513, EPI_ISL_2119522, EPI_ISL_2119525, EPI_ISL_2119528, EPI_ISL_2119546, EPI_ISL_2119556, EPI_ISL_2119562, EPI_ISL_2119564, EPI_ISL_2119567, EPI_ISL_2119576, EPI_ISL_2119577, EPI_ISL_2119589, EPI_ISL_2119593, EPI_ISL_2119597, EPI_ISL_2119610, EPI_ISL_2119625, EPI_ISL_2119635, EPI_ISL_2119642, EPI_ISL_2119652, EPI_ISL_2119654, EPI_ISL_2119658, EPI_ISL_2119661, EPI_ISL_2119662, EPI_ISL_2119664, EPI_ISL_2119671, EPI_ISL_2119680, EPI_ISL_2119682, EPI_ISL_2119692, EPI_ISL_2119699, EPI_ISL_2119712, EPI_ISL_2119717, EPI_ISL_2119724, EPI_ISL_2119741, EPI_ISL_2119745, EPI_ISL_2119748, EPI_ISL_2119749, EPI_ISL_2119753, EPI_ISL_2119754, EPI_ISL_2119778, EPI_ISL_2119782, EPI_ISL_2119792, EPI_ISL_2119803, EPI_ISL_2119805, EPI_ISL_2119819, EPI_ISL_2119825, EPI_ISL_2119828, EPI_ISL_2119832, EPI_ISL_2119836, EPI_ISL_2119841, EPI_ISL_2119848, EPI_ISL_2119858, EPI_ISL_2119863, EPI_ISL_2119864, EPI_ISL_2119870, EPI_ISL_2119912, EPI_ISL_2119918, EPI_ISL_2119921, EPI_ISL_2119924, EPI_ISL_2119925, EPI_ISL_2119928, EPI_ISL_2119930, EPI_ISL_2119933, EPI_ISL_2119934, EPI_ISL_2119938, EPI_ISL_2119939, EPI_ISL_2119941, EPI_ISL_2119950, EPI_ISL_2119964, EPI_ISL_2119967, EPI_ISL_2119968, EPI_ISL_2119981, EPI_ISL_2119984, EPI_ISL_2120005, EPI_ISL_2120010, EPI_ISL_2120016, EPI_ISL_2120021, EPI_ISL_2120024, EPI_ISL_2120025, EPI_ISL_2120030, EPI_ISL_2120033, EPI_ISL_2120035, EPI_ISL_2120039, EPI_ISL_2120046, EPI_ISL_2120049, EPI_ISL_2120066, EPI_ISL_2120067, EPI_ISL_2120075, EPI_ISL_2120078, EPI_ISL_2120098, EPI_ISL_2120116, EPI_ISL_2120120, EPI_ISL_2120126, EPI_ISL_2120133, EPI_ISL_2120140, EPI_ISL_2120148, EPI_ISL_2120154, EPI_ISL_2120165, EPI_ISL_2120181, EPI_ISL_2120209, EPI_ISL_2120210, EPI_ISL_2120218, EPI_ISL_2120222, EPI_ISL_2120224, EPI_ISL_2120226, EPI_ISL_2120229, EPI_ISL_2120230, EPI_ISL_2120238, EPI_ISL_2120245, EPI_ISL_2120247, EPI_ISL_2120250, EPI_ISL_2120253, EPI_ISL_2120259, EPI_ISL_2120262, EPI_ISL_2120267, EPI_ISL_2120280, EPI_ISL_2120282, EPI_ISL_2120290, EPI_ISL_2120295, EPI_ISL_2120296, EPI_ISL_2120307, EPI_ISL_2120319, EPI_ISL_2120322, EPI_ISL_2120324, EPI_ISL_2120326, EPI_ISL_2120339, EPI_ISL_2120343, EPI_ISL_2120345, EPI_ISL_2120362, EPI_ISL_2120365, EPI_ISL_2120373, EPI_ISL_2120381, EPI_ISL_2120394, EPI_ISL_2120395, EPI_ISL_2120406, EPI_ISL_2120407, EPI_ISL_2120418, EPI_ISL_2120422, EPI_ISL_2120431, EPI_ISL_2120436, EPI_ISL_2120439, EPI_ISL_2120440, EPI_ISL_2120443, EPI_ISL_2120448, EPI_ISL_2120466, EPI_ISL_2120472, EPI_ISL_2120474, EPI_ISL_2120484, EPI_ISL_2120497, EPI_ISL_2120508, EPI_ISL_2120515, EPI_ISL_2120520, EPI_ISL_2120528, EPI_ISL_2120532, EPI_ISL_2120535, EPI_ISL_2120549, EPI_ISL_2120557, EPI_ISL_2120564, EPI_ISL_2120566, EPI_ISL_2120570, EPI_ISL_2120597, EPI_ISL_2120598, EPI_ISL_2120601, EPI_ISL_2120616, EPI_ISL_2120622, EPI_ISL_2120631, EPI_ISL_2120634, EPI_ISL_2120636, EPI_ISL_2120638, EPI_ISL_2120646, EPI_ISL_2120649, EPI_ISL_2120654, EPI_ISL_2120657, EPI_ISL_2120690, EPI_ISL_2120703, EPI_ISL_2120708, EPI_ISL_2120717, EPI_ISL_2120720, EPI_ISL_2120723, EPI_ISL_2120733, EPI_ISL_2120735, EPI_ISL_2120736, EPI_ISL_2120738, EPI_ISL_2120739, EPI_ISL_2120740, EPI_ISL_2120753, EPI_ISL_2120770, EPI_ISL_2120773, EPI_ISL_2120774, EPI_ISL_2120775, EPI_ISL_2120781, EPI_ISL_2120787, EPI_ISL_2120790, EPI_ISL_2120798, EPI_ISL_2120806, EPI_ISL_2120808, EPI_ISL_2120814, EPI_ISL_2120816, EPI_ISL_2120827, EPI_ISL_2120833, EPI_ISL_2120840, EPI_ISL_2120842, EPI_ISL_2120846, EPI_ISL_2120848, EPI_ISL_2120849, EPI_ISL_2120858, EPI_ISL_2120871, EPI_ISL_2120873, EPI_ISL_2120874, EPI_ISL_2120876, EPI_ISL_2120880, EPI_ISL_2120883, EPI_ISL_2120886, EPI_ISL_2120892, EPI_ISL_2120898, EPI_ISL_2120899, EPI_ISL_2120904, EPI_ISL_2120907, EPI_ISL_2120908, EPI_ISL_2120922, EPI_ISL_2120928, EPI_ISL_2120949, EPI_ISL_2120954, EPI_ISL_2120966, EPI_ISL_2120969, EPI_ISL_2120971, EPI_ISL_2120980, EPI_ISL_2120983, EPI_ISL_2120986, EPI_ISL_2120987, EPI_ISL_2120996, EPI_ISL_2121004, EPI_ISL_2121016, EPI_ISL_2121020, EPI_ISL_2121028, EPI_ISL_2121034, EPI_ISL_2121035, EPI_ISL_2121038, EPI_ISL_2121039, EPI_ISL_2121042, EPI_ISL_2121049, EPI_ISL_2121053, EPI_ISL_2121055, EPI_ISL_2121058, EPI_ISL_2121061, EPI_ISL_2121063, EPI_ISL_2121064, EPI_ISL_2121067, EPI_ISL_2121070, EPI_ISL_2121073, EPI_ISL_2121075, EPI_ISL_2121081, EPI_ISL_2121084, EPI_ISL_2121085, EPI_ISL_2121086, EPI_ISL_2121088, EPI_ISL_2121089, EPI_ISL_2121091, EPI_ISL_2121094, EPI_ISL_2121099, EPI_ISL_2121102, EPI_ISL_2121105, EPI_ISL_2121110, EPI_ISL_2121113, EPI_ISL_2121119, EPI_ISL_2121120, EPI_ISL_2121125, EPI_ISL_2121126, EPI_ISL_2121128, EPI_ISL_2121129, EPI_ISL_2121131, EPI_ISL_2121132, EPI_ISL_2121134, EPI_ISL_2121135, EPI_ISL_2121139, EPI_ISL_2121143, EPI_ISL_2121144, EPI_ISL_2121146, EPI_ISL_2121147, EPI_ISL_2121149, EPI_ISL_2121154, EPI_ISL_2121160, EPI_ISL_2121163, EPI_ISL_2121169, EPI_ISL_2121171, EPI_ISL_2121172, EPI_ISL_2121174, EPI_ISL_2121175, EPI_ISL_2121179, EPI_ISL_2121180, EPI_ISL_2121185, EPI_ISL_2121186, EPI_ISL_2121190, EPI_ISL_2121192, EPI_ISL_2121193, EPI_ISL_2121199, EPI_ISL_2121203, EPI_ISL_2121206, EPI_ISL_2121207, EPI_ISL_2121213, EPI_ISL_2121216, EPI_ISL_2121218, EPI_ISL_2121219, EPI_ISL_2121221, EPI_ISL_2121224, EPI_ISL_2121225, EPI_ISL_2121227, EPI_ISL_2121228, EPI_ISL_2121231, EPI_ISL_2121234, EPI_ISL_2121235, EPI_ISL_2121244, EPI_ISL_2121249, EPI_ISL_2121250, EPI_ISL_2121252, EPI_ISL_2121258, EPI_ISL_2121261, EPI_ISL_2121262, EPI_ISL_2121268, EPI_ISL_2121269, EPI_ISL_2121271, EPI_ISL_2121274, EPI_ISL_2121279, EPI_ISL_2121280, EPI_ISL_2121281, EPI_ISL_2121283, EPI_ISL_2121285, EPI_ISL_2121288, EPI_ISL_2121290, EPI_ISL_2121297, EPI_ISL_2121299, EPI_ISL_2121302, EPI_ISL_2121303, EPI_ISL_2121310, EPI_ISL_2121319, EPI_ISL_2121321, EPI_ISL_2121322, EPI_ISL_2121324, EPI_ISL_2121325, EPI_ISL_2121327, EPI_ISL_2121330, EPI_ISL_2121331, EPI_ISL_2121334, EPI_ISL_2121337, EPI_ISL_2121343, EPI_ISL_2121344, EPI_ISL_2121349, EPI_ISL_2121352, EPI_ISL_2121353, EPI_ISL_2121359, EPI_ISL_2121361, EPI_ISL_2121362, EPI_ISL_2121365, EPI_ISL_2121367, EPI_ISL_2121368, EPI_ISL_2121378, EPI_ISL_2121380, EPI_ISL_2121381, EPI_ISL_2121383, EPI_ISL_2121386, EPI_ISL_2121388, EPI_ISL_2121390, EPI_ISL_2121391, EPI_ISL_2121394, EPI_ISL_2121396, EPI_ISL_2121405, EPI_ISL_2121407, EPI_ISL_2121408, EPI_ISL_2121410, EPI_ISL_2121414, EPI_ISL_2121419, EPI_ISL_2121428, EPI_ISL_2121431, EPI_ISL_2121434, EPI_ISL_2121470, EPI_ISL_2121476, EPI_ISL_2121478, EPI_ISL_2121487, EPI_ISL_2121490, EPI_ISL_2121492, EPI_ISL_2121496, EPI_ISL_2121503, EPI_ISL_2121504, EPI_ISL_2121511, EPI_ISL_2121528, EPI_ISL_2121530, EPI_ISL_2121532, EPI_ISL_2121535, EPI_ISL_2121543, EPI_ISL_2121545, EPI_ISL_2121548, EPI_ISL_2121551, EPI_ISL_2121552, EPI_ISL_2121554, EPI_ISL_2121555, EPI_ISL_2121559, EPI_ISL_2121561, EPI_ISL_2121563, EPI_ISL_2121564, EPI_ISL_2121566, EPI_ISL_2121569, EPI_ISL_2121571, EPI_ISL_2121579, EPI_ISL_2121585, EPI_ISL_2121604, EPI_ISL_2121606, EPI_ISL_2121620, EPI_ISL_2121623, EPI_ISL_2121637, EPI_ISL_2121640, EPI_ISL_2121642, EPI_ISL_2121654, EPI_ISL_2121656, EPI_ISL_2121658, EPI_ISL_2121663, EPI_ISL_2121673, EPI_ISL_2121674, EPI_ISL_2121682, EPI_ISL_2121694, EPI_ISL_2121699, EPI_ISL_2121702, EPI_ISL_2121705, EPI_ISL_2121713, EPI_ISL_2121721, EPI_ISL_2121722, EPI_ISL_2121724, EPI_ISL_2121727, EPI_ISL_2121729, EPI_ISL_2121732, EPI_ISL_2121737, EPI_ISL_2121738, EPI_ISL_2121740, EPI_ISL_2121741, EPI_ISL_2121745, EPI_ISL_2121747, EPI_ISL_2121751, EPI_ISL_2121765, EPI_ISL_2121777, EPI_ISL_2121780, EPI_ISL_2121787, EPI_ISL_2121788, EPI_ISL_2121791, EPI_ISL_2121795, EPI_ISL_2121799, EPI_ISL_2121809, EPI_ISL_2121813, EPI_ISL_2121816, EPI_ISL_2121817, EPI_ISL_2121825, EPI_ISL_2121830, EPI_ISL_2121831, EPI_ISL_2121840, EPI_ISL_2121843, EPI_ISL_2121844, EPI_ISL_2121849, EPI_ISL_2121869, EPI_ISL_2121878, EPI_ISL_2121881, EPI_ISL_2121885, EPI_ISL_2121894, EPI_ISL_2121896, EPI_ISL_2121917, EPI_ISL_2121923, EPI_ISL_2121925, EPI_ISL_2121928, EPI_ISL_2121950, EPI_ISL_2121954, EPI_ISL_2121965, EPI_ISL_2121968, EPI_ISL_2121970, EPI_ISL_2121971, EPI_ISL_2121973, EPI_ISL_2121979, EPI_ISL_2121982, EPI_ISL_2121985, EPI_ISL_2121986, EPI_ISL_2121994, EPI_ISL_2121999, EPI_ISL_2122013, EPI_ISL_2122016, EPI_ISL_2122021, EPI_ISL_2122027, EPI_ISL_2122044, EPI_ISL_2122048, EPI_ISL_2122055, EPI_ISL_2122073, EPI_ISL_2122078, EPI_ISL_2122092, EPI_ISL_2122097, EPI_ISL_2122098, EPI_ISL_2122120, EPI_ISL_2122123, EPI_ISL_2122143, EPI_ISL_2122146, EPI_ISL_2122152, EPI_ISL_2122153, EPI_ISL_2122156, EPI_ISL_2122169, EPI_ISL_2122170, EPI_ISL_2122174, EPI_ISL_2122178, EPI_ISL_2122186, EPI_ISL_2122192, EPI_ISL_2122213, EPI_ISL_2122220, EPI_ISL_2122223, EPI_ISL_2122224, EPI_ISL_2122226, EPI_ISL_2122231, EPI_ISL_2122238, EPI_ISL_2122243, EPI_ISL_2122250, EPI_ISL_2122253, EPI_ISL_2122256, EPI_ISL_2122278, EPI_ISL_2122281, EPI_ISL_2122290, EPI_ISL_2122292, EPI_ISL_2122293, EPI_ISL_2122295, EPI_ISL_2122300, EPI_ISL_2122306, EPI_ISL_2122314, EPI_ISL_2122316, EPI_ISL_2122317, EPI_ISL_2122320, EPI_ISL_2122337, EPI_ISL_2122338, EPI_ISL_2122340, EPI_ISL_2122346, EPI_ISL_2122357, EPI_ISL_2122359, EPI_ISL_2122362, EPI_ISL_2122371, EPI_ISL_2122372, EPI_ISL_2122374, EPI_ISL_2122377, EPI_ISL_2122392, EPI_ISL_2122394, EPI_ISL_2122395, EPI_ISL_2122406, EPI_ISL_2122410, EPI_ISL_2122415, EPI_ISL_2122418, EPI_ISL_2122423, EPI_ISL_2122426, EPI_ISL_2122428, EPI_ISL_2122437, EPI_ISL_2122447, EPI_ISL_2122450, EPI_ISL_2122452, EPI_ISL_2122453, EPI_ISL_2122459, EPI_ISL_2122465, EPI_ISL_2122467, EPI_ISL_2122476, EPI_ISL_2122479, EPI_ISL_2122480, EPI_ISL_2122482, EPI_ISL_2122485, EPI_ISL_2122494, EPI_ISL_2122496, EPI_ISL_2122507, EPI_ISL_2122512, EPI_ISL_2122520, EPI_ISL_2122521, EPI_ISL_2122543, EPI_ISL_2122554, EPI_ISL_2122559, EPI_ISL_2122566, EPI_ISL_2122569, EPI_ISL_2122571, EPI_ISL_2122573, EPI_ISL_2122576, EPI_ISL_2122578, EPI_ISL_2122579, EPI_ISL_2122581, EPI_ISL_2122584, EPI_ISL_2122588, EPI_ISL_2122594, EPI_ISL_2122596, EPI_ISL_2122603, EPI_ISL_2122612, EPI_ISL_2122619, EPI_ISL_2122626, EPI_ISL_2122627, EPI_ISL_2122633, EPI_ISL_2122635, EPI_ISL_2122641, EPI_ISL_2122642, EPI_ISL_2122649, EPI_ISL_2122652, EPI_ISL_2122656, EPI_ISL_2122659, EPI_ISL_2122669, EPI_ISL_2122674, EPI_ISL_2122693, EPI_ISL_2122695, EPI_ISL_2137923, EPI_ISL_2137953, EPI_ISL_2138005, EPI_ISL_2138033, EPI_ISL_2138050, EPI_ISL_2138165, EPI_ISL_2138185, EPI_ISL_2138270, EPI_ISL_2138295, EPI_ISL_2138701

see above | Lighthouse Lab in Glasgow | Wellcome Sanger Institute for the COVID-19 Genomics UK (COG-UK) Consortium | Anna Dominiczak and Alex Alderton; Carol Clugston; Cordelia Langford; David Gray; David K. Jackson; Dominic Kwiatkowski; Ewan Harrison; Harper VanSteenhouse; Ian Johnston; Jeffrey Barrett; John Sillitoe on behalf of the Wellcome Sanger Institute COVID-19 Surveillance Team; Roberto Amato; Sonia Goncalves; Yumi Kasai

EPI_ISL_2004620, EPI_ISL_2004621, EPI_ISL_2004622, EPI_ISL_2004623, EPI_ISL_2004624, EPI_ISL_2004626, EPI_ISL_2004628, EPI_ISL_2004629, EPI_ISL_2004630, EPI_ISL_2004631, EPI_ISL_2004634, EPI_ISL_2004636, EPI_ISL_2004638, EPI_ISL_2004640, EPI_ISL_2004642, EPI_ISL_2004645, EPI_ISL_2004647, EPI_ISL_2004649, EPI_ISL_2004650, EPI_ISL_2004654, EPI_ISL_2004655, EPI_ISL_2004656, EPI_ISL_2004659, EPI_ISL_2004661, EPI_ISL_2004663, EPI_ISL_2004664, EPI_ISL_2004665, EPI_ISL_2004666, EPI_ISL_2004669, EPI_ISL_2004672, EPI_ISL_2004678, EPI_ISL_2004681, EPI_ISL_2004683, EPI_ISL_2004684, EPI_ISL_2004686, EPI_ISL_2004689, EPI_ISL_2004690, EPI_ISL_2004692, EPI_ISL_2004696, EPI_ISL_2004698, EPI_ISL_2004699, EPI_ISL_2004700, EPI_ISL_2004705, EPI_ISL_2004706, EPI_ISL_2004707, EPI_ISL_2004708, EPI_ISL_2004710, EPI_ISL_2004712, EPI_ISL_2004714, EPI_ISL_2004718, EPI_ISL_2004719, EPI_ISL_2004720, EPI_ISL_2004722, EPI_ISL_2004723, EPI_ISL_2004727, EPI_ISL_2004730, EPI_ISL_2004731, EPI_ISL_2004733, EPI_ISL_2004736, EPI_ISL_2004737, EPI_ISL_2004739, EPI_ISL_2004740, EPI_ISL_2004741, EPI_ISL_2004742, EPI_ISL_2004746, EPI_ISL_2004750,

EPI_ISL_2004756, EPI_ISL_2004760, EPI_ISL_2004761, EPI_ISL_2004763, EPI_ISL_2004765, EPI_ISL_2004766, EPI_ISL_2004770, EPI_ISL_2004771, EPI_ISL_2004772, EPI_ISL_2004776, EPI_ISL_2004777, EPI_ISL_2004778, EPI_ISL_2004780, EPI_ISL_2004781, EPI_ISL_2004782, EPI_ISL_2004784, EPI_ISL_2004785, EPI_ISL_2004786, EPI_ISL_2004791, EPI_ISL_2004793, EPI_ISL_2004797, EPI_ISL_2004800,
EPI_ISL_2004801, EPI_ISL_2004802, EPI_ISL_2004803, EPI_ISL_2004808, EPI_ISL_2004809, EPI_ISL_2004819, EPI_ISL_2004820, EPI_ISL_2004822, EPI_ISL_2004825, EPI_ISL_2004826, EPI_ISL_2004827, EPI_ISL_2004828, EPI_ISL_2004829, EPI_ISL_2004831, EPI_ISL_2004832, EPI_ISL_2004835, EPI_ISL_2004838, EPI_ISL_2004839, EPI_ISL_2004843, EPI_ISL_2004848, EPI_ISL_2004851, EPI_ISL_2004853,
EPI_ISL_2004855, EPI_ISL_2004856, EPI_ISL_2004857, EPI_ISL_2004858, EPI_ISL_2004859, EPI_ISL_2004861, EPI_ISL_2004863, EPI_ISL_2004867, EPI_ISL_2004870, EPI_ISL_2004872, EPI_ISL_2004874, EPI_ISL_2004875, EPI_ISL_2004879, EPI_ISL_2005811, EPI_ISL_2005812, EPI_ISL_2005813, EPI_ISL_2005815, EPI_ISL_2005817, EPI_ISL_2005819, EPI_ISL_2005822, EPI_ISL_2005825, EPI_ISL_2005826,
EPI_ISL_2005827, EPI_ISL_2005829, EPI_ISL_2005830, EPI_ISL_2005831, EPI_ISL_2005832, EPI_ISL_2005836, EPI_ISL_2005840, EPI_ISL_2005848, EPI_ISL_2005849, EPI_ISL_2005853, EPI_ISL_2005854, EPI_ISL_2005855, EPI_ISL_2005857, EPI_ISL_2005859, EPI_ISL_2005860, EPI_ISL_2005861, EPI_ISL_2005864, EPI_ISL_2005870, EPI_ISL_2005872, EPI_ISL_2005873, EPI_ISL_2005874, EPI_ISL_2005880,
EPI_ISL_2005883, EPI_ISL_2005885, EPI_ISL_2005888, EPI_ISL_2005892, EPI_ISL_2005897, EPI_ISL_2005899, EPI_ISL_2005903, EPI_ISL_2005907, EPI_ISL_2005908, EPI_ISL_2005909, EPI_ISL_2005915, EPI_ISL_2005916, EPI_ISL_2005917, EPI_ISL_2005920, EPI_ISL_2005921, EPI_ISL_2005922, EPI_ISL_2005929, EPI_ISL_2005933, EPI_ISL_2005934, EPI_ISL_2005935, EPI_ISL_2005937, EPI_ISL_2005942,
EPI_ISL_2005943, EPI_ISL_2005944, EPI_ISL_2005948, EPI_ISL_2005949, EPI_ISL_2005950, EPI_ISL_2005951, EPI_ISL_2005952, EPI_ISL_2005956, EPI_ISL_2005963, EPI_ISL_2005966, EPI_ISL_2005971, EPI_ISL_2005972, EPI_ISL_2005973, EPI_ISL_2005974, EPI_ISL_2005975, EPI_ISL_2005977, EPI_ISL_2005978, EPI_ISL_2005980, EPI_ISL_2005981, EPI_ISL_2005983, EPI_ISL_2005987, EPI_ISL_2005990,
EPI_ISL_2005995, EPI_ISL_2005997, EPI_ISL_2005999, EPI_ISL_2006001, EPI_ISL_2006004, EPI_ISL_2006005, EPI_ISL_2006009, EPI_ISL_2006011, EPI_ISL_2006014, EPI_ISL_2006017, EPI_ISL_2006019, EPI_ISL_2006020, EPI_ISL_2006022, EPI_ISL_2006023, EPI_ISL_2006025, EPI_ISL_2006028, EPI_ISL_2006030, EPI_ISL_2006033, EPI_ISL_2006037, EPI_ISL_2006038, EPI_ISL_2006041,
EPI_ISL_2006042, EPI_ISL_2006046, EPI_ISL_2006048, EPI_ISL_2006049, EPI_ISL_2006056, EPI_ISL_2006060, EPI_ISL_2006063, EPI_ISL_2006064, EPI_ISL_2006065, EPI_ISL_2006066, EPI_ISL_2006069, EPI_ISL_2006070, EPI_ISL_2021069, EPI_ISL_2021114, EPI_ISL_2021143, EPI_ISL_2021161, EPI_ISL_2021189, EPI_ISL_2021229, EPI_ISL_2021253, EPI_ISL_2021368, EPI_ISL_2021382, EPI_ISL_2021383,
EPI_ISL_2021387, EPI_ISL_2021392, EPI_ISL_2021398, EPI_ISL_2021399, EPI_ISL_2021401, EPI_ISL_2021405, EPI_ISL_2021415, EPI_ISL_2021419, EPI_ISL_2021421, EPI_ISL_2021422, EPI_ISL_2021425, EPI_ISL_2021427, EPI_ISL_2021430, EPI_ISL_2021434, EPI_ISL_2021439, EPI_ISL_2021444, EPI_ISL_2021448, EPI_ISL_2021453, EPI_ISL_2021461, EPI_ISL_2021462, EPI_ISL_2021465, EPI_ISL_2021467,
EPI_ISL_2021477, EPI_ISL_2021478, EPI_ISL_2021483, EPI_ISL_2021492, EPI_ISL_2021494, EPI_ISL_2021496, EPI_ISL_2021508, EPI_ISL_2021509, EPI_ISL_2021511, EPI_ISL_2021536, EPI_ISL_2021541, EPI_ISL_2021544, EPI_ISL_2021546, EPI_ISL_2021548, EPI_ISL_2021549, EPI_ISL_2021558, EPI_ISL_2021575, EPI_ISL_2021576, EPI_ISL_2021593, EPI_ISL_2021601, EPI_ISL_2021603, EPI_ISL_2021610,
EPI_ISL_2021612, EPI_ISL_2021613, EPI_ISL_2021614, EPI_ISL_2021620, EPI_ISL_2021622, EPI_ISL_2021625, EPI_ISL_2021635, EPI_ISL_2021636, EPI_ISL_2021638, EPI_ISL_2021654, EPI_ISL_2021658, EPI_ISL_2021659, EPI_ISL_2021661, EPI_ISL_2021663, EPI_ISL_2021665, EPI_ISL_2021671, EPI_ISL_2021675, EPI_ISL_2021679, EPI_ISL_2021683, EPI_ISL_2021690, EPI_ISL_2021755, EPI_ISL_2021823,
EPI_ISL_2021849, EPI_ISL_2021850, EPI_ISL_2021851, EPI_ISL_2021854, EPI_ISL_2021856, EPI_ISL_2021862, EPI_ISL_2021868, EPI_ISL_2021872, EPI_ISL_2021874, EPI_ISL_2021877, EPI_ISL_2021878, EPI_ISL_2021879, EPI_ISL_2021881, EPI_ISL_2021884, EPI_ISL_2021887, EPI_ISL_2021893, EPI_ISL_2021894, EPI_ISL_2021896, EPI_ISL_2021897, EPI_ISL_2021898, EPI_ISL_2021902, EPI_ISL_2021910,
EPI_ISL_2021914, EPI_ISL_2021917, EPI_ISL_2021922, EPI_ISL_2021926, EPI_ISL_2021930, EPI_ISL_2021931, EPI_ISL_2021932, EPI_ISL_2021939, EPI_ISL_2021940, EPI_ISL_2021944, EPI_ISL_2021947, EPI_ISL_2021948, EPI_ISL_2021951, EPI_ISL_2021955, EPI_ISL_2021957, EPI_ISL_2021958, EPI_ISL_2021962, EPI_ISL_2021965, EPI_ISL_2021969, EPI_ISL_2021972, EPI_ISL_2021973, EPI_ISL_2021974,
EPI_ISL_2021978, EPI_ISL_2021981, EPI_ISL_2021983, EPI_ISL_2021990, EPI_ISL_2021991, EPI_ISL_2021995, EPI_ISL_2021996, EPI_ISL_2021998, EPI_ISL_2022006, EPI_ISL_2022008, EPI_ISL_2022013, EPI_ISL_2022014, EPI_ISL_2022015, EPI_ISL_2022016, EPI_ISL_2022019, EPI_ISL_2022022, EPI_ISL_2022025, EPI_ISL_2022026, EPI_ISL_2022028, EPI_ISL_2022031, EPI_ISL_2022032, EPI_ISL_2022036,
EPI_ISL_2022039, EPI_ISL_2022043, EPI_ISL_2022044, EPI_ISL_2022045, EPI_ISL_2022046, EPI_ISL_2022048, EPI_ISL_2022049, EPI_ISL_2022050, EPI_ISL_2022054, EPI_ISL_2022059, EPI_ISL_2022060, EPI_ISL_2022065, EPI_ISL_2022071, EPI_ISL_2022082, EPI_ISL_2022084, EPI_ISL_2022090, EPI_ISL_2022094, EPI_ISL_2022095, EPI_ISL_2022102, EPI_ISL_2022103, EPI_ISL_2022104,
EPI_ISL_2022105, EPI_ISL_2022106, EPI_ISL_2022107, EPI_ISL_2022108, EPI_ISL_2022110, EPI_ISL_2022113, EPI_ISL_2022116, EPI_ISL_2022122, EPI_ISL_2022123, EPI_ISL_2022125, EPI_ISL_2022126, EPI_ISL_2022128, EPI_ISL_2022130, EPI_ISL_2022131, EPI_ISL_2022134, EPI_ISL_2022137, EPI_ISL_2022141, EPI_ISL_2022146, EPI_ISL_2022148, EPI_ISL_2022151, EPI_ISL_2022155, EPI_ISL_2022157,
EPI_ISL_2022158, EPI_ISL_2022159, EPI_ISL_2022161, EPI_ISL_2022167, EPI_ISL_2022169, EPI_ISL_2022170, EPI_ISL_2022173, EPI_ISL_2022174, EPI_ISL_2022175, EPI_ISL_2022176, EPI_ISL_2022177, EPI_ISL_2022180, EPI_ISL_2022186, EPI_ISL_2022187, EPI_ISL_2022191, EPI_ISL_2022194, EPI_ISL_2022199, EPI_ISL_2022200, EPI_ISL_2022201, EPI_ISL_2022202, EPI_ISL_2022203, EPI_ISL_2022207,
EPI_ISL_2022213, EPI_ISL_2022224, EPI_ISL_2022231, EPI_ISL_2022236, EPI_ISL_2022238, EPI_ISL_2022239, EPI_ISL_2022240, EPI_ISL_2022241, EPI_ISL_2022242, EPI_ISL_2022244, EPI_ISL_2022250, EPI_ISL_2022251, EPI_ISL_2022256, EPI_ISL_2022258, EPI_ISL_2022262, EPI_ISL_2022263, EPI_ISL_2022264, EPI_ISL_2022265, EPI_ISL_2022272, EPI_ISL_2022277, EPI_ISL_2022279, EPI_ISL_2022280,
EPI_ISL_2022281, EPI_ISL_2022285, EPI_ISL_2022286, EPI_ISL_2022288, EPI_ISL_2022290, EPI_ISL_2022295, EPI_ISL_2022296, EPI_ISL_2022299, EPI_ISL_2022304, EPI_ISL_2022305, EPI_ISL_2022306, EPI_ISL_2022308, EPI_ISL_2022317, EPI_ISL_2022319, EPI_ISL_2022327, EPI_ISL_2022330, EPI_ISL_2022332, EPI_ISL_2022337, EPI_ISL_2022342, EPI_ISL_2022343, EPI_ISL_2022348, EPI_ISL_2022351,
EPI_ISL_2022352, EPI_ISL_2022356, EPI_ISL_2022358, EPI_ISL_2022359, EPI_ISL_2022364, EPI_ISL_2022367, EPI_ISL_2022369, EPI_ISL_2022373, EPI_ISL_2022374, EPI_ISL_2022375, EPI_ISL_2022376, EPI_ISL_2022381, EPI_ISL_2022383, EPI_ISL_2022387, EPI_ISL_2022388, EPI_ISL_2022397, EPI_ISL_2022398, EPI_ISL_2022399, EPI_ISL_2022400, EPI_ISL_2022408, EPI_ISL_2022410, EPI_ISL_2022415,
EPI_ISL_2022419, EPI_ISL_2022420, EPI_ISL_2022423, EPI_ISL_2022428, EPI_ISL_2022429, EPI_ISL_2022437, EPI_ISL_2022542, EPI_ISL_2022577, EPI_ISL_2022579, EPI_ISL_2022584, EPI_ISL_2022902, EPI_ISL_2022912, EPI_ISL_2022913, EPI_ISL_2022922, EPI_ISL_2080476, EPI_ISL_2080478, EPI_ISL_2080480, EPI_ISL_2091794, EPI_ISL_2091795, EPI_ISL_2091796, EPI_ISL_2091797, EPI_ISL_2091798,
EPI_ISL_2091799, EPI_ISL_2091802, EPI_ISL_2091803, EPI_ISL_2091805, EPI_ISL_2091806, EPI_ISL_2091807, EPI_ISL_2091809, EPI_ISL_2091811, EPI_ISL_2091813, EPI_ISL_2091815, EPI_ISL_2091817, EPI_ISL_2091818, EPI_ISL_2091820, EPI_ISL_2091821, EPI_ISL_2091822, EPI_ISL_2091826, EPI_ISL_2091832, EPI_ISL_2091834, EPI_ISL_2091835, EPI_ISL_2091837, EPI_ISL_2091838, EPI_ISL_2091839,
EPI_ISL_2091840, EPI_ISL_2091843, EPI_ISL_2091848, EPI_ISL_2091849, EPI_ISL_2091850, EPI_ISL_2091853, EPI_ISL_2091855, EPI_ISL_2091857, EPI_ISL_2091860, EPI_ISL_2091861, EPI_ISL_2091862, EPI_ISL_2091867, EPI_ISL_2091871, EPI_ISL_2091872, EPI_ISL_2091873, EPI_ISL_2091875, EPI_ISL_2091876, EPI_ISL_2091879, EPI_ISL_2091882, EPI_ISL_2091883, EPI_ISL_2091886, EPI_ISL_2091887,
EPI_ISL_2091890, EPI_ISL_2091895, EPI_ISL_2091899, EPI_ISL_2091900, EPI_ISL_2091901, EPI_ISL_2091902, EPI_ISL_2091903, EPI_ISL_2091906, EPI_ISL_2091908, EPI_ISL_2091909, EPI_ISL_2091912, EPI_ISL_2091913, EPI_ISL_2091914, EPI_ISL_2091916, EPI_ISL_2091918, EPI_ISL_2091920, EPI_ISL_2091923, EPI_ISL_2091925, EPI_ISL_2091926, EPI_ISL_2091927, EPI_ISL_2091928, EPI_ISL_2091932,
EPI_ISL_2091933, EPI_ISL_2091935, EPI_ISL_2091938, EPI_ISL_2091939, EPI_ISL_2091941, EPI_ISL_2091942, EPI_ISL_2091944, EPI_ISL_2091946, EPI_ISL_2091948, EPI_ISL_2091949, EPI_ISL_2091950, EPI_ISL_2091952, EPI_ISL_2091953, EPI_ISL_2091956, EPI_ISL_2091957, EPI_ISL_2091960, EPI_ISL_2091963, EPI_ISL_2091969, EPI_ISL_2091971, EPI_ISL_2091972, EPI_ISL_2091973, EPI_ISL_2091976,
EPI_ISL_2091981, EPI_ISL_2091984, EPI_ISL_2091986, EPI_ISL_2091988, EPI_ISL_2091989, EPI_ISL_2091990, EPI_ISL_2091991, EPI_ISL_2091993, EPI_ISL_2091994, EPI_ISL_2091997, EPI_ISL_2092000, EPI_ISL_2092004, EPI_ISL_2092007, EPI_ISL_2092009, EPI_ISL_2092012, EPI_ISL_2092014, EPI_ISL_2092015, EPI_ISL_2092017, EPI_ISL_2092018, EPI_ISL_2092021, EPI_ISL_2092024, EPI_ISL_2092025,
EPI_ISL_2092027, EPI_ISL_2092028, EPI_ISL_2092029, EPI_ISL_2092030, EPI_ISL_2092031, EPI_ISL_2092032, EPI_ISL_2092033, EPI_ISL_2092035, EPI_ISL_2092036, EPI_ISL_2092037, EPI_ISL_2092041, EPI_ISL_2092043, EPI_ISL_2092044, EPI_ISL_2092045, EPI_ISL_2092051, EPI_ISL_2092053, EPI_ISL_2092056, EPI_ISL_2092062, EPI_ISL_2092063, EPI_ISL_2092064, EPI_ISL_2092065, EPI_ISL_2092066,
EPI_ISL_2092067, EPI_ISL_2092068, EPI_ISL_2092069, EPI_ISL_2092070, EPI_ISL_2092071, EPI_ISL_2092073, EPI_ISL_2092074, EPI_ISL_2092077, EPI_ISL_2092078, EPI_ISL_2092079, EPI_ISL_2092080, EPI_ISL_2092081, EPI_ISL_2092086, EPI_ISL_2092087, EPI_ISL_2092088, EPI_ISL_2092089, EPI_ISL_2092090, EPI_ISL_2092093, EPI_ISL_2092094, EPI_ISL_2092095, EPI_ISL_2092097, EPI_ISL_2092098,
EPI_ISL_2092101, EPI_ISL_2092675, EPI_ISL_2117515, EPI_ISL_2117516, EPI_ISL_2117523, EPI_ISL_2117529, EPI_ISL_2117538, EPI_ISL_2117542, EPI_ISL_2117544, EPI_ISL_2117549, EPI_ISL_2117556, EPI_ISL_2117559, EPI_ISL_2117562, EPI_ISL_2117566, EPI_ISL_2117572, EPI_ISL_2117582, EPI_ISL_2117584, EPI_ISL_2117585, EPI_ISL_2117586, EPI_ISL_2117591, EPI_ISL_2117592, EPI_ISL_2117598,
EPI_ISL_2117601, EPI_ISL_2117606, EPI_ISL_2117607, EPI_ISL_2117608, EPI_ISL_2117611, EPI_ISL_2117613, EPI_ISL_2117615, EPI_ISL_2117617, EPI_ISL_2117629, EPI_ISL_2117630, EPI_ISL_2117635, EPI_ISL_2117636, EPI_ISL_2117638, EPI_ISL_2117641, EPI_ISL_2117656, EPI_ISL_2117664, EPI_ISL_2117676, EPI_ISL_2117678, EPI_ISL_2117689, EPI_ISL_2117700, EPI_ISL_2117705, EPI_ISL_2117706,
EPI_ISL_2117709, EPI_ISL_2117711, EPI_ISL_2117722, EPI_ISL_2117725, EPI_ISL_2117728, EPI_ISL_2117730, EPI_ISL_2117733, EPI_ISL_2117737, EPI_ISL_2117739, EPI_ISL_2117740, EPI_ISL_2117743, EPI_ISL_2117747, EPI_ISL_2117755, EPI_ISL_2117783, EPI_ISL_2117785, EPI_ISL_2117788, EPI_ISL_2117790, EPI_ISL_2117791, EPI_ISL_2117793, EPI_ISL_2117797, EPI_ISL_2117800, EPI_ISL_2117801,
EPI_ISL_2117803, EPI_ISL_2117812, EPI_ISL_2117816, EPI_ISL_2117822, EPI_ISL_2117825, EPI_ISL_2117835, EPI_ISL_2117840, EPI_ISL_2117846, EPI_ISL_2117848, EPI_ISL_2117850, EPI_ISL_2117859, EPI_ISL_2117862, EPI_ISL_2117867, EPI_ISL_2117868, EPI_ISL_2117874, EPI_ISL_2117876, EPI_ISL_2117882, EPI_ISL_2117886, EPI_ISL_2117888, EPI_ISL_2117894, EPI_ISL_2117897, EPI_ISL_2117898,
EPI_ISL_2117901, EPI_ISL_2117905, EPI_ISL_2117910, EPI_ISL_2117913, EPI_ISL_2117916, EPI_ISL_2117918, EPI_ISL_2117924, EPI_ISL_2117928, EPI_ISL_2117936, EPI_ISL_2117942, EPI_ISL_2117943, EPI_ISL_2117952, EPI_ISL_2117955, EPI_ISL_2117960, EPI_ISL_2117961, EPI_ISL_2117964, EPI_ISL_2117972, EPI_ISL_2117975, EPI_ISL_2117983, EPI_ISL_2117987, EPI_ISL_2117991, EPI_ISL_2118000,
EPI_ISL_2118007, EPI_ISL_2118014, EPI_ISL_2118019, EPI_ISL_2118023, EPI_ISL_2118026, EPI_ISL_2118032, EPI_ISL_2118035, EPI_ISL_2118036, EPI_ISL_2118040, EPI_ISL_2118045, EPI_ISL_2118050, EPI_ISL_2118058, EPI_ISL_2118084, EPI_ISL_2118089, EPI_ISL_2118093, EPI_ISL_2118096, EPI_ISL_2118097, EPI_ISL_2118113, EPI_ISL_2118116, EPI_ISL_2118118, EPI_ISL_2118119, EPI_ISL_2118122,
EPI_ISL_2118123, EPI_ISL_2118140, EPI_ISL_2118148, EPI_ISL_2118155, EPI_ISL_2118156, EPI_ISL_2118161, EPI_ISL_2118164, EPI_ISL_2118168, EPI_ISL_2118179, EPI_ISL_2118182, EPI_ISL_2118183, EPI_ISL_2118186, EPI_ISL_2118188, EPI_ISL_2118189, EPI_ISL_2118197, EPI_ISL_2118201, EPI_ISL_2118205, EPI_ISL_2118206, EPI_ISL_2118208, EPI_ISL_2118210, EPI_ISL_2118214, EPI_ISL_2118215,
EPI_ISL_2118217, EPI_ISL_2118223, EPI_ISL_2118225, EPI_ISL_2118226, EPI_ISL_2118228, EPI_ISL_2118229, EPI_ISL_2118231, EPI_ISL_2118234, EPI_ISL_2118258, EPI_ISL_2118259, EPI_ISL_2118311, EPI_ISL_2118316, EPI_ISL_2118324, EPI_ISL_2118325, EPI_ISL_2118338, EPI_ISL_2118339, EPI_ISL_2118343, EPI_ISL_2118345, EPI_ISL_2118356, EPI_ISL_2118359, EPI_ISL_2118363, EPI_ISL_2118368,
EPI_ISL_2118370, EPI_ISL_2118382, EPI_ISL_2118390, EPI_ISL_2118401, EPI_ISL_2118402, EPI_ISL_2118406, EPI_ISL_2118407, EPI_ISL_2118410, EPI_ISL_2118413, EPI_ISL_2118425, EPI_ISL_2118435, EPI_ISL_2118436, EPI_ISL_2118438, EPI_ISL_2118440, EPI_ISL_2118450, EPI_ISL_2118452, EPI_ISL_2118453, EPI_ISL_2118459, EPI_ISL_2118463, EPI_ISL_2118466, EPI_ISL_2118468, EPI_ISL_2118499,
EPI_ISL_2118503, EPI_ISL_2118507, EPI_ISL_2118519, EPI_ISL_2118526, EPI_ISL_2118549, EPI_ISL_2118557, EPI_ISL_2118562, EPI_ISL_2118570, EPI_ISL_2118572, EPI_ISL_2118576, EPI_ISL_2118578, EPI_ISL_2118583, EPI_ISL_2118588, EPI_ISL_2118592, EPI_ISL_2118603, EPI_ISL_2118605, EPI_ISL_2118607, EPI_ISL_2118616, EPI_ISL_2118619, EPI_ISL_2118621, EPI_ISL_2118627, EPI_ISL_2118633,
EPI_ISL_2118640, EPI_ISL_2118647, EPI_ISL_2118651, EPI_ISL_2118664, EPI_ISL_2118665, EPI_ISL_2118681, EPI_ISL_2118695, EPI_ISL_2118702, EPI_ISL_2118706, EPI_ISL_2118707, EPI_ISL_2118715, EPI_ISL_2118719, EPI_ISL_2118727, EPI_ISL_2118736, EPI_ISL_2118747, EPI_ISL_2118753, EPI_ISL_2118754, EPI_ISL_2118759, EPI_ISL_2118760, EPI_ISL_2118767, EPI_ISL_2118772, EPI_ISL_2118782,
EPI_ISL_2118783, EPI_ISL_2118789, EPI_ISL_2118794, EPI_ISL_2118798, EPI_ISL_2118800, EPI_ISL_2118801, EPI_ISL_2118804, EPI_ISL_2118809, EPI_ISL_2118810, EPI_ISL_2118820, EPI_ISL_2118823, EPI_ISL_2118825, EPI_ISL_2118826, EPI_ISL_2118834, EPI_ISL_2118837, EPI_ISL_2118838, EPI_ISL_2118840, EPI_ISL_2118841, EPI_ISL_2118846, EPI_ISL_2118847, EPI_ISL_2118854, EPI_ISL_2118858,
EPI_ISL_2118859, EPI_ISL_2118861, EPI_ISL_2118862, EPI_ISL_2118866, EPI_ISL_2118870, EPI_ISL_2118871, EPI_ISL_2118874, EPI_ISL_2118879, EPI_ISL_2118880, EPI_ISL_2118883, EPI_ISL_2118888, EPI_ISL_2118892, EPI_ISL_2118894, EPI_ISL_2118896, EPI_ISL_2118899, EPI_ISL_2118900, EPI_ISL_2118902, EPI_ISL_2118905, EPI_ISL_2118906, EPI_ISL_2118909, EPI_ISL_2118911, EPI_ISL_2118914,
EPI_ISL_2118917, EPI_ISL_2118919, EPI_ISL_2118920, EPI_ISL_2118921, EPI_ISL_2118925, EPI_ISL_2118927, EPI_ISL_2118929, EPI_ISL_2118931, EPI_ISL_2118932, EPI_ISL_2118934, EPI_ISL_2118936, EPI_ISL_2118938, EPI_ISL_2118941, EPI_ISL_2118942, EPI_ISL_2118947, EPI_ISL_2118949, EPI_ISL_2118950, EPI_ISL_2118952, EPI_ISL_2118953, EPI_ISL_2118956, EPI_ISL_2118959, EPI_ISL_2118962,
EPI_ISL_2118964, EPI_ISL_2118966, EPI_ISL_2118969, EPI_ISL_2118972, EPI_ISL_2118974, EPI_ISL_2118975, EPI_ISL_2118976, EPI_ISL_2118977, EPI_ISL_2118980, EPI_ISL_2118982, EPI_ISL_2118986, EPI_ISL_2118994, EPI_ISL_2118995, EPI_ISL_2118996, EPI_ISL_2119000, EPI_ISL_2119001, EPI_ISL_2119003, EPI_ISL_2119007, EPI_ISL_2119011, EPI_ISL_2119012, EPI_ISL_2119018, EPI_ISL_2119021,
EPI_ISL_2119025, EPI_ISL_2119028, EPI_ISL_2119030, EPI_ISL_2119031, EPI_ISL_2119033, EPI_ISL_2119034, EPI_ISL_2119036, EPI_ISL_2119037, EPI_ISL_2119046, EPI_ISL_2119080, EPI_ISL_2119085, EPI_ISL_2119092, EPI_ISL_2119094, EPI_ISL_2119095, EPI_ISL_2119103, EPI_ISL_2119105, EPI_ISL_2119106, EPI_ISL_2119107, EPI_ISL_2119109, EPI_ISL_2119110, EPI_ISL_2119112, EPI_ISL_2119115,
EPI_ISL_2119116, EPI_ISL_2119118, EPI_ISL_2119119, EPI_ISL_2119125, EPI_ISL_2119131, EPI_ISL_2119136, EPI_ISL_2119139, EPI_ISL_2119144, EPI_ISL_2119148, EPI_ISL_2119154, EPI_ISL_2119157, EPI_ISL_2119161, EPI_ISL_2119168, EPI_ISL_2119171, EPI_ISL_2119174, EPI_ISL_2119175, EPI_ISL_2119177, EPI_ISL_2119178, EPI_ISL_2119179, EPI_ISL_2119181, EPI_ISL_2119186, EPI_ISL_2119188,
EPI_ISL_2119189, EPI_ISL_2119191, EPI_ISL_2119193, EPI_ISL_2119194, EPI_ISL_2119197, EPI_ISL_2119200, EPI_ISL_2119202, EPI_ISL_2119203, EPI_ISL_2119208, EPI_ISL_2119213, EPI_ISL_2119214, EPI_ISL_2119219, EPI_ISL_2119223, EPI_ISL_2119225, EPI_ISL_2119228, EPI_ISL_2119231, EPI_ISL_2119233, EPI_ISL_2119235, EPI_ISL_2119238, EPI_ISL_2119240, EPI_ISL_2119242, EPI_ISL_2119246,
EPI_ISL_2119252, EPI_ISL_2119254, EPI_ISL_2119255, EPI_ISL_2119258, EPI_ISL_2119261, EPI_ISL_2119263, EPI_ISL_2119270, EPI_ISL_2119273, EPI_ISL_2119281, EPI_ISL_2119283, EPI_ISL_2119292, EPI_ISL_2119296, EPI_ISL_2119298, EPI_ISL_2119303, EPI_ISL_2119304, EPI_ISL_2119306, EPI_ISL_2119307, EPI_ISL_2119312, EPI_ISL_2119314, EPI_ISL_2119317, EPI_ISL_2119319, EPI_ISL_2119322,
EPI_ISL_2119325, EPI_ISL_2119328, EPI_ISL_2119332, EPI_ISL_2119333, EPI_ISL_2119335, EPI_ISL_2119339, EPI_ISL_2119341, EPI_ISL_2119344, EPI_ISL_2119348, EPI_ISL_2119352, EPI_ISL_2119356, EPI_ISL_2119358, EPI_ISL_2119361, EPI_ISL_2119363, EPI_ISL_2119367, EPI_ISL_2119368, EPI_ISL_2119371, EPI_ISL_2119373, EPI_ISL_2119376, EPI_ISL_2119379, EPI_ISL_2119381, EPI_ISL_2119383,
EPI_ISL_2119385, EPI_ISL_2119390, EPI_ISL_2119393, EPI_ISL_2119397, EPI_ISL_2119400, EPI_ISL_2119402, EPI_ISL_2119405, EPI_ISL_2119408, EPI_ISL_2119411, EPI_ISL_2119412, EPI_ISL_2119413, EPI_ISL_2119414, EPI_ISL_2119417, EPI_ISL_2119419, EPI_ISL_2119422, EPI_ISL_2119427, EPI_ISL_2119434, EPI_ISL_2119437, EPI_ISL_2119447, EPI_ISL_2119450, EPI_ISL_2119453,
EPI_ISL_2119454, EPI_ISL_2119456, EPI_ISL_2119457, EPI_ISL_2119458, EPI_ISL_2119465, EPI_ISL_2119467, EPI_ISL_2119468, EPI_ISL_2119477, EPI_ISL_2119479, EPI_ISL_2119489, EPI_ISL_2119492, EPI_ISL_2119495, EPI_ISL_2119497, EPI_ISL_2119500, EPI_ISL_2119503, EPI_ISL_2119506, EPI_ISL_2119507, EPI_ISL_2119509, EPI_ISL_2119510, EPI_ISL_2119515, EPI_ISL_2119517, EPI_ISL_2119520,
EPI_ISL_2119523, EPI_ISL_2119526, EPI_ISL_2119529, EPI_ISL_2119531, EPI_ISL_2119532, EPI_ISL_2119535, EPI_ISL_2119545, EPI_ISL_2119548, EPI_ISL_2119549, EPI_ISL_2119552, EPI_ISL_2119557, EPI_ISL_2119565, EPI_ISL_2119572, EPI_ISL_2119579, EPI_ISL_2119580, EPI_ISL_2119590, EPI_ISL_2119596, EPI_ISL_2119601, EPI_ISL_2119602, EPI_ISL_2119604, EPI_ISL_2119606, EPI_ISL_2119607,
EPI_ISL_2119612, EPI_ISL_2119613, EPI_ISL_2119615, EPI_ISL_2119617, EPI_ISL_2119618, EPI_ISL_2119630, EPI_ISL_2119641, EPI_ISL_2119644, EPI_ISL_2119647, EPI_ISL_2119649, EPI_ISL_2119655, EPI_ISL_2119657, EPI_ISL_2119666, EPI_ISL_2119667, EPI_ISL_2119669, EPI_ISL_2119672, EPI_ISL_2119674, EPI_ISL_2119676, EPI_ISL_2119677, EPI_ISL_2119685, EPI_ISL_2119686, EPI_ISL_2119688,
EPI_ISL_2119689, EPI_ISL_2119694, EPI_ISL_2119696, EPI_ISL_2119697, EPI_ISL_2119701, EPI_ISL_2119702, EPI_ISL_2119704, EPI_ISL_2119706, EPI_ISL_2119707, EPI_ISL_2119718, EPI_ISL_2119720, EPI_ISL_2119723, EPI_ISL_2119727, EPI_ISL_2119731, EPI_ISL_2119732, EPI_ISL_2119735, EPI_ISL_2119740, EPI_ISL_2119743, EPI_ISL_2119746, EPI_ISL_2119751, EPI_ISL_2119756, EPI_ISL_2119757,
EPI_ISL_2119759, EPI_ISL_2119761, EPI_ISL_2119764, EPI_ISL_2119767, EPI_ISL_2119770, EPI_ISL_2119774, EPI_ISL_2119777, EPI_ISL_2119784, EPI_ISL_2119786, EPI_ISL_2119787, EPI_ISL_2119789, EPI_ISL_2119790, EPI_ISL_2119795, EPI_ISL_2119797, EPI_ISL_2119798, EPI_ISL_2119800, EPI_ISL_2119801, EPI_ISL_2119806, EPI_ISL_2119808, EPI_ISL_2119809, EPI_ISL_2119811, EPI_ISL_2119812,
EPI_ISL_2119814, EPI_ISL_2119816, EPI_ISL_2119817, EPI_ISL_2119820, EPI_ISL_2119823, EPI_ISL_2119826, EPI_ISL_2119830, EPI_ISL_2119833, EPI_ISL_2119839, EPI_ISL_2119843, EPI_ISL_2119844, EPI_ISL_2119846, EPI_ISL_2119851, EPI_ISL_2119854, EPI_ISL_2119857, EPI_ISL_2119861, EPI_ISL_2119865, EPI_ISL_2119869, EPI_ISL_2119875, EPI_ISL_2119904, EPI_ISL_2119906, EPI_ISL_2119907,
EPI_ISL_2119910, EPI_ISL_2119911, EPI_ISL_2119914, EPI_ISL_2119917, EPI_ISL_2119922, EPI_ISL_2119927, EPI_ISL_2119931, EPI_ISL_2119936, EPI_ISL_2119942, EPI_ISL_2119945, EPI_ISL_2119946, EPI_ISL_2119948, EPI_ISL_2119952, EPI_ISL_2119954, EPI_ISL_2119955, EPI_ISL_2119957, EPI_ISL_2119960, EPI_ISL_2119963, EPI_ISL_2119965, EPI_ISL_2119970, EPI_ISL_2119972, EPI_ISL_2119973,
EPI_ISL_2119974, EPI_ISL_2119975, EPI_ISL_2119976, EPI_ISL_2119977, EPI_ISL_2119979, EPI_ISL_2119980, EPI_ISL_2119982, EPI_ISL_2119985, EPI_ISL_2119986, EPI_ISL_2119987, EPI_ISL_2119990, EPI_ISL_2119991, EPI_ISL_2119993, EPI_ISL_2119996, EPI_ISL_2119997, EPI_ISL_2120000, EPI_ISL_2120002, EPI_ISL_2120003, EPI_ISL_2120006, EPI_ISL_2120008, EPI_ISL_2120009, EPI_ISL_2120013,
EPI_ISL_2120027, EPI_ISL_2120028, EPI_ISL_2120036, EPI_ISL_2120037, EPI_ISL_2120040, EPI_ISL_2120042, EPI_ISL_2120045, EPI_ISL_2120048, EPI_ISL_2120052, EPI_ISL_2120053, EPI_ISL_2120054, EPI_ISL_2120057, EPI_ISL_2120059, EPI_ISL_2120061, EPI_ISL_2120062, EPI_ISL_2120064, EPI_ISL_2120068, EPI_ISL_2120071, EPI_ISL_2120074, EPI_ISL_2120077, EPI_ISL_2120080, EPI_ISL_2120082,
EPI_ISL_2120085, EPI_ISL_2120088, EPI_ISL_2120089, EPI_ISL_2120091, EPI_ISL_2120094, EPI_ISL_2120095, EPI_ISL_2120096, EPI_ISL_2120100, EPI_ISL_2120101, EPI_ISL_2120103, EPI_ISL_2120104, EPI_ISL_2120107, EPI_ISL_2120109, EPI_ISL_2120110, EPI_ISL_2120112, EPI_ISL_2120113, EPI_ISL_2120114, EPI_ISL_2120115, EPI_ISL_2120117, EPI_ISL_2120119, EPI_ISL_2120123, EPI_ISL_2120132,
EPI_ISL_2120135, EPI_ISL_2120138, EPI_ISL_2120141, EPI_ISL_2120143, EPI_ISL_2120146, EPI_ISL_2120150, EPI_ISL_2120152, EPI_ISL_2120155, EPI_ISL_2120157, EPI_ISL_2120158, EPI_ISL_2120160, EPI_ISL_2120163, EPI_ISL_2120166, EPI_ISL_2120168, EPI_ISL_2120171, EPI_ISL_2120172, EPI_ISL_2120173, EPI_ISL_2120176, EPI_ISL_2120179, EPI_ISL_2120182, EPI_ISL_2120184, EPI_ISL_2120185,
EPI_ISL_2120188, EPI_ISL_2120189, EPI_ISL_2120191, EPI_ISL_2120192, EPI_ISL_2120193, EPI_ISL_2120195, EPI_ISL_2120196, EPI_ISL_2120198, EPI_ISL_2120202, EPI_ISL_2120206, EPI_ISL_2120207, EPI_ISL_2120211, EPI_ISL_2120213, EPI_ISL_2120214, EPI_ISL_2120216, EPI_ISL_2120217, EPI_ISL_2120220, EPI_ISL_2120221, EPI_ISL_2120227, EPI_ISL_2120232, EPI_ISL_2120236, EPI_ISL_2120239,
EPI_ISL_2120240, EPI_ISL_2120242, EPI_ISL_2120244, EPI_ISL_2120252, EPI_ISL_2120254, EPI_ISL_2120256, EPI_ISL_2120257, EPI_ISL_2120264, EPI_ISL_2120268, EPI_ISL_2120270, EPI_ISL_2120273, EPI_ISL_2120274, EPI_ISL_2120276, EPI_ISL_2120277, EPI_ISL_2120279, EPI_ISL_2120283, EPI_ISL_2120285, EPI_ISL_2120286, EPI_ISL_2120287, EPI_ISL_2120289, EPI_ISL_2120292, EPI_ISL_2120293,
EPI_ISL_2120298, EPI_ISL_2120299, EPI_ISL_2120301, EPI_ISL_2120305, EPI_ISL_2120310, EPI_ISL_2120311, EPI_ISL_2120313, EPI_ISL_2120314, EPI_ISL_2120316, EPI_ISL_2120317, EPI_ISL_2120320, EPI_ISL_2120323, EPI_ISL_2120328, EPI_ISL_2120329, EPI_ISL_2120331, EPI_ISL_2120334, EPI_ISL_2120337, EPI_ISL_2120340, EPI_ISL_2120342, EPI_ISL_2120346, EPI_ISL_2120349, EPI_ISL_2120350,
EPI_ISL_2120352, EPI_ISL_2120353, EPI_ISL_2120355, EPI_ISL_2120357, EPI_ISL_2120358, EPI_ISL_2120360, EPI_ISL_2120367, EPI_ISL_2120369, EPI_ISL_2120371, EPI_ISL_2120376, EPI_ISL_2120379, EPI_ISL_2120384, EPI_ISL_2120388, EPI_ISL_2120390, EPI_ISL_2120391, EPI_ISL_2120393, EPI_ISL_2120397, EPI_ISL_2120400, EPI_ISL_2120402, EPI_ISL_2120405, EPI_ISL_2120409,
EPI_ISL_2120410, EPI_ISL_2120412, EPI_ISL_2120413, EPI_ISL_2120417, EPI_ISL_2120419, EPI_ISL_2120420, EPI_ISL_2120423, EPI_ISL_2120426, EPI_ISL_2120428, EPI_ISL_2120430, EPI_ISL_2120433, EPI_ISL_2120437, EPI_ISL_2120442, EPI_ISL_2120446, EPI_ISL_2120449, EPI_ISL_2120450, EPI_ISL_2120452, EPI_ISL_2120457, EPI_ISL_2120471, EPI_ISL_2120476, EPI_ISL_2120477, EPI_ISL_2120487,
EPI_ISL_2120491, EPI_ISL_2120503, EPI_ISL_2120514, EPI_ISL_2120522, EPI_ISL_2120526, EPI_ISL_2120538, EPI_ISL_2120541, EPI_ISL_2120545, EPI_ISL_2120560, EPI_ISL_2120578, EPI_ISL_2120584, EPI_ISL_2120585, EPI_ISL_2120590, EPI_ISL_2120594, EPI_ISL_2120605, EPI_ISL_2120607, EPI_ISL_2120608, EPI_ISL_2120610, EPI_ISL_2120613, EPI_ISL_2120614, EPI_ISL_2120628, EPI_ISL_2120630,
EPI_ISL_2120642, EPI_ISL_2120643, EPI_ISL_2120652, EPI_ISL_2120655, EPI_ISL_2120695, EPI_ISL_2120697, EPI_ISL_2120700, EPI_ISL_2120713, EPI_ISL_2120724, EPI_ISL_2120726, EPI_ISL_2120745, EPI_ISL_2120746, EPI_ISL_2120751, EPI_ISL_2120754, EPI_ISL_2120756, EPI_ISL_2120760, EPI_ISL_2120761, EPI_ISL_2120776, EPI_ISL_2120782, EPI_ISL_2120791, EPI_ISL_2120793, EPI_ISL_2120795,
EPI_ISL_2120800, EPI_ISL_2120802, EPI_ISL_2120812, EPI_ISL_2120828, EPI_ISL_2120836, EPI_ISL_2120837, EPI_ISL_2120839, EPI_ISL_2120852, EPI_ISL_2120861, EPI_ISL_2120866, EPI_ISL_2120868, EPI_ISL_2120877, EPI_ISL_2120882, EPI_ISL_2120891, EPI_ISL_2120902, EPI_ISL_2120905, EPI_ISL_2120910, EPI_ISL_2120915, EPI_ISL_2120921, EPI_ISL_2120926, EPI_ISL_2120929,
EPI_ISL_2120932, EPI_ISL_2120943, EPI_ISL_2120947, EPI_ISL_2120948, EPI_ISL_2120951, EPI_ISL_2120956, EPI_ISL_2120959, EPI_ISL_2120965, EPI_ISL_2120968, EPI_ISL_2120979, EPI_ISL_2120992, EPI_ISL_2121005, EPI_ISL_2121007, EPI_ISL_2121010, EPI_ISL_2121011, EPI_ISL_2121013, EPI_ISL_2121021, EPI_ISL_2121023, EPI_ISL_2121025, EPI_ISL_2121026, EPI_ISL_2121031, EPI_ISL_2121032,
EPI_ISL_2121037, EPI_ISL_2121041, EPI_ISL_2121046, EPI_ISL_2121047, EPI_ISL_2121050, EPI_ISL_2121056, EPI_ISL_2121057, EPI_ISL_2121060, EPI_ISL_2121066, EPI_ISL_2121069, EPI_ISL_2121072, EPI_ISL_2121076, EPI_ISL_2121078, EPI_ISL_2121079, EPI_ISL_2121082, EPI_ISL_2121092, EPI_ISL_2121097, EPI_ISL_2121098, EPI_ISL_2121101, EPI_ISL_2121104, EPI_ISL_2121107,
EPI_ISL_2121108, EPI_ISL_2121111, EPI_ISL_2121114, EPI_ISL_2121116, EPI_ISL_2121117, EPI_ISL_2121122, EPI_ISL_2121123, EPI_ISL_2121137, EPI_ISL_2121138, EPI_ISL_2121141, EPI_ISL_2121150, EPI_ISL_2121151, EPI_ISL_2121156, EPI_ISL_2121157, EPI_ISL_2121159, EPI_ISL_2121162, EPI_ISL_2121165, EPI_ISL_2121166, EPI_ISL_2121168, EPI_ISL_2121177, EPI_ISL_2121182, EPI_ISL_2121183,
EPI_ISL_2121195, EPI_ISL_2121196, EPI_ISL_2121198, EPI_ISL_2121201, EPI_ISL_2121204, EPI_ISL_2121209, EPI_ISL_2121210, EPI_ISL_2121215, EPI_ISL_2121222, EPI_ISL_2121230, EPI_ISL_2121232, EPI_ISL_2121237, EPI_ISL_2121239, EPI_ISL_2121240, EPI_ISL_2121242, EPI_ISL_2121243, EPI_ISL_2121246, EPI_ISL_2121247, EPI_ISL_2121253, EPI_ISL_2121255, EPI_ISL_2121256, EPI_ISL_2121259,
EPI_ISL_2121263, EPI_ISL_2121265, EPI_ISL_2121266, EPI_ISL_2121273, EPI_ISL_2121275, EPI_ISL_2121277, EPI_ISL_2121286, EPI_ISL_2121291, EPI_ISL_2121293, EPI_ISL_2121294, EPI_ISL_2121296, EPI_ISL_2121300, EPI_ISL_2121305, EPI_ISL_2121306, EPI_ISL_2121308, EPI_ISL_2121309, EPI_ISL_2121312, EPI_ISL_2121313, EPI_ISL_2121315, EPI_ISL_2121316, EPI_ISL_2121318, EPI_ISL_2121333,
EPI_ISL_2121335, EPI_ISL_2121338, EPI_ISL_2121340, EPI_ISL_2121346, EPI_ISL_2121347, EPI_ISL_2121350, EPI_ISL_2121355, EPI_ISL_2121356, EPI_ISL_2121358, EPI_ISL_2121364, EPI_ISL_2121371, EPI_ISL_2121373, EPI_ISL_2121374, EPI_ISL_2121376, EPI_ISL_2121382, EPI_ISL_2121385, EPI_ISL_2121393, EPI_ISL_2121397, EPI_ISL_2121399, EPI_ISL_2121401, EPI_ISL_2121402, EPI_ISL_2121404,
EPI_ISL_2121413, EPI_ISL_2121416, EPI_ISL_2121417, EPI_ISL_2121420, EPI_ISL_2121422, EPI_ISL_2121425, EPI_ISL_2121426, EPI_ISL_2121429, EPI_ISL_2121432, EPI_ISL_2121436, EPI_ISL_2121437, EPI_ISL_2121469, EPI_ISL_2121472, EPI_ISL_2121473, EPI_ISL_2121475, EPI_ISL_2121479, EPI_ISL_2121481, EPI_ISL_2121483, EPI_ISL_2121484, EPI_ISL_2121489, EPI_ISL_2121493, EPI_ISL_2121495,
EPI_ISL_2121498, EPI_ISL_2121500, EPI_ISL_2121501, EPI_ISL_2121506, EPI_ISL_2121508, EPI_ISL_2121509, EPI_ISL_2121513, EPI_ISL_2121514, EPI_ISL_2121517, EPI_ISL_2121519, EPI_ISL_2121521, EPI_ISL_2121522, EPI_ISL_2121524, EPI_ISL_2121525, EPI_ISL_2121527, EPI_ISL_2121533, EPI_ISL_2121536, EPI_ISL_2121538, EPI_ISL_2121542, EPI_ISL_2121547, EPI_ISL_2121549,
EPI_ISL_2121557, EPI_ISL_2121568, EPI_ISL_2121572, EPI_ISL_2121574, EPI_ISL_2121576, EPI_ISL_2121577, EPI_ISL_2121580, EPI_ISL_2121582, EPI_ISL_2121584, EPI_ISL_2121587, EPI_ISL_2121588, EPI_ISL_2121590, EPI_ISL_2121591, EPI_ISL_2121593, EPI_ISL_2121594, EPI_ISL_2121596, EPI_ISL_2121598, EPI_ISL_2121600, EPI_ISL_2121601, EPI_ISL_2121603, EPI_ISL_2121607, EPI_ISL_2121609,
EPI_ISL_2121611, EPI_ISL_2121613, EPI_ISL_2121614, EPI_ISL_2121615, EPI_ISL_2121617, EPI_ISL_2121618, EPI_ISL_2121622, EPI_ISL_2121625, EPI_ISL_2121626, EPI_ISL_2121628, EPI_ISL_2121629, EPI_ISL_2121631, EPI_ISL_2121632, EPI_ISL_2121634, EPI_ISL_2121636, EPI_ISL_2121639, EPI_ISL_2121645, EPI_ISL_2121647, EPI_ISL_2121648, EPI_ISL_2121650, EPI_ISL_2121651, EPI_ISL_2121653,
EPI_ISL_2121657, EPI_ISL_2121660, EPI_ISL_2121662, EPI_ISL_2121665, EPI_ISL_2121666, EPI_ISL_2121668, EPI_ISL_2121670, EPI_ISL_2121671, EPI_ISL_2121676, EPI_ISL_2121677, EPI_ISL_2121679, EPI_ISL_2121680, EPI_ISL_2121684, EPI_ISL_2121685, EPI_ISL_2121687, EPI_ISL_2121688, EPI_ISL_2121690, EPI_ISL_2121691, EPI_ISL_2121693, EPI_ISL_2121696, EPI_ISL_2121697, EPI_ISL_2121700,
EPI_ISL_2121704, EPI_ISL_2121707, EPI_ISL_2121708, EPI_ISL_2121710, EPI_ISL_2121711, EPI_ISL_2121714, EPI_ISL_2121716, EPI_ISL_2121718, EPI_ISL_2121719, EPI_ISL_2121725, EPI_ISL_2121731, EPI_ISL_2121733, EPI_ISL_2121735, EPI_ISL_2121743, EPI_ISL_2121748, EPI_ISL_2121750, EPI_ISL_2121753, EPI_ISL_2121755, EPI_ISL_2121758, EPI_ISL_2121759, EPI_ISL_2121761,
EPI_ISL_2121762, EPI_ISL_2121764, EPI_ISL_2121767, EPI_ISL_2121769, EPI_ISL_2121770, EPI_ISL_2121772, EPI_ISL_2121773, EPI_ISL_2121775, EPI_ISL_2121778, EPI_ISL_2121781, EPI_ISL_2121782, EPI_ISL_2121784, EPI_ISL_2121785, EPI_ISL_2121790, EPI_ISL_2121793, EPI_ISL_2121796, EPI_ISL_2121798, EPI_ISL_2121801, EPI_ISL_2121802, EPI_ISL_2121804, EPI_ISL_2121806, EPI_ISL_2121807,
EPI_ISL_2121810, EPI_ISL_2121812, EPI_ISL_2121819, EPI_ISL_2121820, EPI_ISL_2121822, EPI_ISL_2121823, EPI_ISL_2121826, EPI_ISL_2121828, EPI_ISL_2121833, EPI_ISL_2121835, EPI_ISL_2121837, EPI_ISL_2121838, EPI_ISL_2121841, EPI_ISL_2121847, EPI_ISL_2121850, EPI_ISL_2121852, EPI_ISL_2121853, EPI_ISL_2121855, EPI_ISL_2121856, EPI_ISL_2121858, EPI_ISL_2121859, EPI_ISL_2121861,
EPI_ISL_2121863, EPI_ISL_2121864, EPI_ISL_2121866, EPI_ISL_2121867, EPI_ISL_2121870, EPI_ISL_2121872, EPI_ISL_2121874, EPI_ISL_2121875, EPI_ISL_2121877, EPI_ISL_2121880, EPI_ISL_2121883, EPI_ISL_2121886, EPI_ISL_2121888, EPI_ISL_2121889, EPI_ISL_2121891, EPI_ISL_2121893, EPI_ISL_2121897, EPI_ISL_2121899, EPI_ISL_2121901, EPI_ISL_2121902, EPI_ISL_2121904, EPI_ISL_2121906,
EPI_ISL_2121907, EPI_ISL_2121909, EPI_ISL_2121911, EPI_ISL_2121912, EPI_ISL_2121914, EPI_ISL_2121915, EPI_ISL_2121918, EPI_ISL_2121920, EPI_ISL_2121922, EPI_ISL_2121927, EPI_ISL_2121930, EPI_ISL_2121931, EPI_ISL_2121933, EPI_ISL_2121934, EPI_ISL_2121936, EPI_ISL_2121938, EPI_ISL_2121939, EPI_ISL_2121941, EPI_ISL_2121942, EPI_ISL_2121944, EPI_ISL_2121946, EPI_ISL_2121947,
EPI_ISL_2121949, EPI_ISL_2121952, EPI_ISL_2121955, EPI_ISL_2121957, EPI_ISL_2121958, EPI_ISL_2121960, EPI_ISL_2121962, EPI_ISL_2121963, EPI_ISL_2121966, EPI_ISL_2121974, EPI_ISL_2121976, EPI_ISL_2121977, EPI_ISL_2121980, EPI_ISL_2121983, EPI_ISL_2121988, EPI_ISL_2121989, EPI_ISL_2121991, EPI_ISL_2121995, EPI_ISL_2121997, EPI_ISL_2122000, EPI_ISL_2122002, EPI_ISL_2122004,
EPI_ISL_2122005, EPI_ISL_2122007, EPI_ISL_2122009, EPI_ISL_2122011, EPI_ISL_2122014, EPI_ISL_2122018, EPI_ISL_2122019, EPI_ISL_2122023, EPI_ISL_2122024, EPI_ISL_2122026, EPI_ISL_2122029, EPI_ISL_2122031, EPI_ISL_2122032, EPI_ISL_2122034, EPI_ISL_2122036, EPI_ISL_2122037, EPI_ISL_2122039, EPI_ISL_2122040, EPI_ISL_2122042, EPI_ISL_2122043, EPI_ISL_2122045, EPI_ISL_2122046,
EPI_ISL_2122050, EPI_ISL_2122051, EPI_ISL_2122053, EPI_ISL_2122056, EPI_ISL_2122058, EPI_ISL_2122059, EPI_ISL_2122060, EPI_ISL_2122062, EPI_ISL_2122063, EPI_ISL_2122065, EPI_ISL_2122066, EPI_ISL_2122068, EPI_ISL_2122069, EPI_ISL_2122071, EPI_ISL_2122074, EPI_ISL_2122076, EPI_ISL_2122079, EPI_ISL_2122080, EPI_ISL_2122082, EPI_ISL_2122084, EPI_ISL_2122085, EPI_ISL_2122086,
EPI_ISL_2122088, EPI_ISL_2122089, EPI_ISL_2122091, EPI_ISL_2122094, EPI_ISL_2122096, EPI_ISL_2122100, EPI_ISL_2122102, EPI_ISL_2122103, EPI_ISL_2122105, EPI_ISL_2122106, EPI_ISL_2122108, EPI_ISL_2122109, EPI_ISL_2122111, EPI_ISL_2122113, EPI_ISL_2122114, EPI_ISL_2122116, EPI_ISL_2122117, EPI_ISL_2122119, EPI_ISL_2122122, EPI_ISL_2122147, EPI_ISL_2122148, EPI_ISL_2122150,
EPI_ISL_2122155, EPI_ISL_2122167, EPI_ISL_2122176, EPI_ISL_2122180, EPI_ISL_2122184, EPI_ISL_2122187, EPI_ISL_2122200, EPI_ISL_2122202, EPI_ISL_2122203, EPI_ISL_2122205, EPI_ISL_2122211, EPI_ISL_2122234, EPI_ISL_2122279, EPI_ISL_2122284, EPI_ISL_2122287, EPI_ISL_2122303, EPI_ISL_2122308, EPI_ISL_2122309, EPI_ISL_2122312, EPI_ISL_2122319, EPI_ISL_2122323, EPI_ISL_2122326,
EPI_ISL_2122331, EPI_ISL_2122332, EPI_ISL_2122351, EPI_ISL_2122369, EPI_ISL_2122388, EPI_ISL_2122402, EPI_ISL_2122468, EPI_ISL_2122470, EPI_ISL_2122473, EPI_ISL_2122484, EPI_ISL_2122487, EPI_ISL_2122497, EPI_ISL_2122515, EPI_ISL_2122518, EPI_ISL_2122525, EPI_ISL_2122532, EPI_ISL_2122533, EPI_ISL_2122535, EPI_ISL_2122537, EPI_ISL_2122551, EPI_ISL_2122562,
EPI_ISL_2122568, EPI_ISL_2122586, EPI_ISL_2122590, EPI_ISL_2122591, EPI_ISL_2122598, EPI_ISL_2122600, EPI_ISL_2122611, EPI_ISL_2122613, EPI_ISL_2122616, EPI_ISL_2122621, EPI_ISL_2122632, EPI_ISL_2122639, EPI_ISL_2122661, EPI_ISL_2122665, EPI_ISL_2122667, EPI_ISL_2122681, EPI_ISL_2122701, EPI_ISL_2122702, EPI_ISL_2122704, EPI_ISL_2137847, EPI_ISL_2137849, EPI_ISL_2137851,
EPI_ISL_2137853, EPI_ISL_2137855, EPI_ISL_2137856, EPI_ISL_2137857, EPI_ISL_2137858, EPI_ISL_2137860, EPI_ISL_2137861, EPI_ISL_2137862, EPI_ISL_2137863, EPI_ISL_2137864, EPI_ISL_2137865, EPI_ISL_2137866, EPI_ISL_2137867, EPI_ISL_2137868, EPI_ISL_2137869, EPI_ISL_2137870, EPI_ISL_2137871, EPI_ISL_2137872, EPI_ISL_2137873, EPI_ISL_2137874, EPI_ISL_2137878,
EPI_ISL_2137880, EPI_ISL_2137881, EPI_ISL_2137883, EPI_ISL_2137884, EPI_ISL_2137886, EPI_ISL_2137889, EPI_ISL_2137892, EPI_ISL_2137895, EPI_ISL_2137897, EPI_ISL_2137901, EPI_ISL_2137902, EPI_ISL_2137905, EPI_ISL_2137906, EPI_ISL_2137908, EPI_ISL_2137909, EPI_ISL_2137912, EPI_ISL_2137913, EPI_ISL_2137914, EPI_ISL_2137915, EPI_ISL_2137916, EPI_ISL_2137917, EPI_ISL_2137918,
EPI_ISL_2137921, EPI_ISL_2137922, EPI_ISL_2137924, EPI_ISL_2137926, EPI_ISL_2137927, EPI_ISL_2137931, EPI_ISL_2137932, EPI_ISL_2137934, EPI_ISL_2137935, EPI_ISL_2137936, EPI_ISL_2137937, EPI_ISL_2137938, EPI_ISL_2137939, EPI_ISL_2137940, EPI_ISL_2137941, EPI_ISL_2137942, EPI_ISL_2137943, EPI_ISL_2137944, EPI_ISL_2137945, EPI_ISL_2137946, EPI_ISL_2137948,
EPI_ISL_2137949, EPI_ISL_2137951, EPI_ISL_2137955, EPI_ISL_2137957, EPI_ISL_2137958, EPI_ISL_2137959, EPI_ISL_2137960, EPI_ISL_2137961, EPI_ISL_2137962, EPI_ISL_2137964, EPI_ISL_2137965, EPI_ISL_2137966, EPI_ISL_2137969, EPI_ISL_2137970, EPI_ISL_2137971, EPI_ISL_2137973, EPI_ISL_2137974, EPI_ISL_2137975, EPI_ISL_2137977, EPI_ISL_2137980, EPI_ISL_2137981, EPI_ISL_2137982,
EPI_ISL_2137984, EPI_ISL_2137985, EPI_ISL_2137986, EPI_ISL_2137987, EPI_ISL_2137988, EPI_ISL_2137989, EPI_ISL_2137991, EPI_ISL_2137992, EPI_ISL_2137994, EPI_ISL_2137995, EPI_ISL_2137996, EPI_ISL_2137999, EPI_ISL_2138007, EPI_ISL_2138008, EPI_ISL_2138009, EPI_ISL_2138010, EPI_ISL_2138011, EPI_ISL_2138013, EPI_ISL_2138014, EPI_ISL_2138015, EPI_ISL_2138016, EPI_ISL_2138017,
EPI_ISL_2138018, EPI_ISL_2138021, EPI_ISL_2138025, EPI_ISL_2138026, EPI_ISL_2138027, EPI_ISL_2138029, EPI_ISL_2138030, EPI_ISL_2138035, EPI_ISL_2138036, EPI_ISL_2138037, EPI_ISL_2138038, EPI_ISL_2138039, EPI_ISL_2138040, EPI_ISL_2138042, EPI_ISL_2138045, EPI_ISL_2138051, EPI_ISL_2138052, EPI_ISL_2138053, EPI_ISL_2138055, EPI_ISL_2138056, EPI_ISL_2138057, EPI_ISL_2138058,
EPI_ISL_2138059, EPI_ISL_2138060, EPI_ISL_2138061, EPI_ISL_2138064, EPI_ISL_2138065, EPI_ISL_2138067, EPI_ISL_2138068, EPI_ISL_2138069, EPI_ISL_2138070, EPI_ISL_2138072, EPI_ISL_2138074, EPI_ISL_2138077, EPI_ISL_2138079, EPI_ISL_2138080, EPI_ISL_2138081, EPI_ISL_2138083, EPI_ISL_2138084, EPI_ISL_2138085, EPI_ISL_2138086, EPI_ISL_2138088, EPI_ISL_2138089, EPI_ISL_2138090,
EPI_ISL_2138091, EPI_ISL_2138095, EPI_ISL_2138097, EPI_ISL_2138099, EPI_ISL_2138100, EPI_ISL_2138101, EPI_ISL_2138102, EPI_ISL_2138103, EPI_ISL_2138104, EPI_ISL_2138105, EPI_ISL_2138108, EPI_ISL_2138110, EPI_ISL_2138113, EPI_ISL_2138114, EPI_ISL_2138115, EPI_ISL_2138116, EPI_ISL_2138117, EPI_ISL_2138118, EPI_ISL_2138119, EPI_ISL_2138121, EPI_ISL_2138123, EPI_ISL_2138124,
EPI_ISL_2138125, EPI_ISL_2138126, EPI_ISL_2138127, EPI_ISL_2138130, EPI_ISL_2138131, EPI_ISL_2138133, EPI_ISL_2138134, EPI_ISL_2138135, EPI_ISL_2138138, EPI_ISL_2138140, EPI_ISL_2138141, EPI_ISL_2138146, EPI_ISL_2138148, EPI_ISL_2138149, EPI_ISL_2138150, EPI_ISL_2138151, EPI_ISL_2138152, EPI_ISL_2138153, EPI_ISL_2138154, EPI_ISL_2138156,
EPI_ISL_2138157, EPI_ISL_2138160, EPI_ISL_2138161, EPI_ISL_2138162, EPI_ISL_2138166, EPI_ISL_2138168, EPI_ISL_2138169, EPI_ISL_2138170, EPI_ISL_2138172, EPI_ISL_2138174, EPI_ISL_2138177, EPI_ISL_2138179, EPI_ISL_2138180, EPI_ISL_2138182, EPI_ISL_2138186, EPI_ISL_2138188, EPI_ISL_2138189, EPI_ISL_2138190, EPI_ISL_2138191, EPI_ISL_2138194, EPI_ISL_2138195, EPI_ISL_2138196,
EPI_ISL_2138197, EPI_ISL_2138199, EPI_ISL_2138200, EPI_ISL_2138201, EPI_ISL_2138202, EPI_ISL_2138203, EPI_ISL_2138204, EPI_ISL_2138207, EPI_ISL_2138209, EPI_ISL_2138211, EPI_ISL_2138212, EPI_ISL_2138216, EPI_ISL_2138217, EPI_ISL_2138218, EPI_ISL_2138220, EPI_ISL_2138221, EPI_ISL_2138222, EPI_ISL_2138223, EPI_ISL_2138227, EPI_ISL_2138228,
EPI_ISL_2138229, EPI_ISL_2138230, EPI_ISL_2138231, EPI_ISL_2138233, EPI_ISL_2138234, EPI_ISL_2138235, EPI_ISL_2138236, EPI_ISL_2138239, EPI_ISL_2138240, EPI_ISL_2138241, EPI_ISL_2138243, EPI_ISL_2138244, EPI_ISL_2138245, EPI_ISL_2138249, EPI_ISL_2138250, EPI_ISL_2138251, EPI_ISL_2138252, EPI_ISL_2138253, EPI_ISL_2138254, EPI_ISL_2138256, EPI_ISL_2138257, EPI_ISL_2138258,
EPI_ISL_2138259, EPI_ISL_2138261, EPI_ISL_2138263, EPI_ISL_2138265, EPI_ISL_2138266, EPI_ISL_2138267, EPI_ISL_2138269, EPI_ISL_2138271, EPI_ISL_2138275, EPI_ISL_2138277, EPI_ISL_2138278, EPI_ISL_2138279, EPI_ISL_2138281, EPI_ISL_2138282, EPI_ISL_2138284, EPI_ISL_2138286, EPI_ISL_2138290, EPI_ISL_2138292, EPI_ISL_2138296, EPI_ISL_2138298, EPI_ISL_2138299, EPI_ISL_2138302,
EPI_ISL_2138303, EPI_ISL_2138304, EPI_ISL_2138305, EPI_ISL_2138308, EPI_ISL_2138309, EPI_ISL_2138310, EPI_ISL_2138312, EPI_ISL_2138313, EPI_ISL_2138314, EPI_ISL_2138316, EPI_ISL_2138317, EPI_ISL_2138320, EPI_ISL_2138323, EPI_ISL_2138324, EPI_ISL_2138326, EPI_ISL_2138327, EPI_ISL_2138329, EPI_ISL_2138330, EPI_ISL_2138332, EPI_ISL_2138333, EPI_ISL_2138334, EPI_ISL_2138335,
EPI_ISL_2138336, EPI_ISL_2138338, EPI_ISL_2138340, EPI_ISL_2138341, EPI_ISL_2138342, EPI_ISL_2138343, EPI_ISL_2138345, EPI_ISL_2138347, EPI_ISL_2138348, EPI_ISL_2138349, EPI_ISL_2138350, EPI_ISL_2138351, EPI_ISL_2138352, EPI_ISL_2138357, EPI_ISL_2138358, EPI_ISL_2138361, EPI_ISL_2138362, EPI_ISL_2138364, EPI_ISL_2138365, EPI_ISL_2138366, EPI_ISL_2138367,
EPI_ISL_2138368, EPI_ISL_2138369, EPI_ISL_2138371, EPI_ISL_2138373, EPI_ISL_2138374, EPI_ISL_2138375, EPI_ISL_2138376, EPI_ISL_2138377, EPI_ISL_2138378, EPI_ISL_2138380, EPI_ISL_2138381, EPI_ISL_2138383, EPI_ISL_2138384, EPI_ISL_2138387, EPI_ISL_2138389, EPI_ISL_2138392, EPI_ISL_2138393, EPI_ISL_2138394, EPI_ISL_2138395, EPI_ISL_2138396, EPI_ISL_2138397, EPI_ISL_2138399,
EPI_ISL_2138400, EPI_ISL_2138401, EPI_ISL_2138403, EPI_ISL_2138404, EPI_ISL_2138405, EPI_ISL_2138410, EPI_ISL_2138412, EPI_ISL_2138413, EPI_ISL_2138417, EPI_ISL_2138419, EPI_ISL_2138421, EPI_ISL_2138422, EPI_ISL_2138423, EPI_ISL_2138424, EPI_ISL_2138426, EPI_ISL_2138427, EPI_ISL_2138432, EPI_ISL_2138433, EPI_ISL_2138435, EPI_ISL_2138437,
EPI_ISL_2138438, EPI_ISL_2138439, EPI_ISL_2138443, EPI_ISL_2138444, EPI_ISL_2138445, EPI_ISL_2138447, EPI_ISL_2138449, EPI_ISL_2138450, EPI_ISL_2138452, EPI_ISL_2138453, EPI_ISL_2138457, EPI_ISL_2138462, EPI_ISL_2138463, EPI_ISL_2138464, EPI_ISL_2138465, EPI_ISL_2138466, EPI_ISL_2138468, EPI_ISL_2138473, EPI_ISL_2138474, EPI_ISL_2138478, EPI_ISL_2138482, EPI_ISL_2138486,
EPI_ISL_2138490, EPI_ISL_2138495, EPI_ISL_2138496, EPI_ISL_2138498, EPI_ISL_2138499, EPI_ISL_2138500, EPI_ISL_2138502, EPI_ISL_2138504, EPI_ISL_2138506, EPI_ISL_2138507, EPI_ISL_2138509, EPI_ISL_2138510, EPI_ISL_2138513, EPI_ISL_2138514, EPI_ISL_2138515, EPI_ISL_2138518, EPI_ISL_2138519, EPI_ISL_2138525, EPI_ISL_2138526, EPI_ISL_2138527, EPI_ISL_2138529, EPI_ISL_2138530,

| | | | |
|---|---|---|---|
| EPI_ISL_2138531, EPI_ISL_2138534, EPI_ISL_2138535, EPI_ISL_2138536, EPI_ISL_2138537, EPI_ISL_2138539, EPI_ISL_2138540, EPI_ISL_2138541, EPI_ISL_2138545, EPI_ISL_2138546, EPI_ISL_2138547, EPI_ISL_2138548, EPI_ISL_2138550, EPI_ISL_2138551, EPI_ISL_2138552, EPI_ISL_2138554, EPI_ISL_2138555, EPI_ISL_2138556, EPI_ISL_2138557, EPI_ISL_2138564, EPI_ISL_2138566, EPI_ISL_2138567, EPI_ISL_2138570, EPI_ISL_2138573, EPI_ISL_2138574, EPI_ISL_2138575, EPI_ISL_2138576, EPI_ISL_2138581, EPI_ISL_2138585, EPI_ISL_2138588, EPI_ISL_2138590, EPI_ISL_2138591, EPI_ISL_2138592, EPI_ISL_2138593, EPI_ISL_2138595, EPI_ISL_2138598, EPI_ISL_2138599, EPI_ISL_2138605, EPI_ISL_2138611, EPI_ISL_2138613, EPI_ISL_2138614, EPI_ISL_2138615, EPI_ISL_2138618, EPI_ISL_2138621, EPI_ISL_2138623, EPI_ISL_2138625, EPI_ISL_2138626, EPI_ISL_2138627, EPI_ISL_2138628, EPI_ISL_2138629, EPI_ISL_2138630, EPI_ISL_2138631, EPI_ISL_2138632, EPI_ISL_2138634, EPI_ISL_2138637, EPI_ISL_2138638, EPI_ISL_2138639, EPI_ISL_2138640, EPI_ISL_2138642, EPI_ISL_2138643, EPI_ISL_2138645, EPI_ISL_2138646, EPI_ISL_2138648, EPI_ISL_2138651, EPI_ISL_2138652, EPI_ISL_2138654, EPI_ISL_2138655, EPI_ISL_2138656, EPI_ISL_2138657, EPI_ISL_2138658, EPI_ISL_2138659, EPI_ISL_2138660, EPI_ISL_2138661, EPI_ISL_2138662, EPI_ISL_2138664, EPI_ISL_2138666, EPI_ISL_2138667, EPI_ISL_2138668, EPI_ISL_2138669, EPI_ISL_2138671, EPI_ISL_2138672, EPI_ISL_2138673, EPI_ISL_2138676, EPI_ISL_2138677, EPI_ISL_2138679, EPI_ISL_2138680, EPI_ISL_2138682, EPI_ISL_2138685, EPI_ISL_2138687, EPI_ISL_2138690, EPI_ISL_2138695, EPI_ISL_2138696, EPI_ISL_2138699, EPI_ISL_2138700, EPI_ISL_2138702, EPI_ISL_2138703, EPI_ISL_2138704, EPI_ISL_2138706, EPI_ISL_2138707, EPI_ISL_2138708, EPI_ISL_2138709, EPI_ISL_2138711, EPI_ISL_2138712, EPI_ISL_2138714, EPI_ISL_2138716 | | | |
| see above | Lighthouse Lab in Milton Keynes | Wellcome Sanger Institute for the COVID-19 Genomics UK (COG-UK) Consortium | Cordelia Langford; David K. Jackson; Dominic Kwiatkowski; Ewan Harrison; Ian Johnston; Jeffrey Barrett; John Sillitoe on behalf of the Wellcome Sanger Institute COVID-19 Surveillance Team; Roberto Amato; Sonia Goncalves; The Lighthouse Lab in Milton Keynes and Alex Alderton |
| EPI_ISL_1975041, EPI_ISL_1975042, EPI_ISL_1975044, EPI_ISL_1975045, EPI_ISL_1975047, EPI_ISL_1975049, EPI_ISL_2125988, EPI_ISL_2125989, EPI_ISL_2125990, EPI_ISL_2125992, EPI_ISL_2125993, EPI_ISL_2125995, EPI_ISL_2125997 | | | |
| see above | Liverpool Clinical Laboratories | COVID-19 Genomics UK (COG-UK) Consortium | A Alrezaihi; Alessandro Gerada; Alistair Darby; Angela Cowell; Anita Lucaci; Anu Chawla; Cassie Olateju; Catherine Hartley; Charlotte Nelson; Ecaterina Vamos; Elaine O'Toole; Eleanor G Bentley; Ghada T Shawli; Hermione Webster; Isabel García-Dorival; James Johnson; James P Stewart; Jenifer Manson; Joanne Watts; Jones Benjamin; Jordan J Clark; Julian Hiscox; L Luu; Lucille Rainbow; M Almsaud; Margaret Hughes; Mark Whitehead; Matthew Gemmell; Miren Iturriza-Gomara; Muhannad Alruwaili; N.P Randle; Neil Swainston; PKF Gilmore; Parul Sharma; Rebekah Penrice-Randal; Richard Eccles; Richard Gregory; Sam Haldenby; Steve Paterson; Stuart D Armstrong; Trevor Ian Robinson; Ximeng Han |
| EPI_ISL_2126181, EPI_ISL_2126184, EPI_ISL_2126186, EPI_ISL_2127047, EPI_ISL_2127048, EPI_ISL_2127049, EPI_ISL_2127050, EPI_ISL_2127052, EPI_ISL_2127054, EPI_ISL_2127299, EPI_ISL_2127301, EPI_ISL_2127303, EPI_ISL_2127305, EPI_ISL_2127534, EPI_ISL_2127536, EPI_ISL_2127538, EPI_ISL_2127540, EPI_ISL_2127543, EPI_ISL_2127546, EPI_ISL_2127548, EPI_ISL_2127550, EPI_ISL_2127552, EPI_ISL_2127553, EPI_ISL_2127556, EPI_ISL_2127557, EPI_ISL_2127559, EPI_ISL_2127561, EPI_ISL_2127563, EPI_ISL_2127565, EPI_ISL_2127567, EPI_ISL_2127568, EPI_ISL_2127570, EPI_ISL_2127572, EPI_ISL_2127574, EPI_ISL_2127576, EPI_ISL_2127578, EPI_ISL_2127580, EPI_ISL_2127581, EPI_ISL_2127583, EPI_ISL_2127585, EPI_ISL_2127586, EPI_ISL_2127588, EPI_ISL_2127590, EPI_ISL_2127592 | | | |
| see above | Northumbria University / South Tees Hospitals NHS Foundation Trust / North Cumbria Integrated Care NHS Foundation Trust / North Tees and Hartlepool NHS Foundation Trust / Newcastle Hospitals NHS Foundation Trust | COVID-19 Genomics UK (COG-UK) Consortium | Andrew Nelson; Brendan Payne; Clive Graham; Darren L Smith; Debra Padgett; Edward Barton; Emma Swindells; Garren Scott; Gary Black; Gary Eltringham; Giles S Holt; Greg R Young; Jane Greenaway; Jennifer Collins; John Allan; Joshua Loh; Lynn Dover; Matthew Bashton; Mohammad A Tariq; Paul Baker; Sarah Essex; Steve Liggett; Wen C Yew; Yusri Taha |
| EPI_ISL_1979272, EPI_ISL_1979311, EPI_ISL_1979313, EPI_ISL_1979315, EPI_ISL_1979317, EPI_ISL_1979326, EPI_ISL_1979338, EPI_ISL_1979493, EPI_ISL_1979497, EPI_ISL_1979498, EPI_ISL_1979500, EPI_ISL_1979503, EPI_ISL_1979504, EPI_ISL_1979512, EPI_ISL_1979515, EPI_ISL_1979517, EPI_ISL_1979518, EPI_ISL_1979520, EPI_ISL_1979572, EPI_ISL_1979602, EPI_ISL_1979605, EPI_ISL_1979610, EPI_ISL_1979612, EPI_ISL_1979616, EPI_ISL_1979618, EPI_ISL_1979620, EPI_ISL_1979623, EPI_ISL_1979624, EPI_ISL_1979626, EPI_ISL_1979632, EPI_ISL_1979634, EPI_ISL_1979635, EPI_ISL_1979637, EPI_ISL_1979640, EPI_ISL_1979643, EPI_ISL_1979646, EPI_ISL_1979648, EPI_ISL_1979652, EPI_ISL_1979700, EPI_ISL_1979701, EPI_ISL_1979706, EPI_ISL_1979709, EPI_ISL_1979713, EPI_ISL_1979714, EPI_ISL_1979715, EPI_ISL_1979723, EPI_ISL_1979725, EPI_ISL_1979726, EPI_ISL_1979728, EPI_ISL_1979736, EPI_ISL_1979741 | | | |
| see above | Originating lab: Wales Specialist Virology Centre Sequencing lab: Pathogen Genomics Unit | Public Health Wales Microbiology Cardiff Wales Specialist Virology Centre | Alec Birchley; Alexander Adams; Amy Gaskin; Angela Marchbank; Bree Gatica-Wilcox; Catherine Moore; Jason Coombes; Joanne Watkins; Joel Southgate; Johnathan Evans; Laura Gifford; Lauren Gilbert; Lee Graham; Malorie Perry; Matthew Bull; Nicole Pacchiarini; Sally Corden; Sara Kumziene-Summerhayes; Sara Rey; Sarah Taylor; Simon Cottrell; Sophie Jones; Tom Connor |
| EPI_ISL_1978959, EPI_ISL_1978960, EPI_ISL_2127604, EPI_ISL_2127606, EPI_ISL_2127608, EPI_ISL_2127610, EPI_ISL_2127611, EPI_ISL_2127613, EPI_ISL_2127615, EPI_ISL_2127617, EPI_ISL_2127619, EPI_ISL_2127621, EPI_ISL_2127623, EPI_ISL_2127624 | | | |
| see above | Queens Medical Centre, Clinical Microbiology Department / DeepSeq Nottingham | COVID-19 Genomics UK (COG-UK) Consortium | Christopher Moore; Emily Park; Fei Sang; Gemma Clark; Hannah Howson-Wells; Jack Hill; Jayasree Dey; Johnny Debebe; Jonathan Ball; Joseph Chappell; Manjinder Khakh; Matthew Carlisle; Matthew Loose; Michelle M Lister; Nadine Holmes; Patrick McClure; Theocharis Tsoleridis; Timothy Byaruhanga; Vicki M Fleming; Victoria Wright; Wendy Smith |
| EPI_ISL_2004374, EPI_ISL_2004436, EPI_ISL_2004473, EPI_ISL_2004476, EPI_ISL_2004485, EPI_ISL_2004494, EPI_ISL_2004545, EPI_ISL_2004584, EPI_ISL_2021372, EPI_ISL_2021378, EPI_ISL_2021523, EPI_ISL_2021572, EPI_ISL_2021573, EPI_ISL_2021574, EPI_ISL_2021639, EPI_ISL_2021714, EPI_ISL_2021721, EPI_ISL_2021722, EPI_ISL_2021740, EPI_ISL_2021745, EPI_ISL_2021750, EPI_ISL_2021751, EPI_ISL_2021759, EPI_ISL_2021763, EPI_ISL_2021768, EPI_ISL_2021776, EPI_ISL_2021788, EPI_ISL_2021801, EPI_ISL_2021803, EPI_ISL_2021815, EPI_ISL_2022530, EPI_ISL_2022563, EPI_ISL_2091824, EPI_ISL_2091846, EPI_ISL_2091975, EPI_ISL_2091985, EPI_ISL_2091992, EPI_ISL_2092016, EPI_ISL_2092019, EPI_ISL_2092042, EPI_ISL_2092083, EPI_ISL_2092112, EPI_ISL_2092114, EPI_ISL_2092118, EPI_ISL_2092120, EPI_ISL_2092140, EPI_ISL_2092141, EPI_ISL_2092143, EPI_ISL_2092152, EPI_ISL_2092156, EPI_ISL_2092162, EPI_ISL_2092163, EPI_ISL_2092167, EPI_ISL_2092171, EPI_ISL_2092181, EPI_ISL_2092183, EPI_ISL_2092188, EPI_ISL_2092193, EPI_ISL_2092195, EPI_ISL_2092200, EPI_ISL_2092202, EPI_ISL_2092203, EPI_ISL_2092214, EPI_ISL_2092222, EPI_ISL_2092223, EPI_ISL_2092233, EPI_ISL_2092239, EPI_ISL_2092240, EPI_ISL_2092243, EPI_ISL_2092245, EPI_ISL_2092256, EPI_ISL_2092259, EPI_ISL_2092264, EPI_ISL_2092265, EPI_ISL_2092266, EPI_ISL_2092270, EPI_ISL_2092272, EPI_ISL_2092273, EPI_ISL_2092278, EPI_ISL_2092280, EPI_ISL_2092296, EPI_ISL_2092312, EPI_ISL_2092338, EPI_ISL_2092339, EPI_ISL_2092341, EPI_ISL_2092342, EPI_ISL_2092347, EPI_ISL_2092362, EPI_ISL_2092365, EPI_ISL_2092373, EPI_ISL_2092380, EPI_ISL_2092401, EPI_ISL_2092406, EPI_ISL_2092411, EPI_ISL_2092414, EPI_ISL_2092416, EPI_ISL_2092417, EPI_ISL_2092419, EPI_ISL_2092425, EPI_ISL_2092428, EPI_ISL_2092438, EPI_ISL_2092440, EPI_ISL_2092441, EPI_ISL_2092442, EPI_ISL_2092451, EPI_ISL_2092459, EPI_ISL_2122125, EPI_ISL_2122126, EPI_ISL_2122142, EPI_ISL_2122144, EPI_ISL_2122159, EPI_ISL_2122162, EPI_ISL_2122164, EPI_ISL_2122171, EPI_ISL_2122197, EPI_ISL_2122198, EPI_ISL_2122232, EPI_ISL_2122237, EPI_ISL_2122297, EPI_ISL_2122304, EPI_ISL_2122366, EPI_ISL_2122380, EPI_ISL_2122405, EPI_ISL_2122416, EPI_ISL_2122471, EPI_ISL_2122474, EPI_ISL_2122488, EPI_ISL_2122491, EPI_ISL_2122502, EPI_ISL_2122503, EPI_ISL_2122505, EPI_ISL_2122513, EPI_ISL_2122550, EPI_ISL_2122556, EPI_ISL_2122572, EPI_ISL_2122575, EPI_ISL_2122607, EPI_ISL_2122618, EPI_ISL_2122625, EPI_ISL_2122636, EPI_ISL_2122658, EPI_ISL_2122691, EPI_ISL_2122692, EPI_ISL_2122696 | | | |
| see above | Randox Laboratories | Wellcome Sanger Institute for the COVID-19 Genomics UK (COG-UK) Consortium | Cordelia Langford; David K. Jackson; Dominic Kwiatkowski; Ewan Harrison; Ian Johnston; Jeffrey Barrett; John Sillitoe on behalf of the Wellcome Sanger Institute COVID-19 Surveillance Team; Randox Laboratories and Alex Alderton; Roberto Amato; Sonia Goncalves |
| EPI_ISL_1975115, EPI_ISL_1975118, EPI_ISL_1975122, EPI_ISL_1975130, EPI_ISL_1975133, EPI_ISL_1975171, EPI_ISL_1975179, EPI_ISL_1975180, EPI_ISL_2126130, EPI_ISL_2126132, EPI_ISL_2126136, EPI_ISL_2126138, EPI_ISL_2126140, EPI_ISL_2126142, EPI_ISL_2126145, EPI_ISL_2126147, EPI_ISL_2126149, EPI_ISL_2126151, EPI_ISL_2126152, EPI_ISL_2126154, EPI_ISL_2126155, EPI_ISL_2126157, EPI_ISL_2126159, EPI_ISL_2126161, EPI_ISL_2126163, EPI_ISL_2126164, EPI_ISL_2126166, EPI_ISL_2126168, EPI_ISL_2126170, EPI_ISL_2126171, EPI_ISL_2126173, EPI_ISL_2126175, EPI_ISL_2126177 | | | |
| see above | University College London, Great Ormond Street Hospital for Children NHS Foundation Trust, Imperial College Healthcare NHS Trust | COVID-19 Genomics UK (COG-UK) Consortium | Alison Holmes; Charlotte Williams; Helena Tutill; Jacqueline Findlay; James Price; Judith Breuer; Julianne Brown; Kathryn Harris; Leysa Forrest; Mark Kristiansen; Paola Niola; Paola Resende Silva; Patricia Dyal; Paul Randell; Rachel Williams; Samuel Weeks; Sergi Castellano; Sunando Roy; Tony Brooks; Yasmin Panchbhaya |
| EPI_ISL_2125426, EPI_ISL_2125428, EPI_ISL_2125430, EPI_ISL_2125432, EPI_ISL_2125434, EPI_ISL_2125436, EPI_ISL_2125438, EPI_ISL_2125439, EPI_ISL_2125441, EPI_ISL_2125443, EPI_ISL_2125444, EPI_ISL_2125446, EPI_ISL_2125447, EPI_ISL_2125449 | | | |
| see above | University of Birmingham | COVID-19 Genomics UK (COG-UK) Consortium | Alex Richter; Andrew Bosworth; Andrew Bosworth. Queen Elizabeth Hospital: Anna Casey; Andrew D Beggs PHE Heartlands Lab: Husam Osman; Charlotte Poxon; Institute of Microbiology; Joanne Stockton; Josh Quick; Kasun Wanigasooriya; Mike Kidd; Nicholas Loman. University of Birmingham Testing Laboratory: Celina M Whalley; Oliver Pickles; Radoslaw Poplawski; Samuel Nicholls; University of Birmingham: Claire McMurray; Will Rowe |
| EPI_ISL_2125669, EPI_ISL_2125671, EPI_ISL_2125673, EPI_ISL_2125675, EPI_ISL_2125677, EPI_ISL_2125678, EPI_ISL_2125680, EPI_ISL_2125682, EPI_ISL_2125684, EPI_ISL_2125686, EPI_ISL_2125687, EPI_ISL_2125719, EPI_ISL_2125720, EPI_ISL_2125722, EPI_ISL_2125724, EPI_ISL_2125726, EPI_ISL_2125728, EPI_ISL_2125740, EPI_ISL_2125742, EPI_ISL_2125744, EPI_ISL_2125745, EPI_ISL_2125748, EPI_ISL_2125749, EPI_ISL_2125753, EPI_ISL_2125755, EPI_ISL_2125756, EPI_ISL_2125758, EPI_ISL_2125888, EPI_ISL_2125890, EPI_ISL_2125891 | | | |
| see above | Virology Department, Royal Infirmary of Edinburgh, NHS Lothian / School of Biological Sciences, University of Edinburgh | COVID-19 Genomics UK (COG-UK) Consortium | Colquhoun R; Cotton S; Dewar R; Gallagher A; Hill V; Jackson B; Maloney D; McCrone JT; McHugh M; O'Toole Á; Rambaut A; Rooke S; Scher E; Templeton K; Yu X |
| EPI_ISL_1980356, EPI_ISL_1980368, EPI_ISL_1980375, EPI_ISL_1980404, EPI_ISL_1980417, EPI_ISL_1980422, EPI_ISL_1980423, EPI_ISL_1980441, EPI_ISL_2128460, EPI_ISL_2128463, EPI_ISL_2128467, EPI_ISL_2128469, EPI_ISL_2128471, EPI_ISL_2128474, EPI_ISL_2128481, EPI_ISL_2128491, EPI_ISL_2128495, EPI_ISL_2128502 | | | |
| see above | Virology Department, Sheffield Teaching Hospitals NHS Foundation Trust/Department of Infection, Immunity and Cardiovascular Disease, The Medical School, University of Sheffield | COVID-19 Genomics UK (COG-UK) Consortium | Adri Angyal; Alex Keeley; Benjamin Lindsey; Cariad Evans; Danielle Groves; Dave Partridge; Katie Johnson; Laura Carrilero; Luke Green; Matthew Parker; Matthew Wyles; Mehmet Yavuz; Mohammad Raza; Nikki Smith; Paul Parsons; Rachel Tucker; Rebecca Brown; Thushan de Silva |
| EPI_ISL_2127728, EPI_ISL_2127729, EPI_ISL_2127730, EPI_ISL_2127731, EPI_ISL_2127732, EPI_ISL_2127733, EPI_ISL_2127734, EPI_ISL_2127735, EPI_ISL_2127736, EPI_ISL_2127738, EPI_ISL_2127739, EPI_ISL_2127741, EPI_ISL_2127743, EPI_ISL_2127745, EPI_ISL_2127746, EPI_ISL_2127748, EPI_ISL_2127750, EPI_ISL_2127752, EPI_ISL_2127753, EPI_ISL_2127755, EPI_ISL_2127757, EPI_ISL_2127759, EPI_ISL_2127761, EPI_ISL_2127763, EPI_ISL_2127764, EPI_ISL_2127765, EPI_ISL_2127767, EPI_ISL_2127768, EPI_ISL_2127770, EPI_ISL_2127772, EPI_ISL_2127773, EPI_ISL_2127774, EPI_ISL_2127776, EPI_ISL_2127777, EPI_ISL_2127778, EPI_ISL_2127780, EPI_ISL_2127782, EPI_ISL_2127784, EPI_ISL_2127785, EPI_ISL_2127787, EPI_ISL_2127789, EPI_ISL_2127791, EPI_ISL_2127792, EPI_ISL_2127795, EPI_ISL_2127797, EPI_ISL_2127798, EPI_ISL_2127800, EPI_ISL_2127802, EPI_ISL_2127804, EPI_ISL_2127805, EPI_ISL_2127807, EPI_ISL_2127809, EPI_ISL_2127811, EPI_ISL_2127812, EPI_ISL_2127814, EPI_ISL_2127815, EPI_ISL_2127816, EPI_ISL_2127818, EPI_ISL_2127820, EPI_ISL_2127822, EPI_ISL_2127823, EPI_ISL_2127825, EPI_ISL_2127827, EPI_ISL_2127829, EPI_ISL_2127831, EPI_ISL_2127833, EPI_ISL_2127835, EPI_ISL_2127837, EPI_ISL_2127839, EPI_ISL_2127841, EPI_ISL_2127842, EPI_ISL_2127845, EPI_ISL_2127847, EPI_ISL_2127849, EPI_ISL_2127851, EPI_ISL_2127852, EPI_ISL_2127854, EPI_ISL_2127858, EPI_ISL_2127860, EPI_ISL_2127861, EPI_ISL_2127863, EPI_ISL_2127865, EPI_ISL_2127867, EPI_ISL_2127869, EPI_ISL_2127871, EPI_ISL_2127873, EPI_ISL_2127874, EPI_ISL_2127880, EPI_ISL_2127883 | | | |
| see above | Wales Specialist Virology Centre Sequencing lab: Pathogen Genomics Unit | Public Health Wales Microbiology Cardiff Wales Specialist Virology Centre | Alec Birchley; Alexander Adams; Amy Gaskin; Angela Marchbank; Bree Gatica-Wilcox; Catherine Moore; Jason Coombes; Joanne Watkins; Joel Southgate; Johnathan Evans; Laura Gifford; Lauren Gilbert; Lee Graham; Malorie Perry; Matthew Bull; Nicole Pacchiarini; Sally Corden; Sara Kumziene-Summerhayes; Sara Rey; Sarah Taylor; Simon Cottrell; Sophie Jones; Tom Connor |
| EPI_ISL_2125502 | West of Scotland Specialist Virology Centre, NHSGGC / MRC-University of Glasgow Centre for Virus Research | COVID-19 Genomics UK (COG-UK) Consortium | Alasdair MacLean; Alice Broos; Ana da Silva Filipe; Antonia Ho; Daniel Mair; David L Robertson; Emma Thomson; Guy Mollett; Ioulia Tsatsani; James Shepherd; Jenna Nichols; Joseph Hughes; Kathy Li; Kathy Smollett; Kyriaki Nomikou; Lily Tong; Matthew Holden; Natasha Johnson; Rachel Blacow; Richard Orton; Rory Gunson; Sarah McDonald; Sharif Shaaban; Sreenu Vattipally; Stephen Carmichael |

We gratefully acknowledge the following Authors from the Originating laboratories responsible for obtaining the specimens, as well as the Submitting laboratories where the genome data were generated and shared via GISAID, on which this research is based.

All Submitters of data may be contacted directly via www.gisaid.org

Authors are sorted alphabetically.

| Accession ID | Originating Laboratory | Submitting Laboratory | Authors |
|---|---|---|---|
| EPI_ISL_2784848, EPI_ISL_2875529, EPI_ISL_2875530, EPI_ISL_2875727, EPI_ISL_2875789 | Aegis Sciences Corporation | Centers for Disease Control and Prevention Division of Viral Diseases, Pathogen Discovery | Adrian Paskey; Alec Vest; Benjamin Rambo-Martin; Christopher Gulvick; Clinton R. Paden; Cyndi Clark; Dakota Howard; Darlene Wagner; Dhwani Batra; Dillon Nall; Duncan MacCannell; Ethan Sanders; Holly Houdeshell; Jason Caravas; Kara Moser; Matthew Hardison; Matthew Schmerer; Ola Kvalvaag; Patrick Campbell; Peter W. Cook; Rob Case; Scott Sammons; Shatavia Morrison; Shaun Westlund; Vikramsinha Ghorpade; Yvette Unoarumhi |
| EPI_ISL_2758414, EPI_ISL_2758415, EPI_ISL_2758417, EPI_ISL_2758472 | Alameda County Public Health Lab | Chan-Zuckerberg Biohub | CZB Cliahub Consortium |
| EPI_ISL_2893401, EPI_ISL_2893403, EPI_ISL_2893413 | Arizona State University | Arizona State University | Efrem S. Lim; Joshua LaBaer; Joy M. Blain; LaRinda A. Holland; Matthew F. Smith; Nicholas J. Mellor; Peter T. Skidmore; Rabia Maqsood; Valerie Harris; Vel Murugan |
| EPI_ISL_2931211 | Arkansas Children's Hospital | Center for Global Health, University of New Mexico Health Sciences Center | Ashton Ingold; Bobby Boyanton; Darrell Dinwiddie; Daryl Domman; Joshua L. Kennedy; Kurt Schwalm; Valerie Morley |
| EPI_ISL_2939088 | Arkansas Department of Health | Center for Global Health, University of New Mexico Health Sciences Center | Amanda Novack; Darrell Dinwiddie; Daryl Domman; Dirk Haselow; Joshua L. Kennedy; Kurt Schwalm; Valerie Morley |
| EPI_ISL_2868462 | BOSTON HEART DIAGNOSTICS CORP | Wadsworth Center, New York State Department of Health | Alexis Russell; Catharine Prussing; Daryl M. Lamson; Erasmus Schneider; Erica Lasek-Nesselquist; John Kelly; Jonathan Plitnick; Kirsten St. George; Matthew Shudt; Melissa A Leisner; Navjot Singh |
| EPI_ISL_2886488 | Bozeman Health Deaconess Hospital | Wiedenheft lab, Montana State University | Aidan McVay; Anna Nemudraia; Artem Nemudryi; Calvin Cicha; Helen Lee; Tanner Wiegand; and Blake Wiedenheft |
| EPI_ISL_2782401, EPI_ISL_2782432, EPI_ISL_2782455, EPI_ISL_2782491, EPI_ISL_2782495, EPI_ISL_2782510, EPI_ISL_2782526, EPI_ISL_2782544, EPI_ISL_2782552, EPI_ISL_2782585, EPI_ISL_2782589, EPI_ISL_2782591, EPI_ISL_2782611, EPI_ISL_2782632, EPI_ISL_2782633, EPI_ISL_2782701, EPI_ISL_2782761, EPI_ISL_2782801, EPI_ISL_2782955, EPI_ISL_2782977, EPI_ISL_2783092, EPI_ISL_2783125, EPI_ISL_2783134, EPI_ISL_2783203, EPI_ISL_2783211, EPI_ISL_2783389, EPI_ISL_2783451, EPI_ISL_2783464, EPI_ISL_2783490, EPI_ISL_2783491, EPI_ISL_2783510, EPI_ISL_2796547, EPI_ISL_2796550, EPI_ISL_2796556, EPI_ISL_2796575, EPI_ISL_2796580, EPI_ISL_2796597, EPI_ISL_2796616, EPI_ISL_2796643, EPI_ISL_2796651, EPI_ISL_2796663, EPI_ISL_2796669, EPI_ISL_2796694, EPI_ISL_2796695, EPI_ISL_2796707, EPI_ISL_2796721, EPI_ISL_2796725, EPI_ISL_2796737, EPI_ISL_2796740, EPI_ISL_2796781, EPI_ISL_2796836, EPI_ISL_2796866, EPI_ISL_2796870, EPI_ISL_2796871, EPI_ISL_2796949, EPI_ISL_2796958, EPI_ISL_2796964, EPI_ISL_2797000, EPI_ISL_2797027, EPI_ISL_2797088, EPI_ISL_2797089, EPI_ISL_2797092, EPI_ISL_2797156, EPI_ISL_2797181, EPI_ISL_2797190, EPI_ISL_2797217, EPI_ISL_2797238, EPI_ISL_2797258, EPI_ISL_2797318, EPI_ISL_2797424, EPI_ISL_2797448, EPI_ISL_2797482, EPI_ISL_2797489, EPI_ISL_2797496, EPI_ISL_2797498, EPI_ISL_2797531, EPI_ISL_2797550, EPI_ISL_2797555, EPI_ISL_2797588, EPI_ISL_2797656, EPI_ISL_2797668, EPI_ISL_2797674, EPI_ISL_2797707, EPI_ISL_2797708, EPI_ISL_2797729, EPI_ISL_2797780, EPI_ISL_2797786, EPI_ISL_2797793, EPI_ISL_2797845, EPI_ISL_2797861, EPI_ISL_2797863, EPI_ISL_2797891, EPI_ISL_2797917, EPI_ISL_2797920, EPI_ISL_2797927, EPI_ISL_2797995, EPI_ISL_2798004, EPI_ISL_2798057, EPI_ISL_2798083, EPI_ISL_2798116, EPI_ISL_2798123, EPI_ISL_2798124, EPI_ISL_2798143, EPI_ISL_2825147, EPI_ISL_2825148, EPI_ISL_2825152, EPI_ISL_2825153, EPI_ISL_2825161, EPI_ISL_2825175, EPI_ISL_2825215, EPI_ISL_2825216, EPI_ISL_2825218, EPI_ISL_2825225, EPI_ISL_2825227, EPI_ISL_2825233, EPI_ISL_2825241, EPI_ISL_2825260, EPI_ISL_2825270, EPI_ISL_2825291, EPI_ISL_2825421, EPI_ISL_2825454, EPI_ISL_2825469, EPI_ISL_2825473, EPI_ISL_2825554, EPI_ISL_2825609, EPI_ISL_2825611, EPI_ISL_2825647, EPI_ISL_2825674, EPI_ISL_2825683, EPI_ISL_2825700, EPI_ISL_2825705, EPI_ISL_2825710, EPI_ISL_2825719, EPI_ISL_2825798, EPI_ISL_2825828, EPI_ISL_2825829, EPI_ISL_2825830, EPI_ISL_2825852, EPI_ISL_2825863, EPI_ISL_2825884, EPI_ISL_2825893, EPI_ISL_2825910, EPI_ISL_2825918, EPI_ISL_2825935, EPI_ISL_2825942, EPI_ISL_2825948, EPI_ISL_2825954, EPI_ISL_2825961, EPI_ISL_2825996, EPI_ISL_2826005, EPI_ISL_2826024, EPI_ISL_2826027, EPI_ISL_2826028, EPI_ISL_2826031, EPI_ISL_2826045, EPI_ISL_2826049, EPI_ISL_2826055, EPI_ISL_2826056, EPI_ISL_2826069, EPI_ISL_2826071, EPI_ISL_2826084, EPI_ISL_2826097, EPI_ISL_2826099, EPI_ISL_2826104, EPI_ISL_2826105, EPI_ISL_2826120, EPI_ISL_2826129, EPI_ISL_2826137, EPI_ISL_2826151, EPI_ISL_2826160, EPI_ISL_2826163, EPI_ISL_2826169, EPI_ISL_2826173, EPI_ISL_2826181, EPI_ISL_2826183, EPI_ISL_2826202, EPI_ISL_2826214, EPI_ISL_2826221, EPI_ISL_2826251, EPI_ISL_2826265, EPI_ISL_2826273, EPI_ISL_2826281, EPI_ISL_2826282, EPI_ISL_2826292, EPI_ISL_2826295, EPI_ISL_2826314, EPI_ISL_2826330, EPI_ISL_2826339, EPI_ISL_2826345, EPI_ISL_2826357, EPI_ISL_2826363, EPI_ISL_2826364, EPI_ISL_2826368, EPI_ISL_2826377, EPI_ISL_2826395, EPI_ISL_2826397, EPI_ISL_2826401, EPI_ISL_2826412, EPI_ISL_2826416, EPI_ISL_2826437, EPI_ISL_2826444, EPI_ISL_2826504, EPI_ISL_2826517, EPI_ISL_2826537, EPI_ISL_2826543, EPI_ISL_2826568, EPI_ISL_2826624, EPI_ISL_2826640, EPI_ISL_2826664, EPI_ISL_2826692, EPI_ISL_2826700, EPI_ISL_2826705, EPI_ISL_2826708, EPI_ISL_2826711, EPI_ISL_2826724, EPI_ISL_2826738, EPI_ISL_2826746, EPI_ISL_2826750, EPI_ISL_2826751, EPI_ISL_2826752, EPI_ISL_2826754, EPI_ISL_2826769, EPI_ISL_2826814, EPI_ISL_2826831, EPI_ISL_2826882, EPI_ISL_2826892, EPI_ISL_2826907, EPI_ISL_2853622, EPI_ISL_2853636, EPI_ISL_2853695, EPI_ISL_2853764, EPI_ISL_2853819, EPI_ISL_2853836, EPI_ISL_2853936, EPI_ISL_2854000, EPI_ISL_2854002, EPI_ISL_2854977, EPI_ISL_2854985, EPI_ISL_2855000, EPI_ISL_2855016, EPI_ISL_2855066, EPI_ISL_2855104, EPI_ISL_2855129, EPI_ISL_2855184, EPI_ISL_2855193, EPI_ISL_2855215, EPI_ISL_2855273, EPI_ISL_2855296, EPI_ISL_2855304, EPI_ISL_2855363, EPI_ISL_2855424, EPI_ISL_2855461, EPI_ISL_2855482, EPI_ISL_2855507, EPI_ISL_2855541, EPI_ISL_2855560, EPI_ISL_2855580, EPI_ISL_2855595, EPI_ISL_2855638, EPI_ISL_2855692, EPI_ISL_2855696, EPI_ISL_2855710, EPI_ISL_2855768, EPI_ISL_2855837, EPI_ISL_2855839, EPI_ISL_2858358, EPI_ISL_2858551, EPI_ISL_2858565, EPI_ISL_2881997, EPI_ISL_2882071, EPI_ISL_2882074, EPI_ISL_2882119, EPI_ISL_2882179, EPI_ISL_2882180, EPI_ISL_2882209, EPI_ISL_2882225, EPI_ISL_2882269, EPI_ISL_2882279, EPI_ISL_2882378, EPI_ISL_2882421, EPI_ISL_2882467, EPI_ISL_2882470, EPI_ISL_2882485, EPI_ISL_2882494, EPI_ISL_2882504, EPI_ISL_2882507, EPI_ISL_2882512, EPI_ISL_2882521, EPI_ISL_2882528, EPI_ISL_2882532, EPI_ISL_2882533, EPI_ISL_2882554, EPI_ISL_2882562, EPI_ISL_2882563, EPI_ISL_2882576, EPI_ISL_2882579, EPI_ISL_2882607, EPI_ISL_2882643, EPI_ISL_2882670, EPI_ISL_2882671, EPI_ISL_2882731, EPI_ISL_2882751, EPI_ISL_2882816, EPI_ISL_2882867, EPI_ISL_2883038, EPI_ISL_2883060, EPI_ISL_2889861, EPI_ISL_2890039, EPI_ISL_2890052, EPI_ISL_2890111, EPI_ISL_2890124, EPI_ISL_2890126, EPI_ISL_2890150, EPI_ISL_2890153, EPI_ISL_2890199, EPI_ISL_2890211, EPI_ISL_2890285, EPI_ISL_2890286, EPI_ISL_2890293, EPI_ISL_2890391, EPI_ISL_2890426, EPI_ISL_2890492, EPI_ISL_2890520, EPI_ISL_2890522, EPI_ISL_2890524, EPI_ISL_2890537, EPI_ISL_2890682, EPI_ISL_2890740, EPI_ISL_2890860, EPI_ISL_2891048, EPI_ISL_2891107, EPI_ISL_2891113, EPI_ISL_2891114, EPI_ISL_2891169, EPI_ISL_2891173, EPI_ISL_2891176, EPI_ISL_2891308, EPI_ISL_2891311, EPI_ISL_2891314, EPI_ISL_2891317, EPI_ISL_2891343, EPI_ISL_2891346, EPI_ISL_2891407, EPI_ISL_2891417, EPI_ISL_2891510, EPI_ISL_2891520, EPI_ISL_2891538, EPI_ISL_2891570, EPI_ISL_2891706, EPI_ISL_2891954, EPI_ISL_2892386, EPI_ISL_2921716, EPI_ISL_2921739, EPI_ISL_2922265, EPI_ISL_2923306, EPI_ISL_2923377, EPI_ISL_2923412, EPI_ISL_2923588, EPI_ISL_2923590, EPI_ISL_2923720, EPI_ISL_2934103, EPI_ISL_2934110, EPI_ISL_2934127, EPI_ISL_2934169, EPI_ISL_2934173, EPI_ISL_2934196, EPI_ISL_2934240, EPI_ISL_2934255, EPI_ISL_2934296, EPI_ISL_2934308, EPI_ISL_2934316, EPI_ISL_2934337, EPI_ISL_2934355, EPI_ISL_2934363, EPI_ISL_2934380, EPI_ISL_2934382, EPI_ISL_2934384, EPI_ISL_2934390, EPI_ISL_2934406, EPI_ISL_2934413, EPI_ISL_2934423, EPI_ISL_2934434, EPI_ISL_2934455, EPI_ISL_2934463, EPI_ISL_2934469, EPI_ISL_2934474, EPI_ISL_2934480, EPI_ISL_2934485, EPI_ISL_2934488, EPI_ISL_2934490, EPI_ISL_2934491, EPI_ISL_2934499, EPI_ISL_2934501, EPI_ISL_2934502, EPI_ISL_2934509, EPI_ISL_2934515, EPI_ISL_2934516, EPI_ISL_2934521, EPI_ISL_2934526, EPI_ISL_2934528, EPI_ISL_2934529, EPI_ISL_2934534, EPI_ISL_2934538, EPI_ISL_2934546, EPI_ISL_2934559 | | | |
| see above | CDPH VBL | California Department of Public Health | CDPH-COVIDNet; SwabSeq/UCLA; UCI Genome Sciences Center/GHTF; UCLA Technology Center for Genomics & Bioinformatics; UCSF Center for Advanced Technology |
| EPI_ISL_2776230, EPI_ISL_2776231, EPI_ISL_2776257, EPI_ISL_2778116, EPI_ISL_2778128, EPI_ISL_2778139, EPI_ISL_2778175, EPI_ISL_2778178, EPI_ISL_2778179, EPI_ISL_2778188, EPI_ISL_2778190, EPI_ISL_2778193, EPI_ISL_2778209, EPI_ISL_2778210, EPI_ISL_2778249, EPI_ISL_2778256, EPI_ISL_2778307, EPI_ISL_2778313, EPI_ISL_2778314, EPI_ISL_2778319, EPI_ISL_2778335, EPI_ISL_2778382, EPI_ISL_2778389, EPI_ISL_2778393, EPI_ISL_2778406, EPI_ISL_2778483, EPI_ISL_2778593, EPI_ISL_2778595, EPI_ISL_2778596, EPI_ISL_2778599, EPI_ISL_2778601, EPI_ISL_2778609, EPI_ISL_2778647, EPI_ISL_2778657, EPI_ISL_2778661, EPI_ISL_2778663, EPI_ISL_2778706, EPI_ISL_2778707, EPI_ISL_2778720, EPI_ISL_2778747, EPI_ISL_2778752, EPI_ISL_2778759, EPI_ISL_2778760, EPI_ISL_2778784, EPI_ISL_2778785, EPI_ISL_2778818, EPI_ISL_2778842, EPI_ISL_2778855, EPI_ISL_2778871, EPI_ISL_2778885, EPI_ISL_2778948, EPI_ISL_2778950, EPI_ISL_2778954, EPI_ISL_2837353, EPI_ISL_2837364, EPI_ISL_2837368, EPI_ISL_2837593, EPI_ISL_2837610, EPI_ISL_2837697, EPI_ISL_2837816, EPI_ISL_2837819, EPI_ISL_2837873, EPI_ISL_2837906, EPI_ISL_2837928, EPI_ISL_2837931, EPI_ISL_2837954, EPI_ISL_2838013 | | | |
| see above | California Department of Public Health Valencia Branch Laboratory (CDPH VBL) | California Department of Public Health | Arias Lab at UCSB; CDPH-COVIDNet; UCLA Technology Center for Genomics & Bioinformatics |
| EPI_ISL_2941578, EPI_ISL_2941609 | Colorado Department of Public Health and Environment | Colorado Department of Public Health and Environment | Alexandria Rossheim; Diana Ir; Emily A. Travanty; Laura Bankers; Mandy Waters; Michael A. Martin; Molly C. Hetherington-Rauth; Sarah Elizabeth Totten; Shannon R. Matzinger |
| EPI_ISL_2783576, EPI_ISL_2783691, EPI_ISL_2783695, EPI_ISL_2783832, EPI_ISL_2783870, EPI_ISL_2941933, EPI_ISL_2942173 | | | |
| see above | Curative Labs | Curative Labs | Elias L. Salfati; Eugenia Khorosheva; George Way; J.Cesar Ignacio-Espinoza; Janet Chen; Mikhail Hanewich-Hollatz; Nabjot Sandhu; Sophia Quasem; Vladimir Slepnev; Zhiyi Xie |
| EPI_ISL_2854087, EPI_ISL_2876497, EPI_ISL_2876498, EPI_ISL_2876509, EPI_ISL_2876512 | DC Public Health Lab/ Dept. of Forensic Sciences | DC Public Health Lab/ Dept. of Forensic Sciences | Brittany Hamilton; Connie Maza; David Payne; Elizabeth Zelaya; Janis Doss; Jocelyn Hauser; Monica Mann; Sarah Scott; Scott Nguyen |
| EPI_ISL_2790697, EPI_ISL_2790702, EPI_ISL_2790706, EPI_ISL_2790708, EPI_ISL_2790709, EPI_ISL_2790710, EPI_ISL_2790711, EPI_ISL_2790712, EPI_ISL_2790715 | | | |
| see above | Emory Molecular Diagnostics Laboratory, Emory Healthcare | Genomics and Discovery, Respiratory Viruses Branch, Division of Viral Diseases, Centers for Disease Control and Prevention | Adam Retchless; Ahmed Babiker; Anna Kelleher; Anne Piantadosi; Brian Lynch; Clinton R. Paden; Dhwani Batra; Haibin Wang; Han Jia Justin Ng; Jasmine Padilla; Jing Zhang; Justin Lee; Krista Queen; Mark Burroughs; Mili Sheth; Morgan Davis; Peter Cook; Rachel Marine; Sarah Nobles; Suxiang Tong; Tara Coalter; Yan Li; Ying Tao |
| EPI_ISL_2836594 | Florida Bureau of Public Health Laboratories | Florida Bureau of Public Health Laboratories | Jason Blanton; Sarah Schmedes |
| EPI_ISL_2785525, EPI_ISL_2785684, EPI_ISL_2785728, EPI_ISL_2873457 | Fulgent Genetics | Centers for Disease Control and Prevention Division of Viral Diseases, Pathogen Discovery | Adrian Paskey; Becky Tsai; Benafsh Sapra; Benjamin Rambo-Martin; Christopher Gulvick; Clinton R. Paden; Dakota Howard; Darlene Wagner; Dhwani Batra; Doreen Ng; Duncan MacCannell; Harry Gao; James Xie; Jason Caravas; John Gao; Joseph Fierro; Kara Moser; Matthew Schmerer; Mickey Li; Peter W. Cook; Scott Sammons; Shatavia Morrison; Yan Meng; Yvette Unoarumhi |
| EPI_ISL_2800952, EPI_ISL_2800953, EPI_ISL_2800962, EPI_ISL_2800987, EPI_ISL_2801012, EPI_ISL_2801025, EPI_ISL_2801032, EPI_ISL_2801101, EPI_ISL_2801107, EPI_ISL_2801120, EPI_ISL_2801121, EPI_ISL_2801123, EPI_ISL_2801127, EPI_ISL_2801130, EPI_ISL_2801168, EPI_ISL_2801170, EPI_ISL_2801184, EPI_ISL_2801195, EPI_ISL_2801211, EPI_ISL_2801212, EPI_ISL_2801213, EPI_ISL_2801251, EPI_ISL_2801261, EPI_ISL_2801277 | | | |
| see above | Illinois Department of Public Health | Gagnon Lab, Southern Illinois University | Keith Gagnon |
| EPI_ISL_2884891, EPI_ISL_2884910, EPI_ISL_2884911, EPI_ISL_2884914, EPI_ISL_2884915, EPI_ISL_2884916, EPI_ISL_2884917, EPI_ISL_2884919, EPI_ISL_2884932, EPI_ISL_2884934, EPI_ISL_2884935, EPI_ISL_2884936, EPI_ISL_2884937, EPI_ISL_2884939, EPI_ISL_2884940, EPI_ISL_2884945, EPI_ISL_2884947, EPI_ISL_2884948, EPI_ISL_2884951, EPI_ISL_2884952, EPI_ISL_2884953, EPI_ISL_2884954, EPI_ISL_2884955, EPI_ISL_2884956, EPI_ISL_2884957, EPI_ISL_2884959, EPI_ISL_2884961, EPI_ISL_2884962, EPI_ISL_2884963, EPI_ISL_2884965, EPI_ISL_2884966, EPI_ISL_2884967, EPI_ISL_2884968, EPI_ISL_2884975, EPI_ISL_2884976, EPI_ISL_2884977, EPI_ISL_2884978, EPI_ISL_2884980, EPI_ISL_2884983, EPI_ISL_2884984, EPI_ISL_2884986, EPI_ISL_2884990, EPI_ISL_2884991, EPI_ISL_2884992, EPI_ISL_2884997, EPI_ISL_2885000, EPI_ISL_2885002, EPI_ISL_2885004, EPI_ISL_2885005, EPI_ISL_2885006, EPI_ISL_2885007, EPI_ISL_2885009, EPI_ISL_2885010, EPI_ISL_2885011, EPI_ISL_2885017, EPI_ISL_2885020, EPI_ISL_2885023, EPI_ISL_2885028, EPI_ISL_2885029, EPI_ISL_2885030, EPI_ISL_2885032, EPI_ISL_2885033, EPI_ISL_2885034, EPI_ISL_2885035, EPI_ISL_2885038, EPI_ISL_2885039, EPI_ISL_2885042, EPI_ISL_2885044, EPI_ISL_2885046, EPI_ISL_2885047, EPI_ISL_2885048, EPI_ISL_2885049, EPI_ISL_2885052, EPI_ISL_2885054, EPI_ISL_2885055, EPI_ISL_2885058, EPI_ISL_2885059, EPI_ISL_2885060, EPI_ISL_2885062, EPI_ISL_2885068, EPI_ISL_2885070, EPI_ISL_2885071, EPI_ISL_2885075, EPI_ISL_2885078, EPI_ISL_2885080, EPI_ISL_2885084, EPI_ISL_2885086, EPI_ISL_2885088, EPI_ISL_2885089, EPI_ISL_2885092, EPI_ISL_2885096, EPI_ISL_2885099, EPI_ISL_2885104, EPI_ISL_2885105, EPI_ISL_2885106, EPI_ISL_2885107, EPI_ISL_2885114, EPI_ISL_2885115, EPI_ISL_2885116, EPI_ISL_2885117, EPI_ISL_2885119, EPI_ISL_2885120, EPI_ISL_2885121, EPI_ISL_2885124, EPI_ISL_2885125, EPI_ISL_2885128, EPI_ISL_2885129, EPI_ISL_2885130, EPI_ISL_2885133, EPI_ISL_2885134, EPI_ISL_2885138, EPI_ISL_2885139, EPI_ISL_2885142, EPI_ISL_2885146, EPI_ISL_2885155, EPI_ISL_2885168, EPI_ISL_2885172 | | | |
| see above | Johns Hopkins Hospital Department of Pathology | Johns Hopkins Hospital Department of Pathology | Adannaya Amadi; C. Paul Morris; Chun Huai Luo; Heba H. Mostafa; Matthew Schwartz; Nicholas Gallagher |
| EPI_ISL_2789912, EPI_ISL_2789927, EPI_ISL_2893678 | Kansas Health and Environmental Lab | Kansas Health and Environmental Lab | Ben Olsen; Jonathan Barnell; Katherine Wiggins; Mike Grose; and Phil Adam |
| EPI_ISL_2872951, EPI_ISL_2873018, | Laboratory Corporation of America | Centers for Disease Control and Prevention Division of Viral | Adrian Paskey; Amanda Douglas; Amanda Suchanek; Andrea Throop; Ayla Burns; Benjamin Rambo-Martin; Bobbi Croy; Brian Krueger; Brian Norvell; Christopher Gulvick; Christos Petropoulos; Clinton R. Paden; Craig Lukasik; Dakota Howard; Darlene Wagner; Debbie Boles; Dhwani Batra; Duncan MacCannell; Eyad Almasri; Goran Stevovic; Howard Engler; Hrushikesh Deshmukh; Jake Humphrey; Jana Schroth; Jason Caravas; Joe Voshell; John Pruitt; Jonathan Meltzer; Jonathan Williams; Kara Moser; Kimberly Wagner; Lax Iyer; Lisa Pfefferle; Lyndon Tilson; Manoj Jain; Marcia Eisenberg; Mary Ann |

| | | | |
|---|---|---|---|
| EPI_ISL_2926267, EPI_ISL_2926609 | | Diseases, Pathogen Discovery | Cristobal; Mary Williamson; Matthew Robinson; Matthew Schmerer; Michael Levandoski; Mike Sapeta; Mindy Nye; Minoo Agarwal; Mohan Kolli; Nuthawin Charoensri; Oren Cohen; Peter W. Cook; Prashant Gupta; Qian Zeng; Rama Ghatti; Scott Parker; Scott Ryan; Scott Sammons; Shatavia Morrison; Stanley Letovsky; Steven Ragan; Suresh Babu Selvaraju; Susan Countryman; Susan Hicks; Suzanne Dale; Thomas Urban; Tim Kuphal; Tricia Zwiefelhofer; Vincent Drouillon; Yvette Unoarumhi |
| EPI_ISL_2835673 | Laboratory Medicine, University of Washington | Laboratory Medicine, University of Washington | Baker; Bakhash; Ellis, S.; Greninger, A.; Huang; Jerome; K.R.; M.-L.; Mills, M.; N.R.; Roychoudhury, P.; S.M.; Shrestha, L.; Xie, H. |
| EPI_ISL_2790076, EPI_ISL_2955570 | MD PHL | MD PHL | Maryland Department of Health Laboratories Administration |
| EPI_ISL_2802371, EPI_ISL_2802612, EPI_ISL_2802804, EPI_ISL_2802817 | Maine Health and Environmental Testing Laboratory | Tewhey Lab, The Jackson Laboratory | Barter, M.; Dewey, H.; H. and Tewhey, R.; Iosue, F.; Lynch, R.; Matluk, N.; Munger |
| EPI_ISL_2834696, EPI_ISL_2834697 | Michigan State University Veterinary Diagnostic Laboratory | Diagnostic Virology Laboratory, National Veterinary Services Laboratories, USDA1920 Dayton Avenue, Ames, IA 50010, USA | Emily R. Love; Kerrie M. Franzen; Mary L. Killian; Mia Torchetti; Suelee Robbe-Austerman; Tod P. Stuber |
| EPI_ISL_2834612 | Montana DPHHS Public Health Laboratory | Center for Global Health, University of New Mexico Health Sciences Center | Carrie Biskupiak; Darrell Dinwiddie; Daryl Domman; Deborah Gibson; Joy Ritter; Kurt Schwalm; Valerie Morley |
| EPI_ISL_2941774 | Montana Public Health Laboratory | Montana Public Health Laboratory | Carrie Biskupiak; Deborah Gibson; Joy Ritter; Michael Dills; Michelle Mozer |
| EPI_ISL_2955357 | Monterey County Public Health Laboratory | Monterey County Public Health Laboratory | Monterey County Public Health Laboratory |
| EPI_ISL_2801498, EPI_ISL_2942193 | Nevada State Public Health Laboratory | Nevada State Public Health Laboratory | Andrew Gorzalski; Mark Pandori |
| EPI_ISL_2776403, EPI_ISL_2776408, EPI_ISL_2776409, EPI_ISL_2776417, EPI_ISL_2858815, EPI_ISL_2858817, EPI_ISL_2876426, EPI_ISL_2876427, EPI_ISL_2876428, EPI_ISL_2876430, EPI_ISL_2876431, EPI_ISL_2876434, EPI_ISL_2876437, EPI_ISL_2876438, EPI_ISL_2876449, EPI_ISL_2876454, EPI_ISL_2876468, EPI_ISL_2876472, EPI_ISL_2876485 | | | |
| see above | North Dakota Department of Health, Public Health Laboratory | North Dakota Department of Health, Public Health Laboratory | Lisa Wingerter |
| EPI_ISL_2839527, EPI_ISL_2839532, EPI_ISL_2839554, EPI_ISL_2841622, EPI_ISL_2841628 | Notre Dame COVID Response and Testing Laboratory | Notre Dame Genomics & Bioinformatics Core Facility | Joseph Sarro; Melissa Stephens; Michael Pfrender |
| EPI_ISL_2838580, EPI_ISL_2838624, EPI_ISL_2838651, EPI_ISL_2838655, EPI_ISL_2838719, EPI_ISL_2838780, EPI_ISL_2839022, EPI_ISL_2839051, EPI_ISL_2839062 | | | |
| see above | OHSU Lab Services Molecular Microbiology Lab | Ginkgo Bioworks Clinical Laboratory | '[UsersoroakBox SyncPHI_SARS-COV-2_Sample_Sheet_infoGISAID_Submission_Infometa_master_2021.xlsx]release20_final'!D:AC; 26; FALSE); VLOOKUP(C144; VLOOKUP(C226; VLOOKUP(C246; VLOOKUP(C266; VLOOKUP(C280; VLOOKUP(C315; VLOOKUP(C346; VLOOKUP(C347; VLOOKUP(C349 |
| EPI_ISL_2838567, EPI_ISL_2838724, EPI_ISL_2838899, EPI_ISL_2842723 | OHSU Lab Services Molecular Microbiology Lab | Oregon SARS-CoV-2 Genome Sequencing Center | '[UsersoroakBox SyncPHI_SARS-COV-2_Sample_Sheet_infoGISAID_Submission_Infometa_master_2021.xlsx]release20_final'!D:AC; 26; FALSE); VLOOKUP(C18; VLOOKUP(C56; VLOOKUP(C57; VLOOKUP(C8 |
| EPI_ISL_2895821, EPI_ISL_2895842, EPI_ISL_2895866, EPI_ISL_2895997, EPI_ISL_2896104 | OHSU-MM Lab | OHSU-MM Lab | Rebecca O'Gara |
| EPI_ISL_2811925, EPI_ISL_2811934, EPI_ISL_2812015, EPI_ISL_2812024 | OmniPathology Laboratory | Los Angeles County Public Health Laboratories | P. Hemarajata et al. |
| EPI_ISL_2758432 | Orange County Public Health Lab | Chan-Zuckerberg Biohub | CZB Cliahub Consortium |
| EPI_ISL_2790752, EPI_ISL_2790753, EPI_ISL_2790754, EPI_ISL_2790755, EPI_ISL_2790756, EPI_ISL_2790757, EPI_ISL_2790758, EPI_ISL_2790759, EPI_ISL_2790761, EPI_ISL_2839145, EPI_ISL_2839146, EPI_ISL_2839147, EPI_ISL_2839148, EPI_ISL_2839149, EPI_ISL_2839150, EPI_ISL_2839151, EPI_ISL_2839153, EPI_ISL_2839157, EPI_ISL_2839160, EPI_ISL_2839162, EPI_ISL_2933951, EPI_ISL_2933953, EPI_ISL_2933954, EPI_ISL_2933956, EPI_ISL_2933959, EPI_ISL_2933960, EPI_ISL_2933961, EPI_ISL_2933962 | | | |
| see above | Oregon State Public Health Laboratory | Oregon State Public Health Laboratory | Eugene Yeboah; John Fontana and Shane Sevey; Laura Tsaknaridis; Rafia Razzaque; Vanda Makris |
| EPI_ISL_2827190, EPI_ISL_2827225, EPI_ISL_2827254, EPI_ISL_2827301, EPI_ISL_2827334, EPI_ISL_2827361, EPI_ISL_2827391, EPI_ISL_2827393, EPI_ISL_2827417, EPI_ISL_2827448, EPI_ISL_2827526, EPI_ISL_2827546, EPI_ISL_2827556, EPI_ISL_2827586, EPI_ISL_2827607, EPI_ISL_2827609, EPI_ISL_2827623, EPI_ISL_2827673, EPI_ISL_2827697, EPI_ISL_2827712, EPI_ISL_2827730, EPI_ISL_2923826, EPI_ISL_2923829, EPI_ISL_2923859, EPI_ISL_2923871, EPI_ISL_2923900, EPI_ISL_2923973, EPI_ISL_2923975, EPI_ISL_2923996, EPI_ISL_2923999 | | | |
| see above | Pandemic Response Lab - NYC | Pandemic Response Lab, R&D | Cybill del Castillo; Dylan Law; Haiping Hao; Henry Lee; Isabel Fernandez Escapa; Jon Laurent; Melissa Hopkins; Michael Hammerling; Pradeep Bugga; Shinyoung Clair Kang; Sol Rey; William Ward |
| EPI_ISL_2993488 | Pathology, UAB | Pathology, UAB | E.J.; Hendrickson; Hernandez, J.; Leal; Lefkowitz; Moates, D.; R.C.; S.M. |
| EPI_ISL_2835243, EPI_ISL_2835251 | Rhode Island State Health Laboratory | Rhode Island State Health Laboratory | Ewa King; Kristin Carpenter-Azevedo; Richard C. Huard |
| EPI_ISL_2787245 | SC Dept of Health and Env. Control-Bureau of Laboratories | Centers for Disease Control and Prevention Division of Viral Diseases, Pathogen Discovery | Alison Laufer Halpin; Ben L. Rambo-Martin; Clinton R. Paden; Dakota Howard; Darlene Wagner; Dave Wentworth; Dhwani Batra; Jasmine Padilla; Justin Lee; Katie Dillon; Krista Queen; Kristen Knipe; Kristine Lacek; Mark Burroughs; Matthew Schmerer; Mili Sheth; Peter Cook; Sam Shepard; Sarah Nobles; Shoshona Le; Suxiang Tong; Vivien Dugan; Yvette Unoarumhi |
| EPI_ISL_2920360, EPI_ISL_2920365, EPI_ISL_2920370, EPI_ISL_2920391 | San Diego County Public Health Laboratory | Andersen lab at Scripps Research | Brett Austin; Jovan Shephard; SEARCH Alliance San Diego with Ashleigh Murphy |
| EPI_ISL_2834957, EPI_ISL_2840492 | San Diego County Public Health Laboratory | San Diego County Public Health Laboratory | Ashleigh Murphy Schafer; Brett Austin |
| EPI_ISL_2779556, EPI_ISL_2779557, EPI_ISL_2779559, EPI_ISL_2779564, EPI_ISL_2779583 | Santa Clara County Public Health Laboratory | Santa Clara County Public Health Laboratory | Santa Clara County Public Health Department |
| EPI_ISL_2920287, EPI_ISL_2920330, EPI_ISL_2920418 | Sharp HealthCare Laboratory | Andersen lab at Scripps Research | Art Mendoza; Cathy Woerle; Jacquelyn Berumen; Liam McGinnis; Omid Bakhtar; SEARCH Alliance San Diego with Aaron Harding |
| EPI_ISL_2769187, EPI_ISL_2769207, EPI_ISL_2769209, EPI_ISL_2769213, EPI_ISL_2769224, EPI_ISL_2769228, EPI_ISL_2769229, EPI_ISL_2769233, EPI_ISL_2769246, EPI_ISL_2769253, EPI_ISL_2769258, EPI_ISL_2769280, EPI_ISL_2769285, EPI_ISL_2769291, EPI_ISL_2769302, EPI_ISL_2769304, EPI_ISL_2769310, EPI_ISL_2769314, EPI_ISL_2769317, EPI_ISL_2769330, EPI_ISL_2769331, EPI_ISL_2769335, EPI_ISL_2769341, EPI_ISL_2769344, EPI_ISL_2769348, EPI_ISL_2769353, EPI_ISL_2769354, EPI_ISL_2919295, EPI_ISL_2919303, EPI_ISL_2919313, EPI_ISL_2919324, EPI_ISL_2919327, EPI_ISL_2919329, EPI_ISL_2919331, EPI_ISL_2919332, EPI_ISL_2919335, EPI_ISL_2919336, EPI_ISL_2919347, EPI_ISL_2919348, EPI_ISL_2919351, EPI_ISL_2919361, EPI_ISL_2919362, EPI_ISL_2919370, EPI_ISL_2919384, EPI_ISL_2919389, EPI_ISL_2919390, EPI_ISL_2919396, EPI_ISL_2919398, EPI_ISL_2919402, EPI_ISL_2919404, EPI_ISL_2919409, EPI_ISL_2919417, EPI_ISL_2919431, EPI_ISL_2919432, EPI_ISL_2919439, EPI_ISL_2919444, EPI_ISL_2919464, EPI_ISL_2919466, EPI_ISL_2919467, EPI_ISL_2919468, EPI_ISL_2919474, EPI_ISL_2919478, EPI_ISL_2919487, EPI_ISL_2919494, EPI_ISL_2919495, EPI_ISL_2919502, EPI_ISL_2919504, EPI_ISL_2919507, EPI_ISL_2919514, EPI_ISL_2919518, EPI_ISL_2919535, EPI_ISL_2919536, EPI_ISL_2919545, EPI_ISL_2919548, EPI_ISL_2919553, EPI_ISL_2919567, EPI_ISL_2919572, EPI_ISL_2919581, EPI_ISL_2919607, EPI_ISL_2919623, EPI_ISL_2919625, EPI_ISL_2919633, EPI_ISL_2919638, EPI_ISL_2919640, EPI_ISL_2919685, EPI_ISL_2919696, EPI_ISL_2919698, EPI_ISL_2919700, EPI_ISL_2919705, EPI_ISL_2919710, EPI_ISL_2919715, EPI_ISL_2919716, EPI_ISL_2919720, EPI_ISL_2919728, EPI_ISL_2919731, EPI_ISL_2919732, EPI_ISL_2919750, EPI_ISL_2919755, EPI_ISL_2919760, EPI_ISL_2919765, EPI_ISL_2919775, EPI_ISL_2919782, EPI_ISL_2919785, EPI_ISL_2919787, EPI_ISL_2919791, EPI_ISL_2919793, EPI_ISL_2919795, EPI_ISL_2919812, EPI_ISL_2919813, EPI_ISL_2919822, EPI_ISL_2919823, EPI_ISL_2919830, EPI_ISL_2919831, EPI_ISL_2919911, EPI_ISL_2919912, EPI_ISL_2919916, EPI_ISL_2919927, EPI_ISL_2919928, EPI_ISL_2919930, EPI_ISL_2919935, EPI_ISL_2919947, EPI_ISL_2919989, EPI_ISL_2919999, EPI_ISL_2920002, EPI_ISL_2920005, EPI_ISL_2920035, EPI_ISL_2920037 | | | |
| see above | TGen North | TGen North | "Jolene Bowers; Brett Van Tassel; Chris French; Darrin Lemmer; Dave Engelthaler"; Hayley Yaglom; Heather Centner |
| EPI_ISL_2758863, EPI_ISL_2758932 | The Ohio State University Applied Microbiology Services Laboratory | The Ohio State University Applied Microbiology Services Laboratory | Seth A. Faith PhD |
| EPI_ISL_2791392 | UMGC | UMGC | David Xing; Gregg Thomas; Jeffrey Good; Nathanael Herrera; Tim Thurman |
| EPI_ISL_2775680, EPI_ISL_2775790, EPI_ISL_2775858, EPI_ISL_2775859 | UPMC Clinical Microbiology Laboratory | Microbial Genome Sequencing Center; Microbial Genomic Epidemiology Laboratory, University of Pittsburgh | Daniel J. Snyder; Jane W. Marsh; Kady D. Waggle; Lee H. Harrison; Marissa P. Griffith; Vatsala R. Srinivasa; Vaughn S. Cooper |
| EPI_ISL_2773872, EPI_ISL_2773892, EPI_ISL_2773912, EPI_ISL_2773913, EPI_ISL_2773914, EPI_ISL_2773916, EPI_ISL_2773917, EPI_ISL_2773918, EPI_ISL_2773919, EPI_ISL_2773920, EPI_ISL_2773921, EPI_ISL_2773922, EPI_ISL_2773923, EPI_ISL_2773924, EPI_ISL_2773925, EPI_ISL_2773939, EPI_ISL_2773940, EPI_ISL_2773941, EPI_ISL_2773942, EPI_ISL_2773943, EPI_ISL_2773944, EPI_ISL_2790445, EPI_ISL_2790458, EPI_ISL_2790459, EPI_ISL_2790461, EPI_ISL_2790614, EPI_ISL_2790615, EPI_ISL_2791037, EPI_ISL_2791039, EPI_ISL_2791040, EPI_ISL_2791041, EPI_ISL_2791042, EPI_ISL_2791043, EPI_ISL_2791044, EPI_ISL_2791045, EPI_ISL_2791068, EPI_ISL_2791069, EPI_ISL_2791075, EPI_ISL_2791076, EPI_ISL_2791077, EPI_ISL_2791079, EPI_ISL_2791117, EPI_ISL_2791118, EPI_ISL_2791119, EPI_ISL_2791120, EPI_ISL_2791121, EPI_ISL_2791122, EPI_ISL_2791123, EPI_ISL_2791124, EPI_ISL_2791125, EPI_ISL_2791126, EPI_ISL_2791127, EPI_ISL_2791128, EPI_ISL_2791129, EPI_ISL_2791147, EPI_ISL_2791148, EPI_ISL_2791195, EPI_ISL_2791214, EPI_ISL_2791215, EPI_ISL_2791216, EPI_ISL_2791217, EPI_ISL_2791218, EPI_ISL_2803004, EPI_ISL_2864201, EPI_ISL_2864202, EPI_ISL_2864218 | | | |
| see above | UW Virology Lab | UW Virology Lab | Alexander Greninger; Hong Xie; Keith R Jerome; Lasata Shrestha; Margaret Mills; Meei-Li Huang; Michelle Lin; Nathan Breit; Noah Baker; Noah R. Baker; Patrick Mathias; Pavitra Roychoudhury; Ricardo Perez; Robert J. Livingston; Sean Ellis; Shah Mohamed Bakhash; Tien V. Nguyen |
| EPI_ISL_2858782 | University of Mississippi Medical Center, Department of Pathology | University of Mississippi Medical Center, Molecular and Genomics | Ashley C. Johnson; D. Ashley Robinson; Ithiel J. Frame; Krishna K. Ayyalasomayajula; Michael R. Garrett; Wenjie Wu |

| | | | |
|---|---|---|---|
| | | Core Facility | |
| EPI_ISL_2855951, EPI_ISL_2856112, EPI_ISL_2856182, EPI_ISL_2858104, EPI_ISL_2858126 | Utah Public Health Laboratory | Utah Public Health Laboratory | Erin L. Young; Kelly F. Oakeson; Olinto Linares-Perdomo; Tara Gallagher |
| EPI_ISL_3049039, EPI_ISL_3049047, EPI_ISL_3049051, EPI_ISL_3049057, EPI_ISL_3049058, EPI_ISL_3049061, EPI_ISL_3049068, EPI_ISL_3049070, EPI_ISL_3049071, EPI_ISL_3049086, EPI_ISL_3049087 | | | |
| see above | Utah Public Health Laboratory, Utah Public Health Laboratory Infectious Disease submission group | Utah Public Health Laboratory, Utah Public Health Laboratory Infectious Disease submission group | Linares-Perdomo, O.; Oakeson, K.; Young, E. |
| EPI_ISL_2834689, EPI_ISL_2834690, EPI_ISL_2834691, EPI_ISL_2834693, EPI_ISL_2834694, EPI_ISL_2834695, EPI_ISL_2834937, EPI_ISL_2834938, EPI_ISL_2834939, EPI_ISL_2834940, EPI_ISL_2834941, EPI_ISL_2834942, EPI_ISL_2834943, EPI_ISL_2834944, EPI_ISL_2834945, EPI_ISL_2896207, EPI_ISL_2896208, EPI_ISL_2896209, EPI_ISL_2896210, EPI_ISL_2896211, EPI_ISL_2896212, EPI_ISL_2896213 | | | |
| see above | Utah Veterinary Diagnostic Laboratory | Diagnostic Virology Laboratory, National Veterinary Services Laboratories, USDA1920 Dayton Avenue, Ames, IA 50010, USA | Emily R. Love; Kerrie M. Franzen; Mary L. Killian; Mia Torchetti; Suelee Robbe-Austerman; Tod P. Stuber |
| EPI_ISL_2931129 | Vanderbilt University Medical Center | Genomics and Discovery, Respiratory Viruses Branch, Division of Viral Diseases, Centers for Disease Control and Prevention | Adam Retchless; Anna Kelleher; Anna Uehara; Brian Lynch; Clinton R. Paden; Dhwani Batra; Haibin Wang; Han Jia Justin Ng; Jasmine Padilla; Jing Zhang; Justin Lee; Krista Queen; Mark Burroughs; Mili Sheth; Morgan Davis; Peter Cook; Rachel Marine; Sarah Nobles; Suxiang Tong; Tara Coalter; Yan Li; Ying Tao |
| EPI_ISL_3050735 | Virus Ecology, NIH | Virus Ecology, NIH | Banakis, S.; Bushmaker, C.; Bushmaker, T.; Chung, M.; Ghedin, E.; Holbrook; M.G.; Munster; Roder, A.; V.J.; Van Doremalen, N.; Wang, W. |
| EPI_ISL_2896203, EPI_ISL_2896204, EPI_ISL_2896205, EPI_ISL_2896206 | Washington Animal Disease Diagnostic Laboratory | Diagnostic Virology Laboratory, National Veterinary Services Laboratories, USDA1920 Dayton Avenue, Ames, IA 50010, USA | Emily R. Love; Kerrie M. Franzen; Mary L. Killian; Mia Torchetti; Suelee Robbe-Austerman; Tod P. Stuber |
| EPI_ISL_2834698, EPI_ISL_2834699, EPI_ISL_2834700, EPI_ISL_2834701, EPI_ISL_2834702 | Wisconsin Veterinary Diagnostic Laboratory | Diagnostic Virology Laboratory, National Veterinary Services Laboratories, USDA1920 Dayton Avenue, Ames, IA 50010, USA | Emily R. Love; Kerrie M. Franzen; Mary L. Killian; Mia Torchetti; Suelee Robbe-Austerman; Tod P. Stuber |
| EPI_ISL_2775504, EPI_ISL_2775509, EPI_ISL_2775513, EPI_ISL_2775515, EPI_ISL_2775525, EPI_ISL_2775535, EPI_ISL_2775536, EPI_ISL_2775537, EPI_ISL_2775540, EPI_ISL_2775627, EPI_ISL_2775673, EPI_ISL_2776132, EPI_ISL_2833829, EPI_ISL_2833957, EPI_ISL_2833973, EPI_ISL_2834999, EPI_ISL_2842734, EPI_ISL_2842755, EPI_ISL_2859423, EPI_ISL_2876545, EPI_ISL_2885583, EPI_ISL_2920168, EPI_ISL_2920169, EPI_ISL_2920192, EPI_ISL_2924020, EPI_ISL_2924054, EPI_ISL_2925633, EPI_ISL_2925642, EPI_ISL_2931223, EPI_ISL_2931632, EPI_ISL_2931665, EPI_ISL_2931685, EPI_ISL_2941830, EPI_ISL_2943099, EPI_ISL_2943105 | | | |
| see above | Wyoming Public Health Laboratory | Wyoming Public Health Laboratory | Ashley Norberg; Brian Dominguez; Cari Sloma; Channing Weber; Chayse Rowley; Elliot Thomasson; Jim Mildenberger; Marley Goetz; Robert Petit; Sam Britz; Taylor Fearing; and Rob Christensen |

We gratefully acknowledge the following Authors from the Originating laboratories responsible for obtaining the specimens, as well as the Submitting laboratories where the genome data were generated and shared via GISAID, on which this research is based.

All Submitters of data may be contacted directly via www.gisaid.org

Authors are sorted alphabetically.

| Accession ID | Originating Laboratory | Submitting Laboratory | Authors |
| --- | --- | --- | --- |
| EPI_ISL_2394601, EPI_ISL_2394668, EPI_ISL_2394703, EPI_ISL_2394825 | Health Services Laboratories | Wellcome Sanger Institute for the COVID-19 Genomics UK (COG-UK) Consortium | Cordelia Langford; David K. Jackson; Dominic Kwiatkowski; Ewan Harrison; Health Services Laboratories and Alex Alderton; Ian Johnston; Jeffrey Barrett; John Sillitoe on behalf of the Wellcome Sanger Institute COVID-19 Surveillance Team; Roberto Amato; Sonia Goncalves |
| EPI_ISL_2392201, EPI_ISL_2392204, EPI_ISL_2392205, EPI_ISL_2392206, EPI_ISL_2392207, EPI_ISL_2392212, EPI_ISL_2392217, EPI_ISL_2392226, EPI_ISL_2392231, EPI_ISL_2392240, EPI_ISL_2392241, EPI_ISL_2392246, EPI_ISL_2392248, EPI_ISL_2392251, EPI_ISL_2392255, EPI_ISL_2392259, EPI_ISL_2392269, EPI_ISL_2392283, EPI_ISL_2392284, EPI_ISL_2392289, EPI_ISL_2392290, EPI_ISL_2392301, EPI_ISL_2392306, EPI_ISL_2392317, EPI_ISL_2392354, EPI_ISL_2392358, EPI_ISL_2392361, EPI_ISL_2392363, EPI_ISL_2392370, EPI_ISL_2392379, EPI_ISL_2392386, EPI_ISL_2392396, EPI_ISL_2392406, EPI_ISL_2392416, EPI_ISL_2392417, EPI_ISL_2392419, EPI_ISL_2392424, EPI_ISL_2392432, EPI_ISL_2392437, EPI_ISL_2392453, EPI_ISL_2392466, EPI_ISL_2392488, EPI_ISL_2392528, EPI_ISL_2392530, EPI_ISL_2392539, EPI_ISL_2392544, EPI_ISL_2392546, EPI_ISL_2392566, EPI_ISL_2392630, EPI_ISL_2392650, EPI_ISL_2392655, EPI_ISL_2392667, EPI_ISL_2392688, EPI_ISL_2392698, EPI_ISL_2392714, EPI_ISL_2392730, EPI_ISL_2392736, EPI_ISL_2392750, EPI_ISL_2392754, EPI_ISL_2392952, EPI_ISL_2392955, EPI_ISL_2392966, EPI_ISL_2392967, EPI_ISL_2392968, EPI_ISL_2392976, EPI_ISL_2392983, EPI_ISL_2392986, EPI_ISL_2392989, EPI_ISL_2392998, EPI_ISL_2393000, EPI_ISL_2393008, EPI_ISL_2393011, EPI_ISL_2393026, EPI_ISL_2393030, EPI_ISL_2393035, EPI_ISL_2393038, EPI_ISL_2393045, EPI_ISL_2393052, EPI_ISL_2393072, EPI_ISL_2393076, EPI_ISL_2393078, EPI_ISL_2393083, EPI_ISL_2393092, EPI_ISL_2393095, EPI_ISL_2393099, EPI_ISL_2393106, EPI_ISL_2393118, EPI_ISL_2393120, EPI_ISL_2393128, EPI_ISL_2393136, EPI_ISL_2393160, EPI_ISL_2393162, EPI_ISL_2393166, EPI_ISL_2393167, EPI_ISL_2393173, EPI_ISL_2393193, EPI_ISL_2393196, EPI_ISL_2393209, EPI_ISL_2393210, EPI_ISL_2393212, EPI_ISL_2393247, EPI_ISL_2393248, EPI_ISL_2393256, EPI_ISL_2393257, EPI_ISL_2393270, EPI_ISL_2393283, EPI_ISL_2393306, EPI_ISL_2393307, EPI_ISL_2393316, EPI_ISL_2393322, EPI_ISL_2393325, EPI_ISL_2393344, EPI_ISL_2393361, EPI_ISL_2393380, EPI_ISL_2393385, EPI_ISL_2393386, EPI_ISL_2393388, EPI_ISL_2393391, EPI_ISL_2393399, EPI_ISL_2393404, EPI_ISL_2393409, EPI_ISL_2393415, EPI_ISL_2393427, EPI_ISL_2393443, EPI_ISL_2393535, EPI_ISL_2393547, EPI_ISL_2393580, EPI_ISL_2393596, EPI_ISL_2393609, EPI_ISL_2393617, EPI_ISL_2393632, EPI_ISL_2393637, EPI_ISL_2393643, EPI_ISL_2393648, EPI_ISL_2393675, EPI_ISL_2393698, EPI_ISL_2393706, EPI_ISL_2393720, EPI_ISL_2393732, EPI_ISL_2393744, EPI_ISL_2393760, EPI_ISL_2393774, EPI_ISL_2393777, EPI_ISL_2393782, EPI_ISL_2393788, EPI_ISL_2393793, EPI_ISL_2393797, EPI_ISL_2393811, EPI_ISL_2393825, EPI_ISL_2393852, EPI_ISL_2393862, EPI_ISL_2393866, EPI_ISL_2393878, EPI_ISL_2393920, EPI_ISL_2393927, EPI_ISL_2393936, EPI_ISL_2393947, EPI_ISL_2393952, EPI_ISL_2393964, EPI_ISL_2393987, EPI_ISL_2393989, EPI_ISL_2393990, EPI_ISL_2394005, EPI_ISL_2394028, EPI_ISL_2394037, EPI_ISL_2394038, EPI_ISL_2394044, EPI_ISL_2394049, EPI_ISL_2394057, EPI_ISL_2394078, EPI_ISL_2394084, EPI_ISL_2394096, EPI_ISL_2394100, EPI_ISL_2394101, EPI_ISL_2394103, EPI_ISL_2394105, EPI_ISL_2394108, EPI_ISL_2394118, EPI_ISL_2394130, EPI_ISL_2394138, EPI_ISL_2394139, EPI_ISL_2394153, EPI_ISL_2394163, EPI_ISL_2394164, EPI_ISL_2394167, EPI_ISL_2394171, EPI_ISL_2394199, EPI_ISL_2394203, EPI_ISL_2394220, EPI_ISL_2394221, EPI_ISL_2394222, EPI_ISL_2394229, EPI_ISL_2394263, EPI_ISL_2394292, EPI_ISL_2394293, EPI_ISL_2394298, EPI_ISL_2394338, EPI_ISL_2394344, EPI_ISL_2394345, EPI_ISL_2394347, EPI_ISL_2394352, EPI_ISL_2394367, EPI_ISL_2394380, EPI_ISL_2394386, EPI_ISL_2394391, EPI_ISL_2394401, EPI_ISL_2394412, EPI_ISL_2394416, EPI_ISL_2394429, EPI_ISL_2394434, EPI_ISL_2394440, EPI_ISL_2394472, EPI_ISL_2394510, EPI_ISL_2394525, EPI_ISL_2394532, EPI_ISL_2394541, EPI_ISL_2394545, EPI_ISL_2394562, EPI_ISL_2394564, EPI_ISL_2394575, EPI_ISL_2394579, EPI_ISL_2394587, EPI_ISL_2394594, EPI_ISL_2394599, EPI_ISL_2394621, EPI_ISL_2394627, EPI_ISL_2394630, EPI_ISL_2394646, EPI_ISL_2394656, EPI_ISL_2394657, EPI_ISL_2394666, EPI_ISL_2394675, EPI_ISL_2394709, EPI_ISL_2394712, EPI_ISL_2394743, EPI_ISL_2394745, EPI_ISL_2394752, EPI_ISL_2394770, EPI_ISL_2394771, EPI_ISL_2394775, EPI_ISL_2394777, EPI_ISL_2394790, EPI_ISL_2394796, EPI_ISL_2394798, EPI_ISL_2394801, EPI_ISL_2394812, EPI_ISL_2394822, EPI_ISL_2394827, EPI_ISL_2394837, EPI_ISL_2394842, EPI_ISL_2394854, EPI_ISL_2394855, EPI_ISL_2394862, EPI_ISL_2394864, EPI_ISL_2394897, EPI_ISL_2394915, EPI_ISL_2394921, EPI_ISL_2394935, EPI_ISL_2394941, EPI_ISL_2394942, EPI_ISL_2394968, EPI_ISL_2394989, EPI_ISL_2394994, EPI_ISL_2395006, EPI_ISL_2395021, EPI_ISL_2395024, EPI_ISL_2395027, EPI_ISL_2395068, EPI_ISL_2395087, EPI_ISL_2395092, EPI_ISL_2395094, EPI_ISL_2395106, EPI_ISL_2395107, EPI_ISL_2395125, EPI_ISL_2395127, EPI_ISL_2395135, EPI_ISL_2395136, EPI_ISL_2395144, EPI_ISL_2395145, EPI_ISL_2395149, EPI_ISL_2395160, EPI_ISL_2395214, EPI_ISL_2395220, EPI_ISL_2395233, EPI_ISL_2395237, EPI_ISL_2395241, EPI_ISL_2395244, EPI_ISL_2395256, EPI_ISL_2395265, EPI_ISL_2395267, EPI_ISL_2395275, EPI_ISL_2395294, EPI_ISL_2395306, EPI_ISL_2395312, EPI_ISL_2395317, EPI_ISL_2395322, EPI_ISL_2395368, EPI_ISL_2395381, EPI_ISL_2395400, EPI_ISL_2395456, EPI_ISL_2395478, EPI_ISL_2395491, EPI_ISL_2395587, EPI_ISL_2395675, EPI_ISL_2396029, EPI_ISL_2396030, EPI_ISL_2396032, EPI_ISL_2396035, EPI_ISL_2396046, EPI_ISL_2396062, EPI_ISL_2396064, EPI_ISL_2396075, EPI_ISL_2396076, EPI_ISL_2396079, EPI_ISL_2396082, EPI_ISL_2396083, EPI_ISL_2396107, EPI_ISL_2396115, EPI_ISL_2396125, EPI_ISL_2396127, EPI_ISL_2396128, EPI_ISL_2396133, EPI_ISL_2396137, EPI_ISL_2396141, EPI_ISL_2396142, EPI_ISL_2396143, EPI_ISL_2396146, EPI_ISL_2396147, EPI_ISL_2396157, EPI_ISL_2396161, EPI_ISL_2396164, EPI_ISL_2396165, EPI_ISL_2396169, EPI_ISL_2396175, EPI_ISL_2396176, EPI_ISL_2396177, EPI_ISL_2396179, EPI_ISL_2396184, EPI_ISL_2396185, EPI_ISL_2396195, EPI_ISL_2396200, EPI_ISL_2396203, EPI_ISL_2396205, EPI_ISL_2396210, EPI_ISL_2396212, EPI_ISL_2396217, EPI_ISL_2396223, EPI_ISL_2396227, EPI_ISL_2396232, EPI_ISL_2396233, EPI_ISL_2396237, EPI_ISL_2396238, EPI_ISL_2396248, EPI_ISL_2396253, EPI_ISL_2396256, EPI_ISL_2396259, EPI_ISL_2396263, EPI_ISL_2396270, EPI_ISL_2396273, EPI_ISL_2396274, EPI_ISL_2396276, EPI_ISL_2396278, EPI_ISL_2396280, EPI_ISL_2396282, EPI_ISL_2396290, EPI_ISL_2396296, EPI_ISL_2396297, EPI_ISL_2396302, EPI_ISL_2396305, EPI_ISL_2396309, EPI_ISL_2396314, EPI_ISL_2396319, EPI_ISL_2396321, EPI_ISL_2396322, EPI_ISL_2396324, EPI_ISL_2396334, EPI_ISL_2396336, EPI_ISL_2396342, EPI_ISL_2396344, EPI_ISL_2396798, EPI_ISL_2396809, EPI_ISL_2396817 | | | |
| see above | Lighthouse Lab in Alderley Park | Wellcome Sanger Institute for the COVID-19 Genomics UK (COG-UK) Consortium | Cordelia Langford; David K. Jackson; Dominic Kwiatkowski; Ewan Harrison; Ian Johnston; Jacquelyn Wynn; Jeffrey Barrett; John Sillitoe on behalf of the Wellcome Sanger Institute COVID-19 Surveillance Team; Mairead Hyland; Roberto Amato; Sonia Goncalves; The Lighthouse Lab in Alderley Park and Alex Alderton |
| EPI_ISL_2392593, EPI_ISL_2392626, EPI_ISL_2392710, EPI_ISL_2392716, EPI_ISL_2392785, EPI_ISL_2392972, EPI_ISL_2392995, EPI_ISL_2393114, EPI_ISL_2393555, EPI_ISL_2393663, EPI_ISL_2393683, EPI_ISL_2393701, EPI_ISL_2393773, EPI_ISL_2393798, EPI_ISL_2393824, EPI_ISL_2393847, EPI_ISL_2394045, EPI_ISL_2394069, EPI_ISL_2394094, EPI_ISL_2394316, EPI_ISL_2394417, EPI_ISL_2394462, EPI_ISL_2394467, EPI_ISL_2394893, EPI_ISL_2394991, EPI_ISL_2395109, EPI_ISL_2395150, EPI_ISL_2395273, EPI_ISL_2395309, EPI_ISL_2395337, EPI_ISL_2395385, EPI_ISL_2395406, EPI_ISL_2395416, EPI_ISL_2395424, EPI_ISL_2395431, EPI_ISL_2395448, EPI_ISL_2395454, EPI_ISL_2395483, EPI_ISL_2395531, EPI_ISL_2395537, EPI_ISL_2395539, EPI_ISL_2395558, EPI_ISL_2395586, EPI_ISL_2395602, EPI_ISL_2395606, EPI_ISL_2395650, EPI_ISL_2395660, EPI_ISL_2395661, EPI_ISL_2395667, EPI_ISL_2395685, EPI_ISL_2395690, EPI_ISL_2395692, EPI_ISL_2395694, EPI_ISL_2395695, EPI_ISL_2395701, EPI_ISL_2395708, EPI_ISL_2395709, EPI_ISL_2395711, EPI_ISL_2395715, EPI_ISL_2395722, EPI_ISL_2395728, EPI_ISL_2395729, EPI_ISL_2395730, EPI_ISL_2395745, EPI_ISL_2395747, EPI_ISL_2395752, EPI_ISL_2395758, EPI_ISL_2395761, EPI_ISL_2395764, EPI_ISL_2395766, EPI_ISL_2395771, EPI_ISL_2395773, EPI_ISL_2395775, EPI_ISL_2395787, EPI_ISL_2395788, EPI_ISL_2395792, EPI_ISL_2395793, EPI_ISL_2395794, EPI_ISL_2395800, EPI_ISL_2395806, EPI_ISL_2395808, EPI_ISL_2395810, EPI_ISL_2395816, EPI_ISL_2395823, EPI_ISL_2395826, EPI_ISL_2395829, EPI_ISL_2395838, EPI_ISL_2395841, EPI_ISL_2395842, EPI_ISL_2395846, EPI_ISL_2395855, EPI_ISL_2395859, EPI_ISL_2395865, EPI_ISL_2395866, EPI_ISL_2395867, EPI_ISL_2395871, EPI_ISL_2395873, EPI_ISL_2395879, EPI_ISL_2395885, EPI_ISL_2395888, EPI_ISL_2395894, EPI_ISL_2395896, EPI_ISL_2395900, EPI_ISL_2395903, EPI_ISL_2395906, EPI_ISL_2395911, EPI_ISL_2395913, EPI_ISL_2395914, EPI_ISL_2395916, EPI_ISL_2395919, EPI_ISL_2395921, EPI_ISL_2395925, EPI_ISL_2395931, EPI_ISL_2395932, EPI_ISL_2395960, EPI_ISL_2395962, EPI_ISL_2395970, EPI_ISL_2395980, EPI_ISL_2395993, EPI_ISL_2395997, EPI_ISL_2396000, EPI_ISL_2396001, EPI_ISL_2396004, EPI_ISL_2396008, EPI_ISL_2396345, EPI_ISL_2396346, EPI_ISL_2396347, EPI_ISL_2396348, EPI_ISL_2396349, EPI_ISL_2396350, EPI_ISL_2396351, EPI_ISL_2396352, EPI_ISL_2396353, EPI_ISL_2396354, EPI_ISL_2396356, EPI_ISL_2396359, EPI_ISL_2396361, EPI_ISL_2396362, EPI_ISL_2396363, EPI_ISL_2396364, EPI_ISL_2396367, EPI_ISL_2396368, EPI_ISL_2396369, EPI_ISL_2396371, EPI_ISL_2396372, EPI_ISL_2396373, EPI_ISL_2396374, EPI_ISL_2396376, EPI_ISL_2396378, EPI_ISL_2396379, EPI_ISL_2396381, EPI_ISL_2396382, EPI_ISL_2396383, EPI_ISL_2396384, EPI_ISL_2396385, EPI_ISL_2396386, EPI_ISL_2396387, EPI_ISL_2396391, EPI_ISL_2396393, EPI_ISL_2396395, EPI_ISL_2396396, EPI_ISL_2396398, EPI_ISL_2396400, EPI_ISL_2396402, EPI_ISL_2396403, EPI_ISL_2396405, EPI_ISL_2396409, EPI_ISL_2396410, EPI_ISL_2396411, EPI_ISL_2396413, EPI_ISL_2396415, EPI_ISL_2396416, EPI_ISL_2396417, EPI_ISL_2396419, EPI_ISL_2396420, EPI_ISL_2396421, EPI_ISL_2396423, EPI_ISL_2396426, EPI_ISL_2396429, EPI_ISL_2396430, EPI_ISL_2396432, EPI_ISL_2396433, EPI_ISL_2396434, EPI_ISL_2396435, EPI_ISL_2396436, EPI_ISL_2396437, EPI_ISL_2396438, EPI_ISL_2396439, EPI_ISL_2396440, EPI_ISL_2396441, EPI_ISL_2396442, EPI_ISL_2396443, EPI_ISL_2396444, EPI_ISL_2396445, EPI_ISL_2396446, EPI_ISL_2396447, EPI_ISL_2396448, EPI_ISL_2396449, EPI_ISL_2396450, EPI_ISL_2396451, EPI_ISL_2396453, EPI_ISL_2396455, EPI_ISL_2396458, EPI_ISL_2396459, EPI_ISL_2396460, EPI_ISL_2396461, EPI_ISL_2396463, EPI_ISL_2396465, EPI_ISL_2396467, EPI_ISL_2396468, EPI_ISL_2396469, EPI_ISL_2396473, EPI_ISL_2396474, EPI_ISL_2396475, EPI_ISL_2396478, EPI_ISL_2396479, EPI_ISL_2396480, EPI_ISL_2396481, EPI_ISL_2396482, EPI_ISL_2396483, EPI_ISL_2396485, EPI_ISL_2396486, EPI_ISL_2396487, EPI_ISL_2396488, EPI_ISL_2396492, EPI_ISL_2396494, EPI_ISL_2396497, EPI_ISL_2396498, EPI_ISL_2396499, EPI_ISL_2396500, EPI_ISL_2396501, EPI_ISL_2396502, EPI_ISL_2396504, EPI_ISL_2396505, EPI_ISL_2396506, EPI_ISL_2396507, EPI_ISL_2396510, EPI_ISL_2396511, EPI_ISL_2396512, EPI_ISL_2396513, EPI_ISL_2396523, EPI_ISL_2396526, EPI_ISL_2396530, EPI_ISL_2396533, EPI_ISL_2396537, EPI_ISL_2396538, EPI_ISL_2396539, EPI_ISL_2396541, EPI_ISL_2396542, EPI_ISL_2396547, EPI_ISL_2396548, EPI_ISL_2396551, EPI_ISL_2396552, EPI_ISL_2396556, EPI_ISL_2396564, EPI_ISL_2396565, EPI_ISL_2396566, EPI_ISL_2396571, EPI_ISL_2396572, EPI_ISL_2396579, EPI_ISL_2396584, EPI_ISL_2396587, EPI_ISL_2396591, EPI_ISL_2396595, EPI_ISL_2396596, EPI_ISL_2396599, EPI_ISL_2396601, EPI_ISL_2396610, EPI_ISL_2396614, EPI_ISL_2396615, EPI_ISL_2396621, EPI_ISL_2396625, EPI_ISL_2396627, EPI_ISL_2396630, EPI_ISL_2396632, EPI_ISL_2396640, EPI_ISL_2396641, EPI_ISL_2396645, EPI_ISL_2396661, EPI_ISL_2396672, EPI_ISL_2396673, EPI_ISL_2396674, EPI_ISL_2396676, EPI_ISL_2396678, EPI_ISL_2396685, EPI_ISL_2396692, EPI_ISL_2396693, EPI_ISL_2396694, EPI_ISL_2396699, EPI_ISL_2396716, EPI_ISL_2396717, EPI_ISL_2396721, EPI_ISL_2396724, EPI_ISL_2396731, EPI_ISL_2396732, EPI_ISL_2396733, EPI_ISL_2396736, EPI_ISL_2396739, EPI_ISL_2396748, EPI_ISL_2396750, EPI_ISL_2396755, EPI_ISL_2396757, EPI_ISL_2396758, EPI_ISL_2396759, EPI_ISL_2396760, EPI_ISL_2396764, EPI_ISL_2396767, EPI_ISL_2396770, EPI_ISL_2396773, EPI_ISL_2396774, EPI_ISL_2396775, EPI_ISL_2396776, EPI_ISL_2396777, EPI_ISL_2396780, EPI_ISL_2396781, EPI_ISL_2396782, EPI_ISL_2396783, EPI_ISL_2396784, EPI_ISL_2396785, EPI_ISL_2396788, EPI_ISL_2396789, EPI_ISL_2396793, EPI_ISL_2396795, EPI_ISL_2396796 | | | |
| see above | Lighthouse Lab in Glasgow | Wellcome Sanger Institute for the COVID-19 Genomics UK (COG-UK) Consortium | Anna Dominiczak and Alex Alderton; Carol Clugston; Cordelia Langford; David Gray; David K. Jackson; Dominic Kwiatkowski; Ewan Harrison; Harper VanSteenhouse; Ian Johnston; Jeffrey Barrett; John Sillitoe on behalf of the Wellcome Sanger Institute COVID-19 Surveillance Team; Roberto Amato; Sonia Goncalves; Yumi Kasai |
| EPI_ISL_2392249, EPI_ISL_2392279, EPI_ISL_2392323, EPI_ISL_2392420, EPI_ISL_2392442, EPI_ISL_2392522, EPI_ISL_2392592, EPI_ISL_2392595, EPI_ISL_2392618, EPI_ISL_2392627, EPI_ISL_2392641, EPI_ISL_2392705, EPI_ISL_2392737, EPI_ISL_2392759, EPI_ISL_2392771, EPI_ISL_2392780, EPI_ISL_2392781, EPI_ISL_2392782, EPI_ISL_2392794, EPI_ISL_2392810, EPI_ISL_2392817, EPI_ISL_2392818, EPI_ISL_2392822, EPI_ISL_2392832, EPI_ISL_2392835, EPI_ISL_2392839, EPI_ISL_2392851, EPI_ISL_2392852, EPI_ISL_2392855, EPI_ISL_2392859, EPI_ISL_2392861, EPI_ISL_2392879, EPI_ISL_2392898, EPI_ISL_2392906, EPI_ISL_2392909, EPI_ISL_2392917, EPI_ISL_2392922, EPI_ISL_2392929, EPI_ISL_2392935, EPI_ISL_2392936, EPI_ISL_2392937, EPI_ISL_2392942, EPI_ISL_2393141, EPI_ISL_2393142, EPI_ISL_2393152, EPI_ISL_2393191, EPI_ISL_2393205, EPI_ISL_2393208, EPI_ISL_2393214, EPI_ISL_2393236, EPI_ISL_2393249, EPI_ISL_2393267, EPI_ISL_2393296, EPI_ISL_2393303, EPI_ISL_2393315, EPI_ISL_2393317, EPI_ISL_2393324, EPI_ISL_2393337, EPI_ISL_2393345, EPI_ISL_2393383, EPI_ISL_2393384, EPI_ISL_2393393, EPI_ISL_2393394, EPI_ISL_2393413, EPI_ISL_2393425, EPI_ISL_2393430, EPI_ISL_2393437, EPI_ISL_2393439, EPI_ISL_2393475, EPI_ISL_2393486, EPI_ISL_2393537, EPI_ISL_2393559, EPI_ISL_2393565, EPI_ISL_2393566, EPI_ISL_2393569, EPI_ISL_2393592, EPI_ISL_2393599, EPI_ISL_2393620, EPI_ISL_2393621, EPI_ISL_2393666, EPI_ISL_2393703, EPI_ISL_2393710, EPI_ISL_2393716, EPI_ISL_2393719, EPI_ISL_2393726, EPI_ISL_2393727, EPI_ISL_2393746, EPI_ISL_2393763, EPI_ISL_2393765, EPI_ISL_2393786, EPI_ISL_2393787, EPI_ISL_2393826, EPI_ISL_2393843, EPI_ISL_2393849, EPI_ISL_2393861, EPI_ISL_2393865, EPI_ISL_2393886, EPI_ISL_2393889, EPI_ISL_2393896, EPI_ISL_2393898, EPI_ISL_2393903, EPI_ISL_2393913, EPI_ISL_2393916, EPI_ISL_2393917, EPI_ISL_2393929, EPI_ISL_2393935, EPI_ISL_2393939, EPI_ISL_2393940, EPI_ISL_2393944, EPI_ISL_2393946, EPI_ISL_2393954, EPI_ISL_2393969, EPI_ISL_2393977, EPI_ISL_2394016, EPI_ISL_2394026, EPI_ISL_2394876, EPI_ISL_2394895, EPI_ISL_2394901, EPI_ISL_2394903, EPI_ISL_2394927, EPI_ISL_2394930, EPI_ISL_2394936, EPI_ISL_2394937, EPI_ISL_2394944, EPI_ISL_2394953, EPI_ISL_2394978, EPI_ISL_2395001, EPI_ISL_2395008, EPI_ISL_2395010, EPI_ISL_2395013, EPI_ISL_2395017, EPI_ISL_2395020, EPI_ISL_2395026, EPI_ISL_2395029, EPI_ISL_2395034, EPI_ISL_2395347, EPI_ISL_2395349, EPI_ISL_2395351, EPI_ISL_2395355, EPI_ISL_2395369, EPI_ISL_2395370, EPI_ISL_2395371, EPI_ISL_2395373, EPI_ISL_2395384, EPI_ISL_2395395, EPI_ISL_2395397, EPI_ISL_2395412, EPI_ISL_2395415, EPI_ISL_2395421, EPI_ISL_2395434, EPI_ISL_2395443, EPI_ISL_2395445, EPI_ISL_2395447, EPI_ISL_2395452, EPI_ISL_2395479, EPI_ISL_2395487, EPI_ISL_2395489, EPI_ISL_2395508, EPI_ISL_2395512, EPI_ISL_2395523, EPI_ISL_2395534, EPI_ISL_2395540, EPI_ISL_2395542, EPI_ISL_2395550, EPI_ISL_2395565, EPI_ISL_2395570, EPI_ISL_2395574, EPI_ISL_2395577, EPI_ISL_2395582, EPI_ISL_2395583, EPI_ISL_2395585, EPI_ISL_2395590, EPI_ISL_2395591, EPI_ISL_2395593, EPI_ISL_2395619, EPI_ISL_2395630, EPI_ISL_2395644, EPI_ISL_2395655, EPI_ISL_2395663, EPI_ISL_2395673, EPI_ISL_2395684, EPI_ISL_2395686, EPI_ISL_2395698, EPI_ISL_2395699, EPI_ISL_2395706, EPI_ISL_2395710, EPI_ISL_2395713, EPI_ISL_2395725, EPI_ISL_2395735, EPI_ISL_2395746, EPI_ISL_2395750, EPI_ISL_2395768, EPI_ISL_2395778, EPI_ISL_2395783, EPI_ISL_2395785, EPI_ISL_2395790, EPI_ISL_2395802, EPI_ISL_2395805, EPI_ISL_2395809, EPI_ISL_2395819, EPI_ISL_2395852, EPI_ISL_2395861, EPI_ISL_2395887, EPI_ISL_2395890, EPI_ISL_2395891, EPI_ISL_2395892, EPI_ISL_2395901, EPI_ISL_2395904, EPI_ISL_2395910, EPI_ISL_2395920, EPI_ISL_2395927, EPI_ISL_2395942, EPI_ISL_2395963, EPI_ISL_2395979, EPI_ISL_2395998, EPI_ISL_2396007, EPI_ISL_2396011, EPI_ISL_2396014, EPI_ISL_2396586, EPI_ISL_2396810, EPI_ISL_2396818 | | | |
| see above | Lighthouse Lab in Milton Keynes | Wellcome Sanger Institute for the COVID-19 Genomics UK (COG-UK) Consortium | Cordelia Langford; David K. Jackson; Dominic Kwiatkowski; Ewan Harrison; Ian Johnston; Jeffrey Barrett; John Sillitoe on behalf of the Wellcome Sanger Institute COVID-19 Surveillance Team; Roberto Amato; Sonia Goncalves; The Lighthouse Lab in Milton Keynes and Alex Alderton |
| EPI_ISL_2393159, EPI_ISL_2393189, EPI_ISL_2393211, EPI_ISL_2393215, EPI_ISL_2393219, EPI_ISL_2393228, EPI_ISL_2393240, EPI_ISL_2393243, EPI_ISL_2393244, EPI_ISL_2393245, EPI_ISL_2393279, EPI_ISL_2393293, EPI_ISL_2393304, EPI_ISL_2393342, EPI_ISL_2393347, EPI_ISL_2393353, EPI_ISL_2393368, EPI_ISL_2393406 | | | |
| see above | Lighthouse Laboratory Plymouth | Wellcome Sanger Institute for the COVID-19 Genomics UK (COG-UK) Consortium | Cordelia Langford; David K. Jackson; Dominic Kwiatkowski; Ewan Harrison; Ian Johnston; Jeffrey Barrett; John Sillitoe on behalf of the Wellcome Sanger Institute COVID-19 Surveillance Team; Lighthouse Laboratory Plymouth and Alex Alderton; Roberto Amato; Sonia Goncalves |
| EPI_ISL_2393158, EPI_ISL_2393185, EPI_ISL_2393229, EPI_ISL_2393273, EPI_ISL_2393287, EPI_ISL_2393298, EPI_ISL_2393336, EPI_ISL_2393449, EPI_ISL_2393471, EPI_ISL_2396707, EPI_ISL_2396711, EPI_ISL_2396718, EPI_ISL_2396800, EPI_ISL_2396801, EPI_ISL_2396802, EPI_ISL_2396805, EPI_ISL_2396806, EPI_ISL_2396807, EPI_ISL_2396808, EPI_ISL_2396811, EPI_ISL_2396819, EPI_ISL_2396821, EPI_ISL_2396822, EPI_ISL_2396823, EPI_ISL_2396824, EPI_ISL_2396825, EPI_ISL_2396826, EPI_ISL_2396827, EPI_ISL_2396829, EPI_ISL_2396830, EPI_ISL_2396831, EPI_ISL_2396832, EPI_ISL_2396833, EPI_ISL_2396835, EPI_ISL_2396836, EPI_ISL_2396837, EPI_ISL_2396839, EPI_ISL_2396840, EPI_ISL_2396841, EPI_ISL_2396842, EPI_ISL_2396843, EPI_ISL_2396844, EPI_ISL_2396845, EPI_ISL_2396846, EPI_ISL_2396847, EPI_ISL_2396848, EPI_ISL_2396849, EPI_ISL_2396850, EPI_ISL_2396852, EPI_ISL_2396853, EPI_ISL_2396854, EPI_ISL_2396855, EPI_ISL_2396856, EPI_ISL_2396857, EPI_ISL_2396858, EPI_ISL_2396859, EPI_ISL_2396860, EPI_ISL_2396861, EPI_ISL_2396862, EPI_ISL_2396863 | | | |
| see above | Randox Laboratories | Wellcome Sanger Institute for the COVID-19 Genomics UK (COG-UK) Consortium | Cordelia Langford; David K. Jackson; Dominic Kwiatkowski; Ewan Harrison; Ian Johnston; Jeffrey Barrett; John Sillitoe on behalf of the Wellcome Sanger Institute COVID-19 Surveillance Team; Randox Laboratories and Alex Alderton; Roberto Amato; Sonia Goncalves |

We gratefully acknowledge the following Authors from the Originating laboratories responsible for obtaining the specimens, as well as the Submitting laboratories where the genome data were generated and shared via GISAID, on which this research is based.

All Submitters of data may be contacted directly via www.gisaid.org

Authors are sorted alphabetically.

| Accession ID | Originating Laboratory | Submitting Laboratory | Authors |
|---|---|---|---|
| EPI_ISL_2785045 | Aegis Sciences Corporation | Centers for Disease Control and Prevention Division of Viral Diseases, Pathogen Discovery | Adrian Paskey; Alec Vest; Benjamin Rambo-Martin; Christopher Gulvick; Clinton R. Paden; Cyndi Clark; Dakota Howard; Darlene Wagner; Dhwani Batra; Dillon Nall; Duncan MacCannell; Ethan Sanders; Holly Houdeshell; Jason Caravas; Kara Moser; Matthew Hardison; Matthew Schmerer; Ola Kvalvaag; Patrick Campbell; Peter W. Cook; Rob Case; Scott Sammons; Shatavia Morrison; Shaun Westlund; Vikramsinha Ghorpade; Yvette Unoarumhi |
| EPI_ISL_2758471 | Alameda County Public Health Lab | Chan-Zuckerberg Biohub | CZB Cliahub Consortium |
| EPI_ISL_2782391, EPI_ISL_2782393, EPI_ISL_2782394, EPI_ISL_2782400, EPI_ISL_2782403, EPI_ISL_2782431, EPI_ISL_2782439, EPI_ISL_2782441, EPI_ISL_2782445, EPI_ISL_2782448, EPI_ISL_2782461, EPI_ISL_2782473, EPI_ISL_2782475, EPI_ISL_2782476, EPI_ISL_2782480, EPI_ISL_2782481, EPI_ISL_2782484, EPI_ISL_2782486, EPI_ISL_2782488, EPI_ISL_2782499, EPI_ISL_2782504, EPI_ISL_2782505, EPI_ISL_2782528, EPI_ISL_2782539, EPI_ISL_2782554, EPI_ISL_2782557, EPI_ISL_2782564, EPI_ISL_2782565, EPI_ISL_2782570, EPI_ISL_2782573, EPI_ISL_2782576, EPI_ISL_2782583, EPI_ISL_2782587, EPI_ISL_2782596, EPI_ISL_2782597, EPI_ISL_2782604, EPI_ISL_2782609, EPI_ISL_2782615, EPI_ISL_2782619, EPI_ISL_2782620, EPI_ISL_2782621, EPI_ISL_2782623, EPI_ISL_2782624, EPI_ISL_2782634, EPI_ISL_2782635, EPI_ISL_2782637, EPI_ISL_2782639, EPI_ISL_2782641, EPI_ISL_2782655, EPI_ISL_2782660, EPI_ISL_2782662, EPI_ISL_2782675, EPI_ISL_2782679, EPI_ISL_2782690, EPI_ISL_2782691, EPI_ISL_2782697, EPI_ISL_2782703, EPI_ISL_2782705, EPI_ISL_2782706, EPI_ISL_2782707, EPI_ISL_2782710, EPI_ISL_2782717, EPI_ISL_2782718, EPI_ISL_2782719, EPI_ISL_2782721, EPI_ISL_2782724, EPI_ISL_2782726, EPI_ISL_2782738, EPI_ISL_2782739, EPI_ISL_2782755, EPI_ISL_2782772, EPI_ISL_2782779, EPI_ISL_2782781, EPI_ISL_2782784, EPI_ISL_2782792, EPI_ISL_2782795, EPI_ISL_2782807, EPI_ISL_2782810, EPI_ISL_2782823, EPI_ISL_2782825, EPI_ISL_2782826, EPI_ISL_2782835, EPI_ISL_2782837, EPI_ISL_2782844, EPI_ISL_2782849, EPI_ISL_2782855, EPI_ISL_2782860, EPI_ISL_2782867, EPI_ISL_2782872, EPI_ISL_2782888, EPI_ISL_2782895, EPI_ISL_2782896, EPI_ISL_2782905, EPI_ISL_2782923, EPI_ISL_2782924, EPI_ISL_2782925, EPI_ISL_2782930, EPI_ISL_2782940, EPI_ISL_2782941, EPI_ISL_2782948, EPI_ISL_2782951, EPI_ISL_2782959, EPI_ISL_2782963, EPI_ISL_2782965, EPI_ISL_2782966, EPI_ISL_2782968, EPI_ISL_2782969, EPI_ISL_2782976, EPI_ISL_2782979, EPI_ISL_2782999, EPI_ISL_2783000, EPI_ISL_2783003, EPI_ISL_2783010, EPI_ISL_2783012, EPI_ISL_2783024, EPI_ISL_2783041, EPI_ISL_2783042, EPI_ISL_2783060, EPI_ISL_2783061, EPI_ISL_2783062, EPI_ISL_2783077, EPI_ISL_2783085, EPI_ISL_2783087, EPI_ISL_2783096, EPI_ISL_2783099, EPI_ISL_2783105, EPI_ISL_2783106, EPI_ISL_2783108, EPI_ISL_2783109, EPI_ISL_2783112, EPI_ISL_2783118, EPI_ISL_2783119, EPI_ISL_2783127, EPI_ISL_2783137, EPI_ISL_2783140, EPI_ISL_2783155, EPI_ISL_2783159, EPI_ISL_2783164, EPI_ISL_2783167, EPI_ISL_2783168, EPI_ISL_2783176, EPI_ISL_2783177, EPI_ISL_2783181, EPI_ISL_2783188, EPI_ISL_2783194, EPI_ISL_2783222, EPI_ISL_2783244, EPI_ISL_2783254, EPI_ISL_2783259, EPI_ISL_2783261, EPI_ISL_2783269, EPI_ISL_2783275, EPI_ISL_2783278, EPI_ISL_2783280, EPI_ISL_2783284, EPI_ISL_2783295, EPI_ISL_2783300, EPI_ISL_2783308, EPI_ISL_2783310, EPI_ISL_2783312, EPI_ISL_2783326, EPI_ISL_2783341, EPI_ISL_2783347, EPI_ISL_2783358, EPI_ISL_2783361, EPI_ISL_2783362, EPI_ISL_2783365, EPI_ISL_2783372, EPI_ISL_2783374, EPI_ISL_2783379, EPI_ISL_2783383, EPI_ISL_2783392, EPI_ISL_2783393, EPI_ISL_2783396, EPI_ISL_2783406, EPI_ISL_2783408, EPI_ISL_2783409, EPI_ISL_2783427, EPI_ISL_2783432, EPI_ISL_2783435, EPI_ISL_2783440, EPI_ISL_2783445, EPI_ISL_2783454, EPI_ISL_2783460, EPI_ISL_2783463, EPI_ISL_2783466, EPI_ISL_2783468, EPI_ISL_2783469, EPI_ISL_2783471, EPI_ISL_2783473, EPI_ISL_2783474, EPI_ISL_2783477, EPI_ISL_2783480, EPI_ISL_2783482, EPI_ISL_2783484, EPI_ISL_2783488, EPI_ISL_2783499, EPI_ISL_2783501, EPI_ISL_2783503, EPI_ISL_2783504, EPI_ISL_2783506, EPI_ISL_2783507, EPI_ISL_2783508, EPI_ISL_2796532, EPI_ISL_2796534, EPI_ISL_2796540, EPI_ISL_2796541, EPI_ISL_2796542, EPI_ISL_2796544, EPI_ISL_2796545, EPI_ISL_2796548, EPI_ISL_2796564, EPI_ISL_2796570, EPI_ISL_2796577, EPI_ISL_2796596, EPI_ISL_2796600, EPI_ISL_2796603, EPI_ISL_2796604, EPI_ISL_2796608, EPI_ISL_2796613, EPI_ISL_2796614, EPI_ISL_2796615, EPI_ISL_2796617, EPI_ISL_2796618, EPI_ISL_2796622, EPI_ISL_2796634, EPI_ISL_2796635, EPI_ISL_2796647, EPI_ISL_2796648, EPI_ISL_2796652, EPI_ISL_2796654, EPI_ISL_2796656, EPI_ISL_2796658, EPI_ISL_2796662, EPI_ISL_2796665, EPI_ISL_2796672, EPI_ISL_2796688, EPI_ISL_2796689, EPI_ISL_2796693, EPI_ISL_2796696, EPI_ISL_2796697, EPI_ISL_2796700, EPI_ISL_2796703, EPI_ISL_2796712, EPI_ISL_2796718, EPI_ISL_2796728, EPI_ISL_2796731, EPI_ISL_2796736, EPI_ISL_2796738, EPI_ISL_2796741, EPI_ISL_2796742, EPI_ISL_2796743, EPI_ISL_2796745, EPI_ISL_2796751, EPI_ISL_2796767, EPI_ISL_2796768, EPI_ISL_2796769, EPI_ISL_2796770, EPI_ISL_2796771, EPI_ISL_2796783, EPI_ISL_2796787, EPI_ISL_2796792, EPI_ISL_2796795, EPI_ISL_2796799, EPI_ISL_2796806, EPI_ISL_2796808, EPI_ISL_2796810, EPI_ISL_2796817, EPI_ISL_2796821, EPI_ISL_2796822, EPI_ISL_2796827, EPI_ISL_2796847, EPI_ISL_2796869, EPI_ISL_2796872, EPI_ISL_2796875, EPI_ISL_2796876, EPI_ISL_2796877, EPI_ISL_2796878, EPI_ISL_2796880, EPI_ISL_2796886, EPI_ISL_2796889, EPI_ISL_2796890, EPI_ISL_2796897, EPI_ISL_2796900, EPI_ISL_2796902, EPI_ISL_2796906, EPI_ISL_2796908, EPI_ISL_2796913, EPI_ISL_2796917, EPI_ISL_2796919, EPI_ISL_2796921, EPI_ISL_2796922, EPI_ISL_2796931, EPI_ISL_2796945, EPI_ISL_2796948, EPI_ISL_2796950, EPI_ISL_2796956, EPI_ISL_2796959, EPI_ISL_2796961, EPI_ISL_2796962, EPI_ISL_2796965, EPI_ISL_2796968, EPI_ISL_2796973, EPI_ISL_2796974, EPI_ISL_2796975, EPI_ISL_2796981, EPI_ISL_2796988, EPI_ISL_2797004, EPI_ISL_2797006, EPI_ISL_2797014, EPI_ISL_2797016, EPI_ISL_2797021, EPI_ISL_2797023, EPI_ISL_2797031, EPI_ISL_2797033, EPI_ISL_2797037, EPI_ISL_2797040, EPI_ISL_2797054, EPI_ISL_2797057, EPI_ISL_2797069, EPI_ISL_2797084, EPI_ISL_2797085, EPI_ISL_2797103, EPI_ISL_2797105, EPI_ISL_2797121, EPI_ISL_2797122, EPI_ISL_2797125, EPI_ISL_2797130, EPI_ISL_2797131, EPI_ISL_2797160, EPI_ISL_2797161, EPI_ISL_2797166, EPI_ISL_2797171, EPI_ISL_2797172, EPI_ISL_2797176, EPI_ISL_2797201, EPI_ISL_2797202, EPI_ISL_2797211, EPI_ISL_2797212, EPI_ISL_2797214, EPI_ISL_2797215, EPI_ISL_2797228, EPI_ISL_2797235, EPI_ISL_2797236, EPI_ISL_2797241, EPI_ISL_2797250, EPI_ISL_2797282, EPI_ISL_2797285, EPI_ISL_2797288, EPI_ISL_2797295, EPI_ISL_2797297, EPI_ISL_2797302, EPI_ISL_2797303, EPI_ISL_2797305, EPI_ISL_2797307, EPI_ISL_2797337, EPI_ISL_2797356, EPI_ISL_2797365, EPI_ISL_2797387, EPI_ISL_2797391, EPI_ISL_2797402, EPI_ISL_2797404, EPI_ISL_2797414, EPI_ISL_2797420, EPI_ISL_2797423, EPI_ISL_2797429, EPI_ISL_2797441, EPI_ISL_2797445, EPI_ISL_2797446, EPI_ISL_2797456, EPI_ISL_2797459, EPI_ISL_2797461, EPI_ISL_2797464, EPI_ISL_2797466, EPI_ISL_2797470, EPI_ISL_2797472, EPI_ISL_2797483, EPI_ISL_2797493, EPI_ISL_2797495, EPI_ISL_2797503, EPI_ISL_2797506, EPI_ISL_2797520, EPI_ISL_2797521, EPI_ISL_2797527, EPI_ISL_2797535, EPI_ISL_2797536, EPI_ISL_2797538, EPI_ISL_2797541, EPI_ISL_2797543, EPI_ISL_2797547, EPI_ISL_2797548, EPI_ISL_2797549, EPI_ISL_2797551, EPI_ISL_2797559, EPI_ISL_2797566, EPI_ISL_2797570, EPI_ISL_2797572, EPI_ISL_2797573, EPI_ISL_2797576, EPI_ISL_2797578, EPI_ISL_2797584, EPI_ISL_2797586, EPI_ISL_2797587, EPI_ISL_2797594, EPI_ISL_2797608, EPI_ISL_2797610, EPI_ISL_2797615, EPI_ISL_2797628, EPI_ISL_2797630, EPI_ISL_2797631, EPI_ISL_2797638, EPI_ISL_2797639, EPI_ISL_2797647, EPI_ISL_2797654, EPI_ISL_2797655, EPI_ISL_2797658, EPI_ISL_2797662, EPI_ISL_2797665, EPI_ISL_2797691, EPI_ISL_2797701, EPI_ISL_2797702, EPI_ISL_2797710, EPI_ISL_2797713, EPI_ISL_2797714, EPI_ISL_2797721, EPI_ISL_2797726, EPI_ISL_2797732, EPI_ISL_2797733, EPI_ISL_2797742, EPI_ISL_2797744, EPI_ISL_2797751, EPI_ISL_2797754, EPI_ISL_2797763, EPI_ISL_2797766, EPI_ISL_2797782, EPI_ISL_2797785, EPI_ISL_2797792, EPI_ISL_2797794, EPI_ISL_2797798, EPI_ISL_2797805, EPI_ISL_2797813, EPI_ISL_2797816, EPI_ISL_2797826, EPI_ISL_2797834, EPI_ISL_2797838, EPI_ISL_2797850, EPI_ISL_2797856, EPI_ISL_2797868, EPI_ISL_2797873, EPI_ISL_2797875, EPI_ISL_2797877, EPI_ISL_2797880, EPI_ISL_2797882, EPI_ISL_2797885, EPI_ISL_2797892, EPI_ISL_2797900, EPI_ISL_2797909, EPI_ISL_2797914, EPI_ISL_2797929, EPI_ISL_2797935, EPI_ISL_2797940, EPI_ISL_2797942, EPI_ISL_2797943, EPI_ISL_2797957, EPI_ISL_2797958, EPI_ISL_2797960, EPI_ISL_2797971, EPI_ISL_2797980, EPI_ISL_2797981, EPI_ISL_2797985, EPI_ISL_2797992, EPI_ISL_2798009, EPI_ISL_2798020, EPI_ISL_2798022, EPI_ISL_2798023, EPI_ISL_2798027, EPI_ISL_2798029, EPI_ISL_2798031, EPI_ISL_2798051, EPI_ISL_2798053, EPI_ISL_2798054, EPI_ISL_2798064, EPI_ISL_2798065, EPI_ISL_2798068, EPI_ISL_2798070, EPI_ISL_2798073, EPI_ISL_2798079, EPI_ISL_2798082, EPI_ISL_2798088, EPI_ISL_2798098, EPI_ISL_2798131, EPI_ISL_2798133, EPI_ISL_2798136, EPI_ISL_2798144, EPI_ISL_2798156, EPI_ISL_2798176 | | | |
| see above | CDPH VBL | California Department of Public Health | CDPH-COVIDNet; UCI Genome Sciences Center/GHTF; UCSF Center for Advanced Technology |
| EPI_ISL_2776227, EPI_ISL_2776239, EPI_ISL_2776244, EPI_ISL_2776256, EPI_ISL_2776260, EPI_ISL_2776267, EPI_ISL_2776271, EPI_ISL_2776272, EPI_ISL_2776277, EPI_ISL_2776279, EPI_ISL_2776285, EPI_ISL_2776286, EPI_ISL_2776293, EPI_ISL_2776294, EPI_ISL_2776298, EPI_ISL_2776303, EPI_ISL_2776304, EPI_ISL_2776307, EPI_ISL_2776310, EPI_ISL_2776312, EPI_ISL_2776313, EPI_ISL_2776315, EPI_ISL_2776319, EPI_ISL_2776322, EPI_ISL_2776326, EPI_ISL_2776333, EPI_ISL_2776339, EPI_ISL_2776343, EPI_ISL_2776344, EPI_ISL_2776349, EPI_ISL_2776350, EPI_ISL_2776356, EPI_ISL_2776358, EPI_ISL_2776363, EPI_ISL_2776367, EPI_ISL_2778105, EPI_ISL_2778106, EPI_ISL_2778108, EPI_ISL_2778111, EPI_ISL_2778118, EPI_ISL_2778121, EPI_ISL_2778124, EPI_ISL_2778134, EPI_ISL_2778142, EPI_ISL_2778146, EPI_ISL_2778151, EPI_ISL_2778155, EPI_ISL_2778177, EPI_ISL_2778181, EPI_ISL_2778186, EPI_ISL_2778194, EPI_ISL_2778195, EPI_ISL_2778198, EPI_ISL_2778199, EPI_ISL_2778200, EPI_ISL_2778201, EPI_ISL_2778202, EPI_ISL_2778211, EPI_ISL_2778212, EPI_ISL_2778226, EPI_ISL_2778230, EPI_ISL_2778234, EPI_ISL_2778235, EPI_ISL_2778241, EPI_ISL_2778244, EPI_ISL_2778252, EPI_ISL_2778257, EPI_ISL_2778274, EPI_ISL_2778283, EPI_ISL_2778285, EPI_ISL_2778290, EPI_ISL_2778301, EPI_ISL_2778308, EPI_ISL_2778309, EPI_ISL_2778311, EPI_ISL_2778322, EPI_ISL_2778337, EPI_ISL_2778366, EPI_ISL_2778370, EPI_ISL_2778372, EPI_ISL_2778375, EPI_ISL_2778381, EPI_ISL_2778384, EPI_ISL_2778387, EPI_ISL_2778390, EPI_ISL_2778392, EPI_ISL_2778397, EPI_ISL_2778398, EPI_ISL_2778399, EPI_ISL_2778401, EPI_ISL_2778403, EPI_ISL_2778421, EPI_ISL_2778422, EPI_ISL_2778424, EPI_ISL_2778427, EPI_ISL_2778441, EPI_ISL_2778444, EPI_ISL_2778452, EPI_ISL_2778453, EPI_ISL_2778471, EPI_ISL_2778476, EPI_ISL_2778485, EPI_ISL_2778487, EPI_ISL_2778494, EPI_ISL_2778496, EPI_ISL_2778578, EPI_ISL_2778579, EPI_ISL_2778582, EPI_ISL_2778583, EPI_ISL_2778584, EPI_ISL_2778603, EPI_ISL_2778612, EPI_ISL_2778616, EPI_ISL_2778619, EPI_ISL_2778623, EPI_ISL_2778624, EPI_ISL_2778631, EPI_ISL_2778692, EPI_ISL_2778744, EPI_ISL_2778828 | | | |
| see above | California Department of Public Health Valencia Branch Laboratory (CDPH VBL) | California Department of Public Health | Arias Lab at UCSB; CDPH-COVIDNet; UCLA Technology Center for Genomics & Bioinformatics |
| EPI_ISL_2758456 | Contra Costa County Public Health Lab | Chan-Zuckerberg Biohub | CZB Cliahub Consortium |
| EPI_ISL_2758597, EPI_ISL_2758599 | County of San Luis Obispo Public Health Laboratory | County of San Luis Obispo Public Health Laboratory | Frances Sidhu; Katrina Erwin |
| EPI_ISL_2783620 | Curative Labs | Curative Labs | Elias L. Salfati; Eugenia Khorosheva; George Way; J.Cesar Ignacio-Espinoza; Janet Chen; Mikhail Hanewich-Hollatz; Nabjot Sandhu; Sophia Quasem; Vladimir Slepnev; Zhiyi Xie |
| EPI_ISL_2758427, EPI_ISL_2758428 | Humboldt County Public Health Laboratory | Chan-Zuckerberg Biohub | CZB Cliahub Consortium |
| EPI_ISL_2801477 | IVY3 Central Lab, Vanderbilt University Medical Center | Lauring Lab, University of Michigan, Department of Microbiology and Immunology | Valesano |
| EPI_ISL_2800948, EPI_ISL_2800950, EPI_ISL_2800954, EPI_ISL_2800967, EPI_ISL_2800986, EPI_ISL_2800996, EPI_ISL_2800998, EPI_ISL_2801006, EPI_ISL_2801007, EPI_ISL_2801008, EPI_ISL_2801013, EPI_ISL_2801014, EPI_ISL_2801033, EPI_ISL_2801102, EPI_ISL_2801108, EPI_ISL_2801131, EPI_ISL_2801155, EPI_ISL_2801171, EPI_ISL_2801187, EPI_ISL_2801193, EPI_ISL_2801196, EPI_ISL_2801197, EPI_ISL_2801199, EPI_ISL_2801204, EPI_ISL_2801205, EPI_ISL_2801214, EPI_ISL_2801215, EPI_ISL_2801216, EPI_ISL_2801217, EPI_ISL_2801218, EPI_ISL_2801219, EPI_ISL_2801220, EPI_ISL_2801222, EPI_ISL_2801229, EPI_ISL_2801235, EPI_ISL_2801237, EPI_ISL_2801241, EPI_ISL_2801242, EPI_ISL_2801244, EPI_ISL_2801248, EPI_ISL_2801254, EPI_ISL_2801255, EPI_ISL_2801257, EPI_ISL_2801258, EPI_ISL_2801259, EPI_ISL_2801260, EPI_ISL_2801276, EPI_ISL_2801279, EPI_ISL_2801284, EPI_ISL_2801289, EPI_ISL_2801291, EPI_ISL_2801292, EPI_ISL_2801294, EPI_ISL_2801296, EPI_ISL_2801300, EPI_ISL_2801301, EPI_ISL_2801302 | | | |
| see above | Illinois Department of Public Health | Gagnon Lab, Southern Illinois University | Keith Gagnon |
| EPI_ISL_2774565, EPI_ISL_2774566 | Los Angeles County Public Health Laboratories | Los Angeles County Public Health Laboratories | P. Hemarajata et al. |
| EPI_ISL_2790067, EPI_ISL_2790069, EPI_ISL_2790070, EPI_ISL_2790071, EPI_ISL_2790072, EPI_ISL_2790077, EPI_ISL_2790080 | | | |
| see above | MD PHL | MD PHL | Maryland Department of Health Laboratories Administration |
| EPI_ISL_2802202, EPI_ISL_2802203, EPI_ISL_2802207, EPI_ISL_2802215, EPI_ISL_2802218, EPI_ISL_2802219, EPI_ISL_2802221, EPI_ISL_2802222, EPI_ISL_2802246, EPI_ISL_2802249, EPI_ISL_2802251, EPI_ISL_2802252, EPI_ISL_2802266, EPI_ISL_2802272, EPI_ISL_2802273, EPI_ISL_2802274, EPI_ISL_2802276, EPI_ISL_2802280, EPI_ISL_2802288, EPI_ISL_2802289, EPI_ISL_2802290, EPI_ISL_2802291, EPI_ISL_2802384, EPI_ISL_2802397, EPI_ISL_2802634, EPI_ISL_2802686 | | | |
| see above | Maine Health and Environmental Testing Laboratory | Tewhey Lab, The Jackson Laboratory | Barter, M.; Dewey, H.; H. and Tewhey, R.; Iosue, F.; Lynch, R.; Matluk, N.; Munger |
| EPI_ISL_2776388, EPI_ISL_2776389, EPI_ISL_2776390, EPI_ISL_2776391, EPI_ISL_2776392, EPI_ISL_2776393, EPI_ISL_2776394, EPI_ISL_2776395, EPI_ISL_2776396, EPI_ISL_2776397, EPI_ISL_2776399, EPI_ISL_2776400, EPI_ISL_2776401, EPI_ISL_2776402, EPI_ISL_2776404, EPI_ISL_2776405, EPI_ISL_2776406, EPI_ISL_2776407, EPI_ISL_2776410, EPI_ISL_2776411, EPI_ISL_2776412, EPI_ISL_2776413, EPI_ISL_2776415, EPI_ISL_2776416, EPI_ISL_2776418, EPI_ISL_2776419, EPI_ISL_2776420, EPI_ISL_2776421, EPI_ISL_2776422, EPI_ISL_2776423, EPI_ISL_2776424, EPI_ISL_2776425, EPI_ISL_2776426, EPI_ISL_2776427, EPI_ISL_2776428, EPI_ISL_2776429 | | | |
| see above | North Dakota Department of Health, Public Health Laboratory | North Dakota Department of Health, Public Health Laboratory | Lisa Wingerter |
| EPI_ISL_2758416, EPI_ISL_2758421, EPI_ISL_2758433, EPI_ISL_2758491 | Orange County Public Health Lab | Chan-Zuckerberg Biohub | CZB Cliahub Consortium |
| EPI_ISL_2758413, EPI_ISL_2758418, EPI_ISL_2758419, EPI_ISL_2758501 | Santa Clara County Public Health Laboratory | Chan-Zuckerberg Biohub | CZB Cliahub Consortium |
| EPI_ISL_2779551, EPI_ISL_2779555, EPI_ISL_2779562, EPI_ISL_2779574, EPI_ISL_2779575, EPI_ISL_2779585, EPI_ISL_2779589, EPI_ISL_2779590, EPI_ISL_2779594, EPI_ISL_2779607, EPI_ISL_2779611, EPI_ISL_2779612 | | | |
| see above | Santa Clara County Public Health Laboratory | Santa Clara County Public Health Laboratory | Santa Clara County Public Health Department |
| EPI_ISL_2800691, EPI_ISL_2800698, EPI_ISL_2800700, EPI_ISL_2800702 | State Testing Facility | Altius Institute for Biomedical Research | Alex Isner; Alex Nguyen; Amanda Gale; Audra Johnson; Clem Green; Daniel Bates; Eric Thorland; Jacob Rodriguez; Jean Robinston; Jemma Nelson; Jessica Kunder; John Stamatoyannopoulos; Joshua Richards; Julia Wald; Kneshay Harper; Lauren Mitchell; Mark Frerker; Matt Hartman; Michael Buckley; Muhammad Halimun; Rebecca Bruders; Sadie Patraw; Sofia Olsson; Tobias Ragoczy |
| EPI_ISL_2769159, EPI_ISL_2769160, EPI_ISL_2769162, EPI_ISL_2769164, EPI_ISL_2769172, EPI_ISL_2769183, EPI_ISL_2769193, EPI_ISL_2769196, EPI_ISL_2769206, EPI_ISL_2769208, EPI_ISL_2769214, EPI_ISL_2769221, EPI_ISL_2769255, EPI_ISL_2769269, EPI_ISL_2769271, EPI_ISL_2769272, EPI_ISL_2769273, EPI_ISL_2769274, EPI_ISL_2769281, EPI_ISL_2769282, EPI_ISL_2769283, EPI_ISL_2769308, EPI_ISL_2769312, EPI_ISL_2769328, EPI_ISL_2769329, EPI_ISL_2769347, EPI_ISL_2769351 | | | |
| see above | TGen North | TGen North | "Jolene Bowers; Brett Van Tassel; Chris French; Darrin Lemmer; Dave Engelthaler"; Hayley Yaglom; Heather Centner |
| EPI_ISL_2758842, EPI_ISL_2758873, EPI_ISL_2758874, EPI_ISL_2758904 | The Ohio State University Applied Microbiology Services Laboratory | The Ohio State University Applied Microbiology Services Laboratory | Seth A. Faith PhD |
| EPI_ISL_2791381, EPI_ISL_2791382, EPI_ISL_2791383, EPI_ISL_2791385, EPI_ISL_2791386, EPI_ISL_2791387, EPI_ISL_2791388, EPI_ISL_2791389, EPI_ISL_2791390, EPI_ISL_2791391, EPI_ISL_2791395, EPI_ISL_2791398, EPI_ISL_2791400, EPI_ISL_2791401, EPI_ISL_2791404, EPI_ISL_2791407, EPI_ISL_2791408, EPI_ISL_2791410, EPI_ISL_2791412, EPI_ISL_2791414, EPI_ISL_2791415, EPI_ISL_2791416, EPI_ISL_2791417, EPI_ISL_2791418, EPI_ISL_2791421, EPI_ISL_2791422, EPI_ISL_2791423, EPI_ISL_2791424, EPI_ISL_2791425, EPI_ISL_2791426, EPI_ISL_2791427, EPI_ISL_2791429, EPI_ISL_2791430, EPI_ISL_2791432, EPI_ISL_2791434, EPI_ISL_2791435, EPI_ISL_2791436, EPI_ISL_2791437, EPI_ISL_2791438, EPI_ISL_2791441, EPI_ISL_2791442, EPI_ISL_2791444, EPI_ISL_2791446, EPI_ISL_2791448, EPI_ISL_2791451, EPI_ISL_2791452, EPI_ISL_2791453, EPI_ISL_2791454 | | | |
| see above | UMGC | UMGC | David Xing; Gregg Thomas; Jeffrey Good; Nathanael Herrera; Tim Thurman |
| EPI_ISL_2775678, EPI_ISL_2775679, EPI_ISL_2775681, EPI_ISL_2775682, EPI_ISL_2775683, EPI_ISL_2775685, EPI_ISL_2775687, EPI_ISL_2775688, EPI_ISL_2775689, EPI_ISL_2775690, EPI_ISL_2775691, EPI_ISL_2775692, EPI_ISL_2775693, EPI_ISL_2775694, EPI_ISL_2775695, EPI_ISL_2775696, EPI_ISL_2775697, EPI_ISL_2775699, EPI_ISL_2775702, EPI_ISL_2775707, EPI_ISL_2775709, EPI_ISL_2775711, EPI_ISL_2775715, EPI_ISL_2775716, EPI_ISL_2775717, EPI_ISL_2775718, EPI_ISL_2775719, EPI_ISL_2775721, EPI_ISL_2775723, EPI_ISL_2775724, EPI_ISL_2775725, EPI_ISL_2775726, EPI_ISL_2775727, EPI_ISL_2775728, EPI_ISL_2775729, EPI_ISL_2775731, EPI_ISL_2775733, EPI_ISL_2775735, EPI_ISL_2775736, EPI_ISL_2775737, EPI_ISL_2775738, EPI_ISL_2775743, EPI_ISL_2775746, EPI_ISL_2775747, EPI_ISL_2775751, EPI_ISL_2775752, EPI_ISL_2775756, EPI_ISL_2775760, EPI_ISL_2775761, EPI_ISL_2775764, EPI_ISL_2775765, EPI_ISL_2775768, EPI_ISL_2775769, EPI_ISL_2775774, EPI_ISL_2775776, EPI_ISL_2775778, EPI_ISL_2775789, EPI_ISL_2775795, EPI_ISL_2775806, EPI_ISL_2775868, EPI_ISL_2775869, EPI_ISL_2775870, EPI_ISL_2775871, EPI_ISL_2775872, EPI_ISL_2775873, EPI_ISL_2775874, EPI_ISL_2775875 | | | |
| see above | UPMC Clinical Microbiology Laboratory | Microbial Genome Sequencing Center; Microbial Genomic Epidemiology Laboratory, University of Pittsburgh | Daniel J. Snyder; Jane W. Marsh; Kady D. Waggle; Lee H. Harrison; Marissa P. Griffith; Vatsala R. Srinivasa; Vaughn S. Cooper |
| EPI_ISL_2773884, EPI_ISL_2773885, EPI_ISL_2773886, EPI_ISL_2773887, EPI_ISL_2773888, EPI_ISL_2773889, EPI_ISL_2773890, EPI_ISL_2773891, EPI_ISL_2784711, EPI_ISL_2784712, EPI_ISL_2784713, EPI_ISL_2790460, EPI_ISL_2790556, EPI_ISL_2790565, EPI_ISL_2791116, EPI_ISL_2791175, EPI_ISL_2791176, EPI_ISL_2791177, EPI_ISL_2791178, EPI_ISL_2791179, EPI_ISL_2791180, EPI_ISL_2791181, EPI_ISL_2791182, EPI_ISL_2791183, EPI_ISL_2791185, EPI_ISL_2791186, EPI_ISL_2791187, EPI_ISL_2791211 | | | |
| see above | UW Virology Lab | UW Virology Lab | Alexander Greninger; Hong Xie; Keith R Jerome; Lasata Shrestha; Margaret Mills; Meei-Li Huang; Noah Baker; Pavitra Roychoudhury; Sean Ellis; Shah Mohamed Bakhash |
| EPI_ISL_2828623, EPI_ISL_2828624 | Utah Public Health Laboratory, Utah Public Health Laboratory Infectious Disease submission group | Utah Public Health Laboratory, Utah Public Health Laboratory Infectious Disease submission group | E.L.; Gallagher, T.; K.F.; Oakeson; Park, S.; Young |

| | | | |
|---|---|---|---|
| EPI_ISL_2772719 | Vanderbilt University Medical Center | Genomics and Discovery, Respiratory Viruses Branch, Division of Viral Diseases, Centers for Disease Control and Prevention | Adam Retchless; Anna Kelleher; Anna Uehara; Brian Lynch; Clinton R. Paden; Dhwani Batra; Haibin Wang; Han Jia Justin Ng; Jasmine Padilla; Jing Zhang; Justin Lee; Krista Queen; Mark Burroughs; Mili Sheth; Morgan Davis; Peter Cook; Rachel Marine; Sarah Nobles; Suxiang Tong; Tara Coalter; Yan Li; Ying Tao |
| EPI_ISL_2775502, EPI_ISL_2775503, EPI_ISL_2775506, EPI_ISL_2775508, EPI_ISL_2775511, EPI_ISL_2775512, EPI_ISL_2775514, EPI_ISL_2775517, EPI_ISL_2775519, EPI_ISL_2775520, EPI_ISL_2775526, EPI_ISL_2775538, EPI_ISL_2775542, EPI_ISL_2775543, EPI_ISL_2775544, EPI_ISL_2775545, EPI_ISL_2775549, EPI_ISL_2775550, EPI_ISL_2775552, EPI_ISL_2775553, EPI_ISL_2775554, EPI_ISL_2775558, EPI_ISL_2775559, EPI_ISL_2775561, EPI_ISL_2775562, EPI_ISL_2775571, EPI_ISL_2775573, EPI_ISL_2775574, EPI_ISL_2775575, EPI_ISL_2775576, EPI_ISL_2775595, EPI_ISL_2775614, EPI_ISL_2775615, EPI_ISL_2775616, EPI_ISL_2775617, EPI_ISL_2775619, EPI_ISL_2775620, EPI_ISL_2775621, EPI_ISL_2775623, EPI_ISL_2775624, EPI_ISL_2775626, EPI_ISL_2775632, EPI_ISL_2775633, EPI_ISL_2775635, EPI_ISL_2775636, EPI_ISL_2775637, EPI_ISL_2775640, EPI_ISL_2775672, EPI_ISL_2776125, EPI_ISL_2776126, EPI_ISL_2776127, EPI_ISL_2776128, EPI_ISL_2776130, EPI_ISL_2776133, EPI_ISL_2776135, EPI_ISL_2776136, EPI_ISL_2801535, EPI_ISL_2801536, EPI_ISL_2801539, EPI_ISL_2801541, EPI_ISL_2801542, EPI_ISL_2801543, EPI_ISL_2801544, EPI_ISL_2801545, EPI_ISL_2801546, EPI_ISL_2801548, EPI_ISL_2801549, EPI_ISL_2801551, EPI_ISL_2810754, EPI_ISL_2810756, EPI_ISL_2810758, EPI_ISL_2810759, EPI_ISL_2810766, EPI_ISL_2810769, EPI_ISL_2810770, EPI_ISL_2810771, EPI_ISL_2810773, EPI_ISL_2810774, EPI_ISL_2810785 | | | |
| see above | Wyoming Public Health Laboratory | Wyoming Public Health Laboratory | Ashley Norberg; Brian Dominguez; Cari Sloma; Channing Weber; Chayse Rowley; Elliot Thomasson; Jim Mildenberger; Marley Goetz; Robert Petit; Sam Britz; Taylor Fearing; and Rob Christensen |

We gratefully acknowledge the following Authors from the Originating laboratories responsible for obtaining the specimens, as well as the Submitting laboratories where the genome data were generated and shared via GISAID, on which this research is based.

All Submitters of data may be contacted directly via www.gisaid.org

Authors are sorted alphabetically.

| Accession ID | Originating Laboratory | Submitting Laboratory | Authors |
|---|---|---|---|
| EPI_ISL_2621099, EPI_ISL_2621100, EPI_ISL_2621103, EPI_ISL_2621134, EPI_ISL_2621136, EPI_ISL_2621144 | 2 Military Hospital wc MAA | NHLS/UCT | Arash Iranzadeh; Bruna Galvao; Carolyn Williamson; Deelan Doolabh; Diana Hardie; Gert Marais; Innocent Mudau; Lynn Tyers; Marvin Hsiao; Stephen Korsman |
| EPI_ISL_2547411, EPI_ISL_2547413, EPI_ISL_2547421, EPI_ISL_2547422 | 2 Military Hospital wc MAA | National Health Laboratory Service/University of Cape Town (NHLS/UCT) | Arash Iranzadeh; Bruna Galvao; Carolyn Williamson; Deelan Doolabh; Diana Hardie; Gert Marais; Innocent Mudau; Lynn Tyers; Marvin Hsiao; Stephen Korsman |
| EPI_ISL_2621060 | Alma CDC wc AHC | NHLS/UCT | Arash Iranzadeh; Bruna Galvao; Carolyn Williamson; Deelan Doolabh; Diana Hardie; Gert Marais; Innocent Mudau; Lynn Tyers; Marvin Hsiao; Stephen Korsman |
| EPI_ISL_2710306, EPI_ISL_2710307, EPI_ISL_2710309, EPI_ISL_2710310, EPI_ISL_2710311, EPI_ISL_2710317, EPI_ISL_2710321, EPI_ISL_2710322 | | | |
| see above | BARC / Lancet | KRISP, KZN Research Innovation and Sequencing Platform | Giandhari Jennifer; Naidoo Yeshnee; Pillay Sureshnee; San James; Sisonke; Tegally Houriiyah; Tshabuila Derek; Wilkinson Eduan; Yajna Ramphal; de Oliveira Tulio |
| EPI_ISL_2695506 | Bertha gxowa hospital | National Institute for Communicable Diseases of the National Health Laboratory Service | Amoako DG; Bhiman JN; Everatt J; Ismail A; Mahlangu B; Mnguni A; Mohale T; Ntuli N; Scheepers C |
| EPI_ISL_2694629, EPI_ISL_2694630, EPI_ISL_2694631, EPI_ISL_2694633, EPI_ISL_2694635, EPI_ISL_2694643, EPI_ISL_2694646, EPI_ISL_2694647, EPI_ISL_2694652, EPI_ISL_2694653, EPI_ISL_2694655, EPI_ISL_2694656, EPI_ISL_2694660, EPI_ISL_2694661, EPI_ISL_2694662, EPI_ISL_2694668, EPI_ISL_2694673, EPI_ISL_2694674, EPI_ISL_2694675, EPI_ISL_2694676, EPI_ISL_2709880, EPI_ISL_2709883, EPI_ISL_2709884, EPI_ISL_2709897, EPI_ISL_2709898, EPI_ISL_2709900, EPI_ISL_2709901, EPI_ISL_2709902, EPI_ISL_2709906, EPI_ISL_2709908, EPI_ISL_2709909, EPI_ISL_2709911, EPI_ISL_2709917, EPI_ISL_2709919, EPI_ISL_2709921, EPI_ISL_2709922, EPI_ISL_2709924, EPI_ISL_2709926, EPI_ISL_2709927, EPI_ISL_2709930, EPI_ISL_2709931, EPI_ISL_2709933, EPI_ISL_2709934, EPI_ISL_2709936, EPI_ISL_2709938, EPI_ISL_2709940, EPI_ISL_2709942 | | | |
| see above | CHRIS HANI BARAGWANATH | National Institute for Communicable Diseases of the National Health Laboratory Service | Amoako DG; Bhiman JN; Everatt J; Ismail A; Mahlangu B; Mnguni A; Mohale T; Ntuli N; Scheepers C |
| EPI_ISL_2582812, EPI_ISL_2582815, EPI_ISL_2582818, EPI_ISL_2582822, EPI_ISL_2582825, EPI_ISL_2662469, EPI_ISL_2662470, EPI_ISL_2662473, EPI_ISL_2662474, EPI_ISL_2662492, EPI_ISL_2662494, EPI_ISL_2662495, EPI_ISL_2662496, EPI_ISL_2662497, EPI_ISL_2662498, EPI_ISL_2662499, EPI_ISL_2662500, EPI_ISL_2662501, EPI_ISL_2662502, EPI_ISL_2662503, EPI_ISL_2662504, EPI_ISL_2662505, EPI_ISL_2662506, EPI_ISL_2662507, EPI_ISL_2662509, EPI_ISL_2662510, EPI_ISL_2662554, EPI_ISL_2662555, EPI_ISL_2662618, EPI_ISL_2662619, EPI_ISL_2662620, EPI_ISL_2662621, EPI_ISL_2662626, EPI_ISL_2662627, EPI_ISL_2662631, EPI_ISL_2662632, EPI_ISL_2662636, EPI_ISL_2662638, EPI_ISL_2662639, EPI_ISL_2662641, EPI_ISL_2662642, EPI_ISL_2662643, EPI_ISL_2662644, EPI_ISL_2662645, EPI_ISL_2662648, EPI_ISL_2662649, EPI_ISL_2662651, EPI_ISL_2662654, EPI_ISL_2662655, EPI_ISL_2662656, EPI_ISL_2662657, EPI_ISL_2662658, EPI_ISL_2662660 | | | |
| see above | CHRIS HANI BARAGWANATH LABORATORY | National Institute for Communicable Diseases of the National Health Laboratory Service | Amoako DG; Bhiman JN; Everatt J; Ismail A; Mahlangu B; Mnguni A; Mohale T; Ntuli N; Scheepers C |
| EPI_ISL_2547365 | Calvinia Hospital | National Health Laboratory Service/University of Cape Town (NHLS/UCT) | Arash Iranzadeh; Bruna Galvao; Carolyn Williamson; Deelan Doolabh; Diana Hardie; Gert Marais; Innocent Mudau; Lynn Tyers; Marvin Hsiao; Stephen Korsman |
| EPI_ISL_2547356, EPI_ISL_2547357, EPI_ISL_2547358, EPI_ISL_2547359, EPI_ISL_2547360, EPI_ISL_2547361, EPI_ISL_2547362 | | | |
| see above | Calvinia RSC Clinic | National Health Laboratory Service/University of Cape Town (NHLS/UCT) | Arash Iranzadeh; Bruna Galvao; Carolyn Williamson; Deelan Doolabh; Diana Hardie; Gert Marais; Innocent Mudau; Lynn Tyers; Marvin Hsiao; Stephen Korsman |
| EPI_ISL_2621065, EPI_ISL_2621068, EPI_ISL_2621070, EPI_ISL_2621073, EPI_ISL_2621075, EPI_ISL_2621076, EPI_ISL_2621079, EPI_ISL_2621092, EPI_ISL_2621093 | | | |
| see above | Conville CDC wc CVC | NHLS/UCT | Arash Iranzadeh; Bruna Galvao; Carolyn Williamson; Deelan Doolabh; Diana Hardie; Gert Marais; Innocent Mudau; Lynn Tyers; Marvin Hsiao; Stephen Korsman |
| EPI_ISL_2621143 | D'Almeida Clinic wc DAL | NHLS/UCT | Arash Iranzadeh; Bruna Galvao; Carolyn Williamson; Deelan Doolabh; Diana Hardie; Gert Marais; Innocent Mudau; Lynn Tyers; Marvin Hsiao; Stephen Korsman |
| EPI_ISL_2582809, EPI_ISL_2662511, EPI_ISL_2662512, EPI_ISL_2662513, EPI_ISL_2662514, EPI_ISL_2662515, EPI_ISL_2662516, EPI_ISL_2662517, EPI_ISL_2662518, EPI_ISL_2662519, EPI_ISL_2662520, EPI_ISL_2662521, EPI_ISL_2662522, EPI_ISL_2662525, EPI_ISL_2662527, EPI_ISL_2662528, EPI_ISL_2662538, EPI_ISL_2662540, EPI_ISL_2662541, EPI_ISL_2662542, EPI_ISL_2662543, EPI_ISL_2662544, EPI_ISL_2662545, EPI_ISL_2662546, EPI_ISL_2662547, EPI_ISL_2662548, EPI_ISL_2662550, EPI_ISL_2662551 | | | |
| see above | DR GEORGE MUKHARI LABORATORY | National Institute for Communicable Diseases of the National Health Laboratory Service | Amoako DG; Bhiman JN; Everatt J; Ismail A; Mahlangu B; Mnguni A; Mohale T; Ntuli N; Scheepers C |
| EPI_ISL_2695801, EPI_ISL_2695802, EPI_ISL_2695804, EPI_ISL_2695805, EPI_ISL_2695806, EPI_ISL_2695809, EPI_ISL_2695810, EPI_ISL_2695849, EPI_ISL_2695850, EPI_ISL_2695851, EPI_ISL_2695852, EPI_ISL_2695854, EPI_ISL_2695856, EPI_ISL_2695857, EPI_ISL_2695858, EPI_ISL_2695859, EPI_ISL_2695861, EPI_ISL_2695862, EPI_ISL_2695863, EPI_ISL_2695864, EPI_ISL_2695865, EPI_ISL_2695866, EPI_ISL_2695867, EPI_ISL_2695868, EPI_ISL_2695869, EPI_ISL_2695870, EPI_ISL_2695871, EPI_ISL_2695872, EPI_ISL_2695873 | | | |
| see above | Division of Medical Virology, National Health Laboratory Service (NHLS), Tygerberg Hospital / Stellenbosch University | Division of Medical Virology, National Health Laboratory Service (NHLS), Tygerberg Hospital / Stellenbosch University | Bronwyn Kleinhans; Gert van Zyl; San Emmanual James; Susan Engelbrecht; Tania Stander; Tongai Maponga; Tulio de Oliveira; Wolfgang Preiser |
| EPI_ISL_2621146, EPI_ISL_2621147 | Dysselsdorp Clinic wc DDC | NHLS/UCT | Arash Iranzadeh; Bruna Galvao; Carolyn Williamson; Deelan Doolabh; Diana Hardie; Gert Marais; Innocent Mudau; Lynn Tyers; Marvin Hsiao; Stephen Korsman |
| EPI_ISL_2547341 | Garies PHC Clinic | National Health Laboratory Service/University of Cape Town (NHLS/UCT) | Arash Iranzadeh; Bruna Galvao; Carolyn Williamson; Deelan Doolabh; Diana Hardie; Gert Marais; Innocent Mudau; Lynn Tyers; Marvin Hsiao; Stephen Korsman |
| EPI_ISL_2621072, EPI_ISL_2621137 | George Hospital wc GRH | NHLS/UCT | Arash Iranzadeh; Bruna Galvao; Carolyn Williamson; Deelan Doolabh; Diana Hardie; Gert Marais; Innocent Mudau; Lynn Tyers; Marvin Hsiao; Stephen Korsman |
| EPI_ISL_2547426 | Great Brak River Clinic wc GBC | National Health Laboratory Service/University of Cape Town (NHLS/UCT) | Arash Iranzadeh; Bruna Galvao; Carolyn Williamson; Deelan Doolabh; Diana Hardie; Gert Marais; Innocent Mudau; Lynn Tyers; Marvin Hsiao; Stephen Korsman |
| EPI_ISL_2621062 | Green Point NHLS | NHLS/UCT | Arash Iranzadeh; Bruna Galvao; Carolyn Williamson; Deelan Doolabh; Diana Hardie; Gert Marais; Innocent Mudau; Lynn Tyers; Marvin Hsiao; Stephen Korsman |
| EPI_ISL_2621054, EPI_ISL_2621098, EPI_ISL_2621102, EPI_ISL_2621105, EPI_ISL_2621108, EPI_ISL_2621109, EPI_ISL_2621110, EPI_ISL_2621111, EPI_ISL_2621112, EPI_ISL_2621113, EPI_ISL_2621114, EPI_ISL_2621115, EPI_ISL_2621116, EPI_ISL_2621117, EPI_ISL_2621118, EPI_ISL_2621119, EPI_ISL_2621120, EPI_ISL_2621122, EPI_ISL_2621123, EPI_ISL_2621124, EPI_ISL_2621126, EPI_ISL_2621127, EPI_ISL_2621128, EPI_ISL_2621129, EPI_ISL_2621130, EPI_ISL_2621131, EPI_ISL_2621132, EPI_ISL_2621133 | | | |
| see above | Groote Schuur Hospital wc GSH | NHLS/UCT | Arash Iranzadeh; Bruna Galvao; Carolyn Williamson; Deelan Doolabh; Diana Hardie; Gert Marais; Innocent Mudau; Lynn Tyers; Marvin Hsiao; Stephen Korsman |
| EPI_ISL_2547373, EPI_ISL_2547374, EPI_ISL_2547375, EPI_ISL_2547376, EPI_ISL_2547377, EPI_ISL_2547378, EPI_ISL_2547379, EPI_ISL_2547380, EPI_ISL_2547381, EPI_ISL_2547382, EPI_ISL_2547383, EPI_ISL_2547384, EPI_ISL_2547385, EPI_ISL_2547386, EPI_ISL_2547388, EPI_ISL_2547389, EPI_ISL_2547390, EPI_ISL_2547391, EPI_ISL_2547392, EPI_ISL_2547393, EPI_ISL_2547394, EPI_ISL_2547395, EPI_ISL_2547396, EPI_ISL_2547397, EPI_ISL_2547398, EPI_ISL_2547399, EPI_ISL_2547400, EPI_ISL_2547401, EPI_ISL_2547402, EPI_ISL_2547403, EPI_ISL_2547404, EPI_ISL_2547405, EPI_ISL_2547406, EPI_ISL_2547407, EPI_ISL_2547408, EPI_ISL_2547418, EPI_ISL_2547419, EPI_ISL_2547424, EPI_ISL_2547425 | | | |
| see above | Groote Schuur Hospital wc GSH | National Health Laboratory Service/University of Cape Town (NHLS/UCT) | Arash Iranzadeh; Bruna Galvao; Carolyn Williamson; Deelan Doolabh; Diana Hardie; Gert Marais; Innocent Mudau; Lynn Tyers; Marvin Hsiao; Stephen Korsman |
| EPI_ISL_2695590, EPI_ISL_2695595, EPI_ISL_2695601, EPI_ISL_2695602, EPI_ISL_2695604, EPI_ISL_2695606, EPI_ISL_2695732, EPI_ISL_2695739, EPI_ISL_2695744, EPI_ISL_2709863, EPI_ISL_2709872, EPI_ISL_2709875, EPI_ISL_2709878, EPI_ISL_2709885 | | | |
| see above | HELEN JOSEPH | National Institute for Communicable Diseases of the National Health Laboratory Service | Amoako DG; Bhiman JN; Everatt J; Ismail A; Mahlangu B; Mnguni A; Mohale T; Ntuli N; Scheepers C |
| EPI_ISL_2547372 | Helderberg Hospital wc HHH | National Health Laboratory Service/University of Cape Town (NHLS/UCT) | Arash Iranzadeh; Bruna Galvao; Carolyn Williamson; Deelan Doolabh; Diana Hardie; Gert Marais; Innocent Mudau; Lynn Tyers; Marvin Hsiao; Stephen Korsman |
| EPI_ISL_2547352 | Hondeklipbaai Clinic | National Health Laboratory Service/University of Cape Town (NHLS/UCT) | Arash Iranzadeh; Bruna Galvao; Carolyn Williamson; Deelan Doolabh; Diana Hardie; Gert Marais; Innocent Mudau; Lynn Tyers; Marvin Hsiao; Stephen Korsman |
| EPI_ISL_2621096, EPI_ISL_2621107 | Hornlee Clinic wc HLC | NHLS/UCT | Arash Iranzadeh; Bruna Galvao; Carolyn Williamson; Deelan Doolabh; Diana Hardie; Gert Marais; Innocent Mudau; Lynn Tyers; Marvin Hsiao; Stephen Korsman |
| EPI_ISL_2547427 | Hornlee Clinic wc HLC | National Health Laboratory Service/University of Cape Town (NHLS/UCT) | Arash Iranzadeh; Bruna Galvao; Carolyn Williamson; Deelan Doolabh; Diana Hardie; Gert Marais; Innocent Mudau; Lynn Tyers; Marvin Hsiao; Stephen Korsman |
| EPI_ISL_2547340 | Joe Slovo Hospital | National Health Laboratory Service/University of Cape Town (National Health Laboratory Service/University of Cape Town (NHLS/UCT)) | Arash Iranzadeh; Bruna Galvao; Carolyn Williamson; Deelan Doolabh; Diana Hardie; Gert Marais; Innocent Mudau; Lynn Tyers; Marvin Hsiao; Stephen Korsman |
| EPI_ISL_2582848, EPI_ISL_2582852, EPI_ISL_2582855, EPI_ISL_2582858, EPI_ISL_2582862, EPI_ISL_2582865, EPI_ISL_2582868, EPI_ISL_2582870, EPI_ISL_2582872, EPI_ISL_2582876, EPI_ISL_2582879, EPI_ISL_2582882, EPI_ISL_2582885, EPI_ISL_2582888, EPI_ISL_2582891, EPI_ISL_2582894, EPI_ISL_2582897, EPI_ISL_2582900, EPI_ISL_2582907, EPI_ISL_2582909, EPI_ISL_2582912, EPI_ISL_2582915, EPI_ISL_2582922, EPI_ISL_2582924, EPI_ISL_2582928, EPI_ISL_2582931, EPI_ISL_2582937, EPI_ISL_2582940, EPI_ISL_2582943, EPI_ISL_2582950, EPI_ISL_2582953, EPI_ISL_2582956, EPI_ISL_2582960, EPI_ISL_2582963, EPI_ISL_2582968, EPI_ISL_2582974, EPI_ISL_2582977, EPI_ISL_2582980, EPI_ISL_2662665, EPI_ISL_2662666, EPI_ISL_2662667, EPI_ISL_2662668, EPI_ISL_2662669, EPI_ISL_2662670, EPI_ISL_2662671, EPI_ISL_2662672, EPI_ISL_2662673, EPI_ISL_2662674, EPI_ISL_2662675, EPI_ISL_2662676, EPI_ISL_2662677, EPI_ISL_2662678, EPI_ISL_2662679, EPI_ISL_2662680, EPI_ISL_2662681, EPI_ISL_2662682, EPI_ISL_2662683, EPI_ISL_2662684, EPI_ISL_2662685, EPI_ISL_2662686, EPI_ISL_2662687, EPI_ISL_2662689, EPI_ISL_2662690, EPI_ISL_2662691, EPI_ISL_2662692, EPI_ISL_2662693, EPI_ISL_2662694, EPI_ISL_2662695, EPI_ISL_2662696, EPI_ISL_2662697, EPI_ISL_2662698, EPI_ISL_2662699, EPI_ISL_2662700, EPI_ISL_2662701, EPI_ISL_2662702, EPI_ISL_2662703, EPI_ISL_2662704, EPI_ISL_2662705, EPI_ISL_2662706, EPI_ISL_2662707, EPI_ISL_2695701, EPI_ISL_2695702, EPI_ISL_2695703, EPI_ISL_2695704, EPI_ISL_2695705, EPI_ISL_2695706, EPI_ISL_2695707, EPI_ISL_2695708, EPI_ISL_2695709, EPI_ISL_2695710, EPI_ISL_2695711, EPI_ISL_2695712, EPI_ISL_2695713, EPI_ISL_2695714, EPI_ISL_2695715, EPI_ISL_2695716, EPI_ISL_2695717, EPI_ISL_2695718, EPI_ISL_2695719, EPI_ISL_2695720, EPI_ISL_2695722, EPI_ISL_2695723, EPI_ISL_2695724 | | | |
| see above | KIMBERLEY LABORATORY | National Institute for Communicable Diseases of the National Health Laboratory Service | Amoako DG; Bhiman JN; Everatt J; Ismail A; Mahlangu B; Mnguni A; Mohale T; Ntuli N; Scheepers C |
| EPI_ISL_2694614, EPI_ISL_2694615, EPI_ISL_2695512 | KOPANONG | National Institute for Communicable Diseases of the National Health Laboratory Service | Amoako DG; Bhiman JN; Everatt J; Ismail A; Mahlangu B; Mnguni A; Mohale T; Ntuli N; Scheepers C |
| EPI_ISL_2582990, EPI_ISL_2582993, EPI_ISL_2582996, EPI_ISL_2582999, EPI_ISL_2583001, EPI_ISL_2662663, EPI_ISL_2662664, EPI_ISL_2695760, EPI_ISL_2695761 | | | |
| see above | KOPANONG LABORATORY | National Institute for Communicable Diseases of the National Health Laboratory Service | Amoako DG; Bhiman JN; Everatt J; Ismail A; Mahlangu B; Mnguni A; Mohale T; Ntuli N; Scheepers C |
| EPI_ISL_2688547, EPI_ISL_2688548 | KRISP, KZn Research Innovation and Sequencing Platform | KRISP, KZn Research Innovation and Sequencing Platform | Giandhari Jennifer; Naidoo Yeshnee; Pillay Sureshnee; San James; Tegally Houriiyah; Tshabuila Derek; Wilkinson Eduan; Yajna Ramphal; de Oliveira Tulio |
| EPI_ISL_2621097, EPI_ISL_2621145 | Knysna CDC wc WLC | NHLS/UCT | Arash Iranzadeh; Bruna Galvao; Carolyn Williamson; Deelan Doolabh; Diana Hardie; Gert Marais; Innocent Mudau; Lynn Tyers; Marvin Hsiao; Stephen Korsman |
| EPI_ISL_2547415 | Knysna Hospital wc KNY | National Health Laboratory Service/University of Cape Town (NHLS/UCT) | Arash Iranzadeh; Bruna Galvao; Carolyn Williamson; Deelan Doolabh; Diana Hardie; Gert Marais; Innocent Mudau; Lynn Tyers; Marvin Hsiao; Stephen Korsman |

| | | | |
|---|---|---|---|
| EPI_ISL_2547368, EPI_ISL_2547370 | Kuruman Hospital | National Health Laboratory Service/University of Cape Town (NHLS/UCT) | Arash Iranzadeh; Bruna Galvao; Carolyn Williamson; Deelan Doolabh; Diana Hardie; Gert Marais; Innocent Mudau; Lynn Tyers; Marvin Hsiao; Stephen Korsman |
| EPI_ISL_2547366 | Lutzville Clinic wc LVP | National Health Laboratory Service/University of Cape Town (NHLS/UCT) | Arash Iranzadeh; Bruna Galvao; Carolyn Williamson; Deelan Doolabh; Diana Hardie; Gert Marais; Innocent Mudau; Lynn Tyers; Marvin Hsiao; Stephen Korsman |
| EPI_ISL_2547344 | Matjieskloof Clinic | National Health Laboratory Service/University of Cape Town (NHLS/UCT) | Arash Iranzadeh; Bruna Galvao; Carolyn Williamson; Deelan Doolabh; Diana Hardie; Gert Marais; Innocent Mudau; Lynn Tyers; Marvin Hsiao; Stephen Korsman |
| EPI_ISL_2547354 | Max Shapiro Clinic | National Health Laboratory Service/University of Cape Town (NHLS/UCT) | Arash Iranzadeh; Bruna Galvao; Carolyn Williamson; Deelan Doolabh; Diana Hardie; Gert Marais; Innocent Mudau; Lynn Tyers; Marvin Hsiao; Stephen Korsman |
| EPI_ISL_2621106 | Mitchells Plain Hospital wc MPH | NHLS/UCT | Arash Iranzadeh; Bruna Galvao; Carolyn Williamson; Deelan Doolabh; Diana Hardie; Gert Marais; Innocent Mudau; Lynn Tyers; Marvin Hsiao; Stephen Korsman |
| EPI_ISL_2710251, EPI_ISL_2710254, EPI_ISL_2710263, EPI_ISL_2710276, EPI_ISL_2710281, EPI_ISL_2710285, EPI_ISL_2710289, EPI_ISL_2710291, EPI_ISL_2710293, EPI_ISL_2710304, EPI_ISL_2710305, EPI_ISL_2726856, EPI_ISL_2770441, EPI_ISL_2770442, EPI_ISL_2770443, EPI_ISL_2770446, EPI_ISL_2770448, EPI_ISL_2770449, EPI_ISL_2770454, EPI_ISL_2770455, EPI_ISL_2770456, EPI_ISL_2770470, EPI_ISL_2770471, EPI_ISL_2770474, EPI_ISL_2770475, EPI_ISL_2770476, EPI_ISL_2770478, EPI_ISL_2770481, EPI_ISL_2770482, EPI_ISL_2770483, EPI_ISL_2770485, EPI_ISL_2770487 | | | |
| see above | NHLS Charlotte Maxeke Johannesburg Academic Hospital and the University of the Witwatersrand | KRISP, KZN Research Innovation and Sequencing Platform | Bulelani Manene; Florette Treurnicht; Giandhari Jennifer; Kathleen Subramoney; Naidoo Yeshnee; Pillay Sureshnee; San James; Sisonke; Tegally Houriiyah; Tshabuila Derek; Wilkinson Eduan; Yajna Ramphal; de Oliveira Tulio |
| EPI_ISL_2693040, EPI_ISL_2693044, EPI_ISL_2693049, EPI_ISL_2693051, EPI_ISL_2693052, EPI_ISL_2693054, EPI_ISL_2693057, EPI_ISL_2693060, EPI_ISL_2693070, EPI_ISL_2693072, EPI_ISL_2693074, EPI_ISL_2693075 | | | |
| see above | NHLS_VIRO | KRISP, KZN Research Innovation and Sequencing Platform | Giandhari Jennifer; Naidoo Yeshnee; Pillay Sureshnee; San James; Tegally Houriiyah; Tshabuila Derek; Wilkinson Eduan; Yajna Ramphal; de Oliveira Tulio |
| EPI_ISL_2727382, EPI_ISL_2727385, EPI_ISL_2727390 | NHLS_VIRO | KRISP, KZn Research Innovation and Sequencing Platform | Giandhari Jennifer; Naidoo Yeshnee; Pillay Sureshnee; San James; Tegally Houriiyah; Tshabuila Derek; Wilkinson Eduan; Yajna Ramphal; de Oliveira Tulio |
| EPI_ISL_2693824, EPI_ISL_2693826, EPI_ISL_2693828, EPI_ISL_2693829, EPI_ISL_2693839, EPI_ISL_2693840, EPI_ISL_2693843, EPI_ISL_2693849, EPI_ISL_2693850, EPI_ISL_2693852, EPI_ISL_2693853, EPI_ISL_2693855, EPI_ISL_2693856, EPI_ISL_2693857, EPI_ISL_2693858, EPI_ISL_2693859, EPI_ISL_2693860, EPI_ISL_2693861, EPI_ISL_2693862, EPI_ISL_2693863, EPI_ISL_2693867, EPI_ISL_2693869, EPI_ISL_2693870, EPI_ISL_2693871, EPI_ISL_2693872, EPI_ISL_2693873, EPI_ISL_2693874, EPI_ISL_2693881, EPI_ISL_2693883, EPI_ISL_2693884, EPI_ISL_2693889, EPI_ISL_2693892, EPI_ISL_2693893 | | | |
| see above | NHLS_VIRO Eastern Cape | KRISP, KZN Research Innovation and Sequencing Platform | Giandhari Jennifer; Naidoo Yeshnee; Pillay Sureshnee; San James; Tegally Houriiyah; Tshabuila Derek; Wilkinson Eduan; Yajna Ramphal; de Oliveira Tulio |
| EPI_ISL_2727175, EPI_ISL_2727176, EPI_ISL_2727178, EPI_ISL_2727179, EPI_ISL_2727180, EPI_ISL_2727186, EPI_ISL_2727187, EPI_ISL_2727189, EPI_ISL_2727191, EPI_ISL_2727192, EPI_ISL_2727193, EPI_ISL_2727194, EPI_ISL_2727195, EPI_ISL_2727196, EPI_ISL_2727197, EPI_ISL_2727198, EPI_ISL_2727199, EPI_ISL_2727201, EPI_ISL_2727203, EPI_ISL_2727204, EPI_ISL_2727205, EPI_ISL_2727206, EPI_ISL_2727208, EPI_ISL_2727212, EPI_ISL_2727214, EPI_ISL_2727215, EPI_ISL_2727216, EPI_ISL_2727220, EPI_ISL_2727221, EPI_ISL_2727222, EPI_ISL_2727224, EPI_ISL_2727226, EPI_ISL_2727228, EPI_ISL_2727230 | | | |
| see above | NHLS_VIRO Eastern Cape | KRISP, KZn Research Innovation and Sequencing Platform | Emmanuel SJ; Giandhari J; Khan S; Laguda-Akingba O; Lessells R; Mdlalose K; Pillay S; Tegally H; Wilkinson E; York D; de Oliveira T |
| EPI_ISL_2547343 | Nababeep Hospital | National Health Laboratory Service/University of Cape Town (NHLS/UCT) | Arash Iranzadeh; Bruna Galvao; Carolyn Williamson; Deelan Doolabh; Diana Hardie; Gert Marais; Innocent Mudau; Lynn Tyers; Marvin Hsiao; Stephen Korsman |
| EPI_ISL_2617186, EPI_ISL_2617187, EPI_ISL_2617188, EPI_ISL_2617189, EPI_ISL_2617190, EPI_ISL_2617191, EPI_ISL_2617192, EPI_ISL_2617193, EPI_ISL_2617194, EPI_ISL_2617195, EPI_ISL_2617196, EPI_ISL_2617197, EPI_ISL_2617198, EPI_ISL_2617199, EPI_ISL_2617200, EPI_ISL_2617201, EPI_ISL_2617202, EPI_ISL_2617203, EPI_ISL_2617204, EPI_ISL_2617205, EPI_ISL_2617206, EPI_ISL_2617207, EPI_ISL_2617208, EPI_ISL_2617209, EPI_ISL_2617210, EPI_ISL_2617211, EPI_ISL_2617212, EPI_ISL_2617213, EPI_ISL_2617214, EPI_ISL_2617215, EPI_ISL_2688532, EPI_ISL_2688534, EPI_ISL_2688535, EPI_ISL_2688537, EPI_ISL_2688538, EPI_ISL_2688539, EPI_ISL_2688540, EPI_ISL_2688541, EPI_ISL_2688542, EPI_ISL_2688543, EPI_ISL_2688544, EPI_ISL_2688545, EPI_ISL_2688549, EPI_ISL_2688551, EPI_ISL_2688552, EPI_ISL_2688553, EPI_ISL_2688557, EPI_ISL_2688559, EPI_ISL_2688560, EPI_ISL_2688561, EPI_ISL_2688562, EPI_ISL_2688563, EPI_ISL_2688565, EPI_ISL_2688568, EPI_ISL_2688569, EPI_ISL_2688570, EPI_ISL_2688571, EPI_ISL_2688572, EPI_ISL_2688573, EPI_ISL_2688574, EPI_ISL_2688575 | | | |
| see above | National Health Laboratory Service, South Africa | KRISP, KZn Research Innovation and Sequencing Platform | Giandhari Jennifer; Naidoo Yeshnee; Pillay Sureshnee; San James; Tegally Houriiyah; Tshabuila Derek; Wilkinson Eduan; Yajna Ramphal; de Oliveira Tulio |
| EPI_ISL_2662708, EPI_ISL_2662710, EPI_ISL_2662711, EPI_ISL_2662712, EPI_ISL_2662714, EPI_ISL_2662716, EPI_ISL_2662719, EPI_ISL_2662720, EPI_ISL_2662722, EPI_ISL_2662723, EPI_ISL_2662724, EPI_ISL_2662725 | | | |
| see above | National Institute for Communicable Diseases of the National Health Laboratory Service | National Institute for Communicable Diseases of the National Health Laboratory Service | Amoako DG; Bhiman JN; Everatt J; Ismail A; Mahlangu B; Mnguni A; Mohale T; Ntuli N; Scheepers C |
| EPI_ISL_2547420 | Nelspoort Clinic wc NPP | National Health Laboratory Service/University of Cape Town (NHLS/UCT) | Arash Iranzadeh; Bruna Galvao; Carolyn Williamson; Deelan Doolabh; Diana Hardie; Gert Marais; Innocent Mudau; Lynn Tyers; Marvin Hsiao; Stephen Korsman |
| EPI_ISL_2770489 | Netcare Kingsway | KRISP, KZN Research Innovation and Sequencing Platform | Giandhari Jennifer; Naidoo Yeshnee; Pillay Sureshnee; San James; Tegally Houriiyah; Tshabuila Derek; Wilkinson Eduan; Yajna Ramphal; de Oliveira Tulio |
| EPI_ISL_2693062 | Netcare Kingsway Hospital | KRISP, KZN Research Innovation and Sequencing Platform | Giandhari Jennifer; Naidoo Yeshnee; Pillay Sureshnee; San James; Tegally Houriiyah; Tshabuila Derek; Wilkinson Eduan; Yajna Ramphal; de Oliveira Tulio |
| EPI_ISL_2547342, EPI_ISL_2547345, EPI_ISL_2547351, EPI_ISL_2547353 | Okiep CHC | National Health Laboratory Service/University of Cape Town (NHLS/UCT) | Arash Iranzadeh; Bruna Galvao; Carolyn Williamson; Deelan Doolabh; Diana Hardie; Gert Marais; Innocent Mudau; Lynn Tyers; Marvin Hsiao; Stephen Korsman |
| EPI_ISL_2695584, EPI_ISL_2695585 | PATHCARE LABORATORY | National Institute for Communicable Diseases of the National Health Laboratory Service | Amoako DG; Bhiman JN; Everatt J; Ismail A; Mahlangu B; Mnguni A; Mohale T; Ntuli N; Scheepers C |
| EPI_ISL_2695541, EPI_ISL_2695542, EPI_ISL_2695543, EPI_ISL_2695544, EPI_ISL_2695545, EPI_ISL_2695546, EPI_ISL_2695547, EPI_ISL_2695552, EPI_ISL_2695553, EPI_ISL_2695554, EPI_ISL_2695555, EPI_ISL_2695558, EPI_ISL_2695559, EPI_ISL_2695560, EPI_ISL_2695561, EPI_ISL_2695563, EPI_ISL_2695566, EPI_ISL_2695567, EPI_ISL_2695568, EPI_ISL_2695570, EPI_ISL_2695571, EPI_ISL_2695572, EPI_ISL_2695575, EPI_ISL_2695577, EPI_ISL_2695579, EPI_ISL_2695582, EPI_ISL_2709849, EPI_ISL_2709850, EPI_ISL_2709851, EPI_ISL_2709852, EPI_ISL_2709861, EPI_ISL_2709862, EPI_ISL_2709871, EPI_ISL_2709916 | | | |
| see above | POLOKWANE MANKWENG HOSPITAL | National Institute for Communicable Diseases of the National Health Laboratory Service | Amoako DG; Bhiman JN; Everatt J; Ismail A; Mahlangu B; Mnguni A; Mohale T; Ntuli N; Scheepers C |
| EPI_ISL_2582702, EPI_ISL_2582707, EPI_ISL_2582710, EPI_ISL_2582717, EPI_ISL_2582720 | PORT ELIZABETH LABORATORY | National Institute for Communicable Diseases of the National Health Laboratory Service | Amoako DG; Bhiman JN; Everatt J; Ismail A; Mahlangu B; Mnguni A; Mohale T; Ntuli N; Scheepers C |
| EPI_ISL_2694593, EPI_ISL_2694601, EPI_ISL_2694603, EPI_ISL_2694604, EPI_ISL_2694608, EPI_ISL_2695811, EPI_ISL_2695812, EPI_ISL_2695813, EPI_ISL_2695814, EPI_ISL_2695815, EPI_ISL_2695816, EPI_ISL_2695824, EPI_ISL_2695826, EPI_ISL_2695827, EPI_ISL_2695828, EPI_ISL_2695830, EPI_ISL_2695831, EPI_ISL_2695832, EPI_ISL_2695833, EPI_ISL_2695835, EPI_ISL_2695837, EPI_ISL_2695838, EPI_ISL_2695839, EPI_ISL_2695840, EPI_ISL_2695848 | | | |
| see above | PathCare, Cape Town | Division of Medical Virology, National Health Laboratory Service (NHLS), Tygerberg Hospital / Stellenbosch University | Bronwyn Kleinhans; Gert van Zyl; Houriiyah Tegally; Jean Maritz; Petra Raimond; San Emmanual James; San Emmanuel James; Sisonke; Susan Engelbrecht; Tania Stander; Tongai Maponga; Tulio de Oliveira; Wolfgang Preiser |
| EPI_ISL_2621080 | Plettenberg Bay Clinic wc PLC | NHLS/UCT | Arash Iranzadeh; Bruna Galvao; Carolyn Williamson; Deelan Doolabh; Diana Hardie; Gert Marais; Innocent Mudau; Lynn Tyers; Marvin Hsiao; Stephen Korsman |
| EPI_ISL_2547371 | Postdene Clinic | National Health Laboratory Service/University of Cape Town (NHLS/UCT) | Arash Iranzadeh; Bruna Galvao; Carolyn Williamson; Deelan Doolabh; Diana Hardie; Gert Marais; Innocent Mudau; Lynn Tyers; Marvin Hsiao; Stephen Korsman |
| EPI_ISL_2582639, EPI_ISL_2582643, EPI_ISL_2582651, EPI_ISL_2582659, EPI_ISL_2582665, EPI_ISL_2582668, EPI_ISL_2582674, EPI_ISL_2582680, EPI_ISL_2582684, EPI_ISL_2582687, EPI_ISL_2582693, EPI_ISL_2582698, EPI_ISL_2582827, EPI_ISL_2582833, EPI_ISL_2582837, EPI_ISL_2662475, EPI_ISL_2695607, EPI_ISL_2695608, EPI_ISL_2695609, EPI_ISL_2695611, EPI_ISL_2695612, EPI_ISL_2695613, EPI_ISL_2695615, EPI_ISL_2695617, EPI_ISL_2695620, EPI_ISL_2695621, EPI_ISL_2695624, EPI_ISL_2695754, EPI_ISL_2695755, EPI_ISL_2695762, EPI_ISL_2695763, EPI_ISL_2695764, EPI_ISL_2695765, EPI_ISL_2695768, EPI_ISL_2695769, EPI_ISL_2695770, EPI_ISL_2695771 | | | |
| see above | ROB FERREIRA LABORATORY | National Institute for Communicable Diseases of the National Health Laboratory Service | Amoako DG; Bhiman JN; Everatt J; Ismail A; Mahlangu B; Mnguni A; Mohale T; Ntuli N; Scheepers C |
| EPI_ISL_2547346, EPI_ISL_2547347, EPI_ISL_2547348, EPI_ISL_2547349, EPI_ISL_2547350 | Sanddrift nc Clinic | National Health Laboratory Service/University of Cape Town (NHLS/UCT) | Arash Iranzadeh; Bruna Galvao; Carolyn Williamson; Deelan Doolabh; Diana Hardie; Gert Marais; Innocent Mudau; Lynn Tyers; Marvin Hsiao; Stephen Korsman |
| EPI_ISL_2621121 | Sedgefield Clinic wc SGE | NHLS/UCT | Arash Iranzadeh; Bruna Galvao; Carolyn Williamson; Deelan Doolabh; Diana Hardie; Gert Marais; Innocent Mudau; Lynn Tyers; Marvin Hsiao; Stephen Korsman |
| EPI_ISL_2694612, EPI_ISL_2694618, EPI_ISL_2694620, EPI_ISL_2694622, EPI_ISL_2694624, EPI_ISL_2694625, EPI_ISL_2694632, EPI_ISL_2694681, EPI_ISL_2694682, EPI_ISL_2694683, EPI_ISL_2694684, EPI_ISL_2694694, EPI_ISL_2694695, EPI_ISL_2694697, EPI_ISL_2694703, EPI_ISL_2694704, EPI_ISL_2694705, EPI_ISL_2694706, EPI_ISL_2694707, EPI_ISL_2694708, EPI_ISL_2694710, EPI_ISL_2694712, EPI_ISL_2694713, EPI_ISL_2694714, EPI_ISL_2694717, EPI_ISL_2694720, EPI_ISL_2694721, EPI_ISL_2694722, EPI_ISL_2695504, EPI_ISL_2695507, EPI_ISL_2695509, EPI_ISL_2695510, EPI_ISL_2695511, EPI_ISL_2695519, EPI_ISL_2695521, EPI_ISL_2695525, EPI_ISL_2695528 | | | |
| see above | TAMBO MEMORIAL | National Institute for Communicable Diseases of the National Health Laboratory Service | Amoako DG; Bhiman JN; Everatt J; Ismail A; Mahlangu B; Mnguni A; Mohale T; Ntuli N; Scheepers C |
| EPI_ISL_2662486, EPI_ISL_2662487, EPI_ISL_2662489, EPI_ISL_2662490, EPI_ISL_2662491, EPI_ISL_2662556, EPI_ISL_2662558, EPI_ISL_2662559, EPI_ISL_2662560, EPI_ISL_2662561, EPI_ISL_2662562, EPI_ISL_2662563, EPI_ISL_2662565, EPI_ISL_2662566, EPI_ISL_2662567, EPI_ISL_2662568, EPI_ISL_2662570, EPI_ISL_2662571, EPI_ISL_2662572, EPI_ISL_2662573, EPI_ISL_2662574, EPI_ISL_2662575, EPI_ISL_2662577, EPI_ISL_2662578, EPI_ISL_2662579, EPI_ISL_2662580, EPI_ISL_2662581, EPI_ISL_2662582, EPI_ISL_2662584, EPI_ISL_2662585, EPI_ISL_2662586, EPI_ISL_2662587, EPI_ISL_2662588, EPI_ISL_2662590, EPI_ISL_2662591, EPI_ISL_2662592, EPI_ISL_2662593, EPI_ISL_2662594, EPI_ISL_2662596, EPI_ISL_2662597, EPI_ISL_2662598, EPI_ISL_2662601, EPI_ISL_2662602, EPI_ISL_2662603, EPI_ISL_2662604, EPI_ISL_2662608, EPI_ISL_2662609, EPI_ISL_2662610, EPI_ISL_2662612, EPI_ISL_2662613, EPI_ISL_2662614, EPI_ISL_2662615, EPI_ISL_2662617, EPI_ISL_2695775, EPI_ISL_2695776, EPI_ISL_2695778, EPI_ISL_2695779, EPI_ISL_2695784, EPI_ISL_2709943, EPI_ISL_2709949, EPI_ISL_2709953, EPI_ISL_2709956, EPI_ISL_2709957, EPI_ISL_2709959, EPI_ISL_2709960, EPI_ISL_2709962, EPI_ISL_2709963, EPI_ISL_2709964, EPI_ISL_2709966, EPI_ISL_2709968, EPI_ISL_2709969, EPI_ISL_2709972, EPI_ISL_2709976, EPI_ISL_2709979, EPI_ISL_2709981, EPI_ISL_2709982, EPI_ISL_2709983, EPI_ISL_2709987, EPI_ISL_2709988, EPI_ISL_2709989, EPI_ISL_2709992, EPI_ISL_2709993 | | | |
| see above | TAMBO MEMORIAL LABORATORY | National Institute for Communicable Diseases of the National Health Laboratory Service | Amoako DG; Bhiman JN; Everatt J; Ismail A; Mahlangu B; Mnguni A; Mohale T; Ntuli N; Scheepers C |
| EPI_ISL_2621074, EPI_ISL_2621078, EPI_ISL_2621081 | Thembalethu CDC wc THC | NHLS/UCT | Arash Iranzadeh; Bruna Galvao; Carolyn Williamson; Deelan Doolabh; Diana Hardie; Gert Marais; Innocent Mudau; Lynn Tyers; Marvin Hsiao; Stephen Korsman |
| EPI_ISL_2662476, EPI_ISL_2662477, EPI_ISL_2662478, EPI_ISL_2662479, EPI_ISL_2662480, EPI_ISL_2662481, EPI_ISL_2662482, EPI_ISL_2662483, EPI_ISL_2662484, EPI_ISL_2662485, EPI_ISL_2662529, EPI_ISL_2662530, EPI_ISL_2662531, EPI_ISL_2662533, EPI_ISL_2662536, EPI_ISL_2662537 | | | |
| see above | VERMAAK LABORATORY | National Institute for Communicable Diseases of the National Health Laboratory Service | Amoako DG; Bhiman JN; Everatt J; Ismail A; Mahlangu B; Mnguni A; Mohale T; Ntuli N; Scheepers C |
| EPI_ISL_2582723, EPI_ISL_2582726, EPI_ISL_2582730, EPI_ISL_2582733, EPI_ISL_2582736, EPI_ISL_2582739, EPI_ISL_2582742, EPI_ISL_2582746, EPI_ISL_2582748, EPI_ISL_2582749, EPI_ISL_2582752, EPI_ISL_2582754, EPI_ISL_2582757, EPI_ISL_2582765, EPI_ISL_2582772, EPI_ISL_2582774, EPI_ISL_2582781, EPI_ISL_2582786, EPI_ISL_2582790, EPI_ISL_2582792, EPI_ISL_2582794, EPI_ISL_2582797, EPI_ISL_2582800, EPI_ISL_2582803 | | | |
| see above | Vermaak & Partners Laboratory | National Institute for Communicable Diseases of the National Health Laboratory Service | Amoako DG; Bhiman JN; Everatt J; Ismail A; Mahlangu B; Mnguni A; Mohale T; Ntuli N; Scheepers C |
| EPI_ISL_2621061, EPI_ISL_2621101, EPI_ISL_2621104 | Victoria Hospital wc VHW | NHLS/UCT | Arash Iranzadeh; Bruna Galvao; Carolyn Williamson; Deelan Doolabh; Diana Hardie; Gert Marais; Innocent Mudau; Lynn Tyers; Marvin Hsiao; Stephen Korsman |
| EPI_ISL_2547363 | Vredendal Hospital wc VRE | National Health Laboratory Service/University of Cape Town (NHLS/UCT) | Arash Iranzadeh; Bruna Galvao; Carolyn Williamson; Deelan Doolabh; Diana Hardie; Gert Marais; Innocent Mudau; Lynn Tyers; Marvin Hsiao; Stephen Korsman |
| EPI_ISL_2547367 | Vredendal North Clinic wc VDN | National Health Laboratory Service/University of Cape Town (NHLS/UCT) | Arash Iranzadeh; Bruna Galvao; Carolyn Williamson; Deelan Doolabh; Diana Hardie; Gert Marais; Innocent Mudau; Lynn Tyers; Marvin Hsiao; Stephen Korsman |
| EPI_ISL_2547364 | Worcester Hospital wc WOC | National Health Laboratory Service/University of Cape Town (NHLS/UCT) | Arash Iranzadeh; Bruna Galvao; Carolyn Williamson; Deelan Doolabh; Diana Hardie; Gert Marais; Innocent Mudau; Lynn Tyers; Marvin Hsiao; Stephen Korsman |
| EPI_ISL_2727240, EPI_ISL_2727244, EPI_ISL_2727246, EPI_ISL_2727251, EPI_ISL_2727253, EPI_ISL_2727256, EPI_ISL_2727257, EPI_ISL_2727258, EPI_ISL_2727259, EPI_ISL_2727260, EPI_ISL_2727266, EPI_ISL_2727268, EPI_ISL_2727269, EPI_ISL_2727270, EPI_ISL_2727272, EPI_ISL_2727274, EPI_ISL_2727279, EPI_ISL_2727280, EPI_ISL_2727282, EPI_ISL_2727283, EPI_ISL_2727285, EPI_ISL_2727286, EPI_ISL_2727288, EPI_ISL_2727289, EPI_ISL_2727290 | | | |
| see above | ZARV/NHLS, Department Medical Virology, University of Pretoria | KRISP, KZN Research Innovation and Sequencing Platform | Adriano Mendes; Amy Strydom; Emmanuel SJ; Giandhari J; Lessells R; Micheala Davids; Naidoo Y; Pillay S; Ramphal U; Sim Mayaphi and Marietjie Venter; Tegally H; Wilkinson E; de Oliveira T |

EPI_ISL_2727331, EPI_ISL_2727332, EPI_ISL_2727333, EPI_ISL_2727334, EPI_ISL_2727336, EPI_ISL_2727337, EPI_ISL_2727338, EPI_ISL_2727339, EPI_ISL_2727340, EPI_ISL_2727341, EPI_ISL_2727342, EPI_ISL_2727344, EPI_ISL_2727345, EPI_ISL_2727346, EPI_ISL_2727348, EPI_ISL_2727350, EPI_ISL_2727356, EPI_ISL_2727368, EPI_ISL_2727372

| | | | |
|---|---|---|---|
| see above | ZARV/NHLS, Department Medical Virology, University of Pretoria | KRISP, KZn Research Innovation and Sequencing Platform | Adriano Mendes; Amy Strydom; Emmanuel SJ; Giandhari J; Lessells R; Micheala Davids; Naidoo Y; Pillay S; Ramphal U; Sim Mayaphi and Marietjie Venter; Tegally H; Wilkinson E; de Oliveira T |

We gratefully acknowledge the following Authors from the Originating laboratories responsible for obtaining the specimens, as well as the Submitting laboratories where the genome data were generated and shared via GISAID, on which this research is based.

All Submitters of data may be contacted directly via www.gisaid.org

Authors are sorted alphabetically.

| Accession ID | Originating Laboratory | Submitting Laboratory | Authors |
|---|---|---|---|
| EPI_ISL_2785301, EPI_ISL_2785302 | Aegis Sciences Corporation | Centers for Disease Control and Prevention Division of Viral Diseases, Pathogen Discovery | Adrian Paskey; Alec Vest; Benjamin Rambo-Martin; Christopher Gulvick; Clinton R. Paden; Cyndi Clark; Dakota Howard; Darlene Wagner; Dhwani Batra; Dillon Nall; Duncan MacCannell; Ethan Sanders; Holly Houdeshell; Jason Caravas; Kara Moser; Matthew Hardison; Matthew Schmerer; Ola Kvalvaag; Patrick Campbell; Peter W. Cook; Rob Case; Scott Sammons; Shatavia Morrison; Shaun Westlund; Vikramsinha Ghorpade; Yvette Unoarumhi |
| EPI_ISL_2758444 | Alameda County Public Health Lab | Chan-Zuckerberg Biohub | CZB Cliahub Consortium |
| EPI_ISL_2758441, EPI_ISL_2758464, EPI_ISL_2758466, EPI_ISL_2758477 | CA DPH Viral and Rickettsial Disease Laboratory | Chan-Zuckerberg Biohub | CZB Cliahub Consortium |
| EPI_ISL_2757841, EPI_ISL_2757847, EPI_ISL_2757848, EPI_ISL_2782387, EPI_ISL_2782392, EPI_ISL_2782406, EPI_ISL_2782410, EPI_ISL_2782430, EPI_ISL_2782437, EPI_ISL_2782447, EPI_ISL_2782464, EPI_ISL_2782517, EPI_ISL_2782532, EPI_ISL_2782534, EPI_ISL_2782535, EPI_ISL_2782588, EPI_ISL_2782595, EPI_ISL_2782606, EPI_ISL_2782607, EPI_ISL_2782608, EPI_ISL_2782612, EPI_ISL_2782617, EPI_ISL_2782636, EPI_ISL_2782645, EPI_ISL_2782659, EPI_ISL_2782661, EPI_ISL_2782664, EPI_ISL_2782683, EPI_ISL_2782687, EPI_ISL_2782692, EPI_ISL_2782709, EPI_ISL_2782711, EPI_ISL_2782712, EPI_ISL_2782716, EPI_ISL_2782727, EPI_ISL_2782728, EPI_ISL_2782730, EPI_ISL_2782731, EPI_ISL_2782736, EPI_ISL_2782737, EPI_ISL_2782741, EPI_ISL_2782746, EPI_ISL_2782747, EPI_ISL_2782749, EPI_ISL_2782750, EPI_ISL_2782751, EPI_ISL_2782752, EPI_ISL_2782753, EPI_ISL_2782759, EPI_ISL_2782764, EPI_ISL_2782768, EPI_ISL_2782770, EPI_ISL_2782791, EPI_ISL_2782794, EPI_ISL_2782796, EPI_ISL_2782799, EPI_ISL_2782821, EPI_ISL_2782830, EPI_ISL_2782833, EPI_ISL_2782834, EPI_ISL_2782838, EPI_ISL_2782840, EPI_ISL_2782842, EPI_ISL_2782843, EPI_ISL_2782846, EPI_ISL_2782847, EPI_ISL_2782851, EPI_ISL_2782852, EPI_ISL_2782857, EPI_ISL_2782859, EPI_ISL_2782861, EPI_ISL_2782863, EPI_ISL_2782865, EPI_ISL_2782869, EPI_ISL_2782870, EPI_ISL_2782873, EPI_ISL_2782880, EPI_ISL_2782882, EPI_ISL_2782885, EPI_ISL_2782886, EPI_ISL_2782892, EPI_ISL_2782893, EPI_ISL_2782897, EPI_ISL_2782899, EPI_ISL_2782902, EPI_ISL_2782906, EPI_ISL_2782907, EPI_ISL_2782908, EPI_ISL_2782911, EPI_ISL_2782913, EPI_ISL_2782937, EPI_ISL_2782949, EPI_ISL_2782956, EPI_ISL_2782961, EPI_ISL_2782967, EPI_ISL_2782971, EPI_ISL_2782972, EPI_ISL_2782973, EPI_ISL_2782978, EPI_ISL_2782984, EPI_ISL_2782986, EPI_ISL_2782989, EPI_ISL_2782992, EPI_ISL_2782995, EPI_ISL_2782997, EPI_ISL_2783001, EPI_ISL_2783015, EPI_ISL_2783018, EPI_ISL_2783026, EPI_ISL_2783035, EPI_ISL_2783036, EPI_ISL_2783038, EPI_ISL_2783043, EPI_ISL_2783047, EPI_ISL_2783052, EPI_ISL_2783054, EPI_ISL_2783055, EPI_ISL_2783058, EPI_ISL_2783059, EPI_ISL_2783068, EPI_ISL_2783071, EPI_ISL_2783083, EPI_ISL_2783095, EPI_ISL_2783097, EPI_ISL_2783102, EPI_ISL_2783111, EPI_ISL_2783122, EPI_ISL_2783132, EPI_ISL_2783135, EPI_ISL_2783138, EPI_ISL_2783144, EPI_ISL_2783145, EPI_ISL_2783146, EPI_ISL_2783147, EPI_ISL_2783151, EPI_ISL_2783158, EPI_ISL_2783161, EPI_ISL_2783162, EPI_ISL_2783170, EPI_ISL_2783172, EPI_ISL_2783173, EPI_ISL_2783174, EPI_ISL_2783179, EPI_ISL_2783213, EPI_ISL_2783216, EPI_ISL_2783217, EPI_ISL_2783218, EPI_ISL_2783220, EPI_ISL_2783221, EPI_ISL_2783226, EPI_ISL_2783231, EPI_ISL_2783232, EPI_ISL_2783233, EPI_ISL_2783234, EPI_ISL_2783236, EPI_ISL_2783237, EPI_ISL_2783239, EPI_ISL_2783246, EPI_ISL_2783247, EPI_ISL_2783248, EPI_ISL_2783250, EPI_ISL_2783251, EPI_ISL_2783252, EPI_ISL_2783256, EPI_ISL_2783270, EPI_ISL_2783276, EPI_ISL_2783277, EPI_ISL_2783291, EPI_ISL_2783293, EPI_ISL_2783301, EPI_ISL_2783302, EPI_ISL_2783303, EPI_ISL_2783307, EPI_ISL_2783309, EPI_ISL_2783316, EPI_ISL_2783318, EPI_ISL_2783329, EPI_ISL_2783333, EPI_ISL_2783337, EPI_ISL_2783357, EPI_ISL_2783359, EPI_ISL_2783373, EPI_ISL_2783380, EPI_ISL_2783387, EPI_ISL_2783395, EPI_ISL_2783402, EPI_ISL_2783405, EPI_ISL_2783407, EPI_ISL_2783411, EPI_ISL_2783417, EPI_ISL_2783420, EPI_ISL_2783426, EPI_ISL_2783428, EPI_ISL_2783431, EPI_ISL_2783433, EPI_ISL_2783436, EPI_ISL_2783437, EPI_ISL_2783438, EPI_ISL_2783442, EPI_ISL_2783444, EPI_ISL_2783448, EPI_ISL_2783450, EPI_ISL_2783452, EPI_ISL_2783453, EPI_ISL_2783457, EPI_ISL_2783493, EPI_ISL_2783494, EPI_ISL_2783500, EPI_ISL_2783505, EPI_ISL_2783509 | | | |
| see above | CDPH VBL | California Department of Public Health | CDPH-COVIDNet; UCI Genome Sciences Center/GHTF; UCSF Center for Advanced Technology |
| EPI_ISL_2776222, EPI_ISL_2776229, EPI_ISL_2776238, EPI_ISL_2776241, EPI_ISL_2776246, EPI_ISL_2776252, EPI_ISL_2776274, EPI_ISL_2776287, EPI_ISL_2776290, EPI_ISL_2776296, EPI_ISL_2776302, EPI_ISL_2776311, EPI_ISL_2776321, EPI_ISL_2776332, EPI_ISL_2776335, EPI_ISL_2776351, EPI_ISL_2776352, EPI_ISL_2776357, EPI_ISL_2776359, EPI_ISL_2776362, EPI_ISL_2776366, EPI_ISL_2776368, EPI_ISL_2778104, EPI_ISL_2778110, EPI_ISL_2778112, EPI_ISL_2778123, EPI_ISL_2778125, EPI_ISL_2778129, EPI_ISL_2778138, EPI_ISL_2778143, EPI_ISL_2778147, EPI_ISL_2778150, EPI_ISL_2778169, EPI_ISL_2778171, EPI_ISL_2778180, EPI_ISL_2778192, EPI_ISL_2778205, EPI_ISL_2778224, EPI_ISL_2778227, EPI_ISL_2778228, EPI_ISL_2778229, EPI_ISL_2778231, EPI_ISL_2778245, EPI_ISL_2778247, EPI_ISL_2778272, EPI_ISL_2778279, EPI_ISL_2778287, EPI_ISL_2778288, EPI_ISL_2778289, EPI_ISL_2778299, EPI_ISL_2778302, EPI_ISL_2778303, EPI_ISL_2778327, EPI_ISL_2778330, EPI_ISL_2778334, EPI_ISL_2778340, EPI_ISL_2778342, EPI_ISL_2778348, EPI_ISL_2778358, EPI_ISL_2778361, EPI_ISL_2778367, EPI_ISL_2778368, EPI_ISL_2778373, EPI_ISL_2778374, EPI_ISL_2778376, EPI_ISL_2778377, EPI_ISL_2778378, EPI_ISL_2778391, EPI_ISL_2778395, EPI_ISL_2778414, EPI_ISL_2778416, EPI_ISL_2778417, EPI_ISL_2778419, EPI_ISL_2778443, EPI_ISL_2778448, EPI_ISL_2778449, EPI_ISL_2778456, EPI_ISL_2778458, EPI_ISL_2778461, EPI_ISL_2778463, EPI_ISL_2778465, EPI_ISL_2778469, EPI_ISL_2778473, EPI_ISL_2778474, EPI_ISL_2778477, EPI_ISL_2778480, EPI_ISL_2778481, EPI_ISL_2778486, EPI_ISL_2778495, EPI_ISL_2778497, EPI_ISL_2778500, EPI_ISL_2778502, EPI_ISL_2778503, EPI_ISL_2778506, EPI_ISL_2778516, EPI_ISL_2778518, EPI_ISL_2778519, EPI_ISL_2778520, EPI_ISL_2778521, EPI_ISL_2778525, EPI_ISL_2778528, EPI_ISL_2778543, EPI_ISL_2778545, EPI_ISL_2778552, EPI_ISL_2778555, EPI_ISL_2778559, EPI_ISL_2778562, EPI_ISL_2778564, EPI_ISL_2778565, EPI_ISL_2778568, EPI_ISL_2778569, EPI_ISL_2778572, EPI_ISL_2778573, EPI_ISL_2778575, EPI_ISL_2778576, EPI_ISL_2778577, EPI_ISL_2778580, EPI_ISL_2778581, EPI_ISL_2778597, EPI_ISL_2778604, EPI_ISL_2778606, EPI_ISL_2778607, EPI_ISL_2778615, EPI_ISL_2778620, EPI_ISL_2778622, EPI_ISL_2778627, EPI_ISL_2778628, EPI_ISL_2778630, EPI_ISL_2778639, EPI_ISL_2778643, EPI_ISL_2778653, EPI_ISL_2778660, EPI_ISL_2778667, EPI_ISL_2778674, EPI_ISL_2778677, EPI_ISL_2778681, EPI_ISL_2778687, EPI_ISL_2778698, EPI_ISL_2778699, EPI_ISL_2778700, EPI_ISL_2778701, EPI_ISL_2778714, EPI_ISL_2778722, EPI_ISL_2778726, EPI_ISL_2778728, EPI_ISL_2778731, EPI_ISL_2778741, EPI_ISL_2778753, EPI_ISL_2778754, EPI_ISL_2778765, EPI_ISL_2778766, EPI_ISL_2778769, EPI_ISL_2778773, EPI_ISL_2778781, EPI_ISL_2778782, EPI_ISL_2778805, EPI_ISL_2778809, EPI_ISL_2778810, EPI_ISL_2778827, EPI_ISL_2778837, EPI_ISL_2778851, EPI_ISL_2778854, EPI_ISL_2778882, EPI_ISL_2778890, EPI_ISL_2778915, EPI_ISL_2778945 | | | |
| see above | California Department of Public Health Valencia Branch Laboratory (CDPH VBL) | California Department of Public Health | Arias Lab at UCSB; CDPH-COVIDNet; UCLA Technology Center for Genomics & Bioinformatics |
| EPI_ISL_2758596 | County of San Luis Obispo Public Health Laboratory | County of San Luis Obispo Public Health Laboratory | Frances Sidhu; Katrina Erwin |
| EPI_ISL_2785635, EPI_ISL_2785658 | Fulgent Genetics | Centers for Disease Control and Prevention Division of Viral Diseases, Pathogen Discovery | Adrian Paskey; Becky Tsai; Benafsh Sapra; Benjamin Rambo-Martin; Christopher Gulvick; Clinton R. Paden; Dakota Howard; Darlene Wagner; Dhwani Batra; Doreen Ng; Duncan MacCannell; Harry Gao; James Xie; Jason Caravas; John Gao; Joseph Fierro; Kara Moser; Matthew Schmerer; Mickey Li; Peter W. Cook; Scott Sammons; Shatavia Morrison; Yan Meng; Yvette Unoarumhi |
| EPI_ISL_2716877, EPI_ISL_2716881, EPI_ISL_2716882, EPI_ISL_2716884, EPI_ISL_2716887, EPI_ISL_2754038, EPI_ISL_2754039, EPI_ISL_2754042, EPI_ISL_2759079, EPI_ISL_2759084, EPI_ISL_2774519, EPI_ISL_2774521, EPI_ISL_2774523 | | | |
| see above | Monterey County Public Health Laboratory | Monterey County Public Health Laboratory | Monterey County Public Health Laboratory |
| EPI_ISL_2753974 | PAMF Palo Alto | Santa Clara County Public Health Laboratory | Santa Clara County Public Health Department |
| EPI_ISL_2725518, EPI_ISL_2725524, EPI_ISL_2758429, EPI_ISL_2758492, EPI_ISL_2758502, EPI_ISL_2758515 | Santa Clara County Public Health Laboratory | Chan-Zuckerberg Biohub | CZB Cliahub Consortium |
| EPI_ISL_2779561, EPI_ISL_2779566, EPI_ISL_2779568, EPI_ISL_2779569, EPI_ISL_2779577, EPI_ISL_2779586, EPI_ISL_2779591, EPI_ISL_2779592, EPI_ISL_2779593, EPI_ISL_2779598, EPI_ISL_2779608, EPI_ISL_2779610 | | | |
| see above | Santa Clara County Public Health Laboratory | Santa Clara County Public Health Laboratory | Santa Clara County Public Health Department |
| EPI_ISL_2774576, EPI_ISL_2774578, EPI_ISL_2774579 | Santa Clara Valley Medical Center | Santa Clara County Public Health Laboratory | Santa Clara County Public Health Department |

We gratefully acknowledge the following Authors from the Originating laboratories responsible for obtaining the specimens, as well as the Submitting laboratories where the genome data were generated and shared via GISAID, on which this research is based.

All Submitters of data may be contacted directly via www.gisaid.org

Authors are sorted alphabetically.

| Accession ID | Originating Laboratory | Submitting Laboratory | Authors |
|---|---|---|---|
| EPI_ISL_2758448, EPI_ISL_2758465, EPI_ISL_2758489, EPI_ISL_2758520 | CA DPH Viral and Rickettsial Disease Laboratory | Chan-Zuckerberg Biohub | CZB Cliahub Consortium |
| EPI_ISL_2757838, EPI_ISL_2757839, EPI_ISL_2757844, EPI_ISL_2757846, EPI_ISL_2757849, EPI_ISL_2782396, EPI_ISL_2782404, EPI_ISL_2782408, EPI_ISL_2782420, EPI_ISL_2782429, EPI_ISL_2782443, EPI_ISL_2782451, EPI_ISL_2782452, EPI_ISL_2782456, EPI_ISL_2782457, EPI_ISL_2782458, EPI_ISL_2782463, EPI_ISL_2782474, EPI_ISL_2782483, EPI_ISL_2782494, EPI_ISL_2782524, EPI_ISL_2782531, EPI_ISL_2782541, EPI_ISL_2782548, EPI_ISL_2782549, EPI_ISL_2782550, EPI_ISL_2782555, EPI_ISL_2782567, EPI_ISL_2782571, EPI_ISL_2782581, EPI_ISL_2782599, EPI_ISL_2782602, EPI_ISL_2782603, EPI_ISL_2782625, EPI_ISL_2782627, EPI_ISL_2782628, EPI_ISL_2782643, EPI_ISL_2782644, EPI_ISL_2782647, EPI_ISL_2782649, EPI_ISL_2782654, EPI_ISL_2782656, EPI_ISL_2782658, EPI_ISL_2782663, EPI_ISL_2782665, EPI_ISL_2782666, EPI_ISL_2782668, EPI_ISL_2782671, EPI_ISL_2782672, EPI_ISL_2782673, EPI_ISL_2782677, EPI_ISL_2782678, EPI_ISL_2782684, EPI_ISL_2782685, EPI_ISL_2782693, EPI_ISL_2782699, EPI_ISL_2782713, EPI_ISL_2782715, EPI_ISL_2782720, EPI_ISL_2782722, EPI_ISL_2782723, EPI_ISL_2782734, EPI_ISL_2782735, EPI_ISL_2782742, EPI_ISL_2782745, EPI_ISL_2782748, EPI_ISL_2782756, EPI_ISL_2782763, EPI_ISL_2782765, EPI_ISL_2782771, EPI_ISL_2782773, EPI_ISL_2782775, EPI_ISL_2782783, EPI_ISL_2782785, EPI_ISL_2782787, EPI_ISL_2782797, EPI_ISL_2782798, EPI_ISL_2782805, EPI_ISL_2782806, EPI_ISL_2782808, EPI_ISL_2782809, EPI_ISL_2782811, EPI_ISL_2782812, EPI_ISL_2782813, EPI_ISL_2782814, EPI_ISL_2782815, EPI_ISL_2782816, EPI_ISL_2782819, EPI_ISL_2782820, EPI_ISL_2782822, EPI_ISL_2782824, EPI_ISL_2782827, EPI_ISL_2782829, EPI_ISL_2782831, EPI_ISL_2782836, EPI_ISL_2782839, EPI_ISL_2782845, EPI_ISL_2782850, EPI_ISL_2782854, EPI_ISL_2782856, EPI_ISL_2782858, EPI_ISL_2782862, EPI_ISL_2782868, EPI_ISL_2782871, EPI_ISL_2782876, EPI_ISL_2782878, EPI_ISL_2782881, EPI_ISL_2782887, EPI_ISL_2782898, EPI_ISL_2782900, EPI_ISL_2782901, EPI_ISL_2782904, EPI_ISL_2782909, EPI_ISL_2782910, EPI_ISL_2782916, EPI_ISL_2782917, EPI_ISL_2782919, EPI_ISL_2782921, EPI_ISL_2782929, EPI_ISL_2782931, EPI_ISL_2782932, EPI_ISL_2782935, EPI_ISL_2782936, EPI_ISL_2782943, EPI_ISL_2782946, EPI_ISL_2782950, EPI_ISL_2782952, EPI_ISL_2782953, EPI_ISL_2782954, EPI_ISL_2782957, EPI_ISL_2782958, EPI_ISL_2782960, EPI_ISL_2782962, EPI_ISL_2782970, EPI_ISL_2782975, EPI_ISL_2782980, EPI_ISL_2782982, EPI_ISL_2782983, EPI_ISL_2782985, EPI_ISL_2782994, EPI_ISL_2783004, EPI_ISL_2783017, EPI_ISL_2783019, EPI_ISL_2783022, EPI_ISL_2783023, EPI_ISL_2783025, EPI_ISL_2783027, EPI_ISL_2783028, EPI_ISL_2783030, EPI_ISL_2783031, EPI_ISL_2783034, EPI_ISL_2783037, EPI_ISL_2783040, EPI_ISL_2783049, EPI_ISL_2783050, EPI_ISL_2783051, EPI_ISL_2783053, EPI_ISL_2783056, EPI_ISL_2783063, EPI_ISL_2783064, EPI_ISL_2783066, EPI_ISL_2783067, EPI_ISL_2783072, EPI_ISL_2783073, EPI_ISL_2783074, EPI_ISL_2783075, EPI_ISL_2783078, EPI_ISL_2783079, EPI_ISL_2783080, EPI_ISL_2783082, EPI_ISL_2783086, EPI_ISL_2783088, EPI_ISL_2783090, EPI_ISL_2783093, EPI_ISL_2783094, EPI_ISL_2783098, EPI_ISL_2783103, EPI_ISL_2783107, EPI_ISL_2783110, EPI_ISL_2783116, EPI_ISL_2783120, EPI_ISL_2783123, EPI_ISL_2783124, EPI_ISL_2783128, EPI_ISL_2783129, EPI_ISL_2783130, EPI_ISL_2783131, EPI_ISL_2783133, EPI_ISL_2783139, EPI_ISL_2783141, EPI_ISL_2783142, EPI_ISL_2783143, EPI_ISL_2783152, EPI_ISL_2783153, EPI_ISL_2783154, EPI_ISL_2783156, EPI_ISL_2783157, EPI_ISL_2783169, EPI_ISL_2783175, EPI_ISL_2783182, EPI_ISL_2783183, EPI_ISL_2783186, EPI_ISL_2783187, EPI_ISL_2783189, EPI_ISL_2783192, EPI_ISL_2783193, EPI_ISL_2783195, EPI_ISL_2783197, EPI_ISL_2783198, EPI_ISL_2783200, EPI_ISL_2783202, EPI_ISL_2783206, EPI_ISL_2783207, EPI_ISL_2783208, EPI_ISL_2783210, EPI_ISL_2783214, EPI_ISL_2783215, EPI_ISL_2783219, EPI_ISL_2783223, EPI_ISL_2783224, EPI_ISL_2783225, EPI_ISL_2783227, EPI_ISL_2783229, EPI_ISL_2783230, EPI_ISL_2783238, EPI_ISL_2783242, EPI_ISL_2783245, EPI_ISL_2783249, EPI_ISL_2783253, EPI_ISL_2783266, EPI_ISL_2783267, EPI_ISL_2783271, EPI_ISL_2783272, EPI_ISL_2783281, EPI_ISL_2783286, EPI_ISL_2783313, EPI_ISL_2783315, EPI_ISL_2783325, EPI_ISL_2783330, EPI_ISL_2783331, EPI_ISL_2783332, EPI_ISL_2783335, EPI_ISL_2783338, EPI_ISL_2783340, EPI_ISL_2783342, EPI_ISL_2783343, EPI_ISL_2783346, EPI_ISL_2783348, EPI_ISL_2783349, EPI_ISL_2783350, EPI_ISL_2783351, EPI_ISL_2783352, EPI_ISL_2783353, EPI_ISL_2783354, EPI_ISL_2783355, EPI_ISL_2783356, EPI_ISL_2783363, EPI_ISL_2783364, EPI_ISL_2783367, EPI_ISL_2783368, EPI_ISL_2783369, EPI_ISL_2783370, EPI_ISL_2783371, EPI_ISL_2783376, EPI_ISL_2783378, EPI_ISL_2783381, EPI_ISL_2783382, EPI_ISL_2783384, EPI_ISL_2783386, EPI_ISL_2783388, EPI_ISL_2783394, EPI_ISL_2783397, EPI_ISL_2783398, EPI_ISL_2783403, EPI_ISL_2783413, EPI_ISL_2783421, EPI_ISL_2783423, EPI_ISL_2783424, EPI_ISL_2783429, EPI_ISL_2783430, EPI_ISL_2783434, EPI_ISL_2783446, EPI_ISL_2783456, EPI_ISL_2783458, EPI_ISL_2783459, EPI_ISL_2783465, EPI_ISL_2783467, EPI_ISL_2783475, EPI_ISL_2783478, EPI_ISL_2783487, EPI_ISL_2783495, EPI_ISL_2783496 | | | |
| see above | CDPH VBL | California Department of Public Health | CDPH-COVIDNet; UCI Genome Sciences Center/GHTF; UCSF Center for Advanced Technology |
| EPI_ISL_2776221, EPI_ISL_2776223, EPI_ISL_2776224, EPI_ISL_2776226, EPI_ISL_2776234, EPI_ISL_2776235, EPI_ISL_2776237, EPI_ISL_2776240, EPI_ISL_2776245, EPI_ISL_2776247, EPI_ISL_2776249, EPI_ISL_2776250, EPI_ISL_2776251, EPI_ISL_2776254, EPI_ISL_2776255, EPI_ISL_2776258, EPI_ISL_2776261, EPI_ISL_2776262, EPI_ISL_2776266, EPI_ISL_2776268, EPI_ISL_2776269, EPI_ISL_2776273, EPI_ISL_2776275, EPI_ISL_2776280, EPI_ISL_2776281, EPI_ISL_2776282, EPI_ISL_2776283, EPI_ISL_2776288, EPI_ISL_2776289, EPI_ISL_2776291, EPI_ISL_2776297, EPI_ISL_2776299, EPI_ISL_2776309, EPI_ISL_2776317, EPI_ISL_2776318, EPI_ISL_2776320, EPI_ISL_2776323, EPI_ISL_2776328, EPI_ISL_2776330, EPI_ISL_2776331, EPI_ISL_2776337, EPI_ISL_2776347, EPI_ISL_2776348, EPI_ISL_2776354, EPI_ISL_2776360, EPI_ISL_2776361, EPI_ISL_2776364, EPI_ISL_2776365, EPI_ISL_2778109, EPI_ISL_2778113, EPI_ISL_2778114, EPI_ISL_2778115, EPI_ISL_2778117, EPI_ISL_2778120, EPI_ISL_2778126, EPI_ISL_2778127, EPI_ISL_2778130, EPI_ISL_2778132, EPI_ISL_2778136, EPI_ISL_2778144, EPI_ISL_2778145, EPI_ISL_2778148, EPI_ISL_2778149, EPI_ISL_2778153, EPI_ISL_2778154, EPI_ISL_2778156, EPI_ISL_2778157, EPI_ISL_2778158, EPI_ISL_2778159, EPI_ISL_2778160, EPI_ISL_2778161, EPI_ISL_2778162, EPI_ISL_2778163, EPI_ISL_2778164, EPI_ISL_2778165, EPI_ISL_2778166, EPI_ISL_2778167, EPI_ISL_2778168, EPI_ISL_2778170, EPI_ISL_2778172, EPI_ISL_2778173, EPI_ISL_2778174, EPI_ISL_2778176, EPI_ISL_2778184, EPI_ISL_2778189, EPI_ISL_2778191, EPI_ISL_2778196, EPI_ISL_2778203, EPI_ISL_2778204, EPI_ISL_2778207, EPI_ISL_2778213, EPI_ISL_2778215, EPI_ISL_2778216, EPI_ISL_2778217, EPI_ISL_2778218, EPI_ISL_2778222, EPI_ISL_2778223, EPI_ISL_2778233, EPI_ISL_2778237, EPI_ISL_2778240, EPI_ISL_2778242, EPI_ISL_2778243, EPI_ISL_2778246, EPI_ISL_2778250, EPI_ISL_2778251, EPI_ISL_2778254, EPI_ISL_2778258, EPI_ISL_2778259, EPI_ISL_2778260, EPI_ISL_2778261, EPI_ISL_2778262, EPI_ISL_2778264, EPI_ISL_2778265, EPI_ISL_2778266, EPI_ISL_2778267, EPI_ISL_2778268, EPI_ISL_2778269, EPI_ISL_2778270, EPI_ISL_2778271, EPI_ISL_2778273, EPI_ISL_2778276, EPI_ISL_2778277, EPI_ISL_2778278, EPI_ISL_2778280, EPI_ISL_2778281, EPI_ISL_2778282, EPI_ISL_2778292, EPI_ISL_2778293, EPI_ISL_2778294, EPI_ISL_2778295, EPI_ISL_2778296, EPI_ISL_2778297, EPI_ISL_2778298, EPI_ISL_2778304, EPI_ISL_2778310, EPI_ISL_2778315, EPI_ISL_2778316, EPI_ISL_2778317, EPI_ISL_2778320, EPI_ISL_2778321, EPI_ISL_2778323, EPI_ISL_2778324, EPI_ISL_2778325, EPI_ISL_2778326, EPI_ISL_2778328, EPI_ISL_2778331, EPI_ISL_2778332, EPI_ISL_2778333, EPI_ISL_2778336, EPI_ISL_2778339, EPI_ISL_2778341, EPI_ISL_2778347, EPI_ISL_2778349, EPI_ISL_2778350, EPI_ISL_2778352, EPI_ISL_2778353, EPI_ISL_2778357, EPI_ISL_2778359, EPI_ISL_2778371, EPI_ISL_2778380, EPI_ISL_2778383, EPI_ISL_2778385, EPI_ISL_2778388, EPI_ISL_2778394, EPI_ISL_2778396, EPI_ISL_2778402, EPI_ISL_2778404, EPI_ISL_2778405, EPI_ISL_2778407, EPI_ISL_2778408, EPI_ISL_2778409, EPI_ISL_2778410, EPI_ISL_2778412, EPI_ISL_2778413, EPI_ISL_2778420, EPI_ISL_2778428, EPI_ISL_2778430, EPI_ISL_2778431, EPI_ISL_2778432, EPI_ISL_2778433, EPI_ISL_2778436, EPI_ISL_2778437, EPI_ISL_2778438, EPI_ISL_2778440, EPI_ISL_2778445, EPI_ISL_2778446, EPI_ISL_2778447, EPI_ISL_2778451, EPI_ISL_2778454, EPI_ISL_2778455, EPI_ISL_2778459, EPI_ISL_2778460, EPI_ISL_2778464, EPI_ISL_2778467, EPI_ISL_2778468, EPI_ISL_2778470, EPI_ISL_2778472, EPI_ISL_2778475, EPI_ISL_2778479, EPI_ISL_2778482, EPI_ISL_2778484, EPI_ISL_2778489, EPI_ISL_2778492, EPI_ISL_2778493, EPI_ISL_2778498, EPI_ISL_2778499, EPI_ISL_2778501, EPI_ISL_2778504, EPI_ISL_2778505, EPI_ISL_2778507, EPI_ISL_2778508, EPI_ISL_2778510, EPI_ISL_2778514, EPI_ISL_2778515, EPI_ISL_2778522, EPI_ISL_2778526, EPI_ISL_2778530, EPI_ISL_2778531, EPI_ISL_2778533, EPI_ISL_2778534, EPI_ISL_2778541, EPI_ISL_2778542, EPI_ISL_2778544, EPI_ISL_2778548, EPI_ISL_2778550, EPI_ISL_2778551, EPI_ISL_2778557, EPI_ISL_2778558, EPI_ISL_2778561, EPI_ISL_2778563, EPI_ISL_2778566, EPI_ISL_2778570, EPI_ISL_2778571, EPI_ISL_2778574, EPI_ISL_2778585, EPI_ISL_2778586, EPI_ISL_2778589, EPI_ISL_2778590, EPI_ISL_2778598, EPI_ISL_2778600, EPI_ISL_2778602, EPI_ISL_2778605, EPI_ISL_2778611, EPI_ISL_2778613, EPI_ISL_2778614, EPI_ISL_2778617, EPI_ISL_2778621, EPI_ISL_2778625, EPI_ISL_2778626, EPI_ISL_2778632, EPI_ISL_2778633, EPI_ISL_2778634, EPI_ISL_2778635, EPI_ISL_2778637, EPI_ISL_2778638, EPI_ISL_2778642, EPI_ISL_2778644, EPI_ISL_2778648, EPI_ISL_2778649, EPI_ISL_2778650, EPI_ISL_2778651, EPI_ISL_2778652, EPI_ISL_2778654, EPI_ISL_2778655, EPI_ISL_2778662, EPI_ISL_2778665, EPI_ISL_2778666, EPI_ISL_2778668, EPI_ISL_2778672, EPI_ISL_2778673, EPI_ISL_2778675, EPI_ISL_2778678, EPI_ISL_2778679, EPI_ISL_2778680, EPI_ISL_2778684, EPI_ISL_2778693, EPI_ISL_2778697, EPI_ISL_2778710, EPI_ISL_2778718, EPI_ISL_2778719, EPI_ISL_2778721, EPI_ISL_2778723, EPI_ISL_2778724, EPI_ISL_2778725, EPI_ISL_2778727, EPI_ISL_2778729, EPI_ISL_2778732, EPI_ISL_2778733, EPI_ISL_2778734, EPI_ISL_2778742, EPI_ISL_2778746, EPI_ISL_2778755, EPI_ISL_2778756, EPI_ISL_2778757, EPI_ISL_2778770, EPI_ISL_2778776, EPI_ISL_2778804, EPI_ISL_2778815, EPI_ISL_2778861, EPI_ISL_2778864, EPI_ISL_2778865, EPI_ISL_2778869, EPI_ISL_2778878, EPI_ISL_2778905, EPI_ISL_2778908, EPI_ISL_2778909, EPI_ISL_2778927, EPI_ISL_2778940, EPI_ISL_2778947 | | | |
| see above | California Department of Public Health Valencia Branch Laboratory (CDPH VBL) | California Department of Public Health | Arias Lab at UCSB; CDPH-COVIDNet; UCLA Technology Center for Genomics & Bioinformatics |
| EPI_ISL_2758459, EPI_ISL_2758468, EPI_ISL_2758469, EPI_ISL_2758480, EPI_ISL_2758481 | Contra Costa County Public Health Lab | Chan-Zuckerberg Biohub | CZB Cliahub Consortium |
| EPI_ISL_2758474, EPI_ISL_2758482 | County of San Luis Obispo Public Health Laboratory | Chan-Zuckerberg Biohub | CZB Cliahub Consortium |
| EPI_ISL_2758595 | County of San Luis Obispo Public Health Laboratory | County of San Luis Obispo Public Health Laboratory | Frances Sidhu; Katrina Erwin |
| EPI_ISL_2758453, EPI_ISL_2758460, EPI_ISL_2758473 | Humboldt County Public Health Laboratory | Chan-Zuckerberg Biohub | CZB Cliahub Consortium |
| EPI_ISL_2753968, EPI_ISL_2753971 | Kaiser San Jose | Santa Clara County Public Health Laboratory | Santa Clara County Public Health Department |
| EPI_ISL_2774563, EPI_ISL_2774570, EPI_ISL_2774571, EPI_ISL_2774573, EPI_ISL_2774574 | Los Angeles County Public Health Laboratories | Los Angeles County Public Health Laboratories | P. Hemarajata et al. |
| EPI_ISL_2716876, EPI_ISL_2716879, EPI_ISL_2716883, EPI_ISL_2716885 | Monterey County Public Health Laboratory | Monterey County Public Health Laboratory | Monterey County Public Health Laboratory |
| EPI_ISL_2758434, EPI_ISL_2758435 | Orange County Public Health Lab | Chan-Zuckerberg Biohub | CZB Cliahub Consortium |
| EPI_ISL_2758420, EPI_ISL_2758430, EPI_ISL_2758451 | Santa Clara County Public Health Laboratory | Chan-Zuckerberg Biohub | CZB Cliahub Consortium |
| EPI_ISL_2779560, EPI_ISL_2779567, EPI_ISL_2779570, EPI_ISL_2779571, EPI_ISL_2779572, EPI_ISL_2779573, EPI_ISL_2779581, EPI_ISL_2779588, EPI_ISL_2779595, EPI_ISL_2779597, EPI_ISL_2779601, EPI_ISL_2779602, EPI_ISL_2779604, EPI_ISL_2779605 | | | |
| see above | Santa Clara County Public Health Laboratory | Santa Clara County Public Health Laboratory | Santa Clara County Public Health Department |
| EPI_ISL_2772509 | UCLA Clinical Micro Lab | Los Angeles County Public Health Laboratories | P. Hemarajata et al. |
| EPI_ISL_2774567 | USC Clinical Lab | Los Angeles County Public Health Laboratories | P. Hemarajata et al. |

We gratefully acknowledge the following Authors from the Originating laboratories responsible for obtaining the specimens, as well as the Submitting laboratories where the genome data were generated and shared via GISAID, on which this research is based.

All Submitters of data may be contacted directly via www.gisaid.org

Authors are sorted alphabetically.

| Accession ID | Originating Laboratory | Submitting Laboratory | Authors |
|---|---|---|---|
| EPI_ISL_2650809 | A.S.L. NOVARA - ISTITUTO S.S. TRINITAEUR - BORGOMANERO - S.S. LABORATORIO ANALISI CHIMICO CLINICHE | Fondazione del Piemonte per l'Oncologia IRCCS | Antonino Sottile; Giorgio Giardina; Paola Marino; Silvia Brossa |
| EPI_ISL_2978723 | A.S.L. NOVARA-BORGOMANERO | Fondazione del Piemonte per l'Oncologia IRCCS | Antonino Sottile; Giorgio Giardina; Paola Marino; Silvia Brossa |
| EPI_ISL_2720511, EPI_ISL_2721308 | A.S.L. TO4 | Fondazione del Piemonte per l'Oncologia IRCCS | Antonino Sottile; Giorgio Giardina; Paola Marino; Silvia Brossa |
| EPI_ISL_2720402 | A.S.L. VCO | Fondazione del Piemonte per l'Oncologia IRCCS | Antonino Sottile; Giorgio Giardina; Paola Marino; Silvia Brossa |
| EPI_ISL_2650871 | A.S.L. VERBANO-CUSIO-OSSOLA - OSPEDALE UNICO PLURISEDE - POLIAMBULATORIO OSPEDALIERO P.O. VERBANIA | Fondazione del Piemonte per l'Oncologia IRCCS | Antonino Sottile; Giorgio Giardina; Paola Marino; Silvia Brossa |
| EPI_ISL_2720404 | A.S.L. VERCELLI | Fondazione del Piemonte per l'Oncologia IRCCS | Antonino Sottile; Giorgio Giardina; Paola Marino; Silvia Brossa |
| EPI_ISL_2975147, EPI_ISL_2975151 | AULSS 2 Marca Trevigiana | Istituto Zooprofilattico Sperimentale delle Venezie | Adelaide Milani; Alessia Schivo; Alice Fusaro; Ambra Pastori; Annalisa Salviato; Antonia Ricci; Calogero Terregino; Edoardo Giussani; Elisa Palumbo; Erika Giorgia Quaranta; Isabella Monne; Luca Tassoni |
| EPI_ISL_2727152 | AZDelta | AZ Delta Medical Laboratories in Roeselare, Belgium | Dieter De Smet; Geert Martens; Merijn Vanhee; on behalf of AZ Delta COVID-19 Genomics core (member of Genomic surveillance of SARS-CoV-2 in Belgium network) |
| EPI_ISL_2528220, EPI_ISL_2600220, EPI_ISL_2600245, EPI_ISL_2687044, EPI_ISL_2687176 | Aegis Sciences Corporation | Centers for Disease Control and Prevention Division of Viral Diseases, Pathogen Discovery | Adrian Paskey; Alec Vest; Benjamin Rambo-Martin; Christopher Gulvick; Clinton R. Paden; Cyndi Clark; Dakota Howard; Darlene Wagner; Dhwani Batra; Dillon Nall; Duncan MacCannell; Ethan Sanders; Holly Houdeshell; Jason Caravas; Kara Moser; Matthew Hardison; Matthew Schmerer; Ola Kvalvaag; Patrick Campbell; Peter W. Cook; Rob Case; Scott Sammons; Shatavia Morrison; Shaun Westlund; Vikramsinha Ghorpade; Yvette Unoarumhi |
| EPI_ISL_2547409 | Alma CDC wc AHC | National Health Laboratory Service/University of Cape Town (NHLS/UCT) | Arash Iranzadeh; Bruna Galvao; Carolyn Williamson; Deelan Doolabh; Diana Hardie; Gert Marais; Innocent Mudau; Lynn Tyers; Marvin Hsiao; Stephen Korsman |
| EPI_ISL_2691703 | Analytica | Institute of Medical Virology | Alexandra Trkola; Annette Audigé; Cyril Shah; Gabriela Ziltener; Guido Bloemberg; Jon Huder; Jürg Böni; Kevin Steiner; Maria Grünberg; Maryam Zaheri; Michael Huber; Riccarda Capaul; Stefan Schmutz; Verena Kufner |
| EPI_ISL_2864430 | Arcispedale Santa Maria Nuova, Autoimmunità, Allergologia e Biotecnologie Innovative | Istituto Zooprofilattico Sperimentale della Lombardia e dell'Emilia Romagna (IZSLER), Risk Analysis and Genomic Epidemiology Unit | Alessandro Zerbini; Erika Scaltriti; Ilaria Menozzi; Lucia Belloni; Marina Morganti; Stefania Croci; Stefano Pongolini |
| EPI_ISL_3123113 | Austrian Agency for Health and Food Safety (AGES) | Bergthaler laboratory, CeMM Research Center for Molecular Medicine of the Austrian Academy of Sciences | Andreas Bergthaler; Anna Schedl; Bekir Erguner; Benedikt Agerer; Christoph Bock; Fabian Amman; Jan Laine; Lukas Endler; Maelle Le Moing; Martin Senekowitsch; Matthew Thornton; Michael Schuster; Petr Triska; Thomas Penz |
| EPI_ISL_2622164, EPI_ISL_2622165, EPI_ISL_2622166, EPI_ISL_2622167, EPI_ISL_2622168, EPI_ISL_2622169, EPI_ISL_2622170, EPI_ISL_2622171, EPI_ISL_2622172, EPI_ISL_2622173, EPI_ISL_2622174, EPI_ISL_2622175 | | | |
| see above | Azienda Ospedaliera Pugliese Ciaccio di Catanzaro SOC Microbiologia e Virologia | Azienda Ospedaliera Pugliese Ciaccio di Catanzaro SOC Microbiologia e Virologia | Rossana Tallerico Federica Pasceri Marco De Fazio Ilenia Talotta Giuseppina Panduri Pasquale Minchella |
| EPI_ISL_2604147 | Azienda Ospedaliera Terni | Istituto Zooprofilattico Sperimentale dell'Abruzzo e Molise "G. Caporale" | Ancora M; Calistri P; Cammà C; Curini V; Delli Compagni E; Di Domenico M; Di Pasquale A; Lorusso A; Mangone I; Marcacci M; Palumbo M; Puglia I; Rinaldi A; Savini G; Scaccetti A; Scialabba S |
| EPI_ISL_2864115, EPI_ISL_2864124 | Azienda Sanitaria Locale di Piacenza - Presidio Ospedaliero - Laboratorio di Microbiologia | Istituto Zooprofilattico Sperimentale della Lombardia e dell'Emilia Romagna (IZSLER), Risk Analysis and Genomic Epidemiology Unit | Erika Scaltriti; Ilaria Menozzi; Lo Cascio Giuliana; Marina Morganti; Peroni Irene; Schiavo Roberta; Stefano Pongolini |
| EPI_ISL_2522960, EPI_ISL_2523254, EPI_ISL_2525006, EPI_ISL_2528024, EPI_ISL_2654106, EPI_ISL_2654226, EPI_ISL_2654738, EPI_ISL_2654944, EPI_ISL_2654949, EPI_ISL_2654951, EPI_ISL_2654990, EPI_ISL_2655020, EPI_ISL_2655731, EPI_ISL_2655915, EPI_ISL_2656238, EPI_ISL_2656240, EPI_ISL_2656241, EPI_ISL_2656310, EPI_ISL_2656657, EPI_ISL_2656785, EPI_ISL_2718650, EPI_ISL_2718858, EPI_ISL_2719916, EPI_ISL_2720249, EPI_ISL_2720267, EPI_ISL_2720309, EPI_ISL_2720322, EPI_ISL_2720584, EPI_ISL_2720585, EPI_ISL_2720807, EPI_ISL_2720823, EPI_ISL_2720989, EPI_ISL_2721136, EPI_ISL_2721372, EPI_ISL_2721422, EPI_ISL_2721426, EPI_ISL_2721453, EPI_ISL_2721826, EPI_ISL_2721840, EPI_ISL_2721869, EPI_ISL_2721907, EPI_ISL_2721997 | | | |
| see above | BCCDC Public Health Laboratory | BCCDC Public Health Laboratory | Ana Pacagnella; Corrinne Ng; Dan Fornika; John Tyson; Kim Macdonald; Kimia Kamelian; Linda Hoang; Loretta Janz; Mel Krajden; Prystajecky Natalie; Robert Azana; Shannon Russell |
| EPI_ISL_2933444, EPI_ISL_2933445, EPI_ISL_2933446 | BIOMNIS LYON | CNR Virus des Infections Respiratoires - France SUD | Antonin Bal; Bruno Lina; Gregory Destras; Gwendolyne Burfin; Hadrien Regue; Laurence Josset; Martine Valette; Quentin Semanas |
| EPI_ISL_2957149 | Bayerisches Landesamt für Gesundheit und Lebensmittelsicherheit (LGL) | Robert Koch Institute | |
| EPI_ISL_2615960, EPI_ISL_2615963, EPI_ISL_2615964, EPI_ISL_2615965, EPI_ISL_2615968, EPI_ISL_2615969, EPI_ISL_2615972 | | | |
| see above | Berkeley Medical Center | WVU and Marshall University Combined Genomics Core Facilities | James Denvir; Peter Perrotta; Peter Stoilov; Ryan Percifield; Wesley Kimble |
| EPI_ISL_2795724 | Biogroup Bio Lam-LCD Saint-Denis | Department of Virology, Henri Mondor University Hospital, Assistance Publique Hôpitaux de Paris, Université Paris-Est Créteil, INSERM U955 | Alexandre Soulier; Christophe Rodriguez; Elisabeth Trawinski; Guillaume Gricourt; Jean-Michel Pawlotsky; Melissa N'Debi; Slim Fourati; Vanessa Demontant |
| EPI_ISL_3144965, EPI_ISL_3144967, EPI_ISL_3144979, EPI_ISL_3145450, EPI_ISL_3145455, EPI_ISL_3146533 | BioneXt Lab | Laboratoire national de sante, Microbiology, Microbial Genomics Platform | Anke Wienecke-Baldacchino; Catherine Ragimbeau; Elodie Solarino; Fatu Djabi; Jessica Tapp; Lise Pignon; Raoul Salmon; Tamir Abdelrahman; Thibault Ferrandon; Virginie Jover |
| EPI_ISL_2504547, EPI_ISL_2504551, EPI_ISL_2504553, EPI_ISL_2504556, EPI_ISL_2504558, EPI_ISL_2504572, EPI_ISL_2504576 | | | |
| see above | Biotechnology Division, NCDC Delhi | NCDC Delhi, Biotechnology Division | Hema Gogia; Hemlata Lall; Kalaiarasan Ponnusamy; Mahesh S Dhar; Manoj K Singh; Meena Datta; Partha Rakshit; Preeti Madan; Priyanka Singh; Radhakrishnan V. S; Robin Marwal; Sandhya Kabra; Sujeet K Singh; Uma Sharma |
| EPI_ISL_2547410 | Brandwacht Sat Clinic wc BBC | National Health Laboratory Service/University of Cape Town (NHLS/UCT) | Arash Iranzadeh; Bruna Galvao; Carolyn Williamson; Deelan Doolabh; Diana Hardie; Gert Marais; Innocent Mudau; Lynn Tyers; Marvin Hsiao; Stephen Korsman |
| EPI_ISL_2536143 | Bundeswehr Institute of Microbiology | Bundeswehr Institute of Microbiology | Alexandra Rehn; Enrico Georgi; Malena Bestehorn-Willmann; Markus Antwerpen; Mathias Walter; Mike Pillukat; Roman Wölfel; Sabine Zange |
| EPI_ISL_2779290 | Busia border point | USAMRD-A, Basic Science Laboratory | Alan Lemtudo; Beth Mutai; Brian Andika; Carol Kifude; Clement Masakwe; Eric Muthanje; Esther Omuseni; Faith Sigei; Gathii Kimita; George Awinda; John Waitumbi; Josphat Nyataya; Rachel Githii; Rehema Liyai; Stephen Ochola |
| EPI_ISL_2758539 | CA DPH Viral and Rickettsial Disease Laboratory | Chan-Zuckerberg Biohub | CZB Cliahub Consortium |
| EPI_ISL_2842012 | CAP MANLLEU | Banc de Sang i Teixits | Carlos Hobeich; Francisco Vidal; Irene Corrales; Lorena Ramírez; Maria Glòria Soria; Natália Comes; Nina Borràs; Noemí Gonzalez; Silvia Sauleda |
| EPI_ISL_2881948, EPI_ISL_2934213 | CDPH VBL | California Department of Public Health | CDPH-COVIDNet; SwabSeq/UCLA |
| EPI_ISL_2761765, | CENTOGENE Frankfurt Laboratory: Niederlassung | Robert Koch Institute | |

| | | | |
|---|---|---|---|
| EPI_ISL_2845498, EPI_ISL_2845501 | Industriepark Höchst | | |
| EPI_ISL_3153690 | CERBALLIANCE | UMR PIMIT | Dr Camille Lebarbenchon; Dr David A Wilkinson; Dr Patrick Mavingui; Magali Turpin; Marie-Alice Simbi |
| EPI_ISL_2894087 | CERBALLIANCE - LAMAT | CNR Virus des Infections Respiratoires - France SUD | Antonin Bal; Bruno Lina; Gregory Destras; Gwendolyne Burfin; Hadrien Regue; Laurence Josset; Martine Valette; Quentin Semanas |
| EPI_ISL_2791368 | CH.INTERCOMMUNAL DE CRETEIL | Department of Virology, Henri Mondor University Hospital, Assistance Publique Hôpitaux de Paris, Université Paris-Est Créteil, INSERM U955 | Alexandre Soulier; Christophe Rodriguez; Elisabeth Trawinski; Guillaume Gricourt; Jean-Michel Pawlotsky; Melissa N'Debi; Slim Fourati; Vanessa Demontant |
| EPI_ISL_2695344 | CHEDV | Instituto Nacional de Saude (INSA) | Borges et al |
| EPI_ISL_2989108 | CHLC | Instituto Nacional de Saude (INSA) | Borges et al |
| EPI_ISL_2824971 | CHU Coimbra | Instituto Nacional de Saude (INSA) and Faculdade de Medicina da Universidade de Coimbra | Borges et al |
| EPI_ISL_2791352 | CHU DE DIJON | Department of Virology, Henri Mondor University Hospital, Assistance Publique Hôpitaux de Paris, Université Paris-Est Créteil, INSERM U955 | Alexandre Soulier; Christophe Rodriguez; Elisabeth Trawinski; Guillaume Gricourt; Jean-Michel Pawlotsky; Melissa N'Debi; Slim Fourati; Vanessa Demontant |
| EPI_ISL_2566670 | CHU NIMES | CHU NIMES | Agathe Boudet; Marie-josée Carles; Sophie Bravo; Stephan Robin |
| EPI_ISL_2709248, EPI_ISL_2709250 | CHU Pontchaillou | CHU Pontchaillou | DE TAYRAC Marie; DENOUAL Florent; ETCHEVERRY Amandine; FEBREAU Christine; GALIBERT Marie Dominique; GROLHIER Claire; JAGLINE Steven; PRONIER Charlotte; SASSI Mohamed; THIBAULT Vincent |
| EPI_ISL_2567487 | CHU ST ETIENNE HOPITAL NORD | CNR Virus des Infections Respiratoires - France SUD | Antonin Bal; Bruno Lina; Gregory Destras; Gwendolyne Burfin; Hadrien Regue; Laurence Josset; Martine Valette; Quentin Semanas |
| EPI_ISL_2510045 | CHUGA-IBP-laboratoire de Virologie | IBP-laboratoire de virologie | Alban Caporossi; Anne Signori -Schmuck; Anne-Karen Faure; Aurélie Truffot; Benjamin Nemoz; Hugo Jardin; Julien Andréani; Julien Lupo; Léa Ponderand; Pascal Poignard; Patrice Morand; Raphaële Germi; Sylvie Larrat |
| EPI_ISL_2637460 | CL. DU SUD THIAIS | Department of Virology, Henri Mondor University Hospital, Assistance Publique Hôpitaux de Paris, Université Paris-Est Créteil, INSERM U955 | Alexandre Soulier; Christophe Rodriguez; Elisabeth Trawinski; Guillaume Gricourt; Jean-Michel Pawlotsky; Melissa N'Debi; Slim Fourati; Vanessa Demontant |
| EPI_ISL_2516569, EPI_ISL_2637249, EPI_ISL_2637266 | CLILAB | Microbiology Department | Aida Gonzalez-Diaz; Carmen Ardanuy; Jordi Camara; Jordi Niubó; Laura Calatayud; M Angeles Domínguez; Miguel Fernandez-Huerta; Sara Marti |
| EPI_ISL_2502671, EPI_ISL_2502672 | CQRC_QUALITY CONTROL CHEMICAL BIOLOGICAL RISK_AOOR Villa Sofia Cervello Palermo | CQRC_QUALITY CONTROL CHEMICAL BIOLOGICAL RISK_AOOR Villa Sofia Cervello Palermo | Brunacci, G.; Contino, F.; Di Gaudio, F. |
| EPI_ISL_2892537 | California Department of Public Health | California Department of Public Health | CDPH IDLB COVIDNet et al |
| EPI_ISL_2778797 | California Department of Public Health Valencia Branch Laboratory (CDPH VBL) | California Department of Public Health | CDPH-COVIDNet; UCLA Technology Center for Genomics & Bioinformatics |
| EPI_ISL_3120377, EPI_ISL_3120428, EPI_ISL_3120449, EPI_ISL_3120550, EPI_ISL_3120567, EPI_ISL_3120581, EPI_ISL_3120597, EPI_ISL_3120599, EPI_ISL_3120611, EPI_ISL_3120620, EPI_ISL_3120638 | | | |
| see above | Carl-Thiem-Klinikum Cottbus - Zentrum für Laboratoriumsmedizin Mikrobiologie und Krankenhaushygiene | Robert Koch Institute | |
| EPI_ISL_2688721, EPI_ISL_2688781, EPI_ISL_2688784, EPI_ISL_2688802, EPI_ISL_2688803 | Centre Hospitalier Universitaire de Rouen Laboratoire de Virologie | Centre Hospitalier Universitaire de Rouen Laboratoire de Virologie | Alice Moisan; Fabienne De Oliveira; Marie Leoz |
| EPI_ISL_3074072 | City of Milwaukee Health Department Laboratory | City of Milwaukee Health Department Laboratory | Amy Bauer; Jennifer Lentz; Manjeet Khubbar; Samantha Scott; Sanjib Bhattacharyya |
| EPI_ISL_3089258, EPI_ISL_3089259, EPI_ISL_3089260 | Colorado Department of Public Health and Environment | Colorado Department of Public Health and Environment | Alexandria Rossheim; Diana Ir; Emily A. Travanty; Laura Bankers; Mandy Waters; Michael Martin; Molly C. Hetherington-Rauth; Sarah Elizabeth Totten; Shannon R. Matzinger |
| EPI_ISL_2658698 | Contra Costa County Public Health Lab | Chan-Zuckerberg Biohub | CZB Cliahub Consortium |
| EPI_ISL_2636153 | Corona-Testzentrum ifp Institut für Produktqualität GmbH | Robert Koch Institute | |
| EPI_ISL_2755177 | Curative Labs | Curative Labs | Elias L. Salfati; Eugenia Khorosheva; George Way; J.Cesar Ignacio-Espinoza; Janet Chen; Mikhail Hanewich-Hollatz; Nabjot Sandhu; Sophia Quasem; Vladimir Slepnev; Zhiyi Xie |
| EPI_ISL_2928050, EPI_ISL_2928056 | Curative Labs | New Mexico Department of Health Scientific Laboratory | Anastacia Griego-Fisher; D'eldra Malone; Ellie Johnson; Jennifer Benoit; Keila Gutierrez; Linda Salazar; Ratheesh Rajan |
| EPI_ISL_2621141 | D'Almeida Clinic wc DAL | NHLS/UCT | Arash Iranzadeh; Bruna Galvao; Carolyn Williamson; Deelan Doolabh; Diana Hardie; Gert Marais; Innocent Mudau; Lynn Tyers; Marvin Hsiao; Stephen Korsman |
| EPI_ISL_2725458, EPI_ISL_2725465, EPI_ISL_2854076, EPI_ISL_2854083, EPI_ISL_2854088, EPI_ISL_2876508, EPI_ISL_2930630, EPI_ISL_3010136 | | | |
| see above | DC Public Health Lab/ Dept. of Forensic Sciences | DC Public Health Lab/ Dept. of Forensic Sciences | Brittany Hamilton; Connie Maza; David Payne; Elizabeth Zelaya; Janis Doss; Jocelyn Hauser; Monica Mann; Sarah Scott; Scott Nguyen |
| EPI_ISL_2567347, EPI_ISL_2629386 | DYOMEDEA-LABORATOIRE DE LA SAUVEGARDE | CNR Virus des Infections Respiratoires - France SUD | Antonin Bal; Bruno Lina; Gregory Destras; Gwendolyne Burfin; Hadrien Regue; Laurence Josset; Martine Valette; Quentin Semanas |
| EPI_ISL_2693666 | Demy Labs, Abuja | National Reference Laboratory, Nigeria Centre for Disease Control | Anthony Ahumibe; Catherine Okoi; Chimaobi Chukwu; Dr Chikwe Ihekweazu; Dr Ndodo Nnaemeka; Dr Omoare Adesuyi; Grace Esebanmen; Nwando Mba; Olusola Anuoluwapo Akanbi |
| EPI_ISL_2693668 | Demy Labs, Abuja | National Reference Laboratory, Nigeria Centre for Disease Control | Anthony Ahumibe; Catherine Okoi; Chimaobi Chukwu; Dr Chikwe Ihekweazu; Dr Ndodo Nnaemeka; Dr Omoare Adesuyi; Grace Esebanmen; Nwando Mba; Olusola Anuoluwapo Akanbi |
| EPI_ISL_2604709, EPI_ISL_2609066 | Department of Virology and Immunology, University of Helsinki and Helsinki University Hospital, Huslab Finland | Department of Virology, Faculty of Medicine, University of Helsinki, Helsinki, Finland | Essi Korhonen; Hanna Jarva; Hanna Liimatainen; Hannimari Kallio-Kokko; Harri Kangas; Hussein Alburkat; Jenni Virtanen; Maija Lappalainen; Maija Suvanto; Olli Vapalahti; Pekka Ellonen; Phuoc Truong; Ravi Kant; Sari Hannula; Satu Kurkela; Teemu Smura |
| EPI_ISL_2887349 | Division of Emerging Infectious Diseases, Bureau of Infectious Diseases Diagnosis Control, Korea Disease Control and Prevention Agency | Division of Emerging Infectious Diseases, Bureau of Infectious Diseases Diagnosis Control, Korea Disease Control and Prevention Agency | Ae Kyung Park; Chae Young Lee; Eun-Jin Kim; Heui Man Kim; Il-Hwan Kim; Jeong-Ah Kim; Jeong-Min Kim |
| EPI_ISL_2695860 | Division of Medical Virology, National Health Laboratory Service (NHLS), Tygerberg Hospital / Stellenbosch University | Division of Medical Virology, National Health Laboratory Service (NHLS), Tygerberg Hospital / Stellenbosch University | Bronwyn Kleinhans; Gert van Zyl; San Emmanual James; Susan Engelbrecht; Tania Stander; Tongai Maponga; Tulio de Oliveira; Wolfgang Preiser |
| EPI_ISL_2610614 | Dutch COVID-19 response team | National Institute for Public Health and the Environment (RIVM) | Adam Meijer; AnneMarie van den Brandt; Annelies Kroneman; Bas van der Veer; Chantal Reusken; Dennis Schmitz; Dirk Eggink; Euníce Then; Florian Zwagemaker; Harry Vennema; Jeroen Cremer; Karim Hajji; Kim Freriks; Linda van de Nes; Lisa Wijsman; Lynn Aarts; Melissa van Tuil; Ryanne Jaarsma; Sanne Bos; Sharon van den Brink; on behalf of the national COVID-19 response team |
| EPI_ISL_2938480, EPI_ISL_2938481, EPI_ISL_2938483, EPI_ISL_2938486, EPI_ISL_2938498, EPI_ISL_2938512, EPI_ISL_2938522, EPI_ISL_2938541, EPI_ISL_2938559, EPI_ISL_2938584, EPI_ISL_2938586, EPI_ISL_2938641, EPI_ISL_2938642, EPI_ISL_2938683, EPI_ISL_2938688, EPI_ISL_2938690, EPI_ISL_2938707, EPI_ISL_2938722, EPI_ISL_2938740, EPI_ISL_2938761, EPI_ISL_2938834, EPI_ISL_2938836, EPI_ISL_2938837, EPI_ISL_2938852, EPI_ISL_2938858, EPI_ISL_2938894, EPI_ISL_2938948, EPI_ISL_2938953, EPI_ISL_2938963, EPI_ISL_2938964, EPI_ISL_2938966, EPI_ISL_2939132, EPI_ISL_2939135, EPI_ISL_2939139, EPI_ISL_2939141, EPI_ISL_2939155, EPI_ISL_2939158, EPI_ISL_2939160, EPI_ISL_2939162, EPI_ISL_2939165, EPI_ISL_2939172, EPI_ISL_2939173, EPI_ISL_2939174, EPI_ISL_2939175, EPI_ISL_2939176, EPI_ISL_2939177, EPI_ISL_2939184, EPI_ISL_2939185, EPI_ISL_2939189, EPI_ISL_2939190, EPI_ISL_2939193, EPI_ISL_2939196, EPI_ISL_2939198, EPI_ISL_2939200, EPI_ISL_2939203, EPI_ISL_2939207, EPI_ISL_2939208, EPI_ISL_2939211, EPI_ISL_2939241, EPI_ISL_2939246, EPI_ISL_2939255, EPI_ISL_2939256, EPI_ISL_2939258, EPI_ISL_2939287, EPI_ISL_2939310, EPI_ISL_2939318, EPI_ISL_2939325, EPI_ISL_2939328, EPI_ISL_2939332, EPI_ISL_2939337, EPI_ISL_2939430, EPI_ISL_2939432, EPI_ISL_2939453, EPI_ISL_2939457, EPI_ISL_2939460, EPI_ISL_2939461, EPI_ISL_2939468, EPI_ISL_2939469, EPI_ISL_2939470, EPI_ISL_2939478, EPI_ISL_2939509, EPI_ISL_2939520, EPI_ISL_2939524, EPI_ISL_2939527, EPI_ISL_2939529, EPI_ISL_2939536, EPI_ISL_2939537, EPI_ISL_2939541, EPI_ISL_2939544, EPI_ISL_2939545, EPI_ISL_2939550, EPI_ISL_2939551, EPI_ISL_2939553, EPI_ISL_2939554, EPI_ISL_2939555, EPI_ISL_2939563, EPI_ISL_2939564, EPI_ISL_2939620, EPI_ISL_2939635, EPI_ISL_2939653, EPI_ISL_2939654, EPI_ISL_2939655, EPI_ISL_2939663, EPI_ISL_2939701, EPI_ISL_2939702, EPI_ISL_2939708, EPI_ISL_2939709, EPI_ISL_2939712, EPI_ISL_2939713, EPI_ISL_2939714, EPI_ISL_2939716, EPI_ISL_2939717, EPI_ISL_2939718, EPI_ISL_2939722, EPI_ISL_2939728, EPI_ISL_2939729, EPI_ISL_2939730, EPI_ISL_2940059, EPI_ISL_2940062, EPI_ISL_2940063, EPI_ISL_2940415, EPI_ISL_2940423, EPI_ISL_2940429, EPI_ISL_2940451, EPI_ISL_2940452, EPI_ISL_2940456, EPI_ISL_2940472, EPI_ISL_2940482, EPI_ISL_2940488, EPI_ISL_2940515, EPI_ISL_2940516, EPI_ISL_2940523, EPI_ISL_2940600, EPI_ISL_2940601, EPI_ISL_2940641, EPI_ISL_2940797, EPI_ISL_2940799, EPI_ISL_2940823, EPI_ISL_2940834, EPI_ISL_2940853, EPI_ISL_2940855, EPI_ISL_2940858, EPI_ISL_2940864, EPI_ISL_2940869, EPI_ISL_2940873, EPI_ISL_2940882, EPI_ISL_2940886, EPI_ISL_2940892, EPI_ISL_2940895, EPI_ISL_2940896, EPI_ISL_2940899, EPI_ISL_2940904, EPI_ISL_2940908, EPI_ISL_2940909, EPI_ISL_2940912, EPI_ISL_2940916, EPI_ISL_2940922, EPI_ISL_2940925, EPI_ISL_2940930, EPI_ISL_2940932, EPI_ISL_2940933, EPI_ISL_2940934, EPI_ISL_2940940, EPI_ISL_2940952, EPI_ISL_2940953, EPI_ISL_2940956, EPI_ISL_2941005, EPI_ISL_2941021, EPI_ISL_2941025, EPI_ISL_2941030, EPI_ISL_2941035, EPI_ISL_2941038, EPI_ISL_2941039, EPI_ISL_2941040, EPI_ISL_2941044, EPI_ISL_2941045, EPI_ISL_2941046, EPI_ISL_2941047, EPI_ISL_2941048, EPI_ISL_2941049, EPI_ISL_2941057, EPI_ISL_2941069, EPI_ISL_2941075, EPI_ISL_2941077, EPI_ISL_2941080, EPI_ISL_2941082, EPI_ISL_2941085, EPI_ISL_2941093, EPI_ISL_2941101, EPI_ISL_2941102, EPI_ISL_2941106, EPI_ISL_2941110, EPI_ISL_2941111, EPI_ISL_2941113, EPI_ISL_2941116, EPI_ISL_2941117, EPI_ISL_2941120, EPI_ISL_2941125, | | | |

| | | | |
|---|---|---|---|
| EPI_ISL_2941126, EPI_ISL_2941167, EPI_ISL_2941169, EPI_ISL_2941171, EPI_ISL_2941178, EPI_ISL_2941189, EPI_ISL_2941191, EPI_ISL_2941223, EPI_ISL_2941228, EPI_ISL_2941235, EPI_ISL_2941243, EPI_ISL_2941245, EPI_ISL_2941254, EPI_ISL_2941262, EPI_ISL_2941295, EPI_ISL_2941317, EPI_ISL_2941338, EPI_ISL_2941348, EPI_ISL_2941356, EPI_ISL_2941367, EPI_ISL_2941380, EPI_ISL_2941408, EPI_ISL_2941412, EPI_ISL_2941433, EPI_ISL_2941495, EPI_ISL_3116934, EPI_ISL_3117237, EPI_ISL_3117440, EPI_ISL_3117441, EPI_ISL_3117442 | | | |
| see above | Edmonton Provincial Lab | Public Health Agency of Canada (PHAC) National Microbiology Laboratory | Buss; Croxen M; Deo A; Dieu P; E; Ferrato C; Gill K; Khan F; Koleva P; Li V; Lloyd C; Lynch T; Ma R; Murphy S; Pabbaraju K; Shokoples S; Thayer J; Tipples G; Whitehouse M; Wong A; Yu C; Zelyas N |
| EPI_ISL_2835994 | Epidemiology of Microbial Diseases, Yale School of Public Health | Epidemiology of Microbial Diseases, Yale School of Public Health | A.E.; A.F.; Alpert, T.; Breban; Brito; C.B.; C.C.; Fauver; Grubaugh; I.M.; Iwasaki, A.; J.E.; J.R.; Kalinich; Landry; Lucas, C.; M.E.; M.I.; M.L.; N.D.; Ott; Petrone; Rothman; Vogels; Watkins |
| EPI_ISL_2634508 | Eurofins LifeCodexx GmbH | Robert Koch Institute | |
| EPI_ISL_2693096, EPI_ISL_2693100, EPI_ISL_2693101, EPI_ISL_2693102, EPI_ISL_2693106, EPI_ISL_2693108, EPI_ISL_2693112, EPI_ISL_2803749, EPI_ISL_2803750, EPI_ISL_2803755 | | | |
| see above | Everight Diagnostic Laboratory | National Reference Laboratory, Nigeria Centre for Disease Control | Anthony Ahumibe; Catherine Okoi; Chimaobi Chukwu; Dr Chikwe Ihekweazu; Dr Ndodo Nnaemeka; Dr Omoare Adesuyi; Grace Esebanmen; Nwando Mba; Olusola Anuoluwapo Akanbi |
| EPI_ISL_2803754 | Everight Diagnostic Laboratory | National Reference Laboratory, Nigeria Centre for Disease Control Durumi III, Gaduwa, Abuja, Nigeria | Anthony Ahumibe; Catherine Okoi; Chimaobi Chukwu; Dr Chikwe Ihekweazu; Dr Ndodo Nnaemeka; Dr Omoare Adesuyi; Grace Esebanmen; Nwando Mba; Olusola Anuoluwapo Akanbi |
| EPI_ISL_2505980, EPI_ISL_2507908 | Fimlab Laboratoriot Oy Tampere | Expert Microbiology, National Institute for Health and Welfare | Carita Savolainen-Kopra; Erika Lindh; Haider al-Hello; Jani Halkilahti; Kirsi Liitsola; Niina Ikonen; Olli Vapalahti; Pekka Ellonen; Phuoc Truong; Päivi Laurila; Ravi Kant; Sari Hannula; Soile Blomqvist; Teemu Smura |
| EPI_ISL_2620903, EPI_ISL_2620905, EPI_ISL_2621505 | Fondazione IRCCS Ca' Granda Ospedale Maggiore Policlinico | Fondazione IRCCS Ca' Granda Ospedale Maggiore Policlinico | Ferruccio Ceriotti; Sara Uceda Renteria |
| EPI_ISL_2620733, EPI_ISL_2620734 | Functional Genomics and Proteomics Laboratory (FGPL), Department of Genetic Engineering and Biotechnology, University of Chittagong | Virology Laboratory, International Centre for Diarrhoeal Disease Research, Bangladesh (ICDDR,B) | Amit Datta; Amlan Bhattacharjee; Dr. Laila Khaleda; Dr. Mohammad Al-Forkan; Dr. Mohammad Enayet Hossain; Dr. Mustafizur Rahman; Dr. Sujat Paul; Inzamamul Ismail Shawon; Md. Abdur Rahman; Md. Jibran Alam; Miftah Mushfique; Mojnu Miah; Rahee Hasan Chowdhury; Rashedul Hasan; Shuva Das |
| EPI_ISL_2637406, EPI_ISL_2637565 | GH A.CHENEVIER-H.MONDOR | Department of Virology, Henri Mondor University Hospital, Assistance Publique Hôpitaux de Paris, Université Paris-Est Créteil, INSERM U955 | Alexandre Soulier; Christophe Rodriguez; Elisabeth Trawinski; Guillaume Gricourt; Jean-Michel Pawlotsky; Melissa N'Debi; Slim Fourati; Vanessa Demontant |
| EPI_ISL_2779313, EPI_ISL_2779314, EPI_ISL_2779315, EPI_ISL_2779316, EPI_ISL_2779349 | GK Prisons Dispensary Kapsabet | USAMRD-A, Basic Science Laboratory | Alan Lemtudo; Beth Mutai; Brian Andika; Carol Kifude; Clement Masakwe; Eric Muthanje; Esther Omuseni; Faith Sigei; Gathii Kimita; George Awinda; John Waitumbi; Josphat Nyataya; Rachel Githii; Rehema Liyai; Stephen Ochola |
| EPI_ISL_2536018, EPI_ISL_2602043, EPI_ISL_2602307, EPI_ISL_2602311 | Gravity Diagnostics, LLC | Gravity Diagnostics, LLC | Gravity Diagnostics |
| EPI_ISL_2621142 | Great Brak River Clinic wc GBC | NHLS/UCT | Arash Iranzadeh; Bruna Galvao; Carolyn Williamson; Deelan Doolabh; Diana Hardie; Gert Marais; Innocent Mudau; Lynn Tyers; Marvin Hsiao; Stephen Korsman |
| EPI_ISL_2895268 | H Braga | Instituto Nacional de Saude (INSA) | Borges et al |
| EPI_ISL_2629010, EPI_ISL_2629033, EPI_ISL_2629097, EPI_ISL_2629122, EPI_ISL_2629154 | HEGP - Laboratoire de Virologie | HEGP - Laboratoire de Virologie | David Veyer; Hélène Péré; Julien Puech; Maxime Wack; Nicolas Robillard; Sandrine Imbeaud |
| EPI_ISL_2523657 | HELIX LLC | WHO National Influenza Centre Russian Federation | Andrey Komissarov; Artem Fadeev; Daria Danilenko; Dmitry Lioznov; Elena Nabieva; Georgii Bazykin; Kirill Varchenko; Ksenia Safina; Kseniya Komissarova; Maria Pisareva; Maria Timofeeva; Mikhail Bakaev; Nikita Yolshin; Oula Mansour; Tamila Musaeva; Veronika Eder |
| EPI_ISL_2534411, EPI_ISL_2534430 | HOSPITAL UNIVERSITARIO SON ESPASES | HOSPITAL UNIVERSITARIO SON ESPASES | Hospital Universitario Son Espases; Servicio de Microbiología |
| EPI_ISL_2598867 | Helix/Illumina | Centers for Disease Control and Prevention Division of Viral Diseases, Pathogen Discovery | Adrian Paskey; Alexandre Bolze; Ary Ascencio; Benjamin Rambo-Martin; Brad Sickler; Charlotte Rivera-Garcia; Christine Tran; Christopher Gulvick; Clinton R. Paden; Dakota Howard; Darlene Wagner; David Becker; Dhwani Batra; Duncan MacCannell; Efren Sandoval; Eileen de Feo; Elizabeth Cirulli; Eric Allen; Geraint Levan; James Lu; Jan Antico; Jason Caravas; Jason Nguyen; Jimmy Ramirez; Jingtao Liu; Kara Moser; Kelly Schiabor Barrett; Kim Gietzen; Magnus Isaksson; Marc Laurent; Matthew Schmerer; Matthew Tolentino; Nicole L. Washington; Peter W. Cook; Phil Febbo; Ryan Cho; Scott Sammons; Shannon Wickline; Shatavia Morrison; Sherry Wang; Simon White; Tyler Cassens; William Lee; Yvette Unoarumhi |
| EPI_ISL_2531819, EPI_ISL_2531823, EPI_ISL_2532189, EPI_ISL_2535268, EPI_ISL_2535273, EPI_ISL_2535479, EPI_ISL_2535482, EPI_ISL_2709260, EPI_ISL_2790926 | | | |
| see above | Hospital | National Reference Center for Viruses of Respiratory Infections, Institut Pasteur, Paris | Alexandra Ducancelle; Angela Brisebarre; Camille Capel; Christophe Malabat; CléMence Guillaume; Corinne Maufrais; Damien Mornico; Emmanuelle Permal; Eric Farfour; Etienne Simon-Lorière; Frédéric Lemoine; JéRome Guinard; Laurence Courdavault; Louise Lefrançois; Marion Barbet; Maud Vanpeene; Méline Bizard; Olivia Raulin; Pascale Martres; Sylvie Behillil; Sylvie Van der Werf; Victoire Baillet; Vincent Enouf |
| EPI_ISL_3147285 | Hospital Center Luxembourg | Laboratoire national de sante, Microbiology, Microbial Genomics Platform | Anke Wienecke-Baldacchino; Catherine Ragimbeau; Elodie Solarino; Fatu Djabi; Jean-Hugues Francois; Jessica Tapp; Lise Pignon; Michel Kohnen; Raoul Salmon; Tamir Abdelrahman; Virginie Jover |
| EPI_ISL_2598345 | Hospital General Universitario de Alicante - Instituto de Investigación Sanitaria y Biomédica de Alicante | SeqCOVID-SPAIN consortium/IBV(CSIC) | Carmen Molina Pardines and SeqCOVID-SPAIN consortium; Maripaz Ventero Martín |
| EPI_ISL_2877271, EPI_ISL_2877272, EPI_ISL_2877273, EPI_ISL_2877274, EPI_ISL_2877275, EPI_ISL_2877276, EPI_ISL_2877278 | | | |
| see above | Hospital Universitari Arnau de Vilanova de Lleida | SeqCOVID-SPAIN consortium/IBV(CSIC) | Alba Bellés Bellés; Albert Bernet Sánchez; Eric López González; Iván Prats Sánchez; Mercè García González and SeqCOVID-SPAIN consortium |
| EPI_ISL_2516503 | Hospital Universitari Bellvitge | Microbiology Department | Aida Gonzalez-Diaz; Carmen Ardanuy; Jordi Camara; Jordi Niubó; Laura Calatayud; M Angeles Domínguez; Miguel Fernandez-Huerta; Sara Marti |
| EPI_ISL_2652057 | Hospital Universitari Joan XXIII | Hospital Universitari Joan XXIII | Carla Martín; Clara Benavent; Cristina Gutiérrez; Ester Picó; Gemma Recio; Margarida Terrón; Natalia Bastón |
| EPI_ISL_2828219 | Hospital Universitari Vall d'Hebron - Vall d'Hebron Institut de Recerca | Hospital Universitari Vall d'Hebron - Vall d'Hebron Institut de Recerca | Alejandra González-Sánchez; Andrés Antón; Ariadna Rando; Carla Castillo; Cristina Andrés; Damir Garcia-Cehic; Josep Quer; Juliana Esperalba; Maria Carmen Martin; Maria Gema Codina; Maria Piñana; Tomàs Pumarola |
| EPI_ISL_2597670, EPI_ISL_3022638, EPI_ISL_3022693, EPI_ISL_3022916, EPI_ISL_3023034, EPI_ISL_3023129, EPI_ISL_3023133 | | | |
| see above | Hospital of the University of Pennsylvania Molecular Pathology Lab | Bushman Lab - University of Pennsylvania | Aoife M. Roche; Arupa Ganguly; Ayannah S. Fitzgerald; Brendan Kelly; Jevon Graham-Wooten; John K. Everett; Kyle Rodino; Layla A. Khatib; Mike Feldman; Ronald G. Collman and Frederic Bushan; Samantha A. Whiteside; Scott Sherrill-Mix; Shantan Reddy; Young Hwang |
| EPI_ISL_2637513, EPI_ISL_2791327 | Hôpital Avicenne | Department of Virology, Henri Mondor University Hospital, Assistance Publique Hôpitaux de Paris, Université Paris-Est Créteil, INSERM U955 | Alexandre Soulier; Christophe Rodriguez; Elisabeth Trawinski; Guillaume Gricourt; Jean-Michel Pawlotsky; Melissa N'Debi; Slim Fourati; Vanessa Demontant |
| EPI_ISL_2688693 | Hôpital Jacques Monod | Centre Hospitalier Universitaire de Rouen Laboratoire de Virologie | Alice Moisan; Fabienne De Oliveira; Marie Leoz |
| EPI_ISL_2637575 | Hôpital Saint-Louis | Department of Virology, Henri Mondor University Hospital, Assistance Publique Hôpitaux de Paris, Université Paris-Est Créteil, INSERM U955 | Alexandre Soulier; Christophe Rodriguez; Elisabeth Trawinski; Guillaume Gricourt; Jean-Michel Pawlotsky; Melissa N'Debi; Slim Fourati; Vanessa Demontant |
| EPI_ISL_2546049, EPI_ISL_2546061, EPI_ISL_2546128, EPI_ISL_2546152, EPI_ISL_2546166, EPI_ISL_2546421, EPI_ISL_2546441, EPI_ISL_2546460, EPI_ISL_2546475, EPI_ISL_2546476, EPI_ISL_2546745 | | | |
| see above | ICMR-National Institute of Virology - INSACOG | NIV Influenza | Dr. Varsha Potdar and NIC Team |
| EPI_ISL_2878848, EPI_ISL_2878868, EPI_ISL_2878886, EPI_ISL_2878890, EPI_ISL_2878915, EPI_ISL_2879120, EPI_ISL_2879144, EPI_ISL_2881084 | | | |
| see above | INSACOG Surveillance | INSACOG at CSIR Institute of Genomics and Integrative Biology | INSACOG |
| EPI_ISL_3019106 | INSACOG-Mizoram | National Institute of Biomedical Genomics – INSACOG | Arindam Maitra; Gracy Laldinmawii; N Senthil Kumar; Nidhan Kumar Biswas; Saumitra Das; Sreedhar Chinnaswamy; Swagnik Roy |
| EPI_ISL_2503157, EPI_ISL_2503216, EPI_ISL_2503549, EPI_ISL_2503550, EPI_ISL_2503556, EPI_ISL_2503557, EPI_ISL_2503566, EPI_ISL_2503569, EPI_ISL_2548609, EPI_ISL_2548796, EPI_ISL_2548830, EPI_ISL_2549040 | | | |
| see above | INSACOG-WB | National Institute of Biomedical Genomics – INSACOG | Ajay Chakraborti; Arindam Maitra; Bhaswati Bandyopadhyay; Nidhan Kumar Biswas; Saumitra Das; Sreedhar Chinnaswamy; Tamal Ghosh |
| EPI_ISL_2672210, | INSP, Bamako | Malaria Research and Training | Abdoulaye Djimde; Amadou Daou; Antoine Dara; Demba Koita; Ibrehima Guindo |

| | | | |
|---|---|---|---|
| EPI_ISL_2672216 | | Center-Parasito | |
| EPI_ISL_2598286 | IVY3 Central Lab, Vanderbilt University Medical Center | Lauring Lab, University of Michigan, Department of Microbiology and Immunology | Valesano |
| EPI_ISL_2933108 | IZSM | IZSM-U.O.C. Virologia | Antonio Limone; Claudio de Martinis; Esterina De Carlo; Giovanna Fusco; Lorena Cardillo; Maurizio Viscardi |
| EPI_ISL_3010501, EPI_ISL_3010530 | Idaho Bureau of Laboratories | Boise VA Medical Center, PALMS | Aimee Ceniseros; Cheri Lamb McFarlane; Christian Loera; Dr. Christopher Ball; Emily Bartlette; James Razor; Matthew Burns; Ying Pei |
| EPI_ISL_2536480, EPI_ISL_2692429, EPI_ISL_3015611 | Incyte Diagnostics Spokane | Seattle Flu Study | Amanda Adler; Barry R. Lutz; Benjamin Pelle; Brian Hiatt; Caitlin R. Wolf; Chris D. Frazar; Deborah A. Nickerson; Elisabeth Brandstetter; Erica Ryke; Geoff Melly; Helen Y. Chu; Janet A. Englund; Jay Shendure; Jover Lee; Kairsten Fay; Kirsten Lacombe; Lea M. Starita; Mark J. Rieder; Matthew Richardson; Matthew Thompson; Melissa Truong; Michael Boeckh; Michael Famulare; Misja Ilcisin; Peter D. Han; Philip Dykema; Romesh Gautom; Scott Lindquist; Thomas R. Sibley; Trevor Bedford |
| EPI_ISL_2567624, EPI_ISL_2567626 | Infectious Disease Hospital | National Reference Laboratory, Nigeria Centre for Disease Control | Anthony Ahumibe; Catherine Okoi; Chimaobi Chukwu; Dr Chikwe Ihekweazu; Dr Ndodo Nnaemeka; Dr Omoare Adesuyi; Grace Esebanmen; Nwando Mba; Olusola Anuoluwapo Akanbi |
| EPI_ISL_2686891, EPI_ISL_2709505, EPI_ISL_2786670 | Infinity Biologix | Centers for Disease Control and Prevention Division of Viral Diseases, Pathogen Discovery | Adrian Paskey; Benjamin Rambo-Martin; Chirayu Goswami; Christian Bixby; Christopher Gulvick; Clinton R. Paden; Dakota Howard; Darlene Wagner; Dhwani Batra; Duncan MacCannell; Jason Caravas; Jonathan Schultz; Kara Moser; Matthew Schmerer; Peter W. Cook; Robin Grimwood; Russ Hager; Scott Sammons; Shatavia Morrison; Yihe Wang; Yvette Unoarumhi |
| EPI_ISL_2774041 | Institute of Epidemiology, Disease Control and Research (IEDCR) | IEDCR-ideSHi-icddr,b | Firdausi Qadri; Hassan Afrad; Manjur Hossain Khan; Sadia Rahman; Tahmina Shirin |
| EPI_ISL_2893229, EPI_ISL_2893234, EPI_ISL_2893235, EPI_ISL_2893236 | Instituto Nacional de Investigação em Saúde | CERI, Centre for Epidemic Response and Innovation, Stellenbosch Univeristy & KRISP, KZN Research Innovation and Sequencing Platform | Afonso P; David K; Emmanuel SJ; Freitas RH; Giandhari J; Inglês L; Lutucuta S; Miranda J; Morais J; Mufinda M; Naidoo Y; Neto Z; Paulo A Carralero RR Paixão JP; Pereira A; Pillay S; Tegally H; Wilkinson E; de Oliveira T |
| EPI_ISL_2609502, EPI_ISL_2609522, EPI_ISL_2609528 | Instituto Nacional de Investigação em Saúde | KRISP, KZN Research Innovation and Sequencing Platform | Afonso P; David K; Emmanuel SJ; Freitas RH; Giandhari J; Inglês L; Lutucuta S; Miranda J; Morais J; Mufinda M; Naidoo Y; Neto Z; Paulo A Carralero RR Paixão JP; Pereira A; Pillay S; Tegally H; Wilkinson E; de Oliveira T |
| EPI_ISL_2976195 | Interpath Laboratories - Boise ID | Idaho Bureau of Laboratories | "R. Beukelman; Aimee Ceniseros; Christian Loera; Christopher Ball"; Matthew Charles Burns; Robert L. Voermans |
| EPI_ISL_2620892, EPI_ISL_2873816, EPI_ISL_2873817, EPI_ISL_2873819, EPI_ISL_2873823, EPI_ISL_2873826, EPI_ISL_2887851, EPI_ISL_2887862 | | | |
| see above | Iressef Genomics lab | IRESSEF | Abdou PADANE; Ambroise AHOUIDI; Aminata DIA; Aminata MBOUP; Astou Gaye GAYE; Barada CISSE; Birahim Piere NDIAYE; Diabou Diagne; Gora LO; Khadim GUEYE; Moustapha MBOW; Nafisatou LEYE; Ndeye Coumba Toure KANE; Papa Alassane DIAW; Samba Ndiour; Seni Ndiaye; Souleymane MBOUP; Yacine DIA |
| EPI_ISL_2589880 | Islab, Etelä-Savon aluelaboratorio | Expert Microbiology, National Institute for Health and Welfare | Carita Savolainen-Kopra; Erika Lindh; Haider al-Hello; Jani Halkilahti; Kirsi Liitsola; Niina Ikonen; Olli Vapalahti; Pekka Ellonen; Phuoc Truong; Päivi Laurila; Ravi Kant; Sari Hannula; Soile Blomqvist; Teemu Smura |
| EPI_ISL_2505234, EPI_ISL_2590053, EPI_ISL_2590282, EPI_ISL_2590284, EPI_ISL_2590376, EPI_ISL_2591505, EPI_ISL_2591528, EPI_ISL_2591532, EPI_ISL_2591542, EPI_ISL_2591553, EPI_ISL_2591630, EPI_ISL_2591636 | | | |
| see above | Islab, Pohjois-Savon aluelaboratorio | Expert Microbiology, National Institute for Health and Welfare | Carita Savolainen-Kopra; Erika Lindh; Haider al-Hello; Jani Halkilahti; Kirsi Liitsola; Niina Ikonen; Olli Vapalahti; Pekka Ellonen; Phuoc Truong; Päivi Laurila; Ravi Kant; Sari Hannula; Soile Blomqvist; Teemu Smura |
| EPI_ISL_2636159 | Israel Central Virology laboratory | Israel National Consortium for SARS-CoV-2 sequencing | Dana Bar-Ilan; Efrat Dahan Bucris; Efrat Glick-Saar; Ella Mendelson; Gideon Rechavi; Michal Mandelboim; Miranda Geva; Neta Zuckerman; Omri Nayshool; Oran Erster; Orna Mor |
| EPI_ISL_2725308, EPI_ISL_2725351, EPI_ISL_2725353 | Istituto Zooprofilattico Sperimentale del Mezzogiorno - Azienda Ospedaliera "Pugliese Ciaccio" di Catanzaro | Telethon Institute of Genetics and Medicine (TIGEM) | Antonio Grimaldi Patrizia Annunziata Francesco Panariello Biancamaria Pierri Claudia Tiberio Teresa Giuliano Valentina Bouche Chiara Colantuono Maria Concetta Cuomo Denise Di Concilio Lucio Di Filippo Anna Manfredi Pasquale Minchella Marcello Salvi Antonio Limone Luigi Atripaldi Pellegrino Cerino Andrea Ballabio Davide Cacchiarelli |
| EPI_ISL_2615967, EPI_ISL_2615970, EPI_ISL_2615971 | J.W. Ruby Memorial Hospital | WVU and Marshall University Combined Genomics Core Facilities | James Denvir; Peter Perrotta; Peter Stoilov; Ryan Percifield; Wesley Kimble |
| EPI_ISL_2858251 | Jaber Quarantine | Virology Lab, Jaber Al Ahmad Hospital | Adil Afridi; Dr. Ebaa Al-Awadhi; Dr. Zahrah Buhamad; Haroon Masih |
| EPI_ISL_3031386 | KCRH KEMRI/DEID Surveillance Site | USAMRD-A, Basic Science Laboratory | Alan Lemtudo; Beth Mutai; Brian Andika; Carol Kifude; Clement Masakwe; Eric Muthanje; Esther Omuseni; Faith Sigei; Gathii Kimita; George Awinda; John Waitumbi; Josphat Nyataya; Rachel Githii; Rehema Liyai; Stephen Ochola |
| EPI_ISL_2602995, EPI_ISL_2603003, EPI_ISL_2603009, EPI_ISL_2603026, EPI_ISL_2603108, EPI_ISL_3049731 | KEMRI-Wellcome Trust Research Programme,Kilifi | KEMRI-Wellcome Trust Research Programme,Kilifi | ; Githinji G.; Matoke D.; Mburu M.W.; Mohamed K.S.; Onyango C.; Thiongo K.; de Laurent Z. |
| EPI_ISL_2779323 | KEMRI/DEID Surveillance Site (Isebani Sub-County Hospital) | USAMRD-A, Basic Science Laboratory | Alan Lemtudo; Beth Mutai; Brian Andika; Carol Kifude; Clement Masakwe; Eric Muthanje; Esther Omuseni; Faith Sigei; Gathii Kimita; George Awinda; John Waitumbi; Josphat Nyataya; Rachel Githii; Rehema Liyai; Stephen Ochola |
| EPI_ISL_2779284 | KEMRI/DEID Surveillance Site (Mtongwe Navy Base Hospital) | USAMRD-A, Basic Science Laboratory | Alan Lemtudo; Beth Mutai; Brian Andika; Carol Kifude; Clement Masakwe; Eric Muthanje; Esther Omuseni; Faith Sigei; Gathii Kimita; George Awinda; John Waitumbi; Josphat Nyataya; Rachel Githii; Rehema Liyai; Stephen Ochola |
| EPI_ISL_2725600, EPI_ISL_2726488, EPI_ISL_2832423, EPI_ISL_2832458, EPI_ISL_2832700, EPI_ISL_2832903, EPI_ISL_2833052, EPI_ISL_2833204, EPI_ISL_2833207, EPI_ISL_2834794, EPI_ISL_2964858 | | | |
| see above | KU Leuven, Rega Institute, Clinical and Epidemiological Virology | KU Leuven, Rega Institute, Clinical and Epidemiological Virology | Bert Vanmechelen; Joan Marti-Carerras; Piet Maes; Tony Wawina-Bokalanga |
| EPI_ISL_2629562 | LABM DU PRE - LABOMAINE | CNR Virus des Infections Respiratoires - France SUD | Antonin Bal; Bruno Lina; Gregory Destras; Gwendolyne Burfin; Hadrien Regue; Laurence Josset; Martine Valette; Quentin Semanas |
| EPI_ISL_2933347 | LABORATOIRE CERBALLIANCE PLT VILLON | CNR Virus des Infections Respiratoires - France SUD | Antonin Bal; Bruno Lina; Gregory Destras; Gwendolyne Burfin; Hadrien Regue; Laurence Josset; Martine Valette; Quentin Semanas |
| EPI_ISL_2676580 | LABORATOIRE DE BIOLOGIE MEDICALE | CNR Virus des Infections Respiratoires - France SUD | Antonin Bal; Bruno Lina; Gregory Destras; Gwendolyne Burfin; Hadrien Regue; Laurence Josset; Martine Valette; Quentin Semanas |
| EPI_ISL_2893697 | LABOSUD | GenBio | Gwladys SEVRIN |
| EPI_ISL_2795986 | LAC UCoimbra | Instituto Nacional de Saude (INSA) and Faculdade de Medicina da Universidade de Coimbra | Borges et al |
| EPI_ISL_2957002 | LADR Zentrallabor DR. Kramer & Kollegen Geesthacht | Robert Koch Institute | |
| EPI_ISL_2544805, EPI_ISL_2558066, EPI_ISL_2558082, EPI_ISL_2558084 | LBM ALPHABIO, Marseille | LBM ALPHABIO, Marseille | Vincent GARCIA |
| EPI_ISL_2885901 | LBM Bonhoure | Cerba lab | John Doe; e.g. Jane Doe |
| EPI_ISL_2651265 | LBM Saint-Benoit | Laboratoire de virologie, CNR arbovirus Associé, Chu de la Réunion | Etienne Frumence; Marie-Christine Jaffar Bandjee; Nicolas Traversier; Sabrina Petit Genet |
| EPI_ISL_2567399, EPI_ISL_2567499, EPI_ISL_2567503, EPI_ISL_2676536 | LBM UNIBIO VALENTIN | CNR Virus des Infections Respiratoires - France SUD | Antonin Bal; Bruno Lina; Gregory Destras; Gwendolyne Burfin; Hadrien Regue; Laurence Josset; Martine Valette; Quentin Semanas |
| EPI_ISL_2617525 | Lab Genomik Solidaritas Indonesia | National Institute of Health Research and Development | Arie Ardiansyah Nugraha; Hana Apsari Pawestri; Hartanti Dian Ikawati; Kartika Dewi Puspa; Krisna Pangesti; Meutia Kumaheri; Nelly Puspandari; Subangkit; Triyani Soekarso; Vivi Setiawaty |
| EPI_ISL_2725219, EPI_ISL_2725247, EPI_ISL_2725296 | Lab. Microbiologia e Virologia Cotugno A.O. dei Colli - Istituto Zooprofilattico Sperimentale del Mezzogiorno | Telethon Institute of Genetics and Medicine (TIGEM) | Antonio Grimaldi Patrizia Annunziata Francesco Panariello Biancamaria Pierri Claudia Tiberio Teresa Giuliano Valentina Bouche Chiara Colantuono Maria Concetta Cuomo Denise Di Concilio Lucio Di Filippo Anna Manfredi Marcello Salvi Antonio Limone Luigi Atripaldi Pellegrino Cerino Andrea Ballabio Davide Cacchiarelli |
| EPI_ISL_2532289, EPI_ISL_2532407, EPI_ISL_2532431, EPI_ISL_2532432, EPI_ISL_2532455, EPI_ISL_2535263, EPI_ISL_2612607, EPI_ISL_2627951, EPI_ISL_2628003, EPI_ISL_2835750, EPI_ISL_2835751, EPI_ISL_2835753, EPI_ISL_2835754, EPI_ISL_2835759, EPI_ISL_2835814, EPI_ISL_2835855 | | | |
| see above | Labo Analyses Med | National Reference Center for Viruses of Respiratory Infections, Institut Pasteur, Paris | Angela Brisebarre; Arditi Cocco; BéNéDicte Roquebert; Camille Capel; Christophe Malabat; Corinne Maufrais; Etienne Simon-Lorière; Franck Ennouchi; Frédéric Lemoine; GréGoire Potiron; Louise Lefrançois; Marion Barbet; Maud Vanpeene; Méline Bizard; Pierre-Yves Leonard; Sabine Trombert; Sylvie Behillil; Sylvie Van der Werf; Vincent Enouf; Vincent Vieillefond |
| EPI_ISL_2694831 | Labor Berlin Charité Vivantes GmbH / Institut für | Charité Universitätsmedizin Berlin, | Barbara Mühlemann; Christian Drosten; Christine Stephan; Peter Menzel; Rolf Schwarzer; Terry Jones; Victor M Corman |

| | | | |
|---|---|---|---|
| | Virologie | Institut für Virologie/Labor Berlin | |
| EPI_ISL_2635844, EPI_ISL_2635848, EPI_ISL_2845216, EPI_ISL_2845233 | Labor Dr. Heidrich & Kollegen MVZ GmbH Hamburg | Robert Koch Institute | |
| EPI_ISL_2631829, EPI_ISL_2631830, EPI_ISL_2631835, EPI_ISL_2845285 | Labor Dr. Wisplinghoff - Köln | Robert Koch Institute | |
| EPI_ISL_2761711, EPI_ISL_2761725 | Labor Mönchengladbach MVZ Dr. Stein + Kollegen GbR | Robert Koch Institute | |
| EPI_ISL_2678767 | Labor ZOTZ\|KLIMAS; MVZ Düsseldorf-Centrum | Robert Koch Institute | |
| EPI_ISL_2632037, EPI_ISL_2632085, EPI_ISL_2633440, EPI_ISL_2633443, EPI_ISL_2636890, EPI_ISL_2636900, EPI_ISL_2761928 | | | |
| see above | Laborarztpraxis Osnabrück | Robert Koch Institute | |
| EPI_ISL_2967031, EPI_ISL_2967032, EPI_ISL_3133652, EPI_ISL_3133653 | Laboratoire National de Santé Publique du Cameroun | Pathogen Genomics Lab, National Institute for Biomedical Research (INRB) | Amuri Aziza; Andrew Rambaut; Catherine Pratt; Eddy Kinganda-Lusamaki; Edith Nkwembe; Emmanuel Lokilo Lofiko; Francisca Muyembe Mawete; Gabriel Kabamba; Ian Goodfellow; Jean Claude Makangara; Jean-Jacques Muyembe Tamfum; Josh Quick; Marie Claire Okomo; Matthias Pauthner; Michael Wiley; Nick Loman; Placide Mbala-Kingebeni; Raphael Lumembe; Steve Ahuka-Mundeke; Trevor Bedford |
| EPI_ISL_3129580 | Laboratoire Professeur Daniel GAHOUMA (LPDG) | Centre de Recherches Médicales de Lambaréné (CERMEL) | Ayong More; Bertrand Lell; Davy Leger Mouangala; Elvyre Mbongo-Nkama; Emilio Skarwan; Georgelin Nguema Ondo; Guy Stéphane Padzys; Gédéon Prince Manouana; Jean Bernard Lekana-Douki; Joël-Fleury Djoba Siawaya and Ayola Akim Adegnika; Kevine Zang Ella; Ludovic Mewono; Moustapha Nzamba Maloum; Noé Patrick Mbondoukwe; Rodrigue Bikangui; Rodrigue Mintsa Nguema; Sam O'neilla Oye Bingono; Samira Zoa Assoumou; Sandrine Zeh Nfor; Srinivas reddy Pallerla; Steffen Borrmann; Thirumalaisamy P. Velavan |
| EPI_ISL_2989549, EPI_ISL_2989550, EPI_ISL_3021521, EPI_ISL_3021522, EPI_ISL_3143949, EPI_ISL_3143950, EPI_ISL_3144011, EPI_ISL_3144430, EPI_ISL_3144431 | | | |
| see above | Laboratoire de santé publique du Québec | Laboratoire de santé publique du Québec | Guillaume Bourque; Ioannis Ragoussis; Jesse Shapiro; Mark Lathrop and Michel Roger on behalf of the CoVSeQ research group; Mark Lathrop and Michel Roger on behalf of the CoVSeQ research group (http://covseq.ca/researchgroup); Sandrine Moreira |
| EPI_ISL_2932532, EPI_ISL_2932544, EPI_ISL_2932554, EPI_ISL_2932555, EPI_ISL_2932558, EPI_ISL_2932560, EPI_ISL_2932571, EPI_ISL_2932573, EPI_ISL_2932576, EPI_ISL_2932577, EPI_ISL_2932580, EPI_ISL_2932582, EPI_ISL_2958662, EPI_ISL_2958664 | | | |
| see above | Laboratoire des Fièvres Hémorragiques Virales du Benin | Institut für Virologie - Institute of Virology - Charite | Andres Moreira-Soto; Anges Yadouleton; Anna-Lena Sander; Benjamin Hounkpatin and Jan Felix Drexler; Benjamin Hounkpatin and Jan Felix Drexler; Carine Tchibozo; Christian Drosten; Dossou Ange; Eclou Sedjro; Edmilson F de Oliveira Filho; Gildas Hounkanrin; Hinson Fidelia; Keke K. René; Mamoudou Harouna Djingarey; Melchior A. Joël Aïssi; Michael Nagel; Olfert Landt; Praise Adewumi; Salifou Sourakatou; Victor Max Corman; Wendy Karen Jo; Yvette Badou |
| EPI_ISL_2536216 | Laboratorio HUB -Azienda Ospedaliero Universitaria - AOU – Cagliari | Laboratorio SPOKE Biologia Molecolare -Azienda Ospedaliero Universitaria - AOU – Cagliari | Alessandra Scano; Ferdinando Coghe; Germano Orrù; Miriam Loddo; Riccardo Cappai; Sara Fais; Valentina Medda |
| EPI_ISL_2650008, EPI_ISL_2650434, EPI_ISL_2650442, EPI_ISL_2820494, EPI_ISL_2886198, EPI_ISL_2886200 | Laboratorio di Riferimento Regionale della Sicilia Occidentale per l'Emergenza COVID-19 | Laboratorio di Riferimento Regionale della Sicilia Occidentale per l'Emergenza COVID-19 | Carmelo Massimo Maida; Claudio Costantino; Daniela Di Naro; Fabio Tramuto; Francesco Vitale; Giorgio Graziano; Giulia Randazzo; Vincenzo Restivo; Walter Mazzucco |
| EPI_ISL_2611495, EPI_ISL_2872942, EPI_ISL_2927684 | Laboratory Corporation of America | Centers for Disease Control and Prevention Division of Viral Diseases, Pathogen Discovery | Adrian Paskey; Amanda Douglas; Amanda Suchanek; Andrea Throop; Ayla Burns; Benjamin Rambo-Martin; Bobbi Croy; Brian Krueger; Brian Norvell; Christopher Gulvick; Christos Petropoulos; Clinton R. Paden; Craig Lukasik; Dakota Howard; Darlene Wagner; Debbie Boles; Dhwani Batra; Duncan MacCannell; Eyad Almasri; Goran Stevovic; Howard Engler; Hrushikesh Deshmukh; Jake Humphrey; Jana Schroth; Jason Caravas; Joe Voshell; John Pruitt; Jonathan Meltzer; Jonathan Williams; Kara Moser; Kimberly Wagner; Lax Iyer; Lisa Pfefferle; Lyndon Tilson; Manoj Jain; Marcia Eisenberg; Mary Ann Cristobal; Mary Williamson; Matthew Robinson; Matthew Schmerer; Michael Levandoski; Mike Sapeta; Mindy Nye; Minoo Agarwal; Mohan Kolli; Nuthawin Charoensri; Oren Cohen; Peter W. Cook; Prashant Gupta; Qian Zeng; Rama Ghatti; Scott Parker; Scott Ryan; Scott Sammons; Shatavia Morrison; Stanley Letovsky; Steven Ragan; Suresh Babu Selvaraju; Susan Countryman; Susan Hicks; Suzanne Dale; Thomas Urban; Tim Kuphal; Tricia Zwiefelhofer; Vincent Drouillon; Yvette Unoarumhi |
| EPI_ISL_2696361, EPI_ISL_2696813 | Laboratory, Gravity Diagnostics | Laboratory, Gravity Diagnostics | Diagnostics, G. |
| EPI_ISL_2636903, EPI_ISL_2636907 | Labormedizin Darmstadt | Robert Koch Institute | |
| EPI_ISL_2544771 | MD PHL | MD PHL | Maryland Department of Health Laboratories Administration |
| EPI_ISL_2595752, EPI_ISL_2595840, EPI_ISL_2597048, EPI_ISL_2597094, EPI_ISL_2597134, EPI_ISL_2597152, EPI_ISL_2597160, EPI_ISL_2597189, EPI_ISL_2659660, EPI_ISL_2659663, EPI_ISL_2659677, EPI_ISL_2659701, EPI_ISL_2843618, EPI_ISL_3054109, EPI_ISL_3054246, EPI_ISL_3054681, EPI_ISL_3054756 | | | |
| see above | MEPHI, Aix Marseille University | MEPHI, Aix Marseille University | Anthony LEVASSEUR |
| EPI_ISL_2567445, EPI_ISL_3130887, EPI_ISL_3130889, EPI_ISL_3130894 | MIRIALIS CLUSES BECHET | CNR Virus des Infections Respiratoires - France SUD | Antonin Bal; Bruno Lina; Gregory Destras; Gwendolyne Burfin; Hadrien Regue; Laurence Josset; Martine Valette; Quentin Semanas |
| EPI_ISL_2566572, EPI_ISL_2566575, EPI_ISL_2566584, EPI_ISL_2566585, EPI_ISL_2566589, EPI_ISL_2686207, EPI_ISL_2686209, EPI_ISL_2686210, EPI_ISL_2686211, EPI_ISL_2686214, EPI_ISL_2686215, EPI_ISL_2686221, EPI_ISL_2686223, EPI_ISL_2686224, EPI_ISL_2686228, EPI_ISL_2686229, EPI_ISL_2686230, EPI_ISL_3060277, EPI_ISL_3060278 | | | |
| see above | MKK Mühlenkreiskliniken, Johannes Wesling Klinikum Minden | Hannover Medical School, Institute of Virology | Lars Steinbrück |
| EPI_ISL_2557309, EPI_ISL_2557459 | MOH - Jaber Al-Ahmad Hospital (Innovation Research Laboratory) | MOH - Jaber Al-Ahmad Hospital (Innovation Research Laboratory) | Mohammad Alghounaim; Salman Al-Sabah |
| EPI_ISL_2690456, EPI_ISL_2690457, EPI_ISL_2690459 | MRC/UVRI & LSHTM Uganda Research Unit, Central Public Health Laboratories, Rakai Health Sciences Program | MRC/UVRI & LSHTM Uganda Research Unit, Central Public Health Laboratories | Dan Lule Bugembe; Hellen Nansumba; Isaac Sseeewanyana; Matthew Cotten; My V.T. Phan; Patrick Semanda; Pontiano Kaleebu; Susan Nabadda |
| EPI_ISL_2690465, EPI_ISL_2690466, EPI_ISL_2690467, EPI_ISL_2690468, EPI_ISL_2690469, EPI_ISL_2690470, EPI_ISL_2690471, EPI_ISL_2690472, EPI_ISL_2690473, EPI_ISL_2690474, EPI_ISL_2690475, EPI_ISL_2690476, EPI_ISL_2690477, EPI_ISL_2690478 | | | |
| see above | MRC/UVRI & LSHTM Uganda Research Unit, Central Public Health Laboratories, Rakai Health Sciences Program | MRC/UVRI & LSHTM Uganda Research Unit, Rakai Health Sciences Program | Charles Ssuuna; Dan Lule Bugembe; Matthew Cotten; My V.T. Phan; Pontiano Kaleebu; Ronald Moses Galiwango; Steven J Reynolds |
| EPI_ISL_2941569, EPI_ISL_2958626, EPI_ISL_2958632 | MRCG at LSHTM Genomics lab | MRCG at LSHTM Genomics lab | Abdoulie Kanteh; Abdul Karim sesay; Bakary Sanyang; Dabiri Damilari; Jarra Manneh; Mariama Kujabi |
| EPI_ISL_2636898 | MVZ Dr. Eberhard & Partner Dortmund | Robert Koch Institute | |
| EPI_ISL_2762285 | MVZ Labor Dr. Fenner und Kollegen (Standort Hamburg) | Robert Koch Institute | |
| EPI_ISL_2678695 | MVZ Labor Dr. Limbach & Kollegen GbR | Robert Koch Institute | |
| EPI_ISL_2631941, EPI_ISL_2631960, EPI_ISL_2631965, EPI_ISL_2631988, EPI_ISL_2845315 | MVZ Labor Krone GbR | Robert Koch Institute | |
| EPI_ISL_2632877 | MVZ Medizinisches Labor Hannover GmbH | Robert Koch Institute | |
| EPI_ISL_2631766, EPI_ISL_2631775, EPI_ISL_2631779, EPI_ISL_2635841, EPI_ISL_2636491 | MVZ für Laboratoriumsmedizin und Mikrobiologie Koblenz-Mittelrhein (Labor Koblenz) | Robert Koch Institute | |
| EPI_ISL_2661118, EPI_ISL_2661132, EPI_ISL_2661141, EPI_ISL_2774635, EPI_ISL_2774680, EPI_ISL_2775131, EPI_ISL_3060702 | | | |
| see above | Mapmygenome | CSIR-Centre for Cellular and Molecular Biology-INSACOG | ; Amareshwar Vodapalli; Ara Sreenivas; Archana Bharadwaj Siva; B Himasri; Divya Tej Sowpati; Jandhyala Sai Krishna; Karthik Bharadwaj Tallapaka; Lamuk Zaveri; Onkar Kulkarni; Payel Mukherjee; Priya Nurkurthy; Rakesh K Mishra; Shreekant Verma; Sofia Banu; Sumedha Avadhanula; Tulasi Nagabandi; Valli Nagalakshmi Undamatla; Vidhyadhari Methuku |
| EPI_ISL_3011660, EPI_ISL_3011810, EPI_ISL_3011877, EPI_ISL_3012142 | Maryland Genomics, Institute for Genome Sciences, University of Maryland School of Medicine | Maryland Genomics, Institute for Genome Sciences, University of Maryland School of Medicine | Claire M; Fraser; Hazen; Holly; Humphrys; Ivette; Jacques; Jonathan; Kranthi; Lim; Lisa D; Luke J; Mike; Ott; Ravel; Roussey; Sadzewicz; Sandra; Santana-Cruz; Tallon; Tracy; Vavikolanu |
| EPI_ISL_2635787 | Med. Labor Prof. Schenk Dr. Ansorge & Kollegen | Robert Koch Institute | |
| EPI_ISL_2636094 | Medizinisch-Diagnostisches Labor Kempten allgäulab | Robert Koch Institute | |
| EPI_ISL_2632026, EPI_ISL_2632102, EPI_ISL_2633436 | Medizinisches Labor Wahl Lüdenscheid | Robert Koch Institute | |

| | | | |
|---|---|---|---|
| EPI_ISL_2533755 | Michael E. Debakey VA Medical Center | Genomics and Discovery, Respiratory Viruses Branch, Division of Viral Diseases, Centers for Disease Control and Prevention | Adam Retchless; Anna Kelleher; Anna Uehara; Brian Lynch; Clinton R. Paden; Dhwani Batra; Haibin Wang; Han Jia Justin Ng; Jasmine Padilla; Jing Zhang; Justin Lee; Krista Queen; Mark Burroughs; Mili Sheth; Morgan Davis; Peter Cook; Rachel Marine; Sarah Nobles; Suxiang Tong; Tara Coalter; Yan Li; Ying Tao |
| EPI_ISL_2955984 | Microbiology Department & Molecular Biology CORE \| CDB \| Hospital Clinic \| Barcelona | Microbiology Department & Molecular Biology CORE \| CDB \| Hospital Clinic \| Barcelona | Andrea Vergara Gomez; Jose Luis Villanueva Cañas; Miguel Julian Martínez Yoldi; Mª Angeles Marcos Maeso; Mª del Mar Mosquera Gutierrez |
| EPI_ISL_2650188, EPI_ISL_2650252, EPI_ISL_2650289, EPI_ISL_2671454, EPI_ISL_2681581, EPI_ISL_2718558, EPI_ISL_2832181, EPI_ISL_2926136, EPI_ISL_2926184 | | | |
| see above | Microbiology Department, Laboratori Clínic Metropolitana Nord. Hospital Universitari Germans Trias i Pujol | Can Ruti SARS-CoV-2 Sequencing Hub (HUGTiP/IrsiCaixa/IGTP) | Alba Sánchez; Alexia París; Anna Not; Antoni E Bordoy; Bonaventura Clotet; Cristina Casañ; Cristina Esteban; David Panisello; Francesc Catala-Moll; Gemma Clara; Ignacio Blanco; Laia Soler; Lauro Sumoy; Marc Noguera-Julian; Maria Casadellà; Mariona Parera; Mercedes Guerrero; Montserrat Giménez; Pere-Joan Cardona; Pilar Armengol; Roger Paredes; Verónica Saludes; and Elisa Martró on behalf of the Can Ruti SARS-CoV-2 Sequencing Hub |
| EPI_ISL_2774095 | Microbiology Department, University Hospital Donostia | Microbiology Department, University Hospital Donostia | Cilla G.; Gomez M; Marimon JM; Martin-Peñaranda T; Montes M; Piñeiro L; Sorarrain A |
| EPI_ISL_2629188 | Microbiology Division, South Carolina Department of Health and Environmental Control | Microbiology Division, South Carolina Department of Health and Environmental Control | Flores; Freeman; H.V.; J.S. |
| EPI_ISL_2812246 | Microvida | Microvida | Jaco J. Verweij; Joep J. J. M. Stohr; Suzan D. Pas |
| EPI_ISL_2535627 | Ministry of Health Hospitals | Institute of Health and Community Medicine | Chan Chia Jui; Chua Hock Hin; David Perera; Ooi Mong How; Tonnii Sia Loong Loong; Wong Jyn Shan; Wong Kiing Aik |
| EPI_ISL_2936192, EPI_ISL_2936656, EPI_ISL_2936679, EPI_ISL_2936705, EPI_ISL_2984781, EPI_ISL_2987741, EPI_ISL_2987868, EPI_ISL_3032542, EPI_ISL_3032551, EPI_ISL_3032553, EPI_ISL_3032561, EPI_ISL_3032567, EPI_ISL_3032568, EPI_ISL_3032573, EPI_ISL_3032637, EPI_ISL_3063292, EPI_ISL_3063355, EPI_ISL_3063398 | | | |
| see above | Ministry of Health Turkey | Ministry of Health Turkey | Fatma Bayrakdar; Gulay Korukluoglu; Suleyman Yalcin; Yasemin Cosgun |
| EPI_ISL_2603857, EPI_ISL_2604185, EPI_ISL_2604187 | Ministry of Health, Jaber Al-Ahmad Hospital | Virology Unit, Department of Microbiology, Faculty of Medicine, Kuwait University | Anfal Al-Adwani; Ebaa Al-Awadhi; Hussain Safar; Nada Madi |
| EPI_ISL_3142727, EPI_ISL_3142936 | Molecular & Genomic Pathology Laboratory, Thomas Jefferson University Hospital | Molecular & Genomic Pathology Laboratory, Thomas Jefferson University Hospital | Nitika Badjatia; Run Jin; Shannon Mahoney; Zi-xuan Wang |
| EPI_ISL_2718542 | Molecular Diagnostics Pathology Department Mater Dei Hospital Malta | Molecular Diagnostics Pathology Department Mater Dei Hospital Malta | C Cilia; G Zahra; L Grech; M Briffa; R Borg |
| EPI_ISL_3101069, EPI_ISL_3101079, EPI_ISL_3101353 | Molecular diagnostic laboratory of Federal Budget Institution of Science "Central Research Institute of Epidemiology" of The Federal Service on Customers' Rights Protection and Human Well-being Surveillance | Group of Genomics and Postgenomic Technologies of Central Research Institute of Epidemiology | Akimkin V.G.; Berlina Y.Y.; Bulanenko V.P.; Cherkashina A.S.; Dohoyan A.Y.; Golubeva A.G.; Kaptelova V.V.; Kondrasheva L.Y.; Korneenko E.V.; Nadtoka M.I.; Saenko S.S.; Samoilov A.E.; Shipulina O.Y.; Sinitsyn S.O.; Smirnova Y.S.; Solovieva E.D.; Speranskaya A.S.; Tivanova E.V.; Valdohina A.V.; Zotova M.I.; Zuev S.N. |
| EPI_ISL_2816385, EPI_ISL_2964419 | N.F. Gamaleya Research Center for Epidemiology and Microbiology | WHO National Influenza Centre Russian Federation | Alexander Gintsburg; Alexey Shchetinin; Alina Odintsova; Andrei Botikov; Andrei Pochtovyi; Andrei Siniavin; Andrey Komissarov; Anna Kovyrshina; Artem Fadeev; Artem Tkachuk; Daria Danilenko; Denis Kleymenov; Denis Logunov; Dmitry Lioznov; Dmitry Shcheblyakov; Elena Mazunina; Elena Nabieva; Elena Shidlovskaya; Elizaveta Divisenko; Evgeniia Bykonia; Georgii Bazykin; Inna Dolzhikova; Kirill Varchenko; Ksenia Safina; Kseniya Komissarova; Liubov Popova; Ludmila Vasilchenko; Maria Nikiforova; Maria Pisareva; Mikhail Bakaev; Nadezhda Kuznetsova; Nikita Yolshin; Oula Mansour; Tamila Musaeva; Veronika Eder; Vladimir Gushchin |
| EPI_ISL_2842671 | NJDOH, Public Health and Environmental Laboratories | NJ_PHEL | Allison Roder; Byeong Jeong; Chelsea San Filippo; Dana Woell; Jacquelyn Deverell; Lindsey Bodnar; Ryan Pachucki; Shiv K. Verma |
| EPI_ISL_2753948 | NYU Langone Health | Departments of Pathology and Medicine, New York University School of Medicine | Adriana Heguy; Christian Marier; Dacia Dimartino; Emily Guzman; Gael Westby; Guiqing Wang; Paolo Cotzia; Paul Zappile; Peter Meyn; Sitharam Ramaswami; Yutong Zhang |
| EPI_ISL_2504395, EPI_ISL_2504494, EPI_ISL_2504496, EPI_ISL_2504499, EPI_ISL_2505074, EPI_ISL_2505094, EPI_ISL_2555557, EPI_ISL_2555621, EPI_ISL_2556360, EPI_ISL_2556371, EPI_ISL_2556383, EPI_ISL_2556451, EPI_ISL_2556452, EPI_ISL_2556455, EPI_ISL_2556459, EPI_ISL_2556464, EPI_ISL_2556466, EPI_ISL_2556469, EPI_ISL_2556470, EPI_ISL_2556472, EPI_ISL_2556490, EPI_ISL_2556536, EPI_ISL_2556609, EPI_ISL_2556713, EPI_ISL_2556718, EPI_ISL_2556914, EPI_ISL_2556924, EPI_ISL_2556928, EPI_ISL_2556935, EPI_ISL_2556939, EPI_ISL_2556943, EPI_ISL_2556945, EPI_ISL_2556949, EPI_ISL_2556955, EPI_ISL_2556956, EPI_ISL_2556961, EPI_ISL_2556962, EPI_ISL_2955658, EPI_ISL_2955692 | | | |
| see above | National Centre for Disease Control (NCDC) Biotechnology Division, Delhi | NCDC Delhi, Biotechnology Division INSACOG | Hema Gogia; Hemlata Lall; Kalaiarasan Ponnusamy; Mahesh S Dhar; Manoj K Singh; Meena Datta; Partha Rakshit; Preeti Madan; Priyanka Singh; Radhakrishnan V. S; Robin Marwal; Sandhya Kabra; Sujeet K Singh; Uma Sharma |
| EPI_ISL_2535814, EPI_ISL_2535822, EPI_ISL_2873841, EPI_ISL_2873846 | National Influenza Centre | National Influenza Centre | ; Benjamiin B. Lindsey; Benjamin H. Foulkes; Dennis Laryea; Ernest Asiedu; Franklin Asiedu-Bekoe; Gordon Awandare; Ivy A. Asante; Joseph Oliver-Commey; Joyce Ngoi; Linda Boatemaa; Lorreta Kwasah; Mathew D. Parker; Michael Marks; Mildred Adusei-Poku; Sharon Hsu; Thushan I de Silva; William K. Ampofo |
| EPI_ISL_2693104, EPI_ISL_2821075, EPI_ISL_2821081, EPI_ISL_2821257, EPI_ISL_2821326, EPI_ISL_2821509, EPI_ISL_2821510, EPI_ISL_2821669 | | | |
| see above | National Reference Laboratory, Nigeria Centre for Disease Control | National Reference Laboratory, Nigeria Centre for Disease Control | Anthony Ahumibe; Catherine Okoi; Chimaobi Chukwu; Dr Chikwe Ihekweazu; Dr Ndodo Nnaemeka; Dr Omoare Adesuyi; Grace Esebanmen; Nwando Mba; Olusola Anuoluwapo Akanbi |
| EPI_ISL_2821076 | National Reference Laboratory, Nigeria Centre for Disease Control | National Reference Laboratory, Nigeria Centre for Disease Control Durumi III, Gaduwa, Abuja, Nigeria | Anthony Ahumibe; Catherine Okoi; Chimaobi Chukwu; Dr Chikwe Ihekweazu; Dr Ndodo Nnaemeka; Dr Omoare Adesuyi; Grace Esebanmen; Nwando Mba; Olusola Anuoluwapo Akanbi |
| EPI_ISL_2533168, EPI_ISL_2533440, EPI_ISL_2533640, EPI_ISL_2533669, EPI_ISL_2601319, EPI_ISL_2601361, EPI_ISL_2710647, EPI_ISL_2710724, EPI_ISL_2710759, EPI_ISL_2821086 | | | |
| see above | National Virus Reference Laboratory | National Virus Reference Laboratory | Calum Walsh; Charlene Bennet; Charlene Bennett; Cillian F De Gascun; Fiona Crispie; Gabriel Gonzalez; Jonathan Dean; Matthew McCabe; Michael Carr; Paul Cotter; Zoe Yandle |
| EPI_ISL_2632240, EPI_ISL_2632816, EPI_ISL_2632817, EPI_ISL_2632818, EPI_ISL_2632819, EPI_ISL_2632821, EPI_ISL_2632823 | | | |
| see above | Niedersächsisches Landesgesundheitsamt (NLGA) | Robert Koch Institute | |
| EPI_ISL_2565887, EPI_ISL_2565888, EPI_ISL_2565891, EPI_ISL_2565893, EPI_ISL_2565896, EPI_ISL_2565897, EPI_ISL_2565898 | | | |
| see above | Nigeria Centre for Disease Control (NCDC) | Africa Centre for Excellence for Genomics of Infectious Diseases (ACEGID), Redeemer's University | A.T.; Abechi; Ajogbasile; Akano; C.A.; C.T.; Eromon; F.V.; Folarin, O.; Happi; I.B.; J.N.; J.U.; K.O.; Kayode; Nosamiefan, I.; Oguzie; Olawoye; Olumade; Oluniyi; P.E.; P.S.; T.J.; Ugwu; Uwanibe |
| EPI_ISL_3100116 | OLVZ Aalst | OLVZ Aalst | Astrid Holderbeke |
| EPI_ISL_2651238 | OSPEDALE "Giovanni Paolo II" | AOU Policlinico "G. Rodolico- S. Marco" PO G. Rodolico and Biometec UNICT | Stefani and Scalia |
| EPI_ISL_2840677 | OSPEDALE CIVILE TERAMO - CENTRO TRASFUSIONALE | Istituto Zooprofilattico Sperimentale dell'Abruzzo e Molise "G. Caporale" | Ancora M; Calistri P; Cammà C; Caporale M; Curini V; Delli Compagni E; Di Domenico M; Di Lollo Valeria; Di Pasquale A; Lorusso A; Mangone I; Marcacci M; Puglia I; Rinaldi A; Savini G; Scialabba S |
| EPI_ISL_2747989, EPI_ISL_2748655, EPI_ISL_2771492 | Ohio Department of Health Laboratory | Ohio Department of Health Laboratory | Allison Black; Brent Lee; Caitlin McDonnell; Eric Brandt; Erica Leasure; Glen McGillivary; Heather Blankenship; Holmes; Jade Mowery; Jennifer; Kelsey Florek; Keoni Omura; Kirtana Ramadugu; Quanta Brown; Stephanie Mccracken; Tyler Payne; and Tammy Bannerman |
| EPI_ISL_2641661 | Ospedale Regionale Umberto Parini | U.O. Genomica - Istituto Zooprofilattico Sperimentale del Piemonte, Liguria e Valle d'Aosta | Simone Peletto |
| EPI_ISL_3032284 | Ospedale Santissima Annunziata | Istituto Zooprofilattico Sperimentale della Puglia e della Basilicata | Bianco A.; Capozzi L.; Del Sambro L.; Difato L.; Giannico A; Parisi A.; Ridolfi D.; Simone D.; Tampoia M. |
| EPI_ISL_3008169, EPI_ISL_3008170, EPI_ISL_3031167 | Ostfold Hospital Trust - Kalnes, Centre for Laboratory Medicine, Section for gene technology and infection serology | Norwegian Institute of Public Health, Department of Virology | Atiya R Ali; Debech Nadia; Engebretsen Serina Beate; Garcia Llorente Ignacio; Hilde Elshaug; Hilde Vollan; Jon Bråte; Kamilla Heddeland Instefjord; Karoline Bragstad; Kathrine Stene-Johansen; Line Victoria Moen; Marie Paulsen Madsen; Olav Hungnes; Pedersen Benedikte Nevjen; Rasmus Riis Kopperud |
| EPI_ISL_2547416 | Pacaltsdorp Clinic wc PAC | National Health Laboratory Service/University of Cape Town (NHLS/UCT) | Arash Iranzadeh; Bruna Galvao; Carolyn Williamson; Deelan Doolabh; Diana Hardie; Gert Marais; Innocent Mudau; Lynn Tyers; Marvin Hsiao; Stephen Korsman |
| EPI_ISL_2993463 | Pathology, UAB | Pathology, UAB | E.J.; Hendrickson; Hernandez, J.; Leal; Lefkowitz; Moates, D.; R.C.; S.M. |
| EPI_ISL_3046916 | Pennsylvania Department of Health Bureau of Laboratories | Pennsylvania Department of Health Bureau of Laboratories | Dongxiang Xia |
| EPI_ISL_2779318 | Port Health Clinic, Malaba border point | USAMRD-A, Basic Science Laboratory | Alan Lemtudo; Beth Mutai; Brian Andika; Carol Kifude; Clement Masakwe; Eric Muthanje; Esther Omuseni; Faith Sigei; Gathii Kimita; George Awinda; John Waitumbi; Josphat Nyataya; Rachel Githii; Rehema Liyai; Stephen Ochola |
| EPI_ISL_2859961 | Prime Care Alpha Covid-19 Testing Laboratory | Philippine Genome Center | Alethea R. de Guzman; Anna Ong-Lim; Arianne A. Zamora; Benedict A. Maralit; Carlo M. Lapid; Celia Carlos; Devon Ray Pacial; Diomedes A. Carino; Edsel Maurice Salvaña; El King D. Morado; Elcid Aaron R. Pangilinan; Eva Maria Cutiongco-de la Paz; Francis A. Tablizo; Henrietta Marie |

| | | | |
|---|---|---|---|
| | | | Rodriguez; Jaime C. Montoya; Jan Michael C. Yap; Jarvin E. Nipales; Jo-Hannah S. Llames; John Q. Wong; Joshua Gregor A. Dizon; Juan Antonio R. Magalang; Karol Sophia Agape R. Padilla; Kenneth M. Kim; Kris P. Punayan; Krisitna Patriz Dela Cruz; Lindsay Claire D.L. Carandang; Ma. Exanil Plantig; Marc Edsel C. Ayes; Maria Rosario Singh-Vergeire and Cynthia P. Saloma; Maria Sofia L. Yangzon; Marielle M Gamboa; Marissa Alejandria; Nina Francesca Bustamante; Razel Nikka M. Hao; Renato Jacinto Q. Mantaring; Rianna Patricia S. Cruz; Sheila Mae M. Araiza; Yvonne Valerie Austria; Zipporah Mariebelle R. Enriquez; Zyrel V. Mollejon |
| EPI_ISL_2551373 | Providence Oregon Regional Laboratories | Providence St. Joseph Health Molecular Genomics Laboratory | Alexa K Dowdell; Brian D Piening; Carlo B Bifulco; Fred L Robinson; Mary Campbell; Rogan Rattray |
| EPI_ISL_2698849 | Public Health Laboratory, Minnesota Department of Health | University of Minnesota Genomics Center | Corbin Dirkx; Daryl M. Gohl; Jaquelyn Kuriger-Laber; John Garbe; and Sean Wang |
| EPI_ISL_2547557, EPI_ISL_2547590, EPI_ISL_2547591, EPI_ISL_2547593, EPI_ISL_2548091, EPI_ISL_2548116, EPI_ISL_2630188, EPI_ISL_2630231, EPI_ISL_2630243, EPI_ISL_2630311, EPI_ISL_2630681, EPI_ISL_2630682, EPI_ISL_2630683, EPI_ISL_2630785, EPI_ISL_2630817, EPI_ISL_2630819, EPI_ISL_2630902, EPI_ISL_2630929, EPI_ISL_2713486, EPI_ISL_2713663, EPI_ISL_2713765, EPI_ISL_2713848, EPI_ISL_2714269, EPI_ISL_2714272, EPI_ISL_2714469, EPI_ISL_2714527, EPI_ISL_2822351, EPI_ISL_2822604, EPI_ISL_2823275, EPI_ISL_2823330, EPI_ISL_2823331, EPI_ISL_2823342, EPI_ISL_2823351, EPI_ISL_2823352, EPI_ISL_2823354, EPI_ISL_2823355, EPI_ISL_2823358, EPI_ISL_2823447, EPI_ISL_2922317, EPI_ISL_2922695, EPI_ISL_3012301, EPI_ISL_3089954 | | | |
| see above | Public Health Ontario Laboratory | Public Health Ontario Laboratory | Aimin Li; Alireza Eshaghi; Andre Villegas; Ashleigh Sullivan; Christine Frantz; Dean Maxwell; Esha Joshi; Jared Simpson; Jennifer L Guthrie; Jonathan B Gubbay; Karthikeyan Sivaraman; Lawrence Heisler; Matthew Watson; Michael CY Li; Michael Laszloffy; Nahuel Fittipaldi; Philip Banh; Richard de Borja; Samir N Patel; Sandeep Nagra; Sandra Zittermann; Sarah Teatero; Vanessa G Allen; Yao Chen; Yogi Sundaravadanam |
| EPI_ISL_2860636, EPI_ISL_2860637, EPI_ISL_2860638, EPI_ISL_2860639, EPI_ISL_2860640, EPI_ISL_2860641, EPI_ISL_2860642, EPI_ISL_2860643, EPI_ISL_2860644, EPI_ISL_2860645, EPI_ISL_2860647 | | | |
| see above | Public Health Reference Laboratory | Erasmus Medical Center | Anne van der Linden; Annemiek van der Eijk; Bas Oude Munnink; Corine GeurtsvanKessel; David Nieuwenhuijse; Emmanuelle Munger; Irina Chestakova; Marion Koopmans; Marjan Boter; Omar Elahmer-Abdulla Bashein- Rahma Algheriani-Ahlam Alarif; Reina Sikkema; Richard Molenkamp; on behalf of the Dutch national COVID-19 response team. |
| EPI_ISL_2615961, EPI_ISL_2758356 | QLabs | WVU and Marshall University Combined Genomics Core Facilities | James Denvir; Peter Perrotta; Peter Stoilov; Ryan Percifield; Wesley Kimble |
| EPI_ISL_2869102, EPI_ISL_2869107, EPI_ISL_2929442, EPI_ISL_2929559 | Quest Diagnostics Incorporated | Centers for Disease Control and Prevention Division of Viral Diseases, Pathogen Discovery | A. Gerasimova; A. Perez; Adrian Paskey; B. Anderson; Benjamin Rambo-Martin; Christopher Gulvick; Clinton R. Paden; Dakota Howard; Darlene Wagner; Dhwani Batra; Duncan MacCannell; F. Lacbawan; I. A. Shlyakhter; Jason Caravas; K.E. Livingston; Kara Moser; L.E. Bernstein; M. Hua; Matthew Schmerer; P. Tanpaiboon; Peter W. Cook; R. M. Kagan; R. Owen; R. V. Rolando; S. H. Rosenthal; Scott Sammons; Shatavia Morrison; Y. Liu; Yvette Unoarumhi |
| EPI_ISL_3153576, EPI_ISL_3154311, EPI_ISL_3154312 | REUNILAB | UMR PIMIT | Dr Camille Lebarbenchon; Dr David A Wilkinson; Dr Patrick Mavingui; Magali Turpin; Marie-Alice Simbi |
| EPI_ISL_2695767 | ROB FERREIRA LABORATORY | National Institute for Communicable Diseases of the National Health Laboratory Service | Amoako DG; Bhiman JN; Everatt J; Ismail A; Mahlangu B; Mnguni A; Mohale T; Ntuli N; Scheepers C |
| EPI_ISL_3138822 | RS Kartika Pulomas | National Institute of Health Research and Development | Arie Ardiansyah Nugraha; Fajar Nur Sulistiyohadi; Hana Apsari Pawestri; Hartanti Dian Ikawati; Kartika Dewi Puspa; Krisna Pangesti; Nelly Puspandari; Subangkit; Triyani Soekarso; Vivi Setiawaty |
| EPI_ISL_2854721 | RSUP Dr. Hasan Sadikin Bandung | National Institute of Health Research and Development | ; Arie Ardiansyah Nugraha; Hana Apsari Pawestri; Hartanti Dian Ikawati; Kartika Dewi Puspa; Krisna Pangesti; Nelly Puspandari; Subangkit; Triyani Soekarso; Vivi Setiawaty |
| EPI_ISL_3046655, EPI_ISL_3046658 | Rami Kantor lab | Rami Kantor lab | Ewa King; Josephine Darpolor; Mark Howison; Rami Kantor; Richard Huard; Vlad Novitsky |
| EPI_ISL_2733023, EPI_ISL_2733024, EPI_ISL_2734687, EPI_ISL_2734689, EPI_ISL_2734693, EPI_ISL_2734700, EPI_ISL_2734729, EPI_ISL_2734750, EPI_ISL_2734794, EPI_ISL_2734808, EPI_ISL_2734812, EPI_ISL_2734844, EPI_ISL_2734857, EPI_ISL_2734858, EPI_ISL_2734859, EPI_ISL_2734860, EPI_ISL_2734862, EPI_ISL_2734864, EPI_ISL_2734865, EPI_ISL_2734873, EPI_ISL_2735065, EPI_ISL_2735066, EPI_ISL_2735116, EPI_ISL_2737315, EPI_ISL_2739031, EPI_ISL_2739448, EPI_ISL_2739807, EPI_ISL_2740076, EPI_ISL_2740077, EPI_ISL_2740172, EPI_ISL_2740523, EPI_ISL_2740557, EPI_ISL_2740573, EPI_ISL_2740636, EPI_ISL_2740740, EPI_ISL_2740746, EPI_ISL_2740781, EPI_ISL_2740863, EPI_ISL_2740874, EPI_ISL_2741192, EPI_ISL_2741208, EPI_ISL_2741290, EPI_ISL_2741299, EPI_ISL_2741344, EPI_ISL_2741348, EPI_ISL_2741395, EPI_ISL_2741454, EPI_ISL_2742001, EPI_ISL_2742041, EPI_ISL_2742088, EPI_ISL_2742124, EPI_ISL_2742717, EPI_ISL_2748273, EPI_ISL_2748464, EPI_ISL_2748481, EPI_ISL_2748522, EPI_ISL_2748532, EPI_ISL_2748587, EPI_ISL_2748590, EPI_ISL_2748644, EPI_ISL_2748645, EPI_ISL_2748842, EPI_ISL_2748857, EPI_ISL_2748971, EPI_ISL_2748991, EPI_ISL_2749011, EPI_ISL_2749041, EPI_ISL_2749094, EPI_ISL_2749116, EPI_ISL_2749130, EPI_ISL_2749202, EPI_ISL_2749227, EPI_ISL_2749274, EPI_ISL_2749323, EPI_ISL_2749806, EPI_ISL_2818301, EPI_ISL_2900072, EPI_ISL_2904319, EPI_ISL_3004182, EPI_ISL_3004260, EPI_ISL_3081003 | | | |
| see above | Respiratory Virus Unit, Microbiology Services Colindale, Public Health England | COVID-19 Genomics UK (COG-UK) Consortium | PHE Covid Sequencing Team |
| EPI_ISL_2604855 | Rhode Island State Health Laboratories | Rhode Island State Health Laboratories | Ewa King; Kristin Carpenter-Azevedo; Richard C. Huard |
| EPI_ISL_2603892, EPI_ISL_2835252 | Rhode Island State Health Laboratory | Rhode Island State Health Laboratory | Ewa King; Kristin Carpenter-Azevedo; Richard C. Huard |
| EPI_ISL_2649124, EPI_ISL_2649497, EPI_ISL_2791578, EPI_ISL_2791829, EPI_ISL_2791835, EPI_ISL_2791913 | SARS-CoV-2 Sequencing Castilla y Leon-Spain Consortium | SARS-CoV-2 Sequencing Castilla y Leon-Spain Consortium | Antonio Orduña-Domingo; Carlos Fuster Foz; Carmen Aldea-Mansilla; Carmen Gimeno Crespo; David Abad; Gregoria Megías Lobón; Jose María Eiros Bouza; Laura Sánchez de Prada; M. Isabel Fernandez-Natal; Marta Dominguez-Gil; Marta Hernandez; María Antonia García Castro; Mª Fe Brezmes-Valdivieso; Noelia Arenal Andrés; Silvia Rojo; Sonsoles Garcinuño Pérez |
| EPI_ISL_2621740 | SARS-CoV-2 testing team, National Institute of Infectious Diseases | Pathogen Genomics Center, National Institute of Infectious Diseases | Hazuka Y Furihata; Hiromizu Takahashi; Kentaro Itokawa; Makoto Kuroda; Masanori Hashino; Masumichi Saito; Naomi Nojiri; Nozomu Hanaoka; Rina Tanaka; Sana Uchikoba; Tsuguto Fujimoto; Tsuyoshi Sekizuka |
| EPI_ISL_2585440, EPI_ISL_2587459 | SK-Roy Romanow Provincial Laboratory | National Microbiology Laboratory (NML) | Amanda Lang; Anna Majer; Anneliese Landgraff; CanCOGeN's metadata curation team; Darian Hole; Elsie Grudeski; Gary Van Domselaar; Grace Seo; Jennifer Tanner; Jessica Minion; Kirsten Biggar; Madison Chapel; Morag Graham; Natalie Knox; Nathalie Bastien; Philip Mabon; Public Health Agency of Canada CanCOGeN team; Rachel DePaulo; Rhiannon Huzarewich; Russell Mandes; Ryan McDonald; Shari Tyson; Timothy Booth; Yan Li |
| EPI_ISL_2642063 | SYNLAB Eesti OÜ | Department of Microbiology, Institute of Biomedicine and Translational Medicine, University of Tartu | Aare Abroi; Andrio Lahesaare; Arina Shablinskaja; Dagmar Hoidmets; Ene-Ly Jõgeda; Eveli Kallas; Heiki Niglas; Irja Lutsar; Kai Truusalu; Kaisa Truus; Katrin Kaarna; Kristi Huik; Liidia Dotsenko; Lili Azin Milani; Mari-Anne Härma; Mats Hansen; Meri Pauskar; Olga Sadikova; Paul Naaber; Radko Avi; Taavi Päll; Tuuli Reisberg; Ulvi Gerst Talas |
| EPI_ISL_2956176, EPI_ISL_2956177 | SYNLAB MVZ Trier | Robert Koch Institute | |
| EPI_ISL_2800601, EPI_ISL_2800602 | Seattle University Athletics | Altius Institute for Biomedical Research | Alex Isner; Alex Nguyen; Amanda Gale; Audra Johnson; Clem Green; Daniel Bates; Eric Thorland; Jacob Rodriguez; Jean Robinston; Jemma Nelson; Jessica Kunder; John Stamatoyannopoulos; Joshua Richards; Julia Wald; Kneshay Harper; Lauren Mitchell; Mark Frerker; Matt Hartman; Michael Buckley; Muhammad Halimun; Rebecca Bruders; Sadie Patraw; Sofia Olsson; Tobias Ragoczy |
| EPI_ISL_2621140 | Sedgefield Clinic wc SGE | NHLS/UCT | Arash Iranzadeh; Bruna Galvao; Carolyn Williamson; Deelan Doolabh; Diana Hardie; Gert Marais; Innocent Mudau; Lynn Tyers; Marvin Hsiao; Stephen Korsman |
| EPI_ISL_2709318, EPI_ISL_2709339 | Servicio Microbiología Hospital La Paz | Servicio Microbiología Hospital La Paz | Elie Dahdouh; Fernando Lázaro; Jesús Mingorance Cruz; Rubén Cáceres |
| EPI_ISL_2516675 | Servicio de Microbiología Clínica (Complejo Hospitalario de Navarra, Pamplona) | Centro de Secuenciación NASERTIC | Ana Miqueleiz; Ana Navascués; Carmen Ezpeleta Baquedano |
| EPI_ISL_2545360, EPI_ISL_2545382, EPI_ISL_2545412 | Servicio de Microbiología, Laboratori Clínic Metropolitana Nord. Hospital Universitari Germans Trias i Pujol. Institut d'Investigació en Ciències de la Salut Germans Trias i Pujol (IGTP) | SeqCOVID-SPAIN consortium/IBV(CSIC) | Adrián Antuori; Anabel Fernández; Anna Not; Antoni E. Bordoy; Cristina Casañ and SeqCOVID-SPAIN consortium; Elisa Martró; Nona Romaní; Verónica Saludes |
| EPI_ISL_2627527, EPI_ISL_2627530 | Servicio de Microbiología. Hospital Universitario Doctor Peset | SeqCOVID-SPAIN consortium/IBV(CSIC) | José Miguel Nogueira Coito and SeqCOVID-SPAIN consortium; Juan Alberola Enguídanos; Juan José Camarena Miñana; Rosa González Pellicer |
| EPI_ISL_3087845 | Servizo de Microbioloxía. Complexo Hospitalario de Santiago de Compostela | Servizo de Microbioloxía. Complexo Hospitalario de Santiago de Compostela | Amparo Coira; Antonio Aguilera; Carlos García_Riestra; Daniel Navarro; Gema Barbeito; José Llovo; Laura Millán; Manuela Hernández; María Luisa Pérez_del_Molino; Mercedes Treviño; Rocío trastoy; Teresa Lopez_Valiño; Xosé Costa |
| EPI_ISL_2567026 | Shamir Medical Center (Asaf Harofe) | Shamir Medical Center (Asaf Harofe) | Abu Hamad Ramzia; Adina Bar Chaim; Anna Vishnevsky; Chen Weiner; Nir Rainy; Patricia Benveniste-Lekovitz; Reut Sorek Abramovich; Yevgeni Yegorov |
| EPI_ISL_2761997 | Sonic - Dr. Staber & Kollegen GmbH München | Robert Koch Institute | |
| EPI_ISL_2928015, EPI_ISL_2928018, EPI_ISL_2928020, EPI_ISL_2928021, EPI_ISL_2928022, EPI_ISL_2928027 | South Sudan Ministry of Health, WHO South Sudan, MRC/UVRI & LSHTM Uganda Research Unit | MRC/UVRI & LSHTM Uganda Research Unit, South Sudan Ministry of Health, WHO South Sudan | Abe G. Abias; Dan Lule Bugembe; Dennis Kenyi Lodiongo; James Ayei; John Rumunu; Joseph Francis Wamala; Juma John HM; Lul Lojok Deng; Matthew Cotten; My V.T. Phan; Pontiano Kaleebu; Richard Lino Loro Lako; Sudhir Bunga |
| EPI_ISL_2659060 | Stanford Clinical Virology Laboratory | Santa Clara County Public Health Laboratory | Santa Clara County Public Health Department |
| EPI_ISL_3050279 | Stanford Health Care | Stanford University School of Medicine, Clinical Virology Laboratory | Becky Jiang; Bernadette Troung; Daniel Solis; Fiona Yamamoto; James Zehnder; Malaya K. Sahoo; Mamdouh Sibai; Nathan Hammond; Selamawit Bihon; and Benjamin A. Pinsky |
| EPI_ISL_2677473 | Synlab Haut de France | UMR 8199/1283 EGID | Derhourhi Mehdi |
| EPI_ISL_2662595, EPI_ISL_2662607 | TAMBO MEMORIAL LABORATORY | National Institute for Communicable Diseases of the National Health Laboratory Service | Amoako DG; Bhiman JN; Everatt J; Ismail A; Mahlangu B; Mnguni A; Mohale T; Ntuli N; Scheepers C |
| EPI_ISL_2978670 | U.O. Microbiologia Laboratorio Unico Centro Servizi | U.O. Microbiologia Laboratorio | Francesca Taddei; Giorgio Dirani; Giulia Gatti; Ilaria Poggianti; Silvia Zannoli; Vittorio Sambri |

| | | | |
|---|---|---|---|
| | AUSL della Romagna | Unico Centro Servizi AUSL della Romagna | |
| EPI_ISL_3060014 | UAB "Rezus.lt" | Institute of Biotechnology, Life Sciences Center, Vilnius University | Albertas Timinskas; Alma Gedvilaite; Danguole Ziogiene; Emilija Vasiliunaite; Milda Norkiene |
| EPI_ISL_2878091 | UAB Medicina practica laboratorija | Institute of Biotechnology, Life Sciences Center, Vilnius University | Albertas Timinskas; Alma Gedvilaite; Danguole Ziogiene; Emilija Vasiliunaite; Milda Norkiene |
| EPI_ISL_2989737, EPI_ISL_2989738 | UF Health Shands Hospital - Core Laboratory | Salemi Lab, University of Florida | Cash M; Lauzardo M; Magalis BR; Mavian C; Riva A; Salemi M |
| EPI_ISL_2824973 | ULS Norte Alentejano | Instituto Nacional de Saude (INSA) and Faculdade de Medicina da Universidade de Coimbra | Borges et al |
| EPI_ISL_2888010 | UMC Utrecht | UMC Utrecht | Annemarie Wensing; Anniek Tanja; Pascal Mantiri; Rob Schuurman |
| EPI_ISL_2628710, EPI_ISL_2646019, EPI_ISL_2646083 | UNILABS | Instituto Nacional de Saude (INSA) | Borges et al |
| EPI_ISL_2796141 | UNILABS | Instituto Nacional de Saude (INSA) and Instituto Gulbenkian de Ciencia (IGC) | Borges et al |
| EPI_ISL_2820963 | UOC Microbiologia e Virologia, Azienda Ospedaliera Universitaria Senese, Siena, Italy | Dipartimento di Biotecnologie Mediche | Claudia Gandolfo; David Pinzauti; Francesco Santoro; Gabriele Anichini; Gianni Gori Savellini; Gianni Pozzi; Maria Grazia Cusi |
| EPI_ISL_3025001, EPI_ISL_3025042, EPI_ISL_3025255 | UVA Clinical Microbiology Lab | Mathers Lab | Amy J. Mathers; Hardik I. Parikh; Katie E. Barry |
| EPI_ISL_2618555 | UW Virology Lab | UW Virology Lab | Alexander Greninger; Hong Xie; Keith R Jerome; Lasata Shrestha; Meei-Li Huang; Noah R. Baker; Pavitra Roychoudhury; Sean Ellis; Shah Mohamed Bakhash; Tien V. Nguyen |
| EPI_ISL_2897527 | Unilabs | Karolinska University Hospital | Annelie Bjerkner; Isak Sylvin; Jan Albert; Karolina Ininbergs; Lina Guerra Blomqvist; Lynda Eneh; Martin Ekman; Martina Wahlund; Robert Dyrdak; Sandra Broddesson; Tanja Normark; Tobias Allander; Valtteri Wirta; Zhibing Yun |
| EPI_ISL_2681467 | University Hospitals of Geneva, Laboratory of Virology | HUG, Laboratory of Virology and the Health2030 Genome Center | Ana Rita Goncalves; Deborah Penet; Emmanouil Dermitzakis; Henri Pegeot; Ioannis Xenarios; Keith Harshman; Laurent Kaiser; Lorenzo Cerutti; Melyssa Elies; Samuel Cordey |
| EPI_ISL_3014274, EPI_ISL_3014357 | Università Federico II - Dipartimento di scienze mediche traslazionali - Napoli | TIGEM | Antonio Grimaldi Patrizia Annunziata Francesco Panariello Teresa Giuliano Michele Cennamo Valentina Bouche Chiara Colantuono Lucio Di Filippo Mariano Fiorenza Anna Manfredi Marcello Salvi Giuseppe Portella Andrea Ballabio Davide Cacchiarelli |
| EPI_ISL_2600757, EPI_ISL_2600763, EPI_ISL_2600764, EPI_ISL_2600765, EPI_ISL_2600776, EPI_ISL_3030459 | Università degli Studi di Perugia | Istituto Zooprofilattico Sperimentale dell'Abruzzo e Molise "G. Caporale" | Ancora M; Biagetti M; Calistri P; Camilloni B; Cammà C; Curini V; Delli Compagni E; Di Domenico M; Di Pasquale A; Giammarioli M; Lorusso A; Mangone I; Marcacci M; Mencacci A; Puglia I; Rinaldi A; Savini G; Scialabba S |
| EPI_ISL_2845651 | Universitätsklinikum Köln; Institut für Virologie | Robert Koch Institute | |
| EPI_ISL_2975130 | Usansolo-Galdakao University Hospital | Cruces University Hospital | Ana Belén de la Hoz; Ana Gual-de-Torrella; Izaskun Alejo-Cancho; Mikel Gallego |
| EPI_ISL_2677769, EPI_ISL_2677789, EPI_ISL_2678990, EPI_ISL_2856059, EPI_ISL_2857237, EPI_ISL_2857239 | Utah Public Health Laboratory | Utah Public Health Laboratory | Erin L. Young; Kelly F. Oakeson; Olinto Linares-Perdomo; Tara Gallagher |
| EPI_ISL_2966637, EPI_ISL_2966642, EPI_ISL_2966643, EPI_ISL_2966648, EPI_ISL_2966650, EPI_ISL_2968543, EPI_ISL_2968689, EPI_ISL_3086917 | | | |
| see above | Viral Respiratory Lab, National Institute for Biomedical Research (INRB) | Pathogen Sequencing Lab, National Institute for Biomedical Research (INRB) | Allison Black; Amuri Aziza; Andrew Rambaut; Catherine Pratt; Eddy Kinganda-Lusamaki; Edith Nkwembe; Emmanuel Lokilo Lofiko; Francisca Muyembe Mawete; Gabriel Kabamba; Ian Goodfellow; James Hadfield; Jean Claude Makangara; Jean-Jacques Muyembe Tamfum; Josh Quick; Kristian Andersen; Matthias Pauthner; Michael Wiley; Nick Loman; Placide Mbala-Kingebeni; Raphaël Lumembe; Steve Ahuka-Mundeke; Trevor Bedford |
| EPI_ISL_2776049, EPI_ISL_3025856 | Virginia Division of Consolidated Laboratory Services | Virginia Division of Consolidated Laboratory Services | Virginia Division of Consolidated Laboratory Services |
| EPI_ISL_2884442 | Virology Laboratory, Scientific Department, Army Medical Center | Virology Laboratory, Scientific Department, Army Medical Center | Anella Monte; Anna Anselmo; Antonella Fortunato; Filippo Molinari; Florigio Lista; Francesco Giordani; Giancarlo Petralito; Giandomenico Cerreto; Giulia Campoli; Lucia Nicosia; Marzia Cavalli; Riccardo De Sanctis; Rossella Brandi; Silvia Fillo; Vanessa Vera Fain |
| EPI_ISL_3115349, EPI_ISL_3115354, EPI_ISL_3115360 | Virology laboratory, University of Maiduguri Teaching Hospital | Africa Centre for Excellence for Genomics of Infectious Diseases (ACEGID), Redeemer's University | A.T.; Abechi; Ajogbasile; Akano; C.A.; C.T.; Eromon; F.V.; Folarin, O.; Happi; I.B.; J.N.; J.U.; K.O.; Kayode; Nosamiefan, I.; Oguzie; Olawoye; Olumade; Oluniyi; P.E.; P.S.; T.J.; Ugwu; Uwanibe |
| EPI_ISL_2836853, EPI_ISL_2836862, EPI_ISL_2836864, EPI_ISL_2836868, EPI_ISL_2836882, EPI_ISL_2836886, EPI_ISL_2836890, EPI_ISL_2836892, EPI_ISL_2836893, EPI_ISL_2836895, EPI_ISL_2836900, EPI_ISL_2836909, EPI_ISL_2836914, EPI_ISL_2836961, EPI_ISL_2836964 | | | |
| see above | WACCBIP, University of Ghana, Accra, Ghana | WACCBIP, University of Ghana, Volta Road, Legon-Accra, Ghana | Collins M. Morang'a; Dominic S.Y. Amuzu; Evelyn B. Quansah; Evelyn Y. Bonney; Frederick Tei-Maya; Ivy A. Asante; Joe K. Mutungi; John K. Odoom; Joseph H.K. Bonney; Joyce M. Ngoi; Lawrence Ofori-Boadu; Lucas N. Amenga-Etego and Gordon A. Awandare; Mildred Adusei-Poku; Nicaise T. Ndam; Peter K Quashie; Philip M. Soglo; Samirah Saiid; Vincent Appiah; Violette V. M'cormack; William K. Ampofo; Yaw Bediako |
| EPI_ISL_2615962, EPI_ISL_2615966 | WV Labs | WVU and Marshall University Combined Genomics Core Facilities | James Denvir; Peter Perrotta; Peter Stoilov; Ryan Percifield; Wesley Kimble |
| EPI_ISL_2622322 | West of Scotland Specialist Virology Centre, NHSGGC / MRC-University of Glasgow Centre for Virus Research | COVID-19 Genomics UK (COG-UK) Consortium | Alasdair MacLean; Alice Broos; Ana da Silva Filipe; Antonia Ho; Daniel Mair; David L Robertson; Emma Thomson; Guy Mollett; Ioulia Tsatsani; James Shepherd; Jenna Nichols; Joseph Hughes; Kathy Li; Kathy Smollett; Kyriaki Nomikou; Lily Tong; Matthew Holden; Natasha Johnson; Rachel Blacow; Richard Orton; Rory Gunson; Sarah McDonald; Sharif Shaaban; Sreenu Vattipally; Stephen Carmichael |
| EPI_ISL_2598669 | Yale Clinical Virology Lab | Grubaugh Lab - Yale School of Public Health | Anderson Brito; Anne Wyllie; Chaney Kalinich; Chantal Vogels; Isabel Ott; Joseph Fauver; Mallery Breban; Mary Petrone; Nathan Grubaugh; Tara Alpert |
| EPI_ISL_2727349 | ZARV/NHLS, Department Medical Virology, University of Pretoria | KRISP, KZn Research Innovation and Sequencing Platform | Adriano Mendes; Amy Strydom; Emmanuel SJ; Giandhari J; Lessells R; Micheala Davids; Naidoo Y; Pillay S; Ramphal U; Sim Mayaphi and Marietjie Venter; Tegally H; Wilkinson E; de Oliveira T |
| EPI_ISL_2561989, EPI_ISL_2613699, EPI_ISL_2613713, EPI_ISL_2622043 | cerballiance-IDF | Cerba lab | Aude Lessenne; Bénédicte Roquebert; Emmanuel Lecorche; Kader Merah; Laura Verdurme; Patrice Herisson; Sabine Trombert-Paolantoni; Stéphanie Haïm-Boukobza; Thierry Collin |
| EPI_ISL_2562007 | cerballiance-cote d'azur | Cerba lab | Bénédicte Roquebert; Emmanuel Lecorche; Laura Verdurme; Sabine Trombert-Paolantoni; Stéphanie Haïm-Boukobza |
| EPI_ISL_2637400, EPI_ISL_2637499, EPI_ISL_2637517, EPI_ISL_2637521 | laboratoire Belle Epine | Department of Virology, Henri Mondor University Hospital, Assistance Publique Hôpitaux de Paris, Université Paris-Est Créteil, INSERM U955 | Alexandre Soulier; Christophe Rodriguez; Elisabeth Trawinski; Guillaume Gricourt; Jean-Michel Pawlotsky; Melissa N'Debi; Slim Fourati; Vanessa Demontant |

We gratefully acknowledge the following Authors from the Originating laboratories responsible for obtaining the specimens, as well as the Submitting laboratories where the genome data were generated and shared via GISAID, on which this research is based.

All Submitters of data may be contacted directly via www.gisaid.org

Authors are sorted alphabetically.

| Accession ID | Originating Laboratory | Submitting Laboratory | Authors |
|---|---|---|---|
| EPI_ISL_2422313, EPI_ISL_2627000, EPI_ISL_2627158, EPI_ISL_2627159, EPI_ISL_2727104, EPI_ISL_2727113 | ALBANY MEDICAL CENTER | Wadsworth Center, New York State Department of Health | Alexis Russell; Catharine Prussing; Daryl M. Lamson; Erasmus Schneider; Erica Lasek-Nesselquist; John Kelly; Jonathan Plitnick; Kirsten St. George; Matthew Shudt; Melissa A Leisner; Navjot Singh |
| EPI_ISL_2421084, EPI_ISL_2421192, EPI_ISL_2421194, EPI_ISL_2421195, EPI_ISL_2421232, EPI_ISL_2421234, EPI_ISL_2421235, EPI_ISL_2421236, EPI_ISL_2421237, EPI_ISL_2421245, EPI_ISL_2421248, EPI_ISL_2448309, EPI_ISL_2868475, EPI_ISL_2868543, EPI_ISL_2868544 | | | |
| see above | BOSTON HEART DIAGNOSTICS CORP | Wadsworth Center, New York State Department of Health | Alexis Russell; Catharine Prussing; Daryl M. Lamson; Erasmus Schneider; Erica Lasek-Nesselquist; John Kelly; Jonathan Plitnick; Kirsten St. George; Matthew Shudt; Melissa A Leisner; Navjot Singh |
| EPI_ISL_2500657, EPI_ISL_2500662, EPI_ISL_2500674, EPI_ISL_2500683, EPI_ISL_2500684, EPI_ISL_2500697, EPI_ISL_2500714, EPI_ISL_2500715, EPI_ISL_2500717, EPI_ISL_2500721, EPI_ISL_2500726, EPI_ISL_2500730, EPI_ISL_2500732, EPI_ISL_2500735, EPI_ISL_2500740, EPI_ISL_2500743, EPI_ISL_2500746, EPI_ISL_2500755, EPI_ISL_2500761, EPI_ISL_2500766, EPI_ISL_2500767, EPI_ISL_2500771, EPI_ISL_2500780, EPI_ISL_2500782, EPI_ISL_2500787, EPI_ISL_2500788, EPI_ISL_2500789, EPI_ISL_2500790, EPI_ISL_2500793, EPI_ISL_2500794, EPI_ISL_2500795, EPI_ISL_2500801, EPI_ISL_2500807, EPI_ISL_2500808, EPI_ISL_2500811, EPI_ISL_2500816, EPI_ISL_2500817, EPI_ISL_2500822, EPI_ISL_2500825, EPI_ISL_2500830, EPI_ISL_2500843, EPI_ISL_2500844 | | | |
| see above | Clinical Microbiology Laboratory, NewYork Presbyterian Hospital/Columbia University Irving Medical Center | Uhlemann Laboratory, Columbia University Irving Medical Center | Anne Kelley; Anne-Catrin Uhlemann; Maya Tagliavia; Medini K. Annavajhala |
| EPI_ISL_2868665 | Columbia University Irving Medical Center | Wadsworth Center, New York State Department of Health | Alexis Russell; Catharine Prussing; Daryl M. Lamson; Erasmus Schneider; Erica Lasek-Nesselquist; John Kelly; Jonathan Plitnick; Kirsten St. George; Matthew Shudt; Melissa A Leisner; Navjot Singh |
| EPI_ISL_2400431 | DOHMH PHL | New York City Public Health Laboratory | Jade Wang; et al. |
| EPI_ISL_2466697 | Department of Homeless Services | New York City Public Health Laboratory | Jade Wang; et al. |
| EPI_ISL_2662902 | Erie County Public Health (ECPHL) | University at Buffalo Genomics and Bioinformatics Core | Alyssa Pohlman; Amanda Boccolucci; Brandon Marzullo; Donald Yergeau; Jennifer Surtees; Jonathan Bard; Natalie Lamb; Norma Nowak |
| EPI_ISL_3372616 | Genesis Diagnostics | New York City Public Health Laboratory | Jade Wang; et al. |
| EPI_ISL_2662931 | Kaleida Health Laboratories | University at Buffalo Genomics and Bioinformatics Core | Alyssa Pohlman; Amanda Boccolucci; Brandon Marzullo; Donald Yergeau; Jennifer Surtees; Jonathan Bard; Natalie Lamb; Norma Nowak |
| EPI_ISL_2627144, EPI_ISL_2868661, EPI_ISL_2868663, EPI_ISL_2868664, EPI_ISL_2868666, EPI_ISL_2868667, EPI_ISL_2868671, EPI_ISL_2868672, EPI_ISL_2868673, EPI_ISL_2868674 | | | |
| see above | MONTEFIORE MEDICAL CENTER LABORATORIES | Wadsworth Center, New York State Department of Health | Alexis Russell; Catharine Prussing; Daryl M. Lamson; Erasmus Schneider; Erica Lasek-Nesselquist; John Kelly; Jonathan Plitnick; Kirsten St. George; Matthew Shudt; Melissa A Leisner; Navjot Singh |
| EPI_ISL_2400448, EPI_ISL_2400450, EPI_ISL_2400453, EPI_ISL_2400458, EPI_ISL_2400459, EPI_ISL_3046588, EPI_ISL_3046593, EPI_ISL_3046594, EPI_ISL_3046596, EPI_ISL_3046597, EPI_ISL_3046598, EPI_ISL_3046599, EPI_ISL_3046602, EPI_ISL_3046604, EPI_ISL_3046607, EPI_ISL_3060281, EPI_ISL_3060282, EPI_ISL_3060286, EPI_ISL_3060290, EPI_ISL_3060294, EPI_ISL_3060295, EPI_ISL_3060302 | | | |
| see above | Molecular Biology Laboratory, Westchester Medical Center | Genomics Core Laboratory, New York Medical College | Changhong Yin; Jian Zhuge; Utsav Pandey; Weihua Huang |
| EPI_ISL_2421044, EPI_ISL_2421221, EPI_ISL_2627166, EPI_ISL_2627168, EPI_ISL_2627172, EPI_ISL_2627173 | NORTHWELL HEALTH LABORATORIES | Wadsworth Center, New York State Department of Health | Alexis Russell; Catharine Prussing; Daryl M. Lamson; Erasmus Schneider; Erica Lasek-Nesselquist; John Kelly; Jonathan Plitnick; Kirsten St. George; Matthew Shudt; Melissa A Leisner; Navjot Singh |
| EPI_ISL_2452050, EPI_ISL_2452054, EPI_ISL_2452057, EPI_ISL_2452063, EPI_ISL_2452064, EPI_ISL_2452067, EPI_ISL_2452069, EPI_ISL_2452071 | | | |
| see above | NYC Department of Health and Mental Hygiene | Centers for Disease Control and Prevention Division of Viral Diseases, Pathogen Discovery | Alison Laufer Halpin; Ben L. Rambo-Martin; Clinton R. Paden; Dakota Howard; Darlene Wagner; Dave Wentworth; Dhwani Batra; Jasmine Padilla; Justin Lee; Katie Dillon; Krista Queen; Kristen Knipe; Kristine Lacek; Mark Burroughs; Matthew Schmerer; Mili Sheth; Peter Cook; Sam Shepard; Sarah Nobles; Shoshona Le; Suxiang Tong; Vivien Dugan; Yvette Unoarumhi |
| EPI_ISL_2400382, EPI_ISL_2427405, EPI_ISL_2427418, EPI_ISL_2427419, EPI_ISL_2427421, EPI_ISL_2427422, EPI_ISL_2427434, EPI_ISL_2427447, EPI_ISL_2427448, EPI_ISL_2427451, EPI_ISL_2427457, EPI_ISL_2427473, EPI_ISL_2427478, EPI_ISL_2427485, EPI_ISL_2427488, EPI_ISL_2427501, EPI_ISL_2443070, EPI_ISL_2458171, EPI_ISL_2458188, EPI_ISL_2458190, EPI_ISL_2458191, EPI_ISL_2458194, EPI_ISL_2458203, EPI_ISL_2458205, EPI_ISL_2458207, EPI_ISL_2458208, EPI_ISL_2663516, EPI_ISL_2663517, EPI_ISL_2663534, EPI_ISL_2663537, EPI_ISL_2663548, EPI_ISL_2663550, EPI_ISL_2753945 | | | |
| see above | NYU Langone Health | Departments of Pathology and Medicine, New York University School of Medicine | Adriana Heguy; Christian Marier; Dacia Dimartino; Emily Guzman; Gael Westby; Guiqing Wang; Paolo Cotzia; Paul Zappile; Peter Meyn; Sitharam Ramaswami; Yutong Zhang |
| EPI_ISL_2400428, EPI_ISL_2466700 | OCME Office Of Chief Medical Examiner | New York City Public Health Laboratory | Jade Wang; et al. |
| EPI_ISL_2446054, EPI_ISL_2446056, EPI_ISL_2446067, EPI_ISL_2446073, EPI_ISL_2446079, EPI_ISL_2446081, EPI_ISL_2446096, EPI_ISL_2631368, EPI_ISL_2631373, EPI_ISL_2631377, EPI_ISL_2631394, EPI_ISL_2633530, EPI_ISL_2633663, EPI_ISL_2633874, EPI_ISL_2633996, EPI_ISL_2634024, EPI_ISL_2634123, EPI_ISL_2634154, EPI_ISL_2634183, EPI_ISL_2634214, EPI_ISL_2634229, EPI_ISL_2634252, EPI_ISL_2634279, EPI_ISL_2634280, EPI_ISL_2634282, EPI_ISL_2634313, EPI_ISL_2634346, EPI_ISL_2634377, EPI_ISL_2634388, EPI_ISL_2634393, EPI_ISL_2634402, EPI_ISL_2634523, EPI_ISL_2634527, EPI_ISL_2634544, EPI_ISL_2634598, EPI_ISL_2634633, EPI_ISL_2634696, EPI_ISL_2634708, EPI_ISL_2634739, EPI_ISL_2634843, EPI_ISL_2634870, EPI_ISL_2634894, EPI_ISL_2634907, EPI_ISL_2634920, EPI_ISL_2715384, EPI_ISL_2715397, EPI_ISL_2715400, EPI_ISL_2715408, EPI_ISL_2715420, EPI_ISL_2715423, EPI_ISL_2715428, EPI_ISL_2715432, EPI_ISL_2715445, EPI_ISL_2715447, EPI_ISL_2715448, EPI_ISL_2715450, EPI_ISL_2715452, EPI_ISL_2715463, EPI_ISL_2715466, EPI_ISL_2715470, EPI_ISL_2715475, EPI_ISL_2715479, EPI_ISL_2715490, EPI_ISL_2715493, EPI_ISL_2715494, EPI_ISL_2715506, EPI_ISL_2715529, EPI_ISL_2715538, EPI_ISL_2715539, EPI_ISL_2715544, EPI_ISL_2715556, EPI_ISL_2715566, EPI_ISL_2715571, EPI_ISL_2715580, EPI_ISL_2715584, EPI_ISL_2715599, EPI_ISL_2715600, EPI_ISL_2715624, EPI_ISL_2715626, EPI_ISL_2715628, EPI_ISL_2715637, EPI_ISL_2715643, EPI_ISL_2715650, EPI_ISL_2715680, EPI_ISL_2715693, EPI_ISL_2715697, EPI_ISL_2715723, EPI_ISL_2715734, EPI_ISL_2715737, EPI_ISL_2715743, EPI_ISL_2715748, EPI_ISL_2715756, EPI_ISL_2715769, EPI_ISL_2715770, EPI_ISL_2715775, EPI_ISL_2715780, EPI_ISL_2715786, EPI_ISL_2715800, EPI_ISL_2715803, EPI_ISL_2715813, EPI_ISL_2715822, EPI_ISL_2715839, EPI_ISL_2715842, EPI_ISL_2715843, EPI_ISL_2715846, EPI_ISL_2715862, EPI_ISL_2715863, EPI_ISL_2715867, EPI_ISL_2715872, EPI_ISL_2715880, EPI_ISL_2715889, EPI_ISL_2715895, EPI_ISL_2715896, EPI_ISL_2715903, EPI_ISL_2715916, EPI_ISL_2715921, EPI_ISL_2715922, EPI_ISL_2715931, EPI_ISL_2715944, EPI_ISL_2715951, EPI_ISL_2715961, EPI_ISL_2715969, EPI_ISL_2715972, EPI_ISL_2715973, EPI_ISL_2715975, EPI_ISL_2715989, EPI_ISL_2716011, EPI_ISL_2716013, EPI_ISL_2716018, EPI_ISL_2716025, EPI_ISL_2716026, EPI_ISL_2716032, EPI_ISL_2716039, EPI_ISL_2716040, EPI_ISL_2716041, EPI_ISL_2716045, EPI_ISL_2716046, EPI_ISL_2716051, EPI_ISL_2716065, EPI_ISL_2716068, EPI_ISL_2716071, EPI_ISL_2716077, EPI_ISL_2716080, EPI_ISL_2716082, EPI_ISL_2716084, EPI_ISL_2716092, EPI_ISL_2716111, EPI_ISL_2716124, EPI_ISL_2716146, EPI_ISL_2716148, EPI_ISL_2716153, EPI_ISL_2716159, EPI_ISL_2716164, EPI_ISL_2716165, EPI_ISL_2716169, EPI_ISL_2716178, EPI_ISL_2716183, EPI_ISL_2716185, EPI_ISL_2716187, EPI_ISL_2716202, EPI_ISL_2716205, EPI_ISL_2716206, EPI_ISL_2827235, EPI_ISL_2827517, EPI_ISL_2964958, EPI_ISL_2964960, EPI_ISL_2964961, EPI_ISL_2964965, EPI_ISL_2964966, EPI_ISL_2964967, EPI_ISL_2964968, EPI_ISL_2964969, EPI_ISL_2964971, EPI_ISL_2964974, EPI_ISL_2964976, EPI_ISL_2964979, EPI_ISL_2964983, EPI_ISL_2964984, EPI_ISL_2964985, EPI_ISL_2964986, EPI_ISL_2964994, EPI_ISL_2964995, EPI_ISL_2964996, EPI_ISL_2964997, EPI_ISL_2964998, EPI_ISL_2965000, EPI_ISL_2965004, EPI_ISL_2965005, EPI_ISL_2965010, EPI_ISL_2965012, EPI_ISL_2965015, EPI_ISL_2965017, EPI_ISL_2965028, EPI_ISL_2965029, EPI_ISL_2965031, EPI_ISL_2965032, EPI_ISL_2965033, EPI_ISL_2965034, EPI_ISL_2965039, EPI_ISL_2965041, EPI_ISL_2965043, EPI_ISL_2965046, EPI_ISL_2965048, EPI_ISL_2965050, EPI_ISL_2965051, EPI_ISL_2965052, EPI_ISL_2965053, EPI_ISL_2965054, EPI_ISL_2965057, EPI_ISL_2965058, EPI_ISL_2965059, EPI_ISL_2965063, EPI_ISL_2965064, EPI_ISL_2965065, EPI_ISL_2965066, EPI_ISL_2965069, EPI_ISL_2965070, EPI_ISL_2965071, EPI_ISL_2965072, EPI_ISL_2965074, EPI_ISL_2965075, EPI_ISL_2965078, EPI_ISL_2965082, EPI_ISL_2965083, EPI_ISL_2965084, EPI_ISL_2965085, EPI_ISL_2965086, EPI_ISL_2965087, EPI_ISL_2965088, EPI_ISL_2965089, EPI_ISL_2965090, EPI_ISL_2965092, EPI_ISL_2965093, EPI_ISL_2965094, EPI_ISL_2965095, EPI_ISL_2965096, EPI_ISL_2965097, EPI_ISL_2965100, EPI_ISL_2965103, EPI_ISL_2965104, EPI_ISL_2965105, EPI_ISL_2965127, EPI_ISL_2965128, EPI_ISL_2965129, EPI_ISL_2965130, EPI_ISL_2965131, EPI_ISL_2965132, EPI_ISL_2965133, EPI_ISL_2965134, EPI_ISL_2965135, EPI_ISL_2965136, EPI_ISL_2965145, EPI_ISL_2965146, EPI_ISL_2965147, EPI_ISL_2965148, EPI_ISL_2965149, EPI_ISL_2965156, EPI_ISL_2965157, EPI_ISL_2965158, EPI_ISL_2965159, EPI_ISL_2965160, EPI_ISL_2965161, EPI_ISL_2965163, EPI_ISL_2965168, EPI_ISL_2965170, EPI_ISL_2965171, EPI_ISL_2965172, EPI_ISL_2965180, EPI_ISL_2965181, EPI_ISL_2965183, EPI_ISL_2965184, EPI_ISL_2965187, EPI_ISL_2965188, EPI_ISL_2965191, EPI_ISL_2965192, EPI_ISL_2965193, EPI_ISL_2965194, EPI_ISL_2965195, EPI_ISL_2965196, EPI_ISL_2965198, EPI_ISL_2965204, EPI_ISL_2965205, EPI_ISL_2965206, EPI_ISL_2965207, EPI_ISL_2965208, EPI_ISL_2965209, EPI_ISL_2965210, EPI_ISL_2965211, EPI_ISL_2965212, EPI_ISL_2965213, EPI_ISL_2965214, EPI_ISL_2965215, EPI_ISL_2965216, EPI_ISL_2965217, EPI_ISL_2965230, EPI_ISL_2965231, EPI_ISL_2965233, EPI_ISL_2965234, EPI_ISL_2965235, EPI_ISL_2965236, EPI_ISL_2965237, EPI_ISL_2965238, EPI_ISL_2965239, EPI_ISL_2965242, EPI_ISL_2965243, EPI_ISL_2965244, EPI_ISL_2965245, EPI_ISL_2965246, EPI_ISL_2965248, EPI_ISL_2965251, EPI_ISL_2965252, EPI_ISL_2965253, EPI_ISL_2965254, EPI_ISL_2965256, EPI_ISL_2965258, EPI_ISL_2965274, EPI_ISL_2965275, EPI_ISL_2965279, EPI_ISL_2965296, EPI_ISL_2965297, EPI_ISL_2965301, EPI_ISL_2965306, EPI_ISL_2965309, EPI_ISL_2965312, EPI_ISL_2965315, EPI_ISL_2965324, EPI_ISL_2965329, EPI_ISL_2965331, EPI_ISL_2965334, EPI_ISL_2965335, EPI_ISL_2965347, EPI_ISL_2965359, EPI_ISL_2965371, EPI_ISL_2965372, EPI_ISL_2965378, EPI_ISL_2965379, EPI_ISL_2965381, EPI_ISL_2965385, EPI_ISL_2965386, EPI_ISL_2965389, EPI_ISL_2965391, EPI_ISL_2965398, EPI_ISL_2965400, EPI_ISL_2965401, EPI_ISL_2965410, EPI_ISL_2965415, EPI_ISL_2965418, EPI_ISL_2965422, EPI_ISL_2965424, EPI_ISL_2965427, EPI_ISL_2965434, EPI_ISL_2965442, EPI_ISL_2965448, EPI_ISL_2965468, EPI_ISL_2965472, EPI_ISL_2965476, EPI_ISL_2965479, EPI_ISL_2965482, EPI_ISL_2965483, EPI_ISL_2965484, EPI_ISL_2965496, EPI_ISL_2965500, EPI_ISL_2965516, EPI_ISL_2965526, EPI_ISL_2965527, EPI_ISL_2965531, EPI_ISL_2965532, EPI_ISL_2965533, EPI_ISL_3010727, EPI_ISL_3010748 | | | |
| see above | Pandemic Response Lab - NYC | Pandemic Response Lab, R&D | Cybill del Castillo; Dylan Law; Haiping Hao; Henry Lee; Isabel Fernandez Escapa; Jon Laurent; Katharine Nelson; Melissa Hopkins; Michael Hammerling; Pradeep Bugga; Shinyoung Clair Kang; Sol Rey; William Ward |
| EPI_ISL_2421046, EPI_ISL_2422315 | STONY BROOK UNIVERSITY HOSPITAL | Wadsworth Center, New York State Department of Health | Alexis Russell; Catharine Prussing; Daryl M. Lamson; Erasmus Schneider; Erica Lasek-Nesselquist; John Kelly; Jonathan Plitnick; Kirsten St. George; Matthew Shudt; Melissa A Leisner; Navjot Singh |
| EPI_ISL_2421063, EPI_ISL_2422289, EPI_ISL_2422307, EPI_ISL_2422312, EPI_ISL_2868262, EPI_ISL_2868542, EPI_ISL_3030935, EPI_ISL_3030939, EPI_ISL_3030940, EPI_ISL_3030942, EPI_ISL_3030944, EPI_ISL_3031080 | | | |
| see above | SUNY UPSTATE MEDICAL UNIVERSITY | Wadsworth Center, New York State Department of Health | Alexis Russell; Catharine Prussing; Daryl M. Lamson; Erasmus Schneider; Erica Lasek-Nesselquist; John Kelly; Jonathan Plitnick; Kirsten St. George; Matthew Shudt; Melissa A Leisner; Navjot Singh |
| EPI_ISL_2421249, EPI_ISL_2421252 | Saratoga Hospital Laboratory | Wadsworth Center, New York State Department of Health | Alexis Russell; Catharine Prussing; Daryl M. Lamson; Erasmus Schneider; Erica Lasek-Nesselquist; John Kelly; Jonathan Plitnick; Kirsten St. George; Matthew Shudt; Melissa A Leisner; Navjot Singh |
| EPI_ISL_2466691, EPI_ISL_2466692, EPI_ISL_2466693, EPI_ISL_2466694, EPI_ISL_2466695, EPI_ISL_2466696 | Sherman Abrams Laboratory | New York City Public Health Laboratory | Jade Wang; et al. |
| EPI_ISL_2576919, EPI_ISL_2576924 | Sonic Reference Laboratory | Sonic Reference Laboratory | "Robert Moller; Bijaya Dhakal; Cora Lahey; Ellen Pizarek"; John Woody; Krishna Maddula |
| EPI_ISL_2421164, EPI_ISL_2421170, EPI_ISL_2421176, EPI_ISL_2421181, EPI_ISL_2421187, EPI_ISL_2421188, EPI_ISL_2626931, EPI_ISL_2626932, EPI_ISL_2626937, EPI_ISL_2626939, EPI_ISL_2626940, EPI_ISL_2626953, EPI_ISL_2626958, EPI_ISL_2627117, EPI_ISL_2627121, EPI_ISL_2627131, EPI_ISL_2868577, EPI_ISL_2868579, EPI_ISL_2868620, EPI_ISL_2868623, EPI_ISL_3030946 | | | |
| see above | TEMPUS LABS INC | Wadsworth Center, New York State Department of Health | Alexis Russell; Catharine Prussing; Daryl M. Lamson; Erasmus Schneider; Erica Lasek-Nesselquist; John Kelly; Jonathan Plitnick; Kirsten St. George; Matthew Shudt; Melissa A Leisner; Navjot Singh |
| EPI_ISL_2885704, EPI_ISL_2885710, EPI_ISL_2885728, EPI_ISL_2920232, EPI_ISL_2987193, EPI_ISL_3087184 | URMC LABS | URMC LABS | Andrew Cameron; Dwight Hardy; Ghinwa Dumyati; Kelly DeLary; Stepan Voljehn |
| EPI_ISL_2421150, EPI_ISL_2421152, EPI_ISL_2422333, EPI_ISL_2422343, EPI_ISL_2422344, EPI_ISL_2626979, EPI_ISL_2626987, EPI_ISL_2627031, EPI_ISL_2627064, EPI_ISL_2627071, EPI_ISL_2627073, EPI_ISL_2627074, EPI_ISL_2627075, EPI_ISL_2868238, EPI_ISL_2868265, EPI_ISL_2868275, EPI_ISL_2868279, EPI_ISL_2868281, EPI_ISL_2868282, EPI_ISL_2868288, EPI_ISL_2868293, EPI_ISL_2868304, EPI_ISL_2868307, EPI_ISL_2868309, EPI_ISL_2868466, EPI_ISL_2868469, EPI_ISL_2868472, EPI_ISL_2868477, EPI_ISL_2868522, EPI_ISL_2868531, EPI_ISL_2868532, EPI_ISL_2868533, EPI_ISL_2868534, EPI_ISL_2868535, EPI_ISL_2868538, EPI_ISL_2868539, EPI_ISL_2868540, EPI_ISL_2868541, EPI_ISL_2868556, EPI_ISL_2868559, EPI_ISL_2919190, EPI_ISL_2919195, EPI_ISL_3030965, EPI_ISL_3030978, EPI_ISL_3030981, EPI_ISL_3030987, EPI_ISL_3030991, EPI_ISL_3031004, EPI_ISL_3031005, EPI_ISL_3031232, EPI_ISL_3031244, EPI_ISL_3031247, EPI_ISL_3031258, EPI_ISL_3148991 | | | |
| see above | URMC LABS | Wadsworth Center, New York State Department of Health | Alexis Russell; Catharine Prussing; Daryl M. Lamson; Erasmus Schneider; Erica Lasek-Nesselquist; John Kelly; Jonathan Plitnick; Kirsten St. George; Matthew Shudt; Melissa A Leisner; Navjot Singh |
| EPI_ISL_2421107, EPI_ISL_2627138, EPI_ISL_3148943, EPI_ISL_3148946 | WESTCHESTER MEDICAL CENTER | Wadsworth Center, New York State Department of Health | Alexis Russell; Catharine Prussing; Daryl M. Lamson; Erasmus Schneider; Erica Lasek-Nesselquist; John Kelly; Jonathan Plitnick; Kirsten St. George; Matthew Shudt; Melissa A Leisner; Navjot Singh |

We gratefully acknowledge the following Authors from the Originating laboratories responsible for obtaining the specimens, as well as the Submitting laboratories where the genome data were generated and shared via GISAID, on which this research is based.

All Submitters of data may be contacted directly via www.gisaid.org

Authors are sorted alphabetically.

| Accession ID | Originating Laboratory | Submitting Laboratory | Authors |
|---|---|---|---|
| EPI_ISL_2775309 | AP SSO | CSIR-Centre for Cellular and Molecular Biology-INSACOG | Amareshwar Vodapalli; Ara Sreenivas; Archana Bharadwaj Siva; B Himasri; Divya Tej Sowpati; Jandhyala Sai Krishna; Karthik Bharadwaj Tallapaka; Lamuk Zaveri; Onkar Kulkarni; Payel Mukherjee; Priya Nurkurthy; Rakesh K Mishra; Shreekant Verma; Sofia Banu; Sumedha Avadhanula; Tulasi Nagabandi; Valli Nagalakshmi Undamatla; Vidhyadhari Methuku |
| EPI_ISL_2728298, EPI_ISL_2728299, EPI_ISL_2728301, EPI_ISL_2728448, EPI_ISL_2728449, EPI_ISL_2728450, EPI_ISL_2728451, EPI_ISL_2728461, EPI_ISL_2728462, EPI_ISL_2728464, EPI_ISL_2728467, EPI_ISL_2728468, EPI_ISL_2728470, EPI_ISL_2728474, EPI_ISL_2728523, EPI_ISL_2842195, EPI_ISL_2842196, EPI_ISL_2842200, EPI_ISL_2842223, EPI_ISL_2842226, EPI_ISL_2842229, EPI_ISL_2842233, EPI_ISL_2842234, EPI_ISL_2842240, EPI_ISL_2842241, EPI_ISL_2842270, EPI_ISL_2842274, EPI_ISL_2842277, EPI_ISL_2842278 | | | |
| see above | BBMP Urban | INSACOG-KA, NIMHANS | Ananthapadmanabha Kotambail; Anita S Desai; Anson Kunjumon George; Chetan G K; Chitra Pattabiraman; Darshan Sreenivas; Ellango Ramasamy; Gautham Arunachal Udupi; Mahesh Kumar.C.S; Sony Sharma; V Ravi |
| EPI_ISL_2545214, EPI_ISL_2545220 | BBMP Urban PHC | INSACOG-KA, NIMHANS | Ananthapadmanabha Kotambail; Anita S Desai; Anson Kunjumon George; Chetan G K; Chitra Pattabiraman; Darshan Sreenivas; Gautham Arunachal Udupi; Pramada Prasad; V Ravi |
| EPI_ISL_2504583 | Biotechnology Division, NCDC Delhi | NCDC Delhi, Biotechnology Division | Hema Gogia; Hemlata Lall; Kalaiarasan Ponnusamy; Mahesh S Dhar; Manoj K Singh; Meena Datta; Partha Rakshit; Preeti Madan; Priyanka Singh; Radhakrishnan V. S; Robin Marwal; Sandhya Kabra; Sujeet K Singh; Uma Sharma |
| EPI_ISL_2661048, EPI_ISL_2661078, EPI_ISL_2661202, EPI_ISL_2661230, EPI_ISL_2661252, EPI_ISL_2661266, EPI_ISL_2661277, EPI_ISL_2661278, EPI_ISL_2774842, EPI_ISL_2774843, EPI_ISL_2774845, EPI_ISL_2774848, EPI_ISL_2774853, EPI_ISL_2774854, EPI_ISL_2774855, EPI_ISL_2774856, EPI_ISL_2774857, EPI_ISL_2774894, EPI_ISL_2774895, EPI_ISL_2774896, EPI_ISL_2774900, EPI_ISL_2774904, EPI_ISL_2774907, EPI_ISL_2775078, EPI_ISL_2775107, EPI_ISL_2775110, EPI_ISL_2775111, EPI_ISL_2775113, EPI_ISL_2775115, EPI_ISL_2775118, EPI_ISL_2775120, EPI_ISL_2775121 | | | |
| see above | CSIR-Centre for Cellular and Molecular Biology | CSIR-Centre for Cellular and Molecular Biology-INSACOG | Amareshwar Vodapalli; Ara Sreenivas; Archana Bharadwaj Siva; B Himasri; Divya Tej Sowpati; Jandhyala Sai Krishna; Karthik Bharadwaj Tallapaka; Lamuk Zaveri; Onkar Kulkarni; Payel Mukherjee; Priya Nurkurthy; Rakesh K Mishra; Shreekant Verma; Sofia Banu; Sumedha Avadhanula; Tulasi Nagabandi; Valli Nagalakshmi Undamatla; Vidhyadhari Methuku |
| EPI_ISL_2728350, EPI_ISL_2728358, EPI_ISL_2728382, EPI_ISL_2728387, EPI_ISL_2728407, EPI_ISL_2728411, EPI_ISL_2842432 | | | |
| see above | DSU VIJAYAPURA | INSACOG-KA, NIMHANS | Ananthapadmanabha Kotambail; Anita S Desai; Anson Kunjumon George; Chetan G K; Chitra Pattabiraman; Darshan Sreenivas; Ellango Ramasamy; Gautham Arunachal Udupi; Mahesh Kumar.C.S; Sony Sharma; V Ravi |
| EPI_ISL_2521785, EPI_ISL_2521786, EPI_ISL_2521787, EPI_ISL_2521788, EPI_ISL_2521801, EPI_ISL_2521822, EPI_ISL_2521824, EPI_ISL_2521825, EPI_ISL_2521835, EPI_ISL_2521867, EPI_ISL_2521879, EPI_ISL_2521897, EPI_ISL_2521916, EPI_ISL_2521952, EPI_ISL_2521953, EPI_ISL_2521954 | | | |
| see above | Dr S raju, Director of public heath and preventive medicine | inStem NCBS - INSACOG | Uma Ramakrishnan Dasaradhi Palakodeti Aswin SaiNarain |
| EPI_ISL_2723774, EPI_ISL_2723789, EPI_ISL_2723793, EPI_ISL_2723846, EPI_ISL_2723899, EPI_ISL_2724022, EPI_ISL_2724053, EPI_ISL_2724241, EPI_ISL_2724242, EPI_ISL_2724246, EPI_ISL_2724274, EPI_ISL_2724282 | | | |
| see above | Dr S raju, Director of public heath and preventive medicine | inStem NCBS – INSACOG | Uma Ramakrishnan Dasaradhi Palakodeti Aswin SaiNarain |
| EPI_ISL_2723816 | Dr. Madhusudhan, MBBD, MD, ICMR VRDL | inStem NCBS – INSACOG | Uma Ramakrishnan Dasaradhi Palakodeti Aswin SaiNarain |
| EPI_ISL_2530111, EPI_ISL_2530113, EPI_ISL_2530120, EPI_ISL_2530122, EPI_ISL_2530124, EPI_ISL_2530126, EPI_ISL_2530128, EPI_ISL_2530129, EPI_ISL_2530131, EPI_ISL_2530136, EPI_ISL_2530137, EPI_ISL_2530139, EPI_ISL_2530141, EPI_ISL_2530144, EPI_ISL_2530146, EPI_ISL_2530149, EPI_ISL_2530151, EPI_ISL_2530154, EPI_ISL_2530155, EPI_ISL_2530157, EPI_ISL_2530158, EPI_ISL_2530165, EPI_ISL_2530167, EPI_ISL_2530170, EPI_ISL_2530172, EPI_ISL_2530176, EPI_ISL_2530177, EPI_ISL_2530180, EPI_ISL_2530182, EPI_ISL_2530183, EPI_ISL_2530187, EPI_ISL_2530188, EPI_ISL_2530190, EPI_ISL_2530216, EPI_ISL_2530238, EPI_ISL_2530261, EPI_ISL_2530267, EPI_ISL_2530272, EPI_ISL_2530273, EPI_ISL_2530275, EPI_ISL_2530276, EPI_ISL_2530278, EPI_ISL_2530279, EPI_ISL_2530281, EPI_ISL_2530283, EPI_ISL_2530285, EPI_ISL_2530286, EPI_ISL_2530295, EPI_ISL_2530313, EPI_ISL_2530317, EPI_ISL_2530321, EPI_ISL_2530324, EPI_ISL_2530326, EPI_ISL_2530330, EPI_ISL_2530334, EPI_ISL_2530336, EPI_ISL_2530337, EPI_ISL_2530339, EPI_ISL_2530340, EPI_ISL_2530344, EPI_ISL_2530349, EPI_ISL_2530353, EPI_ISL_2530357, EPI_ISL_2530360, EPI_ISL_2530364, EPI_ISL_2530366, EPI_ISL_2530367, EPI_ISL_2530369, EPI_ISL_2530371, EPI_ISL_2530376, EPI_ISL_2600953, EPI_ISL_2600954, EPI_ISL_2600955, EPI_ISL_2600956, EPI_ISL_2600957, EPI_ISL_2600960, EPI_ISL_2600962 | | | |
| see above | Hematopathology Laboratory, ACTREC, TMC | Hematopathology Laboratory, ACTREC, TMC | ACTREC; Hematopathology Laboratory |
| EPI_ISL_2555685, EPI_ISL_2555686, EPI_ISL_2555688, EPI_ISL_2555691, EPI_ISL_2555694, EPI_ISL_2555698, EPI_ISL_2555700, EPI_ISL_2555703, EPI_ISL_2555704, EPI_ISL_2555707, EPI_ISL_2555721, EPI_ISL_2555726, EPI_ISL_2555731 | | | |
| see above | ICMR-National Institute of Virology | NCDC Delhi, Biotechnology Division | A Walimbe; A.Awhale; A.Titkare; B.Apoorva; G.Divekar; H.Kengale; Hema Gogia; Hemlata Lall; K P.Shinde; K.Iyengar; K.Korabu; Kalaiarasan Ponnusamy; M L Choudhary; M.Das; Mahesh S Dhar; Manoj K Singh; Meena Datta; Partha Rakshit; Preeti Madan; Priyanka Singh; R.Verma; Radhakrishnan V. S; Robin Marwal; S Jadhav; S.Bhorekar; S.Jadhav; S.Shelkande; Sandhya Kabra; Sujeet K Singh; T.Sanjeevi; U.Saha; Uma Sharma; V Malik; V.Autude; V.Vipat; Z. Sayyed |
| EPI_ISL_2545994, EPI_ISL_2545996, EPI_ISL_2545997, EPI_ISL_2545998, EPI_ISL_2546000, EPI_ISL_2546001, EPI_ISL_2546002, EPI_ISL_2546004, EPI_ISL_2546006, EPI_ISL_2546009, EPI_ISL_2546010, EPI_ISL_2546011, EPI_ISL_2546012, EPI_ISL_2546014, EPI_ISL_2546016, EPI_ISL_2546017, EPI_ISL_2546019, EPI_ISL_2546020, EPI_ISL_2546022, EPI_ISL_2546024, EPI_ISL_2546025, EPI_ISL_2546026, EPI_ISL_2546027, EPI_ISL_2546028, EPI_ISL_2546029, EPI_ISL_2546030, EPI_ISL_2546031, EPI_ISL_2546032, EPI_ISL_2546033, EPI_ISL_2546035, EPI_ISL_2546037, EPI_ISL_2546038, EPI_ISL_2546039, EPI_ISL_2546040, EPI_ISL_2546041, EPI_ISL_2546042, EPI_ISL_2546043, EPI_ISL_2546044, EPI_ISL_2546045, EPI_ISL_2546047, EPI_ISL_2546050, EPI_ISL_2546051, EPI_ISL_2546052, EPI_ISL_2546054, EPI_ISL_2546055, EPI_ISL_2546056, EPI_ISL_2546057, EPI_ISL_2546058, EPI_ISL_2546059, EPI_ISL_2546065, EPI_ISL_2546067, EPI_ISL_2546069, EPI_ISL_2546070, EPI_ISL_2546071, EPI_ISL_2546072, EPI_ISL_2546073, EPI_ISL_2546074, EPI_ISL_2546075, EPI_ISL_2546077, EPI_ISL_2546079, EPI_ISL_2546080, EPI_ISL_2546081, EPI_ISL_2546082, EPI_ISL_2546083, EPI_ISL_2546084, EPI_ISL_2546086, EPI_ISL_2546087, EPI_ISL_2546088, EPI_ISL_2546090, EPI_ISL_2546091, EPI_ISL_2546092, EPI_ISL_2546093, EPI_ISL_2546094, EPI_ISL_2546095, EPI_ISL_2546096, EPI_ISL_2546097, EPI_ISL_2546098, EPI_ISL_2546100, EPI_ISL_2546103, EPI_ISL_2546105, EPI_ISL_2546108, EPI_ISL_2546109, EPI_ISL_2546113, EPI_ISL_2546117, EPI_ISL_2546118, EPI_ISL_2546121, EPI_ISL_2546124, EPI_ISL_2546125, EPI_ISL_2546126, EPI_ISL_2546130, EPI_ISL_2546135, EPI_ISL_2546137, EPI_ISL_2546138, EPI_ISL_2546139, EPI_ISL_2546140, EPI_ISL_2546141, EPI_ISL_2546142, EPI_ISL_2546143, EPI_ISL_2546144, EPI_ISL_2546146, EPI_ISL_2546147, EPI_ISL_2546148, EPI_ISL_2546149, EPI_ISL_2546153, EPI_ISL_2546155, EPI_ISL_2546159, EPI_ISL_2546160, EPI_ISL_2546161, EPI_ISL_2546165, EPI_ISL_2546167, EPI_ISL_2546168, EPI_ISL_2546169, EPI_ISL_2546170, EPI_ISL_2546171, EPI_ISL_2546172, EPI_ISL_2546174, EPI_ISL_2546175, EPI_ISL_2546176, EPI_ISL_2546177, EPI_ISL_2546178, EPI_ISL_2546179, EPI_ISL_2546181, EPI_ISL_2546182, EPI_ISL_2546184, EPI_ISL_2546185, EPI_ISL_2546186, EPI_ISL_2546188, EPI_ISL_2546189, EPI_ISL_2546190, EPI_ISL_2546191, EPI_ISL_2546192, EPI_ISL_2546193, EPI_ISL_2546194, EPI_ISL_2546195, EPI_ISL_2546196, EPI_ISL_2546197, EPI_ISL_2546198, EPI_ISL_2546199, EPI_ISL_2546200, EPI_ISL_2546204, EPI_ISL_2546205, EPI_ISL_2546206, EPI_ISL_2546207, EPI_ISL_2546208, EPI_ISL_2546209, EPI_ISL_2546210, EPI_ISL_2546211, EPI_ISL_2546217, EPI_ISL_2546218, EPI_ISL_2546219, EPI_ISL_2546220, EPI_ISL_2546223, EPI_ISL_2546226, EPI_ISL_2546227, EPI_ISL_2546234, EPI_ISL_2546235, EPI_ISL_2546236, EPI_ISL_2546237, EPI_ISL_2546238, EPI_ISL_2546239, EPI_ISL_2546253, EPI_ISL_2546255, EPI_ISL_2546257, EPI_ISL_2546262, EPI_ISL_2546263, EPI_ISL_2546264, EPI_ISL_2546265, EPI_ISL_2546267, EPI_ISL_2546269, EPI_ISL_2546270, EPI_ISL_2546273, EPI_ISL_2546274, EPI_ISL_2546280, EPI_ISL_2546282, EPI_ISL_2546285, EPI_ISL_2546286, EPI_ISL_2546287, EPI_ISL_2546290, EPI_ISL_2546292, EPI_ISL_2546293, EPI_ISL_2546299, EPI_ISL_2546300, EPI_ISL_2546302, EPI_ISL_2546303, EPI_ISL_2546306, EPI_ISL_2546307, EPI_ISL_2546309, EPI_ISL_2546322, EPI_ISL_2546323, EPI_ISL_2546326, EPI_ISL_2546328, EPI_ISL_2546329, EPI_ISL_2546332, EPI_ISL_2546334, EPI_ISL_2546335, EPI_ISL_2546338, EPI_ISL_2546340, EPI_ISL_2546341, EPI_ISL_2546342, EPI_ISL_2546343, EPI_ISL_2546344, EPI_ISL_2546351, EPI_ISL_2546352, EPI_ISL_2546353, EPI_ISL_2546354, EPI_ISL_2546358, EPI_ISL_2546361, EPI_ISL_2546363, EPI_ISL_2546369, EPI_ISL_2546370, EPI_ISL_2546371, EPI_ISL_2546373, EPI_ISL_2546376, EPI_ISL_2546377, EPI_ISL_2546378, EPI_ISL_2546379, EPI_ISL_2546382, EPI_ISL_2546383, EPI_ISL_2546384, EPI_ISL_2546385, EPI_ISL_2546386, EPI_ISL_2546390, EPI_ISL_2546393, EPI_ISL_2546398, EPI_ISL_2546399, EPI_ISL_2546400, EPI_ISL_2546408, EPI_ISL_2546409, EPI_ISL_2546412, EPI_ISL_2546415, EPI_ISL_2546420, EPI_ISL_2546424, EPI_ISL_2546426, EPI_ISL_2546427, EPI_ISL_2546433, EPI_ISL_2546436, EPI_ISL_2546445, EPI_ISL_2546446, EPI_ISL_2546448, EPI_ISL_2546449, EPI_ISL_2546450, EPI_ISL_2546451, EPI_ISL_2546454, EPI_ISL_2546455, EPI_ISL_2546456, EPI_ISL_2546457, EPI_ISL_2546458, EPI_ISL_2546463, EPI_ISL_2546469, EPI_ISL_2546470, EPI_ISL_2546479, EPI_ISL_2546480, EPI_ISL_2546481, EPI_ISL_2546484, EPI_ISL_2546488, EPI_ISL_2546491, EPI_ISL_2546492, EPI_ISL_2546494, EPI_ISL_2546498, EPI_ISL_2546503, EPI_ISL_2546507, EPI_ISL_2546513, EPI_ISL_2546517, EPI_ISL_2546521, EPI_ISL_2546525, EPI_ISL_2546531, EPI_ISL_2546533, EPI_ISL_2546535, EPI_ISL_2546538, EPI_ISL_2546545, EPI_ISL_2546551, EPI_ISL_2546554, EPI_ISL_2546557, EPI_ISL_2546560, EPI_ISL_2546561, EPI_ISL_2546571, EPI_ISL_2546573, EPI_ISL_2546574, EPI_ISL_2546579, EPI_ISL_2546588, EPI_ISL_2546590, EPI_ISL_2546608, EPI_ISL_2546618, EPI_ISL_2546621, EPI_ISL_2546626, EPI_ISL_2546629, EPI_ISL_2546637, EPI_ISL_2546651, EPI_ISL_2546663, EPI_ISL_2546666, EPI_ISL_2546671, EPI_ISL_2546672, EPI_ISL_2546674, EPI_ISL_2546675, EPI_ISL_2546676, EPI_ISL_2546678, EPI_ISL_2546679, EPI_ISL_2546681, EPI_ISL_2546688, EPI_ISL_2546706, EPI_ISL_2546714, EPI_ISL_2546723, EPI_ISL_2546810, EPI_ISL_2546837, EPI_ISL_2546876, EPI_ISL_2547121 | | | |
| see above | ICMR-National Institute of Virology - INSACOG | NIV Influenza | Dr. Varsha Potdar and NIC Team |
| EPI_ISL_2503823, EPI_ISL_2503824, EPI_ISL_2503825, EPI_ISL_2503829, EPI_ISL_2503830, EPI_ISL_2503837 | INSACOG-ARUNACHAL PRADESH | National Institute of Biomedical Genomics – INSACOG | Arindam Maitra; Lobsang Jampa; Mika Umpo; Nidhan Kumar Biswas; Saumitra Das; Sreedhar Chinnaswamy; Yompe Kamki |
| EPI_ISL_2547430, EPI_ISL_2547431, EPI_ISL_2547432, EPI_ISL_2547433, EPI_ISL_2547434, EPI_ISL_2547435, EPI_ISL_2547437, EPI_ISL_2547438, EPI_ISL_2547442, EPI_ISL_2547443, EPI_ISL_2547449, EPI_ISL_2547452, EPI_ISL_2547458, EPI_ISL_2547462, EPI_ISL_2547463, EPI_ISL_2547472, EPI_ISL_2547473, EPI_ISL_2547481, EPI_ISL_2547484, EPI_ISL_2547508, EPI_ISL_2547510 | | | |
| see above | INSACOG-MEGHALAYA | National Institute of Biomedical Genomics – INSACOG | Arindam Maitra; E Marak; E Shadap; Nidhan Kumar Biswas; R. Lyngdoh; Saumitra Das; Sreedhar Chinnaswamy |
| EPI_ISL_2675354, EPI_ISL_2675376 | INSACOG-NAGALAND | National Institute of Biomedical Genomics – INSACOG | Arindam Maitra; Chenole Keppen; Nidhan Kumar Biswas; Saumitra Das; Sreedhar Chinnaswamy; Vinotsole Khamo; Wetetsho Kapfo |
| EPI_ISL_2503147, EPI_ISL_2503149, EPI_ISL_2503150, EPI_ISL_2503152, EPI_ISL_2503158, EPI_ISL_2503163, EPI_ISL_2503164, EPI_ISL_2503168, EPI_ISL_2503177, EPI_ISL_2503178, EPI_ISL_2503179, EPI_ISL_2503180, EPI_ISL_2503181, EPI_ISL_2503182, EPI_ISL_2503183, EPI_ISL_2503184, EPI_ISL_2503187, EPI_ISL_2503192, EPI_ISL_2503196, EPI_ISL_2503197, EPI_ISL_2503198, EPI_ISL_2503200, EPI_ISL_2503201, EPI_ISL_2503202, EPI_ISL_2503203, EPI_ISL_2503205, EPI_ISL_2503209, EPI_ISL_2503210, EPI_ISL_2503212, EPI_ISL_2503215, EPI_ISL_2503217, EPI_ISL_2503218, EPI_ISL_2503219, EPI_ISL_2503220, EPI_ISL_2503221, EPI_ISL_2503222, EPI_ISL_2503223, EPI_ISL_2503224, EPI_ISL_2503228, EPI_ISL_2503229, EPI_ISL_2503230, EPI_ISL_2503231, EPI_ISL_2503232, EPI_ISL_2503233, EPI_ISL_2503234, EPI_ISL_2503237, EPI_ISL_2503239, EPI_ISL_2503240, EPI_ISL_2503242, EPI_ISL_2503244, EPI_ISL_2503245, EPI_ISL_2503247, EPI_ISL_2503248, EPI_ISL_2503249, EPI_ISL_2503250, EPI_ISL_2503251, EPI_ISL_2503252, EPI_ISL_2503254, EPI_ISL_2503255, EPI_ISL_2503256, EPI_ISL_2503258, EPI_ISL_2503259, EPI_ISL_2503260, EPI_ISL_2503264, EPI_ISL_2503266, EPI_ISL_2503267, EPI_ISL_2503268, EPI_ISL_2503269, EPI_ISL_2503271, EPI_ISL_2503272, EPI_ISL_2503275, EPI_ISL_2503276, EPI_ISL_2503278, EPI_ISL_2503279, EPI_ISL_2503280, EPI_ISL_2503281, EPI_ISL_2503282, EPI_ISL_2503283, EPI_ISL_2503284, EPI_ISL_2503285, EPI_ISL_2503286, EPI_ISL_2503292, EPI_ISL_2503293, EPI_ISL_2503297, EPI_ISL_2503299, EPI_ISL_2503302, EPI_ISL_2503303, EPI_ISL_2503306, EPI_ISL_2503309, EPI_ISL_2503310, EPI_ISL_2503311, EPI_ISL_2503313, EPI_ISL_2503316, EPI_ISL_2503317, EPI_ISL_2503318, EPI_ISL_2503319, EPI_ISL_2503322, EPI_ISL_2503323, EPI_ISL_2503324, EPI_ISL_2503325, EPI_ISL_2503328, EPI_ISL_2503331, EPI_ISL_2503332, EPI_ISL_2503333, EPI_ISL_2503334, EPI_ISL_2503336, EPI_ISL_2503337, EPI_ISL_2503338, EPI_ISL_2503340, EPI_ISL_2503344, EPI_ISL_2503345, EPI_ISL_2503347, EPI_ISL_2503352, EPI_ISL_2503354, EPI_ISL_2503355, EPI_ISL_2503356, EPI_ISL_2503357, EPI_ISL_2503360, EPI_ISL_2503361, EPI_ISL_2503364, EPI_ISL_2503365, EPI_ISL_2503366, EPI_ISL_2503368, EPI_ISL_2503370, EPI_ISL_2503374, EPI_ISL_2503375, EPI_ISL_2503376, EPI_ISL_2503378, EPI_ISL_2503379, EPI_ISL_2503380, EPI_ISL_2503381, EPI_ISL_2503383, EPI_ISL_2503384, EPI_ISL_2503385, EPI_ISL_2503386, EPI_ISL_2503387, EPI_ISL_2503388, EPI_ISL_2503389, EPI_ISL_2503390, EPI_ISL_2503391, EPI_ISL_2503392, EPI_ISL_2503393, EPI_ISL_2503394, EPI_ISL_2503396, EPI_ISL_2503398, EPI_ISL_2503400, EPI_ISL_2503403, EPI_ISL_2503404, EPI_ISL_2503405, EPI_ISL_2503406, EPI_ISL_2503408, EPI_ISL_2503410, EPI_ISL_2503413, EPI_ISL_2503414, EPI_ISL_2503416, EPI_ISL_2503417, EPI_ISL_2503419, EPI_ISL_2503420, EPI_ISL_2503421, EPI_ISL_2503422, EPI_ISL_2503425, EPI_ISL_2503427, EPI_ISL_2503428, EPI_ISL_2503442, EPI_ISL_2503449, EPI_ISL_2503451, EPI_ISL_2503454, EPI_ISL_2503457, EPI_ISL_2503458, EPI_ISL_2503462, EPI_ISL_2503464, EPI_ISL_2503466, EPI_ISL_2503469, EPI_ISL_2503470, EPI_ISL_2503472, EPI_ISL_2503473, EPI_ISL_2503475, EPI_ISL_2503476, EPI_ISL_2503477, EPI_ISL_2503478, EPI_ISL_2503479, EPI_ISL_2503481, EPI_ISL_2503482, EPI_ISL_2503483, EPI_ISL_2503484, EPI_ISL_2503486, EPI_ISL_2503487, EPI_ISL_2503489, EPI_ISL_2503490, EPI_ISL_2503491, EPI_ISL_2503492, EPI_ISL_2503494, EPI_ISL_2503495, EPI_ISL_2503498, EPI_ISL_2503505, EPI_ISL_2503511, EPI_ISL_2503512, EPI_ISL_2503513, EPI_ISL_2503514, EPI_ISL_2503518, EPI_ISL_2503531, EPI_ISL_2503533, EPI_ISL_2503534, EPI_ISL_2503537, EPI_ISL_2503539, EPI_ISL_2503540, EPI_ISL_2503542, EPI_ISL_2503543, EPI_ISL_2503544, EPI_ISL_2503545, EPI_ISL_2503546, EPI_ISL_2503560, EPI_ISL_2503570, EPI_ISL_2503573, EPI_ISL_2503574, EPI_ISL_2503575, EPI_ISL_2503576, EPI_ISL_2503582, EPI_ISL_2503583, EPI_ISL_2503587, EPI_ISL_2503589, EPI_ISL_2503594, EPI_ISL_2503598, EPI_ISL_2503600, EPI_ISL_2503602, EPI_ISL_2503605, EPI_ISL_2503607, EPI_ISL_2503612, EPI_ISL_2503613, EPI_ISL_2503617, EPI_ISL_2503621, EPI_ISL_2503625, EPI_ISL_2503626, EPI_ISL_2503630, EPI_ISL_2503631, EPI_ISL_2503633, EPI_ISL_2503634, EPI_ISL_2503639, EPI_ISL_2503642, EPI_ISL_2503643, EPI_ISL_2503644, EPI_ISL_2503645, EPI_ISL_2503646, EPI_ISL_2503647, EPI_ISL_2503648, EPI_ISL_2503649, EPI_ISL_2503650, EPI_ISL_2503651, EPI_ISL_2503652, EPI_ISL_2503655, EPI_ISL_2503660, EPI_ISL_2503661, EPI_ISL_2503668, EPI_ISL_2503669, EPI_ISL_2503670, EPI_ISL_2503671, EPI_ISL_2503676, EPI_ISL_2503678, EPI_ISL_2503679, EPI_ISL_2503680, EPI_ISL_2503682, EPI_ISL_2503689, EPI_ISL_2503690, EPI_ISL_2503691, EPI_ISL_2503693, EPI_ISL_2503695, EPI_ISL_2503696, EPI_ISL_2503697, EPI_ISL_2503698, EPI_ISL_2503699, EPI_ISL_2503700, EPI_ISL_2503702, EPI_ISL_2503703, EPI_ISL_2503705, EPI_ISL_2503706, EPI_ISL_2503713, EPI_ISL_2503714, EPI_ISL_2503718, EPI_ISL_2503726, EPI_ISL_2503727, EPI_ISL_2503728, EPI_ISL_2503729, EPI_ISL_2503730, EPI_ISL_2503731, EPI_ISL_2503734, EPI_ISL_2503736, EPI_ISL_2503737, EPI_ISL_2503738, EPI_ISL_2503740, EPI_ISL_2503745, EPI_ISL_2503746, EPI_ISL_2503748, EPI_ISL_2503760, EPI_ISL_2503761, EPI_ISL_2503768, EPI_ISL_2503770, EPI_ISL_2503773, EPI_ISL_2503778, EPI_ISL_2503780, EPI_ISL_2503781, EPI_ISL_2503782, EPI_ISL_2503783, EPI_ISL_2503784, EPI_ISL_2503785, EPI_ISL_2503786, EPI_ISL_2503787, EPI_ISL_2503788, EPI_ISL_2503789, EPI_ISL_2503795, EPI_ISL_2503796, EPI_ISL_2503798, EPI_ISL_2503799, EPI_ISL_2503801, EPI_ISL_2503804, EPI_ISL_2503807, EPI_ISL_2503808, EPI_ISL_2503813, EPI_ISL_2503814, EPI_ISL_2503817, EPI_ISL_2503820, EPI_ISL_2548374, EPI_ISL_2548379, EPI_ISL_2548444, EPI_ISL_2548501, EPI_ISL_2548502, EPI_ISL_2548527, EPI_ISL_2548529, EPI_ISL_2548530, EPI_ISL_2548577, EPI_ISL_2548585, EPI_ISL_2548599, EPI_ISL_2548611, EPI_ISL_2548617, EPI_ISL_2548621, EPI_ISL_2548624, EPI_ISL_2548625, EPI_ISL_2548626, EPI_ISL_2548628, EPI_ISL_2548629, EPI_ISL_2548637, EPI_ISL_2548639, EPI_ISL_2548640, EPI_ISL_2548642, EPI_ISL_2548643, EPI_ISL_2548644, EPI_ISL_2548648, EPI_ISL_2548651, EPI_ISL_2548652, EPI_ISL_2548658, EPI_ISL_2548662, EPI_ISL_2548665, EPI_ISL_2548666, EPI_ISL_2548667, EPI_ISL_2548668, EPI_ISL_2548674, EPI_ISL_2548681, EPI_ISL_2548682, EPI_ISL_2548683, EPI_ISL_2548687, EPI_ISL_2548688, EPI_ISL_2548689, EPI_ISL_2548691, EPI_ISL_2548692, EPI_ISL_2548695, EPI_ISL_2548696, EPI_ISL_2548697, EPI_ISL_2548703, EPI_ISL_2548705, EPI_ISL_2548707, EPI_ISL_2548712, EPI_ISL_2548713, EPI_ISL_2548714, EPI_ISL_2548715, EPI_ISL_2548716, EPI_ISL_2548718, EPI_ISL_2548719, EPI_ISL_2548720, EPI_ISL_2548721, EPI_ISL_2548722, EPI_ISL_2548723, EPI_ISL_2548726, EPI_ISL_2548727, EPI_ISL_2548729, EPI_ISL_2548730, EPI_ISL_2548734, EPI_ISL_2548735, EPI_ISL_2548738, EPI_ISL_2548746, EPI_ISL_2548750, EPI_ISL_2548754, EPI_ISL_2548755, EPI_ISL_2548756, EPI_ISL_2548757, EPI_ISL_2548763, EPI_ISL_2548771, EPI_ISL_2548772, EPI_ISL_2548775, EPI_ISL_2548781, EPI_ISL_2548788, EPI_ISL_2548793, EPI_ISL_2548795, EPI_ISL_2548797, EPI_ISL_2548799, EPI_ISL_2548804, EPI_ISL_2548805, EPI_ISL_2548807, EPI_ISL_2548814, EPI_ISL_2548816, EPI_ISL_2548822, EPI_ISL_2548829, EPI_ISL_2548831, EPI_ISL_2548856, EPI_ISL_2548869, EPI_ISL_2548872, EPI_ISL_2548879, EPI_ISL_2548896, EPI_ISL_2548899, EPI_ISL_2548916, EPI_ISL_2548941, EPI_ISL_2548968, EPI_ISL_2548987, EPI_ISL_2548988, EPI_ISL_2549001, EPI_ISL_2549002, EPI_ISL_2549011, EPI_ISL_2549020, EPI_ISL_2549035, EPI_ISL_2549044, EPI_ISL_2549045, EPI_ISL_2549046, EPI_ISL_2549047, EPI_ISL_2549049, EPI_ISL_2549050, EPI_ISL_2549051, EPI_ISL_2549052, EPI_ISL_2675908, EPI_ISL_2675909, EPI_ISL_2675944, EPI_ISL_2675945, EPI_ISL_2675969, EPI_ISL_2675972, EPI_ISL_2675982, EPI_ISL_2676010, EPI_ISL_2676013, EPI_ISL_2676025, EPI_ISL_2676026, EPI_ISL_2676043, EPI_ISL_2676045, EPI_ISL_2860948, EPI_ISL_2860957, EPI_ISL_2860960, EPI_ISL_2860962, EPI_ISL_2860966, EPI_ISL_2860968, EPI_ISL_2860973, EPI_ISL_2860974, EPI_ISL_2860977 | | | |
| see above | INSACOG-WB | National Institute of Biomedical Genomics – INSACOG | Ajay Chakraborti; Arindam Maitra; Bhaswati Bandyopadhyay; Nidhan Kumar Biswas; Saumitra Das; Sreedhar Chinnaswamy; Tamal Ghosh |
| EPI_ISL_2549258, EPI_ISL_2549262, EPI_ISL_2549264, EPI_ISL_2549269 | Indian Council of Medical Research-National Institute of Virology, Microbial Containment Complex | Indian Council of Medical Research-National Institute of Virology, Microbial Containment Complex | Pragya D. Yadav |
| EPI_ISL_2545200, EPI_ISL_2545202, EPI_ISL_2545205, EPI_ISL_2545210, EPI_ISL_2842217, EPI_ISL_2842220, EPI_ISL_2842221 | | | |
| see above | KEMPEGOWDA INTERNATIONAL AIRPORT | INSACOG-KA, NIMHANS | Ananthapadmanabha Kotambail; Anita S Desai; Anson Kunjumon George; Chetan G K; Chitra Pattabiraman; Darshan Sreenivas; Ellango Ramasamy; Gautham Arunachal Udupi; Mahesh Kumar.C.S; Pramada Prasad; Sony Sharma; V Ravi |
| EPI_ISL_2774661, EPI_ISL_2774671, EPI_ISL_2775138 | Mapmygenome | CSIR-Centre for Cellular and Molecular Biology-INSACOG | Amareshwar Vodapalli; Ara Sreenivas; Archana Bharadwaj Siva; B Himasri; Divya Tej Sowpati; Jandhyala Sai Krishna; Karthik Bharadwaj Tallapaka; Lamuk Zaveri; Onkar Kulkarni; Payel Mukherjee; Priya Nurkurthy; Rakesh K Mishra; Shreekant Verma; Sofia Banu; Sumedha Avadhanula; Tulasi Nagabandi; Valli Nagalakshmi Undamatla; Vidhyadhari Methuku |
| EPI_ISL_2509947, EPI_ISL_2509948, EPI_ISL_2509956 | N.H.L Municipal Medical College, Ahmedabad | Gujarat Biotechnology Research Centre | Chaitanya Joshi; Dinesh Kumar; Janvi Raval; Jayshri Pethani; Madhvi Joshi; Nitesh Shah; Nitin Savaliya; Ramesh Pandit; Sonal Sharma; Twinkle Soni; Umang Mishra; Zarna Patel; Zuber Saiyed |
| EPI_ISL_2651330, EPI_ISL_2728429, EPI_ISL_2728445, EPI_ISL_2728453, EPI_ISL_2728454, EPI_ISL_2842205, EPI_ISL_2842207, EPI_ISL_2842208, EPI_ISL_2842243, EPI_ISL_2842244, EPI_ISL_2842245, EPI_ISL_2842282, EPI_ISL_2842283, EPI_ISL_2842284 | | | |

| | | | |
|---|---|---|---|
| see above | NIMHANS | INSACOG-KA, NIMHANS | Ananthapadmanabha Kotambail; Anita S Desai; Anson Kunjumon George; Chetan G K; Chitra Pattabiraman; Darshan Sreenivas; Ellango Ramasamy; Gautham Arunachal Udupi; Mahesh Kumar.C.S; Pramada Prasad; Sony Sharma; V Ravi |
| EPI_ISL_2504332, EPI_ISL_2504333, EPI_ISL_2504346, EPI_ISL_2504397, EPI_ISL_2504399, EPI_ISL_2504454, EPI_ISL_2504486, EPI_ISL_2504831, EPI_ISL_2504838, EPI_ISL_2504843, EPI_ISL_2504845, EPI_ISL_2504847, EPI_ISL_2504850, EPI_ISL_2504851, EPI_ISL_2504855, EPI_ISL_2504856, EPI_ISL_2504857, EPI_ISL_2504862, EPI_ISL_2504969, EPI_ISL_2504970, EPI_ISL_2505035, EPI_ISL_2505037, EPI_ISL_2505101, EPI_ISL_2505143, EPI_ISL_2505144, EPI_ISL_2555547, EPI_ISL_2555548, EPI_ISL_2555556, EPI_ISL_2555560, EPI_ISL_2555573, EPI_ISL_2555598, EPI_ISL_2555601, EPI_ISL_2555602, EPI_ISL_2555604, EPI_ISL_2555616, EPI_ISL_2555641, EPI_ISL_2555647, EPI_ISL_2555649, EPI_ISL_2555734, EPI_ISL_2555776, EPI_ISL_2555823, EPI_ISL_2555828, EPI_ISL_2555838, EPI_ISL_2555839, EPI_ISL_2555844, EPI_ISL_2555848, EPI_ISL_2555850, EPI_ISL_2555875, EPI_ISL_2555876, EPI_ISL_2555923, EPI_ISL_2555958, EPI_ISL_2555973, EPI_ISL_2555986, EPI_ISL_2555990, EPI_ISL_2556042, EPI_ISL_2556045, EPI_ISL_2556063, EPI_ISL_2556065, EPI_ISL_2556069, EPI_ISL_2556084, EPI_ISL_2556121, EPI_ISL_2556129, EPI_ISL_2556131, EPI_ISL_2556136, EPI_ISL_2556137, EPI_ISL_2556138, EPI_ISL_2556142, EPI_ISL_2556146, EPI_ISL_2556152, EPI_ISL_2556168, EPI_ISL_2556174, EPI_ISL_2556191, EPI_ISL_2556193, EPI_ISL_2556206, EPI_ISL_2556210, EPI_ISL_2556212, EPI_ISL_2556214, EPI_ISL_2556226, EPI_ISL_2556227, EPI_ISL_2556234, EPI_ISL_2556241, EPI_ISL_2556244, EPI_ISL_2556247, EPI_ISL_2556249, EPI_ISL_2556264, EPI_ISL_2556265, EPI_ISL_2556281, EPI_ISL_2556285, EPI_ISL_2556286, EPI_ISL_2556288, EPI_ISL_2556337, EPI_ISL_2556370, EPI_ISL_2556417, EPI_ISL_2556423, EPI_ISL_2556433, EPI_ISL_2556435, EPI_ISL_2556444, EPI_ISL_2556476, EPI_ISL_2556510, EPI_ISL_2556511, EPI_ISL_2556522, EPI_ISL_2556577, EPI_ISL_2556591, EPI_ISL_2556596, EPI_ISL_2556712, EPI_ISL_2556717, EPI_ISL_2556749, EPI_ISL_2556751, EPI_ISL_2556753, EPI_ISL_2556755, EPI_ISL_2556756, EPI_ISL_2556834, EPI_ISL_2556840, EPI_ISL_2556844, EPI_ISL_2556846, EPI_ISL_2556854, EPI_ISL_2556855, EPI_ISL_2556856, EPI_ISL_2556861, EPI_ISL_2556863, EPI_ISL_2556868, EPI_ISL_2556879, EPI_ISL_2556880, EPI_ISL_2556883, EPI_ISL_2556899, EPI_ISL_2556903, EPI_ISL_2556905, EPI_ISL_2556910, EPI_ISL_2556936, EPI_ISL_2556975, EPI_ISL_2556977, EPI_ISL_2556984, EPI_ISL_2556991, EPI_ISL_2556992, EPI_ISL_2556996, EPI_ISL_2556997, EPI_ISL_2556998, EPI_ISL_2557012, EPI_ISL_2557019, EPI_ISL_2557020, EPI_ISL_2557024, EPI_ISL_2557025 | | | |
| see above | National Centre for Disease Control (NCDC) Biotechnology Division, Delhi | NCDC Delhi, Biotechnology Division INSACOG | Hema Gogia; Hemlata Lall; Kalaiarasan Ponnusamy; Mahesh S Dhar; Manoj K Singh; Meena Datta; Partha Rakshit; Preeti Madan; Priyanka Singh; Radhakrishnan V. S; Robin Marwal; Sandhya Kabra; Sujeet K Singh; Uma Sharma |
| EPI_ISL_2660817, EPI_ISL_2660853, EPI_ISL_2660893 | PASTEUR MOLECULAR LABORATORY - INSACOG MEGHALAYA | CSIR-Centre for Cellular and Molecular Biology-INSACOG | Amareshwar Vodapalli; Ara Sreenivas; Archana Bharadwaj Siva; B Himasri; Divya Tej Sowpati; E Marak; E Shadap; Karthik Bharadwaj Tallapaka; Lamuk Zaveri; Onkar Kulkarni; Payel Mukherjee; Priya Nurkurthy; R. Lyngdoh; Rakesh K Mishra; Shreekant Verma; Sofia Banu; Sumedha Avadhanula; Tulasi Nagabandi; Valli Nagalakshmi Undamatla; Vidhyadhari Methuku |
| EPI_ISL_2728457, EPI_ISL_2728483, EPI_ISL_2728497, EPI_ISL_2842252, EPI_ISL_2842255, EPI_ISL_2842288 | RAILWAY HOSPITAL | INSACOG-KA, NIMHANS | Ananthapadmanabha Kotambail; Anita S Desai; Anson Kunjumon George; Chetan G K; Chitra Pattabiraman; Darshan Sreenivas; Ellango Ramasamy; Gautham Arunachal Udupi; Mahesh Kumar.C.S; Sony Sharma; V Ravi |
| EPI_ISL_2661442 | RUHS College of Medical Sciences | CSIR-Centre for Cellular and Molecular Biology - INSACOG | Amareshwar Vodapalli; Ara Sreenivas; Archana Bharadwaj Siva; B Himasri; Divya Tej Sowpati; Dr. Gaurav Dalela; Dr. Jitendra Panda; Dr. Nilofer Khayyam; Dr. Raja Babu Panwar; Dr. Rajeev Gupta; Dr. Ramesh Sharma; Dr. Sonali Sharma; Dr. Sudhanshu Kacker; Dr. Vaseem Naheed Baig; Jandhyala Sai Krishna; Karthik Bharadwaj Tallapaka; Lamuk Zaveri; Onkar Kulkarni; Payel Mukherjee; Priya Nurkurthy; Rakesh K Mishra; Shreekant Verma; Sofia Banu; Sumedha Avadhanula; Tulasi Nagabandi; Valli Nagalakshmi Undamatla; Vidhyadhari Methuku |
| EPI_ISL_2710000, EPI_ISL_2710002, EPI_ISL_2710007 | Regional VRDL,Dibrugarh | Regional VRDL,Dibrugarh | A Jakharia; B Borkakoty; C Bhattacharya; D Biswas; K Narain; Mandakini Das Sarma; R Hazarika |
| EPI_ISL_2728491, EPI_ISL_2728524 | SANJAY GANDHI INSTITUTE OF TRAUMA AND ORTHOPAEDICS | INSACOG-KA, NIMHANS | Ananthapadmanabha Kotambail; Anita S Desai; Anson Kunjumon George; Chetan G K; Chitra Pattabiraman; Darshan Sreenivas; Ellango Ramasamy; Gautham Arunachal Udupi; Mahesh Kumar.C.S; Sony Sharma; V Ravi |

We gratefully acknowledge the following Authors from the Originating laboratories responsible for obtaining the specimens, as well as the Submitting laboratories where the genome data were generated and shared via GISAID, on which this research is based.

All Submitters of data may be contacted directly via www.gisaid.org

Authors are sorted alphabetically.

| Accession ID | Originating Laboratory | Submitting Laboratory | Authors |
|---|---|---|---|
| EPI_ISL_2774724, EPI_ISL_2774725, EPI_ISL_2774730, EPI_ISL_2774732, EPI_ISL_2774733, EPI_ISL_2774734, EPI_ISL_2774735, EPI_ISL_2774736, EPI_ISL_2774737, EPI_ISL_2774738, EPI_ISL_2774739, EPI_ISL_2774740, EPI_ISL_2774741, EPI_ISL_2774742, EPI_ISL_2774743, EPI_ISL_2774744, EPI_ISL_2774745, EPI_ISL_2774747, EPI_ISL_2774748, EPI_ISL_2774750, EPI_ISL_2774751, EPI_ISL_2774753, EPI_ISL_2774754, EPI_ISL_2774755, EPI_ISL_2774756, EPI_ISL_2774757, EPI_ISL_2774758, EPI_ISL_2774759, EPI_ISL_2774762, EPI_ISL_2774765, EPI_ISL_2774767, EPI_ISL_2774769, EPI_ISL_2774770, EPI_ISL_2774772, EPI_ISL_2774774, EPI_ISL_2774776, EPI_ISL_2774777, EPI_ISL_2774778, EPI_ISL_2774779, EPI_ISL_2774780, EPI_ISL_2774781, EPI_ISL_2774783, EPI_ISL_2774784, EPI_ISL_2774785, EPI_ISL_2774790, EPI_ISL_2774791, EPI_ISL_2774792, EPI_ISL_2774793, EPI_ISL_2774795, EPI_ISL_2774796, EPI_ISL_2774798, EPI_ISL_2774799, EPI_ISL_2774800, EPI_ISL_2774802, EPI_ISL_2774804, EPI_ISL_2774805, EPI_ISL_2774808, EPI_ISL_2774811, EPI_ISL_2774812, EPI_ISL_2774816, EPI_ISL_2774819, EPI_ISL_2774820, EPI_ISL_2774821, EPI_ISL_2774822, EPI_ISL_2774823, EPI_ISL_2774827, EPI_ISL_2774828, EPI_ISL_2774832, EPI_ISL_2775288, EPI_ISL_2775289, EPI_ISL_2775293, EPI_ISL_2775294, EPI_ISL_2775297, EPI_ISL_2775301, EPI_ISL_2775302, EPI_ISL_2775303, EPI_ISL_2775305, EPI_ISL_2775314, EPI_ISL_2775315, EPI_ISL_2775326, EPI_ISL_2775333, EPI_ISL_2775334, EPI_ISL_2775336, EPI_ISL_2775338, EPI_ISL_2775339, EPI_ISL_2775341, EPI_ISL_2775342, EPI_ISL_2775344, EPI_ISL_2775345, EPI_ISL_2775350, EPI_ISL_2775351, EPI_ISL_2775353, EPI_ISL_2775354, EPI_ISL_2775355, EPI_ISL_2775357, EPI_ISL_2775358, EPI_ISL_2775359, EPI_ISL_2775360, EPI_ISL_2775363, EPI_ISL_2775365, EPI_ISL_2775369, EPI_ISL_2775371, EPI_ISL_2775376, EPI_ISL_2775381, EPI_ISL_2775383, EPI_ISL_2775389, EPI_ISL_2789349, EPI_ISL_2789350 | | | |
| see above | AP SSO | CSIR-Centre for Cellular and Molecular Biology-INSACOG | Amareshwar Vodapalli; Ara Sreenivas; Archana Bharadwaj Siva; B Himasri; Divya Tej Sowpati; Jandhyala Sai Krishna; Karthik Bharadwaj Tallapaka; Lamuk Zaveri; Onkar Kulkarni; Payel Mukherjee; Priya Nurkurthy; Rakesh K Mishra; Shreekant Verma; Sofia Banu; Sumedha Avadhanula; Tulasi Nagabandi; Valli Nagalakshmi Undamatla; Vidhyadhari Methuku |
| EPI_ISL_2842119, EPI_ISL_2842122, EPI_ISL_2842123, EPI_ISL_2842124, EPI_ISL_2842130, EPI_ISL_2842145, EPI_ISL_2842158, EPI_ISL_2842159, EPI_ISL_2842175, EPI_ISL_2842184, EPI_ISL_2842198, EPI_ISL_2842225, EPI_ISL_2842228, EPI_ISL_2842236, EPI_ISL_2842238, EPI_ISL_2842258, EPI_ISL_2842265, EPI_ISL_2842271, EPI_ISL_2842272, EPI_ISL_2842275, EPI_ISL_2842279, EPI_ISL_2842292, EPI_ISL_2842293, EPI_ISL_2842298, EPI_ISL_2842299, EPI_ISL_2842302, EPI_ISL_2842307, EPI_ISL_2842309, EPI_ISL_2842311, EPI_ISL_2842319, EPI_ISL_2842322, EPI_ISL_2842324, EPI_ISL_2842325, EPI_ISL_2842326, EPI_ISL_2842327, EPI_ISL_2842329, EPI_ISL_2842330, EPI_ISL_2842331, EPI_ISL_2842332, EPI_ISL_2842333, EPI_ISL_2842334, EPI_ISL_2842336, EPI_ISL_2842338, EPI_ISL_2842339, EPI_ISL_2842340, EPI_ISL_2842341, EPI_ISL_2842343, EPI_ISL_2842344, EPI_ISL_2842345, EPI_ISL_2842346, EPI_ISL_2842348, EPI_ISL_2842349, EPI_ISL_2842351, EPI_ISL_2842356, EPI_ISL_2842357, EPI_ISL_2842358, EPI_ISL_2842359, EPI_ISL_2842360, EPI_ISL_2842361, EPI_ISL_2842363, EPI_ISL_2842364, EPI_ISL_2842365, EPI_ISL_2842367, EPI_ISL_2842369, EPI_ISL_2842370, EPI_ISL_2842371, EPI_ISL_2842372, EPI_ISL_2842375, EPI_ISL_2842377, EPI_ISL_2842378, EPI_ISL_2842381, EPI_ISL_2842382, EPI_ISL_2842383, EPI_ISL_2842384, EPI_ISL_2842385, EPI_ISL_2842386, EPI_ISL_2842387, EPI_ISL_2842389, EPI_ISL_2842390, EPI_ISL_2842391, EPI_ISL_2842393, EPI_ISL_2842394, EPI_ISL_2842399, EPI_ISL_2842410, EPI_ISL_2842438, EPI_ISL_2842439, EPI_ISL_2842442, EPI_ISL_2842446, EPI_ISL_2842447, EPI_ISL_2842449, EPI_ISL_2842450, EPI_ISL_2842451, EPI_ISL_2842454, EPI_ISL_2842456, EPI_ISL_2842457 | | | |
| see above | BBMP Urban | INSACOG-KA, NIMHANS | Ananthapadmanabha Kotambail; Anita S Desai; Anson Kunjumon George; Chetan G K; Chitra Pattabiraman; Darshan Sreenivas; Ellango Ramasamy; Gautham Arunachal Udupi; Mahesh Kumar.C.S; Sony Sharma; V Ravi |
| EPI_ISL_2774839, EPI_ISL_2774844, EPI_ISL_2774858, EPI_ISL_2774868, EPI_ISL_2774878, EPI_ISL_2774881, EPI_ISL_2774882, EPI_ISL_2774897, EPI_ISL_2774899, EPI_ISL_2774901, EPI_ISL_2774902, EPI_ISL_2774903, EPI_ISL_2774909, EPI_ISL_2774913, EPI_ISL_2774915, EPI_ISL_2774917, EPI_ISL_2774918, EPI_ISL_2774920, EPI_ISL_2774921, EPI_ISL_2774924, EPI_ISL_2774925, EPI_ISL_2774927, EPI_ISL_2774931, EPI_ISL_2774932, EPI_ISL_2774933, EPI_ISL_2774934, EPI_ISL_2774935, EPI_ISL_2774939, EPI_ISL_2774943, EPI_ISL_2774947, EPI_ISL_2774949, EPI_ISL_2774950, EPI_ISL_2774955, EPI_ISL_2774956, EPI_ISL_2774957, EPI_ISL_2774968, EPI_ISL_2774971, EPI_ISL_2774974, EPI_ISL_2774977, EPI_ISL_2774978, EPI_ISL_2774980, EPI_ISL_2774981, EPI_ISL_2774982, EPI_ISL_2774983, EPI_ISL_2774987, EPI_ISL_2774991, EPI_ISL_2774993, EPI_ISL_2774994, EPI_ISL_2774995, EPI_ISL_2774996, EPI_ISL_2774997, EPI_ISL_2774998, EPI_ISL_2774999, EPI_ISL_2775004, EPI_ISL_2775007, EPI_ISL_2775008, EPI_ISL_2775009, EPI_ISL_2775011, EPI_ISL_2775017, EPI_ISL_2775019, EPI_ISL_2775023, EPI_ISL_2775025, EPI_ISL_2775027, EPI_ISL_2775034, EPI_ISL_2775035, EPI_ISL_2775041, EPI_ISL_2775044, EPI_ISL_2775047, EPI_ISL_2775048, EPI_ISL_2775050, EPI_ISL_2775052, EPI_ISL_2775054, EPI_ISL_2775055, EPI_ISL_2775056, EPI_ISL_2775057, EPI_ISL_2775059, EPI_ISL_2775060, EPI_ISL_2775062, EPI_ISL_2775063, EPI_ISL_2775064, EPI_ISL_2775065, EPI_ISL_2775069, EPI_ISL_2775070, EPI_ISL_2775072, EPI_ISL_2775074, EPI_ISL_2775075, EPI_ISL_2775076, EPI_ISL_2775077, EPI_ISL_2775079, EPI_ISL_2775083, EPI_ISL_2775085, EPI_ISL_2775086, EPI_ISL_2775088, EPI_ISL_2775094, EPI_ISL_2775097, EPI_ISL_2775099, EPI_ISL_2775103, EPI_ISL_2775104, EPI_ISL_2775105, EPI_ISL_2775109, EPI_ISL_2775112, EPI_ISL_2775114, EPI_ISL_2775116, EPI_ISL_2775117, EPI_ISL_2775119, EPI_ISL_2775150, EPI_ISL_2775160, EPI_ISL_2775166, EPI_ISL_2775167, EPI_ISL_2775168, EPI_ISL_2775169, EPI_ISL_2775172, EPI_ISL_2775176, EPI_ISL_2775179, EPI_ISL_2775180, EPI_ISL_2775181, EPI_ISL_2775182, EPI_ISL_2775186, EPI_ISL_2775188, EPI_ISL_2775192, EPI_ISL_2775193, EPI_ISL_2775194, EPI_ISL_2775195, EPI_ISL_2775198, EPI_ISL_2775205, EPI_ISL_2775207, EPI_ISL_2775209, EPI_ISL_2775210, EPI_ISL_2775212, EPI_ISL_2775217, EPI_ISL_2775218, EPI_ISL_2775221, EPI_ISL_2775222, EPI_ISL_2775223, EPI_ISL_2775224, EPI_ISL_2775225, EPI_ISL_2775227, EPI_ISL_2775236, EPI_ISL_2775237, EPI_ISL_2775239, EPI_ISL_2775240, EPI_ISL_2775241, EPI_ISL_2775242, EPI_ISL_2775243, EPI_ISL_2775244, EPI_ISL_2775246, EPI_ISL_2775247, EPI_ISL_2775248, EPI_ISL_2775250, EPI_ISL_2775253, EPI_ISL_2775257, EPI_ISL_2775258, EPI_ISL_2775260, EPI_ISL_2775261, EPI_ISL_2775262, EPI_ISL_2775263, EPI_ISL_2775265, EPI_ISL_2775272, EPI_ISL_2775275, EPI_ISL_2775281, EPI_ISL_2775287 | | | |
| see above | CSIR-Centre for Cellular and Molecular Biology | CSIR-Centre for Cellular and Molecular Biology-INSACOG | ; Amareshwar Vodapalli; Ara Sreenivas; Archana Bharadwaj Siva; B Himasri; Divya Tej Sowpati; Jandhyala Sai Krishna; Karthik Bharadwaj Tallapaka; Lamuk Zaveri; Onkar Kulkarni; Payel Mukherjee; Priya Nurkurthy; Rakesh K Mishra; Shreekant Verma; Sofia Banu; Sumedha Avadhanula; Tulasi Nagabandi; Valli Nagalakshmi Undamatla; Vidhyadhari Methuku |
| EPI_ISL_2842477 | DSU CHAMRAJANAGARA | INSACOG-KA, NIMHANS | Ananthapadmanabha Kotambail; Anita S Desai; Anson Kunjumon George; Chetan G K; Chitra Pattabiraman; Darshan Sreenivas; Ellango Ramasamy; Gautham Arunachal Udupi; Mahesh Kumar.C.S; Sony Sharma; V Ravi |
| EPI_ISL_2842491, EPI_ISL_2842493, EPI_ISL_2842494, EPI_ISL_2842496, EPI_ISL_2842497, EPI_ISL_2842499 | DSU HASSAN | INSACOG-KA, NIMHANS | Ananthapadmanabha Kotambail; Anita S Desai; Anson Kunjumon George; Chetan G K; Chitra Pattabiraman; Darshan Sreenivas; Ellango Ramasamy; Gautham Arunachal Udupi; Mahesh Kumar.C.S; Sony Sharma; V Ravi |
| EPI_ISL_2842482, EPI_ISL_2842483, EPI_ISL_2842484, EPI_ISL_2842490 | DSU KALABURAGI | INSACOG-KA, NIMHANS | Ananthapadmanabha Kotambail; Anita S Desai; Anson Kunjumon George; Chetan G K; Chitra Pattabiraman; Darshan Sreenivas; Ellango Ramasamy; Gautham Arunachal Udupi; Mahesh Kumar.C.S; Sony Sharma; V Ravi |
| EPI_ISL_2842478, EPI_ISL_2842479, EPI_ISL_2842480 | DSU KODAGU | INSACOG-KA, NIMHANS | Ananthapadmanabha Kotambail; Anita S Desai; Anson Kunjumon George; Chetan G K; Chitra Pattabiraman; Darshan Sreenivas; Ellango Ramasamy; Gautham Arunachal Udupi; Mahesh Kumar.C.S; Sony Sharma; V Ravi |
| EPI_ISL_2842104, EPI_ISL_2842501, EPI_ISL_2842502 | DSU KOLAR | INSACOG-KA, NIMHANS | Ananthapadmanabha Kotambail; Anita S Desai; Anson Kunjumon George; Chetan G K; Chitra Pattabiraman; Darshan Sreenivas; Ellango Ramasamy; Gautham Arunachal Udupi; Mahesh Kumar.C.S; Sony Sharma; V Ravi |
| EPI_ISL_2842473, EPI_ISL_2842474 | DSU MANDYA | INSACOG-KA, NIMHANS | Ananthapadmanabha Kotambail; Anita S Desai; Anson Kunjumon George; Chetan G K; Chitra Pattabiraman; Darshan Sreenivas; Ellango Ramasamy; Gautham Arunachal Udupi; Mahesh Kumar.C.S; Sony Sharma; V Ravi |
| EPI_ISL_2842467, EPI_ISL_2842469, EPI_ISL_2842470, EPI_ISL_2842471, EPI_ISL_2842472 | DSU MYSURU | INSACOG-KA, NIMHANS | Ananthapadmanabha Kotambail; Anita S Desai; Anson Kunjumon George; Chetan G K; Chitra Pattabiraman; Darshan Sreenivas; Ellango Ramasamy; Gautham Arunachal Udupi; Mahesh Kumar.C.S; Sony Sharma; V Ravi |
| EPI_ISL_2842431, EPI_ISL_2842433, EPI_ISL_2842434, EPI_ISL_2842436 | DSU VIJAYAPURA | INSACOG-KA, NIMHANS | Ananthapadmanabha Kotambail; Anita S Desai; Anson Kunjumon George; Chetan G K; Chitra Pattabiraman; Darshan Sreenivas; Ellango Ramasamy; Gautham Arunachal Udupi; Mahesh Kumar.C.S; Sony Sharma; V Ravi |
| EPI_ISL_2812579, EPI_ISL_2812580, EPI_ISL_2812581, EPI_ISL_2812582, EPI_ISL_2812583, EPI_ISL_2812584, EPI_ISL_2812585, EPI_ISL_2812586, EPI_ISL_2812587, EPI_ISL_2812588, EPI_ISL_2812589, EPI_ISL_2812590, EPI_ISL_2812591, EPI_ISL_2812592, EPI_ISL_2812593, EPI_ISL_2812594, EPI_ISL_2812595, EPI_ISL_2812597, EPI_ISL_2812598, EPI_ISL_2812599, EPI_ISL_2812600, EPI_ISL_2812601, EPI_ISL_2812602, EPI_ISL_2812603, EPI_ISL_2812604, EPI_ISL_2812605, EPI_ISL_2812606, EPI_ISL_2812607, EPI_ISL_2812608, EPI_ISL_2812609, EPI_ISL_2812610, EPI_ISL_2812611, EPI_ISL_2812612, EPI_ISL_2812613, EPI_ISL_2812614, EPI_ISL_2812615, EPI_ISL_2812616, EPI_ISL_2812617, EPI_ISL_2812618, EPI_ISL_2812619, EPI_ISL_2812620, EPI_ISL_2812621, EPI_ISL_2812622, EPI_ISL_2812623, EPI_ISL_2812624, EPI_ISL_2812625, EPI_ISL_2812626, EPI_ISL_2812627, EPI_ISL_2812628, EPI_ISL_2812630, EPI_ISL_2812631, EPI_ISL_2812632, EPI_ISL_2812633, EPI_ISL_2812634, EPI_ISL_2812635, EPI_ISL_2812636, EPI_ISL_2812637, EPI_ISL_2812638, EPI_ISL_2812639, EPI_ISL_2812640, EPI_ISL_2812641, EPI_ISL_2812642, EPI_ISL_2812643, EPI_ISL_2812644, EPI_ISL_2812645, EPI_ISL_2812646 | | | |
| see above | INSACOG-MANIPUR | National Institute of Biomedical Genomics – INSACOG | Arindam Maitra; Khumanthem Ranjana; Nanaocha Sharma; Nidhan Kumar Biswas; R K Manglem Singh; Saumitra Das; Sreedhar Chinnaswamy |
| EPI_ISL_2860930, EPI_ISL_2860931, EPI_ISL_2860933, EPI_ISL_2860934, EPI_ISL_2860935, EPI_ISL_2860938, EPI_ISL_2860941, EPI_ISL_2860942, EPI_ISL_2860943, EPI_ISL_2860946, EPI_ISL_2860947, EPI_ISL_2860949, EPI_ISL_2860950, EPI_ISL_2860951, EPI_ISL_2860952, EPI_ISL_2860953, EPI_ISL_2860954, EPI_ISL_2860955, EPI_ISL_2860958, EPI_ISL_2860961, EPI_ISL_2860963, EPI_ISL_2860965, EPI_ISL_2860967, EPI_ISL_2860969, EPI_ISL_2860971, EPI_ISL_2860972, EPI_ISL_2860975, EPI_ISL_2860976, EPI_ISL_2860979, EPI_ISL_2860980, EPI_ISL_2860981, EPI_ISL_2860982, EPI_ISL_2860983, EPI_ISL_2860984, EPI_ISL_2860985, EPI_ISL_2860987, EPI_ISL_2860990, EPI_ISL_2860991, EPI_ISL_2860992, EPI_ISL_2860993, EPI_ISL_2860994, EPI_ISL_2860995, EPI_ISL_2860996, EPI_ISL_2860997, EPI_ISL_2860998, EPI_ISL_2860999, EPI_ISL_2861002, EPI_ISL_2861003, EPI_ISL_2861004, EPI_ISL_2861005, EPI_ISL_2861009 | | | |
| see above | INSACOG-WB | National Institute of Biomedical Genomics – INSACOG | Ajay Chakraborti; Arindam Maitra; Bhaswati Bandyopadhyay; Nidhan Kumar Biswas; Saumitra Das; Sreedhar Chinnaswamy; Tamal Ghosh |
| EPI_ISL_2842169, EPI_ISL_2842379 | JAYADEVA H | INSACOG-KA, NIMHANS | Ananthapadmanabha Kotambail; Anita S Desai; Anson Kunjumon George; Chetan G K; Chitra Pattabiraman; Darshan Sreenivas; Ellango Ramasamy; Gautham Arunachal Udupi; Mahesh Kumar.C.S; Sony Sharma; V Ravi |
| EPI_ISL_2842216, EPI_ISL_2842218, EPI_ISL_2842219 | KEMPEGOWDA INTERNATIONAL AIRPORT | INSACOG-KA, NIMHANS | Ananthapadmanabha Kotambail; Anita S Desai; Anson Kunjumon George; Chetan G K; Chitra Pattabiraman; Darshan Sreenivas; Ellango Ramasamy; Gautham Arunachal Udupi; Mahesh Kumar.C.S; Sony Sharma; V Ravi |
| EPI_ISL_2811394, EPI_ISL_2811395 | MAX Hospital, Delhi | CSIR-IGIB | Jayanthi Shastri; Nirjhar Chaterjee; Priti Devi; Priyanka Mehta; Rajesh Pandey; Rajit Shah; Sachee Agrawal; Swapneil Parikh; Veena Aggarwal |
| EPI_ISL_2774679, EPI_ISL_2774696, EPI_ISL_2774698, EPI_ISL_2774707, EPI_ISL_2774708, EPI_ISL_2774713, EPI_ISL_2774714, EPI_ISL_2775128 | | | |
| see above | Mapmygenome | CSIR-Centre for Cellular and Molecular Biology-INSACOG | ; Amareshwar Vodapalli; Ara Sreenivas; Archana Bharadwaj Siva; B Himasri; Divya Tej Sowpati; Jandhyala Sai Krishna; Karthik Bharadwaj Tallapaka; Lamuk Zaveri; Onkar Kulkarni; Payel Mukherjee; Priya Nurkurthy; Rakesh K Mishra; Shreekant Verma; Sofia Banu; Sumedha Avadhanula; Tulasi Nagabandi; Valli Nagalakshmi Undamatla; Vidhyadhari Methuku |
| EPI_ISL_2842101, EPI_ISL_2842132, EPI_ISL_2842133, EPI_ISL_2842171, EPI_ISL_2842182, EPI_ISL_2842210, EPI_ISL_2842211, EPI_ISL_2842213, EPI_ISL_2842246, EPI_ISL_2842328, EPI_ISL_2842347, EPI_ISL_2842355, EPI_ISL_2842396 | | | |
| see above | NIMHANS | INSACOG-KA, NIMHANS | Ananthapadmanabha Kotambail; Anita S Desai; Anson Kunjumon George; Chetan G K; Chitra Pattabiraman; Darshan Sreenivas; Ellango Ramasamy; Gautham Arunachal Udupi; Mahesh Kumar.C.S; Sony Sharma; V Ravi |
| EPI_ISL_2811888 | National Centre For Cell Science | National Centre For Cell Science – INSACOG | Ajay Pillai; Dhiraj Paul; Girish R Nair; Harshada Ghode; INSACOG Consortium team; Kalpesh Chandrakant Bavkar; Kartik Jayant Deopujari; Manoj Kumar Bhat; Mitali Inamdar; Shivang P. Bhanushali; Yogesh Shouche |
| EPI_ISL_2821077, EPI_ISL_2821078, EPI_ISL_2821079, EPI_ISL_2821080 | National Institute of High Security Animal Diseases | National Institute of High Security Animal Diseases | Anamika Mishra; Ashutosh Aasdev; Ashwin A Raut; Chandan K Dubey; Dipesh K Nayak; Meghna Tripathi; Naveen Kumar; Pradeep N Gandhale; Sandeep Bhatia; Sushil Tripathi; Vijendra P Singh |
| EPI_ISL_2842118, EPI_ISL_2842120, EPI_ISL_2842127, EPI_ISL_2842167, EPI_ISL_2842168, EPI_ISL_2842178, EPI_ISL_2842248, EPI_ISL_2842249, EPI_ISL_2842253, EPI_ISL_2842285, EPI_ISL_2842289, EPI_ISL_2842314, EPI_ISL_2842374, EPI_ISL_2842397, EPI_ISL_2842437, EPI_ISL_2842440, EPI_ISL_2842441, EPI_ISL_2842444, EPI_ISL_2842448, EPI_ISL_2842452, EPI_ISL_2842455 | | | |
| see above | RAILWAY HOSPITAL | INSACOG-KA, NIMHANS | Ananthapadmanabha Kotambail; Anita S Desai; Anson Kunjumon George; Chetan G K; Chitra Pattabiraman; Darshan Sreenivas; Ellango Ramasamy; Gautham Arunachal Udupi; Mahesh Kumar.C.S; Sony Sharma; V Ravi |
| EPI_ISL_2842392 | SRI JAYADEVA INSTITUTE OF CARDIOLOGY | INSACOG-KA, NIMHANS | Ananthapadmanabha Kotambail; Anita S Desai; Anson Kunjumon George; Chetan G K; Chitra Pattabiraman; Darshan Sreenivas; Ellango Ramasamy; Gautham Arunachal Udupi; Mahesh Kumar.C.S; Sony Sharma; V Ravi |
| EPI_ISL_2842458, EPI_ISL_2842460, EPI_ISL_2842465, EPI_ISL_2842466 | VICTORIA HOSPITAL | INSACOG-KA, NIMHANS | Ananthapadmanabha Kotambail; Anita S Desai; Anson Kunjumon George; Chetan G K; Chitra Pattabiraman; Darshan Sreenivas; Ellango Ramasamy; Gautham Arunachal Udupi; Mahesh Kumar.C.S; Sony Sharma; V Ravi |
| EPI_ISL_2842134, EPI_ISL_2842135, EPI_ISL_2842144 | VRDL VIJAYAPURA | INSACOG-KA, NIMHANS | Ananthapadmanabha Kotambail; Anita S Desai; Anson Kunjumon George; Chetan G K; Chitra Pattabiraman; Darshan Sreenivas; Ellango Ramasamy; Gautham Arunachal Udupi; Mahesh Kumar.C.S; Sony Sharma; V Ravi |

We gratefully acknowledge the following Authors from the Originating laboratories responsible for obtaining the specimens, as well as the Submitting laboratories where the genome data were generated and shared via GISAID, on which this research is based.

All Submitters of data may be contacted directly via www.gisaid.org

Authors are sorted alphabetically.

| Accession ID | Originating Laboratory | Submitting Laboratory | Authors |
|---|---|---|---|
| EPI_ISL_2231661, EPI_ISL_2231833 | AP SSO | CSIR-Centre for Cellular and Molecular Biology-INSACOG | Amareshwar Vodapalli; Ara Sreenivas; Archana Bharadwaj Siva; B Himasri; Blessy B John; Divya Tej Sowpati; Karthik Bharadwaj Tallapaka; Lamuk Zaveri; Payel Mukherjee; Rakesh K Mishra; Sharath Chandra Thota; Shreekant Verma; Sofia Banu; Tulasi Nagabandi; Valli Nagalakshmi Undamatla; Viswagithe S L |
| EPI_ISL_2426250 | All India Institute of Medical Sciences, Delhi | CSIR Institute of Genomics and Integrative Biology | Abhinav Jain; Afra Shamnath; Anjali Bajaj; Arvinden VR; Bani Jolly; COVID CBNAAT CORE GROUP; Kiran Bala; Mercy Rophina; Mohamed Imran; Mohit Kumar Divakar; Nayer Jamshed; Praveen Aggarwal; Rahul C. Bhoyar; Rama Chaudhry; Randeep Guleria; Ritu Gupta; Sridhar Sivasubbu; Subrata Sinha; Urvashi B Singh; Vigneshwar Senthivel; Vinod Scaria |
| EPI_ISL_2304119 | Armed Forces Medical College Pune | National Centre For Cell Science – INSACOG | Ajay Pillai; Dhiraj Paul; INSACOG Consortium team; Manoj Kumar Bhat; Mitali Inamdar; Mohak P Gujare; Shivang P. Bhanushali; Yogesh Shouche |
| EPI_ISL_2400383 | BJ Medical College and Civil Hospital, Ahmedabad | Gujarat Biotechnology Research Centre | Chaitanya Joshi; Dinesh Kumar; Janvi Raval; Madhvi Joshi; Nitesh Shah; Nitin Savaliya; Pranay Shah; Ramesh Pandit; Sonal Sharma; Twinkle Soni; Umang Mishra; Zarna Patel; Zuber Saiyed |
| EPI_ISL_2342865 | BRLSABVM Government Medical College, Rajnandgaon, Chhattisgarh | Institute of Life Sciences - INSACOG | Ajay Parida; Amol M. Kanampalliwar; Arup Ghosh; Atimukta Jha; INSACOG Consortium; Omprakash Shiriwas; Punit Prasad; Rajeeb Swain; Rupesh Dash; Safal Walia; Sana Fatma; Shifu Aggarwal; Sunil K. Raghav |
| EPI_ISL_1838377, EPI_ISL_1838380, EPI_ISL_1838545, EPI_ISL_1838552, EPI_ISL_1838553, EPI_ISL_1838655, EPI_ISL_1838772, EPI_ISL_2157045, EPI_ISL_2157048, EPI_ISL_2373285, EPI_ISL_2774861 | | | |
| see above | CSIR-Centre for Cellular and Molecular Biology | CSIR-Centre for Cellular and Molecular Biology-INSACOG | Amareshwar Vodapalli; Ara Sreenivas; Archana Bharadwaj Siva; B Himasri; Blessy B John; Divya Tej Sowpati; Jandhyala Sai Krishna; Karthik Bharadwaj Tallapaka; Lamuk Zaveri; Onkar Kulkarni; Payel Mukherjee; Priya Nurkurthy; Rakesh K Mishra; Sharath Chandra Thota; Shreekant Verma; Sofia Banu; Sumedha Avadhanula; Tulasi Nagabandi; Valli Nagalakshmi Undamatla; Vidhyadhari Methuku; Viswagithe S L |
| EPI_ISL_2441377, EPI_ISL_2441378 | CSIR-National Environmental Engineering Research Institute | CSIR-Centre for Cellular and Molecular Biology-INSACOG | Amareshwar Vodapalli; Ara Sreenivas; Archana Bharadwaj Siva; B Himasri; Divya Tej Sowpati; Karthik Bharadwaj Tallapaka; Krishna Khairnar; Lamuk Zaveri; Onkar Kulkarni; Payel Mukherjee; Priya Nurkurthy; Rakesh K Mishra; Shreekant Verma; Sofia Banu; Sumedha Avadhanula; Tulasi Nagabandi; Valli Nagalakshmi Undamatla; Vidhyadhari Methuku |
| EPI_ISL_1663495 | Capital hospital, Bhubaneswar | Institute of Life Sciences - INSACOG | Ajay Parida; Amol M. Kanampalliwar; Arup Ghosh; Atimukta Jha; INSACOG Consortium; Punit Prasad; Rajeeb Swain; Rupesh Dash; Safal Walia; Shifu Aggarwal; Sunil K. Raghav |
| EPI_ISL_2724202 | Dr S raju, Director of public heath and preventive medicine | inStem NCBS – INSACOG | Uma Ramakrishnan Dasaradhi Palakodeti Aswin SaiNarain |
| EPI_ISL_2600959, EPI_ISL_2600961, EPI_ISL_2600963 | Hematopathology Laboratory, ACTREC, TMC | Hematopathology Laboratory, ACTREC, TMC | ACTREC; Hematopathology Laboratory |
| EPI_ISL_2555665, EPI_ISL_2555669, EPI_ISL_2555673, EPI_ISL_2555693, EPI_ISL_2555699, EPI_ISL_2555702, EPI_ISL_2555712, EPI_ISL_2555718, EPI_ISL_2555719, EPI_ISL_2555722, EPI_ISL_2555723, EPI_ISL_2555727, EPI_ISL_2555729, EPI_ISL_2555730 | | | |
| see above | ICMR-National Institute of Virology | NCDC Delhi, Biotechnology Division | A Walimbe; A.Awhale; A.Titkare; B.Apoorva; G.Divekar; H.Kengale; Hema Gogia; Hemlata Lall; K P.Shinde; K.Iyengar; K.Korabu; Kalaiarasan Ponnusamy; M L Choudhary; M.Das; Mahesh S Dhar; Manoj K Singh; Meena Datta; Partha Rakshit; Preeti Madan; Priyanka Singh; R.Verma; Radhakrishnan V. S; Robin Marwal; S Jadhav; S.Bhorekar; S.Jadhav; S.Shelkande; Sandhya Kabra; Sujeet K Singh; T.Sanjeevi; U.Saha; Uma Sharma; V Malik; V.Autude; V.Vipat; Z. Sayyed |
| EPI_ISL_1703659, EPI_ISL_1703672, EPI_ISL_1703689, EPI_ISL_1703819, EPI_ISL_1703842, EPI_ISL_1703843, EPI_ISL_1703860, EPI_ISL_1703866, EPI_ISL_1703891, EPI_ISL_1704392, EPI_ISL_1704407, EPI_ISL_1704466, EPI_ISL_1704467, EPI_ISL_1704471, EPI_ISL_1704472, EPI_ISL_1704474, EPI_ISL_1704519, EPI_ISL_1704520, EPI_ISL_1704526, EPI_ISL_1704531, EPI_ISL_1704535, EPI_ISL_1704537, EPI_ISL_1704541, EPI_ISL_1704623, EPI_ISL_1928408, EPI_ISL_1928411, EPI_ISL_1928427, EPI_ISL_1928430, EPI_ISL_1928432, EPI_ISL_1928444, EPI_ISL_1928459, EPI_ISL_1928510, EPI_ISL_1928524, EPI_ISL_1970498, EPI_ISL_1970532, EPI_ISL_2131539, EPI_ISL_2546036, EPI_ISL_2546076, EPI_ISL_2546101, EPI_ISL_2546120, EPI_ISL_2546162, EPI_ISL_2546212, EPI_ISL_2546283, EPI_ISL_2546284, EPI_ISL_2546288, EPI_ISL_2546316, EPI_ISL_2546317, EPI_ISL_2546325, EPI_ISL_2546331, EPI_ISL_2546333, EPI_ISL_2546339, EPI_ISL_2546345, EPI_ISL_2546346, EPI_ISL_2546347, EPI_ISL_2546372, EPI_ISL_2546391, EPI_ISL_2546392, EPI_ISL_2546395, EPI_ISL_2546402, EPI_ISL_2546404, EPI_ISL_2546405, EPI_ISL_2546406, EPI_ISL_2546423, EPI_ISL_2546428, EPI_ISL_2546429, EPI_ISL_2546430, EPI_ISL_2546440, EPI_ISL_2546442, EPI_ISL_2546478, EPI_ISL_2546482, EPI_ISL_2546483, EPI_ISL_2546485, EPI_ISL_2546701, EPI_ISL_2546862 | | | |
| see above | ICMR-National Institute of Virology - INSACOG | NIV Influenza | Dr. Varsha Potdar; Dr. Varsha Potdar and NIC Team |
| EPI_ISL_2878727, EPI_ISL_2878878, EPI_ISL_2878881, EPI_ISL_2878898, EPI_ISL_2878930, EPI_ISL_2878982, EPI_ISL_2878986, EPI_ISL_2878994, EPI_ISL_2878995, EPI_ISL_2878997, EPI_ISL_2879001, EPI_ISL_2879002, EPI_ISL_2879006, EPI_ISL_2879011, EPI_ISL_2879086, EPI_ISL_2879087, EPI_ISL_2879088, EPI_ISL_2879143, EPI_ISL_2879145, EPI_ISL_2879158, EPI_ISL_2879310, EPI_ISL_2879316, EPI_ISL_2879323, EPI_ISL_2879325, EPI_ISL_2879389, EPI_ISL_2879414, EPI_ISL_2879648, EPI_ISL_2880056, EPI_ISL_2880796, EPI_ISL_2880808, EPI_ISL_2880830, EPI_ISL_2881025 | | | |
| see above | INSACOG Surveillance | INSACOG at CSIR Institute of Genomics and Integrative Biology | INSACOG |
| EPI_ISL_2433266 | INSACOG-Tripura | National Institute of Biomedical Genomics – INSACOG | Apurba Sarkar; Arindam Maitra; Nidhan Kumar Biswas; Saikat Majumder; Saumitra Das; Sreedhar Chinnaswamy; Tapan Majumdar |
| EPI_ISL_2132715, EPI_ISL_2132718 | INSACOG-WB | National Institute of Biomedical Genomics – INSACOG | Ajay Chakraborti; Arindam Maitra; Bhaswati Bandyopadhyay; Nidhan Kumar Biswas; Saumitra Das; Sreedhar Chinnaswamy; Tamal Ghosh |
| EPI_ISL_2497905 | Indian Council of Medical Research-National Institute of Virology, Microbial Containment Complex | Indian Council of Medical Research-National Institute of Virology, Microbial Containment Complex | Pragya D Yadav |
| EPI_ISL_1663546 | MGM Medical College, Jamshedpur | Institute of Life Sciences - INSACOG | Ajay Parida; Amol M. Kanampalliwar; Arup Ghosh; Atimukta Jha; INSACOG Consortium; Punit Prasad; Rajeeb Swain; Rupesh Dash; Safal Walia; Shifu Aggarwal; Sunil K. Raghav |
| EPI_ISL_2373311 | Mapmygenome | CSIR-Centre for Cellular and Molecular Biology-INSACOG | Amareshwar Vodapalli; Ara Sreenivas; Archana Bharadwaj Siva; B Himasri; Blessy B John; Divya Tej Sowpati; Karthik Bharadwaj Tallapaka; Lamuk Zaveri; Payel Mukherjee; Rakesh K Mishra; Sharath Chandra Thota; Shreekant Verma; Sofia Banu; Tulasi Nagabandi; Valli Nagalakshmi Undamatla; Viswagithe S L |
| EPI_ISL_3006801, EPI_ISL_3006802 | Medini Rai Medical College and Hospital, Medininagar, Palamu | Institute of Life Sciences - INSACOG | Ajay Parida; Amol M. Kanampalliwar; Arup Ghosh; Atimukta Jha; INSACOG Consortium; Omprakash Shiriwas; Punit Prasad; Rajeeb Swain; Rupesh Dash; Safal Walia; Sana Fatma; Shifu Aggarwal; Sunil K. Raghav |
| EPI_ISL_1939891 | NCCS | inStem NCBS - INSACOG | Uma Ramakrishnan Dasaradhi Palakodeti Aswin SaiNarain |
| EPI_ISL_1663380, EPI_ISL_1663409, EPI_ISL_1663418, EPI_ISL_1663421, EPI_ISL_2342916, EPI_ISL_2342925, EPI_ISL_2342933, EPI_ISL_2342949, EPI_ISL_2342962, EPI_ISL_2342965 | | | |
| see above | NCCS, Pune | Institute of Life Sciences - INSACOG | Ajay Parida; Amol M. Kanampalliwar; Arup Ghosh; Atimukta Jha; INSACOG Consortium; Omprakash Shiriwas; Punit Prasad; Rajeeb Swain; Rupesh Dash; Safal Walia; Sana Fatma; Shifu Aggarwal; Sunil K. Raghav |
| EPI_ISL_3162117 | NHL Municipal Medical College, Ahmedbad | Gujarat Biotechnology Research Centre | Chaitanya Joshi; Dinesh Kumar; Janvi Raval; Jayshri Pethani; Madhvi Joshi; Nitesh Shah; Nitin Savaliya; Nitin Shukla; Ramesh Pandit; Sonal Sharma; Twinkle Soni; Umang Mishra; Zarna Patel; Zuber Saiyed |
| EPI_ISL_1415101, EPI_ISL_1415164, EPI_ISL_1415165, EPI_ISL_1415172, EPI_ISL_1415181, EPI_ISL_1415203, EPI_ISL_1415233, EPI_ISL_1415276, EPI_ISL_1415277, EPI_ISL_1415278, EPI_ISL_1415286, EPI_ISL_1415317, EPI_ISL_1415318, EPI_ISL_1415319, EPI_ISL_1415356, EPI_ISL_1415357, EPI_ISL_1415358, EPI_ISL_1415386, EPI_ISL_1415387, EPI_ISL_2162387, EPI_ISL_2162400, EPI_ISL_2162418, EPI_ISL_2162467, EPI_ISL_2304130, EPI_ISL_2332720, EPI_ISL_2332785 | | | |
| see above | National Centre For Cell Science | National Centre For Cell Science – INSACOG | Ajay Pillai; Dhiraj Paul; INSACOG Consortium team; Manoj Kumar Bhat; Mitali Inamdar; Mohak P Gujare; Shivang P. Bhanushali; Sonal Manik Chavan; Yogesh Shouche; Yogesh Shouche. |
| EPI_ISL_2382991, EPI_ISL_2382993 | National Centre For Cell Science | National Centre For Cell Science-INSACOG | Ajay Pillai; Dhiraj Paul; INSACOG Consortium team; Manoj Kumar Bhat; Mitali Inamdar; Mohak P Gujare; Shivang P. Bhanushali; Yogesh Shouche |
| EPI_ISL_2107031, EPI_ISL_2107058 | National Centre For Cell Science – INSACOG | National Centre For Cell Science | Ajay Pillai; Dhiraj Paul; INSACOG Consortium team; Manoj Kumar Bhat; Mitali Inamdar; Mohak P Gujare; Shivang P. Bhanushali; Yogesh Shouche |
| EPI_ISL_2502084, EPI_ISL_2502086, EPI_ISL_2502177, EPI_ISL_2502225, EPI_ISL_2502228, EPI_ISL_2502242, EPI_ISL_2502276, EPI_ISL_2502292, EPI_ISL_2502302, EPI_ISL_2504327, EPI_ISL_2504337, EPI_ISL_2504338, EPI_ISL_2504916, EPI_ISL_2555595, EPI_ISL_2555597, EPI_ISL_2555791, EPI_ISL_2555864, EPI_ISL_2555919, EPI_ISL_2555935, EPI_ISL_2555939, EPI_ISL_2556009, EPI_ISL_2556051, EPI_ISL_2556278, EPI_ISL_2556516, EPI_ISL_2556750, EPI_ISL_2556860 | | | |
| see above | National Centre for Disease Control (NCDC) Biotechnology Division, Delhi | NCDC Delhi, Biotechnology Division INSACOG | Hema Gogia; Hemlata Lall; Kalaiarasan Ponnusamy; Mahesh S Dhar; Manoj K Singh; Meena Datta; Partha Rakshit; Preeti Madan; Priyanka Singh; Radhakrishnan V. S; Robin Marwal; Sandhya Kabra; Sujeet K Singh; Uma Sharma |
| EPI_ISL_2967210 | Sri Ganga Ram Hospital, New Delhi, India | NCDC Delhi, Biotechnology Division | Hema Gogia; Hemlata Lall; Kalaiarasan Ponnusamy; Mahesh S Dhar; Manoj K Singh; Meena Datta; Partha Rakshit; Preeti Madan; Priyanka Singh; Radhakrishnan V. S; Robin Marwal; Sandhya Kabra; Sujeet K Singh; Uma Sharma |
| EPI_ISL_2023383 | The Foundation for Medical Research | The Foundation for Medical Research | Ambreen Shaikh; Ayan Mandal; Grishma Patel; Kayzad Nilgiriwala; Nerges Mistry; Smriti Vaswani; Swapneil Parikh |
| EPI_ISL_1838050, EPI_ISL_1838052, EPI_ISL_1838057, EPI_ISL_1838097, EPI_ISL_1838131, EPI_ISL_1838345 | The National Centre for Cell Science | CSIR-Centre for Cellular and Molecular Biology-INSACOG | Ajay Pillai; Amareshwar Vodapalli; Ara Sreenivas; Archana Bharadwaj Siva; B Himasri; Blessy B John; Dhiraj Paul; Divya Tej Sowpati; INSACOG Consortium team; Karthik Bharadwaj Tallapaka; Lamuk Zaveri; Manoj Kumar Bhat; Mitali Inamdar; Mohak P Gujare; Onkar Kulkarni; Payel Mukherjee; Rakesh K Mishra; Sharath Chandra Thota; Shivang P. Bhanushali; Shreekant Verma; Sofia Banu; Sonal Manik Chavan; Tulasi Nagabandi; Valli Nagalakshmi Undamatla; Viswagithe S L; Yogesh Shouche |

We gratefully acknowledge the following Authors from the Originating laboratories responsible for obtaining the specimens, as well as the Submitting laboratories where the genome data were generated and shared via GISAID, on which this research is based.

All Submitters of data may be contacted directly via www.gisaid.org

Authors are sorted alphabetically.

| Accession ID | Originating Laboratory | Submitting Laboratory | Authors |
|---|---|---|---|
| EPI_ISL_3050652, EPI_ISL_3050653, EPI_ISL_3050654, EPI_ISL_3050655, EPI_ISL_3050656, EPI_ISL_3050657, EPI_ISL_3050658, EPI_ISL_3050659, EPI_ISL_3050660, EPI_ISL_3050661, EPI_ISL_3050662, EPI_ISL_3050663, EPI_ISL_3050664, EPI_ISL_3050665, EPI_ISL_3050666, EPI_ISL_3050668, EPI_ISL_3050669, EPI_ISL_3050670, EPI_ISL_3050672, EPI_ISL_3050673, EPI_ISL_3050675, EPI_ISL_3050676, EPI_ISL_3050677, EPI_ISL_3050678, EPI_ISL_3050679, EPI_ISL_3050680, EPI_ISL_3050681, EPI_ISL_3050683, EPI_ISL_3050684, EPI_ISL_3050685, EPI_ISL_3050686, EPI_ISL_3050687, EPI_ISL_3050688, EPI_ISL_3050689, EPI_ISL_3050690, EPI_ISL_3050692, EPI_ISL_3050693, EPI_ISL_3050694, EPI_ISL_3050696, EPI_ISL_3050698, EPI_ISL_3050699, EPI_ISL_3050702, EPI_ISL_3050703, EPI_ISL_3050705, EPI_ISL_3050706, EPI_ISL_3050707, EPI_ISL_3050708, EPI_ISL_3050709, EPI_ISL_3050710, EPI_ISL_3050711, EPI_ISL_3050712, EPI_ISL_3050713, EPI_ISL_3050714, EPI_ISL_3050715, EPI_ISL_3050716, EPI_ISL_3050717, EPI_ISL_3050719, EPI_ISL_3050721, EPI_ISL_3050722, EPI_ISL_3050724, EPI_ISL_3050726, EPI_ISL_3050727, EPI_ISL_3050728, EPI_ISL_3050729, EPI_ISL_3050730, EPI_ISL_3050731 | | | |
| see above | Dirección regional de salud del Callao (DIRESA-CALLAO) | Centro de Investigaciones Tecnológicas, Biomédicas y Medioambientales (CITBM) | B; Huaman; J. Alarcon; M. Cuellar; M. Ramirez; M. Sovero |
| EPI_ISL_2791488, EPI_ISL_2791489, EPI_ISL_2791490 | Gatuario Veterinary Clinic | Faculty of Veterinary Medicine, Universidad Peruana Cayetano Heredia | Francesca Schiaffino |
| EPI_ISL_3020167, EPI_ISL_3020169, EPI_ISL_3020172, EPI_ISL_3020176, EPI_ISL_3020179, EPI_ISL_3020186, EPI_ISL_3020197, EPI_ISL_3020205, EPI_ISL_3020210, EPI_ISL_3020212, EPI_ISL_3020216, EPI_ISL_3020220, EPI_ISL_3020223 | | | |
| see above | Hospital Carlos Alberto Seguín Escobedo - EsSalud | Laboratorio de Genómica Microbiana, Universidad Peruana Cayetano Heredia | Alejandra Dávila-Barclay; Diego Cuicapuza; Guillermo Salvatierra; Janet Huancachoque; Luis González; Pablo Tsukayama; Pedro E. Romero; Pool Marcos |
| EPI_ISL_3020243, EPI_ISL_3020246, EPI_ISL_3020247, EPI_ISL_3020248, EPI_ISL_3020249, EPI_ISL_3020250, EPI_ISL_3020251, EPI_ISL_3020252, EPI_ISL_3020253, EPI_ISL_3020254, EPI_ISL_3020255, EPI_ISL_3020256, EPI_ISL_3020257, EPI_ISL_3020258, EPI_ISL_3020259, EPI_ISL_3020260, EPI_ISL_3020261, EPI_ISL_3020262, EPI_ISL_3020263, EPI_ISL_3020264, EPI_ISL_3020265, EPI_ISL_3020266, EPI_ISL_3020267, EPI_ISL_3020268, EPI_ISL_3020269, EPI_ISL_3020270, EPI_ISL_3020271, EPI_ISL_3020272 | | | |
| see above | Instituto de Medicina Tropical Alexander Von Humboldt, Sede Cusco | Laboratorio de Genómica Microbiana, Universidad Peruana Cayetano Heredia | Alejandra Dávila-Barclay; Diego Cuicapuza; Guillermo Salvatierra; Janet Huancachoque; Luis González; Pablo Tsukayama; Pedro E. Romero; Pool Marcos |
| EPI_ISL_3020225, EPI_ISL_3020226, EPI_ISL_3020227, EPI_ISL_3020228, EPI_ISL_3020229, EPI_ISL_3020230, EPI_ISL_3020231, EPI_ISL_3020232, EPI_ISL_3020233, EPI_ISL_3020234, EPI_ISL_3020235, EPI_ISL_3020236, EPI_ISL_3020238, EPI_ISL_3020239, EPI_ISL_3020240, EPI_ISL_3020241, EPI_ISL_3020242, EPI_ISL_3020274, EPI_ISL_3020275, EPI_ISL_3020276, EPI_ISL_3020277, EPI_ISL_3020278 | | | |
| see above | Laboratorio de Genómica Microbiana, Universidad Peruana Cayetano Heredia | Laboratorio de Genómica Microbiana, Universidad Peruana Cayetano Heredia | Alejandra Dávila-Barclay; Diego Cuicapuza; Guillermo Salvatierra; Janet Huancachoque; Luis González; Pablo Tsukayama; Pedro E. Romero; Pool Marcos |
| EPI_ISL_2536706, EPI_ISL_2536707, EPI_ISL_2536708, EPI_ISL_2536709, EPI_ISL_2536710, EPI_ISL_2536711, EPI_ISL_2536712, EPI_ISL_2536713, EPI_ISL_2536714, EPI_ISL_2536717, EPI_ISL_2536718, EPI_ISL_2536719, EPI_ISL_2536720, EPI_ISL_2536721, EPI_ISL_2536722, EPI_ISL_2536723, EPI_ISL_2536725, EPI_ISL_2536726, EPI_ISL_2536727, EPI_ISL_2536728, EPI_ISL_2536729, EPI_ISL_2536730, EPI_ISL_2536731, EPI_ISL_2536732, EPI_ISL_2536733, EPI_ISL_2536734, EPI_ISL_2536735, EPI_ISL_2536736, EPI_ISL_2536737, EPI_ISL_2536738, EPI_ISL_2536739, EPI_ISL_2536740, EPI_ISL_2536742, EPI_ISL_2536743, EPI_ISL_2536744, EPI_ISL_2536745, EPI_ISL_2536746, EPI_ISL_2536747, EPI_ISL_2536748, EPI_ISL_2536749, EPI_ISL_2536750, EPI_ISL_2536751, EPI_ISL_2536752, EPI_ISL_2536753, EPI_ISL_2536754, EPI_ISL_2536755, EPI_ISL_2536756, EPI_ISL_2536757, EPI_ISL_2536758, EPI_ISL_2536759, EPI_ISL_2536760, EPI_ISL_2536761, EPI_ISL_2536762, EPI_ISL_2536764, EPI_ISL_2536765, EPI_ISL_2536766, EPI_ISL_2536767, EPI_ISL_2536768, EPI_ISL_2536769, EPI_ISL_2536770, EPI_ISL_2536771, EPI_ISL_2536772, EPI_ISL_2536773, EPI_ISL_2536774, EPI_ISL_2536775, EPI_ISL_2536776, EPI_ISL_2536777, EPI_ISL_2536778, EPI_ISL_2536779, EPI_ISL_2536780, EPI_ISL_2536781, EPI_ISL_2536783, EPI_ISL_2536784, EPI_ISL_2536785, EPI_ISL_2536786, EPI_ISL_2536787, EPI_ISL_2536788, EPI_ISL_2536789, EPI_ISL_2536791, EPI_ISL_2536792, EPI_ISL_2536793, EPI_ISL_2536794, EPI_ISL_2536795, EPI_ISL_2536796, EPI_ISL_2536797, EPI_ISL_2536798, EPI_ISL_2536799, EPI_ISL_2536800, EPI_ISL_2536801, EPI_ISL_2536803, EPI_ISL_2536804, EPI_ISL_2536805, EPI_ISL_2536806, EPI_ISL_2536807, EPI_ISL_2536808, EPI_ISL_2536809, EPI_ISL_2536810, EPI_ISL_2536811, EPI_ISL_2536812, EPI_ISL_2536813, EPI_ISL_2536814, EPI_ISL_2536815, EPI_ISL_2536816, EPI_ISL_2536817, EPI_ISL_2536818, EPI_ISL_2536819, EPI_ISL_2536820, EPI_ISL_2536821, EPI_ISL_2536822, EPI_ISL_2536823, EPI_ISL_2536824, EPI_ISL_2536825, EPI_ISL_2536826, EPI_ISL_2536827, EPI_ISL_2536828, EPI_ISL_2536831, EPI_ISL_2536832, EPI_ISL_2536833, EPI_ISL_2536834, EPI_ISL_2536835, EPI_ISL_2536836, EPI_ISL_2536837, EPI_ISL_2536838, EPI_ISL_2536839, EPI_ISL_2536841, EPI_ISL_2536842, EPI_ISL_2536843, EPI_ISL_2536844, EPI_ISL_2536845, EPI_ISL_2536846, EPI_ISL_2536847, EPI_ISL_2536849, EPI_ISL_2536850, EPI_ISL_2536851, EPI_ISL_2536852, EPI_ISL_2536853, EPI_ISL_2536855, EPI_ISL_2536856, EPI_ISL_2536857, EPI_ISL_2536858, EPI_ISL_2536859, EPI_ISL_2536860, EPI_ISL_2536861, EPI_ISL_2536862, EPI_ISL_2536863, EPI_ISL_2536865, EPI_ISL_2536867, EPI_ISL_2536868, EPI_ISL_2536869, EPI_ISL_2536870, EPI_ISL_2536871, EPI_ISL_2536872, EPI_ISL_2536873, EPI_ISL_2536874, EPI_ISL_2536875, EPI_ISL_2536876, EPI_ISL_2536877, EPI_ISL_2536878, EPI_ISL_2536879, EPI_ISL_2536880, EPI_ISL_2536881, EPI_ISL_2536882, EPI_ISL_2536883, EPI_ISL_2536884, EPI_ISL_2536885, EPI_ISL_2536886, EPI_ISL_2536887, EPI_ISL_2536888, EPI_ISL_2536889, EPI_ISL_2536890, EPI_ISL_2536891, EPI_ISL_2536892, EPI_ISL_2844696, EPI_ISL_2844697, EPI_ISL_2844698, EPI_ISL_2844699, EPI_ISL_2844701, EPI_ISL_2844702, EPI_ISL_2844703, EPI_ISL_2844704, EPI_ISL_2844705, EPI_ISL_2844706, EPI_ISL_2844707, EPI_ISL_2844709, EPI_ISL_2844710, EPI_ISL_2844711, EPI_ISL_2844712, EPI_ISL_2844713, EPI_ISL_2844714, EPI_ISL_2844715, EPI_ISL_2844716, EPI_ISL_2844717, EPI_ISL_2844718, EPI_ISL_2844719, EPI_ISL_2844720, EPI_ISL_2844721, EPI_ISL_2844723, EPI_ISL_2844724, EPI_ISL_2844725, EPI_ISL_2844726, EPI_ISL_2844727, EPI_ISL_2844728, EPI_ISL_2844729, EPI_ISL_2844730, EPI_ISL_2844731, EPI_ISL_2844732, EPI_ISL_2844734, EPI_ISL_2844735, EPI_ISL_2844736, EPI_ISL_2844737, EPI_ISL_2844738, EPI_ISL_2844739, EPI_ISL_2844740, EPI_ISL_2844743, EPI_ISL_2844744, EPI_ISL_2844745, EPI_ISL_2844746, EPI_ISL_2844747, EPI_ISL_2844748, EPI_ISL_2844749, EPI_ISL_2844750, EPI_ISL_2844752, EPI_ISL_2844753, EPI_ISL_2844754, EPI_ISL_2844755, EPI_ISL_2844756, EPI_ISL_2844759, EPI_ISL_2844761, EPI_ISL_2844762, EPI_ISL_2844763, EPI_ISL_2844764, EPI_ISL_2844765, EPI_ISL_2844766, EPI_ISL_2844767, EPI_ISL_2844768, EPI_ISL_2844769, EPI_ISL_2844770, EPI_ISL_2844771, EPI_ISL_2844772, EPI_ISL_2844773, EPI_ISL_2844774, EPI_ISL_2844775, EPI_ISL_2844776, EPI_ISL_2844777, EPI_ISL_2844778, EPI_ISL_2844779, EPI_ISL_2844780, EPI_ISL_2844781, EPI_ISL_2844782, EPI_ISL_2844783, EPI_ISL_2844784, EPI_ISL_2921267, EPI_ISL_2921268, EPI_ISL_2921269, EPI_ISL_2921270, EPI_ISL_2921271, EPI_ISL_2921272, EPI_ISL_2921273, EPI_ISL_2921274, EPI_ISL_2921275, EPI_ISL_2921276, EPI_ISL_2921277, EPI_ISL_2921278, EPI_ISL_2921279, EPI_ISL_2921280, EPI_ISL_2921281, EPI_ISL_2921282, EPI_ISL_2921283, EPI_ISL_2921284, EPI_ISL_2921285, EPI_ISL_2921286, EPI_ISL_2921287, EPI_ISL_2921288, EPI_ISL_2921289, EPI_ISL_2921290, EPI_ISL_2921291, EPI_ISL_2921292, EPI_ISL_2921293, EPI_ISL_2921294, EPI_ISL_2921295, EPI_ISL_2921296, EPI_ISL_2921297, EPI_ISL_2921298, EPI_ISL_2921299, EPI_ISL_2921300, EPI_ISL_2921301, EPI_ISL_2921302, EPI_ISL_2921303, EPI_ISL_2921304, EPI_ISL_2921305, EPI_ISL_2921307, EPI_ISL_2921308, EPI_ISL_2921309, EPI_ISL_2921310, EPI_ISL_2921311, EPI_ISL_2921312, EPI_ISL_2921313, EPI_ISL_2921314, EPI_ISL_2921315, EPI_ISL_2921316, EPI_ISL_2921317, EPI_ISL_2921318, EPI_ISL_2921319, EPI_ISL_2921320, EPI_ISL_2921321, EPI_ISL_2921322, EPI_ISL_2921323, EPI_ISL_2921324, EPI_ISL_2921325, EPI_ISL_2921326, EPI_ISL_2921327, EPI_ISL_2921328, EPI_ISL_2921329, EPI_ISL_2921330, EPI_ISL_2921331, EPI_ISL_2921332, EPI_ISL_2921333, EPI_ISL_2921334, EPI_ISL_2921335, EPI_ISL_2921336, EPI_ISL_2921337, EPI_ISL_2921338, EPI_ISL_2921339, EPI_ISL_2921340, EPI_ISL_2921341, EPI_ISL_2921342, EPI_ISL_2921343, EPI_ISL_2921344, EPI_ISL_2921345, EPI_ISL_2921346, EPI_ISL_2921347, EPI_ISL_2921348, EPI_ISL_2921349, EPI_ISL_2921350, EPI_ISL_2921351, EPI_ISL_2921352, EPI_ISL_2921353, EPI_ISL_2921354, EPI_ISL_2921355, EPI_ISL_2921356, EPI_ISL_2921357, EPI_ISL_2921358, EPI_ISL_2921359, EPI_ISL_2921360, EPI_ISL_2921361, EPI_ISL_2921362, EPI_ISL_2921363, EPI_ISL_2921364, EPI_ISL_2921365, EPI_ISL_2921366, EPI_ISL_2921367, EPI_ISL_2921368, EPI_ISL_2921369, EPI_ISL_2921370, EPI_ISL_2921371, EPI_ISL_2921372, EPI_ISL_2921374, EPI_ISL_2921375, EPI_ISL_2921376, EPI_ISL_2921377, EPI_ISL_2921378, EPI_ISL_2921379, EPI_ISL_2921380, EPI_ISL_2921381, EPI_ISL_2921382, EPI_ISL_2921383, EPI_ISL_2921384, EPI_ISL_2921385, EPI_ISL_2921386, EPI_ISL_2921387, EPI_ISL_2921388, EPI_ISL_2921389, EPI_ISL_2921390, EPI_ISL_2921391, EPI_ISL_2921392, EPI_ISL_2921393, EPI_ISL_2921394, EPI_ISL_2921395, EPI_ISL_2921396, EPI_ISL_2921397, EPI_ISL_2921398, EPI_ISL_2921399, EPI_ISL_2921401, EPI_ISL_2921402, EPI_ISL_2921403, EPI_ISL_2921404, EPI_ISL_2921405, EPI_ISL_2921406, EPI_ISL_2921407, EPI_ISL_2921408, EPI_ISL_2921409, EPI_ISL_2921410, EPI_ISL_2921411, EPI_ISL_2921412, EPI_ISL_2921413, EPI_ISL_2921414, EPI_ISL_2921416, EPI_ISL_2921432, EPI_ISL_2921439, EPI_ISL_2921440, EPI_ISL_2921441, EPI_ISL_2921442, EPI_ISL_2921443, EPI_ISL_2921444, EPI_ISL_2921445, EPI_ISL_2921446, EPI_ISL_2921447, EPI_ISL_2921448, EPI_ISL_2921451, EPI_ISL_2921452, EPI_ISL_2921453, EPI_ISL_2921454, EPI_ISL_2921455, EPI_ISL_2921456, EPI_ISL_2921457, EPI_ISL_2921458, EPI_ISL_2921459, EPI_ISL_2921460, EPI_ISL_2921462, EPI_ISL_2921463, EPI_ISL_2921464, EPI_ISL_2921465, EPI_ISL_2921466, EPI_ISL_2921467, EPI_ISL_2921468, EPI_ISL_2921469, EPI_ISL_2921470, EPI_ISL_2921471, EPI_ISL_2921472, EPI_ISL_2921473, EPI_ISL_2921474, EPI_ISL_2921475, EPI_ISL_2921476, EPI_ISL_2921477, EPI_ISL_2921479, EPI_ISL_2921480, EPI_ISL_2921481, EPI_ISL_2921482, EPI_ISL_2921483, EPI_ISL_2921484, EPI_ISL_2921485, EPI_ISL_2921486, EPI_ISL_2921487, EPI_ISL_2921488, EPI_ISL_2921489, EPI_ISL_2921490, EPI_ISL_2921491, EPI_ISL_2921492, EPI_ISL_2921493, EPI_ISL_2921494, EPI_ISL_2921495, EPI_ISL_2921496, EPI_ISL_2921497, EPI_ISL_2921498, EPI_ISL_2921499, EPI_ISL_2921500, EPI_ISL_2921501, EPI_ISL_2921502, EPI_ISL_2921503, EPI_ISL_2921504, EPI_ISL_2921505, EPI_ISL_2921506, EPI_ISL_2921507, EPI_ISL_2921508, EPI_ISL_2921509, EPI_ISL_2921510, EPI_ISL_2921511, EPI_ISL_2921512, EPI_ISL_2921513, EPI_ISL_2921514, EPI_ISL_2921515, EPI_ISL_2921516, EPI_ISL_2921517, EPI_ISL_2921518, EPI_ISL_2921519, EPI_ISL_2921520, EPI_ISL_2921521, EPI_ISL_2921522, EPI_ISL_2921523, EPI_ISL_2921524, EPI_ISL_2921525, EPI_ISL_2921526, EPI_ISL_2921527, EPI_ISL_2921528, EPI_ISL_2921529, EPI_ISL_2921530, EPI_ISL_2921531, EPI_ISL_2921532, EPI_ISL_3023377, EPI_ISL_3023379, EPI_ISL_3023380, EPI_ISL_3023381, EPI_ISL_3023382, EPI_ISL_3023383, EPI_ISL_3023384, EPI_ISL_3023385, EPI_ISL_3023386, EPI_ISL_3023387, EPI_ISL_3023388, EPI_ISL_3023390, EPI_ISL_3023391, EPI_ISL_3023394, EPI_ISL_3023400, EPI_ISL_3023403, EPI_ISL_3023404, EPI_ISL_3023405, EPI_ISL_3023420, EPI_ISL_3023423, EPI_ISL_3023426, EPI_ISL_3023428, EPI_ISL_3023429, EPI_ISL_3023430, EPI_ISL_3023431, EPI_ISL_3023432, EPI_ISL_3023433, EPI_ISL_3023434, EPI_ISL_3023435, EPI_ISL_3023436, EPI_ISL_3023437, EPI_ISL_3023438, EPI_ISL_3023441, EPI_ISL_3023442, EPI_ISL_3023445, EPI_ISL_3023447, EPI_ISL_3023448, EPI_ISL_3023449, EPI_ISL_3023450, EPI_ISL_3023451, EPI_ISL_3023452, EPI_ISL_3023453, EPI_ISL_3023454, EPI_ISL_3023455, EPI_ISL_3023457, EPI_ISL_3023458, EPI_ISL_3023460, EPI_ISL_3023461, EPI_ISL_3023462, EPI_ISL_3023463, EPI_ISL_3023465, EPI_ISL_3023466, EPI_ISL_3023467, EPI_ISL_3023468, EPI_ISL_3023469, EPI_ISL_3023470, EPI_ISL_3023471, EPI_ISL_3023472, EPI_ISL_3023473, EPI_ISL_3023475, EPI_ISL_3023476, EPI_ISL_3023480, EPI_ISL_3023481, EPI_ISL_3023482, EPI_ISL_3023483, EPI_ISL_3023484, EPI_ISL_3023485, EPI_ISL_3023486, EPI_ISL_3023487, EPI_ISL_3023488, EPI_ISL_3023490, EPI_ISL_3023491, EPI_ISL_3023492, EPI_ISL_3023494, EPI_ISL_3023495, EPI_ISL_3023496, EPI_ISL_3023497, EPI_ISL_3023498, EPI_ISL_3023499, EPI_ISL_3023500, EPI_ISL_3023503, EPI_ISL_3023505, EPI_ISL_3023506, EPI_ISL_3023507, EPI_ISL_3023508, EPI_ISL_3023509, EPI_ISL_3023510, EPI_ISL_3023511, EPI_ISL_3023512, EPI_ISL_3023513, EPI_ISL_3023514, EPI_ISL_3023516, EPI_ISL_3023517, EPI_ISL_3023518, EPI_ISL_3023519, EPI_ISL_3023520, EPI_ISL_3023521, EPI_ISL_3023522, EPI_ISL_3023525, EPI_ISL_3023527, EPI_ISL_3023528, EPI_ISL_3023529, EPI_ISL_3023530, EPI_ISL_3023532, EPI_ISL_3023534, EPI_ISL_3023535, EPI_ISL_3023537, EPI_ISL_3023538, EPI_ISL_3023539, EPI_ISL_3023540, EPI_ISL_3023541, EPI_ISL_3023542, EPI_ISL_3023543, EPI_ISL_3023544, EPI_ISL_3023545, EPI_ISL_3023546, EPI_ISL_3023547, EPI_ISL_3023548, EPI_ISL_3023549, EPI_ISL_3023550, EPI_ISL_3023551, EPI_ISL_3023552, EPI_ISL_3023553, EPI_ISL_3023554, EPI_ISL_3023555, EPI_ISL_3023556, EPI_ISL_3023557, EPI_ISL_3023558, EPI_ISL_3023559, EPI_ISL_3023560, EPI_ISL_3023561, EPI_ISL_3023563, EPI_ISL_3023564, EPI_ISL_3023565, EPI_ISL_3023569, EPI_ISL_3023570, EPI_ISL_3023571, EPI_ISL_3023573, EPI_ISL_3023574, EPI_ISL_3023576, EPI_ISL_3023577, EPI_ISL_3023578, EPI_ISL_3023580, EPI_ISL_3023583, EPI_ISL_3023586, EPI_ISL_3023589, EPI_ISL_3023591, EPI_ISL_3023592, EPI_ISL_3023594, EPI_ISL_3023595, EPI_ISL_3023596, EPI_ISL_3023597, EPI_ISL_3023598, EPI_ISL_3023600, EPI_ISL_3023601, EPI_ISL_3023602, EPI_ISL_3023603, EPI_ISL_3023604, EPI_ISL_3023605, EPI_ISL_3023609, EPI_ISL_3023610, EPI_ISL_3023611, EPI_ISL_3023612, EPI_ISL_3023613, EPI_ISL_3023614, EPI_ISL_3023615, EPI_ISL_3023617, EPI_ISL_3023619, EPI_ISL_3023620, EPI_ISL_3023621, EPI_ISL_3023622, EPI_ISL_3023623, EPI_ISL_3023624, EPI_ISL_3023625, EPI_ISL_3023626, EPI_ISL_3023627, EPI_ISL_3023628, EPI_ISL_3023629, EPI_ISL_3023630, EPI_ISL_3023633, EPI_ISL_3023634, EPI_ISL_3023635, EPI_ISL_3023636, EPI_ISL_3023638, EPI_ISL_3023639, EPI_ISL_3023641, EPI_ISL_3023643, EPI_ISL_3023644, EPI_ISL_3023646, EPI_ISL_3023647 | | | |
| see above | Laboratorio de Referencial Nacional de Virus Respiratorios | Laboratorio de Referencial Nacional de Virus Respiratorios | Carlos Padilla Rojas; Henri Bailon Calderon; Iris Silva Molina; Joseph Huayra Niquen; Lely Solari Zerpa; Luis Barcena Flores; Marco Galarza Perez; Nancy Rojas Serrano; Omar Caceres Rey; Orson Mestanza Millones; Priscila Lope Pari; Sandra Morales Ruiz; Steve Acedo Lazo; Veronica Hurtado Vela |

We gratefully acknowledge the following Authors from the Originating laboratories responsible for obtaining the specimens, as well as the Submitting laboratories where the genome data were generated and shared via GISAID, on which this research is based.

All Submitters of data may be contacted directly via www.gisaid.org

Authors are sorted alphabetically.

| Accession ID | Originating Laboratory | Submitting Laboratory | Authors |
|---|---|---|---|
| EPI_ISL_2756427, EPI_ISL_2756428, EPI_ISL_2756429, EPI_ISL_2756430, EPI_ISL_2756431, EPI_ISL_2756433, EPI_ISL_2756434, EPI_ISL_2756435, EPI_ISL_2756436, EPI_ISL_2756440, EPI_ISL_2756441, EPI_ISL_2756443, EPI_ISL_2756445, EPI_ISL_2756446, EPI_ISL_2756447, EPI_ISL_2756449, EPI_ISL_2756454, EPI_ISL_2756455, EPI_ISL_2756456, EPI_ISL_2756457, EPI_ISL_2756459, EPI_ISL_2756461, EPI_ISL_2756462, EPI_ISL_2756463, EPI_ISL_2756464, EPI_ISL_2756465, EPI_ISL_2756466, EPI_ISL_2756467, EPI_ISL_2756468, EPI_ISL_2756470, EPI_ISL_2756471, EPI_ISL_2756472, EPI_ISL_2756473, EPI_ISL_2756474, EPI_ISL_2756475, EPI_ISL_2756476, EPI_ISL_2756479, EPI_ISL_2756480, EPI_ISL_2756482, EPI_ISL_2756485, EPI_ISL_2756486, EPI_ISL_2756487, EPI_ISL_2756488, EPI_ISL_2756489, EPI_ISL_2756490, EPI_ISL_2756491, EPI_ISL_2756492, EPI_ISL_2756493 | | | |
| see above | Instituto Adolfo Lutz Central | Instituto Adolfo Lutz, Interdiciplinary Procedures Center, Strategic Laboratory | Caio Vinicius Dias Lopes; Claudia Regina Gonçalves; Claudio Tavares Sacchi; Erica Valessa Ramos Gomes; Karoline Rodrigues Campos; Leonardo Jose Tadeu de Araujo |
| EPI_ISL_3418671, EPI_ISL_3418673, EPI_ISL_3418674, EPI_ISL_3418676, EPI_ISL_3418678, EPI_ISL_3418679, EPI_ISL_3418680, EPI_ISL_3418681, EPI_ISL_3418682, EPI_ISL_3418683, EPI_ISL_3418684 | | | |
| see above | Instituto Carlos Chagas - ICC, FIOCRUZ Parana | Instituto Carlos Chagas - ICC, FIOCRUZ Parana | A.A.; A.M.; A.R.; Aguiar; Albrecht, L.; Alves; Avila; Balsanelli, E.; Becker, G.; Blanes, L.; Dallagiovanna, B.; Debur; E.M.; F.K.; F.O.; Faoro, H.; Graef, T.; H.G.; I.N.; L.G.; L.R.; M.M.; M.O.; Marchini; Md.C.; Morello; Nardeli; Oliveira; P.C.; Passetti, F.; Pedrosa; Resende; Riediger; S.C.; Schemberger; Suzukawa; V.A.; Zanette, D.; de Baura; de Souza; dos Santos |
| EPI_ISL_2758988, EPI_ISL_2758992, EPI_ISL_2758994, EPI_ISL_2758995, EPI_ISL_2758997, EPI_ISL_2759001, EPI_ISL_2759002, EPI_ISL_2759004, EPI_ISL_2759006, EPI_ISL_2759011, EPI_ISL_2759012, EPI_ISL_2759014, EPI_ISL_2759016, EPI_ISL_2759017, EPI_ISL_2759020, EPI_ISL_2759021, EPI_ISL_2759022, EPI_ISL_2759024, EPI_ISL_2759025, EPI_ISL_2759026, EPI_ISL_2759028, EPI_ISL_2759033, EPI_ISL_2759035, EPI_ISL_2759037, EPI_ISL_2759041, EPI_ISL_2759045, EPI_ISL_2759046, EPI_ISL_2759047, EPI_ISL_2759050, EPI_ISL_2759051, EPI_ISL_2759052, EPI_ISL_2759054, EPI_ISL_2759056, EPI_ISL_2759059, EPI_ISL_2759061, EPI_ISL_2759063, EPI_ISL_2759065, EPI_ISL_2759066, EPI_ISL_2759068, EPI_ISL_2775421 | | | |
| see above | Instituto de Biologia Molecular do Paraná (LAC) | Instituto Carlos Chagas - Fiocruz | Alessandra De Melo Aguiar; Andreia Akemi Suzukawa; Andréa Rodrigues Ávila; Bruno Dallagiovanna; Dalila Zanette; Eduardo Balsanelli; Emanuel Maltempi de Souza; Fabio Passetti; Fabricio Klerynton Marchini; Fábio de Oliveira Pedrosa; Guilherme Becker; Helisson Faoro; Hellen Geremias dos Santos; Irina Nastassja Riediger; Letusa Albrecht; Lucas Blanes; Luis Gustavo Morello; Lysangela Ronalte Alves; Maria do Carmo Debur; Mauro de Medeiros Oliveira; Michelle Orane Schemberger; Paola Cristina Resende; Sheila Cristina Nardeli; Tiago Gräf; Valter Antônio de Baura |
| EPI_ISL_2761936 | LACEN do Mato Grosso do Sul | Instituto Adolfo Lutz, Interdiciplinary Procedures Center, Strategic Laboratory | Caio Vinicius Dias Lopes; Claudia Regina Gonçalves; Claudio Tavares Sacchi; Erica Valessa Ramos Gomes; Karoline Rodrigues Campos; Leonardo Jose Tadeu de Araujo |
| EPI_ISL_2758940, EPI_ISL_2758942, EPI_ISL_2758943, EPI_ISL_2758945, EPI_ISL_2758947, EPI_ISL_2758948, EPI_ISL_2758949, EPI_ISL_2758950, EPI_ISL_2758951, EPI_ISL_2758952, EPI_ISL_2758953, EPI_ISL_2758957, EPI_ISL_2758958, EPI_ISL_2758961, EPI_ISL_2758962, EPI_ISL_2758963, EPI_ISL_2758964, EPI_ISL_2758965, EPI_ISL_2758966, EPI_ISL_2758967, EPI_ISL_2758968, EPI_ISL_2758970, EPI_ISL_2758971, EPI_ISL_2758972, EPI_ISL_2758975, EPI_ISL_2758979, EPI_ISL_2758981, EPI_ISL_2758982, EPI_ISL_2758989, EPI_ISL_2758991, EPI_ISL_2758993, EPI_ISL_2758996, EPI_ISL_2758998, EPI_ISL_2758999, EPI_ISL_2759003, EPI_ISL_2759007, EPI_ISL_2759009, EPI_ISL_2759010, EPI_ISL_2759013, EPI_ISL_2759015, EPI_ISL_2759018, EPI_ISL_2759019, EPI_ISL_2759023, EPI_ISL_2759027, EPI_ISL_2759029, EPI_ISL_2759031, EPI_ISL_2759032, EPI_ISL_2759036, EPI_ISL_2759038, EPI_ISL_2759039, EPI_ISL_2759042, EPI_ISL_2759043, EPI_ISL_2759044, EPI_ISL_2759053, EPI_ISL_2759055, EPI_ISL_2759058, EPI_ISL_2759060, EPI_ISL_2759064, EPI_ISL_2759067, EPI_ISL_2759069, EPI_ISL_2775390, EPI_ISL_2775391, EPI_ISL_2775393, EPI_ISL_2775394, EPI_ISL_2775395, EPI_ISL_2775399, EPI_ISL_2775400, EPI_ISL_2775403, EPI_ISL_2775404, EPI_ISL_2775405, EPI_ISL_2775407, EPI_ISL_2775410, EPI_ISL_2775411, EPI_ISL_2775412, EPI_ISL_2775413, EPI_ISL_2775414, EPI_ISL_2775415, EPI_ISL_2775416, EPI_ISL_2775417, EPI_ISL_2775418, EPI_ISL_2775422, EPI_ISL_2775487 | | | |
| see above | Laboratorio Central de Saude Publica do Estado do Parana (Instituto de Biologia Molecular do Paraná (LAC)EN-PR) | Instituto Carlos Chagas - Fiocruz | Alessandra De Melo Aguiar; Andreia Akemi Suzukawa; Andréa Rodrigues Ávila; Bruno Dallagiovanna; Dalila Zanette; Eduardo Balsanelli; Emanuel Maltempi de Souza; Fabio Passetti; Fabricio Klerynton Marchini; Fábio de Oliveira Pedrosa; Guilherme Becker; Helisson Faoro; Hellen Geremias dos Santos; Irina Nastassja Riediger; Letusa Albrecht; Lucas Blanes; Luis Gustavo Morello; Lysangela Ronalte Alves; Maria do Carmo Debur; Mauro de Medeiros Oliveira; Michelle Orane Schemberger; Paola Cristina Resende; Sheila Cristina Nardeli; Tiago Gräf; Valter Antônio de Baura |
| EPI_ISL_2777363, EPI_ISL_2777364, EPI_ISL_2777373, EPI_ISL_2777374, EPI_ISL_2777375, EPI_ISL_2777376, EPI_ISL_2777377, EPI_ISL_2777378, EPI_ISL_2777380, EPI_ISL_2777381, EPI_ISL_2777382, EPI_ISL_2777383, EPI_ISL_2777384, EPI_ISL_2777385, EPI_ISL_2777388, EPI_ISL_2777393, EPI_ISL_2777395, EPI_ISL_2777396, EPI_ISL_2777397, EPI_ISL_2777398, EPI_ISL_2777399, EPI_ISL_2777400, EPI_ISL_2777401, EPI_ISL_2777402, EPI_ISL_2777403, EPI_ISL_2777404, EPI_ISL_2777405, EPI_ISL_2777406, EPI_ISL_2777407, EPI_ISL_2777408, EPI_ISL_2777409, EPI_ISL_2777410, EPI_ISL_2777411, EPI_ISL_2777412, EPI_ISL_2777413, EPI_ISL_2777414, EPI_ISL_2777415, EPI_ISL_2777416, EPI_ISL_2777417, EPI_ISL_2777418, EPI_ISL_2777419, EPI_ISL_2777420, EPI_ISL_2777421, EPI_ISL_2777422, EPI_ISL_2777423, EPI_ISL_2777424, EPI_ISL_2777425, EPI_ISL_2777426, EPI_ISL_2777427, EPI_ISL_2777428, EPI_ISL_2777429, EPI_ISL_2777430, EPI_ISL_2777431, EPI_ISL_2777432, EPI_ISL_2777435, EPI_ISL_2777453, EPI_ISL_2777454, EPI_ISL_2777455, EPI_ISL_2777456, EPI_ISL_2777457, EPI_ISL_2777458, EPI_ISL_2777459, EPI_ISL_2777460, EPI_ISL_2777461, EPI_ISL_2777462, EPI_ISL_2777463, EPI_ISL_2777464, EPI_ISL_2777465, EPI_ISL_2777466, EPI_ISL_2777467, EPI_ISL_2777468, EPI_ISL_2777469, EPI_ISL_2777470, EPI_ISL_2777471, EPI_ISL_2777472, EPI_ISL_2777473, EPI_ISL_2777474, EPI_ISL_2777475, EPI_ISL_2777476, EPI_ISL_2777477, EPI_ISL_2777478, EPI_ISL_2777479, EPI_ISL_2777480, EPI_ISL_2777481, EPI_ISL_2777482, EPI_ISL_2777483, EPI_ISL_2777484, EPI_ISL_2777485, EPI_ISL_2777486, EPI_ISL_2777487, EPI_ISL_2777488, EPI_ISL_2777489, EPI_ISL_2777491, EPI_ISL_2777492, EPI_ISL_2777493, EPI_ISL_2777494, EPI_ISL_2777995, EPI_ISL_2777996, EPI_ISL_2777997, EPI_ISL_2777998, EPI_ISL_2777999, EPI_ISL_2778000, EPI_ISL_2778001, EPI_ISL_2778003 | | | |
| see above | Laboratorio de Ecologia de Doencas Transmissiveis na Amazonia, Instituto Leonidas e Maria Deane - Fiocruz Amazonia | Laboratorio de Ecologia de Doencas Transmissiveis na Amazonia, Instituto Leonidas e Maria Deane - Fiocruz Amazonia | André Corado; Debora Duarte; Felipe Naveca; Fernanda Nascimento; George Silva; Karina Pessoa; Luciana Gonçalves; Maria Júlia Brandão; Matilde Mejía; Michele Jesus; Valdinete Nascimento; Victor Souza; Ágatha Costa |
| EPI_ISL_2777235, EPI_ISL_2777236, EPI_ISL_2777237, EPI_ISL_2777238, EPI_ISL_2777239, EPI_ISL_2777240, EPI_ISL_2777241, EPI_ISL_2777242, EPI_ISL_2777243, EPI_ISL_2777244, EPI_ISL_2777245, EPI_ISL_2777246, EPI_ISL_2777247, EPI_ISL_2777248, EPI_ISL_2777249, EPI_ISL_2777250, EPI_ISL_2777251, EPI_ISL_2777252, EPI_ISL_2777253, EPI_ISL_2777254, EPI_ISL_2777255, EPI_ISL_2777256, EPI_ISL_2777257, EPI_ISL_2777258, EPI_ISL_2777259, EPI_ISL_2777260, EPI_ISL_2777261, EPI_ISL_2777262, EPI_ISL_2777263, EPI_ISL_2777264, EPI_ISL_2777265, EPI_ISL_2777266, EPI_ISL_2777267, EPI_ISL_2777268, EPI_ISL_2777269, EPI_ISL_2777270, EPI_ISL_2777271, EPI_ISL_2777272, EPI_ISL_2777273, EPI_ISL_2777274, EPI_ISL_2777275, EPI_ISL_2777276, EPI_ISL_2777277, EPI_ISL_2777278, EPI_ISL_2777279, EPI_ISL_2777280, EPI_ISL_2777281, EPI_ISL_2777282, EPI_ISL_2777283, EPI_ISL_2777284, EPI_ISL_2777285, EPI_ISL_2777290, EPI_ISL_2777292, EPI_ISL_2777293, EPI_ISL_2777295, EPI_ISL_2777297, EPI_ISL_2777299, EPI_ISL_2777301, EPI_ISL_2777302, EPI_ISL_2777304, EPI_ISL_2777306, EPI_ISL_2777309, EPI_ISL_2777313, EPI_ISL_2777318, EPI_ISL_2777320, EPI_ISL_2777321, EPI_ISL_2777324, EPI_ISL_2777327, EPI_ISL_2777328, EPI_ISL_2777330, EPI_ISL_2777331, EPI_ISL_2777333, EPI_ISL_2777334, EPI_ISL_2777340, EPI_ISL_2777345, EPI_ISL_2777346, EPI_ISL_2777347, EPI_ISL_2777348, EPI_ISL_2777356, EPI_ISL_2777357, EPI_ISL_2777358, EPI_ISL_2777495, EPI_ISL_2777496, EPI_ISL_2777498, EPI_ISL_2777499, EPI_ISL_2777501, EPI_ISL_2777502, EPI_ISL_2777503, EPI_ISL_2777504, EPI_ISL_2777505, EPI_ISL_2777506, EPI_ISL_2777507, EPI_ISL_2777508, EPI_ISL_2777509, EPI_ISL_2777510, EPI_ISL_2777511, EPI_ISL_2777512, EPI_ISL_2777513, EPI_ISL_2777515, EPI_ISL_2777516, EPI_ISL_2777517, EPI_ISL_2777518, EPI_ISL_2777519, EPI_ISL_2777520, EPI_ISL_2777521, EPI_ISL_2777522, EPI_ISL_2777524, EPI_ISL_2777525, EPI_ISL_2777526, EPI_ISL_2777527, EPI_ISL_2777528, EPI_ISL_2777530, EPI_ISL_2777532, EPI_ISL_2777533, EPI_ISL_2777534, EPI_ISL_2777535, EPI_ISL_2777536, EPI_ISL_2777537, EPI_ISL_2777538, EPI_ISL_2777539, EPI_ISL_2777540, EPI_ISL_2777541, EPI_ISL_2777542, EPI_ISL_2777543, EPI_ISL_2777544, EPI_ISL_2777545, EPI_ISL_2777546, EPI_ISL_2777549, EPI_ISL_2777551, EPI_ISL_2777552, EPI_ISL_2777553, EPI_ISL_2777554, EPI_ISL_2777556, EPI_ISL_2777557, EPI_ISL_2777558, EPI_ISL_2777559, EPI_ISL_2777560, EPI_ISL_2777561, EPI_ISL_2777562, EPI_ISL_2777563, EPI_ISL_2777564, EPI_ISL_2777565, EPI_ISL_2777566, EPI_ISL_2777567, EPI_ISL_2777569, EPI_ISL_2777570, EPI_ISL_2777571, EPI_ISL_2777572, EPI_ISL_2777573, EPI_ISL_2777574, EPI_ISL_2777575, EPI_ISL_2777576, EPI_ISL_2777581, EPI_ISL_2777584, EPI_ISL_2777585, EPI_ISL_2777586, EPI_ISL_2777588, EPI_ISL_2777589, EPI_ISL_2777591, EPI_ISL_2777593, EPI_ISL_2777599, EPI_ISL_2777600, EPI_ISL_2777601, EPI_ISL_2777603, EPI_ISL_2777604, EPI_ISL_2777605, EPI_ISL_2777606, EPI_ISL_2777609, EPI_ISL_2777612, EPI_ISL_2777615, EPI_ISL_2777616, EPI_ISL_2777618, EPI_ISL_2777620, EPI_ISL_2777621, EPI_ISL_2777622, EPI_ISL_2777624, EPI_ISL_2777627, EPI_ISL_2777628, EPI_ISL_2777630, EPI_ISL_2777631, EPI_ISL_2777632, EPI_ISL_2777634, EPI_ISL_2777635, EPI_ISL_2777636, EPI_ISL_2777637, EPI_ISL_2777638, EPI_ISL_2777639, EPI_ISL_2777641, EPI_ISL_2777642, EPI_ISL_2777643, EPI_ISL_2777645, EPI_ISL_2777646, EPI_ISL_2777647, EPI_ISL_2777651, EPI_ISL_2777654, EPI_ISL_2777655, EPI_ISL_2777657, EPI_ISL_2777658, EPI_ISL_2777659, EPI_ISL_2777662, EPI_ISL_2777668, EPI_ISL_2777669, EPI_ISL_2777670, EPI_ISL_2777675, EPI_ISL_2777676, EPI_ISL_2777683, EPI_ISL_2777686, EPI_ISL_2777687, EPI_ISL_2777690, EPI_ISL_2777691, EPI_ISL_2777692, EPI_ISL_2777693, EPI_ISL_2777696, EPI_ISL_2777697, EPI_ISL_2777698, EPI_ISL_2777699, EPI_ISL_2777700, EPI_ISL_2777701, EPI_ISL_2777702, EPI_ISL_2777703, EPI_ISL_2777704, EPI_ISL_2777705, EPI_ISL_2777706, EPI_ISL_2777707, EPI_ISL_2777708, EPI_ISL_2777709, EPI_ISL_2777710, EPI_ISL_2777711, EPI_ISL_2777713, EPI_ISL_2777714, EPI_ISL_2777715, EPI_ISL_2777716, EPI_ISL_2777717, EPI_ISL_2777718, EPI_ISL_2777719, EPI_ISL_2777720, EPI_ISL_2777721, EPI_ISL_2777722, EPI_ISL_2777723, EPI_ISL_2777724, EPI_ISL_2777725, EPI_ISL_2777726, EPI_ISL_2777727, EPI_ISL_2777728, EPI_ISL_2777729, EPI_ISL_2777730, EPI_ISL_2777731, EPI_ISL_2777732, EPI_ISL_2777733, EPI_ISL_2777734, EPI_ISL_2777735, EPI_ISL_2777736, EPI_ISL_2777737, EPI_ISL_2777738, EPI_ISL_2777739, EPI_ISL_2777740, EPI_ISL_2777742, EPI_ISL_2777743, EPI_ISL_2777744, EPI_ISL_2777745, EPI_ISL_2777746, EPI_ISL_2777747, EPI_ISL_2777748, EPI_ISL_2777749, EPI_ISL_2777750, EPI_ISL_2777751, EPI_ISL_2777752, EPI_ISL_2777754, EPI_ISL_2777755, EPI_ISL_2777757, EPI_ISL_2777758, EPI_ISL_2777759, EPI_ISL_2777762, EPI_ISL_2777763, EPI_ISL_2777764, EPI_ISL_2777765, EPI_ISL_2777766, EPI_ISL_2777767, EPI_ISL_2777768, EPI_ISL_2777769, EPI_ISL_2777771, EPI_ISL_2777772, EPI_ISL_2777775, EPI_ISL_2777776, EPI_ISL_2777779, EPI_ISL_2777780, EPI_ISL_2777781, EPI_ISL_2777782, EPI_ISL_2777783, EPI_ISL_2777784, EPI_ISL_2777785, EPI_ISL_2777786, EPI_ISL_2777787, EPI_ISL_2777788, EPI_ISL_2777789, EPI_ISL_2777790, EPI_ISL_2777791, EPI_ISL_2777792, EPI_ISL_2777793, EPI_ISL_2777794, EPI_ISL_2777795, EPI_ISL_2777796, EPI_ISL_2777797, EPI_ISL_2777798, EPI_ISL_2777799, EPI_ISL_2777800, EPI_ISL_2777801, EPI_ISL_2777802, EPI_ISL_2777803, EPI_ISL_2777804, EPI_ISL_2777805, EPI_ISL_2777806, EPI_ISL_2777807, EPI_ISL_2777808, EPI_ISL_2777811, EPI_ISL_2777812, EPI_ISL_2777815, EPI_ISL_2777816, EPI_ISL_2777817, EPI_ISL_2777818, EPI_ISL_2777819, EPI_ISL_2777820, EPI_ISL_2777821, EPI_ISL_2777823, EPI_ISL_2777827, EPI_ISL_2777828, EPI_ISL_2777829, EPI_ISL_2777830, EPI_ISL_2777833, EPI_ISL_2777834, EPI_ISL_2777837, EPI_ISL_2777838, EPI_ISL_2777839, EPI_ISL_2777840, EPI_ISL_2777841, EPI_ISL_2777842, EPI_ISL_2777843, EPI_ISL_2777844, EPI_ISL_2777845, EPI_ISL_2777847, EPI_ISL_2777848, EPI_ISL_2777849, EPI_ISL_2777850, EPI_ISL_2777855, EPI_ISL_2777857, EPI_ISL_2777858, EPI_ISL_2777859, EPI_ISL_2777861, EPI_ISL_2777862, EPI_ISL_2777863, EPI_ISL_2777864, EPI_ISL_2777867, EPI_ISL_2777868, EPI_ISL_2777870, EPI_ISL_2777871, EPI_ISL_2777872, EPI_ISL_2777873, EPI_ISL_2777874, EPI_ISL_2777875, EPI_ISL_2777876, EPI_ISL_2777877, EPI_ISL_2777878, EPI_ISL_2777879, EPI_ISL_2777880, EPI_ISL_2777881, EPI_ISL_2777882, EPI_ISL_2777883, EPI_ISL_2777884, EPI_ISL_2777885, EPI_ISL_2777886, EPI_ISL_2777888, EPI_ISL_2777889, EPI_ISL_2777892, EPI_ISL_2777894, EPI_ISL_2777896, EPI_ISL_2777897, EPI_ISL_2777898, EPI_ISL_2777899, EPI_ISL_2777900, EPI_ISL_2777903, EPI_ISL_2777911, EPI_ISL_2777913, EPI_ISL_2777915, EPI_ISL_2777916, EPI_ISL_2777917, EPI_ISL_2777919, EPI_ISL_2777922, EPI_ISL_2777923, EPI_ISL_2777932, EPI_ISL_2777934, EPI_ISL_2777935, EPI_ISL_2777936, EPI_ISL_2777938, EPI_ISL_2777939, EPI_ISL_2777940, EPI_ISL_2777948, EPI_ISL_2777950, EPI_ISL_2777952, EPI_ISL_2777954, EPI_ISL_2777955, EPI_ISL_2777962, EPI_ISL_2777963, EPI_ISL_2777964, EPI_ISL_2777965, EPI_ISL_2777980, EPI_ISL_2777986, EPI_ISL_2777987, EPI_ISL_2777988, EPI_ISL_2777992, EPI_ISL_2777993, EPI_ISL_2777994, EPI_ISL_2778002, EPI_ISL_2778004, EPI_ISL_2778005, EPI_ISL_2778006, EPI_ISL_2778007 | | | |
| see above | Laboratório Central de Saúde Pública do Amazonas - LACEN-AM | Laboratorio de Ecologia de Doencas Transmissiveis na Amazonia, Instituto Leonidas e Maria Deane - Fiocruz Amazonia | André Corado; Debora Duarte; Felipe Naveca; Fernanda Nascimento; George Silva; Karina Pessoa; Luciana Gonçalves; Maria Júlia Brandão; Matilde Mejía; Michele Jesus; Valdinete Nascimento; Victor Souza; Ágatha Costa |
| EPI_ISL_2790137, EPI_ISL_2790138, EPI_ISL_2790139, EPI_ISL_2790140, EPI_ISL_2790142, EPI_ISL_2790143, EPI_ISL_2790144, EPI_ISL_2790145, EPI_ISL_2790146, EPI_ISL_2790147, EPI_ISL_2790148, EPI_ISL_2790149, EPI_ISL_2790150, EPI_ISL_2790151, EPI_ISL_2790152, EPI_ISL_2790153, EPI_ISL_2790154, EPI_ISL_2790155, EPI_ISL_2790156, EPI_ISL_2790157, EPI_ISL_2790158, EPI_ISL_2790159, EPI_ISL_2790160, EPI_ISL_2790161, EPI_ISL_2790162, EPI_ISL_2790163, EPI_ISL_2790164, EPI_ISL_2790165, EPI_ISL_2790166, EPI_ISL_2790167, EPI_ISL_2790168, EPI_ISL_2790169, EPI_ISL_2790170, EPI_ISL_2790171, EPI_ISL_2790172, EPI_ISL_2790173, EPI_ISL_2790174 | | | |
| see above | Laboratório de Biotecnologia | Laboratory of Respiratory | Alice Sampaio Rocha; Ana Carolina Mendonca; Anna Carolina Paixao; Elisa Cavalcante Pereira; Felipe Allan da Silva Costa; Fernando Motta; Jayme Augusto de Souza Neto; Leonardo Nazario de Moraes; Luciana Appolinario; Marilda Siqueira on behalf of the Fiocruz COVID-19 Genomic Surveillance Network; Paola Resende; Patricia Akemi Assato; Rejane Maria Tommasini; Renata Serrano Lopes; Taina Venas |

| | | | |
|---|---|---|---|
| | Aplicada (LBA) - Laboratório de Biologia Molecular - Hospital das Clínicas, Faculdade de Medicina de Botucatu, Departamento de Bioprocessos e Biotecnologia - Faculdade de Ciências Agronômicas, UNESP – Botucatu/SP | Viruses and Measles, Oswaldo Cruz Institute, FIOCRUZ | |
| EPI_ISL_2758715, EPI_ISL_2758723, EPI_ISL_2758728, EPI_ISL_2758732, EPI_ISL_2758738, EPI_ISL_2758741 | UEL | IPEC Guarapuava | NAPI-Genômica (Novos Arranjo de Pesquisa e Inovação em Genômica): Ademar Dantas da Cunha Júnior Adriano Ferrasa Adriano Mondini Aldo Przybysz Alessandra Lourenço Cecchini Armani Alex Sandro Jorge Alexandra Ivo de Medeiros Alexandre Maller Aline Cristina Batista Rodrigues Johann Ana Lucia Ferreira Ana Marisa Fusco Almeida Anderson Joel Martino Andrade André Luís Laforga Vanzela Andrea Duarte Doetzer Andrea Name Colado Simao Andressa Pereira de Souza Anelisa Ramão Angelica Beate Winter Boldt Anna Hermínia Castro Gomes de Amorim Anna Silvia Penteado Setti da Rocha Antonio Camilo da Silva Filho Antonio Stabelini Neto Arthur Hirata Bertachi Barbara Mendes Paz Chao Betty Cristiane Kuhn Bruno Ambrozio Galindo Bruno Ribeiro Cruz Camilla Reginatto De Pierri Carla Fredrichsen Moya Araujo Carla Fredrichsen Moya Araujo Carlos Alberto Oliveira de Biagi Junior Carlos Augusto Nassar Carlos Eduardo Buss Carlos Gilberto Carlotti Junior Carlos Henrique Schneider Carolina Panis Carolina Weigert Galvão Caroline de Jesus Coelho Donha Caroline Guisantes de Salvo Toni Caryna Eurich Mazur Catiuscie Cabreira da Silva Tortorella Celso F. D. Doliveira Cesar Luiz Boguszewski Christiane Pienna Soares Chung Man Chin Claudia Moro Cleverson Busso Cristiane Cominetti Daiane Priscila Simão-Silva Dalila Luciola Zanette Daniel de Paula Daniel de Paula Daniel Rech Daniela Fiori Gradia Daniela Pretti da Cunha Tirapelli Daniela Viganó Zanoti Jeronymo Daniele Ukan Danielle Malheiros Ferreira Danielle Venturini Deborah Catharine de Assis Leite Deivid Calebe de Souza Dennis Armando Bertolini Edenir Inez Pamero Edna Maria Vissoci Reiche Edson Roberto Arpini Miguel Eduardo José de Almeida Araújo Eliana Carolina Vespero Eliandro Reis Tavares Elza Kimura Grimshaw Emanuel Maltempi de Souza Emanuele Cristina Gustani Buss Émerson Carraro Emiliana Cristina Melo ENILze Maria de Souza Fonseca Ribeiro Enilze Maria de Souza Fonseca Ribeiro Erika Izumi Erika Seki Kioshima Cotica Evani Marques Pereira Fabio Negretti Fábio Rodrigues Ferreira Seiva Felipe Dunin dos Santos Felipe Tuon Fernanda Andreia Rosa Fernanda Cestaro Prado Cortez Fernanda Ivanski Fernanda Maris Peria Flavia Regina Oliveira de Barros Franciele Aní Caovilla Follador Franciele Mara Lucca Zanardo Bohm Francinete Ramos Campos Fulviana Silva Nishiyama GABRIEL RIBEIRO CORDEIRO Gabriela Datsch Bennemann Gisele Santos de Oliveira Glaucio Valdameri Glauco Akelinghton Freire Vitiello Glauco Vieira Miranda Glaura Scantamburlo ALves Fernandes Guilherme Ferreira Silveira Gustavo Bianchini Porfírio Gustavo Lenci Marques Hélito Volpato Hildebrando Masshiro Nagai Huei Diana Lee Ilce Mara de Syllos Cólus Iris Rabinovich Israel Gomy Jackson Kawakami Jacques Duílio Brancher Jaime Luís Lopes Rocha Jaqueline Carvalho de Oliveira Jean Henrique da Silva Rodrigues Jean Leandro dos Santos Jeane Eliete Laguila Visentainer João Paulo Bianchi Ximenez Joaquim Manoel da Silva Jociani Ascari Joel Donazzolo Jorge Luis Maria Ruiz Jose Knopfholz José Luis da Conceição Silva José Sebastião dos Santos Joseane Carla Schabarum Juliana Cheleski Wiggers Juliana Mara Serpeloni Juliana Morini Küpper Cardoso Perseguini Karen Brajão de Oliveira Karin Braun Prado Karine Aparecida de Lima Katiany Rizzieri Caleffi Ferracioli Katiuscia de Oliveira Francisco Gabriel Kelvinson Fernandes Viana Larissa Beatriz Cossalter Larissa Danielle Bahls Pinto Laurival Antonio Vilas Boas Léia Carolina Lucio Líbero Mezzadri Neto Ligia Carla Faccin Galhardi Lirane Elize Defante Ferreto Luciana Furlaneto Maia Luciana Oliveira de Fariña Luciana Reis Azevedo Alanis Luciane Regina Cavalli Lucy Megumi Yamauchi Lioni Luis Paulo Gomes Mascarenhas Luis Paulo Gomes Mascarenhas Luís Paulo Mascarenhas Lupe Furtado Alle Lyvia Regina Biagi Silva Bertachi Mara Antonia Ramos Costa Mara L. Cordeiro Marcela Maria Birolim Marcelo Ricardo Vicari Marcia Edilaine Lopes Consolaro Marcia Holsbach Beltrame Marcia Regina Eches Perugini Marcos Abdo Arbex Marcos Pileggi MARCOS TADEU GRZELCZAK Marcus Peikriszwili Tartaruga Maria Angelica Ehara Watanabe Maria Antonia Ramos Costa Maria Claudia Gross Maria José Soares Mendes Giannini Maria Leandra Terencio Maria Lúcia Bonfleur Maria Luiza Guimarães de Oliveira Maria Luiza Petzl-Erler Mariana Abe Vicente Cavagnari Marina Kimiko Kadowaki Marise Fonseca dos Santos Marla Karine Amarante Maurício Turkiewicz Mauro Antonio Alves Castro Michel Rodrigo Zambrano Passarini Michele Potrich Michelle Orane Schemberger Milena Massumi Kozonoe Mônica Degraf Cavallin Monica Tereza Suldofski Mucio Luiz de Assis Cirino Nadia Graciele Krohn Najeh Maissar Khalil Nédia de Castilhos Ghisi Neide Tomimura Costa Neiva Leite Neyva Maria Lopes Romeiro Patrícia Amâncio da Rosa Patricia Dayane Carvalho Schaker Patricia Oehlmeyer Nassar Patrícia Savio de Araújo-Souza Patricia Silva Lucio Paulo Henrique Couto Souza Paulo Roberto Donadio Percy Nohama Quirino Alves de Lima Neto Rafael Deminice Rafael dos Santos Bezerra Raquel Alves dos Santos Renan Manozzo Galante Renata Ernlund Freitas de Macedo Rita de Cássia Garcia Simão Roberta Losi Guembarovski Roberto H. Herai Roberto Rosati Rodrigo Ferreira Rodrigo Rodrigues Matiello Rogério Neri Shinsato Rogério Pincela Mateus Rosane Aparecida Ribeiro Rosilene Fressatti Cardoso Rosilene Fressatti Cardoso Sandra Mara Guse Scós Venske Selene Elifio Esposito Sérgio Ossamu Ioshii Silvana Giuliatti Silvia Mara de Souza Halick Silvio Henrique Maia de Almeida Simone Neumann Wendt Spencer Luiz Marques Payão Stefan Wolanski Negrão Stephane Janaina de Moura Escobar Sueli Fumie Yamada Ogatta SUELI PERCIO QUINAIA Taciane Finatto Tatiana Mayumi Veiga Iriyoda Tayza Katelline Danilau Ostroski Tony Alexander Hild Valeria Valente Vanessa Nascimento Kozak Vanessa Santos Sotomaior Victor Breno Pedrosa Victoria Zeghbi Cochenski Borba Vivian Rotuno Moure Valdameri Wander Rogerio Pavanelli Weber Cláudio Francisco Nunes da Silva Willian Augusto de Melo Yohandra Reyes Torres |

We gratefully acknowledge the following Authors from the Originating laboratories responsible for obtaining the specimens, as well as the Submitting laboratories where the genome data were generated and shared via GISAID, on which this research is based.

All Submitters of data may be contacted directly via www.gisaid.org

Authors are sorted alphabetically.

| Accession ID | Originating Laboratory | Submitting Laboratory | Authors |
|---|---|---|---|
| EPI_ISL_2551525, EPI_ISL_2551526, EPI_ISL_2551527 | Belo Horizonte center-south emergency care unit - UPA-BH | Laboratório de Virologia Clínica e Molecular | Alex Fiorini; Ana Paula Salles Fernandes; Bruna Larotonda Telezynski; Danielle Bruna Leal Oliveira; Edison Luiz Durigon; Erick Gustavo Dorlass; Flavio Fonseca e Santuza Teixeira; Guilherme Pereira Scagion; Helena Perez Coelho; Hugo Sato; Karine Lima Lourenço; Luciano Matsumiya Thomazelli; Renata Peixoto; Rubens Daniel Miserani Magalhães; Tatiana Ometto |
| EPI_ISL_2698107 | Biology, UFLA, Universidade Federal de Lavras | Biology, UFLA | Barcante, J.; Cherem, J.; Fernandes, G.; Luciano, P.; Melo, D.; Pylro, V. |
| EPI_ISL_2445444 | CENTRO DE SAUDE DE IBIRAREMA | Instituto Butantan | Antonio Jorge Martins; Claudia Renata dos Santos Barros; David Schlesinger; Debora Botequio Moretti; Dimas Tadeu Covas; Elaine Cristina Marqueze; Elaine Vieira Santos; Evandra Strazza Rodrigues; Heidge Fukumasu; Jayme Augusto de Souza-Neto; José Salvatore Leister Patané; Luiz Alcantara; Luiz Lehmann Coutinho; Maria Carolina Elias; Maurício Lacerda Nogueira; Rafael dos Santos Bezerra; Raul Machado Neto; Rejane Maria Tommasini Grotto; Ricardo Haddad; Sandra Coccuzzo Sampaio Vessoni; Simone Kashima; Svetoslav Nanev Slavov; Vincent Louis Viala |
| EPI_ISL_2612346, EPI_ISL_2612350, EPI_ISL_2612360 | Centro de Infectologia Charles Mérieux/ Laboratório Rodolphe Mérieux, FUNDHACRE | Bioinformatics Laboratory / LNCC | Alessandra P Lamarca; Alexandra L Gerber; Ana Paula de C Guimarães; Ana Tereza R Vasconcelos; Andreas Stocker; Cirley Maria de Oliveira Lobato; Douglas Terra Machado; Luiz Fellype Alves de Souza; Luiz G P de Almeida; Ronaldo da Silva F Jr |
| EPI_ISL_2491690, EPI_ISL_2491692, EPI_ISL_2491719, EPI_ISL_2491720, EPI_ISL_2491764, EPI_ISL_2491765, EPI_ISL_2491766, EPI_ISL_2491767, EPI_ISL_2491768, EPI_ISL_2491769, EPI_ISL_2491770, EPI_ISL_2491771, EPI_ISL_2491773, EPI_ISL_2491774, EPI_ISL_2491775, EPI_ISL_2491776, EPI_ISL_2491777, EPI_ISL_2491778, EPI_ISL_2491779, EPI_ISL_2491781, EPI_ISL_2491782, EPI_ISL_2491783, EPI_ISL_2491784, EPI_ISL_2491785, EPI_ISL_2491786, EPI_ISL_2491787, EPI_ISL_2491788, EPI_ISL_2491789 | | | |
| see above | Centro de Pesquisa Gonçalo Moniz (CPqGM - FIOCRUZ/BA) | Laboratory of Respiratory Viruses and Measles, Oswaldo Cruz Institute, FIOCRUZ | Alice Sampaio Rocha; Ana Carolina Mendonca; Anna Carolina Paixao; Camila I. de Oliveira; Elisa Cavalcante Pereira; Fernando Motta; Luciana Appolinario; Marilda Siqueira on behalf of the Fiocruz COVID-19 Genomic Surveillance Network; Paola Resende; Renata Serrano Lopes; Ricardo Khouri; Taina Venas |
| EPI_ISL_2551534 | Hospital Infantil Menino Jesus | Laboratório de Virologia Clínica e Molecular | Alex Fiorini; Ana Paula Salles Fernandes; Bruna Larotonda Telezynski; Danielle Bruna Leal Oliveira; Edison Luiz Durigon; Erick Gustavo Dorlass; Flavio Fonseca e Santuza Teixeira; Guilherme Pereira Scagion; Helena Perez Coelho; Hugo Sato; Karine Lima Lourenço; Luciano Matsumiya Thomazelli; Renata Peixoto; Rubens Daniel Miserani Magalhães; Tatiana Ometto |
| EPI_ISL_2614548 | IAL Presidente Prudente | Instituto Adolfo Lutz, Interdiciplinary Procedures Center, Strategic Laboratory | Caio Vinicius Dias Lopes; Claudia Regina Gonçalves; Claudio Tavares Sacchi; Erica Valessa Ramos Gomes; Karoline Rodrigues Campos; Leonardo Jose Tadeu de Araujo |
| EPI_ISL_2614543 | IdenGene Medicina Diagnostica SA | Instituto Adolfo Lutz, Interdiciplinary Procedures Center, Strategic Laboratory | Caio Vinicius Dias Lopes; Claudia Regina Gonçalves; Claudio Tavares Sacchi; Erica Valessa Ramos Gomes; Karoline Rodrigues Campos; Leonardo Jose Tadeu de Araujo |
| EPI_ISL_2614513 | Instituto Adolfo Lutz Central | Instituto Adolfo Lutz, Interdiciplinary Procedures Center, Strategic Laboratory | Caio Vinicius Dias Lopes; Claudia Regina Gonçalves; Claudio Tavares Sacchi; Erica Valessa Ramos Gomes; Karoline Rodrigues Campos; Leonardo Jose Tadeu de Araujo |
| EPI_ISL_2614607 | Instituto Biologico | Instituto Adolfo Lutz, Interdiciplinary Procedures Center, Strategic Laboratory | Caio Vinicius Dias Lopes; Claudia Regina Gonçalves; Claudio Tavares Sacchi; Erica Valessa Ramos Gomes; Karoline Rodrigues Campos; Leonardo Jose Tadeu de Araujo |
| EPI_ISL_2425435, EPI_ISL_2425439, EPI_ISL_2425440, EPI_ISL_2425441 | Instituto Butantan | Instituto de Medicina Tropical de Sao Paulo | Brazil-UK Centre for Arbovirus Discovery Diagnosis Genomics and Epidemiology (CADDE) Genomic Network - Instituto de Medicina Tropical |
| EPI_ISL_2614382, EPI_ISL_2614383 | Instituto Estadual do Cerebro Paulo Niemeyer | Laboratory of Respiratory Viruses and Measles, Oswaldo Cruz Institute, FIOCRUZ | Alice Sampaio Rocha; Ana Carolina Mendonca; Anna Carolina Paixao; Carlos Azevedo; Elisa Cavalcante Pereira; Fernando Motta; Luciana Appolinario; Marilda Siqueira on behalf of the Fiocruz COVID-19 Genomic Surveillance Network; Paola Resende; Renata Serrano Lopes; Taina Venas |
| EPI_ISL_2617877 | LABCOVID_HCPA | LABRESIS_HCPA | Barth AL; Martins AF; Monteiro F; Volpato F; Wink PL; Zavascki AP |
| EPI_ISL_2488778, EPI_ISL_2488791 | LACEN - Laboratório Central de Saúde Pública de Pernambuco | Evandro Chagas Institute | A.M.; Barbagelata; E.C.; E.M.A.; Ferreira; J.A.; Junior; K.C.; L.C.; L.S.; M.C.; P.S.; Pinheiro; Santos; Silva; Sousa; Sousa Junior; W.D.C.; da Silva |
| EPI_ISL_2488777 | LACEN - Laboratório Central de Saúde Pública do Amapá | Evandro Chagas Institute | A.M.; Barbagelata; E.C.; E.M.A.; Ferreira; J.A.; Junior; K.C.; L.C.; L.S.; M.C.; P.S.; Pinheiro; Santos; Silva; Sousa; Sousa Junior; W.D.C.; da Silva |
| EPI_ISL_2488771 | LACEN - Laboratório Central de Saúde Pública do Maranhao | Evandro Chagas Institute | A.M.; Barbagelata; E.C.; E.M.A.; Ferreira; J.A.; Junior; K.C.; L.C.; L.S.; M.C.; P.S.; Pinheiro; Santos; Silva; Sousa; Sousa Junior; W.D.C.; da Silva |
| EPI_ISL_2488797, EPI_ISL_2488798 | LACEN - Laboratório Central de Saúde Pública do Rio Grande do Norte | Evandro Chagas Institute | A.M.; Barbagelata; E.C.; E.M.A.; Ferreira; J.A.; Junior; K.C.; L.C.; L.S.; M.C.; P.S.; Pinheiro; Santos; Silva; Sousa; Sousa Junior; W.D.C.; da Silva |
| EPI_ISL_2557347, EPI_ISL_2557349 | Laboratorio Central de Saude Publica do Estado de Minas Gerais (LACEN/MG) | Laboratory of Respiratory Viruses and Measles, Oswaldo Cruz Institute, FIOCRUZ | Alice Sampaio Rocha; Ana Carolina Mendonca; Andre Felipe Leal Bernardes; Anna Carolina Paixao; Elisa Cavalcante Pereira; Fernando Motta; Luciana Appolinario; Marilda Siqueira on behalf of the Fiocruz COVID-19 Genomic Surveillance Network; Paola Resende; Renata Serrano Lopes; Taina Venas |
| EPI_ISL_2645825, EPI_ISL_2645833, EPI_ISL_2645834, EPI_ISL_2645835, EPI_ISL_2645836, EPI_ISL_2645837, EPI_ISL_2645839, EPI_ISL_2645841, EPI_ISL_2645843, EPI_ISL_2645846, EPI_ISL_2645858, EPI_ISL_2645859, EPI_ISL_2645870, EPI_ISL_2645881, EPI_ISL_2645882, EPI_ISL_2645885, EPI_ISL_2645909, EPI_ISL_2645917, EPI_ISL_2645927, EPI_ISL_2645928, EPI_ISL_2645929 | | | |
| see above | Laboratorio Central de Saude Publica do Estado do Para (LACEN/PA) | Laboratory of Respiratory Viruses and Measles, Oswaldo Cruz Institute, FIOCRUZ | Alice Sampaio Rocha; Ana Carolina Mendonca; Anna Carolina Paixao; Elisa Cavalcante Pereira; Fernando Motta; Luciana Appolinario; Marilda Siqueira on behalf of the Fiocruz COVID-19 Genomic Surveillance Network; Paola Resende; Renata Serrano Lopes; Taina Venas; Valnete Andrade |
| EPI_ISL_2385479, EPI_ISL_2534924 | Laboratorio Central Noel Nutels | Bioinformatics Laboratory / LNCC | Alessandra P Lamarca; Alexandra L Gerber; Amilcar Tanuri; Ana Paula de C Guimaraes; Ana Tereza R Vasconcelos; Andrea Cony Cavalcanti; Caio Luiz Pereira Ribeiro; Cassia Alves; Cintia Policarpo; Claudia Maria Braga de Mello; Cristiane Gomes da Silva; Diana Mariani; Douglas Terra Machado; Flavio Dias da Silva; Gleidson da Silva de Oliveira; Leandro Magalhaes de Souza; Liliane Cavalcante; Luiz G P de Almeida; Marcio Henrique de Oliveira Garcia; Mario Sergio Ribeiro; Ronaldo da Silva F Jr; Silvia Carvalho |
| EPI_ISL_2491732, EPI_ISL_2491733, EPI_ISL_2491734, EPI_ISL_2491735, EPI_ISL_2491736, EPI_ISL_2491737, EPI_ISL_2491741, EPI_ISL_2491742, EPI_ISL_2491744, EPI_ISL_2491745, EPI_ISL_2491746, EPI_ISL_2491747, EPI_ISL_2491748, EPI_ISL_2491749, EPI_ISL_2491750, EPI_ISL_2491752, EPI_ISL_2491753, EPI_ISL_2491754 | | | |
| see above | Laboratorio Central de Saude Publica do Estado da Bahia (LACEN/BA) | Laboratory of Respiratory Viruses and Measles, Oswaldo Cruz Institute, FIOCRUZ | Alex Pauvolid-Corrêa; Alice Sampaio Rocha; Ana Beatriz Machado Lima; Ana Carolina Mendonca; Anna Carolina Paixao; Elisa Cavalcante Pereira; Felicidade Pereira; Fernando Motta; Luciana Appolinario; Marilda Siqueira on behalf of the Fiocruz COVID-19 Genomic Surveillance Network; Mia Ferreira de Araujo; Paola Resende; Renata Serrano Lopes; Taina Venas |
| EPI_ISL_2536311 | Laboratorio Central de Saude Publica do Estado da Paraiba (LACEN-PB) | Laboratory of Respiratory Viruses and Measles, Oswaldo Cruz Institute, FIOCRUZ | Alice Sampaio Rocha; Ana Carolina Mendonca; Anna Carolina Paixao; Dalane Loudal Florentino Teixeira; Elisa Cavalcante Pereira; Fernando Motta; Joao Felipe Bezerra; Luciana Appolinario; Marilda Siqueira on behalf of the Fiocruz COVID-19 Genomic Surveillance Network; Paola Resende; Renata Serrano Lopes; Taina Venas |
| EPI_ISL_2443555, EPI_ISL_2645635, EPI_ISL_2645636, EPI_ISL_2645640, EPI_ISL_2645641, EPI_ISL_2645642, EPI_ISL_2645643, EPI_ISL_2645644, EPI_ISL_2645646, EPI_ISL_2645647, EPI_ISL_2645649, EPI_ISL_2645650, EPI_ISL_2645651, EPI_ISL_2645652, EPI_ISL_2645653, EPI_ISL_2645655, EPI_ISL_2645665, EPI_ISL_2645667, EPI_ISL_2645670, EPI_ISL_2645672, EPI_ISL_2645673, EPI_ISL_2645675, EPI_ISL_2645676, EPI_ISL_2645677, EPI_ISL_2645678, EPI_ISL_2645679, EPI_ISL_2645681, EPI_ISL_2645682, EPI_ISL_2645684, EPI_ISL_2645686, EPI_ISL_2645687, EPI_ISL_2645693, EPI_ISL_2645697, EPI_ISL_2645699, EPI_ISL_2645704, EPI_ISL_2645705, EPI_ISL_2645712, EPI_ISL_2645714, EPI_ISL_2645716, EPI_ISL_2645717, EPI_ISL_2645719, EPI_ISL_2645720, EPI_ISL_2645721, EPI_ISL_2645723, EPI_ISL_2645725, EPI_ISL_2645726, EPI_ISL_2645727, EPI_ISL_2645728, EPI_ISL_2645730, EPI_ISL_2645731, EPI_ISL_2645735, EPI_ISL_2645737 | | | |
| see above | Laboratorio Central de Saude Publica do Estado de Alagoas (LACEN/AL) | Laboratory of Respiratory Viruses and Measles, Oswaldo Cruz Institute, FIOCRUZ | Alice Sampaio Rocha; Ana Carolina Mendonca; Anderson Brandao Leite; Anna Carolina Paixao; Elisa Cavalcante Pereira; Fernando Motta; Luciana Appolinario; Marilda Siqueira on behalf of the Fiocruz COVID-19 Genomic Surveillance Network; Paola Resende; Renata Serrano Lopes; Taina Venas |
| EPI_ISL_2645514, EPI_ISL_2645516, EPI_ISL_2645545, EPI_ISL_2645547, EPI_ISL_2645552, EPI_ISL_2645555, EPI_ISL_2645562, EPI_ISL_2645566, EPI_ISL_2645567, EPI_ISL_2645571, EPI_ISL_2645573, EPI_ISL_2645575, EPI_ISL_2645576, EPI_ISL_2645577, EPI_ISL_2645580, EPI_ISL_2645581, EPI_ISL_2645582, EPI_ISL_2645585, EPI_ISL_2645587, EPI_ISL_2645590, EPI_ISL_2645591, EPI_ISL_2645592, EPI_ISL_2645596, EPI_ISL_2645597, EPI_ISL_2645598, EPI_ISL_2645600, EPI_ISL_2645601, EPI_ISL_2645602, EPI_ISL_2645603, EPI_ISL_2645608, EPI_ISL_2645609, EPI_ISL_2645610, EPI_ISL_2645611, EPI_ISL_2645612, EPI_ISL_2645615, EPI_ISL_2645616, EPI_ISL_2645617, EPI_ISL_2645618, EPI_ISL_2645619, EPI_ISL_2645620, EPI_ISL_2645629, EPI_ISL_2645631, EPI_ISL_2645632 | | | |
| see above | Laboratorio Central de Saude Publica do Estado do Espirito Santo (LACEN/ES) | Laboratory of Respiratory Viruses and Measles, Oswaldo Cruz Institute, FIOCRUZ | Alice Sampaio Rocha; Ana Carolina Mendonca; Anna Carolina Paixao; Eilisa Cavalcante Pereira; Fernando Motta; Luciana Appolinario; Marilda Siqueira on behalf of the Fiocruz COVID-19 Genomic Surveillance Network; Paola Resende; Renata Serrano Lopes; Rodrigo Ribeiro Rodrigues; Taina Venas |
| EPI_ISL_2443674, EPI_ISL_2466125, EPI_ISL_2466137 | Laboratorio Central de Saude Publica do Estado do Rio Grande do Sul (LACEN-RS) | Laboratory of Respiratory Viruses and Measles, Oswaldo Cruz Institute, FIOCRUZ | Alice Sampaio Rocha; Ana Carolina Mendonca; Anderson Brandao Leite; Anna Carolina Paixao; Elisa Cavalcante Pereira; Fernando Motta; Luciana Appolinario; Marilda Siqueira on behalf of the Fiocruz COVID-19 Genomic Surveillance Network; Paola Resende; Renata Serrano Lopes; Richard Salvato; Taina Venas; Tatiana Schaffer Gregianini |
| EPI_ISL_2543759, EPI_ISL_2544839, EPI_ISL_2544840, EPI_ISL_2544843, EPI_ISL_2544844, EPI_ISL_2544845, EPI_ISL_2544846, EPI_ISL_2544847, EPI_ISL_2544850, EPI_ISL_2544852, EPI_ISL_2544854, EPI_ISL_2544855, EPI_ISL_2544859, EPI_ISL_2544860, EPI_ISL_2544861, EPI_ISL_2544863, EPI_ISL_2544865, EPI_ISL_2544866, EPI_ISL_2544867, EPI_ISL_2544868, EPI_ISL_2544869, EPI_ISL_2544871, EPI_ISL_2544872, EPI_ISL_2544875, EPI_ISL_2544883, EPI_ISL_2544886, EPI_ISL_2544898 | | | |
| see above | Laboratorio de Pesquisa em Virologia, FAMERP, SJRP | Laboratorio de Pesquisa em Virologia, FAMERP, SJRP | Cecília Artico Banho; Cíntia Bittar; Fábio Sossai Possebon; Guilherme Campos; Helena Lage Ferreira; Jorge A. Petroli Marchesi; João Pessoa Araújo Jr.; Leila Sabrina Ullmann; Lívia Sacchetto; Maisa C. Pereira Parra; Marília Moraes; Maurício L. Nogueira.; Paula Rahal; Paulo Inacio da Costa |
| EPI_ISL_2614070, EPI_ISL_2614071, EPI_ISL_2614073, EPI_ISL_2614075, EPI_ISL_2614079, EPI_ISL_2614084, EPI_ISL_2614085, EPI_ISL_2614086 | | | |
| see above | Laboratory of Molecular Virology, Federal University of Rio de Janeito, UFRJ | Laboratory of Respiratory Viruses and Measles, Oswaldo Cruz Institute, FIOCRUZ | Alice Sampaio Rocha; Amilcar Tanuri; Ana Carolina Mendonca; Anna Carolina Paixao; Elisa Cavalcante Pereira; Fernando Motta; Luciana Appolinario; Marilda Siqueira on behalf of the Fiocruz COVID-19 Genomic Surveillance Network; Paola Resende; Renata Serrano Lopes; Taina Venas |
| EPI_ISL_2443578, EPI_ISL_2443579, EPI_ISL_2443580, EPI_ISL_2443608, EPI_ISL_2443633, EPI_ISL_2443663, EPI_ISL_2443664, EPI_ISL_2443666, EPI_ISL_2443667, EPI_ISL_2443668, EPI_ISL_2443669, EPI_ISL_2443670, EPI_ISL_2443671, EPI_ISL_2466122, EPI_ISL_2466123, EPI_ISL_2466124, EPI_ISL_2557385, EPI_ISL_2603478, EPI_ISL_2603479, EPI_ISL_2603480, EPI_ISL_2603481, EPI_ISL_2603482, | | | |

| | | | |
|---|---|---|---|
| EPI_ISL_2603483, EPI_ISL_2603484, EPI_ISL_2603485, EPI_ISL_2603486, EPI_ISL_2603488, EPI_ISL_2603490, EPI_ISL_2603491, EPI_ISL_2603492, EPI_ISL_2603496, EPI_ISL_2603497, EPI_ISL_2614096, EPI_ISL_2614097, EPI_ISL_2614098, EPI_ISL_2614100, EPI_ISL_2614102, EPI_ISL_2614105, EPI_ISL_2614106, EPI_ISL_2614107, EPI_ISL_2614108, EPI_ISL_2614109, EPI_ISL_2614110, EPI_ISL_2614111, EPI_ISL_2614112, EPI_ISL_2614113, EPI_ISL_2614114, EPI_ISL_2614115, EPI_ISL_2614116, EPI_ISL_2614117, EPI_ISL_2614118, EPI_ISL_2614119, EPI_ISL_2614120, EPI_ISL_2614121, EPI_ISL_2614122, EPI_ISL_2614123, EPI_ISL_2614124, EPI_ISL_2614125, EPI_ISL_2614126, EPI_ISL_2614127, EPI_ISL_2614128, EPI_ISL_2614129, EPI_ISL_2614130, EPI_ISL_2614131, EPI_ISL_2614132, EPI_ISL_2614133, EPI_ISL_2614134, EPI_ISL_2614135, EPI_ISL_2614136, EPI_ISL_2614137, EPI_ISL_2614138, EPI_ISL_2614139, EPI_ISL_2614140, EPI_ISL_2614141, EPI_ISL_2614142, EPI_ISL_2614143, EPI_ISL_2614145, EPI_ISL_2614146, EPI_ISL_2614147, EPI_ISL_2614148, EPI_ISL_2614149, EPI_ISL_2614150, EPI_ISL_2614151, EPI_ISL_2614152, EPI_ISL_2614153, EPI_ISL_2614154, EPI_ISL_2614155, EPI_ISL_2614157, EPI_ISL_2614158, EPI_ISL_2614159, EPI_ISL_2614161, EPI_ISL_2614162, EPI_ISL_2614163, EPI_ISL_2614164, EPI_ISL_2614165, EPI_ISL_2614166, EPI_ISL_2614167, EPI_ISL_2614168, EPI_ISL_2614170, EPI_ISL_2614171, EPI_ISL_2614172, EPI_ISL_2614173, EPI_ISL_2614174, EPI_ISL_2614175, EPI_ISL_2614176, EPI_ISL_2614177, EPI_ISL_2614178, EPI_ISL_2614179, EPI_ISL_2614180, EPI_ISL_2614182, EPI_ISL_2614184, EPI_ISL_2614185, EPI_ISL_2614186, EPI_ISL_2614187, EPI_ISL_2614188, EPI_ISL_2614189, EPI_ISL_2614190, EPI_ISL_2614192, EPI_ISL_2614193, EPI_ISL_2614194, EPI_ISL_2614195, EPI_ISL_2614198, EPI_ISL_2614199, EPI_ISL_2614200, EPI_ISL_2614201, EPI_ISL_2614202, EPI_ISL_2614203, EPI_ISL_2614204, EPI_ISL_2614205, EPI_ISL_2614206, EPI_ISL_2614208, EPI_ISL_2614209, EPI_ISL_2614211, EPI_ISL_2614213, EPI_ISL_2614215, EPI_ISL_2614216, EPI_ISL_2614219, EPI_ISL_2614227, EPI_ISL_2614230, EPI_ISL_2614235, EPI_ISL_2614236, EPI_ISL_2614275, EPI_ISL_2614278, EPI_ISL_2614293, EPI_ISL_2614296, EPI_ISL_2614299, EPI_ISL_2614311, EPI_ISL_2614315 | | | |
| see above | Laboratory of Respiratory Viruses and Measles, Oswaldo Cruz Institute, FIOCRUZ | Laboratory of Respiratory Viruses and Measles, Oswaldo Cruz Institute, FIOCRUZ | Alice Sampaio Rocha; Amilcar Tanuri; Ana Carolina Mendonca; Anna Carolina Paixao; Elisa Cavalcante Pereira; Fernando Motta; Luciana Appolinario; Marilda Siqueira on behalf of the Fiocruz COVID-19 Genomic Surveillance Network; Paola Resende; Renata Serrano Lopes; Taina Venas |
| EPI_ISL_2466139, EPI_ISL_2466140, EPI_ISL_2466141, EPI_ISL_2466142, EPI_ISL_2466143, EPI_ISL_2466144, EPI_ISL_2466145, EPI_ISL_2466204, EPI_ISL_2466207, EPI_ISL_2466219, EPI_ISL_2466221, EPI_ISL_2466224, EPI_ISL_2466226, EPI_ISL_2466227, EPI_ISL_2466228 | | | |
| see above | Laboratório de Biologia Molecular de Doenças Infecciosas e do Câncer (LADIC - UFRN) | Laboratory of Respiratory Viruses and Measles, Oswaldo Cruz Institute, FIOCRUZ | Alice Sampaio Rocha; Ana Carolina Mendonca; Anna Carolina Paixao; Elisa Cavalcante Pereira; Fernando Motta; Josélio Araújo; Luciana Appolinario; Marilda Siqueira on behalf of the Fiocruz COVID-19 Genomic Surveillance Network; Paola Resende; Renata Serrano Lopes; Taina Venas |
| EPI_ISL_2399431, EPI_ISL_2399432, EPI_ISL_2399433, EPI_ISL_2399434, EPI_ISL_2399435, EPI_ISL_2399437, EPI_ISL_2431855 | | | |
| see above | Laboratório de Microbiologia Molecular - Universidade FEEVALE | Molecular Microbiology Laboratory | Alana Witt Hansen; Fernando Rosado Spilki; Flávio Silveira; Fágner Henrique Heldt; Juliana Schons Gularte; Juliane Deise Fleck; Mariana Soares da Silva; Matheus Nunes Weber; Meriane Demoliner; Micheli Filippi.; Paula Rodrigues de Almeida |
| EPI_ISL_2629595, EPI_ISL_2629596, EPI_ISL_2629597, EPI_ISL_2629598, EPI_ISL_2629600, EPI_ISL_2629601, EPI_ISL_2629603, EPI_ISL_2629604, EPI_ISL_2629606, EPI_ISL_2629607, EPI_ISL_2629609, EPI_ISL_2629610, EPI_ISL_2629611, EPI_ISL_2629612, EPI_ISL_2629613, EPI_ISL_2629615, EPI_ISL_2629616, EPI_ISL_2629617, EPI_ISL_2629618, EPI_ISL_2629620, EPI_ISL_2629621, EPI_ISL_2629622, EPI_ISL_2629623, EPI_ISL_2629625, EPI_ISL_2629626, EPI_ISL_2629627, EPI_ISL_2629628, EPI_ISL_2629629, EPI_ISL_2629630, EPI_ISL_2629631, EPI_ISL_2629632, EPI_ISL_2629633, EPI_ISL_2629635, EPI_ISL_2629636, EPI_ISL_2629637, EPI_ISL_2629638, EPI_ISL_2629639, EPI_ISL_2629641, EPI_ISL_2629644, EPI_ISL_2629645, EPI_ISL_2629646, EPI_ISL_2629647, EPI_ISL_2629648, EPI_ISL_2629649, EPI_ISL_2629650, EPI_ISL_2629651, EPI_ISL_2629652, EPI_ISL_2629653, EPI_ISL_2629654, EPI_ISL_2629655, EPI_ISL_2629656, EPI_ISL_2629657, EPI_ISL_2629658, EPI_ISL_2629660, EPI_ISL_2629661, EPI_ISL_2629663, EPI_ISL_2629664, EPI_ISL_2629665, EPI_ISL_2629666, EPI_ISL_2629667, EPI_ISL_2629668, EPI_ISL_2629669, EPI_ISL_2629670, EPI_ISL_2629672, EPI_ISL_2629673, EPI_ISL_2629674, EPI_ISL_2629675, EPI_ISL_2629676, EPI_ISL_2629677, EPI_ISL_2629678, EPI_ISL_2629680, EPI_ISL_2629681, EPI_ISL_2629683, EPI_ISL_2629684, EPI_ISL_2629685, EPI_ISL_2629686, EPI_ISL_2629687, EPI_ISL_2629688, EPI_ISL_2629689, EPI_ISL_2629690, EPI_ISL_2629691, EPI_ISL_2629692, EPI_ISL_2629693, EPI_ISL_2629694, EPI_ISL_2629695, EPI_ISL_2629696, EPI_ISL_2629697, EPI_ISL_2629698, EPI_ISL_2629699, EPI_ISL_2629700, EPI_ISL_2629701, EPI_ISL_2629703, EPI_ISL_2629704, EPI_ISL_2629708, EPI_ISL_2629710, EPI_ISL_2629712, EPI_ISL_2629713, EPI_ISL_2629722, EPI_ISL_2629723, EPI_ISL_2629725, EPI_ISL_2629726, EPI_ISL_2629728, EPI_ISL_2629729, EPI_ISL_2629735, EPI_ISL_2629737, EPI_ISL_2629744, EPI_ISL_2629759, EPI_ISL_2629760, EPI_ISL_2629822, EPI_ISL_2629832 | | | |
| see above | Laboratório de Virologia Molecular - Universidade Federal do Rio de Janeiro | Laboratório de Virologia Molecular - Universidade Federal do Rio de Janeiro | ; Alice Laschuk Herlinger; Amilcar Tanuri; André Felipe Andrade dos Santos; Carolina Moreira Voloch; Cássia Cristina Alves Gonçalves; Diana Mariani; Débora Souza Faffe; Filipe Romero Rebello Moreira; Francine Bittencourt Schiffler; Isabela de Carvalho Leitão; Marcelo Calado de Paula Tôrres; Matheus Augusto Calvano Cosentino; Mirela D'arc; Orlando da Costa Ferreira Junior; Rafael Mello Galliez; Raíssa Mirella dos Santos Cunha da Costa; Renato Santana de Aguiar; Terezinha Marta Pereira Pinto Castineiras; Thamiris dos Santos Miranda; Átila Duque Rossi |
| EPI_ISL_2614361 | Labortorio Central de Saude Publica do Estado do Rio de Janeiro (LACEN/RJ) | Laboratory of Respiratory Viruses and Measles, Oswaldo Cruz Institute, FIOCRUZ | Alice Sampaio Rocha; Ana Carolina Mendonca; Andrea Cony Cavalcanti; Anna Carolina Paixao; Elisa Cavalcante Pereira; Fernando Motta; Luciana Appolinario; Marilda Siqueira on behalf of the Fiocruz COVID-19 Genomic Surveillance Network; Paola Resende; Renata Serrano Lopes; Taina Venas |
| EPI_ISL_2443531 | Lboratorio Central de Saude Publica do Estado do Parana (LACEN/PR) | Laboratory of Respiratory Viruses and Measles, Oswaldo Cruz Institute, FIOCRUZ | Alice Sampaio Rocha; Ana Carolina Mendonca; Anna Carolina Paixao; Elisa Cavalcante Pereira; Fernando Motta; Irina Riediger; Luciana Appolinario; Marilda Siqueira on behalf of the Fiocruz COVID-19 Genomic Surveillance Network; Paola Resende; Renata Serrano Lopes; Taina Venas |
| EPI_ISL_2535214 | Unidade de apoio ao diagnostico da COVID - UNADIG | Bioinformatics Laboratory / LNCC | Alessandra P Lamarca; Alexandra L Gerber; Amilcar Tanuri; Ana Paula de C Guimaraes; Ana Tereza R Vasconcelos; Andrea Cony Cavalcanti; Caio Luiz Pereira Ribeiro; Cassia Alves; Cintia Policarpo; Claudia Maria Braga de Mello; Cristiane Gomes da Silva; Diana Mariani; Douglas Terra Machado; Flavio Dias da Silva; Gleidson da Silva de Oliveira; Leandro Magalhaes de Souza; Liliane Cavalcante; Luiz G P de Almeida; Marcio Henrique de Oliveira Garcia; Mario Sergio Ribeiro; Ronaldo da Silva F Jr; Silvia Carvalho |

We gratefully acknowledge the following Authors from the Originating laboratories responsible for obtaining the specimens, as well as the Submitting laboratories where the genome data were generated and shared via GISAID, on which this research is based.

All Submitters of data may be contacted directly via www.gisaid.org

Authors are sorted alphabetically.

| Accession ID | Originating Laboratory | Submitting Laboratory | Authors |
|---|---|---|---|
| EPI_ISL_2859280 | Adela Serra TY Memorial Center | Philippine Genome Center | Alethea R. de Guzman; Anna Ong-Lim; Arianne A. Zamora; Benedict A. Maralit; Carlo M. Lapid; Celia Carlos; Devon Ray Pacial; Diomedes A. Carino; Edsel Maurice Salvaña; El King D. Morado; Elcid Aaron R. Pangilinan; Eva Maria Cutiongco-de la Paz; Francis A. Tablizo; Henrietta Marie Rodriguez; Jaime C. Montoya; Jan Michael C. Yap; Jarvin E. Nipales; Jo-Hannah S. Llames; John Q. Wong; Joshua Gregor A. Dizon; Juan Antonio R. Magalang; Karol Sophia Agape R. Padilla; Kenneth M. Kim; Kris P. Punayan; Krisitna Patriz Dela Cruz; Lindsay Claire D.L. Carandang; Ma. Exanil Plantig; Marc Edsel C. Ayes; Maria Rosario Singh-Vergeire and Cynthia P. Saloma; Maria Sofia L. Yangzon; Marielle M Gamboa; Marissa Alejandria; Nina Francesca Bustamante; Razel Nikka M. Hao; Renato Jacinto Q. Mantaring; Rianna Patricia S. Cruz; Sheila Mae M. Araiza; Yvonne Valerie Austria; Zipporah Mariebelle R. Enriquez; Zyrel V. Mollejon |
| EPI_ISL_2181026, EPI_ISL_2370650 | Aegis Sciences Corporation | Centers for Disease Control and Prevention Division of Viral Diseases, Pathogen Discovery | Adrian Paskey; Alec Vest; Benjamin Rambo-Martin; Christopher Gulvick; Clinton R. Paden; Cyndi Clark; Dakota Howard; Darlene Wagner; Dhwani Batra; Dillon Nall; Duncan MacCannell; Ethan Sanders; Holly Houdeshell; Jason Caravas; Kara Moser; Matthew Hardison; Matthew Schmerer; Ola Kvalvaag; Patrick Campbell; Peter W. Cook; Rob Case; Scott Sammons; Shatavia Morrison; Shaun Westlund; Vikramsinha Ghorpade; Yvette Unoarumhi |
| EPI_ISL_2156599, EPI_ISL_2188702, EPI_ISL_2188812 | Baguio General Hospital Medical Center (BGHMC) | Philippine Genome Center | Alethea R. de Guzman; Anna Ong-Lim; Arianne A. Zamora; Asia Louisa U. Chong; Benedict A. Maralit; Candice Francheska B. Tambaoan; Carlo M. Lapid; Celia Carlos; Devon Ray Pacial; Edsel Maurice Salvaña; El King D. Morado; Elcid Aaron R. Pangilinan; Eva Maria Cutiongco-de la Paz; Francis A. Tablizo; Irish Coleen A. Asin; Jaime C. Montoya; Jan Michael C. Yap; Jo-Hannah S. Llames; John Q. Wong; Joshua Gregor A. Dizon; Juan Antonio R. Magalang; Karol Sophia Agape R. Padilla; Kenneth M. Kim; Kris P. Punayan; Marc Edsel C. Ayes; Marc Jerrone R. Castro; Maria Rosario Singh-Vergeire and Cynthia P. Saloma; Maria Sofia L. Yangzon; Marissa Alejandria; Razel Nikka M. Hao; Renato Jacinto Q. Mantaring; Rianna Patricia S. Cruz; Sheila Mae M. Araiza |
| EPI_ISL_1213504, EPI_ISL_1213506, EPI_ISL_1213508, EPI_ISL_1213509, EPI_ISL_1213511, EPI_ISL_1213513, EPI_ISL_1213514, EPI_ISL_1213516, EPI_ISL_1213518, EPI_ISL_1213519, EPI_ISL_1213521, EPI_ISL_1213523, EPI_ISL_1213524, EPI_ISL_1213526, EPI_ISL_1213528, EPI_ISL_1213530, EPI_ISL_1213531, EPI_ISL_2091460, EPI_ISL_2156475, EPI_ISL_2156481 | | | |
| see above | Bohol Containerized PCR Laboratory | Philippine Genome Center | Alethea R. de Guzman; Anna Ong-Lim; Arianne A. Zamora; Asia Louisa U. Chong; Benedict A. Maralit; Candice Francheska B. Tambaoan; Carlo M. Lapid; Celia Carlos; Devon Ray Pacial; Edsel Maurice Salvaña; El King D. Morado; Elcid Aaron R. Pangilinan; Eva Maria Cutiongco-de la Paz; Francis A. Tablizo; Irish Coleen A. Asin; Jaime C. Montoya; Jan Michael C. Yap; Jo-Hannah S. Llames; John Q. Wong; Joshua Gregor A. Dizon; Juan Antonio R. Magalang; Karol Sophia Agape R. Padilla; Kenneth M. Kim; Kris P. Punayan; Marc Edsel C. Ayes; Marc Jerrone R. Castro; Maria Rosario Singh-Vergeire and Cynthia P. Saloma; Maria Sofia L. Yangzon; Marissa Alejandria; Razel Nikka M. Hao; Rianna Patricia S. Cruz; Sheila Mae M. Araiza |
| EPI_ISL_2535620, EPI_ISL_2535683, EPI_ISL_2535781 | Borneo Medical Centre | Institute of Health and Community Medicine | Chan Chia Jui; Chua Hock Hin; David Perera; Ooi Mong How; Tonnii Sia Loong Loong; Wong Jyn Shan; Wong Kiing Aik |
| EPI_ISL_2156467, EPI_ISL_2156468, EPI_ISL_2188870, EPI_ISL_2188906, EPI_ISL_2189046, EPI_ISL_2189233, EPI_ISL_2189252, EPI_ISL_2189273, EPI_ISL_2189296, EPI_ISL_2189380, EPI_ISL_2189493 | | | |
| see above | Butuan Medical Center | Philippine Genome Center | Alethea R. de Guzman; Anna Ong-Lim; Arianne A. Zamora; Asia Louisa U. Chong; Benedict A. Maralit; Candice Francheska B. Tambaoan; Carlo M. Lapid; Celia Carlos; Devon Ray Pacial; Edsel Maurice Salvaña; El King D. Morado; Elcid Aaron R. Pangilinan; Eva Maria Cutiongco-de la Paz; Francis A. Tablizo; Irish Coleen A. Asin; Jaime C. Montoya; Jan Michael C. Yap; Jo-Hannah S. Llames; John Q. Wong; Joshua Gregor A. Dizon; Juan Antonio R. Magalang; Karol Sophia Agape R. Padilla; Kenneth M. Kim; Kris P. Punayan; Marc Edsel C. Ayes; Marc Jerrone R. Castro; Maria Rosario Singh-Vergeire and Cynthia P. Saloma; Maria Sofia L. Yangzon; Marissa Alejandria; Razel Nikka M. Hao; Renato Jacinto Q. Mantaring; Rianna Patricia S. Cruz; Sheila Mae M. Araiza |
| EPI_ISL_2761769 | CENTOGENE Frankfurt Laboratory: Niederlassung Industriepark Höchst | Robert Koch Institute | |
| EPI_ISL_1122426, EPI_ISL_1122427, EPI_ISL_1122428, EPI_ISL_1122429, EPI_ISL_1122430, EPI_ISL_1122431, EPI_ISL_1122432, EPI_ISL_1122433, EPI_ISL_1122434, EPI_ISL_1122435, EPI_ISL_1122436, EPI_ISL_1122437, EPI_ISL_1122438, EPI_ISL_1122439, EPI_ISL_1122440, EPI_ISL_1122441, EPI_ISL_1122442, EPI_ISL_1122443, EPI_ISL_1122444, EPI_ISL_1122445, EPI_ISL_1122446, EPI_ISL_1122447, EPI_ISL_1122448, EPI_ISL_1122449, EPI_ISL_1122450, EPI_ISL_1122451, EPI_ISL_1122452, EPI_ISL_1122453, EPI_ISL_1122454, EPI_ISL_1213533, EPI_ISL_1213535, EPI_ISL_1213537, EPI_ISL_1213539, EPI_ISL_1213540, EPI_ISL_1213542, EPI_ISL_1213544, EPI_ISL_1213546, EPI_ISL_1213547, EPI_ISL_1213549, EPI_ISL_1213551, EPI_ISL_1213552, EPI_ISL_1213554, EPI_ISL_1213556, EPI_ISL_1213558, EPI_ISL_1213559, EPI_ISL_1213561, EPI_ISL_1219702, EPI_ISL_2091458, EPI_ISL_2091461, EPI_ISL_2105320, EPI_ISL_2105327, EPI_ISL_2105379, EPI_ISL_2105468, EPI_ISL_2105476, EPI_ISL_2105477, EPI_ISL_2105479, EPI_ISL_2105485, EPI_ISL_2105492, EPI_ISL_2105495, EPI_ISL_2105496, EPI_ISL_2105500, EPI_ISL_2105501, EPI_ISL_2105503, EPI_ISL_2105505, EPI_ISL_2105526, EPI_ISL_2105527, EPI_ISL_2105560, EPI_ISL_2155587, EPI_ISL_2155590, EPI_ISL_2155919, EPI_ISL_2155923, EPI_ISL_2155925, EPI_ISL_2155930, EPI_ISL_2155936, EPI_ISL_2155939, EPI_ISL_2155940, EPI_ISL_2156124, EPI_ISL_2156125, EPI_ISL_2156126, EPI_ISL_2156127, EPI_ISL_2156128 | | | |
| see above | Cebu TB Reference Laboratory | Philippine Genome Center | Alethea R. de Guzman; Anna Ong-Lim; Arianne A. Zamora; Asia Louisa U. Chong; Benedict A. Maralit; Candice Francheska B. Tambaoan; Carlo M. Lapid; Celia Carlos; Devon Ray Pacial; Edsel Maurice Salvaña; El King D. Morado; Eva Maria Cutiongco-de la Paz; Francis A. Tablizo; Irish Coleen A. Asin; Jaime C. Montoya; Jan Michael C. Yap; Jo-Hannah S. Llames; John Q. Wong; Joshua Gregor A. Dizon; Juan Antonio R. Magalang; Karol Sophia Agape R. Padilla; Kenneth M. Kim; Kris P. Punayan; Marc Edsel C. Ayes; Marc Jerrone R. Castro; Maria Rosario Singh-Vergeire and Cynthia P. Saloma; Maria Sofia L. Yangzon; Marissa Alejandria; Razel Nikka M. Hao; Rianna Patricia S. Cruz; Sheila Mae M. Araiza |
| EPI_ISL_2105547 | Chinese General Hospital | Philippine Genome Center | Alethea R. de Guzman; Anna Ong-Lim; Arianne A. Zamora; Asia Louisa U. Chong; Benedict A. Maralit; Candice Francheska B. Tambaoan; Carlo M. Lapid; Celia Carlos; Devon Ray Pacial; Edsel Maurice Salvaña; El King D. Morado; Eva Maria Cutiongco-de la Paz; Francis A. Tablizo; Irish Coleen A. Asin; Jaime C. Montoya; Jan Michael C. Yap; Jo-Hannah S. Llames; John Q. Wong; Joshua Gregor A. Dizon; Juan Antonio R. Magalang; Karol Sophia Agape R. Padilla; Kenneth M. Kim; Kris P. Punayan; Marc Edsel C. Ayes; Marc Jerrone R. Castro; Maria Rosario Singh-Vergeire and Cynthia P. Saloma; Maria Sofia L. Yangzon; Marissa Alejandria; Razel Nikka M. Hao; Rianna Patricia S. Cruz; Sheila Mae M. Araiza |
| EPI_ISL_1213563, EPI_ISL_2859372 | Cotabato Regional and Medical Center | Philippine Genome Center | Alethea R. de Guzman; Anna Ong-Lim; Arianne A. Zamora; Asia Louisa U. Chong; Benedict A. Maralit; Carlo M. Lapid; Celia Carlos; Devon Ray Pacial; Diomedes A. Carino; Edsel Maurice Salvaña; El King D. Morado; Elcid Aaron R. Pangilinan; Eva Maria Cutiongco-de la Paz; Francis A. Tablizo; Henrietta Marie Rodriguez; Irish Coleen A. Asin; Jaime C. Montoya; Jan Michael C. Yap; Jarvin E. Nipales; Jo-Hannah S. Llames; John Q. Wong; Joshua Gregor A. Dizon; Juan Antonio R. Magalang; Karol Sophia Agape R. Padilla; Kenneth M. Kim; Kris P. Punayan; Krisitna Patriz Dela Cruz; Lindsay Claire D.L. Carandang; Ma. Exanil Plantig; Marc Edsel C. Ayes; Marc Jerrone R. Castro; Maria Rosario Singh-Vergeire and Cynthia P. Saloma; Maria Sofia L. Yangzon; Marielle M Gamboa; Marissa Alejandria; Nina Francesca Bustamante; Razel Nikka M. Hao; Renato Jacinto Q. Mantaring; Rianna Patricia S. Cruz; Sheila Mae M. Araiza; Yvonne Valerie Austria; Zipporah Mariebelle R. Enriquez; Zyrel V. Mollejon |
| EPI_ISL_2930802 | Department of Clinical Microbiology | GIGA Medical Genomics | Bouchra Boujemla; Claire Gourzonès; Cécile Meex; Keith Durkin; Laurent Gillet; Maria Artesi; Marie-Pierre Hayette; Nadine Cambisano; Nathalie Renotte; Olivier Ek; Sébastien Bontems; Vincent Bours |
| EPI_ISL_2860802 | Department of Health Technology and Informatics, The Hong Kong Polytechnic University | Department of Health Technology and Informatics, The Hong Kong Polytechnic University | Alan Ka-Lun Wu; Alex Yat-Man Ho; Barry Kin-Chung Wong; Chloe Toi-Mei Chan; David Ho-Keung Shum; Denise Sze-Hang Wong; Gilman Kit-Hang Siu; Hiu-Yin Lao; Hoi-Ching Jim; Ivan Tak-Fai Wong; Jake Siu-Lun Leung; Kam-Tong Yip; Kenneth Siu-Sing Leung; Kingsley King-Gee Tam; Kitty Sau-Chun Fung; Kristine Luk; Lam-Kwong Lee; Miranda Chong-Yee Yau; Sandy Ka-Yee Chau; Shea Ping Yip; Tak-Lun Que; Timothy Ting-Leung Ng; Wing Cheong Yam; Wing-Hei Lo; Wing-Kin To; Yvette Wai-Man Lai |
| EPI_ISL_1073934, EPI_ISL_1323166 | Department of Medical Microbiology - section Molde, Molde Hospital | Norwegian Institute of Public Health, Department of Virology | Atiya R Ali; Debech Nadia; Engebretsen Serina Beate; Engebretsen Serina Beate Atiya R Ali; Garcia Llorente Ignacio; Hilde Elshaug; Hilde Vollan; Jon Bråte; Kamilla Heddeland Instefjord; Karoline Bragstad; Kathrine Stene-Johansen; Marie Paulsen Madsen; Olav Hungnes; Pedersen Benedikte Nevjen; Rasmus Riis Kopperud |
| EPI_ISL_2423558 | Department of Microbiology, The University of Hong Kong | Department of Microbiology, The University of Hong Kong | Kelvin K.W. To; Kwok-Yung Yuen |
| EPI_ISL_1660475 | Department of Microbiology, The University of Hong Kong | The University of Hong Kong Department of Microbiology | Kelvin K.W. To; Kwok-Yung Yuen |
| EPI_ISL_3135014 | Department of Virology and Immunology, University of Helsinki and Helsinki University Hospital, Huslab Finland | Department of Virology, Faculty of Medicine, University of Helsinki, Helsinki, Finland | Essi Korhonen; Hanna Jarva; Hanna Liimatainen; Hannimari Kallio-Kokko; Harri Kangas; Hussein Alburkat; Jenni Virtanen; Maija Lappalainen; Maija Suvanto; Olli Vapalahti; Pekka Ellonen; Phuoc Truong; Ravi Kant; Sari Hannula; Satu Kurkela; Teemu Smura |
| EPI_ISL_1647347, EPI_ISL_3026635 | Division of Emerging Infectious Diseases, Bureau of Infectious Diseases Diagnosis Control, Korea Disease Control and Prevention Agency | Division of Emerging Infectious Diseases, Bureau of Infectious Diseases Diagnosis Control, Korea Disease Control and Prevention Agency | Ae Kyung Park; Chae Young Lee; Eun-Jin Kim; Heui Man Kim; Il-Hwan Kim; Jeong-Ah Kim; Jeong-Min Kim; Jin Sun No |
| EPI_ISL_1522133, EPI_ISL_1522134, EPI_ISL_1597109, EPI_ISL_1597203, EPI_ISL_1792918, EPI_ISL_1792919, EPI_ISL_2476340 | | | |
| see above | Dutch COVID-19 response team | National Institute for Public Health and the Environment (RIVM) | Adam Meijer; AnneMarie van den Brandt; Annelies Kroneman; Bas van der Veer; Chantal Reusken; Dennis Schmitz; Dirk Eggink; Euníce Then; Florian Zwagemaker; Harry Vennema; James Groot; Jeroen Cremer; Jolienke Hardeman; Karim Hajji; Kim Freriks; Linda van de Nes; Lisa Wijsman; Lynn Aarts; Melissa van Tuil; Robert Kohl; Ryanne Jaarsma; Sanne Bos; Sharon van den Brink; Sjoerd Kuiling; on behalf of the national COVID-19 response team |
| EPI_ISL_2859373 | Eastern Visayas Regional Covid-19 Testing Center | Philippine Genome Center | Alethea R. de Guzman; Anna Ong-Lim; Arianne A. Zamora; Benedict A. Maralit; Carlo M. Lapid; Celia Carlos; Devon Ray Pacial; Diomedes A. Carino; Edsel Maurice Salvaña; El King D. Morado; Elcid Aaron R. Pangilinan; Eva Maria Cutiongco-de la Paz; Francis A. Tablizo; Henrietta Marie Rodriguez; Jaime C. Montoya; Jan Michael C. Yap; Jarvin E. Nipales; Jo-Hannah S. Llames; John Q. Wong; Joshua Gregor A. Dizon; Juan Antonio R. Magalang; Karol Sophia Agape R. Padilla; Kenneth M. Kim; Kris P. Punayan; Krisitna Patriz Dela Cruz; Lindsay Claire D.L. Carandang; Ma. Exanil Plantig; Marc Edsel C. Ayes; Maria Rosario Singh-Vergeire and Cynthia P. Saloma; Maria Sofia L. Yangzon; Marielle M Gamboa; Marissa Alejandria; Nina Francesca Bustamante; Razel Nikka M. Hao; Renato Jacinto Q. Mantaring; Rianna Patricia S. Cruz; Sheila Mae M. Araiza; Yvonne Valerie Austria; Zipporah Mariebelle R. Enriquez; Zyrel V. Mollejon |
| EPI_ISL_1571348 | Eurofins LifeCodexx GmbH | Robert Koch Institute | |
| EPI_ISL_2188852 | First Aide Diagnostic Center | Philippine Genome Center | Alethea R. de Guzman; Anna Ong-Lim; Arianne A. Zamora; Asia Louisa U. Chong; Benedict A. Maralit; Candice Francheska B. Tambaoan; Carlo M. Lapid; Celia Carlos; Devon Ray Pacial; Edsel Maurice Salvaña; El King D. Morado; Elcid Aaron R. Pangilinan; Eva Maria Cutiongco-de la Paz; Francis A. Tablizo; Irish Coleen A. Asin; Jaime C. Montoya; Jan Michael C. Yap; Jo-Hannah S. Llames; John Q. Wong; Joshua Gregor A. Dizon; Juan Antonio R. Magalang; Karol Sophia Agape R. Padilla; Kenneth M. Kim; Kris P. Punayan; Marc Edsel C. Ayes; Maria Rosario Singh-Vergeire and Cynthia P. Saloma; Maria Sofia L. Yangzon; Marissa Alejandria; Razel Nikka M. Hao; Renato Jacinto Q. Mantaring; Rianna Patricia S. Cruz; Sheila Mae M. Araiza |
| EPI_ISL_1525595, EPI_ISL_1614114 | Fulgent Genetics | Centers for Disease Control and Prevention Division of Viral Diseases, Pathogen Discovery | Adrian Paskey; Becky Tsai; Benafsh Sapra; Benjamin Rambo-Martin; Christopher Gulvick; Clinton R. Paden; Dakota Howard; Darlene Wagner; Dhwani Batra; Doreen Ng; Duncan MacCannell; Harry Gao; James Xie; Jason Caravas; John Gao; Joseph Fierro; Kara Moser; Matthew Schmerer; Mickey Li; Peter W. Cook; Scott Sammons; Shatavia Morrison; Yan Meng; Yvette Unoarumhi |
| EPI_ISL_3133822 | Furst Medical Laboratory | Norwegian Institute of Public Health, Department of Virology | Atiya R Ali; Debech Nadia; Engebretsen Serina Beate; Garcia Llorente Ignacio; Hilde Elshaug; Hilde Vollan; Jon Bråte; Kamilla Heddeland Instefjord; Karoline Bragstad; Kathrine Stene-Johansen; Line Victoria Moen; Marie Paulsen Madsen; Olav Hungnes; Pedersen Benedikte Nevjen; Rasmus Riis Kopperud |
| EPI_ISL_1213565, EPI_ISL_1213566, EPI_ISL_2091459 | Hi-Precision Diagnostic Center (QC) | Philippine Genome Center | Alethea R. de Guzman; Anna Ong-Lim; Arianne A. Zamora; Asia Louisa U. Chong; Benedict A. Maralit; Candice Francheska B. Tambaoan; Carlo M. Lapid; Celia Carlos; Devon Ray Pacial; Edsel Maurice Salvaña; El King D. Morado; Eva Maria Cutiongco-de la Paz; Francis A. Tablizo; Irish Coleen A. Asin; Jaime C. Montoya; Jan Michael C. Yap; Jo-Hannah S. Llames; John Q. Wong; Joshua Gregor A. Dizon; Juan Antonio R. Magalang; Karol Sophia Agape R. Padilla; Kenneth M. Kim; Kris P. Punayan; Marc Edsel C. Ayes; Marc Jerrone R. Castro; Maria Rosario Singh-Vergeire and Cynthia P. Saloma; Maria Sofia L. Yangzon; Marissa Alejandria; Razel Nikka M. Hao; Rianna Patricia S. Cruz; Sheila Mae M. Araiza |
| EPI_ISL_1122457 | Hi-Precision Diagnostics | Philippine Genome Center | Alethea R. de Guzman; Anna Ong-Lim; Arianne A. Zamora; Asia Louisa U. Chong; Benedict A. Maralit; Candice Francheska B. Tambaoan; Carlo M. Lapid; Celia Carlos; Devon Ray Pacial; Edsel Maurice Salvaña; El King D. Morado; Eva Maria Cutiongco-de la Paz; Francis A. Tablizo; Irish Coleen A. Asin; Jaime C. Montoya; Jan Michael C. Yap; Jo-Hannah S. Llames; John Q. Wong; Joshua Gregor A. Dizon; Juan Antonio R. Magalang; Karol Sophia Agape R. Padilla; Kenneth M. Kim; Kris P. Punayan; Marc Edsel C. Ayes; Marc Jerrone R. Castro; Maria Rosario Singh-Vergeire and Cynthia P. Saloma; Maria Sofia L. Yangzon; Marissa Alejandria; Razel Nikka M. Hao; Rianna Patricia S. Cruz; Sheila Mae M. Araiza |
| EPI_ISL_1914574, EPI_ISL_1963594, EPI_ISL_1963595, EPI_ISL_1963603, EPI_ISL_1963631, EPI_ISL_1963636 | Hong Kong Department of Health | School of Public Health, The University of Hong Kong | Daniel K.W. Chu; Dominic N.C. Tsang; Haogao Gu; Leo L.M. Poon; Malik Peiris; Tong Zhang |
| EPI_ISL_2492563, | Instituto Nacional de Investigação | CERI, Centre for Epidemic Response | Afonso P; David K; Emmanuel SJ; Freitas RH; Giandhari J; Inglês L; Lutucuta S; Miranda J; Morais J; Mufinda M; Naidoo Y; Neto Z; Paulo A Carralero RR Paixão JP; Pereira A; Pillay S; Tegally H; Wilkinson E; de Oliveira T |

| | | | |
|---|---|---|---|
| EPI_ISL_2492682 | em Saúde | and Innvoation, Stellenbosch University and KRISP, KZN Research Innovation and Sequencing Platform, UKZN. | |
| EPI_ISL_2455312 | Johns Hopkins Hospital Department of Pathology | Johns Hopkins Hospital Department of Pathology | Adannaya Amadi; C. Paul Morris; Chun Huai Luo; Heba H. Mostafa; Matthew Schwartz; Nicholas Gallagher |
| EPI_ISL_3085785 | Karolinska University Hospital Solna | Karolinska University Hospital | Annelie Bjerkner; Isak Sylvin; Jan Albert; Karolina Ininbergs; Lina Guerra Blomqvist; Lynda Eneh; Martin Ekman; Martina Wahlund; Robert Dyrdak; Sandra Broddesson; Tanja Normark; Tobias Allander; Valtteri Wirta; Zhibing Yun |
| EPI_ISL_1315326 | LabPLUS | Institute of Environmental Science and Research (ESR) | Anja Werno; Antje van der Linden; Arlo Upton; Chris Mansell; David Hammer; Dragana Drinkovic; Erasmus Smit; Gary McAuliffe; Hana Sofia Andersson; Hermes Perez; James Ussher; Jill Sherwood; Jing Wang; Joep de Ligt; Josh Freeman; Julia Howard; Juliet Elvy; Lauren Jelly; Mary DeAlmeida; Matt Blakiston; Matt Storey; Matthew Rogers; Max Bloomfield; Michael Addidle; Michelle Balm; Muhammad Faisal; Nikki Freed; Olin Silander; Olivia Stroeven; Rachel Boyle; Sally Roberts; SallyAnn Harbison; Sarah Jefferies; Sharmini Muttaiyah; Susan Morpeth; Susan Taylor; Timothy Blackmore; Vani Sathyendran; Veronica Playle; Virginia Hope; Xiaoyun Ren |
| EPI_ISL_2635559 | Labor Becker & Kollegen (Standort München) | Robert Koch Institute | |
| EPI_ISL_2481469, EPI_ISL_2927544, EPI_ISL_2927545 | Laboratory Corporation of America | Centers for Disease Control and Prevention Division of Viral Diseases, Pathogen Discovery | Adrian Paskey; Amanda Douglas; Amanda Suchanek; Andrea Throop; Ayla Burns; Benjamin Rambo-Martin; Bobbi Croy; Brian Krueger; Brian Norvell; Christopher Gulvick; Christos Petropoulos; Clinton R. Paden; Craig Lukasik; Dakota Howard; Darlene Wagner; Debbie Boles; Dhwani Batra; Duncan MacCannell; Eyad Almasri; Goran Stevovic; Howard Engler; Hrushikesh Deshmukh; Jake Humphrey; Jana Schroth; Jason Caravas; Joe Voshell; John Pruitt; Jonathan Meltzer; Jonathan Williams; Kara Moser; Kimberly Wagner; Lax Iyer; Lisa Pfefferle; Lyndon Tilson; Manoj Jain; Marcia Eisenberg; Mary Ann Cristobal; Mary Williamson; Matthew Robinson; Matthew Schmerer; Michael Levandoski; Mike Sapeta; Mindy Nye; Minoo Agarwal; Mohan Kolli; Nuthawin Charoensri; Oren Cohen; Peter W. Cook; Prashant Gupta; Qian Zeng; Rama Ghatti; Scott Parker; Scott Ryan; Scott Sammons; Shatavia Morrison; Stanley Letovsky; Steven Ragan; Suresh Babu Selvaraju; Susan Countryman; Susan Hicks; Suzanne Dale; Thomas Urban; Tim Kuphal; Tricia Zwiefelhofer; Vincent Drouillon; Yvette Unoarumhi |
| EPI_ISL_2188206 | Laguna Provincial Hospital - San Pablo District Hospital | Philippine Genome Center | Alethea R. de Guzman; Anna Ong-Lim; Arianne A. Zamora; Asia Louisa U. Chong; Benedict A. Maralit; Candice Francheska B. Tambaoan; Carlo M. Lapid; Celia Carlos; Devon Ray Pacial; Edsel Maurice Salvaña; Elcid Aaron R. Pangilinan; Eva Maria Cutiongco-de la Paz; Francis A. Tablizo; Irish Coleen A. Asin; Jaime C. Montoya; Jan Michael C. Yap; Jo-Hannah S. Llames; John Q. Wong; Joshua Gregor A. Dizon; Juan Antonio R. Magalang; Karol Sophia Agape R. Padilla; Kenneth M. Kim; Kris P. Punayan; Marc Edsel C. Ayes; Marc Jerrone R. Castro; Maria Rosario Singh-Vergeire and Cynthia P. Saloma; Maria Sofia L. Yangzon; Marissa Alejandria; Razel Nikka M. Hao; Rianna Patricia S. Cruz; Sheila Mae M. Araiza |
| EPI_ISL_1244897, EPI_ISL_1256377, EPI_ISL_1256430, EPI_ISL_1275649, EPI_ISL_1333537 | Lighthouse Lab in Cambridge | Wellcome Sanger Institute for the COVID-19 Genomics UK (COG-UK) Consortium | Cordelia Langford; David K. Jackson; Dominic Kwiatkowski; Ewan Harrison; Ian Johnston; Jeffrey Barrett; John Sillitoe on behalf of the Wellcome Sanger Institute COVID-19 Surveillance Team; Rob Howes; Roberto Amato; Sonia Goncalves; The Lighthouse Lab in Cambridge and Alex Alderton |
| EPI_ISL_1986837, EPI_ISL_2352773 | Lighthouse Lab in Milton Keynes | Wellcome Sanger Institute for the COVID-19 Genomics UK (COG-UK) Consortium | Cordelia Langford; David K. Jackson; Dominic Kwiatkowski; Ewan Harrison; Ian Johnston; Jeffrey Barrett; John Sillitoe on behalf of the Wellcome Sanger Institute COVID-19 Surveillance Team; Roberto Amato; Sonia Goncalves; The Lighthouse Lab in Milton Keynes and Alex Alderton |
| EPI_ISL_1213568 | Lung Center of the Philippines (LCP) | Philippine Genome Center | Alethea R. de Guzman; Anna Ong-Lim; Arianne A. Zamora; Asia Louisa U. Chong; Benedict A. Maralit; Candice Francheska B. Tambaoan; Carlo M. Lapid; Celia Carlos; Devon Ray Pacial; Edsel Maurice Salvaña; El King D. Morado; Eva Maria Cutiongco-de la Paz; Francis A. Tablizo; Irish Coleen A. Asin; Jaime C. Montoya; Jan Michael C. Yap; Jo-Hannah S. Llames; John Q. Wong; Joshua Gregor A. Dizon; Juan Antonio R. Magalang; Karol Sophia Agape R. Padilla; Kenneth M. Kim; Kris P. Punayan; Marc Edsel C. Ayes; Marc Jerrone R. Castro; Maria Rosario Singh-Vergeire and Cynthia P. Saloma; Maria Sofia L. Yangzon; Marissa Alejandria; Razel Nikka M. Hao; Rianna Patricia S. Cruz; Sheila Mae M. Araiza |
| EPI_ISL_1153668 | MVZ Labor Dr. Fenner und Kollegen (Standort Hamburg) | Robert Koch Institute | |
| EPI_ISL_1570731, EPI_ISL_1643403 | MVZ Labor Dr. Limbach & Kollegen GbR | Robert Koch Institute | |
| EPI_ISL_1122456, EPI_ISL_1213569, EPI_ISL_2105475, EPI_ISL_2105558 | Manila Doctors Hospital | Philippine Genome Center | Alethea R. de Guzman; Anna Ong-Lim; Arianne A. Zamora; Asia Louisa U. Chong; Benedict A. Maralit; Candice Francheska B. Tambaoan; Carlo M. Lapid; Celia Carlos; Devon Ray Pacial; Edsel Maurice Salvaña; El King D. Morado; Eva Maria Cutiongco-de la Paz; Francis A. Tablizo; Irish Coleen A. Asin; Jaime C. Montoya; Jan Michael C. Yap; Jo-Hannah S. Llames; John Q. Wong; Joshua Gregor A. Dizon; Juan Antonio R. Magalang; Karol Sophia Agape R. Padilla; Kenneth M. Kim; Kris P. Punayan; Marc Edsel C. Ayes; Marc Jerrone R. Castro; Maria Rosario Singh-Vergeire and Cynthia P. Saloma; Maria Sofia L. Yangzon; Marissa Alejandria; Razel Nikka M. Hao; Rianna Patricia S. Cruz; Sheila Mae M. Araiza |
| EPI_ISL_2189441, EPI_ISL_2860180 | Mary Mediatrix Medical Center | Philippine Genome Center | Alethea R. de Guzman; Anna Ong-Lim; Arianne A. Zamora; Asia Louisa U. Chong; Benedict A. Maralit; Candice Francheska B. Tambaoan; Carlo M. Lapid; Celia Carlos; Devon Ray Pacial; Diomedes A. Carino; Edsel Maurice Salvaña; El King D. Morado; Elcid Aaron R. Pangilinan; Eva Maria Cutiongco-de la Paz; Francis A. Tablizo; Henrietta Marie Rodriguez; Irish Coleen A. Asin; Jaime C. Montoya; Jan Michael C. Yap; Jarvin E. Nipales; Jo-Hannah S. Llames; John Q. Wong; Joshua Gregor A. Dizon; Juan Antonio R. Magalang; Karol Sophia Agape R. Padilla; Kenneth M. Kim; Kris P. Punayan; Krisitna Patriz Dela Cruz; Lindsay Claire D.L. Carandang; Ma. Exanil Plantig; Marc Edsel C. Ayes; Maria Rosario Singh-Vergeire and Cynthia P. Saloma; Maria Sofia L. Yangzon; Marielle M Gamboa; Marissa Alejandria; Nina Francesca Bustamante; Razel Nikka M. Hao; Renato Jacinto Q. Mantaring; Rianna Patricia S. Cruz; Sheila Mae M. Araiza; Yvonne Valerie Austria; Zipporah Mariebelle R. Enriquez; Zyrel V. Mollejon |
| EPI_ISL_1315325, EPI_ISL_1469105 | Middlemore Hospital | Institute of Environmental Science and Research (ESR) | Anja Werno; Antje van der Linden; Arlo Upton; Chris Mansell; David Hammer; Dragana Drinkovic; Erasmus Smit; Gary McAuliffe; Hana Sofia Andersson; Hermes Perez; James Ussher; Jill Sherwood; Jing Wang; Joep de Ligt; Josh Freeman; Julia Howard; Juliet Elvy; Lauren Jelly; Mary DeAlmeida; Matt Blakiston; Matt Storey; Matthew Rogers; Max Bloomfield; Michael Addidle; Michelle Balm; Muhammad Faisal; Nikki Freed; Olin Silander; Olivia Stroeven; Rachel Boyle; Sally Roberts; SallyAnn Harbison; Sarah Jefferies; Sharmini Muttaiyah; Susan Morpeth; Susan Taylor; Timothy Blackmore; Vani Sathyendran; Veronica Playle; Virginia Hope; Xiaoyun Ren |
| EPI_ISL_2535610, EPI_ISL_2535650, EPI_ISL_2535665, EPI_ISL_2535667, EPI_ISL_2535668, EPI_ISL_2535773, EPI_ISL_2535774 | | | |
| see above | Ministry of Health Hospitals | Institute of Health and Community Medicine | Chan Chia Jui; Chua Hock Hin; David Perera; Ooi Mong How; Tonnii Sia Loong Loong; Wong Jyn Shan; Wong Kiing Aik |
| EPI_ISL_2171098 | National Kidney and Transplant Institute (NKTI) | Philippine Genome Center | Alethea R. de Guzman; Anna Ong-Lim; Arianne A. Zamora; Asia Louisa U. Chong; Benedict A. Maralit; Candice Francheska B. Tambaoan; Carlo M. Lapid; Celia Carlos; Devon Ray Pacial; Edsel Maurice Salvaña; El King D. Morado; Elcid Aaron R. Pangilinan; Eva Maria Cutiongco-de la Paz; Francis A. Tablizo; Irish Coleen A. Asin; Jaime C. Montoya; Jan Michael C. Yap; Jo-Hannah S. Llames; John Q. Wong; Joshua Gregor A. Dizon; Juan Antonio R. Magalang; Karol Sophia Agape R. Padilla; Kenneth M. Kim; Kris P. Punayan; Marc Edsel C. Ayes; Marc Jerrone R. Castro; Maria Rosario Singh-Vergeire and Cynthia P. Saloma; Maria Sofia L. Yangzon; Marissa Alejandria; Razel Nikka M. Hao; Rianna Patricia S. Cruz; Sheila Mae M. Araiza |
| EPI_ISL_1652091, EPI_ISL_2349790, EPI_ISL_2508713 | National Public Health Laboratory, National Centre for Infectious Diseases | National Public Health Laboratory, National Centre for Infectious Diseases | Grace Jie Yin Ngan; Lin Cui; Raymond Tzer Pin Lin; Royce Ang; Tze Minn Mak; Zhenyang Zhou |
| EPI_ISL_1213571, EPI_ISL_1213573 | Negros Oriental Provincial Hospital | Philippine Genome Center | Alethea R. de Guzman; Anna Ong-Lim; Arianne A. Zamora; Asia Louisa U. Chong; Benedict A. Maralit; Candice Francheska B. Tambaoan; Carlo M. Lapid; Celia Carlos; Devon Ray Pacial; Edsel Maurice Salvaña; El King D. Morado; Eva Maria Cutiongco-de la Paz; Francis A. Tablizo; Irish Coleen A. Asin; Jaime C. Montoya; Jan Michael C. Yap; Jo-Hannah S. Llames; John Q. Wong; Joshua Gregor A. Dizon; Juan Antonio R. Magalang; Karol Sophia Agape R. Padilla; Kenneth M. Kim; Kris P. Punayan; Marc Edsel C. Ayes; Marc Jerrone R. Castro; Maria Rosario Singh-Vergeire and Cynthia P. Saloma; Maria Sofia L. Yangzon; Marissa Alejandria; Razel Nikka M. Hao; Rianna Patricia S. Cruz; Sheila Mae M. Araiza |
| EPI_ISL_2189152, EPI_ISL_2189187, EPI_ISL_2189350 | Northern Mindanao TB Regional Center | Philippine Genome Center | Alethea R. de Guzman; Anna Ong-Lim; Arianne A. Zamora; Asia Louisa U. Chong; Benedict A. Maralit; Candice Francheska B. Tambaoan; Carlo M. Lapid; Celia Carlos; Devon Ray Pacial; Edsel Maurice Salvaña; El King D. Morado; Elcid Aaron R. Pangilinan; Eva Maria Cutiongco-de la Paz; Francis A. Tablizo; Irish Coleen A. Asin; Jaime C. Montoya; Jan Michael C. Yap; Jo-Hannah S. Llames; John Q. Wong; Joshua Gregor A. Dizon; Juan Antonio R. Magalang; Karol Sophia Agape R. Padilla; Kenneth M. Kim; Kris P. Punayan; Marc Edsel C. Ayes; Maria Rosario Singh-Vergeire and Cynthia P. Saloma; Maria Sofia L. Yangzon; Marissa Alejandria; Razel Nikka M. Hao; Renato Jacinto Q. Mantaring; Rianna Patricia S. Cruz; Sheila Mae M. Araiza |
| EPI_ISL_2188053, EPI_ISL_2188249 | OSPITAL NG MUNTINLUPA | Philippine Genome Center | Alethea R. de Guzman; Anna Ong-Lim; Arianne A. Zamora; Asia Louisa U. Chong; Benedict A. Maralit; Candice Francheska B. Tambaoan; Carlo M. Lapid; Celia Carlos; Devon Ray Pacial; Edsel Maurice Salvaña; El King D. Morado; Elcid Aaron R. Pangilinan; Eva Maria Cutiongco-de la Paz; Francis A. Tablizo; Irish Coleen A. Asin; Jaime C. Montoya; Jan Michael C. Yap; Jo-Hannah S. Llames; John Q. Wong; Joshua Gregor A. Dizon; Juan Antonio R. Magalang; Karol Sophia Agape R. Padilla; Kenneth M. Kim; Kris P. Punayan; Marc Edsel C. Ayes; Marc Jerrone R. Castro; Maria Rosario Singh-Vergeire and Cynthia P. Saloma; Maria Sofia L. Yangzon; Marissa Alejandria; Razel Nikka M. Hao; Rianna Patricia S. Cruz; Sheila Mae M. Araiza |
| EPI_ISL_2859906 | Ospital ng Imus | Philippine Genome Center | Alethea R. de Guzman; Anna Ong-Lim; Arianne A. Zamora; Benedict A. Maralit; Carlo M. Lapid; Celia Carlos; Devon Ray Pacial; Diomedes A. Carino; Edsel Maurice Salvaña; El King D. Morado; Elcid Aaron R. Pangilinan; Eva Maria Cutiongco-de la Paz; Francis A. Tablizo; Henrietta Marie Rodriguez; Jaime C. Montoya; Jan Michael C. Yap; Jarvin E. Nipales; Jo-Hannah S. Llames; John Q. Wong; Joshua Gregor A. Dizon; Juan Antonio R. Magalang; Karol Sophia Agape R. Padilla; Kenneth M. Kim; Kris P. Punayan; Krisitna Patriz Dela Cruz; Lindsay Claire D.L. Carandang; Ma. Exanil Plantig; Marc Edsel C. Ayes; Maria Rosario Singh-Vergeire and Cynthia P. Saloma; Maria Sofia L. Yangzon; Marielle M Gamboa; Marissa Alejandria; Nina Francesca Bustamante; Razel Nikka M. Hao; Renato Jacinto Q. Mantaring; Rianna Patricia S. Cruz; Sheila Mae M. Araiza; Yvonne Valerie Austria; Zipporah Mariebelle R. Enriquez; Zyrel V. Mollejon |
| EPI_ISL_1213574, EPI_ISL_1213576, EPI_ISL_2105474, EPI_ISL_2105478, EPI_ISL_2156284 | PHILIPPINE AIRPORT DIAGNOSTIC LABORATORY | Philippine Genome Center | Alethea R. de Guzman; Anna Ong-Lim; Arianne A. Zamora; Asia Louisa U. Chong; Benedict A. Maralit; Candice Francheska B. Tambaoan; Carlo M. Lapid; Celia Carlos; Devon Ray Pacial; Edsel Maurice Salvaña; El King D. Morado; Elcid Aaron R. Pangilinan; Eva Maria Cutiongco-de la Paz; Francis A. Tablizo; Irish Coleen A. Asin; Jaime C. Montoya; Jan Michael C. Yap; Jo-Hannah S. Llames; John Q. Wong; Joshua Gregor A. Dizon; Juan Antonio R. Magalang; Karol Sophia Agape R. Padilla; Kenneth M. Kim; Kris P. Punayan; Marc Edsel C. Ayes; Marc Jerrone R. Castro; Maria Rosario Singh-Vergeire and Cynthia P. Saloma; Maria Sofia L. Yangzon; Marissa Alejandria; Razel Nikka M. Hao; Rianna Patricia S. Cruz; Sheila Mae M. Araiza |
| EPI_ISL_2156490 | Philippine Airport Diagnostic Laboratory | Philippine Genome Center | Alethea R. de Guzman; Anna Ong-Lim; Arianne A. Zamora; Asia Louisa U. Chong; Benedict A. Maralit; Candice Francheska B. Tambaoan; Carlo M. Lapid; Celia Carlos; Devon Ray Pacial; Edsel Maurice Salvaña; El King D. Morado; Elcid Aaron R. Pangilinan; Eva Maria Cutiongco-de la Paz; Francis A. Tablizo; Irish Coleen A. Asin; Jaime C. Montoya; Jan Michael C. Yap; Jo-Hannah S. Llames; John Q. Wong; Joshua Gregor A. Dizon; Juan Antonio R. Magalang; Karol Sophia Agape R. Padilla; Kenneth M. Kim; Kris P. Punayan; Marc Edsel C. Ayes; Marc Jerrone R. Castro; Maria Rosario Singh-Vergeire and Cynthia P. Saloma; Maria Sofia L. Yangzon; Marissa Alejandria; Razel Nikka M. Hao; Rianna Patricia S. Cruz; Sheila Mae M. Araiza |
| EPI_ISL_2105472, EPI_ISL_2189095 | Philippine Red Cross - National Blood Center | Philippine Genome Center | Alethea R. de Guzman; Anna Ong-Lim; Arianne A. Zamora; Asia Louisa U. Chong; Benedict A. Maralit; Candice Francheska B. Tambaoan; Carlo M. Lapid; Celia Carlos; Devon Ray Pacial; Edsel Maurice Salvaña; El King D. Morado; Elcid Aaron R. Pangilinan; Eva Maria Cutiongco-de la Paz; Francis A. Tablizo; Irish Coleen A. Asin; Jaime C. Montoya; Jan Michael C. Yap; Jo-Hannah S. Llames; John Q. Wong; Joshua Gregor A. Dizon; Juan Antonio R. Magalang; Karol Sophia Agape R. Padilla; Kenneth M. Kim; Kris P. Punayan; Marc Edsel C. Ayes; Marc Jerrone R. Castro; Maria Rosario Singh-Vergeire and Cynthia P. Saloma; Maria Sofia L. Yangzon; Marissa Alejandria; Razel Nikka M. Hao; Renato Jacinto Q. Mantaring; Rianna Patricia S. Cruz; Sheila Mae M. Araiza |
| EPI_ISL_1213578, EPI_ISL_2156334, EPI_ISL_2156391, EPI_ISL_2171148, EPI_ISL_2171174, EPI_ISL_2171237, EPI_ISL_2171239, EPI_ISL_2188075, EPI_ISL_2188699, EPI_ISL_2188811, EPI_ISL_2188826, EPI_ISL_2188853, EPI_ISL_2188892, EPI_ISL_2188938, EPI_ISL_2189035, EPI_ISL_2189124, EPI_ISL_2189194, EPI_ISL_2859599, EPI_ISL_2859908, EPI_ISL_2860178, EPI_ISL_2860179, EPI_ISL_2860181 | | | |
| see above | Philippine Red Cross - Port Area | Philippine Genome Center | Alethea R. de Guzman; Anna Ong-Lim; Arianne A. Zamora; Asia Louisa U. Chong; Benedict A. Maralit; Candice Francheska B. Tambaoan; Carlo M. Lapid; Celia Carlos; Devon Ray Pacial; Diomedes A. Carino; Edsel Maurice Salvaña; El King D. Morado; Elcid Aaron R. Pangilinan; Eva Maria Cutiongco-de la Paz; Francis A. Tablizo; Henrietta Marie Rodriguez; Irish Coleen A. Asin; Jaime C. Montoya; Jan Michael C. Yap; Jarvin E. Nipales; Jo-Hannah S. Llames; John Q. Wong; Joshua Gregor A. Dizon; Juan Antonio R. Magalang; Karol Sophia Agape R. Padilla; Kenneth M. Kim; Kris P. Punayan; Krisitna Patriz Dela Cruz; Lindsay Claire D.L. Carandang; Ma. Exanil Plantig; Marc Edsel C. Ayes; Marc Jerrone R. Castro; Maria Rosario Singh-Vergeire and Cynthia P. Saloma; Maria Sofia L. Yangzon; Marielle M Gamboa; Marissa Alejandria; Nina Francesca Bustamante; Razel Nikka M. Hao; Renato Jacinto Q. Mantaring; Rianna Patricia S. Cruz; Sheila Mae M. Araiza; Yvonne Valerie Austria; Zipporah Mariebelle R. Enriquez; Zyrel V. Mollejon |
| EPI_ISL_1213580, EPI_ISL_2156294, EPI_ISL_2188199, EPI_ISL_2188697 | Philippine Red Cross Logistics and Multipurpose Center | Philippine Genome Center | Alethea R. de Guzman; Anna Ong-Lim; Arianne A. Zamora; Asia Louisa U. Chong; Benedict A. Maralit; Candice Francheska B. Tambaoan; Carlo M. Lapid; Celia Carlos; Devon Ray Pacial; Edsel Maurice Salvaña; El King D. Morado; Elcid Aaron R. Pangilinan; Eva Maria Cutiongco-de la Paz; Francis A. Tablizo; Irish Coleen A. Asin; Jaime C. Montoya; Jan Michael C. Yap; Jo-Hannah S. Llames; John Q. Wong; Joshua Gregor A. Dizon; Juan Antonio R. Magalang; Karol Sophia Agape R. Padilla; Kenneth M. Kim; Kris P. Punayan; Marc Edsel C. Ayes; Marc Jerrone R. Castro; Maria Rosario Singh-Vergeire and Cynthia P. Saloma; Maria Sofia L. Yangzon; Marissa Alejandria; Razel Nikka M. Hao; Renato Jacinto Q. Mantaring; Rianna Patricia S. Cruz; Sheila Mae M. Araiza |
| EPI_ISL_2501732, EPI_ISL_2501733, EPI_ISL_2501734, EPI_ISL_2501735 | Premier Laboratory Services | BioInfoExperts LLC | Cheryl Metzler; David Nolan; Rebecca Rose; Susanna Lamers; Valerie Murray |
| EPI_ISL_1213581 | Prime Care Alpha Covid-19 Testing Laboratory | Philippine Genome Center | Alethea R. de Guzman; Anna Ong-Lim; Arianne A. Zamora; Asia Louisa U. Chong; Benedict A. Maralit; Candice Francheska B. Tambaoan; Carlo M. Lapid; Celia Carlos; Devon Ray Pacial; Edsel Maurice Salvaña; El King D. Morado; Eva Maria Cutiongco-de la Paz; Francis A. Tablizo; Irish Coleen A. Asin; Jaime C. Montoya; Jan Michael C. Yap; Jo-Hannah S. Llames; John Q. Wong; Joshua Gregor A. Dizon; Juan Antonio R. Magalang; Karol Sophia Agape R. Padilla; Kenneth M. Kim; Kris P. Punayan; Marc Edsel C. Ayes; Marc Jerrone R. Castro; Maria Rosario Singh-Vergeire and Cynthia P. Saloma; Maria Sofia L. Yangzon; Marissa Alejandria; Razel Nikka M. Hao; Rianna Patricia S. Cruz; Sheila Mae M. Araiza |
| EPI_ISL_2859862 | Qualimed Hospital Sta. Rosa | Philippine Genome Center | Alethea R. de Guzman; Anna Ong-Lim; Arianne A. Zamora; Benedict A. Maralit; Carlo M. Lapid; Celia Carlos; Devon Ray Pacial; Diomedes A. Carino; Edsel Maurice Salvaña; El King D. Morado; Elcid Aaron R. Pangilinan; Eva Maria Cutiongco-de la Paz; Francis A. Tablizo; Henrietta Marie Rodriguez; Jaime C. Montoya; Jan Michael C. Yap; Jarvin E. Nipales; Jo-Hannah S. Llames; John Q. Wong; Joshua Gregor A. Dizon; Juan Antonio R. Magalang; Karol Sophia Agape R. Padilla; Kenneth M. Kim; Kris P. Punayan; Krisitna Patriz Dela Cruz; Lindsay Claire D.L. Carandang; Ma. Exanil Plantig; Marc Edsel C. Ayes; Maria Rosario Singh-Vergeire and Cynthia P. Saloma; Maria Sofia L. Yangzon; Marielle M Gamboa; Marissa Alejandria; Nina Francesca Bustamante; Razel Nikka M. Hao; Renato Jacinto Q. Mantaring; Rianna Patricia S. Cruz; Sheila Mae M. Araiza; Yvonne Valerie Austria; Zipporah Mariebelle R. Enriquez; Zyrel V. Mollejon |

| | | | |
|---|---|---|---|
| EPI_ISL_1273082, EPI_ISL_1306121, EPI_ISL_2661519 | Queensland Health Forensic and Scientific Services | Queensland Health Forensic and Scientific Services | Son Nguyen |
| EPI_ISL_2156354, EPI_ISL_2156357, EPI_ISL_2156359, EPI_ISL_2188079, EPI_ISL_2188756, EPI_ISL_2188848 | Research Institute for Tropical Medicine, Inc. (RITM) | Philippine Genome Center | Alethea R. de Guzman; Anna Ong-Lim; Arianne A. Zamora; Asia Louisa U. Chong; Benedict A. Maralit; Candice Francheska B. Tambaoan; Carlo M. Lapid; Celia Carlos; Devon Ray Pacial; Edsel Maurice Salvaña; El King D. Morado; Elcid Aaron R. Pangilinan; Eva Maria Cutiongco-de la Paz; Francis A. Tablizo; Irish Coleen A. Asin; Jaime C. Montoya; Jan Michael C. Yap; Jo-Hannah S. Llames; John Q. Wong; Joshua Gregor A. Dizon; Juan Antonio R. Magalang; Karol Sophia Agape R. Padilla; Kenneth M. Kim; Kris P. Punayan; Marc Edsel C. Ayes; Marc Jerrone R. Castro; Maria Rosario Singh-Vergeire and Cynthia P. Saloma; Maria Sofia L. Yangzon; Marissa Alejandria; Razel Nikka M. Hao; Renato Jacinto Q. Mantaring; Rianna Patricia S. Cruz; Sheila Mae M. Araiza |
| EPI_ISL_1229178 | SA Pathology | SA Pathology | Chuan Kok Lim; Geoff Higgins; Ivan Bastian; Julien Soubrier; Karin Kassahn; Lex Leong; Mark Turra; Song Gao |
| EPI_ISL_1198832, EPI_ISL_1425714, EPI_ISL_1425715, EPI_ISL_2253110, EPI_ISL_2828258, EPI_ISL_3076715 | SARS-CoV-2 testing team, National Institute of Infectious Diseases | Pathogen Genomics Center, National Institute of Infectious Diseases | Chang-Kweng Lim; Daisuke Kobayashi; Eri Nakayama; Hazuka Y Furihata; Hiromizu Takahashi; Hussein H. Aly; Kentaro Itokawa; Kousho Wakae; Makoto Kuroda; Masanori Hashino; Masumichi Saito; Motohiko Ogawa; Naomi Nojiri; Nozomu Hanaoka; Rina Tanaka; Sana Uchikoba; Shigeru Tanjima; Takahiro Maeki; Takanobu Kato; Tsuguto Fujimoto; Tsuyoshi Sekizuka |
| EPI_ISL_2883244, EPI_ISL_2883487 | SK-Roy Romanow Provincial Laboratory | National Microbiology Laboratory (NML) | Alanna Senecal; Amanda Lang; Anna Majer; Anneliese Landgraff; CanCOGeN's metadata curation team; Darian Hole; Elsie Grudeski; Gary Van Domselaar; Grace Seo; Jennifer Tanner; Jessica Minion; Kara Loos; Keith MacKenzie; Kirsten Biggar; Madison Chapel; Meredith Faires; Morag Graham; Natalie Knox; Nathalie Bastien; Philip Mabon; Public Health Agency of Canada CanCOGeN team; Rachel DePaulo; Rhiannon Huzarewich; Russell Mandes; Ryan McDonald; Shari Tyson; Timothy Booth; Yan Li |
| EPI_ISL_2930742, EPI_ISL_2930789 | SYNLAB | GIGA Medical Genomics | Bouchra Boujemla; Claire Gourzonès; Cécile Meex; Keith Durkin; Laurent Gillet; Maria Artesi; Marie-Pierre Hayette; Nadine Cambisano; Nathalie Renotte; Olivier Ek; Sébastien Bontems; Vincent Bours |
| EPI_ISL_2171206 | South Super Highway Molecular Diagnostic Laboratory | Philippine Genome Center | Alethea R. de Guzman; Anna Ong-Lim; Arianne A. Zamora; Asia Louisa U. Chong; Benedict A. Maralit; Candice Francheska B. Tambaoan; Carlo M. Lapid; Celia Carlos; Devon Ray Pacial; Edsel Maurice Salvaña; El King D. Morado; Elcid Aaron R. Pangilinan; Eva Maria Cutiongco-de la Paz; Francis A. Tablizo; Irish Coleen A. Asin; Jaime C. Montoya; Jan Michael C. Yap; Jo-Hannah S. Llames; John Q. Wong; Joshua Gregor A. Dizon; Juan Antonio R. Magalang; Karol Sophia Agape R. Padilla; Kenneth M. Kim; Kris P. Punayan; Marc Edsel C. Ayes; Marc Jerrone R. Castro; Maria Rosario Singh-Vergeire and Cynthia P. Saloma; Maria Sofia L. Yangzon; Marissa Alejandria; Razel Nikka M. Hao; Rianna Patricia S. Cruz; Sheila Mae M. Araiza |
| EPI_ISL_1122455 | Southern Philippines Medical Center | Philippine Genome Center | Alethea R. de Guzman; Anna Ong-Lim; Arianne A. Zamora; Asia Louisa U. Chong; Benedict A. Maralit; Candice Francheska B. Tambaoan; Carlo M. Lapid; Celia Carlos; Devon Ray Pacial; Edsel Maurice Salvaña; El King D. Morado; Eva Maria Cutiongco-de la Paz; Francis A. Tablizo; Irish Coleen A. Asin; Jaime C. Montoya; Jan Michael C. Yap; Jo-Hannah S. Llames; John Q. Wong; Joshua Gregor A. Dizon; Juan Antonio R. Magalang; Karol Sophia Agape R. Padilla; Kenneth M. Kim; Kris P. Punayan; Marc Edsel C. Ayes; Marc Jerrone R. Castro; Maria Rosario Singh-Vergeire and Cynthia P. Saloma; Maria Sofia L. Yangzon; Marissa Alejandria; Razel Nikka M. Hao; Rianna Patricia S. Cruz; Sheila Mae M. Araiza |
| EPI_ISL_3156962 | Swedish national genomic surveillance program of SARS-CoV-2 | The Public Health Agency of Sweden | Alma Brolund; Maria Lind Karlberg; Maximilian Riess; Swedish national genomic surveillance program of SARS-CoV-2 |
| EPI_ISL_2105462, EPI_ISL_2105489, EPI_ISL_2156263, EPI_ISL_2156264 | THE LORD'S GRACE MEDICAL AND INDUSTRIAL CLINIC | Philippine Genome Center | Alethea R. de Guzman; Anna Ong-Lim; Arianne A. Zamora; Asia Louisa U. Chong; Benedict A. Maralit; Candice Francheska B. Tambaoan; Carlo M. Lapid; Celia Carlos; Devon Ray Pacial; Edsel Maurice Salvaña; El King D. Morado; Elcid Aaron R. Pangilinan; Eva Maria Cutiongco-de la Paz; Francis A. Tablizo; Irish Coleen A. Asin; Jaime C. Montoya; Jan Michael C. Yap; Jo-Hannah S. Llames; John Q. Wong; Joshua Gregor A. Dizon; Juan Antonio R. Magalang; Karol Sophia Agape R. Padilla; Kenneth M. Kim; Kris P. Punayan; Marc Edsel C. Ayes; Marc Jerrone R. Castro; Maria Rosario Singh-Vergeire and Cynthia P. Saloma; Maria Sofia L. Yangzon; Marissa Alejandria; Razel Nikka M. Hao; Rianna Patricia S. Cruz; Sheila Mae M. Araiza |
| EPI_ISL_1213583 | Teresita Jalandoni Provincial Hospital | Philippine Genome Center | Alethea R. de Guzman; Anna Ong-Lim; Arianne A. Zamora; Asia Louisa U. Chong; Benedict A. Maralit; Candice Francheska B. Tambaoan; Carlo M. Lapid; Celia Carlos; Devon Ray Pacial; Edsel Maurice Salvaña; El King D. Morado; Eva Maria Cutiongco-de la Paz; Francis A. Tablizo; Irish Coleen A. Asin; Jaime C. Montoya; Jan Michael C. Yap; Jo-Hannah S. Llames; John Q. Wong; Joshua Gregor A. Dizon; Juan Antonio R. Magalang; Karol Sophia Agape R. Padilla; Kenneth M. Kim; Kris P. Punayan; Marc Edsel C. Ayes; Marc Jerrone R. Castro; Maria Rosario Singh-Vergeire and Cynthia P. Saloma; Maria Sofia L. Yangzon; Marissa Alejandria; Razel Nikka M. Hao; Rianna Patricia S. Cruz; Sheila Mae M. Araiza |
| EPI_ISL_1122458, EPI_ISL_1213585, EPI_ISL_1213586, EPI_ISL_1219701, EPI_ISL_2189090, EPI_ISL_2189463 | The Lord's Grace Medical and Industrial Clinic | Philippine Genome Center | Alethea R. de Guzman; Anna Ong-Lim; Arianne A. Zamora; Asia Louisa U. Chong; Benedict A. Maralit; Candice Francheska B. Tambaoan; Carlo M. Lapid; Celia Carlos; Devon Ray Pacial; Edsel Maurice Salvaña; El King D. Morado; Elcid Aaron R. Pangilinan; Eva Maria Cutiongco-de la Paz; Francis A. Tablizo; Irish Coleen A. Asin; Jaime C. Montoya; Jan Michael C. Yap; Jo-Hannah S. Llames; John Q. Wong; Joshua Gregor A. Dizon; Juan Antonio R. Magalang; Karol Sophia Agape R. Padilla; Kenneth M. Kim; Kris P. Punayan; Marc Edsel C. Ayes; Marc Jerrone R. Castro; Maria Rosario Singh-Vergeire and Cynthia P. Saloma; Maria Sofia L. Yangzon; Marissa Alejandria; Razel Nikka M. Hao; Renato Jacinto Q. Mantaring; Rianna Patricia S. Cruz; Sheila Mae M. Araiza |
| EPI_ISL_2189408, EPI_ISL_2860175, EPI_ISL_2860176, EPI_ISL_2860177 | The Medical City | Philippine Genome Center | Alethea R. de Guzman; Anna Ong-Lim; Arianne A. Zamora; Asia Louisa U. Chong; Benedict A. Maralit; Candice Francheska B. Tambaoan; Carlo M. Lapid; Celia Carlos; Devon Ray Pacial; Diomedes A. Carino; Edsel Maurice Salvaña; El King D. Morado; Elcid Aaron R. Pangilinan; Eva Maria Cutiongco-de la Paz; Francis A. Tablizo; Henrietta Marie Rodriguez; Irish Coleen A. Asin; Jaime C. Montoya; Jan Michael C. Yap; Jarvin E. Nipales; Jo-Hannah S. Llames; John Q. Wong; Joshua Gregor A. Dizon; Juan Antonio R. Magalang; Karol Sophia Agape R. Padilla; Kenneth M. Kim; Kris P. Punayan; Krisitna Patriz Dela Cruz; Lindsay Claire D.L. Carandang; Ma. Exanil Plantig; Marc Edsel C. Ayes; Maria Rosario Singh-Vergeire and Cynthia P. Saloma; Maria Sofia L. Yangzon; Marielle M Gamboa; Marissa Alejandria; Nina Francesca Bustamante; Razel Nikka M. Hao; Renato Jacinto Q. Mantaring; Rianna Patricia S. Cruz; Sheila Mae M. Araiza; Yvonne Valerie Austria; Zipporah Mariebelle R. Enriquez; Zyrel V. Mollejon |
| EPI_ISL_1213588 | The Medical City - Ortigas | Philippine Genome Center | Alethea R. de Guzman; Anna Ong-Lim; Arianne A. Zamora; Asia Louisa U. Chong; Benedict A. Maralit; Candice Francheska B. Tambaoan; Carlo M. Lapid; Celia Carlos; Devon Ray Pacial; Edsel Maurice Salvaña; El King D. Morado; Eva Maria Cutiongco-de la Paz; Francis A. Tablizo; Irish Coleen A. Asin; Jaime C. Montoya; Jan Michael C. Yap; Jo-Hannah S. Llames; John Q. Wong; Joshua Gregor A. Dizon; Juan Antonio R. Magalang; Karol Sophia Agape R. Padilla; Kenneth M. Kim; Kris P. Punayan; Marc Edsel C. Ayes; Marc Jerrone R. Castro; Maria Rosario Singh-Vergeire and Cynthia P. Saloma; Maria Sofia L. Yangzon; Marissa Alejandria; Razel Nikka M. Hao; Rianna Patricia S. Cruz; Sheila Mae M. Araiza |
| EPI_ISL_2188058, EPI_ISL_2188167, EPI_ISL_2188203 | The Medical City -Â Ortigas | Philippine Genome Center | Alethea R. de Guzman; Anna Ong-Lim; Arianne A. Zamora; Asia Louisa U. Chong; Benedict A. Maralit; Candice Francheska B. Tambaoan; Carlo M. Lapid; Celia Carlos; Devon Ray Pacial; Edsel Maurice Salvaña; El King D. Morado; Elcid Aaron R. Pangilinan; Eva Maria Cutiongco-de la Paz; Francis A. Tablizo; Irish Coleen A. Asin; Jaime C. Montoya; Jan Michael C. Yap; Jo-Hannah S. Llames; John Q. Wong; Joshua Gregor A. Dizon; Juan Antonio R. Magalang; Karol Sophia Agape R. Padilla; Kenneth M. Kim; Kris P. Punayan; Marc Edsel C. Ayes; Marc Jerrone R. Castro; Maria Rosario Singh-Vergeire and Cynthia P. Saloma; Maria Sofia L. Yangzon; Marissa Alejandria; Razel Nikka M. Hao; Rianna Patricia S. Cruz; Sheila Mae M. Araiza |
| EPI_ISL_2104833, EPI_ISL_2161918, EPI_ISL_2333059 | UW Virology Lab | UW Virology Lab | Alexander Greninger; Hong Xie; Keith R Jerome; Lasata Shrestha; Meei-Li Huang; Michelle Lin; Nathan Breit; Noah R. Baker; Patrick Mathias; Pavitra Roychoudhury; Ricardo Perez; Robert J. Livingston; Sean Ellis; Shah Mohamed Bakhash; Tien V. Nguyen |
| EPI_ISL_2188103 | University of the Philippines National Institutes of Health (UP NIH) | Philippine Genome Center | Alethea R. de Guzman; Anna Ong-Lim; Arianne A. Zamora; Asia Louisa U. Chong; Benedict A. Maralit; Candice Francheska B. Tambaoan; Carlo M. Lapid; Celia Carlos; Devon Ray Pacial; Edsel Maurice Salvaña; El King D. Morado; Elcid Aaron R. Pangilinan; Eva Maria Cutiongco-de la Paz; Francis A. Tablizo; Irish Coleen A. Asin; Jaime C. Montoya; Jan Michael C. Yap; Jo-Hannah S. Llames; John Q. Wong; Joshua Gregor A. Dizon; Juan Antonio R. Magalang; Karol Sophia Agape R. Padilla; Kenneth M. Kim; Kris P. Punayan; Marc Edsel C. Ayes; Marc Jerrone R. Castro; Maria Rosario Singh-Vergeire and Cynthia P. Saloma; Maria Sofia L. Yangzon; Marissa Alejandria; Razel Nikka M. Hao; Rianna Patricia S. Cruz; Sheila Mae M. Araiza |
| EPI_ISL_1570318, EPI_ISL_1722848 | UniversitÃ¤tsklinikum Heidelberg | Robert Koch Institute | |
| EPI_ISL_1354944, EPI_ISL_1354960, EPI_ISL_1354990 | Universitätsklinikum Heidelberg | Robert Koch Institute | |
| EPI_ISL_2229455 | Virginia Division of Consolidated Laboratory Services | Virginia Division of Consolidated Laboratory Services | Virginia DCLS |
| EPI_ISL_2859279 | Zamboanga City Medical Center GeneXpert Laboratory | Philippine Genome Center | Alethea R. de Guzman; Anna Ong-Lim; Arianne A. Zamora; Benedict A. Maralit; Carlo M. Lapid; Celia Carlos; Devon Ray Pacial; Diomedes A. Carino; Edsel Maurice Salvaña; El King D. Morado; Elcid Aaron R. Pangilinan; Eva Maria Cutiongco-de la Paz; Francis A. Tablizo; Henrietta Marie Rodriguez; Jaime C. Montoya; Jan Michael C. Yap; Jarvin E. Nipales; Jo-Hannah S. Llames; John Q. Wong; Joshua Gregor A. Dizon; Juan Antonio R. Magalang; Karol Sophia Agape R. Padilla; Kenneth M. Kim; Kris P. Punayan; Krisitna Patriz Dela Cruz; Lindsay Claire D.L. Carandang; Ma. Exanil Plantig; Marc Edsel C. Ayes; Maria Rosario Singh-Vergeire and Cynthia P. Saloma; Maria Sofia L. Yangzon; Marielle M Gamboa; Marissa Alejandria; Nina Francesca Bustamante; Razel Nikka M. Hao; Renato Jacinto Q. Mantaring; Rianna Patricia S. Cruz; Sheila Mae M. Araiza; Yvonne Valerie Austria; Zipporah Mariebelle R. Enriquez; Zyrel V. Mollejon |
| EPI_ISL_2860218 | Zamboanga City Medical Center-Department of Agriculture Satellite Laboratory for COVID-19 Testing | Philippine Genome Center | Alethea R. de Guzman; Anna Ong-Lim; Arianne A. Zamora; Benedict A. Maralit; Carlo M. Lapid; Celia Carlos; Devon Ray Pacial; Diomedes A. Carino; Edsel Maurice Salvaña; El King D. Morado; Elcid Aaron R. Pangilinan; Eva Maria Cutiongco-de la Paz; Francis A. Tablizo; Henrietta Marie Rodriguez; Jaime C. Montoya; Jan Michael C. Yap; Jarvin E. Nipales; Jo-Hannah S. Llames; John Q. Wong; Joshua Gregor A. Dizon; Juan Antonio R. Magalang; Karol Sophia Agape R. Padilla; Kenneth M. Kim; Kris P. Punayan; Krisitna Patriz Dela Cruz; Lindsay Claire D.L. Carandang; Ma. Exanil Plantig; Marc Edsel C. Ayes; Maria Rosario Singh-Vergeire and Cynthia P. Saloma; Maria Sofia L. Yangzon; Marielle M Gamboa; Marissa Alejandria; Nina Francesca Bustamante; Razel Nikka M. Hao; Renato Jacinto Q. Mantaring; Rianna Patricia S. Cruz; Sheila Mae M. Araiza; Yvonne Valerie Austria; Zipporah Mariebelle R. Enriquez; Zyrel V. Mollejon |
| EPI_ISL_2859287, EPI_ISL_2859289 | Zamboanga Del Sur Medical Center PCR Laboratory | Philippine Genome Center | Alethea R. de Guzman; Anna Ong-Lim; Arianne A. Zamora; Benedict A. Maralit; Carlo M. Lapid; Celia Carlos; Devon Ray Pacial; Diomedes A. Carino; Edsel Maurice Salvaña; El King D. Morado; Elcid Aaron R. Pangilinan; Eva Maria Cutiongco-de la Paz; Francis A. Tablizo; Henrietta Marie Rodriguez; Jaime C. Montoya; Jan Michael C. Yap; Jarvin E. Nipales; Jo-Hannah S. Llames; John Q. Wong; Joshua Gregor A. Dizon; Juan Antonio R. Magalang; Karol Sophia Agape R. Padilla; Kenneth M. Kim; Kris P. Punayan; Krisitna Patriz Dela Cruz; Lindsay Claire D.L. Carandang; Ma. Exanil Plantig; Marc Edsel C. Ayes; Maria Rosario Singh-Vergeire and Cynthia P. Saloma; Maria Sofia L. Yangzon; Marielle M Gamboa; Marissa Alejandria; Nina Francesca Bustamante; Razel Nikka M. Hao; Renato Jacinto Q. Mantaring; Rianna Patricia S. Cruz; Sheila Mae M. Araiza; Yvonne Valerie Austria; Zipporah Mariebelle R. Enriquez; Zyrel V. Mollejon |

We gratefully acknowledge the following Authors from the Originating laboratories responsible for obtaining the specimens, as well as the Submitting laboratories where the genome data were generated and shared via GISAID, on which this research is based.

All Submitters of data may be contacted directly via www.gisaid.org

Authors are sorted alphabetically.

| Accession ID | Originating Laboratory | Submitting Laboratory | Authors |
|---|---|---|---|
| EPI_ISL_2713072 | ADPH | ADPH | Adrianna Walker |
| EPI_ISL_2718842 | BCCDC Public Health Laboratory | BCCDC Public Health Laboratory | Ana Pacagnella; Corrinne Ng; Dan Fornika; John Tyson; Kim Macdonald; Kimia Kamelian; Linda Hoang; Loretta Janz; Mel Krajden; Prystajecky Natalie; Robert Azana; Shannon Russell |
| EPI_ISL_2797327, EPI_ISL_2825616, EPI_ISL_2825645, EPI_ISL_2882986, EPI_ISL_2883019, EPI_ISL_2889897, EPI_ISL_2890889, EPI_ISL_2891103, EPI_ISL_2922030, EPI_ISL_2923591 | | | |
| see above | CDPH VBL | California Department of Public Health | CDPH-COVIDNet; UCLA Technology Center for Genomics & Bioinformatics; UCSF Center for Advanced Technology |
| EPI_ISL_2837366, EPI_ISL_2837369 | California Department of Public Health Valencia Branch Laboratory (CDPH VBL) | California Department of Public Health | CDPH-COVIDNet |
| EPI_ISL_2760812, EPI_ISL_2760814, EPI_ISL_2760815, EPI_ISL_2760817, EPI_ISL_2760819, EPI_ISL_2760820, EPI_ISL_2760824, EPI_ISL_2760825, EPI_ISL_2760827, EPI_ISL_2760828, EPI_ISL_2760832, EPI_ISL_2760833, EPI_ISL_2760837, EPI_ISL_2764039, EPI_ISL_2764040, EPI_ISL_2764041, EPI_ISL_2764042, EPI_ISL_2764043, EPI_ISL_2764044, EPI_ISL_2764045, EPI_ISL_2764046, EPI_ISL_2764047, EPI_ISL_2764048, EPI_ISL_2764049, EPI_ISL_2768844, EPI_ISL_2768845, EPI_ISL_2768846 | | | |
| see above | Chiba Prefectural Institute of Public Health | Pathogen Genomics Center, National Institute of Infectious Diseases | Kentaro Itokawa; Makoto Kuroda; Masanori Hashino; Rina Tanaka; Tsuyoshi Sekizuka |
| EPI_ISL_2627831, EPI_ISL_2627843, EPI_ISL_2657779 | Croatian Institute of Public Health | Croatian Institute of Public Health | Irena Tabain; Ivana Ferenčak |
| EPI_ISL_2674604, EPI_ISL_2723738 | Curative | New Mexico Department of Health Scientific Laboratory | Anastacia Griego-Fisher; D'eldra Malone; Ellie Johnson; Jennifer Benoit; Keila Gutierrez; Linda Salazar; Ratheesh Rajan |
| EPI_ISL_2810810, EPI_ISL_2811254, EPI_ISL_2811255, EPI_ISL_2811256, EPI_ISL_2811257 | Department of Infectious Diseases, Kobe Institute of Health | Department of Infectious Diseases, Kobe Institute of Health | Kentaro Itokawa; Makoto Kuroda; Masanori Hashino; Noriko Nakanishi; Rina Tanaka; Ryohei Nomoto; Tomotada Iwamoto; Tsuyoshi Sekizuka |
| EPI_ISL_2836002 | Epidemiology of Microbial Diseases, Yale School of Public Health | Epidemiology of Microbial Diseases, Yale School of Public Health | A.E.; A.F.; Alpert, T.; Breban; Brito; C.B.; C.C.; Fauver; Grubaugh; I.M.; Iwasaki, A.; J.E.; J.R.; Kalinich; Landry; Lucas, C.; M.E.; M.I.; M.L.; N.D.; Ott; Petrone; Rothman; Vogels; Watkins |
| EPI_ISL_2662912 | Erie County Public Health (ECPHL) | University at Buffalo Genomics and Bioinformatics Core | Alyssa Pohlman; Amanda Boccolucci; Brandon Marzullo; Donald Yergeau; Jennifer Surtees; Jonathan Bard; Natalie Lamb; Norma Nowak |
| EPI_ISL_2836352 | Florida Bureau of Public Health Laboratories | Florida Bureau of Public Health Laboratories | Jason Blanton; Sarah Schmedes |
| EPI_ISL_2772733, EPI_ISL_2772751, EPI_ISL_2772755, EPI_ISL_2772790, EPI_ISL_2772791, EPI_ISL_2772837, EPI_ISL_2772838, EPI_ISL_2772875, EPI_ISL_2772876, EPI_ISL_2772916, EPI_ISL_2773064, EPI_ISL_2773100, EPI_ISL_2773101, EPI_ISL_2773113, EPI_ISL_2773114, EPI_ISL_2773115, EPI_ISL_2773175, EPI_ISL_2773176, EPI_ISL_2773177, EPI_ISL_2773178, EPI_ISL_2773179, EPI_ISL_2773180, EPI_ISL_2773181, EPI_ISL_2773182, EPI_ISL_2773183, EPI_ISL_2773184, EPI_ISL_2773185, EPI_ISL_2773186, EPI_ISL_2773187, EPI_ISL_2773188, EPI_ISL_2773189, EPI_ISL_2773207, EPI_ISL_2773208, EPI_ISL_2773313, EPI_ISL_2773332, EPI_ISL_2773417 | | | |
| see above | Fukuoka Institute of Health and Environmental Sciences | Fukuoka Institute of Health and Environmental Sciences | Akira Ohishi; Carl Yuki; Chiharu Katamune; Hiroaki Shigemura; Kentaro Itokawa; Makoto Kuroda; Masanori Hashino; Mitsuhiro Hamasaki; Rina Tanaka; Saori Ueda; Takayuki Kobayashi; Tsuyoshi Sekizuka; Yoshiki Etoh; Yuki Ashizuka |
| EPI_ISL_2812187, EPI_ISL_2812190, EPI_ISL_2812191, EPI_ISL_2812192, EPI_ISL_2812193, EPI_ISL_2812194, EPI_ISL_2812195, EPI_ISL_2812196, EPI_ISL_2812197, EPI_ISL_2812198, EPI_ISL_2812199, EPI_ISL_2812200, EPI_ISL_2812201, EPI_ISL_2812202, EPI_ISL_2812203, EPI_ISL_2812204, EPI_ISL_2812205, EPI_ISL_2812206, EPI_ISL_2812207, EPI_ISL_2812208, EPI_ISL_2812209, EPI_ISL_2812210, EPI_ISL_2812211, EPI_ISL_2812212, EPI_ISL_2812213, EPI_ISL_2812214, EPI_ISL_2812215, EPI_ISL_2812216, EPI_ISL_2812217, EPI_ISL_2812218, EPI_ISL_2812219, EPI_ISL_2812220, EPI_ISL_2812221, EPI_ISL_2812222, EPI_ISL_2812226, EPI_ISL_2812227, EPI_ISL_2812228, EPI_ISL_2812229, EPI_ISL_2812230 | | | |
| see above | Fukushima Prefectural Institute of Public Health | Fukushima Prefectural Institute of Public Health | Kadzuhiro Kitakawa; Kazunori Suzuki; Kentaro Itokawa; Makoto Kuroda; Masanori Hashino; Naoko Kashiwabara; Rie Suzuki; Rina Tanaka; Shohei Fujita; Tsuyoshi Sekizuka |
| EPI_ISL_2621731, EPI_ISL_2657371 | Genome Analysis Center, Yamanashi Central Hospital | Genome Analysis Center, Yamanashi Central Hospital | Yosuke Hirotsu |
| EPI_ISL_2759127, EPI_ISL_2759132 | Genome Information Research Center, Research Institute for Microbial Diseases, Osaka University | Genome Information Research Center, Research Institute for Microbial Diseases, Osaka University | Daisuke Motooka; Hiroya Oki; Kentaro Tanaka; and Shota Nakamura |
| EPI_ISL_2788648, EPI_ISL_2788651, EPI_ISL_2788652, EPI_ISL_2788653, EPI_ISL_2788656, EPI_ISL_2788657, EPI_ISL_2788658, EPI_ISL_2788667, EPI_ISL_2788668, EPI_ISL_2788680, EPI_ISL_2788690, EPI_ISL_2788705, EPI_ISL_2788706 | | | |
| see above | Gunma Prefectural Institute of Public Health and Environmental Sciences | Gunma Prefectural Institute of Public Health and Environmental Sciences | Daisuke Shinoda; Hiroyuki Tsukagoshi; Kentaro Itokawa; Makoto Kuroda; Mariko Saito; Masanori Hashino; Rina Tanaka; Tsuyoshi Sekizuka |
| EPI_ISL_2773702, EPI_ISL_2773703, EPI_ISL_2773719, EPI_ISL_2773720, EPI_ISL_2773732, EPI_ISL_2773734, EPI_ISL_2773735, EPI_ISL_2773736, EPI_ISL_2773737, EPI_ISL_2773738, EPI_ISL_2773741, EPI_ISL_2773742, EPI_ISL_2773746, EPI_ISL_2773747, EPI_ISL_2773749, EPI_ISL_2773750, EPI_ISL_2773751, EPI_ISL_2773752, EPI_ISL_2773754, EPI_ISL_2773756, EPI_ISL_2773762, EPI_ISL_2773764, EPI_ISL_2773766, EPI_ISL_2773768, EPI_ISL_2773769 | | | |
| see above | Hiroshima Prefectural Technology Research Institute, Public Health and Environment Center | Hiroshima Prefectural Technology Research Institute, Public Health and Environment Center | Akie Nakashima; Ayano Ito; Hiroko Akita; Kanako Masuda; Kentaro Itokawa; Makoto Kuroda; Mana Suei; Masanori Hashino; Naoki Shigemoto; Rina Tanaka; Shinichi Takao; Syuhei Ikeda; Takahiro Hiratsuka; Tsuyoshi Sekizuka; Yukie Shimazu |
| EPI_ISL_2884711, EPI_ISL_2884727, EPI_ISL_2884774 | Hokkaido Institute of Public Health | Hokkaido Institute of Public Health | Kazuhiro Okubo; Kentaro Itokawa; Makoto Kuroda; Masanori Hashino; Rina Tanaka; Tsuyoshi Sekizuka; Yuta Ohno |
| EPI_ISL_2835848 | Hospital | National Reference Center for Viruses of Respiratory Infections, Institut Pasteur, Paris | Angela Brisebarre; Camille Capel; Christophe Malabat; Corinne Maufrais; Etienne Simon-Lorière; Evelyne Schvoerer; Frédéric Lemoine; Louise Lefrançois; Marion Barbet; Maud Vanpeene; Méline Bizard; Sylvie Behillil; Sylvie Van der Werf; Vincent Enouf |
| EPI_ISL_2860651, EPI_ISL_2860670 | Hyogo Prefectural Institute of Public Health science | Pathogen Genomics Center, National Institute of Infectious Diseases | Denshi Takai; Etsuko Saito; Kenichi Ogita; Kentaro Itokawa; Kyoko Suzuki; Makoto Kuroda; Masanori Hashino; Masatsugu Chikahira; Miki Ogi; Rina Tanaka; Tomohiro Oshibe; Tsuyoshi Sekizuka; Yumi Akiyama |
| EPI_ISL_2786787, EPI_ISL_2786788, EPI_ISL_2786789, EPI_ISL_2786790, EPI_ISL_2786791, EPI_ISL_2786792, EPI_ISL_2786793, EPI_ISL_2786794, EPI_ISL_2786795, EPI_ISL_2786796, EPI_ISL_2786797, EPI_ISL_2786798, EPI_ISL_2786799, EPI_ISL_2786800, EPI_ISL_2786801, EPI_ISL_2786802, EPI_ISL_2786803, EPI_ISL_2786804, EPI_ISL_2786805, EPI_ISL_2786806, EPI_ISL_2786807, EPI_ISL_2786808, EPI_ISL_2786809, EPI_ISL_2786810, EPI_ISL_2786811, EPI_ISL_2786812, EPI_ISL_2786813, EPI_ISL_2786814, EPI_ISL_2786815, EPI_ISL_2786816, EPI_ISL_2786817, EPI_ISL_2786818, EPI_ISL_2786819, EPI_ISL_2786820, EPI_ISL_2786821, EPI_ISL_2786822, EPI_ISL_2786823, EPI_ISL_2786824, EPI_ISL_2786825, EPI_ISL_2786826, EPI_ISL_2786827, EPI_ISL_2786828, EPI_ISL_2786830, EPI_ISL_2786831, EPI_ISL_2786832, EPI_ISL_2786833, EPI_ISL_2786834, EPI_ISL_2786835, EPI_ISL_2786836, EPI_ISL_2786837, EPI_ISL_2786838, EPI_ISL_2786839, EPI_ISL_2786840, EPI_ISL_2786841, EPI_ISL_2786842, EPI_ISL_2786843, EPI_ISL_2786844, EPI_ISL_2786845, EPI_ISL_2786846, EPI_ISL_2786847, EPI_ISL_2786848, EPI_ISL_2786849, EPI_ISL_2786850, EPI_ISL_2786851, EPI_ISL_2786852, EPI_ISL_2786853, EPI_ISL_2786854, EPI_ISL_2786855, EPI_ISL_2786856, EPI_ISL_2786857, EPI_ISL_2786858, EPI_ISL_2786859, EPI_ISL_2786860, EPI_ISL_2786861, EPI_ISL_2786862, EPI_ISL_2786863, EPI_ISL_2786864, EPI_ISL_2786865, EPI_ISL_2786866, EPI_ISL_2786867, EPI_ISL_2786868, EPI_ISL_2786869, EPI_ISL_2786870, EPI_ISL_2786871, EPI_ISL_2786872, EPI_ISL_2786873, EPI_ISL_2786874, EPI_ISL_2786875, EPI_ISL_2786876, EPI_ISL_2786877, EPI_ISL_2786878, EPI_ISL_2786879, EPI_ISL_2786880, EPI_ISL_2786881 | | | |
| see above | Ibaraki Prefectural Institute of Public Health | Ibaraki Prefectural Institute of Public Health | Keiko Goto; Kentaro Itokawa; Makoto Kuroda; Masanori Hashino; Rina Tanaka; Tsuyoshi Sekizuka |
| EPI_ISL_2671890, EPI_ISL_2671894, EPI_ISL_2960339, EPI_ISL_2964598, EPI_ISL_2964605, EPI_ISL_2964610, EPI_ISL_2964617 | | | |
| see above | Juntendo University Hospital | Juntendo University Hospital | Koji Tsuchiya; Koji Tuchiya; Yoko Tabe; Yoshie Hosaka |
| EPI_ISL_2787104, EPI_ISL_2787106 | Kagawa Prefectural Research Institute for Environmental Sciences and Public Health | Pathogen Genomics Center, National Institute of Infectious Diseases | Kenji Kuwahara; Kentaro Itokawa; Makoto Kuroda; Masanori Hashino; Mayumi Arizuka; Rina Tanaka; Tsuyoshi Sekizuka; Yukari Terajima |
| EPI_ISL_2842531, EPI_ISL_2842533, EPI_ISL_2842534, EPI_ISL_2842535, EPI_ISL_2842536, EPI_ISL_2842537, EPI_ISL_2842538, EPI_ISL_2842539, EPI_ISL_2842540, EPI_ISL_2842541, EPI_ISL_2842542, EPI_ISL_2842543, EPI_ISL_2842545, EPI_ISL_2842550, EPI_ISL_2842551, EPI_ISL_2842552, EPI_ISL_2842553, EPI_ISL_2842554, EPI_ISL_2842555, EPI_ISL_2842556, EPI_ISL_2842557, EPI_ISL_2842558, EPI_ISL_2842562 | | | |
| see above | Kagoshima Prefectural Institute for Environmental Research and Public Health | Kagoshima Prefectural Institute for Environmental Research and Public Health | Kentaro Itokawa; Makoto Kuroda; Masanori Hashino; Rina Tanaka; Tsuyoshi Sekizuka; Yuka Hamada |
| EPI_ISL_2898188, EPI_ISL_2898193, EPI_ISL_2898316, EPI_ISL_2898321, EPI_ISL_2898386, EPI_ISL_2898471, EPI_ISL_2898564, EPI_ISL_2898587, EPI_ISL_2898639, EPI_ISL_2898829, EPI_ISL_2898997, EPI_ISL_2899000 | | | |
| see above | Karolinska University Hospital Solna | Karolinska University Hospital | Annelie Bjerkner; Isak Sylvin; Jan Albert; Karolina Ininbergs; Lina Guerra Blomqvist; Lynda Eneh; Martin Ekman; Martina Wahlund; Robert Dyrdak; Sandra Broddesson; Tanja Normark; Tobias Allander; Valtteri Wirta; Zhibing Yun |
| EPI_ISL_2839615, EPI_ISL_2839616, EPI_ISL_2839618, EPI_ISL_2839619, EPI_ISL_2839653 | Kumamoto City Medical Association PCR Center | Sato Laboratory, Division of Genomics and Transcriptomics, The Joint Research Center for Human Retrovirus Infection, Kumamoto University | Fumiya Ishikawa; Ryota Irie; Samiul Alam Rajib; Yorifumi Sato |
| EPI_ISL_2635752, EPI_ISL_2636181 | Labor Mönchengladbach MVZ Dr. Stein + Kollegen GbR | Robert Koch Institute | |
| EPI_ISL_2690298, EPI_ISL_2691076 | MSK Microbiology Lab | MSK Microbiology Lab | Esther Babady; Krupa Jani; Tracy McMillen |
| EPI_ISL_2646770, EPI_ISL_2647840 | Mako Medical | Centers for Disease Control and Prevention Division of Viral Diseases, Pathogen Discovery | Adrian Paskey; Benjamin Rambo-Martin; Christopher Gulvick; Clinton R. Paden; Dakota Howard; Darlene Wagner; Dhwani Batra; Duncan MacCannell; Jason Caravas; Kara Moser; Lauren Moon; Matthew Schmerer; Matthew Tugwell; Peter W. Cook; Scott Sammons; Shatavia Morrison; Yvette Unoarumhi |
| EPI_ISL_3011653, EPI_ISL_3011680, EPI_ISL_3011721, EPI_ISL_3011736, EPI_ISL_3011744, EPI_ISL_3011897, EPI_ISL_3012111 | | | |
| see above | Maryland Genomics, Institute for Genome Sciences, University of Maryland School of Medicine | Maryland Genomics, Institute for Genome Sciences, University of Maryland School of Medicine | Claire M; Fraser; Hazen; Holly; Humphrys; Ivette; Jacques; Jonathan; Kranthi; Lim; Lisa D; Luke J; Mike; Ott; Ravel; Roussey; Sadzewicz; Sandra; Santana-Cruz; Tallon; Tracy; Vavikolanu |
| EPI_ISL_2841491, EPI_ISL_2841492, EPI_ISL_2841493, EPI_ISL_2841494, EPI_ISL_2841495, EPI_ISL_2841496, EPI_ISL_2841498, EPI_ISL_2841499, EPI_ISL_2841500, EPI_ISL_2841501, EPI_ISL_2841502, EPI_ISL_2841503, EPI_ISL_2841504, EPI_ISL_2841505 | | | |
| see above | Medical Microbiology and Zoology Section, Okinawa Prefectural Institute of Health and Environment | Pathogen Genomics Center, National Institute of Infectious Diseases | Hisako Kyan; Kentaro Itokawa; Makoto Kuroda; Masanori Hashino; Minoru Nidaira; Noriyuki Maeshiro; Rina Tanaka; Taketoshi Takara; Tetsuya Kakita; Tsuyoshi Kudeken; Tsuyoshi Sekizuka; Yumani Kuba |
| EPI_ISL_2984554 | Ministry of Health Turkey | Ministry of Health Turkey | Fatma Bayrakdar; Gulay Korukluoglu; Suleyman Yalcin; Yasemin Cosgun |
| EPI_ISL_2842674 | NJDOH, Public Health and Environmental Laboratories | NJ_PHEL | Allison Roder; Byeong Jeong; Chelsea San Filippo; Dana Woell; Jacquelyn Deverell; Lindsey Bodnar; Ryan Pachucki; Shiv K. Verma |
| EPI_ISL_2764056, EPI_ISL_2764057, EPI_ISL_2764058, EPI_ISL_2764059, EPI_ISL_2764060, EPI_ISL_2764061, EPI_ISL_2764062, EPI_ISL_2764063, EPI_ISL_2764064, EPI_ISL_2764065, EPI_ISL_2764066, EPI_ISL_2764067, EPI_ISL_2764068, EPI_ISL_2764069, EPI_ISL_2764070, EPI_ISL_2764071, EPI_ISL_2764072, EPI_ISL_2764073, EPI_ISL_2764074, EPI_ISL_2764075, EPI_ISL_2764076, EPI_ISL_2764077, EPI_ISL_2764078, EPI_ISL_2764079, EPI_ISL_2764080, EPI_ISL_2764081, EPI_ISL_2764082, EPI_ISL_2764083, EPI_ISL_2764084, EPI_ISL_2764085, EPI_ISL_2764086, EPI_ISL_2764087, EPI_ISL_2764088, EPI_ISL_2764089, EPI_ISL_2768427, EPI_ISL_2768428, EPI_ISL_2768429, EPI_ISL_2768430, EPI_ISL_2768431, EPI_ISL_2768432, EPI_ISL_2768433, EPI_ISL_2768434, EPI_ISL_2768435, EPI_ISL_2768436, EPI_ISL_2768437, EPI_ISL_2768438, EPI_ISL_2768439, EPI_ISL_2768440, EPI_ISL_2768441, EPI_ISL_2768442, EPI_ISL_2768443, EPI_ISL_2768444, EPI_ISL_2768445, EPI_ISL_2768446, EPI_ISL_2768447, EPI_ISL_2768448, EPI_ISL_2768449, EPI_ISL_2768450, EPI_ISL_2768451, EPI_ISL_2768452, EPI_ISL_2768453, EPI_ISL_2768454, EPI_ISL_2768455, EPI_ISL_2768456, EPI_ISL_2768457, EPI_ISL_2768458, EPI_ISL_2768459, EPI_ISL_2768460, EPI_ISL_2768461, EPI_ISL_2768462, EPI_ISL_2768463, EPI_ISL_2768464 | | | |
| see above | Niigata Prefectural Institute of Public Health and Environmental Sciences | Pathogen Genomics Center, National Institute of Infectious Diseases | Kentaro Itokawa; Makoto Kuroda; Masanori Hashino; Rina Tanaka; Tsuyoshi Sekizuka |

| | | | |
|---|---|---|---|
| EPI_ISL_2764050, EPI_ISL_2764051, EPI_ISL_2764052, EPI_ISL_2764053, EPI_ISL_2764054, EPI_ISL_2764055 | Osaka Institute of Public Health, Morinomiya Center | Pathogen Genomics Center, National Institute of Infectious Diseases | Kentaro Itokawa; Makoto Kuroda; Masanori Hashino; Rina Tanaka; Tsuyoshi Sekizuka |
| EPI_ISL_2663782, EPI_ISL_2663795 | Osaka University Hospital | Genome Information Research Center, Research Institute for Microbial Diseases, Osaka University | Daisuke Motooka; Haruhiko Hirata; Hiroshi Ogura; Hiroya Oki; Hisatake Matsumoto; Kentaro Tanaka; Shota Nakamura; Takeshi Ebihara; Yasuhiro Kato; and Atsushi Kumanogoh |
| EPI_ISL_2633633, EPI_ISL_2715543, EPI_ISL_2715915, EPI_ISL_2716143, EPI_ISL_2827332, EPI_ISL_2964973, EPI_ISL_2965469 | | | |
| see above | Pandemic Response Lab - NYC | Pandemic Response Lab, R&D | Cybill del Castillo; Dylan Law; Haiping Hao; Henry Lee; Isabel Fernandez Escapa; Jon Laurent; Katharine Nelson; Melissa Hopkins; Michael Hammerling; Pradeep Bugga; Shinyoung Clair Kang; Sol Rey; William Ward |
| EPI_ISL_2760080, EPI_ISL_2760082, EPI_ISL_2760202, EPI_ISL_2760204, EPI_ISL_2760205, EPI_ISL_2760208, EPI_ISL_2760210, EPI_ISL_2760284, EPI_ISL_2760286, EPI_ISL_2760287, EPI_ISL_2760289, EPI_ISL_2760290, EPI_ISL_2760292, EPI_ISL_2760294, EPI_ISL_2760295, EPI_ISL_2760297, EPI_ISL_2760299, EPI_ISL_2760300, EPI_ISL_2760302, EPI_ISL_2760303, EPI_ISL_2760304, EPI_ISL_2760305, EPI_ISL_2760306, EPI_ISL_2760307, EPI_ISL_2760309, EPI_ISL_2760311, EPI_ISL_2760312, EPI_ISL_2760314, EPI_ISL_2760316, EPI_ISL_2760318, EPI_ISL_2760319, EPI_ISL_2760321, EPI_ISL_2760323, EPI_ISL_2760325, EPI_ISL_2760326, EPI_ISL_2760327, EPI_ISL_2760328, EPI_ISL_2760329, EPI_ISL_2760333, EPI_ISL_2760335, EPI_ISL_2760337, EPI_ISL_2760338, EPI_ISL_2760340, EPI_ISL_2760342, EPI_ISL_2760343, EPI_ISL_2760345, EPI_ISL_2760346, EPI_ISL_2760348, EPI_ISL_2760355, EPI_ISL_2760357, EPI_ISL_2760358, EPI_ISL_2760360, EPI_ISL_2760361, EPI_ISL_2760363, EPI_ISL_2760365, EPI_ISL_2760367, EPI_ISL_2760369, EPI_ISL_2760370, EPI_ISL_2760372, EPI_ISL_2760375, EPI_ISL_2760377, EPI_ISL_2760379, EPI_ISL_2760382, EPI_ISL_2760384, EPI_ISL_2760388, EPI_ISL_2760391, EPI_ISL_2760393, EPI_ISL_2760395, EPI_ISL_2760397, EPI_ISL_2760398, EPI_ISL_2760400, EPI_ISL_2760401, EPI_ISL_2760403, EPI_ISL_2760406, EPI_ISL_2760410, EPI_ISL_2760411, EPI_ISL_2760415, EPI_ISL_2760416, EPI_ISL_2760421, EPI_ISL_2760425, EPI_ISL_2760427, EPI_ISL_2760429, EPI_ISL_2760430, EPI_ISL_2760432, EPI_ISL_2760434, EPI_ISL_2760435, EPI_ISL_2760437, EPI_ISL_2760442, EPI_ISL_2760443, EPI_ISL_2760445, EPI_ISL_2760446, EPI_ISL_2760448, EPI_ISL_2760450, EPI_ISL_2760453, EPI_ISL_2760456, EPI_ISL_2760459, EPI_ISL_2760461, EPI_ISL_2760462, EPI_ISL_2760464, EPI_ISL_2760465, EPI_ISL_2760805, EPI_ISL_2760807, EPI_ISL_2760809, EPI_ISL_2760810, EPI_ISL_2760845, EPI_ISL_2760847, EPI_ISL_2760848, EPI_ISL_2760850, EPI_ISL_2760851, EPI_ISL_2760853, EPI_ISL_2760855, EPI_ISL_2760856, EPI_ISL_2760858, EPI_ISL_2760860, EPI_ISL_2760861, EPI_ISL_2760863, EPI_ISL_2760865, EPI_ISL_2760866, EPI_ISL_2760868, EPI_ISL_2760869, EPI_ISL_2762924, EPI_ISL_2762925, EPI_ISL_2762926, EPI_ISL_2762928, EPI_ISL_2762929, EPI_ISL_2762930, EPI_ISL_2762931, EPI_ISL_2762934, EPI_ISL_2762935, EPI_ISL_2762936, EPI_ISL_2762937, EPI_ISL_2762938, EPI_ISL_2762939, EPI_ISL_2762940, EPI_ISL_2762941, EPI_ISL_2762943, EPI_ISL_2762944, EPI_ISL_2762945, EPI_ISL_2762946, EPI_ISL_2762947, EPI_ISL_2762948, EPI_ISL_2762949, EPI_ISL_2762950, EPI_ISL_2762951, EPI_ISL_2762952, EPI_ISL_2762953, EPI_ISL_2762954, EPI_ISL_2762955, EPI_ISL_2762956, EPI_ISL_2762957, EPI_ISL_2762958, EPI_ISL_2762959, EPI_ISL_2762960, EPI_ISL_2762962, EPI_ISL_2762963, EPI_ISL_2762964, EPI_ISL_2762965, EPI_ISL_2762966, EPI_ISL_2762967, EPI_ISL_2762968, EPI_ISL_2762970, EPI_ISL_2762971, EPI_ISL_2762972, EPI_ISL_2762973, EPI_ISL_2762974, EPI_ISL_2762975, EPI_ISL_2762976, EPI_ISL_2762977, EPI_ISL_2762978, EPI_ISL_2762979, EPI_ISL_2762980, EPI_ISL_2762981, EPI_ISL_2762982, EPI_ISL_2762983, EPI_ISL_2762984, EPI_ISL_2762985, EPI_ISL_2762986, EPI_ISL_2762987, EPI_ISL_2762988, EPI_ISL_2762989, EPI_ISL_2762991, EPI_ISL_2762992, EPI_ISL_2762993, EPI_ISL_2762994, EPI_ISL_2762995, EPI_ISL_2762996, EPI_ISL_2762997, EPI_ISL_2762998, EPI_ISL_2762999, EPI_ISL_2763000, EPI_ISL_2763001, EPI_ISL_2763002, EPI_ISL_2763003, EPI_ISL_2763004, EPI_ISL_2763005, EPI_ISL_2763006, EPI_ISL_2763007, EPI_ISL_2763008, EPI_ISL_2763009, EPI_ISL_2763010, EPI_ISL_2763106, EPI_ISL_2763107, EPI_ISL_2763108, EPI_ISL_2763110, EPI_ISL_2763111, EPI_ISL_2763112, EPI_ISL_2763113, EPI_ISL_2763114, EPI_ISL_2763115, EPI_ISL_2763116, EPI_ISL_2763117, EPI_ISL_2763118, EPI_ISL_2763119, EPI_ISL_2763120, EPI_ISL_2763121, EPI_ISL_2763122, EPI_ISL_2763123, EPI_ISL_2763124, EPI_ISL_2763125, EPI_ISL_2763126, EPI_ISL_2763127, EPI_ISL_2763128, EPI_ISL_2763129, EPI_ISL_2763130, EPI_ISL_2763131, EPI_ISL_2763132, EPI_ISL_2763133, EPI_ISL_2763134, EPI_ISL_2763135, EPI_ISL_2763136, EPI_ISL_2763137, EPI_ISL_2763138, EPI_ISL_2763139, EPI_ISL_2763140, EPI_ISL_2763141, EPI_ISL_2763142, EPI_ISL_2763143, EPI_ISL_2763144, EPI_ISL_2763145, EPI_ISL_2763146, EPI_ISL_2763147, EPI_ISL_2763148, EPI_ISL_2763149, EPI_ISL_2763150, EPI_ISL_2763151, EPI_ISL_2763962, EPI_ISL_2763963, EPI_ISL_2763964, EPI_ISL_2763965, EPI_ISL_2763966, EPI_ISL_2763967, EPI_ISL_2763968, EPI_ISL_2763969, EPI_ISL_2763971, EPI_ISL_2763972, EPI_ISL_2763973, EPI_ISL_2763974, EPI_ISL_2763975, EPI_ISL_2763976, EPI_ISL_2763977, EPI_ISL_2763978, EPI_ISL_2763979, EPI_ISL_2763980, EPI_ISL_2763981, EPI_ISL_2763982, EPI_ISL_2763983, EPI_ISL_2763984, EPI_ISL_2763985, EPI_ISL_2763986, EPI_ISL_2763988, EPI_ISL_2763990, EPI_ISL_2763991, EPI_ISL_2763992, EPI_ISL_2763993, EPI_ISL_2763994, EPI_ISL_2763995, EPI_ISL_2763996, EPI_ISL_2763997, EPI_ISL_2763998, EPI_ISL_2763999, EPI_ISL_2764000, EPI_ISL_2764001, EPI_ISL_2764002, EPI_ISL_2764003, EPI_ISL_2764004, EPI_ISL_2764005, EPI_ISL_2764006, EPI_ISL_2764007, EPI_ISL_2764008, EPI_ISL_2764009, EPI_ISL_2764010, EPI_ISL_2764012, EPI_ISL_2764013, EPI_ISL_2764014, EPI_ISL_2764015, EPI_ISL_2764016, EPI_ISL_2764019, EPI_ISL_2764020, EPI_ISL_2764021, EPI_ISL_2764022, EPI_ISL_2764023, EPI_ISL_2764024, EPI_ISL_2764025, EPI_ISL_2764026, EPI_ISL_2764027, EPI_ISL_2764028, EPI_ISL_2764029, EPI_ISL_2764030, EPI_ISL_2764031, EPI_ISL_2764032, EPI_ISL_2764033, EPI_ISL_2764035, EPI_ISL_2764036, EPI_ISL_2764037, EPI_ISL_2764038, EPI_ISL_2764091, EPI_ISL_2764092, EPI_ISL_2764093, EPI_ISL_2764094, EPI_ISL_2764095, EPI_ISL_2764096, EPI_ISL_2764097, EPI_ISL_2764098, EPI_ISL_2764099, EPI_ISL_2764100, EPI_ISL_2764949, EPI_ISL_2764951, EPI_ISL_2764952, EPI_ISL_2764953, EPI_ISL_2764955, EPI_ISL_2768232, EPI_ISL_2768244, EPI_ISL_2768247, EPI_ISL_2768249, EPI_ISL_2768250, EPI_ISL_2768251, EPI_ISL_2768252, EPI_ISL_2768253, EPI_ISL_2768254, EPI_ISL_2768256, EPI_ISL_2768260, EPI_ISL_2768263, EPI_ISL_2768267, EPI_ISL_2768271, EPI_ISL_2768275, EPI_ISL_2768288, EPI_ISL_2768297, EPI_ISL_2768306, EPI_ISL_2768308, EPI_ISL_2768312, EPI_ISL_2768316, EPI_ISL_2768317, EPI_ISL_2768319, EPI_ISL_2768320, EPI_ISL_2768321, EPI_ISL_2768325, EPI_ISL_2768327, EPI_ISL_2768328, EPI_ISL_2768330, EPI_ISL_2768331, EPI_ISL_2768332, EPI_ISL_2768333, EPI_ISL_2768334, EPI_ISL_2768335, EPI_ISL_2768337, EPI_ISL_2768339, EPI_ISL_2768340, EPI_ISL_2768342, EPI_ISL_2768343, EPI_ISL_2768344, EPI_ISL_2768346, EPI_ISL_2768347, EPI_ISL_2768348, EPI_ISL_2768349, EPI_ISL_2768350, EPI_ISL_2768351, EPI_ISL_2768352, EPI_ISL_2768353, EPI_ISL_2768354, EPI_ISL_2768356, EPI_ISL_2768358, EPI_ISL_2768360, EPI_ISL_2768361, EPI_ISL_2768362, EPI_ISL_2768365, EPI_ISL_2768367, EPI_ISL_2768369, EPI_ISL_2768373, EPI_ISL_2768374, EPI_ISL_2768376, EPI_ISL_2768377, EPI_ISL_2768378, EPI_ISL_2768379, EPI_ISL_2768380, EPI_ISL_2768381, EPI_ISL_2768382, EPI_ISL_2768383, EPI_ISL_2768384, EPI_ISL_2768385, EPI_ISL_2768386, EPI_ISL_2768387, EPI_ISL_2768388, EPI_ISL_2768389, EPI_ISL_2768390, EPI_ISL_2768391, EPI_ISL_2768392, EPI_ISL_2768393, EPI_ISL_2768394, EPI_ISL_2768399, EPI_ISL_2768405, EPI_ISL_2768407, EPI_ISL_2768408, EPI_ISL_2768410, EPI_ISL_2768412, EPI_ISL_2768413, EPI_ISL_2768417, EPI_ISL_2768847, EPI_ISL_2768848, EPI_ISL_2768853, EPI_ISL_2768855, EPI_ISL_2768856, EPI_ISL_2768857, EPI_ISL_2768867, EPI_ISL_2768871, EPI_ISL_2768872, EPI_ISL_2768873, EPI_ISL_2768875, EPI_ISL_2768876, EPI_ISL_2768878, EPI_ISL_2768881, EPI_ISL_2768887, EPI_ISL_2768888, EPI_ISL_2768889, EPI_ISL_2768890, EPI_ISL_2768891, EPI_ISL_2768896, EPI_ISL_2768900, EPI_ISL_2768902, EPI_ISL_2768904, EPI_ISL_2768911, EPI_ISL_2768913, EPI_ISL_2768916, EPI_ISL_2768917, EPI_ISL_2768919, EPI_ISL_2768925, EPI_ISL_2768930, EPI_ISL_2768931, EPI_ISL_2768932, EPI_ISL_2768933, EPI_ISL_2768934, EPI_ISL_2768935, EPI_ISL_2768936, EPI_ISL_2768937, EPI_ISL_2768938, EPI_ISL_2768939, EPI_ISL_2768940, EPI_ISL_2768941, EPI_ISL_2768942, EPI_ISL_2768943, EPI_ISL_2768944, EPI_ISL_2768945, EPI_ISL_2768946, EPI_ISL_2768947, EPI_ISL_2768948, EPI_ISL_2768949, EPI_ISL_2768950, EPI_ISL_2768951, EPI_ISL_2768952, EPI_ISL_2768953, EPI_ISL_2768954, EPI_ISL_2768955, EPI_ISL_2768956, EPI_ISL_2768957, EPI_ISL_2768958, EPI_ISL_2768959, EPI_ISL_2768960, EPI_ISL_2768961, EPI_ISL_2768962, EPI_ISL_2768963, EPI_ISL_2768964, EPI_ISL_2768965, EPI_ISL_2768966, EPI_ISL_2768967, EPI_ISL_2768968, EPI_ISL_2768969, EPI_ISL_2768970, EPI_ISL_2768971, EPI_ISL_2768972, EPI_ISL_2768973, EPI_ISL_2768974, EPI_ISL_2768975, EPI_ISL_2768976, EPI_ISL_2768977, EPI_ISL_2768978, EPI_ISL_2768979, EPI_ISL_2768980, EPI_ISL_2768981, EPI_ISL_2768982, EPI_ISL_2768983, EPI_ISL_2768984, EPI_ISL_2768985, EPI_ISL_2768986, EPI_ISL_2768987, EPI_ISL_2768988, EPI_ISL_2768990, EPI_ISL_2768991, EPI_ISL_2768992, EPI_ISL_2768993, EPI_ISL_2768994, EPI_ISL_2768995, EPI_ISL_2768996, EPI_ISL_2768997, EPI_ISL_2768998, EPI_ISL_2768999, EPI_ISL_2769000, EPI_ISL_2769001, EPI_ISL_2769002, EPI_ISL_2769004, EPI_ISL_2769005, EPI_ISL_2769006, EPI_ISL_2769007, EPI_ISL_2769008, EPI_ISL_2769009, EPI_ISL_2769010, EPI_ISL_2769011, EPI_ISL_2769012, EPI_ISL_2769013, EPI_ISL_2769014, EPI_ISL_2769015, EPI_ISL_2769016, EPI_ISL_2769017, EPI_ISL_2770259, EPI_ISL_2770260, EPI_ISL_2770267, EPI_ISL_2770275, EPI_ISL_2770288, EPI_ISL_2770300, EPI_ISL_2770329, EPI_ISL_2770341, EPI_ISL_2770684, EPI_ISL_2770686, EPI_ISL_2770691, EPI_ISL_2770692, EPI_ISL_2770693, EPI_ISL_2770694, EPI_ISL_2770695, EPI_ISL_2770697, EPI_ISL_2770699, EPI_ISL_2770701, EPI_ISL_2770706, EPI_ISL_2770715, EPI_ISL_2770723, EPI_ISL_2770725, EPI_ISL_2770731, EPI_ISL_2770732, EPI_ISL_2770746, EPI_ISL_2770747, EPI_ISL_2770748, EPI_ISL_2770749, EPI_ISL_2770751, EPI_ISL_2770755, EPI_ISL_2770756, EPI_ISL_2770757, EPI_ISL_2771526, EPI_ISL_2771533, EPI_ISL_2771562, EPI_ISL_2771569, EPI_ISL_2772115, EPI_ISL_2772120, EPI_ISL_2772122, EPI_ISL_2772125, EPI_ISL_2772126, EPI_ISL_2772129, EPI_ISL_2772167, EPI_ISL_2772170, EPI_ISL_2772173, EPI_ISL_2772194, EPI_ISL_2773981, EPI_ISL_2773982, EPI_ISL_2773983, EPI_ISL_2773984, EPI_ISL_2773985, EPI_ISL_2932276, EPI_ISL_2932292, EPI_ISL_2932293, EPI_ISL_2932294, EPI_ISL_2932309, EPI_ISL_2932310, EPI_ISL_2932311, EPI_ISL_2932312, EPI_ISL_2932313, EPI_ISL_2932314, EPI_ISL_2932315 | | | |
| see above | Pathogen Genomics Center, National Institute of Infectious Diseases | Pathogen Genomics Center, National Institute of Infectious Diseases | Kentaro Itokawa; Makoto Kuroda; Masanori Hashino; Rina Tanaka; Tsuyoshi Sekizuka |
| EPI_ISL_2993470 | Pathology, UAB | Pathology, UAB | E.J.; Hendrickson; Hernandez, J.; Leal; Lefkowitz; Moates, D.; R.C.; S.M. |
| EPI_ISL_2759154, EPI_ISL_2759378, EPI_ISL_2759379, EPI_ISL_2759380, EPI_ISL_2759408, EPI_ISL_2759416, EPI_ISL_2759552, EPI_ISL_2759569, EPI_ISL_2759575 | | | |
| see above | Population Medicine and Diagnostic Sciences, Cornell University | Population Medicine and Diagnostic Sciences, Cornell University | Anderson; B.D.; Caserta; Cronk; D.G.; Diel; L.C.; Laverack, M.; Mitchell; P.K.; Plocharczyk, E.; R.R.; Venugopalan, R. |
| EPI_ISL_2697775 | Public Health Laboratory, Minnesota Department of Health | University of Minnesota Genomics Center | Corbin Dirkx; Daryl M. Gohl; Jaquelyn Kuriger-Laber; John Garbe; and Sean Wang |
| EPI_ISL_2740505, EPI_ISL_2748878 | Respiratory Virus Unit, Microbiology Services Colindale, Public Health England | COVID-19 Genomics UK (COG-UK) Consortium | PHE Covid Sequencing Team |
| EPI_ISL_2835244 | Rhode Island State Health Laboratory | Rhode Island State Health Laboratory | Ewa King; Kristin Carpenter-Azevedo; Richard C. Huard |
| EPI_ISL_2763863, EPI_ISL_2768597, EPI_ISL_2769827, EPI_ISL_2771992, EPI_ISL_2771993, EPI_ISL_2771994, EPI_ISL_2776444, EPI_ISL_2776445, EPI_ISL_2776446, EPI_ISL_2776447, EPI_ISL_2776448, EPI_ISL_2776625, EPI_ISL_2776628, EPI_ISL_2776629, EPI_ISL_2776666, EPI_ISL_2776667, EPI_ISL_2776668, EPI_ISL_2776669, EPI_ISL_2776670, EPI_ISL_2776671, EPI_ISL_2776672, EPI_ISL_2776673, EPI_ISL_2776674, EPI_ISL_2776675, EPI_ISL_2776676, EPI_ISL_2776677, EPI_ISL_2776689, EPI_ISL_2776704, EPI_ISL_2776705, EPI_ISL_2776729, EPI_ISL_2776733, EPI_ISL_2776737, EPI_ISL_2776784, EPI_ISL_2776785, EPI_ISL_2776788, EPI_ISL_2776789, EPI_ISL_2776790, EPI_ISL_2776791, EPI_ISL_2776792, EPI_ISL_2776793, EPI_ISL_2776794, EPI_ISL_2776795, EPI_ISL_2776801, EPI_ISL_2776833, EPI_ISL_2776836, EPI_ISL_2776837, EPI_ISL_2776872, EPI_ISL_2776876, EPI_ISL_2777102, EPI_ISL_2777103, EPI_ISL_2777126 | | | |
| see above | SARS-CoV-2 testing team, National Institute of Infectious Diseases | Pathogen Genomics Center, National Institute of Infectious Diseases | Hazuka Y Furihata; Hiromizu Takahashi; Kentaro Itokawa; Makoto Kuroda; Masanori Hashino; Masumichi Saito; Naomi Nojiri; Nozomu Hanaoka; Rina Tanaka; Tsuguto Fujimoto; Tsuyoshi Sekizuka |
| EPI_ISL_2868676, EPI_ISL_2868677 | SUNY UPSTATE MEDICAL UNIVERSITY | Wadsworth Center, New York State Department of Health | Alexis Russell; Catharine Prussing; Daryl M. Lamson; Erasmus Schneider; Erica Lasek-Nesselquist; John Kelly; Jonathan Plitnick; Kirsten St. George; Matthew Shudt; Melissa A Leisner; Navjot Singh |
| EPI_ISL_2812046, EPI_ISL_2812047, EPI_ISL_2812053, EPI_ISL_2812057, EPI_ISL_2812058, EPI_ISL_2812065, EPI_ISL_2812067, EPI_ISL_2812084, EPI_ISL_2812091, EPI_ISL_2812092, EPI_ISL_2812093 | | | |
| see above | Saitama Prefectural Institute of Public Health | Saitama Prefectural Institute of Public Health | Eri Aonuma; Hayato Ehara; Kentaro Itokawa; Makoto Kuroda; Masanori Hashino; Rina Tanaka; Satoshi Daisaki; Tsuyoshi Sekizuka |
| EPI_ISL_2659021, EPI_ISL_2659081 | Santa Clara Valley Medical Center | Santa Clara County Public Health Laboratory | Santa Clara County Public Health Department |
| EPI_ISL_2812029 | Sapporo City Institute of Public Health | Sapporo City Institute of Public Health | Asami Ohnishi; Kentaro Itokawa; Makoto Kuroda; Masanori Hashino; Masayuki Kikuchi; Rie Shimazaki; Rina Tanaka; Tsuyoshi Sekizuka; Yusuke Oguchi |
| EPI_ISL_2736362 | Seattle Flu Study | Seattle Flu Study | Amanda Adler; Barry R. Lutz; Benjamin Pelle; Caitlin R. Wolf; Chris D. Frazar; Deborah A. Nickerson; Elisabeth Brandstetter; Erica Ryke; Helen Y. Chu; Janet A. Englund; Jay Shendure; Jover Lee; Kairsten Fay; Kirsten Lacombe; Lea M. Starita; Mark J. Rieder; Matthew Richardson; Matthew Thompson; Melissa Truong; Michael Boeckh; Michael Famulare; Misja Ilcisin; Peter D. Han; Thomas R. Sibley; Trevor Bedford |
| EPI_ISL_2885182, EPI_ISL_2885192 | Shimane Prefectural Institute of Public Health and Environmental Science | Shimane Prefectural Institute of Public Health and Environmental Science | Aiko Fukuma; Kentaro Itokawa; Makoto Kuroda; Masanori Hashino; Naoki Fujisawa; Rie Onishi; Rina Tanaka; Tsuyoshi Sekizuka; Yurie Kanba |
| EPI_ISL_2716624, EPI_ISL_2716625, EPI_ISL_2716626, EPI_ISL_2716627, EPI_ISL_2716628, EPI_ISL_2716629, EPI_ISL_2716630, EPI_ISL_2716631, EPI_ISL_2716632, EPI_ISL_2716633, EPI_ISL_2716634, EPI_ISL_2716635, EPI_ISL_2716636, EPI_ISL_2716637, EPI_ISL_2716638, EPI_ISL_2716639 | | | |
| see above | State Key Laboratory of Pathogen and Biosecurity, Beijing Institute of Microbiology and Epidemiology | State Key Laboratory of Pathogen and Biosecurity, Beijing Institute of Microbiology and Epidemiology | Campbell, A.; Campbell, J.; Gevao; Harding, D.; Lin, L.; Liu, Y.; P.F.; P.S.; Rogers, J.; Sahr; Wurie, I.; Zhang, J. |
| EPI_ISL_2618870, EPI_ISL_2618874, EPI_ISL_2618937, EPI_ISL_2619755, EPI_ISL_2792544, EPI_ISL_2792802, EPI_ISL_2792830, EPI_ISL_2792912, EPI_ISL_2792987, EPI_ISL_2793009, EPI_ISL_2793094, EPI_ISL_2795151, EPI_ISL_2889076, EPI_ISL_2980128 | | | |
| see above | Swedish national genomic surveillance program of SARS-CoV-2 | The Public Health Agency of Sweden | Alma Brolund; Maria Lind Karlberg; Maximilian Riess; Swedish national genomic surveillance program of SARS-CoV-2 |
| EPI_ISL_2631415, EPI_ISL_2631417, EPI_ISL_2631424, EPI_ISL_2631431, EPI_ISL_2631434, EPI_ISL_2631436, EPI_ISL_2631437, EPI_ISL_2631438 | | | |
| see above | The Caribbean Public Health Agency | Carrington Lab, Department of PreClinical Sciences, Faculty of Medical Sciences, The University of the West Indies, St Augustine Campus | Anushka Ramjag; Arianne Brown-Jordan; Avery Hinds; Christine V. F. Carrington; Christopher Oura; Gabriel Escobar; Karla Georges; Naresh Nandram; Nikita S. D. Sahadeo; Nuno Faria; Oliver Pybus; Risha Singh; Roshan Parasram; Sarah Hill; SueMin Nathaniel; Vernie Ramkissoon |
| EPI_ISL_2621687, EPI_ISL_2621696, EPI_ISL_2621698, EPI_ISL_2621704, EPI_ISL_2621706, EPI_ISL_2621707, EPI_ISL_2621708, EPI_ISL_2621709, EPI_ISL_2621710, EPI_ISL_2621711, EPI_ISL_2621712, EPI_ISL_2621713, EPI_ISL_2621714, EPI_ISL_2621715, EPI_ISL_2621716, EPI_ISL_2621717, EPI_ISL_2621718, EPI_ISL_2621719, EPI_ISL_2621721, EPI_ISL_2621722, EPI_ISL_2621723, EPI_ISL_2621724, EPI_ISL_2626664, EPI_ISL_2626667, EPI_ISL_2626670, EPI_ISL_2626672, EPI_ISL_2626673, EPI_ISL_2626674, EPI_ISL_2626675, EPI_ISL_2626676, EPI_ISL_2626677, EPI_ISL_2626678, EPI_ISL_2626679, EPI_ISL_2626680, EPI_ISL_2626681, EPI_ISL_2626682, EPI_ISL_2626683, EPI_ISL_2626685, EPI_ISL_2626686 | | | |
| see above | The Caribbean Public Health Agency | Carrington Lab, Department of PreClinical Sciences, Faculty of Medical Sciences, The University of the West Indies | Anushka Ramjag; Arianne Brown-Jordan; Avery Hinds; Christine V. F. Carrington; Christopher Oura; Gabriel Escobar; Karla Georges; Naresh Nandram; Nikita S. D. Sahadeo; Nuno Faria; Oliver Pybus; Risha Singh; Roshan Parasram; Sarah Hill; SueMin Nathaniel; Vernie Ramkissoon |
| EPI_ISL_2617922, EPI_ISL_2617924, EPI_ISL_2617925, EPI_ISL_2617926, EPI_ISL_2617927, EPI_ISL_2617929, EPI_ISL_2617930, EPI_ISL_2617931, EPI_ISL_2617932, EPI_ISL_2617933, EPI_ISL_2617934, EPI_ISL_2617936, EPI_ISL_2617938, EPI_ISL_2617940, EPI_ISL_2617941, EPI_ISL_2649745, EPI_ISL_2649746, EPI_ISL_2649747, EPI_ISL_2649748, EPI_ISL_2649749, EPI_ISL_2649779, EPI_ISL_2649780, EPI_ISL_2649781, EPI_ISL_2649782, EPI_ISL_2649785, EPI_ISL_2649786, EPI_ISL_2649787, EPI_ISL_2649789, EPI_ISL_2649791, EPI_ISL_2649792, EPI_ISL_2649793, EPI_ISL_2649794, EPI_ISL_2649795, EPI_ISL_2649796, EPI_ISL_2658371, EPI_ISL_2658372, EPI_ISL_2658373, EPI_ISL_2658376, EPI_ISL_2658379, EPI_ISL_2658381, EPI_ISL_2658383, EPI_ISL_2658384, EPI_ISL_2658385, EPI_ISL_2658386, EPI_ISL_2658387, EPI_ISL_2658388, EPI_ISL_2658391, EPI_ISL_2658394, EPI_ISL_2658399, EPI_ISL_2658402, EPI_ISL_2658403, EPI_ISL_2658404, EPI_ISL_2658405, EPI_ISL_2658411, EPI_ISL_2678139, EPI_ISL_2678186, EPI_ISL_2716585, EPI_ISL_2716592 | | | |
| see above | The Caribbean Public Health Agency | Carrington Lab, Department of Preclinical Sciences, Faculty of Medical Sciences, The University of the West Indies, St Augustine Campus | Anushka Ramjag; Arianne Brown-Jordan; Avery Hinds; Christine V. F. Carrington; Christopher Oura; Gabriel Escobar; Karla Georges; Naresh Nandram; Nikita S. D. Sahadeo; Nuno Faria; Oliver Pybus; Risha Singh; Roshan Parasram; Sarah Hill; SueMin Nathaniel; Vernie Ramkissoon |
| EPI_ISL_2631430 | The Trinidad and Tobago Public Health Laboratory | Carrington Lab, Department of PreClinical Sciences, Faculty of Medical Sciences, The University of the West Indies, St Augustine Campus | Anushka Ramjag; Arianne Brown-Jordan; Avery Hinds; Christine V. F. Carrington; Christopher Oura; Gabriel Escobar; Karla Georges; Naresh Nandram; Nikita S. D. Sahadeo; Nuno Faria; Oliver Pybus; Risha Singh; Roshan Parasram; Sarah Hill; SueMin Nathaniel; Vernie Ramkissoon |
| EPI_ISL_2658353 | The Trinidad and Tobago Public Health Laboratory | Carrington Lab, Department of PreClinical Sciences, Faculty of Medical Sciences, The University of the West Indies, St Augustine Campus | Anushka Ramjag; Arianne Brown-Jordan; Avery Hinds; Christine V. F. Carrington; Christopher Oura; Gabriel Escobar; Karla Georges; Naresh Nandram; Nikita S. D. Sahadeo; Nuno Faria; Oliver Pybus; Risha Singh; Roshan Parasram; Sarah Hill; SueMin Nathaniel; Vernie Ramkissoon |
| EPI_ISL_2786936 | Toyama Institute of Health | Toyama Institute of Health | Emiko Igarashi; Hideki Tani; Kazunori Oishi; Kentaro Itokawa; Makoto Kuroda; Masae Itamochi; Masanori Hashino; Noriko Inasaki; Rina Tanaka; Shunsuke Yazawa; Sumiyo Hasegawa; Tsuyoshi Sekizuka; Yumiko Saga |
| EPI_ISL_2990203 | UF Health Shands Hospital - Core Laboratory | Salemi Lab, University of Florida | Cash M; Lauzardo M; Magalis BR; Mavian C; Riva A; Salemi M |
| EPI_ISL_2868659, EPI_ISL_2868675 | URMC LABS | Wadsworth Center, New York State Department of Health | Alexis Russell; Catharine Prussing; Daryl M. Lamson; Erasmus Schneider; Erica Lasek-Nesselquist; John Kelly; Jonathan Plitnick; Kirsten St. George; Matthew Shudt; Melissa A Leisner; Navjot Singh |
| EPI_ISL_2960258 | USA Health Care Authority | Synergy Laboratories | Megan Cornwell |
| EPI_ISL_2840072, EPI_ISL_2899489 | Unilabs | Karolinska University Hospital | Annelie Bjerkner; Isak Sylvin; Jan Albert; Karolina Ininbergs; Lina Guerra Blomqvist; Lynda Eneh; Martin Ekman; Martina Wahlund; Robert Dyrdak; Sandra Broddesson; Tanja Normark; Tobias Allander; Valtteri Wirta; Zhibing Yun |
| EPI_ISL_2658901 | University Hospital for Infectious Diseases | Croatian Institute of Public Health | Irena Tabain; Ivana Ferenčak |
| EPI_ISL_2989602, EPI_ISL_2991220, EPI_ISL_2991241 | University of Florida Health Pathology Laboratories | Salemi Lab, University of Florida | Cash M; Lauzardo M; Magalis BR; Mavian C; Riva A; Salemi M |
| EPI_ISL_2677775 | Utah Public Health Laboratory | Utah Public Health Laboratory | Erin L. Young; Kelly F. Oakeson; Tara Gallagher |
| EPI_ISL_2788084, EPI_ISL_2788090 | VA-Division of Consolidated Laboratory Services | Centers for Disease Control and Prevention Division of Viral Diseases, Pathogen Discovery | Alison Laufer Halpin; Ben L. Rambo-Martin; Clinton R. Paden; Dakota Howard; Darlene Wagner; Dave Wentworth; Dhwani Batra; Jasmine Padilla; Justin Lee; Katie Dillon; Krista Queen; Kristen Knipe; Kristine Lacek; Mark Burroughs; Matthew Schmerer; Mili Sheth; Peter Cook; Sam Shepard; Sarah Nobles; Shoshona Le; Suxiang Tong; Vivien Dugan; Yvette Unoarumhi |

EPI_ISL_2841513, EPI_ISL_2841514, EPI_ISL_2841515, EPI_ISL_2841516, EPI_ISL_2841517, EPI_ISL_2841518, EPI_ISL_2841519, EPI_ISL_2841520, EPI_ISL_2841521, EPI_ISL_2841522, EPI_ISL_2841523, EPI_ISL_2841524, EPI_ISL_2841525, EPI_ISL_2841526, EPI_ISL_2841527, EPI_ISL_2841528, EPI_ISL_2841529, EPI_ISL_2841530

| | | | |
|---|---|---|---|
| see above | Yamagata Prefectural Institute of Public Health | Pathogen Genomics Center, National Institute of Infectious Diseases | Akinobu Wada; Hideyuki Shinohara; Junji Seto; Katsumi Mizuta; Kenichi Komabayashi; Kentaro Itokawa; Makoto Kuroda; Masanori Hashino; Midori Taruishi; Mika Sampei; Naomi Ogawa; Rina Tanaka; Tsuyoshi Sekizuka; Waka Tanaka; Yoko Aoki; Yoko Ikeda; Yoshitaka Ishida |

EPI_ISL_2859474, EPI_ISL_2859476, EPI_ISL_2859478, EPI_ISL_2859479, EPI_ISL_2859481, EPI_ISL_2859482, EPI_ISL_2859483, EPI_ISL_2859484, EPI_ISL_2859486, EPI_ISL_2859487, EPI_ISL_2859489, EPI_ISL_2859490

| | | | |
|---|---|---|---|
| see above | Yamaguchi Prefectural Institute of Public Health and Environment | Yamaguchi Prefectural Institute of Public Health and Environment | Kanako Kawasaki; Kentaro Itokawa; Makoto Kuroda; Masanori Hashino; Mitushiro Kameyama; Rina Tanaka; Tsuyoshi Sekizuka |

We gratefully acknowledge the following Authors from the Originating laboratories responsible for obtaining the specimens, as well as the Submitting laboratories where the genome data were generated and shared via GISAID, on which this research is based.

All Submitters of data may be contacted directly via www.gisaid.org

Authors are sorted alphabetically.

| Accession ID | Originating Laboratory | Submitting Laboratory | Authors |
|---|---|---|---|
| EPI_ISL_2304313 | Alameda County Public Health Department | Alameda County Public Health Department | Kevin Libuit; Kristna Hsieh; Maria Elma |
| EPI_ISL_454607, EPI_ISL_454608, EPI_ISL_454609, EPI_ISL_454610, EPI_ISL_454611, EPI_ISL_454612, EPI_ISL_454613, EPI_ISL_468357, EPI_ISL_468358, EPI_ISL_468359, EPI_ISL_468360, EPI_ISL_468361, EPI_ISL_468362, EPI_ISL_468363, EPI_ISL_468364, EPI_ISL_468365, EPI_ISL_468366, EPI_ISL_468367, EPI_ISL_468368, EPI_ISL_468369, EPI_ISL_468370, EPI_ISL_468371, EPI_ISL_468372, EPI_ISL_468373, EPI_ISL_468374, EPI_ISL_468375, EPI_ISL_468376, EPI_ISL_468377, EPI_ISL_468378, EPI_ISL_468379, EPI_ISL_468380, EPI_ISL_468381, EPI_ISL_468382, EPI_ISL_468383, EPI_ISL_468384, EPI_ISL_468385, EPI_ISL_468386, EPI_ISL_468387, EPI_ISL_625500, EPI_ISL_625501, EPI_ISL_625502, EPI_ISL_625503, EPI_ISL_625504, EPI_ISL_625505, EPI_ISL_625506, EPI_ISL_625507, EPI_ISL_625508, EPI_ISL_625509, EPI_ISL_625510, EPI_ISL_625511, EPI_ISL_625512, EPI_ISL_625513, EPI_ISL_625514, EPI_ISL_625515, EPI_ISL_625516, EPI_ISL_625517, EPI_ISL_625518, EPI_ISL_625519, EPI_ISL_625520, EPI_ISL_625521, EPI_ISL_625522, EPI_ISL_625523, EPI_ISL_625524, EPI_ISL_625525, EPI_ISL_625526, EPI_ISL_625527, EPI_ISL_625528, EPI_ISL_625529, EPI_ISL_625530, EPI_ISL_625531, EPI_ISL_625532, EPI_ISL_625533, EPI_ISL_625534, EPI_ISL_625535, EPI_ISL_625536, EPI_ISL_625537, EPI_ISL_625538, EPI_ISL_625539, EPI_ISL_625540, EPI_ISL_625541, EPI_ISL_625542, EPI_ISL_625543, EPI_ISL_625544, EPI_ISL_625545, EPI_ISL_625546, EPI_ISL_625611, EPI_ISL_625612, EPI_ISL_625613, EPI_ISL_625614, EPI_ISL_625615, EPI_ISL_625616, EPI_ISL_625617, EPI_ISL_625618, EPI_ISL_625619, EPI_ISL_625620, EPI_ISL_625621, EPI_ISL_625622, EPI_ISL_672014, EPI_ISL_672023, EPI_ISL_672024, EPI_ISL_672029, EPI_ISL_672032, EPI_ISL_672036, EPI_ISL_672038, EPI_ISL_672039, EPI_ISL_672040, EPI_ISL_672045, EPI_ISL_672050, EPI_ISL_672053, EPI_ISL_672054, EPI_ISL_672060, EPI_ISL_672062, EPI_ISL_672064, EPI_ISL_672066, EPI_ISL_672076, EPI_ISL_672078, EPI_ISL_672082, EPI_ISL_672086, EPI_ISL_672091, EPI_ISL_672092, EPI_ISL_672095, EPI_ISL_672097, EPI_ISL_672113, EPI_ISL_672305, EPI_ISL_672306, EPI_ISL_672307, EPI_ISL_672308, EPI_ISL_672309, EPI_ISL_672310, EPI_ISL_672311, EPI_ISL_672312, EPI_ISL_672313, EPI_ISL_672314, EPI_ISL_672315, EPI_ISL_672316, EPI_ISL_672317, EPI_ISL_672318, EPI_ISL_672319, EPI_ISL_672320, EPI_ISL_672321, EPI_ISL_672322, EPI_ISL_672323, EPI_ISL_672324, EPI_ISL_672325, EPI_ISL_672326, EPI_ISL_672327, EPI_ISL_672328, EPI_ISL_672329, EPI_ISL_672330, EPI_ISL_672331, EPI_ISL_672332, EPI_ISL_672333, EPI_ISL_672334, EPI_ISL_672335, EPI_ISL_672336, EPI_ISL_672337, EPI_ISL_672338, EPI_ISL_672339, EPI_ISL_672340, EPI_ISL_672341, EPI_ISL_672342, EPI_ISL_672343, EPI_ISL_672344, EPI_ISL_672345, EPI_ISL_672346, EPI_ISL_672347, EPI_ISL_672348, EPI_ISL_672349, EPI_ISL_672350, EPI_ISL_672351, EPI_ISL_672352, EPI_ISL_672353, EPI_ISL_672439, EPI_ISL_672440, EPI_ISL_672441, EPI_ISL_672442, EPI_ISL_672443, EPI_ISL_672444, EPI_ISL_672445, EPI_ISL_672446, EPI_ISL_672447, EPI_ISL_672448, EPI_ISL_672449, EPI_ISL_672450, EPI_ISL_672451, EPI_ISL_672452, EPI_ISL_672453, EPI_ISL_672454, EPI_ISL_672455, EPI_ISL_672456, EPI_ISL_672457, EPI_ISL_672458, EPI_ISL_672459, EPI_ISL_672460, EPI_ISL_672461, EPI_ISL_672462, EPI_ISL_672463, EPI_ISL_672464, EPI_ISL_672465, EPI_ISL_672466, EPI_ISL_672467, EPI_ISL_672468, EPI_ISL_672469, EPI_ISL_672470, EPI_ISL_672471, EPI_ISL_672472, EPI_ISL_672473, EPI_ISL_672474, EPI_ISL_672475, EPI_ISL_672476, EPI_ISL_738509, EPI_ISL_738510, EPI_ISL_738513, EPI_ISL_738514, EPI_ISL_738519, EPI_ISL_738522, EPI_ISL_738523, EPI_ISL_738524, EPI_ISL_738527, EPI_ISL_738528, EPI_ISL_738531, EPI_ISL_738535, EPI_ISL_738537, EPI_ISL_738538, EPI_ISL_738540, EPI_ISL_738545, EPI_ISL_738547, EPI_ISL_738553, EPI_ISL_738554, EPI_ISL_738561, EPI_ISL_738562, EPI_ISL_738564, EPI_ISL_738565, EPI_ISL_738572, EPI_ISL_738573, EPI_ISL_738577, EPI_ISL_738580, EPI_ISL_738584, EPI_ISL_738585, EPI_ISL_738587, EPI_ISL_738592, EPI_ISL_738604, EPI_ISL_738605, EPI_ISL_738606, EPI_ISL_738607, EPI_ISL_738609, EPI_ISL_738613, EPI_ISL_738614, EPI_ISL_738615, EPI_ISL_738616, EPI_ISL_738619, EPI_ISL_738622, EPI_ISL_738623, EPI_ISL_738625, EPI_ISL_738626, EPI_ISL_738627, EPI_ISL_738633, EPI_ISL_738642, EPI_ISL_738648, EPI_ISL_738654, EPI_ISL_738656, EPI_ISL_738659, EPI_ISL_738665, EPI_ISL_738668, EPI_ISL_738671, EPI_ISL_738674, EPI_ISL_738675, EPI_ISL_738676, EPI_ISL_738677, EPI_ISL_738678, EPI_ISL_738679, EPI_ISL_738680, EPI_ISL_738686, EPI_ISL_738687, EPI_ISL_738692, EPI_ISL_738699, EPI_ISL_738711, EPI_ISL_738715, EPI_ISL_738721, EPI_ISL_738724, EPI_ISL_738743, EPI_ISL_738744, EPI_ISL_738745, EPI_ISL_738748, EPI_ISL_738749, EPI_ISL_738751, EPI_ISL_738753, EPI_ISL_738760, EPI_ISL_738766, EPI_ISL_738767, EPI_ISL_738768, EPI_ISL_738773, EPI_ISL_738774, EPI_ISL_738775, EPI_ISL_738776, EPI_ISL_738781, EPI_ISL_738783, EPI_ISL_738788, EPI_ISL_738792, EPI_ISL_738793, EPI_ISL_738796, EPI_ISL_738797, EPI_ISL_738800, EPI_ISL_738804, EPI_ISL_738813, EPI_ISL_738814, EPI_ISL_738817, EPI_ISL_738819, EPI_ISL_738821, EPI_ISL_738822, EPI_ISL_738825, EPI_ISL_738828, EPI_ISL_738831, EPI_ISL_738833, EPI_ISL_738835, EPI_ISL_738837, EPI_ISL_738838, EPI_ISL_738843, EPI_ISL_738846, EPI_ISL_738847, EPI_ISL_738848, EPI_ISL_738858, EPI_ISL_738868, EPI_ISL_738876, EPI_ISL_738881, EPI_ISL_738886, EPI_ISL_738891, EPI_ISL_738892, EPI_ISL_738896, EPI_ISL_738904, EPI_ISL_738914, EPI_ISL_738919, EPI_ISL_738932, EPI_ISL_738934, EPI_ISL_738940, EPI_ISL_738945, EPI_ISL_738950, EPI_ISL_738957, EPI_ISL_738963, EPI_ISL_738974, EPI_ISL_738976, EPI_ISL_738982, EPI_ISL_738983, EPI_ISL_738986, EPI_ISL_738992, EPI_ISL_739003, EPI_ISL_739018, EPI_ISL_739023, EPI_ISL_739029, EPI_ISL_739033, EPI_ISL_739035, EPI_ISL_739046, EPI_ISL_739047, EPI_ISL_739049, EPI_ISL_739054, EPI_ISL_739056, EPI_ISL_739057, EPI_ISL_739059, EPI_ISL_739062, EPI_ISL_739064, EPI_ISL_739073, EPI_ISL_739075, EPI_ISL_739077, EPI_ISL_739082, EPI_ISL_739083, EPI_ISL_739084, EPI_ISL_739088, EPI_ISL_739093, EPI_ISL_739096, EPI_ISL_739100, EPI_ISL_739109, EPI_ISL_739110, EPI_ISL_739111, EPI_ISL_739113, EPI_ISL_739115, EPI_ISL_739118, EPI_ISL_739120, EPI_ISL_739124, EPI_ISL_739127, EPI_ISL_739134, EPI_ISL_739135, EPI_ISL_739136, EPI_ISL_739137, EPI_ISL_739140, EPI_ISL_739144, EPI_ISL_739149, EPI_ISL_739150, EPI_ISL_739158, EPI_ISL_739160, EPI_ISL_739161, EPI_ISL_739163, EPI_ISL_739164, EPI_ISL_739171, EPI_ISL_739173, EPI_ISL_739174, EPI_ISL_739178, EPI_ISL_739179, EPI_ISL_739189, EPI_ISL_739190, EPI_ISL_739192, EPI_ISL_739196, EPI_ISL_739200, EPI_ISL_739201, EPI_ISL_739202, EPI_ISL_739206, EPI_ISL_739207, EPI_ISL_739208, EPI_ISL_739212, EPI_ISL_739213, EPI_ISL_739217, EPI_ISL_739218, EPI_ISL_739220, EPI_ISL_739226, EPI_ISL_739229, EPI_ISL_739231, EPI_ISL_739233, EPI_ISL_739236, EPI_ISL_739241, EPI_ISL_739243, EPI_ISL_739244, EPI_ISL_739257, EPI_ISL_739261, EPI_ISL_739262, EPI_ISL_739268, EPI_ISL_739270, EPI_ISL_739280, EPI_ISL_739283, EPI_ISL_739285, EPI_ISL_739286, EPI_ISL_739288, EPI_ISL_739290, EPI_ISL_739294, EPI_ISL_739300, EPI_ISL_739302, EPI_ISL_739303, EPI_ISL_739306, EPI_ISL_739307, EPI_ISL_739308, EPI_ISL_739309, EPI_ISL_739312, EPI_ISL_739323, EPI_ISL_739324, EPI_ISL_739326, EPI_ISL_739328, EPI_ISL_739333, EPI_ISL_739337, EPI_ISL_739338, EPI_ISL_739340, EPI_ISL_739349, EPI_ISL_739356, EPI_ISL_739358, EPI_ISL_739361, EPI_ISL_739368, EPI_ISL_739378, EPI_ISL_739383, EPI_ISL_739385, EPI_ISL_739387, EPI_ISL_739391, EPI_ISL_739395, EPI_ISL_739406, EPI_ISL_739407, EPI_ISL_739408, EPI_ISL_739422, EPI_ISL_739424, EPI_ISL_739426, EPI_ISL_739427, EPI_ISL_739434, EPI_ISL_739435, EPI_ISL_739439, EPI_ISL_739444, EPI_ISL_739446, EPI_ISL_739448, EPI_ISL_739449, EPI_ISL_739454, EPI_ISL_739459, EPI_ISL_739463, EPI_ISL_739468, EPI_ISL_739470, EPI_ISL_739472, EPI_ISL_739473, EPI_ISL_739483, EPI_ISL_739484, EPI_ISL_739495, EPI_ISL_739503, EPI_ISL_739506, EPI_ISL_739507, EPI_ISL_739511, EPI_ISL_739515, EPI_ISL_739516, EPI_ISL_739518, EPI_ISL_739519, EPI_ISL_739522, EPI_ISL_739523, EPI_ISL_739526, EPI_ISL_739528, EPI_ISL_739535, EPI_ISL_739538, EPI_ISL_739541, EPI_ISL_739544, EPI_ISL_739551, EPI_ISL_739552, EPI_ISL_739555, EPI_ISL_739563, EPI_ISL_739565, EPI_ISL_739569, EPI_ISL_739572, EPI_ISL_739575, EPI_ISL_739576, EPI_ISL_739578, EPI_ISL_739583, EPI_ISL_739587, EPI_ISL_739590, EPI_ISL_739598, EPI_ISL_739600, EPI_ISL_739601, EPI_ISL_739604, EPI_ISL_739610, EPI_ISL_739616, EPI_ISL_739619, EPI_ISL_739622, EPI_ISL_739623, EPI_ISL_739631, EPI_ISL_739634, EPI_ISL_739638, EPI_ISL_739640, EPI_ISL_739643, EPI_ISL_739644, EPI_ISL_739648, EPI_ISL_739652, EPI_ISL_739656, EPI_ISL_1137082, EPI_ISL_1137083, EPI_ISL_1137084, EPI_ISL_1137085, EPI_ISL_1137086, EPI_ISL_1137087, EPI_ISL_1137088, EPI_ISL_1137089, EPI_ISL_1137090, EPI_ISL_1137091, EPI_ISL_1137092, EPI_ISL_1137093, EPI_ISL_1137094, EPI_ISL_1137095, EPI_ISL_1137096, EPI_ISL_1137097, EPI_ISL_1137098, EPI_ISL_1137099, EPI_ISL_1137100, EPI_ISL_1137101, EPI_ISL_1137102, EPI_ISL_1137103, EPI_ISL_1137104, EPI_ISL_1137105, EPI_ISL_1137106, EPI_ISL_1137107, EPI_ISL_1137108, EPI_ISL_1137109, EPI_ISL_1137110, EPI_ISL_1137111, EPI_ISL_1137112, EPI_ISL_1137113, EPI_ISL_1137114, EPI_ISL_1137115, EPI_ISL_1137116, EPI_ISL_1137117, EPI_ISL_1137118, EPI_ISL_1137119, EPI_ISL_1137120, EPI_ISL_1137121, EPI_ISL_1137122, EPI_ISL_1137123, EPI_ISL_1137124, EPI_ISL_1137125, EPI_ISL_1137126, EPI_ISL_1137127, EPI_ISL_1137128, EPI_ISL_1137129, EPI_ISL_1137130, EPI_ISL_1137131, EPI_ISL_1137132, EPI_ISL_1137133, EPI_ISL_1137134, EPI_ISL_1137135, EPI_ISL_1137136, EPI_ISL_1137137, EPI_ISL_1137138, EPI_ISL_1137139, EPI_ISL_1137140, EPI_ISL_1137141, EPI_ISL_1137142, EPI_ISL_1137143, EPI_ISL_1137144, EPI_ISL_1137145, EPI_ISL_1137146, EPI_ISL_1137147, EPI_ISL_1137148, EPI_ISL_1137149, EPI_ISL_1137150, EPI_ISL_1137151, EPI_ISL_1137152, EPI_ISL_1137153, EPI_ISL_1137154, EPI_ISL_1137155, EPI_ISL_1137156, EPI_ISL_1137157, EPI_ISL_1137158, EPI_ISL_1137159, EPI_ISL_1137160, EPI_ISL_1137161, EPI_ISL_1137162, EPI_ISL_1137163, EPI_ISL_1137164, EPI_ISL_1137165, EPI_ISL_1137166, EPI_ISL_1137167, EPI_ISL_1137168, EPI_ISL_1137169, EPI_ISL_1477262, EPI_ISL_1477269, EPI_ISL_1477335, EPI_ISL_1477446, EPI_ISL_1477447, EPI_ISL_1477462, EPI_ISL_1477466, EPI_ISL_1477468, EPI_ISL_1477483, EPI_ISL_1477487, EPI_ISL_1477489, EPI_ISL_1477491, EPI_ISL_1477497, EPI_ISL_1477499, EPI_ISL_1477520, EPI_ISL_1477545, EPI_ISL_1477546, EPI_ISL_1477547, EPI_ISL_1477554, EPI_ISL_1477557, EPI_ISL_1477565, EPI_ISL_1477569, EPI_ISL_1477584, EPI_ISL_1477594, EPI_ISL_1477595, EPI_ISL_1477605, EPI_ISL_1477617, EPI_ISL_1477627, EPI_ISL_1477630, EPI_ISL_1477632, EPI_ISL_1477639, EPI_ISL_1477647, EPI_ISL_1477649, EPI_ISL_1477657, EPI_ISL_1701120, EPI_ISL_1701123, EPI_ISL_1701124, EPI_ISL_1701132, EPI_ISL_1701188, EPI_ISL_1701213, EPI_ISL_1701230, EPI_ISL_1701233, EPI_ISL_1701235, EPI_ISL_1701245, EPI_ISL_1701247, EPI_ISL_1701302, EPI_ISL_1924616, EPI_ISL_2658579, EPI_ISL_2658603, EPI_ISL_2658620, EPI_ISL_2658631, EPI_ISL_2658634, EPI_ISL_2658638, EPI_ISL_2658654, EPI_ISL_2658658, EPI_ISL_2658664, EPI_ISL_2658666, EPI_ISL_2658682, EPI_ISL_2658687, EPI_ISL_2658697, EPI_ISL_2658712, EPI_ISL_2658734, EPI_ISL_2659582, EPI_ISL_2659653, EPI_ISL_2659654, EPI_ISL_2758414, EPI_ISL_2758415, EPI_ISL_2758417, EPI_ISL_2758463 | | | |
| see above | Alameda County Public Health Lab | Chan-Zuckerberg Biohub | CZB Cliahub Consortium |
| EPI_ISL_414648, EPI_ISL_416457 | Andersen Lab, The Scripps Research Institute | Andersen Lab, The Scripps Research Institute | Catie Anderson; Emily Spender; Karthik Gangavarapu; Kristian Andersen; Laura Nicholson; Mark Zeller; Raphaelle Klitting; Refugio Robles-Sikisaka; Sarah Topol |
| EPI_ISL_429991, EPI_ISL_430016 | Andersen lab at Scripps Research | Andersen lab at Scripps Research | SEARCH Alliance San Diego |
| EPI_ISL_884866, EPI_ISL_884867, EPI_ISL_884868, EPI_ISL_884869, EPI_ISL_884870, EPI_ISL_1924583, EPI_ISL_1924584, EPI_ISL_1924585, EPI_ISL_1924586, EPI_ISL_1924587, EPI_ISL_1924588, EPI_ISL_1924589, EPI_ISL_1924591, EPI_ISL_1924594, EPI_ISL_1924595, EPI_ISL_1924597, EPI_ISL_1924598, EPI_ISL_1924599, EPI_ISL_1924601, EPI_ISL_1924603, EPI_ISL_1924606, EPI_ISL_1924607, EPI_ISL_1924608, EPI_ISL_1924609, EPI_ISL_1924610, EPI_ISL_1924611, EPI_ISL_1924612, EPI_ISL_1924613, EPI_ISL_1924614, EPI_ISL_1924615, EPI_ISL_1924617, EPI_ISL_1924620, EPI_ISL_1924621, EPI_ISL_1924622, EPI_ISL_1924623, EPI_ISL_1924624, EPI_ISL_1924625, EPI_ISL_1924626, EPI_ISL_1924627, EPI_ISL_1924628 | | | |
| see above | CA DPH Viral and Rickettsial Disease Laboratory | Chan-Zuckerberg Biohub | CZB Cliahub Consortium |
| EPI_ISL_576177 | CA, CDPH, Viral and Rickettsial Disease Laboratory | Pathogen Discovery, Respiratory Viruses Branch, Division of Viral Diseases, Centers for Disease Control and Prevention | Anna Uehara; Brian Lynch; Clinton R. Paden; Haibin Wang; Jing Zhang; Krista Queen; Peter Cook; Suxiang Tong; Yan Li; Ying Tao |
| EPI_ISL_535662, EPI_ISL_535663, EPI_ISL_535664, EPI_ISL_535665, EPI_ISL_535666, EPI_ISL_535667, EPI_ISL_535668, EPI_ISL_535669, EPI_ISL_535670, EPI_ISL_535671, EPI_ISL_535672, EPI_ISL_535673, EPI_ISL_535674, EPI_ISL_535675, EPI_ISL_535676, EPI_ISL_535677, EPI_ISL_535678, EPI_ISL_535679, EPI_ISL_535680, EPI_ISL_535681, EPI_ISL_535682, EPI_ISL_535683, EPI_ISL_535684, EPI_ISL_535685, EPI_ISL_535686, EPI_ISL_535687, EPI_ISL_535688, EPI_ISL_535689, EPI_ISL_535690, EPI_ISL_535691, EPI_ISL_535692, EPI_ISL_535693, EPI_ISL_535694, EPI_ISL_535695, EPI_ISL_535696, EPI_ISL_535697, EPI_ISL_535698, EPI_ISL_535699, EPI_ISL_535700, EPI_ISL_535701, EPI_ISL_535702, EPI_ISL_535703, EPI_ISL_535704, EPI_ISL_535705, EPI_ISL_535706, EPI_ISL_535707, EPI_ISL_535708, EPI_ISL_535709, EPI_ISL_535710, EPI_ISL_535711, EPI_ISL_535712, EPI_ISL_535713, EPI_ISL_535714, EPI_ISL_535715 | | | |
| see above | CDPH, Microbial Diseases Laboratory | Pathogen Discovery, Respiratory Viruses Branch, Division of Viral Diseases, Centers for Disease Control and Prevention | Anna Uehara; Brian Lynch; Clinton R. Paden; Haibin Wang; Jing Zhang; Krista Queen; Rachel Marine; Suxiang Tong; Yan Li; Ying Tao |
| EPI_ISL_408009, EPI_ISL_408010 | California Department of Health | Pathogen Discovery, Respiratory Viruses Branch, Division of Viral Diseases, Centers for Dieases Control and Prevention | Anna Uehara; Brett L. Whitaker; Brian Lynch; Clinton Paden; Janna' R. Murray; Jing Zhang; Krista Queen; Lijuan Wang; Senthil Kumar K. Sakthivel; Shifaq Kamili; Stephen Lindstrom; Susan I. Gerber; Suxiang Tong; Xiaoyan Lu; Yan Li; Ying Tao |
| EPI_ISL_408008 | California Department of Health | Pathogen Discovery, Respiratory Viruses Branch, Division of Viral Diseases, Centers for Disease Control and Prevention | Anna Uehara; Brett L. Whitaker; Brian Lynch; Clinton Paden; Janna' R. Murray; Jing Zhang; Krista Queen; Lijuan Wang; Senthil Kumar K. Sakthivel; Shifaq Kamili; Stephen Lindstrom; Susan I. Gerber; Suxiang Tong; Xiaoyan Lu; Yan Li; Ying Tao |
| EPI_ISL_515894, EPI_ISL_515895, EPI_ISL_515896, EPI_ISL_515897, EPI_ISL_515898, EPI_ISL_515899, EPI_ISL_515900, EPI_ISL_515901, EPI_ISL_515902, EPI_ISL_515903, EPI_ISL_515904, EPI_ISL_515905, EPI_ISL_515906, EPI_ISL_515907, EPI_ISL_515908, EPI_ISL_515909, EPI_ISL_515910, EPI_ISL_515911, EPI_ISL_515912, EPI_ISL_515913, EPI_ISL_515914, EPI_ISL_515915, EPI_ISL_515916, EPI_ISL_515917, EPI_ISL_515918, EPI_ISL_515919, EPI_ISL_515920, EPI_ISL_515921, EPI_ISL_515922, EPI_ISL_515923, EPI_ISL_515924, EPI_ISL_515925, EPI_ISL_515926, EPI_ISL_515927, EPI_ISL_515928, EPI_ISL_515929, EPI_ISL_754915, EPI_ISL_754916, EPI_ISL_754917, EPI_ISL_754918, EPI_ISL_754919, EPI_ISL_754920, EPI_ISL_754921, EPI_ISL_754922, EPI_ISL_754923, EPI_ISL_754925, EPI_ISL_754926, EPI_ISL_754927, EPI_ISL_754928, EPI_ISL_754929, EPI_ISL_754930, EPI_ISL_754931, EPI_ISL_754932, EPI_ISL_754933, EPI_ISL_754934, EPI_ISL_754935, EPI_ISL_754936, EPI_ISL_754937, EPI_ISL_754938, EPI_ISL_754939, EPI_ISL_754940, EPI_ISL_754941, EPI_ISL_754943, EPI_ISL_754944, EPI_ISL_754945, EPI_ISL_754946, EPI_ISL_754947, EPI_ISL_754950, EPI_ISL_754951, EPI_ISL_754952, EPI_ISL_754953, EPI_ISL_754954, EPI_ISL_754956, EPI_ISL_754958, EPI_ISL_754959, EPI_ISL_754960, EPI_ISL_754961, EPI_ISL_754962, EPI_ISL_754963, EPI_ISL_754964, EPI_ISL_754968, EPI_ISL_754969, EPI_ISL_754970, EPI_ISL_754972, EPI_ISL_754974, EPI_ISL_754975, EPI_ISL_754976, EPI_ISL_754977, EPI_ISL_754978, EPI_ISL_754979, EPI_ISL_754980, EPI_ISL_754981, EPI_ISL_754984, EPI_ISL_754985, EPI_ISL_754986, EPI_ISL_754987, EPI_ISL_754988, EPI_ISL_754989, EPI_ISL_754990, EPI_ISL_754991, EPI_ISL_754992, EPI_ISL_754994, EPI_ISL_754995, EPI_ISL_754996, EPI_ISL_754998, EPI_ISL_754999, EPI_ISL_755001, EPI_ISL_755004, EPI_ISL_755006, EPI_ISL_755008, EPI_ISL_755011, EPI_ISL_755012, EPI_ISL_755016, EPI_ISL_755017, EPI_ISL_755018, EPI_ISL_755019, EPI_ISL_755020, EPI_ISL_755022, EPI_ISL_755023, EPI_ISL_755025, EPI_ISL_755027, EPI_ISL_755028, EPI_ISL_755029, EPI_ISL_755030, EPI_ISL_755031, EPI_ISL_755032, EPI_ISL_755033, EPI_ISL_755034, EPI_ISL_755035, EPI_ISL_755036, EPI_ISL_755037, EPI_ISL_755038, EPI_ISL_755039, EPI_ISL_755040, EPI_ISL_755041, EPI_ISL_755042, EPI_ISL_755043, EPI_ISL_755044, EPI_ISL_755045, EPI_ISL_755046, EPI_ISL_755047, EPI_ISL_755048, EPI_ISL_755049, EPI_ISL_755050, EPI_ISL_755051, EPI_ISL_755052, EPI_ISL_755053, EPI_ISL_755054, EPI_ISL_755055, EPI_ISL_755056, EPI_ISL_755057, EPI_ISL_755058, EPI_ISL_755059, EPI_ISL_755060, EPI_ISL_755061, EPI_ISL_755065, EPI_ISL_755066, EPI_ISL_755081, EPI_ISL_2233912, EPI_ISL_2233913, EPI_ISL_2233938, EPI_ISL_2233939, EPI_ISL_2233940, EPI_ISL_2233941, EPI_ISL_2233942, EPI_ISL_2233972, EPI_ISL_2233973, EPI_ISL_2233974, EPI_ISL_2233975, EPI_ISL_2233976, EPI_ISL_2233977, EPI_ISL_2233978, EPI_ISL_2233979, EPI_ISL_2233996, EPI_ISL_2233997, EPI_ISL_2233998, EPI_ISL_2233999, EPI_ISL_2234000, EPI_ISL_2234024, EPI_ISL_2234025, EPI_ISL_2234027 | | | |
| see above | California Department of Public Health | California Department of Public Health | CDPH IDLB COVIDNet; CDPH IDLB COVIDNet et al |
| EPI_ISL_413930 | California Department of Public Health | Chiu Laboratory UCSF-Abbott Viral Diagnostics and Discovery Center University of California, San Francisco | Alicia Sotomayor-Gonzalez; Allan Gopez; Chao-Yang Pan; Debra A. Wadford; Guixia Yu; Hugo Guevara; Scot Federman; Steve Miller; Wei Gu; Xianding Deng; and Charles Y. Chiu |
| EPI_ISL_413557, EPI_ISL_413558, EPI_ISL_413559, EPI_ISL_413561, EPI_ISL_413922, EPI_ISL_413924, EPI_ISL_413925, EPI_ISL_413926, EPI_ISL_413928, EPI_ISL_413931, EPI_ISL_417322, EPI_ISL_417323, EPI_ISL_417324, EPI_ISL_417325, EPI_ISL_417326, EPI_ISL_417327, EPI_ISL_417328, EPI_ISL_417329, EPI_ISL_429875, EPI_ISL_429876, EPI_ISL_429881, EPI_ISL_984389, EPI_ISL_984390, EPI_ISL_984409, EPI_ISL_984490, EPI_ISL_984491, EPI_ISL_984492, EPI_ISL_984493, EPI_ISL_984494, EPI_ISL_984495, EPI_ISL_984496, EPI_ISL_984497, EPI_ISL_984498, EPI_ISL_984499, EPI_ISL_984500, EPI_ISL_984501, EPI_ISL_984502, EPI_ISL_984503, EPI_ISL_984504, EPI_ISL_984505, EPI_ISL_984506, EPI_ISL_984507, EPI_ISL_984508, EPI_ISL_984509, EPI_ISL_984510, EPI_ISL_984511, EPI_ISL_984512, EPI_ISL_984513, EPI_ISL_984514, EPI_ISL_984515, EPI_ISL_984516, EPI_ISL_984517, EPI_ISL_984518, EPI_ISL_984519, EPI_ISL_984520, EPI_ISL_984521, EPI_ISL_984522, EPI_ISL_984523, EPI_ISL_984524, EPI_ISL_984525, EPI_ISL_984526, EPI_ISL_984527, EPI_ISL_984528, EPI_ISL_984529, EPI_ISL_984530, EPI_ISL_984531, EPI_ISL_984532, EPI_ISL_984533, EPI_ISL_984534, EPI_ISL_984535, EPI_ISL_984536, EPI_ISL_984537, EPI_ISL_984538, EPI_ISL_984539, EPI_ISL_984540, EPI_ISL_984541, EPI_ISL_984542, EPI_ISL_984543, EPI_ISL_984544, EPI_ISL_984545, EPI_ISL_984546, EPI_ISL_984547, EPI_ISL_984548, EPI_ISL_984549, EPI_ISL_984550, EPI_ISL_984551, EPI_ISL_984552, EPI_ISL_984553, EPI_ISL_984554, EPI_ISL_984555, EPI_ISL_984556, EPI_ISL_984557, EPI_ISL_984558, EPI_ISL_984559 | | | |
| see above | California Department of Public Health | Chiu Laboratory, University of California, San Francisco | Candace Wang; Chao-Yang Pan; Charles Chiu; Debra A. Wadford; Debra Wadford; Hugo Guevara; Jill Hacker; Scot Federman; Venice Servellita; Wei Gu; Xianding (Wayne) Deng; Xianding Deng; and Charles Y. Chiu |
| EPI_ISL_406034, EPI_ISL_406036, EPI_ISL_410044, EPI_ISL_411954, EPI_ISL_411955 | California Department of Public Health | Pathogen Discovery, Respiratory Viruses Branch, Division of Viral Diseases, Centers for Dieases Control and Prevention | Anna Uehara; Brett L. Whitaker; Brian Lynch; Clinton R. Paden; Haibin Wang; Janna' R. Murray; Jing Zhang; Krista Queen; Lijuan Wang; Senthil Kumar K. Sakthivel; Shifaq Kamili; Stephen Lindstrom; Susan I. Gerber; Suxiang Tong; Xiaoyan Lu; Yan Li; Ying Tao |
| EPI_ISL_412862, EPI_ISL_419554, EPI_ISL_594458 | California Department of Public Health | Pathogen Discovery, Respiratory Viruses Branch, Division of Viral Diseases, Centers for Disease Control and Prevention | Anna Uehara; Brett L. Whitaker; Brian Lynch; Clinton Paden; Clinton R. Paden; Haibin Wang; Janna' R. Murray; Jasmine Padilla; Jing Zhang; Julu Bhatnagar; Justin Lee; Krista Queen; Lijuan Wang; Senthil Kumar K. Sakthivel; Shifaq Kamili; Stephen Lindstrom; Susan I. Gerber; Suxiang Tong; Xiaoyan Lu; Yan Li; Ying Tao |
| EPI_ISL_418865 | California Department of Public Health | University of California, San Francisco | Chao-Yang Pan; Debra A. Wadford; Hugo Guevara; Scot Federman; Wei Gu; Xianding Deng; and Charles Y. Chiu |
| EPI_ISL_885128, EPI_ISL_912187, EPI_ISL_912188, EPI_ISL_912189, EPI_ISL_912190, EPI_ISL_912191, EPI_ISL_912192, EPI_ISL_912193, EPI_ISL_912194, EPI_ISL_1234947, EPI_ISL_1234951, EPI_ISL_1234964, EPI_ISL_1235027, EPI_ISL_1235035, EPI_ISL_1701619, EPI_ISL_2536197, EPI_ISL_2536198 | | | |
| see above | California Institute of Technology | Chan-Zuckerberg Biohub | CZB Cliahub Consortium |
| EPI_ISL_467809, EPI_ISL_475574, EPI_ISL_475575, EPI_ISL_475576, EPI_ISL_475577, EPI_ISL_475578, EPI_ISL_475579, EPI_ISL_475580, EPI_ISL_475581, EPI_ISL_475582, EPI_ISL_475583, EPI_ISL_475584, EPI_ISL_475585, EPI_ISL_475586, EPI_ISL_475587, EPI_ISL_475588, EPI_ISL_475589, EPI_ISL_475590, EPI_ISL_475591, EPI_ISL_475592, EPI_ISL_475593, EPI_ISL_475594, EPI_ISL_475595, EPI_ISL_475596, EPI_ISL_475597, EPI_ISL_475598, EPI_ISL_475599, EPI_ISL_475600, EPI_ISL_475601, EPI_ISL_475602, EPI_ISL_475603, EPI_ISL_475604, EPI_ISL_475605, EPI_ISL_475606, EPI_ISL_475607, EPI_ISL_475608, EPI_ISL_475609, EPI_ISL_475610, EPI_ISL_475611, EPI_ISL_475612, EPI_ISL_475613, EPI_ISL_475614, EPI_ISL_475615, EPI_ISL_475616, EPI_ISL_475617, EPI_ISL_475618, EPI_ISL_475619, EPI_ISL_475620, EPI_ISL_475621, EPI_ISL_475622, EPI_ISL_475623, EPI_ISL_475624, EPI_ISL_475625, EPI_ISL_475626, EPI_ISL_475627, EPI_ISL_475628, EPI_ISL_475629, EPI_ISL_475630, EPI_ISL_475631, EPI_ISL_475632, EPI_ISL_475633, EPI_ISL_475634, EPI_ISL_475635, EPI_ISL_475636, EPI_ISL_475637, EPI_ISL_475638, EPI_ISL_475639, EPI_ISL_475640, EPI_ISL_475641, EPI_ISL_475642, EPI_ISL_475643, EPI_ISL_475644, EPI_ISL_475645, EPI_ISL_475646, EPI_ISL_475647, EPI_ISL_475648, EPI_ISL_475649, EPI_ISL_475650, EPI_ISL_475651, EPI_ISL_475652, EPI_ISL_475653, EPI_ISL_475654, EPI_ISL_475655, EPI_ISL_475656, EPI_ISL_475657, EPI_ISL_475658, EPI_ISL_475659, EPI_ISL_475660, EPI_ISL_475661, EPI_ISL_475662, EPI_ISL_475663, EPI_ISL_475664, EPI_ISL_475665, EPI_ISL_475666, EPI_ISL_475667, EPI_ISL_475668, EPI_ISL_475669, EPI_ISL_475670, EPI_ISL_475671, EPI_ISL_475672, EPI_ISL_475673, EPI_ISL_475674, EPI_ISL_475675, EPI_ISL_475676, EPI_ISL_475677, EPI_ISL_475678, EPI_ISL_475679, EPI_ISL_475680, EPI_ISL_475681, EPI_ISL_475682, EPI_ISL_475683, EPI_ISL_475684, EPI_ISL_475685, EPI_ISL_475686, EPI_ISL_475687, EPI_ISL_475688, EPI_ISL_475689, EPI_ISL_475690, EPI_ISL_475691, EPI_ISL_475692, EPI_ISL_475693, EPI_ISL_475694, EPI_ISL_475695, EPI_ISL_475696, EPI_ISL_475697, EPI_ISL_475698, EPI_ISL_475699, EPI_ISL_475700, EPI_ISL_475701, EPI_ISL_475702, EPI_ISL_475703, EPI_ISL_475704, EPI_ISL_475705, EPI_ISL_475706, EPI_ISL_475707, EPI_ISL_475708, EPI_ISL_475709, EPI_ISL_475710, EPI_ISL_475711, EPI_ISL_475712, EPI_ISL_475713, EPI_ISL_475714, EPI_ISL_475715, EPI_ISL_475716 | | | |
| see above | Cedars-Sinai Medical Center, Department of Pathology & Laboratory Medicine, Molecular Pathology Laboratory | Cedars-Sinai Medical Center, Molecular Pathology Laboratory of Department of Pathology & Laboratory Medicine and Genomic Core | Brian Davis; Eric Vail; Jasmine T Plummer; Jean Lopategui; Jianbo Song; John Paul Govindavari; Jong Taek Kim; Stephanie Chen; Wenjuan Zhang |
| EPI_ISL_417330, EPI_ISL_417331, EPI_ISL_417332, EPI_ISL_429877, EPI_ISL_429878, EPI_ISL_977910, EPI_ISL_977911, EPI_ISL_977912, EPI_ISL_977913, EPI_ISL_977914, EPI_ISL_977915, EPI_ISL_977916, EPI_ISL_977917 | | | |
| see above | Chiu Laboratory, University of California, San Francisco | Chiu Laboratory, University of California, San Francisco | Candace Wang; Charles Chiu; Debra Wadford; Jill Hacker; Scot Federman; Venice Servellita; Wei Gu; Xianding (Wayne) Deng; Xianding Deng; and Charles Y. Chiu |
| EPI_ISL_753324, EPI_ISL_1295564, EPI_ISL_1295565, EPI_ISL_1295566, EPI_ISL_1295567, EPI_ISL_1295568, EPI_ISL_1295569, EPI_ISL_1295570, EPI_ISL_1295571, EPI_ISL_1295572, EPI_ISL_1295573, EPI_ISL_1295574, EPI_ISL_1295575, EPI_ISL_1295576, EPI_ISL_1295577, EPI_ISL_1295578, EPI_ISL_1295579, EPI_ISL_1295580, EPI_ISL_1295581, EPI_ISL_1295582, EPI_ISL_1295583, EPI_ISL_1295584 | | | |
| see above | Clinical virology Laboratory, Children's Hospital Los Angeles | Center for Personalized Medicine, Children's Hospital Los Angeles | Alex Ryutov; Alexander R. Judkins; Ananthanarayanan Govindarajan; David Ruble; Dejerianne Ostrow; Dennis Maglinte; Gai et al; Jaclyn A. Biegel; Jennifer Dien Bard; Jennifer Han; Lishuang Shen; Moiz Bootwalla; Timothy J. Triche; Utsav Pandey; Xiaowu Gai |

| | | | |
|---|---|---|---|
| EPI_ISL_1017617, EPI_ISL_1017618 | Connecticut Veterans' Affairs Hospital | Grubaugh Lab - Yale School of Public Health | Chantal Vogels; Danielle Plank; Isabel Ott; Joseph Fauver; Mary Petrone; Nathan Grubaugh; Shaili Gupta; Tara Alpert |
| EPI_ISL_672488, EPI_ISL_672489, EPI_ISL_1924604, EPI_ISL_2758533 | Contra Costa County Public Health Lab | Chan-Zuckerberg Biohub | CZB Cliahub Consortium |
| EPI_ISL_468615, EPI_ISL_468616, EPI_ISL_468617, EPI_ISL_468618, EPI_ISL_468619, EPI_ISL_468620, EPI_ISL_468621, EPI_ISL_468622, EPI_ISL_468623, EPI_ISL_468624, EPI_ISL_468625, EPI_ISL_468626, EPI_ISL_468627, EPI_ISL_468628, EPI_ISL_468629, EPI_ISL_468630, EPI_ISL_468631, EPI_ISL_468632, EPI_ISL_468633, EPI_ISL_468634, EPI_ISL_468635, EPI_ISL_468636, EPI_ISL_468637, EPI_ISL_468638, EPI_ISL_468639, EPI_ISL_468640, EPI_ISL_468641, EPI_ISL_468642, EPI_ISL_468643, EPI_ISL_468644, EPI_ISL_468645, EPI_ISL_468646, EPI_ISL_468647, EPI_ISL_468648, EPI_ISL_468649, EPI_ISL_468650, EPI_ISL_468651, EPI_ISL_468652, EPI_ISL_468653, EPI_ISL_468654, EPI_ISL_468655, EPI_ISL_548491, EPI_ISL_548492, EPI_ISL_548495, EPI_ISL_548499, EPI_ISL_548501, EPI_ISL_548506, EPI_ISL_548507, EPI_ISL_548508, EPI_ISL_548510, EPI_ISL_548515, EPI_ISL_548516, EPI_ISL_548520, EPI_ISL_548524, EPI_ISL_548527, EPI_ISL_548528, EPI_ISL_548531, EPI_ISL_548534, EPI_ISL_548535, EPI_ISL_548537, EPI_ISL_548538, EPI_ISL_548539, EPI_ISL_548540, EPI_ISL_548545, EPI_ISL_548548, EPI_ISL_548549, EPI_ISL_548552, EPI_ISL_548554, EPI_ISL_548556, EPI_ISL_548557, EPI_ISL_548558, EPI_ISL_548561, EPI_ISL_548564, EPI_ISL_548565, EPI_ISL_548566, EPI_ISL_548567, EPI_ISL_548569, EPI_ISL_548572, EPI_ISL_548573, EPI_ISL_548574, EPI_ISL_548575, EPI_ISL_548577, EPI_ISL_548579 | | | |
| see above | Contra Costa Public Health Lab | Chan-Zuckerberg Biohub | CZB Cliahub Consortium |
| EPI_ISL_454635 | County Of San Luis Obispo Public Health Laboratory | Chan-Zuckerberg Biohub | CZB Cliahub Consortium |
| EPI_ISL_468388, EPI_ISL_468389, EPI_ISL_468390, EPI_ISL_468391, EPI_ISL_468392, EPI_ISL_468393, EPI_ISL_468394, EPI_ISL_468395, EPI_ISL_468396, EPI_ISL_468397, EPI_ISL_468398, EPI_ISL_468399, EPI_ISL_468400, EPI_ISL_468401, EPI_ISL_468402, EPI_ISL_468403, EPI_ISL_468404, EPI_ISL_468405, EPI_ISL_468406, EPI_ISL_468407, EPI_ISL_468408, EPI_ISL_468409, EPI_ISL_468410, EPI_ISL_468411, EPI_ISL_468412, EPI_ISL_468413, EPI_ISL_468414, EPI_ISL_468415, EPI_ISL_468416, EPI_ISL_468417, EPI_ISL_468418, EPI_ISL_468419, EPI_ISL_468420, EPI_ISL_468421, EPI_ISL_468422, EPI_ISL_468423, EPI_ISL_468424, EPI_ISL_468425, EPI_ISL_468426, EPI_ISL_468427, EPI_ISL_468428, EPI_ISL_468429, EPI_ISL_468430, EPI_ISL_468431, EPI_ISL_468432, EPI_ISL_468433, EPI_ISL_468434, EPI_ISL_468435, EPI_ISL_468436, EPI_ISL_468437, EPI_ISL_548265, EPI_ISL_548268, EPI_ISL_548269, EPI_ISL_548280, EPI_ISL_548281, EPI_ISL_548284, EPI_ISL_548286, EPI_ISL_548288, EPI_ISL_548292, EPI_ISL_548294, EPI_ISL_548295, EPI_ISL_548304, EPI_ISL_548311, EPI_ISL_548316, EPI_ISL_548322, EPI_ISL_548325, EPI_ISL_548329, EPI_ISL_548330, EPI_ISL_548335, EPI_ISL_548336, EPI_ISL_548347, EPI_ISL_548350, EPI_ISL_548351, EPI_ISL_548357, EPI_ISL_548359, EPI_ISL_548360, EPI_ISL_548361, EPI_ISL_548365, EPI_ISL_548366, EPI_ISL_548369, EPI_ISL_548370, EPI_ISL_548391, EPI_ISL_548425, EPI_ISL_548428, EPI_ISL_548436, EPI_ISL_548453, EPI_ISL_548457, EPI_ISL_548493, EPI_ISL_548502, EPI_ISL_548504, EPI_ISL_548517, EPI_ISL_548519, EPI_ISL_548521, EPI_ISL_548532, EPI_ISL_548542, EPI_ISL_548544, EPI_ISL_548560, EPI_ISL_548603, EPI_ISL_548631, EPI_ISL_548644, EPI_ISL_548663, EPI_ISL_548671, EPI_ISL_625547, EPI_ISL_625548, EPI_ISL_625549, EPI_ISL_625623, EPI_ISL_625624, EPI_ISL_625625, EPI_ISL_625626, EPI_ISL_672028, EPI_ISL_672068, EPI_ISL_672087, EPI_ISL_672354, EPI_ISL_672355, EPI_ISL_672356, EPI_ISL_672357, EPI_ISL_672358, EPI_ISL_672359, EPI_ISL_738890, EPI_ISL_738990, EPI_ISL_739017, EPI_ISL_739038, EPI_ISL_739095, EPI_ISL_739102, EPI_ISL_739363, EPI_ISL_739492, EPI_ISL_739588, EPI_ISL_1235029, EPI_ISL_1477436, EPI_ISL_1477454, EPI_ISL_1477530, EPI_ISL_1477564, EPI_ISL_1477634, EPI_ISL_1529147, EPI_ISL_1529225, EPI_ISL_1529246, EPI_ISL_1529248, EPI_ISL_1529343, EPI_ISL_2497545, EPI_ISL_2497547, EPI_ISL_2497550, EPI_ISL_2497551, EPI_ISL_2497552, EPI_ISL_2497642, EPI_ISL_2497643, EPI_ISL_2497682, EPI_ISL_2497683, EPI_ISL_2497699, EPI_ISL_2497716, EPI_ISL_2497717, EPI_ISL_2497722, EPI_ISL_2497725, EPI_ISL_2497730, EPI_ISL_2497741, EPI_ISL_2497742, EPI_ISL_2497746, EPI_ISL_2497751, EPI_ISL_2497766, EPI_ISL_2497769, EPI_ISL_2497772, EPI_ISL_2497780, EPI_ISL_2497782, EPI_ISL_2497786, EPI_ISL_2497791, EPI_ISL_2497792, EPI_ISL_2497795, EPI_ISL_2497797, EPI_ISL_2497800, EPI_ISL_2497823, EPI_ISL_2497834, EPI_ISL_2497836, EPI_ISL_2497839, EPI_ISL_2497840, EPI_ISL_2497842, EPI_ISL_2500027, EPI_ISL_2500040, EPI_ISL_2500041, EPI_ISL_2500045, EPI_ISL_2500046, EPI_ISL_2500055, EPI_ISL_2500056, EPI_ISL_2500065, EPI_ISL_2500071, EPI_ISL_2500088, EPI_ISL_2500091, EPI_ISL_2500095, EPI_ISL_2500101, EPI_ISL_2500103, EPI_ISL_2500105, EPI_ISL_2500107, EPI_ISL_2500114, EPI_ISL_2500117, EPI_ISL_2500119, EPI_ISL_2500123, EPI_ISL_2500124, EPI_ISL_2500128, EPI_ISL_2500141, EPI_ISL_2500161, EPI_ISL_2500165, EPI_ISL_2500167, EPI_ISL_2500172, EPI_ISL_2500175, EPI_ISL_2500176, EPI_ISL_2500191, EPI_ISL_2500197, EPI_ISL_2500198, EPI_ISL_2500215, EPI_ISL_2658613, EPI_ISL_2658632, EPI_ISL_2658678, EPI_ISL_2659606, EPI_ISL_2758543, EPI_ISL_2758545, EPI_ISL_2758546 | | | |
| see above | County of San Luis Obispo Public Health Laboratory | Chan-Zuckerberg Biohub | CZB Cliahub Consortium |
| EPI_ISL_2758595, EPI_ISL_2758596 | County of San Luis Obispo Public Health Laboratory | County of San Luis Obispo Public Health Laboratory | Frances Sidhu; Katrina Erwin |
| EPI_ISL_437043, EPI_ISL_437044, EPI_ISL_437045, EPI_ISL_437046, EPI_ISL_437047, EPI_ISL_437048, EPI_ISL_437049, EPI_ISL_437050, EPI_ISL_437051, EPI_ISL_437052, EPI_ISL_437053, EPI_ISL_437054, EPI_ISL_437055, EPI_ISL_437056, EPI_ISL_437057, EPI_ISL_437058, EPI_ISL_437059, EPI_ISL_437060, EPI_ISL_437061, EPI_ISL_437062, EPI_ISL_437063, EPI_ISL_437064, EPI_ISL_437065, EPI_ISL_437066, EPI_ISL_437067, EPI_ISL_437068, EPI_ISL_437069, EPI_ISL_437070, EPI_ISL_437071, EPI_ISL_437072, EPI_ISL_437073, EPI_ISL_437074, EPI_ISL_437075, EPI_ISL_437076, EPI_ISL_437077, EPI_ISL_437078, EPI_ISL_437079, EPI_ISL_437080, EPI_ISL_437081, EPI_ISL_437082, EPI_ISL_437083, EPI_ISL_437084, EPI_ISL_437085, EPI_ISL_437086, EPI_ISL_437087, EPI_ISL_437088, EPI_ISL_444023, EPI_ISL_444024, EPI_ISL_444025, EPI_ISL_444026 | | | |
| see above | County of Santa Clara Public Health | Chan-Zuckerberg Biohub | CZB Cliahub Consortium |
| EPI_ISL_436641, EPI_ISL_436642, EPI_ISL_436643, EPI_ISL_436644, EPI_ISL_436645, EPI_ISL_436646, EPI_ISL_436647, EPI_ISL_436648, EPI_ISL_436649, EPI_ISL_436650, EPI_ISL_436651, EPI_ISL_436652, EPI_ISL_436653, EPI_ISL_436654, EPI_ISL_436655, EPI_ISL_436656, EPI_ISL_436657, EPI_ISL_436658, EPI_ISL_436659, EPI_ISL_436660, EPI_ISL_436661, EPI_ISL_436662, EPI_ISL_436663, EPI_ISL_436664, EPI_ISL_436665, EPI_ISL_436666, EPI_ISL_436667, EPI_ISL_436668, EPI_ISL_436669, EPI_ISL_436670, EPI_ISL_436671, EPI_ISL_436672, EPI_ISL_436673, EPI_ISL_436674, EPI_ISL_436675, EPI_ISL_436676, EPI_ISL_436677, EPI_ISL_436678, EPI_ISL_436679, EPI_ISL_436680, EPI_ISL_436681, EPI_ISL_436682, EPI_ISL_436683, EPI_ISL_454653, EPI_ISL_454654, EPI_ISL_454655, EPI_ISL_454656, EPI_ISL_454657, EPI_ISL_454658, EPI_ISL_454659, EPI_ISL_454660, EPI_ISL_454661, EPI_ISL_454662, EPI_ISL_454663, EPI_ISL_454664, EPI_ISL_454665, EPI_ISL_454666, EPI_ISL_454667, EPI_ISL_454668, EPI_ISL_454669, EPI_ISL_454670, EPI_ISL_454671, EPI_ISL_454672, EPI_ISL_454673, EPI_ISL_454674, EPI_ISL_454675, EPI_ISL_454676, EPI_ISL_454677, EPI_ISL_454678, EPI_ISL_454679, EPI_ISL_454680, EPI_ISL_454681, EPI_ISL_454682, EPI_ISL_454683, EPI_ISL_454684, EPI_ISL_454685, EPI_ISL_454686, EPI_ISL_454687, EPI_ISL_454688, EPI_ISL_454689, EPI_ISL_468345, EPI_ISL_468346, EPI_ISL_468347, EPI_ISL_468348, EPI_ISL_468349, EPI_ISL_468350, EPI_ISL_468351, EPI_ISL_468352, EPI_ISL_468353, EPI_ISL_468354, EPI_ISL_468355, EPI_ISL_468356, EPI_ISL_486115, EPI_ISL_486116, EPI_ISL_486117, EPI_ISL_486118, EPI_ISL_486119, EPI_ISL_513773, EPI_ISL_513774, EPI_ISL_513775, EPI_ISL_513776, EPI_ISL_513777, EPI_ISL_513778, EPI_ISL_513779, EPI_ISL_513780, EPI_ISL_513781, EPI_ISL_513782, EPI_ISL_513783, EPI_ISL_513784, EPI_ISL_513785, EPI_ISL_513786, EPI_ISL_513787, EPI_ISL_513788, EPI_ISL_548264, EPI_ISL_548266, EPI_ISL_548267, EPI_ISL_548277, EPI_ISL_548279, EPI_ISL_548282, EPI_ISL_548290, EPI_ISL_548299, EPI_ISL_548302, EPI_ISL_548303, EPI_ISL_548307, EPI_ISL_548310, EPI_ISL_548314, EPI_ISL_548319, EPI_ISL_548320, EPI_ISL_548321, EPI_ISL_548324, EPI_ISL_548327, EPI_ISL_548331, EPI_ISL_548333, EPI_ISL_548337, EPI_ISL_548338, EPI_ISL_548339, EPI_ISL_548340, EPI_ISL_548341, EPI_ISL_548342, EPI_ISL_548343, EPI_ISL_548344, EPI_ISL_548345, EPI_ISL_548371, EPI_ISL_548379, EPI_ISL_548384, EPI_ISL_548385, EPI_ISL_548388, EPI_ISL_548392, EPI_ISL_548394, EPI_ISL_548395, EPI_ISL_548396, EPI_ISL_548399, EPI_ISL_548401, EPI_ISL_548403, EPI_ISL_548407, EPI_ISL_548415, EPI_ISL_548417, EPI_ISL_548419, EPI_ISL_548420, EPI_ISL_548421, EPI_ISL_548422, EPI_ISL_548423, EPI_ISL_548426, EPI_ISL_548429, EPI_ISL_548432, EPI_ISL_548433, EPI_ISL_548434, EPI_ISL_548437, EPI_ISL_548446, EPI_ISL_548448, EPI_ISL_548451, EPI_ISL_548466, EPI_ISL_548468, EPI_ISL_548470, EPI_ISL_548472, EPI_ISL_548478, EPI_ISL_548482, EPI_ISL_548494, EPI_ISL_548496, EPI_ISL_548497, EPI_ISL_548498, EPI_ISL_548500, EPI_ISL_548503, EPI_ISL_548509, EPI_ISL_548511, EPI_ISL_548512, EPI_ISL_548514, EPI_ISL_548518, EPI_ISL_548523, EPI_ISL_548525, EPI_ISL_548526, EPI_ISL_548529, EPI_ISL_548530, EPI_ISL_548533, EPI_ISL_548536, EPI_ISL_548541, EPI_ISL_548543, EPI_ISL_548546, EPI_ISL_548547, EPI_ISL_548550, EPI_ISL_548551, EPI_ISL_548553, EPI_ISL_548559, EPI_ISL_548562, EPI_ISL_548563, EPI_ISL_548568, EPI_ISL_548570, EPI_ISL_548576, EPI_ISL_548578, EPI_ISL_548582, EPI_ISL_548583, EPI_ISL_548584, EPI_ISL_548588, EPI_ISL_548593, EPI_ISL_548595, EPI_ISL_548596, EPI_ISL_548597, EPI_ISL_548598, EPI_ISL_548600, EPI_ISL_548602, EPI_ISL_548604, EPI_ISL_548605, EPI_ISL_548606, EPI_ISL_548607, EPI_ISL_548610, EPI_ISL_548612, EPI_ISL_548616, EPI_ISL_548617, EPI_ISL_548618, EPI_ISL_548619, EPI_ISL_548621, EPI_ISL_548622, EPI_ISL_548624, EPI_ISL_548629, EPI_ISL_548630, EPI_ISL_548632, EPI_ISL_548633, EPI_ISL_548636, EPI_ISL_548639, EPI_ISL_548642, EPI_ISL_548643, EPI_ISL_548645, EPI_ISL_548647, EPI_ISL_548649, EPI_ISL_548650, EPI_ISL_548652, EPI_ISL_548655, EPI_ISL_548656, EPI_ISL_548657, EPI_ISL_548658, EPI_ISL_548659, EPI_ISL_548661, EPI_ISL_548664, EPI_ISL_548669, EPI_ISL_548670, EPI_ISL_548672, EPI_ISL_548673, EPI_ISL_548676, EPI_ISL_548678, EPI_ISL_548679, EPI_ISL_582851, EPI_ISL_582852, EPI_ISL_582853, EPI_ISL_582854, EPI_ISL_582855, EPI_ISL_582856, EPI_ISL_582857, EPI_ISL_582858, EPI_ISL_582859, EPI_ISL_582860, EPI_ISL_582861, EPI_ISL_582862, EPI_ISL_582863, EPI_ISL_582864, EPI_ISL_582865, EPI_ISL_582866, EPI_ISL_582867, EPI_ISL_582868, EPI_ISL_582869, EPI_ISL_582870, EPI_ISL_582871, EPI_ISL_582872, EPI_ISL_582873, EPI_ISL_582874, EPI_ISL_582875, EPI_ISL_582876, EPI_ISL_582877, EPI_ISL_582878, EPI_ISL_582879, EPI_ISL_582880, EPI_ISL_582881, EPI_ISL_582882, EPI_ISL_582883, EPI_ISL_582884, EPI_ISL_582885, EPI_ISL_582886, EPI_ISL_582887, EPI_ISL_582888, EPI_ISL_582889, EPI_ISL_582890, EPI_ISL_582891, EPI_ISL_582892, EPI_ISL_582893, EPI_ISL_582894, EPI_ISL_582895, EPI_ISL_582896, EPI_ISL_582897, EPI_ISL_582898, EPI_ISL_582899, EPI_ISL_582900, EPI_ISL_582901, EPI_ISL_582902, EPI_ISL_582903, EPI_ISL_582904, EPI_ISL_582905, EPI_ISL_582906, EPI_ISL_582907, EPI_ISL_582908, EPI_ISL_582909, EPI_ISL_582910, EPI_ISL_582911, EPI_ISL_582912, EPI_ISL_582913, EPI_ISL_582914, EPI_ISL_582915, EPI_ISL_582916, EPI_ISL_582917, EPI_ISL_582918, EPI_ISL_582919, EPI_ISL_582920, EPI_ISL_582921, EPI_ISL_582922, EPI_ISL_582923, EPI_ISL_582924, EPI_ISL_582925, EPI_ISL_582926, EPI_ISL_582927, EPI_ISL_582928, EPI_ISL_582929, EPI_ISL_582930, EPI_ISL_582931, EPI_ISL_582932, EPI_ISL_582933, EPI_ISL_582934, EPI_ISL_582935, EPI_ISL_582936, EPI_ISL_582937, EPI_ISL_582938, EPI_ISL_582939, EPI_ISL_582940, EPI_ISL_582941, EPI_ISL_582942, EPI_ISL_582943, EPI_ISL_582944, EPI_ISL_582945, EPI_ISL_582946, EPI_ISL_582947, EPI_ISL_582948, EPI_ISL_582949, EPI_ISL_582950, EPI_ISL_582951, EPI_ISL_582952, EPI_ISL_582953, EPI_ISL_582954, EPI_ISL_582955, EPI_ISL_582956, EPI_ISL_582957, EPI_ISL_582958 | | | |
| see above | County of Santa Clara Public Health Department | Chan-Zuckerberg Biohub | CZB Cliahub Consortium |
| EPI_ISL_529151, EPI_ISL_529152, EPI_ISL_529153, EPI_ISL_529154 | Department of Immunology, The Scripps Research Institute | Andersen lab at Scripps Research | Austin; B. with SEARCH Alliance San Diego; Basler, T.; I. with SEARCH Alliance San Diego; Mchardy; Quigley, M.; Shephard, J.; Stefanski, E. |
| EPI_ISL_524435, EPI_ISL_524436 | Department of Immunology, The Scripps Research Institute | Department of Immunology, The Scripps Research Institute | Bell, A.; Drouin, A.; Fusco, D.; Garry, R.; Genemaras, K.; J.H.; Melnik, L.; Pride, D.; S.D.; SEARCH Alliance; Sabino-Santos, G.; Shin; Smither, A.; Snarski, P. |
| EPI_ISL_1811648, EPI_ISL_1811657, EPI_ISL_1811663, EPI_ISL_1811667, EPI_ISL_1811679, EPI_ISL_1811791, EPI_ISL_1811794, EPI_ISL_1811851, EPI_ISL_1811869, EPI_ISL_1811932, EPI_ISL_1812039, EPI_ISL_1812139, EPI_ISL_1812230, EPI_ISL_1812367, EPI_ISL_1812381, EPI_ISL_1812412, EPI_ISL_1812553, EPI_ISL_1812558, EPI_ISL_1812671, EPI_ISL_1812757, EPI_ISL_1812777, EPI_ISL_1812925, EPI_ISL_1812963, EPI_ISL_1813054, EPI_ISL_1813177, EPI_ISL_1813475, EPI_ISL_1813508, EPI_ISL_1813630, EPI_ISL_1815039, EPI_ISL_1816081, EPI_ISL_1816545, EPI_ISL_1816760, EPI_ISL_1816792, EPI_ISL_1816832, EPI_ISL_1816856, EPI_ISL_1816858, EPI_ISL_1816874, EPI_ISL_1816897, EPI_ISL_1816900, EPI_ISL_1816912 | | | |
| see above | EXCITE Lab | Andersen lab at Scripps Research | Abigail Schnapper; Angela Scioscia; Chip Schooley; David Pride; Helena Tubb; Natasha Martin Cheryl Anderson; Sawyer Farmer; Sharon Reed; Sydney Morgan + Marni Jacobs + Celestine Magallanes + Holly Valentine + Louise C. Laurent + SEARCH; Tommy Valles + SEARCH |
| EPI_ISL_672423, EPI_ISL_672424, EPI_ISL_672425 | Fresno County Public Health Laboratory | Chan-Zuckerberg Biohub | CZB Cliahub Consortium |
| EPI_ISL_454636, EPI_ISL_454637, EPI_ISL_454638, EPI_ISL_454639, EPI_ISL_454640, EPI_ISL_454641, EPI_ISL_468438, EPI_ISL_468439, EPI_ISL_468440, EPI_ISL_468441, EPI_ISL_468442, EPI_ISL_468443, EPI_ISL_468444, EPI_ISL_468445, EPI_ISL_468446, EPI_ISL_468447, EPI_ISL_468448, EPI_ISL_468449, EPI_ISL_468450, EPI_ISL_468451, EPI_ISL_468452, EPI_ISL_468453, EPI_ISL_468454, EPI_ISL_468455, EPI_ISL_468456, EPI_ISL_468457, EPI_ISL_468458, EPI_ISL_468459, EPI_ISL_468460, EPI_ISL_468461, EPI_ISL_486280, EPI_ISL_486281, EPI_ISL_486282, EPI_ISL_486283, EPI_ISL_486284, EPI_ISL_486285, EPI_ISL_486286, EPI_ISL_513840, EPI_ISL_513841, EPI_ISL_513842, EPI_ISL_513843, EPI_ISL_513844, EPI_ISL_513845, EPI_ISL_513846, EPI_ISL_513847, EPI_ISL_513848, EPI_ISL_513849, EPI_ISL_513850, EPI_ISL_513851, EPI_ISL_513852, EPI_ISL_513853, EPI_ISL_513854, EPI_ISL_513855, EPI_ISL_583046, EPI_ISL_583047, EPI_ISL_583048, EPI_ISL_583049, EPI_ISL_583050, EPI_ISL_583051, EPI_ISL_583052, EPI_ISL_583053, EPI_ISL_583054, EPI_ISL_583055, EPI_ISL_583056, EPI_ISL_583057, EPI_ISL_583058, EPI_ISL_583059, EPI_ISL_583060, EPI_ISL_583061, EPI_ISL_583062, EPI_ISL_583063, EPI_ISL_583064, EPI_ISL_583065, EPI_ISL_583066, EPI_ISL_583067, EPI_ISL_583068, EPI_ISL_583069, EPI_ISL_583070, EPI_ISL_583071, EPI_ISL_583072, EPI_ISL_583073, EPI_ISL_583074, EPI_ISL_583075, EPI_ISL_583076, EPI_ISL_583077, EPI_ISL_583078, EPI_ISL_583079, EPI_ISL_583080, EPI_ISL_583081, EPI_ISL_583082, EPI_ISL_583083, EPI_ISL_583084, EPI_ISL_583085, EPI_ISL_583086, EPI_ISL_583087, EPI_ISL_583088, EPI_ISL_583089, EPI_ISL_583090, EPI_ISL_583091, EPI_ISL_583092, EPI_ISL_583093, EPI_ISL_583094, EPI_ISL_583095, EPI_ISL_583096, EPI_ISL_583097, EPI_ISL_583098, EPI_ISL_583099, EPI_ISL_583100, EPI_ISL_583101, EPI_ISL_583102, EPI_ISL_583103, EPI_ISL_583104, EPI_ISL_583105, EPI_ISL_583106, EPI_ISL_583107, EPI_ISL_583108, EPI_ISL_583109, EPI_ISL_583110, EPI_ISL_583111, EPI_ISL_583112, EPI_ISL_583113, EPI_ISL_583114, EPI_ISL_583115, EPI_ISL_583116, EPI_ISL_583117, EPI_ISL_583118, EPI_ISL_583119, EPI_ISL_583120, EPI_ISL_583121, EPI_ISL_583122, EPI_ISL_583123, EPI_ISL_583124, EPI_ISL_583125, EPI_ISL_583126, EPI_ISL_583127, EPI_ISL_583128, EPI_ISL_583129, EPI_ISL_583130, EPI_ISL_583131, EPI_ISL_583132, EPI_ISL_583133, EPI_ISL_583134, EPI_ISL_583135, EPI_ISL_583136, EPI_ISL_583137, EPI_ISL_583138, EPI_ISL_583139, EPI_ISL_583140, EPI_ISL_583141, EPI_ISL_583142, EPI_ISL_583143, EPI_ISL_583144, EPI_ISL_583145, EPI_ISL_583146, EPI_ISL_583147, EPI_ISL_583148, EPI_ISL_583149, EPI_ISL_583150, EPI_ISL_583151, EPI_ISL_583152, EPI_ISL_583153, EPI_ISL_583154, EPI_ISL_583155, EPI_ISL_583156, EPI_ISL_583157, EPI_ISL_583158, EPI_ISL_583159, EPI_ISL_583160, EPI_ISL_583161, EPI_ISL_583162, EPI_ISL_583163, EPI_ISL_583164, EPI_ISL_583165, EPI_ISL_583166, EPI_ISL_583167, EPI_ISL_583168, EPI_ISL_583169, EPI_ISL_583170, EPI_ISL_738530, EPI_ISL_738756, EPI_ISL_738778, EPI_ISL_738785, EPI_ISL_738879, EPI_ISL_738951, EPI_ISL_738961, EPI_ISL_738988, EPI_ISL_739011, EPI_ISL_739086, EPI_ISL_739148, EPI_ISL_739177, EPI_ISL_739225, EPI_ISL_739264, EPI_ISL_739297, EPI_ISL_739311, EPI_ISL_739366, EPI_ISL_739430, EPI_ISL_739489, EPI_ISL_739533, EPI_ISL_739558, EPI_ISL_739571, EPI_ISL_1924619, EPI_ISL_2658689, EPI_ISL_2658704, EPI_ISL_2659205, EPI_ISL_2659209, EPI_ISL_2659260, EPI_ISL_2659346, EPI_ISL_2659371, EPI_ISL_2659385, EPI_ISL_2659403, EPI_ISL_2659405, EPI_ISL_2659408, EPI_ISL_2659414, EPI_ISL_2659461, EPI_ISL_2659486, EPI_ISL_2659487, EPI_ISL_2659492, EPI_ISL_2659500, EPI_ISL_2659549, EPI_ISL_2659591, EPI_ISL_2659638, EPI_ISL_2758407, EPI_ISL_2758408, EPI_ISL_2758409 | | | |
| see above | Humboldt County Public Health Laboratory | Chan-Zuckerberg Biohub | CZB Cliahub Consortium |
| EPI_ISL_1233894, EPI_ISL_2604244, EPI_ISL_2604245 | Infectious Diseases, Chan-Zuckerberg Biohub Covidtracker | Infectious Diseases, Chan-Zuckerberg Biohub Covidtracker | CZB Cliahub Consortium |
| EPI_ISL_884307, EPI_ISL_884308, EPI_ISL_884309, EPI_ISL_884310, EPI_ISL_884311, EPI_ISL_884312, EPI_ISL_884313, EPI_ISL_884314, EPI_ISL_884315 | | | |
| see above | Infectious Diseases, Quest Diagnostics | Infectious Diseases, Quest Diagnostics | Anderson, B.; Bernstein; D.F.; Gerasimova, A.; Hua, M.; K.E.; Kagan; L.E.; Lacbawan, F.; Liu, Y.; Livingston; Owen, R.; R.M.; Rosenthal; S.H.; Shalhout |
| EPI_ISL_649152 | Infectious diseases, Chan Zuckerberg Biohub | Infectious diseases, Chan Zuckerberg Biohub | Bhatt; K.D. |
| EPI_ISL_1512783, EPI_ISL_1512787, EPI_ISL_1512802, EPI_ISL_1512803, EPI_ISL_1512804, EPI_ISL_1512818, EPI_ISL_1512819, EPI_ISL_1512864, EPI_ISL_1512878, EPI_ISL_1512963, EPI_ISL_1702182, EPI_ISL_2013304, EPI_ISL_2013305, EPI_ISL_2013317, EPI_ISL_2013335 | | | |
| see above | Infinity Biologix | Centers for Disease Control and Prevention Division of Viral Diseases, Pathogen Discovery | Adrian Paskey; Benjamin Rambo-Martin; Chirayu Goswami; Christian Bixby; Christopher Gulvick; Clinton R. Paden; Dakota Howard; Darlene Wagner; Dhwani Batra; Duncan MacCannell; Jason Caravas; Jonathan Schultz; Kara Moser; Matthew Schmerer; Peter W. Cook; Robin Grimwood; Russ Hager; Scott Sammons; Shatavia Morrison; Yihe Wang; Yvette Unoarumhi |
| EPI_ISL_467946, EPI_ISL_467947, EPI_ISL_467948, EPI_ISL_467949, EPI_ISL_495122, EPI_ISL_495123, EPI_ISL_495124, EPI_ISL_495125, EPI_ISL_495126, EPI_ISL_495127, EPI_ISL_495128, EPI_ISL_495129, EPI_ISL_495130, EPI_ISL_495131, EPI_ISL_495132, EPI_ISL_495133, EPI_ISL_495134, EPI_ISL_495135, EPI_ISL_495136, EPI_ISL_495137, EPI_ISL_495138, EPI_ISL_495139, EPI_ISL_495140, EPI_ISL_495141, EPI_ISL_495142, EPI_ISL_495143, EPI_ISL_495144, EPI_ISL_495145, EPI_ISL_495146, EPI_ISL_495147, EPI_ISL_495148, EPI_ISL_495149, EPI_ISL_495150, EPI_ISL_495151, EPI_ISL_495152, EPI_ISL_495153, EPI_ISL_495154, EPI_ISL_495155, EPI_ISL_495156, EPI_ISL_495157, EPI_ISL_495158, EPI_ISL_495159, EPI_ISL_495160, EPI_ISL_511805, EPI_ISL_511806, EPI_ISL_511807, EPI_ISL_511808, EPI_ISL_511809, EPI_ISL_511810, EPI_ISL_511811, EPI_ISL_511812, EPI_ISL_511813, EPI_ISL_511814, EPI_ISL_511815, EPI_ISL_511816, EPI_ISL_511817, EPI_ISL_511818, EPI_ISL_511819, EPI_ISL_511820, EPI_ISL_511821, EPI_ISL_511822, EPI_ISL_511823, EPI_ISL_511824, EPI_ISL_511825, EPI_ISL_511826, EPI_ISL_511827, EPI_ISL_511828, EPI_ISL_511829, EPI_ISL_511830, EPI_ISL_511831, EPI_ISL_511832, EPI_ISL_511833, EPI_ISL_511834, EPI_ISL_511835, EPI_ISL_511836, EPI_ISL_511837, EPI_ISL_511838, EPI_ISL_511839, EPI_ISL_511840, EPI_ISL_511841, EPI_ISL_511842, EPI_ISL_511843, EPI_ISL_511844, EPI_ISL_511845, EPI_ISL_511846, EPI_ISL_511847, EPI_ISL_511848, EPI_ISL_511849, EPI_ISL_511850, EPI_ISL_511851, EPI_ISL_511862, EPI_ISL_569887, EPI_ISL_569888, EPI_ISL_569889, EPI_ISL_569890, EPI_ISL_569891, EPI_ISL_569892, EPI_ISL_569893, EPI_ISL_569894, EPI_ISL_569895, EPI_ISL_569896, EPI_ISL_569897, EPI_ISL_569898, EPI_ISL_569899, EPI_ISL_569900, EPI_ISL_569901, EPI_ISL_569902, EPI_ISL_569903, EPI_ISL_569904, EPI_ISL_569905, EPI_ISL_569906, EPI_ISL_569907, EPI_ISL_569908, EPI_ISL_569909, EPI_ISL_569910, EPI_ISL_569911, EPI_ISL_569912, EPI_ISL_569913, EPI_ISL_569914, EPI_ISL_569915, EPI_ISL_569916, EPI_ISL_569917, EPI_ISL_569918, EPI_ISL_569919, EPI_ISL_569920, EPI_ISL_569921, EPI_ISL_569922, EPI_ISL_569923, EPI_ISL_569924, EPI_ISL_569925, EPI_ISL_569926, EPI_ISL_569927, EPI_ISL_569928, EPI_ISL_569929, EPI_ISL_569930, EPI_ISL_569931, EPI_ISL_569932, EPI_ISL_569933, EPI_ISL_569934, EPI_ISL_569935, EPI_ISL_569936, EPI_ISL_569937, EPI_ISL_569938, EPI_ISL_569939, EPI_ISL_569940, EPI_ISL_569941, EPI_ISL_569942, EPI_ISL_569943, EPI_ISL_569944, EPI_ISL_569945, EPI_ISL_569946, EPI_ISL_569947, EPI_ISL_569948, EPI_ISL_576518, EPI_ISL_576519, EPI_ISL_576520, EPI_ISL_576521, EPI_ISL_576522, EPI_ISL_576523, EPI_ISL_576524, EPI_ISL_576525, EPI_ISL_576526, EPI_ISL_576527, EPI_ISL_576528, EPI_ISL_576529, EPI_ISL_576530, EPI_ISL_576531, EPI_ISL_576532, EPI_ISL_576533, EPI_ISL_576534, EPI_ISL_576535, EPI_ISL_576536, EPI_ISL_576537, EPI_ISL_576538, EPI_ISL_576539, EPI_ISL_576540, EPI_ISL_576541, EPI_ISL_576542, EPI_ISL_576543, EPI_ISL_576544, EPI_ISL_576545, EPI_ISL_576546, EPI_ISL_576547, EPI_ISL_576548, EPI_ISL_576549, EPI_ISL_576550, EPI_ISL_576551, EPI_ISL_576552, EPI_ISL_576553, EPI_ISL_576554, EPI_ISL_576555, EPI_ISL_576556, EPI_ISL_677821, EPI_ISL_677822, EPI_ISL_677823, EPI_ISL_677824, EPI_ISL_677825, EPI_ISL_677826, EPI_ISL_677827, EPI_ISL_677828, EPI_ISL_677829, EPI_ISL_677830, EPI_ISL_677831, EPI_ISL_677832, EPI_ISL_677833, EPI_ISL_677834, EPI_ISL_677835, EPI_ISL_677836, EPI_ISL_677837, EPI_ISL_677838, EPI_ISL_677839, EPI_ISL_677840, EPI_ISL_677841, EPI_ISL_677842, EPI_ISL_677843, EPI_ISL_677844, EPI_ISL_677845, EPI_ISL_677846, EPI_ISL_677847, EPI_ISL_677848, EPI_ISL_677849, EPI_ISL_677850, EPI_ISL_677851, EPI_ISL_677852, EPI_ISL_677853, EPI_ISL_677854, EPI_ISL_677855, EPI_ISL_677856, EPI_ISL_677857, EPI_ISL_677858, EPI_ISL_677859, EPI_ISL_677860, EPI_ISL_677861, EPI_ISL_677862, EPI_ISL_677863, EPI_ISL_677864, EPI_ISL_677865, EPI_ISL_677866, EPI_ISL_677867, EPI_ISL_677868, EPI_ISL_677869, EPI_ISL_677870, EPI_ISL_677871, EPI_ISL_677872, EPI_ISL_677873, EPI_ISL_677874, EPI_ISL_677875, EPI_ISL_677876, EPI_ISL_677877, EPI_ISL_677878, EPI_ISL_677879, EPI_ISL_677880, EPI_ISL_677881, EPI_ISL_677882, EPI_ISL_677883, EPI_ISL_677884, EPI_ISL_677885, EPI_ISL_677886, EPI_ISL_677887, EPI_ISL_677888, EPI_ISL_677889, EPI_ISL_677890, EPI_ISL_677891, EPI_ISL_677892, EPI_ISL_677893, EPI_ISL_677894, EPI_ISL_677895, EPI_ISL_677896, EPI_ISL_677897, EPI_ISL_677898, EPI_ISL_677899, EPI_ISL_677900, EPI_ISL_677901, EPI_ISL_677902, EPI_ISL_677903, EPI_ISL_677904, EPI_ISL_677905, EPI_ISL_677906, EPI_ISL_677907, EPI_ISL_754857, EPI_ISL_754858, EPI_ISL_754859, EPI_ISL_754860, EPI_ISL_754861, EPI_ISL_754862, EPI_ISL_754863, EPI_ISL_754864, EPI_ISL_754865, EPI_ISL_754866, EPI_ISL_754867, EPI_ISL_754868, EPI_ISL_754869, EPI_ISL_754870, EPI_ISL_754871, EPI_ISL_754872, EPI_ISL_754873, EPI_ISL_754874, EPI_ISL_754875, EPI_ISL_754876, EPI_ISL_754877, EPI_ISL_754878, EPI_ISL_754879, EPI_ISL_754880, EPI_ISL_754881, EPI_ISL_754882, EPI_ISL_754883, EPI_ISL_754884, EPI_ISL_754885, EPI_ISL_754886, EPI_ISL_754887, EPI_ISL_754888, EPI_ISL_754889, EPI_ISL_754890, EPI_ISL_754891, EPI_ISL_754892, EPI_ISL_754893, EPI_ISL_754894, EPI_ISL_754895, EPI_ISL_754896, EPI_ISL_754897, EPI_ISL_754898, EPI_ISL_754899, EPI_ISL_754900, EPI_ISL_754901, EPI_ISL_756312, EPI_ISL_756313, EPI_ISL_756314, EPI_ISL_756315, EPI_ISL_756316, EPI_ISL_756317, EPI_ISL_756318, EPI_ISL_756319, EPI_ISL_756320, EPI_ISL_756321, EPI_ISL_756322, EPI_ISL_756323, EPI_ISL_756324, EPI_ISL_756325, EPI_ISL_756326, EPI_ISL_756327, EPI_ISL_756328, EPI_ISL_756329, EPI_ISL_756330, EPI_ISL_756331, EPI_ISL_756332, EPI_ISL_756333, EPI_ISL_756334, EPI_ISL_756335, EPI_ISL_756336, EPI_ISL_756337, EPI_ISL_756338, EPI_ISL_756339, EPI_ISL_756340, EPI_ISL_756341, EPI_ISL_756342, EPI_ISL_756343, EPI_ISL_756344, EPI_ISL_756345, EPI_ISL_756346, EPI_ISL_756347, EPI_ISL_756348, EPI_ISL_756349, EPI_ISL_756350, EPI_ISL_756351, EPI_ISL_756352, EPI_ISL_756353, EPI_ISL_756354, EPI_ISL_756355, EPI_ISL_756356, EPI_ISL_1041034, EPI_ISL_1041035, EPI_ISL_1041036, EPI_ISL_1041037, EPI_ISL_1041038, EPI_ISL_1041039, EPI_ISL_1041040, EPI_ISL_1041041, EPI_ISL_1041042, EPI_ISL_1041043, EPI_ISL_1041044, EPI_ISL_1041045, EPI_ISL_1041046, EPI_ISL_1041047, EPI_ISL_1041048, EPI_ISL_1041049, EPI_ISL_1041050, EPI_ISL_1041051, EPI_ISL_1041052, EPI_ISL_1041053, EPI_ISL_1041056, EPI_ISL_1041057, EPI_ISL_1041058, EPI_ISL_1041059, EPI_ISL_1041060, EPI_ISL_1041061, EPI_ISL_1041062, EPI_ISL_1041063, EPI_ISL_1041064, EPI_ISL_1041065, EPI_ISL_1041066, EPI_ISL_1041067, EPI_ISL_1041068, EPI_ISL_1041069, EPI_ISL_1041070, EPI_ISL_1041071, EPI_ISL_1041072, EPI_ISL_1041073, EPI_ISL_1041074, EPI_ISL_1041075, EPI_ISL_1041076, EPI_ISL_1041077, EPI_ISL_1041078, EPI_ISL_1041079, EPI_ISL_1041080, EPI_ISL_1041081, EPI_ISL_1041082, EPI_ISL_1041083, EPI_ISL_1041084, EPI_ISL_1041085, EPI_ISL_1041086, EPI_ISL_1041087, EPI_ISL_1041088, EPI_ISL_1041089, EPI_ISL_1041090, EPI_ISL_1041091, EPI_ISL_1041092, EPI_ISL_1041093, EPI_ISL_1041094, EPI_ISL_1041095, EPI_ISL_1041096, EPI_ISL_1041097, EPI_ISL_1041098, EPI_ISL_1041099, EPI_ISL_1041100, EPI_ISL_1041101, EPI_ISL_1041102, EPI_ISL_1041103, EPI_ISL_1041104, EPI_ISL_1041105, EPI_ISL_1041106, EPI_ISL_1041107, EPI_ISL_1041181, EPI_ISL_1041182, EPI_ISL_1041183, EPI_ISL_1041184, EPI_ISL_1041185, EPI_ISL_1041186, EPI_ISL_1041187, EPI_ISL_1041188, EPI_ISL_1041189, EPI_ISL_1041190, EPI_ISL_1041191, EPI_ISL_1041192, EPI_ISL_1041193, EPI_ISL_1041194, EPI_ISL_1712486, EPI_ISL_1712493, EPI_ISL_1712494, EPI_ISL_1712500, EPI_ISL_1712507, EPI_ISL_1712509, EPI_ISL_1712511, EPI_ISL_1712512, EPI_ISL_1712514, EPI_ISL_1712517, EPI_ISL_1712518, EPI_ISL_1712519 | | | |
| see above | Innovative Genomics Institute, UC Berkeley | Innovative Genomics Institute, UC Berkeley | Haridha Shivram; Holly Gildea; John Boyle; Justin Choi; Liana Lareau; Phil Frankino; Shana McDevitt; Stacia Wyman |
| EPI_ISL_466648 | Innovative Genomics Institute, UCB | Innovative Genomics Institute, UCB | Haridha Shivram; Justin Choi; Liana Lareau; Shana McDevitt; Stacia Wyman |
| EPI_ISL_694046, EPI_ISL_707693, EPI_ISL_707694, EPI_ISL_707695, EPI_ISL_707883, EPI_ISL_707884, EPI_ISL_707885, EPI_ISL_707886, EPI_ISL_707887, EPI_ISL_707888, EPI_ISL_707889, EPI_ISL_707903, EPI_ISL_707904, EPI_ISL_707905, EPI_ISL_707906, EPI_ISL_707907, EPI_ISL_707908, EPI_ISL_707909, EPI_ISL_707910, EPI_ISL_707911, EPI_ISL_707912, EPI_ISL_707913, EPI_ISL_707914, EPI_ISL_707915, EPI_ISL_707916, EPI_ISL_707917, EPI_ISL_707918, EPI_ISL_707919, EPI_ISL_707920, EPI_ISL_707921, EPI_ISL_707922, EPI_ISL_707923, EPI_ISL_707924, EPI_ISL_707925, EPI_ISL_707926, EPI_ISL_707927, EPI_ISL_707929, EPI_ISL_707930, EPI_ISL_707931, EPI_ISL_707932, EPI_ISL_708182, EPI_ISL_708183, EPI_ISL_708505, EPI_ISL_708506, EPI_ISL_708507, EPI_ISL_708528, EPI_ISL_708724, EPI_ISL_708725, EPI_ISL_708726, EPI_ISL_708727, EPI_ISL_708728, EPI_ISL_708729, EPI_ISL_708730, EPI_ISL_708731, EPI_ISL_708732, EPI_ISL_708733, EPI_ISL_708734, EPI_ISL_710099, EPI_ISL_710100, EPI_ISL_710101, EPI_ISL_710102, EPI_ISL_710103, EPI_ISL_710104, EPI_ISL_710105, EPI_ISL_710106, EPI_ISL_710107, EPI_ISL_710108, EPI_ISL_710109, EPI_ISL_710110, EPI_ISL_710111, EPI_ISL_710112, EPI_ISL_710113, EPI_ISL_710114, EPI_ISL_710115, EPI_ISL_710116, EPI_ISL_710117, EPI_ISL_710118, EPI_ISL_710416, EPI_ISL_710417, EPI_ISL_710418, EPI_ISL_710419, EPI_ISL_710420, EPI_ISL_710421, EPI_ISL_710422, EPI_ISL_710423, EPI_ISL_710425, EPI_ISL_710426, EPI_ISL_710429, EPI_ISL_710430, EPI_ISL_710431, EPI_ISL_710432, EPI_ISL_710433, EPI_ISL_735374, EPI_ISL_735375, EPI_ISL_735376, EPI_ISL_735377, EPI_ISL_735378, EPI_ISL_735379, EPI_ISL_735380, EPI_ISL_735381, EPI_ISL_735382, EPI_ISL_735383, EPI_ISL_1497543 | | | |

| | | | |
|---|---|---|---|
| see above | Los Angeles County Public Health Laboratories | Los Angeles County Public Health Laboratories | P. Hemarajata et al.; Peera Hemarajata et al. |
| EPI_ISL_1501809 | Lurie Children's Hospital of Chicago | Northwestern University - Ozer Lab | Egon A. Ozer; Judd F. Hultquist; Lacy M. Simons; Larry K. Kociolek; Michael G. Ison; Ramon Lorenzo-Redondo; Taylor J. Dean; William J. Muller; Xiaotian; Zheng |
| EPI_ISL_648545, EPI_ISL_648546, EPI_ISL_648547, EPI_ISL_648548, EPI_ISL_648549, EPI_ISL_648550, EPI_ISL_648551, EPI_ISL_648552, EPI_ISL_648553, EPI_ISL_648554, EPI_ISL_672019, EPI_ISL_672022, EPI_ISL_672026, EPI_ISL_672037, EPI_ISL_672041, EPI_ISL_672046, EPI_ISL_672059, EPI_ISL_672081, EPI_ISL_672098, EPI_ISL_672099, EPI_ISL_672100, EPI_ISL_672101, EPI_ISL_672102, EPI_ISL_672394, EPI_ISL_672395, EPI_ISL_672396, EPI_ISL_672397, EPI_ISL_672398, EPI_ISL_672399, EPI_ISL_672400, EPI_ISL_672401, EPI_ISL_672402, EPI_ISL_672403, EPI_ISL_672404, EPI_ISL_672405, EPI_ISL_672406, EPI_ISL_672407, EPI_ISL_672408, EPI_ISL_672409, EPI_ISL_672410, EPI_ISL_672411, EPI_ISL_672412, EPI_ISL_672413, EPI_ISL_672414, EPI_ISL_672415, EPI_ISL_672416, EPI_ISL_672417, EPI_ISL_672418, EPI_ISL_672419, EPI_ISL_672420, EPI_ISL_672421, EPI_ISL_672422, EPI_ISL_672502, EPI_ISL_672503, EPI_ISL_672506, EPI_ISL_672507, EPI_ISL_672508, EPI_ISL_672509, EPI_ISL_672510, EPI_ISL_672511, EPI_ISL_672512, EPI_ISL_672513, EPI_ISL_672514, EPI_ISL_672515, EPI_ISL_672516, EPI_ISL_672517, EPI_ISL_672518, EPI_ISL_672519, EPI_ISL_672520, EPI_ISL_672521, EPI_ISL_672522, EPI_ISL_672523, EPI_ISL_672524, EPI_ISL_672525, EPI_ISL_672526, EPI_ISL_672527, EPI_ISL_672528, EPI_ISL_672529, EPI_ISL_672530, EPI_ISL_738666, EPI_ISL_738707, EPI_ISL_738708, EPI_ISL_738964, EPI_ISL_739061, EPI_ISL_739133, EPI_ISL_739197, EPI_ISL_739390, EPI_ISL_739429, EPI_ISL_739539, EPI_ISL_2658670, EPI_ISL_2658720, EPI_ISL_2659203 | | | |
| see above | Madera County Department of Public Health | Chan-Zuckerberg Biohub | CZB Cliahub Consortium |
| EPI_ISL_583198 | Marin County Public Health Department | Chan-Zuckerberg Biohub | CZB Cliahub Consortium |
| EPI_ISL_2259242 | Martin Luther King Jr. Community Hospital | Los Angeles County Public Health Laboratories | P. Hemarajata et al. |
| EPI_ISL_500532, EPI_ISL_500533, EPI_ISL_500534 | Mayo Clinic Laboratories | University of Washington Virology Lab | Alexander Greninger; Amin Addetia; Hong Xie; Keith R Jerome; Lasata Shrestha; Meei-Li Huang; Pavitra Roychoudhury; Truong Nguyen; Victoria M Rachleff |
| EPI_ISL_1810946, EPI_ISL_2006728, EPI_ISL_2009448, EPI_ISL_2009450, EPI_ISL_2009505 | Moderna Inc. | Moderna Inc. | Groves Dixon; Rolando Pajon; Yamuna D. Paila; Yamuna Paila |
| EPI_ISL_444455 | Molecular Infectious Disease | Molecular Infectious Disease | Anderson; B.P.; Gerasimova, A.; Kagan; R.M. and Owen, R.; Rosenthal; S.H. |
| EPI_ISL_738506, EPI_ISL_739101, EPI_ISL_739329, EPI_ISL_739336, EPI_ISL_739364, EPI_ISL_739372, EPI_ISL_739546 | | | |
| see above | Monterey County Public Health Lab | Chan-Zuckerberg Biohub | CZB Cliahub Consortium |
| EPI_ISL_1263207, EPI_ISL_1263208, EPI_ISL_1263209, EPI_ISL_1263210, EPI_ISL_2308283 | Monterey County Public Health Laboratory | Monterey County Public Health Laboratory | Monterey County Public Health Laboratory |
| EPI_ISL_625559, EPI_ISL_625560, EPI_ISL_625561, EPI_ISL_625562, EPI_ISL_625563, EPI_ISL_625564, EPI_ISL_625565, EPI_ISL_625566, EPI_ISL_625567, EPI_ISL_625568, EPI_ISL_625569, EPI_ISL_625570, EPI_ISL_625571, EPI_ISL_625572, EPI_ISL_625573, EPI_ISL_625574, EPI_ISL_625575, EPI_ISL_625576, EPI_ISL_625577, EPI_ISL_625578, EPI_ISL_625579, EPI_ISL_625580, EPI_ISL_625581, EPI_ISL_625582, EPI_ISL_625583, EPI_ISL_625584, EPI_ISL_625585, EPI_ISL_625586, EPI_ISL_625587, EPI_ISL_625588, EPI_ISL_625589, EPI_ISL_625590, EPI_ISL_625591, EPI_ISL_625592, EPI_ISL_625593 | | | |
| see above | NaN | Chan-Zuckerberg Biohub | CZB Cliahub Consortium |
| EPI_ISL_738512, EPI_ISL_738541, EPI_ISL_738754, EPI_ISL_738777, EPI_ISL_738820, EPI_ISL_738836, EPI_ISL_738870, EPI_ISL_738897, EPI_ISL_738911, EPI_ISL_738952, EPI_ISL_738959, EPI_ISL_739169, EPI_ISL_739181, EPI_ISL_739223, EPI_ISL_739251, EPI_ISL_739301, EPI_ISL_739315, EPI_ISL_739325, EPI_ISL_739339, EPI_ISL_739469, EPI_ISL_739471, EPI_ISL_739474, EPI_ISL_739514, EPI_ISL_739557, EPI_ISL_739628, EPI_ISL_739635, EPI_ISL_739645, EPI_ISL_2659565 | | | |
| see above | Napa-Solano-Yolo- Marin County (NSYM) Public Health Laboratories | Chan-Zuckerberg Biohub | CZB Cliahub Consortium |
| EPI_ISL_444994, EPI_ISL_1624239, EPI_ISL_1624240, EPI_ISL_1624241, EPI_ISL_1624242, EPI_ISL_1624243, EPI_ISL_1624244, EPI_ISL_1624245 | | | |
| see above | Naval Health Research Center | Naval Medical Research Center Biological Defense Research Directorate | Adrian Paskey; Chris Myers; Dessiree Pena-Gomez; Ewell Hollis; Francisco Malagon Bautista; Kimberly Bishop-Lilly; Kyle Long; Lindsay Glang; Logan Voegtly; Melinda Balansay-Ames; Nathaniel Christy; Regina Cer; Roger Pan |
| EPI_ISL_475232 | Nebraska Public Health Laboratory | UNMC COVID-19 Response Team | UNMC COVID-19 Response Team |
| EPI_ISL_525803 | OHSU Lab Services Molecular Microbiology Lab | Ginkgo Bioworks Clinical Laboratory | Alec J. Hirsch; Alex Plocik; Andrew C. Adey; Becky Schilling; Benjamin N. Bimber; Birgitte Simen; Brendan L. O'Connell; Brian J. O'Roak; Daniel N. Streblow; Guang Fan; James McGann; Jim Griffin; Keith Robison; Malaika Mckenzie-Bennett; Michelle Spencer; Rebecca Littlefield; Ruth V. Nichols; William B. Messer |
| EPI_ISL_525804, EPI_ISL_525805, EPI_ISL_667088, EPI_ISL_667300, EPI_ISL_667417, EPI_ISL_667424 | OHSU Lab Services Molecular Microbiology Lab | Oregon SARS-CoV-2 Genome Sequencing Center | Alec J. Hirsch; Andrew C. Adey; Benjamin N. Bimber; Brendan L. O'Connell; Brian J. O'Roak; Daniel N. Streblow; Donna Hansel; Guang Fan; Ruth V. Nichols; Sally Grindstaff; William B. Messer |
| EPI_ISL_2620802, EPI_ISL_2620803, EPI_ISL_2620804, EPI_ISL_2620805, EPI_ISL_2620806, EPI_ISL_2620807, EPI_ISL_2620808, EPI_ISL_2620809, EPI_ISL_2620810, EPI_ISL_2620811, EPI_ISL_2620812, EPI_ISL_2620813, EPI_ISL_2620814, EPI_ISL_2620815, EPI_ISL_2620816, EPI_ISL_2620817, EPI_ISL_2620818, EPI_ISL_2620819, EPI_ISL_2620820, EPI_ISL_2620821, EPI_ISL_2620822, EPI_ISL_2620830, EPI_ISL_2620831, EPI_ISL_2620832, EPI_ISL_2620833, EPI_ISL_2620834, EPI_ISL_2620835, EPI_ISL_2620836, EPI_ISL_2620837, EPI_ISL_2620838, EPI_ISL_2620839, EPI_ISL_2620840, EPI_ISL_2620841, EPI_ISL_2620842, EPI_ISL_2620843, EPI_ISL_2620844, EPI_ISL_2620845, EPI_ISL_2620846, EPI_ISL_2620847, EPI_ISL_2620848, EPI_ISL_2620849, EPI_ISL_2811918, EPI_ISL_2811919, EPI_ISL_2811923, EPI_ISL_2811924, EPI_ISL_2811925, EPI_ISL_2811926, EPI_ISL_2811927, EPI_ISL_2811928, EPI_ISL_2811929, EPI_ISL_2811930, EPI_ISL_2811931, EPI_ISL_2811932, EPI_ISL_2811933, EPI_ISL_2811934, EPI_ISL_2811935, EPI_ISL_2811936, EPI_ISL_2811937, EPI_ISL_2811938, EPI_ISL_2811960, EPI_ISL_2812015, EPI_ISL_2812016, EPI_ISL_2812017, EPI_ISL_2812018, EPI_ISL_2812019, EPI_ISL_2812020, EPI_ISL_2812021, EPI_ISL_2812022, EPI_ISL_2812023, EPI_ISL_2812024, EPI_ISL_2812025, EPI_ISL_2821256, EPI_ISL_2894570, EPI_ISL_3029852, EPI_ISL_3029865, EPI_ISL_3029866, EPI_ISL_3029867, EPI_ISL_3029873, EPI_ISL_3029874, EPI_ISL_3029876, EPI_ISL_3029877, EPI_ISL_3029878, EPI_ISL_3029879 | | | |
| see above | OmniPathology Laboratory | Los Angeles County Public Health Laboratories | P. Hemarajata et al. |
| EPI_ISL_2502592, EPI_ISL_2502596, EPI_ISL_2502597, EPI_ISL_2502598, EPI_ISL_2502599, EPI_ISL_2502600 | Operation Infectious Diseases, Naval Health Research Center | Operation Infectious Diseases, Naval Health Research Center | A.E.; Balansay-Ames; C.A.; Fierro, M.; Iniguez-Stevens, E.; Kriner, P.; M.S.; Moser, K.; Myers; Pan; Phippard; R.S.; R.W.; Underwood |
| EPI_ISL_1714785, EPI_ISL_1714786, EPI_ISL_1714787, EPI_ISL_1714788, EPI_ISL_1714789, EPI_ISL_1714790, EPI_ISL_1714791, EPI_ISL_1714792, EPI_ISL_1714793, EPI_ISL_1714794, EPI_ISL_1714795 | | | |
| see above | Operational Infectious Diseases, Naval Health Research Center | Operational Infectious Diseases, Naval Health Research Center | A.E.; Balansay-Ames; C.A.; Fierro, M.; Iniguez-Stevens, E.; Kriner, P.; M.S.; Moser, K.; Myers; Pan; Phippard; R.S.; R.W.; Underwood; Vo, S. |
| EPI_ISL_468592, EPI_ISL_468593, EPI_ISL_468594, EPI_ISL_468595, EPI_ISL_468596, EPI_ISL_468597, EPI_ISL_468598, EPI_ISL_468599, EPI_ISL_468600, EPI_ISL_468601, EPI_ISL_468602, EPI_ISL_468603, EPI_ISL_468604, EPI_ISL_468605, EPI_ISL_468606, EPI_ISL_468607, EPI_ISL_468608, EPI_ISL_468609, EPI_ISL_468610, EPI_ISL_468611, EPI_ISL_468612, EPI_ISL_468613, EPI_ISL_468614, EPI_ISL_513707, EPI_ISL_513708, EPI_ISL_513709, EPI_ISL_513710, EPI_ISL_513711, EPI_ISL_513712, EPI_ISL_513713, EPI_ISL_513714, EPI_ISL_513715, EPI_ISL_513716, EPI_ISL_513717, EPI_ISL_513718, EPI_ISL_513719, EPI_ISL_513720, EPI_ISL_513721, EPI_ISL_513722, EPI_ISL_513723, EPI_ISL_513724, EPI_ISL_513725, EPI_ISL_513726, EPI_ISL_513727, EPI_ISL_513728, EPI_ISL_513729, EPI_ISL_513730, EPI_ISL_513731, EPI_ISL_513732, EPI_ISL_513733, EPI_ISL_513734, EPI_ISL_513735, EPI_ISL_513736, EPI_ISL_513737, EPI_ISL_513738, EPI_ISL_513739, EPI_ISL_513740, EPI_ISL_513741, EPI_ISL_513742, EPI_ISL_513743, EPI_ISL_513744, EPI_ISL_513745, EPI_ISL_513746, EPI_ISL_513747, EPI_ISL_513748, EPI_ISL_513749, EPI_ISL_513750, EPI_ISL_513751, EPI_ISL_513752, EPI_ISL_513753, EPI_ISL_513754, EPI_ISL_513755, EPI_ISL_513756, EPI_ISL_513757, EPI_ISL_513758, EPI_ISL_513759, EPI_ISL_513760, EPI_ISL_513761, EPI_ISL_513762, EPI_ISL_513763, EPI_ISL_513764, EPI_ISL_513765, EPI_ISL_513766, EPI_ISL_513767, EPI_ISL_513768, EPI_ISL_513769, EPI_ISL_513770, EPI_ISL_513771, EPI_ISL_513772, EPI_ISL_582975, EPI_ISL_582976, EPI_ISL_582977, EPI_ISL_582978, EPI_ISL_582979, EPI_ISL_582980, EPI_ISL_582981, EPI_ISL_582982, EPI_ISL_582983, EPI_ISL_582984, EPI_ISL_582985, EPI_ISL_582986, EPI_ISL_582987, EPI_ISL_582988, EPI_ISL_582989, EPI_ISL_582990, EPI_ISL_582991, EPI_ISL_582992, EPI_ISL_582993, EPI_ISL_582994, EPI_ISL_582995, EPI_ISL_582996, EPI_ISL_582997, EPI_ISL_582998, EPI_ISL_582999, EPI_ISL_583000, EPI_ISL_583001, EPI_ISL_583002, EPI_ISL_583003, EPI_ISL_583004, EPI_ISL_583005, EPI_ISL_583006, EPI_ISL_583007, EPI_ISL_583008, EPI_ISL_583009, EPI_ISL_583010, EPI_ISL_583011, EPI_ISL_583012, EPI_ISL_583013, EPI_ISL_583014, EPI_ISL_583015, EPI_ISL_583016, EPI_ISL_583017, EPI_ISL_583018, EPI_ISL_583019, EPI_ISL_583020, EPI_ISL_583021, EPI_ISL_583022, EPI_ISL_583023, EPI_ISL_583024, EPI_ISL_583025, EPI_ISL_583026, EPI_ISL_583027, EPI_ISL_583028, EPI_ISL_583029, EPI_ISL_583030, EPI_ISL_583031, EPI_ISL_583032, EPI_ISL_583033, EPI_ISL_583034, EPI_ISL_583035, EPI_ISL_583036, EPI_ISL_583037, EPI_ISL_583038, EPI_ISL_583039, EPI_ISL_583040, EPI_ISL_583041, EPI_ISL_583042, EPI_ISL_583043, EPI_ISL_583044, EPI_ISL_583045, EPI_ISL_625627, EPI_ISL_625628, EPI_ISL_625629, EPI_ISL_625630, EPI_ISL_625631, EPI_ISL_625632, EPI_ISL_625633, EPI_ISL_625634, EPI_ISL_625635, EPI_ISL_625636, EPI_ISL_625637, EPI_ISL_625638, EPI_ISL_625639, EPI_ISL_625640, EPI_ISL_625641, EPI_ISL_625642, EPI_ISL_625643, EPI_ISL_625644, EPI_ISL_625645, EPI_ISL_625646, EPI_ISL_625647, EPI_ISL_625648, EPI_ISL_625649, EPI_ISL_625650, EPI_ISL_625651, EPI_ISL_625652, EPI_ISL_625653, EPI_ISL_625654, EPI_ISL_625655, EPI_ISL_625656, EPI_ISL_625657, EPI_ISL_625658, EPI_ISL_625659, EPI_ISL_648496, EPI_ISL_648497, EPI_ISL_648498, EPI_ISL_648499, EPI_ISL_648500, EPI_ISL_648501, EPI_ISL_648502, EPI_ISL_648503, EPI_ISL_648504, EPI_ISL_648505, EPI_ISL_648506, EPI_ISL_648507, EPI_ISL_648508, EPI_ISL_648509, EPI_ISL_648510, EPI_ISL_648511, EPI_ISL_648512, EPI_ISL_648513, EPI_ISL_648514, EPI_ISL_648515, EPI_ISL_648516, EPI_ISL_648517, EPI_ISL_648518, EPI_ISL_648519, EPI_ISL_648520, EPI_ISL_648521, EPI_ISL_648522, EPI_ISL_648523, EPI_ISL_648524, EPI_ISL_648525, EPI_ISL_648526, EPI_ISL_648527, EPI_ISL_648528, EPI_ISL_648529, EPI_ISL_648530, EPI_ISL_672075, EPI_ISL_672084, EPI_ISL_672103, EPI_ISL_672109, EPI_ISL_672110, EPI_ISL_672360, EPI_ISL_672361, EPI_ISL_672362, EPI_ISL_672363, EPI_ISL_672364, EPI_ISL_672365, EPI_ISL_672366, EPI_ISL_672367, EPI_ISL_672368, EPI_ISL_672369, EPI_ISL_672370, EPI_ISL_672371, EPI_ISL_672372, EPI_ISL_672373, EPI_ISL_672374, EPI_ISL_672477, EPI_ISL_672478, EPI_ISL_672479, EPI_ISL_672480, EPI_ISL_955692, EPI_ISL_1924582, EPI_ISL_1924590, EPI_ISL_1924593, EPI_ISL_1924596, EPI_ISL_1924602, EPI_ISL_1924629, EPI_ISL_2103035, EPI_ISL_2658614, EPI_ISL_2658625, EPI_ISL_2658637, EPI_ISL_2658656, EPI_ISL_2658659, EPI_ISL_2658691, EPI_ISL_2658721, EPI_ISL_2658740, EPI_ISL_2659206, EPI_ISL_2659474, EPI_ISL_2659540, EPI_ISL_2659547, EPI_ISL_2659553, EPI_ISL_2659622, EPI_ISL_2758410, EPI_ISL_2758411, EPI_ISL_2758425, EPI_ISL_2758431, EPI_ISL_2758432, EPI_ISL_2758439 | | | |
| see above | Orange County Public Health Lab | Chan-Zuckerberg Biohub | CZB Cliahub Consortium |
| EPI_ISL_486120, EPI_ISL_486121, EPI_ISL_486122, EPI_ISL_486123, EPI_ISL_486124, EPI_ISL_486125, EPI_ISL_486126, EPI_ISL_486127, EPI_ISL_486128, EPI_ISL_486129, EPI_ISL_486130, EPI_ISL_486131, EPI_ISL_486132, EPI_ISL_486133, EPI_ISL_486134, EPI_ISL_486135, EPI_ISL_486136, EPI_ISL_486137, EPI_ISL_486138, EPI_ISL_486139, EPI_ISL_486140, EPI_ISL_486141, EPI_ISL_486142, EPI_ISL_486143, EPI_ISL_486144, EPI_ISL_486145, EPI_ISL_486146, EPI_ISL_486147, EPI_ISL_486148, EPI_ISL_486149, EPI_ISL_486150, EPI_ISL_486151, EPI_ISL_486152, EPI_ISL_486153, EPI_ISL_486154, EPI_ISL_486155, EPI_ISL_486156, EPI_ISL_486157, EPI_ISL_486158, EPI_ISL_486159, EPI_ISL_486160, EPI_ISL_486161, EPI_ISL_486162, EPI_ISL_486163, EPI_ISL_486164, EPI_ISL_486165, EPI_ISL_486166, EPI_ISL_486167, EPI_ISL_486168, EPI_ISL_486169, EPI_ISL_486170, EPI_ISL_486171, EPI_ISL_486172, EPI_ISL_486173, EPI_ISL_486174, EPI_ISL_486175, EPI_ISL_486176, EPI_ISL_486177, EPI_ISL_486178, EPI_ISL_486179, EPI_ISL_486180, EPI_ISL_486181, EPI_ISL_486182, EPI_ISL_486183, EPI_ISL_486184, EPI_ISL_486185, EPI_ISL_486186, EPI_ISL_486187, EPI_ISL_486188, EPI_ISL_486189, EPI_ISL_486190, EPI_ISL_486191, EPI_ISL_486192, EPI_ISL_486193, EPI_ISL_486194, EPI_ISL_486195, EPI_ISL_486196, EPI_ISL_486197, EPI_ISL_486198, EPI_ISL_486199, EPI_ISL_486200, EPI_ISL_486201, EPI_ISL_486202, EPI_ISL_486203, EPI_ISL_486204, EPI_ISL_486205, EPI_ISL_486206, EPI_ISL_486207, EPI_ISL_486208, EPI_ISL_486209, EPI_ISL_486210, EPI_ISL_486211, EPI_ISL_486212, EPI_ISL_486213, EPI_ISL_486214, EPI_ISL_486215, EPI_ISL_486216, EPI_ISL_486217, EPI_ISL_486218, EPI_ISL_486219, EPI_ISL_486220, EPI_ISL_486221, EPI_ISL_486222, EPI_ISL_486223, EPI_ISL_486224, EPI_ISL_486225, EPI_ISL_486226, EPI_ISL_486227, EPI_ISL_486228, EPI_ISL_486229, EPI_ISL_486230, EPI_ISL_486231, EPI_ISL_486232, EPI_ISL_486233, EPI_ISL_486234, EPI_ISL_486235, EPI_ISL_486236, EPI_ISL_486237, EPI_ISL_486238, EPI_ISL_486239, EPI_ISL_486240, EPI_ISL_486241, EPI_ISL_486242, EPI_ISL_486243, EPI_ISL_486244, EPI_ISL_486245, EPI_ISL_486246, EPI_ISL_486247, EPI_ISL_486248, EPI_ISL_486249, EPI_ISL_486250, EPI_ISL_486251, EPI_ISL_486252, EPI_ISL_486253, EPI_ISL_486254, EPI_ISL_486255, EPI_ISL_486256, EPI_ISL_486257, EPI_ISL_486258, EPI_ISL_486259, EPI_ISL_486260, EPI_ISL_486261, EPI_ISL_486262, EPI_ISL_486263, EPI_ISL_486264, EPI_ISL_486265, EPI_ISL_486266, EPI_ISL_486267, EPI_ISL_486268, EPI_ISL_486269, EPI_ISL_486270, EPI_ISL_486271, EPI_ISL_486272, EPI_ISL_486273, EPI_ISL_486274, EPI_ISL_486275, EPI_ISL_486276, EPI_ISL_486277, EPI_ISL_486278, EPI_ISL_486279, EPI_ISL_513789, EPI_ISL_513790, EPI_ISL_513791, EPI_ISL_513792, EPI_ISL_513793, EPI_ISL_513794, EPI_ISL_513795, EPI_ISL_513796, EPI_ISL_513797, EPI_ISL_513798, EPI_ISL_513799, EPI_ISL_513800, EPI_ISL_513801, EPI_ISL_513802, EPI_ISL_513803, EPI_ISL_513804, EPI_ISL_513805, EPI_ISL_513806, EPI_ISL_513807, EPI_ISL_513808, EPI_ISL_513809, EPI_ISL_513810, EPI_ISL_513811, EPI_ISL_513812, EPI_ISL_513813, EPI_ISL_513814, EPI_ISL_513815, EPI_ISL_513816, EPI_ISL_513817, EPI_ISL_513818, EPI_ISL_513819, EPI_ISL_513820, EPI_ISL_513821, EPI_ISL_513822, EPI_ISL_513823, EPI_ISL_513824, EPI_ISL_513825, EPI_ISL_513826, EPI_ISL_513827, EPI_ISL_513828, EPI_ISL_513829, EPI_ISL_513830, EPI_ISL_513831, EPI_ISL_513832, EPI_ISL_513833, EPI_ISL_513834, EPI_ISL_513835, EPI_ISL_513836, EPI_ISL_513837, EPI_ISL_513838, EPI_ISL_513839, EPI_ISL_548270, EPI_ISL_548271, EPI_ISL_548272, EPI_ISL_548273, EPI_ISL_548274, EPI_ISL_548275, EPI_ISL_548276, EPI_ISL_548278, EPI_ISL_548283, EPI_ISL_548285, EPI_ISL_548287, EPI_ISL_548289, EPI_ISL_548291, EPI_ISL_548293, EPI_ISL_548296, EPI_ISL_548297, EPI_ISL_548298, EPI_ISL_548300, EPI_ISL_548301, EPI_ISL_548305, EPI_ISL_548306, EPI_ISL_548308, EPI_ISL_548309, EPI_ISL_548312, EPI_ISL_548313, EPI_ISL_548315, EPI_ISL_548317, EPI_ISL_548318, EPI_ISL_548323, EPI_ISL_548326, EPI_ISL_548328, EPI_ISL_548332, EPI_ISL_548334, EPI_ISL_548373, EPI_ISL_548375, EPI_ISL_548377, EPI_ISL_548378, EPI_ISL_548381, EPI_ISL_548383, EPI_ISL_548386, EPI_ISL_548387, EPI_ISL_548389, EPI_ISL_548393, EPI_ISL_548400, EPI_ISL_548404, EPI_ISL_548405, EPI_ISL_548408, EPI_ISL_548411, EPI_ISL_548412, EPI_ISL_548413, EPI_ISL_548416, EPI_ISL_548418, EPI_ISL_548424, EPI_ISL_548431, EPI_ISL_548438, EPI_ISL_548439, EPI_ISL_548440, EPI_ISL_548441, EPI_ISL_548444, EPI_ISL_548445, EPI_ISL_548449, EPI_ISL_548454, EPI_ISL_548456, EPI_ISL_548460, EPI_ISL_548462, EPI_ISL_548463, EPI_ISL_548464, EPI_ISL_548465, EPI_ISL_548467, EPI_ISL_548469, EPI_ISL_548484, EPI_ISL_548485, EPI_ISL_548488, EPI_ISL_548580, EPI_ISL_548581, EPI_ISL_548585, EPI_ISL_548586, EPI_ISL_548587, EPI_ISL_548589, EPI_ISL_548590, EPI_ISL_548591, EPI_ISL_548592, EPI_ISL_548594, EPI_ISL_548601, EPI_ISL_548608, EPI_ISL_548609, EPI_ISL_548611, EPI_ISL_548613, EPI_ISL_548614, EPI_ISL_548615, EPI_ISL_548620, EPI_ISL_548623, EPI_ISL_548625, EPI_ISL_548627, EPI_ISL_548628, EPI_ISL_548634, EPI_ISL_548635, EPI_ISL_548637, EPI_ISL_548638, EPI_ISL_548640, EPI_ISL_548641, EPI_ISL_548646, EPI_ISL_548648, EPI_ISL_548651, EPI_ISL_548653, EPI_ISL_548654, EPI_ISL_548660, EPI_ISL_548662, EPI_ISL_548665, EPI_ISL_548666, EPI_ISL_548667, EPI_ISL_548668, EPI_ISL_548674, EPI_ISL_548675, EPI_ISL_548677 | | | |
| see above | Orange County Public Health Laboratory | Chan-Zuckerberg Biohub | CZB Cliahub Consortium |
| EPI_ISL_2443713, EPI_ISL_2443714, EPI_ISL_2443715, EPI_ISL_2443716, EPI_ISL_2443717, EPI_ISL_2443718, EPI_ISL_2443719, EPI_ISL_2443720, EPI_ISL_2443721, EPI_ISL_2443722, EPI_ISL_2443723, EPI_ISL_2443724, EPI_ISL_2443725, EPI_ISL_2443726, EPI_ISL_2443727, EPI_ISL_2443728, EPI_ISL_2443729, EPI_ISL_2443730, EPI_ISL_2443731, EPI_ISL_2443732, EPI_ISL_2443733, EPI_ISL_2443734, EPI_ISL_2443735, EPI_ISL_2443736, EPI_ISL_2443737, EPI_ISL_2443738, EPI_ISL_2443739, EPI_ISL_2443740, EPI_ISL_2443741, EPI_ISL_2443742, EPI_ISL_2443743, EPI_ISL_2443744, EPI_ISL_2443745, EPI_ISL_2443746, EPI_ISL_2443747, EPI_ISL_2443748, EPI_ISL_2443749, EPI_ISL_2443750, EPI_ISL_2443751, EPI_ISL_2443752, EPI_ISL_2443753, EPI_ISL_2443754, EPI_ISL_2443755, EPI_ISL_2443756, EPI_ISL_2443757, EPI_ISL_2443758, EPI_ISL_2443759, EPI_ISL_2443760, EPI_ISL_2443761, EPI_ISL_2443762, EPI_ISL_2443763, EPI_ISL_2443764, EPI_ISL_2443765, EPI_ISL_2443766, EPI_ISL_2443767, EPI_ISL_2443768, EPI_ISL_2443769, EPI_ISL_2443770, EPI_ISL_2443771, EPI_ISL_2443772, EPI_ISL_2443773, EPI_ISL_2443774, EPI_ISL_2443775, EPI_ISL_2443776, EPI_ISL_2443777 | | | |
| see above | Orange County Public Health Laboratory | University of California, Irvine | Amanda N. Pinski; Ilhem Messaoudi |
| EPI_ISL_490210, EPI_ISL_490211, EPI_ISL_490212, EPI_ISL_490213, EPI_ISL_490214, EPI_ISL_490215, EPI_ISL_490216, EPI_ISL_490217, EPI_ISL_490218, EPI_ISL_490219, EPI_ISL_490220, EPI_ISL_490221, EPI_ISL_490222, EPI_ISL_490223 | | | |
| see above | Quest Diagnostics | Q Squared Solutions - QRTP facility | Victor J Weigman |
| EPI_ISL_454693, EPI_ISL_467811, EPI_ISL_467812, EPI_ISL_467813, EPI_ISL_467814, EPI_ISL_467815, EPI_ISL_467816, EPI_ISL_467817, EPI_ISL_467818, EPI_ISL_467819, EPI_ISL_467820, EPI_ISL_467821, EPI_ISL_467822, EPI_ISL_467823, EPI_ISL_467824, EPI_ISL_467825, EPI_ISL_467826, EPI_ISL_467827, EPI_ISL_467828, EPI_ISL_467829, EPI_ISL_467830, EPI_ISL_467831, EPI_ISL_467832, EPI_ISL_467833, EPI_ISL_467834, EPI_ISL_467835, EPI_ISL_467836, EPI_ISL_467837, EPI_ISL_467838, EPI_ISL_467839, EPI_ISL_467840, EPI_ISL_467841, EPI_ISL_467842, EPI_ISL_467843, EPI_ISL_468560, EPI_ISL_468561, EPI_ISL_468562, EPI_ISL_468563, EPI_ISL_468564, EPI_ISL_468565, EPI_ISL_468566, EPI_ISL_468567, EPI_ISL_468568, EPI_ISL_468569, EPI_ISL_468570, EPI_ISL_494505, EPI_ISL_494506, EPI_ISL_494507, EPI_ISL_494508, EPI_ISL_494509, EPI_ISL_498697, EPI_ISL_498698, EPI_ISL_498699, EPI_ISL_498700, EPI_ISL_498701, EPI_ISL_498702, EPI_ISL_498703, EPI_ISL_498704, EPI_ISL_571020, EPI_ISL_571021, EPI_ISL_571022, EPI_ISL_571023, EPI_ISL_571024, EPI_ISL_571025, EPI_ISL_571026, EPI_ISL_571156, EPI_ISL_571157, EPI_ISL_571158, EPI_ISL_571159, EPI_ISL_571160, EPI_ISL_571161, EPI_ISL_571162, EPI_ISL_571163, EPI_ISL_571164, EPI_ISL_571165, EPI_ISL_571166, EPI_ISL_571167, EPI_ISL_571168, EPI_ISL_571169, EPI_ISL_571170, EPI_ISL_571171, EPI_ISL_571172, EPI_ISL_571173, EPI_ISL_571174, EPI_ISL_571175, EPI_ISL_571176, EPI_ISL_571177, EPI_ISL_571268, EPI_ISL_571269, EPI_ISL_571270, EPI_ISL_571271, EPI_ISL_571272, EPI_ISL_571273, EPI_ISL_571274, EPI_ISL_571275, EPI_ISL_571276, EPI_ISL_571277, EPI_ISL_571278, EPI_ISL_571279, EPI_ISL_571280, EPI_ISL_571281, EPI_ISL_571282, EPI_ISL_571283, EPI_ISL_571284, EPI_ISL_571285, EPI_ISL_571410, EPI_ISL_571411, EPI_ISL_571412, EPI_ISL_571413, EPI_ISL_571414, EPI_ISL_571415, EPI_ISL_571416, EPI_ISL_571417, EPI_ISL_571418, EPI_ISL_571419, EPI_ISL_571420, EPI_ISL_571421, EPI_ISL_571422, EPI_ISL_571423, EPI_ISL_571484, EPI_ISL_571485, EPI_ISL_571486, EPI_ISL_571487, EPI_ISL_571488, EPI_ISL_571489, EPI_ISL_571490, EPI_ISL_571491, EPI_ISL_571492, EPI_ISL_571493, EPI_ISL_571494, EPI_ISL_571495, EPI_ISL_571496, EPI_ISL_571497, EPI_ISL_571498, EPI_ISL_571499, EPI_ISL_571500, EPI_ISL_571501, EPI_ISL_571502, EPI_ISL_571503, EPI_ISL_571504, EPI_ISL_571505, EPI_ISL_571506, EPI_ISL_571507, EPI_ISL_571508, EPI_ISL_571573, EPI_ISL_571574, EPI_ISL_571575, EPI_ISL_571576, EPI_ISL_571577, EPI_ISL_571578, EPI_ISL_571579, EPI_ISL_571580, EPI_ISL_571581, EPI_ISL_571582, EPI_ISL_571583, EPI_ISL_571584, EPI_ISL_571585, EPI_ISL_571586, EPI_ISL_571587, EPI_ISL_571588, EPI_ISL_571589, EPI_ISL_571590, EPI_ISL_571591, EPI_ISL_571694, EPI_ISL_571695, EPI_ISL_571696, EPI_ISL_571697, EPI_ISL_571698, EPI_ISL_571699, EPI_ISL_571700, EPI_ISL_571701, EPI_ISL_571702, EPI_ISL_571703, EPI_ISL_571704, EPI_ISL_571705, EPI_ISL_571706, EPI_ISL_571707, EPI_ISL_571708, EPI_ISL_571709, EPI_ISL_571710, EPI_ISL_571711, EPI_ISL_571712, EPI_ISL_571713, EPI_ISL_571714, EPI_ISL_571715, EPI_ISL_571804, EPI_ISL_571805, EPI_ISL_571806, EPI_ISL_571807, EPI_ISL_571808, EPI_ISL_571809, EPI_ISL_571810, EPI_ISL_571811, EPI_ISL_571812, EPI_ISL_571813, EPI_ISL_571814, EPI_ISL_571815, EPI_ISL_571816, EPI_ISL_571817, EPI_ISL_571818, EPI_ISL_571819, EPI_ISL_571820, EPI_ISL_571917, EPI_ISL_571918, EPI_ISL_571919, EPI_ISL_571920, EPI_ISL_571921, EPI_ISL_571922, EPI_ISL_571923, EPI_ISL_571924, EPI_ISL_572055, EPI_ISL_572056, EPI_ISL_572057, EPI_ISL_572058, EPI_ISL_572059, EPI_ISL_572060, EPI_ISL_572061, EPI_ISL_572062, EPI_ISL_572063, EPI_ISL_572064, EPI_ISL_572065, EPI_ISL_572066, EPI_ISL_572067, EPI_ISL_572068, EPI_ISL_572069, EPI_ISL_572070, EPI_ISL_572071, EPI_ISL_572072, EPI_ISL_572073, EPI_ISL_572074, EPI_ISL_572075, EPI_ISL_572076, EPI_ISL_572077, EPI_ISL_572078, EPI_ISL_572079, EPI_ISL_572080, EPI_ISL_572081, EPI_ISL_572082, EPI_ISL_572083, EPI_ISL_572084, EPI_ISL_572085, EPI_ISL_572086, EPI_ISL_572087, EPI_ISL_572088, EPI_ISL_572089, EPI_ISL_572090, EPI_ISL_572091, EPI_ISL_572092, EPI_ISL_572093, EPI_ISL_572094, EPI_ISL_572095, EPI_ISL_572096, EPI_ISL_603761, EPI_ISL_603762, EPI_ISL_603763, EPI_ISL_603764, EPI_ISL_603765, EPI_ISL_603766, EPI_ISL_603767, EPI_ISL_603768, EPI_ISL_603769, EPI_ISL_603770, EPI_ISL_603771, EPI_ISL_603772, EPI_ISL_603773, EPI_ISL_603774, EPI_ISL_603775, EPI_ISL_603776, EPI_ISL_603777, EPI_ISL_603778, EPI_ISL_603779, EPI_ISL_603780, EPI_ISL_603781, EPI_ISL_603782, EPI_ISL_603783, EPI_ISL_603784, EPI_ISL_603785, EPI_ISL_603786, EPI_ISL_603787, EPI_ISL_603788, EPI_ISL_603789, EPI_ISL_603790, EPI_ISL_603791, EPI_ISL_603792, EPI_ISL_603793, EPI_ISL_603794, EPI_ISL_603795, EPI_ISL_603796, EPI_ISL_603797, EPI_ISL_603798, EPI_ISL_603799, EPI_ISL_603800, EPI_ISL_603801, EPI_ISL_603802, EPI_ISL_603803, EPI_ISL_603883, EPI_ISL_603884, EPI_ISL_603885, EPI_ISL_603886, EPI_ISL_603887, EPI_ISL_603888, EPI_ISL_603889, EPI_ISL_603890, EPI_ISL_603891, EPI_ISL_603892, EPI_ISL_603893, EPI_ISL_603894, EPI_ISL_603895, EPI_ISL_603896, EPI_ISL_603897, EPI_ISL_603898, EPI_ISL_603899, EPI_ISL_603900, EPI_ISL_603901, EPI_ISL_603902, EPI_ISL_603903, EPI_ISL_603904, EPI_ISL_603905, EPI_ISL_603906, EPI_ISL_603907, EPI_ISL_603908, EPI_ISL_603909, EPI_ISL_603910, EPI_ISL_603911, EPI_ISL_603912, EPI_ISL_603913, EPI_ISL_603914, EPI_ISL_603915, EPI_ISL_603916, EPI_ISL_603917, EPI_ISL_603918, EPI_ISL_603919, EPI_ISL_603920, EPI_ISL_603921, EPI_ISL_603922, EPI_ISL_603923, EPI_ISL_603924, EPI_ISL_603925, EPI_ISL_603926, EPI_ISL_603927, EPI_ISL_603928, EPI_ISL_603929, EPI_ISL_603930, EPI_ISL_603931, EPI_ISL_603932, EPI_ISL_603933, EPI_ISL_603934, EPI_ISL_604017, EPI_ISL_604018, EPI_ISL_604019, EPI_ISL_604020, EPI_ISL_604098, EPI_ISL_604099, EPI_ISL_604100, EPI_ISL_604101, EPI_ISL_604102, EPI_ISL_604103, EPI_ISL_604104, EPI_ISL_604185, EPI_ISL_604186, EPI_ISL_604187, EPI_ISL_604188, EPI_ISL_604189, EPI_ISL_604190, EPI_ISL_604191, EPI_ISL_604192, EPI_ISL_604193, EPI_ISL_604194, EPI_ISL_604195, EPI_ISL_604196, EPI_ISL_604197, EPI_ISL_604198, EPI_ISL_604199, EPI_ISL_604200, EPI_ISL_604201, EPI_ISL_604202, EPI_ISL_604203, EPI_ISL_604204, EPI_ISL_604205, EPI_ISL_604206, EPI_ISL_604207, EPI_ISL_604208, EPI_ISL_604209, EPI_ISL_604210, | | | |

EPI_ISL_604211, EPI_ISL_604212, EPI_ISL_604213, EPI_ISL_604214, EPI_ISL_604215, EPI_ISL_604216, EPI_ISL_604217, EPI_ISL_604218, EPI_ISL_604219, EPI_ISL_604293, EPI_ISL_604294, EPI_ISL_604295, EPI_ISL_604296, EPI_ISL_604297, EPI_ISL_604298, EPI_ISL_604299, EPI_ISL_604300, EPI_ISL_604301, EPI_ISL_604302, EPI_ISL_604303, EPI_ISL_604304, EPI_ISL_604305, EPI_ISL_604306, EPI_ISL_604307, EPI_ISL_604308, EPI_ISL_604309, EPI_ISL_604310, EPI_ISL_604311, EPI_ISL_604312, EPI_ISL_604313, EPI_ISL_604314, EPI_ISL_604315, EPI_ISL_604316, EPI_ISL_604317, EPI_ISL_604318, EPI_ISL_604319, EPI_ISL_604320, EPI_ISL_604321, EPI_ISL_604322, EPI_ISL_604323, EPI_ISL_604324, EPI_ISL_604325, EPI_ISL_604393, EPI_ISL_604394, EPI_ISL_604395, EPI_ISL_604396, EPI_ISL_604397, EPI_ISL_604398, EPI_ISL_604399, EPI_ISL_604400, EPI_ISL_604401, EPI_ISL_604402, EPI_ISL_604403, EPI_ISL_604404, EPI_ISL_604405, EPI_ISL_604406, EPI_ISL_604407, EPI_ISL_604408, EPI_ISL_604409, EPI_ISL_604410, EPI_ISL_604411, EPI_ISL_604412, EPI_ISL_604413, EPI_ISL_604414, EPI_ISL_604415, EPI_ISL_604416, EPI_ISL_604417, EPI_ISL_604418, EPI_ISL_604419, EPI_ISL_604420, EPI_ISL_604421, EPI_ISL_604422, EPI_ISL_604423, EPI_ISL_604424, EPI_ISL_604425, EPI_ISL_604426, EPI_ISL_604427, EPI_ISL_604428, EPI_ISL_604429, EPI_ISL_604430, EPI_ISL_604431, EPI_ISL_604432, EPI_ISL_604433, EPI_ISL_604434, EPI_ISL_604435, EPI_ISL_604436, EPI_ISL_604437, EPI_ISL_604438, EPI_ISL_604439, EPI_ISL_604440, EPI_ISL_604441, EPI_ISL_604442, EPI_ISL_604443, EPI_ISL_604444, EPI_ISL_604445, EPI_ISL_604446, EPI_ISL_604505, EPI_ISL_604506, EPI_ISL_604507, EPI_ISL_604508, EPI_ISL_604509, EPI_ISL_604510, EPI_ISL_604511, EPI_ISL_604512, EPI_ISL_604513, EPI_ISL_604514, EPI_ISL_604515, EPI_ISL_604628, EPI_ISL_604629, EPI_ISL_604630, EPI_ISL_604631, EPI_ISL_604632, EPI_ISL_604633, EPI_ISL_604634, EPI_ISL_604635, EPI_ISL_604636, EPI_ISL_604637, EPI_ISL_604638, EPI_ISL_604639, EPI_ISL_604640, EPI_ISL_604641, EPI_ISL_604642, EPI_ISL_604643, EPI_ISL_604644, EPI_ISL_604645, EPI_ISL_604646, EPI_ISL_604647, EPI_ISL_604648, EPI_ISL_604649, EPI_ISL_604650, EPI_ISL_604651, EPI_ISL_604652, EPI_ISL_604653, EPI_ISL_604654, EPI_ISL_604655, EPI_ISL_604656, EPI_ISL_604657, EPI_ISL_604658, EPI_ISL_604659, EPI_ISL_604660, EPI_ISL_604661, EPI_ISL_604662, EPI_ISL_604663, EPI_ISL_604664, EPI_ISL_604665, EPI_ISL_604666, EPI_ISL_604667, EPI_ISL_604668, EPI_ISL_604669, EPI_ISL_604670, EPI_ISL_604671, EPI_ISL_604672, EPI_ISL_604673, EPI_ISL_604674, EPI_ISL_604675, EPI_ISL_604676, EPI_ISL_604677, EPI_ISL_604678, EPI_ISL_604679, EPI_ISL_604680, EPI_ISL_604681, EPI_ISL_604682, EPI_ISL_604683, EPI_ISL_604743, EPI_ISL_604744, EPI_ISL_604745, EPI_ISL_604746, EPI_ISL_604747, EPI_ISL_604748, EPI_ISL_604749, EPI_ISL_604750, EPI_ISL_604751, EPI_ISL_604752, EPI_ISL_604753, EPI_ISL_604754, EPI_ISL_604755, EPI_ISL_604756, EPI_ISL_604757, EPI_ISL_604758, EPI_ISL_604759, EPI_ISL_604760, EPI_ISL_604761, EPI_ISL_604762, EPI_ISL_604763, EPI_ISL_604764, EPI_ISL_604765, EPI_ISL_604766, EPI_ISL_604767, EPI_ISL_604768, EPI_ISL_604769, EPI_ISL_604770, EPI_ISL_604771, EPI_ISL_604772, EPI_ISL_604874, EPI_ISL_604875, EPI_ISL_604876, EPI_ISL_604877, EPI_ISL_604878, EPI_ISL_604879, EPI_ISL_604880, EPI_ISL_604881, EPI_ISL_604882, EPI_ISL_604883, EPI_ISL_604884, EPI_ISL_604885, EPI_ISL_604886, EPI_ISL_604887, EPI_ISL_604888, EPI_ISL_604889, EPI_ISL_604890, EPI_ISL_604891, EPI_ISL_604892, EPI_ISL_604893, EPI_ISL_604894, EPI_ISL_604895, EPI_ISL_604896, EPI_ISL_604897, EPI_ISL_604898, EPI_ISL_604899, EPI_ISL_604900, EPI_ISL_604901, EPI_ISL_604902, EPI_ISL_604903, EPI_ISL_604904, EPI_ISL_604905, EPI_ISL_604906, EPI_ISL_604907, EPI_ISL_604908, EPI_ISL_604909, EPI_ISL_604910, EPI_ISL_604911, EPI_ISL_604912, EPI_ISL_604913, EPI_ISL_604914, EPI_ISL_604915, EPI_ISL_604916, EPI_ISL_604917, EPI_ISL_604918, EPI_ISL_604993, EPI_ISL_604994, EPI_ISL_604995, EPI_ISL_604996, EPI_ISL_604997, EPI_ISL_604998, EPI_ISL_604999, EPI_ISL_605000, EPI_ISL_605001, EPI_ISL_605002, EPI_ISL_605003, EPI_ISL_605004, EPI_ISL_605005, EPI_ISL_605006, EPI_ISL_605007, EPI_ISL_605008, EPI_ISL_605009, EPI_ISL_605010, EPI_ISL_605011, EPI_ISL_605012, EPI_ISL_605013, EPI_ISL_605014, EPI_ISL_605015, EPI_ISL_605016, EPI_ISL_605017, EPI_ISL_605018, EPI_ISL_605019, EPI_ISL_605020, EPI_ISL_605021, EPI_ISL_605022, EPI_ISL_845657, EPI_ISL_845658, EPI_ISL_845659, EPI_ISL_845660, EPI_ISL_845661, EPI_ISL_876827, EPI_ISL_876828, EPI_ISL_876882, EPI_ISL_876887, EPI_ISL_876888, EPI_ISL_876894, EPI_ISL_876905, EPI_ISL_876910, EPI_ISL_876913, EPI_ISL_876926, EPI_ISL_876930, EPI_ISL_876936, EPI_ISL_876950, EPI_ISL_876953, EPI_ISL_937027, EPI_ISL_937028, EPI_ISL_937029, EPI_ISL_937030, EPI_ISL_937031, EPI_ISL_937032, EPI_ISL_937033, EPI_ISL_937034

| | | | |
|---|---|---|---|
| see above | Quest Diagnostics | Quest Diagnostics | Anderson; Anderson, B.; B.P.; Bernstein; D.F.; Gerasimova, A.; Grover, D.; Hua, M.; I.A.; K.E.; Kagan; L.E.; Lacbawan, F.; Liu Y.; Livingston; Owen, R.; Perez, A.; R.M.; R.M. and Owen, R.; Rosenthal; S.H.; Shalhout; Shlyakhter; Tanpaiboon, P. |

EPI_ISL_429990, EPI_ISL_435473, EPI_ISL_435474, EPI_ISL_445118, EPI_ISL_450190, EPI_ISL_467976, EPI_ISL_467981, EPI_ISL_467984, EPI_ISL_494570, EPI_ISL_878637, EPI_ISL_878640, EPI_ISL_878643, EPI_ISL_878645, EPI_ISL_878651, EPI_ISL_878653, EPI_ISL_878656, EPI_ISL_878664, EPI_ISL_878669, EPI_ISL_878671, EPI_ISL_878677, EPI_ISL_878679, EPI_ISL_878687, EPI_ISL_878690, EPI_ISL_878693, EPI_ISL_878710

| | | | |
|---|---|---|---|
| see above | Rady's Childrens Hospital | Andersen lab at Scripps Research | Christina Clarke; David Dimmock; Denise Malicki; Kathryn Bouic; Linda Luo; Michelle Vanderpool; SEARCH Alliance San Diego; SEARCH Alliance San Diego with Christina Clarke; SEARCH Alliance San Diego with Nanda Radamchar; Teresa Mueller |
| EPI_ISL_2313551, EPI_ISL_2313553 | Respiratory Viruses Branch, Centers for Disease Control and Prevention | Respiratory Viruses Branch, Centers for Disease Control and Prevention | Batra, D.; Bixby, C.; C.R.; Caravas, J.; Cook; Goswami, C.; Grimwood, R.; Gulvick, C.; Hager, R.; Howard, D.; MacCannell, D.; Morrison, S.; Moser, K.; P.W.; Paden; Paskey, A.; Rambo-Martin, B.; Sammons, S.; Schmerer, M.; Schultz, J.; Unoarumhi, Y.; Wagner, D.; Wang, Y. |

EPI_ISL_583173, EPI_ISL_583174, EPI_ISL_583175, EPI_ISL_583176, EPI_ISL_583177, EPI_ISL_583178, EPI_ISL_583179, EPI_ISL_583180, EPI_ISL_583181, EPI_ISL_583182, EPI_ISL_583183, EPI_ISL_583184, EPI_ISL_583185, EPI_ISL_583186, EPI_ISL_583187, EPI_ISL_583188, EPI_ISL_583189

| | | | |
|---|---|---|---|
| see above | San Bernardino County Public Health Lab | Chan-Zuckerberg Biohub | CZB Cliahub Consortium |

EPI_ISL_467950, EPI_ISL_467951, EPI_ISL_467952, EPI_ISL_467953, EPI_ISL_467954, EPI_ISL_467955, EPI_ISL_467956, EPI_ISL_467957, EPI_ISL_467958, EPI_ISL_467959, EPI_ISL_467960, EPI_ISL_467961, EPI_ISL_467962, EPI_ISL_467963, EPI_ISL_467964, EPI_ISL_467965, EPI_ISL_467966, EPI_ISL_467968, EPI_ISL_467969, EPI_ISL_467970, EPI_ISL_467971, EPI_ISL_467973, EPI_ISL_467974, EPI_ISL_467975, EPI_ISL_467977, EPI_ISL_467978, EPI_ISL_467979, EPI_ISL_467980, EPI_ISL_467982, EPI_ISL_467983, EPI_ISL_483158, EPI_ISL_483159, EPI_ISL_483160, EPI_ISL_483161, EPI_ISL_483162, EPI_ISL_483163, EPI_ISL_483164, EPI_ISL_483531, EPI_ISL_483532, EPI_ISL_483533, EPI_ISL_483534, EPI_ISL_483535, EPI_ISL_483536, EPI_ISL_483537, EPI_ISL_483538, EPI_ISL_483539, EPI_ISL_483540, EPI_ISL_483541, EPI_ISL_494372, EPI_ISL_494373, EPI_ISL_494374, EPI_ISL_494375, EPI_ISL_494376, EPI_ISL_494377, EPI_ISL_494378, EPI_ISL_494379, EPI_ISL_494380, EPI_ISL_494381, EPI_ISL_494382, EPI_ISL_494383, EPI_ISL_494384, EPI_ISL_494385, EPI_ISL_494386, EPI_ISL_494387, EPI_ISL_494388, EPI_ISL_494389, EPI_ISL_494390, EPI_ISL_494391, EPI_ISL_494392, EPI_ISL_494393, EPI_ISL_494394, EPI_ISL_494395, EPI_ISL_494396, EPI_ISL_494397, EPI_ISL_494398, EPI_ISL_494399, EPI_ISL_494400, EPI_ISL_494401, EPI_ISL_494402, EPI_ISL_494403, EPI_ISL_494404, EPI_ISL_494405, EPI_ISL_494406, EPI_ISL_494407, EPI_ISL_494408, EPI_ISL_494409, EPI_ISL_494410, EPI_ISL_494411, EPI_ISL_494412, EPI_ISL_494413, EPI_ISL_494414, EPI_ISL_494415, EPI_ISL_494416, EPI_ISL_494417, EPI_ISL_494418, EPI_ISL_494419, EPI_ISL_494420, EPI_ISL_494421, EPI_ISL_494422, EPI_ISL_494423, EPI_ISL_494424, EPI_ISL_494425, EPI_ISL_494426, EPI_ISL_494427, EPI_ISL_494428, EPI_ISL_494429, EPI_ISL_494430, EPI_ISL_494431, EPI_ISL_494432, EPI_ISL_494433, EPI_ISL_494434, EPI_ISL_494435, EPI_ISL_494436, EPI_ISL_494437, EPI_ISL_494438, EPI_ISL_494439, EPI_ISL_494440, EPI_ISL_494441, EPI_ISL_494442, EPI_ISL_494443, EPI_ISL_494444, EPI_ISL_494445, EPI_ISL_494446, EPI_ISL_494447, EPI_ISL_494448, EPI_ISL_494449, EPI_ISL_494450, EPI_ISL_494451, EPI_ISL_494452, EPI_ISL_494453, EPI_ISL_494454, EPI_ISL_494455, EPI_ISL_494456, EPI_ISL_494457, EPI_ISL_494458, EPI_ISL_494459, EPI_ISL_494460, EPI_ISL_494461, EPI_ISL_494462, EPI_ISL_494463, EPI_ISL_494464, EPI_ISL_494465, EPI_ISL_494466, EPI_ISL_494467, EPI_ISL_494468, EPI_ISL_494469, EPI_ISL_494470, EPI_ISL_494471, EPI_ISL_494472, EPI_ISL_494473, EPI_ISL_494474, EPI_ISL_494475, EPI_ISL_494476, EPI_ISL_494477, EPI_ISL_494478, EPI_ISL_494479, EPI_ISL_494480, EPI_ISL_494481, EPI_ISL_494482, EPI_ISL_494483, EPI_ISL_494484, EPI_ISL_494485, EPI_ISL_494486, EPI_ISL_494487, EPI_ISL_494488, EPI_ISL_494489, EPI_ISL_494490, EPI_ISL_494491, EPI_ISL_494492, EPI_ISL_494493, EPI_ISL_494494, EPI_ISL_494495, EPI_ISL_494496, EPI_ISL_494497, EPI_ISL_494498, EPI_ISL_494499, EPI_ISL_494500, EPI_ISL_494501, EPI_ISL_494502, EPI_ISL_494503, EPI_ISL_494568, EPI_ISL_494569, EPI_ISL_494571, EPI_ISL_494572, EPI_ISL_494573, EPI_ISL_494574, EPI_ISL_494575, EPI_ISL_494576, EPI_ISL_494577, EPI_ISL_494578, EPI_ISL_494579, EPI_ISL_494580, EPI_ISL_494581, EPI_ISL_494582, EPI_ISL_494583, EPI_ISL_494584, EPI_ISL_494585, EPI_ISL_494586, EPI_ISL_494587, EPI_ISL_494588, EPI_ISL_494589, EPI_ISL_494590, EPI_ISL_494591, EPI_ISL_494592, EPI_ISL_494593, EPI_ISL_494594, EPI_ISL_494595, EPI_ISL_494620, EPI_ISL_494621, EPI_ISL_494622, EPI_ISL_494623, EPI_ISL_494624, EPI_ISL_494625, EPI_ISL_494626, EPI_ISL_494627, EPI_ISL_494628, EPI_ISL_494629, EPI_ISL_494630, EPI_ISL_494631, EPI_ISL_494632, EPI_ISL_494633, EPI_ISL_494634, EPI_ISL_494714, EPI_ISL_494715, EPI_ISL_494716, EPI_ISL_494717, EPI_ISL_494718, EPI_ISL_494719, EPI_ISL_494720, EPI_ISL_494721, EPI_ISL_494722, EPI_ISL_494723, EPI_ISL_494724, EPI_ISL_494725, EPI_ISL_494726, EPI_ISL_494727, EPI_ISL_494728, EPI_ISL_494729, EPI_ISL_494730, EPI_ISL_494731, EPI_ISL_494732, EPI_ISL_494733, EPI_ISL_494734, EPI_ISL_494735, EPI_ISL_494736, EPI_ISL_494737, EPI_ISL_494738, EPI_ISL_494739, EPI_ISL_494740, EPI_ISL_494741, EPI_ISL_494742, EPI_ISL_494743, EPI_ISL_494744, EPI_ISL_494745, EPI_ISL_494746, EPI_ISL_512158, EPI_ISL_512159, EPI_ISL_512160, EPI_ISL_512161, EPI_ISL_512162, EPI_ISL_512163, EPI_ISL_512164, EPI_ISL_512165, EPI_ISL_512166, EPI_ISL_512167, EPI_ISL_512168, EPI_ISL_512169, EPI_ISL_512170, EPI_ISL_512171, EPI_ISL_512172, EPI_ISL_512173, EPI_ISL_512174, EPI_ISL_512175, EPI_ISL_512176, EPI_ISL_512177, EPI_ISL_512178, EPI_ISL_512179, EPI_ISL_512180, EPI_ISL_512181, EPI_ISL_512182, EPI_ISL_512183, EPI_ISL_512184, EPI_ISL_512185, EPI_ISL_512186, EPI_ISL_512187, EPI_ISL_512188, EPI_ISL_512189, EPI_ISL_512190, EPI_ISL_512191, EPI_ISL_512192, EPI_ISL_512193, EPI_ISL_512194, EPI_ISL_512195, EPI_ISL_512196, EPI_ISL_512197, EPI_ISL_512198, EPI_ISL_512199, EPI_ISL_512200, EPI_ISL_512201, EPI_ISL_512202, EPI_ISL_512203, EPI_ISL_512204, EPI_ISL_512205, EPI_ISL_512206, EPI_ISL_512207, EPI_ISL_512208, EPI_ISL_512209, EPI_ISL_512210, EPI_ISL_512211, EPI_ISL_512212, EPI_ISL_512213, EPI_ISL_512214, EPI_ISL_512215, EPI_ISL_512216, EPI_ISL_512217, EPI_ISL_512218, EPI_ISL_512219, EPI_ISL_512220, EPI_ISL_512221, EPI_ISL_512222, EPI_ISL_512223, EPI_ISL_512224, EPI_ISL_512225, EPI_ISL_512226, EPI_ISL_512227, EPI_ISL_512228, EPI_ISL_512229, EPI_ISL_512230, EPI_ISL_512231, EPI_ISL_512232, EPI_ISL_512233, EPI_ISL_512234, EPI_ISL_512235, EPI_ISL_512236, EPI_ISL_512237, EPI_ISL_512238, EPI_ISL_512239, EPI_ISL_512240, EPI_ISL_512241, EPI_ISL_512242, EPI_ISL_512243, EPI_ISL_512244, EPI_ISL_512245, EPI_ISL_512246, EPI_ISL_512247, EPI_ISL_512248, EPI_ISL_512249, EPI_ISL_512250, EPI_ISL_512251, EPI_ISL_512252, EPI_ISL_512253, EPI_ISL_512254, EPI_ISL_512255, EPI_ISL_512256, EPI_ISL_512257, EPI_ISL_512258, EPI_ISL_512259, EPI_ISL_512260, EPI_ISL_512261, EPI_ISL_512262, EPI_ISL_512263, EPI_ISL_512264, EPI_ISL_512265, EPI_ISL_512266, EPI_ISL_512267, EPI_ISL_512268, EPI_ISL_512269, EPI_ISL_512270, EPI_ISL_512271, EPI_ISL_512272, EPI_ISL_512273, EPI_ISL_635300, EPI_ISL_635301, EPI_ISL_635302, EPI_ISL_635303, EPI_ISL_635304, EPI_ISL_635305, EPI_ISL_635306, EPI_ISL_635307, EPI_ISL_635308, EPI_ISL_635309, EPI_ISL_635310, EPI_ISL_635311, EPI_ISL_635312, EPI_ISL_635313, EPI_ISL_635314, EPI_ISL_635315, EPI_ISL_635316, EPI_ISL_635317, EPI_ISL_635318, EPI_ISL_635319, EPI_ISL_635320, EPI_ISL_635321, EPI_ISL_635322, EPI_ISL_635323, EPI_ISL_635324, EPI_ISL_635325, EPI_ISL_635326, EPI_ISL_635327, EPI_ISL_635328, EPI_ISL_635329, EPI_ISL_635330, EPI_ISL_635331, EPI_ISL_635332, EPI_ISL_635333, EPI_ISL_635334, EPI_ISL_635335, EPI_ISL_635336, EPI_ISL_635337, EPI_ISL_635338, EPI_ISL_635339, EPI_ISL_635340, EPI_ISL_635341, EPI_ISL_635342, EPI_ISL_635343, EPI_ISL_635344, EPI_ISL_635345, EPI_ISL_635346, EPI_ISL_635347, EPI_ISL_635348, EPI_ISL_635349, EPI_ISL_635350, EPI_ISL_635351, EPI_ISL_635352, EPI_ISL_635353, EPI_ISL_635354, EPI_ISL_635355, EPI_ISL_635356, EPI_ISL_635357, EPI_ISL_635358, EPI_ISL_635359, EPI_ISL_635360, EPI_ISL_635361, EPI_ISL_635362, EPI_ISL_635363, EPI_ISL_635364, EPI_ISL_635365, EPI_ISL_635366, EPI_ISL_635367, EPI_ISL_635368, EPI_ISL_635369, EPI_ISL_635370, EPI_ISL_635371, EPI_ISL_635372, EPI_ISL_635373, EPI_ISL_635374, EPI_ISL_635375, EPI_ISL_635376, EPI_ISL_635377, EPI_ISL_635378, EPI_ISL_635379, EPI_ISL_635380, EPI_ISL_635381, EPI_ISL_635382, EPI_ISL_635383, EPI_ISL_635384, EPI_ISL_635385, EPI_ISL_635386, EPI_ISL_635387, EPI_ISL_635388, EPI_ISL_635389, EPI_ISL_635390, EPI_ISL_635391, EPI_ISL_635392, EPI_ISL_635393, EPI_ISL_635394, EPI_ISL_635395, EPI_ISL_635396, EPI_ISL_635397, EPI_ISL_635398, EPI_ISL_635399, EPI_ISL_635400, EPI_ISL_635401, EPI_ISL_635402, EPI_ISL_635403, EPI_ISL_635404, EPI_ISL_635405, EPI_ISL_635406, EPI_ISL_635407, EPI_ISL_635408, EPI_ISL_635409, EPI_ISL_635410, EPI_ISL_635411, EPI_ISL_635412, EPI_ISL_635413, EPI_ISL_635414, EPI_ISL_635415, EPI_ISL_635416, EPI_ISL_635417, EPI_ISL_635418, EPI_ISL_635419, EPI_ISL_635420, EPI_ISL_635421, EPI_ISL_635422, EPI_ISL_635423, EPI_ISL_635424, EPI_ISL_635425, EPI_ISL_635426, EPI_ISL_635427, EPI_ISL_635428, EPI_ISL_635429, EPI_ISL_635430, EPI_ISL_635431, EPI_ISL_635432, EPI_ISL_635433, EPI_ISL_635434, EPI_ISL_635435, EPI_ISL_635436, EPI_ISL_635437, EPI_ISL_635438, EPI_ISL_635439, EPI_ISL_635440, EPI_ISL_635441, EPI_ISL_635442, EPI_ISL_635443, EPI_ISL_635444, EPI_ISL_635445, EPI_ISL_635446, EPI_ISL_635447, EPI_ISL_635448, EPI_ISL_635449, EPI_ISL_635450, EPI_ISL_635451, EPI_ISL_635452, EPI_ISL_635453, EPI_ISL_635454, EPI_ISL_635455, EPI_ISL_635456, EPI_ISL_635457, EPI_ISL_635458, EPI_ISL_635459, EPI_ISL_635460, EPI_ISL_635461, EPI_ISL_635462, EPI_ISL_635463, EPI_ISL_635464, EPI_ISL_635465, EPI_ISL_635466, EPI_ISL_635467, EPI_ISL_635468, EPI_ISL_635469, EPI_ISL_635470, EPI_ISL_635471, EPI_ISL_635472, EPI_ISL_635473, EPI_ISL_635474, EPI_ISL_635475, EPI_ISL_635476, EPI_ISL_635477, EPI_ISL_635478, EPI_ISL_635577, EPI_ISL_635578, EPI_ISL_635579, EPI_ISL_635580, EPI_ISL_635581, EPI_ISL_635582, EPI_ISL_635583, EPI_ISL_635584, EPI_ISL_635585, EPI_ISL_635586, EPI_ISL_635587, EPI_ISL_635588, EPI_ISL_635589, EPI_ISL_635590, EPI_ISL_635591, EPI_ISL_635592, EPI_ISL_635593, EPI_ISL_635594, EPI_ISL_635595, EPI_ISL_635596, EPI_ISL_635597, EPI_ISL_635598, EPI_ISL_635599, EPI_ISL_635600, EPI_ISL_635601, EPI_ISL_635602, EPI_ISL_635603, EPI_ISL_635604, EPI_ISL_635605, EPI_ISL_635606, EPI_ISL_635607, EPI_ISL_635608, EPI_ISL_635609, EPI_ISL_635610, EPI_ISL_635611, EPI_ISL_635612, EPI_ISL_635613, EPI_ISL_635614, EPI_ISL_635615, EPI_ISL_635616, EPI_ISL_635617, EPI_ISL_635618, EPI_ISL_635619, EPI_ISL_635620, EPI_ISL_635621, EPI_ISL_635622, EPI_ISL_635623, EPI_ISL_635624, EPI_ISL_635625, EPI_ISL_635626, EPI_ISL_635627, EPI_ISL_635628, EPI_ISL_635629, EPI_ISL_635630, EPI_ISL_635631, EPI_ISL_635632, EPI_ISL_635633, EPI_ISL_635634, EPI_ISL_635635, EPI_ISL_635636, EPI_ISL_635637, EPI_ISL_635638, EPI_ISL_635639, EPI_ISL_635640, EPI_ISL_635641, EPI_ISL_635642, EPI_ISL_635643, EPI_ISL_635644, EPI_ISL_635645, EPI_ISL_635646, EPI_ISL_635647, EPI_ISL_635648, EPI_ISL_635649, EPI_ISL_635650, EPI_ISL_635651, EPI_ISL_635652, EPI_ISL_635653, EPI_ISL_635654, EPI_ISL_635655, EPI_ISL_635656, EPI_ISL_635657, EPI_ISL_635658, EPI_ISL_635659, EPI_ISL_635660, EPI_ISL_635661, EPI_ISL_635662, EPI_ISL_635663, EPI_ISL_635664, EPI_ISL_635665, EPI_ISL_635666, EPI_ISL_635667, EPI_ISL_635668, EPI_ISL_635669, EPI_ISL_635670, EPI_ISL_635671, EPI_ISL_635672, EPI_ISL_635673, EPI_ISL_635674, EPI_ISL_635675, EPI_ISL_635676, EPI_ISL_635677, EPI_ISL_635678, EPI_ISL_635679, EPI_ISL_635680, EPI_ISL_635681, EPI_ISL_635682, EPI_ISL_635683, EPI_ISL_635684, EPI_ISL_635685, EPI_ISL_635686, EPI_ISL_635687, EPI_ISL_635688, EPI_ISL_635689, EPI_ISL_635690, EPI_ISL_635691, EPI_ISL_635692, EPI_ISL_635693, EPI_ISL_635694, EPI_ISL_635695, EPI_ISL_635696, EPI_ISL_635697, EPI_ISL_635698, EPI_ISL_635699, EPI_ISL_635700, EPI_ISL_635701, EPI_ISL_635702, EPI_ISL_635703, EPI_ISL_635704, EPI_ISL_635705, EPI_ISL_635706, EPI_ISL_635707, EPI_ISL_635708, EPI_ISL_635709, EPI_ISL_635710, EPI_ISL_635711, EPI_ISL_635712, EPI_ISL_635713, EPI_ISL_635714, EPI_ISL_635715, EPI_ISL_635716, EPI_ISL_635717, EPI_ISL_635718, EPI_ISL_635719, EPI_ISL_635720, EPI_ISL_635721, EPI_ISL_635722, EPI_ISL_635723, EPI_ISL_635724, EPI_ISL_635725, EPI_ISL_635726, EPI_ISL_635727, EPI_ISL_635728, EPI_ISL_635729, EPI_ISL_635730, EPI_ISL_635731, EPI_ISL_635732, EPI_ISL_635733, EPI_ISL_635734, EPI_ISL_635735, EPI_ISL_635736, EPI_ISL_635737, EPI_ISL_635738, EPI_ISL_635739, EPI_ISL_635740, EPI_ISL_635741, EPI_ISL_635742, EPI_ISL_635743, EPI_ISL_635744, EPI_ISL_635745, EPI_ISL_635746, EPI_ISL_635747, EPI_ISL_635748, EPI_ISL_635749, EPI_ISL_635750, EPI_ISL_635751, EPI_ISL_635752, EPI_ISL_635753, EPI_ISL_635754, EPI_ISL_635755, EPI_ISL_635756, EPI_ISL_635757, EPI_ISL_635758, EPI_ISL_635759, EPI_ISL_635760, EPI_ISL_635761, EPI_ISL_635762, EPI_ISL_635763, EPI_ISL_635764, EPI_ISL_635765, EPI_ISL_635766, EPI_ISL_635767, EPI_ISL_635768, EPI_ISL_635769, EPI_ISL_635770, EPI_ISL_635771, EPI_ISL_635772, EPI_ISL_635773, EPI_ISL_635774, EPI_ISL_635775, EPI_ISL_635776, EPI_ISL_635783, EPI_ISL_635784, EPI_ISL_635785, EPI_ISL_635786, EPI_ISL_635787, EPI_ISL_635788, EPI_ISL_635789, EPI_ISL_635790, EPI_ISL_635791, EPI_ISL_635792, EPI_ISL_635793, EPI_ISL_635794, EPI_ISL_635795, EPI_ISL_635796, EPI_ISL_635797, EPI_ISL_635798, EPI_ISL_635799, EPI_ISL_635800, EPI_ISL_635801, EPI_ISL_635802, EPI_ISL_635803, EPI_ISL_635804, EPI_ISL_635805, EPI_ISL_635806, EPI_ISL_635807, EPI_ISL_635808, EPI_ISL_635809, EPI_ISL_635810, EPI_ISL_635811, EPI_ISL_635812, EPI_ISL_635813, EPI_ISL_635814, EPI_ISL_635815, EPI_ISL_635816, EPI_ISL_635817, EPI_ISL_635818, EPI_ISL_635819, EPI_ISL_635820, EPI_ISL_635821, EPI_ISL_635822, EPI_ISL_635823, EPI_ISL_635824, EPI_ISL_635825, EPI_ISL_635826, EPI_ISL_635827, EPI_ISL_635828, EPI_ISL_635829, EPI_ISL_635830, EPI_ISL_635831, EPI_ISL_635832, EPI_ISL_635833, EPI_ISL_635834, EPI_ISL_635835, EPI_ISL_635836, EPI_ISL_635837, EPI_ISL_635838, EPI_ISL_635839, EPI_ISL_635840, EPI_ISL_635841, EPI_ISL_635842, EPI_ISL_635843, EPI_ISL_635844, EPI_ISL_635845, EPI_ISL_635846, EPI_ISL_635847, EPI_ISL_635848, EPI_ISL_635849, EPI_ISL_635850, EPI_ISL_635851, EPI_ISL_635852, EPI_ISL_635853, EPI_ISL_635854, EPI_ISL_635855, EPI_ISL_635856, EPI_ISL_635857, EPI_ISL_635858, EPI_ISL_635859, EPI_ISL_635860, EPI_ISL_635861, EPI_ISL_635862, EPI_ISL_635863, EPI_ISL_635864, EPI_ISL_635865, EPI_ISL_635866, EPI_ISL_635867, EPI_ISL_635868, EPI_ISL_635869, EPI_ISL_635870, EPI_ISL_635871, EPI_ISL_635872, EPI_ISL_635873, EPI_ISL_635874, EPI_ISL_635875, EPI_ISL_635876, EPI_ISL_635877, EPI_ISL_635878, EPI_ISL_635879, EPI_ISL_635880, EPI_ISL_635881, EPI_ISL_635882, EPI_ISL_635883, EPI_ISL_635884, EPI_ISL_635885, EPI_ISL_635886, EPI_ISL_635887, EPI_ISL_635888, EPI_ISL_635889, EPI_ISL_635890, EPI_ISL_635891, EPI_ISL_635892, EPI_ISL_635893, EPI_ISL_635894, EPI_ISL_635895, EPI_ISL_635896, EPI_ISL_635897, EPI_ISL_635898, EPI_ISL_635899, EPI_ISL_635900, EPI_ISL_635901, EPI_ISL_635902, EPI_ISL_635903, EPI_ISL_635904, EPI_ISL_635905, EPI_ISL_635906, EPI_ISL_635907, EPI_ISL_635908, EPI_ISL_635909, EPI_ISL_635910, EPI_ISL_635911, EPI_ISL_635912, EPI_ISL_635913, EPI_ISL_635914, EPI_ISL_635915, EPI_ISL_635916, EPI_ISL_635917, EPI_ISL_635918, EPI_ISL_635919, EPI_ISL_635920, EPI_ISL_635921, EPI_ISL_635922, EPI_ISL_635923, EPI_ISL_635924, EPI_ISL_635925, EPI_ISL_635926, EPI_ISL_635927, EPI_ISL_635928, EPI_ISL_635929, EPI_ISL_635930, EPI_ISL_635931, EPI_ISL_635932, EPI_ISL_635933, EPI_ISL_635934, EPI_ISL_635935, EPI_ISL_635936, EPI_ISL_635937, EPI_ISL_635938, EPI_ISL_635939, EPI_ISL_635940, EPI_ISL_635941, EPI_ISL_635942, EPI_ISL_635943, EPI_ISL_635944, EPI_ISL_635945, EPI_ISL_635946, EPI_ISL_635947, EPI_ISL_635948, EPI_ISL_635949, EPI_ISL_635950, EPI_ISL_635951, EPI_ISL_635952, EPI_ISL_635953, EPI_ISL_635954, EPI_ISL_635955, EPI_ISL_635956, EPI_ISL_635957, EPI_ISL_635958, EPI_ISL_635959, EPI_ISL_635960, EPI_ISL_635961, EPI_ISL_635962, EPI_ISL_635963, EPI_ISL_635964, EPI_ISL_635965, EPI_ISL_635966, EPI_ISL_635967, EPI_ISL_635968, EPI_ISL_635969, EPI_ISL_635970, EPI_ISL_635971, EPI_ISL_635972, EPI_ISL_635973, EPI_ISL_635974, EPI_ISL_635975, EPI_ISL_635976, EPI_ISL_635977, EPI_ISL_635978, EPI_ISL_635979, EPI_ISL_635980, EPI_ISL_635981, EPI_ISL_635982, EPI_ISL_635983, EPI_ISL_635984, EPI_ISL_635985, EPI_ISL_635986, EPI_ISL_635987, EPI_ISL_635988, EPI_ISL_635989, EPI_ISL_635990, EPI_ISL_635991, EPI_ISL_635992, EPI_ISL_635993, EPI_ISL_635994, EPI_ISL_635995, EPI_ISL_635996, EPI_ISL_635997, EPI_ISL_635998, EPI_ISL_635999, EPI_ISL_636000, EPI_ISL_636001, EPI_ISL_636002, EPI_ISL_636003, EPI_ISL_636004, EPI_ISL_636005, EPI_ISL_636006, EPI_ISL_636007, EPI_ISL_636008, EPI_ISL_636009, EPI_ISL_636010, EPI_ISL_636011, EPI_ISL_636012, EPI_ISL_636013, EPI_ISL_636014, EPI_ISL_636015, EPI_ISL_636016, EPI_ISL_636017, EPI_ISL_636018, EPI_ISL_636019, EPI_ISL_636020, EPI_ISL_636021, EPI_ISL_636022, EPI_ISL_636023, EPI_ISL_636024, EPI_ISL_636025, EPI_ISL_636026, EPI_ISL_636027, EPI_ISL_636028, EPI_ISL_636029, EPI_ISL_636030, EPI_ISL_636031, EPI_ISL_636032, EPI_ISL_636033, EPI_ISL_636034, EPI_ISL_636035, EPI_ISL_636036, EPI_ISL_636037, EPI_ISL_636038, EPI_ISL_636039, EPI_ISL_636040, EPI_ISL_636041, EPI_ISL_636042, EPI_ISL_636043, EPI_ISL_636044, EPI_ISL_636045, EPI_ISL_636046, EPI_ISL_636047, EPI_ISL_636048, EPI_ISL_636049, EPI_ISL_636050, EPI_ISL_636051, EPI_ISL_636052, EPI_ISL_636053, EPI_ISL_636054, EPI_ISL_636055, EPI_ISL_636056, EPI_ISL_636057, EPI_ISL_636058, EPI_ISL_636059, EPI_ISL_636060, EPI_ISL_636061, EPI_ISL_636062, EPI_ISL_636063, EPI_ISL_636064, EPI_ISL_636065, EPI_ISL_636066, EPI_ISL_636067, EPI_ISL_636068, EPI_ISL_636069, EPI_ISL_636070, EPI_ISL_636071, EPI_ISL_636072, EPI_ISL_636073, EPI_ISL_636074, EPI_ISL_636075, EPI_ISL_636076, EPI_ISL_636077, EPI_ISL_636078, EPI_ISL_636079, EPI_ISL_636080, EPI_ISL_636081, EPI_ISL_636082, EPI_ISL_636083, EPI_ISL_636084, EPI_ISL_636085, EPI_ISL_636086, EPI_ISL_636087, EPI_ISL_636088, EPI_ISL_636089, EPI_ISL_636090, EPI_ISL_636091, EPI_ISL_636092, EPI_ISL_636093, EPI_ISL_636094, EPI_ISL_636095, EPI_ISL_636096, EPI_ISL_636097, EPI_ISL_636098, EPI_ISL_636099, EPI_ISL_636100, EPI_ISL_636101, EPI_ISL_636102, EPI_ISL_636103, EPI_ISL_636104, EPI_ISL_636105, EPI_ISL_636106, EPI_ISL_636107, EPI_ISL_636108, EPI_ISL_636109, EPI_ISL_636110, EPI_ISL_636111, EPI_ISL_636112, EPI_ISL_636121, EPI_ISL_636122, EPI_ISL_636123, EPI_ISL_636124, EPI_ISL_636125, EPI_ISL_636126, EPI_ISL_636127, EPI_ISL_636128, EPI_ISL_636129, EPI_ISL_636130, EPI_ISL_636131, EPI_ISL_636132, EPI_ISL_636133, EPI_ISL_636134, EPI_ISL_636135, EPI_ISL_636136, EPI_ISL_636137, EPI_ISL_636138, EPI_ISL_636139, EPI_ISL_636140, EPI_ISL_636141, EPI_ISL_636142, EPI_ISL_636143, EPI_ISL_636144, EPI_ISL_636145, EPI_ISL_636146, EPI_ISL_636147, EPI_ISL_636148, EPI_ISL_636149, EPI_ISL_636150, EPI_ISL_636151, EPI_ISL_636152, EPI_ISL_636153, EPI_ISL_636154, EPI_ISL_636155, EPI_ISL_636156, EPI_ISL_636157, EPI_ISL_636158, EPI_ISL_636159, EPI_ISL_636160, EPI_ISL_636161, EPI_ISL_636162, EPI_ISL_636163, EPI_ISL_636164, EPI_ISL_636165, EPI_ISL_636166, EPI_ISL_636167, EPI_ISL_636168, EPI_ISL_636169, EPI_ISL_636170, EPI_ISL_636171, EPI_ISL_636172, EPI_ISL_636173, EPI_ISL_636174, EPI_ISL_636175, EPI_ISL_636176, EPI_ISL_636177, EPI_ISL_636178, EPI_ISL_636179, EPI_ISL_636180, EPI_ISL_636181, EPI_ISL_636182, EPI_ISL_636183, EPI_ISL_636184, EPI_ISL_636185, EPI_ISL_636186, EPI_ISL_636187, EPI_ISL_636188, EPI_ISL_636189, EPI_ISL_636190, EPI_ISL_636191, EPI_ISL_636192, EPI_ISL_636193, EPI_ISL_636194, EPI_ISL_636195, EPI_ISL_636196, EPI_ISL_636197, EPI_ISL_636198, EPI_ISL_636199, EPI_ISL_636200, EPI_ISL_636201, EPI_ISL_636202, EPI_ISL_636203, EPI_ISL_636204, EPI_ISL_636205, EPI_ISL_636206, EPI_ISL_636207, EPI_ISL_636208, EPI_ISL_636209, EPI_ISL_636210, EPI_ISL_636211, EPI_ISL_636212, EPI_ISL_636213, EPI_ISL_636214, EPI_ISL_636215, EPI_ISL_636216, EPI_ISL_636217, EPI_ISL_636218, EPI_ISL_636219, EPI_ISL_636220, EPI_ISL_636221, EPI_ISL_636223, EPI_ISL_636224, EPI_ISL_636225, EPI_ISL_636226, EPI_ISL_636227, EPI_ISL_636228, EPI_ISL_636229, EPI_ISL_636230, EPI_ISL_636231, EPI_ISL_636232, EPI_ISL_636233, EPI_ISL_636234, EPI_ISL_636235, EPI_ISL_636236, EPI_ISL_636237, EPI_ISL_636238, EPI_ISL_636239, EPI_ISL_636240, EPI_ISL_636241, EPI_ISL_636242, EPI_ISL_636243, EPI_ISL_636244, EPI_ISL_636245, EPI_ISL_636246, EPI_ISL_636247, EPI_ISL_636248, EPI_ISL_636249, EPI_ISL_636250, EPI_ISL_636251, EPI_ISL_636252, EPI_ISL_636253, EPI_ISL_636254, EPI_ISL_636255, EPI_ISL_636256, EPI_ISL_636257, EPI_ISL_636258, EPI_ISL_636259, EPI_ISL_636260, EPI_ISL_636261, EPI_ISL_636262, EPI_ISL_636263, EPI_ISL_636264, EPI_ISL_636265, EPI_ISL_636266, EPI_ISL_648823, EPI_ISL_648824, EPI_ISL_648825, EPI_ISL_648826, EPI_ISL_648827, EPI_ISL_648828, EPI_ISL_648829, EPI_ISL_648830, EPI_ISL_648831, EPI_ISL_648832, EPI_ISL_648833, EPI_ISL_648834, EPI_ISL_648835, EPI_ISL_648836, EPI_ISL_648837, EPI_ISL_648838, EPI_ISL_648839, EPI_ISL_648840, EPI_ISL_648841, EPI_ISL_648842, EPI_ISL_648843, EPI_ISL_648844, EPI_ISL_648845, EPI_ISL_648846, EPI_ISL_648847, EPI_ISL_648848, EPI_ISL_648849, EPI_ISL_648850, EPI_ISL_648851, EPI_ISL_648852, EPI_ISL_648853, EPI_ISL_648854, EPI_ISL_648855, EPI_ISL_648856, EPI_ISL_648857, EPI_ISL_648858, EPI_ISL_648859, EPI_ISL_648860, EPI_ISL_648861, EPI_ISL_648862, EPI_ISL_648863, EPI_ISL_648864, EPI_ISL_648865, EPI_ISL_648866, EPI_ISL_648867, EPI_ISL_648868, EPI_ISL_648869, EPI_ISL_648870, EPI_ISL_648871, EPI_ISL_648872, EPI_ISL_648873, EPI_ISL_648874, EPI_ISL_648875, EPI_ISL_648876, EPI_ISL_648877, EPI_ISL_648878, EPI_ISL_648879, EPI_ISL_648880, EPI_ISL_648881, EPI_ISL_648882, EPI_ISL_648883, EPI_ISL_648884, EPI_ISL_648885, EPI_ISL_648886, EPI_ISL_648887, EPI_ISL_648888, EPI_ISL_648889, EPI_ISL_648890, EPI_ISL_648891, EPI_ISL_648892, EPI_ISL_648893, EPI_ISL_648894, EPI_ISL_648895, EPI_ISL_648896, EPI_ISL_648897, EPI_ISL_648898, EPI_ISL_648899, EPI_ISL_648900, EPI_ISL_648901, EPI_ISL_648902, EPI_ISL_648903, EPI_ISL_648904, EPI_ISL_648905, EPI_ISL_648906, EPI_ISL_648907, EPI_ISL_648908, EPI_ISL_648909, EPI_ISL_648910, EPI_ISL_648911, EPI_ISL_648912, EPI_ISL_648913, EPI_ISL_648914, EPI_ISL_648915, EPI_ISL_648916, EPI_ISL_648917, EPI_ISL_648918, EPI_ISL_648919, EPI_ISL_648920, EPI_ISL_648921, EPI_ISL_648922, EPI_ISL_648923, EPI_ISL_648924, EPI_ISL_648925, EPI_ISL_648926, EPI_ISL_648927, EPI_ISL_648928, EPI_ISL_648929, EPI_ISL_648930, EPI_ISL_648931, EPI_ISL_648932, EPI_ISL_648933, EPI_ISL_648934, EPI_ISL_648935, EPI_ISL_648936, EPI_ISL_648937, EPI_ISL_648938, EPI_ISL_648939, EPI_ISL_648940, EPI_ISL_648941, EPI_ISL_648942, EPI_ISL_648943, EPI_ISL_648944, EPI_ISL_648945, EPI_ISL_648946, EPI_ISL_648947, EPI_ISL_648948, EPI_ISL_648949, EPI_ISL_648950, EPI_ISL_648951, EPI_ISL_648952, EPI_ISL_648953, EPI_ISL_648954, EPI_ISL_648955, EPI_ISL_648956, EPI_ISL_648957, EPI_ISL_648958, EPI_ISL_648959, EPI_ISL_648960, EPI_ISL_648961, EPI_ISL_648962, EPI_ISL_648963, EPI_ISL_648964, EPI_ISL_648965, EPI_ISL_648966, EPI_ISL_648967, EPI_ISL_648968, EPI_ISL_648969, EPI_ISL_648970, EPI_ISL_648971, EPI_ISL_648972, EPI_ISL_648973, EPI_ISL_648974, EPI_ISL_648975, EPI_ISL_648976, EPI_ISL_648977, EPI_ISL_648978, EPI_ISL_648979, EPI_ISL_648980, EPI_ISL_648981, EPI_ISL_648982, EPI_ISL_648983, EPI_ISL_648984, EPI_ISL_648985, EPI_ISL_648986, EPI_ISL_648987, EPI_ISL_648988, EPI_ISL_648989, EPI_ISL_648990, EPI_ISL_648991, EPI_ISL_648992, EPI_ISL_648993, EPI_ISL_648994, EPI_ISL_648995, EPI_ISL_648996, EPI_ISL_648997, EPI_ISL_648998, EPI_ISL_648999, EPI_ISL_649000, EPI_ISL_649001, EPI_ISL_649002, EPI_ISL_649003, EPI_ISL_649004, EPI_ISL_649005, EPI_ISL_649006, EPI_ISL_649007, EPI_ISL_649008, EPI_ISL_649009, EPI_ISL_649010, EPI_ISL_649011, EPI_ISL_649012, EPI_ISL_649013, EPI_ISL_649014, EPI_ISL_649015, EPI_ISL_649016, EPI_ISL_649017, EPI_ISL_649018, EPI_ISL_649019, EPI_ISL_649020, EPI_ISL_649021, EPI_ISL_649022, EPI_ISL_649023, EPI_ISL_649024, EPI_ISL_649025, EPI_ISL_649026, EPI_ISL_649027, EPI_ISL_649028, EPI_ISL_649029, EPI_ISL_649030, EPI_ISL_649031, EPI_ISL_649032, EPI_ISL_649033, EPI_ISL_649034, EPI_ISL_649035, EPI_ISL_649036, EPI_ISL_649037, EPI_ISL_649038, EPI_ISL_649039, EPI_ISL_649040, EPI_ISL_649041, EPI_ISL_649042, EPI_ISL_649043, EPI_ISL_649044, EPI_ISL_649045, EPI_ISL_649046, EPI_ISL_649047, EPI_ISL_649048, EPI_ISL_649049, EPI_ISL_649050, EPI_ISL_649051, EPI_ISL_649052, EPI_ISL_649053, EPI_ISL_649054, EPI_ISL_649055, EPI_ISL_649056, EPI_ISL_666965, EPI_ISL_666966, EPI_ISL_666967, EPI_ISL_666968, EPI_ISL_666969, EPI_ISL_666970, EPI_ISL_666971, EPI_ISL_666974, EPI_ISL_666975, EPI_ISL_666976, EPI_ISL_666977, EPI_ISL_666978, EPI_ISL_666979, EPI_ISL_666980, EPI_ISL_666981, EPI_ISL_666982, EPI_ISL_666983, EPI_ISL_666984, EPI_ISL_666985, EPI_ISL_666986, EPI_ISL_666987, EPI_ISL_666988, EPI_ISL_666989, EPI_ISL_666990, EPI_ISL_666991, EPI_ISL_666992, EPI_ISL_666993, EPI_ISL_666994, EPI_ISL_666995, EPI_ISL_666996, EPI_ISL_666997, EPI_ISL_666998, EPI_ISL_666999, EPI_ISL_667000, EPI_ISL_667001, EPI_ISL_667002, EPI_ISL_667003, EPI_ISL_667004, EPI_ISL_667005, EPI_ISL_667006, EPI_ISL_667007, EPI_ISL_667008, EPI_ISL_667009, EPI_ISL_667010, EPI_ISL_667011, EPI_ISL_667012, EPI_ISL_667013, EPI_ISL_667014, EPI_ISL_667015, EPI_ISL_667016, EPI_ISL_667017, EPI_ISL_667018, EPI_ISL_667019, EPI_ISL_667020, EPI_ISL_667021, EPI_ISL_667022, EPI_ISL_667023, EPI_ISL_667024, EPI_ISL_667025, EPI_ISL_667026, EPI_ISL_667027, EPI_ISL_667028, EPI_ISL_667029, EPI_ISL_667030, EPI_ISL_667031, EPI_ISL_667033, EPI_ISL_667034, EPI_ISL_667035, EPI_ISL_667036, EPI_ISL_667037, EPI_ISL_667038, EPI_ISL_667039, EPI_ISL_667040, EPI_ISL_667041, EPI_ISL_667044, EPI_ISL_667045, EPI_ISL_667046, EPI_ISL_667047, EPI_ISL_667048, EPI_ISL_667049, EPI_ISL_667050, EPI_ISL_667051, EPI_ISL_667052, EPI_ISL_667053, EPI_ISL_667056, EPI_ISL_667062, EPI_ISL_667063, EPI_ISL_730085, EPI_ISL_730087, EPI_ISL_730088, EPI_ISL_730089, EPI_ISL_730090, EPI_ISL_730091, EPI_ISL_730092, EPI_ISL_730093, EPI_ISL_730094, EPI_ISL_730095, EPI_ISL_730096, EPI_ISL_730097, EPI_ISL_730098, EPI_ISL_730099, EPI_ISL_730100, EPI_ISL_730101, EPI_ISL_730102, EPI_ISL_730103, EPI_ISL_730104, EPI_ISL_730105, EPI_ISL_730106, EPI_ISL_730107, EPI_ISL_730108, EPI_ISL_730109, EPI_ISL_730110, EPI_ISL_730111, EPI_ISL_730112, EPI_ISL_730113, EPI_ISL_730114, EPI_ISL_730115, EPI_ISL_730116, EPI_ISL_730117, EPI_ISL_730118, EPI_ISL_730119, EPI_ISL_730120, EPI_ISL_730121, EPI_ISL_730122, EPI_ISL_730123, EPI_ISL_730124, EPI_ISL_730125, EPI_ISL_730126, EPI_ISL_730163, EPI_ISL_730165, EPI_ISL_730170, EPI_ISL_730172, EPI_ISL_730173, EPI_ISL_730180, EPI_ISL_730181, EPI_ISL_730184, EPI_ISL_730185, EPI_ISL_730192, EPI_ISL_730318, EPI_ISL_730321, EPI_ISL_730323, EPI_ISL_730326, EPI_ISL_730327, EPI_ISL_730330, EPI_ISL_730331, EPI_ISL_730333, EPI_ISL_730334, EPI_ISL_730335, EPI_ISL_730337, EPI_ISL_730339, EPI_ISL_730341, EPI_ISL_730343, EPI_ISL_730344, EPI_ISL_730345, EPI_ISL_730347, EPI_ISL_730348, EPI_ISL_1794655, EPI_ISL_1794661, EPI_ISL_1794664, EPI_ISL_1794672, EPI_ISL_1794677, EPI_ISL_1794682, EPI_ISL_1969287, EPI_ISL_1969288, EPI_ISL_1969289, EPI_ISL_1969290, EPI_ISL_1969291, EPI_ISL_1969292, EPI_ISL_1969294, EPI_ISL_1969295, EPI_ISL_1969296, EPI_ISL_1969297, EPI_ISL_1969298, EPI_ISL_1969299, EPI_ISL_1969300, EPI_ISL_1969301, EPI_ISL_1969302, EPI_ISL_1969303, EPI_ISL_1969304, EPI_ISL_1969306, EPI_ISL_1969307, EPI_ISL_1969308, EPI_ISL_1969309, EPI_ISL_1969310, EPI_ISL_1969311, EPI_ISL_1969371, EPI_ISL_1969372, EPI_ISL_1969377, EPI_ISL_1969378, EPI_ISL_1969379, EPI_ISL_1969383, EPI_ISL_1969384, EPI_ISL_1969385, EPI_ISL_1969390, EPI_ISL_1969391, EPI_ISL_1969392, EPI_ISL_1969396, EPI_ISL_1969397, EPI_ISL_1969398, EPI_ISL_1969401, EPI_ISL_1969402, EPI_ISL_1969403, EPI_ISL_1969404, EPI_ISL_1969407, EPI_ISL_1969408, EPI_ISL_1969409, EPI_ISL_1969412, EPI_ISL_1969413, EPI_ISL_1969414, EPI_ISL_1969415, EPI_ISL_1969419, EPI_ISL_1969420, EPI_ISL_1969421, EPI_ISL_1969422, EPI_ISL_1969425, EPI_ISL_1969426, EPI_ISL_1969427, EPI_ISL_1969428, EPI_ISL_1969431, EPI_ISL_1969432, EPI_ISL_1969433, EPI_ISL_1969435, EPI_ISL_1969436, EPI_ISL_1969497, EPI_ISL_1969498, EPI_ISL_1969503, EPI_ISL_1969504, EPI_ISL_1969505, EPI_ISL_1969506, EPI_ISL_2231475, EPI_ISL_2231478, EPI_ISL_2231485, EPI_ISL_2329468, EPI_ISL_2329473, EPI_ISL_2329482, EPI_ISL_2329483, EPI_ISL_2329489, EPI_ISL_2329490, EPI_ISL_2329495, EPI_ISL_2329496, EPI_ISL_2329500, EPI_ISL_2329501, EPI_ISL_2329506, EPI_ISL_2329507, EPI_ISL_2329513, EPI_ISL_2329520, EPI_ISL_2329528, EPI_ISL_2329541, EPI_ISL_2329545, EPI_ISL_2329546, EPI_ISL_2329552, EPI_ISL_2329553, EPI_ISL_2329554, EPI_ISL_2329558, EPI_ISL_2329559, EPI_ISL_2329560, EPI_ISL_2329564, EPI_ISL_2329565, EPI_ISL_2329566, EPI_ISL_2329569, EPI_ISL_2329570, EPI_ISL_2329571, EPI_ISL_2329572, EPI_ISL_2329577, EPI_ISL_2329578, EPI_ISL_2329579, EPI_ISL_2329582, EPI_ISL_2329583, EPI_ISL_2329587, EPI_ISL_2329588, EPI_ISL_2329591, EPI_ISL_2329592, EPI_ISL_2329593, EPI_ISL_2329594, EPI_ISL_2329598, EPI_ISL_2329599, EPI_ISL_2329604, EPI_ISL_2329606, EPI_ISL_2628313, EPI_ISL_2628340, EPI_ISL_2628344, EPI_ISL_2628351, EPI_ISL_2628353, EPI_ISL_2628354, EPI_ISL_2628438, EPI_ISL_2628442, EPI_ISL_2628447, EPI_ISL_2628449, EPI_ISL_2628453, EPI_ISL_2628455, EPI_ISL_2628456, EPI_ISL_2628459, EPI_ISL_2628464, EPI_ISL_2628469, EPI_ISL_2628470, EPI_ISL_2628472, EPI_ISL_2628473, EPI_ISL_2628475, EPI_ISL_2628477, EPI_ISL_2628479, EPI_ISL_2628480, EPI_ISL_2628482, EPI_ISL_2628483, EPI_ISL_2628485, EPI_ISL_2628487, EPI_ISL_2628489, EPI_ISL_2628490, EPI_ISL_2628491, EPI_ISL_2628492, EPI_ISL_2628493, EPI_ISL_2628495, EPI_ISL_2628508, EPI_ISL_2628509, EPI_ISL_2628511, EPI_ISL_2628548, EPI_ISL_2628554, EPI_ISL_2628559, EPI_ISL_2628560, EPI_ISL_2628564, EPI_ISL_2628575, EPI_ISL_2628579, EPI_ISL_2628581, EPI_ISL_2628582, EPI_ISL_2628586, EPI_ISL_2628587, EPI_ISL_2628593, EPI_ISL_2628594, EPI_ISL_2712781, EPI_ISL_2712794, EPI_ISL_2712802, EPI_ISL_2712833, EPI_ISL_2712837, EPI_ISL_2920365, EPI_ISL_2920370

| | | | |
|---|---|---|---|
| see above | San Diego County Public Health Laboratory | Andersen lab at Scripps Research | Brett Austin; Ellen Stefanski; Ian Mchardy; Jovan Shephard; SEARCH Alliance San Diego with Ashleigh Murphy; SEARCH Alliance San Diego with Michael Quigley; SEARCH Alliance San Diego with Tracy Basler |

EPI_ISL_513856, EPI_ISL_513857, EPI_ISL_513858, EPI_ISL_513859, EPI_ISL_513860, EPI_ISL_513861, EPI_ISL_513862, EPI_ISL_513863, EPI_ISL_513864, EPI_ISL_513865, EPI_ISL_513866, EPI_ISL_513867, EPI_ISL_513868, EPI_ISL_513869, EPI_ISL_513870, EPI_ISL_513871, EPI_ISL_513872, EPI_ISL_513873, EPI_ISL_513874, EPI_ISL_513875, EPI_ISL_513876, EPI_ISL_513877, EPI_ISL_513878, EPI_ISL_513879, EPI_ISL_513880, EPI_ISL_513881, EPI_ISL_513882, EPI_ISL_583190, EPI_ISL_583191, EPI_ISL_583192, EPI_ISL_583193, EPI_ISL_583194, EPI_ISL_583195, EPI_ISL_583196, EPI_ISL_583197, EPI_ISL_672077, EPI_ISL_672375, EPI_ISL_672376, EPI_ISL_672377, EPI_ISL_672378, EPI_ISL_672379, EPI_ISL_672380, EPI_ISL_672381, EPI_ISL_672382, EPI_ISL_672383, EPI_ISL_672384, EPI_ISL_672385, EPI_ISL_2658652

| | | | |
|---|---|---|---|
| see above | San Francisco Public Health Laboratory | Chan-Zuckerberg Biohub | CZB Cliahub Consortium |

EPI_ISL_468506, EPI_ISL_468507, EPI_ISL_468508, EPI_ISL_468509, EPI_ISL_468510, EPI_ISL_468511, EPI_ISL_468512, EPI_ISL_468513, EPI_ISL_468514, EPI_ISL_468515, EPI_ISL_468516, EPI_ISL_468517, EPI_ISL_468518, EPI_ISL_468519, EPI_ISL_468520, EPI_ISL_468521, EPI_ISL_468522, EPI_ISL_468523, EPI_ISL_468524, EPI_ISL_468525, EPI_ISL_468526, EPI_ISL_468527, EPI_ISL_468528, EPI_ISL_468529,

EPI_ISL_468530, EPI_ISL_468531, EPI_ISL_468532, EPI_ISL_468533, EPI_ISL_468534, EPI_ISL_468535, EPI_ISL_468536, EPI_ISL_468537, EPI_ISL_468538, EPI_ISL_468539, EPI_ISL_468540, EPI_ISL_468541, EPI_ISL_468542, EPI_ISL_468543, EPI_ISL_468544, EPI_ISL_468545, EPI_ISL_468546, EPI_ISL_468547, EPI_ISL_468548, EPI_ISL_468549, EPI_ISL_468550, EPI_ISL_468551, EPI_ISL_468552, EPI_ISL_468553, EPI_ISL_468554, EPI_ISL_468555, EPI_ISL_468556, EPI_ISL_468557, EPI_ISL_468558, EPI_ISL_468559, EPI_ISL_486287, EPI_ISL_486288, EPI_ISL_486289, EPI_ISL_486290, EPI_ISL_486291, EPI_ISL_486292, EPI_ISL_486293, EPI_ISL_486294, EPI_ISL_486295, EPI_ISL_486296, EPI_ISL_486297, EPI_ISL_486298, EPI_ISL_486299, EPI_ISL_486300, EPI_ISL_486301, EPI_ISL_486302, EPI_ISL_486303, EPI_ISL_486304, EPI_ISL_486305, EPI_ISL_486306, EPI_ISL_486307, EPI_ISL_486308, EPI_ISL_486309, EPI_ISL_486310, EPI_ISL_486311, EPI_ISL_486312, EPI_ISL_486313, EPI_ISL_486314, EPI_ISL_486315, EPI_ISL_486316, EPI_ISL_486317, EPI_ISL_486318, EPI_ISL_486319, EPI_ISL_486320, EPI_ISL_486321, EPI_ISL_486322, EPI_ISL_486323, EPI_ISL_486324, EPI_ISL_486325, EPI_ISL_486326, EPI_ISL_486327, EPI_ISL_486328, EPI_ISL_486329, EPI_ISL_486330, EPI_ISL_486331, EPI_ISL_486332, EPI_ISL_486333, EPI_ISL_486334, EPI_ISL_486335, EPI_ISL_486336, EPI_ISL_486337, EPI_ISL_486338, EPI_ISL_625660, EPI_ISL_1924618

| | | | |
|---|---|---|---|
| see above | San Joaquin County Public Health Lab | Chan-Zuckerberg Biohub | CZB Cliahub Consortium |

EPI_ISL_582959, EPI_ISL_582960, EPI_ISL_582961, EPI_ISL_582962, EPI_ISL_582963, EPI_ISL_582964, EPI_ISL_582965, EPI_ISL_582966, EPI_ISL_582967, EPI_ISL_582968, EPI_ISL_582969, EPI_ISL_582970, EPI_ISL_582971, EPI_ISL_582972, EPI_ISL_582973, EPI_ISL_582974

| | | | |
|---|---|---|---|
| see above | San Luis Obispo Public Health Department | Chan-Zuckerberg Biohub | CZB Cliahub Consortium |

EPI_ISL_417317, EPI_ISL_417318, EPI_ISL_417319, EPI_ISL_417320, EPI_ISL_417321, EPI_ISL_429879, EPI_ISL_429880, EPI_ISL_435580, EPI_ISL_435581, EPI_ISL_435582, EPI_ISL_435583, EPI_ISL_435584, EPI_ISL_435585, EPI_ISL_435586, EPI_ISL_435587, EPI_ISL_435588, EPI_ISL_435589, EPI_ISL_435590, EPI_ISL_435591, EPI_ISL_435592, EPI_ISL_435593, EPI_ISL_435594, EPI_ISL_435595, EPI_ISL_435596, EPI_ISL_435597, EPI_ISL_435598, EPI_ISL_435599, EPI_ISL_435600, EPI_ISL_435601, EPI_ISL_435602, EPI_ISL_435603, EPI_ISL_435604, EPI_ISL_435605, EPI_ISL_435606, EPI_ISL_435607, EPI_ISL_435608, EPI_ISL_435609, EPI_ISL_435610, EPI_ISL_435611, EPI_ISL_435612, EPI_ISL_435613, EPI_ISL_435614, EPI_ISL_435615, EPI_ISL_435616, EPI_ISL_435617, EPI_ISL_435618, EPI_ISL_435619, EPI_ISL_435620, EPI_ISL_435621, EPI_ISL_435622, EPI_ISL_435623, EPI_ISL_435624, EPI_ISL_435625, EPI_ISL_435626, EPI_ISL_435627, EPI_ISL_435628, EPI_ISL_435629, EPI_ISL_435630, EPI_ISL_435631, EPI_ISL_435632, EPI_ISL_435633, EPI_ISL_435634, EPI_ISL_435635, EPI_ISL_435636, EPI_ISL_435637, EPI_ISL_435638, EPI_ISL_435639, EPI_ISL_435640, EPI_ISL_435641, EPI_ISL_435642, EPI_ISL_435643, EPI_ISL_435644, EPI_ISL_435645, EPI_ISL_435646, EPI_ISL_435647, EPI_ISL_435648, EPI_ISL_435649, EPI_ISL_435650, EPI_ISL_435651, EPI_ISL_435652, EPI_ISL_435653, EPI_ISL_435654, EPI_ISL_435655, EPI_ISL_435656, EPI_ISL_435657, EPI_ISL_435658, EPI_ISL_435659, EPI_ISL_435660, EPI_ISL_435661, EPI_ISL_435662, EPI_ISL_435663, EPI_ISL_435664, EPI_ISL_435665, EPI_ISL_435666, EPI_ISL_435667, EPI_ISL_435668, EPI_ISL_435669, EPI_ISL_435670, EPI_ISL_435671, EPI_ISL_435672, EPI_ISL_435673, EPI_ISL_671764, EPI_ISL_671765, EPI_ISL_671766, EPI_ISL_671767, EPI_ISL_671768, EPI_ISL_672663

| | | | |
|---|---|---|---|
| see above | Santa Clara County Public Health Department | Chiu Laboratory, University of California, San Francisco | Brandon Bonin; Debra A. Wadford; Elsa Villarino; Scot Federman; Wei Gu; Xianding Deng; and Charles Y. Chiu |

EPI_ISL_625478, EPI_ISL_625479, EPI_ISL_625480, EPI_ISL_625481, EPI_ISL_625482, EPI_ISL_625483, EPI_ISL_625484, EPI_ISL_625485, EPI_ISL_625486, EPI_ISL_625487, EPI_ISL_625488, EPI_ISL_625489, EPI_ISL_625490, EPI_ISL_625491, EPI_ISL_625492, EPI_ISL_625493, EPI_ISL_625494, EPI_ISL_625495, EPI_ISL_625496, EPI_ISL_625497, EPI_ISL_625498, EPI_ISL_625499, EPI_ISL_625594, EPI_ISL_625595, EPI_ISL_625596, EPI_ISL_625597, EPI_ISL_625598, EPI_ISL_625599, EPI_ISL_625600, EPI_ISL_625601, EPI_ISL_625602, EPI_ISL_625603, EPI_ISL_625604, EPI_ISL_625605, EPI_ISL_625606, EPI_ISL_625607, EPI_ISL_625608, EPI_ISL_625609, EPI_ISL_625610, EPI_ISL_648381, EPI_ISL_648382, EPI_ISL_648383, EPI_ISL_648384, EPI_ISL_648385, EPI_ISL_648386, EPI_ISL_648387, EPI_ISL_648388, EPI_ISL_648389, EPI_ISL_648390, EPI_ISL_648391, EPI_ISL_648392, EPI_ISL_648393, EPI_ISL_648394, EPI_ISL_648395, EPI_ISL_648396, EPI_ISL_648397, EPI_ISL_648398, EPI_ISL_648399, EPI_ISL_648400, EPI_ISL_648401, EPI_ISL_648402, EPI_ISL_648403, EPI_ISL_648404, EPI_ISL_648405, EPI_ISL_648406, EPI_ISL_648407, EPI_ISL_648408, EPI_ISL_648409, EPI_ISL_648410, EPI_ISL_648411, EPI_ISL_648412, EPI_ISL_648413, EPI_ISL_648414, EPI_ISL_648415, EPI_ISL_648416, EPI_ISL_648417, EPI_ISL_648418, EPI_ISL_648419, EPI_ISL_648420, EPI_ISL_648421, EPI_ISL_648422, EPI_ISL_648423, EPI_ISL_648424, EPI_ISL_648425, EPI_ISL_648426, EPI_ISL_648427, EPI_ISL_648428, EPI_ISL_648429, EPI_ISL_648430, EPI_ISL_648431, EPI_ISL_648432, EPI_ISL_648433, EPI_ISL_648434, EPI_ISL_648435, EPI_ISL_648436, EPI_ISL_648437, EPI_ISL_648438, EPI_ISL_648439, EPI_ISL_648440, EPI_ISL_648441, EPI_ISL_648442, EPI_ISL_648443, EPI_ISL_648444, EPI_ISL_648445, EPI_ISL_648446, EPI_ISL_648447, EPI_ISL_648448, EPI_ISL_648449, EPI_ISL_648450, EPI_ISL_648451, EPI_ISL_648452, EPI_ISL_648453, EPI_ISL_648454, EPI_ISL_648455, EPI_ISL_648456, EPI_ISL_648457, EPI_ISL_648458, EPI_ISL_648459, EPI_ISL_648460, EPI_ISL_648461, EPI_ISL_648462, EPI_ISL_648463, EPI_ISL_648464, EPI_ISL_648465, EPI_ISL_648466, EPI_ISL_648467, EPI_ISL_648468, EPI_ISL_648469, EPI_ISL_648470, EPI_ISL_648471, EPI_ISL_648472, EPI_ISL_648473, EPI_ISL_648474, EPI_ISL_648475, EPI_ISL_648476, EPI_ISL_648477, EPI_ISL_648478, EPI_ISL_648479, EPI_ISL_648480, EPI_ISL_648481, EPI_ISL_648482, EPI_ISL_648483, EPI_ISL_648484, EPI_ISL_648485, EPI_ISL_648486, EPI_ISL_648487, EPI_ISL_648488, EPI_ISL_648489, EPI_ISL_648490, EPI_ISL_648491, EPI_ISL_648492, EPI_ISL_648493, EPI_ISL_648494, EPI_ISL_648495, EPI_ISL_648542, EPI_ISL_660108, EPI_ISL_672013, EPI_ISL_672031, EPI_ISL_672033, EPI_ISL_672035, EPI_ISL_672043, EPI_ISL_672044, EPI_ISL_672052, EPI_ISL_672057, EPI_ISL_672065, EPI_ISL_672069, EPI_ISL_672070, EPI_ISL_672072, EPI_ISL_672073, EPI_ISL_672080, EPI_ISL_672085, EPI_ISL_672088, EPI_ISL_672089, EPI_ISL_672104, EPI_ISL_672106, EPI_ISL_672107, EPI_ISL_672108, EPI_ISL_672111, EPI_ISL_672275, EPI_ISL_672276, EPI_ISL_672277, EPI_ISL_672278, EPI_ISL_672279, EPI_ISL_672280, EPI_ISL_672281, EPI_ISL_672282, EPI_ISL_672283, EPI_ISL_672284, EPI_ISL_672285, EPI_ISL_672286, EPI_ISL_672287, EPI_ISL_672288, EPI_ISL_672289, EPI_ISL_672290, EPI_ISL_672291, EPI_ISL_672292, EPI_ISL_672293, EPI_ISL_672294, EPI_ISL_672295, EPI_ISL_672296, EPI_ISL_672297, EPI_ISL_672298, EPI_ISL_672299, EPI_ISL_672300, EPI_ISL_672301, EPI_ISL_672302, EPI_ISL_672303, EPI_ISL_672304, EPI_ISL_672429, EPI_ISL_672430, EPI_ISL_672431, EPI_ISL_672432, EPI_ISL_672433, EPI_ISL_739024, EPI_ISL_739249, EPI_ISL_739402, EPI_ISL_739414, EPI_ISL_739554, EPI_ISL_739615, EPI_ISL_853416, EPI_ISL_853546, EPI_ISL_853547, EPI_ISL_853548, EPI_ISL_853549, EPI_ISL_853550, EPI_ISL_853551, EPI_ISL_853552, EPI_ISL_853553, EPI_ISL_853554, EPI_ISL_853555, EPI_ISL_853556, EPI_ISL_853557, EPI_ISL_853558, EPI_ISL_853559, EPI_ISL_979145, EPI_ISL_979146, EPI_ISL_979147, EPI_ISL_979148, EPI_ISL_979149, EPI_ISL_979150, EPI_ISL_979151, EPI_ISL_979152, EPI_ISL_979153, EPI_ISL_979154, EPI_ISL_979155, EPI_ISL_979156, EPI_ISL_979157, EPI_ISL_979158, EPI_ISL_1093053, EPI_ISL_1093054, EPI_ISL_1093055, EPI_ISL_1093056, EPI_ISL_1093057, EPI_ISL_1093058, EPI_ISL_1093059, EPI_ISL_1093060, EPI_ISL_1093061, EPI_ISL_1093062, EPI_ISL_1093063, EPI_ISL_1093064, EPI_ISL_1093065, EPI_ISL_1093066, EPI_ISL_1093067, EPI_ISL_1093068, EPI_ISL_1924600, EPI_ISL_2658572, EPI_ISL_2658573, EPI_ISL_2658574, EPI_ISL_2658576, EPI_ISL_2658585, EPI_ISL_2658611, EPI_ISL_2658612, EPI_ISL_2658623, EPI_ISL_2658628, EPI_ISL_2658650, EPI_ISL_2658675, EPI_ISL_2658703, EPI_ISL_2658711, EPI_ISL_2658725, EPI_ISL_2658728, EPI_ISL_2658733, EPI_ISL_2658743, EPI_ISL_2659218, EPI_ISL_2659227, EPI_ISL_2659230, EPI_ISL_2659241, EPI_ISL_2659250, EPI_ISL_2659268, EPI_ISL_2659299, EPI_ISL_2659302, EPI_ISL_2659307, EPI_ISL_2659313, EPI_ISL_2659317, EPI_ISL_2659320, EPI_ISL_2659358, EPI_ISL_2659360, EPI_ISL_2659363, EPI_ISL_2659367, EPI_ISL_2659370, EPI_ISL_2659397, EPI_ISL_2659406, EPI_ISL_2659415, EPI_ISL_2659420, EPI_ISL_2659424, EPI_ISL_2659456, EPI_ISL_2659471, EPI_ISL_2659472, EPI_ISL_2659473, EPI_ISL_2659478, EPI_ISL_2659483, EPI_ISL_2659485, EPI_ISL_2659490, EPI_ISL_2659494, EPI_ISL_2659511, EPI_ISL_2659523, EPI_ISL_2659530, EPI_ISL_2659535, EPI_ISL_2659536, EPI_ISL_2659564, EPI_ISL_2659569, EPI_ISL_2659571, EPI_ISL_2659574, EPI_ISL_2659575, EPI_ISL_2659581, EPI_ISL_2659587, EPI_ISL_2659590, EPI_ISL_2659602, EPI_ISL_2659605, EPI_ISL_2659616, EPI_ISL_2659631, EPI_ISL_2659632, EPI_ISL_2659633, EPI_ISL_2659639, EPI_ISL_2659647, EPI_ISL_2659649, EPI_ISL_2758412, EPI_ISL_2758423, EPI_ISL_2758424, EPI_ISL_2758542, EPI_ISL_2758547, EPI_ISL_2758548

| | | | |
|---|---|---|---|
| see above | Santa Clara County Public Health Laboratory | Chan-Zuckerberg Biohub | CZB Cliahub Consortium |

EPI_ISL_806543, EPI_ISL_806545, EPI_ISL_812115, EPI_ISL_812116, EPI_ISL_812119, EPI_ISL_812120, EPI_ISL_812121, EPI_ISL_1920837, EPI_ISL_1920838, EPI_ISL_1920844, EPI_ISL_1920845, EPI_ISL_1920849, EPI_ISL_1920850, EPI_ISL_1920853, EPI_ISL_1920864, EPI_ISL_1920870, EPI_ISL_1920878, EPI_ISL_1920886, EPI_ISL_1920890, EPI_ISL_1920897, EPI_ISL_2779610

| | | | |
|---|---|---|---|
| see above | Santa Clara County Public Health Laboratory | Santa Clara County Public Health Laboratory | Santa CLara County Public Health Department; Santa Clara Co Public Health Department; Santa Clara County Public Health Department |

EPI_ISL_437549, EPI_ISL_437550, EPI_ISL_437551, EPI_ISL_437552, EPI_ISL_437553, EPI_ISL_437554, EPI_ISL_437555, EPI_ISL_437556, EPI_ISL_437557, EPI_ISL_437558, EPI_ISL_437559, EPI_ISL_437560, EPI_ISL_437561, EPI_ISL_437562, EPI_ISL_437563, EPI_ISL_437564, EPI_ISL_437565, EPI_ISL_437566, EPI_ISL_437567, EPI_ISL_437568, EPI_ISL_437569, EPI_ISL_437570, EPI_ISL_437571, EPI_ISL_437572, EPI_ISL_437573, EPI_ISL_437574, EPI_ISL_437575, EPI_ISL_437576, EPI_ISL_437577, EPI_ISL_437578, EPI_ISL_437579, EPI_ISL_437580, EPI_ISL_437581, EPI_ISL_437582, EPI_ISL_437583, EPI_ISL_437584, EPI_ISL_437585, EPI_ISL_437586, EPI_ISL_437587, EPI_ISL_437588, EPI_ISL_437589, EPI_ISL_437590, EPI_ISL_437591, EPI_ISL_437592, EPI_ISL_437593, EPI_ISL_437594, EPI_ISL_437595, EPI_ISL_437596, EPI_ISL_437597, EPI_ISL_437598, EPI_ISL_437599, EPI_ISL_437600, EPI_ISL_445164, EPI_ISL_445165, EPI_ISL_445166, EPI_ISL_445167, EPI_ISL_445168, EPI_ISL_450191, EPI_ISL_450192, EPI_ISL_458237, EPI_ISL_458238, EPI_ISL_458239, EPI_ISL_458240, EPI_ISL_458241, EPI_ISL_458242, EPI_ISL_458243, EPI_ISL_458244, EPI_ISL_458245, EPI_ISL_458246, EPI_ISL_458247, EPI_ISL_458248, EPI_ISL_458249, EPI_ISL_458250, EPI_ISL_458251, EPI_ISL_458252, EPI_ISL_458253, EPI_ISL_458254, EPI_ISL_458255, EPI_ISL_458256, EPI_ISL_458257, EPI_ISL_458258, EPI_ISL_458259, EPI_ISL_458260, EPI_ISL_458261, EPI_ISL_458262, EPI_ISL_458263, EPI_ISL_458264, EPI_ISL_458265, EPI_ISL_458266, EPI_ISL_458267, EPI_ISL_458268, EPI_ISL_458269, EPI_ISL_458270, EPI_ISL_458271, EPI_ISL_458272, EPI_ISL_458273, EPI_ISL_458274, EPI_ISL_458275, EPI_ISL_458276, EPI_ISL_458277, EPI_ISL_458278, EPI_ISL_458279, EPI_ISL_458280, EPI_ISL_458281, EPI_ISL_458282, EPI_ISL_458283, EPI_ISL_458284, EPI_ISL_467967, EPI_ISL_467972, EPI_ISL_494596, EPI_ISL_494597, EPI_ISL_494598, EPI_ISL_494599, EPI_ISL_494600, EPI_ISL_494601, EPI_ISL_494602, EPI_ISL_494603, EPI_ISL_494604, EPI_ISL_494605, EPI_ISL_494606, EPI_ISL_494607, EPI_ISL_494608, EPI_ISL_494609, EPI_ISL_494610, EPI_ISL_494611, EPI_ISL_494612, EPI_ISL_494613, EPI_ISL_494614, EPI_ISL_494615, EPI_ISL_494616, EPI_ISL_494617, EPI_ISL_494618, EPI_ISL_494619, EPI_ISL_494635, EPI_ISL_494636, EPI_ISL_494637, EPI_ISL_494638, EPI_ISL_494639, EPI_ISL_494640, EPI_ISL_494641, EPI_ISL_494642, EPI_ISL_494643, EPI_ISL_494644, EPI_ISL_494645, EPI_ISL_494646, EPI_ISL_494647, EPI_ISL_494648, EPI_ISL_494649, EPI_ISL_494650, EPI_ISL_494651, EPI_ISL_494652, EPI_ISL_494653, EPI_ISL_494654, EPI_ISL_494655, EPI_ISL_494656, EPI_ISL_494657, EPI_ISL_494658, EPI_ISL_494659, EPI_ISL_494660, EPI_ISL_494661, EPI_ISL_494662, EPI_ISL_494663, EPI_ISL_494664, EPI_ISL_494665, EPI_ISL_494666, EPI_ISL_494667, EPI_ISL_494668, EPI_ISL_494669, EPI_ISL_494670, EPI_ISL_494671, EPI_ISL_494672, EPI_ISL_494673, EPI_ISL_494674, EPI_ISL_494675, EPI_ISL_494676, EPI_ISL_494677, EPI_ISL_494678, EPI_ISL_494679, EPI_ISL_494680, EPI_ISL_494681, EPI_ISL_494682, EPI_ISL_494683, EPI_ISL_494684, EPI_ISL_494685, EPI_ISL_494686, EPI_ISL_494687, EPI_ISL_494688, EPI_ISL_494689, EPI_ISL_494690, EPI_ISL_494691, EPI_ISL_494692, EPI_ISL_494693, EPI_ISL_494694, EPI_ISL_494695, EPI_ISL_494696, EPI_ISL_494697, EPI_ISL_494698, EPI_ISL_494699, EPI_ISL_494700, EPI_ISL_494701, EPI_ISL_494702, EPI_ISL_494703, EPI_ISL_494704, EPI_ISL_494705, EPI_ISL_494706, EPI_ISL_494707, EPI_ISL_494708, EPI_ISL_494709, EPI_ISL_494710, EPI_ISL_494711, EPI_ISL_494712, EPI_ISL_494713, EPI_ISL_730137, EPI_ISL_730138, EPI_ISL_730139, EPI_ISL_730140, EPI_ISL_730141, EPI_ISL_730142, EPI_ISL_730143, EPI_ISL_730144, EPI_ISL_730145, EPI_ISL_730146, EPI_ISL_730147, EPI_ISL_730148, EPI_ISL_730149, EPI_ISL_730150, EPI_ISL_730151, EPI_ISL_730152, EPI_ISL_730153, EPI_ISL_730154, EPI_ISL_730155, EPI_ISL_730156, EPI_ISL_730157, EPI_ISL_730158, EPI_ISL_730159, EPI_ISL_730160

| | | | |
|---|---|---|---|
| see above | Scripps Medical Laboratory | Andersen lab at Scripps Research | Brett Austin; Ellen Stefanski; Ian Mchardy; Jovan Shephard; SEARCH Alliance San Diego with Michael Quigley; SEARCH Alliance San Diego with Tracy Basler |
| EPI_ISL_2835566 | Sharp HealthCare Laboratory | Andersen lab at Scripps Research | Art Mendoza; Cathy Woerle; Jacquelyn Berumen; Liam McGinnis; Omid Bakhtar; SEARCH Alliance San Diego with Aaron Harding |

EPI_ISL_450445, EPI_ISL_450446, EPI_ISL_450447, EPI_ISL_450448, EPI_ISL_450449, EPI_ISL_450450, EPI_ISL_450451, EPI_ISL_450452, EPI_ISL_450453, EPI_ISL_450454, EPI_ISL_450455, EPI_ISL_450456, EPI_ISL_450457, EPI_ISL_450458, EPI_ISL_450459, EPI_ISL_450460, EPI_ISL_450461, EPI_ISL_450462, EPI_ISL_450463, EPI_ISL_450464, EPI_ISL_450465, EPI_ISL_450466, EPI_ISL_450467, EPI_ISL_450468, EPI_ISL_450469, EPI_ISL_450470, EPI_ISL_450471, EPI_ISL_450472, EPI_ISL_450473, EPI_ISL_450474, EPI_ISL_450475, EPI_ISL_450476, EPI_ISL_450477, EPI_ISL_450478, EPI_ISL_450479, EPI_ISL_450480, EPI_ISL_450481, EPI_ISL_476767, EPI_ISL_476768, EPI_ISL_476769, EPI_ISL_476770, EPI_ISL_476771, EPI_ISL_476772, EPI_ISL_476773, EPI_ISL_476774, EPI_ISL_476775, EPI_ISL_476776, EPI_ISL_476777, EPI_ISL_476778, EPI_ISL_476779, EPI_ISL_476780, EPI_ISL_476781, EPI_ISL_476782, EPI_ISL_476783, EPI_ISL_476784, EPI_ISL_476785, EPI_ISL_476786, EPI_ISL_476787, EPI_ISL_476788, EPI_ISL_476789, EPI_ISL_476790, EPI_ISL_476791, EPI_ISL_476792, EPI_ISL_476793, EPI_ISL_476794

| | | | |
|---|---|---|---|
| see above | Stanford clinical virology lab | Chan-Zuckerberg Biohub | Benjamin Pinksy; Carlos Bustamante; Euan Ashley; Hannah N. DeJong; Jason Andrews; John Gorzynski; Katharine Walter; Manuel Rivas; Matthew T. Wheeler; Victoria N. Parikh; with CZB Cliahub Consortium |

EPI_ISL_672017, EPI_ISL_672027, EPI_ISL_672030, EPI_ISL_672042, EPI_ISL_672047, EPI_ISL_672049, EPI_ISL_672051, EPI_ISL_672055, EPI_ISL_672056, EPI_ISL_672058, EPI_ISL_672061, EPI_ISL_672063, EPI_ISL_672071, EPI_ISL_672074, EPI_ISL_672079, EPI_ISL_672083, EPI_ISL_672090, EPI_ISL_672093, EPI_ISL_672094, EPI_ISL_672096, EPI_ISL_672105, EPI_ISL_672112, EPI_ISL_672114, EPI_ISL_672115, EPI_ISL_672116, EPI_ISL_672117, EPI_ISL_672118, EPI_ISL_672119, EPI_ISL_672120, EPI_ISL_672121, EPI_ISL_672122, EPI_ISL_672123, EPI_ISL_672124, EPI_ISL_672125, EPI_ISL_672126, EPI_ISL_672127, EPI_ISL_672128, EPI_ISL_672129, EPI_ISL_672130, EPI_ISL_672131, EPI_ISL_672132, EPI_ISL_672133, EPI_ISL_672134, EPI_ISL_672135, EPI_ISL_672136, EPI_ISL_672137, EPI_ISL_672138, EPI_ISL_672139, EPI_ISL_672140, EPI_ISL_672141, EPI_ISL_672142, EPI_ISL_672143, EPI_ISL_672144, EPI_ISL_672145, EPI_ISL_672146, EPI_ISL_672147, EPI_ISL_672148, EPI_ISL_672149, EPI_ISL_672150, EPI_ISL_672151, EPI_ISL_672152, EPI_ISL_672153, EPI_ISL_672154, EPI_ISL_672155, EPI_ISL_672156, EPI_ISL_672157, EPI_ISL_672158, EPI_ISL_672159, EPI_ISL_672160, EPI_ISL_672161, EPI_ISL_672162, EPI_ISL_672163, EPI_ISL_672164, EPI_ISL_672165, EPI_ISL_672166, EPI_ISL_672167, EPI_ISL_672168, EPI_ISL_672169, EPI_ISL_672170, EPI_ISL_672171, EPI_ISL_672172, EPI_ISL_672173, EPI_ISL_672174, EPI_ISL_672175, EPI_ISL_672176, EPI_ISL_672177, EPI_ISL_672178, EPI_ISL_672179, EPI_ISL_672180, EPI_ISL_672181, EPI_ISL_672182, EPI_ISL_672183, EPI_ISL_672184, EPI_ISL_672185, EPI_ISL_672186, EPI_ISL_672187, EPI_ISL_672188, EPI_ISL_672189, EPI_ISL_672190, EPI_ISL_672191, EPI_ISL_672192, EPI_ISL_672193, EPI_ISL_672194, EPI_ISL_672195, EPI_ISL_672196, EPI_ISL_672197, EPI_ISL_672198, EPI_ISL_672199, EPI_ISL_672200, EPI_ISL_672201, EPI_ISL_672202, EPI_ISL_672203, EPI_ISL_672204, EPI_ISL_672205, EPI_ISL_672206, EPI_ISL_672207, EPI_ISL_672208, EPI_ISL_672209, EPI_ISL_672210, EPI_ISL_672211, EPI_ISL_672212, EPI_ISL_672213, EPI_ISL_672214, EPI_ISL_672215, EPI_ISL_672216, EPI_ISL_672217, EPI_ISL_672218, EPI_ISL_672219, EPI_ISL_672220, EPI_ISL_672221, EPI_ISL_672222, EPI_ISL_672223, EPI_ISL_672224, EPI_ISL_672225, EPI_ISL_672226, EPI_ISL_672227, EPI_ISL_672228, EPI_ISL_672229, EPI_ISL_672230, EPI_ISL_672231, EPI_ISL_672232, EPI_ISL_672233, EPI_ISL_672234, EPI_ISL_672235, EPI_ISL_672236, EPI_ISL_672237, EPI_ISL_672238, EPI_ISL_672239, EPI_ISL_672240, EPI_ISL_672241, EPI_ISL_672242, EPI_ISL_672243, EPI_ISL_672244, EPI_ISL_672245, EPI_ISL_672246, EPI_ISL_672247, EPI_ISL_672248, EPI_ISL_672249, EPI_ISL_672250, EPI_ISL_672251, EPI_ISL_672252, EPI_ISL_672253, EPI_ISL_672254, EPI_ISL_672255, EPI_ISL_672256, EPI_ISL_672257, EPI_ISL_672258, EPI_ISL_672259, EPI_ISL_672260, EPI_ISL_672261, EPI_ISL_672262, EPI_ISL_672263, EPI_ISL_672264, EPI_ISL_672265, EPI_ISL_672266, EPI_ISL_672267, EPI_ISL_672268, EPI_ISL_672269, EPI_ISL_672270, EPI_ISL_672271, EPI_ISL_672272, EPI_ISL_672273, EPI_ISL_672274, EPI_ISL_856974, EPI_ISL_856975, EPI_ISL_856976, EPI_ISL_856977, EPI_ISL_856978, EPI_ISL_856979, EPI_ISL_856980, EPI_ISL_856981, EPI_ISL_856982, EPI_ISL_856983, EPI_ISL_856984, EPI_ISL_856985, EPI_ISL_856986, EPI_ISL_856987, EPI_ISL_856988, EPI_ISL_856989, EPI_ISL_856990, EPI_ISL_856991, EPI_ISL_856992, EPI_ISL_856993, EPI_ISL_856994, EPI_ISL_856995, EPI_ISL_856996, EPI_ISL_856997, EPI_ISL_856998, EPI_ISL_856999, EPI_ISL_857000, EPI_ISL_857001, EPI_ISL_857002, EPI_ISL_857003, EPI_ISL_857004, EPI_ISL_857005, EPI_ISL_857006, EPI_ISL_857007, EPI_ISL_857008, EPI_ISL_857009, EPI_ISL_857010, EPI_ISL_857011, EPI_ISL_857012, EPI_ISL_857013, EPI_ISL_857014, EPI_ISL_857015, EPI_ISL_857016, EPI_ISL_857017, EPI_ISL_857018, EPI_ISL_857019, EPI_ISL_857020, EPI_ISL_857021, EPI_ISL_857022, EPI_ISL_857023, EPI_ISL_857024, EPI_ISL_857025, EPI_ISL_857026, EPI_ISL_857027, EPI_ISL_857028, EPI_ISL_857029, EPI_ISL_857030, EPI_ISL_857031, EPI_ISL_857032, EPI_ISL_857033, EPI_ISL_857034, EPI_ISL_857035, EPI_ISL_857036, EPI_ISL_857037, EPI_ISL_857038, EPI_ISL_857039

| | | | |
|---|---|---|---|
| see above | The Ashley Laboratory, Stanford University | Chan-Zuckerberg Biohub | CZB Cliahub Consortium |

EPI_ISL_583171, EPI_ISL_583172, EPI_ISL_648531, EPI_ISL_648532, EPI_ISL_648533, EPI_ISL_648534, EPI_ISL_648535, EPI_ISL_648536, EPI_ISL_648537, EPI_ISL_648538, EPI_ISL_648539, EPI_ISL_648540, EPI_ISL_648541, EPI_ISL_739119, EPI_ISL_739193, EPI_ISL_739639

| | | | |
|---|---|---|---|
| see above | Tulare County Public Health Lab | Chan-Zuckerberg Biohub | CZB Cliahub Consortium |
| EPI_ISL_548363, EPI_ISL_548599, EPI_ISL_548626 | Tuolumne County Public Health | Chan-Zuckerberg Biohub | CZB Cliahub Consortium |

EPI_ISL_849928, EPI_ISL_849929, EPI_ISL_849930, EPI_ISL_849931, EPI_ISL_849932, EPI_ISL_849933, EPI_ISL_849934, EPI_ISL_849935, EPI_ISL_849936, EPI_ISL_849937, EPI_ISL_849938, EPI_ISL_849939, EPI_ISL_849940, EPI_ISL_849941, EPI_ISL_849942, EPI_ISL_849943, EPI_ISL_849946, EPI_ISL_849947, EPI_ISL_849949, EPI_ISL_849950

| | | | |
|---|---|---|---|
| see above | UC Davis- Department of Pathology and Laboratory Medicine | Chan-Zuckerberg Biohub | CZB Cliahub Consortium |

EPI_ISL_445094, EPI_ISL_445095, EPI_ISL_445096, EPI_ISL_445097, EPI_ISL_445098, EPI_ISL_445099, EPI_ISL_445100, EPI_ISL_445101, EPI_ISL_445102, EPI_ISL_445103, EPI_ISL_445104, EPI_ISL_445105, EPI_ISL_445106, EPI_ISL_445107, EPI_ISL_445108, EPI_ISL_445109, EPI_ISL_445110, EPI_ISL_445111, EPI_ISL_445112, EPI_ISL_445113, EPI_ISL_445114, EPI_ISL_445115, EPI_ISL_445116, EPI_ISL_445117, EPI_ISL_483165, EPI_ISL_483166, EPI_ISL_483167, EPI_ISL_483168, EPI_ISL_483169, EPI_ISL_483170, EPI_ISL_483171, EPI_ISL_483172, EPI_ISL_483173, EPI_ISL_483174, EPI_ISL_483175, EPI_ISL_483176, EPI_ISL_483177, EPI_ISL_483178, EPI_ISL_483179, EPI_ISL_483180, EPI_ISL_483181, EPI_ISL_483182, EPI_ISL_483183, EPI_ISL_483184, EPI_ISL_483185, EPI_ISL_483186, EPI_ISL_483187, EPI_ISL_483188, EPI_ISL_483189, EPI_ISL_483190, EPI_ISL_483191, EPI_ISL_483192, EPI_ISL_483193, EPI_ISL_483194, EPI_ISL_483195, EPI_ISL_483196, EPI_ISL_483197, EPI_ISL_483198, EPI_ISL_483199, EPI_ISL_483200, EPI_ISL_483201, EPI_ISL_483202, EPI_ISL_483203, EPI_ISL_483204, EPI_ISL_483205, EPI_ISL_483206, EPI_ISL_483207, EPI_ISL_483208, EPI_ISL_483209, EPI_ISL_483210, EPI_ISL_483211, EPI_ISL_483212, EPI_ISL_483213, EPI_ISL_483214, EPI_ISL_483215, EPI_ISL_483216, EPI_ISL_483217, EPI_ISL_483218, EPI_ISL_483219, EPI_ISL_483220, EPI_ISL_483221, EPI_ISL_483222, EPI_ISL_483223, EPI_ISL_483224, EPI_ISL_483225, EPI_ISL_483226, EPI_ISL_483227, EPI_ISL_483228, EPI_ISL_483229, EPI_ISL_483230, EPI_ISL_483231, EPI_ISL_483232, EPI_ISL_483233, EPI_ISL_483234, EPI_ISL_483235, EPI_ISL_483236, EPI_ISL_483237, EPI_ISL_483238, EPI_ISL_483239, EPI_ISL_483240, EPI_ISL_483241, EPI_ISL_483242, EPI_ISL_483243, EPI_ISL_483244, EPI_ISL_483245, EPI_ISL_483246, EPI_ISL_483247, EPI_ISL_483248, EPI_ISL_483249, EPI_ISL_483250, EPI_ISL_483251, EPI_ISL_483252, EPI_ISL_483253, EPI_ISL_483254, EPI_ISL_483255, EPI_ISL_483256, EPI_ISL_483257, EPI_ISL_483258, EPI_ISL_483259, EPI_ISL_483260, EPI_ISL_483261, EPI_ISL_483262, EPI_ISL_483263, EPI_ISL_483264, EPI_ISL_483265, EPI_ISL_483266, EPI_ISL_483267, EPI_ISL_483268, EPI_ISL_483269, EPI_ISL_483270, EPI_ISL_483271, EPI_ISL_483272, EPI_ISL_483273, EPI_ISL_483274, EPI_ISL_483275, EPI_ISL_483276, EPI_ISL_483277, EPI_ISL_483278, EPI_ISL_483279, EPI_ISL_483280, EPI_ISL_483281, EPI_ISL_483282, EPI_ISL_483283, EPI_ISL_483284, EPI_ISL_483285, EPI_ISL_483286, EPI_ISL_483287, EPI_ISL_483288, EPI_ISL_483289, EPI_ISL_483290, EPI_ISL_483291, EPI_ISL_483292, EPI_ISL_483293, EPI_ISL_483294, EPI_ISL_483295, EPI_ISL_483296, EPI_ISL_483297, EPI_ISL_483298, EPI_ISL_483299, EPI_ISL_483300, EPI_ISL_483301, EPI_ISL_483302, EPI_ISL_483303, EPI_ISL_483304, EPI_ISL_483305, EPI_ISL_483306, EPI_ISL_483307, EPI_ISL_483308, EPI_ISL_483309, EPI_ISL_483310, EPI_ISL_483311, EPI_ISL_483312, EPI_ISL_483313, EPI_ISL_483314, EPI_ISL_483315, EPI_ISL_483316, EPI_ISL_483317, EPI_ISL_483318, EPI_ISL_483319, EPI_ISL_483320, EPI_ISL_483321, EPI_ISL_483322, EPI_ISL_483323, EPI_ISL_483324, EPI_ISL_483325, EPI_ISL_483326, EPI_ISL_483327, EPI_ISL_483328, EPI_ISL_483329, EPI_ISL_483330, EPI_ISL_483331, EPI_ISL_483332, EPI_ISL_483333, EPI_ISL_483334, EPI_ISL_483335, EPI_ISL_483336, EPI_ISL_483337, EPI_ISL_483338, EPI_ISL_483339, EPI_ISL_483340, EPI_ISL_483341, EPI_ISL_483342, EPI_ISL_483343, EPI_ISL_483344, EPI_ISL_483345, EPI_ISL_483346, EPI_ISL_483347, EPI_ISL_483348, EPI_ISL_483349, EPI_ISL_483350, EPI_ISL_483351, EPI_ISL_483352, EPI_ISL_483353, EPI_ISL_483354, EPI_ISL_483355, EPI_ISL_483356, EPI_ISL_483357, EPI_ISL_483358, EPI_ISL_483359, EPI_ISL_483360, EPI_ISL_483361, EPI_ISL_483362, EPI_ISL_483363, EPI_ISL_483364, EPI_ISL_483365, EPI_ISL_483366, EPI_ISL_483367, EPI_ISL_483368, EPI_ISL_483369, EPI_ISL_483370, EPI_ISL_483371, EPI_ISL_483372, EPI_ISL_483373, EPI_ISL_483374, EPI_ISL_483375, EPI_ISL_483376, EPI_ISL_483377, EPI_ISL_483378, EPI_ISL_483379, EPI_ISL_483380, EPI_ISL_483381, EPI_ISL_483382, EPI_ISL_483383, EPI_ISL_483384, EPI_ISL_483385, EPI_ISL_483386, EPI_ISL_483387, EPI_ISL_483388, EPI_ISL_483389, EPI_ISL_483390, EPI_ISL_483391, EPI_ISL_483392, EPI_ISL_483393, EPI_ISL_483394, EPI_ISL_483395, EPI_ISL_483396, EPI_ISL_483397, EPI_ISL_483403, EPI_ISL_483404, EPI_ISL_483405, EPI_ISL_483406, EPI_ISL_483407, EPI_ISL_483408, EPI_ISL_483409, EPI_ISL_483410, EPI_ISL_483411, EPI_ISL_483412, EPI_ISL_483413, EPI_ISL_483414, EPI_ISL_483415, EPI_ISL_483416, EPI_ISL_483417, EPI_ISL_483418, EPI_ISL_483419, EPI_ISL_483420, EPI_ISL_483421, EPI_ISL_483422, EPI_ISL_483423, EPI_ISL_483424, EPI_ISL_483425, EPI_ISL_483426, EPI_ISL_483427, EPI_ISL_483428, EPI_ISL_483429, EPI_ISL_483430, EPI_ISL_483431, EPI_ISL_483432, EPI_ISL_483433, EPI_ISL_483434, EPI_ISL_483435, EPI_ISL_483436, EPI_ISL_483437, EPI_ISL_483438, EPI_ISL_483439, EPI_ISL_483440, EPI_ISL_483441, EPI_ISL_483442, EPI_ISL_483443, EPI_ISL_483444, EPI_ISL_483445, EPI_ISL_483446, EPI_ISL_483447, EPI_ISL_483449, EPI_ISL_483450, EPI_ISL_483451, EPI_ISL_483452, EPI_ISL_483453, EPI_ISL_483454, EPI_ISL_483455, EPI_ISL_483456, EPI_ISL_483457, EPI_ISL_483458, EPI_ISL_483459, EPI_ISL_483460, EPI_ISL_483461, EPI_ISL_483462, EPI_ISL_483463, EPI_ISL_483464, EPI_ISL_483465, EPI_ISL_483466, EPI_ISL_483467, EPI_ISL_483468, EPI_ISL_483469, EPI_ISL_483470, EPI_ISL_483471, EPI_ISL_483472, EPI_ISL_483473, EPI_ISL_483474, EPI_ISL_483475, EPI_ISL_483476, EPI_ISL_483477, EPI_ISL_483478, EPI_ISL_483479, EPI_ISL_483480, EPI_ISL_483481, EPI_ISL_483482, EPI_ISL_483483, EPI_ISL_483484, EPI_ISL_483485, EPI_ISL_483486, EPI_ISL_483487, EPI_ISL_483488, EPI_ISL_483489, EPI_ISL_483490, EPI_ISL_483491, EPI_ISL_483492, EPI_ISL_483493, EPI_ISL_483494, EPI_ISL_483495, EPI_ISL_483496, EPI_ISL_483497, EPI_ISL_483498, EPI_ISL_483499, EPI_ISL_483500, EPI_ISL_483501, EPI_ISL_483502, EPI_ISL_483503, EPI_ISL_483504, EPI_ISL_483505, EPI_ISL_483506, EPI_ISL_483507, EPI_ISL_483508, EPI_ISL_483509, EPI_ISL_483510, EPI_ISL_483511, EPI_ISL_483512, EPI_ISL_483513, EPI_ISL_483514, EPI_ISL_483515, EPI_ISL_483516, EPI_ISL_483517, EPI_ISL_483518, EPI_ISL_483519, EPI_ISL_483520, EPI_ISL_483521, EPI_ISL_483522, EPI_ISL_483523, EPI_ISL_483524, EPI_ISL_483525, EPI_ISL_483526, EPI_ISL_483527, EPI_ISL_483528, EPI_ISL_483529, EPI_ISL_483530

| | | | |
|---|---|---|---|
| see above | UC San Diego Center for Advanced Laboratory Medicine | Andersen lab at Scripps Research | Ji H Shin; SEARCH Alliance San Diego with David Pride |

EPI_ISL_537467, EPI_ISL_537468, EPI_ISL_537469, EPI_ISL_537470, EPI_ISL_537471, EPI_ISL_537472, EPI_ISL_537473, EPI_ISL_537474, EPI_ISL_537475, EPI_ISL_537476, EPI_ISL_537477, EPI_ISL_537478, EPI_ISL_537479, EPI_ISL_537480, EPI_ISL_537481, EPI_ISL_537482, EPI_ISL_537483, EPI_ISL_537484, EPI_ISL_537485, EPI_ISL_537486, EPI_ISL_537487, EPI_ISL_537488, EPI_ISL_537489, EPI_ISL_537490, EPI_ISL_537491, EPI_ISL_537492, EPI_ISL_537493, EPI_ISL_537494, EPI_ISL_537495, EPI_ISL_537496, EPI_ISL_537497, EPI_ISL_537498, EPI_ISL_537499, EPI_ISL_537500, EPI_ISL_537501, EPI_ISL_537502, EPI_ISL_537503, EPI_ISL_537504, EPI_ISL_537505, EPI_ISL_537506, EPI_ISL_537507, EPI_ISL_537508, EPI_ISL_537509, EPI_ISL_537510, EPI_ISL_537511, EPI_ISL_537512, EPI_ISL_537513, EPI_ISL_537514, EPI_ISL_537515, EPI_ISL_537516, EPI_ISL_537517, EPI_ISL_537518, EPI_ISL_537519, EPI_ISL_537520, EPI_ISL_537521, EPI_ISL_537522, EPI_ISL_537523, EPI_ISL_537524, EPI_ISL_537525, EPI_ISL_537526, EPI_ISL_537527, EPI_ISL_537528, EPI_ISL_537529, EPI_ISL_537530, EPI_ISL_537531, EPI_ISL_537532, EPI_ISL_537533, EPI_ISL_537534, EPI_ISL_537535, EPI_ISL_537536, EPI_ISL_537537, EPI_ISL_537538, EPI_ISL_537539, EPI_ISL_537540, EPI_ISL_537541, EPI_ISL_537542, EPI_ISL_537543, EPI_ISL_537544, EPI_ISL_537545, EPI_ISL_537546, EPI_ISL_537547, EPI_ISL_537548, EPI_ISL_537549, EPI_ISL_537550, EPI_ISL_537551, EPI_ISL_537552, EPI_ISL_537553, EPI_ISL_537554, EPI_ISL_537555, EPI_ISL_537556, EPI_ISL_537557, EPI_ISL_537558, EPI_ISL_537559, EPI_ISL_537560, EPI_ISL_537561, EPI_ISL_537562, EPI_ISL_537563, EPI_ISL_537564, EPI_ISL_537565, EPI_ISL_537566, EPI_ISL_537567, EPI_ISL_537568, EPI_ISL_537569, EPI_ISL_537570, EPI_ISL_537571, EPI_ISL_537572, EPI_ISL_537573, EPI_ISL_537574, EPI_ISL_537575, EPI_ISL_537576, EPI_ISL_537577, EPI_ISL_537578, EPI_ISL_537579, EPI_ISL_537580, EPI_ISL_537581, EPI_ISL_537582, EPI_ISL_537583, EPI_ISL_537584, EPI_ISL_537585, EPI_ISL_537586, EPI_ISL_537587, EPI_ISL_537588, EPI_ISL_537589, EPI_ISL_537590, EPI_ISL_537591, EPI_ISL_537592, EPI_ISL_537593, EPI_ISL_537594, EPI_ISL_537595, EPI_ISL_537596, EPI_ISL_537597, EPI_ISL_537598, EPI_ISL_537599, EPI_ISL_537600, EPI_ISL_537601, EPI_ISL_537602, EPI_ISL_537603, EPI_ISL_537604, EPI_ISL_537605, EPI_ISL_537606, EPI_ISL_537607

| | | | |
|---|---|---|---|
| see above | UCLA Pathology Clinical Microbiology Lab | Kruglyak Lab | Guo et al. |

EPI_ISL_1469864, EPI_ISL_1469865, EPI_ISL_1469884, EPI_ISL_1469885, EPI_ISL_1469886, EPI_ISL_1469888, EPI_ISL_1469892, EPI_ISL_1469893, EPI_ISL_1469898, EPI_ISL_1469902, EPI_ISL_1469905, EPI_ISL_1469906, EPI_ISL_1469914, EPI_ISL_1469929, EPI_ISL_1921119, EPI_ISL_1921121, EPI_ISL_1921129, EPI_ISL_1921133, EPI_ISL_1921136, EPI_ISL_1921138, EPI_ISL_1921140, EPI_ISL_1921149, EPI_ISL_1921150, EPI_ISL_1921160, EPI_ISL_1921161, EPI_ISL_1921163, EPI_ISL_1921164, EPI_ISL_2031792, EPI_ISL_2031812, EPI_ISL_2031816, EPI_ISL_2031818, EPI_ISL_2031820, EPI_ISL_2031821, EPI_ISL_2031826, EPI_ISL_2031828, EPI_ISL_2031833, EPI_ISL_2031835, EPI_ISL_2031840, EPI_ISL_2031841, EPI_ISL_2031844, EPI_ISL_2081235, EPI_ISL_2081243, EPI_ISL_2081251, EPI_ISL_2081252, EPI_ISL_2081258, EPI_ISL_2081259, EPI_ISL_2081263, EPI_ISL_2081264, EPI_ISL_2081267, EPI_ISL_2081268, EPI_ISL_2081273, EPI_ISL_2081291, EPI_ISL_2081539, EPI_ISL_2081545, EPI_ISL_2081553, EPI_ISL_2081554, EPI_ISL_2081555, EPI_ISL_2081556, EPI_ISL_2081559, EPI_ISL_2081564, EPI_ISL_2081565, EPI_ISL_2081603, EPI_ISL_2082939, EPI_ISL_2082940, EPI_ISL_2082941, EPI_ISL_2082946, EPI_ISL_2082953, EPI_ISL_2082954, EPI_ISL_2082955, EPI_ISL_2082956, EPI_ISL_2082979, EPI_ISL_2082980, EPI_ISL_2082988, EPI_ISL_2082989, EPI_ISL_2082990, EPI_ISL_2082993, EPI_ISL_2083004, EPI_ISL_2083005, EPI_ISL_2083011, EPI_ISL_2083012, EPI_ISL_2083013, EPI_ISL_2083014, EPI_ISL_2179219, EPI_ISL_2179254, EPI_ISL_2179255, EPI_ISL_2179256, EPI_ISL_2179257, EPI_ISL_2179258, EPI_ISL_2179259, EPI_ISL_2179260, EPI_ISL_2179263, EPI_ISL_2295468, EPI_ISL_2295478, EPI_ISL_2295479, EPI_ISL_2295484, EPI_ISL_2295487, EPI_ISL_2295488, EPI_ISL_2295489, EPI_ISL_2295496, EPI_ISL_2295507, EPI_ISL_2295515, EPI_ISL_2295516, EPI_ISL_2378761, EPI_ISL_2378814, EPI_ISL_2494652, EPI_ISL_2494653, EPI_ISL_2494654, EPI_ISL_2494666, EPI_ISL_2494667, EPI_ISL_2494668, EPI_ISL_2494674

| | | | |
|---|---|---|---|
| see above | UCSC Genomics Institute | UCSC Genomics Institute | A. Marm Kilpatrick; Angie Hinrichs; Beth Shapiro; Bryan Thornlow; Ciara Wanket; David Haussler; Eric Beraut; Hugh Olsen; Ikenna Anigbogu; Isabel Bjork; Jakob McBroome; Jeremy Sanford; Joshua Kapp; Mark Akeson; Maximilian |

| | | | |
|---|---|---|---|
| | | | Haeussler; Michael Stone; Miten Jain; Molly Cassatt-Johnstone; Namrita Dhillon; Russell Corbett-Detig; Terren Chang; Yatish Turakhia |
| EPI_ISL_755137, EPI_ISL_755197, EPI_ISL_755215 | UCSD EXCITE lab | Andersen lab at Scripps Research | SEARCH Alliance San Diego |
| EPI_ISL_417931, EPI_ISL_417932, EPI_ISL_417933, EPI_ISL_417935, EPI_ISL_417937, EPI_ISL_417938, EPI_ISL_417939, EPI_ISL_428990, EPI_ISL_428991, EPI_ISL_428992, EPI_ISL_428993, EPI_ISL_428994, EPI_ISL_428995, EPI_ISL_428996, EPI_ISL_428997, EPI_ISL_428998, EPI_ISL_428999, EPI_ISL_429000, EPI_ISL_429001, EPI_ISL_429002, EPI_ISL_429003, EPI_ISL_429004, EPI_ISL_429005, EPI_ISL_429006, EPI_ISL_429007, EPI_ISL_429008, EPI_ISL_429009, EPI_ISL_429010, EPI_ISL_429011, EPI_ISL_429012, EPI_ISL_429013, EPI_ISL_429014, EPI_ISL_429015, EPI_ISL_429016, EPI_ISL_429017, EPI_ISL_429018, EPI_ISL_429019, EPI_ISL_429020, EPI_ISL_429021, EPI_ISL_429022, EPI_ISL_429023, EPI_ISL_429024, EPI_ISL_429025, EPI_ISL_429026, EPI_ISL_429027, EPI_ISL_429028, EPI_ISL_429029, EPI_ISL_429030, EPI_ISL_429031, EPI_ISL_429032, EPI_ISL_429033, EPI_ISL_429034, EPI_ISL_429035, EPI_ISL_429036, EPI_ISL_429037, EPI_ISL_429038, EPI_ISL_429039, EPI_ISL_429040, EPI_ISL_429041, EPI_ISL_429042, EPI_ISL_429043, EPI_ISL_429044, EPI_ISL_429045, EPI_ISL_429046, EPI_ISL_429047, EPI_ISL_429048, EPI_ISL_429049, EPI_ISL_429050, EPI_ISL_429051, EPI_ISL_429052, EPI_ISL_429053, EPI_ISL_429054, EPI_ISL_429055, EPI_ISL_429056, EPI_ISL_429057, EPI_ISL_429058, EPI_ISL_429059, EPI_ISL_429060, EPI_ISL_429061, EPI_ISL_429062, EPI_ISL_429063, EPI_ISL_429064, EPI_ISL_429065, EPI_ISL_429066, EPI_ISL_429067, EPI_ISL_429068, EPI_ISL_429069, EPI_ISL_429070, EPI_ISL_429071, EPI_ISL_429072, EPI_ISL_429073, EPI_ISL_430791, EPI_ISL_430792, EPI_ISL_444051, EPI_ISL_444052, EPI_ISL_444053, EPI_ISL_444054, EPI_ISL_444055, EPI_ISL_444056, EPI_ISL_444057, EPI_ISL_444058, EPI_ISL_444059, EPI_ISL_444060, EPI_ISL_444061, EPI_ISL_444062, EPI_ISL_444063, EPI_ISL_444064, EPI_ISL_444065, EPI_ISL_444066, EPI_ISL_444067, EPI_ISL_444068, EPI_ISL_444069, EPI_ISL_444070, EPI_ISL_444071, EPI_ISL_444072, EPI_ISL_444073, EPI_ISL_444074, EPI_ISL_444075, EPI_ISL_444076, EPI_ISL_444077, EPI_ISL_444078, EPI_ISL_445169, EPI_ISL_445170, EPI_ISL_445171, EPI_ISL_445172, EPI_ISL_445173, EPI_ISL_445174, EPI_ISL_445175, EPI_ISL_445176, EPI_ISL_445177, EPI_ISL_445178, EPI_ISL_445179, EPI_ISL_445180, EPI_ISL_445181, EPI_ISL_445182, EPI_ISL_454614, EPI_ISL_454615, EPI_ISL_454616, EPI_ISL_454617, EPI_ISL_454618, EPI_ISL_454619, EPI_ISL_454620, EPI_ISL_454621, EPI_ISL_454622, EPI_ISL_454623, EPI_ISL_454624, EPI_ISL_454625, EPI_ISL_454626, EPI_ISL_454627, EPI_ISL_454628, EPI_ISL_454629, EPI_ISL_454630, EPI_ISL_454631, EPI_ISL_454632, EPI_ISL_454633, EPI_ISL_454634, EPI_ISL_486339, EPI_ISL_486340, EPI_ISL_486341, EPI_ISL_486342, EPI_ISL_486343, EPI_ISL_486344, EPI_ISL_486345, EPI_ISL_486346, EPI_ISL_486347, EPI_ISL_486348, EPI_ISL_486349, EPI_ISL_486350, EPI_ISL_486351, EPI_ISL_486352, EPI_ISL_486353, EPI_ISL_486354, EPI_ISL_486355, EPI_ISL_486356, EPI_ISL_486357, EPI_ISL_486358, EPI_ISL_486359, EPI_ISL_486360, EPI_ISL_486361, EPI_ISL_486362, EPI_ISL_486363, EPI_ISL_486364, EPI_ISL_486365, EPI_ISL_513883, EPI_ISL_513884, EPI_ISL_513885, EPI_ISL_513886, EPI_ISL_513887, EPI_ISL_513888, EPI_ISL_513889, EPI_ISL_513890, EPI_ISL_513891, EPI_ISL_513892, EPI_ISL_513893, EPI_ISL_513894, EPI_ISL_513895, EPI_ISL_513896, EPI_ISL_513897, EPI_ISL_513898, EPI_ISL_513899, EPI_ISL_513900, EPI_ISL_513901, EPI_ISL_513902, EPI_ISL_513903, EPI_ISL_513904, EPI_ISL_513905, EPI_ISL_513906, EPI_ISL_513907, EPI_ISL_513908, EPI_ISL_513909, EPI_ISL_513910, EPI_ISL_516723, EPI_ISL_516724, EPI_ISL_516725, EPI_ISL_516726, EPI_ISL_516727, EPI_ISL_516728, EPI_ISL_516729, EPI_ISL_516730, EPI_ISL_516731, EPI_ISL_516732, EPI_ISL_516733, EPI_ISL_516734, EPI_ISL_516735, EPI_ISL_516736, EPI_ISL_516737, EPI_ISL_516738, EPI_ISL_516739, EPI_ISL_516740, EPI_ISL_516741, EPI_ISL_516742, EPI_ISL_516743, EPI_ISL_516744, EPI_ISL_516745, EPI_ISL_516746, EPI_ISL_516747, EPI_ISL_548505, EPI_ISL_548513, EPI_ISL_548522, EPI_ISL_548555, EPI_ISL_548571, EPI_ISL_583199, EPI_ISL_583200, EPI_ISL_583201, EPI_ISL_583202, EPI_ISL_583203, EPI_ISL_583204, EPI_ISL_583205, EPI_ISL_583206, EPI_ISL_583207, EPI_ISL_583208, EPI_ISL_583209, EPI_ISL_583210, EPI_ISL_583211, EPI_ISL_583212, EPI_ISL_583213, EPI_ISL_583214, EPI_ISL_583215, EPI_ISL_583216, EPI_ISL_583217, EPI_ISL_583218, EPI_ISL_583219, EPI_ISL_583220, EPI_ISL_583221, EPI_ISL_583222, EPI_ISL_583223, EPI_ISL_583224, EPI_ISL_583225, EPI_ISL_583226, EPI_ISL_583227, EPI_ISL_583228, EPI_ISL_583229, EPI_ISL_583230, EPI_ISL_583231, EPI_ISL_625550, EPI_ISL_625551, EPI_ISL_625552, EPI_ISL_625553, EPI_ISL_625554, EPI_ISL_625555, EPI_ISL_625556, EPI_ISL_625557, EPI_ISL_625558, EPI_ISL_625661, EPI_ISL_625662, EPI_ISL_625663, EPI_ISL_625664, EPI_ISL_625665, EPI_ISL_625666, EPI_ISL_625667, EPI_ISL_625668, EPI_ISL_625669, EPI_ISL_625670, EPI_ISL_648543, EPI_ISL_648544, EPI_ISL_672015, EPI_ISL_672016, EPI_ISL_672018, EPI_ISL_672020, EPI_ISL_672021, EPI_ISL_672025, EPI_ISL_672386, EPI_ISL_672387, EPI_ISL_672388, EPI_ISL_672389, EPI_ISL_672390, EPI_ISL_672391, EPI_ISL_672392, EPI_ISL_672393, EPI_ISL_672426, EPI_ISL_672427, EPI_ISL_672428, EPI_ISL_672490, EPI_ISL_672491, EPI_ISL_672492, EPI_ISL_672493, EPI_ISL_672494, EPI_ISL_672495, EPI_ISL_672496, EPI_ISL_672497, EPI_ISL_672498, EPI_ISL_672499, EPI_ISL_672500, EPI_ISL_672501, EPI_ISL_738544, EPI_ISL_739004, EPI_ISL_739203, EPI_ISL_739295, EPI_ISL_739613, EPI_ISL_739632, EPI_ISL_739633, EPI_ISL_2658587, EPI_ISL_2658660, EPI_ISL_2658661, EPI_ISL_2659468, EPI_ISL_2659537, EPI_ISL_2941520 | | | |
| see above | UCSF Clinical Microbiology Laboratory | Chan-Zuckerberg Biohub | Amy Kistler; Angela Detweiler; Angela Pisco; CZB Cliahub Consortium; Chaz Langelier; Chunyu Zhao; David Dynerman; G. Renuka Kumar; Gloria Castaneda; Jack Kamm; Josh Batson; Lucy Li; Lusajo Mwakibete; Maira Phelps; Michelle Tan; Norma Neff; Olga Botvinnik; Samantha Hao; Shaun Arevalo; Steve Miller |
| EPI_ISL_450232, EPI_ISL_450233, EPI_ISL_450234, EPI_ISL_450235, EPI_ISL_450236, EPI_ISL_450237, EPI_ISL_450238, EPI_ISL_450239, EPI_ISL_450240 | | | |
| see above | UCSF Clinical Microbiology Laboratory | Chiu Laboratory, University of California, San Francisco | Scot Federman; Wei Gu; Xianding Deng; and Charles Y. Chiu |
| EPI_ISL_765994, EPI_ISL_765995, EPI_ISL_765996, EPI_ISL_765997, EPI_ISL_766015, EPI_ISL_939619, EPI_ISL_939620, EPI_ISL_939621, EPI_ISL_939622, EPI_ISL_939623, EPI_ISL_939624, EPI_ISL_939625, EPI_ISL_1005491, EPI_ISL_1005492, EPI_ISL_1005493, EPI_ISL_1005494, EPI_ISL_1005558, EPI_ISL_1040167, EPI_ISL_1040168, EPI_ISL_1040169, EPI_ISL_1040170, EPI_ISL_1040171, EPI_ISL_1040172, EPI_ISL_1040173, EPI_ISL_1040174, EPI_ISL_1040175, EPI_ISL_1040176, EPI_ISL_1040177, EPI_ISL_1040178, EPI_ISL_1040179, EPI_ISL_1040180, EPI_ISL_1040427, EPI_ISL_1040428, EPI_ISL_1040429, EPI_ISL_1040430, EPI_ISL_1040431, EPI_ISL_1040432, EPI_ISL_1040433, EPI_ISL_1040434, EPI_ISL_1040435, EPI_ISL_1040436, EPI_ISL_1040437, EPI_ISL_1192367, EPI_ISL_1201558, EPI_ISL_2405186, EPI_ISL_2405187, EPI_ISL_2405188, EPI_ISL_2405189, EPI_ISL_2405190, EPI_ISL_2405191, EPI_ISL_2405192, EPI_ISL_2405193, EPI_ISL_2405194, EPI_ISL_2405195, EPI_ISL_2405196, EPI_ISL_2405197, EPI_ISL_2405198, EPI_ISL_2405199, EPI_ISL_2405200, EPI_ISL_2424167, EPI_ISL_2424174, EPI_ISL_2424176, EPI_ISL_2424184, EPI_ISL_2424185, EPI_ISL_2424186, EPI_ISL_2424187, EPI_ISL_2424213, EPI_ISL_2424214, EPI_ISL_2424215, EPI_ISL_2424216, EPI_ISL_2424217, EPI_ISL_2424218, EPI_ISL_2424219, EPI_ISL_2424220, EPI_ISL_2424221, EPI_ISL_2424222, EPI_ISL_2424223, EPI_ISL_2424224, EPI_ISL_2424225, EPI_ISL_2424226, EPI_ISL_2424227, EPI_ISL_2424228, EPI_ISL_2424229, EPI_ISL_2424230, EPI_ISL_2424231, EPI_ISL_2424232, EPI_ISL_2424233, EPI_ISL_2617827, EPI_ISL_2617828, EPI_ISL_2617829, EPI_ISL_2617830, EPI_ISL_2617831, EPI_ISL_2617832, EPI_ISL_2617833, EPI_ISL_2617834, EPI_ISL_2617835, EPI_ISL_2617836, EPI_ISL_2617837, EPI_ISL_2617838, EPI_ISL_2617839, EPI_ISL_2617840, EPI_ISL_2617841, EPI_ISL_2617842, EPI_ISL_2617843, EPI_ISL_2617844, EPI_ISL_2617845, EPI_ISL_2617846, EPI_ISL_2617847, EPI_ISL_2617848, EPI_ISL_2617849, EPI_ISL_2617850, EPI_ISL_2617851, EPI_ISL_2617852, EPI_ISL_2617853, EPI_ISL_2617855, EPI_ISL_2617856, EPI_ISL_2617857, EPI_ISL_2617858, EPI_ISL_2617859, EPI_ISL_2617860, EPI_ISL_2617861, EPI_ISL_2617862, EPI_ISL_2617863, EPI_ISL_2617864, EPI_ISL_2617865, EPI_ISL_2617866, EPI_ISL_2617867, EPI_ISL_2617868, EPI_ISL_2617869, EPI_ISL_2617870, EPI_ISL_2617871, EPI_ISL_2617872, EPI_ISL_2617873, EPI_ISL_2617874, EPI_ISL_2617960, EPI_ISL_2617961, EPI_ISL_2617962, EPI_ISL_2617963, EPI_ISL_2617964, EPI_ISL_2617965, EPI_ISL_2617966, EPI_ISL_2620795, EPI_ISL_2620796, EPI_ISL_2620797, EPI_ISL_2620798, EPI_ISL_2620799, EPI_ISL_2620800 | | | |
| see above | USC Clinical Lab | Los Angeles County Public Health Laboratories | P. Hemarajata el al.; P. Hemarajata et al. |
| EPI_ISL_2790456, EPI_ISL_2790457, EPI_ISL_2790578, EPI_ISL_2791153 | UW Virology Lab | UW Virology Lab | Alexander Greninger; Hong Xie; Keith R Jerome; Lasata Shrestha; Margaret Mills; Meei-Li Huang; Noah Baker; Pavitra Roychoudhury; Sean Ellis; Shah Mohamed Bakhash |
| EPI_ISL_812311, EPI_ISL_812312, EPI_ISL_812530, EPI_ISL_812537, EPI_ISL_812687 | United States Air Force School of Aerospace Medicine | United States Air Force School of Aerospace Medicine | Amanda Javorina; Anthony Fries; Clarise Starr; Elizabeth Macias; Jennifer Meyer; Sarah Purves; William Gruner |
| EPI_ISL_447887, EPI_ISL_447888, EPI_ISL_447889, EPI_ISL_447890, EPI_ISL_447891, EPI_ISL_447892, EPI_ISL_447893, EPI_ISL_447894, EPI_ISL_447895, EPI_ISL_447896, EPI_ISL_548374, EPI_ISL_548380, EPI_ISL_548382, EPI_ISL_548397, EPI_ISL_548414, EPI_ISL_548435, EPI_ISL_548447, EPI_ISL_548452, EPI_ISL_548458, EPI_ISL_548474, EPI_ISL_548477, EPI_ISL_548479, EPI_ISL_548486, EPI_ISL_548490 | | | |
| see above | University of California, Davis | Chan-Zuckerberg Biohub | CZB Cliahub Consortium |
| EPI_ISL_450701, EPI_ISL_484981, EPI_ISL_491300, EPI_ISL_491301, EPI_ISL_491302, EPI_ISL_491303, EPI_ISL_509799, EPI_ISL_536668, EPI_ISL_536678 | | | |
| see above | University of Wisconsin-Madison AIDS Vaccine Research Laboratories | University of Wisconsin-Madison AIDS Vaccine Research Laboratories | Gage Moreno; Katarina Braun; et al. AIDS Vaccine Research Laboratories |
| EPI_ISL_468462, EPI_ISL_468463, EPI_ISL_468464, EPI_ISL_468465, EPI_ISL_468466, EPI_ISL_468467, EPI_ISL_468468, EPI_ISL_468469, EPI_ISL_468470, EPI_ISL_468471, EPI_ISL_468472, EPI_ISL_468473, EPI_ISL_468474, EPI_ISL_468475, EPI_ISL_468476, EPI_ISL_468477, EPI_ISL_468478, EPI_ISL_468479, EPI_ISL_468480, EPI_ISL_468481, EPI_ISL_468482, EPI_ISL_468483, EPI_ISL_468484, EPI_ISL_468485, EPI_ISL_468486, EPI_ISL_468487, EPI_ISL_468488, EPI_ISL_468489, EPI_ISL_468490, EPI_ISL_468491, EPI_ISL_468492, EPI_ISL_468493, EPI_ISL_468494, EPI_ISL_468495, EPI_ISL_468496, EPI_ISL_468497, EPI_ISL_468498, EPI_ISL_468499, EPI_ISL_468500, EPI_ISL_468501, EPI_ISL_468502, EPI_ISL_468503, EPI_ISL_468504, EPI_ISL_468505, EPI_ISL_548346, EPI_ISL_548348, EPI_ISL_548349, EPI_ISL_548352, EPI_ISL_548353, EPI_ISL_548354, EPI_ISL_548355, EPI_ISL_548356, EPI_ISL_548358, EPI_ISL_548362, EPI_ISL_548364, EPI_ISL_548367, EPI_ISL_548368, EPI_ISL_548372, EPI_ISL_548376, EPI_ISL_548390, EPI_ISL_548398, EPI_ISL_548402, EPI_ISL_548406, EPI_ISL_548409, EPI_ISL_548410, EPI_ISL_548427, EPI_ISL_548430, EPI_ISL_548442, EPI_ISL_548443, EPI_ISL_548450, EPI_ISL_548455, EPI_ISL_548459, EPI_ISL_548461, EPI_ISL_548471, EPI_ISL_548473, EPI_ISL_548475, EPI_ISL_548476, EPI_ISL_548480, EPI_ISL_548481, EPI_ISL_548483, EPI_ISL_548487, EPI_ISL_548489, EPI_ISL_1924592, EPI_ISL_2659465 | | | |
| see above | Ventura County Public Health Lab | Chan-Zuckerberg Biohub | CZB Cliahub Consortium |
| EPI_ISL_2101779, EPI_ISL_2101780, EPI_ISL_2101781, EPI_ISL_2101782, EPI_ISL_2101786, EPI_ISL_2101799, EPI_ISL_2101802, EPI_ISL_2101803, EPI_ISL_2101804, EPI_ISL_2101805, EPI_ISL_2101806, EPI_ISL_2101807, EPI_ISL_2101808, EPI_ISL_2101809, EPI_ISL_2101810, EPI_ISL_2101811, EPI_ISL_2101812, EPI_ISL_2101813, EPI_ISL_2101814, EPI_ISL_2101815, EPI_ISL_2101816, EPI_ISL_2101817, EPI_ISL_2101818, EPI_ISL_2101819, EPI_ISL_2101820, EPI_ISL_2101821, EPI_ISL_2101822, EPI_ISL_2101823, EPI_ISL_2101824, EPI_ISL_2101825, EPI_ISL_2101826, EPI_ISL_2101827, EPI_ISL_2101828, EPI_ISL_2101829, EPI_ISL_2101830, EPI_ISL_2101831, EPI_ISL_2101832, EPI_ISL_2101833, EPI_ISL_2101834, EPI_ISL_2101835, EPI_ISL_2101836, EPI_ISL_2101837, EPI_ISL_2101838, EPI_ISL_2101839, EPI_ISL_2101840, EPI_ISL_2101841, EPI_ISL_2101842, EPI_ISL_2101843, EPI_ISL_2101844, EPI_ISL_2101845, EPI_ISL_2101846, EPI_ISL_2101847, EPI_ISL_2101848, EPI_ISL_2101849, EPI_ISL_2101850, EPI_ISL_2101851, EPI_ISL_2101852, EPI_ISL_2101853, EPI_ISL_2101854, EPI_ISL_2101855, EPI_ISL_2101856, EPI_ISL_2101857, EPI_ISL_2101858, EPI_ISL_2101859, EPI_ISL_2101860, EPI_ISL_2101861, EPI_ISL_2101862, EPI_ISL_2101863, EPI_ISL_2101864, EPI_ISL_2101865, EPI_ISL_2101866, EPI_ISL_2101867, EPI_ISL_2101868, EPI_ISL_2246783, EPI_ISL_2246784, EPI_ISL_2246785, EPI_ISL_2246786 | | | |
| see above | WLA VAMC | Pathogen Discovery, Respiratory Viruses Branch, Division of Viral Diseases, Centers for Disease Control and Prevention | Adam Retchless; Anna Kelleher; Anna Uehara; Brian Lynch; Clinton R. Paden; Dhwani Batra; Haibin Wang; Han Jia Justin Ng; Jasmine Padilla; Jing Zhang; Justin Lee; Krista Queen; Mark Burroughs; Mili Sheth; Morgan Davis; Peter Cook; Rachel Marine; Sarah Nobles; Suxiang Tong; Tara Coalter; Yan Li; Ying Tao |
| EPI_ISL_497341 | Washington State Department of Health | Seattle Flu Study | Amanda Adler; Barry R. Lutz; Benjamin Pelle; Brian Hiatt; Caitlin R. Wolf; Chris D. Frazar; Deborah A. Nickerson; Elisabeth Brandstetter; Geoff Melly; Helen Y. Chu; Janet A. Englund; Jay Shendure; Jover Lee; Kairsten Fay; Kirsten Lacombe; Lea M. Starita; Mark J. Rieder; Matthew Richardson; Matthew Thompson; Melissa Truong; Michael Boeckh; Michael Famulare; Misja Ilcisin; Peter D. Han; Philip Dykema; Romesh Gautom; Scott Lindquist; Thomas R. Sibley; Trevor Bedford |
| EPI_ISL_450484, EPI_ISL_450485, EPI_ISL_450486, EPI_ISL_450487 | unknown | Data Science | C.J.; Carroll; Cohen; Miller; N.K.; S.H.; T.D.; Tran |
| EPI_ISL_447886 | unknown | Pathogen Discovery | Anna Uehara; Clinton R. Paden; Haibin Wang; Jing Zhang; Julu Bhatnagar; Krista Queen; Suxiang Tong; Yan Li; Ying Tao |

We gratefully acknowledge the following Authors from the Originating laboratories responsible for obtaining the specimens, as well as the Submitting laboratories where the genome data were generated and shared via GISAID, on which this research is based.

All Submitters of data may be contacted directly via www.gisaid.org

Authors are sorted alphabetically.

| Accession ID | Originating Laboratory | Submitting Laboratory | Authors |
|---|---|---|---|
| EPI_ISL_1225963, EPI_ISL_1225964, EPI_ISL_1225965, EPI_ISL_1225966 | "CDPH, Viral and Rickettsial Disease Laboratory" | Genomics and Discovery, Respiratory Viruses Branch, Division of Viral Diseases, Centers for Disease Control and Prevention | Anna Montmayeur; Anna Uehara; Ben L. Rambo-Martin; Clinton R. Paden; Dhwani Batra; Haibin Wang; Jasmine Padilla; Jing Zhang; Justin Lee; Katie Dillon; Krista Queen; Kristen Knipe; Kristine Lacek; Lori Rowe; Mark Burroughs; Matthew Schmerer; Mili Sheth; Peter W. Cook; Rachel Marine; Sam Shepard; Sarah Nobles; Shoshona Le; Suxiang Tong; Yan Li; Ying Tao |

EPI_ISL_1093547, EPI_ISL_1093548, EPI_ISL_1093549, EPI_ISL_1093550, EPI_ISL_1093553, EPI_ISL_1093554, EPI_ISL_1093555, EPI_ISL_1093556, EPI_ISL_1093563, EPI_ISL_1093565, EPI_ISL_1093566, EPI_ISL_1093568, EPI_ISL_1093571, EPI_ISL_1093573, EPI_ISL_1093574, EPI_ISL_1093575, EPI_ISL_1093576, EPI_ISL_1093577, EPI_ISL_1093578, EPI_ISL_1093579, EPI_ISL_1093580, EPI_ISL_1093581, EPI_ISL_1093582, EPI_ISL_1093583, EPI_ISL_1093584, EPI_ISL_1093585, EPI_ISL_1093586, EPI_ISL_1093587, EPI_ISL_1093588, EPI_ISL_1093589, EPI_ISL_1093590, EPI_ISL_1093591, EPI_ISL_1093592

| | | | |
|---|---|---|---|
| see above | Alameda County Public Health Department | Alameda County Public Health Department | Kevin Libuit; Kristna Hsieh; Maria Elma |

EPI_ISL_738629, EPI_ISL_738802, EPI_ISL_738895, EPI_ISL_738925, EPI_ISL_739090, EPI_ISL_739209, EPI_ISL_739215, EPI_ISL_739351, EPI_ISL_739606, EPI_ISL_955386, EPI_ISL_955387, EPI_ISL_955388, EPI_ISL_955389, EPI_ISL_955390, EPI_ISL_1027296, EPI_ISL_1027303, EPI_ISL_1027307, EPI_ISL_1027308, EPI_ISL_1027309, EPI_ISL_1027323, EPI_ISL_1027324, EPI_ISL_1027330, EPI_ISL_1027333, EPI_ISL_1027336, EPI_ISL_1027341, EPI_ISL_1027345, EPI_ISL_1027348, EPI_ISL_1027361, EPI_ISL_1027363, EPI_ISL_1027365, EPI_ISL_1027445, EPI_ISL_1027446, EPI_ISL_1027447, EPI_ISL_1027448, EPI_ISL_1027449, EPI_ISL_1027450, EPI_ISL_1027451, EPI_ISL_1027452, EPI_ISL_1027453, EPI_ISL_1027454, EPI_ISL_1027455, EPI_ISL_1027456, EPI_ISL_1027457, EPI_ISL_1027458, EPI_ISL_1027459, EPI_ISL_1027460, EPI_ISL_1027461, EPI_ISL_1027462, EPI_ISL_1027463, EPI_ISL_1027464, EPI_ISL_1027465, EPI_ISL_1027466, EPI_ISL_1027467, EPI_ISL_1027468, EPI_ISL_1027469, EPI_ISL_1027470, EPI_ISL_1027471, EPI_ISL_1027472, EPI_ISL_1027473, EPI_ISL_1027543, EPI_ISL_1027560, EPI_ISL_1184624, EPI_ISL_1184625, EPI_ISL_1184626, EPI_ISL_1184627, EPI_ISL_1184628, EPI_ISL_1184629, EPI_ISL_1184630, EPI_ISL_1184631, EPI_ISL_1184632, EPI_ISL_1184633, EPI_ISL_1184634, EPI_ISL_1184635, EPI_ISL_1184636, EPI_ISL_1184637, EPI_ISL_1184638, EPI_ISL_1184639, EPI_ISL_1184640, EPI_ISL_1184641, EPI_ISL_1184642, EPI_ISL_1184643, EPI_ISL_1184644, EPI_ISL_1184645, EPI_ISL_1184646, EPI_ISL_1184647, EPI_ISL_1184648, EPI_ISL_1184649, EPI_ISL_1184650, EPI_ISL_1184651, EPI_ISL_1184652, EPI_ISL_1184653, EPI_ISL_1184654, EPI_ISL_1184655, EPI_ISL_1184656, EPI_ISL_1184657, EPI_ISL_1184658, EPI_ISL_1184659, EPI_ISL_1184660, EPI_ISL_1184661, EPI_ISL_1184662, EPI_ISL_1184663, EPI_ISL_1184664, EPI_ISL_1184665, EPI_ISL_1184666, EPI_ISL_1184667, EPI_ISL_1184668, EPI_ISL_1184669, EPI_ISL_1184670, EPI_ISL_1184671, EPI_ISL_1184672, EPI_ISL_1184673, EPI_ISL_1184674, EPI_ISL_1184675, EPI_ISL_1184676, EPI_ISL_1184677, EPI_ISL_1184678, EPI_ISL_1184679, EPI_ISL_1184680, EPI_ISL_1184861, EPI_ISL_1184862, EPI_ISL_1184863, EPI_ISL_1184864, EPI_ISL_1184865, EPI_ISL_1184866, EPI_ISL_1184867, EPI_ISL_1184868, EPI_ISL_1184869, EPI_ISL_1184870, EPI_ISL_1184871, EPI_ISL_1184872, EPI_ISL_1184873, EPI_ISL_1184874, EPI_ISL_1184875, EPI_ISL_1184876, EPI_ISL_1184877, EPI_ISL_1184878, EPI_ISL_1184879, EPI_ISL_1184880, EPI_ISL_1184881, EPI_ISL_1184882, EPI_ISL_1184883, EPI_ISL_1184884, EPI_ISL_1184885, EPI_ISL_1184886, EPI_ISL_1184887, EPI_ISL_1184888, EPI_ISL_1184889, EPI_ISL_1184890, EPI_ISL_1184891, EPI_ISL_1184892, EPI_ISL_1184893, EPI_ISL_1184894, EPI_ISL_1184895, EPI_ISL_1184896, EPI_ISL_1184897, EPI_ISL_1184898, EPI_ISL_1184899, EPI_ISL_1184900, EPI_ISL_1184901, EPI_ISL_1184902, EPI_ISL_1184903, EPI_ISL_1184904, EPI_ISL_1184905, EPI_ISL_1184906, EPI_ISL_1184907, EPI_ISL_1184908, EPI_ISL_1184909, EPI_ISL_1184910, EPI_ISL_1184911, EPI_ISL_1184912, EPI_ISL_1184913, EPI_ISL_1184914, EPI_ISL_1184915, EPI_ISL_1184916, EPI_ISL_1184917, EPI_ISL_1184918, EPI_ISL_1184919, EPI_ISL_1184920, EPI_ISL_1184921, EPI_ISL_1184922, EPI_ISL_1234542, EPI_ISL_1234543, EPI_ISL_1234562, EPI_ISL_1234564, EPI_ISL_1234565, EPI_ISL_1234566, EPI_ISL_1234567, EPI_ISL_1234568, EPI_ISL_1234574, EPI_ISL_1234597, EPI_ISL_1234600, EPI_ISL_1234603, EPI_ISL_1234604, EPI_ISL_1234611, EPI_ISL_1234612, EPI_ISL_1234617, EPI_ISL_1234621, EPI_ISL_1234637, EPI_ISL_1234640, EPI_ISL_1234646, EPI_ISL_1234648, EPI_ISL_1234661, EPI_ISL_1234665, EPI_ISL_1234666, EPI_ISL_1234670, EPI_ISL_1234672, EPI_ISL_1234676, EPI_ISL_1234677, EPI_ISL_1234679, EPI_ISL_1234683, EPI_ISL_1234698, EPI_ISL_1234701, EPI_ISL_1234716, EPI_ISL_1234718, EPI_ISL_1234720, EPI_ISL_1234723, EPI_ISL_1234732, EPI_ISL_1234739, EPI_ISL_1234742, EPI_ISL_1234743, EPI_ISL_1234751, EPI_ISL_1234755, EPI_ISL_1234758, EPI_ISL_1234762, EPI_ISL_1234765, EPI_ISL_1234767, EPI_ISL_1234773, EPI_ISL_1234776, EPI_ISL_1234779, EPI_ISL_1234782, EPI_ISL_1234785, EPI_ISL_1234790, EPI_ISL_1234795, EPI_ISL_1234802, EPI_ISL_1234808, EPI_ISL_1234809, EPI_ISL_1234815, EPI_ISL_1234819, EPI_ISL_1234820, EPI_ISL_1234827, EPI_ISL_1234829, EPI_ISL_1234830, EPI_ISL_1234832, EPI_ISL_1234834, EPI_ISL_1234835, EPI_ISL_1234839, EPI_ISL_1234844, EPI_ISL_1234848, EPI_ISL_1234858, EPI_ISL_1234860, EPI_ISL_1234862, EPI_ISL_1234870, EPI_ISL_1234872, EPI_ISL_1234873, EPI_ISL_1234877, EPI_ISL_1234892, EPI_ISL_1234894, EPI_ISL_1234895, EPI_ISL_1234900, EPI_ISL_1234901, EPI_ISL_1234904, EPI_ISL_1234907, EPI_ISL_1234920, EPI_ISL_1234921, EPI_ISL_1234924, EPI_ISL_1234932, EPI_ISL_1234933, EPI_ISL_1234939, EPI_ISL_1234941, EPI_ISL_1234950, EPI_ISL_1234952, EPI_ISL_1234953, EPI_ISL_1234956, EPI_ISL_1234959, EPI_ISL_1234968, EPI_ISL_1234969, EPI_ISL_1234979, EPI_ISL_1234982, EPI_ISL_1234986, EPI_ISL_1234987, EPI_ISL_1234990, EPI_ISL_1234995, EPI_ISL_1235006, EPI_ISL_1235013, EPI_ISL_1235021, EPI_ISL_1235022, EPI_ISL_1235025, EPI_ISL_1235028, EPI_ISL_1235032, EPI_ISL_1235042, EPI_ISL_1235048, EPI_ISL_1235054, EPI_ISL_1235059, EPI_ISL_1235075, EPI_ISL_1235077, EPI_ISL_1235083, EPI_ISL_1235091, EPI_ISL_1235093, EPI_ISL_1235094, EPI_ISL_1235098, EPI_ISL_1235099, EPI_ISL_1235102, EPI_ISL_1235111, EPI_ISL_1235115, EPI_ISL_1235116, EPI_ISL_1235122, EPI_ISL_1235124, EPI_ISL_1235141, EPI_ISL_1235142, EPI_ISL_1235147, EPI_ISL_1235149, EPI_ISL_1235158, EPI_ISL_1235165, EPI_ISL_1235171, EPI_ISL_1235175, EPI_ISL_1235178, EPI_ISL_1235181, EPI_ISL_1235183, EPI_ISL_1235185, EPI_ISL_1235188, EPI_ISL_1235194, EPI_ISL_1235198, EPI_ISL_1235210, EPI_ISL_1235213, EPI_ISL_1235217, EPI_ISL_1235219, EPI_ISL_1235223, EPI_ISL_1235230, EPI_ISL_1235235, EPI_ISL_1235241, EPI_ISL_1235243, EPI_ISL_1235244, EPI_ISL_1235249, EPI_ISL_1235255, EPI_ISL_1235256, EPI_ISL_1235263, EPI_ISL_1235270, EPI_ISL_1235276, EPI_ISL_1235279, EPI_ISL_1235281, EPI_ISL_1235289, EPI_ISL_1235292, EPI_ISL_1235294, EPI_ISL_1235299, EPI_ISL_1235303, EPI_ISL_1235308, EPI_ISL_1235309, EPI_ISL_1235312, EPI_ISL_1235313, EPI_ISL_1235314, EPI_ISL_1235318, EPI_ISL_1235335, EPI_ISL_1235338, EPI_ISL_1235339, EPI_ISL_1235359, EPI_ISL_1235368, EPI_ISL_1235369, EPI_ISL_1235380, EPI_ISL_1235387, EPI_ISL_1235388, EPI_ISL_1235391, EPI_ISL_1235392, EPI_ISL_1235404, EPI_ISL_1235405, EPI_ISL_1235406, EPI_ISL_1235407, EPI_ISL_1235414, EPI_ISL_1235440, EPI_ISL_1235448, EPI_ISL_1235459, EPI_ISL_1235471, EPI_ISL_1235477, EPI_ISL_1235480, EPI_ISL_1235482, EPI_ISL_1235485, EPI_ISL_1235486, EPI_ISL_1235492, EPI_ISL_1235495, EPI_ISL_1235497, EPI_ISL_1235499, EPI_ISL_1235503, EPI_ISL_1235504, EPI_ISL_1235505, EPI_ISL_1235512, EPI_ISL_1235516, EPI_ISL_1235520, EPI_ISL_1235524, EPI_ISL_1235529, EPI_ISL_1235532, EPI_ISL_1235537, EPI_ISL_1235543, EPI_ISL_1235550, EPI_ISL_1235552, EPI_ISL_1235554, EPI_ISL_1235557, EPI_ISL_1235559, EPI_ISL_1235562, EPI_ISL_1235564, EPI_ISL_1235566, EPI_ISL_1235567, EPI_ISL_1235573, EPI_ISL_1235574, EPI_ISL_1235575, EPI_ISL_1235579, EPI_ISL_1235580, EPI_ISL_1235586, EPI_ISL_1235589, EPI_ISL_1235590, EPI_ISL_1235592, EPI_ISL_1235597, EPI_ISL_1235598, EPI_ISL_1235599, EPI_ISL_1235605, EPI_ISL_1235610, EPI_ISL_1477170, EPI_ISL_1477175, EPI_ISL_1477178, EPI_ISL_1477199, EPI_ISL_1477216, EPI_ISL_1477228, EPI_ISL_1477243, EPI_ISL_1477244, EPI_ISL_1477251, EPI_ISL_1477252, EPI_ISL_1477259, EPI_ISL_1477260, EPI_ISL_1477268, EPI_ISL_1477270, EPI_ISL_1477271, EPI_ISL_1477279, EPI_ISL_1477287, EPI_ISL_1477288, EPI_ISL_1477292, EPI_ISL_1477293, EPI_ISL_1477295, EPI_ISL_1477301, EPI_ISL_1477308, EPI_ISL_1477309, EPI_ISL_1477313, EPI_ISL_1477318, EPI_ISL_1477322, EPI_ISL_1477328, EPI_ISL_1477329, EPI_ISL_1477337, EPI_ISL_1477343, EPI_ISL_1477352, EPI_ISL_1477359, EPI_ISL_1477364, EPI_ISL_1477367, EPI_ISL_1477371, EPI_ISL_1477373, EPI_ISL_1477377, EPI_ISL_1477380, EPI_ISL_1477382, EPI_ISL_1477385, EPI_ISL_1477388, EPI_ISL_1477405, EPI_ISL_1477406, EPI_ISL_1477410, EPI_ISL_1477411, EPI_ISL_1477413, EPI_ISL_1477414, EPI_ISL_1477415, EPI_ISL_1477432, EPI_ISL_1477433, EPI_ISL_1477435, EPI_ISL_1477668, EPI_ISL_1477669, EPI_ISL_1477671, EPI_ISL_1477672, EPI_ISL_1477674, EPI_ISL_1477683, EPI_ISL_1477684, EPI_ISL_1477686, EPI_ISL_1477688, EPI_ISL_1477691, EPI_ISL_1477696, EPI_ISL_1477701, EPI_ISL_1477702, EPI_ISL_1477703, EPI_ISL_1477708, EPI_ISL_1477710, EPI_ISL_1477712, EPI_ISL_1477720, EPI_ISL_1477733, EPI_ISL_1477735, EPI_ISL_1477741, EPI_ISL_1477747, EPI_ISL_1477751, EPI_ISL_1477753, EPI_ISL_1477754, EPI_ISL_1477757, EPI_ISL_1477758, EPI_ISL_1477766, EPI_ISL_1477771, EPI_ISL_1477772, EPI_ISL_1477773, EPI_ISL_1477775, EPI_ISL_1477779, EPI_ISL_1477783, EPI_ISL_1477784, EPI_ISL_1477791, EPI_ISL_1477801, EPI_ISL_1477804, EPI_ISL_1477805, EPI_ISL_1477807, EPI_ISL_1477809, EPI_ISL_1477820, EPI_ISL_1477823, EPI_ISL_1477824, EPI_ISL_1477825, EPI_ISL_1477828, EPI_ISL_1477832, EPI_ISL_1477841, EPI_ISL_1477847, EPI_ISL_1477857, EPI_ISL_1477858, EPI_ISL_1477862, EPI_ISL_1477865, EPI_ISL_1477880, EPI_ISL_1477886, EPI_ISL_1477887, EPI_ISL_1477889, EPI_ISL_1477896, EPI_ISL_1477899, EPI_ISL_1477900, EPI_ISL_1477904, EPI_ISL_1477905, EPI_ISL_1477906, EPI_ISL_1477914, EPI_ISL_1477915, EPI_ISL_1477919, EPI_ISL_1477922, EPI_ISL_1477928, EPI_ISL_1477932, EPI_ISL_1477933, EPI_ISL_1477936, EPI_ISL_1477941, EPI_ISL_1477942, EPI_ISL_1477945, EPI_ISL_1477948, EPI_ISL_1663927, EPI_ISL_1663928, EPI_ISL_1663935, EPI_ISL_1663938, EPI_ISL_1663947, EPI_ISL_1663956, EPI_ISL_1663958, EPI_ISL_1663959, EPI_ISL_1663960, EPI_ISL_1663969, EPI_ISL_1663972, EPI_ISL_1663986, EPI_ISL_1663990, EPI_ISL_1664003, EPI_ISL_1664006, EPI_ISL_1664010, EPI_ISL_1664015, EPI_ISL_1664021, EPI_ISL_1664037, EPI_ISL_1664039, EPI_ISL_1664040, EPI_ISL_1664051, EPI_ISL_1664052, EPI_ISL_1664056, EPI_ISL_1664065, EPI_ISL_1664090, EPI_ISL_1664101, EPI_ISL_1664106, EPI_ISL_1701046, EPI_ISL_1701057, EPI_ISL_1701065, EPI_ISL_1701071, EPI_ISL_1701072, EPI_ISL_1701074, EPI_ISL_1701078, EPI_ISL_1701080, EPI_ISL_1701081, EPI_ISL_1701082, EPI_ISL_1701083, EPI_ISL_1701084, EPI_ISL_1701087, EPI_ISL_1701090, EPI_ISL_1701091, EPI_ISL_1701092, EPI_ISL_1701099, EPI_ISL_1701104, EPI_ISL_1701106, EPI_ISL_1701115, EPI_ISL_1701119, EPI_ISL_1701127, EPI_ISL_1701135, EPI_ISL_1701137, EPI_ISL_1701147, EPI_ISL_1701148, EPI_ISL_1701150, EPI_ISL_1701155, EPI_ISL_1701156, EPI_ISL_1701157, EPI_ISL_1701161, EPI_ISL_1701162, EPI_ISL_1701167, EPI_ISL_1701171, EPI_ISL_1701172, EPI_ISL_1701174, EPI_ISL_1701175, EPI_ISL_1701176, EPI_ISL_1701177, EPI_ISL_1701178, EPI_ISL_1701179, EPI_ISL_1701180, EPI_ISL_1701194, EPI_ISL_1701205, EPI_ISL_1701210, EPI_ISL_1701212, EPI_ISL_1701214, EPI_ISL_1701217, EPI_ISL_1701218, EPI_ISL_1701222, EPI_ISL_1701224, EPI_ISL_1701228, EPI_ISL_1701234, EPI_ISL_1701249, EPI_ISL_1701250, EPI_ISL_1701267, EPI_ISL_1701273, EPI_ISL_1701274, EPI_ISL_1701277, EPI_ISL_1701285, EPI_ISL_1701289, EPI_ISL_1701291, EPI_ISL_1701293, EPI_ISL_1701295, EPI_ISL_1701299, EPI_ISL_1701301, EPI_ISL_1701303, EPI_ISL_1701306, EPI_ISL_1701307, EPI_ISL_1701397, EPI_ISL_1701403, EPI_ISL_1701407, EPI_ISL_1701409, EPI_ISL_1701415, EPI_ISL_1701416, EPI_ISL_1701417, EPI_ISL_1701418, EPI_ISL_1701421, EPI_ISL_1701424, EPI_ISL_1701427, EPI_ISL_1701432, EPI_ISL_1701434, EPI_ISL_1701436, EPI_ISL_1701442, EPI_ISL_1701443, EPI_ISL_1701449, EPI_ISL_1701452, EPI_ISL_1701453, EPI_ISL_1701455, EPI_ISL_1701459, EPI_ISL_1701463, EPI_ISL_1701466, EPI_ISL_1701470, EPI_ISL_1701475, EPI_ISL_1701485, EPI_ISL_1701487, EPI_ISL_1701488, EPI_ISL_1701489, EPI_ISL_1701492, EPI_ISL_1701499, EPI_ISL_1701503, EPI_ISL_1701506, EPI_ISL_1701508, EPI_ISL_1701512, EPI_ISL_1701514, EPI_ISL_1701515, EPI_ISL_1701521, EPI_ISL_1701522, EPI_ISL_1701524, EPI_ISL_1701529, EPI_ISL_1701535, EPI_ISL_1701546, EPI_ISL_1701547, EPI_ISL_1701551, EPI_ISL_1701552, EPI_ISL_1701553, EPI_ISL_1701556, EPI_ISL_1701561, EPI_ISL_1701563, EPI_ISL_1701570, EPI_ISL_1701572, EPI_ISL_1701576, EPI_ISL_1701578, EPI_ISL_1701580, EPI_ISL_1701585, EPI_ISL_1701592, EPI_ISL_1701606, EPI_ISL_1701610, EPI_ISL_1701613, EPI_ISL_1701621, EPI_ISL_1701622, EPI_ISL_1701632, EPI_ISL_1701633, EPI_ISL_1701635, EPI_ISL_1701642, EPI_ISL_1701644, EPI_ISL_1701647, EPI_ISL_1701649, EPI_ISL_1701650, EPI_ISL_1701651, EPI_ISL_1701652, EPI_ISL_1701655, EPI_ISL_2658568, EPI_ISL_2658571, EPI_ISL_2658577, EPI_ISL_2658583, EPI_ISL_2658596, EPI_ISL_2658599, EPI_ISL_2658626, EPI_ISL_2658671, EPI_ISL_2658690, EPI_ISL_2658692, EPI_ISL_2658693, EPI_ISL_2658706, EPI_ISL_2658713, EPI_ISL_2658739, EPI_ISL_2659215, EPI_ISL_2659220, EPI_ISL_2659224, EPI_ISL_2659229, EPI_ISL_2659232, EPI_ISL_2659235, EPI_ISL_2659272, EPI_ISL_2659311, EPI_ISL_2659324, EPI_ISL_2659325, EPI_ISL_2659328, EPI_ISL_2659332, EPI_ISL_2659355, EPI_ISL_2659356, EPI_ISL_2659377, EPI_ISL_2659390, EPI_ISL_2659404, EPI_ISL_2659407, EPI_ISL_2659417, EPI_ISL_2659419, EPI_ISL_2659423, EPI_ISL_2659428, EPI_ISL_2659450, EPI_ISL_2659455, EPI_ISL_2659480, EPI_ISL_2659482, EPI_ISL_2659484, EPI_ISL_2659505, EPI_ISL_2659508, EPI_ISL_2659519, EPI_ISL_2659534, EPI_ISL_2659588, EPI_ISL_2659595, EPI_ISL_2659611, EPI_ISL_2659614, EPI_ISL_2659628, EPI_ISL_2659636, EPI_ISL_2659641, EPI_ISL_2725519, EPI_ISL_2758443, EPI_ISL_2758454, EPI_ISL_2758455, EPI_ISL_2758470, EPI_ISL_2758471, EPI_ISL_2758472

| | | | |
|---|---|---|---|
| see above | Alameda County Public Health Lab | Chan-Zuckerberg Biohub | CZB Cliahub Consortium |

EPI_ISL_1184809, EPI_ISL_1184810, EPI_ISL_1184811, EPI_ISL_1184812, EPI_ISL_1184813, EPI_ISL_1184814, EPI_ISL_1184815, EPI_ISL_1184816, EPI_ISL_1184817, EPI_ISL_1184818, EPI_ISL_1185289, EPI_ISL_1185290, EPI_ISL_1185293, EPI_ISL_1185295, EPI_ISL_1185296, EPI_ISL_1185297, EPI_ISL_1185299, EPI_ISL_1185303, EPI_ISL_1185307, EPI_ISL_1185310, EPI_ISL_1185311, EPI_ISL_1185314, EPI_ISL_1185316, EPI_ISL_1185317, EPI_ISL_1185320, EPI_ISL_1185321, EPI_ISL_1185327, EPI_ISL_1185330, EPI_ISL_1185331, EPI_ISL_1185332, EPI_ISL_1185333, EPI_ISL_1185334, EPI_ISL_1185335, EPI_ISL_1185336, EPI_ISL_1185337, EPI_ISL_1185338, EPI_ISL_1185339, EPI_ISL_1185341, EPI_ISL_1185342, EPI_ISL_1185343, EPI_ISL_1185344, EPI_ISL_1185347, EPI_ISL_1185348, EPI_ISL_1185349, EPI_ISL_1185350, EPI_ISL_1185354, EPI_ISL_1185366, EPI_ISL_1185370, EPI_ISL_1185371, EPI_ISL_1185373, EPI_ISL_1185374, EPI_ISL_1185375, EPI_ISL_1185377, EPI_ISL_1185378, EPI_ISL_1185379, EPI_ISL_1185380, EPI_ISL_1185381, EPI_ISL_1185384, EPI_ISL_1185386, EPI_ISL_1185389, EPI_ISL_1185390, EPI_ISL_1185391, EPI_ISL_1185392, EPI_ISL_1185396, EPI_ISL_1185397, EPI_ISL_1185398, EPI_ISL_1185400, EPI_ISL_1185402, EPI_ISL_1185403, EPI_ISL_1185404, EPI_ISL_1185405, EPI_ISL_1185406, EPI_ISL_1185411, EPI_ISL_1185415, EPI_ISL_1185416, EPI_ISL_1185418, EPI_ISL_1185419, EPI_ISL_1185420, EPI_ISL_1185421, EPI_ISL_1185423, EPI_ISL_1185426, EPI_ISL_1185429, EPI_ISL_1185433, EPI_ISL_1185434, EPI_ISL_1185435, EPI_ISL_1185437, EPI_ISL_1185441, EPI_ISL_1185442, EPI_ISL_1185444, EPI_ISL_1185445, EPI_ISL_1185446, EPI_ISL_1185450, EPI_ISL_1185451, EPI_ISL_1185453, EPI_ISL_1185458, EPI_ISL_1185462, EPI_ISL_1185464, EPI_ISL_1185465, EPI_ISL_1185467, EPI_ISL_1185470, EPI_ISL_1185472, EPI_ISL_1185473, EPI_ISL_1185477, EPI_ISL_1185478, EPI_ISL_1185482, EPI_ISL_1185483, EPI_ISL_1185484, EPI_ISL_1185485, EPI_ISL_1185490, EPI_ISL_1185492, EPI_ISL_1185494, EPI_ISL_1185495, EPI_ISL_1185500, EPI_ISL_1185506, EPI_ISL_1185508, EPI_ISL_1185510, EPI_ISL_1185511, EPI_ISL_1185512, EPI_ISL_1185513, EPI_ISL_1185514, EPI_ISL_1185515, EPI_ISL_1185520, EPI_ISL_1185522, EPI_ISL_1185523, EPI_ISL_1185524, EPI_ISL_1185525, EPI_ISL_1185527, EPI_ISL_1185530, EPI_ISL_1185531, EPI_ISL_1185532, EPI_ISL_2659296, EPI_ISL_2659387, EPI_ISL_2659524, EPI_ISL_2659593, EPI_ISL_2659604, EPI_ISL_2659642, EPI_ISL_2758440, EPI_ISL_2758441

| | | | |
|---|---|---|---|
| see above | CA DPH Viral and Rickettsial Disease Laboratory | Chan-Zuckerberg Biohub | CZB Cliahub Consortium |

EPI_ISL_2826026, EPI_ISL_2826027, EPI_ISL_2826028, EPI_ISL_2826029, EPI_ISL_2826030, EPI_ISL_2826031, EPI_ISL_2826032, EPI_ISL_2826033, EPI_ISL_2826034, EPI_ISL_2826035, EPI_ISL_2826036, EPI_ISL_2826037, EPI_ISL_2826038, EPI_ISL_2826039, EPI_ISL_2826040, EPI_ISL_2826041, EPI_ISL_2826042, EPI_ISL_2826043, EPI_ISL_2826044, EPI_ISL_2826045, EPI_ISL_2826046, EPI_ISL_2826047, EPI_ISL_2826048, EPI_ISL_2826049, EPI_ISL_2826050, EPI_ISL_2826051, EPI_ISL_2826052, EPI_ISL_2826053, EPI_ISL_2826054, EPI_ISL_2826055, EPI_ISL_2826056, EPI_ISL_2826057, EPI_ISL_2826058, EPI_ISL_2826059, EPI_ISL_2826060, EPI_ISL_2826061, EPI_ISL_2826062, EPI_ISL_2826063, EPI_ISL_2826064, EPI_ISL_2826065, EPI_ISL_2826066, EPI_ISL_2826067, EPI_ISL_2826068, EPI_ISL_2826069, EPI_ISL_2826070, EPI_ISL_2826071, EPI_ISL_2826072, EPI_ISL_2826073, EPI_ISL_2826074, EPI_ISL_2826075, EPI_ISL_2826076, EPI_ISL_2826077, EPI_ISL_2826078, EPI_ISL_2826079, EPI_ISL_2826080, EPI_ISL_2826081, EPI_ISL_2826082, EPI_ISL_2826083, EPI_ISL_2826084, EPI_ISL_2826085, EPI_ISL_2826086, EPI_ISL_2826087, EPI_ISL_2826088, EPI_ISL_2826089, EPI_ISL_2826090, EPI_ISL_2826091, EPI_ISL_2826092, EPI_ISL_2826093, EPI_ISL_2826094, EPI_ISL_2826095, EPI_ISL_2826096, EPI_ISL_2826097, EPI_ISL_2826098, EPI_ISL_2826099, EPI_ISL_2826100, EPI_ISL_2826101, EPI_ISL_2826102, EPI_ISL_2826103, EPI_ISL_2826104, EPI_ISL_2826105, EPI_ISL_2826106, EPI_ISL_2826107, EPI_ISL_2826108, EPI_ISL_2826109, EPI_ISL_2826110, EPI_ISL_2826111, EPI_ISL_2826112, EPI_ISL_2826113, EPI_ISL_2826114, EPI_ISL_2826115, EPI_ISL_2826116, EPI_ISL_2826117, EPI_ISL_2826118, EPI_ISL_2826119, EPI_ISL_2826120, EPI_ISL_2826121, EPI_ISL_2826122, EPI_ISL_2826123, EPI_ISL_2826124, EPI_ISL_2826125, EPI_ISL_2826126, EPI_ISL_2826127, EPI_ISL_2826128, EPI_ISL_2826129, EPI_ISL_2826130, EPI_ISL_2826131, EPI_ISL_2826132, EPI_ISL_2826133, EPI_ISL_2826134, EPI_ISL_2826135, EPI_ISL_2826136, EPI_ISL_2826137, EPI_ISL_2826138, EPI_ISL_2826139, EPI_ISL_2826140, EPI_ISL_2826141, EPI_ISL_2826142, EPI_ISL_2826143, EPI_ISL_2826144, EPI_ISL_2826145, EPI_ISL_2826146, EPI_ISL_2826147, EPI_ISL_2826148, EPI_ISL_2826149, EPI_ISL_2826150, EPI_ISL_2826151, EPI_ISL_2826152, EPI_ISL_2826153, EPI_ISL_2826154, EPI_ISL_2826155, EPI_ISL_2826156, EPI_ISL_2826157, EPI_ISL_2826158, EPI_ISL_2826159, EPI_ISL_2826160, EPI_ISL_2826161, EPI_ISL_2826162, EPI_ISL_2826163, EPI_ISL_2826164, EPI_ISL_2826165, EPI_ISL_2826166, EPI_ISL_2826167, EPI_ISL_2826168, EPI_ISL_2826169, EPI_ISL_2826170, EPI_ISL_2826171, EPI_ISL_2826172, EPI_ISL_2826173, EPI_ISL_2934103, EPI_ISL_2934105, EPI_ISL_2934107, EPI_ISL_2934108, EPI_ISL_2934113, EPI_ISL_2934118, EPI_ISL_2934134, EPI_ISL_2934151, EPI_ISL_2934152, EPI_ISL_2934153, EPI_ISL_2934162, EPI_ISL_2934166, EPI_ISL_2934178, EPI_ISL_2934192, EPI_ISL_2934195, EPI_ISL_2934197, EPI_ISL_2934199, EPI_ISL_2934200, EPI_ISL_2934205, EPI_ISL_2934209, EPI_ISL_2934211, EPI_ISL_2934215, EPI_ISL_2934224, EPI_ISL_2934225, EPI_ISL_2934239, EPI_ISL_2934241, EPI_ISL_2934246, EPI_ISL_2934252, EPI_ISL_2934255, EPI_ISL_2934272, EPI_ISL_2934274, EPI_ISL_2934282, EPI_ISL_2934286, EPI_ISL_2934304, EPI_ISL_2934306, EPI_ISL_2934318, EPI_ISL_2934321, EPI_ISL_2934326, EPI_ISL_2934327, EPI_ISL_2934331, EPI_ISL_2934332, EPI_ISL_2934346, EPI_ISL_2934352, EPI_ISL_2934355, EPI_ISL_2934372, EPI_ISL_2934378, EPI_ISL_2934383, EPI_ISL_2934384, EPI_ISL_2934400, EPI_ISL_2934413, EPI_ISL_2934414, EPI_ISL_2934420, EPI_ISL_2934431, EPI_ISL_2934440, EPI_ISL_2934442, EPI_ISL_2934450, EPI_ISL_2934462, EPI_ISL_2934464, EPI_ISL_2934469, EPI_ISL_2934479, EPI_ISL_2934486, EPI_ISL_2934489, EPI_ISL_2934497, EPI_ISL_2934498, EPI_ISL_2934504, EPI_ISL_2934512, EPI_ISL_2934520, EPI_ISL_2934526, EPI_ISL_2934527, EPI_ISL_2934529, EPI_ISL_2934531, EPI_ISL_2934532, EPI_ISL_2934540, EPI_ISL_2934543, EPI_ISL_2934545, EPI_ISL_2934552, EPI_ISL_2934559, EPI_ISL_3067067, EPI_ISL_3067068, EPI_ISL_3067069, EPI_ISL_3067070, EPI_ISL_3067071, EPI_ISL_3067072, EPI_ISL_3067073, EPI_ISL_3067074, EPI_ISL_3067075, EPI_ISL_3067076, EPI_ISL_3067078, EPI_ISL_3067079, EPI_ISL_3067081, EPI_ISL_3067082, EPI_ISL_3067083, EPI_ISL_3067084, EPI_ISL_3067085, EPI_ISL_3067087, EPI_ISL_3067088, EPI_ISL_3067089, EPI_ISL_3067090, EPI_ISL_3067091, EPI_ISL_3067092, EPI_ISL_3067093, EPI_ISL_3067094, EPI_ISL_3067095, EPI_ISL_3067097, EPI_ISL_3067099, EPI_ISL_3067100, EPI_ISL_3067101, EPI_ISL_3067102, EPI_ISL_3067103, EPI_ISL_3067104, EPI_ISL_3067105, EPI_ISL_3067107, EPI_ISL_3067108, EPI_ISL_3067109, EPI_ISL_3067110, EPI_ISL_3067111, EPI_ISL_3067112, EPI_ISL_3067113, EPI_ISL_3067114, EPI_ISL_3067115, EPI_ISL_3067117, EPI_ISL_3067119, EPI_ISL_3067120, EPI_ISL_3067121, EPI_ISL_3067122, EPI_ISL_3067123, EPI_ISL_3067124, EPI_ISL_3067125, EPI_ISL_3067127, EPI_ISL_3067128, EPI_ISL_3067129, EPI_ISL_3067131, EPI_ISL_3067132, EPI_ISL_3067133, EPI_ISL_3067134, EPI_ISL_3067136, EPI_ISL_3067137, EPI_ISL_3067138, EPI_ISL_3067139, EPI_ISL_3067141, EPI_ISL_3067143, EPI_ISL_3067144, EPI_ISL_3067145, EPI_ISL_3067146, EPI_ISL_3067147, EPI_ISL_3067148, EPI_ISL_3067149, EPI_ISL_3067151, EPI_ISL_3067152, EPI_ISL_3067153, EPI_ISL_3067154, EPI_ISL_3067155, EPI_ISL_3067156, EPI_ISL_3067157, EPI_ISL_3067158, EPI_ISL_3067160, EPI_ISL_3067161, EPI_ISL_3067162, EPI_ISL_3067163, EPI_ISL_3067164, EPI_ISL_3067165, EPI_ISL_3067167, EPI_ISL_3067168, EPI_ISL_3067170, EPI_ISL_3067171, EPI_ISL_3067172, EPI_ISL_3067174, EPI_ISL_3067175, EPI_ISL_3067177, EPI_ISL_3067178, EPI_ISL_3067179, EPI_ISL_3067180, EPI_ISL_3067181, EPI_ISL_3067182, EPI_ISL_3067183, EPI_ISL_3067184, EPI_ISL_3067185, EPI_ISL_3067186, EPI_ISL_3067187, EPI_ISL_3067188, EPI_ISL_3067189, EPI_ISL_3067190, EPI_ISL_3067191, EPI_ISL_3067192, EPI_ISL_3067194, EPI_ISL_3067195, EPI_ISL_3067196, EPI_ISL_3067197, EPI_ISL_3067198, EPI_ISL_3067199, EPI_ISL_3067200, EPI_ISL_3067201, EPI_ISL_3067202, EPI_ISL_3067203, EPI_ISL_3067204, EPI_ISL_3067205, EPI_ISL_3067206, EPI_ISL_3067207, EPI_ISL_3067208, EPI_ISL_3067209, EPI_ISL_3067210, EPI_ISL_3067211, EPI_ISL_3067212, EPI_ISL_3067213, EPI_ISL_3067214, EPI_ISL_3067215, EPI_ISL_3067216, EPI_ISL_3067217, EPI_ISL_3067220, EPI_ISL_3067221, EPI_ISL_3067222, EPI_ISL_3067223, EPI_ISL_3067226, EPI_ISL_3067227, EPI_ISL_3067229, EPI_ISL_3067230, EPI_ISL_3067231, EPI_ISL_3067232, EPI_ISL_3067233, EPI_ISL_3067234, EPI_ISL_3067236, EPI_ISL_3067238, EPI_ISL_3067239, EPI_ISL_3067240, EPI_ISL_3067241, EPI_ISL_3067242, EPI_ISL_3067243, EPI_ISL_3067244, EPI_ISL_3067245, EPI_ISL_3067246, EPI_ISL_3067247, EPI_ISL_3067248, EPI_ISL_3067249, EPI_ISL_3067251, EPI_ISL_3067252, EPI_ISL_3067254, EPI_ISL_3067255, EPI_ISL_3067256, EPI_ISL_3067257, EPI_ISL_3067258, EPI_ISL_3067259, EPI_ISL_3067261, EPI_ISL_3067262, EPI_ISL_3067263, EPI_ISL_3067264, EPI_ISL_3067266, EPI_ISL_3067267, EPI_ISL_3067268, EPI_ISL_3067270, EPI_ISL_3067271, EPI_ISL_3067272, EPI_ISL_3067273, EPI_ISL_3067274, EPI_ISL_3067275, EPI_ISL_3067276, EPI_ISL_3067278, EPI_ISL_3067279, EPI_ISL_3067280, EPI_ISL_3067281, EPI_ISL_3067283, EPI_ISL_3067284, EPI_ISL_3067285, EPI_ISL_3067286, EPI_ISL_3067287, EPI_ISL_3067288, EPI_ISL_3067289, EPI_ISL_3067291, EPI_ISL_3067292, EPI_ISL_3067293, EPI_ISL_3067294, EPI_ISL_3067295, EPI_ISL_3067297, EPI_ISL_3067298, EPI_ISL_3067299, EPI_ISL_3067300, EPI_ISL_3067301, EPI_ISL_3067302, EPI_ISL_3067303, EPI_ISL_3067304, EPI_ISL_3067305, EPI_ISL_3067306, EPI_ISL_3067307, EPI_ISL_3067308, EPI_ISL_3067309, EPI_ISL_3067310, EPI_ISL_3067312, EPI_ISL_3067313, EPI_ISL_3067315, EPI_ISL_3067316, EPI_ISL_3067317, EPI_ISL_3067318, EPI_ISL_3067319, EPI_ISL_3067320, EPI_ISL_3067321, EPI_ISL_3067322, EPI_ISL_3067323, EPI_ISL_3067324, EPI_ISL_3067325, EPI_ISL_3067326, EPI_ISL_3067327, EPI_ISL_3067328, EPI_ISL_3067329, EPI_ISL_3067330, EPI_ISL_3067331, EPI_ISL_3067332, EPI_ISL_3067333, EPI_ISL_3067334, EPI_ISL_3067335, EPI_ISL_3067336, EPI_ISL_3067337, EPI_ISL_3067338, EPI_ISL_3067339, EPI_ISL_3067340, EPI_ISL_3067341, EPI_ISL_3067342, EPI_ISL_3067343, EPI_ISL_3067344, EPI_ISL_3067345, EPI_ISL_3067346, EPI_ISL_3067347, EPI_ISL_3067348, EPI_ISL_3067349, EPI_ISL_3067351, EPI_ISL_3067352, EPI_ISL_3067354, EPI_ISL_3067355, EPI_ISL_3067357, EPI_ISL_3067358, EPI_ISL_3067359, EPI_ISL_3067360, EPI_ISL_3067361

| | | | |
|---|---|---|---|
| see above | CDPH VBL | California Department of Public Health | CDPH-COVIDNet; SwabSeq/UCLA; UCI Genome Sciences Center/GHTF |

EPI_ISL_2968797, EPI_ISL_2968798, EPI_ISL_2968803, EPI_ISL_2968810, EPI_ISL_2968811, EPI_ISL_2968812, EPI_ISL_2968814, EPI_ISL_2968815, EPI_ISL_2968816, EPI_ISL_2968817, EPI_ISL_2968818, EPI_ISL_2968819, EPI_ISL_2968820, EPI_ISL_2968821, EPI_ISL_2968822, EPI_ISL_2968823, EPI_ISL_2968824, EPI_ISL_2968825, EPI_ISL_2968826, EPI_ISL_2968827, EPI_ISL_2968828, EPI_ISL_2968829, EPI_ISL_2968830, EPI_ISL_2968831, EPI_ISL_2968832, EPI_ISL_2968833, EPI_ISL_2968834, EPI_ISL_2968835, EPI_ISL_2968836, EPI_ISL_2968837, EPI_ISL_2968838, EPI_ISL_2968839, EPI_ISL_2968840, EPI_ISL_2968841, EPI_ISL_2968842, EPI_ISL_2968843, EPI_ISL_2968844, EPI_ISL_2968845, EPI_ISL_2968846, EPI_ISL_2968847, EPI_ISL_2968848, EPI_ISL_2968849, EPI_ISL_2968850, EPI_ISL_2968851, EPI_ISL_2968852, EPI_ISL_2968853, EPI_ISL_2968854, EPI_ISL_2968855, EPI_ISL_2968856, EPI_ISL_2968857, EPI_ISL_2968858, EPI_ISL_2968859, EPI_ISL_2968860, EPI_ISL_2968861, EPI_ISL_2968862, EPI_ISL_2968863, EPI_ISL_2968864, EPI_ISL_2968865, EPI_ISL_2968866, EPI_ISL_2968867, EPI_ISL_2968868, EPI_ISL_2968869, EPI_ISL_2968870, EPI_ISL_2968871, EPI_ISL_2968872, EPI_ISL_2968873, EPI_ISL_2968874, EPI_ISL_2968875, EPI_ISL_2968876, EPI_ISL_2968877, EPI_ISL_2968878, EPI_ISL_2968879, EPI_ISL_2968880, EPI_ISL_2968881, EPI_ISL_2968882, EPI_ISL_2968883, EPI_ISL_2968884, EPI_ISL_2968885, EPI_ISL_2968886, EPI_ISL_2968887, EPI_ISL_2968888, EPI_ISL_2968889, EPI_ISL_2968890, EPI_ISL_2968891, EPI_ISL_2968892, EPI_ISL_2968893, EPI_ISL_2968894, EPI_ISL_2968895, EPI_ISL_2968896, EPI_ISL_2968897, EPI_ISL_2968898, EPI_ISL_2968899, EPI_ISL_2968900, EPI_ISL_2968901, EPI_ISL_2968902, EPI_ISL_2968903, EPI_ISL_2968904, EPI_ISL_2968905, EPI_ISL_2968906, EPI_ISL_2968907, EPI_ISL_2968908, EPI_ISL_2968909, EPI_ISL_2968910, EPI_ISL_2968911, EPI_ISL_2968912, EPI_ISL_2968913, EPI_ISL_2968914, EPI_ISL_2968915, EPI_ISL_2968916, EPI_ISL_2968917, EPI_ISL_2968918, EPI_ISL_2968919, EPI_ISL_2968920, EPI_ISL_2968921, EPI_ISL_2968922, EPI_ISL_2968923, EPI_ISL_2968924, EPI_ISL_2968925, EPI_ISL_2968926, EPI_ISL_2968927, EPI_ISL_2968928, EPI_ISL_2968929, EPI_ISL_2968930, EPI_ISL_2968931, EPI_ISL_2968932, EPI_ISL_2968933, EPI_ISL_2968934, EPI_ISL_2968935, EPI_ISL_2968936, EPI_ISL_2968937, EPI_ISL_2968938, EPI_ISL_2968939, EPI_ISL_2968940, EPI_ISL_2968941, EPI_ISL_2968942, EPI_ISL_2968943, EPI_ISL_2968944, EPI_ISL_2968945, EPI_ISL_2968946, EPI_ISL_2968947, EPI_ISL_2968948, EPI_ISL_2968949, EPI_ISL_2968950, EPI_ISL_2968951, EPI_ISL_2968952, EPI_ISL_2968953, EPI_ISL_2968954, EPI_ISL_2968955, EPI_ISL_2968956, EPI_ISL_2968957, EPI_ISL_2968958, EPI_ISL_2968959, EPI_ISL_2968960, EPI_ISL_2968961, EPI_ISL_2968962, EPI_ISL_2968963, EPI_ISL_2968964, EPI_ISL_2968965, EPI_ISL_2968966, EPI_ISL_2968967, EPI_ISL_2968968, EPI_ISL_2968969, EPI_ISL_2968970, EPI_ISL_2968971, EPI_ISL_2968972, EPI_ISL_2968973, EPI_ISL_2968974, EPI_ISL_2968975, EPI_ISL_2968976, EPI_ISL_2968977, EPI_ISL_2968978, EPI_ISL_2968979, EPI_ISL_2968980, EPI_ISL_2968981, EPI_ISL_2968982, EPI_ISL_2968983, EPI_ISL_2968984, EPI_ISL_2968985, EPI_ISL_2968986, EPI_ISL_2968987, EPI_ISL_2968988, EPI_ISL_2968989, EPI_ISL_2968990, EPI_ISL_2968991, EPI_ISL_2968992, EPI_ISL_2968993, EPI_ISL_2968994, EPI_ISL_2968995, EPI_ISL_2968996, EPI_ISL_2968997, EPI_ISL_2968998, EPI_ISL_2968999, EPI_ISL_2969000, EPI_ISL_2969001, EPI_ISL_2969002, EPI_ISL_2969003, EPI_ISL_2969004, EPI_ISL_2969005, EPI_ISL_2969006, EPI_ISL_2969007, EPI_ISL_2969008, EPI_ISL_2969009, EPI_ISL_2969010, EPI_ISL_2969011, EPI_ISL_2969012, EPI_ISL_2969013, EPI_ISL_2969014, EPI_ISL_2969015, EPI_ISL_2969016, EPI_ISL_2969017, EPI_ISL_2969018, EPI_ISL_2969019, EPI_ISL_2969020, EPI_ISL_2969021, EPI_ISL_2969022, EPI_ISL_2969023, EPI_ISL_2969024, EPI_ISL_2969025, EPI_ISL_2969026, EPI_ISL_2969027, EPI_ISL_2969028, EPI_ISL_2969029, EPI_ISL_2969030

| | | | |
|---|---|---|---|
| see above | CDPH Valencia Branch Laboratory | California Department of Public Health | CDPH-COVIDNet; Priva Path Labs Inc. |

EPI_ISL_754914, EPI_ISL_754924, EPI_ISL_754942, EPI_ISL_754948, EPI_ISL_754949, EPI_ISL_754955, EPI_ISL_754957, EPI_ISL_754965, EPI_ISL_754966, EPI_ISL_754967, EPI_ISL_754971, EPI_ISL_754973, EPI_ISL_754982, EPI_ISL_754983, EPI_ISL_754993, EPI_ISL_754997, EPI_ISL_755000, EPI_ISL_755002, EPI_ISL_755003, EPI_ISL_755005, EPI_ISL_755007, EPI_ISL_755009, EPI_ISL_755010, EPI_ISL_755013, EPI_ISL_755014, EPI_ISL_755015, EPI_ISL_755021, EPI_ISL_755024, EPI_ISL_755026, EPI_ISL_755062, EPI_ISL_755063, EPI_ISL_755064, EPI_ISL_755067, EPI_ISL_755068, EPI_ISL_755069, EPI_ISL_755070, EPI_ISL_755071, EPI_ISL_755072, EPI_ISL_755073, EPI_ISL_755074, EPI_ISL_755075, EPI_ISL_755076, EPI_ISL_755077, EPI_ISL_755078, EPI_ISL_755079, EPI_ISL_755080, EPI_ISL_755082, EPI_ISL_755083, EPI_ISL_755084, EPI_ISL_755085, EPI_ISL_755086, EPI_ISL_755087, EPI_ISL_755088, EPI_ISL_755089, EPI_ISL_755090, EPI_ISL_755091, EPI_ISL_755092, EPI_ISL_755093, EPI_ISL_755094, EPI_ISL_755095, EPI_ISL_755096, EPI_ISL_755097, EPI_ISL_755098, EPI_ISL_755099, EPI_ISL_755100, EPI_ISL_755101, EPI_ISL_755102, EPI_ISL_755103, EPI_ISL_755104, EPI_ISL_755105, EPI_ISL_755106, EPI_ISL_755107, EPI_ISL_755108, EPI_ISL_755109, EPI_ISL_755110, EPI_ISL_755111, EPI_ISL_755112, EPI_ISL_755113, EPI_ISL_755114, EPI_ISL_824390, EPI_ISL_824391, EPI_ISL_824392, EPI_ISL_824393, EPI_ISL_824394, EPI_ISL_824395, EPI_ISL_824396, EPI_ISL_824397, EPI_ISL_824398, EPI_ISL_824399, EPI_ISL_824400, EPI_ISL_824401, EPI_ISL_824402, EPI_ISL_824403, EPI_ISL_824404, EPI_ISL_824405, EPI_ISL_824406, EPI_ISL_2233910, EPI_ISL_2233911, EPI_ISL_2233914, EPI_ISL_2233915, EPI_ISL_2233916, EPI_ISL_2233917, EPI_ISL_2233918, EPI_ISL_2233919, EPI_ISL_2233920, EPI_ISL_2233921, EPI_ISL_2233922, EPI_ISL_2233923, EPI_ISL_2233924, EPI_ISL_2233925, EPI_ISL_2233926, EPI_ISL_2233927, EPI_ISL_2233928, EPI_ISL_2233929, EPI_ISL_2233930, EPI_ISL_2233931, EPI_ISL_2233935, EPI_ISL_2233936, EPI_ISL_2233943, EPI_ISL_2233944, EPI_ISL_2233945, EPI_ISL_2233946, EPI_ISL_2233947, EPI_ISL_2233948, EPI_ISL_2233949, EPI_ISL_2233950, EPI_ISL_2233951, EPI_ISL_2233952, EPI_ISL_2233953, EPI_ISL_2233954, EPI_ISL_2233955, EPI_ISL_2233956, EPI_ISL_2233957, EPI_ISL_2233958, EPI_ISL_2233959, EPI_ISL_2233960, EPI_ISL_2233961, EPI_ISL_2233962, EPI_ISL_2233963, EPI_ISL_2233964,

EPI_ISL_2233965, EPI_ISL_2233966, EPI_ISL_2233967, EPI_ISL_2233968, EPI_ISL_2233969, EPI_ISL_2233970, EPI_ISL_2233971, EPI_ISL_2233980, EPI_ISL_2233981, EPI_ISL_2233982, EPI_ISL_2233986, EPI_ISL_2233992, EPI_ISL_2233994, EPI_ISL_2234001, EPI_ISL_2234002, EPI_ISL_2234003, EPI_ISL_2234004, EPI_ISL_2234005, EPI_ISL_2234006, EPI_ISL_2234007, EPI_ISL_2234008, EPI_ISL_2234009, EPI_ISL_2234010, EPI_ISL_2234011, EPI_ISL_2234012, EPI_ISL_2234013, EPI_ISL_2234014, EPI_ISL_2234015, EPI_ISL_2234016, EPI_ISL_2234017, EPI_ISL_2234018, EPI_ISL_2234019, EPI_ISL_2234020, EPI_ISL_2234026, EPI_ISL_2234028, EPI_ISL_2234037, EPI_ISL_2234038, EPI_ISL_2234039, EPI_ISL_2234070, EPI_ISL_2234071, EPI_ISL_2234072, EPI_ISL_2234073, EPI_ISL_2234074, EPI_ISL_2234075, EPI_ISL_2234076, EPI_ISL_2234077, EPI_ISL_2234078, EPI_ISL_2234079, EPI_ISL_2234080, EPI_ISL_2234126, EPI_ISL_2234127, EPI_ISL_2234128, EPI_ISL_2234185, EPI_ISL_2234186, EPI_ISL_2234187, EPI_ISL_2234188, EPI_ISL_2234189, EPI_ISL_2234190, EPI_ISL_2234191, EPI_ISL_2234192, EPI_ISL_2234193, EPI_ISL_2234206, EPI_ISL_2234207, EPI_ISL_2234208, EPI_ISL_2234265, EPI_ISL_2234266, EPI_ISL_2234297, EPI_ISL_2234298, EPI_ISL_2234299, EPI_ISL_2234300, EPI_ISL_2234301, EPI_ISL_2234302, EPI_ISL_2234303, EPI_ISL_2234304, EPI_ISL_2234305, EPI_ISL_2234306, EPI_ISL_2234307, EPI_ISL_2234308, EPI_ISL_2234309, EPI_ISL_2234310, EPI_ISL_2892408, EPI_ISL_2892551

| | | | |
|---|---|---|---|
| see above | California Department of Public Health | California Department of Public Health | CDPH IDLB COVIDNet; CDPH IDLB COVIDNet et al |

EPI_ISL_755940, EPI_ISL_755941, EPI_ISL_755942, EPI_ISL_847516, EPI_ISL_847518, EPI_ISL_847519, EPI_ISL_847520, EPI_ISL_847521, EPI_ISL_847522, EPI_ISL_847523, EPI_ISL_847524, EPI_ISL_847525, EPI_ISL_847526, EPI_ISL_847527, EPI_ISL_847528, EPI_ISL_847529, EPI_ISL_847530, EPI_ISL_847531, EPI_ISL_847532, EPI_ISL_847533, EPI_ISL_847534, EPI_ISL_847535, EPI_ISL_847537, EPI_ISL_847538, EPI_ISL_847539, EPI_ISL_847541, EPI_ISL_847542, EPI_ISL_847543, EPI_ISL_847544, EPI_ISL_847547, EPI_ISL_847548, EPI_ISL_847549, EPI_ISL_847550, EPI_ISL_847551, EPI_ISL_847552, EPI_ISL_847554, EPI_ISL_847555, EPI_ISL_847556, EPI_ISL_847557, EPI_ISL_847558, EPI_ISL_847559, EPI_ISL_847560, EPI_ISL_847561, EPI_ISL_847562, EPI_ISL_847565, EPI_ISL_847566, EPI_ISL_847567, EPI_ISL_847569, EPI_ISL_847570, EPI_ISL_847571, EPI_ISL_847572, EPI_ISL_847573, EPI_ISL_847574, EPI_ISL_847575, EPI_ISL_847576, EPI_ISL_847577, EPI_ISL_847578, EPI_ISL_847580, EPI_ISL_847581, EPI_ISL_847582, EPI_ISL_847583, EPI_ISL_847584, EPI_ISL_847585, EPI_ISL_847586, EPI_ISL_847587, EPI_ISL_847588, EPI_ISL_847595, EPI_ISL_847596, EPI_ISL_847597, EPI_ISL_847598, EPI_ISL_847599, EPI_ISL_847600, EPI_ISL_847601, EPI_ISL_847602, EPI_ISL_847603, EPI_ISL_847604, EPI_ISL_847605, EPI_ISL_847606, EPI_ISL_847607, EPI_ISL_847608, EPI_ISL_847609, EPI_ISL_847610, EPI_ISL_847611, EPI_ISL_847612, EPI_ISL_847613, EPI_ISL_847614, EPI_ISL_847615, EPI_ISL_847616, EPI_ISL_847617, EPI_ISL_847618, EPI_ISL_847623, EPI_ISL_847624, EPI_ISL_847625, EPI_ISL_847626, EPI_ISL_847627, EPI_ISL_847628, EPI_ISL_847629, EPI_ISL_847630, EPI_ISL_847631, EPI_ISL_847632, EPI_ISL_847633, EPI_ISL_847634, EPI_ISL_847635, EPI_ISL_847636, EPI_ISL_847637, EPI_ISL_847638, EPI_ISL_847639, EPI_ISL_847645, EPI_ISL_847646, EPI_ISL_847647, EPI_ISL_847648, EPI_ISL_847649, EPI_ISL_847650, EPI_ISL_847651, EPI_ISL_847652, EPI_ISL_847653, EPI_ISL_847654, EPI_ISL_847655, EPI_ISL_847656, EPI_ISL_847657, EPI_ISL_847658, EPI_ISL_847659, EPI_ISL_847660, EPI_ISL_847661, EPI_ISL_847662, EPI_ISL_847663, EPI_ISL_847664, EPI_ISL_847665, EPI_ISL_847666, EPI_ISL_847667, EPI_ISL_847668, EPI_ISL_847669, EPI_ISL_847670, EPI_ISL_847671, EPI_ISL_847672, EPI_ISL_847673, EPI_ISL_847674, EPI_ISL_847675, EPI_ISL_847676, EPI_ISL_847677, EPI_ISL_847678, EPI_ISL_847679, EPI_ISL_847680, EPI_ISL_847681, EPI_ISL_847682, EPI_ISL_847683, EPI_ISL_847684, EPI_ISL_847685, EPI_ISL_847686, EPI_ISL_847687, EPI_ISL_847688, EPI_ISL_847689, EPI_ISL_847690, EPI_ISL_847691, EPI_ISL_847692, EPI_ISL_847693, EPI_ISL_847694, EPI_ISL_847695, EPI_ISL_847696, EPI_ISL_847697, EPI_ISL_847698, EPI_ISL_847699, EPI_ISL_847700, EPI_ISL_847701, EPI_ISL_847702, EPI_ISL_847703, EPI_ISL_847704, EPI_ISL_847705, EPI_ISL_847706, EPI_ISL_847707, EPI_ISL_847708, EPI_ISL_847709, EPI_ISL_847710, EPI_ISL_847711, EPI_ISL_847712, EPI_ISL_847713, EPI_ISL_847714, EPI_ISL_847715, EPI_ISL_847717, EPI_ISL_847718, EPI_ISL_847719, EPI_ISL_847721, EPI_ISL_847722, EPI_ISL_847723, EPI_ISL_847724, EPI_ISL_847725, EPI_ISL_847726, EPI_ISL_847727, EPI_ISL_847728, EPI_ISL_847729, EPI_ISL_847732, EPI_ISL_847733, EPI_ISL_847734, EPI_ISL_847735, EPI_ISL_847737, EPI_ISL_847739, EPI_ISL_847740, EPI_ISL_847745, EPI_ISL_847748, EPI_ISL_847749, EPI_ISL_847750, EPI_ISL_847751, EPI_ISL_847752, EPI_ISL_847753, EPI_ISL_847754, EPI_ISL_847755, EPI_ISL_847756, EPI_ISL_847757, EPI_ISL_847760, EPI_ISL_847761, EPI_ISL_847762, EPI_ISL_847763, EPI_ISL_847764, EPI_ISL_847765, EPI_ISL_847766, EPI_ISL_847767, EPI_ISL_847768, EPI_ISL_847769, EPI_ISL_847770, EPI_ISL_847771, EPI_ISL_847772, EPI_ISL_847773, EPI_ISL_847774, EPI_ISL_847775, EPI_ISL_847776, EPI_ISL_847777, EPI_ISL_847778, EPI_ISL_847779, EPI_ISL_847780, EPI_ISL_847781, EPI_ISL_847783, EPI_ISL_847784, EPI_ISL_847785, EPI_ISL_847786, EPI_ISL_847787, EPI_ISL_847788, EPI_ISL_847789, EPI_ISL_847790, EPI_ISL_847791, EPI_ISL_847792, EPI_ISL_847793, EPI_ISL_847794, EPI_ISL_847795, EPI_ISL_847796, EPI_ISL_847797, EPI_ISL_847798, EPI_ISL_847799, EPI_ISL_847800, EPI_ISL_847801, EPI_ISL_847802, EPI_ISL_847803, EPI_ISL_847804, EPI_ISL_847805, EPI_ISL_847806, EPI_ISL_847807, EPI_ISL_847808, EPI_ISL_847809, EPI_ISL_847810, EPI_ISL_847811, EPI_ISL_847812, EPI_ISL_847813, EPI_ISL_847814, EPI_ISL_847815, EPI_ISL_847816, EPI_ISL_847817, EPI_ISL_847818, EPI_ISL_855389, EPI_ISL_855390, EPI_ISL_855391, EPI_ISL_855392, EPI_ISL_855393, EPI_ISL_855394, EPI_ISL_855395, EPI_ISL_855396, EPI_ISL_855397, EPI_ISL_855398, EPI_ISL_977721, EPI_ISL_977722, EPI_ISL_977723, EPI_ISL_977724, EPI_ISL_977725, EPI_ISL_977726, EPI_ISL_977727, EPI_ISL_977728, EPI_ISL_977729, EPI_ISL_977730, EPI_ISL_977731, EPI_ISL_977732, EPI_ISL_977733, EPI_ISL_977734, EPI_ISL_977735, EPI_ISL_977736, EPI_ISL_977737, EPI_ISL_977738, EPI_ISL_977739, EPI_ISL_977740, EPI_ISL_977741, EPI_ISL_977742, EPI_ISL_977745, EPI_ISL_977746, EPI_ISL_977747, EPI_ISL_977748, EPI_ISL_977749, EPI_ISL_977750, EPI_ISL_977751, EPI_ISL_977752, EPI_ISL_977753, EPI_ISL_977754, EPI_ISL_977755, EPI_ISL_977757, EPI_ISL_977758, EPI_ISL_977759, EPI_ISL_977760, EPI_ISL_977761, EPI_ISL_977762, EPI_ISL_977763, EPI_ISL_977764, EPI_ISL_977765, EPI_ISL_977766, EPI_ISL_977767, EPI_ISL_977768, EPI_ISL_977769, EPI_ISL_977770, EPI_ISL_977771, EPI_ISL_977772, EPI_ISL_977773, EPI_ISL_977774, EPI_ISL_977775, EPI_ISL_977776, EPI_ISL_977777, EPI_ISL_977778, EPI_ISL_977779, EPI_ISL_977780, EPI_ISL_977781, EPI_ISL_977782, EPI_ISL_977783, EPI_ISL_977784, EPI_ISL_977785, EPI_ISL_977786, EPI_ISL_977787, EPI_ISL_977788, EPI_ISL_977789, EPI_ISL_977790, EPI_ISL_977791, EPI_ISL_977792, EPI_ISL_977793, EPI_ISL_977794, EPI_ISL_977795, EPI_ISL_977796, EPI_ISL_977797, EPI_ISL_977798, EPI_ISL_977799, EPI_ISL_977800, EPI_ISL_977801, EPI_ISL_977802, EPI_ISL_977803, EPI_ISL_977804, EPI_ISL_977805, EPI_ISL_977806, EPI_ISL_977807, EPI_ISL_977808, EPI_ISL_977809, EPI_ISL_977810, EPI_ISL_977811, EPI_ISL_977812, EPI_ISL_977813, EPI_ISL_977814, EPI_ISL_977815, EPI_ISL_977816, EPI_ISL_977817, EPI_ISL_977818, EPI_ISL_977819, EPI_ISL_977820, EPI_ISL_977821, EPI_ISL_977822, EPI_ISL_977823, EPI_ISL_977824, EPI_ISL_977825, EPI_ISL_977826, EPI_ISL_977827, EPI_ISL_977828, EPI_ISL_977829, EPI_ISL_977830, EPI_ISL_977831, EPI_ISL_977832, EPI_ISL_977833, EPI_ISL_977834, EPI_ISL_977835, EPI_ISL_977836, EPI_ISL_977837, EPI_ISL_977838, EPI_ISL_977839, EPI_ISL_977840, EPI_ISL_977841, EPI_ISL_977842, EPI_ISL_977843, EPI_ISL_977844, EPI_ISL_977845, EPI_ISL_977846, EPI_ISL_977847, EPI_ISL_977848, EPI_ISL_977849, EPI_ISL_977850, EPI_ISL_977851, EPI_ISL_977864, EPI_ISL_977865, EPI_ISL_977866, EPI_ISL_977867, EPI_ISL_977869, EPI_ISL_977878, EPI_ISL_978976, EPI_ISL_978977, EPI_ISL_978978, EPI_ISL_978979, EPI_ISL_978980, EPI_ISL_978981, EPI_ISL_978982, EPI_ISL_978983, EPI_ISL_984318, EPI_ISL_984319, EPI_ISL_984320, EPI_ISL_984321, EPI_ISL_984322, EPI_ISL_984323, EPI_ISL_984324, EPI_ISL_984325, EPI_ISL_984326, EPI_ISL_984327, EPI_ISL_984328, EPI_ISL_984329, EPI_ISL_984330, EPI_ISL_984331, EPI_ISL_984333, EPI_ISL_984334, EPI_ISL_984335, EPI_ISL_984336, EPI_ISL_984337, EPI_ISL_984338, EPI_ISL_984339, EPI_ISL_984340, EPI_ISL_984341, EPI_ISL_984342, EPI_ISL_984343, EPI_ISL_984344, EPI_ISL_984345, EPI_ISL_984346, EPI_ISL_984348, EPI_ISL_984349, EPI_ISL_984350, EPI_ISL_984351, EPI_ISL_984352, EPI_ISL_984353, EPI_ISL_984355, EPI_ISL_984356, EPI_ISL_984357, EPI_ISL_984359, EPI_ISL_984361, EPI_ISL_984362, EPI_ISL_984365, EPI_ISL_984366, EPI_ISL_984367, EPI_ISL_984368, EPI_ISL_984369, EPI_ISL_984370, EPI_ISL_984375, EPI_ISL_984376, EPI_ISL_984377, EPI_ISL_984378, EPI_ISL_984380, EPI_ISL_984384, EPI_ISL_984385, EPI_ISL_984386, EPI_ISL_984391, EPI_ISL_984393, EPI_ISL_984394, EPI_ISL_984395, EPI_ISL_984396, EPI_ISL_984397, EPI_ISL_984398, EPI_ISL_984399, EPI_ISL_984400, EPI_ISL_984401, EPI_ISL_984402, EPI_ISL_984403, EPI_ISL_984404, EPI_ISL_984405, EPI_ISL_984406, EPI_ISL_984407, EPI_ISL_984408, EPI_ISL_984410, EPI_ISL_984411, EPI_ISL_984412, EPI_ISL_984413, EPI_ISL_984414, EPI_ISL_984415, EPI_ISL_984416, EPI_ISL_984417, EPI_ISL_984418, EPI_ISL_984420, EPI_ISL_984421, EPI_ISL_984422, EPI_ISL_984423, EPI_ISL_984424, EPI_ISL_984425, EPI_ISL_984426, EPI_ISL_984427, EPI_ISL_984428, EPI_ISL_984429, EPI_ISL_984430, EPI_ISL_984431, EPI_ISL_984432, EPI_ISL_984433, EPI_ISL_984434, EPI_ISL_984435, EPI_ISL_984437, EPI_ISL_984439, EPI_ISL_984440, EPI_ISL_984441, EPI_ISL_984442, EPI_ISL_984443, EPI_ISL_984444, EPI_ISL_984560, EPI_ISL_984561, EPI_ISL_984562, EPI_ISL_984563, EPI_ISL_984564, EPI_ISL_984565, EPI_ISL_984566, EPI_ISL_984567, EPI_ISL_984568, EPI_ISL_984569, EPI_ISL_984570, EPI_ISL_984571, EPI_ISL_984572, EPI_ISL_984573, EPI_ISL_984574, EPI_ISL_984575, EPI_ISL_984576, EPI_ISL_984577, EPI_ISL_984578, EPI_ISL_984579, EPI_ISL_984580, EPI_ISL_984581, EPI_ISL_984582, EPI_ISL_984583, EPI_ISL_984584, EPI_ISL_984585, EPI_ISL_984586, EPI_ISL_984587, EPI_ISL_984588, EPI_ISL_984589, EPI_ISL_984590, EPI_ISL_984591, EPI_ISL_984592, EPI_ISL_984593, EPI_ISL_984594, EPI_ISL_984595, EPI_ISL_984596, EPI_ISL_984597, EPI_ISL_984598, EPI_ISL_984599, EPI_ISL_984600, EPI_ISL_984601, EPI_ISL_984602, EPI_ISL_984603, EPI_ISL_984604, EPI_ISL_984605, EPI_ISL_984606, EPI_ISL_984607, EPI_ISL_984608, EPI_ISL_984609, EPI_ISL_984610, EPI_ISL_984611, EPI_ISL_984612, EPI_ISL_984613

| | | | |
|---|---|---|---|
| see above | California Department of Public Health | Chiu Laboratory, University of California, San Francisco | Brian Bushnell; Candace Wang; Charles Chiu; Debra Wadford; Jill Hacker; Scot Federman; Venice Servellita; Xianding (Wayne) Deng |
| EPI_ISL_2778104 | California Department of Public Health Valencia Branch Laboratory (CDPH VBL) | California Department of Public Health | CDPH-COVIDNet; UCLA Technology Center for Genomics & Bioinformatics |

EPI_ISL_885129, EPI_ISL_885130, EPI_ISL_885131, EPI_ISL_885132, EPI_ISL_885133, EPI_ISL_885134, EPI_ISL_885135, EPI_ISL_885136, EPI_ISL_885137, EPI_ISL_885138, EPI_ISL_1235014, EPI_ISL_1235056, EPI_ISL_1701638

| | | | |
|---|---|---|---|
| see above | California Institute of Technology | Chan-Zuckerberg Biohub | CZB Cliahub Consortium |

EPI_ISL_824555, EPI_ISL_824556, EPI_ISL_824557, EPI_ISL_824558, EPI_ISL_824559, EPI_ISL_824560, EPI_ISL_824561, EPI_ISL_824562, EPI_ISL_824563, EPI_ISL_824564, EPI_ISL_824565, EPI_ISL_824566, EPI_ISL_824567, EPI_ISL_824568, EPI_ISL_824569, EPI_ISL_824570, EPI_ISL_824571, EPI_ISL_824572, EPI_ISL_824573, EPI_ISL_824574, EPI_ISL_824575, EPI_ISL_824576, EPI_ISL_824577, EPI_ISL_824578, EPI_ISL_824579, EPI_ISL_824580, EPI_ISL_824581, EPI_ISL_824582, EPI_ISL_824583, EPI_ISL_824584, EPI_ISL_824585, EPI_ISL_824586, EPI_ISL_824587, EPI_ISL_824588, EPI_ISL_824589, EPI_ISL_824590, EPI_ISL_824591, EPI_ISL_824592, EPI_ISL_824593, EPI_ISL_824594, EPI_ISL_824595, EPI_ISL_824596, EPI_ISL_824597, EPI_ISL_824598, EPI_ISL_824599, EPI_ISL_824600, EPI_ISL_824601, EPI_ISL_824602, EPI_ISL_824603, EPI_ISL_824604, EPI_ISL_824605, EPI_ISL_824606, EPI_ISL_824607, EPI_ISL_824608, EPI_ISL_824609, EPI_ISL_824610, EPI_ISL_824611, EPI_ISL_824612, EPI_ISL_824613, EPI_ISL_824614, EPI_ISL_824615, EPI_ISL_824616, EPI_ISL_824618, EPI_ISL_824619, EPI_ISL_824620, EPI_ISL_824621, EPI_ISL_824622, EPI_ISL_824623, EPI_ISL_824624, EPI_ISL_824625, EPI_ISL_824626, EPI_ISL_824627, EPI_ISL_824628, EPI_ISL_824629, EPI_ISL_824630, EPI_ISL_824631, EPI_ISL_824632, EPI_ISL_824633, EPI_ISL_824634, EPI_ISL_824635, EPI_ISL_824636, EPI_ISL_824637, EPI_ISL_824638, EPI_ISL_824639, EPI_ISL_824640, EPI_ISL_824641, EPI_ISL_824642, EPI_ISL_824643, EPI_ISL_824644, EPI_ISL_824645, EPI_ISL_824646, EPI_ISL_824647, EPI_ISL_824648, EPI_ISL_824649, EPI_ISL_824650, EPI_ISL_824651, EPI_ISL_824652, EPI_ISL_824653, EPI_ISL_824654, EPI_ISL_824655, EPI_ISL_824656, EPI_ISL_824658, EPI_ISL_824659, EPI_ISL_824660, EPI_ISL_824661, EPI_ISL_824662, EPI_ISL_824663, EPI_ISL_824664, EPI_ISL_824665, EPI_ISL_824666, EPI_ISL_824667, EPI_ISL_824668, EPI_ISL_824669, EPI_ISL_824670, EPI_ISL_824671, EPI_ISL_824672, EPI_ISL_824673, EPI_ISL_824674, EPI_ISL_824675, EPI_ISL_824676, EPI_ISL_824677, EPI_ISL_824678, EPI_ISL_824679, EPI_ISL_824680, EPI_ISL_824681, EPI_ISL_824682, EPI_ISL_824683, EPI_ISL_824684, EPI_ISL_824685, EPI_ISL_824686, EPI_ISL_824687, EPI_ISL_824688, EPI_ISL_824689, EPI_ISL_824690, EPI_ISL_824691, EPI_ISL_824692, EPI_ISL_824693, EPI_ISL_824694, EPI_ISL_824695, EPI_ISL_824696, EPI_ISL_824697, EPI_ISL_824698, EPI_ISL_824699, EPI_ISL_824700, EPI_ISL_824701, EPI_ISL_824702, EPI_ISL_824703, EPI_ISL_824704, EPI_ISL_824705, EPI_ISL_824706, EPI_ISL_824707, EPI_ISL_824708, EPI_ISL_824709, EPI_ISL_824710, EPI_ISL_824711, EPI_ISL_824712, EPI_ISL_824713, EPI_ISL_824714, EPI_ISL_824715, EPI_ISL_824716, EPI_ISL_824717, EPI_ISL_824718, EPI_ISL_824719, EPI_ISL_824720, EPI_ISL_824721, EPI_ISL_824722, EPI_ISL_824723, EPI_ISL_824724, EPI_ISL_824725, EPI_ISL_824726, EPI_ISL_824727, EPI_ISL_824728, EPI_ISL_824729, EPI_ISL_824730, EPI_ISL_824731, EPI_ISL_824732, EPI_ISL_824733, EPI_ISL_824734, EPI_ISL_824735, EPI_ISL_824736, EPI_ISL_824737, EPI_ISL_824738, EPI_ISL_824739, EPI_ISL_824740, EPI_ISL_824741

| | | | |
|---|---|---|---|
| see above | Cedars-Sinai Medical Center, Department of Pathology & Laboratory Medicine, Molecular Pathology Laboratory | Cedars-Sinai Medical Center, Molecular Pathology Laboratory of Department of Pathology & Laboratory Medicine and Genomic Core | Brian Davis; Eric Vail; Jasmine T Plummer; Jorge Mario Sincuir Martinez; Stephanie Chen; Wenjuan Zhang |

EPI_ISL_847515, EPI_ISL_847517, EPI_ISL_847536, EPI_ISL_847540, EPI_ISL_847545, EPI_ISL_847546, EPI_ISL_847553, EPI_ISL_847563, EPI_ISL_847579, EPI_ISL_847619, EPI_ISL_847620, EPI_ISL_847621, EPI_ISL_847622, EPI_ISL_847640, EPI_ISL_847641, EPI_ISL_847642, EPI_ISL_847643, EPI_ISL_847644, EPI_ISL_847716, EPI_ISL_847720, EPI_ISL_847731, EPI_ISL_847736, EPI_ISL_847738, EPI_ISL_847741, EPI_ISL_847742, EPI_ISL_847744, EPI_ISL_847746, EPI_ISL_847747, EPI_ISL_847782, EPI_ISL_855399, EPI_ISL_855400, EPI_ISL_855401, EPI_ISL_977885, EPI_ISL_977886, EPI_ISL_977887, EPI_ISL_977888, EPI_ISL_977889, EPI_ISL_977890, EPI_ISL_977891, EPI_ISL_977892, EPI_ISL_977893, EPI_ISL_977894, EPI_ISL_977895, EPI_ISL_977896, EPI_ISL_977897, EPI_ISL_977898, EPI_ISL_977899, EPI_ISL_977900, EPI_ISL_977901, EPI_ISL_977902, EPI_ISL_977903, EPI_ISL_977904, EPI_ISL_977905, EPI_ISL_977906, EPI_ISL_977907, EPI_ISL_977908, EPI_ISL_977909, EPI_ISL_977981, EPI_ISL_977982, EPI_ISL_977983, EPI_ISL_977985, EPI_ISL_977986, EPI_ISL_977987, EPI_ISL_977988, EPI_ISL_977989, EPI_ISL_977990, EPI_ISL_977991, EPI_ISL_977992, EPI_ISL_977993, EPI_ISL_977994, EPI_ISL_977995, EPI_ISL_977996, EPI_ISL_977997, EPI_ISL_977998, EPI_ISL_977999, EPI_ISL_978000, EPI_ISL_978001, EPI_ISL_978002, EPI_ISL_978003, EPI_ISL_978004, EPI_ISL_978005, EPI_ISL_978006, EPI_ISL_978007, EPI_ISL_978008, EPI_ISL_978009, EPI_ISL_978010, EPI_ISL_978011, EPI_ISL_978012, EPI_ISL_978013, EPI_ISL_978014, EPI_ISL_978015, EPI_ISL_978016, EPI_ISL_978017, EPI_ISL_978018, EPI_ISL_978019, EPI_ISL_978020, EPI_ISL_978021, EPI_ISL_978022, EPI_ISL_978023, EPI_ISL_978024, EPI_ISL_978025, EPI_ISL_978026, EPI_ISL_978027, EPI_ISL_978028, EPI_ISL_978029, EPI_ISL_978030, EPI_ISL_978031, EPI_ISL_978032, EPI_ISL_978033, EPI_ISL_978034, EPI_ISL_978035, EPI_ISL_978036, EPI_ISL_978037, EPI_ISL_978038, EPI_ISL_978039, EPI_ISL_978040, EPI_ISL_978041, EPI_ISL_978042, EPI_ISL_978043, EPI_ISL_978044, EPI_ISL_978045, EPI_ISL_978046, EPI_ISL_978047, EPI_ISL_978048, EPI_ISL_978049, EPI_ISL_978050, EPI_ISL_978051, EPI_ISL_978052, EPI_ISL_978053, EPI_ISL_978054, EPI_ISL_978055, EPI_ISL_978056, EPI_ISL_978057, EPI_ISL_978058, EPI_ISL_978059, EPI_ISL_978060, EPI_ISL_978061, EPI_ISL_978062, EPI_ISL_978070, EPI_ISL_978071, EPI_ISL_978072, EPI_ISL_978073, EPI_ISL_978074, EPI_ISL_978119, EPI_ISL_978120, EPI_ISL_978121, EPI_ISL_978122, EPI_ISL_978123, EPI_ISL_978129, EPI_ISL_978130, EPI_ISL_978131, EPI_ISL_978160, EPI_ISL_978161, EPI_ISL_978162, EPI_ISL_978163, EPI_ISL_978164, EPI_ISL_978165, EPI_ISL_978166, EPI_ISL_978167, EPI_ISL_978168, EPI_ISL_978955, EPI_ISL_978956, EPI_ISL_978957, EPI_ISL_978958, EPI_ISL_978959, EPI_ISL_978960, EPI_ISL_978967, EPI_ISL_978968, EPI_ISL_978969, EPI_ISL_978970, EPI_ISL_978971, EPI_ISL_978972

| | | | |
|---|---|---|---|
| see above | Chiu Laboratory, University of California, San Francisco | Chiu Laboratory, University of California, San Francisco | Brian Bushnell; Candace Wang; Charles Chiu; Debra Wadford; Jill Hacker; Scot Federman; Venice Servellita; Xianding (Wayne) Deng |

EPI_ISL_753262, EPI_ISL_753263, EPI_ISL_753264, EPI_ISL_753265, EPI_ISL_753266, EPI_ISL_753267, EPI_ISL_753268, EPI_ISL_753269, EPI_ISL_753270, EPI_ISL_753271, EPI_ISL_753272, EPI_ISL_753273, EPI_ISL_753274, EPI_ISL_753275, EPI_ISL_753276, EPI_ISL_753277, EPI_ISL_753278, EPI_ISL_753279, EPI_ISL_753280, EPI_ISL_753281, EPI_ISL_753282, EPI_ISL_753283, EPI_ISL_753284, EPI_ISL_753285, EPI_ISL_753286, EPI_ISL_753287, EPI_ISL_753288, EPI_ISL_753289, EPI_ISL_753290, EPI_ISL_753291, EPI_ISL_753292, EPI_ISL_753293, EPI_ISL_753294, EPI_ISL_753295, EPI_ISL_753296, EPI_ISL_753297, EPI_ISL_753298, EPI_ISL_753299, EPI_ISL_753300, EPI_ISL_753301, EPI_ISL_753302, EPI_ISL_753303, EPI_ISL_753304, EPI_ISL_753305, EPI_ISL_753306, EPI_ISL_753307, EPI_ISL_753308, EPI_ISL_753309, EPI_ISL_753310, EPI_ISL_753311, EPI_ISL_753312, EPI_ISL_753313, EPI_ISL_753314, EPI_ISL_753315, EPI_ISL_753316, EPI_ISL_753317, EPI_ISL_753318, EPI_ISL_753319, EPI_ISL_753320, EPI_ISL_753321, EPI_ISL_753322, EPI_ISL_753323, EPI_ISL_753325, EPI_ISL_753326, EPI_ISL_753327, EPI_ISL_753328, EPI_ISL_753329, EPI_ISL_753330, EPI_ISL_753331, EPI_ISL_753332, EPI_ISL_753333, EPI_ISL_753334, EPI_ISL_753335, EPI_ISL_753336, EPI_ISL_753337, EPI_ISL_753338, EPI_ISL_753339, EPI_ISL_753340, EPI_ISL_753341, EPI_ISL_753342, EPI_ISL_753343, EPI_ISL_753344, EPI_ISL_753345, EPI_ISL_753346, EPI_ISL_753347, EPI_ISL_753348, EPI_ISL_753349, EPI_ISL_753350, EPI_ISL_753351, EPI_ISL_753352, EPI_ISL_753353, EPI_ISL_753354, EPI_ISL_753355, EPI_ISL_753356, EPI_ISL_753357, EPI_ISL_753358, EPI_ISL_753359, EPI_ISL_753360, EPI_ISL_753361, EPI_ISL_753362, EPI_ISL_753363, EPI_ISL_753364, EPI_ISL_753365, EPI_ISL_753366, EPI_ISL_753367, EPI_ISL_753368, EPI_ISL_753369, EPI_ISL_753370, EPI_ISL_753371, EPI_ISL_753372, EPI_ISL_753373, EPI_ISL_753374, EPI_ISL_753375, EPI_ISL_753376, EPI_ISL_753377, EPI_ISL_753378, EPI_ISL_753379, EPI_ISL_753380, EPI_ISL_753381, EPI_ISL_753382, EPI_ISL_753383, EPI_ISL_753384, EPI_ISL_753385, EPI_ISL_753386, EPI_ISL_753387, EPI_ISL_753388, EPI_ISL_753389, EPI_ISL_753390, EPI_ISL_753391, EPI_ISL_753392, EPI_ISL_753393, EPI_ISL_753394, EPI_ISL_753395, EPI_ISL_753396, EPI_ISL_753397, EPI_ISL_753398, EPI_ISL_753399, EPI_ISL_753400, EPI_ISL_753401, EPI_ISL_753402, EPI_ISL_753403, EPI_ISL_753404, EPI_ISL_753405, EPI_ISL_753406, EPI_ISL_753407, EPI_ISL_753408, EPI_ISL_753409, EPI_ISL_753410, EPI_ISL_753411, EPI_ISL_753412, EPI_ISL_753413, EPI_ISL_753414, EPI_ISL_753415, EPI_ISL_753416, EPI_ISL_753417, EPI_ISL_753418, EPI_ISL_753419, EPI_ISL_753420, EPI_ISL_753421, EPI_ISL_753422, EPI_ISL_753423, EPI_ISL_753424, EPI_ISL_753425, EPI_ISL_753426, EPI_ISL_753427, EPI_ISL_753428, EPI_ISL_753429, EPI_ISL_753430, EPI_ISL_753431, EPI_ISL_753432, EPI_ISL_753433, EPI_ISL_753434, EPI_ISL_753435, EPI_ISL_753436, EPI_ISL_753437, EPI_ISL_753438, EPI_ISL_753439, EPI_ISL_753440, EPI_ISL_753441, EPI_ISL_753442, EPI_ISL_753443, EPI_ISL_753444, EPI_ISL_753445, EPI_ISL_753446, EPI_ISL_753447, EPI_ISL_753448, EPI_ISL_753449, EPI_ISL_753450, EPI_ISL_753451, EPI_ISL_753452, EPI_ISL_753453, EPI_ISL_753454, EPI_ISL_753455, EPI_ISL_753456, EPI_ISL_753457, EPI_ISL_753458, EPI_ISL_753459, EPI_ISL_753460, EPI_ISL_753461, EPI_ISL_753462, EPI_ISL_753463, EPI_ISL_753464, EPI_ISL_753465, EPI_ISL_753466, EPI_ISL_753467, EPI_ISL_753468, EPI_ISL_753469, EPI_ISL_753470, EPI_ISL_753471, EPI_ISL_753472, EPI_ISL_753473, EPI_ISL_753474, EPI_ISL_753475, EPI_ISL_753476, EPI_ISL_753477, EPI_ISL_753478, EPI_ISL_753479, EPI_ISL_753480, EPI_ISL_753481, EPI_ISL_753482, EPI_ISL_753483, EPI_ISL_753484, EPI_ISL_753485, EPI_ISL_753486, EPI_ISL_753487, EPI_ISL_753488, EPI_ISL_753489, EPI_ISL_753490, EPI_ISL_753491, EPI_ISL_753492, EPI_ISL_753493, EPI_ISL_753494, EPI_ISL_753495, EPI_ISL_753496, EPI_ISL_753497, EPI_ISL_753498, EPI_ISL_753499, EPI_ISL_753500, EPI_ISL_753501, EPI_ISL_753502, EPI_ISL_753503, EPI_ISL_753504, EPI_ISL_753505, EPI_ISL_753506, EPI_ISL_753507, EPI_ISL_753508, EPI_ISL_753509, EPI_ISL_753510, EPI_ISL_753511, EPI_ISL_753512, EPI_ISL_753513, EPI_ISL_753514, EPI_ISL_753515, EPI_ISL_753516, EPI_ISL_753517, EPI_ISL_753518, EPI_ISL_753519, EPI_ISL_753520, EPI_ISL_753521, EPI_ISL_753522, EPI_ISL_753523, EPI_ISL_753524, EPI_ISL_753525, EPI_ISL_753526, EPI_ISL_753527, EPI_ISL_753528, EPI_ISL_753529, EPI_ISL_753530, EPI_ISL_753531, EPI_ISL_753532, EPI_ISL_753533, EPI_ISL_753534, EPI_ISL_753535, EPI_ISL_753536, EPI_ISL_753537, EPI_ISL_753538, EPI_ISL_753539, EPI_ISL_753540, EPI_ISL_753541, EPI_ISL_753542, EPI_ISL_753543, EPI_ISL_753544, EPI_ISL_753545, EPI_ISL_753546, EPI_ISL_753547, EPI_ISL_753548, EPI_ISL_753549, EPI_ISL_753550, EPI_ISL_753551, EPI_ISL_753552, EPI_ISL_753553, EPI_ISL_753554, EPI_ISL_753555, EPI_ISL_753556, EPI_ISL_753557, EPI_ISL_753558, EPI_ISL_753559, EPI_ISL_753560, EPI_ISL_753561, EPI_ISL_753562, EPI_ISL_753563, EPI_ISL_753564, EPI_ISL_753565, EPI_ISL_753566, EPI_ISL_753567, EPI_ISL_753568, EPI_ISL_753569, EPI_ISL_753570, EPI_ISL_753571, EPI_ISL_753572, EPI_ISL_753573, EPI_ISL_753574, EPI_ISL_753575, EPI_ISL_753576, EPI_ISL_753577, EPI_ISL_753578, EPI_ISL_753580, EPI_ISL_753581, EPI_ISL_753582, EPI_ISL_753583, EPI_ISL_753584, EPI_ISL_753585, EPI_ISL_753586, EPI_ISL_753587, EPI_ISL_753588, EPI_ISL_753589, EPI_ISL_753590, EPI_ISL_753591, EPI_ISL_753592, EPI_ISL_753593, EPI_ISL_753594, EPI_ISL_753595, EPI_ISL_753596, EPI_ISL_753597, EPI_ISL_753598, EPI_ISL_753599, EPI_ISL_753600, EPI_ISL_753601, EPI_ISL_753602, EPI_ISL_753603, EPI_ISL_753604, EPI_ISL_753605, EPI_ISL_753606, EPI_ISL_753607, EPI_ISL_753608, EPI_ISL_753609, EPI_ISL_753610, EPI_ISL_753611, EPI_ISL_753612, EPI_ISL_753613, EPI_ISL_753614, EPI_ISL_753615, EPI_ISL_753616, EPI_ISL_753617, EPI_ISL_753618, EPI_ISL_753619, EPI_ISL_753620, EPI_ISL_753621, EPI_ISL_753622, EPI_ISL_753623, EPI_ISL_753624, EPI_ISL_753625, EPI_ISL_753626, EPI_ISL_753627, EPI_ISL_753628, EPI_ISL_753629, EPI_ISL_753630, EPI_ISL_753631, EPI_ISL_753632, EPI_ISL_753633, EPI_ISL_753634, EPI_ISL_753635, EPI_ISL_753636, EPI_ISL_753637, EPI_ISL_753638, EPI_ISL_753639, EPI_ISL_753640, EPI_ISL_753641, EPI_ISL_753642, EPI_ISL_753643, EPI_ISL_753644, EPI_ISL_753645, EPI_ISL_753646, EPI_ISL_753647, EPI_ISL_753648, EPI_ISL_753649, EPI_ISL_753650, EPI_ISL_753651, EPI_ISL_753652, EPI_ISL_753653, EPI_ISL_753654, EPI_ISL_753655, EPI_ISL_753656, EPI_ISL_753657, EPI_ISL_753658, EPI_ISL_753659, EPI_ISL_753660, EPI_ISL_753661, EPI_ISL_753662, EPI_ISL_753663, EPI_ISL_753664, EPI_ISL_753665, EPI_ISL_753666, EPI_ISL_753667, EPI_ISL_753668, EPI_ISL_753669, EPI_ISL_753670, EPI_ISL_753671, EPI_ISL_753672, EPI_ISL_753673, EPI_ISL_753674, EPI_ISL_753675, EPI_ISL_753676, EPI_ISL_753677, EPI_ISL_753678, EPI_ISL_753679, EPI_ISL_753680, EPI_ISL_753681, EPI_ISL_753682, EPI_ISL_753683, EPI_ISL_753684, EPI_ISL_753685, EPI_ISL_753686, EPI_ISL_753687, EPI_ISL_753688, EPI_ISL_753689, EPI_ISL_753690, EPI_ISL_753691, EPI_ISL_753692

| | | | |
|---|---|---|---|
| see above | Clinical virology Laboratory, Children's Hospital Los Angeles | Center for Personalized Medicine, Children's Hospital Los Angeles | Gai et al |

EPI_ISL_739399, EPI_ISL_739480, EPI_ISL_1477965, EPI_ISL_1478006, EPI_ISL_1478018, EPI_ISL_1478025, EPI_ISL_1478044, EPI_ISL_1478045, EPI_ISL_1478061, EPI_ISL_1478070, EPI_ISL_1478074, EPI_ISL_1478080, EPI_ISL_1478118, EPI_ISL_1478136, EPI_ISL_1478179, EPI_ISL_1478195, EPI_ISL_1478198, EPI_ISL_1529168, EPI_ISL_1529237, EPI_ISL_1529255, EPI_ISL_1529256, EPI_ISL_1529268, EPI_ISL_1529270, EPI_ISL_1529279, EPI_ISL_1529285, EPI_ISL_1529292, EPI_ISL_1529293, EPI_ISL_1529311, EPI_ISL_1701066, EPI_ISL_1701073, EPI_ISL_1701093, EPI_ISL_1701168, EPI_ISL_1701189, EPI_ISL_1701219, EPI_ISL_1701264, EPI_ISL_1701268, EPI_ISL_1701408, EPI_ISL_1701419, EPI_ISL_1701460, EPI_ISL_1701461, EPI_ISL_1701464, EPI_ISL_1701468, EPI_ISL_1701476, EPI_ISL_1701490, EPI_ISL_1701519, EPI_ISL_1701531, EPI_ISL_1701539, EPI_ISL_1701540, EPI_ISL_1701557, EPI_ISL_1701562, EPI_ISL_1701569, EPI_ISL_1701582, EPI_ISL_2102802, EPI_ISL_2102870, EPI_ISL_2102889, EPI_ISL_2102925, EPI_ISL_2102926, EPI_ISL_2102938, EPI_ISL_2102953, EPI_ISL_2102971, EPI_ISL_2103044, EPI_ISL_2103096, EPI_ISL_2228263, EPI_ISL_2228274, EPI_ISL_2228297, EPI_ISL_2228311, EPI_ISL_2228367, EPI_ISL_2228420, EPI_ISL_2497553, EPI_ISL_2497554, EPI_ISL_2497676, EPI_ISL_2497680, EPI_ISL_2497695, EPI_ISL_2497696, EPI_ISL_2497701, EPI_ISL_2497702, EPI_ISL_2497703, EPI_ISL_2497705, EPI_ISL_2497706, EPI_ISL_2497711, EPI_ISL_2497712, EPI_ISL_2497713, EPI_ISL_2497715, EPI_ISL_2497718, EPI_ISL_2497720, EPI_ISL_2497723, EPI_ISL_2497726, EPI_ISL_2497727, EPI_ISL_2497728, EPI_ISL_2497729, EPI_ISL_2497731, EPI_ISL_2497732, EPI_ISL_2497735, EPI_ISL_2497736, EPI_ISL_2497738, EPI_ISL_2497739, EPI_ISL_2497740, EPI_ISL_2497744, EPI_ISL_2497745, EPI_ISL_2497747, EPI_ISL_2497748, EPI_ISL_2497750, EPI_ISL_2497753, EPI_ISL_2497754, EPI_ISL_2497755, EPI_ISL_2497758, EPI_ISL_2497760, EPI_ISL_2497761, EPI_ISL_2497763, EPI_ISL_2497764, EPI_ISL_2497765, EPI_ISL_2497767, EPI_ISL_2497770, EPI_ISL_2497771, EPI_ISL_2497773, EPI_ISL_2497775, EPI_ISL_2497776, EPI_ISL_2497777, EPI_ISL_2497778, EPI_ISL_2497779, EPI_ISL_2497781, EPI_ISL_2497784, EPI_ISL_2497785, EPI_ISL_2497787, EPI_ISL_2497789, EPI_ISL_2497790, EPI_ISL_2497796, EPI_ISL_2497799, EPI_ISL_2497802, EPI_ISL_2497803, EPI_ISL_2497804, EPI_ISL_2497805, EPI_ISL_2497806, EPI_ISL_2497807, EPI_ISL_2497808, EPI_ISL_2497809, EPI_ISL_2497810, EPI_ISL_2497814, EPI_ISL_2497815, EPI_ISL_2497817, EPI_ISL_2497818, EPI_ISL_2497819, EPI_ISL_2497820, EPI_ISL_2497824, EPI_ISL_2497825, EPI_ISL_2497826, EPI_ISL_2497828, EPI_ISL_2497829, EPI_ISL_2497830, EPI_ISL_2497831, EPI_ISL_2497832, EPI_ISL_2497833, EPI_ISL_2497838, EPI_ISL_2497844, EPI_ISL_2497846, EPI_ISL_2658635, EPI_ISL_2658636, EPI_ISL_2659252, EPI_ISL_2659289, EPI_ISL_2659337

| | | | |
|---|---|---|---|
| see above | County of San Luis Obispo Public Health Laboratory | Chan-Zuckerberg Biohub | CZB Cliahub Consortium |

EPI_ISL_2758597, EPI_ISL_2758598, EPI_ISL_2758599, EPI_ISL_2758600, EPI_ISL_2758601, EPI_ISL_2758602, EPI_ISL_2758603, EPI_ISL_2758604, EPI_ISL_2758605, EPI_ISL_2758606

| | | | |
|---|---|---|---|
| see above | County of San Luis Obispo Public Health Laboratory | County of San Luis Obispo Public Health Laboratory | Frances Sidhu; Katrina Erwin |

EPI_ISL_1811644, EPI_ISL_1811645, EPI_ISL_1811655, EPI_ISL_1811660, EPI_ISL_1811664, EPI_ISL_1811668, EPI_ISL_1811672, EPI_ISL_1811673, EPI_ISL_1811677, EPI_ISL_1811684, EPI_ISL_1811709, EPI_ISL_1811713, EPI_ISL_1811716, EPI_ISL_1811723, EPI_ISL_1811734, EPI_ISL_1811790, EPI_ISL_1811792, EPI_ISL_1811793, EPI_ISL_1811812, EPI_ISL_1811824, EPI_ISL_1811835, EPI_ISL_1811867, EPI_ISL_1811891, EPI_ISL_1811940, EPI_ISL_1811966, EPI_ISL_1812007, EPI_ISL_1812013, EPI_ISL_1812033, EPI_ISL_1812047, EPI_ISL_1812073, EPI_ISL_1812159, EPI_ISL_1812178, EPI_ISL_1812199, EPI_ISL_1812217, EPI_ISL_1812261, EPI_ISL_1812271, EPI_ISL_1812281, EPI_ISL_1812282, EPI_ISL_1812291, EPI_ISL_1812330, EPI_ISL_1812361, EPI_ISL_1812363, EPI_ISL_1812371, EPI_ISL_1812448, EPI_ISL_1812449, EPI_ISL_1812492, EPI_ISL_1812495, EPI_ISL_1812520, EPI_ISL_1812590, EPI_ISL_1812638, EPI_ISL_1812646, EPI_ISL_1812683, EPI_ISL_1812758, EPI_ISL_1812766, EPI_ISL_1812784, EPI_ISL_1812798, EPI_ISL_1812799, EPI_ISL_1812810, EPI_ISL_1812826, EPI_ISL_1812854, EPI_ISL_1812859, EPI_ISL_1812900, EPI_ISL_1812905, EPI_ISL_1812910, EPI_ISL_1812917, EPI_ISL_1812929, EPI_ISL_1812955, EPI_ISL_1812981, EPI_ISL_1812987, EPI_ISL_1812994, EPI_ISL_1813002, EPI_ISL_1813040, EPI_ISL_1813042, EPI_ISL_1813059, EPI_ISL_1813063, EPI_ISL_1813079, EPI_ISL_1813093, EPI_ISL_1813097, EPI_ISL_1813113, EPI_ISL_1813134, EPI_ISL_1813139, EPI_ISL_1813145, EPI_ISL_1813304, EPI_ISL_1813306, EPI_ISL_1813340, EPI_ISL_1813342, EPI_ISL_1813377, EPI_ISL_1813405, EPI_ISL_1813409, EPI_ISL_1813425, EPI_ISL_1813430, EPI_ISL_1813497, EPI_ISL_1813500, EPI_ISL_1813512, EPI_ISL_1813600, EPI_ISL_1813711, EPI_ISL_1813783, EPI_ISL_1813806, EPI_ISL_1813851, EPI_ISL_1813925, EPI_ISL_1813975, EPI_ISL_1814026, EPI_ISL_1814049, EPI_ISL_1814055, EPI_ISL_1814237, EPI_ISL_1814460, EPI_ISL_1814601, EPI_ISL_1814637, EPI_ISL_1814649, EPI_ISL_1814744, EPI_ISL_1814766, EPI_ISL_1814815, EPI_ISL_1815137, EPI_ISL_1815209, EPI_ISL_1815351, EPI_ISL_1815380, EPI_ISL_1815495, EPI_ISL_1815515, EPI_ISL_1815519, EPI_ISL_1815566, EPI_ISL_1815594, EPI_ISL_1815677, EPI_ISL_1815748, EPI_ISL_1815764, EPI_ISL_1815789, EPI_ISL_1816066, EPI_ISL_1816079, EPI_ISL_1816123, EPI_ISL_1816127, EPI_ISL_1816197, EPI_ISL_1816220, EPI_ISL_1816264, EPI_ISL_1816265, EPI_ISL_1816270, EPI_ISL_1816277, EPI_ISL_1816290, EPI_ISL_1816297, EPI_ISL_1816405, EPI_ISL_1816489, EPI_ISL_1816556, EPI_ISL_1816583, EPI_ISL_1816597, EPI_ISL_1816633, EPI_ISL_1816659, EPI_ISL_1816674, EPI_ISL_1816742, EPI_ISL_1816750, EPI_ISL_1816761, EPI_ISL_1816764, EPI_ISL_1816808, EPI_ISL_1816821, EPI_ISL_1816825, EPI_ISL_1816830, EPI_ISL_1816831, EPI_ISL_1816833, EPI_ISL_1816834, EPI_ISL_1816852, EPI_ISL_1816859, EPI_ISL_1816860, EPI_ISL_1816865, EPI_ISL_1816871, EPI_ISL_1816873, EPI_ISL_1816876, EPI_ISL_1816878, EPI_ISL_1816880, EPI_ISL_1816882, EPI_ISL_1816895, EPI_ISL_1816896, EPI_ISL_1816899, EPI_ISL_1816901, EPI_ISL_1816908

| | | | |
|---|---|---|---|
| see above | EXCITE Lab | Andersen lab at Scripps Research | Abigail Schnapper; Angela Scioscia; Chip Schooley; David Pride; Helena Tubb; Natasha Martin Cheryl Anderson; Sawyer Farmer; Sharon Reed; Sydney Morgan + Marni Jacobs + Celestine Magallanes + Holly Valentine + Louise C. Laurent + SEARCH; Tommy Valles + SEARCH |

| | | | |
|---|---|---|---|
| EPI_ISL_1235371, EPI_ISL_1235408, EPI_ISL_1235413, EPI_ISL_1235422, EPI_ISL_1235560, EPI_ISL_1235607 | Fresno County Public Health Laboratory | Chan-Zuckerberg Biohub | CZB Cliahub Consortium |
| EPI_ISL_955261, EPI_ISL_955263, EPI_ISL_1005583, EPI_ISL_1005584, EPI_ISL_1005585, EPI_ISL_1005586, EPI_ISL_1005587, EPI_ISL_1005588, EPI_ISL_1005589, EPI_ISL_1005590, EPI_ISL_1005591, EPI_ISL_1005592, EPI_ISL_1005593, EPI_ISL_1005594, EPI_ISL_1005595, EPI_ISL_1005596, EPI_ISL_1005597, EPI_ISL_1005598, EPI_ISL_1005599, EPI_ISL_1005600, EPI_ISL_1005601, EPI_ISL_1005602, EPI_ISL_1005603, EPI_ISL_1005604, EPI_ISL_1005605, EPI_ISL_1005606, EPI_ISL_1005607, EPI_ISL_1005608, EPI_ISL_1005609, EPI_ISL_1005610, EPI_ISL_1005611, EPI_ISL_1005612, EPI_ISL_1005613, EPI_ISL_1005614, EPI_ISL_1005615, EPI_ISL_1005616, EPI_ISL_1005617, EPI_ISL_1005618, EPI_ISL_1005619, EPI_ISL_1005620, EPI_ISL_1005621, EPI_ISL_1005622, EPI_ISL_1005623, EPI_ISL_1005624, EPI_ISL_1005625, EPI_ISL_1005626, EPI_ISL_1005627, EPI_ISL_1005628, EPI_ISL_1005629, EPI_ISL_1005630, EPI_ISL_1005631, EPI_ISL_1005632, EPI_ISL_1005633, EPI_ISL_1005634, EPI_ISL_1005635, EPI_ISL_1005636, EPI_ISL_1005637, EPI_ISL_1005638, EPI_ISL_1005639, EPI_ISL_1005640, EPI_ISL_1005641, EPI_ISL_1005642, EPI_ISL_1005643, EPI_ISL_1005644, EPI_ISL_1005645, EPI_ISL_1005646, EPI_ISL_1005647, EPI_ISL_1005648, EPI_ISL_1005649, EPI_ISL_1005650, EPI_ISL_1005651, EPI_ISL_1005652, EPI_ISL_1005653, EPI_ISL_1005654, EPI_ISL_1027178, EPI_ISL_1027179, EPI_ISL_1027180, EPI_ISL_1027181, EPI_ISL_1027182, EPI_ISL_1027183, EPI_ISL_1027184, EPI_ISL_1027185, EPI_ISL_1027186, EPI_ISL_1027187, EPI_ISL_1027188, EPI_ISL_1027189, EPI_ISL_1027190, EPI_ISL_1027191, EPI_ISL_1027192, EPI_ISL_1027193, EPI_ISL_1027194, EPI_ISL_1027195, EPI_ISL_1027196, EPI_ISL_1027197, EPI_ISL_1027198, EPI_ISL_1027199, EPI_ISL_1027200, EPI_ISL_1027201, EPI_ISL_1027202, EPI_ISL_1027203, EPI_ISL_1027204, EPI_ISL_1027205, EPI_ISL_1027206, EPI_ISL_1027207, EPI_ISL_1027208, EPI_ISL_1027209, EPI_ISL_1027210, EPI_ISL_1027211, EPI_ISL_1027212, EPI_ISL_1027213, EPI_ISL_1027214, EPI_ISL_1027215, EPI_ISL_1027216, EPI_ISL_1027217, EPI_ISL_1027218, EPI_ISL_1027219, EPI_ISL_1027220, EPI_ISL_1027221, EPI_ISL_1027222, EPI_ISL_1027223, EPI_ISL_1027224, EPI_ISL_1027225, EPI_ISL_1027226, EPI_ISL_1027227, EPI_ISL_1027228, EPI_ISL_1027229, EPI_ISL_1027230, EPI_ISL_1027231, EPI_ISL_1027232, EPI_ISL_1027233, EPI_ISL_1027234, EPI_ISL_1027235, EPI_ISL_1027236, EPI_ISL_1027237, EPI_ISL_1027238, EPI_ISL_1027239, EPI_ISL_1027240, EPI_ISL_1027241, EPI_ISL_1027242, EPI_ISL_1027243, EPI_ISL_1027244, EPI_ISL_1027245, EPI_ISL_1027246, EPI_ISL_1027247, EPI_ISL_3067433 | | | |
| see above | Fulgent Genetics | Fulgent Genetics | Becky Tsai; Benafsh Sapra; Doreen Ng; Harry Gao; James Xie; John Gao; Joseph Fierro; Mickey Li; Yan Meng |
| EPI_ISL_755594, EPI_ISL_755595, EPI_ISL_755596, EPI_ISL_755597, EPI_ISL_755602, EPI_ISL_755613, EPI_ISL_802604, EPI_ISL_802605, EPI_ISL_802607, EPI_ISL_802608, EPI_ISL_802609, EPI_ISL_802610, EPI_ISL_802611, EPI_ISL_802612, EPI_ISL_802613, EPI_ISL_802614, EPI_ISL_802615, EPI_ISL_802616, EPI_ISL_802617, EPI_ISL_802618, EPI_ISL_802619, EPI_ISL_802620, EPI_ISL_802621, EPI_ISL_802622, EPI_ISL_802623, EPI_ISL_802624, EPI_ISL_802657, EPI_ISL_802658, EPI_ISL_802659, EPI_ISL_802660, EPI_ISL_802661, EPI_ISL_802662, EPI_ISL_802663, EPI_ISL_802664, EPI_ISL_802665, EPI_ISL_802720 | | | |
| see above | Helix/Illumina | Genomics and Discovery, Respiratory Viruses Branch, Division of Viral Diseases, Centers for Disease Control and Prevention | Alexandre Bolze; Ary Ascencio; Ben L. Rambo-Martin Eileen de Feo; Brad Sickler; Charlotte Rivera-Garcia; Christine Tran; Clinton R. Paden; David Becker; Dhwani Batra; Duncan MacCannell; Efren Sandoval; Eileen de Feo; Elizabeth Cirulli; Eric Allen; Geraint Levan; James Lu; Jan Antico; Jason Nguyen; Jimmy Ramirez; Jingtao Liu; Kelly Schiabor Barrett; Kim Gietzen; Magnus Isaksson; Marc Laurent; Matthew Tolentino; Nicole L. Washington; Peter W. Cook; Phil Febbo; Ryan Cho; Shannon Wickline; Sherry Wang; Simon White; Summer Galloway; Suxiang Tong; Tyler Cassens; William Lee |
| EPI_ISL_738504, EPI_ISL_738505, EPI_ISL_738539, EPI_ISL_738563, EPI_ISL_738600, EPI_ISL_738647, EPI_ISL_738649, EPI_ISL_738651, EPI_ISL_738742, EPI_ISL_738759, EPI_ISL_738779, EPI_ISL_738786, EPI_ISL_738795, EPI_ISL_738801, EPI_ISL_738806, EPI_ISL_738807, EPI_ISL_738809, EPI_ISL_738810, EPI_ISL_738827, EPI_ISL_738832, EPI_ISL_738844, EPI_ISL_738849, EPI_ISL_738852, EPI_ISL_738854, EPI_ISL_738865, EPI_ISL_738875, EPI_ISL_738902, EPI_ISL_738912, EPI_ISL_738913, EPI_ISL_738917, EPI_ISL_738918, EPI_ISL_738923, EPI_ISL_738929, EPI_ISL_738938, EPI_ISL_738947, EPI_ISL_738953, EPI_ISL_738958, EPI_ISL_738969, EPI_ISL_738980, EPI_ISL_738985, EPI_ISL_738999, EPI_ISL_739000, EPI_ISL_739001, EPI_ISL_739007, EPI_ISL_739032, EPI_ISL_739043, EPI_ISL_739051, EPI_ISL_739052, EPI_ISL_739079, EPI_ISL_739106, EPI_ISL_739112, EPI_ISL_739114, EPI_ISL_739123, EPI_ISL_739126, EPI_ISL_739131, EPI_ISL_739155, EPI_ISL_739159, EPI_ISL_739165, EPI_ISL_739170, EPI_ISL_739183, EPI_ISL_739194, EPI_ISL_739198, EPI_ISL_739210, EPI_ISL_739228, EPI_ISL_739235, EPI_ISL_739240, EPI_ISL_739242, EPI_ISL_739250, EPI_ISL_739269, EPI_ISL_739272, EPI_ISL_739274, EPI_ISL_739287, EPI_ISL_739292, EPI_ISL_739299, EPI_ISL_739313, EPI_ISL_739318, EPI_ISL_739330, EPI_ISL_739367, EPI_ISL_739389, EPI_ISL_739403, EPI_ISL_739405, EPI_ISL_739411, EPI_ISL_739417, EPI_ISL_739418, EPI_ISL_739425, EPI_ISL_739433, EPI_ISL_739436, EPI_ISL_739442, EPI_ISL_739447, EPI_ISL_739452, EPI_ISL_739461, EPI_ISL_739475, EPI_ISL_739499, EPI_ISL_739525, EPI_ISL_739527, EPI_ISL_739530, EPI_ISL_739531, EPI_ISL_739542, EPI_ISL_739553, EPI_ISL_739585, EPI_ISL_739608, EPI_ISL_739611, EPI_ISL_739621, EPI_ISL_739651, EPI_ISL_955496, EPI_ISL_955497, EPI_ISL_955498, EPI_ISL_955499, EPI_ISL_955500, EPI_ISL_955501, EPI_ISL_955502, EPI_ISL_955503, EPI_ISL_955504, EPI_ISL_955505, EPI_ISL_955506, EPI_ISL_955507, EPI_ISL_955508, EPI_ISL_955509, EPI_ISL_955510, EPI_ISL_955511, EPI_ISL_955512, EPI_ISL_955513, EPI_ISL_955514, EPI_ISL_955515, EPI_ISL_955516, EPI_ISL_955517, EPI_ISL_955518, EPI_ISL_955519, EPI_ISL_955520, EPI_ISL_955521, EPI_ISL_955522, EPI_ISL_955523, EPI_ISL_955524, EPI_ISL_955525, EPI_ISL_955526, EPI_ISL_955527, EPI_ISL_955528, EPI_ISL_955529, EPI_ISL_955530, EPI_ISL_955531, EPI_ISL_955532, EPI_ISL_955533, EPI_ISL_955534, EPI_ISL_955535, EPI_ISL_955536, EPI_ISL_955537, EPI_ISL_955538, EPI_ISL_955539, EPI_ISL_955540, EPI_ISL_955541, EPI_ISL_955542, EPI_ISL_955543, EPI_ISL_955544, EPI_ISL_955545, EPI_ISL_955546, EPI_ISL_955547, EPI_ISL_955548, EPI_ISL_955549, EPI_ISL_955550, EPI_ISL_955551, EPI_ISL_955552, EPI_ISL_955553, EPI_ISL_955554, EPI_ISL_955555, EPI_ISL_955556, EPI_ISL_955557, EPI_ISL_955558, EPI_ISL_955559, EPI_ISL_955560, EPI_ISL_955693, EPI_ISL_955694, EPI_ISL_955695, EPI_ISL_955696, EPI_ISL_955697, EPI_ISL_955698, EPI_ISL_955699, EPI_ISL_955700, EPI_ISL_955701, EPI_ISL_955702, EPI_ISL_955703, EPI_ISL_955704, EPI_ISL_955705, EPI_ISL_955706, EPI_ISL_955707, EPI_ISL_955708, EPI_ISL_955709, EPI_ISL_955710, EPI_ISL_955711, EPI_ISL_955712, EPI_ISL_955713, EPI_ISL_955714, EPI_ISL_955715, EPI_ISL_955716, EPI_ISL_955717, EPI_ISL_955718, EPI_ISL_955719, EPI_ISL_955720, EPI_ISL_955721, EPI_ISL_955722, EPI_ISL_955723, EPI_ISL_955724, EPI_ISL_955725, EPI_ISL_955726, EPI_ISL_955727, EPI_ISL_955728, EPI_ISL_955729, EPI_ISL_955731, EPI_ISL_955732, EPI_ISL_955733, EPI_ISL_955734, EPI_ISL_955735, EPI_ISL_955739, EPI_ISL_955740, EPI_ISL_955741, EPI_ISL_955743, EPI_ISL_955744, EPI_ISL_955748, EPI_ISL_955749, EPI_ISL_979162, EPI_ISL_979163, EPI_ISL_979164, EPI_ISL_979165, EPI_ISL_979166, EPI_ISL_979167, EPI_ISL_979168, EPI_ISL_979169, EPI_ISL_979170, EPI_ISL_979171, EPI_ISL_979172, EPI_ISL_979173, EPI_ISL_979174, EPI_ISL_979175, EPI_ISL_979176, EPI_ISL_979177, EPI_ISL_979178, EPI_ISL_979179, EPI_ISL_979180, EPI_ISL_979181, EPI_ISL_979182, EPI_ISL_979183, EPI_ISL_979184, EPI_ISL_979185, EPI_ISL_979186, EPI_ISL_979187, EPI_ISL_979188, EPI_ISL_979189, EPI_ISL_979190, EPI_ISL_979191, EPI_ISL_979192, EPI_ISL_979193, EPI_ISL_979194, EPI_ISL_979195, EPI_ISL_979196, EPI_ISL_979197, EPI_ISL_979198, EPI_ISL_979199, EPI_ISL_979200, EPI_ISL_979201, EPI_ISL_979202, EPI_ISL_979203, EPI_ISL_979204, EPI_ISL_979205, EPI_ISL_979206, EPI_ISL_979207, EPI_ISL_979208, EPI_ISL_979209, EPI_ISL_979210, EPI_ISL_979211, EPI_ISL_979212, EPI_ISL_979213, EPI_ISL_979214, EPI_ISL_979215, EPI_ISL_979216, EPI_ISL_979217, EPI_ISL_979218, EPI_ISL_979219, EPI_ISL_979220, EPI_ISL_979221, EPI_ISL_979222, EPI_ISL_979223, EPI_ISL_979224, EPI_ISL_979225, EPI_ISL_979226, EPI_ISL_979227, EPI_ISL_979228, EPI_ISL_979229, EPI_ISL_979230, EPI_ISL_979231, EPI_ISL_979232, EPI_ISL_979233, EPI_ISL_979234, EPI_ISL_979235, EPI_ISL_979236, EPI_ISL_979237, EPI_ISL_979238, EPI_ISL_979239, EPI_ISL_979240, EPI_ISL_979241, EPI_ISL_979242, EPI_ISL_979243, EPI_ISL_979244, EPI_ISL_979245, EPI_ISL_979710, EPI_ISL_979711, EPI_ISL_979712, EPI_ISL_979713, EPI_ISL_979714, EPI_ISL_979715, EPI_ISL_979716, EPI_ISL_979717, EPI_ISL_979718, EPI_ISL_979719, EPI_ISL_979720, EPI_ISL_979721, EPI_ISL_979722, EPI_ISL_979723, EPI_ISL_979724, EPI_ISL_979725, EPI_ISL_979726, EPI_ISL_979727, EPI_ISL_979728, EPI_ISL_979729, EPI_ISL_979730, EPI_ISL_979731, EPI_ISL_979732, EPI_ISL_979733, EPI_ISL_979734, EPI_ISL_979735, EPI_ISL_979736, EPI_ISL_979737, EPI_ISL_979738, EPI_ISL_979739, EPI_ISL_979740, EPI_ISL_979741, EPI_ISL_979742, EPI_ISL_979743, EPI_ISL_979744, EPI_ISL_979745, EPI_ISL_979746, EPI_ISL_979747, EPI_ISL_979748, EPI_ISL_979749, EPI_ISL_979750, EPI_ISL_979751, EPI_ISL_979752, EPI_ISL_979753, EPI_ISL_979754, EPI_ISL_979755, EPI_ISL_979756, EPI_ISL_979757, EPI_ISL_979758, EPI_ISL_979759, EPI_ISL_979760, EPI_ISL_979761, EPI_ISL_979762, EPI_ISL_979763, EPI_ISL_979764, EPI_ISL_979765, EPI_ISL_979766, EPI_ISL_979767, EPI_ISL_979768, EPI_ISL_979769, EPI_ISL_979770, EPI_ISL_979771, EPI_ISL_979772, EPI_ISL_979773, EPI_ISL_979774, EPI_ISL_979775, EPI_ISL_979776, EPI_ISL_979777, EPI_ISL_979778, EPI_ISL_979779, EPI_ISL_979780, EPI_ISL_979781, EPI_ISL_979782, EPI_ISL_979783, EPI_ISL_979784, EPI_ISL_979785, EPI_ISL_979786, EPI_ISL_979787, EPI_ISL_979788, EPI_ISL_979789, EPI_ISL_979790, EPI_ISL_979791, EPI_ISL_979792, EPI_ISL_979793, EPI_ISL_979794, EPI_ISL_979795, EPI_ISL_979796, EPI_ISL_979797, EPI_ISL_1027535, EPI_ISL_1027536, EPI_ISL_1027537, EPI_ISL_1027538, EPI_ISL_1027539, EPI_ISL_1027540, EPI_ISL_1027541, EPI_ISL_1027561, EPI_ISL_1030499, EPI_ISL_2658604, EPI_ISL_2658662, EPI_ISL_2658668, EPI_ISL_2659213, EPI_ISL_2659251, EPI_ISL_2659259, EPI_ISL_2659308, EPI_ISL_2659362, EPI_ISL_2659384, EPI_ISL_2758426, EPI_ISL_2758427, EPI_ISL_2758428 | | | |
| see above | Humboldt County Public Health Laboratory | Chan-Zuckerberg Biohub | CZB Cliahub Consortium |
| EPI_ISL_1081170 | IEH Laboratories & Consulting Group | ID Genomics | Curtis Fowler; Evgeni Sokurenko; Hao Geng; Hemantha Kulakesara; Kai Deng; Kirthi Kutumbaka; Lydia Larson; Mansour Samadpour; Veronika Tchesnokova |
| EPI_ISL_1233893, EPI_ISL_2253173, EPI_ISL_2604247 | Infectious Diseases, Chan-Zuckerberg Biohub Covidtracker | Infectious Diseases, Chan-Zuckerberg Biohub Covidtracker | CZB Cliahub Consortium |
| EPI_ISL_1512788, EPI_ISL_1512859, EPI_ISL_1512861 | Infinity Biologix | Centers for Disease Control and Prevention Division of Viral Diseases, Pathogen Discovery | Adrian Paskey; Benjamin Rambo-Martin; Chirayu Goswami; Christian Bixby; Christopher Gulvick; Clinton R. Paden; Dakota Howard; Darlene Wagner; Dhwani Batra; Duncan MacCannell; Jason Caravas; Jonathan Schultz; Kara Moser; Matthew Schmerer; Peter W. Cook; Robin Grimwood; Russ Hager; Scott Sammons; Shatavia Morrison; Yihe Wang; Yvette Unoarumhi |
| EPI_ISL_918178, EPI_ISL_918184, EPI_ISL_918186, EPI_ISL_918193, EPI_ISL_918197, EPI_ISL_918202, EPI_ISL_918205, EPI_ISL_918206, EPI_ISL_918207, EPI_ISL_918208, EPI_ISL_918209, EPI_ISL_918210, EPI_ISL_918211, EPI_ISL_918212, EPI_ISL_918213, EPI_ISL_918214, EPI_ISL_918215, EPI_ISL_918216, EPI_ISL_918217, EPI_ISL_918218, EPI_ISL_918219, EPI_ISL_918220, EPI_ISL_918221, EPI_ISL_918222, EPI_ISL_918223, EPI_ISL_918224, EPI_ISL_918225, EPI_ISL_918228, EPI_ISL_918233, EPI_ISL_918239, EPI_ISL_918240, EPI_ISL_918241, EPI_ISL_918242, EPI_ISL_918244, EPI_ISL_918248, EPI_ISL_918250, EPI_ISL_918261, EPI_ISL_955126, EPI_ISL_955128, EPI_ISL_980974, EPI_ISL_1041055, EPI_ISL_1049262, EPI_ISL_1049263, EPI_ISL_1049264, EPI_ISL_1049265, EPI_ISL_1049266, EPI_ISL_1049267, EPI_ISL_1049268, EPI_ISL_1049269, EPI_ISL_1049270, EPI_ISL_1049271, EPI_ISL_1049272, EPI_ISL_1049273, EPI_ISL_1049274, EPI_ISL_1049275, EPI_ISL_1049276, EPI_ISL_1049277, EPI_ISL_1049278, EPI_ISL_1049279, EPI_ISL_1049280, EPI_ISL_1049281, EPI_ISL_1049282, EPI_ISL_1049283, EPI_ISL_1049284, EPI_ISL_1049285, EPI_ISL_1049286, EPI_ISL_1049287, EPI_ISL_1049288, EPI_ISL_1049289, EPI_ISL_1049290, EPI_ISL_1049291, EPI_ISL_1049292, EPI_ISL_1049293, EPI_ISL_1049294, EPI_ISL_1049295, EPI_ISL_1049296, EPI_ISL_1049297, EPI_ISL_1049298, EPI_ISL_1049299, EPI_ISL_1049300, EPI_ISL_1049301, EPI_ISL_1049302, EPI_ISL_1049303, EPI_ISL_1049304, EPI_ISL_1049305, EPI_ISL_1049306, EPI_ISL_1049307, EPI_ISL_1049308, EPI_ISL_1049309, EPI_ISL_1049310, EPI_ISL_1049311, EPI_ISL_1049312, EPI_ISL_1049313, EPI_ISL_1049314, EPI_ISL_1049315, EPI_ISL_1049316, EPI_ISL_1049317, EPI_ISL_1049318, EPI_ISL_1049319, EPI_ISL_1049320, EPI_ISL_1049321, EPI_ISL_1049322, EPI_ISL_1049323, EPI_ISL_1049324, EPI_ISL_1049325, EPI_ISL_1049326, EPI_ISL_1049327, EPI_ISL_1049328, EPI_ISL_1049329, EPI_ISL_1049330, EPI_ISL_1049331, EPI_ISL_1049332, EPI_ISL_1049333, EPI_ISL_1049334, EPI_ISL_1049335, EPI_ISL_1049336, EPI_ISL_1049337, EPI_ISL_1712457, EPI_ISL_1712458, EPI_ISL_1712459, EPI_ISL_1712460, EPI_ISL_1712461, EPI_ISL_1712480, EPI_ISL_1712481, EPI_ISL_1712482, EPI_ISL_1712483, EPI_ISL_1712484, EPI_ISL_1712485, EPI_ISL_1712487, EPI_ISL_1712488, EPI_ISL_1712489, EPI_ISL_1712490, EPI_ISL_1712491, EPI_ISL_1712492, EPI_ISL_1712495, EPI_ISL_1712496, EPI_ISL_1712497, EPI_ISL_1712498, EPI_ISL_1712499, EPI_ISL_1712501, EPI_ISL_1712502, EPI_ISL_1712503, EPI_ISL_1712504, EPI_ISL_1712505, EPI_ISL_1712506, EPI_ISL_1712508, EPI_ISL_1712510, EPI_ISL_1712513, EPI_ISL_1712515, EPI_ISL_1712516, EPI_ISL_1712520, EPI_ISL_1712521, EPI_ISL_1712522 | | | |
| see above | Innovative Genomics Institute, UC Berkeley | Innovative Genomics Institute, UC Berkeley | Alison Ciling; Haridha Shivram; Holly Gildea; John Boyle; Justin Choi; Liana Lareau; Netravathi Krishnappa; Phil Frankino; Shana McDevitt; Stacia Wyman |
| EPI_ISL_1113761, EPI_ISL_1121043 | Kashi Clinical Laboratory | Kashi Clinical Laboratory | Jenefer DeKoning; Zahra Kashi |
| EPI_ISL_887678, EPI_ISL_887696, EPI_ISL_887698, EPI_ISL_887762, EPI_ISL_887766, EPI_ISL_887779, EPI_ISL_887831, EPI_ISL_887859, EPI_ISL_887890, EPI_ISL_887891, EPI_ISL_887901, EPI_ISL_887903, EPI_ISL_887953, EPI_ISL_887960, EPI_ISL_887971, EPI_ISL_887994, EPI_ISL_888015, EPI_ISL_888018, EPI_ISL_888019, EPI_ISL_888024, EPI_ISL_888030, EPI_ISL_888057, EPI_ISL_888066, EPI_ISL_888116, EPI_ISL_888117, EPI_ISL_888122, EPI_ISL_888151, EPI_ISL_888163, EPI_ISL_888199, EPI_ISL_888221, EPI_ISL_888225, EPI_ISL_888241, EPI_ISL_888272, EPI_ISL_888274, EPI_ISL_888325, EPI_ISL_888330, EPI_ISL_888334, EPI_ISL_888373, EPI_ISL_888382, EPI_ISL_888454, EPI_ISL_888486, EPI_ISL_888547, EPI_ISL_888567, EPI_ISL_888582 | | | |
| see above | Labcorp | Genomics and Discovery, Respiratory Viruses Branch, Division of Viral Diseases, Centers for Disease Control and Prevention | ; Amanda Douglas; Amanda Suchanek; Andrea Throop; Ayla Burns; Ben L. Rambo-Martin; Bobbi Croy; Brian Krueger; Brian Norvell; Christos Petropoulos; Clinton R. Paden; Craig Lukasik; Debbie Boles; Dhwani Batra; Duncan MacCannell; Eyad Almasri; Goran Stevovic; Howard Engler; Hrushikesh Deshmukh; Jake Humphrey; Jana Schroth; Joe Voshell; John Pruitt; Jonathan Meltzer; Jonathan Williams; Kimberly Wagner; Lax Iyer; Lyndon Tilson; Manoj Jain; Marcia Eisenberg; Mary Ann Cristobal; Mary Williamson; Michael Levandoski; Mike Sapeta; Mindy Nye; Minoo Agarwal; Mohan Kolli; Nuthawin Charoensri; Oren Cohen; Peter W. Cook; Prashant Gupta; Qian Zeng; Rama Ghatti; Scott Parker; Scott Ryan; Stanley Letovsky; Steven Ragan; Summer Galloway; Suresh Babu Selvaraju; Susan Countryman; Susan Hicks; Suxiang Tong; Suzanne Dale; Thomas Urban; Tim Kuphal; Tricia Zwiefelhofer; Vincent Drouillon |
| EPI_ISL_1027773, EPI_ISL_1027789, EPI_ISL_1027838, EPI_ISL_1027839, EPI_ISL_1027912, EPI_ISL_1027916, EPI_ISL_1027917, EPI_ISL_1027936, EPI_ISL_1028029, EPI_ISL_1028053, EPI_ISL_1028145, EPI_ISL_1028202, EPI_ISL_1028241, EPI_ISL_1028294, EPI_ISL_1028347, EPI_ISL_1028348, EPI_ISL_1028381, EPI_ISL_1028412, EPI_ISL_1028448, EPI_ISL_1028516, EPI_ISL_1028580, EPI_ISL_1028610, EPI_ISL_1028679, EPI_ISL_1028731, EPI_ISL_1028800, EPI_ISL_1028822, EPI_ISL_1028825, EPI_ISL_1028826, EPI_ISL_1028864, EPI_ISL_1028866, EPI_ISL_1028867, EPI_ISL_1028873, EPI_ISL_1028874, EPI_ISL_1028886, EPI_ISL_1028899, EPI_ISL_1028941, EPI_ISL_1028956, EPI_ISL_1028957, EPI_ISL_1028965, EPI_ISL_1028998, EPI_ISL_1029035, EPI_ISL_1029068, EPI_ISL_1029069, EPI_ISL_1029079, EPI_ISL_1029274, EPI_ISL_1029285, EPI_ISL_1029300, EPI_ISL_1029301, EPI_ISL_1029303, EPI_ISL_1029305, EPI_ISL_1029306, EPI_ISL_1029307, EPI_ISL_1029308, EPI_ISL_1029309, EPI_ISL_1029310, EPI_ISL_1029311, EPI_ISL_1029312, EPI_ISL_1029313, EPI_ISL_1029314, EPI_ISL_1029315, EPI_ISL_1029316, EPI_ISL_1029317, EPI_ISL_1029318, EPI_ISL_1029319, EPI_ISL_1029320, EPI_ISL_1029321, EPI_ISL_1029322, EPI_ISL_1029323, EPI_ISL_1029324, EPI_ISL_1029325, EPI_ISL_1029326, EPI_ISL_1029414, EPI_ISL_1029415, EPI_ISL_1029416, EPI_ISL_1029417, EPI_ISL_1029418, EPI_ISL_1029419, EPI_ISL_1029420, EPI_ISL_1029421, EPI_ISL_1029422, EPI_ISL_1029423, EPI_ISL_1029532, EPI_ISL_1029579, EPI_ISL_1029580, EPI_ISL_1029581, EPI_ISL_1029721 | | | |
| see above | Laboratory Corporation of America | Respiratory Viruses Branch, Division of Viral Diseases, Centers for Disease Control and Prevention | Ben L. Rambo-Martin; Clinton R. Paden; Dakota Howard; Dhwani Batra; Duncan MacCannell; Peter W. Cook; Suxiang Tong |
| EPI_ISL_710424, EPI_ISL_710427, EPI_ISL_710428, EPI_ISL_737236, EPI_ISL_737237, EPI_ISL_737238, EPI_ISL_737239, EPI_ISL_737240, EPI_ISL_737241, EPI_ISL_737242, EPI_ISL_737243, EPI_ISL_737244, EPI_ISL_737245, EPI_ISL_737246, EPI_ISL_737247, EPI_ISL_737248, EPI_ISL_737249, EPI_ISL_737250, EPI_ISL_737251, EPI_ISL_737252, EPI_ISL_737253, EPI_ISL_737254, EPI_ISL_737255, EPI_ISL_737256, EPI_ISL_737257, EPI_ISL_737258, EPI_ISL_737259, EPI_ISL_737260, EPI_ISL_737261, EPI_ISL_754130, EPI_ISL_754131, EPI_ISL_754132, EPI_ISL_765998, EPI_ISL_765999, EPI_ISL_766000, EPI_ISL_766001, EPI_ISL_766002, EPI_ISL_766003, EPI_ISL_766004, EPI_ISL_766005, EPI_ISL_766006, EPI_ISL_766007, EPI_ISL_766008, EPI_ISL_766009, EPI_ISL_766010, EPI_ISL_766011, EPI_ISL_766012, EPI_ISL_766013, EPI_ISL_766014, EPI_ISL_792655, EPI_ISL_792656, EPI_ISL_792657, EPI_ISL_792658, EPI_ISL_792659, EPI_ISL_792660, EPI_ISL_792661, EPI_ISL_792662, EPI_ISL_792663, EPI_ISL_792664, EPI_ISL_792665, EPI_ISL_792666, EPI_ISL_792667, EPI_ISL_792668, EPI_ISL_792669, EPI_ISL_792670, EPI_ISL_792671, EPI_ISL_792672, EPI_ISL_792673, EPI_ISL_792674, EPI_ISL_792675, EPI_ISL_792676, EPI_ISL_792677, EPI_ISL_792678, EPI_ISL_861698, EPI_ISL_861699, EPI_ISL_903367, EPI_ISL_903368, EPI_ISL_903369, EPI_ISL_903370, EPI_ISL_915438, EPI_ISL_1503992, EPI_ISL_1503995, EPI_ISL_1707677, EPI_ISL_1707678, EPI_ISL_1707679, EPI_ISL_2031979, EPI_ISL_2031980, EPI_ISL_2031981, EPI_ISL_2031982, EPI_ISL_2031983, EPI_ISL_2031984, EPI_ISL_2031985, EPI_ISL_2031986, EPI_ISL_2031987, EPI_ISL_2031988, EPI_ISL_2031989, EPI_ISL_2031990, EPI_ISL_2031991, EPI_ISL_2031992, EPI_ISL_2031993, EPI_ISL_2031994, EPI_ISL_2031995, EPI_ISL_2031996, EPI_ISL_2031997, EPI_ISL_2106182, EPI_ISL_2259246 | | | |
| see above | Los Angeles County Public Health Laboratories | Los Angeles County Public Health Laboratories | Hemarajata; P. Hemarajata et al.; P. and Green, N. |
| EPI_ISL_832105, EPI_ISL_832106, EPI_ISL_832107, EPI_ISL_832108, EPI_ISL_832109, EPI_ISL_832110, EPI_ISL_832111, EPI_ISL_832112, EPI_ISL_832113, EPI_ISL_832114, EPI_ISL_832115, EPI_ISL_832116, EPI_ISL_832117, EPI_ISL_832118, EPI_ISL_832119, EPI_ISL_832120, EPI_ISL_1303483 | | | |
| see above | MD Laboratories | Los Angeles County Public Health Laboratories | P. Hemarajata et al. |
| EPI_ISL_672048, EPI_ISL_672067, EPI_ISL_672504, EPI_ISL_672505, EPI_ISL_738916, EPI_ISL_739172, EPI_ISL_739393, EPI_ISL_739567, EPI_ISL_2658745, EPI_ISL_2659365 | | | |
| see above | Madera County Department of Public Health | Chan-Zuckerberg Biohub | CZB Cliahub Consortium |
| EPI_ISL_2006747, EPI_ISL_2006753, EPI_ISL_2006754, EPI_ISL_2006758, EPI_ISL_2006761, EPI_ISL_2006774, EPI_ISL_2006779, EPI_ISL_2006780, EPI_ISL_2006789, EPI_ISL_2006790, EPI_ISL_2009412, EPI_ISL_2009495, EPI_ISL_2009496, EPI_ISL_2011702 | | | |
| see above | Moderna Inc. | Moderna Inc. | Groves Dixon; Rolando Pajon; Yamuna Paila |
| EPI_ISL_738658 | Monterey County Public Health Lab | Chan-Zuckerberg Biohub | CZB Cliahub Consortium |
| EPI_ISL_1263211, EPI_ISL_1263212, EPI_ISL_1263213, EPI_ISL_1263214, EPI_ISL_1263215, EPI_ISL_1263216, EPI_ISL_1263217, EPI_ISL_1263218, EPI_ISL_1263219, EPI_ISL_1263220, EPI_ISL_1263221, EPI_ISL_1263222, EPI_ISL_1263223, EPI_ISL_1263224, EPI_ISL_1263225, EPI_ISL_1263226, EPI_ISL_2308284, EPI_ISL_2308285, EPI_ISL_2716874 | | | |
| see above | Monterey County Public Health Laboratory | Monterey County Public Health Laboratory | Monterey County Public Health Laboratory |
| EPI_ISL_1171051, EPI_ISL_2839054 | OHSU Lab Services Molecular Microbiology Lab | Ginkgo Bioworks Clinical Laboratory | '[UsersoroakBox SyncPHI_SARS-COV-2_Sample_Sheet_infoGISAID_Submission_Infometa_master_2021.xlsx]release20_final'!D:AC; 26; Alec J. Hirsch; Alex Plocik; Amanda Riccardi; Andrew C. Adey; Becky Schilling; Benjamin N. Bimber; Birgitte Simen; Brendan L. O'Connell; Brian J. O'Roak; Daniel N. Streblow; Donna Hansel; FALSE); Guang Fan; James McGann; Jim Griffin; Kayla Carter; Keith Robison; Malaika Mckenzie-Bennett; Martha Pierson; Michelle Spencer; Rebecca Littlefield; Ruth V. Nichols; Sally Grindstaff; VLOOKUP(C299; William B. Messer; Xuan Qin |
| EPI_ISL_832675, EPI_ISL_832758 | OHSU Lab Services Molecular Microbiology Lab | Oregon SARS-CoV-2 Genome Sequencing Center | Alec J. Hirsch; Andrew C. Adey; Benjamin N. Bimber; Brendan L. O'Connell; Brian J. O'Roak; Daniel N. Streblow; Donna Hansel; Guang Fan; Ruth V. Nichols; Sally Grindstaff; William B. Messer |
| EPI_ISL_792679 | Olive View Medical Center | Los Angeles County Public Health | P. Hemarajata et al. |

| | | | |
|---|---|---|---|
| | | Laboratories | |
| EPI_ISL_2620823, EPI_ISL_2811980 | OmniPathology Laboratory | Los Angeles County Public Health Laboratories | P. Hemarajata et al. |
| EPI_ISL_2502593, EPI_ISL_2502594, EPI_ISL_2502595, EPI_ISL_2502601, EPI_ISL_2502602, EPI_ISL_2502603, EPI_ISL_2502604, EPI_ISL_2502605, EPI_ISL_2502606, EPI_ISL_2502607, EPI_ISL_2502608, EPI_ISL_2502609, EPI_ISL_2502610, EPI_ISL_2502611, EPI_ISL_2566362, EPI_ISL_2566363, EPI_ISL_2566364, EPI_ISL_2566366, EPI_ISL_2566367 | | | |
| see above | Operation Infectious Diseases, Naval Health Research Center | Operation Infectious Diseases, Naval Health Research Center | A.E.; Balansay-Ames; C.A.; Fierro, M.; Iniguez-Stevens, E.; Kriner, P.; M.S.; Moser, K.; Myers; Pan; Phippard; R.S.; R.W.; Underwood |
| EPI_ISL_672481, EPI_ISL_672482, EPI_ISL_672483, EPI_ISL_672484, EPI_ISL_672485, EPI_ISL_672486, EPI_ISL_672487, EPI_ISL_738503, EPI_ISL_738507, EPI_ISL_738515, EPI_ISL_738518, EPI_ISL_738529, EPI_ISL_738548, EPI_ISL_738559, EPI_ISL_738569, EPI_ISL_738570, EPI_ISL_738571, EPI_ISL_738588, EPI_ISL_738594, EPI_ISL_738599, EPI_ISL_738602, EPI_ISL_738603, EPI_ISL_738620, EPI_ISL_738621, EPI_ISL_738624, EPI_ISL_738630, EPI_ISL_738632, EPI_ISL_738634, EPI_ISL_738635, EPI_ISL_738636, EPI_ISL_738637, EPI_ISL_738638, EPI_ISL_738639, EPI_ISL_738640, EPI_ISL_738641, EPI_ISL_738646, EPI_ISL_738655, EPI_ISL_738664, EPI_ISL_738669, EPI_ISL_738688, EPI_ISL_738693, EPI_ISL_738694, EPI_ISL_738703, EPI_ISL_738704, EPI_ISL_738709, EPI_ISL_738722, EPI_ISL_738729, EPI_ISL_738731, EPI_ISL_738732, EPI_ISL_738733, EPI_ISL_738734, EPI_ISL_738735, EPI_ISL_738736, EPI_ISL_738737, EPI_ISL_738738, EPI_ISL_738739, EPI_ISL_738740, EPI_ISL_738741, EPI_ISL_738752, EPI_ISL_738755, EPI_ISL_738757, EPI_ISL_738758, EPI_ISL_738761, EPI_ISL_738762, EPI_ISL_738782, EPI_ISL_738784, EPI_ISL_738787, EPI_ISL_738790, EPI_ISL_738799, EPI_ISL_738808, EPI_ISL_738811, EPI_ISL_738815, EPI_ISL_738816, EPI_ISL_738818, EPI_ISL_738823, EPI_ISL_738824, EPI_ISL_738826, EPI_ISL_738829, EPI_ISL_738830, EPI_ISL_738840, EPI_ISL_738841, EPI_ISL_738845, EPI_ISL_738850, EPI_ISL_738856, EPI_ISL_738857, EPI_ISL_738860, EPI_ISL_738862, EPI_ISL_738863, EPI_ISL_738866, EPI_ISL_738869, EPI_ISL_738871, EPI_ISL_738872, EPI_ISL_738873, EPI_ISL_738874, EPI_ISL_738878, EPI_ISL_738880, EPI_ISL_738882, EPI_ISL_738884, EPI_ISL_738885, EPI_ISL_738888, EPI_ISL_738893, EPI_ISL_738899, EPI_ISL_738900, EPI_ISL_738905, EPI_ISL_738907, EPI_ISL_738908, EPI_ISL_738921, EPI_ISL_738928, EPI_ISL_738930, EPI_ISL_738935, EPI_ISL_738937, EPI_ISL_738939, EPI_ISL_738944, EPI_ISL_738946, EPI_ISL_738949, EPI_ISL_738954, EPI_ISL_738966, EPI_ISL_738968, EPI_ISL_738970, EPI_ISL_738973, EPI_ISL_738975, EPI_ISL_738978, EPI_ISL_738981, EPI_ISL_738989, EPI_ISL_738991, EPI_ISL_738993, EPI_ISL_738995, EPI_ISL_739002, EPI_ISL_739013, EPI_ISL_739016, EPI_ISL_739028, EPI_ISL_739034, EPI_ISL_739042, EPI_ISL_739044, EPI_ISL_739048, EPI_ISL_739055, EPI_ISL_739063, EPI_ISL_739069, EPI_ISL_739071, EPI_ISL_739072, EPI_ISL_739078, EPI_ISL_739081, EPI_ISL_739089, EPI_ISL_739092, EPI_ISL_739094, EPI_ISL_739097, EPI_ISL_739099, EPI_ISL_739105, EPI_ISL_739108, EPI_ISL_739116, EPI_ISL_739125, EPI_ISL_739132, EPI_ISL_739139, EPI_ISL_739142, EPI_ISL_739143, EPI_ISL_739151, EPI_ISL_739154, EPI_ISL_739156, EPI_ISL_739162, EPI_ISL_739175, EPI_ISL_739176, EPI_ISL_739182, EPI_ISL_739185, EPI_ISL_739191, EPI_ISL_739199, EPI_ISL_739205, EPI_ISL_739211, EPI_ISL_739216, EPI_ISL_739222, EPI_ISL_739227, EPI_ISL_739230, EPI_ISL_739238, EPI_ISL_739239, EPI_ISL_739245, EPI_ISL_739255, EPI_ISL_739258, EPI_ISL_739259, EPI_ISL_739265, EPI_ISL_739266, EPI_ISL_739267, EPI_ISL_739271, EPI_ISL_739276, EPI_ISL_739279, EPI_ISL_739289, EPI_ISL_739291, EPI_ISL_739296, EPI_ISL_739304, EPI_ISL_739314, EPI_ISL_739322, EPI_ISL_739331, EPI_ISL_739334, EPI_ISL_739341, EPI_ISL_739342, EPI_ISL_739345, EPI_ISL_739347, EPI_ISL_739350, EPI_ISL_739353, EPI_ISL_739359, EPI_ISL_739362, EPI_ISL_739369, EPI_ISL_739373, EPI_ISL_739375, EPI_ISL_739379, EPI_ISL_739384, EPI_ISL_739386, EPI_ISL_739396, EPI_ISL_739397, EPI_ISL_739401, EPI_ISL_739410, EPI_ISL_739412, EPI_ISL_739420, EPI_ISL_739421, EPI_ISL_739423, EPI_ISL_739428, EPI_ISL_739432, EPI_ISL_739437, EPI_ISL_739438, EPI_ISL_739441, EPI_ISL_739445, EPI_ISL_739450, EPI_ISL_739455, EPI_ISL_739460, EPI_ISL_739462, EPI_ISL_739464, EPI_ISL_739465, EPI_ISL_739477, EPI_ISL_739482, EPI_ISL_739486, EPI_ISL_739488, EPI_ISL_739490, EPI_ISL_739493, EPI_ISL_739494, EPI_ISL_739496, EPI_ISL_739497, EPI_ISL_739500, EPI_ISL_739513, EPI_ISL_739517, EPI_ISL_739524, EPI_ISL_739529, EPI_ISL_739532, EPI_ISL_739534, EPI_ISL_739540, EPI_ISL_739545, EPI_ISL_739547, EPI_ISL_739556, EPI_ISL_739559, EPI_ISL_739566, EPI_ISL_739568, EPI_ISL_739570, EPI_ISL_739573, EPI_ISL_739577, EPI_ISL_739581, EPI_ISL_739582, EPI_ISL_739586, EPI_ISL_739591, EPI_ISL_739592, EPI_ISL_739595, EPI_ISL_739599, EPI_ISL_739607, EPI_ISL_739609, EPI_ISL_739614, EPI_ISL_739620, EPI_ISL_739625, EPI_ISL_739627, EPI_ISL_739636, EPI_ISL_739637, EPI_ISL_739641, EPI_ISL_739646, EPI_ISL_739654, EPI_ISL_739658, EPI_ISL_885052, EPI_ISL_885053, EPI_ISL_885054, EPI_ISL_885055, EPI_ISL_885056, EPI_ISL_885057, EPI_ISL_885058, EPI_ISL_885059, EPI_ISL_885060, EPI_ISL_885061, EPI_ISL_885062, EPI_ISL_885063, EPI_ISL_885064, EPI_ISL_885065, EPI_ISL_885066, EPI_ISL_885067, EPI_ISL_885068, EPI_ISL_885069, EPI_ISL_885070, EPI_ISL_885071, EPI_ISL_885072, EPI_ISL_885073, EPI_ISL_885074, EPI_ISL_885075, EPI_ISL_885076, EPI_ISL_885077, EPI_ISL_885078, EPI_ISL_885079, EPI_ISL_885080, EPI_ISL_885081, EPI_ISL_885082, EPI_ISL_885083, EPI_ISL_885084, EPI_ISL_885085, EPI_ISL_885086, EPI_ISL_885087, EPI_ISL_885088, EPI_ISL_885089, EPI_ISL_885090, EPI_ISL_885091, EPI_ISL_885092, EPI_ISL_885093, EPI_ISL_885094, EPI_ISL_885095, EPI_ISL_885096, EPI_ISL_885097, EPI_ISL_885098, EPI_ISL_885099, EPI_ISL_885100, EPI_ISL_885101, EPI_ISL_885102, EPI_ISL_885103, EPI_ISL_885104, EPI_ISL_885105, EPI_ISL_885106, EPI_ISL_885107, EPI_ISL_885108, EPI_ISL_885109, EPI_ISL_885110, EPI_ISL_885111, EPI_ISL_885112, EPI_ISL_885113, EPI_ISL_885114, EPI_ISL_885115, EPI_ISL_885116, EPI_ISL_885117, EPI_ISL_885118, EPI_ISL_885119, EPI_ISL_885120, EPI_ISL_885121, EPI_ISL_885122, EPI_ISL_885123, EPI_ISL_885124, EPI_ISL_885125, EPI_ISL_885126, EPI_ISL_885127, EPI_ISL_955628, EPI_ISL_955629, EPI_ISL_955630, EPI_ISL_955631, EPI_ISL_955632, EPI_ISL_955633, EPI_ISL_955634, EPI_ISL_955635, EPI_ISL_955636, EPI_ISL_955637, EPI_ISL_955638, EPI_ISL_955639, EPI_ISL_955640, EPI_ISL_955641, EPI_ISL_955642, EPI_ISL_955643, EPI_ISL_955644, EPI_ISL_955645, EPI_ISL_955646, EPI_ISL_955647, EPI_ISL_955648, EPI_ISL_955649, EPI_ISL_955650, EPI_ISL_955651, EPI_ISL_955652, EPI_ISL_955653, EPI_ISL_955654, EPI_ISL_955655, EPI_ISL_955656, EPI_ISL_955657, EPI_ISL_955658, EPI_ISL_955659, EPI_ISL_955660, EPI_ISL_955661, EPI_ISL_955662, EPI_ISL_955663, EPI_ISL_955664, EPI_ISL_955665, EPI_ISL_955666, EPI_ISL_955667, EPI_ISL_955668, EPI_ISL_955669, EPI_ISL_955670, EPI_ISL_955671, EPI_ISL_955672, EPI_ISL_955673, EPI_ISL_955674, EPI_ISL_955675, EPI_ISL_955676, EPI_ISL_955677, EPI_ISL_955678, EPI_ISL_955679, EPI_ISL_955680, EPI_ISL_955681, EPI_ISL_955682, EPI_ISL_955683, EPI_ISL_955684, EPI_ISL_955685, EPI_ISL_955686, EPI_ISL_955687, EPI_ISL_955688, EPI_ISL_955689, EPI_ISL_955690, EPI_ISL_955691, EPI_ISL_979643, EPI_ISL_979644, EPI_ISL_979645, EPI_ISL_979646, EPI_ISL_979647, EPI_ISL_979648, EPI_ISL_979649, EPI_ISL_979650, EPI_ISL_979651, EPI_ISL_979652, EPI_ISL_979653, EPI_ISL_979654, EPI_ISL_979655, EPI_ISL_979656, EPI_ISL_979657, EPI_ISL_979658, EPI_ISL_979659, EPI_ISL_979660, EPI_ISL_979661, EPI_ISL_979662, EPI_ISL_979663, EPI_ISL_979664, EPI_ISL_979665, EPI_ISL_979666, EPI_ISL_979667, EPI_ISL_979668, EPI_ISL_979669, EPI_ISL_979670, EPI_ISL_979671, EPI_ISL_979672, EPI_ISL_979673, EPI_ISL_979674, EPI_ISL_979675, EPI_ISL_979676, EPI_ISL_979677, EPI_ISL_979678, EPI_ISL_979679, EPI_ISL_979680, EPI_ISL_979681, EPI_ISL_979682, EPI_ISL_979683, EPI_ISL_979684, EPI_ISL_979685, EPI_ISL_979686, EPI_ISL_979687, EPI_ISL_979688, EPI_ISL_979689, EPI_ISL_1027294, EPI_ISL_1027321, EPI_ISL_1027329, EPI_ISL_1027331, EPI_ISL_1027335, EPI_ISL_1027339, EPI_ISL_1027340, EPI_ISL_1027344, EPI_ISL_1027350, EPI_ISL_1027351, EPI_ISL_1027359, EPI_ISL_1027378, EPI_ISL_1027379, EPI_ISL_1027380, EPI_ISL_1027381, EPI_ISL_1027382, EPI_ISL_1027383, EPI_ISL_1027384, EPI_ISL_1027385, EPI_ISL_1027386, EPI_ISL_1027387, EPI_ISL_1027388, EPI_ISL_1027389, EPI_ISL_1027390, EPI_ISL_1027391, EPI_ISL_1027392, EPI_ISL_1027393, EPI_ISL_1027394, EPI_ISL_1027395, EPI_ISL_1027396, EPI_ISL_1027397, EPI_ISL_1027398, EPI_ISL_1027399, EPI_ISL_1027400, EPI_ISL_1027401, EPI_ISL_1027402, EPI_ISL_1027403, EPI_ISL_1027404, EPI_ISL_1027405, EPI_ISL_1027406, EPI_ISL_1027407, EPI_ISL_1027408, EPI_ISL_1027409, EPI_ISL_1027410, EPI_ISL_1027411, EPI_ISL_1027412, EPI_ISL_1027413, EPI_ISL_1027414, EPI_ISL_1027415, EPI_ISL_1027416, EPI_ISL_1027417, EPI_ISL_1027418, EPI_ISL_1027419, EPI_ISL_1027420, EPI_ISL_1027421, EPI_ISL_1027422, EPI_ISL_1027423, EPI_ISL_1027424, EPI_ISL_1027425, EPI_ISL_1027426, EPI_ISL_1027427, EPI_ISL_1027428, EPI_ISL_1027429, EPI_ISL_1027430, EPI_ISL_1027431, EPI_ISL_1027432, EPI_ISL_1027433, EPI_ISL_1027434, EPI_ISL_1027435, EPI_ISL_1027436, EPI_ISL_1030290, EPI_ISL_1030291, EPI_ISL_1030292, EPI_ISL_1030293, EPI_ISL_1030294, EPI_ISL_1030295, EPI_ISL_1030296, EPI_ISL_1030297, EPI_ISL_1030298, EPI_ISL_1030299, EPI_ISL_1030308, EPI_ISL_1030312, EPI_ISL_1030444, EPI_ISL_1030445, EPI_ISL_1030446, EPI_ISL_1030447, EPI_ISL_1030448, EPI_ISL_1030449, EPI_ISL_1030450, EPI_ISL_1030451, EPI_ISL_1030452, EPI_ISL_1030453, EPI_ISL_1030454, EPI_ISL_1030455, EPI_ISL_1030456, EPI_ISL_1030457, EPI_ISL_1030458, EPI_ISL_1030459, EPI_ISL_1030460, EPI_ISL_1030461, EPI_ISL_1030462, EPI_ISL_1030463, EPI_ISL_1030464, EPI_ISL_1030465, EPI_ISL_1030466, EPI_ISL_1030467, EPI_ISL_1030468, EPI_ISL_1030469, EPI_ISL_1030470, EPI_ISL_1030471, EPI_ISL_1030472, EPI_ISL_1030473, EPI_ISL_1030474, EPI_ISL_1030475, EPI_ISL_1030476, EPI_ISL_1030477, EPI_ISL_1030478, EPI_ISL_1030479, EPI_ISL_1030480, EPI_ISL_1030481, EPI_ISL_1030482, EPI_ISL_1030483, EPI_ISL_1030484, EPI_ISL_1030485, EPI_ISL_1030486, EPI_ISL_1030487, EPI_ISL_1030488, EPI_ISL_1030489, EPI_ISL_1030490, EPI_ISL_1030491, EPI_ISL_1030492, EPI_ISL_1030493, EPI_ISL_1030494, EPI_ISL_1030495, EPI_ISL_1030496, EPI_ISL_1030497, EPI_ISL_1030498, EPI_ISL_1184681, EPI_ISL_1184682, EPI_ISL_1184683, EPI_ISL_1184684, EPI_ISL_1184685, EPI_ISL_1184686, EPI_ISL_1184687, EPI_ISL_1184688, EPI_ISL_1184689, EPI_ISL_1184690, EPI_ISL_1184691, EPI_ISL_1184692, EPI_ISL_1184693, EPI_ISL_1184694, EPI_ISL_1184695, EPI_ISL_1184696, EPI_ISL_1184697, EPI_ISL_1184698, EPI_ISL_1184699, EPI_ISL_1184700, EPI_ISL_1184701, EPI_ISL_1184702, EPI_ISL_1184703, EPI_ISL_1184704, EPI_ISL_1184705, EPI_ISL_1184706, EPI_ISL_1184707, EPI_ISL_1184708, EPI_ISL_1184709, EPI_ISL_1184710, EPI_ISL_1184711, EPI_ISL_1184712, EPI_ISL_1184713, EPI_ISL_1184714, EPI_ISL_1184715, EPI_ISL_1184716, EPI_ISL_1184717, EPI_ISL_1184718, EPI_ISL_1184719, EPI_ISL_1184720, EPI_ISL_1184721, EPI_ISL_1184722, EPI_ISL_1184723, EPI_ISL_1184724, EPI_ISL_1184725, EPI_ISL_1184726, EPI_ISL_1184727, EPI_ISL_1184728, EPI_ISL_1184729, EPI_ISL_1184730, EPI_ISL_1184731, EPI_ISL_1184732, EPI_ISL_1184733, EPI_ISL_1184734, EPI_ISL_1184735, EPI_ISL_1184736, EPI_ISL_1184737, EPI_ISL_1184738, EPI_ISL_1184739, EPI_ISL_1184740, EPI_ISL_1184741, EPI_ISL_1184742, EPI_ISL_1184743, EPI_ISL_1184744, EPI_ISL_1184745, EPI_ISL_1184746, EPI_ISL_1184747, EPI_ISL_1184748, EPI_ISL_1184749, EPI_ISL_1184750, EPI_ISL_1184751, EPI_ISL_1184752, EPI_ISL_1184753, EPI_ISL_1184754, EPI_ISL_1184755, EPI_ISL_1184756, EPI_ISL_1184757, EPI_ISL_1184758, EPI_ISL_1184759, EPI_ISL_1184760, EPI_ISL_1184761, EPI_ISL_1184762, EPI_ISL_1184763, EPI_ISL_1184764, EPI_ISL_1184765, EPI_ISL_1184766, EPI_ISL_1184767, EPI_ISL_1184768, EPI_ISL_1184769, EPI_ISL_1184770, EPI_ISL_1184771, EPI_ISL_1184772, EPI_ISL_1184773, EPI_ISL_1184774, EPI_ISL_1184775, EPI_ISL_1184776, EPI_ISL_1184777, EPI_ISL_1184778, EPI_ISL_1184779, EPI_ISL_1184780, EPI_ISL_1184781, EPI_ISL_1184782, EPI_ISL_1184783, EPI_ISL_1184784, EPI_ISL_1184785, EPI_ISL_1184786, EPI_ISL_1184787, EPI_ISL_1184788, EPI_ISL_1184789, EPI_ISL_1184790, EPI_ISL_1184791, EPI_ISL_1184792, EPI_ISL_1184793, EPI_ISL_1184794, EPI_ISL_1184795, EPI_ISL_1184796, EPI_ISL_1184797, EPI_ISL_1184798, EPI_ISL_1184799, EPI_ISL_1184800, EPI_ISL_1184801, EPI_ISL_1184802, EPI_ISL_1184803, EPI_ISL_1184804, EPI_ISL_1184805, EPI_ISL_1184806, EPI_ISL_1184807, EPI_ISL_1184808, EPI_ISL_1184967, EPI_ISL_1184968, EPI_ISL_1184969, EPI_ISL_1184970, EPI_ISL_1184971, EPI_ISL_1184972, EPI_ISL_1184973, EPI_ISL_1184974, EPI_ISL_1184975, EPI_ISL_1184976, EPI_ISL_1184977, EPI_ISL_1184978, EPI_ISL_1184979, EPI_ISL_1184980, EPI_ISL_1184981, EPI_ISL_1184982, EPI_ISL_1184983, EPI_ISL_1184984, EPI_ISL_1184985, EPI_ISL_1184986, EPI_ISL_1184987, EPI_ISL_1184988, EPI_ISL_1184989, EPI_ISL_1184990, EPI_ISL_1184991, EPI_ISL_1184992, EPI_ISL_1184993, EPI_ISL_1184994, EPI_ISL_1184995, EPI_ISL_1184996, EPI_ISL_1184997, EPI_ISL_1184998, EPI_ISL_1184999, EPI_ISL_1185000, EPI_ISL_1185001, EPI_ISL_1185002, EPI_ISL_1185003, EPI_ISL_1185004, EPI_ISL_1185005, EPI_ISL_1185006, EPI_ISL_1185007, EPI_ISL_1185008, EPI_ISL_1185009, EPI_ISL_1185010, EPI_ISL_1185011, EPI_ISL_1185012, EPI_ISL_1185013, EPI_ISL_1185014, EPI_ISL_1185015, EPI_ISL_1701400, EPI_ISL_1701426, EPI_ISL_1701431, EPI_ISL_1701450, EPI_ISL_1701465, EPI_ISL_1701478, EPI_ISL_1701483, EPI_ISL_1701491, EPI_ISL_1701511, EPI_ISL_1701513, EPI_ISL_1701533, EPI_ISL_1701537, EPI_ISL_1701543, EPI_ISL_1701545, EPI_ISL_1701586, EPI_ISL_1701599, EPI_ISL_1701611, EPI_ISL_1701616, EPI_ISL_1701623, EPI_ISL_1701629, EPI_ISL_1701657, EPI_ISL_2103006, EPI_ISL_2103025, EPI_ISL_2103028, EPI_ISL_2103029, EPI_ISL_2103036, EPI_ISL_2103067, EPI_ISL_2103081, EPI_ISL_2103085, EPI_ISL_2103086, EPI_ISL_2103094, EPI_ISL_2103100, EPI_ISL_2103114, EPI_ISL_2103115, EPI_ISL_2103122, EPI_ISL_2103134, EPI_ISL_2103152, EPI_ISL_2103158, EPI_ISL_2103159, EPI_ISL_2103178, EPI_ISL_2103187, EPI_ISL_2658580, EPI_ISL_2658602, EPI_ISL_2658647, EPI_ISL_2658669, EPI_ISL_2658716, EPI_ISL_2658718, EPI_ISL_2658723, EPI_ISL_2659217, EPI_ISL_2659262, EPI_ISL_2659270, EPI_ISL_2659276, EPI_ISL_2659278, EPI_ISL_2659287, EPI_ISL_2659291, EPI_ISL_2659398, EPI_ISL_2659418, EPI_ISL_2659462, EPI_ISL_2659550, EPI_ISL_2659561, EPI_ISL_2659603, EPI_ISL_2659640, EPI_ISL_2659644, EPI_ISL_2758416, EPI_ISL_2758421, EPI_ISL_2758422, EPI_ISL_2758434, EPI_ISL_2758435, EPI_ISL_2758491 | | | |
| see above | Orange County Public Health Lab | Chan-Zuckerberg Biohub | CZB Cliahub Consortium |
| EPI_ISL_2928151 | Orange County Public Health Laboratory | Orange County Public Health Laboratory | Angelica Torres; Julia Wolfe |
| EPI_ISL_1017051, EPI_ISL_1017059, EPI_ISL_1017060, EPI_ISL_1017061, EPI_ISL_1017062, EPI_ISL_1017063, EPI_ISL_1017065, EPI_ISL_1017097, EPI_ISL_1017098, EPI_ISL_1017099, EPI_ISL_1017108, EPI_ISL_1017109, EPI_ISL_1017110, EPI_ISL_1017111, EPI_ISL_1017116, EPI_ISL_1017117, EPI_ISL_1017126, EPI_ISL_1017127, EPI_ISL_1017128, EPI_ISL_1017129, EPI_ISL_1017130, EPI_ISL_1017131, EPI_ISL_1017132, EPI_ISL_1017147, EPI_ISL_1017148, EPI_ISL_1017149, EPI_ISL_1017150, EPI_ISL_1017151, EPI_ISL_1017152, EPI_ISL_1017153, EPI_ISL_1017154, EPI_ISL_1017155, EPI_ISL_1017156, EPI_ISL_1017157, EPI_ISL_1017158, EPI_ISL_1017159, EPI_ISL_1017160, EPI_ISL_2551187, EPI_ISL_2551188, EPI_ISL_2551189, EPI_ISL_2551190, EPI_ISL_2551191, EPI_ISL_2551192, EPI_ISL_2551193, EPI_ISL_2551194, EPI_ISL_2551195, EPI_ISL_2551196, EPI_ISL_2551197, EPI_ISL_2551198, EPI_ISL_2551199, EPI_ISL_2551200, EPI_ISL_2551201, EPI_ISL_2551202, EPI_ISL_2551203, EPI_ISL_2551204, EPI_ISL_2551205, EPI_ISL_2551206, EPI_ISL_2551207, EPI_ISL_2551208, EPI_ISL_2551209, EPI_ISL_2551210, EPI_ISL_2551211, EPI_ISL_2551212, EPI_ISL_2551213, EPI_ISL_2551214, EPI_ISL_2551215, EPI_ISL_2551216, EPI_ISL_2551217, EPI_ISL_2551218, EPI_ISL_2551219, EPI_ISL_2551220, EPI_ISL_2551221 | | | |
| see above | Providence Saint Joseph Medical Center - Burbank | Providence St. Joseph Health Molecular Genomics Laboratory | Alexa K Dowdell; Brian D Piening; Carlo B Bifulco; Fred L Robinson; Mary Campbell; Rogan Rattray |
| EPI_ISL_1017058, EPI_ISL_1017080, EPI_ISL_1017081, EPI_ISL_1017082, EPI_ISL_1017083, EPI_ISL_1017084, EPI_ISL_1017085, EPI_ISL_1017086, EPI_ISL_1017087, EPI_ISL_1017088, EPI_ISL_1017089, EPI_ISL_1017091, EPI_ISL_1017092, EPI_ISL_1017093, EPI_ISL_1017094, EPI_ISL_1017095, EPI_ISL_1017106, EPI_ISL_1017118, EPI_ISL_1017119, EPI_ISL_1017133, EPI_ISL_1017134, EPI_ISL_1017135, EPI_ISL_1017136, EPI_ISL_1017137, EPI_ISL_1017138, EPI_ISL_1017139, EPI_ISL_1017140, EPI_ISL_1017141, EPI_ISL_1017142, EPI_ISL_1017143, EPI_ISL_1017144, EPI_ISL_1017145, EPI_ISL_1017146 | | | |
| see above | Providence St. Joseph Health Orange - Chapman | Providence St. Joseph Health Molecular Genomics Laboratory | Alexa K Dowdell; Brian D Piening; Carlo B Bifulco; Fred L Robinson; Mary Campbell |
| EPI_ISL_802996, EPI_ISL_802997, EPI_ISL_802998, EPI_ISL_802999, EPI_ISL_803000, EPI_ISL_803001, EPI_ISL_803002, EPI_ISL_803003, EPI_ISL_803004, EPI_ISL_803005, EPI_ISL_803006, EPI_ISL_803007, EPI_ISL_803008, EPI_ISL_803009, EPI_ISL_803010, EPI_ISL_803011, EPI_ISL_803012, EPI_ISL_803013, EPI_ISL_803014, EPI_ISL_803015, EPI_ISL_803016, EPI_ISL_803017, EPI_ISL_803018, EPI_ISL_803019, EPI_ISL_803020, EPI_ISL_803021, EPI_ISL_803022, EPI_ISL_803023, EPI_ISL_803024, EPI_ISL_803025, EPI_ISL_803026, EPI_ISL_803027, EPI_ISL_803028, EPI_ISL_803029, EPI_ISL_803030, EPI_ISL_803031, EPI_ISL_803032, EPI_ISL_803033, EPI_ISL_803034, EPI_ISL_803035, EPI_ISL_803036, EPI_ISL_803037, EPI_ISL_803038, EPI_ISL_803039, EPI_ISL_803040, EPI_ISL_803041, EPI_ISL_803042, EPI_ISL_803043, EPI_ISL_803044, EPI_ISL_803045, EPI_ISL_803046, EPI_ISL_803047, EPI_ISL_803048, EPI_ISL_803049, EPI_ISL_803050, EPI_ISL_803051, EPI_ISL_803052, EPI_ISL_803053, EPI_ISL_803054, EPI_ISL_803055, EPI_ISL_803056, EPI_ISL_803057, EPI_ISL_803058, EPI_ISL_803059, EPI_ISL_803060, EPI_ISL_803061, EPI_ISL_803062, EPI_ISL_803063, EPI_ISL_803064, EPI_ISL_803065, EPI_ISL_803066, EPI_ISL_803067, EPI_ISL_803068, EPI_ISL_803069, EPI_ISL_803070, EPI_ISL_803071, EPI_ISL_803072, EPI_ISL_803073, EPI_ISL_803074, EPI_ISL_803075, EPI_ISL_803076, EPI_ISL_803077, EPI_ISL_803078, EPI_ISL_803079, EPI_ISL_803080, EPI_ISL_803081, EPI_ISL_803082, EPI_ISL_803083, EPI_ISL_803084, EPI_ISL_803085, EPI_ISL_803086, EPI_ISL_803087, EPI_ISL_803088, EPI_ISL_803089, EPI_ISL_803090, EPI_ISL_803091, EPI_ISL_803092, EPI_ISL_803093, EPI_ISL_803094, EPI_ISL_803095, EPI_ISL_803096, EPI_ISL_803097, EPI_ISL_855016, EPI_ISL_855019, EPI_ISL_855023, EPI_ISL_855027, EPI_ISL_855030, EPI_ISL_855031, EPI_ISL_855033, EPI_ISL_855034, EPI_ISL_855037, EPI_ISL_855038, EPI_ISL_855039, EPI_ISL_855041, EPI_ISL_855046, EPI_ISL_855047, EPI_ISL_855049, EPI_ISL_855050, EPI_ISL_855052, EPI_ISL_855053, EPI_ISL_855054, EPI_ISL_855057, EPI_ISL_855059, EPI_ISL_855060, EPI_ISL_855061, EPI_ISL_855062, EPI_ISL_855063, EPI_ISL_877186, EPI_ISL_877188 | | | |
| see above | Quest Diagnostics | Quest Diagnostics | Anderson, B.; Bernstein; D.F.; Gerasimova, A.; Hua, M.; I.A.; K.E.; Kagan; L.E.; Lacbawan, F.; Liu Y.; Livingston; Owen, R.; Perez, A.; R.M.; Rosenthal; S.H.; Shalhout; Shlyakhter; Tanpaiboon, P. |
| EPI_ISL_878648, EPI_ISL_878658, EPI_ISL_878661, EPI_ISL_878666, EPI_ISL_878674, EPI_ISL_878682, EPI_ISL_878684, EPI_ISL_878697, EPI_ISL_878700, EPI_ISL_878702, EPI_ISL_878705, EPI_ISL_878707, EPI_ISL_878713, EPI_ISL_878716, EPI_ISL_878719, EPI_ISL_878721, EPI_ISL_878724, EPI_ISL_878726, EPI_ISL_878729, EPI_ISL_878732, EPI_ISL_878736, EPI_ISL_878739, EPI_ISL_878742, EPI_ISL_878747, EPI_ISL_878755, EPI_ISL_878757, EPI_ISL_878760, EPI_ISL_878763, EPI_ISL_878765, EPI_ISL_878768, EPI_ISL_878771, EPI_ISL_878773, EPI_ISL_878776, EPI_ISL_878779, EPI_ISL_878781, EPI_ISL_878786, EPI_ISL_878789, EPI_ISL_878791, EPI_ISL_878797, EPI_ISL_880172, EPI_ISL_880173, EPI_ISL_880174, EPI_ISL_880175, EPI_ISL_880180, EPI_ISL_880181, EPI_ISL_880184, EPI_ISL_880185, EPI_ISL_880186, EPI_ISL_880187, EPI_ISL_880188, EPI_ISL_880189, EPI_ISL_880190, EPI_ISL_880191, EPI_ISL_880193, EPI_ISL_880194, EPI_ISL_880195, EPI_ISL_880196, EPI_ISL_880197, EPI_ISL_880198, EPI_ISL_880199, EPI_ISL_880200 | | | |
| see above | Rady's Childrens Hospital | Andersen lab at Scripps Research | Christina Clarke; David Dimmock; Denise Malicki; Kathryn Bouic; Linda Luo; SEARCH Alliance San Diego with Nanda Radamchar; Teresa Mueller |
| EPI_ISL_738567, EPI_ISL_738574, EPI_ISL_738575, EPI_ISL_738576, EPI_ISL_738591, EPI_ISL_738660, EPI_ISL_738662, EPI_ISL_738712, EPI_ISL_738713, EPI_ISL_738714, EPI_ISL_738723, EPI_ISL_738725, EPI_ISL_738727, EPI_ISL_738770, EPI_ISL_738851, EPI_ISL_738877, EPI_ISL_738901, EPI_ISL_738977, EPI_ISL_739014, EPI_ISL_739022, EPI_ISL_739316, EPI_ISL_739348, EPI_ISL_739355, EPI_ISL_739394, EPI_ISL_739466, EPI_ISL_739467, EPI_ISL_739478, EPI_ISL_739564, EPI_ISL_739630, EPI_ISL_739655, EPI_ISL_850019, EPI_ISL_850020, EPI_ISL_850021, EPI_ISL_850022, EPI_ISL_850023, EPI_ISL_850024, EPI_ISL_850025, EPI_ISL_850026, EPI_ISL_850027, EPI_ISL_850028, EPI_ISL_850029, EPI_ISL_850030, EPI_ISL_850031, EPI_ISL_850032, EPI_ISL_850033, EPI_ISL_850034, EPI_ISL_850035, EPI_ISL_850036, EPI_ISL_850037, EPI_ISL_850038, EPI_ISL_850039, EPI_ISL_850040, EPI_ISL_850041, EPI_ISL_850042, EPI_ISL_850043, EPI_ISL_850044, EPI_ISL_850045, EPI_ISL_850046, EPI_ISL_850047, EPI_ISL_850048, EPI_ISL_850049, EPI_ISL_850050, EPI_ISL_850051, EPI_ISL_850052, EPI_ISL_850053, EPI_ISL_850054, EPI_ISL_850055, EPI_ISL_850056, EPI_ISL_850057, EPI_ISL_850058, EPI_ISL_850059, EPI_ISL_850060, EPI_ISL_850061, EPI_ISL_850062, EPI_ISL_850063, EPI_ISL_850064, EPI_ISL_850065, EPI_ISL_850066, EPI_ISL_850067, EPI_ISL_850068, EPI_ISL_850069, EPI_ISL_850070, EPI_ISL_850071, EPI_ISL_850072, EPI_ISL_850073, EPI_ISL_850074, EPI_ISL_850075, EPI_ISL_850076, EPI_ISL_850077, EPI_ISL_850078, EPI_ISL_850079, EPI_ISL_850080, EPI_ISL_850081, EPI_ISL_850082, EPI_ISL_850083, EPI_ISL_850084, EPI_ISL_850085, EPI_ISL_850086, EPI_ISL_850087, EPI_ISL_850088, EPI_ISL_850089, EPI_ISL_850090, EPI_ISL_850091, EPI_ISL_850092, EPI_ISL_850093, EPI_ISL_850094, EPI_ISL_850095, EPI_ISL_850096, EPI_ISL_850097, EPI_ISL_850098, EPI_ISL_850099, EPI_ISL_850100, EPI_ISL_850101, EPI_ISL_850102, EPI_ISL_850103, EPI_ISL_850104, EPI_ISL_850105, EPI_ISL_850106, EPI_ISL_850107, EPI_ISL_850108, EPI_ISL_850109, EPI_ISL_850110, EPI_ISL_850111, EPI_ISL_850112, EPI_ISL_850113, EPI_ISL_850114, EPI_ISL_850115, EPI_ISL_850116, EPI_ISL_850117, EPI_ISL_850118, EPI_ISL_850119, EPI_ISL_850120, EPI_ISL_850121, EPI_ISL_850122, EPI_ISL_850123, EPI_ISL_850124, EPI_ISL_850125, EPI_ISL_850126, EPI_ISL_850127, EPI_ISL_850128, EPI_ISL_850129, EPI_ISL_850130, EPI_ISL_850131, EPI_ISL_850132, EPI_ISL_850133, EPI_ISL_850134, EPI_ISL_850135, EPI_ISL_850136, EPI_ISL_850137, EPI_ISL_850138, EPI_ISL_850139, EPI_ISL_850140, EPI_ISL_850141, EPI_ISL_850142, EPI_ISL_850143, EPI_ISL_850144, EPI_ISL_850145, EPI_ISL_850146, EPI_ISL_850147, EPI_ISL_850148, EPI_ISL_850149, EPI_ISL_850150, EPI_ISL_850151, EPI_ISL_850152, EPI_ISL_850153, EPI_ISL_850154, EPI_ISL_850155, EPI_ISL_850156, EPI_ISL_850157, EPI_ISL_850158, EPI_ISL_850159, EPI_ISL_850160, EPI_ISL_850161, EPI_ISL_850162, EPI_ISL_850163, EPI_ISL_850164, EPI_ISL_850165, EPI_ISL_850166, EPI_ISL_850167, EPI_ISL_850168, EPI_ISL_850169, EPI_ISL_850170, EPI_ISL_850171, EPI_ISL_850172, EPI_ISL_850173, EPI_ISL_850174, EPI_ISL_850175, EPI_ISL_850176, EPI_ISL_850177, EPI_ISL_850178, EPI_ISL_850179 | | | |
| see above | Renegade | Chan-Zuckerberg Biohub | CZB Cliahub Consortium |
| EPI_ISL_2313619, EPI_ISL_2313621 | Respiratory Viruses Branch, Centers for Disease Control and Prevention | Respiratory Viruses Branch, Centers for Disease Control and Prevention | Batra, D.; Bixby, C.; C.R.; Caravas, J.; Cook; Goswami, C.; Grimwood, R.; Gulvick, C.; Hager, R.; Howard, D.; MacCannell, D.; Morrison, S.; Moser, K.; P.W.; Paden; Paskey, A.; Rambo-Martin, B.; Sammons, S.; Schmerer, M.; Schultz, J.; Unoarumhi, Y.; Wagner, D.; Wang, Y. |
| EPI_ISL_666972, EPI_ISL_666973, EPI_ISL_667042, EPI_ISL_667043, EPI_ISL_667054, EPI_ISL_667055, EPI_ISL_667057, EPI_ISL_667058, EPI_ISL_667059, EPI_ISL_667060, EPI_ISL_667061, EPI_ISL_667064, EPI_ISL_667065, EPI_ISL_667066, EPI_ISL_667067, EPI_ISL_667068, EPI_ISL_667069, EPI_ISL_667070, EPI_ISL_667071, EPI_ISL_667072, EPI_ISL_667073, EPI_ISL_667074, EPI_ISL_667075, EPI_ISL_667076, EPI_ISL_667077, EPI_ISL_667078, EPI_ISL_730086, EPI_ISL_730161, EPI_ISL_730162, EPI_ISL_730164, EPI_ISL_730166, EPI_ISL_730167, EPI_ISL_730168, EPI_ISL_730169, EPI_ISL_730171, EPI_ISL_730174, EPI_ISL_730175, EPI_ISL_730176, EPI_ISL_730177, EPI_ISL_730178, EPI_ISL_730179, EPI_ISL_730182, EPI_ISL_730183, EPI_ISL_730186, EPI_ISL_730187, EPI_ISL_730188, EPI_ISL_730189, EPI_ISL_730190, EPI_ISL_730191, EPI_ISL_730193, EPI_ISL_730194, EPI_ISL_730195, EPI_ISL_730196, EPI_ISL_730291, EPI_ISL_730292, EPI_ISL_730293, EPI_ISL_730294, EPI_ISL_730295, EPI_ISL_730296, EPI_ISL_730297, EPI_ISL_730298, EPI_ISL_730299, EPI_ISL_730300, EPI_ISL_730301, EPI_ISL_730302, EPI_ISL_730303, EPI_ISL_730304, EPI_ISL_730305, EPI_ISL_730306, EPI_ISL_730307, EPI_ISL_730308, EPI_ISL_730309, EPI_ISL_730310, EPI_ISL_730311, EPI_ISL_730312, EPI_ISL_730313, EPI_ISL_730314, EPI_ISL_730315, EPI_ISL_730316, EPI_ISL_730317, EPI_ISL_730319, EPI_ISL_730320, EPI_ISL_730322, EPI_ISL_730324, EPI_ISL_730325, EPI_ISL_730328, EPI_ISL_730329, EPI_ISL_730332, EPI_ISL_730336, EPI_ISL_730338, EPI_ISL_730340, EPI_ISL_730342, EPI_ISL_730346, EPI_ISL_730349, EPI_ISL_730350, EPI_ISL_730351, EPI_ISL_730352, EPI_ISL_730353, EPI_ISL_730354, EPI_ISL_730355, EPI_ISL_730356, EPI_ISL_730357, EPI_ISL_730358, EPI_ISL_730359, EPI_ISL_730360, EPI_ISL_730361, EPI_ISL_730362, EPI_ISL_730363, EPI_ISL_730364, EPI_ISL_730365, EPI_ISL_730366, EPI_ISL_730367, EPI_ISL_730368, EPI_ISL_730369, EPI_ISL_730370, EPI_ISL_730371, EPI_ISL_755115, EPI_ISL_755116, EPI_ISL_755117, EPI_ISL_755119, EPI_ISL_755130, EPI_ISL_755132, EPI_ISL_755134, EPI_ISL_755135, EPI_ISL_755136, EPI_ISL_755138, EPI_ISL_755139, EPI_ISL_755269, EPI_ISL_755270, EPI_ISL_755271, EPI_ISL_755272, EPI_ISL_755273, EPI_ISL_755274, EPI_ISL_755275, EPI_ISL_755276, EPI_ISL_755277, EPI_ISL_755278, EPI_ISL_755279, EPI_ISL_755280, EPI_ISL_755281, EPI_ISL_755282, EPI_ISL_755283, EPI_ISL_755284, EPI_ISL_755285, EPI_ISL_755286, EPI_ISL_755287, EPI_ISL_755288, EPI_ISL_755289, EPI_ISL_755290, EPI_ISL_755291, EPI_ISL_755292, EPI_ISL_755293, EPI_ISL_755294, EPI_ISL_755295, EPI_ISL_755296, EPI_ISL_755297, EPI_ISL_755298, EPI_ISL_755299, EPI_ISL_755300, EPI_ISL_878152, EPI_ISL_878155, EPI_ISL_878157, EPI_ISL_878159, EPI_ISL_878162, EPI_ISL_878165, EPI_ISL_878167, EPI_ISL_878170, EPI_ISL_878172, EPI_ISL_878175, EPI_ISL_878178, EPI_ISL_878180, EPI_ISL_878183, EPI_ISL_878186, EPI_ISL_878189, EPI_ISL_878191, EPI_ISL_878194, EPI_ISL_878197, EPI_ISL_878199, EPI_ISL_878202, EPI_ISL_878205, EPI_ISL_878207, EPI_ISL_878209, EPI_ISL_878215, EPI_ISL_878217, EPI_ISL_878221, EPI_ISL_878223, EPI_ISL_878229, EPI_ISL_878232, EPI_ISL_878234, EPI_ISL_878237, EPI_ISL_878239, EPI_ISL_878242, EPI_ISL_878245, EPI_ISL_878248, EPI_ISL_878250, EPI_ISL_878253, EPI_ISL_878256, EPI_ISL_878258, EPI_ISL_878260, EPI_ISL_878263, EPI_ISL_878266, EPI_ISL_878268, EPI_ISL_878271, EPI_ISL_878274, EPI_ISL_878276, EPI_ISL_878279, EPI_ISL_878281, EPI_ISL_878284, EPI_ISL_878287, EPI_ISL_878290, EPI_ISL_878292, EPI_ISL_878295, EPI_ISL_878298, EPI_ISL_878303, EPI_ISL_878305, EPI_ISL_878308, EPI_ISL_878311, EPI_ISL_878314, EPI_ISL_878316, EPI_ISL_878319, EPI_ISL_878322, EPI_ISL_878325, EPI_ISL_878327, EPI_ISL_878330, EPI_ISL_878333, EPI_ISL_878336, EPI_ISL_878338, EPI_ISL_878341, EPI_ISL_878342, EPI_ISL_878343, EPI_ISL_878344, EPI_ISL_878345, EPI_ISL_878346, EPI_ISL_878347, EPI_ISL_878348, EPI_ISL_878349, EPI_ISL_878350, EPI_ISL_878351, EPI_ISL_878352, EPI_ISL_878353, EPI_ISL_878354, EPI_ISL_878355, EPI_ISL_878356, EPI_ISL_878357, EPI_ISL_878358, EPI_ISL_878359, EPI_ISL_878360, EPI_ISL_878361, EPI_ISL_878362, EPI_ISL_878363, EPI_ISL_878364, EPI_ISL_878365, EPI_ISL_878366, EPI_ISL_878368, EPI_ISL_878369, EPI_ISL_878370, EPI_ISL_878371, EPI_ISL_878372, EPI_ISL_878373, EPI_ISL_878374, EPI_ISL_878376, EPI_ISL_878377, EPI_ISL_878378, EPI_ISL_878379, EPI_ISL_878380, EPI_ISL_878381, EPI_ISL_878382, EPI_ISL_878383, EPI_ISL_878384, EPI_ISL_878385, EPI_ISL_878386, EPI_ISL_878387, EPI_ISL_878388, EPI_ISL_878389, EPI_ISL_878391, EPI_ISL_878392, EPI_ISL_878393, EPI_ISL_878394, EPI_ISL_878395, EPI_ISL_878396, EPI_ISL_878397, EPI_ISL_878398, EPI_ISL_878399, EPI_ISL_878402, EPI_ISL_878403, EPI_ISL_878405, EPI_ISL_878406, EPI_ISL_878407, EPI_ISL_878411, EPI_ISL_878413, EPI_ISL_878416, EPI_ISL_878418, EPI_ISL_878421, EPI_ISL_878423, EPI_ISL_878426, EPI_ISL_878429, EPI_ISL_878431, EPI_ISL_878434, EPI_ISL_878584, EPI_ISL_878587, EPI_ISL_878589, EPI_ISL_878592, EPI_ISL_878595, EPI_ISL_878597, EPI_ISL_878600, EPI_ISL_878603, EPI_ISL_878605, EPI_ISL_878608, EPI_ISL_878611, EPI_ISL_878613, EPI_ISL_878616, EPI_ISL_878619, EPI_ISL_878624, EPI_ISL_878629, EPI_ISL_878632, EPI_ISL_878635, EPI_ISL_879924, EPI_ISL_879926, EPI_ISL_879929, EPI_ISL_879932, EPI_ISL_879934, EPI_ISL_879937, EPI_ISL_879939, EPI_ISL_879942, EPI_ISL_879945, EPI_ISL_879947, EPI_ISL_879953, EPI_ISL_879956, EPI_ISL_879959, EPI_ISL_879961, EPI_ISL_879964, EPI_ISL_879967, EPI_ISL_879969, EPI_ISL_879972, EPI_ISL_879975, EPI_ISL_879980, EPI_ISL_879982, EPI_ISL_879985, EPI_ISL_879987, EPI_ISL_879990, EPI_ISL_879993, EPI_ISL_879995, EPI_ISL_879998, EPI_ISL_880000, EPI_ISL_880003, EPI_ISL_880006, EPI_ISL_880009, EPI_ISL_880011, EPI_ISL_880014, EPI_ISL_880017, EPI_ISL_880019, EPI_ISL_880022, EPI_ISL_880024, EPI_ISL_880027, EPI_ISL_880029, EPI_ISL_880032, EPI_ISL_880034, EPI_ISL_962609, EPI_ISL_962610, EPI_ISL_962612, EPI_ISL_962613, EPI_ISL_962614, EPI_ISL_962615, EPI_ISL_962616, EPI_ISL_962617, EPI_ISL_962766, EPI_ISL_962767, EPI_ISL_962768, EPI_ISL_962769, EPI_ISL_962770, EPI_ISL_962771, EPI_ISL_962772, EPI_ISL_962773, EPI_ISL_962774, EPI_ISL_962775, EPI_ISL_962776, EPI_ISL_962777, EPI_ISL_962778, EPI_ISL_962779, EPI_ISL_962780, EPI_ISL_962781, EPI_ISL_962782, EPI_ISL_962783, EPI_ISL_962784, EPI_ISL_962785, EPI_ISL_962786, EPI_ISL_962787, EPI_ISL_962788, EPI_ISL_962789, EPI_ISL_962790, EPI_ISL_962791, EPI_ISL_962792, EPI_ISL_962793, EPI_ISL_962794, EPI_ISL_962795, EPI_ISL_962796, EPI_ISL_962797, EPI_ISL_962798, EPI_ISL_962799, EPI_ISL_962800, EPI_ISL_962801, EPI_ISL_962802, EPI_ISL_962803, EPI_ISL_962804, EPI_ISL_962805, EPI_ISL_962806, EPI_ISL_962807, EPI_ISL_962808, EPI_ISL_962809, EPI_ISL_962810, EPI_ISL_962811, EPI_ISL_1081457, EPI_ISL_1081458, EPI_ISL_1081619, EPI_ISL_1081620, EPI_ISL_1081621, EPI_ISL_1081622, EPI_ISL_1081623, EPI_ISL_1081624, EPI_ISL_1081625, EPI_ISL_1081626, EPI_ISL_1081627, EPI_ISL_1366747, EPI_ISL_1531942, EPI_ISL_1531943, EPI_ISL_1531944, EPI_ISL_1531945, EPI_ISL_1531946, EPI_ISL_1531947, EPI_ISL_1531948, EPI_ISL_1531949, EPI_ISL_1531950, EPI_ISL_1531951, EPI_ISL_1531952, EPI_ISL_1531953, EPI_ISL_1531954, EPI_ISL_1531955, EPI_ISL_1531956, EPI_ISL_1531957, EPI_ISL_1531958, EPI_ISL_1531959, EPI_ISL_1531960, EPI_ISL_1531961, EPI_ISL_1531962, EPI_ISL_1531963, EPI_ISL_1531964, EPI_ISL_1531965, EPI_ISL_1531966, EPI_ISL_1531967, EPI_ISL_1531968, EPI_ISL_1531969, EPI_ISL_1531970, EPI_ISL_1531971, EPI_ISL_1531972, EPI_ISL_1531973, EPI_ISL_1531974, EPI_ISL_1531975, EPI_ISL_1531976, EPI_ISL_1531977, EPI_ISL_1531978, EPI_ISL_1531979, EPI_ISL_1531980, EPI_ISL_1531981, EPI_ISL_1531982, EPI_ISL_1531983, EPI_ISL_1531984, EPI_ISL_1531985, EPI_ISL_1531986, EPI_ISL_1531987, EPI_ISL_1531988, EPI_ISL_1531989, EPI_ISL_1531990, EPI_ISL_1531991, EPI_ISL_1531992, EPI_ISL_1531993, EPI_ISL_1531994, EPI_ISL_1531995, EPI_ISL_1531996, EPI_ISL_1531997, EPI_ISL_1531998, EPI_ISL_1531999, EPI_ISL_1532000, EPI_ISL_1532001, EPI_ISL_1532002, EPI_ISL_1532003, EPI_ISL_1532004, EPI_ISL_1532005, EPI_ISL_1532006, EPI_ISL_1532007, EPI_ISL_1532008, EPI_ISL_1532009, EPI_ISL_1532010, EPI_ISL_1532011, EPI_ISL_1532012, EPI_ISL_1532013, EPI_ISL_1532014, EPI_ISL_1532015, EPI_ISL_1532016, EPI_ISL_1532017, EPI_ISL_1532018, EPI_ISL_1532019, EPI_ISL_1532020, EPI_ISL_1532021, EPI_ISL_1532022, EPI_ISL_1532023, EPI_ISL_1532024, EPI_ISL_1532025, EPI_ISL_1532026, EPI_ISL_1532027, EPI_ISL_1532028, EPI_ISL_1532029, EPI_ISL_1532030, EPI_ISL_1532031, EPI_ISL_1532032, EPI_ISL_1532033, EPI_ISL_1532034, EPI_ISL_1532035, EPI_ISL_1532036, EPI_ISL_1532037, EPI_ISL_1532038, EPI_ISL_1532039, EPI_ISL_1532040, EPI_ISL_1532041, EPI_ISL_1532042, EPI_ISL_1532043, EPI_ISL_1532044, EPI_ISL_1532045, EPI_ISL_1532046, EPI_ISL_1532047, EPI_ISL_1532048, EPI_ISL_1532049, EPI_ISL_1532050, EPI_ISL_1532051, EPI_ISL_1532052, EPI_ISL_1532053, EPI_ISL_1532054, EPI_ISL_1532055, EPI_ISL_1532056, EPI_ISL_1532057, EPI_ISL_1532058, | | | |

| | | | |
|---|---|---|---|
| EPI_ISL_1532059, EPI_ISL_1532060, EPI_ISL_1532061, EPI_ISL_1532062, EPI_ISL_1532063, EPI_ISL_1532064, EPI_ISL_1794620, EPI_ISL_1794627, EPI_ISL_1794629, EPI_ISL_1794634, EPI_ISL_1794636, EPI_ISL_1794641, EPI_ISL_1794645, EPI_ISL_1794650, EPI_ISL_1794656, EPI_ISL_1794657, EPI_ISL_1794658, EPI_ISL_1794659, EPI_ISL_1794660, EPI_ISL_1794662, EPI_ISL_1794663, EPI_ISL_1794665, EPI_ISL_1794666, EPI_ISL_1794667, EPI_ISL_1794669, EPI_ISL_1794671, EPI_ISL_1794673, EPI_ISL_1794678, EPI_ISL_1794680, EPI_ISL_1794683, EPI_ISL_1794684, EPI_ISL_1794939, EPI_ISL_1794940, EPI_ISL_1794941, EPI_ISL_1794943, EPI_ISL_1794945, EPI_ISL_1794946, EPI_ISL_1794947, EPI_ISL_1794951, EPI_ISL_1794955, EPI_ISL_1794956, EPI_ISL_1794960, EPI_ISL_1794962, EPI_ISL_1794963, EPI_ISL_1794964, EPI_ISL_1794967, EPI_ISL_1794968, EPI_ISL_1794971, EPI_ISL_1794972, EPI_ISL_1794976, EPI_ISL_1794977, EPI_ISL_1794978, EPI_ISL_1794980, EPI_ISL_1794981, EPI_ISL_1794982, EPI_ISL_1969282, EPI_ISL_1969284, EPI_ISL_1969285, EPI_ISL_1969286, EPI_ISL_1969293, EPI_ISL_1969305, EPI_ISL_1969319, EPI_ISL_1969320, EPI_ISL_1969321, EPI_ISL_1969322, EPI_ISL_1969323, EPI_ISL_1969451, EPI_ISL_1969452, EPI_ISL_1969453, EPI_ISL_1969454, EPI_ISL_1969455, EPI_ISL_1969456, EPI_ISL_1969457, EPI_ISL_1969458, EPI_ISL_1969459, EPI_ISL_1969460, EPI_ISL_1969461, EPI_ISL_1969462, EPI_ISL_1969463, EPI_ISL_1969464, EPI_ISL_1969465, EPI_ISL_1969466, EPI_ISL_1969467, EPI_ISL_1969468, EPI_ISL_1969469, EPI_ISL_1969470, EPI_ISL_1969471, EPI_ISL_1969472, EPI_ISL_1969473, EPI_ISL_1969474, EPI_ISL_1969475, EPI_ISL_1969476, EPI_ISL_1969477, EPI_ISL_1969478, EPI_ISL_1969479, EPI_ISL_1969480, EPI_ISL_1969481, EPI_ISL_1969482, EPI_ISL_1969483, EPI_ISL_1969484, EPI_ISL_1969485, EPI_ISL_1969486, EPI_ISL_1969487, EPI_ISL_1969488, EPI_ISL_1969489, EPI_ISL_1969490, EPI_ISL_1969491, EPI_ISL_1969492, EPI_ISL_1969493, EPI_ISL_1969494, EPI_ISL_1969495, EPI_ISL_1969496, EPI_ISL_1969501, EPI_ISL_1969502, EPI_ISL_2231039, EPI_ISL_2231040, EPI_ISL_2231042, EPI_ISL_2231043, EPI_ISL_2231045, EPI_ISL_2231048, EPI_ISL_2231052, EPI_ISL_2231053, EPI_ISL_2231060, EPI_ISL_2231065, EPI_ISL_2231067, EPI_ISL_2231069, EPI_ISL_2231070, EPI_ISL_2231071, EPI_ISL_2231077, EPI_ISL_2231078, EPI_ISL_2231081, EPI_ISL_2231083, EPI_ISL_2231084, EPI_ISL_2231086, EPI_ISL_2231087, EPI_ISL_2231089, EPI_ISL_2231173, EPI_ISL_2231193, EPI_ISL_2231201, EPI_ISL_2231202, EPI_ISL_2231203, EPI_ISL_2231207, EPI_ISL_2231208, EPI_ISL_2231209, EPI_ISL_2231210, EPI_ISL_2231211, EPI_ISL_2231212, EPI_ISL_2231213, EPI_ISL_2231214, EPI_ISL_2231215, EPI_ISL_2231216, EPI_ISL_2231217, EPI_ISL_2231218, EPI_ISL_2231219, EPI_ISL_2231220, EPI_ISL_2231221, EPI_ISL_2231222, EPI_ISL_2231223, EPI_ISL_2231224, EPI_ISL_2231225, EPI_ISL_2231226, EPI_ISL_2231227, EPI_ISL_2231228, EPI_ISL_2231229, EPI_ISL_2231230, EPI_ISL_2231231, EPI_ISL_2231232, EPI_ISL_2231233, EPI_ISL_2231234, EPI_ISL_2231235, EPI_ISL_2231236, EPI_ISL_2231237, EPI_ISL_2231238, EPI_ISL_2231239, EPI_ISL_2231240, EPI_ISL_2231241, EPI_ISL_2231242, EPI_ISL_2231243, EPI_ISL_2231244, EPI_ISL_2231245, EPI_ISL_2231246, EPI_ISL_2231247, EPI_ISL_2231248, EPI_ISL_2231249, EPI_ISL_2231250, EPI_ISL_2231251, EPI_ISL_2231252, EPI_ISL_2231253, EPI_ISL_2231254, EPI_ISL_2231255, EPI_ISL_2231256, EPI_ISL_2231257, EPI_ISL_2231258, EPI_ISL_2231259, EPI_ISL_2231260, EPI_ISL_2231261, EPI_ISL_2231262, EPI_ISL_2231263, EPI_ISL_2231264, EPI_ISL_2231265, EPI_ISL_2231266, EPI_ISL_2231267, EPI_ISL_2231268, EPI_ISL_2231269, EPI_ISL_2231436, EPI_ISL_2231437, EPI_ISL_2231438, EPI_ISL_2231439, EPI_ISL_2231440, EPI_ISL_2231441, EPI_ISL_2231443, EPI_ISL_2231444, EPI_ISL_2231445, EPI_ISL_2231446, EPI_ISL_2231447, EPI_ISL_2231448, EPI_ISL_2231449, EPI_ISL_2231450, EPI_ISL_2231451, EPI_ISL_2231452, EPI_ISL_2231453, EPI_ISL_2231454, EPI_ISL_2231455, EPI_ISL_2231456, EPI_ISL_2231457, EPI_ISL_2231458, EPI_ISL_2231459, EPI_ISL_2231460, EPI_ISL_2231461, EPI_ISL_2329335, EPI_ISL_2329337, EPI_ISL_2329338, EPI_ISL_2329340, EPI_ISL_2329341, EPI_ISL_2329342, EPI_ISL_2329343, EPI_ISL_2329344, EPI_ISL_2329346, EPI_ISL_2329347, EPI_ISL_2329348, EPI_ISL_2329349, EPI_ISL_2329350, EPI_ISL_2329352, EPI_ISL_2329353, EPI_ISL_2329354, EPI_ISL_2329355, EPI_ISL_2329356, EPI_ISL_2329359, EPI_ISL_2329360, EPI_ISL_2329361, EPI_ISL_2329362, EPI_ISL_2329363, EPI_ISL_2329366, EPI_ISL_2329367, EPI_ISL_2329368, EPI_ISL_2329369, EPI_ISL_2329372, EPI_ISL_2329373, EPI_ISL_2329374, EPI_ISL_2329375, EPI_ISL_2329376, EPI_ISL_2329377, EPI_ISL_2329378, EPI_ISL_2329379, EPI_ISL_2329380, EPI_ISL_2329382, EPI_ISL_2329383, EPI_ISL_2329384, EPI_ISL_2329385, EPI_ISL_2329387, EPI_ISL_2329388, EPI_ISL_2329389, EPI_ISL_2329390, EPI_ISL_2329391, EPI_ISL_2329394, EPI_ISL_2329395, EPI_ISL_2329396, EPI_ISL_2329397, EPI_ISL_2329398, EPI_ISL_2329401, EPI_ISL_2329402, EPI_ISL_2329403, EPI_ISL_2329404, EPI_ISL_2329405, EPI_ISL_2329406, EPI_ISL_2329469, EPI_ISL_2329474, EPI_ISL_2329478, EPI_ISL_2329484, EPI_ISL_2329491, EPI_ISL_2329497, EPI_ISL_2329502, EPI_ISL_2329508, EPI_ISL_2329514, EPI_ISL_2329515, EPI_ISL_2329521, EPI_ISL_2329535, EPI_ISL_2422573, EPI_ISL_2422576, EPI_ISL_2422580, EPI_ISL_2422584, EPI_ISL_2422587, EPI_ISL_2422590, EPI_ISL_2422592, EPI_ISL_2422596, EPI_ISL_2422599, EPI_ISL_2422602, EPI_ISL_2422606, EPI_ISL_2422609, EPI_ISL_2422613, EPI_ISL_2422616, EPI_ISL_2422620, EPI_ISL_2422623, EPI_ISL_2422626, EPI_ISL_2422629, EPI_ISL_2422631, EPI_ISL_2422632, EPI_ISL_2422633, EPI_ISL_2422634, EPI_ISL_2422635, EPI_ISL_2422636, EPI_ISL_2422637, EPI_ISL_2422638, EPI_ISL_2422639, EPI_ISL_2422640, EPI_ISL_2422641, EPI_ISL_2422642, EPI_ISL_2422643, EPI_ISL_2422644, EPI_ISL_2422645, EPI_ISL_2422646, EPI_ISL_2422647, EPI_ISL_2422648, EPI_ISL_2422649, EPI_ISL_2422651, EPI_ISL_2422652, EPI_ISL_2422653, EPI_ISL_2422654, EPI_ISL_2422655, EPI_ISL_2422656, EPI_ISL_2422657, EPI_ISL_2422658, EPI_ISL_2422659, EPI_ISL_2422660, EPI_ISL_2422661, EPI_ISL_2422662, EPI_ISL_2422663, EPI_ISL_2422664, EPI_ISL_2422665, EPI_ISL_2422666, EPI_ISL_2422667, EPI_ISL_2422668, EPI_ISL_2422897, EPI_ISL_2422898, EPI_ISL_2422903, EPI_ISL_2422909, EPI_ISL_2422910, EPI_ISL_2422911, EPI_ISL_2422916, EPI_ISL_2422917, EPI_ISL_2422923, EPI_ISL_2422924, EPI_ISL_2422925, EPI_ISL_2422928, EPI_ISL_2422929, EPI_ISL_2422930, EPI_ISL_2422935, EPI_ISL_2422936, EPI_ISL_2422937, EPI_ISL_2422943, EPI_ISL_2422944, EPI_ISL_2422951, EPI_ISL_2422952, EPI_ISL_2422958, EPI_ISL_2422961, EPI_ISL_2422963, EPI_ISL_2422969, EPI_ISL_2422970, EPI_ISL_2628314, EPI_ISL_2628318, EPI_ISL_2628322, EPI_ISL_2628327, EPI_ISL_2628330, EPI_ISL_2628335, EPI_ISL_2628436, EPI_ISL_2628437, EPI_ISL_2628441, EPI_ISL_2628445, EPI_ISL_2628448, EPI_ISL_2628450, EPI_ISL_2628451, EPI_ISL_2628454, EPI_ISL_2628458, EPI_ISL_2628462, EPI_ISL_2628463, EPI_ISL_2628467, EPI_ISL_2628468, EPI_ISL_2628474, EPI_ISL_2628507, EPI_ISL_2628549, EPI_ISL_2628561, EPI_ISL_2628565, EPI_ISL_2628570, EPI_ISL_2628597, EPI_ISL_2628603, EPI_ISL_2628615, EPI_ISL_2628684, EPI_ISL_2628689, EPI_ISL_2628695, EPI_ISL_2628699, EPI_ISL_2628703, EPI_ISL_2628707, EPI_ISL_2628711, EPI_ISL_2628735, EPI_ISL_2712771, EPI_ISL_2712773, EPI_ISL_2712774, EPI_ISL_2712776, EPI_ISL_2712778, EPI_ISL_2712782, EPI_ISL_2712784, EPI_ISL_2712788, EPI_ISL_2712790, EPI_ISL_2712795, EPI_ISL_2712797, EPI_ISL_2712798, EPI_ISL_2712803, EPI_ISL_2712804, EPI_ISL_2712805, EPI_ISL_2712808, EPI_ISL_2712809, EPI_ISL_2712810, EPI_ISL_2712811, EPI_ISL_2712812, EPI_ISL_2712815, EPI_ISL_2712816, EPI_ISL_2712817, EPI_ISL_2712818, EPI_ISL_2712821, EPI_ISL_2712822, EPI_ISL_2712823, EPI_ISL_2712824, EPI_ISL_2712825, EPI_ISL_2712828, EPI_ISL_2712829, EPI_ISL_2712830, EPI_ISL_2712834, EPI_ISL_2712835, EPI_ISL_2712838, EPI_ISL_2712840, EPI_ISL_2920336, EPI_ISL_2920337, EPI_ISL_2920342, EPI_ISL_2920343, EPI_ISL_2920346, EPI_ISL_2920352, EPI_ISL_2920356, EPI_ISL_2920362, EPI_ISL_2920366, EPI_ISL_2920371, EPI_ISL_2920376, EPI_ISL_2920387, EPI_ISL_2920388, EPI_ISL_2920394, EPI_ISL_2920395 | | | |
| see above | San Diego County Public Health Laboratory | Andersen lab at Scripps Research | Brett Austin; Jovan Shephard; SEARCH Alliance San Diego with Ashleigh Murphy; SEARCH Alliance San Diego with Tracy Basler |
| EPI_ISL_2840492, EPI_ISL_2840544, EPI_ISL_2840583, EPI_ISL_2840584, EPI_ISL_2840590, EPI_ISL_2840865, EPI_ISL_2840866, EPI_ISL_2840923 | | | |
| see above | San Diego County Public Health Laboratory | San Diego County Public Health Laboratory | Ashleigh Murphy Schafer; Brett Austin |
| EPI_ISL_739603 | San Francisco Public Health Laboratory | Chan-Zuckerberg Biohub | CZB Cliahub Consortium |
| EPI_ISL_672434, EPI_ISL_672435, EPI_ISL_672436, EPI_ISL_672437, EPI_ISL_672438, EPI_ISL_738500, EPI_ISL_738501, EPI_ISL_738502, EPI_ISL_738508, EPI_ISL_738511, EPI_ISL_738516, EPI_ISL_738517, EPI_ISL_738520, EPI_ISL_738521, EPI_ISL_738525, EPI_ISL_738526, EPI_ISL_738532, EPI_ISL_738533, EPI_ISL_738534, EPI_ISL_738536, EPI_ISL_738542, EPI_ISL_738543, EPI_ISL_738546, EPI_ISL_738549, EPI_ISL_738550, EPI_ISL_738551, EPI_ISL_738552, EPI_ISL_738555, EPI_ISL_738556, EPI_ISL_738557, EPI_ISL_738558, EPI_ISL_738560, EPI_ISL_738566, EPI_ISL_738568, EPI_ISL_738578, EPI_ISL_738579, EPI_ISL_738581, EPI_ISL_738582, EPI_ISL_738583, EPI_ISL_738586, EPI_ISL_738589, EPI_ISL_738590, EPI_ISL_738593, EPI_ISL_738595, EPI_ISL_738596, EPI_ISL_738597, EPI_ISL_738598, EPI_ISL_738601, EPI_ISL_738608, EPI_ISL_738610, EPI_ISL_738611, EPI_ISL_738612, EPI_ISL_738617, EPI_ISL_738618, EPI_ISL_738628, EPI_ISL_738631, EPI_ISL_738643, EPI_ISL_738644, EPI_ISL_738645, EPI_ISL_738650, EPI_ISL_738652, EPI_ISL_738653, EPI_ISL_738657, EPI_ISL_738661, EPI_ISL_738663, EPI_ISL_738667, EPI_ISL_738670, EPI_ISL_738672, EPI_ISL_738673, EPI_ISL_738681, EPI_ISL_738682, EPI_ISL_738683, EPI_ISL_738684, EPI_ISL_738685, EPI_ISL_738689, EPI_ISL_738690, EPI_ISL_738691, EPI_ISL_738695, EPI_ISL_738696, EPI_ISL_738697, EPI_ISL_738698, EPI_ISL_738700, EPI_ISL_738701, EPI_ISL_738702, EPI_ISL_738705, EPI_ISL_738706, EPI_ISL_738710, EPI_ISL_738716, EPI_ISL_738717, EPI_ISL_738718, EPI_ISL_738719, EPI_ISL_738720, EPI_ISL_738726, EPI_ISL_738728, EPI_ISL_738730, EPI_ISL_738746, EPI_ISL_738747, EPI_ISL_738750, EPI_ISL_738763, EPI_ISL_738764, EPI_ISL_738765, EPI_ISL_738769, EPI_ISL_738771, EPI_ISL_738772, EPI_ISL_738780, EPI_ISL_738789, EPI_ISL_738791, EPI_ISL_738794, EPI_ISL_738798, EPI_ISL_738803, EPI_ISL_738805, EPI_ISL_738812, EPI_ISL_738834, EPI_ISL_738839, EPI_ISL_738842, EPI_ISL_738853, EPI_ISL_738855, EPI_ISL_738859, EPI_ISL_738861, EPI_ISL_738864, EPI_ISL_738867, EPI_ISL_738883, EPI_ISL_738887, EPI_ISL_738889, EPI_ISL_738894, EPI_ISL_738898, EPI_ISL_738903, EPI_ISL_738906, EPI_ISL_738909, EPI_ISL_738910, EPI_ISL_738915, EPI_ISL_738920, EPI_ISL_738922, EPI_ISL_738924, EPI_ISL_738926, EPI_ISL_738927, EPI_ISL_738931, EPI_ISL_738933, EPI_ISL_738936, EPI_ISL_738941, EPI_ISL_738942, EPI_ISL_738943, EPI_ISL_738948, EPI_ISL_738955, EPI_ISL_738956, EPI_ISL_738960, EPI_ISL_738962, EPI_ISL_738965, EPI_ISL_738967, EPI_ISL_738971, EPI_ISL_738972, EPI_ISL_738979, EPI_ISL_738984, EPI_ISL_738987, EPI_ISL_738994, EPI_ISL_738996, EPI_ISL_738997, EPI_ISL_738998, EPI_ISL_739005, EPI_ISL_739006, EPI_ISL_739008, EPI_ISL_739009, EPI_ISL_739010, EPI_ISL_739012, EPI_ISL_739015, EPI_ISL_739019, EPI_ISL_739020, EPI_ISL_739021, EPI_ISL_739025, EPI_ISL_739026, EPI_ISL_739027, EPI_ISL_739030, EPI_ISL_739031, EPI_ISL_739036, EPI_ISL_739037, EPI_ISL_739039, EPI_ISL_739040, EPI_ISL_739041, EPI_ISL_739045, EPI_ISL_739050, EPI_ISL_739053, EPI_ISL_739058, EPI_ISL_739060, EPI_ISL_739065, EPI_ISL_739066, EPI_ISL_739067, EPI_ISL_739068, EPI_ISL_739070, EPI_ISL_739074, EPI_ISL_739076, EPI_ISL_739080, EPI_ISL_739085, EPI_ISL_739087, EPI_ISL_739098, EPI_ISL_739103, EPI_ISL_739104, EPI_ISL_739107, EPI_ISL_739117, EPI_ISL_739121, EPI_ISL_739122, EPI_ISL_739128, EPI_ISL_739129, EPI_ISL_739130, EPI_ISL_739138, EPI_ISL_739141, EPI_ISL_739145, EPI_ISL_739146, EPI_ISL_739147, EPI_ISL_739152, EPI_ISL_739153, EPI_ISL_739157, EPI_ISL_739166, EPI_ISL_739167, EPI_ISL_739168, EPI_ISL_739180, EPI_ISL_739184, EPI_ISL_739186, EPI_ISL_739187, EPI_ISL_739188, EPI_ISL_739195, EPI_ISL_739204, EPI_ISL_739214, EPI_ISL_739219, EPI_ISL_739221, EPI_ISL_739224, EPI_ISL_739232, EPI_ISL_739234, EPI_ISL_739237, EPI_ISL_739246, EPI_ISL_739247, EPI_ISL_739248, EPI_ISL_739252, EPI_ISL_739253, EPI_ISL_739254, EPI_ISL_739256, EPI_ISL_739260, EPI_ISL_739263, EPI_ISL_739273, EPI_ISL_739275, EPI_ISL_739277, EPI_ISL_739278, EPI_ISL_739281, EPI_ISL_739282, EPI_ISL_739284, EPI_ISL_739293, EPI_ISL_739298, EPI_ISL_739305, EPI_ISL_739310, EPI_ISL_739317, EPI_ISL_739319, EPI_ISL_739320, EPI_ISL_739321, EPI_ISL_739327, EPI_ISL_739332, EPI_ISL_739335, EPI_ISL_739343, EPI_ISL_739344, EPI_ISL_739346, EPI_ISL_739352, EPI_ISL_739354, EPI_ISL_739357, EPI_ISL_739360, EPI_ISL_739365, EPI_ISL_739370, EPI_ISL_739371, EPI_ISL_739374, EPI_ISL_739376, EPI_ISL_739377, EPI_ISL_739380, EPI_ISL_739381, EPI_ISL_739382, EPI_ISL_739388, EPI_ISL_739392, EPI_ISL_739398, EPI_ISL_739400, EPI_ISL_739404, EPI_ISL_739409, EPI_ISL_739413, EPI_ISL_739415, EPI_ISL_739416, EPI_ISL_739419, EPI_ISL_739431, EPI_ISL_739440, EPI_ISL_739443, EPI_ISL_739451, EPI_ISL_739453, EPI_ISL_739456, EPI_ISL_739457, EPI_ISL_739458, EPI_ISL_739476, EPI_ISL_739479, EPI_ISL_739481, EPI_ISL_739485, EPI_ISL_739487, EPI_ISL_739491, EPI_ISL_739498, EPI_ISL_739501, EPI_ISL_739502, EPI_ISL_739504, EPI_ISL_739505, EPI_ISL_739508, EPI_ISL_739509, EPI_ISL_739510, EPI_ISL_739512, EPI_ISL_739520, EPI_ISL_739521, EPI_ISL_739536, EPI_ISL_739537, EPI_ISL_739543, EPI_ISL_739548, EPI_ISL_739549, EPI_ISL_739550, EPI_ISL_739560, EPI_ISL_739561, EPI_ISL_739562, EPI_ISL_739574, EPI_ISL_739579, EPI_ISL_739580, EPI_ISL_739584, EPI_ISL_739589, EPI_ISL_739593, EPI_ISL_739594, EPI_ISL_739596, EPI_ISL_739597, EPI_ISL_739602, EPI_ISL_739605, EPI_ISL_739612, EPI_ISL_739617, EPI_ISL_739618, EPI_ISL_739624, EPI_ISL_739626, EPI_ISL_739629, EPI_ISL_739642, EPI_ISL_739647, EPI_ISL_739649, EPI_ISL_739650, EPI_ISL_739653, EPI_ISL_739657, EPI_ISL_849953, EPI_ISL_849954, EPI_ISL_849955, EPI_ISL_849956, EPI_ISL_849957, EPI_ISL_849959, EPI_ISL_849960, EPI_ISL_849961, EPI_ISL_849962, EPI_ISL_849963, EPI_ISL_849964, EPI_ISL_849965, EPI_ISL_849966, EPI_ISL_849967, EPI_ISL_849968, EPI_ISL_849969, EPI_ISL_849970, EPI_ISL_849971, EPI_ISL_849972, EPI_ISL_849973, EPI_ISL_849974, EPI_ISL_849975, EPI_ISL_849976, EPI_ISL_849977, EPI_ISL_849978, EPI_ISL_849979, EPI_ISL_849980, EPI_ISL_849981, EPI_ISL_849982, EPI_ISL_849983, EPI_ISL_849984, EPI_ISL_849985, EPI_ISL_849986, EPI_ISL_849987, EPI_ISL_849988, EPI_ISL_849989, EPI_ISL_849990, EPI_ISL_849991, EPI_ISL_849992, EPI_ISL_849993, EPI_ISL_849994, EPI_ISL_849995, EPI_ISL_849996, EPI_ISL_849997, EPI_ISL_849998, EPI_ISL_849999, EPI_ISL_850000, EPI_ISL_850001, EPI_ISL_850002, EPI_ISL_850003, EPI_ISL_850004, EPI_ISL_850005, EPI_ISL_850006, EPI_ISL_850007, EPI_ISL_850008, EPI_ISL_850009, EPI_ISL_850010, EPI_ISL_850011, EPI_ISL_850012, EPI_ISL_850013, EPI_ISL_850014, EPI_ISL_850015, EPI_ISL_850016, EPI_ISL_850017, EPI_ISL_850018, EPI_ISL_853417, EPI_ISL_853418, EPI_ISL_853419, EPI_ISL_853420, EPI_ISL_853421, EPI_ISL_853422, EPI_ISL_853423, EPI_ISL_853424, EPI_ISL_853425, EPI_ISL_853426, EPI_ISL_853427, EPI_ISL_853428, EPI_ISL_853429, EPI_ISL_853430, EPI_ISL_853431, EPI_ISL_853432, EPI_ISL_853433, EPI_ISL_853434, EPI_ISL_853435, EPI_ISL_853436, EPI_ISL_853437, EPI_ISL_853438, EPI_ISL_853439, EPI_ISL_853440, EPI_ISL_853441, EPI_ISL_853442, EPI_ISL_853443, EPI_ISL_853444, EPI_ISL_853445, EPI_ISL_853446, EPI_ISL_853447, EPI_ISL_853448, EPI_ISL_853449, EPI_ISL_853450, EPI_ISL_853451, EPI_ISL_853452, EPI_ISL_853453, EPI_ISL_853454, EPI_ISL_853455, EPI_ISL_853456, EPI_ISL_853457, EPI_ISL_853458, EPI_ISL_853459, EPI_ISL_853460, EPI_ISL_853461, EPI_ISL_853462, EPI_ISL_853463, EPI_ISL_853464, EPI_ISL_853465, EPI_ISL_853466, EPI_ISL_853467, EPI_ISL_853468, EPI_ISL_853469, EPI_ISL_853470, EPI_ISL_853471, EPI_ISL_853472, EPI_ISL_853473, EPI_ISL_853474, EPI_ISL_853475, EPI_ISL_853476, EPI_ISL_853477, EPI_ISL_853478, EPI_ISL_853479, EPI_ISL_853480, EPI_ISL_853481, EPI_ISL_853482, EPI_ISL_853483, EPI_ISL_853484, EPI_ISL_853485, EPI_ISL_853486, EPI_ISL_853487, EPI_ISL_853488, EPI_ISL_853489, EPI_ISL_853490, EPI_ISL_853491, EPI_ISL_853492, EPI_ISL_853493, EPI_ISL_853494, EPI_ISL_853495, EPI_ISL_853496, EPI_ISL_853497, EPI_ISL_853498, EPI_ISL_853499, EPI_ISL_853500, EPI_ISL_853501, EPI_ISL_853502, EPI_ISL_853503, EPI_ISL_853504, EPI_ISL_853505, EPI_ISL_853506, EPI_ISL_853507, EPI_ISL_853508, EPI_ISL_853509, EPI_ISL_853510, EPI_ISL_853511, EPI_ISL_853512, EPI_ISL_853513, EPI_ISL_853514, EPI_ISL_853515, EPI_ISL_853516, EPI_ISL_853517, EPI_ISL_853518, EPI_ISL_853519, EPI_ISL_853520, EPI_ISL_853521, EPI_ISL_853522, EPI_ISL_853523, EPI_ISL_853524, EPI_ISL_853525, EPI_ISL_853526, EPI_ISL_853527, EPI_ISL_853528, EPI_ISL_853529, EPI_ISL_853530, EPI_ISL_853531, EPI_ISL_853532, EPI_ISL_853533, EPI_ISL_853534, EPI_ISL_853535, EPI_ISL_853536, EPI_ISL_853537, EPI_ISL_853538, EPI_ISL_853539, EPI_ISL_853540, EPI_ISL_853541, EPI_ISL_853542, EPI_ISL_853543, EPI_ISL_853544, EPI_ISL_853545, EPI_ISL_853560, EPI_ISL_853561, EPI_ISL_853562, EPI_ISL_853563, EPI_ISL_853564, EPI_ISL_853565, EPI_ISL_884871, EPI_ISL_884872, EPI_ISL_884873, EPI_ISL_884874, EPI_ISL_884875, EPI_ISL_884876, EPI_ISL_884877, EPI_ISL_884878, EPI_ISL_884879, EPI_ISL_884880, EPI_ISL_884881, EPI_ISL_884882, EPI_ISL_884883, EPI_ISL_884884, EPI_ISL_884885, EPI_ISL_884886, EPI_ISL_884887, EPI_ISL_884888, EPI_ISL_884889, EPI_ISL_884890, EPI_ISL_884891, EPI_ISL_884892, EPI_ISL_884893, EPI_ISL_884894, EPI_ISL_884895, EPI_ISL_884896, EPI_ISL_884897, EPI_ISL_884898, EPI_ISL_884899, EPI_ISL_884900, EPI_ISL_884901, EPI_ISL_884902, EPI_ISL_884903, EPI_ISL_884904, EPI_ISL_884905, EPI_ISL_884906, EPI_ISL_884907, EPI_ISL_884908, EPI_ISL_884909, EPI_ISL_884910, EPI_ISL_884911, EPI_ISL_884912, EPI_ISL_884913, EPI_ISL_884914, EPI_ISL_884915, EPI_ISL_884916, EPI_ISL_884917, EPI_ISL_884918, EPI_ISL_884919, EPI_ISL_884920, EPI_ISL_884921, EPI_ISL_884922, EPI_ISL_884923, EPI_ISL_884924, EPI_ISL_884925, EPI_ISL_884926, EPI_ISL_884927, EPI_ISL_884928, EPI_ISL_884929, EPI_ISL_884930, EPI_ISL_884931, EPI_ISL_884932, EPI_ISL_884933, EPI_ISL_884934, EPI_ISL_884935, EPI_ISL_884936, EPI_ISL_884937, EPI_ISL_884938, EPI_ISL_884939, EPI_ISL_884940, EPI_ISL_884941, EPI_ISL_884942, EPI_ISL_884943, EPI_ISL_884944, EPI_ISL_884945, EPI_ISL_884946, EPI_ISL_884947, EPI_ISL_884948, EPI_ISL_884949, EPI_ISL_884950, EPI_ISL_884951, EPI_ISL_884952, EPI_ISL_884953, EPI_ISL_884954, EPI_ISL_884955, EPI_ISL_884956, EPI_ISL_884957, EPI_ISL_884958, EPI_ISL_884959, EPI_ISL_884960, EPI_ISL_884961, EPI_ISL_884962, EPI_ISL_884963, EPI_ISL_884964, EPI_ISL_884965, EPI_ISL_884966, EPI_ISL_884967, EPI_ISL_884968, EPI_ISL_884969, EPI_ISL_884970, EPI_ISL_884971, EPI_ISL_884972, EPI_ISL_884973, EPI_ISL_884974, EPI_ISL_884975, EPI_ISL_884976, EPI_ISL_884977, EPI_ISL_884978, EPI_ISL_884979, EPI_ISL_884980, EPI_ISL_884981, EPI_ISL_884982, EPI_ISL_884983, EPI_ISL_884984, EPI_ISL_884985, EPI_ISL_884986, EPI_ISL_884987, EPI_ISL_884988, EPI_ISL_884989, EPI_ISL_884990, EPI_ISL_884991, EPI_ISL_884992, EPI_ISL_884993, EPI_ISL_884994, EPI_ISL_884995, EPI_ISL_884996, EPI_ISL_884997, EPI_ISL_884998, EPI_ISL_884999, EPI_ISL_885000, EPI_ISL_885001, EPI_ISL_885002, EPI_ISL_885003, EPI_ISL_885004, EPI_ISL_885005, EPI_ISL_885006, EPI_ISL_885007, EPI_ISL_885008, EPI_ISL_885009, EPI_ISL_885010, EPI_ISL_885011, EPI_ISL_885012, EPI_ISL_885013, EPI_ISL_885018, EPI_ISL_885051, EPI_ISL_955353, EPI_ISL_955354, EPI_ISL_955355, EPI_ISL_955356, EPI_ISL_955357, EPI_ISL_955358, EPI_ISL_955359, EPI_ISL_955360, EPI_ISL_955361, EPI_ISL_955362, EPI_ISL_955363, EPI_ISL_955364, EPI_ISL_955365, EPI_ISL_955366, EPI_ISL_955367, EPI_ISL_955369, EPI_ISL_955370, EPI_ISL_955371, EPI_ISL_955372, EPI_ISL_955373, EPI_ISL_955374, EPI_ISL_955375, EPI_ISL_955376, EPI_ISL_955377, EPI_ISL_955378, EPI_ISL_955379, EPI_ISL_955380, EPI_ISL_955381, EPI_ISL_955382, EPI_ISL_955383, EPI_ISL_955384, EPI_ISL_955385, EPI_ISL_955561, EPI_ISL_955562, EPI_ISL_955563, EPI_ISL_955564, EPI_ISL_955565, EPI_ISL_955566, EPI_ISL_955567, EPI_ISL_955568, EPI_ISL_955569, EPI_ISL_955570, EPI_ISL_955571, EPI_ISL_955572, EPI_ISL_955573, EPI_ISL_955574, EPI_ISL_955575, EPI_ISL_955576, EPI_ISL_955577, EPI_ISL_955578, EPI_ISL_955579, EPI_ISL_955580, EPI_ISL_955581, EPI_ISL_955582, EPI_ISL_955583, EPI_ISL_955584, EPI_ISL_955585, EPI_ISL_955586, EPI_ISL_955587, EPI_ISL_955588, EPI_ISL_955589, EPI_ISL_955590, EPI_ISL_955591, EPI_ISL_955592, EPI_ISL_955593, EPI_ISL_955594, EPI_ISL_955595, EPI_ISL_955596, EPI_ISL_955597, EPI_ISL_955598, EPI_ISL_955599, EPI_ISL_955600, EPI_ISL_955601, EPI_ISL_955602, EPI_ISL_955603, EPI_ISL_955604, EPI_ISL_955605, EPI_ISL_955606, EPI_ISL_955607, EPI_ISL_955608, EPI_ISL_955609, EPI_ISL_955610, EPI_ISL_955611, EPI_ISL_955612, EPI_ISL_955613, EPI_ISL_955614, EPI_ISL_955615, EPI_ISL_955616, EPI_ISL_955617, EPI_ISL_955618, EPI_ISL_955619, EPI_ISL_955620, EPI_ISL_955621, EPI_ISL_955622, EPI_ISL_955623, EPI_ISL_955624, EPI_ISL_955625, EPI_ISL_955626, EPI_ISL_955627, EPI_ISL_978989, EPI_ISL_978990, EPI_ISL_978991, EPI_ISL_978992, EPI_ISL_978993, EPI_ISL_978994, EPI_ISL_978995, EPI_ISL_978996, EPI_ISL_978997, EPI_ISL_978998, EPI_ISL_978999, EPI_ISL_979000, EPI_ISL_979001, EPI_ISL_979002, EPI_ISL_979003, EPI_ISL_979004, EPI_ISL_979005, EPI_ISL_979006, EPI_ISL_979007, EPI_ISL_979008, EPI_ISL_979009, EPI_ISL_979010, EPI_ISL_979011, EPI_ISL_979012, EPI_ISL_979013, EPI_ISL_979014, EPI_ISL_979015, EPI_ISL_979016, EPI_ISL_979017, EPI_ISL_979018, EPI_ISL_979019, EPI_ISL_979020, EPI_ISL_979021, EPI_ISL_979022, EPI_ISL_979023, EPI_ISL_979024, EPI_ISL_979025, EPI_ISL_979026, EPI_ISL_979027, EPI_ISL_979028, EPI_ISL_979029, EPI_ISL_979030, EPI_ISL_979031, EPI_ISL_979032, EPI_ISL_979033, EPI_ISL_979034, EPI_ISL_979035, EPI_ISL_979036, EPI_ISL_979037, EPI_ISL_979038, EPI_ISL_979039, EPI_ISL_979040, EPI_ISL_979041, EPI_ISL_979042, EPI_ISL_979043, EPI_ISL_979044, EPI_ISL_979045, EPI_ISL_979046, EPI_ISL_979047, EPI_ISL_979048, EPI_ISL_979049, EPI_ISL_979050, EPI_ISL_979051, EPI_ISL_979052, EPI_ISL_979053, EPI_ISL_979054, EPI_ISL_979055, EPI_ISL_979060, EPI_ISL_979545, EPI_ISL_979546, EPI_ISL_979547, EPI_ISL_979548, EPI_ISL_979549, EPI_ISL_979550, EPI_ISL_979551, EPI_ISL_979552, EPI_ISL_979553, EPI_ISL_979554, EPI_ISL_979555, EPI_ISL_979556, EPI_ISL_979557, EPI_ISL_979558, EPI_ISL_979559, EPI_ISL_979560, EPI_ISL_979561, EPI_ISL_979562, EPI_ISL_979563, EPI_ISL_979564, EPI_ISL_979565, EPI_ISL_979566, EPI_ISL_979567, EPI_ISL_979568, EPI_ISL_979569, EPI_ISL_979570, EPI_ISL_979571, EPI_ISL_979572, EPI_ISL_979573, EPI_ISL_979574, EPI_ISL_979575, EPI_ISL_979576, EPI_ISL_979577, EPI_ISL_979578, EPI_ISL_979579, EPI_ISL_979580, EPI_ISL_979581, EPI_ISL_979582, EPI_ISL_979583, EPI_ISL_979584, EPI_ISL_979585, EPI_ISL_979586, EPI_ISL_979587, EPI_ISL_979588, EPI_ISL_979589, EPI_ISL_979590, EPI_ISL_979591, EPI_ISL_979592, EPI_ISL_979593, EPI_ISL_979594, EPI_ISL_979595, EPI_ISL_979596, EPI_ISL_979597, EPI_ISL_979598, EPI_ISL_979599, EPI_ISL_979600, EPI_ISL_979601, EPI_ISL_979602, EPI_ISL_979603, EPI_ISL_979604, EPI_ISL_979605, EPI_ISL_979606, EPI_ISL_979607, EPI_ISL_979608, EPI_ISL_979609, EPI_ISL_979610, EPI_ISL_979611, EPI_ISL_979612, EPI_ISL_979613, EPI_ISL_979614, EPI_ISL_979615, EPI_ISL_979616, EPI_ISL_979617, EPI_ISL_979618, EPI_ISL_979619, EPI_ISL_979620, EPI_ISL_979621, EPI_ISL_979622, EPI_ISL_979623, EPI_ISL_979624, EPI_ISL_979629, EPI_ISL_979630, EPI_ISL_979631, EPI_ISL_979632, EPI_ISL_979633, EPI_ISL_1027293, EPI_ISL_1027306, EPI_ISL_1027312, EPI_ISL_1027314, EPI_ISL_1027315, EPI_ISL_1027317, EPI_ISL_1027319, EPI_ISL_1027320, EPI_ISL_1027325, EPI_ISL_1027326, EPI_ISL_1027327, EPI_ISL_1027332, EPI_ISL_1027337, EPI_ISL_1027338, EPI_ISL_1027342, EPI_ISL_1027343, EPI_ISL_1027347, EPI_ISL_1027355, EPI_ISL_1027357, EPI_ISL_1027362, EPI_ISL_1027364, EPI_ISL_1027437, EPI_ISL_1027438, EPI_ISL_1027439, EPI_ISL_1027440, EPI_ISL_1027441, EPI_ISL_1027442, EPI_ISL_1027443, EPI_ISL_1027444, EPI_ISL_1027474, EPI_ISL_1027477, EPI_ISL_1027478, EPI_ISL_1027481, EPI_ISL_1027483, EPI_ISL_1027485, EPI_ISL_1027488, EPI_ISL_1027492, EPI_ISL_1027493, EPI_ISL_1027495, EPI_ISL_1027497, EPI_ISL_1027500, EPI_ISL_1027508, EPI_ISL_1027509, EPI_ISL_1027510, EPI_ISL_1027511, EPI_ISL_1027512, EPI_ISL_1027514, EPI_ISL_1027516, EPI_ISL_1027519, EPI_ISL_1027522, EPI_ISL_1027524, EPI_ISL_1027527, EPI_ISL_1027529, EPI_ISL_1027533, EPI_ISL_1027534, EPI_ISL_1027542, EPI_ISL_1027544, EPI_ISL_1027545, EPI_ISL_1027546, EPI_ISL_1027547, EPI_ISL_1027548, EPI_ISL_1027549, EPI_ISL_1027550, EPI_ISL_1027551, EPI_ISL_1027552, EPI_ISL_1027553, EPI_ISL_1027554, EPI_ISL_1027555, EPI_ISL_1027556, EPI_ISL_1027557, EPI_ISL_1027558, EPI_ISL_1027559, EPI_ISL_1030252, EPI_ISL_1030253, EPI_ISL_1030254, EPI_ISL_1030255, EPI_ISL_1030256, EPI_ISL_1030257, EPI_ISL_1030258, EPI_ISL_1030259, EPI_ISL_1030260, EPI_ISL_1030261, EPI_ISL_1030262, EPI_ISL_1030263, EPI_ISL_1030264, EPI_ISL_1030265, EPI_ISL_1030266, EPI_ISL_1030267, EPI_ISL_1030268, EPI_ISL_1030270, EPI_ISL_1030271, EPI_ISL_1030272, EPI_ISL_1030273, EPI_ISL_1030274, EPI_ISL_1030275, EPI_ISL_1030276, EPI_ISL_1030277, EPI_ISL_1030278, EPI_ISL_1030279, EPI_ISL_1030280, EPI_ISL_1030283, EPI_ISL_1030284, EPI_ISL_1030285, EPI_ISL_1030286, EPI_ISL_1030287, EPI_ISL_1030288, EPI_ISL_1030289, EPI_ISL_1030300, EPI_ISL_1030301, EPI_ISL_1030304, EPI_ISL_1030305, EPI_ISL_1030391, EPI_ISL_1030392, EPI_ISL_1030393, EPI_ISL_1030394, EPI_ISL_1030395, EPI_ISL_1030396, EPI_ISL_1030397, EPI_ISL_1030398, EPI_ISL_1030399, EPI_ISL_1030400, EPI_ISL_1030401, EPI_ISL_1030402, EPI_ISL_1030403, EPI_ISL_1030404, EPI_ISL_1030405, EPI_ISL_1030406, EPI_ISL_1030407, EPI_ISL_1030408, EPI_ISL_1030409, EPI_ISL_1030410, EPI_ISL_1030411, EPI_ISL_1030412, EPI_ISL_1030413, EPI_ISL_1030414, EPI_ISL_1030415, EPI_ISL_1030416, EPI_ISL_1030417, EPI_ISL_1030418, EPI_ISL_1030419, EPI_ISL_1030420, EPI_ISL_1030421, EPI_ISL_1030422, EPI_ISL_1030423, EPI_ISL_1030424, EPI_ISL_1030425, EPI_ISL_1030426, EPI_ISL_1030427, EPI_ISL_1030428, EPI_ISL_1030429, EPI_ISL_1030430, EPI_ISL_1030431, EPI_ISL_1030432, EPI_ISL_1030433, EPI_ISL_1030434, EPI_ISL_1030435, EPI_ISL_1030436, EPI_ISL_1030437, EPI_ISL_1030438, EPI_ISL_1030439, EPI_ISL_1030440, EPI_ISL_1030441, EPI_ISL_1030442, EPI_ISL_1030443, EPI_ISL_1030500, EPI_ISL_1030501, EPI_ISL_1030502, EPI_ISL_1184583, EPI_ISL_1184584, EPI_ISL_1184585, EPI_ISL_1184586, EPI_ISL_1184587, EPI_ISL_1184588, EPI_ISL_1184589, EPI_ISL_1184590, EPI_ISL_1184591, EPI_ISL_1184592, EPI_ISL_1184593, EPI_ISL_1184594, EPI_ISL_1184595, EPI_ISL_1184596, EPI_ISL_1184597, EPI_ISL_1184598, EPI_ISL_1184599, EPI_ISL_1184600, EPI_ISL_1184601, EPI_ISL_1184602, EPI_ISL_1184603, EPI_ISL_1184604, EPI_ISL_1184605, EPI_ISL_1184606, EPI_ISL_1184607, EPI_ISL_1184608, EPI_ISL_1184609, EPI_ISL_1184610, EPI_ISL_1184611, EPI_ISL_1184612, EPI_ISL_1184613, EPI_ISL_1184614, EPI_ISL_1184615, EPI_ISL_1184616, EPI_ISL_1184617, EPI_ISL_1184618, EPI_ISL_1184619, EPI_ISL_1184620, EPI_ISL_1184621, EPI_ISL_1184622, EPI_ISL_1184623, EPI_ISL_1184827, EPI_ISL_1184828, EPI_ISL_1184829, EPI_ISL_1184830, EPI_ISL_1184831, EPI_ISL_1184832, EPI_ISL_1184833, EPI_ISL_1184834, EPI_ISL_1184835, EPI_ISL_1184836, EPI_ISL_1184837, EPI_ISL_1184838, EPI_ISL_1184839, EPI_ISL_1184840, EPI_ISL_1184841, EPI_ISL_1184842, EPI_ISL_1184843, EPI_ISL_1184844, EPI_ISL_1184845, EPI_ISL_1184846, EPI_ISL_1184847, EPI_ISL_1184848, EPI_ISL_1184849, EPI_ISL_1184850, EPI_ISL_1184851, EPI_ISL_1184852, EPI_ISL_1184853, EPI_ISL_1184854, EPI_ISL_1184855, EPI_ISL_1184856, EPI_ISL_1184857, EPI_ISL_1184858, EPI_ISL_1184859, EPI_ISL_1184860, EPI_ISL_2658605, EPI_ISL_2658606, EPI_ISL_2658607, EPI_ISL_2658608, EPI_ISL_2658617, EPI_ISL_2658624, EPI_ISL_2658627, EPI_ISL_2658633, EPI_ISL_2658648, EPI_ISL_2658655, EPI_ISL_2658663, EPI_ISL_2658673, EPI_ISL_2658677, EPI_ISL_2658679, EPI_ISL_2658688, EPI_ISL_2658694, EPI_ISL_2658696, EPI_ISL_2658714, EPI_ISL_2658726, EPI_ISL_2658742, EPI_ISL_2658744, EPI_ISL_2659199, EPI_ISL_2659201, EPI_ISL_2659208, EPI_ISL_2659214, EPI_ISL_2659219, EPI_ISL_2659221, EPI_ISL_2659222, EPI_ISL_2659225, EPI_ISL_2659234, EPI_ISL_2659236, EPI_ISL_2659237, EPI_ISL_2659239, EPI_ISL_2659242, EPI_ISL_2659243, EPI_ISL_2659244, EPI_ISL_2659246, EPI_ISL_2659247, EPI_ISL_2659248, EPI_ISL_2659249, EPI_ISL_2659253, EPI_ISL_2659254, EPI_ISL_2659256, EPI_ISL_2659263, EPI_ISL_2659264, EPI_ISL_2659269, EPI_ISL_2659273, EPI_ISL_2659274, EPI_ISL_2659275, EPI_ISL_2659281, EPI_ISL_2659284, EPI_ISL_2659285, EPI_ISL_2659286, EPI_ISL_2659288, EPI_ISL_2659290, EPI_ISL_2659292, EPI_ISL_2659293, EPI_ISL_2659298, EPI_ISL_2659301, EPI_ISL_2659314, EPI_ISL_2659315, EPI_ISL_2659323, EPI_ISL_2659326, EPI_ISL_2659327, EPI_ISL_2659329, EPI_ISL_2659335, EPI_ISL_2659340, EPI_ISL_2659345, EPI_ISL_2659350, EPI_ISL_2659352, EPI_ISL_2659355, EPI_ISL_2659364, EPI_ISL_2659366, EPI_ISL_2659374, EPI_ISL_2659375, EPI_ISL_2659376, EPI_ISL_2659378, EPI_ISL_2659379, EPI_ISL_2659381, EPI_ISL_2659382, EPI_ISL_2659383, EPI_ISL_2659388, EPI_ISL_2659389, EPI_ISL_2659391, EPI_ISL_2659394, EPI_ISL_2659395, EPI_ISL_2659399, EPI_ISL_2659401, EPI_ISL_2659402, EPI_ISL_2659410, EPI_ISL_2659411, EPI_ISL_2659413, EPI_ISL_2659422, EPI_ISL_2659425, EPI_ISL_2659426, EPI_ISL_2659427, EPI_ISL_2659429, EPI_ISL_2659432, EPI_ISL_2659433, EPI_ISL_2659435, EPI_ISL_2659437, EPI_ISL_2659439, EPI_ISL_2659440, EPI_ISL_2659441, EPI_ISL_2659442, EPI_ISL_2659443, EPI_ISL_2659444, EPI_ISL_2659446, EPI_ISL_2659447, EPI_ISL_2659448, EPI_ISL_2659451, EPI_ISL_2659453, EPI_ISL_2659454, EPI_ISL_2659458, EPI_ISL_2659459, EPI_ISL_2659460, EPI_ISL_2659466, EPI_ISL_2659470, EPI_ISL_2659475, EPI_ISL_2659476, EPI_ISL_2659479, EPI_ISL_2659491, EPI_ISL_2659493, EPI_ISL_2659496, EPI_ISL_2659498, EPI_ISL_2659499, EPI_ISL_2659501, EPI_ISL_2659506, EPI_ISL_2659510, EPI_ISL_2659512, EPI_ISL_2659513, EPI_ISL_2659515, EPI_ISL_2659516, EPI_ISL_2659518, EPI_ISL_2659520, EPI_ISL_2659525, EPI_ISL_2659527, EPI_ISL_2659528, EPI_ISL_2659531, EPI_ISL_2659532, EPI_ISL_2659541, EPI_ISL_2659542, EPI_ISL_2659543, EPI_ISL_2659545, EPI_ISL_2659546, EPI_ISL_2659555, EPI_ISL_2659559, EPI_ISL_2659563, EPI_ISL_2659567, EPI_ISL_2659572, EPI_ISL_2659573, EPI_ISL_2659577, EPI_ISL_2659580, EPI_ISL_2659583, EPI_ISL_2659584, EPI_ISL_2659585, EPI_ISL_2659589, EPI_ISL_2659594, EPI_ISL_2659598, EPI_ISL_2659599, EPI_ISL_2659600, EPI_ISL_2659607, EPI_ISL_2659610, EPI_ISL_2659613, EPI_ISL_2659615, EPI_ISL_2659617, EPI_ISL_2659618, EPI_ISL_2659619, EPI_ISL_2659621, EPI_ISL_2659623, EPI_ISL_2659625, EPI_ISL_2659626, EPI_ISL_2659627, EPI_ISL_2659629, EPI_ISL_2659634, EPI_ISL_2659637, EPI_ISL_2659643, EPI_ISL_2659645, EPI_ISL_2659646, EPI_ISL_2659648, EPI_ISL_2659651, EPI_ISL_2659655, EPI_ISL_2659656, EPI_ISL_2758413, EPI_ISL_2758418, EPI_ISL_2758419, EPI_ISL_2758420, EPI_ISL_2758429, EPI_ISL_2758430 | | | |
| see above | Santa Clara County Public Health Laboratory | Chan-Zuckerberg Biohub | CZB Cliahub Consortium |
| EPI_ISL_812350, EPI_ISL_831648, EPI_ISL_831662, EPI_ISL_831687, EPI_ISL_831893, EPI_ISL_831894, EPI_ISL_831895, EPI_ISL_831896, EPI_ISL_831897, EPI_ISL_831899, EPI_ISL_832066, EPI_ISL_832068, EPI_ISL_832075, EPI_ISL_832076, EPI_ISL_832375, EPI_ISL_832376, EPI_ISL_832377, EPI_ISL_832379, EPI_ISL_832380, EPI_ISL_832381, EPI_ISL_832382, EPI_ISL_832383, EPI_ISL_832388, EPI_ISL_832391, EPI_ISL_832392, EPI_ISL_832394, EPI_ISL_832395, EPI_ISL_832396, EPI_ISL_832398, EPI_ISL_832399, EPI_ISL_832400, EPI_ISL_832401, EPI_ISL_832402, EPI_ISL_832403, EPI_ISL_832404, EPI_ISL_845544, EPI_ISL_914646, EPI_ISL_1081192, EPI_ISL_1137637, EPI_ISL_1169590, EPI_ISL_1169591, EPI_ISL_1203818, EPI_ISL_1920833, EPI_ISL_1920834, EPI_ISL_1920835, EPI_ISL_1920839, EPI_ISL_1920840, EPI_ISL_1920841, EPI_ISL_1920842, EPI_ISL_1920846, EPI_ISL_1920848, EPI_ISL_1920851, EPI_ISL_1920852, EPI_ISL_1920854, EPI_ISL_1920855, EPI_ISL_1920856, EPI_ISL_1920857, EPI_ISL_1920858, EPI_ISL_1920859, EPI_ISL_1920860, EPI_ISL_1920862, EPI_ISL_1920863, EPI_ISL_1920865, EPI_ISL_1920866, EPI_ISL_1920868, EPI_ISL_1920869, EPI_ISL_1920871, EPI_ISL_1920872, EPI_ISL_1920873, EPI_ISL_1920874, EPI_ISL_1920876, EPI_ISL_1920877, EPI_ISL_1920879, EPI_ISL_1920880, EPI_ISL_1920881, EPI_ISL_1920882, EPI_ISL_1920884, EPI_ISL_1920885, EPI_ISL_1920887, EPI_ISL_1920888, EPI_ISL_1920889, EPI_ISL_1920891, EPI_ISL_1920892, EPI_ISL_1920893, EPI_ISL_1920894, EPI_ISL_1920895, EPI_ISL_1920896, EPI_ISL_1920898, EPI_ISL_1920899, EPI_ISL_1920900, EPI_ISL_1920901 | | | |
| see above | Santa Clara County Public Health Laboratory | Santa Clara County Public Health Laboratory | Santa Clara County Public Health Department |
| EPI_ISL_1794577, EPI_ISL_1794578, EPI_ISL_1794579, EPI_ISL_1794580, EPI_ISL_1794581, EPI_ISL_1794582, EPI_ISL_1794583, EPI_ISL_1794584, EPI_ISL_1794585, EPI_ISL_1794586, EPI_ISL_1794587, EPI_ISL_1794588, EPI_ISL_1794589, EPI_ISL_1794590, EPI_ISL_1794591, EPI_ISL_1794592, EPI_ISL_1794593, EPI_ISL_1794594, EPI_ISL_1794595, EPI_ISL_1794596, EPI_ISL_1794597, EPI_ISL_1794598, EPI_ISL_1794599, EPI_ISL_1794600, EPI_ISL_1794601, EPI_ISL_1794602, EPI_ISL_1794603, EPI_ISL_1794604, EPI_ISL_1794605, EPI_ISL_1794606, EPI_ISL_1794607, EPI_ISL_1794608, EPI_ISL_1794609, EPI_ISL_1794610, EPI_ISL_1794611, EPI_ISL_1794612, EPI_ISL_1794613, EPI_ISL_1794614, EPI_ISL_1794615, EPI_ISL_1794787, EPI_ISL_1794788, EPI_ISL_1794789, EPI_ISL_1794790, EPI_ISL_1794791, EPI_ISL_1794792, EPI_ISL_1794793, EPI_ISL_1794794, EPI_ISL_1794795, EPI_ISL_1794796, EPI_ISL_1794797, EPI_ISL_1794798, EPI_ISL_1794799, EPI_ISL_1794800, EPI_ISL_1794801, EPI_ISL_1794802, EPI_ISL_1794803, EPI_ISL_1794804, EPI_ISL_1794805, EPI_ISL_1794806, EPI_ISL_1794807, EPI_ISL_1794808, EPI_ISL_1794809, EPI_ISL_1794810, EPI_ISL_1794811, EPI_ISL_1794812, EPI_ISL_1794813, EPI_ISL_1794814, EPI_ISL_1794815, EPI_ISL_1794816, EPI_ISL_1794817, EPI_ISL_1794818, EPI_ISL_1794819, EPI_ISL_1794820, EPI_ISL_1794821, EPI_ISL_1794822, EPI_ISL_1794823, EPI_ISL_1794824, EPI_ISL_1794825, EPI_ISL_1794826, EPI_ISL_1794827, EPI_ISL_1794828, EPI_ISL_1794829, EPI_ISL_1794830, EPI_ISL_1794831, EPI_ISL_1794832, EPI_ISL_1794833, EPI_ISL_1794834, EPI_ISL_1794835, EPI_ISL_1794836, EPI_ISL_1794837, EPI_ISL_1794838, EPI_ISL_1969271, EPI_ISL_1969272, EPI_ISL_1969273, EPI_ISL_1969274, EPI_ISL_1969275, EPI_ISL_1969276, EPI_ISL_1969277, EPI_ISL_1969278, EPI_ISL_1969279, EPI_ISL_1969280, EPI_ISL_1969281, EPI_ISL_1969314, EPI_ISL_1969327, EPI_ISL_1969328, EPI_ISL_1969329, EPI_ISL_1969330, EPI_ISL_1969331, EPI_ISL_1969332, EPI_ISL_1969333, EPI_ISL_1969334, EPI_ISL_1969335, EPI_ISL_1969336, EPI_ISL_1969337, EPI_ISL_1969338, EPI_ISL_1969339, EPI_ISL_1969340, EPI_ISL_1969341, EPI_ISL_1969342, EPI_ISL_1969343, EPI_ISL_1969344, EPI_ISL_1969345, EPI_ISL_1969346, EPI_ISL_1969347, EPI_ISL_1969348, EPI_ISL_1969349, EPI_ISL_1969350, EPI_ISL_1969351, EPI_ISL_1969352, EPI_ISL_1969353, EPI_ISL_1969354, EPI_ISL_1969355, EPI_ISL_1969356, EPI_ISL_1969357, EPI_ISL_1969358, EPI_ISL_1969359, EPI_ISL_1969360, EPI_ISL_1969361, EPI_ISL_1969362, EPI_ISL_1969363, EPI_ISL_1969364, EPI_ISL_1969365, EPI_ISL_1969366, EPI_ISL_1969367, EPI_ISL_1969368, EPI_ISL_1969369, EPI_ISL_1969370 | | | |
| see above | Scripps Medical Laboratory | Andersen lab at Scripps Research | Ellen Stefanski; Ian Mchardy; SEARCH Alliance San Diego with Michael Quigley |

| | | | |
|---|---|---|---|
| EPI_ISL_1794854, EPI_ISL_1794855, EPI_ISL_1794856, EPI_ISL_1794857, EPI_ISL_1794858, EPI_ISL_1794859, EPI_ISL_1794860, EPI_ISL_1794861, EPI_ISL_1794862, EPI_ISL_1794863, EPI_ISL_1794864, EPI_ISL_1794865, EPI_ISL_1794866, EPI_ISL_1794867, EPI_ISL_1794868, EPI_ISL_1794869, EPI_ISL_1794870, EPI_ISL_1794871, EPI_ISL_1794872, EPI_ISL_1794873, EPI_ISL_1794874, EPI_ISL_1794875, EPI_ISL_1794876, EPI_ISL_1794877, EPI_ISL_1794878, EPI_ISL_1794879, EPI_ISL_1794880, EPI_ISL_1794881, EPI_ISL_1794882, EPI_ISL_1794884, EPI_ISL_1794919, EPI_ISL_1794920, EPI_ISL_1794921, EPI_ISL_1794922, EPI_ISL_1794923, EPI_ISL_1794924, EPI_ISL_1794926, EPI_ISL_1794927, EPI_ISL_1794928, EPI_ISL_1794929, EPI_ISL_1794930, EPI_ISL_1794931, EPI_ISL_1794932, EPI_ISL_1794933, EPI_ISL_1969315, EPI_ISL_1969316, EPI_ISL_1969317, EPI_ISL_1969318, EPI_ISL_1969554, EPI_ISL_1969555, EPI_ISL_1969556, EPI_ISL_1969557, EPI_ISL_1969558, EPI_ISL_1969559, EPI_ISL_1969560, EPI_ISL_1969561, EPI_ISL_1969562, EPI_ISL_1969602, EPI_ISL_1969603, EPI_ISL_1969604, EPI_ISL_1969605, EPI_ISL_1969607, EPI_ISL_1969611, EPI_ISL_1969612, EPI_ISL_1969613, EPI_ISL_1969614, EPI_ISL_1969615, EPI_ISL_1969616, EPI_ISL_1969617, EPI_ISL_1969618, EPI_ISL_1969619, EPI_ISL_1969620, EPI_ISL_1969621, EPI_ISL_1969622, EPI_ISL_1969623, EPI_ISL_1969624, EPI_ISL_1969625, EPI_ISL_1969626, EPI_ISL_1969627, EPI_ISL_1969628, EPI_ISL_1969629, EPI_ISL_1969630, EPI_ISL_1969631, EPI_ISL_1969632, EPI_ISL_1969633, EPI_ISL_1969634, EPI_ISL_1969635, EPI_ISL_1969636, EPI_ISL_1969637, EPI_ISL_1969638, EPI_ISL_1969639, EPI_ISL_1969640, EPI_ISL_1969641, EPI_ISL_1969642, EPI_ISL_1969643, EPI_ISL_1969644, EPI_ISL_1969645, EPI_ISL_1969646, EPI_ISL_1969647, EPI_ISL_1969648, EPI_ISL_1969649, EPI_ISL_1969650, EPI_ISL_1969651, EPI_ISL_1969652, EPI_ISL_1969653, EPI_ISL_1969654, EPI_ISL_1969655, EPI_ISL_1969656, EPI_ISL_1969657, EPI_ISL_1969658, EPI_ISL_1969659, EPI_ISL_1969660, EPI_ISL_1969661, EPI_ISL_1969662, EPI_ISL_1969663, EPI_ISL_1969664, EPI_ISL_1969665, EPI_ISL_1969666, EPI_ISL_2231044, EPI_ISL_2231046, EPI_ISL_2231047, EPI_ISL_2231049, EPI_ISL_2231054, EPI_ISL_2231056, EPI_ISL_2231057, EPI_ISL_2231058, EPI_ISL_2231059, EPI_ISL_2231062, EPI_ISL_2231064, EPI_ISL_2231068, EPI_ISL_2231073, EPI_ISL_2231074, EPI_ISL_2231076, EPI_ISL_2231079, EPI_ISL_2231080, EPI_ISL_2231085, EPI_ISL_2231088, EPI_ISL_2231154, EPI_ISL_2231175, EPI_ISL_2231186, EPI_ISL_2231187, EPI_ISL_2231188, EPI_ISL_2231189, EPI_ISL_2231194, EPI_ISL_2231196, EPI_ISL_2231197, EPI_ISL_2231270, EPI_ISL_2231271, EPI_ISL_2231272, EPI_ISL_2231273, EPI_ISL_2231274, EPI_ISL_2231275, EPI_ISL_2231276, EPI_ISL_2231277, EPI_ISL_2231278, EPI_ISL_2231279, EPI_ISL_2231280, EPI_ISL_2231281, EPI_ISL_2231282, EPI_ISL_2231283, EPI_ISL_2231285, EPI_ISL_2231286, EPI_ISL_2231287, EPI_ISL_2231288, EPI_ISL_2231289, EPI_ISL_2231290, EPI_ISL_2231291, EPI_ISL_2231292, EPI_ISL_2231293, EPI_ISL_2231294, EPI_ISL_2231295, EPI_ISL_2231296, EPI_ISL_2231297, EPI_ISL_2231298, EPI_ISL_2231299, EPI_ISL_2231300, EPI_ISL_2231301, EPI_ISL_2231302, EPI_ISL_2231303, EPI_ISL_2231304, EPI_ISL_2231305, EPI_ISL_2231306, EPI_ISL_2231307, EPI_ISL_2231308, EPI_ISL_2231309, EPI_ISL_2231310, EPI_ISL_2231311, EPI_ISL_2231312, EPI_ISL_2231313, EPI_ISL_2231314, EPI_ISL_2231315, EPI_ISL_2231316, EPI_ISL_2231317, EPI_ISL_2231528, EPI_ISL_2231531, EPI_ISL_2231532, EPI_ISL_2231533, EPI_ISL_2231534, EPI_ISL_2231535, EPI_ISL_2231536, EPI_ISL_2231537, EPI_ISL_2231538, EPI_ISL_2231542, EPI_ISL_2231543, EPI_ISL_2231544, EPI_ISL_2231546, EPI_ISL_2231548, EPI_ISL_2231549, EPI_ISL_2231550, EPI_ISL_2231551, EPI_ISL_2231552, EPI_ISL_2231553, EPI_ISL_2231554, EPI_ISL_2231555, EPI_ISL_2231556, EPI_ISL_2231557, EPI_ISL_2231558, EPI_ISL_2231559, EPI_ISL_2231560, EPI_ISL_2231561, EPI_ISL_2231562, EPI_ISL_2231563, EPI_ISL_2231564, EPI_ISL_2329286, EPI_ISL_2329287, EPI_ISL_2329288, EPI_ISL_2329289, EPI_ISL_2329290, EPI_ISL_2329291, EPI_ISL_2329292, EPI_ISL_2329293, EPI_ISL_2329294, EPI_ISL_2329295, EPI_ISL_2329296, EPI_ISL_2329297, EPI_ISL_2329298, EPI_ISL_2329299, EPI_ISL_2329300, EPI_ISL_2329301, EPI_ISL_2329302, EPI_ISL_2329303, EPI_ISL_2329304, EPI_ISL_2329305, EPI_ISL_2329306, EPI_ISL_2329307, EPI_ISL_2329308, EPI_ISL_2329309, EPI_ISL_2329310, EPI_ISL_2329311, EPI_ISL_2329312, EPI_ISL_2329313, EPI_ISL_2329314, EPI_ISL_2329315, EPI_ISL_2329316, EPI_ISL_2329317, EPI_ISL_2329318, EPI_ISL_2329319, EPI_ISL_2329320, EPI_ISL_2329321, EPI_ISL_2329322, EPI_ISL_2329323, EPI_ISL_2329324, EPI_ISL_2329325, EPI_ISL_2329326, EPI_ISL_2329327, EPI_ISL_2329328, EPI_ISL_2329329, EPI_ISL_2329330, EPI_ISL_2329331, EPI_ISL_2329332, EPI_ISL_2329333, EPI_ISL_2329334 | | | |
| see above | Sharp HealthCare Laboratory | Andersen lab at Scripps Research | Art Mendoza; Cathy Woerle; Jacquelyn Berumen; Liam McGinnis; Omid Bakhtar; SEARCH Alliance San Diego with Aaron Harding |
| EPI_ISL_1379719, EPI_ISL_1379720, EPI_ISL_1379721, EPI_ISL_1379722, EPI_ISL_1379723, EPI_ISL_1379724, EPI_ISL_1379725, EPI_ISL_1379726, EPI_ISL_1379731, EPI_ISL_1379733, EPI_ISL_1379751, EPI_ISL_1379813, EPI_ISL_1379824, EPI_ISL_1379829, EPI_ISL_1379830, EPI_ISL_1379832, EPI_ISL_1379833, EPI_ISL_1379834, EPI_ISL_1379838, EPI_ISL_1379841, EPI_ISL_1379842, EPI_ISL_1379845, EPI_ISL_1379846, EPI_ISL_1379861, EPI_ISL_1379878, EPI_ISL_1379881, EPI_ISL_1424067, EPI_ISL_1424068, EPI_ISL_1424069, EPI_ISL_1424070, EPI_ISL_1424071, EPI_ISL_1424095, EPI_ISL_2009050, EPI_ISL_2009051, EPI_ISL_2009052, EPI_ISL_2009070, EPI_ISL_2009071, EPI_ISL_3050261, EPI_ISL_3050262, EPI_ISL_3050263 | | | |
| see above | Stanford Health Care | Stanford University School of Medicine, Clinical Virology Laboratory | Becky Jiang; Bernadette Troung; ChunHong Huang; Daniel Solis; Fiona Yamamoto; Fumiko Yamamoto; James Zehnder; Malaya K. Sahoo; Mamdouh Sibai; Michelle Vergese; Michelle Verghese; Nathan Hammond; Selamawit Bihon; and Benjamin A. Pinsky |
| EPI_ISL_849944, EPI_ISL_849945, EPI_ISL_849948, EPI_ISL_849951, EPI_ISL_849952, EPI_ISL_955337, EPI_ISL_955338, EPI_ISL_955339, EPI_ISL_955340, EPI_ISL_955341, EPI_ISL_955342, EPI_ISL_955343, EPI_ISL_955344, EPI_ISL_955345, EPI_ISL_955346, EPI_ISL_955347, EPI_ISL_955348, EPI_ISL_955349, EPI_ISL_955350, EPI_ISL_955351, EPI_ISL_955352 | | | |
| see above | UC Davis- Department of Pathology and Laboratory Medicine | Chan-Zuckerberg Biohub | CZB Cliahub Consortium |
| EPI_ISL_754110, EPI_ISL_754111, EPI_ISL_754112, EPI_ISL_754113, EPI_ISL_754114, EPI_ISL_754115, EPI_ISL_754116, EPI_ISL_754117, EPI_ISL_754118, EPI_ISL_754119, EPI_ISL_754120, EPI_ISL_754121, EPI_ISL_754122, EPI_ISL_754123, EPI_ISL_765987, EPI_ISL_765988, EPI_ISL_765989, EPI_ISL_765990, EPI_ISL_765991, EPI_ISL_765992, EPI_ISL_765993, EPI_ISL_962506, EPI_ISL_962507, EPI_ISL_962508, EPI_ISL_962514, EPI_ISL_962876 | | | |
| see above | UCLA Clinical Micro Lab | Los Angeles County Public Health Laboratories | P. Hemarajata et al. |
| EPI_ISL_1469854, EPI_ISL_1469855, EPI_ISL_1469856, EPI_ISL_1469857, EPI_ISL_1469858, EPI_ISL_1469859, EPI_ISL_1469860, EPI_ISL_1469861, EPI_ISL_1469862, EPI_ISL_1469863, EPI_ISL_1469866, EPI_ISL_1469867, EPI_ISL_1469868, EPI_ISL_1469869, EPI_ISL_1469870, EPI_ISL_1469871, EPI_ISL_1469872, EPI_ISL_1469873, EPI_ISL_1469874, EPI_ISL_1469887, EPI_ISL_1469889, EPI_ISL_1469890, EPI_ISL_1469891, EPI_ISL_1469894, EPI_ISL_1469895, EPI_ISL_1469896, EPI_ISL_1469897, EPI_ISL_1469899, EPI_ISL_1469900, EPI_ISL_1469901, EPI_ISL_1469903, EPI_ISL_1469904, EPI_ISL_1469907, EPI_ISL_1469908, EPI_ISL_1469909, EPI_ISL_1469910, EPI_ISL_1469911, EPI_ISL_1469912, EPI_ISL_1469913, EPI_ISL_1469915, EPI_ISL_1469916, EPI_ISL_1469917, EPI_ISL_1469918, EPI_ISL_1469919, EPI_ISL_1469920, EPI_ISL_1469921, EPI_ISL_1469922, EPI_ISL_1469923, EPI_ISL_1469924, EPI_ISL_1469925, EPI_ISL_1469926, EPI_ISL_1469927, EPI_ISL_1469928, EPI_ISL_1469930, EPI_ISL_1469931, EPI_ISL_1469932, EPI_ISL_1469933, EPI_ISL_1469934, EPI_ISL_1469935, EPI_ISL_1469936, EPI_ISL_1921085, EPI_ISL_1921086, EPI_ISL_1921087, EPI_ISL_1921088, EPI_ISL_1921089, EPI_ISL_1921090, EPI_ISL_1921091, EPI_ISL_1921092, EPI_ISL_1921093, EPI_ISL_1921094, EPI_ISL_1921095, EPI_ISL_1921096, EPI_ISL_1921097, EPI_ISL_1921098, EPI_ISL_1921099, EPI_ISL_1921100, EPI_ISL_1921101, EPI_ISL_1921102, EPI_ISL_1921103, EPI_ISL_1921104, EPI_ISL_1921105, EPI_ISL_1921106, EPI_ISL_1921107, EPI_ISL_1921108, EPI_ISL_1921109, EPI_ISL_1921110, EPI_ISL_1921111, EPI_ISL_1921116, EPI_ISL_1921117, EPI_ISL_1921118, EPI_ISL_1921120, EPI_ISL_1921122, EPI_ISL_1921123, EPI_ISL_1921124, EPI_ISL_1921125, EPI_ISL_1921126, EPI_ISL_1921127, EPI_ISL_1921130, EPI_ISL_1921131, EPI_ISL_1921132, EPI_ISL_1921134, EPI_ISL_1921135, EPI_ISL_1921137, EPI_ISL_1921139, EPI_ISL_1921141, EPI_ISL_1921142, EPI_ISL_1921143, EPI_ISL_1921144, EPI_ISL_1921145, EPI_ISL_1921147, EPI_ISL_1921148, EPI_ISL_1921151, EPI_ISL_1921152, EPI_ISL_1921153, EPI_ISL_1921154, EPI_ISL_1921155, EPI_ISL_1921156, EPI_ISL_1921157, EPI_ISL_1921158, EPI_ISL_1921159, EPI_ISL_1921162, EPI_ISL_2031787, EPI_ISL_2031788, EPI_ISL_2031789, EPI_ISL_2031790, EPI_ISL_2031791, EPI_ISL_2031793, EPI_ISL_2031794, EPI_ISL_2031795, EPI_ISL_2031796, EPI_ISL_2031797, EPI_ISL_2031798, EPI_ISL_2031799, EPI_ISL_2031800, EPI_ISL_2031801, EPI_ISL_2031802, EPI_ISL_2031803, EPI_ISL_2031804, EPI_ISL_2031805, EPI_ISL_2031806, EPI_ISL_2031807, EPI_ISL_2031808, EPI_ISL_2031809, EPI_ISL_2031810, EPI_ISL_2031811, EPI_ISL_2031813, EPI_ISL_2031814, EPI_ISL_2031815, EPI_ISL_2031817, EPI_ISL_2031819, EPI_ISL_2031822, EPI_ISL_2031823, EPI_ISL_2031824, EPI_ISL_2031825, EPI_ISL_2031827, EPI_ISL_2031829, EPI_ISL_2031830, EPI_ISL_2031831, EPI_ISL_2031832, EPI_ISL_2031834, EPI_ISL_2031836, EPI_ISL_2031837, EPI_ISL_2031838, EPI_ISL_2031839, EPI_ISL_2031842, EPI_ISL_2031843, EPI_ISL_2031845, EPI_ISL_2031846, EPI_ISL_2031847, EPI_ISL_2031848, EPI_ISL_2031849, EPI_ISL_2031850, EPI_ISL_2031851, EPI_ISL_2031852, EPI_ISL_2031853, EPI_ISL_2031854, EPI_ISL_2031855, EPI_ISL_2031856, EPI_ISL_2031857, EPI_ISL_2081230, EPI_ISL_2081236, EPI_ISL_2081240, EPI_ISL_2081241, EPI_ISL_2081242, EPI_ISL_2081244, EPI_ISL_2081245, EPI_ISL_2081247, EPI_ISL_2081248, EPI_ISL_2081249, EPI_ISL_2081250, EPI_ISL_2081253, EPI_ISL_2081254, EPI_ISL_2081255, EPI_ISL_2081256, EPI_ISL_2081257, EPI_ISL_2081260, EPI_ISL_2081261, EPI_ISL_2081262, EPI_ISL_2081265, EPI_ISL_2081266, EPI_ISL_2081269, EPI_ISL_2081270, EPI_ISL_2081271, EPI_ISL_2081272, EPI_ISL_2081274, EPI_ISL_2081275, EPI_ISL_2081276, EPI_ISL_2081277, EPI_ISL_2081278, EPI_ISL_2081279, EPI_ISL_2081280, EPI_ISL_2081281, EPI_ISL_2081282, EPI_ISL_2081283, EPI_ISL_2081284, EPI_ISL_2081285, EPI_ISL_2081286, EPI_ISL_2081287, EPI_ISL_2081288, EPI_ISL_2081289, EPI_ISL_2081290, EPI_ISL_2081292, EPI_ISL_2081293, EPI_ISL_2081294, EPI_ISL_2081295, EPI_ISL_2081296, EPI_ISL_2081297, EPI_ISL_2081298, EPI_ISL_2081299, EPI_ISL_2081300, EPI_ISL_2081301, EPI_ISL_2081302, EPI_ISL_2081303, EPI_ISL_2081304, EPI_ISL_2081305, EPI_ISL_2081306, EPI_ISL_2081307, EPI_ISL_2081308, EPI_ISL_2081309, EPI_ISL_2081530, EPI_ISL_2081531, EPI_ISL_2081532, EPI_ISL_2081534, EPI_ISL_2081535, EPI_ISL_2081536, EPI_ISL_2081538, EPI_ISL_2081542, EPI_ISL_2081543, EPI_ISL_2081544, EPI_ISL_2081546, EPI_ISL_2081548, EPI_ISL_2081549, EPI_ISL_2081550, EPI_ISL_2081557, EPI_ISL_2081558, EPI_ISL_2081560, EPI_ISL_2081561, EPI_ISL_2081562, EPI_ISL_2081563, EPI_ISL_2081566, EPI_ISL_2081567, EPI_ISL_2081568, EPI_ISL_2081569, EPI_ISL_2081570, EPI_ISL_2081571, EPI_ISL_2081572, EPI_ISL_2081573, EPI_ISL_2081574, EPI_ISL_2081575, EPI_ISL_2081576, EPI_ISL_2081577, EPI_ISL_2081578, EPI_ISL_2081579, EPI_ISL_2081580, EPI_ISL_2081581, EPI_ISL_2081582, EPI_ISL_2081583, EPI_ISL_2081584, EPI_ISL_2081585, EPI_ISL_2081586, EPI_ISL_2081587, EPI_ISL_2081588, EPI_ISL_2081589, EPI_ISL_2081590, EPI_ISL_2081591, EPI_ISL_2081592, EPI_ISL_2081593, EPI_ISL_2081594, EPI_ISL_2081595, EPI_ISL_2081596, EPI_ISL_2081597, EPI_ISL_2081598, EPI_ISL_2081599, EPI_ISL_2081600, EPI_ISL_2081601, EPI_ISL_2081602, EPI_ISL_2081604, EPI_ISL_2081605, EPI_ISL_2081606, EPI_ISL_2081607, EPI_ISL_2081608, EPI_ISL_2081609, EPI_ISL_2081610, EPI_ISL_2081611, EPI_ISL_2081612, EPI_ISL_2081613, EPI_ISL_2082936, EPI_ISL_2082937, EPI_ISL_2082938, EPI_ISL_2082943, EPI_ISL_2082944, EPI_ISL_2082945, EPI_ISL_2082947, EPI_ISL_2082948, EPI_ISL_2082949, EPI_ISL_2082950, EPI_ISL_2082951, EPI_ISL_2082952, EPI_ISL_2082957, EPI_ISL_2082958, EPI_ISL_2082959, EPI_ISL_2082960, EPI_ISL_2082961, EPI_ISL_2082962, EPI_ISL_2082963, EPI_ISL_2082964, EPI_ISL_2082965, EPI_ISL_2082966, EPI_ISL_2082967, EPI_ISL_2082968, EPI_ISL_2082969, EPI_ISL_2082970, EPI_ISL_2082971, EPI_ISL_2082972, EPI_ISL_2082973, EPI_ISL_2082974, EPI_ISL_2082975, EPI_ISL_2082976, EPI_ISL_2082977, EPI_ISL_2082978, EPI_ISL_2082981, EPI_ISL_2082982, EPI_ISL_2082983, EPI_ISL_2082984, EPI_ISL_2082985, EPI_ISL_2082986, EPI_ISL_2082987, EPI_ISL_2082991, EPI_ISL_2082992, EPI_ISL_2082994, EPI_ISL_2082995, EPI_ISL_2082996, EPI_ISL_2082997, EPI_ISL_2082998, EPI_ISL_2082999, EPI_ISL_2083000, EPI_ISL_2083001, EPI_ISL_2083003, EPI_ISL_2083006, EPI_ISL_2083007, EPI_ISL_2083008, EPI_ISL_2083009, EPI_ISL_2083015, EPI_ISL_2179191, EPI_ISL_2179192, EPI_ISL_2179193, EPI_ISL_2179194, EPI_ISL_2179195, EPI_ISL_2179196, EPI_ISL_2179197, EPI_ISL_2179198, EPI_ISL_2179199, EPI_ISL_2179200, EPI_ISL_2179201, EPI_ISL_2179202, EPI_ISL_2179203, EPI_ISL_2179204, EPI_ISL_2179205, EPI_ISL_2179206, EPI_ISL_2179207, EPI_ISL_2179208, EPI_ISL_2179209, EPI_ISL_2179210, EPI_ISL_2179211, EPI_ISL_2179212, EPI_ISL_2179213, EPI_ISL_2179214, EPI_ISL_2179215, EPI_ISL_2179216, EPI_ISL_2179220, EPI_ISL_2179221, EPI_ISL_2179222, EPI_ISL_2179223, EPI_ISL_2179224, EPI_ISL_2179225, EPI_ISL_2179226, EPI_ISL_2179227, EPI_ISL_2179228, EPI_ISL_2179229, EPI_ISL_2179230, EPI_ISL_2179231, EPI_ISL_2179232, EPI_ISL_2179233, EPI_ISL_2179234, EPI_ISL_2179235, EPI_ISL_2179236, EPI_ISL_2179237, EPI_ISL_2179238, EPI_ISL_2179239, EPI_ISL_2179240, EPI_ISL_2179241, EPI_ISL_2179242, EPI_ISL_2179243, EPI_ISL_2179244, EPI_ISL_2179245, EPI_ISL_2179246, EPI_ISL_2179247, EPI_ISL_2179248, EPI_ISL_2179249, EPI_ISL_2179250, EPI_ISL_2179251, EPI_ISL_2179252, EPI_ISL_2179253, EPI_ISL_2179261, EPI_ISL_2179262, EPI_ISL_2179264, EPI_ISL_2179265, EPI_ISL_2179266, EPI_ISL_2179267, EPI_ISL_2179268, EPI_ISL_2179269, EPI_ISL_2179273, EPI_ISL_2179274, EPI_ISL_2295453, EPI_ISL_2295454, EPI_ISL_2295455, EPI_ISL_2295458, EPI_ISL_2295459, EPI_ISL_2295460, EPI_ISL_2295461, EPI_ISL_2295462, EPI_ISL_2295463, EPI_ISL_2295464, EPI_ISL_2295465, EPI_ISL_2295466, EPI_ISL_2295467, EPI_ISL_2295469, EPI_ISL_2295470, EPI_ISL_2295471, EPI_ISL_2295472, EPI_ISL_2295473, EPI_ISL_2295474, EPI_ISL_2295475, EPI_ISL_2295476, EPI_ISL_2295477, EPI_ISL_2295480, EPI_ISL_2295481, EPI_ISL_2295482, EPI_ISL_2295483, EPI_ISL_2295485, EPI_ISL_2295486, EPI_ISL_2295490, EPI_ISL_2295491, EPI_ISL_2295492, EPI_ISL_2295493, EPI_ISL_2295494, EPI_ISL_2295495, EPI_ISL_2295497, EPI_ISL_2295498, EPI_ISL_2295499, EPI_ISL_2295500, EPI_ISL_2295501, EPI_ISL_2295502, EPI_ISL_2295503, EPI_ISL_2295504, EPI_ISL_2295505, EPI_ISL_2295506, EPI_ISL_2295508, EPI_ISL_2295509, EPI_ISL_2295510, EPI_ISL_2295511, EPI_ISL_2295512, EPI_ISL_2295513, EPI_ISL_2295514, EPI_ISL_2295517, EPI_ISL_2295518, EPI_ISL_2295519, EPI_ISL_2295520, EPI_ISL_2295521, EPI_ISL_2295522, EPI_ISL_2295523, EPI_ISL_2295524, EPI_ISL_2295525, EPI_ISL_2295526, EPI_ISL_2295527, EPI_ISL_2295528, EPI_ISL_2295529, EPI_ISL_2295530, EPI_ISL_2295531, EPI_ISL_2295532, EPI_ISL_2295533, EPI_ISL_2341412, EPI_ISL_2341413, EPI_ISL_2378758, EPI_ISL_2378759, EPI_ISL_2378762, EPI_ISL_2378763, EPI_ISL_2378764, EPI_ISL_2378765, EPI_ISL_2378767, EPI_ISL_2378770, EPI_ISL_2378771, EPI_ISL_2378773, EPI_ISL_2378774, EPI_ISL_2378775, EPI_ISL_2378776, EPI_ISL_2378780, EPI_ISL_2378783, EPI_ISL_2378784, EPI_ISL_2378789, EPI_ISL_2378792, EPI_ISL_2378793, EPI_ISL_2378794, EPI_ISL_2378795, EPI_ISL_2378796, EPI_ISL_2378797, EPI_ISL_2378798, EPI_ISL_2378802, EPI_ISL_2378803, EPI_ISL_2378804, EPI_ISL_2378806, EPI_ISL_2378807, EPI_ISL_2378808, EPI_ISL_2378812, EPI_ISL_2378813, EPI_ISL_2378815, EPI_ISL_2378816, EPI_ISL_2378817, EPI_ISL_2378818, EPI_ISL_2378819, EPI_ISL_2378820, EPI_ISL_2494636, EPI_ISL_2494637, EPI_ISL_2494638, EPI_ISL_2494639, EPI_ISL_2494640, EPI_ISL_2494645, EPI_ISL_2494646, EPI_ISL_2494647, EPI_ISL_2494651, EPI_ISL_2494655, EPI_ISL_2494656, EPI_ISL_2494657, EPI_ISL_2494658, EPI_ISL_2494659, EPI_ISL_2494660, EPI_ISL_2494661, EPI_ISL_2494662, EPI_ISL_2494663, EPI_ISL_2494664, EPI_ISL_2494665, EPI_ISL_2494669, EPI_ISL_2494670, EPI_ISL_2494671, EPI_ISL_2494672, EPI_ISL_2494673, EPI_ISL_2494678, EPI_ISL_2494690, EPI_ISL_2494691, EPI_ISL_2494699, EPI_ISL_2494700, EPI_ISL_2494701, EPI_ISL_2494702, EPI_ISL_2494703, EPI_ISL_2494704, EPI_ISL_2494705, EPI_ISL_2494706, EPI_ISL_2494707, EPI_ISL_2494708, EPI_ISL_2494709 | | | |
| see above | UCSC Genomics Institute | UCSC Genomics Institute | A. Marm Kilpatrick; Angie Hinrichs; Beth Shapiro; Bryan Thornlow; Ciara Wanket; David Haussler; Eric Beraut; Hugh Olsen; Ikenna Anigbogu; Isabel Bjork; Jakob McBroome; Jeremy Sanford; Joshua Kapp; Mark Akeson; Maximilian Haeussler; Michael Stone; Miten Jain; Molly Cassatt-Johnstone; Namrita Dhillon; Russell Corbett-Detig; Terren Chang; Yatish Turakhia |
| EPI_ISL_751801 | UCSD EXCITE | Andersen lab at Scripps Research | SEARCH Alliance San Diego |
| EPI_ISL_1301442, EPI_ISL_1301443 | UCSD EXCITE | NBACC | Aaron Phillips; Adam Bazinet; Amanda Graham; Brian Holland; David Miller; Denise Freeburger; Elyse Abbott; Gregory Williams; Jason Yolitz; Jennifer Biryukov; Jordan Bohannon; Karla Garcia; Katie Beck; M.J. Rosovitz; Melissa Krause; Michael Schuit; Paul Dabisch; Sean Lovett; Shanna Ratnesar-Shumate; Stewart Wood; Victoria Wahl; Wade Weaver |
| EPI_ISL_755133, EPI_ISL_755140, EPI_ISL_755146, EPI_ISL_755147, EPI_ISL_755148, EPI_ISL_755149, EPI_ISL_755150, EPI_ISL_755151, EPI_ISL_755152, EPI_ISL_755153, EPI_ISL_755154, EPI_ISL_755155, EPI_ISL_755156, EPI_ISL_755157, EPI_ISL_755158, EPI_ISL_755159, EPI_ISL_755160, EPI_ISL_755161, EPI_ISL_755162, EPI_ISL_755163, EPI_ISL_755164, EPI_ISL_755165, EPI_ISL_755166, EPI_ISL_755167, EPI_ISL_755168, EPI_ISL_755169, EPI_ISL_755170, EPI_ISL_755171, EPI_ISL_755172, EPI_ISL_755173, EPI_ISL_755174, EPI_ISL_755175, EPI_ISL_755176, EPI_ISL_755177, EPI_ISL_755178, EPI_ISL_755179, EPI_ISL_755180, EPI_ISL_755181, EPI_ISL_755182, EPI_ISL_755183, EPI_ISL_755184, EPI_ISL_755185, EPI_ISL_755186, EPI_ISL_755187, EPI_ISL_755188, EPI_ISL_755189, EPI_ISL_755190, EPI_ISL_755191, EPI_ISL_755192, EPI_ISL_755193, EPI_ISL_755194, EPI_ISL_755195, EPI_ISL_755196, EPI_ISL_755198, EPI_ISL_755199, EPI_ISL_755200, EPI_ISL_755201, EPI_ISL_755202, EPI_ISL_755203, EPI_ISL_755204, EPI_ISL_755205, EPI_ISL_755206, EPI_ISL_755207, EPI_ISL_755208, EPI_ISL_755209, EPI_ISL_755216, EPI_ISL_755217, EPI_ISL_755218, EPI_ISL_755219, EPI_ISL_755220, EPI_ISL_755221, EPI_ISL_755222, EPI_ISL_755223, EPI_ISL_755224, EPI_ISL_755225, EPI_ISL_755226, EPI_ISL_755227, EPI_ISL_755228, EPI_ISL_755229, EPI_ISL_755230, EPI_ISL_755231 | | | |
| see above | UCSD EXCITE lab | Andersen lab at Scripps Research | SEARCH Alliance San Diego |
| EPI_ISL_849926 | UCSF Clinical Microbiology Laboratory | Chan-Zuckerberg Biohub | CZB Cliahub Consortium |
| EPI_ISL_754124, EPI_ISL_754125, EPI_ISL_754126, EPI_ISL_754127, EPI_ISL_754128, EPI_ISL_754129, EPI_ISL_754133, EPI_ISL_754134, EPI_ISL_754135, EPI_ISL_754390 | | | |
| see above | USC Clinical Lab | Los Angeles County Public Health Laboratories | P. Hemarajata et al. |
| EPI_ISL_831700, EPI_ISL_831737, EPI_ISL_831784, EPI_ISL_831816, EPI_ISL_831840, EPI_ISL_831849, EPI_ISL_831864, EPI_ISL_831880 | | | |
| see above | United States Air Force School of Aerospace Medicine | United States Air Force School of Aerospace Medicine | Amanda Javorina; Anthony Fries; Clarise Starr; Elizabeth Macias; Jennifer Meyer; Sarah Purves; William Gruner |
| EPI_ISL_2295290, EPI_ISL_2295291, EPI_ISL_2295292, EPI_ISL_2295293, EPI_ISL_2295294, EPI_ISL_2295295, EPI_ISL_2295296, EPI_ISL_2295297, EPI_ISL_2295298, EPI_ISL_2295299, EPI_ISL_2295300, EPI_ISL_2295301, EPI_ISL_2295302, EPI_ISL_2295303, EPI_ISL_2295304, EPI_ISL_2295305, EPI_ISL_2295306, EPI_ISL_2295307, EPI_ISL_2295308, EPI_ISL_2295309, EPI_ISL_2295310, EPI_ISL_2295311, EPI_ISL_2295312, EPI_ISL_2295313, EPI_ISL_2295314, EPI_ISL_2295315, EPI_ISL_2295316, EPI_ISL_2295317, EPI_ISL_2295318, EPI_ISL_2295319, EPI_ISL_2295320, EPI_ISL_2295321, EPI_ISL_2295322, EPI_ISL_2295323, EPI_ISL_2295324, EPI_ISL_2295325, EPI_ISL_2295326, EPI_ISL_2295327, EPI_ISL_2295328, EPI_ISL_2295329, EPI_ISL_2295330, EPI_ISL_2295331, EPI_ISL_2295332, EPI_ISL_2295333, EPI_ISL_2295334, EPI_ISL_2295335, EPI_ISL_2295336, EPI_ISL_2295337, EPI_ISL_2295338, EPI_ISL_2295339, EPI_ISL_2295340, EPI_ISL_2295341, EPI_ISL_2295342, EPI_ISL_2295343, EPI_ISL_2295344, EPI_ISL_2295345, EPI_ISL_2295346, EPI_ISL_2295347, EPI_ISL_2295348, EPI_ISL_2295349, EPI_ISL_2295350, EPI_ISL_2295381, EPI_ISL_2295383, EPI_ISL_2322841, EPI_ISL_2322842, EPI_ISL_2322844 | | | |
| see above | WLA VAMC | Genomics and Discovery, Respiratory Viruses Branch, Division of Viral Diseases, Centers for Disease Control and Prevention | Adam Retchless; Anna Kelleher; Anna Uehara; Brian Lynch; Clinton R. Paden; Dhwani Batra; Haibin Wang; Han Jia Justin Ng; Jasmine Padilla; Jing Zhang; Justin Lee; Krista Queen; Mark Burroughs; Mili Sheth; Morgan Davis; Peter Cook; Rachel Marine; Sarah Nobles; Suxiang Tong; Tara Coalter; Yan Li; Ying Tao |
| EPI_ISL_849577 | Washington State Department of Health | Seattle Flu Study | Amanda Adler; Barry R. Lutz; Benjamin Pelle; Brian Hiatt; Caitlin R. Wolf; Chris D. Frazar; Deborah A. Nickerson; Elisabeth Brandstetter; Geoff Melly; Helen Y. Chu; Janet A. Englund; Jay Shendure; Jover Lee; Kairsten Fay; Kirsten Lacombe; Lea M. Starita; Mark J. Rieder; Matthew Richardson; Matthew Thompson; Melissa Truong; Michael Boeckh; Michael Famulare; Misja Ilcisin; Peter D. Han; Philip Dykema; Romesh Gautom; Scott Lindquist; Thomas R. Sibley; Trevor Bedford |
| EPI_ISL_977984 | ` | Chiu Laboratory, University of California, San Francisco | Candace Wang; Charles Chiu; Debra Wadford; Jill Hacker; Venice Servellita; Xianding (Wayne) Deng |

We gratefully acknowledge the following Authors from the Originating laboratories responsible for obtaining the specimens, as well as the Submitting laboratories where the genome data were generated and shared via GISAID, on which this research is based.

All Submitters of data may be contacted directly via www.gisaid.org

Authors are sorted alphabetically.

| Accession ID | Originating Laboratory | Submitting Laboratory | Authors |
|---|---|---|---|
| EPI_ISL_1225958, EPI_ISL_1225959, EPI_ISL_1225960, EPI_ISL_1225961, EPI_ISL_1225962, EPI_ISL_1225967, EPI_ISL_1225968, EPI_ISL_1225969, EPI_ISL_1225970, EPI_ISL_1225971, EPI_ISL_1225972, EPI_ISL_1225973, EPI_ISL_1225974 | | | |
| see above | "CDPH, Viral and Rickettsial Disease Laboratory" | Genomics and Discovery, Respiratory Viruses Branch, Division of Viral Diseases, Centers for Disease Control and Prevention | Anna Montmayeur; Anna Uehara; Ben L. Rambo-Martin; Clinton R. Paden; Dhwani Batra; Haibin Wang; Jasmine Padilla; Jing Zhang; Justin Lee; Katie Dillon; Krista Queen; Kristen Knipe; Kristine Lacek; Lori Rowe; Mark Burroughs; Matthew Schmerer; Mili Sheth; Peter W. Cook; Rachel Marine; Sam Shepard; Sarah Nobles; Shoshona Le; Suxiang Tong; Yan Li; Ying Tao |
| EPI_ISL_1093564, EPI_ISL_1093567, EPI_ISL_1093570, EPI_ISL_1093572 | Alameda County Public Health Department | Alameda County Public Health Department | Kevin Libuit; Kristna Hsieh; Maria Elma |
| EPI_ISL_955391, EPI_ISL_955392, EPI_ISL_955393, EPI_ISL_955394, EPI_ISL_955395, EPI_ISL_955396, EPI_ISL_955397, EPI_ISL_955398, EPI_ISL_955399, EPI_ISL_955400, EPI_ISL_955401, EPI_ISL_955402, EPI_ISL_955403, EPI_ISL_955404, EPI_ISL_955405, EPI_ISL_955406, EPI_ISL_955407, EPI_ISL_955408, EPI_ISL_955409, EPI_ISL_955410, EPI_ISL_955411, EPI_ISL_955412, EPI_ISL_955413, EPI_ISL_955414, EPI_ISL_955415, EPI_ISL_955416, EPI_ISL_955417, EPI_ISL_955418, EPI_ISL_955419, EPI_ISL_955420, EPI_ISL_955421, EPI_ISL_955422, EPI_ISL_955423, EPI_ISL_955424, EPI_ISL_955425, EPI_ISL_955426, EPI_ISL_1027479, EPI_ISL_1027568, EPI_ISL_1027578, EPI_ISL_1027582, EPI_ISL_1184923, EPI_ISL_1184924, EPI_ISL_1184925, EPI_ISL_1184926, EPI_ISL_1184927, EPI_ISL_1184928, EPI_ISL_1184929, EPI_ISL_1184930, EPI_ISL_1184931, EPI_ISL_1184932, EPI_ISL_1184933, EPI_ISL_1184934, EPI_ISL_1184935, EPI_ISL_1184936, EPI_ISL_1184937, EPI_ISL_1184938, EPI_ISL_1184939, EPI_ISL_1184940, EPI_ISL_1184941, EPI_ISL_1184942, EPI_ISL_1184943, EPI_ISL_1184944, EPI_ISL_1184945, EPI_ISL_1184946, EPI_ISL_1184947, EPI_ISL_1184948, EPI_ISL_1184949, EPI_ISL_1184950, EPI_ISL_1184951, EPI_ISL_1184952, EPI_ISL_1184953, EPI_ISL_1184954, EPI_ISL_1184955, EPI_ISL_1184956, EPI_ISL_1184957, EPI_ISL_1184958, EPI_ISL_1184959, EPI_ISL_1184960, EPI_ISL_1184961, EPI_ISL_1184962, EPI_ISL_1184963, EPI_ISL_1184964, EPI_ISL_1184965, EPI_ISL_1184966, EPI_ISL_1185027, EPI_ISL_1185028, EPI_ISL_1185031, EPI_ISL_1185033, EPI_ISL_1185034, EPI_ISL_1185035, EPI_ISL_1185036, EPI_ISL_1185037, EPI_ISL_1185038, EPI_ISL_1185041, EPI_ISL_1185047, EPI_ISL_1185054, EPI_ISL_1185056, EPI_ISL_1185057, EPI_ISL_1185066, EPI_ISL_1185071, EPI_ISL_1185073, EPI_ISL_1185082, EPI_ISL_1185085, EPI_ISL_1185087, EPI_ISL_1185094, EPI_ISL_1185095, EPI_ISL_1185100, EPI_ISL_1185102, EPI_ISL_1185105, EPI_ISL_1185109, EPI_ISL_1185117, EPI_ISL_1185121, EPI_ISL_1185124, EPI_ISL_1185126, EPI_ISL_1185128, EPI_ISL_1185130, EPI_ISL_1185136, EPI_ISL_1185142, EPI_ISL_1185157, EPI_ISL_1185161, EPI_ISL_1185162, EPI_ISL_1185164, EPI_ISL_1185172, EPI_ISL_1185176, EPI_ISL_1185179, EPI_ISL_1185187, EPI_ISL_1185198, EPI_ISL_1185199, EPI_ISL_1185204, EPI_ISL_1185207, EPI_ISL_1185215, EPI_ISL_1185222, EPI_ISL_1185229, EPI_ISL_1185232, EPI_ISL_1185243, EPI_ISL_1185250, EPI_ISL_1185258, EPI_ISL_1185259, EPI_ISL_1185262, EPI_ISL_1185271, EPI_ISL_1185281, EPI_ISL_1185283, EPI_ISL_1185286, EPI_ISL_1185305, EPI_ISL_1185313, EPI_ISL_1185315, EPI_ISL_1185322, EPI_ISL_1185351, EPI_ISL_1185356, EPI_ISL_1185357, EPI_ISL_1185358, EPI_ISL_1185362, EPI_ISL_1185363, EPI_ISL_1185364, EPI_ISL_1185367, EPI_ISL_1185368, EPI_ISL_1185372, EPI_ISL_1185376, EPI_ISL_1185383, EPI_ISL_1185388, EPI_ISL_1185393, EPI_ISL_1185401, EPI_ISL_1185414, EPI_ISL_1185422, EPI_ISL_1185427, EPI_ISL_1185430, EPI_ISL_1185431, EPI_ISL_1185447, EPI_ISL_1185449, EPI_ISL_1185452, EPI_ISL_1185456, EPI_ISL_1185457, EPI_ISL_1185459, EPI_ISL_1185461, EPI_ISL_1185466, EPI_ISL_1185475, EPI_ISL_1185480, EPI_ISL_1185491, EPI_ISL_1185493, EPI_ISL_1185496, EPI_ISL_1185499, EPI_ISL_1185501, EPI_ISL_1185502, EPI_ISL_1185503, EPI_ISL_1185517, EPI_ISL_1185529, EPI_ISL_1185533, EPI_ISL_1185534, EPI_ISL_2658737, EPI_ISL_2659245, EPI_ISL_2659267, EPI_ISL_2659331, EPI_ISL_2659372, EPI_ISL_2659416, EPI_ISL_2659544, EPI_ISL_2758444, EPI_ISL_2758445, EPI_ISL_2758446, EPI_ISL_2758447 | | | |
| see above | Alameda County Public Health Lab | Chan-Zuckerberg Biohub | CZB Cliahub Consortium |
| EPI_ISL_833405, EPI_ISL_833406, EPI_ISL_833407, EPI_ISL_833408, EPI_ISL_833409, EPI_ISL_833410, EPI_ISL_833411, EPI_ISL_833412 | | | |
| see above | Beverly Hospital | Los Angeles County Public Health Laboratories | P. Hemarajata et al. |
| EPI_ISL_1184819, EPI_ISL_1184820, EPI_ISL_1184821, EPI_ISL_1184822, EPI_ISL_1184823, EPI_ISL_1184824, EPI_ISL_1185016, EPI_ISL_1185017, EPI_ISL_1185018, EPI_ISL_1185019, EPI_ISL_1185020, EPI_ISL_1185021, EPI_ISL_1185022, EPI_ISL_1185023, EPI_ISL_1185024, EPI_ISL_1185025, EPI_ISL_1185030, EPI_ISL_1185042, EPI_ISL_1185050, EPI_ISL_1185052, EPI_ISL_1185059, EPI_ISL_1185060, EPI_ISL_1185061, EPI_ISL_1185063, EPI_ISL_1185064, EPI_ISL_1185067, EPI_ISL_1185068, EPI_ISL_1185069, EPI_ISL_1185077, EPI_ISL_1185078, EPI_ISL_1185079, EPI_ISL_1185080, EPI_ISL_1185084, EPI_ISL_1185088, EPI_ISL_1185089, EPI_ISL_1185090, EPI_ISL_1185096, EPI_ISL_1185099, EPI_ISL_1185103, EPI_ISL_1185119, EPI_ISL_1185122, EPI_ISL_1185132, EPI_ISL_1185133, EPI_ISL_1185135, EPI_ISL_1185138, EPI_ISL_1185139, EPI_ISL_1185144, EPI_ISL_1185147, EPI_ISL_1185150, EPI_ISL_1185153, EPI_ISL_1185155, EPI_ISL_1185156, EPI_ISL_1185158, EPI_ISL_1185166, EPI_ISL_1185168, EPI_ISL_1185169, EPI_ISL_1185173, EPI_ISL_1185177, EPI_ISL_1185178, EPI_ISL_1185182, EPI_ISL_1185184, EPI_ISL_1185186, EPI_ISL_1185188, EPI_ISL_1185190, EPI_ISL_1185192, EPI_ISL_1185193, EPI_ISL_1185195, EPI_ISL_1185196, EPI_ISL_1185202, EPI_ISL_1185203, EPI_ISL_1185205, EPI_ISL_1185208, EPI_ISL_1185210, EPI_ISL_1185211, EPI_ISL_1185213, EPI_ISL_1185220, EPI_ISL_1185223, EPI_ISL_1185226, EPI_ISL_1185230, EPI_ISL_1185244, EPI_ISL_1185246, EPI_ISL_1185247, EPI_ISL_1185248, EPI_ISL_1185249, EPI_ISL_1185253, EPI_ISL_1185254, EPI_ISL_1185256, EPI_ISL_1185260, EPI_ISL_1185264, EPI_ISL_1185265, EPI_ISL_1185268, EPI_ISL_1185270, EPI_ISL_1185275, EPI_ISL_1185276, EPI_ISL_1185279, EPI_ISL_1185282, EPI_ISL_1234544, EPI_ISL_1234545, EPI_ISL_1234550, EPI_ISL_1234551, EPI_ISL_1234554, EPI_ISL_1234557, EPI_ISL_1234571, EPI_ISL_1234575, EPI_ISL_1234576, EPI_ISL_1234581, EPI_ISL_1234582, EPI_ISL_1234589, EPI_ISL_1234591, EPI_ISL_1234592, EPI_ISL_1234593, EPI_ISL_1234594, EPI_ISL_1234595, EPI_ISL_1234599, EPI_ISL_1234601, EPI_ISL_1234605, EPI_ISL_1234608, EPI_ISL_1234619, EPI_ISL_1234620, EPI_ISL_1234623, EPI_ISL_1234624, EPI_ISL_1234625, EPI_ISL_1234627, EPI_ISL_1234633, EPI_ISL_1234634, EPI_ISL_1234635, EPI_ISL_1234638, EPI_ISL_1234639, EPI_ISL_1234644, EPI_ISL_1234649, EPI_ISL_1234650, EPI_ISL_1234651, EPI_ISL_1234652, EPI_ISL_1234656, EPI_ISL_1234658, EPI_ISL_1234663, EPI_ISL_1234668, EPI_ISL_1234669, EPI_ISL_1234673, EPI_ISL_1234681, EPI_ISL_1234695, EPI_ISL_1234699, EPI_ISL_1234700, EPI_ISL_1234703, EPI_ISL_1234704, EPI_ISL_1234712, EPI_ISL_1234713, EPI_ISL_1234714, EPI_ISL_1234715, EPI_ISL_1234717, EPI_ISL_1234719, EPI_ISL_1234722, EPI_ISL_1234730, EPI_ISL_1234731, EPI_ISL_1234733, EPI_ISL_1234734, EPI_ISL_1234735, EPI_ISL_1234740, EPI_ISL_1234747, EPI_ISL_1234748, EPI_ISL_1234750, EPI_ISL_1234752, EPI_ISL_1234753, EPI_ISL_1234754, EPI_ISL_1234761, EPI_ISL_1234766, EPI_ISL_1234771, EPI_ISL_1234774, EPI_ISL_1234778, EPI_ISL_1234780, EPI_ISL_1234781, EPI_ISL_1234784, EPI_ISL_1235060, EPI_ISL_1235063, EPI_ISL_1235068, EPI_ISL_1235070, EPI_ISL_1235074, EPI_ISL_1235079, EPI_ISL_1235084, EPI_ISL_1235092, EPI_ISL_1235095, EPI_ISL_1235097, EPI_ISL_1235104, EPI_ISL_1235107, EPI_ISL_1235112, EPI_ISL_1235118, EPI_ISL_1235119, EPI_ISL_1235121, EPI_ISL_1235123, EPI_ISL_1235125, EPI_ISL_1235126, EPI_ISL_1235127, EPI_ISL_1235128, EPI_ISL_1235132, EPI_ISL_1235133, EPI_ISL_1235135, EPI_ISL_1235136, EPI_ISL_1235138, EPI_ISL_1235140, EPI_ISL_1235144, EPI_ISL_1235148, EPI_ISL_1235150, EPI_ISL_1235152, EPI_ISL_1235162, EPI_ISL_1235176, EPI_ISL_1235182, EPI_ISL_1235186, EPI_ISL_1235190, EPI_ISL_1235192, EPI_ISL_1235193, EPI_ISL_1235195, EPI_ISL_1235197, EPI_ISL_1235199, EPI_ISL_1235203, EPI_ISL_1235206, EPI_ISL_1235212, EPI_ISL_1235220, EPI_ISL_1235222, EPI_ISL_1235226, EPI_ISL_1235229, EPI_ISL_1235232, EPI_ISL_1235234, EPI_ISL_1235236, EPI_ISL_1235238, EPI_ISL_1235242, EPI_ISL_1235248, EPI_ISL_1235251, EPI_ISL_1235252, EPI_ISL_1235254, EPI_ISL_1235269, EPI_ISL_1235272, EPI_ISL_1235273, EPI_ISL_1235274, EPI_ISL_1235282, EPI_ISL_1235285, EPI_ISL_1235287, EPI_ISL_1235290, EPI_ISL_1235291, EPI_ISL_1235295, EPI_ISL_1235298, EPI_ISL_1235307, EPI_ISL_1235310, EPI_ISL_1235317, EPI_ISL_1235322, EPI_ISL_1235346, EPI_ISL_1235351, EPI_ISL_1235354, EPI_ISL_1477438, EPI_ISL_1477439, EPI_ISL_1477444, EPI_ISL_1477449, EPI_ISL_1477452, EPI_ISL_1477453, EPI_ISL_1477457, EPI_ISL_1477458, EPI_ISL_1477459, EPI_ISL_1477460, EPI_ISL_1477463, EPI_ISL_1477465, EPI_ISL_1477471, EPI_ISL_1477473, EPI_ISL_1477476, EPI_ISL_1477477, EPI_ISL_1477478, EPI_ISL_1477481, EPI_ISL_1477482, EPI_ISL_1477488, EPI_ISL_1477493, EPI_ISL_1477500, EPI_ISL_1477502, EPI_ISL_1477503, EPI_ISL_1477504, EPI_ISL_1477506, EPI_ISL_1477508, EPI_ISL_1477511, EPI_ISL_1477514, EPI_ISL_1477515, EPI_ISL_1477517, EPI_ISL_1477521, EPI_ISL_1477525, EPI_ISL_1477531, EPI_ISL_1477532, EPI_ISL_1477533, EPI_ISL_1477536, EPI_ISL_1477537, EPI_ISL_1477539, EPI_ISL_1477543, EPI_ISL_1477544, EPI_ISL_1477551, EPI_ISL_1477555, EPI_ISL_1477561, EPI_ISL_1477562, EPI_ISL_1477571, EPI_ISL_1477577, EPI_ISL_1477587, EPI_ISL_1477588, EPI_ISL_1477589, EPI_ISL_1477593, EPI_ISL_1477597, EPI_ISL_1477600, EPI_ISL_1477601, EPI_ISL_1477604, EPI_ISL_1477610, EPI_ISL_1477612, EPI_ISL_1477613, EPI_ISL_1477614, EPI_ISL_1477615, EPI_ISL_1477619, EPI_ISL_1477623, EPI_ISL_1477624, EPI_ISL_1477631, EPI_ISL_1477635, EPI_ISL_1477641, EPI_ISL_1477645, EPI_ISL_1477646, EPI_ISL_1477650, EPI_ISL_1477656, EPI_ISL_1477658, EPI_ISL_1477659, EPI_ISL_1477660, EPI_ISL_1477665, EPI_ISL_1477666, EPI_ISL_1477667, EPI_ISL_1477673, EPI_ISL_1477678, EPI_ISL_1477680, EPI_ISL_1477687, EPI_ISL_1477692, EPI_ISL_1477693, EPI_ISL_1477694, EPI_ISL_1477700, EPI_ISL_1477705, EPI_ISL_1477707, EPI_ISL_1477709, EPI_ISL_1477711, EPI_ISL_1477715, EPI_ISL_1477717, EPI_ISL_1477718, EPI_ISL_1477719, EPI_ISL_1477721, EPI_ISL_1477723, EPI_ISL_1477725, EPI_ISL_1477728, EPI_ISL_1477736, EPI_ISL_1477740, EPI_ISL_1477742, EPI_ISL_1477756, EPI_ISL_1477759, EPI_ISL_1477763, EPI_ISL_1477769, EPI_ISL_1477777, EPI_ISL_1477778, EPI_ISL_1477786, EPI_ISL_1477787, EPI_ISL_1477792, EPI_ISL_1477796, EPI_ISL_1477798, EPI_ISL_1477803, EPI_ISL_1477808, EPI_ISL_1477811, EPI_ISL_1477812, EPI_ISL_1477819, EPI_ISL_1477830, EPI_ISL_1477831, EPI_ISL_1477837, EPI_ISL_1477838, EPI_ISL_1477840, EPI_ISL_1477848, EPI_ISL_1477852, EPI_ISL_1477853, EPI_ISL_1477855, EPI_ISL_1477868, EPI_ISL_1477873, EPI_ISL_1477875, EPI_ISL_1477877, EPI_ISL_1477883, EPI_ISL_1477884, EPI_ISL_1477890, EPI_ISL_1477891, EPI_ISL_1477897, EPI_ISL_1477901, EPI_ISL_1477903, EPI_ISL_1477908, EPI_ISL_1477913, EPI_ISL_1477917, EPI_ISL_1477920, EPI_ISL_1477921, EPI_ISL_1477924, EPI_ISL_1477929, EPI_ISL_1477934, EPI_ISL_1477938, EPI_ISL_1477940, EPI_ISL_1477949, EPI_ISL_2658595, EPI_ISL_2658719, EPI_ISL_2659280, EPI_ISL_2659306, EPI_ISL_2659318, EPI_ISL_2659333, EPI_ISL_2659369, EPI_ISL_2659445, EPI_ISL_2659495, EPI_ISL_2659592, EPI_ISL_2659612, EPI_ISL_2659620, EPI_ISL_2758442, EPI_ISL_2758448, EPI_ISL_2758449, EPI_ISL_2758464, EPI_ISL_2758465, EPI_ISL_2758466, EPI_ISL_2758467 | | | |
| see above | CA DPH Viral and Rickettsial Disease Laboratory | Chan-Zuckerberg Biohub | CZB Cliahub Consortium |
| EPI_ISL_1225949, EPI_ISL_1225950, EPI_ISL_1225951, EPI_ISL_1225952, EPI_ISL_1225953, EPI_ISL_1225954, EPI_ISL_1225955, EPI_ISL_1225956, EPI_ISL_1225957 | | | |
| see above | CA-Los Angeles County Public Health Laboratory | Genomics and Discovery, Respiratory Viruses Branch, Division of Viral Diseases, Centers for Disease Control and Prevention | Anna Montmayeur; Anna Uehara; Ben L. Rambo-Martin; Clinton R. Paden; Dhwani Batra; Haibin Wang; Jasmine Padilla; Jing Zhang; Justin Lee; Katie Dillon; Krista Queen; Kristen Knipe; Kristine Lacek; Lori Rowe; Mark Burroughs; Matthew Schmerer; Mili Sheth; Peter W. Cook; Rachel Marine; Sam Shepard; Sarah Nobles; Shoshona Le; Suxiang Tong; Yan Li; Ying Tao |
| EPI_ISL_1094362, EPI_ISL_1094363, EPI_ISL_1094560, EPI_ISL_1094562, EPI_ISL_1094563, EPI_ISL_1094566, EPI_ISL_1094567, EPI_ISL_1094568, EPI_ISL_1094569, EPI_ISL_1094571 | | | |
| see above | CA-Los Angeles County Public Health Laboratory | Respiratory Viruses Branch, Division of Viral Diseases, Centers for Disease Control and Prevention | Anna Montmayeur; Anna Uehara; Ben L. Rambo-Martin; Clinton R. Paden; Dhwani Batra; Haibin Wang; Jasmine Padilla; Jing Zhang; Justin Lee; Krista Queen; Lori Rowe; Mark Burroughs; Mili Sheth; Peter W. Cook; Rachel Marine; Sarah Nobles; Suxiang Tong; Yan Li; Ying Tao |
| EPI_ISL_2782383, EPI_ISL_2782386, EPI_ISL_2782388, EPI_ISL_2782390, EPI_ISL_2782393, EPI_ISL_2782394, EPI_ISL_2782396, EPI_ISL_2782397, EPI_ISL_2782400, EPI_ISL_2782403, EPI_ISL_2782406, EPI_ISL_2782408, EPI_ISL_2782409, EPI_ISL_2782410, EPI_ISL_2782418, EPI_ISL_2782421, EPI_ISL_2782422, EPI_ISL_2782434, EPI_ISL_2782436, EPI_ISL_2782439, EPI_ISL_2782447, EPI_ISL_2782452, EPI_ISL_2782468, EPI_ISL_2782476, EPI_ISL_2782480, EPI_ISL_2782481, EPI_ISL_2782486, EPI_ISL_2782487, EPI_ISL_2782489, EPI_ISL_2782491, EPI_ISL_2782493, EPI_ISL_2782495, EPI_ISL_2782504, EPI_ISL_2782506, EPI_ISL_2782511, EPI_ISL_2782514, EPI_ISL_2782520, EPI_ISL_2782521, EPI_ISL_2782524, EPI_ISL_2782526, EPI_ISL_2782539, EPI_ISL_2782541, EPI_ISL_2782542, EPI_ISL_2782546, EPI_ISL_2782547, EPI_ISL_2782550, EPI_ISL_2782553, EPI_ISL_2782559, EPI_ISL_2782560, EPI_ISL_2782569, EPI_ISL_2782576, EPI_ISL_2782580, EPI_ISL_2782586, EPI_ISL_2782602, EPI_ISL_2782603, EPI_ISL_2782605, EPI_ISL_2782610, EPI_ISL_2782611, EPI_ISL_2782612, EPI_ISL_2782613, EPI_ISL_2782614, EPI_ISL_2782615, EPI_ISL_2782616, EPI_ISL_2782617, EPI_ISL_2782618, EPI_ISL_2782619, EPI_ISL_2782620, EPI_ISL_2782621, EPI_ISL_2782622, EPI_ISL_2782623, EPI_ISL_2782624, EPI_ISL_2782625, EPI_ISL_2782626, EPI_ISL_2782627, EPI_ISL_2782628, EPI_ISL_2782629, EPI_ISL_2782630, EPI_ISL_2782631, EPI_ISL_2782632, EPI_ISL_2782633, EPI_ISL_2782634, EPI_ISL_2782635, EPI_ISL_2782636, EPI_ISL_2782637, EPI_ISL_2782638, EPI_ISL_2782639, EPI_ISL_2782640, EPI_ISL_2782641, EPI_ISL_2782642, EPI_ISL_2782643, EPI_ISL_2782644, EPI_ISL_2782645, EPI_ISL_2782646, EPI_ISL_2782647, EPI_ISL_2782648, EPI_ISL_2782649, EPI_ISL_2782650, EPI_ISL_2782651, EPI_ISL_2782652, EPI_ISL_2782653, EPI_ISL_2782654, EPI_ISL_2782655, EPI_ISL_2782656, EPI_ISL_2782657, EPI_ISL_2782658, EPI_ISL_2782659, EPI_ISL_2782660, EPI_ISL_2782661, EPI_ISL_2782662, EPI_ISL_2782663, EPI_ISL_2782664, EPI_ISL_2782665, EPI_ISL_2782666, EPI_ISL_2782667, EPI_ISL_2782668, EPI_ISL_2782669, EPI_ISL_2782670, EPI_ISL_2782671, EPI_ISL_2782672, EPI_ISL_2782673, EPI_ISL_2782674, EPI_ISL_2782675, EPI_ISL_2782676, EPI_ISL_2782677, EPI_ISL_2782678, EPI_ISL_2782679, EPI_ISL_2782680, EPI_ISL_2782681, EPI_ISL_2782682, EPI_ISL_2782683, EPI_ISL_2782684, EPI_ISL_2782685, EPI_ISL_2782686, EPI_ISL_2782687, EPI_ISL_2782688, EPI_ISL_2782689, EPI_ISL_2782690, EPI_ISL_2782691, EPI_ISL_2782692, EPI_ISL_2782693, EPI_ISL_2782694, EPI_ISL_2782695, EPI_ISL_2782696, EPI_ISL_2782697, EPI_ISL_2782698, EPI_ISL_2782699, EPI_ISL_2782700, EPI_ISL_2782701, EPI_ISL_2782702, EPI_ISL_2782703, EPI_ISL_2782704, EPI_ISL_2782705, EPI_ISL_2782706, EPI_ISL_2782707, EPI_ISL_2782708, EPI_ISL_2782709, EPI_ISL_2782710, EPI_ISL_2782711, EPI_ISL_2782712, EPI_ISL_2782713, EPI_ISL_2782714, EPI_ISL_2782715, EPI_ISL_2782716, EPI_ISL_2782717, EPI_ISL_2782718, EPI_ISL_2782719, EPI_ISL_2782720, EPI_ISL_2782721, EPI_ISL_2782722, EPI_ISL_2782723, EPI_ISL_2782724, EPI_ISL_2782725, EPI_ISL_2782726, EPI_ISL_2782727, EPI_ISL_2782728, EPI_ISL_2782729, EPI_ISL_2782730, EPI_ISL_2782731, EPI_ISL_2782732, EPI_ISL_2782733, EPI_ISL_2782734, EPI_ISL_2782735, EPI_ISL_2782736, EPI_ISL_2782737, EPI_ISL_2782738, EPI_ISL_2782739, EPI_ISL_2782740, EPI_ISL_2782741, EPI_ISL_2782742, EPI_ISL_2782743, EPI_ISL_2782744, EPI_ISL_2782745, EPI_ISL_2782746, EPI_ISL_2782747, EPI_ISL_2782748, EPI_ISL_2782749, EPI_ISL_2782750, EPI_ISL_2782751, EPI_ISL_2782752, EPI_ISL_2782753, EPI_ISL_2782754, EPI_ISL_2782755, EPI_ISL_2782756, EPI_ISL_2782757, EPI_ISL_2782758, EPI_ISL_2782759, EPI_ISL_2782760, EPI_ISL_2782761, EPI_ISL_2782762, EPI_ISL_2782763, EPI_ISL_2782764, EPI_ISL_2782765, EPI_ISL_2782766, EPI_ISL_2782767, EPI_ISL_2782768, EPI_ISL_2782769, EPI_ISL_2782770, EPI_ISL_2782771, EPI_ISL_2782772, EPI_ISL_2782773, EPI_ISL_2782774, EPI_ISL_2782775, EPI_ISL_2782776, EPI_ISL_2782777, EPI_ISL_2782778, EPI_ISL_2782779, EPI_ISL_2782780, EPI_ISL_2782781, EPI_ISL_2782782, EPI_ISL_2782783, EPI_ISL_2782784, EPI_ISL_2782785, EPI_ISL_2782786, EPI_ISL_2782787, EPI_ISL_2782788, EPI_ISL_2782789, EPI_ISL_2782790, EPI_ISL_2782791, EPI_ISL_2782792, EPI_ISL_2782793, EPI_ISL_2782794, EPI_ISL_2782795, EPI_ISL_2783244, EPI_ISL_2783245, EPI_ISL_2783246, EPI_ISL_2783247, EPI_ISL_2783248, EPI_ISL_2783249, EPI_ISL_2783250, EPI_ISL_2783251, EPI_ISL_2783252, EPI_ISL_2783253, EPI_ISL_2783254, EPI_ISL_2783255, EPI_ISL_2783256, EPI_ISL_2783257, EPI_ISL_2783258, EPI_ISL_2783259, EPI_ISL_2783260, EPI_ISL_2783261, EPI_ISL_2783262, EPI_ISL_2783263, EPI_ISL_2783264, EPI_ISL_2783265, EPI_ISL_2783266, EPI_ISL_2783267, EPI_ISL_2783268, EPI_ISL_2783269, EPI_ISL_2783270, EPI_ISL_2783271, EPI_ISL_2783272, EPI_ISL_2783273, EPI_ISL_2783274, EPI_ISL_2783275, EPI_ISL_2783276, EPI_ISL_2783277, EPI_ISL_2783278, EPI_ISL_2783279, EPI_ISL_2783280, EPI_ISL_2783281, EPI_ISL_2783282, EPI_ISL_2783283, EPI_ISL_2783284, EPI_ISL_2783285, EPI_ISL_2783286, EPI_ISL_2783287, EPI_ISL_2783288, EPI_ISL_2783289, EPI_ISL_2783290, EPI_ISL_2783291, EPI_ISL_2783292, EPI_ISL_2783293, EPI_ISL_2783294, EPI_ISL_2783295, EPI_ISL_2783296, EPI_ISL_2783297, EPI_ISL_2783298, EPI_ISL_2783299, EPI_ISL_2783300, EPI_ISL_2783301, EPI_ISL_2783302, EPI_ISL_2783303, EPI_ISL_2783304, EPI_ISL_2783305, EPI_ISL_2783306, EPI_ISL_2783307, EPI_ISL_2783308, EPI_ISL_2783309, EPI_ISL_2783310, EPI_ISL_2783311, EPI_ISL_2783312, EPI_ISL_2783313, EPI_ISL_2783314, EPI_ISL_2783315, EPI_ISL_2783316, EPI_ISL_2783317, EPI_ISL_2783318, EPI_ISL_2783319, EPI_ISL_2783320, EPI_ISL_2783321, EPI_ISL_2783322, EPI_ISL_2783323, EPI_ISL_2783324, EPI_ISL_2783325, EPI_ISL_2783326, EPI_ISL_2783327, EPI_ISL_2783328, EPI_ISL_2783329, EPI_ISL_2783330, EPI_ISL_2783331, EPI_ISL_2783332, EPI_ISL_2783333, EPI_ISL_2783334, EPI_ISL_2783335, EPI_ISL_2783336, EPI_ISL_2783337, EPI_ISL_2783338, EPI_ISL_2783339, EPI_ISL_2783340, EPI_ISL_2783341, EPI_ISL_2783342, EPI_ISL_2783343, EPI_ISL_2783344, EPI_ISL_2783345, EPI_ISL_2783346, EPI_ISL_2783347, EPI_ISL_2783348, EPI_ISL_2783349, EPI_ISL_2783350, EPI_ISL_2783351, EPI_ISL_2783352, EPI_ISL_2783353, EPI_ISL_2783354, EPI_ISL_2783355, EPI_ISL_2783356, EPI_ISL_2783357, EPI_ISL_2783358, EPI_ISL_2783359, EPI_ISL_2783360, EPI_ISL_2783361, EPI_ISL_2783362, EPI_ISL_2783363, EPI_ISL_2783364, EPI_ISL_2783365, EPI_ISL_2783366, EPI_ISL_2783367, EPI_ISL_2783368, EPI_ISL_2783369, EPI_ISL_2783370, EPI_ISL_2783371, EPI_ISL_2783372, EPI_ISL_2783373, EPI_ISL_2783374, EPI_ISL_2783375, EPI_ISL_2783376, EPI_ISL_2783377, EPI_ISL_2783378, EPI_ISL_2783379, EPI_ISL_2783380, EPI_ISL_2783381, EPI_ISL_2783382, EPI_ISL_2783383, EPI_ISL_2783384, EPI_ISL_2783385, EPI_ISL_2783386, EPI_ISL_2783387, EPI_ISL_2783388, EPI_ISL_2783389, EPI_ISL_2783390, EPI_ISL_2783391, EPI_ISL_2783392, EPI_ISL_2783393, EPI_ISL_2783394, EPI_ISL_2783395, EPI_ISL_2783396, EPI_ISL_2783397, EPI_ISL_2783398, EPI_ISL_2783399, EPI_ISL_2783400, EPI_ISL_2783401, EPI_ISL_2783402, EPI_ISL_2783403, EPI_ISL_2783404, EPI_ISL_2783405, EPI_ISL_2783406, EPI_ISL_2783407, EPI_ISL_2783408, EPI_ISL_2783409, EPI_ISL_2783410, EPI_ISL_2783411, EPI_ISL_2783412, EPI_ISL_2783413, EPI_ISL_2783414, EPI_ISL_2783415, EPI_ISL_2783416, EPI_ISL_2783417, EPI_ISL_2783418, EPI_ISL_2783419, EPI_ISL_2783420, EPI_ISL_2783421, EPI_ISL_2783422, EPI_ISL_2783423, EPI_ISL_2783424, EPI_ISL_2783425, EPI_ISL_2783426, EPI_ISL_2783427, EPI_ISL_2783428, EPI_ISL_2783429, EPI_ISL_2783430, EPI_ISL_2783431, EPI_ISL_2783432, EPI_ISL_2783433, EPI_ISL_2783434, EPI_ISL_2783435, EPI_ISL_2783436, EPI_ISL_2783437, EPI_ISL_2783438, EPI_ISL_2783439, EPI_ISL_2783440, EPI_ISL_2783441, EPI_ISL_2783442, EPI_ISL_2783443, EPI_ISL_2783444, EPI_ISL_2783445, EPI_ISL_2783446, EPI_ISL_2783447, EPI_ISL_2783448, EPI_ISL_2783449, EPI_ISL_2783450, EPI_ISL_2783451, EPI_ISL_2783452, EPI_ISL_2783453, EPI_ISL_2783454, EPI_ISL_2783455, EPI_ISL_2783456, EPI_ISL_2783457, EPI_ISL_2783458, EPI_ISL_2783459, EPI_ISL_2783460, EPI_ISL_2783461, EPI_ISL_2783462, EPI_ISL_2783463, EPI_ISL_2783464, EPI_ISL_2783465, EPI_ISL_2783466, EPI_ISL_2783467, EPI_ISL_2783468, EPI_ISL_2783469, EPI_ISL_2783470, EPI_ISL_2783471, EPI_ISL_2783472, EPI_ISL_2783473, EPI_ISL_2783474, EPI_ISL_2783475, EPI_ISL_2783476, EPI_ISL_2783477, EPI_ISL_2783478, EPI_ISL_2783479, EPI_ISL_2783480, EPI_ISL_2783481, EPI_ISL_2783482, EPI_ISL_2783483, EPI_ISL_2783484, EPI_ISL_2783485, EPI_ISL_2783486, EPI_ISL_2783487, EPI_ISL_2783488, EPI_ISL_2783489, EPI_ISL_2783490, EPI_ISL_2783491, EPI_ISL_2783492, EPI_ISL_2783493, EPI_ISL_2783494, EPI_ISL_2783495, EPI_ISL_2783496, EPI_ISL_2783497, EPI_ISL_2783498, EPI_ISL_2783499, EPI_ISL_2783500, EPI_ISL_2783501, EPI_ISL_2783502, EPI_ISL_2783503, EPI_ISL_2783504, EPI_ISL_2783505, EPI_ISL_2783506, EPI_ISL_2783507, EPI_ISL_2783508, EPI_ISL_2783509, EPI_ISL_2783510, EPI_ISL_2796532, EPI_ISL_2796533, EPI_ISL_2796534, EPI_ISL_2796535, EPI_ISL_2796536, EPI_ISL_2796537, EPI_ISL_2796538, EPI_ISL_2796539, EPI_ISL_2796540, EPI_ISL_2796541, EPI_ISL_2796542, EPI_ISL_2796543, EPI_ISL_2796544, EPI_ISL_2796545, EPI_ISL_2796546, EPI_ISL_2796547, EPI_ISL_2796548, EPI_ISL_2796549, EPI_ISL_2796550, EPI_ISL_2796551, EPI_ISL_2796552, EPI_ISL_2796553, EPI_ISL_2796554, EPI_ISL_2796555, EPI_ISL_2796556, EPI_ISL_2796557, EPI_ISL_2796558, EPI_ISL_2796559, EPI_ISL_2796560, EPI_ISL_2796561, EPI_ISL_2796562, EPI_ISL_2796563, EPI_ISL_2796564, EPI_ISL_2796565, EPI_ISL_2796566, EPI_ISL_2796567, EPI_ISL_2796568, EPI_ISL_2796569, EPI_ISL_2796570, EPI_ISL_2796571, EPI_ISL_2796572, EPI_ISL_2796573, EPI_ISL_2796574, EPI_ISL_2796575, EPI_ISL_2796576, EPI_ISL_2796577, EPI_ISL_2796578, EPI_ISL_2796579, EPI_ISL_2796580, EPI_ISL_2796581, EPI_ISL_2796582, EPI_ISL_2796583, EPI_ISL_2796584, EPI_ISL_2796585, EPI_ISL_2796586, EPI_ISL_2796587, EPI_ISL_2796588, EPI_ISL_2796589, EPI_ISL_2796590, EPI_ISL_2796591, EPI_ISL_2796592, EPI_ISL_2796593, EPI_ISL_2796594, EPI_ISL_2796595, EPI_ISL_2796596, EPI_ISL_2796597, EPI_ISL_2796598, EPI_ISL_2796599, EPI_ISL_2796600, EPI_ISL_2796601, EPI_ISL_2796602, EPI_ISL_2796603, EPI_ISL_2796604, EPI_ISL_2796605, EPI_ISL_2796606, EPI_ISL_2796607, EPI_ISL_2796608, EPI_ISL_2796609, EPI_ISL_2796610, EPI_ISL_2796611, EPI_ISL_2796612, EPI_ISL_2796613, EPI_ISL_2796614, EPI_ISL_2796615, EPI_ISL_2796616, EPI_ISL_2796617, EPI_ISL_2796618, EPI_ISL_2796619, EPI_ISL_2796620, EPI_ISL_2796621, EPI_ISL_2796622, EPI_ISL_2796623, EPI_ISL_2796624, EPI_ISL_2796625, EPI_ISL_2796626, EPI_ISL_2796627, EPI_ISL_2796628, EPI_ISL_2796629, EPI_ISL_2796630, EPI_ISL_2796631, EPI_ISL_2796632, EPI_ISL_2796633, EPI_ISL_2796634, EPI_ISL_2796635, EPI_ISL_2796636, EPI_ISL_2796637, EPI_ISL_2796638, EPI_ISL_2796639, EPI_ISL_2796640, EPI_ISL_2796641, EPI_ISL_2796642, EPI_ISL_2796643, EPI_ISL_2796644, EPI_ISL_2796645, EPI_ISL_2796646, EPI_ISL_2796647, EPI_ISL_2796648, EPI_ISL_2796649, EPI_ISL_2796650, EPI_ISL_2796651, EPI_ISL_2796652, EPI_ISL_2796653, EPI_ISL_2796654, EPI_ISL_2796655, EPI_ISL_2796656, EPI_ISL_2796657, EPI_ISL_2796658, EPI_ISL_2796659, EPI_ISL_2796660, EPI_ISL_2796661, EPI_ISL_2796662, EPI_ISL_2796663, EPI_ISL_2796664, EPI_ISL_2796665, EPI_ISL_2796666, EPI_ISL_2796667, EPI_ISL_2796668, EPI_ISL_2796669, EPI_ISL_2796670, EPI_ISL_2796671, EPI_ISL_2796672, EPI_ISL_2796673, EPI_ISL_2796674, EPI_ISL_2796675, EPI_ISL_2796676, EPI_ISL_2796677, EPI_ISL_2796678, EPI_ISL_2796679, EPI_ISL_2796680, EPI_ISL_2796681, EPI_ISL_2796682, EPI_ISL_2796683, EPI_ISL_2796684, EPI_ISL_2796685, EPI_ISL_2796686, EPI_ISL_2796687, EPI_ISL_2796688, EPI_ISL_2796689, EPI_ISL_2796690, EPI_ISL_2796691, EPI_ISL_2796692, EPI_ISL_2796693, EPI_ISL_2796694, EPI_ISL_2796695, EPI_ISL_2796696, EPI_ISL_2796697, EPI_ISL_2796698, EPI_ISL_2796699, EPI_ISL_2796700, EPI_ISL_2796701, EPI_ISL_2796702, EPI_ISL_2796703, EPI_ISL_2796704, EPI_ISL_2796705, EPI_ISL_2796706, EPI_ISL_2796707, EPI_ISL_2796708, EPI_ISL_2796709, EPI_ISL_2796711, EPI_ISL_2796712, EPI_ISL_2796713, EPI_ISL_2796714, EPI_ISL_2796715, EPI_ISL_2796716, EPI_ISL_2796717, EPI_ISL_2796718, EPI_ISL_2796719, EPI_ISL_2796720, EPI_ISL_2796721, EPI_ISL_2796722, EPI_ISL_2796723, EPI_ISL_2796724, EPI_ISL_2796725, EPI_ISL_2796726, EPI_ISL_2796727, EPI_ISL_2796728, EPI_ISL_2796729, EPI_ISL_2796730, EPI_ISL_2796731, EPI_ISL_2796732, EPI_ISL_2796733, EPI_ISL_2796734, EPI_ISL_2796735, EPI_ISL_2796736, EPI_ISL_2796737, EPI_ISL_2796738, EPI_ISL_2796739, EPI_ISL_2796740, EPI_ISL_2796741, EPI_ISL_2796742, EPI_ISL_2796743, EPI_ISL_2797383, EPI_ISL_2797384, EPI_ISL_2797385, EPI_ISL_2797386, EPI_ISL_2797387, EPI_ISL_2797388, EPI_ISL_2797389, EPI_ISL_2797390, EPI_ISL_2797391, EPI_ISL_2797392, EPI_ISL_2797393, EPI_ISL_2797394, EPI_ISL_2797395, EPI_ISL_2797396, EPI_ISL_2797397, EPI_ISL_2797398, EPI_ISL_2797399, EPI_ISL_2797400, EPI_ISL_2797401, EPI_ISL_2797402, EPI_ISL_2797403, EPI_ISL_2797404, EPI_ISL_2797405, EPI_ISL_2797406, EPI_ISL_2797407, EPI_ISL_2797408, EPI_ISL_2797409, EPI_ISL_2797410, EPI_ISL_2797411, EPI_ISL_2797412, EPI_ISL_2797413, EPI_ISL_2797414, EPI_ISL_2797415, EPI_ISL_2797416, EPI_ISL_2797417, EPI_ISL_2797418, EPI_ISL_2797419, EPI_ISL_2797420, EPI_ISL_2797421, EPI_ISL_2797422, EPI_ISL_2797423, EPI_ISL_2797424, EPI_ISL_2797425, EPI_ISL_2797426, EPI_ISL_2797427, EPI_ISL_2797428, EPI_ISL_2797429, EPI_ISL_2797430, EPI_ISL_2797431, EPI_ISL_2797432, EPI_ISL_2797433, EPI_ISL_2797434, EPI_ISL_2797435, EPI_ISL_2797436, EPI_ISL_2797437, EPI_ISL_2797438, EPI_ISL_2797439, EPI_ISL_2797440, EPI_ISL_2797441, EPI_ISL_2797442, EPI_ISL_2797443, EPI_ISL_2797444, EPI_ISL_2797445, EPI_ISL_2797446, EPI_ISL_2797447, EPI_ISL_2797448, EPI_ISL_2797449, EPI_ISL_2797450, EPI_ISL_2797451, EPI_ISL_2797452, EPI_ISL_2797453, EPI_ISL_2797454, EPI_ISL_2797455, EPI_ISL_2797456, EPI_ISL_2797457, EPI_ISL_2797458, EPI_ISL_2797459, EPI_ISL_2797460, EPI_ISL_2797461, EPI_ISL_2797462, EPI_ISL_2797463, EPI_ISL_2797464, EPI_ISL_2797465, EPI_ISL_2797466, EPI_ISL_2797467, EPI_ISL_2797468, EPI_ISL_2797469, EPI_ISL_2797470, EPI_ISL_2797471, EPI_ISL_2797472, EPI_ISL_2797473, EPI_ISL_2797474, EPI_ISL_2797475, EPI_ISL_2797476, EPI_ISL_2797477, EPI_ISL_2797478, EPI_ISL_2797479, EPI_ISL_2797480, EPI_ISL_2797481, EPI_ISL_2797482, EPI_ISL_2797483, EPI_ISL_2797484, EPI_ISL_2797485, EPI_ISL_2797486, EPI_ISL_2797487, EPI_ISL_2797488, EPI_ISL_2797489, EPI_ISL_2797490, EPI_ISL_2797491, EPI_ISL_2797492, EPI_ISL_2797493, EPI_ISL_2797494, EPI_ISL_2797495, EPI_ISL_2797496, EPI_ISL_2797497, EPI_ISL_2797498, | | | |

| | | | |
|---|---|---|---|
| EPI_ISL_2797499, EPI_ISL_2797500, EPI_ISL_2797501, EPI_ISL_2797502, EPI_ISL_2797503, EPI_ISL_2797504, EPI_ISL_2797505, EPI_ISL_2797506, EPI_ISL_2797507, EPI_ISL_2797508, EPI_ISL_2797509, EPI_ISL_2797510, EPI_ISL_2797511, EPI_ISL_2797512, EPI_ISL_2797513, EPI_ISL_2797514, EPI_ISL_2797515, EPI_ISL_2797516, EPI_ISL_2797517, EPI_ISL_2797518, EPI_ISL_2797519, EPI_ISL_2797520, EPI_ISL_2797521, EPI_ISL_2797522, EPI_ISL_2797523, EPI_ISL_2797524, EPI_ISL_2797525, EPI_ISL_2797526, EPI_ISL_2797527, EPI_ISL_2797528, EPI_ISL_2797529, EPI_ISL_2797530, EPI_ISL_2797531, EPI_ISL_2797532, EPI_ISL_2797533, EPI_ISL_2797534, EPI_ISL_2797535, EPI_ISL_2797536, EPI_ISL_2797537, EPI_ISL_2797538, EPI_ISL_2797539, EPI_ISL_2797540, EPI_ISL_2797541, EPI_ISL_2797542, EPI_ISL_2797543, EPI_ISL_2797544, EPI_ISL_2797545, EPI_ISL_2797546, EPI_ISL_2797547, EPI_ISL_2797548, EPI_ISL_2797549, EPI_ISL_2797550, EPI_ISL_2797551, EPI_ISL_2797552, EPI_ISL_2797553, EPI_ISL_2797554, EPI_ISL_2797555, EPI_ISL_2797556, EPI_ISL_2797557, EPI_ISL_2797558, EPI_ISL_2797559, EPI_ISL_2797560, EPI_ISL_2797561, EPI_ISL_2797562, EPI_ISL_2797563, EPI_ISL_2797564, EPI_ISL_2797565, EPI_ISL_2797566, EPI_ISL_2797567, EPI_ISL_2797568, EPI_ISL_2797569, EPI_ISL_2797570, EPI_ISL_2797571, EPI_ISL_2797572, EPI_ISL_2797573, EPI_ISL_2797574, EPI_ISL_2797575, EPI_ISL_2797576, EPI_ISL_2797577, EPI_ISL_2797578, EPI_ISL_2797579, EPI_ISL_2797580, EPI_ISL_2797581, EPI_ISL_2797582, EPI_ISL_2797583, EPI_ISL_2797584, EPI_ISL_2797585, EPI_ISL_2797586, EPI_ISL_2797587, EPI_ISL_2797588, EPI_ISL_2797589, EPI_ISL_2797590, EPI_ISL_2976558, EPI_ISL_2976559, EPI_ISL_2976560, EPI_ISL_2976561, EPI_ISL_2976562, EPI_ISL_2976563, EPI_ISL_2976564, EPI_ISL_2976565, EPI_ISL_2976566, EPI_ISL_2976567, EPI_ISL_2976568, EPI_ISL_2976569, EPI_ISL_2976570, EPI_ISL_2976571, EPI_ISL_2976572, EPI_ISL_2976573, EPI_ISL_2976574, EPI_ISL_2976575, EPI_ISL_2976576, EPI_ISL_2976577, EPI_ISL_2976578, EPI_ISL_2976579, EPI_ISL_2976580, EPI_ISL_2976581, EPI_ISL_2976582, EPI_ISL_2976583, EPI_ISL_2976584, EPI_ISL_2976585, EPI_ISL_2976586, EPI_ISL_2976587, EPI_ISL_2976588, EPI_ISL_2976589, EPI_ISL_2976590, EPI_ISL_2976591, EPI_ISL_2976592, EPI_ISL_2976593, EPI_ISL_2976594, EPI_ISL_2976595, EPI_ISL_2976596, EPI_ISL_2976597, EPI_ISL_2976598, EPI_ISL_2976599, EPI_ISL_2976600, EPI_ISL_2976601, EPI_ISL_2976602, EPI_ISL_2976603, EPI_ISL_2976604, EPI_ISL_2976605, EPI_ISL_2976606, EPI_ISL_2976607, EPI_ISL_2976608, EPI_ISL_2976609, EPI_ISL_2976610, EPI_ISL_2976611, EPI_ISL_2976612, EPI_ISL_2976613, EPI_ISL_2976614, EPI_ISL_2976615, EPI_ISL_2976616, EPI_ISL_2976617, EPI_ISL_2976618, EPI_ISL_2976619, EPI_ISL_2976620, EPI_ISL_2976621, EPI_ISL_2976622, EPI_ISL_2976623, EPI_ISL_2976624, EPI_ISL_2976625, EPI_ISL_2976626, EPI_ISL_2976627, EPI_ISL_2976628, EPI_ISL_2976629, EPI_ISL_2976630, EPI_ISL_2976631, EPI_ISL_2976632, EPI_ISL_2976633, EPI_ISL_2976634, EPI_ISL_2976635, EPI_ISL_2976636, EPI_ISL_2976637, EPI_ISL_2976638, EPI_ISL_2976639, EPI_ISL_2976640, EPI_ISL_2976641, EPI_ISL_2976642, EPI_ISL_2976643, EPI_ISL_2976644, EPI_ISL_2976645, EPI_ISL_2976646, EPI_ISL_2976647, EPI_ISL_2976648, EPI_ISL_2976649, EPI_ISL_2976650, EPI_ISL_2976651, EPI_ISL_2976652, EPI_ISL_2976654, EPI_ISL_2976655, EPI_ISL_2976656, EPI_ISL_2976657, EPI_ISL_2976658, EPI_ISL_2976659, EPI_ISL_2976660, EPI_ISL_2976661, EPI_ISL_2976662, EPI_ISL_2976663, EPI_ISL_2976664, EPI_ISL_2976665, EPI_ISL_2976667, EPI_ISL_2976668, EPI_ISL_2976669, EPI_ISL_2976670, EPI_ISL_2976671, EPI_ISL_2976672, EPI_ISL_2976673, EPI_ISL_2976674, EPI_ISL_2976675, EPI_ISL_2976676, EPI_ISL_2976677, EPI_ISL_2976678, EPI_ISL_2976679, EPI_ISL_2976680, EPI_ISL_2976681, EPI_ISL_2976682, EPI_ISL_2976683, EPI_ISL_2976684, EPI_ISL_2976685, EPI_ISL_2976686, EPI_ISL_2976687, EPI_ISL_2976688, EPI_ISL_2976690, EPI_ISL_2976693, EPI_ISL_2976695, EPI_ISL_2976696, EPI_ISL_2976697, EPI_ISL_2976698, EPI_ISL_2976700, EPI_ISL_2976702, EPI_ISL_2976703, EPI_ISL_2976705, EPI_ISL_2976706, EPI_ISL_2976707, EPI_ISL_2976708, EPI_ISL_2976709, EPI_ISL_2976710, EPI_ISL_2976712, EPI_ISL_2976714, EPI_ISL_2976715, EPI_ISL_2976716, EPI_ISL_2976717, EPI_ISL_2976718, EPI_ISL_2976719, EPI_ISL_2976720, EPI_ISL_2976721, EPI_ISL_2976724, EPI_ISL_2976725, EPI_ISL_2976727, EPI_ISL_2976728, EPI_ISL_2976730, EPI_ISL_2976731, EPI_ISL_2976732, EPI_ISL_2976733, EPI_ISL_2976734, EPI_ISL_2976735, EPI_ISL_2976736, EPI_ISL_2976737, EPI_ISL_2976738, EPI_ISL_2976739, EPI_ISL_2976740, EPI_ISL_2976741, EPI_ISL_2976743, EPI_ISL_2976744, EPI_ISL_2976745, EPI_ISL_2976746, EPI_ISL_2976747, EPI_ISL_2976748, EPI_ISL_2976749, EPI_ISL_2976750, EPI_ISL_2976752, EPI_ISL_2976754, EPI_ISL_2976758, EPI_ISL_2976761, EPI_ISL_2976762, EPI_ISL_2976764, EPI_ISL_2976765, EPI_ISL_2976766, EPI_ISL_2976767, EPI_ISL_2976768, EPI_ISL_2976769, EPI_ISL_2976770, EPI_ISL_2976771, EPI_ISL_2976773, EPI_ISL_2976774, EPI_ISL_2976775, EPI_ISL_2976776, EPI_ISL_2976777, EPI_ISL_2976778, EPI_ISL_2976779, EPI_ISL_2976780, EPI_ISL_2976781, EPI_ISL_2976782, EPI_ISL_2976783, EPI_ISL_2976784, EPI_ISL_2976785, EPI_ISL_2976786, EPI_ISL_2976787, EPI_ISL_2976788, EPI_ISL_2976789, EPI_ISL_2976790, EPI_ISL_2976791, EPI_ISL_2976792, EPI_ISL_2976793, EPI_ISL_2976794, EPI_ISL_2976795, EPI_ISL_2976796, EPI_ISL_2976797, EPI_ISL_2976798, EPI_ISL_2976799, EPI_ISL_2976800, EPI_ISL_2976802, EPI_ISL_2976803, EPI_ISL_2976804, EPI_ISL_2976807, EPI_ISL_2976808, EPI_ISL_2976809, EPI_ISL_2976810, EPI_ISL_2976811, EPI_ISL_2976812, EPI_ISL_2976813, EPI_ISL_2976814, EPI_ISL_2976815, EPI_ISL_2976816, EPI_ISL_2976817, EPI_ISL_2976818, EPI_ISL_2976819, EPI_ISL_2976821, EPI_ISL_2976822, EPI_ISL_2976823, EPI_ISL_2976824, EPI_ISL_2976825, EPI_ISL_2976826, EPI_ISL_2976827, EPI_ISL_2976828, EPI_ISL_2976829, EPI_ISL_2976830, EPI_ISL_2976831, EPI_ISL_2976832, EPI_ISL_2976833, EPI_ISL_2976835, EPI_ISL_2976837, EPI_ISL_2976838, EPI_ISL_2976839, EPI_ISL_2976840, EPI_ISL_2976841, EPI_ISL_2976842, EPI_ISL_2976843, EPI_ISL_2976844, EPI_ISL_2976845, EPI_ISL_2976846, EPI_ISL_2976847, EPI_ISL_2976848, EPI_ISL_2976849, EPI_ISL_2976850, EPI_ISL_2976851, EPI_ISL_2976852, EPI_ISL_2976853, EPI_ISL_2976854, EPI_ISL_2976855, EPI_ISL_2976856, EPI_ISL_2976857, EPI_ISL_2976858, EPI_ISL_2976859, EPI_ISL_2976860, EPI_ISL_2976861, EPI_ISL_2976862, EPI_ISL_2976863, EPI_ISL_2976864, EPI_ISL_2976865, EPI_ISL_2976866, EPI_ISL_2976867, EPI_ISL_2976868, EPI_ISL_2976869, EPI_ISL_2976870, EPI_ISL_2976871, EPI_ISL_2976873, EPI_ISL_2976874, EPI_ISL_2976876, EPI_ISL_2976877, EPI_ISL_2976878, EPI_ISL_2976879, EPI_ISL_2976880, EPI_ISL_2976881, EPI_ISL_2976882, EPI_ISL_2976883, EPI_ISL_2976884, EPI_ISL_2976885, EPI_ISL_2976886, EPI_ISL_2976887, EPI_ISL_2976888, EPI_ISL_2976889, EPI_ISL_2976890, EPI_ISL_2976891, EPI_ISL_2976892, EPI_ISL_2976893, EPI_ISL_2976894, EPI_ISL_2976895, EPI_ISL_2976896, EPI_ISL_2976897, EPI_ISL_2976898, EPI_ISL_2976899, EPI_ISL_2976900, EPI_ISL_2976901, EPI_ISL_2976902, EPI_ISL_2976904, EPI_ISL_2976905, EPI_ISL_2976906, EPI_ISL_2976908, EPI_ISL_2976909, EPI_ISL_2976910, EPI_ISL_2976911, EPI_ISL_2976912, EPI_ISL_2976913, EPI_ISL_2976915, EPI_ISL_2976916, EPI_ISL_2976917, EPI_ISL_2976918, EPI_ISL_2976919, EPI_ISL_2976920, EPI_ISL_2976921, EPI_ISL_2976922, EPI_ISL_2976923, EPI_ISL_2976924, EPI_ISL_2976925, EPI_ISL_2976926, EPI_ISL_2976927, EPI_ISL_2976928, EPI_ISL_2976929, EPI_ISL_2976930, EPI_ISL_2976931, EPI_ISL_2977475, EPI_ISL_2977476, EPI_ISL_2977477, EPI_ISL_2977478, EPI_ISL_2977479, EPI_ISL_2977480, EPI_ISL_2977481, EPI_ISL_2977482, EPI_ISL_2977483, EPI_ISL_2977484, EPI_ISL_2977485, EPI_ISL_2977486, EPI_ISL_2977487, EPI_ISL_2977488, EPI_ISL_2977489, EPI_ISL_2977490, EPI_ISL_2977491, EPI_ISL_2977492, EPI_ISL_2977493, EPI_ISL_2977494, EPI_ISL_2977495, EPI_ISL_2977496, EPI_ISL_2977497, EPI_ISL_2977498, EPI_ISL_2977499, EPI_ISL_2977500, EPI_ISL_2977501, EPI_ISL_2977502, EPI_ISL_2977503, EPI_ISL_2977504, EPI_ISL_2977505, EPI_ISL_2977506, EPI_ISL_2977507, EPI_ISL_2977508, EPI_ISL_2977509, EPI_ISL_2977510, EPI_ISL_2977511, EPI_ISL_2977512, EPI_ISL_2977513, EPI_ISL_2977514, EPI_ISL_2977515, EPI_ISL_2977516, EPI_ISL_2977517, EPI_ISL_2977518, EPI_ISL_2977519, EPI_ISL_2977520, EPI_ISL_2977521, EPI_ISL_2977522, EPI_ISL_2977523, EPI_ISL_2977524, EPI_ISL_2977525, EPI_ISL_2977526, EPI_ISL_2977527, EPI_ISL_2977528, EPI_ISL_2977529, EPI_ISL_2977530, EPI_ISL_2977531, EPI_ISL_2977532, EPI_ISL_2977533, EPI_ISL_2977534, EPI_ISL_2977535, EPI_ISL_2977536, EPI_ISL_2977537, EPI_ISL_2977538, EPI_ISL_2977539, EPI_ISL_2977540, EPI_ISL_2977541, EPI_ISL_2977542, EPI_ISL_2977544, EPI_ISL_2977545, EPI_ISL_2977547, EPI_ISL_2977548, EPI_ISL_2977550, EPI_ISL_2977551, EPI_ISL_2977553, EPI_ISL_2977554, EPI_ISL_2977555, EPI_ISL_2977556, EPI_ISL_2977557, EPI_ISL_2977559, EPI_ISL_2977560, EPI_ISL_2977561, EPI_ISL_2977563, EPI_ISL_2977564, EPI_ISL_2977567, EPI_ISL_2977568, EPI_ISL_2977569, EPI_ISL_2977572, EPI_ISL_2977573, EPI_ISL_2977575, EPI_ISL_2977576, EPI_ISL_2977577, EPI_ISL_2977578, EPI_ISL_2977582, EPI_ISL_2977583, EPI_ISL_2977584, EPI_ISL_2977585, EPI_ISL_2977586, EPI_ISL_2977587, EPI_ISL_2977588, EPI_ISL_2977589, EPI_ISL_2977590, EPI_ISL_2977591, EPI_ISL_2977592, EPI_ISL_2977593, EPI_ISL_2977594, EPI_ISL_2977595, EPI_ISL_2977599, EPI_ISL_2977600, EPI_ISL_2977602, EPI_ISL_2977603, EPI_ISL_2977604, EPI_ISL_2977605, EPI_ISL_2977606, EPI_ISL_2977607, EPI_ISL_2977608, EPI_ISL_2977609, EPI_ISL_2977610, EPI_ISL_2977611, EPI_ISL_2977612, EPI_ISL_2977613, EPI_ISL_2977614, EPI_ISL_2977615, EPI_ISL_2977616, EPI_ISL_2977617, EPI_ISL_2977618, EPI_ISL_2977619, EPI_ISL_2977620, EPI_ISL_2977621, EPI_ISL_2977622, EPI_ISL_2977623, EPI_ISL_2977626, EPI_ISL_2977627, EPI_ISL_2977629, EPI_ISL_2977630, EPI_ISL_2977631, EPI_ISL_2977632, EPI_ISL_2977633, EPI_ISL_2977634, EPI_ISL_2977635, EPI_ISL_2977636, EPI_ISL_2977637, EPI_ISL_2977638, EPI_ISL_2977641, EPI_ISL_2977642, EPI_ISL_2977643, EPI_ISL_2977644, EPI_ISL_2977645, EPI_ISL_2977646, EPI_ISL_2977647, EPI_ISL_2977648, EPI_ISL_2977649, EPI_ISL_2977650, EPI_ISL_2977651, EPI_ISL_2977652, EPI_ISL_2977653, EPI_ISL_2977654, EPI_ISL_2977655, EPI_ISL_2977656, EPI_ISL_2977657, EPI_ISL_2977658, EPI_ISL_2977659, EPI_ISL_2977660, EPI_ISL_2977663, EPI_ISL_2977665, EPI_ISL_2977666, EPI_ISL_2977669, EPI_ISL_2977670, EPI_ISL_2977671, EPI_ISL_2977672, EPI_ISL_2977673, EPI_ISL_2977674, EPI_ISL_2977675, EPI_ISL_2977676, EPI_ISL_2977677, EPI_ISL_2977678, EPI_ISL_2977679, EPI_ISL_2977680, EPI_ISL_2977681, EPI_ISL_2977682, EPI_ISL_2977683, EPI_ISL_2977684, EPI_ISL_2977685, EPI_ISL_2977686, EPI_ISL_2977687, EPI_ISL_2977688, EPI_ISL_2977689, EPI_ISL_2977690, EPI_ISL_2977691, EPI_ISL_2977692, EPI_ISL_2977693, EPI_ISL_2977694, EPI_ISL_2977695, EPI_ISL_2977696, EPI_ISL_2977698, EPI_ISL_2977699, EPI_ISL_2977700, EPI_ISL_2977701, EPI_ISL_2977702, EPI_ISL_2977703, EPI_ISL_2977704, EPI_ISL_2977705, EPI_ISL_2977706, EPI_ISL_2977707, EPI_ISL_2977708, EPI_ISL_2977709, EPI_ISL_2977710, EPI_ISL_2977711, EPI_ISL_2977712, EPI_ISL_2977713, EPI_ISL_2977714, EPI_ISL_2977715, EPI_ISL_2977716, EPI_ISL_2977717, EPI_ISL_2977718, EPI_ISL_2977719, EPI_ISL_2977720, EPI_ISL_2977721, EPI_ISL_2977722, EPI_ISL_2977723, EPI_ISL_2977724, EPI_ISL_2977725, EPI_ISL_2977727, EPI_ISL_2977728, EPI_ISL_2977730, EPI_ISL_2977732, EPI_ISL_2977734, EPI_ISL_2977735, EPI_ISL_2977736, EPI_ISL_2977737, EPI_ISL_2977738, EPI_ISL_2977739, EPI_ISL_2977740, EPI_ISL_2977742, EPI_ISL_2977743, EPI_ISL_2977744, EPI_ISL_2977745, EPI_ISL_2977746, EPI_ISL_2977747, EPI_ISL_2977748, EPI_ISL_2977749, EPI_ISL_2977750, EPI_ISL_2977751, EPI_ISL_2977752, EPI_ISL_2977753, EPI_ISL_2977754, EPI_ISL_2977755, EPI_ISL_2977756, EPI_ISL_2977757, EPI_ISL_2977758, EPI_ISL_2977759, EPI_ISL_2977760, EPI_ISL_2977761, EPI_ISL_2977762, EPI_ISL_2977763, EPI_ISL_2977764, EPI_ISL_2977765, EPI_ISL_2977766, EPI_ISL_2977767, EPI_ISL_2977768, EPI_ISL_2977769, EPI_ISL_2977770, EPI_ISL_2977773, EPI_ISL_2977774, EPI_ISL_2977775, EPI_ISL_2977776, EPI_ISL_2977777, EPI_ISL_2977778, EPI_ISL_2977779, EPI_ISL_2977782, EPI_ISL_2977783, EPI_ISL_2977784, EPI_ISL_2977785, EPI_ISL_2977786, EPI_ISL_2977787, EPI_ISL_2977788, EPI_ISL_2977789, EPI_ISL_2977790, EPI_ISL_2977791, EPI_ISL_2977792, EPI_ISL_2977793, EPI_ISL_2977794, EPI_ISL_2977795, EPI_ISL_2977796, EPI_ISL_2977797, EPI_ISL_2977798, EPI_ISL_2977799, EPI_ISL_2977800, EPI_ISL_2977801, EPI_ISL_2977802, EPI_ISL_2977803, EPI_ISL_2977804, EPI_ISL_2977805, EPI_ISL_2977806, EPI_ISL_2977807, EPI_ISL_2977808, EPI_ISL_2977809, EPI_ISL_2977810, EPI_ISL_2977811, EPI_ISL_2977812, EPI_ISL_2977813, EPI_ISL_2977814, EPI_ISL_2977815, EPI_ISL_2977816, EPI_ISL_2977817, EPI_ISL_2977818, EPI_ISL_2977819, EPI_ISL_2977820, EPI_ISL_3063821, EPI_ISL_3063822, EPI_ISL_3063823, EPI_ISL_3063824, EPI_ISL_3063825, EPI_ISL_3063826, EPI_ISL_3063827, EPI_ISL_3063828, EPI_ISL_3063829, EPI_ISL_3063830, EPI_ISL_3063831, EPI_ISL_3063832, EPI_ISL_3063833, EPI_ISL_3063834, EPI_ISL_3063835, EPI_ISL_3063836, EPI_ISL_3063837, EPI_ISL_3063838, EPI_ISL_3063839, EPI_ISL_3063840, EPI_ISL_3063841, EPI_ISL_3063842, EPI_ISL_3063843, EPI_ISL_3063844, EPI_ISL_3063845, EPI_ISL_3063846, EPI_ISL_3063847, EPI_ISL_3063848, EPI_ISL_3063849, EPI_ISL_3063850, EPI_ISL_3063851, EPI_ISL_3063852, EPI_ISL_3063853, EPI_ISL_3063854, EPI_ISL_3063855, EPI_ISL_3063856, EPI_ISL_3063857, EPI_ISL_3063858, EPI_ISL_3063859, EPI_ISL_3063860, EPI_ISL_3063861, EPI_ISL_3063862, EPI_ISL_3063863, EPI_ISL_3063864, EPI_ISL_3063865, EPI_ISL_3063866, EPI_ISL_3063867, EPI_ISL_3063868, EPI_ISL_3063869, EPI_ISL_3063870, EPI_ISL_3063871, EPI_ISL_3063872, EPI_ISL_3063873, EPI_ISL_3063874, EPI_ISL_3063875, EPI_ISL_3063876, EPI_ISL_3063877, EPI_ISL_3063878, EPI_ISL_3063879, EPI_ISL_3063880, EPI_ISL_3063881, EPI_ISL_3063882, EPI_ISL_3063883, EPI_ISL_3063884, EPI_ISL_3063885, EPI_ISL_3063886, EPI_ISL_3063887, EPI_ISL_3063888, EPI_ISL_3063889, EPI_ISL_3063890, EPI_ISL_3063891, EPI_ISL_3063892, EPI_ISL_3063893, EPI_ISL_3063894, EPI_ISL_3063895, EPI_ISL_3063896, EPI_ISL_3063897, EPI_ISL_3063898, EPI_ISL_3063899, EPI_ISL_3063900, EPI_ISL_3063901, EPI_ISL_3063902, EPI_ISL_3063903, EPI_ISL_3063904, EPI_ISL_3063905, EPI_ISL_3063906, EPI_ISL_3063907, EPI_ISL_3063908, EPI_ISL_3063909, EPI_ISL_3063910, EPI_ISL_3063911, EPI_ISL_3063912, EPI_ISL_3063913, EPI_ISL_3063914, EPI_ISL_3063915, EPI_ISL_3063916, EPI_ISL_3063917, EPI_ISL_3063918, EPI_ISL_3063919, EPI_ISL_3063920, EPI_ISL_3063921, EPI_ISL_3063922, EPI_ISL_3063923, EPI_ISL_3063924, EPI_ISL_3063925, EPI_ISL_3063926, EPI_ISL_3063927, EPI_ISL_3063928, EPI_ISL_3063929, EPI_ISL_3063930, EPI_ISL_3063931, EPI_ISL_3063932, EPI_ISL_3063933, EPI_ISL_3063934, EPI_ISL_3063935, EPI_ISL_3063936, EPI_ISL_3063937, EPI_ISL_3063938, EPI_ISL_3063939, EPI_ISL_3063940, EPI_ISL_3063941, EPI_ISL_3063942, EPI_ISL_3063943, EPI_ISL_3063944, EPI_ISL_3063945, EPI_ISL_3063946, EPI_ISL_3063947, EPI_ISL_3063948, EPI_ISL_3063949, EPI_ISL_3063950, EPI_ISL_3063951, EPI_ISL_3063952, EPI_ISL_3063953, EPI_ISL_3063954, EPI_ISL_3063955, EPI_ISL_3063956, EPI_ISL_3063957, EPI_ISL_3063958, EPI_ISL_3063959, EPI_ISL_3063960, EPI_ISL_3063961, EPI_ISL_3063962, EPI_ISL_3063963, EPI_ISL_3063964, EPI_ISL_3063965, EPI_ISL_3063966, EPI_ISL_3063967, EPI_ISL_3063968, EPI_ISL_3063969, EPI_ISL_3063970, EPI_ISL_3063971, EPI_ISL_3063972, EPI_ISL_3063973, EPI_ISL_3063974, EPI_ISL_3063975, EPI_ISL_3063976, EPI_ISL_3063977, EPI_ISL_3063978, EPI_ISL_3063979, EPI_ISL_3063980, EPI_ISL_3063981, EPI_ISL_3063982, EPI_ISL_3063983, EPI_ISL_3063984, EPI_ISL_3063985, EPI_ISL_3063986, EPI_ISL_3063987, EPI_ISL_3063988, EPI_ISL_3063989, EPI_ISL_3063990, EPI_ISL_3063991, EPI_ISL_3063992, EPI_ISL_3063993, EPI_ISL_3063994, EPI_ISL_3063995, EPI_ISL_3063996, EPI_ISL_3063997, EPI_ISL_3063998, EPI_ISL_3063999, EPI_ISL_3064000, EPI_ISL_3064001, EPI_ISL_3064002, EPI_ISL_3064003, EPI_ISL_3064004, EPI_ISL_3064005, EPI_ISL_3064006, EPI_ISL_3064007, EPI_ISL_3064008, EPI_ISL_3064009, EPI_ISL_3064010, EPI_ISL_3064011, EPI_ISL_3064012, EPI_ISL_3064013, EPI_ISL_3064014, EPI_ISL_3064015, EPI_ISL_3064016, EPI_ISL_3064017, EPI_ISL_3064018, EPI_ISL_3064019, EPI_ISL_3064020, EPI_ISL_3064021, EPI_ISL_3064022, EPI_ISL_3064023, EPI_ISL_3064024, EPI_ISL_3064025, EPI_ISL_3064026, EPI_ISL_3064027, EPI_ISL_3064028, EPI_ISL_3064029, EPI_ISL_3064030, EPI_ISL_3064031, EPI_ISL_3064032, EPI_ISL_3064033, EPI_ISL_3064034, EPI_ISL_3064035, EPI_ISL_3064036, EPI_ISL_3064037, EPI_ISL_3064038, EPI_ISL_3064039, EPI_ISL_3064040, EPI_ISL_3064041, EPI_ISL_3064042, EPI_ISL_3064043, EPI_ISL_3064044, EPI_ISL_3064045, EPI_ISL_3064046, EPI_ISL_3064047, EPI_ISL_3064048, EPI_ISL_3064049, EPI_ISL_3064050, EPI_ISL_3064051, EPI_ISL_3064052, EPI_ISL_3064053, EPI_ISL_3064054, EPI_ISL_3064055, EPI_ISL_3064056, EPI_ISL_3064057, EPI_ISL_3064058, EPI_ISL_3064059, EPI_ISL_3064060, EPI_ISL_3064061, EPI_ISL_3064062, EPI_ISL_3064063, EPI_ISL_3064064, EPI_ISL_3064065, EPI_ISL_3064066, EPI_ISL_3064067, EPI_ISL_3064068, EPI_ISL_3064069, EPI_ISL_3064070, EPI_ISL_3064071, EPI_ISL_3064072, EPI_ISL_3064073, EPI_ISL_3064074, EPI_ISL_3064075, EPI_ISL_3064076, EPI_ISL_3064077, EPI_ISL_3064078, EPI_ISL_3064079, EPI_ISL_3064080, EPI_ISL_3064081, EPI_ISL_3064082, EPI_ISL_3064083, EPI_ISL_3064084, EPI_ISL_3064085, EPI_ISL_3064086, EPI_ISL_3064087, EPI_ISL_3064088, EPI_ISL_3064089, EPI_ISL_3064090, EPI_ISL_3064091, EPI_ISL_3064092, EPI_ISL_3064093, EPI_ISL_3064094, EPI_ISL_3064095, EPI_ISL_3064096, EPI_ISL_3064097, EPI_ISL_3064098, EPI_ISL_3064099, EPI_ISL_3064100, EPI_ISL_3064101, EPI_ISL_3064102, EPI_ISL_3064103, EPI_ISL_3064104, EPI_ISL_3064105, EPI_ISL_3064106, EPI_ISL_3064107, EPI_ISL_3064108, EPI_ISL_3064109, EPI_ISL_3064110, EPI_ISL_3064111, EPI_ISL_3064112, EPI_ISL_3064113, EPI_ISL_3064114, EPI_ISL_3064115, EPI_ISL_3064116, EPI_ISL_3064117, EPI_ISL_3064118, EPI_ISL_3064119, EPI_ISL_3064120, EPI_ISL_3064121, EPI_ISL_3064122, EPI_ISL_3064123, EPI_ISL_3064124, EPI_ISL_3064125, EPI_ISL_3064126, EPI_ISL_3064127, EPI_ISL_3067077, EPI_ISL_3067080, EPI_ISL_3067086, EPI_ISL_3067096, EPI_ISL_3067098, EPI_ISL_3067106, EPI_ISL_3067116, EPI_ISL_3067118, EPI_ISL_3067126, EPI_ISL_3067130, EPI_ISL_3067135, EPI_ISL_3067140, EPI_ISL_3067142, EPI_ISL_3067150, EPI_ISL_3067159, EPI_ISL_3067166, EPI_ISL_3067169, EPI_ISL_3067173, EPI_ISL_3067176, EPI_ISL_3067193, EPI_ISL_3067218, EPI_ISL_3067219, EPI_ISL_3067224, EPI_ISL_3067225, EPI_ISL_3067228, EPI_ISL_3067235, EPI_ISL_3067237, EPI_ISL_3067250, EPI_ISL_3067253, EPI_ISL_3067260, EPI_ISL_3067265, EPI_ISL_3067269, EPI_ISL_3067277, EPI_ISL_3067282, EPI_ISL_3067290, EPI_ISL_3067296, EPI_ISL_3067311, EPI_ISL_3067314, EPI_ISL_3067350, EPI_ISL_3067353, EPI_ISL_3067356 | | | |
| see above | CDPH VBL | California Department of Public Health | CDPH-COVIDNet; UCI Genome Sciences Center/GHTF; UCSD EXCITE lab; UCSF Center for Advanced Technology |
| EPI_ISL_2968768, EPI_ISL_2968769, EPI_ISL_2968770, EPI_ISL_2968771, EPI_ISL_2968772, EPI_ISL_2968773, EPI_ISL_2968774, EPI_ISL_2968775, EPI_ISL_2968776, EPI_ISL_2968777, EPI_ISL_2968778, EPI_ISL_2968779, EPI_ISL_2968780, EPI_ISL_2968781, EPI_ISL_2968782, EPI_ISL_2968783, EPI_ISL_2968784, EPI_ISL_2968785, EPI_ISL_2968786, EPI_ISL_2968787, EPI_ISL_2968788, EPI_ISL_2968789, EPI_ISL_2968790, EPI_ISL_2968791, EPI_ISL_2968792, EPI_ISL_2968793, EPI_ISL_2968794, EPI_ISL_2968795, EPI_ISL_2968796, EPI_ISL_2968799, EPI_ISL_2968800, EPI_ISL_2968801, EPI_ISL_2968802, EPI_ISL_2968804, EPI_ISL_2968805, EPI_ISL_2968806, EPI_ISL_2968807, EPI_ISL_2968808, EPI_ISL_2968809, EPI_ISL_2968813, EPI_ISL_2969031, EPI_ISL_2969032, EPI_ISL_2969033, EPI_ISL_2969034, EPI_ISL_2969035, EPI_ISL_2969036, EPI_ISL_2969037, EPI_ISL_2969038, EPI_ISL_2969039, EPI_ISL_2969040, EPI_ISL_2969041, EPI_ISL_2969042, EPI_ISL_2969043, EPI_ISL_2969044, EPI_ISL_2969045, EPI_ISL_2969046, EPI_ISL_2969047, EPI_ISL_2969048, EPI_ISL_2969049, EPI_ISL_2969050, EPI_ISL_2969051, EPI_ISL_2969052, EPI_ISL_2969053, EPI_ISL_2969054, EPI_ISL_2969055, EPI_ISL_2969056, EPI_ISL_2969057, EPI_ISL_2969058, EPI_ISL_2969059, EPI_ISL_2969060, EPI_ISL_2969061, EPI_ISL_2969062, EPI_ISL_2969063, EPI_ISL_2969064, EPI_ISL_2969065, EPI_ISL_2969066, EPI_ISL_2969067, EPI_ISL_2969068, EPI_ISL_2969069, EPI_ISL_2969070, EPI_ISL_2969071, EPI_ISL_2969072, EPI_ISL_2969073, EPI_ISL_2969074, EPI_ISL_2969075, EPI_ISL_2969076, EPI_ISL_2969077, EPI_ISL_2969078, EPI_ISL_2969079, EPI_ISL_2969080, EPI_ISL_2969081, EPI_ISL_2969082, EPI_ISL_2969083, EPI_ISL_2969084, EPI_ISL_2969085, EPI_ISL_2969086, EPI_ISL_2969087, EPI_ISL_2969088, EPI_ISL_2969089, EPI_ISL_2969090, EPI_ISL_2969091, EPI_ISL_2969092, EPI_ISL_2969093, EPI_ISL_2969094, EPI_ISL_2969095, EPI_ISL_2969096, EPI_ISL_2969097, EPI_ISL_2969098, EPI_ISL_2969099, EPI_ISL_2969100, EPI_ISL_2969101, EPI_ISL_2969102, EPI_ISL_2969103, EPI_ISL_2969104, EPI_ISL_2969105, EPI_ISL_2969106, EPI_ISL_2969107, EPI_ISL_2969108, EPI_ISL_2969109, EPI_ISL_2969110, EPI_ISL_2969111, EPI_ISL_2969112, EPI_ISL_2969113, EPI_ISL_2969114, EPI_ISL_2969115, EPI_ISL_2969116, EPI_ISL_2969117, EPI_ISL_2969118, EPI_ISL_2969119, EPI_ISL_2969120, EPI_ISL_2969121, EPI_ISL_2969122, EPI_ISL_2969123, EPI_ISL_2969124, EPI_ISL_2969125, EPI_ISL_2969126, EPI_ISL_2969127, EPI_ISL_2969128, EPI_ISL_2969129, EPI_ISL_2969130, EPI_ISL_2969131, EPI_ISL_2969132, EPI_ISL_2969133, EPI_ISL_2969134, EPI_ISL_2969135, EPI_ISL_2969136, EPI_ISL_2969137, EPI_ISL_2969138, EPI_ISL_2969139, EPI_ISL_2969140, EPI_ISL_2969141, EPI_ISL_2969142, EPI_ISL_2969143, EPI_ISL_2969144, EPI_ISL_2969145, EPI_ISL_2969146, EPI_ISL_2969147, EPI_ISL_2969148, EPI_ISL_2969149, EPI_ISL_2969150, EPI_ISL_2969151, EPI_ISL_2969152, EPI_ISL_2969153, EPI_ISL_2969154, EPI_ISL_2969155, EPI_ISL_2969156, EPI_ISL_2969157, EPI_ISL_2969158, EPI_ISL_2969159, EPI_ISL_2969160, EPI_ISL_2969161, EPI_ISL_2969162, EPI_ISL_2969163, EPI_ISL_2969164, EPI_ISL_2969165, EPI_ISL_2969166, EPI_ISL_2969167, EPI_ISL_2969168, EPI_ISL_2969169, EPI_ISL_2969170, EPI_ISL_2969171, EPI_ISL_2969172, EPI_ISL_2969173, EPI_ISL_2969174, EPI_ISL_2969175, EPI_ISL_2969176, EPI_ISL_2969177, EPI_ISL_2969178, EPI_ISL_2969179, EPI_ISL_2969180, EPI_ISL_2969181, EPI_ISL_2969182, EPI_ISL_2969183, EPI_ISL_2969184, EPI_ISL_2969185, EPI_ISL_2969186, EPI_ISL_2969187, EPI_ISL_2969188, EPI_ISL_2969189, EPI_ISL_2969190, EPI_ISL_2969191, EPI_ISL_2969192, EPI_ISL_2969193, EPI_ISL_2969194, EPI_ISL_2969195, EPI_ISL_2969196, EPI_ISL_2969197, EPI_ISL_2969198, EPI_ISL_2969199, EPI_ISL_2969200, EPI_ISL_2969201, EPI_ISL_2969202, EPI_ISL_2969203, EPI_ISL_2969204, EPI_ISL_2969205, EPI_ISL_2969206, EPI_ISL_2969207, EPI_ISL_2969208, EPI_ISL_2969209, EPI_ISL_2969210, EPI_ISL_2969211, EPI_ISL_2969212, EPI_ISL_2969213, EPI_ISL_2969214, EPI_ISL_2969215, EPI_ISL_2969216, EPI_ISL_2969217, EPI_ISL_2969218, EPI_ISL_2969219, EPI_ISL_2969220, EPI_ISL_2969221, EPI_ISL_2969222, EPI_ISL_2969223, EPI_ISL_2969224, EPI_ISL_2969225, EPI_ISL_2969226, EPI_ISL_2969227, EPI_ISL_2969228, EPI_ISL_2969229, EPI_ISL_2969230, EPI_ISL_2969231, EPI_ISL_2969232, EPI_ISL_2969233, EPI_ISL_2969234, EPI_ISL_2969235, EPI_ISL_2969236, EPI_ISL_2969237, EPI_ISL_2969238, EPI_ISL_2969239, EPI_ISL_2969240, EPI_ISL_2969241, EPI_ISL_2969242, EPI_ISL_2969243, EPI_ISL_2969244, EPI_ISL_2969245, EPI_ISL_2969246, EPI_ISL_2969247, EPI_ISL_2969248, EPI_ISL_2969249, EPI_ISL_2969250, EPI_ISL_2969251, EPI_ISL_2969252, EPI_ISL_2969253, EPI_ISL_2969254, EPI_ISL_2969255, EPI_ISL_2969256, EPI_ISL_2969257, EPI_ISL_2969258, EPI_ISL_2969259, EPI_ISL_2969260, EPI_ISL_2969261, EPI_ISL_2969262, EPI_ISL_2969263, EPI_ISL_2969264, EPI_ISL_2969265, EPI_ISL_2969266, EPI_ISL_2969267, EPI_ISL_2969268, EPI_ISL_2969269, EPI_ISL_2969270, EPI_ISL_2969271, EPI_ISL_2969272, EPI_ISL_2969273, EPI_ISL_2969274, EPI_ISL_2969275, EPI_ISL_2969276, EPI_ISL_2969277, EPI_ISL_2969278, EPI_ISL_2969279, EPI_ISL_2969280, EPI_ISL_2969281, EPI_ISL_2969282, EPI_ISL_2969283, EPI_ISL_2969284, EPI_ISL_2969285, EPI_ISL_2969286, EPI_ISL_2969287, EPI_ISL_2969288, EPI_ISL_2969289, EPI_ISL_2969290, EPI_ISL_2969291, EPI_ISL_2969292, EPI_ISL_2969293, EPI_ISL_2969294, EPI_ISL_2969295, EPI_ISL_2969296, EPI_ISL_2969297, EPI_ISL_2969298, EPI_ISL_2969299, EPI_ISL_2969300, EPI_ISL_2969301, EPI_ISL_2969302, EPI_ISL_2969303, EPI_ISL_2969304, EPI_ISL_2969305, EPI_ISL_2969306, EPI_ISL_2969307, EPI_ISL_2969308, EPI_ISL_2969309, EPI_ISL_2969310, EPI_ISL_2969311, EPI_ISL_2969312, EPI_ISL_2969313, EPI_ISL_2969314, EPI_ISL_2969315, EPI_ISL_2969316, EPI_ISL_2969317, EPI_ISL_2969318, EPI_ISL_2969319, EPI_ISL_2969320, EPI_ISL_2969321, EPI_ISL_2969322, EPI_ISL_2969323, EPI_ISL_2969324, EPI_ISL_2969325, EPI_ISL_2969326, EPI_ISL_2969327, EPI_ISL_2969328, EPI_ISL_2969329, EPI_ISL_2969330, EPI_ISL_2969331, EPI_ISL_2969332, EPI_ISL_2969333, EPI_ISL_2969334, EPI_ISL_2969335, EPI_ISL_2969336, EPI_ISL_2969337, EPI_ISL_2969338, EPI_ISL_2969339, EPI_ISL_2969340, EPI_ISL_2969341, EPI_ISL_2969342, EPI_ISL_2969343, EPI_ISL_2969344, EPI_ISL_2969345, EPI_ISL_2969346, EPI_ISL_2969347, EPI_ISL_2969348, EPI_ISL_2969349, EPI_ISL_2969350, EPI_ISL_2969351, EPI_ISL_2969352, EPI_ISL_2969353, EPI_ISL_2969354, EPI_ISL_2969355, EPI_ISL_2969356, EPI_ISL_2969357, EPI_ISL_2969358, EPI_ISL_2969359, EPI_ISL_2969360, EPI_ISL_2969361, EPI_ISL_2969362, EPI_ISL_2969363, EPI_ISL_2969364, EPI_ISL_2969365, EPI_ISL_2969366, EPI_ISL_2969367, EPI_ISL_2969368, EPI_ISL_2969369, EPI_ISL_2969370, EPI_ISL_2969371, EPI_ISL_2969372, EPI_ISL_2969373, EPI_ISL_2969374, EPI_ISL_2969375, EPI_ISL_2969376, EPI_ISL_2969377, EPI_ISL_2969378, EPI_ISL_2969379, EPI_ISL_2969380, EPI_ISL_2969381, EPI_ISL_2969382, EPI_ISL_2969383, EPI_ISL_2969384, EPI_ISL_2969385, EPI_ISL_2969386, EPI_ISL_2969387, EPI_ISL_2969388, EPI_ISL_2969389, EPI_ISL_2969390, EPI_ISL_2969391, EPI_ISL_2969392, EPI_ISL_2969393, EPI_ISL_2969394, EPI_ISL_2969395, EPI_ISL_2969396, EPI_ISL_2969397, EPI_ISL_2969398, EPI_ISL_2969399, EPI_ISL_2969400, EPI_ISL_2969401, EPI_ISL_2969402, EPI_ISL_2969403, EPI_ISL_2969404, EPI_ISL_2969405, EPI_ISL_2969406, EPI_ISL_2969407, EPI_ISL_2969408, EPI_ISL_2969409, EPI_ISL_2969410, EPI_ISL_2969411, EPI_ISL_2969412, EPI_ISL_2969413, EPI_ISL_2969414, EPI_ISL_2969415, EPI_ISL_2969416, EPI_ISL_2969417, EPI_ISL_2969418, EPI_ISL_2969419, EPI_ISL_2969420, EPI_ISL_2969421, EPI_ISL_2969422, EPI_ISL_2969423, EPI_ISL_2969424, EPI_ISL_2969425, EPI_ISL_2969426, EPI_ISL_2969427, EPI_ISL_2969428, EPI_ISL_2969429, EPI_ISL_2969430, EPI_ISL_2969431, EPI_ISL_2969432, EPI_ISL_2969433, EPI_ISL_2969434, EPI_ISL_2969435, EPI_ISL_2969436, EPI_ISL_2969437, EPI_ISL_2969438, EPI_ISL_2969439, EPI_ISL_2969440, EPI_ISL_2969441, EPI_ISL_2969442, EPI_ISL_2969443, EPI_ISL_2969444, EPI_ISL_2969445, EPI_ISL_2969446, EPI_ISL_2969447, EPI_ISL_2969448, EPI_ISL_2969449, EPI_ISL_2969450, EPI_ISL_2969451, EPI_ISL_2969452, EPI_ISL_2969453, EPI_ISL_2969454, EPI_ISL_2969455, EPI_ISL_2969456, EPI_ISL_2969457, EPI_ISL_2969458, EPI_ISL_2969459, EPI_ISL_2969460, EPI_ISL_2969461, EPI_ISL_2969462, EPI_ISL_2969463, EPI_ISL_2969464, EPI_ISL_2969465, EPI_ISL_2969466, EPI_ISL_2969467, EPI_ISL_2969468, EPI_ISL_2969469, EPI_ISL_2969470, EPI_ISL_2969471, EPI_ISL_2969472, EPI_ISL_2969473, EPI_ISL_2969474, EPI_ISL_2969475, EPI_ISL_2969476, EPI_ISL_2969477, EPI_ISL_2969478, EPI_ISL_2969479, EPI_ISL_2969480, EPI_ISL_2969481, EPI_ISL_2969482, EPI_ISL_2969483, EPI_ISL_2969484, EPI_ISL_2969485, EPI_ISL_2969486, EPI_ISL_2969487, EPI_ISL_2969488, EPI_ISL_2969489, EPI_ISL_2969490, EPI_ISL_2969491, EPI_ISL_2969492, EPI_ISL_2969493, EPI_ISL_2969494, EPI_ISL_2969495, EPI_ISL_2969496, EPI_ISL_2969497, EPI_ISL_2969498, EPI_ISL_2969499, EPI_ISL_2969500, EPI_ISL_2969501, EPI_ISL_2969502, EPI_ISL_2969503, EPI_ISL_2969504, EPI_ISL_2969505, EPI_ISL_2969506, EPI_ISL_2969507, EPI_ISL_2969508, EPI_ISL_2969509, EPI_ISL_2969510, EPI_ISL_2969511, EPI_ISL_2969512, EPI_ISL_2969513, EPI_ISL_2969514, EPI_ISL_2969515, EPI_ISL_2969516, EPI_ISL_2969517, EPI_ISL_2969518, EPI_ISL_2969519 | | | |
| see above | CDPH Valencia Branch Laboratory | California Department of Public Health | CDPH-COVIDNet; Priva Path Labs Inc. |
| EPI_ISL_1798774, EPI_ISL_1798785, EPI_ISL_1798795 | CDPH, Viral and Rickettsial Disease Laboratory | Centers for Disease Control and Prevention Division of Viral Diseases, Pathogen Discovery | Alison Laufer Halpin; Ben L. Rambo-Martin; Clinton R. Paden; Dakota Howard; Darlene Wagner; Dave Wentworth; Dhwani Batra; Jasmine Padilla; Justin Lee; Katie Dillon; Krista Queen; Kristen Knipe; Kristine Lacek; Mark Burroughs; Matthew Schmerer; Mili Sheth; Peter Cook; Sam Shepard; Sarah Nobles; Shoshona Le; Suxiang Tong; Vivien Dugan; Yvette Unoarumhi |
| EPI_ISL_1094224, EPI_ISL_1094583, EPI_ISL_1094584, EPI_ISL_1094586, EPI_ISL_1094587, EPI_ISL_1094589, EPI_ISL_1095191, EPI_ISL_1095193, EPI_ISL_1095194, EPI_ISL_1095195, EPI_ISL_1095198, EPI_ISL_1095199 | | | |
| see above | CDPH, Viral and Rickettsial Disease Laboratory | Respiratory Viruses Branch, Division of Viral Diseases, Centers for Disease Control and Prevention | Anna Montmayeur; Anna Uehara; Ben L. Rambo-Martin; Clinton R. Paden; Dhwani Batra; Haibin Wang; Jasmine Padilla; Jing Zhang; Justin Lee; Krista Queen; Lori Rowe; Mark Burroughs; Mili Sheth; Peter W. Cook; Rachel Marine; Sarah Nobles; Suxiang Tong; Yan Li; Ying Tao |
| EPI_ISL_2233932, EPI_ISL_2233933, EPI_ISL_2233934, EPI_ISL_2233937, EPI_ISL_2233983, EPI_ISL_2233984, EPI_ISL_2233985, EPI_ISL_2233987, EPI_ISL_2233988, EPI_ISL_2233989, EPI_ISL_2233990, EPI_ISL_2233991, EPI_ISL_2233993, EPI_ISL_2233995, EPI_ISL_2234021, EPI_ISL_2234022, EPI_ISL_2234023, EPI_ISL_2234029, EPI_ISL_2234030, EPI_ISL_2234035, EPI_ISL_2234040, EPI_ISL_2234041, EPI_ISL_2234042, EPI_ISL_2234043, EPI_ISL_2234044, EPI_ISL_2234045, EPI_ISL_2234046, EPI_ISL_2234047, EPI_ISL_2234048, EPI_ISL_2234066, EPI_ISL_2234068, EPI_ISL_2234081, EPI_ISL_2234082, EPI_ISL_2234083, EPI_ISL_2234089, EPI_ISL_2234158, EPI_ISL_2234159, EPI_ISL_2234160, EPI_ISL_2234161, EPI_ISL_2234162, EPI_ISL_2234171, EPI_ISL_2234172, EPI_ISL_2234173, EPI_ISL_2234174, EPI_ISL_2234175, EPI_ISL_2234176, EPI_ISL_2234177, EPI_ISL_2234178, EPI_ISL_2234179, EPI_ISL_2234180, EPI_ISL_2234181, EPI_ISL_2234182, EPI_ISL_2234183, EPI_ISL_2234184, EPI_ISL_2234264, EPI_ISL_2234321, EPI_ISL_2892541, EPI_ISL_2892542, EPI_ISL_2892543, EPI_ISL_2892545, EPI_ISL_2892546 | | | |
| see above | California Department of Public Health | California Department of Public Health | CDPH IDLB COVIDNet et al |
| EPI_ISL_847564, EPI_ISL_847568, EPI_ISL_847589, EPI_ISL_847590, EPI_ISL_847591, EPI_ISL_847592, EPI_ISL_847593, EPI_ISL_847594, EPI_ISL_847758, EPI_ISL_847759, EPI_ISL_984332, EPI_ISL_984347, EPI_ISL_984354, EPI_ISL_984358, EPI_ISL_984360, EPI_ISL_984364, EPI_ISL_984371, EPI_ISL_984372, EPI_ISL_984373, EPI_ISL_984374, EPI_ISL_984379, EPI_ISL_984381, EPI_ISL_984382, EPI_ISL_984383, EPI_ISL_984387, EPI_ISL_984388, EPI_ISL_984392, EPI_ISL_984419, EPI_ISL_984436, EPI_ISL_984438, EPI_ISL_984445, EPI_ISL_984446, EPI_ISL_984447, EPI_ISL_984448, EPI_ISL_984449, EPI_ISL_984450, EPI_ISL_984451, EPI_ISL_984452, EPI_ISL_984453, EPI_ISL_984454, EPI_ISL_984455, EPI_ISL_984456, EPI_ISL_984457, EPI_ISL_984458, EPI_ISL_984459, EPI_ISL_984460, EPI_ISL_984461, EPI_ISL_984462, EPI_ISL_984463, EPI_ISL_984464, EPI_ISL_984465, EPI_ISL_984466, EPI_ISL_984467, EPI_ISL_984468, EPI_ISL_984469, EPI_ISL_984470, EPI_ISL_984471, EPI_ISL_984472, EPI_ISL_984473, EPI_ISL_984474, EPI_ISL_984475, EPI_ISL_984476, EPI_ISL_984477, EPI_ISL_984478, EPI_ISL_984479, EPI_ISL_984480, EPI_ISL_984481, EPI_ISL_984482, EPI_ISL_984483, EPI_ISL_984484, EPI_ISL_984485, EPI_ISL_984486, EPI_ISL_984487, EPI_ISL_984488, EPI_ISL_984489 | | | |
| see above | California Department of Public Health | Chiu Laboratory, University of California, San Francisco | Brian Bushnell; Candace Wang; Charles Chiu; Debra Wadford; Jill Hacker; Scot Federman; Venice Servellita; Xianding (Wayne) Deng |
| EPI_ISL_2776221, EPI_ISL_2776222, EPI_ISL_2776223, EPI_ISL_2776224, EPI_ISL_2776225, EPI_ISL_2776226, EPI_ISL_2776227, EPI_ISL_2776228, EPI_ISL_2776229, EPI_ISL_2776230, EPI_ISL_2776231, EPI_ISL_2776232, EPI_ISL_2776233, EPI_ISL_2776234, EPI_ISL_2776235, EPI_ISL_2776236, EPI_ISL_2776237, EPI_ISL_2776238, EPI_ISL_2776239, EPI_ISL_2776240, EPI_ISL_2776241, EPI_ISL_2776242, EPI_ISL_2776243, EPI_ISL_2776244, EPI_ISL_2776245, EPI_ISL_2776246, EPI_ISL_2776247, EPI_ISL_2776248, EPI_ISL_2776249, EPI_ISL_2776250, EPI_ISL_2776251, EPI_ISL_2776252, EPI_ISL_2776253, EPI_ISL_2776254, EPI_ISL_2776255, EPI_ISL_2776256, EPI_ISL_2776257, EPI_ISL_2776258, EPI_ISL_2776259, EPI_ISL_2776260, EPI_ISL_2776261, EPI_ISL_2776262, EPI_ISL_2776263, EPI_ISL_2776264, EPI_ISL_2776265, EPI_ISL_2776266, EPI_ISL_2776267, EPI_ISL_2776268, EPI_ISL_2776269, EPI_ISL_2776270, EPI_ISL_2776271, EPI_ISL_2776272, EPI_ISL_2776273, EPI_ISL_2776274, EPI_ISL_2776275, EPI_ISL_2776276, EPI_ISL_2776277, EPI_ISL_2776278, EPI_ISL_2776279, EPI_ISL_2776280, EPI_ISL_2776281, EPI_ISL_2776282, EPI_ISL_2776283, EPI_ISL_2776284, EPI_ISL_2776285, EPI_ISL_2776286, EPI_ISL_2776287, EPI_ISL_2776288, EPI_ISL_2776289, EPI_ISL_2776290, EPI_ISL_2776291, EPI_ISL_2776292, EPI_ISL_2776293, EPI_ISL_2776294, EPI_ISL_2776295, EPI_ISL_2776296, EPI_ISL_2776297, EPI_ISL_2776298, EPI_ISL_2776299, EPI_ISL_2776300, EPI_ISL_2776301, EPI_ISL_2776302, EPI_ISL_2776303, EPI_ISL_2776304, EPI_ISL_2776305, EPI_ISL_2776306, EPI_ISL_2776307, EPI_ISL_2776308, EPI_ISL_2776309, EPI_ISL_2776310, EPI_ISL_2776311, EPI_ISL_2776312, EPI_ISL_2776313, EPI_ISL_2776314, EPI_ISL_2776315, EPI_ISL_2776316, EPI_ISL_2776317, EPI_ISL_2776318, EPI_ISL_2776319, EPI_ISL_2776320, EPI_ISL_2776321, EPI_ISL_2776322, EPI_ISL_2776323, EPI_ISL_2776324, EPI_ISL_2776325, EPI_ISL_2776326, EPI_ISL_2776327, EPI_ISL_2776328, EPI_ISL_2776329, EPI_ISL_2776330, EPI_ISL_2776331, EPI_ISL_2776332, EPI_ISL_2776333, EPI_ISL_2776334, EPI_ISL_2776335, EPI_ISL_2776336, EPI_ISL_2776337, EPI_ISL_2776338, EPI_ISL_2776339, EPI_ISL_2776340, EPI_ISL_2776341, EPI_ISL_2776342, EPI_ISL_2776343, EPI_ISL_2776344, EPI_ISL_2776345, EPI_ISL_2776346, EPI_ISL_2776347, EPI_ISL_2776348, EPI_ISL_2776349, EPI_ISL_2776350, EPI_ISL_2776351, EPI_ISL_2776352, EPI_ISL_2776353, EPI_ISL_2776354, EPI_ISL_2776355, EPI_ISL_2776356, EPI_ISL_2776357, EPI_ISL_2776358, EPI_ISL_2776359, EPI_ISL_2776360, EPI_ISL_2776361, EPI_ISL_2776362, EPI_ISL_2776363, EPI_ISL_2776364, EPI_ISL_2776365, EPI_ISL_2776366, EPI_ISL_2776367, EPI_ISL_2776368, EPI_ISL_2778106, EPI_ISL_2778107, EPI_ISL_2778115 | | | |
| see above | California Department | California Department of Public | Arias Lab at UCSB; CDPH-COVIDNet; UCLA Technology Center for Genomics & Bioinformatics |

| | | | |
|---|---|---|---|
| | of Public Health Valencia Branch Laboratory (CDPH VBL) | Health | |
| EPI_ISL_1234867, EPI_ISL_1234978, EPI_ISL_1235055, EPI_ISL_1701456 | California Institute of Technology | Chan-Zuckerberg Biohub | CZB Cliahub Consortium |
| EPI_ISL_847730, EPI_ISL_847743, EPI_ISL_977918, EPI_ISL_977919, EPI_ISL_977920, EPI_ISL_977921, EPI_ISL_977922, EPI_ISL_977923, EPI_ISL_977924, EPI_ISL_977925, EPI_ISL_977926, EPI_ISL_977927, EPI_ISL_977928, EPI_ISL_977929, EPI_ISL_977930, EPI_ISL_977931, EPI_ISL_977932, EPI_ISL_977933, EPI_ISL_977934, EPI_ISL_977935, EPI_ISL_977936, EPI_ISL_977937, EPI_ISL_977938, EPI_ISL_977939, EPI_ISL_977940, EPI_ISL_977941, EPI_ISL_977942, EPI_ISL_977943, EPI_ISL_977944, EPI_ISL_977945, EPI_ISL_977946, EPI_ISL_977947, EPI_ISL_977948, EPI_ISL_977949, EPI_ISL_977950, EPI_ISL_977951, EPI_ISL_977952, EPI_ISL_977953, EPI_ISL_977954, EPI_ISL_977955, EPI_ISL_977956, EPI_ISL_977957, EPI_ISL_977958, EPI_ISL_977959, EPI_ISL_977960, EPI_ISL_977961, EPI_ISL_977962, EPI_ISL_977963, EPI_ISL_977964, EPI_ISL_977965, EPI_ISL_977966, EPI_ISL_977967, EPI_ISL_977968, EPI_ISL_977969, EPI_ISL_977970, EPI_ISL_977971, EPI_ISL_977972, EPI_ISL_977973, EPI_ISL_977974, EPI_ISL_977975, EPI_ISL_977976, EPI_ISL_977977, EPI_ISL_977978, EPI_ISL_977979, EPI_ISL_977980, EPI_ISL_978063, EPI_ISL_978064, EPI_ISL_978065, EPI_ISL_978066, EPI_ISL_978067, EPI_ISL_978068, EPI_ISL_978069, EPI_ISL_978075, EPI_ISL_978076, EPI_ISL_978077, EPI_ISL_978078, EPI_ISL_978079, EPI_ISL_978080, EPI_ISL_978081, EPI_ISL_978082, EPI_ISL_978083, EPI_ISL_978084, EPI_ISL_978085, EPI_ISL_978086, EPI_ISL_978087, EPI_ISL_978088, EPI_ISL_978089, EPI_ISL_978090, EPI_ISL_978091, EPI_ISL_978092, EPI_ISL_978093, EPI_ISL_978094, EPI_ISL_978095, EPI_ISL_978096, EPI_ISL_978097, EPI_ISL_978098, EPI_ISL_978099, EPI_ISL_978100, EPI_ISL_978101, EPI_ISL_978102, EPI_ISL_978103, EPI_ISL_978104, EPI_ISL_978105, EPI_ISL_978106, EPI_ISL_978107, EPI_ISL_978108, EPI_ISL_978109, EPI_ISL_978110, EPI_ISL_978111, EPI_ISL_978112, EPI_ISL_978113, EPI_ISL_978114, EPI_ISL_978115, EPI_ISL_978116, EPI_ISL_978117, EPI_ISL_978118, EPI_ISL_978124, EPI_ISL_978125, EPI_ISL_978126, EPI_ISL_978127, EPI_ISL_978128, EPI_ISL_978132, EPI_ISL_978133, EPI_ISL_978134, EPI_ISL_978135, EPI_ISL_978136, EPI_ISL_978137, EPI_ISL_978138, EPI_ISL_978139, EPI_ISL_978140, EPI_ISL_978141, EPI_ISL_978142, EPI_ISL_978143, EPI_ISL_978144, EPI_ISL_978145, EPI_ISL_978146, EPI_ISL_978147, EPI_ISL_978148, EPI_ISL_978149, EPI_ISL_978150, EPI_ISL_978151, EPI_ISL_978152, EPI_ISL_978153, EPI_ISL_978154, EPI_ISL_978155, EPI_ISL_978156, EPI_ISL_978157, EPI_ISL_978158, EPI_ISL_978159, EPI_ISL_978961, EPI_ISL_978962, EPI_ISL_978963, EPI_ISL_978964, EPI_ISL_978965, EPI_ISL_978966, EPI_ISL_978973 | | | |
| see above | Chiu Laboratory, University of California, San Francisco | Chiu Laboratory, University of California, San Francisco | Brian Bushnell; Candace Wang; Charles Chiu; Debra Wadford; Jill Hacker; Scot Federman; Venice Servellita; Xianding (Wayne) Deng |
| EPI_ISL_833413 | City of Hope Comprehensive Cancer Center | Los Angeles County Public Health Laboratories | P. Hemarajata et al. |
| EPI_ISL_1030269, EPI_ISL_1030282, EPI_ISL_1030302, EPI_ISL_1030303, EPI_ISL_1030306, EPI_ISL_1030307, EPI_ISL_1030309, EPI_ISL_1030311, EPI_ISL_1030329, EPI_ISL_1030331, EPI_ISL_1030332, EPI_ISL_1030333, EPI_ISL_1030334, EPI_ISL_1030335, EPI_ISL_1030336, EPI_ISL_1030337, EPI_ISL_1030338, EPI_ISL_1030339, EPI_ISL_1030340, EPI_ISL_1030341, EPI_ISL_1030342, EPI_ISL_1030343, EPI_ISL_1030344, EPI_ISL_1030345, EPI_ISL_1030346, EPI_ISL_1030347, EPI_ISL_1030348, EPI_ISL_1030349, EPI_ISL_1030350, EPI_ISL_1030351, EPI_ISL_1030352, EPI_ISL_1030353, EPI_ISL_1030354, EPI_ISL_1030355, EPI_ISL_1030356, EPI_ISL_1030357, EPI_ISL_1030358, EPI_ISL_1030359, EPI_ISL_1030360, EPI_ISL_1030361, EPI_ISL_1030362, EPI_ISL_1030363, EPI_ISL_1030364, EPI_ISL_1030365, EPI_ISL_1030366, EPI_ISL_1030367, EPI_ISL_1030368, EPI_ISL_1030369, EPI_ISL_1030370, EPI_ISL_1030371, EPI_ISL_1030372, EPI_ISL_1030373, EPI_ISL_1030374, EPI_ISL_1030375, EPI_ISL_1030376, EPI_ISL_1030377, EPI_ISL_1030378, EPI_ISL_1030379, EPI_ISL_1030380, EPI_ISL_1030381, EPI_ISL_1030382, EPI_ISL_1030383, EPI_ISL_1030384, EPI_ISL_1030385, EPI_ISL_1030386, EPI_ISL_1030387, EPI_ISL_1030388, EPI_ISL_1030389, EPI_ISL_1030390, EPI_ISL_1234538, EPI_ISL_1234540, EPI_ISL_1234546, EPI_ISL_1234552, EPI_ISL_1234553, EPI_ISL_1234555, EPI_ISL_1234556, EPI_ISL_1234558, EPI_ISL_1234559, EPI_ISL_1234563, EPI_ISL_1234569, EPI_ISL_1234570, EPI_ISL_1234572, EPI_ISL_1234573, EPI_ISL_1234577, EPI_ISL_1234579, EPI_ISL_1234580, EPI_ISL_1234583, EPI_ISL_1234585, EPI_ISL_1234587, EPI_ISL_1234590, EPI_ISL_1234596, EPI_ISL_1234598, EPI_ISL_1234602, EPI_ISL_1234607, EPI_ISL_1234610, EPI_ISL_1234614, EPI_ISL_1234615, EPI_ISL_1234616, EPI_ISL_1234618, EPI_ISL_1234622, EPI_ISL_1234626, EPI_ISL_1234628, EPI_ISL_1234629, EPI_ISL_1234630, EPI_ISL_1234631, EPI_ISL_1234632, EPI_ISL_1234636, EPI_ISL_1234642, EPI_ISL_1234643, EPI_ISL_1234647, EPI_ISL_1234654, EPI_ISL_1234655, EPI_ISL_1234659, EPI_ISL_1234662, EPI_ISL_1234664, EPI_ISL_1234674, EPI_ISL_1234675, EPI_ISL_1234680, EPI_ISL_1234682, EPI_ISL_1234684, EPI_ISL_1234685, EPI_ISL_1234686, EPI_ISL_1234688, EPI_ISL_1234690, EPI_ISL_1234691, EPI_ISL_1234692, EPI_ISL_1234693, EPI_ISL_1234696, EPI_ISL_1234697, EPI_ISL_1234702, EPI_ISL_1234705, EPI_ISL_1234707, EPI_ISL_1234708, EPI_ISL_1234709, EPI_ISL_1234710, EPI_ISL_1234726, EPI_ISL_1234727, EPI_ISL_1234736, EPI_ISL_1234738, EPI_ISL_1234741, EPI_ISL_1234745, EPI_ISL_1234746, EPI_ISL_1234749, EPI_ISL_1234756, EPI_ISL_1234757, EPI_ISL_1234763, EPI_ISL_1234764, EPI_ISL_1234768, EPI_ISL_1234770, EPI_ISL_1234772, EPI_ISL_1234775, EPI_ISL_1234777, EPI_ISL_1234783, EPI_ISL_1235080, EPI_ISL_1235088, EPI_ISL_1235106, EPI_ISL_1235146, EPI_ISL_1235157, EPI_ISL_1235159, EPI_ISL_1235184, EPI_ISL_1235209, EPI_ISL_1235221, EPI_ISL_1235240, EPI_ISL_1235278, EPI_ISL_1235321, EPI_ISL_1235330, EPI_ISL_1235334, EPI_ISL_1477746, EPI_ISL_1477760, EPI_ISL_1477794, EPI_ISL_1477821, EPI_ISL_1477833, EPI_ISL_1477867, EPI_ISL_1477950, EPI_ISL_1477952, EPI_ISL_1477953, EPI_ISL_1477960, EPI_ISL_1477972, EPI_ISL_1477977, EPI_ISL_1477990, EPI_ISL_1477993, EPI_ISL_1478002, EPI_ISL_1478003, EPI_ISL_1478008, EPI_ISL_1478011, EPI_ISL_1478017, EPI_ISL_1478022, EPI_ISL_1478032, EPI_ISL_1478034, EPI_ISL_1478035, EPI_ISL_1478036, EPI_ISL_1478037, EPI_ISL_1478039, EPI_ISL_1478040, EPI_ISL_1478043, EPI_ISL_1478053, EPI_ISL_1478057, EPI_ISL_1478072, EPI_ISL_1478078, EPI_ISL_1478088, EPI_ISL_1478099, EPI_ISL_1478108, EPI_ISL_1478111, EPI_ISL_1478122, EPI_ISL_1478125, EPI_ISL_1478126, EPI_ISL_1478131, EPI_ISL_1478147, EPI_ISL_1478154, EPI_ISL_1478180, EPI_ISL_1478187, EPI_ISL_1478188, EPI_ISL_1478193, EPI_ISL_1478196, EPI_ISL_1478197, EPI_ISL_1478199, EPI_ISL_1663923, EPI_ISL_1663926, EPI_ISL_1663930, EPI_ISL_1663934, EPI_ISL_1663936, EPI_ISL_1663937, EPI_ISL_1663940, EPI_ISL_1663943, EPI_ISL_1663946, EPI_ISL_1663948, EPI_ISL_1663950, EPI_ISL_1663951, EPI_ISL_1663953, EPI_ISL_1663954, EPI_ISL_1663955, EPI_ISL_1663957, EPI_ISL_1663962, EPI_ISL_1663964, EPI_ISL_1663968, EPI_ISL_1663970, EPI_ISL_1663971, EPI_ISL_1663974, EPI_ISL_1663975, EPI_ISL_1663976, EPI_ISL_1663977, EPI_ISL_1663983, EPI_ISL_1663985, EPI_ISL_1663987, EPI_ISL_1663989, EPI_ISL_1663992, EPI_ISL_1663994, EPI_ISL_1663995, EPI_ISL_1663996, EPI_ISL_1663999, EPI_ISL_1664005, EPI_ISL_1664008, EPI_ISL_1664014, EPI_ISL_1664024, EPI_ISL_1664026, EPI_ISL_1664030, EPI_ISL_1664032, EPI_ISL_1664035, EPI_ISL_1664038, EPI_ISL_1664045, EPI_ISL_1664046, EPI_ISL_1664047, EPI_ISL_1664049, EPI_ISL_1664050, EPI_ISL_1664053, EPI_ISL_1664054, EPI_ISL_1664057, EPI_ISL_1664058, EPI_ISL_1664060, EPI_ISL_1664063, EPI_ISL_1664068, EPI_ISL_1664069, EPI_ISL_1664071, EPI_ISL_1664072, EPI_ISL_1664081, EPI_ISL_1664082, EPI_ISL_1664084, EPI_ISL_1664085, EPI_ISL_1664087, EPI_ISL_1664088, EPI_ISL_1664096, EPI_ISL_1664097, EPI_ISL_1664098, EPI_ISL_1664100, EPI_ISL_1664102, EPI_ISL_1664103, EPI_ISL_1664104, EPI_ISL_1664105, EPI_ISL_1664108, EPI_ISL_1664109, EPI_ISL_1664110, EPI_ISL_1664111, EPI_ISL_1664112, EPI_ISL_2102522, EPI_ISL_2102524, EPI_ISL_2102526, EPI_ISL_2102534, EPI_ISL_2102538, EPI_ISL_2102541, EPI_ISL_2102543, EPI_ISL_2102555, EPI_ISL_2102569, EPI_ISL_2102579, EPI_ISL_2102588, EPI_ISL_2102590, EPI_ISL_2102591, EPI_ISL_2102746, EPI_ISL_2102758, EPI_ISL_2102761, EPI_ISL_2102764, EPI_ISL_2102769, EPI_ISL_2102782, EPI_ISL_2102788, EPI_ISL_2102795, EPI_ISL_2102796, EPI_ISL_2102804, EPI_ISL_2102815, EPI_ISL_2102817, EPI_ISL_2102828, EPI_ISL_2102836, EPI_ISL_2102840, EPI_ISL_2102841, EPI_ISL_2102858, EPI_ISL_2102863, EPI_ISL_2102868, EPI_ISL_2102880, EPI_ISL_2102885, EPI_ISL_2102894, EPI_ISL_2102896, EPI_ISL_2102901, EPI_ISL_2102907, EPI_ISL_2102908, EPI_ISL_2102909, EPI_ISL_2102916, EPI_ISL_2102917, EPI_ISL_2102918, EPI_ISL_2102922, EPI_ISL_2102927, EPI_ISL_2102931, EPI_ISL_2102936, EPI_ISL_2102939, EPI_ISL_2102949, EPI_ISL_2102951, EPI_ISL_2102970, EPI_ISL_2102975, EPI_ISL_2102985, EPI_ISL_2102990, EPI_ISL_2103049, EPI_ISL_2103195, EPI_ISL_2658644, EPI_ISL_2658722, EPI_ISL_2658741, EPI_ISL_2659202, EPI_ISL_2659226, EPI_ISL_2659271, EPI_ISL_2659344, EPI_ISL_2659347, EPI_ISL_2659386, EPI_ISL_2659412, EPI_ISL_2659529, EPI_ISL_2659538, EPI_ISL_2659554, EPI_ISL_2659560, EPI_ISL_2659596, EPI_ISL_2659597, EPI_ISL_2659601, EPI_ISL_2725520, EPI_ISL_2725521, EPI_ISL_2725522, EPI_ISL_2725523, EPI_ISL_2758436, EPI_ISL_2758456 | | | |
| see above | Contra Costa County Public Health Lab | Chan-Zuckerberg Biohub | CZB Cliahub Consortium |
| EPI_ISL_979159, EPI_ISL_979160, EPI_ISL_979161, EPI_ISL_2758474 | County of San Luis Obispo Public Health Laboratory | Chan-Zuckerberg Biohub | CZB Cliahub Consortium |
| EPI_ISL_1195946, EPI_ISL_1255102 | Curative Inc | Curative Inc | Katie Bond; Vladimir Slepnev |
| EPI_ISL_1093603, EPI_ISL_1093606, EPI_ISL_1093607, EPI_ISL_1093609, EPI_ISL_1093615, EPI_ISL_1093616, EPI_ISL_1093617, EPI_ISL_1093618, EPI_ISL_1093623, EPI_ISL_1093626, EPI_ISL_1093628, EPI_ISL_1093635, EPI_ISL_1093637, EPI_ISL_1093639, EPI_ISL_1093640, EPI_ISL_1093643, EPI_ISL_1093647, EPI_ISL_1093648, EPI_ISL_1093649, EPI_ISL_1093657, EPI_ISL_1093662, EPI_ISL_1093663, EPI_ISL_1093665, EPI_ISL_1093669, EPI_ISL_1093671, EPI_ISL_1093672, EPI_ISL_1093674, EPI_ISL_1093677, EPI_ISL_1093679, EPI_ISL_1093682, EPI_ISL_1093686, EPI_ISL_1093687, EPI_ISL_1093697, EPI_ISL_1093702, EPI_ISL_1093703, EPI_ISL_1093705, EPI_ISL_1093720, EPI_ISL_1093724, EPI_ISL_1093725, EPI_ISL_1093733, EPI_ISL_1093739, EPI_ISL_1093743, EPI_ISL_1093749, EPI_ISL_1093753, EPI_ISL_1093755, EPI_ISL_1093756, EPI_ISL_1093758, EPI_ISL_1093761, EPI_ISL_1093762, EPI_ISL_1093763, EPI_ISL_1093766, EPI_ISL_1093768, EPI_ISL_1093769, EPI_ISL_1093771, EPI_ISL_1093776, EPI_ISL_1093779, EPI_ISL_1093781, EPI_ISL_1093784, EPI_ISL_1093791, EPI_ISL_1093792, EPI_ISL_1093796, EPI_ISL_1093798, EPI_ISL_1093799, EPI_ISL_1093800, EPI_ISL_1093801, EPI_ISL_1093807, EPI_ISL_1093810, EPI_ISL_1093813, EPI_ISL_1093817, EPI_ISL_1093819, EPI_ISL_1093821, EPI_ISL_1093823, EPI_ISL_1093825, EPI_ISL_1093826, EPI_ISL_1093827, EPI_ISL_1093832, EPI_ISL_1093833, EPI_ISL_1093834, EPI_ISL_1093840, EPI_ISL_1093841, EPI_ISL_1093844, EPI_ISL_1093845, EPI_ISL_1093847, EPI_ISL_1093860, EPI_ISL_1093861, EPI_ISL_1093863, EPI_ISL_1093864, EPI_ISL_1093867, EPI_ISL_1093869, EPI_ISL_1093871, EPI_ISL_1093875, EPI_ISL_1093876, EPI_ISL_1093878, EPI_ISL_1093880, EPI_ISL_1093881, EPI_ISL_1093883, EPI_ISL_1093888, EPI_ISL_1093890, EPI_ISL_1093891, EPI_ISL_1093893, EPI_ISL_1093894, EPI_ISL_1093902, EPI_ISL_1093916, EPI_ISL_1093918, EPI_ISL_1093920, EPI_ISL_1093923, EPI_ISL_1093926, EPI_ISL_1093932, EPI_ISL_1093935, EPI_ISL_1093936, EPI_ISL_1093939, EPI_ISL_1093943, EPI_ISL_1093946, EPI_ISL_1093947, EPI_ISL_1093948, EPI_ISL_1093950, EPI_ISL_1093951, EPI_ISL_1093952, EPI_ISL_1093958, EPI_ISL_1093959, EPI_ISL_1093960, EPI_ISL_1093962, EPI_ISL_1093966, EPI_ISL_1093970, EPI_ISL_1093973, EPI_ISL_1093974, EPI_ISL_1093976, EPI_ISL_1093979, EPI_ISL_1093981, EPI_ISL_1093988, EPI_ISL_1093989, EPI_ISL_1093991, EPI_ISL_1093994, EPI_ISL_1094000, EPI_ISL_1094003, EPI_ISL_1094005, EPI_ISL_1094006, EPI_ISL_1094009, EPI_ISL_1094011, EPI_ISL_1094012, EPI_ISL_1094013, EPI_ISL_1094014, EPI_ISL_1094015, EPI_ISL_1094017, EPI_ISL_1094018, EPI_ISL_1094019, EPI_ISL_1094020, EPI_ISL_1094021, EPI_ISL_1094022, EPI_ISL_1094027, EPI_ISL_1094034, EPI_ISL_1094035, EPI_ISL_1094038, EPI_ISL_1094039, EPI_ISL_1094040, EPI_ISL_1094046, EPI_ISL_1094047, EPI_ISL_1094049, EPI_ISL_1094050, EPI_ISL_1094051, EPI_ISL_1094052, EPI_ISL_1094054, EPI_ISL_1094055, EPI_ISL_1094061, EPI_ISL_1094062, EPI_ISL_1094063, EPI_ISL_1094065, EPI_ISL_1094067, EPI_ISL_1094068, EPI_ISL_1094069, EPI_ISL_1094071, EPI_ISL_1094072, EPI_ISL_1094074, EPI_ISL_1094075, EPI_ISL_1094081, EPI_ISL_1094082, EPI_ISL_1094086, EPI_ISL_1094090, EPI_ISL_1094091, EPI_ISL_1094095, EPI_ISL_1094097, EPI_ISL_1094099, EPI_ISL_1094103, EPI_ISL_1094107, EPI_ISL_1094108, EPI_ISL_1094109, EPI_ISL_1094111, EPI_ISL_1094113, EPI_ISL_1094115, EPI_ISL_1094116, EPI_ISL_1094117, EPI_ISL_1094119, EPI_ISL_1094120, EPI_ISL_1094121, EPI_ISL_1094122, EPI_ISL_1094129, EPI_ISL_1094130, EPI_ISL_1094134, EPI_ISL_1094136, EPI_ISL_1094140, EPI_ISL_1094143, EPI_ISL_1094144, EPI_ISL_1094145, EPI_ISL_1094151, EPI_ISL_1094152, EPI_ISL_1094153, EPI_ISL_1094156, EPI_ISL_1094158, EPI_ISL_1094161, EPI_ISL_1094162, EPI_ISL_1094163, EPI_ISL_1094166, EPI_ISL_1094167, EPI_ISL_1094168, EPI_ISL_1094170, EPI_ISL_1094172, EPI_ISL_1094173, EPI_ISL_1094179, EPI_ISL_1094180, EPI_ISL_1094183, EPI_ISL_1094184, EPI_ISL_1094187, EPI_ISL_1094190, EPI_ISL_1094191, EPI_ISL_1098800, EPI_ISL_1098801, EPI_ISL_1098802, EPI_ISL_1098803, EPI_ISL_1098804, EPI_ISL_1098805, EPI_ISL_1098807, EPI_ISL_1098808, EPI_ISL_1098809, EPI_ISL_1098810, EPI_ISL_1098811, EPI_ISL_1098812, EPI_ISL_1098813, EPI_ISL_1098815, EPI_ISL_1098817, EPI_ISL_1098819, EPI_ISL_1098820, EPI_ISL_1098822, EPI_ISL_1098824, EPI_ISL_1098828, EPI_ISL_1318698, EPI_ISL_1318700, EPI_ISL_1318703, EPI_ISL_1318706, EPI_ISL_1318717, EPI_ISL_1318719, EPI_ISL_1318723, EPI_ISL_1318724, EPI_ISL_1318725, EPI_ISL_1318732, EPI_ISL_1318734, EPI_ISL_1318735, EPI_ISL_1318737, EPI_ISL_1318749, EPI_ISL_1318752, EPI_ISL_1318756, EPI_ISL_1318765, EPI_ISL_1318766, EPI_ISL_1318768, EPI_ISL_1318772, EPI_ISL_1318774, EPI_ISL_1318778, EPI_ISL_1318784, EPI_ISL_1318785, EPI_ISL_1318788, EPI_ISL_1318792, EPI_ISL_1318795, EPI_ISL_1318797, EPI_ISL_1318800, EPI_ISL_1318801, EPI_ISL_1318807, EPI_ISL_1318809, EPI_ISL_1318814, EPI_ISL_1318815, EPI_ISL_1318816, EPI_ISL_1318822, EPI_ISL_1318824, EPI_ISL_1318825, EPI_ISL_1318835, EPI_ISL_1318837, EPI_ISL_1318843, EPI_ISL_1318849, EPI_ISL_1318854, EPI_ISL_1318860, EPI_ISL_1318864, EPI_ISL_1318865, EPI_ISL_1318872, EPI_ISL_1318881, EPI_ISL_1318887, EPI_ISL_1318892, EPI_ISL_1318894, EPI_ISL_1318895, EPI_ISL_1318897, EPI_ISL_1318899, EPI_ISL_1318909, EPI_ISL_1318910, EPI_ISL_1318913, EPI_ISL_1318922, EPI_ISL_1318923, EPI_ISL_1318931, EPI_ISL_1318932, EPI_ISL_1318939, EPI_ISL_1318940, EPI_ISL_1318941, EPI_ISL_1318942, EPI_ISL_1318944, EPI_ISL_1318949, EPI_ISL_1318950, EPI_ISL_1318959, EPI_ISL_1318964, EPI_ISL_1318965, EPI_ISL_1318966, EPI_ISL_1318968, EPI_ISL_1318973, EPI_ISL_1318974, EPI_ISL_1318975, EPI_ISL_1318977, EPI_ISL_1318980, EPI_ISL_1318983, EPI_ISL_1318984, EPI_ISL_1318985, EPI_ISL_1347778, EPI_ISL_1347779, EPI_ISL_1347780, EPI_ISL_1347781, EPI_ISL_1347782, EPI_ISL_1347784, EPI_ISL_1349077 | | | |
| see above | DeRisi Lab, University of California, San Francisco | DeRisi Lab, University of California, San Francisco | Amy Kistler; Anthea Mitchell; Carina Marquez; Diane Havlir; Diane Jones; Douglas Black; Eric Chow; Gabriel Chamie; Genay Pilarowski; IDseq Team; Jackie Martinez; James Peng; Jamin Liu; Jon Jacobo; Lucy Li; Luis Rubio; Manu Vanaerschot; Matthew Laurie; Maya Petersen; Patrick Ayscue; Sabrina Mann; Sara Sunshine; Susana Rojas; Susy Rojas; Valerie Tulier-Laiwa; and Joseph DeRisi |
| EPI_ISL_1811691, EPI_ISL_1811694, EPI_ISL_1811699, EPI_ISL_1811700, EPI_ISL_1811704, EPI_ISL_1811728, EPI_ISL_1811756, EPI_ISL_1811757, EPI_ISL_1811768, EPI_ISL_1811774, EPI_ISL_1811781, EPI_ISL_1811798, EPI_ISL_1811800, EPI_ISL_1811823, EPI_ISL_1811838, EPI_ISL_1811839, EPI_ISL_1811842, EPI_ISL_1811844, EPI_ISL_1811846, EPI_ISL_1811856, EPI_ISL_1811859, EPI_ISL_1811860, EPI_ISL_1811861, EPI_ISL_1811863, EPI_ISL_1811864, EPI_ISL_1811865, EPI_ISL_1811866, EPI_ISL_1811868, EPI_ISL_1811870, EPI_ISL_1811874, EPI_ISL_1811876, EPI_ISL_1811877, EPI_ISL_1811878, EPI_ISL_1811879, EPI_ISL_1811880, EPI_ISL_1811881, EPI_ISL_1811882, EPI_ISL_1811886, EPI_ISL_1811888, EPI_ISL_1811890, EPI_ISL_1811893, EPI_ISL_1811894, EPI_ISL_1811895, EPI_ISL_1811896, EPI_ISL_1811898, EPI_ISL_1811899, EPI_ISL_1811900, EPI_ISL_1811901, EPI_ISL_1811903, EPI_ISL_1811904, EPI_ISL_1811905, EPI_ISL_1811906, EPI_ISL_1811907, EPI_ISL_1811908, EPI_ISL_1811909, EPI_ISL_1811911, EPI_ISL_1811912, EPI_ISL_1811913, EPI_ISL_1811915, EPI_ISL_1811916, EPI_ISL_1811917, EPI_ISL_1811918, EPI_ISL_1811919, EPI_ISL_1811920, EPI_ISL_1811921, EPI_ISL_1811923, EPI_ISL_1811925, EPI_ISL_1811928, EPI_ISL_1811930, EPI_ISL_1811933, EPI_ISL_1811934, EPI_ISL_1811935, EPI_ISL_1811937, EPI_ISL_1811938, EPI_ISL_1811939, EPI_ISL_1811941, EPI_ISL_1811942, EPI_ISL_1811943, EPI_ISL_1811944, EPI_ISL_1811945, EPI_ISL_1811946, EPI_ISL_1811947, EPI_ISL_1811948, EPI_ISL_1811949, EPI_ISL_1811950, EPI_ISL_1811951, EPI_ISL_1811952, EPI_ISL_1811953, EPI_ISL_1811954, EPI_ISL_1811955, EPI_ISL_1811956, EPI_ISL_1811957, EPI_ISL_1811958, EPI_ISL_1811959, EPI_ISL_1811960, EPI_ISL_1811961, EPI_ISL_1811962, EPI_ISL_1811963, EPI_ISL_1811964, EPI_ISL_1811968, EPI_ISL_1811970, EPI_ISL_1811971, EPI_ISL_1811972, EPI_ISL_1811974, EPI_ISL_1811975, EPI_ISL_1811976, EPI_ISL_1811977, EPI_ISL_1811978, EPI_ISL_1811981, EPI_ISL_1811982, EPI_ISL_1811984, EPI_ISL_1811985, EPI_ISL_1811986, EPI_ISL_1811987, EPI_ISL_1811989, EPI_ISL_1811990, EPI_ISL_1811993, EPI_ISL_1811995, EPI_ISL_1811997, EPI_ISL_1811998, EPI_ISL_1811999, EPI_ISL_1812003, EPI_ISL_1812004, EPI_ISL_1812005, EPI_ISL_1812006, EPI_ISL_1812009, EPI_ISL_1812014, EPI_ISL_1812018, EPI_ISL_1812020, EPI_ISL_1812022, EPI_ISL_1812023, EPI_ISL_1812024, EPI_ISL_1812025, EPI_ISL_1812026, EPI_ISL_1812027, EPI_ISL_1812034, EPI_ISL_1812037, EPI_ISL_1812038, EPI_ISL_1812040, EPI_ISL_1812041, EPI_ISL_1812048, EPI_ISL_1812053, EPI_ISL_1812054, EPI_ISL_1812055, EPI_ISL_1812056, EPI_ISL_1812058, EPI_ISL_1812059, EPI_ISL_1812063, EPI_ISL_1812064, EPI_ISL_1812066, EPI_ISL_1812067, EPI_ISL_1812069, EPI_ISL_1812070, EPI_ISL_1812072, EPI_ISL_1812078, EPI_ISL_1812080, EPI_ISL_1812081, EPI_ISL_1812082, EPI_ISL_1812083, EPI_ISL_1812084, EPI_ISL_1812085, EPI_ISL_1812088, EPI_ISL_1812089, EPI_ISL_1812091, EPI_ISL_1812092, EPI_ISL_1812093, EPI_ISL_1812094, EPI_ISL_1812096, EPI_ISL_1812101, EPI_ISL_1812103, EPI_ISL_1812107, EPI_ISL_1812108, EPI_ISL_1812110, EPI_ISL_1812111, EPI_ISL_1812113, EPI_ISL_1812115, EPI_ISL_1812118, EPI_ISL_1812124, EPI_ISL_1812125, EPI_ISL_1812126, EPI_ISL_1812129, EPI_ISL_1812132, EPI_ISL_1812133, EPI_ISL_1812135, EPI_ISL_1812136, EPI_ISL_1812138, EPI_ISL_1812140, EPI_ISL_1812142, EPI_ISL_1812143, EPI_ISL_1812146, EPI_ISL_1812147, EPI_ISL_1812148, EPI_ISL_1812149, EPI_ISL_1812151, EPI_ISL_1812152, EPI_ISL_1812153, EPI_ISL_1812158, EPI_ISL_1812161, EPI_ISL_1812164, EPI_ISL_1812165, EPI_ISL_1812167, EPI_ISL_1812169, EPI_ISL_1812172, EPI_ISL_1812173, EPI_ISL_1812174, EPI_ISL_1812175, EPI_ISL_1812176, EPI_ISL_1812177, EPI_ISL_1812179, EPI_ISL_1812186, EPI_ISL_1812189, EPI_ISL_1812190, EPI_ISL_1812194, EPI_ISL_1812195, EPI_ISL_1812196, EPI_ISL_1812200, EPI_ISL_1812202, EPI_ISL_1812207, EPI_ISL_1812208, EPI_ISL_1812210, EPI_ISL_1812212, EPI_ISL_1812213, EPI_ISL_1812214, EPI_ISL_1812215, EPI_ISL_1812220, EPI_ISL_1812221, EPI_ISL_1812222, EPI_ISL_1812223, EPI_ISL_1812224, EPI_ISL_1812226, EPI_ISL_1812232, EPI_ISL_1812233, EPI_ISL_1812234, EPI_ISL_1812237, EPI_ISL_1812241, EPI_ISL_1812243, EPI_ISL_1812244, EPI_ISL_1812245, EPI_ISL_1812246, EPI_ISL_1812249, EPI_ISL_1812250, EPI_ISL_1812251, EPI_ISL_1812254, EPI_ISL_1812255, EPI_ISL_1812256, EPI_ISL_1812258, EPI_ISL_1812259, EPI_ISL_1812260, EPI_ISL_1812262, EPI_ISL_1812263, EPI_ISL_1812264, EPI_ISL_1812266, EPI_ISL_1812269, EPI_ISL_1812270, EPI_ISL_1812272, EPI_ISL_1812273, EPI_ISL_1812274, EPI_ISL_1812277, EPI_ISL_1812278, EPI_ISL_1812280, EPI_ISL_1812283, EPI_ISL_1812289, EPI_ISL_1812293, EPI_ISL_1812297, EPI_ISL_1812298, EPI_ISL_1812299, EPI_ISL_1812300, EPI_ISL_1812302, EPI_ISL_1812304, EPI_ISL_1812305, EPI_ISL_1812308, EPI_ISL_1812309, EPI_ISL_1812315, EPI_ISL_1812316, EPI_ISL_1812321, EPI_ISL_1812322, EPI_ISL_1812333, EPI_ISL_1812334, EPI_ISL_1812335, EPI_ISL_1812339, EPI_ISL_1812340, EPI_ISL_1812342, EPI_ISL_1812344, EPI_ISL_1812345, EPI_ISL_1812346, EPI_ISL_1812347, EPI_ISL_1812350, EPI_ISL_1812353, EPI_ISL_1812355, EPI_ISL_1812358, EPI_ISL_1812359, EPI_ISL_1812366, EPI_ISL_1812370, EPI_ISL_1812374, EPI_ISL_1812375, EPI_ISL_1812380, EPI_ISL_1812383, EPI_ISL_1812386, EPI_ISL_1812394, EPI_ISL_1812396, EPI_ISL_1812397, EPI_ISL_1812401, EPI_ISL_1812409, EPI_ISL_1812410, EPI_ISL_1812413, EPI_ISL_1812416, EPI_ISL_1812418, EPI_ISL_1812419, EPI_ISL_1812425, EPI_ISL_1812426, EPI_ISL_1812428, EPI_ISL_1812435, EPI_ISL_1812437, EPI_ISL_1812438, EPI_ISL_1812440, EPI_ISL_1812442, EPI_ISL_1812444, EPI_ISL_1812450, EPI_ISL_1812451, EPI_ISL_1812452, EPI_ISL_1812455, EPI_ISL_1812461, EPI_ISL_1812462, EPI_ISL_1812466, EPI_ISL_1812469, EPI_ISL_1812473, EPI_ISL_1812476, EPI_ISL_1812479, EPI_ISL_1812483, EPI_ISL_1812484, EPI_ISL_1812485, EPI_ISL_1812493, EPI_ISL_1812494, EPI_ISL_1812501, EPI_ISL_1812502, EPI_ISL_1812503, EPI_ISL_1812504, EPI_ISL_1812510, EPI_ISL_1812511, EPI_ISL_1812513, EPI_ISL_1812514, EPI_ISL_1812516, EPI_ISL_1812519, EPI_ISL_1812523, EPI_ISL_1812528, EPI_ISL_1812529, EPI_ISL_1812532, EPI_ISL_1812543, EPI_ISL_1812547, EPI_ISL_1812551, EPI_ISL_1812555, EPI_ISL_1812557, EPI_ISL_1812560, EPI_ISL_1812565, EPI_ISL_1812570, EPI_ISL_1812572, EPI_ISL_1812573, EPI_ISL_1812575, EPI_ISL_1812580, EPI_ISL_1812581, EPI_ISL_1812584, EPI_ISL_1812585, EPI_ISL_1812588, EPI_ISL_1812594, EPI_ISL_1812595, EPI_ISL_1812596, EPI_ISL_1812600, EPI_ISL_1812602, EPI_ISL_1812605, EPI_ISL_1812607, EPI_ISL_1812609, EPI_ISL_1812616, EPI_ISL_1812617, EPI_ISL_1812619, EPI_ISL_1812620, EPI_ISL_1812622, EPI_ISL_1812624, EPI_ISL_1812630, EPI_ISL_1812633, EPI_ISL_1812635, EPI_ISL_1812636, EPI_ISL_1812645, EPI_ISL_1812652, EPI_ISL_1812654, EPI_ISL_1812656, EPI_ISL_1812659, EPI_ISL_1812662, EPI_ISL_1812663, EPI_ISL_1812664, EPI_ISL_1812667, EPI_ISL_1812672, EPI_ISL_1812678, EPI_ISL_1812680, EPI_ISL_1812681, EPI_ISL_1812685, EPI_ISL_1812690, EPI_ISL_1812700, EPI_ISL_1812706, EPI_ISL_1812708, EPI_ISL_1812710, EPI_ISL_1812711, EPI_ISL_1812713, EPI_ISL_1812715, EPI_ISL_1812717, EPI_ISL_1812718, EPI_ISL_1812719, EPI_ISL_1812721, EPI_ISL_1812725, EPI_ISL_1812726, EPI_ISL_1812730, EPI_ISL_1812734, EPI_ISL_1812738, EPI_ISL_1812747, EPI_ISL_1812748, EPI_ISL_1812754, EPI_ISL_1812761, EPI_ISL_1812770, EPI_ISL_1812772, EPI_ISL_1812773, EPI_ISL_1812774, EPI_ISL_1812778, EPI_ISL_1812783, EPI_ISL_1812786, EPI_ISL_1812787, EPI_ISL_1812788, EPI_ISL_1812789, EPI_ISL_1812791, EPI_ISL_1812793, EPI_ISL_1812794, EPI_ISL_1812808, EPI_ISL_1812809, EPI_ISL_1812813, EPI_ISL_1812814, EPI_ISL_1812823, EPI_ISL_1812824, EPI_ISL_1812830, EPI_ISL_1812832, EPI_ISL_1812834, EPI_ISL_1812847, EPI_ISL_1812848, EPI_ISL_1812849, EPI_ISL_1812851, EPI_ISL_1812855, EPI_ISL_1812856, EPI_ISL_1812862, EPI_ISL_1812863, EPI_ISL_1812867, EPI_ISL_1812870, EPI_ISL_1812884, EPI_ISL_1812888, EPI_ISL_1812894, EPI_ISL_1812898, EPI_ISL_1812899, EPI_ISL_1812902, EPI_ISL_1812903, EPI_ISL_1812904, EPI_ISL_1812907, EPI_ISL_1812909, EPI_ISL_1812911, EPI_ISL_1812912, EPI_ISL_1812920, EPI_ISL_1812928, EPI_ISL_1812932, EPI_ISL_1812933, EPI_ISL_1812939, EPI_ISL_1812941, EPI_ISL_1812947, EPI_ISL_1812948, EPI_ISL_1812949, EPI_ISL_1812950, EPI_ISL_1812952, EPI_ISL_1812954, EPI_ISL_1812958, EPI_ISL_1812961, EPI_ISL_1812965, EPI_ISL_1812968, EPI_ISL_1812971, EPI_ISL_1812972, EPI_ISL_1812975, EPI_ISL_1812976, EPI_ISL_1812979, EPI_ISL_1812980, EPI_ISL_1812982, EPI_ISL_1812984, EPI_ISL_1812991, EPI_ISL_1813000, EPI_ISL_1813003, EPI_ISL_1813007, EPI_ISL_1813011, EPI_ISL_1813012, EPI_ISL_1813013, EPI_ISL_1813014, EPI_ISL_1813015, EPI_ISL_1813017, EPI_ISL_1813023, EPI_ISL_1813029, EPI_ISL_1813033, EPI_ISL_1813034, EPI_ISL_1813036, EPI_ISL_1813037, EPI_ISL_1813039, EPI_ISL_1813044, EPI_ISL_1813049, EPI_ISL_1813050, EPI_ISL_1813052, EPI_ISL_1813053, EPI_ISL_1813056, EPI_ISL_1813057, EPI_ISL_1813061, EPI_ISL_1813062, EPI_ISL_1813066, EPI_ISL_1813069, EPI_ISL_1813077, EPI_ISL_1813080, EPI_ISL_1813082, EPI_ISL_1813084, EPI_ISL_1813085, EPI_ISL_1813088, EPI_ISL_1813090, EPI_ISL_1813091, EPI_ISL_1813099, EPI_ISL_1813100, EPI_ISL_1813103, EPI_ISL_1813105, EPI_ISL_1813108, EPI_ISL_1813110, EPI_ISL_1813112, EPI_ISL_1813114, EPI_ISL_1813118, EPI_ISL_1813121, EPI_ISL_1813122, EPI_ISL_1813124, EPI_ISL_1813125, EPI_ISL_1813128, EPI_ISL_1813129, EPI_ISL_1813131, EPI_ISL_1813132, EPI_ISL_1813135, EPI_ISL_1813136, EPI_ISL_1813140, EPI_ISL_1813142, EPI_ISL_1813143, EPI_ISL_1813147, EPI_ISL_1813155, EPI_ISL_1813156, EPI_ISL_1813159, EPI_ISL_1813160, EPI_ISL_1813163, EPI_ISL_1813167, EPI_ISL_1813169, EPI_ISL_1813170, EPI_ISL_1813171, EPI_ISL_1813173, EPI_ISL_1813183, EPI_ISL_1813187, EPI_ISL_1813188, EPI_ISL_1813190, EPI_ISL_1813191, EPI_ISL_1813196, EPI_ISL_1813197, EPI_ISL_1813200, EPI_ISL_1813203, EPI_ISL_1813205, EPI_ISL_1813206, EPI_ISL_1813207, EPI_ISL_1813210, EPI_ISL_1813213, EPI_ISL_1813216, EPI_ISL_1813221, EPI_ISL_1813225, EPI_ISL_1813227, EPI_ISL_1813228, EPI_ISL_1813229, EPI_ISL_1813230, EPI_ISL_1813232, EPI_ISL_1813233, EPI_ISL_1813234, EPI_ISL_1813239, EPI_ISL_1813243, EPI_ISL_1813244, EPI_ISL_1813247, EPI_ISL_1813251, EPI_ISL_1813252, EPI_ISL_1813256, EPI_ISL_1813259, EPI_ISL_1813262, EPI_ISL_1813265, EPI_ISL_1813266, EPI_ISL_1813267, EPI_ISL_1813270, EPI_ISL_1813273, EPI_ISL_1813276, EPI_ISL_1813279, EPI_ISL_1813280, EPI_ISL_1813281, EPI_ISL_1813287, EPI_ISL_1813288, EPI_ISL_1813291, EPI_ISL_1813292, EPI_ISL_1813293, EPI_ISL_1813298, EPI_ISL_1813300, EPI_ISL_1813303, EPI_ISL_1813310, EPI_ISL_1813312, EPI_ISL_1813313, EPI_ISL_1813322, EPI_ISL_1813323, EPI_ISL_1813324, EPI_ISL_1813325, EPI_ISL_1813329, EPI_ISL_1813331, EPI_ISL_1813337, EPI_ISL_1813338, EPI_ISL_1813343, EPI_ISL_1813345, EPI_ISL_1813347, EPI_ISL_1813350, EPI_ISL_1813353, EPI_ISL_1813359, EPI_ISL_1813361, EPI_ISL_1813363, EPI_ISL_1813369, EPI_ISL_1813378, EPI_ISL_1813382, EPI_ISL_1813386, EPI_ISL_1813390, EPI_ISL_1813394, EPI_ISL_1813395, EPI_ISL_1813396, EPI_ISL_1813398, EPI_ISL_1813399, EPI_ISL_1813401, EPI_ISL_1813403, EPI_ISL_1813404, EPI_ISL_1813408, EPI_ISL_1813412, EPI_ISL_1813413, EPI_ISL_1813414, EPI_ISL_1813416, EPI_ISL_1813420, EPI_ISL_1813421, EPI_ISL_1813426, EPI_ISL_1813427, EPI_ISL_1813434, EPI_ISL_1813435, EPI_ISL_1813442, EPI_ISL_1813444, EPI_ISL_1813446, EPI_ISL_1813450, EPI_ISL_1813451, EPI_ISL_1813452, EPI_ISL_1813454, EPI_ISL_1813456, EPI_ISL_1813457, EPI_ISL_1813458, EPI_ISL_1813460, EPI_ISL_1813462, EPI_ISL_1813465, EPI_ISL_1813470, EPI_ISL_1813473, EPI_ISL_1813474, EPI_ISL_1813483, EPI_ISL_1813486, EPI_ISL_1813495, EPI_ISL_1813501, EPI_ISL_1813506, EPI_ISL_1813511, EPI_ISL_1813520, EPI_ISL_1813526, EPI_ISL_1813532, EPI_ISL_1813534, EPI_ISL_1813536, EPI_ISL_1813537, EPI_ISL_1813538, EPI_ISL_1813539, EPI_ISL_1813540, EPI_ISL_1813544, EPI_ISL_1813553, EPI_ISL_1813554, EPI_ISL_1813560, EPI_ISL_1813562, EPI_ISL_1813567, EPI_ISL_1813568, EPI_ISL_1813571, EPI_ISL_1813574, EPI_ISL_1813583, EPI_ISL_1813584, EPI_ISL_1813587, EPI_ISL_1813588, EPI_ISL_1813589, EPI_ISL_1813590, EPI_ISL_1813592, EPI_ISL_1813593, EPI_ISL_1813595, EPI_ISL_1813596, EPI_ISL_1813597, EPI_ISL_1813599, EPI_ISL_1813605, EPI_ISL_1813608, EPI_ISL_1813610, EPI_ISL_1813611, EPI_ISL_1813615, EPI_ISL_1813616, EPI_ISL_1813624, EPI_ISL_1813626, EPI_ISL_1813633, EPI_ISL_1813638, EPI_ISL_1813644, EPI_ISL_1813647, EPI_ISL_1813653, EPI_ISL_1813654, EPI_ISL_1813657, EPI_ISL_1813660, EPI_ISL_1813665, EPI_ISL_1813677, EPI_ISL_1813682, EPI_ISL_1813687, EPI_ISL_1813690, EPI_ISL_1813691, EPI_ISL_1813696, EPI_ISL_1813697, EPI_ISL_1813700, EPI_ISL_1813702, EPI_ISL_1813713, EPI_ISL_1813720, EPI_ISL_1813721, EPI_ISL_1813723, EPI_ISL_1813727, EPI_ISL_1813728, EPI_ISL_1813731, EPI_ISL_1813732, EPI_ISL_1813739, EPI_ISL_1813744, EPI_ISL_1813745, EPI_ISL_1813746, EPI_ISL_1813752, EPI_ISL_1813753, EPI_ISL_1813755, EPI_ISL_1813758, EPI_ISL_1813759, EPI_ISL_1813764, EPI_ISL_1813766, EPI_ISL_1813767, EPI_ISL_1813784, EPI_ISL_1813789, EPI_ISL_1813791, EPI_ISL_1813793, EPI_ISL_1813798, EPI_ISL_1813800, EPI_ISL_1813801, EPI_ISL_1813807, EPI_ISL_1813808, EPI_ISL_1813809, EPI_ISL_1813815, EPI_ISL_1813817, EPI_ISL_1813824, EPI_ISL_1813834, EPI_ISL_1813843, EPI_ISL_1813846, EPI_ISL_1813860, EPI_ISL_1813861, EPI_ISL_1813862, EPI_ISL_1813868, EPI_ISL_1813873, EPI_ISL_1813874, EPI_ISL_1813876, EPI_ISL_1813881, EPI_ISL_1813886, EPI_ISL_1813887, EPI_ISL_1813892, EPI_ISL_1813894, EPI_ISL_1813897, EPI_ISL_1813903, EPI_ISL_1813908, EPI_ISL_1813910, EPI_ISL_1813934, EPI_ISL_1813938, EPI_ISL_1813939, EPI_ISL_1813947, EPI_ISL_1813949, EPI_ISL_1813953, EPI_ISL_1813954, EPI_ISL_1813962, EPI_ISL_1813964, EPI_ISL_1813971, EPI_ISL_1813977, EPI_ISL_1813979, EPI_ISL_1813981, EPI_ISL_1813988, EPI_ISL_1813990, EPI_ISL_1813991, EPI_ISL_1813992, EPI_ISL_1813995, EPI_ISL_1814000, EPI_ISL_1814006, EPI_ISL_1814007, EPI_ISL_1814008, EPI_ISL_1814012, EPI_ISL_1814018, EPI_ISL_1814020, EPI_ISL_1814022, EPI_ISL_1814031, EPI_ISL_1814045, EPI_ISL_1814050, EPI_ISL_1814052, EPI_ISL_1814054, EPI_ISL_1814059, EPI_ISL_1814061, EPI_ISL_1814072, EPI_ISL_1814075, EPI_ISL_1814077, EPI_ISL_1814078, EPI_ISL_1814091, EPI_ISL_1814094, EPI_ISL_1814107, EPI_ISL_1814108, EPI_ISL_1814111, EPI_ISL_1814113, EPI_ISL_1814116, EPI_ISL_1814118, EPI_ISL_1814120, EPI_ISL_1814125, EPI_ISL_1814126, EPI_ISL_1814127, EPI_ISL_1814132, EPI_ISL_1814146, EPI_ISL_1814148, EPI_ISL_1814151, EPI_ISL_1814159, EPI_ISL_1814160, EPI_ISL_1814162, EPI_ISL_1814179, EPI_ISL_1814180, EPI_ISL_1814186, EPI_ISL_1814193, EPI_ISL_1814198, EPI_ISL_1814202, EPI_ISL_1814209, EPI_ISL_1814211, EPI_ISL_1814213, EPI_ISL_1814235, EPI_ISL_1814238, EPI_ISL_1814239, EPI_ISL_1814240, EPI_ISL_1814245, EPI_ISL_1814249, EPI_ISL_1814258, EPI_ISL_1814267, EPI_ISL_1814274, EPI_ISL_1814280, EPI_ISL_1814285, EPI_ISL_1814289, EPI_ISL_1814294, EPI_ISL_1814303, EPI_ISL_1814304, EPI_ISL_1814319, EPI_ISL_1814337, EPI_ISL_1814347, EPI_ISL_1814358, EPI_ISL_1814367, EPI_ISL_1814370, EPI_ISL_1814375, EPI_ISL_1814376, EPI_ISL_1814379, EPI_ISL_1814380, EPI_ISL_1814387, EPI_ISL_1814393, EPI_ISL_1814400, EPI_ISL_1814403, EPI_ISL_1814405, EPI_ISL_1814409, EPI_ISL_1814428, EPI_ISL_1814448, EPI_ISL_1814450, EPI_ISL_1814454, EPI_ISL_1814458, EPI_ISL_1814477, EPI_ISL_1814487, EPI_ISL_1814491, EPI_ISL_1814492, EPI_ISL_1814501, EPI_ISL_1814513, EPI_ISL_1814520, EPI_ISL_1814526, EPI_ISL_1814528, EPI_ISL_1814529, EPI_ISL_1814531, EPI_ISL_1814535, EPI_ISL_1814542, EPI_ISL_1814554, EPI_ISL_1814560, EPI_ISL_1814564, EPI_ISL_1814565, EPI_ISL_1814567, EPI_ISL_1814571, EPI_ISL_1814580, EPI_ISL_1814581, EPI_ISL_1814587, EPI_ISL_1814595, EPI_ISL_1814599, EPI_ISL_1814603, | | | |

EPI_ISL_1814608, EPI_ISL_1814615, EPI_ISL_1814616, EPI_ISL_1814640, EPI_ISL_1814647, EPI_ISL_1814651, EPI_ISL_1814660, EPI_ISL_1814671, EPI_ISL_1814679, EPI_ISL_1814690, EPI_ISL_1814691, EPI_ISL_1814693, EPI_ISL_1814696, EPI_ISL_1814700, EPI_ISL_1814701, EPI_ISL_1814707, EPI_ISL_1814724, EPI_ISL_1814728, EPI_ISL_1814729, EPI_ISL_1814735, EPI_ISL_1814750, EPI_ISL_1814753, EPI_ISL_1814776, EPI_ISL_1814777, EPI_ISL_1814778, EPI_ISL_1814807, EPI_ISL_1814821, EPI_ISL_1814838, EPI_ISL_1814841, EPI_ISL_1814854, EPI_ISL_1814857, EPI_ISL_1814858, EPI_ISL_1814862, EPI_ISL_1814867, EPI_ISL_1814873, EPI_ISL_1814886, EPI_ISL_1814902, EPI_ISL_1814910, EPI_ISL_1814912, EPI_ISL_1814919, EPI_ISL_1814928, EPI_ISL_1814935, EPI_ISL_1814940, EPI_ISL_1814943, EPI_ISL_1814946, EPI_ISL_1814952, EPI_ISL_1814955, EPI_ISL_1814956, EPI_ISL_1814961, EPI_ISL_1814963, EPI_ISL_1814967, EPI_ISL_1814973, EPI_ISL_1814974, EPI_ISL_1814980, EPI_ISL_1814991, EPI_ISL_1815001, EPI_ISL_1815011, EPI_ISL_1815022, EPI_ISL_1815028, EPI_ISL_1815030, EPI_ISL_1815033, EPI_ISL_1815036, EPI_ISL_1815041, EPI_ISL_1815045, EPI_ISL_1815056, EPI_ISL_1815057, EPI_ISL_1815067, EPI_ISL_1815086, EPI_ISL_1815094, EPI_ISL_1815099, EPI_ISL_1815100, EPI_ISL_1815101, EPI_ISL_1815116, EPI_ISL_1815134, EPI_ISL_1815144, EPI_ISL_1815149, EPI_ISL_1815150, EPI_ISL_1815156, EPI_ISL_1815176, EPI_ISL_1815179, EPI_ISL_1815202, EPI_ISL_1815219, EPI_ISL_1815221, EPI_ISL_1815223, EPI_ISL_1815224, EPI_ISL_1815225, EPI_ISL_1815234, EPI_ISL_1815240, EPI_ISL_1815241, EPI_ISL_1815248, EPI_ISL_1815260, EPI_ISL_1815261, EPI_ISL_1815263, EPI_ISL_1815268, EPI_ISL_1815276, EPI_ISL_1815284, EPI_ISL_1815285, EPI_ISL_1815306, EPI_ISL_1815321, EPI_ISL_1815324, EPI_ISL_1815331, EPI_ISL_1815332, EPI_ISL_1815342, EPI_ISL_1815344, EPI_ISL_1815369, EPI_ISL_1815374, EPI_ISL_1815407, EPI_ISL_1815411, EPI_ISL_1815418, EPI_ISL_1815456, EPI_ISL_1815468, EPI_ISL_1815512, EPI_ISL_1815543, EPI_ISL_1815545, EPI_ISL_1815547, EPI_ISL_1815568, EPI_ISL_1815569, EPI_ISL_1815586, EPI_ISL_1815592, EPI_ISL_1815609, EPI_ISL_1815612, EPI_ISL_1815621, EPI_ISL_1815625, EPI_ISL_1815630, EPI_ISL_1815645, EPI_ISL_1815648, EPI_ISL_1815652, EPI_ISL_1815688, EPI_ISL_1815693, EPI_ISL_1815707, EPI_ISL_1815710, EPI_ISL_1815717, EPI_ISL_1815719, EPI_ISL_1815724, EPI_ISL_1815734, EPI_ISL_1815737, EPI_ISL_1815738, EPI_ISL_1815750, EPI_ISL_1815756, EPI_ISL_1815800, EPI_ISL_1815801, EPI_ISL_1815807, EPI_ISL_1815817, EPI_ISL_1815826, EPI_ISL_1815834, EPI_ISL_1815841, EPI_ISL_1815844, EPI_ISL_1815877, EPI_ISL_1815886, EPI_ISL_1815888, EPI_ISL_1815889, EPI_ISL_1815896, EPI_ISL_1815906, EPI_ISL_1815919, EPI_ISL_1815927, EPI_ISL_1815930, EPI_ISL_1815941, EPI_ISL_1815960, EPI_ISL_1815965, EPI_ISL_1815969, EPI_ISL_1815975, EPI_ISL_1815977, EPI_ISL_1815978, EPI_ISL_1815981, EPI_ISL_1816006, EPI_ISL_1816009, EPI_ISL_1816017, EPI_ISL_1816033, EPI_ISL_1816063, EPI_ISL_1816068, EPI_ISL_1816074, EPI_ISL_1816076, EPI_ISL_1816082, EPI_ISL_1816086, EPI_ISL_1816094, EPI_ISL_1816152, EPI_ISL_1816155, EPI_ISL_1816182, EPI_ISL_1816198, EPI_ISL_1816199, EPI_ISL_1816230, EPI_ISL_1816231, EPI_ISL_1816251, EPI_ISL_1816282, EPI_ISL_1816286, EPI_ISL_1816318, EPI_ISL_1816332, EPI_ISL_1816340, EPI_ISL_1816347, EPI_ISL_1816370, EPI_ISL_1816379, EPI_ISL_1816408, EPI_ISL_1816413, EPI_ISL_1816419, EPI_ISL_1816421, EPI_ISL_1816445, EPI_ISL_1816474, EPI_ISL_1816480, EPI_ISL_1816492, EPI_ISL_1816497, EPI_ISL_1816501, EPI_ISL_1816504, EPI_ISL_1816517, EPI_ISL_1816518, EPI_ISL_1816536, EPI_ISL_1816539, EPI_ISL_1816558, EPI_ISL_1816562, EPI_ISL_1816582, EPI_ISL_1816585, EPI_ISL_1816588, EPI_ISL_1816596, EPI_ISL_1816620, EPI_ISL_1816635, EPI_ISL_1816657, EPI_ISL_1816658, EPI_ISL_1816660, EPI_ISL_1816662, EPI_ISL_1816668, EPI_ISL_1816710, EPI_ISL_1816730, EPI_ISL_1816747, EPI_ISL_1816766, EPI_ISL_1816771, EPI_ISL_1816775, EPI_ISL_1816784, EPI_ISL_1816788, EPI_ISL_1816817, EPI_ISL_1816836, EPI_ISL_1816845, EPI_ISL_1816846, EPI_ISL_1816850, EPI_ISL_1816863, EPI_ISL_1816868, EPI_ISL_1816888, EPI_ISL_1816889, EPI_ISL_1816898, EPI_ISL_1816910, EPI_ISL_1816914

| | | | |
|---|---|---|---|
| see above | EXCITE Lab | Andersen lab at Scripps Research | Abigail Schnapper; Alexandre Bolze; Alice Summerfield; Angela Scioscia; Araz Majnoonian; Ashkan Hassani; Brent Brainard+ SEARCH; Celena Andrade; Charlotte Rivera-Garcia; Cheryl Anderson; Chip Schooley; Christine Mitchell + SEARCH; David Becker; David Pride; Efren Sandoval; Elizabeth Cirulli; Fitri Carrissa Wijaya; Francisco Tanudjaja; Fred Wu; Geraint Levan; Greg Humphrey; Helena Tubb; J. Joelle Donofrio-Odmann; James Lu + SEARCH; Jason Nguyen; Jimmy Ramirez; Kelly Schiabor Barrett; Laura Kohn; Magnus Isaksson; Marc Laurent; Marlene Flores; Megan Nguyen; Natasha Martin; Natasha Martin Cheryl Anderson; Nicole L Washington; Patricia Gonzalez-Zuniga; Rebecca Fielding-Miller; Rodolfo Salido; Ryan Cho; Sawyer Farmer; Sharon Reed; Sherry Wang; Simon White; Smruthi Karthikeyan; Sydney Morgan + Marni Jacobs + Celestine Magallanes + Holly Valentine + Louise C. Laurent + SEARCH; Tommy Valles + SEARCH; Tyler Cassens; Victor Cantu; Vinton Omaleki; William Lee |

EPI_ISL_977120, EPI_ISL_977121, EPI_ISL_977122, EPI_ISL_977123, EPI_ISL_977124, EPI_ISL_977125, EPI_ISL_977126, EPI_ISL_977127, EPI_ISL_977128, EPI_ISL_977129, EPI_ISL_977130, EPI_ISL_977131, EPI_ISL_977132, EPI_ISL_977133, EPI_ISL_977134, EPI_ISL_977138, EPI_ISL_977139, EPI_ISL_977140, EPI_ISL_977141, EPI_ISL_977142, EPI_ISL_977143

| | | | |
|---|---|---|---|
| see above | Flow Health | Infectious Disease Program, Broad Institute of Harvard and MIT | Adams, G.; B.L.; B.W.; Bauer, M.; Birren; Carter, A.; Chaluvadi, S.; D.J.; DeRuff, K.; Gallagher, G.; Gladden-Young, A.; J.E.; K.J.; Lagerborg, K.; Lemieux; Loreth, C.; MacInnis; Normandin, E.; P.C.; Park; Reilly, S.; Rudy, M.; Siddle; Smole, S.; Tomkins-Tinch, C.; and Sabeti |

EPI_ISL_1010191, EPI_ISL_1010192, EPI_ISL_1010193, EPI_ISL_1010194, EPI_ISL_1010195, EPI_ISL_1010196, EPI_ISL_1010197, EPI_ISL_1010198, EPI_ISL_1010199, EPI_ISL_1010200, EPI_ISL_1010201, EPI_ISL_1010202, EPI_ISL_1010203, EPI_ISL_1010204, EPI_ISL_1010205, EPI_ISL_1010206, EPI_ISL_1010207, EPI_ISL_1010208, EPI_ISL_1010209, EPI_ISL_1010210, EPI_ISL_1010211, EPI_ISL_1010212, EPI_ISL_1010213, EPI_ISL_1010214, EPI_ISL_1010215, EPI_ISL_1010216, EPI_ISL_1010217, EPI_ISL_1010218, EPI_ISL_1010219, EPI_ISL_1010220, EPI_ISL_1010221, EPI_ISL_1010222, EPI_ISL_1010223, EPI_ISL_1010224, EPI_ISL_1010225, EPI_ISL_1010226, EPI_ISL_1010227, EPI_ISL_1010228, EPI_ISL_1010229, EPI_ISL_1010230, EPI_ISL_1010231, EPI_ISL_1010232, EPI_ISL_1010233, EPI_ISL_1010234, EPI_ISL_1010235, EPI_ISL_1010236, EPI_ISL_1010237, EPI_ISL_1010238, EPI_ISL_1010239, EPI_ISL_1010240, EPI_ISL_1010241, EPI_ISL_1010242, EPI_ISL_1010243, EPI_ISL_1010244, EPI_ISL_1010245, EPI_ISL_1010246, EPI_ISL_1010247, EPI_ISL_1010248, EPI_ISL_1010249, EPI_ISL_1010250, EPI_ISL_1010251, EPI_ISL_1010252, EPI_ISL_1010253, EPI_ISL_1010254, EPI_ISL_1010255, EPI_ISL_1010256, EPI_ISL_1010257, EPI_ISL_1010258, EPI_ISL_1010259, EPI_ISL_1010260, EPI_ISL_1010261, EPI_ISL_1010262, EPI_ISL_1010263, EPI_ISL_1010264, EPI_ISL_1010265, EPI_ISL_1010266, EPI_ISL_1010267, EPI_ISL_1032946

| | | | |
|---|---|---|---|
| see above | Fulgent Genetics | Fulgent Genetics | Becky Tsai; Benafsh Sapra; Doreen Ng; Harry Gao; James Xie; John Gao; Joseph Fierro; Mickey Li; Yan Meng |

EPI_ISL_2626475, EPI_ISL_2626476, EPI_ISL_2626477, EPI_ISL_2626478, EPI_ISL_2626479, EPI_ISL_2626480, EPI_ISL_2626481, EPI_ISL_2626482, EPI_ISL_2626483, EPI_ISL_2626484

| | | | |
|---|---|---|---|
| see above | Helix | Helix | Andrew dei Rossi; Celina Andrade; Chelsea Andrews; Dana Wyman; David Becker; Efren Sandoval; Francisco Tanudjaja; Geraint Levan; Henrique Machado; James Lu; Jason Nguyen; Jimmy Ramirez; Justin Eagles; Magnus Isaksson; Marc Laurent; Sherry Wang; Shishi Luo; Simon White; Tyler Cassens; William Lee |

EPI_ISL_850690, EPI_ISL_850691, EPI_ISL_850693, EPI_ISL_850695, EPI_ISL_850696, EPI_ISL_850697, EPI_ISL_850698, EPI_ISL_850699, EPI_ISL_850700, EPI_ISL_850701, EPI_ISL_850702, EPI_ISL_850703, EPI_ISL_850704, EPI_ISL_850710, EPI_ISL_850735, EPI_ISL_850765, EPI_ISL_850781, EPI_ISL_850784, EPI_ISL_850788, EPI_ISL_850790, EPI_ISL_850791, EPI_ISL_850792, EPI_ISL_850793, EPI_ISL_850794, EPI_ISL_850795, EPI_ISL_850796, EPI_ISL_850797, EPI_ISL_850798, EPI_ISL_850799, EPI_ISL_850800, EPI_ISL_850801, EPI_ISL_850802, EPI_ISL_850803, EPI_ISL_850804, EPI_ISL_850805, EPI_ISL_850806, EPI_ISL_850807, EPI_ISL_850808, EPI_ISL_850809, EPI_ISL_850811, EPI_ISL_850812, EPI_ISL_850813, EPI_ISL_850814, EPI_ISL_850815, EPI_ISL_850816

| | | | |
|---|---|---|---|
| see above | Helix / Illumina | Genomics and Discovery, Respiratory Viruses Branch, Division of Viral Diseases, Centers for Disease Control and Prevention | Alexandre Bolze; Ary Ascencio; Ben L. Rambo-Martin Eileen de Feo; Brad Sickler; Charlotte Rivera-Garcia; Christine Tran; Clinton R. Paden; David Becker; Dhwani Batra; Duncan MacCannell; Efren Sandoval; Elizabeth Cirulli; Eric Allen; Geraint Levan; James Lu; Jan Antico; Jason Nguyen; Jimmy Ramirez; Jingtao Liu; Kelly Schiabor Barrett; Kim Gietzen; Magnus Isaksson; Marc Laurent; Matthew Tolentino; Nicole L. Washington; Peter W. Cook; Phil Febbo; Ryan Cho; Shannon Wickline; Sherry Wang; Simon White; Summer Galloway; Suxiang Tong; Tyler Cassens; William Lee |

EPI_ISL_802601, EPI_ISL_802625, EPI_ISL_802626, EPI_ISL_802627, EPI_ISL_802628, EPI_ISL_802652, EPI_ISL_802666, EPI_ISL_802667, EPI_ISL_802668, EPI_ISL_850517, EPI_ISL_850518, EPI_ISL_850519, EPI_ISL_850520, EPI_ISL_850521, EPI_ISL_850522, EPI_ISL_850523, EPI_ISL_850524, EPI_ISL_850525, EPI_ISL_850527, EPI_ISL_850528, EPI_ISL_850588, EPI_ISL_850631, EPI_ISL_850632, EPI_ISL_850633, EPI_ISL_850634, EPI_ISL_850635, EPI_ISL_850637, EPI_ISL_850638, EPI_ISL_850953, EPI_ISL_850954, EPI_ISL_850956, EPI_ISL_850957, EPI_ISL_850958, EPI_ISL_850959, EPI_ISL_851000, EPI_ISL_851001, EPI_ISL_851002, EPI_ISL_851003, EPI_ISL_851004, EPI_ISL_851005, EPI_ISL_851006, EPI_ISL_851011, EPI_ISL_851013, EPI_ISL_851021, EPI_ISL_851022, EPI_ISL_851023, EPI_ISL_851025, EPI_ISL_851032, EPI_ISL_876634, EPI_ISL_876635, EPI_ISL_876641, EPI_ISL_876644, EPI_ISL_876645, EPI_ISL_876658, EPI_ISL_876661, EPI_ISL_876670, EPI_ISL_876671, EPI_ISL_876672, EPI_ISL_876673, EPI_ISL_876674, EPI_ISL_876675, EPI_ISL_876676

| | | | |
|---|---|---|---|
| see above | Helix/Illumina | Genomics and Discovery, Respiratory Viruses Branch, Division of Viral Diseases, Centers for Disease Control and Prevention | ; Alexandre Bolze; Ary Ascencio; Ben L. Rambo-Martin; Ben L. Rambo-Martin Eileen de Feo; Brad Sickler; Charlotte Rivera-Garcia; Christine Tran; Clinton R. Paden; David Becker; Dhwani Batra; Duncan MacCannell; Efren Sandoval; Eileen de Feo; Elizabeth Cirulli; Eric Allen; Geraint Levan; James Lu; Jan Antico; Jason Nguyen; Jimmy Ramirez; Jingtao Liu; Kelly Schiabor Barrett; Kim Gietzen; Magnus Isaksson; Marc Laurent; Matthew Tolentino; Nicole L. Washington; Peter W. Cook; Phil Febbo; Ryan Cho; Shannon Wickline; Sherry Wang; Simon White; Summer Galloway; Suxiang Tong; Tyler Cassens; William Lee |

EPI_ISL_966497, EPI_ISL_966498, EPI_ISL_966499, EPI_ISL_966500, EPI_ISL_966501, EPI_ISL_966505, EPI_ISL_966532, EPI_ISL_966534, EPI_ISL_966535, EPI_ISL_966536, EPI_ISL_966537, EPI_ISL_966538, EPI_ISL_966545, EPI_ISL_966583, EPI_ISL_966584, EPI_ISL_966585, EPI_ISL_966586, EPI_ISL_966587, EPI_ISL_966588, EPI_ISL_966590, EPI_ISL_967175, EPI_ISL_967193, EPI_ISL_967199, EPI_ISL_967211, EPI_ISL_967214, EPI_ISL_967218, EPI_ISL_967243, EPI_ISL_967257, EPI_ISL_967259, EPI_ISL_967305, EPI_ISL_967318, EPI_ISL_967324, EPI_ISL_967346, EPI_ISL_967347, EPI_ISL_967354, EPI_ISL_967355, EPI_ISL_967361, EPI_ISL_967376, EPI_ISL_967390, EPI_ISL_967392, EPI_ISL_967403, EPI_ISL_967407, EPI_ISL_967413, EPI_ISL_967418, EPI_ISL_967425, EPI_ISL_967426, EPI_ISL_967427, EPI_ISL_967428, EPI_ISL_967429, EPI_ISL_967430, EPI_ISL_967505, EPI_ISL_967506, EPI_ISL_967514, EPI_ISL_967517, EPI_ISL_967700, EPI_ISL_967710, EPI_ISL_967719, EPI_ISL_967726, EPI_ISL_967736, EPI_ISL_967748, EPI_ISL_967775, EPI_ISL_967777, EPI_ISL_967779, EPI_ISL_967780, EPI_ISL_967813, EPI_ISL_967817, EPI_ISL_967818, EPI_ISL_967819, EPI_ISL_967820, EPI_ISL_967849, EPI_ISL_978622, EPI_ISL_978623, EPI_ISL_978624, EPI_ISL_978630, EPI_ISL_978640, EPI_ISL_978642, EPI_ISL_978643, EPI_ISL_978644, EPI_ISL_978646, EPI_ISL_978648, EPI_ISL_978718, EPI_ISL_978727, EPI_ISL_978730, EPI_ISL_978733, EPI_ISL_978734, EPI_ISL_1036687, EPI_ISL_1036698, EPI_ISL_1036699, EPI_ISL_1036700, EPI_ISL_1036707, EPI_ISL_1036709, EPI_ISL_1036712, EPI_ISL_1036714, EPI_ISL_1036716, EPI_ISL_1036719

| | | | |
|---|---|---|---|
| see above | Helix/Illumina | Respiratory Viruses Branch, Division of Viral Diseases, Centers for Disease Control and Prevention | ; Alexandre Bolze; Ary Ascencio; Ben L. Rambo-Martin; Brad Sickler; Charlotte Rivera-Garcia; Christine Tran; Clinton R. Paden; Dakota Howard; David Becker; Dhwani Batra; Duncan MacCannell; Efren Sandoval; Eileen de Feo; Elizabeth Cirulli; Eric Allen; Geraint Levan; James Lu; Jan Antico; Jason Nguyen; Jimmy Ramirez; Jingtao Liu; Kelly Schiabor Barrett; Kim Gietzen; Magnus Isaksson; Marc Laurent; Matthew Tolentino; Nicole L. Washington; Peter W. Cook; Phil Febbo; Ryan Cho; Shannon Wickline; Sherry Wang; Simon White; Summer Galloway; Suxiang Tong; Tyler Cassens; William Lee |

EPI_ISL_955730, EPI_ISL_955736, EPI_ISL_955737, EPI_ISL_955738, EPI_ISL_955742, EPI_ISL_955745, EPI_ISL_955746, EPI_ISL_955747, EPI_ISL_955750, EPI_ISL_955751, EPI_ISL_955752, EPI_ISL_955753, EPI_ISL_955754, EPI_ISL_955755, EPI_ISL_955756, EPI_ISL_955757, EPI_ISL_955758, EPI_ISL_955759, EPI_ISL_955760, EPI_ISL_955761, EPI_ISL_955762, EPI_ISL_955763, EPI_ISL_955764, EPI_ISL_955765, EPI_ISL_955766, EPI_ISL_955767, EPI_ISL_955768, EPI_ISL_955769, EPI_ISL_955770, EPI_ISL_955771, EPI_ISL_955772, EPI_ISL_955773, EPI_ISL_955774, EPI_ISL_955775, EPI_ISL_955776, EPI_ISL_955777, EPI_ISL_1027476, EPI_ISL_1027517, EPI_ISL_1185287, EPI_ISL_1185291, EPI_ISL_1185292, EPI_ISL_1185294, EPI_ISL_1185298, EPI_ISL_1185300, EPI_ISL_1185301, EPI_ISL_1185302, EPI_ISL_1185304, EPI_ISL_1185306, EPI_ISL_1185308, EPI_ISL_1185309, EPI_ISL_1185312, EPI_ISL_1185318, EPI_ISL_1185319, EPI_ISL_1185323, EPI_ISL_1185324, EPI_ISL_1185325, EPI_ISL_1185326, EPI_ISL_1185328, EPI_ISL_1185329, EPI_ISL_1185340, EPI_ISL_1185345, EPI_ISL_1185346, EPI_ISL_1185352, EPI_ISL_1185353, EPI_ISL_1185355, EPI_ISL_1185359, EPI_ISL_1185360, EPI_ISL_1185361, EPI_ISL_1185365, EPI_ISL_1185369, EPI_ISL_1185382, EPI_ISL_1185385, EPI_ISL_1185394, EPI_ISL_1185395, EPI_ISL_1185399, EPI_ISL_1185407, EPI_ISL_1185408, EPI_ISL_1185409, EPI_ISL_1185410, EPI_ISL_1185412, EPI_ISL_1185413, EPI_ISL_1185417, EPI_ISL_1185424, EPI_ISL_1185425, EPI_ISL_1185428, EPI_ISL_1185432, EPI_ISL_1185436, EPI_ISL_1185438, EPI_ISL_1185439, EPI_ISL_1185440, EPI_ISL_1185448, EPI_ISL_1185454, EPI_ISL_1185455, EPI_ISL_1185460, EPI_ISL_1185463, EPI_ISL_1185468, EPI_ISL_1185469, EPI_ISL_1185471, EPI_ISL_1185474, EPI_ISL_1185476, EPI_ISL_1185479, EPI_ISL_1185481, EPI_ISL_1185486, EPI_ISL_1185487, EPI_ISL_1185488, EPI_ISL_1185489, EPI_ISL_1185497, EPI_ISL_1185498, EPI_ISL_1185504, EPI_ISL_1185505, EPI_ISL_1185507, EPI_ISL_1185509, EPI_ISL_1185516, EPI_ISL_1185518, EPI_ISL_1185519, EPI_ISL_1185521, EPI_ISL_1185526, EPI_ISL_1185528, EPI_ISL_1234789, EPI_ISL_1234793, EPI_ISL_1234794, EPI_ISL_1234799, EPI_ISL_1234807, EPI_ISL_1234826, EPI_ISL_1234831, EPI_ISL_1234833, EPI_ISL_1234836, EPI_ISL_1234871, EPI_ISL_1234882, EPI_ISL_1234908, EPI_ISL_1234930, EPI_ISL_1234949, EPI_ISL_1234955, EPI_ISL_1235002, EPI_ISL_1235011, EPI_ISL_1235012, EPI_ISL_1235023, EPI_ISL_1235030, EPI_ISL_2658600, EPI_ISL_2658710, EPI_ISL_2659265, EPI_ISL_2659316, EPI_ISL_2659319, EPI_ISL_2659336, EPI_ISL_2659338, EPI_ISL_2659359, EPI_ISL_2659380, EPI_ISL_2659421, EPI_ISL_2659557, EPI_ISL_2659609, EPI_ISL_2659635

| | | | |
|---|---|---|---|
| see above | Humboldt County Public Health Laboratory | Chan-Zuckerberg Biohub | CZB Cliahub Consortium |
| EPI_ISL_1081337, EPI_ISL_1081339, EPI_ISL_1081341 | IEH Laboratories and Consulting Group | IEH and ID Genomics | Curtis Fowler; Evgeni Sokurenko; Hao Geng; Hemantha Kulakesara; Kai Deng; Kirthi Kutumbaka; Lydia Larson; Mansour Samadpour; Veronika Tchesnokova |
| EPI_ISL_2253171 | Infectious Diseases, Chan-Zuckerberg Biohub Covidtracker | Infectious Diseases, Chan-Zuckerberg Biohub Covidtracker | CZB Cliahub Consortium |

EPI_ISL_906920, EPI_ISL_906921, EPI_ISL_906922, EPI_ISL_906923, EPI_ISL_906924, EPI_ISL_906925, EPI_ISL_906926, EPI_ISL_906927, EPI_ISL_906928, EPI_ISL_906929, EPI_ISL_906930, EPI_ISL_906931, EPI_ISL_906932, EPI_ISL_906933, EPI_ISL_906934, EPI_ISL_906935, EPI_ISL_906936, EPI_ISL_906937, EPI_ISL_906938, EPI_ISL_906939, EPI_ISL_906940, EPI_ISL_906941, EPI_ISL_906942, EPI_ISL_906943, EPI_ISL_906944, EPI_ISL_906945, EPI_ISL_906946, EPI_ISL_906947, EPI_ISL_906948, EPI_ISL_906949, EPI_ISL_906950, EPI_ISL_906951, EPI_ISL_906952, EPI_ISL_906953, EPI_ISL_906954, EPI_ISL_906955, EPI_ISL_906956, EPI_ISL_906957, EPI_ISL_906958, EPI_ISL_906959, EPI_ISL_906960, EPI_ISL_906961, EPI_ISL_906962, EPI_ISL_906963, EPI_ISL_906964, EPI_ISL_906965, EPI_ISL_906966, EPI_ISL_906967, EPI_ISL_906968, EPI_ISL_906969, EPI_ISL_906970, EPI_ISL_906971, EPI_ISL_906972, EPI_ISL_906973, EPI_ISL_906974, EPI_ISL_906975, EPI_ISL_906976, EPI_ISL_906977, EPI_ISL_906978, EPI_ISL_906979, EPI_ISL_906980, EPI_ISL_906981, EPI_ISL_906982, EPI_ISL_906983, EPI_ISL_906984, EPI_ISL_906985, EPI_ISL_906986, EPI_ISL_906987, EPI_ISL_906988, EPI_ISL_906989, EPI_ISL_906990, EPI_ISL_906991, EPI_ISL_906992, EPI_ISL_906993, EPI_ISL_906994, EPI_ISL_906995, EPI_ISL_906996, EPI_ISL_906997, EPI_ISL_906998, EPI_ISL_906999, EPI_ISL_907000, EPI_ISL_907001, EPI_ISL_907002, EPI_ISL_907003, EPI_ISL_907004, EPI_ISL_907005, EPI_ISL_907006, EPI_ISL_907007, EPI_ISL_907008, EPI_ISL_907009, EPI_ISL_907010, EPI_ISL_907011, EPI_ISL_907012, EPI_ISL_907013, EPI_ISL_907014, EPI_ISL_907015, EPI_ISL_907016, EPI_ISL_907017, EPI_ISL_907018, EPI_ISL_907019, EPI_ISL_907020, EPI_ISL_907021, EPI_ISL_907022, EPI_ISL_907023, EPI_ISL_907024, EPI_ISL_907025, EPI_ISL_907026, EPI_ISL_907027, EPI_ISL_907028, EPI_ISL_907029, EPI_ISL_907030, EPI_ISL_907031, EPI_ISL_907032, EPI_ISL_907033, EPI_ISL_907034, EPI_ISL_907035, EPI_ISL_907036, EPI_ISL_907037, EPI_ISL_907038, EPI_ISL_907039, EPI_ISL_907040, EPI_ISL_907041, EPI_ISL_907042, EPI_ISL_907043, EPI_ISL_907044, EPI_ISL_907045, EPI_ISL_907046, EPI_ISL_907047, EPI_ISL_907048, EPI_ISL_907049, EPI_ISL_907050, EPI_ISL_907051, EPI_ISL_907052, EPI_ISL_907053, EPI_ISL_907054, EPI_ISL_907055, EPI_ISL_907056, EPI_ISL_907057, EPI_ISL_907058, EPI_ISL_907059, EPI_ISL_907060, EPI_ISL_907061, EPI_ISL_907062, EPI_ISL_907063, EPI_ISL_1233901, EPI_ISL_1233902, EPI_ISL_1233903, EPI_ISL_1233919, EPI_ISL_1233920

| | | | |
|---|---|---|---|
| see above | Infectious Diseases, Quest Diagnostics | Infectious Diseases, Quest Diagnostics | Anderson, B.; Bernstein; D.F.; Gerasimova, A.; Hua, M.; I.A.; K.E.; Kagan; L.E.; Lacbawan, F.; Liu, Y.; Livingston; Owen, R.; Perez, A.; R.M.; Rosenthal; S.H.; Shalhout; Shlyakhter; Tanpaiboon, P. |

EPI_ISL_918177, EPI_ISL_918179, EPI_ISL_918180, EPI_ISL_918181, EPI_ISL_918182, EPI_ISL_918183, EPI_ISL_918185, EPI_ISL_918187, EPI_ISL_918188, EPI_ISL_918189, EPI_ISL_918190, EPI_ISL_918194, EPI_ISL_918195, EPI_ISL_918196, EPI_ISL_918199, EPI_ISL_918200, EPI_ISL_918201, EPI_ISL_918203, EPI_ISL_918204, EPI_ISL_918226, EPI_ISL_918227, EPI_ISL_918229, EPI_ISL_918230, EPI_ISL_918231, EPI_ISL_918234, EPI_ISL_918235, EPI_ISL_918236, EPI_ISL_918237, EPI_ISL_918238, EPI_ISL_918243, EPI_ISL_918245, EPI_ISL_918246, EPI_ISL_918247, EPI_ISL_918249, EPI_ISL_918251, EPI_ISL_918252, EPI_ISL_918253, EPI_ISL_918254, EPI_ISL_918255, EPI_ISL_918256, EPI_ISL_918257, EPI_ISL_918258, EPI_ISL_918259, EPI_ISL_918260, EPI_ISL_918262, EPI_ISL_955127, EPI_ISL_955129, EPI_ISL_980845, EPI_ISL_980852, EPI_ISL_980854, EPI_ISL_980855, EPI_ISL_980859, EPI_ISL_980897, EPI_ISL_980898, EPI_ISL_980899, EPI_ISL_980902, EPI_ISL_980905, EPI_ISL_980916, EPI_ISL_980920, EPI_ISL_980921, EPI_ISL_980922, EPI_ISL_980930, EPI_ISL_980933, EPI_ISL_980935, EPI_ISL_980936, EPI_ISL_980937, EPI_ISL_980938, EPI_ISL_980939, EPI_ISL_980975, EPI_ISL_980976, EPI_ISL_980977, EPI_ISL_980978, EPI_ISL_980979, EPI_ISL_980980, EPI_ISL_980981, EPI_ISL_980982, EPI_ISL_980983, EPI_ISL_984662, EPI_ISL_1712450, EPI_ISL_1712451, EPI_ISL_1712452, EPI_ISL_1712453, EPI_ISL_1712454, EPI_ISL_1712455, EPI_ISL_1712456

| | | | |
|---|---|---|---|
| see above | Innovative Genomics Institute, UC Berkeley | Innovative Genomics Institute, UC Berkeley | Haridha Shivram; Holly Gildea; John Boyle; Justin Choi; Liana Lareau; Phil Frankino; Shana McDevitt; Stacia Wyman |

EPI_ISL_886303, EPI_ISL_886330, EPI_ISL_886348, EPI_ISL_886351, EPI_ISL_886352, EPI_ISL_886354, EPI_ISL_886358, EPI_ISL_886463, EPI_ISL_886465, EPI_ISL_886557, EPI_ISL_886720, EPI_ISL_886730, EPI_ISL_886865, EPI_ISL_886907, EPI_ISL_887035, EPI_ISL_887641, EPI_ISL_887729, EPI_ISL_887995, EPI_ISL_888059, EPI_ISL_888522, EPI_ISL_888583

| | | | |
|---|---|---|---|
| see above | Labcorp | Genomics and Discovery, Respiratory Viruses Branch, Division of Viral Diseases, Centers for Disease Control and Prevention | ; Amanda Douglas; Amanda Suchanek; Andrea Throop; Ayla Burns; Ben L. Rambo-Martin; Bobbi Croy; Brian Krueger; Brian Norvell; Christos Petropoulos; Clinton R. Paden; Craig Lukasik; Debbie Boles; Dhwani Batra; Duncan MacCannell; Eyad Almasri; Goran Stevovic; Howard Engler; Hrushikesh Deshmukh; Jake Humphrey; Jana Schroth; Joe Voshell; John Pruitt; Jonathan Meltzer; Jonathan Williams; Kimberly Wagner; Lax Iyer; Lyndon Tilson; Manoj Jain; Marcia Eisenberg; Mary Ann Cristobal; Mary Williamson; Michael Levandoski; Mike Sapeta; Mindy Nye; Minoo Agarwal; Mohan Kolli; Nuthawin Charoensri; Oren Cohen; Peter W. Cook; Prashant Gupta; Qian Zeng; Rama Ghatti; Scott Parker; Scott Ryan; Stanley Letovsky; Steven Ragan; Summer Galloway; Suresh Babu Selvaraju; Susan Countryman; Susan Hicks; Suxiang Tong; Suzanne Dale; Thomas Urban; Tim Kuphal; Tricia Zwiefelhofer; Vincent Drouillon |

EPI_ISL_1021343, EPI_ISL_1021367, EPI_ISL_1021373, EPI_ISL_1021374, EPI_ISL_1021380, EPI_ISL_1021382, EPI_ISL_1021397, EPI_ISL_1021402, EPI_ISL_1021403, EPI_ISL_1021415, EPI_ISL_1021416, EPI_ISL_1021445, EPI_ISL_1021448, EPI_ISL_1021462, EPI_ISL_1021468, EPI_ISL_1021526, EPI_ISL_1021529, EPI_ISL_1021554, EPI_ISL_1021559, EPI_ISL_1021579, EPI_ISL_1021612, EPI_ISL_1021616, EPI_ISL_1021617, EPI_ISL_1021622, EPI_ISL_1021623, EPI_ISL_1021624, EPI_ISL_1021646, EPI_ISL_1021647, EPI_ISL_1021648, EPI_ISL_1021651, EPI_ISL_1021652, EPI_ISL_1021665, EPI_ISL_1021671, EPI_ISL_1021680, EPI_ISL_1021696, EPI_ISL_1021697, EPI_ISL_1021698, EPI_ISL_1021723, EPI_ISL_1021724, EPI_ISL_1021794, EPI_ISL_1021795, EPI_ISL_1021810, EPI_ISL_1021814, EPI_ISL_1021815, EPI_ISL_1021823, EPI_ISL_1021826, EPI_ISL_1021827, EPI_ISL_1021830, EPI_ISL_1021834, EPI_ISL_1021837, EPI_ISL_1021838, EPI_ISL_1021841, EPI_ISL_1021853, EPI_ISL_1021860, EPI_ISL_1021861, EPI_ISL_1021862, EPI_ISL_1021871, EPI_ISL_1021877, EPI_ISL_1021892, EPI_ISL_1021897, EPI_ISL_1021899, EPI_ISL_1021906, EPI_ISL_1021907, EPI_ISL_1021908, EPI_ISL_1021914, EPI_ISL_1021915, EPI_ISL_1021923, EPI_ISL_1021926, EPI_ISL_1021932, EPI_ISL_1021933, EPI_ISL_1021934, EPI_ISL_1021935, EPI_ISL_1021936, EPI_ISL_1021937, EPI_ISL_1021961, EPI_ISL_1021964, EPI_ISL_1021975, EPI_ISL_1021978, EPI_ISL_1021987, EPI_ISL_1021998, EPI_ISL_1022003, EPI_ISL_1022007, EPI_ISL_1022009, EPI_ISL_1022018, EPI_ISL_1022019, EPI_ISL_1022020, EPI_ISL_1022025, EPI_ISL_1022028, EPI_ISL_1022029, EPI_ISL_1022036, EPI_ISL_1022040, EPI_ISL_1022045, EPI_ISL_1022052, EPI_ISL_1022055, EPI_ISL_1022057, EPI_ISL_1022060, EPI_ISL_1022064, EPI_ISL_1022065, EPI_ISL_1022067, EPI_ISL_1022068, EPI_ISL_1022070, EPI_ISL_1022072, EPI_ISL_1022073, EPI_ISL_1022079, EPI_ISL_1022083, EPI_ISL_1022084, EPI_ISL_1022086, EPI_ISL_1022090, EPI_ISL_1022091, EPI_ISL_1022093, EPI_ISL_1022094, EPI_ISL_1022095, EPI_ISL_1022099, EPI_ISL_1022101, EPI_ISL_1022103, EPI_ISL_1022104, EPI_ISL_1022105, EPI_ISL_1022106, EPI_ISL_1022109, EPI_ISL_1022110, EPI_ISL_1022116, EPI_ISL_1022120, EPI_ISL_1022124, EPI_ISL_1022125, EPI_ISL_1022126, EPI_ISL_1022127, EPI_ISL_1022128, EPI_ISL_1022138, EPI_ISL_1022139, EPI_ISL_1022140, EPI_ISL_1022141, EPI_ISL_1022159, EPI_ISL_1022160, EPI_ISL_1022161, EPI_ISL_1022162, EPI_ISL_1022163, EPI_ISL_1022164, EPI_ISL_1022169, EPI_ISL_1022174, EPI_ISL_1022188, EPI_ISL_1022221, EPI_ISL_1022222, EPI_ISL_1022225, EPI_ISL_1022226, EPI_ISL_1022232, EPI_ISL_1022250, EPI_ISL_1022255, EPI_ISL_1022267, EPI_ISL_1022295, EPI_ISL_1022326, EPI_ISL_1022369, EPI_ISL_1022375, EPI_ISL_1022380, EPI_ISL_1022393, EPI_ISL_1022394, EPI_ISL_1022407, EPI_ISL_1022419, EPI_ISL_1025966, EPI_ISL_1025999, EPI_ISL_1026019, EPI_ISL_1026025, EPI_ISL_1026056, EPI_ISL_1026151, EPI_ISL_1026171, EPI_ISL_1026183, EPI_ISL_1026340, EPI_ISL_1026355, EPI_ISL_1026469, EPI_ISL_1026495, EPI_ISL_1026564, EPI_ISL_1026638, EPI_ISL_1026728, EPI_ISL_1026760, EPI_ISL_1026809, EPI_ISL_1027752, EPI_ISL_1027888, EPI_ISL_1028135, EPI_ISL_1028205, EPI_ISL_1028413, EPI_ISL_1028426, EPI_ISL_1028552, EPI_ISL_1028634, EPI_ISL_1028641, EPI_ISL_1028751, EPI_ISL_1029767, EPI_ISL_1029768, EPI_ISL_1029774, EPI_ISL_1029775, EPI_ISL_1029801, EPI_ISL_1029866

| | | | |
|---|---|---|---|
| see above | Laboratory Corporation of America | Respiratory Viruses Branch, Division of Viral Diseases, Centers for Disease Control and Prevention | Ben L. Rambo-Martin; Clinton R. Paden; Dakota Howard; Dhwani Batra; Duncan MacCannell; Peter W. Cook; Suxiang Tong |

EPI_ISL_2988733, EPI_ISL_2988734, EPI_ISL_2988735, EPI_ISL_2988736, EPI_ISL_2988737, EPI_ISL_2988738, EPI_ISL_2988739, EPI_ISL_2988740, EPI_ISL_2988741, EPI_ISL_2988742, EPI_ISL_2988743, EPI_ISL_2988744, EPI_ISL_2988745, EPI_ISL_2988746, EPI_ISL_2988747, EPI_ISL_2988748, EPI_ISL_2988749, EPI_ISL_2988750, EPI_ISL_2988751, EPI_ISL_2988752, EPI_ISL_2988753, EPI_ISL_2988754, EPI_ISL_2988755, EPI_ISL_2988756, EPI_ISL_2988757, EPI_ISL_2988758, EPI_ISL_2988759, EPI_ISL_2988760, EPI_ISL_2988761, EPI_ISL_2988762, EPI_ISL_2988763, EPI_ISL_2988764, EPI_ISL_2988765, EPI_ISL_2988766, EPI_ISL_2988767, EPI_ISL_2988768, EPI_ISL_2988769, EPI_ISL_2988770, EPI_ISL_2988771, EPI_ISL_2988772, EPI_ISL_2988773, EPI_ISL_2988774, EPI_ISL_2988775, EPI_ISL_2988776, EPI_ISL_2988777, EPI_ISL_2988778, EPI_ISL_2988779, EPI_ISL_2988780, EPI_ISL_2988781, EPI_ISL_2988782, EPI_ISL_2988783, EPI_ISL_2988784, EPI_ISL_2988785, EPI_ISL_2988786, EPI_ISL_2988787, EPI_ISL_2988788, EPI_ISL_2988789, EPI_ISL_2988790, EPI_ISL_2988791, EPI_ISL_2988792, EPI_ISL_2988793, EPI_ISL_2988794, EPI_ISL_2988795, EPI_ISL_2988796, EPI_ISL_2988797, EPI_ISL_2988798, EPI_ISL_2988799, EPI_ISL_2988800, EPI_ISL_2988801, EPI_ISL_2988802, EPI_ISL_2988803, EPI_ISL_2988804

| | | | |
|---|---|---|---|
| see above | Los Angeles County Public Health Laboratories | Andersen lab at Scripps Research | H. Rivas; P. Hemarajata; SEARCH Alliance San Diego with N. Green; T. Burleson et al. |
| EPI_ISL_861686, EPI_ISL_861687, EPI_ISL_861688, EPI_ISL_861689, EPI_ISL_861690, EPI_ISL_861691, EPI_ISL_861692, EPI_ISL_861693, EPI_ISL_861694, EPI_ISL_861695, EPI_ISL_861696, EPI_ISL_861697, EPI_ISL_903371, EPI_ISL_903372, EPI_ISL_917936, EPI_ISL_917938, EPI_ISL_917941, EPI_ISL_939652, EPI_ISL_1005487, EPI_ISL_1018060, EPI_ISL_1018061, EPI_ISL_1018062, EPI_ISL_2964661, EPI_ISL_2964662, EPI_ISL_2964663, EPI_ISL_2964664, EPI_ISL_2964665, EPI_ISL_2964667, EPI_ISL_2964668, EPI_ISL_2964669, EPI_ISL_2964670 | | | |
| see above | Los Angeles County Public Health Laboratories | Los Angeles County Public Health Laboratories | Green, N.; Hemarajata, P.; P. Hemarajata et al. |
| EPI_ISL_833417, EPI_ISL_833418, EPI_ISL_833419, EPI_ISL_833420, EPI_ISL_833421, EPI_ISL_833422, EPI_ISL_833423, EPI_ISL_833424, EPI_ISL_833425, EPI_ISL_833426, EPI_ISL_833427, EPI_ISL_833490, EPI_ISL_833491, EPI_ISL_861700, EPI_ISL_861701, EPI_ISL_861702, EPI_ISL_861703, EPI_ISL_862121, EPI_ISL_882963, EPI_ISL_882964, EPI_ISL_882965, EPI_ISL_882966, EPI_ISL_882985, EPI_ISL_882986, EPI_ISL_882987, EPI_ISL_882988, EPI_ISL_882989, EPI_ISL_882990, EPI_ISL_882991, EPI_ISL_882992, EPI_ISL_882993, EPI_ISL_882994, EPI_ISL_882995, EPI_ISL_882996, EPI_ISL_892249, EPI_ISL_892250, EPI_ISL_892251, EPI_ISL_892252, EPI_ISL_892253, EPI_ISL_892254, EPI_ISL_892255, EPI_ISL_892256, EPI_ISL_892257, EPI_ISL_892258, EPI_ISL_892259, EPI_ISL_892260, EPI_ISL_892261, EPI_ISL_892262, EPI_ISL_892263, EPI_ISL_892264, EPI_ISL_892265, EPI_ISL_892266, EPI_ISL_902925, EPI_ISL_903373, EPI_ISL_903374, EPI_ISL_905765, EPI_ISL_905766, EPI_ISL_905767, EPI_ISL_905768, EPI_ISL_905769, EPI_ISL_905770, EPI_ISL_905771, EPI_ISL_905772, EPI_ISL_906047, EPI_ISL_906048, EPI_ISL_915434, EPI_ISL_915435, EPI_ISL_915436, EPI_ISL_915437, EPI_ISL_915439, EPI_ISL_917944, EPI_ISL_917945, EPI_ISL_917946, EPI_ISL_917947, EPI_ISL_917948, EPI_ISL_917949, EPI_ISL_917950, EPI_ISL_939651, EPI_ISL_939653, EPI_ISL_962515, EPI_ISL_962516, EPI_ISL_962847, EPI_ISL_1005555, EPI_ISL_1015733, EPI_ISL_1098628, EPI_ISL_1098646 | | | |
| see above | MD Laboratories | Los Angeles County Public Health Laboratories | P. Hemarajata et al. |
| EPI_ISL_2009465, EPI_ISL_2009467 | Moderna Inc. | Moderna Inc. | Groves Dixon; Rolando Pajon; Yamuna Paila |
| EPI_ISL_930791, EPI_ISL_930792, EPI_ISL_930794, EPI_ISL_930796, EPI_ISL_930797, EPI_ISL_930800, EPI_ISL_930804, EPI_ISL_930805, EPI_ISL_930830, EPI_ISL_930833, EPI_ISL_930845, EPI_ISL_930847, EPI_ISL_930848, EPI_ISL_930851, EPI_ISL_930852, EPI_ISL_968069, EPI_ISL_968070, EPI_ISL_968071, EPI_ISL_968072, EPI_ISL_968073, EPI_ISL_968074, EPI_ISL_968075, EPI_ISL_968076, EPI_ISL_968079, EPI_ISL_1263227, EPI_ISL_1263229, EPI_ISL_1664599, EPI_ISL_1664600, EPI_ISL_1664601, EPI_ISL_1664602, EPI_ISL_1664603, EPI_ISL_1664604, EPI_ISL_1664605, EPI_ISL_1700696, EPI_ISL_1700697, EPI_ISL_1700698, EPI_ISL_1700699, EPI_ISL_2754036, EPI_ISL_2754038, EPI_ISL_2754039, EPI_ISL_2754041, EPI_ISL_2759084, EPI_ISL_2774522, EPI_ISL_2955354, EPI_ISL_2955355, EPI_ISL_2955357, EPI_ISL_2955358, EPI_ISL_2955359, EPI_ISL_2955360, EPI_ISL_2955361, EPI_ISL_2955362, EPI_ISL_3065506 | | | |
| see above | Monterey County Public Health Laboratory | Monterey County Public Health Laboratory | ; C. Anaya et al.; Monterey County Public Health Laboratory |
| EPI_ISL_832204 | Northwest Pathology | UW Virology Lab | Alexander Greninger; Hong Xie; Keith R Jerome; Lasata Shrestha; Meei-Li Huang; Michelle Lin; Pavitra Roychoudhury |
| EPI_ISL_936990 | Northwestern Memorial Hospital | Ozer Lab | Chad J. Achenbach; Egon A. Ozer; Judd F. Hultquist; Lacy M. Simons; Lawrence J. Jennings; Michael G. Ison; Ramon Lorenzo-Redondo |
| EPI_ISL_2839062 | OHSU Lab Services Molecular Microbiology Lab | Ginkgo Bioworks Clinical Laboratory | '[UsersoroakBox SyncPHI_SARS-COV-2_Sample_Sheet_infoGISAID_Submission_Infometa_master_2021.xlsx]release20_final'!D:AC; 26; FALSE); VLOOKUP(C315 |
| EPI_ISL_1122131, EPI_ISL_1541732, EPI_ISL_1541768, EPI_ISL_1541889 | OHSU Lab Services Molecular Microbiology Lab | Oregon SARS-CoV-2 Genome Sequencing Center | Alec J. Hirsch; Andrew C. Adey; Benjamin N. Bimber; Brendan L. O'Connell; Brian J. O'Roak; Daniel N. Streblow; Donna Hansel; Guang Fan; Kayla Carter; Qin; Ruth V. Nichols; Sally Grindstaff; Sonia Acharya; William B. Messer; Xuan; Xuan Qin |
| EPI_ISL_3029875, EPI_ISL_3029880, EPI_ISL_3029881, EPI_ISL_3029882, EPI_ISL_3029883, EPI_ISL_3029884, EPI_ISL_3029885, EPI_ISL_3029886, EPI_ISL_3029887, EPI_ISL_3029888, EPI_ISL_3029889, EPI_ISL_3029890, EPI_ISL_3029891, EPI_ISL_3029892, EPI_ISL_3029893, EPI_ISL_3029894, EPI_ISL_3029895, EPI_ISL_3029896, EPI_ISL_3029897 | | | |
| see above | Olive View Medical Center | Los Angeles County Public Health Laboratories | P. Hemarajata et al. |
| EPI_ISL_2502612, EPI_ISL_2502613, EPI_ISL_2502614, EPI_ISL_2566368, EPI_ISL_2566369, EPI_ISL_2566370, EPI_ISL_2566371, EPI_ISL_2566372, EPI_ISL_2566373, EPI_ISL_2566374, EPI_ISL_2566375, EPI_ISL_2566376, EPI_ISL_2566377, EPI_ISL_2566378, EPI_ISL_2566379 | | | |
| see above | Operation Infectious Diseases, Naval Health Research Center | Operation Infectious Diseases, Naval Health Research Center | A.E.; Balansay-Ames; C.A.; Fierro, M.; Iniguez-Stevens, E.; Kriner, P.; M.S.; Moser, K.; Myers; Pan; Phippard; R.S.; R.W.; Underwood |
| EPI_ISL_849922, EPI_ISL_955427, EPI_ISL_955428, EPI_ISL_955429, EPI_ISL_955430, EPI_ISL_955431, EPI_ISL_955432, EPI_ISL_955433, EPI_ISL_955434, EPI_ISL_955435, EPI_ISL_955436, EPI_ISL_955437, EPI_ISL_955438, EPI_ISL_955439, EPI_ISL_955440, EPI_ISL_955441, EPI_ISL_955442, EPI_ISL_955443, EPI_ISL_955444, EPI_ISL_955445, EPI_ISL_955446, EPI_ISL_955447, EPI_ISL_955448, EPI_ISL_955449, EPI_ISL_955450, EPI_ISL_955451, EPI_ISL_955452, EPI_ISL_955453, EPI_ISL_955454, EPI_ISL_955455, EPI_ISL_955456, EPI_ISL_955457, EPI_ISL_955458, EPI_ISL_955459, EPI_ISL_955460, EPI_ISL_955461, EPI_ISL_955462, EPI_ISL_955463, EPI_ISL_955464, EPI_ISL_955465, EPI_ISL_955466, EPI_ISL_955467, EPI_ISL_955468, EPI_ISL_955469, EPI_ISL_955470, EPI_ISL_955471, EPI_ISL_955472, EPI_ISL_955473, EPI_ISL_955474, EPI_ISL_955475, EPI_ISL_955476, EPI_ISL_955477, EPI_ISL_955478, EPI_ISL_955479, EPI_ISL_955480, EPI_ISL_955481, EPI_ISL_955482, EPI_ISL_955483, EPI_ISL_955484, EPI_ISL_955485, EPI_ISL_955486, EPI_ISL_955487, EPI_ISL_955488, EPI_ISL_955489, EPI_ISL_955490, EPI_ISL_955491, EPI_ISL_955492, EPI_ISL_955493, EPI_ISL_955494, EPI_ISL_955495, EPI_ISL_979690, EPI_ISL_979691, EPI_ISL_979692, EPI_ISL_979693, EPI_ISL_979694, EPI_ISL_979695, EPI_ISL_979696, EPI_ISL_979697, EPI_ISL_979698, EPI_ISL_979699, EPI_ISL_979700, EPI_ISL_979701, EPI_ISL_979702, EPI_ISL_979703, EPI_ISL_979704, EPI_ISL_979705, EPI_ISL_979706, EPI_ISL_979707, EPI_ISL_979708, EPI_ISL_979709, EPI_ISL_1027487, EPI_ISL_1027503, EPI_ISL_1027504, EPI_ISL_1027531, EPI_ISL_1027532, EPI_ISL_1027573, EPI_ISL_1027583, EPI_ISL_1027584, EPI_ISL_1027587, EPI_ISL_1027588, EPI_ISL_1027589, EPI_ISL_1027590, EPI_ISL_1027591, EPI_ISL_1027592, EPI_ISL_1027593, EPI_ISL_1027594, EPI_ISL_1027595, EPI_ISL_1027596, EPI_ISL_1027597, EPI_ISL_1027598, EPI_ISL_1027599, EPI_ISL_1027600, EPI_ISL_1027601, EPI_ISL_1027602, EPI_ISL_1027603, EPI_ISL_1027604, EPI_ISL_1027605, EPI_ISL_1027606, EPI_ISL_1027607, EPI_ISL_1027608, EPI_ISL_1027609, EPI_ISL_1027610, EPI_ISL_1185032, EPI_ISL_1185039, EPI_ISL_1185040, EPI_ISL_1185043, EPI_ISL_1185045, EPI_ISL_1185048, EPI_ISL_1185049, EPI_ISL_1185055, EPI_ISL_1185058, EPI_ISL_1185062, EPI_ISL_1185070, EPI_ISL_1185075, EPI_ISL_1185076, EPI_ISL_1185093, EPI_ISL_1185098, EPI_ISL_1185101, EPI_ISL_1185104, EPI_ISL_1185106, EPI_ISL_1185107, EPI_ISL_1185108, EPI_ISL_1185112, EPI_ISL_1185113, EPI_ISL_1185114, EPI_ISL_1185115, EPI_ISL_1185118, EPI_ISL_1185123, EPI_ISL_1185125, EPI_ISL_1185127, EPI_ISL_1185131, EPI_ISL_1185134, EPI_ISL_1185140, EPI_ISL_1185143, EPI_ISL_1185145, EPI_ISL_1185146, EPI_ISL_1185148, EPI_ISL_1185149, EPI_ISL_1185152, EPI_ISL_1185154, EPI_ISL_1185159, EPI_ISL_1185160, EPI_ISL_1185163, EPI_ISL_1185165, EPI_ISL_1185167, EPI_ISL_1185171, EPI_ISL_1185174, EPI_ISL_1185175, EPI_ISL_1185180, EPI_ISL_1185189, EPI_ISL_1185194, EPI_ISL_1185201, EPI_ISL_1185206, EPI_ISL_1185209, EPI_ISL_1185212, EPI_ISL_1185214, EPI_ISL_1185216, EPI_ISL_1185217, EPI_ISL_1185221, EPI_ISL_1185224, EPI_ISL_1185225, EPI_ISL_1185227, EPI_ISL_1185228, EPI_ISL_1185231, EPI_ISL_1185233, EPI_ISL_1185235, EPI_ISL_1185236, EPI_ISL_1185237, EPI_ISL_1185238, EPI_ISL_1185240, EPI_ISL_1185241, EPI_ISL_1185242, EPI_ISL_1185257, EPI_ISL_1185261, EPI_ISL_1185263, EPI_ISL_1185266, EPI_ISL_1185269, EPI_ISL_1185272, EPI_ISL_1185274, EPI_ISL_1185278, EPI_ISL_1185280, EPI_ISL_2103061, EPI_ISL_2103110, EPI_ISL_2658639, EPI_ISL_2658643, EPI_ISL_2658702, EPI_ISL_2658730, EPI_ISL_2659304, EPI_ISL_2659330, EPI_ISL_2659341, EPI_ISL_2659488, EPI_ISL_2659650, EPI_ISL_2758433 | | | |
| see above | Orange County Public Health Lab | Chan-Zuckerberg Biohub | CZB Cliahub Consortium |
| EPI_ISL_1017052, EPI_ISL_1017075, EPI_ISL_1017076, EPI_ISL_1017077, EPI_ISL_1017078, EPI_ISL_1017079 | Providence St. Joseph Health Orange - Chapman | Providence St. Joseph Health Molecular Genomics Laboratory | Alexa K Dowdell; Brian D Piening; Carlo B Bifulco; Fred L Robinson; Mary Campbell |
| EPI_ISL_854810, EPI_ISL_854811, EPI_ISL_854812, EPI_ISL_854813, EPI_ISL_854814, EPI_ISL_854815, EPI_ISL_854816, EPI_ISL_854817, EPI_ISL_854818, EPI_ISL_854819, EPI_ISL_854820, EPI_ISL_854821, EPI_ISL_854822, EPI_ISL_854823, EPI_ISL_854824, EPI_ISL_854825, EPI_ISL_854826, EPI_ISL_854827, EPI_ISL_854828, EPI_ISL_854829, EPI_ISL_854830, EPI_ISL_854831, EPI_ISL_854832, EPI_ISL_854833, EPI_ISL_854834, EPI_ISL_854835, EPI_ISL_854836, EPI_ISL_854837, EPI_ISL_854838, EPI_ISL_854839, EPI_ISL_854840, EPI_ISL_854841, EPI_ISL_855015, EPI_ISL_855017, EPI_ISL_855018, EPI_ISL_855020, EPI_ISL_855021, EPI_ISL_855022, EPI_ISL_855024, EPI_ISL_855025, EPI_ISL_855026, EPI_ISL_855028, EPI_ISL_855029, EPI_ISL_855032, EPI_ISL_855035, EPI_ISL_855036, EPI_ISL_855040, EPI_ISL_855042, EPI_ISL_855043, EPI_ISL_855044, EPI_ISL_855045, EPI_ISL_855048, EPI_ISL_855051, EPI_ISL_855055, EPI_ISL_855056, EPI_ISL_855058, EPI_ISL_855064, EPI_ISL_855065, EPI_ISL_855066, EPI_ISL_855067, EPI_ISL_855068, EPI_ISL_855069, EPI_ISL_855070, EPI_ISL_855071, EPI_ISL_855072, EPI_ISL_855073, EPI_ISL_855074, EPI_ISL_855075, EPI_ISL_855076, EPI_ISL_855077, EPI_ISL_855078, EPI_ISL_855079, EPI_ISL_855080, EPI_ISL_877037, EPI_ISL_877038, EPI_ISL_877039, EPI_ISL_877040, EPI_ISL_877041, EPI_ISL_877042, EPI_ISL_877043, EPI_ISL_877044, EPI_ISL_877045, EPI_ISL_877046, EPI_ISL_877047, EPI_ISL_877048, EPI_ISL_877049, EPI_ISL_877050, EPI_ISL_877053, EPI_ISL_877054, EPI_ISL_877055, EPI_ISL_877056, EPI_ISL_877057, EPI_ISL_877058, EPI_ISL_877059, EPI_ISL_877060, EPI_ISL_877061, EPI_ISL_877062, EPI_ISL_877063, EPI_ISL_877064, EPI_ISL_877065, EPI_ISL_877066, EPI_ISL_877067, EPI_ISL_877068, EPI_ISL_877069, EPI_ISL_877070, EPI_ISL_877071, EPI_ISL_877072, EPI_ISL_877073, EPI_ISL_877074, EPI_ISL_877075, EPI_ISL_877076, EPI_ISL_877077, EPI_ISL_877078, EPI_ISL_877079, EPI_ISL_877080, EPI_ISL_877081, EPI_ISL_877082, EPI_ISL_877083, EPI_ISL_877084, EPI_ISL_877085, EPI_ISL_877086, EPI_ISL_877087, EPI_ISL_877088, EPI_ISL_877089, EPI_ISL_877090, EPI_ISL_877091, EPI_ISL_877092, EPI_ISL_877093, EPI_ISL_877094, EPI_ISL_877095, EPI_ISL_877096, EPI_ISL_877097, EPI_ISL_877098, EPI_ISL_877099, EPI_ISL_877100, EPI_ISL_877101, EPI_ISL_877102, EPI_ISL_877103, EPI_ISL_877104, EPI_ISL_877105, EPI_ISL_877106, EPI_ISL_877107, EPI_ISL_877108, EPI_ISL_877109, EPI_ISL_877110, EPI_ISL_877111, EPI_ISL_877112, EPI_ISL_877113, EPI_ISL_877114, EPI_ISL_877115, EPI_ISL_877116, EPI_ISL_877117, EPI_ISL_877118, EPI_ISL_877187, EPI_ISL_877197, EPI_ISL_903390, EPI_ISL_903391, EPI_ISL_903392, EPI_ISL_903394, EPI_ISL_903395, EPI_ISL_903396, EPI_ISL_903397, EPI_ISL_903398, EPI_ISL_903399, EPI_ISL_903400, EPI_ISL_903401, EPI_ISL_903402, EPI_ISL_903403, EPI_ISL_903404, EPI_ISL_903405, EPI_ISL_903407, EPI_ISL_903408, EPI_ISL_903409, EPI_ISL_903410, EPI_ISL_903411, EPI_ISL_903412, EPI_ISL_903413, EPI_ISL_903414, EPI_ISL_903415, EPI_ISL_903416, EPI_ISL_903417, EPI_ISL_903418, EPI_ISL_903419, EPI_ISL_903420, EPI_ISL_903421, EPI_ISL_903422, EPI_ISL_903423, EPI_ISL_903424, EPI_ISL_903425, EPI_ISL_903426, EPI_ISL_903427, EPI_ISL_903428, EPI_ISL_903429, EPI_ISL_903430, EPI_ISL_903431, EPI_ISL_903432, EPI_ISL_903433, EPI_ISL_903434, EPI_ISL_903435, EPI_ISL_903436, EPI_ISL_903437, EPI_ISL_903438, EPI_ISL_903439, EPI_ISL_903440, EPI_ISL_903441, EPI_ISL_903442, EPI_ISL_903443, EPI_ISL_903444, EPI_ISL_903445, EPI_ISL_903446, EPI_ISL_903447, EPI_ISL_903448, EPI_ISL_903449, EPI_ISL_903450, EPI_ISL_903451, EPI_ISL_903452, EPI_ISL_903453, EPI_ISL_903454, EPI_ISL_903455, EPI_ISL_903456, EPI_ISL_903457, EPI_ISL_903458, EPI_ISL_903459, EPI_ISL_903460, EPI_ISL_903461, EPI_ISL_903462, EPI_ISL_903463, EPI_ISL_903464, EPI_ISL_903465, EPI_ISL_903466, EPI_ISL_903467, EPI_ISL_903468, EPI_ISL_903469, EPI_ISL_903470, EPI_ISL_903471, EPI_ISL_903472, EPI_ISL_903473, EPI_ISL_903474, EPI_ISL_903475, EPI_ISL_903476, EPI_ISL_903477, EPI_ISL_903478, EPI_ISL_903479, EPI_ISL_903480, EPI_ISL_903481, EPI_ISL_903482, EPI_ISL_903483, EPI_ISL_903484, EPI_ISL_903485, EPI_ISL_903486, EPI_ISL_903487, EPI_ISL_903488, EPI_ISL_903489, EPI_ISL_903490, EPI_ISL_903491, EPI_ISL_903492, EPI_ISL_903493, EPI_ISL_903494, EPI_ISL_903495, EPI_ISL_903496, EPI_ISL_903497, EPI_ISL_903498, EPI_ISL_903499, EPI_ISL_903500, EPI_ISL_903501, EPI_ISL_903502, EPI_ISL_903503, EPI_ISL_903504, EPI_ISL_903505, EPI_ISL_903506, EPI_ISL_903507, EPI_ISL_903508, EPI_ISL_903509, EPI_ISL_903510, EPI_ISL_903511, EPI_ISL_903512, EPI_ISL_903513, EPI_ISL_903514, EPI_ISL_903515, EPI_ISL_903516, EPI_ISL_903517, EPI_ISL_903518, EPI_ISL_903519, EPI_ISL_903520, EPI_ISL_903521, EPI_ISL_903522, EPI_ISL_903523, EPI_ISL_903524, EPI_ISL_903525, EPI_ISL_904127, EPI_ISL_904128, EPI_ISL_904129, EPI_ISL_904130, EPI_ISL_906308, EPI_ISL_906309, EPI_ISL_906310, EPI_ISL_906311, EPI_ISL_906312, EPI_ISL_906313, EPI_ISL_906314, EPI_ISL_906315, EPI_ISL_906316, EPI_ISL_906317, EPI_ISL_906318, EPI_ISL_906319, EPI_ISL_906320, EPI_ISL_906321, EPI_ISL_906322, EPI_ISL_906323, EPI_ISL_906324, EPI_ISL_906325, EPI_ISL_906326, EPI_ISL_906327, EPI_ISL_906328, EPI_ISL_906329, EPI_ISL_906330, EPI_ISL_906331, EPI_ISL_906332, EPI_ISL_906333, EPI_ISL_906334, EPI_ISL_906335, EPI_ISL_906336, EPI_ISL_906337, EPI_ISL_906338, EPI_ISL_906339, EPI_ISL_906340, EPI_ISL_906341, EPI_ISL_906342, EPI_ISL_906343, EPI_ISL_906344, EPI_ISL_906345, EPI_ISL_906346, EPI_ISL_906347, EPI_ISL_906348, EPI_ISL_906349, EPI_ISL_906350, EPI_ISL_906351, EPI_ISL_906352, EPI_ISL_906353, EPI_ISL_906354, EPI_ISL_906355, EPI_ISL_906356, EPI_ISL_906357, EPI_ISL_906358, EPI_ISL_906359, EPI_ISL_906360, EPI_ISL_906361, EPI_ISL_906362, EPI_ISL_906363, EPI_ISL_906364, EPI_ISL_906365, EPI_ISL_906366, EPI_ISL_906367, EPI_ISL_906368, EPI_ISL_906369, EPI_ISL_906370, EPI_ISL_906371, EPI_ISL_906372, EPI_ISL_906373, EPI_ISL_906374, EPI_ISL_906375, EPI_ISL_906377, EPI_ISL_906378, EPI_ISL_906379, EPI_ISL_906380, EPI_ISL_906381, EPI_ISL_906382, EPI_ISL_906383, EPI_ISL_906384, EPI_ISL_906385, EPI_ISL_906386, EPI_ISL_906387, EPI_ISL_906388, EPI_ISL_906389, EPI_ISL_906390, EPI_ISL_906391, EPI_ISL_906392, EPI_ISL_906393, EPI_ISL_906394, EPI_ISL_906395, EPI_ISL_906396, EPI_ISL_906397, EPI_ISL_906398, EPI_ISL_906399, EPI_ISL_906400, EPI_ISL_906401, EPI_ISL_906402, EPI_ISL_906403, EPI_ISL_906404, EPI_ISL_906405, EPI_ISL_906406, EPI_ISL_906407, EPI_ISL_906408, EPI_ISL_906409, EPI_ISL_906410, EPI_ISL_906411, EPI_ISL_906412, EPI_ISL_906413, EPI_ISL_906414, EPI_ISL_906415, EPI_ISL_906416, EPI_ISL_906417, EPI_ISL_906418, EPI_ISL_906419, EPI_ISL_906420, EPI_ISL_906421, EPI_ISL_906422, EPI_ISL_906423, EPI_ISL_911704, EPI_ISL_911705, EPI_ISL_911706, EPI_ISL_914890, EPI_ISL_914891, EPI_ISL_914892, EPI_ISL_914893, EPI_ISL_914894, EPI_ISL_914895, EPI_ISL_914896, EPI_ISL_914897, EPI_ISL_914898, EPI_ISL_914899, EPI_ISL_914900, EPI_ISL_914901, EPI_ISL_914902, EPI_ISL_914903, EPI_ISL_914904, EPI_ISL_914905, EPI_ISL_914906, EPI_ISL_914907, EPI_ISL_914908, EPI_ISL_914909, EPI_ISL_914910, EPI_ISL_914911, EPI_ISL_914912, EPI_ISL_914913, EPI_ISL_914914, EPI_ISL_914915, EPI_ISL_914916, EPI_ISL_914917, EPI_ISL_914918, EPI_ISL_914919, EPI_ISL_914920, EPI_ISL_914921, EPI_ISL_914922, EPI_ISL_914923, EPI_ISL_914924, EPI_ISL_914925, EPI_ISL_914926, EPI_ISL_914927, EPI_ISL_914928, EPI_ISL_914929, EPI_ISL_914930, EPI_ISL_914931, EPI_ISL_914932, EPI_ISL_914933, EPI_ISL_914934, EPI_ISL_914935, EPI_ISL_914936, EPI_ISL_914937, EPI_ISL_914938, EPI_ISL_914939, EPI_ISL_914940, EPI_ISL_914941, EPI_ISL_914942, EPI_ISL_914943, EPI_ISL_914944, EPI_ISL_914945, EPI_ISL_914946, EPI_ISL_914947, EPI_ISL_914948, EPI_ISL_914949, EPI_ISL_914950, EPI_ISL_914951, EPI_ISL_914952, EPI_ISL_914953, EPI_ISL_914954, EPI_ISL_914955, EPI_ISL_914956, EPI_ISL_914957, EPI_ISL_914958, EPI_ISL_914959, EPI_ISL_914960, EPI_ISL_914961, EPI_ISL_914962, EPI_ISL_914963, EPI_ISL_914964, EPI_ISL_914965, EPI_ISL_914966, EPI_ISL_914967, EPI_ISL_914968, EPI_ISL_914969, EPI_ISL_915196, EPI_ISL_915197, EPI_ISL_915198, EPI_ISL_915199, EPI_ISL_915200, EPI_ISL_915201, EPI_ISL_915202, EPI_ISL_915203, EPI_ISL_915204, EPI_ISL_915205, EPI_ISL_915206, EPI_ISL_915207, EPI_ISL_915208, EPI_ISL_915209, EPI_ISL_915210, EPI_ISL_915211, EPI_ISL_915212, EPI_ISL_915213, EPI_ISL_915214, EPI_ISL_915215, EPI_ISL_915216, EPI_ISL_915217, EPI_ISL_915218, EPI_ISL_915219, EPI_ISL_915220, EPI_ISL_915221, EPI_ISL_915222, EPI_ISL_915223, EPI_ISL_915224, EPI_ISL_915225, EPI_ISL_915226, EPI_ISL_915227, EPI_ISL_915228, EPI_ISL_915229, EPI_ISL_915230, EPI_ISL_915231, EPI_ISL_915232, EPI_ISL_915233, EPI_ISL_915234, EPI_ISL_915235, EPI_ISL_915236, EPI_ISL_915237, EPI_ISL_915238, EPI_ISL_915239, EPI_ISL_915240, EPI_ISL_915241, EPI_ISL_915242, EPI_ISL_915243, EPI_ISL_915244, EPI_ISL_915245, EPI_ISL_915246, EPI_ISL_915247, EPI_ISL_915248, EPI_ISL_915249, EPI_ISL_915250, EPI_ISL_915251, EPI_ISL_915252, EPI_ISL_915253, EPI_ISL_915254, EPI_ISL_915255, EPI_ISL_915256, EPI_ISL_915257, EPI_ISL_915258, EPI_ISL_915259, EPI_ISL_915260, EPI_ISL_915261, EPI_ISL_915262, EPI_ISL_915263, EPI_ISL_915264, EPI_ISL_915265, EPI_ISL_915266, EPI_ISL_915267, EPI_ISL_915268, EPI_ISL_915269, EPI_ISL_915270, EPI_ISL_915271, EPI_ISL_915272, EPI_ISL_915273, EPI_ISL_915274, EPI_ISL_915275, EPI_ISL_915276, EPI_ISL_915277, EPI_ISL_915278, EPI_ISL_915279, EPI_ISL_915280, EPI_ISL_915281, EPI_ISL_915282, EPI_ISL_915283, EPI_ISL_915284, EPI_ISL_915285, EPI_ISL_915286, EPI_ISL_915287, EPI_ISL_915288, EPI_ISL_915289, EPI_ISL_915290, EPI_ISL_915291, EPI_ISL_915292, EPI_ISL_915293, EPI_ISL_915294, EPI_ISL_915295, EPI_ISL_915296, EPI_ISL_915297, EPI_ISL_915298, EPI_ISL_915299, EPI_ISL_915300, EPI_ISL_915301, EPI_ISL_915302, EPI_ISL_915303, EPI_ISL_915304, EPI_ISL_915305, EPI_ISL_915306, EPI_ISL_915307, EPI_ISL_925137, EPI_ISL_925138, EPI_ISL_925139, EPI_ISL_925140, EPI_ISL_937121, EPI_ISL_937122, EPI_ISL_937123, EPI_ISL_937124, EPI_ISL_937125 | | | |
| see above | Quest Diagnostics | Quest Diagnostics | Anderson, B.; Bernstein; D.F.; Gerasimova, A.; Hua, M.; I.A.; K.E.; Kagan; L.E.; Lacbawan, F.; Liu Y.; Livingston; Owen, R.; Perez, A.; R.M.; Rosenthal; S.H.; Shalhout; Shlyakhter; Tanpaiboon, P. |
| EPI_ISL_1091089 | Quest Diagnostics Incorporated | Respiratory Viruses Branch, Division of Viral Diseases, Centers for Disease Control and Prevention | A. Gerasimova; A. Perez; B. Anderson; Ben L. Rambo-Martin; Clinton R. Paden; Dakota Howard; Dhwani Batra; Duncan MacCannell; F. Lacbawan; I. A. Shlyakhter; K.E. Livingston; L.E. Bernstein; M. Hua; P. Tanpaiboon; Peter W. Cook; R. M. Kagan; R. Owen; R. V. Rolando; S. H. Rosenthal; Suxiang Tong; Y. Liu |
| EPI_ISL_878695, EPI_ISL_878734, EPI_ISL_878744, EPI_ISL_878750, EPI_ISL_878752, EPI_ISL_878784, EPI_ISL_878794, EPI_ISL_880171, EPI_ISL_880176, EPI_ISL_880177, EPI_ISL_880178, EPI_ISL_880179, EPI_ISL_880182, EPI_ISL_880183, EPI_ISL_880192, EPI_ISL_1081571, EPI_ISL_1081573, EPI_ISL_1081574, EPI_ISL_1081576, EPI_ISL_1081577, EPI_ISL_1081578, EPI_ISL_1081580, EPI_ISL_1081582, EPI_ISL_1081583, EPI_ISL_1081584, EPI_ISL_1081585, EPI_ISL_1081586, EPI_ISL_1081587, EPI_ISL_1081588, EPI_ISL_1081589, EPI_ISL_1081590, EPI_ISL_1081591, EPI_ISL_1081592, EPI_ISL_1081595, EPI_ISL_1081596, EPI_ISL_1081597 | | | |
| see above | Rady's Childrens Hospital | Andersen lab at Scripps Research | Christina Clarke; David Dimmock; Denise Malicki; Kathryn Bouic; Linda Luo; SEARCH Alliance San Diego with Nanda Radamchar; Teresa Mueller |
| EPI_ISL_884582, EPI_ISL_884584, EPI_ISL_884586, EPI_ISL_884590, EPI_ISL_884638, EPI_ISL_884641, EPI_ISL_884642, EPI_ISL_884643, EPI_ISL_884644, EPI_ISL_884645, EPI_ISL_884646, EPI_ISL_884647, EPI_ISL_884648, EPI_ISL_884650, EPI_ISL_884653, EPI_ISL_884656, EPI_ISL_884658, EPI_ISL_884663, EPI_ISL_884665, EPI_ISL_884690, EPI_ISL_884695, EPI_ISL_884704, EPI_ISL_884729, EPI_ISL_884730, EPI_ISL_884733, EPI_ISL_884735, EPI_ISL_884737, EPI_ISL_884739, EPI_ISL_884741, EPI_ISL_884742, EPI_ISL_884743, EPI_ISL_884746, EPI_ISL_884823, EPI_ISL_1233929 | | | |
| see above | Respiratory Viruses Branch, Centers for Disease Control and Prevention | Respiratory Viruses Branch, Centers for Disease Control and Prevention | Allen, E.; Antico, J.; Ascencio, A.; B.L.; Barret; Batra, D.; Becker, D.; Bolze, A.; C.R.; Cassens, T.; Cho, R.; Cirulli, E.; Cook; Febbo, P.; Galloway, S.; Gietzen, K.; Isaksson, M.; K.S.; Laurent, M.; Lee, W.; Levan, G.; Liu, J.; Lu, J.; MacCannell, D.; N.L.; Nguyen, J.; P.W.; Paden; Rambo-Martin; Ramirez, J.; Rivera-Garcia, C.; Sandoval, E.; Sickler, B.; Tolentino, M.; Tong, S.; Tran, C.; Wang, S.; Washington; White, S.; Wickline, S.; de Feo, E. |
| EPI_ISL_833394, EPI_ISL_833395, EPI_ISL_833396, EPI_ISL_833397, EPI_ISL_833398, EPI_ISL_833399, EPI_ISL_833400, EPI_ISL_833401, EPI_ISL_833402, EPI_ISL_833403, EPI_ISL_833404, EPI_ISL_833489 | | | |
| see above | Saint Francis Medical | Los Angeles County Public Health | P. Hemarajata et al. |

| | Center | Laboratories | |
|---|---|---|---|
| EPI_ISL_862723, EPI_ISL_878622, EPI_ISL_878627, EPI_ISL_962611, EPI_ISL_1081459, EPI_ISL_1081460, EPI_ISL_1081461, EPI_ISL_1081462, EPI_ISL_1081463, EPI_ISL_1081464, EPI_ISL_1081465, EPI_ISL_1081466, EPI_ISL_1081467, EPI_ISL_1081468, EPI_ISL_1081469, EPI_ISL_1081470, EPI_ISL_1081471, EPI_ISL_1081742, EPI_ISL_1081743, EPI_ISL_1081744, EPI_ISL_1081745, EPI_ISL_1081746, EPI_ISL_1081747, EPI_ISL_1081748, EPI_ISL_1081749, EPI_ISL_1081750, EPI_ISL_1081751, EPI_ISL_1081752, EPI_ISL_1081753, EPI_ISL_1081754, EPI_ISL_1081755, EPI_ISL_1081756, EPI_ISL_1081757, EPI_ISL_1081758, EPI_ISL_1081759, EPI_ISL_1081760, EPI_ISL_1081761, EPI_ISL_1081762, EPI_ISL_1081763, EPI_ISL_1081764, EPI_ISL_1081765, EPI_ISL_1081766, EPI_ISL_1081767, EPI_ISL_1081768, EPI_ISL_1081769, EPI_ISL_1081770, EPI_ISL_1081771, EPI_ISL_1081772, EPI_ISL_1081773, EPI_ISL_1081774, EPI_ISL_1081775, EPI_ISL_1081776, EPI_ISL_1081777, EPI_ISL_1081778, EPI_ISL_1081779, EPI_ISL_1081780, EPI_ISL_1081781, EPI_ISL_1081782, EPI_ISL_1081783, EPI_ISL_1081785, EPI_ISL_1081786, EPI_ISL_1081788, EPI_ISL_1081789, EPI_ISL_1081790, EPI_ISL_1081791, EPI_ISL_1366377, EPI_ISL_1366378, EPI_ISL_1366748, EPI_ISL_1406045, EPI_ISL_1468837, EPI_ISL_1468959, EPI_ISL_1794668, EPI_ISL_1794670, EPI_ISL_1794674, EPI_ISL_1794675, EPI_ISL_1794676, EPI_ISL_1794679, EPI_ISL_1794681, EPI_ISL_1794983, EPI_ISL_1794984, EPI_ISL_1794985, EPI_ISL_1794986, EPI_ISL_1794988, EPI_ISL_1794989, EPI_ISL_1794990, EPI_ISL_1794991, EPI_ISL_1794992, EPI_ISL_1794993, EPI_ISL_1794994, EPI_ISL_1794995, EPI_ISL_1794998, EPI_ISL_1794999, EPI_ISL_1795000, EPI_ISL_1795003, EPI_ISL_1795004, EPI_ISL_1795005, EPI_ISL_1795008, EPI_ISL_1795009, EPI_ISL_1795010, EPI_ISL_1795013, EPI_ISL_1795014, EPI_ISL_1795015, EPI_ISL_1795016, EPI_ISL_1795018, EPI_ISL_1795019, EPI_ISL_1795020, EPI_ISL_1795021, EPI_ISL_1795024, EPI_ISL_1795027, EPI_ISL_1795028, EPI_ISL_1795029, EPI_ISL_1795030, EPI_ISL_1795031, EPI_ISL_1795035, EPI_ISL_1795036, EPI_ISL_1795037, EPI_ISL_1795039, EPI_ISL_1795040, EPI_ISL_1795041, EPI_ISL_1795042, EPI_ISL_1969324, EPI_ISL_1969325, EPI_ISL_1969326, EPI_ISL_2231038, EPI_ISL_2231050, EPI_ISL_2231204, EPI_ISL_2231462, EPI_ISL_2231463, EPI_ISL_2231464, EPI_ISL_2231465, EPI_ISL_2231468, EPI_ISL_2231469, EPI_ISL_2231470, EPI_ISL_2231471, EPI_ISL_2231473, EPI_ISL_2231474, EPI_ISL_2231477, EPI_ISL_2231480, EPI_ISL_2231481, EPI_ISL_2231482, EPI_ISL_2231483, EPI_ISL_2231484, EPI_ISL_2231487, EPI_ISL_2231488, EPI_ISL_2231489, EPI_ISL_2231490, EPI_ISL_2231491, EPI_ISL_2231492, EPI_ISL_2231493, EPI_ISL_2231495, EPI_ISL_2231496, EPI_ISL_2231497, EPI_ISL_2231500, EPI_ISL_2231501, EPI_ISL_2231504, EPI_ISL_2231505, EPI_ISL_2231506, EPI_ISL_2231507, EPI_ISL_2231510, EPI_ISL_2231511, EPI_ISL_2231512, EPI_ISL_2231513, EPI_ISL_2231514, EPI_ISL_2231518, EPI_ISL_2231519, EPI_ISL_2231520, EPI_ISL_2231521, EPI_ISL_2231522, EPI_ISL_2329336, EPI_ISL_2329339, EPI_ISL_2329345, EPI_ISL_2329351, EPI_ISL_2329358, EPI_ISL_2329365, EPI_ISL_2329371, EPI_ISL_2329386, EPI_ISL_2329392, EPI_ISL_2329393, EPI_ISL_2329399, EPI_ISL_2329400, EPI_ISL_2329407, EPI_ISL_2329408, EPI_ISL_2329466, EPI_ISL_2329467, EPI_ISL_2329471, EPI_ISL_2329472, EPI_ISL_2329475, EPI_ISL_2329476, EPI_ISL_2329477, EPI_ISL_2329480, EPI_ISL_2329481, EPI_ISL_2329485, EPI_ISL_2329486, EPI_ISL_2329487, EPI_ISL_2329488, EPI_ISL_2329493, EPI_ISL_2329494, EPI_ISL_2329498, EPI_ISL_2329499, EPI_ISL_2329504, EPI_ISL_2329505, EPI_ISL_2329510, EPI_ISL_2329511, EPI_ISL_2329512, EPI_ISL_2329516, EPI_ISL_2329517, EPI_ISL_2329518, EPI_ISL_2329519, EPI_ISL_2329524, EPI_ISL_2329525, EPI_ISL_2329526, EPI_ISL_2329527, EPI_ISL_2329531, EPI_ISL_2329532, EPI_ISL_2329533, EPI_ISL_2329534, EPI_ISL_2422720, EPI_ISL_2422721, EPI_ISL_2422722, EPI_ISL_2422723, EPI_ISL_2422724, EPI_ISL_2422725, EPI_ISL_2422726, EPI_ISL_2422727, EPI_ISL_2422728, EPI_ISL_2422729, EPI_ISL_2422730, EPI_ISL_2422731, EPI_ISL_2422732, EPI_ISL_2422733, EPI_ISL_2422734, EPI_ISL_2422735, EPI_ISL_2422736, EPI_ISL_2422737, EPI_ISL_2422738, EPI_ISL_2422739, EPI_ISL_2422740, EPI_ISL_2422741, EPI_ISL_2422742, EPI_ISL_2422743, EPI_ISL_2422744, EPI_ISL_2422745, EPI_ISL_2422746, EPI_ISL_2422747, EPI_ISL_2422748, EPI_ISL_2422749, EPI_ISL_2422750, EPI_ISL_2422751, EPI_ISL_2422752, EPI_ISL_2422753, EPI_ISL_2422754, EPI_ISL_2422755, EPI_ISL_2422756, EPI_ISL_2422757, EPI_ISL_2422758, EPI_ISL_2422759, EPI_ISL_2422760, EPI_ISL_2422761, EPI_ISL_2422762, EPI_ISL_2422763, EPI_ISL_2422764, EPI_ISL_2422765, EPI_ISL_2422766, EPI_ISL_2422767, EPI_ISL_2422768, EPI_ISL_2422769, EPI_ISL_2422770, EPI_ISL_2422771, EPI_ISL_2422772, EPI_ISL_2422773, EPI_ISL_2422774, EPI_ISL_2422775, EPI_ISL_2422776, EPI_ISL_2422777, EPI_ISL_2422778, EPI_ISL_2422779, EPI_ISL_2422780, EPI_ISL_2422781, EPI_ISL_2422782, EPI_ISL_2422899, EPI_ISL_2422900, EPI_ISL_2422901, EPI_ISL_2422902, EPI_ISL_2422904, EPI_ISL_2422905, EPI_ISL_2422906, EPI_ISL_2422907, EPI_ISL_2422908, EPI_ISL_2422912, EPI_ISL_2422913, EPI_ISL_2422914, EPI_ISL_2422915, EPI_ISL_2422918, EPI_ISL_2422919, EPI_ISL_2422920, EPI_ISL_2422921, EPI_ISL_2422922, EPI_ISL_2422926, EPI_ISL_2422927, EPI_ISL_2422931, EPI_ISL_2422932, EPI_ISL_2422933, EPI_ISL_2422934, EPI_ISL_2422938, EPI_ISL_2422939, EPI_ISL_2422940, EPI_ISL_2422941, EPI_ISL_2422942, EPI_ISL_2422945, EPI_ISL_2422946, EPI_ISL_2422947, EPI_ISL_2422948, EPI_ISL_2422949, EPI_ISL_2422950, EPI_ISL_2422953, EPI_ISL_2422954, EPI_ISL_2422955, EPI_ISL_2422956, EPI_ISL_2422957, EPI_ISL_2422959, EPI_ISL_2422960, EPI_ISL_2422962, EPI_ISL_2422964, EPI_ISL_2422965, EPI_ISL_2422966, EPI_ISL_2422967, EPI_ISL_2422968, EPI_ISL_2422971, EPI_ISL_2422972, EPI_ISL_2422973, EPI_ISL_2422974, EPI_ISL_2422975, EPI_ISL_2628311, EPI_ISL_2628312, EPI_ISL_2628315, EPI_ISL_2628316, EPI_ISL_2628317, EPI_ISL_2628319, EPI_ISL_2628320, EPI_ISL_2628321, EPI_ISL_2628323, EPI_ISL_2628324, EPI_ISL_2628325, EPI_ISL_2628326, EPI_ISL_2628328, EPI_ISL_2628331, EPI_ISL_2628332, EPI_ISL_2628333, EPI_ISL_2628336, EPI_ISL_2628337, EPI_ISL_2628338, EPI_ISL_2628339, EPI_ISL_2628341, EPI_ISL_2628342, EPI_ISL_2628343, EPI_ISL_2628345, EPI_ISL_2628347, EPI_ISL_2628348, EPI_ISL_2628349, EPI_ISL_2628352, EPI_ISL_2628355, EPI_ISL_2628551, EPI_ISL_2628552, EPI_ISL_2628553, EPI_ISL_2628555, EPI_ISL_2628556, EPI_ISL_2628557, EPI_ISL_2628558, EPI_ISL_2628562, EPI_ISL_2628563, EPI_ISL_2628566, EPI_ISL_2628567, EPI_ISL_2628568, EPI_ISL_2628569, EPI_ISL_2628571, EPI_ISL_2628572, EPI_ISL_2628573, EPI_ISL_2628574, EPI_ISL_2628576, EPI_ISL_2628577, EPI_ISL_2628578, EPI_ISL_2628580, EPI_ISL_2628584, EPI_ISL_2628585, EPI_ISL_2628589, EPI_ISL_2628590, EPI_ISL_2628591, EPI_ISL_2628592, EPI_ISL_2628595, EPI_ISL_2628596, EPI_ISL_2628599, EPI_ISL_2628600, EPI_ISL_2628601, EPI_ISL_2628602, EPI_ISL_2628604, EPI_ISL_2628692, EPI_ISL_2628715, EPI_ISL_2628719, EPI_ISL_2712770, EPI_ISL_2712772, EPI_ISL_2712775, EPI_ISL_2712777, EPI_ISL_2712779, EPI_ISL_2712780, EPI_ISL_2712783, EPI_ISL_2712785, EPI_ISL_2712786, EPI_ISL_2712787, EPI_ISL_2712789, EPI_ISL_2712791, EPI_ISL_2712792, EPI_ISL_2712793, EPI_ISL_2712796, EPI_ISL_2712799, EPI_ISL_2712800, EPI_ISL_2712801, EPI_ISL_2712806, EPI_ISL_2712807, EPI_ISL_2712813, EPI_ISL_2712814, EPI_ISL_2712819, EPI_ISL_2712820, EPI_ISL_2712826, EPI_ISL_2712827, EPI_ISL_2712832, EPI_ISL_2712836, EPI_ISL_2712839, EPI_ISL_2712841, EPI_ISL_2920333, EPI_ISL_2920334, EPI_ISL_2920335, EPI_ISL_2920338, EPI_ISL_2920339, EPI_ISL_2920340, EPI_ISL_2920341, EPI_ISL_2920344, EPI_ISL_2920345, EPI_ISL_2920347, EPI_ISL_2920348, EPI_ISL_2920349, EPI_ISL_2920350, EPI_ISL_2920351, EPI_ISL_2920353, EPI_ISL_2920354, EPI_ISL_2920355, EPI_ISL_2920357, EPI_ISL_2920358, EPI_ISL_2920359, EPI_ISL_2920360, EPI_ISL_2920361, EPI_ISL_2920363, EPI_ISL_2920364, EPI_ISL_2920367, EPI_ISL_2920368, EPI_ISL_2920369, EPI_ISL_2920372, EPI_ISL_2920373, EPI_ISL_2920374, EPI_ISL_2920375, EPI_ISL_2920377, EPI_ISL_2920378, EPI_ISL_2920379, EPI_ISL_2920380, EPI_ISL_2920381, EPI_ISL_2920382, EPI_ISL_2920383, EPI_ISL_2920384, EPI_ISL_2920385, EPI_ISL_2920386, EPI_ISL_2920389, EPI_ISL_2920390, EPI_ISL_2920391, EPI_ISL_2920392, EPI_ISL_2920393, EPI_ISL_2920396 | | | |
| see above | San Diego County Public Health Laboratory | Andersen lab at Scripps Research | Brett Austin; Jovan Shephard; SEARCH Alliance San Diego with Ashleigh Murphy; SEARCH Alliance San Diego with Tracy Basler |
| EPI_ISL_2840591 | San Diego County Public Health Laboratory | San Diego County Public Health Laboratory | Ashleigh Murphy Schafer; Brett Austin |
| EPI_ISL_885015, EPI_ISL_885016, EPI_ISL_885017, EPI_ISL_885019, EPI_ISL_885020, EPI_ISL_885021, EPI_ISL_885022, EPI_ISL_885023, EPI_ISL_885024, EPI_ISL_885025, EPI_ISL_885026, EPI_ISL_885027, EPI_ISL_885028, EPI_ISL_885029, EPI_ISL_885030, EPI_ISL_885031, EPI_ISL_885032, EPI_ISL_885033, EPI_ISL_885034, EPI_ISL_885035, EPI_ISL_885036, EPI_ISL_885037, EPI_ISL_885038, EPI_ISL_885039, EPI_ISL_885040, EPI_ISL_885041, EPI_ISL_885042, EPI_ISL_885043, EPI_ISL_885044, EPI_ISL_885045, EPI_ISL_885046, EPI_ISL_885047, EPI_ISL_885048, EPI_ISL_885049, EPI_ISL_885050, EPI_ISL_979056, EPI_ISL_979057, EPI_ISL_979058, EPI_ISL_979059, EPI_ISL_979061, EPI_ISL_979062, EPI_ISL_979063, EPI_ISL_979064, EPI_ISL_979065, EPI_ISL_979066, EPI_ISL_979067, EPI_ISL_979068, EPI_ISL_979069, EPI_ISL_979070, EPI_ISL_979071, EPI_ISL_979072, EPI_ISL_979073, EPI_ISL_979074, EPI_ISL_979075, EPI_ISL_979076, EPI_ISL_979077, EPI_ISL_979078, EPI_ISL_979079, EPI_ISL_979080, EPI_ISL_979081, EPI_ISL_979082, EPI_ISL_979083, EPI_ISL_979084, EPI_ISL_979085, EPI_ISL_979086, EPI_ISL_979087, EPI_ISL_979088, EPI_ISL_979089, EPI_ISL_979090, EPI_ISL_979091, EPI_ISL_979092, EPI_ISL_979093, EPI_ISL_979094, EPI_ISL_979095, EPI_ISL_979096, EPI_ISL_979097, EPI_ISL_979098, EPI_ISL_979099, EPI_ISL_979100, EPI_ISL_979101, EPI_ISL_979102, EPI_ISL_979103, EPI_ISL_979104, EPI_ISL_979105, EPI_ISL_979106, EPI_ISL_979107, EPI_ISL_979108, EPI_ISL_979109, EPI_ISL_979110, EPI_ISL_979111, EPI_ISL_979112, EPI_ISL_979113, EPI_ISL_979114, EPI_ISL_979115, EPI_ISL_979116, EPI_ISL_979117, EPI_ISL_979118, EPI_ISL_979119, EPI_ISL_979120, EPI_ISL_979121, EPI_ISL_979122, EPI_ISL_979123, EPI_ISL_979124, EPI_ISL_979125, EPI_ISL_979126, EPI_ISL_979127, EPI_ISL_979128, EPI_ISL_979129, EPI_ISL_979130, EPI_ISL_979131, EPI_ISL_979132, EPI_ISL_979133, EPI_ISL_979134, EPI_ISL_979135, EPI_ISL_979136, EPI_ISL_979137, EPI_ISL_979138, EPI_ISL_979139, EPI_ISL_979140, EPI_ISL_979141, EPI_ISL_979142, EPI_ISL_979143, EPI_ISL_979144, EPI_ISL_979625, EPI_ISL_979626, EPI_ISL_979627, EPI_ISL_979628, EPI_ISL_979634, EPI_ISL_979635, EPI_ISL_979636, EPI_ISL_979637, EPI_ISL_1027295, EPI_ISL_1027297, EPI_ISL_1027298, EPI_ISL_1027299, EPI_ISL_1027300, EPI_ISL_1027301, EPI_ISL_1027302, EPI_ISL_1027304, EPI_ISL_1027305, EPI_ISL_1027310, EPI_ISL_1027311, EPI_ISL_1027313, EPI_ISL_1027316, EPI_ISL_1027318, EPI_ISL_1027322, EPI_ISL_1027328, EPI_ISL_1027334, EPI_ISL_1027346, EPI_ISL_1027349, EPI_ISL_1027352, EPI_ISL_1027353, EPI_ISL_1027354, EPI_ISL_1027356, EPI_ISL_1027358, EPI_ISL_1027360, EPI_ISL_1027368, EPI_ISL_1027369, EPI_ISL_1027370, EPI_ISL_1027371, EPI_ISL_1027372, EPI_ISL_1027373, EPI_ISL_1027374, EPI_ISL_1027375, EPI_ISL_1027376, EPI_ISL_1027377, EPI_ISL_1027475, EPI_ISL_1027480, EPI_ISL_1027482, EPI_ISL_1027484, EPI_ISL_1027486, EPI_ISL_1027489, EPI_ISL_1027490, EPI_ISL_1027491, EPI_ISL_1027494, EPI_ISL_1027496, EPI_ISL_1027498, EPI_ISL_1027499, EPI_ISL_1027501, EPI_ISL_1027502, EPI_ISL_1027505, EPI_ISL_1027506, EPI_ISL_1027513, EPI_ISL_1027515, EPI_ISL_1027518, EPI_ISL_1027520, EPI_ISL_1027521, EPI_ISL_1027523, EPI_ISL_1027525, EPI_ISL_1027526, EPI_ISL_1027528, EPI_ISL_1027562, EPI_ISL_1027563, EPI_ISL_1027564, EPI_ISL_1027565, EPI_ISL_1027566, EPI_ISL_1027567, EPI_ISL_1027569, EPI_ISL_1027570, EPI_ISL_1027571, EPI_ISL_1027572, EPI_ISL_1027574, EPI_ISL_1027575, EPI_ISL_1027576, EPI_ISL_1027577, EPI_ISL_1027579, EPI_ISL_1027580, EPI_ISL_1027581, EPI_ISL_1027585, EPI_ISL_1027586, EPI_ISL_1030330, EPI_ISL_1185151, EPI_ISL_1701405, EPI_ISL_1701406, EPI_ISL_1701410, EPI_ISL_1701414, EPI_ISL_1701467, EPI_ISL_1701469, EPI_ISL_1701482, EPI_ISL_1701502, EPI_ISL_1701504, EPI_ISL_1701505, EPI_ISL_1701517, EPI_ISL_1701520, EPI_ISL_1701525, EPI_ISL_1701530, EPI_ISL_1701536, EPI_ISL_1701541, EPI_ISL_1701542, EPI_ISL_1701559, EPI_ISL_1701574, EPI_ISL_1701579, EPI_ISL_1701581, EPI_ISL_1701602, EPI_ISL_1701628, EPI_ISL_1701636, EPI_ISL_1701639, EPI_ISL_1701654, EPI_ISL_1701656, EPI_ISL_2658591, EPI_ISL_2658640, EPI_ISL_2658649, EPI_ISL_2658695, EPI_ISL_2659216, EPI_ISL_2659303, EPI_ISL_2659431 | | | |
| see above | Santa Clara County Public Health Laboratory | Chan-Zuckerberg Biohub | CZB Cliahub Consortium |
| EPI_ISL_832079, EPI_ISL_832080, EPI_ISL_832081, EPI_ISL_832090, EPI_ISL_832095, EPI_ISL_832097, EPI_ISL_884297, EPI_ISL_884298, EPI_ISL_884299, EPI_ISL_884300, EPI_ISL_884301, EPI_ISL_884302, EPI_ISL_914641, EPI_ISL_914642, EPI_ISL_914644, EPI_ISL_936493, EPI_ISL_1060547, EPI_ISL_1137631, EPI_ISL_1137632, EPI_ISL_1137633, EPI_ISL_1137634, EPI_ISL_1137635, EPI_ISL_1137636, EPI_ISL_1137638, EPI_ISL_1137639, EPI_ISL_1137640, EPI_ISL_1137641, EPI_ISL_1137642, EPI_ISL_1137643, EPI_ISL_1137644, EPI_ISL_1137645, EPI_ISL_1137648, EPI_ISL_1169579, EPI_ISL_1169581, EPI_ISL_1169582, EPI_ISL_1169583, EPI_ISL_1169584, EPI_ISL_1169585, EPI_ISL_1169586, EPI_ISL_1169587, EPI_ISL_1169588, EPI_ISL_1169589, EPI_ISL_1169592, EPI_ISL_1169593, EPI_ISL_1169594, EPI_ISL_1169595, EPI_ISL_1169596, EPI_ISL_1169597, EPI_ISL_1169598, EPI_ISL_1222758, EPI_ISL_1235639, EPI_ISL_1235641, EPI_ISL_1406164, EPI_ISL_1406167, EPI_ISL_1920836, EPI_ISL_1920843, EPI_ISL_1920847, EPI_ISL_1920875, EPI_ISL_1920883, EPI_ISL_2631190, EPI_ISL_2631191, EPI_ISL_2631192, EPI_ISL_2631193, EPI_ISL_2631194, EPI_ISL_2631195, EPI_ISL_2631196, EPI_ISL_2631197, EPI_ISL_2631198, EPI_ISL_2631199, EPI_ISL_2631200, EPI_ISL_2631201, EPI_ISL_2659052, EPI_ISL_2659053, EPI_ISL_2659054, EPI_ISL_2659055, EPI_ISL_2659056, EPI_ISL_2779551, EPI_ISL_2779552, EPI_ISL_2779553, EPI_ISL_2779554, EPI_ISL_2779555, EPI_ISL_2779556, EPI_ISL_2779557, EPI_ISL_2779558, EPI_ISL_2779559, EPI_ISL_2779560, EPI_ISL_2779561, EPI_ISL_2779562, EPI_ISL_2779563, EPI_ISL_2779564, EPI_ISL_2779565, EPI_ISL_2779566, EPI_ISL_2779567, EPI_ISL_2779568, EPI_ISL_2779569, EPI_ISL_2779570, EPI_ISL_2779571, EPI_ISL_2779572, EPI_ISL_2779573, EPI_ISL_2779574, EPI_ISL_2779575, EPI_ISL_2779576, EPI_ISL_2779577, EPI_ISL_2779578, EPI_ISL_2779579, EPI_ISL_2779580, EPI_ISL_2779581, EPI_ISL_2779582, EPI_ISL_2779583, EPI_ISL_2779584, EPI_ISL_2779585, EPI_ISL_2779586, EPI_ISL_2779587, EPI_ISL_2779588, EPI_ISL_2779589, EPI_ISL_2779590, EPI_ISL_2779591, EPI_ISL_2779592, EPI_ISL_2779593, EPI_ISL_2779594, EPI_ISL_2779595, EPI_ISL_2779596, EPI_ISL_2779597, EPI_ISL_2779598, EPI_ISL_2779599, EPI_ISL_2779600, EPI_ISL_2779601, EPI_ISL_2779602, EPI_ISL_2779603, EPI_ISL_2779604, EPI_ISL_2779605, EPI_ISL_2779606, EPI_ISL_2779607, EPI_ISL_2779608, EPI_ISL_2779609, EPI_ISL_2779611, EPI_ISL_2779612, EPI_ISL_2779613 | | | |
| see above | Santa Clara County Public Health Laboratory | Santa Clara County Public Health Laboratory | Santa Clara County Public Health Department |
| EPI_ISL_2230760, EPI_ISL_2230761, EPI_ISL_2230762, EPI_ISL_2230763, EPI_ISL_2230764, EPI_ISL_2230765, EPI_ISL_2230766, EPI_ISL_2230767, EPI_ISL_2230768, EPI_ISL_2230769, EPI_ISL_2230770, EPI_ISL_2230771 | | | |
| see above | Santa Clara County public Health Laboratory | Santa Clara County public Health Laboratory | Santa Clara County Public Health Department |
| EPI_ISL_880201, EPI_ISL_880202, EPI_ISL_880203, EPI_ISL_880204, EPI_ISL_880205, EPI_ISL_880206, EPI_ISL_880207, EPI_ISL_880208, EPI_ISL_880209, EPI_ISL_880212, EPI_ISL_880215, EPI_ISL_880218, EPI_ISL_880220, EPI_ISL_880223, EPI_ISL_880225, EPI_ISL_880228, EPI_ISL_880231, EPI_ISL_880233, EPI_ISL_880236, EPI_ISL_880239, EPI_ISL_880241, EPI_ISL_880244, EPI_ISL_880246, EPI_ISL_880249, EPI_ISL_880251, EPI_ISL_880253, EPI_ISL_880256, EPI_ISL_880259, EPI_ISL_880261, EPI_ISL_880263, EPI_ISL_880266, EPI_ISL_880268, EPI_ISL_880271, EPI_ISL_880274, EPI_ISL_880277, EPI_ISL_880279, EPI_ISL_880282, EPI_ISL_880284, EPI_ISL_880287, EPI_ISL_880290, EPI_ISL_880292, EPI_ISL_880294, EPI_ISL_880297, EPI_ISL_880300, EPI_ISL_880303, EPI_ISL_880305, EPI_ISL_880308, EPI_ISL_880310, EPI_ISL_880313, EPI_ISL_880316, EPI_ISL_880319, EPI_ISL_880321, EPI_ISL_880324, EPI_ISL_880326, EPI_ISL_880329, EPI_ISL_962585, EPI_ISL_962586, EPI_ISL_962587, EPI_ISL_962588, EPI_ISL_962589, EPI_ISL_962590, EPI_ISL_962591, EPI_ISL_962592, EPI_ISL_962593, EPI_ISL_962594, EPI_ISL_962595, EPI_ISL_962596, EPI_ISL_962597, EPI_ISL_962598, EPI_ISL_962599, EPI_ISL_962600, EPI_ISL_962601, EPI_ISL_962618, EPI_ISL_962619, EPI_ISL_962620, EPI_ISL_962621, EPI_ISL_962622, EPI_ISL_962623, EPI_ISL_962624, EPI_ISL_962625, EPI_ISL_962626, EPI_ISL_962627, EPI_ISL_962628, EPI_ISL_962629, EPI_ISL_962630, EPI_ISL_962631, EPI_ISL_962632, EPI_ISL_962633, EPI_ISL_962634, EPI_ISL_962635, EPI_ISL_962636, EPI_ISL_962637, EPI_ISL_962638, EPI_ISL_962639, EPI_ISL_962640, EPI_ISL_962641, EPI_ISL_962642, EPI_ISL_962643, EPI_ISL_962644, EPI_ISL_962645, EPI_ISL_962646, EPI_ISL_962647, EPI_ISL_962648, EPI_ISL_962649, EPI_ISL_1081558, EPI_ISL_1081628, EPI_ISL_1081629, EPI_ISL_1081630, EPI_ISL_1081631, EPI_ISL_1081632, EPI_ISL_1081633, EPI_ISL_1081634, EPI_ISL_1081635, EPI_ISL_1081636, EPI_ISL_1081637, EPI_ISL_1081638, EPI_ISL_1081639, EPI_ISL_1081640, EPI_ISL_1081641, EPI_ISL_1081642, EPI_ISL_1081643, EPI_ISL_1081644, EPI_ISL_1081645, EPI_ISL_1081646, EPI_ISL_1081647, EPI_ISL_1081648, EPI_ISL_1081649, EPI_ISL_1081650, EPI_ISL_1081651, EPI_ISL_1081652, EPI_ISL_1081653, EPI_ISL_1081654, EPI_ISL_1081655, EPI_ISL_1081656, EPI_ISL_1081657, EPI_ISL_1081658, EPI_ISL_1081659, EPI_ISL_1081660, EPI_ISL_1081661, EPI_ISL_1081662, EPI_ISL_1081663, EPI_ISL_1081664, EPI_ISL_1081665, EPI_ISL_1081666, EPI_ISL_1081667, EPI_ISL_1081668, EPI_ISL_1081669, EPI_ISL_1081670, EPI_ISL_1081671, EPI_ISL_1081672, EPI_ISL_1081673, EPI_ISL_1081674, EPI_ISL_1081675, EPI_ISL_1081676, EPI_ISL_1081677, EPI_ISL_1081678, EPI_ISL_1081679, EPI_ISL_1081680, EPI_ISL_1081681, EPI_ISL_1081682, EPI_ISL_1081683, EPI_ISL_1081684, EPI_ISL_1081685, EPI_ISL_1081686, EPI_ISL_1081687, EPI_ISL_1081688, EPI_ISL_1081689, EPI_ISL_1081690, EPI_ISL_1081691, EPI_ISL_1081692, EPI_ISL_1081693, EPI_ISL_1469043, EPI_ISL_1469044 | | | |
| see above | Scripps Medical Laboratory | Andersen lab at Scripps Research | Ellen Stefanski; Ian Mchardy; SEARCH Alliance San Diego with Michael Quigley |
| EPI_ISL_880037, EPI_ISL_880040, EPI_ISL_880042, EPI_ISL_880045, EPI_ISL_880048, EPI_ISL_880050, EPI_ISL_880053, EPI_ISL_880056, EPI_ISL_880058, EPI_ISL_880061, EPI_ISL_880063, EPI_ISL_880066, EPI_ISL_880069, EPI_ISL_880071, EPI_ISL_880074, EPI_ISL_880077, EPI_ISL_880079, EPI_ISL_880082, EPI_ISL_880085, EPI_ISL_880087, EPI_ISL_880090, EPI_ISL_880092, EPI_ISL_880095, EPI_ISL_880098, EPI_ISL_880101, EPI_ISL_880103, EPI_ISL_880106, EPI_ISL_880109, EPI_ISL_880111, EPI_ISL_880114, EPI_ISL_880117, EPI_ISL_880119, EPI_ISL_880122, EPI_ISL_880125, EPI_ISL_880128, EPI_ISL_880130, EPI_ISL_880133, EPI_ISL_880135, EPI_ISL_880138, EPI_ISL_880140, EPI_ISL_880141, EPI_ISL_880142, EPI_ISL_880143, EPI_ISL_880144, EPI_ISL_880145, EPI_ISL_880146, EPI_ISL_880147, EPI_ISL_880148, EPI_ISL_880149, EPI_ISL_880150, EPI_ISL_880151, EPI_ISL_880152, EPI_ISL_880153, EPI_ISL_880154, EPI_ISL_880155, EPI_ISL_880156, EPI_ISL_880157, EPI_ISL_880158, EPI_ISL_880159, EPI_ISL_880160, EPI_ISL_880161, EPI_ISL_880162, EPI_ISL_880163, EPI_ISL_880164, EPI_ISL_880165, EPI_ISL_880166, EPI_ISL_880167, EPI_ISL_880168, EPI_ISL_880169, EPI_ISL_880170, EPI_ISL_962602, EPI_ISL_962603, EPI_ISL_962604, EPI_ISL_962605, EPI_ISL_962606, EPI_ISL_962607, EPI_ISL_962608, EPI_ISL_962650, EPI_ISL_962651, EPI_ISL_962652, EPI_ISL_962653, EPI_ISL_962654, EPI_ISL_962655, EPI_ISL_962656, EPI_ISL_962657, EPI_ISL_962658, EPI_ISL_962659, EPI_ISL_962660, EPI_ISL_962661, EPI_ISL_962664, EPI_ISL_962665, EPI_ISL_962666, EPI_ISL_962667, EPI_ISL_962668, EPI_ISL_962669, EPI_ISL_962670, EPI_ISL_962671, EPI_ISL_962672, EPI_ISL_962673, EPI_ISL_962674, EPI_ISL_962675, EPI_ISL_962676, EPI_ISL_962677, EPI_ISL_962678, EPI_ISL_962679, EPI_ISL_962680, EPI_ISL_962681, EPI_ISL_962682, EPI_ISL_962683, EPI_ISL_962684, EPI_ISL_962685, EPI_ISL_962686, EPI_ISL_962687, EPI_ISL_962688, EPI_ISL_962689, EPI_ISL_962690, EPI_ISL_962691, EPI_ISL_962692, EPI_ISL_962693, EPI_ISL_962694, EPI_ISL_962695, EPI_ISL_962696, EPI_ISL_962697, EPI_ISL_962698, EPI_ISL_962699, EPI_ISL_962700, EPI_ISL_962701, EPI_ISL_962702, EPI_ISL_962703, EPI_ISL_962704, EPI_ISL_962705, EPI_ISL_962706, EPI_ISL_962707, EPI_ISL_962708, EPI_ISL_962709, EPI_ISL_962710, EPI_ISL_962711, EPI_ISL_962712, EPI_ISL_962713, EPI_ISL_962714, EPI_ISL_962715, EPI_ISL_962716, EPI_ISL_962717, EPI_ISL_962718, EPI_ISL_962719, EPI_ISL_962720, EPI_ISL_962721, EPI_ISL_962722, EPI_ISL_962723, EPI_ISL_962724, EPI_ISL_962725, EPI_ISL_962726, EPI_ISL_962727, EPI_ISL_962728, EPI_ISL_962729, EPI_ISL_962730, EPI_ISL_962731, EPI_ISL_962732, EPI_ISL_962733, EPI_ISL_962734, EPI_ISL_962735, EPI_ISL_962736, EPI_ISL_962737, EPI_ISL_962738, EPI_ISL_962739, EPI_ISL_962740, EPI_ISL_962741, EPI_ISL_962742, EPI_ISL_962743, EPI_ISL_962744, EPI_ISL_962745, EPI_ISL_962746, EPI_ISL_962747, EPI_ISL_962748, EPI_ISL_962749, EPI_ISL_962750, EPI_ISL_962751, EPI_ISL_962752, EPI_ISL_962753, EPI_ISL_962754, EPI_ISL_962755, EPI_ISL_962756, EPI_ISL_962757, EPI_ISL_962758, EPI_ISL_962759, EPI_ISL_962760, EPI_ISL_962761, EPI_ISL_962762, EPI_ISL_962763, EPI_ISL_962764, EPI_ISL_962765, EPI_ISL_1081511, EPI_ISL_1081522, EPI_ISL_1081540, EPI_ISL_1081568, EPI_ISL_1081569, EPI_ISL_1081570, EPI_ISL_1794883, EPI_ISL_2329409, EPI_ISL_2329410, EPI_ISL_2329411, EPI_ISL_2329412, EPI_ISL_2329413, EPI_ISL_2329414, EPI_ISL_2329416, EPI_ISL_2329417, EPI_ISL_2329419, EPI_ISL_2329420, EPI_ISL_2329421, EPI_ISL_2329422, EPI_ISL_2329423, EPI_ISL_2329424, EPI_ISL_2329425, EPI_ISL_2329426, EPI_ISL_2329448, EPI_ISL_2329449, EPI_ISL_2329465, EPI_ISL_2422669, EPI_ISL_2422670, EPI_ISL_2422671, EPI_ISL_2422672, EPI_ISL_2422673, EPI_ISL_2422674, EPI_ISL_2422675, EPI_ISL_2422676, EPI_ISL_2422677, EPI_ISL_2422678, EPI_ISL_2422679, EPI_ISL_2422680, EPI_ISL_2422681, EPI_ISL_2422682, EPI_ISL_2422683, EPI_ISL_2422684, EPI_ISL_2422685, EPI_ISL_2422686, EPI_ISL_2422687, EPI_ISL_2422688, EPI_ISL_2422689, EPI_ISL_2422690, EPI_ISL_2422691, EPI_ISL_2422692, EPI_ISL_2422693, EPI_ISL_2422694, EPI_ISL_2422695, EPI_ISL_2422696, EPI_ISL_2422697, EPI_ISL_2422698, EPI_ISL_2422699, EPI_ISL_2422700, EPI_ISL_2422701, EPI_ISL_2422702, EPI_ISL_2422703, EPI_ISL_2422704, EPI_ISL_2422705, EPI_ISL_2422706, EPI_ISL_2422707, EPI_ISL_2422708, EPI_ISL_2422709, EPI_ISL_2422710, EPI_ISL_2422711, EPI_ISL_2422712, EPI_ISL_2422713, EPI_ISL_2422714, EPI_ISL_2422715, EPI_ISL_2422716, EPI_ISL_2422717, EPI_ISL_2422718, EPI_ISL_2422719, EPI_ISL_2422783, EPI_ISL_2422784, EPI_ISL_2422785, EPI_ISL_2422786, EPI_ISL_2422787, EPI_ISL_2422788, EPI_ISL_2422789, EPI_ISL_2422790, EPI_ISL_2422791, EPI_ISL_2422792, EPI_ISL_2422793, EPI_ISL_2422794, EPI_ISL_2422795, EPI_ISL_2422796, EPI_ISL_2422797, EPI_ISL_2422798, EPI_ISL_2422799, EPI_ISL_2422800, EPI_ISL_2422801, EPI_ISL_2422802, EPI_ISL_2422803, EPI_ISL_2422804, EPI_ISL_2422805, EPI_ISL_2422806, EPI_ISL_2422807, EPI_ISL_2422808, EPI_ISL_2422809, EPI_ISL_2422810, EPI_ISL_2422811, EPI_ISL_2422812, EPI_ISL_2422813, EPI_ISL_2422814, EPI_ISL_2422828, EPI_ISL_2422829, EPI_ISL_2422830, EPI_ISL_2422834, EPI_ISL_2422835, EPI_ISL_2422836, EPI_ISL_2422837, EPI_ISL_2422838, EPI_ISL_2422839, EPI_ISL_2422840, EPI_ISL_2422841, EPI_ISL_2422842, EPI_ISL_2422843, EPI_ISL_2422844, EPI_ISL_2422845, EPI_ISL_2422846, EPI_ISL_2422847, EPI_ISL_2422848, EPI_ISL_2422849, EPI_ISL_2422850, EPI_ISL_2422851, EPI_ISL_2422852, EPI_ISL_2422853, EPI_ISL_2422854, EPI_ISL_2422855, EPI_ISL_2422856, EPI_ISL_2422857, EPI_ISL_2422858, EPI_ISL_2422859, EPI_ISL_2422860, EPI_ISL_2422861, EPI_ISL_2422862, EPI_ISL_2422863, EPI_ISL_2422864, EPI_ISL_2422865, EPI_ISL_2422866, EPI_ISL_2422867, EPI_ISL_2422868, EPI_ISL_2422869, EPI_ISL_2422870, EPI_ISL_2422871, EPI_ISL_2422872, EPI_ISL_2422873, EPI_ISL_2422874, EPI_ISL_2422875, EPI_ISL_2422876, EPI_ISL_2422877, EPI_ISL_2422878, EPI_ISL_2422879, EPI_ISL_2422880, EPI_ISL_2422881, EPI_ISL_2422882, EPI_ISL_2422883, EPI_ISL_2422884, EPI_ISL_2422885, EPI_ISL_2422886, EPI_ISL_2422887, EPI_ISL_2422888, EPI_ISL_2422889, EPI_ISL_2422890, EPI_ISL_2422891, EPI_ISL_2422892, EPI_ISL_2422893, EPI_ISL_2422894, EPI_ISL_2422895, EPI_ISL_2422896, EPI_ISL_2628356, EPI_ISL_2628357, EPI_ISL_2628358, EPI_ISL_2628359, EPI_ISL_2628360, EPI_ISL_2628361, EPI_ISL_2628362, EPI_ISL_2628363, EPI_ISL_2628364, EPI_ISL_2628365, EPI_ISL_2628366, EPI_ISL_2628367, EPI_ISL_2628368, EPI_ISL_2628369, EPI_ISL_2628370, EPI_ISL_2628371, EPI_ISL_2628372, EPI_ISL_2628373, EPI_ISL_2628374, EPI_ISL_2628375, EPI_ISL_2628376, EPI_ISL_2628377, EPI_ISL_2628378, EPI_ISL_2628379, EPI_ISL_2628396, EPI_ISL_2628397, EPI_ISL_2628398, EPI_ISL_2628399, EPI_ISL_2628400, EPI_ISL_2628401, EPI_ISL_2628402, EPI_ISL_2628403, EPI_ISL_2628404, EPI_ISL_2628405, EPI_ISL_2628406, EPI_ISL_2628407, EPI_ISL_2628408, EPI_ISL_2628409, EPI_ISL_2628410, EPI_ISL_2628411, EPI_ISL_2628412, EPI_ISL_2628413, EPI_ISL_2628414, EPI_ISL_2628415, EPI_ISL_2628416, EPI_ISL_2628417, EPI_ISL_2628418, EPI_ISL_2628419, EPI_ISL_2628420, EPI_ISL_2628421, EPI_ISL_2628422, EPI_ISL_2628423, EPI_ISL_2628424, EPI_ISL_2628425, EPI_ISL_2628426, EPI_ISL_2628427, EPI_ISL_2628428, EPI_ISL_2628429, EPI_ISL_2628430, EPI_ISL_2628431, EPI_ISL_2628432, EPI_ISL_2628433, EPI_ISL_2628434, EPI_ISL_2628435, EPI_ISL_2628512, EPI_ISL_2628513, EPI_ISL_2628514, EPI_ISL_2628515, EPI_ISL_2628516, EPI_ISL_2628517, EPI_ISL_2628518, EPI_ISL_2628519, EPI_ISL_2628520, EPI_ISL_2628521, EPI_ISL_2628522, EPI_ISL_2628523, EPI_ISL_2628524, EPI_ISL_2628525, EPI_ISL_2628526, EPI_ISL_2628527, EPI_ISL_2628528, EPI_ISL_2628529, EPI_ISL_2628530, EPI_ISL_2628531, EPI_ISL_2628532, EPI_ISL_2628533, EPI_ISL_2628534, EPI_ISL_2628535, EPI_ISL_2712456, EPI_ISL_2712461, EPI_ISL_2712462, EPI_ISL_2712464, EPI_ISL_2712465, EPI_ISL_2712466, EPI_ISL_2712467, EPI_ISL_2712468, EPI_ISL_2712469, EPI_ISL_2712470, EPI_ISL_2712472, EPI_ISL_2712473, EPI_ISL_2712474, EPI_ISL_2712475, EPI_ISL_2712476, EPI_ISL_2712477, EPI_ISL_2712478, EPI_ISL_2712480, EPI_ISL_2712481, EPI_ISL_2712483, EPI_ISL_2712484, EPI_ISL_2712485, EPI_ISL_2712486, EPI_ISL_2712487, EPI_ISL_2712488, EPI_ISL_2712489, EPI_ISL_2712490, EPI_ISL_2712492, EPI_ISL_2712493, EPI_ISL_2712494, EPI_ISL_2712495, EPI_ISL_2712496, EPI_ISL_2712497, EPI_ISL_2712498, EPI_ISL_2712499, EPI_ISL_2712500, EPI_ISL_2712501, EPI_ISL_2712502, EPI_ISL_2712503, EPI_ISL_2712504, EPI_ISL_2712505, EPI_ISL_2712506, EPI_ISL_2712507, EPI_ISL_2712508, EPI_ISL_2712509, EPI_ISL_2712510, EPI_ISL_2712511, EPI_ISL_2712512, EPI_ISL_2712513, EPI_ISL_2712514, EPI_ISL_2712515, EPI_ISL_2712516, EPI_ISL_2712517, EPI_ISL_2712518, EPI_ISL_2712521, EPI_ISL_2712522, EPI_ISL_2712523, EPI_ISL_2712524, EPI_ISL_2712525, EPI_ISL_2712526, EPI_ISL_2712527, EPI_ISL_2712528, EPI_ISL_2712529, EPI_ISL_2712530, EPI_ISL_2712531, EPI_ISL_2712532, EPI_ISL_2712533, EPI_ISL_2712534, EPI_ISL_2712556, EPI_ISL_2712557, EPI_ISL_2712558, EPI_ISL_2712559, EPI_ISL_2712560, EPI_ISL_2712561, EPI_ISL_2712562, EPI_ISL_2712563, EPI_ISL_2712564, EPI_ISL_2712565, EPI_ISL_2712566, EPI_ISL_2712567, EPI_ISL_2712568, EPI_ISL_2712569, EPI_ISL_2712570, EPI_ISL_2712571, EPI_ISL_2712572, EPI_ISL_2712573, EPI_ISL_2712574, EPI_ISL_2712575, EPI_ISL_2712576, EPI_ISL_2712577, EPI_ISL_2712578, EPI_ISL_2712579, EPI_ISL_2712580, EPI_ISL_2712581, EPI_ISL_2712582, EPI_ISL_2712583, EPI_ISL_2712584, EPI_ISL_2712585, EPI_ISL_2712586, EPI_ISL_2712587, EPI_ISL_2712588, EPI_ISL_2712589, EPI_ISL_2712590, EPI_ISL_2712591, EPI_ISL_2712592, EPI_ISL_2712593, EPI_ISL_2712594, EPI_ISL_2712595, EPI_ISL_2712596, EPI_ISL_2712597, EPI_ISL_2712598, EPI_ISL_2712599, EPI_ISL_2712600, EPI_ISL_2712601, EPI_ISL_2712602, EPI_ISL_2712603, EPI_ISL_2712604, EPI_ISL_2712605, EPI_ISL_2712606, EPI_ISL_2712608, EPI_ISL_2712609, EPI_ISL_2712610, EPI_ISL_2712611, EPI_ISL_2712612 | | | |
| see above | Sharp HealthCare Laboratory | Andersen lab at Scripps Research | Art Mendoza; Cathy Woerle; Jacquelyn Berumen; Liam McGinnis; Omid Bakhtar; SEARCH Alliance San Diego with Aaron Harding |
| EPI_ISL_1508963 | Stanford Clinical Virology Laboratory | Santa Clara County Public Health Laboratory | Santa Clara County Public Health Department |
| EPI_ISL_1335872 | Stanford Health Care | Clinical Virology Laboratory, Stanford University School or Medicine | ChunHong Huang; Daniel Solis; Fumiko Yamamoto; James Zehnder; Malaya K. Sahoo; Mamdouh Sibai; Michelle Vergese; and Benjamin A. Pinsky |
| EPI_ISL_1364488, EPI_ISL_1364489, EPI_ISL_1364490, EPI_ISL_1364491, EPI_ISL_1364492, EPI_ISL_1364493, EPI_ISL_1364494, EPI_ISL_1364495, EPI_ISL_1364496, EPI_ISL_1364497, EPI_ISL_1364498, EPI_ISL_1364499, EPI_ISL_1364500, EPI_ISL_1364501, EPI_ISL_1364502, EPI_ISL_1364503, EPI_ISL_1364504, EPI_ISL_1364505, EPI_ISL_1364508, EPI_ISL_1379727, EPI_ISL_1379728, EPI_ISL_1379729, EPI_ISL_1379730, EPI_ISL_1379732, EPI_ISL_1379734, EPI_ISL_1379735, EPI_ISL_1379736, EPI_ISL_1379737, EPI_ISL_1379738, EPI_ISL_1379739, EPI_ISL_1379740, EPI_ISL_1379741, EPI_ISL_1379742, EPI_ISL_1379743, EPI_ISL_1379744, EPI_ISL_1379745, EPI_ISL_1379746, EPI_ISL_1379747, EPI_ISL_1379748, EPI_ISL_1379749, EPI_ISL_1379750, EPI_ISL_1379752, EPI_ISL_1379753, EPI_ISL_1379754, EPI_ISL_1379755, EPI_ISL_1379756, EPI_ISL_1379757, EPI_ISL_1379758, EPI_ISL_1379759, EPI_ISL_1379760, EPI_ISL_1379761, EPI_ISL_1379762, EPI_ISL_1379763, EPI_ISL_1379764, EPI_ISL_1379765, EPI_ISL_1379766, EPI_ISL_1379767, EPI_ISL_1379768, EPI_ISL_1379769, EPI_ISL_1379770, EPI_ISL_1379771, EPI_ISL_1379772, EPI_ISL_1379773, EPI_ISL_1379774, EPI_ISL_1379775, EPI_ISL_1379776, EPI_ISL_1379777, EPI_ISL_1379779, EPI_ISL_1379783, EPI_ISL_1379785, EPI_ISL_1379786, EPI_ISL_1379787, EPI_ISL_1379788, EPI_ISL_1379794, EPI_ISL_1379795, EPI_ISL_1379805, EPI_ISL_1379806, EPI_ISL_1379808, EPI_ISL_1379814, EPI_ISL_1379818, EPI_ISL_1379819, EPI_ISL_1379825, EPI_ISL_1379828, EPI_ISL_1379837, EPI_ISL_1379839, EPI_ISL_1379840, EPI_ISL_1379843, EPI_ISL_1379844, | | | |

| | | | |
|---|---|---|---|
| EPI_ISL_1379849, EPI_ISL_1379850, EPI_ISL_1379851, EPI_ISL_1379852, EPI_ISL_1379855, EPI_ISL_1379856, EPI_ISL_1379860, EPI_ISL_1379864, EPI_ISL_1379867, EPI_ISL_1379868, EPI_ISL_1379871, EPI_ISL_1379872, EPI_ISL_1379873, EPI_ISL_1379875, EPI_ISL_1379876, EPI_ISL_1379879, EPI_ISL_1379880, EPI_ISL_1424072, EPI_ISL_1424073, EPI_ISL_1424074, EPI_ISL_1424075, EPI_ISL_1424076, EPI_ISL_1424077, EPI_ISL_1424078, EPI_ISL_1424079, EPI_ISL_1424080, EPI_ISL_1424082, EPI_ISL_1424084, EPI_ISL_1424088, EPI_ISL_1424089, EPI_ISL_1424092, EPI_ISL_1424093, EPI_ISL_1424094, EPI_ISL_1424096, EPI_ISL_1424097, EPI_ISL_1424100, EPI_ISL_1424101, EPI_ISL_1424102, EPI_ISL_1424104, EPI_ISL_1424106, EPI_ISL_2009053, EPI_ISL_3050264, EPI_ISL_3050265, EPI_ISL_3050266 | | | |
| see above | Stanford Health Care | Stanford University School of Medicine, Clinical Virology Laboratory | Becky Jiang; Bernadette Troung; ChunHong Huang; Daniel Solis; Fiona Yamamoto; Fumiko Yamamoto; James Zehnder; Malaya K. Sahoo; Mamdouh Sibai; Michelle Vergese; Michelle Verghese; Nathan Hammond; Selamawit Bihon; and Benjamin A. Pinsky |
| EPI_ISL_845836 | TGen North | TGen North | Ashlyn Pfeiffer; Chris French; Darrin Lemmer; Dave Engelthaler; Hayley Yaglom; Jolene Bowers; Megan Folkerts; The Arizona COVID Genomics Union (ACGU) |
| EPI_ISL_833432 | Torrance Memorial Medical Center | Los Angeles County Public Health Laboratories | P. Hemarajata et al. |
| EPI_ISL_2324441, EPI_ISL_2324442, EPI_ISL_2324444, EPI_ISL_2341048, EPI_ISL_2341051, EPI_ISL_2341057, EPI_ISL_2341067, EPI_ISL_2341179, EPI_ISL_2341180, EPI_ISL_2341181, EPI_ISL_2341182, EPI_ISL_2341183, EPI_ISL_2341184, EPI_ISL_2341185, EPI_ISL_2341186, EPI_ISL_2341187, EPI_ISL_2341188, EPI_ISL_2341189, EPI_ISL_2341190, EPI_ISL_2341191, EPI_ISL_2341192, EPI_ISL_2341193, EPI_ISL_2341194, EPI_ISL_2341195, EPI_ISL_2341196, EPI_ISL_2341197, EPI_ISL_2341198, EPI_ISL_2341199, EPI_ISL_2341200, EPI_ISL_2341201, EPI_ISL_2341202, EPI_ISL_2341206, EPI_ISL_2341207, EPI_ISL_2341208, EPI_ISL_2341209, EPI_ISL_2341210, EPI_ISL_2341211, EPI_ISL_2341212, EPI_ISL_2341213, EPI_ISL_2341214, EPI_ISL_2341215, EPI_ISL_2341216, EPI_ISL_2341217, EPI_ISL_2341218, EPI_ISL_2341219, EPI_ISL_2341220, EPI_ISL_2341221, EPI_ISL_2341222, EPI_ISL_2341223, EPI_ISL_2341224, EPI_ISL_2341225, EPI_ISL_2341226, EPI_ISL_2341227, EPI_ISL_2341228, EPI_ISL_2341230, EPI_ISL_2341231, EPI_ISL_2341232, EPI_ISL_2341233, EPI_ISL_2341234, EPI_ISL_2341235, EPI_ISL_2341236, EPI_ISL_2341237, EPI_ISL_2341238, EPI_ISL_2341239, EPI_ISL_2341240, EPI_ISL_2341241, EPI_ISL_2341242, EPI_ISL_2341260, EPI_ISL_2341261, EPI_ISL_2341262, EPI_ISL_2341263, EPI_ISL_2341308, EPI_ISL_2341309, EPI_ISL_2341310, EPI_ISL_2341311, EPI_ISL_2341312, EPI_ISL_2341313, EPI_ISL_2341314, EPI_ISL_2600374, EPI_ISL_3049195, EPI_ISL_3049196, EPI_ISL_3049197, EPI_ISL_3049201, EPI_ISL_3049206, EPI_ISL_3049214, EPI_ISL_3049215, EPI_ISL_3049216, EPI_ISL_3049217, EPI_ISL_3049218, EPI_ISL_3049219, EPI_ISL_3049220, EPI_ISL_3049221, EPI_ISL_3049222, EPI_ISL_3049223, EPI_ISL_3049224, EPI_ISL_3049225, EPI_ISL_3049226, EPI_ISL_3049227, EPI_ISL_3049234, EPI_ISL_3049235, EPI_ISL_3049236, EPI_ISL_3049237, EPI_ISL_3049238, EPI_ISL_3049239, EPI_ISL_3049240, EPI_ISL_3049241, EPI_ISL_3049242, EPI_ISL_3049243, EPI_ISL_3049244, EPI_ISL_3049245, EPI_ISL_3049246, EPI_ISL_3049247, EPI_ISL_3049248, EPI_ISL_3049249, EPI_ISL_3049250, EPI_ISL_3049251, EPI_ISL_3049252, EPI_ISL_3049253, EPI_ISL_3049254, EPI_ISL_3049255 | | | |
| see above | Tulare County Public Health Laboratory | Tulare County Public Health Laboratory | Denise L Lopez; Joshua Kootstra |
| EPI_ISL_794641, EPI_ISL_794642, EPI_ISL_794643, EPI_ISL_794644, EPI_ISL_794645, EPI_ISL_794646, EPI_ISL_794647, EPI_ISL_794648, EPI_ISL_794670, EPI_ISL_833428, EPI_ISL_833429, EPI_ISL_833430, EPI_ISL_833431, EPI_ISL_861704, EPI_ISL_861705, EPI_ISL_861706, EPI_ISL_862122, EPI_ISL_862123, EPI_ISL_882967, EPI_ISL_882968, EPI_ISL_882969, EPI_ISL_882970, EPI_ISL_882971, EPI_ISL_882972, EPI_ISL_882973, EPI_ISL_882974, EPI_ISL_882975, EPI_ISL_882976, EPI_ISL_882977, EPI_ISL_882978, EPI_ISL_882979, EPI_ISL_882980, EPI_ISL_882981, EPI_ISL_882982, EPI_ISL_882983, EPI_ISL_882984, EPI_ISL_892267, EPI_ISL_892268, EPI_ISL_905773, EPI_ISL_905774, EPI_ISL_905775, EPI_ISL_905776, EPI_ISL_905777, EPI_ISL_905778, EPI_ISL_905779, EPI_ISL_905780, EPI_ISL_905781, EPI_ISL_905782, EPI_ISL_905783, EPI_ISL_905784, EPI_ISL_905785, EPI_ISL_905786, EPI_ISL_905787, EPI_ISL_905788, EPI_ISL_905789, EPI_ISL_905790, EPI_ISL_905791, EPI_ISL_962509, EPI_ISL_962510, EPI_ISL_962511, EPI_ISL_962512, EPI_ISL_962521, EPI_ISL_1040053, EPI_ISL_1706603, EPI_ISL_2259228, EPI_ISL_2617959 | | | |
| see above | UCLA Clinical Micro Lab | Los Angeles County Public Health Laboratories | P. Hemarajata et al. |
| EPI_ISL_1405599, EPI_ISL_1405600, EPI_ISL_1405601, EPI_ISL_1405602, EPI_ISL_1405603, EPI_ISL_1405604, EPI_ISL_1405605, EPI_ISL_1405606, EPI_ISL_1405607, EPI_ISL_1405608, EPI_ISL_1405609, EPI_ISL_1405610, EPI_ISL_1405611, EPI_ISL_1405612, EPI_ISL_1405613, EPI_ISL_1405614, EPI_ISL_1405615, EPI_ISL_1405616, EPI_ISL_1405617, EPI_ISL_1405618, EPI_ISL_1405619, EPI_ISL_1405620, EPI_ISL_1405621, EPI_ISL_1405622, EPI_ISL_1405623, EPI_ISL_1405624, EPI_ISL_1405625, EPI_ISL_1405626, EPI_ISL_1405627, EPI_ISL_1405628, EPI_ISL_1405629, EPI_ISL_1405630, EPI_ISL_1405631, EPI_ISL_1405632, EPI_ISL_1405633, EPI_ISL_1405634, EPI_ISL_1405635, EPI_ISL_1405636, EPI_ISL_1405637, EPI_ISL_1405638, EPI_ISL_1405639, EPI_ISL_1405640, EPI_ISL_1405641, EPI_ISL_1405814, EPI_ISL_1405815, EPI_ISL_1405816, EPI_ISL_1405817, EPI_ISL_1405818, EPI_ISL_1405819, EPI_ISL_1405820, EPI_ISL_1405821, EPI_ISL_1405822, EPI_ISL_1405823, EPI_ISL_1405824, EPI_ISL_1405825, EPI_ISL_1405826, EPI_ISL_1405827, EPI_ISL_1405828, EPI_ISL_1405829, EPI_ISL_1405830, EPI_ISL_1405831, EPI_ISL_1405832, EPI_ISL_1405833, EPI_ISL_1405834, EPI_ISL_1405835, EPI_ISL_1405836, EPI_ISL_1405837, EPI_ISL_1405838, EPI_ISL_1405839, EPI_ISL_1405840, EPI_ISL_1405841, EPI_ISL_1405842, EPI_ISL_1405843, EPI_ISL_1405844, EPI_ISL_1405845, EPI_ISL_1405846, EPI_ISL_1405847, EPI_ISL_1405848, EPI_ISL_1405849, EPI_ISL_1405850, EPI_ISL_1405851, EPI_ISL_1405852, EPI_ISL_1405853, EPI_ISL_1405854, EPI_ISL_1405855, EPI_ISL_1405856, EPI_ISL_1405857, EPI_ISL_1405858, EPI_ISL_1405859, EPI_ISL_1405860, EPI_ISL_1405861, EPI_ISL_1405862, EPI_ISL_1405863, EPI_ISL_1405864, EPI_ISL_1405865, EPI_ISL_1405866, EPI_ISL_1405867, EPI_ISL_1405868, EPI_ISL_1405869, EPI_ISL_1405870, EPI_ISL_1405871, EPI_ISL_1405872, EPI_ISL_1405873, EPI_ISL_1405874, EPI_ISL_1405875, EPI_ISL_1405876, EPI_ISL_1405877, EPI_ISL_1405878, EPI_ISL_1405879, EPI_ISL_1405880, EPI_ISL_1405881, EPI_ISL_1405882, EPI_ISL_1405883, EPI_ISL_1405884, EPI_ISL_1405885, EPI_ISL_1405886, EPI_ISL_1405887, EPI_ISL_1405888, EPI_ISL_1405889, EPI_ISL_1405890, EPI_ISL_1405891, EPI_ISL_1405892, EPI_ISL_1405893, EPI_ISL_1405894, EPI_ISL_1405895, EPI_ISL_1405896, EPI_ISL_1405897, EPI_ISL_1469120, EPI_ISL_1469121, EPI_ISL_2378757, EPI_ISL_2378760, EPI_ISL_2378768, EPI_ISL_2378769, EPI_ISL_2378772, EPI_ISL_2378777, EPI_ISL_2378779, EPI_ISL_2378781, EPI_ISL_2378782, EPI_ISL_2378785, EPI_ISL_2378786, EPI_ISL_2378787, EPI_ISL_2378788, EPI_ISL_2378790, EPI_ISL_2378791, EPI_ISL_2378799, EPI_ISL_2378800, EPI_ISL_2378801, EPI_ISL_2378805, EPI_ISL_2378809, EPI_ISL_2378810, EPI_ISL_2378811, EPI_ISL_2378821, EPI_ISL_2378822, EPI_ISL_2378823, EPI_ISL_2494641, EPI_ISL_2494642, EPI_ISL_2494643, EPI_ISL_2494644, EPI_ISL_2494648, EPI_ISL_2494649, EPI_ISL_2494650, EPI_ISL_2494679, EPI_ISL_2494680 | | | |
| see above | UCSC Genomics Institute | UCSC Genomics Institute | A. Marm Kilpatrick; Angie Hinrichs; Beth Shapiro; Bryan Thornlow; Ciara Wanket; David Haussler; Eric Beraut; Hugh Olsen; Ikenna Anigbogu; Isabel Bjork; Jakob McBroome; Jeremy Sanford; Joshua Kapp; Mark Akeson; Maximilian Haeussler; Michael Stone; Miten Jain; Molly Cassatt-Johnstone; Namrita Dhillon; Russell Corbett-Detig; Terren Chang; Yatish Turakhia |
| EPI_ISL_849923, EPI_ISL_849924, EPI_ISL_849925, EPI_ISL_849927 | UCSF Clinical Microbiology Laboratory | Chan-Zuckerberg Biohub | CZB Cliahub Consortium |
| EPI_ISL_794635, EPI_ISL_794636, EPI_ISL_794637, EPI_ISL_794638, EPI_ISL_794639, EPI_ISL_794640, EPI_ISL_794669, EPI_ISL_833414, EPI_ISL_833415, EPI_ISL_833416 | | | |
| see above | USC Clinical Lab | Los Angeles County Public Health Laboratories | P. Hemarajata et al. |
| EPI_ISL_2295351, EPI_ISL_2295352, EPI_ISL_2295353, EPI_ISL_2295354, EPI_ISL_2295355, EPI_ISL_2295356, EPI_ISL_2295357, EPI_ISL_2295358, EPI_ISL_2295359, EPI_ISL_2295360, EPI_ISL_2295361, EPI_ISL_2295362, EPI_ISL_2295382, EPI_ISL_2295384 | | | |
| see above | WLA VAMC | Genomics and Discovery, Respiratory Viruses Branch, Division of Viral Diseases, Centers for Disease Control and Prevention | Adam Retchless; Anna Kelleher; Anna Uehara; Brian Lynch; Clinton R. Paden; Dhwani Batra; Haibin Wang; Han Jia Justin Ng; Jasmine Padilla; Jing Zhang; Justin Lee; Krista Queen; Mark Burroughs; Mili Sheth; Morgan Davis; Peter Cook; Rachel Marine; Sarah Nobles; Suxiang Tong; Tara Coalter; Yan Li; Ying Tao |

We gratefully acknowledge the following Authors from the Originating laboratories responsible for obtaining the specimens, as well as the Submitting laboratories where the genome data were generated and shared via GISAID, on which this research is based.

All Submitters of data may be contacted directly via www.gisaid.org

Authors are sorted alphabetically.

| Accession ID | Originating Laboratory | Submitting Laboratory | Authors |
|---|---|---|---|
| EPI_ISL_1226237, EPI_ISL_1226238, EPI_ISL_1226239, EPI_ISL_1226240, EPI_ISL_1226246, EPI_ISL_1226249, EPI_ISL_1226250, EPI_ISL_1226251, EPI_ISL_1226252, EPI_ISL_1226255, EPI_ISL_1226256, EPI_ISL_1226257, EPI_ISL_1226258, EPI_ISL_1226266, EPI_ISL_1226267, EPI_ISL_1226268, EPI_ISL_1226269, EPI_ISL_1226271, EPI_ISL_1226272, EPI_ISL_1226274, EPI_ISL_1226275 | | | |
| see above | "CDPH, Viral and Rickettsial Disease Laboratory" | Genomics and Discovery, Respiratory Viruses Branch, Division of Viral Diseases, Centers for Disease Control and Prevention | Anna Montmayeur; Anna Uehara; Ben L. Rambo-Martin; Clinton R. Paden; Dhwani Batra; Haibin Wang; Jasmine Padilla; Jing Zhang; Justin Lee; Katie Dillon; Krista Queen; Kristen Knipe; Kristine Lacek; Lori Rowe; Mark Burroughs; Matthew Schmerer; Mili Sheth; Peter W. Cook; Rachel Marine; Sam Shepard; Sarah Nobles; Shoshona Le; Suxiang Tong; Yan Li; Ying Tao |
| EPI_ISL_1093551, EPI_ISL_1093569, EPI_ISL_1093595, EPI_ISL_1093596, EPI_ISL_1093597, EPI_ISL_1093598, EPI_ISL_1196780, EPI_ISL_1205471, EPI_ISL_1205472, EPI_ISL_1205475, EPI_ISL_1205478, EPI_ISL_1205479, EPI_ISL_1373135, EPI_ISL_1373136, EPI_ISL_1373137, EPI_ISL_1373138, EPI_ISL_1373139, EPI_ISL_1373140, EPI_ISL_1373141, EPI_ISL_1373142, EPI_ISL_1373143, EPI_ISL_1373144, EPI_ISL_1373145, EPI_ISL_1373146, EPI_ISL_1373147, EPI_ISL_1373148, EPI_ISL_1373149, EPI_ISL_1373150, EPI_ISL_1373151, EPI_ISL_1373152, EPI_ISL_1373153, EPI_ISL_1373154, EPI_ISL_1373155, EPI_ISL_1373156, EPI_ISL_1373157, EPI_ISL_1373158, EPI_ISL_1373159, EPI_ISL_1373160, EPI_ISL_1576992, EPI_ISL_1576993, EPI_ISL_1576994, EPI_ISL_1576995, EPI_ISL_1576996, EPI_ISL_1576997, EPI_ISL_1576998, EPI_ISL_2304330 | | | |
| see above | Alameda County Public Health Department | Alameda County Public Health Department | Kevin Libuit; Kristina Hsieh; Kristna Hsieh; Maria Elma |
| EPI_ISL_1235361, EPI_ISL_1235363, EPI_ISL_1235364, EPI_ISL_1235365, EPI_ISL_1235366, EPI_ISL_1235374, EPI_ISL_1235376, EPI_ISL_1235385, EPI_ISL_1235386, EPI_ISL_1235389, EPI_ISL_1235390, EPI_ISL_1235393, EPI_ISL_1235397, EPI_ISL_1235398, EPI_ISL_1235399, EPI_ISL_1235402, EPI_ISL_1235421, EPI_ISL_1235423, EPI_ISL_1235425, EPI_ISL_1235426, EPI_ISL_1235432, EPI_ISL_1235433, EPI_ISL_1235437, EPI_ISL_1235443, EPI_ISL_1235446, EPI_ISL_1235447, EPI_ISL_1235456, EPI_ISL_1235457, EPI_ISL_1235458, EPI_ISL_1235460, EPI_ISL_1235462, EPI_ISL_1235463, EPI_ISL_1235464, EPI_ISL_1235465, EPI_ISL_1235475, EPI_ISL_1235476, EPI_ISL_1235481, EPI_ISL_1235489, EPI_ISL_1235493, EPI_ISL_1235494, EPI_ISL_1235500, EPI_ISL_1235510, EPI_ISL_1235513, EPI_ISL_1235514, EPI_ISL_1235517, EPI_ISL_1235523, EPI_ISL_1235527, EPI_ISL_1235538, EPI_ISL_1235548, EPI_ISL_1235553, EPI_ISL_1235555, EPI_ISL_1235561, EPI_ISL_1235565, EPI_ISL_1235568, EPI_ISL_1235569, EPI_ISL_1235571, EPI_ISL_1235576, EPI_ISL_1235577, EPI_ISL_1235584, EPI_ISL_1235585, EPI_ISL_1235587, EPI_ISL_1235594, EPI_ISL_1235595, EPI_ISL_1235603, EPI_ISL_1235609, EPI_ISL_1235611, EPI_ISL_2658569, EPI_ISL_2658618, EPI_ISL_2659261, EPI_ISL_2659266, EPI_ISL_2659449 | | | |
| see above | Alameda County Public Health Lab | Chan-Zuckerberg Biohub | CZB Cliahub Consortium |
| EPI_ISL_1015897 | Altius Institute for Biomedical Sciences | Seattle Flu Study | Alex Nguyen; Amanda Adler; Andrew Meuser; Barry R. Lutz; Benjamin Pelle; Caitlin R. Wolf; Chris D. Frazar; Clem Green; Daniel Bates; Deborah A. Nickerson; Elisabeth Brandstetter; Erica Ryke; Hannah Petersen; Helen Y. Chu; Jacob Rodriguez; Janet A. Englund; Jay Shendure; Jessica Halow; John Stamatoyannopoulos; Joshua Richards; Jover Lee; Julia Wald; Kairsten Fay; Kirsten Lacombe; Kneshay Harper; Lea M. Starita; Mark J. Rieder; Matt Hartman; Matthew Richardson; Matthew Thompson; Melissa Truong; Michael Boeckh; Michael Famulare; Misja Ilcisin; Muhammad Halimun; Olivia Waltner; Peter D. Han; Rebecca Bruders; Ryan Alexander; Sadie Patraw; Sofia Olsson; Stephanie DeBaun; Thomas R. Sibley; Tobias Ragoczy; Trevor Bedford; Truong Nguyen |
| EPI_ISL_1235360, EPI_ISL_1235367, EPI_ISL_1235372, EPI_ISL_1235373, EPI_ISL_1235375, EPI_ISL_1235377, EPI_ISL_1235378, EPI_ISL_1235381, EPI_ISL_1235383, EPI_ISL_1235384, EPI_ISL_1235400, EPI_ISL_1235401, EPI_ISL_1235403, EPI_ISL_1235410, EPI_ISL_1235416, EPI_ISL_1235417, EPI_ISL_1235418, EPI_ISL_1235419, EPI_ISL_1235424, EPI_ISL_1235427, EPI_ISL_1235434, EPI_ISL_1235435, EPI_ISL_1235438, EPI_ISL_1235441, EPI_ISL_1235442, EPI_ISL_1235450, EPI_ISL_1235451, EPI_ISL_1235455, EPI_ISL_1235468, EPI_ISL_1235469, EPI_ISL_1235472, EPI_ISL_1235473, EPI_ISL_1235478, EPI_ISL_1235488, EPI_ISL_1235490, EPI_ISL_1235496, EPI_ISL_1235498, EPI_ISL_1235501, EPI_ISL_1235502, EPI_ISL_1235506, EPI_ISL_1235507, EPI_ISL_1235509, EPI_ISL_1235511, EPI_ISL_1235515, EPI_ISL_1235521, EPI_ISL_1235522, EPI_ISL_1235525, EPI_ISL_1235526, EPI_ISL_1235528, EPI_ISL_1235539, EPI_ISL_1235540, EPI_ISL_1235544, EPI_ISL_1235545, EPI_ISL_1235546, EPI_ISL_1235547, EPI_ISL_1235549, EPI_ISL_1235558, EPI_ISL_1235570, EPI_ISL_1235572, EPI_ISL_1235578, EPI_ISL_1235582, EPI_ISL_1235588, EPI_ISL_1235591, EPI_ISL_1235596, EPI_ISL_1235600, EPI_ISL_1235604, EPI_ISL_1235606, EPI_ISL_1235608, EPI_ISL_1477168, EPI_ISL_1477179, EPI_ISL_1477184, EPI_ISL_1477187, EPI_ISL_1477193, EPI_ISL_1477196, EPI_ISL_1477201, EPI_ISL_1477202, EPI_ISL_1477204, EPI_ISL_1477205, EPI_ISL_1477209, EPI_ISL_1477214, EPI_ISL_1477221, EPI_ISL_1477222, EPI_ISL_1477223, EPI_ISL_1477231, EPI_ISL_1477232, EPI_ISL_1477237, EPI_ISL_1477238, EPI_ISL_1477241, EPI_ISL_1477242, EPI_ISL_1477245, EPI_ISL_1477246, EPI_ISL_1477248, EPI_ISL_1477250, EPI_ISL_1477256, EPI_ISL_1477265, EPI_ISL_1477266, EPI_ISL_1477283, EPI_ISL_1477285, EPI_ISL_1477286, EPI_ISL_1477291, EPI_ISL_1477297, EPI_ISL_1477300, EPI_ISL_1477302, EPI_ISL_1477303, EPI_ISL_1477304, EPI_ISL_1477307, EPI_ISL_1477310, EPI_ISL_1477315, EPI_ISL_1477316, EPI_ISL_1477319, EPI_ISL_1477321, EPI_ISL_1477323, EPI_ISL_1477324, EPI_ISL_1477331, EPI_ISL_1477333, EPI_ISL_1477336, EPI_ISL_1477338, EPI_ISL_1477344, EPI_ISL_1477345, EPI_ISL_1477347, EPI_ISL_1477353, EPI_ISL_1477356, EPI_ISL_1477357, EPI_ISL_1477361, EPI_ISL_1477362, EPI_ISL_1477363, EPI_ISL_1477369, EPI_ISL_1477372, EPI_ISL_1477374, EPI_ISL_1477378, EPI_ISL_1477386, EPI_ISL_1477398, EPI_ISL_1477407, EPI_ISL_1477416, EPI_ISL_1477418, EPI_ISL_1477419, EPI_ISL_1477422, EPI_ISL_1477423, EPI_ISL_1477424, EPI_ISL_1477425, EPI_ISL_1477427, EPI_ISL_1477428, EPI_ISL_1477429, EPI_ISL_1477434, EPI_ISL_1477563, EPI_ISL_1477580, EPI_ISL_1477607, EPI_ISL_1821316, EPI_ISL_1821318, EPI_ISL_1821321, EPI_ISL_1821322, EPI_ISL_1821325, EPI_ISL_1821334, EPI_ISL_1821335, EPI_ISL_1821336, EPI_ISL_1821343, EPI_ISL_1821347, EPI_ISL_1821348, EPI_ISL_1821349, EPI_ISL_1821351, EPI_ISL_1821352, EPI_ISL_1821353, EPI_ISL_1821370, EPI_ISL_1821393, EPI_ISL_1821395, EPI_ISL_1821400, EPI_ISL_1821401, EPI_ISL_1821406, EPI_ISL_1821407, EPI_ISL_1821409, EPI_ISL_1821411, EPI_ISL_1821415, EPI_ISL_1821416, EPI_ISL_1821417, EPI_ISL_1821418, EPI_ISL_1821420, EPI_ISL_1821421, EPI_ISL_1821422, EPI_ISL_1821423, EPI_ISL_1821425, EPI_ISL_1821429, EPI_ISL_1821430, EPI_ISL_1821431, EPI_ISL_1821432, EPI_ISL_1821433, EPI_ISL_1821435, EPI_ISL_1821436, EPI_ISL_1821438, EPI_ISL_1821439, EPI_ISL_1821441, EPI_ISL_1821444, EPI_ISL_1821446, EPI_ISL_1821448, EPI_ISL_1821449, EPI_ISL_1821450, EPI_ISL_1821451, EPI_ISL_1821452, EPI_ISL_1821453, EPI_ISL_1821456, EPI_ISL_1821457, EPI_ISL_1821459, EPI_ISL_1821462, EPI_ISL_1821464, EPI_ISL_1821465, EPI_ISL_1821467, EPI_ISL_1821469, EPI_ISL_1821471, EPI_ISL_1821472, EPI_ISL_2102523, EPI_ISL_2102529, EPI_ISL_2102530, EPI_ISL_2102531, EPI_ISL_2102532, EPI_ISL_2102535, EPI_ISL_2102536, EPI_ISL_2102544, EPI_ISL_2102545, EPI_ISL_2102547, EPI_ISL_2102548, EPI_ISL_2102549, EPI_ISL_2102553, EPI_ISL_2102554, EPI_ISL_2102556, EPI_ISL_2102557, EPI_ISL_2102558, EPI_ISL_2102559, EPI_ISL_2102560, EPI_ISL_2102563, EPI_ISL_2102565, EPI_ISL_2102567, EPI_ISL_2102571, EPI_ISL_2102574, EPI_ISL_2102576, EPI_ISL_2102577, EPI_ISL_2102578, EPI_ISL_2102581, EPI_ISL_2102582, EPI_ISL_2102583, EPI_ISL_2102589, EPI_ISL_2102593, EPI_ISL_2102594, EPI_ISL_2102596, EPI_ISL_2102598, EPI_ISL_2103149, EPI_ISL_2228248, EPI_ISL_2228250, EPI_ISL_2228253, EPI_ISL_2228254, EPI_ISL_2228258, EPI_ISL_2228260, EPI_ISL_2228261, EPI_ISL_2228268, EPI_ISL_2228269, EPI_ISL_2228272, EPI_ISL_2228273, EPI_ISL_2228276, EPI_ISL_2228278, EPI_ISL_2228279, EPI_ISL_2228281, EPI_ISL_2228287, EPI_ISL_2228300, EPI_ISL_2228302, EPI_ISL_2228306, EPI_ISL_2228307, EPI_ISL_2228313, EPI_ISL_2228316, EPI_ISL_2228318, EPI_ISL_2228327, EPI_ISL_2228329, EPI_ISL_2228332, EPI_ISL_2228337, EPI_ISL_2228340, EPI_ISL_2228345, EPI_ISL_2228346, EPI_ISL_2228348, EPI_ISL_2228349, EPI_ISL_2228350, EPI_ISL_2228353, EPI_ISL_2228355, EPI_ISL_2228356, EPI_ISL_2228358, EPI_ISL_2228360, EPI_ISL_2228361, EPI_ISL_2228369, EPI_ISL_2228370, EPI_ISL_2228374, EPI_ISL_2228376, EPI_ISL_2228380, EPI_ISL_2228382, EPI_ISL_2228385, EPI_ISL_2228389, EPI_ISL_2228391, EPI_ISL_2228402, EPI_ISL_2228405, EPI_ISL_2228406, EPI_ISL_2228410, EPI_ISL_2228414, EPI_ISL_2228418, EPI_ISL_2228421, EPI_ISL_2228426, EPI_ISL_2658665, EPI_ISL_2658701, EPI_ISL_2659204, EPI_ISL_2659305, EPI_ISL_2659334, EPI_ISL_2659481, EPI_ISL_2659502, EPI_ISL_2659556, EPI_ISL_2758488, EPI_ISL_2758489, EPI_ISL_2758519, EPI_ISL_2758520 | | | |
| see above | CA DPH Viral and Rickettsial Disease Laboratory | Chan-Zuckerberg Biohub | CZB Cliahub Consortium |
| EPI_ISL_1094325, EPI_ISL_1094361, EPI_ISL_1094559, EPI_ISL_1094561, EPI_ISL_1094564, EPI_ISL_1094565, EPI_ISL_1094570 | | | |
| see above | CA-Los Angeles County Public Health Laboratory | Respiratory Viruses Branch, Division of Viral Diseases, Centers for Disease Control and Prevention | Anna Montmayeur; Anna Uehara; Ben L. Rambo-Martin; Clinton R. Paden; Dhwani Batra; Haibin Wang; Jasmine Padilla; Jing Zhang; Justin Lee; Krista Queen; Lori Rowe; Mark Burroughs; Mili Sheth; Peter W. Cook; Rachel Marine; Sarah Nobles; Suxiang Tong; Yan Li; Ying Tao |
| EPI_ISL_2757838, EPI_ISL_2757839, EPI_ISL_2757840, EPI_ISL_2757841, EPI_ISL_2757842, EPI_ISL_2757843, EPI_ISL_2757844, EPI_ISL_2757845, EPI_ISL_2757846, EPI_ISL_2757847, EPI_ISL_2757848, EPI_ISL_2757849, EPI_ISL_2782384, EPI_ISL_2782385, EPI_ISL_2782387, EPI_ISL_2782389, EPI_ISL_2782391, EPI_ISL_2782392, EPI_ISL_2782398, EPI_ISL_2782399, EPI_ISL_2782401, EPI_ISL_2782402, EPI_ISL_2782404, EPI_ISL_2782405, EPI_ISL_2782407, EPI_ISL_2782412, EPI_ISL_2782413, EPI_ISL_2782415, EPI_ISL_2782417, EPI_ISL_2782419, EPI_ISL_2782423, EPI_ISL_2782425, EPI_ISL_2782426, EPI_ISL_2782430, EPI_ISL_2782431, EPI_ISL_2782432, EPI_ISL_2782433, EPI_ISL_2782435, EPI_ISL_2782437, EPI_ISL_2782440, EPI_ISL_2782442, EPI_ISL_2782443, EPI_ISL_2782445, EPI_ISL_2782446, EPI_ISL_2782448, EPI_ISL_2782449, EPI_ISL_2782450, EPI_ISL_2782453, EPI_ISL_2782454, EPI_ISL_2782455, EPI_ISL_2782456, EPI_ISL_2782457, EPI_ISL_2782458, EPI_ISL_2782459, EPI_ISL_2782460, EPI_ISL_2782461, EPI_ISL_2782462, EPI_ISL_2782464, EPI_ISL_2782465, EPI_ISL_2782466, EPI_ISL_2782467, EPI_ISL_2782469, EPI_ISL_2782470, EPI_ISL_2782471, EPI_ISL_2782472, EPI_ISL_2782473, EPI_ISL_2782475, EPI_ISL_2782477, EPI_ISL_2782478, EPI_ISL_2782479, EPI_ISL_2782482, EPI_ISL_2782483, EPI_ISL_2782484, EPI_ISL_2782485, EPI_ISL_2782488, EPI_ISL_2782492, EPI_ISL_2782494, EPI_ISL_2782496, EPI_ISL_2782497, EPI_ISL_2782498, EPI_ISL_2782499, EPI_ISL_2782500, EPI_ISL_2782501, EPI_ISL_2782502, EPI_ISL_2782503, EPI_ISL_2782505, EPI_ISL_2782507, EPI_ISL_2782515, EPI_ISL_2782516, EPI_ISL_2782517, EPI_ISL_2782518, EPI_ISL_2782519, EPI_ISL_2782522, EPI_ISL_2782523, EPI_ISL_2782525, EPI_ISL_2782527, EPI_ISL_2782528, EPI_ISL_2782529, EPI_ISL_2782531, EPI_ISL_2782532, EPI_ISL_2782533, EPI_ISL_2782535, EPI_ISL_2782536, EPI_ISL_2782538, EPI_ISL_2782540, EPI_ISL_2782543, EPI_ISL_2782544, EPI_ISL_2782551, EPI_ISL_2782552, EPI_ISL_2782554, EPI_ISL_2782556, EPI_ISL_2782557, EPI_ISL_2782558, EPI_ISL_2782561, EPI_ISL_2782562, EPI_ISL_2782563, EPI_ISL_2782564, EPI_ISL_2782565, EPI_ISL_2782567, EPI_ISL_2782568, EPI_ISL_2782572, EPI_ISL_2782573, EPI_ISL_2782575, EPI_ISL_2782577, EPI_ISL_2782578, EPI_ISL_2782581, EPI_ISL_2782582, EPI_ISL_2782584, EPI_ISL_2782585, EPI_ISL_2782587, EPI_ISL_2782589, EPI_ISL_2782590, EPI_ISL_2782591, EPI_ISL_2782596, EPI_ISL_2782597, EPI_ISL_2782598, EPI_ISL_2782599, EPI_ISL_2782604, EPI_ISL_2782606, EPI_ISL_2782607, EPI_ISL_2782609, EPI_ISL_2782796, EPI_ISL_2782797, EPI_ISL_2782798, EPI_ISL_2782799, EPI_ISL_2782800, EPI_ISL_2782801, EPI_ISL_2782802, EPI_ISL_2782803, EPI_ISL_2782804, EPI_ISL_2782805, EPI_ISL_2782806, EPI_ISL_2782807, EPI_ISL_2782808, EPI_ISL_2782809, EPI_ISL_2782810, EPI_ISL_2782811, EPI_ISL_2782812, EPI_ISL_2782813, EPI_ISL_2782814, EPI_ISL_2782815, EPI_ISL_2782816, EPI_ISL_2782817, EPI_ISL_2782818, EPI_ISL_2782819, EPI_ISL_2782820, EPI_ISL_2782821, EPI_ISL_2782822, EPI_ISL_2782823, EPI_ISL_2782824, EPI_ISL_2782825, EPI_ISL_2782826, EPI_ISL_2782827, EPI_ISL_2782828, EPI_ISL_2782829, EPI_ISL_2782830, EPI_ISL_2782831, EPI_ISL_2782832, EPI_ISL_2782833, EPI_ISL_2782834, EPI_ISL_2782835, EPI_ISL_2782836, EPI_ISL_2782837, EPI_ISL_2782838, EPI_ISL_2782839, EPI_ISL_2782840, EPI_ISL_2782841, EPI_ISL_2782842, EPI_ISL_2782843, EPI_ISL_2782844, EPI_ISL_2782845, EPI_ISL_2782846, EPI_ISL_2782847, EPI_ISL_2782848, EPI_ISL_2782849, EPI_ISL_2782850, EPI_ISL_2782851, EPI_ISL_2782852, EPI_ISL_2782853, EPI_ISL_2782854, EPI_ISL_2782855, EPI_ISL_2782856, EPI_ISL_2782857, EPI_ISL_2782858, EPI_ISL_2782859, EPI_ISL_2782860, EPI_ISL_2782861, EPI_ISL_2782862, EPI_ISL_2782863, EPI_ISL_2782864, EPI_ISL_2782865, EPI_ISL_2782866, EPI_ISL_2782867, EPI_ISL_2782868, EPI_ISL_2782869, EPI_ISL_2782870, EPI_ISL_2782871, EPI_ISL_2782872, EPI_ISL_2782873, EPI_ISL_2782874, EPI_ISL_2782875, EPI_ISL_2782876, EPI_ISL_2782877, EPI_ISL_2782878, EPI_ISL_2782879, EPI_ISL_2782880, EPI_ISL_2782881, EPI_ISL_2782882, EPI_ISL_2782883, EPI_ISL_2782884, EPI_ISL_2782885, EPI_ISL_2782886, EPI_ISL_2782887, EPI_ISL_2782888, EPI_ISL_2782889, EPI_ISL_2782890, EPI_ISL_2782891, EPI_ISL_2782892, EPI_ISL_2782893, EPI_ISL_2782894, EPI_ISL_2782895, EPI_ISL_2782896, EPI_ISL_2782897, EPI_ISL_2782898, EPI_ISL_2782899, EPI_ISL_2782900, EPI_ISL_2782901, EPI_ISL_2782902, EPI_ISL_2782903, EPI_ISL_2782904, EPI_ISL_2782905, EPI_ISL_2782906, EPI_ISL_2782907, EPI_ISL_2782908, EPI_ISL_2782909, EPI_ISL_2782910, EPI_ISL_2782911, EPI_ISL_2782912, EPI_ISL_2782913, EPI_ISL_2782914, EPI_ISL_2782915, EPI_ISL_2782916, EPI_ISL_2782917, EPI_ISL_2782918, EPI_ISL_2782919, EPI_ISL_2782920, EPI_ISL_2782921, EPI_ISL_2782922, EPI_ISL_2782923, EPI_ISL_2782924, EPI_ISL_2782925, EPI_ISL_2782926, EPI_ISL_2782927, EPI_ISL_2782928, EPI_ISL_2782929, EPI_ISL_2782930, EPI_ISL_2782931, EPI_ISL_2782932, EPI_ISL_2782933, EPI_ISL_2782934, EPI_ISL_2782935, EPI_ISL_2782936, EPI_ISL_2782937, EPI_ISL_2782938, EPI_ISL_2782939, EPI_ISL_2782940, EPI_ISL_2782941, EPI_ISL_2782942, EPI_ISL_2782943, EPI_ISL_2782944, EPI_ISL_2782945, EPI_ISL_2782946, EPI_ISL_2782947, EPI_ISL_2782948, EPI_ISL_2782949, EPI_ISL_2782950, EPI_ISL_2782951, EPI_ISL_2782952, EPI_ISL_2782953, EPI_ISL_2782954, EPI_ISL_2782955, EPI_ISL_2782956, EPI_ISL_2782957, EPI_ISL_2782958, EPI_ISL_2782959, EPI_ISL_2782960, EPI_ISL_2782961, EPI_ISL_2782962, EPI_ISL_2782963, EPI_ISL_2782964, EPI_ISL_2782965, EPI_ISL_2782966, EPI_ISL_2782967, EPI_ISL_2782968, EPI_ISL_2782969, EPI_ISL_2782970, EPI_ISL_2782971, EPI_ISL_2782972, EPI_ISL_2782973, EPI_ISL_2782974, EPI_ISL_2782975, EPI_ISL_2782976, EPI_ISL_2782977, EPI_ISL_2782978, EPI_ISL_2782979, EPI_ISL_2782980, EPI_ISL_2782981, EPI_ISL_2782982, EPI_ISL_2782983, EPI_ISL_2782984, EPI_ISL_2782985, EPI_ISL_2782986, EPI_ISL_2782987, EPI_ISL_2782988, EPI_ISL_2782989, EPI_ISL_2782990, EPI_ISL_2782991, EPI_ISL_2782992, EPI_ISL_2782993, EPI_ISL_2782994, EPI_ISL_2782995, EPI_ISL_2782996, EPI_ISL_2782997, EPI_ISL_2782998, EPI_ISL_2782999, EPI_ISL_2783000, EPI_ISL_2783001, EPI_ISL_2783002, EPI_ISL_2783003, EPI_ISL_2783004, EPI_ISL_2783005, EPI_ISL_2783006, EPI_ISL_2783007, EPI_ISL_2783008, EPI_ISL_2783009, EPI_ISL_2783010, EPI_ISL_2783011, EPI_ISL_2783012, EPI_ISL_2783013, EPI_ISL_2783014, EPI_ISL_2783015, EPI_ISL_2783016, EPI_ISL_2783017, EPI_ISL_2783018, EPI_ISL_2783019, EPI_ISL_2783020, EPI_ISL_2783021, EPI_ISL_2783022, EPI_ISL_2783023, EPI_ISL_2783024, EPI_ISL_2783025, EPI_ISL_2783026, EPI_ISL_2783027, EPI_ISL_2783028, EPI_ISL_2783029, EPI_ISL_2783030, EPI_ISL_2783031, EPI_ISL_2783032, EPI_ISL_2783033, EPI_ISL_2783034, EPI_ISL_2783035, EPI_ISL_2783039, EPI_ISL_2783040, EPI_ISL_2783041, EPI_ISL_2783042, EPI_ISL_2783043, EPI_ISL_2783044, EPI_ISL_2783045, EPI_ISL_2783046, EPI_ISL_2783047, EPI_ISL_2783048, EPI_ISL_2783049, EPI_ISL_2783050, EPI_ISL_2783051, EPI_ISL_2783052, EPI_ISL_2783053, EPI_ISL_2783054, EPI_ISL_2783055, EPI_ISL_2783058, EPI_ISL_2783060, EPI_ISL_2783061, EPI_ISL_2783062, EPI_ISL_2783068, EPI_ISL_2783070, EPI_ISL_2783071, EPI_ISL_2783072, EPI_ISL_2783073, EPI_ISL_2783074, EPI_ISL_2783075, EPI_ISL_2783076, EPI_ISL_2783077, EPI_ISL_2783078, EPI_ISL_2783079, EPI_ISL_2783080, EPI_ISL_2783081, EPI_ISL_2783082, EPI_ISL_2783084, EPI_ISL_2783085, EPI_ISL_2783086, EPI_ISL_2783087, EPI_ISL_2783089, EPI_ISL_2783090, EPI_ISL_2783091, EPI_ISL_2783092, EPI_ISL_2783093, EPI_ISL_2783094, EPI_ISL_2783095, EPI_ISL_2783096, EPI_ISL_2783097, EPI_ISL_2783098, EPI_ISL_2783099, EPI_ISL_2783100, EPI_ISL_2783101, EPI_ISL_2783102, EPI_ISL_2783103, EPI_ISL_2783104, EPI_ISL_2783105, EPI_ISL_2783106, EPI_ISL_2783107, EPI_ISL_2783108, EPI_ISL_2783109, EPI_ISL_2783110, EPI_ISL_2783111, EPI_ISL_2783112, EPI_ISL_2783113, EPI_ISL_2783114, EPI_ISL_2783115, EPI_ISL_2783116, EPI_ISL_2783117, EPI_ISL_2783118, EPI_ISL_2783119, EPI_ISL_2783120, EPI_ISL_2783121, EPI_ISL_2783122, EPI_ISL_2783123, EPI_ISL_2783124, EPI_ISL_2783125, EPI_ISL_2783126, EPI_ISL_2783127, EPI_ISL_2783128, EPI_ISL_2783129, EPI_ISL_2783130, EPI_ISL_2783131, EPI_ISL_2783132, EPI_ISL_2783133, EPI_ISL_2783134, EPI_ISL_2783135, EPI_ISL_2783136, EPI_ISL_2783137, EPI_ISL_2783138, EPI_ISL_2783139, EPI_ISL_2783140, EPI_ISL_2783141, EPI_ISL_2783142, EPI_ISL_2783143, EPI_ISL_2783144, EPI_ISL_2783145, EPI_ISL_2783146, EPI_ISL_2783147, EPI_ISL_2783148, EPI_ISL_2783149, EPI_ISL_2783150, EPI_ISL_2783151, EPI_ISL_2783152, EPI_ISL_2783153, EPI_ISL_2783154, EPI_ISL_2783155, EPI_ISL_2783156, EPI_ISL_2783157, EPI_ISL_2783158, EPI_ISL_2783159, EPI_ISL_2783160, EPI_ISL_2783161, EPI_ISL_2783162, EPI_ISL_2783163, EPI_ISL_2783164, EPI_ISL_2783165, EPI_ISL_2783166, EPI_ISL_2783167, EPI_ISL_2783168, EPI_ISL_2783169, EPI_ISL_2783170, EPI_ISL_2783171, EPI_ISL_2783172, EPI_ISL_2783173, EPI_ISL_2783174, EPI_ISL_2783175, EPI_ISL_2783176, EPI_ISL_2783177, EPI_ISL_2783178, EPI_ISL_2783179, EPI_ISL_2783180, EPI_ISL_2783181, EPI_ISL_2783182, EPI_ISL_2783183, EPI_ISL_2783184, EPI_ISL_2783185, EPI_ISL_2783186, EPI_ISL_2783187, EPI_ISL_2783188, EPI_ISL_2783189, EPI_ISL_2783190, EPI_ISL_2783191, EPI_ISL_2783192, EPI_ISL_2783193, EPI_ISL_2783194, EPI_ISL_2783195, EPI_ISL_2783196, EPI_ISL_2783197, EPI_ISL_2796744, EPI_ISL_2796745, EPI_ISL_2796746, EPI_ISL_2796747, EPI_ISL_2796748, EPI_ISL_2796749, EPI_ISL_2796750, EPI_ISL_2796751, EPI_ISL_2796752, EPI_ISL_2796753, EPI_ISL_2796754, EPI_ISL_2796755, EPI_ISL_2796756, EPI_ISL_2796757, EPI_ISL_2796758, EPI_ISL_2796759, EPI_ISL_2796760, EPI_ISL_2796761, EPI_ISL_2796762, EPI_ISL_2796763, EPI_ISL_2796764, EPI_ISL_2796765, EPI_ISL_2796766, EPI_ISL_2796767, EPI_ISL_2796768, EPI_ISL_2796769, EPI_ISL_2796770, EPI_ISL_2796771, EPI_ISL_2796772, EPI_ISL_2796773, EPI_ISL_2796774, EPI_ISL_2796775, EPI_ISL_2796776, EPI_ISL_2796777, EPI_ISL_2796778, EPI_ISL_2796779, EPI_ISL_2796780, EPI_ISL_2796781, EPI_ISL_2796782, EPI_ISL_2796783, EPI_ISL_2796784, EPI_ISL_2796785, EPI_ISL_2796786, EPI_ISL_2796787, EPI_ISL_2796788, EPI_ISL_2796789, EPI_ISL_2796790, EPI_ISL_2796791, EPI_ISL_2796792, EPI_ISL_2796793, EPI_ISL_2796794, EPI_ISL_2796795, EPI_ISL_2796796, EPI_ISL_2796797, EPI_ISL_2796798, EPI_ISL_2796799, EPI_ISL_2796800, EPI_ISL_2796801, EPI_ISL_2796802, EPI_ISL_2796803, EPI_ISL_2796804, EPI_ISL_2796805, EPI_ISL_2796806, EPI_ISL_2796807, EPI_ISL_2796808, EPI_ISL_2796809, EPI_ISL_2796810, EPI_ISL_2796811, EPI_ISL_2796812, EPI_ISL_2796813, EPI_ISL_2796814, EPI_ISL_2796815, EPI_ISL_2796816, EPI_ISL_2796817, EPI_ISL_2796818, EPI_ISL_2796819, EPI_ISL_2796820, EPI_ISL_2796821, EPI_ISL_2796822, EPI_ISL_2796823, EPI_ISL_2796824, EPI_ISL_2796825, EPI_ISL_2796826, EPI_ISL_2796827, EPI_ISL_2796828, EPI_ISL_2796829, EPI_ISL_2796830, EPI_ISL_2796831, EPI_ISL_2796832, EPI_ISL_2796833, EPI_ISL_2796834, EPI_ISL_2796835, EPI_ISL_2796836, EPI_ISL_2796837, EPI_ISL_2796838, EPI_ISL_2796839, EPI_ISL_2796840, EPI_ISL_2796841, EPI_ISL_2796842, EPI_ISL_2796843, EPI_ISL_2796844, EPI_ISL_2796845, EPI_ISL_2796846, EPI_ISL_2796847, EPI_ISL_2796848, EPI_ISL_2796849, EPI_ISL_2796850, EPI_ISL_2796851, EPI_ISL_2796852, EPI_ISL_2796853, EPI_ISL_2796854, EPI_ISL_2796855, EPI_ISL_2796856, EPI_ISL_2796857, EPI_ISL_2796858, EPI_ISL_2796859, EPI_ISL_2796860, EPI_ISL_2796861, EPI_ISL_2796862, EPI_ISL_2796863, EPI_ISL_2796864, EPI_ISL_2796865, EPI_ISL_2796866, EPI_ISL_2796867, EPI_ISL_2796868, EPI_ISL_2796869, EPI_ISL_2796870, EPI_ISL_2796871, EPI_ISL_2796872, EPI_ISL_2796873, EPI_ISL_2796874, EPI_ISL_2796875, EPI_ISL_2796876, EPI_ISL_2796877, EPI_ISL_2796878, EPI_ISL_2796879, EPI_ISL_2796880, EPI_ISL_2796881, EPI_ISL_2796882, EPI_ISL_2796883, EPI_ISL_2796884, EPI_ISL_2796885, EPI_ISL_2796886, EPI_ISL_2796887, EPI_ISL_2796888, EPI_ISL_2796889, EPI_ISL_2796890, EPI_ISL_2796891, EPI_ISL_2796892, EPI_ISL_2796893, EPI_ISL_2796894, EPI_ISL_2796895, EPI_ISL_2796896, EPI_ISL_2796897, EPI_ISL_2796898, EPI_ISL_2796899, EPI_ISL_2796900, EPI_ISL_2796901, EPI_ISL_2796902, EPI_ISL_2796903, EPI_ISL_2796904, EPI_ISL_2796905, EPI_ISL_2796906, EPI_ISL_2796907, EPI_ISL_2796908, EPI_ISL_2796909, EPI_ISL_2796910, EPI_ISL_2796911, EPI_ISL_2796912, EPI_ISL_2796913, EPI_ISL_2796914, EPI_ISL_2796915, EPI_ISL_2796916, EPI_ISL_2796917, EPI_ISL_2796918, EPI_ISL_2796919, EPI_ISL_2796920, EPI_ISL_2796921, EPI_ISL_2796922, EPI_ISL_2796923, EPI_ISL_2796924, EPI_ISL_2796925, EPI_ISL_2796926, EPI_ISL_2796927, EPI_ISL_2796928, EPI_ISL_2796929, EPI_ISL_2796930, EPI_ISL_2796931, EPI_ISL_2796932, EPI_ISL_2796933, EPI_ISL_2796934, EPI_ISL_2796935, EPI_ISL_2796936, EPI_ISL_2796937, EPI_ISL_2796938, EPI_ISL_2796939, EPI_ISL_2796940, EPI_ISL_2796941, EPI_ISL_2796942, EPI_ISL_2796943, EPI_ISL_2796944, EPI_ISL_2796945, EPI_ISL_2796946, EPI_ISL_2796947, EPI_ISL_2796948, EPI_ISL_2796949, EPI_ISL_2796950, EPI_ISL_2796951, EPI_ISL_2796952, EPI_ISL_2796953, EPI_ISL_2796954, EPI_ISL_2796955, EPI_ISL_2796956, EPI_ISL_2796957, EPI_ISL_2796958, EPI_ISL_2796959, EPI_ISL_2796960, EPI_ISL_2796961, EPI_ISL_2796962, EPI_ISL_2796963, EPI_ISL_2796964, EPI_ISL_2796965, EPI_ISL_2796966, EPI_ISL_2796967, EPI_ISL_2796968, EPI_ISL_2796969, EPI_ISL_2796970, EPI_ISL_2796971, EPI_ISL_2796972, EPI_ISL_2796973, EPI_ISL_2796974, EPI_ISL_2796975, EPI_ISL_2796976, EPI_ISL_2796977, EPI_ISL_2796978, EPI_ISL_2796979, EPI_ISL_2796980, EPI_ISL_2796981, EPI_ISL_2796982, EPI_ISL_2796983, EPI_ISL_2796984, EPI_ISL_2796985, EPI_ISL_2796986, EPI_ISL_2796987, EPI_ISL_2796988, EPI_ISL_2796989, EPI_ISL_2796990, EPI_ISL_2796991, EPI_ISL_2796992, EPI_ISL_2796993, EPI_ISL_2796994, EPI_ISL_2796995, EPI_ISL_2796996, EPI_ISL_2796997, EPI_ISL_2796998, EPI_ISL_2796999, EPI_ISL_2797000, EPI_ISL_2797001, EPI_ISL_2797002, EPI_ISL_2797003, EPI_ISL_2797004, EPI_ISL_2797005, EPI_ISL_2797006, EPI_ISL_2797007, EPI_ISL_2797008, EPI_ISL_2797009, EPI_ISL_2797010, EPI_ISL_2797011, EPI_ISL_2797012, EPI_ISL_2797013, EPI_ISL_2797014, EPI_ISL_2797015, EPI_ISL_2797016, EPI_ISL_2797017, EPI_ISL_2797018, EPI_ISL_2797019, EPI_ISL_2797020, EPI_ISL_2797021, EPI_ISL_2797022, EPI_ISL_2797023, EPI_ISL_2797024, EPI_ISL_2797025, EPI_ISL_2797026, EPI_ISL_2797027, EPI_ISL_2797028, EPI_ISL_2797029, EPI_ISL_2797030, EPI_ISL_2797031, EPI_ISL_2797032, EPI_ISL_2797033, EPI_ISL_2797034, EPI_ISL_2797035, EPI_ISL_2797036, EPI_ISL_2797037, EPI_ISL_2797038, EPI_ISL_2797039, EPI_ISL_2797040, EPI_ISL_2797041, EPI_ISL_2797042, EPI_ISL_2797043, EPI_ISL_2797044, EPI_ISL_2797045, EPI_ISL_2797046, EPI_ISL_2797047, EPI_ISL_2797048, EPI_ISL_2797049, EPI_ISL_2797050, EPI_ISL_2797051, EPI_ISL_2797052, EPI_ISL_2797053, EPI_ISL_2797054, EPI_ISL_2797055, EPI_ISL_2797056, EPI_ISL_2797057, EPI_ISL_2797058, EPI_ISL_2797059, EPI_ISL_2797060, EPI_ISL_2797061, EPI_ISL_2797062, EPI_ISL_2797063, EPI_ISL_2797064, EPI_ISL_2797065, EPI_ISL_2797066, EPI_ISL_2797067, EPI_ISL_2797068, EPI_ISL_2797069, EPI_ISL_2797070, EPI_ISL_2797071, EPI_ISL_2797072, EPI_ISL_2797073, EPI_ISL_2797074, EPI_ISL_2797075, EPI_ISL_2797076, EPI_ISL_2797077, EPI_ISL_2797078, EPI_ISL_2797079, EPI_ISL_2797080, EPI_ISL_2797081, EPI_ISL_2797082, EPI_ISL_2797084, EPI_ISL_2797085, EPI_ISL_2797086, EPI_ISL_2797087, EPI_ISL_2797088, EPI_ISL_2797089, EPI_ISL_2797090, EPI_ISL_2797091, EPI_ISL_2797092, EPI_ISL_2797093, EPI_ISL_2797094, EPI_ISL_2797095, EPI_ISL_2797096, EPI_ISL_2797097, EPI_ISL_2797098, EPI_ISL_2797099, EPI_ISL_2797101, EPI_ISL_2797102, EPI_ISL_2797103, EPI_ISL_2797105, EPI_ISL_2797107, EPI_ISL_2797108, EPI_ISL_2797110, EPI_ISL_2797111, EPI_ISL_2797112, EPI_ISL_2797113, EPI_ISL_2797116, EPI_ISL_2797117, EPI_ISL_2797119, EPI_ISL_2797120, EPI_ISL_2797122, EPI_ISL_2797123, EPI_ISL_2797124, EPI_ISL_2797125, EPI_ISL_2797126, EPI_ISL_2797145, EPI_ISL_2797147, EPI_ISL_2797148, EPI_ISL_2797149, EPI_ISL_2797150, EPI_ISL_2797152, EPI_ISL_2797153, EPI_ISL_2797155, EPI_ISL_2797157, EPI_ISL_2797158, EPI_ISL_2797159, EPI_ISL_2797160, EPI_ISL_2797162, EPI_ISL_2797163, EPI_ISL_2797164, EPI_ISL_2797165, EPI_ISL_2797166, EPI_ISL_2797167, EPI_ISL_2797168, EPI_ISL_2797169, EPI_ISL_2797170, EPI_ISL_2797171, EPI_ISL_2797172, EPI_ISL_2797173, EPI_ISL_2797174, EPI_ISL_2797175, EPI_ISL_2797176, EPI_ISL_2797177, EPI_ISL_2797178, EPI_ISL_2797179, EPI_ISL_2797180, EPI_ISL_2797182, EPI_ISL_2797183, EPI_ISL_2797184, EPI_ISL_2797185, EPI_ISL_2797186, EPI_ISL_2797187, EPI_ISL_2797188, EPI_ISL_2797189, EPI_ISL_2797190, EPI_ISL_2797191, EPI_ISL_2797192, EPI_ISL_2797193, EPI_ISL_2797194, EPI_ISL_2797195, EPI_ISL_2797196, EPI_ISL_2797197, EPI_ISL_2797198, EPI_ISL_2797199, EPI_ISL_2797200, EPI_ISL_2797201, EPI_ISL_2797202, EPI_ISL_2797203, EPI_ISL_2797204, EPI_ISL_2797205, EPI_ISL_2797206, EPI_ISL_2797207, EPI_ISL_2797208, EPI_ISL_2797209, EPI_ISL_2797210, EPI_ISL_2797211, EPI_ISL_2797212, EPI_ISL_2797213, EPI_ISL_2797214, EPI_ISL_2797215, EPI_ISL_2797216, EPI_ISL_2797217, EPI_ISL_2797218, EPI_ISL_2797219, EPI_ISL_2797220, EPI_ISL_2797221, EPI_ISL_2797222, EPI_ISL_2797223, EPI_ISL_2797224, EPI_ISL_2797225, EPI_ISL_2797226, EPI_ISL_2797227, EPI_ISL_2797228, EPI_ISL_2797229, EPI_ISL_2797230, EPI_ISL_2797231, EPI_ISL_2797232, EPI_ISL_2797233, EPI_ISL_2797234, EPI_ISL_2797235, EPI_ISL_2797236, EPI_ISL_2797237, EPI_ISL_2797238, EPI_ISL_2797239, EPI_ISL_2797240, EPI_ISL_2797241, EPI_ISL_2797242, EPI_ISL_2797243, EPI_ISL_2797244, EPI_ISL_2797245, EPI_ISL_2797246, EPI_ISL_2797247, EPI_ISL_2797248, EPI_ISL_2797249, EPI_ISL_2797250, EPI_ISL_2797251, EPI_ISL_2797252, EPI_ISL_2797253, EPI_ISL_2797254, EPI_ISL_2797255, EPI_ISL_2797256, EPI_ISL_2797257, EPI_ISL_2797258, EPI_ISL_2797259, EPI_ISL_2797260, EPI_ISL_2797261, EPI_ISL_2797262, EPI_ISL_2797263, EPI_ISL_2797264, EPI_ISL_2797265, EPI_ISL_2797266, EPI_ISL_2797267, EPI_ISL_2797268, EPI_ISL_2797269, EPI_ISL_2797270, EPI_ISL_2797271, EPI_ISL_2797272, EPI_ISL_2797273, EPI_ISL_2797274, EPI_ISL_2797275, EPI_ISL_2797276, EPI_ISL_2797277, EPI_ISL_2797278, EPI_ISL_2797279, EPI_ISL_2797280, EPI_ISL_2797281, EPI_ISL_2797282, EPI_ISL_2797283, EPI_ISL_2797284, EPI_ISL_2797285, EPI_ISL_2797286, EPI_ISL_2797287, EPI_ISL_2797288, EPI_ISL_2797289, EPI_ISL_2797290, EPI_ISL_2797291, EPI_ISL_2797292, EPI_ISL_2797293, EPI_ISL_2797294, EPI_ISL_2797295, EPI_ISL_2797296, EPI_ISL_2797297, EPI_ISL_2797298, EPI_ISL_2797299, EPI_ISL_2797300, EPI_ISL_2797301, EPI_ISL_2797302, EPI_ISL_2797303, EPI_ISL_2797304, EPI_ISL_2797305, EPI_ISL_2797306, EPI_ISL_2797307, EPI_ISL_2797308, EPI_ISL_2797309, EPI_ISL_2797310, EPI_ISL_2797311, EPI_ISL_2797312, EPI_ISL_2797313, EPI_ISL_2797314, EPI_ISL_2797591, EPI_ISL_2797592, EPI_ISL_2797593, EPI_ISL_2797594, EPI_ISL_2797595, EPI_ISL_2797596, EPI_ISL_2797597, EPI_ISL_2797598, EPI_ISL_2797599, EPI_ISL_2797600, EPI_ISL_2797601, EPI_ISL_2797602, EPI_ISL_2797603, EPI_ISL_2797604, EPI_ISL_2797605, EPI_ISL_2797606, EPI_ISL_2797607, EPI_ISL_2797608, EPI_ISL_2797609, EPI_ISL_2797610, EPI_ISL_2797611, EPI_ISL_2797612, EPI_ISL_2797613, EPI_ISL_2797614, EPI_ISL_2797615, EPI_ISL_2797616, EPI_ISL_2797617, EPI_ISL_2797618, EPI_ISL_2797619, EPI_ISL_2797620, EPI_ISL_2797621, EPI_ISL_2797622, EPI_ISL_2797623, EPI_ISL_2797624, EPI_ISL_2797625, EPI_ISL_2797626, EPI_ISL_2797627, EPI_ISL_2797628, EPI_ISL_2797629, EPI_ISL_2797630, EPI_ISL_2797631, EPI_ISL_2797632, EPI_ISL_2797633, EPI_ISL_2797634, EPI_ISL_2797635, EPI_ISL_2797636, EPI_ISL_2797637, EPI_ISL_2797638, EPI_ISL_2797639, EPI_ISL_2797640, EPI_ISL_2797641, EPI_ISL_2797642, EPI_ISL_2797643, EPI_ISL_2797644, EPI_ISL_2797645, EPI_ISL_2797646, EPI_ISL_2797647, EPI_ISL_2797648, EPI_ISL_2797649, EPI_ISL_2797650, EPI_ISL_2797651, EPI_ISL_2797652, EPI_ISL_2797653, EPI_ISL_2797654, EPI_ISL_2797655, EPI_ISL_2797656, EPI_ISL_2797657, EPI_ISL_2797658, EPI_ISL_2797659, EPI_ISL_2797660, EPI_ISL_2797661, EPI_ISL_2797662, EPI_ISL_2797663, EPI_ISL_2797664, EPI_ISL_2797665, EPI_ISL_2797666, EPI_ISL_2797667, | | | |

EPI_ISL_2797668, EPI_ISL_2797669, EPI_ISL_2797670, EPI_ISL_2797671, EPI_ISL_2797672, EPI_ISL_2797673, EPI_ISL_2797674, EPI_ISL_2797675, EPI_ISL_2797676, EPI_ISL_2797677, EPI_ISL_2797678, EPI_ISL_2797679, EPI_ISL_2797680, EPI_ISL_2797681, EPI_ISL_2797682, EPI_ISL_2797683, EPI_ISL_2797684, EPI_ISL_2797685, EPI_ISL_2797686, EPI_ISL_2797687, EPI_ISL_2797688, EPI_ISL_2797689, EPI_ISL_2797690, EPI_ISL_2797691, EPI_ISL_2797692, EPI_ISL_2797693, EPI_ISL_2797694, EPI_ISL_2797695, EPI_ISL_2797696, EPI_ISL_2797697, EPI_ISL_2797698, EPI_ISL_2797699, EPI_ISL_2797700, EPI_ISL_2797701, EPI_ISL_2797702, EPI_ISL_2797703, EPI_ISL_2797704, EPI_ISL_2797705, EPI_ISL_2797706, EPI_ISL_2797707, EPI_ISL_2797708, EPI_ISL_2797709, EPI_ISL_2797710, EPI_ISL_2797711, EPI_ISL_2797712, EPI_ISL_2797713, EPI_ISL_2797714, EPI_ISL_2797715, EPI_ISL_2797716, EPI_ISL_2797717, EPI_ISL_2797718, EPI_ISL_2797719, EPI_ISL_2797720, EPI_ISL_2797721, EPI_ISL_2797722, EPI_ISL_2797723, EPI_ISL_2797724, EPI_ISL_2797725, EPI_ISL_2797726, EPI_ISL_2797727, EPI_ISL_2797728, EPI_ISL_2797729, EPI_ISL_2797730, EPI_ISL_2797731, EPI_ISL_2797732, EPI_ISL_2797733, EPI_ISL_2797734, EPI_ISL_2797735, EPI_ISL_2797736, EPI_ISL_2797737, EPI_ISL_2797738, EPI_ISL_2797739, EPI_ISL_2797740, EPI_ISL_2797741, EPI_ISL_2797742, EPI_ISL_2797743, EPI_ISL_2797744, EPI_ISL_2797745, EPI_ISL_2797746, EPI_ISL_2797747, EPI_ISL_2797748, EPI_ISL_2797749, EPI_ISL_2797750, EPI_ISL_2797751, EPI_ISL_2797752, EPI_ISL_2797753, EPI_ISL_2797754, EPI_ISL_2797755, EPI_ISL_2797756, EPI_ISL_2797757, EPI_ISL_2797758, EPI_ISL_2797759, EPI_ISL_2797760, EPI_ISL_2797761, EPI_ISL_2797762, EPI_ISL_2797763, EPI_ISL_2797764, EPI_ISL_2797765, EPI_ISL_2797766, EPI_ISL_2797767, EPI_ISL_2797768, EPI_ISL_2797769, EPI_ISL_2797770, EPI_ISL_2797771, EPI_ISL_2797772, EPI_ISL_2797773, EPI_ISL_2797774, EPI_ISL_2797775, EPI_ISL_2797776, EPI_ISL_2797777, EPI_ISL_2797778, EPI_ISL_2797779, EPI_ISL_2797780, EPI_ISL_2797781, EPI_ISL_2797782, EPI_ISL_2797783, EPI_ISL_2797784, EPI_ISL_2797785, EPI_ISL_2797786, EPI_ISL_2797787, EPI_ISL_2797788, EPI_ISL_2797789, EPI_ISL_2797790, EPI_ISL_2797791, EPI_ISL_2797792, EPI_ISL_2797793, EPI_ISL_2797794, EPI_ISL_2797795, EPI_ISL_2797796, EPI_ISL_2797797, EPI_ISL_2797798, EPI_ISL_2797799, EPI_ISL_2797800, EPI_ISL_2797801, EPI_ISL_2797802, EPI_ISL_2797803, EPI_ISL_2797804, EPI_ISL_2797805, EPI_ISL_2797806, EPI_ISL_2797807, EPI_ISL_2797808, EPI_ISL_2797809, EPI_ISL_2797810, EPI_ISL_2797811, EPI_ISL_2797812, EPI_ISL_2797813, EPI_ISL_2797814, EPI_ISL_2797815, EPI_ISL_2797816, EPI_ISL_2797817, EPI_ISL_2797818, EPI_ISL_2797819, EPI_ISL_2797820, EPI_ISL_2797821, EPI_ISL_2797822, EPI_ISL_2797823, EPI_ISL_2797824, EPI_ISL_2797825, EPI_ISL_2797826, EPI_ISL_2797827, EPI_ISL_2797828, EPI_ISL_2797829, EPI_ISL_2797830, EPI_ISL_2797831, EPI_ISL_2797832, EPI_ISL_2797833, EPI_ISL_2797834, EPI_ISL_2797835, EPI_ISL_2797836, EPI_ISL_2797837, EPI_ISL_2797838, EPI_ISL_2797839, EPI_ISL_2797840, EPI_ISL_2797841, EPI_ISL_2797842, EPI_ISL_2797843, EPI_ISL_2797844, EPI_ISL_2797845, EPI_ISL_2797846, EPI_ISL_2797847, EPI_ISL_2797848, EPI_ISL_2797849, EPI_ISL_2797850, EPI_ISL_2797851, EPI_ISL_2797852, EPI_ISL_2797853, EPI_ISL_2797854, EPI_ISL_2797855, EPI_ISL_2797856, EPI_ISL_2797857, EPI_ISL_2797858, EPI_ISL_2797859, EPI_ISL_2797860, EPI_ISL_2797861, EPI_ISL_2797862, EPI_ISL_2797863, EPI_ISL_2797864, EPI_ISL_2797865, EPI_ISL_2797866, EPI_ISL_2797867, EPI_ISL_2797868, EPI_ISL_2797869, EPI_ISL_2797870, EPI_ISL_2797871, EPI_ISL_2797872, EPI_ISL_2797873, EPI_ISL_2797874, EPI_ISL_2797875, EPI_ISL_2797876, EPI_ISL_2797877, EPI_ISL_2797878, EPI_ISL_2797879, EPI_ISL_2797880, EPI_ISL_2797881, EPI_ISL_2797882, EPI_ISL_2797883, EPI_ISL_2797884, EPI_ISL_2797885, EPI_ISL_2797886, EPI_ISL_2797887, EPI_ISL_2797888, EPI_ISL_2797889, EPI_ISL_2797890, EPI_ISL_2797891, EPI_ISL_2797892, EPI_ISL_2797893, EPI_ISL_2797894, EPI_ISL_2797895, EPI_ISL_2797896, EPI_ISL_2797897, EPI_ISL_2797898, EPI_ISL_2797899, EPI_ISL_2797900, EPI_ISL_2797901, EPI_ISL_2797902, EPI_ISL_2797903, EPI_ISL_2797904, EPI_ISL_2797906, EPI_ISL_2797907, EPI_ISL_2797908, EPI_ISL_2797909, EPI_ISL_2797910, EPI_ISL_2797911, EPI_ISL_2797912, EPI_ISL_2797913, EPI_ISL_2797914, EPI_ISL_2797915, EPI_ISL_2797916, EPI_ISL_2797917, EPI_ISL_2797918, EPI_ISL_2797921, EPI_ISL_2797923, EPI_ISL_2797924, EPI_ISL_2797926, EPI_ISL_2797927, EPI_ISL_2797932, EPI_ISL_2797935, EPI_ISL_2797936, EPI_ISL_2797937, EPI_ISL_2797940, EPI_ISL_2797941, EPI_ISL_2797943, EPI_ISL_2797956, EPI_ISL_2797957, EPI_ISL_2797961, EPI_ISL_2797962, EPI_ISL_2797963, EPI_ISL_2797964, EPI_ISL_2797965, EPI_ISL_2797966, EPI_ISL_2797967, EPI_ISL_2797968, EPI_ISL_2797969, EPI_ISL_2797970, EPI_ISL_2797971, EPI_ISL_2797972, EPI_ISL_2797973, EPI_ISL_2797974, EPI_ISL_2797975, EPI_ISL_2797976, EPI_ISL_2797977, EPI_ISL_2797978, EPI_ISL_2797979, EPI_ISL_2797980, EPI_ISL_2797981, EPI_ISL_2797982, EPI_ISL_2797983, EPI_ISL_2797984, EPI_ISL_2797985, EPI_ISL_2797986, EPI_ISL_2797987, EPI_ISL_2797988, EPI_ISL_2797989, EPI_ISL_2797990, EPI_ISL_2797991, EPI_ISL_2797992, EPI_ISL_2797993, EPI_ISL_2797994, EPI_ISL_2797995, EPI_ISL_2797996, EPI_ISL_2797997, EPI_ISL_2797998, EPI_ISL_2797999, EPI_ISL_2798000, EPI_ISL_2798001, EPI_ISL_2798002, EPI_ISL_2798003, EPI_ISL_2798004, EPI_ISL_2798005, EPI_ISL_2798006, EPI_ISL_2798007, EPI_ISL_2798008, EPI_ISL_2798009, EPI_ISL_2798010, EPI_ISL_2798011, EPI_ISL_2798012, EPI_ISL_2798013, EPI_ISL_2798014, EPI_ISL_2798015, EPI_ISL_2798016, EPI_ISL_2798017, EPI_ISL_2798018, EPI_ISL_2798019, EPI_ISL_2798020, EPI_ISL_2798021, EPI_ISL_2798022, EPI_ISL_2798023, EPI_ISL_2798024, EPI_ISL_2798025, EPI_ISL_2798026, EPI_ISL_2798027, EPI_ISL_2798028, EPI_ISL_2798029, EPI_ISL_2798030, EPI_ISL_2798031, EPI_ISL_2798032, EPI_ISL_2798033, EPI_ISL_2798034, EPI_ISL_2798035, EPI_ISL_2798036, EPI_ISL_2798037, EPI_ISL_2798038, EPI_ISL_2798039, EPI_ISL_2798040, EPI_ISL_2798041, EPI_ISL_2798042, EPI_ISL_2798043, EPI_ISL_2798044, EPI_ISL_2798045, EPI_ISL_2798046, EPI_ISL_2798047, EPI_ISL_2798048, EPI_ISL_2798049, EPI_ISL_2798050, EPI_ISL_2798051, EPI_ISL_2798052, EPI_ISL_2798053, EPI_ISL_2798054, EPI_ISL_2798055, EPI_ISL_2798056, EPI_ISL_2798057, EPI_ISL_2798058, EPI_ISL_2798059, EPI_ISL_2798060, EPI_ISL_2798061, EPI_ISL_2798062, EPI_ISL_2798063, EPI_ISL_2798064, EPI_ISL_2798065, EPI_ISL_2798066, EPI_ISL_2798067, EPI_ISL_2798068, EPI_ISL_2798069, EPI_ISL_2798070, EPI_ISL_2798071, EPI_ISL_2798072, EPI_ISL_2798073, EPI_ISL_2798074, EPI_ISL_2798075, EPI_ISL_2798076, EPI_ISL_2798077, EPI_ISL_2798078, EPI_ISL_2798079, EPI_ISL_2798080, EPI_ISL_2798081, EPI_ISL_2798082, EPI_ISL_2798083, EPI_ISL_2798084, EPI_ISL_2798085, EPI_ISL_2798086, EPI_ISL_2798087, EPI_ISL_2798088, EPI_ISL_2798089, EPI_ISL_2798090, EPI_ISL_2798091, EPI_ISL_2798092, EPI_ISL_2798093, EPI_ISL_2798094, EPI_ISL_2798095, EPI_ISL_2798096, EPI_ISL_2798097, EPI_ISL_2798098, EPI_ISL_2798099, EPI_ISL_2798100, EPI_ISL_2798101, EPI_ISL_2798102, EPI_ISL_2825138, EPI_ISL_2825139, EPI_ISL_2825140, EPI_ISL_2825141, EPI_ISL_2825142, EPI_ISL_2825143, EPI_ISL_2825144, EPI_ISL_2825145, EPI_ISL_2825146, EPI_ISL_2825147, EPI_ISL_2825148, EPI_ISL_2825149, EPI_ISL_2825150, EPI_ISL_2825151, EPI_ISL_2825152, EPI_ISL_2825153, EPI_ISL_2825154, EPI_ISL_2825155, EPI_ISL_2825156, EPI_ISL_2825157, EPI_ISL_2825158, EPI_ISL_2825159, EPI_ISL_2825160, EPI_ISL_2825161, EPI_ISL_2825162, EPI_ISL_2825163, EPI_ISL_2825164, EPI_ISL_2825165, EPI_ISL_2825166, EPI_ISL_2825167, EPI_ISL_2825168, EPI_ISL_2825169, EPI_ISL_2825170, EPI_ISL_2825171, EPI_ISL_2825172, EPI_ISL_2825173, EPI_ISL_2825174, EPI_ISL_2825175, EPI_ISL_2825176, EPI_ISL_2825177, EPI_ISL_2825178, EPI_ISL_2825179, EPI_ISL_2825180, EPI_ISL_2825181, EPI_ISL_2825182, EPI_ISL_2825183, EPI_ISL_2825184, EPI_ISL_2825185, EPI_ISL_2825186, EPI_ISL_2825187, EPI_ISL_2825188, EPI_ISL_2825189, EPI_ISL_2825190, EPI_ISL_2825191, EPI_ISL_2825192, EPI_ISL_2825193, EPI_ISL_2825194, EPI_ISL_2825195, EPI_ISL_2825196, EPI_ISL_2825197, EPI_ISL_2825198, EPI_ISL_2825199, EPI_ISL_2825200, EPI_ISL_2825201, EPI_ISL_2825202, EPI_ISL_2825203, EPI_ISL_2825204, EPI_ISL_2825205, EPI_ISL_2825206, EPI_ISL_2825207, EPI_ISL_2825208, EPI_ISL_2825209, EPI_ISL_2825210, EPI_ISL_2825211, EPI_ISL_2825212, EPI_ISL_2825213, EPI_ISL_2825214, EPI_ISL_2825215, EPI_ISL_2825216, EPI_ISL_2825217, EPI_ISL_2825218, EPI_ISL_2825219, EPI_ISL_2825220, EPI_ISL_2825221, EPI_ISL_2825222, EPI_ISL_2825223, EPI_ISL_2825224, EPI_ISL_2825225, EPI_ISL_2825226, EPI_ISL_2825227, EPI_ISL_2825228, EPI_ISL_2825229, EPI_ISL_2825230, EPI_ISL_2825231, EPI_ISL_2825232, EPI_ISL_2825233, EPI_ISL_2825234, EPI_ISL_2825235, EPI_ISL_2825236, EPI_ISL_2825237, EPI_ISL_2825238, EPI_ISL_2825239, EPI_ISL_2825240, EPI_ISL_2825241, EPI_ISL_2825242, EPI_ISL_2825243, EPI_ISL_2825244, EPI_ISL_2825245, EPI_ISL_2825246, EPI_ISL_2825247, EPI_ISL_2825248, EPI_ISL_2825249, EPI_ISL_2825250, EPI_ISL_2825251, EPI_ISL_2825252, EPI_ISL_2825253, EPI_ISL_2825254, EPI_ISL_2825255, EPI_ISL_2825257, EPI_ISL_2825258, EPI_ISL_2825259, EPI_ISL_2825260, EPI_ISL_2825261, EPI_ISL_2825262, EPI_ISL_2825264, EPI_ISL_2825265, EPI_ISL_2825266, EPI_ISL_2825267, EPI_ISL_2825270, EPI_ISL_2825271, EPI_ISL_2825275, EPI_ISL_2825276, EPI_ISL_2825278, EPI_ISL_2825279, EPI_ISL_2825280, EPI_ISL_2825281, EPI_ISL_2825282, EPI_ISL_2825283, EPI_ISL_2825284, EPI_ISL_2825285, EPI_ISL_2825286, EPI_ISL_2825287, EPI_ISL_2825288, EPI_ISL_2825289, EPI_ISL_2825290, EPI_ISL_2825291, EPI_ISL_2825292, EPI_ISL_2825293, EPI_ISL_2825294, EPI_ISL_2825295, EPI_ISL_2825297, EPI_ISL_2825298, EPI_ISL_2825299, EPI_ISL_2826174, EPI_ISL_2826175, EPI_ISL_2826176, EPI_ISL_2826177, EPI_ISL_2826178, EPI_ISL_2826179, EPI_ISL_2826180, EPI_ISL_2826181, EPI_ISL_2826182, EPI_ISL_2826183, EPI_ISL_2826184, EPI_ISL_2826185, EPI_ISL_2826186, EPI_ISL_2826187, EPI_ISL_2826188, EPI_ISL_2826189, EPI_ISL_2826190, EPI_ISL_2826191, EPI_ISL_2826192, EPI_ISL_2826193, EPI_ISL_2826194, EPI_ISL_2826195, EPI_ISL_2826196, EPI_ISL_2826197, EPI_ISL_2826198, EPI_ISL_2826199, EPI_ISL_2826200, EPI_ISL_2826201, EPI_ISL_2826202, EPI_ISL_2826203, EPI_ISL_2826204, EPI_ISL_2826205, EPI_ISL_2826206, EPI_ISL_2826207, EPI_ISL_2826208, EPI_ISL_2826209, EPI_ISL_2826210, EPI_ISL_2826211, EPI_ISL_2826212, EPI_ISL_2826213, EPI_ISL_2826214, EPI_ISL_2826215, EPI_ISL_2826216, EPI_ISL_2826217, EPI_ISL_2826218, EPI_ISL_2826219, EPI_ISL_2826220, EPI_ISL_2826221, EPI_ISL_2826222, EPI_ISL_2826223, EPI_ISL_2826224, EPI_ISL_2826225, EPI_ISL_2826226, EPI_ISL_2826227, EPI_ISL_2826228, EPI_ISL_2826229, EPI_ISL_2826230, EPI_ISL_2826231, EPI_ISL_2826232, EPI_ISL_2826233, EPI_ISL_2826234, EPI_ISL_2826235, EPI_ISL_2826236, EPI_ISL_2826237, EPI_ISL_2826238, EPI_ISL_2826239, EPI_ISL_2826240, EPI_ISL_2826241, EPI_ISL_2826242, EPI_ISL_2826243, EPI_ISL_2826244, EPI_ISL_2826245, EPI_ISL_2826246, EPI_ISL_2826247, EPI_ISL_2826248, EPI_ISL_2826249, EPI_ISL_2826250, EPI_ISL_2826251, EPI_ISL_2826252, EPI_ISL_2826253, EPI_ISL_2826254, EPI_ISL_2826255, EPI_ISL_2826256, EPI_ISL_2826257, EPI_ISL_2826258, EPI_ISL_2826259, EPI_ISL_2826260, EPI_ISL_2826261, EPI_ISL_2826262, EPI_ISL_2826263, EPI_ISL_2826264, EPI_ISL_2826265, EPI_ISL_2826266, EPI_ISL_2826267, EPI_ISL_2826268, EPI_ISL_2826269, EPI_ISL_2826270, EPI_ISL_2826271, EPI_ISL_2826272, EPI_ISL_2826273, EPI_ISL_2826274, EPI_ISL_2826275, EPI_ISL_2826276, EPI_ISL_2826277, EPI_ISL_2826278, EPI_ISL_2826279, EPI_ISL_2826280, EPI_ISL_2826281, EPI_ISL_2826282, EPI_ISL_2826283, EPI_ISL_2826284, EPI_ISL_2826285, EPI_ISL_2826286, EPI_ISL_2826287, EPI_ISL_2826288, EPI_ISL_2826289, EPI_ISL_2826290, EPI_ISL_2826291, EPI_ISL_2826292, EPI_ISL_2826293, EPI_ISL_2826294, EPI_ISL_2826295, EPI_ISL_2826296, EPI_ISL_2826297, EPI_ISL_2826298, EPI_ISL_2826299, EPI_ISL_2826300, EPI_ISL_2826301, EPI_ISL_2826302, EPI_ISL_2826303, EPI_ISL_2826304, EPI_ISL_2826305, EPI_ISL_2826306, EPI_ISL_2826307, EPI_ISL_2826308, EPI_ISL_2826309, EPI_ISL_2826310, EPI_ISL_2826311, EPI_ISL_2826312, EPI_ISL_2826313, EPI_ISL_2826314, EPI_ISL_2826315, EPI_ISL_2826316, EPI_ISL_2826317, EPI_ISL_2826318, EPI_ISL_2826319, EPI_ISL_2826320, EPI_ISL_2826321, EPI_ISL_2826322, EPI_ISL_2826323, EPI_ISL_2826324, EPI_ISL_2826325, EPI_ISL_2826326, EPI_ISL_2858254, EPI_ISL_2858255, EPI_ISL_2858256, EPI_ISL_2858358, EPI_ISL_2858370, EPI_ISL_2858394, EPI_ISL_2858445, EPI_ISL_2858446, EPI_ISL_2858458, EPI_ISL_2858493, EPI_ISL_2858569, EPI_ISL_2934125, EPI_ISL_2934126, EPI_ISL_2934127, EPI_ISL_2934129, EPI_ISL_2934132, EPI_ISL_2934139, EPI_ISL_2934143, EPI_ISL_2934144, EPI_ISL_2934150, EPI_ISL_2934155, EPI_ISL_2934156, EPI_ISL_2934158, EPI_ISL_2934159, EPI_ISL_2934160, EPI_ISL_2934167, EPI_ISL_2934169, EPI_ISL_2934173, EPI_ISL_2934191, EPI_ISL_2934193, EPI_ISL_2934204, EPI_ISL_2934210, EPI_ISL_2934216, EPI_ISL_2934230, EPI_ISL_2934232, EPI_ISL_2934240, EPI_ISL_2934244, EPI_ISL_2934247, EPI_ISL_2934250, EPI_ISL_2934256, EPI_ISL_2934258, EPI_ISL_2934263, EPI_ISL_2934267, EPI_ISL_2934268, EPI_ISL_2934269, EPI_ISL_2934273, EPI_ISL_2934279, EPI_ISL_2934283, EPI_ISL_2934288, EPI_ISL_2934289, EPI_ISL_2934293, EPI_ISL_2934295, EPI_ISL_2934296, EPI_ISL_2934305, EPI_ISL_2934307, EPI_ISL_2934308, EPI_ISL_2934309, EPI_ISL_2934324, EPI_ISL_2934325, EPI_ISL_2934334, EPI_ISL_2934336, EPI_ISL_2934337, EPI_ISL_2934344, EPI_ISL_2934348, EPI_ISL_2934353, EPI_ISL_2934361, EPI_ISL_2934363, EPI_ISL_2934366, EPI_ISL_2934373, EPI_ISL_2934393, EPI_ISL_2934403, EPI_ISL_2934404, EPI_ISL_2934412, EPI_ISL_2934423, EPI_ISL_2934425, EPI_ISL_2934430, EPI_ISL_2934434, EPI_ISL_2934437, EPI_ISL_2934443, EPI_ISL_2934447, EPI_ISL_2934451, EPI_ISL_2934459, EPI_ISL_2934468, EPI_ISL_2934470, EPI_ISL_2934472, EPI_ISL_2934480, EPI_ISL_2934490, EPI_ISL_2934491, EPI_ISL_2934499, EPI_ISL_2934513, EPI_ISL_2934517, EPI_ISL_2934522, EPI_ISL_2934523, EPI_ISL_2934534, EPI_ISL_2934538, EPI_ISL_2934544, EPI_ISL_2934562, EPI_ISL_2934564, EPI_ISL_2934565, EPI_ISL_2934567, EPI_ISL_2976653, EPI_ISL_2976666, EPI_ISL_2976689, EPI_ISL_2976691, EPI_ISL_2976692, EPI_ISL_2976694, EPI_ISL_2976699, EPI_ISL_2976701, EPI_ISL_2976704, EPI_ISL_2976711, EPI_ISL_2976713, EPI_ISL_2976722, EPI_ISL_2976723, EPI_ISL_2976726, EPI_ISL_2976729, EPI_ISL_2976742, EPI_ISL_2976751, EPI_ISL_2976753, EPI_ISL_2976755, EPI_ISL_2976756, EPI_ISL_2976757, EPI_ISL_2976759, EPI_ISL_2976760, EPI_ISL_2976763, EPI_ISL_2976772, EPI_ISL_2976801, EPI_ISL_2976805, EPI_ISL_2976806, EPI_ISL_2976820, EPI_ISL_2976834, EPI_ISL_2976836, EPI_ISL_2976872, EPI_ISL_2976875, EPI_ISL_2976903, EPI_ISL_2976907, EPI_ISL_2976914, EPI_ISL_2977543, EPI_ISL_2977546, EPI_ISL_2977549, EPI_ISL_2977552, EPI_ISL_2977558, EPI_ISL_2977562, EPI_ISL_2977565, EPI_ISL_2977566, EPI_ISL_2977570, EPI_ISL_2977571, EPI_ISL_2977574, EPI_ISL_2977579, EPI_ISL_2977580, EPI_ISL_2977581, EPI_ISL_2977596, EPI_ISL_2977597, EPI_ISL_2977598, EPI_ISL_2977601, EPI_ISL_2977624, EPI_ISL_2977625, EPI_ISL_2977628, EPI_ISL_2977639, EPI_ISL_2977640, EPI_ISL_2977661, EPI_ISL_2977662, EPI_ISL_2977664, EPI_ISL_2977667, EPI_ISL_2977668, EPI_ISL_2977697, EPI_ISL_2977726, EPI_ISL_2977729, EPI_ISL_2977731, EPI_ISL_2977733, EPI_ISL_2977741, EPI_ISL_2977771, EPI_ISL_2977772, EPI_ISL_2977780, EPI_ISL_2977781, EPI_ISL_2977821, EPI_ISL_2977822, EPI_ISL_2977823, EPI_ISL_2977824, EPI_ISL_2977825, EPI_ISL_2977826, EPI_ISL_2977827, EPI_ISL_2977828, EPI_ISL_2977829, EPI_ISL_2977830, EPI_ISL_2977831, EPI_ISL_2977832, EPI_ISL_2977833, EPI_ISL_2977834, EPI_ISL_2977835, EPI_ISL_2977836, EPI_ISL_2977837, EPI_ISL_2977838, EPI_ISL_2977839, EPI_ISL_2977840, EPI_ISL_2977841, EPI_ISL_2977842, EPI_ISL_2977843, EPI_ISL_2977844, EPI_ISL_2977845, EPI_ISL_2977846, EPI_ISL_2977847, EPI_ISL_2977848, EPI_ISL_2977849, EPI_ISL_2977850, EPI_ISL_2977851, EPI_ISL_2977852, EPI_ISL_2977853, EPI_ISL_2977854, EPI_ISL_2977855, EPI_ISL_2977856, EPI_ISL_2977857, EPI_ISL_2977858, EPI_ISL_2977859, EPI_ISL_2977860, EPI_ISL_2977861, EPI_ISL_2977862, EPI_ISL_2977863, EPI_ISL_2977864, EPI_ISL_2977865, EPI_ISL_2977866, EPI_ISL_2977867, EPI_ISL_2977868, EPI_ISL_2977869, EPI_ISL_2977870, EPI_ISL_2977871, EPI_ISL_2977872, EPI_ISL_2977873, EPI_ISL_2977874, EPI_ISL_2977875, EPI_ISL_2977876, EPI_ISL_2977877, EPI_ISL_2977878, EPI_ISL_2977879, EPI_ISL_2977880, EPI_ISL_2977881, EPI_ISL_2977882, EPI_ISL_2977883, EPI_ISL_2977884, EPI_ISL_2977885, EPI_ISL_2977886, EPI_ISL_2977887, EPI_ISL_2977888, EPI_ISL_2977889, EPI_ISL_2977890, EPI_ISL_2977891, EPI_ISL_2977892, EPI_ISL_2977893, EPI_ISL_2977894, EPI_ISL_2977895, EPI_ISL_2977896, EPI_ISL_2977897, EPI_ISL_2977898, EPI_ISL_2977899, EPI_ISL_2977900, EPI_ISL_2977901, EPI_ISL_2977902, EPI_ISL_2977903, EPI_ISL_2977904, EPI_ISL_2977905, EPI_ISL_2977906, EPI_ISL_2977907, EPI_ISL_2977908, EPI_ISL_2977909, EPI_ISL_2977910, EPI_ISL_2977911, EPI_ISL_2977912, EPI_ISL_2977913, EPI_ISL_2977914, EPI_ISL_2977915, EPI_ISL_2977916, EPI_ISL_2977917, EPI_ISL_2977918, EPI_ISL_2977919, EPI_ISL_2977920, EPI_ISL_2977921, EPI_ISL_2977922, EPI_ISL_2977923, EPI_ISL_2977924, EPI_ISL_2977925, EPI_ISL_2977926, EPI_ISL_2977927, EPI_ISL_2977928, EPI_ISL_2977929, EPI_ISL_2977930, EPI_ISL_2977931, EPI_ISL_2977932, EPI_ISL_2977933, EPI_ISL_2977934, EPI_ISL_2977935, EPI_ISL_2977936, EPI_ISL_2977937, EPI_ISL_2977938, EPI_ISL_2977939, EPI_ISL_2977940, EPI_ISL_2977941, EPI_ISL_2977942, EPI_ISL_2977943, EPI_ISL_2977944, EPI_ISL_2977945, EPI_ISL_2977946, EPI_ISL_2977947, EPI_ISL_2977948, EPI_ISL_2977949, EPI_ISL_2977950, EPI_ISL_2977951, EPI_ISL_2977952, EPI_ISL_2977953, EPI_ISL_2977954, EPI_ISL_2977955, EPI_ISL_2977956, EPI_ISL_2977957, EPI_ISL_2977958, EPI_ISL_2977959, EPI_ISL_2977960, EPI_ISL_2977961, EPI_ISL_2977962, EPI_ISL_2977963, EPI_ISL_2977964, EPI_ISL_2977965, EPI_ISL_2977966, EPI_ISL_2977967, EPI_ISL_2977968, EPI_ISL_2977969, EPI_ISL_2977970, EPI_ISL_2977971, EPI_ISL_2977972, EPI_ISL_2977973, EPI_ISL_2977974, EPI_ISL_2977975, EPI_ISL_2977976, EPI_ISL_2977977, EPI_ISL_2977978, EPI_ISL_2977979, EPI_ISL_2977980, EPI_ISL_2977981, EPI_ISL_2977982, EPI_ISL_2977983, EPI_ISL_2977984, EPI_ISL_2977985, EPI_ISL_2977986, EPI_ISL_2977987, EPI_ISL_2977988, EPI_ISL_2977989, EPI_ISL_2977990, EPI_ISL_2977991, EPI_ISL_2977992, EPI_ISL_2977993, EPI_ISL_2977994, EPI_ISL_2977995, EPI_ISL_2977996, EPI_ISL_2977997, EPI_ISL_2977998, EPI_ISL_2977999, EPI_ISL_2978000, EPI_ISL_2978001, EPI_ISL_2978002, EPI_ISL_2978003, EPI_ISL_2978004, EPI_ISL_2978005, EPI_ISL_2978006, EPI_ISL_2978007, EPI_ISL_2978008, EPI_ISL_2978009, EPI_ISL_2978010, EPI_ISL_2978011, EPI_ISL_2978012, EPI_ISL_2978013, EPI_ISL_2978014, EPI_ISL_2978015, EPI_ISL_2978016, EPI_ISL_2978017, EPI_ISL_2978018, EPI_ISL_2978019, EPI_ISL_2978020, EPI_ISL_2978021, EPI_ISL_2978022, EPI_ISL_2978023, EPI_ISL_2978024, EPI_ISL_2978025, EPI_ISL_2978026, EPI_ISL_2978027, EPI_ISL_2978028, EPI_ISL_2978029, EPI_ISL_2978030, EPI_ISL_2978031, EPI_ISL_2978032, EPI_ISL_2978033, EPI_ISL_2978034, EPI_ISL_2978035, EPI_ISL_2978036, EPI_ISL_2978037, EPI_ISL_2978038, EPI_ISL_2978039, EPI_ISL_2978040, EPI_ISL_2978041, EPI_ISL_2978042, EPI_ISL_2978043, EPI_ISL_2978044, EPI_ISL_2978045, EPI_ISL_2978046, EPI_ISL_2978047, EPI_ISL_2978048, EPI_ISL_2978049, EPI_ISL_2978050, EPI_ISL_2978051, EPI_ISL_2978052, EPI_ISL_2978053, EPI_ISL_2978054, EPI_ISL_2978055, EPI_ISL_2978056, EPI_ISL_2978057, EPI_ISL_2978058, EPI_ISL_2978059, EPI_ISL_2978060, EPI_ISL_2978061, EPI_ISL_2978062, EPI_ISL_2978063, EPI_ISL_2978064, EPI_ISL_2978065, EPI_ISL_2978066, EPI_ISL_2978067, EPI_ISL_2978068, EPI_ISL_2978069, EPI_ISL_2978070, EPI_ISL_2978071, EPI_ISL_2978072, EPI_ISL_2978073, EPI_ISL_2978074, EPI_ISL_2978075, EPI_ISL_2978076, EPI_ISL_2978077, EPI_ISL_2978078, EPI_ISL_2978079, EPI_ISL_2978080, EPI_ISL_2978081, EPI_ISL_2978082, EPI_ISL_2978083, EPI_ISL_2978084, EPI_ISL_2978085, EPI_ISL_2978086, EPI_ISL_2978087, EPI_ISL_2978088, EPI_ISL_2978089, EPI_ISL_2978090, EPI_ISL_2978091, EPI_ISL_2978092, EPI_ISL_2978093, EPI_ISL_2978094, EPI_ISL_2978095, EPI_ISL_2978096, EPI_ISL_2978097, EPI_ISL_2978098, EPI_ISL_2978099, EPI_ISL_2978100, EPI_ISL_2978101, EPI_ISL_2978102, EPI_ISL_2978103, EPI_ISL_2978104, EPI_ISL_2978105, EPI_ISL_2978106, EPI_ISL_2978107, EPI_ISL_2978108, EPI_ISL_2978109, EPI_ISL_2978110, EPI_ISL_2978111, EPI_ISL_2978112, EPI_ISL_2978113, EPI_ISL_2978114, EPI_ISL_2978115, EPI_ISL_2978116, EPI_ISL_2978117, EPI_ISL_2978118, EPI_ISL_2978119, EPI_ISL_2978120, EPI_ISL_2978121, EPI_ISL_2978122, EPI_ISL_2978123, EPI_ISL_2978124, EPI_ISL_2978125, EPI_ISL_2978126, EPI_ISL_2978127, EPI_ISL_2978128, EPI_ISL_2978129, EPI_ISL_2978130, EPI_ISL_2978131, EPI_ISL_2978132, EPI_ISL_2978133, EPI_ISL_2978134, EPI_ISL_2978135, EPI_ISL_2978136, EPI_ISL_2978137, EPI_ISL_2978138, EPI_ISL_2978139, EPI_ISL_2978140, EPI_ISL_2978141, EPI_ISL_2978142, EPI_ISL_2978143, EPI_ISL_2978144, EPI_ISL_2978145, EPI_ISL_2978146

| | | | |
|---|---|---|---|
| see above | CDPH VBL | California Department of Public Health | CDPH-COVIDNet; SwabSeq/UCLA; UCLA Technology Center for Genomics & Bioinformatics; UCSD EXCITE lab; UCSF Center for Advanced Technology |
| EPI_ISL_1094536 | CDPH, Microbial Diseases Laboratory | Respiratory Viruses Branch, Division of Viral Diseases, Centers for Disease Control and Prevention | Anna Montmayeur; Anna Uehara; Ben L. Rambo-Martin; Clinton R. Paden; Dhwani Batra; Haibin Wang; Jasmine Padilla; Jing Zhang; Justin Lee; Krista Queen; Lori Rowe; Mark Burroughs; Mili Sheth; Peter W. Cook; Rachel Marine; Sarah Nobles; Suxiang Tong; Yan Li; Ying Tao |

EPI_ISL_1373839, EPI_ISL_1373845, EPI_ISL_1373852, EPI_ISL_1373857, EPI_ISL_1373859, EPI_ISL_1373866, EPI_ISL_1373868, EPI_ISL_1373871, EPI_ISL_1373874

| | | | |
|---|---|---|---|
| see above | CDPH, Viral and Rickettsial Disease Laboratory | Genomics and Discovery, Respiratory Viruses Branch, Division of Viral Diseases, Centers for Disease Control and Prevention | Anna Kelleher; Anna Uehara; Brian Lynch; Clinton R. Paden; Haibin Wang; Han Jia Justin Ng; Jing Zhang; Krista Queen; Peter Cook; Suxiang Tong; Yan Li; Ying Tao |

EPI_ISL_1094216, EPI_ISL_1094233, EPI_ISL_1094268, EPI_ISL_1094348, EPI_ISL_1094577, EPI_ISL_1094579, EPI_ISL_1094580, EPI_ISL_1094581, EPI_ISL_1094585, EPI_ISL_1094588, EPI_ISL_1095190, EPI_ISL_1095192, EPI_ISL_1095196, EPI_ISL_1095197, EPI_ISL_1095200, EPI_ISL_1095201, EPI_ISL_1095203

| | | | |
|---|---|---|---|
| see above | CDPH, Viral and Rickettsial Disease Laboratory | Respiratory Viruses Branch, Division of Viral Diseases, Centers for Disease Control and Prevention | Anna Montmayeur; Anna Uehara; Ben L. Rambo-Martin; Clinton R. Paden; Dhwani Batra; Haibin Wang; Jasmine Padilla; Jing Zhang; Justin Lee; Krista Queen; Lori Rowe; Mark Burroughs; Mili Sheth; Peter W. Cook; Rachel Marine; Sarah Nobles; Suxiang Tong; Yan Li; Ying Tao |

EPI_ISL_2234031, EPI_ISL_2234032, EPI_ISL_2234033, EPI_ISL_2234034, EPI_ISL_2234036, EPI_ISL_2234049, EPI_ISL_2234050, EPI_ISL_2234051, EPI_ISL_2234052, EPI_ISL_2234053, EPI_ISL_2234054, EPI_ISL_2234055, EPI_ISL_2234056, EPI_ISL_2234057, EPI_ISL_2234058, EPI_ISL_2234059, EPI_ISL_2234060, EPI_ISL_2234061, EPI_ISL_2234062, EPI_ISL_2234063, EPI_ISL_2234064, EPI_ISL_2234065, EPI_ISL_2234067, EPI_ISL_2234069, EPI_ISL_2234084, EPI_ISL_2234085, EPI_ISL_2234086, EPI_ISL_2234087, EPI_ISL_2234088, EPI_ISL_2234090, EPI_ISL_2234091, EPI_ISL_2234092, EPI_ISL_2234093, EPI_ISL_2234094, EPI_ISL_2234095, EPI_ISL_2234096, EPI_ISL_2234097, EPI_ISL_2234120, EPI_ISL_2234129, EPI_ISL_2234130, EPI_ISL_2234139, EPI_ISL_2234141, EPI_ISL_2234147, EPI_ISL_2234148, EPI_ISL_2234149, EPI_ISL_2234150, EPI_ISL_2234151, EPI_ISL_2234152, EPI_ISL_2234153, EPI_ISL_2234154, EPI_ISL_2234155, EPI_ISL_2234156, EPI_ISL_2234163, EPI_ISL_2234194, EPI_ISL_2234317, EPI_ISL_2234318, EPI_ISL_2234319, EPI_ISL_2234320, EPI_ISL_2234322, EPI_ISL_2234323, EPI_ISL_2234324, EPI_ISL_2234325, EPI_ISL_2234327, EPI_ISL_2234328, EPI_ISL_2234329, EPI_ISL_2234330, EPI_ISL_2234355, EPI_ISL_2234356, EPI_ISL_2537394, EPI_ISL_2537405, EPI_ISL_2537406, EPI_ISL_2537420, EPI_ISL_2892544

| | | | |
|---|---|---|---|
| see above | California Department of Public Health | California Department of Public Health | CDPH IDLB COVIDNet et al |

EPI_ISL_2778105, EPI_ISL_2778108, EPI_ISL_2778109, EPI_ISL_2778110, EPI_ISL_2778111, EPI_ISL_2778112, EPI_ISL_2778113, EPI_ISL_2778114, EPI_ISL_2778116, EPI_ISL_2778117, EPI_ISL_2778118, EPI_ISL_2778119, EPI_ISL_2778120, EPI_ISL_2778121, EPI_ISL_2778122, EPI_ISL_2778123, EPI_ISL_2778124, EPI_ISL_2778125, EPI_ISL_2778126, EPI_ISL_2778127, EPI_ISL_2778128, EPI_ISL_2778129, EPI_ISL_2778130, EPI_ISL_2778131, EPI_ISL_2778132, EPI_ISL_2778133, EPI_ISL_2778134, EPI_ISL_2778135, EPI_ISL_2778136, EPI_ISL_2778137, EPI_ISL_2778138, EPI_ISL_2778139, EPI_ISL_2778140, EPI_ISL_2778141, EPI_ISL_2778142, EPI_ISL_2778143, EPI_ISL_2778144, EPI_ISL_2778145, EPI_ISL_2778146, EPI_ISL_2778147, EPI_ISL_2778148, EPI_ISL_2778149, EPI_ISL_2778150, EPI_ISL_2778151, EPI_ISL_2778152, EPI_ISL_2778153, EPI_ISL_2778154, EPI_ISL_2778155, EPI_ISL_2778156, EPI_ISL_2778157, EPI_ISL_2778158, EPI_ISL_2778159, EPI_ISL_2778160, EPI_ISL_2778161, EPI_ISL_2778162, EPI_ISL_2778163, EPI_ISL_2778164, EPI_ISL_2778165, EPI_ISL_2778166, EPI_ISL_2778167, EPI_ISL_2778168, EPI_ISL_2778169, EPI_ISL_2778170, EPI_ISL_2778171, EPI_ISL_2778172, EPI_ISL_2778173, EPI_ISL_2778174, EPI_ISL_2778175, EPI_ISL_2778176, EPI_ISL_2778177, EPI_ISL_2778178, EPI_ISL_2778179, EPI_ISL_2778180, EPI_ISL_2778181, EPI_ISL_2778182, EPI_ISL_2778183, EPI_ISL_2778184, EPI_ISL_2778185, EPI_ISL_2778186, EPI_ISL_2778187, EPI_ISL_2778188, EPI_ISL_2778189, EPI_ISL_2778190, EPI_ISL_2778191, EPI_ISL_2778192, EPI_ISL_2778193, EPI_ISL_2778194, EPI_ISL_2778195, EPI_ISL_2778196, EPI_ISL_2778197, EPI_ISL_2778198, EPI_ISL_2778199, EPI_ISL_2778200, EPI_ISL_2778201, EPI_ISL_2778202, EPI_ISL_2778203, EPI_ISL_2778204, EPI_ISL_2778205, EPI_ISL_2778206, EPI_ISL_2778207, EPI_ISL_2778208, EPI_ISL_2778209, EPI_ISL_2778210, EPI_ISL_2778211, EPI_ISL_2778212, EPI_ISL_2778214, EPI_ISL_2778216, EPI_ISL_2778217, EPI_ISL_2778218, EPI_ISL_2778219, EPI_ISL_2778222, EPI_ISL_2778223, EPI_ISL_2778224, EPI_ISL_2778225, EPI_ISL_2778226, EPI_ISL_2778229, EPI_ISL_2778230, EPI_ISL_2778231, EPI_ISL_2778233, EPI_ISL_2778234, EPI_ISL_2778235, EPI_ISL_2778236, EPI_ISL_2778237, EPI_ISL_2778238, EPI_ISL_2778241, EPI_ISL_2778242, EPI_ISL_2778243, EPI_ISL_2778244, EPI_ISL_2778245, EPI_ISL_2778246, EPI_ISL_2778247, EPI_ISL_2778248, EPI_ISL_2778249, EPI_ISL_2778250, EPI_ISL_2778251, EPI_ISL_2778252, EPI_ISL_2778253, EPI_ISL_2778254, EPI_ISL_2778255, EPI_ISL_2778256, EPI_ISL_2778257, EPI_ISL_2778258, EPI_ISL_2778259, EPI_ISL_2778260, EPI_ISL_2778261, EPI_ISL_2778262, EPI_ISL_2778263, EPI_ISL_2778264, EPI_ISL_2778265, EPI_ISL_2778266, EPI_ISL_2778267, EPI_ISL_2778268, EPI_ISL_2778269, EPI_ISL_2778270, EPI_ISL_2837323, EPI_ISL_2837405, EPI_ISL_2837406, EPI_ISL_2837407, EPI_ISL_2837408, EPI_ISL_2837409, EPI_ISL_2837431, EPI_ISL_2837445, EPI_ISL_2837512, EPI_ISL_2837513, EPI_ISL_2837569, EPI_ISL_2837570, EPI_ISL_2837571, EPI_ISL_2837696, EPI_ISL_2837697, EPI_ISL_2837715, EPI_ISL_2837749, EPI_ISL_2837750, EPI_ISL_2837751, EPI_ISL_2837752, EPI_ISL_2837802, EPI_ISL_2837803, EPI_ISL_2837804, EPI_ISL_2837893, EPI_ISL_2837894, EPI_ISL_2837895, EPI_ISL_2837954, EPI_ISL_2838012, EPI_ISL_2838013, EPI_ISL_2838014, EPI_ISL_2838022, EPI_ISL_2838023

| | | | |
|---|---|---|---|
| see above | California Department of Public Health Valencia Branch Laboratory (CDPH VBL) | California Department of Public Health | CDPH-COVIDNet; UCLA Technology Center for Genomics & Bioinformatics |

EPI_ISL_1234845, EPI_ISL_1234885, EPI_ISL_1234915, EPI_ISL_1234992, EPI_ISL_1235026, EPI_ISL_1235036, EPI_ISL_1701620

| | | | |
|---|---|---|---|
| see above | California Institute of Technology | Chan-Zuckerberg Biohub | CZB Cliahub Consortium |

EPI_ISL_1824296, EPI_ISL_1824297, EPI_ISL_1824298, EPI_ISL_1824299, EPI_ISL_1824300, EPI_ISL_1824301, EPI_ISL_1824302, EPI_ISL_1824303, EPI_ISL_1824304, EPI_ISL_1824305, EPI_ISL_1824306, EPI_ISL_1824307, EPI_ISL_1824308, EPI_ISL_1824309, EPI_ISL_1824310, EPI_ISL_1824316, EPI_ISL_1824317, EPI_ISL_1824319

| | | | |
|---|---|---|---|
| see above | Chiu Laboratory, University of California, San Francisco | Chiu Laboratory, University of California, San Francisco | Alicia Sotomayor-Gonzalez; Candace Wang; Charles Chiu; Daniel Wong; Venice Servellita; Yueyuan Zhang |

EPI_ISL_1824720, EPI_ISL_1824721, EPI_ISL_1824722, EPI_ISL_1824723, EPI_ISL_1824724, EPI_ISL_1824725, EPI_ISL_1824726, EPI_ISL_1824727, EPI_ISL_1824728, EPI_ISL_1824729, EPI_ISL_1824730, EPI_ISL_1824731, EPI_ISL_1824732, EPI_ISL_1824733, EPI_ISL_1824734, EPI_ISL_1824735, EPI_ISL_1824736, EPI_ISL_1824737, EPI_ISL_1824738, EPI_ISL_1824739, EPI_ISL_1824740, EPI_ISL_1824741, EPI_ISL_1824742, EPI_ISL_1824743, EPI_ISL_1824744, EPI_ISL_1824745, EPI_ISL_1824746, EPI_ISL_1824747, EPI_ISL_1824748, EPI_ISL_1824749, EPI_ISL_1824750, EPI_ISL_1824751, EPI_ISL_1824752, EPI_ISL_1824753, EPI_ISL_1824754, EPI_ISL_1824755, EPI_ISL_1824756, EPI_ISL_1824757, EPI_ISL_1824759, EPI_ISL_1824760, EPI_ISL_1824761, EPI_ISL_1824762, EPI_ISL_1824763, EPI_ISL_1824764, EPI_ISL_1824765, EPI_ISL_1824766, EPI_ISL_1824767, EPI_ISL_1824768, EPI_ISL_1824769, EPI_ISL_1824770, EPI_ISL_1824771, EPI_ISL_1824772, EPI_ISL_1824773, EPI_ISL_1824774, EPI_ISL_1824775, EPI_ISL_1824776, EPI_ISL_1824777, EPI_ISL_1824778, EPI_ISL_1824779, EPI_ISL_1824780

| | | | |
|---|---|---|---|
| see above | Color Genomics | Chiu Laboratory, University of California, San Francisco | Alicia Sotomayor-Gonzalez; Alicia Zhou; Candace Wang; Charles Chiu; Daniel Wong; Erika Torres; Madyson Wanner; Scott Topper; Venice Servellita; Yueyuan Zhang |

EPI_ISL_1030281, EPI_ISL_1030310, EPI_ISL_1030313, EPI_ISL_1030314, EPI_ISL_1030315, EPI_ISL_1030316, EPI_ISL_1030317, EPI_ISL_1030318, EPI_ISL_1030319, EPI_ISL_1030320, EPI_ISL_1030321, EPI_ISL_1030322, EPI_ISL_1030323, EPI_ISL_1030324, EPI_ISL_1030325, EPI_ISL_1030326, EPI_ISL_1030327, EPI_ISL_1030328, EPI_ISL_1235061, EPI_ISL_1235062, EPI_ISL_1235064, EPI_ISL_1235065,

EPI_ISL_1235066, EPI_ISL_1235067, EPI_ISL_1235069, EPI_ISL_1235071, EPI_ISL_1235072, EPI_ISL_1235073, EPI_ISL_1235076, EPI_ISL_1235078, EPI_ISL_1235081, EPI_ISL_1235082, EPI_ISL_1235085, EPI_ISL_1235086, EPI_ISL_1235087, EPI_ISL_1235089, EPI_ISL_1235090, EPI_ISL_1235096, EPI_ISL_1235100, EPI_ISL_1235101, EPI_ISL_1235103, EPI_ISL_1235105, EPI_ISL_1235108, EPI_ISL_1235109, EPI_ISL_1235110, EPI_ISL_1235113, EPI_ISL_1235114, EPI_ISL_1235117, EPI_ISL_1235120, EPI_ISL_1235129, EPI_ISL_1235130, EPI_ISL_1235131, EPI_ISL_1235134, EPI_ISL_1235137, EPI_ISL_1235139, EPI_ISL_1235143, EPI_ISL_1235145, EPI_ISL_1235151, EPI_ISL_1235153, EPI_ISL_1235154, EPI_ISL_1235155, EPI_ISL_1235156, EPI_ISL_1235160, EPI_ISL_1235161, EPI_ISL_1235163, EPI_ISL_1235164, EPI_ISL_1235166, EPI_ISL_1235167, EPI_ISL_1235168, EPI_ISL_1235169, EPI_ISL_1235170, EPI_ISL_1235172, EPI_ISL_1235173, EPI_ISL_1235174, EPI_ISL_1235177, EPI_ISL_1235179, EPI_ISL_1235180, EPI_ISL_1235187, EPI_ISL_1235189, EPI_ISL_1235191, EPI_ISL_1235196, EPI_ISL_1235200, EPI_ISL_1235201, EPI_ISL_1235202, EPI_ISL_1235204, EPI_ISL_1235205, EPI_ISL_1235207, EPI_ISL_1235208, EPI_ISL_1235211, EPI_ISL_1235214, EPI_ISL_1235215, EPI_ISL_1235216, EPI_ISL_1235218, EPI_ISL_1235224, EPI_ISL_1235225, EPI_ISL_1235227, EPI_ISL_1235228, EPI_ISL_1235231, EPI_ISL_1235233, EPI_ISL_1235237, EPI_ISL_1235239, EPI_ISL_1235245, EPI_ISL_1235246, EPI_ISL_1235247, EPI_ISL_1235250, EPI_ISL_1235253, EPI_ISL_1235257, EPI_ISL_1235258, EPI_ISL_1235259, EPI_ISL_1235260, EPI_ISL_1235261, EPI_ISL_1235262, EPI_ISL_1235264, EPI_ISL_1235265, EPI_ISL_1235266, EPI_ISL_1235267, EPI_ISL_1235268, EPI_ISL_1235271, EPI_ISL_1235275, EPI_ISL_1235277, EPI_ISL_1235280, EPI_ISL_1235283, EPI_ISL_1235284, EPI_ISL_1235286, EPI_ISL_1235288, EPI_ISL_1235293, EPI_ISL_1235296, EPI_ISL_1235297, EPI_ISL_1235300, EPI_ISL_1235301, EPI_ISL_1235302, EPI_ISL_1235304, EPI_ISL_1235305, EPI_ISL_1235306, EPI_ISL_1235311, EPI_ISL_1235315, EPI_ISL_1235316, EPI_ISL_1235319, EPI_ISL_1235320, EPI_ISL_1235323, EPI_ISL_1235324, EPI_ISL_1235325, EPI_ISL_1235326, EPI_ISL_1235327, EPI_ISL_1235328, EPI_ISL_1235329, EPI_ISL_1235331, EPI_ISL_1235332, EPI_ISL_1235333, EPI_ISL_1235336, EPI_ISL_1235337, EPI_ISL_1235340, EPI_ISL_1235341, EPI_ISL_1235342, EPI_ISL_1235343, EPI_ISL_1235344, EPI_ISL_1235345, EPI_ISL_1235347, EPI_ISL_1235348, EPI_ISL_1235349, EPI_ISL_1235350, EPI_ISL_1235352, EPI_ISL_1235353, EPI_ISL_1235355, EPI_ISL_1235356, EPI_ISL_1235357, EPI_ISL_1235358, EPI_ISL_1477697, EPI_ISL_1477737, EPI_ISL_1477774, EPI_ISL_1477781, EPI_ISL_1477802, EPI_ISL_1477844, EPI_ISL_1477930, EPI_ISL_1477946, EPI_ISL_1478091, EPI_ISL_1529103, EPI_ISL_1529144, EPI_ISL_1529149, EPI_ISL_1529163, EPI_ISL_1529164, EPI_ISL_1529165, EPI_ISL_1529166, EPI_ISL_1529171, EPI_ISL_1529177, EPI_ISL_1529181, EPI_ISL_1529186, EPI_ISL_1529188, EPI_ISL_1529194, EPI_ISL_1529195, EPI_ISL_1529196, EPI_ISL_1529204, EPI_ISL_1529210, EPI_ISL_1529211, EPI_ISL_1529212, EPI_ISL_1529216, EPI_ISL_1529217, EPI_ISL_1529227, EPI_ISL_1529228, EPI_ISL_1529229, EPI_ISL_1529236, EPI_ISL_1529239, EPI_ISL_1529250, EPI_ISL_1529253, EPI_ISL_1529261, EPI_ISL_1529269, EPI_ISL_1529271, EPI_ISL_1529272, EPI_ISL_1529275, EPI_ISL_1529282, EPI_ISL_1529296, EPI_ISL_1529297, EPI_ISL_1529298, EPI_ISL_1529301, EPI_ISL_1529302, EPI_ISL_1529304, EPI_ISL_1529305, EPI_ISL_1529310, EPI_ISL_1529320, EPI_ISL_1529328, EPI_ISL_1529330, EPI_ISL_1529331, EPI_ISL_1529334, EPI_ISL_1529340, EPI_ISL_1529346, EPI_ISL_1529348, EPI_ISL_1529350, EPI_ISL_1529352, EPI_ISL_1529353, EPI_ISL_1529354, EPI_ISL_1529355, EPI_ISL_1529365, EPI_ISL_2102745, EPI_ISL_2102757, EPI_ISL_2102759, EPI_ISL_2102760, EPI_ISL_2102762, EPI_ISL_2102767, EPI_ISL_2102770, EPI_ISL_2102792, EPI_ISL_2102800, EPI_ISL_2102801, EPI_ISL_2102814, EPI_ISL_2102821, EPI_ISL_2102833, EPI_ISL_2102856, EPI_ISL_2102886, EPI_ISL_2102919, EPI_ISL_2102930, EPI_ISL_2102937, EPI_ISL_2102952, EPI_ISL_2102961, EPI_ISL_2102973, EPI_ISL_2102981, EPI_ISL_2103005, EPI_ISL_2103007, EPI_ISL_2103017, EPI_ISL_2103022, EPI_ISL_2103039, EPI_ISL_2103043, EPI_ISL_2103052, EPI_ISL_2103070, EPI_ISL_2103078, EPI_ISL_2103080, EPI_ISL_2103099, EPI_ISL_2103102, EPI_ISL_2103105, EPI_ISL_2103111, EPI_ISL_2103113, EPI_ISL_2103136, EPI_ISL_2103143, EPI_ISL_2103156, EPI_ISL_2103157, EPI_ISL_2103179, EPI_ISL_2103185, EPI_ISL_2103186, EPI_ISL_2103196, EPI_ISL_2658575, EPI_ISL_2658594, EPI_ISL_2658619, EPI_ISL_2658653, EPI_ISL_2658698, EPI_ISL_2658717, EPI_ISL_2659339, EPI_ISL_2659396, EPI_ISL_2659409, EPI_ISL_2659568, EPI_ISL_2758457, EPI_ISL_2758458, EPI_ISL_2758459, EPI_ISL_2758479, EPI_ISL_2758480, EPI_ISL_2758481

| | | | |
|---|---|---|---|
| see above | Contra Costa County Public Health Lab | Chan-Zuckerberg Biohub | CZB Cliahub Consortium |

EPI_ISL_1027366, EPI_ISL_1027367, EPI_ISL_1185288, EPI_ISL_1185387, EPI_ISL_1701471, EPI_ISL_1701604, EPI_ISL_2659297, EPI_ISL_2659504

| | | | |
|---|---|---|---|
| see above | County of San Luis Obispo Public Health Laboratory | Chan-Zuckerberg Biohub | CZB Cliahub Consortium |

EPI_ISL_1093602, EPI_ISL_1093604, EPI_ISL_1093605, EPI_ISL_1093608, EPI_ISL_1093610, EPI_ISL_1093611, EPI_ISL_1093612, EPI_ISL_1093613, EPI_ISL_1093614, EPI_ISL_1093619, EPI_ISL_1093620, EPI_ISL_1093621, EPI_ISL_1093622, EPI_ISL_1093624, EPI_ISL_1093625, EPI_ISL_1093627, EPI_ISL_1093629, EPI_ISL_1093630, EPI_ISL_1093631, EPI_ISL_1093632, EPI_ISL_1093633, EPI_ISL_1093634, EPI_ISL_1093636, EPI_ISL_1093638, EPI_ISL_1093641, EPI_ISL_1093642, EPI_ISL_1093644, EPI_ISL_1093645, EPI_ISL_1093646, EPI_ISL_1093650, EPI_ISL_1093651, EPI_ISL_1093652, EPI_ISL_1093653, EPI_ISL_1093654, EPI_ISL_1093655, EPI_ISL_1093656, EPI_ISL_1093658, EPI_ISL_1093659, EPI_ISL_1093660, EPI_ISL_1093661, EPI_ISL_1093664, EPI_ISL_1093666, EPI_ISL_1093667, EPI_ISL_1093668, EPI_ISL_1093670, EPI_ISL_1093673, EPI_ISL_1093675, EPI_ISL_1093676, EPI_ISL_1093678, EPI_ISL_1093680, EPI_ISL_1093681, EPI_ISL_1093683, EPI_ISL_1093684, EPI_ISL_1093685, EPI_ISL_1093688, EPI_ISL_1093689, EPI_ISL_1093690, EPI_ISL_1093691, EPI_ISL_1093692, EPI_ISL_1093693, EPI_ISL_1093694, EPI_ISL_1093695, EPI_ISL_1093696, EPI_ISL_1093698, EPI_ISL_1093699, EPI_ISL_1093700, EPI_ISL_1093701, EPI_ISL_1093704, EPI_ISL_1093706, EPI_ISL_1093707, EPI_ISL_1093708, EPI_ISL_1093709, EPI_ISL_1093710, EPI_ISL_1093711, EPI_ISL_1093712, EPI_ISL_1093713, EPI_ISL_1093714, EPI_ISL_1093715, EPI_ISL_1093716, EPI_ISL_1093717, EPI_ISL_1093718, EPI_ISL_1093719, EPI_ISL_1093721, EPI_ISL_1093722, EPI_ISL_1093723, EPI_ISL_1093726, EPI_ISL_1093727, EPI_ISL_1093728, EPI_ISL_1093729, EPI_ISL_1093730, EPI_ISL_1093731, EPI_ISL_1093732, EPI_ISL_1093734, EPI_ISL_1093735, EPI_ISL_1093736, EPI_ISL_1093737, EPI_ISL_1093738, EPI_ISL_1093740, EPI_ISL_1093741, EPI_ISL_1093742, EPI_ISL_1093744, EPI_ISL_1093745, EPI_ISL_1093746, EPI_ISL_1093747, EPI_ISL_1093748, EPI_ISL_1093750, EPI_ISL_1093751, EPI_ISL_1093752, EPI_ISL_1093754, EPI_ISL_1093757, EPI_ISL_1093759, EPI_ISL_1093760, EPI_ISL_1093764, EPI_ISL_1093765, EPI_ISL_1093767, EPI_ISL_1093770, EPI_ISL_1093772, EPI_ISL_1093773, EPI_ISL_1093774, EPI_ISL_1093775, EPI_ISL_1093777, EPI_ISL_1093778, EPI_ISL_1093780, EPI_ISL_1093782, EPI_ISL_1093783, EPI_ISL_1093785, EPI_ISL_1093786, EPI_ISL_1093787, EPI_ISL_1093788, EPI_ISL_1093789, EPI_ISL_1093790, EPI_ISL_1093793, EPI_ISL_1093794, EPI_ISL_1093795, EPI_ISL_1093797, EPI_ISL_1093802, EPI_ISL_1093803, EPI_ISL_1093804, EPI_ISL_1093805, EPI_ISL_1093806, EPI_ISL_1093808, EPI_ISL_1093809, EPI_ISL_1093811, EPI_ISL_1093812, EPI_ISL_1093814, EPI_ISL_1093815, EPI_ISL_1093816, EPI_ISL_1093818, EPI_ISL_1093820, EPI_ISL_1093822, EPI_ISL_1093824, EPI_ISL_1093828, EPI_ISL_1093829, EPI_ISL_1093830, EPI_ISL_1093831, EPI_ISL_1093835, EPI_ISL_1093836, EPI_ISL_1093837, EPI_ISL_1093838, EPI_ISL_1093839, EPI_ISL_1093842, EPI_ISL_1093843, EPI_ISL_1093846, EPI_ISL_1093848, EPI_ISL_1093849, EPI_ISL_1093850, EPI_ISL_1093851, EPI_ISL_1093852, EPI_ISL_1093853, EPI_ISL_1093854, EPI_ISL_1093855, EPI_ISL_1093856, EPI_ISL_1093857, EPI_ISL_1093858, EPI_ISL_1093859, EPI_ISL_1093862, EPI_ISL_1093865, EPI_ISL_1093866, EPI_ISL_1093868, EPI_ISL_1093870, EPI_ISL_1093872, EPI_ISL_1093873, EPI_ISL_1093874, EPI_ISL_1093877, EPI_ISL_1093879, EPI_ISL_1093882, EPI_ISL_1093884, EPI_ISL_1093885, EPI_ISL_1093886, EPI_ISL_1093887, EPI_ISL_1093889, EPI_ISL_1093892, EPI_ISL_1093895, EPI_ISL_1093896, EPI_ISL_1093897, EPI_ISL_1093898, EPI_ISL_1093899, EPI_ISL_1093900, EPI_ISL_1093901, EPI_ISL_1093903, EPI_ISL_1093904, EPI_ISL_1093905, EPI_ISL_1093906, EPI_ISL_1093907, EPI_ISL_1093908, EPI_ISL_1093909, EPI_ISL_1093910, EPI_ISL_1093911, EPI_ISL_1093912, EPI_ISL_1093913, EPI_ISL_1093914, EPI_ISL_1093915, EPI_ISL_1093917, EPI_ISL_1093919, EPI_ISL_1093921, EPI_ISL_1093922, EPI_ISL_1093924, EPI_ISL_1093925, EPI_ISL_1093927, EPI_ISL_1093928, EPI_ISL_1093929, EPI_ISL_1093930, EPI_ISL_1093931, EPI_ISL_1093933, EPI_ISL_1093934, EPI_ISL_1093937, EPI_ISL_1093938, EPI_ISL_1093940, EPI_ISL_1093941, EPI_ISL_1093942, EPI_ISL_1093944, EPI_ISL_1093945, EPI_ISL_1093949, EPI_ISL_1093953, EPI_ISL_1093954, EPI_ISL_1093955, EPI_ISL_1093956, EPI_ISL_1093957, EPI_ISL_1093961, EPI_ISL_1093963, EPI_ISL_1093964, EPI_ISL_1093965, EPI_ISL_1093967, EPI_ISL_1093968, EPI_ISL_1093969, EPI_ISL_1093971, EPI_ISL_1093972, EPI_ISL_1093975, EPI_ISL_1093977, EPI_ISL_1093978, EPI_ISL_1093980, EPI_ISL_1093982, EPI_ISL_1093983, EPI_ISL_1093984, EPI_ISL_1093985, EPI_ISL_1093986, EPI_ISL_1093987, EPI_ISL_1093990, EPI_ISL_1093992, EPI_ISL_1093993, EPI_ISL_1093995, EPI_ISL_1093996, EPI_ISL_1093997, EPI_ISL_1093998, EPI_ISL_1093999, EPI_ISL_1094001, EPI_ISL_1094002, EPI_ISL_1094004, EPI_ISL_1094007, EPI_ISL_1094008, EPI_ISL_1094010, EPI_ISL_1094016, EPI_ISL_1094023, EPI_ISL_1094024, EPI_ISL_1094025, EPI_ISL_1094026, EPI_ISL_1094028, EPI_ISL_1094029, EPI_ISL_1094030, EPI_ISL_1094031, EPI_ISL_1094032, EPI_ISL_1094033, EPI_ISL_1094036, EPI_ISL_1094037, EPI_ISL_1094041, EPI_ISL_1094042, EPI_ISL_1094043, EPI_ISL_1094044, EPI_ISL_1094045, EPI_ISL_1094048, EPI_ISL_1094053, EPI_ISL_1094056, EPI_ISL_1094057, EPI_ISL_1094058, EPI_ISL_1094059, EPI_ISL_1094060, EPI_ISL_1094064, EPI_ISL_1094066, EPI_ISL_1094070, EPI_ISL_1094073, EPI_ISL_1094076, EPI_ISL_1094077, EPI_ISL_1094078, EPI_ISL_1094079, EPI_ISL_1094080, EPI_ISL_1094083, EPI_ISL_1094084, EPI_ISL_1094085, EPI_ISL_1094087, EPI_ISL_1094088, EPI_ISL_1094089, EPI_ISL_1094092, EPI_ISL_1094093, EPI_ISL_1094094, EPI_ISL_1094096, EPI_ISL_1094098, EPI_ISL_1094100, EPI_ISL_1094101, EPI_ISL_1094102, EPI_ISL_1094104, EPI_ISL_1094105, EPI_ISL_1094106, EPI_ISL_1094110, EPI_ISL_1094112, EPI_ISL_1094114, EPI_ISL_1094118, EPI_ISL_1094123, EPI_ISL_1094124, EPI_ISL_1094125, EPI_ISL_1094126, EPI_ISL_1094127, EPI_ISL_1094128, EPI_ISL_1094131, EPI_ISL_1094132, EPI_ISL_1094133, EPI_ISL_1094135, EPI_ISL_1094137, EPI_ISL_1094138, EPI_ISL_1094139, EPI_ISL_1094141, EPI_ISL_1094142, EPI_ISL_1094146, EPI_ISL_1094147, EPI_ISL_1094148, EPI_ISL_1094149, EPI_ISL_1094150, EPI_ISL_1094154, EPI_ISL_1094155, EPI_ISL_1094157, EPI_ISL_1094159, EPI_ISL_1094160, EPI_ISL_1094164, EPI_ISL_1094165, EPI_ISL_1094169, EPI_ISL_1094171, EPI_ISL_1094174, EPI_ISL_1094175, EPI_ISL_1094176, EPI_ISL_1094177, EPI_ISL_1094178, EPI_ISL_1094181, EPI_ISL_1094182, EPI_ISL_1094185, EPI_ISL_1094186, EPI_ISL_1094188, EPI_ISL_1094189, EPI_ISL_1098806, EPI_ISL_1098814, EPI_ISL_1098816, EPI_ISL_1098818, EPI_ISL_1098821, EPI_ISL_1098823, EPI_ISL_1098825, EPI_ISL_1098826, EPI_ISL_1098827, EPI_ISL_1098829, EPI_ISL_1318707, EPI_ISL_1318710, EPI_ISL_1318713, EPI_ISL_1318716, EPI_ISL_1318722, EPI_ISL_1318729, EPI_ISL_1318733, EPI_ISL_1318738, EPI_ISL_1318739, EPI_ISL_1318742, EPI_ISL_1318747, EPI_ISL_1318759, EPI_ISL_1318762, EPI_ISL_1318770, EPI_ISL_1318771, EPI_ISL_1318773, EPI_ISL_1318782, EPI_ISL_1318790, EPI_ISL_1318793, EPI_ISL_1318794, EPI_ISL_1318813, EPI_ISL_1318819, EPI_ISL_1318821, EPI_ISL_1318827, EPI_ISL_1318828, EPI_ISL_1318838, EPI_ISL_1318839, EPI_ISL_1318844, EPI_ISL_1318850, EPI_ISL_1318858, EPI_ISL_1318859, EPI_ISL_1318870, EPI_ISL_1318877, EPI_ISL_1318882, EPI_ISL_1318883, EPI_ISL_1318884, EPI_ISL_1318888, EPI_ISL_1318891, EPI_ISL_1318903, EPI_ISL_1318912, EPI_ISL_1318914, EPI_ISL_1318925, EPI_ISL_1318935, EPI_ISL_1318938, EPI_ISL_1318947, EPI_ISL_1318948, EPI_ISL_1318955, EPI_ISL_1318960, EPI_ISL_1318961, EPI_ISL_1318962, EPI_ISL_1318967, EPI_ISL_1318978, EPI_ISL_1347783, EPI_ISL_1347786, EPI_ISL_1347787, EPI_ISL_1347788, EPI_ISL_1349087

| | | | |
|---|---|---|---|
| see above | DeRisi Lab, University of California, San Francisco | DeRisi Lab, University of California, San Francisco | Amy Kistler; Anthea Mitchell; Carina Marquez; Diane Havlir; Diane Jones; Douglas Black; Eric Chow; Gabriel Chamie; Genay Pilarowski; IDseq Team; Jackie Martinez; James Peng; Jamin Liu; Jon Jacobo; Lucy Li; Luis Rubio; Manu Vanaerschot; Matthew Laurie; Maya Petersen; Patrick Ayscue; Sabrina Mann; Sara Sunshine; Susana Rojas; Susy Rojas; Valerie Tulier-Laiwa; and Joseph DeRisi |

EPI_ISL_1811656, EPI_ISL_1811666, EPI_ISL_1811676, EPI_ISL_1811687, EPI_ISL_1811707, EPI_ISL_1811724, EPI_ISL_1811731, EPI_ISL_1811735, EPI_ISL_1811752, EPI_ISL_1811765, EPI_ISL_1811767, EPI_ISL_1811783, EPI_ISL_1811834, EPI_ISL_1811841, EPI_ISL_1811857, EPI_ISL_1811871, EPI_ISL_1811883, EPI_ISL_1811884, EPI_ISL_1811892, EPI_ISL_1811910, EPI_ISL_1811922, EPI_ISL_1811926, EPI_ISL_1811967, EPI_ISL_1811979, EPI_ISL_1811983, EPI_ISL_1811988, EPI_ISL_1811991, EPI_ISL_1812000, EPI_ISL_1812002, EPI_ISL_1812008, EPI_ISL_1812010, EPI_ISL_1812028, EPI_ISL_1812030, EPI_ISL_1812032, EPI_ISL_1812035, EPI_ISL_1812046, EPI_ISL_1812057, EPI_ISL_1812061, EPI_ISL_1812095, EPI_ISL_1812097, EPI_ISL_1812105, EPI_ISL_1812109, EPI_ISL_1812112, EPI_ISL_1812128, EPI_ISL_1812141, EPI_ISL_1812144, EPI_ISL_1812155, EPI_ISL_1812156, EPI_ISL_1812162, EPI_ISL_1812166, EPI_ISL_1812168, EPI_ISL_1812182, EPI_ISL_1812184, EPI_ISL_1812193, EPI_ISL_1812205, EPI_ISL_1812209, EPI_ISL_1812218, EPI_ISL_1812225, EPI_ISL_1812227, EPI_ISL_1812228, EPI_ISL_1812235, EPI_ISL_1812236, EPI_ISL_1812240, EPI_ISL_1812247, EPI_ISL_1812248, EPI_ISL_1812253, EPI_ISL_1812267, EPI_ISL_1812279, EPI_ISL_1812285, EPI_ISL_1812288, EPI_ISL_1812296, EPI_ISL_1812301, EPI_ISL_1812303, EPI_ISL_1812306, EPI_ISL_1812307, EPI_ISL_1812313, EPI_ISL_1812318, EPI_ISL_1812319, EPI_ISL_1812320, EPI_ISL_1812325, EPI_ISL_1812331, EPI_ISL_1812332, EPI_ISL_1812337, EPI_ISL_1812351, EPI_ISL_1812356, EPI_ISL_1812362, EPI_ISL_1812369, EPI_ISL_1812376, EPI_ISL_1812377, EPI_ISL_1812399, EPI_ISL_1812403, EPI_ISL_1812405, EPI_ISL_1812415, EPI_ISL_1812421, EPI_ISL_1812422, EPI_ISL_1812429, EPI_ISL_1812431, EPI_ISL_1812432, EPI_ISL_1812433, EPI_ISL_1812439, EPI_ISL_1812443, EPI_ISL_1812454, EPI_ISL_1812456, EPI_ISL_1812457, EPI_ISL_1812459, EPI_ISL_1812463, EPI_ISL_1812467, EPI_ISL_1812470, EPI_ISL_1812471, EPI_ISL_1812474, EPI_ISL_1812475, EPI_ISL_1812478, EPI_ISL_1812488, EPI_ISL_1812490, EPI_ISL_1812496, EPI_ISL_1812498, EPI_ISL_1812499, EPI_ISL_1812500, EPI_ISL_1812505, EPI_ISL_1812507, EPI_ISL_1812508, EPI_ISL_1812515, EPI_ISL_1812521, EPI_ISL_1812527, EPI_ISL_1812539, EPI_ISL_1812544, EPI_ISL_1812545, EPI_ISL_1812546, EPI_ISL_1812554, EPI_ISL_1812561, EPI_ISL_1812563, EPI_ISL_1812567, EPI_ISL_1812574, EPI_ISL_1812583, EPI_ISL_1812589, EPI_ISL_1812591, EPI_ISL_1812593, EPI_ISL_1812606, EPI_ISL_1812610, EPI_ISL_1812613, EPI_ISL_1812618, EPI_ISL_1812631, EPI_ISL_1812643, EPI_ISL_1812644, EPI_ISL_1812649, EPI_ISL_1812658, EPI_ISL_1812666, EPI_ISL_1812669, EPI_ISL_1812676, EPI_ISL_1812679, EPI_ISL_1812693, EPI_ISL_1812694, EPI_ISL_1812697, EPI_ISL_1812698, EPI_ISL_1812701, EPI_ISL_1812704, EPI_ISL_1812707, EPI_ISL_1812709, EPI_ISL_1812714, EPI_ISL_1812720, EPI_ISL_1812723, EPI_ISL_1812735, EPI_ISL_1812737, EPI_ISL_1812740, EPI_ISL_1812741, EPI_ISL_1812742, EPI_ISL_1812743, EPI_ISL_1812744, EPI_ISL_1812750, EPI_ISL_1812752, EPI_ISL_1812759, EPI_ISL_1812762, EPI_ISL_1812767, EPI_ISL_1812776, EPI_ISL_1812779, EPI_ISL_1812780, EPI_ISL_1812782, EPI_ISL_1812785, EPI_ISL_1812802, EPI_ISL_1812805, EPI_ISL_1812806, EPI_ISL_1812807, EPI_ISL_1812818, EPI_ISL_1812819, EPI_ISL_1812821, EPI_ISL_1812822, EPI_ISL_1812831, EPI_ISL_1812835, EPI_ISL_1812836, EPI_ISL_1812839, EPI_ISL_1812843, EPI_ISL_1812845, EPI_ISL_1812871, EPI_ISL_1812873, EPI_ISL_1812875, EPI_ISL_1812876, EPI_ISL_1812880, EPI_ISL_1812883, EPI_ISL_1812885, EPI_ISL_1812889, EPI_ISL_1812895, EPI_ISL_1812906, EPI_ISL_1812908, EPI_ISL_1812916, EPI_ISL_1812923, EPI_ISL_1812924, EPI_ISL_1812927, EPI_ISL_1812930, EPI_ISL_1812938, EPI_ISL_1812943, EPI_ISL_1812944, EPI_ISL_1812945, EPI_ISL_1812951, EPI_ISL_1812953, EPI_ISL_1812957, EPI_ISL_1812962, EPI_ISL_1812966, EPI_ISL_1812967, EPI_ISL_1812973, EPI_ISL_1812977, EPI_ISL_1812986, EPI_ISL_1812988, EPI_ISL_1812989, EPI_ISL_1812992, EPI_ISL_1812995, EPI_ISL_1812997, EPI_ISL_1813006, EPI_ISL_1813010, EPI_ISL_1813020, EPI_ISL_1813021, EPI_ISL_1813024, EPI_ISL_1813026, EPI_ISL_1813027, EPI_ISL_1813031, EPI_ISL_1813032, EPI_ISL_1813046, EPI_ISL_1813055, EPI_ISL_1813060, EPI_ISL_1813065, EPI_ISL_1813067, EPI_ISL_1813071, EPI_ISL_1813073, EPI_ISL_1813083, EPI_ISL_1813086, EPI_ISL_1813087, EPI_ISL_1813089, EPI_ISL_1813092, EPI_ISL_1813095, EPI_ISL_1813106, EPI_ISL_1813107, EPI_ISL_1813111, EPI_ISL_1813115, EPI_ISL_1813120, EPI_ISL_1813133, EPI_ISL_1813141, EPI_ISL_1813146, EPI_ISL_1813150, EPI_ISL_1813152, EPI_ISL_1813154, EPI_ISL_1813161, EPI_ISL_1813164, EPI_ISL_1813174, EPI_ISL_1813175, EPI_ISL_1813179, EPI_ISL_1813180, EPI_ISL_1813181, EPI_ISL_1813184, EPI_ISL_1813185, EPI_ISL_1813186, EPI_ISL_1813193, EPI_ISL_1813194, EPI_ISL_1813208, EPI_ISL_1813209, EPI_ISL_1813222, EPI_ISL_1813224, EPI_ISL_1813226, EPI_ISL_1813235, EPI_ISL_1813242, EPI_ISL_1813250, EPI_ISL_1813258, EPI_ISL_1813260, EPI_ISL_1813263, EPI_ISL_1813268, EPI_ISL_1813269, EPI_ISL_1813272, EPI_ISL_1813275, EPI_ISL_1813282, EPI_ISL_1813283, EPI_ISL_1813294, EPI_ISL_1813297, EPI_ISL_1813299, EPI_ISL_1813302, EPI_ISL_1813308, EPI_ISL_1813309, EPI_ISL_1813315, EPI_ISL_1813317, EPI_ISL_1813318, EPI_ISL_1813330, EPI_ISL_1813332, EPI_ISL_1813335, EPI_ISL_1813339, EPI_ISL_1813351, EPI_ISL_1813358, EPI_ISL_1813360, EPI_ISL_1813362, EPI_ISL_1813373, EPI_ISL_1813374, EPI_ISL_1813375, EPI_ISL_1813381, EPI_ISL_1813383, EPI_ISL_1813388, EPI_ISL_1813389, EPI_ISL_1813392, EPI_ISL_1813393, EPI_ISL_1813400, EPI_ISL_1813402, EPI_ISL_1813422, EPI_ISL_1813423, EPI_ISL_1813431, EPI_ISL_1813436, EPI_ISL_1813437, EPI_ISL_1813438, EPI_ISL_1813439, EPI_ISL_1813441, EPI_ISL_1813443, EPI_ISL_1813449, EPI_ISL_1813455, EPI_ISL_1813467, EPI_ISL_1813479, EPI_ISL_1813481, EPI_ISL_1813488, EPI_ISL_1813489, EPI_ISL_1813492, EPI_ISL_1813504, EPI_ISL_1813507, EPI_ISL_1813510, EPI_ISL_1813513, EPI_ISL_1813514, EPI_ISL_1813515, EPI_ISL_1813516, EPI_ISL_1813517, EPI_ISL_1813518, EPI_ISL_1813519, EPI_ISL_1813523, EPI_ISL_1813525, EPI_ISL_1813535, EPI_ISL_1813545, EPI_ISL_1813548, EPI_ISL_1813552, EPI_ISL_1813556, EPI_ISL_1813559, EPI_ISL_1813563, EPI_ISL_1813564, EPI_ISL_1813565, EPI_ISL_1813573, EPI_ISL_1813575, EPI_ISL_1813578, EPI_ISL_1813586, EPI_ISL_1813598, EPI_ISL_1813607, EPI_ISL_1813612, EPI_ISL_1813620, EPI_ISL_1813621, EPI_ISL_1813627, EPI_ISL_1813629, EPI_ISL_1813632, EPI_ISL_1813639, EPI_ISL_1813641, EPI_ISL_1813643, EPI_ISL_1813645, EPI_ISL_1813649, EPI_ISL_1813650, EPI_ISL_1813652, EPI_ISL_1813658, EPI_ISL_1813659, EPI_ISL_1813661, EPI_ISL_1813663, EPI_ISL_1813664, EPI_ISL_1813672, EPI_ISL_1813674, EPI_ISL_1813676, EPI_ISL_1813683, EPI_ISL_1813684, EPI_ISL_1813686, EPI_ISL_1813688, EPI_ISL_1813689, EPI_ISL_1813694, EPI_ISL_1813695, EPI_ISL_1813699, EPI_ISL_1813704, EPI_ISL_1813710, EPI_ISL_1813716, EPI_ISL_1813718, EPI_ISL_1813737, EPI_ISL_1813738, EPI_ISL_1813740, EPI_ISL_1813743, EPI_ISL_1813748, EPI_ISL_1813749, EPI_ISL_1813762, EPI_ISL_1813763, EPI_ISL_1813765, EPI_ISL_1813768, EPI_ISL_1813770, EPI_ISL_1813773, EPI_ISL_1813774, EPI_ISL_1813776, EPI_ISL_1813777, EPI_ISL_1813780, EPI_ISL_1813782, EPI_ISL_1813788, EPI_ISL_1813794, EPI_ISL_1813797, EPI_ISL_1813799, EPI_ISL_1813803, EPI_ISL_1813804, EPI_ISL_1813811, EPI_ISL_1813812, EPI_ISL_1813816, EPI_ISL_1813820, EPI_ISL_1813823, EPI_ISL_1813825, EPI_ISL_1813826, EPI_ISL_1813830, EPI_ISL_1813831, EPI_ISL_1813832, EPI_ISL_1813833, EPI_ISL_1813839, EPI_ISL_1813841, EPI_ISL_1813847, EPI_ISL_1813848, EPI_ISL_1813852, EPI_ISL_1813853, EPI_ISL_1813856, EPI_ISL_1813858, EPI_ISL_1813867, EPI_ISL_1813869, EPI_ISL_1813871, EPI_ISL_1813875, EPI_ISL_1813880, EPI_ISL_1813883, EPI_ISL_1813885, EPI_ISL_1813888, EPI_ISL_1813889, EPI_ISL_1813895, EPI_ISL_1813898, EPI_ISL_1813902, EPI_ISL_1813911, EPI_ISL_1813913, EPI_ISL_1813918, EPI_ISL_1813919, EPI_ISL_1813920, EPI_ISL_1813927, EPI_ISL_1813928, EPI_ISL_1813930, EPI_ISL_1813933, EPI_ISL_1813940, EPI_ISL_1813941, EPI_ISL_1813944, EPI_ISL_1813946, EPI_ISL_1813950, EPI_ISL_1813951, EPI_ISL_1813955, EPI_ISL_1813959, EPI_ISL_1813963, EPI_ISL_1813965, EPI_ISL_1813966, EPI_ISL_1813968, EPI_ISL_1813969, EPI_ISL_1813972, EPI_ISL_1813980, EPI_ISL_1813983, EPI_ISL_1813985, EPI_ISL_1813998, EPI_ISL_1813999, EPI_ISL_1814010, EPI_ISL_1814013, EPI_ISL_1814015, EPI_ISL_1814023, EPI_ISL_1814025, EPI_ISL_1814028, EPI_ISL_1814030, EPI_ISL_1814033, EPI_ISL_1814034, EPI_ISL_1814035, EPI_ISL_1814039, EPI_ISL_1814041, EPI_ISL_1814048, EPI_ISL_1814051, EPI_ISL_1814064, EPI_ISL_1814067, EPI_ISL_1814069, EPI_ISL_1814070, EPI_ISL_1814074, EPI_ISL_1814079, EPI_ISL_1814081, EPI_ISL_1814083, EPI_ISL_1814085, EPI_ISL_1814086, EPI_ISL_1814088, EPI_ISL_1814090, EPI_ISL_1814093, EPI_ISL_1814097, EPI_ISL_1814099, EPI_ISL_1814102, EPI_ISL_1814112, EPI_ISL_1814114, EPI_ISL_1814115, EPI_ISL_1814128, EPI_ISL_1814129, EPI_ISL_1814137, EPI_ISL_1814141, EPI_ISL_1814142, EPI_ISL_1814143, EPI_ISL_1814144, EPI_ISL_1814155, EPI_ISL_1814156, EPI_ISL_1814157, EPI_ISL_1814163, EPI_ISL_1814165, EPI_ISL_1814166, EPI_ISL_1814167, EPI_ISL_1814168, EPI_ISL_1814171, EPI_ISL_1814175, EPI_ISL_1814176, EPI_ISL_1814177, EPI_ISL_1814181, EPI_ISL_1814183, EPI_ISL_1814189, EPI_ISL_1814195, EPI_ISL_1814200, EPI_ISL_1814201, EPI_ISL_1814205, EPI_ISL_1814206, EPI_ISL_1814219, EPI_ISL_1814223, EPI_ISL_1814224, EPI_ISL_1814225, EPI_ISL_1814242, EPI_ISL_1814243, EPI_ISL_1814246, EPI_ISL_1814247, EPI_ISL_1814248, EPI_ISL_1814250, EPI_ISL_1814251, EPI_ISL_1814253, EPI_ISL_1814256, EPI_ISL_1814257, EPI_ISL_1814264, EPI_ISL_1814265, EPI_ISL_1814266, EPI_ISL_1814270, EPI_ISL_1814272, EPI_ISL_1814273, EPI_ISL_1814275, EPI_ISL_1814277, EPI_ISL_1814282, EPI_ISL_1814284, EPI_ISL_1814292, EPI_ISL_1814293, EPI_ISL_1814295, EPI_ISL_1814308, EPI_ISL_1814311, EPI_ISL_1814316, EPI_ISL_1814327, EPI_ISL_1814328, EPI_ISL_1814331, EPI_ISL_1814340, EPI_ISL_1814342, EPI_ISL_1814348, EPI_ISL_1814349, EPI_ISL_1814350, EPI_ISL_1814351, EPI_ISL_1814352, EPI_ISL_1814355, EPI_ISL_1814363, EPI_ISL_1814365, EPI_ISL_1814366, EPI_ISL_1814368, EPI_ISL_1814369, EPI_ISL_1814372, EPI_ISL_1814373, EPI_ISL_1814377, EPI_ISL_1814389, EPI_ISL_1814390, EPI_ISL_1814406, EPI_ISL_1814408, EPI_ISL_1814418, EPI_ISL_1814420, EPI_ISL_1814422, EPI_ISL_1814423, EPI_ISL_1814425, EPI_ISL_1814429, EPI_ISL_1814431, EPI_ISL_1814433, EPI_ISL_1814436, EPI_ISL_1814437, EPI_ISL_1814438, EPI_ISL_1814445, EPI_ISL_1814451, EPI_ISL_1814456, EPI_ISL_1814464, EPI_ISL_1814466, EPI_ISL_1814468, EPI_ISL_1814469, EPI_ISL_1814470, EPI_ISL_1814471, EPI_ISL_1814475, EPI_ISL_1814478, EPI_ISL_1814480, EPI_ISL_1814486, EPI_ISL_1814489, EPI_ISL_1814496, EPI_ISL_1814499, EPI_ISL_1814502, EPI_ISL_1814503, EPI_ISL_1814506, EPI_ISL_1814507, EPI_ISL_1814508, EPI_ISL_1814509, EPI_ISL_1814511, EPI_ISL_1814516, EPI_ISL_1814521, EPI_ISL_1814530, EPI_ISL_1814534, EPI_ISL_1814543, EPI_ISL_1814548, EPI_ISL_1814550, EPI_ISL_1814562, EPI_ISL_1814563, EPI_ISL_1814576, EPI_ISL_1814577, EPI_ISL_1814578, EPI_ISL_1814579, EPI_ISL_1814585, EPI_ISL_1814596, EPI_ISL_1814607, EPI_ISL_1814609, EPI_ISL_1814610, EPI_ISL_1814613, EPI_ISL_1814618, EPI_ISL_1814619, EPI_ISL_1814620, EPI_ISL_1814621, EPI_ISL_1814622, EPI_ISL_1814627, EPI_ISL_1814636, EPI_ISL_1814639, EPI_ISL_1814644, EPI_ISL_1814645, EPI_ISL_1814646, EPI_ISL_1814650, EPI_ISL_1814653, EPI_ISL_1814657, EPI_ISL_1814659, EPI_ISL_1814661, EPI_ISL_1814664, EPI_ISL_1814665, EPI_ISL_1814668, EPI_ISL_1814675, EPI_ISL_1814678, EPI_ISL_1814681, EPI_ISL_1814688, EPI_ISL_1814692, EPI_ISL_1814694, EPI_ISL_1814702, EPI_ISL_1814703, EPI_ISL_1814706, EPI_ISL_1814708, EPI_ISL_1814709, EPI_ISL_1814719, EPI_ISL_1814722, EPI_ISL_1814725, EPI_ISL_1814741, EPI_ISL_1814752, EPI_ISL_1814754, EPI_ISL_1814760, EPI_ISL_1814761, EPI_ISL_1814767, EPI_ISL_1814768, EPI_ISL_1814769, EPI_ISL_1814770, EPI_ISL_1814780, EPI_ISL_1814784, EPI_ISL_1814785, EPI_ISL_1814786, EPI_ISL_1814787, EPI_ISL_1814789, EPI_ISL_1814790, EPI_ISL_1814793, EPI_ISL_1814796, EPI_ISL_1814799, EPI_ISL_1814803, EPI_ISL_1814804, EPI_ISL_1814813, EPI_ISL_1814818, EPI_ISL_1814819, EPI_ISL_1814820, EPI_ISL_1814822, EPI_ISL_1814827, EPI_ISL_1814828, EPI_ISL_1814832, EPI_ISL_1814835, EPI_ISL_1814843, EPI_ISL_1814849, EPI_ISL_1814853, EPI_ISL_1814860, EPI_ISL_1814863, EPI_ISL_1814864, EPI_ISL_1814874, EPI_ISL_1814880, EPI_ISL_1814882, EPI_ISL_1814885, EPI_ISL_1814887, EPI_ISL_1814889, EPI_ISL_1814897, EPI_ISL_1814899, EPI_ISL_1814901, EPI_ISL_1814903, EPI_ISL_1814909, EPI_ISL_1814916, EPI_ISL_1814920, EPI_ISL_1814923, EPI_ISL_1814927, EPI_ISL_1814929, EPI_ISL_1814931, EPI_ISL_1814932, EPI_ISL_1814933, EPI_ISL_1814937, EPI_ISL_1814944, EPI_ISL_1814945, EPI_ISL_1814949, EPI_ISL_1814958, EPI_ISL_1814965, EPI_ISL_1814968, EPI_ISL_1814969, EPI_ISL_1814970, EPI_ISL_1814972, EPI_ISL_1814976, EPI_ISL_1814982, EPI_ISL_1814983, EPI_ISL_1814985, EPI_ISL_1814987, EPI_ISL_1814990, EPI_ISL_1814993, EPI_ISL_1814994, EPI_ISL_1814996, EPI_ISL_1814999, EPI_ISL_1815005, EPI_ISL_1815010, EPI_ISL_1815012, EPI_ISL_1815013, EPI_ISL_1815014, EPI_ISL_1815019, EPI_ISL_1815020, EPI_ISL_1815023, EPI_ISL_1815027, EPI_ISL_1815029, EPI_ISL_1815032, EPI_ISL_1815034, EPI_ISL_1815038, EPI_ISL_1815042, EPI_ISL_1815043, EPI_ISL_1815044, EPI_ISL_1815047, EPI_ISL_1815049, EPI_ISL_1815052, EPI_ISL_1815053, EPI_ISL_1815058, EPI_ISL_1815064, EPI_ISL_1815068, EPI_ISL_1815069, EPI_ISL_1815073, EPI_ISL_1815077, EPI_ISL_1815078, EPI_ISL_1815079, EPI_ISL_1815080, EPI_ISL_1815081, EPI_ISL_1815090, EPI_ISL_1815093, EPI_ISL_1815095, EPI_ISL_1815097, EPI_ISL_1815105, EPI_ISL_1815107, EPI_ISL_1815111, EPI_ISL_1815113, EPI_ISL_1815119, EPI_ISL_1815122, EPI_ISL_1815123, EPI_ISL_1815125, EPI_ISL_1815127, EPI_ISL_1815128, EPI_ISL_1815130, EPI_ISL_1815131, EPI_ISL_1815141, EPI_ISL_1815145, EPI_ISL_1815152, EPI_ISL_1815153, EPI_ISL_1815163, EPI_ISL_1815167, EPI_ISL_1815168, EPI_ISL_1815173, EPI_ISL_1815177, EPI_ISL_1815181, EPI_ISL_1815182, EPI_ISL_1815184, EPI_ISL_1815185, EPI_ISL_1815186, EPI_ISL_1815187, EPI_ISL_1815188, EPI_ISL_1815190, EPI_ISL_1815193, EPI_ISL_1815196, EPI_ISL_1815199, EPI_ISL_1815204, EPI_ISL_1815205, EPI_ISL_1815208, EPI_ISL_1815210, EPI_ISL_1815211, EPI_ISL_1815217, EPI_ISL_1815222, EPI_ISL_1815232, EPI_ISL_1815233, EPI_ISL_1815236, EPI_ISL_1815238, EPI_ISL_1815239, EPI_ISL_1815244, EPI_ISL_1815245, EPI_ISL_1815246, EPI_ISL_1815247, EPI_ISL_1815249, EPI_ISL_1815262, EPI_ISL_1815265, EPI_ISL_1815269, EPI_ISL_1815275, EPI_ISL_1815277, EPI_ISL_1815279, EPI_ISL_1815280, EPI_ISL_1815282, EPI_ISL_1815289, EPI_ISL_1815292, EPI_ISL_1815294, EPI_ISL_1815297, EPI_ISL_1815299, EPI_ISL_1815300, EPI_ISL_1815302, EPI_ISL_1815308, EPI_ISL_1815311, EPI_ISL_1815312, EPI_ISL_1815313, EPI_ISL_1815314, EPI_ISL_1815316, EPI_ISL_1815317, EPI_ISL_1815319, EPI_ISL_1815323, EPI_ISL_1815325, EPI_ISL_1815334, EPI_ISL_1815338, EPI_ISL_1815345, EPI_ISL_1815349, EPI_ISL_1815352, EPI_ISL_1815353, EPI_ISL_1815355, EPI_ISL_1815358, EPI_ISL_1815364, EPI_ISL_1815365, EPI_ISL_1815370, EPI_ISL_1815372, EPI_ISL_1815377, EPI_ISL_1815381, EPI_ISL_1815397, EPI_ISL_1815401, EPI_ISL_1815403, EPI_ISL_1815410, EPI_ISL_1815412, EPI_ISL_1815414, EPI_ISL_1815416, EPI_ISL_1815419, EPI_ISL_1815421, EPI_ISL_1815423, EPI_ISL_1815424, EPI_ISL_1815426, EPI_ISL_1815433, EPI_ISL_1815434, EPI_ISL_1815436, EPI_ISL_1815437, EPI_ISL_1815438, EPI_ISL_1815440, EPI_ISL_1815442, EPI_ISL_1815447, EPI_ISL_1815448, EPI_ISL_1815459, EPI_ISL_1815461, EPI_ISL_1815476, EPI_ISL_1815477, EPI_ISL_1815479, EPI_ISL_1815480, EPI_ISL_1815481, EPI_ISL_1815482, EPI_ISL_1815483, EPI_ISL_1815484, EPI_ISL_1815487, EPI_ISL_1815488, EPI_ISL_1815492, EPI_ISL_1815494, EPI_ISL_1815498, EPI_ISL_1815499, EPI_ISL_1815500, EPI_ISL_1815503, EPI_ISL_1815508, EPI_ISL_1815511, EPI_ISL_1815516, EPI_ISL_1815522, EPI_ISL_1815524, EPI_ISL_1815527, EPI_ISL_1815528, EPI_ISL_1815531, EPI_ISL_1815532, EPI_ISL_1815549, EPI_ISL_1815552, EPI_ISL_1815555, EPI_ISL_1815556, EPI_ISL_1815557, EPI_ISL_1815560, EPI_ISL_1815561, EPI_ISL_1815570, EPI_ISL_1815571, EPI_ISL_1815581, EPI_ISL_1815583, EPI_ISL_1815589, EPI_ISL_1815595, EPI_ISL_1815596, EPI_ISL_1815597, EPI_ISL_1815608, EPI_ISL_1815610, EPI_ISL_1815611, EPI_ISL_1815627, EPI_ISL_1815629, EPI_ISL_1815633, EPI_ISL_1815637, EPI_ISL_1815646, EPI_ISL_1815649, EPI_ISL_1815651, EPI_ISL_1815655, EPI_ISL_1815659, EPI_ISL_1815661, EPI_ISL_1815666, EPI_ISL_1815675, EPI_ISL_1815678, EPI_ISL_1815679, EPI_ISL_1815680, EPI_ISL_1815684, EPI_ISL_1815685, EPI_ISL_1815695, EPI_ISL_1815697, EPI_ISL_1815699, EPI_ISL_1815704, EPI_ISL_1815711, EPI_ISL_1815716, EPI_ISL_1815723, EPI_ISL_1815730, EPI_ISL_1815731, EPI_ISL_1815732, EPI_ISL_1815742, EPI_ISL_1815746, EPI_ISL_1815752, EPI_ISL_1815766, EPI_ISL_1815772, EPI_ISL_1815773, EPI_ISL_1815777, EPI_ISL_1815779, EPI_ISL_1815787, EPI_ISL_1815791, EPI_ISL_1815795, EPI_ISL_1815796, EPI_ISL_1815799, EPI_ISL_1815803, EPI_ISL_1815804, EPI_ISL_1815814, EPI_ISL_1815816, EPI_ISL_1815819, EPI_ISL_1815820, EPI_ISL_1815824, EPI_ISL_1815829, EPI_ISL_1815835, EPI_ISL_1815836, EPI_ISL_1815837, EPI_ISL_1815839, EPI_ISL_1815840, EPI_ISL_1815842, EPI_ISL_1815843, EPI_ISL_1815853, EPI_ISL_1815858, EPI_ISL_1815859, EPI_ISL_1815860, EPI_ISL_1815862, EPI_ISL_1815866, EPI_ISL_1815867, EPI_ISL_1815883, EPI_ISL_1815898, EPI_ISL_1815899, EPI_ISL_1815903, EPI_ISL_1815909, EPI_ISL_1815913, EPI_ISL_1815916, EPI_ISL_1815918, EPI_ISL_1815920, EPI_ISL_1815925, EPI_ISL_1815928, EPI_ISL_1815929, EPI_ISL_1815934, EPI_ISL_1815935, EPI_ISL_1815939, EPI_ISL_1815943, EPI_ISL_1815944, EPI_ISL_1815945, EPI_ISL_1815947, EPI_ISL_1815949, EPI_ISL_1815950, EPI_ISL_1815952, EPI_ISL_1815956, EPI_ISL_1815958, EPI_ISL_1815966, EPI_ISL_1815968, EPI_ISL_1815974, EPI_ISL_1815979, EPI_ISL_1815983, EPI_ISL_1815984, EPI_ISL_1815986, EPI_ISL_1815989, EPI_ISL_1815990, EPI_ISL_1815992, EPI_ISL_1815995, EPI_ISL_1815997, EPI_ISL_1815999, EPI_ISL_1816004, EPI_ISL_1816005, EPI_ISL_1816007, EPI_ISL_1816008, EPI_ISL_1816015, EPI_ISL_1816018, EPI_ISL_1816021, EPI_ISL_1816024, EPI_ISL_1816026, EPI_ISL_1816034, EPI_ISL_1816035, EPI_ISL_1816036, EPI_ISL_1816039, EPI_ISL_1816041, EPI_ISL_1816045, EPI_ISL_1816049, EPI_ISL_1816050, EPI_ISL_1816052, EPI_ISL_1816059, EPI_ISL_1816067, EPI_ISL_1816072, EPI_ISL_1816073, EPI_ISL_1816077, EPI_ISL_1816078, EPI_ISL_1816080, EPI_ISL_1816083, EPI_ISL_1816088, EPI_ISL_1816108, EPI_ISL_1816118, EPI_ISL_1816120, EPI_ISL_1816125, EPI_ISL_1816128, EPI_ISL_1816129, EPI_ISL_1816132, EPI_ISL_1816134, EPI_ISL_1816135, EPI_ISL_1816137, EPI_ISL_1816138, EPI_ISL_1816140, EPI_ISL_1816142, EPI_ISL_1816147, EPI_ISL_1816153, EPI_ISL_1816156, EPI_ISL_1816161, EPI_ISL_1816165, EPI_ISL_1816168, EPI_ISL_1816171, EPI_ISL_1816172, EPI_ISL_1816175, EPI_ISL_1816177, EPI_ISL_1816178, EPI_ISL_1816179, EPI_ISL_1816181, EPI_ISL_1816185, EPI_ISL_1816195, EPI_ISL_1816208, EPI_ISL_1816216, EPI_ISL_1816218, EPI_ISL_1816227, EPI_ISL_1816233, EPI_ISL_1816237, EPI_ISL_1816238, EPI_ISL_1816242, EPI_ISL_1816243, EPI_ISL_1816244, EPI_ISL_1816248, EPI_ISL_1816249, EPI_ISL_1816252, EPI_ISL_1816254, EPI_ISL_1816255, EPI_ISL_1816262, EPI_ISL_1816272, EPI_ISL_1816276, EPI_ISL_1816281, EPI_ISL_1816283, EPI_ISL_1816287, EPI_ISL_1816291, EPI_ISL_1816294, EPI_ISL_1816296, EPI_ISL_1816298, EPI_ISL_1816301, EPI_ISL_1816303, EPI_ISL_1816304, EPI_ISL_1816305, EPI_ISL_1816313, EPI_ISL_1816315, EPI_ISL_1816317, EPI_ISL_1816321, EPI_ISL_1816323, EPI_ISL_1816334, EPI_ISL_1816336, EPI_ISL_1816337, EPI_ISL_1816341, EPI_ISL_1816343, EPI_ISL_1816344, EPI_ISL_1816345, EPI_ISL_1816350, EPI_ISL_1816352, EPI_ISL_1816353, EPI_ISL_1816358, EPI_ISL_1816359, EPI_ISL_1816360, EPI_ISL_1816362, EPI_ISL_1816364, EPI_ISL_1816365, EPI_ISL_1816367, EPI_ISL_1816368, EPI_ISL_1816372, EPI_ISL_1816374, EPI_ISL_1816376, EPI_ISL_1816378, EPI_ISL_1816382, EPI_ISL_1816386, EPI_ISL_1816387, EPI_ISL_1816389, EPI_ISL_1816390, EPI_ISL_1816393, EPI_ISL_1816395, EPI_ISL_1816396, EPI_ISL_1816397, EPI_ISL_1816406, EPI_ISL_1816407, EPI_ISL_1816410, EPI_ISL_1816417, EPI_ISL_1816422, EPI_ISL_1816424, EPI_ISL_1816427, EPI_ISL_1816431, EPI_ISL_1816434, EPI_ISL_1816439, EPI_ISL_1816441, EPI_ISL_1816442, EPI_ISL_1816452, EPI_ISL_1816453, EPI_ISL_1816455, EPI_ISL_1816456, EPI_ISL_1816457, EPI_ISL_1816460, EPI_ISL_1816462, EPI_ISL_1816466, EPI_ISL_1816468, EPI_ISL_1816476, EPI_ISL_1816477, EPI_ISL_1816479, EPI_ISL_1816485, EPI_ISL_1816486, EPI_ISL_1816490, EPI_ISL_1816491, EPI_ISL_1816495, EPI_ISL_1816513, EPI_ISL_1816514, EPI_ISL_1816538, EPI_ISL_1816543, EPI_ISL_1816546, EPI_ISL_1816549, EPI_ISL_1816554, EPI_ISL_1816555, EPI_ISL_1816561, EPI_ISL_1816564, EPI_ISL_1816565, EPI_ISL_1816566, EPI_ISL_1816570, EPI_ISL_1816573, EPI_ISL_1816579, EPI_ISL_1816580, EPI_ISL_1816581, EPI_ISL_1816586, EPI_ISL_1816589, EPI_ISL_1816590, EPI_ISL_1816591, EPI_ISL_1816593, EPI_ISL_1816595, EPI_ISL_1816600, EPI_ISL_1816601, EPI_ISL_1816606, EPI_ISL_1816607, EPI_ISL_1816617, EPI_ISL_1816623, EPI_ISL_1816625, EPI_ISL_1816626, EPI_ISL_1816630, EPI_ISL_1816631, EPI_ISL_1816638, EPI_ISL_1816645, EPI_ISL_1816646, EPI_ISL_1816647, EPI_ISL_1816651, EPI_ISL_1816653, EPI_ISL_1816654, EPI_ISL_1816656, EPI_ISL_1816663, EPI_ISL_1816664, EPI_ISL_1816667, EPI_ISL_1816672, EPI_ISL_1816673, EPI_ISL_1816676, EPI_ISL_1816677, EPI_ISL_1816682, EPI_ISL_1816683, EPI_ISL_1816684, EPI_ISL_1816685, EPI_ISL_1816686, EPI_ISL_1816689, EPI_ISL_1816691, EPI_ISL_1816692, EPI_ISL_1816696, EPI_ISL_1816697, EPI_ISL_1816698, EPI_ISL_1816701, EPI_ISL_1816702, EPI_ISL_1816703, EPI_ISL_1816706, EPI_ISL_1816709, EPI_ISL_1816713, EPI_ISL_1816715, EPI_ISL_1816720, EPI_ISL_1816721, EPI_ISL_1816724, EPI_ISL_1816726, EPI_ISL_1816728, EPI_ISL_1816729, EPI_ISL_1816731, EPI_ISL_1816732, EPI_ISL_1816735, EPI_ISL_1816738, EPI_ISL_1816739, EPI_ISL_1816740, EPI_ISL_1816743, EPI_ISL_1816745, EPI_ISL_1816746, EPI_ISL_1816749, EPI_ISL_1816751, EPI_ISL_1816757, EPI_ISL_1816758, EPI_ISL_1816762, EPI_ISL_1816765, EPI_ISL_1816768, EPI_ISL_1816769, EPI_ISL_1816777, EPI_ISL_1816778, EPI_ISL_1816779, EPI_ISL_1816781, EPI_ISL_1816782, EPI_ISL_1816789, EPI_ISL_1816790, EPI_ISL_1816791, EPI_ISL_1816801, EPI_ISL_1816803, EPI_ISL_1816804, EPI_ISL_1816810, EPI_ISL_1816812, EPI_ISL_1816814, EPI_ISL_1816816, EPI_ISL_1816820, EPI_ISL_1816822, EPI_ISL_1816823, EPI_ISL_1816826, EPI_ISL_1816837, EPI_ISL_1816844, EPI_ISL_1816847, EPI_ISL_1816851, EPI_ISL_1816854, EPI_ISL_1816857, EPI_ISL_1816862, EPI_ISL_1816866, EPI_ISL_1816869, EPI_ISL_1816870, EPI_ISL_1816877, EPI_ISL_1816883, EPI_ISL_1816890, EPI_ISL_1816906

| | | | |
|---|---|---|---|
| see above | EXCITE Lab | Andersen lab at Scripps Research | Abigail Schnapper; Alexandre Bolze; Alice Summerfield; Angela Scioscia; Araz Majnoonian; Ashkan Hassani; Brent Brainard+ SEARCH; Celena Andrade; Charlotte Rivera-Garcia; Cheryl Anderson; Chip Schooley; Christine Mitchell + SEARCH; David Becker; David Pride; Efren Sandoval; Elizabeth Cirulli; Fitri Carrissa Wijaya; Francisco Tanudjaja; Fred Wu; Geraint Levan; Greg Humphrey; Helena Tubb; J. Joelle Donofrio-Odmann; James Lu + SEARCH; Jason Nguyen; Jimmy Ramirez; Kelly Schiabor Barrett; Laura Kohn; Magnus Isaksson; Marc Laurent; Marlene Flores; Megan Nguyen; Natasha Martin; Natasha Martin Cheryl Anderson; Nicole L Washington; Patricia Gonzalez-Zuniga; Rebecca Fielding-Miller; Rodolfo Salido; Ryan Cho; Sawyer Farmer; Sharon Reed; Sherry Wang; Simon White; Smruthi Karthikeyan; Sydney Morgan + Marni Jacobs + Celestine Magallanes + Holly Valentine + Louise C. Laurent + SEARCH; Tommy Valles + SEARCH; Tyler Cassens; Victor Cantu; Vinton Omaleki; William Lee |

EPI_ISL_917943, EPI_ISL_1097739 — Flow Diagnostics Lab — Los Angeles County Public Health Laboratories

EPI_ISL_977135, EPI_ISL_977136, EPI_ISL_977137, EPI_ISL_977144, EPI_ISL_1253713, EPI_ISL_1253714, EPI_ISL_1253715, EPI_ISL_1253716, EPI_ISL_1253717, EPI_ISL_1253718

see above — Flow Health — Infectious Disease Program, Broad Institute of Harvard and MIT — Adams, G.; B.L.; B.W.; Bauer, M.; Birren; Carter, A.; Chaluvadi, S.; D.J.; DeRuff, K.; Gallagher, G.; Gladden-Young, A.; J.E.; K.J.; Lagerborg, K.; Lemieux; Loreth, C.; MacInnis; Normandin, E.; P.C.; Park; Reilly, S.; Rudy, M.; Siddle; Smole, S.; Tomkins-Tinch, C.; and Sabeti

EPI_ISL_984753, EPI_ISL_984754, EPI_ISL_984948, EPI_ISL_1009868, EPI_ISL_1009869, EPI_ISL_1009870, EPI_ISL_1009871, EPI_ISL_1009872, EPI_ISL_1009873, EPI_ISL_1009874, EPI_ISL_1009875, EPI_ISL_1009876, EPI_ISL_1009877, EPI_ISL_1009878, EPI_ISL_1009879, EPI_ISL_1009880, EPI_ISL_1009881, EPI_ISL_1009882, EPI_ISL_1009883, EPI_ISL_1009884, EPI_ISL_1009885, EPI_ISL_1009886,
EPI_ISL_1009887, EPI_ISL_1009888, EPI_ISL_1009889, EPI_ISL_1009890, EPI_ISL_1009891, EPI_ISL_1009892, EPI_ISL_1009893, EPI_ISL_1009894, EPI_ISL_1009895, EPI_ISL_1009896, EPI_ISL_1009897, EPI_ISL_1009898, EPI_ISL_1009899, EPI_ISL_1009900, EPI_ISL_1009901, EPI_ISL_1009902, EPI_ISL_1009903, EPI_ISL_1009904, EPI_ISL_1009905, EPI_ISL_1009906, EPI_ISL_1009907, EPI_ISL_1009908,
EPI_ISL_1009909, EPI_ISL_1009910, EPI_ISL_1009911, EPI_ISL_1009912, EPI_ISL_1009913, EPI_ISL_1009914, EPI_ISL_1009915, EPI_ISL_1009916, EPI_ISL_1009917, EPI_ISL_1009918, EPI_ISL_1009919, EPI_ISL_1009920, EPI_ISL_1009921, EPI_ISL_1009922, EPI_ISL_1009923, EPI_ISL_1009924, EPI_ISL_1009925, EPI_ISL_1009926, EPI_ISL_1009927, EPI_ISL_1009928, EPI_ISL_1009929, EPI_ISL_1009930,
EPI_ISL_1009931, EPI_ISL_1009932, EPI_ISL_1009933, EPI_ISL_1009934, EPI_ISL_1009935, EPI_ISL_1009936, EPI_ISL_1009937, EPI_ISL_1009938, EPI_ISL_1009939, EPI_ISL_1009940, EPI_ISL_1009941, EPI_ISL_1009942, EPI_ISL_1009943, EPI_ISL_1009944, EPI_ISL_1009945, EPI_ISL_1009946, EPI_ISL_1009947, EPI_ISL_1009948, EPI_ISL_1009949, EPI_ISL_1009950, EPI_ISL_1009951, EPI_ISL_1009952,
EPI_ISL_1009953, EPI_ISL_1009954, EPI_ISL_1009955, EPI_ISL_1009956, EPI_ISL_1009957, EPI_ISL_1009958, EPI_ISL_1009959, EPI_ISL_1009960, EPI_ISL_1009961, EPI_ISL_1009962, EPI_ISL_1009963, EPI_ISL_1009964, EPI_ISL_1009965, EPI_ISL_1009966, EPI_ISL_1009967, EPI_ISL_1009968, EPI_ISL_1009969, EPI_ISL_1009970, EPI_ISL_1009971, EPI_ISL_1009972, EPI_ISL_1009973, EPI_ISL_1009974,
EPI_ISL_1009975, EPI_ISL_1009976, EPI_ISL_1009977, EPI_ISL_1009978, EPI_ISL_1009979, EPI_ISL_1009980, EPI_ISL_1009981, EPI_ISL_1009982, EPI_ISL_1009983, EPI_ISL_1009984, EPI_ISL_1009985, EPI_ISL_1009986, EPI_ISL_1009987, EPI_ISL_1009988, EPI_ISL_1009989, EPI_ISL_1009990, EPI_ISL_1009991, EPI_ISL_1009992, EPI_ISL_1009993, EPI_ISL_1009994, EPI_ISL_1009995, EPI_ISL_1009996,
EPI_ISL_1009997, EPI_ISL_1009998, EPI_ISL_1009999, EPI_ISL_1010000, EPI_ISL_1010001, EPI_ISL_1010002, EPI_ISL_1010003, EPI_ISL_1010004, EPI_ISL_1010005, EPI_ISL_1010006, EPI_ISL_1010007, EPI_ISL_1010008, EPI_ISL_1010009, EPI_ISL_1010010, EPI_ISL_1010011, EPI_ISL_1010012, EPI_ISL_1010013, EPI_ISL_1010014, EPI_ISL_1010015, EPI_ISL_1010016, EPI_ISL_1010017, EPI_ISL_1010018,
EPI_ISL_1010019, EPI_ISL_1010020, EPI_ISL_1010021, EPI_ISL_1010022, EPI_ISL_1010023, EPI_ISL_1010024, EPI_ISL_1010025, EPI_ISL_1010026, EPI_ISL_1010027, EPI_ISL_1010028, EPI_ISL_1010029, EPI_ISL_1010030, EPI_ISL_1010031, EPI_ISL_1010032, EPI_ISL_1010033, EPI_ISL_1010034, EPI_ISL_1010035, EPI_ISL_1010036, EPI_ISL_1010037, EPI_ISL_1010038, EPI_ISL_1010039, EPI_ISL_1010040,
EPI_ISL_1010041, EPI_ISL_1010042, EPI_ISL_1010043, EPI_ISL_1010044, EPI_ISL_1010045, EPI_ISL_1010046, EPI_ISL_1010047, EPI_ISL_1010048, EPI_ISL_1010049, EPI_ISL_1010050, EPI_ISL_1010051, EPI_ISL_1010052, EPI_ISL_1010053, EPI_ISL_1010054, EPI_ISL_1010055, EPI_ISL_1010056, EPI_ISL_1010057, EPI_ISL_1010058, EPI_ISL_1010059, EPI_ISL_1010060, EPI_ISL_1010061, EPI_ISL_1010062,
EPI_ISL_1010063, EPI_ISL_1010064, EPI_ISL_1010065, EPI_ISL_1010066, EPI_ISL_1010067, EPI_ISL_1010068, EPI_ISL_1010069, EPI_ISL_1010070, EPI_ISL_1010071, EPI_ISL_1010072, EPI_ISL_1010073, EPI_ISL_1010074, EPI_ISL_1010075, EPI_ISL_1010076, EPI_ISL_1010077, EPI_ISL_1010078, EPI_ISL_1010079, EPI_ISL_1010080, EPI_ISL_1010081, EPI_ISL_1010082, EPI_ISL_1010083, EPI_ISL_1010084,
EPI_ISL_1010085, EPI_ISL_1010086, EPI_ISL_1010087, EPI_ISL_1010088, EPI_ISL_1010089, EPI_ISL_1010090, EPI_ISL_1010091, EPI_ISL_1010092, EPI_ISL_1010093, EPI_ISL_1010094, EPI_ISL_1010095, EPI_ISL_1010096, EPI_ISL_1010097, EPI_ISL_1010098, EPI_ISL_1010099, EPI_ISL_1010100, EPI_ISL_1010101, EPI_ISL_1010102, EPI_ISL_1010103, EPI_ISL_1010104, EPI_ISL_1010105, EPI_ISL_1010106,
EPI_ISL_1010107, EPI_ISL_1010108, EPI_ISL_1010109, EPI_ISL_1010110, EPI_ISL_1010111, EPI_ISL_1010112, EPI_ISL_1010113, EPI_ISL_1010114, EPI_ISL_1010115, EPI_ISL_1010116, EPI_ISL_1010117, EPI_ISL_1010118, EPI_ISL_1010119, EPI_ISL_1010120, EPI_ISL_1010121, EPI_ISL_1010122, EPI_ISL_1010123, EPI_ISL_1010124, EPI_ISL_1010125, EPI_ISL_1010126, EPI_ISL_1010127, EPI_ISL_1010128,
EPI_ISL_1010129, EPI_ISL_1010130, EPI_ISL_1010131, EPI_ISL_1010132, EPI_ISL_1010133, EPI_ISL_1010134, EPI_ISL_1010135, EPI_ISL_1010136, EPI_ISL_1010137, EPI_ISL_1010138, EPI_ISL_1010139, EPI_ISL_1010140, EPI_ISL_1010141, EPI_ISL_1010142, EPI_ISL_1010143, EPI_ISL_1010144, EPI_ISL_1010145, EPI_ISL_1010146, EPI_ISL_1010147, EPI_ISL_1010148, EPI_ISL_1010149, EPI_ISL_1010150,
EPI_ISL_1010151, EPI_ISL_1010152, EPI_ISL_1010153, EPI_ISL_1010154, EPI_ISL_1010155, EPI_ISL_1010156, EPI_ISL_1010157, EPI_ISL_1010158, EPI_ISL_1010159, EPI_ISL_1010160, EPI_ISL_1010161, EPI_ISL_1010162, EPI_ISL_1010163, EPI_ISL_1010164, EPI_ISL_1010165, EPI_ISL_1010166, EPI_ISL_1010167, EPI_ISL_1010168, EPI_ISL_1010169, EPI_ISL_1010170, EPI_ISL_1010171, EPI_ISL_1010172,
EPI_ISL_1010173, EPI_ISL_1010174, EPI_ISL_1010175, EPI_ISL_1010176, EPI_ISL_1010177, EPI_ISL_1010178, EPI_ISL_1010179, EPI_ISL_1010180, EPI_ISL_1010181, EPI_ISL_1010182, EPI_ISL_1010183, EPI_ISL_1010184, EPI_ISL_1010185, EPI_ISL_1010186, EPI_ISL_1010187, EPI_ISL_1010188, EPI_ISL_1010189, EPI_ISL_1010190, EPI_ISL_1010268, EPI_ISL_1010269, EPI_ISL_1010270, EPI_ISL_1010271,
EPI_ISL_1010272, EPI_ISL_1010273, EPI_ISL_1010274, EPI_ISL_1010275, EPI_ISL_1010276, EPI_ISL_1010277, EPI_ISL_1010278, EPI_ISL_1010279, EPI_ISL_1010280, EPI_ISL_1010281, EPI_ISL_1010282, EPI_ISL_1010283, EPI_ISL_1010284, EPI_ISL_1010285, EPI_ISL_1010286, EPI_ISL_1010287, EPI_ISL_1010288, EPI_ISL_1010289, EPI_ISL_1010290, EPI_ISL_1010291, EPI_ISL_1010292, EPI_ISL_1010293,
EPI_ISL_1010294, EPI_ISL_1010295, EPI_ISL_1010296, EPI_ISL_1010297, EPI_ISL_1010298, EPI_ISL_1010299, EPI_ISL_1010300, EPI_ISL_1010301, EPI_ISL_1010302, EPI_ISL_1010303, EPI_ISL_1010304, EPI_ISL_1010305, EPI_ISL_1010306, EPI_ISL_1010307, EPI_ISL_1010309, EPI_ISL_1010310, EPI_ISL_1010311, EPI_ISL_1010312, EPI_ISL_1010313, EPI_ISL_1010314, EPI_ISL_1010315, EPI_ISL_1010316,
EPI_ISL_1010317, EPI_ISL_1010318, EPI_ISL_1010319, EPI_ISL_1010320, EPI_ISL_1010321, EPI_ISL_1010322, EPI_ISL_1010323, EPI_ISL_1010324, EPI_ISL_1010325, EPI_ISL_1010326, EPI_ISL_1010327, EPI_ISL_1010328, EPI_ISL_1010329, EPI_ISL_1010330, EPI_ISL_1010331, EPI_ISL_1010332, EPI_ISL_1010333, EPI_ISL_1010334, EPI_ISL_1010335, EPI_ISL_1010336, EPI_ISL_1010337, EPI_ISL_1010338,
EPI_ISL_1010339, EPI_ISL_1010340, EPI_ISL_1010341, EPI_ISL_1010342, EPI_ISL_1010343, EPI_ISL_1010344, EPI_ISL_1010345, EPI_ISL_1010346, EPI_ISL_1010347, EPI_ISL_1010348, EPI_ISL_1010349, EPI_ISL_1010350, EPI_ISL_1010351, EPI_ISL_1010352, EPI_ISL_1010353, EPI_ISL_1010354, EPI_ISL_1010355, EPI_ISL_1010356, EPI_ISL_1010357, EPI_ISL_1010358, EPI_ISL_1010359, EPI_ISL_1010360,
EPI_ISL_1010361, EPI_ISL_1010362, EPI_ISL_1010363, EPI_ISL_1010364, EPI_ISL_1010365, EPI_ISL_1010366, EPI_ISL_1010367, EPI_ISL_1010368, EPI_ISL_1010369, EPI_ISL_1010370, EPI_ISL_1010371, EPI_ISL_1010372, EPI_ISL_1010373, EPI_ISL_1010374, EPI_ISL_1010375, EPI_ISL_1010376, EPI_ISL_1010377, EPI_ISL_1010378, EPI_ISL_1010379, EPI_ISL_1010380, EPI_ISL_1010381, EPI_ISL_1010382,
EPI_ISL_1010383, EPI_ISL_1010384, EPI_ISL_1010385, EPI_ISL_1010386, EPI_ISL_1010387, EPI_ISL_1010388, EPI_ISL_1010389, EPI_ISL_1010390, EPI_ISL_1010391, EPI_ISL_1010392, EPI_ISL_1010393, EPI_ISL_1010394, EPI_ISL_1010395, EPI_ISL_1010396, EPI_ISL_1010397, EPI_ISL_1010398, EPI_ISL_1010399, EPI_ISL_1010400, EPI_ISL_1010401, EPI_ISL_1010402, EPI_ISL_1010403, EPI_ISL_1010404,
EPI_ISL_1010405, EPI_ISL_1010406, EPI_ISL_1010407, EPI_ISL_1010408, EPI_ISL_1010409, EPI_ISL_1010410, EPI_ISL_1010411, EPI_ISL_1010412, EPI_ISL_1010413, EPI_ISL_1010414, EPI_ISL_1010415, EPI_ISL_1010416, EPI_ISL_1010417, EPI_ISL_1010418, EPI_ISL_1010419, EPI_ISL_1010420, EPI_ISL_1010421, EPI_ISL_1010422, EPI_ISL_1010423, EPI_ISL_1010424, EPI_ISL_1010425, EPI_ISL_1010426,
EPI_ISL_1010427, EPI_ISL_1010428, EPI_ISL_1010429, EPI_ISL_1010430, EPI_ISL_1010431, EPI_ISL_1010432, EPI_ISL_1010433, EPI_ISL_1010434, EPI_ISL_1010435, EPI_ISL_1010436, EPI_ISL_1010437, EPI_ISL_1010438, EPI_ISL_1010439, EPI_ISL_1010440, EPI_ISL_1010441, EPI_ISL_1010442, EPI_ISL_1010443, EPI_ISL_1010444, EPI_ISL_1010445, EPI_ISL_1010446, EPI_ISL_1010447, EPI_ISL_1010448,
EPI_ISL_1010449, EPI_ISL_1010450, EPI_ISL_1010451, EPI_ISL_1010452, EPI_ISL_1010453, EPI_ISL_1010454, EPI_ISL_1010455, EPI_ISL_1010456, EPI_ISL_1010457, EPI_ISL_1010458, EPI_ISL_1010459, EPI_ISL_1010460, EPI_ISL_1010461, EPI_ISL_1010462, EPI_ISL_1010463, EPI_ISL_1010464, EPI_ISL_1010465, EPI_ISL_1010466, EPI_ISL_1010467, EPI_ISL_1010468, EPI_ISL_1010469, EPI_ISL_1010470,
EPI_ISL_1010471, EPI_ISL_1010472, EPI_ISL_1010473, EPI_ISL_1010474, EPI_ISL_1010475, EPI_ISL_1010476, EPI_ISL_1010477, EPI_ISL_1010478, EPI_ISL_1010479, EPI_ISL_1010480, EPI_ISL_1010481, EPI_ISL_1010482, EPI_ISL_1010483, EPI_ISL_1010484, EPI_ISL_1010485, EPI_ISL_1010486, EPI_ISL_1010487, EPI_ISL_1010488, EPI_ISL_1010489, EPI_ISL_1010490, EPI_ISL_1010491, EPI_ISL_1010492,
EPI_ISL_1010493, EPI_ISL_1010494, EPI_ISL_1010495, EPI_ISL_1010496, EPI_ISL_1010497, EPI_ISL_1010498, EPI_ISL_1010499, EPI_ISL_1010500, EPI_ISL_1010501, EPI_ISL_1010502, EPI_ISL_1010503, EPI_ISL_1010504, EPI_ISL_1010505, EPI_ISL_1010506, EPI_ISL_1010507, EPI_ISL_1010508, EPI_ISL_1010509, EPI_ISL_1010510, EPI_ISL_1010511, EPI_ISL_1010512, EPI_ISL_1010513, EPI_ISL_1010514,
EPI_ISL_1010515, EPI_ISL_1010516, EPI_ISL_1010517, EPI_ISL_1010518, EPI_ISL_1010519, EPI_ISL_1010520, EPI_ISL_1010521, EPI_ISL_1010522, EPI_ISL_1010523, EPI_ISL_1010524, EPI_ISL_1010525, EPI_ISL_1010526, EPI_ISL_1010527, EPI_ISL_1010528, EPI_ISL_1010529, EPI_ISL_1010530, EPI_ISL_1010531, EPI_ISL_1010532, EPI_ISL_1010533, EPI_ISL_1010534, EPI_ISL_1010535, EPI_ISL_1010536,
EPI_ISL_1010537, EPI_ISL_1010538, EPI_ISL_1010539, EPI_ISL_1010540, EPI_ISL_1010541, EPI_ISL_1010542, EPI_ISL_1010543, EPI_ISL_1010544, EPI_ISL_1010545, EPI_ISL_1010546, EPI_ISL_1010547, EPI_ISL_1010548, EPI_ISL_1010549, EPI_ISL_1010550, EPI_ISL_1010551, EPI_ISL_1010552, EPI_ISL_1010553, EPI_ISL_1010554, EPI_ISL_1010555, EPI_ISL_1010556, EPI_ISL_1010557, EPI_ISL_1010558,
EPI_ISL_1010559, EPI_ISL_1010560, EPI_ISL_1010561, EPI_ISL_1010562, EPI_ISL_1010563, EPI_ISL_1010564, EPI_ISL_1010565, EPI_ISL_1010566, EPI_ISL_1010567, EPI_ISL_1010568, EPI_ISL_1010569, EPI_ISL_1010570, EPI_ISL_1010571, EPI_ISL_1010572, EPI_ISL_1010573, EPI_ISL_1010574, EPI_ISL_1010575, EPI_ISL_1010576, EPI_ISL_1010577, EPI_ISL_1010578, EPI_ISL_1010579, EPI_ISL_1010580,
EPI_ISL_1010581, EPI_ISL_1010582, EPI_ISL_1010583, EPI_ISL_1010584, EPI_ISL_1010585, EPI_ISL_1010586, EPI_ISL_1010587, EPI_ISL_1010588, EPI_ISL_1010589, EPI_ISL_1010590, EPI_ISL_1010591, EPI_ISL_1010592, EPI_ISL_1010593, EPI_ISL_1010594, EPI_ISL_1010595, EPI_ISL_1010596, EPI_ISL_1010597, EPI_ISL_1010598, EPI_ISL_1010599, EPI_ISL_1010600, EPI_ISL_1010601, EPI_ISL_1010602,
EPI_ISL_1010603, EPI_ISL_1010604, EPI_ISL_1010605, EPI_ISL_1010606, EPI_ISL_1010607, EPI_ISL_1010608, EPI_ISL_1010609, EPI_ISL_1010610, EPI_ISL_1010611, EPI_ISL_1010612, EPI_ISL_1010613, EPI_ISL_1010614, EPI_ISL_1010615, EPI_ISL_1010616, EPI_ISL_1010617, EPI_ISL_1010618, EPI_ISL_1010619, EPI_ISL_1010620, EPI_ISL_1010621, EPI_ISL_1010622, EPI_ISL_1010623, EPI_ISL_1010624,
EPI_ISL_1010625, EPI_ISL_1010626, EPI_ISL_1010627, EPI_ISL_1010628, EPI_ISL_1010629, EPI_ISL_1010630, EPI_ISL_1010631, EPI_ISL_1010632, EPI_ISL_1010633, EPI_ISL_1010634, EPI_ISL_1010635, EPI_ISL_1010636, EPI_ISL_1010637, EPI_ISL_1010638, EPI_ISL_1010639, EPI_ISL_1010640, EPI_ISL_1010641, EPI_ISL_1010642, EPI_ISL_1010643, EPI_ISL_1010644, EPI_ISL_1010645, EPI_ISL_1010646,
EPI_ISL_1010647, EPI_ISL_1010648, EPI_ISL_1010649, EPI_ISL_1010650, EPI_ISL_1010651, EPI_ISL_1010652, EPI_ISL_1010653, EPI_ISL_1010654, EPI_ISL_1010655, EPI_ISL_1010656, EPI_ISL_1010657, EPI_ISL_1010658, EPI_ISL_1010659, EPI_ISL_1010660, EPI_ISL_1010661, EPI_ISL_1010662, EPI_ISL_1010663, EPI_ISL_1010664, EPI_ISL_1010665, EPI_ISL_1010666, EPI_ISL_1010667, EPI_ISL_1010668,
EPI_ISL_1010669, EPI_ISL_1010670, EPI_ISL_1010671, EPI_ISL_1010672, EPI_ISL_1010673, EPI_ISL_1010674, EPI_ISL_1010675, EPI_ISL_1010676, EPI_ISL_1010677, EPI_ISL_1010678, EPI_ISL_1010679, EPI_ISL_1010680, EPI_ISL_1010681, EPI_ISL_1010682, EPI_ISL_1010683, EPI_ISL_1010684, EPI_ISL_1010685, EPI_ISL_1010686, EPI_ISL_1010687, EPI_ISL_1010688, EPI_ISL_1010689, EPI_ISL_1010690,
EPI_ISL_1010691, EPI_ISL_1010692, EPI_ISL_1010693, EPI_ISL_1010694, EPI_ISL_1010695, EPI_ISL_1010696, EPI_ISL_1010697, EPI_ISL_1010698, EPI_ISL_1010737, EPI_ISL_1010738, EPI_ISL_1010739, EPI_ISL_1010740, EPI_ISL_1010741, EPI_ISL_1010742, EPI_ISL_1010743, EPI_ISL_1010744, EPI_ISL_1010745, EPI_ISL_1010746, EPI_ISL_1010747, EPI_ISL_1010748, EPI_ISL_1010749, EPI_ISL_1010750,
EPI_ISL_1010751, EPI_ISL_1010752, EPI_ISL_1010753, EPI_ISL_1010754, EPI_ISL_1010755, EPI_ISL_1010756, EPI_ISL_1010757, EPI_ISL_1010758, EPI_ISL_1010759, EPI_ISL_1010760, EPI_ISL_1010761, EPI_ISL_1010762, EPI_ISL_1010763, EPI_ISL_1010764, EPI_ISL_1010765, EPI_ISL_1010766, EPI_ISL_1010767, EPI_ISL_1010768, EPI_ISL_1010769, EPI_ISL_1010770, EPI_ISL_1010771, EPI_ISL_1010772,
EPI_ISL_1010773, EPI_ISL_1010774, EPI_ISL_1010775, EPI_ISL_1010776, EPI_ISL_1010777, EPI_ISL_1010778, EPI_ISL_1010779, EPI_ISL_1010780, EPI_ISL_1010781, EPI_ISL_1010782, EPI_ISL_1010783, EPI_ISL_1010784, EPI_ISL_1010785, EPI_ISL_1010786, EPI_ISL_1010787, EPI_ISL_1010788, EPI_ISL_1010789, EPI_ISL_1010790, EPI_ISL_1010791, EPI_ISL_1010792, EPI_ISL_1010793, EPI_ISL_1010794,
EPI_ISL_1010795, EPI_ISL_1010796, EPI_ISL_1010797, EPI_ISL_1010798, EPI_ISL_1010799, EPI_ISL_1010800, EPI_ISL_1010801, EPI_ISL_1010802, EPI_ISL_1010803, EPI_ISL_1010804, EPI_ISL_1010805, EPI_ISL_1010806, EPI_ISL_1010807, EPI_ISL_1010808, EPI_ISL_1010809, EPI_ISL_1010810, EPI_ISL_1010811, EPI_ISL_1010812, EPI_ISL_1010813, EPI_ISL_1010814, EPI_ISL_1010815, EPI_ISL_1010816,
EPI_ISL_1010817, EPI_ISL_1010818, EPI_ISL_1010819, EPI_ISL_1010820, EPI_ISL_1010821, EPI_ISL_1010822, EPI_ISL_1010823, EPI_ISL_1010824, EPI_ISL_1010825, EPI_ISL_1010826, EPI_ISL_1010827, EPI_ISL_1010828, EPI_ISL_1010829, EPI_ISL_1010830, EPI_ISL_1010831, EPI_ISL_1010832, EPI_ISL_1010833, EPI_ISL_1010834, EPI_ISL_1010835, EPI_ISL_1010836, EPI_ISL_1010837, EPI_ISL_1010838,
EPI_ISL_1010839, EPI_ISL_1010840, EPI_ISL_1010841, EPI_ISL_1010842, EPI_ISL_1010843, EPI_ISL_1010844, EPI_ISL_1010845, EPI_ISL_1010846, EPI_ISL_1010847, EPI_ISL_1010848, EPI_ISL_1010849, EPI_ISL_1010850, EPI_ISL_1010851, EPI_ISL_1010852, EPI_ISL_1010853, EPI_ISL_1010854, EPI_ISL_1010855, EPI_ISL_1010856, EPI_ISL_1010857, EPI_ISL_1010858, EPI_ISL_1010859, EPI_ISL_1010860,
EPI_ISL_1010861, EPI_ISL_1010862, EPI_ISL_1010863, EPI_ISL_1010864, EPI_ISL_1010865, EPI_ISL_1010866, EPI_ISL_1010867, EPI_ISL_1010868, EPI_ISL_1010869, EPI_ISL_1010870, EPI_ISL_1010871, EPI_ISL_1010872, EPI_ISL_1010873, EPI_ISL_1010874, EPI_ISL_1010875, EPI_ISL_1010876, EPI_ISL_1010877, EPI_ISL_1010878, EPI_ISL_1010879, EPI_ISL_1010880, EPI_ISL_1010881, EPI_ISL_1010882,
EPI_ISL_1010883, EPI_ISL_1010884, EPI_ISL_1010885, EPI_ISL_1010886, EPI_ISL_1010887, EPI_ISL_1010888, EPI_ISL_1010889, EPI_ISL_1010890, EPI_ISL_1010891, EPI_ISL_1010892, EPI_ISL_1010893, EPI_ISL_1010894, EPI_ISL_1010895, EPI_ISL_1010896, EPI_ISL_1010897, EPI_ISL_1010898, EPI_ISL_1010899, EPI_ISL_1010900, EPI_ISL_1010901, EPI_ISL_1010902, EPI_ISL_1010903, EPI_ISL_1010904,
EPI_ISL_1010905, EPI_ISL_1010906, EPI_ISL_1010907, EPI_ISL_1010908, EPI_ISL_1010909, EPI_ISL_1010910, EPI_ISL_1010911, EPI_ISL_1010912, EPI_ISL_1010913, EPI_ISL_1010914, EPI_ISL_1010915, EPI_ISL_1010916, EPI_ISL_1010917, EPI_ISL_1010918, EPI_ISL_1010919, EPI_ISL_1010920, EPI_ISL_1010921, EPI_ISL_1010922, EPI_ISL_1010923, EPI_ISL_1010924, EPI_ISL_1010925, EPI_ISL_1010926,
EPI_ISL_1010927, EPI_ISL_1010928, EPI_ISL_1010929, EPI_ISL_1010930, EPI_ISL_1010931, EPI_ISL_1010932, EPI_ISL_1010933, EPI_ISL_1010934, EPI_ISL_1010935, EPI_ISL_1010936, EPI_ISL_1010937, EPI_ISL_1010938, EPI_ISL_1010939, EPI_ISL_1010940, EPI_ISL_1010941, EPI_ISL_1010942, EPI_ISL_1010943, EPI_ISL_1010944, EPI_ISL_1010945, EPI_ISL_1010946, EPI_ISL_1010947, EPI_ISL_1010948,
EPI_ISL_1010949, EPI_ISL_1010950, EPI_ISL_1010951, EPI_ISL_1010952, EPI_ISL_1010953, EPI_ISL_1010954, EPI_ISL_1010955, EPI_ISL_1010956, EPI_ISL_1010957, EPI_ISL_1010958, EPI_ISL_1010959, EPI_ISL_1010960, EPI_ISL_1010961, EPI_ISL_1010962, EPI_ISL_1010963, EPI_ISL_1010964, EPI_ISL_1010965, EPI_ISL_1010966, EPI_ISL_1010967, EPI_ISL_1010968, EPI_ISL_1010969, EPI_ISL_1010970,
EPI_ISL_1010971, EPI_ISL_1010972, EPI_ISL_1010973, EPI_ISL_1010974, EPI_ISL_1010975, EPI_ISL_1010976, EPI_ISL_1010977, EPI_ISL_1010978, EPI_ISL_1010979, EPI_ISL_1010980, EPI_ISL_1010981, EPI_ISL_1010982, EPI_ISL_1010983, EPI_ISL_1010984, EPI_ISL_1010985, EPI_ISL_1010986, EPI_ISL_1010987, EPI_ISL_1010988, EPI_ISL_1010989, EPI_ISL_1010990, EPI_ISL_1010991, EPI_ISL_1010992,
EPI_ISL_1010993, EPI_ISL_1010994, EPI_ISL_1010995, EPI_ISL_1010996, EPI_ISL_1010997, EPI_ISL_1010998, EPI_ISL_1010999, EPI_ISL_1011000, EPI_ISL_1011001, EPI_ISL_1011002, EPI_ISL_1011003, EPI_ISL_1011004, EPI_ISL_1011005, EPI_ISL_1011006, EPI_ISL_1011007, EPI_ISL_1011008, EPI_ISL_1011009, EPI_ISL_1011010, EPI_ISL_1011011, EPI_ISL_1011012, EPI_ISL_1011013, EPI_ISL_1011014,
EPI_ISL_1011015, EPI_ISL_1011016, EPI_ISL_1011017, EPI_ISL_1011018, EPI_ISL_1011019, EPI_ISL_1011020, EPI_ISL_1011021, EPI_ISL_1011022, EPI_ISL_1011023, EPI_ISL_1011024, EPI_ISL_1011025, EPI_ISL_1011026, EPI_ISL_1011027, EPI_ISL_1011028, EPI_ISL_1011029, EPI_ISL_1011030, EPI_ISL_1011031, EPI_ISL_1011032, EPI_ISL_1011033, EPI_ISL_1011034, EPI_ISL_1011035, EPI_ISL_1011036,
EPI_ISL_1011037, EPI_ISL_1011038, EPI_ISL_1011039, EPI_ISL_1011040, EPI_ISL_1011041, EPI_ISL_1011042, EPI_ISL_1011043, EPI_ISL_1011044, EPI_ISL_1011045, EPI_ISL_1011046, EPI_ISL_1011047, EPI_ISL_1011048, EPI_ISL_1011049, EPI_ISL_1011050, EPI_ISL_1011051, EPI_ISL_1011052, EPI_ISL_1011053, EPI_ISL_1011054, EPI_ISL_1011055, EPI_ISL_1011056, EPI_ISL_1011057, EPI_ISL_1011058,
EPI_ISL_1011059, EPI_ISL_1011060, EPI_ISL_1011061, EPI_ISL_1011062, EPI_ISL_1011063, EPI_ISL_1011064, EPI_ISL_1011065, EPI_ISL_1011066, EPI_ISL_1011067, EPI_ISL_1011068, EPI_ISL_1011069, EPI_ISL_1011070, EPI_ISL_1011071, EPI_ISL_1011072, EPI_ISL_1011073, EPI_ISL_1011074, EPI_ISL_1011075, EPI_ISL_1011076, EPI_ISL_1011077, EPI_ISL_1011078, EPI_ISL_1011079, EPI_ISL_1011080,
EPI_ISL_1011081, EPI_ISL_1011082, EPI_ISL_1011083, EPI_ISL_1011084, EPI_ISL_1011085, EPI_ISL_1011086, EPI_ISL_1011087, EPI_ISL_1011088, EPI_ISL_1011089, EPI_ISL_1011090, EPI_ISL_1011091, EPI_ISL_1011092, EPI_ISL_1011093, EPI_ISL_1011094, EPI_ISL_1011095, EPI_ISL_1011096, EPI_ISL_1011097, EPI_ISL_1011098, EPI_ISL_1011099, EPI_ISL_1011100, EPI_ISL_1011101, EPI_ISL_1011102,
EPI_ISL_1011103, EPI_ISL_1011104, EPI_ISL_1011105, EPI_ISL_1011106, EPI_ISL_1011107, EPI_ISL_1011108, EPI_ISL_1011109, EPI_ISL_1011110, EPI_ISL_1011111, EPI_ISL_1011112, EPI_ISL_1011113, EPI_ISL_1011114, EPI_ISL_1011115, EPI_ISL_1011116, EPI_ISL_1011117, EPI_ISL_1011118, EPI_ISL_1011119, EPI_ISL_1011120, EPI_ISL_1011121, EPI_ISL_1011122, EPI_ISL_1011123, EPI_ISL_1011124,
EPI_ISL_1011125, EPI_ISL_1011126, EPI_ISL_1011127, EPI_ISL_1011128, EPI_ISL_1011129, EPI_ISL_1011130, EPI_ISL_1011131, EPI_ISL_1011132, EPI_ISL_1011133, EPI_ISL_1011134, EPI_ISL_1011135, EPI_ISL_1011136, EPI_ISL_1011137, EPI_ISL_1011138, EPI_ISL_1011139, EPI_ISL_1011140, EPI_ISL_1011141, EPI_ISL_1011142, EPI_ISL_1011143, EPI_ISL_1011144, EPI_ISL_1011145, EPI_ISL_1011146,
EPI_ISL_1011147, EPI_ISL_1011148, EPI_ISL_1011149, EPI_ISL_1011150, EPI_ISL_1011151, EPI_ISL_1011152, EPI_ISL_1011153, EPI_ISL_1011154, EPI_ISL_1011155, EPI_ISL_1011156, EPI_ISL_1011157, EPI_ISL_1011158, EPI_ISL_1011159, EPI_ISL_1011160, EPI_ISL_1011161, EPI_ISL_1011162, EPI_ISL_1011163, EPI_ISL_1011164, EPI_ISL_1011165, EPI_ISL_1011166, EPI_ISL_1011167, EPI_ISL_1011168,
EPI_ISL_1011169, EPI_ISL_1011170, EPI_ISL_1011171, EPI_ISL_1011172, EPI_ISL_1011173, EPI_ISL_1011174, EPI_ISL_1011175, EPI_ISL_1011176, EPI_ISL_1011177, EPI_ISL_1011178, EPI_ISL_1011179, EPI_ISL_1011180, EPI_ISL_1011181, EPI_ISL_1011182, EPI_ISL_1011183, EPI_ISL_1011184, EPI_ISL_1011185, EPI_ISL_1011186, EPI_ISL_1011187, EPI_ISL_1011188, EPI_ISL_1011189, EPI_ISL_1011190,
EPI_ISL_1011191, EPI_ISL_1011192, EPI_ISL_1011193, EPI_ISL_1011194, EPI_ISL_1011195, EPI_ISL_1011196, EPI_ISL_1011197, EPI_ISL_1011198, EPI_ISL_1011199, EPI_ISL_1011200, EPI_ISL_1011201, EPI_ISL_1011202, EPI_ISL_1011203, EPI_ISL_1011204, EPI_ISL_1011205, EPI_ISL_1011206, EPI_ISL_1011207, EPI_ISL_1011208, EPI_ISL_1011209, EPI_ISL_1011210, EPI_ISL_1011211, EPI_ISL_1011212,
EPI_ISL_1011213, EPI_ISL_1011214, EPI_ISL_1011215, EPI_ISL_1011216, EPI_ISL_1011217, EPI_ISL_1011218, EPI_ISL_1011219, EPI_ISL_1011220, EPI_ISL_1011221, EPI_ISL_1011222, EPI_ISL_1011223, EPI_ISL_1011224, EPI_ISL_1011225, EPI_ISL_1011226, EPI_ISL_1011227, EPI_ISL_1011228, EPI_ISL_1011229, EPI_ISL_1011230, EPI_ISL_1011231, EPI_ISL_1011232, EPI_ISL_1011233, EPI_ISL_1011234,
EPI_ISL_1011235, EPI_ISL_1011236, EPI_ISL_1011237, EPI_ISL_1011238, EPI_ISL_1011239, EPI_ISL_1011240, EPI_ISL_1011241, EPI_ISL_1011242, EPI_ISL_1011243, EPI_ISL_1011244, EPI_ISL_1011245, EPI_ISL_1011246, EPI_ISL_1011247, EPI_ISL_1011248, EPI_ISL_1011249, EPI_ISL_1011250, EPI_ISL_1011251, EPI_ISL_1011252, EPI_ISL_1011253, EPI_ISL_1011254, EPI_ISL_1011255, EPI_ISL_1011256,
EPI_ISL_1011257, EPI_ISL_1011258, EPI_ISL_1011259, EPI_ISL_1011260, EPI_ISL_1011261, EPI_ISL_1011262, EPI_ISL_1011263, EPI_ISL_1011264, EPI_ISL_1011265, EPI_ISL_1011266, EPI_ISL_1011267, EPI_ISL_1011268, EPI_ISL_1011269, EPI_ISL_1011270, EPI_ISL_1011271, EPI_ISL_1011272, EPI_ISL_1011273, EPI_ISL_1011274, EPI_ISL_1011275, EPI_ISL_1011276, EPI_ISL_1011277, EPI_ISL_1011278,
EPI_ISL_1011279, EPI_ISL_1011280, EPI_ISL_1011281, EPI_ISL_1011282, EPI_ISL_1011283, EPI_ISL_1011284, EPI_ISL_1011285, EPI_ISL_1011286, EPI_ISL_1011287, EPI_ISL_1011288, EPI_ISL_1011289, EPI_ISL_1011290, EPI_ISL_1011291, EPI_ISL_1011292, EPI_ISL_1011293, EPI_ISL_1011294, EPI_ISL_1011295, EPI_ISL_1011296, EPI_ISL_1011297, EPI_ISL_1011298, EPI_ISL_1011299, EPI_ISL_1011300,
EPI_ISL_1011301, EPI_ISL_1011302, EPI_ISL_1011303, EPI_ISL_1011304, EPI_ISL_1011305, EPI_ISL_1011306, EPI_ISL_1011307, EPI_ISL_1011308, EPI_ISL_1011309, EPI_ISL_1011310, EPI_ISL_1011311, EPI_ISL_1011312, EPI_ISL_1011313, EPI_ISL_1011314, EPI_ISL_1011315, EPI_ISL_1011316, EPI_ISL_1011317, EPI_ISL_1011318, EPI_ISL_1011319, EPI_ISL_1011320, EPI_ISL_1011321, EPI_ISL_1011322,
EPI_ISL_1011323, EPI_ISL_1011324, EPI_ISL_1011325, EPI_ISL_1011326, EPI_ISL_1011327, EPI_ISL_1011328, EPI_ISL_1011329, EPI_ISL_1011330, EPI_ISL_1011331, EPI_ISL_1011332, EPI_ISL_1011333, EPI_ISL_1011334, EPI_ISL_1011335, EPI_ISL_1011336, EPI_ISL_1011337, EPI_ISL_1011338, EPI_ISL_1011339, EPI_ISL_1011340, EPI_ISL_1011341, EPI_ISL_1011342, EPI_ISL_1011343, EPI_ISL_1011344,
EPI_ISL_1011345, EPI_ISL_1011346, EPI_ISL_1011347, EPI_ISL_1011348, EPI_ISL_1011349, EPI_ISL_1011350, EPI_ISL_1011351, EPI_ISL_1011352, EPI_ISL_1011353, EPI_ISL_1011354, EPI_ISL_1011355, EPI_ISL_1011356, EPI_ISL_1011357, EPI_ISL_1011358, EPI_ISL_1011359, EPI_ISL_1011360, EPI_ISL_1011361, EPI_ISL_1011362, EPI_ISL_1011363, EPI_ISL_1011364, EPI_ISL_1011365, EPI_ISL_1011366,
EPI_ISL_1011367, EPI_ISL_1011368, EPI_ISL_1011369, EPI_ISL_1011370, EPI_ISL_1011371, EPI_ISL_1011372, EPI_ISL_1011373, EPI_ISL_1011374, EPI_ISL_1011375, EPI_ISL_1011376, EPI_ISL_1011377, EPI_ISL_1011378, EPI_ISL_1011379, EPI_ISL_1011380, EPI_ISL_1011381, EPI_ISL_1011382, EPI_ISL_1011383, EPI_ISL_1011384, EPI_ISL_1011385, EPI_ISL_1011386, EPI_ISL_1011387, EPI_ISL_1011388,
EPI_ISL_1011389, EPI_ISL_1011390, EPI_ISL_1011391, EPI_ISL_1011392, EPI_ISL_1011393, EPI_ISL_1011394, EPI_ISL_1011395, EPI_ISL_1011396, EPI_ISL_1011397, EPI_ISL_1011398, EPI_ISL_1011399, EPI_ISL_1011400, EPI_ISL_1011401, EPI_ISL_1011402, EPI_ISL_1011403, EPI_ISL_1011404, EPI_ISL_1011405, EPI_ISL_1011406, EPI_ISL_1011407, EPI_ISL_1011408, EPI_ISL_1011409, EPI_ISL_1011410,
EPI_ISL_1011411, EPI_ISL_1011412, EPI_ISL_1011413, EPI_ISL_1011414, EPI_ISL_1011415, EPI_ISL_1011416, EPI_ISL_1011417, EPI_ISL_1011418, EPI_ISL_1011419, EPI_ISL_1011420, EPI_ISL_1011421, EPI_ISL_1011422, EPI_ISL_1011423, EPI_ISL_1011424, EPI_ISL_1011425, EPI_ISL_1011426, EPI_ISL_1011427, EPI_ISL_1011428, EPI_ISL_1011429, EPI_ISL_1011430, EPI_ISL_1011431, EPI_ISL_1011432,
EPI_ISL_1011433, EPI_ISL_1011434, EPI_ISL_1011435, EPI_ISL_1011436, EPI_ISL_1011437, EPI_ISL_1011438, EPI_ISL_1011439, EPI_ISL_1011440, EPI_ISL_1011441, EPI_ISL_1011442, EPI_ISL_1011443, EPI_ISL_1011444, EPI_ISL_1011445, EPI_ISL_1011446, EPI_ISL_1011447, EPI_ISL_1011448, EPI_ISL_1011449, EPI_ISL_1011450, EPI_ISL_1011451, EPI_ISL_1011452, EPI_ISL_1011453, EPI_ISL_1011454,
EPI_ISL_1011455, EPI_ISL_1011456, EPI_ISL_1011457, EPI_ISL_1011458, EPI_ISL_1011459, EPI_ISL_1011460, EPI_ISL_1011461, EPI_ISL_1011462, EPI_ISL_1011463, EPI_ISL_1011464, EPI_ISL_1011465, EPI_ISL_1011466, EPI_ISL_1011467, EPI_ISL_1011468, EPI_ISL_1011469, EPI_ISL_1011470, EPI_ISL_1011471, EPI_ISL_1011472, EPI_ISL_1011473, EPI_ISL_1011474, EPI_ISL_1011475, EPI_ISL_1011476,
EPI_ISL_1011477, EPI_ISL_1011478, EPI_ISL_1011479, EPI_ISL_1011480, EPI_ISL_1011481, EPI_ISL_1011482, EPI_ISL_1011483, EPI_ISL_1011484, EPI_ISL_1011485, EPI_ISL_1011486, EPI_ISL_1011487, EPI_ISL_1011488, EPI_ISL_1011489, EPI_ISL_1011490, EPI_ISL_1011491, EPI_ISL_1011492, EPI_ISL_1011493, EPI_ISL_1011494, EPI_ISL_1011495, EPI_ISL_1011496, EPI_ISL_1011497, EPI_ISL_1011498,
EPI_ISL_1011499, EPI_ISL_1011500, EPI_ISL_1011501, EPI_ISL_1011502, EPI_ISL_1011503, EPI_ISL_1011504, EPI_ISL_1011505, EPI_ISL_1011506, EPI_ISL_1011507, EPI_ISL_1011508, EPI_ISL_1026945, EPI_ISL_1026946, EPI_ISL_1026947, EPI_ISL_1026948, EPI_ISL_1026949, EPI_ISL_1026950, EPI_ISL_1026951, EPI_ISL_1027046, EPI_ISL_1027074, EPI_ISL_1027080, EPI_ISL_1027086, EPI_ISL_1027099,
EPI_ISL_1027100, EPI_ISL_1027103, EPI_ISL_1027107, EPI_ISL_1027124, EPI_ISL_1027147, EPI_ISL_1027177, EPI_ISL_1032947, EPI_ISL_1032948, EPI_ISL_1032949, EPI_ISL_1032950, EPI_ISL_1032952, EPI_ISL_1032953, EPI_ISL_1032954, EPI_ISL_1032955, EPI_ISL_1032956, EPI_ISL_1032957, EPI_ISL_1032958, EPI_ISL_1032959, EPI_ISL_1032960, EPI_ISL_1032961, EPI_ISL_1032962, EPI_ISL_1032963,
EPI_ISL_1032964, EPI_ISL_1032965, EPI_ISL_1032966, EPI_ISL_1032967, EPI_ISL_1032968, EPI_ISL_1032969, EPI_ISL_1032970, EPI_ISL_1032971, EPI_ISL_1032972, EPI_ISL_1032973, EPI_ISL_1032974, EPI_ISL_1032975, EPI_ISL_1032976, EPI_ISL_1032977, EPI_ISL_1032978, EPI_ISL_1032979, EPI_ISL_1032980, EPI_ISL_1032981, EPI_ISL_1032982, EPI_ISL_1032983, EPI_ISL_1032984, EPI_ISL_1032985,
EPI_ISL_1032986, EPI_ISL_1032987, EPI_ISL_1032988, EPI_ISL_1032989, EPI_ISL_1032990, EPI_ISL_1032991, EPI_ISL_1032992, EPI_ISL_1032993, EPI_ISL_1032994, EPI_ISL_1032995, EPI_ISL_1032996, EPI_ISL_1032997, EPI_ISL_1032998, EPI_ISL_1032999, EPI_ISL_1033000, EPI_ISL_1033001, EPI_ISL_1033002, EPI_ISL_1033003, EPI_ISL_1033004, EPI_ISL_1033005, EPI_ISL_1033006, EPI_ISL_1033007,
EPI_ISL_1033008, EPI_ISL_1033009, EPI_ISL_1033010, EPI_ISL_1033011, EPI_ISL_1033012, EPI_ISL_1033013, EPI_ISL_1033014, EPI_ISL_1033015, EPI_ISL_1033016, EPI_ISL_1033017, EPI_ISL_1033018, EPI_ISL_1033019, EPI_ISL_1033020, EPI_ISL_1033021, EPI_ISL_1033022, EPI_ISL_1033023, EPI_ISL_1068402, EPI_ISL_1068406, EPI_ISL_1068413, EPI_ISL_1068438, EPI_ISL_1068447, EPI_ISL_1068691,
EPI_ISL_1068694, EPI_ISL_1068706, EPI_ISL_1068707, EPI_ISL_1068710, EPI_ISL_1068723, EPI_ISL_1068724, EPI_ISL_1068725, EPI_ISL_1068726, EPI_ISL_1069085, EPI_ISL_1069125, EPI_ISL_1069126, EPI_ISL_1069127, EPI_ISL_1069128, EPI_ISL_1069129, EPI_ISL_1069130, EPI_ISL_1069131, EPI_ISL_1069132, EPI_ISL_1069133, EPI_ISL_1069134, EPI_ISL_1069135, EPI_ISL_1069136, EPI_ISL_1069137,
EPI_ISL_1069138, EPI_ISL_1069139, EPI_ISL_1069140, EPI_ISL_1069141, EPI_ISL_1069142, EPI_ISL_1069143, EPI_ISL_1069144, EPI_ISL_1069145, EPI_ISL_1069146, EPI_ISL_1069147, EPI_ISL_1069148, EPI_ISL_1069149, EPI_ISL_1069150, EPI_ISL_1069151, EPI_ISL_1069152, EPI_ISL_1095645, EPI_ISL_1095646, EPI_ISL_1095647, EPI_ISL_1095649, EPI_ISL_1095650, EPI_ISL_1095651, EPI_ISL_1095654,
EPI_ISL_1095655, EPI_ISL_1095656, EPI_ISL_1095657, EPI_ISL_1095658, EPI_ISL_1095659, EPI_ISL_1095660, EPI_ISL_1095661, EPI_ISL_1095662, EPI_ISL_1095664, EPI_ISL_1095665, EPI_ISL_1095666, EPI_ISL_1095669, EPI_ISL_1095670, EPI_ISL_1095671, EPI_ISL_1095672, EPI_ISL_1095673, EPI_ISL_1095674, EPI_ISL_1095675, EPI_ISL_1095676, EPI_ISL_1095678, EPI_ISL_1095679, EPI_ISL_1095681,
EPI_ISL_1095682, EPI_ISL_1095683, EPI_ISL_1095685, EPI_ISL_1095687, EPI_ISL_1095688, EPI_ISL_1095689, EPI_ISL_1095691, EPI_ISL_1095694, EPI_ISL_1095695, EPI_ISL_1095698, EPI_ISL_1095699, EPI_ISL_1095700, EPI_ISL_1095701, EPI_ISL_1095702, EPI_ISL_1095703, EPI_ISL_1095705, EPI_ISL_1095706, EPI_ISL_1095707, EPI_ISL_1095708, EPI_ISL_1132915, EPI_ISL_1132916, EPI_ISL_1132917,
EPI_ISL_1132918, EPI_ISL_1132919, EPI_ISL_1132920, EPI_ISL_1132921, EPI_ISL_1132922, EPI_ISL_1132923, EPI_ISL_1132924, EPI_ISL_1132925, EPI_ISL_1132926, EPI_ISL_1132927, EPI_ISL_1132928, EPI_ISL_1132929, EPI_ISL_1132930, EPI_ISL_1132931, EPI_ISL_1132932, EPI_ISL_1132933, EPI_ISL_1132934, EPI_ISL_1132935, EPI_ISL_1132936, EPI_ISL_1132937, EPI_ISL_1132938, EPI_ISL_1132939,
EPI_ISL_1132940, EPI_ISL_1132941, EPI_ISL_1132942, EPI_ISL_1132943, EPI_ISL_1132944, EPI_ISL_1132945, EPI_ISL_1132946, EPI_ISL_1132947, EPI_ISL_1132948, EPI_ISL_1132949, EPI_ISL_1132950, EPI_ISL_1132951, EPI_ISL_1132952, EPI_ISL_1132953, EPI_ISL_1132954, EPI_ISL_1132955, EPI_ISL_1132956, EPI_ISL_1132957, EPI_ISL_1132958, EPI_ISL_1132959, EPI_ISL_1132960, EPI_ISL_1132961,
EPI_ISL_1132962, EPI_ISL_1132963, EPI_ISL_1132964, EPI_ISL_1132965, EPI_ISL_1132966, EPI_ISL_1132967, EPI_ISL_1132968, EPI_ISL_1132969, EPI_ISL_1132970, EPI_ISL_1134840, EPI_ISL_1134841, EPI_ISL_1134842, EPI_ISL_1134843, EPI_ISL_1134844, EPI_ISL_1134845, EPI_ISL_1134846, EPI_ISL_1134847, EPI_ISL_1134848, EPI_ISL_1134849, EPI_ISL_1134850, EPI_ISL_1134851, EPI_ISL_1134852,
EPI_ISL_1134853, EPI_ISL_1134854, EPI_ISL_1134855, EPI_ISL_1134856, EPI_ISL_1134857, EPI_ISL_1134858, EPI_ISL_1134859, EPI_ISL_1134860, EPI_ISL_1134861, EPI_ISL_1134862, EPI_ISL_1134863, EPI_ISL_1134864, EPI_ISL_1134865, EPI_ISL_1134866, EPI_ISL_1134867, EPI_ISL_1134868, EPI_ISL_1134869, EPI_ISL_1134870, EPI_ISL_1134871, EPI_ISL_1134872, EPI_ISL_1134873, EPI_ISL_1134874,
EPI_ISL_1134875, EPI_ISL_1134876, EPI_ISL_1134877, EPI_ISL_1134878, EPI_ISL_1134879, EPI_ISL_1134880, EPI_ISL_1134881, EPI_ISL_1134882, EPI_ISL_1134883, EPI_ISL_1134884, EPI_ISL_1134885, EPI_ISL_1134886, EPI_ISL_1134887, EPI_ISL_1134888, EPI_ISL_1134889, EPI_ISL_1134890, EPI_ISL_1134891, EPI_ISL_1134892, EPI_ISL_1134893, EPI_ISL_1134894, EPI_ISL_1134895, EPI_ISL_1134896,
EPI_ISL_1134897, EPI_ISL_1134898, EPI_ISL_1134899, EPI_ISL_1134900, EPI_ISL_1134901, EPI_ISL_1134902, EPI_ISL_1134903, EPI_ISL_1172047, EPI_ISL_1172048, EPI_ISL_1172049, EPI_ISL_1172050, EPI_ISL_1172051, EPI_ISL_1172052, EPI_ISL_1172053, EPI_ISL_1172054, EPI_ISL_1172055, EPI_ISL_1172056, EPI_ISL_1172057, EPI_ISL_1172058, EPI_ISL_1172060, EPI_ISL_1172061, EPI_ISL_1172062,
EPI_ISL_1172063, EPI_ISL_1172064, EPI_ISL_1172065, EPI_ISL_1172066, EPI_ISL_1172068, EPI_ISL_1172069, EPI_ISL_1172070, EPI_ISL_1172071, EPI_ISL_1172078, EPI_ISL_1172079, EPI_ISL_1172088, EPI_ISL_1172089, EPI_ISL_1172090, EPI_ISL_1172097, EPI_ISL_1172099, EPI_ISL_1172100, EPI_ISL_1195948, EPI_ISL_1195949, EPI_ISL_1251260

see above — Fulgent Genetics — Fulgent Genetics — Becky Tsai; Benafsh Sapra; Doreen Ng; Harry Gao; James Xie; John Gao; Joseph Fierro; Mickey Li; Yan Meng

EPI_ISL_1508978, EPI_ISL_1532118, EPI_ISL_1532120, EPI_ISL_1532121, EPI_ISL_1532122 — Good Samaritan Hospital — Santa Clara County Public Health Laboratory — Santa Clara County Public Health Department

EPI_ISL_1088591, EPI_ISL_1088654, EPI_ISL_1088663, EPI_ISL_1088727, EPI_ISL_1088774, EPI_ISL_1088777, EPI_ISL_1088822, EPI_ISL_1088841, EPI_ISL_1088867, EPI_ISL_1088891, EPI_ISL_1088907, EPI_ISL_1088933, EPI_ISL_1088956, EPI_ISL_1088965, EPI_ISL_1088974, EPI_ISL_1088979, EPI_ISL_1089007, EPI_ISL_1089266, EPI_ISL_1089267, EPI_ISL_1089269, EPI_ISL_1089272, EPI_ISL_1089273,
EPI_ISL_1089274, EPI_ISL_1089275, EPI_ISL_1089279, EPI_ISL_1089280, EPI_ISL_1089284, EPI_ISL_1089285, EPI_ISL_1089288, EPI_ISL_1089289, EPI_ISL_1089290, EPI_ISL_1089291, EPI_ISL_1089293, EPI_ISL_1089297, EPI_ISL_1089298, EPI_ISL_1089299, EPI_ISL_1089302, EPI_ISL_1089304, EPI_ISL_1089305, EPI_ISL_1089308, EPI_ISL_1089310, EPI_ISL_1089311, EPI_ISL_1089312, EPI_ISL_1089314,
EPI_ISL_1089316, EPI_ISL_1089318, EPI_ISL_1089319, EPI_ISL_1089320, EPI_ISL_1089321

see above — Helix / Illumina — Respiratory Viruses Branch, Division of Viral Diseases, Centers for Disease Control and Prevention — Alexandre Bolze; Ary Ascencio; Ben L. Rambo-Martin; Brad Sickler; Charlotte Rivera-Garcia; Christine Tran; Clinton R. Paden; Dakota Howard; David Becker; Dhwani Batra; Duncan MacCannell; Efren Sandoval; Eileen de Feo; Elizabeth Cirulli; Eric Allen; Geraint Levan; James Lu; Jan Antico; Jason Nguyen; Jimmy Ramirez; Jingtao Liu; Kelly Schiabor Barrett; Kim Gietzen; Magnus Isaksson; Marc Laurent; Matthew Tolentino; Nicole L. Washington; Peter W. Cook; Phil Febbo; Ryan Cho; Shannon Wickline; Sherry Wang; Simon White; Summer Galloway; Suxiang Tong; Tyler Cassens; William Lee

EPI_ISL_966556, EPI_ISL_966561, EPI_ISL_966562, EPI_ISL_966563, EPI_ISL_966567, EPI_ISL_966568, EPI_ISL_966570, EPI_ISL_966571, EPI_ISL_966603, EPI_ISL_966608, EPI_ISL_966615, EPI_ISL_966622, EPI_ISL_966624, EPI_ISL_966625, EPI_ISL_966626, EPI_ISL_966627, EPI_ISL_966628, EPI_ISL_966630, EPI_ISL_966632, EPI_ISL_966633, EPI_ISL_966636, EPI_ISL_966637, EPI_ISL_966638, EPI_ISL_966640,
EPI_ISL_966657, EPI_ISL_966665, EPI_ISL_966670, EPI_ISL_966672, EPI_ISL_966680, EPI_ISL_966683, EPI_ISL_966686, EPI_ISL_966694, EPI_ISL_966702, EPI_ISL_966720, EPI_ISL_966725, EPI_ISL_966729, EPI_ISL_966852, EPI_ISL_966853, EPI_ISL_966855, EPI_ISL_966857, EPI_ISL_966858, EPI_ISL_966860, EPI_ISL_966861, EPI_ISL_966862, EPI_ISL_966910, EPI_ISL_966912, EPI_ISL_966921, EPI_ISL_966922,
EPI_ISL_966923, EPI_ISL_966924, EPI_ISL_966925, EPI_ISL_966926, EPI_ISL_966927, EPI_ISL_966928, EPI_ISL_966942, EPI_ISL_966974, EPI_ISL_966984, EPI_ISL_966985, EPI_ISL_966986, EPI_ISL_966991, EPI_ISL_966992, EPI_ISL_966993, EPI_ISL_966995, EPI_ISL_967000, EPI_ISL_967046, EPI_ISL_967047, EPI_ISL_967048, EPI_ISL_967057, EPI_ISL_967058, EPI_ISL_967060, EPI_ISL_967061, EPI_ISL_967062,
EPI_ISL_967065, EPI_ISL_967072, EPI_ISL_967073, EPI_ISL_967081, EPI_ISL_967099, EPI_ISL_967104, EPI_ISL_967107, EPI_ISL_967113, EPI_ISL_967116, EPI_ISL_967117, EPI_ISL_967431, EPI_ISL_967432, EPI_ISL_967433, EPI_ISL_967437, EPI_ISL_967439, EPI_ISL_967451, EPI_ISL_967454, EPI_ISL_967456, EPI_ISL_967460, EPI_ISL_967464, EPI_ISL_967465, EPI_ISL_967487, EPI_ISL_967488, EPI_ISL_967489,
EPI_ISL_967490, EPI_ISL_967492, EPI_ISL_967493, EPI_ISL_967494, EPI_ISL_967495, EPI_ISL_978557, EPI_ISL_978561, EPI_ISL_978562, EPI_ISL_978564, EPI_ISL_978565, EPI_ISL_978568, EPI_ISL_978569, EPI_ISL_978581, EPI_ISL_978585, EPI_ISL_978586, EPI_ISL_978587, EPI_ISL_978589, EPI_ISL_978597, EPI_ISL_978609, EPI_ISL_978618, EPI_ISL_1016510, EPI_ISL_1016513, EPI_ISL_1016518,

| | | | |
|---|---|---|---|
| EPI_ISL_1016528, EPI_ISL_1016532, EPI_ISL_1016538, EPI_ISL_1016545, EPI_ISL_1016549, EPI_ISL_1016554, EPI_ISL_1016555, EPI_ISL_1016578, EPI_ISL_1016592, EPI_ISL_1016599, EPI_ISL_1016603, EPI_ISL_1016604, EPI_ISL_1016606, EPI_ISL_1016608, EPI_ISL_1016609, EPI_ISL_1016610, EPI_ISL_1016611, EPI_ISL_1016612, EPI_ISL_1016618, EPI_ISL_1016621, EPI_ISL_1016622, EPI_ISL_1016623, EPI_ISL_1016647, EPI_ISL_1016652, EPI_ISL_1016654, EPI_ISL_1016657, EPI_ISL_1016658, EPI_ISL_1016746, EPI_ISL_1016786, EPI_ISL_1016804, EPI_ISL_1016833, EPI_ISL_1016834, EPI_ISL_1081049, EPI_ISL_1081059, EPI_ISL_1081063, EPI_ISL_1081083 | | | |
| see above | Helix/Illumina | Respiratory Viruses Branch, Division of Viral Diseases, Centers for Disease Control and Prevention | ; Alexandre Bolze; Ary Ascencio; Ben L. Rambo-Martin; Brad Sickler; Charlotte Rivera-Garcia; Christine Tran; Clinton R. Paden; Dakota Howard; David Becker; Dhwani Batra; Duncan MacCannell; Efren Sandoval; Eileen de Feo; Elizabeth Cirulli; Eric Allen; Geraint Levan; James Lu; Jan Antico; Jason Nguyen; Jimmy Ramirez; Jingtao Liu; Kelly Schiabor Barrett; Kim Gietzen; Magnus Isaksson; Marc Laurent; Matthew Tolentino; Nicole L. Washington; Peter W. Cook; Phil Febbo; Ryan Cho; Shannon Wickline; Sherry Wang; Simon White; Summer Galloway; Suxiang Tong; Tyler Cassens; William Lee |
| EPI_ISL_1234786, EPI_ISL_1234788, EPI_ISL_1234792, EPI_ISL_1234801, EPI_ISL_1234803, EPI_ISL_1234805, EPI_ISL_1234811, EPI_ISL_1234814, EPI_ISL_1234816, EPI_ISL_1234817, EPI_ISL_1234821, EPI_ISL_1234822, EPI_ISL_1234823, EPI_ISL_1234824, EPI_ISL_1234852, EPI_ISL_1234853, EPI_ISL_1234856, EPI_ISL_1234859, EPI_ISL_1234861, EPI_ISL_1234863, EPI_ISL_1234868, EPI_ISL_1234874, EPI_ISL_1234888, EPI_ISL_1234889, EPI_ISL_1234890, EPI_ISL_1234893, EPI_ISL_1234896, EPI_ISL_1234898, EPI_ISL_1234903, EPI_ISL_1234905, EPI_ISL_1234906, EPI_ISL_1234910, EPI_ISL_1234911, EPI_ISL_1234912, EPI_ISL_1234914, EPI_ISL_1234916, EPI_ISL_1234922, EPI_ISL_1234927, EPI_ISL_1234928, EPI_ISL_1234931, EPI_ISL_1234934, EPI_ISL_1234936, EPI_ISL_1234948, EPI_ISL_1234960, EPI_ISL_1234963, EPI_ISL_1234966, EPI_ISL_1234971, EPI_ISL_1234974, EPI_ISL_1234980, EPI_ISL_1234981, EPI_ISL_1234984, EPI_ISL_1234989, EPI_ISL_1234998, EPI_ISL_1235001, EPI_ISL_1235003, EPI_ISL_1235004, EPI_ISL_1235005, EPI_ISL_1235033, EPI_ISL_1235037, EPI_ISL_1235039, EPI_ISL_1235040, EPI_ISL_1235045, EPI_ISL_1235046, EPI_ISL_1235053, EPI_ISL_1477169, EPI_ISL_1477176, EPI_ISL_1477180, EPI_ISL_1477208, EPI_ISL_1477227, EPI_ISL_1477234, EPI_ISL_1477255, EPI_ISL_1477264, EPI_ISL_1477272, EPI_ISL_1477284, EPI_ISL_1477290, EPI_ISL_1477294, EPI_ISL_1477306, EPI_ISL_1477348, EPI_ISL_1477375, EPI_ISL_1477399, EPI_ISL_1477417, EPI_ISL_2658582, EPI_ISL_2658646, EPI_ISL_2659310, EPI_ISL_2758452, EPI_ISL_2758453 | | | |
| see above | Humboldt County Public Health Laboratory | Chan-Zuckerberg Biohub | CZB Cliahub Consortium |
| EPI_ISL_1081346, EPI_ISL_1081347 | IEH Laboratories and Consulting Group | IEH and ID Genomics | Curtis Fowler; Evgeni Sokurenko; Hao Geng; Hemantha Kulakesara; Kai Deng; Kirthi Kutumbaka; Lydia Larson; Mansour Samadpour; Veronika Tchesnokova |
| EPI_ISL_2253166, EPI_ISL_2604248 | Infectious Diseases, Chan-Zuckerberg Biohub Covidtracker | Infectious Diseases, Chan-Zuckerberg Biohub Covidtracker | CZB Cliahub Consortium |
| EPI_ISL_918191, EPI_ISL_918192, EPI_ISL_980840, EPI_ISL_980841, EPI_ISL_980842, EPI_ISL_980843, EPI_ISL_980844, EPI_ISL_980846, EPI_ISL_980847, EPI_ISL_980848, EPI_ISL_980849, EPI_ISL_980850, EPI_ISL_980851, EPI_ISL_980853, EPI_ISL_980856, EPI_ISL_980857, EPI_ISL_980858, EPI_ISL_980860, EPI_ISL_980861, EPI_ISL_980862, EPI_ISL_980863, EPI_ISL_980864, EPI_ISL_980865, EPI_ISL_980866, EPI_ISL_980867, EPI_ISL_980868, EPI_ISL_980869, EPI_ISL_980870, EPI_ISL_980871, EPI_ISL_980872, EPI_ISL_980873, EPI_ISL_980874, EPI_ISL_980875, EPI_ISL_980876, EPI_ISL_980877, EPI_ISL_980878, EPI_ISL_980879, EPI_ISL_980880, EPI_ISL_980881, EPI_ISL_980882, EPI_ISL_980883, EPI_ISL_980884, EPI_ISL_980885, EPI_ISL_980886, EPI_ISL_980887, EPI_ISL_980888, EPI_ISL_980889, EPI_ISL_980890, EPI_ISL_980891, EPI_ISL_980892, EPI_ISL_980893, EPI_ISL_980894, EPI_ISL_980895, EPI_ISL_980896, EPI_ISL_980900, EPI_ISL_980901, EPI_ISL_980903, EPI_ISL_980904, EPI_ISL_980906, EPI_ISL_980907, EPI_ISL_980908, EPI_ISL_980909, EPI_ISL_980910, EPI_ISL_980911, EPI_ISL_980912, EPI_ISL_980913, EPI_ISL_980914, EPI_ISL_980915, EPI_ISL_980917, EPI_ISL_980918, EPI_ISL_980919, EPI_ISL_980923, EPI_ISL_980924, EPI_ISL_980926, EPI_ISL_980927, EPI_ISL_980928, EPI_ISL_980929, EPI_ISL_980931, EPI_ISL_980932, EPI_ISL_980934, EPI_ISL_980940, EPI_ISL_980941, EPI_ISL_980942, EPI_ISL_980943, EPI_ISL_980944, EPI_ISL_980945, EPI_ISL_980946, EPI_ISL_980947, EPI_ISL_980948, EPI_ISL_980949, EPI_ISL_980950, EPI_ISL_980951, EPI_ISL_980952, EPI_ISL_980953, EPI_ISL_980954, EPI_ISL_980955, EPI_ISL_980956, EPI_ISL_980957, EPI_ISL_980958, EPI_ISL_980959, EPI_ISL_980960, EPI_ISL_980961, EPI_ISL_980962, EPI_ISL_980963, EPI_ISL_980964, EPI_ISL_980965, EPI_ISL_980966, EPI_ISL_980967, EPI_ISL_980968, EPI_ISL_980969, EPI_ISL_980970, EPI_ISL_980971, EPI_ISL_980972, EPI_ISL_980973, EPI_ISL_980984, EPI_ISL_980985, EPI_ISL_980986, EPI_ISL_980987, EPI_ISL_980988, EPI_ISL_980989, EPI_ISL_980990, EPI_ISL_980991, EPI_ISL_980992, EPI_ISL_980993, EPI_ISL_980994, EPI_ISL_980995, EPI_ISL_980996, EPI_ISL_980997, EPI_ISL_980998, EPI_ISL_980999, EPI_ISL_981000, EPI_ISL_981001, EPI_ISL_981002, EPI_ISL_981003, EPI_ISL_984659, EPI_ISL_984660, EPI_ISL_984661, EPI_ISL_1060041, EPI_ISL_1060043, EPI_ISL_1060044, EPI_ISL_1060045, EPI_ISL_1060053, EPI_ISL_1060054, EPI_ISL_1060057, EPI_ISL_1060058, EPI_ISL_1060060, EPI_ISL_1060068, EPI_ISL_1060075, EPI_ISL_1060076, EPI_ISL_1060078, EPI_ISL_1060079, EPI_ISL_1060085, EPI_ISL_1060096, EPI_ISL_1060098, EPI_ISL_1060112, EPI_ISL_1060113, EPI_ISL_1060114, EPI_ISL_1060119, EPI_ISL_1060121, EPI_ISL_1060130, EPI_ISL_1060131, EPI_ISL_1060135, EPI_ISL_1060138, EPI_ISL_1060143, EPI_ISL_1060145, EPI_ISL_1060147, EPI_ISL_1060150, EPI_ISL_1060151, EPI_ISL_1060152, EPI_ISL_1060156, EPI_ISL_1060157, EPI_ISL_1060159, EPI_ISL_1060162, EPI_ISL_1060175, EPI_ISL_1060176, EPI_ISL_1060189, EPI_ISL_1060190, EPI_ISL_1060191, EPI_ISL_1081288, EPI_ISL_1081289, EPI_ISL_1081290, EPI_ISL_1712441, EPI_ISL_1712442, EPI_ISL_1712443, EPI_ISL_1712444, EPI_ISL_1712445, EPI_ISL_1712446, EPI_ISL_1712447, EPI_ISL_1712448, EPI_ISL_1712449 | | | |
| see above | Innovative Genomics Institute, UC Berkeley | Innovative Genomics Institute, UC Berkeley | Alison Ciling; Haridha Shivram; Holly Gildea; John Boyle; Justin Choi; Liana Lareau; Netravathi Krishnappa; Phil Frankino; Shana McDevitt; Stacia Wyman |
| EPI_ISL_1532141, EPI_ISL_1532142, EPI_ISL_1532143 | Kaiser San Jose | Santa Clara County Public Health Laboratory | Santa Clara County Public Health Department |
| EPI_ISL_1664228, EPI_ISL_1664229, EPI_ISL_1664230, EPI_ISL_1664231, EPI_ISL_1664232, EPI_ISL_1664233, EPI_ISL_1664234, EPI_ISL_1664237, EPI_ISL_1664238, EPI_ISL_1664240, EPI_ISL_1664244, EPI_ISL_1664245, EPI_ISL_1664247, EPI_ISL_1664248, EPI_ISL_1664249, EPI_ISL_1664250, EPI_ISL_1664251, EPI_ISL_1664252, EPI_ISL_1664253 | | | |
| see above | Kaiser San Jose | Santa Clara County public Health Laboratory | Santa Clara County Public Health Department |
| EPI_ISL_1193289, EPI_ISL_1193290 | Laboratory Corporation of America | Centers for Disease Control and Prevention Division of Viral Diseases, Pathogen Discovery | Amanda Douglas; Amanda Suchanek; Andrea Throop; Ayla Burns; Ben L. Rambo-Martin; Bobbi Croy; Brian Krueger; Brian Norvell; Christos Petropoulos; Clinton R. Paden; Craig Lukasik; Dakota Howard; Dhwani Batra; Duncan MacCannell; Eyad Almasri Debbie Boles; Goran Stevovic; Howard Engler; Hrushikesh Deshmukh; Jake Humphrey; Jana Schroth; Joe Voshell; John Pruitt; Jonathan Meltzer; Jonathan Williams; Kimberly Wagner; Lax Iyer; Lyndon Tilson; Manoj Jain; Marcia Eisenberg; Mary Ann Cristobal; Mary Williamson; Michael Levandoski; Mike Sapeta; Mindy Nye; Minoo Agarwal; Mohan Kolli; Nuthawin Charoensri; Oren Cohen; Peter W. Cook; Prashant Gupta; Qian Zeng; Rama Ghatti; Scott Parker; Scott Ryan; Stanley Letovsky; Steven Ragan; Suresh Babu Selvaraju; Susan Countryman; Susan Hicks; Suxiang Tong; Suzanne Dale; Thomas Urban; Tim Kuphal; Tricia Zwiefelhofer; Vincent Drouillon |
| EPI_ISL_1020770, EPI_ISL_1020784, EPI_ISL_1020807, EPI_ISL_1020853, EPI_ISL_1020857, EPI_ISL_1020859, EPI_ISL_1020877, EPI_ISL_1020882, EPI_ISL_1020885, EPI_ISL_1020890, EPI_ISL_1020899, EPI_ISL_1020944, EPI_ISL_1020966, EPI_ISL_1020967, EPI_ISL_1020977, EPI_ISL_1021008, EPI_ISL_1021045, EPI_ISL_1021049, EPI_ISL_1021060, EPI_ISL_1021063, EPI_ISL_1021086, EPI_ISL_1021095, EPI_ISL_1021099, EPI_ISL_1021101, EPI_ISL_1021129, EPI_ISL_1021130, EPI_ISL_1021131, EPI_ISL_1021139, EPI_ISL_1021173, EPI_ISL_1021245, EPI_ISL_1021246, EPI_ISL_1021280, EPI_ISL_1021308, EPI_ISL_1021309, EPI_ISL_1021310, EPI_ISL_1021311, EPI_ISL_1021312, EPI_ISL_1021313, EPI_ISL_1021314, EPI_ISL_1021341, EPI_ISL_1030654, EPI_ISL_1030691, EPI_ISL_1030709, EPI_ISL_1030972, EPI_ISL_1031067, EPI_ISL_1031072, EPI_ISL_1031132, EPI_ISL_1031536, EPI_ISL_1031697, EPI_ISL_1031798, EPI_ISL_1031799, EPI_ISL_1031803, EPI_ISL_1032409, EPI_ISL_1032411, EPI_ISL_1032412, EPI_ISL_1032413, EPI_ISL_1032527, EPI_ISL_1032528, EPI_ISL_1032542, EPI_ISL_1036992, EPI_ISL_1037031, EPI_ISL_1037106, EPI_ISL_1037109, EPI_ISL_1037112, EPI_ISL_1037115, EPI_ISL_1037126, EPI_ISL_1037144, EPI_ISL_1037147, EPI_ISL_1037180, EPI_ISL_1037222, EPI_ISL_1037249, EPI_ISL_1037316, EPI_ISL_1037373, EPI_ISL_1037404, EPI_ISL_1037417, EPI_ISL_1037423, EPI_ISL_1037448, EPI_ISL_1037473, EPI_ISL_1037515, EPI_ISL_1037559, EPI_ISL_1037576, EPI_ISL_1037578, EPI_ISL_1037612, EPI_ISL_1037639, EPI_ISL_1037651, EPI_ISL_1037660, EPI_ISL_1037674, EPI_ISL_1037682, EPI_ISL_1037709, EPI_ISL_1037740, EPI_ISL_1037747, EPI_ISL_1037757, EPI_ISL_1037760, EPI_ISL_1037762, EPI_ISL_1037763, EPI_ISL_1037781, EPI_ISL_1037782, EPI_ISL_1037783, EPI_ISL_1037806, EPI_ISL_1037874, EPI_ISL_1037875, EPI_ISL_1037886, EPI_ISL_1037888, EPI_ISL_1037918, EPI_ISL_1037938, EPI_ISL_1037993, EPI_ISL_1038066, EPI_ISL_1038085, EPI_ISL_1038148, EPI_ISL_1038150, EPI_ISL_1038181, EPI_ISL_1038243, EPI_ISL_1038244, EPI_ISL_1038245, EPI_ISL_1038246, EPI_ISL_1038275, EPI_ISL_1038283, EPI_ISL_1038351, EPI_ISL_1038362, EPI_ISL_1038488, EPI_ISL_1038489, EPI_ISL_1038673, EPI_ISL_1038674, EPI_ISL_1038675, EPI_ISL_1038676, EPI_ISL_1038677, EPI_ISL_1038678, EPI_ISL_1038679, EPI_ISL_1038680, EPI_ISL_1038681, EPI_ISL_1038682, EPI_ISL_1038683, EPI_ISL_1038684, EPI_ISL_1038685, EPI_ISL_1038686, EPI_ISL_1038687, EPI_ISL_1038689, EPI_ISL_1038690, EPI_ISL_1038691, EPI_ISL_1038692, EPI_ISL_1038693, EPI_ISL_1038694, EPI_ISL_1038695, EPI_ISL_1038769, EPI_ISL_1081267 | | | |
| see above | Laboratory Corporation of America | Respiratory Viruses Branch, Division of Viral Diseases, Centers for Disease Control and Prevention | Alexandre Bolze; Ary Ascencio; Ben L. Rambo-Martin; Brad Sickler; Charlotte Rivera-Garcia; Christine Tran; Clinton R. Paden; Dakota Howard; David Becker; Dhwani Batra; Duncan MacCannell; Efren Sandoval; Eileen de Feo; Elizabeth Cirulli; Eric Allen; Geraint Levan; James Lu; Jan Antico; Jason Nguyen; Jimmy Ramirez; Jingtao Liu; Kelly Schiabor Barrett; Kim Gietzen; Magnus Isaksson; Marc Laurent; Matthew Tolentino; Nicole L. Washington; Peter W. Cook; Phil Febbo; Ryan Cho; Shannon Wickline; Sherry Wang; Simon White; Summer Galloway; Suxiang Tong; Tyler Cassens; William Lee |
| EPI_ISL_917951, EPI_ISL_917952, EPI_ISL_917953, EPI_ISL_917955, EPI_ISL_939609, EPI_ISL_939610, EPI_ISL_939611, EPI_ISL_939612, EPI_ISL_939613, EPI_ISL_939614, EPI_ISL_939615, EPI_ISL_939616, EPI_ISL_939617, EPI_ISL_939618, EPI_ISL_962848, EPI_ISL_962849, EPI_ISL_962850, EPI_ISL_962851, EPI_ISL_962852, EPI_ISL_962853, EPI_ISL_962854, EPI_ISL_962855, EPI_ISL_962856, EPI_ISL_962857, EPI_ISL_962858, EPI_ISL_962859, EPI_ISL_962860, EPI_ISL_962861, EPI_ISL_962862, EPI_ISL_962863, EPI_ISL_962864, EPI_ISL_962865, EPI_ISL_962866, EPI_ISL_962867, EPI_ISL_1005483, EPI_ISL_1005484, EPI_ISL_1005485, EPI_ISL_1005486, EPI_ISL_1005488, EPI_ISL_1005489, EPI_ISL_1005490, EPI_ISL_1005508, EPI_ISL_1005509, EPI_ISL_1005510, EPI_ISL_1040052, EPI_ISL_1040425, EPI_ISL_1040426, EPI_ISL_1097725, EPI_ISL_1097726, EPI_ISL_1097727, EPI_ISL_1097728, EPI_ISL_1097729, EPI_ISL_1097730, EPI_ISL_1097731, EPI_ISL_1097732, EPI_ISL_1097733, EPI_ISL_1097734, EPI_ISL_1097735, EPI_ISL_1097736, EPI_ISL_1097737, EPI_ISL_1097740, EPI_ISL_1097741, EPI_ISL_1097743, EPI_ISL_2774563, EPI_ISL_2774564, EPI_ISL_2774565, EPI_ISL_2774566, EPI_ISL_2774570, EPI_ISL_2774571, EPI_ISL_2894593, EPI_ISL_2894594, EPI_ISL_2964666 | | | |
| see above | Los Angeles County Public Health Laboratories | Los Angeles County Public Health Laboratories | P. Hemarajata et al. |
| EPI_ISL_930788, EPI_ISL_930789, EPI_ISL_968077, EPI_ISL_968078, EPI_ISL_1263230, EPI_ISL_1263231, EPI_ISL_1263232, EPI_ISL_1664606, EPI_ISL_1664607, EPI_ISL_1664608, EPI_ISL_1664609, EPI_ISL_1664610, EPI_ISL_1664611, EPI_ISL_1664612, EPI_ISL_1664613, EPI_ISL_1664614, EPI_ISL_1664615, EPI_ISL_1664616, EPI_ISL_2308282, EPI_ISL_2308286, EPI_ISL_2308287, EPI_ISL_2308288, EPI_ISL_2308354, EPI_ISL_2308355, EPI_ISL_2308356, EPI_ISL_2308357, EPI_ISL_2308358, EPI_ISL_2308359, EPI_ISL_2308360, EPI_ISL_2308361, EPI_ISL_2308362, EPI_ISL_2308363, EPI_ISL_2308364, EPI_ISL_2308365, EPI_ISL_2308366, EPI_ISL_2716876, EPI_ISL_2716877, EPI_ISL_2716878, EPI_ISL_2716879, EPI_ISL_2716880, EPI_ISL_2716881, EPI_ISL_2716882, EPI_ISL_2716883, EPI_ISL_2716884, EPI_ISL_2716885, EPI_ISL_2716886, EPI_ISL_2716887, EPI_ISL_2716888, EPI_ISL_2754037, EPI_ISL_2774519, EPI_ISL_2774520, EPI_ISL_2774521, EPI_ISL_2774523, EPI_ISL_2955356 | | | |
| see above | Monterey County Public Health Laboratory | Monterey County Public Health Laboratory | ; C. Anaya et al.; Monterey County Public Health Laboratory |
| EPI_ISL_882997, EPI_ISL_882998 | Motion Picture Television Fund Hospital Lab | Los Angeles County Public Health Laboratories | P. Hemarajata et al. |
| EPI_ISL_1541297, EPI_ISL_1541555 | OHSU Lab Services Molecular Microbiology Lab | Oregon SARS-CoV-2 Genome Sequencing Center | Alec J. Hirsch; Andrew C. Adey; Benjamin N. Bimber; Brendan L. O'Connell; Brian J. O'Roak; Daniel N. Streblow; Donna Hansel; Guang Fan; Kayla Carter; Ruth V. Nichols; Sally Grindstaff; Sonia Acharya; William B. Messer; Xuan Qin |
| EPI_ISL_1005541, EPI_ISL_1005556 | Olive View Medical Center | Los Angeles County Public Health Laboratories | P. Hemarajata et al. |
| EPI_ISL_1027507, EPI_ISL_1027530, EPI_ISL_1027611, EPI_ISL_1027612, EPI_ISL_1027613, EPI_ISL_1027614, EPI_ISL_1027615, EPI_ISL_1027616, EPI_ISL_1027617, EPI_ISL_1027618, EPI_ISL_1027619, EPI_ISL_1027620, EPI_ISL_1027621, EPI_ISL_1027622, EPI_ISL_1027623, EPI_ISL_1027624, EPI_ISL_1027625, EPI_ISL_1027626, EPI_ISL_1027627, EPI_ISL_1234797, EPI_ISL_1234804, EPI_ISL_1234825, EPI_ISL_1234828, EPI_ISL_1234842, EPI_ISL_1234869, EPI_ISL_1234887, EPI_ISL_1234918, EPI_ISL_1234923, EPI_ISL_1234929, EPI_ISL_1234937, EPI_ISL_1234940, EPI_ISL_1234942, EPI_ISL_1234944, EPI_ISL_1234970, EPI_ISL_1235007, EPI_ISL_1235009, EPI_ISL_1235016, EPI_ISL_1235041, EPI_ISL_1235044, EPI_ISL_1235052, EPI_ISL_1235057, EPI_ISL_1701566, EPI_ISL_1701571, EPI_ISL_2659240, EPI_ISL_2659430, EPI_ISL_2659578 | | | |
| see above | Orange County Public Health Lab | Chan-Zuckerberg Biohub | CZB Cliahub Consortium |
| EPI_ISL_917954 | Pomona Valley Hospital Medical Center | Los Angeles County Public Health Laboratories | P. Hemarajata et al. |
| EPI_ISL_2551231, EPI_ISL_2551232, EPI_ISL_2551233, EPI_ISL_2551234, EPI_ISL_2551235, EPI_ISL_2551236, EPI_ISL_2551237, EPI_ISL_2551242, EPI_ISL_2551243, EPI_ISL_2551244, EPI_ISL_2551245, EPI_ISL_2551246, EPI_ISL_2551247, EPI_ISL_2551248, EPI_ISL_2551249, EPI_ISL_2551250, EPI_ISL_2551251 | | | |
| see above | Providence St. Joseph Health Orange - Chapman | Providence St. Joseph Health Molecular Genomics Laboratory | Alexa K Dowdell; Brian D Piening; Carlo B Bifulco; Fred L Robinson; Mary Campbell; Rogan Rattray |
| EPI_ISL_1194381, EPI_ISL_1194383, EPI_ISL_1194402, EPI_ISL_1194403, EPI_ISL_1194404 | Quest Diagnostics Incorporated | Centers for Disease Control and Prevention Division of Viral Diseases, Pathogen Discovery | A. Gerasimova; A. Perez; B. Anderson; Ben L. Rambo-Martin; Clinton R. Paden; Dakota Howard; Dhwani Batra; Duncan MacCannell; F. Lacbawan; I. A. Shlyakhter; K.E. Livingston; L.E. Bernstein; M. Hua; P. Tanpaiboon; Peter W. Cook; R. M. Kagan; R. Owen; R. V. Rolando; S. H. Rosenthal; Suxiang Tong; Y. Liu |
| EPI_ISL_1086225, EPI_ISL_1086232, EPI_ISL_1086236, EPI_ISL_1086244, EPI_ISL_1086245, EPI_ISL_1086252, EPI_ISL_1086270, EPI_ISL_1086277, EPI_ISL_1086280, EPI_ISL_1086292, EPI_ISL_1086301, EPI_ISL_1086331, EPI_ISL_1086368, EPI_ISL_1086378, EPI_ISL_1086380, EPI_ISL_1086382, EPI_ISL_1086383, EPI_ISL_1086384, EPI_ISL_1086402, EPI_ISL_1086494, EPI_ISL_1086500, EPI_ISL_1086503, EPI_ISL_1086506, EPI_ISL_1086508, EPI_ISL_1086510, EPI_ISL_1086512, EPI_ISL_1086518, EPI_ISL_1086523, EPI_ISL_1086525, EPI_ISL_1086526, EPI_ISL_1086537, EPI_ISL_1086539, EPI_ISL_1086541, EPI_ISL_1086542, EPI_ISL_1086543, EPI_ISL_1086546, EPI_ISL_1086548, EPI_ISL_1086549, EPI_ISL_1086557, EPI_ISL_1086558, EPI_ISL_1086566, EPI_ISL_1086568, EPI_ISL_1086571, EPI_ISL_1086576, EPI_ISL_1086579, EPI_ISL_1086585, EPI_ISL_1086586, EPI_ISL_1086587, EPI_ISL_1086589, EPI_ISL_1086598, EPI_ISL_1086605, EPI_ISL_1086607, EPI_ISL_1086609, EPI_ISL_1086615, EPI_ISL_1086620, EPI_ISL_1086621, EPI_ISL_1086622, EPI_ISL_1086628, EPI_ISL_1086632, EPI_ISL_1086633, EPI_ISL_1086637, EPI_ISL_1086639, EPI_ISL_1086644, EPI_ISL_1086648, EPI_ISL_1086649, EPI_ISL_1086650, EPI_ISL_1086657, EPI_ISL_1086658, EPI_ISL_1086665, EPI_ISL_1086666, EPI_ISL_1086667, EPI_ISL_1086668, EPI_ISL_1086670, EPI_ISL_1086671, EPI_ISL_1086674, EPI_ISL_1086677, EPI_ISL_1086679, EPI_ISL_1086681, EPI_ISL_1086682, EPI_ISL_1086684, EPI_ISL_1086685, EPI_ISL_1086688, EPI_ISL_1086689, EPI_ISL_1086690, EPI_ISL_1086693, EPI_ISL_1086694, EPI_ISL_1086697, EPI_ISL_1086699, EPI_ISL_1086700, EPI_ISL_1086701, EPI_ISL_1086703, EPI_ISL_1086709, EPI_ISL_1086710, EPI_ISL_1086712, EPI_ISL_1086715, EPI_ISL_1086716, EPI_ISL_1086719, EPI_ISL_1086721, EPI_ISL_1086722, EPI_ISL_1086725, EPI_ISL_1086726, EPI_ISL_1086728, EPI_ISL_1086729, EPI_ISL_1086730, EPI_ISL_1086731, EPI_ISL_1086733, EPI_ISL_1086735, EPI_ISL_1086736, EPI_ISL_1086738, EPI_ISL_1086739, EPI_ISL_1086741, EPI_ISL_1086742, EPI_ISL_1086745, EPI_ISL_1086747, EPI_ISL_1086749, EPI_ISL_1086750, EPI_ISL_1086752, EPI_ISL_1086756, EPI_ISL_1086759, EPI_ISL_1086760, EPI_ISL_1086761, EPI_ISL_1086762, EPI_ISL_1086765, EPI_ISL_1086767, EPI_ISL_1086769, EPI_ISL_1086770, EPI_ISL_1086771, EPI_ISL_1086772, EPI_ISL_1086774, EPI_ISL_1086775, EPI_ISL_1086777, EPI_ISL_1086778, EPI_ISL_1086779, EPI_ISL_1086780, EPI_ISL_1086783, EPI_ISL_1086784, EPI_ISL_1086785, EPI_ISL_1086787, EPI_ISL_1086792, EPI_ISL_1086793, EPI_ISL_1086796, EPI_ISL_1086797, EPI_ISL_1086799, EPI_ISL_1086801, EPI_ISL_1086802, EPI_ISL_1086803, EPI_ISL_1086804, EPI_ISL_1086805, EPI_ISL_1086806, EPI_ISL_1086808, EPI_ISL_1086809, EPI_ISL_1086810, EPI_ISL_1086811, EPI_ISL_1086813, EPI_ISL_1086814, EPI_ISL_1086815, EPI_ISL_1086816, EPI_ISL_1086819, EPI_ISL_1086820, EPI_ISL_1086822, EPI_ISL_1087439, EPI_ISL_1087517, EPI_ISL_1087525, EPI_ISL_1087541, EPI_ISL_1087552, EPI_ISL_1087628, EPI_ISL_1087633, EPI_ISL_1087634, EPI_ISL_1087636, EPI_ISL_1087651, EPI_ISL_1087655, EPI_ISL_1087656, EPI_ISL_1087659, EPI_ISL_1087661, EPI_ISL_1087663, EPI_ISL_1087667, EPI_ISL_1087669, EPI_ISL_1087675, EPI_ISL_1087679, EPI_ISL_1087684, EPI_ISL_1087693, EPI_ISL_1087727, EPI_ISL_1087755, EPI_ISL_1087760, EPI_ISL_1087768, EPI_ISL_1087782, EPI_ISL_1087786, EPI_ISL_1087788, EPI_ISL_1087789, EPI_ISL_1087793, EPI_ISL_1087794, EPI_ISL_1087799, EPI_ISL_1087800, EPI_ISL_1087813, EPI_ISL_1087814, EPI_ISL_1087817, EPI_ISL_1087819, EPI_ISL_1087824, EPI_ISL_1087825, EPI_ISL_1087826, EPI_ISL_1087827, EPI_ISL_1087828, EPI_ISL_1087829, EPI_ISL_1087831, EPI_ISL_1087834, EPI_ISL_1087835, EPI_ISL_1087837, EPI_ISL_1087838, EPI_ISL_1087840, EPI_ISL_1087842, EPI_ISL_1087845, EPI_ISL_1087847, EPI_ISL_1087850, EPI_ISL_1087852, EPI_ISL_1087853, EPI_ISL_1087854, EPI_ISL_1087855, EPI_ISL_1087856, EPI_ISL_1087857, EPI_ISL_1087858, EPI_ISL_1087859, EPI_ISL_1087860, EPI_ISL_1087895, EPI_ISL_1087896, EPI_ISL_1087897, EPI_ISL_1087898, EPI_ISL_1087900, EPI_ISL_1087903, EPI_ISL_1087905, EPI_ISL_1087906, EPI_ISL_1087909, EPI_ISL_1087912, EPI_ISL_1087917, EPI_ISL_1087918, EPI_ISL_1087919, EPI_ISL_1087921, EPI_ISL_1087925, EPI_ISL_1087930, EPI_ISL_1087932, EPI_ISL_1087933, EPI_ISL_1087934, EPI_ISL_1087935, EPI_ISL_1087943, EPI_ISL_1087945, EPI_ISL_1087948, EPI_ISL_1087949, EPI_ISL_1087950, EPI_ISL_1087951, EPI_ISL_1087952, EPI_ISL_1087954, EPI_ISL_1087957, EPI_ISL_1087959, EPI_ISL_1087976, EPI_ISL_1088080, EPI_ISL_1088087, EPI_ISL_1088107, EPI_ISL_1088117, EPI_ISL_1088123, EPI_ISL_1088140, EPI_ISL_1088142, EPI_ISL_1088143, EPI_ISL_1088152, EPI_ISL_1088170, EPI_ISL_1088188, EPI_ISL_1088192, EPI_ISL_1088211, EPI_ISL_1088216, EPI_ISL_1088232, EPI_ISL_1088381, EPI_ISL_1088382, EPI_ISL_1088383, EPI_ISL_1088385, EPI_ISL_1088386, EPI_ISL_1088390, EPI_ISL_1088393, EPI_ISL_1088395, EPI_ISL_1088398, EPI_ISL_1088399, EPI_ISL_1088400, EPI_ISL_1088402, EPI_ISL_1088403, EPI_ISL_1088404, EPI_ISL_1088409, EPI_ISL_1088410, EPI_ISL_1088411, EPI_ISL_1088412, EPI_ISL_1088413, EPI_ISL_1088414, EPI_ISL_1088415, EPI_ISL_1088416, EPI_ISL_1088417, EPI_ISL_1088419, EPI_ISL_1088420, EPI_ISL_1088422, EPI_ISL_1088423, EPI_ISL_1088425, EPI_ISL_1088426, EPI_ISL_1088428, EPI_ISL_1088429, EPI_ISL_1088430, EPI_ISL_1088431, EPI_ISL_1088432, EPI_ISL_1088434, EPI_ISL_1088437, EPI_ISL_1088438, EPI_ISL_1088439, EPI_ISL_1088440, EPI_ISL_1088441, EPI_ISL_1088442, EPI_ISL_1088443, EPI_ISL_1088444, EPI_ISL_1088445, EPI_ISL_1088447, EPI_ISL_1088448, EPI_ISL_1088453, EPI_ISL_1088455, EPI_ISL_1088456, EPI_ISL_1088457, EPI_ISL_1088458, EPI_ISL_1088460, EPI_ISL_1088462, EPI_ISL_1088464, EPI_ISL_1088466, EPI_ISL_1088467, EPI_ISL_1088469, EPI_ISL_1088471, EPI_ISL_1088472, EPI_ISL_1088474, EPI_ISL_1088475, EPI_ISL_1088477, | | | |

| | | | |
|---|---|---|---|
| EPI_ISL_1088478, EPI_ISL_1088479, EPI_ISL_1088480, EPI_ISL_1088481, EPI_ISL_1088482, EPI_ISL_1088483, EPI_ISL_1088488, EPI_ISL_1088489, EPI_ISL_1088490, EPI_ISL_1088491, EPI_ISL_1088493, EPI_ISL_1088494, EPI_ISL_1088495, EPI_ISL_1088496, EPI_ISL_1088497, EPI_ISL_1088499, EPI_ISL_1088501, EPI_ISL_1088502, EPI_ISL_1088503, EPI_ISL_1088505, EPI_ISL_1088506, EPI_ISL_1088508, EPI_ISL_1088509, EPI_ISL_1088511, EPI_ISL_1088512, EPI_ISL_1088513, EPI_ISL_1088514, EPI_ISL_1088516, EPI_ISL_1088517, EPI_ISL_1088518, EPI_ISL_1088520, EPI_ISL_1088521, EPI_ISL_1088522, EPI_ISL_1088523, EPI_ISL_1088525, EPI_ISL_1088528, EPI_ISL_1088529, EPI_ISL_1088530, EPI_ISL_1088532, EPI_ISL_1088533, EPI_ISL_1088534, EPI_ISL_1088537, EPI_ISL_1088538, EPI_ISL_1088539, EPI_ISL_1088541, EPI_ISL_1088542, EPI_ISL_1088543, EPI_ISL_1088545, EPI_ISL_1088547, EPI_ISL_1088548, EPI_ISL_1088549, EPI_ISL_1090697 | | | |
| see above | Quest Diagnostics Incorporated | Respiratory Viruses Branch, Division of Viral Diseases, Centers for Disease Control and Prevention | A. Gerasimova; A. Perez; B. Anderson; Ben L. Rambo-Martin; Clinton R. Paden; Dakota Howard; Dhwani Batra; Duncan MacCannell; F. Lacbawan; I. A. Shlyakhter; K.E. Livingston; L.E. Bernstein; M. Hua; P. Tanpaiboon; Peter W. Cook; R. M. Kagan; R. Owen; R. V. Rolando; S. H. Rosenthal; Suxiang Tong; Y. Liu |
| EPI_ISL_1295796, EPI_ISL_1295797, EPI_ISL_1295801, EPI_ISL_1295803, EPI_ISL_1295804, EPI_ISL_1295805, EPI_ISL_1295806, EPI_ISL_1295808, EPI_ISL_1295810, EPI_ISL_1295811, EPI_ISL_1295814, EPI_ISL_1794771, EPI_ISL_1794774, EPI_ISL_1794775, EPI_ISL_1794780 | | | |
| see above | Rady Children's Hospital - San Diego | Andersen lab at Scripps Research | Christina Clarke; David Dimmock; Denise Malicki; Kathryn Bouic; Linda Luo; SEARCH Alliance San Diego with Nanda Radamchar; Teresa Mueller |
| EPI_ISL_1081572, EPI_ISL_1081575, EPI_ISL_1081579, EPI_ISL_1081581, EPI_ISL_1081593, EPI_ISL_1081594 | Rady's Childrens Hospital | Andersen lab at Scripps Research | Christina Clarke; David Dimmock; Denise Malicki; Kathryn Bouic; Linda Luo; SEARCH Alliance San Diego with Nanda Radamchar; Teresa Mueller |
| EPI_ISL_1081472, EPI_ISL_1081784, EPI_ISL_1081787, EPI_ISL_1185961, EPI_ISL_1185967, EPI_ISL_1185969, EPI_ISL_1185971, EPI_ISL_1185976, EPI_ISL_1185981, EPI_ISL_1185985, EPI_ISL_1185989, EPI_ISL_1185991, EPI_ISL_1185992, EPI_ISL_1185993, EPI_ISL_1185994, EPI_ISL_1185995, EPI_ISL_1185996, EPI_ISL_1185997, EPI_ISL_1185998, EPI_ISL_1185999, EPI_ISL_1186000, EPI_ISL_1186001, EPI_ISL_1186002, EPI_ISL_1186003, EPI_ISL_1186004, EPI_ISL_1186005, EPI_ISL_1186006, EPI_ISL_1186007, EPI_ISL_1186008, EPI_ISL_1186009, EPI_ISL_1186010, EPI_ISL_1186011, EPI_ISL_1186012, EPI_ISL_1186013, EPI_ISL_1186014, EPI_ISL_1186015, EPI_ISL_1186016, EPI_ISL_1186017, EPI_ISL_1186018, EPI_ISL_1186019, EPI_ISL_1186020, EPI_ISL_1186021, EPI_ISL_1186022, EPI_ISL_1468876, EPI_ISL_1468989, EPI_ISL_1468994, EPI_ISL_1468999, EPI_ISL_1469000, EPI_ISL_1469004, EPI_ISL_1469008, EPI_ISL_1469009, EPI_ISL_1469012, EPI_ISL_1469020, EPI_ISL_1794621, EPI_ISL_1794635, EPI_ISL_1794638 | | | |
| see above | San Diego County Public Health Laboratory | Andersen lab at Scripps Research | Brett Austin; Jovan Shephard; SEARCH Alliance San Diego with Tracy Basler |
| EPI_ISL_2841616, EPI_ISL_2841617, EPI_ISL_2841630, EPI_ISL_2841631, EPI_ISL_2841633, EPI_ISL_2841634 | San Diego County Public Health Laboratory | San Diego County Public Health Laboratory | Ashleigh Murphy Schafer; Brett Austin |
| EPI_ISL_1701040, EPI_ISL_1701041, EPI_ISL_1701050, EPI_ISL_1701052, EPI_ISL_1701061, EPI_ISL_1701069, EPI_ISL_1701086, EPI_ISL_1701096, EPI_ISL_1701117, EPI_ISL_1701142, EPI_ISL_1701204, EPI_ISL_1701208, EPI_ISL_1701211, EPI_ISL_1701236, EPI_ISL_1701262, EPI_ISL_1701280, EPI_ISL_1701281, EPI_ISL_1701290, EPI_ISL_2659295 | | | |
| see above | San Joaquin County Public Health Lab | Chan-Zuckerberg Biohub | CZB Cliahub Consortium |
| EPI_ISL_979638, EPI_ISL_979639, EPI_ISL_979640, EPI_ISL_979641, EPI_ISL_979642, EPI_ISL_1185029, EPI_ISL_1185044, EPI_ISL_1185046, EPI_ISL_1185051, EPI_ISL_1185053, EPI_ISL_1185065, EPI_ISL_1185072, EPI_ISL_1185074, EPI_ISL_1185081, EPI_ISL_1185083, EPI_ISL_1185086, EPI_ISL_1185091, EPI_ISL_1185092, EPI_ISL_1185097, EPI_ISL_1185110, EPI_ISL_1185111, EPI_ISL_1185116, EPI_ISL_1185120, EPI_ISL_1185129, EPI_ISL_1185137, EPI_ISL_1185141, EPI_ISL_1185170, EPI_ISL_1185181, EPI_ISL_1185183, EPI_ISL_1185185, EPI_ISL_1185191, EPI_ISL_1185197, EPI_ISL_1185200, EPI_ISL_1185218, EPI_ISL_1185219, EPI_ISL_1185234, EPI_ISL_1185239, EPI_ISL_1185251, EPI_ISL_1185252, EPI_ISL_1185255, EPI_ISL_1185267, EPI_ISL_1185273, EPI_ISL_1185277, EPI_ISL_1185284, EPI_ISL_1234537, EPI_ISL_1234539, EPI_ISL_1234541, EPI_ISL_1234547, EPI_ISL_1234548, EPI_ISL_1234549, EPI_ISL_1234560, EPI_ISL_1234561, EPI_ISL_1234578, EPI_ISL_1234586, EPI_ISL_1234588, EPI_ISL_1234606, EPI_ISL_1234609, EPI_ISL_1234613, EPI_ISL_1234641, EPI_ISL_1234645, EPI_ISL_1234653, EPI_ISL_1234657, EPI_ISL_1234660, EPI_ISL_1234667, EPI_ISL_1234671, EPI_ISL_1234678, EPI_ISL_1234689, EPI_ISL_1234694, EPI_ISL_1234706, EPI_ISL_1234711, EPI_ISL_1234721, EPI_ISL_1234724, EPI_ISL_1234725, EPI_ISL_1234728, EPI_ISL_1234729, EPI_ISL_1234737, EPI_ISL_1234744, EPI_ISL_1234759, EPI_ISL_1234760, EPI_ISL_1234769, EPI_ISL_1234787, EPI_ISL_1234798, EPI_ISL_1234800, EPI_ISL_1234810, EPI_ISL_1234812, EPI_ISL_1234818, EPI_ISL_1234840, EPI_ISL_1234841, EPI_ISL_1234843, EPI_ISL_1234846, EPI_ISL_1234849, EPI_ISL_1234850, EPI_ISL_1234851, EPI_ISL_1234854, EPI_ISL_1234857, EPI_ISL_1234865, EPI_ISL_1234866, EPI_ISL_1234875, EPI_ISL_1234876, EPI_ISL_1234878, EPI_ISL_1234879, EPI_ISL_1234880, EPI_ISL_1234881, EPI_ISL_1234883, EPI_ISL_1234884, EPI_ISL_1234886, EPI_ISL_1234897, EPI_ISL_1234899, EPI_ISL_1234902, EPI_ISL_1234917, EPI_ISL_1234919, EPI_ISL_1234925, EPI_ISL_1234935, EPI_ISL_1234946, EPI_ISL_1234957, EPI_ISL_1234961, EPI_ISL_1234962, EPI_ISL_1234972, EPI_ISL_1234973, EPI_ISL_1234975, EPI_ISL_1234983, EPI_ISL_1234985, EPI_ISL_1234991, EPI_ISL_1234993, EPI_ISL_1234996, EPI_ISL_1234999, EPI_ISL_1235000, EPI_ISL_1235010, EPI_ISL_1235015, EPI_ISL_1235017, EPI_ISL_1235018, EPI_ISL_1235019, EPI_ISL_1235020, EPI_ISL_1235024, EPI_ISL_1235034, EPI_ISL_1235043, EPI_ISL_1235047, EPI_ISL_1235049, EPI_ISL_1235050, EPI_ISL_1235051, EPI_ISL_1235362, EPI_ISL_1235370, EPI_ISL_1235394, EPI_ISL_1235452, EPI_ISL_1235487, EPI_ISL_1235518, EPI_ISL_1235583, EPI_ISL_1235601, EPI_ISL_1477958, EPI_ISL_1477961, EPI_ISL_1477963, EPI_ISL_1477967, EPI_ISL_1477979, EPI_ISL_1477983, EPI_ISL_1478012, EPI_ISL_1478013, EPI_ISL_1478019, EPI_ISL_1478031, EPI_ISL_1478038, EPI_ISL_1478081, EPI_ISL_1478084, EPI_ISL_1478087, EPI_ISL_1478092, EPI_ISL_1478102, EPI_ISL_1478103, EPI_ISL_1478129, EPI_ISL_1478153, EPI_ISL_1478170, EPI_ISL_2658578, EPI_ISL_2658584, EPI_ISL_2658590, EPI_ISL_2658629, EPI_ISL_2658667, EPI_ISL_2658715, EPI_ISL_2658727, EPI_ISL_2659238, EPI_ISL_2659373, EPI_ISL_2659463, EPI_ISL_2659576, EPI_ISL_2758437, EPI_ISL_2758438, EPI_ISL_2758450, EPI_ISL_2758451 | | | |
| see above | Santa Clara County Public Health Laboratory | Chan-Zuckerberg Biohub | CZB Cliahub Consortium |
| EPI_ISL_902730, EPI_ISL_902731, EPI_ISL_902732, EPI_ISL_902733, EPI_ISL_902734, EPI_ISL_914638, EPI_ISL_914639, EPI_ISL_914640, EPI_ISL_914643, EPI_ISL_1001005, EPI_ISL_1001006, EPI_ISL_1001007, EPI_ISL_1001008, EPI_ISL_1001009, EPI_ISL_1001010, EPI_ISL_1001011, EPI_ISL_1001012, EPI_ISL_1001013, EPI_ISL_1001014, EPI_ISL_1001015, EPI_ISL_1001016, EPI_ISL_1001017, EPI_ISL_1001018, EPI_ISL_1001019, EPI_ISL_1001020, EPI_ISL_1001021, EPI_ISL_1001022, EPI_ISL_1001023, EPI_ISL_1001024, EPI_ISL_1001025, EPI_ISL_1001026, EPI_ISL_1001027, EPI_ISL_1001028, EPI_ISL_1001029, EPI_ISL_1001030, EPI_ISL_1001031, EPI_ISL_1009655, EPI_ISL_1009657, EPI_ISL_1009659, EPI_ISL_1060543, EPI_ISL_1060544, EPI_ISL_1060545, EPI_ISL_1060546, EPI_ISL_1060548, EPI_ISL_1060549, EPI_ISL_1060551, EPI_ISL_1060552, EPI_ISL_1060553, EPI_ISL_1060554, EPI_ISL_1060555, EPI_ISL_1060556, EPI_ISL_1169580, EPI_ISL_1169599, EPI_ISL_1335804, EPI_ISL_1335808, EPI_ISL_1335809, EPI_ISL_1335810, EPI_ISL_1335811, EPI_ISL_1335827, EPI_ISL_1335828, EPI_ISL_1335829, EPI_ISL_1335830, EPI_ISL_1335831, EPI_ISL_1335832, EPI_ISL_1335833, EPI_ISL_1335834, EPI_ISL_1335835, EPI_ISL_1335836, EPI_ISL_1335837, EPI_ISL_1335838, EPI_ISL_1335839, EPI_ISL_1335840, EPI_ISL_1335841, EPI_ISL_1335842, EPI_ISL_1335843, EPI_ISL_1335844, EPI_ISL_1335845, EPI_ISL_1335846, EPI_ISL_1335847, EPI_ISL_1335848, EPI_ISL_1335849, EPI_ISL_1335850, EPI_ISL_1335851, EPI_ISL_1335852, EPI_ISL_1335856, EPI_ISL_1335857, EPI_ISL_1335858, EPI_ISL_1335859, EPI_ISL_1335860, EPI_ISL_1335862, EPI_ISL_1335863, EPI_ISL_1335865, EPI_ISL_1335867, EPI_ISL_1406156, EPI_ISL_1406157, EPI_ISL_1406158, EPI_ISL_1406159, EPI_ISL_1406160, EPI_ISL_1406161, EPI_ISL_1406170, EPI_ISL_1406171, EPI_ISL_1406172, EPI_ISL_1406173, EPI_ISL_1406174, EPI_ISL_1406175, EPI_ISL_1406176 | | | |
| see above | Santa Clara County Public Health Laboratory | Santa Clara County Public Health Laboratory | Santa Clara County Public Health Department |
| EPI_ISL_1081550, EPI_ISL_1081551, EPI_ISL_1081552, EPI_ISL_1081553, EPI_ISL_1081554, EPI_ISL_1081555, EPI_ISL_1081556, EPI_ISL_1081557, EPI_ISL_1081559, EPI_ISL_1081560, EPI_ISL_1081561, EPI_ISL_1081562, EPI_ISL_1081563, EPI_ISL_1081564, EPI_ISL_1081565, EPI_ISL_1081566, EPI_ISL_1081567, EPI_ISL_1081598, EPI_ISL_1081599, EPI_ISL_1081600, EPI_ISL_1081601, EPI_ISL_1081602, EPI_ISL_1081603, EPI_ISL_1081604, EPI_ISL_1081605, EPI_ISL_1081606, EPI_ISL_1081607, EPI_ISL_1081608, EPI_ISL_1081609, EPI_ISL_1081610, EPI_ISL_1081611, EPI_ISL_1081612, EPI_ISL_1081613, EPI_ISL_1081614, EPI_ISL_1081615, EPI_ISL_1081616, EPI_ISL_1081617, EPI_ISL_1081618, EPI_ISL_1081694, EPI_ISL_1081695, EPI_ISL_1081696, EPI_ISL_1081697, EPI_ISL_1081698, EPI_ISL_1081699, EPI_ISL_1081700, EPI_ISL_1081701, EPI_ISL_1081702, EPI_ISL_1081703, EPI_ISL_1081704, EPI_ISL_1081705, EPI_ISL_1081706, EPI_ISL_1081707, EPI_ISL_1081708, EPI_ISL_1081709, EPI_ISL_1081710, EPI_ISL_1081711, EPI_ISL_1081712, EPI_ISL_1081713, EPI_ISL_1081714, EPI_ISL_1081715, EPI_ISL_1081716, EPI_ISL_1081717, EPI_ISL_1081718, EPI_ISL_1081719, EPI_ISL_1081720, EPI_ISL_1081721, EPI_ISL_1081722, EPI_ISL_1081723, EPI_ISL_1081724, EPI_ISL_1081725, EPI_ISL_1081726, EPI_ISL_1081727, EPI_ISL_1081728, EPI_ISL_1081729, EPI_ISL_1081730, EPI_ISL_1081731, EPI_ISL_1081732, EPI_ISL_1081733, EPI_ISL_1081734, EPI_ISL_1081735, EPI_ISL_1081736, EPI_ISL_1081737, EPI_ISL_1081738, EPI_ISL_1081739, EPI_ISL_1081740, EPI_ISL_1081741, EPI_ISL_1185854, EPI_ISL_1185855, EPI_ISL_1185856, EPI_ISL_1185857, EPI_ISL_1185858, EPI_ISL_1185859, EPI_ISL_1185860, EPI_ISL_1185861, EPI_ISL_1185862, EPI_ISL_1185863, EPI_ISL_1185864, EPI_ISL_1185865, EPI_ISL_1185866, EPI_ISL_1185867, EPI_ISL_1185868, EPI_ISL_1185869, EPI_ISL_1185870, EPI_ISL_1185871, EPI_ISL_1185872, EPI_ISL_1185873, EPI_ISL_1185874, EPI_ISL_1185875, EPI_ISL_1185876, EPI_ISL_1185877, EPI_ISL_1185878, EPI_ISL_1185879, EPI_ISL_1185880, EPI_ISL_1185881, EPI_ISL_1185882, EPI_ISL_1185883, EPI_ISL_1185884, EPI_ISL_1185885, EPI_ISL_1185886, EPI_ISL_1185887, EPI_ISL_1185888, EPI_ISL_1185889, EPI_ISL_1185890, EPI_ISL_1366282, EPI_ISL_1366285, EPI_ISL_1366320, EPI_ISL_1366379, EPI_ISL_1366380, EPI_ISL_1366381, EPI_ISL_1366382, EPI_ISL_1366383, EPI_ISL_1366384, EPI_ISL_1366385, EPI_ISL_1366386, EPI_ISL_1366387, EPI_ISL_1366388, EPI_ISL_1366389, EPI_ISL_1366390, EPI_ISL_1366391, EPI_ISL_1366392, EPI_ISL_1366393, EPI_ISL_1366394, EPI_ISL_1366395, EPI_ISL_1366396, EPI_ISL_1366397, EPI_ISL_1366398, EPI_ISL_1366399, EPI_ISL_1366400, EPI_ISL_1366401, EPI_ISL_1366402, EPI_ISL_1366403, EPI_ISL_1366404, EPI_ISL_1366405, EPI_ISL_1366406, EPI_ISL_1366407, EPI_ISL_1366408, EPI_ISL_1366409 | | | |
| see above | Scripps Medical Laboratory | Andersen lab at Scripps Research | Ellen Stefanski; Ian Mchardy; SEARCH Alliance San Diego with Michael Quigley |
| EPI_ISL_1081373, EPI_ISL_1081374, EPI_ISL_1081375, EPI_ISL_1081376, EPI_ISL_1081377, EPI_ISL_1081378, EPI_ISL_1081379, EPI_ISL_1081380, EPI_ISL_1081381, EPI_ISL_1081382, EPI_ISL_1081383, EPI_ISL_1081384, EPI_ISL_1081385, EPI_ISL_1081386, EPI_ISL_1081387, EPI_ISL_1081388, EPI_ISL_1081389, EPI_ISL_1081390, EPI_ISL_1081391, EPI_ISL_1081392, EPI_ISL_1081393, EPI_ISL_1081394, EPI_ISL_1081395, EPI_ISL_1081396, EPI_ISL_1081397, EPI_ISL_1081398, EPI_ISL_1081399, EPI_ISL_1081400, EPI_ISL_1081401, EPI_ISL_1081402, EPI_ISL_1081403, EPI_ISL_1081404, EPI_ISL_1081405, EPI_ISL_1081406, EPI_ISL_1081407, EPI_ISL_1081408, EPI_ISL_1081409, EPI_ISL_1081410, EPI_ISL_1081411, EPI_ISL_1081412, EPI_ISL_1081413, EPI_ISL_1081414, EPI_ISL_1081415, EPI_ISL_1081417, EPI_ISL_1081418, EPI_ISL_1081419, EPI_ISL_1081420, EPI_ISL_1081421, EPI_ISL_1081422, EPI_ISL_1081496, EPI_ISL_1081497, EPI_ISL_1081498, EPI_ISL_1081499, EPI_ISL_1081500, EPI_ISL_1081501, EPI_ISL_1081502, EPI_ISL_1081503, EPI_ISL_1081504, EPI_ISL_1081505, EPI_ISL_1081507, EPI_ISL_1081508, EPI_ISL_1081509, EPI_ISL_1081510, EPI_ISL_1081512, EPI_ISL_1081513, EPI_ISL_1081514, EPI_ISL_1081515, EPI_ISL_1081516, EPI_ISL_1081517, EPI_ISL_1081518, EPI_ISL_1081519, EPI_ISL_1081520, EPI_ISL_1081521, EPI_ISL_1081523, EPI_ISL_1081524, EPI_ISL_1081525, EPI_ISL_1081526, EPI_ISL_1081527, EPI_ISL_1081528, EPI_ISL_1081529, EPI_ISL_1081530, EPI_ISL_1081531, EPI_ISL_1081532, EPI_ISL_1081533, EPI_ISL_1081534, EPI_ISL_1081535, EPI_ISL_1081536, EPI_ISL_1081537, EPI_ISL_1081538, EPI_ISL_1081539, EPI_ISL_1081541, EPI_ISL_1081542, EPI_ISL_1081543, EPI_ISL_1081545, EPI_ISL_1081546, EPI_ISL_1081547, EPI_ISL_1081548, EPI_ISL_1081549, EPI_ISL_1185891, EPI_ISL_1185892, EPI_ISL_1185893, EPI_ISL_1185894, EPI_ISL_1185895, EPI_ISL_1185896, EPI_ISL_1185897, EPI_ISL_1185898, EPI_ISL_1185899, EPI_ISL_1185900, EPI_ISL_1185901, EPI_ISL_1185902, EPI_ISL_1185903, EPI_ISL_1185904, EPI_ISL_1185905, EPI_ISL_1185906, EPI_ISL_1185907, EPI_ISL_1185908, EPI_ISL_1185909, EPI_ISL_1185910, EPI_ISL_1185911, EPI_ISL_1185912, EPI_ISL_1185913, EPI_ISL_1185914, EPI_ISL_1185915, EPI_ISL_1185916, EPI_ISL_1185917, EPI_ISL_1185918, EPI_ISL_1185919, EPI_ISL_1185920, EPI_ISL_1185921, EPI_ISL_1185922, EPI_ISL_1185923, EPI_ISL_1185924, EPI_ISL_1185925, EPI_ISL_1185926, EPI_ISL_1185927, EPI_ISL_1185928, EPI_ISL_1185930, EPI_ISL_1185932, EPI_ISL_1185933, EPI_ISL_1185937, EPI_ISL_1185938, EPI_ISL_1185939, EPI_ISL_1185940, EPI_ISL_1185941, EPI_ISL_1185942, EPI_ISL_1185943, EPI_ISL_1185944, EPI_ISL_1185945, EPI_ISL_1185946, EPI_ISL_1185947, EPI_ISL_1185948, EPI_ISL_1185949, EPI_ISL_1185950, EPI_ISL_1185951, EPI_ISL_1185952, EPI_ISL_1185953, EPI_ISL_1185954, EPI_ISL_1185955, EPI_ISL_1185956, EPI_ISL_1185957, EPI_ISL_1185958, EPI_ISL_1185959, EPI_ISL_1295819, EPI_ISL_1295823, EPI_ISL_1295824, EPI_ISL_1295825, EPI_ISL_1295826, EPI_ISL_1295827, EPI_ISL_1295828, EPI_ISL_1295829, EPI_ISL_1295830, EPI_ISL_1295831, EPI_ISL_1295832, EPI_ISL_1295833, EPI_ISL_1295834, EPI_ISL_1295835, EPI_ISL_1295836, EPI_ISL_1295838, EPI_ISL_1295839, EPI_ISL_1295840, EPI_ISL_1295841, EPI_ISL_1295842, EPI_ISL_1295844, EPI_ISL_1295845, EPI_ISL_1295846, EPI_ISL_1295847, EPI_ISL_1295848, EPI_ISL_1295849, EPI_ISL_1295850, EPI_ISL_1295851, EPI_ISL_1295852, EPI_ISL_1295853, EPI_ISL_1295854, EPI_ISL_1295855, EPI_ISL_1295856, EPI_ISL_1295857, EPI_ISL_1295858, EPI_ISL_1295859, EPI_ISL_1295860, EPI_ISL_1295861, EPI_ISL_1295862, EPI_ISL_1295863, EPI_ISL_1295864, EPI_ISL_1295865, EPI_ISL_1295866, EPI_ISL_1295867, EPI_ISL_1295868, EPI_ISL_1295869, EPI_ISL_1295870, EPI_ISL_1295871, EPI_ISL_1295872, EPI_ISL_1295873, EPI_ISL_1295874, EPI_ISL_1295875, EPI_ISL_1295876, EPI_ISL_1295877, EPI_ISL_1295878, EPI_ISL_1295879, EPI_ISL_1295880, EPI_ISL_1295881, EPI_ISL_1295882, EPI_ISL_1295883, EPI_ISL_1295884, EPI_ISL_1295885, EPI_ISL_1295886, EPI_ISL_1295887, EPI_ISL_1295888, EPI_ISL_1295889, EPI_ISL_1295890, EPI_ISL_1295891, EPI_ISL_1295892, EPI_ISL_1295893, EPI_ISL_1295894, EPI_ISL_1295895, EPI_ISL_1295896, EPI_ISL_1295897, EPI_ISL_1295898, EPI_ISL_1295899, EPI_ISL_1295900, EPI_ISL_1366280, EPI_ISL_1366293, EPI_ISL_1366294, EPI_ISL_1366295, EPI_ISL_1366296, EPI_ISL_1366316, EPI_ISL_1366324, EPI_ISL_1366335, EPI_ISL_1366336, EPI_ISL_1366337, EPI_ISL_1366338, EPI_ISL_1366339, EPI_ISL_1366340, EPI_ISL_1366341, EPI_ISL_1366342, EPI_ISL_1366343, EPI_ISL_1366344, EPI_ISL_1366345, EPI_ISL_1366346, EPI_ISL_1366347, EPI_ISL_1366348, EPI_ISL_1366349, EPI_ISL_1366350, EPI_ISL_1366351, EPI_ISL_1366352, EPI_ISL_1366353, EPI_ISL_1366354, EPI_ISL_1366355, EPI_ISL_1366356, EPI_ISL_1366357, EPI_ISL_1366358, EPI_ISL_1366359, EPI_ISL_1366360, EPI_ISL_1366361, EPI_ISL_1366362, EPI_ISL_1366363, EPI_ISL_1366364, EPI_ISL_1366365, EPI_ISL_1366366, EPI_ISL_1366367, EPI_ISL_1366368, EPI_ISL_1366369, EPI_ISL_1366370, EPI_ISL_1366371, EPI_ISL_1366372, EPI_ISL_1366373, EPI_ISL_1366374, EPI_ISL_1366375, EPI_ISL_1366376, EPI_ISL_2628545, EPI_ISL_2628546, EPI_ISL_2628547, EPI_ISL_2712451, EPI_ISL_2712452, EPI_ISL_2712453, EPI_ISL_2712454, EPI_ISL_2712455, EPI_ISL_2712457, EPI_ISL_2712458, EPI_ISL_2712459, EPI_ISL_2712460, EPI_ISL_2712463, EPI_ISL_2712471, EPI_ISL_2712491, EPI_ISL_2835511, EPI_ISL_2835512, EPI_ISL_2835513, EPI_ISL_2835514, EPI_ISL_2835515, EPI_ISL_2835516, EPI_ISL_2835517, EPI_ISL_2835518, EPI_ISL_2835519, EPI_ISL_2835520, EPI_ISL_2835521, EPI_ISL_2835522, EPI_ISL_2835523, EPI_ISL_2835524, EPI_ISL_2835525, EPI_ISL_2835526, EPI_ISL_2835527, EPI_ISL_2835528, EPI_ISL_2835529, EPI_ISL_2835530, EPI_ISL_2835531, EPI_ISL_2835532, EPI_ISL_2835533, EPI_ISL_2835534, EPI_ISL_2835535, EPI_ISL_2835536, EPI_ISL_2835537, EPI_ISL_2835538, EPI_ISL_2835539, EPI_ISL_2835541, EPI_ISL_2835542, EPI_ISL_2835543, EPI_ISL_2835546, EPI_ISL_2835547, EPI_ISL_2835548, EPI_ISL_2835549, EPI_ISL_2835550, EPI_ISL_2835551, EPI_ISL_2920284, EPI_ISL_2920285, EPI_ISL_2920286, EPI_ISL_2920287, EPI_ISL_2920288, EPI_ISL_2920289, EPI_ISL_2920290, EPI_ISL_2920291, EPI_ISL_2920292, EPI_ISL_2920293, EPI_ISL_2920294, EPI_ISL_2920295, EPI_ISL_2920296, EPI_ISL_2920297, EPI_ISL_2920298, EPI_ISL_2920299, EPI_ISL_2920300, EPI_ISL_2920302, EPI_ISL_2920303, EPI_ISL_2920304, EPI_ISL_2920305, EPI_ISL_2920306, EPI_ISL_2920307, EPI_ISL_2920308, EPI_ISL_2920309, EPI_ISL_2920310, EPI_ISL_2920311, EPI_ISL_2920312, EPI_ISL_2920313, EPI_ISL_2920314, EPI_ISL_2920315, EPI_ISL_2920316, EPI_ISL_2920317, EPI_ISL_2920318, EPI_ISL_2920319, EPI_ISL_2920320, EPI_ISL_2920321, EPI_ISL_2920322, EPI_ISL_2920323, EPI_ISL_2920324, EPI_ISL_2920325, EPI_ISL_2920326, EPI_ISL_2920327, EPI_ISL_2920328, EPI_ISL_2920329, EPI_ISL_2920330, EPI_ISL_2920331, EPI_ISL_2920332, EPI_ISL_2920398, EPI_ISL_2920399, EPI_ISL_2920400, EPI_ISL_2920401, EPI_ISL_2920402, EPI_ISL_2920403, EPI_ISL_2920404, EPI_ISL_2920407, EPI_ISL_2920410, EPI_ISL_2920412, EPI_ISL_2920413, EPI_ISL_2920414, EPI_ISL_2920415, EPI_ISL_2920416, EPI_ISL_2920417, EPI_ISL_2920418, EPI_ISL_2920419, EPI_ISL_2920420, EPI_ISL_2920421, EPI_ISL_2920422, EPI_ISL_2920423, EPI_ISL_2920424, EPI_ISL_2920425, EPI_ISL_2920426, EPI_ISL_2920427, EPI_ISL_2920428, EPI_ISL_2920429, EPI_ISL_2920430, EPI_ISL_2920431, EPI_ISL_2920444, EPI_ISL_2920445, EPI_ISL_2920446, EPI_ISL_2920447, EPI_ISL_2920448, EPI_ISL_2920449, EPI_ISL_2920450, EPI_ISL_2920451, EPI_ISL_2920452, EPI_ISL_2920453, EPI_ISL_2920454, EPI_ISL_2988805, EPI_ISL_2988806, EPI_ISL_2988807, EPI_ISL_2988808, EPI_ISL_2988809, EPI_ISL_2988810, EPI_ISL_2988811, EPI_ISL_2988812, EPI_ISL_2988813, EPI_ISL_2988814, EPI_ISL_2988815, EPI_ISL_2988816, EPI_ISL_2988817, EPI_ISL_2988818, EPI_ISL_2988819, EPI_ISL_2988820, EPI_ISL_2988821, EPI_ISL_2988822, EPI_ISL_2988823, EPI_ISL_2988824, EPI_ISL_2988825, EPI_ISL_2988826, EPI_ISL_2988827, EPI_ISL_2988828, EPI_ISL_2988829, EPI_ISL_2988830, EPI_ISL_2988831, EPI_ISL_2988832, EPI_ISL_2988833, EPI_ISL_2988834, EPI_ISL_2988835, EPI_ISL_2988837, EPI_ISL_2988838, EPI_ISL_2988839 | | | |
| see above | Sharp HealthCare Laboratory | Andersen lab at Scripps Research | Art Mendoza; Cathy Woerle; Jacquelyn Berumen; Liam McGinnis; Omid Bakhtar; SEARCH Alliance San Diego with Aaron Harding |
| EPI_ISL_1483795 | Stanford Clinical Virology Laboratory | Santa Clara County Public Health Laboratory | Santa Clara County Public Health Department |
| EPI_ISL_1335871 | Stanford Health Care | Clinical Virology Laboratory, Stanford University School or Medicine | ChunHong Huang; Daniel Solis; Fumiko Yamamoto; James Zehnder; Malaya K. Sahoo; Mamdouh Sibai; Michelle Vergese; and Benjamin A. Pinsky |
| EPI_ISL_1364506, EPI_ISL_1364507, EPI_ISL_1364509, EPI_ISL_1364510, EPI_ISL_1364511, EPI_ISL_1364512, EPI_ISL_1364513, EPI_ISL_1364514, EPI_ISL_1364515, EPI_ISL_1364516, EPI_ISL_1364518, EPI_ISL_1364519, EPI_ISL_1364520, EPI_ISL_1379778, EPI_ISL_1379780, EPI_ISL_1379781, EPI_ISL_1379782, EPI_ISL_1379784, EPI_ISL_1379789, EPI_ISL_1379790, EPI_ISL_1379791, EPI_ISL_1379792, EPI_ISL_1379793, EPI_ISL_1379796, EPI_ISL_1379797, EPI_ISL_1379798, EPI_ISL_1379799, EPI_ISL_1379800, EPI_ISL_1379801, EPI_ISL_1379802, EPI_ISL_1379803, EPI_ISL_1379804, EPI_ISL_1379807, EPI_ISL_1379809, EPI_ISL_1379810, EPI_ISL_1379811, EPI_ISL_1379812, EPI_ISL_1379815, EPI_ISL_1379816, EPI_ISL_1379817, EPI_ISL_1379820, EPI_ISL_1379821, EPI_ISL_1379822, EPI_ISL_1379823, EPI_ISL_1379826, EPI_ISL_1379827, EPI_ISL_1379831, EPI_ISL_1379835, EPI_ISL_1379836, EPI_ISL_1379847, EPI_ISL_1379848, EPI_ISL_1379853, EPI_ISL_1379857, EPI_ISL_1379858, EPI_ISL_1379859, EPI_ISL_1379862, EPI_ISL_1379863, EPI_ISL_1379865, EPI_ISL_1379866, EPI_ISL_1379869, EPI_ISL_1379870, EPI_ISL_1379874, EPI_ISL_1379877, EPI_ISL_1379882, EPI_ISL_1379883, EPI_ISL_1379884, EPI_ISL_1379885, EPI_ISL_1379886, EPI_ISL_1379887, EPI_ISL_1424066, EPI_ISL_1424081, EPI_ISL_1424083, EPI_ISL_1424085, EPI_ISL_1424086, EPI_ISL_1424087, EPI_ISL_1424090, EPI_ISL_1424091, EPI_ISL_1424098, EPI_ISL_1424099, EPI_ISL_1424103, EPI_ISL_1424105, EPI_ISL_1424107, EPI_ISL_1424108, EPI_ISL_1424109, EPI_ISL_1424110, EPI_ISL_1424111, EPI_ISL_1424112, EPI_ISL_1424113, EPI_ISL_1424114, EPI_ISL_1424115, EPI_ISL_1424126, EPI_ISL_1424127, EPI_ISL_2009049, EPI_ISL_2009054, EPI_ISL_2009056, EPI_ISL_2009057, EPI_ISL_2027140, EPI_ISL_2027147, EPI_ISL_2027153, EPI_ISL_2027157, EPI_ISL_2027160, EPI_ISL_2027164, EPI_ISL_2027202, EPI_ISL_2027205, EPI_ISL_2027208, EPI_ISL_2027212, EPI_ISL_2027215, EPI_ISL_2027218, EPI_ISL_2027221, EPI_ISL_2027225, EPI_ISL_2027228, EPI_ISL_3050267, EPI_ISL_3050284, EPI_ISL_3050287, EPI_ISL_3050288, EPI_ISL_3050290 | | | |
| see above | Stanford Health Care | Stanford University School of Medicine, Clinical Virology Laboratory | Becky Jiang; Bernadette Troung; ChunHong Huang; Daniel Solis; Fiona Yamamoto; Fumiko Yamamoto; James Zehnder; Malaya K. Sahoo; Mamdouh Sibai; Michelle Vergese; Michelle Verghese; Nathan Hammond; Selamawit Bihon; and Benjamin A. Pinsky |
| EPI_ISL_2171674, EPI_ISL_2324440, EPI_ISL_2324443, EPI_ISL_2341049, EPI_ISL_2341053, EPI_ISL_2341056, EPI_ISL_2341074, EPI_ISL_2341075, EPI_ISL_2341076, EPI_ISL_2341078, EPI_ISL_2341079, EPI_ISL_2341080, EPI_ISL_2341087, EPI_ISL_2341144, EPI_ISL_2341145, EPI_ISL_2341146, EPI_ISL_2341147, EPI_ISL_2341148, EPI_ISL_2341149, EPI_ISL_2341155, EPI_ISL_2341156, EPI_ISL_2341157, EPI_ISL_2341158, EPI_ISL_2341159, EPI_ISL_2341160, EPI_ISL_2341161, EPI_ISL_2341162, EPI_ISL_2341163, EPI_ISL_2341164, EPI_ISL_2341165, EPI_ISL_2341166, EPI_ISL_2341167, EPI_ISL_2341168, EPI_ISL_2341169, EPI_ISL_2341170, EPI_ISL_2341171, EPI_ISL_2341172, EPI_ISL_2341173, EPI_ISL_2341174, EPI_ISL_2341177, EPI_ISL_2341178, EPI_ISL_2341253, EPI_ISL_2341254, EPI_ISL_2341255, EPI_ISL_2341256, EPI_ISL_2341257, EPI_ISL_2341258, EPI_ISL_2341268, EPI_ISL_2341269, EPI_ISL_2341270, EPI_ISL_2341271, EPI_ISL_2341272, EPI_ISL_2341273, EPI_ISL_2341274, EPI_ISL_2341275, EPI_ISL_2341276, EPI_ISL_2341277, EPI_ISL_2341278, EPI_ISL_2341279, EPI_ISL_2341280, EPI_ISL_2341281, EPI_ISL_2341282, EPI_ISL_2341283, EPI_ISL_2341284, EPI_ISL_2341285, EPI_ISL_2341286, EPI_ISL_2341287, EPI_ISL_2341288, EPI_ISL_2341289, EPI_ISL_2341290, EPI_ISL_2341291, EPI_ISL_2341292, EPI_ISL_2341293, EPI_ISL_2341294, EPI_ISL_2341295, EPI_ISL_2341296, EPI_ISL_2341297, EPI_ISL_2341298, EPI_ISL_2341299, EPI_ISL_2341300, EPI_ISL_2341301, EPI_ISL_2341302, EPI_ISL_2341303, EPI_ISL_2341304, EPI_ISL_2341305, EPI_ISL_2341306, EPI_ISL_2341307, EPI_ISL_3049199, | | | |

| | | | |
|---|---|---|---|
| EPI_ISL_3049203, EPI_ISL_3049204, EPI_ISL_3049205 | | | |
| see above | Tulare County Public Health Laboratory | Tulare County Public Health Laboratory | Denise L Lopez; Joshua Kootstra |
| EPI_ISL_1009813, EPI_ISL_1009827 | UC Davis Genome Center and Healthy Davis Together | UC Davis Genome Center | Sam Diaz-Muñoz and AJ Campbell |
| EPI_ISL_962505, EPI_ISL_962513, EPI_ISL_962833, EPI_ISL_962834, EPI_ISL_962835, EPI_ISL_962836, EPI_ISL_962837, EPI_ISL_962838, EPI_ISL_962839, EPI_ISL_962840, EPI_ISL_962841, EPI_ISL_962842, EPI_ISL_962843, EPI_ISL_962844, EPI_ISL_962845, EPI_ISL_962846, EPI_ISL_962879, EPI_ISL_1005511, EPI_ISL_1005512, EPI_ISL_1005513, EPI_ISL_1005514, EPI_ISL_1005515, EPI_ISL_1005516, EPI_ISL_1005517, EPI_ISL_1005518, EPI_ISL_1005519, EPI_ISL_1005520, EPI_ISL_1005521, EPI_ISL_1005542, EPI_ISL_1005543, EPI_ISL_1005544, EPI_ISL_1005545, EPI_ISL_1005546, EPI_ISL_1005547, EPI_ISL_1005548, EPI_ISL_1005549, EPI_ISL_1005550, EPI_ISL_1005551, EPI_ISL_1005552, EPI_ISL_1005553, EPI_ISL_1005554, EPI_ISL_1098629, EPI_ISL_1191129, EPI_ISL_1191130, EPI_ISL_1706604 | | | |
| see above | UCLA Clinical Micro Lab | Los Angeles County Public Health Laboratories | P. Hemarajata et al. |
| EPI_ISL_1405642, EPI_ISL_1405643, EPI_ISL_1405644, EPI_ISL_1405645, EPI_ISL_1405646, EPI_ISL_1405647, EPI_ISL_1405648, EPI_ISL_1405649, EPI_ISL_1405650, EPI_ISL_1405651, EPI_ISL_1405652, EPI_ISL_1405653, EPI_ISL_1405654, EPI_ISL_1405655, EPI_ISL_1405656, EPI_ISL_1405657, EPI_ISL_1405658, EPI_ISL_1405659, EPI_ISL_1405660, EPI_ISL_1405661, EPI_ISL_1405662, EPI_ISL_1405663, EPI_ISL_1405664, EPI_ISL_1405665, EPI_ISL_1405666, EPI_ISL_1405667, EPI_ISL_1405668, EPI_ISL_1405669, EPI_ISL_1405670, EPI_ISL_1405671, EPI_ISL_2494675, EPI_ISL_2494676, EPI_ISL_2494677, EPI_ISL_2494681, EPI_ISL_2494682, EPI_ISL_2494683, EPI_ISL_2494684, EPI_ISL_2494685, EPI_ISL_2494686, EPI_ISL_2494687, EPI_ISL_2494688, EPI_ISL_2494689, EPI_ISL_2494692, EPI_ISL_2494693, EPI_ISL_2494694, EPI_ISL_2494695, EPI_ISL_2494696, EPI_ISL_2494697, EPI_ISL_2494698 | | | |
| see above | UCSC Genomics Institute | UCSC Genomics Institute | A. Marm Kilpatrick; Angie Hinrichs; Beth Shapiro; Bryan Thornlow; Ciara Wanket; David Haussler; Eric Beraut; Hugh Olsen; Ikenna Anigbogu; Isabel Bjork; Jakob McBroome; Jeremy Sanford; Joshua Kapp; Mark Akeson; Maximilian Haeussler; Michael Stone; Miten Jain; Molly Cassatt-Johnstone; Namrita Dhillon; Russell Corbett-Detig; Terren Chang; Yatish Turakhia |
| EPI_ISL_1005495, EPI_ISL_1005496, EPI_ISL_1005497, EPI_ISL_1005498, EPI_ISL_1005499, EPI_ISL_1005500, EPI_ISL_1005501, EPI_ISL_1005502, EPI_ISL_1005503, EPI_ISL_1005504, EPI_ISL_1005505, EPI_ISL_1005506, EPI_ISL_1005507, EPI_ISL_1005522, EPI_ISL_1005523, EPI_ISL_1005524, EPI_ISL_1005525, EPI_ISL_1005526, EPI_ISL_1005527, EPI_ISL_1005528, EPI_ISL_1005529, EPI_ISL_1005530, EPI_ISL_1005531, EPI_ISL_1005532, EPI_ISL_1005533, EPI_ISL_1005534, EPI_ISL_1005535, EPI_ISL_1508930 | | | |
| see above | USC Clinical Lab | Los Angeles County Public Health Laboratories | P. Hemarajata et al. |
| EPI_ISL_962517, EPI_ISL_962518, EPI_ISL_962519, EPI_ISL_962520 | VA Medical Center | Los Angeles County Public Health Laboratories | P. Hemarajata et al. |
| EPI_ISL_2295363, EPI_ISL_2295364, EPI_ISL_2295365, EPI_ISL_2295366, EPI_ISL_2295367, EPI_ISL_2295368, EPI_ISL_2295369, EPI_ISL_2295370, EPI_ISL_2295371, EPI_ISL_2295372, EPI_ISL_2295373, EPI_ISL_2295374, EPI_ISL_2295375 | | | |
| see above | WLA VAMC | Genomics and Discovery, Respiratory Viruses Branch, Division of Viral Diseases, Centers for Disease Control and Prevention | Adam Retchless; Anna Kelleher; Anna Uehara; Brian Lynch; Clinton R. Paden; Dhwani Batra; Haibin Wang; Han Jia Justin Ng; Jasmine Padilla; Jing Zhang; Justin Lee; Krista Queen; Mark Burroughs; Mili Sheth; Morgan Davis; Peter Cook; Rachel Marine; Sarah Nobles; Suxiang Tong; Tara Coalter; Yan Li; Ying Tao |

We gratefully acknowledge the following Authors from the Originating laboratories responsible for obtaining the specimens, as well as the Submitting laboratories where the genome data were generated and shared via GISAID, on which this research is based.

All Submitters of data may be contacted directly via www.gisaid.org

Authors are sorted alphabetically.

| Accession ID | Originating Laboratory | Submitting Laboratory | Authors |
|---|---|---|---|
| EPI_ISL_1318635, EPI_ISL_1318637, EPI_ISL_1318638, EPI_ISL_1318643, EPI_ISL_1318645, EPI_ISL_1318646, EPI_ISL_1318647, EPI_ISL_1318649, EPI_ISL_1318651, EPI_ISL_1318653, EPI_ISL_1318654, EPI_ISL_1318659, EPI_ISL_1318660, EPI_ISL_1318663, EPI_ISL_1318664, EPI_ISL_1318665, EPI_ISL_1446781, EPI_ISL_1446784, EPI_ISL_1446791, EPI_ISL_1446794 | | | |
| see above | "CDPH, Viral and Rickettsial Disease Laboratory" | Centers for Disease Control and Prevention Division of Viral Diseases, Pathogen Discovery | Alison Laufer Halpin; Anna Montmayeur; Anna Uehara; Ben L. Rambo-Martin; Clinton R. Paden; Dakota Howard; Darlene Wagner; Dave Wentworth; Dhwani Batra; Haibin Wang; Jasmine Padilla; Jing Zhang; Justin Lee; Katie Dillon; Krista Queen; Kristen Knipe; Kristine Lacek; Lori Rowe; Mark Burroughs; Matthew Schmerer; Mili Sheth; Peter Cook; Peter W. Cook; Rachel Marine; Sam Shepard; Sarah Nobles; Shoshona Le; Suxiang Tong; Vivien Dugan; Yan Li; Ying Tao; Yvette Unoarumhi |
| EPI_ISL_1226241, EPI_ISL_1226242, EPI_ISL_1226243, EPI_ISL_1226244, EPI_ISL_1226245, EPI_ISL_1226247, EPI_ISL_1226248, EPI_ISL_1226249, EPI_ISL_1226250, EPI_ISL_1226253, EPI_ISL_1226254, EPI_ISL_1226258, EPI_ISL_1226259, EPI_ISL_1226260, EPI_ISL_1226261, EPI_ISL_1226262, EPI_ISL_1226263, EPI_ISL_1226264, EPI_ISL_1226265, EPI_ISL_1226270, EPI_ISL_1226273 | | | |
| see above | "CDPH, Viral and Rickettsial Disease Laboratory" | Genomics and Discovery, Respiratory Viruses Branch, Division of Viral Diseases, Centers for Disease Control and Prevention | Anna Montmayeur; Anna Uehara; Ben L. Rambo-Martin; Clinton R. Paden; Dhwani Batra; Haibin Wang; Jasmine Padilla; Jing Zhang; Justin Lee; Katie Dillon; Krista Queen; Kristen Knipe; Kristine Lacek; Lori Rowe; Mark Burroughs; Matthew Schmerer; Mili Sheth; Peter W. Cook; Rachel Marine; Sam Shepard; Sarah Nobles; Shoshona Le; Suxiang Tong; Yan Li; Ying Tao |
| EPI_ISL_1093546, EPI_ISL_1093552, EPI_ISL_1093557, EPI_ISL_1093558, EPI_ISL_1093559, EPI_ISL_1093560, EPI_ISL_1093561, EPI_ISL_1093562, EPI_ISL_1093593, EPI_ISL_1093594, EPI_ISL_1093599, EPI_ISL_1093600, EPI_ISL_1093601, EPI_ISL_1196775, EPI_ISL_1196776, EPI_ISL_1196777, EPI_ISL_1196778, EPI_ISL_1196779, EPI_ISL_1196780, EPI_ISL_1196781, EPI_ISL_1196782, EPI_ISL_1196783, EPI_ISL_1196784, EPI_ISL_1196785, EPI_ISL_1196786, EPI_ISL_1196787, EPI_ISL_1196788, EPI_ISL_1196789, EPI_ISL_1196790, EPI_ISL_1196791, EPI_ISL_1205472, EPI_ISL_1205475, EPI_ISL_1205476, EPI_ISL_1205477, EPI_ISL_1205480, EPI_ISL_1205481, EPI_ISL_1205482 | | | |
| see above | Alameda County Public Health Department | Alameda County Public Health Department | Kevin Libuit; Kristna Hsieh; Maria Elma |
| EPI_ISL_1364729 | Arizona State University | Arizona State University | Efrem S. Lim; Emily A. Kaelin; Joshua LaBaer; Joy M. Blain; Kristina Buss; LaRinda A. Holland; Neal W Woodbury; Nicholas J. Mellor; Peter T. Skidmore; Rabia Maqsood; Valerie Harris; Vel Murugan |
| EPI_ISL_1235360, EPI_ISL_1235367, EPI_ISL_1235372, EPI_ISL_1235373, EPI_ISL_1235375, EPI_ISL_1235377, EPI_ISL_1235378, EPI_ISL_1235381, EPI_ISL_1235383, EPI_ISL_1235384, EPI_ISL_1235400, EPI_ISL_1235401, EPI_ISL_1235403, EPI_ISL_1235410, EPI_ISL_1235416, EPI_ISL_1235418, EPI_ISL_1235419, EPI_ISL_1235424, EPI_ISL_1235427, EPI_ISL_1235434, EPI_ISL_1235435, EPI_ISL_1235438, EPI_ISL_1235441, EPI_ISL_1235442, EPI_ISL_1235450, EPI_ISL_1235451, EPI_ISL_1235455, EPI_ISL_1235468, EPI_ISL_1235469, EPI_ISL_1235472, EPI_ISL_1235473, EPI_ISL_1235478, EPI_ISL_1235488, EPI_ISL_1235490, EPI_ISL_1235496, EPI_ISL_1235498, EPI_ISL_1235501, EPI_ISL_1235502, EPI_ISL_1235506, EPI_ISL_1235507, EPI_ISL_1235509, EPI_ISL_1235511, EPI_ISL_1235515, EPI_ISL_1235521, EPI_ISL_1235522, EPI_ISL_1235525, EPI_ISL_1235526, EPI_ISL_1235528, EPI_ISL_1235539, EPI_ISL_1235540, EPI_ISL_1235544, EPI_ISL_1235545, EPI_ISL_1235546, EPI_ISL_1235547, EPI_ISL_1235549, EPI_ISL_1235551, EPI_ISL_1235558, EPI_ISL_1235570, EPI_ISL_1235572, EPI_ISL_1235578, EPI_ISL_1235582, EPI_ISL_1235588, EPI_ISL_1235591, EPI_ISL_1235596, EPI_ISL_1235600, EPI_ISL_1235602, EPI_ISL_1235604, EPI_ISL_1235606, EPI_ISL_1235608, EPI_ISL_1477167, EPI_ISL_1477168, EPI_ISL_1477171, EPI_ISL_1477177, EPI_ISL_1477179, EPI_ISL_1477181, EPI_ISL_1477182, EPI_ISL_1477184, EPI_ISL_1477185, EPI_ISL_1477186, EPI_ISL_1477187, EPI_ISL_1477188, EPI_ISL_1477190, EPI_ISL_1477191, EPI_ISL_1477192, EPI_ISL_1477193, EPI_ISL_1477194, EPI_ISL_1477196, EPI_ISL_1477197, EPI_ISL_1477198, EPI_ISL_1477200, EPI_ISL_1477201, EPI_ISL_1477202, EPI_ISL_1477203, EPI_ISL_1477204, EPI_ISL_1477205, EPI_ISL_1477209, EPI_ISL_1477210, EPI_ISL_1477211, EPI_ISL_1477212, EPI_ISL_1477213, EPI_ISL_1477214, EPI_ISL_1477215, EPI_ISL_1477217, EPI_ISL_1477219, EPI_ISL_1477221, EPI_ISL_1477222, EPI_ISL_1477223, EPI_ISL_1477225, EPI_ISL_1477229, EPI_ISL_1477230, EPI_ISL_1477231, EPI_ISL_1477232, EPI_ISL_1477233, EPI_ISL_1477235, EPI_ISL_1477237, EPI_ISL_1477238, EPI_ISL_1477240, EPI_ISL_1477241, EPI_ISL_1477242, EPI_ISL_1477245, EPI_ISL_1477246, EPI_ISL_1477248, EPI_ISL_1477250, EPI_ISL_1477253, EPI_ISL_1477254, EPI_ISL_1477256, EPI_ISL_1477258, EPI_ISL_1477261, EPI_ISL_1477263, EPI_ISL_1477265, EPI_ISL_1477266, EPI_ISL_1477267, EPI_ISL_1477273, EPI_ISL_1477274, EPI_ISL_1477275, EPI_ISL_1477278, EPI_ISL_1477281, EPI_ISL_1477283, EPI_ISL_1477285, EPI_ISL_1477286, EPI_ISL_1477289, EPI_ISL_1477291, EPI_ISL_1477297, EPI_ISL_1477298, EPI_ISL_1477300, EPI_ISL_1477302, EPI_ISL_1477303, EPI_ISL_1477304, EPI_ISL_1477305, EPI_ISL_1477307, EPI_ISL_1477310, EPI_ISL_1477311, EPI_ISL_1477314, EPI_ISL_1477315, EPI_ISL_1477316, EPI_ISL_1477317, EPI_ISL_1477319, EPI_ISL_1477320, EPI_ISL_1477321, EPI_ISL_1477323, EPI_ISL_1477324, EPI_ISL_1477327, EPI_ISL_1477330, EPI_ISL_1477331, EPI_ISL_1477333, EPI_ISL_1477336, EPI_ISL_1477338, EPI_ISL_1477342, EPI_ISL_1477344, EPI_ISL_1477345, EPI_ISL_1477347, EPI_ISL_1477349, EPI_ISL_1477350, EPI_ISL_1477351, EPI_ISL_1477353, EPI_ISL_1477356, EPI_ISL_1477357, EPI_ISL_1477358, EPI_ISL_1477360, EPI_ISL_1477361, EPI_ISL_1477362, EPI_ISL_1477363, EPI_ISL_1477365, EPI_ISL_1477366, EPI_ISL_1477368, EPI_ISL_1477369, EPI_ISL_1477370, EPI_ISL_1477372, EPI_ISL_1477374, EPI_ISL_1477378, EPI_ISL_1477383, EPI_ISL_1477384, EPI_ISL_1477386, EPI_ISL_1477387, EPI_ISL_1477390, EPI_ISL_1477391, EPI_ISL_1477392, EPI_ISL_1477393, EPI_ISL_1477395, EPI_ISL_1477398, EPI_ISL_1477400, EPI_ISL_1477402, EPI_ISL_1477403, EPI_ISL_1477404, EPI_ISL_1477407, EPI_ISL_1477409, EPI_ISL_1477416, EPI_ISL_1477418, EPI_ISL_1477419, EPI_ISL_1477420, EPI_ISL_1477421, EPI_ISL_1477422, EPI_ISL_1477423, EPI_ISL_1477424, EPI_ISL_1477425, EPI_ISL_1477426, EPI_ISL_1477427, EPI_ISL_1477428, EPI_ISL_1477429, EPI_ISL_1477431, EPI_ISL_1477434, EPI_ISL_1477437, EPI_ISL_1477440, EPI_ISL_1477442, EPI_ISL_1477448, EPI_ISL_1477450, EPI_ISL_1477451, EPI_ISL_1477456, EPI_ISL_1477464, EPI_ISL_1477467, EPI_ISL_1477469, EPI_ISL_1477470, EPI_ISL_1477472, EPI_ISL_1477474, EPI_ISL_1477475, EPI_ISL_1477479, EPI_ISL_1477480, EPI_ISL_1477485, EPI_ISL_1477490, EPI_ISL_1477492, EPI_ISL_1477505, EPI_ISL_1477509, EPI_ISL_1477512, EPI_ISL_1477513, EPI_ISL_1477516, EPI_ISL_1477522, EPI_ISL_1477523, EPI_ISL_1477524, EPI_ISL_1477526, EPI_ISL_1477527, EPI_ISL_1477528, EPI_ISL_1477538, EPI_ISL_1477542, EPI_ISL_1477548, EPI_ISL_1477549, EPI_ISL_1477553, EPI_ISL_1477556, EPI_ISL_1477559, EPI_ISL_1477560, EPI_ISL_1477563, EPI_ISL_1477567, EPI_ISL_1477568, EPI_ISL_1477570, EPI_ISL_1477573, EPI_ISL_1477574, EPI_ISL_1477576, EPI_ISL_1477579, EPI_ISL_1477580, EPI_ISL_1477583, EPI_ISL_1477585, EPI_ISL_1477586, EPI_ISL_1477591, EPI_ISL_1477592, EPI_ISL_1477596, EPI_ISL_1477599, EPI_ISL_1477602, EPI_ISL_1477606, EPI_ISL_1477607, EPI_ISL_1477608, EPI_ISL_1477609, EPI_ISL_1477616, EPI_ISL_1477618, EPI_ISL_1477620, EPI_ISL_1477621, EPI_ISL_1477625, EPI_ISL_1477628, EPI_ISL_1477629, EPI_ISL_1477633, EPI_ISL_1477636, EPI_ISL_1477638, EPI_ISL_1477640, EPI_ISL_1477642, EPI_ISL_1477643, EPI_ISL_1477648, EPI_ISL_1477651, EPI_ISL_1477652, EPI_ISL_1477653, EPI_ISL_1477654, EPI_ISL_1477661, EPI_ISL_1477662, EPI_ISL_1529100, EPI_ISL_1529105, EPI_ISL_1529111, EPI_ISL_1529112, EPI_ISL_1529114, EPI_ISL_1529115, EPI_ISL_1529116, EPI_ISL_1529117, EPI_ISL_1529120, EPI_ISL_1529125, EPI_ISL_1529126, EPI_ISL_1529139, EPI_ISL_1529140, EPI_ISL_1529143, EPI_ISL_1529145, EPI_ISL_1529146, EPI_ISL_1529150, EPI_ISL_1529153, EPI_ISL_1529157, EPI_ISL_1529159, EPI_ISL_1529160, EPI_ISL_1529170, EPI_ISL_1529172, EPI_ISL_1529174, EPI_ISL_1529178, EPI_ISL_1529182, EPI_ISL_1529187, EPI_ISL_1529189, EPI_ISL_1529192, EPI_ISL_1529197, EPI_ISL_1529198, EPI_ISL_1529199, EPI_ISL_1529200, EPI_ISL_1529203, EPI_ISL_1529214, EPI_ISL_1529215, EPI_ISL_1529219, EPI_ISL_1529222, EPI_ISL_1529223, EPI_ISL_1529230, EPI_ISL_1529231, EPI_ISL_1529233, EPI_ISL_1529242, EPI_ISL_1529243, EPI_ISL_1529244, EPI_ISL_1529249, EPI_ISL_1529254, EPI_ISL_1529257, EPI_ISL_1529265, EPI_ISL_1529266, EPI_ISL_1529274, EPI_ISL_1529276, EPI_ISL_1529277, EPI_ISL_1529280, EPI_ISL_1529281, EPI_ISL_1529286, EPI_ISL_1529288, EPI_ISL_1529290, EPI_ISL_1529291, EPI_ISL_1529294, EPI_ISL_1529299, EPI_ISL_1529306, EPI_ISL_1529312, EPI_ISL_1529313, EPI_ISL_1529314, EPI_ISL_1529319, EPI_ISL_1529321, EPI_ISL_1529324, EPI_ISL_1529325, EPI_ISL_1529326, EPI_ISL_1529332, EPI_ISL_1529333, EPI_ISL_1529335, EPI_ISL_1529337, EPI_ISL_1529357, EPI_ISL_1529359, EPI_ISL_1529360, EPI_ISL_1529361, EPI_ISL_1529362, EPI_ISL_1529363, EPI_ISL_1529366, EPI_ISL_1529369, EPI_ISL_1663921, EPI_ISL_1663922, EPI_ISL_1663924, EPI_ISL_1663929, EPI_ISL_1663932, EPI_ISL_1663941, EPI_ISL_1663942, EPI_ISL_1663944, EPI_ISL_1663949, EPI_ISL_1663961, EPI_ISL_1663965, EPI_ISL_1663966, EPI_ISL_1663967, EPI_ISL_1663973, EPI_ISL_1663978, EPI_ISL_1663981, EPI_ISL_1663988, EPI_ISL_1663991, EPI_ISL_1663997, EPI_ISL_1663998, EPI_ISL_1664000, EPI_ISL_1664001, EPI_ISL_1664002, EPI_ISL_1664007, EPI_ISL_1664011, EPI_ISL_1664012, EPI_ISL_1664017, EPI_ISL_1664018, EPI_ISL_1664019, EPI_ISL_1664022, EPI_ISL_1664023, EPI_ISL_1664027, EPI_ISL_1664029, EPI_ISL_1664031, EPI_ISL_1664033, EPI_ISL_1664034, EPI_ISL_1664041, EPI_ISL_1664042, EPI_ISL_1664044, EPI_ISL_1664055, EPI_ISL_1664059, EPI_ISL_1664061, EPI_ISL_1664066, EPI_ISL_1664067, EPI_ISL_1664074, EPI_ISL_1664075, EPI_ISL_1664076, EPI_ISL_1664077, EPI_ISL_1664078, EPI_ISL_1664092, EPI_ISL_1664099, EPI_ISL_1821423, EPI_ISL_1821457, EPI_ISL_2102571, EPI_ISL_2658665, EPI_ISL_2658686, EPI_ISL_2658731, EPI_ISL_2659204, EPI_ISL_2659211, EPI_ISL_2659258, EPI_ISL_2659282, EPI_ISL_2659305, EPI_ISL_2659464, EPI_ISL_2659509, EPI_ISL_2659652, EPI_ISL_2725517 | | | |
| see above | CA DPH Viral and Rickettsial Disease Laboratory | Chan-Zuckerberg Biohub | CZB Cliahub Consortium |
| EPI_ISL_1272876, EPI_ISL_1272877, EPI_ISL_1272880, EPI_ISL_1272881, EPI_ISL_1272882, EPI_ISL_1272883, EPI_ISL_1272884, EPI_ISL_1272885, EPI_ISL_1272887, EPI_ISL_1272889, EPI_ISL_1272890, EPI_ISL_1272892, EPI_ISL_1272893, EPI_ISL_1447049 | | | |
| see above | CA-Los Angeles County Public Health Laboratory | Centers for Disease Control and Prevention Division of Viral Diseases, Pathogen Discovery | Alison Laufer Halpin; Anna Montmayeur; Anna Uehara; Ben L. Rambo-Martin; Clinton R. Paden; Dakota Howard; Darlene Wagner; Dave Wentworth; Dhwani Batra; Haibin Wang; Jasmine Padilla; Jing Zhang; Justin Lee; Katie Dillon; Krista Queen; Kristen Knipe; Kristine Lacek; Lori Rowe; Mark Burroughs; Matthew Schmerer; Mili Sheth; Peter Cook; Peter W. Cook; Rachel Marine; Sam Shepard; Sarah Nobles; Shoshona Le; Suxiang Tong; Vivien Dugan; Yan Li; Ying Tao; Yvette Unoarumhi |
| EPI_ISL_1226169, EPI_ISL_1226170, EPI_ISL_1226171, EPI_ISL_1226172, EPI_ISL_1226173, EPI_ISL_1226174, EPI_ISL_1226175, EPI_ISL_1226176 | | | |
| see above | CA-Los Angeles County Public Health Laboratory | Genomics and Discovery, Respiratory Viruses Branch, Division of Viral Diseases, Centers for Disease Control and Prevention | Anna Montmayeur; Anna Uehara; Ben L. Rambo-Martin; Clinton R. Paden; Dhwani Batra; Haibin Wang; Jasmine Padilla; Jing Zhang; Justin Lee; Katie Dillon; Krista Queen; Kristen Knipe; Kristine Lacek; Lori Rowe; Mark Burroughs; Matthew Schmerer; Mili Sheth; Peter W. Cook; Rachel Marine; Sam Shepard; Sarah Nobles; Shoshona Le; Suxiang Tong; Yan Li; Ying Tao |
| EPI_ISL_431140, EPI_ISL_431141, EPI_ISL_2782391, EPI_ISL_2782401, EPI_ISL_2782402, EPI_ISL_2782413, EPI_ISL_2782420, EPI_ISL_2782426, EPI_ISL_2782429, EPI_ISL_2782438, EPI_ISL_2782441, EPI_ISL_2782443, EPI_ISL_2782451, EPI_ISL_2782454, EPI_ISL_2782458, EPI_ISL_2782463, EPI_ISL_2782464, EPI_ISL_2782467, EPI_ISL_2782471, EPI_ISL_2782472, EPI_ISL_2782474, EPI_ISL_2782485, EPI_ISL_2782492, EPI_ISL_2782494, EPI_ISL_2782502, EPI_ISL_2782505, EPI_ISL_2782510, EPI_ISL_2782518, EPI_ISL_2782528, EPI_ISL_2782531, EPI_ISL_2782532, EPI_ISL_2782534, EPI_ISL_2782545, EPI_ISL_2782548, EPI_ISL_2782549, EPI_ISL_2782554, EPI_ISL_2782555, EPI_ISL_2782558, EPI_ISL_2782562, EPI_ISL_2782563, EPI_ISL_2782571, EPI_ISL_2782575, EPI_ISL_2782577, EPI_ISL_2782578, EPI_ISL_2782591, EPI_ISL_2783036, EPI_ISL_2783037, EPI_ISL_2783038, EPI_ISL_2783039, EPI_ISL_2783040, EPI_ISL_2783041, EPI_ISL_2783042, EPI_ISL_2783043, EPI_ISL_2783044, EPI_ISL_2783045, EPI_ISL_2783046, EPI_ISL_2783047, EPI_ISL_2783048, EPI_ISL_2783049, EPI_ISL_2783050, EPI_ISL_2783051, EPI_ISL_2783052, EPI_ISL_2783053, EPI_ISL_2783054, EPI_ISL_2783055, EPI_ISL_2783056, EPI_ISL_2783057, EPI_ISL_2783058, EPI_ISL_2783059, EPI_ISL_2783060, EPI_ISL_2783061, EPI_ISL_2783062, EPI_ISL_2783063, EPI_ISL_2783064, EPI_ISL_2783065, EPI_ISL_2783066, EPI_ISL_2783067, EPI_ISL_2783068, EPI_ISL_2783069, EPI_ISL_2783070, EPI_ISL_2783071, EPI_ISL_2783072, EPI_ISL_2783073, EPI_ISL_2783074, EPI_ISL_2783075, EPI_ISL_2783076, EPI_ISL_2783077, EPI_ISL_2783078, EPI_ISL_2783080, EPI_ISL_2783081, EPI_ISL_2783082, EPI_ISL_2783083, EPI_ISL_2783084, EPI_ISL_2783085, EPI_ISL_2783086, EPI_ISL_2783087, EPI_ISL_2783088, EPI_ISL_2783089, EPI_ISL_2783090, EPI_ISL_2783093, EPI_ISL_2783101, EPI_ISL_2783103, EPI_ISL_2783122, EPI_ISL_2783172, EPI_ISL_2783173, EPI_ISL_2783176, EPI_ISL_2783185, EPI_ISL_2783186, EPI_ISL_2783188, EPI_ISL_2783189, EPI_ISL_2797075, EPI_ISL_2797076, EPI_ISL_2797077, EPI_ISL_2797078, EPI_ISL_2797079, EPI_ISL_2797080, EPI_ISL_2797081, EPI_ISL_2797082, EPI_ISL_2797083, EPI_ISL_2797084, EPI_ISL_2797085, EPI_ISL_2797086, EPI_ISL_2797087, EPI_ISL_2797088, EPI_ISL_2797089, EPI_ISL_2797090, EPI_ISL_2797091, EPI_ISL_2797092, EPI_ISL_2797093, EPI_ISL_2797094, EPI_ISL_2797095, EPI_ISL_2797096, EPI_ISL_2797097, EPI_ISL_2797098, EPI_ISL_2797099, EPI_ISL_2797100, EPI_ISL_2797101, EPI_ISL_2797102, EPI_ISL_2797103, EPI_ISL_2797104, EPI_ISL_2797105, EPI_ISL_2797106, EPI_ISL_2797107, EPI_ISL_2797108, EPI_ISL_2797109, EPI_ISL_2797110, EPI_ISL_2797111, EPI_ISL_2797112, EPI_ISL_2797113, EPI_ISL_2797114, EPI_ISL_2797115, EPI_ISL_2797116, EPI_ISL_2797117, EPI_ISL_2797118, EPI_ISL_2797119, EPI_ISL_2797120, EPI_ISL_2797121, EPI_ISL_2797122, EPI_ISL_2797123, EPI_ISL_2797124, EPI_ISL_2797125, EPI_ISL_2797126, EPI_ISL_2797127, EPI_ISL_2797128, EPI_ISL_2797129, EPI_ISL_2797130, EPI_ISL_2797131, EPI_ISL_2797132, EPI_ISL_2797133, EPI_ISL_2797134, EPI_ISL_2797135, EPI_ISL_2797136, EPI_ISL_2797137, EPI_ISL_2797138, EPI_ISL_2797139, EPI_ISL_2797140, EPI_ISL_2797141, EPI_ISL_2797142, EPI_ISL_2797143, EPI_ISL_2797144, EPI_ISL_2797145, EPI_ISL_2797146, EPI_ISL_2797147, EPI_ISL_2797148, EPI_ISL_2797149, EPI_ISL_2797150, EPI_ISL_2797151, EPI_ISL_2797152, EPI_ISL_2797153, EPI_ISL_2797154, EPI_ISL_2797155, EPI_ISL_2797156, EPI_ISL_2797157, EPI_ISL_2797158, EPI_ISL_2797159, EPI_ISL_2797160, EPI_ISL_2797161, EPI_ISL_2797162, EPI_ISL_2797163, EPI_ISL_2797164, EPI_ISL_2797165, EPI_ISL_2797166, EPI_ISL_2797167, EPI_ISL_2797168, EPI_ISL_2797169, EPI_ISL_2797170, EPI_ISL_2797171, EPI_ISL_2797172, EPI_ISL_2797173, EPI_ISL_2797174, EPI_ISL_2797175, EPI_ISL_2797178, EPI_ISL_2797179, EPI_ISL_2797180, EPI_ISL_2797181, EPI_ISL_2797182, EPI_ISL_2797183, EPI_ISL_2797184, EPI_ISL_2797185, EPI_ISL_2797186, EPI_ISL_2797187, EPI_ISL_2797189, EPI_ISL_2797191, EPI_ISL_2797201, EPI_ISL_2797203, EPI_ISL_2797207, EPI_ISL_2797219, EPI_ISL_2797220, EPI_ISL_2797225, EPI_ISL_2797226, EPI_ISL_2797232, EPI_ISL_2797235, EPI_ISL_2797237, EPI_ISL_2797239, EPI_ISL_2797250, EPI_ISL_2797273, EPI_ISL_2797275, EPI_ISL_2797278, EPI_ISL_2797303, EPI_ISL_2797305, EPI_ISL_2797314, EPI_ISL_2797889, EPI_ISL_2797890, EPI_ISL_2797891, EPI_ISL_2797892, EPI_ISL_2797893, EPI_ISL_2797894, EPI_ISL_2797895, EPI_ISL_2797896, EPI_ISL_2797897, EPI_ISL_2797898, EPI_ISL_2797899, EPI_ISL_2797900, EPI_ISL_2797901, EPI_ISL_2797902, EPI_ISL_2797903, EPI_ISL_2797904, EPI_ISL_2797905, EPI_ISL_2797906, EPI_ISL_2797907, EPI_ISL_2797908, EPI_ISL_2797909, EPI_ISL_2797910, EPI_ISL_2797911, EPI_ISL_2797912, EPI_ISL_2797913, EPI_ISL_2797914, EPI_ISL_2797915, EPI_ISL_2797916, EPI_ISL_2797917, EPI_ISL_2797918, EPI_ISL_2797919, EPI_ISL_2797920, EPI_ISL_2797921, EPI_ISL_2797922, EPI_ISL_2797924, EPI_ISL_2797925, EPI_ISL_2797926, EPI_ISL_2797928, EPI_ISL_2797929, EPI_ISL_2797930, EPI_ISL_2797931, EPI_ISL_2797932, EPI_ISL_2797933, EPI_ISL_2797934, EPI_ISL_2797935, EPI_ISL_2797936, EPI_ISL_2797937, EPI_ISL_2797938, EPI_ISL_2797939, EPI_ISL_2797940, EPI_ISL_2797941, EPI_ISL_2797942, EPI_ISL_2797943, EPI_ISL_2797944, EPI_ISL_2797945, EPI_ISL_2797946, EPI_ISL_2797947, EPI_ISL_2797948, EPI_ISL_2797949, EPI_ISL_2797950, EPI_ISL_2797951, EPI_ISL_2797952, EPI_ISL_2797953, EPI_ISL_2797954, EPI_ISL_2797955, EPI_ISL_2797956, EPI_ISL_2797957, EPI_ISL_2797958, EPI_ISL_2797959, EPI_ISL_2797960, EPI_ISL_2797961, EPI_ISL_2797962, EPI_ISL_2797963, EPI_ISL_2797964, EPI_ISL_2797965, EPI_ISL_2797966, EPI_ISL_2797968, EPI_ISL_2797969, EPI_ISL_2797970, EPI_ISL_2797971, EPI_ISL_2797972, EPI_ISL_2797973, EPI_ISL_2797974, EPI_ISL_2797975, EPI_ISL_2797976, EPI_ISL_2797977, EPI_ISL_2797978, EPI_ISL_2797979, EPI_ISL_2797980, EPI_ISL_2797981, EPI_ISL_2797982, EPI_ISL_2797983, EPI_ISL_2797984, EPI_ISL_2797989, EPI_ISL_2797994, EPI_ISL_2798001, EPI_ISL_2798008, EPI_ISL_2798009, EPI_ISL_2798021, EPI_ISL_2798027, EPI_ISL_2798028, EPI_ISL_2798029, EPI_ISL_2798030, EPI_ISL_2798031, EPI_ISL_2798043, EPI_ISL_2798048, EPI_ISL_2798055, EPI_ISL_2798062, EPI_ISL_2798064, EPI_ISL_2798065, EPI_ISL_2798066, EPI_ISL_2798068, EPI_ISL_2798070, EPI_ISL_2825247, EPI_ISL_2825248, EPI_ISL_2825249, EPI_ISL_2825250, EPI_ISL_2825251, EPI_ISL_2825252, EPI_ISL_2825253, EPI_ISL_2825254, EPI_ISL_2825255, EPI_ISL_2825256, EPI_ISL_2825257, EPI_ISL_2825258, EPI_ISL_2825259, EPI_ISL_2825260, EPI_ISL_2825261, EPI_ISL_2825262, EPI_ISL_2825263, EPI_ISL_2825264, EPI_ISL_2825265, EPI_ISL_2825266, EPI_ISL_2825268, EPI_ISL_2825269, EPI_ISL_2825270, EPI_ISL_2825271, EPI_ISL_2825272, EPI_ISL_2825273, EPI_ISL_2825274, EPI_ISL_2825275, EPI_ISL_2825276, EPI_ISL_2825284, EPI_ISL_2825285, EPI_ISL_2825286, EPI_ISL_2825291, EPI_ISL_2825300, EPI_ISL_2825301, EPI_ISL_2825302, EPI_ISL_2825303, EPI_ISL_2825304, EPI_ISL_2825305, EPI_ISL_2825306, EPI_ISL_2825307, EPI_ISL_2825308, EPI_ISL_2825309, EPI_ISL_2825310, EPI_ISL_2825311, EPI_ISL_2825312, EPI_ISL_2825313, EPI_ISL_2825315, EPI_ISL_2825316, EPI_ISL_2825317, EPI_ISL_2825318, EPI_ISL_2825319, EPI_ISL_2825320, EPI_ISL_2825321, EPI_ISL_2825322, EPI_ISL_2825323, EPI_ISL_2825324, EPI_ISL_2825325, EPI_ISL_2825326, EPI_ISL_2825327, EPI_ISL_2825328, EPI_ISL_2825329, EPI_ISL_2825330, EPI_ISL_2825332, EPI_ISL_2825333, EPI_ISL_2825334, EPI_ISL_2825335, EPI_ISL_2825336, EPI_ISL_2825337, EPI_ISL_2825338, EPI_ISL_2825339, EPI_ISL_2825340, EPI_ISL_2825341, EPI_ISL_2825342, EPI_ISL_2825343, EPI_ISL_2825344, EPI_ISL_2825345, EPI_ISL_2825346, EPI_ISL_2825347, EPI_ISL_2825349, EPI_ISL_2825350, EPI_ISL_2825351, EPI_ISL_2825352, EPI_ISL_2825353, EPI_ISL_2825354, EPI_ISL_2825355, EPI_ISL_2825356, EPI_ISL_2825357, EPI_ISL_2825358, EPI_ISL_2825359, EPI_ISL_2825360, EPI_ISL_2825361, EPI_ISL_2825362, EPI_ISL_2825363, EPI_ISL_2825364, EPI_ISL_2825366, EPI_ISL_2825367, EPI_ISL_2825368, EPI_ISL_2825369, EPI_ISL_2825370, EPI_ISL_2825371, EPI_ISL_2825372, EPI_ISL_2825373, EPI_ISL_2825374, EPI_ISL_2825375, EPI_ISL_2825376, EPI_ISL_2825377, EPI_ISL_2825378, EPI_ISL_2825379, EPI_ISL_2825380, EPI_ISL_2825381, EPI_ISL_2825382, EPI_ISL_2825384, EPI_ISL_2825385, EPI_ISL_2825387, EPI_ISL_2825389, EPI_ISL_2825392, EPI_ISL_2825393, EPI_ISL_2825394, EPI_ISL_2825395, EPI_ISL_2825396, EPI_ISL_2825397, EPI_ISL_2825398, EPI_ISL_2825399, EPI_ISL_2825400, EPI_ISL_2825402, EPI_ISL_2825403, EPI_ISL_2825404, EPI_ISL_2825405, EPI_ISL_2825406, EPI_ISL_2825407, EPI_ISL_2825408, EPI_ISL_2825409, EPI_ISL_2825411, EPI_ISL_2825412, EPI_ISL_2825418, EPI_ISL_2825422, EPI_ISL_2825423, EPI_ISL_2825424, EPI_ISL_2825425, EPI_ISL_2825439, EPI_ISL_2826307, EPI_ISL_2826308, EPI_ISL_2826309, EPI_ISL_2826310, EPI_ISL_2826311, EPI_ISL_2826312, EPI_ISL_2826317, EPI_ISL_2826326, EPI_ISL_2826327, EPI_ISL_2826328, EPI_ISL_2826329, EPI_ISL_2826330, EPI_ISL_2826331, EPI_ISL_2826332, EPI_ISL_2826333, EPI_ISL_2826334, EPI_ISL_2826335, EPI_ISL_2826336, EPI_ISL_2826337, EPI_ISL_2826338, EPI_ISL_2826339, EPI_ISL_2826340, EPI_ISL_2826341, EPI_ISL_2826342, EPI_ISL_2826343, EPI_ISL_2826344, EPI_ISL_2826345, EPI_ISL_2826346, EPI_ISL_2826347, EPI_ISL_2826348, EPI_ISL_2826349, EPI_ISL_2826350, EPI_ISL_2826351, EPI_ISL_2826352, EPI_ISL_2826353, EPI_ISL_2826354, EPI_ISL_2826355, EPI_ISL_2826356, EPI_ISL_2826357, EPI_ISL_2826358, EPI_ISL_2826359, EPI_ISL_2826360, EPI_ISL_2826361, EPI_ISL_2826362, EPI_ISL_2826363, EPI_ISL_2826364, EPI_ISL_2826365, EPI_ISL_2826366, EPI_ISL_2826367, EPI_ISL_2826368, EPI_ISL_2826369, EPI_ISL_2826370, EPI_ISL_2826371, EPI_ISL_2826372, EPI_ISL_2826373, EPI_ISL_2826374, EPI_ISL_2826375, EPI_ISL_2826376, EPI_ISL_2826377, EPI_ISL_2826378, EPI_ISL_2826379, EPI_ISL_2826380, EPI_ISL_2826381, EPI_ISL_2826382, EPI_ISL_2826383, EPI_ISL_2826384, EPI_ISL_2826385, EPI_ISL_2826386, EPI_ISL_2826387, EPI_ISL_2826388, EPI_ISL_2826389, EPI_ISL_2826390, EPI_ISL_2826391, EPI_ISL_2826392, EPI_ISL_2826393, EPI_ISL_2826394, EPI_ISL_2826395, EPI_ISL_2826396, EPI_ISL_2826397, EPI_ISL_2826398, EPI_ISL_2826399, EPI_ISL_2826400, EPI_ISL_2826401, EPI_ISL_2826402, EPI_ISL_2826403, EPI_ISL_2826404, EPI_ISL_2826405, EPI_ISL_2826406, EPI_ISL_2826407, EPI_ISL_2826408, EPI_ISL_2826409, EPI_ISL_2826410, EPI_ISL_2826411, EPI_ISL_2826412, EPI_ISL_2826413, EPI_ISL_2826414, EPI_ISL_2826415, EPI_ISL_2826416, EPI_ISL_2826417, EPI_ISL_2826418, EPI_ISL_2826419, EPI_ISL_2826420, EPI_ISL_2826421, EPI_ISL_2826422, EPI_ISL_2826423, EPI_ISL_2826424, EPI_ISL_2826425, EPI_ISL_2826426, EPI_ISL_2826427, EPI_ISL_2826428, EPI_ISL_2826429, EPI_ISL_2826430, EPI_ISL_2826431, EPI_ISL_2826432, EPI_ISL_2826433, EPI_ISL_2826434, EPI_ISL_2826435, EPI_ISL_2826436, EPI_ISL_2826437, EPI_ISL_2826438, EPI_ISL_2826439, EPI_ISL_2826440, EPI_ISL_2826441, EPI_ISL_2826442, EPI_ISL_2826443, EPI_ISL_2826444, EPI_ISL_2826445, EPI_ISL_2826446, EPI_ISL_2826447, EPI_ISL_2826448, EPI_ISL_2826449, EPI_ISL_2826450, EPI_ISL_2826451, EPI_ISL_2826452, EPI_ISL_2826453, EPI_ISL_2826454, EPI_ISL_2826455, EPI_ISL_2826456, EPI_ISL_2826457, EPI_ISL_2826458, EPI_ISL_2826459, EPI_ISL_2826460, EPI_ISL_2826461, EPI_ISL_2826462, EPI_ISL_2826463, EPI_ISL_2826464, EPI_ISL_2826465, EPI_ISL_2826466, EPI_ISL_2826467, EPI_ISL_2826468, EPI_ISL_2826469, EPI_ISL_2826470, EPI_ISL_2826471, EPI_ISL_2826472, EPI_ISL_2826473, EPI_ISL_2826474, EPI_ISL_2826475, EPI_ISL_2826476, EPI_ISL_2826477, EPI_ISL_2826478, EPI_ISL_2826479, EPI_ISL_2826480, EPI_ISL_2826481, EPI_ISL_2826482, EPI_ISL_2826483, EPI_ISL_2826484, EPI_ISL_2826485, EPI_ISL_2826486, EPI_ISL_2826487, EPI_ISL_2826488, EPI_ISL_2826489, EPI_ISL_2826490, EPI_ISL_2826491, EPI_ISL_2826492, EPI_ISL_2826493, EPI_ISL_2826494, EPI_ISL_2826495, EPI_ISL_2826496, EPI_ISL_2826497, EPI_ISL_2826498, EPI_ISL_2826499, EPI_ISL_2826500, EPI_ISL_2826501, EPI_ISL_2826502, EPI_ISL_2826503, EPI_ISL_2826504, EPI_ISL_2826505, EPI_ISL_2826506, EPI_ISL_2826507, EPI_ISL_2826508, EPI_ISL_2826509, EPI_ISL_2826510, EPI_ISL_2826511, EPI_ISL_2826512, EPI_ISL_2826513, EPI_ISL_2826514, EPI_ISL_2826515, EPI_ISL_2826516, EPI_ISL_2826517, EPI_ISL_2826518, EPI_ISL_2826519, EPI_ISL_2826520, EPI_ISL_2826521, EPI_ISL_2826522, EPI_ISL_2826523, EPI_ISL_2826524, EPI_ISL_2826525, EPI_ISL_2826526, EPI_ISL_2826528, EPI_ISL_2826529, EPI_ISL_2826530, EPI_ISL_2826531, EPI_ISL_2826532, EPI_ISL_2826533, EPI_ISL_2826534, EPI_ISL_2826535, EPI_ISL_2826536, EPI_ISL_2826537, EPI_ISL_2826538, EPI_ISL_2826539, EPI_ISL_2826540, EPI_ISL_2826541, EPI_ISL_2826542, EPI_ISL_2826543, EPI_ISL_2826544, EPI_ISL_2826545, EPI_ISL_2826546, EPI_ISL_2826547, EPI_ISL_2826548, EPI_ISL_2826549, EPI_ISL_2826550, EPI_ISL_2826551, EPI_ISL_2826552, EPI_ISL_2826553, EPI_ISL_2826554, EPI_ISL_2826555, EPI_ISL_2826556, EPI_ISL_2826557, EPI_ISL_2826558, EPI_ISL_2826559, EPI_ISL_2826560, EPI_ISL_2826561, EPI_ISL_2826562, EPI_ISL_2826563, EPI_ISL_2826564, EPI_ISL_2826565, EPI_ISL_2826566, EPI_ISL_2826567, EPI_ISL_2826568, EPI_ISL_2826569, EPI_ISL_2826570, EPI_ISL_2826571, EPI_ISL_2826572, EPI_ISL_2826573, EPI_ISL_2826574, EPI_ISL_2826575, EPI_ISL_2826576, EPI_ISL_2826577, EPI_ISL_2826578, EPI_ISL_2826579, EPI_ISL_2826580, EPI_ISL_2826581, EPI_ISL_2826582, EPI_ISL_2826585, EPI_ISL_2826586, EPI_ISL_2826589, EPI_ISL_2826590, EPI_ISL_2826591, EPI_ISL_2826592, EPI_ISL_2826593, EPI_ISL_2826594, EPI_ISL_2826595, EPI_ISL_2826596, EPI_ISL_2826597, EPI_ISL_2826598, EPI_ISL_2826599, EPI_ISL_2826600, EPI_ISL_2826601, EPI_ISL_2826602, EPI_ISL_2826603, EPI_ISL_2826604, EPI_ISL_2826605, EPI_ISL_2826606, EPI_ISL_2826607, EPI_ISL_2826608, EPI_ISL_2826609, EPI_ISL_2826610, EPI_ISL_2826611, EPI_ISL_2826612, EPI_ISL_2826613, EPI_ISL_2826614, EPI_ISL_2826615, EPI_ISL_2826616, EPI_ISL_2826617, EPI_ISL_2826618, EPI_ISL_2826619, EPI_ISL_2826620, EPI_ISL_2826621, EPI_ISL_2826622, EPI_ISL_2826623, EPI_ISL_2826624, EPI_ISL_2826625, EPI_ISL_2826626, EPI_ISL_2826627, EPI_ISL_2826628, EPI_ISL_2826629, EPI_ISL_2826630, EPI_ISL_2853588, EPI_ISL_2853589, EPI_ISL_2853590, EPI_ISL_2853591, EPI_ISL_2853592, EPI_ISL_2853593, EPI_ISL_2853594, EPI_ISL_2853595, EPI_ISL_2853596, EPI_ISL_2853597, EPI_ISL_2853598, EPI_ISL_2853599, EPI_ISL_2853600, EPI_ISL_2853601, EPI_ISL_2853602, EPI_ISL_2853603, EPI_ISL_2853604, EPI_ISL_2853605, EPI_ISL_2853606, EPI_ISL_2853607, EPI_ISL_2853608, EPI_ISL_2853609, EPI_ISL_2853610, EPI_ISL_2853611, EPI_ISL_2853612, EPI_ISL_2853613, EPI_ISL_2853614, EPI_ISL_2853615, EPI_ISL_2853616, EPI_ISL_2853617, EPI_ISL_2853618, EPI_ISL_2853619, EPI_ISL_2853620, EPI_ISL_2853621, EPI_ISL_2853622, EPI_ISL_2853623, EPI_ISL_2853624, EPI_ISL_2853625, EPI_ISL_2853626, EPI_ISL_2853627, EPI_ISL_2853628, EPI_ISL_2853629, EPI_ISL_2853630, EPI_ISL_2853631, EPI_ISL_2853632, EPI_ISL_2853633, EPI_ISL_2853634, EPI_ISL_2853635, EPI_ISL_2853636, EPI_ISL_2853637, EPI_ISL_2853638, EPI_ISL_2853639, EPI_ISL_2853640, EPI_ISL_2853641, EPI_ISL_2853642, EPI_ISL_2853643, EPI_ISL_2853644, EPI_ISL_2853645, EPI_ISL_2853646, EPI_ISL_2853647, EPI_ISL_2853648, EPI_ISL_2853649, EPI_ISL_2853650, EPI_ISL_2853651, EPI_ISL_2853652, EPI_ISL_2853653, EPI_ISL_2853654, EPI_ISL_2853655, EPI_ISL_2853656, EPI_ISL_2853657, EPI_ISL_2853658, EPI_ISL_2853659, EPI_ISL_2853660, EPI_ISL_2853661, EPI_ISL_2853662, EPI_ISL_2853663, EPI_ISL_2853664, EPI_ISL_2853665, EPI_ISL_2853666, EPI_ISL_2853667, EPI_ISL_2853668, EPI_ISL_2853669, EPI_ISL_2853670, EPI_ISL_2853671, EPI_ISL_2853672, EPI_ISL_2853673, EPI_ISL_2853674, EPI_ISL_2853675, EPI_ISL_2853676, EPI_ISL_2853677, EPI_ISL_2853678, EPI_ISL_2853679, EPI_ISL_2853680, EPI_ISL_2853681, EPI_ISL_2853682, EPI_ISL_2853683, EPI_ISL_2853684, EPI_ISL_2853685, EPI_ISL_2853686, EPI_ISL_2853687, EPI_ISL_2853688, EPI_ISL_2853689, EPI_ISL_2853690, EPI_ISL_2853691, EPI_ISL_2853692, EPI_ISL_2853693, EPI_ISL_2853694, EPI_ISL_2853695, EPI_ISL_2853696, EPI_ISL_2853697, EPI_ISL_2853698, EPI_ISL_2853699, EPI_ISL_2853700, EPI_ISL_2853701, EPI_ISL_2853702, EPI_ISL_2853703, EPI_ISL_2853704, EPI_ISL_2853705, EPI_ISL_2853706, EPI_ISL_2853707, EPI_ISL_2853708, EPI_ISL_2853709, EPI_ISL_2853710, EPI_ISL_2853711, EPI_ISL_2853712, EPI_ISL_2853713, EPI_ISL_2853714, EPI_ISL_2853715, EPI_ISL_2853716, EPI_ISL_2853717, EPI_ISL_2853718, EPI_ISL_2853719, EPI_ISL_2853720, EPI_ISL_2853721, EPI_ISL_2853722, EPI_ISL_2853723, EPI_ISL_2853724, EPI_ISL_2853725, EPI_ISL_2853726, EPI_ISL_2853727, EPI_ISL_2853728, EPI_ISL_2853729, EPI_ISL_2853730, EPI_ISL_2853731, EPI_ISL_2853732, EPI_ISL_2853733, EPI_ISL_2853734, EPI_ISL_2853735, EPI_ISL_2853736, EPI_ISL_2853737, EPI_ISL_2853738, EPI_ISL_2853739, EPI_ISL_2853740, EPI_ISL_2853741, EPI_ISL_2853742, EPI_ISL_2853743, EPI_ISL_2853744, EPI_ISL_2853745, EPI_ISL_2853746, EPI_ISL_2853747, EPI_ISL_2853748, EPI_ISL_2853749, EPI_ISL_2853750, EPI_ISL_2853751, EPI_ISL_2853752, EPI_ISL_2853753, | | | |

EPI_ISL_2853754, EPI_ISL_2853755, EPI_ISL_2853756, EPI_ISL_2853757, EPI_ISL_2853758, EPI_ISL_2853759, EPI_ISL_2853760, EPI_ISL_2853761, EPI_ISL_2853762, EPI_ISL_2853763, EPI_ISL_2853764, EPI_ISL_2853765, EPI_ISL_2853766, EPI_ISL_2853767, EPI_ISL_2853768, EPI_ISL_2853769, EPI_ISL_2853770, EPI_ISL_2853771, EPI_ISL_2853772, EPI_ISL_2853773, EPI_ISL_2853774, EPI_ISL_2853775, EPI_ISL_2853776, EPI_ISL_2853777, EPI_ISL_2853778, EPI_ISL_2853779, EPI_ISL_2853780, EPI_ISL_2853781, EPI_ISL_2853782, EPI_ISL_2853783, EPI_ISL_2853784, EPI_ISL_2853785, EPI_ISL_2853786, EPI_ISL_2853787, EPI_ISL_2853788, EPI_ISL_2853789, EPI_ISL_2853790, EPI_ISL_2853791, EPI_ISL_2853792, EPI_ISL_2853793, EPI_ISL_2853794, EPI_ISL_2853795, EPI_ISL_2853796, EPI_ISL_2853797, EPI_ISL_2853798, EPI_ISL_2853799, EPI_ISL_2853800, EPI_ISL_2853801, EPI_ISL_2853802, EPI_ISL_2853803, EPI_ISL_2853804, EPI_ISL_2853805, EPI_ISL_2853806, EPI_ISL_2853807, EPI_ISL_2853808, EPI_ISL_2853809, EPI_ISL_2853810, EPI_ISL_2853811, EPI_ISL_2853812, EPI_ISL_2853813, EPI_ISL_2853814, EPI_ISL_2853815, EPI_ISL_2853816, EPI_ISL_2853817, EPI_ISL_2853818, EPI_ISL_2853819, EPI_ISL_2853820, EPI_ISL_2853821, EPI_ISL_2853822, EPI_ISL_2853823, EPI_ISL_2853824, EPI_ISL_2853825, EPI_ISL_2853826, EPI_ISL_2853827, EPI_ISL_2853828, EPI_ISL_2853829, EPI_ISL_2853830, EPI_ISL_2853831, EPI_ISL_2853832, EPI_ISL_2853833, EPI_ISL_2853834, EPI_ISL_2853835, EPI_ISL_2853836, EPI_ISL_2853837, EPI_ISL_2853838, EPI_ISL_2853839, EPI_ISL_2853840, EPI_ISL_2853841, EPI_ISL_2853842, EPI_ISL_2853843, EPI_ISL_2853844, EPI_ISL_2853845, EPI_ISL_2853846, EPI_ISL_2853847, EPI_ISL_2853848, EPI_ISL_2853849, EPI_ISL_2853850, EPI_ISL_2853851, EPI_ISL_2853852, EPI_ISL_2853853, EPI_ISL_2853854, EPI_ISL_2853855, EPI_ISL_2853856, EPI_ISL_2853857, EPI_ISL_2853858, EPI_ISL_2853859, EPI_ISL_2853860, EPI_ISL_2853861, EPI_ISL_2853862, EPI_ISL_2853863, EPI_ISL_2853864, EPI_ISL_2853865, EPI_ISL_2853866, EPI_ISL_2853867, EPI_ISL_2853868, EPI_ISL_2853869, EPI_ISL_2853870, EPI_ISL_2853871, EPI_ISL_2853872, EPI_ISL_2853873, EPI_ISL_2853874, EPI_ISL_2853875, EPI_ISL_2853876, EPI_ISL_2853877, EPI_ISL_2853878, EPI_ISL_2853879, EPI_ISL_2853880, EPI_ISL_2853881, EPI_ISL_2853882, EPI_ISL_2853883, EPI_ISL_2853884, EPI_ISL_2853885, EPI_ISL_2853886, EPI_ISL_2853887, EPI_ISL_2853888, EPI_ISL_2853889, EPI_ISL_2853890, EPI_ISL_2853891, EPI_ISL_2853892, EPI_ISL_2853893, EPI_ISL_2853894, EPI_ISL_2853895, EPI_ISL_2853896, EPI_ISL_2853897, EPI_ISL_2853898, EPI_ISL_2853899, EPI_ISL_2853900, EPI_ISL_2853901, EPI_ISL_2853902, EPI_ISL_2853903, EPI_ISL_2853904, EPI_ISL_2853905, EPI_ISL_2853906, EPI_ISL_2853907, EPI_ISL_2853908, EPI_ISL_2853909, EPI_ISL_2853910, EPI_ISL_2853911, EPI_ISL_2853912, EPI_ISL_2853913, EPI_ISL_2853914, EPI_ISL_2853915, EPI_ISL_2853916, EPI_ISL_2853917, EPI_ISL_2853918, EPI_ISL_2853919, EPI_ISL_2853920, EPI_ISL_2853921, EPI_ISL_2853922, EPI_ISL_2853923, EPI_ISL_2853924, EPI_ISL_2853925, EPI_ISL_2853926, EPI_ISL_2853927, EPI_ISL_2853928, EPI_ISL_2853929, EPI_ISL_2853930, EPI_ISL_2853931, EPI_ISL_2853932, EPI_ISL_2853933, EPI_ISL_2853934, EPI_ISL_2853935, EPI_ISL_2853936, EPI_ISL_2853937, EPI_ISL_2853938, EPI_ISL_2853939, EPI_ISL_2853940, EPI_ISL_2853941, EPI_ISL_2853942, EPI_ISL_2853943, EPI_ISL_2853944, EPI_ISL_2853945, EPI_ISL_2853946, EPI_ISL_2853947, EPI_ISL_2853948, EPI_ISL_2853949, EPI_ISL_2853950, EPI_ISL_2853951, EPI_ISL_2853952, EPI_ISL_2853953, EPI_ISL_2853954, EPI_ISL_2853955, EPI_ISL_2853956, EPI_ISL_2853957, EPI_ISL_2853958, EPI_ISL_2853959, EPI_ISL_2853960, EPI_ISL_2853961, EPI_ISL_2853962, EPI_ISL_2853963, EPI_ISL_2853964, EPI_ISL_2853965, EPI_ISL_2853966, EPI_ISL_2853967, EPI_ISL_2853968, EPI_ISL_2853969, EPI_ISL_2853970, EPI_ISL_2853971, EPI_ISL_2853972, EPI_ISL_2853973, EPI_ISL_2853974, EPI_ISL_2853975, EPI_ISL_2853976, EPI_ISL_2853977, EPI_ISL_2853978, EPI_ISL_2853979, EPI_ISL_2853980, EPI_ISL_2853981, EPI_ISL_2853982, EPI_ISL_2853983, EPI_ISL_2853984, EPI_ISL_2853985, EPI_ISL_2853986, EPI_ISL_2853987, EPI_ISL_2853988, EPI_ISL_2853989, EPI_ISL_2853990, EPI_ISL_2853991, EPI_ISL_2853992, EPI_ISL_2853993, EPI_ISL_2853994, EPI_ISL_2853995, EPI_ISL_2853996, EPI_ISL_2853997, EPI_ISL_2853998, EPI_ISL_2853999, EPI_ISL_2854000, EPI_ISL_2854001, EPI_ISL_2854002, EPI_ISL_2854003, EPI_ISL_2854004, EPI_ISL_2854005, EPI_ISL_2854006, EPI_ISL_2854007, EPI_ISL_2854008, EPI_ISL_2854009, EPI_ISL_2854010, EPI_ISL_2854011, EPI_ISL_2854012, EPI_ISL_2854013, EPI_ISL_2854014, EPI_ISL_2854015, EPI_ISL_2854016, EPI_ISL_2854017, EPI_ISL_2854018, EPI_ISL_2854019, EPI_ISL_2854020, EPI_ISL_2854021, EPI_ISL_2854022, EPI_ISL_2854023, EPI_ISL_2854024, EPI_ISL_2854025, EPI_ISL_2854026, EPI_ISL_2854027, EPI_ISL_2854028, EPI_ISL_2854029, EPI_ISL_2854030, EPI_ISL_2854031, EPI_ISL_2854032, EPI_ISL_2854033, EPI_ISL_2854034, EPI_ISL_2854035, EPI_ISL_2854036, EPI_ISL_2854037, EPI_ISL_2854038, EPI_ISL_2854039, EPI_ISL_2854040, EPI_ISL_2854041, EPI_ISL_2854042, EPI_ISL_2854043, EPI_ISL_2854044, EPI_ISL_2854045, EPI_ISL_2854046, EPI_ISL_2854047, EPI_ISL_2854977, EPI_ISL_2854978, EPI_ISL_2854979, EPI_ISL_2854980, EPI_ISL_2854981, EPI_ISL_2854982, EPI_ISL_2854983, EPI_ISL_2854984, EPI_ISL_2854985, EPI_ISL_2854986, EPI_ISL_2854987, EPI_ISL_2854988, EPI_ISL_2854989, EPI_ISL_2854990, EPI_ISL_2854991, EPI_ISL_2854997, EPI_ISL_2854998, EPI_ISL_2854999, EPI_ISL_2855009, EPI_ISL_2855010, EPI_ISL_2855011, EPI_ISL_2855012, EPI_ISL_2855013, EPI_ISL_2855014, EPI_ISL_2855017, EPI_ISL_2855018, EPI_ISL_2855019, EPI_ISL_2855020, EPI_ISL_2855021, EPI_ISL_2855022, EPI_ISL_2855023, EPI_ISL_2855024, EPI_ISL_2855025, EPI_ISL_2855026, EPI_ISL_2855027, EPI_ISL_2855028, EPI_ISL_2855029, EPI_ISL_2855030, EPI_ISL_2855031, EPI_ISL_2855032, EPI_ISL_2855033, EPI_ISL_2855034, EPI_ISL_2855035, EPI_ISL_2855036, EPI_ISL_2855037, EPI_ISL_2855038, EPI_ISL_2855039, EPI_ISL_2855040, EPI_ISL_2855041, EPI_ISL_2855042, EPI_ISL_2855043, EPI_ISL_2855046, EPI_ISL_2855047, EPI_ISL_2855048, EPI_ISL_2855049, EPI_ISL_2855050, EPI_ISL_2855051, EPI_ISL_2855052, EPI_ISL_2855053, EPI_ISL_2855054, EPI_ISL_2855055, EPI_ISL_2855057, EPI_ISL_2855061, EPI_ISL_2855062, EPI_ISL_2855065, EPI_ISL_2855066, EPI_ISL_2855067, EPI_ISL_2855069, EPI_ISL_2855070, EPI_ISL_2855071, EPI_ISL_2855072, EPI_ISL_2855073, EPI_ISL_2855075, EPI_ISL_2855076, EPI_ISL_2855077, EPI_ISL_2855078, EPI_ISL_2855080, EPI_ISL_2855084, EPI_ISL_2855085, EPI_ISL_2855089, EPI_ISL_2855090, EPI_ISL_2855093, EPI_ISL_2855095, EPI_ISL_2855096, EPI_ISL_2855097, EPI_ISL_2855098, EPI_ISL_2855099, EPI_ISL_2855100, EPI_ISL_2855101, EPI_ISL_2855102, EPI_ISL_2855103, EPI_ISL_2855104, EPI_ISL_2855105, EPI_ISL_2855110, EPI_ISL_2855111, EPI_ISL_2855113, EPI_ISL_2855114, EPI_ISL_2855115, EPI_ISL_2855116, EPI_ISL_2855117, EPI_ISL_2855118, EPI_ISL_2855119, EPI_ISL_2855120, EPI_ISL_2855122, EPI_ISL_2855123, EPI_ISL_2855124, EPI_ISL_2855125, EPI_ISL_2855126, EPI_ISL_2855127, EPI_ISL_2855128, EPI_ISL_2855129, EPI_ISL_2855130, EPI_ISL_2855131, EPI_ISL_2855132, EPI_ISL_2855133, EPI_ISL_2855134, EPI_ISL_2855135, EPI_ISL_2855136, EPI_ISL_2855138, EPI_ISL_2855139, EPI_ISL_2855143, EPI_ISL_2855144, EPI_ISL_2855145, EPI_ISL_2855146, EPI_ISL_2855147, EPI_ISL_2855148, EPI_ISL_2855149, EPI_ISL_2855150, EPI_ISL_2855151, EPI_ISL_2855153, EPI_ISL_2855155, EPI_ISL_2855156, EPI_ISL_2855157, EPI_ISL_2855158, EPI_ISL_2855159, EPI_ISL_2855160, EPI_ISL_2855161, EPI_ISL_2855162, EPI_ISL_2855163, EPI_ISL_2855164, EPI_ISL_2855165, EPI_ISL_2855166, EPI_ISL_2855167, EPI_ISL_2855168, EPI_ISL_2855169, EPI_ISL_2855170, EPI_ISL_2855171, EPI_ISL_2855172, EPI_ISL_2855173, EPI_ISL_2855181, EPI_ISL_2855182, EPI_ISL_2855183, EPI_ISL_2855184, EPI_ISL_2855185, EPI_ISL_2855186, EPI_ISL_2855187, EPI_ISL_2855190, EPI_ISL_2855191, EPI_ISL_2855192, EPI_ISL_2855193, EPI_ISL_2855194, EPI_ISL_2855195, EPI_ISL_2855196, EPI_ISL_2855197, EPI_ISL_2855198, EPI_ISL_2855199, EPI_ISL_2855200, EPI_ISL_2855201, EPI_ISL_2855202, EPI_ISL_2855203, EPI_ISL_2855205, EPI_ISL_2855208, EPI_ISL_2855209, EPI_ISL_2855212, EPI_ISL_2855215, EPI_ISL_2855216, EPI_ISL_2855217, EPI_ISL_2855218, EPI_ISL_2855219, EPI_ISL_2855220, EPI_ISL_2855223, EPI_ISL_2855224, EPI_ISL_2855225, EPI_ISL_2855226, EPI_ISL_2855227, EPI_ISL_2855228, EPI_ISL_2855231, EPI_ISL_2855232, EPI_ISL_2855233, EPI_ISL_2855234, EPI_ISL_2855235, EPI_ISL_2855237, EPI_ISL_2855238, EPI_ISL_2855239, EPI_ISL_2855243, EPI_ISL_2855244, EPI_ISL_2855245, EPI_ISL_2855246, EPI_ISL_2855248, EPI_ISL_2855249, EPI_ISL_2855250, EPI_ISL_2855251, EPI_ISL_2855253, EPI_ISL_2855254, EPI_ISL_2855255, EPI_ISL_2855256, EPI_ISL_2855260, EPI_ISL_2855261, EPI_ISL_2855262, EPI_ISL_2855263, EPI_ISL_2855264, EPI_ISL_2855265, EPI_ISL_2855266, EPI_ISL_2855267, EPI_ISL_2855268, EPI_ISL_2855269, EPI_ISL_2855270, EPI_ISL_2855271, EPI_ISL_2855272, EPI_ISL_2855273, EPI_ISL_2855274, EPI_ISL_2855275, EPI_ISL_2855276, EPI_ISL_2855277, EPI_ISL_2855280, EPI_ISL_2855284, EPI_ISL_2855285, EPI_ISL_2855286, EPI_ISL_2855287, EPI_ISL_2855288, EPI_ISL_2855289, EPI_ISL_2855290, EPI_ISL_2855291, EPI_ISL_2855292, EPI_ISL_2855293, EPI_ISL_2855294, EPI_ISL_2855295, EPI_ISL_2855297, EPI_ISL_2855298, EPI_ISL_2855299, EPI_ISL_2855300, EPI_ISL_2855301, EPI_ISL_2855302, EPI_ISL_2855303, EPI_ISL_2855304, EPI_ISL_2855305, EPI_ISL_2855306, EPI_ISL_2855307, EPI_ISL_2855308, EPI_ISL_2855309, EPI_ISL_2855311, EPI_ISL_2855312, EPI_ISL_2855313, EPI_ISL_2855314, EPI_ISL_2855315, EPI_ISL_2855316, EPI_ISL_2855317, EPI_ISL_2855318, EPI_ISL_2855319, EPI_ISL_2855320, EPI_ISL_2855321, EPI_ISL_2855322, EPI_ISL_2855323, EPI_ISL_2855324, EPI_ISL_2855326, EPI_ISL_2855327, EPI_ISL_2855328, EPI_ISL_2855329, EPI_ISL_2855330, EPI_ISL_2855331, EPI_ISL_2855332, EPI_ISL_2855333, EPI_ISL_2855334, EPI_ISL_2855335, EPI_ISL_2855336, EPI_ISL_2855337, EPI_ISL_2855338, EPI_ISL_2855339, EPI_ISL_2855340, EPI_ISL_2855341, EPI_ISL_2855342, EPI_ISL_2855343, EPI_ISL_2855344, EPI_ISL_2855345, EPI_ISL_2855346, EPI_ISL_2855347, EPI_ISL_2855348, EPI_ISL_2855349, EPI_ISL_2855350, EPI_ISL_2855351, EPI_ISL_2855352, EPI_ISL_2855353, EPI_ISL_2855354, EPI_ISL_2855355, EPI_ISL_2855356, EPI_ISL_2855358, EPI_ISL_2855359, EPI_ISL_2855360, EPI_ISL_2855361, EPI_ISL_2855362, EPI_ISL_2855363, EPI_ISL_2855364, EPI_ISL_2855365, EPI_ISL_2855366, EPI_ISL_2855367, EPI_ISL_2855368, EPI_ISL_2855369, EPI_ISL_2855371, EPI_ISL_2855372, EPI_ISL_2855373, EPI_ISL_2855374, EPI_ISL_2855375, EPI_ISL_2855376, EPI_ISL_2855377, EPI_ISL_2855378, EPI_ISL_2855380, EPI_ISL_2855382, EPI_ISL_2855383, EPI_ISL_2855384, EPI_ISL_2855386, EPI_ISL_2855387, EPI_ISL_2855388, EPI_ISL_2855389, EPI_ISL_2855391, EPI_ISL_2855392, EPI_ISL_2855394, EPI_ISL_2855395, EPI_ISL_2855396, EPI_ISL_2855397, EPI_ISL_2855398, EPI_ISL_2855399, EPI_ISL_2855400, EPI_ISL_2855401, EPI_ISL_2855402, EPI_ISL_2855403, EPI_ISL_2855404, EPI_ISL_2855405, EPI_ISL_2855406, EPI_ISL_2855407, EPI_ISL_2855408, EPI_ISL_2855412, EPI_ISL_2855413, EPI_ISL_2855414, EPI_ISL_2855417, EPI_ISL_2855418, EPI_ISL_2855419, EPI_ISL_2855420, EPI_ISL_2855424, EPI_ISL_2855429, EPI_ISL_2855433, EPI_ISL_2855434, EPI_ISL_2855435, EPI_ISL_2855436, EPI_ISL_2855437, EPI_ISL_2855438, EPI_ISL_2855439, EPI_ISL_2855440, EPI_ISL_2855441, EPI_ISL_2855442, EPI_ISL_2855443, EPI_ISL_2855445, EPI_ISL_2855446, EPI_ISL_2855447, EPI_ISL_2855448, EPI_ISL_2855449, EPI_ISL_2855450, EPI_ISL_2855451, EPI_ISL_2855452, EPI_ISL_2855453, EPI_ISL_2855454, EPI_ISL_2855455, EPI_ISL_2855456, EPI_ISL_2855457, EPI_ISL_2855458, EPI_ISL_2855459, EPI_ISL_2855460, EPI_ISL_2855461, EPI_ISL_2855462, EPI_ISL_2855464, EPI_ISL_2855465, EPI_ISL_2855466, EPI_ISL_2855467, EPI_ISL_2855468, EPI_ISL_2855469, EPI_ISL_2855471, EPI_ISL_2855472, EPI_ISL_2855473, EPI_ISL_2855474, EPI_ISL_2855475, EPI_ISL_2855476, EPI_ISL_2855477, EPI_ISL_2855478, EPI_ISL_2855479, EPI_ISL_2855480, EPI_ISL_2855481, EPI_ISL_2855482, EPI_ISL_2855483, EPI_ISL_2855484, EPI_ISL_2855485, EPI_ISL_2855486, EPI_ISL_2855487, EPI_ISL_2855488, EPI_ISL_2855489, EPI_ISL_2855494, EPI_ISL_2855495, EPI_ISL_2855496, EPI_ISL_2855497, EPI_ISL_2855498, EPI_ISL_2855499, EPI_ISL_2855500, EPI_ISL_2855501, EPI_ISL_2855502, EPI_ISL_2855503, EPI_ISL_2855504, EPI_ISL_2855505, EPI_ISL_2855506, EPI_ISL_2855507, EPI_ISL_2855508, EPI_ISL_2855509, EPI_ISL_2855510, EPI_ISL_2855511, EPI_ISL_2855512, EPI_ISL_2855513, EPI_ISL_2855514, EPI_ISL_2855515, EPI_ISL_2855516, EPI_ISL_2855517, EPI_ISL_2855518, EPI_ISL_2855519, EPI_ISL_2855520, EPI_ISL_2855521, EPI_ISL_2855522, EPI_ISL_2855525, EPI_ISL_2855528, EPI_ISL_2855529, EPI_ISL_2855530, EPI_ISL_2855531, EPI_ISL_2855532, EPI_ISL_2855533, EPI_ISL_2855534, EPI_ISL_2855535, EPI_ISL_2855536, EPI_ISL_2855537, EPI_ISL_2855538, EPI_ISL_2855539, EPI_ISL_2855541, EPI_ISL_2855542, EPI_ISL_2855545, EPI_ISL_2855547, EPI_ISL_2855548, EPI_ISL_2855549, EPI_ISL_2855550, EPI_ISL_2855551, EPI_ISL_2855552, EPI_ISL_2855553, EPI_ISL_2855554, EPI_ISL_2855555, EPI_ISL_2855556, EPI_ISL_2855557, EPI_ISL_2855558, EPI_ISL_2855559, EPI_ISL_2855560, EPI_ISL_2855563, EPI_ISL_2855564, EPI_ISL_2855565, EPI_ISL_2855566, EPI_ISL_2855567, EPI_ISL_2855568, EPI_ISL_2855571, EPI_ISL_2855572, EPI_ISL_2855573, EPI_ISL_2855574, EPI_ISL_2855575, EPI_ISL_2855576, EPI_ISL_2855577, EPI_ISL_2855578, EPI_ISL_2855579, EPI_ISL_2855580, EPI_ISL_2855581, EPI_ISL_2855582, EPI_ISL_2855583, EPI_ISL_2855584, EPI_ISL_2855585, EPI_ISL_2855586, EPI_ISL_2855589, EPI_ISL_2855590, EPI_ISL_2855591, EPI_ISL_2855592, EPI_ISL_2855594, EPI_ISL_2855595, EPI_ISL_2855596, EPI_ISL_2855598, EPI_ISL_2855599, EPI_ISL_2855600, EPI_ISL_2855601, EPI_ISL_2855602, EPI_ISL_2855603, EPI_ISL_2855606, EPI_ISL_2855608, EPI_ISL_2855609, EPI_ISL_2855621, EPI_ISL_2855622, EPI_ISL_2855623, EPI_ISL_2855624, EPI_ISL_2855625, EPI_ISL_2855626, EPI_ISL_2855627, EPI_ISL_2855628, EPI_ISL_2855629, EPI_ISL_2855630, EPI_ISL_2855631, EPI_ISL_2855632, EPI_ISL_2855633, EPI_ISL_2855634, EPI_ISL_2855635, EPI_ISL_2855636, EPI_ISL_2855637, EPI_ISL_2855638, EPI_ISL_2855639, EPI_ISL_2855644, EPI_ISL_2855645, EPI_ISL_2855646, EPI_ISL_2855647, EPI_ISL_2855648, EPI_ISL_2855649, EPI_ISL_2855651, EPI_ISL_2855652, EPI_ISL_2855653, EPI_ISL_2855654, EPI_ISL_2855655, EPI_ISL_2855656, EPI_ISL_2855657, EPI_ISL_2855659, EPI_ISL_2855666, EPI_ISL_2855667, EPI_ISL_2855668, EPI_ISL_2855669, EPI_ISL_2855670, EPI_ISL_2855671, EPI_ISL_2855672, EPI_ISL_2855673, EPI_ISL_2855674, EPI_ISL_2855675, EPI_ISL_2855676, EPI_ISL_2855677, EPI_ISL_2855678, EPI_ISL_2855679, EPI_ISL_2855687, EPI_ISL_2855688, EPI_ISL_2855690, EPI_ISL_2855691, EPI_ISL_2855692, EPI_ISL_2855693, EPI_ISL_2855694, EPI_ISL_2855695, EPI_ISL_2855696, EPI_ISL_2855697, EPI_ISL_2855698, EPI_ISL_2855699, EPI_ISL_2855700, EPI_ISL_2855701, EPI_ISL_2855703, EPI_ISL_2855704, EPI_ISL_2855705, EPI_ISL_2855706, EPI_ISL_2855707, EPI_ISL_2855708, EPI_ISL_2855709, EPI_ISL_2855710, EPI_ISL_2855711, EPI_ISL_2855712, EPI_ISL_2855717, EPI_ISL_2855718, EPI_ISL_2855719, EPI_ISL_2855720, EPI_ISL_2855721, EPI_ISL_2855730, EPI_ISL_2855734, EPI_ISL_2855735, EPI_ISL_2855737, EPI_ISL_2855738, EPI_ISL_2855743, EPI_ISL_2855745, EPI_ISL_2855747, EPI_ISL_2855748, EPI_ISL_2855749, EPI_ISL_2855750, EPI_ISL_2855751, EPI_ISL_2855752, EPI_ISL_2855753, EPI_ISL_2855754, EPI_ISL_2855755, EPI_ISL_2855756, EPI_ISL_2855757, EPI_ISL_2855758, EPI_ISL_2855759, EPI_ISL_2855760, EPI_ISL_2855761, EPI_ISL_2855762, EPI_ISL_2855767, EPI_ISL_2855768, EPI_ISL_2855769, EPI_ISL_2855770, EPI_ISL_2855771, EPI_ISL_2855773, EPI_ISL_2855774, EPI_ISL_2855775, EPI_ISL_2855776, EPI_ISL_2855777, EPI_ISL_2855780, EPI_ISL_2855781, EPI_ISL_2855782, EPI_ISL_2855783, EPI_ISL_2855784, EPI_ISL_2855794, EPI_ISL_2855795, EPI_ISL_2855796, EPI_ISL_2855797, EPI_ISL_2855798, EPI_ISL_2855799, EPI_ISL_2855800, EPI_ISL_2855801, EPI_ISL_2855803, EPI_ISL_2855804, EPI_ISL_2855805, EPI_ISL_2855806, EPI_ISL_2855807, EPI_ISL_2855808, EPI_ISL_2855809, EPI_ISL_2855810, EPI_ISL_2855811, EPI_ISL_2855812, EPI_ISL_2855813, EPI_ISL_2855814, EPI_ISL_2855815, EPI_ISL_2855816, EPI_ISL_2855817, EPI_ISL_2855818, EPI_ISL_2855819, EPI_ISL_2855820, EPI_ISL_2855821, EPI_ISL_2855822, EPI_ISL_2855823, EPI_ISL_2855824, EPI_ISL_2855829, EPI_ISL_2855830, EPI_ISL_2855831, EPI_ISL_2855832, EPI_ISL_2855833, EPI_ISL_2855834, EPI_ISL_2855835, EPI_ISL_2855836, EPI_ISL_2855837, EPI_ISL_2855838, EPI_ISL_2855839, EPI_ISL_2855840, EPI_ISL_2855841, EPI_ISL_2855842, EPI_ISL_2855843, EPI_ISL_2855844, EPI_ISL_2855845, EPI_ISL_2855846, EPI_ISL_2855847, EPI_ISL_2855848, EPI_ISL_2855849, EPI_ISL_2855850, EPI_ISL_2855851, EPI_ISL_2858254, EPI_ISL_2858255, EPI_ISL_2858256, EPI_ISL_2858281, EPI_ISL_2858329, EPI_ISL_2858340, EPI_ISL_2858370, EPI_ISL_2858394, EPI_ISL_2858447, EPI_ISL_2858458, EPI_ISL_2858473, EPI_ISL_2858474, EPI_ISL_2858475, EPI_ISL_2858476, EPI_ISL_2858477, EPI_ISL_2858478, EPI_ISL_2858479, EPI_ISL_2858480, EPI_ISL_2858481, EPI_ISL_2858493, EPI_ISL_2858521, EPI_ISL_2858522, EPI_ISL_2858523, EPI_ISL_2858524, EPI_ISL_2858525, EPI_ISL_2858526, EPI_ISL_2858527, EPI_ISL_2858528, EPI_ISL_2858529, EPI_ISL_2858530, EPI_ISL_2858531, EPI_ISL_2858543, EPI_ISL_2858544, EPI_ISL_2858545, EPI_ISL_2858546, EPI_ISL_2858547, EPI_ISL_2858548, EPI_ISL_2858549, EPI_ISL_2858550, EPI_ISL_2858551, EPI_ISL_2858552, EPI_ISL_2858553, EPI_ISL_2858554, EPI_ISL_2881858, EPI_ISL_2881859, EPI_ISL_2881860, EPI_ISL_2881861, EPI_ISL_2881862, EPI_ISL_2881863, EPI_ISL_2881864, EPI_ISL_2881865, EPI_ISL_2881866, EPI_ISL_2881869, EPI_ISL_2881875, EPI_ISL_2881877, EPI_ISL_2881878, EPI_ISL_2881879, EPI_ISL_2881880, EPI_ISL_2881881, EPI_ISL_2881885, EPI_ISL_2881886, EPI_ISL_2881893, EPI_ISL_2881896, EPI_ISL_2881906, EPI_ISL_2881907, EPI_ISL_2881908, EPI_ISL_2881909, EPI_ISL_2881911, EPI_ISL_2881913, EPI_ISL_2881914, EPI_ISL_2881915, EPI_ISL_2881917, EPI_ISL_2881918, EPI_ISL_2881919, EPI_ISL_2881920, EPI_ISL_2881921, EPI_ISL_2881922, EPI_ISL_2881923, EPI_ISL_2881924, EPI_ISL_2881925, EPI_ISL_2881926, EPI_ISL_2881927, EPI_ISL_2881928, EPI_ISL_2881929, EPI_ISL_2881930, EPI_ISL_2881931, EPI_ISL_2881932, EPI_ISL_2881933, EPI_ISL_2881934, EPI_ISL_2881935, EPI_ISL_2881936, EPI_ISL_2881937, EPI_ISL_2881938, EPI_ISL_2881939, EPI_ISL_2881940, EPI_ISL_2881942, EPI_ISL_2881947, EPI_ISL_2881948, EPI_ISL_2881949, EPI_ISL_2881950, EPI_ISL_2881951, EPI_ISL_2881952, EPI_ISL_2881957, EPI_ISL_2881966, EPI_ISL_2881967, EPI_ISL_2881968, EPI_ISL_2881969, EPI_ISL_2881970, EPI_ISL_2881971, EPI_ISL_2881972, EPI_ISL_2881995, EPI_ISL_2881996, EPI_ISL_2881997, EPI_ISL_2881998, EPI_ISL_2881999, EPI_ISL_2882000, EPI_ISL_2882005, EPI_ISL_2882017, EPI_ISL_2882028, EPI_ISL_2882029, EPI_ISL_2882030, EPI_ISL_2882031, EPI_ISL_2882032, EPI_ISL_2882033, EPI_ISL_2882034, EPI_ISL_2882035, EPI_ISL_2882036, EPI_ISL_2882037, EPI_ISL_2882038, EPI_ISL_2882043, EPI_ISL_2882044, EPI_ISL_2882045, EPI_ISL_2882046, EPI_ISL_2882047, EPI_ISL_2882048, EPI_ISL_2882049, EPI_ISL_2882050, EPI_ISL_2882051, EPI_ISL_2882052, EPI_ISL_2882053, EPI_ISL_2882054, EPI_ISL_2882055, EPI_ISL_2882056, EPI_ISL_2882057, EPI_ISL_2882058, EPI_ISL_2882059, EPI_ISL_2882060, EPI_ISL_2882061, EPI_ISL_2882062, EPI_ISL_2882064, EPI_ISL_2882065, EPI_ISL_2882066, EPI_ISL_2882067, EPI_ISL_2882068, EPI_ISL_2882069, EPI_ISL_2882070, EPI_ISL_2882071, EPI_ISL_2882072, EPI_ISL_2882073, EPI_ISL_2882074, EPI_ISL_2882075, EPI_ISL_2882076, EPI_ISL_2882077, EPI_ISL_2882078, EPI_ISL_2882079, EPI_ISL_2882080, EPI_ISL_2882081, EPI_ISL_2882082, EPI_ISL_2882083, EPI_ISL_2882084, EPI_ISL_2882085, EPI_ISL_2882086, EPI_ISL_2882087, EPI_ISL_2882088, EPI_ISL_2882091, EPI_ISL_2882092, EPI_ISL_2882093, EPI_ISL_2882094, EPI_ISL_2882095, EPI_ISL_2882096, EPI_ISL_2882097, EPI_ISL_2882098, EPI_ISL_2882099, EPI_ISL_2882101, EPI_ISL_2882102, EPI_ISL_2882103, EPI_ISL_2882104, EPI_ISL_2882105, EPI_ISL_2882106, EPI_ISL_2882107, EPI_ISL_2882108, EPI_ISL_2882109, EPI_ISL_2882110, EPI_ISL_2882111, EPI_ISL_2882112, EPI_ISL_2882114, EPI_ISL_2882115, EPI_ISL_2882116, EPI_ISL_2882117, EPI_ISL_2882118, EPI_ISL_2882119, EPI_ISL_2882120, EPI_ISL_2882121, EPI_ISL_2882122, EPI_ISL_2882123, EPI_ISL_2882124, EPI_ISL_2882125, EPI_ISL_2882126, EPI_ISL_2882127, EPI_ISL_2882131, EPI_ISL_2882132, EPI_ISL_2882133, EPI_ISL_2882134, EPI_ISL_2882135, EPI_ISL_2882137, EPI_ISL_2882139, EPI_ISL_2882140, EPI_ISL_2882141, EPI_ISL_2882142, EPI_ISL_2882143, EPI_ISL_2882144, EPI_ISL_2882145, EPI_ISL_2882146, EPI_ISL_2882150, EPI_ISL_2882151, EPI_ISL_2882152, EPI_ISL_2882153, EPI_ISL_2882154, EPI_ISL_2882172, EPI_ISL_2882173, EPI_ISL_2882174, EPI_ISL_2882175, EPI_ISL_2882176, EPI_ISL_2882179, EPI_ISL_2882180, EPI_ISL_2882181, EPI_ISL_2882182, EPI_ISL_2882183, EPI_ISL_2882184, EPI_ISL_2882185, EPI_ISL_2882186, EPI_ISL_2882187, EPI_ISL_2882188, EPI_ISL_2882190, EPI_ISL_2882191, EPI_ISL_2882192, EPI_ISL_2882194, EPI_ISL_2882195, EPI_ISL_2882196, EPI_ISL_2882197, EPI_ISL_2882198, EPI_ISL_2882199, EPI_ISL_2882200, EPI_ISL_2882201, EPI_ISL_2882202, EPI_ISL_2882203, EPI_ISL_2882207, EPI_ISL_2882208, EPI_ISL_2882209, EPI_ISL_2882210, EPI_ISL_2882211, EPI_ISL_2882212, EPI_ISL_2882213, EPI_ISL_2882214, EPI_ISL_2882215, EPI_ISL_2882216, EPI_ISL_2882217, EPI_ISL_2882218, EPI_ISL_2882219, EPI_ISL_2882220, EPI_ISL_2882221, EPI_ISL_2882222, EPI_ISL_2882223, EPI_ISL_2882224, EPI_ISL_2882225, EPI_ISL_2882226, EPI_ISL_2882227, EPI_ISL_2882228, EPI_ISL_2882229, EPI_ISL_2882230, EPI_ISL_2882231, EPI_ISL_2882232, EPI_ISL_2882233, EPI_ISL_2882234, EPI_ISL_2882235, EPI_ISL_2882236, EPI_ISL_2882237, EPI_ISL_2882238, EPI_ISL_2882239, EPI_ISL_2882240, EPI_ISL_2882241, EPI_ISL_2882242, EPI_ISL_2882243, EPI_ISL_2882244, EPI_ISL_2882246, EPI_ISL_2882248, EPI_ISL_2882249, EPI_ISL_2882250, EPI_ISL_2882251, EPI_ISL_2882252, EPI_ISL_2882253, EPI_ISL_2882254, EPI_ISL_2882255, EPI_ISL_2882256, EPI_ISL_2882257, EPI_ISL_2882258, EPI_ISL_2882259, EPI_ISL_2882260, EPI_ISL_2882261, EPI_ISL_2882262, EPI_ISL_2882263, EPI_ISL_2882264, EPI_ISL_2882265, EPI_ISL_2882266, EPI_ISL_2882267, EPI_ISL_2882268, EPI_ISL_2882276, EPI_ISL_2882277, EPI_ISL_2882279, EPI_ISL_2882280, EPI_ISL_2882283, EPI_ISL_2882284, EPI_ISL_2882285, EPI_ISL_2882286, EPI_ISL_2882287, EPI_ISL_2882288, EPI_ISL_2882289, EPI_ISL_2882290, EPI_ISL_2882293, EPI_ISL_2882294, EPI_ISL_2882295, EPI_ISL_2882296, EPI_ISL_2882298, EPI_ISL_2882303, EPI_ISL_2882304, EPI_ISL_2882305, EPI_ISL_2882306, EPI_ISL_2882307, EPI_ISL_2882308, EPI_ISL_2882309, EPI_ISL_2882310, EPI_ISL_2882311, EPI_ISL_2882312, EPI_ISL_2882313, EPI_ISL_2882314, EPI_ISL_2882317, EPI_ISL_2882318, EPI_ISL_2882320, EPI_ISL_2882321, EPI_ISL_2882322, EPI_ISL_2882323, EPI_ISL_2882324, EPI_ISL_2882325, EPI_ISL_2882326, EPI_ISL_2882328, EPI_ISL_2882332, EPI_ISL_2882333, EPI_ISL_2882334, EPI_ISL_2882335, EPI_ISL_2882336, EPI_ISL_2882337, EPI_ISL_2882338, EPI_ISL_2882339, EPI_ISL_2882340, EPI_ISL_2882341, EPI_ISL_2882342, EPI_ISL_2882346, EPI_ISL_2882347, EPI_ISL_2882348, EPI_ISL_2882349, EPI_ISL_2882350, EPI_ISL_2882351, EPI_ISL_2882352, EPI_ISL_2882353, EPI_ISL_2882354, EPI_ISL_2882355, EPI_ISL_2882356, EPI_ISL_2882357, EPI_ISL_2882358, EPI_ISL_2882359, EPI_ISL_2882360, EPI_ISL_2882361, EPI_ISL_2882366, EPI_ISL_2882367, EPI_ISL_2882368, EPI_ISL_2882369, EPI_ISL_2882370, EPI_ISL_2882371, EPI_ISL_2882372, EPI_ISL_2882373, EPI_ISL_2882374, EPI_ISL_2882380, EPI_ISL_2882381, EPI_ISL_2882382, EPI_ISL_2882383, EPI_ISL_2882384, EPI_ISL_2882385, EPI_ISL_2882386, EPI_ISL_2882387, EPI_ISL_2882388, EPI_ISL_2882389, EPI_ISL_2882392, EPI_ISL_2882393, EPI_ISL_2882394, EPI_ISL_2882395, EPI_ISL_2882396, EPI_ISL_2882397, EPI_ISL_2882398, EPI_ISL_2882399, EPI_ISL_2882400, EPI_ISL_2882401, EPI_ISL_2882402, EPI_ISL_2882403, EPI_ISL_2882404, EPI_ISL_2882405, EPI_ISL_2882406, EPI_ISL_2882407, EPI_ISL_2882408, EPI_ISL_2882409, EPI_ISL_2882410, EPI_ISL_2882411, EPI_ISL_2882412, EPI_ISL_2882413, EPI_ISL_2882414, EPI_ISL_2882415, EPI_ISL_2882419, EPI_ISL_2882420, EPI_ISL_2882421, EPI_ISL_2882422, EPI_ISL_2882423, EPI_ISL_2882424, EPI_ISL_2882425, EPI_ISL_2882426, EPI_ISL_2882427, EPI_ISL_2882429, EPI_ISL_2882430, EPI_ISL_2882433, EPI_ISL_2882434, EPI_ISL_2882435, EPI_ISL_2882436, EPI_ISL_2882437, EPI_ISL_2882438, EPI_ISL_2882439, EPI_ISL_2882441, EPI_ISL_2882444, EPI_ISL_2882445, EPI_ISL_2882446, EPI_ISL_2882447, EPI_ISL_2882448, EPI_ISL_2882449, EPI_ISL_2882450, EPI_ISL_2882451, EPI_ISL_2882452, EPI_ISL_2882453, EPI_ISL_2882456, EPI_ISL_2882457, EPI_ISL_2882458, EPI_ISL_2882459, EPI_ISL_2882460, EPI_ISL_2882461, EPI_ISL_2882462, EPI_ISL_2882463, EPI_ISL_2882464, EPI_ISL_2882465, EPI_ISL_2882466, EPI_ISL_2882467, EPI_ISL_2882468, EPI_ISL_2882469, EPI_ISL_2882470, EPI_ISL_2882471, EPI_ISL_2882472, EPI_ISL_2882473, EPI_ISL_2882474, EPI_ISL_2882478, EPI_ISL_2882479, EPI_ISL_2882480, EPI_ISL_2882490, EPI_ISL_2882492, EPI_ISL_2882493, EPI_ISL_2882494, EPI_ISL_2882495, EPI_ISL_2882496, EPI_ISL_2882497, EPI_ISL_2882498, EPI_ISL_2882499, EPI_ISL_2882500, EPI_ISL_2882501, EPI_ISL_2882502, EPI_ISL_2882503, EPI_ISL_2882504, EPI_ISL_2882505, EPI_ISL_2882506, EPI_ISL_2882507, EPI_ISL_2882508, EPI_ISL_2882509, EPI_ISL_2882510, EPI_ISL_2882511, EPI_ISL_2882512, EPI_ISL_2882513, EPI_ISL_2882514, EPI_ISL_2882515, EPI_ISL_2882516, EPI_ISL_2882517, EPI_ISL_2882518, EPI_ISL_2882519, EPI_ISL_2882520, EPI_ISL_2882521, EPI_ISL_2882522, EPI_ISL_2882523, EPI_ISL_2882524, EPI_ISL_2882525, EPI_ISL_2882526, EPI_ISL_2882527, EPI_ISL_2882528, EPI_ISL_2882529, EPI_ISL_2882531, EPI_ISL_2882532, EPI_ISL_2882533, EPI_ISL_2882534, EPI_ISL_2882535, EPI_ISL_2882539, EPI_ISL_2882542, EPI_ISL_2882543, EPI_ISL_2882544, EPI_ISL_2882547, EPI_ISL_2882548, EPI_ISL_2882549, EPI_ISL_2882550, EPI_ISL_2882551, EPI_ISL_2882552, EPI_ISL_2882553, EPI_ISL_2882554, EPI_ISL_2882555, EPI_ISL_2882556, EPI_ISL_2882557, EPI_ISL_2882558, EPI_ISL_2882559, EPI_ISL_2882560, EPI_ISL_2882561, EPI_ISL_2882562, EPI_ISL_2882563, EPI_ISL_2882564, EPI_ISL_2882565, EPI_ISL_2882566, EPI_ISL_2882567, EPI_ISL_2882568, EPI_ISL_2882569, EPI_ISL_2882570, EPI_ISL_2882571, EPI_ISL_2882572, EPI_ISL_2882573, EPI_ISL_2882574, EPI_ISL_2882575, EPI_ISL_2882576, EPI_ISL_2882577, EPI_ISL_2882578, EPI_ISL_2882579, EPI_ISL_2882580, EPI_ISL_2882581, EPI_ISL_2882582, EPI_ISL_2882583, EPI_ISL_2882586, EPI_ISL_2882594, EPI_ISL_2882596, EPI_ISL_2882597, EPI_ISL_2882598, EPI_ISL_2882599, EPI_ISL_2882600, EPI_ISL_2882601, EPI_ISL_2882602, EPI_ISL_2882603, EPI_ISL_2882604, EPI_ISL_2882605, EPI_ISL_2882606, EPI_ISL_2882607, EPI_ISL_2882608, EPI_ISL_2882609, EPI_ISL_2882610, EPI_ISL_2882611, EPI_ISL_2882612, EPI_ISL_2882613, EPI_ISL_2882614, EPI_ISL_2882615, EPI_ISL_2882616, EPI_ISL_2882617, EPI_ISL_2882618, EPI_ISL_2882619, EPI_ISL_2882620, EPI_ISL_2882621, EPI_ISL_2882622, EPI_ISL_2882623, EPI_ISL_2882624, EPI_ISL_2882626, EPI_ISL_2882629, EPI_ISL_2882630, EPI_ISL_2882631, EPI_ISL_2882633, EPI_ISL_2882634, EPI_ISL_2882639, EPI_ISL_2882640, EPI_ISL_2882641, EPI_ISL_2882643, EPI_ISL_2882644, EPI_ISL_2882645, EPI_ISL_2882646, EPI_ISL_2882647, EPI_ISL_2882650, EPI_ISL_2882651, EPI_ISL_2882652, EPI_ISL_2882653, EPI_ISL_2882654, EPI_ISL_2882656, EPI_ISL_2882657, EPI_ISL_2882658, EPI_ISL_2882659, EPI_ISL_2882660, EPI_ISL_2882661, EPI_ISL_2882662, EPI_ISL_2882663, EPI_ISL_2882664, EPI_ISL_2882665, EPI_ISL_2882666, EPI_ISL_2882667, EPI_ISL_2882668, EPI_ISL_2882669, EPI_ISL_2882670, EPI_ISL_2882671, EPI_ISL_2882672, EPI_ISL_2882673, EPI_ISL_2882674, EPI_ISL_2882675, EPI_ISL_2882676, EPI_ISL_2882677, EPI_ISL_2882678, EPI_ISL_2882679, EPI_ISL_2882680, EPI_ISL_2882681, EPI_ISL_2882682, EPI_ISL_2882683, EPI_ISL_2882684, EPI_ISL_2882685, EPI_ISL_2882686, EPI_ISL_2882687, EPI_ISL_2882690, EPI_ISL_2882691, EPI_ISL_2882692, EPI_ISL_2882693, EPI_ISL_2882694, EPI_ISL_2882695, EPI_ISL_2882696, EPI_ISL_2882697, EPI_ISL_2882698, EPI_ISL_2882699, EPI_ISL_2882700, EPI_ISL_2882701, EPI_ISL_2882702, EPI_ISL_2882703, EPI_ISL_2882708, EPI_ISL_2882709, EPI_ISL_2882710, EPI_ISL_2882711, EPI_ISL_2882712, EPI_ISL_2882713, EPI_ISL_2882714, EPI_ISL_2882715, EPI_ISL_2882716, EPI_ISL_2882717, EPI_ISL_2882719, EPI_ISL_2882720, EPI_ISL_2882721, EPI_ISL_2882722, EPI_ISL_2882723, EPI_ISL_2882724, EPI_ISL_2882725, EPI_ISL_2882726, EPI_ISL_2882727, EPI_ISL_2882728, EPI_ISL_2882729, EPI_ISL_2882730, EPI_ISL_2882731, EPI_ISL_2889858, EPI_ISL_2889860, EPI_ISL_2889867, EPI_ISL_2889882, EPI_ISL_2889883, EPI_ISL_2889884, EPI_ISL_2889893, EPI_ISL_2889911, EPI_ISL_2889912, EPI_ISL_2889913, EPI_ISL_2889922, EPI_ISL_2889925, EPI_ISL_2889926, EPI_ISL_2889927, EPI_ISL_2889928, EPI_ISL_2889929, EPI_ISL_2889930, EPI_ISL_2889931, EPI_ISL_2889932, EPI_ISL_2889945, EPI_ISL_2889946, EPI_ISL_2889961, EPI_ISL_2889970, EPI_ISL_2889971, EPI_ISL_2889993, EPI_ISL_2889994, EPI_ISL_2889996, EPI_ISL_2889997, EPI_ISL_2889998, EPI_ISL_2890007, EPI_ISL_2890009, EPI_ISL_2890010, EPI_ISL_2890011, EPI_ISL_2890012, EPI_ISL_2890054, EPI_ISL_2890060, EPI_ISL_2890063, EPI_ISL_2890064, EPI_ISL_2890066, EPI_ISL_2890067, EPI_ISL_2890068, EPI_ISL_2890069, EPI_ISL_2890070, EPI_ISL_2890076, EPI_ISL_2890086, EPI_ISL_2890087, EPI_ISL_2890088, EPI_ISL_2890089, EPI_ISL_2890090, EPI_ISL_2890103, EPI_ISL_2890104, EPI_ISL_2890127, EPI_ISL_2890129, EPI_ISL_2890130, EPI_ISL_2890131, EPI_ISL_2890132, EPI_ISL_2890133, EPI_ISL_2890137, EPI_ISL_2890147, EPI_ISL_2890148, EPI_ISL_2890150, EPI_ISL_2890151, EPI_ISL_2890152, EPI_ISL_2890153, EPI_ISL_2890178, EPI_ISL_2890179, EPI_ISL_2890183, EPI_ISL_2890223, EPI_ISL_2890224, EPI_ISL_2890238, EPI_ISL_2890246, EPI_ISL_2890247, EPI_ISL_2890248, EPI_ISL_2890258, EPI_ISL_2890267, EPI_ISL_2890268, EPI_ISL_2890269, EPI_ISL_2890270, EPI_ISL_2890271, EPI_ISL_2890283, EPI_ISL_2890284, EPI_ISL_2890285, EPI_ISL_2890286, EPI_ISL_2890287, EPI_ISL_2890288, EPI_ISL_2890294, EPI_ISL_2890295, EPI_ISL_2890296, EPI_ISL_2890305, EPI_ISL_2890320, EPI_ISL_2890321, EPI_ISL_2890322, EPI_ISL_2890325, EPI_ISL_2890326, EPI_ISL_2890327, EPI_ISL_2890328, EPI_ISL_2890329, EPI_ISL_2890338, EPI_ISL_2890343, EPI_ISL_2890344, EPI_ISL_2890345, EPI_ISL_2890363, EPI_ISL_2890401, EPI_ISL_2890405, EPI_ISL_2890407, EPI_ISL_2890426, EPI_ISL_2890440, EPI_ISL_2890441, EPI_ISL_2890445, EPI_ISL_2890450, EPI_ISL_2890474, EPI_ISL_2890475, EPI_ISL_2890476, EPI_ISL_2890483, EPI_ISL_2890486, EPI_ISL_2890487, EPI_ISL_2890489, EPI_ISL_2890516, EPI_ISL_2890517, EPI_ISL_2890519, EPI_ISL_2890520, EPI_ISL_2890521, EPI_ISL_2890522, EPI_ISL_2890523, EPI_ISL_2890524, EPI_ISL_2890527, EPI_ISL_2890537, EPI_ISL_2890538, EPI_ISL_2890539, EPI_ISL_2890540, EPI_ISL_2890541, EPI_ISL_2890554, EPI_ISL_2890555, EPI_ISL_2890556, EPI_ISL_2890562, EPI_ISL_2890563, EPI_ISL_2890564, EPI_ISL_2890565, EPI_ISL_2890566, EPI_ISL_2890567, EPI_ISL_2890568, EPI_ISL_2890569, EPI_ISL_2890571, EPI_ISL_2890578, EPI_ISL_2890579, EPI_ISL_2890610, EPI_ISL_2890614, EPI_ISL_2890615, EPI_ISL_2890632, EPI_ISL_2890634, EPI_ISL_2890635, EPI_ISL_2890636, EPI_ISL_2890637, EPI_ISL_2890643, EPI_ISL_2890654, EPI_ISL_2890655, EPI_ISL_2890656, EPI_ISL_2890657, EPI_ISL_2890658, EPI_ISL_2890662, EPI_ISL_2890663, EPI_ISL_2890670, EPI_ISL_2890671, EPI_ISL_2890672, EPI_ISL_2890673, EPI_ISL_2890674, EPI_ISL_2890675, EPI_ISL_2890676, EPI_ISL_2890686, EPI_ISL_2890717, EPI_ISL_2890718, EPI_ISL_2890725, EPI_ISL_2890726, EPI_ISL_2890727, EPI_ISL_2890728, EPI_ISL_2890729, EPI_ISL_2890809, EPI_ISL_2890810, EPI_ISL_2890812, EPI_ISL_2890813, EPI_ISL_2890814, EPI_ISL_2890815, EPI_ISL_2890816, EPI_ISL_2890817, EPI_ISL_2890818, EPI_ISL_2890819, EPI_ISL_2890830, EPI_ISL_2890840, EPI_ISL_2890841, EPI_ISL_2890842, EPI_ISL_2890843, EPI_ISL_2890853, EPI_ISL_2890854, EPI_ISL_2890874, EPI_ISL_2890875, EPI_ISL_2890876, EPI_ISL_2890877, EPI_ISL_2890885, EPI_ISL_2890935, EPI_ISL_2890936, EPI_ISL_2890937, EPI_ISL_2890938, EPI_ISL_2890939, EPI_ISL_2890940, EPI_ISL_2890944, EPI_ISL_2890954, EPI_ISL_2890955, EPI_ISL_2890956, EPI_ISL_2890957, EPI_ISL_2890958, EPI_ISL_2890959, EPI_ISL_2890960, EPI_ISL_2890961, EPI_ISL_2890962, EPI_ISL_2890963, EPI_ISL_2890964, EPI_ISL_2890965, EPI_ISL_2890966, EPI_ISL_2890967, EPI_ISL_2890971, EPI_ISL_2890973, EPI_ISL_2890975, EPI_ISL_2890980, EPI_ISL_2890981, EPI_ISL_2890991, EPI_ISL_2890992, EPI_ISL_2891012, EPI_ISL_2891013, EPI_ISL_2891014, EPI_ISL_2891015, EPI_ISL_2891016, EPI_ISL_2891017, EPI_ISL_2891018, EPI_ISL_2891019, EPI_ISL_2891044, EPI_ISL_2891046, EPI_ISL_2891048, EPI_ISL_2891049, EPI_ISL_2891058, EPI_ISL_2891059, EPI_ISL_2891079, EPI_ISL_2891080, EPI_ISL_2891081, EPI_ISL_2891082, EPI_ISL_2891084, EPI_ISL_2891110, EPI_ISL_2891111, EPI_ISL_2891134, EPI_ISL_2891135, EPI_ISL_2891137, EPI_ISL_2891138, EPI_ISL_2891139, EPI_ISL_2891140, EPI_ISL_2891141, EPI_ISL_2891142, EPI_ISL_2891145, EPI_ISL_2891146, EPI_ISL_2891147, EPI_ISL_2891153, EPI_ISL_2891176, EPI_ISL_2891177, EPI_ISL_2891178, EPI_ISL_2891179, EPI_ISL_2891193, EPI_ISL_2891194, EPI_ISL_2891195, EPI_ISL_2891196, EPI_ISL_2891197, EPI_ISL_2891199, EPI_ISL_2891200, EPI_ISL_2891201, EPI_ISL_2891202, EPI_ISL_2891203, EPI_ISL_2891204, EPI_ISL_2891205, EPI_ISL_2891206, EPI_ISL_2891207, EPI_ISL_2891208, EPI_ISL_2891209, EPI_ISL_2891210, EPI_ISL_2891226, EPI_ISL_2891227, EPI_ISL_2891228, EPI_ISL_2891253, EPI_ISL_2891255, EPI_ISL_2891256, EPI_ISL_2891257, EPI_ISL_2891267, EPI_ISL_2891274, EPI_ISL_2891275, EPI_ISL_2891276, EPI_ISL_2891277, EPI_ISL_2891278, EPI_ISL_2891281, EPI_ISL_2891282, EPI_ISL_2891299, EPI_ISL_2891300, EPI_ISL_2891301, EPI_ISL_2891302, EPI_ISL_2891303, EPI_ISL_2891304, EPI_ISL_2891305, EPI_ISL_2891306, EPI_ISL_2891307, EPI_ISL_2891308, EPI_ISL_2891309, EPI_ISL_2891313, EPI_ISL_2891321, EPI_ISL_2891322, EPI_ISL_2891323, EPI_ISL_2891324, EPI_ISL_2891325, EPI_ISL_2891326, EPI_ISL_2891327, EPI_ISL_2891328, EPI_ISL_2891329, EPI_ISL_2891330, EPI_ISL_2891331, EPI_ISL_2891337, EPI_ISL_2891338, EPI_ISL_2891339, EPI_ISL_2891340, EPI_ISL_2891341, EPI_ISL_2891342, EPI_ISL_2891343, EPI_ISL_2891344, EPI_ISL_2891345, EPI_ISL_2891346, EPI_ISL_2891347, EPI_ISL_2891351, EPI_ISL_2891352, EPI_ISL_2891404, EPI_ISL_2891405, EPI_ISL_2891406, EPI_ISL_2891407, EPI_ISL_2891415, EPI_ISL_2891416, EPI_ISL_2891428, EPI_ISL_2891429, EPI_ISL_2891430, EPI_ISL_2891432, EPI_ISL_2891433, EPI_ISL_2891452, EPI_ISL_2891460, EPI_ISL_2891461, EPI_ISL_2891462, EPI_ISL_2891481, EPI_ISL_2891494, EPI_ISL_2891506, EPI_ISL_2891507, EPI_ISL_2891508, EPI_ISL_2891510, EPI_ISL_2891512, EPI_ISL_2891513, EPI_ISL_2891514, EPI_ISL_2891524, EPI_ISL_2891525, EPI_ISL_2891526, EPI_ISL_2891527, EPI_ISL_2891528, EPI_ISL_2891531, EPI_ISL_2891532, EPI_ISL_2891533, EPI_ISL_2891534, EPI_ISL_2891536, EPI_ISL_2891537, EPI_ISL_2891538, EPI_ISL_2891550, EPI_ISL_2891552, EPI_ISL_2891590, EPI_ISL_2891598, EPI_ISL_2891606, EPI_ISL_2891629, EPI_ISL_2891630, EPI_ISL_2891636, EPI_ISL_2891638, EPI_ISL_2891639, EPI_ISL_2891641, EPI_ISL_2891642, EPI_ISL_2891675, EPI_ISL_2891678, EPI_ISL_2891679, EPI_ISL_2891691, EPI_ISL_2891692, EPI_ISL_2891693, EPI_ISL_2892376, EPI_ISL_2892378, EPI_ISL_2892381, EPI_ISL_2892382, EPI_ISL_2892394, EPI_ISL_2892395, EPI_ISL_2892396, EPI_ISL_2934097, EPI_ISL_2934100, EPI_ISL_2934101, EPI_ISL_2934104, EPI_ISL_2934109, EPI_ISL_2934115, EPI_ISL_2934116, EPI_ISL_2934117, EPI_ISL_2934121, EPI_ISL_2934123, EPI_ISL_2934124, EPI_ISL_2934130, EPI_ISL_2934136, EPI_ISL_2934140, EPI_ISL_2934147, EPI_ISL_2934148, EPI_ISL_2934149, EPI_ISL_2934154, EPI_ISL_2934161, EPI_ISL_2934164, EPI_ISL_2934170, EPI_ISL_2934171, EPI_ISL_2934176, EPI_ISL_2934179, EPI_ISL_2934181, EPI_ISL_2934183, EPI_ISL_2934184, EPI_ISL_2934185, EPI_ISL_2934186, EPI_ISL_2934188, EPI_ISL_2934189, EPI_ISL_2934196, EPI_ISL_2934201, EPI_ISL_2934203, EPI_ISL_2934206, EPI_ISL_2934212, EPI_ISL_2934213, EPI_ISL_2934217, EPI_ISL_2934219, EPI_ISL_2934221, EPI_ISL_2934222, EPI_ISL_2934227, EPI_ISL_2934229, EPI_ISL_2934233, EPI_ISL_2934237, EPI_ISL_2934242, EPI_ISL_2934245, EPI_ISL_2934248, EPI_ISL_2934249, EPI_ISL_2934253, EPI_ISL_2934254, EPI_ISL_2934259, EPI_ISL_2934260, EPI_ISL_2934264, EPI_ISL_2934265, EPI_ISL_2934270, EPI_ISL_2934271, EPI_ISL_2934278, EPI_ISL_2934280, EPI_ISL_2934281, EPI_ISL_2934284, EPI_ISL_2934290, EPI_ISL_2934291, EPI_ISL_2934298, EPI_ISL_2934299, EPI_ISL_2934300, EPI_ISL_2934302, EPI_ISL_2934311, EPI_ISL_2934312, EPI_ISL_2934316, EPI_ISL_2934320, EPI_ISL_2934322, EPI_ISL_2934324, EPI_ISL_2934333, EPI_ISL_2934338, EPI_ISL_2934341, EPI_ISL_2934347, EPI_ISL_2934348, EPI_ISL_2934350, EPI_ISL_2934358, EPI_ISL_2934362, EPI_ISL_2934364, EPI_ISL_2934365, EPI_ISL_2934367, EPI_ISL_2934369, EPI_ISL_2934374, EPI_ISL_2934375, EPI_ISL_2934377, EPI_ISL_2934379, EPI_ISL_2934381, EPI_ISL_2934385, EPI_ISL_2934388, EPI_ISL_2934389, EPI_ISL_2934397, EPI_ISL_2934398, EPI_ISL_2934401, EPI_ISL_2934406, EPI_ISL_2934407, EPI_ISL_2934408, EPI_ISL_2934410, EPI_ISL_2934415, EPI_ISL_2934416, EPI_ISL_2934421, EPI_ISL_2934426, EPI_ISL_2934428, EPI_ISL_2934432, EPI_ISL_2934436, EPI_ISL_2934438, EPI_ISL_2934439, EPI_ISL_2934441, EPI_ISL_2934448, EPI_ISL_2934449, EPI_ISL_2934453, EPI_ISL_2934454, EPI_ISL_2934455, EPI_ISL_2934456, EPI_ISL_2934458, EPI_ISL_2934461, EPI_ISL_2934466, EPI_ISL_2934467, EPI_ISL_2934473, EPI_ISL_2934474, EPI_ISL_2934475, EPI_ISL_2934478, EPI_ISL_2934481, EPI_ISL_2934482, EPI_ISL_2934483, EPI_ISL_2934494, EPI_ISL_2934496, EPI_ISL_2934502, EPI_ISL_2934503, EPI_ISL_2934506, EPI_ISL_2934508, EPI_ISL_2934511, EPI_ISL_2934514, EPI_ISL_2934519, EPI_ISL_2934533, EPI_ISL_2934535, EPI_ISL_2934536, EPI_ISL_2934538, EPI_ISL_2934547, EPI_ISL_2934551, EPI_ISL_2934557

| | | | |
|---|---|---|---|
| see above | CDPH VBL | California Department of Public Health | CDPH-COVIDNet; SwabSeq/UCLA; UCLA Technology Center for Genomics & Bioinformatics; UCSF Center for Advanced Technology |

EPI_ISL_1373726, EPI_ISL_1373729, EPI_ISL_1373741, EPI_ISL_1373770, EPI_ISL_1373814, EPI_ISL_1373840, EPI_ISL_1373841, EPI_ISL_1373842, EPI_ISL_1373843, EPI_ISL_1373844, EPI_ISL_1373846, EPI_ISL_1373848, EPI_ISL_1373849, EPI_ISL_1373851, EPI_ISL_1373855, EPI_ISL_1373856, EPI_ISL_1373860, EPI_ISL_1373861, EPI_ISL_1373864, EPI_ISL_1373867, EPI_ISL_1373869, EPI_ISL_1373870, EPI_ISL_1373873, EPI_ISL_1373875, EPI_ISL_1373876, EPI_ISL_1373878

| | | | |
|---|---|---|---|
| see above | CDPH, Viral and Rickettsial Disease Laboratory | Genomics and Discovery, Respiratory Viruses Branch, Division of Viral Diseases, Centers for Disease Control and Prevention | Anna Kelleher; Anna Uehara; Brian Lynch; Clinton R. Paden; Haibin Wang; Han Jia Justin Ng; Jing Zhang; Krista Queen; Peter Cook; Suxiang Tong; Yan Li; Ying Tao |
| EPI_ISL_1483240 | CDPH, Viral and Rickettsial Disease Laboratory | Pathogen Discovery, Respiratory Viruses Branch, Division of Viral Diseases, Centers for Disease Control | Anna Kelleher; Anna Uehara; Brian Lynch; Clinton R. Paden; Haibin Wang; Han Jia Justin Ng; Jing Zhang; Krista Queen; Peter Cook; Suxiang Tong; Yan Li; Ying Tao |

| | | | |
|---|---|---|---|
| | | and Prevention | |
| EPI_ISL_2234098, EPI_ISL_2234102, EPI_ISL_2234108, EPI_ISL_2234109, EPI_ISL_2234110, EPI_ISL_2234111, EPI_ISL_2234112, EPI_ISL_2234113, EPI_ISL_2234114, EPI_ISL_2234115, EPI_ISL_2234116, EPI_ISL_2234117, EPI_ISL_2234118, EPI_ISL_2234119, EPI_ISL_2234121, EPI_ISL_2234122, EPI_ISL_2234123, EPI_ISL_2234124, EPI_ISL_2234125, EPI_ISL_2234130, EPI_ISL_2234131, EPI_ISL_2234132, EPI_ISL_2234133, EPI_ISL_2234134, EPI_ISL_2234135, EPI_ISL_2234136, EPI_ISL_2234137, EPI_ISL_2234138, EPI_ISL_2234140, EPI_ISL_2234142, EPI_ISL_2234145, EPI_ISL_2234157, EPI_ISL_2234164, EPI_ISL_2234165, EPI_ISL_2234196, EPI_ISL_2234216, EPI_ISL_2234217, EPI_ISL_2234218, EPI_ISL_2234219, EPI_ISL_2234220, EPI_ISL_2234221, EPI_ISL_2234311, EPI_ISL_2234327, EPI_ISL_2234328, EPI_ISL_2234329, EPI_ISL_2234330, EPI_ISL_2537388, EPI_ISL_2537389, EPI_ISL_2537390, EPI_ISL_2537391, EPI_ISL_2537392 | | | |
| see above | California Department of Public Health | California Department of Public Health | CDPH IDLB COVIDNet et al |
| EPI_ISL_2778213, EPI_ISL_2778214, EPI_ISL_2778215, EPI_ISL_2778216, EPI_ISL_2778217, EPI_ISL_2778218, EPI_ISL_2778219, EPI_ISL_2778220, EPI_ISL_2778221, EPI_ISL_2778222, EPI_ISL_2778223, EPI_ISL_2778224, EPI_ISL_2778225, EPI_ISL_2778226, EPI_ISL_2778227, EPI_ISL_2778228, EPI_ISL_2778229, EPI_ISL_2778230, EPI_ISL_2778231, EPI_ISL_2778232, EPI_ISL_2778233, EPI_ISL_2778234, EPI_ISL_2778235, EPI_ISL_2778236, EPI_ISL_2778237, EPI_ISL_2778238, EPI_ISL_2778239, EPI_ISL_2778240, EPI_ISL_2778241, EPI_ISL_2778242, EPI_ISL_2778243, EPI_ISL_2778244, EPI_ISL_2778245, EPI_ISL_2778261, EPI_ISL_2778269, EPI_ISL_2778271, EPI_ISL_2778272, EPI_ISL_2778273, EPI_ISL_2778274, EPI_ISL_2778275, EPI_ISL_2778276, EPI_ISL_2778277, EPI_ISL_2778278, EPI_ISL_2778279, EPI_ISL_2778280, EPI_ISL_2778281, EPI_ISL_2778282, EPI_ISL_2778283, EPI_ISL_2778284, EPI_ISL_2778285, EPI_ISL_2778286, EPI_ISL_2778287, EPI_ISL_2778288, EPI_ISL_2778289, EPI_ISL_2778290, EPI_ISL_2778291, EPI_ISL_2778292, EPI_ISL_2778293, EPI_ISL_2778294, EPI_ISL_2778295, EPI_ISL_2778296, EPI_ISL_2778297, EPI_ISL_2778298, EPI_ISL_2778299, EPI_ISL_2778300, EPI_ISL_2778301, EPI_ISL_2778302, EPI_ISL_2778303, EPI_ISL_2778304, EPI_ISL_2778305, EPI_ISL_2778306, EPI_ISL_2778307, EPI_ISL_2778308, EPI_ISL_2778309, EPI_ISL_2778310, EPI_ISL_2778311, EPI_ISL_2778312, EPI_ISL_2778313, EPI_ISL_2778314, EPI_ISL_2778315, EPI_ISL_2778316, EPI_ISL_2778317, EPI_ISL_2778318, EPI_ISL_2778319, EPI_ISL_2778320, EPI_ISL_2778321, EPI_ISL_2778322, EPI_ISL_2778323, EPI_ISL_2778324, EPI_ISL_2778325, EPI_ISL_2778326, EPI_ISL_2778327, EPI_ISL_2778328, EPI_ISL_2778329, EPI_ISL_2778330, EPI_ISL_2778331, EPI_ISL_2778332, EPI_ISL_2778333, EPI_ISL_2778334, EPI_ISL_2778335, EPI_ISL_2778336, EPI_ISL_2778337, EPI_ISL_2778338, EPI_ISL_2778339, EPI_ISL_2778340, EPI_ISL_2778341, EPI_ISL_2778342, EPI_ISL_2778343, EPI_ISL_2778344, EPI_ISL_2778345, EPI_ISL_2778346, EPI_ISL_2778347, EPI_ISL_2778351, EPI_ISL_2778352, EPI_ISL_2778353, EPI_ISL_2778354, EPI_ISL_2778355, EPI_ISL_2778356, EPI_ISL_2778357, EPI_ISL_2778358, EPI_ISL_2778359, EPI_ISL_2778360, EPI_ISL_2778361, EPI_ISL_2778362, EPI_ISL_2778363, EPI_ISL_2778364, EPI_ISL_2778365, EPI_ISL_2778366, EPI_ISL_2778367, EPI_ISL_2778368, EPI_ISL_2778369, EPI_ISL_2778370, EPI_ISL_2778371, EPI_ISL_2778372, EPI_ISL_2778373, EPI_ISL_2778374, EPI_ISL_2778375, EPI_ISL_2778376, EPI_ISL_2778377, EPI_ISL_2778378, EPI_ISL_2778391, EPI_ISL_2778402, EPI_ISL_2837571, EPI_ISL_2837751, EPI_ISL_2837752 | | | |
| see above | California Department of Public Health Valencia Branch Laboratory (CDPH VBL) | California Department of Public Health | CDPH-COVIDNet; UCLA Technology Center for Genomics & Bioinformatics |
| EPI_ISL_1234813, EPI_ISL_1234891, EPI_ISL_1234967, EPI_ISL_1234994, EPI_ISL_1701473 | California Institute of Technology | Chan-Zuckerberg Biohub | CZB Cliahub Consortium |
| EPI_ISL_1824301, EPI_ISL_1824302, EPI_ISL_1824303, EPI_ISL_1824304, EPI_ISL_1824305, EPI_ISL_1824311, EPI_ISL_1824312, EPI_ISL_1824313, EPI_ISL_1824314, EPI_ISL_1824315, EPI_ISL_1824316, EPI_ISL_1824317, EPI_ISL_1824319, EPI_ISL_1824321, EPI_ISL_1824322, EPI_ISL_1824323, EPI_ISL_1824324, EPI_ISL_1824325, EPI_ISL_1824326, EPI_ISL_1824327, EPI_ISL_1824328, EPI_ISL_1824329, EPI_ISL_1824330, EPI_ISL_1824331, EPI_ISL_1824332, EPI_ISL_1824333, EPI_ISL_1824334, EPI_ISL_1824335, EPI_ISL_1824340, EPI_ISL_1824341, EPI_ISL_1824342, EPI_ISL_1824345, EPI_ISL_1824365 | | | |
| see above | Chiu Laboratory, University of California, San Francisco | Chiu Laboratory, University of California, San Francisco | Alicia Sotomayor-Gonzalez; Candace Wang; Charles Chiu; Daniel Wong; Venice Servellita; Yueyuan Zhang |
| EPI_ISL_1824781, EPI_ISL_1824782, EPI_ISL_1824783, EPI_ISL_1824784, EPI_ISL_1824785, EPI_ISL_1824786, EPI_ISL_1824787, EPI_ISL_1824788, EPI_ISL_1824789, EPI_ISL_1824790, EPI_ISL_1824791, EPI_ISL_1824792, EPI_ISL_1824793, EPI_ISL_1824794, EPI_ISL_1824795, EPI_ISL_1824796, EPI_ISL_1824797, EPI_ISL_1824798, EPI_ISL_1824799, EPI_ISL_1824800, EPI_ISL_1824801, EPI_ISL_1824802, EPI_ISL_1824803, EPI_ISL_1824804, EPI_ISL_1824805, EPI_ISL_1824806, EPI_ISL_1824807, EPI_ISL_1824808, EPI_ISL_1824809, EPI_ISL_1824810, EPI_ISL_1824811, EPI_ISL_1824812, EPI_ISL_1824813, EPI_ISL_1824814, EPI_ISL_1824815, EPI_ISL_1824816, EPI_ISL_1824817, EPI_ISL_1824818, EPI_ISL_1824819, EPI_ISL_1824820, EPI_ISL_1824821, EPI_ISL_1824822, EPI_ISL_1824823, EPI_ISL_1824824, EPI_ISL_1824825, EPI_ISL_1824826, EPI_ISL_1824827, EPI_ISL_1824828, EPI_ISL_1824829, EPI_ISL_1824830, EPI_ISL_1824831, EPI_ISL_1824832, EPI_ISL_1824833, EPI_ISL_1824834, EPI_ISL_1824835, EPI_ISL_1824836, EPI_ISL_1824837, EPI_ISL_1824838, EPI_ISL_1824839, EPI_ISL_1824840, EPI_ISL_1824841, EPI_ISL_1824842, EPI_ISL_1824843, EPI_ISL_1824844, EPI_ISL_1824845, EPI_ISL_1824846, EPI_ISL_1824847, EPI_ISL_1824848, EPI_ISL_1824849, EPI_ISL_1824850, EPI_ISL_1824851, EPI_ISL_1824852, EPI_ISL_1824853, EPI_ISL_1824854, EPI_ISL_1824855, EPI_ISL_1824856, EPI_ISL_1824857, EPI_ISL_1824858, EPI_ISL_1824859, EPI_ISL_1824860, EPI_ISL_1824861, EPI_ISL_1824862, EPI_ISL_1824863, EPI_ISL_1824864, EPI_ISL_1824865, EPI_ISL_1824866, EPI_ISL_1824867, EPI_ISL_1824868, EPI_ISL_1824869, EPI_ISL_1824870, EPI_ISL_1824871, EPI_ISL_1824872, EPI_ISL_1824873, EPI_ISL_1824874, EPI_ISL_1824875, EPI_ISL_1824876, EPI_ISL_1824877, EPI_ISL_1824878, EPI_ISL_1824879, EPI_ISL_1824880, EPI_ISL_1824881, EPI_ISL_1824882, EPI_ISL_1824883, EPI_ISL_1824884, EPI_ISL_1824885, EPI_ISL_1824886, EPI_ISL_1824887, EPI_ISL_1824888, EPI_ISL_1824889, EPI_ISL_1824890, EPI_ISL_1824891, EPI_ISL_1824892, EPI_ISL_1824893, EPI_ISL_1824894, EPI_ISL_1824895, EPI_ISL_1824896, EPI_ISL_1824897, EPI_ISL_1824898, EPI_ISL_1824899, EPI_ISL_1824900, EPI_ISL_1824901, EPI_ISL_1824902, EPI_ISL_1824903, EPI_ISL_1824904, EPI_ISL_1824905, EPI_ISL_1824906, EPI_ISL_1824907, EPI_ISL_1824908, EPI_ISL_1824909, EPI_ISL_1824910, EPI_ISL_1824911, EPI_ISL_1824912, EPI_ISL_1824913, EPI_ISL_1824914, EPI_ISL_1824915, EPI_ISL_1824916 | | | |
| see above | Color Genomics | Chiu Laboratory, University of California, San Francisco | Alicia Sotomayor-Gonzalez; Alicia Zhou; Candace Wang; Charles Chiu; Daniel Wong; Erika Torres; Madyson Wanner; Scott Topper; Venice Servellita; Yueyuan Zhang |
| EPI_ISL_1235065, EPI_ISL_1235086, EPI_ISL_1235101, EPI_ISL_1235151, EPI_ISL_1235227, EPI_ISL_1235231, EPI_ISL_1235336, EPI_ISL_1235340, EPI_ISL_1235341, EPI_ISL_1477716, EPI_ISL_1477732, EPI_ISL_1477738, EPI_ISL_1477743, EPI_ISL_1477781, EPI_ISL_1477802, EPI_ISL_1477813, EPI_ISL_1477826, EPI_ISL_1477827, EPI_ISL_1477829, EPI_ISL_1477835, EPI_ISL_1477849, EPI_ISL_1477851, EPI_ISL_1477861, EPI_ISL_1477866, EPI_ISL_1477869, EPI_ISL_1477910, EPI_ISL_1477918, EPI_ISL_1477927, EPI_ISL_1477939, EPI_ISL_1477956, EPI_ISL_1477957, EPI_ISL_1477966, EPI_ISL_1477980, EPI_ISL_1477995, EPI_ISL_1477998, EPI_ISL_1477999, EPI_ISL_1478000, EPI_ISL_1478005, EPI_ISL_1478007, EPI_ISL_1478020, EPI_ISL_1478023, EPI_ISL_1478047, EPI_ISL_1478049, EPI_ISL_1478052, EPI_ISL_1478069, EPI_ISL_1478086, EPI_ISL_1478090, EPI_ISL_1478091, EPI_ISL_1478098, EPI_ISL_1478101, EPI_ISL_1478120, EPI_ISL_1478128, EPI_ISL_1478133, EPI_ISL_1478139, EPI_ISL_1478141, EPI_ISL_1478145, EPI_ISL_1478155, EPI_ISL_1478157, EPI_ISL_1478160, EPI_ISL_1478181, EPI_ISL_1478182, EPI_ISL_1478189, EPI_ISL_2102996, EPI_ISL_2103003, EPI_ISL_2103010, EPI_ISL_2103011, EPI_ISL_2103012, EPI_ISL_2103013, EPI_ISL_2103016, EPI_ISL_2103024, EPI_ISL_2103030, EPI_ISL_2103037, EPI_ISL_2103040, EPI_ISL_2103041, EPI_ISL_2103048, EPI_ISL_2103054, EPI_ISL_2103055, EPI_ISL_2103056, EPI_ISL_2103057, EPI_ISL_2103060, EPI_ISL_2103063, EPI_ISL_2103065, EPI_ISL_2103068, EPI_ISL_2103069, EPI_ISL_2103072, EPI_ISL_2103076, EPI_ISL_2103077, EPI_ISL_2103079, EPI_ISL_2103082, EPI_ISL_2103084, EPI_ISL_2103087, EPI_ISL_2103101, EPI_ISL_2103104, EPI_ISL_2103108, EPI_ISL_2103117, EPI_ISL_2103125, EPI_ISL_2103127, EPI_ISL_2103131, EPI_ISL_2103141, EPI_ISL_2103145, EPI_ISL_2103150, EPI_ISL_2103154, EPI_ISL_2103164, EPI_ISL_2103168, EPI_ISL_2103170, EPI_ISL_2103171, EPI_ISL_2103174, EPI_ISL_2103175, EPI_ISL_2103182, EPI_ISL_2103183, EPI_ISL_2103184, EPI_ISL_2103191, EPI_ISL_2103198, EPI_ISL_2228251, EPI_ISL_2228257, EPI_ISL_2228427, EPI_ISL_2659255, EPI_ISL_2659552, EPI_ISL_2758459, EPI_ISL_2758469 | | | |
| see above | Contra Costa County Public Health Lab | Chan-Zuckerberg Biohub | CZB Cliahub Consortium |
| EPI_ISL_1185288, EPI_ISL_1185443, EPI_ISL_1234938, EPI_ISL_1529232, EPI_ISL_1701088, EPI_ISL_1701548, EPI_ISL_1701660, EPI_ISL_1821326, EPI_ISL_2102855, EPI_ISL_2102943, EPI_ISL_2103091, EPI_ISL_2103188, EPI_ISL_2659517 | | | |
| see above | County of San Luis Obispo Public Health Laboratory | Chan-Zuckerberg Biohub | CZB Cliahub Consortium |
| EPI_ISL_1318699, EPI_ISL_1318701, EPI_ISL_1318702, EPI_ISL_1318705, EPI_ISL_1318709, EPI_ISL_1318712, EPI_ISL_1318713, EPI_ISL_1318715, EPI_ISL_1318716, EPI_ISL_1318720, EPI_ISL_1318721, EPI_ISL_1318730, EPI_ISL_1318731, EPI_ISL_1318733, EPI_ISL_1318736, EPI_ISL_1318738, EPI_ISL_1318739, EPI_ISL_1318741, EPI_ISL_1318742, EPI_ISL_1318743, EPI_ISL_1318745, EPI_ISL_1318746, EPI_ISL_1318750, EPI_ISL_1318751, EPI_ISL_1318754, EPI_ISL_1318755, EPI_ISL_1318757, EPI_ISL_1318758, EPI_ISL_1318759, EPI_ISL_1318761, EPI_ISL_1318763, EPI_ISL_1318764, EPI_ISL_1318769, EPI_ISL_1318770, EPI_ISL_1318773, EPI_ISL_1318776, EPI_ISL_1318779, EPI_ISL_1318780, EPI_ISL_1318781, EPI_ISL_1318782, EPI_ISL_1318783, EPI_ISL_1318789, EPI_ISL_1318791, EPI_ISL_1318794, EPI_ISL_1318798, EPI_ISL_1318799, EPI_ISL_1318804, EPI_ISL_1318806, EPI_ISL_1318808, EPI_ISL_1318810, EPI_ISL_1318811, EPI_ISL_1318818, EPI_ISL_1318819, EPI_ISL_1318821, EPI_ISL_1318826, EPI_ISL_1318827, EPI_ISL_1318828, EPI_ISL_1318829, EPI_ISL_1318830, EPI_ISL_1318831, EPI_ISL_1318833, EPI_ISL_1318834, EPI_ISL_1318840, EPI_ISL_1318841, EPI_ISL_1318842, EPI_ISL_1318846, EPI_ISL_1318847, EPI_ISL_1318848, EPI_ISL_1318850, EPI_ISL_1318851, EPI_ISL_1318852, EPI_ISL_1318853, EPI_ISL_1318855, EPI_ISL_1318858, EPI_ISL_1318861, EPI_ISL_1318862, EPI_ISL_1318863, EPI_ISL_1318866, EPI_ISL_1318867, EPI_ISL_1318868, EPI_ISL_1318869, EPI_ISL_1318873, EPI_ISL_1318874, EPI_ISL_1318876, EPI_ISL_1318879, EPI_ISL_1318880, EPI_ISL_1318884, EPI_ISL_1318885, EPI_ISL_1318888, EPI_ISL_1318889, EPI_ISL_1318893, EPI_ISL_1318901, EPI_ISL_1318902, EPI_ISL_1318904, EPI_ISL_1318905, EPI_ISL_1318907, EPI_ISL_1318911, EPI_ISL_1318915, EPI_ISL_1318916, EPI_ISL_1318919, EPI_ISL_1318920, EPI_ISL_1318921, EPI_ISL_1318924, EPI_ISL_1318925, EPI_ISL_1318926, EPI_ISL_1318929, EPI_ISL_1318935, EPI_ISL_1318936, EPI_ISL_1318946, EPI_ISL_1318948, EPI_ISL_1318954, EPI_ISL_1318957, EPI_ISL_1318958, EPI_ISL_1318962, EPI_ISL_1318963, EPI_ISL_1318969, EPI_ISL_1318971, EPI_ISL_1318972, EPI_ISL_1318978, EPI_ISL_1318981, EPI_ISL_1318982, EPI_ISL_1318986, EPI_ISL_1318987, EPI_ISL_1347783, EPI_ISL_1347785, EPI_ISL_1347786, EPI_ISL_1347787, EPI_ISL_1347788, EPI_ISL_1349075, EPI_ISL_1349080, EPI_ISL_1349082, EPI_ISL_1349084, EPI_ISL_1349089, EPI_ISL_1349091 | | | |
| see above | DeRisi Lab, University of California, San Francisco | DeRisi Lab, University of California, San Francisco | Amy Kistler; Anthea Mitchell; Carina Marquez; Diane Havlir; Diane Jones; Douglas Black; Eric Chow; Gabriel Chamie; Genay Pilarowski; IDseq Team; Jackie Martinez; James Peng; Jamin Liu; Jon Jacobo; Lucy Li; Luis Rubio; Manu Vanaerschot; Matthew Laurie; Maya Petersen; Patrick Ayscue; Sabrina Mann; Sara Sunshine; Susana Rojas; Susy Rojas; Valerie Tulier-Laiwa; and Joseph DeRisi |
| EPI_ISL_1811642, EPI_ISL_1811643, EPI_ISL_1811647, EPI_ISL_1811649, EPI_ISL_1811650, EPI_ISL_1811651, EPI_ISL_1811652, EPI_ISL_1811656, EPI_ISL_1811659, EPI_ISL_1811670, EPI_ISL_1811671, EPI_ISL_1811674, EPI_ISL_1811675, EPI_ISL_1811678, EPI_ISL_1811681, EPI_ISL_1811682, EPI_ISL_1811683, EPI_ISL_1811686, EPI_ISL_1811688, EPI_ISL_1811690, EPI_ISL_1811692, EPI_ISL_1811698, EPI_ISL_1811701, EPI_ISL_1811703, EPI_ISL_1811705, EPI_ISL_1811708, EPI_ISL_1811710, EPI_ISL_1811711, EPI_ISL_1811712, EPI_ISL_1811714, EPI_ISL_1811715, EPI_ISL_1811717, EPI_ISL_1811718, EPI_ISL_1811719, EPI_ISL_1811720, EPI_ISL_1811721, EPI_ISL_1811722, EPI_ISL_1811725, EPI_ISL_1811726, EPI_ISL_1811730, EPI_ISL_1811733, EPI_ISL_1811736, EPI_ISL_1811737, EPI_ISL_1811738, EPI_ISL_1811739, EPI_ISL_1811743, EPI_ISL_1811744, EPI_ISL_1811745, EPI_ISL_1811746, EPI_ISL_1811747, EPI_ISL_1811748, EPI_ISL_1811749, EPI_ISL_1811750, EPI_ISL_1811751, EPI_ISL_1811753, EPI_ISL_1811754, EPI_ISL_1811755, EPI_ISL_1811758, EPI_ISL_1811759, EPI_ISL_1811760, EPI_ISL_1811761, EPI_ISL_1811762, EPI_ISL_1811764, EPI_ISL_1811770, EPI_ISL_1811772, EPI_ISL_1811773, EPI_ISL_1811775, EPI_ISL_1811776, EPI_ISL_1811777, EPI_ISL_1811778, EPI_ISL_1811779, EPI_ISL_1811780, EPI_ISL_1811782, EPI_ISL_1811784, EPI_ISL_1811785, EPI_ISL_1811786, EPI_ISL_1811787, EPI_ISL_1811788, EPI_ISL_1811789, EPI_ISL_1811795, EPI_ISL_1811796, EPI_ISL_1811799, EPI_ISL_1811801, EPI_ISL_1811802, EPI_ISL_1811803, EPI_ISL_1811804, EPI_ISL_1811805, EPI_ISL_1811806, EPI_ISL_1811808, EPI_ISL_1811809, EPI_ISL_1811810, EPI_ISL_1811811, EPI_ISL_1811813, EPI_ISL_1811814, EPI_ISL_1811815, EPI_ISL_1811816, EPI_ISL_1811818, EPI_ISL_1811819, EPI_ISL_1811820, EPI_ISL_1811822, EPI_ISL_1811826, EPI_ISL_1811827, EPI_ISL_1811830, EPI_ISL_1811832, EPI_ISL_1811833, EPI_ISL_1811837, EPI_ISL_1811840, EPI_ISL_1811843, EPI_ISL_1811845, EPI_ISL_1811848, EPI_ISL_1811849, EPI_ISL_1811850, EPI_ISL_1811852, EPI_ISL_1811854, EPI_ISL_1811855, EPI_ISL_1811862, EPI_ISL_1811872, EPI_ISL_1811873, EPI_ISL_1811875, EPI_ISL_1811885, EPI_ISL_1811887, EPI_ISL_1811902, EPI_ISL_1811927, EPI_ISL_1811929, EPI_ISL_1811931, EPI_ISL_1811936, EPI_ISL_1811965, EPI_ISL_1811969, EPI_ISL_1811996, EPI_ISL_1812001, EPI_ISL_1812017, EPI_ISL_1812019, EPI_ISL_1812021, EPI_ISL_1812031, EPI_ISL_1812036, EPI_ISL_1812043, EPI_ISL_1812045, EPI_ISL_1812049, EPI_ISL_1812052, EPI_ISL_1812065, EPI_ISL_1812068, EPI_ISL_1812074, EPI_ISL_1812075, EPI_ISL_1812076, EPI_ISL_1812098, EPI_ISL_1812100, EPI_ISL_1812102, EPI_ISL_1812104, EPI_ISL_1812120, EPI_ISL_1812121, EPI_ISL_1812123, EPI_ISL_1812130, EPI_ISL_1812145, EPI_ISL_1812150, EPI_ISL_1812157, EPI_ISL_1812163, EPI_ISL_1812180, EPI_ISL_1812185, EPI_ISL_1812192, EPI_ISL_1812197, EPI_ISL_1812198, EPI_ISL_1812203, EPI_ISL_1812204, EPI_ISL_1812206, EPI_ISL_1812238, EPI_ISL_1812239, EPI_ISL_1812252, EPI_ISL_1812257, EPI_ISL_1812268, EPI_ISL_1812275, EPI_ISL_1812284, EPI_ISL_1812286, EPI_ISL_1812287, EPI_ISL_1812294, EPI_ISL_1812295, EPI_ISL_1812311, EPI_ISL_1812323, EPI_ISL_1812327, EPI_ISL_1812328, EPI_ISL_1812338, EPI_ISL_1812341, EPI_ISL_1812349, EPI_ISL_1812360, EPI_ISL_1812364, EPI_ISL_1812373, EPI_ISL_1812378, EPI_ISL_1812382, EPI_ISL_1812388, EPI_ISL_1812389, EPI_ISL_1812390, EPI_ISL_1812391, EPI_ISL_1812393, EPI_ISL_1812395, EPI_ISL_1812411, EPI_ISL_1812414, EPI_ISL_1812420, EPI_ISL_1812424, EPI_ISL_1812427, EPI_ISL_1812433, EPI_ISL_1812436, EPI_ISL_1812445, EPI_ISL_1812446, EPI_ISL_1812447, EPI_ISL_1812458, EPI_ISL_1812464, EPI_ISL_1812465, EPI_ISL_1812472, EPI_ISL_1812480, EPI_ISL_1812481, EPI_ISL_1812486, EPI_ISL_1812487, EPI_ISL_1812506, EPI_ISL_1812509, EPI_ISL_1812512, EPI_ISL_1812518, EPI_ISL_1812522, EPI_ISL_1812524, EPI_ISL_1812525, EPI_ISL_1812526, EPI_ISL_1812531, EPI_ISL_1812533, EPI_ISL_1812534, EPI_ISL_1812536, EPI_ISL_1812537, EPI_ISL_1812548, EPI_ISL_1812549, EPI_ISL_1812550, EPI_ISL_1812556, EPI_ISL_1812559, EPI_ISL_1812566, EPI_ISL_1812569, EPI_ISL_1812574, EPI_ISL_1812577, EPI_ISL_1812578, EPI_ISL_1812579, EPI_ISL_1812582, EPI_ISL_1812586, EPI_ISL_1812597, EPI_ISL_1812599, EPI_ISL_1812601, EPI_ISL_1812608, EPI_ISL_1812614, EPI_ISL_1812615, EPI_ISL_1812621, EPI_ISL_1812623, EPI_ISL_1812625, EPI_ISL_1812627, EPI_ISL_1812628, EPI_ISL_1812629, EPI_ISL_1812632, EPI_ISL_1812639, EPI_ISL_1812640, EPI_ISL_1812642, EPI_ISL_1812647, EPI_ISL_1812648, EPI_ISL_1812650, EPI_ISL_1812651, EPI_ISL_1812657, EPI_ISL_1812660, EPI_ISL_1812661, EPI_ISL_1812665, EPI_ISL_1812673, EPI_ISL_1812677, EPI_ISL_1812682, EPI_ISL_1812684, EPI_ISL_1812686, EPI_ISL_1812688, EPI_ISL_1812691, EPI_ISL_1812692, EPI_ISL_1812696, EPI_ISL_1812702, EPI_ISL_1812703, EPI_ISL_1812705, EPI_ISL_1812712, EPI_ISL_1812724, EPI_ISL_1812727, EPI_ISL_1812728, EPI_ISL_1812729, EPI_ISL_1812731, EPI_ISL_1812733, EPI_ISL_1812736, EPI_ISL_1812745, EPI_ISL_1812749, EPI_ISL_1812751, EPI_ISL_1812752, EPI_ISL_1812755, EPI_ISL_1812764, EPI_ISL_1812769, EPI_ISL_1812775, EPI_ISL_1812795, EPI_ISL_1812797, EPI_ISL_1812800, EPI_ISL_1812812, EPI_ISL_1812815, EPI_ISL_1812816, EPI_ISL_1812820, EPI_ISL_1812827, EPI_ISL_1812829, EPI_ISL_1812833, EPI_ISL_1812844, EPI_ISL_1812846, EPI_ISL_1812850, EPI_ISL_1812857, EPI_ISL_1812864, EPI_ISL_1812865, EPI_ISL_1812866, EPI_ISL_1812868, EPI_ISL_1812869, EPI_ISL_1812872, EPI_ISL_1812881, EPI_ISL_1812886, EPI_ISL_1812891, EPI_ISL_1812893, EPI_ISL_1812901, EPI_ISL_1812913, EPI_ISL_1812918, EPI_ISL_1812919, EPI_ISL_1812926, EPI_ISL_1812931, EPI_ISL_1812934, EPI_ISL_1812935, EPI_ISL_1812936, EPI_ISL_1812937, EPI_ISL_1812942, EPI_ISL_1812956, EPI_ISL_1812959, EPI_ISL_1812964, EPI_ISL_1812969, EPI_ISL_1812983, EPI_ISL_1812986, EPI_ISL_1812996, EPI_ISL_1812997, EPI_ISL_1812999, EPI_ISL_1813001, EPI_ISL_1813004, EPI_ISL_1813005, EPI_ISL_1813016, EPI_ISL_1813018, EPI_ISL_1813022, EPI_ISL_1813025, EPI_ISL_1813026, EPI_ISL_1813028, EPI_ISL_1813035, EPI_ISL_1813048, EPI_ISL_1813058, EPI_ISL_1813064, EPI_ISL_1813068, EPI_ISL_1813072, EPI_ISL_1813074, EPI_ISL_1813076, EPI_ISL_1813078, EPI_ISL_1813083, EPI_ISL_1813094, EPI_ISL_1813096, EPI_ISL_1813098, EPI_ISL_1813102, EPI_ISL_1813107, EPI_ISL_1813116, EPI_ISL_1813117, EPI_ISL_1813123, EPI_ISL_1813133, EPI_ISL_1813137, EPI_ISL_1813146, EPI_ISL_1813148, EPI_ISL_1813165, EPI_ISL_1813176, EPI_ISL_1813182, EPI_ISL_1813185, EPI_ISL_1813189, EPI_ISL_1813195, EPI_ISL_1813199, EPI_ISL_1813204, EPI_ISL_1813212, EPI_ISL_1813214, EPI_ISL_1813215, EPI_ISL_1813218, EPI_ISL_1813220, EPI_ISL_1813224, EPI_ISL_1813231, EPI_ISL_1813240, EPI_ISL_1813245, EPI_ISL_1813249, EPI_ISL_1813253, EPI_ISL_1813254, EPI_ISL_1813269, EPI_ISL_1813271, EPI_ISL_1813274, EPI_ISL_1813283, EPI_ISL_1813284, EPI_ISL_1813285, EPI_ISL_1813286, EPI_ISL_1813289, EPI_ISL_1813294, EPI_ISL_1813295, EPI_ISL_1813299, EPI_ISL_1813301, EPI_ISL_1813305, EPI_ISL_1813307, EPI_ISL_1813314, EPI_ISL_1813316, EPI_ISL_1813319, EPI_ISL_1813320, EPI_ISL_1813327, EPI_ISL_1813328, EPI_ISL_1813334, EPI_ISL_1813344, EPI_ISL_1813348, EPI_ISL_1813352, EPI_ISL_1813357, EPI_ISL_1813365, EPI_ISL_1813368, EPI_ISL_1813370, EPI_ISL_1813372, EPI_ISL_1813376, EPI_ISL_1813380, EPI_ISL_1813384, EPI_ISL_1813400, EPI_ISL_1813406, EPI_ISL_1813407, EPI_ISL_1813410, EPI_ISL_1813411, EPI_ISL_1813415, EPI_ISL_1813429, EPI_ISL_1813431, EPI_ISL_1813433, EPI_ISL_1813443, EPI_ISL_1813461, EPI_ISL_1813463, EPI_ISL_1813464, EPI_ISL_1813466, EPI_ISL_1813468, EPI_ISL_1813469, EPI_ISL_1813472, EPI_ISL_1813484, EPI_ISL_1813487, EPI_ISL_1813489, EPI_ISL_1813499, EPI_ISL_1813502, EPI_ISL_1813505, EPI_ISL_1813509, EPI_ISL_1813522, EPI_ISL_1813523, EPI_ISL_1813524, EPI_ISL_1813529, EPI_ISL_1813541, EPI_ISL_1813550, EPI_ISL_1813556, EPI_ISL_1813557, EPI_ISL_1813566, EPI_ISL_1813569, EPI_ISL_1813572, EPI_ISL_1813576, EPI_ISL_1813580, EPI_ISL_1813585, EPI_ISL_1813602, EPI_ISL_1813603, EPI_ISL_1813613, EPI_ISL_1813614, EPI_ISL_1813619, EPI_ISL_1813620, EPI_ISL_1813622, EPI_ISL_1813623, EPI_ISL_1813625, EPI_ISL_1813631, EPI_ISL_1813635, EPI_ISL_1813637, EPI_ISL_1813641, EPI_ISL_1813646, EPI_ISL_1813651, EPI_ISL_1813656, EPI_ISL_1813667, EPI_ISL_1813670, EPI_ISL_1813679, EPI_ISL_1813685, EPI_ISL_1813692, EPI_ISL_1813695, EPI_ISL_1813698, EPI_ISL_1813708, EPI_ISL_1813709, EPI_ISL_1813712, EPI_ISL_1813715, EPI_ISL_1813717, EPI_ISL_1813719, EPI_ISL_1813724, EPI_ISL_1813730, EPI_ISL_1813733, EPI_ISL_1813736, EPI_ISL_1813741, EPI_ISL_1813742, EPI_ISL_1813747, EPI_ISL_1813754, EPI_ISL_1813757, EPI_ISL_1813768, EPI_ISL_1813769, EPI_ISL_1813772, EPI_ISL_1813775, EPI_ISL_1813781, EPI_ISL_1813785, EPI_ISL_1813796, EPI_ISL_1813805, EPI_ISL_1813810, EPI_ISL_1813813, EPI_ISL_1813819, EPI_ISL_1813820, EPI_ISL_1813828, EPI_ISL_1813835, EPI_ISL_1813836, EPI_ISL_1813838, EPI_ISL_1813844, EPI_ISL_1813848, EPI_ISL_1813854, EPI_ISL_1813869, EPI_ISL_1813872, EPI_ISL_1813879, EPI_ISL_1813882, EPI_ISL_1813883, EPI_ISL_1813890, EPI_ISL_1813900, EPI_ISL_1813907, EPI_ISL_1813909, EPI_ISL_1813911, EPI_ISL_1813914, EPI_ISL_1813915, EPI_ISL_1813916, EPI_ISL_1813917, EPI_ISL_1813923, EPI_ISL_1813926, EPI_ISL_1813929, EPI_ISL_1813931, EPI_ISL_1813932, EPI_ISL_1813935, EPI_ISL_1813936, EPI_ISL_1813937, EPI_ISL_1813941, EPI_ISL_1813942, EPI_ISL_1813958, EPI_ISL_1813960, EPI_ISL_1813961, EPI_ISL_1813967, EPI_ISL_1813970, EPI_ISL_1813978, EPI_ISL_1813989, EPI_ISL_1813993, EPI_ISL_1813996, EPI_ISL_1813999, EPI_ISL_1814001, EPI_ISL_1814002, EPI_ISL_1814004, EPI_ISL_1814005, EPI_ISL_1814016, EPI_ISL_1814017, EPI_ISL_1814021, EPI_ISL_1814024, EPI_ISL_1814027, EPI_ISL_1814028, EPI_ISL_1814029, EPI_ISL_1814030, EPI_ISL_1814032, EPI_ISL_1814037, EPI_ISL_1814038, EPI_ISL_1814040, EPI_ISL_1814041, EPI_ISL_1814042, EPI_ISL_1814044, EPI_ISL_1814047, EPI_ISL_1814051, EPI_ISL_1814057, EPI_ISL_1814058, EPI_ISL_1814062, EPI_ISL_1814068, EPI_ISL_1814073, EPI_ISL_1814080, EPI_ISL_1814084, EPI_ISL_1814092, EPI_ISL_1814095, EPI_ISL_1814096, EPI_ISL_1814100, EPI_ISL_1814103, EPI_ISL_1814104, EPI_ISL_1814105, EPI_ISL_1814109, EPI_ISL_1814110, EPI_ISL_1814119, EPI_ISL_1814124, EPI_ISL_1814133, EPI_ISL_1814134, EPI_ISL_1814136, EPI_ISL_1814138, EPI_ISL_1814139, EPI_ISL_1814149, EPI_ISL_1814158, EPI_ISL_1814161, EPI_ISL_1814164, EPI_ISL_1814165, EPI_ISL_1814169, EPI_ISL_1814171, EPI_ISL_1814172, EPI_ISL_1814174, EPI_ISL_1814176, EPI_ISL_1814177, EPI_ISL_1814178, EPI_ISL_1814182, EPI_ISL_1814184, EPI_ISL_1814185, EPI_ISL_1814187, EPI_ISL_1814188, EPI_ISL_1814189, EPI_ISL_1814190, EPI_ISL_1814191, EPI_ISL_1814192, EPI_ISL_1814194, EPI_ISL_1814196, EPI_ISL_1814203, EPI_ISL_1814204, EPI_ISL_1814205, EPI_ISL_1814206, EPI_ISL_1814208, EPI_ISL_1814215, EPI_ISL_1814216, EPI_ISL_1814217, EPI_ISL_1814219, EPI_ISL_1814220, EPI_ISL_1814222, EPI_ISL_1814223, EPI_ISL_1814224, EPI_ISL_1814228, EPI_ISL_1814229, EPI_ISL_1814230, EPI_ISL_1814231, EPI_ISL_1814232, EPI_ISL_1814233, EPI_ISL_1814236, EPI_ISL_1814241, EPI_ISL_1814244, EPI_ISL_1814253, EPI_ISL_1814254, EPI_ISL_1814255, EPI_ISL_1814256, EPI_ISL_1814262, EPI_ISL_1814263, EPI_ISL_1814268, EPI_ISL_1814271, EPI_ISL_1814281, EPI_ISL_1814283, EPI_ISL_1814288, EPI_ISL_1814291, EPI_ISL_1814296, EPI_ISL_1814299, EPI_ISL_1814300, EPI_ISL_1814302, EPI_ISL_1814305, EPI_ISL_1814307, EPI_ISL_1814309, EPI_ISL_1814317, EPI_ISL_1814321, EPI_ISL_1814322, EPI_ISL_1814323, EPI_ISL_1814328, EPI_ISL_1814339, EPI_ISL_1814340, EPI_ISL_1814343, EPI_ISL_1814344, EPI_ISL_1814356, EPI_ISL_1814371, EPI_ISL_1814374, EPI_ISL_1814382, EPI_ISL_1814383, EPI_ISL_1814388, EPI_ISL_1814391, EPI_ISL_1814394, EPI_ISL_1814395, EPI_ISL_1814399, EPI_ISL_1814404, EPI_ISL_1814410, EPI_ISL_1814411, EPI_ISL_1814412, EPI_ISL_1814413, EPI_ISL_1814416, EPI_ISL_1814419, EPI_ISL_1814421, EPI_ISL_1814424, EPI_ISL_1814426, EPI_ISL_1814429, EPI_ISL_1814430, EPI_ISL_1814432, EPI_ISL_1814435, EPI_ISL_1814437, EPI_ISL_1814440, EPI_ISL_1814441, EPI_ISL_1814442, EPI_ISL_1814444, EPI_ISL_1814447, EPI_ISL_1814452, EPI_ISL_1814453, EPI_ISL_1814455, EPI_ISL_1814459, EPI_ISL_1814467, EPI_ISL_1814472, EPI_ISL_1814473, EPI_ISL_1814474, EPI_ISL_1814476, EPI_ISL_1814478, EPI_ISL_1814479, EPI_ISL_1814481, EPI_ISL_1814482, EPI_ISL_1814485, EPI_ISL_1814490, EPI_ISL_1814495, EPI_ISL_1814497, EPI_ISL_1814500, EPI_ISL_1814505, EPI_ISL_1814510, EPI_ISL_1814512, EPI_ISL_1814514, EPI_ISL_1814516, EPI_ISL_1814517, EPI_ISL_1814518, EPI_ISL_1814523, EPI_ISL_1814530, EPI_ISL_1814532, EPI_ISL_1814533, EPI_ISL_1814538, EPI_ISL_1814544, EPI_ISL_1814546, EPI_ISL_1814551, EPI_ISL_1814555, EPI_ISL_1814559, EPI_ISL_1814566, EPI_ISL_1814568, EPI_ISL_1814573, EPI_ISL_1814575, EPI_ISL_1814578, EPI_ISL_1814584, EPI_ISL_1814586, EPI_ISL_1814588, EPI_ISL_1814589, EPI_ISL_1814590, EPI_ISL_1814591, EPI_ISL_1814592, EPI_ISL_1814593, EPI_ISL_1814594, EPI_ISL_1814604, EPI_ISL_1814606, EPI_ISL_1814611, EPI_ISL_1814614, EPI_ISL_1814617, EPI_ISL_1814624, EPI_ISL_1814626, EPI_ISL_1814629, EPI_ISL_1814633, EPI_ISL_1814638, EPI_ISL_1814639, EPI_ISL_1814643, EPI_ISL_1814646, EPI_ISL_1814648, EPI_ISL_1814654, EPI_ISL_1814663, EPI_ISL_1814666, EPI_ISL_1814667, EPI_ISL_1814672, EPI_ISL_1814676, EPI_ISL_1814677, EPI_ISL_1814680, EPI_ISL_1814685, EPI_ISL_1814695, EPI_ISL_1814698, EPI_ISL_1814699, EPI_ISL_1814705, EPI_ISL_1814706, EPI_ISL_1814708, EPI_ISL_1814712, EPI_ISL_1814713, EPI_ISL_1814714, EPI_ISL_1814716, EPI_ISL_1814719, EPI_ISL_1814723, EPI_ISL_1814726, EPI_ISL_1814727, EPI_ISL_1814730, EPI_ISL_1814731, EPI_ISL_1814734, EPI_ISL_1814737, EPI_ISL_1814738, EPI_ISL_1814740, EPI_ISL_1814748, EPI_ISL_1814749, EPI_ISL_1814755, EPI_ISL_1814756, EPI_ISL_1814758, EPI_ISL_1814759, EPI_ISL_1814764, EPI_ISL_1814772, EPI_ISL_1814774, EPI_ISL_1814779, EPI_ISL_1814782, EPI_ISL_1814791, EPI_ISL_1814797, EPI_ISL_1814800, EPI_ISL_1814801, EPI_ISL_1814802, EPI_ISL_1814805, EPI_ISL_1814808, EPI_ISL_1814809, EPI_ISL_1814810, EPI_ISL_1814817, EPI_ISL_1814823, EPI_ISL_1814824, EPI_ISL_1814829, EPI_ISL_1814831, EPI_ISL_1814837, EPI_ISL_1814842, EPI_ISL_1814843, EPI_ISL_1814852, EPI_ISL_1814865, EPI_ISL_1814866, EPI_ISL_1814869, EPI_ISL_1814871, EPI_ISL_1814875, EPI_ISL_1814876, EPI_ISL_1814884, EPI_ISL_1814888, EPI_ISL_1814890, EPI_ISL_1814897, EPI_ISL_1814898, EPI_ISL_1814900, EPI_ISL_1814904, EPI_ISL_1814907, EPI_ISL_1814908, EPI_ISL_1814911, EPI_ISL_1814913, EPI_ISL_1814915, EPI_ISL_1814921, EPI_ISL_1814922, EPI_ISL_1814925, EPI_ISL_1814932, EPI_ISL_1814934, EPI_ISL_1814936, EPI_ISL_1814938, EPI_ISL_1814941, EPI_ISL_1814947, EPI_ISL_1814948, EPI_ISL_1814951, EPI_ISL_1814959, EPI_ISL_1814960, EPI_ISL_1814964, EPI_ISL_1814966, EPI_ISL_1814975, EPI_ISL_1814977, EPI_ISL_1814978, EPI_ISL_1814982, EPI_ISL_1814988, EPI_ISL_1814990, EPI_ISL_1814992, EPI_ISL_1814995, EPI_ISL_1814997, EPI_ISL_1815003, EPI_ISL_1815004, EPI_ISL_1815006, EPI_ISL_1815007, EPI_ISL_1815009, EPI_ISL_1815015, EPI_ISL_1815016, EPI_ISL_1815021, EPI_ISL_1815023, EPI_ISL_1815024, EPI_ISL_1815025, EPI_ISL_1815032, EPI_ISL_1815037, EPI_ISL_1815046, EPI_ISL_1815049, EPI_ISL_1815050, EPI_ISL_1815051, EPI_ISL_1815052, EPI_ISL_1815054, EPI_ISL_1815059, EPI_ISL_1815061, EPI_ISL_1815065, EPI_ISL_1815066, EPI_ISL_1815071, EPI_ISL_1815072, EPI_ISL_1815076, EPI_ISL_1815078, EPI_ISL_1815080, EPI_ISL_1815082, EPI_ISL_1815083, EPI_ISL_1815085, EPI_ISL_1815087, EPI_ISL_1815089, EPI_ISL_1815096, EPI_ISL_1815098, EPI_ISL_1815102, EPI_ISL_1815104, EPI_ISL_1815105, EPI_ISL_1815106, EPI_ISL_1815117, EPI_ISL_1815120, EPI_ISL_1815131, EPI_ISL_1815132, EPI_ISL_1815136, EPI_ISL_1815138, EPI_ISL_1815143, EPI_ISL_1815146, EPI_ISL_1815148, EPI_ISL_1815153, EPI_ISL_1815154, EPI_ISL_1815171, EPI_ISL_1815174, EPI_ISL_1815177, EPI_ISL_1815185, EPI_ISL_1815187, EPI_ISL_1815189, EPI_ISL_1815192, EPI_ISL_1815195, EPI_ISL_1815196, EPI_ISL_1815199, EPI_ISL_1815210, EPI_ISL_1815214, EPI_ISL_1815215, EPI_ISL_1815218, EPI_ISL_1815226, EPI_ISL_1815227, EPI_ISL_1815228, EPI_ISL_1815229, EPI_ISL_1815242, EPI_ISL_1815243, EPI_ISL_1815249, EPI_ISL_1815250, EPI_ISL_1815252, EPI_ISL_1815254, EPI_ISL_1815256, EPI_ISL_1815258, EPI_ISL_1815259, EPI_ISL_1815262, EPI_ISL_1815266, EPI_ISL_1815272, EPI_ISL_1815281, EPI_ISL_1815286, EPI_ISL_1815291, EPI_ISL_1815295, EPI_ISL_1815300, EPI_ISL_1815302, EPI_ISL_1815304, EPI_ISL_1815310, EPI_ISL_1815311, EPI_ISL_1815313, EPI_ISL_1815322, EPI_ISL_1815323, EPI_ISL_1815327, EPI_ISL_1815328, EPI_ISL_1815330, EPI_ISL_1815339, EPI_ISL_1815343, EPI_ISL_1815347, EPI_ISL_1815348, EPI_ISL_1815353, EPI_ISL_1815356, EPI_ISL_1815360, EPI_ISL_1815361, EPI_ISL_1815362, EPI_ISL_1815366, EPI_ISL_1815367, EPI_ISL_1815376, EPI_ISL_1815379, EPI_ISL_1815382, EPI_ISL_1815384, EPI_ISL_1815389, EPI_ISL_1815394, EPI_ISL_1815398, EPI_ISL_1815400, EPI_ISL_1815406, EPI_ISL_1815408, EPI_ISL_1815409, EPI_ISL_1815413, EPI_ISL_1815417, EPI_ISL_1815425, EPI_ISL_1815427, EPI_ISL_1815428, EPI_ISL_1815430, EPI_ISL_1815431, EPI_ISL_1815432, EPI_ISL_1815437, EPI_ISL_1815439, EPI_ISL_1815443, EPI_ISL_1815444, EPI_ISL_1815445, EPI_ISL_1815458, EPI_ISL_1815462, EPI_ISL_1815469, EPI_ISL_1815470, EPI_ISL_1815472, EPI_ISL_1815478, EPI_ISL_1815480, EPI_ISL_1815481, EPI_ISL_1815485, EPI_ISL_1815489, EPI_ISL_1815490, EPI_ISL_1815494, EPI_ISL_1815497, EPI_ISL_1815498, EPI_ISL_1815504, EPI_ISL_1815505, EPI_ISL_1815518, EPI_ISL_1815520, EPI_ISL_1815523, EPI_ISL_1815525, EPI_ISL_1815529, EPI_ISL_1815536, EPI_ISL_1815537, EPI_ISL_1815539, EPI_ISL_1815540, EPI_ISL_1815541, EPI_ISL_1815542, | | | |

| | | | |
|---|---|---|---|
| EPI_ISL_1815551, EPI_ISL_1815553, EPI_ISL_1815556, EPI_ISL_1815558, EPI_ISL_1815563, EPI_ISL_1815565, EPI_ISL_1815567, EPI_ISL_1815572, EPI_ISL_1815580, EPI_ISL_1815585, EPI_ISL_1815587, EPI_ISL_1815591, EPI_ISL_1815598, EPI_ISL_1815599, EPI_ISL_1815600, EPI_ISL_1815601, EPI_ISL_1815602, EPI_ISL_1815613, EPI_ISL_1815615, EPI_ISL_1815620, EPI_ISL_1815626, EPI_ISL_1815632, EPI_ISL_1815636, EPI_ISL_1815637, EPI_ISL_1815640, EPI_ISL_1815642, EPI_ISL_1815643, EPI_ISL_1815644, EPI_ISL_1815650, EPI_ISL_1815655, EPI_ISL_1815658, EPI_ISL_1815662, EPI_ISL_1815668, EPI_ISL_1815674, EPI_ISL_1815676, EPI_ISL_1815682, EPI_ISL_1815685, EPI_ISL_1815686, EPI_ISL_1815687, EPI_ISL_1815689, EPI_ISL_1815690, EPI_ISL_1815692, EPI_ISL_1815694, EPI_ISL_1815695, EPI_ISL_1815696, EPI_ISL_1815701, EPI_ISL_1815703, EPI_ISL_1815709, EPI_ISL_1815715, EPI_ISL_1815718, EPI_ISL_1815721, EPI_ISL_1815722, EPI_ISL_1815723, EPI_ISL_1815726, EPI_ISL_1815729, EPI_ISL_1815733, EPI_ISL_1815736, EPI_ISL_1815739, EPI_ISL_1815740, EPI_ISL_1815745, EPI_ISL_1815755, EPI_ISL_1815758, EPI_ISL_1815763, EPI_ISL_1815767, EPI_ISL_1815769, EPI_ISL_1815771, EPI_ISL_1815779, EPI_ISL_1815780, EPI_ISL_1815783, EPI_ISL_1815790, EPI_ISL_1815797, EPI_ISL_1815798, EPI_ISL_1815802, EPI_ISL_1815805, EPI_ISL_1815815, EPI_ISL_1815822, EPI_ISL_1815827, EPI_ISL_1815828, EPI_ISL_1815830, EPI_ISL_1815831, EPI_ISL_1815837, EPI_ISL_1815845, EPI_ISL_1815854, EPI_ISL_1815871, EPI_ISL_1815872, EPI_ISL_1815874, EPI_ISL_1815876, EPI_ISL_1815880, EPI_ISL_1815881, EPI_ISL_1815882, EPI_ISL_1815884, EPI_ISL_1815885, EPI_ISL_1815891, EPI_ISL_1815892, EPI_ISL_1815894, EPI_ISL_1815895, EPI_ISL_1815900, EPI_ISL_1815907, EPI_ISL_1815908, EPI_ISL_1815911, EPI_ISL_1815912, EPI_ISL_1815917, EPI_ISL_1815922, EPI_ISL_1815923, EPI_ISL_1815926, EPI_ISL_1815933, EPI_ISL_1815936, EPI_ISL_1815940, EPI_ISL_1815950, EPI_ISL_1815952, EPI_ISL_1815953, EPI_ISL_1815954, EPI_ISL_1815955, EPI_ISL_1815961, EPI_ISL_1815962, EPI_ISL_1815964, EPI_ISL_1815967, EPI_ISL_1815971, EPI_ISL_1815982, EPI_ISL_1815987, EPI_ISL_1815994, EPI_ISL_1815996, EPI_ISL_1815998, EPI_ISL_1816000, EPI_ISL_1816011, EPI_ISL_1816012, EPI_ISL_1816014, EPI_ISL_1816025, EPI_ISL_1816027, EPI_ISL_1816029, EPI_ISL_1816031, EPI_ISL_1816032, EPI_ISL_1816034, EPI_ISL_1816038, EPI_ISL_1816040, EPI_ISL_1816042, EPI_ISL_1816044, EPI_ISL_1816048, EPI_ISL_1816050, EPI_ISL_1816056, EPI_ISL_1816061, EPI_ISL_1816069, EPI_ISL_1816089, EPI_ISL_1816097, EPI_ISL_1816102, EPI_ISL_1816103, EPI_ISL_1816110, EPI_ISL_1816111, EPI_ISL_1816115, EPI_ISL_1816119, EPI_ISL_1816120, EPI_ISL_1816124, EPI_ISL_1816145, EPI_ISL_1816148, EPI_ISL_1816149, EPI_ISL_1816151, EPI_ISL_1816157, EPI_ISL_1816158, EPI_ISL_1816159, EPI_ISL_1816160, EPI_ISL_1816164, EPI_ISL_1816167, EPI_ISL_1816171, EPI_ISL_1816173, EPI_ISL_1816184, EPI_ISL_1816187, EPI_ISL_1816188, EPI_ISL_1816189, EPI_ISL_1816190, EPI_ISL_1816191, EPI_ISL_1816193, EPI_ISL_1816194, EPI_ISL_1816200, EPI_ISL_1816201, EPI_ISL_1816207, EPI_ISL_1816215, EPI_ISL_1816219, EPI_ISL_1816221, EPI_ISL_1816223, EPI_ISL_1816224, EPI_ISL_1816225, EPI_ISL_1816229, EPI_ISL_1816232, EPI_ISL_1816233, EPI_ISL_1816235, EPI_ISL_1816240, EPI_ISL_1816250, EPI_ISL_1816257, EPI_ISL_1816259, EPI_ISL_1816268, EPI_ISL_1816273, EPI_ISL_1816279, EPI_ISL_1816288, EPI_ISL_1816289, EPI_ISL_1816292, EPI_ISL_1816295, EPI_ISL_1816315, EPI_ISL_1816325, EPI_ISL_1816327, EPI_ISL_1816328, EPI_ISL_1816329, EPI_ISL_1816333, EPI_ISL_1816338, EPI_ISL_1816357, EPI_ISL_1816380, EPI_ISL_1816383, EPI_ISL_1816403, EPI_ISL_1816404, EPI_ISL_1816412, EPI_ISL_1816416, EPI_ISL_1816418, EPI_ISL_1816425, EPI_ISL_1816429, EPI_ISL_1816430, EPI_ISL_1816432, EPI_ISL_1816435, EPI_ISL_1816437, EPI_ISL_1816438, EPI_ISL_1816446, EPI_ISL_1816448, EPI_ISL_1816449, EPI_ISL_1816451, EPI_ISL_1816463, EPI_ISL_1816471, EPI_ISL_1816483, EPI_ISL_1816488, EPI_ISL_1816496, EPI_ISL_1816499, EPI_ISL_1816506, EPI_ISL_1816509, EPI_ISL_1816510, EPI_ISL_1816524, EPI_ISL_1816528, EPI_ISL_1816529, EPI_ISL_1816531, EPI_ISL_1816532, EPI_ISL_1816534, EPI_ISL_1816544, EPI_ISL_1816546, EPI_ISL_1816547, EPI_ISL_1816550, EPI_ISL_1816557, EPI_ISL_1816563, EPI_ISL_1816571, EPI_ISL_1816575, EPI_ISL_1816576, EPI_ISL_1816578, EPI_ISL_1816592, EPI_ISL_1816599, EPI_ISL_1816602, EPI_ISL_1816604, EPI_ISL_1816605, EPI_ISL_1816614, EPI_ISL_1816618, EPI_ISL_1816621, EPI_ISL_1816622, EPI_ISL_1816628, EPI_ISL_1816641, EPI_ISL_1816643, EPI_ISL_1816649, EPI_ISL_1816655, EPI_ISL_1816661, EPI_ISL_1816670, EPI_ISL_1816679, EPI_ISL_1816688, EPI_ISL_1816690, EPI_ISL_1816693, EPI_ISL_1816705, EPI_ISL_1816707, EPI_ISL_1816712, EPI_ISL_1816714, EPI_ISL_1816717, EPI_ISL_1816744, EPI_ISL_1816753, EPI_ISL_1816755, EPI_ISL_1816762, EPI_ISL_1816763, EPI_ISL_1816779, EPI_ISL_1816798, EPI_ISL_1816806, EPI_ISL_1816809, EPI_ISL_1816828, EPI_ISL_1816854 | | | |
| see above | EXCITE Lab | Andersen lab at Scripps Research | Abigail Schnapper; Alexandre Bolze; Alice Summerfield; Angela Scioscia; Celena Andrade; Charlotte Rivera-Garcia; Cheryl Anderson; Chip Schooley; Christine Mitchell + SEARCH; David Becker; David Pride; Efren Sandoval; Elizabeth Cirulli; Francisco Tanudjaja; Fred Wu; Geraint Levan; Greg Humphrey; Helena Tubb; James Lu + SEARCH; Jason Nguyen; Jimmy Ramirez; Kelly Schiabor Barrett; Magnus Isaksson; Marc Laurent; Natasha Martin; Natasha Martin Cheryl Anderson; Nicole L Washington; Ryan Cho; Sawyer Farmer; Sharon Reed; Sherry Wang; Simon White; Smruthi Karthikeyan; Sydney Morgan + Marni Jacobs + Celestine Magallanes + Holly Valentine + Louise C. Laurent + SEARCH; Tommy Valles + SEARCH; Tyler Cassens; William Lee |
| EPI_ISL_1098636, EPI_ISL_1098641, EPI_ISL_1192366, EPI_ISL_1192369, EPI_ISL_1197059, EPI_ISL_1198005 | Flow Diagnostics Lab | Los Angeles County Public Health Laboratories | P. Hemarajata et al. |
| EPI_ISL_1253710, EPI_ISL_1253711, EPI_ISL_1253712, EPI_ISL_1254012 | Flow Health | Infectious Disease Program, Broad Institute of Harvard and MIT | Adams, G.; B.L.; B.W.; Bauer, M.; Birren; Carter, A.; Chaluvadi, S.; D.J.; DeRuff, K.; Gallagher, G.; Gladden-Young, A.; J.E.; K.J.; Lagerborg, K.; Lemieux; Loreth, C.; MacInnis; Normandin, E.; P.C.; Park; Reilly, S.; Rudy, M.; Siddle; Smole, S.; Tomkins-Tinch, C.; and Sabeti |
| EPI_ISL_1026866, EPI_ISL_1026867, EPI_ISL_1026868, EPI_ISL_1026869, EPI_ISL_1026870, EPI_ISL_1026871, EPI_ISL_1026872, EPI_ISL_1026873, EPI_ISL_1026874, EPI_ISL_1026875, EPI_ISL_1026876, EPI_ISL_1026877, EPI_ISL_1026878, EPI_ISL_1026879, EPI_ISL_1026880, EPI_ISL_1026881, EPI_ISL_1026882, EPI_ISL_1026883, EPI_ISL_1026884, EPI_ISL_1026885, EPI_ISL_1026886, EPI_ISL_1026887, EPI_ISL_1026888, EPI_ISL_1026889, EPI_ISL_1026890, EPI_ISL_1026891, EPI_ISL_1026892, EPI_ISL_1026893, EPI_ISL_1026894, EPI_ISL_1026895, EPI_ISL_1026896, EPI_ISL_1026897, EPI_ISL_1026898, EPI_ISL_1026899, EPI_ISL_1026900, EPI_ISL_1026901, EPI_ISL_1026902, EPI_ISL_1026903, EPI_ISL_1026904, EPI_ISL_1026905, EPI_ISL_1026906, EPI_ISL_1026907, EPI_ISL_1026908, EPI_ISL_1026909, EPI_ISL_1026910, EPI_ISL_1026911, EPI_ISL_1026912, EPI_ISL_1026913, EPI_ISL_1026914, EPI_ISL_1026915, EPI_ISL_1026916, EPI_ISL_1026917, EPI_ISL_1026918, EPI_ISL_1026919, EPI_ISL_1026920, EPI_ISL_1026921, EPI_ISL_1026922, EPI_ISL_1026923, EPI_ISL_1026924, EPI_ISL_1026925, EPI_ISL_1026926, EPI_ISL_1026927, EPI_ISL_1026928, EPI_ISL_1026929, EPI_ISL_1026930, EPI_ISL_1026931, EPI_ISL_1026932, EPI_ISL_1026933, EPI_ISL_1026934, EPI_ISL_1026935, EPI_ISL_1026936, EPI_ISL_1026937, EPI_ISL_1026938, EPI_ISL_1026939, EPI_ISL_1026940, EPI_ISL_1026941, EPI_ISL_1026942, EPI_ISL_1026943, EPI_ISL_1026944, EPI_ISL_1026945, EPI_ISL_1026946, EPI_ISL_1026947, EPI_ISL_1026948, EPI_ISL_1026949, EPI_ISL_1026950, EPI_ISL_1026951, EPI_ISL_1026952, EPI_ISL_1026953, EPI_ISL_1026954, EPI_ISL_1026955, EPI_ISL_1026956, EPI_ISL_1026957, EPI_ISL_1026958, EPI_ISL_1026959, EPI_ISL_1026960, EPI_ISL_1026961, EPI_ISL_1026962, EPI_ISL_1026963, EPI_ISL_1026964, EPI_ISL_1026965, EPI_ISL_1026966, EPI_ISL_1026967, EPI_ISL_1026968, EPI_ISL_1026969, EPI_ISL_1026970, EPI_ISL_1026971, EPI_ISL_1026972, EPI_ISL_1026973, EPI_ISL_1026974, EPI_ISL_1026975, EPI_ISL_1026976, EPI_ISL_1026977, EPI_ISL_1026978, EPI_ISL_1026979, EPI_ISL_1026980, EPI_ISL_1026981, EPI_ISL_1026982, EPI_ISL_1026983, EPI_ISL_1026984, EPI_ISL_1026985, EPI_ISL_1026986, EPI_ISL_1026987, EPI_ISL_1026988, EPI_ISL_1026989, EPI_ISL_1026990, EPI_ISL_1026991, EPI_ISL_1026992, EPI_ISL_1026993, EPI_ISL_1026994, EPI_ISL_1026995, EPI_ISL_1026996, EPI_ISL_1026997, EPI_ISL_1026998, EPI_ISL_1026999, EPI_ISL_1027000, EPI_ISL_1027001, EPI_ISL_1027002, EPI_ISL_1027003, EPI_ISL_1027004, EPI_ISL_1027005, EPI_ISL_1027006, EPI_ISL_1027007, EPI_ISL_1027008, EPI_ISL_1027009, EPI_ISL_1027010, EPI_ISL_1027011, EPI_ISL_1027012, EPI_ISL_1027013, EPI_ISL_1027014, EPI_ISL_1027015, EPI_ISL_1027016, EPI_ISL_1027017, EPI_ISL_1027018, EPI_ISL_1027019, EPI_ISL_1027020, EPI_ISL_1027021, EPI_ISL_1027022, EPI_ISL_1027023, EPI_ISL_1027024, EPI_ISL_1027025, EPI_ISL_1027026, EPI_ISL_1027028, EPI_ISL_1027029, EPI_ISL_1027030, EPI_ISL_1027031, EPI_ISL_1027032, EPI_ISL_1027033, EPI_ISL_1027034, EPI_ISL_1027035, EPI_ISL_1027037, EPI_ISL_1027038, EPI_ISL_1027039, EPI_ISL_1027040, EPI_ISL_1027041, EPI_ISL_1027042, EPI_ISL_1027043, EPI_ISL_1027044, EPI_ISL_1027045, EPI_ISL_1027046, EPI_ISL_1027047, EPI_ISL_1027048, EPI_ISL_1027049, EPI_ISL_1027050, EPI_ISL_1027051, EPI_ISL_1027052, EPI_ISL_1027053, EPI_ISL_1027054, EPI_ISL_1027055, EPI_ISL_1027056, EPI_ISL_1027057, EPI_ISL_1027058, EPI_ISL_1027059, EPI_ISL_1027060, EPI_ISL_1027061, EPI_ISL_1027062, EPI_ISL_1027063, EPI_ISL_1027064, EPI_ISL_1027065, EPI_ISL_1027066, EPI_ISL_1027067, EPI_ISL_1027068, EPI_ISL_1027069, EPI_ISL_1027070, EPI_ISL_1027071, EPI_ISL_1027072, EPI_ISL_1027073, EPI_ISL_1027074, EPI_ISL_1027075, EPI_ISL_1027076, EPI_ISL_1027077, EPI_ISL_1027078, EPI_ISL_1027079, EPI_ISL_1027080, EPI_ISL_1027081, EPI_ISL_1027082, EPI_ISL_1027083, EPI_ISL_1027084, EPI_ISL_1027086, EPI_ISL_1027087, EPI_ISL_1027088, EPI_ISL_1027089, EPI_ISL_1027090, EPI_ISL_1027091, EPI_ISL_1027092, EPI_ISL_1027093, EPI_ISL_1027094, EPI_ISL_1027095, EPI_ISL_1027096, EPI_ISL_1027097, EPI_ISL_1027098, EPI_ISL_1027099, EPI_ISL_1027100, EPI_ISL_1027101, EPI_ISL_1027102, EPI_ISL_1027103, EPI_ISL_1027104, EPI_ISL_1027105, EPI_ISL_1027106, EPI_ISL_1027107, EPI_ISL_1027108, EPI_ISL_1027109, EPI_ISL_1027110, EPI_ISL_1027111, EPI_ISL_1027112, EPI_ISL_1027113, EPI_ISL_1027114, EPI_ISL_1027115, EPI_ISL_1027116, EPI_ISL_1027117, EPI_ISL_1027118, EPI_ISL_1027119, EPI_ISL_1027120, EPI_ISL_1027121, EPI_ISL_1027122, EPI_ISL_1027123, EPI_ISL_1027124, EPI_ISL_1027125, EPI_ISL_1027126, EPI_ISL_1027127, EPI_ISL_1027128, EPI_ISL_1027129, EPI_ISL_1027130, EPI_ISL_1027131, EPI_ISL_1027132, EPI_ISL_1027133, EPI_ISL_1027134, EPI_ISL_1027135, EPI_ISL_1027136, EPI_ISL_1027137, EPI_ISL_1027138, EPI_ISL_1027139, EPI_ISL_1027140, EPI_ISL_1027141, EPI_ISL_1027142, EPI_ISL_1027143, EPI_ISL_1027144, EPI_ISL_1027145, EPI_ISL_1027146, EPI_ISL_1027147, EPI_ISL_1027148, EPI_ISL_1027149, EPI_ISL_1027150, EPI_ISL_1027151, EPI_ISL_1027152, EPI_ISL_1027153, EPI_ISL_1027154, EPI_ISL_1027155, EPI_ISL_1027156, EPI_ISL_1027157, EPI_ISL_1027158, EPI_ISL_1027159, EPI_ISL_1027160, EPI_ISL_1027161, EPI_ISL_1027162, EPI_ISL_1027163, EPI_ISL_1027164, EPI_ISL_1027165, EPI_ISL_1027166, EPI_ISL_1027167, EPI_ISL_1027168, EPI_ISL_1027169, EPI_ISL_1027170, EPI_ISL_1027171, EPI_ISL_1027172, EPI_ISL_1027173, EPI_ISL_1027174, EPI_ISL_1027175, EPI_ISL_1027176, EPI_ISL_1027177, EPI_ISL_1033008, EPI_ISL_1033009, EPI_ISL_1033010, EPI_ISL_1033011, EPI_ISL_1033012, EPI_ISL_1033013, EPI_ISL_1033014, EPI_ISL_1033015, EPI_ISL_1033017, EPI_ISL_1033018, EPI_ISL_1033019, EPI_ISL_1033020, EPI_ISL_1068395, EPI_ISL_1068396, EPI_ISL_1068397, EPI_ISL_1068398, EPI_ISL_1068399, EPI_ISL_1068400, EPI_ISL_1068401, EPI_ISL_1068402, EPI_ISL_1068403, EPI_ISL_1068404, EPI_ISL_1068405, EPI_ISL_1068406, EPI_ISL_1068407, EPI_ISL_1068408, EPI_ISL_1068409, EPI_ISL_1068410, EPI_ISL_1068411, EPI_ISL_1068412, EPI_ISL_1068413, EPI_ISL_1068414, EPI_ISL_1068415, EPI_ISL_1068416, EPI_ISL_1068417, EPI_ISL_1068418, EPI_ISL_1068419, EPI_ISL_1068420, EPI_ISL_1068421, EPI_ISL_1068422, EPI_ISL_1068423, EPI_ISL_1068424, EPI_ISL_1068425, EPI_ISL_1068426, EPI_ISL_1068427, EPI_ISL_1068428, EPI_ISL_1068429, EPI_ISL_1068430, EPI_ISL_1068431, EPI_ISL_1068432, EPI_ISL_1068433, EPI_ISL_1068434, EPI_ISL_1068435, EPI_ISL_1068436, EPI_ISL_1068437, EPI_ISL_1068438, EPI_ISL_1068439, EPI_ISL_1068440, EPI_ISL_1068441, EPI_ISL_1068442, EPI_ISL_1068443, EPI_ISL_1068444, EPI_ISL_1068445, EPI_ISL_1068446, EPI_ISL_1068447, EPI_ISL_1068448, EPI_ISL_1068449, EPI_ISL_1068450, EPI_ISL_1068451, EPI_ISL_1068452, EPI_ISL_1068453, EPI_ISL_1068454, EPI_ISL_1068455, EPI_ISL_1068456, EPI_ISL_1068457, EPI_ISL_1068458, EPI_ISL_1068459, EPI_ISL_1068460, EPI_ISL_1068461, EPI_ISL_1068462, EPI_ISL_1068463, EPI_ISL_1068464, EPI_ISL_1068465, EPI_ISL_1068466, EPI_ISL_1068467, EPI_ISL_1068468, EPI_ISL_1068469, EPI_ISL_1068470, EPI_ISL_1068471, EPI_ISL_1068472, EPI_ISL_1068473, EPI_ISL_1068474, EPI_ISL_1068475, EPI_ISL_1068476, EPI_ISL_1068477, EPI_ISL_1068478, EPI_ISL_1068479, EPI_ISL_1068480, EPI_ISL_1068481, EPI_ISL_1068482, EPI_ISL_1068483, EPI_ISL_1068484, EPI_ISL_1068485, EPI_ISL_1068486, EPI_ISL_1068487, EPI_ISL_1068488, EPI_ISL_1068489, EPI_ISL_1068490, EPI_ISL_1068491, EPI_ISL_1068492, EPI_ISL_1068493, EPI_ISL_1068494, EPI_ISL_1068495, EPI_ISL_1068496, EPI_ISL_1068497, EPI_ISL_1068498, EPI_ISL_1068499, EPI_ISL_1068500, EPI_ISL_1068501, EPI_ISL_1068502, EPI_ISL_1068503, EPI_ISL_1068504, EPI_ISL_1068505, EPI_ISL_1068506, EPI_ISL_1068507, EPI_ISL_1068508, EPI_ISL_1068509, EPI_ISL_1068510, EPI_ISL_1068511, EPI_ISL_1068512, EPI_ISL_1068513, EPI_ISL_1068514, EPI_ISL_1068515, EPI_ISL_1068516, EPI_ISL_1068517, EPI_ISL_1068518, EPI_ISL_1068519, EPI_ISL_1068520, EPI_ISL_1068521, EPI_ISL_1068522, EPI_ISL_1068523, EPI_ISL_1068524, EPI_ISL_1068525, EPI_ISL_1068526, EPI_ISL_1068527, EPI_ISL_1068528, EPI_ISL_1068529, EPI_ISL_1068530, EPI_ISL_1068531, EPI_ISL_1068532, EPI_ISL_1068533, EPI_ISL_1068534, EPI_ISL_1068535, EPI_ISL_1068536, EPI_ISL_1068537, EPI_ISL_1068538, EPI_ISL_1068539, EPI_ISL_1068540, EPI_ISL_1068541, EPI_ISL_1068542, EPI_ISL_1068543, EPI_ISL_1068544, EPI_ISL_1068545, EPI_ISL_1068546, EPI_ISL_1068547, EPI_ISL_1068548, EPI_ISL_1068549, EPI_ISL_1068550, EPI_ISL_1068551, EPI_ISL_1068552, EPI_ISL_1068553, EPI_ISL_1068554, EPI_ISL_1068555, EPI_ISL_1068556, EPI_ISL_1068557, EPI_ISL_1068558, EPI_ISL_1068559, EPI_ISL_1068560, EPI_ISL_1068561, EPI_ISL_1068562, EPI_ISL_1068563, EPI_ISL_1068564, EPI_ISL_1068565, EPI_ISL_1068566, EPI_ISL_1068567, EPI_ISL_1068568, EPI_ISL_1068569, EPI_ISL_1068570, EPI_ISL_1068571, EPI_ISL_1068572, EPI_ISL_1068573, EPI_ISL_1068574, EPI_ISL_1068575, EPI_ISL_1068576, EPI_ISL_1068577, EPI_ISL_1068578, EPI_ISL_1068579, EPI_ISL_1068580, EPI_ISL_1068581, EPI_ISL_1068582, EPI_ISL_1068583, EPI_ISL_1068584, EPI_ISL_1068585, EPI_ISL_1068586, EPI_ISL_1068587, EPI_ISL_1068588, EPI_ISL_1068589, EPI_ISL_1068590, EPI_ISL_1068591, EPI_ISL_1068592, EPI_ISL_1068594, EPI_ISL_1068595, EPI_ISL_1068596, EPI_ISL_1068597, EPI_ISL_1068598, EPI_ISL_1068599, EPI_ISL_1068600, EPI_ISL_1068601, EPI_ISL_1068602, EPI_ISL_1068603, EPI_ISL_1068604, EPI_ISL_1068605, EPI_ISL_1068606, EPI_ISL_1068607, EPI_ISL_1068608, EPI_ISL_1068609, EPI_ISL_1068610, EPI_ISL_1068611, EPI_ISL_1068612, EPI_ISL_1068613, EPI_ISL_1068614, EPI_ISL_1068615, EPI_ISL_1068616, EPI_ISL_1068617, EPI_ISL_1068618, EPI_ISL_1068619, EPI_ISL_1068620, EPI_ISL_1068621, EPI_ISL_1068622, EPI_ISL_1068623, EPI_ISL_1068624, EPI_ISL_1068625, EPI_ISL_1068626, EPI_ISL_1068627, EPI_ISL_1068628, EPI_ISL_1068629, EPI_ISL_1068630, EPI_ISL_1068631, EPI_ISL_1068632, EPI_ISL_1068633, EPI_ISL_1068634, EPI_ISL_1068635, EPI_ISL_1068636, EPI_ISL_1068637, EPI_ISL_1068638, EPI_ISL_1068639, EPI_ISL_1068641, EPI_ISL_1068642, EPI_ISL_1068643, EPI_ISL_1068644, EPI_ISL_1068645, EPI_ISL_1068646, EPI_ISL_1068647, EPI_ISL_1068648, EPI_ISL_1068649, EPI_ISL_1068650, EPI_ISL_1068651, EPI_ISL_1068652, EPI_ISL_1068653, EPI_ISL_1068654, EPI_ISL_1068655, EPI_ISL_1068656, EPI_ISL_1068657, EPI_ISL_1068658, EPI_ISL_1068659, EPI_ISL_1068660, EPI_ISL_1068661, EPI_ISL_1068662, EPI_ISL_1068663, EPI_ISL_1068664, EPI_ISL_1068665, EPI_ISL_1068666, EPI_ISL_1068667, EPI_ISL_1068668, EPI_ISL_1068669, EPI_ISL_1068670, EPI_ISL_1068671, EPI_ISL_1068672, EPI_ISL_1068673, EPI_ISL_1068674, EPI_ISL_1068675, EPI_ISL_1068676, EPI_ISL_1068677, EPI_ISL_1068678, EPI_ISL_1068679, EPI_ISL_1068680, EPI_ISL_1068681, EPI_ISL_1068683, EPI_ISL_1068684, EPI_ISL_1068685, EPI_ISL_1068686, EPI_ISL_1068687, EPI_ISL_1068688, EPI_ISL_1068689, EPI_ISL_1068690, EPI_ISL_1068691, EPI_ISL_1068692, EPI_ISL_1068693, EPI_ISL_1068694, EPI_ISL_1068695, EPI_ISL_1068696, EPI_ISL_1068697, EPI_ISL_1068698, EPI_ISL_1068699, EPI_ISL_1068700, EPI_ISL_1068701, EPI_ISL_1068702, EPI_ISL_1068703, EPI_ISL_1068704, EPI_ISL_1068705, EPI_ISL_1068706, EPI_ISL_1068707, EPI_ISL_1068708, EPI_ISL_1068709, EPI_ISL_1068710, EPI_ISL_1068711, EPI_ISL_1068712, EPI_ISL_1068713, EPI_ISL_1068714, EPI_ISL_1068715, EPI_ISL_1068716, EPI_ISL_1068717, EPI_ISL_1068718, EPI_ISL_1068719, EPI_ISL_1068720, EPI_ISL_1068721, EPI_ISL_1068722, EPI_ISL_1068723, EPI_ISL_1068724, EPI_ISL_1068725, EPI_ISL_1068726, EPI_ISL_1068727, EPI_ISL_1068728, EPI_ISL_1068729, EPI_ISL_1068730, EPI_ISL_1068731, EPI_ISL_1068732, EPI_ISL_1068733, EPI_ISL_1068734, EPI_ISL_1068735, EPI_ISL_1068736, EPI_ISL_1068737, EPI_ISL_1068738, EPI_ISL_1068739, EPI_ISL_1068740, EPI_ISL_1068741, EPI_ISL_1068742, EPI_ISL_1068743, EPI_ISL_1068744, EPI_ISL_1068745, EPI_ISL_1068746, EPI_ISL_1068747, EPI_ISL_1068748, EPI_ISL_1068749, EPI_ISL_1068750, EPI_ISL_1068751, EPI_ISL_1068752, EPI_ISL_1068753, EPI_ISL_1068754, EPI_ISL_1068755, EPI_ISL_1068756, EPI_ISL_1068757, EPI_ISL_1068758, EPI_ISL_1068759, EPI_ISL_1068760, EPI_ISL_1068761, EPI_ISL_1068762, EPI_ISL_1068763, EPI_ISL_1068764, EPI_ISL_1068765, EPI_ISL_1068766, EPI_ISL_1068767, EPI_ISL_1068768, EPI_ISL_1068769, EPI_ISL_1068770, EPI_ISL_1068771, EPI_ISL_1068772, EPI_ISL_1068773, EPI_ISL_1068774, EPI_ISL_1068775, EPI_ISL_1068776, EPI_ISL_1068777, EPI_ISL_1068778, EPI_ISL_1068779, EPI_ISL_1068780, EPI_ISL_1068781, EPI_ISL_1068782, EPI_ISL_1068783, EPI_ISL_1068784, EPI_ISL_1068785, EPI_ISL_1068786, EPI_ISL_1068787, EPI_ISL_1068788, EPI_ISL_1068789, EPI_ISL_1068790, EPI_ISL_1068791, EPI_ISL_1068792, EPI_ISL_1068793, EPI_ISL_1068794, EPI_ISL_1068795, EPI_ISL_1068796, EPI_ISL_1068797, EPI_ISL_1068798, EPI_ISL_1068799, EPI_ISL_1068800, EPI_ISL_1068801, EPI_ISL_1068802, EPI_ISL_1068803, EPI_ISL_1068804, EPI_ISL_1068805, EPI_ISL_1068806, EPI_ISL_1068807, EPI_ISL_1068808, EPI_ISL_1068809, EPI_ISL_1068810, EPI_ISL_1068811, EPI_ISL_1068812, EPI_ISL_1068813, EPI_ISL_1068814, EPI_ISL_1068815, EPI_ISL_1068816, EPI_ISL_1068817, EPI_ISL_1068818, EPI_ISL_1068819, EPI_ISL_1068820, EPI_ISL_1068821, EPI_ISL_1068822, EPI_ISL_1068823, EPI_ISL_1068824, EPI_ISL_1068825, EPI_ISL_1068826, EPI_ISL_1068827, EPI_ISL_1068828, EPI_ISL_1068829, EPI_ISL_1068830, EPI_ISL_1068831, EPI_ISL_1068832, EPI_ISL_1068833, EPI_ISL_1068834, EPI_ISL_1068835, EPI_ISL_1068836, EPI_ISL_1068837, EPI_ISL_1068838, EPI_ISL_1068839, EPI_ISL_1068840, EPI_ISL_1068841, EPI_ISL_1068842, EPI_ISL_1068843, EPI_ISL_1068844, EPI_ISL_1068845, EPI_ISL_1068846, EPI_ISL_1068847, EPI_ISL_1068848, EPI_ISL_1068852, EPI_ISL_1068853, EPI_ISL_1068854, EPI_ISL_1068855, EPI_ISL_1068856, EPI_ISL_1068857, EPI_ISL_1068858, EPI_ISL_1068859, EPI_ISL_1068860, EPI_ISL_1068861, EPI_ISL_1068862, EPI_ISL_1068863, EPI_ISL_1068864, EPI_ISL_1068865, EPI_ISL_1068866, EPI_ISL_1068867, EPI_ISL_1068868, EPI_ISL_1068869, EPI_ISL_1068870, EPI_ISL_1068871, EPI_ISL_1068872, EPI_ISL_1068873, EPI_ISL_1068874, EPI_ISL_1068875, EPI_ISL_1068876, EPI_ISL_1068877, EPI_ISL_1068878, EPI_ISL_1068879, EPI_ISL_1068880, EPI_ISL_1068881, EPI_ISL_1068882, EPI_ISL_1068883, EPI_ISL_1068884, EPI_ISL_1068885, EPI_ISL_1068886, EPI_ISL_1068887, EPI_ISL_1068888, EPI_ISL_1068889, EPI_ISL_1068890, EPI_ISL_1068891, EPI_ISL_1068892, EPI_ISL_1068893, EPI_ISL_1068894, EPI_ISL_1068895, EPI_ISL_1068896, EPI_ISL_1068897, EPI_ISL_1068898, EPI_ISL_1068899, EPI_ISL_1068900, EPI_ISL_1068901, EPI_ISL_1068902, EPI_ISL_1068903, EPI_ISL_1068904, EPI_ISL_1068905, EPI_ISL_1068906, EPI_ISL_1068907, EPI_ISL_1068908, EPI_ISL_1068909, EPI_ISL_1068910, EPI_ISL_1068911, EPI_ISL_1068912, EPI_ISL_1068913, EPI_ISL_1068914, EPI_ISL_1068915, EPI_ISL_1068916, EPI_ISL_1068917, EPI_ISL_1068918, EPI_ISL_1068919, EPI_ISL_1068920, EPI_ISL_1068921, EPI_ISL_1068922, EPI_ISL_1068923, EPI_ISL_1068924, EPI_ISL_1068925, EPI_ISL_1068926, EPI_ISL_1068930, EPI_ISL_1068931, EPI_ISL_1068932, EPI_ISL_1068933, EPI_ISL_1068934, EPI_ISL_1068935, EPI_ISL_1068936, EPI_ISL_1068937, EPI_ISL_1068938, EPI_ISL_1068939, EPI_ISL_1068940, EPI_ISL_1068941, EPI_ISL_1068942, EPI_ISL_1068943, EPI_ISL_1068944, EPI_ISL_1068945, EPI_ISL_1068946, EPI_ISL_1068947, EPI_ISL_1068948, EPI_ISL_1068949, EPI_ISL_1068950, EPI_ISL_1068951, EPI_ISL_1068952, EPI_ISL_1068953, EPI_ISL_1068954, EPI_ISL_1068955, EPI_ISL_1068956, EPI_ISL_1068957, EPI_ISL_1068958, EPI_ISL_1068959, EPI_ISL_1068960, EPI_ISL_1068961, EPI_ISL_1068962, EPI_ISL_1068963, EPI_ISL_1068964, EPI_ISL_1068965, EPI_ISL_1068966, EPI_ISL_1068967, EPI_ISL_1068968, EPI_ISL_1068969, EPI_ISL_1068970, EPI_ISL_1068971, EPI_ISL_1068972, EPI_ISL_1068973, EPI_ISL_1068974, EPI_ISL_1068975, EPI_ISL_1068976, EPI_ISL_1068977, EPI_ISL_1068978, EPI_ISL_1068979, EPI_ISL_1068980, EPI_ISL_1068981, EPI_ISL_1068982, EPI_ISL_1068983, EPI_ISL_1068984, EPI_ISL_1068985, EPI_ISL_1068986, EPI_ISL_1068987, EPI_ISL_1068988, EPI_ISL_1068989, EPI_ISL_1068990, EPI_ISL_1068991, EPI_ISL_1068992, EPI_ISL_1068993, EPI_ISL_1068994, EPI_ISL_1068995, EPI_ISL_1069000, EPI_ISL_1069001, EPI_ISL_1069002, EPI_ISL_1069003, EPI_ISL_1069004, EPI_ISL_1069005, EPI_ISL_1069006, EPI_ISL_1069007, EPI_ISL_1069008, EPI_ISL_1069009, EPI_ISL_1069010, EPI_ISL_1069011, EPI_ISL_1069012, EPI_ISL_1069013, EPI_ISL_1069014, EPI_ISL_1069015, EPI_ISL_1069016, EPI_ISL_1069017, EPI_ISL_1069018, EPI_ISL_1069019, EPI_ISL_1069020, EPI_ISL_1069021, EPI_ISL_1069022, EPI_ISL_1069023, EPI_ISL_1069024, EPI_ISL_1069025, EPI_ISL_1069026, EPI_ISL_1069027, EPI_ISL_1069028, EPI_ISL_1069029, EPI_ISL_1069030, EPI_ISL_1069031, EPI_ISL_1069032, EPI_ISL_1069033, EPI_ISL_1069034, EPI_ISL_1069035, EPI_ISL_1069036, EPI_ISL_1069037, EPI_ISL_1069038, EPI_ISL_1069039, EPI_ISL_1069040, EPI_ISL_1069041, EPI_ISL_1069042, EPI_ISL_1069043, EPI_ISL_1069044, EPI_ISL_1069045, EPI_ISL_1069046, EPI_ISL_1069047, EPI_ISL_1069048, EPI_ISL_1069049, EPI_ISL_1069050, EPI_ISL_1069051, EPI_ISL_1069052, EPI_ISL_1069053, EPI_ISL_1069054, EPI_ISL_1069055, EPI_ISL_1069056, EPI_ISL_1069057, EPI_ISL_1069058, EPI_ISL_1069059, EPI_ISL_1069060, EPI_ISL_1069061, EPI_ISL_1069062, EPI_ISL_1069063, EPI_ISL_1069064, EPI_ISL_1069065, EPI_ISL_1069066, EPI_ISL_1069067, EPI_ISL_1069068, EPI_ISL_1069069, EPI_ISL_1069070, EPI_ISL_1069071, EPI_ISL_1069072, EPI_ISL_1069073, EPI_ISL_1069074, EPI_ISL_1069075, EPI_ISL_1069085, EPI_ISL_1069086, EPI_ISL_1069087, EPI_ISL_1069088, EPI_ISL_1069089, EPI_ISL_1069090, EPI_ISL_1069091, EPI_ISL_1069092, EPI_ISL_1069093, EPI_ISL_1069094, EPI_ISL_1069095, EPI_ISL_1069096, EPI_ISL_1069097, EPI_ISL_1069098, EPI_ISL_1069099, EPI_ISL_1069100, EPI_ISL_1069101, EPI_ISL_1069102, EPI_ISL_1069103, EPI_ISL_1069104, EPI_ISL_1069105, EPI_ISL_1069106, EPI_ISL_1069107, EPI_ISL_1069108, EPI_ISL_1069109, EPI_ISL_1069110, EPI_ISL_1069111, EPI_ISL_1069112, EPI_ISL_1069113, EPI_ISL_1069114, EPI_ISL_1069115, EPI_ISL_1069116, EPI_ISL_1069117, EPI_ISL_1069118, EPI_ISL_1069119, EPI_ISL_1069120, EPI_ISL_1069121, EPI_ISL_1069122, EPI_ISL_1069123, EPI_ISL_1069124, EPI_ISL_1069125, EPI_ISL_1069126, EPI_ISL_1069131, EPI_ISL_1069132, EPI_ISL_1069133, EPI_ISL_1069135, EPI_ISL_1069136, EPI_ISL_1069137, EPI_ISL_1069138, EPI_ISL_1069139, EPI_ISL_1069140, EPI_ISL_1069141, EPI_ISL_1069142, EPI_ISL_1069143, EPI_ISL_1069144, EPI_ISL_1069148, EPI_ISL_1069150, EPI_ISL_1095645, EPI_ISL_1095646, EPI_ISL_1095647, EPI_ISL_1095648, EPI_ISL_1095649, EPI_ISL_1095650, EPI_ISL_1095651, EPI_ISL_1095652, EPI_ISL_1095653, EPI_ISL_1095654, EPI_ISL_1095655, EPI_ISL_1095656, EPI_ISL_1095657, EPI_ISL_1095658, EPI_ISL_1095659, EPI_ISL_1095660, EPI_ISL_1095661, EPI_ISL_1095662, EPI_ISL_1095663, EPI_ISL_1095664, EPI_ISL_1095665, EPI_ISL_1095666, EPI_ISL_1095667, EPI_ISL_1095668, EPI_ISL_1095669, EPI_ISL_1095670, EPI_ISL_1095671, EPI_ISL_1095672, EPI_ISL_1095673, EPI_ISL_1095674, EPI_ISL_1095675, EPI_ISL_1095676, EPI_ISL_1095677, EPI_ISL_1095678, EPI_ISL_1095679, EPI_ISL_1095680, EPI_ISL_1095681, EPI_ISL_1095682, EPI_ISL_1095683, EPI_ISL_1095684, EPI_ISL_1095685, EPI_ISL_1095686, EPI_ISL_1095687, EPI_ISL_1095688, EPI_ISL_1095689, EPI_ISL_1095690, EPI_ISL_1095691, EPI_ISL_1095692, EPI_ISL_1095693, EPI_ISL_1095694, EPI_ISL_1095695, EPI_ISL_1095696, EPI_ISL_1095697, EPI_ISL_1095698, EPI_ISL_1095699, EPI_ISL_1095700, EPI_ISL_1095701, EPI_ISL_1095702, EPI_ISL_1095703, EPI_ISL_1095704, EPI_ISL_1095705, EPI_ISL_1095706, EPI_ISL_1095707, EPI_ISL_1095708, EPI_ISL_1095709, EPI_ISL_1095710, EPI_ISL_1095711, EPI_ISL_1095712, EPI_ISL_1095713, EPI_ISL_1095714, EPI_ISL_1095715, EPI_ISL_1095716, EPI_ISL_1095717, EPI_ISL_1095718, EPI_ISL_1095719, EPI_ISL_1095720, EPI_ISL_1095721, EPI_ISL_1095722, EPI_ISL_1095723, EPI_ISL_1095724, EPI_ISL_1095725, EPI_ISL_1095726, EPI_ISL_1095727, EPI_ISL_1095728, EPI_ISL_1095729, EPI_ISL_1095730, EPI_ISL_1095731, EPI_ISL_1095732, EPI_ISL_1095733, EPI_ISL_1095734, EPI_ISL_1095735, EPI_ISL_1095736, EPI_ISL_1095737, EPI_ISL_1095738, EPI_ISL_1095739, EPI_ISL_1095740, EPI_ISL_1095741, EPI_ISL_1095742, EPI_ISL_1095743, EPI_ISL_1095744, EPI_ISL_1095745, EPI_ISL_1095746, EPI_ISL_1095747, EPI_ISL_1095748, EPI_ISL_1095749, EPI_ISL_1095750, EPI_ISL_1095751, EPI_ISL_1095752, EPI_ISL_1095753, EPI_ISL_1095754, EPI_ISL_1095755, EPI_ISL_1095756, EPI_ISL_1095757, EPI_ISL_1095758, EPI_ISL_1095759, EPI_ISL_1095760, EPI_ISL_1095761, EPI_ISL_1095762, EPI_ISL_1095763, EPI_ISL_1095764, EPI_ISL_1095765, EPI_ISL_1095766, EPI_ISL_1095767, EPI_ISL_1095768, EPI_ISL_1095769, EPI_ISL_1095770, EPI_ISL_1095771, EPI_ISL_1095772, EPI_ISL_1095773, EPI_ISL_1095774, EPI_ISL_1095775, EPI_ISL_1095776, EPI_ISL_1095777, EPI_ISL_1095778, EPI_ISL_1095779, EPI_ISL_1095780, EPI_ISL_1095781, EPI_ISL_1095782, EPI_ISL_1095783, EPI_ISL_1095784, EPI_ISL_1095785, EPI_ISL_1095786, EPI_ISL_1095787, EPI_ISL_1095788, EPI_ISL_1095789, EPI_ISL_1095790, EPI_ISL_1095791, EPI_ISL_1095792, EPI_ISL_1095793, EPI_ISL_1095794, EPI_ISL_1095795, EPI_ISL_1095796, EPI_ISL_1095797, EPI_ISL_1095798, EPI_ISL_1095799, EPI_ISL_1095800, EPI_ISL_1095801, EPI_ISL_1095802, EPI_ISL_1095803, EPI_ISL_1095804, EPI_ISL_1095805, EPI_ISL_1095806, EPI_ISL_1095807, EPI_ISL_1095808, EPI_ISL_1095809, EPI_ISL_1095810, EPI_ISL_1095811, EPI_ISL_1095812, EPI_ISL_1095813, EPI_ISL_1095814, EPI_ISL_1095815, EPI_ISL_1095816, EPI_ISL_1095817, EPI_ISL_1095818, EPI_ISL_1095819, EPI_ISL_1095820, EPI_ISL_1095821, EPI_ISL_1095822, EPI_ISL_1095823, EPI_ISL_1095824, EPI_ISL_1095825, EPI_ISL_1095826, EPI_ISL_1095827, EPI_ISL_1095828, EPI_ISL_1095829, EPI_ISL_1095830, EPI_ISL_1095831, EPI_ISL_1095832, EPI_ISL_1095833, EPI_ISL_1095834, EPI_ISL_1095835, EPI_ISL_1095836, EPI_ISL_1095837, EPI_ISL_1095838, EPI_ISL_1095839, EPI_ISL_1095840, EPI_ISL_1095841, EPI_ISL_1095842, EPI_ISL_1095843, EPI_ISL_1095844, EPI_ISL_1095845, EPI_ISL_1095846, EPI_ISL_1095847, EPI_ISL_1095848, EPI_ISL_1095849, EPI_ISL_1095850, EPI_ISL_1095851, EPI_ISL_1095852, EPI_ISL_1095853, EPI_ISL_1095854, EPI_ISL_1095855, EPI_ISL_1095856, EPI_ISL_1095857, EPI_ISL_1095858, EPI_ISL_1095859, EPI_ISL_1095860, EPI_ISL_1095861, EPI_ISL_1095862, EPI_ISL_1095863, EPI_ISL_1095864, EPI_ISL_1095865, EPI_ISL_1095866, EPI_ISL_1095867, EPI_ISL_1095868, EPI_ISL_1095869, EPI_ISL_1095870, EPI_ISL_1095871, EPI_ISL_1095872, EPI_ISL_1095873, EPI_ISL_1095874, EPI_ISL_1095875, EPI_ISL_1095876, EPI_ISL_1095877, EPI_ISL_1095878, EPI_ISL_1095879, EPI_ISL_1095880, EPI_ISL_1095881, EPI_ISL_1095882, EPI_ISL_1095883, EPI_ISL_1095884, EPI_ISL_1095885, EPI_ISL_1095886, EPI_ISL_1095887, EPI_ISL_1095888, EPI_ISL_1095889, EPI_ISL_1095890, EPI_ISL_1095891, EPI_ISL_1095892, EPI_ISL_1095893, EPI_ISL_1095894, EPI_ISL_1095895, EPI_ISL_1095896, EPI_ISL_1095897, EPI_ISL_1095898, EPI_ISL_1095899, EPI_ISL_1095900, EPI_ISL_1095901, EPI_ISL_1095902, EPI_ISL_1095903, EPI_ISL_1095904, EPI_ISL_1095905, EPI_ISL_1095906, EPI_ISL_1095907, EPI_ISL_1095908, EPI_ISL_1095909, EPI_ISL_1095910, EPI_ISL_1095911, EPI_ISL_1095912, EPI_ISL_1095914, EPI_ISL_1095915, EPI_ISL_1095916, EPI_ISL_1095917, EPI_ISL_1095918, EPI_ISL_1095919, EPI_ISL_1095920, EPI_ISL_1095921, EPI_ISL_1095922, EPI_ISL_1095923, EPI_ISL_1095924, EPI_ISL_1095925, EPI_ISL_1095926, EPI_ISL_1095927, EPI_ISL_1095928, EPI_ISL_1095929, EPI_ISL_1095930, EPI_ISL_1095931, EPI_ISL_1095932, EPI_ISL_1095933, EPI_ISL_1095934, EPI_ISL_1095935, EPI_ISL_1095936, EPI_ISL_1095937, EPI_ISL_1095938, EPI_ISL_1095939, EPI_ISL_1095940, EPI_ISL_1095941, EPI_ISL_1095942, EPI_ISL_1095943, EPI_ISL_1095944, EPI_ISL_1095945, EPI_ISL_1095946, EPI_ISL_1095947, EPI_ISL_1095948, EPI_ISL_1095949, EPI_ISL_1095950, EPI_ISL_1095951, EPI_ISL_1095952, EPI_ISL_1095953, EPI_ISL_1095954, EPI_ISL_1095955, EPI_ISL_1095956, EPI_ISL_1095957, EPI_ISL_1095958, EPI_ISL_1095959, EPI_ISL_1095960, EPI_ISL_1095961, EPI_ISL_1095962, EPI_ISL_1095963, EPI_ISL_1095964, EPI_ISL_1095965, EPI_ISL_1095966, EPI_ISL_1095967, EPI_ISL_1095968, EPI_ISL_1095969, EPI_ISL_1095970, EPI_ISL_1095971, EPI_ISL_1095972, EPI_ISL_1095973, EPI_ISL_1095974, EPI_ISL_1095975, EPI_ISL_1095976, EPI_ISL_1095977, EPI_ISL_1095978, EPI_ISL_1095979, EPI_ISL_1095980, EPI_ISL_1095981, EPI_ISL_1095982, EPI_ISL_1095983, EPI_ISL_1095984, EPI_ISL_1095985, EPI_ISL_1095986, EPI_ISL_1095987, EPI_ISL_1095988, EPI_ISL_1095989, EPI_ISL_1095990, EPI_ISL_1095991, EPI_ISL_1095992, EPI_ISL_1095993, EPI_ISL_1095994, EPI_ISL_1095995, EPI_ISL_1095996, EPI_ISL_1095997, EPI_ISL_1095998, EPI_ISL_1095999, EPI_ISL_1096000, EPI_ISL_1096001, EPI_ISL_1096002, EPI_ISL_1096003, EPI_ISL_1096004, EPI_ISL_1096005, EPI_ISL_1096006, EPI_ISL_1096007, EPI_ISL_1096008, EPI_ISL_1096009, EPI_ISL_1096010, EPI_ISL_1096011, EPI_ISL_1096012, EPI_ISL_1096013, EPI_ISL_1096014, EPI_ISL_1096015, EPI_ISL_1096016, EPI_ISL_1096017, EPI_ISL_1096018, EPI_ISL_1096019, EPI_ISL_1096020, EPI_ISL_1096021, EPI_ISL_1096022, EPI_ISL_1096023, EPI_ISL_1096024, EPI_ISL_1096025, EPI_ISL_1096026, EPI_ISL_1096027, EPI_ISL_1096028, EPI_ISL_1096029, EPI_ISL_1132845, EPI_ISL_1132846, EPI_ISL_1132847, EPI_ISL_1132848, EPI_ISL_1132849, EPI_ISL_1132850, EPI_ISL_1132851, EPI_ISL_1132852, EPI_ISL_1132853, EPI_ISL_1132854, EPI_ISL_1132855, EPI_ISL_1132856, EPI_ISL_1132857, EPI_ISL_1132858, EPI_ISL_1132859, EPI_ISL_1132860, EPI_ISL_1132861, EPI_ISL_1132862, EPI_ISL_1132863, EPI_ISL_1132864, EPI_ISL_1132865, EPI_ISL_1132866, EPI_ISL_1132867, EPI_ISL_1132868, EPI_ISL_1132869, EPI_ISL_1132870, EPI_ISL_1132871, EPI_ISL_1132872, EPI_ISL_1132873, EPI_ISL_1132874, EPI_ISL_1132875, EPI_ISL_1132876, EPI_ISL_1132877, EPI_ISL_1132878, EPI_ISL_1132879, EPI_ISL_1132880, EPI_ISL_1132881, EPI_ISL_1132882, EPI_ISL_1132883, EPI_ISL_1132884, EPI_ISL_1132885, EPI_ISL_1132886, EPI_ISL_1132887, EPI_ISL_1132888, EPI_ISL_1132889, EPI_ISL_1132890, EPI_ISL_1132891, EPI_ISL_1132892, EPI_ISL_1132893, EPI_ISL_1132894, EPI_ISL_1132895, EPI_ISL_1132896, EPI_ISL_1132897, EPI_ISL_1132898, EPI_ISL_1132899, EPI_ISL_1132900, EPI_ISL_1132901, EPI_ISL_1132902, EPI_ISL_1132903, EPI_ISL_1132904, EPI_ISL_1132905, EPI_ISL_1132906, EPI_ISL_1132907, EPI_ISL_1132908, EPI_ISL_1132909, EPI_ISL_1132910, EPI_ISL_1132911, EPI_ISL_1132912, EPI_ISL_1132913, EPI_ISL_1132914, EPI_ISL_1132915, EPI_ISL_1132916, EPI_ISL_1132917, EPI_ISL_1132918, EPI_ISL_1132919, EPI_ISL_1132920, EPI_ISL_1132921, EPI_ISL_1132922, EPI_ISL_1132923, EPI_ISL_1132924, EPI_ISL_1132925, EPI_ISL_1132926, EPI_ISL_1132927, EPI_ISL_1132928, EPI_ISL_1132929, EPI_ISL_1132930, EPI_ISL_1132931, EPI_ISL_1132932, EPI_ISL_1132933, EPI_ISL_1132934, EPI_ISL_1132935, EPI_ISL_1132936, EPI_ISL_1132937, EPI_ISL_1132938, EPI_ISL_1132939, EPI_ISL_1132940, EPI_ISL_1132941, EPI_ISL_1132942, EPI_ISL_1132943, EPI_ISL_1132944, EPI_ISL_1132945, EPI_ISL_1132946, EPI_ISL_1132947, EPI_ISL_1132948, EPI_ISL_1132949, EPI_ISL_1132950, EPI_ISL_1132951, EPI_ISL_1132952, EPI_ISL_1132953, EPI_ISL_1132954, EPI_ISL_1132955, EPI_ISL_1132956, EPI_ISL_1132957, EPI_ISL_1132958, EPI_ISL_1132959, EPI_ISL_1132960, EPI_ISL_1132961, EPI_ISL_1132962, EPI_ISL_1132963, EPI_ISL_1132964, EPI_ISL_1132965, EPI_ISL_1132966, EPI_ISL_1132967, EPI_ISL_1132968, EPI_ISL_1132969, EPI_ISL_1132970, EPI_ISL_1134840, EPI_ISL_1134841, EPI_ISL_1134842, EPI_ISL_1134843, EPI_ISL_1134844, EPI_ISL_1134845, EPI_ISL_1134846, EPI_ISL_1134847, EPI_ISL_1134848, EPI_ISL_1134849, EPI_ISL_1134850, EPI_ISL_1134851, EPI_ISL_1134852, EPI_ISL_1134853, EPI_ISL_1134854, EPI_ISL_1134855, EPI_ISL_1134856, EPI_ISL_1134857, EPI_ISL_1134858, EPI_ISL_1134859, EPI_ISL_1134860, EPI_ISL_1134861, EPI_ISL_1134862, EPI_ISL_1134863, EPI_ISL_1134864, EPI_ISL_1134865, EPI_ISL_1134866, EPI_ISL_1134867, EPI_ISL_1134868, EPI_ISL_1134869, EPI_ISL_1134870, EPI_ISL_1134871, EPI_ISL_1134872, EPI_ISL_1134873, EPI_ISL_1134874, EPI_ISL_1134875, EPI_ISL_1134876, EPI_ISL_1134877, EPI_ISL_1134878, EPI_ISL_1134879, EPI_ISL_1134880, EPI_ISL_1134881, EPI_ISL_1134882, EPI_ISL_1134883, EPI_ISL_1134884, EPI_ISL_1134885, EPI_ISL_1134886, EPI_ISL_1134887, EPI_ISL_1134888, EPI_ISL_1134889, EPI_ISL_1134890, EPI_ISL_1134891, EPI_ISL_1134892, EPI_ISL_1134893, EPI_ISL_1134894, EPI_ISL_1134895, EPI_ISL_1134896, EPI_ISL_1134897, EPI_ISL_1134898, EPI_ISL_1134899, EPI_ISL_1134900, EPI_ISL_1134901, EPI_ISL_1134902, EPI_ISL_1134903, EPI_ISL_1164360, EPI_ISL_1164361, EPI_ISL_1164362, EPI_ISL_1164363, EPI_ISL_1164364, EPI_ISL_1164365, EPI_ISL_1164366, EPI_ISL_1164367, EPI_ISL_1164368, EPI_ISL_1164369, EPI_ISL_1164370, EPI_ISL_1164371, EPI_ISL_1164372, EPI_ISL_1164373, EPI_ISL_1164374, EPI_ISL_1164375, EPI_ISL_1164376, EPI_ISL_1164377, EPI_ISL_1164378, EPI_ISL_1164379, EPI_ISL_1164380, EPI_ISL_1164381, EPI_ISL_1164382, EPI_ISL_1164383, EPI_ISL_1164384, EPI_ISL_1164385, EPI_ISL_1164386, EPI_ISL_1164387, EPI_ISL_1164388, EPI_ISL_1164389, EPI_ISL_1164390, EPI_ISL_1164391, EPI_ISL_1164392, EPI_ISL_1164393, EPI_ISL_1164394, EPI_ISL_1164395, EPI_ISL_1164396, EPI_ISL_1164397, EPI_ISL_1164398, EPI_ISL_1164399, EPI_ISL_1164400, EPI_ISL_1164401, EPI_ISL_1164402, EPI_ISL_1164403, EPI_ISL_1164404, EPI_ISL_1164405, EPI_ISL_1164406, EPI_ISL_1164407, EPI_ISL_1164408, EPI_ISL_1164409, EPI_ISL_1164410, EPI_ISL_1164411, EPI_ISL_1164412, EPI_ISL_1164413, EPI_ISL_1164414, EPI_ISL_1164415, EPI_ISL_1164416, EPI_ISL_1164417, EPI_ISL_1164418, EPI_ISL_1164419, EPI_ISL_1164420, EPI_ISL_1164421, EPI_ISL_1164422, EPI_ISL_1164423, EPI_ISL_1164424, EPI_ISL_1164425, EPI_ISL_1164426, EPI_ISL_1164427, EPI_ISL_1164428, EPI_ISL_1164429, EPI_ISL_1164430, EPI_ISL_1164431, EPI_ISL_1164432, EPI_ISL_1164433, EPI_ISL_1164434, EPI_ISL_1164435, EPI_ISL_1164436, EPI_ISL_1164437, EPI_ISL_1164438, EPI_ISL_1172046, EPI_ISL_1172047, EPI_ISL_1172048, EPI_ISL_1172049, EPI_ISL_1172050, EPI_ISL_1172051, EPI_ISL_1172052, EPI_ISL_1172053, EPI_ISL_1172054, EPI_ISL_1172055, EPI_ISL_1172056, EPI_ISL_1172057, EPI_ISL_1172058, EPI_ISL_1172059, EPI_ISL_1172060, EPI_ISL_1172061, EPI_ISL_1172062, EPI_ISL_1172063, EPI_ISL_1172064, EPI_ISL_1172065, EPI_ISL_1172066, EPI_ISL_1172067, EPI_ISL_1172068, EPI_ISL_1172069, EPI_ISL_1172070, EPI_ISL_1172071, EPI_ISL_1172072, EPI_ISL_1172073, EPI_ISL_1172074, EPI_ISL_1172075, EPI_ISL_1172076, EPI_ISL_1172077, EPI_ISL_1172078, EPI_ISL_1172079, EPI_ISL_1172080, EPI_ISL_1172081, EPI_ISL_1172082, EPI_ISL_1172083, EPI_ISL_1172084, EPI_ISL_1172085, EPI_ISL_1172086, EPI_ISL_1172087, EPI_ISL_1172088, EPI_ISL_1172089, EPI_ISL_1172090, EPI_ISL_1172091, EPI_ISL_1172092, EPI_ISL_1172093, EPI_ISL_1172094, EPI_ISL_1172095, EPI_ISL_1172096, EPI_ISL_1172097, EPI_ISL_1172098, EPI_ISL_1172099, EPI_ISL_1172100, EPI_ISL_1172101, EPI_ISL_1172102, EPI_ISL_1172103, EPI_ISL_1172104, EPI_ISL_1172105, EPI_ISL_1172106, EPI_ISL_1172107, EPI_ISL_1172108, EPI_ISL_1172109, EPI_ISL_1195947, EPI_ISL_1251260, EPI_ISL_1252050 | | | |
| see above | Fulgent Genetics | Fulgent Genetics | Becky Tsai; Benafsh Sapra; Doreen Ng; Harry Gao; James Xie; John Gao; Joseph Fierro; Mickey Li; Yan Meng |

| | | | |
|---|---|---|---|
| EPI_ISL_1508979, EPI_ISL_1508980, EPI_ISL_1508981, EPI_ISL_1508982, EPI_ISL_1508983, EPI_ISL_1508987 | Good Samaritan Hospital | Santa Clara County Public Health Laboratory | Santa Clara County Public Health Department |
| EPI_ISL_1088585, EPI_ISL_1088593, EPI_ISL_1088595, EPI_ISL_1088599, EPI_ISL_1088600, EPI_ISL_1088601, EPI_ISL_1088603, EPI_ISL_1088605, EPI_ISL_1088606, EPI_ISL_1088610, EPI_ISL_1088611, EPI_ISL_1088616, EPI_ISL_1088617, EPI_ISL_1088625, EPI_ISL_1088627, EPI_ISL_1088628, EPI_ISL_1088630, EPI_ISL_1088633, EPI_ISL_1088635, EPI_ISL_1088640, EPI_ISL_1088652, EPI_ISL_1088655, EPI_ISL_1088661, EPI_ISL_1088666, EPI_ISL_1088677, EPI_ISL_1088678, EPI_ISL_1088681, EPI_ISL_1088684, EPI_ISL_1088689, EPI_ISL_1088690, EPI_ISL_1088696, EPI_ISL_1088698, EPI_ISL_1088703, EPI_ISL_1088706, EPI_ISL_1088715, EPI_ISL_1088720, EPI_ISL_1088725, EPI_ISL_1088726, EPI_ISL_1088730, EPI_ISL_1088751, EPI_ISL_1088755, EPI_ISL_1088762, EPI_ISL_1088763, EPI_ISL_1088765, EPI_ISL_1088772, EPI_ISL_1088791, EPI_ISL_1088793, EPI_ISL_1088816, EPI_ISL_1088819, EPI_ISL_1088820, EPI_ISL_1088828, EPI_ISL_1088833, EPI_ISL_1088835, EPI_ISL_1088850, EPI_ISL_1088851, EPI_ISL_1088853, EPI_ISL_1088865, EPI_ISL_1088871, EPI_ISL_1088874, EPI_ISL_1088880, EPI_ISL_1088887, EPI_ISL_1088900, EPI_ISL_1088901, EPI_ISL_1088903, EPI_ISL_1088904, EPI_ISL_1088913, EPI_ISL_1088914, EPI_ISL_1088933, EPI_ISL_1088956, EPI_ISL_1088967, EPI_ISL_1088974, EPI_ISL_1088979, EPI_ISL_1088995, EPI_ISL_1088997, EPI_ISL_1088999, EPI_ISL_1089000, EPI_ISL_1089001, EPI_ISL_1089004, EPI_ISL_1089007, EPI_ISL_1089012, EPI_ISL_1089027, EPI_ISL_1089050, EPI_ISL_1089053, EPI_ISL_1089054, EPI_ISL_1089062, EPI_ISL_1089063, EPI_ISL_1089069, EPI_ISL_1089074, EPI_ISL_1089075, EPI_ISL_1089080, EPI_ISL_1089086, EPI_ISL_1089090, EPI_ISL_1089101, EPI_ISL_1089108, EPI_ISL_1089117, EPI_ISL_1089118, EPI_ISL_1089127, EPI_ISL_1089136, EPI_ISL_1089138, EPI_ISL_1089148, EPI_ISL_1089152, EPI_ISL_1089153, EPI_ISL_1089154, EPI_ISL_1089155, EPI_ISL_1089157, EPI_ISL_1089159, EPI_ISL_1089171, EPI_ISL_1089173, EPI_ISL_1089185, EPI_ISL_1089197, EPI_ISL_1089198, EPI_ISL_1089207, EPI_ISL_1089212, EPI_ISL_1089214, EPI_ISL_1089215, EPI_ISL_1089216, EPI_ISL_1089221, EPI_ISL_1089222, EPI_ISL_1089239, EPI_ISL_1089244, EPI_ISL_1089258, EPI_ISL_1089260, EPI_ISL_1089262, EPI_ISL_1089429, EPI_ISL_1089430, EPI_ISL_1089433 | | | |
| see above | Helix / Illumina | Respiratory Viruses Branch, Division of Viral Diseases, Centers for Disease Control and Prevention | Alexandre Bolze; Ary Ascencio; Ben L. Rambo-Martin; Brad Sickler; Charlotte Rivera-Garcia; Christine Tran; Clinton R. Paden; Dakota Howard; David Becker; Dhwani Batra; Duncan MacCannell; Efren Sandoval; Eileen de Feo; Elizabeth Cirulli; Eric Allen; Geraint Levan; James Lu; Jan Antico; Jason Nguyen; Jimmy Ramirez; Jingtao Liu; Kelly Schiabor Barrett; Kim Gietzen; Magnus Isaksson; Marc Laurent; Matthew Tolentino; Nicole L. Washington; Peter W. Cook; Phil Febbo; Ryan Cho; Shannon Wickline; Sherry Wang; Simon White; Summer Galloway; Suxiang Tong; Tyler Cassens; William Lee |
| EPI_ISL_1194743, EPI_ISL_1194746, EPI_ISL_1194785, EPI_ISL_1194808, EPI_ISL_1194818, EPI_ISL_1194828, EPI_ISL_1262278, EPI_ISL_1262280, EPI_ISL_1262284, EPI_ISL_1262291, EPI_ISL_1262300, EPI_ISL_1262306, EPI_ISL_1262336, EPI_ISL_1262338, EPI_ISL_1262339, EPI_ISL_1262347, EPI_ISL_1262353, EPI_ISL_1262360, EPI_ISL_1262379, EPI_ISL_1262381, EPI_ISL_1262385, EPI_ISL_1262393, EPI_ISL_1262404, EPI_ISL_1262414, EPI_ISL_1267505, EPI_ISL_1267513, EPI_ISL_1267530, EPI_ISL_1267537, EPI_ISL_1267538, EPI_ISL_1267542, EPI_ISL_1267545, EPI_ISL_1267548, EPI_ISL_1267551, EPI_ISL_1267556, EPI_ISL_1267557, EPI_ISL_1267558, EPI_ISL_1267559, EPI_ISL_1267568, EPI_ISL_1267569, EPI_ISL_1267575, EPI_ISL_1267582, EPI_ISL_1267589, EPI_ISL_1267621, EPI_ISL_1267627, EPI_ISL_1267629, EPI_ISL_1267631, EPI_ISL_1267635, EPI_ISL_1267636, EPI_ISL_1267640, EPI_ISL_1267642, EPI_ISL_1267652, EPI_ISL_1267657, EPI_ISL_1267661, EPI_ISL_1267667, EPI_ISL_1267680, EPI_ISL_1267681, EPI_ISL_1267689, EPI_ISL_1267695, EPI_ISL_1267704, EPI_ISL_1267746, EPI_ISL_1267758, EPI_ISL_1267767, EPI_ISL_1267776, EPI_ISL_1267790, EPI_ISL_1267797, EPI_ISL_1267798, EPI_ISL_1267800, EPI_ISL_1267809, EPI_ISL_1267815, EPI_ISL_1267820, EPI_ISL_1267821, EPI_ISL_1267833, EPI_ISL_1267839, EPI_ISL_1267844, EPI_ISL_1267845, EPI_ISL_1267851, EPI_ISL_1267876, EPI_ISL_1268073, EPI_ISL_1268075, EPI_ISL_1268085, EPI_ISL_1268086, EPI_ISL_1268096, EPI_ISL_1268098, EPI_ISL_1268099, EPI_ISL_1268110, EPI_ISL_1268111, EPI_ISL_1268113, EPI_ISL_1268119, EPI_ISL_1268138, EPI_ISL_1268146, EPI_ISL_1268147, EPI_ISL_1268150, EPI_ISL_1268173, EPI_ISL_1268174, EPI_ISL_1268180, EPI_ISL_1268181, EPI_ISL_1268185, EPI_ISL_1268186, EPI_ISL_1268190, EPI_ISL_1268197, EPI_ISL_1268199, EPI_ISL_1268201, EPI_ISL_1268231, EPI_ISL_1268260, EPI_ISL_1268272, EPI_ISL_1268276, EPI_ISL_1268286, EPI_ISL_1268344, EPI_ISL_1268404, EPI_ISL_1268413, EPI_ISL_1268434, EPI_ISL_1268436, EPI_ISL_1268440, EPI_ISL_1268442, EPI_ISL_1268443, EPI_ISL_1268473, EPI_ISL_1268485, EPI_ISL_1268534, EPI_ISL_1269039, EPI_ISL_1269109, EPI_ISL_1269132, EPI_ISL_1269135, EPI_ISL_1269139, EPI_ISL_1269148, EPI_ISL_1269151, EPI_ISL_1269152, EPI_ISL_1298206, EPI_ISL_1336965, EPI_ISL_1336973, EPI_ISL_1336975, EPI_ISL_1337062, EPI_ISL_1337211, EPI_ISL_1391777, EPI_ISL_1391778, EPI_ISL_1391782, EPI_ISL_1391808, EPI_ISL_1391814, EPI_ISL_1493792 | | | |
| see above | Helix/Illumina | Centers for Disease Control and Prevention Division of Viral Diseases, Pathogen Discovery | Adrian Paskey; Alexandre Bolze; Ary Ascencio; Ben L. Rambo-Martin; Benjamin Rambo-Martin; Brad Sickler; Charlotte Rivera-Garcia; Christine Tran; Christopher Gulvick; Clinton R. Paden; Dakota Howard; Darlene Wagner; David Becker; Dhwani Batra; Duncan MacCannell; Efren Sandoval; Eileen de Feo; Elizabeth Cirulli; Eric Allen; Geraint Levan; James Lu; Jan Antico; Jason Caravas; Jason Nguyen; Jimmy Ramirez; Jingtao Liu; Kara Moser; Kelly Schiabor Barrett; Kim Gietzen; Magnus Isaksson; Marc Laurent; Matthew Schmerer; Matthew Tolentino; Nicole L. Washington; Peter W. Cook; Phil Febbo; Ryan Cho; Scott Sammons; Shannon Wickline; Shatavia Morrison; Sherry Wang; Simon White; Summer Galloway; Suxiang Tong; Tyler Cassens; William Lee; Yvette Unoarumhi |
| EPI_ISL_1016513, EPI_ISL_1016518, EPI_ISL_1016538, EPI_ISL_1016545, EPI_ISL_1016575, EPI_ISL_1081049, EPI_ISL_1081063, EPI_ISL_1110663, EPI_ISL_1133793, EPI_ISL_1133796, EPI_ISL_1133802, EPI_ISL_1133805, EPI_ISL_1133822, EPI_ISL_1133824, EPI_ISL_1133832, EPI_ISL_1133838, EPI_ISL_1133842, EPI_ISL_1133858, EPI_ISL_1133859, EPI_ISL_1133862, EPI_ISL_1133866, EPI_ISL_1133876, EPI_ISL_1133878, EPI_ISL_1133884, EPI_ISL_1133887, EPI_ISL_1133891, EPI_ISL_1133892, EPI_ISL_1133894, EPI_ISL_1133895, EPI_ISL_1133896, EPI_ISL_1133899, EPI_ISL_1133901, EPI_ISL_1133912, EPI_ISL_1133915, EPI_ISL_1133916, EPI_ISL_1133917, EPI_ISL_1133929, EPI_ISL_1133937, EPI_ISL_1133944, EPI_ISL_1133945, EPI_ISL_1133954, EPI_ISL_1133960, EPI_ISL_1133964, EPI_ISL_1133965, EPI_ISL_1133969, EPI_ISL_1133972, EPI_ISL_1133975, EPI_ISL_1133980, EPI_ISL_1133981, EPI_ISL_1133984, EPI_ISL_1133985, EPI_ISL_1133987, EPI_ISL_1133996, EPI_ISL_1134002, EPI_ISL_1134005, EPI_ISL_1134011, EPI_ISL_1134019, EPI_ISL_1134041, EPI_ISL_1134048, EPI_ISL_1134056, EPI_ISL_1134058, EPI_ISL_1134063, EPI_ISL_1134065, EPI_ISL_1134070, EPI_ISL_1134073, EPI_ISL_1134085, EPI_ISL_1134099, EPI_ISL_1134109, EPI_ISL_1134114, EPI_ISL_1134121, EPI_ISL_1134127, EPI_ISL_1134145, EPI_ISL_1134146, EPI_ISL_1134147, EPI_ISL_1134148, EPI_ISL_1134154, EPI_ISL_1134157, EPI_ISL_1134180, EPI_ISL_1134182, EPI_ISL_1134193, EPI_ISL_1134260, EPI_ISL_1134272, EPI_ISL_1134276, EPI_ISL_1134279, EPI_ISL_1134280, EPI_ISL_1134283, EPI_ISL_1134291, EPI_ISL_1134292, EPI_ISL_1134297, EPI_ISL_1134299, EPI_ISL_1134305, EPI_ISL_1134306, EPI_ISL_1134307, EPI_ISL_1134312, EPI_ISL_1134314, EPI_ISL_1134315, EPI_ISL_1134323, EPI_ISL_1134326, EPI_ISL_1134332, EPI_ISL_1134338, EPI_ISL_1134345, EPI_ISL_1134351, EPI_ISL_1134369, EPI_ISL_1134395, EPI_ISL_1134404, EPI_ISL_1134407, EPI_ISL_1134412, EPI_ISL_1134432, EPI_ISL_1134458, EPI_ISL_1134463, EPI_ISL_1134497, EPI_ISL_1134504, EPI_ISL_1134531, EPI_ISL_1134539, EPI_ISL_1134557, EPI_ISL_1134563, EPI_ISL_1134568, EPI_ISL_1134569, EPI_ISL_1134570, EPI_ISL_1134574, EPI_ISL_1134580, EPI_ISL_1134582, EPI_ISL_1134585, EPI_ISL_1134648, EPI_ISL_1134649, EPI_ISL_1134654, EPI_ISL_1134656, EPI_ISL_1134664, EPI_ISL_1134670, EPI_ISL_1134673, EPI_ISL_1134685, EPI_ISL_1134687, EPI_ISL_1134695, EPI_ISL_1134714, EPI_ISL_1134729, EPI_ISL_1134732, EPI_ISL_1134740, EPI_ISL_1134751, EPI_ISL_1134754, EPI_ISL_1134767, EPI_ISL_1134785, EPI_ISL_1134818, EPI_ISL_1134829 | | | |
| see above | Helix/Illumina | Respiratory Viruses Branch, Division of Viral Diseases, Centers for Disease Control and Prevention | Alexandre Bolze; Ary Ascencio; Ben L. Rambo-Martin; Brad Sickler; Charlotte Rivera-Garcia; Christine Tran; Clinton R. Paden; Dakota Howard; David Becker; Dhwani Batra; Duncan MacCannell; Efren Sandoval; Eileen de Feo; Elizabeth Cirulli; Eric Allen; Geraint Levan; James Lu; Jan Antico; Jason Nguyen; Jimmy Ramirez; Jingtao Liu; Kelly Schiabor Barrett; Kim Gietzen; Magnus Isaksson; Marc Laurent; Matthew Tolentino; Nicole L. Washington; Peter W. Cook; Phil Febbo; Ryan Cho; Shannon Wickline; Sherry Wang; Simon White; Summer Galloway; Suxiang Tong; Tyler Cassens; William Lee |
| EPI_ISL_1477174, EPI_ISL_1477189, EPI_ISL_1477207, EPI_ISL_1477218, EPI_ISL_1477220, EPI_ISL_1477224, EPI_ISL_1477226, EPI_ISL_1477236, EPI_ISL_1477239, EPI_ISL_1477249, EPI_ISL_1477257, EPI_ISL_1477264, EPI_ISL_1477294, EPI_ISL_1477312, EPI_ISL_1477326, EPI_ISL_1477334, EPI_ISL_1477340, EPI_ISL_1477381, EPI_ISL_1477389, EPI_ISL_1477394, EPI_ISL_1477396, EPI_ISL_1477397, EPI_ISL_1477401, EPI_ISL_1477412, EPI_ISL_1477430, EPI_ISL_2659309, EPI_ISL_2659526, EPI_ISL_2758460 | | | |
| see above | Humboldt County Public Health Laboratory | Chan-Zuckerberg Biohub | CZB Cliahub Consortium |
| EPI_ISL_1060040, EPI_ISL_1060042, EPI_ISL_1060046, EPI_ISL_1060047, EPI_ISL_1060048, EPI_ISL_1060049, EPI_ISL_1060050, EPI_ISL_1060051, EPI_ISL_1060052, EPI_ISL_1060053, EPI_ISL_1060055, EPI_ISL_1060056, EPI_ISL_1060057, EPI_ISL_1060058, EPI_ISL_1060059, EPI_ISL_1060061, EPI_ISL_1060062, EPI_ISL_1060063, EPI_ISL_1060064, EPI_ISL_1060065, EPI_ISL_1060066, EPI_ISL_1060067, EPI_ISL_1060069, EPI_ISL_1060070, EPI_ISL_1060071, EPI_ISL_1060072, EPI_ISL_1060073, EPI_ISL_1060074, EPI_ISL_1060075, EPI_ISL_1060077, EPI_ISL_1060079, EPI_ISL_1060080, EPI_ISL_1060081, EPI_ISL_1060082, EPI_ISL_1060083, EPI_ISL_1060084, EPI_ISL_1060085, EPI_ISL_1060086, EPI_ISL_1060087, EPI_ISL_1060088, EPI_ISL_1060089, EPI_ISL_1060090, EPI_ISL_1060091, EPI_ISL_1060092, EPI_ISL_1060093, EPI_ISL_1060094, EPI_ISL_1060095, EPI_ISL_1060097, EPI_ISL_1060099, EPI_ISL_1060100, EPI_ISL_1060101, EPI_ISL_1060102, EPI_ISL_1060103, EPI_ISL_1060104, EPI_ISL_1060105, EPI_ISL_1060106, EPI_ISL_1060107, EPI_ISL_1060108, EPI_ISL_1060109, EPI_ISL_1060110, EPI_ISL_1060111, EPI_ISL_1060115, EPI_ISL_1060116, EPI_ISL_1060117, EPI_ISL_1060118, EPI_ISL_1060120, EPI_ISL_1060122, EPI_ISL_1060123, EPI_ISL_1060124, EPI_ISL_1060125, EPI_ISL_1060126, EPI_ISL_1060128, EPI_ISL_1060129, EPI_ISL_1060131, EPI_ISL_1060132, EPI_ISL_1060133, EPI_ISL_1060134, EPI_ISL_1060136, EPI_ISL_1060139, EPI_ISL_1060140, EPI_ISL_1060141, EPI_ISL_1060142, EPI_ISL_1060143, EPI_ISL_1060144, EPI_ISL_1060146, EPI_ISL_1060147, EPI_ISL_1060148, EPI_ISL_1060149, EPI_ISL_1060153, EPI_ISL_1060154, EPI_ISL_1060155, EPI_ISL_1060158, EPI_ISL_1060159, EPI_ISL_1060160, EPI_ISL_1060161, EPI_ISL_1060162, EPI_ISL_1060163, EPI_ISL_1060164, EPI_ISL_1060165, EPI_ISL_1060166, EPI_ISL_1060167, EPI_ISL_1060168, EPI_ISL_1060169, EPI_ISL_1060170, EPI_ISL_1060171, EPI_ISL_1060172, EPI_ISL_1060173, EPI_ISL_1060174, EPI_ISL_1060177, EPI_ISL_1060178, EPI_ISL_1060179, EPI_ISL_1060180, EPI_ISL_1060181, EPI_ISL_1060182, EPI_ISL_1060183, EPI_ISL_1060184, EPI_ISL_1060185, EPI_ISL_1060186, EPI_ISL_1060187, EPI_ISL_1060188, EPI_ISL_1060189, EPI_ISL_1060190, EPI_ISL_1060191, EPI_ISL_1081284, EPI_ISL_1081285, EPI_ISL_1081286, EPI_ISL_1081287, EPI_ISL_1081288 | | | |
| see above | Innovative Genomics Institute, UC Berkeley | Innovative Genomics Institute, UC Berkeley | Alison Ciling; Haridha Shivram; Liana Lareau; Netravathi Krishnappa; Phil Frankino; Stacia Wyman |
| EPI_ISL_1664235, EPI_ISL_1664236, EPI_ISL_1664239, EPI_ISL_1664241, EPI_ISL_1664242, EPI_ISL_1664243, EPI_ISL_1664246, EPI_ISL_1664247 | | | |
| see above | Kaiser San Jose | Santa Clara County public Health Laboratory | Santa Clara County Public Health Department |
| EPI_ISL_1221355, EPI_ISL_1221386, EPI_ISL_1221388, EPI_ISL_1221810, EPI_ISL_1221811, EPI_ISL_1291297, EPI_ISL_1291298 | | | |
| see above | Laboratory Corporation of America | Centers for Disease Control and Prevention Division of Viral Diseases, Pathogen Discovery | Amanda Douglas; Amanda Suchanek; Andrea Throop; Ayla Burns; Ben L. Rambo-Martin; Bobbi Croy; Brian Krueger; Brian Norvell; Christos Petropoulos; Clinton R. Paden; Craig Lukasik; Dakota Howard; Debbie Boles; Dhwani Batra; Duncan MacCannell; Eyad Almasri; Eyad Almasri Debbie Boles; Goran Stevovic; Howard Engler; Hrushikesh Deshmukh; Jake Humphrey; Jana Schroth; Joe Voshell; John Pruitt; Jonathan Meltzer; Jonathan Williams; Kimberly Wagner; Lax Iyer; Lyndon Tilson; Manoj Jain; Marcia Eisenberg; Mary Ann Cristobal; Mary Williamson; Michael Levandoski; Mike Sapeta; Mindy Nye; Minoo Agarwal; Mohan Kolli; Nuthawin Charoensri; Oren Cohen; Peter W. Cook; Prashant Gupta; Qian Zeng; Rama Ghatti; Scott Parker; Scott Ryan; Stanley Letovsky; Steven Ragan; Suresh Babu Selvaraju; Susan Countryman; Susan Hicks; Suxiang Tong; Suzanne Dale; Thomas Urban; Tim Kuphal; Tricia Zwiefelhofer; Vincent Drouillon |
| EPI_ISL_1160013, EPI_ISL_1160036, EPI_ISL_1160061, EPI_ISL_1160078, EPI_ISL_1160082, EPI_ISL_1160113, EPI_ISL_1160210, EPI_ISL_1160222, EPI_ISL_1160278, EPI_ISL_1160329, EPI_ISL_1160354, EPI_ISL_1160367, EPI_ISL_1160373, EPI_ISL_1160390, EPI_ISL_1160411, EPI_ISL_1160445, EPI_ISL_1160453, EPI_ISL_1160497, EPI_ISL_1160508, EPI_ISL_1160511, EPI_ISL_1160527, EPI_ISL_1160560, EPI_ISL_1160583, EPI_ISL_1160660, EPI_ISL_1160799, EPI_ISL_1160807, EPI_ISL_1160886, EPI_ISL_1160923, EPI_ISL_1160924, EPI_ISL_1161290, EPI_ISL_1161291, EPI_ISL_1161335, EPI_ISL_1161343, EPI_ISL_1161344, EPI_ISL_1161345, EPI_ISL_1161346, EPI_ISL_1161347, EPI_ISL_1161351, EPI_ISL_1161352, EPI_ISL_1161353, EPI_ISL_1161358, EPI_ISL_1161359, EPI_ISL_1161360, EPI_ISL_1161419, EPI_ISL_1161420, EPI_ISL_1161422, EPI_ISL_1161598, EPI_ISL_1161599, EPI_ISL_1161600, EPI_ISL_1161601, EPI_ISL_1161602, EPI_ISL_1161603, EPI_ISL_1161691, EPI_ISL_1161709, EPI_ISL_1161710, EPI_ISL_1161711, EPI_ISL_1161761, EPI_ISL_1161923, EPI_ISL_1161943, EPI_ISL_1161960, EPI_ISL_1161970, EPI_ISL_1161997, EPI_ISL_1161998, EPI_ISL_1162020, EPI_ISL_1162050, EPI_ISL_1162096, EPI_ISL_1162130, EPI_ISL_1162136, EPI_ISL_1162293, EPI_ISL_1162312, EPI_ISL_1162342, EPI_ISL_1162494, EPI_ISL_1162497, EPI_ISL_1162539, EPI_ISL_1162542, EPI_ISL_1162553, EPI_ISL_1162561, EPI_ISL_1162562, EPI_ISL_1162604, EPI_ISL_1162624, EPI_ISL_1162640, EPI_ISL_1162641, EPI_ISL_1162687, EPI_ISL_1162754, EPI_ISL_1162783, EPI_ISL_1162839, EPI_ISL_1162937, EPI_ISL_1162938, EPI_ISL_1162939, EPI_ISL_1162940, EPI_ISL_1162976, EPI_ISL_1162984, EPI_ISL_1163192, EPI_ISL_1163423, EPI_ISL_1163424, EPI_ISL_1163425, EPI_ISL_1163426, EPI_ISL_1163427, EPI_ISL_1163428, EPI_ISL_1163429, EPI_ISL_1163430, EPI_ISL_1163431, EPI_ISL_1163432, EPI_ISL_1163433, EPI_ISL_1163434, EPI_ISL_1163435, EPI_ISL_1163436, EPI_ISL_1163437, EPI_ISL_1163438 | | | |
| see above | Laboratory Corporation of America | Respiratory Viruses Branch, Division of Viral Diseases, Centers for Disease Control and Prevention | Amanda Douglas; Amanda Suchanek; Andrea Throop; Ayla Burns; Ben L. Rambo-Martin; Bobbi Croy; Brian Krueger; Brian Norvell; Christos Petropoulos; Clinton R. Paden; Craig Lukasik; Dakota Howard; Dhwani Batra; Duncan MacCannell; Eyad Almasri Debbie Boles; Goran Stevovic; Howard Engler; Hrushikesh Deshmukh; Jake Humphrey; Jana Schroth; Joe Voshell; John Pruitt; Jonathan Meltzer; Jonathan Williams; Kimberly Wagner; Lax Iyer; Lyndon Tilson; Manoj Jain; Marcia Eisenberg; Mary Ann Cristobal; Mary Williamson; Michael Levandoski; Mike Sapeta; Mindy Nye; Minoo Agarwal; Mohan Kolli; Nuthawin Charoensri; Oren Cohen; Peter W. Cook; Prashant Gupta; Qian Zeng; Rama Ghatti; Scott Parker; Scott Ryan; Stanley Letovsky; Steven Ragan; Suresh Babu Selvaraju; Susan Countryman; Susan Hicks; Suxiang Tong; Suzanne Dale; Thomas Urban; Tim Kuphal; Tricia Zwiefelhofer; Vincent Drouillon |
| EPI_ISL_1005557 | Los Angeles Community Hospital | Los Angeles County Public Health Laboratories | P. Hemarajata et al. |
| EPI_ISL_1097724, EPI_ISL_1097740, EPI_ISL_1097741, EPI_ISL_1098639, EPI_ISL_1098640, EPI_ISL_2620780, EPI_ISL_2620781, EPI_ISL_2620782, EPI_ISL_2620784, EPI_ISL_2620785, EPI_ISL_2620786, EPI_ISL_2620787, EPI_ISL_2620788, EPI_ISL_2620789, EPI_ISL_2620790, EPI_ISL_2620791, EPI_ISL_2620792, EPI_ISL_2620793, EPI_ISL_2620794, EPI_ISL_2620801, EPI_ISL_2620824, EPI_ISL_2620825, EPI_ISL_2811983, EPI_ISL_2894585 | | | |
| see above | Los Angeles County Public Health Laboratories | Los Angeles County Public Health Laboratories | P. Hemarajata et al. |
| EPI_ISL_1197043, EPI_ISL_1197050, EPI_ISL_1197055, EPI_ISL_1197056, EPI_ISL_1197057, EPI_ISL_1303439, EPI_ISL_1303447 | | | |
| see above | Martin Luther King Jr. Community Hospital | Los Angeles County Public Health Laboratories | P. Hemarajata et al. |
| EPI_ISL_1263228, EPI_ISL_1263233, EPI_ISL_1263234, EPI_ISL_1664617, EPI_ISL_1700695, EPI_ISL_1921880, EPI_ISL_1921881, EPI_ISL_1921882, EPI_ISL_1921883, EPI_ISL_1921884, EPI_ISL_1921885, EPI_ISL_1921886, EPI_ISL_1921887, EPI_ISL_1921888, EPI_ISL_2308296, EPI_ISL_2308297, EPI_ISL_2308299, EPI_ISL_2308353, EPI_ISL_2308367, EPI_ISL_2308368, EPI_ISL_2754042, EPI_ISL_2759079 | | | |
| see above | Monterey County Public Health Laboratory | Monterey County Public Health Laboratory | Monterey County Public Health Laboratory |
| EPI_ISL_2603592, EPI_ISL_2603593 | NIAID - IRF Frederick, NIH | NIAID - IRF Frederick, NIH | Georgia, S.; Kocher, G. |
| EPI_ISL_1005556 | Olive View Medical Center | Los Angeles County Public Health Laboratories | P. Hemarajata et al. |
| EPI_ISL_1234791, EPI_ISL_1234828, EPI_ISL_1234838, EPI_ISL_1234847, EPI_ISL_1234855, EPI_ISL_1234864, EPI_ISL_1234909, EPI_ISL_1234926, EPI_ISL_1234943, EPI_ISL_1234945, EPI_ISL_1234954, EPI_ISL_1234958, EPI_ISL_1234965, EPI_ISL_1234977, EPI_ISL_1234997, EPI_ISL_1235008, EPI_ISL_1235009, EPI_ISL_1235058, EPI_ISL_1477443, EPI_ISL_1477611, EPI_ISL_1477663, EPI_ISL_1701438, EPI_ISL_1701565, EPI_ISL_1701589 | | | |
| see above | Orange County Public Health Lab | Chan-Zuckerberg Biohub | CZB Cliahub Consortium |
| EPI_ISL_1912975 | Poplar Healthcare | Colleen B. Jonsson | Colleen Jonsson; Jacob Nichols; Joythi Parvathareddy; Mariah Taylor; Surekha Surendranathan; Walter Reichard |
| EPI_ISL_2551226, EPI_ISL_2551227, EPI_ISL_2551228, EPI_ISL_2551229, EPI_ISL_2551230, EPI_ISL_2551238, EPI_ISL_2551239, EPI_ISL_2551241, EPI_ISL_2551252, EPI_ISL_2551253 | | | |
| see above | Providence St. Joseph Health Orange - Chapman | Providence St. Joseph Health Molecular Genomics Laboratory | Alexa K Dowdell; Brian D Piening; Carlo B Bifulco; Fred L Robinson; Mary Campbell; Rogan Rattray |
| EPI_ISL_1159233, EPI_ISL_1159244, EPI_ISL_1159250, EPI_ISL_1159262, EPI_ISL_1159268, EPI_ISL_1159269, EPI_ISL_1159274, EPI_ISL_1159275, EPI_ISL_1159287, EPI_ISL_1159288, EPI_ISL_1159291, EPI_ISL_1159293, EPI_ISL_1159294, EPI_ISL_1159311, EPI_ISL_1159319, EPI_ISL_1159321, EPI_ISL_1159324, EPI_ISL_1159325, EPI_ISL_1159332, EPI_ISL_1193374, EPI_ISL_1193375, EPI_ISL_1193380, EPI_ISL_1193385, EPI_ISL_1193386, EPI_ISL_1193390, EPI_ISL_1193395, EPI_ISL_1193405, EPI_ISL_1193411, EPI_ISL_1193413, EPI_ISL_1193414, EPI_ISL_1193415, EPI_ISL_1193416, EPI_ISL_1193417, EPI_ISL_1193419, EPI_ISL_1193421, EPI_ISL_1193422, EPI_ISL_1193426, EPI_ISL_1193430, EPI_ISL_1193433, EPI_ISL_1193434, EPI_ISL_1193435, EPI_ISL_1193439, EPI_ISL_1193460, EPI_ISL_1193464, EPI_ISL_1193466, EPI_ISL_1193471, EPI_ISL_1193472, EPI_ISL_1193481, EPI_ISL_1193482, EPI_ISL_1193483, EPI_ISL_1193484, EPI_ISL_1193486, EPI_ISL_1193487, EPI_ISL_1193490, EPI_ISL_1193491, EPI_ISL_1193492, EPI_ISL_1193493, EPI_ISL_1193495, EPI_ISL_1193497, EPI_ISL_1193498, EPI_ISL_1193501, EPI_ISL_1193503, EPI_ISL_1193504, EPI_ISL_1193505, EPI_ISL_1193511, EPI_ISL_1193512, EPI_ISL_1193514, EPI_ISL_1193515, EPI_ISL_1193518, EPI_ISL_1193520, EPI_ISL_1193523, EPI_ISL_1193524, EPI_ISL_1193525, EPI_ISL_1193526, EPI_ISL_1193527, EPI_ISL_1193530, EPI_ISL_1193534, EPI_ISL_1193535, EPI_ISL_1193538, EPI_ISL_1193552, EPI_ISL_1193557, EPI_ISL_1193559, EPI_ISL_1193562, EPI_ISL_1193570, EPI_ISL_1193571, EPI_ISL_1193576, EPI_ISL_1193585, EPI_ISL_1193601, EPI_ISL_1193607, EPI_ISL_1193628, EPI_ISL_1193647, EPI_ISL_1193653, EPI_ISL_1193780, EPI_ISL_1193803, EPI_ISL_1193810, EPI_ISL_1193831, EPI_ISL_1193862, EPI_ISL_1193863, EPI_ISL_1193865, EPI_ISL_1193877, EPI_ISL_1193892, EPI_ISL_1193914, EPI_ISL_1193916, EPI_ISL_1193917, EPI_ISL_1193918, EPI_ISL_1193919, EPI_ISL_1193920, EPI_ISL_1193921, EPI_ISL_1193923, EPI_ISL_1193924, EPI_ISL_1193927, EPI_ISL_1193928, EPI_ISL_1193929, EPI_ISL_1193931, EPI_ISL_1193932, EPI_ISL_1193933, EPI_ISL_1193934, EPI_ISL_1193935, EPI_ISL_1193939, EPI_ISL_1193942, EPI_ISL_1193945, EPI_ISL_1193946, EPI_ISL_1193947, EPI_ISL_1193950, EPI_ISL_1193951, EPI_ISL_1193952, EPI_ISL_1193954, EPI_ISL_1193956, EPI_ISL_1193957, EPI_ISL_1193958, EPI_ISL_1193959, EPI_ISL_1193960, | | | |

| | | | |
|---|---|---|---|
| EPI_ISL_1193961, EPI_ISL_1193962, EPI_ISL_1193963, EPI_ISL_1193964, EPI_ISL_1193966, EPI_ISL_1193967, EPI_ISL_1193968, EPI_ISL_1193969, EPI_ISL_1193970, EPI_ISL_1193971, EPI_ISL_1193976, EPI_ISL_1193977, EPI_ISL_1193978, EPI_ISL_1193979, EPI_ISL_1193982, EPI_ISL_1193984, EPI_ISL_1193989, EPI_ISL_1193991, EPI_ISL_1193992, EPI_ISL_1193994, EPI_ISL_1193995, EPI_ISL_1193996, EPI_ISL_1193998, EPI_ISL_1194000, EPI_ISL_1194002, EPI_ISL_1194003, EPI_ISL_1194004, EPI_ISL_1194005, EPI_ISL_1194006, EPI_ISL_1194007, EPI_ISL_1194008, EPI_ISL_1194009, EPI_ISL_1194012, EPI_ISL_1194014, EPI_ISL_1194016, EPI_ISL_1194017, EPI_ISL_1194020, EPI_ISL_1194021, EPI_ISL_1194022, EPI_ISL_1194023, EPI_ISL_1194024, EPI_ISL_1194025, EPI_ISL_1194027, EPI_ISL_1194030, EPI_ISL_1194031, EPI_ISL_1194033, EPI_ISL_1194034, EPI_ISL_1194036, EPI_ISL_1194037, EPI_ISL_1194038, EPI_ISL_1194039, EPI_ISL_1194040, EPI_ISL_1194041, EPI_ISL_1194042, EPI_ISL_1194074, EPI_ISL_1194113, EPI_ISL_1194154, EPI_ISL_1194172, EPI_ISL_1194173, EPI_ISL_1194175, EPI_ISL_1194186, EPI_ISL_1194195, EPI_ISL_1194196, EPI_ISL_1194198, EPI_ISL_1194200, EPI_ISL_1194202, EPI_ISL_1194207, EPI_ISL_1194214, EPI_ISL_1194227, EPI_ISL_1194236, EPI_ISL_1194244, EPI_ISL_1194245, EPI_ISL_1194247, EPI_ISL_1194258, EPI_ISL_1194263, EPI_ISL_1194270, EPI_ISL_1194276, EPI_ISL_1194281, EPI_ISL_1194283, EPI_ISL_1194284, EPI_ISL_1194287, EPI_ISL_1194291, EPI_ISL_1194295, EPI_ISL_1194297, EPI_ISL_1194304, EPI_ISL_1194307, EPI_ISL_1194315, EPI_ISL_1194316, EPI_ISL_1194323, EPI_ISL_1194324, EPI_ISL_1194343, EPI_ISL_1194345, EPI_ISL_1194420, EPI_ISL_1194488, EPI_ISL_1220375, EPI_ISL_1220429, EPI_ISL_1220430, EPI_ISL_1220596, EPI_ISL_1220601, EPI_ISL_1220611, EPI_ISL_1220629, EPI_ISL_1220636, EPI_ISL_1220660, EPI_ISL_1220661, EPI_ISL_1220662, EPI_ISL_1220672, EPI_ISL_1220687, EPI_ISL_1220739, EPI_ISL_1220754, EPI_ISL_1220778, EPI_ISL_1220781, EPI_ISL_1220792, EPI_ISL_1220796, EPI_ISL_1220801, EPI_ISL_1220806, EPI_ISL_1220832, EPI_ISL_1220833, EPI_ISL_1220834, EPI_ISL_1220860, EPI_ISL_1291400, EPI_ISL_1291420, EPI_ISL_1291433, EPI_ISL_1291438, EPI_ISL_1291451, EPI_ISL_1291480, EPI_ISL_1291482, EPI_ISL_1291541, EPI_ISL_1298235, EPI_ISL_1335915 | | | |
| see above | Quest Diagnostics Incorporated | Centers for Disease Control and Prevention Division of Viral Diseases, Pathogen Discovery | A. Gerasimova; A. Perez; B. Anderson; Ben L. Rambo-Martin; Clinton R. Paden; Dakota Howard; Dhwani Batra; Duncan MacCannell; F. Lacbawan; I. A. Shlyakhter; K.E. Livingston; L.E. Bernstein; M. Hua; P. Tanpaiboon; Peter W. Cook; R. M. Kagan; R. Owen; R. V. Rolando; S. H. Rosenthal; Suxiang Tong; Y. Liu |
| EPI_ISL_1087439, EPI_ISL_1088079, EPI_ISL_1088080, EPI_ISL_1088083, EPI_ISL_1088087, EPI_ISL_1088094, EPI_ISL_1088107, EPI_ISL_1088111, EPI_ISL_1088115, EPI_ISL_1088117, EPI_ISL_1088123, EPI_ISL_1088124, EPI_ISL_1088140, EPI_ISL_1088142, EPI_ISL_1088152, EPI_ISL_1088170, EPI_ISL_1088188, EPI_ISL_1088192, EPI_ISL_1088196, EPI_ISL_1088210, EPI_ISL_1088211, EPI_ISL_1088216, EPI_ISL_1088231, EPI_ISL_1088239, EPI_ISL_1088271, EPI_ISL_1088299, EPI_ISL_1088333, EPI_ISL_1088354, EPI_ISL_1090291, EPI_ISL_1090327, EPI_ISL_1090351, EPI_ISL_1090370, EPI_ISL_1090397, EPI_ISL_1090426, EPI_ISL_1090496, EPI_ISL_1090520, EPI_ISL_1090522, EPI_ISL_1090559, EPI_ISL_1090562, EPI_ISL_1090565, EPI_ISL_1090568, EPI_ISL_1090595, EPI_ISL_1090618, EPI_ISL_1090653, EPI_ISL_1090672, EPI_ISL_1090674, EPI_ISL_1090679, EPI_ISL_1090684, EPI_ISL_1090694, EPI_ISL_1090702, EPI_ISL_1090713, EPI_ISL_1090714, EPI_ISL_1090715, EPI_ISL_1090716, EPI_ISL_1090717, EPI_ISL_1090718, EPI_ISL_1090719, EPI_ISL_1090721, EPI_ISL_1090723, EPI_ISL_1090724, EPI_ISL_1090725, EPI_ISL_1090726, EPI_ISL_1090727, EPI_ISL_1090728, EPI_ISL_1090730, EPI_ISL_1090731, EPI_ISL_1090733, EPI_ISL_1090758, EPI_ISL_1090782, EPI_ISL_1090842, EPI_ISL_1090847, EPI_ISL_1090849, EPI_ISL_1090865, EPI_ISL_1090880, EPI_ISL_1090881, EPI_ISL_1090950, EPI_ISL_1090951, EPI_ISL_1090952, EPI_ISL_1090953, EPI_ISL_1090955, EPI_ISL_1090956, EPI_ISL_1090957, EPI_ISL_1090959, EPI_ISL_1090960, EPI_ISL_1090961, EPI_ISL_1090962, EPI_ISL_1090963, EPI_ISL_1090964, EPI_ISL_1090965, EPI_ISL_1090981, EPI_ISL_1090982, EPI_ISL_1090983, EPI_ISL_1090984, EPI_ISL_1090985, EPI_ISL_1090986, EPI_ISL_1090987, EPI_ISL_1090988, EPI_ISL_1090990, EPI_ISL_1090991, EPI_ISL_1090992, EPI_ISL_1090993, EPI_ISL_1090994, EPI_ISL_1090995, EPI_ISL_1090997, EPI_ISL_1090998, EPI_ISL_1090999, EPI_ISL_1091000, EPI_ISL_1091001, EPI_ISL_1091002, EPI_ISL_1091003, EPI_ISL_1091004, EPI_ISL_1091005, EPI_ISL_1091011, EPI_ISL_1091012, EPI_ISL_1091037, EPI_ISL_1091040, EPI_ISL_1091073, EPI_ISL_1091078, EPI_ISL_1113902, EPI_ISL_1114075, EPI_ISL_1114137, EPI_ISL_1139223, EPI_ISL_1139297, EPI_ISL_1139322, EPI_ISL_1139381, EPI_ISL_1139383, EPI_ISL_1139384, EPI_ISL_1139385, EPI_ISL_1139387, EPI_ISL_1139388, EPI_ISL_1139389, EPI_ISL_1139390, EPI_ISL_1139391, EPI_ISL_1139392, EPI_ISL_1139393, EPI_ISL_1139394, EPI_ISL_1139395, EPI_ISL_1139396, EPI_ISL_1139397, EPI_ISL_1139398, EPI_ISL_1139399, EPI_ISL_1139400, EPI_ISL_1139401, EPI_ISL_1139404, EPI_ISL_1139405, EPI_ISL_1139407, EPI_ISL_1139408, EPI_ISL_1139409, EPI_ISL_1139410, EPI_ISL_1139413, EPI_ISL_1139415, EPI_ISL_1139416, EPI_ISL_1139417, EPI_ISL_1139419, EPI_ISL_1139420, EPI_ISL_1139421, EPI_ISL_1139425, EPI_ISL_1139428, EPI_ISL_1139429, EPI_ISL_1139480, EPI_ISL_1139491, EPI_ISL_1139503, EPI_ISL_1139505, EPI_ISL_1139506, EPI_ISL_1139811, EPI_ISL_1139812, EPI_ISL_1139817, EPI_ISL_1139819, EPI_ISL_1139820, EPI_ISL_1139821, EPI_ISL_1139822, EPI_ISL_1139823, EPI_ISL_1139824, EPI_ISL_1139825, EPI_ISL_1139826, EPI_ISL_1139827, EPI_ISL_1139828, EPI_ISL_1139831, EPI_ISL_1139832, EPI_ISL_1139833, EPI_ISL_1139834, EPI_ISL_1139835, EPI_ISL_1139836, EPI_ISL_1139837, EPI_ISL_1139838, EPI_ISL_1139839, EPI_ISL_1139840, EPI_ISL_1139841, EPI_ISL_1139842, EPI_ISL_1139843, EPI_ISL_1139844, EPI_ISL_1139845, EPI_ISL_1139846, EPI_ISL_1139847, EPI_ISL_1139848, EPI_ISL_1139849, EPI_ISL_1139850, EPI_ISL_1139851, EPI_ISL_1139852, EPI_ISL_1139853, EPI_ISL_1139855, EPI_ISL_1139856, EPI_ISL_1139857, EPI_ISL_1139858, EPI_ISL_1139859, EPI_ISL_1139860, EPI_ISL_1139861, EPI_ISL_1139862, EPI_ISL_1139863, EPI_ISL_1139864, EPI_ISL_1139865, EPI_ISL_1139866, EPI_ISL_1139867, EPI_ISL_1139868, EPI_ISL_1139869, EPI_ISL_1139870, EPI_ISL_1139871, EPI_ISL_1139872, EPI_ISL_1139873, EPI_ISL_1139874, EPI_ISL_1139875, EPI_ISL_1139876, EPI_ISL_1139877, EPI_ISL_1139878, EPI_ISL_1139879, EPI_ISL_1139880, EPI_ISL_1139881, EPI_ISL_1139882, EPI_ISL_1139883, EPI_ISL_1139884, EPI_ISL_1139885, EPI_ISL_1139886, EPI_ISL_1139887, EPI_ISL_1139888, EPI_ISL_1139889, EPI_ISL_1139890, EPI_ISL_1139891, EPI_ISL_1139892, EPI_ISL_1139893, EPI_ISL_1139894, EPI_ISL_1139895, EPI_ISL_1139896, EPI_ISL_1139898, EPI_ISL_1139899, EPI_ISL_1139900, EPI_ISL_1139901, EPI_ISL_1139902, EPI_ISL_1139903, EPI_ISL_1139904, EPI_ISL_1139905, EPI_ISL_1139906, EPI_ISL_1139907, EPI_ISL_1139908, EPI_ISL_1139909, EPI_ISL_1139910, EPI_ISL_1139911, EPI_ISL_1139912, EPI_ISL_1139913, EPI_ISL_1139914, EPI_ISL_1139915, EPI_ISL_1139916, EPI_ISL_1139917, EPI_ISL_1139918, EPI_ISL_1139919, EPI_ISL_1139920, EPI_ISL_1139921, EPI_ISL_1139922, EPI_ISL_1139923, EPI_ISL_1139924, EPI_ISL_1139925, EPI_ISL_1139926, EPI_ISL_1139927, EPI_ISL_1139928, EPI_ISL_1139929, EPI_ISL_1139930, EPI_ISL_1139931, EPI_ISL_1139932, EPI_ISL_1139933, EPI_ISL_1139934, EPI_ISL_1139935, EPI_ISL_1139936, EPI_ISL_1139937, EPI_ISL_1139938, EPI_ISL_1139939, EPI_ISL_1139940, EPI_ISL_1139941, EPI_ISL_1139942, EPI_ISL_1139943, EPI_ISL_1139944, EPI_ISL_1139945, EPI_ISL_1139946, EPI_ISL_1139947, EPI_ISL_1139948, EPI_ISL_1139949, EPI_ISL_1139950, EPI_ISL_1139951, EPI_ISL_1139952, EPI_ISL_1139953, EPI_ISL_1139954, EPI_ISL_1139955, EPI_ISL_1139956, EPI_ISL_1139957, EPI_ISL_1139958, EPI_ISL_1139959, EPI_ISL_1139960, EPI_ISL_1139961, EPI_ISL_1139962, EPI_ISL_1139963, EPI_ISL_1139964, EPI_ISL_1139965, EPI_ISL_1139966, EPI_ISL_1139967, EPI_ISL_1139968, EPI_ISL_1139969, EPI_ISL_1139970, EPI_ISL_1139971, EPI_ISL_1139972, EPI_ISL_1139973, EPI_ISL_1139974, EPI_ISL_1139975, EPI_ISL_1139976, EPI_ISL_1139977, EPI_ISL_1139978, EPI_ISL_1139979, EPI_ISL_1139980, EPI_ISL_1139981, EPI_ISL_1139982, EPI_ISL_1139983, EPI_ISL_1139984, EPI_ISL_1139985, EPI_ISL_1139986, EPI_ISL_1139987, EPI_ISL_1139988, EPI_ISL_1139989, EPI_ISL_1139990, EPI_ISL_1139991, EPI_ISL_1139992, EPI_ISL_1139993, EPI_ISL_1139994, EPI_ISL_1139995, EPI_ISL_1139996, EPI_ISL_1139997, EPI_ISL_1139998, EPI_ISL_1139999, EPI_ISL_1140000, EPI_ISL_1140001, EPI_ISL_1140002, EPI_ISL_1140003, EPI_ISL_1140004, EPI_ISL_1140005, EPI_ISL_1140006, EPI_ISL_1140007, EPI_ISL_1140008, EPI_ISL_1140009, EPI_ISL_1140010, EPI_ISL_1140011, EPI_ISL_1140012, EPI_ISL_1140013, EPI_ISL_1140014, EPI_ISL_1140015, EPI_ISL_1140016, EPI_ISL_1140017, EPI_ISL_1140018, EPI_ISL_1140019, EPI_ISL_1140020, EPI_ISL_1140021, EPI_ISL_1140022, EPI_ISL_1140023, EPI_ISL_1140024, EPI_ISL_1140025, EPI_ISL_1140026, EPI_ISL_1140027, EPI_ISL_1140028, EPI_ISL_1140029, EPI_ISL_1140030, EPI_ISL_1140031, EPI_ISL_1140032, EPI_ISL_1140033, EPI_ISL_1140034, EPI_ISL_1140035, EPI_ISL_1140036, EPI_ISL_1140037, EPI_ISL_1140038, EPI_ISL_1140039, EPI_ISL_1140040, EPI_ISL_1140041, EPI_ISL_1140042, EPI_ISL_1140043, EPI_ISL_1140044, EPI_ISL_1140045, EPI_ISL_1140046, EPI_ISL_1158931, EPI_ISL_1158988, EPI_ISL_1159006, EPI_ISL_1159009, EPI_ISL_1159067, EPI_ISL_1159409, EPI_ISL_1159437, EPI_ISL_1159549, EPI_ISL_1159579, EPI_ISL_1159591, EPI_ISL_1159600, EPI_ISL_1159603, EPI_ISL_1159620, EPI_ISL_1159623, EPI_ISL_1159643, EPI_ISL_1159646, EPI_ISL_1159653, EPI_ISL_1159659, EPI_ISL_1159674, EPI_ISL_1293334, EPI_ISL_1293335, EPI_ISL_1293340 | | | |
| see above | Quest Diagnostics Incorporated | Respiratory Viruses Branch, Division of Viral Diseases, Centers for Disease Control and Prevention | A. Gerasimova; A. Perez; B. Anderson; Ben L. Rambo-Martin; Clinton R. Paden; Dakota Howard; Dhwani Batra; Duncan MacCannell; F. Lacbawan; I. A. Shlyakhter; K.E. Livingston; L.E. Bernstein; M. Hua; P. Tanpaiboon; Peter W. Cook; R. M. Kagan; R. Owen; R. V. Rolando; S. H. Rosenthal; Suxiang Tong; Y. Liu |
| EPI_ISL_1295798, EPI_ISL_1295799, EPI_ISL_1295800, EPI_ISL_1295802, EPI_ISL_1295807, EPI_ISL_1295811, EPI_ISL_1295813, EPI_ISL_1794773 | | | |
| see above | Rady Children's Hospital - San Diego | Andersen lab at Scripps Research | Christina Clarke; David Dimmock; Denise Malicki; Kathryn Bouic; Linda Luo; SEARCH Alliance San Diego with Nanda Radamchar; Teresa Mueller |
| EPI_ISL_1233880, EPI_ISL_1233889 | Respiratory Viruses Branch, Centers for Disease Control and Prevention | Respiratory Viruses Branch, Centers for Disease Control and Prevention | Allen, E.; Antico, J.; Ascencio, A.; B.L.; Barrett; Batra, D.; Becker, D.; Bolze, A.; C.R.; Cassens, T.; Cho, R.; Cirulli, E.; Cook; D.T.; Febbo, P.; Galloway, S.; Gietzen, K.; Howard; Isaksson, M.; K.S.; Laurent, M.; Lee, W.; Levan, G.; Liu, J.; Lu, J.; MacCannell, D.; N.L.; Nguyen, J.; P.W.; Paden; Rambo-Martin; Ramirez, J.; Rivera-Garcia, C.; Sandoval, E.; Sickler, B.; Tolentino, M.; Tong, S.; Tran, C.; Wang, S.; Washington; White, S.; Wickline, S.; de Feo, E. |
| EPI_ISL_1185960, EPI_ISL_1185961, EPI_ISL_1185962, EPI_ISL_1185963, EPI_ISL_1185964, EPI_ISL_1185965, EPI_ISL_1185966, EPI_ISL_1185967, EPI_ISL_1185968, EPI_ISL_1185969, EPI_ISL_1185970, EPI_ISL_1185971, EPI_ISL_1185972, EPI_ISL_1185973, EPI_ISL_1185974, EPI_ISL_1185975, EPI_ISL_1185976, EPI_ISL_1185977, EPI_ISL_1185978, EPI_ISL_1185979, EPI_ISL_1185980, EPI_ISL_1185981, EPI_ISL_1185982, EPI_ISL_1185984, EPI_ISL_1185985, EPI_ISL_1185986, EPI_ISL_1185987, EPI_ISL_1185988, EPI_ISL_1185989, EPI_ISL_1185990, EPI_ISL_1186016, EPI_ISL_1468841, EPI_ISL_1468847, EPI_ISL_1468854, EPI_ISL_1468876, EPI_ISL_1468906, EPI_ISL_1468907, EPI_ISL_1468908, EPI_ISL_1468909, EPI_ISL_1468950, EPI_ISL_1468953, EPI_ISL_1468979, EPI_ISL_1468980, EPI_ISL_1468983, EPI_ISL_1468984, EPI_ISL_1468986, EPI_ISL_1468987, EPI_ISL_1468990, EPI_ISL_1468995, EPI_ISL_1468996, EPI_ISL_1469001, EPI_ISL_1469002, EPI_ISL_1469005, EPI_ISL_1469013, EPI_ISL_1469016, EPI_ISL_1469020, EPI_ISL_1469021, EPI_ISL_1469022, EPI_ISL_1794618, EPI_ISL_1794628, EPI_ISL_1794635, EPI_ISL_1794639, EPI_ISL_1794643, EPI_ISL_1794648, EPI_ISL_1794653, EPI_ISL_1969255, EPI_ISL_1969260, EPI_ISL_1969262, EPI_ISL_1969265, EPI_ISL_1969270 | | | |
| see above | San Diego County Public Health Laboratory | Andersen lab at Scripps Research | Brett Austin; Jovan Shephard; SEARCH Alliance San Diego with Tracy Basler |
| EPI_ISL_1701079 | San Joaquin County Public Health Lab | Chan-Zuckerberg Biohub | CZB Cliahub Consortium |
| EPI_ISL_1234578, EPI_ISL_1234584, EPI_ISL_1234687, EPI_ISL_1234800, EPI_ISL_1234810, EPI_ISL_1234812, EPI_ISL_1234837, EPI_ISL_1234880, EPI_ISL_1234913, EPI_ISL_1234976, EPI_ISL_1234985, EPI_ISL_1234988, EPI_ISL_1234999, EPI_ISL_1235031, EPI_ISL_1235038, EPI_ISL_1235043, EPI_ISL_1235362, EPI_ISL_1235370, EPI_ISL_1235379, EPI_ISL_1235382, EPI_ISL_1235394, EPI_ISL_1235395, EPI_ISL_1235396, EPI_ISL_1235409, EPI_ISL_1235411, EPI_ISL_1235412, EPI_ISL_1235415, EPI_ISL_1235420, EPI_ISL_1235428, EPI_ISL_1235429, EPI_ISL_1235431, EPI_ISL_1235436, EPI_ISL_1235439, EPI_ISL_1235444, EPI_ISL_1235445, EPI_ISL_1235449, EPI_ISL_1235452, EPI_ISL_1235453, EPI_ISL_1235454, EPI_ISL_1235461, EPI_ISL_1235466, EPI_ISL_1235467, EPI_ISL_1235470, EPI_ISL_1235474, EPI_ISL_1235479, EPI_ISL_1235483, EPI_ISL_1235484, EPI_ISL_1235487, EPI_ISL_1235491, EPI_ISL_1235508, EPI_ISL_1235518, EPI_ISL_1235519, EPI_ISL_1235530, EPI_ISL_1235531, EPI_ISL_1235533, EPI_ISL_1235534, EPI_ISL_1235535, EPI_ISL_1235536, EPI_ISL_1235541, EPI_ISL_1235542, EPI_ISL_1235556, EPI_ISL_1235563, EPI_ISL_1235581, EPI_ISL_1235593, EPI_ISL_1235601, EPI_ISL_1235612, EPI_ISL_1477441, EPI_ISL_1477445, EPI_ISL_1477455, EPI_ISL_1477461, EPI_ISL_1477484, EPI_ISL_1477486, EPI_ISL_1477494, EPI_ISL_1477496, EPI_ISL_1477501, EPI_ISL_1477510, EPI_ISL_1477518, EPI_ISL_1477529, EPI_ISL_1477534, EPI_ISL_1477535, EPI_ISL_1477540, EPI_ISL_1477541, EPI_ISL_1477552, EPI_ISL_1477558, EPI_ISL_1477566, EPI_ISL_1477575, EPI_ISL_1477578, EPI_ISL_1477581, EPI_ISL_1477590, EPI_ISL_1477598, EPI_ISL_1477603, EPI_ISL_1477622, EPI_ISL_1477637, EPI_ISL_1477655, EPI_ISL_1477664, EPI_ISL_1477670, EPI_ISL_1477677, EPI_ISL_1477681, EPI_ISL_1477682, EPI_ISL_1477690, EPI_ISL_1477695, EPI_ISL_1477699, EPI_ISL_1477706, EPI_ISL_1477722, EPI_ISL_1477726, EPI_ISL_1477727, EPI_ISL_1477729, EPI_ISL_1477730, EPI_ISL_1477731, EPI_ISL_1477734, EPI_ISL_1477745, EPI_ISL_1477749, EPI_ISL_1477750, EPI_ISL_1477752, EPI_ISL_1477762, EPI_ISL_1477767, EPI_ISL_1477768, EPI_ISL_1477776, EPI_ISL_1477780, EPI_ISL_1477793, EPI_ISL_1477797, EPI_ISL_1477799, EPI_ISL_1477800, EPI_ISL_1477814, EPI_ISL_1477817, EPI_ISL_1477818, EPI_ISL_1477822, EPI_ISL_1477836, EPI_ISL_1477839, EPI_ISL_1477842, EPI_ISL_1477845, EPI_ISL_1477846, EPI_ISL_1477850, EPI_ISL_1477854, EPI_ISL_1477856, EPI_ISL_1477859, EPI_ISL_1477860, EPI_ISL_1477863, EPI_ISL_1477871, EPI_ISL_1477874, EPI_ISL_1477878, EPI_ISL_1477879, EPI_ISL_1477881, EPI_ISL_1477882, EPI_ISL_1477885, EPI_ISL_1477888, EPI_ISL_1477892, EPI_ISL_1477893, EPI_ISL_1477895, EPI_ISL_1477902, EPI_ISL_1477907, EPI_ISL_1477909, EPI_ISL_1477912, EPI_ISL_1477923, EPI_ISL_1477925, EPI_ISL_1477926, EPI_ISL_1477935, EPI_ISL_1477937, EPI_ISL_1477943, EPI_ISL_1477974, EPI_ISL_1477982, EPI_ISL_1478028, EPI_ISL_1478046, EPI_ISL_1478048, EPI_ISL_1478050, EPI_ISL_1478059, EPI_ISL_1478089, EPI_ISL_1478095, EPI_ISL_1478097, EPI_ISL_1478123, EPI_ISL_1478124, EPI_ISL_1478149, EPI_ISL_1478152, EPI_ISL_1478161, EPI_ISL_1478171, EPI_ISL_1478192, EPI_ISL_1478200, EPI_ISL_1529101, EPI_ISL_1529102, EPI_ISL_1529110, EPI_ISL_1529113, EPI_ISL_1529119, EPI_ISL_1529138, EPI_ISL_1529151, EPI_ISL_1529152, EPI_ISL_1529173, EPI_ISL_1529206, EPI_ISL_1529207, EPI_ISL_1529218, EPI_ISL_1529226, EPI_ISL_1529241, EPI_ISL_1529252, EPI_ISL_1529258, EPI_ISL_1529295, EPI_ISL_1529344, EPI_ISL_1529347, EPI_ISL_1529351, EPI_ISL_1821317, EPI_ISL_1821328, EPI_ISL_1821344, EPI_ISL_1821346, EPI_ISL_2102805, EPI_ISL_2102812, EPI_ISL_2102878, EPI_ISL_2102892, EPI_ISL_2102895, EPI_ISL_2102904, EPI_ISL_2658622, EPI_ISL_2658630, EPI_ISL_2658645, EPI_ISL_2658657, EPI_ISL_2658736, EPI_ISL_2659223, EPI_ISL_2659228, EPI_ISL_2659321, EPI_ISL_2659342, EPI_ISL_2659351, EPI_ISL_2659354, EPI_ISL_2659368, EPI_ISL_2659373, EPI_ISL_2659436, EPI_ISL_2659438, EPI_ISL_2659489, EPI_ISL_2659558, EPI_ISL_2659576, EPI_ISL_2758475 | | | |
| see above | Santa Clara County Public Health Laboratory | Chan-Zuckerberg Biohub | CZB Cliahub Consortium |
| EPI_ISL_1001005, EPI_ISL_1001006, EPI_ISL_1001007, EPI_ISL_1001008, EPI_ISL_1001009, EPI_ISL_1001032, EPI_ISL_1001033, EPI_ISL_1001034, EPI_ISL_1001035, EPI_ISL_1001036, EPI_ISL_1001037, EPI_ISL_1001038, EPI_ISL_1001039, EPI_ISL_1001040, EPI_ISL_1009659, EPI_ISL_1009660, EPI_ISL_1009661, EPI_ISL_1009662, EPI_ISL_1009663, EPI_ISL_1009664, EPI_ISL_1060544, EPI_ISL_1060550, EPI_ISL_1137646, EPI_ISL_1291743, EPI_ISL_1291745, EPI_ISL_1335805, EPI_ISL_1335806, EPI_ISL_1335807, EPI_ISL_1335812, EPI_ISL_1335813, EPI_ISL_1335814, EPI_ISL_1335815, EPI_ISL_1335816, EPI_ISL_1335817, EPI_ISL_1335818, EPI_ISL_1335819, EPI_ISL_1335820, EPI_ISL_1335821, EPI_ISL_1335822, EPI_ISL_1335823, EPI_ISL_1335824, EPI_ISL_1335825, EPI_ISL_1335826, EPI_ISL_1335853, EPI_ISL_1335854, EPI_ISL_1335855, EPI_ISL_1335861, EPI_ISL_1335864, EPI_ISL_1335866, EPI_ISL_1406162, EPI_ISL_1406163 | | | |
| see above | Santa Clara County Public Health Laboratory | Santa Clara County Public Health Laboratory | Santa Clara County Public Health Department |
| EPI_ISL_1366275, EPI_ISL_1366278, EPI_ISL_1366297, EPI_ISL_1366318, EPI_ISL_1366320, EPI_ISL_1366322, EPI_ISL_1366403, EPI_ISL_1366404, EPI_ISL_1366410, EPI_ISL_1366411, EPI_ISL_1366412, EPI_ISL_1366413, EPI_ISL_1366414, EPI_ISL_1366415, EPI_ISL_1366416, EPI_ISL_1366417, EPI_ISL_1366418, EPI_ISL_1366419, EPI_ISL_1366420, EPI_ISL_1366421, EPI_ISL_1366422, EPI_ISL_1366423, EPI_ISL_1468947, EPI_ISL_1469047, EPI_ISL_1469048, EPI_ISL_1469049, EPI_ISL_1469050 | | | |
| see above | Scripps Medical Laboratory | Andersen lab at Scripps Research | Ellen Stefanski; Ian Mchardy; SEARCH Alliance San Diego with Michael Quigley |
| EPI_ISL_1295646, EPI_ISL_1295647, EPI_ISL_1295648, EPI_ISL_1295649, EPI_ISL_1295650, EPI_ISL_1295651, EPI_ISL_1295652, EPI_ISL_1295653, EPI_ISL_1295654, EPI_ISL_1295655, EPI_ISL_1295656, EPI_ISL_1295657, EPI_ISL_1295658, EPI_ISL_1295659, EPI_ISL_1295660, EPI_ISL_1295661, EPI_ISL_1295662, EPI_ISL_1295663, EPI_ISL_1295664, EPI_ISL_1295665, EPI_ISL_1295666, EPI_ISL_1295667, EPI_ISL_1295668, EPI_ISL_1295669, EPI_ISL_1295670, EPI_ISL_1295672, EPI_ISL_1295673, EPI_ISL_1295674, EPI_ISL_1295675, EPI_ISL_1295677, EPI_ISL_1295678, EPI_ISL_1295679, EPI_ISL_1295680, EPI_ISL_1295681, EPI_ISL_1295682, EPI_ISL_1295683, EPI_ISL_1295684, EPI_ISL_1295685, EPI_ISL_1295686, EPI_ISL_1295687, EPI_ISL_1295688, EPI_ISL_1295689, EPI_ISL_1295690, EPI_ISL_1295692, EPI_ISL_1295693, EPI_ISL_1295694, EPI_ISL_1295695, EPI_ISL_1295696, EPI_ISL_1295697, EPI_ISL_1295698, EPI_ISL_1295699, EPI_ISL_1295700, EPI_ISL_1295701, EPI_ISL_1295702, EPI_ISL_1295703, EPI_ISL_1295704, EPI_ISL_1295705, EPI_ISL_1295706, EPI_ISL_1295707, EPI_ISL_1295708, EPI_ISL_1295709, EPI_ISL_1295710, EPI_ISL_1295711, EPI_ISL_1295712, EPI_ISL_1295713, EPI_ISL_1295714, EPI_ISL_1295715, EPI_ISL_1295716, EPI_ISL_1295717, EPI_ISL_1295718, EPI_ISL_1295719, EPI_ISL_1295720, EPI_ISL_1295721, EPI_ISL_1295722, EPI_ISL_1295723, EPI_ISL_1295724, EPI_ISL_1295725, EPI_ISL_1295726, EPI_ISL_1295727, EPI_ISL_1295728, EPI_ISL_1295729, EPI_ISL_1295731, EPI_ISL_1295732, EPI_ISL_1295733, EPI_ISL_1295734, EPI_ISL_1295735, EPI_ISL_1295737, EPI_ISL_1295778, EPI_ISL_1295782, EPI_ISL_1295787, EPI_ISL_1295789, EPI_ISL_1295790, EPI_ISL_1295818, EPI_ISL_1295820, EPI_ISL_1295821, EPI_ISL_1295822, EPI_ISL_1295824, EPI_ISL_1366288, EPI_ISL_1366289, EPI_ISL_1366290, EPI_ISL_1366291, EPI_ISL_1366292, EPI_ISL_1366298, EPI_ISL_1366299, EPI_ISL_1366300, EPI_ISL_1366306, EPI_ISL_1366307, EPI_ISL_1366323, EPI_ISL_1366424, EPI_ISL_1366425, EPI_ISL_1366426, EPI_ISL_1366427, EPI_ISL_1366428, EPI_ISL_1366429, EPI_ISL_1366430, EPI_ISL_1366431, EPI_ISL_1366432, EPI_ISL_1366433, EPI_ISL_1366434, EPI_ISL_1366435, EPI_ISL_1366436, EPI_ISL_1366437, EPI_ISL_1366438, EPI_ISL_1366439, EPI_ISL_1366440, EPI_ISL_1366441, EPI_ISL_1366442, EPI_ISL_1366443, EPI_ISL_1366444, EPI_ISL_1366445, EPI_ISL_1366446, EPI_ISL_1366447, EPI_ISL_1366448, EPI_ISL_1366449, EPI_ISL_1366450, EPI_ISL_1366451, EPI_ISL_1366452, EPI_ISL_1366453, EPI_ISL_1366454, EPI_ISL_1366455, EPI_ISL_1366456, EPI_ISL_1366457, EPI_ISL_1366475, EPI_ISL_1366477 | | | |
| see above | Sharp HealthCare Laboratory | Andersen lab at Scripps Research | Art Mendoza; Cathy Woerle; Jacquelyn Berumen; Liam McGinnis; Omid Bakhtar; SEARCH Alliance San Diego with Aaron Harding |
| EPI_ISL_1483796, EPI_ISL_1483798 | Stanford Clinical Virology Laboratory | Santa Clara County Public Health Laboratory | Santa Clara County Public Health Department |
| EPI_ISL_1364517, EPI_ISL_1364521, EPI_ISL_1364522, EPI_ISL_1364523, EPI_ISL_1364524, EPI_ISL_1364525, EPI_ISL_1364526, EPI_ISL_1379854, EPI_ISL_1379887, EPI_ISL_1379888, EPI_ISL_1424124, EPI_ISL_1424128, EPI_ISL_1424129, EPI_ISL_2009055, EPI_ISL_2009057, EPI_ISL_2027144, EPI_ISL_2027150, EPI_ISL_2027171, EPI_ISL_2027177, EPI_ISL_2027178, EPI_ISL_2027183, EPI_ISL_2027185, EPI_ISL_2027187, EPI_ISL_2027191, EPI_ISL_2027195, EPI_ISL_2027198 | | | |
| see above | Stanford Health Care | Stanford University School of Medicine, Clinical Virology Laboratory | Becky Jiang; ChunHong Huang; Daniel Solis; Fumiko Yamamoto; James Zehnder; Malaya K. Sahoo; Mamdouh Sibai; Michelle Verghese; and Benjamin A. Pinsky |
| EPI_ISL_2341046, EPI_ISL_2341052, EPI_ISL_2341070, EPI_ISL_2341073, EPI_ISL_2341086, EPI_ISL_2341088, EPI_ISL_2341089, EPI_ISL_2341090, EPI_ISL_2341091, EPI_ISL_2341092, EPI_ISL_2341093, EPI_ISL_2341094, EPI_ISL_2341095, EPI_ISL_2341096, EPI_ISL_2341097, EPI_ISL_2341098, EPI_ISL_2341133, EPI_ISL_2341134, EPI_ISL_2341135, EPI_ISL_2341136, EPI_ISL_2341137, EPI_ISL_2341138, EPI_ISL_2341139, EPI_ISL_2341140, EPI_ISL_2341141, EPI_ISL_2341142, EPI_ISL_2341143, EPI_ISL_2341247, EPI_ISL_2341248, EPI_ISL_2341249, EPI_ISL_2341250, EPI_ISL_2341251, EPI_ISL_2341252, EPI_ISL_3049198, EPI_ISL_3049202 | | | |
| | Tulare County Public Health Laboratory | Tulare County Public Health Laboratory | Denise L Lopez; Joshua Kootstra |
| EPI_ISL_1040054, EPI_ISL_1040055, EPI_ISL_1040056, EPI_ISL_1040057, EPI_ISL_1040058, EPI_ISL_1040059, EPI_ISL_1040060, EPI_ISL_1040061, EPI_ISL_1040062, EPI_ISL_1040063, EPI_ISL_1040064, EPI_ISL_1040065, EPI_ISL_1040066, EPI_ISL_1040067, EPI_ISL_1040068, EPI_ISL_1040069, EPI_ISL_1040070, EPI_ISL_1040071, EPI_ISL_1097742, EPI_ISL_1098609, EPI_ISL_1098610, EPI_ISL_1098611, EPI_ISL_1098612, EPI_ISL_1098613, EPI_ISL_1098614, EPI_ISL_1098615, EPI_ISL_1098616, EPI_ISL_1098618, EPI_ISL_1098619, EPI_ISL_1098620, EPI_ISL_1098621, EPI_ISL_1098622, EPI_ISL_1098623, EPI_ISL_1098624, EPI_ISL_1098625, EPI_ISL_1098626, EPI_ISL_1098627, EPI_ISL_1098631, EPI_ISL_1191110, EPI_ISL_1191111, EPI_ISL_1191112, EPI_ISL_1191113, EPI_ISL_1191114, EPI_ISL_1191117, EPI_ISL_1191118, EPI_ISL_1191119, EPI_ISL_1191120, EPI_ISL_1191121, EPI_ISL_1191122, EPI_ISL_1191124, EPI_ISL_1191130, EPI_ISL_1191131 | | | |
| see above | UCLA Clinical Micro Lab | Los Angeles County Public Health Laboratories | P. Hemarajata et al. |
| EPI_ISL_1402586, EPI_ISL_1402587, EPI_ISL_1402588, EPI_ISL_1405671, EPI_ISL_1405672, EPI_ISL_1405673, EPI_ISL_1405674, EPI_ISL_1405675, EPI_ISL_1405676, EPI_ISL_1405677, EPI_ISL_1405678, EPI_ISL_1405679 | | | |
| see above | UCSC Genomics Institute | UCSC Genomics Institute | A. Marm Kilpatrick; Angie Hinrichs; Beth Shapiro; Bryan Thornlow; David Haussler; Eric Beraut; Hugh Olsen; Ikenna Anigbogu; Isabel Bjork; Jakob McBroome; Jeremy Sanford; Joshua Kapp; Mark Akeson; Maximilian Haeussler; Michael Stone; Miten Jain; Molly Cassatt-Johnstone; Namrita Dhillon; Russell Corbett-Detig; Terren Chang; Yatish Turakhia |
| EPI_ISL_1040051, EPI_ISL_1197042, EPI_ISL_1201559, | USC Clinical Lab | Los Angeles County Public Health Laboratories | P. Hemarajata et al. |

| | | | |
|---|---|---|---|
| EPI_ISL_1201560, EPI_ISL_1303438 | | | |
| EPI_ISL_2295376, EPI_ISL_2295377, EPI_ISL_2295385, EPI_ISL_2295386 | WLA VAMC | Genomics and Discovery, Respiratory Viruses Branch, Division of Viral Diseases, Centers for Disease Control and Prevention | Adam Retchless; Anna Kelleher; Anna Uehara; Brian Lynch; Clinton R. Paden; Dhwani Batra; Haibin Wang; Han Jia Justin Ng; Jasmine Padilla; Jing Zhang; Justin Lee; Krista Queen; Mark Burroughs; Mili Sheth; Morgan Davis; Peter Cook; Rachel Marine; Sarah Nobles; Suxiang Tong; Tara Coalter; Yan Li; Ying Tao |

We gratefully acknowledge the following Authors from the Originating laboratories responsible for obtaining the specimens, as well as the Submitting laboratories where the genome data were generated and shared via GISAID, on which this research is based.

All Submitters of data may be contacted directly via www.gisaid.org

Authors are sorted alphabetically.

| Accession ID | Originating Laboratory | Submitting Laboratory | Authors |
|---|---|---|---|
| EPI_ISL_1318634, EPI_ISL_1318636, EPI_ISL_1318639, EPI_ISL_1318640, EPI_ISL_1318641, EPI_ISL_1318642, EPI_ISL_1318644, EPI_ISL_1318648, EPI_ISL_1318650, EPI_ISL_1318652, EPI_ISL_1318655, EPI_ISL_1318656, EPI_ISL_1318657, EPI_ISL_1318661, EPI_ISL_1318662, EPI_ISL_1318666, EPI_ISL_1318667, EPI_ISL_1446773, EPI_ISL_1446774, EPI_ISL_1446775, EPI_ISL_1446776, EPI_ISL_1446777, EPI_ISL_1446778, EPI_ISL_1446779, EPI_ISL_1446780, EPI_ISL_1446782, EPI_ISL_1446783, EPI_ISL_1446785, EPI_ISL_1446786, EPI_ISL_1446788, EPI_ISL_1446789, EPI_ISL_1446792, EPI_ISL_1446793, EPI_ISL_1446795, EPI_ISL_1446796, EPI_ISL_1446797, EPI_ISL_1446798, EPI_ISL_1446799, EPI_ISL_1446800, EPI_ISL_1446801, EPI_ISL_1446802, EPI_ISL_1446840, EPI_ISL_1446841, EPI_ISL_1446842, EPI_ISL_1446843, EPI_ISL_1446844, EPI_ISL_1446845, EPI_ISL_1447018, EPI_ISL_1678147, EPI_ISL_1678152, EPI_ISL_1678153, EPI_ISL_1678155, EPI_ISL_1678314 | | | |
| see above | "CDPH, Viral and Rickettsial Disease Laboratory" | Centers for Disease Control and Prevention Division of Viral Diseases, Pathogen Discovery | Alison Laufer Halpin; Anna Montmayeur; Anna Uehara; Ben L. Rambo-Martin; Clinton R. Paden; Dakota Howard; Darlene Wagner; Dave Wentworth; Dhwani Batra; Haibin Wang; Jasmine Padilla; Jing Zhang; Justin Lee; Katie Dillon; Krista Queen; Kristen Knipe; Kristine Lacek; Lori Rowe; Mark Burroughs; Matthew Schmerer; Mili Sheth; Peter Cook; Peter W. Cook; Rachel Marine; Sam Shepard; Sarah Nobles; Shoshona Le; Suxiang Tong; Vivien Dugan; Yan Li; Ying Tao; Yvette Unoarumhi |
| EPI_ISL_1491516, EPI_ISL_1491517, EPI_ISL_1491522 | "CDPH, Viral and Rickettsial Disease Laboratory" | Genomics and Discovery, Respiratory Viruses Branch, Division of Viral Diseases, Centers for Disease Control and Prevention | Anna Kelleher; Anna Uehara; Brian Lynch; Clinton R. Paden; Haibin Wang; Han Jia Justin Ng; Jing Zhang; Krista Queen; Peter Cook; Suxiang Tong; Yan Li; Ying Tao |
| EPI_ISL_1445279, EPI_ISL_1445280, EPI_ISL_1445289, EPI_ISL_1445292, EPI_ISL_1445293, EPI_ISL_1445294, EPI_ISL_1445384, EPI_ISL_1445425, EPI_ISL_1445426, EPI_ISL_1445427, EPI_ISL_1445429, EPI_ISL_1445430, EPI_ISL_1445431, EPI_ISL_1445434, EPI_ISL_1445454, EPI_ISL_1445455, EPI_ISL_1445461, EPI_ISL_1445462, EPI_ISL_1445468, EPI_ISL_1445472, EPI_ISL_1445491, EPI_ISL_1445506, EPI_ISL_1445508, EPI_ISL_1445509, EPI_ISL_1445511, EPI_ISL_1445537, EPI_ISL_1445538, EPI_ISL_1445539, EPI_ISL_1445540, EPI_ISL_1445541, EPI_ISL_1445542, EPI_ISL_1445544, EPI_ISL_1445545, EPI_ISL_1445546, EPI_ISL_1445547, EPI_ISL_1445548, EPI_ISL_1445550, EPI_ISL_1445551, EPI_ISL_1445552, EPI_ISL_1445554, EPI_ISL_1445555, EPI_ISL_1445556, EPI_ISL_1445557, EPI_ISL_1445558, EPI_ISL_1445559, EPI_ISL_1445560, EPI_ISL_1445561, EPI_ISL_1445562, EPI_ISL_1445565, EPI_ISL_1445566, EPI_ISL_1445568, EPI_ISL_1445569, EPI_ISL_1445573, EPI_ISL_1445584, EPI_ISL_1445593, EPI_ISL_1445595, EPI_ISL_1445598, EPI_ISL_1445605, EPI_ISL_1445608, EPI_ISL_1445609, EPI_ISL_1445610, EPI_ISL_1445613, EPI_ISL_1445615, EPI_ISL_1445616, EPI_ISL_1445620, EPI_ISL_1445622, EPI_ISL_1445627, EPI_ISL_1445651, EPI_ISL_1445652, EPI_ISL_1445653, EPI_ISL_1445654, EPI_ISL_1445655, EPI_ISL_1445656, EPI_ISL_1445657, EPI_ISL_1445658, EPI_ISL_1445659, EPI_ISL_1445660, EPI_ISL_1445671, EPI_ISL_1445673, EPI_ISL_1445675, EPI_ISL_1445685, EPI_ISL_1445690, EPI_ISL_1445691, EPI_ISL_1445694, EPI_ISL_1445695, EPI_ISL_1445703, EPI_ISL_1445709, EPI_ISL_1445720, EPI_ISL_1445725, EPI_ISL_1445731, EPI_ISL_1445735, EPI_ISL_1445739, EPI_ISL_1445754, EPI_ISL_1445764, EPI_ISL_1445769, EPI_ISL_1445774, EPI_ISL_1445775, EPI_ISL_1445776, EPI_ISL_1445789, EPI_ISL_1445790, EPI_ISL_1445791, EPI_ISL_1445796, EPI_ISL_1445797, EPI_ISL_1445798, EPI_ISL_1445799, EPI_ISL_1445800, EPI_ISL_1445801, EPI_ISL_1445802, EPI_ISL_1445803, EPI_ISL_1445805, EPI_ISL_1445806, EPI_ISL_1445809, EPI_ISL_1445815, EPI_ISL_1445816, EPI_ISL_1445817, EPI_ISL_1445818, EPI_ISL_1445819, EPI_ISL_1445820, EPI_ISL_1445821, EPI_ISL_1445835, EPI_ISL_1445843, EPI_ISL_1445845, EPI_ISL_1445858, EPI_ISL_1445871, EPI_ISL_1445904, EPI_ISL_1445913, EPI_ISL_1445931, EPI_ISL_1445932, EPI_ISL_1445940, EPI_ISL_1445941, EPI_ISL_1445942, EPI_ISL_1445950, EPI_ISL_1445959, EPI_ISL_1446032, EPI_ISL_1446035, EPI_ISL_1446066, EPI_ISL_1446083, EPI_ISL_1446084, EPI_ISL_1446090, EPI_ISL_1446091, EPI_ISL_1446092, EPI_ISL_1446096, EPI_ISL_1446097, EPI_ISL_1446110, EPI_ISL_1446116, EPI_ISL_1446118, EPI_ISL_1446122, EPI_ISL_1446123, EPI_ISL_1446132, EPI_ISL_1446147, EPI_ISL_1446162, EPI_ISL_1446179, EPI_ISL_1446181, EPI_ISL_1446192, EPI_ISL_1479498, EPI_ISL_1479512, EPI_ISL_1479518, EPI_ISL_1479587, EPI_ISL_1479590, EPI_ISL_1479591, EPI_ISL_1479608, EPI_ISL_1479613, EPI_ISL_1479825, EPI_ISL_1479828, EPI_ISL_1479830, EPI_ISL_1479834, EPI_ISL_1479842, EPI_ISL_1493502, EPI_ISL_1493503, EPI_ISL_1493535, EPI_ISL_1493550, EPI_ISL_1493570, EPI_ISL_1493571, EPI_ISL_1550455, EPI_ISL_1550465, EPI_ISL_1550466, EPI_ISL_1550492, EPI_ISL_1550516, EPI_ISL_1550519, EPI_ISL_1550521, EPI_ISL_1550522, EPI_ISL_1550550, EPI_ISL_1550565, EPI_ISL_1550669, EPI_ISL_1550670, EPI_ISL_1550690, EPI_ISL_1550739, EPI_ISL_1550742, EPI_ISL_1550743, EPI_ISL_1560009, EPI_ISL_1560010, EPI_ISL_1560011, EPI_ISL_1560018, EPI_ISL_1560037, EPI_ISL_1560106, EPI_ISL_1560123, EPI_ISL_1560137, EPI_ISL_1560142, EPI_ISL_1560150, EPI_ISL_1560177, EPI_ISL_1560179, EPI_ISL_1560255, EPI_ISL_1561202, EPI_ISL_1561249, EPI_ISL_1561298, EPI_ISL_1561328, EPI_ISL_1561330, EPI_ISL_1561335, EPI_ISL_1561342, EPI_ISL_1561345, EPI_ISL_1561366, EPI_ISL_1561374, EPI_ISL_1561400, EPI_ISL_1561404, EPI_ISL_1561541, EPI_ISL_1561717, EPI_ISL_1561719, EPI_ISL_1561788, EPI_ISL_1561794, EPI_ISL_1561802, EPI_ISL_1561803, EPI_ISL_1561804, EPI_ISL_1561805, EPI_ISL_1561806, EPI_ISL_1561809, EPI_ISL_1561810, EPI_ISL_1561811, EPI_ISL_1561817, EPI_ISL_1561818, EPI_ISL_1561819, EPI_ISL_1561828, EPI_ISL_1561866, EPI_ISL_1561870, EPI_ISL_1561891, EPI_ISL_1561893, EPI_ISL_1561902, EPI_ISL_1561904, EPI_ISL_1561907, EPI_ISL_1561918, EPI_ISL_1561926, EPI_ISL_1561927, EPI_ISL_1562412, EPI_ISL_1562554, EPI_ISL_1562567, EPI_ISL_1562601, EPI_ISL_1562631, EPI_ISL_1562812, EPI_ISL_1562820, EPI_ISL_1563608, EPI_ISL_2185633, EPI_ISL_2185669, EPI_ISL_2185680, EPI_ISL_2185681, EPI_ISL_2185682, EPI_ISL_2185683, EPI_ISL_2185704, EPI_ISL_2185708, EPI_ISL_2185714, EPI_ISL_2185716, EPI_ISL_2185736, EPI_ISL_2185752, EPI_ISL_2185754, EPI_ISL_2185755, EPI_ISL_2185756, EPI_ISL_2185757, EPI_ISL_2185760, EPI_ISL_2185762, EPI_ISL_2185764, EPI_ISL_2185780, EPI_ISL_2185782, EPI_ISL_2185783, EPI_ISL_2185784, EPI_ISL_2185785, EPI_ISL_2185786, EPI_ISL_2185847, EPI_ISL_2185848, EPI_ISL_2185853, EPI_ISL_2185875, EPI_ISL_2185877, EPI_ISL_2185878, EPI_ISL_2185942, EPI_ISL_2186008, EPI_ISL_2186035, EPI_ISL_2186043, EPI_ISL_2241714, EPI_ISL_2241731, EPI_ISL_2241745, EPI_ISL_2241793, EPI_ISL_2241796, EPI_ISL_2241797, EPI_ISL_2241799, EPI_ISL_2241821, EPI_ISL_2241832, EPI_ISL_2241833, EPI_ISL_2241967, EPI_ISL_2241968, EPI_ISL_2242543, EPI_ISL_2242605, EPI_ISL_2242606, EPI_ISL_2242616, EPI_ISL_2242643, EPI_ISL_2242658, EPI_ISL_2242660, EPI_ISL_2242662, EPI_ISL_2242664, EPI_ISL_2242900, EPI_ISL_2242916, EPI_ISL_2243064, EPI_ISL_2243146, EPI_ISL_2243149, EPI_ISL_2243152, EPI_ISL_2243166, EPI_ISL_2243169, EPI_ISL_2243172, EPI_ISL_2243173, EPI_ISL_2243174, EPI_ISL_2243177, EPI_ISL_2243181, EPI_ISL_2243187, EPI_ISL_2243188, EPI_ISL_2243190, EPI_ISL_2243209, EPI_ISL_2243210, EPI_ISL_2243213, EPI_ISL_2243216, EPI_ISL_2243226, EPI_ISL_2243227, EPI_ISL_2243228, EPI_ISL_2243229, EPI_ISL_2243237, EPI_ISL_2243249, EPI_ISL_2243257, EPI_ISL_2243276, EPI_ISL_2243279, EPI_ISL_2243281, EPI_ISL_2243282, EPI_ISL_2243284, EPI_ISL_2243285, EPI_ISL_2243286, EPI_ISL_2243287, EPI_ISL_2243288, EPI_ISL_2243289, EPI_ISL_2243290, EPI_ISL_2243291, EPI_ISL_2243293, EPI_ISL_2243298, EPI_ISL_2243312, EPI_ISL_2243314, EPI_ISL_2243325, EPI_ISL_2243329, EPI_ISL_2243336, EPI_ISL_2243340, EPI_ISL_2243347, EPI_ISL_2243348, EPI_ISL_2243349, EPI_ISL_2243350, EPI_ISL_2243359, EPI_ISL_2243361, EPI_ISL_2243376, EPI_ISL_2243380, EPI_ISL_2243382, EPI_ISL_2243383, EPI_ISL_2243384, EPI_ISL_2243385, EPI_ISL_2243386, EPI_ISL_2243387, EPI_ISL_2243388, EPI_ISL_2243391, EPI_ISL_2243398, EPI_ISL_2243399, EPI_ISL_2243441, EPI_ISL_2243450, EPI_ISL_2243451, EPI_ISL_2243481, EPI_ISL_2243488, EPI_ISL_2243527, EPI_ISL_2243538, EPI_ISL_2243547, EPI_ISL_2243557, EPI_ISL_2243558, EPI_ISL_2243559, EPI_ISL_2243560, EPI_ISL_2243572, EPI_ISL_2243573, EPI_ISL_2243574, EPI_ISL_2243575, EPI_ISL_2243576, EPI_ISL_2243577, EPI_ISL_2243578, EPI_ISL_2243579, EPI_ISL_2243580, EPI_ISL_2243581, EPI_ISL_2243582, EPI_ISL_2243583, EPI_ISL_2243584, EPI_ISL_2243585, EPI_ISL_2243586, EPI_ISL_2243587, EPI_ISL_2243609, EPI_ISL_2243610, EPI_ISL_2243612, EPI_ISL_2243628, EPI_ISL_2243631, EPI_ISL_2243661, EPI_ISL_2243673, EPI_ISL_2243680, EPI_ISL_2243717, EPI_ISL_2243719, EPI_ISL_2243722, EPI_ISL_2243747, EPI_ISL_2243765, EPI_ISL_2243766, EPI_ISL_2243777, EPI_ISL_2243778, EPI_ISL_2243779, EPI_ISL_2243807, EPI_ISL_2243810, EPI_ISL_2243813, EPI_ISL_2243814, EPI_ISL_2243815, EPI_ISL_2243828, EPI_ISL_2243859, EPI_ISL_2243860, EPI_ISL_2243869, EPI_ISL_2243875, EPI_ISL_2243889, EPI_ISL_2243901, EPI_ISL_2243910, EPI_ISL_2243926, EPI_ISL_2243936, EPI_ISL_2243962, EPI_ISL_2243980, EPI_ISL_2243981, EPI_ISL_2244042, EPI_ISL_2244090, EPI_ISL_2244135, EPI_ISL_2244293, EPI_ISL_2244294, EPI_ISL_2244309 | | | |
| see above | Aegis Sciences Corporation | Centers for Disease Control and Prevention Division of Viral Diseases, Pathogen Discovery | Adrian Paskey; Alec Vest; Benjamin Rambo-Martin; Christopher Gulvick; Clinton R. Paden; Cyndi Clark; Dakota Howard; Darlene Wagner; Dhwani Batra; Dillon Nall; Duncan MacCannell; Ethan Sanders; Holly Houdeshell; Jason Caravas; Kara Moser; Matthew Hardison; Matthew Schmerer; Ola Kvalvaag; Patrick Campbell; Peter W. Cook; Rob Case; Scott Sammons; Shatavia Morrison; Shaun Westlund; Vikramsinha Ghorpade; Yvette Unoarumhi |
| EPI_ISL_1205473, EPI_ISL_1205474, EPI_ISL_1205483, EPI_ISL_1205484, EPI_ISL_1575114, EPI_ISL_1575120, EPI_ISL_1575121, EPI_ISL_1575122, EPI_ISL_1575123, EPI_ISL_1575124, EPI_ISL_1576999, EPI_ISL_1577000, EPI_ISL_1577001, EPI_ISL_1577002, EPI_ISL_2304307, EPI_ISL_2304322, EPI_ISL_2304328, EPI_ISL_2304331, EPI_ISL_2304332, EPI_ISL_2304333, EPI_ISL_2304334, EPI_ISL_2304335 | | | |
| see above | Alameda County Public Health Department | Alameda County Public Health Department | Kevin Libuit; Kristina Hsieh; Kristna Hsieh; Maria Elma |
| EPI_ISL_1222640 | Altius Institute | Seattle Flu Study | Alex Nguyen; Amanda Adler; Andrew Meuser; Barry R. Lutz; Benjamin Pelle; Caitlin R. Wolf; Chris D. Frazar; Clem Green; Daniel Bates; Deborah A. Nickerson; Elisabeth Brandstetter; Erica Ryke; Hannah Petersen; Helen Y. Chu; Jacob Rodriguez; Janet A. Englund; Jay Shendure; Jessica Halow; John Stamatoyannopoulos; Joshua Richards; Jover Lee; Julia Wald; Kairsten Fay; Kirsten Lacombe; Kneshay Harper; Lea M. Starita; Mark J. Rieder; Matt Hartman; Matthew Richardson; Matthew Thompson; Melissa Truong; Michael Boeckh; Michael Famulare; Misja Ilcisin; Muhammad Halimun; Olivia Waltner; Peter D. Han; Rebecca Bruders; Ryan Alexander; Sadie Patraw; Sofia Olsson; Stephanie DeBaun; Thomas R. Sibley; Tobias Ragoczy; Trevor Bedford; Truong Nguyen |
| EPI_ISL_1114336 | Altius Institute for Biomedical Sciences | Seattle Flu Study | Alex Nguyen; Amanda Adler; Andrew Meuser; Barry R. Lutz; Benjamin Pelle; Caitlin R. Wolf; Chris D. Frazar; Clem Green; Daniel Bates; Deborah A. Nickerson; Elisabeth Brandstetter; Erica Ryke; Hannah Petersen; Helen Y. Chu; Jacob Rodriguez; Janet A. Englund; Jay Shendure; Jessica Halow; John Stamatoyannopoulos; Joshua Richards; Jover Lee; Julia Wald; Kairsten Fay; Kirsten Lacombe; Kneshay Harper; Lea M. Starita; Mark J. Rieder; Matt Hartman; Matthew Richardson; Matthew Thompson; Melissa Truong; Michael Boeckh; Michael Famulare; Misja Ilcisin; Muhammad Halimun; Olivia Waltner; Peter D. Han; Rebecca Bruders; Ryan Alexander; Sadie Patraw; Sofia Olsson; Stephanie DeBaun; Thomas R. Sibley; Tobias Ragoczy; Trevor Bedford; Truong Nguyen |
| EPI_ISL_1477959, EPI_ISL_1477964, EPI_ISL_1477968, EPI_ISL_1477969, EPI_ISL_1477971, EPI_ISL_1477978, EPI_ISL_1477984, EPI_ISL_1477985, EPI_ISL_1477988, EPI_ISL_1477989, EPI_ISL_1477994, EPI_ISL_1477996, EPI_ISL_1478001, EPI_ISL_1478004, EPI_ISL_1478014, EPI_ISL_1478016, EPI_ISL_1478021, EPI_ISL_1478027, EPI_ISL_1478029, EPI_ISL_1478033, EPI_ISL_1478042, EPI_ISL_1478051, EPI_ISL_1478056, EPI_ISL_1478060, EPI_ISL_1478064, EPI_ISL_1478068, EPI_ISL_1478071, EPI_ISL_1478073, EPI_ISL_1478076, EPI_ISL_1478079, EPI_ISL_1478085, EPI_ISL_1478100, EPI_ISL_1478104, EPI_ISL_1478109, EPI_ISL_1478112, EPI_ISL_1478113, EPI_ISL_1478114, EPI_ISL_1478115, EPI_ISL_1478116, EPI_ISL_1478119, EPI_ISL_1478127, EPI_ISL_1478130, EPI_ISL_1478132, EPI_ISL_1478137, EPI_ISL_1478142, EPI_ISL_1478144, EPI_ISL_1478146, EPI_ISL_1478148, EPI_ISL_1478151, EPI_ISL_1478158, EPI_ISL_1478162, EPI_ISL_1478166, EPI_ISL_1478167, EPI_ISL_1478168, EPI_ISL_1478173, EPI_ISL_1478178, EPI_ISL_1478183, EPI_ISL_1478184, EPI_ISL_1478185, EPI_ISL_1478191, EPI_ISL_1478194, EPI_ISL_1478201, EPI_ISL_1529104, EPI_ISL_1529118, EPI_ISL_1529130, EPI_ISL_1529132, EPI_ISL_1529136, EPI_ISL_1529141, EPI_ISL_1529142, EPI_ISL_1529156, EPI_ISL_1529167, EPI_ISL_1529169, EPI_ISL_1529175, EPI_ISL_1529180, EPI_ISL_1529184, EPI_ISL_1529201, EPI_ISL_1529220, EPI_ISL_1529260, EPI_ISL_1529264, EPI_ISL_1529278, EPI_ISL_1529289, EPI_ISL_1529303, EPI_ISL_1529307, EPI_ISL_1529315, EPI_ISL_1529338, EPI_ISL_1529339, EPI_ISL_1529356, EPI_ISL_1701042, EPI_ISL_1701043, EPI_ISL_1701044, EPI_ISL_1701045, EPI_ISL_1701047, EPI_ISL_1701048, EPI_ISL_1701049, EPI_ISL_1701051, EPI_ISL_1701053, EPI_ISL_1701054, EPI_ISL_1701055, EPI_ISL_1701056, EPI_ISL_1701058, EPI_ISL_1701059, EPI_ISL_1701060, EPI_ISL_1701062, EPI_ISL_1701063, EPI_ISL_1701064, EPI_ISL_1701067, EPI_ISL_1701068, EPI_ISL_1701070, EPI_ISL_1701075, EPI_ISL_1701076, EPI_ISL_1701077, EPI_ISL_1701085, EPI_ISL_1701089, EPI_ISL_1701094, EPI_ISL_1701095, EPI_ISL_1701097, EPI_ISL_1701098, EPI_ISL_1701100, EPI_ISL_1701101, EPI_ISL_1701102, EPI_ISL_1701103, EPI_ISL_1701105, EPI_ISL_1701107, EPI_ISL_1701108, EPI_ISL_1701109, EPI_ISL_1701110, EPI_ISL_1701111, EPI_ISL_1701112, EPI_ISL_1701113, EPI_ISL_1701114, EPI_ISL_1701116, EPI_ISL_1701118, EPI_ISL_1701121, EPI_ISL_1701122, EPI_ISL_1701125, EPI_ISL_1701126, EPI_ISL_1701128, EPI_ISL_1701129, EPI_ISL_1701130, EPI_ISL_1701131, EPI_ISL_1701133, EPI_ISL_1701134, EPI_ISL_1701138, EPI_ISL_1701139, EPI_ISL_1701140, EPI_ISL_1701141, EPI_ISL_1701143, EPI_ISL_1701144, EPI_ISL_1701145, EPI_ISL_1701146, EPI_ISL_1701149, EPI_ISL_1701151, EPI_ISL_1701152, EPI_ISL_1701153, EPI_ISL_1701154, EPI_ISL_1701158, EPI_ISL_1701159, EPI_ISL_1701160, EPI_ISL_1701163, EPI_ISL_1701164, EPI_ISL_1701165, EPI_ISL_1701166, EPI_ISL_1701169, EPI_ISL_1701170, EPI_ISL_1701173, EPI_ISL_1701181, EPI_ISL_1701182, EPI_ISL_1701183, EPI_ISL_1701184, EPI_ISL_1701185, EPI_ISL_1701186, EPI_ISL_1701187, EPI_ISL_1701190, EPI_ISL_1701191, EPI_ISL_1701192, EPI_ISL_1701193, EPI_ISL_1701195, EPI_ISL_1701196, EPI_ISL_1701197, EPI_ISL_1701198, EPI_ISL_1701199, EPI_ISL_1701200, EPI_ISL_1701202, EPI_ISL_1701203, EPI_ISL_1701206, EPI_ISL_1701207, EPI_ISL_1701209, EPI_ISL_1701215, EPI_ISL_1701216, EPI_ISL_1701220, EPI_ISL_1701221, EPI_ISL_1701223, EPI_ISL_1701225, EPI_ISL_1701226, EPI_ISL_1701227, EPI_ISL_1701229, EPI_ISL_1701231, EPI_ISL_1701232, EPI_ISL_1701237, EPI_ISL_1701238, EPI_ISL_1701239, EPI_ISL_1701240, EPI_ISL_1701241, EPI_ISL_1701242, EPI_ISL_1701243, EPI_ISL_1701244, EPI_ISL_1701246, EPI_ISL_1701248, EPI_ISL_1701252, EPI_ISL_1701253, EPI_ISL_1701254, EPI_ISL_1701255, EPI_ISL_1701256, EPI_ISL_1701257, EPI_ISL_1701258, EPI_ISL_1701259, EPI_ISL_1701260, EPI_ISL_1701261, EPI_ISL_1701263, EPI_ISL_1701265, EPI_ISL_1701269, EPI_ISL_1701270, EPI_ISL_1701271, EPI_ISL_1701272, EPI_ISL_1701275, EPI_ISL_1701276, EPI_ISL_1701278, EPI_ISL_1701279, EPI_ISL_1701282, EPI_ISL_1701283, EPI_ISL_1701284, EPI_ISL_1701286, EPI_ISL_1701287, EPI_ISL_1701288, EPI_ISL_1701292, EPI_ISL_1701294, EPI_ISL_1701296, EPI_ISL_1701298, EPI_ISL_1701300, EPI_ISL_1701304, EPI_ISL_1701305, EPI_ISL_1701308, EPI_ISL_1701309, EPI_ISL_1701310, EPI_ISL_2658621, EPI_ISL_2659207, EPI_ISL_2659233, EPI_ISL_2659279, EPI_ISL_2659507, EPI_ISL_2758476, EPI_ISL_2758477, EPI_ISL_2758478, EPI_ISL_2758483, EPI_ISL_2758484, EPI_ISL_2758485, EPI_ISL_2758486 | | | |
| see above | CA DPH Viral and Rickettsial Disease Laboratory | Chan-Zuckerberg Biohub | CZB Cliahub Consortium |
| EPI_ISL_1272873, EPI_ISL_1272874, EPI_ISL_1272875, EPI_ISL_1272878, EPI_ISL_1272879, EPI_ISL_1272886, EPI_ISL_1272888, EPI_ISL_1272891, EPI_ISL_1272894, EPI_ISL_1272895, EPI_ISL_1318377, EPI_ISL_1318378, EPI_ISL_1318379, EPI_ISL_1318380, EPI_ISL_1446714, EPI_ISL_1446715, EPI_ISL_1447050, EPI_ISL_1447051, EPI_ISL_1678280, EPI_ISL_1678281, EPI_ISL_1678284, EPI_ISL_1678285 | | | |
| see above | CA-Los Angeles County Public Health Laboratory | Centers for Disease Control and Prevention Division of Viral Diseases, Pathogen Discovery | Alison Laufer Halpin; Anna Montmayeur; Anna Uehara; Ben L. Rambo-Martin; Clinton R. Paden; Dakota Howard; Darlene Wagner; Dave Wentworth; Dhwani Batra; Haibin Wang; Jasmine Padilla; Jing Zhang; Justin Lee; Katie Dillon; Krista Queen; Kristen Knipe; Kristine Lacek; Lori Rowe; Mark Burroughs; Matthew Schmerer; Mili Sheth; Peter Cook; Peter W. Cook; Rachel Marine; Sam Shepard; Sarah Nobles; Shoshona Le; Suxiang Tong; Vivien Dugan; Yan Li; Ying Tao; Yvette Unoarumhi |
| EPI_ISL_431142, EPI_ISL_431143, EPI_ISL_2825386, EPI_ISL_2825388, EPI_ISL_2825390, EPI_ISL_2825391, EPI_ISL_2825410, EPI_ISL_2825413, EPI_ISL_2825414, EPI_ISL_2825415, EPI_ISL_2825416, EPI_ISL_2825417, EPI_ISL_2825420, EPI_ISL_2825421, EPI_ISL_2825426, EPI_ISL_2825427, EPI_ISL_2825428, EPI_ISL_2825429, EPI_ISL_2825430, EPI_ISL_2825431, EPI_ISL_2825432, EPI_ISL_2825433, EPI_ISL_2825434, EPI_ISL_2825435, EPI_ISL_2825436, EPI_ISL_2825438, EPI_ISL_2825440, EPI_ISL_2825441, EPI_ISL_2825442, EPI_ISL_2825443, EPI_ISL_2825444, EPI_ISL_2825445, EPI_ISL_2825446, EPI_ISL_2825447, EPI_ISL_2825448, EPI_ISL_2825449, EPI_ISL_2825450, EPI_ISL_2825451, EPI_ISL_2825452, EPI_ISL_2825453, EPI_ISL_2825454, EPI_ISL_2825456, EPI_ISL_2825457, EPI_ISL_2825458, EPI_ISL_2825459, EPI_ISL_2825460, EPI_ISL_2825461, EPI_ISL_2825462, EPI_ISL_2825463, EPI_ISL_2825464, EPI_ISL_2825465, EPI_ISL_2825466, EPI_ISL_2825467, EPI_ISL_2825468, EPI_ISL_2825469, EPI_ISL_2825470, EPI_ISL_2825471, EPI_ISL_2825472, EPI_ISL_2825473, EPI_ISL_2825475, EPI_ISL_2825476, EPI_ISL_2825477, EPI_ISL_2825478, EPI_ISL_2825479, EPI_ISL_2825480, EPI_ISL_2825481, EPI_ISL_2825482, EPI_ISL_2825483, EPI_ISL_2825484, EPI_ISL_2825485, EPI_ISL_2825486, EPI_ISL_2825487, EPI_ISL_2825488, EPI_ISL_2825489, EPI_ISL_2825490, EPI_ISL_2825491, EPI_ISL_2825493, EPI_ISL_2825494, EPI_ISL_2825495, EPI_ISL_2825496, EPI_ISL_2825497, EPI_ISL_2825498, EPI_ISL_2825499, EPI_ISL_2825500, EPI_ISL_2825501, EPI_ISL_2825502, EPI_ISL_2825503, EPI_ISL_2825504, EPI_ISL_2825505, EPI_ISL_2825506, EPI_ISL_2825507, EPI_ISL_2825508, EPI_ISL_2825509, EPI_ISL_2825511, EPI_ISL_2825512, EPI_ISL_2825513, EPI_ISL_2825514, EPI_ISL_2825515, EPI_ISL_2825516, EPI_ISL_2825517, EPI_ISL_2825518, EPI_ISL_2825519, EPI_ISL_2825520, EPI_ISL_2825521, EPI_ISL_2825522, EPI_ISL_2825523, EPI_ISL_2825524, EPI_ISL_2825525, EPI_ISL_2825526, EPI_ISL_2825527, EPI_ISL_2825529, EPI_ISL_2825530, EPI_ISL_2825531, EPI_ISL_2825532, EPI_ISL_2825533, EPI_ISL_2825534, EPI_ISL_2825535, EPI_ISL_2825536, EPI_ISL_2825537, EPI_ISL_2825538, EPI_ISL_2825539, EPI_ISL_2825540, EPI_ISL_2825541, EPI_ISL_2825542, EPI_ISL_2825543, EPI_ISL_2825544, EPI_ISL_2825545, EPI_ISL_2825547, EPI_ISL_2825548, EPI_ISL_2825549, EPI_ISL_2825550, EPI_ISL_2825551, EPI_ISL_2825552, EPI_ISL_2825553, EPI_ISL_2825554, EPI_ISL_2825555, EPI_ISL_2825556, EPI_ISL_2825557, EPI_ISL_2825558, EPI_ISL_2825559, EPI_ISL_2825560, EPI_ISL_2825561, EPI_ISL_2825562, EPI_ISL_2825563, EPI_ISL_2825564, EPI_ISL_2825566, EPI_ISL_2825567, EPI_ISL_2825568, EPI_ISL_2825569, EPI_ISL_2825570, EPI_ISL_2825571, EPI_ISL_2825572, EPI_ISL_2825573, EPI_ISL_2825574, EPI_ISL_2825575, EPI_ISL_2825576, EPI_ISL_2825577, EPI_ISL_2825578, EPI_ISL_2825579, EPI_ISL_2825580, EPI_ISL_2825581, EPI_ISL_2825582, EPI_ISL_2825584, EPI_ISL_2825585, EPI_ISL_2825586, EPI_ISL_2825587, EPI_ISL_2825588, EPI_ISL_2825589, EPI_ISL_2825590, EPI_ISL_2825591, EPI_ISL_2825592, EPI_ISL_2825593, EPI_ISL_2825594, EPI_ISL_2825595, EPI_ISL_2825596, EPI_ISL_2825597, EPI_ISL_2825598, EPI_ISL_2825599, EPI_ISL_2825600, EPI_ISL_2825602, EPI_ISL_2825603, EPI_ISL_2825604, EPI_ISL_2825605, EPI_ISL_2825606, EPI_ISL_2825607, EPI_ISL_2825608, EPI_ISL_2825609, EPI_ISL_2825610, EPI_ISL_2825611, EPI_ISL_2825612, EPI_ISL_2825613, EPI_ISL_2825614, EPI_ISL_2825615, EPI_ISL_2825616, EPI_ISL_2825617, EPI_ISL_2825618, EPI_ISL_2825620, EPI_ISL_2825621, EPI_ISL_2825622, EPI_ISL_2825623, EPI_ISL_2825624, EPI_ISL_2825625, EPI_ISL_2825626, EPI_ISL_2825627, EPI_ISL_2825628, EPI_ISL_2825629, EPI_ISL_2825630, EPI_ISL_2825631, EPI_ISL_2825632, EPI_ISL_2825633, EPI_ISL_2825634, EPI_ISL_2825635, EPI_ISL_2825636, EPI_ISL_2825637, EPI_ISL_2825638, EPI_ISL_2825639, EPI_ISL_2825640, EPI_ISL_2825641, EPI_ISL_2825642, EPI_ISL_2825643, EPI_ISL_2825644, EPI_ISL_2825645, EPI_ISL_2825646, EPI_ISL_2825647, EPI_ISL_2825648, EPI_ISL_2825649, EPI_ISL_2825650, EPI_ISL_2825651, EPI_ISL_2825652, EPI_ISL_2825653, EPI_ISL_2825654, EPI_ISL_2825655, EPI_ISL_2825656, EPI_ISL_2825657, EPI_ISL_2825658, EPI_ISL_2825659, EPI_ISL_2825660, EPI_ISL_2825661, EPI_ISL_2825662, EPI_ISL_2825663, EPI_ISL_2825664, EPI_ISL_2825665, EPI_ISL_2825666, EPI_ISL_2825667, EPI_ISL_2825668, EPI_ISL_2825669, EPI_ISL_2825670, EPI_ISL_2825671, EPI_ISL_2825672, EPI_ISL_2825673, EPI_ISL_2825674, EPI_ISL_2825675, EPI_ISL_2825676, EPI_ISL_2825677, EPI_ISL_2825678, EPI_ISL_2825679, EPI_ISL_2825680, EPI_ISL_2825681, EPI_ISL_2825682, EPI_ISL_2825683, EPI_ISL_2825684, EPI_ISL_2825685, EPI_ISL_2825686, EPI_ISL_2825687, EPI_ISL_2825688, EPI_ISL_2826527, EPI_ISL_2826583, EPI_ISL_2826584, EPI_ISL_2826587, EPI_ISL_2826588, EPI_ISL_2826631, EPI_ISL_2826632, EPI_ISL_2826633, EPI_ISL_2826634, EPI_ISL_2826635, EPI_ISL_2826636, EPI_ISL_2826637, EPI_ISL_2826638, EPI_ISL_2826639, EPI_ISL_2826640, EPI_ISL_2826641, EPI_ISL_2826642, EPI_ISL_2826643, EPI_ISL_2826644, EPI_ISL_2826645, EPI_ISL_2826646, EPI_ISL_2826647, EPI_ISL_2826648, EPI_ISL_2826649, EPI_ISL_2826650, EPI_ISL_2826651, EPI_ISL_2826652, EPI_ISL_2826653, EPI_ISL_2826654, EPI_ISL_2826655, EPI_ISL_2826656, EPI_ISL_2826657, EPI_ISL_2826658, EPI_ISL_2826659, EPI_ISL_2826660, EPI_ISL_2826661, EPI_ISL_2826662, EPI_ISL_2826663, EPI_ISL_2826664, EPI_ISL_2826665, EPI_ISL_2826666, EPI_ISL_2826667, EPI_ISL_2826668, EPI_ISL_2826669, EPI_ISL_2826670, EPI_ISL_2826671, EPI_ISL_2826672, EPI_ISL_2826673, EPI_ISL_2826674, EPI_ISL_2826675, EPI_ISL_2826676, EPI_ISL_2826677, EPI_ISL_2826678, EPI_ISL_2826679, EPI_ISL_2826680, EPI_ISL_2826681, EPI_ISL_2826682, EPI_ISL_2826683, EPI_ISL_2826684, EPI_ISL_2826685, EPI_ISL_2826686, EPI_ISL_2826687, EPI_ISL_2826688, EPI_ISL_2826689, EPI_ISL_2826690, EPI_ISL_2826691, EPI_ISL_2826692, EPI_ISL_2826693, EPI_ISL_2826694, EPI_ISL_2826695, EPI_ISL_2826696, EPI_ISL_2826697, EPI_ISL_2826698, EPI_ISL_2826699, EPI_ISL_2826700, EPI_ISL_2826701, EPI_ISL_2826702, EPI_ISL_2826703, EPI_ISL_2826704, EPI_ISL_2826705, EPI_ISL_2826706, EPI_ISL_2826707, EPI_ISL_2826708, EPI_ISL_2826709, EPI_ISL_2826710, EPI_ISL_2826711, EPI_ISL_2826712, EPI_ISL_2826713, EPI_ISL_2826714, EPI_ISL_2826715, EPI_ISL_2826716, EPI_ISL_2826717, EPI_ISL_2826718, EPI_ISL_2826719, EPI_ISL_2826720, EPI_ISL_2826721, EPI_ISL_2826722, EPI_ISL_2826723, EPI_ISL_2826724, EPI_ISL_2826725, EPI_ISL_2826726, EPI_ISL_2826727, EPI_ISL_2826728, EPI_ISL_2826729, EPI_ISL_2826730, EPI_ISL_2826731, EPI_ISL_2826732, EPI_ISL_2826733, EPI_ISL_2826734, EPI_ISL_2826735, EPI_ISL_2826736, EPI_ISL_2826737, EPI_ISL_2826738, EPI_ISL_2826739, EPI_ISL_2826740, EPI_ISL_2826741, EPI_ISL_2826742, EPI_ISL_2826743, EPI_ISL_2826744, EPI_ISL_2826745, EPI_ISL_2826746, EPI_ISL_2826747, EPI_ISL_2826748, EPI_ISL_2826749, EPI_ISL_2826750, EPI_ISL_2826751, EPI_ISL_2826752, EPI_ISL_2826753, EPI_ISL_2826754, EPI_ISL_2826755, EPI_ISL_2826756, EPI_ISL_2826757, EPI_ISL_2826758, EPI_ISL_2826759, EPI_ISL_2826760, EPI_ISL_2826761, EPI_ISL_2826762, EPI_ISL_2826763, EPI_ISL_2826764, EPI_ISL_2826765, EPI_ISL_2826766, EPI_ISL_2826767, EPI_ISL_2826768, EPI_ISL_2826769, EPI_ISL_2826770, EPI_ISL_2826771, EPI_ISL_2826772, EPI_ISL_2826773, EPI_ISL_2826774, EPI_ISL_2826775, EPI_ISL_2826776, EPI_ISL_2826777, EPI_ISL_2826778, EPI_ISL_2826779, EPI_ISL_2826780, EPI_ISL_2826781, EPI_ISL_2826782, EPI_ISL_2826783, EPI_ISL_2826784, EPI_ISL_2826785, EPI_ISL_2826786, EPI_ISL_2826787, EPI_ISL_2826788, EPI_ISL_2826789, EPI_ISL_2826790, EPI_ISL_2826791, EPI_ISL_2826792, EPI_ISL_2826793, EPI_ISL_2826794, EPI_ISL_2826795, EPI_ISL_2826796, EPI_ISL_2826797, EPI_ISL_2826798, EPI_ISL_2826799, EPI_ISL_2826800, EPI_ISL_2826801, EPI_ISL_2826802, EPI_ISL_2826803, EPI_ISL_2826804, EPI_ISL_2826805, EPI_ISL_2826806, EPI_ISL_2826807, EPI_ISL_2826808, EPI_ISL_2826809, EPI_ISL_2826810, EPI_ISL_2826811, EPI_ISL_2826812, EPI_ISL_2826813, EPI_ISL_2826814, EPI_ISL_2826815, EPI_ISL_2826816, EPI_ISL_2826817, EPI_ISL_2826818, EPI_ISL_2826819, EPI_ISL_2826820, EPI_ISL_2826821, EPI_ISL_2826822, EPI_ISL_2826823, EPI_ISL_2826824, EPI_ISL_2826825, EPI_ISL_2826826, EPI_ISL_2826827, EPI_ISL_2826828, EPI_ISL_2826829, EPI_ISL_2826830, EPI_ISL_2826831, EPI_ISL_2826832, EPI_ISL_2826833, EPI_ISL_2826834, EPI_ISL_2826835, EPI_ISL_2826836, EPI_ISL_2854992, EPI_ISL_2854993, EPI_ISL_2854994, EPI_ISL_2854995, EPI_ISL_2854996, EPI_ISL_2855000, EPI_ISL_2855001, EPI_ISL_2855002, EPI_ISL_2855003, EPI_ISL_2855004, EPI_ISL_2855005, EPI_ISL_2855006, EPI_ISL_2855007, EPI_ISL_2855008, EPI_ISL_2855015, EPI_ISL_2855016, EPI_ISL_2855044, EPI_ISL_2855045, EPI_ISL_2855056, EPI_ISL_2855058, EPI_ISL_2855059, EPI_ISL_2855060, EPI_ISL_2855063, EPI_ISL_2855064, EPI_ISL_2855068, EPI_ISL_2855074, EPI_ISL_2855079, EPI_ISL_2855081, EPI_ISL_2855082, EPI_ISL_2855083, EPI_ISL_2855086, EPI_ISL_2855087, EPI_ISL_2855088, EPI_ISL_2855091, EPI_ISL_2855092, EPI_ISL_2855094, EPI_ISL_2855106, EPI_ISL_2855107, EPI_ISL_2855108, EPI_ISL_2855109, EPI_ISL_2855112, EPI_ISL_2855121, EPI_ISL_2855137, EPI_ISL_2855140, EPI_ISL_2855141, EPI_ISL_2855142, EPI_ISL_2855152, EPI_ISL_2855154, EPI_ISL_2855174, EPI_ISL_2855175, EPI_ISL_2855176, EPI_ISL_2855177, EPI_ISL_2855178, EPI_ISL_2855179, EPI_ISL_2855180, EPI_ISL_2855188, EPI_ISL_2855189, EPI_ISL_2855204, EPI_ISL_2855206, EPI_ISL_2855207, EPI_ISL_2855210, EPI_ISL_2855211, EPI_ISL_2855213, EPI_ISL_2855214, EPI_ISL_2855221, EPI_ISL_2855222, EPI_ISL_2855229, EPI_ISL_2855230, EPI_ISL_2855236, EPI_ISL_2855240, EPI_ISL_2855241, EPI_ISL_2855242, EPI_ISL_2855247, EPI_ISL_2855252, EPI_ISL_2855257, EPI_ISL_2855258, EPI_ISL_2855259, EPI_ISL_2855278, EPI_ISL_2855279, EPI_ISL_2855281, EPI_ISL_2855282, EPI_ISL_2855283, EPI_ISL_2855296, EPI_ISL_2855310, EPI_ISL_2855325, EPI_ISL_2855357, EPI_ISL_2855370, EPI_ISL_2855379, EPI_ISL_2855381, EPI_ISL_2855385, EPI_ISL_2855390, EPI_ISL_2855393, EPI_ISL_2855409, EPI_ISL_2855410, EPI_ISL_2855411, EPI_ISL_2855415, EPI_ISL_2855416, EPI_ISL_2855421, EPI_ISL_2855422, EPI_ISL_2855423, EPI_ISL_2855425, EPI_ISL_2855426, EPI_ISL_2855427, EPI_ISL_2855428, EPI_ISL_2855430, EPI_ISL_2855431, EPI_ISL_2855432, EPI_ISL_2855463, EPI_ISL_2855470, EPI_ISL_2855490, EPI_ISL_2855491, EPI_ISL_2855492, EPI_ISL_2855493, EPI_ISL_2855523, EPI_ISL_2855524, EPI_ISL_2855526, EPI_ISL_2855527, EPI_ISL_2855540, EPI_ISL_2855543, EPI_ISL_2855544, EPI_ISL_2855546, EPI_ISL_2855561, EPI_ISL_2855562, EPI_ISL_2855569, EPI_ISL_2855570, EPI_ISL_2855587, EPI_ISL_2855588, EPI_ISL_2855593, EPI_ISL_2855597, EPI_ISL_2855604, EPI_ISL_2855605, EPI_ISL_2855607, EPI_ISL_2855610, EPI_ISL_2855611, EPI_ISL_2855612, EPI_ISL_2855613, EPI_ISL_2855614, EPI_ISL_2855615, EPI_ISL_2855616, EPI_ISL_2855617, EPI_ISL_2855618, EPI_ISL_2855619, EPI_ISL_2855620, EPI_ISL_2855640, EPI_ISL_2855641, EPI_ISL_2855642, EPI_ISL_2855643, EPI_ISL_2855650, EPI_ISL_2855658, EPI_ISL_2855660, EPI_ISL_2855661, EPI_ISL_2855662, EPI_ISL_2855663, EPI_ISL_2855664, EPI_ISL_2855665, EPI_ISL_2855680, EPI_ISL_2855681, EPI_ISL_2855682, EPI_ISL_2855683, EPI_ISL_2855684, EPI_ISL_2855685, EPI_ISL_2855686, EPI_ISL_2855689, EPI_ISL_2855702, EPI_ISL_2855713, EPI_ISL_2855714, EPI_ISL_2855715, EPI_ISL_2855716, EPI_ISL_2855722, EPI_ISL_2855723, EPI_ISL_2855724, EPI_ISL_2855725, EPI_ISL_2855726, EPI_ISL_2855727, EPI_ISL_2855728, EPI_ISL_2855729, EPI_ISL_2855731, EPI_ISL_2855732, EPI_ISL_2855733, EPI_ISL_2855736, EPI_ISL_2855739, EPI_ISL_2855740, EPI_ISL_2855741, EPI_ISL_2855742, | | | |

| | | | |
|---|---|---|---|
| EPI_ISL_2855744, EPI_ISL_2855746, EPI_ISL_2855763, EPI_ISL_2855764, EPI_ISL_2855765, EPI_ISL_2855766, EPI_ISL_2855772, EPI_ISL_2855778, EPI_ISL_2855779, EPI_ISL_2855785, EPI_ISL_2855786, EPI_ISL_2855787, EPI_ISL_2855788, EPI_ISL_2855789, EPI_ISL_2855790, EPI_ISL_2855791, EPI_ISL_2855792, EPI_ISL_2855793, EPI_ISL_2855802, EPI_ISL_2855825, EPI_ISL_2855826, EPI_ISL_2855827, EPI_ISL_2855828, EPI_ISL_2858374, EPI_ISL_2858407, EPI_ISL_2858582, EPI_ISL_2881871, EPI_ISL_2881872, EPI_ISL_2881873, EPI_ISL_2881876, EPI_ISL_2881883, EPI_ISL_2881884, EPI_ISL_2881887, EPI_ISL_2881888, EPI_ISL_2881889, EPI_ISL_2881892, EPI_ISL_2881895, EPI_ISL_2881898, EPI_ISL_2881899, EPI_ISL_2881900, EPI_ISL_2881901, EPI_ISL_2881910, EPI_ISL_2881912, EPI_ISL_2881941, EPI_ISL_2881943, EPI_ISL_2881944, EPI_ISL_2881945, EPI_ISL_2881956, EPI_ISL_2881958, EPI_ISL_2881959, EPI_ISL_2881964, EPI_ISL_2881965, EPI_ISL_2881977, EPI_ISL_2881989, EPI_ISL_2881990, EPI_ISL_2881991, EPI_ISL_2881992, EPI_ISL_2881993, EPI_ISL_2881994, EPI_ISL_2882002, EPI_ISL_2882004, EPI_ISL_2882006, EPI_ISL_2882007, EPI_ISL_2882008, EPI_ISL_2882009, EPI_ISL_2882010, EPI_ISL_2882016, EPI_ISL_2882018, EPI_ISL_2882019, EPI_ISL_2882020, EPI_ISL_2882023, EPI_ISL_2882024, EPI_ISL_2882025, EPI_ISL_2882026, EPI_ISL_2882027, EPI_ISL_2882039, EPI_ISL_2882041, EPI_ISL_2882042, EPI_ISL_2889856, EPI_ISL_2889857, EPI_ISL_2889861, EPI_ISL_2889862, EPI_ISL_2889863, EPI_ISL_2889864, EPI_ISL_2889865, EPI_ISL_2889866, EPI_ISL_2889868, EPI_ISL_2889869, EPI_ISL_2889870, EPI_ISL_2889871, EPI_ISL_2889872, EPI_ISL_2889873, EPI_ISL_2889874, EPI_ISL_2889875, EPI_ISL_2889876, EPI_ISL_2889877, EPI_ISL_2889878, EPI_ISL_2889879, EPI_ISL_2889880, EPI_ISL_2889885, EPI_ISL_2889886, EPI_ISL_2889887, EPI_ISL_2889888, EPI_ISL_2889889, EPI_ISL_2889890, EPI_ISL_2889891, EPI_ISL_2889892, EPI_ISL_2889894, EPI_ISL_2889900, EPI_ISL_2889901, EPI_ISL_2889902, EPI_ISL_2889903, EPI_ISL_2889904, EPI_ISL_2889905, EPI_ISL_2889906, EPI_ISL_2889907, EPI_ISL_2889909, EPI_ISL_2889910, EPI_ISL_2889917, EPI_ISL_2889918, EPI_ISL_2889919, EPI_ISL_2889920, EPI_ISL_2889923, EPI_ISL_2889924, EPI_ISL_2889933, EPI_ISL_2889937, EPI_ISL_2889939, EPI_ISL_2889943, EPI_ISL_2889944, EPI_ISL_2889949, EPI_ISL_2889950, EPI_ISL_2889951, EPI_ISL_2889952, EPI_ISL_2889953, EPI_ISL_2889954, EPI_ISL_2889955, EPI_ISL_2889956, EPI_ISL_2889957, EPI_ISL_2889958, EPI_ISL_2889959, EPI_ISL_2889960, EPI_ISL_2889963, EPI_ISL_2889964, EPI_ISL_2889965, EPI_ISL_2889966, EPI_ISL_2889967, EPI_ISL_2889968, EPI_ISL_2889969, EPI_ISL_2889972, EPI_ISL_2889973, EPI_ISL_2889975, EPI_ISL_2889976, EPI_ISL_2889977, EPI_ISL_2889978, EPI_ISL_2889979, EPI_ISL_2889980, EPI_ISL_2889981, EPI_ISL_2889983, EPI_ISL_2889984, EPI_ISL_2889985, EPI_ISL_2889986, EPI_ISL_2889987, EPI_ISL_2889988, EPI_ISL_2889989, EPI_ISL_2889990, EPI_ISL_2889991, EPI_ISL_2889992, EPI_ISL_2889995, EPI_ISL_2889999, EPI_ISL_2890000, EPI_ISL_2890001, EPI_ISL_2890002, EPI_ISL_2890003, EPI_ISL_2890004, EPI_ISL_2890005, EPI_ISL_2890006, EPI_ISL_2890013, EPI_ISL_2890016, EPI_ISL_2890017, EPI_ISL_2890018, EPI_ISL_2890019, EPI_ISL_2890020, EPI_ISL_2890021, EPI_ISL_2890022, EPI_ISL_2890023, EPI_ISL_2890024, EPI_ISL_2890025, EPI_ISL_2890026, EPI_ISL_2890027, EPI_ISL_2890028, EPI_ISL_2890029, EPI_ISL_2890030, EPI_ISL_2890031, EPI_ISL_2890033, EPI_ISL_2890034, EPI_ISL_2890035, EPI_ISL_2890036, EPI_ISL_2890037, EPI_ISL_2890038, EPI_ISL_2890039, EPI_ISL_2890040, EPI_ISL_2890041, EPI_ISL_2890042, EPI_ISL_2890043, EPI_ISL_2890044, EPI_ISL_2890045, EPI_ISL_2890046, EPI_ISL_2890047, EPI_ISL_2890048, EPI_ISL_2890049, EPI_ISL_2890052, EPI_ISL_2890053, EPI_ISL_2890056, EPI_ISL_2890057, EPI_ISL_2890058, EPI_ISL_2890059, EPI_ISL_2890061, EPI_ISL_2890062, EPI_ISL_2890071, EPI_ISL_2890072, EPI_ISL_2890073, EPI_ISL_2890074, EPI_ISL_2890075, EPI_ISL_2890077, EPI_ISL_2890078, EPI_ISL_2890079, EPI_ISL_2890080, EPI_ISL_2890081, EPI_ISL_2890082, EPI_ISL_2890083, EPI_ISL_2890085, EPI_ISL_2890091, EPI_ISL_2890095, EPI_ISL_2890096, EPI_ISL_2890097, EPI_ISL_2890098, EPI_ISL_2890099, EPI_ISL_2890100, EPI_ISL_2890101, EPI_ISL_2890102, EPI_ISL_2890106, EPI_ISL_2890107, EPI_ISL_2890108, EPI_ISL_2890109, EPI_ISL_2890110, EPI_ISL_2890111, EPI_ISL_2890112, EPI_ISL_2890113, EPI_ISL_2890114, EPI_ISL_2890115, EPI_ISL_2890116, EPI_ISL_2890117, EPI_ISL_2890118, EPI_ISL_2890119, EPI_ISL_2890120, EPI_ISL_2890121, EPI_ISL_2890123, EPI_ISL_2890124, EPI_ISL_2890125, EPI_ISL_2890126, EPI_ISL_2890128, EPI_ISL_2890134, EPI_ISL_2890135, EPI_ISL_2890136, EPI_ISL_2890138, EPI_ISL_2890139, EPI_ISL_2890140, EPI_ISL_2890141, EPI_ISL_2890142, EPI_ISL_2890143, EPI_ISL_2890145, EPI_ISL_2890146, EPI_ISL_2890149, EPI_ISL_2890154, EPI_ISL_2890155, EPI_ISL_2890156, EPI_ISL_2890157, EPI_ISL_2890159, EPI_ISL_2890160, EPI_ISL_2890161, EPI_ISL_2890162, EPI_ISL_2890163, EPI_ISL_2890164, EPI_ISL_2890165, EPI_ISL_2890166, EPI_ISL_2890167, EPI_ISL_2890168, EPI_ISL_2890169, EPI_ISL_2890170, EPI_ISL_2890173, EPI_ISL_2890180, EPI_ISL_2890181, EPI_ISL_2890182, EPI_ISL_2890185, EPI_ISL_2890186, EPI_ISL_2890187, EPI_ISL_2890188, EPI_ISL_2890189, EPI_ISL_2890190, EPI_ISL_2890191, EPI_ISL_2890192, EPI_ISL_2890193, EPI_ISL_2890194, EPI_ISL_2890195, EPI_ISL_2890196, EPI_ISL_2890197, EPI_ISL_2890198, EPI_ISL_2890199, EPI_ISL_2890200, EPI_ISL_2890201, EPI_ISL_2890205, EPI_ISL_2890206, EPI_ISL_2890207, EPI_ISL_2890208, EPI_ISL_2890209, EPI_ISL_2890210, EPI_ISL_2890211, EPI_ISL_2890212, EPI_ISL_2890213, EPI_ISL_2890214, EPI_ISL_2890215, EPI_ISL_2890216, EPI_ISL_2890217, EPI_ISL_2890218, EPI_ISL_2890219, EPI_ISL_2890220, EPI_ISL_2890221, EPI_ISL_2890222, EPI_ISL_2890227, EPI_ISL_2890228, EPI_ISL_2890229, EPI_ISL_2890230, EPI_ISL_2890231, EPI_ISL_2890232, EPI_ISL_2890233, EPI_ISL_2890234, EPI_ISL_2890235, EPI_ISL_2890236, EPI_ISL_2890237, EPI_ISL_2890241, EPI_ISL_2890243, EPI_ISL_2890244, EPI_ISL_2890245, EPI_ISL_2890249, EPI_ISL_2890250, EPI_ISL_2890251, EPI_ISL_2890252, EPI_ISL_2890253, EPI_ISL_2890254, EPI_ISL_2890255, EPI_ISL_2890256, EPI_ISL_2890257, EPI_ISL_2890259, EPI_ISL_2890260, EPI_ISL_2890261, EPI_ISL_2890262, EPI_ISL_2890263, EPI_ISL_2890264, EPI_ISL_2890265, EPI_ISL_2890266, EPI_ISL_2890272, EPI_ISL_2890273, EPI_ISL_2890274, EPI_ISL_2890275, EPI_ISL_2890276, EPI_ISL_2890277, EPI_ISL_2890278, EPI_ISL_2890280, EPI_ISL_2890281, EPI_ISL_2890282, EPI_ISL_2890289, EPI_ISL_2890290, EPI_ISL_2890291, EPI_ISL_2890292, EPI_ISL_2890293, EPI_ISL_2890297, EPI_ISL_2890298, EPI_ISL_2890299, EPI_ISL_2890300, EPI_ISL_2890301, EPI_ISL_2890302, EPI_ISL_2890303, EPI_ISL_2890304, EPI_ISL_2890306, EPI_ISL_2890307, EPI_ISL_2890308, EPI_ISL_2890309, EPI_ISL_2890310, EPI_ISL_2890311, EPI_ISL_2890312, EPI_ISL_2890313, EPI_ISL_2890314, EPI_ISL_2890315, EPI_ISL_2890316, EPI_ISL_2890317, EPI_ISL_2890319, EPI_ISL_2890324, EPI_ISL_2890330, EPI_ISL_2890331, EPI_ISL_2890332, EPI_ISL_2890333, EPI_ISL_2890335, EPI_ISL_2890336, EPI_ISL_2890337, EPI_ISL_2890339, EPI_ISL_2890340, EPI_ISL_2890341, EPI_ISL_2890342, EPI_ISL_2890346, EPI_ISL_2890347, EPI_ISL_2890348, EPI_ISL_2890349, EPI_ISL_2890351, EPI_ISL_2890353, EPI_ISL_2890354, EPI_ISL_2890365, EPI_ISL_2890366, EPI_ISL_2890367, EPI_ISL_2890368, EPI_ISL_2890371, EPI_ISL_2890373, EPI_ISL_2890374, EPI_ISL_2890375, EPI_ISL_2890376, EPI_ISL_2890377, EPI_ISL_2890378, EPI_ISL_2890379, EPI_ISL_2890380, EPI_ISL_2890381, EPI_ISL_2890382, EPI_ISL_2890383, EPI_ISL_2890384, EPI_ISL_2890385, EPI_ISL_2890386, EPI_ISL_2890387, EPI_ISL_2890388, EPI_ISL_2890389, EPI_ISL_2890390, EPI_ISL_2890391, EPI_ISL_2890393, EPI_ISL_2890394, EPI_ISL_2890395, EPI_ISL_2890396, EPI_ISL_2890397, EPI_ISL_2890398, EPI_ISL_2890399, EPI_ISL_2890400, EPI_ISL_2890404, EPI_ISL_2890406, EPI_ISL_2890408, EPI_ISL_2890409, EPI_ISL_2890410, EPI_ISL_2890411, EPI_ISL_2890412, EPI_ISL_2890413, EPI_ISL_2890414, EPI_ISL_2890415, EPI_ISL_2890416, EPI_ISL_2890417, EPI_ISL_2890418, EPI_ISL_2890419, EPI_ISL_2890420, EPI_ISL_2890421, EPI_ISL_2890422, EPI_ISL_2890423, EPI_ISL_2890424, EPI_ISL_2890425, EPI_ISL_2890427, EPI_ISL_2890428, EPI_ISL_2890429, EPI_ISL_2890430, EPI_ISL_2890431, EPI_ISL_2890433, EPI_ISL_2890435, EPI_ISL_2890436, EPI_ISL_2890443, EPI_ISL_2890444, EPI_ISL_2890446, EPI_ISL_2890447, EPI_ISL_2890448, EPI_ISL_2890449, EPI_ISL_2890452, EPI_ISL_2890453, EPI_ISL_2890454, EPI_ISL_2890455, EPI_ISL_2890456, EPI_ISL_2890457, EPI_ISL_2890458, EPI_ISL_2890459, EPI_ISL_2890460, EPI_ISL_2890461, EPI_ISL_2890462, EPI_ISL_2890463, EPI_ISL_2890464, EPI_ISL_2890465, EPI_ISL_2890466, EPI_ISL_2890467, EPI_ISL_2890468, EPI_ISL_2890469, EPI_ISL_2890470, EPI_ISL_2890471, EPI_ISL_2890472, EPI_ISL_2890473, EPI_ISL_2890477, EPI_ISL_2890478, EPI_ISL_2890482, EPI_ISL_2890484, EPI_ISL_2890485, EPI_ISL_2890488, EPI_ISL_2890495, EPI_ISL_2890496, EPI_ISL_2890497, EPI_ISL_2890498, EPI_ISL_2890499, EPI_ISL_2890500, EPI_ISL_2890501, EPI_ISL_2890503, EPI_ISL_2890504, EPI_ISL_2890505, EPI_ISL_2890511, EPI_ISL_2890513, EPI_ISL_2890514, EPI_ISL_2890515, EPI_ISL_2890518, EPI_ISL_2890525, EPI_ISL_2890526, EPI_ISL_2890531, EPI_ISL_2890532, EPI_ISL_2890533, EPI_ISL_2890535, EPI_ISL_2890536, EPI_ISL_2890546, EPI_ISL_2890547, EPI_ISL_2890548, EPI_ISL_2890549, EPI_ISL_2890550, EPI_ISL_2890551, EPI_ISL_2890552, EPI_ISL_2890553, EPI_ISL_2890557, EPI_ISL_2890558, EPI_ISL_2890559, EPI_ISL_2890560, EPI_ISL_2890561, EPI_ISL_2890574, EPI_ISL_2890575, EPI_ISL_2890576, EPI_ISL_2890577, EPI_ISL_2890582, EPI_ISL_2890583, EPI_ISL_2890584, EPI_ISL_2890585, EPI_ISL_2890588, EPI_ISL_2890589, EPI_ISL_2890599, EPI_ISL_2890600, EPI_ISL_2890601, EPI_ISL_2890602, EPI_ISL_2890603, EPI_ISL_2890604, EPI_ISL_2890605, EPI_ISL_2890606, EPI_ISL_2890607, EPI_ISL_2890608, EPI_ISL_2890609, EPI_ISL_2890611, EPI_ISL_2890616, EPI_ISL_2890617, EPI_ISL_2890618, EPI_ISL_2890619, EPI_ISL_2890620, EPI_ISL_2890621, EPI_ISL_2890622, EPI_ISL_2890623, EPI_ISL_2890624, EPI_ISL_2890625, EPI_ISL_2890626, EPI_ISL_2890627, EPI_ISL_2890628, EPI_ISL_2890629, EPI_ISL_2890630, EPI_ISL_2890631, EPI_ISL_2890633, EPI_ISL_2890638, EPI_ISL_2890639, EPI_ISL_2890641, EPI_ISL_2890642, EPI_ISL_2890645, EPI_ISL_2890646, EPI_ISL_2890647, EPI_ISL_2890648, EPI_ISL_2890649, EPI_ISL_2890650, EPI_ISL_2890651, EPI_ISL_2890652, EPI_ISL_2890653, EPI_ISL_2890660, EPI_ISL_2890661, EPI_ISL_2890664, EPI_ISL_2890665, EPI_ISL_2890668, EPI_ISL_2890669, EPI_ISL_2890677, EPI_ISL_2890678, EPI_ISL_2890679, EPI_ISL_2890680, EPI_ISL_2890681, EPI_ISL_2890682, EPI_ISL_2890683, EPI_ISL_2890684, EPI_ISL_2890685, EPI_ISL_2890687, EPI_ISL_2890688, EPI_ISL_2890689, EPI_ISL_2890690, EPI_ISL_2890691, EPI_ISL_2890692, EPI_ISL_2890693, EPI_ISL_2890694, EPI_ISL_2890695, EPI_ISL_2890696, EPI_ISL_2890697, EPI_ISL_2890700, EPI_ISL_2890701, EPI_ISL_2890703, EPI_ISL_2890704, EPI_ISL_2890705, EPI_ISL_2890706, EPI_ISL_2890707, EPI_ISL_2890708, EPI_ISL_2890709, EPI_ISL_2890710, EPI_ISL_2890711, EPI_ISL_2890713, EPI_ISL_2890714, EPI_ISL_2890715, EPI_ISL_2890716, EPI_ISL_2890719, EPI_ISL_2890720, EPI_ISL_2890721, EPI_ISL_2890722, EPI_ISL_2890723, EPI_ISL_2890724, EPI_ISL_2890730, EPI_ISL_2890734, EPI_ISL_2890735, EPI_ISL_2890736, EPI_ISL_2890737, EPI_ISL_2890738, EPI_ISL_2890739, EPI_ISL_2890740, EPI_ISL_2890741, EPI_ISL_2890742, EPI_ISL_2890743, EPI_ISL_2890744, EPI_ISL_2890745, EPI_ISL_2890746, EPI_ISL_2890747, EPI_ISL_2890748, EPI_ISL_2890749, EPI_ISL_2890750, EPI_ISL_2890751, EPI_ISL_2890752, EPI_ISL_2890755, EPI_ISL_2890756, EPI_ISL_2890757, EPI_ISL_2890758, EPI_ISL_2890759, EPI_ISL_2890760, EPI_ISL_2890761, EPI_ISL_2890762, EPI_ISL_2890763, EPI_ISL_2890764, EPI_ISL_2890765, EPI_ISL_2890766, EPI_ISL_2890767, EPI_ISL_2890768, EPI_ISL_2890769, EPI_ISL_2890770, EPI_ISL_2890771, EPI_ISL_2890772, EPI_ISL_2890773, EPI_ISL_2890774, EPI_ISL_2890775, EPI_ISL_2890776, EPI_ISL_2890777, EPI_ISL_2890778, EPI_ISL_2890779, EPI_ISL_2890780, EPI_ISL_2890781, EPI_ISL_2890782, EPI_ISL_2890783, EPI_ISL_2890785, EPI_ISL_2890786, EPI_ISL_2890787, EPI_ISL_2890788, EPI_ISL_2890789, EPI_ISL_2890790, EPI_ISL_2890791, EPI_ISL_2890792, EPI_ISL_2890794, EPI_ISL_2890795, EPI_ISL_2890796, EPI_ISL_2890797, EPI_ISL_2890798, EPI_ISL_2890799, EPI_ISL_2890800, EPI_ISL_2890801, EPI_ISL_2890802, EPI_ISL_2890803, EPI_ISL_2890807, EPI_ISL_2890808, EPI_ISL_2890811, EPI_ISL_2890820, EPI_ISL_2890821, EPI_ISL_2890822, EPI_ISL_2890823, EPI_ISL_2890824, EPI_ISL_2890825, EPI_ISL_2890826, EPI_ISL_2890827, EPI_ISL_2890828, EPI_ISL_2890829, EPI_ISL_2890831, EPI_ISL_2890832, EPI_ISL_2890833, EPI_ISL_2890834, EPI_ISL_2890835, EPI_ISL_2890836, EPI_ISL_2890837, EPI_ISL_2890838, EPI_ISL_2890839, EPI_ISL_2890855, EPI_ISL_2890856, EPI_ISL_2890857, EPI_ISL_2890858, EPI_ISL_2890859, EPI_ISL_2890860, EPI_ISL_2890861, EPI_ISL_2890862, EPI_ISL_2890863, EPI_ISL_2890864, EPI_ISL_2890865, EPI_ISL_2890866, EPI_ISL_2890867, EPI_ISL_2890868, EPI_ISL_2890869, EPI_ISL_2890870, EPI_ISL_2890871, EPI_ISL_2890872, EPI_ISL_2890873, EPI_ISL_2890878, EPI_ISL_2890879, EPI_ISL_2890880, EPI_ISL_2890881, EPI_ISL_2890882, EPI_ISL_2890883, EPI_ISL_2890884, EPI_ISL_2890886, EPI_ISL_2890887, EPI_ISL_2890888, EPI_ISL_2890889, EPI_ISL_2890890, EPI_ISL_2890891, EPI_ISL_2890899, EPI_ISL_2890900, EPI_ISL_2890901, EPI_ISL_2890902, EPI_ISL_2890903, EPI_ISL_2890904, EPI_ISL_2890905, EPI_ISL_2890906, EPI_ISL_2890907, EPI_ISL_2890908, EPI_ISL_2890909, EPI_ISL_2890910, EPI_ISL_2890911, EPI_ISL_2890912, EPI_ISL_2890913, EPI_ISL_2890914, EPI_ISL_2890915, EPI_ISL_2890923, EPI_ISL_2890928, EPI_ISL_2890930, EPI_ISL_2890931, EPI_ISL_2890932, EPI_ISL_2890933, EPI_ISL_2890934, EPI_ISL_2890941, EPI_ISL_2890942, EPI_ISL_2890945, EPI_ISL_2890946, EPI_ISL_2890947, EPI_ISL_2890948, EPI_ISL_2890949, EPI_ISL_2890950, EPI_ISL_2890951, EPI_ISL_2890952, EPI_ISL_2890953, EPI_ISL_2890968, EPI_ISL_2890969, EPI_ISL_2890970, EPI_ISL_2890972, EPI_ISL_2890974, EPI_ISL_2890976, EPI_ISL_2890977, EPI_ISL_2890978, EPI_ISL_2890979, EPI_ISL_2890982, EPI_ISL_2890983, EPI_ISL_2890984, EPI_ISL_2890986, EPI_ISL_2890987, EPI_ISL_2890989, EPI_ISL_2890990, EPI_ISL_2890993, EPI_ISL_2890994, EPI_ISL_2890996, EPI_ISL_2890997, EPI_ISL_2890998, EPI_ISL_2890999, EPI_ISL_2891000, EPI_ISL_2891001, EPI_ISL_2891002, EPI_ISL_2891003, EPI_ISL_2891004, EPI_ISL_2891005, EPI_ISL_2891006, EPI_ISL_2891007, EPI_ISL_2891008, EPI_ISL_2891010, EPI_ISL_2891011, EPI_ISL_2891020, EPI_ISL_2891021, EPI_ISL_2891022, EPI_ISL_2891023, EPI_ISL_2891024, EPI_ISL_2891025, EPI_ISL_2891032, EPI_ISL_2891033, EPI_ISL_2891034, EPI_ISL_2891035, EPI_ISL_2891042, EPI_ISL_2891043, EPI_ISL_2891045, EPI_ISL_2891047, EPI_ISL_2891050, EPI_ISL_2891051, EPI_ISL_2891052, EPI_ISL_2891053, EPI_ISL_2891054, EPI_ISL_2891055, EPI_ISL_2891056, EPI_ISL_2891057, EPI_ISL_2891060, EPI_ISL_2891061, EPI_ISL_2891062, EPI_ISL_2891063, EPI_ISL_2891064, EPI_ISL_2891065, EPI_ISL_2891066, EPI_ISL_2891067, EPI_ISL_2891068, EPI_ISL_2891069, EPI_ISL_2891070, EPI_ISL_2891071, EPI_ISL_2891072, EPI_ISL_2891073, EPI_ISL_2891078, EPI_ISL_2891085, EPI_ISL_2891086, EPI_ISL_2891087, EPI_ISL_2891088, EPI_ISL_2891090, EPI_ISL_2891091, EPI_ISL_2891092, EPI_ISL_2891093, EPI_ISL_2891094, EPI_ISL_2891096, EPI_ISL_2891097, EPI_ISL_2891098, EPI_ISL_2891099, EPI_ISL_2891100, EPI_ISL_2891101, EPI_ISL_2891102, EPI_ISL_2891103, EPI_ISL_2891104, EPI_ISL_2891105, EPI_ISL_2891106, EPI_ISL_2891107, EPI_ISL_2891108, EPI_ISL_2891109, EPI_ISL_2891112, EPI_ISL_2891113, EPI_ISL_2891114, EPI_ISL_2891115, EPI_ISL_2891116, EPI_ISL_2891117, EPI_ISL_2891118, EPI_ISL_2891119, EPI_ISL_2891120, EPI_ISL_2891121, EPI_ISL_2891122, EPI_ISL_2891123, EPI_ISL_2891124, EPI_ISL_2891125, EPI_ISL_2891126, EPI_ISL_2891127, EPI_ISL_2891128, EPI_ISL_2891129, EPI_ISL_2891130, EPI_ISL_2891131, EPI_ISL_2891132, EPI_ISL_2891133, EPI_ISL_2891136, EPI_ISL_2891144, EPI_ISL_2891148, EPI_ISL_2891150, EPI_ISL_2891151, EPI_ISL_2891152, EPI_ISL_2891154, EPI_ISL_2891155, EPI_ISL_2891156, EPI_ISL_2891157, EPI_ISL_2891159, EPI_ISL_2891160, EPI_ISL_2891161, EPI_ISL_2891162, EPI_ISL_2891163, EPI_ISL_2891164, EPI_ISL_2891165, EPI_ISL_2891166, EPI_ISL_2891167, EPI_ISL_2891168, EPI_ISL_2891169, EPI_ISL_2891170, EPI_ISL_2891171, EPI_ISL_2891172, EPI_ISL_2891173, EPI_ISL_2891174, EPI_ISL_2891180, EPI_ISL_2891181, EPI_ISL_2891182, EPI_ISL_2891183, EPI_ISL_2891184, EPI_ISL_2891186, EPI_ISL_2891187, EPI_ISL_2891188, EPI_ISL_2891189, EPI_ISL_2891198, EPI_ISL_2891211, EPI_ISL_2891212, EPI_ISL_2891213, EPI_ISL_2891214, EPI_ISL_2891215, EPI_ISL_2891216, EPI_ISL_2891217, EPI_ISL_2891218, EPI_ISL_2891219, EPI_ISL_2891220, EPI_ISL_2891221, EPI_ISL_2891222, EPI_ISL_2891223, EPI_ISL_2891230, EPI_ISL_2891231, EPI_ISL_2891232, EPI_ISL_2891235, EPI_ISL_2891236, EPI_ISL_2891237, EPI_ISL_2891238, EPI_ISL_2891239, EPI_ISL_2891240, EPI_ISL_2891241, EPI_ISL_2891242, EPI_ISL_2891243, EPI_ISL_2891244, EPI_ISL_2891245, EPI_ISL_2891252, EPI_ISL_2891254, EPI_ISL_2891258, EPI_ISL_2891259, EPI_ISL_2891260, EPI_ISL_2891261, EPI_ISL_2891262, EPI_ISL_2891263, EPI_ISL_2891264, EPI_ISL_2891265, EPI_ISL_2891266, EPI_ISL_2891268, EPI_ISL_2891269, EPI_ISL_2891270, EPI_ISL_2891271, EPI_ISL_2891272, EPI_ISL_2891273, EPI_ISL_2891279, EPI_ISL_2891280, EPI_ISL_2891283, EPI_ISL_2891284, EPI_ISL_2891285, EPI_ISL_2891286, EPI_ISL_2891288, EPI_ISL_2891289, EPI_ISL_2891290, EPI_ISL_2891291, EPI_ISL_2891292, EPI_ISL_2891293, EPI_ISL_2891294, EPI_ISL_2891297, EPI_ISL_2891298, EPI_ISL_2891310, EPI_ISL_2891312, EPI_ISL_2891314, EPI_ISL_2891315, EPI_ISL_2891316, EPI_ISL_2891317, EPI_ISL_2891318, EPI_ISL_2891319, EPI_ISL_2891332, EPI_ISL_2891333, EPI_ISL_2891334, EPI_ISL_2891335, EPI_ISL_2891336, EPI_ISL_2891348, EPI_ISL_2891349, EPI_ISL_2891350, EPI_ISL_2891356, EPI_ISL_2891357, EPI_ISL_2891358, EPI_ISL_2891359, EPI_ISL_2891360, EPI_ISL_2891361, EPI_ISL_2891363, EPI_ISL_2891364, EPI_ISL_2891365, EPI_ISL_2891366, EPI_ISL_2891367, EPI_ISL_2891368, EPI_ISL_2891369, EPI_ISL_2891370, EPI_ISL_2891372, EPI_ISL_2891373, EPI_ISL_2891374, EPI_ISL_2891375, EPI_ISL_2891376, EPI_ISL_2891377, EPI_ISL_2891379, EPI_ISL_2891380, EPI_ISL_2891381, EPI_ISL_2891382, EPI_ISL_2891383, EPI_ISL_2891384, EPI_ISL_2891385, EPI_ISL_2891386, EPI_ISL_2891387, EPI_ISL_2891388, EPI_ISL_2891389, EPI_ISL_2891390, EPI_ISL_2891391, EPI_ISL_2891392, EPI_ISL_2891393, EPI_ISL_2891394, EPI_ISL_2891395, EPI_ISL_2891396, EPI_ISL_2891397, EPI_ISL_2891398, EPI_ISL_2891399, EPI_ISL_2891400, EPI_ISL_2891401, EPI_ISL_2891402, EPI_ISL_2891408, EPI_ISL_2891409, EPI_ISL_2891410, EPI_ISL_2891411, EPI_ISL_2891413, EPI_ISL_2891417, EPI_ISL_2891418, EPI_ISL_2891421, EPI_ISL_2891422, EPI_ISL_2891431, EPI_ISL_2891434, EPI_ISL_2891435, EPI_ISL_2891436, EPI_ISL_2891437, EPI_ISL_2891438, EPI_ISL_2891439, EPI_ISL_2891440, EPI_ISL_2891441, EPI_ISL_2891442, EPI_ISL_2891443, EPI_ISL_2891444, EPI_ISL_2891445, EPI_ISL_2891446, EPI_ISL_2891447, EPI_ISL_2891448, EPI_ISL_2891449, EPI_ISL_2891450, EPI_ISL_2891451, EPI_ISL_2891453, EPI_ISL_2891454, EPI_ISL_2891455, EPI_ISL_2891456, EPI_ISL_2891457, EPI_ISL_2891458, EPI_ISL_2891459, EPI_ISL_2891463, EPI_ISL_2891464, EPI_ISL_2891465, EPI_ISL_2891467, EPI_ISL_2891468, EPI_ISL_2891469, EPI_ISL_2891470, EPI_ISL_2891471, EPI_ISL_2891472, EPI_ISL_2891473, EPI_ISL_2891474, EPI_ISL_2891475, EPI_ISL_2891476, EPI_ISL_2891477, EPI_ISL_2891478, EPI_ISL_2891479, EPI_ISL_2891480, EPI_ISL_2891482, EPI_ISL_2891483, EPI_ISL_2891484, EPI_ISL_2891485, EPI_ISL_2891486, EPI_ISL_2891487, EPI_ISL_2891488, EPI_ISL_2891489, EPI_ISL_2891490, EPI_ISL_2891491, EPI_ISL_2891492, EPI_ISL_2891495, EPI_ISL_2891496, EPI_ISL_2891497, EPI_ISL_2891498, EPI_ISL_2891499, EPI_ISL_2891500, EPI_ISL_2891501, EPI_ISL_2891502, EPI_ISL_2891503, EPI_ISL_2891504, EPI_ISL_2891505, EPI_ISL_2891509, EPI_ISL_2891511, EPI_ISL_2891515, EPI_ISL_2891516, EPI_ISL_2891517, EPI_ISL_2891518, EPI_ISL_2891520, EPI_ISL_2891521, EPI_ISL_2891522, EPI_ISL_2891523, EPI_ISL_2891529, EPI_ISL_2891530, EPI_ISL_2891535, EPI_ISL_2891539, EPI_ISL_2891540, EPI_ISL_2891541, EPI_ISL_2891542, EPI_ISL_2891543, EPI_ISL_2891544, EPI_ISL_2891545, EPI_ISL_2891553, EPI_ISL_2891554, EPI_ISL_2891555, EPI_ISL_2891556, EPI_ISL_2891557, EPI_ISL_2891558, EPI_ISL_2891559, EPI_ISL_2891560, EPI_ISL_2891561, EPI_ISL_2891562, EPI_ISL_2891563, EPI_ISL_2891564, EPI_ISL_2891566, EPI_ISL_2891567, EPI_ISL_2891568, EPI_ISL_2891569, EPI_ISL_2891570, EPI_ISL_2891571, EPI_ISL_2891573, EPI_ISL_2891575, EPI_ISL_2891576, EPI_ISL_2891577, EPI_ISL_2891578, EPI_ISL_2891579, EPI_ISL_2891580, EPI_ISL_2891581, EPI_ISL_2891582, EPI_ISL_2891584, EPI_ISL_2891585, EPI_ISL_2891586, EPI_ISL_2891587, EPI_ISL_2891588, EPI_ISL_2891589, EPI_ISL_2891591, EPI_ISL_2891592, EPI_ISL_2891593, EPI_ISL_2891594, EPI_ISL_2891605, EPI_ISL_2891607, EPI_ISL_2891609, EPI_ISL_2891610, EPI_ISL_2891611, EPI_ISL_2891612, EPI_ISL_2891613, EPI_ISL_2891614, EPI_ISL_2891615, EPI_ISL_2891619, EPI_ISL_2891620, EPI_ISL_2891622, EPI_ISL_2891623, EPI_ISL_2891632, EPI_ISL_2891633, EPI_ISL_2891634, EPI_ISL_2891635, EPI_ISL_2891637, EPI_ISL_2891640, EPI_ISL_2891643, EPI_ISL_2891644, EPI_ISL_2891645, EPI_ISL_2891646, EPI_ISL_2891647, EPI_ISL_2891648, EPI_ISL_2891649, EPI_ISL_2891650, EPI_ISL_2891651, EPI_ISL_2891652, EPI_ISL_2891653, EPI_ISL_2891654, EPI_ISL_2891655, EPI_ISL_2891656, EPI_ISL_2891657, EPI_ISL_2891658, EPI_ISL_2891659, EPI_ISL_2891660, EPI_ISL_2891661, EPI_ISL_2891662, EPI_ISL_2891663, EPI_ISL_2891664, EPI_ISL_2891665, EPI_ISL_2891666, EPI_ISL_2891667, EPI_ISL_2891668, EPI_ISL_2891670, EPI_ISL_2891671, EPI_ISL_2891672, EPI_ISL_2891673, EPI_ISL_2891674, EPI_ISL_2891676, EPI_ISL_2891680, EPI_ISL_2891681, EPI_ISL_2891682, EPI_ISL_2891683, EPI_ISL_2891684, EPI_ISL_2891685, EPI_ISL_2891686, EPI_ISL_2891687, EPI_ISL_2891688, EPI_ISL_2891689, EPI_ISL_2891690, EPI_ISL_2891695, EPI_ISL_2891696, EPI_ISL_2891698, EPI_ISL_2891699, EPI_ISL_2891700, EPI_ISL_2891701, EPI_ISL_2891703, EPI_ISL_2891704, EPI_ISL_2891706, EPI_ISL_2892369, EPI_ISL_2892370, EPI_ISL_2892377, EPI_ISL_2892379, EPI_ISL_2892383, EPI_ISL_2892384, EPI_ISL_2892386, EPI_ISL_2892388, EPI_ISL_2892389, EPI_ISL_2892390, EPI_ISL_2892391, EPI_ISL_2892392, EPI_ISL_2892397, EPI_ISL_2892398, EPI_ISL_2892401, EPI_ISL_2892402, EPI_ISL_2921685, EPI_ISL_2921720, EPI_ISL_2921721, EPI_ISL_2921745, EPI_ISL_2921756, EPI_ISL_2921757, EPI_ISL_2921758, EPI_ISL_2921759, EPI_ISL_2921760, EPI_ISL_2921761, EPI_ISL_2921779, EPI_ISL_2921780, EPI_ISL_2921781, EPI_ISL_2921787, EPI_ISL_2921788, EPI_ISL_2921789, EPI_ISL_2921791, EPI_ISL_2921794, EPI_ISL_2921814, EPI_ISL_2921815, EPI_ISL_2921817, EPI_ISL_2921818, EPI_ISL_2921825, EPI_ISL_2921826, EPI_ISL_2923290, EPI_ISL_2923297, EPI_ISL_2923298, EPI_ISL_2923299, EPI_ISL_2923305, EPI_ISL_2923324, EPI_ISL_2923345, EPI_ISL_2923351, EPI_ISL_2923352, EPI_ISL_2923381, EPI_ISL_2923382, EPI_ISL_2923383, EPI_ISL_2923398, EPI_ISL_2923399, EPI_ISL_2923416, EPI_ISL_2923417, EPI_ISL_2923418, EPI_ISL_2923419, EPI_ISL_2923420, EPI_ISL_2923421, EPI_ISL_2923422, EPI_ISL_2923423, EPI_ISL_2923424, EPI_ISL_2923425, EPI_ISL_2923426, EPI_ISL_2923427, EPI_ISL_2923428, EPI_ISL_2923429, EPI_ISL_2923445, EPI_ISL_2923448, EPI_ISL_2923452, EPI_ISL_2923453, EPI_ISL_2923463, EPI_ISL_2923471, EPI_ISL_2923490, EPI_ISL_2923501, EPI_ISL_2923539, EPI_ISL_2923545, EPI_ISL_2923559, EPI_ISL_2923583, EPI_ISL_2923584, EPI_ISL_2923593, EPI_ISL_2923594, EPI_ISL_2923595, EPI_ISL_2923596, EPI_ISL_2923599, EPI_ISL_2923601, EPI_ISL_2923613, EPI_ISL_2923618, EPI_ISL_2923621, EPI_ISL_2923626, EPI_ISL_2923627, EPI_ISL_2923628, EPI_ISL_2923629, EPI_ISL_2923630, EPI_ISL_2923640, EPI_ISL_2923645, EPI_ISL_2923646, EPI_ISL_2923647, EPI_ISL_2923675, EPI_ISL_2923677, EPI_ISL_2923678, EPI_ISL_2923687, EPI_ISL_2923692, EPI_ISL_2923694, EPI_ISL_2923695, EPI_ISL_2923696, EPI_ISL_2923697, EPI_ISL_2923698, EPI_ISL_2923699, EPI_ISL_2923700, EPI_ISL_2923701, EPI_ISL_2923702, EPI_ISL_2923704, EPI_ISL_2923705, EPI_ISL_2923706, EPI_ISL_2923715, EPI_ISL_2923720, EPI_ISL_2923721, EPI_ISL_2923723, EPI_ISL_2923724, EPI_ISL_2923725, EPI_ISL_2923726, EPI_ISL_2923733, EPI_ISL_2923734, EPI_ISL_2923735, EPI_ISL_2923736, EPI_ISL_2923737, EPI_ISL_2923750, EPI_ISL_2923751, EPI_ISL_2923752, EPI_ISL_2923753, EPI_ISL_2923754, EPI_ISL_2923755, EPI_ISL_2923757, EPI_ISL_2923758, EPI_ISL_2923773, EPI_ISL_2923774, EPI_ISL_2934094, EPI_ISL_2934095, EPI_ISL_2934099, EPI_ISL_2934102, EPI_ISL_2934110, EPI_ISL_2934112, EPI_ISL_2934114, EPI_ISL_2934119, EPI_ISL_2934120, EPI_ISL_2934122, EPI_ISL_2934131, EPI_ISL_2934137, EPI_ISL_2934138, EPI_ISL_2934141, EPI_ISL_2934142, EPI_ISL_2934145, EPI_ISL_2934157, EPI_ISL_2934163, EPI_ISL_2934168, EPI_ISL_2934172, EPI_ISL_2934174, EPI_ISL_2934175, EPI_ISL_2934177, EPI_ISL_2934180, EPI_ISL_2934182, EPI_ISL_2934187, EPI_ISL_2934190, EPI_ISL_2934194, EPI_ISL_2934198, EPI_ISL_2934207, EPI_ISL_2934214, EPI_ISL_2934218, EPI_ISL_2934220, EPI_ISL_2934223, EPI_ISL_2934226, EPI_ISL_2934228, EPI_ISL_2934231, EPI_ISL_2934234, EPI_ISL_2934235, EPI_ISL_2934238, EPI_ISL_2934243, EPI_ISL_2934251, EPI_ISL_2934261, EPI_ISL_2934262, EPI_ISL_2934266, EPI_ISL_2934277, EPI_ISL_2934285, EPI_ISL_2934287, EPI_ISL_2934292, EPI_ISL_2934297, EPI_ISL_2934301, EPI_ISL_2934303, EPI_ISL_2934310, EPI_ISL_2934313, EPI_ISL_2934319, EPI_ISL_2934328, EPI_ISL_2934329, EPI_ISL_2934339, EPI_ISL_2934340, EPI_ISL_2934343, EPI_ISL_2934349, EPI_ISL_2934356, EPI_ISL_2934357, EPI_ISL_2934359, EPI_ISL_2934360, EPI_ISL_2934370, EPI_ISL_2934371, EPI_ISL_2934376, EPI_ISL_2934382, EPI_ISL_2934386, EPI_ISL_2934387, EPI_ISL_2934394, EPI_ISL_2934395, EPI_ISL_2934396, EPI_ISL_2934405, EPI_ISL_2934409, EPI_ISL_2934417, EPI_ISL_2934418, EPI_ISL_2934419, EPI_ISL_2934422, EPI_ISL_2934427, EPI_ISL_2934429, EPI_ISL_2934433, EPI_ISL_2934435, EPI_ISL_2934444, EPI_ISL_2934446, EPI_ISL_2934457, EPI_ISL_2934460, EPI_ISL_2934463, EPI_ISL_2934471, EPI_ISL_2934476, EPI_ISL_2934477, EPI_ISL_2934485, EPI_ISL_2934487, EPI_ISL_2934488, EPI_ISL_2934492, EPI_ISL_2934493, EPI_ISL_2934495, EPI_ISL_2934500, EPI_ISL_2934501, EPI_ISL_2934505, EPI_ISL_2934507, EPI_ISL_2934509, EPI_ISL_2934510, EPI_ISL_2934515, EPI_ISL_2934516, EPI_ISL_2934521, EPI_ISL_2934524, EPI_ISL_2934525, EPI_ISL_2934528, EPI_ISL_2934530, EPI_ISL_2934537, EPI_ISL_2934539, EPI_ISL_2934541, EPI_ISL_2934542, EPI_ISL_2934546, EPI_ISL_2934548, EPI_ISL_2934549, EPI_ISL_2934553, EPI_ISL_2934554, EPI_ISL_2934555, EPI_ISL_2934556, EPI_ISL_2934558, EPI_ISL_2934560, EPI_ISL_2934561, EPI_ISL_2934563, EPI_ISL_2934568 | | | |
| see above | CDPH VBL | California Department of Public Health | CDPH-COVIDNet; SwabSeq/UCLA; UCLA Technology Center for Genomics & Bioinformatics |
| EPI_ISL_1373737, EPI_ISL_1373739, EPI_ISL_1373751, EPI_ISL_1373776, EPI_ISL_1373778, EPI_ISL_1373780, EPI_ISL_1373786, EPI_ISL_1373787, EPI_ISL_1373793, EPI_ISL_1373801, EPI_ISL_1373804, EPI_ISL_1373813, EPI_ISL_1373816, EPI_ISL_1373828, EPI_ISL_1373837 | | | |
| see above | CDPH, Viral and Rickettsial Disease Laboratory | Genomics and Discovery, Respiratory Viruses Branch, Division of Viral Diseases, Centers for Disease Control and Prevention | Anna Kelleher; Anna Uehara; Brian Lynch; Clinton R. Paden; Haibin Wang; Han Jia Justin Ng; Jing Zhang; Krista Queen; Peter Cook; Suxiang Tong; Yan Li; Ying Tao |
| EPI_ISL_1591240, EPI_ISL_1591241, EPI_ISL_1591242, EPI_ISL_1623836, EPI_ISL_1911349, EPI_ISL_1911350, EPI_ISL_1911351, EPI_ISL_1911352, EPI_ISL_1911353, EPI_ISL_1911354, EPI_ISL_1911355, EPI_ISL_1911356, EPI_ISL_1911357, EPI_ISL_1911358, EPI_ISL_1911359, EPI_ISL_1911360, EPI_ISL_1911361, EPI_ISL_1911366, EPI_ISL_1911367, EPI_ISL_1911368 | | | |
| see above | CDPH, Viral and Rickettsial Disease Laboratory | Pathogen Discovery, Respiratory Viruses Branch, Division of Viral Diseases, Centers for Disease Control and Prevention | Adam Retchless; Anna Kelleher; Anna Montmayeur; Anna Uehara; Brian Lynch; Clinton R. Paden; Dhwani Batra; Haibin Wang; Han Jia Justin Ng; Jasmine Padilla; Jing Zhang; Justin Lee; Krista Queen; Mark Burroughs; Mili Sheth; Morgan Davis; Peter Cook; Rachel Marine; Sarah Nobles; Suxiang Tong; Tara Coalter; Yan Li; Ying Tao |
| EPI_ISL_2234099, EPI_ISL_2234100, EPI_ISL_2234101, EPI_ISL_2234103, EPI_ISL_2234104, EPI_ISL_2234105, EPI_ISL_2234106, EPI_ISL_2234107, EPI_ISL_2234143, EPI_ISL_2234144, EPI_ISL_2234146, EPI_ISL_2234166, EPI_ISL_2234167, EPI_ISL_2234168, EPI_ISL_2234169, EPI_ISL_2234170, EPI_ISL_2234195, EPI_ISL_2234197, EPI_ISL_2234198, EPI_ISL_2234199, EPI_ISL_2234200, EPI_ISL_2234201, EPI_ISL_2234202, EPI_ISL_2234203, EPI_ISL_2234204, EPI_ISL_2234205, EPI_ISL_2234209, EPI_ISL_2234210, EPI_ISL_2234211, EPI_ISL_2234212, EPI_ISL_2234213, EPI_ISL_2234214, EPI_ISL_2234215, EPI_ISL_2234222, EPI_ISL_2234223, EPI_ISL_2234224, EPI_ISL_2234225, EPI_ISL_2234226, EPI_ISL_2234227, EPI_ISL_2234228, EPI_ISL_2234229, EPI_ISL_2234230, EPI_ISL_2234231, EPI_ISL_2234232, EPI_ISL_2234233, EPI_ISL_2234234, EPI_ISL_2234235, EPI_ISL_2234236, EPI_ISL_2234237, EPI_ISL_2234238, EPI_ISL_2234239, EPI_ISL_2234240, EPI_ISL_2234242, EPI_ISL_2234244, EPI_ISL_2234245, EPI_ISL_2234246, EPI_ISL_2234247, EPI_ISL_2234256, EPI_ISL_2234257, EPI_ISL_2234262, EPI_ISL_2234267, EPI_ISL_2234268, EPI_ISL_2234281, EPI_ISL_2234287, EPI_ISL_2234288, EPI_ISL_2234289, EPI_ISL_2234290, EPI_ISL_2234291, EPI_ISL_2234292, EPI_ISL_2234293, EPI_ISL_2234294, EPI_ISL_2537395, EPI_ISL_2537396, EPI_ISL_2537397, EPI_ISL_2537398, EPI_ISL_2537399, EPI_ISL_2537435, EPI_ISL_2537438, EPI_ISL_2537439, EPI_ISL_2537470, EPI_ISL_2892454, EPI_ISL_2892538, EPI_ISL_2892547, EPI_ISL_2892566, EPI_ISL_2892567, EPI_ISL_2892568, EPI_ISL_2892569 | | | |
| see above | California Department of Public Health | California Department of Public Health | CDPH IDLB COVIDNet et al |
| EPI_ISL_2778348, EPI_ISL_2778349, EPI_ISL_2778350, EPI_ISL_2778379, EPI_ISL_2778380, EPI_ISL_2778381, EPI_ISL_2778382, EPI_ISL_2778383, EPI_ISL_2778384, EPI_ISL_2778385, EPI_ISL_2778386, EPI_ISL_2778387, EPI_ISL_2778388, EPI_ISL_2778389, EPI_ISL_2778390, EPI_ISL_2778392, EPI_ISL_2778393, EPI_ISL_2778394, EPI_ISL_2778395, EPI_ISL_2778396, EPI_ISL_2778397, EPI_ISL_2778398, EPI_ISL_2778399, EPI_ISL_2778400, EPI_ISL_2778401, EPI_ISL_2778403, EPI_ISL_2778404, EPI_ISL_2778405, EPI_ISL_2778406, EPI_ISL_2778407, EPI_ISL_2778408, EPI_ISL_2778409, EPI_ISL_2778410, EPI_ISL_2778411, EPI_ISL_2778412, EPI_ISL_2778413, EPI_ISL_2778414, EPI_ISL_2778415, EPI_ISL_2778416, EPI_ISL_2778417, EPI_ISL_2778418, EPI_ISL_2778419, EPI_ISL_2778420, EPI_ISL_2778421, EPI_ISL_2778422, EPI_ISL_2778423, EPI_ISL_2778424, EPI_ISL_2778425, EPI_ISL_2778426, EPI_ISL_2778427, EPI_ISL_2778428, EPI_ISL_2778429, EPI_ISL_2778430, EPI_ISL_2778431, EPI_ISL_2778432, EPI_ISL_2778433, EPI_ISL_2778434, EPI_ISL_2778435, EPI_ISL_2778436, EPI_ISL_2778437, EPI_ISL_2778438, EPI_ISL_2778439, EPI_ISL_2778440, EPI_ISL_2778441, EPI_ISL_2778442, EPI_ISL_2778443, EPI_ISL_2778444, EPI_ISL_2778445, EPI_ISL_2778446, EPI_ISL_2778447, EPI_ISL_2778448, EPI_ISL_2778449, EPI_ISL_2778450, EPI_ISL_2778451, EPI_ISL_2778452, EPI_ISL_2778453, EPI_ISL_2778454, EPI_ISL_2778455, EPI_ISL_2778456, EPI_ISL_2778457, EPI_ISL_2778458, EPI_ISL_2778459, EPI_ISL_2778460, EPI_ISL_2778461, EPI_ISL_2778462, EPI_ISL_2778463, EPI_ISL_2778464, EPI_ISL_2778465, EPI_ISL_2778466, EPI_ISL_2778467, EPI_ISL_2778468, EPI_ISL_2778469, EPI_ISL_2778470, EPI_ISL_2778471, EPI_ISL_2778472, EPI_ISL_2778473, EPI_ISL_2778474, EPI_ISL_2778475, EPI_ISL_2778476, EPI_ISL_2778477, EPI_ISL_2778478, EPI_ISL_2778479, EPI_ISL_2778480, EPI_ISL_2778481, EPI_ISL_2778482, EPI_ISL_2778483, EPI_ISL_2778484, EPI_ISL_2778485, EPI_ISL_2778486, EPI_ISL_2778487, EPI_ISL_2778488, EPI_ISL_2778489, EPI_ISL_2778490, EPI_ISL_2778491, EPI_ISL_2778492, EPI_ISL_2778493, EPI_ISL_2778494, EPI_ISL_2778495, EPI_ISL_2778496, EPI_ISL_2778497, EPI_ISL_2778498, EPI_ISL_2778499, EPI_ISL_2778500, EPI_ISL_2778501, EPI_ISL_2778502, EPI_ISL_2778503, EPI_ISL_2778504, EPI_ISL_2778505, EPI_ISL_2778506, EPI_ISL_2778507, EPI_ISL_2778508, EPI_ISL_2778509, EPI_ISL_2778510, EPI_ISL_2778511, EPI_ISL_2778512, EPI_ISL_2778513, EPI_ISL_2778514, EPI_ISL_2778515, EPI_ISL_2778516, EPI_ISL_2778517, EPI_ISL_2778518, EPI_ISL_2778519, EPI_ISL_2778520, EPI_ISL_2778521, EPI_ISL_2778522, EPI_ISL_2778523, EPI_ISL_2778524, EPI_ISL_2778525, EPI_ISL_2778526, EPI_ISL_2778527, EPI_ISL_2778528, EPI_ISL_2778529, EPI_ISL_2778530, EPI_ISL_2778531, EPI_ISL_2778532, EPI_ISL_2778533, EPI_ISL_2778534, EPI_ISL_2778535, EPI_ISL_2778536, EPI_ISL_2778537, EPI_ISL_2778538, EPI_ISL_2778539, EPI_ISL_2778540, EPI_ISL_2778541, EPI_ISL_2778542, EPI_ISL_2778543, EPI_ISL_2778544, EPI_ISL_2778545, EPI_ISL_2778546, EPI_ISL_2778547, EPI_ISL_2778548, EPI_ISL_2778549, EPI_ISL_2778550, EPI_ISL_2778551, EPI_ISL_2778552, EPI_ISL_2778553, EPI_ISL_2778554, EPI_ISL_2778555, EPI_ISL_2778556, EPI_ISL_2778557, EPI_ISL_2778558, EPI_ISL_2778559, EPI_ISL_2778560, EPI_ISL_2778561, EPI_ISL_2778562, EPI_ISL_2778563, EPI_ISL_2778564, EPI_ISL_2778565, EPI_ISL_2778566, EPI_ISL_2778567, EPI_ISL_2778568, EPI_ISL_2778569, EPI_ISL_2778570, EPI_ISL_2778571, EPI_ISL_2778572, EPI_ISL_2778573, EPI_ISL_2778574, EPI_ISL_2778575, EPI_ISL_2778576, EPI_ISL_2778577, EPI_ISL_2778578, EPI_ISL_2778579, EPI_ISL_2778580, EPI_ISL_2778581, EPI_ISL_2778582, EPI_ISL_2778583, EPI_ISL_2778584, EPI_ISL_2778585, EPI_ISL_2778586, EPI_ISL_2778587, EPI_ISL_2778588, EPI_ISL_2778589, EPI_ISL_2778590, EPI_ISL_2778591, EPI_ISL_2778592, EPI_ISL_2778593, EPI_ISL_2778594, EPI_ISL_2778595, EPI_ISL_2778596, EPI_ISL_2778597, EPI_ISL_2778598, EPI_ISL_2778599, EPI_ISL_2778600, EPI_ISL_2778601, EPI_ISL_2778602, EPI_ISL_2778603, EPI_ISL_2778604, EPI_ISL_2778605, EPI_ISL_2778606, EPI_ISL_2778607, EPI_ISL_2778608, EPI_ISL_2778609, EPI_ISL_2778610, EPI_ISL_2778611, EPI_ISL_2778612, EPI_ISL_2778613, EPI_ISL_2778614, EPI_ISL_2778615, EPI_ISL_2778616, EPI_ISL_2778617, EPI_ISL_2778618, EPI_ISL_2778619, EPI_ISL_2778620, EPI_ISL_2778621, EPI_ISL_2778622, EPI_ISL_2778623, EPI_ISL_2778624, EPI_ISL_2778625, EPI_ISL_2778626, EPI_ISL_2778627, EPI_ISL_2778628, EPI_ISL_2778629, EPI_ISL_2778630, EPI_ISL_2778631, EPI_ISL_2778632, EPI_ISL_2778633, EPI_ISL_2778634, EPI_ISL_2778635 | | | |
| see above | California Department of Public Health Valencia Branch Laboratory (CDPH VBL) | California Department of Public Health | CDPH-COVIDNet; UCLA Technology Center for Genomics & Bioinformatics |

| | | | |
|---|---|---|---|
| EPI_ISL_1701495 | California Institute of Technology | Chan-Zuckerberg Biohub | CZB Cliahub Consortium |
| EPI_ISL_1824336, EPI_ISL_1824337, EPI_ISL_1824338, EPI_ISL_1824339, EPI_ISL_1824343, EPI_ISL_1824344, EPI_ISL_1824346, EPI_ISL_1824347, EPI_ISL_1824348, EPI_ISL_1824349, EPI_ISL_1824350, EPI_ISL_1824351, EPI_ISL_1824352, EPI_ISL_1824353, EPI_ISL_1824354, EPI_ISL_1824355, EPI_ISL_1824356, EPI_ISL_1824357, EPI_ISL_1824358, EPI_ISL_1824359, EPI_ISL_1824360, EPI_ISL_1824361, EPI_ISL_1824362, EPI_ISL_1824363, EPI_ISL_1824364, EPI_ISL_1824366, EPI_ISL_1824367, EPI_ISL_1824368, EPI_ISL_1824369, EPI_ISL_1824370, EPI_ISL_1824371, EPI_ISL_1824372, EPI_ISL_1824373, EPI_ISL_1824374, EPI_ISL_1824375, EPI_ISL_1824376, EPI_ISL_1824377, EPI_ISL_1824378, EPI_ISL_1824379, EPI_ISL_1824380, EPI_ISL_1824381, EPI_ISL_1824382, EPI_ISL_1824383, EPI_ISL_1824384, EPI_ISL_1824385, EPI_ISL_1824386, EPI_ISL_1824387, EPI_ISL_1824388, EPI_ISL_1824389, EPI_ISL_1824390, EPI_ISL_1824391, EPI_ISL_1824392 | | | |
| see above | Chiu Laboratory, University of California, San Francisco | Chiu Laboratory, University of California, San Francisco | Alicia Sotomayor-Gonzalez; Candace Wang; Charles Chiu; Daniel Wong; Venice Servellita; Yueyuan Zhang |
| EPI_ISL_1192363 | City of Hope Comprehensive Cancer Center | Los Angeles County Public Health Laboratories | P. Hemarajata et al. |
| EPI_ISL_1824917, EPI_ISL_1824918, EPI_ISL_1824919, EPI_ISL_1824920, EPI_ISL_1824921, EPI_ISL_1824922, EPI_ISL_1824923, EPI_ISL_1824924, EPI_ISL_1824925, EPI_ISL_1824926, EPI_ISL_1824927, EPI_ISL_1824928, EPI_ISL_1824929, EPI_ISL_1824930, EPI_ISL_1824931, EPI_ISL_1824932, EPI_ISL_1824933, EPI_ISL_1824934, EPI_ISL_1824935, EPI_ISL_1824936, EPI_ISL_1824937, EPI_ISL_1824938, EPI_ISL_1824939, EPI_ISL_1824940, EPI_ISL_1824941, EPI_ISL_1824942, EPI_ISL_1824943, EPI_ISL_1824944, EPI_ISL_1824945, EPI_ISL_1824946, EPI_ISL_1824947, EPI_ISL_1824948, EPI_ISL_1824949, EPI_ISL_1824950, EPI_ISL_1824951, EPI_ISL_1824952, EPI_ISL_1824953, EPI_ISL_1824954, EPI_ISL_1824955, EPI_ISL_1824956, EPI_ISL_1824957, EPI_ISL_1824958, EPI_ISL_1824959, EPI_ISL_1824960, EPI_ISL_1824961, EPI_ISL_1824962, EPI_ISL_1824963, EPI_ISL_1824964, EPI_ISL_1824965, EPI_ISL_1824966, EPI_ISL_1824967, EPI_ISL_1824968, EPI_ISL_1824969, EPI_ISL_1824970, EPI_ISL_1824971, EPI_ISL_1824972, EPI_ISL_1824973, EPI_ISL_1824974, EPI_ISL_1824975, EPI_ISL_1824976, EPI_ISL_1824977, EPI_ISL_1824978, EPI_ISL_1824979, EPI_ISL_1824980, EPI_ISL_1824981, EPI_ISL_1824982, EPI_ISL_1824983, EPI_ISL_1824984, EPI_ISL_1824985, EPI_ISL_1824986, EPI_ISL_1824987, EPI_ISL_1824988, EPI_ISL_1824989, EPI_ISL_1824990, EPI_ISL_1824991, EPI_ISL_1824992, EPI_ISL_1824993, EPI_ISL_1824994, EPI_ISL_1824995, EPI_ISL_1824996, EPI_ISL_1824997, EPI_ISL_1824998, EPI_ISL_1824999, EPI_ISL_1825000, EPI_ISL_1825001, EPI_ISL_1825002, EPI_ISL_1825003, EPI_ISL_1825004, EPI_ISL_1825005, EPI_ISL_1825006, EPI_ISL_1825007, EPI_ISL_1825008, EPI_ISL_1825009, EPI_ISL_1825010, EPI_ISL_1825011, EPI_ISL_1825012, EPI_ISL_1825013, EPI_ISL_1825014, EPI_ISL_1825015, EPI_ISL_1825016, EPI_ISL_1825017, EPI_ISL_1825018, EPI_ISL_1825019, EPI_ISL_1825020, EPI_ISL_1825021, EPI_ISL_1825022, EPI_ISL_1825023, EPI_ISL_1825024, EPI_ISL_1825025, EPI_ISL_1825026, EPI_ISL_1825027, EPI_ISL_1825028, EPI_ISL_1825029, EPI_ISL_1825030, EPI_ISL_1825031, EPI_ISL_1825032, EPI_ISL_1825033, EPI_ISL_1825034, EPI_ISL_1825035, EPI_ISL_1825036, EPI_ISL_1825037, EPI_ISL_1825038, EPI_ISL_1825039, EPI_ISL_1825040, EPI_ISL_1825041, EPI_ISL_1825042, EPI_ISL_1825043, EPI_ISL_1825044, EPI_ISL_1825045, EPI_ISL_1825046, EPI_ISL_1825047, EPI_ISL_1825048, EPI_ISL_1825049, EPI_ISL_1825050, EPI_ISL_1825051, EPI_ISL_1825052, EPI_ISL_1825053, EPI_ISL_1825054, EPI_ISL_1825055, EPI_ISL_1825056, EPI_ISL_1825057, EPI_ISL_1825058, EPI_ISL_1825059, EPI_ISL_1825060, EPI_ISL_1825061, EPI_ISL_1825062, EPI_ISL_1825063, EPI_ISL_1825064, EPI_ISL_1825065, EPI_ISL_1825066, EPI_ISL_1825067, EPI_ISL_1825068, EPI_ISL_1825069, EPI_ISL_1825070, EPI_ISL_1825071, EPI_ISL_1825072, EPI_ISL_1825073 | | | |
| see above | Color Genomics | Chiu Laboratory, University of California, San Francisco | Alicia Sotomayor-Gonzalez; Alicia Zhou; Candace Wang; Charles Chiu; Daniel Wong; Erika Torres; Madyson Wanner; Scott Topper; Venice Servellita; Yueyuan Zhang |
| EPI_ISL_1477675, EPI_ISL_1477676, EPI_ISL_1477679, EPI_ISL_1477685, EPI_ISL_1477689, EPI_ISL_1477698, EPI_ISL_1477704, EPI_ISL_1477713, EPI_ISL_1477714, EPI_ISL_1477724, EPI_ISL_1477739, EPI_ISL_1477744, EPI_ISL_1477748, EPI_ISL_1477761, EPI_ISL_1477764, EPI_ISL_1477765, EPI_ISL_1477770, EPI_ISL_1477782, EPI_ISL_1477785, EPI_ISL_1477788, EPI_ISL_1477789, EPI_ISL_1477795, EPI_ISL_1477806, EPI_ISL_1477810, EPI_ISL_1477815, EPI_ISL_1477816, EPI_ISL_1477834, EPI_ISL_1477843, EPI_ISL_1477864, EPI_ISL_1477870, EPI_ISL_1477872, EPI_ISL_1477876, EPI_ISL_1477894, EPI_ISL_1477898, EPI_ISL_1477911, EPI_ISL_1477916, EPI_ISL_1477931, EPI_ISL_1477944, EPI_ISL_1477947, EPI_ISL_1477962, EPI_ISL_1478117, EPI_ISL_1529122, EPI_ISL_1529123, EPI_ISL_1529124, EPI_ISL_1529131, EPI_ISL_1529133, EPI_ISL_1529134, EPI_ISL_1529135, EPI_ISL_1529155, EPI_ISL_1529176, EPI_ISL_1529179, EPI_ISL_1529185, EPI_ISL_1529190, EPI_ISL_1529202, EPI_ISL_1529209, EPI_ISL_1529235, EPI_ISL_1529247, EPI_ISL_1529251, EPI_ISL_1529300, EPI_ISL_1529308, EPI_ISL_1529318, EPI_ISL_1529322, EPI_ISL_1529323, EPI_ISL_1529327, EPI_ISL_1529345, EPI_ISL_1529358, EPI_ISL_1529364, EPI_ISL_1529367, EPI_ISL_1529368, EPI_ISL_1664070, EPI_ISL_1664086, EPI_ISL_1664093, EPI_ISL_2228246, EPI_ISL_2228247, EPI_ISL_2228249, EPI_ISL_2228252, EPI_ISL_2228255, EPI_ISL_2228256, EPI_ISL_2228262, EPI_ISL_2228265, EPI_ISL_2228266, EPI_ISL_2228267, EPI_ISL_2228270, EPI_ISL_2228271, EPI_ISL_2228275, EPI_ISL_2228277, EPI_ISL_2228280, EPI_ISL_2228282, EPI_ISL_2228283, EPI_ISL_2228286, EPI_ISL_2228291, EPI_ISL_2228294, EPI_ISL_2228296, EPI_ISL_2228298, EPI_ISL_2228299, EPI_ISL_2228305, EPI_ISL_2228308, EPI_ISL_2228309, EPI_ISL_2228312, EPI_ISL_2228315, EPI_ISL_2228317, EPI_ISL_2228320, EPI_ISL_2228321, EPI_ISL_2228322, EPI_ISL_2228323, EPI_ISL_2228324, EPI_ISL_2228325, EPI_ISL_2228326, EPI_ISL_2228330, EPI_ISL_2228331, EPI_ISL_2228333, EPI_ISL_2228334, EPI_ISL_2228335, EPI_ISL_2228338, EPI_ISL_2228341, EPI_ISL_2228342, EPI_ISL_2228344, EPI_ISL_2228347, EPI_ISL_2228352, EPI_ISL_2228362, EPI_ISL_2228365, EPI_ISL_2228371, EPI_ISL_2228372, EPI_ISL_2228373, EPI_ISL_2228375, EPI_ISL_2228378, EPI_ISL_2228379, EPI_ISL_2228384, EPI_ISL_2228386, EPI_ISL_2228387, EPI_ISL_2228388, EPI_ISL_2228392, EPI_ISL_2228393, EPI_ISL_2228394, EPI_ISL_2228397, EPI_ISL_2228398, EPI_ISL_2228401, EPI_ISL_2228403, EPI_ISL_2228407, EPI_ISL_2228408, EPI_ISL_2228409, EPI_ISL_2228413, EPI_ISL_2228417, EPI_ISL_2228423, EPI_ISL_2228425, EPI_ISL_2228428, EPI_ISL_2658610, EPI_ISL_2658707, EPI_ISL_2659497, EPI_ISL_2758468 | | | |
| see above | Contra Costa County Public Health Lab | Chan-Zuckerberg Biohub | CZB Cliahub Consortium |
| EPI_ISL_1234806, EPI_ISL_1477550, EPI_ISL_1477626, EPI_ISL_1477992, EPI_ISL_1478174, EPI_ISL_1701201, EPI_ISL_1701297, EPI_ISL_1821359, EPI_ISL_1821367, EPI_ISL_1821408, EPI_ISL_1821447, EPI_ISL_1821468, EPI_ISL_2659393 | | | |
| see above | County of San Luis Obispo Public Health Laboratory | Chan-Zuckerberg Biohub | CZB Cliahub Consortium |
| EPI_ISL_1503991 | Curative Labs | Los Angeles County Public Health Laboratories | P. Hemarajata et al. |
| EPI_ISL_1318697, EPI_ISL_1318704, EPI_ISL_1318708, EPI_ISL_1318711, EPI_ISL_1318714, EPI_ISL_1318718, EPI_ISL_1318726, EPI_ISL_1318727, EPI_ISL_1318728, EPI_ISL_1318740, EPI_ISL_1318744, EPI_ISL_1318748, EPI_ISL_1318753, EPI_ISL_1318760, EPI_ISL_1318767, EPI_ISL_1318775, EPI_ISL_1318777, EPI_ISL_1318786, EPI_ISL_1318787, EPI_ISL_1318796, EPI_ISL_1318802, EPI_ISL_1318803, EPI_ISL_1318805, EPI_ISL_1318812, EPI_ISL_1318817, EPI_ISL_1318820, EPI_ISL_1318823, EPI_ISL_1318832, EPI_ISL_1318836, EPI_ISL_1318845, EPI_ISL_1318856, EPI_ISL_1318857, EPI_ISL_1318871, EPI_ISL_1318875, EPI_ISL_1318878, EPI_ISL_1318886, EPI_ISL_1318890, EPI_ISL_1318896, EPI_ISL_1318898, EPI_ISL_1318900, EPI_ISL_1318906, EPI_ISL_1318908, EPI_ISL_1318917, EPI_ISL_1318918, EPI_ISL_1318927, EPI_ISL_1318928, EPI_ISL_1318930, EPI_ISL_1318933, EPI_ISL_1318934, EPI_ISL_1318937, EPI_ISL_1318943, EPI_ISL_1318945, EPI_ISL_1318951, EPI_ISL_1318952, EPI_ISL_1318953, EPI_ISL_1318956, EPI_ISL_1318970, EPI_ISL_1318976, EPI_ISL_1318979, EPI_ISL_1318988, EPI_ISL_1347789, EPI_ISL_1349078, EPI_ISL_1349085, EPI_ISL_1349093, EPI_ISL_2158084, EPI_ISL_2158087, EPI_ISL_2158088, EPI_ISL_2158092, EPI_ISL_2158094, EPI_ISL_2158095, EPI_ISL_2158096 | | | |
| see above | DeRisi Lab, University of California, San Francisco | DeRisi Lab, University of California, San Francisco | Amy Kistler; Anthea Mitchell; Carina Marquez; Diane Havlir; Diane Jones; Douglas Black; Eric Chow; Gabriel Chamie; Genay Pilarowski; IDseq Team; Jackie Martinez; James Peng; Jamin Liu; Jon Jacobo; Lucy Li; Luis Rubio; Manu Vanaerschot; Matthew Laurie; Maya Petersen; Patrick Ayscue; Sabrina Mann; Sara Sunshine; Susana Rojas; Susy Rojas; Valerie Tulier-Laiwa; and Joseph DeRisi |
| EPI_ISL_1811646, EPI_ISL_1811653, EPI_ISL_1811654, EPI_ISL_1811658, EPI_ISL_1811662, EPI_ISL_1811665, EPI_ISL_1811685, EPI_ISL_1811689, EPI_ISL_1811693, EPI_ISL_1811695, EPI_ISL_1811696, EPI_ISL_1811697, EPI_ISL_1811702, EPI_ISL_1811706, EPI_ISL_1811727, EPI_ISL_1811729, EPI_ISL_1811732, EPI_ISL_1811740, EPI_ISL_1811741, EPI_ISL_1811742, EPI_ISL_1811763, EPI_ISL_1811766, EPI_ISL_1811769, EPI_ISL_1811771, EPI_ISL_1811797, EPI_ISL_1811807, EPI_ISL_1811817, EPI_ISL_1811825, EPI_ISL_1811828, EPI_ISL_1811829, EPI_ISL_1811847, EPI_ISL_1811858, EPI_ISL_1811889, EPI_ISL_1811897, EPI_ISL_1811914, EPI_ISL_1811924, EPI_ISL_1811973, EPI_ISL_1811992, EPI_ISL_1811994, EPI_ISL_1812011, EPI_ISL_1812012, EPI_ISL_1812015, EPI_ISL_1812016, EPI_ISL_1812029, EPI_ISL_1812042, EPI_ISL_1812044, EPI_ISL_1812050, EPI_ISL_1812051, EPI_ISL_1812060, EPI_ISL_1812062, EPI_ISL_1812071, EPI_ISL_1812077, EPI_ISL_1812079, EPI_ISL_1812086, EPI_ISL_1812087, EPI_ISL_1812090, EPI_ISL_1812099, EPI_ISL_1812114, EPI_ISL_1812116, EPI_ISL_1812122, EPI_ISL_1812127, EPI_ISL_1812131, EPI_ISL_1812134, EPI_ISL_1812137, EPI_ISL_1812154, EPI_ISL_1812160, EPI_ISL_1812170, EPI_ISL_1812171, EPI_ISL_1812181, EPI_ISL_1812183, EPI_ISL_1812187, EPI_ISL_1812188, EPI_ISL_1812191, EPI_ISL_1812201, EPI_ISL_1812211, EPI_ISL_1812216, EPI_ISL_1812219, EPI_ISL_1812229, EPI_ISL_1812231, EPI_ISL_1812242, EPI_ISL_1812265, EPI_ISL_1812276, EPI_ISL_1812290, EPI_ISL_1812292, EPI_ISL_1812310, EPI_ISL_1812312, EPI_ISL_1812314, EPI_ISL_1812317, EPI_ISL_1812324, EPI_ISL_1812326, EPI_ISL_1812329, EPI_ISL_1812336, EPI_ISL_1812343, EPI_ISL_1812348, EPI_ISL_1812352, EPI_ISL_1812354, EPI_ISL_1812357, EPI_ISL_1812365, EPI_ISL_1812368, EPI_ISL_1812372, EPI_ISL_1812379, EPI_ISL_1812384, EPI_ISL_1812385, EPI_ISL_1812387, EPI_ISL_1812392, EPI_ISL_1812398, EPI_ISL_1812400, EPI_ISL_1812402, EPI_ISL_1812404, EPI_ISL_1812406, EPI_ISL_1812407, EPI_ISL_1812408, EPI_ISL_1812417, EPI_ISL_1812423, EPI_ISL_1812430, EPI_ISL_1812434, EPI_ISL_1812441, EPI_ISL_1812453, EPI_ISL_1812460, EPI_ISL_1812468, EPI_ISL_1812477, EPI_ISL_1812482, EPI_ISL_1812489, EPI_ISL_1812491, EPI_ISL_1812497, EPI_ISL_1812517, EPI_ISL_1812530, EPI_ISL_1812535, EPI_ISL_1812538, EPI_ISL_1812540, EPI_ISL_1812541, EPI_ISL_1812542, EPI_ISL_1812552, EPI_ISL_1812562, EPI_ISL_1812564, EPI_ISL_1812568, EPI_ISL_1812571, EPI_ISL_1812576, EPI_ISL_1812587, EPI_ISL_1812592, EPI_ISL_1812598, EPI_ISL_1812603, EPI_ISL_1812604, EPI_ISL_1812611, EPI_ISL_1812612, EPI_ISL_1812626, EPI_ISL_1812634, EPI_ISL_1812637, EPI_ISL_1812641, EPI_ISL_1812653, EPI_ISL_1812655, EPI_ISL_1812670, EPI_ISL_1812674, EPI_ISL_1812675, EPI_ISL_1812687, EPI_ISL_1812689, EPI_ISL_1812695, EPI_ISL_1812699, EPI_ISL_1812716, EPI_ISL_1812722, EPI_ISL_1812732, EPI_ISL_1812739, EPI_ISL_1812746, EPI_ISL_1812753, EPI_ISL_1812756, EPI_ISL_1812760, EPI_ISL_1812765, EPI_ISL_1812768, EPI_ISL_1812771, EPI_ISL_1812781, EPI_ISL_1812790, EPI_ISL_1812792, EPI_ISL_1812796, EPI_ISL_1812801, EPI_ISL_1812803, EPI_ISL_1812804, EPI_ISL_1812811, EPI_ISL_1812817, EPI_ISL_1812825, EPI_ISL_1812828, EPI_ISL_1812837, EPI_ISL_1812838, EPI_ISL_1812840, EPI_ISL_1812841, EPI_ISL_1812842, EPI_ISL_1812852, EPI_ISL_1812853, EPI_ISL_1812858, EPI_ISL_1812860, EPI_ISL_1812861, EPI_ISL_1812874, EPI_ISL_1812877, EPI_ISL_1812878, EPI_ISL_1812879, EPI_ISL_1812882, EPI_ISL_1812887, EPI_ISL_1812890, EPI_ISL_1812892, EPI_ISL_1812896, EPI_ISL_1812897, EPI_ISL_1812914, EPI_ISL_1812915, EPI_ISL_1812921, EPI_ISL_1812922, EPI_ISL_1812940, EPI_ISL_1812946, EPI_ISL_1812960, EPI_ISL_1812970, EPI_ISL_1812974, EPI_ISL_1812978, EPI_ISL_1812985, EPI_ISL_1812993, EPI_ISL_1812998, EPI_ISL_1813008, EPI_ISL_1813009, EPI_ISL_1813019, EPI_ISL_1813030, EPI_ISL_1813038, EPI_ISL_1813041, EPI_ISL_1813043, EPI_ISL_1813045, EPI_ISL_1813047, EPI_ISL_1813051, EPI_ISL_1813070, EPI_ISL_1813075, EPI_ISL_1813081, EPI_ISL_1813101, EPI_ISL_1813109, EPI_ISL_1813119, EPI_ISL_1813126, EPI_ISL_1813127, EPI_ISL_1813130, EPI_ISL_1813138, EPI_ISL_1813144, EPI_ISL_1813149, EPI_ISL_1813151, EPI_ISL_1813153, EPI_ISL_1813157, EPI_ISL_1813158, EPI_ISL_1813162, EPI_ISL_1813168, EPI_ISL_1813172, EPI_ISL_1813178, EPI_ISL_1813192, EPI_ISL_1813198, EPI_ISL_1813201, EPI_ISL_1813202, EPI_ISL_1813211, EPI_ISL_1813217, EPI_ISL_1813219, EPI_ISL_1813223, EPI_ISL_1813236, EPI_ISL_1813238, EPI_ISL_1813241, EPI_ISL_1813246, EPI_ISL_1813248, EPI_ISL_1813255, EPI_ISL_1813257, EPI_ISL_1813261, EPI_ISL_1813264, EPI_ISL_1813277, EPI_ISL_1813278, EPI_ISL_1813290, EPI_ISL_1813296, EPI_ISL_1813311, EPI_ISL_1813321, EPI_ISL_1813326, EPI_ISL_1813333, EPI_ISL_1813336, EPI_ISL_1813341, EPI_ISL_1813346, EPI_ISL_1813349, EPI_ISL_1813354, EPI_ISL_1813355, EPI_ISL_1813356, EPI_ISL_1813364, EPI_ISL_1813366, EPI_ISL_1813367, EPI_ISL_1813371, EPI_ISL_1813379, EPI_ISL_1813385, EPI_ISL_1813387, EPI_ISL_1813391, EPI_ISL_1813397, EPI_ISL_1813417, EPI_ISL_1813418, EPI_ISL_1813419, EPI_ISL_1813424, EPI_ISL_1813428, EPI_ISL_1813432, EPI_ISL_1813445, EPI_ISL_1813448, EPI_ISL_1813453, EPI_ISL_1813459, EPI_ISL_1813471, EPI_ISL_1813476, EPI_ISL_1813477, EPI_ISL_1813478, EPI_ISL_1813480, EPI_ISL_1813482, EPI_ISL_1813485, EPI_ISL_1813490, EPI_ISL_1813491, EPI_ISL_1813493, EPI_ISL_1813494, EPI_ISL_1813496, EPI_ISL_1813498, EPI_ISL_1813503, EPI_ISL_1813521, EPI_ISL_1813527, EPI_ISL_1813528, EPI_ISL_1813530, EPI_ISL_1813531, EPI_ISL_1813533, EPI_ISL_1813542, EPI_ISL_1813543, EPI_ISL_1813546, EPI_ISL_1813547, EPI_ISL_1813549, EPI_ISL_1813551, EPI_ISL_1813555, EPI_ISL_1813558, EPI_ISL_1813561, EPI_ISL_1813570, EPI_ISL_1813577, EPI_ISL_1813579, EPI_ISL_1813581, EPI_ISL_1813591, EPI_ISL_1813594, EPI_ISL_1813601, EPI_ISL_1813604, EPI_ISL_1813606, EPI_ISL_1813609, EPI_ISL_1813617, EPI_ISL_1813618, EPI_ISL_1813628, EPI_ISL_1813634, EPI_ISL_1813636, EPI_ISL_1813640, EPI_ISL_1813642, EPI_ISL_1813655, EPI_ISL_1813662, EPI_ISL_1813668, EPI_ISL_1813671, EPI_ISL_1813673, EPI_ISL_1813675, EPI_ISL_1813678, EPI_ISL_1813680, EPI_ISL_1813681, EPI_ISL_1813693, EPI_ISL_1813701, EPI_ISL_1813703, EPI_ISL_1813705, EPI_ISL_1813706, EPI_ISL_1813707, EPI_ISL_1813714, EPI_ISL_1813722, EPI_ISL_1813725, EPI_ISL_1813726, EPI_ISL_1813729, EPI_ISL_1813734, EPI_ISL_1813735, EPI_ISL_1813750, EPI_ISL_1813751, EPI_ISL_1813756, EPI_ISL_1813760, EPI_ISL_1813761, EPI_ISL_1813771, EPI_ISL_1813778, EPI_ISL_1813779, EPI_ISL_1813786, EPI_ISL_1813787, EPI_ISL_1813790, EPI_ISL_1813792, EPI_ISL_1813795, EPI_ISL_1813802, EPI_ISL_1813814, EPI_ISL_1813818, EPI_ISL_1813821, EPI_ISL_1813822, EPI_ISL_1813827, EPI_ISL_1813829, EPI_ISL_1813837, EPI_ISL_1813840, EPI_ISL_1813842, EPI_ISL_1813845, EPI_ISL_1813849, EPI_ISL_1813850, EPI_ISL_1813855, EPI_ISL_1813857, EPI_ISL_1813859, EPI_ISL_1813864, EPI_ISL_1813866, EPI_ISL_1813870, EPI_ISL_1813877, EPI_ISL_1813878, EPI_ISL_1813884, EPI_ISL_1813893, EPI_ISL_1813896, EPI_ISL_1813901, EPI_ISL_1813904, EPI_ISL_1813905, EPI_ISL_1813906, EPI_ISL_1813912, EPI_ISL_1813921, EPI_ISL_1813922, EPI_ISL_1813924, EPI_ISL_1813945, EPI_ISL_1813948, EPI_ISL_1813952, EPI_ISL_1813956, EPI_ISL_1813973, EPI_ISL_1813976, EPI_ISL_1813982, EPI_ISL_1813984, EPI_ISL_1813986, EPI_ISL_1813987, EPI_ISL_1813994, EPI_ISL_1813997, EPI_ISL_1814003, EPI_ISL_1814011, EPI_ISL_1814014, EPI_ISL_1814019, EPI_ISL_1814036, EPI_ISL_1814043, EPI_ISL_1814046, EPI_ISL_1814053, EPI_ISL_1814056, EPI_ISL_1814060, EPI_ISL_1814063, EPI_ISL_1814065, EPI_ISL_1814066, EPI_ISL_1814071, EPI_ISL_1814076, EPI_ISL_1814087, EPI_ISL_1814089, EPI_ISL_1814098, EPI_ISL_1814101, EPI_ISL_1814106, EPI_ISL_1814117, EPI_ISL_1814121, EPI_ISL_1814122, EPI_ISL_1814123, EPI_ISL_1814130, EPI_ISL_1814131, EPI_ISL_1814135, EPI_ISL_1814140, EPI_ISL_1814145, EPI_ISL_1814147, EPI_ISL_1814150, EPI_ISL_1814152, EPI_ISL_1814153, EPI_ISL_1814154, EPI_ISL_1814170, EPI_ISL_1814173, EPI_ISL_1814197, EPI_ISL_1814199, EPI_ISL_1814207, EPI_ISL_1814210, EPI_ISL_1814212, EPI_ISL_1814214, EPI_ISL_1814218, EPI_ISL_1814221, EPI_ISL_1814226, EPI_ISL_1814227, EPI_ISL_1814234, EPI_ISL_1814252, EPI_ISL_1814259, EPI_ISL_1814260, EPI_ISL_1814261, EPI_ISL_1814269, EPI_ISL_1814276, EPI_ISL_1814278, EPI_ISL_1814279, EPI_ISL_1814287, EPI_ISL_1814290, EPI_ISL_1814297, EPI_ISL_1814298, EPI_ISL_1814301, EPI_ISL_1814306, EPI_ISL_1814312, EPI_ISL_1814313, EPI_ISL_1814315, EPI_ISL_1814318, EPI_ISL_1814324, EPI_ISL_1814325, EPI_ISL_1814326, EPI_ISL_1814329, EPI_ISL_1814332, EPI_ISL_1814333, EPI_ISL_1814334, EPI_ISL_1814335, EPI_ISL_1814336, EPI_ISL_1814338, EPI_ISL_1814341, EPI_ISL_1814345, EPI_ISL_1814346, EPI_ISL_1814353, EPI_ISL_1814354, EPI_ISL_1814357, EPI_ISL_1814359, EPI_ISL_1814360, EPI_ISL_1814362, EPI_ISL_1814364, EPI_ISL_1814378, EPI_ISL_1814381, EPI_ISL_1814384, EPI_ISL_1814385, EPI_ISL_1814386, EPI_ISL_1814392, EPI_ISL_1814397, EPI_ISL_1814398, EPI_ISL_1814402, EPI_ISL_1814407, EPI_ISL_1814414, EPI_ISL_1814415, EPI_ISL_1814427, EPI_ISL_1814434, EPI_ISL_1814439, EPI_ISL_1814443, EPI_ISL_1814446, EPI_ISL_1814449, EPI_ISL_1814457, EPI_ISL_1814461, EPI_ISL_1814462, EPI_ISL_1814463, EPI_ISL_1814465, EPI_ISL_1814483, EPI_ISL_1814484, EPI_ISL_1814488, EPI_ISL_1814493, EPI_ISL_1814494, EPI_ISL_1814498, EPI_ISL_1814504, EPI_ISL_1814515, EPI_ISL_1814519, EPI_ISL_1814522, EPI_ISL_1814524, EPI_ISL_1814525, EPI_ISL_1814527, EPI_ISL_1814536, EPI_ISL_1814537, EPI_ISL_1814539, EPI_ISL_1814540, EPI_ISL_1814541, EPI_ISL_1814547, EPI_ISL_1814549, EPI_ISL_1814553, EPI_ISL_1814556, EPI_ISL_1814557, EPI_ISL_1814558, EPI_ISL_1814561, EPI_ISL_1814569, EPI_ISL_1814570, EPI_ISL_1814572, EPI_ISL_1814574, EPI_ISL_1814582, EPI_ISL_1814583, EPI_ISL_1814597, EPI_ISL_1814598, EPI_ISL_1814602, EPI_ISL_1814605, EPI_ISL_1814612, EPI_ISL_1814623, EPI_ISL_1814625, EPI_ISL_1814630, EPI_ISL_1814631, EPI_ISL_1814632, EPI_ISL_1814634, EPI_ISL_1814635, EPI_ISL_1814641, EPI_ISL_1814642, EPI_ISL_1814655, EPI_ISL_1814656, EPI_ISL_1814658, EPI_ISL_1814662, EPI_ISL_1814669, EPI_ISL_1814670, EPI_ISL_1814674, EPI_ISL_1814682, EPI_ISL_1814683, EPI_ISL_1814684, EPI_ISL_1814687, EPI_ISL_1814689, EPI_ISL_1814697, EPI_ISL_1814704, EPI_ISL_1814710, EPI_ISL_1814711, EPI_ISL_1814715, EPI_ISL_1814717, EPI_ISL_1814718, EPI_ISL_1814720, EPI_ISL_1814721, EPI_ISL_1814732, EPI_ISL_1814733, EPI_ISL_1814736, EPI_ISL_1814739, EPI_ISL_1814742, EPI_ISL_1814743, EPI_ISL_1814745, EPI_ISL_1814746, EPI_ISL_1814747, EPI_ISL_1814751, EPI_ISL_1814757, EPI_ISL_1814763, EPI_ISL_1814765, EPI_ISL_1814771, EPI_ISL_1814773, EPI_ISL_1814775, EPI_ISL_1814792, EPI_ISL_1814794, EPI_ISL_1814795, EPI_ISL_1814798, EPI_ISL_1814806, EPI_ISL_1814811, EPI_ISL_1814812, EPI_ISL_1814814, EPI_ISL_1814816, EPI_ISL_1814826, EPI_ISL_1814830, EPI_ISL_1814833, EPI_ISL_1814834, EPI_ISL_1814839, EPI_ISL_1814840, EPI_ISL_1814844, EPI_ISL_1814845, EPI_ISL_1814846, EPI_ISL_1814847, EPI_ISL_1814848, EPI_ISL_1814850, EPI_ISL_1814851, EPI_ISL_1814855, EPI_ISL_1814859, EPI_ISL_1814861, EPI_ISL_1814868, EPI_ISL_1814870, EPI_ISL_1814872, EPI_ISL_1814877, EPI_ISL_1814879, EPI_ISL_1814881, EPI_ISL_1814883, EPI_ISL_1814891, EPI_ISL_1814892, EPI_ISL_1814893, EPI_ISL_1814894, EPI_ISL_1814895, EPI_ISL_1814896, EPI_ISL_1814905, EPI_ISL_1814906, EPI_ISL_1814914, EPI_ISL_1814917, EPI_ISL_1814918, EPI_ISL_1814924, EPI_ISL_1814926, EPI_ISL_1814930, EPI_ISL_1814939, EPI_ISL_1814942, EPI_ISL_1814950, EPI_ISL_1814953, EPI_ISL_1814954, EPI_ISL_1814957, EPI_ISL_1814962, EPI_ISL_1814979, EPI_ISL_1814981, EPI_ISL_1814986, EPI_ISL_1814989, EPI_ISL_1814998, EPI_ISL_1815000, EPI_ISL_1815008, EPI_ISL_1815017, EPI_ISL_1815018, EPI_ISL_1815026, EPI_ISL_1815031, EPI_ISL_1815035, EPI_ISL_1815040, EPI_ISL_1815048, EPI_ISL_1815055, EPI_ISL_1815060, EPI_ISL_1815062, EPI_ISL_1815063, EPI_ISL_1815070, EPI_ISL_1815074, EPI_ISL_1815075, EPI_ISL_1815084, EPI_ISL_1815088, EPI_ISL_1815091, EPI_ISL_1815092, EPI_ISL_1815103, EPI_ISL_1815108, EPI_ISL_1815110, EPI_ISL_1815112, EPI_ISL_1815114, EPI_ISL_1815115, EPI_ISL_1815118, EPI_ISL_1815121, EPI_ISL_1815124, EPI_ISL_1815126, EPI_ISL_1815129, EPI_ISL_1815133, EPI_ISL_1815135, EPI_ISL_1815139, EPI_ISL_1815140, EPI_ISL_1815142, EPI_ISL_1815147, EPI_ISL_1815151, EPI_ISL_1815157, EPI_ISL_1815158, EPI_ISL_1815159, EPI_ISL_1815160, EPI_ISL_1815161, EPI_ISL_1815162, EPI_ISL_1815164, EPI_ISL_1815165, EPI_ISL_1815166, EPI_ISL_1815169, EPI_ISL_1815170, EPI_ISL_1815172, EPI_ISL_1815175, EPI_ISL_1815180, EPI_ISL_1815183, EPI_ISL_1815191, EPI_ISL_1815194, EPI_ISL_1815198, EPI_ISL_1815200, EPI_ISL_1815201, EPI_ISL_1815203, EPI_ISL_1815206, EPI_ISL_1815207, EPI_ISL_1815212, EPI_ISL_1815213, EPI_ISL_1815216, EPI_ISL_1815220, EPI_ISL_1815230, EPI_ISL_1815231, EPI_ISL_1815235, EPI_ISL_1815251, EPI_ISL_1815253, EPI_ISL_1815255, EPI_ISL_1815267, EPI_ISL_1815270, EPI_ISL_1815271, EPI_ISL_1815273, EPI_ISL_1815274, EPI_ISL_1815278, EPI_ISL_1815283, EPI_ISL_1815287, EPI_ISL_1815288, EPI_ISL_1815290, EPI_ISL_1815293, EPI_ISL_1815296, EPI_ISL_1815298, EPI_ISL_1815303, EPI_ISL_1815305, EPI_ISL_1815307, EPI_ISL_1815309, EPI_ISL_1815315, EPI_ISL_1815318, EPI_ISL_1815320, EPI_ISL_1815326, EPI_ISL_1815329, EPI_ISL_1815333, EPI_ISL_1815337, EPI_ISL_1815340, EPI_ISL_1815341, EPI_ISL_1815346, EPI_ISL_1815350, EPI_ISL_1815354, EPI_ISL_1815357, EPI_ISL_1815359, EPI_ISL_1815363, EPI_ISL_1815368, EPI_ISL_1815371, EPI_ISL_1815373, EPI_ISL_1815375, EPI_ISL_1815378, EPI_ISL_1815383, EPI_ISL_1815385, EPI_ISL_1815386, EPI_ISL_1815387, EPI_ISL_1815388, EPI_ISL_1815390, EPI_ISL_1815391, EPI_ISL_1815392, EPI_ISL_1815393, EPI_ISL_1815395, EPI_ISL_1815396, EPI_ISL_1815399, EPI_ISL_1815402, EPI_ISL_1815404, EPI_ISL_1815405, EPI_ISL_1815415, EPI_ISL_1815420, EPI_ISL_1815422, EPI_ISL_1815429, EPI_ISL_1815435, EPI_ISL_1815441, EPI_ISL_1815449, EPI_ISL_1815451, EPI_ISL_1815452, EPI_ISL_1815453, EPI_ISL_1815454, EPI_ISL_1815455, EPI_ISL_1815457, EPI_ISL_1815460, EPI_ISL_1815463, EPI_ISL_1815464, EPI_ISL_1815465, EPI_ISL_1815466, EPI_ISL_1815467, EPI_ISL_1815471, EPI_ISL_1815473, EPI_ISL_1815474, EPI_ISL_1815475, EPI_ISL_1815486, EPI_ISL_1815491, EPI_ISL_1815496, EPI_ISL_1815501, EPI_ISL_1815502, EPI_ISL_1815506, EPI_ISL_1815507, EPI_ISL_1815509, EPI_ISL_1815510, EPI_ISL_1815513, EPI_ISL_1815514, EPI_ISL_1815517, EPI_ISL_1815521, EPI_ISL_1815530, EPI_ISL_1815533, EPI_ISL_1815534, EPI_ISL_1815538, EPI_ISL_1815544, EPI_ISL_1815546, EPI_ISL_1815548, EPI_ISL_1815550, EPI_ISL_1815554, EPI_ISL_1815559, EPI_ISL_1815562, EPI_ISL_1815564, EPI_ISL_1815573, EPI_ISL_1815574, EPI_ISL_1815576, EPI_ISL_1815577, EPI_ISL_1815578, EPI_ISL_1815579, EPI_ISL_1815582, EPI_ISL_1815588, EPI_ISL_1815590, EPI_ISL_1815593, EPI_ISL_1815603, EPI_ISL_1815604, EPI_ISL_1815605, EPI_ISL_1815606, EPI_ISL_1815607, EPI_ISL_1815614, EPI_ISL_1815616, EPI_ISL_1815617, EPI_ISL_1815618, EPI_ISL_1815619, EPI_ISL_1815622, EPI_ISL_1815623, EPI_ISL_1815624, EPI_ISL_1815631, EPI_ISL_1815634, EPI_ISL_1815635, EPI_ISL_1815638, EPI_ISL_1815639, EPI_ISL_1815641, EPI_ISL_1815647, EPI_ISL_1815654, EPI_ISL_1815656, EPI_ISL_1815657, EPI_ISL_1815663, EPI_ISL_1815664, EPI_ISL_1815665, EPI_ISL_1815667, EPI_ISL_1815669, EPI_ISL_1815670, EPI_ISL_1815671, EPI_ISL_1815672, EPI_ISL_1815673, EPI_ISL_1815681, EPI_ISL_1815683, EPI_ISL_1815698, EPI_ISL_1815700, EPI_ISL_1815702, EPI_ISL_1815705, EPI_ISL_1815706, EPI_ISL_1815712, EPI_ISL_1815713, EPI_ISL_1815714, EPI_ISL_1815720, EPI_ISL_1815725, EPI_ISL_1815728, EPI_ISL_1815735, EPI_ISL_1815741, EPI_ISL_1815743, EPI_ISL_1815744, EPI_ISL_1815747, EPI_ISL_1815749, EPI_ISL_1815751, EPI_ISL_1815754, EPI_ISL_1815757, EPI_ISL_1815759, EPI_ISL_1815760, EPI_ISL_1815761, EPI_ISL_1815762, EPI_ISL_1815765, EPI_ISL_1815768, EPI_ISL_1815770, EPI_ISL_1815774, EPI_ISL_1815775, EPI_ISL_1815776, EPI_ISL_1815781, EPI_ISL_1815782, EPI_ISL_1815784, EPI_ISL_1815785, EPI_ISL_1815786, EPI_ISL_1815788, EPI_ISL_1815792, EPI_ISL_1815793, EPI_ISL_1815794, EPI_ISL_1815806, EPI_ISL_1815808, EPI_ISL_1815810, EPI_ISL_1815811, EPI_ISL_1815812, EPI_ISL_1815818, EPI_ISL_1815821, EPI_ISL_1815823, EPI_ISL_1815825, EPI_ISL_1815838, EPI_ISL_1815846, EPI_ISL_1815847, EPI_ISL_1815849, EPI_ISL_1815850, EPI_ISL_1815851, EPI_ISL_1815852, EPI_ISL_1815855, EPI_ISL_1815856, EPI_ISL_1815857, EPI_ISL_1815861, EPI_ISL_1815864, EPI_ISL_1815865, EPI_ISL_1815868, EPI_ISL_1815869, EPI_ISL_1815870, EPI_ISL_1815873, EPI_ISL_1815875, EPI_ISL_1815878, EPI_ISL_1815879, EPI_ISL_1815887, EPI_ISL_1815890, EPI_ISL_1815893, EPI_ISL_1815897, EPI_ISL_1815901, EPI_ISL_1815902, EPI_ISL_1815904, EPI_ISL_1815905, EPI_ISL_1815910, EPI_ISL_1815914, EPI_ISL_1815921, EPI_ISL_1815931, EPI_ISL_1815932, EPI_ISL_1815937, EPI_ISL_1815938, EPI_ISL_1815942, EPI_ISL_1815946, EPI_ISL_1815948, EPI_ISL_1815951, EPI_ISL_1815957, EPI_ISL_1815959, EPI_ISL_1815963, EPI_ISL_1815970, EPI_ISL_1815972, EPI_ISL_1815976, EPI_ISL_1815980, EPI_ISL_1815985, EPI_ISL_1815988, EPI_ISL_1815993, EPI_ISL_1816001, EPI_ISL_1816002, EPI_ISL_1816003, EPI_ISL_1816010, EPI_ISL_1816013, EPI_ISL_1816016, EPI_ISL_1816019, EPI_ISL_1816020, EPI_ISL_1816022, EPI_ISL_1816023, EPI_ISL_1816028, EPI_ISL_1816037, EPI_ISL_1816043, EPI_ISL_1816046, EPI_ISL_1816051, EPI_ISL_1816053, EPI_ISL_1816054, EPI_ISL_1816055, EPI_ISL_1816057, EPI_ISL_1816058, EPI_ISL_1816060, EPI_ISL_1816065, EPI_ISL_1816070, EPI_ISL_1816071, EPI_ISL_1816075, EPI_ISL_1816084, EPI_ISL_1816085, EPI_ISL_1816087, EPI_ISL_1816090, EPI_ISL_1816091, EPI_ISL_1816092, EPI_ISL_1816093, EPI_ISL_1816095, EPI_ISL_1816098, EPI_ISL_1816099, EPI_ISL_1816100, EPI_ISL_1816101, EPI_ISL_1816104, EPI_ISL_1816106, EPI_ISL_1816107, EPI_ISL_1816109, EPI_ISL_1816112, EPI_ISL_1816113, EPI_ISL_1816114, EPI_ISL_1816116, EPI_ISL_1816117, EPI_ISL_1816121, EPI_ISL_1816122, EPI_ISL_1816126, EPI_ISL_1816130, EPI_ISL_1816131, EPI_ISL_1816133, EPI_ISL_1816136, EPI_ISL_1816139, EPI_ISL_1816143, EPI_ISL_1816144, EPI_ISL_1816146, EPI_ISL_1816150, EPI_ISL_1816154, EPI_ISL_1816162, EPI_ISL_1816163, EPI_ISL_1816166, EPI_ISL_1816169, EPI_ISL_1816170, EPI_ISL_1816176, EPI_ISL_1816183, EPI_ISL_1816192, EPI_ISL_1816196, EPI_ISL_1816202, EPI_ISL_1816203, EPI_ISL_1816204, EPI_ISL_1816205, EPI_ISL_1816206, EPI_ISL_1816209, EPI_ISL_1816210, EPI_ISL_1816211, EPI_ISL_1816212, EPI_ISL_1816213, EPI_ISL_1816214, EPI_ISL_1816217, EPI_ISL_1816222, EPI_ISL_1816226, EPI_ISL_1816228, EPI_ISL_1816236, EPI_ISL_1816239, EPI_ISL_1816241, EPI_ISL_1816245, EPI_ISL_1816246, EPI_ISL_1816247, EPI_ISL_1816253, EPI_ISL_1816258, EPI_ISL_1816260, EPI_ISL_1816261, EPI_ISL_1816263, EPI_ISL_1816266, EPI_ISL_1816267, EPI_ISL_1816269, EPI_ISL_1816271, EPI_ISL_1816274, EPI_ISL_1816275, EPI_ISL_1816280, EPI_ISL_1816284, EPI_ISL_1816285, EPI_ISL_1816293, EPI_ISL_1816299, EPI_ISL_1816300, EPI_ISL_1816306, EPI_ISL_1816307, EPI_ISL_1816308, EPI_ISL_1816309, EPI_ISL_1816310, EPI_ISL_1816311, EPI_ISL_1816312, EPI_ISL_1816314, EPI_ISL_1816316, EPI_ISL_1816319, EPI_ISL_1816320, EPI_ISL_1816322, EPI_ISL_1816324, EPI_ISL_1816326, EPI_ISL_1816330, EPI_ISL_1816331, EPI_ISL_1816335, EPI_ISL_1816342, EPI_ISL_1816346, EPI_ISL_1816348, EPI_ISL_1816349, EPI_ISL_1816351, EPI_ISL_1816354, EPI_ISL_1816355, EPI_ISL_1816356, EPI_ISL_1816361, EPI_ISL_1816363, EPI_ISL_1816366, EPI_ISL_1816369, EPI_ISL_1816371, EPI_ISL_1816373, EPI_ISL_1816375, EPI_ISL_1816377, EPI_ISL_1816381, EPI_ISL_1816384, EPI_ISL_1816385, EPI_ISL_1816388, EPI_ISL_1816391, EPI_ISL_1816392, EPI_ISL_1816394, EPI_ISL_1816399, EPI_ISL_1816400, EPI_ISL_1816401, EPI_ISL_1816402, EPI_ISL_1816409, EPI_ISL_1816411, EPI_ISL_1816414, EPI_ISL_1816415, EPI_ISL_1816423, EPI_ISL_1816426, EPI_ISL_1816428, EPI_ISL_1816436, EPI_ISL_1816440, EPI_ISL_1816443, EPI_ISL_1816444, EPI_ISL_1816447, EPI_ISL_1816450, EPI_ISL_1816454, EPI_ISL_1816459, EPI_ISL_1816461, EPI_ISL_1816464, EPI_ISL_1816465, EPI_ISL_1816467, EPI_ISL_1816469, EPI_ISL_1816470, EPI_ISL_1816472, EPI_ISL_1816473, EPI_ISL_1816475, EPI_ISL_1816478, EPI_ISL_1816481, EPI_ISL_1816482, EPI_ISL_1816484, EPI_ISL_1816487, EPI_ISL_1816493, EPI_ISL_1816494, EPI_ISL_1816498, EPI_ISL_1816500, EPI_ISL_1816502, EPI_ISL_1816505, EPI_ISL_1816507, EPI_ISL_1816508, EPI_ISL_1816512, EPI_ISL_1816515, EPI_ISL_1816516, EPI_ISL_1816519, EPI_ISL_1816520, EPI_ISL_1816521, EPI_ISL_1816522, EPI_ISL_1816523, EPI_ISL_1816525, EPI_ISL_1816526, EPI_ISL_1816527, EPI_ISL_1816530, EPI_ISL_1816533, EPI_ISL_1816535, EPI_ISL_1816537, EPI_ISL_1816540, EPI_ISL_1816541, EPI_ISL_1816542, EPI_ISL_1816548, EPI_ISL_1816551, EPI_ISL_1816552, EPI_ISL_1816553, EPI_ISL_1816559, EPI_ISL_1816560, EPI_ISL_1816567, EPI_ISL_1816568, EPI_ISL_1816569, EPI_ISL_1816572, EPI_ISL_1816574, EPI_ISL_1816577, EPI_ISL_1816584, EPI_ISL_1816587, EPI_ISL_1816594, EPI_ISL_1816598, EPI_ISL_1816603, EPI_ISL_1816608, EPI_ISL_1816609, EPI_ISL_1816610, EPI_ISL_1816611, EPI_ISL_1816612, EPI_ISL_1816613, EPI_ISL_1816615, EPI_ISL_1816616, EPI_ISL_1816619, EPI_ISL_1816624, EPI_ISL_1816627, EPI_ISL_1816629, EPI_ISL_1816632, EPI_ISL_1816634, EPI_ISL_1816636, EPI_ISL_1816637, EPI_ISL_1816639, EPI_ISL_1816640, EPI_ISL_1816642, EPI_ISL_1816644, EPI_ISL_1816648, EPI_ISL_1816650, EPI_ISL_1816652, EPI_ISL_1816665, EPI_ISL_1816669, EPI_ISL_1816671, EPI_ISL_1816675, EPI_ISL_1816678, EPI_ISL_1816680, EPI_ISL_1816681, EPI_ISL_1816687, EPI_ISL_1816694, EPI_ISL_1816695, EPI_ISL_1816699, EPI_ISL_1816700, EPI_ISL_1816704, EPI_ISL_1816711, EPI_ISL_1816718, EPI_ISL_1816719, EPI_ISL_1816722, EPI_ISL_1816723, EPI_ISL_1816725, EPI_ISL_1816727, EPI_ISL_1816733, EPI_ISL_1816734, EPI_ISL_1816736, EPI_ISL_1816737, EPI_ISL_1816741, EPI_ISL_1816748, EPI_ISL_1816752, EPI_ISL_1816754, EPI_ISL_1816756, EPI_ISL_1816759, EPI_ISL_1816767, EPI_ISL_1816770, EPI_ISL_1816773, EPI_ISL_1816774, EPI_ISL_1816776, EPI_ISL_1816780, EPI_ISL_1816783, EPI_ISL_1816786, EPI_ISL_1816787, EPI_ISL_1816793, EPI_ISL_1816794, EPI_ISL_1816796, EPI_ISL_1816799, EPI_ISL_1816802, EPI_ISL_1816805, EPI_ISL_1816807, EPI_ISL_1816818, EPI_ISL_1816819, EPI_ISL_1816824, EPI_ISL_1816827, EPI_ISL_1816829, EPI_ISL_1816838, EPI_ISL_1816839, EPI_ISL_1816840, EPI_ISL_1816841, EPI_ISL_1816843, EPI_ISL_1816849, EPI_ISL_1816861, EPI_ISL_1816864, EPI_ISL_1816867, EPI_ISL_1816879, EPI_ISL_1816884, EPI_ISL_1816891, EPI_ISL_1816894, EPI_ISL_1816902, EPI_ISL_1816905 | | | |
| see above | EXCITE Lab | Andersen lab at Scripps Research | Abigail Schnapper; Alexandre Bolze; Alice Summerfield; Angela Scioscia; Araz Majnoonian; Ashkan Hassani; Celena Andrade; Charlotte Rivera-Garcia; Cheryl Anderson; Chip Schooley; Christine Mitchell + SEARCH; David Becker; David Pride; Efren Sandoval; Elizabeth Cirulli; Fitri Carrissa Wijaya; Francisco |

Tanudjaja; Fred Wu; Geraint Levan; Greg Humphrey; Helena Tubb; James Lu + SEARCH; Jason Nguyen; Jimmy Ramirez; Kelly Schiabor Barrett; Laura Kohn; Magnus Isaksson; Marc Laurent; Marlene Flores; Megan Nguyen; Natasha Martin; Natasha Martin Cheryl Anderson; Nicole L Washington; Patricia Gonzalez-Zuniga; Rebecca Fielding-Miller; Rodolfo Salido; Ryan Cho; Sawyer Farmer; Sharon Reed; Sherry Wang; Simon White; Smruthi Karthikeyan; Tommy Valles + SEARCH; Tyler Cassens; Victor Cantu; Vinton Omaleki; William Lee

EPI_ISL_1192370, EPI_ISL_1192371, EPI_ISL_1197060, EPI_ISL_1197061, EPI_ISL_1303426, EPI_ISL_1303474, EPI_ISL_1303475, EPI_ISL_1303476, EPI_ISL_1303477, EPI_ISL_1303478, EPI_ISL_1303479, EPI_ISL_1303484, EPI_ISL_1303485, EPI_ISL_1303486, EPI_ISL_1303487, EPI_ISL_1303488, EPI_ISL_1307636, EPI_ISL_1307637, EPI_ISL_1379978, EPI_ISL_1497542, EPI_ISL_1669342, EPI_ISL_1706605, EPI_ISL_1706606, EPI_ISL_1706607, EPI_ISL_2259250

see above | Flow Diagnostics Lab | Los Angeles County Public Health Laboratories | P. Hemarajata et al.

EPI_ISL_1254009, EPI_ISL_1254010, EPI_ISL_1254011, EPI_ISL_1413763, EPI_ISL_1413766, EPI_ISL_1516746, EPI_ISL_1578329

see above | Flow Health | Infectious Disease Program, Broad Institute of Harvard and MIT | Adams, G.; B.L.; B.W.; Bauer, M.; Birren; Carter, A.; Chaluvadi, S.; D.J.; DeRuff, K.; Gallagher, G.; Gladden-Young, A.; J.E.; K.J.; Lagerborg, K.; Lemieux; Loreth, C.; MacInnis; Normandin, E.; P.C.; Park; Reilly, S.; Rudy, M.; Siddle; Smole, S.; Tomkins-Tinch, C.; and Sabeti

EPI_ISL_1457792, EPI_ISL_1457795, EPI_ISL_1457796, EPI_ISL_1457797, EPI_ISL_1457798, EPI_ISL_1457812, EPI_ISL_1457816, EPI_ISL_1457818, EPI_ISL_1457825, EPI_ISL_1457829, EPI_ISL_1457835, EPI_ISL_1457836, EPI_ISL_1457839, EPI_ISL_1457840, EPI_ISL_1457841, EPI_ISL_1457842, EPI_ISL_1457846, EPI_ISL_1457849, EPI_ISL_1457867, EPI_ISL_1457872, EPI_ISL_1457873, EPI_ISL_1457874, EPI_ISL_1457890, EPI_ISL_1457895, EPI_ISL_1457901, EPI_ISL_1457902, EPI_ISL_1457903, EPI_ISL_1457905, EPI_ISL_1457907, EPI_ISL_1457908, EPI_ISL_1457909, EPI_ISL_1458153, EPI_ISL_1458165, EPI_ISL_1458166, EPI_ISL_1458167, EPI_ISL_1458168, EPI_ISL_1458265, EPI_ISL_1458266, EPI_ISL_1458267, EPI_ISL_1458271, EPI_ISL_1458289, EPI_ISL_1458290, EPI_ISL_1458291, EPI_ISL_1458295, EPI_ISL_1458296, EPI_ISL_1458297, EPI_ISL_1458298, EPI_ISL_1458481, EPI_ISL_1458482, EPI_ISL_1458483, EPI_ISL_1458484, EPI_ISL_1458485, EPI_ISL_1458486, EPI_ISL_1458487, EPI_ISL_1458488, EPI_ISL_1458489, EPI_ISL_1458490, EPI_ISL_1458491, EPI_ISL_1458492, EPI_ISL_1458493, EPI_ISL_1458494, EPI_ISL_1458495, EPI_ISL_1458496, EPI_ISL_1458497, EPI_ISL_1458498, EPI_ISL_1458499, EPI_ISL_1458500, EPI_ISL_1458501, EPI_ISL_1458502, EPI_ISL_1458503, EPI_ISL_1458504, EPI_ISL_1458505, EPI_ISL_1458506, EPI_ISL_1458507, EPI_ISL_1458508, EPI_ISL_1458509, EPI_ISL_1458510, EPI_ISL_1458511, EPI_ISL_1458512, EPI_ISL_1458513, EPI_ISL_1458514, EPI_ISL_1458515, EPI_ISL_1458516, EPI_ISL_1458517, EPI_ISL_1458518, EPI_ISL_1458519, EPI_ISL_1458520, EPI_ISL_1458521, EPI_ISL_1458522, EPI_ISL_1458523, EPI_ISL_1458524, EPI_ISL_1458525, EPI_ISL_1458526, EPI_ISL_1458527, EPI_ISL_1458528, EPI_ISL_1458529, EPI_ISL_1458530, EPI_ISL_1458531, EPI_ISL_1458532, EPI_ISL_1458533, EPI_ISL_1458534, EPI_ISL_1458535, EPI_ISL_1458536, EPI_ISL_1458537, EPI_ISL_1458538, EPI_ISL_1458539, EPI_ISL_1458540, EPI_ISL_1458541, EPI_ISL_1458542, EPI_ISL_1458543, EPI_ISL_1458544, EPI_ISL_1458545, EPI_ISL_1458546, EPI_ISL_1458547, EPI_ISL_1458548, EPI_ISL_1458549, EPI_ISL_1458550, EPI_ISL_1458551, EPI_ISL_1458552, EPI_ISL_1458553, EPI_ISL_1458554, EPI_ISL_1458555, EPI_ISL_1458556, EPI_ISL_1458557, EPI_ISL_1458558, EPI_ISL_1458559, EPI_ISL_1458560, EPI_ISL_1458561, EPI_ISL_1458562, EPI_ISL_1458563, EPI_ISL_1458564, EPI_ISL_1458565, EPI_ISL_1458566, EPI_ISL_1458567, EPI_ISL_1458568, EPI_ISL_1458569, EPI_ISL_1458570, EPI_ISL_1458571, EPI_ISL_1458572, EPI_ISL_1458573, EPI_ISL_1458574, EPI_ISL_1458575, EPI_ISL_1458576, EPI_ISL_1458577, EPI_ISL_1458578, EPI_ISL_1458579, EPI_ISL_1458580, EPI_ISL_1458581, EPI_ISL_1458582, EPI_ISL_1458583, EPI_ISL_1458584, EPI_ISL_1458585, EPI_ISL_1458586, EPI_ISL_1458587, EPI_ISL_1458588, EPI_ISL_1458589, EPI_ISL_1458590, EPI_ISL_1458591, EPI_ISL_1458592, EPI_ISL_1458593, EPI_ISL_1458594, EPI_ISL_1458595, EPI_ISL_1458596, EPI_ISL_1458597, EPI_ISL_1458598, EPI_ISL_1458599, EPI_ISL_1458600, EPI_ISL_1458601, EPI_ISL_1458602, EPI_ISL_1458603, EPI_ISL_1458604, EPI_ISL_1458605, EPI_ISL_1458606, EPI_ISL_1458607, EPI_ISL_1458608, EPI_ISL_1458609, EPI_ISL_1458610, EPI_ISL_1458611, EPI_ISL_1458612, EPI_ISL_1458614, EPI_ISL_1458615, EPI_ISL_1458616, EPI_ISL_1458617, EPI_ISL_1458618, EPI_ISL_1458619, EPI_ISL_1458620, EPI_ISL_1458621, EPI_ISL_1458622, EPI_ISL_1458623, EPI_ISL_1458624, EPI_ISL_1458625, EPI_ISL_1458626, EPI_ISL_1458627, EPI_ISL_1458628, EPI_ISL_1458629, EPI_ISL_1458630, EPI_ISL_1458631, EPI_ISL_1458632, EPI_ISL_1458633, EPI_ISL_1458634, EPI_ISL_1458635, EPI_ISL_1458636, EPI_ISL_1458637, EPI_ISL_1458638, EPI_ISL_1458639, EPI_ISL_1458640, EPI_ISL_1458641, EPI_ISL_1458642, EPI_ISL_1458643, EPI_ISL_1458644, EPI_ISL_1458645, EPI_ISL_1458646, EPI_ISL_1458647, EPI_ISL_1458648, EPI_ISL_1458649, EPI_ISL_1458650, EPI_ISL_1458651, EPI_ISL_1458652, EPI_ISL_1458653, EPI_ISL_1458654, EPI_ISL_1458655, EPI_ISL_1458656, EPI_ISL_1458657, EPI_ISL_1458658, EPI_ISL_1458659, EPI_ISL_1458660, EPI_ISL_1458661, EPI_ISL_1458662, EPI_ISL_1458663, EPI_ISL_1458664, EPI_ISL_1458665, EPI_ISL_1458666, EPI_ISL_1458667, EPI_ISL_1458668, EPI_ISL_1458669, EPI_ISL_1458670, EPI_ISL_1458671, EPI_ISL_1458672, EPI_ISL_1458673, EPI_ISL_1458674, EPI_ISL_1458675, EPI_ISL_1458676, EPI_ISL_1458677, EPI_ISL_1458678, EPI_ISL_1458679, EPI_ISL_1458680, EPI_ISL_1458681, EPI_ISL_1458682, EPI_ISL_1458683, EPI_ISL_1458684, EPI_ISL_1458685, EPI_ISL_1458686, EPI_ISL_1458687, EPI_ISL_1458688, EPI_ISL_1458689, EPI_ISL_1458690, EPI_ISL_1458691, EPI_ISL_1458692, EPI_ISL_1458693, EPI_ISL_1458694, EPI_ISL_1458695, EPI_ISL_1458696, EPI_ISL_1458697, EPI_ISL_1458698, EPI_ISL_1458699, EPI_ISL_1458700, EPI_ISL_1458701, EPI_ISL_1458702, EPI_ISL_1458703, EPI_ISL_1458704, EPI_ISL_1458705, EPI_ISL_1458706, EPI_ISL_1458707, EPI_ISL_1458708, EPI_ISL_1458709, EPI_ISL_1458710, EPI_ISL_1458711, EPI_ISL_1458714, EPI_ISL_1458715, EPI_ISL_1458716, EPI_ISL_1458717, EPI_ISL_1458718, EPI_ISL_1458719, EPI_ISL_1458720, EPI_ISL_1458721, EPI_ISL_1458722, EPI_ISL_1458723, EPI_ISL_1458724, EPI_ISL_1458725, EPI_ISL_1458726, EPI_ISL_1458727, EPI_ISL_1458728, EPI_ISL_1458729, EPI_ISL_1458730, EPI_ISL_1458731, EPI_ISL_1458732, EPI_ISL_1458733, EPI_ISL_1458734, EPI_ISL_1458735, EPI_ISL_1458736, EPI_ISL_1458737, EPI_ISL_1458738, EPI_ISL_1458739, EPI_ISL_1458740, EPI_ISL_1458741, EPI_ISL_1458742, EPI_ISL_1458743, EPI_ISL_1458744, EPI_ISL_1458745, EPI_ISL_1458746, EPI_ISL_1458747, EPI_ISL_1458748, EPI_ISL_1458749, EPI_ISL_1458750, EPI_ISL_1458751, EPI_ISL_1458752, EPI_ISL_1458753, EPI_ISL_1458754, EPI_ISL_1458755, EPI_ISL_1458756, EPI_ISL_1458757, EPI_ISL_1458758, EPI_ISL_1458759, EPI_ISL_1458760, EPI_ISL_1458761, EPI_ISL_1458762, EPI_ISL_1458763, EPI_ISL_1458764, EPI_ISL_1458765, EPI_ISL_1458766, EPI_ISL_1458767, EPI_ISL_1458768, EPI_ISL_1458769, EPI_ISL_1458770, EPI_ISL_1458771, EPI_ISL_1458772, EPI_ISL_1458773, EPI_ISL_1458774, EPI_ISL_1458775, EPI_ISL_1458776, EPI_ISL_1458777, EPI_ISL_1458778, EPI_ISL_1458779, EPI_ISL_1458780, EPI_ISL_1458781, EPI_ISL_1458782, EPI_ISL_1458783, EPI_ISL_1458784, EPI_ISL_1458785, EPI_ISL_1458786, EPI_ISL_1458787, EPI_ISL_1458788, EPI_ISL_1458789, EPI_ISL_1458790, EPI_ISL_1458791, EPI_ISL_1458792, EPI_ISL_1458793, EPI_ISL_1458794, EPI_ISL_1458795, EPI_ISL_1458796, EPI_ISL_1458797, EPI_ISL_1458798, EPI_ISL_1458799, EPI_ISL_1458800, EPI_ISL_1458801, EPI_ISL_1458802, EPI_ISL_1458803, EPI_ISL_1458804, EPI_ISL_1458805, EPI_ISL_1458806, EPI_ISL_1458807, EPI_ISL_1458808, EPI_ISL_1458809, EPI_ISL_1458810, EPI_ISL_1458811, EPI_ISL_1458812, EPI_ISL_1458813, EPI_ISL_1458814, EPI_ISL_1458815, EPI_ISL_1458816, EPI_ISL_1458817, EPI_ISL_1458818, EPI_ISL_1458819, EPI_ISL_1458844, EPI_ISL_1458845, EPI_ISL_1458847, EPI_ISL_1458850, EPI_ISL_1458851, EPI_ISL_1458852, EPI_ISL_1458853, EPI_ISL_1458854, EPI_ISL_1458855, EPI_ISL_1458856, EPI_ISL_1458857, EPI_ISL_1458858, EPI_ISL_1458859, EPI_ISL_1458900, EPI_ISL_1458901, EPI_ISL_1458902, EPI_ISL_1458903, EPI_ISL_1458904, EPI_ISL_1458905, EPI_ISL_1458906, EPI_ISL_1458907, EPI_ISL_1458908, EPI_ISL_1458909, EPI_ISL_1458910, EPI_ISL_1458911, EPI_ISL_1458912, EPI_ISL_1458913, EPI_ISL_1458914, EPI_ISL_1458915, EPI_ISL_1458916, EPI_ISL_1458917, EPI_ISL_1458918, EPI_ISL_1458919, EPI_ISL_1458920, EPI_ISL_1458921, EPI_ISL_1458922, EPI_ISL_1458923, EPI_ISL_1458924, EPI_ISL_1458925, EPI_ISL_1458926, EPI_ISL_1458927, EPI_ISL_1458928, EPI_ISL_1458929, EPI_ISL_1458930, EPI_ISL_1458931, EPI_ISL_1458932, EPI_ISL_1458933, EPI_ISL_1458934, EPI_ISL_1458935, EPI_ISL_1458936, EPI_ISL_1458937, EPI_ISL_1458938, EPI_ISL_1458939, EPI_ISL_1458965, EPI_ISL_1458968, EPI_ISL_1458970, EPI_ISL_1458971, EPI_ISL_1458972, EPI_ISL_1458973, EPI_ISL_1458974, EPI_ISL_1458975, EPI_ISL_1458976, EPI_ISL_1458977, EPI_ISL_1458978, EPI_ISL_1458979, EPI_ISL_1459256, EPI_ISL_1459257, EPI_ISL_1459258, EPI_ISL_1459259, EPI_ISL_1459260, EPI_ISL_1459261, EPI_ISL_1459262, EPI_ISL_1459263, EPI_ISL_1459264, EPI_ISL_1459265, EPI_ISL_1459266, EPI_ISL_1459267, EPI_ISL_1459268, EPI_ISL_1459269, EPI_ISL_1459292, EPI_ISL_1459293, EPI_ISL_1459294, EPI_ISL_1459295, EPI_ISL_1459296, EPI_ISL_1459297, EPI_ISL_1459298, EPI_ISL_1459299, EPI_ISL_1459300, EPI_ISL_1459301, EPI_ISL_1459302, EPI_ISL_1459303, EPI_ISL_1459304, EPI_ISL_1459305, EPI_ISL_1459306, EPI_ISL_1459307, EPI_ISL_1459308, EPI_ISL_1459309, EPI_ISL_1459310, EPI_ISL_1459311, EPI_ISL_1459312, EPI_ISL_1459401, EPI_ISL_1459402, EPI_ISL_1459403, EPI_ISL_1459404, EPI_ISL_1459405, EPI_ISL_1459406, EPI_ISL_1459407, EPI_ISL_1459408, EPI_ISL_1459409, EPI_ISL_1459410, EPI_ISL_1459411, EPI_ISL_1459412, EPI_ISL_1459413, EPI_ISL_1459414, EPI_ISL_1459415, EPI_ISL_1459416, EPI_ISL_1459417, EPI_ISL_1459418, EPI_ISL_1459419, EPI_ISL_1459420, EPI_ISL_1459421, EPI_ISL_1459422, EPI_ISL_1459423, EPI_ISL_1459424, EPI_ISL_1459425, EPI_ISL_1459426, EPI_ISL_1459427, EPI_ISL_1459428, EPI_ISL_1459429, EPI_ISL_1459430, EPI_ISL_1459431, EPI_ISL_1459432, EPI_ISL_1459433, EPI_ISL_1459434, EPI_ISL_1459435, EPI_ISL_1459436, EPI_ISL_1459437, EPI_ISL_1459438, EPI_ISL_1459439, EPI_ISL_1459440, EPI_ISL_1459441, EPI_ISL_1459442, EPI_ISL_1459443, EPI_ISL_1459444, EPI_ISL_1459445, EPI_ISL_1459446, EPI_ISL_1459447, EPI_ISL_1459448, EPI_ISL_1459449, EPI_ISL_1459450, EPI_ISL_1459451, EPI_ISL_1459452, EPI_ISL_1459453, EPI_ISL_1459454, EPI_ISL_1459455, EPI_ISL_1459456, EPI_ISL_1459457, EPI_ISL_1459458, EPI_ISL_1459459, EPI_ISL_1459460, EPI_ISL_1459461, EPI_ISL_1459462, EPI_ISL_1459463, EPI_ISL_1459464, EPI_ISL_1459465, EPI_ISL_1459466, EPI_ISL_1459467, EPI_ISL_1459468, EPI_ISL_1459469, EPI_ISL_1459470, EPI_ISL_1459471, EPI_ISL_1459472, EPI_ISL_1459473, EPI_ISL_1459474, EPI_ISL_1459475, EPI_ISL_1459476, EPI_ISL_1459477, EPI_ISL_1459478, EPI_ISL_1459479, EPI_ISL_1459480, EPI_ISL_1459481, EPI_ISL_1459482, EPI_ISL_1459483, EPI_ISL_1459484, EPI_ISL_1459485, EPI_ISL_1459486, EPI_ISL_1459487, EPI_ISL_1459488, EPI_ISL_1459489, EPI_ISL_1459490, EPI_ISL_1459491, EPI_ISL_1459492, EPI_ISL_1459493, EPI_ISL_1459494, EPI_ISL_1459495, EPI_ISL_1459496, EPI_ISL_1459497, EPI_ISL_1459498, EPI_ISL_1459499, EPI_ISL_1459500, EPI_ISL_1459501, EPI_ISL_1459502, EPI_ISL_1459503, EPI_ISL_1459504, EPI_ISL_1459505, EPI_ISL_1459506, EPI_ISL_1459507, EPI_ISL_1459508, EPI_ISL_1459529, EPI_ISL_1459530, EPI_ISL_1459531, EPI_ISL_1459532, EPI_ISL_1459533, EPI_ISL_1459534, EPI_ISL_1459535, EPI_ISL_1459536, EPI_ISL_1459537, EPI_ISL_1459538, EPI_ISL_1459539, EPI_ISL_1459540, EPI_ISL_1459541, EPI_ISL_1459542, EPI_ISL_1459543, EPI_ISL_1459544, EPI_ISL_1459545, EPI_ISL_1459546, EPI_ISL_1459547, EPI_ISL_1459548, EPI_ISL_1459549, EPI_ISL_1459550, EPI_ISL_1459551, EPI_ISL_1459552, EPI_ISL_1459553, EPI_ISL_1459554, EPI_ISL_1459555, EPI_ISL_1459556, EPI_ISL_1459557, EPI_ISL_1459558, EPI_ISL_1459559, EPI_ISL_1459560, EPI_ISL_1459561, EPI_ISL_1459562, EPI_ISL_1459563, EPI_ISL_1459564, EPI_ISL_1459565, EPI_ISL_1459566, EPI_ISL_1459567, EPI_ISL_1459568, EPI_ISL_1459569, EPI_ISL_1459570, EPI_ISL_1459571, EPI_ISL_1459572, EPI_ISL_1459573, EPI_ISL_1459574, EPI_ISL_1459575, EPI_ISL_1459576, EPI_ISL_1459577, EPI_ISL_1459578, EPI_ISL_1459579, EPI_ISL_1459580, EPI_ISL_1459581, EPI_ISL_1459582, EPI_ISL_1459583, EPI_ISL_1459584, EPI_ISL_1459585, EPI_ISL_1459586, EPI_ISL_1459587, EPI_ISL_1459588, EPI_ISL_1459589, EPI_ISL_1459590, EPI_ISL_1459591, EPI_ISL_1459592, EPI_ISL_1459593, EPI_ISL_1459594, EPI_ISL_1459595, EPI_ISL_1459596, EPI_ISL_1459597, EPI_ISL_1459598, EPI_ISL_1459599, EPI_ISL_1459600, EPI_ISL_1459601, EPI_ISL_1459602, EPI_ISL_1459603, EPI_ISL_1459604, EPI_ISL_1459605, EPI_ISL_1459606, EPI_ISL_1459607, EPI_ISL_1459608, EPI_ISL_1459609, EPI_ISL_1459610, EPI_ISL_1459611, EPI_ISL_1459612, EPI_ISL_1459613, EPI_ISL_1459614, EPI_ISL_1459615, EPI_ISL_1459616, EPI_ISL_1459620, EPI_ISL_1459621, EPI_ISL_1459622, EPI_ISL_1459626, EPI_ISL_1459628, EPI_ISL_1459629, EPI_ISL_1459630, EPI_ISL_1459631, EPI_ISL_1459632, EPI_ISL_1459633, EPI_ISL_1459634, EPI_ISL_1459635, EPI_ISL_1459637, EPI_ISL_1459638, EPI_ISL_1459639, EPI_ISL_1459640, EPI_ISL_1459641, EPI_ISL_1459645, EPI_ISL_1459646, EPI_ISL_1459648, EPI_ISL_1459650, EPI_ISL_1459651, EPI_ISL_1459652, EPI_ISL_1459653, EPI_ISL_1459654, EPI_ISL_1459655, EPI_ISL_1459656, EPI_ISL_1459657, EPI_ISL_1459658, EPI_ISL_1459659, EPI_ISL_1459660, EPI_ISL_1459661, EPI_ISL_1459662, EPI_ISL_1459663, EPI_ISL_1459664, EPI_ISL_1459665, EPI_ISL_1459666, EPI_ISL_1459667, EPI_ISL_1459668, EPI_ISL_1459669, EPI_ISL_1459670, EPI_ISL_1459671, EPI_ISL_1459672, EPI_ISL_1459673, EPI_ISL_1459674, EPI_ISL_1459675, EPI_ISL_1459676, EPI_ISL_1459677, EPI_ISL_1459678, EPI_ISL_1459679, EPI_ISL_1459680, EPI_ISL_1459681, EPI_ISL_1459682, EPI_ISL_1459683, EPI_ISL_1459684, EPI_ISL_1459685, EPI_ISL_1459686, EPI_ISL_1459687, EPI_ISL_1459688, EPI_ISL_1459689, EPI_ISL_1459690, EPI_ISL_1459691, EPI_ISL_1459692, EPI_ISL_1459693, EPI_ISL_1459694, EPI_ISL_1459695, EPI_ISL_1459696, EPI_ISL_1459697, EPI_ISL_1459698, EPI_ISL_1459699, EPI_ISL_1459700, EPI_ISL_1459701, EPI_ISL_1459702, EPI_ISL_1459703, EPI_ISL_1459704, EPI_ISL_1459705, EPI_ISL_1459706, EPI_ISL_1459707, EPI_ISL_1459708, EPI_ISL_1459709, EPI_ISL_1459710, EPI_ISL_1459711, EPI_ISL_1459712, EPI_ISL_1459713, EPI_ISL_1459714, EPI_ISL_1459715, EPI_ISL_1459716, EPI_ISL_1459717, EPI_ISL_1459718, EPI_ISL_1459719, EPI_ISL_1459720, EPI_ISL_1459721, EPI_ISL_1459722, EPI_ISL_1459723, EPI_ISL_1459724, EPI_ISL_1459725, EPI_ISL_1459726, EPI_ISL_1459727, EPI_ISL_1459728, EPI_ISL_1459729, EPI_ISL_1459730, EPI_ISL_1459731, EPI_ISL_1459732, EPI_ISL_1459733, EPI_ISL_1459734, EPI_ISL_1459735, EPI_ISL_1459736, EPI_ISL_1459737, EPI_ISL_1459738, EPI_ISL_1459739, EPI_ISL_1459740, EPI_ISL_1459741, EPI_ISL_1459742, EPI_ISL_1459743, EPI_ISL_1459744, EPI_ISL_1459745, EPI_ISL_1459746, EPI_ISL_1459747, EPI_ISL_1459748, EPI_ISL_1459749, EPI_ISL_1459750, EPI_ISL_1459751, EPI_ISL_1459752, EPI_ISL_1459753, EPI_ISL_1459754, EPI_ISL_1459755, EPI_ISL_1459756, EPI_ISL_1459757, EPI_ISL_1459758, EPI_ISL_1459759, EPI_ISL_1459760, EPI_ISL_1459761, EPI_ISL_1459762, EPI_ISL_1459763, EPI_ISL_1459764, EPI_ISL_1459765, EPI_ISL_1459766, EPI_ISL_1459767, EPI_ISL_1459768, EPI_ISL_1459769, EPI_ISL_1459771, EPI_ISL_1459772, EPI_ISL_1459773, EPI_ISL_1459774, EPI_ISL_1459775, EPI_ISL_1459776, EPI_ISL_1459777, EPI_ISL_1459778, EPI_ISL_1459779, EPI_ISL_1459780, EPI_ISL_1459781, EPI_ISL_1459782, EPI_ISL_1459783, EPI_ISL_1459784, EPI_ISL_1459785, EPI_ISL_1459786, EPI_ISL_1459787, EPI_ISL_1459788, EPI_ISL_1459789, EPI_ISL_1459790, EPI_ISL_1459791, EPI_ISL_1459792, EPI_ISL_1459793, EPI_ISL_1459794, EPI_ISL_1459795, EPI_ISL_1459796, EPI_ISL_1459797, EPI_ISL_1459798, EPI_ISL_1459799, EPI_ISL_1459800, EPI_ISL_1459801, EPI_ISL_1459802, EPI_ISL_1459803, EPI_ISL_1459804, EPI_ISL_1459805, EPI_ISL_1459806, EPI_ISL_1459807, EPI_ISL_1459808, EPI_ISL_1459809, EPI_ISL_1459810, EPI_ISL_1459811, EPI_ISL_1459812, EPI_ISL_1459813, EPI_ISL_1459814, EPI_ISL_1459815, EPI_ISL_1459816, EPI_ISL_1459817, EPI_ISL_1459818, EPI_ISL_1459819, EPI_ISL_1459820, EPI_ISL_1459821, EPI_ISL_1459822, EPI_ISL_1459823, EPI_ISL_1459824, EPI_ISL_1459825, EPI_ISL_1459826, EPI_ISL_1459827, EPI_ISL_1459828, EPI_ISL_1459829, EPI_ISL_1459830, EPI_ISL_1459831, EPI_ISL_1459832, EPI_ISL_1459833, EPI_ISL_1459834, EPI_ISL_1459835, EPI_ISL_1459836, EPI_ISL_1459837, EPI_ISL_1459838, EPI_ISL_1459839, EPI_ISL_1459840, EPI_ISL_1459842, EPI_ISL_1459843, EPI_ISL_1459844, EPI_ISL_1459845, EPI_ISL_1459846, EPI_ISL_1459848, EPI_ISL_1459849, EPI_ISL_1459850, EPI_ISL_1459851, EPI_ISL_1459852, EPI_ISL_1459853, EPI_ISL_1459854, EPI_ISL_1459855, EPI_ISL_1459856, EPI_ISL_1459857, EPI_ISL_1459858, EPI_ISL_1459859, EPI_ISL_1459860, EPI_ISL_1459861, EPI_ISL_1459862, EPI_ISL_1459863, EPI_ISL_1459864, EPI_ISL_1459865, EPI_ISL_1459866, EPI_ISL_1459867, EPI_ISL_1459868, EPI_ISL_1459869, EPI_ISL_1459870, EPI_ISL_1459871, EPI_ISL_1459872, EPI_ISL_1459873, EPI_ISL_1459874, EPI_ISL_1459875, EPI_ISL_1459876, EPI_ISL_1459877, EPI_ISL_1459878, EPI_ISL_1459879, EPI_ISL_1459880, EPI_ISL_1459881, EPI_ISL_1459882, EPI_ISL_1459883, EPI_ISL_1459884, EPI_ISL_1459885, EPI_ISL_1459886, EPI_ISL_1459887, EPI_ISL_1459888, EPI_ISL_1459889, EPI_ISL_1459890, EPI_ISL_1459891, EPI_ISL_1459892, EPI_ISL_1459893, EPI_ISL_1459894, EPI_ISL_1459895, EPI_ISL_1459896, EPI_ISL_1459897, EPI_ISL_1459898, EPI_ISL_1459899, EPI_ISL_1459900, EPI_ISL_1459901, EPI_ISL_1459902, EPI_ISL_1459903, EPI_ISL_1459904, EPI_ISL_1459905, EPI_ISL_1459906, EPI_ISL_1459907, EPI_ISL_1459908, EPI_ISL_1459909, EPI_ISL_1459910, EPI_ISL_1459911, EPI_ISL_1459912, EPI_ISL_1459913, EPI_ISL_1459914, EPI_ISL_1459915, EPI_ISL_1459916, EPI_ISL_1459917, EPI_ISL_1459918, EPI_ISL_1459919, EPI_ISL_1459920, EPI_ISL_1459921, EPI_ISL_1459922, EPI_ISL_1459923, EPI_ISL_1459924, EPI_ISL_1459925, EPI_ISL_1459926, EPI_ISL_1459927, EPI_ISL_1459928, EPI_ISL_1459929, EPI_ISL_1459930, EPI_ISL_1459931, EPI_ISL_1459932, EPI_ISL_1459933, EPI_ISL_1459934, EPI_ISL_1459935, EPI_ISL_1459936, EPI_ISL_1459937, EPI_ISL_1459938, EPI_ISL_1459939, EPI_ISL_1459940, EPI_ISL_1459941, EPI_ISL_1459942, EPI_ISL_1459943, EPI_ISL_1459944, EPI_ISL_1459945, EPI_ISL_1459946, EPI_ISL_1459947, EPI_ISL_1459948, EPI_ISL_1459949, EPI_ISL_1459950, EPI_ISL_1459951, EPI_ISL_1459952, EPI_ISL_1459953, EPI_ISL_1459954, EPI_ISL_1459955, EPI_ISL_1459956, EPI_ISL_1459957, EPI_ISL_1460123, EPI_ISL_1460124, EPI_ISL_1460125, EPI_ISL_1460126, EPI_ISL_1460127, EPI_ISL_1460128, EPI_ISL_1460131, EPI_ISL_1460134, EPI_ISL_1460135, EPI_ISL_1460136, EPI_ISL_1460137, EPI_ISL_1460138, EPI_ISL_1460139, EPI_ISL_1460140, EPI_ISL_1460141, EPI_ISL_1460142, EPI_ISL_1460143, EPI_ISL_1460144, EPI_ISL_1460145, EPI_ISL_1460146, EPI_ISL_1460147, EPI_ISL_1460148, EPI_ISL_1460149, EPI_ISL_1460150, EPI_ISL_1460151, EPI_ISL_1460152, EPI_ISL_1460153, EPI_ISL_1460154, EPI_ISL_1460155, EPI_ISL_1460156, EPI_ISL_1460157, EPI_ISL_1460158, EPI_ISL_1460159, EPI_ISL_1460160, EPI_ISL_1460161, EPI_ISL_1460162, EPI_ISL_1460163, EPI_ISL_1460164, EPI_ISL_1460165, EPI_ISL_1460166, EPI_ISL_1460168, EPI_ISL_1460169, EPI_ISL_1460170, EPI_ISL_1460295, EPI_ISL_1460296, EPI_ISL_1460297, EPI_ISL_1460298, EPI_ISL_1460299, EPI_ISL_1460300, EPI_ISL_1460301, EPI_ISL_1460302, EPI_ISL_1460303, EPI_ISL_1460304, EPI_ISL_1460305, EPI_ISL_1460306, EPI_ISL_1460307, EPI_ISL_1460308, EPI_ISL_1460309, EPI_ISL_1460310, EPI_ISL_1460311, EPI_ISL_1460312, EPI_ISL_1460313, EPI_ISL_1460314, EPI_ISL_1460315, EPI_ISL_1460316, EPI_ISL_1460317, EPI_ISL_1460318, EPI_ISL_1460319, EPI_ISL_1460320, EPI_ISL_1460321, EPI_ISL_1460322, EPI_ISL_1460323, EPI_ISL_1460324, EPI_ISL_1460325, EPI_ISL_1460326, EPI_ISL_1460327, EPI_ISL_1460328, EPI_ISL_1460329, EPI_ISL_1460330, EPI_ISL_1460331, EPI_ISL_1460332, EPI_ISL_1460333, EPI_ISL_1460334, EPI_ISL_1460335, EPI_ISL_1460336, EPI_ISL_1460337, EPI_ISL_1460338, EPI_ISL_1460339, EPI_ISL_1460340, EPI_ISL_1460341, EPI_ISL_1460342, EPI_ISL_1460343, EPI_ISL_1460344, EPI_ISL_1460345, EPI_ISL_1460346, EPI_ISL_1460347, EPI_ISL_1460348, EPI_ISL_1460349, EPI_ISL_1460350, EPI_ISL_1460351, EPI_ISL_1460352, EPI_ISL_1460353, EPI_ISL_1460356, EPI_ISL_1460357, EPI_ISL_1460358, EPI_ISL_1460359, EPI_ISL_1460360, EPI_ISL_1460361, EPI_ISL_1460362, EPI_ISL_1460363, EPI_ISL_1460364, EPI_ISL_1460365, EPI_ISL_1460366, EPI_ISL_1460367, EPI_ISL_1460368, EPI_ISL_1460369, EPI_ISL_1460370, EPI_ISL_1460371, EPI_ISL_1460372, EPI_ISL_1460373, EPI_ISL_1460374, EPI_ISL_1460375, EPI_ISL_1460376, EPI_ISL_1460377, EPI_ISL_1460378, EPI_ISL_1460379, EPI_ISL_1460380, EPI_ISL_1460381, EPI_ISL_1460382, EPI_ISL_1460383, EPI_ISL_1460384, EPI_ISL_1460385, EPI_ISL_1460386, EPI_ISL_1460387, EPI_ISL_1460388, EPI_ISL_1460389, EPI_ISL_1460390, EPI_ISL_1460391, EPI_ISL_1460392, EPI_ISL_1460393, EPI_ISL_1460394, EPI_ISL_1460395, EPI_ISL_1460396, EPI_ISL_1460397, EPI_ISL_1460398, EPI_ISL_1460399, EPI_ISL_1460400, EPI_ISL_1460401, EPI_ISL_1460402, EPI_ISL_1460403, EPI_ISL_1460404, EPI_ISL_1460405, EPI_ISL_1460406, EPI_ISL_1460407, EPI_ISL_1460408, EPI_ISL_1460409, EPI_ISL_1460410, EPI_ISL_1460411, EPI_ISL_1460412, EPI_ISL_1460413, EPI_ISL_1460414, EPI_ISL_1460415, EPI_ISL_1460416, EPI_ISL_1460417, EPI_ISL_1460418, EPI_ISL_1460419, EPI_ISL_1460420, EPI_ISL_1460421, EPI_ISL_1460422, EPI_ISL_1460423, EPI_ISL_1460424, EPI_ISL_1460425, EPI_ISL_1460426, EPI_ISL_1460427, EPI_ISL_1460428, EPI_ISL_1460429, EPI_ISL_1460430, EPI_ISL_1460431, EPI_ISL_1460432, EPI_ISL_1460433, EPI_ISL_1460434, EPI_ISL_1460435, EPI_ISL_1460436, EPI_ISL_1460437, EPI_ISL_1460438, EPI_ISL_1460439, EPI_ISL_1460440, EPI_ISL_1460442, EPI_ISL_1460443, EPI_ISL_1460444, EPI_ISL_1460445, EPI_ISL_1460446, EPI_ISL_1460447, EPI_ISL_1460448, EPI_ISL_1460449, EPI_ISL_1460450, EPI_ISL_1460451, EPI_ISL_1460452, EPI_ISL_1460453, EPI_ISL_1460454, EPI_ISL_1460455, EPI_ISL_1460456, EPI_ISL_1460457, EPI_ISL_1460458, EPI_ISL_1460459, EPI_ISL_1460460, EPI_ISL_1460461, EPI_ISL_1460462, EPI_ISL_1460463, EPI_ISL_1460464, EPI_ISL_1460465, EPI_ISL_1460466, EPI_ISL_1460467, EPI_ISL_1460468, EPI_ISL_1460469, EPI_ISL_1460470, EPI_ISL_1460471, EPI_ISL_1460472, EPI_ISL_1460473, EPI_ISL_1460474, EPI_ISL_1460475, EPI_ISL_1460476, EPI_ISL_1460477, EPI_ISL_1460478, EPI_ISL_1460479, EPI_ISL_1460480, EPI_ISL_1460481, EPI_ISL_1460482, EPI_ISL_1460483, EPI_ISL_1460484, EPI_ISL_1460485, EPI_ISL_1460486, EPI_ISL_1460487, EPI_ISL_1460488, EPI_ISL_1460489, EPI_ISL_1460490, EPI_ISL_1460491, EPI_ISL_1460492, EPI_ISL_1460493, EPI_ISL_1460494, EPI_ISL_1460495, EPI_ISL_1460496, EPI_ISL_1460497, EPI_ISL_1460498, EPI_ISL_1460499, EPI_ISL_1460500, EPI_ISL_1460501, EPI_ISL_1460502, EPI_ISL_1460503, EPI_ISL_1460504, EPI_ISL_1460505, EPI_ISL_1460506, EPI_ISL_1460507, EPI_ISL_1460508, EPI_ISL_1460509, EPI_ISL_1460510, EPI_ISL_1460511, EPI_ISL_1460512, EPI_ISL_1460513, EPI_ISL_1460514, EPI_ISL_1460515, EPI_ISL_1460516, EPI_ISL_1460517, EPI_ISL_1460518, EPI_ISL_1460519, EPI_ISL_1460520, EPI_ISL_1460521, EPI_ISL_1460522, EPI_ISL_1460523, EPI_ISL_1460524, EPI_ISL_1460525, EPI_ISL_1460526, EPI_ISL_1460527, EPI_ISL_1460528, EPI_ISL_1460529, EPI_ISL_1460530, EPI_ISL_1460531, EPI_ISL_1460532, EPI_ISL_1460533, EPI_ISL_1460534, EPI_ISL_1460535, EPI_ISL_1460536, EPI_ISL_1460537, EPI_ISL_1460538, EPI_ISL_1460539, EPI_ISL_1460540, EPI_ISL_1460541, EPI_ISL_1460542, EPI_ISL_1460543, EPI_ISL_1460544, EPI_ISL_1460545, EPI_ISL_1460546, EPI_ISL_1460547, EPI_ISL_1460548, EPI_ISL_1460549, EPI_ISL_1460550, EPI_ISL_1460551, EPI_ISL_1460552, EPI_ISL_1460553, EPI_ISL_1460554, EPI_ISL_1460555, EPI_ISL_1460556, EPI_ISL_1460557, EPI_ISL_1460558, EPI_ISL_1460559, EPI_ISL_1460560, EPI_ISL_1460561, EPI_ISL_1460562, EPI_ISL_1460563, EPI_ISL_1460564, EPI_ISL_1460565, EPI_ISL_1460566, EPI_ISL_1460567, EPI_ISL_1460568, EPI_ISL_1460569, EPI_ISL_1460570, EPI_ISL_1460571, EPI_ISL_1460572, EPI_ISL_1460573, EPI_ISL_1460574, EPI_ISL_1460575, EPI_ISL_1460576, EPI_ISL_1460577, EPI_ISL_1460578, EPI_ISL_1460580, EPI_ISL_1460581, EPI_ISL_1460582, EPI_ISL_1460583, EPI_ISL_1460584, EPI_ISL_1460585, EPI_ISL_1460586, EPI_ISL_1460587, EPI_ISL_1460588, EPI_ISL_1460589, EPI_ISL_1460590, EPI_ISL_1460591, EPI_ISL_1460592, EPI_ISL_1460593, EPI_ISL_1460594, EPI_ISL_1460595, EPI_ISL_1460596, EPI_ISL_1460597, EPI_ISL_1460598, EPI_ISL_1460599, EPI_ISL_1460600, EPI_ISL_1460601, EPI_ISL_1460602, EPI_ISL_1460603, EPI_ISL_1460604, EPI_ISL_1460605, EPI_ISL_1460606, EPI_ISL_1460607, EPI_ISL_1460608, EPI_ISL_1460609, EPI_ISL_1460610, EPI_ISL_1460611, EPI_ISL_1460612, EPI_ISL_1460613, EPI_ISL_1460614, EPI_ISL_1460615, EPI_ISL_1460616, EPI_ISL_1460617, EPI_ISL_1460618, EPI_ISL_1460619, EPI_ISL_1460620, EPI_ISL_1460621, EPI_ISL_1460622, EPI_ISL_1460623, EPI_ISL_1460624, EPI_ISL_1460625, EPI_ISL_1460626, EPI_ISL_1460627, EPI_ISL_1460628, EPI_ISL_1460629, EPI_ISL_1460630, EPI_ISL_1460631, EPI_ISL_1460632, EPI_ISL_1460633, EPI_ISL_1460634, EPI_ISL_1460635, EPI_ISL_1460636, EPI_ISL_1460637, EPI_ISL_1460638, EPI_ISL_1460639, EPI_ISL_1460640, EPI_ISL_1460641, EPI_ISL_1460642, EPI_ISL_1460643, EPI_ISL_1460644, EPI_ISL_1460645, EPI_ISL_1460646, EPI_ISL_1460647, EPI_ISL_1460648, EPI_ISL_1460649, EPI_ISL_1460650, EPI_ISL_1460651, EPI_ISL_1460652, EPI_ISL_1460653, EPI_ISL_1460654, EPI_ISL_1460655, EPI_ISL_1460656, EPI_ISL_1460657, EPI_ISL_1460658, EPI_ISL_1460659, EPI_ISL_1460660, EPI_ISL_1460661, EPI_ISL_1460662, EPI_ISL_1460663, EPI_ISL_1460664, EPI_ISL_1460665, EPI_ISL_1460666, EPI_ISL_1460667, EPI_ISL_1460668, EPI_ISL_1460669, EPI_ISL_1460670, EPI_ISL_1460671, EPI_ISL_1460672, EPI_ISL_1460673, EPI_ISL_1460674, EPI_ISL_1460675, EPI_ISL_1460676, EPI_ISL_1460677, EPI_ISL_1460678, EPI_ISL_1460679, EPI_ISL_1460680, EPI_ISL_1460681, EPI_ISL_1460682, EPI_ISL_1481837, EPI_ISL_1481931, EPI_ISL_1481934, EPI_ISL_1481936, EPI_ISL_1481944, EPI_ISL_1481945, EPI_ISL_1481946, EPI_ISL_1481947, EPI_ISL_1481980, EPI_ISL_1482000, EPI_ISL_1482001, EPI_ISL_1482002, EPI_ISL_1482003, EPI_ISL_1482004, EPI_ISL_1482005, EPI_ISL_1482006, EPI_ISL_1482007, EPI_ISL_1482008, EPI_ISL_1482009, EPI_ISL_1482010, EPI_ISL_1482011, EPI_ISL_1482012, EPI_ISL_1482013, EPI_ISL_1482014, EPI_ISL_1482015, EPI_ISL_1482016, EPI_ISL_1482017, EPI_ISL_1482018, EPI_ISL_1482019, EPI_ISL_1482020, EPI_ISL_1482021, EPI_ISL_1482022, EPI_ISL_1482023, EPI_ISL_1482024, EPI_ISL_1482025, EPI_ISL_1482026, EPI_ISL_1482027, EPI_ISL_1482028, EPI_ISL_1482029, EPI_ISL_1482030, EPI_ISL_1482031, EPI_ISL_1482032, EPI_ISL_1482033, EPI_ISL_1482034, EPI_ISL_1482035, EPI_ISL_1482042, EPI_ISL_1482043, EPI_ISL_1482044, EPI_ISL_1482045, EPI_ISL_1482046, EPI_ISL_1482056, EPI_ISL_1482057, EPI_ISL_1482058, EPI_ISL_1482059, EPI_ISL_1482060, EPI_ISL_1482061, EPI_ISL_1482062, EPI_ISL_1482063, EPI_ISL_1482064, EPI_ISL_1482065, EPI_ISL_1482066, EPI_ISL_1482067, EPI_ISL_1482068, EPI_ISL_1482069, EPI_ISL_1482070, EPI_ISL_1482071, EPI_ISL_1482072, EPI_ISL_1482073, EPI_ISL_1482074, EPI_ISL_1482075, EPI_ISL_1482076, EPI_ISL_1482077, EPI_ISL_1482078, EPI_ISL_1482080, EPI_ISL_1482081, EPI_ISL_1482082, EPI_ISL_1482083, EPI_ISL_1482084, EPI_ISL_1482085, EPI_ISL_1482086, EPI_ISL_1482087, EPI_ISL_1482088, EPI_ISL_1482089, EPI_ISL_1482090, EPI_ISL_1482091, EPI_ISL_1482092, EPI_ISL_1482093, EPI_ISL_1482094, EPI_ISL_1482095, EPI_ISL_1482096, EPI_ISL_1482097, EPI_ISL_1482098, EPI_ISL_1482099, EPI_ISL_1482101, EPI_ISL_1482102, EPI_ISL_1482103, EPI_ISL_1482104, EPI_ISL_1482105, EPI_ISL_1482106, EPI_ISL_1482107, EPI_ISL_1482108, EPI_ISL_1482109, EPI_ISL_1482110, EPI_ISL_1482111, EPI_ISL_1482126, EPI_ISL_1482127, EPI_ISL_1482128, EPI_ISL_1482129, EPI_ISL_1482130, EPI_ISL_1482131, EPI_ISL_1482141, EPI_ISL_1482142, EPI_ISL_1482143, EPI_ISL_1482144, EPI_ISL_1482145, EPI_ISL_1482146, EPI_ISL_1482147, EPI_ISL_1482148, EPI_ISL_1482149, EPI_ISL_1482150, EPI_ISL_1482151, EPI_ISL_1482152, EPI_ISL_1482153, EPI_ISL_1482154, EPI_ISL_1482155, EPI_ISL_1482156, EPI_ISL_1482157, EPI_ISL_1482158, EPI_ISL_1482159, EPI_ISL_1482160, EPI_ISL_1482161, EPI_ISL_1482162, EPI_ISL_1482163, EPI_ISL_1482164, EPI_ISL_1482165, EPI_ISL_1482166, EPI_ISL_1482167, EPI_ISL_1482168, EPI_ISL_1482169, EPI_ISL_1482170, EPI_ISL_1482171, EPI_ISL_1554950, EPI_ISL_1554956, EPI_ISL_1554967, EPI_ISL_1554969, EPI_ISL_1554971, EPI_ISL_1555083, EPI_ISL_1555264, EPI_ISL_1555265, EPI_ISL_1555266, EPI_ISL_1555267, EPI_ISL_1555268, EPI_ISL_1555269, EPI_ISL_1555270, EPI_ISL_1555271, EPI_ISL_1555272, EPI_ISL_1555273, EPI_ISL_1555274, EPI_ISL_1555275, EPI_ISL_1555276, EPI_ISL_1555277, EPI_ISL_1555278, EPI_ISL_1555279, EPI_ISL_1555280, EPI_ISL_1555281, EPI_ISL_1555282, EPI_ISL_1555283, EPI_ISL_1555284, EPI_ISL_1555285, EPI_ISL_1555286, EPI_ISL_1555287, EPI_ISL_1555289, EPI_ISL_1555290, EPI_ISL_1555291, EPI_ISL_1555292, EPI_ISL_1555293, EPI_ISL_1555294, EPI_ISL_1555295, EPI_ISL_1555296, EPI_ISL_1555297, EPI_ISL_1555298, EPI_ISL_1555299, EPI_ISL_1555300, EPI_ISL_1555301, EPI_ISL_1555302, EPI_ISL_1555303, EPI_ISL_1555306, EPI_ISL_1555307, EPI_ISL_1555308, EPI_ISL_1555309, EPI_ISL_1555310, EPI_ISL_1555311, EPI_ISL_1555312, EPI_ISL_1555313, EPI_ISL_1555314, EPI_ISL_1555315, EPI_ISL_1555316, EPI_ISL_1555317, EPI_ISL_1555318, EPI_ISL_1555319, EPI_ISL_1555320, EPI_ISL_1555321, EPI_ISL_1555322, EPI_ISL_1555323, EPI_ISL_1555324, EPI_ISL_1555325, EPI_ISL_1555326, EPI_ISL_1555327, EPI_ISL_1555328, EPI_ISL_1555329, EPI_ISL_1555330, EPI_ISL_1555331, EPI_ISL_1555332, EPI_ISL_1555333, EPI_ISL_1555334, EPI_ISL_1555335, EPI_ISL_1555336, EPI_ISL_1555337, EPI_ISL_1555338, EPI_ISL_1555339, EPI_ISL_1555340, EPI_ISL_1555563, EPI_ISL_1555566, EPI_ISL_1555567, EPI_ISL_1555612, EPI_ISL_1555613, EPI_ISL_1555614, EPI_ISL_1555615, EPI_ISL_1555616, EPI_ISL_1555617, EPI_ISL_1555618, EPI_ISL_1555619, EPI_ISL_1555620, EPI_ISL_1555621, EPI_ISL_1555622, EPI_ISL_1555623, EPI_ISL_1555624, EPI_ISL_1555625, EPI_ISL_1555626, EPI_ISL_1555627, EPI_ISL_1555628, EPI_ISL_1555629, EPI_ISL_1555630, EPI_ISL_1555631, EPI_ISL_1555632, EPI_ISL_1555633, EPI_ISL_1555635, EPI_ISL_1555636, EPI_ISL_1555637, EPI_ISL_1555638, EPI_ISL_1555639, EPI_ISL_1555640, EPI_ISL_1555641, EPI_ISL_1555642, EPI_ISL_1555643, EPI_ISL_1555644, EPI_ISL_1555645, EPI_ISL_1555646, EPI_ISL_1555648, EPI_ISL_1555649, EPI_ISL_1555650, EPI_ISL_1555651, EPI_ISL_1555652, EPI_ISL_1555653, EPI_ISL_1555654, EPI_ISL_1555655, EPI_ISL_1555656, EPI_ISL_1555657, EPI_ISL_1555658, EPI_ISL_1555660, EPI_ISL_1555661, EPI_ISL_1555662, EPI_ISL_1555663, EPI_ISL_1555664, EPI_ISL_1555665, EPI_ISL_1555666, EPI_ISL_1555667, EPI_ISL_1555668, EPI_ISL_1555669, EPI_ISL_1555670, EPI_ISL_1555671, EPI_ISL_1555672, EPI_ISL_1555673, EPI_ISL_1555674, EPI_ISL_1555675, EPI_ISL_1555676, EPI_ISL_1555677, EPI_ISL_1555678, EPI_ISL_1555728, EPI_ISL_1555729, EPI_ISL_1555730, EPI_ISL_1555731, EPI_ISL_1555732, EPI_ISL_1555733, EPI_ISL_1555877, EPI_ISL_1555878, EPI_ISL_1555879, EPI_ISL_1555880, EPI_ISL_1555881, EPI_ISL_1555882, EPI_ISL_1555883, EPI_ISL_1555884, EPI_ISL_1555886, EPI_ISL_1555887, EPI_ISL_1555888, EPI_ISL_1555889, EPI_ISL_1555890, EPI_ISL_1555891, EPI_ISL_1555892, EPI_ISL_1555894, EPI_ISL_1555895, EPI_ISL_1555896, EPI_ISL_1555897, EPI_ISL_1555898, EPI_ISL_1555899, EPI_ISL_1555900, EPI_ISL_1555901, EPI_ISL_1555902, EPI_ISL_1555903, EPI_ISL_1555904, EPI_ISL_1555905, EPI_ISL_1555906, EPI_ISL_1555907, EPI_ISL_1555908, EPI_ISL_1555909, EPI_ISL_1555910, EPI_ISL_1555911, EPI_ISL_1555912, EPI_ISL_1555913, EPI_ISL_1555914, EPI_ISL_1555915, EPI_ISL_1555916, EPI_ISL_1555917, EPI_ISL_1555918, EPI_ISL_1555919, EPI_ISL_1555920, EPI_ISL_1555921, EPI_ISL_1555922, EPI_ISL_1555923, EPI_ISL_1555924, EPI_ISL_1555925, EPI_ISL_1555926, EPI_ISL_1555927, EPI_ISL_1555928, EPI_ISL_1555929, EPI_ISL_1555930, EPI_ISL_1555931, EPI_ISL_1555932, EPI_ISL_1555933, EPI_ISL_1555934, EPI_ISL_1555935, EPI_ISL_1555936, EPI_ISL_1555937, EPI_ISL_1555938, EPI_ISL_1556230, EPI_ISL_1556231, EPI_ISL_1556232, EPI_ISL_1556233, EPI_ISL_1556245, EPI_ISL_1556247, EPI_ISL_1556254, EPI_ISL_1556255, EPI_ISL_1667103, EPI_ISL_1667105, EPI_ISL_1667106, EPI_ISL_1667107, EPI_ISL_1694965, EPI_ISL_1694966, EPI_ISL_1694967, EPI_ISL_1694968

see above | Fulgent Genetics | Centers for Disease Control and Prevention Division of Viral Diseases, Pathogen Discovery | Adrian Paskey; Becky Tsai; Benafsh Sapra; Benjamin Rambo-Martin; Christopher Gulvick; Clinton R. Paden; Dakota Howard; Darlene Wagner; Dhwani Batra; Doreen Ng; Duncan MacCannell; Harry Gao; James Xie; Jason Caravas; John Gao; Joseph Fierro; Kara Moser; Matthew Schmerer; Mickey Li; Peter W. Cook; Scott Sammons; Shatavia Morrison; Yan Meng; Yvette Unoarumhi

EPI_ISL_1231437, EPI_ISL_1231438, EPI_ISL_1231439, EPI_ISL_1231440, EPI_ISL_1231441, EPI_ISL_1231442, EPI_ISL_1231443, EPI_ISL_1231444, EPI_ISL_1231445, EPI_ISL_1231446, EPI_ISL_1231447, EPI_ISL_1231448, EPI_ISL_1231449, EPI_ISL_1231450, EPI_ISL_1231451, EPI_ISL_1231452, EPI_ISL_1231453, EPI_ISL_1231454, EPI_ISL_1231455, EPI_ISL_1231456, EPI_ISL_1231457, EPI_ISL_1231458, EPI_ISL_1231459, EPI_ISL_1231460, EPI_ISL_1231461, EPI_ISL_1231462, EPI_ISL_1231463, EPI_ISL_1231464, EPI_ISL_1231465, EPI_ISL_1231466, EPI_ISL_1231467, EPI_ISL_1231468, EPI_ISL_1231469, EPI_ISL_1231470, EPI_ISL_1231471, EPI_ISL_1231472, EPI_ISL_1231473, EPI_ISL_1231474, EPI_ISL_1231475, EPI_ISL_1231476, EPI_ISL_1231477, EPI_ISL_1231478, EPI_ISL_1231479, EPI_ISL_1231480, EPI_ISL_1231481, EPI_ISL_1231482, EPI_ISL_1231483, EPI_ISL_1231484, EPI_ISL_1231485, EPI_ISL_1231486, EPI_ISL_1231487, EPI_ISL_1231488, EPI_ISL_1231489, EPI_ISL_1231490, EPI_ISL_1231491, EPI_ISL_1231492, EPI_ISL_1231493, EPI_ISL_1231494, EPI_ISL_1231495, EPI_ISL_1231496, EPI_ISL_1231497, EPI_ISL_1231498, EPI_ISL_1231499, EPI_ISL_1231500, EPI_ISL_1231501, EPI_ISL_1231502, EPI_ISL_1231503, EPI_ISL_1231504, EPI_ISL_1231505, EPI_ISL_1231506, EPI_ISL_1231507, EPI_ISL_1231508, EPI_ISL_1231509, EPI_ISL_1231510, EPI_ISL_1231512, EPI_ISL_1231513, EPI_ISL_1231514, EPI_ISL_1231515, EPI_ISL_1231516, EPI_ISL_1231517, EPI_ISL_1231518, EPI_ISL_1231519, EPI_ISL_1231520, EPI_ISL_1231521, EPI_ISL_1231522, EPI_ISL_1231523, EPI_ISL_1231524, EPI_ISL_1231525, EPI_ISL_1231526, EPI_ISL_1231527, EPI_ISL_1231528, EPI_ISL_1231529, EPI_ISL_1231530, EPI_ISL_1231533, EPI_ISL_1231537, EPI_ISL_1231541, EPI_ISL_1231545, EPI_ISL_1231549, EPI_ISL_1231553, EPI_ISL_1231557, EPI_ISL_1231561, EPI_ISL_1231565, EPI_ISL_1231569, EPI_ISL_1231573, EPI_ISL_1231577, EPI_ISL_1231581, EPI_ISL_1231585, EPI_ISL_1231590, EPI_ISL_1231594, EPI_ISL_1231598, EPI_ISL_1231602, EPI_ISL_1231606, EPI_ISL_1231610, EPI_ISL_1231615, EPI_ISL_1231618, EPI_ISL_1231623, EPI_ISL_1231626, EPI_ISL_1231630, EPI_ISL_1231634, EPI_ISL_1231638, EPI_ISL_1231642, EPI_ISL_1231646, EPI_ISL_1231651, EPI_ISL_1231655, EPI_ISL_1231659, EPI_ISL_1231663, EPI_ISL_1231666, EPI_ISL_1231670, EPI_ISL_1231675, EPI_ISL_1231678, EPI_ISL_1231682, EPI_ISL_1231686, EPI_ISL_1231690, EPI_ISL_1231694, EPI_ISL_1231698, EPI_ISL_1231702, EPI_ISL_1231706, EPI_ISL_1231710, EPI_ISL_1231714, EPI_ISL_1231719, EPI_ISL_1231723, EPI_ISL_1231727, EPI_ISL_1231730, EPI_ISL_1231735, EPI_ISL_1231739, EPI_ISL_1231743, EPI_ISL_1231747, EPI_ISL_1231751, EPI_ISL_1231755, EPI_ISL_1231758, EPI_ISL_1231762, EPI_ISL_1231767, EPI_ISL_1231771, EPI_ISL_1231774, EPI_ISL_1231775, EPI_ISL_1231776, EPI_ISL_1231777, EPI_ISL_1231778, EPI_ISL_1231779, EPI_ISL_1231780, EPI_ISL_1231781, EPI_ISL_1231782, EPI_ISL_1231783, EPI_ISL_1231784, EPI_ISL_1231785, EPI_ISL_1231786, EPI_ISL_1231787, EPI_ISL_1231788, EPI_ISL_1231789, EPI_ISL_1231790, EPI_ISL_1231791, EPI_ISL_1231793, EPI_ISL_1231794, EPI_ISL_1231795, EPI_ISL_1231796, EPI_ISL_1231797, EPI_ISL_1231798, EPI_ISL_1231799, EPI_ISL_1231800, EPI_ISL_1231802, EPI_ISL_1231803, EPI_ISL_1231804, EPI_ISL_1231805, EPI_ISL_1231806, EPI_ISL_1231807, EPI_ISL_1231808, EPI_ISL_1231809, EPI_ISL_1231810, EPI_ISL_1231811, EPI_ISL_1231812, EPI_ISL_1231813, EPI_ISL_1231814, EPI_ISL_1231815, EPI_ISL_1231816, EPI_ISL_1231817, EPI_ISL_1231818, EPI_ISL_1231819, EPI_ISL_1231820, EPI_ISL_1231821, EPI_ISL_1231822, EPI_ISL_1231823, EPI_ISL_1231824, EPI_ISL_1231825, EPI_ISL_1231826, EPI_ISL_1231827, EPI_ISL_1231828, EPI_ISL_1231829, EPI_ISL_1231830, EPI_ISL_1231831, EPI_ISL_1231832, EPI_ISL_1231833, EPI_ISL_1231834, EPI_ISL_1231835, EPI_ISL_1231836, EPI_ISL_1231837, EPI_ISL_1231838, EPI_ISL_1231839, EPI_ISL_1231840, EPI_ISL_1231841, EPI_ISL_1231842, EPI_ISL_1231843, EPI_ISL_1231844, EPI_ISL_1231845, EPI_ISL_1231846, EPI_ISL_1231847, EPI_ISL_1231848, EPI_ISL_1231849, EPI_ISL_1231850, EPI_ISL_1231851, EPI_ISL_1231852, EPI_ISL_1231853, EPI_ISL_1231854, EPI_ISL_1231855, EPI_ISL_1231856, EPI_ISL_1231857, EPI_ISL_1231858, EPI_ISL_1231859, EPI_ISL_1231860, EPI_ISL_1231861, EPI_ISL_1231862, EPI_ISL_1231863, EPI_ISL_1231864, EPI_ISL_1231865, EPI_ISL_1231866, EPI_ISL_1231867, EPI_ISL_1231868, EPI_ISL_1231869, EPI_ISL_1231870, EPI_ISL_1231871, EPI_ISL_1231872, EPI_ISL_1231873, EPI_ISL_1231874, EPI_ISL_1231875, EPI_ISL_1231876, EPI_ISL_1231877, EPI_ISL_1231878, EPI_ISL_1231879, EPI_ISL_1231880, EPI_ISL_1231881, EPI_ISL_1231882, EPI_ISL_1231883, EPI_ISL_1231884, EPI_ISL_1231885, EPI_ISL_1231886, EPI_ISL_1231887, EPI_ISL_1231888, EPI_ISL_1231889, EPI_ISL_1231890, EPI_ISL_1231891, EPI_ISL_1231892, EPI_ISL_1231893, EPI_ISL_1231894, EPI_ISL_1231895, EPI_ISL_1231896, EPI_ISL_1231897, EPI_ISL_1231898, EPI_ISL_1231899, EPI_ISL_1231900, EPI_ISL_1231901, EPI_ISL_1231902, EPI_ISL_1231903, EPI_ISL_1231904, EPI_ISL_1231905, EPI_ISL_1231906, EPI_ISL_1231907, EPI_ISL_1231908,

| | | | |
|---|---|---|---|
| EPI_ISL_1231909, EPI_ISL_1231910, EPI_ISL_1231911, EPI_ISL_1231912, EPI_ISL_1231913, EPI_ISL_1232029, EPI_ISL_1232030, EPI_ISL_1232031, EPI_ISL_1232032, EPI_ISL_1232033, EPI_ISL_1232034, EPI_ISL_1232035, EPI_ISL_1232036, EPI_ISL_1232037, EPI_ISL_1232038, EPI_ISL_1232039, EPI_ISL_1232040, EPI_ISL_1232041, EPI_ISL_1232042, EPI_ISL_1232043, EPI_ISL_1232044, EPI_ISL_1232045, EPI_ISL_1232046, EPI_ISL_1232047, EPI_ISL_1232048, EPI_ISL_1232049, EPI_ISL_1232050, EPI_ISL_1232051, EPI_ISL_1232052, EPI_ISL_1232053, EPI_ISL_1232054, EPI_ISL_1232055, EPI_ISL_1232056, EPI_ISL_1232057, EPI_ISL_1232058, EPI_ISL_1232059, EPI_ISL_1232060, EPI_ISL_1232061, EPI_ISL_1232062, EPI_ISL_1232063, EPI_ISL_1232064, EPI_ISL_1232065, EPI_ISL_1232066, EPI_ISL_1232067, EPI_ISL_1232068, EPI_ISL_1232069, EPI_ISL_1232070, EPI_ISL_1232071, EPI_ISL_1232072, EPI_ISL_1232073, EPI_ISL_1232074, EPI_ISL_1232075, EPI_ISL_1232076, EPI_ISL_1232077, EPI_ISL_1232078, EPI_ISL_1232079, EPI_ISL_1232080, EPI_ISL_1232081, EPI_ISL_1232082, EPI_ISL_1232083, EPI_ISL_1232084, EPI_ISL_1232085, EPI_ISL_1232086, EPI_ISL_1232087, EPI_ISL_1232088, EPI_ISL_1232089, EPI_ISL_1232090, EPI_ISL_1232091, EPI_ISL_1232092, EPI_ISL_1232093, EPI_ISL_1232094, EPI_ISL_1232095, EPI_ISL_1232096, EPI_ISL_1232099, EPI_ISL_1232100, EPI_ISL_1232101, EPI_ISL_1232102, EPI_ISL_1232103, EPI_ISL_1232104, EPI_ISL_1232105, EPI_ISL_1232106, EPI_ISL_1232107, EPI_ISL_1232108, EPI_ISL_1232109, EPI_ISL_1232110, EPI_ISL_1232111, EPI_ISL_1232112, EPI_ISL_1232113, EPI_ISL_1232114, EPI_ISL_1232115, EPI_ISL_1232116, EPI_ISL_1232117, EPI_ISL_1232118, EPI_ISL_1232119, EPI_ISL_1232120, EPI_ISL_1232121, EPI_ISL_1232122, EPI_ISL_1232123, EPI_ISL_1232124, EPI_ISL_1232125, EPI_ISL_1232126, EPI_ISL_1232127, EPI_ISL_1232128, EPI_ISL_1232129, EPI_ISL_1232130, EPI_ISL_1232131, EPI_ISL_1232132, EPI_ISL_1232133, EPI_ISL_1232134, EPI_ISL_1232135, EPI_ISL_1232136, EPI_ISL_1232137, EPI_ISL_1232138, EPI_ISL_1232139, EPI_ISL_1232140, EPI_ISL_1232141, EPI_ISL_1232142, EPI_ISL_1232143, EPI_ISL_1232144, EPI_ISL_1232145, EPI_ISL_1232146, EPI_ISL_1232147, EPI_ISL_1232148, EPI_ISL_1232149, EPI_ISL_1232150, EPI_ISL_1232151, EPI_ISL_1232152, EPI_ISL_1232153, EPI_ISL_1232154, EPI_ISL_1232155, EPI_ISL_1232156, EPI_ISL_1232157, EPI_ISL_1232158, EPI_ISL_1232159, EPI_ISL_1232160, EPI_ISL_1232161, EPI_ISL_1232162, EPI_ISL_1232163, EPI_ISL_1232164, EPI_ISL_1232165, EPI_ISL_1232166, EPI_ISL_1232167, EPI_ISL_1232168, EPI_ISL_1232169, EPI_ISL_1232170, EPI_ISL_1232171, EPI_ISL_1232172, EPI_ISL_1232173, EPI_ISL_1232174, EPI_ISL_1232175, EPI_ISL_1232176, EPI_ISL_1232177, EPI_ISL_1232178, EPI_ISL_1232179, EPI_ISL_1232180, EPI_ISL_1232181, EPI_ISL_1232182, EPI_ISL_1232183, EPI_ISL_1232184, EPI_ISL_1232185, EPI_ISL_1232186, EPI_ISL_1232187, EPI_ISL_1232188, EPI_ISL_1232189, EPI_ISL_1232190, EPI_ISL_1232191, EPI_ISL_1232192, EPI_ISL_1251273 | | | |
| see above | Fulgent Genetics | Fulgent Genetics | Becky Tsai; Benafsh Sapra; Doreen Ng; Harry Gao; James Xie; John Gao; Joseph Fierro; Mickey Li; Yan Meng |
| EPI_ISL_1532119 | Good Samaritan Hospital | Santa Clara County Public Health Laboratory | Santa Clara County Public Health Department |
| EPI_ISL_1261900, EPI_ISL_1261904, EPI_ISL_1261907, EPI_ISL_1261921, EPI_ISL_1261932, EPI_ISL_1261951, EPI_ISL_1261969, EPI_ISL_1262005, EPI_ISL_1262219, EPI_ISL_1267892, EPI_ISL_1267897, EPI_ISL_1267898, EPI_ISL_1267901, EPI_ISL_1267903, EPI_ISL_1267904, EPI_ISL_1267906, EPI_ISL_1267911, EPI_ISL_1267912, EPI_ISL_1267913, EPI_ISL_1267916, EPI_ISL_1267920, EPI_ISL_1267971, EPI_ISL_1267978, EPI_ISL_1267993, EPI_ISL_1268000, EPI_ISL_1268012, EPI_ISL_1268017, EPI_ISL_1268038, EPI_ISL_1268039, EPI_ISL_1268048, EPI_ISL_1268049, EPI_ISL_1268273, EPI_ISL_1268312, EPI_ISL_1268339, EPI_ISL_1268363, EPI_ISL_1268378, EPI_ISL_1268509, EPI_ISL_1268544, EPI_ISL_1268549, EPI_ISL_1268554, EPI_ISL_1268575, EPI_ISL_1268599, EPI_ISL_1268603, EPI_ISL_1268604, EPI_ISL_1268606, EPI_ISL_1268634, EPI_ISL_1268640, EPI_ISL_1268744, EPI_ISL_1268748, EPI_ISL_1268842, EPI_ISL_1268866, EPI_ISL_1268870, EPI_ISL_1268897, EPI_ISL_1268901, EPI_ISL_1268909, EPI_ISL_1268915, EPI_ISL_1268917, EPI_ISL_1268931, EPI_ISL_1268935, EPI_ISL_1268953, EPI_ISL_1268955, EPI_ISL_1268991, EPI_ISL_1269016, EPI_ISL_1269023, EPI_ISL_1269059, EPI_ISL_1269094, EPI_ISL_1269141, EPI_ISL_1269194, EPI_ISL_1269196, EPI_ISL_1269198, EPI_ISL_1269205, EPI_ISL_1269207, EPI_ISL_1269214, EPI_ISL_1269219, EPI_ISL_1269230, EPI_ISL_1269239, EPI_ISL_1269270, EPI_ISL_1269278, EPI_ISL_1269312, EPI_ISL_1269313, EPI_ISL_1269332, EPI_ISL_1269532, EPI_ISL_1269554, EPI_ISL_1269557, EPI_ISL_1269576, EPI_ISL_1269588, EPI_ISL_1269593, EPI_ISL_1269596, EPI_ISL_1269606, EPI_ISL_1269609, EPI_ISL_1269621, EPI_ISL_1269630, EPI_ISL_1269648, EPI_ISL_1269673, EPI_ISL_1269676, EPI_ISL_1269697, EPI_ISL_1269716, EPI_ISL_1269727, EPI_ISL_1269730, EPI_ISL_1270144, EPI_ISL_1270145, EPI_ISL_1270147, EPI_ISL_1270161, EPI_ISL_1270174, EPI_ISL_1270184, EPI_ISL_1270207, EPI_ISL_1270246, EPI_ISL_1270253, EPI_ISL_1270273, EPI_ISL_1270279, EPI_ISL_1270442, EPI_ISL_1270443, EPI_ISL_1270448, EPI_ISL_1270451, EPI_ISL_1270459, EPI_ISL_1270461, EPI_ISL_1270462, EPI_ISL_1270464, EPI_ISL_1270469, EPI_ISL_1270470, EPI_ISL_1270471, EPI_ISL_1270473, EPI_ISL_1270474, EPI_ISL_1270475, EPI_ISL_1270478, EPI_ISL_1270480, EPI_ISL_1270504, EPI_ISL_1270663, EPI_ISL_1270668, EPI_ISL_1270669, EPI_ISL_1270703, EPI_ISL_1270706, EPI_ISL_1270708, EPI_ISL_1270727, EPI_ISL_1270746, EPI_ISL_1270749, EPI_ISL_1270761, EPI_ISL_1270767, EPI_ISL_1270770, EPI_ISL_1283377, EPI_ISL_1283443, EPI_ISL_1283456, EPI_ISL_1283639, EPI_ISL_1283692, EPI_ISL_1283707, EPI_ISL_1283742, EPI_ISL_1283831, EPI_ISL_1283862, EPI_ISL_1283864, EPI_ISL_1283882, EPI_ISL_1283906, EPI_ISL_1283935, EPI_ISL_1283952, EPI_ISL_1283990, EPI_ISL_1284006, EPI_ISL_1284051, EPI_ISL_1284264, EPI_ISL_1284385, EPI_ISL_1284410, EPI_ISL_1297927, EPI_ISL_1297941, EPI_ISL_1297944, EPI_ISL_1297951, EPI_ISL_1297955, EPI_ISL_1297959, EPI_ISL_1297975, EPI_ISL_1298016, EPI_ISL_1298067, EPI_ISL_1298069, EPI_ISL_1298074, EPI_ISL_1298077, EPI_ISL_1298080, EPI_ISL_1298084, EPI_ISL_1298098, EPI_ISL_1298111, EPI_ISL_1298114, EPI_ISL_1298122, EPI_ISL_1298168, EPI_ISL_1298171, EPI_ISL_1298172, EPI_ISL_1298182, EPI_ISL_1337010, EPI_ISL_1337017, EPI_ISL_1337027, EPI_ISL_1337029, EPI_ISL_1337039, EPI_ISL_1337040, EPI_ISL_1337068, EPI_ISL_1337071, EPI_ISL_1337073, EPI_ISL_1337095, EPI_ISL_1337109, EPI_ISL_1337132, EPI_ISL_1337138, EPI_ISL_1337188, EPI_ISL_1337399, EPI_ISL_1338057, EPI_ISL_1338070, EPI_ISL_1338072, EPI_ISL_1338074, EPI_ISL_1338085, EPI_ISL_1338087, EPI_ISL_1338091, EPI_ISL_1338097, EPI_ISL_1338101, EPI_ISL_1338122, EPI_ISL_1338124, EPI_ISL_1338130, EPI_ISL_1338133, EPI_ISL_1338140, EPI_ISL_1338156, EPI_ISL_1338166, EPI_ISL_1338170, EPI_ISL_1338194, EPI_ISL_1338204, EPI_ISL_1338210, EPI_ISL_1338212, EPI_ISL_1338226, EPI_ISL_1338230, EPI_ISL_1338233, EPI_ISL_1338245, EPI_ISL_1338289, EPI_ISL_1338307, EPI_ISL_1338333, EPI_ISL_1338394, EPI_ISL_1338397, EPI_ISL_1340060, EPI_ISL_1340061, EPI_ISL_1340068, EPI_ISL_1340083, EPI_ISL_1340084, EPI_ISL_1340088, EPI_ISL_1340090, EPI_ISL_1340096, EPI_ISL_1340124, EPI_ISL_1340129, EPI_ISL_1340225, EPI_ISL_1340249, EPI_ISL_1340251, EPI_ISL_1340254, EPI_ISL_1340258, EPI_ISL_1340260, EPI_ISL_1340264, EPI_ISL_1340265, EPI_ISL_1340275, EPI_ISL_1340301, EPI_ISL_1340305, EPI_ISL_1340308, EPI_ISL_1340337, EPI_ISL_1340339, EPI_ISL_1340368, EPI_ISL_1340373, EPI_ISL_1340377, EPI_ISL_1340384, EPI_ISL_1340447, EPI_ISL_1391729, EPI_ISL_1391750, EPI_ISL_1391839, EPI_ISL_1391843, EPI_ISL_1391848, EPI_ISL_1391853, EPI_ISL_1391864, EPI_ISL_1391865, EPI_ISL_1391888, EPI_ISL_1391895, EPI_ISL_1391905, EPI_ISL_1391919, EPI_ISL_1391929, EPI_ISL_1391934, EPI_ISL_1391939, EPI_ISL_1391943, EPI_ISL_1391949, EPI_ISL_1391962, EPI_ISL_1391982, EPI_ISL_1391991, EPI_ISL_1391995, EPI_ISL_1420835, EPI_ISL_1420925, EPI_ISL_1421655, EPI_ISL_1443819, EPI_ISL_1443827, EPI_ISL_1443831, EPI_ISL_1443835, EPI_ISL_1443842, EPI_ISL_1443846, EPI_ISL_1443865, EPI_ISL_1443918, EPI_ISL_1443932, EPI_ISL_1443938, EPI_ISL_1443950, EPI_ISL_1443965, EPI_ISL_1443968, EPI_ISL_1443973, EPI_ISL_1443981, EPI_ISL_1443984, EPI_ISL_1443997, EPI_ISL_1444004, EPI_ISL_1444005, EPI_ISL_1444006, EPI_ISL_1444015, EPI_ISL_1444022, EPI_ISL_1444024, EPI_ISL_1444030, EPI_ISL_1444041, EPI_ISL_1444046, EPI_ISL_1444064, EPI_ISL_1444074, EPI_ISL_1444092, EPI_ISL_1444098, EPI_ISL_1444100, EPI_ISL_1444188, EPI_ISL_1444190, EPI_ISL_1444192, EPI_ISL_1444200, EPI_ISL_1460798, EPI_ISL_1460801, EPI_ISL_1460802, EPI_ISL_1460831, EPI_ISL_1460835, EPI_ISL_1479333, EPI_ISL_1479353, EPI_ISL_1480009, EPI_ISL_1480066, EPI_ISL_1480190, EPI_ISL_1480238, EPI_ISL_1480260, EPI_ISL_1480271, EPI_ISL_1480341, EPI_ISL_1480451, EPI_ISL_1480513, EPI_ISL_1493758, EPI_ISL_1493886, EPI_ISL_1493898, EPI_ISL_1493921, EPI_ISL_1553291, EPI_ISL_1553301, EPI_ISL_1553931, EPI_ISL_1553965, EPI_ISL_1554030, EPI_ISL_1554035 | | | |
| see above | Helix/Illumina | Centers for Disease Control and Prevention Division of Viral Diseases, Pathogen Discovery | Adrian Paskey; Alexandre Bolze; Ary Ascencio; Ben L. Rambo-Martin; Benjamin Rambo-Martin; Brad Sickler; Charlotte Rivera-Garcia; Christine Tran; Christopher Gulvick; Clinton R. Paden; Dakota Howard; Darlene Wagner; David Becker; Dhwani Batra; Duncan MacCannell; Efren Sandoval; Eileen de Feo; Elizabeth Cirulli; Eric Allen; Geraint Levan; James Lu; Jan Antico; Jason Caravas; Jason Nguyen; Jimmy Ramirez; Jingtao Liu; Kara Moser; Kelly Schiabor Barrett; Kim Gietzen; Magnus Isaksson; Marc Laurent; Matthew Schmerer; Matthew Tolentino; Nicole L. Washington; Peter W. Cook; Phil Febbo; Ryan Cho; Scott Sammons; Shannon Wickline; Shatavia Morrison; Sherry Wang; Simon White; Summer Galloway; Suxiang Tong; Tyler Cassens; William Lee; Yvette Unoarumhi |
| EPI_ISL_1303471 | Henry Mayo Newhall Hospital | Los Angeles County Public Health Laboratories | P. Hemarajata et al. |
| EPI_ISL_1477172, EPI_ISL_1477173, EPI_ISL_1477183, EPI_ISL_1477195, EPI_ISL_1477206, EPI_ISL_1477247, EPI_ISL_1477276, EPI_ISL_1477277, EPI_ISL_1477280, EPI_ISL_1477282, EPI_ISL_1477296, EPI_ISL_1477299, EPI_ISL_1477325, EPI_ISL_1477332, EPI_ISL_1477339, EPI_ISL_1477341, EPI_ISL_1477346, EPI_ISL_1477354, EPI_ISL_1477355, EPI_ISL_1477376, EPI_ISL_1477379, EPI_ISL_1477408, EPI_ISL_1477951, EPI_ISL_1477954, EPI_ISL_1477955, EPI_ISL_1477970, EPI_ISL_1477973, EPI_ISL_1477975, EPI_ISL_1477976, EPI_ISL_1477981, EPI_ISL_1477987, EPI_ISL_1477991, EPI_ISL_1477997, EPI_ISL_1478009, EPI_ISL_1478010, EPI_ISL_1478015, EPI_ISL_1478026, EPI_ISL_1478041, EPI_ISL_1478054, EPI_ISL_1478055, EPI_ISL_1478058, EPI_ISL_1478062, EPI_ISL_1478063, EPI_ISL_1478065, EPI_ISL_1478066, EPI_ISL_1478067, EPI_ISL_1478075, EPI_ISL_1478077, EPI_ISL_1478082, EPI_ISL_1478083, EPI_ISL_1478093, EPI_ISL_1478094, EPI_ISL_1478096, EPI_ISL_1478105, EPI_ISL_1478107, EPI_ISL_1478121, EPI_ISL_1478134, EPI_ISL_1478135, EPI_ISL_1478138, EPI_ISL_1478140, EPI_ISL_1478143, EPI_ISL_1478150, EPI_ISL_1478156, EPI_ISL_1478159, EPI_ISL_1478163, EPI_ISL_1478164, EPI_ISL_1478165, EPI_ISL_1478169, EPI_ISL_1478172, EPI_ISL_1478176, EPI_ISL_1478177, EPI_ISL_1478186, EPI_ISL_1478190, EPI_ISL_1701402, EPI_ISL_1701472, EPI_ISL_1701527, EPI_ISL_1701558, EPI_ISL_1701590, EPI_ISL_1701597, EPI_ISL_1701603, EPI_ISL_1701627, EPI_ISL_2758461, EPI_ISL_2758462, EPI_ISL_2758473 | | | |
| see above | Humboldt County Public Health Laboratory | Chan-Zuckerberg Biohub | CZB Cliahub Consortium |
| EPI_ISL_1192364, EPI_ISL_1192365, EPI_ISL_1497544, EPI_ISL_1497545, EPI_ISL_1497546, EPI_ISL_1497547 | Huntington Memorial Hospital | Los Angeles County Public Health Laboratories | P. Hemarajata et al. |
| EPI_ISL_2253167, EPI_ISL_2253168, EPI_ISL_2253170, EPI_ISL_2253174, EPI_ISL_2253175 | Infectious Diseases, Chan-Zuckerberg Biohub Covidtracker | Infectious Diseases, Chan-Zuckerberg Biohub Covidtracker | CZB Cliahub Consortium |
| EPI_ISL_1712423, EPI_ISL_1712431, EPI_ISL_1712432, EPI_ISL_1712433, EPI_ISL_1712434, EPI_ISL_1712436, EPI_ISL_1712437, EPI_ISL_1712438, EPI_ISL_1712439, EPI_ISL_1712440, EPI_ISL_1712467, EPI_ISL_1712468, EPI_ISL_1712469, EPI_ISL_1712470, EPI_ISL_1712471, EPI_ISL_1712472, EPI_ISL_1712474, EPI_ISL_1712475, EPI_ISL_1712476, EPI_ISL_1712477, EPI_ISL_1712478, EPI_ISL_1712479 | | | |
| see above | Innovative Genomics Institute, UC Berkeley | Innovative Genomics Institute, UC Berkeley | Holly Gildea; John Boyle; Phil Frankino; Stacia Wyman |
| EPI_ISL_1508918 | Kaiser Permanente Downey Medical Center | Los Angeles County Public Health Laboratories | P. Hemarajata et al. |
| EPI_ISL_1508917 | Kaiser Permanente West Los Angeles Medical Center | Los Angeles County Public Health Laboratories | P. Hemarajata et al. |
| EPI_ISL_1508984, EPI_ISL_1508985, EPI_ISL_1508986 | Kaiser Regional | Santa Clara County Public Health Laboratory | Santa Clara County Public Health Department |
| EPI_ISL_1508966, EPI_ISL_1508967, EPI_ISL_1508969, EPI_ISL_1508971, EPI_ISL_1508973, EPI_ISL_1508976, EPI_ISL_1508977 | | | |
| see above | Kaiser Santa Clara | Santa Clara County Public Health Laboratory | Santa Clara County Public Health Department |
| EPI_ISL_1221302, EPI_ISL_1221420, EPI_ISL_1221660, EPI_ISL_1221702, EPI_ISL_1221739, EPI_ISL_1221740, EPI_ISL_1221741, EPI_ISL_1221830, EPI_ISL_1221831, EPI_ISL_1221832, EPI_ISL_1221833, EPI_ISL_1221863, EPI_ISL_1221864, EPI_ISL_1221865, EPI_ISL_1222032, EPI_ISL_1222033, EPI_ISL_1298296, EPI_ISL_1320008, EPI_ISL_1320038, EPI_ISL_1320039, EPI_ISL_1320040, EPI_ISL_1320041, EPI_ISL_1320042, EPI_ISL_1320043, EPI_ISL_1320044, EPI_ISL_1320045, EPI_ISL_1320046, EPI_ISL_1320047, EPI_ISL_1320048, EPI_ISL_1320049, EPI_ISL_1320050, EPI_ISL_1320051, EPI_ISL_1320052, EPI_ISL_1320053, EPI_ISL_1320054, EPI_ISL_1320055, EPI_ISL_1320056, EPI_ISL_1320057, EPI_ISL_1320058, EPI_ISL_1320059, EPI_ISL_1320060, EPI_ISL_1320061, EPI_ISL_1320062, EPI_ISL_1320063, EPI_ISL_1320064, EPI_ISL_1320065, EPI_ISL_1320066, EPI_ISL_1320067, EPI_ISL_1320068, EPI_ISL_1320069, EPI_ISL_1320070, EPI_ISL_1320071, EPI_ISL_1320072, EPI_ISL_1320073, EPI_ISL_1320074, EPI_ISL_1320075, EPI_ISL_1320076, EPI_ISL_1320077, EPI_ISL_1320078, EPI_ISL_1320079, EPI_ISL_1320080, EPI_ISL_1320081, EPI_ISL_1320082, EPI_ISL_1320083, EPI_ISL_1320084, EPI_ISL_1320085, EPI_ISL_1320086, EPI_ISL_1320087, EPI_ISL_1320088, EPI_ISL_1320089, EPI_ISL_1320090, EPI_ISL_1320091, EPI_ISL_1320092, EPI_ISL_1320093, EPI_ISL_1320094, EPI_ISL_1320095, EPI_ISL_1320096, EPI_ISL_1320097, EPI_ISL_1320098, EPI_ISL_1320099, EPI_ISL_1320100, EPI_ISL_1320145, EPI_ISL_1320146, EPI_ISL_1320147, EPI_ISL_1320148, EPI_ISL_1320149, EPI_ISL_1320236, EPI_ISL_1320237, EPI_ISL_1320244, EPI_ISL_1320274, EPI_ISL_1320275, EPI_ISL_1320276, EPI_ISL_1320277, EPI_ISL_1320278, EPI_ISL_1320279, EPI_ISL_1320280, EPI_ISL_1320281, EPI_ISL_1320283, EPI_ISL_1320284, EPI_ISL_1320285, EPI_ISL_1320286, EPI_ISL_1320288, EPI_ISL_1320289, EPI_ISL_1320290, EPI_ISL_1320291, EPI_ISL_1320292, EPI_ISL_1320293, EPI_ISL_1320294, EPI_ISL_1320295, EPI_ISL_1320296, EPI_ISL_1320297, EPI_ISL_1320298, EPI_ISL_1320299, EPI_ISL_1320300, EPI_ISL_1320301, EPI_ISL_1320302, EPI_ISL_1320303, EPI_ISL_1320304, EPI_ISL_1320305, EPI_ISL_1320306, EPI_ISL_1320307, EPI_ISL_1320308, EPI_ISL_1320309, EPI_ISL_1320310, EPI_ISL_1320311, EPI_ISL_1320312, EPI_ISL_1320313, EPI_ISL_1320315, EPI_ISL_1320316, EPI_ISL_1320317, EPI_ISL_1320318, EPI_ISL_1320319, EPI_ISL_1320320, EPI_ISL_1320321, EPI_ISL_1320322, EPI_ISL_1320323, EPI_ISL_1320324, EPI_ISL_1320325, EPI_ISL_1320326, EPI_ISL_1320327, EPI_ISL_1320328, EPI_ISL_1320329, EPI_ISL_1320330, EPI_ISL_1320585, EPI_ISL_1321010, EPI_ISL_1321011, EPI_ISL_1321012, EPI_ISL_1321013, EPI_ISL_1321014, EPI_ISL_1321015, EPI_ISL_1321016, EPI_ISL_1321017, EPI_ISL_1321018, EPI_ISL_1321019, EPI_ISL_1321020, EPI_ISL_1321021, EPI_ISL_1321022, EPI_ISL_1321023, EPI_ISL_1321024, EPI_ISL_1321025, EPI_ISL_1321026, EPI_ISL_1321027, EPI_ISL_1321028, EPI_ISL_1321029, EPI_ISL_1321030, EPI_ISL_1321031, EPI_ISL_1321032, EPI_ISL_1321033, EPI_ISL_1321034, EPI_ISL_1321035, EPI_ISL_1321036, EPI_ISL_1321037, EPI_ISL_1321038, EPI_ISL_1321039, EPI_ISL_1321040, EPI_ISL_1321041, EPI_ISL_1321042, EPI_ISL_1321043, EPI_ISL_1321044, EPI_ISL_1321045, EPI_ISL_1321046, EPI_ISL_1321047, EPI_ISL_1321048, EPI_ISL_1321049, EPI_ISL_1321050, EPI_ISL_1321051, EPI_ISL_1321052, EPI_ISL_1321053, EPI_ISL_1321055, EPI_ISL_1321056, EPI_ISL_1321057, EPI_ISL_1321058, EPI_ISL_1321060, EPI_ISL_1321061, EPI_ISL_1321062, EPI_ISL_1321063, EPI_ISL_1321064, EPI_ISL_1321065, EPI_ISL_1321066, EPI_ISL_1321067, EPI_ISL_1321068, EPI_ISL_1321069, EPI_ISL_1321070, EPI_ISL_1321071, EPI_ISL_1321072, EPI_ISL_1321073, EPI_ISL_1321074, EPI_ISL_1321132, EPI_ISL_1321133, EPI_ISL_1321134, EPI_ISL_1321135, EPI_ISL_1321136, EPI_ISL_1321137, EPI_ISL_1321138, EPI_ISL_1321139, EPI_ISL_1321141, EPI_ISL_1321143, EPI_ISL_1321144, EPI_ISL_1321145, EPI_ISL_1321146, EPI_ISL_1321147, EPI_ISL_1321148, EPI_ISL_1321166, EPI_ISL_1321167, EPI_ISL_1321168, EPI_ISL_1321176, EPI_ISL_1321177, EPI_ISL_1321178, EPI_ISL_1321179, EPI_ISL_1321180, EPI_ISL_1321181, EPI_ISL_1321188, EPI_ISL_1321190, EPI_ISL_1321191, EPI_ISL_1321192, EPI_ISL_1321193, EPI_ISL_1321194, EPI_ISL_1321195, EPI_ISL_1321197, EPI_ISL_1321198, EPI_ISL_1321199, EPI_ISL_1321200, EPI_ISL_1321201, EPI_ISL_1321202, EPI_ISL_1321203, EPI_ISL_1321204, EPI_ISL_1321205, EPI_ISL_1321206, EPI_ISL_1321207, EPI_ISL_1321208, EPI_ISL_1321209, EPI_ISL_1321210, EPI_ISL_1321211, EPI_ISL_1321212, EPI_ISL_1321213, EPI_ISL_1321214, EPI_ISL_1321215, EPI_ISL_1321216, EPI_ISL_1321217, EPI_ISL_1321218, EPI_ISL_1321228, EPI_ISL_1321229, EPI_ISL_1321230, EPI_ISL_1321231, EPI_ISL_1321232, EPI_ISL_1321233, EPI_ISL_1321234, EPI_ISL_1321235, EPI_ISL_1321236, EPI_ISL_1321237, EPI_ISL_1321238, EPI_ISL_1321239, EPI_ISL_1321240, EPI_ISL_1321241, EPI_ISL_1321242, EPI_ISL_1321243, EPI_ISL_1321244, EPI_ISL_1321245, EPI_ISL_1321246, EPI_ISL_1321247, EPI_ISL_1321248, EPI_ISL_1321249, EPI_ISL_1321250, EPI_ISL_1321251, EPI_ISL_1321252, EPI_ISL_1321253, EPI_ISL_1321254, EPI_ISL_1321255, EPI_ISL_1321256, EPI_ISL_1321259, EPI_ISL_1321260, EPI_ISL_1321261, EPI_ISL_1321262, EPI_ISL_1321263, EPI_ISL_1321264, EPI_ISL_1321265, EPI_ISL_1321266, EPI_ISL_1321267, EPI_ISL_1321268, EPI_ISL_1321269, EPI_ISL_1321274, EPI_ISL_1321275, EPI_ISL_1321276, EPI_ISL_1321277, EPI_ISL_1321278, EPI_ISL_1321279, EPI_ISL_1321280, EPI_ISL_1321281, EPI_ISL_1321282, EPI_ISL_1321283, EPI_ISL_1321284, EPI_ISL_1321285, EPI_ISL_1321286, EPI_ISL_1321287, EPI_ISL_1321288, EPI_ISL_1321289, EPI_ISL_1321290, EPI_ISL_1321291, EPI_ISL_1321292, EPI_ISL_1321293, EPI_ISL_1321294, EPI_ISL_1321295, EPI_ISL_1321296, EPI_ISL_1321297, EPI_ISL_1321299, EPI_ISL_1321300, EPI_ISL_1321301, EPI_ISL_1321302, EPI_ISL_1321303, EPI_ISL_1321304, EPI_ISL_1321305, EPI_ISL_1321306, EPI_ISL_1321307, EPI_ISL_1321316, EPI_ISL_1321317, EPI_ISL_1321318, EPI_ISL_1321319, EPI_ISL_1321320, EPI_ISL_1321321, EPI_ISL_1321322, EPI_ISL_1321323, EPI_ISL_1321324, EPI_ISL_1321325, EPI_ISL_1321326, EPI_ISL_1321327, EPI_ISL_1321328, EPI_ISL_1321329, EPI_ISL_1321330, EPI_ISL_1321331, EPI_ISL_1321332, EPI_ISL_1321333, EPI_ISL_1321334, EPI_ISL_1321335, EPI_ISL_1321336, EPI_ISL_1321337, EPI_ISL_1321345, EPI_ISL_1321347, EPI_ISL_1321354, EPI_ISL_1338888, EPI_ISL_1338896, EPI_ISL_1338897, EPI_ISL_1338898, EPI_ISL_1338899, EPI_ISL_1338900, EPI_ISL_1338901, EPI_ISL_1338902, EPI_ISL_1338903, EPI_ISL_1338904, EPI_ISL_1338906, EPI_ISL_1338907, EPI_ISL_1338908, EPI_ISL_1338909, EPI_ISL_1338910, EPI_ISL_1338912, EPI_ISL_1338921, EPI_ISL_1338923, EPI_ISL_1338924, EPI_ISL_1338925, EPI_ISL_1338926, EPI_ISL_1338927, EPI_ISL_1338928, EPI_ISL_1338930, EPI_ISL_1338931, EPI_ISL_1338932, EPI_ISL_1338933, EPI_ISL_1338934, EPI_ISL_1338935, EPI_ISL_1338937, EPI_ISL_1338938, EPI_ISL_1338939, EPI_ISL_1338948, EPI_ISL_1338949, EPI_ISL_1338950, EPI_ISL_1338951, EPI_ISL_1338952, EPI_ISL_1338953, EPI_ISL_1338954, EPI_ISL_1338955, EPI_ISL_1338956, EPI_ISL_1338957, EPI_ISL_1338958, EPI_ISL_1338959, EPI_ISL_1338991, EPI_ISL_1338992, EPI_ISL_1339004, EPI_ISL_1339005, EPI_ISL_1339006, EPI_ISL_1339038, EPI_ISL_1339039, EPI_ISL_1339051, EPI_ISL_1339052, EPI_ISL_1339053, EPI_ISL_1339054, EPI_ISL_1339055, EPI_ISL_1339056, EPI_ISL_1339057, EPI_ISL_1339090, EPI_ISL_1339091, EPI_ISL_1339092, EPI_ISL_1339093, EPI_ISL_1339094, EPI_ISL_1339095, EPI_ISL_1339149, EPI_ISL_1339158, EPI_ISL_1339159, EPI_ISL_1339160, EPI_ISL_1339161, EPI_ISL_1339162, EPI_ISL_1339182, EPI_ISL_1339183, EPI_ISL_1339184, EPI_ISL_1339185, EPI_ISL_1339186, EPI_ISL_1339187, EPI_ISL_1339188, EPI_ISL_1339207, EPI_ISL_1339234, EPI_ISL_1339251, EPI_ISL_1339253, EPI_ISL_1421979, EPI_ISL_1421988, EPI_ISL_1421989, EPI_ISL_1421990, EPI_ISL_1421991, EPI_ISL_1421992, EPI_ISL_1422037, EPI_ISL_1422164, EPI_ISL_1422165, EPI_ISL_1422166, EPI_ISL_1422167, EPI_ISL_1422168, EPI_ISL_1422174, EPI_ISL_1422175, EPI_ISL_1422176, EPI_ISL_1422177, EPI_ISL_1422178, EPI_ISL_1422183, EPI_ISL_1422184, EPI_ISL_1422190, EPI_ISL_1422191, EPI_ISL_1422194, EPI_ISL_1422195, EPI_ISL_1422196, EPI_ISL_1422199, EPI_ISL_1422200, EPI_ISL_1422201, EPI_ISL_1422203, EPI_ISL_1422204, EPI_ISL_1422205, EPI_ISL_1422206, EPI_ISL_1461710, EPI_ISL_1461711, EPI_ISL_1461712, EPI_ISL_1461713, EPI_ISL_1461719, EPI_ISL_1461720, EPI_ISL_1461721, EPI_ISL_1461722, EPI_ISL_1461723, EPI_ISL_1461724, EPI_ISL_1461725, EPI_ISL_1461726, EPI_ISL_1461727, EPI_ISL_1461728, EPI_ISL_1461729, EPI_ISL_1461730, EPI_ISL_1461731, EPI_ISL_1461741, EPI_ISL_1461742, EPI_ISL_1461743, EPI_ISL_1461744, EPI_ISL_1461745, EPI_ISL_1461746, EPI_ISL_1461750, EPI_ISL_1461751, EPI_ISL_1461753, EPI_ISL_1461754, EPI_ISL_1461755, EPI_ISL_1461756, EPI_ISL_1461757, EPI_ISL_1461759, EPI_ISL_1461760, EPI_ISL_1461761, EPI_ISL_1461786, EPI_ISL_1461787, EPI_ISL_1461788, EPI_ISL_1461789, EPI_ISL_1461790, EPI_ISL_1461791, EPI_ISL_1461792, EPI_ISL_1461793, EPI_ISL_1461794, EPI_ISL_1461795, EPI_ISL_1461796, EPI_ISL_1461797, EPI_ISL_1461798, EPI_ISL_1461799, EPI_ISL_1461800, EPI_ISL_1461801, EPI_ISL_1461802, EPI_ISL_1461803, EPI_ISL_1461804, EPI_ISL_1461805, EPI_ISL_1461806, EPI_ISL_1461807, EPI_ISL_1461808, EPI_ISL_1461809, EPI_ISL_1461810, EPI_ISL_1461811, EPI_ISL_1461827, EPI_ISL_1461859, EPI_ISL_1461877, EPI_ISL_1461878, EPI_ISL_1461879, EPI_ISL_1481185, EPI_ISL_1481186, EPI_ISL_1481189, EPI_ISL_1481190, EPI_ISL_1481191, EPI_ISL_1481192, EPI_ISL_1481194, EPI_ISL_1481202, EPI_ISL_1481214, EPI_ISL_1481265, EPI_ISL_1481270, EPI_ISL_1513757, EPI_ISL_1513762, EPI_ISL_1513768, EPI_ISL_1513769, EPI_ISL_1513801, EPI_ISL_1513805, EPI_ISL_1513877, EPI_ISL_1513878, EPI_ISL_1513880, EPI_ISL_1514723, EPI_ISL_1514724, EPI_ISL_1514725, EPI_ISL_1514726, EPI_ISL_1514727, EPI_ISL_1514731, EPI_ISL_1514752, EPI_ISL_1514753, EPI_ISL_1514754, EPI_ISL_1514755, EPI_ISL_1514756, EPI_ISL_1514758, EPI_ISL_1514759, EPI_ISL_1514760, EPI_ISL_1514771, EPI_ISL_1514772, EPI_ISL_1514773, EPI_ISL_1514774, EPI_ISL_1548117, EPI_ISL_1548118, EPI_ISL_1548125, EPI_ISL_1548126, EPI_ISL_1548139, EPI_ISL_1548140, EPI_ISL_1549203, EPI_ISL_1549205 | | | |
| see above | Laboratory Corporation of America | Centers for Disease Control and Prevention Division of Viral Diseases, Pathogen Discovery | Adrian Paskey; Amanda Douglas; Amanda Suchanek; Andrea Throop; Ayla Burns; Ben L. Rambo-Martin; Benjamin Rambo-Martin; Bobbi Croy; Brian Krueger; Brian Norvell; Christopher Gulvick; Christos Petropoulos; Clinton R. Paden; Craig Lukasik; Dakota Howard; Darlene Wagner; Debbie Boles; Dhwani Batra; Duncan MacCannell; Eyad Almasri; Goran Stevovic; Howard Engler; Hrushikesh Deshmukh; Jake Humphrey; Jana Schroth; Jason Caravas; Joe Voshell; John Pruitt; Jonathan Meltzer; Jonathan Williams; Kara Moser; Kimberly Wagner; Lax Iyer; Lyndon Tilson; Manoj Jain; Marcia Eisenberg; Mary Ann Cristobal; Mary Williamson; Matthew Schmerer; Michael Levandoski; Mike Sapeta; Mindy Nye; Minoo Agarwal; Mohan Kolli; Nuthawin Charoensri; Oren Cohen; Peter W. Cook; Prashant Gupta; Qian Zeng; Rama Ghatti; Scott Parker; Scott Ryan; Scott Sammons; Shatavia Morrison; Stanley Letovsky; Steven Ragan; Suresh Babu Selvaraju; Susan Countryman; Susan Hicks; Suxiang Tong; Suzanne Dale; Thomas Urban; Tim Kuphal; Tricia Zwiefelhofer; Vincent Drouillon; Yvette Unoarumhi |
| EPI_ISL_1098632, EPI_ISL_1098633, EPI_ISL_1191109, EPI_ISL_2774573, EPI_ISL_2774574 | Los Angeles County Public Health Laboratories | Los Angeles County Public Health Laboratories | P. Hemarajata et al. |

| | | | |
|---|---|---|---|
| EPI_ISL_1303416, EPI_ISL_1303417, EPI_ISL_1303418, EPI_ISL_1303419, EPI_ISL_1303420, EPI_ISL_1303421, EPI_ISL_1303422, EPI_ISL_1303423, EPI_ISL_1379976, EPI_ISL_1506117, EPI_ISL_1506118, EPI_ISL_1506119 | | | |
| see above | MD Laboratories | Los Angeles County Public Health Laboratories | P. Hemarajata et al. |
| EPI_ISL_1197045, EPI_ISL_1197048, EPI_ISL_1197052, EPI_ISL_1197054, EPI_ISL_1303414, EPI_ISL_1303482 | Martin Luther King Jr. Community Hospital | Los Angeles County Public Health Laboratories | P. Hemarajata et al. |
| EPI_ISL_1263235, EPI_ISL_1664594, EPI_ISL_1664596, EPI_ISL_1664618, EPI_ISL_1664619, EPI_ISL_1921889, EPI_ISL_1921890, EPI_ISL_1921891, EPI_ISL_2308289, EPI_ISL_2308300, EPI_ISL_2308301, EPI_ISL_2308369, EPI_ISL_2716889 | | | |
| see above | Monterey County Public Health Laboratory | Monterey County Public Health Laboratory | Monterey County Public Health Laboratory |
| EPI_ISL_2604866 | NIAID - IRF Frederick, NIH | NIAID - IRF Frederick, NIH | Georgia; S. and Kocher, G. |
| EPI_ISL_1195800, EPI_ISL_1195863 | NYU Langone Health | Departments of Pathology and Medicine, New York University School of Medicine | Adriana Heguy; Christian Marier; Dacia Dimartino; Emily Guzman; Gael Westby; Guiqing Wang; Paolo Cotzia; Paul Zappile; Peter Meyn; Sitharam Ramaswami; Yutong Zhang |
| EPI_ISL_1234796, EPI_ISL_1477495, EPI_ISL_1477498, EPI_ISL_1477572, EPI_ISL_1477582, EPI_ISL_1477986, EPI_ISL_1478024, EPI_ISL_1478030, EPI_ISL_1478106, EPI_ISL_1478110, EPI_ISL_1478175, EPI_ISL_1529158, EPI_ISL_1701399, EPI_ISL_1701430, EPI_ISL_1701646 | | | |
| see above | Orange County Public Health Lab | Chan-Zuckerberg Biohub | CZB Cliahub Consortium |
| EPI_ISL_1303404, EPI_ISL_1303405, EPI_ISL_1303406, EPI_ISL_1303472, EPI_ISL_1303473, EPI_ISL_1497525 | Pomona Valley Hospital Medical Center | Los Angeles County Public Health Laboratories | P. Hemarajata et al. |
| EPI_ISL_2551222, EPI_ISL_2551223, EPI_ISL_2551224, EPI_ISL_2551225, EPI_ISL_2551240 | Providence St. Joseph Health Orange - Chapman | Providence St. Joseph Health Molecular Genomics Laboratory | Alexa K Dowdell; Brian D Piening; Carlo B Bifulco; Fred L Robinson; Mary Campbell; Rogan Rattray |
| EPI_ISL_1159295, EPI_ISL_1159308, EPI_ISL_1193379, EPI_ISL_1193381, EPI_ISL_1193382, EPI_ISL_1193383, EPI_ISL_1193388, EPI_ISL_1193392, EPI_ISL_1193394, EPI_ISL_1193423, EPI_ISL_1193431, EPI_ISL_1193436, EPI_ISL_1193437, EPI_ISL_1193438, EPI_ISL_1193442, EPI_ISL_1193443, EPI_ISL_1193444, EPI_ISL_1193479, EPI_ISL_1193494, EPI_ISL_1193506, EPI_ISL_1193522, EPI_ISL_1193532, EPI_ISL_1193533, EPI_ISL_1193536, EPI_ISL_1193546, EPI_ISL_1193548, EPI_ISL_1193588, EPI_ISL_1193630, EPI_ISL_1193784, EPI_ISL_1193799, EPI_ISL_1193899, EPI_ISL_1193913, EPI_ISL_1193925, EPI_ISL_1193926, EPI_ISL_1193936, EPI_ISL_1193943, EPI_ISL_1193944, EPI_ISL_1193953, EPI_ISL_1193972, EPI_ISL_1193981, EPI_ISL_1193986, EPI_ISL_1194010, EPI_ISL_1194013, EPI_ISL_1194018, EPI_ISL_1194026, EPI_ISL_1194028, EPI_ISL_1194029, EPI_ISL_1194032, EPI_ISL_1194035, EPI_ISL_1194125, EPI_ISL_1194229, EPI_ISL_1194237, EPI_ISL_1194260, EPI_ISL_1194264, EPI_ISL_1194275, EPI_ISL_1194289, EPI_ISL_1194298, EPI_ISL_1194299, EPI_ISL_1194305, EPI_ISL_1194311, EPI_ISL_1194320, EPI_ISL_1194329, EPI_ISL_1194330, EPI_ISL_1194333, EPI_ISL_1194354, EPI_ISL_1194355, EPI_ISL_1194359, EPI_ISL_1194362, EPI_ISL_1220472, EPI_ISL_1220507, EPI_ISL_1220520, EPI_ISL_1220547, EPI_ISL_1220549, EPI_ISL_1220695, EPI_ISL_1220715, EPI_ISL_1220723, EPI_ISL_1220724, EPI_ISL_1220730, EPI_ISL_1220737, EPI_ISL_1220740, EPI_ISL_1220741, EPI_ISL_1220743, EPI_ISL_1220745, EPI_ISL_1220747, EPI_ISL_1220749, EPI_ISL_1220751, EPI_ISL_1220753, EPI_ISL_1220757, EPI_ISL_1220761, EPI_ISL_1220766, EPI_ISL_1220767, EPI_ISL_1220776, EPI_ISL_1220783, EPI_ISL_1220784, EPI_ISL_1220786, EPI_ISL_1220791, EPI_ISL_1220794, EPI_ISL_1220797, EPI_ISL_1220798, EPI_ISL_1220804, EPI_ISL_1220808, EPI_ISL_1220811, EPI_ISL_1220817, EPI_ISL_1220818, EPI_ISL_1220820, EPI_ISL_1220827, EPI_ISL_1220831, EPI_ISL_1220835, EPI_ISL_1220837, EPI_ISL_1220838, EPI_ISL_1220839, EPI_ISL_1220840, EPI_ISL_1220841, EPI_ISL_1220843, EPI_ISL_1220845, EPI_ISL_1220857, EPI_ISL_1220859, EPI_ISL_1220861, EPI_ISL_1252919, EPI_ISL_1252923, EPI_ISL_1252924, EPI_ISL_1252928, EPI_ISL_1252929, EPI_ISL_1252937, EPI_ISL_1252944, EPI_ISL_1252945, EPI_ISL_1252946, EPI_ISL_1252958, EPI_ISL_1252967, EPI_ISL_1252969, EPI_ISL_1252971, EPI_ISL_1252978, EPI_ISL_1252979, EPI_ISL_1252980, EPI_ISL_1252985, EPI_ISL_1252987, EPI_ISL_1252988, EPI_ISL_1252989, EPI_ISL_1252992, EPI_ISL_1252993, EPI_ISL_1253001, EPI_ISL_1253007, EPI_ISL_1253008, EPI_ISL_1253018, EPI_ISL_1253019, EPI_ISL_1253020, EPI_ISL_1253021, EPI_ISL_1253022, EPI_ISL_1253025, EPI_ISL_1253028, EPI_ISL_1253029, EPI_ISL_1253030, EPI_ISL_1253032, EPI_ISL_1253033, EPI_ISL_1253135, EPI_ISL_1253136, EPI_ISL_1253140, EPI_ISL_1253141, EPI_ISL_1253142, EPI_ISL_1253147, EPI_ISL_1253152, EPI_ISL_1253154, EPI_ISL_1253164, EPI_ISL_1253172, EPI_ISL_1253185, EPI_ISL_1253190, EPI_ISL_1253192, EPI_ISL_1253193, EPI_ISL_1253232, EPI_ISL_1253233, EPI_ISL_1253239, EPI_ISL_1253243, EPI_ISL_1253253, EPI_ISL_1253259, EPI_ISL_1253368, EPI_ISL_1253390, EPI_ISL_1253391, EPI_ISL_1253395, EPI_ISL_1253397, EPI_ISL_1253404, EPI_ISL_1253407, EPI_ISL_1253409, EPI_ISL_1253413, EPI_ISL_1253420, EPI_ISL_1253423, EPI_ISL_1253424, EPI_ISL_1253427, EPI_ISL_1253429, EPI_ISL_1253431, EPI_ISL_1253433, EPI_ISL_1253436, EPI_ISL_1253441, EPI_ISL_1253446, EPI_ISL_1253448, EPI_ISL_1253453, EPI_ISL_1253455, EPI_ISL_1253464, EPI_ISL_1253465, EPI_ISL_1253471, EPI_ISL_1253473, EPI_ISL_1253475, EPI_ISL_1253476, EPI_ISL_1253477, EPI_ISL_1253480, EPI_ISL_1253482, EPI_ISL_1253484, EPI_ISL_1253485, EPI_ISL_1253486, EPI_ISL_1253487, EPI_ISL_1253497, EPI_ISL_1253506, EPI_ISL_1253512, EPI_ISL_1253513, EPI_ISL_1253519, EPI_ISL_1253523, EPI_ISL_1253525, EPI_ISL_1253527, EPI_ISL_1253528, EPI_ISL_1253529, EPI_ISL_1253533, EPI_ISL_1253535, EPI_ISL_1253536, EPI_ISL_1254015, EPI_ISL_1254019, EPI_ISL_1254020, EPI_ISL_1254021, EPI_ISL_1254022, EPI_ISL_1254023, EPI_ISL_1254024, EPI_ISL_1254025, EPI_ISL_1254027, EPI_ISL_1254029, EPI_ISL_1254030, EPI_ISL_1254031, EPI_ISL_1254032, EPI_ISL_1254034, EPI_ISL_1254035, EPI_ISL_1254038, EPI_ISL_1254041, EPI_ISL_1254042, EPI_ISL_1254043, EPI_ISL_1254046, EPI_ISL_1254047, EPI_ISL_1254053, EPI_ISL_1254055, EPI_ISL_1254062, EPI_ISL_1254063, EPI_ISL_1254064, EPI_ISL_1254066, EPI_ISL_1254068, EPI_ISL_1254069, EPI_ISL_1254071, EPI_ISL_1254073, EPI_ISL_1254077, EPI_ISL_1254078, EPI_ISL_1254079, EPI_ISL_1254080, EPI_ISL_1254081, EPI_ISL_1254082, EPI_ISL_1254084, EPI_ISL_1254089, EPI_ISL_1254092, EPI_ISL_1254093, EPI_ISL_1254095, EPI_ISL_1254096, EPI_ISL_1254097, EPI_ISL_1254101, EPI_ISL_1254104, EPI_ISL_1254106, EPI_ISL_1254107, EPI_ISL_1254109, EPI_ISL_1254110, EPI_ISL_1254117, EPI_ISL_1254123, EPI_ISL_1254129, EPI_ISL_1254130, EPI_ISL_1254133, EPI_ISL_1254136, EPI_ISL_1254138, EPI_ISL_1254140, EPI_ISL_1254141, EPI_ISL_1254143, EPI_ISL_1254144, EPI_ISL_1254149, EPI_ISL_1254150, EPI_ISL_1254151, EPI_ISL_1254152, EPI_ISL_1254158, EPI_ISL_1254159, EPI_ISL_1254160, EPI_ISL_1254162, EPI_ISL_1254167, EPI_ISL_1254169, EPI_ISL_1254173, EPI_ISL_1254174, EPI_ISL_1254176, EPI_ISL_1254180, EPI_ISL_1254181, EPI_ISL_1254183, EPI_ISL_1254186, EPI_ISL_1254188, EPI_ISL_1254192, EPI_ISL_1254193, EPI_ISL_1254194, EPI_ISL_1254196, EPI_ISL_1254198, EPI_ISL_1254200, EPI_ISL_1254206, EPI_ISL_1254209, EPI_ISL_1254210, EPI_ISL_1254213, EPI_ISL_1254216, EPI_ISL_1254217, EPI_ISL_1254218, EPI_ISL_1254220, EPI_ISL_1254221, EPI_ISL_1254222, EPI_ISL_1254224, EPI_ISL_1254227, EPI_ISL_1254241, EPI_ISL_1254262, EPI_ISL_1254267, EPI_ISL_1254270, EPI_ISL_1254273, EPI_ISL_1254275, EPI_ISL_1254278, EPI_ISL_1254281, EPI_ISL_1254286, EPI_ISL_1254292, EPI_ISL_1254294, EPI_ISL_1254298, EPI_ISL_1254300, EPI_ISL_1254301, EPI_ISL_1254302, EPI_ISL_1254304, EPI_ISL_1254306, EPI_ISL_1254307, EPI_ISL_1254308, EPI_ISL_1254313, EPI_ISL_1254316, EPI_ISL_1254320, EPI_ISL_1254344, EPI_ISL_1254360, EPI_ISL_1254379, EPI_ISL_1254432, EPI_ISL_1254433, EPI_ISL_1254464, EPI_ISL_1254465, EPI_ISL_1254470, EPI_ISL_1254477, EPI_ISL_1254491, EPI_ISL_1265918, EPI_ISL_1265924, EPI_ISL_1265930, EPI_ISL_1265954, EPI_ISL_1265963, EPI_ISL_1265966, EPI_ISL_1265979, EPI_ISL_1266036, EPI_ISL_1266101, EPI_ISL_1266106, EPI_ISL_1266107, EPI_ISL_1266205, EPI_ISL_1266216, EPI_ISL_1266485, EPI_ISL_1266524, EPI_ISL_1266667, EPI_ISL_1266697, EPI_ISL_1266734, EPI_ISL_1266862, EPI_ISL_1267283, EPI_ISL_1267342, EPI_ISL_1267351, EPI_ISL_1267357, EPI_ISL_1267390, EPI_ISL_1267391, EPI_ISL_1267411, EPI_ISL_1267412, EPI_ISL_1267422, EPI_ISL_1267430, EPI_ISL_1267432, EPI_ISL_1267434, EPI_ISL_1267445, EPI_ISL_1267479, EPI_ISL_1267481, EPI_ISL_1291002, EPI_ISL_1291005, EPI_ISL_1291015, EPI_ISL_1291028, EPI_ISL_1291032, EPI_ISL_1291042, EPI_ISL_1291091, EPI_ISL_1291092, EPI_ISL_1291094, EPI_ISL_1291095, EPI_ISL_1291096, EPI_ISL_1291099, EPI_ISL_1291100, EPI_ISL_1291105, EPI_ISL_1291106, EPI_ISL_1291109, EPI_ISL_1291110, EPI_ISL_1291111, EPI_ISL_1291112, EPI_ISL_1291114, EPI_ISL_1291115, EPI_ISL_1291116, EPI_ISL_1291117, EPI_ISL_1291118, EPI_ISL_1291121, EPI_ISL_1291123, EPI_ISL_1291124, EPI_ISL_1291125, EPI_ISL_1291127, EPI_ISL_1291128, EPI_ISL_1291129, EPI_ISL_1291130, EPI_ISL_1291131, EPI_ISL_1291132, EPI_ISL_1291134, EPI_ISL_1291135, EPI_ISL_1291137, EPI_ISL_1291138, EPI_ISL_1291140, EPI_ISL_1291141, EPI_ISL_1291142, EPI_ISL_1291143, EPI_ISL_1291144, EPI_ISL_1291145, EPI_ISL_1291146, EPI_ISL_1291147, EPI_ISL_1291149, EPI_ISL_1291150, EPI_ISL_1291151, EPI_ISL_1291152, EPI_ISL_1291153, EPI_ISL_1291154, EPI_ISL_1291155, EPI_ISL_1291156, EPI_ISL_1291157, EPI_ISL_1291159, EPI_ISL_1291160, EPI_ISL_1291161, EPI_ISL_1291162, EPI_ISL_1291163, EPI_ISL_1291164, EPI_ISL_1291165, EPI_ISL_1291166, EPI_ISL_1291167, EPI_ISL_1291170, EPI_ISL_1291171, EPI_ISL_1291172, EPI_ISL_1291173, EPI_ISL_1291179, EPI_ISL_1291180, EPI_ISL_1291182, EPI_ISL_1291186, EPI_ISL_1291187, EPI_ISL_1291188, EPI_ISL_1297683, EPI_ISL_1297688, EPI_ISL_1297689, EPI_ISL_1297691, EPI_ISL_1297701, EPI_ISL_1297705, EPI_ISL_1297708, EPI_ISL_1297719, EPI_ISL_1297735, EPI_ISL_1297737, EPI_ISL_1297738, EPI_ISL_1297741, EPI_ISL_1297744, EPI_ISL_1297745, EPI_ISL_1297747, EPI_ISL_1297751, EPI_ISL_1297754, EPI_ISL_1297757, EPI_ISL_1297758, EPI_ISL_1297760, EPI_ISL_1314735, EPI_ISL_1335973, EPI_ISL_1335987, EPI_ISL_1335993, EPI_ISL_1335995, EPI_ISL_1335996, EPI_ISL_1336068, EPI_ISL_1336087, EPI_ISL_1336092, EPI_ISL_1336094, EPI_ISL_1336104, EPI_ISL_1336105, EPI_ISL_1336106, EPI_ISL_1336157, EPI_ISL_1339830, EPI_ISL_1339837, EPI_ISL_1339839, EPI_ISL_1339850, EPI_ISL_1339866, EPI_ISL_1339876, EPI_ISL_1339877, EPI_ISL_1339881, EPI_ISL_1339892, EPI_ISL_1339893, EPI_ISL_1339896, EPI_ISL_1339910, EPI_ISL_1339913, EPI_ISL_1339916, EPI_ISL_1339917, EPI_ISL_1339928, EPI_ISL_1339935, EPI_ISL_1339949, EPI_ISL_1339964, EPI_ISL_1340679, EPI_ISL_1367239, EPI_ISL_1367241, EPI_ISL_1367242, EPI_ISL_1367249, EPI_ISL_1367252, EPI_ISL_1367253, EPI_ISL_1367257, EPI_ISL_1367261, EPI_ISL_1367262, EPI_ISL_1367263, EPI_ISL_1367267, EPI_ISL_1367273, EPI_ISL_1367277, EPI_ISL_1367278, EPI_ISL_1367281, EPI_ISL_1367282, EPI_ISL_1367283, EPI_ISL_1367286, EPI_ISL_1367288, EPI_ISL_1367290, EPI_ISL_1367293, EPI_ISL_1367298, EPI_ISL_1367300, EPI_ISL_1367302, EPI_ISL_1367307, EPI_ISL_1367311, EPI_ISL_1367312, EPI_ISL_1367314, EPI_ISL_1367315, EPI_ISL_1367319, EPI_ISL_1367321, EPI_ISL_1367324, EPI_ISL_1367326, EPI_ISL_1367328, EPI_ISL_1391608, EPI_ISL_1422428, EPI_ISL_1422585, EPI_ISL_1422623, EPI_ISL_1422635, EPI_ISL_1422642, EPI_ISL_1422653, EPI_ISL_1422655, EPI_ISL_1422663, EPI_ISL_1422669, EPI_ISL_1422671, EPI_ISL_1422674, EPI_ISL_1422675, EPI_ISL_1422679, EPI_ISL_1422680, EPI_ISL_1422682, EPI_ISL_1422683, EPI_ISL_1422684, EPI_ISL_1422685, EPI_ISL_1422687, EPI_ISL_1422688, EPI_ISL_1422692, EPI_ISL_1422693, EPI_ISL_1422694, EPI_ISL_1422697, EPI_ISL_1422699, EPI_ISL_1422702, EPI_ISL_1422705, EPI_ISL_1422706, EPI_ISL_1422707, EPI_ISL_1422708, EPI_ISL_1422710, EPI_ISL_1422787, EPI_ISL_1422795, EPI_ISL_1422798, EPI_ISL_1444411, EPI_ISL_1444412, EPI_ISL_1444421, EPI_ISL_1444428, EPI_ISL_1444431, EPI_ISL_1444433, EPI_ISL_1444436, EPI_ISL_1444438, EPI_ISL_1444440, EPI_ISL_1444446, EPI_ISL_1444453, EPI_ISL_1444455, EPI_ISL_1444456, EPI_ISL_1444461, EPI_ISL_1444468, EPI_ISL_1444469, EPI_ISL_1444470, EPI_ISL_1444473, EPI_ISL_1444481, EPI_ISL_1444482, EPI_ISL_1444484, EPI_ISL_1444485, EPI_ISL_1444489, EPI_ISL_1444491, EPI_ISL_1444494, EPI_ISL_1444566, EPI_ISL_1444568, EPI_ISL_1444571, EPI_ISL_1444574, EPI_ISL_1444578, EPI_ISL_1444581, EPI_ISL_1444589, EPI_ISL_1444592, EPI_ISL_1444594, EPI_ISL_1444595, EPI_ISL_1444596, EPI_ISL_1444601, EPI_ISL_1444602, EPI_ISL_1444605, EPI_ISL_1444606, EPI_ISL_1444608, EPI_ISL_1444609, EPI_ISL_1444613, EPI_ISL_1444614, EPI_ISL_1444615, EPI_ISL_1444617, EPI_ISL_1444618, EPI_ISL_1444620, EPI_ISL_1444621, EPI_ISL_1444622, EPI_ISL_1444623, EPI_ISL_1444627, EPI_ISL_1444638, EPI_ISL_1444640, EPI_ISL_1444643, EPI_ISL_1444644, EPI_ISL_1444645, EPI_ISL_1444646, EPI_ISL_1444647, EPI_ISL_1444648, EPI_ISL_1444650, EPI_ISL_1444652, EPI_ISL_1444653, EPI_ISL_1444654, EPI_ISL_1444655, EPI_ISL_1444657, EPI_ISL_1444658, EPI_ISL_1444659, EPI_ISL_1444660, EPI_ISL_1444664, EPI_ISL_1444665, EPI_ISL_1444666, EPI_ISL_1444669, EPI_ISL_1480817, EPI_ISL_1480838, EPI_ISL_1480839, EPI_ISL_1480842, EPI_ISL_1480848, EPI_ISL_1480858, EPI_ISL_1480862, EPI_ISL_1493939, EPI_ISL_1493941, EPI_ISL_1493949, EPI_ISL_1493951, EPI_ISL_1493960, EPI_ISL_1493962, EPI_ISL_1493963, EPI_ISL_1493966, EPI_ISL_1493976, EPI_ISL_1493978, EPI_ISL_1493983, EPI_ISL_1494011, EPI_ISL_1494021, EPI_ISL_1551782, EPI_ISL_1551783, EPI_ISL_1551788, EPI_ISL_1551791, EPI_ISL_1551793, EPI_ISL_1551795, EPI_ISL_1551799, EPI_ISL_1551803, EPI_ISL_1551808, EPI_ISL_1551814, EPI_ISL_1551881, EPI_ISL_1551883, EPI_ISL_1551890, EPI_ISL_1551895, EPI_ISL_1551899, EPI_ISL_1551900, EPI_ISL_1551903, EPI_ISL_1551908, EPI_ISL_1551912, EPI_ISL_1551916, EPI_ISL_1551920, EPI_ISL_1551921, EPI_ISL_1551922, EPI_ISL_1551923 | | | |
| see above | Quest Diagnostics Incorporated | Centers for Disease Control and Prevention Division of Viral Diseases, Pathogen Discovery | A. Gerasimova; A. Perez; Adrian Paskey; B. Anderson; Ben L. Rambo-Martin; Benjamin Rambo-Martin; Christopher Gulvick; Clinton R. Paden; Dakota Howard; Darlene Wagner; Dhwani Batra; Duncan MacCannell; F. Lacbawan; I. A. Shlyakhter; Jason Caravas; K.E. Livingston; Kara Moser; L.E. Bernstein; M. Hua; Matthew Schmerer; P. Tanpaiboon; Peter W. Cook; R. M. Kagan; R. Owen; R. V. Rolando; S. H. Rosenthal; Scott Sammons; Shatavia Morrison; Suxiang Tong; Y. Liu; Yvette Unoarumhi |
| EPI_ISL_1139180 | Quest Diagnostics Incorporated | Respiratory Viruses Branch, Division of Viral Diseases, Centers for Disease Control and Prevention | A. Gerasimova; A. Perez; B. Anderson; Ben L. Rambo-Martin; Clinton R. Paden; Dakota Howard; Dhwani Batra; Duncan MacCannell; F. Lacbawan; I. A. Shlyakhter; K.E. Livingston; L.E. Bernstein; M. Hua; P. Tanpaiboon; Peter W. Cook; R. M. Kagan; R. Owen; R. V. Rolando; S. H. Rosenthal; Suxiang Tong; Y. Liu |
| EPI_ISL_1295809, EPI_ISL_1295812, EPI_ISL_1794769, EPI_ISL_1794772, EPI_ISL_1794776, EPI_ISL_1794778, EPI_ISL_1794781 | | | |
| see above | Rady Children's Hospital - San Diego | Andersen lab at Scripps Research | Christina Clarke; David Dimmock; Denise Malicki; Kathryn Bouic; Linda Luo; SEARCH Alliance San Diego with Nanda Radamchar; Teresa Mueller |
| EPI_ISL_1295815, EPI_ISL_1295816, EPI_ISL_1295817, EPI_ISL_1315073, EPI_ISL_1366749, EPI_ISL_1366750, EPI_ISL_1366751, EPI_ISL_1468832, EPI_ISL_1468839, EPI_ISL_1468843, EPI_ISL_1468844, EPI_ISL_1468874, EPI_ISL_1468875, EPI_ISL_1468877, EPI_ISL_1468879, EPI_ISL_1468880, EPI_ISL_1468923, EPI_ISL_1468927, EPI_ISL_1468932, EPI_ISL_1468936, EPI_ISL_1468938, EPI_ISL_1468941, EPI_ISL_1468944, EPI_ISL_1468945, EPI_ISL_1468946, EPI_ISL_1468951, EPI_ISL_1468981, EPI_ISL_1468982, EPI_ISL_1468985, EPI_ISL_1468988, EPI_ISL_1468991, EPI_ISL_1468992, EPI_ISL_1468993, EPI_ISL_1468997, EPI_ISL_1468998, EPI_ISL_1469003, EPI_ISL_1469006, EPI_ISL_1469007, EPI_ISL_1469010, EPI_ISL_1469011, EPI_ISL_1469014, EPI_ISL_1469015, EPI_ISL_1469017, EPI_ISL_1469018, EPI_ISL_1469019, EPI_ISL_1469023, EPI_ISL_1469025, EPI_ISL_1469026, EPI_ISL_1469027, EPI_ISL_1469028, EPI_ISL_1469029, EPI_ISL_1469030, EPI_ISL_1469031, EPI_ISL_1469034, EPI_ISL_1469035, EPI_ISL_1469036, EPI_ISL_1469037, EPI_ISL_1469039, EPI_ISL_1469041, EPI_ISL_1791411, EPI_ISL_1969251, EPI_ISL_1969252, EPI_ISL_1969253, EPI_ISL_1969254, EPI_ISL_1969256, EPI_ISL_1969257, EPI_ISL_1969258, EPI_ISL_1969259, EPI_ISL_1969261, EPI_ISL_1969263, EPI_ISL_1969264, EPI_ISL_1969266, EPI_ISL_1969267, EPI_ISL_1969268, EPI_ISL_1969269 | | | |
| see above | San Diego County Public Health Laboratory | Andersen lab at Scripps Research | Brett Austin; Jovan Shephard; SEARCH Alliance San Diego with Ashleigh Murphy; SEARCH Alliance San Diego with Tracy Basler |
| EPI_ISL_1235430, EPI_ISL_1477507, EPI_ISL_1477519, EPI_ISL_1477644, EPI_ISL_1477755, EPI_ISL_1529106, EPI_ISL_1529107, EPI_ISL_1529108, EPI_ISL_1529109, EPI_ISL_1529121, EPI_ISL_1529137, EPI_ISL_1529154, EPI_ISL_1529162, EPI_ISL_1529183, EPI_ISL_1529191, EPI_ISL_1529193, EPI_ISL_1529205, EPI_ISL_1529221, EPI_ISL_1529224, EPI_ISL_1529234, EPI_ISL_1529238, EPI_ISL_1529240, EPI_ISL_1529245, EPI_ISL_1529259, EPI_ISL_1529262, EPI_ISL_1529263, EPI_ISL_1529267, EPI_ISL_1529273, EPI_ISL_1529283, EPI_ISL_1529287, EPI_ISL_1529309, EPI_ISL_1529316, EPI_ISL_1529317, EPI_ISL_1529349, EPI_ISL_1663931, EPI_ISL_1663933, EPI_ISL_1663939, EPI_ISL_1663963, EPI_ISL_1663979, EPI_ISL_1663980, EPI_ISL_1663982, EPI_ISL_1663993, EPI_ISL_1664004, EPI_ISL_1664016, EPI_ISL_1664025, EPI_ISL_1664036, EPI_ISL_1664043, EPI_ISL_1664062, EPI_ISL_1664073, EPI_ISL_1664079, EPI_ISL_1664080, EPI_ISL_1664083, EPI_ISL_1664089, EPI_ISL_1664094, EPI_ISL_1664095, EPI_ISL_1664107, EPI_ISL_1701401, EPI_ISL_1701413, EPI_ISL_1701439, EPI_ISL_1701440, EPI_ISL_1701445, EPI_ISL_1701457, EPI_ISL_1701462, EPI_ISL_1701477, EPI_ISL_1701480, EPI_ISL_1701481, EPI_ISL_1701496, EPI_ISL_1701498, EPI_ISL_1701526, EPI_ISL_1701532, EPI_ISL_1701544, EPI_ISL_1701550, EPI_ISL_1701554, EPI_ISL_1701564, EPI_ISL_1701567, EPI_ISL_1701573, EPI_ISL_1701575, EPI_ISL_1701583, EPI_ISL_1701596, EPI_ISL_1701598, EPI_ISL_1701609, EPI_ISL_1701612, EPI_ISL_1701625, EPI_ISL_1701630, EPI_ISL_1701631, EPI_ISL_1701640, EPI_ISL_1821338, EPI_ISL_1821342, EPI_ISL_1821350, EPI_ISL_1821398, EPI_ISL_1821414, EPI_ISL_1821426, EPI_ISL_1821428, EPI_ISL_1821445, EPI_ISL_1821460, EPI_ISL_1821470, EPI_ISL_2102742, EPI_ISL_2102744, EPI_ISL_2102768, EPI_ISL_2102779, EPI_ISL_2102803, EPI_ISL_2102810, EPI_ISL_2102813, EPI_ISL_2102825, EPI_ISL_2102831, EPI_ISL_2102851, EPI_ISL_2102859, EPI_ISL_2102862, EPI_ISL_2102865, EPI_ISL_2102883, EPI_ISL_2102898, EPI_ISL_2102899, EPI_ISL_2102913, EPI_ISL_2102914, EPI_ISL_2102920, EPI_ISL_2102923, EPI_ISL_2102941, EPI_ISL_2102959, EPI_ISL_2102963, EPI_ISL_2102966, EPI_ISL_2102967, EPI_ISL_2102969, EPI_ISL_2103021, EPI_ISL_2103089, EPI_ISL_2103148, EPI_ISL_2228364, EPI_ISL_2658588, EPI_ISL_2658642, EPI_ISL_2658672, EPI_ISL_2658709, EPI_ISL_2725518, EPI_ISL_2725524, EPI_ISL_2758487, EPI_ISL_2758492, EPI_ISL_2758502, EPI_ISL_2758515 | | | |
| see above | Santa Clara County Public Health Laboratory | Chan-Zuckerberg Biohub | CZB Cliahub Consortium |
| EPI_ISL_1137647, EPI_ISL_1169600, EPI_ISL_1203817, EPI_ISL_1222757, EPI_ISL_1235633, EPI_ISL_1235634, EPI_ISL_1235635, EPI_ISL_1235636, EPI_ISL_1235637, EPI_ISL_1235638, EPI_ISL_1235640, EPI_ISL_1335868, EPI_ISL_1335869, EPI_ISL_1335870, EPI_ISL_1372415, EPI_ISL_1406165, EPI_ISL_1406166, EPI_ISL_1406168, EPI_ISL_1406169 | | | |
| see above | Santa Clara County Public Health Laboratory | Santa Clara County Public Health Laboratory | Santa Clara County Public Health Department |
| EPI_ISL_1468842, EPI_ISL_1468849, EPI_ISL_1468882, EPI_ISL_1468883, EPI_ISL_1468884, EPI_ISL_1468885, EPI_ISL_1468886, EPI_ISL_1468887, EPI_ISL_1468888, EPI_ISL_1468955, EPI_ISL_1468956, EPI_ISL_1469045, EPI_ISL_1469046, EPI_ISL_1469051, EPI_ISL_1469052, EPI_ISL_1469053, EPI_ISL_1469054, EPI_ISL_1469055, EPI_ISL_1469056, EPI_ISL_1469057, EPI_ISL_1469058, EPI_ISL_1469059, EPI_ISL_1469060 | | | |
| see above | Scripps Medical Laboratory | Andersen lab at Scripps Research | Ellen Stefanski; Ian Mchardy; SEARCH Alliance San Diego with Michael Quigley |
| EPI_ISL_1295736, EPI_ISL_1295738, EPI_ISL_1295739, EPI_ISL_1295740, EPI_ISL_1295741, EPI_ISL_1295742, EPI_ISL_1295743, EPI_ISL_1295744, EPI_ISL_1295745, EPI_ISL_1295746, EPI_ISL_1295747, EPI_ISL_1295748, EPI_ISL_1295749, EPI_ISL_1295750, EPI_ISL_1295751, EPI_ISL_1295752, EPI_ISL_1295753, EPI_ISL_1295754, EPI_ISL_1295755, EPI_ISL_1295756, EPI_ISL_1295757, EPI_ISL_1295758, | | | |

| | | | |
|---|---|---|---|
| EPI_ISL_1295759, EPI_ISL_1295761, EPI_ISL_1295762, EPI_ISL_1295763, EPI_ISL_1295764, EPI_ISL_1295765, EPI_ISL_1295766, EPI_ISL_1295767, EPI_ISL_1295768, EPI_ISL_1295769, EPI_ISL_1295770, EPI_ISL_1295771, EPI_ISL_1295772, EPI_ISL_1295773, EPI_ISL_1295774, EPI_ISL_1295775, EPI_ISL_1295776, EPI_ISL_1295777, EPI_ISL_1295780, EPI_ISL_1295781, EPI_ISL_1295783, EPI_ISL_1295784, EPI_ISL_1295785, EPI_ISL_1295786, EPI_ISL_1295788, EPI_ISL_1295791, EPI_ISL_1295792, EPI_ISL_1295793, EPI_ISL_1295794, EPI_ISL_1295795, EPI_ISL_1366276, EPI_ISL_1366277, EPI_ISL_1366279, EPI_ISL_1366281, EPI_ISL_1366283, EPI_ISL_1366284, EPI_ISL_1366286, EPI_ISL_1366287, EPI_ISL_1366301, EPI_ISL_1366302, EPI_ISL_1366303, EPI_ISL_1366304, EPI_ISL_1366305, EPI_ISL_1366308, EPI_ISL_1366309, EPI_ISL_1366310, EPI_ISL_1366311, EPI_ISL_1366312, EPI_ISL_1366313, EPI_ISL_1366314, EPI_ISL_1366315, EPI_ISL_1366317, EPI_ISL_1366319, EPI_ISL_1366321, EPI_ISL_1366458, EPI_ISL_1366459, EPI_ISL_1366460, EPI_ISL_1366461, EPI_ISL_1366462, EPI_ISL_1366463, EPI_ISL_1366464, EPI_ISL_1366465, EPI_ISL_1366466, EPI_ISL_1366467, EPI_ISL_1366468, EPI_ISL_1366469, EPI_ISL_1366470, EPI_ISL_1366471, EPI_ISL_1366472, EPI_ISL_1366473, EPI_ISL_1366474, EPI_ISL_1366476, EPI_ISL_1366478, EPI_ISL_1366479, EPI_ISL_1366480, EPI_ISL_1366481, EPI_ISL_1366482, EPI_ISL_1366483, EPI_ISL_1366484, EPI_ISL_1366485, EPI_ISL_1366486, EPI_ISL_1366487, EPI_ISL_1366488, EPI_ISL_1366489, EPI_ISL_1366490, EPI_ISL_1366491, EPI_ISL_1366492, EPI_ISL_1366493, EPI_ISL_1366494, EPI_ISL_1366495, EPI_ISL_1366496, EPI_ISL_1468829, EPI_ISL_1468830, EPI_ISL_1468831, EPI_ISL_1468833, EPI_ISL_1468834, EPI_ISL_1468836, EPI_ISL_1468838, EPI_ISL_1468840, EPI_ISL_1468845, EPI_ISL_1468848, EPI_ISL_1468850, EPI_ISL_1468851, EPI_ISL_1468852, EPI_ISL_1468853, EPI_ISL_1468855, EPI_ISL_1468856, EPI_ISL_1468857, EPI_ISL_1468858, EPI_ISL_1468859, EPI_ISL_1468860, EPI_ISL_1468861, EPI_ISL_1468862, EPI_ISL_1468863, EPI_ISL_1468864, EPI_ISL_1468865, EPI_ISL_1468866, EPI_ISL_1468867, EPI_ISL_1468868, EPI_ISL_1468869, EPI_ISL_1468870, EPI_ISL_1468871, EPI_ISL_1468873, EPI_ISL_1468889, EPI_ISL_1468890, EPI_ISL_1468891, EPI_ISL_1468892, EPI_ISL_1468893, EPI_ISL_1468894, EPI_ISL_1468895, EPI_ISL_1468896, EPI_ISL_1468897, EPI_ISL_1468898, EPI_ISL_1468899, EPI_ISL_1468900, EPI_ISL_1468901, EPI_ISL_1468902, EPI_ISL_1468903, EPI_ISL_1468904, EPI_ISL_1468905, EPI_ISL_1468910, EPI_ISL_1468911, EPI_ISL_1468913, EPI_ISL_1468917, EPI_ISL_1468918, EPI_ISL_1468920, EPI_ISL_1468921, EPI_ISL_1468952, EPI_ISL_1468954, EPI_ISL_1468957, EPI_ISL_1468960, EPI_ISL_1468961, EPI_ISL_1468962, EPI_ISL_1468963, EPI_ISL_1468964, EPI_ISL_1468965, EPI_ISL_1468966, EPI_ISL_1468967, EPI_ISL_1468968, EPI_ISL_1468969, EPI_ISL_1468970, EPI_ISL_1468971, EPI_ISL_1468972, EPI_ISL_1468973, EPI_ISL_1468974, EPI_ISL_1468975, EPI_ISL_1468976, EPI_ISL_1468977, EPI_ISL_1468978, EPI_ISL_1469061, EPI_ISL_1469062, EPI_ISL_1469063, EPI_ISL_1469064, EPI_ISL_1469065, EPI_ISL_1469066, EPI_ISL_1469067, EPI_ISL_1469068, EPI_ISL_1469069, EPI_ISL_1469070, EPI_ISL_1469071, EPI_ISL_1469072, EPI_ISL_1469073, EPI_ISL_1469074, EPI_ISL_1469075, EPI_ISL_1469076, EPI_ISL_1469077, EPI_ISL_1469078, EPI_ISL_1469079, EPI_ISL_1469080, EPI_ISL_1469081, EPI_ISL_1469082, EPI_ISL_1469083, EPI_ISL_1469084, EPI_ISL_1469085, EPI_ISL_1469086, EPI_ISL_1469087, EPI_ISL_1531802, EPI_ISL_1531804, EPI_ISL_1531806 | | | |
| see above | Sharp HealthCare Laboratory | Andersen lab at Scripps Research | Art Mendoza; Cathy Woerle; Jacquelyn Berumen; Liam McGinnis; Omid Bakhtar; SEARCH Alliance San Diego with Aaron Harding |
| EPI_ISL_1483797 | Stanford Clinical Virology Laboratory | Santa Clara County Public Health Laboratory | Santa Clara County Public Health Department |
| EPI_ISL_1379889, EPI_ISL_1424116, EPI_ISL_1424117, EPI_ISL_1424118, EPI_ISL_1424119, EPI_ISL_1424120, EPI_ISL_1424121, EPI_ISL_1424122, EPI_ISL_1424123, EPI_ISL_1424125, EPI_ISL_1675223, EPI_ISL_1675224, EPI_ISL_1701679, EPI_ISL_2027133, EPI_ISL_2027136, EPI_ISL_2027138, EPI_ISL_2027167, EPI_ISL_2027174, EPI_ISL_3050259, EPI_ISL_3050260, EPI_ISL_3050268, EPI_ISL_3050269, EPI_ISL_3050271, EPI_ISL_3050273, EPI_ISL_3050274, EPI_ISL_3050275, EPI_ISL_3050292 | | | |
| see above | Stanford Health Care | Stanford University School of Medicine, Clinical Virology Laboratory | Becky Jiang; Bernadette Troung; Bernadette Truong; ChunHong Huang; Daniel Solis; Fiona Yamamoto; Fumiko Yamamoto; Hannah Wang; Jacob Miller; James Zehnder; Kenji O. Mfuh; Malaya K. Sahoo; Mamdouh Sibai; Marilyn Mar; Michelle Verghese; Naomi Iwai; Nathan Hammond; Selamawit Bihon; and Benjamin A. Pinsky |
| EPI_ISL_2341050, EPI_ISL_2341071, EPI_ISL_2341072, EPI_ISL_2341081, EPI_ISL_2341085, EPI_ISL_2341099, EPI_ISL_2341100, EPI_ISL_2341101, EPI_ISL_2341102, EPI_ISL_2341103, EPI_ISL_2341104, EPI_ISL_2341105, EPI_ISL_2341106, EPI_ISL_2341107, EPI_ISL_2341108, EPI_ISL_2341109, EPI_ISL_2341110, EPI_ISL_2341111, EPI_ISL_2341112, EPI_ISL_2341113, EPI_ISL_2341114, EPI_ISL_2341115, EPI_ISL_2341116, EPI_ISL_2341117, EPI_ISL_2341118, EPI_ISL_2341119, EPI_ISL_2341120, EPI_ISL_2341121, EPI_ISL_2341122, EPI_ISL_2341123, EPI_ISL_2341124, EPI_ISL_2341125, EPI_ISL_2341126, EPI_ISL_2341127, EPI_ISL_2341128, EPI_ISL_2341129, EPI_ISL_2341130, EPI_ISL_2341131, EPI_ISL_2341132, EPI_ISL_2341154 | | | |
| see above | Tulare County Public Health Laboratory | Tulare County Public Health Laboratory | Denise L Lopez; Joshua Kootstra |
| EPI_ISL_1191115, EPI_ISL_1191116, EPI_ISL_1191123, EPI_ISL_1191125, EPI_ISL_1191126, EPI_ISL_1191127, EPI_ISL_1191128, EPI_ISL_1197149, EPI_ISL_1303432, EPI_ISL_1303433, EPI_ISL_1303434, EPI_ISL_1303435, EPI_ISL_1303436, EPI_ISL_1303437, EPI_ISL_1303449, EPI_ISL_1303450, EPI_ISL_1303451, EPI_ISL_1303452, EPI_ISL_1303453, EPI_ISL_1303454, EPI_ISL_1303455, EPI_ISL_1303456, EPI_ISL_1303457, EPI_ISL_1303458, EPI_ISL_1303459, EPI_ISL_1303460, EPI_ISL_1303461, EPI_ISL_1379958, EPI_ISL_1379959, EPI_ISL_1379960, EPI_ISL_1379961, EPI_ISL_1379962, EPI_ISL_1379963, EPI_ISL_1379964, EPI_ISL_1379965, EPI_ISL_1379966, EPI_ISL_1379967, EPI_ISL_1379968, EPI_ISL_1379969, EPI_ISL_1379970, EPI_ISL_1379971, EPI_ISL_1379972, EPI_ISL_1379973, EPI_ISL_1379974, EPI_ISL_1379975, EPI_ISL_1497541, EPI_ISL_1506114 | | | |
| see above | UCLA Clinical Micro Lab | Los Angeles County Public Health Laboratories | P. Hemarajata et al. |
| EPI_ISL_1402589, EPI_ISL_1402590, EPI_ISL_1402591, EPI_ISL_1402592, EPI_ISL_1402593, EPI_ISL_1402594, EPI_ISL_1402595, EPI_ISL_1402596, EPI_ISL_1402597, EPI_ISL_1402598, EPI_ISL_1402599, EPI_ISL_1402600, EPI_ISL_1402601, EPI_ISL_1402602, EPI_ISL_1402603, EPI_ISL_1402604, EPI_ISL_1402605, EPI_ISL_1402606, EPI_ISL_1402607, EPI_ISL_1402608, EPI_ISL_1402609, EPI_ISL_1402610, EPI_ISL_1402611, EPI_ISL_1402612, EPI_ISL_1402613, EPI_ISL_1402614, EPI_ISL_1402615, EPI_ISL_1402616, EPI_ISL_1402617, EPI_ISL_1402618, EPI_ISL_1402619, EPI_ISL_1402620, EPI_ISL_1402621, EPI_ISL_1402622, EPI_ISL_1402623, EPI_ISL_1402624, EPI_ISL_1402625, EPI_ISL_1402626, EPI_ISL_1402627, EPI_ISL_1402628, EPI_ISL_1402629, EPI_ISL_1402630, EPI_ISL_1402631, EPI_ISL_1402632, EPI_ISL_1402633, EPI_ISL_1402634, EPI_ISL_1402635, EPI_ISL_1402636, EPI_ISL_1402637, EPI_ISL_1402638, EPI_ISL_1402639, EPI_ISL_1402640, EPI_ISL_1402641, EPI_ISL_1402642, EPI_ISL_1402643, EPI_ISL_1402644, EPI_ISL_1402645, EPI_ISL_1402646, EPI_ISL_1405050, EPI_ISL_1405051, EPI_ISL_1405052, EPI_ISL_1405053, EPI_ISL_1405054, EPI_ISL_1405055 | | | |
| see above | UCSC Genomics Institute | UCSC Genomics Institute | A. Marm Kilpatrick; Angie Hinrichs; Beth Shapiro; Bryan Thornlow; David Haussler; Eric Beraut; Hugh Olsen; Ikenna Anigbogu; Isabel Bjork; Jakob McBroome; Jeremy Sanford; Joshua Kapp; Mark Akeson; Maximilian Haeussler; Michael Stone; Miten Jain; Molly Cassatt-Johnstone; Namrita Dhillon; Russell Corbett-Detig; Terren Chang; Yatish Turakhia |
| EPI_ISL_1192368, EPI_ISL_1197044, EPI_ISL_1197046, EPI_ISL_1197047, EPI_ISL_1197049, EPI_ISL_1197051, EPI_ISL_1197053, EPI_ISL_1197131, EPI_ISL_1197132, EPI_ISL_1197133, EPI_ISL_1197134, EPI_ISL_1197135, EPI_ISL_1197136, EPI_ISL_1197137, EPI_ISL_1197138, EPI_ISL_1197139, EPI_ISL_1197140, EPI_ISL_1197141, EPI_ISL_1197142, EPI_ISL_1197143, EPI_ISL_1197144, EPI_ISL_1197145, EPI_ISL_1197146, EPI_ISL_1197147, EPI_ISL_1197148, EPI_ISL_1303407, EPI_ISL_1303408, EPI_ISL_1303409, EPI_ISL_1303410, EPI_ISL_1303411, EPI_ISL_1303412, EPI_ISL_1303413, EPI_ISL_1303415, EPI_ISL_1303424, EPI_ISL_1303425, EPI_ISL_1303440, EPI_ISL_1303441, EPI_ISL_1303442, EPI_ISL_1303443, EPI_ISL_1303444, EPI_ISL_1303445, EPI_ISL_1303446, EPI_ISL_1303448, EPI_ISL_1303462, EPI_ISL_1303464, EPI_ISL_1303465, EPI_ISL_1303466, EPI_ISL_1303467, EPI_ISL_1303468, EPI_ISL_1303469, EPI_ISL_1303470, EPI_ISL_1303480, EPI_ISL_1303481, EPI_ISL_1303489, EPI_ISL_1303490, EPI_ISL_1303491, EPI_ISL_1303492, EPI_ISL_1303493, EPI_ISL_1303494, EPI_ISL_1303495, EPI_ISL_1303496, EPI_ISL_1303497, EPI_ISL_1303498, EPI_ISL_1379977, EPI_ISL_1497526, EPI_ISL_1497527, EPI_ISL_1497528, EPI_ISL_1497529, EPI_ISL_1497530, EPI_ISL_1497533, EPI_ISL_1497534, EPI_ISL_1497537, EPI_ISL_1497538, EPI_ISL_1503993, EPI_ISL_1503994, EPI_ISL_1504041, EPI_ISL_1508903, EPI_ISL_1508929, EPI_ISL_1508936, EPI_ISL_1508939 | | | |
| see above | USC Clinical Lab | Los Angeles County Public Health Laboratories | P. Hemarajata et al. |
| EPI_ISL_1307638 | VA Medical Center | Los Angeles County Public Health Laboratories | P. Hemarajata et al. |

We gratefully acknowledge the following Authors from the Originating laboratories responsible for obtaining the specimens, as well as the Submitting laboratories where the genome data were generated and shared via GISAID, on which this research is based.

All Submitters of data may be contacted directly via www.gisaid.org

Authors are sorted alphabetically.

| Accession ID | Originating Laboratory | Submitting Laboratory | Authors |
|---|---|---|---|
| EPI_ISL_1678143, EPI_ISL_1678144, EPI_ISL_1678145, EPI_ISL_1678146, EPI_ISL_1678148, EPI_ISL_1678149, EPI_ISL_1678150, EPI_ISL_1678151, EPI_ISL_1678154, EPI_ISL_1678156, EPI_ISL_1678157, EPI_ISL_1678158, EPI_ISL_1678159, EPI_ISL_1678160, EPI_ISL_1678161, EPI_ISL_1678162, EPI_ISL_1678313, EPI_ISL_1678315 | | | |
| see above | "CDPH, Viral and Rickettsial Disease Laboratory" | Centers for Disease Control and Prevention Division of Viral Diseases, Pathogen Discovery | Alison Laufer Halpin; Ben L. Rambo-Martin; Clinton R. Paden; Dakota Howard; Darlene Wagner; Dave Wentworth; Dhwani Batra; Jasmine Padilla; Justin Lee; Katie Dillon; Krista Queen; Kristen Knipe; Kristine Lacek; Mark Burroughs; Matthew Schmerer; Mili Sheth; Peter Cook; Sam Shepard; Sarah Nobles; Shoshona Le; Suxiang Tong; Vivien Dugan; Yvette Unoarumhi |
| EPI_ISL_1479656, EPI_ISL_1479805, EPI_ISL_1491734, EPI_ISL_1491744, EPI_ISL_1491786, EPI_ISL_1491787, EPI_ISL_1491788, EPI_ISL_1491814, EPI_ISL_1491817, EPI_ISL_1491829, EPI_ISL_1491842, EPI_ISL_1491878, EPI_ISL_1491884, EPI_ISL_1491885, EPI_ISL_1491888, EPI_ISL_1491898, EPI_ISL_1491899, EPI_ISL_1491917, EPI_ISL_1491918, EPI_ISL_1491929, EPI_ISL_1491939, EPI_ISL_1491940, EPI_ISL_1513139, EPI_ISL_1513152, EPI_ISL_1513153, EPI_ISL_1513160, EPI_ISL_1513172, EPI_ISL_1513183, EPI_ISL_1513185, EPI_ISL_1513186, EPI_ISL_1513190, EPI_ISL_1513199, EPI_ISL_1513200, EPI_ISL_1513220, EPI_ISL_1513222, EPI_ISL_1513236, EPI_ISL_1513239, EPI_ISL_1513240, EPI_ISL_1513242, EPI_ISL_1513262, EPI_ISL_1513263, EPI_ISL_1513294, EPI_ISL_1513296, EPI_ISL_1513297, EPI_ISL_1513301, EPI_ISL_1513303, EPI_ISL_1513306, EPI_ISL_1513307, EPI_ISL_1513309, EPI_ISL_1513323, EPI_ISL_1513324, EPI_ISL_1513325, EPI_ISL_1513352, EPI_ISL_1513359, EPI_ISL_1513360, EPI_ISL_1513361, EPI_ISL_1513362, EPI_ISL_1513386, EPI_ISL_1513387, EPI_ISL_1513410, EPI_ISL_1513487, EPI_ISL_1513488, EPI_ISL_1513489, EPI_ISL_1513513, EPI_ISL_1513518, EPI_ISL_1513524, EPI_ISL_1513541, EPI_ISL_1513542, EPI_ISL_1513548, EPI_ISL_1513570, EPI_ISL_1513583, EPI_ISL_1513622, EPI_ISL_1513625, EPI_ISL_1513627, EPI_ISL_1525033, EPI_ISL_1525034, EPI_ISL_1525035, EPI_ISL_1525040, EPI_ISL_1525041, EPI_ISL_1525042, EPI_ISL_1525048, EPI_ISL_1525049, EPI_ISL_1525066, EPI_ISL_1525075, EPI_ISL_1525123, EPI_ISL_1525146, EPI_ISL_1525147, EPI_ISL_1525152, EPI_ISL_1525153, EPI_ISL_1525155, EPI_ISL_1525161, EPI_ISL_1525162, EPI_ISL_1525163, EPI_ISL_1525176, EPI_ISL_1525186, EPI_ISL_1525187, EPI_ISL_1525220, EPI_ISL_1550794, EPI_ISL_1550845, EPI_ISL_1550972, EPI_ISL_1550975, EPI_ISL_1550976, EPI_ISL_1550982, EPI_ISL_1550984, EPI_ISL_1551127, EPI_ISL_1551128, EPI_ISL_1551220, EPI_ISL_1551266, EPI_ISL_1551393, EPI_ISL_1551422, EPI_ISL_1551543, EPI_ISL_1551556, EPI_ISL_1551585, EPI_ISL_1551636, EPI_ISL_1559656, EPI_ISL_1559657, EPI_ISL_1559658, EPI_ISL_1559659, EPI_ISL_1559660, EPI_ISL_1559661, EPI_ISL_1559662, EPI_ISL_1559663, EPI_ISL_1559664, EPI_ISL_1559665, EPI_ISL_1559667, EPI_ISL_1559668, EPI_ISL_1559669, EPI_ISL_1559670, EPI_ISL_1559672, EPI_ISL_1559740, EPI_ISL_1559741, EPI_ISL_1559758, EPI_ISL_1559759, EPI_ISL_1559760, EPI_ISL_1559761, EPI_ISL_1559806, EPI_ISL_1559807, EPI_ISL_1559808, EPI_ISL_1559809, EPI_ISL_1559828, EPI_ISL_1559829, EPI_ISL_1559847, EPI_ISL_1559877, EPI_ISL_1559880, EPI_ISL_1559881, EPI_ISL_1559883, EPI_ISL_1559887, EPI_ISL_1559903, EPI_ISL_1560350, EPI_ISL_1560382, EPI_ISL_1560383, EPI_ISL_1560491, EPI_ISL_1560496, EPI_ISL_1560519, EPI_ISL_1560527, EPI_ISL_1560603, EPI_ISL_1560606, EPI_ISL_1560644, EPI_ISL_1560645, EPI_ISL_1560672, EPI_ISL_1560679, EPI_ISL_1560681, EPI_ISL_1560702, EPI_ISL_1560707, EPI_ISL_1560717, EPI_ISL_1560720, EPI_ISL_1560726, EPI_ISL_1560740, EPI_ISL_1560787, EPI_ISL_1560788, EPI_ISL_1560942, EPI_ISL_1560949, EPI_ISL_1561002, EPI_ISL_1561020, EPI_ISL_1561022, EPI_ISL_1561030, EPI_ISL_1561043, EPI_ISL_1561223, EPI_ISL_1561224, EPI_ISL_1561225, EPI_ISL_1561226, EPI_ISL_1561237, EPI_ISL_1561248, EPI_ISL_1561260, EPI_ISL_1561289, EPI_ISL_1561449, EPI_ISL_1561496, EPI_ISL_1561525, EPI_ISL_1561551, EPI_ISL_1561553, EPI_ISL_1561563, EPI_ISL_1561580, EPI_ISL_1561588, EPI_ISL_1561595, EPI_ISL_1561596, EPI_ISL_1561614, EPI_ISL_1561615, EPI_ISL_1561622, EPI_ISL_1561637, EPI_ISL_1561638, EPI_ISL_1561647, EPI_ISL_1561682, EPI_ISL_1561693, EPI_ISL_1561975, EPI_ISL_1561976, EPI_ISL_1561983, EPI_ISL_1561987, EPI_ISL_1561998, EPI_ISL_1562015, EPI_ISL_1562017, EPI_ISL_1562018, EPI_ISL_1562025, EPI_ISL_1562043, EPI_ISL_1562046, EPI_ISL_1562054, EPI_ISL_1562073, EPI_ISL_1562096, EPI_ISL_1562102, EPI_ISL_1562103, EPI_ISL_1562104, EPI_ISL_1562145, EPI_ISL_1562146, EPI_ISL_1562152, EPI_ISL_1562156, EPI_ISL_1562181, EPI_ISL_1562182, EPI_ISL_1562183, EPI_ISL_1562184, EPI_ISL_1562198, EPI_ISL_1562219, EPI_ISL_1562225, EPI_ISL_1562254, EPI_ISL_1562255, EPI_ISL_1562267, EPI_ISL_1562268, EPI_ISL_1562343, EPI_ISL_1562352, EPI_ISL_1562364, EPI_ISL_1562377, EPI_ISL_1562378, EPI_ISL_1562380, EPI_ISL_1562381, EPI_ISL_1562384, EPI_ISL_1562387, EPI_ISL_1562391, EPI_ISL_1562392, EPI_ISL_1562400, EPI_ISL_1562411, EPI_ISL_1562416, EPI_ISL_1562418, EPI_ISL_1562426, EPI_ISL_1562450, EPI_ISL_1562451, EPI_ISL_1562454, EPI_ISL_1562456, EPI_ISL_1562503, EPI_ISL_1562506, EPI_ISL_1562791, EPI_ISL_1562823, EPI_ISL_1562827, EPI_ISL_1562831, EPI_ISL_1562836, EPI_ISL_1562848, EPI_ISL_1562945, EPI_ISL_1562950, EPI_ISL_1562987, EPI_ISL_1562990, EPI_ISL_1563006, EPI_ISL_1563044, EPI_ISL_1563048, EPI_ISL_1563055, EPI_ISL_1563071, EPI_ISL_1563072, EPI_ISL_1563128, EPI_ISL_1563132, EPI_ISL_1563154, EPI_ISL_1563224, EPI_ISL_1563225, EPI_ISL_1563226, EPI_ISL_1563229, EPI_ISL_1563239, EPI_ISL_1563281, EPI_ISL_1563329, EPI_ISL_1563350, EPI_ISL_1563353, EPI_ISL_1563398, EPI_ISL_1563409, EPI_ISL_1563410, EPI_ISL_1563411, EPI_ISL_1563417, EPI_ISL_1563418, EPI_ISL_1563454, EPI_ISL_1563549, EPI_ISL_1563550, EPI_ISL_1563558, EPI_ISL_1563573, EPI_ISL_1563607, EPI_ISL_1648968, EPI_ISL_1649028, EPI_ISL_1649029, EPI_ISL_1649030, EPI_ISL_1649031, EPI_ISL_1649048, EPI_ISL_1649126, EPI_ISL_1649150, EPI_ISL_1649159, EPI_ISL_1649210, EPI_ISL_1649211, EPI_ISL_1649263, EPI_ISL_1649264, EPI_ISL_1649336, EPI_ISL_1649381, EPI_ISL_1649382, EPI_ISL_1649414, EPI_ISL_1649447, EPI_ISL_1649448, EPI_ISL_1649471, EPI_ISL_1649525, EPI_ISL_1649546, EPI_ISL_1649547, EPI_ISL_1649548, EPI_ISL_1649550, EPI_ISL_1649556, EPI_ISL_1649566, EPI_ISL_1649598, EPI_ISL_1649599, EPI_ISL_1649622, EPI_ISL_1649652, EPI_ISL_1649694, EPI_ISL_1649732, EPI_ISL_1649734, EPI_ISL_1649737, EPI_ISL_1649738, EPI_ISL_1649757, EPI_ISL_1649764, EPI_ISL_1649766, EPI_ISL_1649767, EPI_ISL_1649769, EPI_ISL_1649777, EPI_ISL_1649787, EPI_ISL_1649796, EPI_ISL_1649804, EPI_ISL_1649858, EPI_ISL_1649894, EPI_ISL_1649897, EPI_ISL_1649907, EPI_ISL_1649944, EPI_ISL_1650002, EPI_ISL_1650041, EPI_ISL_1650051, EPI_ISL_1650058, EPI_ISL_1650088, EPI_ISL_1650205, EPI_ISL_1650271, EPI_ISL_1650278, EPI_ISL_1650292, EPI_ISL_1650388, EPI_ISL_1650403, EPI_ISL_1650409, EPI_ISL_1650412, EPI_ISL_1650417, EPI_ISL_1650449, EPI_ISL_1650453, EPI_ISL_1650454, EPI_ISL_1650455, EPI_ISL_1650457, EPI_ISL_1650459, EPI_ISL_1650460, EPI_ISL_1650471, EPI_ISL_1650472, EPI_ISL_1650534, EPI_ISL_1650538, EPI_ISL_1650632, EPI_ISL_1650638, EPI_ISL_1650659, EPI_ISL_1650672, EPI_ISL_1650693, EPI_ISL_1650695, EPI_ISL_1650703, EPI_ISL_1650729, EPI_ISL_1650730, EPI_ISL_1650944, EPI_ISL_1650954, EPI_ISL_1650966, EPI_ISL_1650967, EPI_ISL_1651036, EPI_ISL_1651045, EPI_ISL_1651061, EPI_ISL_1651075, EPI_ISL_1651094, EPI_ISL_1651099, EPI_ISL_1651133, EPI_ISL_1651152, EPI_ISL_1651169, EPI_ISL_1651180, EPI_ISL_1651182, EPI_ISL_1667497, EPI_ISL_1667498, EPI_ISL_1667525, EPI_ISL_1667542, EPI_ISL_1667573, EPI_ISL_1667576, EPI_ISL_1667579, EPI_ISL_1667580, EPI_ISL_1667586, EPI_ISL_1667604, EPI_ISL_1667605, EPI_ISL_1667623, EPI_ISL_1667647, EPI_ISL_1667764, EPI_ISL_1667768, EPI_ISL_1667770, EPI_ISL_1667779, EPI_ISL_1667788, EPI_ISL_1667861, EPI_ISL_1667862, EPI_ISL_1667863, EPI_ISL_1667866, EPI_ISL_1667919, EPI_ISL_1667965, EPI_ISL_1667975, EPI_ISL_1667976, EPI_ISL_1667977, EPI_ISL_1667986, EPI_ISL_1667987, EPI_ISL_1668046, EPI_ISL_1668058, EPI_ISL_1668073, EPI_ISL_1668075, EPI_ISL_1668080, EPI_ISL_1668082, EPI_ISL_1668084, EPI_ISL_1668087, EPI_ISL_1668090, EPI_ISL_1668096, EPI_ISL_1668099, EPI_ISL_1668136, EPI_ISL_1668139, EPI_ISL_1668150, EPI_ISL_1668155, EPI_ISL_1668158, EPI_ISL_1668159, EPI_ISL_1668163, EPI_ISL_1668165, EPI_ISL_1668175, EPI_ISL_1668177, EPI_ISL_1668179, EPI_ISL_1668225, EPI_ISL_1668257, EPI_ISL_1668280, EPI_ISL_1668320, EPI_ISL_1668324, EPI_ISL_1668335, EPI_ISL_1685472, EPI_ISL_1685474, EPI_ISL_1685487, EPI_ISL_1685548, EPI_ISL_1685552, EPI_ISL_1685553, EPI_ISL_1685555, EPI_ISL_1685556, EPI_ISL_1685566, EPI_ISL_1685619, EPI_ISL_1685621, EPI_ISL_1685625, EPI_ISL_1685634, EPI_ISL_1685680, EPI_ISL_1685681, EPI_ISL_1685843, EPI_ISL_1685852, EPI_ISL_1685860, EPI_ISL_1685942, EPI_ISL_1685943, EPI_ISL_1686023, EPI_ISL_1686079, EPI_ISL_1686204, EPI_ISL_1686233, EPI_ISL_1686290, EPI_ISL_1686462, EPI_ISL_1686508, EPI_ISL_1686542, EPI_ISL_1686543, EPI_ISL_1686573, EPI_ISL_1686576, EPI_ISL_1686587, EPI_ISL_1686589, EPI_ISL_1686627, EPI_ISL_1686635, EPI_ISL_1686699, EPI_ISL_1686700, EPI_ISL_1686708, EPI_ISL_1686768, EPI_ISL_1686901, EPI_ISL_1686907, EPI_ISL_1686926, EPI_ISL_1686938, EPI_ISL_1686939, EPI_ISL_1686940, EPI_ISL_1686941, EPI_ISL_1686991, EPI_ISL_1686998, EPI_ISL_1687070, EPI_ISL_1687113, EPI_ISL_1687157, EPI_ISL_1687159, EPI_ISL_1687160, EPI_ISL_1687161, EPI_ISL_1687162, EPI_ISL_1687164, EPI_ISL_1687172, EPI_ISL_1687180, EPI_ISL_1687181, EPI_ISL_1687190, EPI_ISL_1687241, EPI_ISL_1687242, EPI_ISL_1687269, EPI_ISL_1687309, EPI_ISL_1687332, EPI_ISL_1687346, EPI_ISL_1687416, EPI_ISL_1687435, EPI_ISL_1687446, EPI_ISL_1687454, EPI_ISL_1687460, EPI_ISL_1687633, EPI_ISL_1687701, EPI_ISL_1687702, EPI_ISL_1687731, EPI_ISL_1687734, EPI_ISL_1687758, EPI_ISL_1687770, EPI_ISL_1687815, EPI_ISL_1687827, EPI_ISL_1687837, EPI_ISL_1687898, EPI_ISL_1687942, EPI_ISL_1687964, EPI_ISL_1687971, EPI_ISL_1687980, EPI_ISL_1687981, EPI_ISL_1687983, EPI_ISL_1687988, EPI_ISL_1687994, EPI_ISL_1688003, EPI_ISL_1688017, EPI_ISL_1688028, EPI_ISL_1688045, EPI_ISL_1688074, EPI_ISL_1688077, EPI_ISL_1688078, EPI_ISL_1688097, EPI_ISL_1688104, EPI_ISL_1688111, EPI_ISL_1688117, EPI_ISL_1688118, EPI_ISL_1688203, EPI_ISL_1688274, EPI_ISL_1688361, EPI_ISL_1688370, EPI_ISL_1688460, EPI_ISL_1688512, EPI_ISL_1688587, EPI_ISL_1688590, EPI_ISL_1688593, EPI_ISL_1688705, EPI_ISL_1688709, EPI_ISL_1688768, EPI_ISL_1688859, EPI_ISL_1688862, EPI_ISL_1688866, EPI_ISL_1689033, EPI_ISL_1689061, EPI_ISL_1689075, EPI_ISL_1689194, EPI_ISL_1689333, EPI_ISL_1689494, EPI_ISL_1689560, EPI_ISL_1689590, EPI_ISL_1689592, EPI_ISL_1689633, EPI_ISL_1689634, EPI_ISL_1689636, EPI_ISL_1689637, EPI_ISL_1689638, EPI_ISL_1689640, EPI_ISL_1689649, EPI_ISL_1689655, EPI_ISL_1689684, EPI_ISL_1689738, EPI_ISL_1689754, EPI_ISL_1689757, EPI_ISL_1689760, EPI_ISL_1689813, EPI_ISL_1689827, EPI_ISL_1689829, EPI_ISL_1689832, EPI_ISL_1689858, EPI_ISL_1689992, EPI_ISL_1690028, EPI_ISL_1690106, EPI_ISL_1690127, EPI_ISL_1690178, EPI_ISL_1690179, EPI_ISL_1690180, EPI_ISL_1690190, EPI_ISL_1690229, EPI_ISL_1690235, EPI_ISL_1690238, EPI_ISL_1690241, EPI_ISL_1695152, EPI_ISL_1695225, EPI_ISL_1695275, EPI_ISL_1695302, EPI_ISL_1695389, EPI_ISL_1695422, EPI_ISL_1695448, EPI_ISL_1695506, EPI_ISL_1695560, EPI_ISL_1701908, EPI_ISL_1701914, EPI_ISL_1701921, EPI_ISL_1701924, EPI_ISL_1701979, EPI_ISL_1701993, EPI_ISL_1702008, EPI_ISL_1702047, EPI_ISL_1702143, EPI_ISL_1702725, EPI_ISL_1702726, EPI_ISL_1702738, EPI_ISL_1702745, EPI_ISL_1702759, EPI_ISL_1702769, EPI_ISL_1702792, EPI_ISL_1702797, EPI_ISL_1702798, EPI_ISL_1702799, EPI_ISL_1702800, EPI_ISL_1702803, EPI_ISL_1702812, EPI_ISL_1702813, EPI_ISL_1702952, EPI_ISL_1735687, EPI_ISL_1735689, EPI_ISL_1735765, EPI_ISL_1735767, EPI_ISL_1735789, EPI_ISL_1735804, EPI_ISL_1735805, EPI_ISL_1735822, EPI_ISL_1735846, EPI_ISL_1735873, EPI_ISL_1735874, EPI_ISL_1735877, EPI_ISL_1735908, EPI_ISL_1735956, EPI_ISL_1735958, EPI_ISL_1735967, EPI_ISL_1735977, EPI_ISL_1736013, EPI_ISL_1736085, EPI_ISL_1736088, EPI_ISL_1736101, EPI_ISL_1736102, EPI_ISL_1736103, EPI_ISL_1736137, EPI_ISL_1736141, EPI_ISL_1736150, EPI_ISL_1736185, EPI_ISL_1736302, EPI_ISL_1736355, EPI_ISL_1736404, EPI_ISL_1736427, EPI_ISL_1736428, EPI_ISL_1736440, EPI_ISL_1736451, EPI_ISL_1736458, EPI_ISL_1736461, EPI_ISL_1736482, EPI_ISL_1736489, EPI_ISL_1736499, EPI_ISL_1736504, EPI_ISL_1736505, EPI_ISL_1736543, EPI_ISL_1736544, EPI_ISL_1736561, EPI_ISL_1736566, EPI_ISL_1736567, EPI_ISL_1736572, EPI_ISL_1736576, EPI_ISL_1736645, EPI_ISL_1736648, EPI_ISL_1736681, EPI_ISL_1736753, EPI_ISL_1736787, EPI_ISL_1736790, EPI_ISL_1736813, EPI_ISL_1736823, EPI_ISL_1736870, EPI_ISL_1736873, EPI_ISL_1736875, EPI_ISL_1736877, EPI_ISL_1736878, EPI_ISL_1737032, EPI_ISL_1737064, EPI_ISL_1737071, EPI_ISL_1737072, EPI_ISL_1737073, EPI_ISL_1737075, EPI_ISL_1737086, EPI_ISL_1737087, EPI_ISL_1737154, EPI_ISL_1737182, EPI_ISL_1737199, EPI_ISL_1737217, EPI_ISL_1737218, EPI_ISL_1737224, EPI_ISL_1737241, EPI_ISL_1737252, EPI_ISL_1737256, EPI_ISL_1737258, EPI_ISL_1737264, EPI_ISL_1737265, EPI_ISL_1737266, EPI_ISL_1737267, EPI_ISL_1737268, EPI_ISL_1737322, EPI_ISL_1737323, EPI_ISL_1737434, EPI_ISL_1737438, EPI_ISL_1737446, EPI_ISL_1737454, EPI_ISL_1737474, EPI_ISL_1737479, EPI_ISL_1737490, EPI_ISL_1737500, EPI_ISL_1737511, EPI_ISL_1737514, EPI_ISL_1737553, EPI_ISL_1737592, EPI_ISL_1737609, EPI_ISL_1738763, EPI_ISL_1753554, EPI_ISL_1753593, EPI_ISL_1753597, EPI_ISL_1834453, EPI_ISL_1834455, EPI_ISL_1834494, EPI_ISL_1834499, EPI_ISL_1834515, EPI_ISL_1834516, EPI_ISL_1834522, EPI_ISL_1834581, EPI_ISL_1834602, EPI_ISL_1834616, EPI_ISL_1834650, EPI_ISL_1834733, EPI_ISL_1834743, EPI_ISL_1834762, EPI_ISL_1834765, EPI_ISL_1834778, EPI_ISL_1834809, EPI_ISL_1834824, EPI_ISL_1834831, EPI_ISL_1834880, EPI_ISL_1834918, EPI_ISL_1834945, EPI_ISL_1834971, EPI_ISL_1835057, EPI_ISL_1835058, EPI_ISL_1835059, EPI_ISL_1835084, EPI_ISL_1835085, EPI_ISL_1835156, EPI_ISL_1835183, EPI_ISL_1835223, EPI_ISL_1835232, EPI_ISL_1835273, EPI_ISL_1835332, EPI_ISL_1835350, EPI_ISL_1835372, EPI_ISL_1835451, EPI_ISL_1835455, EPI_ISL_1835478, EPI_ISL_1835605, EPI_ISL_1835643, EPI_ISL_1835655, EPI_ISL_1835676, EPI_ISL_1835694, EPI_ISL_1835702, EPI_ISL_1835733, EPI_ISL_1835734, EPI_ISL_1835791, EPI_ISL_1835792, EPI_ISL_1835793, EPI_ISL_1835796, EPI_ISL_1835807, EPI_ISL_1835809, EPI_ISL_1835829, EPI_ISL_1835830, EPI_ISL_1835850, EPI_ISL_1835878, EPI_ISL_1835896, EPI_ISL_1835945, EPI_ISL_1835946, EPI_ISL_1835964, EPI_ISL_1835966, EPI_ISL_1836010, EPI_ISL_1836032, EPI_ISL_1836085, EPI_ISL_1836087, EPI_ISL_1836095, EPI_ISL_1836098, EPI_ISL_1836100, EPI_ISL_1836105, EPI_ISL_1836107, EPI_ISL_1836119, EPI_ISL_1836490, EPI_ISL_1836536, EPI_ISL_1837112, EPI_ISL_1837113, EPI_ISL_1837118, EPI_ISL_1837120, EPI_ISL_1837617, EPI_ISL_1837619, EPI_ISL_1837639, EPI_ISL_1837644, EPI_ISL_1837664, EPI_ISL_1900357, EPI_ISL_1925142, EPI_ISL_1925194, EPI_ISL_1925208, EPI_ISL_1925211, EPI_ISL_1925229, EPI_ISL_1925233, EPI_ISL_1925286, EPI_ISL_1925375, EPI_ISL_1925386, EPI_ISL_1925434, EPI_ISL_1925443, EPI_ISL_1925444, EPI_ISL_1925449, EPI_ISL_1925469, EPI_ISL_1925472, EPI_ISL_1925479, EPI_ISL_1925587, EPI_ISL_1925588, EPI_ISL_1995226, EPI_ISL_1995235, EPI_ISL_1996882, EPI_ISL_2185804, EPI_ISL_2185873, EPI_ISL_2185879, EPI_ISL_2185891, EPI_ISL_2185901, EPI_ISL_2185905, EPI_ISL_2185910, EPI_ISL_2185924, EPI_ISL_2185949, EPI_ISL_2185960, EPI_ISL_2185963, EPI_ISL_2185965, EPI_ISL_2185976, EPI_ISL_2185983, EPI_ISL_2185998, EPI_ISL_2186002, EPI_ISL_2186162, EPI_ISL_2186166, EPI_ISL_2242117, EPI_ISL_2242121, EPI_ISL_2242126, EPI_ISL_2242128, EPI_ISL_2242330, EPI_ISL_2242399, EPI_ISL_2242401, EPI_ISL_2242457, EPI_ISL_2242475, EPI_ISL_2242504, EPI_ISL_2242690, EPI_ISL_2242691, EPI_ISL_2242732, EPI_ISL_2242781, EPI_ISL_2242821, EPI_ISL_2242851, EPI_ISL_2242875, EPI_ISL_2242985, EPI_ISL_2244222, EPI_ISL_2244338, EPI_ISL_2244350, EPI_ISL_2244354, EPI_ISL_2244363, EPI_ISL_2244385, EPI_ISL_2244386, EPI_ISL_2244399, EPI_ISL_2244420, EPI_ISL_2244433, EPI_ISL_2244438, EPI_ISL_2244456, EPI_ISL_2244470, EPI_ISL_2244473, EPI_ISL_2244478, EPI_ISL_2244481, EPI_ISL_2244482, EPI_ISL_2244483, EPI_ISL_2244493, EPI_ISL_2244509, EPI_ISL_2244511, EPI_ISL_2244513, EPI_ISL_2244548, EPI_ISL_2244562, EPI_ISL_2244573, EPI_ISL_2244579, EPI_ISL_2244583, EPI_ISL_2244623, EPI_ISL_2244647, EPI_ISL_2244653, EPI_ISL_2244661, EPI_ISL_2244665, EPI_ISL_2244674, EPI_ISL_2244706, EPI_ISL_2244722, EPI_ISL_2244744, EPI_ISL_2244748, EPI_ISL_2244781, EPI_ISL_2244790, EPI_ISL_2244791, EPI_ISL_2244798, EPI_ISL_2244817, EPI_ISL_2244820, EPI_ISL_2244853, EPI_ISL_2244878, EPI_ISL_2421362, EPI_ISL_2421376, EPI_ISL_2421377, EPI_ISL_2421380, EPI_ISL_2421386, EPI_ISL_2421390, EPI_ISL_2421392, EPI_ISL_2421393, EPI_ISL_2421394, EPI_ISL_2421395, EPI_ISL_2421396, EPI_ISL_2421397, EPI_ISL_2421398, EPI_ISL_2421399, EPI_ISL_2421403, EPI_ISL_2421495, EPI_ISL_2421616, EPI_ISL_2421636, EPI_ISL_2421659, EPI_ISL_2421702, EPI_ISL_2421783, EPI_ISL_2421795, EPI_ISL_2421796, EPI_ISL_2421797, EPI_ISL_2421805, EPI_ISL_2421812, EPI_ISL_2442306, EPI_ISL_2442307, EPI_ISL_2442320, EPI_ISL_2482313, EPI_ISL_2482314, EPI_ISL_2482316, EPI_ISL_2482317, EPI_ISL_2482318, EPI_ISL_2482319, EPI_ISL_2482320, EPI_ISL_2482349, EPI_ISL_2482402, EPI_ISL_2482422, EPI_ISL_2600031, EPI_ISL_2600034, EPI_ISL_2600036, EPI_ISL_2600041, EPI_ISL_2600042, EPI_ISL_2600049, EPI_ISL_2600080, EPI_ISL_2600083, EPI_ISL_2600109, EPI_ISL_2600142, EPI_ISL_2600153, EPI_ISL_2600163, EPI_ISL_2649902 | | | |
| see above | Aegis Sciences Corporation | Centers for Disease Control and Prevention Division of Viral Diseases, Pathogen Discovery | Adrian Paskey; Alec Vest; Benjamin Rambo-Martin; Christopher Gulvick; Clinton R. Paden; Cyndi Clark; Dakota Howard; Darlene Wagner; Dhwani Batra; Dillon Nall; Duncan MacCannell; Ethan Sanders; Holly Houdeshell; Jason Caravas; Kara Moser; Matthew Hardison; Matthew Schmerer; Ola Kvalvaag; Patrick Campbell; Peter W. Cook; Rob Case; Scott Sammons; Shatavia Morrison; Shaun Westlund; Vikramsinha Ghorpade; Yvette Unoarumhi |
| EPI_ISL_1575115, EPI_ISL_1575116, EPI_ISL_1575117, EPI_ISL_1575118, EPI_ISL_1575119, EPI_ISL_1575125, EPI_ISL_1575126, EPI_ISL_1576991, EPI_ISL_2304308, EPI_ISL_2304309, EPI_ISL_2304312, EPI_ISL_2304314, EPI_ISL_2304317, EPI_ISL_2304318, EPI_ISL_2304323, EPI_ISL_2304329, EPI_ISL_2304336, EPI_ISL_2304337, EPI_ISL_2304338, EPI_ISL_2304339, EPI_ISL_2304340 | | | |
| see above | Alameda County Public Health Department | Alameda County Public Health Department | Kevin Libuit; Kristina Hsieh; Kristna Hsieh; Maria Elma |
| EPI_ISL_1585085 | Arizona State University | Arizona State University | Efrem S. Lim; Joshua LaBaer; Joy M. Blain; LaRinda A. Holland; Nicholas J. Mellor; Peter T. Skidmore; Rabia Maqsood; Valerie Harris; Vel Murugan |
| EPI_ISL_1853453, EPI_ISL_1853470, EPI_ISL_1853565 | Avellino Labs | Avellino Labs | Milos Pjanic |
| EPI_ISL_1821315, EPI_ISL_1821340, EPI_ISL_1821345, EPI_ISL_1821356, EPI_ISL_1821357, EPI_ISL_1821360, EPI_ISL_1821361, EPI_ISL_1821364, EPI_ISL_1821369, EPI_ISL_1821379, EPI_ISL_1821382, EPI_ISL_1821403, EPI_ISL_1821410, EPI_ISL_1821412, EPI_ISL_1821424, EPI_ISL_1821440, EPI_ISL_1821442, EPI_ISL_1821466, EPI_ISL_2102525, EPI_ISL_2102527, EPI_ISL_2102539, EPI_ISL_2102540, EPI_ISL_2102542, EPI_ISL_2102546, EPI_ISL_2102551, EPI_ISL_2102552, EPI_ISL_2102562, EPI_ISL_2102564, EPI_ISL_2102566, EPI_ISL_2102568, EPI_ISL_2102570, EPI_ISL_2102572, EPI_ISL_2102573, EPI_ISL_2102575, EPI_ISL_2102580, EPI_ISL_2102584, EPI_ISL_2102586, EPI_ISL_2102587, EPI_ISL_2102592, EPI_ISL_2102595, EPI_ISL_2102597, EPI_ISL_2102741, EPI_ISL_2102743, EPI_ISL_2102750, EPI_ISL_2102754, EPI_ISL_2102756, EPI_ISL_2102765, EPI_ISL_2102771, EPI_ISL_2102773, EPI_ISL_2102775, EPI_ISL_2102777, EPI_ISL_2102778, EPI_ISL_2102780, EPI_ISL_2102783, EPI_ISL_2102786, EPI_ISL_2102787, EPI_ISL_2102789, EPI_ISL_2102793, EPI_ISL_2102794, EPI_ISL_2102799, EPI_ISL_2102807, EPI_ISL_2102816, EPI_ISL_2102818, EPI_ISL_2102819, EPI_ISL_2102829, EPI_ISL_2102834, EPI_ISL_2102835, EPI_ISL_2102839, EPI_ISL_2102846, EPI_ISL_2102854, EPI_ISL_2102857, EPI_ISL_2102860, EPI_ISL_2102864, EPI_ISL_2102867, EPI_ISL_2102873, EPI_ISL_2102874, EPI_ISL_2102877, EPI_ISL_2102879, EPI_ISL_2102881, EPI_ISL_2102884, EPI_ISL_2102897, EPI_ISL_2102900, EPI_ISL_2102910, EPI_ISL_2102915, EPI_ISL_2102921, EPI_ISL_2102924, EPI_ISL_2102928, EPI_ISL_2102929, EPI_ISL_2102934, EPI_ISL_2102940, EPI_ISL_2102942, EPI_ISL_2102944, EPI_ISL_2102957, EPI_ISL_2102962, EPI_ISL_2102965, EPI_ISL_2102968, EPI_ISL_2102974, EPI_ISL_2102977, EPI_ISL_2102980, EPI_ISL_2102983, EPI_ISL_2102984, EPI_ISL_2102987, EPI_ISL_2102989, EPI_ISL_2102991, EPI_ISL_2102993, EPI_ISL_2103000, EPI_ISL_2103004, EPI_ISL_2103015, EPI_ISL_2103027, EPI_ISL_2103032, EPI_ISL_2103047, EPI_ISL_2103050, EPI_ISL_2103053, EPI_ISL_2103066, EPI_ISL_2103075, EPI_ISL_2103095, EPI_ISL_2103098, EPI_ISL_2103103, EPI_ISL_2103116, EPI_ISL_2103118, EPI_ISL_2103120, EPI_ISL_2103124, EPI_ISL_2103128, EPI_ISL_2103135, EPI_ISL_2103139, EPI_ISL_2103142, EPI_ISL_2103155, EPI_ISL_2103160, EPI_ISL_2103163, EPI_ISL_2103166, EPI_ISL_2103180, EPI_ISL_2103181, EPI_ISL_2103193, EPI_ISL_2497661, EPI_ISL_2658615, EPI_ISL_2659277, EPI_ISL_2659283, EPI_ISL_2659348, EPI_ISL_2659566, EPI_ISL_2758493, EPI_ISL_2758494, EPI_ISL_2758505, EPI_ISL_2758506, EPI_ISL_2758507, EPI_ISL_2758508, EPI_ISL_2758509, EPI_ISL_2758510, EPI_ISL_2758511, EPI_ISL_2758512, EPI_ISL_2758513 | | | |
| see above | CA DPH Viral and Rickettsial Disease Laboratory | Chan-Zuckerberg Biohub | CZB Cliahub Consortium |
| EPI_ISL_1516361, EPI_ISL_1678085, EPI_ISL_1678086, EPI_ISL_1678087, EPI_ISL_1678088, EPI_ISL_1678089, EPI_ISL_1678090, EPI_ISL_1678091, EPI_ISL_1678092, EPI_ISL_1678093, EPI_ISL_1678094, EPI_ISL_1678095, EPI_ISL_1678276, EPI_ISL_1678277, EPI_ISL_1678278, EPI_ISL_1678279, EPI_ISL_1678282, EPI_ISL_1678283, EPI_ISL_1678286, EPI_ISL_1678287, EPI_ISL_1798830, EPI_ISL_1798832, EPI_ISL_1798834, EPI_ISL_1798836, EPI_ISL_1798839, EPI_ISL_1798841, EPI_ISL_1798843, EPI_ISL_1798844, EPI_ISL_1798846, EPI_ISL_1798848, EPI_ISL_1798850, EPI_ISL_1798853, EPI_ISL_1798855, EPI_ISL_1798857, EPI_ISL_1798858, EPI_ISL_1798860, EPI_ISL_1798862, EPI_ISL_1798864, EPI_ISL_1798866, EPI_ISL_1798868, EPI_ISL_1823713, EPI_ISL_1823714, EPI_ISL_1823715, EPI_ISL_1823716, EPI_ISL_1823717, EPI_ISL_1823718, EPI_ISL_1823719, EPI_ISL_1823720, EPI_ISL_1823721, EPI_ISL_1823722, EPI_ISL_1823723, EPI_ISL_1823724, EPI_ISL_1823725, EPI_ISL_1823726, EPI_ISL_1823727, EPI_ISL_1823728, EPI_ISL_1823729, EPI_ISL_1823730, EPI_ISL_1823731, EPI_ISL_1823732, EPI_ISL_1823733, EPI_ISL_1823734, EPI_ISL_1823735, EPI_ISL_1823736, EPI_ISL_1823737, EPI_ISL_1823738, EPI_ISL_1823739, EPI_ISL_1823740, EPI_ISL_1823741, EPI_ISL_1823742, EPI_ISL_1823743, EPI_ISL_1823744, EPI_ISL_1823745, EPI_ISL_1823746, EPI_ISL_1823747, EPI_ISL_1937813, EPI_ISL_1937820, EPI_ISL_1937823, EPI_ISL_1937825, EPI_ISL_1937827, EPI_ISL_1937829 | | | |
| see above | CA-Los Angeles County Public Health Laboratory | Centers for Disease Control and Prevention Division of Viral Diseases, Pathogen Discovery | Alison Laufer Halpin; Ben L. Rambo-Martin; Clinton R. Paden; Dakota Howard; Darlene Wagner; Dave Wentworth; Dhwani Batra; Jasmine Padilla; Justin Lee; Katie Dillon; Krista Queen; Kristen Knipe; Kristine Lacek; Mark Burroughs; Matthew Schmerer; Mili Sheth; Peter Cook; Sam Shepard; Sarah Nobles; Shoshona Le; Suxiang Tong; Vivien Dugan; Yvette Unoarumhi |
| EPI_ISL_431144, EPI_ISL_2782395, EPI_ISL_2782411, EPI_ISL_2782414, EPI_ISL_2782416, EPI_ISL_2782424, EPI_ISL_2782427, EPI_ISL_2782428, EPI_ISL_2782444, EPI_ISL_2782490, EPI_ISL_2782508, EPI_ISL_2782509, EPI_ISL_2782512, EPI_ISL_2782513, EPI_ISL_2782530, EPI_ISL_2782537, EPI_ISL_2782566, EPI_ISL_2782570, EPI_ISL_2782574, EPI_ISL_2782579, EPI_ISL_2782583, EPI_ISL_2782588, EPI_ISL_2782592, EPI_ISL_2782593, EPI_ISL_2782594, EPI_ISL_2782595, EPI_ISL_2782600, EPI_ISL_2782608, EPI_ISL_2783198, EPI_ISL_2783199, EPI_ISL_2783200, EPI_ISL_2783201, EPI_ISL_2783202, EPI_ISL_2783203, EPI_ISL_2783204, EPI_ISL_2783205, EPI_ISL_2783206, EPI_ISL_2783207, EPI_ISL_2783208, EPI_ISL_2783209, EPI_ISL_2783210, EPI_ISL_2783211, EPI_ISL_2783212, EPI_ISL_2783213, EPI_ISL_2783214, EPI_ISL_2783215, EPI_ISL_2783216, EPI_ISL_2783217, EPI_ISL_2783218, EPI_ISL_2783219, EPI_ISL_2783220, EPI_ISL_2783221, EPI_ISL_2783222, EPI_ISL_2783223, EPI_ISL_2783224, EPI_ISL_2783225, EPI_ISL_2783226, EPI_ISL_2783227, EPI_ISL_2783228, EPI_ISL_2783229, EPI_ISL_2783230, EPI_ISL_2783231, EPI_ISL_2783232, EPI_ISL_2783233, EPI_ISL_2783234, EPI_ISL_2783235, EPI_ISL_2783236, EPI_ISL_2783237, EPI_ISL_2783238, EPI_ISL_2783239, EPI_ISL_2783240, EPI_ISL_2783241, EPI_ISL_2783242, EPI_ISL_2783243, EPI_ISL_2797315, EPI_ISL_2797316, EPI_ISL_2797317, EPI_ISL_2797318, EPI_ISL_2797319, EPI_ISL_2797320, EPI_ISL_2797321, EPI_ISL_2797322, EPI_ISL_2797323, EPI_ISL_2797324, EPI_ISL_2797325, EPI_ISL_2797326, EPI_ISL_2797327, EPI_ISL_2797328, EPI_ISL_2797329, EPI_ISL_2797330, EPI_ISL_2797331, EPI_ISL_2797332, EPI_ISL_2797333, EPI_ISL_2797334, EPI_ISL_2797335, EPI_ISL_2797336, EPI_ISL_2797337, EPI_ISL_2797338, EPI_ISL_2797339, EPI_ISL_2797340, EPI_ISL_2797341, EPI_ISL_2797342, EPI_ISL_2797343, EPI_ISL_2797344, EPI_ISL_2797345, EPI_ISL_2797346, EPI_ISL_2797347, EPI_ISL_2797348, EPI_ISL_2797349, EPI_ISL_2797350, EPI_ISL_2797351, EPI_ISL_2797352, EPI_ISL_2797353, EPI_ISL_2797354, EPI_ISL_2797355, EPI_ISL_2797356, EPI_ISL_2797357, EPI_ISL_2797358, EPI_ISL_2797359, EPI_ISL_2797360, EPI_ISL_2797361, EPI_ISL_2797362, EPI_ISL_2797363, EPI_ISL_2797364, EPI_ISL_2797365, EPI_ISL_2797366, EPI_ISL_2797367, EPI_ISL_2797368, EPI_ISL_2797369, EPI_ISL_2797370, EPI_ISL_2797371, EPI_ISL_2797372, EPI_ISL_2797373, EPI_ISL_2797374, EPI_ISL_2797375, EPI_ISL_2797376, EPI_ISL_2797377, EPI_ISL_2797378, EPI_ISL_2797379, EPI_ISL_2797380, EPI_ISL_2797381, EPI_ISL_2798103, EPI_ISL_2798104, EPI_ISL_2798105, EPI_ISL_2798106, EPI_ISL_2798107, EPI_ISL_2798108, EPI_ISL_2798109, EPI_ISL_2798110, EPI_ISL_2798111, EPI_ISL_2798112, EPI_ISL_2798113, EPI_ISL_2798114, EPI_ISL_2798115, EPI_ISL_2798116, EPI_ISL_2798117, EPI_ISL_2798118, EPI_ISL_2798119, EPI_ISL_2798120, EPI_ISL_2798121, EPI_ISL_2798122, EPI_ISL_2798123, EPI_ISL_2798124, EPI_ISL_2798125, EPI_ISL_2798126, EPI_ISL_2798127, EPI_ISL_2798128, EPI_ISL_2798129, EPI_ISL_2798130, EPI_ISL_2798131, EPI_ISL_2798132, EPI_ISL_2798133, EPI_ISL_2798134, EPI_ISL_2798135, EPI_ISL_2798136, EPI_ISL_2798137, EPI_ISL_2798138, EPI_ISL_2798139, EPI_ISL_2798140, EPI_ISL_2798141, EPI_ISL_2798142, EPI_ISL_2798143, EPI_ISL_2798144, EPI_ISL_2798145, EPI_ISL_2798146, EPI_ISL_2798147, EPI_ISL_2798148, EPI_ISL_2798149, EPI_ISL_2798150, EPI_ISL_2798151, EPI_ISL_2798152, EPI_ISL_2798153, EPI_ISL_2798154, EPI_ISL_2798155, EPI_ISL_2798156, EPI_ISL_2798157, EPI_ISL_2798158, EPI_ISL_2798159, EPI_ISL_2798160, EPI_ISL_2798161, EPI_ISL_2798162, EPI_ISL_2798163, EPI_ISL_2798164, EPI_ISL_2798165, EPI_ISL_2798166, EPI_ISL_2798167, EPI_ISL_2798168, EPI_ISL_2798169, EPI_ISL_2798170, EPI_ISL_2798171, EPI_ISL_2798172, EPI_ISL_2798173, EPI_ISL_2798174, EPI_ISL_2798175, EPI_ISL_2798176, EPI_ISL_2798178, EPI_ISL_2798179, EPI_ISL_2798180, EPI_ISL_2798181, EPI_ISL_2798182, EPI_ISL_2825689, EPI_ISL_2825690, EPI_ISL_2825691, EPI_ISL_2825692, EPI_ISL_2825693, EPI_ISL_2825694, EPI_ISL_2825695, EPI_ISL_2825696, EPI_ISL_2825697, EPI_ISL_2825698, EPI_ISL_2825699, EPI_ISL_2825700, EPI_ISL_2825701, EPI_ISL_2825702, EPI_ISL_2825703, EPI_ISL_2825704, EPI_ISL_2825705, EPI_ISL_2825706, EPI_ISL_2825707, EPI_ISL_2825708, EPI_ISL_2825709, EPI_ISL_2825710, EPI_ISL_2825711, EPI_ISL_2825712, EPI_ISL_2825713, EPI_ISL_2825714, EPI_ISL_2825715, EPI_ISL_2825716, EPI_ISL_2825717, EPI_ISL_2825718, EPI_ISL_2825719, EPI_ISL_2825720, EPI_ISL_2825721, EPI_ISL_2825722, EPI_ISL_2825723, EPI_ISL_2825724, EPI_ISL_2825725, EPI_ISL_2825726, EPI_ISL_2825727, EPI_ISL_2825728, EPI_ISL_2825729, EPI_ISL_2825730, EPI_ISL_2825731, EPI_ISL_2825732, EPI_ISL_2825733, EPI_ISL_2825734, EPI_ISL_2825735, EPI_ISL_2825736, EPI_ISL_2825737, EPI_ISL_2825738, EPI_ISL_2825739, EPI_ISL_2825740, EPI_ISL_2825741, EPI_ISL_2825742, EPI_ISL_2825743, EPI_ISL_2825744, EPI_ISL_2825745, EPI_ISL_2825746, EPI_ISL_2825747, EPI_ISL_2825748, EPI_ISL_2825749, EPI_ISL_2825750, EPI_ISL_2825751, EPI_ISL_2825752, EPI_ISL_2825753, EPI_ISL_2825754, EPI_ISL_2825755, EPI_ISL_2825756, EPI_ISL_2825757, EPI_ISL_2825758, EPI_ISL_2825759, EPI_ISL_2825760, EPI_ISL_2825761, EPI_ISL_2825762, EPI_ISL_2825763, EPI_ISL_2825764, EPI_ISL_2825765, EPI_ISL_2825766, EPI_ISL_2825767, EPI_ISL_2825768, EPI_ISL_2825769, EPI_ISL_2825770, EPI_ISL_2825771, EPI_ISL_2825772, EPI_ISL_2825773, EPI_ISL_2825774, | | | |

EPI_ISL_2825775, EPI_ISL_2825776, EPI_ISL_2825777, EPI_ISL_2825778, EPI_ISL_2825779, EPI_ISL_2825780, EPI_ISL_2825781, EPI_ISL_2825782, EPI_ISL_2825783, EPI_ISL_2825784, EPI_ISL_2825785, EPI_ISL_2825786, EPI_ISL_2825787, EPI_ISL_2825788, EPI_ISL_2825789, EPI_ISL_2825790, EPI_ISL_2825791, EPI_ISL_2825792, EPI_ISL_2825793, EPI_ISL_2825794, EPI_ISL_2825795, EPI_ISL_2825796, EPI_ISL_2825797, EPI_ISL_2825798, EPI_ISL_2825799, EPI_ISL_2825800, EPI_ISL_2825801, EPI_ISL_2825802, EPI_ISL_2825803, EPI_ISL_2825804, EPI_ISL_2825805, EPI_ISL_2825806, EPI_ISL_2825807, EPI_ISL_2825808, EPI_ISL_2825809, EPI_ISL_2826837, EPI_ISL_2826838, EPI_ISL_2826839, EPI_ISL_2826840, EPI_ISL_2826841, EPI_ISL_2826842, EPI_ISL_2826843, EPI_ISL_2826844, EPI_ISL_2826845, EPI_ISL_2826846, EPI_ISL_2826847, EPI_ISL_2826848, EPI_ISL_2826849, EPI_ISL_2826850, EPI_ISL_2826851, EPI_ISL_2826852, EPI_ISL_2826853, EPI_ISL_2826854, EPI_ISL_2826855, EPI_ISL_2826856, EPI_ISL_2826857, EPI_ISL_2826858, EPI_ISL_2826859, EPI_ISL_2826860, EPI_ISL_2826861, EPI_ISL_2826862, EPI_ISL_2826863, EPI_ISL_2826864, EPI_ISL_2826865, EPI_ISL_2826866, EPI_ISL_2826867, EPI_ISL_2826868, EPI_ISL_2826869, EPI_ISL_2826870, EPI_ISL_2826871, EPI_ISL_2826872, EPI_ISL_2826873, EPI_ISL_2826874, EPI_ISL_2826875, EPI_ISL_2826876, EPI_ISL_2826877, EPI_ISL_2826878, EPI_ISL_2826879, EPI_ISL_2826880, EPI_ISL_2826881, EPI_ISL_2826882, EPI_ISL_2826883, EPI_ISL_2826884, EPI_ISL_2826885, EPI_ISL_2826886, EPI_ISL_2826887, EPI_ISL_2826888, EPI_ISL_2826889, EPI_ISL_2826890, EPI_ISL_2826891, EPI_ISL_2826892, EPI_ISL_2826893, EPI_ISL_2826894, EPI_ISL_2826895, EPI_ISL_2826896, EPI_ISL_2826897, EPI_ISL_2826898, EPI_ISL_2826899, EPI_ISL_2826900, EPI_ISL_2826901, EPI_ISL_2826902, EPI_ISL_2826903, EPI_ISL_2826904, EPI_ISL_2826905, EPI_ISL_2826906, EPI_ISL_2826907, EPI_ISL_2826908, EPI_ISL_2826909, EPI_ISL_2826910, EPI_ISL_2826911, EPI_ISL_2858257, EPI_ISL_2858258, EPI_ISL_2858259, EPI_ISL_2858260, EPI_ISL_2858261, EPI_ISL_2858262, EPI_ISL_2858263, EPI_ISL_2858264, EPI_ISL_2858265, EPI_ISL_2858266, EPI_ISL_2858267, EPI_ISL_2858268, EPI_ISL_2858269, EPI_ISL_2858270, EPI_ISL_2858271, EPI_ISL_2858272, EPI_ISL_2858273, EPI_ISL_2858274, EPI_ISL_2858275, EPI_ISL_2858276, EPI_ISL_2858277, EPI_ISL_2858278, EPI_ISL_2858279, EPI_ISL_2858284, EPI_ISL_2858285, EPI_ISL_2858288, EPI_ISL_2858290, EPI_ISL_2858293, EPI_ISL_2858294, EPI_ISL_2858295, EPI_ISL_2858296, EPI_ISL_2858303, EPI_ISL_2858304, EPI_ISL_2858305, EPI_ISL_2858306, EPI_ISL_2858307, EPI_ISL_2858308, EPI_ISL_2858309, EPI_ISL_2858310, EPI_ISL_2858322, EPI_ISL_2858328, EPI_ISL_2858332, EPI_ISL_2858333, EPI_ISL_2858334, EPI_ISL_2858335, EPI_ISL_2858336, EPI_ISL_2858337, EPI_ISL_2858338, EPI_ISL_2858341, EPI_ISL_2858342, EPI_ISL_2858345, EPI_ISL_2858346, EPI_ISL_2858347, EPI_ISL_2858349, EPI_ISL_2858350, EPI_ISL_2858351, EPI_ISL_2858353, EPI_ISL_2858357, EPI_ISL_2858359, EPI_ISL_2858360, EPI_ISL_2858366, EPI_ISL_2858372, EPI_ISL_2858376, EPI_ISL_2858378, EPI_ISL_2858379, EPI_ISL_2858380, EPI_ISL_2858384, EPI_ISL_2858385, EPI_ISL_2858386, EPI_ISL_2858387, EPI_ISL_2858388, EPI_ISL_2858389, EPI_ISL_2858392, EPI_ISL_2858401, EPI_ISL_2858402, EPI_ISL_2858403, EPI_ISL_2858404, EPI_ISL_2858420, EPI_ISL_2858421, EPI_ISL_2858422, EPI_ISL_2858423, EPI_ISL_2858424, EPI_ISL_2858425, EPI_ISL_2858426, EPI_ISL_2858427, EPI_ISL_2858428, EPI_ISL_2858429, EPI_ISL_2858430, EPI_ISL_2858433, EPI_ISL_2858435, EPI_ISL_2858436, EPI_ISL_2858440, EPI_ISL_2858442, EPI_ISL_2858444, EPI_ISL_2858448, EPI_ISL_2858457, EPI_ISL_2858459, EPI_ISL_2858460, EPI_ISL_2858461, EPI_ISL_2858462, EPI_ISL_2858463, EPI_ISL_2858464, EPI_ISL_2858465, EPI_ISL_2858466, EPI_ISL_2858467, EPI_ISL_2858468, EPI_ISL_2858470, EPI_ISL_2858482, EPI_ISL_2858483, EPI_ISL_2858484, EPI_ISL_2858485, EPI_ISL_2858490, EPI_ISL_2858491, EPI_ISL_2858492, EPI_ISL_2858494, EPI_ISL_2858495, EPI_ISL_2858497, EPI_ISL_2858498, EPI_ISL_2858499, EPI_ISL_2858500, EPI_ISL_2858502, EPI_ISL_2858503, EPI_ISL_2858504, EPI_ISL_2858505, EPI_ISL_2858506, EPI_ISL_2858508, EPI_ISL_2858534, EPI_ISL_2858535, EPI_ISL_2858536, EPI_ISL_2858537, EPI_ISL_2858538, EPI_ISL_2858539, EPI_ISL_2858540, EPI_ISL_2858541, EPI_ISL_2858542, EPI_ISL_2858555, EPI_ISL_2858556, EPI_ISL_2858557, EPI_ISL_2858558, EPI_ISL_2858559, EPI_ISL_2858560, EPI_ISL_2858561, EPI_ISL_2858562, EPI_ISL_2858565, EPI_ISL_2858566, EPI_ISL_2858567, EPI_ISL_2858568, EPI_ISL_2858570, EPI_ISL_2858571, EPI_ISL_2858572, EPI_ISL_2858573, EPI_ISL_2858574, EPI_ISL_2858575, EPI_ISL_2858576, EPI_ISL_2858577, EPI_ISL_2858578, EPI_ISL_2858579, EPI_ISL_2858580, EPI_ISL_2858583, EPI_ISL_2858584, EPI_ISL_2858586, EPI_ISL_2858587, EPI_ISL_2881867, EPI_ISL_2881868, EPI_ISL_2881870, EPI_ISL_2881874, EPI_ISL_2881882, EPI_ISL_2881890, EPI_ISL_2881891, EPI_ISL_2881894, EPI_ISL_2881897, EPI_ISL_2881902, EPI_ISL_2881903, EPI_ISL_2881904, EPI_ISL_2881905, EPI_ISL_2881916, EPI_ISL_2881946, EPI_ISL_2881953, EPI_ISL_2881954, EPI_ISL_2881955, EPI_ISL_2881960, EPI_ISL_2881961, EPI_ISL_2881962, EPI_ISL_2881963, EPI_ISL_2881973, EPI_ISL_2881974, EPI_ISL_2881975, EPI_ISL_2881976, EPI_ISL_2881978, EPI_ISL_2881979, EPI_ISL_2881980, EPI_ISL_2881981, EPI_ISL_2881982, EPI_ISL_2881983, EPI_ISL_2881984, EPI_ISL_2881985, EPI_ISL_2881986, EPI_ISL_2881987, EPI_ISL_2881988, EPI_ISL_2882001, EPI_ISL_2882011, EPI_ISL_2882012, EPI_ISL_2882013, EPI_ISL_2882014, EPI_ISL_2882015, EPI_ISL_2882021, EPI_ISL_2882022, EPI_ISL_2882040, EPI_ISL_2882732, EPI_ISL_2882734, EPI_ISL_2882735, EPI_ISL_2882738, EPI_ISL_2882740, EPI_ISL_2882741, EPI_ISL_2882742, EPI_ISL_2882743, EPI_ISL_2882744, EPI_ISL_2882748, EPI_ISL_2882749, EPI_ISL_2882752, EPI_ISL_2882753, EPI_ISL_2882754, EPI_ISL_2882755, EPI_ISL_2882756, EPI_ISL_2882757, EPI_ISL_2882759, EPI_ISL_2882764, EPI_ISL_2882767, EPI_ISL_2882768, EPI_ISL_2882769, EPI_ISL_2882770, EPI_ISL_2882771, EPI_ISL_2882773, EPI_ISL_2882774, EPI_ISL_2882779, EPI_ISL_2882780, EPI_ISL_2882782, EPI_ISL_2882783, EPI_ISL_2882784, EPI_ISL_2882785, EPI_ISL_2882786, EPI_ISL_2882787, EPI_ISL_2882788, EPI_ISL_2882790, EPI_ISL_2882794, EPI_ISL_2882795, EPI_ISL_2882797, EPI_ISL_2882803, EPI_ISL_2882804, EPI_ISL_2882805, EPI_ISL_2882807, EPI_ISL_2882808, EPI_ISL_2882809, EPI_ISL_2882817, EPI_ISL_2882818, EPI_ISL_2882819, EPI_ISL_2882820, EPI_ISL_2882821, EPI_ISL_2882822, EPI_ISL_2882825, EPI_ISL_2882826, EPI_ISL_2882831, EPI_ISL_2882838, EPI_ISL_2882839, EPI_ISL_2882840, EPI_ISL_2882841, EPI_ISL_2882842, EPI_ISL_2882843, EPI_ISL_2882844, EPI_ISL_2882846, EPI_ISL_2882847, EPI_ISL_2882848, EPI_ISL_2882850, EPI_ISL_2882851, EPI_ISL_2882852, EPI_ISL_2882853, EPI_ISL_2882854, EPI_ISL_2882857, EPI_ISL_2882858, EPI_ISL_2882859, EPI_ISL_2882860, EPI_ISL_2882867, EPI_ISL_2882869, EPI_ISL_2882870, EPI_ISL_2882871, EPI_ISL_2882872, EPI_ISL_2882873, EPI_ISL_2882874, EPI_ISL_2882876, EPI_ISL_2882880, EPI_ISL_2882881, EPI_ISL_2882884, EPI_ISL_2882885, EPI_ISL_2882887, EPI_ISL_2882889, EPI_ISL_2882892, EPI_ISL_2882894, EPI_ISL_2882896, EPI_ISL_2882901, EPI_ISL_2882902, EPI_ISL_2882903, EPI_ISL_2882904, EPI_ISL_2882906, EPI_ISL_2882912, EPI_ISL_2882913, EPI_ISL_2882914, EPI_ISL_2882915, EPI_ISL_2882918, EPI_ISL_2882920, EPI_ISL_2882922, EPI_ISL_2882923, EPI_ISL_2882924, EPI_ISL_2882925, EPI_ISL_2882926, EPI_ISL_2882927, EPI_ISL_2882928, EPI_ISL_2882929, EPI_ISL_2882930, EPI_ISL_2882931, EPI_ISL_2882932, EPI_ISL_2882933, EPI_ISL_2882935, EPI_ISL_2882936, EPI_ISL_2882937, EPI_ISL_2882938, EPI_ISL_2882939, EPI_ISL_2882940, EPI_ISL_2882941, EPI_ISL_2882944, EPI_ISL_2882945, EPI_ISL_2882946, EPI_ISL_2882947, EPI_ISL_2882951, EPI_ISL_2882954, EPI_ISL_2882955, EPI_ISL_2882956, EPI_ISL_2882957, EPI_ISL_2882958, EPI_ISL_2882959, EPI_ISL_2882960, EPI_ISL_2882961, EPI_ISL_2882962, EPI_ISL_2882963, EPI_ISL_2882964, EPI_ISL_2882965, EPI_ISL_2882970, EPI_ISL_2882974, EPI_ISL_2882979, EPI_ISL_2882981, EPI_ISL_2882984, EPI_ISL_2882985, EPI_ISL_2882986, EPI_ISL_2882987, EPI_ISL_2882988, EPI_ISL_2882989, EPI_ISL_2882991, EPI_ISL_2882992, EPI_ISL_2882999, EPI_ISL_2883002, EPI_ISL_2883003, EPI_ISL_2883004, EPI_ISL_2883006, EPI_ISL_2883007, EPI_ISL_2883010, EPI_ISL_2883011, EPI_ISL_2883012, EPI_ISL_2883013, EPI_ISL_2883014, EPI_ISL_2883016, EPI_ISL_2883017, EPI_ISL_2883018, EPI_ISL_2883020, EPI_ISL_2883023, EPI_ISL_2883024, EPI_ISL_2883025, EPI_ISL_2883029, EPI_ISL_2883030, EPI_ISL_2883031, EPI_ISL_2883032, EPI_ISL_2883033, EPI_ISL_2883034, EPI_ISL_2883035, EPI_ISL_2883040, EPI_ISL_2883041, EPI_ISL_2883043, EPI_ISL_2883044, EPI_ISL_2883045, EPI_ISL_2883047, EPI_ISL_2883048, EPI_ISL_2883049, EPI_ISL_2883050, EPI_ISL_2883051, EPI_ISL_2883052, EPI_ISL_2883053, EPI_ISL_2883054, EPI_ISL_2883055, EPI_ISL_2883061, EPI_ISL_2883062, EPI_ISL_2883064, EPI_ISL_2883065, EPI_ISL_2883068, EPI_ISL_2883069, EPI_ISL_2883070, EPI_ISL_2883071, EPI_ISL_2889859, EPI_ISL_2889881, EPI_ISL_2889895, EPI_ISL_2889896, EPI_ISL_2889897, EPI_ISL_2889898, EPI_ISL_2889899, EPI_ISL_2889908, EPI_ISL_2889914, EPI_ISL_2889915, EPI_ISL_2889916, EPI_ISL_2889921, EPI_ISL_2889936, EPI_ISL_2889938, EPI_ISL_2889940, EPI_ISL_2889941, EPI_ISL_2889942, EPI_ISL_2889974, EPI_ISL_2889982, EPI_ISL_2890008, EPI_ISL_2890014, EPI_ISL_2890015, EPI_ISL_2890050, EPI_ISL_2890055, EPI_ISL_2890065, EPI_ISL_2890092, EPI_ISL_2890105, EPI_ISL_2890122, EPI_ISL_2890158, EPI_ISL_2890171, EPI_ISL_2890172, EPI_ISL_2890174, EPI_ISL_2890175, EPI_ISL_2890176, EPI_ISL_2890177, EPI_ISL_2890184, EPI_ISL_2890226, EPI_ISL_2890239, EPI_ISL_2890240, EPI_ISL_2890242, EPI_ISL_2890318, EPI_ISL_2890323, EPI_ISL_2890350, EPI_ISL_2890352, EPI_ISL_2890372, EPI_ISL_2890402, EPI_ISL_2890403, EPI_ISL_2890442, EPI_ISL_2890451, EPI_ISL_2890479, EPI_ISL_2890480, EPI_ISL_2890481, EPI_ISL_2890490, EPI_ISL_2890491, EPI_ISL_2890492, EPI_ISL_2890493, EPI_ISL_2890528, EPI_ISL_2890529, EPI_ISL_2890530, EPI_ISL_2890534, EPI_ISL_2890542, EPI_ISL_2890543, EPI_ISL_2890586, EPI_ISL_2890587, EPI_ISL_2890590, EPI_ISL_2890591, EPI_ISL_2890592, EPI_ISL_2890593, EPI_ISL_2890594, EPI_ISL_2890595, EPI_ISL_2890596, EPI_ISL_2890597, EPI_ISL_2890598, EPI_ISL_2890612, EPI_ISL_2890613, EPI_ISL_2890640, EPI_ISL_2890666, EPI_ISL_2890667, EPI_ISL_2890698, EPI_ISL_2890699, EPI_ISL_2890702, EPI_ISL_2890733, EPI_ISL_2890754, EPI_ISL_2890793, EPI_ISL_2890804, EPI_ISL_2890805, EPI_ISL_2890806, EPI_ISL_2890844, EPI_ISL_2890845, EPI_ISL_2890846, EPI_ISL_2890847, EPI_ISL_2890848, EPI_ISL_2890849, EPI_ISL_2890850, EPI_ISL_2890851, EPI_ISL_2890852, EPI_ISL_2890892, EPI_ISL_2890916, EPI_ISL_2890917, EPI_ISL_2890918, EPI_ISL_2890919, EPI_ISL_2890920, EPI_ISL_2890921, EPI_ISL_2890922, EPI_ISL_2890924, EPI_ISL_2890925, EPI_ISL_2890926, EPI_ISL_2890927, EPI_ISL_2890988, EPI_ISL_2890995, EPI_ISL_2891009, EPI_ISL_2891027, EPI_ISL_2891036, EPI_ISL_2891037, EPI_ISL_2891038, EPI_ISL_2891039, EPI_ISL_2891040, EPI_ISL_2891041, EPI_ISL_2891074, EPI_ISL_2891075, EPI_ISL_2891076, EPI_ISL_2891077, EPI_ISL_2891083, EPI_ISL_2891089, EPI_ISL_2891095, EPI_ISL_2891149, EPI_ISL_2891158, EPI_ISL_2891175, EPI_ISL_2891229, EPI_ISL_2891233, EPI_ISL_2891234, EPI_ISL_2891287, EPI_ISL_2891311, EPI_ISL_2891320, EPI_ISL_2891353, EPI_ISL_2891354, EPI_ISL_2891355, EPI_ISL_2891371, EPI_ISL_2891378, EPI_ISL_2891412, EPI_ISL_2891414, EPI_ISL_2891419, EPI_ISL_2891420, EPI_ISL_2891493, EPI_ISL_2891519, EPI_ISL_2891574, EPI_ISL_2891595, EPI_ISL_2891596, EPI_ISL_2891597, EPI_ISL_2891599, EPI_ISL_2891601, EPI_ISL_2891618, EPI_ISL_2891621, EPI_ISL_2891669, EPI_ISL_2891677, EPI_ISL_2891697, EPI_ISL_2891705, EPI_ISL_2892372, EPI_ISL_2892373, EPI_ISL_2892380, EPI_ISL_2892385, EPI_ISL_2892387, EPI_ISL_2921623, EPI_ISL_2921624, EPI_ISL_2921625, EPI_ISL_2921626, EPI_ISL_2921627, EPI_ISL_2921628, EPI_ISL_2921629, EPI_ISL_2921630, EPI_ISL_2921631, EPI_ISL_2921632, EPI_ISL_2921633, EPI_ISL_2921634, EPI_ISL_2921635, EPI_ISL_2921636, EPI_ISL_2921637, EPI_ISL_2921638, EPI_ISL_2921639, EPI_ISL_2921640, EPI_ISL_2921641, EPI_ISL_2921642, EPI_ISL_2921643, EPI_ISL_2921644, EPI_ISL_2921645, EPI_ISL_2921646, EPI_ISL_2921647, EPI_ISL_2921648, EPI_ISL_2921649, EPI_ISL_2921650, EPI_ISL_2921651, EPI_ISL_2921652, EPI_ISL_2921653, EPI_ISL_2921654, EPI_ISL_2921655, EPI_ISL_2921656, EPI_ISL_2921657, EPI_ISL_2921658, EPI_ISL_2921659, EPI_ISL_2921660, EPI_ISL_2921661, EPI_ISL_2921662, EPI_ISL_2921663, EPI_ISL_2921665, EPI_ISL_2921666, EPI_ISL_2921668, EPI_ISL_2921669, EPI_ISL_2921670, EPI_ISL_2921671, EPI_ISL_2921672, EPI_ISL_2921673, EPI_ISL_2921674, EPI_ISL_2921676, EPI_ISL_2921677, EPI_ISL_2921678, EPI_ISL_2921679, EPI_ISL_2921680, EPI_ISL_2921681, EPI_ISL_2921686, EPI_ISL_2921688, EPI_ISL_2921689, EPI_ISL_2921690, EPI_ISL_2921691, EPI_ISL_2921692, EPI_ISL_2921693, EPI_ISL_2921694, EPI_ISL_2921695, EPI_ISL_2921696, EPI_ISL_2921697, EPI_ISL_2921698, EPI_ISL_2921699, EPI_ISL_2921700, EPI_ISL_2921701, EPI_ISL_2921704, EPI_ISL_2921705, EPI_ISL_2921706, EPI_ISL_2921707, EPI_ISL_2921708, EPI_ISL_2921709, EPI_ISL_2921710, EPI_ISL_2921711, EPI_ISL_2921712, EPI_ISL_2921713, EPI_ISL_2921714, EPI_ISL_2921715, EPI_ISL_2921716, EPI_ISL_2921717, EPI_ISL_2921718, EPI_ISL_2921722, EPI_ISL_2921723, EPI_ISL_2921724, EPI_ISL_2921726, EPI_ISL_2921727, EPI_ISL_2921728, EPI_ISL_2921729, EPI_ISL_2921730, EPI_ISL_2921731, EPI_ISL_2921732, EPI_ISL_2921733, EPI_ISL_2921734, EPI_ISL_2921735, EPI_ISL_2921738, EPI_ISL_2921739, EPI_ISL_2921740, EPI_ISL_2921741, EPI_ISL_2921742, EPI_ISL_2921743, EPI_ISL_2921744, EPI_ISL_2921746, EPI_ISL_2921747, EPI_ISL_2921748, EPI_ISL_2921749, EPI_ISL_2921750, EPI_ISL_2921751, EPI_ISL_2921752, EPI_ISL_2921753, EPI_ISL_2921754, EPI_ISL_2921755, EPI_ISL_2921771, EPI_ISL_2921772, EPI_ISL_2921777, EPI_ISL_2921778, EPI_ISL_2921782, EPI_ISL_2921783, EPI_ISL_2921784, EPI_ISL_2921785, EPI_ISL_2921786, EPI_ISL_2921790, EPI_ISL_2921792, EPI_ISL_2921793, EPI_ISL_2921795, EPI_ISL_2921796, EPI_ISL_2921797, EPI_ISL_2921798, EPI_ISL_2921799, EPI_ISL_2921800, EPI_ISL_2921801, EPI_ISL_2921802, EPI_ISL_2921803, EPI_ISL_2921804, EPI_ISL_2921806, EPI_ISL_2921807, EPI_ISL_2921808, EPI_ISL_2921809, EPI_ISL_2921810, EPI_ISL_2921811, EPI_ISL_2921813, EPI_ISL_2921816, EPI_ISL_2921819, EPI_ISL_2921820, EPI_ISL_2921821, EPI_ISL_2921822, EPI_ISL_2921829, EPI_ISL_2921830, EPI_ISL_2921831, EPI_ISL_2921832, EPI_ISL_2921833, EPI_ISL_2921834, EPI_ISL_2921835, EPI_ISL_2921836, EPI_ISL_2921847, EPI_ISL_2921853, EPI_ISL_2921854, EPI_ISL_2921855, EPI_ISL_2921856, EPI_ISL_2921873, EPI_ISL_2921882, EPI_ISL_2921893, EPI_ISL_2921897, EPI_ISL_2921898, EPI_ISL_2921899, EPI_ISL_2921900, EPI_ISL_2921901, EPI_ISL_2921902, EPI_ISL_2921903, EPI_ISL_2921904, EPI_ISL_2921905, EPI_ISL_2921907, EPI_ISL_2921914, EPI_ISL_2921928, EPI_ISL_2921929, EPI_ISL_2921930, EPI_ISL_2921931, EPI_ISL_2921932, EPI_ISL_2921933, EPI_ISL_2921942, EPI_ISL_2921943, EPI_ISL_2921944, EPI_ISL_2921957, EPI_ISL_2921958, EPI_ISL_2921959, EPI_ISL_2921960, EPI_ISL_2921961, EPI_ISL_2921962, EPI_ISL_2921963, EPI_ISL_2921982, EPI_ISL_2921988, EPI_ISL_2921995, EPI_ISL_2922004, EPI_ISL_2922006, EPI_ISL_2922007, EPI_ISL_2922010, EPI_ISL_2922011, EPI_ISL_2922012, EPI_ISL_2922018, EPI_ISL_2922024, EPI_ISL_2922025, EPI_ISL_2922026, EPI_ISL_2922027, EPI_ISL_2922028, EPI_ISL_2922029, EPI_ISL_2922030, EPI_ISL_2922032, EPI_ISL_2922033, EPI_ISL_2922035, EPI_ISL_2922036, EPI_ISL_2922037, EPI_ISL_2922038, EPI_ISL_2922045, EPI_ISL_2922057, EPI_ISL_2922058, EPI_ISL_2922059, EPI_ISL_2922060, EPI_ISL_2922061, EPI_ISL_2922065, EPI_ISL_2922066, EPI_ISL_2922067, EPI_ISL_2922068, EPI_ISL_2922069, EPI_ISL_2922071, EPI_ISL_2922072, EPI_ISL_2922073, EPI_ISL_2922074, EPI_ISL_2922075, EPI_ISL_2922076, EPI_ISL_2922077, EPI_ISL_2922085, EPI_ISL_2922089, EPI_ISL_2922092, EPI_ISL_2922093, EPI_ISL_2922094, EPI_ISL_2922095, EPI_ISL_2922096, EPI_ISL_2922097, EPI_ISL_2922098, EPI_ISL_2922099, EPI_ISL_2922100, EPI_ISL_2922101, EPI_ISL_2922107, EPI_ISL_2922108, EPI_ISL_2922109, EPI_ISL_2922110, EPI_ISL_2922111, EPI_ISL_2922114, EPI_ISL_2922123, EPI_ISL_2922125, EPI_ISL_2922131, EPI_ISL_2922133, EPI_ISL_2922136, EPI_ISL_2922140, EPI_ISL_2922141, EPI_ISL_2922143, EPI_ISL_2922144, EPI_ISL_2922145, EPI_ISL_2922146, EPI_ISL_2922147, EPI_ISL_2922150, EPI_ISL_2922151, EPI_ISL_2922152, EPI_ISL_2922155, EPI_ISL_2922156, EPI_ISL_2922157, EPI_ISL_2922159, EPI_ISL_2922160, EPI_ISL_2922161, EPI_ISL_2922162, EPI_ISL_2922164, EPI_ISL_2922167, EPI_ISL_2922169, EPI_ISL_2922170, EPI_ISL_2922192, EPI_ISL_2922193, EPI_ISL_2922196, EPI_ISL_2922197, EPI_ISL_2922198, EPI_ISL_2922204, EPI_ISL_2922212, EPI_ISL_2922213, EPI_ISL_2922214, EPI_ISL_2922215, EPI_ISL_2922216, EPI_ISL_2922217, EPI_ISL_2922218, EPI_ISL_2922219, EPI_ISL_2922220, EPI_ISL_2922229, EPI_ISL_2922230, EPI_ISL_2922231, EPI_ISL_2922232, EPI_ISL_2922233, EPI_ISL_2922234, EPI_ISL_2922235, EPI_ISL_2922236, EPI_ISL_2922237, EPI_ISL_2922238, EPI_ISL_2922239, EPI_ISL_2922240, EPI_ISL_2922241, EPI_ISL_2922242, EPI_ISL_2922243, EPI_ISL_2922247, EPI_ISL_2922258, EPI_ISL_2922259, EPI_ISL_2922260, EPI_ISL_2922261, EPI_ISL_2922265, EPI_ISL_2922266, EPI_ISL_2922267, EPI_ISL_2922268, EPI_ISL_2922281, EPI_ISL_2922282, EPI_ISL_2922283, EPI_ISL_2922284, EPI_ISL_2922286, EPI_ISL_2922287, EPI_ISL_2922288, EPI_ISL_2922306, EPI_ISL_2922307, EPI_ISL_2922308, EPI_ISL_2922309, EPI_ISL_2922310, EPI_ISL_2922314, EPI_ISL_2922315, EPI_ISL_2922316, EPI_ISL_2923284, EPI_ISL_2923286, EPI_ISL_2923288, EPI_ISL_2923289, EPI_ISL_2923291, EPI_ISL_2923300, EPI_ISL_2923306, EPI_ISL_2923308, EPI_ISL_2923309, EPI_ISL_2923310, EPI_ISL_2923311, EPI_ISL_2923312, EPI_ISL_2923313, EPI_ISL_2923314, EPI_ISL_2923316, EPI_ISL_2923317, EPI_ISL_2923322, EPI_ISL_2923325, EPI_ISL_2923339, EPI_ISL_2923353, EPI_ISL_2923362, EPI_ISL_2923369, EPI_ISL_2923370, EPI_ISL_2923374, EPI_ISL_2923375, EPI_ISL_2923376, EPI_ISL_2923377, EPI_ISL_2923391, EPI_ISL_2923395, EPI_ISL_2923396, EPI_ISL_2923397, EPI_ISL_2923400, EPI_ISL_2923412, EPI_ISL_2923413, EPI_ISL_2923414, EPI_ISL_2923415, EPI_ISL_2923431, EPI_ISL_2923432, EPI_ISL_2923437, EPI_ISL_2923438, EPI_ISL_2923439, EPI_ISL_2923440, EPI_ISL_2923441, EPI_ISL_2923442, EPI_ISL_2923446, EPI_ISL_2923447, EPI_ISL_2923449, EPI_ISL_2923450, EPI_ISL_2923454, EPI_ISL_2923457, EPI_ISL_2923462, EPI_ISL_2923464, EPI_ISL_2923466, EPI_ISL_2923467, EPI_ISL_2923468, EPI_ISL_2923469, EPI_ISL_2923470, EPI_ISL_2923485, EPI_ISL_2923487, EPI_ISL_2923491, EPI_ISL_2923495, EPI_ISL_2923496, EPI_ISL_2923512, EPI_ISL_2923513, EPI_ISL_2923514, EPI_ISL_2923517, EPI_ISL_2923518, EPI_ISL_2923519, EPI_ISL_2923520, EPI_ISL_2923521, EPI_ISL_2923522, EPI_ISL_2923526, EPI_ISL_2923527, EPI_ISL_2923528, EPI_ISL_2923529, EPI_ISL_2923530, EPI_ISL_2923531, EPI_ISL_2923535, EPI_ISL_2923536, EPI_ISL_2923537, EPI_ISL_2923538, EPI_ISL_2923542, EPI_ISL_2923543, EPI_ISL_2923544, EPI_ISL_2923546, EPI_ISL_2923553, EPI_ISL_2923554, EPI_ISL_2923556, EPI_ISL_2923557, EPI_ISL_2923580, EPI_ISL_2923581, EPI_ISL_2923585, EPI_ISL_2923586, EPI_ISL_2923587, EPI_ISL_2923588, EPI_ISL_2923589, EPI_ISL_2923590, EPI_ISL_2923591, EPI_ISL_2923592, EPI_ISL_2923597, EPI_ISL_2923598, EPI_ISL_2923600, EPI_ISL_2923602, EPI_ISL_2923603, EPI_ISL_2923604, EPI_ISL_2923605, EPI_ISL_2923606, EPI_ISL_2923617, EPI_ISL_2923624, EPI_ISL_2923631, EPI_ISL_2923632, EPI_ISL_2923641, EPI_ISL_2923670, EPI_ISL_2923674, EPI_ISL_2923676, EPI_ISL_2923680, EPI_ISL_2923681, EPI_ISL_2923685, EPI_ISL_2923686, EPI_ISL_2923703, EPI_ISL_2923707, EPI_ISL_2923708, EPI_ISL_2923709, EPI_ISL_2923712, EPI_ISL_2923714, EPI_ISL_2923716, EPI_ISL_2923717, EPI_ISL_2923718, EPI_ISL_2923719, EPI_ISL_2923727, EPI_ISL_2923728, EPI_ISL_2923729, EPI_ISL_2923740, EPI_ISL_2923748, EPI_ISL_2923749, EPI_ISL_2923760, EPI_ISL_2923761, EPI_ISL_2923762, EPI_ISL_2923763, EPI_ISL_2923767, EPI_ISL_2923768, EPI_ISL_2923770, EPI_ISL_2923771, EPI_ISL_2923772, EPI_ISL_2934096, EPI_ISL_2934098, EPI_ISL_2934106, EPI_ISL_2934111, EPI_ISL_2934128, EPI_ISL_2934133, EPI_ISL_2934135, EPI_ISL_2934146, EPI_ISL_2934202, EPI_ISL_2934208, EPI_ISL_2934236, EPI_ISL_2934257, EPI_ISL_2934275, EPI_ISL_2934276, EPI_ISL_2934294, EPI_ISL_2934314, EPI_ISL_2934315, EPI_ISL_2934317, EPI_ISL_2934323, EPI_ISL_2934330, EPI_ISL_2934335, EPI_ISL_2934342, EPI_ISL_2934345, EPI_ISL_2934351, EPI_ISL_2934354, EPI_ISL_2934368, EPI_ISL_2934380, EPI_ISL_2934390, EPI_ISL_2934391, EPI_ISL_2934392, EPI_ISL_2934399, EPI_ISL_2934402, EPI_ISL_2934411, EPI_ISL_2934424, EPI_ISL_2934445, EPI_ISL_2934452, EPI_ISL_2934465, EPI_ISL_2934484, EPI_ISL_2934518, EPI_ISL_2934550, EPI_ISL_2934566

| | | | |
|---|---|---|---|
| see above | CDPH VBL | California Department of Public Health | CDPH-COVIDNet; SwabSeq/UCLA; UCLA Technology Center for Genomics & Bioinformatics; UCSF Center for Advanced Technology |

EPI_ISL_1798766, EPI_ISL_1798768, EPI_ISL_1798770, EPI_ISL_1798772, EPI_ISL_1798777, EPI_ISL_1798779, EPI_ISL_1798781, EPI_ISL_1798783, EPI_ISL_1798787, EPI_ISL_1798789, EPI_ISL_1798791, EPI_ISL_1798793, EPI_ISL_1798797, EPI_ISL_1798799, EPI_ISL_1798801, EPI_ISL_2018319, EPI_ISL_2018320, EPI_ISL_2018321, EPI_ISL_2018325

| | | | |
|---|---|---|---|
| see above | CDPH, Viral and Rickettsial Disease Laboratory | Centers for Disease Control and Prevention Division of Viral Diseases, Pathogen Discovery | Alison Laufer Halpin; Ben L. Rambo-Martin; Clinton R. Paden; Dakota Howard; Darlene Wagner; Dave Wentworth; Dhwani Batra; Jasmine Padilla; Justin Lee; Katie Dillon; Krista Queen; Kristen Knipe; Kristine Lacek; Mark Burroughs; Matthew Schmerer; Mili Sheth; Peter Cook; Sam Shepard; Sarah Nobles; Shoshona Le; Suxiang Tong; Vivien Dugan; Yvette Unoarumhi |
| EPI_ISL_2295591 | CDPH, Viral and Rickettsial Disease Laboratory | Genomics and Discovery, Respiratory Viruses Branch, Division of Viral Diseases, Centers for Disease Control and Prevention | Adam Retchless; Anna Kelleher; Anna Uehara; Brian Lynch; Clinton R. Paden; Dhwani Batra; Haibin Wang; Han Jia Justin Ng; Jasmine Padilla; Jing Zhang; Justin Lee; Krista Queen; Mark Burroughs; Mili Sheth; Morgan Davis; Peter Cook; Rachel Marine; Sarah Nobles; Suxiang Tong; Tara Coalter; Yan Li; Ying Tao |

EPI_ISL_1911362, EPI_ISL_1911363, EPI_ISL_1911364, EPI_ISL_1911365, EPI_ISL_2228091, EPI_ISL_2228092, EPI_ISL_2228093, EPI_ISL_2228094, EPI_ISL_2228095, EPI_ISL_2228096, EPI_ISL_2228097, EPI_ISL_2228098, EPI_ISL_2228099

| | | | |
|---|---|---|---|
| see above | CDPH, Viral and Rickettsial Disease Laboratory | Pathogen Discovery, Respiratory Viruses Branch, Division of Viral Diseases, Centers for Disease Control and Prevention | Adam Retchless; Anna Kelleher; Anna Uehara; Brian Lynch; Clinton R. Paden; Dhwani Batra; Haibin Wang; Han Jia Justin Ng; Jasmine Padilla; Jing Zhang; Justin Lee; Krista Queen; Mark Burroughs; Mili Sheth; Morgan Davis; Peter Cook; Rachel Marine; Sarah Nobles; Suxiang Tong; Tara Coalter; Yan Li; Ying Tao |

EPI_ISL_2234241, EPI_ISL_2234243, EPI_ISL_2234248, EPI_ISL_2234249, EPI_ISL_2234250, EPI_ISL_2234251, EPI_ISL_2234252, EPI_ISL_2234253, EPI_ISL_2234254, EPI_ISL_2234255, EPI_ISL_2234258, EPI_ISL_2234259, EPI_ISL_2234260, EPI_ISL_2234261, EPI_ISL_2234263, EPI_ISL_2234269, EPI_ISL_2234270, EPI_ISL_2234271, EPI_ISL_2234272, EPI_ISL_2234273, EPI_ISL_2234274, EPI_ISL_2234275, EPI_ISL_2234276, EPI_ISL_2234277, EPI_ISL_2234278, EPI_ISL_2234279, EPI_ISL_2234280, EPI_ISL_2234282, EPI_ISL_2234283, EPI_ISL_2234284, EPI_ISL_2234285, EPI_ISL_2234286, EPI_ISL_2234295, EPI_ISL_2234296, EPI_ISL_2234312, EPI_ISL_2234313, EPI_ISL_2234314, EPI_ISL_2234315, EPI_ISL_2234316, EPI_ISL_2234326, EPI_ISL_2234331, EPI_ISL_2234332, EPI_ISL_2234333, EPI_ISL_2234334, EPI_ISL_2234335, EPI_ISL_2234336, EPI_ISL_2234337, EPI_ISL_2234338, EPI_ISL_2234339, EPI_ISL_2234340, EPI_ISL_2234341, EPI_ISL_2234342, EPI_ISL_2234343, EPI_ISL_2234344, EPI_ISL_2234345, EPI_ISL_2234346, EPI_ISL_2234347, EPI_ISL_2234348, EPI_ISL_2234349, EPI_ISL_2234350, EPI_ISL_2234351, EPI_ISL_2234352, EPI_ISL_2234353, EPI_ISL_2234354, EPI_ISL_2537393, EPI_ISL_2537400, EPI_ISL_2537401, EPI_ISL_2537402, EPI_ISL_2537403, EPI_ISL_2537404, EPI_ISL_2537407, EPI_ISL_2537408, EPI_ISL_2537409, EPI_ISL_2537410, EPI_ISL_2537411, EPI_ISL_2537412, EPI_ISL_2537413, EPI_ISL_2537414, EPI_ISL_2537415, EPI_ISL_2537416, EPI_ISL_2537417, EPI_ISL_2537418, EPI_ISL_2537419, EPI_ISL_2537421, EPI_ISL_2537422, EPI_ISL_2537423, EPI_ISL_2537424, EPI_ISL_2537425, EPI_ISL_2537426, EPI_ISL_2537428, EPI_ISL_2537429, EPI_ISL_2537436, EPI_ISL_2537437, EPI_ISL_2537445, EPI_ISL_2537446, EPI_ISL_2537448, EPI_ISL_2537449, EPI_ISL_2537450, EPI_ISL_2537455, EPI_ISL_2537456, EPI_ISL_2537465, EPI_ISL_2842805, EPI_ISL_2892455, EPI_ISL_2892491, EPI_ISL_2892492, EPI_ISL_2892493, EPI_ISL_2892494, EPI_ISL_2892495, EPI_ISL_2892505, EPI_ISL_2892537

| | | | |
|---|---|---|---|
| see above | California Department of Public Health | California Department of Public Health | CDPH IDLB COVIDNet et al |

EPI_ISL_2778636, EPI_ISL_2778637, EPI_ISL_2778638, EPI_ISL_2778639, EPI_ISL_2778640, EPI_ISL_2778641, EPI_ISL_2778642, EPI_ISL_2778643, EPI_ISL_2778644, EPI_ISL_2778645, EPI_ISL_2778646, EPI_ISL_2778647, EPI_ISL_2778648, EPI_ISL_2778649, EPI_ISL_2778650, EPI_ISL_2778651, EPI_ISL_2778652, EPI_ISL_2778653, EPI_ISL_2778654, EPI_ISL_2778655, EPI_ISL_2778656, EPI_ISL_2778657, EPI_ISL_2778658, EPI_ISL_2778659, EPI_ISL_2778660, EPI_ISL_2778661, EPI_ISL_2778662, EPI_ISL_2778663, EPI_ISL_2778664, EPI_ISL_2778665, EPI_ISL_2778666, EPI_ISL_2778667, EPI_ISL_2778668, EPI_ISL_2778669, EPI_ISL_2778670, EPI_ISL_2778671, EPI_ISL_2778672, EPI_ISL_2778673, EPI_ISL_2778674, EPI_ISL_2778675, EPI_ISL_2778676, EPI_ISL_2778677, EPI_ISL_2778678, EPI_ISL_2778679, EPI_ISL_2778680, EPI_ISL_2778681, EPI_ISL_2778682, EPI_ISL_2778683, EPI_ISL_2778684, EPI_ISL_2778685, EPI_ISL_2778686, EPI_ISL_2778687, EPI_ISL_2778688, EPI_ISL_2778689, EPI_ISL_2778690, EPI_ISL_2778691, EPI_ISL_2778692, EPI_ISL_2778693, EPI_ISL_2778694, EPI_ISL_2778695, EPI_ISL_2778696, EPI_ISL_2778697, EPI_ISL_2778698, EPI_ISL_2778699, EPI_ISL_2778700, EPI_ISL_2778701, EPI_ISL_2778702, EPI_ISL_2778703, EPI_ISL_2778704, EPI_ISL_2778705, EPI_ISL_2778706, EPI_ISL_2778707, EPI_ISL_2778708, EPI_ISL_2778709, EPI_ISL_2778710, EPI_ISL_2778711, EPI_ISL_2778712, EPI_ISL_2778713, EPI_ISL_2778714, EPI_ISL_2778715, EPI_ISL_2778716, EPI_ISL_2778717, EPI_ISL_2778718, EPI_ISL_2778719, EPI_ISL_2778720, EPI_ISL_2778721, EPI_ISL_2778722, EPI_ISL_2778723, EPI_ISL_2778724, EPI_ISL_2778725, EPI_ISL_2778726, EPI_ISL_2778727, EPI_ISL_2778728, EPI_ISL_2778729, EPI_ISL_2778730, EPI_ISL_2778731, EPI_ISL_2778732, EPI_ISL_2778733, EPI_ISL_2778734, EPI_ISL_2778735, EPI_ISL_2778736, EPI_ISL_2778737, EPI_ISL_2778738, EPI_ISL_2778739, EPI_ISL_2778740, EPI_ISL_2778741, EPI_ISL_2778742, EPI_ISL_2778743, EPI_ISL_2778744, EPI_ISL_2778745, EPI_ISL_2778746, EPI_ISL_2778747, EPI_ISL_2837324, EPI_ISL_2837328, EPI_ISL_2837332, EPI_ISL_2837333, EPI_ISL_2837355, EPI_ISL_2837357, EPI_ISL_2837362, EPI_ISL_2837364, EPI_ISL_2837365, EPI_ISL_2837366, EPI_ISL_2837367, EPI_ISL_2837368, EPI_ISL_2837369, EPI_ISL_2837370, EPI_ISL_2837380, EPI_ISL_2837389, EPI_ISL_2837391, EPI_ISL_2837399, EPI_ISL_2837418, EPI_ISL_2837419, EPI_ISL_2837420, EPI_ISL_2837422, EPI_ISL_2837424, EPI_ISL_2837425, EPI_ISL_2837426, EPI_ISL_2837427, EPI_ISL_2837428, EPI_ISL_2837429, EPI_ISL_2837435, EPI_ISL_2837436, EPI_ISL_2837437, EPI_ISL_2837438, EPI_ISL_2837460, EPI_ISL_2837461, EPI_ISL_2837465, EPI_ISL_2837468, EPI_ISL_2837469, EPI_ISL_2837470, EPI_ISL_2837471, EPI_ISL_2837472, EPI_ISL_2837473, EPI_ISL_2837474, EPI_ISL_2837475, EPI_ISL_2837476, EPI_ISL_2837484, EPI_ISL_2837507, EPI_ISL_2837508, EPI_ISL_2837510, EPI_ISL_2837511, EPI_ISL_2837524, EPI_ISL_2837525, EPI_ISL_2837535, EPI_ISL_2837536, EPI_ISL_2837537, EPI_ISL_2837539, EPI_ISL_2837543, EPI_ISL_2837545, EPI_ISL_2837546, EPI_ISL_2837562, EPI_ISL_2837565, EPI_ISL_2837566, EPI_ISL_2837567, EPI_ISL_2837573, EPI_ISL_2837580, EPI_ISL_2837581, EPI_ISL_2837582, EPI_ISL_2837583, EPI_ISL_2837586, EPI_ISL_2837593, EPI_ISL_2837596, EPI_ISL_2837597, EPI_ISL_2837601, EPI_ISL_2837603, EPI_ISL_2837618, EPI_ISL_2837624, EPI_ISL_2837633, EPI_ISL_2837634, EPI_ISL_2837636, EPI_ISL_2837637, EPI_ISL_2837638, EPI_ISL_2837641, EPI_ISL_2837642, EPI_ISL_2837643, EPI_ISL_2837644, EPI_ISL_2837645, EPI_ISL_2837646, EPI_ISL_2837650, EPI_ISL_2837652, EPI_ISL_2837654, EPI_ISL_2837655, EPI_ISL_2837658, EPI_ISL_2837659, EPI_ISL_2837660, EPI_ISL_2837679, EPI_ISL_2837680, EPI_ISL_2837681, EPI_ISL_2837698, EPI_ISL_2837699, EPI_ISL_2837700, EPI_ISL_2837701, EPI_ISL_2837704, EPI_ISL_2837705, EPI_ISL_2837707, EPI_ISL_2837708, EPI_ISL_2837714, EPI_ISL_2837719, EPI_ISL_2837720, EPI_ISL_2837721, EPI_ISL_2837725, EPI_ISL_2837726, EPI_ISL_2837745, EPI_ISL_2837746, EPI_ISL_2837747, EPI_ISL_2837748, EPI_ISL_2837756, EPI_ISL_2837762, EPI_ISL_2837763, EPI_ISL_2837784, EPI_ISL_2837787, EPI_ISL_2837788, EPI_ISL_2837790, EPI_ISL_2837801, EPI_ISL_2837850, EPI_ISL_2837862, EPI_ISL_2837869, EPI_ISL_2837871, EPI_ISL_2837872, EPI_ISL_2837873, EPI_ISL_2837874, EPI_ISL_2837875, EPI_ISL_2837876, EPI_ISL_2837886, EPI_ISL_2837901, EPI_ISL_2837902, EPI_ISL_2837903, EPI_ISL_2837904, EPI_ISL_2837905, EPI_ISL_2837906, EPI_ISL_2837907, EPI_ISL_2837913, EPI_ISL_2837937, EPI_ISL_2837938, EPI_ISL_2837941, EPI_ISL_2837943, EPI_ISL_2837945, EPI_ISL_2837946, EPI_ISL_2837947, EPI_ISL_2837956, EPI_ISL_2837957, EPI_ISL_2837972, EPI_ISL_2837973, EPI_ISL_2837974, EPI_ISL_2837975, EPI_ISL_2837976, EPI_ISL_2837977, EPI_ISL_2837980, EPI_ISL_2837981, EPI_ISL_2837982, EPI_ISL_2837984, EPI_ISL_2837985, EPI_ISL_2837986, EPI_ISL_2837987, EPI_ISL_2837988, EPI_ISL_2838002, EPI_ISL_2838003, EPI_ISL_2838004, EPI_ISL_2838005, EPI_ISL_2838006, EPI_ISL_2838007, EPI_ISL_2838009, EPI_ISL_2838010, EPI_ISL_2838015, EPI_ISL_2838016, EPI_ISL_2838026, EPI_ISL_2838027, EPI_ISL_2838028, EPI_ISL_2838030, EPI_ISL_2838032, EPI_ISL_2838038, EPI_ISL_2838045

| | | | |
|---|---|---|---|
| see above | California Department of Public Health Valencia Branch Laboratory (CDPH VBL) | California Department of Public Health | CDPH-COVIDNet; UCLA Technology Center for Genomics & Bioinformatics |
| EPI_ISL_1701494 | California Institute of Technology | Chan-Zuckerberg Biohub | CZB Cliahub Consortium |

EPI_ISL_1824393, EPI_ISL_1824394, EPI_ISL_1824395, EPI_ISL_1824396, EPI_ISL_1824397, EPI_ISL_1824398, EPI_ISL_1824399, EPI_ISL_1824400, EPI_ISL_1824401, EPI_ISL_1824402, EPI_ISL_1824403, EPI_ISL_1824404, EPI_ISL_1824405, EPI_ISL_1824406, EPI_ISL_1824407, EPI_ISL_1824408, EPI_ISL_1824409, EPI_ISL_1824410, EPI_ISL_1824411, EPI_ISL_1824412, EPI_ISL_1824413, EPI_ISL_1824414, EPI_ISL_1824415, EPI_ISL_1824416, EPI_ISL_1824417

| | | | |
|---|---|---|---|
| see above | Chiu Laboratory, University of California, San Francisco | Chiu Laboratory, University of California, San Francisco | Alicia Sotomayor-Gonzalez; Candace Wang; Charles Chiu; Daniel Wong; Venice Servellita; Yueyuan Zhang |

| | | | |
|---|---|---|---|
| EPI_ISL_1825074, EPI_ISL_1825075, EPI_ISL_1825076, EPI_ISL_1825078, EPI_ISL_1825079, EPI_ISL_1825080, EPI_ISL_1825081, EPI_ISL_1825082, EPI_ISL_1825083 | | | |
| see above | Color Genomics | Chiu Laboratory, University of California, San Francisco | Alicia Sotomayor-Gonzalez; Alicia Zhou; Candace Wang; Charles Chiu; Daniel Wong; Erika Torres; Madyson Wanner; Scott Topper; Venice Servellita; Yueyuan Zhang |
| EPI_ISL_1664020, EPI_ISL_1664091, EPI_ISL_2102749, EPI_ISL_2102751, EPI_ISL_2102763, EPI_ISL_2102772, EPI_ISL_2102798, EPI_ISL_2102824, EPI_ISL_2102826, EPI_ISL_2102843, EPI_ISL_2102861, EPI_ISL_2102866, EPI_ISL_2102887, EPI_ISL_2102891, EPI_ISL_2102932, EPI_ISL_2102945, EPI_ISL_2102946, EPI_ISL_2102954, EPI_ISL_2102960, EPI_ISL_2102972, EPI_ISL_2102982, EPI_ISL_2102988, EPI_ISL_2103173, EPI_ISL_2341414, EPI_ISL_2341423, EPI_ISL_2341424, EPI_ISL_2341427, EPI_ISL_2341429, EPI_ISL_2341432, EPI_ISL_2341439, EPI_ISL_2341444, EPI_ISL_2341447, EPI_ISL_2341450, EPI_ISL_2341451, EPI_ISL_2341452, EPI_ISL_2341454, EPI_ISL_2341456, EPI_ISL_2341457, EPI_ISL_2341459, EPI_ISL_2341468, EPI_ISL_2341490, EPI_ISL_2341493, EPI_ISL_2341497, EPI_ISL_2341500, EPI_ISL_2341505, EPI_ISL_2341508, EPI_ISL_2341512, EPI_ISL_2341516, EPI_ISL_2341518, EPI_ISL_2341520, EPI_ISL_2341524, EPI_ISL_2341525, EPI_ISL_2341526, EPI_ISL_2341534, EPI_ISL_2341535, EPI_ISL_2341537, EPI_ISL_2341538, EPI_ISL_2341539, EPI_ISL_2341541, EPI_ISL_2341544, EPI_ISL_2341547, EPI_ISL_2341551, EPI_ISL_2341553, EPI_ISL_2341554, EPI_ISL_2341555, EPI_ISL_2341561, EPI_ISL_2341570, EPI_ISL_2341577, EPI_ISL_2341586, EPI_ISL_2341589, EPI_ISL_2341593, EPI_ISL_2341597, EPI_ISL_2341599, EPI_ISL_2341600, EPI_ISL_2341602, EPI_ISL_2341605, EPI_ISL_2341606, EPI_ISL_2341610, EPI_ISL_2341611, EPI_ISL_2341612, EPI_ISL_2341615, EPI_ISL_2341624, EPI_ISL_2341628, EPI_ISL_2341630, EPI_ISL_2341633, EPI_ISL_2341634, EPI_ISL_2341638, EPI_ISL_2341643, EPI_ISL_2341645, EPI_ISL_2341656, EPI_ISL_2341657, EPI_ISL_2341659, EPI_ISL_2341661, EPI_ISL_2341662, EPI_ISL_2341663, EPI_ISL_2497611, EPI_ISL_2497612, EPI_ISL_2497687, EPI_ISL_2497698, EPI_ISL_2497708, EPI_ISL_2497719, EPI_ISL_2497721, EPI_ISL_2497733, EPI_ISL_2497734, EPI_ISL_2497759, EPI_ISL_2497762, EPI_ISL_2497774, EPI_ISL_2497793, EPI_ISL_2758503, EPI_ISL_2758504 | | | |
| see above | Contra Costa County Public Health Lab | Chan-Zuckerberg Biohub | CZB Cliahub Consortium |
| EPI_ISL_1805719, EPI_ISL_1805720, EPI_ISL_1805721, EPI_ISL_1805722, EPI_ISL_1805723, EPI_ISL_1805724, EPI_ISL_1805725, EPI_ISL_1805726, EPI_ISL_1805727, EPI_ISL_1805728, EPI_ISL_1805729, EPI_ISL_1805730, EPI_ISL_1805731, EPI_ISL_1805732, EPI_ISL_1805733, EPI_ISL_1805734, EPI_ISL_1805735, EPI_ISL_1805736, EPI_ISL_1805737, EPI_ISL_1805738, EPI_ISL_1805739, EPI_ISL_1841392, EPI_ISL_1910451, EPI_ISL_1910452, EPI_ISL_1910453, EPI_ISL_1910454, EPI_ISL_1910455, EPI_ISL_1910456, EPI_ISL_1910457, EPI_ISL_1910458, EPI_ISL_1910459, EPI_ISL_1910460, EPI_ISL_1910461, EPI_ISL_1910462, EPI_ISL_1910463, EPI_ISL_1910464, EPI_ISL_1910465, EPI_ISL_1910466, EPI_ISL_1968661 | | | |
| see above | County of San Diego Health and Human Services Agency | Pathogen Discovery, Respiratory Viruses Branch, Division of Viral Diseases, Centers for Disease Control and Prevention | Adam Retchless; Anna Kelleher; Anna Montmayeur; Anna Uehara; Brian Lynch; Clinton R. Paden; Dhwani Batra; Haibin Wang; Han Jia Justin Ng; Jasmine Padilla; Jing Zhang; Justin Lee; Krista Queen; Mark Burroughs; Mili Sheth; Morgan Davis; Peter Cook; Rachel Marine; Sarah Nobles; Suxiang Tong; Tara Coalter; Yan Li; Ying Tao |
| EPI_ISL_1529127, EPI_ISL_1529128, EPI_ISL_1529129, EPI_ISL_1529161, EPI_ISL_1529208, EPI_ISL_1529213, EPI_ISL_1529329, EPI_ISL_1529341, EPI_ISL_1529342, EPI_ISL_1701136, EPI_ISL_1701266, EPI_ISL_1701423, EPI_ISL_1701591, EPI_ISL_1701607, EPI_ISL_1821323, EPI_ISL_1821332, EPI_ISL_1821358, EPI_ISL_1821362, EPI_ISL_1821363, EPI_ISL_1821365, EPI_ISL_1821366, EPI_ISL_1821368, EPI_ISL_1821371, EPI_ISL_1821372, EPI_ISL_1821373, EPI_ISL_1821374, EPI_ISL_1821375, EPI_ISL_1821376, EPI_ISL_1821377, EPI_ISL_1821378, EPI_ISL_1821380, EPI_ISL_1821381, EPI_ISL_1821383, EPI_ISL_1821384, EPI_ISL_1821385, EPI_ISL_1821386, EPI_ISL_1821387, EPI_ISL_1821388, EPI_ISL_1821389, EPI_ISL_1821390, EPI_ISL_1821391, EPI_ISL_1821394, EPI_ISL_1821399, EPI_ISL_1821404, EPI_ISL_1821434, EPI_ISL_1821454, EPI_ISL_1821463, EPI_ISL_2102790, EPI_ISL_2102809, EPI_ISL_2102842, EPI_ISL_2102850, EPI_ISL_2102912, EPI_ISL_2102978, EPI_ISL_2102998, EPI_ISL_2103073, EPI_ISL_2103092, EPI_ISL_2103126, EPI_ISL_2103153, EPI_ISL_2103172, EPI_ISL_2758482, EPI_ISL_2758495, EPI_ISL_2758496, EPI_ISL_2758497, EPI_ISL_2758498, EPI_ISL_2758499, EPI_ISL_2758500 | | | |
| see above | County of San Luis Obispo Public Health Laboratory | Chan-Zuckerberg Biohub | CZB Cliahub Consortium |
| EPI_ISL_2507113, EPI_ISL_2507114, EPI_ISL_2507115, EPI_ISL_2507116, EPI_ISL_2507117, EPI_ISL_2507118, EPI_ISL_2507119, EPI_ISL_2507120, EPI_ISL_2507123, EPI_ISL_2507138, EPI_ISL_2507246, EPI_ISL_2507247, EPI_ISL_2507249, EPI_ISL_2507250, EPI_ISL_2507251, EPI_ISL_2507254, EPI_ISL_2754961, EPI_ISL_2754962, EPI_ISL_2754963, EPI_ISL_2754964, EPI_ISL_2754986, EPI_ISL_2754987, EPI_ISL_2754988, EPI_ISL_2754989, EPI_ISL_2754990, EPI_ISL_2755107, EPI_ISL_2755108, EPI_ISL_2755109, EPI_ISL_2755110, EPI_ISL_2755111, EPI_ISL_2755112, EPI_ISL_2755113, EPI_ISL_2755114, EPI_ISL_2755136, EPI_ISL_2755137, EPI_ISL_2755138, EPI_ISL_2755338 | | | |
| see above | Curative Labs | Curative Labs | Elias L. Salfati; Eugenia Khorosheva; George Way; J.Cesar Ignacio-Espinoza; Janet Chen; Mikhail Hanewich-Hollatz; Nabjot Sandhu; Sophia Quasem; Vladimir Slepnev; Zhiyi Xie |
| EPI_ISL_2158079, EPI_ISL_2158080, EPI_ISL_2158081, EPI_ISL_2158082, EPI_ISL_2158083, EPI_ISL_2158085, EPI_ISL_2158086, EPI_ISL_2158089, EPI_ISL_2158090, EPI_ISL_2158091, EPI_ISL_2158093, EPI_ISL_2158097, EPI_ISL_2158098, EPI_ISL_2158099, EPI_ISL_2158100, EPI_ISL_2158101, EPI_ISL_2158102, EPI_ISL_2158103, EPI_ISL_2158104, EPI_ISL_2158106, EPI_ISL_2158107, EPI_ISL_2178766, EPI_ISL_2178769, EPI_ISL_2178777, EPI_ISL_2178778 | | | |
| see above | DeRisi Lab, University of California, San Francisco | DeRisi Lab, University of California, San Francisco | Amy Kistler; Anthea Mitchell; Carina Marquez; Diane Havlir; Diane Jones; Douglas Black; Eric Chow; Gabriel Chamie; Genay Pilarowski; IDseq Team; Jackie Martinez; James Peng; Jamin Liu; Jon Jacobo; Lucy Li; Luis Rubio; Manu Vanaerschot; Matthew Laurie; Maya Petersen; Patrick Ayscue; Sabrina Mann; Sara Sunshine; Susana Rojas; Susy Rojas; Valerie Tulier-Laiwa; and Joseph DeRisi |
| EPI_ISL_1811669, EPI_ISL_1811831, EPI_ISL_1811836, EPI_ISL_1811853, EPI_ISL_1811980, EPI_ISL_1812119, EPI_ISL_1812763, EPI_ISL_1812990, EPI_ISL_1813104, EPI_ISL_1813166, EPI_ISL_1813237, EPI_ISL_1813440, EPI_ISL_1813447, EPI_ISL_1813582, EPI_ISL_1813648, EPI_ISL_1813666, EPI_ISL_1813669, EPI_ISL_1813863, EPI_ISL_1813865, EPI_ISL_1813891, EPI_ISL_1813899, EPI_ISL_1813943, EPI_ISL_1813957, EPI_ISL_1813974, EPI_ISL_1814009, EPI_ISL_1814082, EPI_ISL_1814286, EPI_ISL_1814310, EPI_ISL_1814314, EPI_ISL_1814320, EPI_ISL_1814330, EPI_ISL_1814361, EPI_ISL_1814396, EPI_ISL_1814401, EPI_ISL_1814545, EPI_ISL_1814552, EPI_ISL_1814600, EPI_ISL_1814628, EPI_ISL_1814652, EPI_ISL_1814673, EPI_ISL_1814686, EPI_ISL_1814762, EPI_ISL_1814781, EPI_ISL_1814783, EPI_ISL_1814788, EPI_ISL_1814825, EPI_ISL_1814836, EPI_ISL_1814856, EPI_ISL_1814878, EPI_ISL_1814971, EPI_ISL_1814984, EPI_ISL_1815002, EPI_ISL_1815109, EPI_ISL_1815155, EPI_ISL_1815178, EPI_ISL_1815197, EPI_ISL_1815237, EPI_ISL_1815257, EPI_ISL_1815264, EPI_ISL_1815301, EPI_ISL_1815335, EPI_ISL_1815336, EPI_ISL_1815446, EPI_ISL_1815450, EPI_ISL_1815493, EPI_ISL_1815526, EPI_ISL_1815535, EPI_ISL_1815575, EPI_ISL_1815584, EPI_ISL_1815653, EPI_ISL_1815660, EPI_ISL_1815691, EPI_ISL_1815708, EPI_ISL_1815727, EPI_ISL_1815753, EPI_ISL_1815778, EPI_ISL_1815809, EPI_ISL_1815813, EPI_ISL_1815832, EPI_ISL_1815833, EPI_ISL_1815848, EPI_ISL_1815863, EPI_ISL_1815915, EPI_ISL_1815924, EPI_ISL_1815973, EPI_ISL_1815991, EPI_ISL_1816047, EPI_ISL_1816062, EPI_ISL_1816064, EPI_ISL_1816096, EPI_ISL_1816105, EPI_ISL_1816141, EPI_ISL_1816174, EPI_ISL_1816180, EPI_ISL_1816186, EPI_ISL_1816234, EPI_ISL_1816256, EPI_ISL_1816278, EPI_ISL_1816302, EPI_ISL_1816339, EPI_ISL_1816398, EPI_ISL_1816420, EPI_ISL_1816433, EPI_ISL_1816458, EPI_ISL_1816503, EPI_ISL_1816511, EPI_ISL_1816666, EPI_ISL_1816708, EPI_ISL_1816716, EPI_ISL_1816772, EPI_ISL_1816785, EPI_ISL_1816795, EPI_ISL_1816797, EPI_ISL_1816811, EPI_ISL_1816813, EPI_ISL_1816835, EPI_ISL_1816842, EPI_ISL_1816848, EPI_ISL_1816855, EPI_ISL_1816875, EPI_ISL_1816881, EPI_ISL_1816887, EPI_ISL_1816892, EPI_ISL_1816893, EPI_ISL_1816903, EPI_ISL_1816904, EPI_ISL_1816907, EPI_ISL_1816909, EPI_ISL_1816911, EPI_ISL_1816913, EPI_ISL_1816915 | | | |
| see above | EXCITE Lab | Andersen lab at Scripps Research | Abigail Schnapper; Alexandre Bolze; Alice Summerfield; Angela Scioscia; Celena Andrade; Charlotte Rivera-Garcia; Cheryl Anderson; Chip Schooley; David Becker; David Pride; Efren Sandoval; Elizabeth Cirulli; Francisco Tanudjaja; Geraint Levan; Greg Humphrey; Helena Tubb; James Lu + SEARCH; Jason Nguyen; Jimmy Ramirez; Kelly Schiabor Barrett; Magnus Isaksson; Marc Laurent; Natasha Martin; Natasha Martin Cheryl Anderson; Nicole L Washington; Ryan Cho; Sawyer Farmer; Sharon Reed; Sherry Wang; Simon White; Smruthi Karthikeyan; Tommy Valles + SEARCH; Tyler Cassens; William Lee |
| EPI_ISL_1483789, EPI_ISL_1483790, EPI_ISL_1483791, EPI_ISL_1483792, EPI_ISL_1483793, EPI_ISL_1483794, EPI_ISL_1483800, EPI_ISL_1483801, EPI_ISL_1532124, EPI_ISL_1532125, EPI_ISL_1532126 | | | |
| see above | El Camino Hospital | Santa Clara County Public Health Laboratory | Santa Clara County Public Health Department |
| EPI_ISL_1664270, EPI_ISL_1664271, EPI_ISL_1664272, EPI_ISL_1664273 | El Camino Hospital | Santa Clara County public Health Laboratory | Santa Clara County Public Health Department |
| EPI_ISL_1669341, EPI_ISL_1669343, EPI_ISL_1669344, EPI_ISL_1669345, EPI_ISL_1669346, EPI_ISL_1669347, EPI_ISL_1697372, EPI_ISL_1707675, EPI_ISL_1707676, EPI_ISL_2031859, EPI_ISL_2031860, EPI_ISL_2259438, EPI_ISL_2260394, EPI_ISL_2260396, EPI_ISL_2391789, EPI_ISL_2391795, EPI_ISL_2391796, EPI_ISL_2391797, EPI_ISL_2392170, EPI_ISL_2620775 | | | |
| see above | Flow Diagnostics Lab | Los Angeles County Public Health Laboratories | P. Hemarajata et al. |
| EPI_ISL_1578136, EPI_ISL_1578139, EPI_ISL_1578144, EPI_ISL_1578153, EPI_ISL_1578157, EPI_ISL_1578162, EPI_ISL_1578176, EPI_ISL_1578180, EPI_ISL_1578215, EPI_ISL_1578299, EPI_ISL_1578327, EPI_ISL_1578328, EPI_ISL_1578330, EPI_ISL_1578331, EPI_ISL_1578333, EPI_ISL_1578336, EPI_ISL_1578338, EPI_ISL_1578339, EPI_ISL_1578451, EPI_ISL_1918992 | | | |
| see above | Flow Health | Infectious Disease Program, Broad Institute of Harvard and MIT | Adams, G.; B.L.; B.W.; Bauer, M.; Birren; Carter, A.; Chaluvadi, S.; D.J.; DeRuff, K.; Gallagher, G.; Gladden-Young, A.; J.E.; K.J.; Lagerborg, K.; Lemieux; Loreth, C.; MacInnis; Normandin, E.; P.C.; Park; Reilly, S.; Rudy, M.; Siddle; Smole, S.; Tomkins-Tinch, C.; and Sabeti |
| EPI_ISL_1457793, EPI_ISL_1457794, EPI_ISL_1457799, EPI_ISL_1457800, EPI_ISL_1457801, EPI_ISL_1457802, EPI_ISL_1457803, EPI_ISL_1457804, EPI_ISL_1457805, EPI_ISL_1457806, EPI_ISL_1457807, EPI_ISL_1457808, EPI_ISL_1457809, EPI_ISL_1457810, EPI_ISL_1457811, EPI_ISL_1457813, EPI_ISL_1457814, EPI_ISL_1457815, EPI_ISL_1457817, EPI_ISL_1457819, EPI_ISL_1457820, EPI_ISL_1457821, EPI_ISL_1457822, EPI_ISL_1457823, EPI_ISL_1457824, EPI_ISL_1457826, EPI_ISL_1457827, EPI_ISL_1457828, EPI_ISL_1457830, EPI_ISL_1457831, EPI_ISL_1457832, EPI_ISL_1457833, EPI_ISL_1457834, EPI_ISL_1457837, EPI_ISL_1457838, EPI_ISL_1457843, EPI_ISL_1457844, EPI_ISL_1457845, EPI_ISL_1457847, EPI_ISL_1457848, EPI_ISL_1457850, EPI_ISL_1457851, EPI_ISL_1457852, EPI_ISL_1457853, EPI_ISL_1457854, EPI_ISL_1457855, EPI_ISL_1457856, EPI_ISL_1457857, EPI_ISL_1457858, EPI_ISL_1457859, EPI_ISL_1457861, EPI_ISL_1457862, EPI_ISL_1457863, EPI_ISL_1457864, EPI_ISL_1457865, EPI_ISL_1457866, EPI_ISL_1457868, EPI_ISL_1457869, EPI_ISL_1457870, EPI_ISL_1457871, EPI_ISL_1457875, EPI_ISL_1457876, EPI_ISL_1457877, EPI_ISL_1457878, EPI_ISL_1457879, EPI_ISL_1457880, EPI_ISL_1457881, EPI_ISL_1457882, EPI_ISL_1457883, EPI_ISL_1457884, EPI_ISL_1457885, EPI_ISL_1457886, EPI_ISL_1457887, EPI_ISL_1457888, EPI_ISL_1457889, EPI_ISL_1457891, EPI_ISL_1457892, EPI_ISL_1457893, EPI_ISL_1457894, EPI_ISL_1457896, EPI_ISL_1457897, EPI_ISL_1457898, EPI_ISL_1457899, EPI_ISL_1457900, EPI_ISL_1457904, EPI_ISL_1457906, EPI_ISL_1457910, EPI_ISL_1457911, EPI_ISL_1457912, EPI_ISL_1457913, EPI_ISL_1457914, EPI_ISL_1457915, EPI_ISL_1457916, EPI_ISL_1457917, EPI_ISL_1457918, EPI_ISL_1457919, EPI_ISL_1457920, EPI_ISL_1457921, EPI_ISL_1457922, EPI_ISL_1457923, EPI_ISL_1457924, EPI_ISL_1457925, EPI_ISL_1457926, EPI_ISL_1457927, EPI_ISL_1457928, EPI_ISL_1457929, EPI_ISL_1457930, EPI_ISL_1457931, EPI_ISL_1457932, EPI_ISL_1457933, EPI_ISL_1457934, EPI_ISL_1457935, EPI_ISL_1457936, EPI_ISL_1457937, EPI_ISL_1457938, EPI_ISL_1457939, EPI_ISL_1457940, EPI_ISL_1457941, EPI_ISL_1457942, EPI_ISL_1457943, EPI_ISL_1457944, EPI_ISL_1457945, EPI_ISL_1457946, EPI_ISL_1457947, EPI_ISL_1457948, EPI_ISL_1457949, EPI_ISL_1457950, EPI_ISL_1457951, EPI_ISL_1457952, EPI_ISL_1457953, EPI_ISL_1457954, EPI_ISL_1457955, EPI_ISL_1457956, EPI_ISL_1457957, EPI_ISL_1457958, EPI_ISL_1457959, EPI_ISL_1457960, EPI_ISL_1457961, EPI_ISL_1457962, EPI_ISL_1457963, EPI_ISL_1457964, EPI_ISL_1457965, EPI_ISL_1457966, EPI_ISL_1457968, EPI_ISL_1457969, EPI_ISL_1457970, EPI_ISL_1457971, EPI_ISL_1457972, EPI_ISL_1457973, EPI_ISL_1457974, EPI_ISL_1457976, EPI_ISL_1457977, EPI_ISL_1457978, EPI_ISL_1457980, EPI_ISL_1457981, EPI_ISL_1457982, EPI_ISL_1457983, EPI_ISL_1457984, EPI_ISL_1457985, EPI_ISL_1457986, EPI_ISL_1457987, EPI_ISL_1457988, EPI_ISL_1457989, EPI_ISL_1457990, EPI_ISL_1457991, EPI_ISL_1457992, EPI_ISL_1457993, EPI_ISL_1457994, EPI_ISL_1457995, EPI_ISL_1457996, EPI_ISL_1457997, EPI_ISL_1457998, EPI_ISL_1457999, EPI_ISL_1458000, EPI_ISL_1458001, EPI_ISL_1458002, EPI_ISL_1458003, EPI_ISL_1458004, EPI_ISL_1458005, EPI_ISL_1458006, EPI_ISL_1458007, EPI_ISL_1458008, EPI_ISL_1458009, EPI_ISL_1458010, EPI_ISL_1458012, EPI_ISL_1458013, EPI_ISL_1458014, EPI_ISL_1458015, EPI_ISL_1458016, EPI_ISL_1458017, EPI_ISL_1458018, EPI_ISL_1458019, EPI_ISL_1458020, EPI_ISL_1458021, EPI_ISL_1458022, EPI_ISL_1458023, EPI_ISL_1458024, EPI_ISL_1458025, EPI_ISL_1458026, EPI_ISL_1458027, EPI_ISL_1458028, EPI_ISL_1458029, EPI_ISL_1458030, EPI_ISL_1458031, EPI_ISL_1458032, EPI_ISL_1458033, EPI_ISL_1458034, EPI_ISL_1458035, EPI_ISL_1458036, EPI_ISL_1458037, EPI_ISL_1458038, EPI_ISL_1458039, EPI_ISL_1458040, EPI_ISL_1458041, EPI_ISL_1458042, EPI_ISL_1458043, EPI_ISL_1458044, EPI_ISL_1458045, EPI_ISL_1458046, EPI_ISL_1458047, EPI_ISL_1458048, EPI_ISL_1458049, EPI_ISL_1458050, EPI_ISL_1458051, EPI_ISL_1458052, EPI_ISL_1458053, EPI_ISL_1458054, EPI_ISL_1458055, EPI_ISL_1458056, EPI_ISL_1458057, EPI_ISL_1458058, EPI_ISL_1458059, EPI_ISL_1458060, EPI_ISL_1458061, EPI_ISL_1458062, EPI_ISL_1458063, EPI_ISL_1458064, EPI_ISL_1458065, EPI_ISL_1458066, EPI_ISL_1458067, EPI_ISL_1458068, EPI_ISL_1458069, EPI_ISL_1458070, EPI_ISL_1458071, EPI_ISL_1458072, EPI_ISL_1458073, EPI_ISL_1458074, EPI_ISL_1458075, EPI_ISL_1458076, EPI_ISL_1458077, EPI_ISL_1458078, EPI_ISL_1458079, EPI_ISL_1458080, EPI_ISL_1458081, EPI_ISL_1458082, EPI_ISL_1458083, EPI_ISL_1458084, EPI_ISL_1458086, EPI_ISL_1458087, EPI_ISL_1458088, EPI_ISL_1458089, EPI_ISL_1458090, EPI_ISL_1458091, EPI_ISL_1458092, EPI_ISL_1458093, EPI_ISL_1458094, EPI_ISL_1458095, EPI_ISL_1458096, EPI_ISL_1458097, EPI_ISL_1458098, EPI_ISL_1458099, EPI_ISL_1458100, EPI_ISL_1458101, EPI_ISL_1458102, EPI_ISL_1458103, EPI_ISL_1458104, EPI_ISL_1458105, EPI_ISL_1458106, EPI_ISL_1458107, EPI_ISL_1458108, EPI_ISL_1458109, EPI_ISL_1458110, EPI_ISL_1458111, EPI_ISL_1458112, EPI_ISL_1458113, EPI_ISL_1458114, EPI_ISL_1458115, EPI_ISL_1458116, EPI_ISL_1458117, EPI_ISL_1458118, EPI_ISL_1458119, EPI_ISL_1458120, EPI_ISL_1458121, EPI_ISL_1458122, EPI_ISL_1458123, EPI_ISL_1458124, EPI_ISL_1458125, EPI_ISL_1458126, EPI_ISL_1458127, EPI_ISL_1458128, EPI_ISL_1458129, EPI_ISL_1458130, EPI_ISL_1458131, EPI_ISL_1458132, EPI_ISL_1458133, EPI_ISL_1458134, EPI_ISL_1458135, EPI_ISL_1458136, EPI_ISL_1458137, EPI_ISL_1458138, EPI_ISL_1458139, EPI_ISL_1458140, EPI_ISL_1458141, EPI_ISL_1458142, EPI_ISL_1458143, EPI_ISL_1458144, EPI_ISL_1458145, EPI_ISL_1458146, EPI_ISL_1458147, EPI_ISL_1458148, EPI_ISL_1458149, EPI_ISL_1458150, EPI_ISL_1458151, EPI_ISL_1458152, EPI_ISL_1458154, EPI_ISL_1458155, EPI_ISL_1458156, EPI_ISL_1458157, EPI_ISL_1458158, EPI_ISL_1458159, EPI_ISL_1458160, EPI_ISL_1458161, EPI_ISL_1458162, EPI_ISL_1458163, EPI_ISL_1458164, EPI_ISL_1458169, EPI_ISL_1458170, EPI_ISL_1458171, EPI_ISL_1458172, EPI_ISL_1458173, EPI_ISL_1458174, EPI_ISL_1458175, EPI_ISL_1458176, EPI_ISL_1458177, EPI_ISL_1458178, EPI_ISL_1458179, EPI_ISL_1458180, EPI_ISL_1458181, EPI_ISL_1458182, EPI_ISL_1458183, EPI_ISL_1458184, EPI_ISL_1458185, EPI_ISL_1458186, EPI_ISL_1458187, EPI_ISL_1458188, EPI_ISL_1458189, EPI_ISL_1458190, EPI_ISL_1458191, EPI_ISL_1458192, EPI_ISL_1458193, EPI_ISL_1458194, EPI_ISL_1458195, EPI_ISL_1458196, EPI_ISL_1458197, EPI_ISL_1458198, EPI_ISL_1458199, EPI_ISL_1458200, EPI_ISL_1458201, EPI_ISL_1458202, EPI_ISL_1458203, EPI_ISL_1458204, EPI_ISL_1458205, EPI_ISL_1458206, EPI_ISL_1458207, EPI_ISL_1458208, EPI_ISL_1458209, EPI_ISL_1458210, EPI_ISL_1458211, EPI_ISL_1458212, EPI_ISL_1458213, EPI_ISL_1458214, EPI_ISL_1458215, EPI_ISL_1458216, EPI_ISL_1458217, EPI_ISL_1458218, EPI_ISL_1458219, EPI_ISL_1458220, EPI_ISL_1458221, EPI_ISL_1458222, EPI_ISL_1458223, EPI_ISL_1458224, EPI_ISL_1458225, EPI_ISL_1458226, EPI_ISL_1458227, EPI_ISL_1458228, EPI_ISL_1458229, EPI_ISL_1458230, EPI_ISL_1458231, EPI_ISL_1458232, EPI_ISL_1458233, EPI_ISL_1458234, EPI_ISL_1458235, EPI_ISL_1458236, EPI_ISL_1458237, EPI_ISL_1458238, EPI_ISL_1458239, EPI_ISL_1458240, EPI_ISL_1458241, EPI_ISL_1458242, EPI_ISL_1458243, EPI_ISL_1458244, EPI_ISL_1458245, EPI_ISL_1458246, EPI_ISL_1458247, EPI_ISL_1458248, EPI_ISL_1458249, EPI_ISL_1458250, EPI_ISL_1458251, EPI_ISL_1458252, EPI_ISL_1458253, EPI_ISL_1458254, EPI_ISL_1458255, EPI_ISL_1458256, EPI_ISL_1458257, EPI_ISL_1458258, EPI_ISL_1458259, EPI_ISL_1458260, EPI_ISL_1458261, EPI_ISL_1458262, EPI_ISL_1458263, EPI_ISL_1458264, EPI_ISL_1458268, EPI_ISL_1458269, EPI_ISL_1458270, EPI_ISL_1458272, EPI_ISL_1458273, EPI_ISL_1458274, EPI_ISL_1458275, EPI_ISL_1458276, EPI_ISL_1458277, EPI_ISL_1458278, EPI_ISL_1458279, EPI_ISL_1458280, EPI_ISL_1458281, EPI_ISL_1458282, EPI_ISL_1458283, EPI_ISL_1458285, EPI_ISL_1458286, EPI_ISL_1458292, EPI_ISL_1458293, EPI_ISL_1458294, EPI_ISL_1458299, EPI_ISL_1458300, EPI_ISL_1458301, EPI_ISL_1458302, EPI_ISL_1458303, EPI_ISL_1458304, EPI_ISL_1458305, EPI_ISL_1458306, EPI_ISL_1458307, EPI_ISL_1458308, EPI_ISL_1458309, EPI_ISL_1458310, EPI_ISL_1458311, EPI_ISL_1458312, EPI_ISL_1458313, EPI_ISL_1458314, EPI_ISL_1458315, EPI_ISL_1458316, EPI_ISL_1458317, EPI_ISL_1458318, EPI_ISL_1458319, EPI_ISL_1458320, EPI_ISL_1458321, EPI_ISL_1458322, EPI_ISL_1458323, EPI_ISL_1458324, EPI_ISL_1458325, EPI_ISL_1458326, EPI_ISL_1458327, EPI_ISL_1458328, EPI_ISL_1458329, EPI_ISL_1458330, EPI_ISL_1458331, EPI_ISL_1458332, EPI_ISL_1458333, EPI_ISL_1458334, EPI_ISL_1458335, EPI_ISL_1458336, EPI_ISL_1458337, EPI_ISL_1458338, EPI_ISL_1458339, EPI_ISL_1458340, EPI_ISL_1458341, EPI_ISL_1458342, EPI_ISL_1458343, EPI_ISL_1458344, EPI_ISL_1458345, EPI_ISL_1458346, EPI_ISL_1458347, EPI_ISL_1458348, EPI_ISL_1458349, EPI_ISL_1458350, EPI_ISL_1458351, EPI_ISL_1458352, EPI_ISL_1458353, EPI_ISL_1458354, EPI_ISL_1458355, EPI_ISL_1458356, EPI_ISL_1458357, EPI_ISL_1458358, EPI_ISL_1458359, EPI_ISL_1458360, EPI_ISL_1458361, EPI_ISL_1458362, EPI_ISL_1458363, EPI_ISL_1458364, EPI_ISL_1458365, EPI_ISL_1458366, EPI_ISL_1458367, EPI_ISL_1458368, EPI_ISL_1458369, EPI_ISL_1458370, EPI_ISL_1458371, EPI_ISL_1458372, EPI_ISL_1458373, EPI_ISL_1458374, EPI_ISL_1458375, EPI_ISL_1458376, EPI_ISL_1458377, EPI_ISL_1458378, EPI_ISL_1458379, EPI_ISL_1458380, EPI_ISL_1458381, EPI_ISL_1458382, EPI_ISL_1458383, EPI_ISL_1458384, EPI_ISL_1458385, EPI_ISL_1458386, EPI_ISL_1458387, EPI_ISL_1458388, EPI_ISL_1458389, EPI_ISL_1458390, EPI_ISL_1458391, EPI_ISL_1458392, EPI_ISL_1458393, EPI_ISL_1458394, EPI_ISL_1458395, EPI_ISL_1458396, EPI_ISL_1458397, EPI_ISL_1458398, EPI_ISL_1458399, EPI_ISL_1458400, EPI_ISL_1458401, EPI_ISL_1458402, EPI_ISL_1458403, EPI_ISL_1458404, EPI_ISL_1458405, EPI_ISL_1458406, EPI_ISL_1458407, EPI_ISL_1458408, EPI_ISL_1458409, EPI_ISL_1458410, EPI_ISL_1458411, EPI_ISL_1458412, EPI_ISL_1458413, EPI_ISL_1458414, EPI_ISL_1458415, EPI_ISL_1458416, EPI_ISL_1458417, EPI_ISL_1458418, EPI_ISL_1459207, EPI_ISL_1459208, EPI_ISL_1459209, EPI_ISL_1459313, EPI_ISL_1459314, EPI_ISL_1459315, EPI_ISL_1459316, EPI_ISL_1459317, EPI_ISL_1459318, EPI_ISL_1459319, EPI_ISL_1459320, EPI_ISL_1459321, EPI_ISL_1459322, EPI_ISL_1459323, EPI_ISL_1459324, EPI_ISL_1459325, EPI_ISL_1459326, EPI_ISL_1459327, EPI_ISL_1459328, EPI_ISL_1459329, EPI_ISL_1459330, EPI_ISL_1459331, EPI_ISL_1459332, EPI_ISL_1459333, EPI_ISL_1459334, EPI_ISL_1459335, EPI_ISL_1459336, EPI_ISL_1459337, EPI_ISL_1459338, EPI_ISL_1459339, EPI_ISL_1459340, EPI_ISL_1459341, EPI_ISL_1459342, EPI_ISL_1459343, EPI_ISL_1459344, EPI_ISL_1459345, EPI_ISL_1459346, EPI_ISL_1459347, EPI_ISL_1459348, EPI_ISL_1459349, EPI_ISL_1459350, EPI_ISL_1459351, EPI_ISL_1459352, EPI_ISL_1459353, EPI_ISL_1459354, EPI_ISL_1459355, EPI_ISL_1459356, EPI_ISL_1459357, EPI_ISL_1459358, EPI_ISL_1459359, EPI_ISL_1459360, EPI_ISL_1459361, EPI_ISL_1459362, EPI_ISL_1459363, EPI_ISL_1459364, EPI_ISL_1459365, EPI_ISL_1459366, EPI_ISL_1459367, EPI_ISL_1459368, EPI_ISL_1459369, EPI_ISL_1459370, EPI_ISL_1459371, EPI_ISL_1459372, EPI_ISL_1459373, EPI_ISL_1459374, EPI_ISL_1459375, EPI_ISL_1459376, EPI_ISL_1459377, EPI_ISL_1459378, EPI_ISL_1459379, EPI_ISL_1459381, EPI_ISL_1459382, EPI_ISL_1459383, EPI_ISL_1459384, EPI_ISL_1459385, EPI_ISL_1459386, EPI_ISL_1459387, EPI_ISL_1459388, EPI_ISL_1459389, EPI_ISL_1459390, EPI_ISL_1459391, EPI_ISL_1459392, EPI_ISL_1459393, EPI_ISL_1459394, EPI_ISL_1459395, EPI_ISL_1459396, EPI_ISL_1459397, EPI_ISL_1459398, EPI_ISL_1459399, EPI_ISL_1459400, EPI_ISL_1459617, EPI_ISL_1459618, EPI_ISL_1459619, EPI_ISL_1459623, EPI_ISL_1459624, EPI_ISL_1459625, EPI_ISL_1459627, EPI_ISL_1459636, EPI_ISL_1459642, EPI_ISL_1459643, EPI_ISL_1459644, EPI_ISL_1459647, EPI_ISL_1459649, EPI_ISL_1459958, EPI_ISL_1459959, EPI_ISL_1459960, EPI_ISL_1459961, EPI_ISL_1459962, EPI_ISL_1459963, EPI_ISL_1459964, EPI_ISL_1459965, EPI_ISL_1459966, EPI_ISL_1459967, EPI_ISL_1459968, EPI_ISL_1459969, EPI_ISL_1459970, EPI_ISL_1459971, EPI_ISL_1459972, EPI_ISL_1459973, EPI_ISL_1459974, EPI_ISL_1459975, EPI_ISL_1459976, EPI_ISL_1459977, EPI_ISL_1459978, EPI_ISL_1459979, EPI_ISL_1459980, EPI_ISL_1459981, EPI_ISL_1459982, EPI_ISL_1459983, EPI_ISL_1459984, EPI_ISL_1459985, EPI_ISL_1459986, EPI_ISL_1459987, EPI_ISL_1459988, EPI_ISL_1459989, EPI_ISL_1459990, EPI_ISL_1459991, EPI_ISL_1459992, EPI_ISL_1459993, EPI_ISL_1459994, EPI_ISL_1459995, EPI_ISL_1459996, EPI_ISL_1459997, EPI_ISL_1459998, EPI_ISL_1459999, EPI_ISL_1460000, EPI_ISL_1460001, EPI_ISL_1460002, EPI_ISL_1460003, EPI_ISL_1460004, EPI_ISL_1460005, EPI_ISL_1460006, EPI_ISL_1460007, EPI_ISL_1460008, EPI_ISL_1460009, EPI_ISL_1460010, EPI_ISL_1460011, EPI_ISL_1460012, EPI_ISL_1460013, EPI_ISL_1460014, EPI_ISL_1460015, EPI_ISL_1460016, EPI_ISL_1460017, EPI_ISL_1460018, EPI_ISL_1460019, EPI_ISL_1460020, EPI_ISL_1460021, EPI_ISL_1460022, EPI_ISL_1460023, EPI_ISL_1460024, EPI_ISL_1460025, EPI_ISL_1460026, EPI_ISL_1460027, EPI_ISL_1460028, EPI_ISL_1460029, EPI_ISL_1460030, EPI_ISL_1460031, EPI_ISL_1460032, EPI_ISL_1460033, EPI_ISL_1460034, EPI_ISL_1460035, EPI_ISL_1460036, EPI_ISL_1460037, EPI_ISL_1460038, EPI_ISL_1460039, EPI_ISL_1460040, EPI_ISL_1460041, EPI_ISL_1460042, EPI_ISL_1460043, EPI_ISL_1460044, EPI_ISL_1460045, EPI_ISL_1460046, EPI_ISL_1460047, EPI_ISL_1460048, EPI_ISL_1460049, EPI_ISL_1460050, EPI_ISL_1460051, EPI_ISL_1460052, EPI_ISL_1460053, EPI_ISL_1460054, EPI_ISL_1460055, EPI_ISL_1460056, EPI_ISL_1460057, EPI_ISL_1460058, EPI_ISL_1460059, EPI_ISL_1460060, EPI_ISL_1460061, EPI_ISL_1460062, EPI_ISL_1460063, EPI_ISL_1460064, EPI_ISL_1460065, EPI_ISL_1460066, EPI_ISL_1460067, EPI_ISL_1460068, EPI_ISL_1460069, EPI_ISL_1460070, EPI_ISL_1460071, EPI_ISL_1460072, EPI_ISL_1460073, EPI_ISL_1460074, EPI_ISL_1460075, EPI_ISL_1460076, EPI_ISL_1460077, EPI_ISL_1460078, EPI_ISL_1460079, EPI_ISL_1460080, EPI_ISL_1460081, EPI_ISL_1460082, EPI_ISL_1460083, EPI_ISL_1460084, EPI_ISL_1460085, EPI_ISL_1460086, EPI_ISL_1460087, EPI_ISL_1460088, EPI_ISL_1460089, EPI_ISL_1460090, EPI_ISL_1460091, EPI_ISL_1460092, EPI_ISL_1460093, EPI_ISL_1460094, EPI_ISL_1460095, EPI_ISL_1460096, EPI_ISL_1460097, EPI_ISL_1460098, EPI_ISL_1460099, EPI_ISL_1460100, EPI_ISL_1460101, EPI_ISL_1460102, EPI_ISL_1460103, EPI_ISL_1460104, EPI_ISL_1460105, EPI_ISL_1460106, EPI_ISL_1460107, EPI_ISL_1460108, EPI_ISL_1460109, EPI_ISL_1460110, EPI_ISL_1460111, EPI_ISL_1460112, EPI_ISL_1460113, EPI_ISL_1460114, EPI_ISL_1460115, EPI_ISL_1460116, EPI_ISL_1460117, EPI_ISL_1460118, EPI_ISL_1460119, EPI_ISL_1460120, EPI_ISL_1460121, EPI_ISL_1460129, EPI_ISL_1460130, EPI_ISL_1460132, EPI_ISL_1460133, EPI_ISL_1460167, EPI_ISL_1460171, EPI_ISL_1460172, EPI_ISL_1460173, EPI_ISL_1460174, EPI_ISL_1460175, EPI_ISL_1460176, EPI_ISL_1460177, EPI_ISL_1460178, EPI_ISL_1460179, EPI_ISL_1460180, EPI_ISL_1460181, EPI_ISL_1460182, EPI_ISL_1460183, EPI_ISL_1460184, EPI_ISL_1460185, EPI_ISL_1460186, EPI_ISL_1460187, EPI_ISL_1460188, EPI_ISL_1460189, EPI_ISL_1460190, EPI_ISL_1460191, EPI_ISL_1460192, EPI_ISL_1460193, EPI_ISL_1460194, EPI_ISL_1460195, EPI_ISL_1460196, EPI_ISL_1460197, EPI_ISL_1460198, EPI_ISL_1460199, EPI_ISL_1460200, EPI_ISL_1460201, EPI_ISL_1460202, EPI_ISL_1460203, EPI_ISL_1460204, EPI_ISL_1460205, EPI_ISL_1460206, EPI_ISL_1460207, EPI_ISL_1460208, EPI_ISL_1460209, EPI_ISL_1460210, EPI_ISL_1460211, EPI_ISL_1460212, EPI_ISL_1460213, EPI_ISL_1460214, EPI_ISL_1460263, EPI_ISL_1460683, EPI_ISL_1460684, EPI_ISL_1460685, EPI_ISL_1460686, EPI_ISL_1460687, EPI_ISL_1460688, EPI_ISL_1460689, EPI_ISL_1460690, EPI_ISL_1460691, EPI_ISL_1460692, EPI_ISL_1460693, EPI_ISL_1460694, EPI_ISL_1460695, EPI_ISL_1460696, EPI_ISL_1460697, EPI_ISL_1460698, EPI_ISL_1460699, EPI_ISL_1460700, EPI_ISL_1460701, EPI_ISL_1460702, EPI_ISL_1460703, EPI_ISL_1460704, EPI_ISL_1460705, EPI_ISL_1460706, EPI_ISL_1460707, EPI_ISL_1460708, EPI_ISL_1460709, EPI_ISL_1460710, EPI_ISL_1460711, EPI_ISL_1460712, EPI_ISL_1460713, EPI_ISL_1460714, EPI_ISL_1460715, EPI_ISL_1460716, EPI_ISL_1460717, EPI_ISL_1460718, EPI_ISL_1460719, EPI_ISL_1460720, EPI_ISL_1460721, EPI_ISL_1460722, EPI_ISL_1460723, EPI_ISL_1460724, EPI_ISL_1460725, EPI_ISL_1460726, EPI_ISL_1460727, EPI_ISL_1460728, EPI_ISL_1460729, EPI_ISL_1481550, EPI_ISL_1481551, EPI_ISL_1481552, EPI_ISL_1481553, EPI_ISL_1481555, EPI_ISL_1481556, EPI_ISL_1481557, EPI_ISL_1481558, EPI_ISL_1481559, EPI_ISL_1481560, EPI_ISL_1481561, EPI_ISL_1481562, EPI_ISL_1481563, EPI_ISL_1481564, EPI_ISL_1481565, EPI_ISL_1481566, EPI_ISL_1481567, EPI_ISL_1481568, EPI_ISL_1481569, EPI_ISL_1481570, EPI_ISL_1481571, EPI_ISL_1481572, EPI_ISL_1481573, EPI_ISL_1481574, EPI_ISL_1481575, EPI_ISL_1481576, EPI_ISL_1481577, EPI_ISL_1481578, EPI_ISL_1481579, EPI_ISL_1481580, EPI_ISL_1481581, EPI_ISL_1481582, EPI_ISL_1481583, EPI_ISL_1481584, EPI_ISL_1481585, EPI_ISL_1481586, EPI_ISL_1481587, EPI_ISL_1481588, EPI_ISL_1481589, EPI_ISL_1481590, EPI_ISL_1481591, EPI_ISL_1481592, EPI_ISL_1481593, EPI_ISL_1481594, EPI_ISL_1481595, EPI_ISL_1481596, EPI_ISL_1481597, EPI_ISL_1481679, EPI_ISL_1481716, EPI_ISL_1481717, EPI_ISL_1481718, EPI_ISL_1481719, EPI_ISL_1481720, EPI_ISL_1481721, EPI_ISL_1481722, EPI_ISL_1481723, EPI_ISL_1481724, EPI_ISL_1481725, EPI_ISL_1481726, EPI_ISL_1481727, EPI_ISL_1481728, EPI_ISL_1481729, EPI_ISL_1481730, EPI_ISL_1481731, EPI_ISL_1481732, EPI_ISL_1481733, EPI_ISL_1481734, EPI_ISL_1481735, EPI_ISL_1481736, EPI_ISL_1481737, EPI_ISL_1481738, EPI_ISL_1481739, EPI_ISL_1481740, EPI_ISL_1481741, EPI_ISL_1481742, EPI_ISL_1481743, EPI_ISL_1481744, EPI_ISL_1481745, EPI_ISL_1481746, EPI_ISL_1481747, EPI_ISL_1481748, EPI_ISL_1481749, EPI_ISL_1481750, EPI_ISL_1481751, EPI_ISL_1481752, EPI_ISL_1481753, EPI_ISL_1481754, EPI_ISL_1481755, EPI_ISL_1481756, EPI_ISL_1481757, EPI_ISL_1481758, EPI_ISL_1481759, EPI_ISL_1481760, EPI_ISL_1481761, EPI_ISL_1481762, EPI_ISL_1481763, EPI_ISL_1481764, EPI_ISL_1481765, EPI_ISL_1481766, EPI_ISL_1481767, EPI_ISL_1481768, EPI_ISL_1481769, EPI_ISL_1481770, EPI_ISL_1481771, EPI_ISL_1481772, EPI_ISL_1481773, EPI_ISL_1481774, EPI_ISL_1481775, EPI_ISL_1481776, EPI_ISL_1481777, EPI_ISL_1481778, EPI_ISL_1481779, EPI_ISL_1481780, EPI_ISL_1481781, EPI_ISL_1481782, EPI_ISL_1481783, EPI_ISL_1481784, EPI_ISL_1481785, EPI_ISL_1481786, EPI_ISL_1481787, EPI_ISL_1481788, EPI_ISL_1481789, EPI_ISL_1481790, EPI_ISL_1481791, EPI_ISL_1481792, EPI_ISL_1481793, EPI_ISL_1481794, EPI_ISL_1481795, EPI_ISL_1481796, EPI_ISL_1481797, EPI_ISL_1481798, EPI_ISL_1481799, EPI_ISL_1481800, EPI_ISL_1481801, EPI_ISL_1481802, EPI_ISL_1481803, EPI_ISL_1481804, EPI_ISL_1481805, EPI_ISL_1481806, EPI_ISL_1481807, EPI_ISL_1481808, | | | |

EPI_ISL_1481809, EPI_ISL_1481810, EPI_ISL_1481811, EPI_ISL_1481812, EPI_ISL_1481813, EPI_ISL_1481814, EPI_ISL_1481815, EPI_ISL_1481816, EPI_ISL_1481817, EPI_ISL_1481818, EPI_ISL_1481819, EPI_ISL_1481901, EPI_ISL_1481904, EPI_ISL_1481905, EPI_ISL_1481906, EPI_ISL_1481907, EPI_ISL_1481908, EPI_ISL_1481909, EPI_ISL_1481910, EPI_ISL_1481911, EPI_ISL_1481912, EPI_ISL_1481913,
EPI_ISL_1481914, EPI_ISL_1481915, EPI_ISL_1481916, EPI_ISL_1481917, EPI_ISL_1481918, EPI_ISL_1481919, EPI_ISL_1481920, EPI_ISL_1481921, EPI_ISL_1481922, EPI_ISL_1481923, EPI_ISL_1481924, EPI_ISL_1481925, EPI_ISL_1481926, EPI_ISL_1481927, EPI_ISL_1481928, EPI_ISL_1481929, EPI_ISL_1481930, EPI_ISL_1481932, EPI_ISL_1481933, EPI_ISL_1481935, EPI_ISL_1481937, EPI_ISL_1481938,
EPI_ISL_1481939, EPI_ISL_1481940, EPI_ISL_1481941, EPI_ISL_1481942, EPI_ISL_1481943, EPI_ISL_1481948, EPI_ISL_1481949, EPI_ISL_1481950, EPI_ISL_1481951, EPI_ISL_1481952, EPI_ISL_1481953, EPI_ISL_1481954, EPI_ISL_1481955, EPI_ISL_1481956, EPI_ISL_1481957, EPI_ISL_1481958, EPI_ISL_1481959, EPI_ISL_1481960, EPI_ISL_1481961, EPI_ISL_1481962, EPI_ISL_1481963, EPI_ISL_1481964,
EPI_ISL_1481965, EPI_ISL_1481966, EPI_ISL_1481967, EPI_ISL_1481968, EPI_ISL_1481969, EPI_ISL_1481970, EPI_ISL_1481971, EPI_ISL_1481972, EPI_ISL_1481973, EPI_ISL_1481974, EPI_ISL_1481975, EPI_ISL_1481976, EPI_ISL_1481977, EPI_ISL_1481978, EPI_ISL_1481979, EPI_ISL_1481981, EPI_ISL_1481982, EPI_ISL_1481983, EPI_ISL_1481984, EPI_ISL_1481985, EPI_ISL_1481986, EPI_ISL_1481987,
EPI_ISL_1481988, EPI_ISL_1481989, EPI_ISL_1481990, EPI_ISL_1481991, EPI_ISL_1481992, EPI_ISL_1481993, EPI_ISL_1481994, EPI_ISL_1481995, EPI_ISL_1481996, EPI_ISL_1481997, EPI_ISL_1482047, EPI_ISL_1482048, EPI_ISL_1482050, EPI_ISL_1482051, EPI_ISL_1482052, EPI_ISL_1482053, EPI_ISL_1482054, EPI_ISL_1482055, EPI_ISL_1482079, EPI_ISL_1482112, EPI_ISL_1482113, EPI_ISL_1482114,
EPI_ISL_1482115, EPI_ISL_1482116, EPI_ISL_1482117, EPI_ISL_1482118, EPI_ISL_1482119, EPI_ISL_1482120, EPI_ISL_1482121, EPI_ISL_1482122, EPI_ISL_1482123, EPI_ISL_1482124, EPI_ISL_1482125, EPI_ISL_1482132, EPI_ISL_1482133, EPI_ISL_1482134, EPI_ISL_1482135, EPI_ISL_1482136, EPI_ISL_1482137, EPI_ISL_1482138, EPI_ISL_1482139, EPI_ISL_1482140, EPI_ISL_1482172, EPI_ISL_1482173,
EPI_ISL_1482174, EPI_ISL_1482175, EPI_ISL_1482176, EPI_ISL_1482194, EPI_ISL_1525267, EPI_ISL_1525268, EPI_ISL_1525269, EPI_ISL_1525270, EPI_ISL_1525271, EPI_ISL_1525272, EPI_ISL_1525273, EPI_ISL_1525274, EPI_ISL_1525275, EPI_ISL_1525276, EPI_ISL_1525277, EPI_ISL_1525278, EPI_ISL_1525279, EPI_ISL_1525280, EPI_ISL_1525281, EPI_ISL_1525282, EPI_ISL_1525283, EPI_ISL_1525284,
EPI_ISL_1525285, EPI_ISL_1525286, EPI_ISL_1525287, EPI_ISL_1525288, EPI_ISL_1525289, EPI_ISL_1525290, EPI_ISL_1525291, EPI_ISL_1525292, EPI_ISL_1525293, EPI_ISL_1525294, EPI_ISL_1525295, EPI_ISL_1525296, EPI_ISL_1525297, EPI_ISL_1525298, EPI_ISL_1525299, EPI_ISL_1525300, EPI_ISL_1525301, EPI_ISL_1525302, EPI_ISL_1525303, EPI_ISL_1525304, EPI_ISL_1525305, EPI_ISL_1525306,
EPI_ISL_1525307, EPI_ISL_1525308, EPI_ISL_1525309, EPI_ISL_1525310, EPI_ISL_1525311, EPI_ISL_1525312, EPI_ISL_1525313, EPI_ISL_1525314, EPI_ISL_1525545, EPI_ISL_1525546, EPI_ISL_1525547, EPI_ISL_1525548, EPI_ISL_1525549, EPI_ISL_1525550, EPI_ISL_1525551, EPI_ISL_1525552, EPI_ISL_1525553, EPI_ISL_1525554, EPI_ISL_1525555, EPI_ISL_1525556, EPI_ISL_1525557, EPI_ISL_1525558,
EPI_ISL_1525559, EPI_ISL_1525560, EPI_ISL_1525561, EPI_ISL_1525562, EPI_ISL_1525563, EPI_ISL_1525564, EPI_ISL_1525565, EPI_ISL_1525566, EPI_ISL_1525567, EPI_ISL_1525568, EPI_ISL_1525569, EPI_ISL_1525570, EPI_ISL_1525571, EPI_ISL_1525572, EPI_ISL_1525573, EPI_ISL_1525574, EPI_ISL_1525575, EPI_ISL_1525576, EPI_ISL_1525577, EPI_ISL_1525578, EPI_ISL_1525579, EPI_ISL_1525580,
EPI_ISL_1525581, EPI_ISL_1525582, EPI_ISL_1525583, EPI_ISL_1525584, EPI_ISL_1525585, EPI_ISL_1525586, EPI_ISL_1525587, EPI_ISL_1525588, EPI_ISL_1525589, EPI_ISL_1525590, EPI_ISL_1525591, EPI_ISL_1525592, EPI_ISL_1525593, EPI_ISL_1525594, EPI_ISL_1525595, EPI_ISL_1525596, EPI_ISL_1525597, EPI_ISL_1525598, EPI_ISL_1525599, EPI_ISL_1525600, EPI_ISL_1525601, EPI_ISL_1525602,
EPI_ISL_1525603, EPI_ISL_1525604, EPI_ISL_1525605, EPI_ISL_1525606, EPI_ISL_1525607, EPI_ISL_1525608, EPI_ISL_1525609, EPI_ISL_1525610, EPI_ISL_1525611, EPI_ISL_1525612, EPI_ISL_1525613, EPI_ISL_1525614, EPI_ISL_1525615, EPI_ISL_1525616, EPI_ISL_1525617, EPI_ISL_1525618, EPI_ISL_1525619, EPI_ISL_1525620, EPI_ISL_1525621, EPI_ISL_1525622, EPI_ISL_1525623, EPI_ISL_1525624,
EPI_ISL_1525625, EPI_ISL_1525626, EPI_ISL_1525627, EPI_ISL_1525628, EPI_ISL_1525629, EPI_ISL_1525630, EPI_ISL_1525631, EPI_ISL_1525632, EPI_ISL_1525633, EPI_ISL_1525634, EPI_ISL_1525635, EPI_ISL_1525636, EPI_ISL_1525637, EPI_ISL_1525638, EPI_ISL_1525639, EPI_ISL_1525640, EPI_ISL_1525641, EPI_ISL_1525642, EPI_ISL_1525643, EPI_ISL_1525644, EPI_ISL_1525645, EPI_ISL_1525646,
EPI_ISL_1525647, EPI_ISL_1525648, EPI_ISL_1525649, EPI_ISL_1525650, EPI_ISL_1525651, EPI_ISL_1525652, EPI_ISL_1525653, EPI_ISL_1525654, EPI_ISL_1525655, EPI_ISL_1525656, EPI_ISL_1525657, EPI_ISL_1525658, EPI_ISL_1525659, EPI_ISL_1525660, EPI_ISL_1525661, EPI_ISL_1525662, EPI_ISL_1525663, EPI_ISL_1525664, EPI_ISL_1525665, EPI_ISL_1525666, EPI_ISL_1525667, EPI_ISL_1525668,
EPI_ISL_1525669, EPI_ISL_1525670, EPI_ISL_1525671, EPI_ISL_1525672, EPI_ISL_1525673, EPI_ISL_1525674, EPI_ISL_1525675, EPI_ISL_1525676, EPI_ISL_1525677, EPI_ISL_1525678, EPI_ISL_1525679, EPI_ISL_1525680, EPI_ISL_1525681, EPI_ISL_1525682, EPI_ISL_1525683, EPI_ISL_1525684, EPI_ISL_1525685, EPI_ISL_1525686, EPI_ISL_1525687, EPI_ISL_1525688, EPI_ISL_1525689, EPI_ISL_1525690,
EPI_ISL_1525691, EPI_ISL_1525692, EPI_ISL_1525693, EPI_ISL_1525694, EPI_ISL_1525695, EPI_ISL_1525696, EPI_ISL_1525697, EPI_ISL_1525698, EPI_ISL_1525699, EPI_ISL_1525700, EPI_ISL_1525701, EPI_ISL_1525702, EPI_ISL_1525703, EPI_ISL_1525704, EPI_ISL_1525705, EPI_ISL_1525706, EPI_ISL_1525707, EPI_ISL_1525708, EPI_ISL_1525709, EPI_ISL_1525710, EPI_ISL_1525711, EPI_ISL_1525712,
EPI_ISL_1525713, EPI_ISL_1525714, EPI_ISL_1525715, EPI_ISL_1525716, EPI_ISL_1525717, EPI_ISL_1525718, EPI_ISL_1525719, EPI_ISL_1525720, EPI_ISL_1525721, EPI_ISL_1525722, EPI_ISL_1525723, EPI_ISL_1525724, EPI_ISL_1525725, EPI_ISL_1525726, EPI_ISL_1525727, EPI_ISL_1525728, EPI_ISL_1525729, EPI_ISL_1525730, EPI_ISL_1525731, EPI_ISL_1525732, EPI_ISL_1525733, EPI_ISL_1525734,
EPI_ISL_1525735, EPI_ISL_1525736, EPI_ISL_1525737, EPI_ISL_1525738, EPI_ISL_1525739, EPI_ISL_1525740, EPI_ISL_1525741, EPI_ISL_1525742, EPI_ISL_1525743, EPI_ISL_1525744, EPI_ISL_1525745, EPI_ISL_1525746, EPI_ISL_1525747, EPI_ISL_1525748, EPI_ISL_1525749, EPI_ISL_1525750, EPI_ISL_1525751, EPI_ISL_1525752, EPI_ISL_1525753, EPI_ISL_1525754, EPI_ISL_1525755, EPI_ISL_1525756,
EPI_ISL_1525757, EPI_ISL_1525758, EPI_ISL_1525759, EPI_ISL_1525760, EPI_ISL_1525761, EPI_ISL_1525762, EPI_ISL_1525763, EPI_ISL_1525764, EPI_ISL_1525765, EPI_ISL_1525766, EPI_ISL_1525767, EPI_ISL_1525768, EPI_ISL_1525769, EPI_ISL_1525770, EPI_ISL_1525771, EPI_ISL_1525772, EPI_ISL_1525773, EPI_ISL_1525774, EPI_ISL_1525775, EPI_ISL_1525776, EPI_ISL_1525777, EPI_ISL_1525778,
EPI_ISL_1525779, EPI_ISL_1525780, EPI_ISL_1525781, EPI_ISL_1525782, EPI_ISL_1525783, EPI_ISL_1525784, EPI_ISL_1525785, EPI_ISL_1525786, EPI_ISL_1525787, EPI_ISL_1525788, EPI_ISL_1525789, EPI_ISL_1525790, EPI_ISL_1525791, EPI_ISL_1525792, EPI_ISL_1525793, EPI_ISL_1525794, EPI_ISL_1525795, EPI_ISL_1525796, EPI_ISL_1525797, EPI_ISL_1525798, EPI_ISL_1525799, EPI_ISL_1554951,
EPI_ISL_1554953, EPI_ISL_1554954, EPI_ISL_1554957, EPI_ISL_1554959, EPI_ISL_1554962, EPI_ISL_1554963, EPI_ISL_1554964, EPI_ISL_1554965, EPI_ISL_1554966, EPI_ISL_1554968, EPI_ISL_1554970, EPI_ISL_1554972, EPI_ISL_1554973, EPI_ISL_1554974, EPI_ISL_1554975, EPI_ISL_1554976, EPI_ISL_1554977, EPI_ISL_1554978, EPI_ISL_1554979, EPI_ISL_1554980, EPI_ISL_1554982, EPI_ISL_1554983,
EPI_ISL_1554984, EPI_ISL_1554985, EPI_ISL_1554986, EPI_ISL_1554987, EPI_ISL_1554988, EPI_ISL_1554989, EPI_ISL_1554990, EPI_ISL_1554991, EPI_ISL_1554992, EPI_ISL_1554993, EPI_ISL_1554994, EPI_ISL_1554995, EPI_ISL_1554996, EPI_ISL_1554997, EPI_ISL_1554998, EPI_ISL_1554999, EPI_ISL_1555000, EPI_ISL_1555001, EPI_ISL_1555002, EPI_ISL_1555003, EPI_ISL_1555004, EPI_ISL_1555005,
EPI_ISL_1555006, EPI_ISL_1555007, EPI_ISL_1555010, EPI_ISL_1555011, EPI_ISL_1555012, EPI_ISL_1555013, EPI_ISL_1555014, EPI_ISL_1555015, EPI_ISL_1555016, EPI_ISL_1555017, EPI_ISL_1555018, EPI_ISL_1555019, EPI_ISL_1555020, EPI_ISL_1555021, EPI_ISL_1555022, EPI_ISL_1555023, EPI_ISL_1555024, EPI_ISL_1555025, EPI_ISL_1555026, EPI_ISL_1555027, EPI_ISL_1555028, EPI_ISL_1555029,
EPI_ISL_1555030, EPI_ISL_1555031, EPI_ISL_1555032, EPI_ISL_1555033, EPI_ISL_1555034, EPI_ISL_1555035, EPI_ISL_1555036, EPI_ISL_1555037, EPI_ISL_1555042, EPI_ISL_1555043, EPI_ISL_1555044, EPI_ISL_1555045, EPI_ISL_1555046, EPI_ISL_1555047, EPI_ISL_1555048, EPI_ISL_1555049, EPI_ISL_1555050, EPI_ISL_1555052, EPI_ISL_1555053, EPI_ISL_1555054, EPI_ISL_1555055, EPI_ISL_1555056,
EPI_ISL_1555057, EPI_ISL_1555058, EPI_ISL_1555059, EPI_ISL_1555060, EPI_ISL_1555061, EPI_ISL_1555062, EPI_ISL_1555063, EPI_ISL_1555064, EPI_ISL_1555065, EPI_ISL_1555066, EPI_ISL_1555067, EPI_ISL_1555068, EPI_ISL_1555069, EPI_ISL_1555070, EPI_ISL_1555071, EPI_ISL_1555072, EPI_ISL_1555073, EPI_ISL_1555074, EPI_ISL_1555075, EPI_ISL_1555076, EPI_ISL_1555077, EPI_ISL_1555078,
EPI_ISL_1555079, EPI_ISL_1555080, EPI_ISL_1555081, EPI_ISL_1555082, EPI_ISL_1555084, EPI_ISL_1555085, EPI_ISL_1555086, EPI_ISL_1555087, EPI_ISL_1555088, EPI_ISL_1555089, EPI_ISL_1555090, EPI_ISL_1555091, EPI_ISL_1555092, EPI_ISL_1555093, EPI_ISL_1555094, EPI_ISL_1555095, EPI_ISL_1555096, EPI_ISL_1555097, EPI_ISL_1555098, EPI_ISL_1555099, EPI_ISL_1555100, EPI_ISL_1555101,
EPI_ISL_1555102, EPI_ISL_1555103, EPI_ISL_1555104, EPI_ISL_1555105, EPI_ISL_1555106, EPI_ISL_1555107, EPI_ISL_1555108, EPI_ISL_1555109, EPI_ISL_1555110, EPI_ISL_1555112, EPI_ISL_1555113, EPI_ISL_1555114, EPI_ISL_1555117, EPI_ISL_1555118, EPI_ISL_1555119, EPI_ISL_1555120, EPI_ISL_1555121, EPI_ISL_1555122, EPI_ISL_1555123, EPI_ISL_1555124, EPI_ISL_1555125, EPI_ISL_1555126,
EPI_ISL_1555127, EPI_ISL_1555128, EPI_ISL_1555129, EPI_ISL_1555130, EPI_ISL_1555131, EPI_ISL_1555132, EPI_ISL_1555133, EPI_ISL_1555134, EPI_ISL_1555135, EPI_ISL_1555136, EPI_ISL_1555137, EPI_ISL_1555138, EPI_ISL_1555139, EPI_ISL_1555140, EPI_ISL_1555141, EPI_ISL_1555142, EPI_ISL_1555143, EPI_ISL_1555144, EPI_ISL_1555145, EPI_ISL_1555146, EPI_ISL_1555147, EPI_ISL_1555148,
EPI_ISL_1555149, EPI_ISL_1555150, EPI_ISL_1555151, EPI_ISL_1555152, EPI_ISL_1555153, EPI_ISL_1555154, EPI_ISL_1555155, EPI_ISL_1555156, EPI_ISL_1555157, EPI_ISL_1555158, EPI_ISL_1555159, EPI_ISL_1555539, EPI_ISL_1555540, EPI_ISL_1555571, EPI_ISL_1555572, EPI_ISL_1555573, EPI_ISL_1555574, EPI_ISL_1555575, EPI_ISL_1555577, EPI_ISL_1555579, EPI_ISL_1555580, EPI_ISL_1555581,
EPI_ISL_1555582, EPI_ISL_1555583, EPI_ISL_1555584, EPI_ISL_1555585, EPI_ISL_1555586, EPI_ISL_1555587, EPI_ISL_1555588, EPI_ISL_1555589, EPI_ISL_1555590, EPI_ISL_1555591, EPI_ISL_1555592, EPI_ISL_1555593, EPI_ISL_1555594, EPI_ISL_1555595, EPI_ISL_1555596, EPI_ISL_1555597, EPI_ISL_1555598, EPI_ISL_1555599, EPI_ISL_1555600, EPI_ISL_1555601, EPI_ISL_1555602, EPI_ISL_1555603,
EPI_ISL_1555604, EPI_ISL_1555605, EPI_ISL_1555606, EPI_ISL_1555607, EPI_ISL_1555608, EPI_ISL_1555609, EPI_ISL_1555610, EPI_ISL_1555611, EPI_ISL_1555647, EPI_ISL_1555679, EPI_ISL_1555680, EPI_ISL_1555681, EPI_ISL_1555682, EPI_ISL_1555683, EPI_ISL_1555684, EPI_ISL_1555685, EPI_ISL_1555686, EPI_ISL_1555687, EPI_ISL_1555688, EPI_ISL_1555689, EPI_ISL_1555690, EPI_ISL_1555691,
EPI_ISL_1555692, EPI_ISL_1555693, EPI_ISL_1555694, EPI_ISL_1555699, EPI_ISL_1555700, EPI_ISL_1555701, EPI_ISL_1555702, EPI_ISL_1555703, EPI_ISL_1555704, EPI_ISL_1555705, EPI_ISL_1555706, EPI_ISL_1555707, EPI_ISL_1555708, EPI_ISL_1555709, EPI_ISL_1555710, EPI_ISL_1555711, EPI_ISL_1555712, EPI_ISL_1555713, EPI_ISL_1555714, EPI_ISL_1555715, EPI_ISL_1555716, EPI_ISL_1555717,
EPI_ISL_1555718, EPI_ISL_1555719, EPI_ISL_1555720, EPI_ISL_1555721, EPI_ISL_1555722, EPI_ISL_1555723, EPI_ISL_1555724, EPI_ISL_1555725, EPI_ISL_1555726, EPI_ISL_1555727, EPI_ISL_1555734, EPI_ISL_1555735, EPI_ISL_1555736, EPI_ISL_1555737, EPI_ISL_1555738, EPI_ISL_1555739, EPI_ISL_1555740, EPI_ISL_1555741, EPI_ISL_1555742, EPI_ISL_1555743, EPI_ISL_1555744, EPI_ISL_1555745,
EPI_ISL_1555746, EPI_ISL_1555747, EPI_ISL_1555748, EPI_ISL_1555749, EPI_ISL_1555939, EPI_ISL_1555940, EPI_ISL_1555941, EPI_ISL_1555942, EPI_ISL_1555943, EPI_ISL_1555944, EPI_ISL_1555945, EPI_ISL_1555946, EPI_ISL_1555947, EPI_ISL_1555948, EPI_ISL_1555949, EPI_ISL_1555950, EPI_ISL_1555951, EPI_ISL_1555952, EPI_ISL_1555953, EPI_ISL_1555954, EPI_ISL_1555955, EPI_ISL_1555956,
EPI_ISL_1555957, EPI_ISL_1555958, EPI_ISL_1555959, EPI_ISL_1555960, EPI_ISL_1555961, EPI_ISL_1555962, EPI_ISL_1555963, EPI_ISL_1555964, EPI_ISL_1555965, EPI_ISL_1555966, EPI_ISL_1555967, EPI_ISL_1555968, EPI_ISL_1556078, EPI_ISL_1556079, EPI_ISL_1556080, EPI_ISL_1556081, EPI_ISL_1556082, EPI_ISL_1556083, EPI_ISL_1556084, EPI_ISL_1556085, EPI_ISL_1556086, EPI_ISL_1556087,
EPI_ISL_1556088, EPI_ISL_1556089, EPI_ISL_1556090, EPI_ISL_1556091, EPI_ISL_1556092, EPI_ISL_1556093, EPI_ISL_1556094, EPI_ISL_1556095, EPI_ISL_1556096, EPI_ISL_1556097, EPI_ISL_1556098, EPI_ISL_1556099, EPI_ISL_1556100, EPI_ISL_1556101, EPI_ISL_1556102, EPI_ISL_1556103, EPI_ISL_1556104, EPI_ISL_1556105, EPI_ISL_1556106, EPI_ISL_1556107, EPI_ISL_1556108, EPI_ISL_1556109,
EPI_ISL_1556110, EPI_ISL_1556111, EPI_ISL_1556112, EPI_ISL_1556113, EPI_ISL_1556114, EPI_ISL_1556115, EPI_ISL_1556116, EPI_ISL_1556117, EPI_ISL_1556119, EPI_ISL_1556197, EPI_ISL_1556198, EPI_ISL_1556199, EPI_ISL_1556200, EPI_ISL_1556202, EPI_ISL_1556203, EPI_ISL_1556204, EPI_ISL_1556205, EPI_ISL_1556206, EPI_ISL_1556207, EPI_ISL_1556208, EPI_ISL_1556209, EPI_ISL_1556210,
EPI_ISL_1556211, EPI_ISL_1556212, EPI_ISL_1556213, EPI_ISL_1556214, EPI_ISL_1556215, EPI_ISL_1556216, EPI_ISL_1556217, EPI_ISL_1556218, EPI_ISL_1556219, EPI_ISL_1556220, EPI_ISL_1556221, EPI_ISL_1556222, EPI_ISL_1556223, EPI_ISL_1556224, EPI_ISL_1556225, EPI_ISL_1556243, EPI_ISL_1556244, EPI_ISL_1556246, EPI_ISL_1556248, EPI_ISL_1556249, EPI_ISL_1556250, EPI_ISL_1556251,
EPI_ISL_1556256, EPI_ISL_1556257, EPI_ISL_1556281, EPI_ISL_1556282, EPI_ISL_1556394, EPI_ISL_1556395, EPI_ISL_1556396, EPI_ISL_1556397, EPI_ISL_1556398, EPI_ISL_1556399, EPI_ISL_1556400, EPI_ISL_1556401, EPI_ISL_1556402, EPI_ISL_1556403, EPI_ISL_1556404, EPI_ISL_1556405, EPI_ISL_1556406, EPI_ISL_1556407, EPI_ISL_1556408, EPI_ISL_1556409, EPI_ISL_1556410, EPI_ISL_1556411,
EPI_ISL_1556412, EPI_ISL_1556413, EPI_ISL_1556414, EPI_ISL_1556415, EPI_ISL_1556416, EPI_ISL_1556417, EPI_ISL_1556418, EPI_ISL_1556419, EPI_ISL_1556875, EPI_ISL_1556876, EPI_ISL_1556877, EPI_ISL_1556878, EPI_ISL_1556879, EPI_ISL_1556880, EPI_ISL_1556881, EPI_ISL_1556882, EPI_ISL_1556883, EPI_ISL_1556884, EPI_ISL_1556885, EPI_ISL_1556886, EPI_ISL_1556887, EPI_ISL_1556888,
EPI_ISL_1556889, EPI_ISL_1556890, EPI_ISL_1556891, EPI_ISL_1556892, EPI_ISL_1556893, EPI_ISL_1556894, EPI_ISL_1556895, EPI_ISL_1556896, EPI_ISL_1556897, EPI_ISL_1556898, EPI_ISL_1556899, EPI_ISL_1556900, EPI_ISL_1556901, EPI_ISL_1556902, EPI_ISL_1556903, EPI_ISL_1556904, EPI_ISL_1556905, EPI_ISL_1556906, EPI_ISL_1556907, EPI_ISL_1556908, EPI_ISL_1556909, EPI_ISL_1556910,
EPI_ISL_1556911, EPI_ISL_1556912, EPI_ISL_1556913, EPI_ISL_1556914, EPI_ISL_1556915, EPI_ISL_1556916, EPI_ISL_1556917, EPI_ISL_1556918, EPI_ISL_1556919, EPI_ISL_1556920, EPI_ISL_1556921, EPI_ISL_1556922, EPI_ISL_1556923, EPI_ISL_1556924, EPI_ISL_1556925, EPI_ISL_1556926, EPI_ISL_1556927, EPI_ISL_1556928, EPI_ISL_1556929, EPI_ISL_1556930, EPI_ISL_1556931, EPI_ISL_1556932,
EPI_ISL_1556933, EPI_ISL_1556934, EPI_ISL_1556935, EPI_ISL_1556936, EPI_ISL_1556937, EPI_ISL_1556938, EPI_ISL_1556939, EPI_ISL_1556940, EPI_ISL_1556941, EPI_ISL_1556942, EPI_ISL_1556943, EPI_ISL_1556944, EPI_ISL_1556945, EPI_ISL_1556946, EPI_ISL_1556947, EPI_ISL_1556948, EPI_ISL_1556949, EPI_ISL_1556950, EPI_ISL_1556951, EPI_ISL_1556952, EPI_ISL_1556953, EPI_ISL_1556954,
EPI_ISL_1556955, EPI_ISL_1556956, EPI_ISL_1556957, EPI_ISL_1556958, EPI_ISL_1556959, EPI_ISL_1556960, EPI_ISL_1556961, EPI_ISL_1556962, EPI_ISL_1556963, EPI_ISL_1556964, EPI_ISL_1556965, EPI_ISL_1556966, EPI_ISL_1556967, EPI_ISL_1556968, EPI_ISL_1556969, EPI_ISL_1556970, EPI_ISL_1556971, EPI_ISL_1556972, EPI_ISL_1556973, EPI_ISL_1556974, EPI_ISL_1556976, EPI_ISL_1556977,
EPI_ISL_1556978, EPI_ISL_1556979, EPI_ISL_1556980, EPI_ISL_1556981, EPI_ISL_1556982, EPI_ISL_1556983, EPI_ISL_1556984, EPI_ISL_1556985, EPI_ISL_1556986, EPI_ISL_1556987, EPI_ISL_1556988, EPI_ISL_1556989, EPI_ISL_1556990, EPI_ISL_1556991, EPI_ISL_1556992, EPI_ISL_1556993, EPI_ISL_1556994, EPI_ISL_1556995, EPI_ISL_1556996, EPI_ISL_1556997, EPI_ISL_1556998, EPI_ISL_1556999,
EPI_ISL_1557000, EPI_ISL_1557001, EPI_ISL_1557002, EPI_ISL_1557003, EPI_ISL_1557004, EPI_ISL_1557005, EPI_ISL_1557006, EPI_ISL_1557007, EPI_ISL_1557008, EPI_ISL_1557009, EPI_ISL_1557010, EPI_ISL_1557011, EPI_ISL_1557012, EPI_ISL_1557013, EPI_ISL_1557014, EPI_ISL_1557015, EPI_ISL_1557016, EPI_ISL_1557017, EPI_ISL_1557018, EPI_ISL_1557019, EPI_ISL_1557020, EPI_ISL_1557021,
EPI_ISL_1557022, EPI_ISL_1557023, EPI_ISL_1557024, EPI_ISL_1557025, EPI_ISL_1557026, EPI_ISL_1557027, EPI_ISL_1557028, EPI_ISL_1557029, EPI_ISL_1557030, EPI_ISL_1557031, EPI_ISL_1557032, EPI_ISL_1557186, EPI_ISL_1612850, EPI_ISL_1612953, EPI_ISL_1612954, EPI_ISL_1612955, EPI_ISL_1612956, EPI_ISL_1612957, EPI_ISL_1612958, EPI_ISL_1612959, EPI_ISL_1612960, EPI_ISL_1612961,
EPI_ISL_1612962, EPI_ISL_1612963, EPI_ISL_1612964, EPI_ISL_1612965, EPI_ISL_1612966, EPI_ISL_1612967, EPI_ISL_1612968, EPI_ISL_1612969, EPI_ISL_1612970, EPI_ISL_1612971, EPI_ISL_1612972, EPI_ISL_1612973, EPI_ISL_1612974, EPI_ISL_1612975, EPI_ISL_1612976, EPI_ISL_1612977, EPI_ISL_1612978, EPI_ISL_1612979, EPI_ISL_1612980, EPI_ISL_1612981, EPI_ISL_1612982, EPI_ISL_1612983,
EPI_ISL_1612984, EPI_ISL_1612985, EPI_ISL_1612986, EPI_ISL_1612987, EPI_ISL_1612988, EPI_ISL_1612989, EPI_ISL_1612990, EPI_ISL_1612991, EPI_ISL_1612992, EPI_ISL_1612993, EPI_ISL_1612994, EPI_ISL_1612995, EPI_ISL_1612996, EPI_ISL_1612997, EPI_ISL_1612998, EPI_ISL_1612999, EPI_ISL_1613000, EPI_ISL_1613001, EPI_ISL_1613002, EPI_ISL_1613003, EPI_ISL_1613004, EPI_ISL_1613005,
EPI_ISL_1613006, EPI_ISL_1613007, EPI_ISL_1613008, EPI_ISL_1613009, EPI_ISL_1613010, EPI_ISL_1613011, EPI_ISL_1613012, EPI_ISL_1613013, EPI_ISL_1613014, EPI_ISL_1613015, EPI_ISL_1613016, EPI_ISL_1613017, EPI_ISL_1613018, EPI_ISL_1613019, EPI_ISL_1613020, EPI_ISL_1613021, EPI_ISL_1613022, EPI_ISL_1613023, EPI_ISL_1613024, EPI_ISL_1613025, EPI_ISL_1613026, EPI_ISL_1613027,
EPI_ISL_1613028, EPI_ISL_1613029, EPI_ISL_1613030, EPI_ISL_1613031, EPI_ISL_1613032, EPI_ISL_1613033, EPI_ISL_1613034, EPI_ISL_1613035, EPI_ISL_1613036, EPI_ISL_1613037, EPI_ISL_1613038, EPI_ISL_1613039, EPI_ISL_1613040, EPI_ISL_1613041, EPI_ISL_1613042, EPI_ISL_1613043, EPI_ISL_1613044, EPI_ISL_1613045, EPI_ISL_1613046, EPI_ISL_1613047, EPI_ISL_1613048, EPI_ISL_1613049,
EPI_ISL_1613050, EPI_ISL_1613051, EPI_ISL_1613052, EPI_ISL_1613053, EPI_ISL_1613054, EPI_ISL_1613055, EPI_ISL_1613056, EPI_ISL_1613057, EPI_ISL_1613058, EPI_ISL_1613059, EPI_ISL_1613060, EPI_ISL_1613061, EPI_ISL_1613062, EPI_ISL_1613063, EPI_ISL_1613064, EPI_ISL_1613065, EPI_ISL_1613066, EPI_ISL_1613067, EPI_ISL_1613068, EPI_ISL_1613069, EPI_ISL_1613070, EPI_ISL_1613071,
EPI_ISL_1613072, EPI_ISL_1613073, EPI_ISL_1613074, EPI_ISL_1613075, EPI_ISL_1613076, EPI_ISL_1613077, EPI_ISL_1613078, EPI_ISL_1613079, EPI_ISL_1613182, EPI_ISL_1613185, EPI_ISL_1613188, EPI_ISL_1613191, EPI_ISL_1613194, EPI_ISL_1613197, EPI_ISL_1613200, EPI_ISL_1613203, EPI_ISL_1613206, EPI_ISL_1613209, EPI_ISL_1613212, EPI_ISL_1613215, EPI_ISL_1613218, EPI_ISL_1613221,
EPI_ISL_1613224, EPI_ISL_1613226, EPI_ISL_1613229, EPI_ISL_1613235, EPI_ISL_1613238, EPI_ISL_1613241, EPI_ISL_1613245, EPI_ISL_1613248, EPI_ISL_1613251, EPI_ISL_1613254, EPI_ISL_1613257, EPI_ISL_1613260, EPI_ISL_1613263, EPI_ISL_1613266, EPI_ISL_1613269, EPI_ISL_1613272, EPI_ISL_1613275, EPI_ISL_1613277, EPI_ISL_1613280, EPI_ISL_1613284, EPI_ISL_1613287, EPI_ISL_1613290,
EPI_ISL_1613292, EPI_ISL_1613295, EPI_ISL_1613298, EPI_ISL_1613301, EPI_ISL_1613304, EPI_ISL_1613307, EPI_ISL_1613310, EPI_ISL_1613313, EPI_ISL_1613316, EPI_ISL_1613319, EPI_ISL_1613321, EPI_ISL_1613324, EPI_ISL_1613327, EPI_ISL_1613331, EPI_ISL_1613334, EPI_ISL_1613339, EPI_ISL_1613342, EPI_ISL_1613345, EPI_ISL_1613348, EPI_ISL_1613351, EPI_ISL_1613354, EPI_ISL_1613357,
EPI_ISL_1613360, EPI_ISL_1613363, EPI_ISL_1613366, EPI_ISL_1613369, EPI_ISL_1613372, EPI_ISL_1613375, EPI_ISL_1613378, EPI_ISL_1613381, EPI_ISL_1613384, EPI_ISL_1613386, EPI_ISL_1613389, EPI_ISL_1613392, EPI_ISL_1613401, EPI_ISL_1613404, EPI_ISL_1613407, EPI_ISL_1613410, EPI_ISL_1613413, EPI_ISL_1613416, EPI_ISL_1613419, EPI_ISL_1613422, EPI_ISL_1613425, EPI_ISL_1613428,
EPI_ISL_1613431, EPI_ISL_1613433, EPI_ISL_1613436, EPI_ISL_1613439, EPI_ISL_1613442, EPI_ISL_1613445, EPI_ISL_1613448, EPI_ISL_1613451, EPI_ISL_1613454, EPI_ISL_1613458, EPI_ISL_1613461, EPI_ISL_1613464, EPI_ISL_1613467, EPI_ISL_1613469, EPI_ISL_1613472, EPI_ISL_1613475, EPI_ISL_1613478, EPI_ISL_1613481, EPI_ISL_1613485, EPI_ISL_1613487, EPI_ISL_1613490, EPI_ISL_1613493,
EPI_ISL_1613496, EPI_ISL_1613499, EPI_ISL_1613502, EPI_ISL_1613505, EPI_ISL_1613508, EPI_ISL_1613511, EPI_ISL_1613514, EPI_ISL_1613517, EPI_ISL_1613520, EPI_ISL_1613522, EPI_ISL_1613529, EPI_ISL_1613532, EPI_ISL_1613534, EPI_ISL_1613538, EPI_ISL_1613540, EPI_ISL_1613543, EPI_ISL_1613546, EPI_ISL_1613549, EPI_ISL_1613552, EPI_ISL_1613555, EPI_ISL_1613558, EPI_ISL_1613562,
EPI_ISL_1613565, EPI_ISL_1613567, EPI_ISL_1613570, EPI_ISL_1613573, EPI_ISL_1613576, EPI_ISL_1613579, EPI_ISL_1613582, EPI_ISL_1613585, EPI_ISL_1613588, EPI_ISL_1613591, EPI_ISL_1613593, EPI_ISL_1613596, EPI_ISL_1613599, EPI_ISL_1613602, EPI_ISL_1613605, EPI_ISL_1613608, EPI_ISL_1613805, EPI_ISL_1613808, EPI_ISL_1613810, EPI_ISL_1613813, EPI_ISL_1613816, EPI_ISL_1613819,
EPI_ISL_1613822, EPI_ISL_1613825, EPI_ISL_1613828, EPI_ISL_1613831, EPI_ISL_1613834, EPI_ISL_1613836, EPI_ISL_1613839, EPI_ISL_1613843, EPI_ISL_1613846, EPI_ISL_1613849, EPI_ISL_1613852, EPI_ISL_1613854, EPI_ISL_1613857, EPI_ISL_1613859, EPI_ISL_1613862, EPI_ISL_1613865, EPI_ISL_1613868, EPI_ISL_1613871, EPI_ISL_1613874, EPI_ISL_1613877, EPI_ISL_1613880, EPI_ISL_1613883,
EPI_ISL_1613886, EPI_ISL_1613889, EPI_ISL_1613891, EPI_ISL_1613894, EPI_ISL_1613897, EPI_ISL_1613900, EPI_ISL_1613902, EPI_ISL_1613905, EPI_ISL_1613908, EPI_ISL_1613911, EPI_ISL_1613914, EPI_ISL_1613916, EPI_ISL_1613919, EPI_ISL_1613922, EPI_ISL_1613925, EPI_ISL_1613928, EPI_ISL_1613931, EPI_ISL_1613934, EPI_ISL_1613937, EPI_ISL_1613938, EPI_ISL_1613939, EPI_ISL_1613940,
EPI_ISL_1613941, EPI_ISL_1613942, EPI_ISL_1613943, EPI_ISL_1613944, EPI_ISL_1613945, EPI_ISL_1613946, EPI_ISL_1613947, EPI_ISL_1613948, EPI_ISL_1613949, EPI_ISL_1613950, EPI_ISL_1613951, EPI_ISL_1613952, EPI_ISL_1613953, EPI_ISL_1613954, EPI_ISL_1613955, EPI_ISL_1613956, EPI_ISL_1613957, EPI_ISL_1613958, EPI_ISL_1613959, EPI_ISL_1613960, EPI_ISL_1613961, EPI_ISL_1613962,
EPI_ISL_1613963, EPI_ISL_1613964, EPI_ISL_1613965, EPI_ISL_1613966, EPI_ISL_1613967, EPI_ISL_1613968, EPI_ISL_1613969, EPI_ISL_1613970, EPI_ISL_1613973, EPI_ISL_1613974, EPI_ISL_1613975, EPI_ISL_1613976, EPI_ISL_1613977, EPI_ISL_1613978, EPI_ISL_1613979, EPI_ISL_1613980, EPI_ISL_1613981, EPI_ISL_1613982, EPI_ISL_1613983, EPI_ISL_1613984, EPI_ISL_1613985, EPI_ISL_1613986,
EPI_ISL_1613987, EPI_ISL_1613988, EPI_ISL_1613989, EPI_ISL_1613990, EPI_ISL_1613991, EPI_ISL_1613992, EPI_ISL_1613993, EPI_ISL_1613994, EPI_ISL_1613995, EPI_ISL_1613996, EPI_ISL_1613997, EPI_ISL_1613998, EPI_ISL_1613999, EPI_ISL_1614000, EPI_ISL_1614001, EPI_ISL_1614002, EPI_ISL_1614003, EPI_ISL_1614004, EPI_ISL_1614005, EPI_ISL_1614006, EPI_ISL_1614007, EPI_ISL_1614008,
EPI_ISL_1614009, EPI_ISL_1614010, EPI_ISL_1614011, EPI_ISL_1614012, EPI_ISL_1614013, EPI_ISL_1614014, EPI_ISL_1614015, EPI_ISL_1614016, EPI_ISL_1614017, EPI_ISL_1614018, EPI_ISL_1614019, EPI_ISL_1614020, EPI_ISL_1614021, EPI_ISL_1614022, EPI_ISL_1614023, EPI_ISL_1614024, EPI_ISL_1614025, EPI_ISL_1614026, EPI_ISL_1614027, EPI_ISL_1614028, EPI_ISL_1614029, EPI_ISL_1614030,
EPI_ISL_1614031, EPI_ISL_1614032, EPI_ISL_1614033, EPI_ISL_1614034, EPI_ISL_1614035, EPI_ISL_1614036, EPI_ISL_1614037, EPI_ISL_1614038, EPI_ISL_1614039, EPI_ISL_1614040, EPI_ISL_1614041, EPI_ISL_1614042, EPI_ISL_1614043, EPI_ISL_1614044, EPI_ISL_1614045, EPI_ISL_1614046, EPI_ISL_1614047, EPI_ISL_1614048, EPI_ISL_1614049, EPI_ISL_1614050, EPI_ISL_1614051, EPI_ISL_1614052,
EPI_ISL_1614053, EPI_ISL_1614054, EPI_ISL_1614055, EPI_ISL_1614056, EPI_ISL_1614057, EPI_ISL_1614058, EPI_ISL_1614059, EPI_ISL_1614060, EPI_ISL_1614061, EPI_ISL_1614062, EPI_ISL_1614063, EPI_ISL_1614064, EPI_ISL_1614065, EPI_ISL_1614066, EPI_ISL_1614067, EPI_ISL_1614068, EPI_ISL_1614069, EPI_ISL_1614070, EPI_ISL_1614071, EPI_ISL_1614072, EPI_ISL_1614073, EPI_ISL_1614074,
EPI_ISL_1614075, EPI_ISL_1614077, EPI_ISL_1614078, EPI_ISL_1614080, EPI_ISL_1614081, EPI_ISL_1614082, EPI_ISL_1614083, EPI_ISL_1614084, EPI_ISL_1614085, EPI_ISL_1614086, EPI_ISL_1614087, EPI_ISL_1614088, EPI_ISL_1614089, EPI_ISL_1614090, EPI_ISL_1614091, EPI_ISL_1614092, EPI_ISL_1614093, EPI_ISL_1614094, EPI_ISL_1614095, EPI_ISL_1614096, EPI_ISL_1614097, EPI_ISL_1614098,
EPI_ISL_1614099, EPI_ISL_1614100, EPI_ISL_1614101, EPI_ISL_1614102, EPI_ISL_1614103, EPI_ISL_1614104, EPI_ISL_1614105, EPI_ISL_1614106, EPI_ISL_1614107, EPI_ISL_1614108, EPI_ISL_1614109, EPI_ISL_1614110, EPI_ISL_1614111, EPI_ISL_1614112, EPI_ISL_1614113, EPI_ISL_1614114, EPI_ISL_1614115, EPI_ISL_1614116, EPI_ISL_1614117, EPI_ISL_1614118, EPI_ISL_1614119, EPI_ISL_1614120,
EPI_ISL_1614121, EPI_ISL_1614122, EPI_ISL_1614123, EPI_ISL_1614124, EPI_ISL_1614125, EPI_ISL_1614126, EPI_ISL_1614127, EPI_ISL_1614128, EPI_ISL_1614129, EPI_ISL_1614130, EPI_ISL_1614131, EPI_ISL_1614132, EPI_ISL_1614133, EPI_ISL_1614134, EPI_ISL_1614135, EPI_ISL_1614136, EPI_ISL_1614137, EPI_ISL_1614138, EPI_ISL_1614139, EPI_ISL_1614140, EPI_ISL_1614141, EPI_ISL_1614142,
EPI_ISL_1614143, EPI_ISL_1614144, EPI_ISL_1614145, EPI_ISL_1614146, EPI_ISL_1614147, EPI_ISL_1614148, EPI_ISL_1614149, EPI_ISL_1614150, EPI_ISL_1614151, EPI_ISL_1614152, EPI_ISL_1614153, EPI_ISL_1614154, EPI_ISL_1614155, EPI_ISL_1614156, EPI_ISL_1614157, EPI_ISL_1614158, EPI_ISL_1614159, EPI_ISL_1614160, EPI_ISL_1614161, EPI_ISL_1614162, EPI_ISL_1614163, EPI_ISL_1614164,
EPI_ISL_1614165, EPI_ISL_1614166, EPI_ISL_1614167, EPI_ISL_1614168, EPI_ISL_1614169, EPI_ISL_1614170, EPI_ISL_1614226, EPI_ISL_1614348, EPI_ISL_1614460, EPI_ISL_1614462, EPI_ISL_1614463, EPI_ISL_1614464, EPI_ISL_1614465, EPI_ISL_1614466, EPI_ISL_1614467, EPI_ISL_1614468, EPI_ISL_1614469, EPI_ISL_1614470, EPI_ISL_1614471, EPI_ISL_1614472, EPI_ISL_1614473, EPI_ISL_1614474,
EPI_ISL_1614475, EPI_ISL_1614476, EPI_ISL_1614477, EPI_ISL_1614478, EPI_ISL_1614479, EPI_ISL_1614480, EPI_ISL_1614481, EPI_ISL_1614482, EPI_ISL_1614483, EPI_ISL_1614484, EPI_ISL_1614485, EPI_ISL_1614486, EPI_ISL_1614487, EPI_ISL_1614488, EPI_ISL_1614489, EPI_ISL_1614490, EPI_ISL_1614491, EPI_ISL_1614492, EPI_ISL_1614493, EPI_ISL_1614494, EPI_ISL_1614495, EPI_ISL_1614496,
EPI_ISL_1614497, EPI_ISL_1614498, EPI_ISL_1614499, EPI_ISL_1614500, EPI_ISL_1614501, EPI_ISL_1614502, EPI_ISL_1614503, EPI_ISL_1614504, EPI_ISL_1614505, EPI_ISL_1614506, EPI_ISL_1614507, EPI_ISL_1614508, EPI_ISL_1614509, EPI_ISL_1614510, EPI_ISL_1614511, EPI_ISL_1614512, EPI_ISL_1614513, EPI_ISL_1614514, EPI_ISL_1614515, EPI_ISL_1614516, EPI_ISL_1614517, EPI_ISL_1614518,
EPI_ISL_1614519, EPI_ISL_1614520, EPI_ISL_1614521, EPI_ISL_1614522, EPI_ISL_1614523, EPI_ISL_1614524, EPI_ISL_1614525, EPI_ISL_1614526, EPI_ISL_1614527, EPI_ISL_1614528, EPI_ISL_1614529, EPI_ISL_1614530, EPI_ISL_1614531, EPI_ISL_1614532, EPI_ISL_1614533, EPI_ISL_1614534, EPI_ISL_1614536, EPI_ISL_1614537, EPI_ISL_1614538, EPI_ISL_1614539, EPI_ISL_1614540, EPI_ISL_1614541,
EPI_ISL_1614542, EPI_ISL_1614543, EPI_ISL_1614544, EPI_ISL_1614545, EPI_ISL_1614546, EPI_ISL_1614547, EPI_ISL_1614548, EPI_ISL_1614549, EPI_ISL_1614550, EPI_ISL_1614551, EPI_ISL_1614552, EPI_ISL_1614553, EPI_ISL_1614554, EPI_ISL_1614555, EPI_ISL_1614556, EPI_ISL_1614557, EPI_ISL_1614558, EPI_ISL_1614559, EPI_ISL_1614560, EPI_ISL_1614561, EPI_ISL_1614562, EPI_ISL_1614563,
EPI_ISL_1614564, EPI_ISL_1614565, EPI_ISL_1614566, EPI_ISL_1614567, EPI_ISL_1614568, EPI_ISL_1614569, EPI_ISL_1614570, EPI_ISL_1614571, EPI_ISL_1614572, EPI_ISL_1614573, EPI_ISL_1614574, EPI_ISL_1614575, EPI_ISL_1614576, EPI_ISL_1614577, EPI_ISL_1614578, EPI_ISL_1614579, EPI_ISL_1614580, EPI_ISL_1614581, EPI_ISL_1614582, EPI_ISL_1614583, EPI_ISL_1614584, EPI_ISL_1614585,
EPI_ISL_1614586, EPI_ISL_1614587, EPI_ISL_1614588, EPI_ISL_1614589, EPI_ISL_1614590, EPI_ISL_1614591, EPI_ISL_1614592, EPI_ISL_1614593, EPI_ISL_1614594, EPI_ISL_1614595, EPI_ISL_1614596, EPI_ISL_1614597, EPI_ISL_1614598, EPI_ISL_1614599, EPI_ISL_1614600, EPI_ISL_1614601, EPI_ISL_1614602, EPI_ISL_1614603, EPI_ISL_1614604, EPI_ISL_1614605, EPI_ISL_1614606, EPI_ISL_1614607,
EPI_ISL_1614608, EPI_ISL_1614609, EPI_ISL_1614610, EPI_ISL_1614611, EPI_ISL_1614612, EPI_ISL_1614613, EPI_ISL_1614614, EPI_ISL_1614615, EPI_ISL_1614616, EPI_ISL_1614617, EPI_ISL_1614618, EPI_ISL_1614619, EPI_ISL_1614620, EPI_ISL_1614621, EPI_ISL_1614622, EPI_ISL_1614623, EPI_ISL_1614624, EPI_ISL_1614625, EPI_ISL_1614626, EPI_ISL_1614627, EPI_ISL_1614628, EPI_ISL_1614629,
EPI_ISL_1614630, EPI_ISL_1614631, EPI_ISL_1614632, EPI_ISL_1614633, EPI_ISL_1614634, EPI_ISL_1614635, EPI_ISL_1614636, EPI_ISL_1614637, EPI_ISL_1614638, EPI_ISL_1614639, EPI_ISL_1614640, EPI_ISL_1614641, EPI_ISL_1614642, EPI_ISL_1614643, EPI_ISL_1614644, EPI_ISL_1614645, EPI_ISL_1614646, EPI_ISL_1614647, EPI_ISL_1614648, EPI_ISL_1614649, EPI_ISL_1614650, EPI_ISL_1614651,
EPI_ISL_1614652, EPI_ISL_1614653, EPI_ISL_1614654, EPI_ISL_1614655, EPI_ISL_1614656, EPI_ISL_1614657, EPI_ISL_1614658, EPI_ISL_1614659, EPI_ISL_1614660, EPI_ISL_1614661, EPI_ISL_1614662, EPI_ISL_1614663, EPI_ISL_1614664, EPI_ISL_1614665, EPI_ISL_1614666, EPI_ISL_1614667, EPI_ISL_1614668, EPI_ISL_1614669, EPI_ISL_1614670, EPI_ISL_1614671, EPI_ISL_1614672, EPI_ISL_1614673,
EPI_ISL_1614674, EPI_ISL_1614675, EPI_ISL_1614676, EPI_ISL_1614677, EPI_ISL_1614678, EPI_ISL_1614679, EPI_ISL_1614680, EPI_ISL_1614681, EPI_ISL_1614682, EPI_ISL_1614683, EPI_ISL_1614684, EPI_ISL_1614685, EPI_ISL_1614686, EPI_ISL_1614687, EPI_ISL_1614688, EPI_ISL_1614689, EPI_ISL_1614690, EPI_ISL_1614691, EPI_ISL_1614692, EPI_ISL_1614693, EPI_ISL_1614694, EPI_ISL_1614695,
EPI_ISL_1614696, EPI_ISL_1614697, EPI_ISL_1614698, EPI_ISL_1614699, EPI_ISL_1614700, EPI_ISL_1614702, EPI_ISL_1614703, EPI_ISL_1614704, EPI_ISL_1614705, EPI_ISL_1614706, EPI_ISL_1614707, EPI_ISL_1614708, EPI_ISL_1614709, EPI_ISL_1614710, EPI_ISL_1614711, EPI_ISL_1614712, EPI_ISL_1614713, EPI_ISL_1614714, EPI_ISL_1614715, EPI_ISL_1614716, EPI_ISL_1614717, EPI_ISL_1614718,
EPI_ISL_1614719, EPI_ISL_1614720, EPI_ISL_1614721, EPI_ISL_1614722, EPI_ISL_1614723, EPI_ISL_1614724, EPI_ISL_1614725, EPI_ISL_1614726, EPI_ISL_1614727, EPI_ISL_1614728, EPI_ISL_1614729, EPI_ISL_1614730, EPI_ISL_1614731, EPI_ISL_1614732, EPI_ISL_1614733, EPI_ISL_1614734, EPI_ISL_1614735, EPI_ISL_1614736, EPI_ISL_1614737, EPI_ISL_1614738, EPI_ISL_1614739, EPI_ISL_1614740,
EPI_ISL_1614741, EPI_ISL_1614742, EPI_ISL_1614743, EPI_ISL_1614745, EPI_ISL_1614746, EPI_ISL_1614747, EPI_ISL_1614748, EPI_ISL_1614749, EPI_ISL_1614750, EPI_ISL_1614751, EPI_ISL_1614752, EPI_ISL_1614753, EPI_ISL_1614754, EPI_ISL_1614755, EPI_ISL_1614756, EPI_ISL_1614757, EPI_ISL_1614760, EPI_ISL_1614761, EPI_ISL_1614762, EPI_ISL_1614763, EPI_ISL_1614764, EPI_ISL_1614765,
EPI_ISL_1614766, EPI_ISL_1614767, EPI_ISL_1614768, EPI_ISL_1614769, EPI_ISL_1614770, EPI_ISL_1614771, EPI_ISL_1614772, EPI_ISL_1614773, EPI_ISL_1614774, EPI_ISL_1614776, EPI_ISL_1614777, EPI_ISL_1614778, EPI_ISL_1614779, EPI_ISL_1614780, EPI_ISL_1614781, EPI_ISL_1614782, EPI_ISL_1614783, EPI_ISL_1614784, EPI_ISL_1614785, EPI_ISL_1614786, EPI_ISL_1614787, EPI_ISL_1614788,
EPI_ISL_1614789, EPI_ISL_1614790, EPI_ISL_1614791, EPI_ISL_1614792, EPI_ISL_1614793, EPI_ISL_1614794, EPI_ISL_1614795, EPI_ISL_1614796, EPI_ISL_1614797, EPI_ISL_1614798, EPI_ISL_1614799, EPI_ISL_1614800, EPI_ISL_1614801, EPI_ISL_1614802, EPI_ISL_1614803, EPI_ISL_1614804, EPI_ISL_1614805, EPI_ISL_1614806, EPI_ISL_1614807, EPI_ISL_1614808, EPI_ISL_1614809, EPI_ISL_1614810,
EPI_ISL_1614811, EPI_ISL_1614812, EPI_ISL_1614813, EPI_ISL_1614814, EPI_ISL_1614815, EPI_ISL_1614816, EPI_ISL_1614817, EPI_ISL_1614818, EPI_ISL_1614819, EPI_ISL_1614820, EPI_ISL_1614821, EPI_ISL_1614822, EPI_ISL_1614823, EPI_ISL_1614824, EPI_ISL_1614825, EPI_ISL_1614826, EPI_ISL_1614827, EPI_ISL_1614828, EPI_ISL_1614829, EPI_ISL_1614830, EPI_ISL_1614831, EPI_ISL_1614832,
EPI_ISL_1614833, EPI_ISL_1614834, EPI_ISL_1614835, EPI_ISL_1614836, EPI_ISL_1614837, EPI_ISL_1614838, EPI_ISL_1614839, EPI_ISL_1614840, EPI_ISL_1614841, EPI_ISL_1614842, EPI_ISL_1614843, EPI_ISL_1614844, EPI_ISL_1614845, EPI_ISL_1614846, EPI_ISL_1614847, EPI_ISL_1614848, EPI_ISL_1614849, EPI_ISL_1614850, EPI_ISL_1614851, EPI_ISL_1614852, EPI_ISL_1614853, EPI_ISL_1614854,
EPI_ISL_1614855, EPI_ISL_1614856, EPI_ISL_1614857, EPI_ISL_1614858, EPI_ISL_1614859, EPI_ISL_1614860, EPI_ISL_1614861, EPI_ISL_1614862, EPI_ISL_1614863, EPI_ISL_1614864, EPI_ISL_1614865, EPI_ISL_1614866, EPI_ISL_1614867, EPI_ISL_1614868, EPI_ISL_1614869, EPI_ISL_1614870, EPI_ISL_1614871, EPI_ISL_1614872, EPI_ISL_1614873, EPI_ISL_1614874, EPI_ISL_1614875, EPI_ISL_1614876,
EPI_ISL_1614877, EPI_ISL_1614878, EPI_ISL_1614879, EPI_ISL_1614880, EPI_ISL_1614881, EPI_ISL_1614882, EPI_ISL_1614883, EPI_ISL_1614884, EPI_ISL_1614885, EPI_ISL_1614886, EPI_ISL_1614887, EPI_ISL_1614888, EPI_ISL_1614889, EPI_ISL_1614890, EPI_ISL_1614891, EPI_ISL_1614893, EPI_ISL_1614895, EPI_ISL_1614898, EPI_ISL_1614900, EPI_ISL_1614901, EPI_ISL_1614902, EPI_ISL_1614903,
EPI_ISL_1614904, EPI_ISL_1665940, EPI_ISL_1666655, EPI_ISL_1666658, EPI_ISL_1666702, EPI_ISL_1666724, EPI_ISL_1666934, EPI_ISL_1667018, EPI_ISL_1667078, EPI_ISL_1667084, EPI_ISL_1667085, EPI_ISL_1667086, EPI_ISL_1667088, EPI_ISL_1667089, EPI_ISL_1667090, EPI_ISL_1667095, EPI_ISL_1667096, EPI_ISL_1667097, EPI_ISL_1667100, EPI_ISL_1667101, EPI_ISL_1667102, EPI_ISL_1667104,
EPI_ISL_1667110, EPI_ISL_1667111, EPI_ISL_1667112, EPI_ISL_1667113, EPI_ISL_1667127, EPI_ISL_1667136, EPI_ISL_1667138, EPI_ISL_1667140, EPI_ISL_1667141, EPI_ISL_1667142, EPI_ISL_1667168, EPI_ISL_1667171, EPI_ISL_1667172, EPI_ISL_1667193, EPI_ISL_1694957, EPI_ISL_1694960, EPI_ISL_1694961, EPI_ISL_1694962, EPI_ISL_1694964, EPI_ISL_1694973, EPI_ISL_1694974, EPI_ISL_1695033,
EPI_ISL_1695036, EPI_ISL_1733806

| | | | |
|---|---|---|---|
| see above | Fulgent Genetics | Centers for Disease Control and Prevention Division of Viral Diseases, Pathogen Discovery | Adrian Paskey; Becky Tsai; Benafsh Sapra; Benjamin Rambo-Martin; Christopher Gulvick; Clinton R. Paden; Dakota Howard; Darlene Wagner; Dhwani Batra; Doreen Ng; Duncan MacCannell; Harry Gao; James Xie; Jason Caravas; John Gao; Joseph Fierro; Kara Moser; Matthew Schmerer; Mickey Li; Peter W. Cook; Scott Sammons; Shatavia Morrison; Yan Meng; Yvette Unoarumhi |
| EPI_ISL_1532123 | Good Samaritan Hospital | Santa Clara County Public Health Laboratory | Santa Clara County Public Health Department |
| EPI_ISL_1664267 | Good Samaritan Hospital | Santa Clara County public Health Laboratory | Santa Clara County Public Health Department |

EPI_ISL_1444227, EPI_ISL_1444242, EPI_ISL_1460853, EPI_ISL_1460917, EPI_ISL_1460959, EPI_ISL_1460975, EPI_ISL_1460981, EPI_ISL_1460985, EPI_ISL_1460997, EPI_ISL_1460998, EPI_ISL_1479226, EPI_ISL_1479271, EPI_ISL_1479275, EPI_ISL_1479283, EPI_ISL_1479285, EPI_ISL_1479425, EPI_ISL_1511861, EPI_ISL_1511862, EPI_ISL_1511867, EPI_ISL_1511871, EPI_ISL_1511872, EPI_ISL_1511875,
EPI_ISL_1511879, EPI_ISL_1511883, EPI_ISL_1511888, EPI_ISL_1511889, EPI_ISL_1511897, EPI_ISL_1511903, EPI_ISL_1511905, EPI_ISL_1511986, EPI_ISL_1511989, EPI_ISL_1512001, EPI_ISL_1512003, EPI_ISL_1512005, EPI_ISL_1512011, EPI_ISL_1512027, EPI_ISL_1512028, EPI_ISL_1512034, EPI_ISL_1512044, EPI_ISL_1512048, EPI_ISL_1512049, EPI_ISL_1512083, EPI_ISL_1512089, EPI_ISL_1512090,
EPI_ISL_1512093, EPI_ISL_1512095, EPI_ISL_1512096, EPI_ISL_1512098, EPI_ISL_1512100, EPI_ISL_1512101, EPI_ISL_1512102, EPI_ISL_1512105, EPI_ISL_1512107, EPI_ISL_1512109, EPI_ISL_1512111, EPI_ISL_1512112, EPI_ISL_1512113, EPI_ISL_1512115, EPI_ISL_1512116, EPI_ISL_1512119, EPI_ISL_1512122, EPI_ISL_1512125, EPI_ISL_1512148, EPI_ISL_1512203, EPI_ISL_1512233, EPI_ISL_1512247,
EPI_ISL_1512248, EPI_ISL_1512249, EPI_ISL_1512252, EPI_ISL_1512253, EPI_ISL_1512255, EPI_ISL_1512259, EPI_ISL_1512266, EPI_ISL_1512267, EPI_ISL_1512268, EPI_ISL_1512314, EPI_ISL_1512317, EPI_ISL_1512319, EPI_ISL_1512321, EPI_ISL_1512322, EPI_ISL_1512324, EPI_ISL_1512326, EPI_ISL_1512329, EPI_ISL_1512331, EPI_ISL_1512335, EPI_ISL_1512349, EPI_ISL_1512351, EPI_ISL_1512352,
EPI_ISL_1512358, EPI_ISL_1512366, EPI_ISL_1512376, EPI_ISL_1512377, EPI_ISL_1512378, EPI_ISL_1512383, EPI_ISL_1512386, EPI_ISL_1512395, EPI_ISL_1512402, EPI_ISL_1512404, EPI_ISL_1512419, EPI_ISL_1512420, EPI_ISL_1512423, EPI_ISL_1512424, EPI_ISL_1512429, EPI_ISL_1512431, EPI_ISL_1512432, EPI_ISL_1512436, EPI_ISL_1512438, EPI_ISL_1512442, EPI_ISL_1512443, EPI_ISL_1512451,
EPI_ISL_1512459, EPI_ISL_1512463, EPI_ISL_1512464, EPI_ISL_1512466, EPI_ISL_1512469, EPI_ISL_1512471, EPI_ISL_1512475, EPI_ISL_1512478, EPI_ISL_1512479, EPI_ISL_1512484, EPI_ISL_1512489, EPI_ISL_1512492, EPI_ISL_1512519, EPI_ISL_1512520, EPI_ISL_1512554, EPI_ISL_1512562, EPI_ISL_1512584, EPI_ISL_1512589, EPI_ISL_1512593, EPI_ISL_1512599, EPI_ISL_1512634, EPI_ISL_1512642,
EPI_ISL_1512652, EPI_ISL_1512664, EPI_ISL_1512667, EPI_ISL_1512672, EPI_ISL_1512686, EPI_ISL_1512707, EPI_ISL_1512718, EPI_ISL_1512734, EPI_ISL_1512735, EPI_ISL_1512736, EPI_ISL_1512742, EPI_ISL_1512744, EPI_ISL_1553418, EPI_ISL_1553432, EPI_ISL_1553447, EPI_ISL_1553474, EPI_ISL_1553500, EPI_ISL_1553518, EPI_ISL_1553595, EPI_ISL_1554197, EPI_ISL_1554200,
EPI_ISL_1554204, EPI_ISL_1554216, EPI_ISL_1554217, EPI_ISL_1554224, EPI_ISL_1554304, EPI_ISL_1554343, EPI_ISL_1554361, EPI_ISL_1554367, EPI_ISL_1554375, EPI_ISL_1554401, EPI_ISL_1554406, EPI_ISL_1554419, EPI_ISL_1554422, EPI_ISL_1554425, EPI_ISL_1554465, EPI_ISL_1554466, EPI_ISL_1554467, EPI_ISL_1554468, EPI_ISL_1554469, EPI_ISL_1554470, EPI_ISL_1554471, EPI_ISL_1554472,

| | | | |
|---|---|---|---|
| EPI_ISL_1554475, EPI_ISL_1554476, EPI_ISL_1554477, EPI_ISL_1554479, EPI_ISL_1554480, EPI_ISL_1554481, EPI_ISL_1554486, EPI_ISL_1554487, EPI_ISL_1554488, EPI_ISL_1554490, EPI_ISL_1554493, EPI_ISL_1554494, EPI_ISL_1554495, EPI_ISL_1554499, EPI_ISL_1554500, EPI_ISL_1554501, EPI_ISL_1554502, EPI_ISL_1554506, EPI_ISL_1554507, EPI_ISL_1554508, EPI_ISL_1554523, EPI_ISL_1554525, EPI_ISL_1554526, EPI_ISL_1554528, EPI_ISL_1554537, EPI_ISL_1554545, EPI_ISL_1554570, EPI_ISL_1554621, EPI_ISL_1554645, EPI_ISL_1554649, EPI_ISL_1554653, EPI_ISL_1554659, EPI_ISL_1554662, EPI_ISL_1554663, EPI_ISL_1554664, EPI_ISL_1554685, EPI_ISL_1554686, EPI_ISL_1554687, EPI_ISL_1554688, EPI_ISL_1554689, EPI_ISL_1554701, EPI_ISL_1554715, EPI_ISL_1554720, EPI_ISL_1554725, EPI_ISL_1554728, EPI_ISL_1554744, EPI_ISL_1554747, EPI_ISL_1554769, EPI_ISL_1554770, EPI_ISL_1554771, EPI_ISL_1554776, EPI_ISL_1554778, EPI_ISL_1554780, EPI_ISL_1554784, EPI_ISL_1554790, EPI_ISL_1554791, EPI_ISL_1554794, EPI_ISL_1554801, EPI_ISL_1554806, EPI_ISL_1554818, EPI_ISL_1554819, EPI_ISL_1554820, EPI_ISL_1554824, EPI_ISL_1554827, EPI_ISL_1554829, EPI_ISL_1554831, EPI_ISL_1554834, EPI_ISL_1554839, EPI_ISL_1554844, EPI_ISL_1554845, EPI_ISL_1554849, EPI_ISL_1554850, EPI_ISL_1554857, EPI_ISL_1554859, EPI_ISL_1554860, EPI_ISL_1554865, EPI_ISL_1554866, EPI_ISL_1554872, EPI_ISL_1575494, EPI_ISL_1575499, EPI_ISL_1575506, EPI_ISL_1575512, EPI_ISL_1575521, EPI_ISL_1575524, EPI_ISL_1575527, EPI_ISL_1575534, EPI_ISL_1575537, EPI_ISL_1575546, EPI_ISL_1575549, EPI_ISL_1575551, EPI_ISL_1575552, EPI_ISL_1575573, EPI_ISL_1575581, EPI_ISL_1575582, EPI_ISL_1575595, EPI_ISL_1575606, EPI_ISL_1575617, EPI_ISL_1575623, EPI_ISL_1575628, EPI_ISL_1575631, EPI_ISL_1575640, EPI_ISL_1575653, EPI_ISL_1575665, EPI_ISL_1575836, EPI_ISL_1575837, EPI_ISL_1575847, EPI_ISL_1575848, EPI_ISL_1575851, EPI_ISL_1575863, EPI_ISL_1575872, EPI_ISL_1575882, EPI_ISL_1575895, EPI_ISL_1575900, EPI_ISL_1575909, EPI_ISL_1575910, EPI_ISL_1575919, EPI_ISL_1575933, EPI_ISL_1575954, EPI_ISL_1575975, EPI_ISL_1575977, EPI_ISL_1575992, EPI_ISL_1575997, EPI_ISL_1576016, EPI_ISL_1576032, EPI_ISL_1576041, EPI_ISL_1576042, EPI_ISL_1576046, EPI_ISL_1576047, EPI_ISL_1576048, EPI_ISL_1576049, EPI_ISL_1576051, EPI_ISL_1576053, EPI_ISL_1576058, EPI_ISL_1576059, EPI_ISL_1576060, EPI_ISL_1576062, EPI_ISL_1576066, EPI_ISL_1576070, EPI_ISL_1576072, EPI_ISL_1576076, EPI_ISL_1576078, EPI_ISL_1576082, EPI_ISL_1576084, EPI_ISL_1576086, EPI_ISL_1576087, EPI_ISL_1576094, EPI_ISL_1576102, EPI_ISL_1576104, EPI_ISL_1576115, EPI_ISL_1576122, EPI_ISL_1576132, EPI_ISL_1576168, EPI_ISL_1576171, EPI_ISL_1576183, EPI_ISL_1576278, EPI_ISL_1576279, EPI_ISL_1576302, EPI_ISL_1576309, EPI_ISL_1576320, EPI_ISL_1576328, EPI_ISL_1576330, EPI_ISL_1576334, EPI_ISL_1576340, EPI_ISL_1576351, EPI_ISL_1576352, EPI_ISL_1576354, EPI_ISL_1576357, EPI_ISL_1576366, EPI_ISL_1576374, EPI_ISL_1576376, EPI_ISL_1576378, EPI_ISL_1576391, EPI_ISL_1576395, EPI_ISL_1576587, EPI_ISL_1576607, EPI_ISL_1576614, EPI_ISL_1576615, EPI_ISL_1576619, EPI_ISL_1576629, EPI_ISL_1576636, EPI_ISL_1576637, EPI_ISL_1576641, EPI_ISL_1576657, EPI_ISL_1576658, EPI_ISL_1576666, EPI_ISL_1576669, EPI_ISL_1576678, EPI_ISL_1576681, EPI_ISL_1576683, EPI_ISL_1576689, EPI_ISL_1576692, EPI_ISL_1576701, EPI_ISL_1576704, EPI_ISL_1576713, EPI_ISL_1576721, EPI_ISL_1576723, EPI_ISL_1576734, EPI_ISL_1576735, EPI_ISL_1576744, EPI_ISL_1576746, EPI_ISL_1576747, EPI_ISL_1576754, EPI_ISL_1576756, EPI_ISL_1576757, EPI_ISL_1580871, EPI_ISL_1580873, EPI_ISL_1580880, EPI_ISL_1580885, EPI_ISL_1580887, EPI_ISL_1580895, EPI_ISL_1580904, EPI_ISL_1580907, EPI_ISL_1580908, EPI_ISL_1580919, EPI_ISL_1580925, EPI_ISL_1580935, EPI_ISL_1580949, EPI_ISL_1581154, EPI_ISL_1581158, EPI_ISL_1581169, EPI_ISL_1581175, EPI_ISL_1581190, EPI_ISL_1581193, EPI_ISL_1581213, EPI_ISL_1581240, EPI_ISL_1581297, EPI_ISL_1581309, EPI_ISL_1581321, EPI_ISL_1581334, EPI_ISL_1581357, EPI_ISL_1581366, EPI_ISL_1581368, EPI_ISL_1581369, EPI_ISL_1581378, EPI_ISL_1581379, EPI_ISL_1581402, EPI_ISL_1581561, EPI_ISL_1581704, EPI_ISL_1581740, EPI_ISL_1581751, EPI_ISL_1581817, EPI_ISL_1592405, EPI_ISL_1592408, EPI_ISL_1592409, EPI_ISL_1592413, EPI_ISL_1592415, EPI_ISL_1592422, EPI_ISL_1592423, EPI_ISL_1592424, EPI_ISL_1592432, EPI_ISL_1592435, EPI_ISL_1592436, EPI_ISL_1592440, EPI_ISL_1592441, EPI_ISL_1592448, EPI_ISL_1592461, EPI_ISL_1592463, EPI_ISL_1592471, EPI_ISL_1592477, EPI_ISL_1592479, EPI_ISL_1592483, EPI_ISL_1592484, EPI_ISL_1592487, EPI_ISL_1592489, EPI_ISL_1592496, EPI_ISL_1592497, EPI_ISL_1592573, EPI_ISL_1592593, EPI_ISL_1592594, EPI_ISL_1592595, EPI_ISL_1592596, EPI_ISL_1592597, EPI_ISL_1592604, EPI_ISL_1592605, EPI_ISL_1592606, EPI_ISL_1592608, EPI_ISL_1592609, EPI_ISL_1592614, EPI_ISL_1592615, EPI_ISL_1592616, EPI_ISL_1592617, EPI_ISL_1592618, EPI_ISL_1592619, EPI_ISL_1592620, EPI_ISL_1592626, EPI_ISL_1592627, EPI_ISL_1592628, EPI_ISL_1592629, EPI_ISL_1592630, EPI_ISL_1592631, EPI_ISL_1592638, EPI_ISL_1592639, EPI_ISL_1592642, EPI_ISL_1592643, EPI_ISL_1592644, EPI_ISL_1592650, EPI_ISL_1592652, EPI_ISL_1592653, EPI_ISL_1592654, EPI_ISL_1592662, EPI_ISL_1592664, EPI_ISL_1592665, EPI_ISL_1592667, EPI_ISL_1592674, EPI_ISL_1592677, EPI_ISL_1592678, EPI_ISL_1592896, EPI_ISL_1592940, EPI_ISL_1593021, EPI_ISL_1593054, EPI_ISL_1593063, EPI_ISL_1593074, EPI_ISL_1593162, EPI_ISL_1593166, EPI_ISL_1593182, EPI_ISL_1593197, EPI_ISL_1615101, EPI_ISL_1615136, EPI_ISL_1615233, EPI_ISL_1615234, EPI_ISL_1615240, EPI_ISL_1615241, EPI_ISL_1615295, EPI_ISL_1615322, EPI_ISL_1615323, EPI_ISL_1615370, EPI_ISL_1615372, EPI_ISL_1615393, EPI_ISL_1615409, EPI_ISL_1615430, EPI_ISL_1615436, EPI_ISL_1615441, EPI_ISL_1615450, EPI_ISL_1615478, EPI_ISL_1615531, EPI_ISL_1615537, EPI_ISL_1615555, EPI_ISL_1678894, EPI_ISL_1678901, EPI_ISL_1678928, EPI_ISL_1678936, EPI_ISL_1678974, EPI_ISL_1678983, EPI_ISL_1678984, EPI_ISL_1679020, EPI_ISL_1679021, EPI_ISL_1679159, EPI_ISL_1679214, EPI_ISL_1679218, EPI_ISL_1679220, EPI_ISL_1679236, EPI_ISL_1679239, EPI_ISL_1679247, EPI_ISL_1679249, EPI_ISL_1679258, EPI_ISL_1679267, EPI_ISL_1679288, EPI_ISL_1679301, EPI_ISL_1679303, EPI_ISL_1679309, EPI_ISL_1679363, EPI_ISL_1679370, EPI_ISL_1679384, EPI_ISL_1679399, EPI_ISL_1679450, EPI_ISL_1679451, EPI_ISL_1679472, EPI_ISL_1679523, EPI_ISL_1679529, EPI_ISL_1679613, EPI_ISL_1679616, EPI_ISL_1679677, EPI_ISL_1679702, EPI_ISL_1679713, EPI_ISL_1679720, EPI_ISL_1679738, EPI_ISL_1679771, EPI_ISL_1679775, EPI_ISL_1679776, EPI_ISL_1679781, EPI_ISL_1679791, EPI_ISL_1679792, EPI_ISL_1679841, EPI_ISL_1679862, EPI_ISL_1679878, EPI_ISL_1679900, EPI_ISL_1679908, EPI_ISL_1679911, EPI_ISL_1679915, EPI_ISL_1679922, EPI_ISL_1680073, EPI_ISL_1680144, EPI_ISL_1680178, EPI_ISL_1690283, EPI_ISL_1690290, EPI_ISL_1690295, EPI_ISL_1690301, EPI_ISL_1690311, EPI_ISL_1690312, EPI_ISL_1690330, EPI_ISL_1690342, EPI_ISL_1690435, EPI_ISL_1690452, EPI_ISL_1690457, EPI_ISL_1690459, EPI_ISL_1690498, EPI_ISL_1690584, EPI_ISL_1690600, EPI_ISL_1690617, EPI_ISL_1690649, EPI_ISL_1690708, EPI_ISL_1690772, EPI_ISL_1690817, EPI_ISL_1690833, EPI_ISL_1690899, EPI_ISL_1690984, EPI_ISL_1691022, EPI_ISL_1691032, EPI_ISL_1691062, EPI_ISL_1691063, EPI_ISL_1691082, EPI_ISL_1691133, EPI_ISL_1691155, EPI_ISL_1691162, EPI_ISL_1691163, EPI_ISL_1691190, EPI_ISL_1691192, EPI_ISL_1691199, EPI_ISL_1691201, EPI_ISL_1691216, EPI_ISL_1691218, EPI_ISL_1691222, EPI_ISL_1691310, EPI_ISL_1691312, EPI_ISL_1691340, EPI_ISL_1691343, EPI_ISL_1694635, EPI_ISL_1694637, EPI_ISL_1694638, EPI_ISL_1694641, EPI_ISL_1694664, EPI_ISL_1695048, EPI_ISL_1701848, EPI_ISL_1701852, EPI_ISL_1701854, EPI_ISL_1702158, EPI_ISL_1734681, EPI_ISL_1734703, EPI_ISL_1734784, EPI_ISL_1734798, EPI_ISL_1734832, EPI_ISL_1734848, EPI_ISL_1734876, EPI_ISL_1734894, EPI_ISL_1734905, EPI_ISL_1734906, EPI_ISL_1734911, EPI_ISL_1734915, EPI_ISL_1734935, EPI_ISL_1734962, EPI_ISL_1734980, EPI_ISL_1734992, EPI_ISL_1735013, EPI_ISL_1735015, EPI_ISL_1735018, EPI_ISL_1735174, EPI_ISL_1735416, EPI_ISL_1735427, EPI_ISL_1735604, EPI_ISL_1735621, EPI_ISL_1735631, EPI_ISL_1735674, EPI_ISL_1753503, EPI_ISL_1797276 | | | |
| see above | Helix/Illumina | Centers for Disease Control and Prevention Division of Viral Diseases, Pathogen Discovery | Adrian Paskey; Alexandre Bolze; Ary Ascencio; Ben L. Rambo-Martin; Benjamin Rambo-Martin; Brad Sickler; Charlotte Rivera-Garcia; Christine Tran; Christopher Gulvick; Clinton R. Paden; Dakota Howard; Darlene Wagner; David Becker; Dhwani Batra; Duncan MacCannell; Efren Sandoval; Eileen de Feo; Elizabeth Cirulli; Eric Allen; Geraint Levan; James Lu; Jan Antico; Jason Caravas; Jason Nguyen; Jimmy Ramirez; Jingtao Liu; Kara Moser; Kelly Schiabor Barrett; Kim Gietzen; Magnus Isaksson; Marc Laurent; Matthew Schmerer; Matthew Tolentino; Nicole L. Washington; Peter W. Cook; Phil Febbo; Ryan Cho; Scott Sammons; Shannon Wickline; Shatavia Morrison; Sherry Wang; Simon White; Summer Galloway; Suxiang Tong; Tyler Cassens; William Lee; Yvette Unoarumhi |
| EPI_ISL_1701396, EPI_ISL_1701398, EPI_ISL_1701404, EPI_ISL_1701411, EPI_ISL_1701412, EPI_ISL_1701420, EPI_ISL_1701422, EPI_ISL_1701428, EPI_ISL_1701429, EPI_ISL_1701433, EPI_ISL_1701435, EPI_ISL_1701441, EPI_ISL_1701444, EPI_ISL_1701446, EPI_ISL_1701447, EPI_ISL_1701448, EPI_ISL_1701451, EPI_ISL_1701454, EPI_ISL_1701458, EPI_ISL_1701474, EPI_ISL_1701479, EPI_ISL_1701484, EPI_ISL_1701493, EPI_ISL_1701497, EPI_ISL_1701500, EPI_ISL_1701507, EPI_ISL_1701509, EPI_ISL_1701534, EPI_ISL_1701538, EPI_ISL_1701549, EPI_ISL_1701555, EPI_ISL_1701560, EPI_ISL_1701568, EPI_ISL_1701577, EPI_ISL_1701584, EPI_ISL_1701587, EPI_ISL_1701588, EPI_ISL_1701594, EPI_ISL_1701595, EPI_ISL_1701600, EPI_ISL_1701601, EPI_ISL_1701605, EPI_ISL_1701608, EPI_ISL_1701614, EPI_ISL_1701617, EPI_ISL_1701618, EPI_ISL_1701624, EPI_ISL_1701637, EPI_ISL_1701643, EPI_ISL_1701658, EPI_ISL_2102533, EPI_ISL_2102585, EPI_ISL_2500031, EPI_ISL_2500032, EPI_ISL_2500043, EPI_ISL_2500049, EPI_ISL_2500077, EPI_ISL_2500109, EPI_ISL_2500122, EPI_ISL_2500132, EPI_ISL_2500188, EPI_ISL_2658700 | | | |
| see above | Humboldt County Public Health Laboratory | Chan-Zuckerberg Biohub | CZB Cliahub Consortium |
| EPI_ISL_1506116, EPI_ISL_2259238, EPI_ISL_2259240 | Huntington Memorial Hospital | Los Angeles County Public Health Laboratories | P. Hemarajata et al. |
| EPI_ISL_2253169, EPI_ISL_2253172, EPI_ISL_2604242, EPI_ISL_2604243, EPI_ISL_2604249 | Infectious Diseases, Chan-Zuckerberg Biohub Covidtracker | Infectious Diseases, Chan-Zuckerberg Biohub Covidtracker | CZB Cliahub Consortium |
| EPI_ISL_1512777, EPI_ISL_1512805, EPI_ISL_1512823, EPI_ISL_1512824, EPI_ISL_1512827, EPI_ISL_1512844, EPI_ISL_1512880, EPI_ISL_1512898, EPI_ISL_1512945, EPI_ISL_1513116, EPI_ISL_1513119, EPI_ISL_1513120, EPI_ISL_1513127, EPI_ISL_1513134, EPI_ISL_1513137, EPI_ISL_1513138, EPI_ISL_1587592, EPI_ISL_1587898, EPI_ISL_1587916, EPI_ISL_1587932, EPI_ISL_1587948, EPI_ISL_1588416, EPI_ISL_1691713, EPI_ISL_1692004, EPI_ISL_1692123, EPI_ISL_1692184, EPI_ISL_1692231, EPI_ISL_1692270, EPI_ISL_1692487, EPI_ISL_1693196, EPI_ISL_1693463, EPI_ISL_1693826, EPI_ISL_1694297, EPI_ISL_1694349, EPI_ISL_1694535, EPI_ISL_1702257, EPI_ISL_1802878, EPI_ISL_1818093, EPI_ISL_2013193, EPI_ISL_2013277, EPI_ISL_2013332 | | | |
| see above | Infinity Biologix | Centers for Disease Control and Prevention Division of Viral Diseases, Pathogen Discovery | Adrian Paskey; Benjamin Rambo-Martin; Chirayu Goswami; Christian Bixby; Christopher Gulvick; Clinton R. Paden; Dakota Howard; Darlene Wagner; Dhwani Batra; Duncan MacCannell; Jason Caravas; Jonathan Schultz; Kara Moser; Matthew Schmerer; Peter W. Cook; Robin Grimwood; Russ Hager; Scott Sammons; Shatavia Morrison; Yihe Wang; Yvette Unoarumhi |
| EPI_ISL_1712422, EPI_ISL_1712424, EPI_ISL_1712425, EPI_ISL_1712426, EPI_ISL_1712427, EPI_ISL_1712429, EPI_ISL_1712430, EPI_ISL_1712462, EPI_ISL_1712463, EPI_ISL_1712464, EPI_ISL_1712465, EPI_ISL_1712466, EPI_ISL_1712523, EPI_ISL_1712524, EPI_ISL_1712525, EPI_ISL_1712526, EPI_ISL_1712527, EPI_ISL_1712528, EPI_ISL_1712529, EPI_ISL_1712530, EPI_ISL_1712531, EPI_ISL_1712532, EPI_ISL_1712533, EPI_ISL_1712534, EPI_ISL_1712535, EPI_ISL_1712536, EPI_ISL_1712537, EPI_ISL_1712542, EPI_ISL_1712543, EPI_ISL_1712544, EPI_ISL_1712545, EPI_ISL_1712547, EPI_ISL_1712548, EPI_ISL_1712549, EPI_ISL_1712550, EPI_ISL_1712551, EPI_ISL_1712552, EPI_ISL_1712553, EPI_ISL_1712554, EPI_ISL_1712555, EPI_ISL_1712556, EPI_ISL_1712557, EPI_ISL_1712558, EPI_ISL_1712559, EPI_ISL_1712560, EPI_ISL_1712561, EPI_ISL_1712562 | | | |
| see above | Innovative Genomics Institute, UC Berkeley | Innovative Genomics Institute, UC Berkeley | Holly Gildea; John Boyle; Phil Frankino; Stacia Wyman |
| EPI_ISL_1508902 | Kaiser Permanente West Los Angeles Medical Center | Los Angeles County Public Health Laboratories | P. Hemarajata et al. |
| EPI_ISL_2753971 | Kaiser San Jose | Santa Clara County Public Health Laboratory | Santa Clara County Public Health Department |
| EPI_ISL_1508968, EPI_ISL_1508970, EPI_ISL_1508972, EPI_ISL_1508974, EPI_ISL_1508975 | Kaiser Santa Clara | Santa Clara County Public Health Laboratory | Santa Clara County Public Health Department |
| EPI_ISL_1664268, EPI_ISL_1664269 | Kaiser Santa Clara | Santa Clara County public Health Laboratory | Santa Clara County Public Health Department |
| EPI_ISL_1513802, EPI_ISL_1513803, EPI_ISL_1513804, EPI_ISL_1513818, EPI_ISL_1513819, EPI_ISL_1513820, EPI_ISL_1513821, EPI_ISL_1513822, EPI_ISL_1513823, EPI_ISL_1513824, EPI_ISL_1513825, EPI_ISL_1513826, EPI_ISL_1513827, EPI_ISL_1513833, EPI_ISL_1513859, EPI_ISL_1513860, EPI_ISL_1513862, EPI_ISL_1513863, EPI_ISL_1513864, EPI_ISL_1513865, EPI_ISL_1513866, EPI_ISL_1513867, EPI_ISL_1513868, EPI_ISL_1513869, EPI_ISL_1513870, EPI_ISL_1513871, EPI_ISL_1513872, EPI_ISL_1513873, EPI_ISL_1513874, EPI_ISL_1513875, EPI_ISL_1513876, EPI_ISL_1513879, EPI_ISL_1513881, EPI_ISL_1513882, EPI_ISL_1513883, EPI_ISL_1513884, EPI_ISL_1513885, EPI_ISL_1513886, EPI_ISL_1513887, EPI_ISL_1513888, EPI_ISL_1513889, EPI_ISL_1513890, EPI_ISL_1513891, EPI_ISL_1513892, EPI_ISL_1513893, EPI_ISL_1513894, EPI_ISL_1513895, EPI_ISL_1513896, EPI_ISL_1513897, EPI_ISL_1513898, EPI_ISL_1513899, EPI_ISL_1513900, EPI_ISL_1513901, EPI_ISL_1513902, EPI_ISL_1513903, EPI_ISL_1513904, EPI_ISL_1513905, EPI_ISL_1513906, EPI_ISL_1513907, EPI_ISL_1513908, EPI_ISL_1513909, EPI_ISL_1514813, EPI_ISL_1514825, EPI_ISL_1514826, EPI_ISL_1514827, EPI_ISL_1514828, EPI_ISL_1514832, EPI_ISL_1514838, EPI_ISL_1514839, EPI_ISL_1514840, EPI_ISL_1514842, EPI_ISL_1514843, EPI_ISL_1514844, EPI_ISL_1514845, EPI_ISL_1514846, EPI_ISL_1514847, EPI_ISL_1514848, EPI_ISL_1514849, EPI_ISL_1514850, EPI_ISL_1514851, EPI_ISL_1514852, EPI_ISL_1514853, EPI_ISL_1514854, EPI_ISL_1514855, EPI_ISL_1514856, EPI_ISL_1514857, EPI_ISL_1514858, EPI_ISL_1514859, EPI_ISL_1514860, EPI_ISL_1514861, EPI_ISL_1514862, EPI_ISL_1514863, EPI_ISL_1514864, EPI_ISL_1514865, EPI_ISL_1514866, EPI_ISL_1514867, EPI_ISL_1514868, EPI_ISL_1514871, EPI_ISL_1514884, EPI_ISL_1514895, EPI_ISL_1514896, EPI_ISL_1514897, EPI_ISL_1514898, EPI_ISL_1514899, EPI_ISL_1514927, EPI_ISL_1514928, EPI_ISL_1514929, EPI_ISL_1514930, EPI_ISL_1514931, EPI_ISL_1514939, EPI_ISL_1514940, EPI_ISL_1514941, EPI_ISL_1514942, EPI_ISL_1514943, EPI_ISL_1514961, EPI_ISL_1514974, EPI_ISL_1514975, EPI_ISL_1514976, EPI_ISL_1514977, EPI_ISL_1514978, EPI_ISL_1514979, EPI_ISL_1514980, EPI_ISL_1514982, EPI_ISL_1514997, EPI_ISL_1514998, EPI_ISL_1514999, EPI_ISL_1515000, EPI_ISL_1515001, EPI_ISL_1515002, EPI_ISL_1515003, EPI_ISL_1515004, EPI_ISL_1515005, EPI_ISL_1515006, EPI_ISL_1515007, EPI_ISL_1515008, EPI_ISL_1515009, EPI_ISL_1515010, EPI_ISL_1515011, EPI_ISL_1515012, EPI_ISL_1515013, EPI_ISL_1515014, EPI_ISL_1515015, EPI_ISL_1515016, EPI_ISL_1515017, EPI_ISL_1515018, EPI_ISL_1515019, EPI_ISL_1515020, EPI_ISL_1515021, EPI_ISL_1515022, EPI_ISL_1515051, EPI_ISL_1515052, EPI_ISL_1515053, EPI_ISL_1515054, EPI_ISL_1515055, EPI_ISL_1515091, EPI_ISL_1515117, EPI_ISL_1515118, EPI_ISL_1515119, EPI_ISL_1515120, EPI_ISL_1515121, EPI_ISL_1515139, EPI_ISL_1515140, EPI_ISL_1515141, EPI_ISL_1515142, EPI_ISL_1515143, EPI_ISL_1515144, EPI_ISL_1515145, EPI_ISL_1515146, EPI_ISL_1515147, EPI_ISL_1515148, EPI_ISL_1515149, EPI_ISL_1515157, EPI_ISL_1515158, EPI_ISL_1515159, EPI_ISL_1515160, EPI_ISL_1515161, EPI_ISL_1515162, EPI_ISL_1515163, EPI_ISL_1515164, EPI_ISL_1515166, EPI_ISL_1515167, EPI_ISL_1515168, EPI_ISL_1515169, EPI_ISL_1515170, EPI_ISL_1515171, EPI_ISL_1515350, EPI_ISL_1515365, EPI_ISL_1515374, EPI_ISL_1515375, EPI_ISL_1515389, EPI_ISL_1515390, EPI_ISL_1515391, EPI_ISL_1515392, EPI_ISL_1515393, EPI_ISL_1515394, EPI_ISL_1515395, EPI_ISL_1515405, EPI_ISL_1515406, EPI_ISL_1515407, EPI_ISL_1515408, EPI_ISL_1515409, EPI_ISL_1515410, EPI_ISL_1515411, EPI_ISL_1515412, EPI_ISL_1515413, EPI_ISL_1515414, EPI_ISL_1515415, EPI_ISL_1515416, EPI_ISL_1515417, EPI_ISL_1515418, EPI_ISL_1515419, EPI_ISL_1515420, EPI_ISL_1515421, EPI_ISL_1515422, EPI_ISL_1515423, EPI_ISL_1515424, EPI_ISL_1515425, EPI_ISL_1515426, EPI_ISL_1515427, EPI_ISL_1515428, EPI_ISL_1515429, EPI_ISL_1515430, EPI_ISL_1515431, EPI_ISL_1515432, EPI_ISL_1515433, EPI_ISL_1515586, EPI_ISL_1515615, EPI_ISL_1515618, EPI_ISL_1515619, EPI_ISL_1515620, EPI_ISL_1515621, EPI_ISL_1515622, EPI_ISL_1515623, EPI_ISL_1515632, EPI_ISL_1515633, EPI_ISL_1515634, EPI_ISL_1515635, EPI_ISL_1515636, EPI_ISL_1515637, EPI_ISL_1515638, EPI_ISL_1515640, EPI_ISL_1515682, EPI_ISL_1515733, EPI_ISL_1515744, EPI_ISL_1549245, EPI_ISL_1549246, EPI_ISL_1549247, EPI_ISL_1549248, EPI_ISL_1549809, EPI_ISL_1549811, EPI_ISL_1549825, EPI_ISL_1549826, EPI_ISL_1549827, EPI_ISL_1549828, EPI_ISL_1549829, EPI_ISL_1549830, EPI_ISL_1549831, EPI_ISL_1549849, EPI_ISL_1549858, EPI_ISL_1549859, EPI_ISL_1549877, EPI_ISL_1549883, EPI_ISL_1549884, EPI_ISL_1549885, EPI_ISL_1549886, EPI_ISL_1549887, EPI_ISL_1549888, EPI_ISL_1549889, EPI_ISL_1549890, EPI_ISL_1549903, EPI_ISL_1549904, EPI_ISL_1549905, EPI_ISL_1549934, EPI_ISL_1549935, EPI_ISL_1549944, EPI_ISL_1549945, EPI_ISL_1549952, EPI_ISL_1550057, EPI_ISL_1550058, EPI_ISL_1550068, EPI_ISL_1550093, EPI_ISL_1550094, EPI_ISL_1550095, EPI_ISL_1550096, EPI_ISL_1550097, EPI_ISL_1550098, EPI_ISL_1550099, EPI_ISL_1550101, EPI_ISL_1550102, EPI_ISL_1550103, EPI_ISL_1550104, EPI_ISL_1550105, EPI_ISL_1550184, EPI_ISL_1550204, EPI_ISL_1550205, EPI_ISL_1550224, EPI_ISL_1550232, EPI_ISL_1550246, EPI_ISL_1550247, EPI_ISL_1550248, EPI_ISL_1550361, EPI_ISL_1612132, EPI_ISL_1612231, EPI_ISL_1612243, EPI_ISL_1612244, EPI_ISL_1612245, EPI_ISL_1612259, EPI_ISL_1612263, EPI_ISL_1612394, EPI_ISL_1612395, EPI_ISL_1612396, EPI_ISL_1612397, EPI_ISL_1612398, EPI_ISL_1612399, EPI_ISL_1612401, EPI_ISL_1612406, EPI_ISL_1612441, EPI_ISL_1612442, EPI_ISL_1612443, EPI_ISL_1612444, EPI_ISL_1612445, EPI_ISL_1612446, EPI_ISL_1612447, EPI_ISL_1612448, EPI_ISL_1612449, EPI_ISL_1612450, EPI_ISL_1612451, EPI_ISL_1612452, EPI_ISL_1612453, EPI_ISL_1612454, EPI_ISL_1612455, EPI_ISL_1612456, EPI_ISL_1612457, EPI_ISL_1612458, EPI_ISL_1612459, EPI_ISL_1612510, EPI_ISL_1612616, EPI_ISL_1612617, EPI_ISL_1612618, EPI_ISL_1612619, EPI_ISL_1612620, EPI_ISL_1612621, EPI_ISL_1612622, EPI_ISL_1612623, EPI_ISL_1612624, EPI_ISL_1612625, EPI_ISL_1612626, EPI_ISL_1612627, EPI_ISL_1612628, EPI_ISL_1612629, EPI_ISL_1612671, EPI_ISL_1612672, EPI_ISL_1612673, EPI_ISL_1612677, EPI_ISL_1612678, EPI_ISL_1612679, EPI_ISL_1612680, EPI_ISL_1612681, EPI_ISL_1612682, EPI_ISL_1612683, EPI_ISL_1612684, EPI_ISL_1612685, EPI_ISL_1612686, EPI_ISL_1612687, EPI_ISL_1612688, EPI_ISL_1612689, EPI_ISL_1612690, EPI_ISL_1612691, EPI_ISL_1612692, EPI_ISL_1612694, EPI_ISL_1612695, EPI_ISL_1612696, EPI_ISL_1612697, EPI_ISL_1612698, EPI_ISL_1612699, EPI_ISL_1612700, EPI_ISL_1612701, EPI_ISL_1612702, EPI_ISL_1612703, EPI_ISL_1612704, EPI_ISL_1612705, EPI_ISL_1612706, EPI_ISL_1612707, EPI_ISL_1612708, EPI_ISL_1612709, EPI_ISL_1612710, EPI_ISL_1612711, EPI_ISL_1612712, EPI_ISL_1612713, EPI_ISL_1612714, EPI_ISL_1612715, EPI_ISL_1612716, EPI_ISL_1612717, EPI_ISL_1612718, EPI_ISL_1612719, EPI_ISL_1612720, EPI_ISL_1612722, EPI_ISL_1612723, EPI_ISL_1612724, EPI_ISL_1612728, EPI_ISL_1612729, EPI_ISL_1612731, EPI_ISL_1612732, EPI_ISL_1612733, EPI_ISL_1612734, EPI_ISL_1612735, EPI_ISL_1612736, EPI_ISL_1612737, EPI_ISL_1612738, EPI_ISL_1612739, EPI_ISL_1612740, EPI_ISL_1612741, EPI_ISL_1612742, EPI_ISL_1612743, EPI_ISL_1612744, EPI_ISL_1612745, EPI_ISL_1612746, EPI_ISL_1612747, EPI_ISL_1612748, EPI_ISL_1612749, EPI_ISL_1612750, EPI_ISL_1612751, EPI_ISL_1612752, EPI_ISL_1612753, EPI_ISL_1612759, EPI_ISL_1612760, EPI_ISL_1612769, EPI_ISL_1612770, EPI_ISL_1612771, EPI_ISL_1612772, EPI_ISL_1612773, EPI_ISL_1612781, EPI_ISL_1612782, EPI_ISL_1612783, EPI_ISL_1612784, EPI_ISL_1612785, EPI_ISL_1612786, EPI_ISL_1612794, EPI_ISL_1612795, EPI_ISL_1612796, EPI_ISL_1612797, EPI_ISL_1612798, EPI_ISL_1612799, EPI_ISL_1612800, EPI_ISL_1612801, EPI_ISL_1612802, EPI_ISL_1612803, EPI_ISL_1612804, EPI_ISL_1612805, EPI_ISL_1684343, EPI_ISL_1684344, EPI_ISL_1684345, EPI_ISL_1684346, EPI_ISL_1684347, EPI_ISL_1684348, EPI_ISL_1684354, EPI_ISL_1684359, EPI_ISL_1684360, EPI_ISL_1684364, EPI_ISL_1684365, EPI_ISL_1684366, EPI_ISL_1684367, EPI_ISL_1684368, EPI_ISL_1684369, EPI_ISL_1684370, EPI_ISL_1684371, EPI_ISL_1684372, EPI_ISL_1684373, EPI_ISL_1684374, EPI_ISL_1684375, EPI_ISL_1684376, EPI_ISL_1684377, EPI_ISL_1684378, EPI_ISL_1684379, EPI_ISL_1684380, EPI_ISL_1684383, EPI_ISL_1684384, EPI_ISL_1684401, EPI_ISL_1684402, EPI_ISL_1684403, EPI_ISL_1684404, EPI_ISL_1684405, EPI_ISL_1684406, EPI_ISL_1684407, EPI_ISL_1684409, EPI_ISL_1684410, EPI_ISL_1684411, EPI_ISL_1684412, EPI_ISL_1684413, EPI_ISL_1684414, EPI_ISL_1684415, EPI_ISL_1684454, EPI_ISL_1684455, EPI_ISL_1684456, EPI_ISL_1684457, EPI_ISL_1684458, EPI_ISL_1684459, EPI_ISL_1684460, EPI_ISL_1684461, EPI_ISL_1684462, EPI_ISL_1684500, EPI_ISL_1684632, EPI_ISL_1684635, EPI_ISL_1684971, EPI_ISL_1684997, EPI_ISL_1695016, EPI_ISL_1695018, EPI_ISL_1695023, EPI_ISL_1798209, EPI_ISL_1798210, EPI_ISL_1798211, EPI_ISL_1798258, EPI_ISL_1798259, EPI_ISL_1798435, EPI_ISL_1798436, EPI_ISL_1798438, EPI_ISL_1798440, EPI_ISL_1798471, EPI_ISL_1798474, EPI_ISL_1798475, EPI_ISL_1798477, EPI_ISL_1798479, EPI_ISL_1798493, EPI_ISL_1798495, EPI_ISL_1798529, EPI_ISL_1798531, EPI_ISL_1798537, EPI_ISL_1798539, EPI_ISL_1798541, EPI_ISL_1798544, EPI_ISL_1798546, EPI_ISL_1798548, EPI_ISL_1798550, EPI_ISL_1798552, EPI_ISL_1798556, EPI_ISL_1798558, EPI_ISL_1798560, EPI_ISL_1798572, EPI_ISL_1798574 | | | |
| see above | Laboratory Corporation of America | Centers for Disease Control and Prevention Division of Viral Diseases, Pathogen Discovery | Adrian Paskey; Amanda Douglas; Amanda Suchanek; Andrea Throop; Ayla Burns; Benjamin Rambo-Martin; Bobbi Croy; Brian Krueger; Brian Norvell; Christopher Gulvick; Christos Petropoulos; Clinton R. Paden; Craig Lukasik; Dakota Howard; Darlene Wagner; Debbie Boles; Dhwani Batra; Duncan MacCannell; Eyad Almasri; Goran Stevovic; Howard Engler; Hrushikesh Deshmukh; Jake Humphrey; Jana Schroth; Jason Caravas; Joe Voshell; John Pruitt; Jonathan Meltzer; Jonathan Williams; Kara Moser; Kimberly Wagner; Lax Iyer; Lyndon Tilson; Manoj Jain; Marcia Eisenberg; Mary Ann Cristobal; Mary Williamson; Matthew Schmerer; Michael Levandoski; Mike Sapeta; Mindy Nye; Minoo Agarwal; Mohan Kolli; Nuthawin Charoensri; Oren Cohen; Peter W. Cook; Prashant Gupta; Qian Zeng; Rama Ghatti; Scott Parker; Scott Ryan; Scott Sammons; Shatavia Morrison; Stanley Letovsky; Steven Ragan; Suresh Babu Selvaraju; Susan Countryman; Susan Hicks; Suzanne Dale; Thomas Urban; Tim Kuphal; Tricia Zwiefelhofer; Vincent Drouillon; Yvette Unoarumhi |
| EPI_ISL_1504027, EPI_ISL_1508909, EPI_ISL_1707686, EPI_ISL_1707687 | Los Angeles County Public Health Laboratories | Los Angeles County Public Health Laboratories | P. Hemarajata et al. |
| EPI_ISL_1502804, EPI_ISL_1502805, EPI_ISL_1502806, EPI_ISL_1502807, EPI_ISL_1502808, EPI_ISL_1502809 | MD Laboratories | Los Angeles County Public Health Laboratories | P. Hemarajata et al. |
| EPI_ISL_1716896 | Martin Luther King Jr. Community Hospital | Los Angeles County Public Health Laboratories | P. Hemarajata et al. |

| | | | |
|---|---|---|---|
| EPI_ISL_1664595, EPI_ISL_1664597, EPI_ISL_1664598, EPI_ISL_1669862, EPI_ISL_1677791, EPI_ISL_1700688, EPI_ISL_1700689, EPI_ISL_1700690, EPI_ISL_1700691, EPI_ISL_1700692, EPI_ISL_2001029, EPI_ISL_2001032, EPI_ISL_2001034, EPI_ISL_2001036, EPI_ISL_2308290, EPI_ISL_2308291, EPI_ISL_2308292, EPI_ISL_2308293, EPI_ISL_2311862, EPI_ISL_2754040 | | | |
| see above | Monterey County Public Health Laboratory | Monterey County Public Health Laboratory | Monterey County Public Health Laboratory |
| EPI_ISL_1529336, EPI_ISL_1701251, EPI_ISL_2103197, EPI_ISL_2758490 | Orange County Public Health Lab | Chan-Zuckerberg Biohub | CZB Cliahub Consortium |
| EPI_ISL_2774580, EPI_ISL_2774581, EPI_ISL_2774582, EPI_ISL_2774583, EPI_ISL_2774584, EPI_ISL_2774585 | PAMF Palo Alto | Santa Clara County Public Health Laboratory | Santa Clara County Public Health Department |
| EPI_ISL_1824248 | Pathology and Laboratory Medicine Institute, Cleveland Clinic, Ohio, USA | Pathology and Laboratory Medicine Institute, Cleveland Clinic, Ohio, USA | Daniel H. Farkas; Daniel Rhoads; David Bosler; David Plunkett; Gary Procop; Jay Brock; Jessica Spildener; Joy Nakitandwe; Kristen McDonnell; Yu-Wei Cheng; Zheng Jin Tu |
| EPI_ISL_1506115, EPI_ISL_1508898, EPI_ISL_1508899, EPI_ISL_1508900, EPI_ISL_1508901 | Pomona Valley Hospital Medical Center | Los Angeles County Public Health Laboratories | P. Hemarajata et al. |
| EPI_ISL_1669340 | Providence Little Company of Mary Medical Center | Los Angeles County Public Health Laboratories | P. Hemarajata et al. |
| EPI_ISL_1391459, EPI_ISL_1391464, EPI_ISL_1391672, EPI_ISL_1391692, EPI_ISL_1444408, EPI_ISL_1444409, EPI_ISL_1444410, EPI_ISL_1444414, EPI_ISL_1444417, EPI_ISL_1444419, EPI_ISL_1444422, EPI_ISL_1444426, EPI_ISL_1444427, EPI_ISL_1444429, EPI_ISL_1444430, EPI_ISL_1444434, EPI_ISL_1444435, EPI_ISL_1444437, EPI_ISL_1444439, EPI_ISL_1444441, EPI_ISL_1444442, EPI_ISL_1444443, EPI_ISL_1444444, EPI_ISL_1444445, EPI_ISL_1444447, EPI_ISL_1444448, EPI_ISL_1444449, EPI_ISL_1444452, EPI_ISL_1444454, EPI_ISL_1444457, EPI_ISL_1444459, EPI_ISL_1444462, EPI_ISL_1444463, EPI_ISL_1444465, EPI_ISL_1444466, EPI_ISL_1444474, EPI_ISL_1444475, EPI_ISL_1444476, EPI_ISL_1444477, EPI_ISL_1444478, EPI_ISL_1444479, EPI_ISL_1444483, EPI_ISL_1444486, EPI_ISL_1444487, EPI_ISL_1444490, EPI_ISL_1444493, EPI_ISL_1444495, EPI_ISL_1444496, EPI_ISL_1444497, EPI_ISL_1444499, EPI_ISL_1444523, EPI_ISL_1444527, EPI_ISL_1444569, EPI_ISL_1444573, EPI_ISL_1444576, EPI_ISL_1444579, EPI_ISL_1444580, EPI_ISL_1444583, EPI_ISL_1444585, EPI_ISL_1444588, EPI_ISL_1444590, EPI_ISL_1444593, EPI_ISL_1444597, EPI_ISL_1444604, EPI_ISL_1444611, EPI_ISL_1444612, EPI_ISL_1444619, EPI_ISL_1444624, EPI_ISL_1444625, EPI_ISL_1444626, EPI_ISL_1444628, EPI_ISL_1444631, EPI_ISL_1444632, EPI_ISL_1444633, EPI_ISL_1444639, EPI_ISL_1444642, EPI_ISL_1444649, EPI_ISL_1444651, EPI_ISL_1444656, EPI_ISL_1444661, EPI_ISL_1444662, EPI_ISL_1444663, EPI_ISL_1444667, EPI_ISL_1444714, EPI_ISL_1460730, EPI_ISL_1460731, EPI_ISL_1460733, EPI_ISL_1460735, EPI_ISL_1460737, EPI_ISL_1460739, EPI_ISL_1460740, EPI_ISL_1460741, EPI_ISL_1460742, EPI_ISL_1460748, EPI_ISL_1460751, EPI_ISL_1460752, EPI_ISL_1460753, EPI_ISL_1460754, EPI_ISL_1460755, EPI_ISL_1460756, EPI_ISL_1460757, EPI_ISL_1460760, EPI_ISL_1460761, EPI_ISL_1460762, EPI_ISL_1460763, EPI_ISL_1460764, EPI_ISL_1460765, EPI_ISL_1460766, EPI_ISL_1460767, EPI_ISL_1460768, EPI_ISL_1460769, EPI_ISL_1460770, EPI_ISL_1460771, EPI_ISL_1460772, EPI_ISL_1460774, EPI_ISL_1460776, EPI_ISL_1460777, EPI_ISL_1460778, EPI_ISL_1460779, EPI_ISL_1460780, EPI_ISL_1460782, EPI_ISL_1460783, EPI_ISL_1460784, EPI_ISL_1479739, EPI_ISL_1479757, EPI_ISL_1479758, EPI_ISL_1479759, EPI_ISL_1479767, EPI_ISL_1479768, EPI_ISL_1479769, EPI_ISL_1479770, EPI_ISL_1479774, EPI_ISL_1479776, EPI_ISL_1479779, EPI_ISL_1479782, EPI_ISL_1479783, EPI_ISL_1479784, EPI_ISL_1479789, EPI_ISL_1479793, EPI_ISL_1479798, EPI_ISL_1479801, EPI_ISL_1479950, EPI_ISL_1491950, EPI_ISL_1511667, EPI_ISL_1511706, EPI_ISL_1511711, EPI_ISL_1511721, EPI_ISL_1511735, EPI_ISL_1525910, EPI_ISL_1525912, EPI_ISL_1525939, EPI_ISL_1525955, EPI_ISL_1525956, EPI_ISL_1525969, EPI_ISL_1525971, EPI_ISL_1525977, EPI_ISL_1525978, EPI_ISL_1525979, EPI_ISL_1525980, EPI_ISL_1525981, EPI_ISL_1525983, EPI_ISL_1525984, EPI_ISL_1525986, EPI_ISL_1525987, EPI_ISL_1525988, EPI_ISL_1525989, EPI_ISL_1525992, EPI_ISL_1525993, EPI_ISL_1525994, EPI_ISL_1525996, EPI_ISL_1525998, EPI_ISL_1525999, EPI_ISL_1526000, EPI_ISL_1526001, EPI_ISL_1526002, EPI_ISL_1526003, EPI_ISL_1526004, EPI_ISL_1526005, EPI_ISL_1526008, EPI_ISL_1526009, EPI_ISL_1526010, EPI_ISL_1526012, EPI_ISL_1526013, EPI_ISL_1526016, EPI_ISL_1526017, EPI_ISL_1526018, EPI_ISL_1526019, EPI_ISL_1526020, EPI_ISL_1526021, EPI_ISL_1526022, EPI_ISL_1526024, EPI_ISL_1526025, EPI_ISL_1526026, EPI_ISL_1526027, EPI_ISL_1526028, EPI_ISL_1526029, EPI_ISL_1526032, EPI_ISL_1526033, EPI_ISL_1526034, EPI_ISL_1526035, EPI_ISL_1526037, EPI_ISL_1526038, EPI_ISL_1526039, EPI_ISL_1526040, EPI_ISL_1526041, EPI_ISL_1526042, EPI_ISL_1526043, EPI_ISL_1526044, EPI_ISL_1526045, EPI_ISL_1526046, EPI_ISL_1526049, EPI_ISL_1526050, EPI_ISL_1526051, EPI_ISL_1526052, EPI_ISL_1526054, EPI_ISL_1526056, EPI_ISL_1526057, EPI_ISL_1526058, EPI_ISL_1526059, EPI_ISL_1526061, EPI_ISL_1526062, EPI_ISL_1526063, EPI_ISL_1526064, EPI_ISL_1526065, EPI_ISL_1526068, EPI_ISL_1526070, EPI_ISL_1551776, EPI_ISL_1551778, EPI_ISL_1551780, EPI_ISL_1551781, EPI_ISL_1551784, EPI_ISL_1551785, EPI_ISL_1551789, EPI_ISL_1551792, EPI_ISL_1551794, EPI_ISL_1551801, EPI_ISL_1551802, EPI_ISL_1551805, EPI_ISL_1551806, EPI_ISL_1551807, EPI_ISL_1551809, EPI_ISL_1551810, EPI_ISL_1551813, EPI_ISL_1551816, EPI_ISL_1551817, EPI_ISL_1551820, EPI_ISL_1551885, EPI_ISL_1551886, EPI_ISL_1551887, EPI_ISL_1551891, EPI_ISL_1551892, EPI_ISL_1551901, EPI_ISL_1551905, EPI_ISL_1551906, EPI_ISL_1551911, EPI_ISL_1551913, EPI_ISL_1551917, EPI_ISL_1551919, EPI_ISL_1552010, EPI_ISL_1552012, EPI_ISL_1552013, EPI_ISL_1552020, EPI_ISL_1552021, EPI_ISL_1552025, EPI_ISL_1552028, EPI_ISL_1552029, EPI_ISL_1552031, EPI_ISL_1552032, EPI_ISL_1552033, EPI_ISL_1552037, EPI_ISL_1552038, EPI_ISL_1552040, EPI_ISL_1552047, EPI_ISL_1552051, EPI_ISL_1552055, EPI_ISL_1552057, EPI_ISL_1552062, EPI_ISL_1552063, EPI_ISL_1552064, EPI_ISL_1552070, EPI_ISL_1552074, EPI_ISL_1552174, EPI_ISL_1552202, EPI_ISL_1552214, EPI_ISL_1552703, EPI_ISL_1552721, EPI_ISL_1552738, EPI_ISL_1552752, EPI_ISL_1552761, EPI_ISL_1552762, EPI_ISL_1552763, EPI_ISL_1552765, EPI_ISL_1552767, EPI_ISL_1552768, EPI_ISL_1552769, EPI_ISL_1552774, EPI_ISL_1552781, EPI_ISL_1552785, EPI_ISL_1552787, EPI_ISL_1552788, EPI_ISL_1552789, EPI_ISL_1552791, EPI_ISL_1552792, EPI_ISL_1552793, EPI_ISL_1552794, EPI_ISL_1552798, EPI_ISL_1552799, EPI_ISL_1552800, EPI_ISL_1552801, EPI_ISL_1552802, EPI_ISL_1552803, EPI_ISL_1552804, EPI_ISL_1552806, EPI_ISL_1552809, EPI_ISL_1552813, EPI_ISL_1552815, EPI_ISL_1552816, EPI_ISL_1552855, EPI_ISL_1559413, EPI_ISL_1559416, EPI_ISL_1559418, EPI_ISL_1559425, EPI_ISL_1559431, EPI_ISL_1559432, EPI_ISL_1559437, EPI_ISL_1559443, EPI_ISL_1559450, EPI_ISL_1559454, EPI_ISL_1559455, EPI_ISL_1559457, EPI_ISL_1559458, EPI_ISL_1559461, EPI_ISL_1559463, EPI_ISL_1559464, EPI_ISL_1559476, EPI_ISL_1559480, EPI_ISL_1559481, EPI_ISL_1559485, EPI_ISL_1559486, EPI_ISL_1559488, EPI_ISL_1559491, EPI_ISL_1559495, EPI_ISL_1559496, EPI_ISL_1559498, EPI_ISL_1559506, EPI_ISL_1559510, EPI_ISL_1559512, EPI_ISL_1559514, EPI_ISL_1559515, EPI_ISL_1559517, EPI_ISL_1559518, EPI_ISL_1559534, EPI_ISL_1559535, EPI_ISL_1559536, EPI_ISL_1559537, EPI_ISL_1559538, EPI_ISL_1559540, EPI_ISL_1559541, EPI_ISL_1559542, EPI_ISL_1559543, EPI_ISL_1559545, EPI_ISL_1559549, EPI_ISL_1559550, EPI_ISL_1559551, EPI_ISL_1559553, EPI_ISL_1559555, EPI_ISL_1559559, EPI_ISL_1559560, EPI_ISL_1559561, EPI_ISL_1559562, EPI_ISL_1559563, EPI_ISL_1559565, EPI_ISL_1559569, EPI_ISL_1559577, EPI_ISL_1559579, EPI_ISL_1559582, EPI_ISL_1559583, EPI_ISL_1559586, EPI_ISL_1581859, EPI_ISL_1581889, EPI_ISL_1581943, EPI_ISL_1581945, EPI_ISL_1581947, EPI_ISL_1581958, EPI_ISL_1581978, EPI_ISL_1582001, EPI_ISL_1582032, EPI_ISL_1582080, EPI_ISL_1582093, EPI_ISL_1582122, EPI_ISL_1582123, EPI_ISL_1582135, EPI_ISL_1582136, EPI_ISL_1582139, EPI_ISL_1582140, EPI_ISL_1582146, EPI_ISL_1582163, EPI_ISL_1622104, EPI_ISL_1648141, EPI_ISL_1648142, EPI_ISL_1648143, EPI_ISL_1648154, EPI_ISL_1648155, EPI_ISL_1648156, EPI_ISL_1648157, EPI_ISL_1648160, EPI_ISL_1648166, EPI_ISL_1648172, EPI_ISL_1648175, EPI_ISL_1648180, EPI_ISL_1648182, EPI_ISL_1648185, EPI_ISL_1648186, EPI_ISL_1648188, EPI_ISL_1648190, EPI_ISL_1648191, EPI_ISL_1648192, EPI_ISL_1648196, EPI_ISL_1648197, EPI_ISL_1648198, EPI_ISL_1648199, EPI_ISL_1648201, EPI_ISL_1648203, EPI_ISL_1648205, EPI_ISL_1648210, EPI_ISL_1648214, EPI_ISL_1648217, EPI_ISL_1648221, EPI_ISL_1648222, EPI_ISL_1648224, EPI_ISL_1648228, EPI_ISL_1648234, EPI_ISL_1648235, EPI_ISL_1648239, EPI_ISL_1648242, EPI_ISL_1648243, EPI_ISL_1648247, EPI_ISL_1648254, EPI_ISL_1648258, EPI_ISL_1648261, EPI_ISL_1648262, EPI_ISL_1648266, EPI_ISL_1648268, EPI_ISL_1648269, EPI_ISL_1648270, EPI_ISL_1648271, EPI_ISL_1648273, EPI_ISL_1648275, EPI_ISL_1648277, EPI_ISL_1648278, EPI_ISL_1648281, EPI_ISL_1648282, EPI_ISL_1648283, EPI_ISL_1648290, EPI_ISL_1648291, EPI_ISL_1648292, EPI_ISL_1648298, EPI_ISL_1648299, EPI_ISL_1648306, EPI_ISL_1648311, EPI_ISL_1648312, EPI_ISL_1648313, EPI_ISL_1648319, EPI_ISL_1648321, EPI_ISL_1648323, EPI_ISL_1648324, EPI_ISL_1648326, EPI_ISL_1648330, EPI_ISL_1648331, EPI_ISL_1648333, EPI_ISL_1648334, EPI_ISL_1648335, EPI_ISL_1648337, EPI_ISL_1648343, EPI_ISL_1648345, EPI_ISL_1648346, EPI_ISL_1648347, EPI_ISL_1648349, EPI_ISL_1648351, EPI_ISL_1648352, EPI_ISL_1648355, EPI_ISL_1648364, EPI_ISL_1648366, EPI_ISL_1648369, EPI_ISL_1648376, EPI_ISL_1648380, EPI_ISL_1648381, EPI_ISL_1648385, EPI_ISL_1648391, EPI_ISL_1648393, EPI_ISL_1648394, EPI_ISL_1648395, EPI_ISL_1648398, EPI_ISL_1648400, EPI_ISL_1648403, EPI_ISL_1648406, EPI_ISL_1648407, EPI_ISL_1648416, EPI_ISL_1648419, EPI_ISL_1648420, EPI_ISL_1648421, EPI_ISL_1648422, EPI_ISL_1648423, EPI_ISL_1648424, EPI_ISL_1648427, EPI_ISL_1648431, EPI_ISL_1648432, EPI_ISL_1648435, EPI_ISL_1648436, EPI_ISL_1648438, EPI_ISL_1648441, EPI_ISL_1648444, EPI_ISL_1648447, EPI_ISL_1648448, EPI_ISL_1648449, EPI_ISL_1648452, EPI_ISL_1648453, EPI_ISL_1648454, EPI_ISL_1648618, EPI_ISL_1648629, EPI_ISL_1648631, EPI_ISL_1648639, EPI_ISL_1648643, EPI_ISL_1648645, EPI_ISL_1648654, EPI_ISL_1648666, EPI_ISL_1648667, EPI_ISL_1648670, EPI_ISL_1648679, EPI_ISL_1648690, EPI_ISL_1648693, EPI_ISL_1648699, EPI_ISL_1648702, EPI_ISL_1648714, EPI_ISL_1648723, EPI_ISL_1648725, EPI_ISL_1648728, EPI_ISL_1648742, EPI_ISL_1648754, EPI_ISL_1648766, EPI_ISL_1648770, EPI_ISL_1648833, EPI_ISL_1648893, EPI_ISL_1668548, EPI_ISL_1668614, EPI_ISL_1668634, EPI_ISL_1668635, EPI_ISL_1668657, EPI_ISL_1668659, EPI_ISL_1668661, EPI_ISL_1668667, EPI_ISL_1668668, EPI_ISL_1668669, EPI_ISL_1668681, EPI_ISL_1668687, EPI_ISL_1668691, EPI_ISL_1668694, EPI_ISL_1668713, EPI_ISL_1668728, EPI_ISL_1668733, EPI_ISL_1668737, EPI_ISL_1668753, EPI_ISL_1668760, EPI_ISL_1668776, EPI_ISL_1668777, EPI_ISL_1668789, EPI_ISL_1668790, EPI_ISL_1668798, EPI_ISL_1668803, EPI_ISL_1668843, EPI_ISL_1668845, EPI_ISL_1668871, EPI_ISL_1668889, EPI_ISL_1668944, EPI_ISL_1668991, EPI_ISL_1669000, EPI_ISL_1669004, EPI_ISL_1669021, EPI_ISL_1669033, EPI_ISL_1669046, EPI_ISL_1669061, EPI_ISL_1669064, EPI_ISL_1669074, EPI_ISL_1669092, EPI_ISL_1669102, EPI_ISL_1669111, EPI_ISL_1669129, EPI_ISL_1669148, EPI_ISL_1669190, EPI_ISL_1669204, EPI_ISL_1669224, EPI_ISL_1669260, EPI_ISL_1669264, EPI_ISL_1669304, EPI_ISL_1691398, EPI_ISL_1691472, EPI_ISL_1691476, EPI_ISL_1691482, EPI_ISL_1691488, EPI_ISL_1694666, EPI_ISL_1694667, EPI_ISL_1694668, EPI_ISL_1694692, EPI_ISL_1694694, EPI_ISL_1694695, EPI_ISL_1695040, EPI_ISL_1695057, EPI_ISL_1695073, EPI_ISL_1695076, EPI_ISL_1695077, EPI_ISL_1695078, EPI_ISL_1695080, EPI_ISL_1695082, EPI_ISL_1695084, EPI_ISL_1695085, EPI_ISL_1695087, EPI_ISL_1695091, EPI_ISL_1695674, EPI_ISL_1695736, EPI_ISL_1702691, EPI_ISL_1738087, EPI_ISL_1738101, EPI_ISL_1738178, EPI_ISL_1738195, EPI_ISL_1738204, EPI_ISL_1738207, EPI_ISL_1738210, EPI_ISL_1738222, EPI_ISL_1738231, EPI_ISL_1738238, EPI_ISL_1738246, EPI_ISL_1738250, EPI_ISL_1738263, EPI_ISL_1738280, EPI_ISL_1738292, EPI_ISL_1738296, EPI_ISL_1738383, EPI_ISL_1738474, EPI_ISL_1738477, EPI_ISL_1738482, EPI_ISL_1738491, EPI_ISL_1738492, EPI_ISL_1738494, EPI_ISL_1738508, EPI_ISL_1738518, EPI_ISL_1738523, EPI_ISL_1738526, EPI_ISL_1738531, EPI_ISL_1738565, EPI_ISL_1738597, EPI_ISL_1738598, EPI_ISL_1738620, EPI_ISL_1738622, EPI_ISL_1738663, EPI_ISL_1738684, EPI_ISL_1738688, EPI_ISL_1738690, EPI_ISL_1738713, EPI_ISL_1738721, EPI_ISL_1738736, EPI_ISL_1738747, EPI_ISL_1753182, EPI_ISL_1753183, EPI_ISL_1753184, EPI_ISL_1753185, EPI_ISL_1753186, EPI_ISL_1753187, EPI_ISL_1753188, EPI_ISL_1753190, EPI_ISL_1753193, EPI_ISL_1753196, EPI_ISL_1753197, EPI_ISL_1753200, EPI_ISL_1753203, EPI_ISL_1753211, EPI_ISL_1753213, EPI_ISL_1753219, EPI_ISL_1753222, EPI_ISL_1753223, EPI_ISL_1753240, EPI_ISL_1753248, EPI_ISL_1753250, EPI_ISL_1753252, EPI_ISL_1753255, EPI_ISL_1753261, EPI_ISL_1753265, EPI_ISL_1753266, EPI_ISL_1753274, EPI_ISL_1753276, EPI_ISL_1753279, EPI_ISL_1753280, EPI_ISL_1753284, EPI_ISL_1753287, EPI_ISL_1753300, EPI_ISL_1753306, EPI_ISL_1753316, EPI_ISL_1753326, EPI_ISL_1753332, EPI_ISL_1753344, EPI_ISL_1753347, EPI_ISL_1753352, EPI_ISL_1753356, EPI_ISL_1753375, EPI_ISL_1753382, EPI_ISL_1753389, EPI_ISL_1753393, EPI_ISL_1753416, EPI_ISL_1753646, EPI_ISL_1753647, EPI_ISL_1753657, EPI_ISL_1761881, EPI_ISL_1761885, EPI_ISL_1761891 | | | |
| see above | Quest Diagnostics Incorporated | Centers for Disease Control and Prevention Division of Viral Diseases, Pathogen Discovery | A. Gerasimova; A. Perez; Adrian Paskey; B. Anderson; Ben L. Rambo-Martin; Benjamin Rambo-Martin; Christopher Gulvick; Clinton R. Paden; Dakota Howard; Darlene Wagner; Dhwani Batra; Duncan MacCannell; F. Lacbawan; I. A. Shlyakhter; Jason Caravas; K.E. Livingston; Kara Moser; L.E. Bernstein; M. Hua; Matthew Schmerer; P. Tanpaiboon; Peter W. Cook; R. M. Kagan; R. Owen; R. V. Rolando; S. H. Rosenthal; Scott Sammons; Shatavia Morrison; Suxiang Tong; Y. Liu; Yvette Unoarumhi |
| EPI_ISL_1794768, EPI_ISL_1794770, EPI_ISL_1794777, EPI_ISL_1794779, EPI_ISL_2628505, EPI_ISL_2628506 | Rady Children's Hospital - San Diego | Andersen lab at Scripps Research | Christina Clarke; David Dimmock; Denise Malicki; Kathryn Bouic; Linda Luo; SEARCH Alliance San Diego with Nanda Radamchar; Teresa Mueller |
| EPI_ISL_2313477, EPI_ISL_2313488, EPI_ISL_2313501, EPI_ISL_2313523, EPI_ISL_2313525, EPI_ISL_2313527, EPI_ISL_2313528, EPI_ISL_2313564, EPI_ISL_2313657, EPI_ISL_2313691, EPI_ISL_2313799, EPI_ISL_2313878 | | | |
| see above | Respiratory Viruses Branch, Centers for Disease Control and Prevention | Respiratory Viruses Branch, Centers for Disease Control and Prevention | Batra, D.; Bixby, C.; C.R.; Caravas, J.; Cook; Goswami, C.; Grimwood, R.; Gulvick, C.; Hager, R.; Howard, D.; MacCannell, D.; Morrison, S.; Moser, K.; P.W.; Paden; Paskey, A.; Rambo-Martin, B.; Sammons, S.; Schmerer, M.; Schultz, J.; Unoarumhi, Y.; Wagner, D.; Wang, Y. |
| EPI_ISL_1468878, EPI_ISL_1468881, EPI_ISL_1468914, EPI_ISL_1468915, EPI_ISL_1468916, EPI_ISL_1468922, EPI_ISL_1468924, EPI_ISL_1468925, EPI_ISL_1468926, EPI_ISL_1468928, EPI_ISL_1468929, EPI_ISL_1468930, EPI_ISL_1468931, EPI_ISL_1468933, EPI_ISL_1468934, EPI_ISL_1468935, EPI_ISL_1468937, EPI_ISL_1468939, EPI_ISL_1468940, EPI_ISL_1468942, EPI_ISL_1468943, EPI_ISL_1468948, EPI_ISL_1468949, EPI_ISL_1469024, EPI_ISL_1469032, EPI_ISL_1469033, EPI_ISL_1469038, EPI_ISL_1469040, EPI_ISL_1469042, EPI_ISL_1791403, EPI_ISL_1791404, EPI_ISL_1791405, EPI_ISL_1791406, EPI_ISL_1791407, EPI_ISL_1791408, EPI_ISL_1791409, EPI_ISL_1791410, EPI_ISL_1791412, EPI_ISL_1791413, EPI_ISL_1791414, EPI_ISL_1791415, EPI_ISL_1791416, EPI_ISL_1791417, EPI_ISL_1791418, EPI_ISL_1791419, EPI_ISL_1791420, EPI_ISL_1791421, EPI_ISL_1791422, EPI_ISL_1791423, EPI_ISL_1791424, EPI_ISL_1791425, EPI_ISL_1791426, EPI_ISL_1791427, EPI_ISL_1791428, EPI_ISL_1791429, EPI_ISL_1791430, EPI_ISL_1791431, EPI_ISL_1791432, EPI_ISL_1791433, EPI_ISL_1791434, EPI_ISL_1791435, EPI_ISL_1791436, EPI_ISL_1791437, EPI_ISL_1791438, EPI_ISL_1791439, EPI_ISL_1791440, EPI_ISL_1791441, EPI_ISL_1791442, EPI_ISL_1791443, EPI_ISL_1791444, EPI_ISL_1791445, EPI_ISL_1791446, EPI_ISL_1791447, EPI_ISL_1791448, EPI_ISL_1791449, EPI_ISL_1791450, EPI_ISL_1791451, EPI_ISL_1791452, EPI_ISL_1791453, EPI_ISL_1791454, EPI_ISL_1791455, EPI_ISL_1791456, EPI_ISL_1794616, EPI_ISL_1794617, EPI_ISL_1794619, EPI_ISL_1794622, EPI_ISL_1794623, EPI_ISL_1794624, EPI_ISL_1794625, EPI_ISL_1794626, EPI_ISL_1794630, EPI_ISL_1794631, EPI_ISL_1794632, EPI_ISL_1794633, EPI_ISL_1794637, EPI_ISL_1794640, EPI_ISL_1794642, EPI_ISL_1794644, EPI_ISL_1794646, EPI_ISL_1794647, EPI_ISL_1794649, EPI_ISL_1794651, EPI_ISL_1794652, EPI_ISL_1794654, EPI_ISL_1794937, EPI_ISL_1794938, EPI_ISL_1794942, EPI_ISL_1794944, EPI_ISL_1794950, EPI_ISL_1794952, EPI_ISL_1794953, EPI_ISL_1794954, EPI_ISL_1794957, EPI_ISL_1794958, EPI_ISL_1794959, EPI_ISL_1794961, EPI_ISL_1794965, EPI_ISL_1794966, EPI_ISL_1794969, EPI_ISL_1794970, EPI_ISL_1794973, EPI_ISL_1794974, EPI_ISL_1794975, EPI_ISL_1794979, EPI_ISL_1794997, EPI_ISL_1795001, EPI_ISL_1795006, EPI_ISL_1795011, EPI_ISL_1795012, EPI_ISL_1795017, EPI_ISL_1795023, EPI_ISL_1795025, EPI_ISL_1795034, EPI_ISL_1969283, EPI_ISL_1969373, EPI_ISL_1969386, EPI_ISL_1969393, EPI_ISL_1969410, EPI_ISL_1969416, EPI_ISL_1969423, EPI_ISL_1969429, EPI_ISL_1969437, EPI_ISL_1969499, EPI_ISL_1969500, EPI_ISL_2329568, EPI_ISL_2329575, EPI_ISL_2423010, EPI_ISL_2628727, EPI_ISL_2628731 | | | |
| see above | San Diego County Public Health Laboratory | Andersen lab at Scripps Research | Brett Austin; Jovan Shephard; SEARCH Alliance San Diego with Ashleigh Murphy; SEARCH Alliance San Diego with Tracy Basler |
| EPI_ISL_1529148, EPI_ISL_1529284, EPI_ISL_1701437, EPI_ISL_1701486, EPI_ISL_1701501, EPI_ISL_1701510, EPI_ISL_1701516, EPI_ISL_1701518, EPI_ISL_1701523, EPI_ISL_1701528, EPI_ISL_1701593, EPI_ISL_1701645, EPI_ISL_1701653, EPI_ISL_1821319, EPI_ISL_1821320, EPI_ISL_1821324, EPI_ISL_1821327, EPI_ISL_1821329, EPI_ISL_1821330, EPI_ISL_1821331, EPI_ISL_1821333, EPI_ISL_1821337, EPI_ISL_1821339, EPI_ISL_1821341, EPI_ISL_1821355, EPI_ISL_1821392, EPI_ISL_1821396, EPI_ISL_1821397, EPI_ISL_1821402, EPI_ISL_1821405, EPI_ISL_1821413, EPI_ISL_1821419, EPI_ISL_1821427, EPI_ISL_1821437, EPI_ISL_1821443, EPI_ISL_1821455, EPI_ISL_1821458, EPI_ISL_1821461, EPI_ISL_2102766, EPI_ISL_2102774, EPI_ISL_2102781, EPI_ISL_2102784, EPI_ISL_2102785, EPI_ISL_2102823, EPI_ISL_2102827, EPI_ISL_2102871, EPI_ISL_2102875, EPI_ISL_2102876, EPI_ISL_2102882, EPI_ISL_2102911, EPI_ISL_2102950, EPI_ISL_2102956, EPI_ISL_2102995, EPI_ISL_2103014, EPI_ISL_2103023, EPI_ISL_2103033, EPI_ISL_2103042, EPI_ISL_2103058, EPI_ISL_2103059, EPI_ISL_2103083, EPI_ISL_2103109, EPI_ISL_2103121, EPI_ISL_2103123, EPI_ISL_2103144, EPI_ISL_2103190, EPI_ISL_2228259, EPI_ISL_2228354, EPI_ISL_2228381, EPI_ISL_2228396, EPI_ISL_2228415, EPI_ISL_2496838, EPI_ISL_2658570, EPI_ISL_2658593, EPI_ISL_2658641, EPI_ISL_2658676, EPI_ISL_2658684, EPI_ISL_2758501 | | | |
| see above | Santa Clara County Public Health Laboratory | Chan-Zuckerberg Biohub | CZB Cliahub Consortium |
| EPI_ISL_1406155, EPI_ISL_1483802, EPI_ISL_1508964, EPI_ISL_1508965, EPI_ISL_1532139, EPI_ISL_1532140, EPI_ISL_1532144, EPI_ISL_1620461, EPI_ISL_1920832 | | | |
| see above | Santa Clara County Public Health Laboratory | Santa Clara County Public Health Laboratory | Santa Clara County Public Health Department |
| EPI_ISL_1664263, EPI_ISL_1664264, EPI_ISL_1664265 | Santa Clara County Public Health Laboratory | Santa Clara County public Health Laboratory | Santa Clara County Public Health Department |
| EPI_ISL_1483799, EPI_ISL_1532127, EPI_ISL_1532128, EPI_ISL_1532129, EPI_ISL_1532130, EPI_ISL_1532131, EPI_ISL_1532132, EPI_ISL_1532133, EPI_ISL_1532134, EPI_ISL_1532135, EPI_ISL_1532136, EPI_ISL_1532137, EPI_ISL_1532138, EPI_ISL_1620456, EPI_ISL_2691219, EPI_ISL_2691220, EPI_ISL_2691221, EPI_ISL_2691222, EPI_ISL_2691223 | | | |
| see above | Santa Clara Valley Medical Center | Santa Clara County Public Health Laboratory | Santa Clara County Public Health Department |
| EPI_ISL_1664276, EPI_ISL_1664277, EPI_ISL_1664278, EPI_ISL_1664279, EPI_ISL_1664280, EPI_ISL_1664281, EPI_ISL_1664282, EPI_ISL_1664283, EPI_ISL_1664284, EPI_ISL_1664285, EPI_ISL_1664286, EPI_ISL_1664287, EPI_ISL_1664288, EPI_ISL_1664289, EPI_ISL_1664290, EPI_ISL_1664291, EPI_ISL_1664292, EPI_ISL_1664293, EPI_ISL_1664294, EPI_ISL_1664295, EPI_ISL_1664296, EPI_ISL_1664297, | | | |

| | | | |
|---|---|---|---|
| EPI_ISL_1664298, EPI_ISL_1664299, EPI_ISL_1913215, EPI_ISL_1913247 | | | |
| see above | Santa Clara Valley Medical Center | Santa Clara County public Health Laboratory | Santa Clara County Public Health Department |
| EPI_ISL_1794721, EPI_ISL_1794722, EPI_ISL_1794723, EPI_ISL_1794724, EPI_ISL_1794725, EPI_ISL_1794726, EPI_ISL_1794727, EPI_ISL_1794728, EPI_ISL_1794729, EPI_ISL_1794730, EPI_ISL_1794731, EPI_ISL_1794732, EPI_ISL_1794733, EPI_ISL_1794734, EPI_ISL_1794735, EPI_ISL_1794736, EPI_ISL_1794737, EPI_ISL_1794738, EPI_ISL_1794739, EPI_ISL_1794740, EPI_ISL_1794741, EPI_ISL_1794839, EPI_ISL_1794842, EPI_ISL_1794845, EPI_ISL_1794846, EPI_ISL_1794847, EPI_ISL_1794848, EPI_ISL_1794850, EPI_ISL_1794851, EPI_ISL_1794852, EPI_ISL_1794853, EPI_ISL_1794934 | | | |
| see above | Scripps Medical Laboratory | Andersen lab at Scripps Research | Ellen Stefanski; Ian Mchardy; SEARCH Alliance San Diego with Michael Quigley |
| EPI_ISL_1468872, EPI_ISL_1468912, EPI_ISL_1531777, EPI_ISL_1531779, EPI_ISL_1531780, EPI_ISL_1531781, EPI_ISL_1531782, EPI_ISL_1531783, EPI_ISL_1531784, EPI_ISL_1531785, EPI_ISL_1531786, EPI_ISL_1531788, EPI_ISL_1531789, EPI_ISL_1531790, EPI_ISL_1531791, EPI_ISL_1531792, EPI_ISL_1531793, EPI_ISL_1531794, EPI_ISL_1531795, EPI_ISL_1531796, EPI_ISL_1531797, EPI_ISL_1531798, EPI_ISL_1531799, EPI_ISL_1531800, EPI_ISL_1531801, EPI_ISL_1531803, EPI_ISL_1531805, EPI_ISL_1794685, EPI_ISL_1794686, EPI_ISL_1794687, EPI_ISL_1794688, EPI_ISL_1794689, EPI_ISL_1794690, EPI_ISL_1794691, EPI_ISL_1794692, EPI_ISL_1794693, EPI_ISL_1794694, EPI_ISL_1794695, EPI_ISL_1794696, EPI_ISL_1794697, EPI_ISL_1794699, EPI_ISL_1794700, EPI_ISL_1794701, EPI_ISL_1794702, EPI_ISL_1794703, EPI_ISL_1794704, EPI_ISL_1794705, EPI_ISL_1794706, EPI_ISL_1794707, EPI_ISL_1794708, EPI_ISL_1794709, EPI_ISL_1794710, EPI_ISL_1794711, EPI_ISL_1794712, EPI_ISL_1794713, EPI_ISL_1794714, EPI_ISL_1794715, EPI_ISL_1794716, EPI_ISL_1794717, EPI_ISL_1794718, EPI_ISL_1794719, EPI_ISL_1794720, EPI_ISL_1794742, EPI_ISL_1794743, EPI_ISL_1794745, EPI_ISL_1794746, EPI_ISL_1794748, EPI_ISL_1794749, EPI_ISL_1794750, EPI_ISL_1794751, EPI_ISL_1794752, EPI_ISL_1794753, EPI_ISL_1794754, EPI_ISL_1794755, EPI_ISL_1794756, EPI_ISL_1794757, EPI_ISL_1794758, EPI_ISL_1794759, EPI_ISL_1794760, EPI_ISL_1794761, EPI_ISL_1794762, EPI_ISL_1794763, EPI_ISL_1794764, EPI_ISL_1794765, EPI_ISL_1794766, EPI_ISL_1794767, EPI_ISL_1794782, EPI_ISL_1794783, EPI_ISL_1794784, EPI_ISL_1794785, EPI_ISL_1794786, EPI_ISL_1794891, EPI_ISL_1794895, EPI_ISL_1794896, EPI_ISL_1794898, EPI_ISL_1794899, EPI_ISL_1794901, EPI_ISL_1794903, EPI_ISL_1794904, EPI_ISL_1794905, EPI_ISL_1794906, EPI_ISL_1794907, EPI_ISL_1794908, EPI_ISL_1794909, EPI_ISL_1794910, EPI_ISL_1794911, EPI_ISL_1794912, EPI_ISL_1794915, EPI_ISL_1969313 | | | |
| see above | Sharp HealthCare Laboratory | Andersen lab at Scripps Research | Art Mendoza; Cathy Woerle; Jacquelyn Berumen; Liam McGinnis; Omid Bakhtar; SEARCH Alliance San Diego with Aaron Harding |
| EPI_ISL_1620455, EPI_ISL_1620457, EPI_ISL_1620458, EPI_ISL_1620459, EPI_ISL_1620460, EPI_ISL_1620462, EPI_ISL_1620463, EPI_ISL_1620464, EPI_ISL_1620465 | | | |
| see above | Stanford Clinical Virology Laboratory | Santa Clara County Public Health Laboratory | Santa Clara County Public Health Department |
| EPI_ISL_1924635, EPI_ISL_1924637, EPI_ISL_1924638, EPI_ISL_1924639, EPI_ISL_1924640, EPI_ISL_1924641 | Stanford Clinical Virology Laboratory | Santa Clara County public Health Laboratory | Santa Clara County Public Health Department |
| EPI_ISL_1675225, EPI_ISL_1675226, EPI_ISL_1701680, EPI_ISL_1701681, EPI_ISL_3050270, EPI_ISL_3050272, EPI_ISL_3050276, EPI_ISL_3050277, EPI_ISL_3050278, EPI_ISL_3050279, EPI_ISL_3050280, EPI_ISL_3050281, EPI_ISL_3050282, EPI_ISL_3050283, EPI_ISL_3050285, EPI_ISL_3050286, EPI_ISL_3050289, EPI_ISL_3050291, EPI_ISL_3050293 | | | |
| see above | Stanford Health Care | Stanford University School of Medicine, Clinical Virology Laboratory | Becky Jiang; Bernadette Troung; Bernadette Truong; Daniel Solis; Fiona Yamamoto; Fumiko Yamamoto; Hannah Wang; Jacob Miller; James Zehnder; Kenji O. Mfuh; Malaya K. Sahoo; Mamdouh Sibai; Marilyn Mar; Michelle Verghese; Naomi Iwai; Nathan Hammond; Selamawit Bihon; and Benjamin A. Pinsky |
| EPI_ISL_1540751 | The Jackson Laboratory | The Jackson Laboratory | Adams M; Bergeron D; Kelly K; Li L; Long J; Omerza G; Renzette N |
| EPI_ISL_2341047, EPI_ISL_2341055, EPI_ISL_2341059, EPI_ISL_2341060, EPI_ISL_2341066, EPI_ISL_2341068, EPI_ISL_2341069, EPI_ISL_2341077, EPI_ISL_2341150, EPI_ISL_2341151, EPI_ISL_2341152, EPI_ISL_2341153, EPI_ISL_2341175, EPI_ISL_2341176, EPI_ISL_2341203, EPI_ISL_2341204, EPI_ISL_2341205, EPI_ISL_2341229, EPI_ISL_2341243, EPI_ISL_2341244, EPI_ISL_3049200 | | | |
| see above | Tulare County Public Health Laboratory | Tulare County Public Health Laboratory | Denise L Lopez; Joshua Kootstra |
| EPI_ISL_1707042, EPI_ISL_1707043, EPI_ISL_1707044, EPI_ISL_1707045, EPI_ISL_1707046, EPI_ISL_1707047, EPI_ISL_1707048, EPI_ISL_1707049, EPI_ISL_1707050, EPI_ISL_1707051, EPI_ISL_1707052, EPI_ISL_1707053, EPI_ISL_1707054, EPI_ISL_1707055, EPI_ISL_1707056, EPI_ISL_1707057, EPI_ISL_1707058, EPI_ISL_1707059, EPI_ISL_1707060, EPI_ISL_1707061, EPI_ISL_1716873, EPI_ISL_1716874, EPI_ISL_1716875, EPI_ISL_1716876, EPI_ISL_2620761 | | | |
| see above | UCLA Clinical Micro Lab | Los Angeles County Public Health Laboratories | P. Hemarajata et al. |
| EPI_ISL_1402647, EPI_ISL_1402648, EPI_ISL_1402649, EPI_ISL_1402650, EPI_ISL_1402651, EPI_ISL_1402652, EPI_ISL_1402653, EPI_ISL_1402654, EPI_ISL_1405056, EPI_ISL_1405057, EPI_ISL_1405058, EPI_ISL_1405059, EPI_ISL_1405060, EPI_ISL_1469852, EPI_ISL_1469853, EPI_ISL_1469875, EPI_ISL_1469876, EPI_ISL_1469877, EPI_ISL_1469878, EPI_ISL_1469879, EPI_ISL_1469880, EPI_ISL_1469881, EPI_ISL_1469882, EPI_ISL_1469883, EPI_ISL_1921080, EPI_ISL_1921081, EPI_ISL_1921082, EPI_ISL_1921083, EPI_ISL_1921084, EPI_ISL_1921112, EPI_ISL_1921113, EPI_ISL_1921114, EPI_ISL_1921115, EPI_ISL_1921128, EPI_ISL_1921146, EPI_ISL_2031773, EPI_ISL_2031781 | | | |
| see above | UCSC Genomics Institute | UCSC Genomics Institute | A. Marm Kilpatrick; Angie Hinrichs; Beth Shapiro; Bryan Thornlow; Ciara Wanket; David Haussler; Eric Beraut; Hugh Olsen; Ikenna Anigbogu; Isabel Bjork; Jakob McBroome; Jeremy Sanford; Joshua Kapp; Mark Akeson; Maximilian Haeussler; Michael Stone; Miten Jain; Molly Cassatt-Johnstone; Namrita Dhillon; Russell Corbett-Detig; Terren Chang; Yatish Turakhia |
| EPI_ISL_1497531, EPI_ISL_1497532, EPI_ISL_1497535, EPI_ISL_1497536, EPI_ISL_1497539, EPI_ISL_1497540, EPI_ISL_1502810, EPI_ISL_1502811, EPI_ISL_1502812, EPI_ISL_1503989, EPI_ISL_1503990, EPI_ISL_1503996, EPI_ISL_1503997, EPI_ISL_1504025, EPI_ISL_1504028, EPI_ISL_1504029, EPI_ISL_1504030, EPI_ISL_1504031, EPI_ISL_1504032, EPI_ISL_1504033, EPI_ISL_1504034, EPI_ISL_1504035, EPI_ISL_1504036, EPI_ISL_1504037, EPI_ISL_1504038, EPI_ISL_1504039, EPI_ISL_1504040, EPI_ISL_1506120, EPI_ISL_1506121, EPI_ISL_1506122, EPI_ISL_1506123, EPI_ISL_1506124, EPI_ISL_1506125, EPI_ISL_1506126, EPI_ISL_1508904, EPI_ISL_1508905, EPI_ISL_1508906, EPI_ISL_1508907, EPI_ISL_1508908, EPI_ISL_1508910, EPI_ISL_1508911, EPI_ISL_1508912, EPI_ISL_1508913, EPI_ISL_1508914, EPI_ISL_1508919, EPI_ISL_1508920, EPI_ISL_1508921, EPI_ISL_1508922, EPI_ISL_1508923, EPI_ISL_1508924, EPI_ISL_1508925, EPI_ISL_1508926, EPI_ISL_1508927, EPI_ISL_1508928, EPI_ISL_1508931, EPI_ISL_1508932, EPI_ISL_1508933, EPI_ISL_1508934, EPI_ISL_1508935, EPI_ISL_1508937, EPI_ISL_1508938, EPI_ISL_1509274, EPI_ISL_1509275, EPI_ISL_1509276, EPI_ISL_1509277, EPI_ISL_1509278, EPI_ISL_1509279, EPI_ISL_1509280, EPI_ISL_1509281, EPI_ISL_1509282, EPI_ISL_1509283, EPI_ISL_1509284, EPI_ISL_1509285, EPI_ISL_1509286, EPI_ISL_1509287, EPI_ISL_1509288, EPI_ISL_1509289, EPI_ISL_1509290, EPI_ISL_1509291, EPI_ISL_1509292, EPI_ISL_1509293, EPI_ISL_1509294, EPI_ISL_1509295, EPI_ISL_1509296, EPI_ISL_1509297, EPI_ISL_1515800, EPI_ISL_1669348, EPI_ISL_1706608, EPI_ISL_1706609, EPI_ISL_1707078, EPI_ISL_1707079, EPI_ISL_1707080, EPI_ISL_1707081, EPI_ISL_1707082, EPI_ISL_1707083, EPI_ISL_1707084, EPI_ISL_1707085, EPI_ISL_1707086, EPI_ISL_1707087, EPI_ISL_1707088, EPI_ISL_1707089, EPI_ISL_1707090, EPI_ISL_1707091, EPI_ISL_1707092, EPI_ISL_1707093, EPI_ISL_1707094, EPI_ISL_1707095, EPI_ISL_1707096, EPI_ISL_1707097, EPI_ISL_1707098, EPI_ISL_1716893, EPI_ISL_1716894, EPI_ISL_1716895, EPI_ISL_1716897, EPI_ISL_1716898, EPI_ISL_2774567 | | | |
| see above | USC Clinical Lab | Los Angeles County Public Health Laboratories | P. Hemarajata et al. |
| EPI_ISL_1504026, EPI_ISL_1506127 | VA Medical Center | Los Angeles County Public Health Laboratories | P. Hemarajata et al. |
| EPI_ISL_2295378 | WLA VAMC | Genomics and Discovery, Respiratory Viruses Branch, Division of Viral Diseases, Centers for Disease Control and Prevention | Adam Retchless; Anna Kelleher; Anna Uehara; Brian Lynch; Clinton R. Paden; Dhwani Batra; Haibin Wang; Han Jia Justin Ng; Jasmine Padilla; Jing Zhang; Justin Lee; Krista Queen; Mark Burroughs; Mili Sheth; Morgan Davis; Peter Cook; Rachel Marine; Sarah Nobles; Suxiang Tong; Tara Coalter; Yan Li; Ying Tao |

We gratefully acknowledge the following Authors from the Originating laboratories responsible for obtaining the specimens, as well as the Submitting laboratories where the genome data were generated and shared via GISAID, on which this research is based.

All Submitters of data may be contacted directly via www.gisaid.org

Authors are sorted alphabetically.

| Accession ID | Originating Laboratory | Submitting Laboratory | Authors |
|---|---|---|---|
| EPI_ISL_2229144, EPI_ISL_2229145, EPI_ISL_2229146, EPI_ISL_2229148, EPI_ISL_2229149, EPI_ISL_2229150, EPI_ISL_2229151, EPI_ISL_2229152, EPI_ISL_2229153, EPI_ISL_2229154, EPI_ISL_2229155, EPI_ISL_2229156, EPI_ISL_2248727, EPI_ISL_2788097, EPI_ISL_2788118 | | | |
| see above | "CDPH, Viral and Rickettsial Disease Laboratory" | Centers for Disease Control and Prevention Division of Viral Diseases, Pathogen Discovery | Alison Laufer Halpin; Ben L. Rambo-Martin; Clinton R. Paden; Dakota Howard; Darlene Wagner; Dave Wentworth; Dhwani Batra; Jasmine Padilla; Justin Lee; Katie Dillon; Krista Queen; Kristen Knipe; Kristine Lacek; Mark Burroughs; Matthew Schmerer; Mili Sheth; Peter Cook; Sam Shepard; Sarah Nobles; Shoshona Le; Suxiang Tong; Vivien Dugan; Yvette Unoarumhi |
| EPI_ISL_1667437, EPI_ISL_1667677, EPI_ISL_1685762, EPI_ISL_1685764, EPI_ISL_1685770, EPI_ISL_1685786, EPI_ISL_1687470, EPI_ISL_1687474, EPI_ISL_1687481, EPI_ISL_1687484, EPI_ISL_1687485, EPI_ISL_1687486, EPI_ISL_1687510, EPI_ISL_1687512, EPI_ISL_1687517, EPI_ISL_1737653, EPI_ISL_1737654, EPI_ISL_1737655, EPI_ISL_1737679, EPI_ISL_1737693, EPI_ISL_1737694, EPI_ISL_1737696, EPI_ISL_1737715, EPI_ISL_1737723, EPI_ISL_1737739, EPI_ISL_1737740, EPI_ISL_1737742, EPI_ISL_1737750, EPI_ISL_1737755, EPI_ISL_1737777, EPI_ISL_1737778, EPI_ISL_1737782, EPI_ISL_1737783, EPI_ISL_1737787, EPI_ISL_1737788, EPI_ISL_1737798, EPI_ISL_1737805, EPI_ISL_1737831, EPI_ISL_1737833, EPI_ISL_1737834, EPI_ISL_1737838, EPI_ISL_1737840, EPI_ISL_1753605, EPI_ISL_1753609, EPI_ISL_1835392, EPI_ISL_1835404, EPI_ISL_1835495, EPI_ISL_1835513, EPI_ISL_1835517, EPI_ISL_1835545, EPI_ISL_1835546, EPI_ISL_1835556, EPI_ISL_1835561, EPI_ISL_1836335, EPI_ISL_1836338, EPI_ISL_1836339, EPI_ISL_1836354, EPI_ISL_1836426, EPI_ISL_1836428, EPI_ISL_1836429, EPI_ISL_1836430, EPI_ISL_1836464, EPI_ISL_1836465, EPI_ISL_1836477, EPI_ISL_1836483, EPI_ISL_1836493, EPI_ISL_1836499, EPI_ISL_1836518, EPI_ISL_1836521, EPI_ISL_1836580, EPI_ISL_1836610, EPI_ISL_1836611, EPI_ISL_1836612, EPI_ISL_1836639, EPI_ISL_1836640, EPI_ISL_1836644, EPI_ISL_1836668, EPI_ISL_1836687, EPI_ISL_1836826, EPI_ISL_1836840, EPI_ISL_1836853, EPI_ISL_1836886, EPI_ISL_1836909, EPI_ISL_1836913, EPI_ISL_1836914, EPI_ISL_1836916, EPI_ISL_1836918, EPI_ISL_1836952, EPI_ISL_1836958, EPI_ISL_1836986, EPI_ISL_1837008, EPI_ISL_1837039, EPI_ISL_1837068, EPI_ISL_1837097, EPI_ISL_1837098, EPI_ISL_1837115, EPI_ISL_1837116, EPI_ISL_1837119, EPI_ISL_1837176, EPI_ISL_1837314, EPI_ISL_1837385, EPI_ISL_1837417, EPI_ISL_1837432, EPI_ISL_1837447, EPI_ISL_1837448, EPI_ISL_1837449, EPI_ISL_1837452, EPI_ISL_1837454, EPI_ISL_1837455, EPI_ISL_1837456, EPI_ISL_1837477, EPI_ISL_1837479, EPI_ISL_1837480, EPI_ISL_1837499, EPI_ISL_1837514, EPI_ISL_1837565, EPI_ISL_1837670, EPI_ISL_1837701, EPI_ISL_1837702, EPI_ISL_1837703, EPI_ISL_1837717, EPI_ISL_1837758, EPI_ISL_1837957, EPI_ISL_1837959, EPI_ISL_1837970, EPI_ISL_1837971, EPI_ISL_1838211, EPI_ISL_1838407, EPI_ISL_1900378, EPI_ISL_1900408, EPI_ISL_1922910, EPI_ISL_1922912, EPI_ISL_1922913, EPI_ISL_1922917, EPI_ISL_1922934, EPI_ISL_1922968, EPI_ISL_1922970, EPI_ISL_1923038, EPI_ISL_1923154, EPI_ISL_1923155, EPI_ISL_1923156, EPI_ISL_1995006, EPI_ISL_1995061, EPI_ISL_1995067, EPI_ISL_1995107, EPI_ISL_1995113, EPI_ISL_1995116, EPI_ISL_1995136, EPI_ISL_1995177, EPI_ISL_1995188, EPI_ISL_1995230, EPI_ISL_1995244, EPI_ISL_1995259, EPI_ISL_1995260, EPI_ISL_1995261, EPI_ISL_1995264, EPI_ISL_1995322, EPI_ISL_1995323, EPI_ISL_1995341, EPI_ISL_1995348, EPI_ISL_1995410, EPI_ISL_1995455, EPI_ISL_1995476, EPI_ISL_1995483, EPI_ISL_1995490, EPI_ISL_1995491, EPI_ISL_1995507, EPI_ISL_1995541, EPI_ISL_1995552, EPI_ISL_1995570, EPI_ISL_1995582, EPI_ISL_1995599, EPI_ISL_1995722, EPI_ISL_1995831, EPI_ISL_1995863, EPI_ISL_1995872, EPI_ISL_1995942, EPI_ISL_1995964, EPI_ISL_1995990, EPI_ISL_1995991, EPI_ISL_1996059, EPI_ISL_1996083, EPI_ISL_1996085, EPI_ISL_1996086, EPI_ISL_1996088, EPI_ISL_1996089, EPI_ISL_1996090, EPI_ISL_1996092, EPI_ISL_1996093, EPI_ISL_1996188, EPI_ISL_1996203, EPI_ISL_1996264, EPI_ISL_1996267, EPI_ISL_1996311, EPI_ISL_1996440, EPI_ISL_1996486, EPI_ISL_1996523, EPI_ISL_1996594, EPI_ISL_1996596, EPI_ISL_1996598, EPI_ISL_1996600, EPI_ISL_1996626, EPI_ISL_1996657, EPI_ISL_1996661, EPI_ISL_1996664, EPI_ISL_1996665, EPI_ISL_1996667, EPI_ISL_1996671, EPI_ISL_1996672, EPI_ISL_1996673, EPI_ISL_1996681, EPI_ISL_1996707, EPI_ISL_1996719, EPI_ISL_1996730, EPI_ISL_1996731, EPI_ISL_1996732, EPI_ISL_1996745, EPI_ISL_1996746, EPI_ISL_1996855, EPI_ISL_1996857, EPI_ISL_1996861, EPI_ISL_1996872, EPI_ISL_1996874, EPI_ISL_1996892, EPI_ISL_1996944, EPI_ISL_1996949, EPI_ISL_1996973, EPI_ISL_1996987, EPI_ISL_1996990, EPI_ISL_1997023, EPI_ISL_1997063, EPI_ISL_1997079, EPI_ISL_1997088, EPI_ISL_1997101, EPI_ISL_1997107, EPI_ISL_1997108, EPI_ISL_1997122, EPI_ISL_1997123, EPI_ISL_1997124, EPI_ISL_1997126, EPI_ISL_1997130, EPI_ISL_1997132, EPI_ISL_1997133, EPI_ISL_1997134, EPI_ISL_1997135, EPI_ISL_1997136, EPI_ISL_1997138, EPI_ISL_1997156, EPI_ISL_1997159, EPI_ISL_1997168, EPI_ISL_1997247, EPI_ISL_1997262, EPI_ISL_1997314, EPI_ISL_1997362, EPI_ISL_1997363, EPI_ISL_1997399, EPI_ISL_1997400, EPI_ISL_1997404, EPI_ISL_1997416, EPI_ISL_1997426, EPI_ISL_1997427, EPI_ISL_1997428, EPI_ISL_1997431, EPI_ISL_1997432, EPI_ISL_1997435, EPI_ISL_1997436, EPI_ISL_1997439, EPI_ISL_1997440, EPI_ISL_1997496, EPI_ISL_1997497, EPI_ISL_1997498, EPI_ISL_1997528, EPI_ISL_1997613, EPI_ISL_1997614, EPI_ISL_1997615, EPI_ISL_1997616, EPI_ISL_1997617, EPI_ISL_1997618, EPI_ISL_1997620, EPI_ISL_1997621, EPI_ISL_1997622, EPI_ISL_1997628, EPI_ISL_1997629, EPI_ISL_1997630, EPI_ISL_1997641, EPI_ISL_1997722, EPI_ISL_1997739, EPI_ISL_1997740, EPI_ISL_1997745, EPI_ISL_1997747, EPI_ISL_1997751, EPI_ISL_1997754, EPI_ISL_1997756, EPI_ISL_1997762, EPI_ISL_1997763, EPI_ISL_1997769, EPI_ISL_1997805, EPI_ISL_1997806, EPI_ISL_1997824, EPI_ISL_1997851, EPI_ISL_1997869, EPI_ISL_1997922, EPI_ISL_1997935, EPI_ISL_1997949, EPI_ISL_1997950, EPI_ISL_1997986, EPI_ISL_1998013, EPI_ISL_1998025, EPI_ISL_1998026, EPI_ISL_1998054, EPI_ISL_1998061, EPI_ISL_1998088, EPI_ISL_1998111, EPI_ISL_1998112, EPI_ISL_1998120, EPI_ISL_1998122, EPI_ISL_1998123, EPI_ISL_1998187, EPI_ISL_1998212, EPI_ISL_1998217, EPI_ISL_1998237, EPI_ISL_1998238, EPI_ISL_1998240, EPI_ISL_1998268, EPI_ISL_1998269, EPI_ISL_1998283, EPI_ISL_1998285, EPI_ISL_1998307, EPI_ISL_1998375, EPI_ISL_1998377, EPI_ISL_1998381, EPI_ISL_1998407, EPI_ISL_1998408, EPI_ISL_1998410, EPI_ISL_1998411, EPI_ISL_1998412, EPI_ISL_1998419, EPI_ISL_1998432, EPI_ISL_1998439, EPI_ISL_1998467, EPI_ISL_1998513, EPI_ISL_1998514, EPI_ISL_1998518, EPI_ISL_1998537, EPI_ISL_1998538, EPI_ISL_1998620, EPI_ISL_1998668, EPI_ISL_1998672, EPI_ISL_1998722, EPI_ISL_1998755, EPI_ISL_1998774, EPI_ISL_1998789, EPI_ISL_1998799, EPI_ISL_1998831, EPI_ISL_1998848, EPI_ISL_1998859, EPI_ISL_1998860, EPI_ISL_1998862, EPI_ISL_1998863, EPI_ISL_1998882, EPI_ISL_1998896, EPI_ISL_1998965, EPI_ISL_1998975, EPI_ISL_1999023, EPI_ISL_1999024, EPI_ISL_1999034, EPI_ISL_1999040, EPI_ISL_1999098, EPI_ISL_1999132, EPI_ISL_1999141, EPI_ISL_1999162, EPI_ISL_1999189, EPI_ISL_1999195, EPI_ISL_1999196, EPI_ISL_1999200, EPI_ISL_1999211, EPI_ISL_1999212, EPI_ISL_1999227, EPI_ISL_1999231, EPI_ISL_1999248, EPI_ISL_1999293, EPI_ISL_1999299, EPI_ISL_1999302, EPI_ISL_1999309, EPI_ISL_1999311, EPI_ISL_1999366, EPI_ISL_1999368, EPI_ISL_1999375, EPI_ISL_1999446, EPI_ISL_1999491, EPI_ISL_1999569, EPI_ISL_1999600, EPI_ISL_1999716, EPI_ISL_1999717, EPI_ISL_1999780, EPI_ISL_1999878, EPI_ISL_1999914, EPI_ISL_2000128, EPI_ISL_2000130, EPI_ISL_2000131, EPI_ISL_2000175, EPI_ISL_2000180, EPI_ISL_2000209, EPI_ISL_2000210, EPI_ISL_2000215, EPI_ISL_2000220, EPI_ISL_2000222, EPI_ISL_2000226, EPI_ISL_2000233, EPI_ISL_2000234, EPI_ISL_2000237, EPI_ISL_2000262, EPI_ISL_2000270, EPI_ISL_2000271, EPI_ISL_2000275, EPI_ISL_2000296, EPI_ISL_2000333, EPI_ISL_2000343, EPI_ISL_2000393, EPI_ISL_2000407, EPI_ISL_2000410, EPI_ISL_2000412, EPI_ISL_2000417, EPI_ISL_2000483, EPI_ISL_2000484, EPI_ISL_2000509, EPI_ISL_2000512, EPI_ISL_2000520, EPI_ISL_2035046, EPI_ISL_2035069, EPI_ISL_2035110, EPI_ISL_2035138, EPI_ISL_2035143, EPI_ISL_2035151, EPI_ISL_2035184, EPI_ISL_2035189, EPI_ISL_2035223, EPI_ISL_2035225, EPI_ISL_2039034, EPI_ISL_2039064, EPI_ISL_2039065, EPI_ISL_2039067, EPI_ISL_2039088, EPI_ISL_2039089, EPI_ISL_2039095, EPI_ISL_2039101, EPI_ISL_2039125, EPI_ISL_2039126, EPI_ISL_2039131, EPI_ISL_2039135, EPI_ISL_2039141, EPI_ISL_2039143, EPI_ISL_2039152, EPI_ISL_2039154, EPI_ISL_2039155, EPI_ISL_2039393, EPI_ISL_2039395, EPI_ISL_2039397, EPI_ISL_2039445, EPI_ISL_2039481, EPI_ISL_2039484, EPI_ISL_2039487, EPI_ISL_2039503, EPI_ISL_2039507, EPI_ISL_2039521, EPI_ISL_2039522, EPI_ISL_2039523, EPI_ISL_2039564, EPI_ISL_2039577, EPI_ISL_2039579, EPI_ISL_2039580, EPI_ISL_2039592, EPI_ISL_2039632, EPI_ISL_2039700, EPI_ISL_2039710, EPI_ISL_2039711, EPI_ISL_2039715, EPI_ISL_2039719, EPI_ISL_2039734, EPI_ISL_2039736, EPI_ISL_2039737, EPI_ISL_2039765, EPI_ISL_2039818, EPI_ISL_2039824, EPI_ISL_2039827, EPI_ISL_2039880, EPI_ISL_2039893, EPI_ISL_2039954, EPI_ISL_2039988, EPI_ISL_2039989, EPI_ISL_2040082, EPI_ISL_2040090, EPI_ISL_2040187, EPI_ISL_2040189, EPI_ISL_2040190, EPI_ISL_2040191, EPI_ISL_2040254, EPI_ISL_2040265, EPI_ISL_2040317, EPI_ISL_2040325, EPI_ISL_2040326, EPI_ISL_2040328, EPI_ISL_2040331, EPI_ISL_2040332, EPI_ISL_2040333, EPI_ISL_2040352, EPI_ISL_2040357, EPI_ISL_2040361, EPI_ISL_2040410, EPI_ISL_2040424, EPI_ISL_2040429, EPI_ISL_2040477, EPI_ISL_2040478, EPI_ISL_2040555, EPI_ISL_2040573, EPI_ISL_2040581, EPI_ISL_2040592, EPI_ISL_2040607, EPI_ISL_2040614, EPI_ISL_2040615, EPI_ISL_2040616, EPI_ISL_2040617, EPI_ISL_2040634, EPI_ISL_2040636, EPI_ISL_2040640, EPI_ISL_2040657, EPI_ISL_2040746, EPI_ISL_2040747, EPI_ISL_2040778, EPI_ISL_2040812, EPI_ISL_2040829, EPI_ISL_2040905, EPI_ISL_2040906, EPI_ISL_2040942, EPI_ISL_2040943, EPI_ISL_2040949, EPI_ISL_2040950, EPI_ISL_2040955, EPI_ISL_2040996, EPI_ISL_2041016, EPI_ISL_2041038, EPI_ISL_2041070, EPI_ISL_2041075, EPI_ISL_2041096, EPI_ISL_2041104, EPI_ISL_2041125, EPI_ISL_2041209, EPI_ISL_2041215, EPI_ISL_2041216, EPI_ISL_2041217, EPI_ISL_2041218, EPI_ISL_2041234, EPI_ISL_2041235, EPI_ISL_2041239, EPI_ISL_2041240, EPI_ISL_2041247, EPI_ISL_2041285, EPI_ISL_2041286, EPI_ISL_2041309, EPI_ISL_2041327, EPI_ISL_2041347, EPI_ISL_2041403, EPI_ISL_2041416, EPI_ISL_2041418, EPI_ISL_2041484, EPI_ISL_2041500, EPI_ISL_2041544, EPI_ISL_2041554, EPI_ISL_2041556, EPI_ISL_2041557, EPI_ISL_2041558, EPI_ISL_2041560, EPI_ISL_2041614, EPI_ISL_2041621, EPI_ISL_2041674, EPI_ISL_2041675, EPI_ISL_2041676, EPI_ISL_2041678, EPI_ISL_2041704, EPI_ISL_2041751, EPI_ISL_2041821, EPI_ISL_2041838, EPI_ISL_2041841, EPI_ISL_2041873, EPI_ISL_2041874, EPI_ISL_2041895, EPI_ISL_2041955, EPI_ISL_2041959, EPI_ISL_2041960, EPI_ISL_2041961, EPI_ISL_2041962, EPI_ISL_2041964, EPI_ISL_2042042, EPI_ISL_2042053, EPI_ISL_2042057, EPI_ISL_2042065, EPI_ISL_2042074, EPI_ISL_2042199, EPI_ISL_2042258, EPI_ISL_2042292, EPI_ISL_2042293, EPI_ISL_2042298, EPI_ISL_2042299, EPI_ISL_2042315, EPI_ISL_2042346, EPI_ISL_2042351, EPI_ISL_2042352, EPI_ISL_2042353, EPI_ISL_2042370, EPI_ISL_2042459, EPI_ISL_2042494, EPI_ISL_2042502, EPI_ISL_2042505, EPI_ISL_2042517, EPI_ISL_2042530, EPI_ISL_2042541, EPI_ISL_2042603, EPI_ISL_2042626, EPI_ISL_2042650, EPI_ISL_2042661, EPI_ISL_2042662, EPI_ISL_2042663, EPI_ISL_2042664, EPI_ISL_2042665, EPI_ISL_2042669, EPI_ISL_2042677, EPI_ISL_2042685, EPI_ISL_2042709, EPI_ISL_2042710, EPI_ISL_2042712, EPI_ISL_2042717, EPI_ISL_2042718, EPI_ISL_2042751, EPI_ISL_2042756, EPI_ISL_2042771, EPI_ISL_2042790, EPI_ISL_2042795, EPI_ISL_2042830, EPI_ISL_2042832, EPI_ISL_2042845, EPI_ISL_2042860, EPI_ISL_2042868, EPI_ISL_2042892, EPI_ISL_2042907, EPI_ISL_2042909, EPI_ISL_2042934, EPI_ISL_2042950, EPI_ISL_2043014, EPI_ISL_2043020, EPI_ISL_2043032, EPI_ISL_2043036, EPI_ISL_2043037, EPI_ISL_2043043, EPI_ISL_2043044, EPI_ISL_2043045, EPI_ISL_2043054, EPI_ISL_2043073, EPI_ISL_2043130, EPI_ISL_2043133, EPI_ISL_2043134, EPI_ISL_2043146, EPI_ISL_2043156, EPI_ISL_2043167, EPI_ISL_2043170, EPI_ISL_2043181, EPI_ISL_2043190, EPI_ISL_2043196, EPI_ISL_2043228, EPI_ISL_2043235, EPI_ISL_2043238, EPI_ISL_2043293, EPI_ISL_2043296, EPI_ISL_2043304, EPI_ISL_2043328, EPI_ISL_2043329, EPI_ISL_2043330, EPI_ISL_2043390, EPI_ISL_2043399, EPI_ISL_2043400, EPI_ISL_2043403, EPI_ISL_2043416, EPI_ISL_2043426, EPI_ISL_2043427, EPI_ISL_2043435, EPI_ISL_2043443, EPI_ISL_2043445, EPI_ISL_2043448, EPI_ISL_2043460, EPI_ISL_2043463, EPI_ISL_2043488, EPI_ISL_2043493, EPI_ISL_2043504, EPI_ISL_2043528, EPI_ISL_2043534, EPI_ISL_2043549, EPI_ISL_2089613, EPI_ISL_2089660, EPI_ISL_2089680, EPI_ISL_2089686, EPI_ISL_2089693, EPI_ISL_2089694, EPI_ISL_2089707, EPI_ISL_2089709, EPI_ISL_2089710, EPI_ISL_2089785, EPI_ISL_2089818, EPI_ISL_2089829, EPI_ISL_2089842, EPI_ISL_2089896, EPI_ISL_2089911, EPI_ISL_2089913, EPI_ISL_2089924, EPI_ISL_2089930, EPI_ISL_2089934, EPI_ISL_2089951, EPI_ISL_2089953, EPI_ISL_2089956, EPI_ISL_2089959, EPI_ISL_2089972, EPI_ISL_2089994, EPI_ISL_2089995, EPI_ISL_2089997, EPI_ISL_2090045, EPI_ISL_2090103, EPI_ISL_2090105, EPI_ISL_2090141, EPI_ISL_2090142, EPI_ISL_2090143, EPI_ISL_2090154, EPI_ISL_2090157, EPI_ISL_2090158, EPI_ISL_2090160, EPI_ISL_2090169, EPI_ISL_2090183, EPI_ISL_2090225, EPI_ISL_2090268, EPI_ISL_2090269, EPI_ISL_2090272, EPI_ISL_2090273, EPI_ISL_2090274, EPI_ISL_2090275, EPI_ISL_2090311, EPI_ISL_2090323, EPI_ISL_2090421, EPI_ISL_2090428, EPI_ISL_2090430, EPI_ISL_2090431, EPI_ISL_2090434, EPI_ISL_2090436, EPI_ISL_2090437, EPI_ISL_2090438, EPI_ISL_2090443, EPI_ISL_2132537, EPI_ISL_2132553, EPI_ISL_2132567, EPI_ISL_2145804, EPI_ISL_2145814, EPI_ISL_2145825, EPI_ISL_2145832, EPI_ISL_2145836, EPI_ISL_2145839, EPI_ISL_2145864, EPI_ISL_2145882, EPI_ISL_2145888, EPI_ISL_2145889, EPI_ISL_2145893, EPI_ISL_2145928, EPI_ISL_2145942, EPI_ISL_2145986, EPI_ISL_2146001, EPI_ISL_2146011, EPI_ISL_2146091, EPI_ISL_2146094, EPI_ISL_2146095, EPI_ISL_2146100, EPI_ISL_2146169, EPI_ISL_2147080, EPI_ISL_2147081, EPI_ISL_2147797, EPI_ISL_2148007, EPI_ISL_2148024, EPI_ISL_2148028, EPI_ISL_2148029, EPI_ISL_2148045, EPI_ISL_2148048, EPI_ISL_2148058, EPI_ISL_2148120, EPI_ISL_2148144, EPI_ISL_2148145, EPI_ISL_2148148, EPI_ISL_2148149, EPI_ISL_2148151, EPI_ISL_2148152, EPI_ISL_2148153, EPI_ISL_2148164, EPI_ISL_2148165, EPI_ISL_2148166, EPI_ISL_2148167, EPI_ISL_2148168, EPI_ISL_2148169, EPI_ISL_2148275, EPI_ISL_2148283, EPI_ISL_2148287, EPI_ISL_2148297, EPI_ISL_2148298, EPI_ISL_2148304, EPI_ISL_2148403, EPI_ISL_2148404, EPI_ISL_2148406, EPI_ISL_2148409, EPI_ISL_2148410, EPI_ISL_2148412, EPI_ISL_2148413, EPI_ISL_2148415, EPI_ISL_2148457, EPI_ISL_2148465, EPI_ISL_2148470, EPI_ISL_2148551, EPI_ISL_2148572, EPI_ISL_2148579, EPI_ISL_2148600, EPI_ISL_2148604, EPI_ISL_2148610, EPI_ISL_2148617, EPI_ISL_2148638, EPI_ISL_2148646, EPI_ISL_2148671, EPI_ISL_2148700, EPI_ISL_2148701, EPI_ISL_2148717, EPI_ISL_2148721, EPI_ISL_2148732, EPI_ISL_2148735, EPI_ISL_2148736, EPI_ISL_2148840, EPI_ISL_2148877, EPI_ISL_2148889, EPI_ISL_2148894, EPI_ISL_2148909, EPI_ISL_2148910, EPI_ISL_2148925, EPI_ISL_2149147, EPI_ISL_2149151, EPI_ISL_2149153, EPI_ISL_2149155, EPI_ISL_2149165, EPI_ISL_2149166, EPI_ISL_2149167, EPI_ISL_2149172, EPI_ISL_2149173, EPI_ISL_2149174, EPI_ISL_2149183, EPI_ISL_2149185, EPI_ISL_2149221, EPI_ISL_2149222, EPI_ISL_2149223, EPI_ISL_2149224, EPI_ISL_2149225, EPI_ISL_2149226, EPI_ISL_2149227, EPI_ISL_2149228, EPI_ISL_2149231, EPI_ISL_2149256, EPI_ISL_2149257, EPI_ISL_2149258, EPI_ISL_2149262, EPI_ISL_2149296, EPI_ISL_2149302, EPI_ISL_2149306, EPI_ISL_2149327, EPI_ISL_2149328, EPI_ISL_2149329, EPI_ISL_2149330, EPI_ISL_2149334, EPI_ISL_2149379, EPI_ISL_2149395, EPI_ISL_2149399, EPI_ISL_2149420, EPI_ISL_2149423, EPI_ISL_2149427, EPI_ISL_2149428, EPI_ISL_2149441, EPI_ISL_2149448, EPI_ISL_2149474, EPI_ISL_2149475, EPI_ISL_2149531, EPI_ISL_2149549, EPI_ISL_2149558, EPI_ISL_2149559, EPI_ISL_2149567, EPI_ISL_2149568, EPI_ISL_2149569, EPI_ISL_2149605, EPI_ISL_2149611, EPI_ISL_2149628, EPI_ISL_2149629, EPI_ISL_2149634, EPI_ISL_2149654, EPI_ISL_2149658, EPI_ISL_2149659, EPI_ISL_2149666, EPI_ISL_2149668, EPI_ISL_2149672, EPI_ISL_2149677, EPI_ISL_2149691, EPI_ISL_2149733, EPI_ISL_2149745, EPI_ISL_2149772, EPI_ISL_2149774, EPI_ISL_2149829, EPI_ISL_2149856, EPI_ISL_2149858, EPI_ISL_2149886, EPI_ISL_2149897, EPI_ISL_2150038, EPI_ISL_2150071, EPI_ISL_2150121, EPI_ISL_2150221, EPI_ISL_2150222, EPI_ISL_2150235, EPI_ISL_2150251, EPI_ISL_2150296, EPI_ISL_2150309, EPI_ISL_2150326, EPI_ISL_2150328, EPI_ISL_2150342, EPI_ISL_2150343, EPI_ISL_2150344, EPI_ISL_2150354, EPI_ISL_2150357, EPI_ISL_2150377, EPI_ISL_2150384, EPI_ISL_2150385, EPI_ISL_2150386, EPI_ISL_2150405, EPI_ISL_2150415, EPI_ISL_2150432, EPI_ISL_2150446, EPI_ISL_2150452, EPI_ISL_2150462, EPI_ISL_2150463, EPI_ISL_2150480, EPI_ISL_2150513, EPI_ISL_2150514, EPI_ISL_2150547, EPI_ISL_2150548, EPI_ISL_2150556, EPI_ISL_2150663, EPI_ISL_2150707, EPI_ISL_2150718, EPI_ISL_2150730, EPI_ISL_2150744, EPI_ISL_2150819, EPI_ISL_2150937, EPI_ISL_2150962, EPI_ISL_2150965, EPI_ISL_2150966, EPI_ISL_2159021, EPI_ISL_2159214, EPI_ISL_2159223, EPI_ISL_2159225, EPI_ISL_2159226, EPI_ISL_2159248, EPI_ISL_2159261, EPI_ISL_2159272, EPI_ISL_2159273, EPI_ISL_2159275, EPI_ISL_2159327, EPI_ISL_2159328, EPI_ISL_2159340, EPI_ISL_2159364, EPI_ISL_2159377, EPI_ISL_2159378, EPI_ISL_2159427, EPI_ISL_2187362, EPI_ISL_2187419, EPI_ISL_2187448, EPI_ISL_2187481, EPI_ISL_2187500, EPI_ISL_2187526, EPI_ISL_2187531, EPI_ISL_2187539, EPI_ISL_2187579, EPI_ISL_2187589, EPI_ISL_2201148, EPI_ISL_2201229, EPI_ISL_2201246, EPI_ISL_2201295, EPI_ISL_2201438, EPI_ISL_2201442, EPI_ISL_2201543, EPI_ISL_2201545, EPI_ISL_2201546, EPI_ISL_2201765, EPI_ISL_2201830, EPI_ISL_2201831, EPI_ISL_2201833, EPI_ISL_2201878, EPI_ISL_2201880, EPI_ISL_2201884, EPI_ISL_2201893, EPI_ISL_2201897, EPI_ISL_2201904, EPI_ISL_2201905, EPI_ISL_2201912, EPI_ISL_2201914, EPI_ISL_2201942, EPI_ISL_2201944, EPI_ISL_2201965, EPI_ISL_2201973, EPI_ISL_2202013, EPI_ISL_2202074, EPI_ISL_2202131, EPI_ISL_2202134, EPI_ISL_2202161, EPI_ISL_2202213, EPI_ISL_2202227, EPI_ISL_2202258, EPI_ISL_2202430, EPI_ISL_2202441, EPI_ISL_2202444, EPI_ISL_2202448, EPI_ISL_2202455, EPI_ISL_2202593, EPI_ISL_2202632, EPI_ISL_2202637, EPI_ISL_2202639, EPI_ISL_2202653, EPI_ISL_2202760, EPI_ISL_2202769, EPI_ISL_2202806, EPI_ISL_2202975, EPI_ISL_2202983, EPI_ISL_2203005, EPI_ISL_2203031, EPI_ISL_2203058, EPI_ISL_2203065, EPI_ISL_2203109, EPI_ISL_2203118, EPI_ISL_2203123, EPI_ISL_2203133, EPI_ISL_2203145, EPI_ISL_2203164, EPI_ISL_2203181, EPI_ISL_2203195, EPI_ISL_2203198, EPI_ISL_2203215, EPI_ISL_2203240, EPI_ISL_2268019, EPI_ISL_2268026, EPI_ISL_2268045, EPI_ISL_2268076, EPI_ISL_2268115, EPI_ISL_2370290, EPI_ISL_2372447, EPI_ISL_2421823, EPI_ISL_2421824, EPI_ISL_2421827, EPI_ISL_2421848, EPI_ISL_2421886, EPI_ISL_2421926, EPI_ISL_2421931, EPI_ISL_2421934, EPI_ISL_2421939, EPI_ISL_2489827, EPI_ISL_2489828, EPI_ISL_2489831, EPI_ISL_2489832, EPI_ISL_2489835, EPI_ISL_2489841, EPI_ISL_2489842, EPI_ISL_2489843, EPI_ISL_2489851, EPI_ISL_2489872, EPI_ISL_2489879, EPI_ISL_2531085, EPI_ISL_2600190, EPI_ISL_2600227, EPI_ISL_2600230, EPI_ISL_2600232, EPI_ISL_2600234, EPI_ISL_2600242, EPI_ISL_2600258, EPI_ISL_2600266, EPI_ISL_2600267, EPI_ISL_2600269, EPI_ISL_2600270, EPI_ISL_2600284, EPI_ISL_2600300 | | | |
| see above | Aegis Sciences Corporation | Centers for Disease Control and Prevention Division of Viral Diseases, Pathogen Discovery | Adrian Paskey; Alec Vest; Benjamin Rambo-Martin; Christopher Gulvick; Clinton R. Paden; Cyndi Clark; Dakota Howard; Darlene Wagner; Dhwani Batra; Dillon Nall; Duncan MacCannell; Ethan Sanders; Holly Houdeshell; Jason Caravas; Kara Moser; Matthew Hardison; Matthew Schmerer; Ola Kvalvaag; Patrick Campbell; Peter W. Cook; Rob Case; Scott Sammons; Shatavia Morrison; Shaun Westlund; Vikramsinha Ghorpade; Yvette Unoarumhi |
| EPI_ISL_2304310, EPI_ISL_2304311, EPI_ISL_2304315, EPI_ISL_2304316, EPI_ISL_2304319, EPI_ISL_2304320, EPI_ISL_2304321, EPI_ISL_2304325, EPI_ISL_2304326, EPI_ISL_2304341, EPI_ISL_2304342, EPI_ISL_2304343, EPI_ISL_2304344 | | | |
| see above | Alameda County Public Health Department | Alameda County Public Health Department | Kevin Libuit; Kristina Hsieh; Kristna Hsieh; Maria Elma |
| EPI_ISL_2102999, EPI_ISL_2103001, EPI_ISL_2103002, EPI_ISL_2103018, EPI_ISL_2103019, EPI_ISL_2103020, EPI_ISL_2103034, EPI_ISL_2103038, EPI_ISL_2103046, EPI_ISL_2103051, EPI_ISL_2103064, EPI_ISL_2103071, EPI_ISL_2103074, EPI_ISL_2103088, EPI_ISL_2103093, EPI_ISL_2103107, EPI_ISL_2103119, EPI_ISL_2103129, EPI_ISL_2103132, EPI_ISL_2103137, EPI_ISL_2103140, EPI_ISL_2103147, EPI_ISL_2103151, EPI_ISL_2103162, EPI_ISL_2103165, EPI_ISL_2103167, EPI_ISL_2103192, EPI_ISL_2103194, EPI_ISL_2341416, EPI_ISL_2341417, EPI_ISL_2341418, EPI_ISL_2341419, EPI_ISL_2341420, EPI_ISL_2341422, EPI_ISL_2341425, EPI_ISL_2341426, EPI_ISL_2341428, EPI_ISL_2341430, EPI_ISL_2341433, EPI_ISL_2341434, EPI_ISL_2341435, EPI_ISL_2341440, EPI_ISL_2341441, EPI_ISL_2341443, EPI_ISL_2341445, EPI_ISL_2341446, EPI_ISL_2341453, EPI_ISL_2341461, EPI_ISL_2341462, EPI_ISL_2341464, EPI_ISL_2341465, EPI_ISL_2341466, EPI_ISL_2341467, EPI_ISL_2341470, EPI_ISL_2341474, EPI_ISL_2341475, EPI_ISL_2341476, EPI_ISL_2341477, EPI_ISL_2341479, EPI_ISL_2341483, EPI_ISL_2341484, EPI_ISL_2341485, EPI_ISL_2341486, EPI_ISL_2341491, EPI_ISL_2341492, EPI_ISL_2341502, EPI_ISL_2341503, EPI_ISL_2341504, EPI_ISL_2341506, EPI_ISL_2341507, EPI_ISL_2341510, EPI_ISL_2341513, EPI_ISL_2341514, EPI_ISL_2341519, EPI_ISL_2341521, EPI_ISL_2341522, EPI_ISL_2341523, EPI_ISL_2341527, EPI_ISL_2341528, EPI_ISL_2341529, EPI_ISL_2341530, EPI_ISL_2341532, EPI_ISL_2341540, EPI_ISL_2341545, EPI_ISL_2341546, EPI_ISL_2341548, EPI_ISL_2341549, EPI_ISL_2341550, EPI_ISL_2341552, EPI_ISL_2341559, EPI_ISL_2341562, EPI_ISL_2341566, EPI_ISL_2341568, EPI_ISL_2341569, EPI_ISL_2341571, EPI_ISL_2341573, EPI_ISL_2341574, EPI_ISL_2341575, EPI_ISL_2341576, EPI_ISL_2341578, EPI_ISL_2341579, EPI_ISL_2341580, EPI_ISL_2341581, EPI_ISL_2341582, EPI_ISL_2341583, EPI_ISL_2341585, EPI_ISL_2341587, EPI_ISL_2341588, EPI_ISL_2341591, EPI_ISL_2341594, EPI_ISL_2341596, EPI_ISL_2341598, EPI_ISL_2341601, EPI_ISL_2341604, EPI_ISL_2341607, EPI_ISL_2341609, EPI_ISL_2341617, EPI_ISL_2341618, EPI_ISL_2341619, EPI_ISL_2341620, EPI_ISL_2341626, EPI_ISL_2341631, EPI_ISL_2341635, EPI_ISL_2341639, EPI_ISL_2341644, EPI_ISL_2341646, EPI_ISL_2341647, EPI_ISL_2341648, EPI_ISL_2341651, EPI_ISL_2341652, EPI_ISL_2341653, EPI_ISL_2341658, EPI_ISL_2341660, EPI_ISL_2500024, EPI_ISL_2500026, EPI_ISL_2500028, EPI_ISL_2500030, EPI_ISL_2500035, EPI_ISL_2500036, EPI_ISL_2500037, EPI_ISL_2500038, EPI_ISL_2500039, EPI_ISL_2500042, EPI_ISL_2500044, EPI_ISL_2500047, EPI_ISL_2500050, EPI_ISL_2500051, EPI_ISL_2500052, EPI_ISL_2500057, EPI_ISL_2500062, EPI_ISL_2500067, EPI_ISL_2500068, EPI_ISL_2500070, EPI_ISL_2500072, EPI_ISL_2500073, EPI_ISL_2500075, EPI_ISL_2500079, EPI_ISL_2500080, EPI_ISL_2500081, EPI_ISL_2500082, EPI_ISL_2500083, EPI_ISL_2500084, EPI_ISL_2500086, EPI_ISL_2500089, EPI_ISL_2500090, EPI_ISL_2500092, EPI_ISL_2500093, EPI_ISL_2500094, EPI_ISL_2500096, EPI_ISL_2500097, EPI_ISL_2500098, EPI_ISL_2500099, EPI_ISL_2500102, EPI_ISL_2500106, EPI_ISL_2500110, EPI_ISL_2500111, EPI_ISL_2500112, EPI_ISL_2500113, EPI_ISL_2500118, EPI_ISL_2500120, EPI_ISL_2500121, EPI_ISL_2500126, EPI_ISL_2500135, EPI_ISL_2500137, EPI_ISL_2500138, EPI_ISL_2500139, EPI_ISL_2500140, EPI_ISL_2500143, EPI_ISL_2500144, EPI_ISL_2500145, EPI_ISL_2500146, EPI_ISL_2500147, EPI_ISL_2500149, EPI_ISL_2500152, EPI_ISL_2500153, EPI_ISL_2500158, EPI_ISL_2500160, EPI_ISL_2500163, EPI_ISL_2500164, EPI_ISL_2500166, EPI_ISL_2500168, EPI_ISL_2500169, EPI_ISL_2500170, EPI_ISL_2500174, EPI_ISL_2500177, EPI_ISL_2500178, EPI_ISL_2500179, EPI_ISL_2500180, EPI_ISL_2500181, EPI_ISL_2500184, EPI_ISL_2500186, EPI_ISL_2500190, EPI_ISL_2500193, EPI_ISL_2500194, EPI_ISL_2500195, EPI_ISL_2500203, EPI_ISL_2500204, EPI_ISL_2500205, EPI_ISL_2500208, EPI_ISL_2500209, EPI_ISL_2500210, EPI_ISL_2500212, EPI_ISL_2500216, EPI_ISL_2658581, EPI_ISL_2658597, EPI_ISL_2658680, EPI_ISL_2658729, EPI_ISL_2659231, EPI_ISL_2659322, EPI_ISL_2659361, EPI_ISL_2659452, EPI_ISL_2659477, EPI_ISL_2659586, EPI_ISL_2659608, EPI_ISL_2758521, EPI_ISL_2758522, EPI_ISL_2758523, EPI_ISL_2758524, EPI_ISL_2758525, EPI_ISL_2758526, EPI_ISL_2758527, EPI_ISL_2758528, EPI_ISL_2758529, EPI_ISL_2758530 | | | |
| see above | CA DPH Viral and Rickettsial Disease Laboratory | Chan-Zuckerberg Biohub | CZB Cliahub Consortium |
| EPI_ISL_1937812, EPI_ISL_1937814, EPI_ISL_1937815, EPI_ISL_1937816, EPI_ISL_1937817, EPI_ISL_1937818, EPI_ISL_1937819, EPI_ISL_1937821, EPI_ISL_1937822, EPI_ISL_1937824, EPI_ISL_1937826, EPI_ISL_1937828, EPI_ISL_2105107, EPI_ISL_2105108, EPI_ISL_2105109, EPI_ISL_2105128, EPI_ISL_2105134, EPI_ISL_2105135, EPI_ISL_2105158, EPI_ISL_2229193, EPI_ISL_2248730 | | | |
| see above | CA-Los Angeles County Public Health Laboratory | Centers for Disease Control and Prevention Division of Viral Diseases, Pathogen Discovery | Alison Laufer Halpin; Ben L. Rambo-Martin; Clinton R. Paden; Dakota Howard; Darlene Wagner; Dave Wentworth; Dhwani Batra; Jasmine Padilla; Justin Lee; Katie Dillon; Krista Queen; Kristen Knipe; Kristine Lacek; Mark Burroughs; Matthew Schmerer; Mili Sheth; Peter Cook; Sam Shepard; Sarah Nobles; Shoshona Le; Suxiang Tong; Vivien Dugan; Yvette Unoarumhi |
| EPI_ISL_2825810, EPI_ISL_2825811, EPI_ISL_2825812, EPI_ISL_2825813, EPI_ISL_2825814, EPI_ISL_2825815, EPI_ISL_2825816, EPI_ISL_2825817, EPI_ISL_2825818, EPI_ISL_2825819, EPI_ISL_2825820, EPI_ISL_2825821, EPI_ISL_2825822, EPI_ISL_2825823, EPI_ISL_2825824, EPI_ISL_2825825, EPI_ISL_2825826, EPI_ISL_2825827, EPI_ISL_2825828, EPI_ISL_2825829, EPI_ISL_2825830, EPI_ISL_2825831, EPI_ISL_2825832, EPI_ISL_2825833, EPI_ISL_2825834, EPI_ISL_2825835, EPI_ISL_2825836, EPI_ISL_2825837, EPI_ISL_2825838, EPI_ISL_2825839, EPI_ISL_2825840, EPI_ISL_2825841, EPI_ISL_2825842, EPI_ISL_2825843, EPI_ISL_2825844, EPI_ISL_2825845, EPI_ISL_2825846, EPI_ISL_2825847, EPI_ISL_2825848, EPI_ISL_2825849, EPI_ISL_2825850, EPI_ISL_2825851, EPI_ISL_2825852, EPI_ISL_2825853, EPI_ISL_2825854, EPI_ISL_2825855, EPI_ISL_2825856, EPI_ISL_2825857, EPI_ISL_2825858, EPI_ISL_2825859, EPI_ISL_2825860, EPI_ISL_2825861, EPI_ISL_2825862, EPI_ISL_2825863, EPI_ISL_2825864, EPI_ISL_2825865, EPI_ISL_2825866, EPI_ISL_2825867, EPI_ISL_2825868, EPI_ISL_2825869, EPI_ISL_2825870, EPI_ISL_2825871, EPI_ISL_2825872, EPI_ISL_2825873, EPI_ISL_2825874, EPI_ISL_2825875, EPI_ISL_2825876, EPI_ISL_2825877, EPI_ISL_2825878, EPI_ISL_2825879, EPI_ISL_2825880, EPI_ISL_2825881, EPI_ISL_2825882, EPI_ISL_2825883, EPI_ISL_2825884, EPI_ISL_2825885, EPI_ISL_2825886, EPI_ISL_2825887, EPI_ISL_2825888, EPI_ISL_2825889, EPI_ISL_2825890, EPI_ISL_2825891, EPI_ISL_2825892, EPI_ISL_2825893, EPI_ISL_2825894, EPI_ISL_2825895, EPI_ISL_2825896, EPI_ISL_2825897, EPI_ISL_2825898, EPI_ISL_2825899, EPI_ISL_2825900, EPI_ISL_2825901, EPI_ISL_2825902, EPI_ISL_2825903, EPI_ISL_2825904, EPI_ISL_2825905, EPI_ISL_2825906, EPI_ISL_2825907, EPI_ISL_2825908, EPI_ISL_2825909, EPI_ISL_2825910, EPI_ISL_2825911, EPI_ISL_2825912, EPI_ISL_2825913, EPI_ISL_2825914, EPI_ISL_2825915, EPI_ISL_2825916, EPI_ISL_2825917, EPI_ISL_2825918, EPI_ISL_2825919, EPI_ISL_2825920, EPI_ISL_2825921, EPI_ISL_2825922, EPI_ISL_2825923, EPI_ISL_2825924, EPI_ISL_2825925, EPI_ISL_2825926, EPI_ISL_2825927, EPI_ISL_2825928, EPI_ISL_2825929, EPI_ISL_2825930, EPI_ISL_2825931, EPI_ISL_2825932, EPI_ISL_2825933, EPI_ISL_2825934, EPI_ISL_2825935, EPI_ISL_2825936, EPI_ISL_2825937, EPI_ISL_2825938, EPI_ISL_2825939, EPI_ISL_2825940, EPI_ISL_2825941, EPI_ISL_2825942, EPI_ISL_2825943, EPI_ISL_2825944, EPI_ISL_2825945, EPI_ISL_2825946, EPI_ISL_2825947, EPI_ISL_2825948, EPI_ISL_2825949, EPI_ISL_2825950, EPI_ISL_2825951, EPI_ISL_2825952, EPI_ISL_2825953, EPI_ISL_2825954, EPI_ISL_2825955, EPI_ISL_2825956, EPI_ISL_2825957, EPI_ISL_2825958, EPI_ISL_2825959, EPI_ISL_2825960, EPI_ISL_2825961, EPI_ISL_2825962, EPI_ISL_2825963, EPI_ISL_2825964, EPI_ISL_2825965, EPI_ISL_2825966, EPI_ISL_2825967, EPI_ISL_2825968, EPI_ISL_2825969, EPI_ISL_2825970, EPI_ISL_2825971, EPI_ISL_2825972, EPI_ISL_2825973, EPI_ISL_2825974, EPI_ISL_2825975, EPI_ISL_2825976, EPI_ISL_2825977, EPI_ISL_2825978, EPI_ISL_2825979, EPI_ISL_2825980, EPI_ISL_2825981, EPI_ISL_2825982, EPI_ISL_2825983, EPI_ISL_2825984, EPI_ISL_2825985, EPI_ISL_2825986, EPI_ISL_2825987, EPI_ISL_2825988, EPI_ISL_2825989, EPI_ISL_2825990, EPI_ISL_2825991, EPI_ISL_2825992, EPI_ISL_2825993, EPI_ISL_2825994, EPI_ISL_2825995, EPI_ISL_2825996, EPI_ISL_2825997, EPI_ISL_2825998, EPI_ISL_2825999, EPI_ISL_2826000, EPI_ISL_2826001, EPI_ISL_2826002, EPI_ISL_2826003, EPI_ISL_2826004, EPI_ISL_2826005, EPI_ISL_2826006, EPI_ISL_2826007, EPI_ISL_2826008, EPI_ISL_2826009, EPI_ISL_2826010, EPI_ISL_2826011, EPI_ISL_2826012, EPI_ISL_2826013, EPI_ISL_2826014, EPI_ISL_2826015, EPI_ISL_2826016, EPI_ISL_2826017, EPI_ISL_2826018, EPI_ISL_2826019, EPI_ISL_2826020, EPI_ISL_2826021, EPI_ISL_2826022, EPI_ISL_2826023, EPI_ISL_2826024, EPI_ISL_2858280, EPI_ISL_2858282, EPI_ISL_2858283, EPI_ISL_2858286, EPI_ISL_2858287, EPI_ISL_2858289, EPI_ISL_2858291, EPI_ISL_2858292, EPI_ISL_2858297, EPI_ISL_2858298, EPI_ISL_2858299, EPI_ISL_2858300, EPI_ISL_2858301, EPI_ISL_2858302, EPI_ISL_2858311, EPI_ISL_2858312, EPI_ISL_2858313, EPI_ISL_2858314, EPI_ISL_2858315, EPI_ISL_2858316, EPI_ISL_2858317, EPI_ISL_2858318, EPI_ISL_2858319, EPI_ISL_2858320, EPI_ISL_2858321, EPI_ISL_2858323, EPI_ISL_2858324, EPI_ISL_2858325, EPI_ISL_2858326, EPI_ISL_2858327, EPI_ISL_2858330, EPI_ISL_2858331, EPI_ISL_2858339, EPI_ISL_2858343, EPI_ISL_2858344, EPI_ISL_2858348, EPI_ISL_2858352, EPI_ISL_2858354, EPI_ISL_2858355, EPI_ISL_2858356, EPI_ISL_2858361, EPI_ISL_2858362, EPI_ISL_2858363, EPI_ISL_2858364, EPI_ISL_2858365, EPI_ISL_2858367, EPI_ISL_2858368, EPI_ISL_2858369, EPI_ISL_2858371, EPI_ISL_2858373, EPI_ISL_2858375, EPI_ISL_2858377, EPI_ISL_2858381, EPI_ISL_2858382, EPI_ISL_2858383, EPI_ISL_2858390, EPI_ISL_2858391, EPI_ISL_2858393, EPI_ISL_2858395, EPI_ISL_2858396, EPI_ISL_2858397, EPI_ISL_2858398, EPI_ISL_2858399, EPI_ISL_2858400, EPI_ISL_2858405, EPI_ISL_2858406, EPI_ISL_2858408, EPI_ISL_2858409, EPI_ISL_2858410, EPI_ISL_2858411, EPI_ISL_2858412, | | | |

EPI_ISL_2858413, EPI_ISL_2858414, EPI_ISL_2858415, EPI_ISL_2858416, EPI_ISL_2858417, EPI_ISL_2858418, EPI_ISL_2858419, EPI_ISL_2858431, EPI_ISL_2858432, EPI_ISL_2858434, EPI_ISL_2858437, EPI_ISL_2858438, EPI_ISL_2858439, EPI_ISL_2858441, EPI_ISL_2858443, EPI_ISL_2858449, EPI_ISL_2858450, EPI_ISL_2858451, EPI_ISL_2858452, EPI_ISL_2858453, EPI_ISL_2858454, EPI_ISL_2858455, EPI_ISL_2858456, EPI_ISL_2858469, EPI_ISL_2858471, EPI_ISL_2858472, EPI_ISL_2858486, EPI_ISL_2858487, EPI_ISL_2858488, EPI_ISL_2858489, EPI_ISL_2858496, EPI_ISL_2858501, EPI_ISL_2858507, EPI_ISL_2858509, EPI_ISL_2858510, EPI_ISL_2858511, EPI_ISL_2858512, EPI_ISL_2858513, EPI_ISL_2858514, EPI_ISL_2858515, EPI_ISL_2858516, EPI_ISL_2858517, EPI_ISL_2858518, EPI_ISL_2858519, EPI_ISL_2858520, EPI_ISL_2858532, EPI_ISL_2858533, EPI_ISL_2858563, EPI_ISL_2858564, EPI_ISL_2858581, EPI_ISL_2858585, EPI_ISL_2882063, EPI_ISL_2882089, EPI_ISL_2882090, EPI_ISL_2882100, EPI_ISL_2882113, EPI_ISL_2882128, EPI_ISL_2882129, EPI_ISL_2882130, EPI_ISL_2882136, EPI_ISL_2882138, EPI_ISL_2882147, EPI_ISL_2882148, EPI_ISL_2882155, EPI_ISL_2882156, EPI_ISL_2882157, EPI_ISL_2882158, EPI_ISL_2882159, EPI_ISL_2882160, EPI_ISL_2882161, EPI_ISL_2882162, EPI_ISL_2882163, EPI_ISL_2882164, EPI_ISL_2882165, EPI_ISL_2882166, EPI_ISL_2882167, EPI_ISL_2882168, EPI_ISL_2882169, EPI_ISL_2882170, EPI_ISL_2882171, EPI_ISL_2882177, EPI_ISL_2882178, EPI_ISL_2882189, EPI_ISL_2882193, EPI_ISL_2882204, EPI_ISL_2882205, EPI_ISL_2882206, EPI_ISL_2882245, EPI_ISL_2882247, EPI_ISL_2882269, EPI_ISL_2882270, EPI_ISL_2882271, EPI_ISL_2882272, EPI_ISL_2882273, EPI_ISL_2882274, EPI_ISL_2882275, EPI_ISL_2882278, EPI_ISL_2882281, EPI_ISL_2882282, EPI_ISL_2882291, EPI_ISL_2882292, EPI_ISL_2882297, EPI_ISL_2882299, EPI_ISL_2882300, EPI_ISL_2882301, EPI_ISL_2882302, EPI_ISL_2882315, EPI_ISL_2882316, EPI_ISL_2882319, EPI_ISL_2882327, EPI_ISL_2882329, EPI_ISL_2882330, EPI_ISL_2882331, EPI_ISL_2882343, EPI_ISL_2882344, EPI_ISL_2882345, EPI_ISL_2882362, EPI_ISL_2882363, EPI_ISL_2882364, EPI_ISL_2882365, EPI_ISL_2882375, EPI_ISL_2882376, EPI_ISL_2882377, EPI_ISL_2882378, EPI_ISL_2882379, EPI_ISL_2882390, EPI_ISL_2882391, EPI_ISL_2882416, EPI_ISL_2882417, EPI_ISL_2882418, EPI_ISL_2882428, EPI_ISL_2882431, EPI_ISL_2882432, EPI_ISL_2882440, EPI_ISL_2882442, EPI_ISL_2882443, EPI_ISL_2882454, EPI_ISL_2882455, EPI_ISL_2882475, EPI_ISL_2882476, EPI_ISL_2882477, EPI_ISL_2882481, EPI_ISL_2882482, EPI_ISL_2882483, EPI_ISL_2882484, EPI_ISL_2882485, EPI_ISL_2882486, EPI_ISL_2882487, EPI_ISL_2882488, EPI_ISL_2882489, EPI_ISL_2882491, EPI_ISL_2882530, EPI_ISL_2882536, EPI_ISL_2882537, EPI_ISL_2882538, EPI_ISL_2882540, EPI_ISL_2882541, EPI_ISL_2882545, EPI_ISL_2882546, EPI_ISL_2882584, EPI_ISL_2882585, EPI_ISL_2882587, EPI_ISL_2882588, EPI_ISL_2882589, EPI_ISL_2882590, EPI_ISL_2882591, EPI_ISL_2882592, EPI_ISL_2882593, EPI_ISL_2882595, EPI_ISL_2882625, EPI_ISL_2882627, EPI_ISL_2882628, EPI_ISL_2882632, EPI_ISL_2882635, EPI_ISL_2882636, EPI_ISL_2882637, EPI_ISL_2882638, EPI_ISL_2882642, EPI_ISL_2882648, EPI_ISL_2882649, EPI_ISL_2882655, EPI_ISL_2882688, EPI_ISL_2882689, EPI_ISL_2882704, EPI_ISL_2882705, EPI_ISL_2882706, EPI_ISL_2882707, EPI_ISL_2882718, EPI_ISL_2882733, EPI_ISL_2882736, EPI_ISL_2882737, EPI_ISL_2882739, EPI_ISL_2882745, EPI_ISL_2882746, EPI_ISL_2882747, EPI_ISL_2882750, EPI_ISL_2882751, EPI_ISL_2882758, EPI_ISL_2882760, EPI_ISL_2882761, EPI_ISL_2882762, EPI_ISL_2882763, EPI_ISL_2882765, EPI_ISL_2882766, EPI_ISL_2882772, EPI_ISL_2882775, EPI_ISL_2882776, EPI_ISL_2882777, EPI_ISL_2882778, EPI_ISL_2882781, EPI_ISL_2882789, EPI_ISL_2882791, EPI_ISL_2882792, EPI_ISL_2882793, EPI_ISL_2882796, EPI_ISL_2882798, EPI_ISL_2882799, EPI_ISL_2882800, EPI_ISL_2882801, EPI_ISL_2882802, EPI_ISL_2882806, EPI_ISL_2882810, EPI_ISL_2882811, EPI_ISL_2882812, EPI_ISL_2882813, EPI_ISL_2882814, EPI_ISL_2882815, EPI_ISL_2882816, EPI_ISL_2882823, EPI_ISL_2882824, EPI_ISL_2882827, EPI_ISL_2882828, EPI_ISL_2882829, EPI_ISL_2882830, EPI_ISL_2882832, EPI_ISL_2882833, EPI_ISL_2882834, EPI_ISL_2882835, EPI_ISL_2882836, EPI_ISL_2882837, EPI_ISL_2882845, EPI_ISL_2882849, EPI_ISL_2882855, EPI_ISL_2882856, EPI_ISL_2882861, EPI_ISL_2882862, EPI_ISL_2882863, EPI_ISL_2882864, EPI_ISL_2882865, EPI_ISL_2882866, EPI_ISL_2882868, EPI_ISL_2882875, EPI_ISL_2882877, EPI_ISL_2882878, EPI_ISL_2882879, EPI_ISL_2882882, EPI_ISL_2882883, EPI_ISL_2882886, EPI_ISL_2882888, EPI_ISL_2882890, EPI_ISL_2882891, EPI_ISL_2882893, EPI_ISL_2882895, EPI_ISL_2882897, EPI_ISL_2882898, EPI_ISL_2882899, EPI_ISL_2882900, EPI_ISL_2882905, EPI_ISL_2882907, EPI_ISL_2882908, EPI_ISL_2882909, EPI_ISL_2882910, EPI_ISL_2882911, EPI_ISL_2882916, EPI_ISL_2882917, EPI_ISL_2882919, EPI_ISL_2882921, EPI_ISL_2882934, EPI_ISL_2882942, EPI_ISL_2882943, EPI_ISL_2882948, EPI_ISL_2882949, EPI_ISL_2882950, EPI_ISL_2882952, EPI_ISL_2882953, EPI_ISL_2882966, EPI_ISL_2882967, EPI_ISL_2882968, EPI_ISL_2882969, EPI_ISL_2882971, EPI_ISL_2882972, EPI_ISL_2882973, EPI_ISL_2882975, EPI_ISL_2882976, EPI_ISL_2882977, EPI_ISL_2882978, EPI_ISL_2882980, EPI_ISL_2882982, EPI_ISL_2882983, EPI_ISL_2882990, EPI_ISL_2882993, EPI_ISL_2882994, EPI_ISL_2882995, EPI_ISL_2882996, EPI_ISL_2882997, EPI_ISL_2882998, EPI_ISL_2883000, EPI_ISL_2883001, EPI_ISL_2883005, EPI_ISL_2883008, EPI_ISL_2883009, EPI_ISL_2883015, EPI_ISL_2883019, EPI_ISL_2883021, EPI_ISL_2883022, EPI_ISL_2883026, EPI_ISL_2883027, EPI_ISL_2883028, EPI_ISL_2883036, EPI_ISL_2883037, EPI_ISL_2883038, EPI_ISL_2883039, EPI_ISL_2883042, EPI_ISL_2883046, EPI_ISL_2883056, EPI_ISL_2883057, EPI_ISL_2883058, EPI_ISL_2883059, EPI_ISL_2883060, EPI_ISL_2883063, EPI_ISL_2883066, EPI_ISL_2883067, EPI_ISL_2883072, EPI_ISL_2883073, EPI_ISL_2889855, EPI_ISL_2889934, EPI_ISL_2889935, EPI_ISL_2889947, EPI_ISL_2889948, EPI_ISL_2889962, EPI_ISL_2890051, EPI_ISL_2890084, EPI_ISL_2890093, EPI_ISL_2890094, EPI_ISL_2890144, EPI_ISL_2890202, EPI_ISL_2890203, EPI_ISL_2890204, EPI_ISL_2890225, EPI_ISL_2890279, EPI_ISL_2890355, EPI_ISL_2890356, EPI_ISL_2890357, EPI_ISL_2890358, EPI_ISL_2890359, EPI_ISL_2890360, EPI_ISL_2890361, EPI_ISL_2890362, EPI_ISL_2890364, EPI_ISL_2890432, EPI_ISL_2890434, EPI_ISL_2890437, EPI_ISL_2890438, EPI_ISL_2890439, EPI_ISL_2890494, EPI_ISL_2890502, EPI_ISL_2890506, EPI_ISL_2890507, EPI_ISL_2890508, EPI_ISL_2890509, EPI_ISL_2890510, EPI_ISL_2890512, EPI_ISL_2890570, EPI_ISL_2890572, EPI_ISL_2890573, EPI_ISL_2890580, EPI_ISL_2890581, EPI_ISL_2890644, EPI_ISL_2890659, EPI_ISL_2890712, EPI_ISL_2890731, EPI_ISL_2890732, EPI_ISL_2890753, EPI_ISL_2890784, EPI_ISL_2890893, EPI_ISL_2890894, EPI_ISL_2890895, EPI_ISL_2890896, EPI_ISL_2890897, EPI_ISL_2890898, EPI_ISL_2890929, EPI_ISL_2890985, EPI_ISL_2891026, EPI_ISL_2891028, EPI_ISL_2891029, EPI_ISL_2891030, EPI_ISL_2891031, EPI_ISL_2891143, EPI_ISL_2891185, EPI_ISL_2891190, EPI_ISL_2891191, EPI_ISL_2891192, EPI_ISL_2891224, EPI_ISL_2891225, EPI_ISL_2891246, EPI_ISL_2891247, EPI_ISL_2891248, EPI_ISL_2891249, EPI_ISL_2891250, EPI_ISL_2891251, EPI_ISL_2891295, EPI_ISL_2891296, EPI_ISL_2891362, EPI_ISL_2891403, EPI_ISL_2891423, EPI_ISL_2891424, EPI_ISL_2891425, EPI_ISL_2891426, EPI_ISL_2891427, EPI_ISL_2891466, EPI_ISL_2891546, EPI_ISL_2891547, EPI_ISL_2891548, EPI_ISL_2891549, EPI_ISL_2891551, EPI_ISL_2891565, EPI_ISL_2891572, EPI_ISL_2891583, EPI_ISL_2891600, EPI_ISL_2891602, EPI_ISL_2891603, EPI_ISL_2891604, EPI_ISL_2891616, EPI_ISL_2891617, EPI_ISL_2891624, EPI_ISL_2891625, EPI_ISL_2891626, EPI_ISL_2891627, EPI_ISL_2891628, EPI_ISL_2891631, EPI_ISL_2891694, EPI_ISL_2891707, EPI_ISL_2891718, EPI_ISL_2891719, EPI_ISL_2891720, EPI_ISL_2891721, EPI_ISL_2891728, EPI_ISL_2891729, EPI_ISL_2891741, EPI_ISL_2891742, EPI_ISL_2891744, EPI_ISL_2891750, EPI_ISL_2891751, EPI_ISL_2891752, EPI_ISL_2891755, EPI_ISL_2891756, EPI_ISL_2891758, EPI_ISL_2891759, EPI_ISL_2891765, EPI_ISL_2891766, EPI_ISL_2891768, EPI_ISL_2891781, EPI_ISL_2891802, EPI_ISL_2891857, EPI_ISL_2891858, EPI_ISL_2891860, EPI_ISL_2891871, EPI_ISL_2891872, EPI_ISL_2891873, EPI_ISL_2891874, EPI_ISL_2891875, EPI_ISL_2891877, EPI_ISL_2891878, EPI_ISL_2891879, EPI_ISL_2891880, EPI_ISL_2891885, EPI_ISL_2891886, EPI_ISL_2891913, EPI_ISL_2891954, EPI_ISL_2891955, EPI_ISL_2891957, EPI_ISL_2891970, EPI_ISL_2891971, EPI_ISL_2891972, EPI_ISL_2891973, EPI_ISL_2891979, EPI_ISL_2891980, EPI_ISL_2891981, EPI_ISL_2891982, EPI_ISL_2891986, EPI_ISL_2891987, EPI_ISL_2891988, EPI_ISL_2891990, EPI_ISL_2891993, EPI_ISL_2891997, EPI_ISL_2891998, EPI_ISL_2891999, EPI_ISL_2892000, EPI_ISL_2892001, EPI_ISL_2892015, EPI_ISL_2892017, EPI_ISL_2892019, EPI_ISL_2892020, EPI_ISL_2892040, EPI_ISL_2892041, EPI_ISL_2892054, EPI_ISL_2892057, EPI_ISL_2892058, EPI_ISL_2892061, EPI_ISL_2892069, EPI_ISL_2892071, EPI_ISL_2892072, EPI_ISL_2892082, EPI_ISL_2892091, EPI_ISL_2892092, EPI_ISL_2892104, EPI_ISL_2892106, EPI_ISL_2892109, EPI_ISL_2892118, EPI_ISL_2892122, EPI_ISL_2892125, EPI_ISL_2892126, EPI_ISL_2892129, EPI_ISL_2892132, EPI_ISL_2892135, EPI_ISL_2892136, EPI_ISL_2892137, EPI_ISL_2892138, EPI_ISL_2892139, EPI_ISL_2892151, EPI_ISL_2892157, EPI_ISL_2892158, EPI_ISL_2892167, EPI_ISL_2892168, EPI_ISL_2892169, EPI_ISL_2892172, EPI_ISL_2892173, EPI_ISL_2892179, EPI_ISL_2892188, EPI_ISL_2892194, EPI_ISL_2892203, EPI_ISL_2892210, EPI_ISL_2892212, EPI_ISL_2892214, EPI_ISL_2892218, EPI_ISL_2892219, EPI_ISL_2892220, EPI_ISL_2892222, EPI_ISL_2892223, EPI_ISL_2892226, EPI_ISL_2892227, EPI_ISL_2892228, EPI_ISL_2892229, EPI_ISL_2892230, EPI_ISL_2892233, EPI_ISL_2892234, EPI_ISL_2892235, EPI_ISL_2892236, EPI_ISL_2892237, EPI_ISL_2892238, EPI_ISL_2892239, EPI_ISL_2892240, EPI_ISL_2892245, EPI_ISL_2892246, EPI_ISL_2892247, EPI_ISL_2892250, EPI_ISL_2892252, EPI_ISL_2892258, EPI_ISL_2892260, EPI_ISL_2892261, EPI_ISL_2892264, EPI_ISL_2892272, EPI_ISL_2892273, EPI_ISL_2892278, EPI_ISL_2892279, EPI_ISL_2892280, EPI_ISL_2892281, EPI_ISL_2892282, EPI_ISL_2892286, EPI_ISL_2892287, EPI_ISL_2892288, EPI_ISL_2892289, EPI_ISL_2892292, EPI_ISL_2892308, EPI_ISL_2892309, EPI_ISL_2892310, EPI_ISL_2892321, EPI_ISL_2892322, EPI_ISL_2892324, EPI_ISL_2892325, EPI_ISL_2892328, EPI_ISL_2892329, EPI_ISL_2892330, EPI_ISL_2892332, EPI_ISL_2892337, EPI_ISL_2892338, EPI_ISL_2892339, EPI_ISL_2892340, EPI_ISL_2892341, EPI_ISL_2892342, EPI_ISL_2892343, EPI_ISL_2892352, EPI_ISL_2892357, EPI_ISL_2892358, EPI_ISL_2892359, EPI_ISL_2892360, EPI_ISL_2892374, EPI_ISL_2892375, EPI_ISL_2892393, EPI_ISL_2921664, EPI_ISL_2921667, EPI_ISL_2921675, EPI_ISL_2921682, EPI_ISL_2921683, EPI_ISL_2921684, EPI_ISL_2921687, EPI_ISL_2921702, EPI_ISL_2921703, EPI_ISL_2921719, EPI_ISL_2921725, EPI_ISL_2921736, EPI_ISL_2921737, EPI_ISL_2921762, EPI_ISL_2921763, EPI_ISL_2921764, EPI_ISL_2921765, EPI_ISL_2921766, EPI_ISL_2921767, EPI_ISL_2921768, EPI_ISL_2921769, EPI_ISL_2921770, EPI_ISL_2921773, EPI_ISL_2921774, EPI_ISL_2921775, EPI_ISL_2921776, EPI_ISL_2921805, EPI_ISL_2921812, EPI_ISL_2921823, EPI_ISL_2921824, EPI_ISL_2921827, EPI_ISL_2921828, EPI_ISL_2921837, EPI_ISL_2921838, EPI_ISL_2921839, EPI_ISL_2921840, EPI_ISL_2921841, EPI_ISL_2921846, EPI_ISL_2921848, EPI_ISL_2921849, EPI_ISL_2921850, EPI_ISL_2921851, EPI_ISL_2921857, EPI_ISL_2921858, EPI_ISL_2921859, EPI_ISL_2921860, EPI_ISL_2921862, EPI_ISL_2921863, EPI_ISL_2921868, EPI_ISL_2921870, EPI_ISL_2921874, EPI_ISL_2921875, EPI_ISL_2921876, EPI_ISL_2921877, EPI_ISL_2921878, EPI_ISL_2921879, EPI_ISL_2921880, EPI_ISL_2921881, EPI_ISL_2921883, EPI_ISL_2921884, EPI_ISL_2921886, EPI_ISL_2921889, EPI_ISL_2921890, EPI_ISL_2921891, EPI_ISL_2921892, EPI_ISL_2921906, EPI_ISL_2921908, EPI_ISL_2921911, EPI_ISL_2921912, EPI_ISL_2921913, EPI_ISL_2921915, EPI_ISL_2921916, EPI_ISL_2921917, EPI_ISL_2921918, EPI_ISL_2921919, EPI_ISL_2921920, EPI_ISL_2921922, EPI_ISL_2921923, EPI_ISL_2921924, EPI_ISL_2921925, EPI_ISL_2921926, EPI_ISL_2921934, EPI_ISL_2921938, EPI_ISL_2921940, EPI_ISL_2921949, EPI_ISL_2921950, EPI_ISL_2921951, EPI_ISL_2921952, EPI_ISL_2921953, EPI_ISL_2921954, EPI_ISL_2921955, EPI_ISL_2921956, EPI_ISL_2921964, EPI_ISL_2921967, EPI_ISL_2921968, EPI_ISL_2921970, EPI_ISL_2921974, EPI_ISL_2921976, EPI_ISL_2921977, EPI_ISL_2921978, EPI_ISL_2921979, EPI_ISL_2921983, EPI_ISL_2921984, EPI_ISL_2921985, EPI_ISL_2921987, EPI_ISL_2921992, EPI_ISL_2921993, EPI_ISL_2921994, EPI_ISL_2922002, EPI_ISL_2922008, EPI_ISL_2922009, EPI_ISL_2922013, EPI_ISL_2922014, EPI_ISL_2922015, EPI_ISL_2922016, EPI_ISL_2922017, EPI_ISL_2922019, EPI_ISL_2922020, EPI_ISL_2922022, EPI_ISL_2922023, EPI_ISL_2922031, EPI_ISL_2922039, EPI_ISL_2922040, EPI_ISL_2922041, EPI_ISL_2922042, EPI_ISL_2922050, EPI_ISL_2922052, EPI_ISL_2922062, EPI_ISL_2922063, EPI_ISL_2922064, EPI_ISL_2922070, EPI_ISL_2922078, EPI_ISL_2922080, EPI_ISL_2922082, EPI_ISL_2922083, EPI_ISL_2922084, EPI_ISL_2922087, EPI_ISL_2922088, EPI_ISL_2922090, EPI_ISL_2922091, EPI_ISL_2922105, EPI_ISL_2922106, EPI_ISL_2922112, EPI_ISL_2922116, EPI_ISL_2922117, EPI_ISL_2922120, EPI_ISL_2922121, EPI_ISL_2922122, EPI_ISL_2922124, EPI_ISL_2922126, EPI_ISL_2922129, EPI_ISL_2922130, EPI_ISL_2922132, EPI_ISL_2922134, EPI_ISL_2922137, EPI_ISL_2922138, EPI_ISL_2922139, EPI_ISL_2922148, EPI_ISL_2922149, EPI_ISL_2922150, EPI_ISL_2922154, EPI_ISL_2922158, EPI_ISL_2922165, EPI_ISL_2922168, EPI_ISL_2922172, EPI_ISL_2922173, EPI_ISL_2922174, EPI_ISL_2922175, EPI_ISL_2922176, EPI_ISL_2922177, EPI_ISL_2922178, EPI_ISL_2922179, EPI_ISL_2922180, EPI_ISL_2922185, EPI_ISL_2922186, EPI_ISL_2922187, EPI_ISL_2922188, EPI_ISL_2922191, EPI_ISL_2922194, EPI_ISL_2922195, EPI_ISL_2922199, EPI_ISL_2922200, EPI_ISL_2922201, EPI_ISL_2922202, EPI_ISL_2922203, EPI_ISL_2922222, EPI_ISL_2922223, EPI_ISL_2922224, EPI_ISL_2922244, EPI_ISL_2922245, EPI_ISL_2922246, EPI_ISL_2922248, EPI_ISL_2922252, EPI_ISL_2922253, EPI_ISL_2922254, EPI_ISL_2922255, EPI_ISL_2922256, EPI_ISL_2922257, EPI_ISL_2922270, EPI_ISL_2922271, EPI_ISL_2922272, EPI_ISL_2922274, EPI_ISL_2922275, EPI_ISL_2922276, EPI_ISL_2922277, EPI_ISL_2922278, EPI_ISL_2922279, EPI_ISL_2922280, EPI_ISL_2922290, EPI_ISL_2922291, EPI_ISL_2922292, EPI_ISL_2922305, EPI_ISL_2922311, EPI_ISL_2922312, EPI_ISL_2922313, EPI_ISL_2923287, EPI_ISL_2923296, EPI_ISL_2923315, EPI_ISL_2923330, EPI_ISL_2923331, EPI_ISL_2923338, EPI_ISL_2923344, EPI_ISL_2923346, EPI_ISL_2923347, EPI_ISL_2923348, EPI_ISL_2923349, EPI_ISL_2923350, EPI_ISL_2923354, EPI_ISL_2923366, EPI_ISL_2923378, EPI_ISL_2923392, EPI_ISL_2923393, EPI_ISL_2923394, EPI_ISL_2923402, EPI_ISL_2923403, EPI_ISL_2923404, EPI_ISL_2923443, EPI_ISL_2923444, EPI_ISL_2923451, EPI_ISL_2923465, EPI_ISL_2923478, EPI_ISL_2923480, EPI_ISL_2923481, EPI_ISL_2923482, EPI_ISL_2923483, EPI_ISL_2923484, EPI_ISL_2923489, EPI_ISL_2923497, EPI_ISL_2923498, EPI_ISL_2923499, EPI_ISL_2923500, EPI_ISL_2923508, EPI_ISL_2923509, EPI_ISL_2923510, EPI_ISL_2923516, EPI_ISL_2923524, EPI_ISL_2923532, EPI_ISL_2923533, EPI_ISL_2923534, EPI_ISL_2923540, EPI_ISL_2923547, EPI_ISL_2923548, EPI_ISL_2923549, EPI_ISL_2923550, EPI_ISL_2923551, EPI_ISL_2923552, EPI_ISL_2923582, EPI_ISL_2923607, EPI_ISL_2923608, EPI_ISL_2923609, EPI_ISL_2923610, EPI_ISL_2923611, EPI_ISL_2923612, EPI_ISL_2923616, EPI_ISL_2923619, EPI_ISL_2923622, EPI_ISL_2923625, EPI_ISL_2923642, EPI_ISL_2923648, EPI_ISL_2923649, EPI_ISL_2923650, EPI_ISL_2923651, EPI_ISL_2923655, EPI_ISL_2923656, EPI_ISL_2923657, EPI_ISL_2923658, EPI_ISL_2923669, EPI_ISL_2923743, EPI_ISL_2923766, EPI_ISL_2976932, EPI_ISL_2976933, EPI_ISL_2976934, EPI_ISL_2976935, EPI_ISL_2976936, EPI_ISL_2976937, EPI_ISL_2976938, EPI_ISL_2976939, EPI_ISL_2976940, EPI_ISL_2976941, EPI_ISL_2976942, EPI_ISL_2976943, EPI_ISL_2976944, EPI_ISL_2976945, EPI_ISL_2976946, EPI_ISL_2976947, EPI_ISL_2976948, EPI_ISL_2976949, EPI_ISL_2976950, EPI_ISL_2976951, EPI_ISL_2976952, EPI_ISL_2976953, EPI_ISL_2976954, EPI_ISL_2976955, EPI_ISL_2976956, EPI_ISL_2976957, EPI_ISL_2976958, EPI_ISL_2976959, EPI_ISL_2976960, EPI_ISL_2976961, EPI_ISL_2976962, EPI_ISL_2976963, EPI_ISL_2976964, EPI_ISL_2976965, EPI_ISL_2976966, EPI_ISL_2976967, EPI_ISL_2976968, EPI_ISL_2976969, EPI_ISL_2976970, EPI_ISL_2976971, EPI_ISL_2976972, EPI_ISL_2976973, EPI_ISL_2976974, EPI_ISL_2976975, EPI_ISL_2976976, EPI_ISL_2976977, EPI_ISL_2976978, EPI_ISL_2976979, EPI_ISL_2976980, EPI_ISL_2976981, EPI_ISL_2976982, EPI_ISL_2976983, EPI_ISL_2976984, EPI_ISL_2976985, EPI_ISL_2976986, EPI_ISL_2976987, EPI_ISL_2976988, EPI_ISL_2976989, EPI_ISL_2976990, EPI_ISL_2976991, EPI_ISL_2976992, EPI_ISL_2976993, EPI_ISL_2976994, EPI_ISL_2976995, EPI_ISL_2976996, EPI_ISL_2976997, EPI_ISL_2976998, EPI_ISL_2976999, EPI_ISL_2977000, EPI_ISL_2977001, EPI_ISL_2977002, EPI_ISL_2977003, EPI_ISL_2977004, EPI_ISL_2977005, EPI_ISL_2977006, EPI_ISL_2977007, EPI_ISL_2977008, EPI_ISL_2977009, EPI_ISL_2977010, EPI_ISL_2977011, EPI_ISL_2977012, EPI_ISL_2977013, EPI_ISL_2977014, EPI_ISL_2977015, EPI_ISL_2977016, EPI_ISL_2977017, EPI_ISL_2977018, EPI_ISL_2977019, EPI_ISL_2977020, EPI_ISL_2977021, EPI_ISL_2977022, EPI_ISL_2977023, EPI_ISL_2977024, EPI_ISL_2977025, EPI_ISL_2977026, EPI_ISL_2977027, EPI_ISL_2977028, EPI_ISL_2977029, EPI_ISL_2977030, EPI_ISL_2977031, EPI_ISL_2977032, EPI_ISL_2977033, EPI_ISL_2977034, EPI_ISL_2977035, EPI_ISL_2977036, EPI_ISL_2977037, EPI_ISL_2977038, EPI_ISL_2977039, EPI_ISL_2977040, EPI_ISL_2977041, EPI_ISL_2977042, EPI_ISL_2977043, EPI_ISL_2977044, EPI_ISL_2977045, EPI_ISL_2977046, EPI_ISL_2977047, EPI_ISL_2977048, EPI_ISL_2977049, EPI_ISL_2977050, EPI_ISL_2977051, EPI_ISL_2977052, EPI_ISL_2977053, EPI_ISL_2977054, EPI_ISL_2977055, EPI_ISL_2977056, EPI_ISL_2977057, EPI_ISL_2977058, EPI_ISL_2977059, EPI_ISL_2977060, EPI_ISL_2977061, EPI_ISL_2977062, EPI_ISL_2977063, EPI_ISL_2977064, EPI_ISL_2977065, EPI_ISL_2977066, EPI_ISL_2977067, EPI_ISL_2977068, EPI_ISL_2977069, EPI_ISL_2977070, EPI_ISL_2977071, EPI_ISL_2977072, EPI_ISL_2977073, EPI_ISL_2977074, EPI_ISL_2977075, EPI_ISL_2977076, EPI_ISL_2977077, EPI_ISL_2977078, EPI_ISL_2977079, EPI_ISL_2977080, EPI_ISL_2977081, EPI_ISL_2977082, EPI_ISL_2977083, EPI_ISL_2977084, EPI_ISL_2977085, EPI_ISL_2977086, EPI_ISL_2977087, EPI_ISL_2977088, EPI_ISL_2977089, EPI_ISL_2977090, EPI_ISL_2977091, EPI_ISL_2977092, EPI_ISL_2977093, EPI_ISL_2977094, EPI_ISL_2977095, EPI_ISL_2977099, EPI_ISL_2977100, EPI_ISL_2977101, EPI_ISL_2977102, EPI_ISL_2977106, EPI_ISL_2977107, EPI_ISL_2977108, EPI_ISL_2977109, EPI_ISL_2977114, EPI_ISL_2977115, EPI_ISL_2977116, EPI_ISL_2977117, EPI_ISL_2977121, EPI_ISL_2977122, EPI_ISL_2977123, EPI_ISL_2977124, EPI_ISL_2977127, EPI_ISL_2977128, EPI_ISL_2977129, EPI_ISL_2977130, EPI_ISL_2977131, EPI_ISL_2977135, EPI_ISL_2977136, EPI_ISL_2977137, EPI_ISL_2977138, EPI_ISL_2977139, EPI_ISL_2977143, EPI_ISL_2977144, EPI_ISL_2977145, EPI_ISL_2977148, EPI_ISL_2977149, EPI_ISL_2977150, EPI_ISL_2977155, EPI_ISL_2977156, EPI_ISL_2977157, EPI_ISL_2977158, EPI_ISL_2977163, EPI_ISL_2977164, EPI_ISL_2977165, EPI_ISL_2977166, EPI_ISL_2977170, EPI_ISL_2977171, EPI_ISL_2977172, EPI_ISL_2977173, EPI_ISL_2978147, EPI_ISL_2978148, EPI_ISL_2978149, EPI_ISL_2978150, EPI_ISL_2978151, EPI_ISL_2978152, EPI_ISL_2978153, EPI_ISL_2978154, EPI_ISL_2978155, EPI_ISL_2978156, EPI_ISL_2978158, EPI_ISL_2978159, EPI_ISL_2978161, EPI_ISL_2978163

| | | | |
|---|---|---|---|
| see above | CDPH VBL | California Department of Public Health | CDPH-COVIDNet; UCLA Technology Center for Genomics & Bioinformatics; UCSD EXCITE lab |

EPI_ISL_2018310, EPI_ISL_2018311, EPI_ISL_2018312, EPI_ISL_2018313, EPI_ISL_2018314, EPI_ISL_2018315, EPI_ISL_2018316, EPI_ISL_2018317, EPI_ISL_2018318, EPI_ISL_2018322, EPI_ISL_2018323, EPI_ISL_2018324, EPI_ISL_2452033

| | | | |
|---|---|---|---|
| see above | CDPH, Viral and Rickettsial Disease Laboratory | Centers for Disease Control and Prevention Division of Viral Diseases, Pathogen Discovery | Alison Laufer Halpin; Ben L. Rambo-Martin; Clinton R. Paden; Dakota Howard; Darlene Wagner; Dave Wentworth; Dhwani Batra; Jasmine Padilla; Justin Lee; Katie Dillon; Krista Queen; Kristen Knipe; Kristine Lacek; Mark Burroughs; Matthew Schmerer; Mili Sheth; Peter Cook; Sam Shepard; Sarah Nobles; Shoshona Le; Suxiang Tong; Vivien Dugan; Yvette Unoarumhi |
| EPI_ISL_2228100 | CDPH, Viral and Rickettsial Disease Laboratory | Pathogen Discovery, Respiratory Viruses Branch, Division of Viral Diseases, Centers for Disease Control and Prevention | Adam Retchless; Anna Kelleher; Anna Uehara; Brian Lynch; Clinton R. Paden; Dhwani Batra; Haibin Wang; Han Jia Justin Ng; Jasmine Padilla; Jing Zhang; Justin Lee; Krista Queen; Mark Burroughs; Mili Sheth; Morgan Davis; Peter Cook; Rachel Marine; Sarah Nobles; Suxiang Tong; Tara Coalter; Yan Li; Ying Tao |
| EPI_ISL_1924630 | CHILDRENS HOSPITAL OF NORCAL | Santa Clara County public Health Laboratory | Santa Clara County Public Health Department |

EPI_ISL_2537427, EPI_ISL_2537430, EPI_ISL_2537431, EPI_ISL_2537432, EPI_ISL_2537433, EPI_ISL_2537434, EPI_ISL_2537440, EPI_ISL_2537441, EPI_ISL_2537442, EPI_ISL_2537443, EPI_ISL_2537444, EPI_ISL_2537447, EPI_ISL_2537451, EPI_ISL_2537452, EPI_ISL_2537453, EPI_ISL_2537454, EPI_ISL_2537457, EPI_ISL_2537458, EPI_ISL_2537459, EPI_ISL_2537460, EPI_ISL_2537461, EPI_ISL_2537462, EPI_ISL_2537463, EPI_ISL_2537464, EPI_ISL_2537466, EPI_ISL_2537467, EPI_ISL_2537468, EPI_ISL_2537469, EPI_ISL_2537471, EPI_ISL_2537472, EPI_ISL_2537473, EPI_ISL_2537474, EPI_ISL_2537475, EPI_ISL_2537476, EPI_ISL_2537477, EPI_ISL_2537478, EPI_ISL_2537479, EPI_ISL_2842806, EPI_ISL_2842807, EPI_ISL_2842808, EPI_ISL_2842809, EPI_ISL_2842810, EPI_ISL_2842817, EPI_ISL_2842819, EPI_ISL_2842820, EPI_ISL_2842821, EPI_ISL_2842822, EPI_ISL_2842823, EPI_ISL_2842824, EPI_ISL_2842825, EPI_ISL_2842826, EPI_ISL_2842827, EPI_ISL_2842828, EPI_ISL_2842829, EPI_ISL_2842830, EPI_ISL_2842831, EPI_ISL_2842833, EPI_ISL_2842834, EPI_ISL_2842835, EPI_ISL_2842836, EPI_ISL_2892403, EPI_ISL_2892404, EPI_ISL_2892412, EPI_ISL_2892414, EPI_ISL_2892420, EPI_ISL_2892421, EPI_ISL_2892422, EPI_ISL_2892423, EPI_ISL_2892424, EPI_ISL_2892425, EPI_ISL_2892426, EPI_ISL_2892427, EPI_ISL_2892428, EPI_ISL_2892429, EPI_ISL_2892430, EPI_ISL_2892431, EPI_ISL_2892432, EPI_ISL_2892433, EPI_ISL_2892434, EPI_ISL_2892435, EPI_ISL_2892437, EPI_ISL_2892438, EPI_ISL_2892456, EPI_ISL_2892457, EPI_ISL_2892496, EPI_ISL_2892497, EPI_ISL_2892498, EPI_ISL_2892499, EPI_ISL_2892500, EPI_ISL_2892501, EPI_ISL_2892502, EPI_ISL_2892506, EPI_ISL_2892511, EPI_ISL_2892524, EPI_ISL_2892530, EPI_ISL_2892539, EPI_ISL_2892548, EPI_ISL_2892549, EPI_ISL_2892570, EPI_ISL_2892571, EPI_ISL_2892572, EPI_ISL_2892573, EPI_ISL_2892574, EPI_ISL_2892575, EPI_ISL_2892577, EPI_ISL_2892587, EPI_ISL_2892588, EPI_ISL_2892589, EPI_ISL_2892590, EPI_ISL_2892591, EPI_ISL_2892592

| | | | |
|---|---|---|---|
| see above | California Department of Public Health | California Department of Public Health | CDPH IDLB COVIDNet et al |

EPI_ISL_2778748, EPI_ISL_2778749, EPI_ISL_2778750, EPI_ISL_2778751, EPI_ISL_2778752, EPI_ISL_2778753, EPI_ISL_2778754, EPI_ISL_2778755, EPI_ISL_2778756, EPI_ISL_2778757, EPI_ISL_2778758, EPI_ISL_2778759, EPI_ISL_2778760, EPI_ISL_2778761, EPI_ISL_2778762, EPI_ISL_2778763, EPI_ISL_2778764, EPI_ISL_2778765, EPI_ISL_2778766, EPI_ISL_2778767, EPI_ISL_2778768, EPI_ISL_2778769, EPI_ISL_2778770, EPI_ISL_2778771, EPI_ISL_2778772, EPI_ISL_2778773, EPI_ISL_2778774, EPI_ISL_2778775, EPI_ISL_2778776, EPI_ISL_2778777, EPI_ISL_2778778, EPI_ISL_2778779, EPI_ISL_2778780, EPI_ISL_2778781, EPI_ISL_2778782, EPI_ISL_2778783, EPI_ISL_2778784, EPI_ISL_2778785, EPI_ISL_2778786, EPI_ISL_2778787, EPI_ISL_2778788, EPI_ISL_2778789, EPI_ISL_2778790, EPI_ISL_2778791, EPI_ISL_2778792, EPI_ISL_2778793, EPI_ISL_2778794, EPI_ISL_2778795, EPI_ISL_2778796, EPI_ISL_2778797, EPI_ISL_2778798, EPI_ISL_2778799, EPI_ISL_2778800, EPI_ISL_2778801, EPI_ISL_2778802, EPI_ISL_2778803, EPI_ISL_2778804, EPI_ISL_2778805, EPI_ISL_2778806, EPI_ISL_2778807, EPI_ISL_2778808, EPI_ISL_2778809, EPI_ISL_2778810, EPI_ISL_2778811, EPI_ISL_2778812, EPI_ISL_2778813, EPI_ISL_2778814, EPI_ISL_2778815, EPI_ISL_2778816, EPI_ISL_2778817, EPI_ISL_2778818, EPI_ISL_2778819, EPI_ISL_2778820, EPI_ISL_2778821, EPI_ISL_2778822, EPI_ISL_2778823, EPI_ISL_2778824, EPI_ISL_2778825, EPI_ISL_2778826, EPI_ISL_2778827, EPI_ISL_2778828, EPI_ISL_2778829, EPI_ISL_2778830, EPI_ISL_2778831, EPI_ISL_2778832, EPI_ISL_2778833, EPI_ISL_2778834, EPI_ISL_2778835, EPI_ISL_2778836, EPI_ISL_2778837, EPI_ISL_2778838, EPI_ISL_2778839, EPI_ISL_2778840, EPI_ISL_2778841, EPI_ISL_2778842, EPI_ISL_2778843, EPI_ISL_2778844, EPI_ISL_2778845, EPI_ISL_2778847, EPI_ISL_2778848, EPI_ISL_2778849, EPI_ISL_2778850, EPI_ISL_2778851, EPI_ISL_2778852, EPI_ISL_2778853, EPI_ISL_2778854, EPI_ISL_2778855, EPI_ISL_2778856, EPI_ISL_2778857, EPI_ISL_2778858, EPI_ISL_2778859, EPI_ISL_2778860, EPI_ISL_2778861, EPI_ISL_2778862, EPI_ISL_2778863, EPI_ISL_2778864, EPI_ISL_2778865, EPI_ISL_2778866, EPI_ISL_2778867, EPI_ISL_2778868, EPI_ISL_2778869, EPI_ISL_2778870, EPI_ISL_2778871, EPI_ISL_2778872, EPI_ISL_2778873, EPI_ISL_2778874, EPI_ISL_2778875, EPI_ISL_2778876, EPI_ISL_2778877, EPI_ISL_2778878, EPI_ISL_2778879, EPI_ISL_2778880, EPI_ISL_2778881, EPI_ISL_2778882, EPI_ISL_2778883, EPI_ISL_2778884, EPI_ISL_2778885, EPI_ISL_2778886, EPI_ISL_2778887, EPI_ISL_2778888, EPI_ISL_2778889, EPI_ISL_2778890, EPI_ISL_2778891, EPI_ISL_2778892, EPI_ISL_2778893, EPI_ISL_2778894, EPI_ISL_2778895, EPI_ISL_2778896, EPI_ISL_2778897, EPI_ISL_2778898, EPI_ISL_2778899, EPI_ISL_2778900, EPI_ISL_2778901, EPI_ISL_2778902, EPI_ISL_2778903, EPI_ISL_2778904, EPI_ISL_2778905, EPI_ISL_2778906, EPI_ISL_2778907, EPI_ISL_2778908, EPI_ISL_2778909, EPI_ISL_2778910, EPI_ISL_2778911, EPI_ISL_2778912, EPI_ISL_2778913, EPI_ISL_2778914, EPI_ISL_2778915, EPI_ISL_2778916, EPI_ISL_2778917, EPI_ISL_2778918, EPI_ISL_2778919, EPI_ISL_2778920, EPI_ISL_2778921, EPI_ISL_2778922, EPI_ISL_2778923, EPI_ISL_2778924, EPI_ISL_2778925, EPI_ISL_2778926, EPI_ISL_2778927, EPI_ISL_2778928, EPI_ISL_2778929, EPI_ISL_2778930, EPI_ISL_2778931, EPI_ISL_2778932, EPI_ISL_2778933, EPI_ISL_2778934, EPI_ISL_2778935, EPI_ISL_2778936, EPI_ISL_2778937, EPI_ISL_2778938, EPI_ISL_2778939, EPI_ISL_2778940, EPI_ISL_2778941, EPI_ISL_2778942, EPI_ISL_2778943, EPI_ISL_2778944, EPI_ISL_2778945, EPI_ISL_2778946, EPI_ISL_2778947, EPI_ISL_2778948, EPI_ISL_2778949, EPI_ISL_2778950, EPI_ISL_2778951, EPI_ISL_2778952, EPI_ISL_2778953, EPI_ISL_2778954, EPI_ISL_2837315, EPI_ISL_2837316, EPI_ISL_2837317, EPI_ISL_2837318, EPI_ISL_2837319, EPI_ISL_2837320, EPI_ISL_2837321, EPI_ISL_2837322, EPI_ISL_2837325, EPI_ISL_2837326, EPI_ISL_2837327, EPI_ISL_2837329, EPI_ISL_2837330, EPI_ISL_2837331, EPI_ISL_2837334, EPI_ISL_2837335, EPI_ISL_2837336, EPI_ISL_2837337, EPI_ISL_2837338, EPI_ISL_2837339, EPI_ISL_2837340, EPI_ISL_2837341, EPI_ISL_2837342, EPI_ISL_2837343, EPI_ISL_2837344, EPI_ISL_2837345, EPI_ISL_2837346, EPI_ISL_2837347, EPI_ISL_2837348, EPI_ISL_2837349, EPI_ISL_2837350, EPI_ISL_2837351, EPI_ISL_2837352, EPI_ISL_2837353, EPI_ISL_2837354, EPI_ISL_2837356, EPI_ISL_2837358, EPI_ISL_2837359, EPI_ISL_2837360, EPI_ISL_2837361, EPI_ISL_2837363, EPI_ISL_2837371, EPI_ISL_2837372, EPI_ISL_2837373, EPI_ISL_2837374, EPI_ISL_2837375, EPI_ISL_2837376, EPI_ISL_2837377, EPI_ISL_2837378, EPI_ISL_2837379, EPI_ISL_2837381, EPI_ISL_2837382, EPI_ISL_2837383, EPI_ISL_2837384, EPI_ISL_2837385, EPI_ISL_2837386, EPI_ISL_2837387, EPI_ISL_2837388, EPI_ISL_2837390, EPI_ISL_2837392, EPI_ISL_2837393, EPI_ISL_2837394, EPI_ISL_2837395, EPI_ISL_2837396, EPI_ISL_2837397, EPI_ISL_2837398, EPI_ISL_2837400, EPI_ISL_2837401, EPI_ISL_2837402, EPI_ISL_2837403, EPI_ISL_2837404, EPI_ISL_2837410, EPI_ISL_2837411, EPI_ISL_2837412, EPI_ISL_2837413, EPI_ISL_2837414, EPI_ISL_2837415, EPI_ISL_2837416, EPI_ISL_2837417, EPI_ISL_2837421, EPI_ISL_2837423, EPI_ISL_2837430, EPI_ISL_2837432, EPI_ISL_2837433, EPI_ISL_2837434, EPI_ISL_2837439, EPI_ISL_2837440, EPI_ISL_2837441, EPI_ISL_2837442, EPI_ISL_2837443, EPI_ISL_2837444, EPI_ISL_2837446, EPI_ISL_2837447, EPI_ISL_2837448, EPI_ISL_2837449, EPI_ISL_2837450, EPI_ISL_2837451, EPI_ISL_2837452, EPI_ISL_2837453, EPI_ISL_2837454, EPI_ISL_2837455, EPI_ISL_2837456, EPI_ISL_2837457, EPI_ISL_2837458, EPI_ISL_2837459, EPI_ISL_2837462, EPI_ISL_2837463, EPI_ISL_2837464, EPI_ISL_2837466, EPI_ISL_2837467, EPI_ISL_2837477, EPI_ISL_2837478, EPI_ISL_2837479, EPI_ISL_2837480, EPI_ISL_2837481, EPI_ISL_2837482, EPI_ISL_2837483, EPI_ISL_2837485, EPI_ISL_2837486, EPI_ISL_2837487, EPI_ISL_2837488, EPI_ISL_2837489, EPI_ISL_2837490, EPI_ISL_2837491, EPI_ISL_2837492, EPI_ISL_2837493, EPI_ISL_2837494, EPI_ISL_2837495, EPI_ISL_2837496, EPI_ISL_2837497, EPI_ISL_2837498, EPI_ISL_2837499, EPI_ISL_2837500, EPI_ISL_2837501, EPI_ISL_2837502, EPI_ISL_2837503, EPI_ISL_2837504, EPI_ISL_2837505, EPI_ISL_2837506, EPI_ISL_2837509, EPI_ISL_2837514, EPI_ISL_2837515, EPI_ISL_2837516, EPI_ISL_2837517, EPI_ISL_2837518, EPI_ISL_2837519, EPI_ISL_2837520, EPI_ISL_2837521, EPI_ISL_2837522, EPI_ISL_2837523, EPI_ISL_2837526, EPI_ISL_2837527, EPI_ISL_2837528, EPI_ISL_2837529, EPI_ISL_2837530, EPI_ISL_2837531, EPI_ISL_2837532, EPI_ISL_2837533, EPI_ISL_2837534, EPI_ISL_2837538, EPI_ISL_2837540, EPI_ISL_2837541, EPI_ISL_2837542, EPI_ISL_2837544, EPI_ISL_2837547, EPI_ISL_2837548, EPI_ISL_2837549, EPI_ISL_2837550, EPI_ISL_2837551, EPI_ISL_2837552, EPI_ISL_2837553, EPI_ISL_2837554, EPI_ISL_2837555, EPI_ISL_2837556, EPI_ISL_2837557, EPI_ISL_2837558, EPI_ISL_2837559, EPI_ISL_2837560, EPI_ISL_2837561, EPI_ISL_2837563, EPI_ISL_2837564, EPI_ISL_2837568, EPI_ISL_2837572, EPI_ISL_2837574, EPI_ISL_2837575, EPI_ISL_2837576, EPI_ISL_2837577, EPI_ISL_2837578, EPI_ISL_2837579, EPI_ISL_2837584, EPI_ISL_2837585, EPI_ISL_2837587, EPI_ISL_2837588, EPI_ISL_2837589, EPI_ISL_2837590, EPI_ISL_2837591, EPI_ISL_2837592, EPI_ISL_2837594, EPI_ISL_2837595, EPI_ISL_2837598, EPI_ISL_2837599, EPI_ISL_2837600, EPI_ISL_2837602, EPI_ISL_2837604, EPI_ISL_2837605, EPI_ISL_2837606, EPI_ISL_2837607, EPI_ISL_2837608, EPI_ISL_2837609, EPI_ISL_2837610, EPI_ISL_2837611, EPI_ISL_2837612, EPI_ISL_2837613, EPI_ISL_2837614, EPI_ISL_2837615, EPI_ISL_2837616, EPI_ISL_2837617, EPI_ISL_2837619, EPI_ISL_2837620, EPI_ISL_2837621, EPI_ISL_2837622, EPI_ISL_2837623, EPI_ISL_2837625, EPI_ISL_2837626, EPI_ISL_2837627, EPI_ISL_2837628, EPI_ISL_2837629, EPI_ISL_2837630, EPI_ISL_2837631, EPI_ISL_2837632, EPI_ISL_2837635, EPI_ISL_2837639, EPI_ISL_2837640, EPI_ISL_2837647, EPI_ISL_2837648, EPI_ISL_2837649, EPI_ISL_2837651, EPI_ISL_2837653, EPI_ISL_2837656, EPI_ISL_2837657, EPI_ISL_2837661, EPI_ISL_2837662, EPI_ISL_2837663, EPI_ISL_2837664, EPI_ISL_2837665, EPI_ISL_2837667, EPI_ISL_2837668, EPI_ISL_2837669, EPI_ISL_2837670, EPI_ISL_2837671, EPI_ISL_2837672, EPI_ISL_2837673, EPI_ISL_2837674, EPI_ISL_2837675, EPI_ISL_2837676, EPI_ISL_2837677, EPI_ISL_2837678, EPI_ISL_2837682, EPI_ISL_2837683, EPI_ISL_2837684, EPI_ISL_2837685, EPI_ISL_2837686, EPI_ISL_2837687, EPI_ISL_2837688, EPI_ISL_2837689, EPI_ISL_2837690, EPI_ISL_2837691, EPI_ISL_2837692, EPI_ISL_2837693, EPI_ISL_2837694, EPI_ISL_2837695, EPI_ISL_2837702, EPI_ISL_2837703, EPI_ISL_2837706, EPI_ISL_2837709, EPI_ISL_2837710, EPI_ISL_2837711, EPI_ISL_2837712, EPI_ISL_2837713, EPI_ISL_2837716, EPI_ISL_2837717, EPI_ISL_2837718, EPI_ISL_2837722, EPI_ISL_2837723, EPI_ISL_2837724, EPI_ISL_2837727, EPI_ISL_2837728, EPI_ISL_2837729, EPI_ISL_2837730, EPI_ISL_2837731, EPI_ISL_2837732, EPI_ISL_2837733, EPI_ISL_2837734, EPI_ISL_2837735, EPI_ISL_2837736, EPI_ISL_2837737, EPI_ISL_2837738, EPI_ISL_2837739, EPI_ISL_2837740, EPI_ISL_2837741, EPI_ISL_2837742, EPI_ISL_2837743, EPI_ISL_2837744, EPI_ISL_2837753, EPI_ISL_2837754, EPI_ISL_2837755, EPI_ISL_2837757, EPI_ISL_2837758, EPI_ISL_2837759, EPI_ISL_2837760, EPI_ISL_2837761, EPI_ISL_2837764, EPI_ISL_2837765, EPI_ISL_2837766, EPI_ISL_2837767, EPI_ISL_2837768, EPI_ISL_2837769, EPI_ISL_2837770, EPI_ISL_2837771, EPI_ISL_2837772, EPI_ISL_2837773, EPI_ISL_2837774, EPI_ISL_2837775, EPI_ISL_2837776, EPI_ISL_2837777, EPI_ISL_2837778, EPI_ISL_2837779, EPI_ISL_2837780, EPI_ISL_2837781, EPI_ISL_2837782, EPI_ISL_2837783, EPI_ISL_2837785, EPI_ISL_2837786, EPI_ISL_2837789, EPI_ISL_2837791, EPI_ISL_2837792, EPI_ISL_2837793, EPI_ISL_2837794, EPI_ISL_2837795, EPI_ISL_2837796, EPI_ISL_2837797, EPI_ISL_2837798, EPI_ISL_2837799, EPI_ISL_2837800, EPI_ISL_2837805, EPI_ISL_2837806, EPI_ISL_2837807, EPI_ISL_2837808, EPI_ISL_2837809, EPI_ISL_2837810, EPI_ISL_2837811, EPI_ISL_2837813, EPI_ISL_2837814, EPI_ISL_2837816, EPI_ISL_2837817, EPI_ISL_2837818, EPI_ISL_2837819, EPI_ISL_2837820, EPI_ISL_2837821, EPI_ISL_2837822, EPI_ISL_2837823, EPI_ISL_2837824, EPI_ISL_2837825, EPI_ISL_2837826, EPI_ISL_2837827, EPI_ISL_2837828, EPI_ISL_2837829, EPI_ISL_2837830, EPI_ISL_2837831, EPI_ISL_2837832, EPI_ISL_2837833, EPI_ISL_2837834, EPI_ISL_2837835, EPI_ISL_2837836, EPI_ISL_2837837, EPI_ISL_2837838, EPI_ISL_2837839, EPI_ISL_2837840, EPI_ISL_2837841, EPI_ISL_2837842, EPI_ISL_2837843, EPI_ISL_2837844, EPI_ISL_2837845, EPI_ISL_2837846, EPI_ISL_2837847, EPI_ISL_2837848, EPI_ISL_2837849, EPI_ISL_2837851, EPI_ISL_2837852, EPI_ISL_2837853, EPI_ISL_2837854, EPI_ISL_2837855, EPI_ISL_2837856, EPI_ISL_2837857, EPI_ISL_2837858, EPI_ISL_2837859, EPI_ISL_2837860, EPI_ISL_2837861, EPI_ISL_2837863, EPI_ISL_2837864, EPI_ISL_2837865, EPI_ISL_2837866, EPI_ISL_2837867, EPI_ISL_2837868, EPI_ISL_2837870, EPI_ISL_2837877, EPI_ISL_2837878, EPI_ISL_2837879, EPI_ISL_2837880, EPI_ISL_2837881, EPI_ISL_2837882, EPI_ISL_2837883, EPI_ISL_2837884, EPI_ISL_2837885, EPI_ISL_2837887, EPI_ISL_2837888, EPI_ISL_2837889, EPI_ISL_2837890, EPI_ISL_2837891, EPI_ISL_2837892, EPI_ISL_2837896, EPI_ISL_2837897, EPI_ISL_2837898, EPI_ISL_2837899, EPI_ISL_2837900, EPI_ISL_2837908, EPI_ISL_2837909, EPI_ISL_2837910, EPI_ISL_2837911, EPI_ISL_2837912, EPI_ISL_2837914, EPI_ISL_2837915, EPI_ISL_2837916, EPI_ISL_2837918, EPI_ISL_2837919, EPI_ISL_2837920, EPI_ISL_2837921, EPI_ISL_2837922, EPI_ISL_2837923, EPI_ISL_2837924, EPI_ISL_2837925, EPI_ISL_2837926, EPI_ISL_2837927, EPI_ISL_2837928, EPI_ISL_2837929, EPI_ISL_2837930, EPI_ISL_2837931, EPI_ISL_2837932, EPI_ISL_2837933, EPI_ISL_2837934, EPI_ISL_2837935, EPI_ISL_2837936, EPI_ISL_2837939, EPI_ISL_2837940, EPI_ISL_2837942, EPI_ISL_2837944, EPI_ISL_2837948, EPI_ISL_2837949, EPI_ISL_2837950, EPI_ISL_2837951, EPI_ISL_2837952, EPI_ISL_2837953, EPI_ISL_2837955, EPI_ISL_2837958, EPI_ISL_2837959, EPI_ISL_2837960, EPI_ISL_2837961, EPI_ISL_2837962, EPI_ISL_2837963, EPI_ISL_2837964, EPI_ISL_2837965, EPI_ISL_2837966, EPI_ISL_2837967, EPI_ISL_2837968, EPI_ISL_2837969, EPI_ISL_2837970, EPI_ISL_2837971, EPI_ISL_2837978, EPI_ISL_2837983, EPI_ISL_2837989, EPI_ISL_2837990, EPI_ISL_2837991, EPI_ISL_2837993, EPI_ISL_2837994, EPI_ISL_2837995, EPI_ISL_2837996, EPI_ISL_2837997, EPI_ISL_2837998, EPI_ISL_2837999, EPI_ISL_2838000, EPI_ISL_2838001, EPI_ISL_2838008, EPI_ISL_2838011, EPI_ISL_2838017, EPI_ISL_2838018, EPI_ISL_2838019, EPI_ISL_2838020, EPI_ISL_2838021, EPI_ISL_2838024, EPI_ISL_2838025, EPI_ISL_2838029, EPI_ISL_2838031, EPI_ISL_2838033, EPI_ISL_2838034, EPI_ISL_2838035, EPI_ISL_2838036, EPI_ISL_2838037, EPI_ISL_2838039, EPI_ISL_2838040, EPI_ISL_2838041, EPI_ISL_2838042, EPI_ISL_2838043, EPI_ISL_2838044, EPI_ISL_2838046, EPI_ISL_2838047

| | | | |
|---|---|---|---|
| see above | California Department of Public Health Valencia Branch Laboratory (CDPH VBL) | California Department of Public Health | CDPH-COVIDNet; UCLA Technology Center for Genomics & Bioinformatics |
| EPI_ISL_2102997, EPI_ISL_2103009, EPI_ISL_2103031, EPI_ISL_2103062, EPI_ISL_2103097, EPI_ISL_2103106, EPI_ISL_2103138, EPI_ISL_2103169, EPI_ISL_2341431, EPI_ISL_2341437, EPI_ISL_2341455, EPI_ISL_2341481, EPI_ISL_2341494, EPI_ISL_2341499, EPI_ISL_2341515, EPI_ISL_2341558, EPI_ISL_2341590, EPI_ISL_2341608, EPI_ISL_2341614, EPI_ISL_2341622, EPI_ISL_2341632, EPI_ISL_2341640, EPI_ISL_2341642, EPI_ISL_2341654, EPI_ISL_2341664, EPI_ISL_2496784, EPI_ISL_2496793, EPI_ISL_2496795, EPI_ISL_2496800, EPI_ISL_2496805, EPI_ISL_2496812, EPI_ISL_2496814, EPI_ISL_2496821, EPI_ISL_2496827, EPI_ISL_2496828, EPI_ISL_2496829, EPI_ISL_2496832, EPI_ISL_2496834, EPI_ISL_2496843, EPI_ISL_2496847, EPI_ISL_2496852, EPI_ISL_2496853, EPI_ISL_2496855, EPI_ISL_2496858, EPI_ISL_2496864, EPI_ISL_2496866, EPI_ISL_2496867, EPI_ISL_2496872, EPI_ISL_2496877, EPI_ISL_2496878, EPI_ISL_2496879, EPI_ISL_2496881, EPI_ISL_2496885, EPI_ISL_2496886, EPI_ISL_2496887, EPI_ISL_2496904, EPI_ISL_2496910, EPI_ISL_2496912, EPI_ISL_2496914, EPI_ISL_2496916, EPI_ISL_2496917, EPI_ISL_2496919, EPI_ISL_2496920, EPI_ISL_2496921, EPI_ISL_2496924, EPI_ISL_2496925, EPI_ISL_2496932, EPI_ISL_2496933, EPI_ISL_2496935, EPI_ISL_2496936, EPI_ISL_2496938, EPI_ISL_2496939, EPI_ISL_2496942, EPI_ISL_2496943, EPI_ISL_2496945, EPI_ISL_2496947, EPI_ISL_2496956, EPI_ISL_2496960, EPI_ISL_2496965, EPI_ISL_2496968, EPI_ISL_2496969, EPI_ISL_2496971, EPI_ISL_2496972, EPI_ISL_2496973, EPI_ISL_2496975, EPI_ISL_2496977, EPI_ISL_2496979, EPI_ISL_2496980, EPI_ISL_2496981, EPI_ISL_2496987, EPI_ISL_2496989, EPI_ISL_2496990, EPI_ISL_2496992, EPI_ISL_2496995, EPI_ISL_2496996, EPI_ISL_2496999, EPI_ISL_2497000, EPI_ISL_2497555, EPI_ISL_2497558, EPI_ISL_2497559, EPI_ISL_2497562, EPI_ISL_2497565, EPI_ISL_2497566, EPI_ISL_2497569, EPI_ISL_2497571, EPI_ISL_2497572, EPI_ISL_2497576, EPI_ISL_2497577, EPI_ISL_2497578, EPI_ISL_2497579, EPI_ISL_2497581, EPI_ISL_2497584, EPI_ISL_2497586, EPI_ISL_2497587, EPI_ISL_2497589, EPI_ISL_2497590, EPI_ISL_2497591, EPI_ISL_2497592, EPI_ISL_2497593, EPI_ISL_2497595, EPI_ISL_2497596, EPI_ISL_2497599, EPI_ISL_2497602, EPI_ISL_2497603, EPI_ISL_2497604, EPI_ISL_2497605, EPI_ISL_2497607, EPI_ISL_2497614, EPI_ISL_2497618, EPI_ISL_2497629, EPI_ISL_2497635, EPI_ISL_2497638, EPI_ISL_2497639, EPI_ISL_2497641, EPI_ISL_2497644, EPI_ISL_2497659, EPI_ISL_2497667, EPI_ISL_2497675, EPI_ISL_2497677, EPI_ISL_2497678, EPI_ISL_2497679, EPI_ISL_2497681, EPI_ISL_2497684, EPI_ISL_2497688, EPI_ISL_2497692, EPI_ISL_2497700, EPI_ISL_2497704, EPI_ISL_2497709, EPI_ISL_2497714, EPI_ISL_2497724, EPI_ISL_2497743, EPI_ISL_2497749, EPI_ISL_2497752, EPI_ISL_2497756, EPI_ISL_2497757, EPI_ISL_2497783, EPI_ISL_2497788, EPI_ISL_2497794, EPI_ISL_2497798, EPI_ISL_2497801, EPI_ISL_2497811, EPI_ISL_2497812, EPI_ISL_2497813, EPI_ISL_2497816, EPI_ISL_2497821, EPI_ISL_2497822, EPI_ISL_2497827, EPI_ISL_2497835, EPI_ISL_2497837, EPI_ISL_2497841, EPI_ISL_2497843, EPI_ISL_2658616, EPI_ISL_2658674, EPI_ISL_2658708, EPI_ISL_2659210, EPI_ISL_2659212, EPI_ISL_2659257, EPI_ISL_2659294, EPI_ISL_2659312, EPI_ISL_2659343, EPI_ISL_2659353, EPI_ISL_2659434, EPI_ISL_2659467, EPI_ISL_2659469, EPI_ISL_2659533, EPI_ISL_2659539, EPI_ISL_2659562, EPI_ISL_2659570, EPI_ISL_2659624, EPI_ISL_2659630, EPI_ISL_2758532 | | | |
| see above | Contra Costa County Public Health Lab | Chan-Zuckerberg Biohub | CZB Cliahub Consortium |
| EPI_ISL_1821354, EPI_ISL_2102747, EPI_ISL_2102748, EPI_ISL_2102755, EPI_ISL_2102776, EPI_ISL_2102811, EPI_ISL_2102820, EPI_ISL_2102830, EPI_ISL_2102837, EPI_ISL_2102844, EPI_ISL_2102845, EPI_ISL_2102848, EPI_ISL_2102849, EPI_ISL_2102869, EPI_ISL_2102890, EPI_ISL_2102893, EPI_ISL_2102902, EPI_ISL_2102905, EPI_ISL_2102933, EPI_ISL_2102947, EPI_ISL_2102955, EPI_ISL_2102958, EPI_ISL_2102976, EPI_ISL_2102994, EPI_ISL_2103008, EPI_ISL_2103026, EPI_ISL_2103045, EPI_ISL_2103090, EPI_ISL_2103112, EPI_ISL_2103130, EPI_ISL_2103133, EPI_ISL_2103161, EPI_ISL_2103176, EPI_ISL_2103189, EPI_ISL_2228293, EPI_ISL_2228339, EPI_ISL_2228357, EPI_ISL_2228390, EPI_ISL_2228400, EPI_ISL_2658601, EPI_ISL_2758514, EPI_ISL_2758518 | | | |
| see above | County of San Luis Obispo Public Health Laboratory | Chan-Zuckerberg Biohub | CZB Cliahub Consortium |
| EPI_ISL_2507097, EPI_ISL_2507098, EPI_ISL_2507099, EPI_ISL_2507100, EPI_ISL_2507101, EPI_ISL_2507105, EPI_ISL_2507109, EPI_ISL_2507111, EPI_ISL_2507124, EPI_ISL_2507126, EPI_ISL_2507129, EPI_ISL_2507130, EPI_ISL_2507131, EPI_ISL_2507135, EPI_ISL_2507140, EPI_ISL_2507146, EPI_ISL_2507147, EPI_ISL_2507149, EPI_ISL_2507150, EPI_ISL_2507152, EPI_ISL_2507153, EPI_ISL_2507157, EPI_ISL_2507158, EPI_ISL_2507161, EPI_ISL_2507162, EPI_ISL_2507167, EPI_ISL_2507169, EPI_ISL_2507170, EPI_ISL_2507171, EPI_ISL_2507174, EPI_ISL_2507176, EPI_ISL_2507177, EPI_ISL_2507178, EPI_ISL_2507179, EPI_ISL_2507184, EPI_ISL_2507193, EPI_ISL_2507195, EPI_ISL_2507197, EPI_ISL_2507198, EPI_ISL_2507199, EPI_ISL_2507200, EPI_ISL_2507202, EPI_ISL_2507208, EPI_ISL_2507210, EPI_ISL_2507211, EPI_ISL_2507213, EPI_ISL_2507214, EPI_ISL_2507217, EPI_ISL_2507221, EPI_ISL_2507224, EPI_ISL_2507225, EPI_ISL_2507228, EPI_ISL_2507231, EPI_ISL_2507232, EPI_ISL_2507236, EPI_ISL_2507238, EPI_ISL_2507241, EPI_ISL_2507242, EPI_ISL_2507248, EPI_ISL_2507253, EPI_ISL_2507261, EPI_ISL_2507262, EPI_ISL_2507265, EPI_ISL_2507271, EPI_ISL_2507279, EPI_ISL_2507282, EPI_ISL_2507292, EPI_ISL_2507293, EPI_ISL_2507303, EPI_ISL_2507321, EPI_ISL_2507323, EPI_ISL_2507324, EPI_ISL_2507335, EPI_ISL_2507340, EPI_ISL_2507356, EPI_ISL_2507360, EPI_ISL_2754965, EPI_ISL_2754966, EPI_ISL_2754967, EPI_ISL_2754968, EPI_ISL_2754969, EPI_ISL_2754970, EPI_ISL_2754971, EPI_ISL_2754972, EPI_ISL_2754973, EPI_ISL_2754974, EPI_ISL_2754975, EPI_ISL_2754976, EPI_ISL_2754977, EPI_ISL_2754978, EPI_ISL_2754979, EPI_ISL_2754980, EPI_ISL_2754981, EPI_ISL_2754982, EPI_ISL_2754983, EPI_ISL_2754984, EPI_ISL_2754985, EPI_ISL_2754991, EPI_ISL_2754992, EPI_ISL_2754993, EPI_ISL_2754994, EPI_ISL_2754995, EPI_ISL_2754996, EPI_ISL_2754997, EPI_ISL_2754998, EPI_ISL_2754999, EPI_ISL_2755000, EPI_ISL_2755001, EPI_ISL_2755002, EPI_ISL_2755003, EPI_ISL_2755004, EPI_ISL_2755005, EPI_ISL_2755006, EPI_ISL_2755007, EPI_ISL_2755008, EPI_ISL_2755009, EPI_ISL_2755010, EPI_ISL_2755011, EPI_ISL_2755012, EPI_ISL_2755013, EPI_ISL_2755014, EPI_ISL_2755015, EPI_ISL_2755016, EPI_ISL_2755017, EPI_ISL_2755018, EPI_ISL_2755019, EPI_ISL_2755020, EPI_ISL_2755021, EPI_ISL_2755022, EPI_ISL_2755023, EPI_ISL_2755024, EPI_ISL_2755025, EPI_ISL_2755026, EPI_ISL_2755027, EPI_ISL_2755028, EPI_ISL_2755029, EPI_ISL_2755030, EPI_ISL_2755031, EPI_ISL_2755032, EPI_ISL_2755033, EPI_ISL_2755034, EPI_ISL_2755035, EPI_ISL_2755036, EPI_ISL_2755037, EPI_ISL_2755038, EPI_ISL_2755039, EPI_ISL_2755040, EPI_ISL_2755041, EPI_ISL_2755042, EPI_ISL_2755043, EPI_ISL_2755044, EPI_ISL_2755045, EPI_ISL_2755046, EPI_ISL_2755047, EPI_ISL_2755048, EPI_ISL_2755049, EPI_ISL_2755050, EPI_ISL_2755051, EPI_ISL_2755052, EPI_ISL_2755053, EPI_ISL_2755054, EPI_ISL_2755055, EPI_ISL_2755056, EPI_ISL_2755058, EPI_ISL_2755059, EPI_ISL_2755060, EPI_ISL_2755061, EPI_ISL_2755062, EPI_ISL_2755063, EPI_ISL_2755064, EPI_ISL_2755065, EPI_ISL_2755066, EPI_ISL_2755067, EPI_ISL_2755068, EPI_ISL_2755069, EPI_ISL_2755070, EPI_ISL_2755071, EPI_ISL_2755072, EPI_ISL_2755073, EPI_ISL_2755074, EPI_ISL_2755075, EPI_ISL_2755076, EPI_ISL_2755077, EPI_ISL_2755078, EPI_ISL_2755079, EPI_ISL_2755080, EPI_ISL_2755081, EPI_ISL_2755082, EPI_ISL_2755083, EPI_ISL_2755085, EPI_ISL_2755086, EPI_ISL_2755087, EPI_ISL_2755088, EPI_ISL_2755089, EPI_ISL_2755090, EPI_ISL_2755091, EPI_ISL_2755092, EPI_ISL_2755093, EPI_ISL_2755094, EPI_ISL_2755095, EPI_ISL_2755096, EPI_ISL_2755097, EPI_ISL_2755098, EPI_ISL_2755099, EPI_ISL_2755101, EPI_ISL_2755102, EPI_ISL_2755103, EPI_ISL_2755104, EPI_ISL_2755105, EPI_ISL_2755106, EPI_ISL_2755115, EPI_ISL_2755116, EPI_ISL_2755117, EPI_ISL_2755118, EPI_ISL_2755119, EPI_ISL_2755121, EPI_ISL_2755122, EPI_ISL_2755123, EPI_ISL_2755124, EPI_ISL_2755125, EPI_ISL_2755126, EPI_ISL_2755127, EPI_ISL_2755128, EPI_ISL_2755129, EPI_ISL_2755130, EPI_ISL_2755131, EPI_ISL_2755132, EPI_ISL_2755133, EPI_ISL_2755134, EPI_ISL_2755135, EPI_ISL_2755139, EPI_ISL_2755140, EPI_ISL_2755145, EPI_ISL_2755146, EPI_ISL_2755147, EPI_ISL_2755148, EPI_ISL_2755149, EPI_ISL_2755150, EPI_ISL_2755151, EPI_ISL_2755167, EPI_ISL_2755168, EPI_ISL_2755169, EPI_ISL_2755170, EPI_ISL_2755171, EPI_ISL_2755172, EPI_ISL_2755173, EPI_ISL_2755174, EPI_ISL_2755175, EPI_ISL_2755176, EPI_ISL_2755177, EPI_ISL_2755178, EPI_ISL_2755179, EPI_ISL_2755200, EPI_ISL_2755201, EPI_ISL_2755203, EPI_ISL_2755204, EPI_ISL_2755205, EPI_ISL_2755206, EPI_ISL_2755207, EPI_ISL_2755208, EPI_ISL_2755209, EPI_ISL_2755210, EPI_ISL_2755211, EPI_ISL_2755212, EPI_ISL_2755213, EPI_ISL_2755214, EPI_ISL_2755215, EPI_ISL_2755231, EPI_ISL_2755232, EPI_ISL_2755233, EPI_ISL_2755234, EPI_ISL_2755235, EPI_ISL_2755236, EPI_ISL_2755237, EPI_ISL_2755238, EPI_ISL_2755239, EPI_ISL_2755264, EPI_ISL_2755265, EPI_ISL_2755266, EPI_ISL_2755267, EPI_ISL_2755301, EPI_ISL_2755302, EPI_ISL_2755303, EPI_ISL_2755339, EPI_ISL_2755340, EPI_ISL_2755374, EPI_ISL_2755375, EPI_ISL_2755376, EPI_ISL_2755377, EPI_ISL_2755416, EPI_ISL_2755417, EPI_ISL_2755418, EPI_ISL_2755419, EPI_ISL_2755420, EPI_ISL_2755421 | | | |
| see above | Curative Labs | Curative Labs | Elias L. Salfati; Eugenia Khorosheva; George Way; J.Cesar Ignacio-Espinoza; Janet Chen; Mikhail Hanewich-Hollatz; Nabjot Sandhu; Sophia Quasem; Vladimir Slepnev; Zhiyi Xie |
| EPI_ISL_2178765, EPI_ISL_2178767, EPI_ISL_2178768, EPI_ISL_2178770, EPI_ISL_2178771, EPI_ISL_2178772, EPI_ISL_2178773, EPI_ISL_2178774, EPI_ISL_2178775, EPI_ISL_2178776, EPI_ISL_2508349, EPI_ISL_2508350, EPI_ISL_2508351, EPI_ISL_2508352, EPI_ISL_2508353, EPI_ISL_2508354, EPI_ISL_2508355, EPI_ISL_2508356, EPI_ISL_2508357, EPI_ISL_2508358 | | | |
| see above | DeRisi Lab, University of California, San Francisco | DeRisi Lab, University of California, San Francisco | Amy Kistler; Anthea Mitchell; Carina Marquez; Diane Havlir; Diane Jones; Douglas Black; Eric Chow; Gabriel Chamie; Genay Pilarowski; IDseq Team; Jackie Martinez; James Peng; Jamin Liu; Jon Jacobo; Lucy Li; Luis Rubio; Manu Vanaerschot; Matthew Laurie; Maya Petersen; Patrick Ayscue; Sabrina Mann; Sara Sunshine; Susana Rojas; Susy Rojas; Valerie Tulier-Laiwa; and Joseph DeRisi |
| EPI_ISL_1913219, EPI_ISL_1913220, EPI_ISL_1913221, EPI_ISL_1913222, EPI_ISL_1913223, EPI_ISL_1913224 | EL CAMINO HOSPITAL | Santa Clara County public Health Laboratory | Santa Clara County Public Health Department |
| EPI_ISL_2659065, EPI_ISL_2659066, EPI_ISL_2659067, EPI_ISL_2659068, EPI_ISL_2659069, EPI_ISL_2659070, EPI_ISL_2659071, EPI_ISL_2659072 | | | |
| see above | El Camino Hospital | Santa Clara County Public Health Laboratory | Santa Clara County Public Health Department |
| EPI_ISL_1664274, EPI_ISL_1824117, EPI_ISL_1824118, EPI_ISL_1824119 | El Camino Hospital | Santa Clara County public Health Laboratory | Santa Clara County Public Health Department |
| EPI_ISL_2391754, EPI_ISL_2391755, EPI_ISL_2391756, EPI_ISL_2391757, EPI_ISL_2391758, EPI_ISL_2391759, EPI_ISL_2391798, EPI_ISL_2391799, EPI_ISL_2391800, EPI_ISL_2391801, EPI_ISL_2391802, EPI_ISL_2391803, EPI_ISL_2391804, EPI_ISL_2391805, EPI_ISL_2391806, EPI_ISL_2392169, EPI_ISL_2392190, EPI_ISL_2399618, EPI_ISL_2405211, EPI_ISL_2405212, EPI_ISL_2424191 | | | |
| see above | Flow Diagnostics Lab | Los Angeles County Public Health Laboratories | P. Hemarajata et al. |
| EPI_ISL_1918991 | Flow Health | Infectious Disease Program, Broad Institute of Harvard and MIT | Adams, G.; B.L.; B.W.; Bauer, M.; Birren; Carter, A.; Chaluvadi, S.; D.J.; DeRuff, K.; Gallagher, G.; Gladden-Young, A.; J.E.; K.J.; Lagerborg, K.; Lemieux; Loreth, C.; MacInnis; Normandin, E.; P.C.; Park; Reilly, S.; Rudy, M.; Siddle; Smole, S.; Tomkins-Tinch, C.; and Sabeti |
| EPI_ISL_1666000, EPI_ISL_1666001, EPI_ISL_1666002, EPI_ISL_1666003, EPI_ISL_1666004, EPI_ISL_1666005, EPI_ISL_1666006, EPI_ISL_1666007, EPI_ISL_1666008, EPI_ISL_1666009, EPI_ISL_1666010, EPI_ISL_1666011, EPI_ISL_1666012, EPI_ISL_1666013, EPI_ISL_1666014, EPI_ISL_1666015, EPI_ISL_1666016, EPI_ISL_1666017, EPI_ISL_1666018, EPI_ISL_1666019, EPI_ISL_1666020, EPI_ISL_1666021, EPI_ISL_1666022, EPI_ISL_1666023, EPI_ISL_1666024, EPI_ISL_1666025, EPI_ISL_1666026, EPI_ISL_1666027, EPI_ISL_1666028, EPI_ISL_1666029, EPI_ISL_1666030, EPI_ISL_1666031, EPI_ISL_1666032, EPI_ISL_1666033, EPI_ISL_1666034, EPI_ISL_1666035, EPI_ISL_1666036, EPI_ISL_1666037, EPI_ISL_1666038, EPI_ISL_1666039, EPI_ISL_1666040, EPI_ISL_1666041, EPI_ISL_1666042, EPI_ISL_1666043, EPI_ISL_1666044, EPI_ISL_1666045, EPI_ISL_1666046, EPI_ISL_1666047, EPI_ISL_1666048, EPI_ISL_1666049, EPI_ISL_1666050, EPI_ISL_1666051, EPI_ISL_1666052, EPI_ISL_1666053, EPI_ISL_1666054, EPI_ISL_1666055, EPI_ISL_1666056, EPI_ISL_1666060, EPI_ISL_1666061, EPI_ISL_1666062, EPI_ISL_1666063, EPI_ISL_1666064, EPI_ISL_1666065, EPI_ISL_1666066, EPI_ISL_1666067, EPI_ISL_1666068, EPI_ISL_1666069, EPI_ISL_1666070, EPI_ISL_1666071, EPI_ISL_1666072, EPI_ISL_1666073, EPI_ISL_1666074, EPI_ISL_1666075, EPI_ISL_1666076, EPI_ISL_1666077, EPI_ISL_1666078, EPI_ISL_1666079, EPI_ISL_1666080, EPI_ISL_1666081, EPI_ISL_1666082, EPI_ISL_1666083, EPI_ISL_1666084, EPI_ISL_1666085, EPI_ISL_1666086, EPI_ISL_1666087, EPI_ISL_1666088, EPI_ISL_1666089, EPI_ISL_1666090, EPI_ISL_1666091, EPI_ISL_1666092, EPI_ISL_1666093, EPI_ISL_1666094, EPI_ISL_1666095, EPI_ISL_1666096, EPI_ISL_1666097, EPI_ISL_1666098, EPI_ISL_1666099, EPI_ISL_1666100, EPI_ISL_1666101, EPI_ISL_1666102, EPI_ISL_1666103, EPI_ISL_1666104, EPI_ISL_1666105, EPI_ISL_1666106, EPI_ISL_1666107, EPI_ISL_1666108, EPI_ISL_1666109, EPI_ISL_1666110, EPI_ISL_1666111, EPI_ISL_1666112, EPI_ISL_1666113, EPI_ISL_1666114, EPI_ISL_1666115, EPI_ISL_1666116, EPI_ISL_1666117, EPI_ISL_1666118, EPI_ISL_1666119, EPI_ISL_1666120, EPI_ISL_1666121, EPI_ISL_1666122, EPI_ISL_1666123, EPI_ISL_1666124, EPI_ISL_1666125, EPI_ISL_1666126, EPI_ISL_1666127, EPI_ISL_1666128, EPI_ISL_1666129, EPI_ISL_1666130, EPI_ISL_1666131, EPI_ISL_1666132, EPI_ISL_1666133, EPI_ISL_1666134, EPI_ISL_1666135, EPI_ISL_1666136, EPI_ISL_1666137, EPI_ISL_1666138, EPI_ISL_1666139, EPI_ISL_1666140, EPI_ISL_1666141, EPI_ISL_1666142, EPI_ISL_1666143, EPI_ISL_1666144, EPI_ISL_1666145, EPI_ISL_1666146, EPI_ISL_1666147, EPI_ISL_1666148, EPI_ISL_1666149, EPI_ISL_1666150, EPI_ISL_1666151, EPI_ISL_1666152, EPI_ISL_1666153, EPI_ISL_1666154, EPI_ISL_1666155, EPI_ISL_1666156, EPI_ISL_1666157, EPI_ISL_1666158, EPI_ISL_1666159, EPI_ISL_1666160, EPI_ISL_1666161, EPI_ISL_1666162, EPI_ISL_1666163, EPI_ISL_1666164, EPI_ISL_1666165, EPI_ISL_1666166, EPI_ISL_1666167, EPI_ISL_1666168, EPI_ISL_1666169, EPI_ISL_1666170, EPI_ISL_1666171, EPI_ISL_1666172, EPI_ISL_1666173, EPI_ISL_1666174, EPI_ISL_1666176, EPI_ISL_1666177, EPI_ISL_1666178, EPI_ISL_1666179, EPI_ISL_1666180, EPI_ISL_1666181, EPI_ISL_1666182, EPI_ISL_1666183, EPI_ISL_1666184, EPI_ISL_1666185, EPI_ISL_1666187, EPI_ISL_1666188, EPI_ISL_1666189, EPI_ISL_1666190, EPI_ISL_1666191, EPI_ISL_1666192, EPI_ISL_1666193, EPI_ISL_1666194, EPI_ISL_1666195, EPI_ISL_1666196, EPI_ISL_1666197, EPI_ISL_1666198, EPI_ISL_1666199, EPI_ISL_1666200, EPI_ISL_1666201, EPI_ISL_1666202, EPI_ISL_1666203, EPI_ISL_1666204, EPI_ISL_1666205, EPI_ISL_1666206, EPI_ISL_1666207, EPI_ISL_1666208, EPI_ISL_1666209, EPI_ISL_1666210, EPI_ISL_1666211, EPI_ISL_1666212, EPI_ISL_1666213, EPI_ISL_1666214, EPI_ISL_1666215, EPI_ISL_1666216, EPI_ISL_1666217, EPI_ISL_1666218, EPI_ISL_1666219, EPI_ISL_1666221, EPI_ISL_1666222, EPI_ISL_1666223, EPI_ISL_1666224, EPI_ISL_1666225, EPI_ISL_1666226, EPI_ISL_1666227, EPI_ISL_1666228, EPI_ISL_1666229, EPI_ISL_1666230, EPI_ISL_1666231, EPI_ISL_1666232, EPI_ISL_1666233, EPI_ISL_1666234, EPI_ISL_1666235, EPI_ISL_1666236, EPI_ISL_1666237, EPI_ISL_1666238, EPI_ISL_1666239, EPI_ISL_1666240, EPI_ISL_1666241, EPI_ISL_1666242, EPI_ISL_1666243, EPI_ISL_1666244, EPI_ISL_1666245, EPI_ISL_1666246, EPI_ISL_1666247, EPI_ISL_1666248, EPI_ISL_1666249, EPI_ISL_1666250, EPI_ISL_1666251, EPI_ISL_1666252, EPI_ISL_1666253, EPI_ISL_1666254, EPI_ISL_1666255, EPI_ISL_1666256, EPI_ISL_1666257, EPI_ISL_1666258, EPI_ISL_1666259, EPI_ISL_1666260, EPI_ISL_1666261, EPI_ISL_1666262, EPI_ISL_1666263, EPI_ISL_1666264, EPI_ISL_1666265, EPI_ISL_1666266, EPI_ISL_1666267, EPI_ISL_1666268, EPI_ISL_1666269, EPI_ISL_1666270, EPI_ISL_1666271, EPI_ISL_1666272, EPI_ISL_1666273, EPI_ISL_1666274, EPI_ISL_1666275, EPI_ISL_1666276, EPI_ISL_1666277, EPI_ISL_1666278, EPI_ISL_1666279, EPI_ISL_1666280, EPI_ISL_1666281, EPI_ISL_1666282, EPI_ISL_1666283, EPI_ISL_1666284, EPI_ISL_1666285, EPI_ISL_1666286, EPI_ISL_1666287, EPI_ISL_1666288, EPI_ISL_1666289, EPI_ISL_1666290, EPI_ISL_1666291, EPI_ISL_1666292, EPI_ISL_1666293, EPI_ISL_1666294, EPI_ISL_1666295, EPI_ISL_1666296, EPI_ISL_1666297, EPI_ISL_1666298, EPI_ISL_1666299, EPI_ISL_1666300, EPI_ISL_1666301, EPI_ISL_1666302, EPI_ISL_1666303, EPI_ISL_1666304, EPI_ISL_1666305, EPI_ISL_1666306, EPI_ISL_1666307, EPI_ISL_1666308, EPI_ISL_1666309, EPI_ISL_1666310, EPI_ISL_1666311, EPI_ISL_1666312, EPI_ISL_1666313, EPI_ISL_1666314, EPI_ISL_1666315, EPI_ISL_1666316, EPI_ISL_1666317, EPI_ISL_1666318, EPI_ISL_1666319, EPI_ISL_1666320, EPI_ISL_1666321, EPI_ISL_1666322, EPI_ISL_1666323, EPI_ISL_1666324, EPI_ISL_1666325, EPI_ISL_1666326, EPI_ISL_1666327, EPI_ISL_1666329, EPI_ISL_1666330, EPI_ISL_1666331, EPI_ISL_1666332, EPI_ISL_1666333, EPI_ISL_1666334, EPI_ISL_1666335, EPI_ISL_1666336, EPI_ISL_1666337, EPI_ISL_1666338, EPI_ISL_1666339, EPI_ISL_1666340, EPI_ISL_1666341, EPI_ISL_1666342, EPI_ISL_1666343, EPI_ISL_1666344, EPI_ISL_1666345, EPI_ISL_1666346, EPI_ISL_1666347, EPI_ISL_1666348, EPI_ISL_1666349, EPI_ISL_1666350, EPI_ISL_1666351, EPI_ISL_1666352, EPI_ISL_1666353, EPI_ISL_1666354, EPI_ISL_1666355, EPI_ISL_1666356, EPI_ISL_1666357, EPI_ISL_1666358, EPI_ISL_1666359, EPI_ISL_1666360, EPI_ISL_1666361, EPI_ISL_1666362, EPI_ISL_1666363, EPI_ISL_1666365, EPI_ISL_1666366, EPI_ISL_1666367, EPI_ISL_1666368, EPI_ISL_1666369, EPI_ISL_1666370, EPI_ISL_1666371, EPI_ISL_1666372, EPI_ISL_1666373, EPI_ISL_1666374, EPI_ISL_1666375, EPI_ISL_1666376, EPI_ISL_1666377, EPI_ISL_1666378, EPI_ISL_1666379, EPI_ISL_1666380, EPI_ISL_1666381, EPI_ISL_1666382, EPI_ISL_1666383, EPI_ISL_1666384, EPI_ISL_1666385, EPI_ISL_1666386, EPI_ISL_1666387, EPI_ISL_1666388, EPI_ISL_1666389, EPI_ISL_1666390, EPI_ISL_1666391, EPI_ISL_1666392, EPI_ISL_1666393, EPI_ISL_1666394, EPI_ISL_1666395, EPI_ISL_1666396, EPI_ISL_1666397, EPI_ISL_1666398, EPI_ISL_1666399, EPI_ISL_1666400, EPI_ISL_1666401, EPI_ISL_1666402, EPI_ISL_1666404, EPI_ISL_1666405, EPI_ISL_1666406, EPI_ISL_1666407, EPI_ISL_1666408, EPI_ISL_1666409, EPI_ISL_1666410, EPI_ISL_1666411, EPI_ISL_1666412, EPI_ISL_1666415, EPI_ISL_1666416, EPI_ISL_1666417, EPI_ISL_1666418, EPI_ISL_1666419, EPI_ISL_1666420, EPI_ISL_1666421, EPI_ISL_1666422, EPI_ISL_1666423, EPI_ISL_1666424, EPI_ISL_1666425, EPI_ISL_1666426, EPI_ISL_1666427, EPI_ISL_1666428, EPI_ISL_1666429, EPI_ISL_1666430, EPI_ISL_1666431, EPI_ISL_1666432, EPI_ISL_1666433, EPI_ISL_1666434, EPI_ISL_1666435, EPI_ISL_1666436, EPI_ISL_1666437, EPI_ISL_1666438, EPI_ISL_1666439, EPI_ISL_1666440, EPI_ISL_1666441, EPI_ISL_1666442, EPI_ISL_1666443, EPI_ISL_1666444, EPI_ISL_1666445, EPI_ISL_1666446, EPI_ISL_1666447, EPI_ISL_1666448, EPI_ISL_1666449, EPI_ISL_1666450, EPI_ISL_1666451, EPI_ISL_1666452, EPI_ISL_1666453, EPI_ISL_1666454, EPI_ISL_1666455, EPI_ISL_1666456, EPI_ISL_1666457, EPI_ISL_1666458, EPI_ISL_1666459, EPI_ISL_1666460, EPI_ISL_1666461, EPI_ISL_1666462, EPI_ISL_1666463, EPI_ISL_1666464, EPI_ISL_1666465, EPI_ISL_1666466, EPI_ISL_1666467, EPI_ISL_1666468, EPI_ISL_1666469, EPI_ISL_1666470, EPI_ISL_1666471, EPI_ISL_1666472, EPI_ISL_1666473, EPI_ISL_1666474, EPI_ISL_1666475, EPI_ISL_1666476, EPI_ISL_1666478, EPI_ISL_1666479, EPI_ISL_1666480, EPI_ISL_1666481, EPI_ISL_1666482, EPI_ISL_1666483, EPI_ISL_1666484, EPI_ISL_1666485, EPI_ISL_1666486, EPI_ISL_1666487, EPI_ISL_1666488, EPI_ISL_1666489, EPI_ISL_1666490, EPI_ISL_1666491, EPI_ISL_1666492, EPI_ISL_1666493, EPI_ISL_1666494, EPI_ISL_1666495, EPI_ISL_1666496, EPI_ISL_1666497, EPI_ISL_1666498, EPI_ISL_1666499, EPI_ISL_1666500, EPI_ISL_1666501, EPI_ISL_1666502, EPI_ISL_1666503, EPI_ISL_1666504, EPI_ISL_1666505, EPI_ISL_1666507, EPI_ISL_1666508, EPI_ISL_1666509, EPI_ISL_1666510, EPI_ISL_1666511, EPI_ISL_1666512, EPI_ISL_1666513, EPI_ISL_1666514, EPI_ISL_1666515, EPI_ISL_1666516, EPI_ISL_1666517, EPI_ISL_1666518, EPI_ISL_1666519, EPI_ISL_1666520, EPI_ISL_1666521, EPI_ISL_1666522, EPI_ISL_1666523, EPI_ISL_1666524, EPI_ISL_1666525, EPI_ISL_1666526, EPI_ISL_1666527, EPI_ISL_1666528, EPI_ISL_1666529, EPI_ISL_1666530, EPI_ISL_1666531, EPI_ISL_1666532, EPI_ISL_1666533, EPI_ISL_1666534, EPI_ISL_1666535, EPI_ISL_1666536, EPI_ISL_1666537, EPI_ISL_1666538, EPI_ISL_1666539, EPI_ISL_1666540, EPI_ISL_1666541, EPI_ISL_1666542, EPI_ISL_1667077, EPI_ISL_1667083, EPI_ISL_1667087, EPI_ISL_1667091, EPI_ISL_1667092, EPI_ISL_1667093, EPI_ISL_1667094, EPI_ISL_1667098, EPI_ISL_1667099, EPI_ISL_1667108, EPI_ISL_1667109, EPI_ISL_1667114, EPI_ISL_1667115, EPI_ISL_1667116, EPI_ISL_1667117, EPI_ISL_1667118, EPI_ISL_1667119, EPI_ISL_1667120, EPI_ISL_1667121, EPI_ISL_1667122, EPI_ISL_1667123, EPI_ISL_1667124, EPI_ISL_1667125, EPI_ISL_1667126, EPI_ISL_1667128, EPI_ISL_1667129, EPI_ISL_1667130, EPI_ISL_1667131, EPI_ISL_1667132, EPI_ISL_1667133, EPI_ISL_1667134, EPI_ISL_1667135, EPI_ISL_1667137, EPI_ISL_1667139, EPI_ISL_1667143, EPI_ISL_1667144, EPI_ISL_1667145, EPI_ISL_1667146, EPI_ISL_1667147, EPI_ISL_1667148, EPI_ISL_1667149, EPI_ISL_1667150, EPI_ISL_1667151, EPI_ISL_1667152, EPI_ISL_1667153, EPI_ISL_1667154, EPI_ISL_1667155, EPI_ISL_1667156, EPI_ISL_1667157, EPI_ISL_1667158, EPI_ISL_1667159, EPI_ISL_1667160, EPI_ISL_1667161, EPI_ISL_1667162, EPI_ISL_1667163, EPI_ISL_1667164, EPI_ISL_1667165, EPI_ISL_1667166, EPI_ISL_1667167, EPI_ISL_1667169, EPI_ISL_1667170, EPI_ISL_1667174, EPI_ISL_1667175, EPI_ISL_1667176, EPI_ISL_1667177, EPI_ISL_1667178, EPI_ISL_1667179, EPI_ISL_1667180, EPI_ISL_1667181, EPI_ISL_1667182, EPI_ISL_1667183, EPI_ISL_1667184, EPI_ISL_1667185, EPI_ISL_1667186, EPI_ISL_1667187, EPI_ISL_1667188, EPI_ISL_1667189, EPI_ISL_1667190, EPI_ISL_1667191, EPI_ISL_1667192, EPI_ISL_1667194, EPI_ISL_1667195, EPI_ISL_1667196, EPI_ISL_1667197, EPI_ISL_1667198, EPI_ISL_1667199, EPI_ISL_1667200, EPI_ISL_1667202, EPI_ISL_1667203, EPI_ISL_1667204, EPI_ISL_1667205, EPI_ISL_1667206, EPI_ISL_1667207, EPI_ISL_1667208, EPI_ISL_1667209, EPI_ISL_1667210, EPI_ISL_1667211, EPI_ISL_1667212, EPI_ISL_1667213, EPI_ISL_1667214, EPI_ISL_1667215, EPI_ISL_1667216, EPI_ISL_1667217, EPI_ISL_1667218, EPI_ISL_1667219, EPI_ISL_1667220, EPI_ISL_1667221, EPI_ISL_1667222, EPI_ISL_1667223, EPI_ISL_1667224, EPI_ISL_1667225, EPI_ISL_1667226, EPI_ISL_1667227, EPI_ISL_1667228, EPI_ISL_1667229, EPI_ISL_1667230, EPI_ISL_1667231, EPI_ISL_1667232, EPI_ISL_1667233, EPI_ISL_1667234, EPI_ISL_1667235, EPI_ISL_1667236, EPI_ISL_1667237, EPI_ISL_1667238, EPI_ISL_1667239, EPI_ISL_1667240, EPI_ISL_1667241, EPI_ISL_1667242, EPI_ISL_1667243, EPI_ISL_1667244, EPI_ISL_1667245, EPI_ISL_1667246, EPI_ISL_1667247, EPI_ISL_1667248, EPI_ISL_1667249, EPI_ISL_1667250, EPI_ISL_1667251, EPI_ISL_1667252, EPI_ISL_1667253, EPI_ISL_1667254, EPI_ISL_1667255, EPI_ISL_1667256, EPI_ISL_1667257, EPI_ISL_1667258, EPI_ISL_1667259, EPI_ISL_1667260, EPI_ISL_1667261, EPI_ISL_1667262, EPI_ISL_1667263, EPI_ISL_1667264, EPI_ISL_1667265, EPI_ISL_1667266, EPI_ISL_1667267, EPI_ISL_1667268, EPI_ISL_1667269, EPI_ISL_1667270, EPI_ISL_1667271, EPI_ISL_1667272, EPI_ISL_1667273, EPI_ISL_1667274, EPI_ISL_1667275, EPI_ISL_1667276, EPI_ISL_1667277, EPI_ISL_1667278, EPI_ISL_1667279, EPI_ISL_1667280, EPI_ISL_1667281, EPI_ISL_1667282, EPI_ISL_1667283, EPI_ISL_1667284, EPI_ISL_1667285, EPI_ISL_1667286, EPI_ISL_1667287, EPI_ISL_1667288, EPI_ISL_1667289, EPI_ISL_1667290, EPI_ISL_1667291, EPI_ISL_1667292, EPI_ISL_1667293, EPI_ISL_1667294, EPI_ISL_1667295, EPI_ISL_1667296, EPI_ISL_1667297, EPI_ISL_1667298, EPI_ISL_1667299, EPI_ISL_1667300, EPI_ISL_1667302, EPI_ISL_1667303, EPI_ISL_1667304, EPI_ISL_1667305, EPI_ISL_1667306, EPI_ISL_1667307, EPI_ISL_1667308, EPI_ISL_1667309, EPI_ISL_1667310, EPI_ISL_1667311, EPI_ISL_1667312, EPI_ISL_1667313, EPI_ISL_1667314, EPI_ISL_1667315, EPI_ISL_1667316, EPI_ISL_1667317, EPI_ISL_1667318, EPI_ISL_1667319, EPI_ISL_1667320, EPI_ISL_1667321, EPI_ISL_1667322, EPI_ISL_1667323, EPI_ISL_1667324, EPI_ISL_1667325, EPI_ISL_1667326, EPI_ISL_1667327, EPI_ISL_1667328, EPI_ISL_1667329, EPI_ISL_1667330, EPI_ISL_1667331, EPI_ISL_1667332, EPI_ISL_1667333, EPI_ISL_1667334, EPI_ISL_1667335, EPI_ISL_1667336, EPI_ISL_1667337, EPI_ISL_1667338, EPI_ISL_1667339, EPI_ISL_1667340, EPI_ISL_1667341, EPI_ISL_1667342, EPI_ISL_1667343, EPI_ISL_1667344, EPI_ISL_1667345, EPI_ISL_1667346, EPI_ISL_1667347, EPI_ISL_1667348, EPI_ISL_1667349, EPI_ISL_1667350, EPI_ISL_1667351, EPI_ISL_1667352, EPI_ISL_1667353, EPI_ISL_1667354, EPI_ISL_1667355, EPI_ISL_1667356, EPI_ISL_1667357, EPI_ISL_1667358, EPI_ISL_1667359, EPI_ISL_1667360, EPI_ISL_1667361, EPI_ISL_1667362, EPI_ISL_1667363, EPI_ISL_1667364, EPI_ISL_1667365, EPI_ISL_1667366, EPI_ISL_1667367, EPI_ISL_1667368, EPI_ISL_1667369, EPI_ISL_1667370, EPI_ISL_1667371, EPI_ISL_1667372, EPI_ISL_1667373, EPI_ISL_1667374, EPI_ISL_1667375, EPI_ISL_1667376, EPI_ISL_1667377, EPI_ISL_1667378, EPI_ISL_1667379, EPI_ISL_1667380, EPI_ISL_1667381, EPI_ISL_1733838, EPI_ISL_1733839, EPI_ISL_1733840, EPI_ISL_1733841, EPI_ISL_1733842, EPI_ISL_1733843, EPI_ISL_1733844, EPI_ISL_1733845, EPI_ISL_1733846, EPI_ISL_1733847, EPI_ISL_1733848, EPI_ISL_1733849, EPI_ISL_1733850, EPI_ISL_1733851, EPI_ISL_1733852, EPI_ISL_1733853, EPI_ISL_1733854, EPI_ISL_1733855, EPI_ISL_1733856, EPI_ISL_1733857, EPI_ISL_1733858, EPI_ISL_1733859, EPI_ISL_1733860, EPI_ISL_1733861, EPI_ISL_1733862, EPI_ISL_1733863, EPI_ISL_1733864, EPI_ISL_1733865, EPI_ISL_1733866, EPI_ISL_1733867, EPI_ISL_1733868, EPI_ISL_1733869, EPI_ISL_1733870, EPI_ISL_1733871, EPI_ISL_1733872, EPI_ISL_1733873, EPI_ISL_1733874, EPI_ISL_1733875, EPI_ISL_1733876, EPI_ISL_1733877, EPI_ISL_1733878, EPI_ISL_1733879, EPI_ISL_1733880, EPI_ISL_1733881, EPI_ISL_1733882, EPI_ISL_1733883, EPI_ISL_1733884, EPI_ISL_1733885, EPI_ISL_1733886, EPI_ISL_1733887, EPI_ISL_1733888, EPI_ISL_1733889, EPI_ISL_1733890, EPI_ISL_1733891, EPI_ISL_1733892, EPI_ISL_1733893, EPI_ISL_1733894, EPI_ISL_1733895, EPI_ISL_1733896, EPI_ISL_1733897, EPI_ISL_1733898, EPI_ISL_1733899, EPI_ISL_1733900, EPI_ISL_1733901, EPI_ISL_1733902, EPI_ISL_1733903, EPI_ISL_1733904, EPI_ISL_1733905, EPI_ISL_1733906, EPI_ISL_1733907, EPI_ISL_1733908, EPI_ISL_1733909, EPI_ISL_1733910, EPI_ISL_1733911, EPI_ISL_1733912, EPI_ISL_1733913, EPI_ISL_1733914, EPI_ISL_1733915, EPI_ISL_1733916, EPI_ISL_1733919, EPI_ISL_1733920, EPI_ISL_1733921, EPI_ISL_1733922, EPI_ISL_1733923, EPI_ISL_1733924, EPI_ISL_1733925, EPI_ISL_1733926, EPI_ISL_1733927, EPI_ISL_1733928, EPI_ISL_1733929, EPI_ISL_1733931, EPI_ISL_1733932, EPI_ISL_1733933, EPI_ISL_1733934, EPI_ISL_1733935, EPI_ISL_1733936, EPI_ISL_1733937, EPI_ISL_1733938, EPI_ISL_1733939, EPI_ISL_1733940, EPI_ISL_1733941, EPI_ISL_1733942, EPI_ISL_1733943, EPI_ISL_1733944, EPI_ISL_1733945, EPI_ISL_1733946, EPI_ISL_1733947, EPI_ISL_1733948, EPI_ISL_1733949, EPI_ISL_1733950, EPI_ISL_1733951, EPI_ISL_1733952, EPI_ISL_1733953, EPI_ISL_1733954, EPI_ISL_1733955, EPI_ISL_1733956, EPI_ISL_1733957, EPI_ISL_1733958, EPI_ISL_1733959, EPI_ISL_1733960, EPI_ISL_1733961, EPI_ISL_1733962, EPI_ISL_1733963, EPI_ISL_1733964, EPI_ISL_1733965, EPI_ISL_1733966, EPI_ISL_1733967, EPI_ISL_1733968, EPI_ISL_1733969, EPI_ISL_1733970, EPI_ISL_1733971, EPI_ISL_1733972, EPI_ISL_1733973, EPI_ISL_1733974, EPI_ISL_1733975, EPI_ISL_1733976, EPI_ISL_1734413, EPI_ISL_1839210, EPI_ISL_1839211, EPI_ISL_1839212, EPI_ISL_1839215, EPI_ISL_1839216, EPI_ISL_1839217, EPI_ISL_1839218, EPI_ISL_1839219, EPI_ISL_1839220, EPI_ISL_1839221, EPI_ISL_1839222, EPI_ISL_1839223, EPI_ISL_1839224, EPI_ISL_1839225, EPI_ISL_1839226, EPI_ISL_1839227, EPI_ISL_1839228, EPI_ISL_1839229, EPI_ISL_1839230, EPI_ISL_1839231, EPI_ISL_1839232, EPI_ISL_1839233, EPI_ISL_1839234, EPI_ISL_1839235, EPI_ISL_1839236, EPI_ISL_1839237, EPI_ISL_1839238, EPI_ISL_1839239, EPI_ISL_1839240, EPI_ISL_1839241, EPI_ISL_1839242, EPI_ISL_1839243, EPI_ISL_1839244, EPI_ISL_1839245, EPI_ISL_1839246, EPI_ISL_1839247, EPI_ISL_1839248, EPI_ISL_1839249, EPI_ISL_1839250, EPI_ISL_1839251, EPI_ISL_1839253, EPI_ISL_1839254, EPI_ISL_1839255, EPI_ISL_1839256, EPI_ISL_1839257, EPI_ISL_1839258, EPI_ISL_1839259, EPI_ISL_1839260, EPI_ISL_1839261, EPI_ISL_1839262, EPI_ISL_1839263, EPI_ISL_1839264, EPI_ISL_1839265, EPI_ISL_1839266, EPI_ISL_1839267, EPI_ISL_1839268, EPI_ISL_1839269, | | | |

EPI_ISL_1839270, EPI_ISL_1839271, EPI_ISL_1839272, EPI_ISL_1839273, EPI_ISL_1839274, EPI_ISL_1839275, EPI_ISL_1839276, EPI_ISL_1839277, EPI_ISL_1839402, EPI_ISL_1839546, EPI_ISL_1839547, EPI_ISL_1839548, EPI_ISL_1839549, EPI_ISL_1839550, EPI_ISL_1839551, EPI_ISL_1839552, EPI_ISL_1839553, EPI_ISL_1839554, EPI_ISL_1839555, EPI_ISL_1839556, EPI_ISL_1839557, EPI_ISL_1839558, EPI_ISL_1839559, EPI_ISL_1839560, EPI_ISL_1839561, EPI_ISL_1839562, EPI_ISL_1839563, EPI_ISL_1839564, EPI_ISL_1839565, EPI_ISL_1839566, EPI_ISL_1839567, EPI_ISL_1839568, EPI_ISL_1839569, EPI_ISL_1839570, EPI_ISL_1839571, EPI_ISL_1839572, EPI_ISL_1839573, EPI_ISL_1839574, EPI_ISL_1839576, EPI_ISL_1839577, EPI_ISL_1839578, EPI_ISL_1839579, EPI_ISL_1839580, EPI_ISL_1839581, EPI_ISL_1839582, EPI_ISL_1839584, EPI_ISL_1839585, EPI_ISL_1839586, EPI_ISL_1839587, EPI_ISL_1839588, EPI_ISL_1839589, EPI_ISL_1839590, EPI_ISL_1839591, EPI_ISL_1839592, EPI_ISL_1839593, EPI_ISL_1839594, EPI_ISL_1839595, EPI_ISL_1839596, EPI_ISL_1839597, EPI_ISL_1839598, EPI_ISL_1839599, EPI_ISL_1839600, EPI_ISL_1839601, EPI_ISL_1839602, EPI_ISL_1839603, EPI_ISL_1839604, EPI_ISL_1839605, EPI_ISL_1839606, EPI_ISL_1839607, EPI_ISL_1839608, EPI_ISL_1839609, EPI_ISL_1839610, EPI_ISL_1839611, EPI_ISL_1839612, EPI_ISL_1839613, EPI_ISL_1839614, EPI_ISL_1839615, EPI_ISL_1839843, EPI_ISL_1839847, EPI_ISL_1839848, EPI_ISL_1839849, EPI_ISL_1839850, EPI_ISL_1839851, EPI_ISL_1839852, EPI_ISL_1839853, EPI_ISL_1839854, EPI_ISL_1839855, EPI_ISL_1839856, EPI_ISL_1839857, EPI_ISL_1839858, EPI_ISL_1839859, EPI_ISL_1839860, EPI_ISL_1839861, EPI_ISL_1839862, EPI_ISL_1839863, EPI_ISL_1839864, EPI_ISL_1839865, EPI_ISL_1839866, EPI_ISL_1839867, EPI_ISL_1839868, EPI_ISL_1839869, EPI_ISL_1839870, EPI_ISL_1839871, EPI_ISL_1839872, EPI_ISL_1839873, EPI_ISL_1839874, EPI_ISL_1839875, EPI_ISL_1839876, EPI_ISL_1839877, EPI_ISL_1839878, EPI_ISL_1839879, EPI_ISL_1839880, EPI_ISL_1839881, EPI_ISL_1839882, EPI_ISL_1839884, EPI_ISL_1839885, EPI_ISL_1839886, EPI_ISL_1839888, EPI_ISL_1839889, EPI_ISL_1839890, EPI_ISL_1839891, EPI_ISL_1839892, EPI_ISL_1839893, EPI_ISL_1839894, EPI_ISL_1839895, EPI_ISL_1839896, EPI_ISL_1839897, EPI_ISL_1839898, EPI_ISL_1839899, EPI_ISL_1839900, EPI_ISL_1839942, EPI_ISL_1839943, EPI_ISL_1839944, EPI_ISL_1839945, EPI_ISL_1839946, EPI_ISL_1839947, EPI_ISL_1839948, EPI_ISL_1839949, EPI_ISL_1839950, EPI_ISL_1839951, EPI_ISL_1839952, EPI_ISL_1839953, EPI_ISL_1839954, EPI_ISL_1839955, EPI_ISL_1839956, EPI_ISL_1839957, EPI_ISL_1839958, EPI_ISL_1839959, EPI_ISL_1839960, EPI_ISL_1839961, EPI_ISL_1839962, EPI_ISL_1839964, EPI_ISL_1839965, EPI_ISL_1839966, EPI_ISL_1839967, EPI_ISL_1839968, EPI_ISL_1839969, EPI_ISL_1839970, EPI_ISL_1839972, EPI_ISL_1839973, EPI_ISL_1839974, EPI_ISL_1839975, EPI_ISL_1839976, EPI_ISL_1839977, EPI_ISL_1839978, EPI_ISL_1839979, EPI_ISL_1839980, EPI_ISL_1839981, EPI_ISL_1839982, EPI_ISL_1839983, EPI_ISL_1839984, EPI_ISL_1839985, EPI_ISL_1839986, EPI_ISL_1839987, EPI_ISL_1839988, EPI_ISL_1839989, EPI_ISL_1839990, EPI_ISL_1839991, EPI_ISL_1839992, EPI_ISL_1839993, EPI_ISL_1839994, EPI_ISL_1839995, EPI_ISL_1839996, EPI_ISL_1839997, EPI_ISL_1839998, EPI_ISL_1839999, EPI_ISL_1840000, EPI_ISL_1840001, EPI_ISL_1840002, EPI_ISL_1840003, EPI_ISL_1840005, EPI_ISL_1840006, EPI_ISL_1840007, EPI_ISL_1840008, EPI_ISL_1840009, EPI_ISL_1840010, EPI_ISL_1840011, EPI_ISL_1840012, EPI_ISL_1840013, EPI_ISL_1840014, EPI_ISL_1840015, EPI_ISL_1840016, EPI_ISL_1840017, EPI_ISL_1840018, EPI_ISL_1840019, EPI_ISL_1840020, EPI_ISL_1840021, EPI_ISL_1840022, EPI_ISL_1840023, EPI_ISL_1840024, EPI_ISL_1840025, EPI_ISL_1840026, EPI_ISL_1840027, EPI_ISL_1840028, EPI_ISL_1840029, EPI_ISL_1840030, EPI_ISL_1840031, EPI_ISL_1840032, EPI_ISL_1840033, EPI_ISL_1840034, EPI_ISL_1840035, EPI_ISL_1840036, EPI_ISL_1840037, EPI_ISL_1840038, EPI_ISL_1840039, EPI_ISL_1840040, EPI_ISL_1840041, EPI_ISL_1840042, EPI_ISL_1840043, EPI_ISL_1840044, EPI_ISL_1840045, EPI_ISL_1840046, EPI_ISL_1840047, EPI_ISL_1840048, EPI_ISL_1840049, EPI_ISL_1840050, EPI_ISL_1840051, EPI_ISL_1840052, EPI_ISL_1840053, EPI_ISL_1840054, EPI_ISL_1840055, EPI_ISL_1840056, EPI_ISL_1840057, EPI_ISL_1840058, EPI_ISL_1840059, EPI_ISL_1840060, EPI_ISL_1840061, EPI_ISL_1840062, EPI_ISL_1840063, EPI_ISL_1840064, EPI_ISL_1840065, EPI_ISL_1840066, EPI_ISL_1840067, EPI_ISL_1840068, EPI_ISL_1840069, EPI_ISL_1840070, EPI_ISL_1840071, EPI_ISL_1840072, EPI_ISL_1840073, EPI_ISL_1840074, EPI_ISL_1840075, EPI_ISL_1840076, EPI_ISL_1840077, EPI_ISL_1840078, EPI_ISL_1840079, EPI_ISL_1840080, EPI_ISL_1840081, EPI_ISL_1840082, EPI_ISL_1840083, EPI_ISL_1840084, EPI_ISL_1840085, EPI_ISL_1840086, EPI_ISL_1840087, EPI_ISL_1840088, EPI_ISL_1840089, EPI_ISL_1840091, EPI_ISL_1840092, EPI_ISL_1840093, EPI_ISL_1840094, EPI_ISL_1840095, EPI_ISL_1840096, EPI_ISL_1840097, EPI_ISL_1840098, EPI_ISL_1840099, EPI_ISL_1840100, EPI_ISL_1840101, EPI_ISL_1840102, EPI_ISL_1840103, EPI_ISL_1840104, EPI_ISL_1840105, EPI_ISL_1840106, EPI_ISL_1840107, EPI_ISL_1840108, EPI_ISL_1840109, EPI_ISL_1840110, EPI_ISL_1840111, EPI_ISL_1840112, EPI_ISL_1840113, EPI_ISL_1840114, EPI_ISL_1840115, EPI_ISL_1840116, EPI_ISL_1840117, EPI_ISL_1840118, EPI_ISL_1840119, EPI_ISL_1840120, EPI_ISL_1840121, EPI_ISL_1840122, EPI_ISL_1840123, EPI_ISL_1840124, EPI_ISL_1840125, EPI_ISL_1992018, EPI_ISL_1992019, EPI_ISL_1992022, EPI_ISL_1992023, EPI_ISL_1992024, EPI_ISL_1992025, EPI_ISL_1992026, EPI_ISL_1992027, EPI_ISL_1992028, EPI_ISL_1992029, EPI_ISL_1992030, EPI_ISL_1992031, EPI_ISL_1992032, EPI_ISL_1992033, EPI_ISL_1992034, EPI_ISL_1992035, EPI_ISL_1992036, EPI_ISL_1992037, EPI_ISL_1992038, EPI_ISL_1992039, EPI_ISL_1992040, EPI_ISL_1992041, EPI_ISL_1992042, EPI_ISL_1992043, EPI_ISL_1992044, EPI_ISL_1992045, EPI_ISL_1992046, EPI_ISL_1992047, EPI_ISL_1992048, EPI_ISL_1992049, EPI_ISL_1992050, EPI_ISL_1992051, EPI_ISL_1992052, EPI_ISL_1992053, EPI_ISL_1992054, EPI_ISL_1992055, EPI_ISL_1992056, EPI_ISL_1992057, EPI_ISL_1992058, EPI_ISL_1992059, EPI_ISL_1992060, EPI_ISL_1992061, EPI_ISL_1992062, EPI_ISL_1992063, EPI_ISL_1992064, EPI_ISL_1992065, EPI_ISL_1992066, EPI_ISL_1992067, EPI_ISL_1992068, EPI_ISL_1992069, EPI_ISL_1992070, EPI_ISL_1992071, EPI_ISL_1992072, EPI_ISL_1992073, EPI_ISL_1992074, EPI_ISL_1992075, EPI_ISL_1992076, EPI_ISL_1992077, EPI_ISL_1992078, EPI_ISL_1992079, EPI_ISL_1992080, EPI_ISL_1992081, EPI_ISL_1992082, EPI_ISL_1992083, EPI_ISL_1992084, EPI_ISL_1992085, EPI_ISL_1992086, EPI_ISL_1992087, EPI_ISL_1992088, EPI_ISL_1992092, EPI_ISL_1992093, EPI_ISL_1992096, EPI_ISL_1992097, EPI_ISL_1992098, EPI_ISL_1992099, EPI_ISL_1992101, EPI_ISL_1992102, EPI_ISL_1992103, EPI_ISL_1992104, EPI_ISL_1992105, EPI_ISL_1992106, EPI_ISL_1992107, EPI_ISL_1992108, EPI_ISL_1992109, EPI_ISL_1992110, EPI_ISL_1992111, EPI_ISL_1992112, EPI_ISL_1992113, EPI_ISL_1992114, EPI_ISL_1992115, EPI_ISL_1992116, EPI_ISL_1992118, EPI_ISL_1992119, EPI_ISL_1992120, EPI_ISL_1992121, EPI_ISL_1992122, EPI_ISL_1992123, EPI_ISL_1992124, EPI_ISL_1992125, EPI_ISL_1992127, EPI_ISL_1992128, EPI_ISL_1992129, EPI_ISL_1992130, EPI_ISL_1992131, EPI_ISL_1992132, EPI_ISL_1992133, EPI_ISL_1992134, EPI_ISL_1992135, EPI_ISL_1992136, EPI_ISL_1992137, EPI_ISL_1992138, EPI_ISL_1992139, EPI_ISL_1992140, EPI_ISL_1992141, EPI_ISL_1992142, EPI_ISL_1992143, EPI_ISL_1992144, EPI_ISL_1992145, EPI_ISL_1992146, EPI_ISL_1992147, EPI_ISL_1992148, EPI_ISL_1992149, EPI_ISL_1992150, EPI_ISL_1992151, EPI_ISL_1992152, EPI_ISL_1992153, EPI_ISL_1992154, EPI_ISL_1992265, EPI_ISL_1992266, EPI_ISL_1992267, EPI_ISL_1992268, EPI_ISL_1992269, EPI_ISL_1992270, EPI_ISL_1992271, EPI_ISL_1992272, EPI_ISL_1992273, EPI_ISL_1992274, EPI_ISL_1992275, EPI_ISL_1992276, EPI_ISL_1992277, EPI_ISL_1992278, EPI_ISL_1992279, EPI_ISL_1992280, EPI_ISL_1992281, EPI_ISL_1992282, EPI_ISL_1992283, EPI_ISL_1992284, EPI_ISL_1992285, EPI_ISL_1992286, EPI_ISL_1992287, EPI_ISL_1992288, EPI_ISL_1992289, EPI_ISL_1992290, EPI_ISL_1992291, EPI_ISL_1992292, EPI_ISL_1992293, EPI_ISL_1992294, EPI_ISL_1992295, EPI_ISL_1992296, EPI_ISL_1992297, EPI_ISL_1992298, EPI_ISL_1992299, EPI_ISL_1992300, EPI_ISL_1992301, EPI_ISL_1992302, EPI_ISL_1992303, EPI_ISL_1992304, EPI_ISL_1992305, EPI_ISL_1992306, EPI_ISL_1992307, EPI_ISL_1992308, EPI_ISL_1992309, EPI_ISL_1992310, EPI_ISL_1992311, EPI_ISL_1992312, EPI_ISL_1992313, EPI_ISL_1992314, EPI_ISL_1992315, EPI_ISL_1992316, EPI_ISL_1992317, EPI_ISL_1992318, EPI_ISL_1992319, EPI_ISL_1992320, EPI_ISL_1992321, EPI_ISL_1992322, EPI_ISL_1992323, EPI_ISL_1992324, EPI_ISL_1992329, EPI_ISL_1992330, EPI_ISL_1992331, EPI_ISL_1992332, EPI_ISL_1992333, EPI_ISL_1992334, EPI_ISL_1992335, EPI_ISL_1992336, EPI_ISL_1992337, EPI_ISL_1992338, EPI_ISL_1992339, EPI_ISL_1992340, EPI_ISL_1992341, EPI_ISL_1992342, EPI_ISL_1992343, EPI_ISL_1992344, EPI_ISL_1992345, EPI_ISL_1992346, EPI_ISL_1992347, EPI_ISL_1992348, EPI_ISL_1992349, EPI_ISL_1992350, EPI_ISL_1992351, EPI_ISL_1992352, EPI_ISL_1992353, EPI_ISL_1992354, EPI_ISL_1992355, EPI_ISL_1992356, EPI_ISL_1992357, EPI_ISL_1992358, EPI_ISL_1992359, EPI_ISL_1992360, EPI_ISL_1992361, EPI_ISL_1992362, EPI_ISL_1992363, EPI_ISL_1992364, EPI_ISL_1992365, EPI_ISL_1992366, EPI_ISL_1992367, EPI_ISL_1992368, EPI_ISL_1992369, EPI_ISL_1992370, EPI_ISL_1992371, EPI_ISL_1992373, EPI_ISL_1992374, EPI_ISL_1992375, EPI_ISL_1992376, EPI_ISL_1992377, EPI_ISL_1992378, EPI_ISL_1992379, EPI_ISL_1992380, EPI_ISL_1992381, EPI_ISL_1992382, EPI_ISL_1992383, EPI_ISL_1992384, EPI_ISL_1992385, EPI_ISL_1992386, EPI_ISL_1992387, EPI_ISL_1992388, EPI_ISL_1992389, EPI_ISL_1992390, EPI_ISL_1992391, EPI_ISL_1992392, EPI_ISL_1992393, EPI_ISL_1992407, EPI_ISL_1992665, EPI_ISL_1992777, EPI_ISL_1992980, EPI_ISL_1992984, EPI_ISL_1992985, EPI_ISL_1992986, EPI_ISL_1992987, EPI_ISL_1992988, EPI_ISL_1992989, EPI_ISL_1992990, EPI_ISL_1992991, EPI_ISL_1992992, EPI_ISL_1992993, EPI_ISL_1992994, EPI_ISL_1992995, EPI_ISL_1992996, EPI_ISL_1992997, EPI_ISL_1992998, EPI_ISL_1992999, EPI_ISL_1993000, EPI_ISL_1993001, EPI_ISL_1993002, EPI_ISL_1993003, EPI_ISL_1993004, EPI_ISL_1993005, EPI_ISL_1993006, EPI_ISL_1993007, EPI_ISL_1993008, EPI_ISL_1993009, EPI_ISL_1993010, EPI_ISL_1993011, EPI_ISL_1993012, EPI_ISL_1993013, EPI_ISL_1993014, EPI_ISL_1993015, EPI_ISL_1993016, EPI_ISL_1993017, EPI_ISL_1993018, EPI_ISL_1993019, EPI_ISL_1993020, EPI_ISL_1993021, EPI_ISL_1993022, EPI_ISL_1993023, EPI_ISL_1993024, EPI_ISL_1993025, EPI_ISL_1993026, EPI_ISL_1993027, EPI_ISL_1993028, EPI_ISL_1993029, EPI_ISL_1993030, EPI_ISL_1993031, EPI_ISL_1993032, EPI_ISL_1993033, EPI_ISL_1993034, EPI_ISL_1993035, EPI_ISL_1993036, EPI_ISL_1993037, EPI_ISL_1993040, EPI_ISL_1993042, EPI_ISL_1993043, EPI_ISL_1993044, EPI_ISL_1993045, EPI_ISL_1993046, EPI_ISL_1993047, EPI_ISL_1993048, EPI_ISL_1993049, EPI_ISL_1993050, EPI_ISL_1993051, EPI_ISL_1993052, EPI_ISL_1993053, EPI_ISL_1993054, EPI_ISL_1993055, EPI_ISL_1993056, EPI_ISL_1993057, EPI_ISL_1993058, EPI_ISL_1993059, EPI_ISL_1993060, EPI_ISL_1993061, EPI_ISL_1993062, EPI_ISL_1993063, EPI_ISL_1993064, EPI_ISL_1993065, EPI_ISL_1993066, EPI_ISL_1993067, EPI_ISL_1993068, EPI_ISL_1993069, EPI_ISL_1993070, EPI_ISL_1993071, EPI_ISL_1993072, EPI_ISL_1993073, EPI_ISL_1993074, EPI_ISL_1993075, EPI_ISL_1993076, EPI_ISL_1993077, EPI_ISL_1993078, EPI_ISL_1993079, EPI_ISL_1993080, EPI_ISL_1993081, EPI_ISL_1993082, EPI_ISL_1993083, EPI_ISL_1993084, EPI_ISL_1993085, EPI_ISL_1993086, EPI_ISL_1993087, EPI_ISL_1993088, EPI_ISL_1993089, EPI_ISL_1993090, EPI_ISL_1993091, EPI_ISL_1993092, EPI_ISL_1993093, EPI_ISL_1993094, EPI_ISL_1993097, EPI_ISL_1993098, EPI_ISL_1993099, EPI_ISL_1993100, EPI_ISL_1993101, EPI_ISL_1993102, EPI_ISL_1993103, EPI_ISL_1993104, EPI_ISL_1993105, EPI_ISL_1993106, EPI_ISL_1993107, EPI_ISL_1993108, EPI_ISL_1993109, EPI_ISL_1993110, EPI_ISL_1993111, EPI_ISL_1993112, EPI_ISL_1993113, EPI_ISL_1993114, EPI_ISL_1993115, EPI_ISL_1993116, EPI_ISL_1993117, EPI_ISL_1993118, EPI_ISL_1993119, EPI_ISL_1993120, EPI_ISL_1993121, EPI_ISL_1993122, EPI_ISL_1993123, EPI_ISL_1993124, EPI_ISL_1993125, EPI_ISL_1993126, EPI_ISL_1993127, EPI_ISL_1993128, EPI_ISL_1993129, EPI_ISL_1993130, EPI_ISL_1993131, EPI_ISL_1993132, EPI_ISL_1993133, EPI_ISL_1993134, EPI_ISL_1993135, EPI_ISL_1993136, EPI_ISL_1993137, EPI_ISL_1993138, EPI_ISL_1993139, EPI_ISL_1993140, EPI_ISL_1993141, EPI_ISL_1993142, EPI_ISL_1993143, EPI_ISL_1993144, EPI_ISL_1993145, EPI_ISL_1993146, EPI_ISL_1993147, EPI_ISL_1993148, EPI_ISL_1993149, EPI_ISL_1993150, EPI_ISL_1993151, EPI_ISL_1993152, EPI_ISL_1993153, EPI_ISL_1993210, EPI_ISL_1993222, EPI_ISL_1993223, EPI_ISL_1993225, EPI_ISL_1993226, EPI_ISL_1993228, EPI_ISL_1993231, EPI_ISL_1993232, EPI_ISL_1993234, EPI_ISL_1993235, EPI_ISL_1993236, EPI_ISL_1993239, EPI_ISL_1993240, EPI_ISL_1993241, EPI_ISL_1993243, EPI_ISL_1993248, EPI_ISL_1993249, EPI_ISL_1993252, EPI_ISL_1993255, EPI_ISL_1993256, EPI_ISL_1993257, EPI_ISL_1993258, EPI_ISL_1993259, EPI_ISL_1993261, EPI_ISL_1993265, EPI_ISL_1993266, EPI_ISL_1993269, EPI_ISL_1993271, EPI_ISL_1993287, EPI_ISL_2034996, EPI_ISL_2035000, EPI_ISL_2037981, EPI_ISL_2038260, EPI_ISL_2038261, EPI_ISL_2038262, EPI_ISL_2038263, EPI_ISL_2038266, EPI_ISL_2038267, EPI_ISL_2038268, EPI_ISL_2038269, EPI_ISL_2038270, EPI_ISL_2038271, EPI_ISL_2038272, EPI_ISL_2038273, EPI_ISL_2038274, EPI_ISL_2038275, EPI_ISL_2038276, EPI_ISL_2038277, EPI_ISL_2038278, EPI_ISL_2038279, EPI_ISL_2038280, EPI_ISL_2038281, EPI_ISL_2038282, EPI_ISL_2038283, EPI_ISL_2038284, EPI_ISL_2038285, EPI_ISL_2038286, EPI_ISL_2038287, EPI_ISL_2038288, EPI_ISL_2038289, EPI_ISL_2038290, EPI_ISL_2038291, EPI_ISL_2038292, EPI_ISL_2038293, EPI_ISL_2038294, EPI_ISL_2038295, EPI_ISL_2038296, EPI_ISL_2038297, EPI_ISL_2038298, EPI_ISL_2038299, EPI_ISL_2038300, EPI_ISL_2038301, EPI_ISL_2038303, EPI_ISL_2038304, EPI_ISL_2038305, EPI_ISL_2038306, EPI_ISL_2038307, EPI_ISL_2038309, EPI_ISL_2038310, EPI_ISL_2038311, EPI_ISL_2038312, EPI_ISL_2038313, EPI_ISL_2038314, EPI_ISL_2038315, EPI_ISL_2038316, EPI_ISL_2038317, EPI_ISL_2038318, EPI_ISL_2038319, EPI_ISL_2038320, EPI_ISL_2038321, EPI_ISL_2038322, EPI_ISL_2038323, EPI_ISL_2038324, EPI_ISL_2038325, EPI_ISL_2038326, EPI_ISL_2038327, EPI_ISL_2038328, EPI_ISL_2038329, EPI_ISL_2038459, EPI_ISL_2038460, EPI_ISL_2038461, EPI_ISL_2038462, EPI_ISL_2038463, EPI_ISL_2038464, EPI_ISL_2038465, EPI_ISL_2038466, EPI_ISL_2038467, EPI_ISL_2038468, EPI_ISL_2038469, EPI_ISL_2038470, EPI_ISL_2038471, EPI_ISL_2038472, EPI_ISL_2038473, EPI_ISL_2038474, EPI_ISL_2038475, EPI_ISL_2038476, EPI_ISL_2038477, EPI_ISL_2038478, EPI_ISL_2038479, EPI_ISL_2038480, EPI_ISL_2038481, EPI_ISL_2038482, EPI_ISL_2038483, EPI_ISL_2038484, EPI_ISL_2038485, EPI_ISL_2038486, EPI_ISL_2038487, EPI_ISL_2038488, EPI_ISL_2038489, EPI_ISL_2038490, EPI_ISL_2038491, EPI_ISL_2038492, EPI_ISL_2038493, EPI_ISL_2038494, EPI_ISL_2038495, EPI_ISL_2038496, EPI_ISL_2038497, EPI_ISL_2038498, EPI_ISL_2038499, EPI_ISL_2038500, EPI_ISL_2038501, EPI_ISL_2038502, EPI_ISL_2038503, EPI_ISL_2038504, EPI_ISL_2038505, EPI_ISL_2038506, EPI_ISL_2038507, EPI_ISL_2038508, EPI_ISL_2038509, EPI_ISL_2038510, EPI_ISL_2038511, EPI_ISL_2038513, EPI_ISL_2038514, EPI_ISL_2038515, EPI_ISL_2038516, EPI_ISL_2038517, EPI_ISL_2038518, EPI_ISL_2038519, EPI_ISL_2038520, EPI_ISL_2038521, EPI_ISL_2038522, EPI_ISL_2038524, EPI_ISL_2038525, EPI_ISL_2038526, EPI_ISL_2038527, EPI_ISL_2038528, EPI_ISL_2038529, EPI_ISL_2038530, EPI_ISL_2038531, EPI_ISL_2038532, EPI_ISL_2038533, EPI_ISL_2038534, EPI_ISL_2038535, EPI_ISL_2038536, EPI_ISL_2038537, EPI_ISL_2038538, EPI_ISL_2038539, EPI_ISL_2038540, EPI_ISL_2038541, EPI_ISL_2038542, EPI_ISL_2038543, EPI_ISL_2038544, EPI_ISL_2038545, EPI_ISL_2038546, EPI_ISL_2038547, EPI_ISL_2038548, EPI_ISL_2038549, EPI_ISL_2038550, EPI_ISL_2038551, EPI_ISL_2038552, EPI_ISL_2038553, EPI_ISL_2038554, EPI_ISL_2038555, EPI_ISL_2038556, EPI_ISL_2038557, EPI_ISL_2038558, EPI_ISL_2038559, EPI_ISL_2038560, EPI_ISL_2038561, EPI_ISL_2038562, EPI_ISL_2038563, EPI_ISL_2038565, EPI_ISL_2038566, EPI_ISL_2038567, EPI_ISL_2038568, EPI_ISL_2038569, EPI_ISL_2038570, EPI_ISL_2038571, EPI_ISL_2038572, EPI_ISL_2038573, EPI_ISL_2038574, EPI_ISL_2038575, EPI_ISL_2038576, EPI_ISL_2038577, EPI_ISL_2038578, EPI_ISL_2038579, EPI_ISL_2038580, EPI_ISL_2038581, EPI_ISL_2038582, EPI_ISL_2038583, EPI_ISL_2038584, EPI_ISL_2038585, EPI_ISL_2038586, EPI_ISL_2038587, EPI_ISL_2038588, EPI_ISL_2038591, EPI_ISL_2038592, EPI_ISL_2038593, EPI_ISL_2038594, EPI_ISL_2038595, EPI_ISL_2038596, EPI_ISL_2038597, EPI_ISL_2038598, EPI_ISL_2038599, EPI_ISL_2038600, EPI_ISL_2038601, EPI_ISL_2038602, EPI_ISL_2038603, EPI_ISL_2038604, EPI_ISL_2038605, EPI_ISL_2038606, EPI_ISL_2038607, EPI_ISL_2038608, EPI_ISL_2038609, EPI_ISL_2038610, EPI_ISL_2038611, EPI_ISL_2038612, EPI_ISL_2038613, EPI_ISL_2038614, EPI_ISL_2038615, EPI_ISL_2038616, EPI_ISL_2038617, EPI_ISL_2038618, EPI_ISL_2038619, EPI_ISL_2038620, EPI_ISL_2038621, EPI_ISL_2038622, EPI_ISL_2038624, EPI_ISL_2038625, EPI_ISL_2038626, EPI_ISL_2038627, EPI_ISL_2038628, EPI_ISL_2038629, EPI_ISL_2038630, EPI_ISL_2038631, EPI_ISL_2038632, EPI_ISL_2038633, EPI_ISL_2038634, EPI_ISL_2038635, EPI_ISL_2038636, EPI_ISL_2038637, EPI_ISL_2038638, EPI_ISL_2038639, EPI_ISL_2038640, EPI_ISL_2038641, EPI_ISL_2038642, EPI_ISL_2038643, EPI_ISL_2038644, EPI_ISL_2038645, EPI_ISL_2038646, EPI_ISL_2038647, EPI_ISL_2038648, EPI_ISL_2038649, EPI_ISL_2038650, EPI_ISL_2038651, EPI_ISL_2038652, EPI_ISL_2038653, EPI_ISL_2038654, EPI_ISL_2038655, EPI_ISL_2038656, EPI_ISL_2038657, EPI_ISL_2038659, EPI_ISL_2038660, EPI_ISL_2038661, EPI_ISL_2038662, EPI_ISL_2038663, EPI_ISL_2038664, EPI_ISL_2038665, EPI_ISL_2038666, EPI_ISL_2038667, EPI_ISL_2038668, EPI_ISL_2038669, EPI_ISL_2038670, EPI_ISL_2038671, EPI_ISL_2038672, EPI_ISL_2038673, EPI_ISL_2038674, EPI_ISL_2038675, EPI_ISL_2038676, EPI_ISL_2038677, EPI_ISL_2038678, EPI_ISL_2038679, EPI_ISL_2038680, EPI_ISL_2038681, EPI_ISL_2038682, EPI_ISL_2038683, EPI_ISL_2038684, EPI_ISL_2038685, EPI_ISL_2038686, EPI_ISL_2038687, EPI_ISL_2038688, EPI_ISL_2038689, EPI_ISL_2038690, EPI_ISL_2038691, EPI_ISL_2038692, EPI_ISL_2038693, EPI_ISL_2038694, EPI_ISL_2038695, EPI_ISL_2038696, EPI_ISL_2038697, EPI_ISL_2038698, EPI_ISL_2038699, EPI_ISL_2038700, EPI_ISL_2038701, EPI_ISL_2038702, EPI_ISL_2038703, EPI_ISL_2038704, EPI_ISL_2038705, EPI_ISL_2038706, EPI_ISL_2038707, EPI_ISL_2038708, EPI_ISL_2038709, EPI_ISL_2038710, EPI_ISL_2038711, EPI_ISL_2038712, EPI_ISL_2038713, EPI_ISL_2038714, EPI_ISL_2038715, EPI_ISL_2038716, EPI_ISL_2038717, EPI_ISL_2038718, EPI_ISL_2038719, EPI_ISL_2038720, EPI_ISL_2038721, EPI_ISL_2038722, EPI_ISL_2038723, EPI_ISL_2038724, EPI_ISL_2038725, EPI_ISL_2038726, EPI_ISL_2038727, EPI_ISL_2038728, EPI_ISL_2038729, EPI_ISL_2038730, EPI_ISL_2038731, EPI_ISL_2038732, EPI_ISL_2038733, EPI_ISL_2038734, EPI_ISL_2038735, EPI_ISL_2038736, EPI_ISL_2038737, EPI_ISL_2038738, EPI_ISL_2038739, EPI_ISL_2038740, EPI_ISL_2038741, EPI_ISL_2038742, EPI_ISL_2038743, EPI_ISL_2038744, EPI_ISL_2038745, EPI_ISL_2038746, EPI_ISL_2038747, EPI_ISL_2038748, EPI_ISL_2038749, EPI_ISL_2038750, EPI_ISL_2038752, EPI_ISL_2038753, EPI_ISL_2038754, EPI_ISL_2038755, EPI_ISL_2038756, EPI_ISL_2038757, EPI_ISL_2038758, EPI_ISL_2038759, EPI_ISL_2038760, EPI_ISL_2038761, EPI_ISL_2038762, EPI_ISL_2038763, EPI_ISL_2038764, EPI_ISL_2038765, EPI_ISL_2038766, EPI_ISL_2038767, EPI_ISL_2038768, EPI_ISL_2038769, EPI_ISL_2038770, EPI_ISL_2038771, EPI_ISL_2038772, EPI_ISL_2038773, EPI_ISL_2038774, EPI_ISL_2038775, EPI_ISL_2038776, EPI_ISL_2038777, EPI_ISL_2038778, EPI_ISL_2038779, EPI_ISL_2038780, EPI_ISL_2038781, EPI_ISL_2038782, EPI_ISL_2038783, EPI_ISL_2038784, EPI_ISL_2038785, EPI_ISL_2038786, EPI_ISL_2038787, EPI_ISL_2038788, EPI_ISL_2038789, EPI_ISL_2038790, EPI_ISL_2038791, EPI_ISL_2038792, EPI_ISL_2038793, EPI_ISL_2038794, EPI_ISL_2038795, EPI_ISL_2038796, EPI_ISL_2038797, EPI_ISL_2038798, EPI_ISL_2043633, EPI_ISL_2043955, EPI_ISL_2043982

| | | | |
|---|---|---|---|
| see above | Fulgent Genetics | Centers for Disease Control and Prevention Division of Viral Diseases, Pathogen Discovery | Adrian Paskey; Becky Tsai; Benafsh Sapra; Benjamin Rambo-Martin; Christopher Gulvick; Clinton R. Paden; Dakota Howard; Darlene Wagner; Dhwani Batra; Doreen Ng; Duncan MacCannell; Harry Gao; James Xie; Jason Caravas; John Gao; Joseph Fierro; Kara Moser; Matthew Schmerer; Mickey Li; Peter W. Cook; Scott Sammons; Shatavia Morrison; Yan Meng; Yvette Unoarumhi |
| EPI_ISL_1707680, EPI_ISL_1707681, EPI_ISL_1707682, EPI_ISL_1707683, EPI_ISL_1707684, EPI_ISL_1707685 | Fulgent Genetics | Los Angeles County Public Health Laboratories | P. Hemarajata et al. |
| EPI_ISL_2404801 | GOOD SAMARITAN HOSPTIAL | Santa Clara County public Health Laboratory | Santa Clara County Public Health Department |
| EPI_ISL_2659073 | Good Samaritan Hospital | Santa Clara County Public Health Laboratory | Santa Clara County Public Health Department |
| EPI_ISL_1824116, EPI_ISL_2967926, EPI_ISL_2967927 | Good Samaritan Hospital | Santa Clara County public Health Laboratory | Santa Clara County Public Health Department |

EPI_ISL_1735217, EPI_ISL_1735227, EPI_ISL_1735228, EPI_ISL_1735238, EPI_ISL_1735291, EPI_ISL_1735300, EPI_ISL_1735323, EPI_ISL_1735429, EPI_ISL_1735438, EPI_ISL_1753510, EPI_ISL_1796139, EPI_ISL_1796218, EPI_ISL_1796229, EPI_ISL_1796231, EPI_ISL_1796237, EPI_ISL_1796241, EPI_ISL_1796250, EPI_ISL_1796251, EPI_ISL_1796259, EPI_ISL_1796275, EPI_ISL_1796285, EPI_ISL_1796341, EPI_ISL_1796428, EPI_ISL_1796435, EPI_ISL_1796454, EPI_ISL_1796474, EPI_ISL_1796487, EPI_ISL_1796500, EPI_ISL_1796595, EPI_ISL_1796613, EPI_ISL_1796621, EPI_ISL_1796630, EPI_ISL_1796639, EPI_ISL_1796641, EPI_ISL_1796708, EPI_ISL_1796718, EPI_ISL_1796853, EPI_ISL_1796869, EPI_ISL_1796882, EPI_ISL_1796894, EPI_ISL_1796897, EPI_ISL_1796911, EPI_ISL_1796922, EPI_ISL_1796977, EPI_ISL_1796992, EPI_ISL_1796993, EPI_ISL_1797102, EPI_ISL_1797124, EPI_ISL_1797134, EPI_ISL_1797265, EPI_ISL_1797277, EPI_ISL_1797299, EPI_ISL_1797335, EPI_ISL_1797371, EPI_ISL_1797385, EPI_ISL_1797392, EPI_ISL_1797395, EPI_ISL_1797416, EPI_ISL_1797425, EPI_ISL_1803526, EPI_ISL_1803535, EPI_ISL_1803554, EPI_ISL_1803592, EPI_ISL_1803593, EPI_ISL_1803614, EPI_ISL_1803627, EPI_ISL_1803663, EPI_ISL_1803669, EPI_ISL_1803673, EPI_ISL_1803674, EPI_ISL_1803676, EPI_ISL_1803694, EPI_ISL_1803697, EPI_ISL_1803704, EPI_ISL_1803709, EPI_ISL_1803722, EPI_ISL_1803729, EPI_ISL_1803815, EPI_ISL_1803820, EPI_ISL_1803964, EPI_ISL_1803987, EPI_ISL_1803990, EPI_ISL_1803997, EPI_ISL_1804000, EPI_ISL_1804083, EPI_ISL_1804094, EPI_ISL_1804095, EPI_ISL_1804103, EPI_ISL_1804108, EPI_ISL_1804122, EPI_ISL_1804135, EPI_ISL_1804143, EPI_ISL_1804169, EPI_ISL_1804179, EPI_ISL_1804191, EPI_ISL_1804202, EPI_ISL_1804213, EPI_ISL_1804256, EPI_ISL_1804518, EPI_ISL_1804524, EPI_ISL_1804534, EPI_ISL_1804535, EPI_ISL_1804554, EPI_ISL_1804564, EPI_ISL_1804568, EPI_ISL_1804583, EPI_ISL_1804589, EPI_ISL_1804601, EPI_ISL_1804611, EPI_ISL_1804625, EPI_ISL_1804643, EPI_ISL_1804676, EPI_ISL_1804681, EPI_ISL_1804702, EPI_ISL_1804713, EPI_ISL_1804722, EPI_ISL_1804729, EPI_ISL_1818008, EPI_ISL_1818009, EPI_ISL_1818028, EPI_ISL_1906970, EPI_ISL_1907014, EPI_ISL_1907023, EPI_ISL_1907031, EPI_ISL_1907068, EPI_ISL_1907098, EPI_ISL_1907130, EPI_ISL_1907133, EPI_ISL_1907221, EPI_ISL_1907252, EPI_ISL_1907255, EPI_ISL_1907262, EPI_ISL_1907283, EPI_ISL_1907384, EPI_ISL_1907395, EPI_ISL_1907424, EPI_ISL_1907455, EPI_ISL_1907456, EPI_ISL_1907464, EPI_ISL_1907469, EPI_ISL_1907475, EPI_ISL_1907480, EPI_ISL_1907492, EPI_ISL_1907493, EPI_ISL_1907513, EPI_ISL_1907520, EPI_ISL_1907521, EPI_ISL_1907531, EPI_ISL_1907544, EPI_ISL_1907546, EPI_ISL_1907679, EPI_ISL_1907692, EPI_ISL_1907702, EPI_ISL_1907745, EPI_ISL_1907785, EPI_ISL_1907797, EPI_ISL_1907831, EPI_ISL_1907832, EPI_ISL_1907851, EPI_ISL_1907860, EPI_ISL_1907873, EPI_ISL_1907875, EPI_ISL_1907879, EPI_ISL_1908068, EPI_ISL_1908103, EPI_ISL_1908114, EPI_ISL_1908138, EPI_ISL_1923183, EPI_ISL_1923209, EPI_ISL_1923235, EPI_ISL_1923287, EPI_ISL_1923355, EPI_ISL_1923393, EPI_ISL_1923399, EPI_ISL_1923415, EPI_ISL_1923426, EPI_ISL_1923446, EPI_ISL_1923453, EPI_ISL_1923459, EPI_ISL_1923638, EPI_ISL_1923639, EPI_ISL_1923650, EPI_ISL_1923711, EPI_ISL_1923743, EPI_ISL_1923763, EPI_ISL_1923768, EPI_ISL_1923784, EPI_ISL_1923785, EPI_ISL_1923793, EPI_ISL_1923832, EPI_ISL_1923833, EPI_ISL_1923847, EPI_ISL_1923848, EPI_ISL_1923861, EPI_ISL_1923926, EPI_ISL_1923936, EPI_ISL_1923979, EPI_ISL_1923990, EPI_ISL_1923999, EPI_ISL_1924035, EPI_ISL_1924060, EPI_ISL_1924080, EPI_ISL_1924110, EPI_ISL_1924143, EPI_ISL_1924231, EPI_ISL_1990551, EPI_ISL_1990555, EPI_ISL_1990560, EPI_ISL_1990590, EPI_ISL_1990594, EPI_ISL_1990613, EPI_ISL_1990620, EPI_ISL_1990678, EPI_ISL_1990723, EPI_ISL_1990836, EPI_ISL_1990848, EPI_ISL_1990859, EPI_ISL_1990889, EPI_ISL_1990901, EPI_ISL_1990902, EPI_ISL_1990913, EPI_ISL_1990918, EPI_ISL_1990924, EPI_ISL_1990932, EPI_ISL_1990943, EPI_ISL_1990954, EPI_ISL_1990957, EPI_ISL_1990975, EPI_ISL_1990986, EPI_ISL_1991010, EPI_ISL_1991022, EPI_ISL_1991061, EPI_ISL_1991073, EPI_ISL_1991087, EPI_ISL_1991100, EPI_ISL_1991113, EPI_ISL_1991134, EPI_ISL_1991160, EPI_ISL_1991162, EPI_ISL_1991269, EPI_ISL_1991300, EPI_ISL_1991301, EPI_ISL_1991308, EPI_ISL_1991487, EPI_ISL_1991501, EPI_ISL_1991513, EPI_ISL_1991524, EPI_ISL_1991545, EPI_ISL_1991725, EPI_ISL_1991734, EPI_ISL_1991740, EPI_ISL_1991745, EPI_ISL_1991748, EPI_ISL_1991758, EPI_ISL_1991784, EPI_ISL_1991785, EPI_ISL_1991805, EPI_ISL_1991824, EPI_ISL_1991826, EPI_ISL_2009725, EPI_ISL_2009845, EPI_ISL_2009856, EPI_ISL_2009887, EPI_ISL_2009896, EPI_ISL_2009907, EPI_ISL_2009917, EPI_ISL_2010005, EPI_ISL_2010015, EPI_ISL_2010020, EPI_ISL_2010062, EPI_ISL_2010175, EPI_ISL_2010179, EPI_ISL_2010181, EPI_ISL_2010186, EPI_ISL_2010192, EPI_ISL_2010201, EPI_ISL_2010209, EPI_ISL_2010210, EPI_ISL_2010215, EPI_ISL_2010217, EPI_ISL_2010229, EPI_ISL_2010237, EPI_ISL_2010273, EPI_ISL_2010282, EPI_ISL_2010316, EPI_ISL_2010413, EPI_ISL_2010427, EPI_ISL_2010497, EPI_ISL_2010552, EPI_ISL_2010553, EPI_ISL_2010560, EPI_ISL_2010561, EPI_ISL_2010589, EPI_ISL_2010714, EPI_ISL_2010747, EPI_ISL_2010758, EPI_ISL_2010778, EPI_ISL_2010785, EPI_ISL_2010796, EPI_ISL_2010797, EPI_ISL_2035278, EPI_ISL_2035285, EPI_ISL_2035287, EPI_ISL_2097374, EPI_ISL_2097388, EPI_ISL_2097391, EPI_ISL_2097395, EPI_ISL_2097422, EPI_ISL_2097430, EPI_ISL_2097469, EPI_ISL_2097496, EPI_ISL_2097624, EPI_ISL_2097638, EPI_ISL_2097693, EPI_ISL_2097702, EPI_ISL_2097845, EPI_ISL_2097898, EPI_ISL_2097899, EPI_ISL_2097966, EPI_ISL_2098005, EPI_ISL_2098011, EPI_ISL_2098029, EPI_ISL_2098049, EPI_ISL_2098077, EPI_ISL_2098095, EPI_ISL_2098106, EPI_ISL_2132607, EPI_ISL_2132613, EPI_ISL_2144118, EPI_ISL_2144141, EPI_ISL_2144340, EPI_ISL_2144371, EPI_ISL_2144401, EPI_ISL_2144411, EPI_ISL_2144413, EPI_ISL_2144520, EPI_ISL_2144583, EPI_ISL_2144601, EPI_ISL_2144619, EPI_ISL_2144655, EPI_ISL_2144747, EPI_ISL_2144793, EPI_ISL_2144803, EPI_ISL_2144843, EPI_ISL_2144929, EPI_ISL_2145000, EPI_ISL_2145463, EPI_ISL_2145501, EPI_ISL_2145576, EPI_ISL_2145685, EPI_ISL_2159490, EPI_ISL_2159518, EPI_ISL_2159520, EPI_ISL_2159539, EPI_ISL_2159541, EPI_ISL_2159686, EPI_ISL_2159774, EPI_ISL_2159787, EPI_ISL_2159794, EPI_ISL_2159795, EPI_ISL_2159806, EPI_ISL_2159807, EPI_ISL_2159811, EPI_ISL_2159812, EPI_ISL_2159815, EPI_ISL_2159817, EPI_ISL_2159823, EPI_ISL_2159830, EPI_ISL_2159839, EPI_ISL_2159842, EPI_ISL_2159853, EPI_ISL_2159872, EPI_ISL_2159891, EPI_ISL_2159899, EPI_ISL_2159910, EPI_ISL_2160089, EPI_ISL_2160094, EPI_ISL_2160105, EPI_ISL_2160115, EPI_ISL_2160116, EPI_ISL_2160125, EPI_ISL_2160131, EPI_ISL_2160156, EPI_ISL_2160224, EPI_ISL_2160231, EPI_ISL_2160327, EPI_ISL_2160335, EPI_ISL_2160348, EPI_ISL_2160360, EPI_ISL_2160375, EPI_ISL_2160481, EPI_ISL_2160490, EPI_ISL_2160514, EPI_ISL_2160572, EPI_ISL_2160576, EPI_ISL_2247687, EPI_ISL_2247690, EPI_ISL_2247842, EPI_ISL_2247899, EPI_ISL_2248330

| | | | |
|---|---|---|---|
| see above | Helix/Illumina | Centers for Disease Control and Prevention Division of Viral Diseases, Pathogen Discovery | Adrian Paskey; Alexandre Bolze; Ary Ascencio; Benjamin Rambo-Martin; Brad Sickler; Charlotte Rivera-Garcia; Christine Tran; Christopher Gulvick; Clinton R. Paden; Dakota Howard; Darlene Wagner; David Becker; Dhwani Batra; Duncan MacCannell; Efren Sandoval; Eileen de Feo; Elizabeth Cirulli; Eric Allen; Geraint Levan; James Lu; Jan Antico; Jason Caravas; Jason Nguyen; Jimmy Ramirez; Jingtao Liu; Kara Moser; Kelly Schiabor Barrett; Kim Gietzen; Magnus Isaksson; Marc Laurent; Matthew Schmerer; Matthew Tolentino; Nicole L. Washington; Peter W. Cook; Phil Febbo; Ryan Cho; Scott Sammons; Shannon Wickline; Shatavia Morrison; Sherry Wang; Simon White; Tyler Cassens; William Lee; Yvette Unoarumhi |

EPI_ISL_2102528, EPI_ISL_2102537, EPI_ISL_2102550, EPI_ISL_2102561, EPI_ISL_2102752, EPI_ISL_2102753, EPI_ISL_2102791, EPI_ISL_2102797, EPI_ISL_2102806, EPI_ISL_2102808, EPI_ISL_2102822, EPI_ISL_2102832, EPI_ISL_2102838, EPI_ISL_2102847, EPI_ISL_2102852, EPI_ISL_2102853, EPI_ISL_2102872, EPI_ISL_2102888, EPI_ISL_2102903, EPI_ISL_2102906, EPI_ISL_2102935, EPI_ISL_2102948, EPI_ISL_2102964, EPI_ISL_2102979, EPI_ISL_2102986, EPI_ISL_2102992, EPI_ISL_2103203, EPI_ISL_2103204, EPI_ISL_2103205, EPI_ISL_2103206, EPI_ISL_2103207, EPI_ISL_2103208, EPI_ISL_2103209, EPI_ISL_2103210, EPI_ISL_2103211, EPI_ISL_2103212, EPI_ISL_2103213, EPI_ISL_2103214, EPI_ISL_2103215, EPI_ISL_2103216, EPI_ISL_2103217, EPI_ISL_2103218, EPI_ISL_2103219, EPI_ISL_2103220, EPI_ISL_2103221, EPI_ISL_2103222, EPI_ISL_2103223, EPI_ISL_2103224, EPI_ISL_2103225, EPI_ISL_2103226, EPI_ISL_2103227, EPI_ISL_2103228, EPI_ISL_2228264, EPI_ISL_2228284, EPI_ISL_2228285, EPI_ISL_2228288, EPI_ISL_2228289, EPI_ISL_2228290, EPI_ISL_2228295, EPI_ISL_2228301, EPI_ISL_2228303, EPI_ISL_2228304, EPI_ISL_2228310, EPI_ISL_2228314, EPI_ISL_2228319, EPI_ISL_2228328, EPI_ISL_2228336, EPI_ISL_2228343, EPI_ISL_2228359, EPI_ISL_2228363, EPI_ISL_2228366, EPI_ISL_2228368, EPI_ISL_2228377, EPI_ISL_2228395, EPI_ISL_2228399, EPI_ISL_2228404, EPI_ISL_2228411, EPI_ISL_2228412, EPI_ISL_2228416, EPI_ISL_2228419, EPI_ISL_2228422, EPI_ISL_2228424, EPI_ISL_2341442, EPI_ISL_2341478, EPI_ISL_2341488, EPI_ISL_2341509, EPI_ISL_2341542, EPI_ISL_2341567, EPI_ISL_2500187, EPI_ISL_2500206, EPI_ISL_2500214, EPI_ISL_2658586, EPI_ISL_2658609, EPI_ISL_2658651, EPI_ISL_2658699, EPI_ISL_2658732, EPI_ISL_2758517, EPI_ISL_2758531

| | | | |
|---|---|---|---|
| see above | Humboldt County Public Health Laboratory | Chan-Zuckerberg Biohub | CZB Cliahub Consortium |
| EPI_ISL_2259237, EPI_ISL_2259241 | Huntington Memorial Hospital | Los Angeles County Public Health Laboratories | P. Hemarajata et al. |
| EPI_ISL_2604241, EPI_ISL_2604250 | Infectious Diseases, Chan-Zuckerberg Biohub Covidtracker | Infectious Diseases, Chan-Zuckerberg Biohub Covidtracker | CZB Cliahub Consortium |

EPI_ISL_1795476, EPI_ISL_1795480, EPI_ISL_1795494, EPI_ISL_1795606, EPI_ISL_2014190, EPI_ISL_2087709, EPI_ISL_2087772, EPI_ISL_2087845, EPI_ISL_2087859, EPI_ISL_2088291, EPI_ISL_2088565, EPI_ISL_2089006, EPI_ISL_2089217, EPI_ISL_2089299, EPI_ISL_2089312, EPI_ISL_2089408, EPI_ISL_2132366, EPI_ISL_2282944, EPI_ISL_2282953, EPI_ISL_2283329, EPI_ISL_2283355, EPI_ISL_2283398,

| | | | |
|---|---|---|---|
| EPI_ISL_2283423, EPI_ISL_2283478, EPI_ISL_2283554, EPI_ISL_2283580, EPI_ISL_2283588, EPI_ISL_2284220 | | | |
| see above | Infinity Biologix | Centers for Disease Control and Prevention Division of Viral Diseases, Pathogen Discovery | Adrian Paskey; Benjamin Rambo-Martin; Chirayu Goswami; Christian Bixby; Christopher Gulvick; Clinton R. Paden; Dakota Howard; Darlene Wagner; Dhwani Batra; Duncan MacCannell; Jason Caravas; Jonathan Schultz; Kara Moser; Matthew Schmerer; Peter W. Cook; Robin Grimwood; Russ Hager; Scott Sammons; Shatavia Morrison; Yihe Wang; Yvette Unoarumhi |
| EPI_ISL_1712538, EPI_ISL_1712539, EPI_ISL_1712540, EPI_ISL_1712541 | Innovative Genomics Institute, UC Berkeley | Innovative Genomics Institute, UC Berkeley | Holly Gildea; John Boyle; Phil Frankino; Stacia Wyman |
| EPI_ISL_1913217, EPI_ISL_1913225 | KAISER SAN JOSE | Santa Clara County public Health Laboratory | Santa Clara County Public Health Department |
| EPI_ISL_2404800 | KAISER SANTA CLARA | Santa Clara County public Health Laboratory | Santa Clara County Public Health Department |
| EPI_ISL_2753991, EPI_ISL_2754001 | Kaiser Regional | Santa Clara County Public Health Laboratory | Santa Clara County Public Health Department |
| EPI_ISL_2753968, EPI_ISL_2753969, EPI_ISL_2753970, EPI_ISL_2753972, EPI_ISL_2753973 | Kaiser San Jose | Santa Clara County Public Health Laboratory | Santa Clara County Public Health Department |
| EPI_ISL_1824111, EPI_ISL_1824112, EPI_ISL_1824113, EPI_ISL_1824114, EPI_ISL_1824115 | Kaiser San Jose | Santa Clara County public Health Laboratory | Santa Clara County Public Health Department |
| EPI_ISL_2658984 | Kaiser Santa Clara | Santa Clara County Public Health Laboratory | Santa Clara County Public Health Department |
| EPI_ISL_1824120 | Kaiser Santa Clara | Santa Clara County public Health Laboratory | Santa Clara County Public Health Department |
| EPI_ISL_1681623, EPI_ISL_1684758, EPI_ISL_1684786, EPI_ISL_1684833, EPI_ISL_1684834, EPI_ISL_1684836, EPI_ISL_1684848, EPI_ISL_1684853, EPI_ISL_1684854, EPI_ISL_1684855, EPI_ISL_1684883, EPI_ISL_1684894, EPI_ISL_1684895, EPI_ISL_1684896, EPI_ISL_1684897, EPI_ISL_1684898, EPI_ISL_1798260, EPI_ISL_1798292, EPI_ISL_1798297, EPI_ISL_1798298, EPI_ISL_1798299, EPI_ISL_1798300, EPI_ISL_1798301, EPI_ISL_1798302, EPI_ISL_1798308, EPI_ISL_1798309, EPI_ISL_1798310, EPI_ISL_1798313, EPI_ISL_1798317, EPI_ISL_1798318, EPI_ISL_1798321, EPI_ISL_1798322, EPI_ISL_1798326, EPI_ISL_1798328, EPI_ISL_1798330, EPI_ISL_1798562, EPI_ISL_1798564, EPI_ISL_1798567, EPI_ISL_1798569, EPI_ISL_1798570, EPI_ISL_1798587, EPI_ISL_1798588, EPI_ISL_1798590, EPI_ISL_1798592, EPI_ISL_1798594, EPI_ISL_1798596, EPI_ISL_1798598, EPI_ISL_1798601, EPI_ISL_1798602, EPI_ISL_1798629, EPI_ISL_1798897, EPI_ISL_1798949, EPI_ISL_1798950, EPI_ISL_1798951, EPI_ISL_1798952, EPI_ISL_1798953, EPI_ISL_1798954, EPI_ISL_1798959, EPI_ISL_1798960, EPI_ISL_1798976, EPI_ISL_1798978, EPI_ISL_1798979, EPI_ISL_1798980, EPI_ISL_1798981, EPI_ISL_1798982, EPI_ISL_1798983, EPI_ISL_1798984, EPI_ISL_1798985, EPI_ISL_1798986, EPI_ISL_1798987, EPI_ISL_1798988, EPI_ISL_1798989, EPI_ISL_1798991, EPI_ISL_1798992, EPI_ISL_1798993, EPI_ISL_1798994, EPI_ISL_1798995, EPI_ISL_1798996, EPI_ISL_1798997, EPI_ISL_1798998, EPI_ISL_1798999, EPI_ISL_1799000, EPI_ISL_1799001, EPI_ISL_1799002, EPI_ISL_1799003, EPI_ISL_1799004, EPI_ISL_1799006, EPI_ISL_1799007, EPI_ISL_1799008, EPI_ISL_1799009, EPI_ISL_1799079, EPI_ISL_1799142, EPI_ISL_1799143, EPI_ISL_1799153, EPI_ISL_1799167, EPI_ISL_1799168, EPI_ISL_1799169, EPI_ISL_1799259, EPI_ISL_1799277, EPI_ISL_1799290, EPI_ISL_1799291, EPI_ISL_1799292, EPI_ISL_1799335, EPI_ISL_1799336, EPI_ISL_1799337, EPI_ISL_1799338, EPI_ISL_1799399, EPI_ISL_1799400, EPI_ISL_1799401, EPI_ISL_1799425, EPI_ISL_1799426, EPI_ISL_1799427, EPI_ISL_1799428, EPI_ISL_1799447, EPI_ISL_1799448, EPI_ISL_1799449, EPI_ISL_1799454, EPI_ISL_1799455, EPI_ISL_1799456, EPI_ISL_1799457, EPI_ISL_1799471, EPI_ISL_1799472, EPI_ISL_1799473, EPI_ISL_1799474, EPI_ISL_1799484, EPI_ISL_1799485, EPI_ISL_1799535, EPI_ISL_1799551, EPI_ISL_1799552, EPI_ISL_1799564, EPI_ISL_1799605, EPI_ISL_1799620, EPI_ISL_1799621, EPI_ISL_1799640, EPI_ISL_1925654, EPI_ISL_1926318, EPI_ISL_1926668, EPI_ISL_1930643, EPI_ISL_1930646, EPI_ISL_1930650, EPI_ISL_1930653, EPI_ISL_1930656, EPI_ISL_1930659, EPI_ISL_1930668, EPI_ISL_1930669, EPI_ISL_1930674, EPI_ISL_1930675, EPI_ISL_1930676, EPI_ISL_1930679, EPI_ISL_1930681, EPI_ISL_1930682, EPI_ISL_1930687, EPI_ISL_1930688, EPI_ISL_1930689, EPI_ISL_1930701, EPI_ISL_1930702, EPI_ISL_1930703, EPI_ISL_1930704, EPI_ISL_1930705, EPI_ISL_1930706, EPI_ISL_1930707, EPI_ISL_1930708, EPI_ISL_1930709, EPI_ISL_1930710, EPI_ISL_1930727, EPI_ISL_1930745, EPI_ISL_1930786, EPI_ISL_1930787, EPI_ISL_1930788, EPI_ISL_1930790, EPI_ISL_1930791, EPI_ISL_1930792, EPI_ISL_1930829, EPI_ISL_1930830, EPI_ISL_1930831, EPI_ISL_1930832, EPI_ISL_1930833, EPI_ISL_1930834, EPI_ISL_1930835, EPI_ISL_1930836, EPI_ISL_1930848, EPI_ISL_1930849, EPI_ISL_1930850, EPI_ISL_1930851, EPI_ISL_1930852, EPI_ISL_1930853, EPI_ISL_1930854, EPI_ISL_1930855, EPI_ISL_1930857, EPI_ISL_1930858, EPI_ISL_1930859, EPI_ISL_1930860, EPI_ISL_1930861, EPI_ISL_1930970, EPI_ISL_1930989, EPI_ISL_1930992, EPI_ISL_1930995, EPI_ISL_1930998, EPI_ISL_1931001, EPI_ISL_1931003, EPI_ISL_1931034, EPI_ISL_1931037, EPI_ISL_1931040, EPI_ISL_1931042, EPI_ISL_1931046, EPI_ISL_1931049, EPI_ISL_1931052, EPI_ISL_1931122, EPI_ISL_1931123, EPI_ISL_1931124, EPI_ISL_1931126, EPI_ISL_1931127, EPI_ISL_1931129, EPI_ISL_1931130, EPI_ISL_1931131, EPI_ISL_1931133, EPI_ISL_1931134, EPI_ISL_1931136, EPI_ISL_1931418, EPI_ISL_1931484, EPI_ISL_1931487, EPI_ISL_1931490, EPI_ISL_1931492, EPI_ISL_1931495, EPI_ISL_1931606, EPI_ISL_1931609, EPI_ISL_1931612, EPI_ISL_1931615, EPI_ISL_1931616, EPI_ISL_1931623, EPI_ISL_1931672, EPI_ISL_1931673, EPI_ISL_1931675, EPI_ISL_1931676, EPI_ISL_1931678, EPI_ISL_1931679, EPI_ISL_1931680, EPI_ISL_1931683, EPI_ISL_1931685, EPI_ISL_1931708, EPI_ISL_1931710, EPI_ISL_1931711, EPI_ISL_1931713, EPI_ISL_1931714, EPI_ISL_1931716, EPI_ISL_1931717, EPI_ISL_1931718, EPI_ISL_1931742, EPI_ISL_1931745, EPI_ISL_1931748, EPI_ISL_1931751, EPI_ISL_1931754, EPI_ISL_1931755, EPI_ISL_1931756, EPI_ISL_1931758, EPI_ISL_1931759, EPI_ISL_1931760, EPI_ISL_1931762, EPI_ISL_1931994, EPI_ISL_1931996, EPI_ISL_1932045, EPI_ISL_1932048, EPI_ISL_1932050, EPI_ISL_1932051, EPI_ISL_1932053, EPI_ISL_1932054, EPI_ISL_2044694, EPI_ISL_2044742, EPI_ISL_2045197, EPI_ISL_2046011, EPI_ISL_2046027, EPI_ISL_2046028, EPI_ISL_2046029, EPI_ISL_2046064, EPI_ISL_2046071, EPI_ISL_2046072, EPI_ISL_2046190, EPI_ISL_2046216, EPI_ISL_2046217, EPI_ISL_2046218, EPI_ISL_2046219, EPI_ISL_2046220, EPI_ISL_2046221, EPI_ISL_2046252, EPI_ISL_2046253, EPI_ISL_2046254, EPI_ISL_2046312, EPI_ISL_2046356, EPI_ISL_2046357, EPI_ISL_2046358, EPI_ISL_2046359, EPI_ISL_2046360, EPI_ISL_2046361, EPI_ISL_2046362, EPI_ISL_2046363, EPI_ISL_2046364, EPI_ISL_2046409, EPI_ISL_2046410, EPI_ISL_2046411, EPI_ISL_2046412, EPI_ISL_2046413, EPI_ISL_2046414, EPI_ISL_2046415, EPI_ISL_2046416, EPI_ISL_2046417, EPI_ISL_2046418, EPI_ISL_2046419, EPI_ISL_2046420, EPI_ISL_2046421, EPI_ISL_2046422, EPI_ISL_2046423, EPI_ISL_2046424, EPI_ISL_2046425, EPI_ISL_2046426, EPI_ISL_2046427, EPI_ISL_2046428, EPI_ISL_2046429, EPI_ISL_2046430, EPI_ISL_2046431, EPI_ISL_2046488, EPI_ISL_2046489, EPI_ISL_2046490, EPI_ISL_2046491, EPI_ISL_2046492, EPI_ISL_2046493, EPI_ISL_2046564, EPI_ISL_2046565, EPI_ISL_2046566, EPI_ISL_2046567, EPI_ISL_2046568, EPI_ISL_2046569, EPI_ISL_2046570, EPI_ISL_2046571, EPI_ISL_2046572, EPI_ISL_2046573, EPI_ISL_2046574, EPI_ISL_2046611, EPI_ISL_2046612, EPI_ISL_2046613, EPI_ISL_2046614, EPI_ISL_2046615, EPI_ISL_2046680, EPI_ISL_2046716, EPI_ISL_2046717, EPI_ISL_2046776, EPI_ISL_2046778, EPI_ISL_2046790, EPI_ISL_2046792, EPI_ISL_2046793, EPI_ISL_2046821 | | | |
| see above | Laboratory Corporation of America | Centers for Disease Control and Prevention Division of Viral Diseases, Pathogen Discovery | Adrian Paskey; Amanda Douglas; Amanda Suchanek; Andrea Throop; Ayla Burns; Benjamin Rambo-Martin; Bobbi Croy; Brian Krueger; Brian Norvell; Christopher Gulvick; Christos Petropoulos; Clinton R. Paden; Craig Lukasik; Dakota Howard; Darlene Wagner; Debbie Boles; Dhwani Batra; Duncan MacCannell; Eyad Almasri; Goran Stevovic; Howard Engler; Hrushikesh Deshmukh; Jake Humphrey; Jana Schroth; Jason Caravas; Joe Voshell; John Pruitt; Jonathan Meltzer; Jonathan Williams; Kara Moser; Kimberly Wagner; Lax Iyer; Lyndon Tilson; Manoj Jain; Marcia Eisenberg; Mary Ann Cristobal; Mary Williamson; Matthew Schmerer; Michael Levandoski; Mike Sapeta; Mindy Nye; Minoo Agarwal; Mohan Kolli; Nuthawin Charoensri; Oren Cohen; Peter W. Cook; Prashant Gupta; Qian Zeng; Rama Ghatti; Scott Parker; Scott Ryan; Scott Sammons; Shatavia Morrison; Stanley Letovsky; Steven Ragan; Suresh Babu Selvaraju; Susan Countryman; Susan Hicks; Suzanne Dale; Thomas Urban; Tim Kuphal; Tricia Zwiefelhofer; Vincent Drouillon; Yvette Unoarumhi |
| EPI_ISL_2106181, EPI_ISL_2259243, EPI_ISL_2259244, EPI_ISL_2259245, EPI_ISL_2259248, EPI_ISL_2617854 | Los Angeles County Public Health Laboratories | Los Angeles County Public Health Laboratories | Hemarajata; P. Hemarajata et al.; P. and Green, N. |
| EPI_ISL_2009392 | Lurie Children's Hospital of Chicago | Northwestern University - Ozer Lab | Egon A. Ozer; Judd F. Hultquist; Lacy M. Simons; Larry K. Kociolek; Michael G. Ison; Ramon Lorenzo-Redondo; Taylor J. Dean; William J. Muller; Xiaotian; Zheng |
| EPI_ISL_1716899, EPI_ISL_2424212, EPI_ISL_2774562 | Martin Luther King Jr. Community Hospital | Los Angeles County Public Health Laboratories | P. Hemarajata et al. |
| EPI_ISL_1700693, EPI_ISL_1700694, EPI_ISL_2001039, EPI_ISL_2308294, EPI_ISL_2308295, EPI_ISL_2308298, EPI_ISL_2759080, EPI_ISL_2759081 | | | |
| see above | Monterey County Public Health Laboratory | Monterey County Public Health Laboratory | Monterey County Public Health Laboratory |
| EPI_ISL_2009353 | Northwestern Memorial Hospital | Northwestern University - Ozer Lab | Chad J. Achenbach; Chao Qi; Egon A. Ozer; Judd F. Hultquist; Lacy M. Simons; Lawrence J. Jennings; Michael G. Ison; Ramon Lorenzo-Redondo; Taylor J. Dean |
| EPI_ISL_2080756 | OHSU Lab Services Molecular Microbiology Lab | Oregon SARS-CoV-2 Genome Sequencing Center | Alec J. Hirsch; Andrew C. Adey; Benjamin N. Bimber; Brendan L. O'Connell; Brian J. O'Roak; Cierra LaBlanc; Daniel N. Streblow; Donna Hansel; Guang Fan; Kayla Carter; Ruth V. Nichols; Sally Grindstaff; Sonia Acharya; William B. Messer; Xuan Qin |
| EPI_ISL_2811962, EPI_ISL_2811963, EPI_ISL_2811964, EPI_ISL_2811965 | Olive View Medical Center | Los Angeles County Public Health Laboratories | P. Hemarajata et al. |
| EPI_ISL_2566342, EPI_ISL_2566360, EPI_ISL_2566361 | Operation Infectious Diseases, Naval Health Research Center | Operation Infectious Diseases, Naval Health Research Center | A.E.; Balansay-Ames; C.A.; Fierro, M.; Iniguez-Stevens, E.; Kriner, P.; M.S.; Moser, K.; Myers; Pan; Phippard; R.S.; R.W.; Underwood |
| EPI_ISL_2228292, EPI_ISL_2228351, EPI_ISL_2228383, EPI_ISL_2758516 | Orange County Public Health Lab | Chan-Zuckerberg Biohub | CZB Cliahub Consortium |
| EPI_ISL_2659104, EPI_ISL_2659105, EPI_ISL_2659106, EPI_ISL_2659107, EPI_ISL_2659108, EPI_ISL_2659109, EPI_ISL_2753974, EPI_ISL_2774586, EPI_ISL_2774587, EPI_ISL_2774588, EPI_ISL_2774589, EPI_ISL_2774590 | | | |
| see above | PAMF Palo Alto | Santa Clara County Public Health Laboratory | Santa Clara County Public Health Department |
| EPI_ISL_2364736 | Pathology and Laboratory Medicine Institute, Cleveland Clinic, Ohio, USA | Pathology and Laboratory Medicine Institute, Cleveland Clinic, Ohio, USA | Concetta Peck; Daniel H. Farkas; Daniel Rhoads; David Bosler; David Plunkett; Gary Procop; Jay Brock; Jennifer Starbuck; Joy Nakitandwe; Kristen McDonnell; Yu-Wei Cheng; Zheng Jin Tu |
| EPI_ISL_2259229, EPI_ISL_2259230, EPI_ISL_2259231, EPI_ISL_2259232, EPI_ISL_2259233, EPI_ISL_2259234, EPI_ISL_2259235, EPI_ISL_2259427, EPI_ISL_2259428, EPI_ISL_2259429, EPI_ISL_2259430, EPI_ISL_2259431, EPI_ISL_2259437, EPI_ISL_2391749, EPI_ISL_2391750, EPI_ISL_2391752, EPI_ISL_2391780, EPI_ISL_2392167 | | | |
| see above | Pomona Valley Hospital Medical Center | Los Angeles County Public Health Laboratories | P. Hemarajata et al. |
| EPI_ISL_2259439 | Providence Little Company of Mary Medical Center | Los Angeles County Public Health Laboratories | P. Hemarajata et al. |
| EPI_ISL_2259251 | Quest Diagnostics | Los Angeles County Public Health Laboratories | P. Hemarajata et al. |
| EPI_ISL_1668641, EPI_ISL_1668729, EPI_ISL_1668781, EPI_ISL_1669009, EPI_ISL_1669080, EPI_ISL_1738064, EPI_ISL_1738069, EPI_ISL_1738123, EPI_ISL_1738329, EPI_ISL_1738401, EPI_ISL_1738467, EPI_ISL_1738481, EPI_ISL_1738487, EPI_ISL_1738497, EPI_ISL_1738506, EPI_ISL_1738507, EPI_ISL_1738511, EPI_ISL_1738515, EPI_ISL_1738524, EPI_ISL_1738532, EPI_ISL_1738536, EPI_ISL_1738544, EPI_ISL_1738547, EPI_ISL_1738550, EPI_ISL_1738562, EPI_ISL_1738583, EPI_ISL_1738599, EPI_ISL_1738600, EPI_ISL_1738606, EPI_ISL_1738630, EPI_ISL_1738641, EPI_ISL_1738652, EPI_ISL_1738662, EPI_ISL_1738739, EPI_ISL_1753033, EPI_ISL_1753034, EPI_ISL_1753035, EPI_ISL_1753038, EPI_ISL_1753041, EPI_ISL_1753045, EPI_ISL_1753062, EPI_ISL_1753063, EPI_ISL_1753068, EPI_ISL_1753070, | | | |

| | | | |
|---|---|---|---|
| EPI_ISL_1753084, EPI_ISL_1753086, EPI_ISL_1753091, EPI_ISL_1753105, EPI_ISL_1753108, EPI_ISL_1753111, EPI_ISL_1753113, EPI_ISL_1753126, EPI_ISL_1753139, EPI_ISL_1753143, EPI_ISL_1753145, EPI_ISL_1753149, EPI_ISL_1753154, EPI_ISL_1753156, EPI_ISL_1753161, EPI_ISL_1753163, EPI_ISL_1753165, EPI_ISL_1753170, EPI_ISL_1753172, EPI_ISL_1753175, EPI_ISL_1753177, EPI_ISL_1753178, EPI_ISL_1753179, EPI_ISL_1753205, EPI_ISL_1753207, EPI_ISL_1753212, EPI_ISL_1753220, EPI_ISL_1753221, EPI_ISL_1753226, EPI_ISL_1753228, EPI_ISL_1753233, EPI_ISL_1753238, EPI_ISL_1753242, EPI_ISL_1753243, EPI_ISL_1753257, EPI_ISL_1753260, EPI_ISL_1753262, EPI_ISL_1753264, EPI_ISL_1753267, EPI_ISL_1753269, EPI_ISL_1753307, EPI_ISL_1753349, EPI_ISL_1753361, EPI_ISL_1753365, EPI_ISL_1753372, EPI_ISL_1753379, EPI_ISL_1753380, EPI_ISL_1753383, EPI_ISL_1753396, EPI_ISL_1753404, EPI_ISL_1753621, EPI_ISL_1753645, EPI_ISL_1753653, EPI_ISL_1753655, EPI_ISL_1753656, EPI_ISL_1761871, EPI_ISL_1761872, EPI_ISL_1761873, EPI_ISL_1761877, EPI_ISL_1761879, EPI_ISL_1761880, EPI_ISL_1761902, EPI_ISL_1761904, EPI_ISL_1761909, EPI_ISL_1761911, EPI_ISL_1761912, EPI_ISL_1761916, EPI_ISL_1761917, EPI_ISL_1761930, EPI_ISL_1761934, EPI_ISL_1761935, EPI_ISL_1761938, EPI_ISL_1761943, EPI_ISL_1761945, EPI_ISL_1761948, EPI_ISL_1761950, EPI_ISL_1761956, EPI_ISL_1761958, EPI_ISL_1761959, EPI_ISL_1761960, EPI_ISL_1761964, EPI_ISL_1761966, EPI_ISL_1761969, EPI_ISL_1761973, EPI_ISL_1761979, EPI_ISL_1761982, EPI_ISL_1761987, EPI_ISL_1761991, EPI_ISL_1761998, EPI_ISL_1762007, EPI_ISL_1762009, EPI_ISL_1762011, EPI_ISL_1762022, EPI_ISL_1762025, EPI_ISL_1762026, EPI_ISL_1797926, EPI_ISL_1798036, EPI_ISL_1798071, EPI_ISL_1798086, EPI_ISL_1798096, EPI_ISL_1798104, EPI_ISL_1798118, EPI_ISL_1798129, EPI_ISL_1798151, EPI_ISL_1798161, EPI_ISL_1798183, EPI_ISL_1804812, EPI_ISL_1804815, EPI_ISL_1804816, EPI_ISL_1804819, EPI_ISL_1818064, EPI_ISL_1818069, EPI_ISL_1818309, EPI_ISL_1818410, EPI_ISL_1834201, EPI_ISL_1834227, EPI_ISL_1834293, EPI_ISL_1834301, EPI_ISL_1834310, EPI_ISL_1834311, EPI_ISL_1834402, EPI_ISL_1840260, EPI_ISL_1840275, EPI_ISL_1840279, EPI_ISL_1840284, EPI_ISL_1840286, EPI_ISL_1840294, EPI_ISL_1840304, EPI_ISL_1840315, EPI_ISL_1840336, EPI_ISL_1840339, EPI_ISL_1840346, EPI_ISL_1840375, EPI_ISL_1840380, EPI_ISL_1840385, EPI_ISL_1840398, EPI_ISL_1840406, EPI_ISL_1900132, EPI_ISL_1900147, EPI_ISL_1900442, EPI_ISL_1900445, EPI_ISL_1900449, EPI_ISL_1900451, EPI_ISL_1905848, EPI_ISL_1905852, EPI_ISL_1905859, EPI_ISL_1905866, EPI_ISL_1905869, EPI_ISL_1905901, EPI_ISL_1905905, EPI_ISL_1905925, EPI_ISL_1905927, EPI_ISL_1906009, EPI_ISL_1906030, EPI_ISL_1906036, EPI_ISL_1906074, EPI_ISL_1906102, EPI_ISL_1906126, EPI_ISL_1906143, EPI_ISL_1906145, EPI_ISL_1906168, EPI_ISL_1906301, EPI_ISL_1906332, EPI_ISL_1906450, EPI_ISL_1906571, EPI_ISL_1906584, EPI_ISL_1906586, EPI_ISL_1906589, EPI_ISL_1906591, EPI_ISL_1906613, EPI_ISL_1906617, EPI_ISL_1906621, EPI_ISL_1906625, EPI_ISL_1906641, EPI_ISL_1906650, EPI_ISL_1906654, EPI_ISL_1906663, EPI_ISL_1906672, EPI_ISL_1906684, EPI_ISL_1906736, EPI_ISL_1906742, EPI_ISL_1906744, EPI_ISL_1906749, EPI_ISL_1906751, EPI_ISL_1906753, EPI_ISL_1906757, EPI_ISL_1906763, EPI_ISL_1906768, EPI_ISL_1906772, EPI_ISL_1906778, EPI_ISL_1906790, EPI_ISL_1906804, EPI_ISL_1906808, EPI_ISL_1906819, EPI_ISL_1906825, EPI_ISL_1906830, EPI_ISL_1906846, EPI_ISL_1906847, EPI_ISL_1906848, EPI_ISL_1906856, EPI_ISL_1906863, EPI_ISL_1906864, EPI_ISL_1906867, EPI_ISL_1906871, EPI_ISL_1906874, EPI_ISL_1906882, EPI_ISL_1924317, EPI_ISL_1924347, EPI_ISL_1924356, EPI_ISL_1924651, EPI_ISL_1924652, EPI_ISL_1924653, EPI_ISL_1924660, EPI_ISL_1924661, EPI_ISL_1924663, EPI_ISL_1924669, EPI_ISL_1924671, EPI_ISL_1924672, EPI_ISL_1924677, EPI_ISL_1924680, EPI_ISL_1924686, EPI_ISL_1924690, EPI_ISL_1924694, EPI_ISL_1924696, EPI_ISL_1924699, EPI_ISL_1924704, EPI_ISL_1924709, EPI_ISL_1924714, EPI_ISL_1924716, EPI_ISL_1924719, EPI_ISL_1924724, EPI_ISL_1924733, EPI_ISL_1924741, EPI_ISL_1924742, EPI_ISL_1924744, EPI_ISL_1924746, EPI_ISL_1924749, EPI_ISL_1924751, EPI_ISL_1924753, EPI_ISL_1924757, EPI_ISL_1924760, EPI_ISL_1924761, EPI_ISL_1924762, EPI_ISL_1924767, EPI_ISL_1924769, EPI_ISL_1924778, EPI_ISL_1924785, EPI_ISL_1924786, EPI_ISL_1924787, EPI_ISL_1924788, EPI_ISL_1924792, EPI_ISL_1924795, EPI_ISL_1924810, EPI_ISL_1924819, EPI_ISL_1924821, EPI_ISL_1924835, EPI_ISL_1924847, EPI_ISL_1924866, EPI_ISL_1924867, EPI_ISL_1924877, EPI_ISL_1924881, EPI_ISL_1924887, EPI_ISL_1924888, EPI_ISL_1924900, EPI_ISL_1924912, EPI_ISL_1935558, EPI_ISL_1935563, EPI_ISL_1935571, EPI_ISL_1935576, EPI_ISL_1935582, EPI_ISL_1942184, EPI_ISL_1942187, EPI_ISL_1942189, EPI_ISL_1942190, EPI_ISL_1942199, EPI_ISL_1942201, EPI_ISL_1942203, EPI_ISL_1942205, EPI_ISL_1942206, EPI_ISL_1942208, EPI_ISL_1942209, EPI_ISL_1942210, EPI_ISL_1942214, EPI_ISL_1989696, EPI_ISL_1989710, EPI_ISL_1989720, EPI_ISL_1989721, EPI_ISL_1989726, EPI_ISL_1989727, EPI_ISL_1989729, EPI_ISL_1989738, EPI_ISL_1989744, EPI_ISL_1989758, EPI_ISL_1989773, EPI_ISL_1989776, EPI_ISL_1989782, EPI_ISL_1989787, EPI_ISL_1989790, EPI_ISL_1989793, EPI_ISL_1989796, EPI_ISL_1989805, EPI_ISL_1989806, EPI_ISL_1989807, EPI_ISL_1989814, EPI_ISL_1989847, EPI_ISL_1989853, EPI_ISL_1989868, EPI_ISL_1989881, EPI_ISL_1989975, EPI_ISL_1989985, EPI_ISL_1989991, EPI_ISL_1990052, EPI_ISL_1990160, EPI_ISL_1990166, EPI_ISL_1990180, EPI_ISL_1990181, EPI_ISL_1990216, EPI_ISL_1990258, EPI_ISL_1990280, EPI_ISL_1990281, EPI_ISL_1990290, EPI_ISL_1990291, EPI_ISL_1990294, EPI_ISL_1990295, EPI_ISL_1990298, EPI_ISL_1990311, EPI_ISL_1990322, EPI_ISL_1990326, EPI_ISL_1990328, EPI_ISL_1990330, EPI_ISL_1990333, EPI_ISL_1990334, EPI_ISL_1990350, EPI_ISL_1990368, EPI_ISL_1990376, EPI_ISL_1990394, EPI_ISL_1990401, EPI_ISL_1990406, EPI_ISL_2010890, EPI_ISL_2010895, EPI_ISL_2010903, EPI_ISL_2010906, EPI_ISL_2010909, EPI_ISL_2010912, EPI_ISL_2010915, EPI_ISL_2010916, EPI_ISL_2010918, EPI_ISL_2010919, EPI_ISL_2010920, EPI_ISL_2010922, EPI_ISL_2010924, EPI_ISL_2010925, EPI_ISL_2010926, EPI_ISL_2010927, EPI_ISL_2010928, EPI_ISL_2010933, EPI_ISL_2010937, EPI_ISL_2010938, EPI_ISL_2010940, EPI_ISL_2010942, EPI_ISL_2010943, EPI_ISL_2010944, EPI_ISL_2010945, EPI_ISL_2010946, EPI_ISL_2010948, EPI_ISL_2010949, EPI_ISL_2010950, EPI_ISL_2010953, EPI_ISL_2010955, EPI_ISL_2010958, EPI_ISL_2010960, EPI_ISL_2010964, EPI_ISL_2010965, EPI_ISL_2010967, EPI_ISL_2010968, EPI_ISL_2010974, EPI_ISL_2010976, EPI_ISL_2010978, EPI_ISL_2010979, EPI_ISL_2010980, EPI_ISL_2010981, EPI_ISL_2010982, EPI_ISL_2010985, EPI_ISL_2010986, EPI_ISL_2010992, EPI_ISL_2010994, EPI_ISL_2010996, EPI_ISL_2010997, EPI_ISL_2010998, EPI_ISL_2011001, EPI_ISL_2011002, EPI_ISL_2011005, EPI_ISL_2011007, EPI_ISL_2011008, EPI_ISL_2011009, EPI_ISL_2011011, EPI_ISL_2011012, EPI_ISL_2011014, EPI_ISL_2011019, EPI_ISL_2011020, EPI_ISL_2011023, EPI_ISL_2011171, EPI_ISL_2011221, EPI_ISL_2034722, EPI_ISL_2034729, EPI_ISL_2034736, EPI_ISL_2034762, EPI_ISL_2034766, EPI_ISL_2034769, EPI_ISL_2034776, EPI_ISL_2034779, EPI_ISL_2034782, EPI_ISL_2035302, EPI_ISL_2035303, EPI_ISL_2035304, EPI_ISL_2035306, EPI_ISL_2035307, EPI_ISL_2035309, EPI_ISL_2035310, EPI_ISL_2035311, EPI_ISL_2035312, EPI_ISL_2035313, EPI_ISL_2035314, EPI_ISL_2035315, EPI_ISL_2035316, EPI_ISL_2035317, EPI_ISL_2035318, EPI_ISL_2035319, EPI_ISL_2035320, EPI_ISL_2035322, EPI_ISL_2046840, EPI_ISL_2046844, EPI_ISL_2046884, EPI_ISL_2046891, EPI_ISL_2046976, EPI_ISL_2047006, EPI_ISL_2047022, EPI_ISL_2047026, EPI_ISL_2047028, EPI_ISL_2047100, EPI_ISL_2047151, EPI_ISL_2047154, EPI_ISL_2047180, EPI_ISL_2047199, EPI_ISL_2047200, EPI_ISL_2047249, EPI_ISL_2090474, EPI_ISL_2090477, EPI_ISL_2090478, EPI_ISL_2090482, EPI_ISL_2090485, EPI_ISL_2090516, EPI_ISL_2090529, EPI_ISL_2090531, EPI_ISL_2090532, EPI_ISL_2090539, EPI_ISL_2090552, EPI_ISL_2090560, EPI_ISL_2090566, EPI_ISL_2090599, EPI_ISL_2098355, EPI_ISL_2098401, EPI_ISL_2132575, EPI_ISL_2132581, EPI_ISL_2132582, EPI_ISL_2132584, EPI_ISL_2132585, EPI_ISL_2132619, EPI_ISL_2133176, EPI_ISL_2133281, EPI_ISL_2133318, EPI_ISL_2133319, EPI_ISL_2133326, EPI_ISL_2133392, EPI_ISL_2133393, EPI_ISL_2133404, EPI_ISL_2133414, EPI_ISL_2133416, EPI_ISL_2133417, EPI_ISL_2133421, EPI_ISL_2133423, EPI_ISL_2133426, EPI_ISL_2133428, EPI_ISL_2133429, EPI_ISL_2133430, EPI_ISL_2133431, EPI_ISL_2133433, EPI_ISL_2133437, EPI_ISL_2133445, EPI_ISL_2133447, EPI_ISL_2133449, EPI_ISL_2133453, EPI_ISL_2133461, EPI_ISL_2133463, EPI_ISL_2133464, EPI_ISL_2133467, EPI_ISL_2133471, EPI_ISL_2133472, EPI_ISL_2133475, EPI_ISL_2133476, EPI_ISL_2133477, EPI_ISL_2133487, EPI_ISL_2133489, EPI_ISL_2133490, EPI_ISL_2133496, EPI_ISL_2133497, EPI_ISL_2133498, EPI_ISL_2133504, EPI_ISL_2133509, EPI_ISL_2133511, EPI_ISL_2133515, EPI_ISL_2133527, EPI_ISL_2133528, EPI_ISL_2133530, EPI_ISL_2133533, EPI_ISL_2133535, EPI_ISL_2133540, EPI_ISL_2133542, EPI_ISL_2133543, EPI_ISL_2133544, EPI_ISL_2133547, EPI_ISL_2133549, EPI_ISL_2133551, EPI_ISL_2133553, EPI_ISL_2133555, EPI_ISL_2133556, EPI_ISL_2133557, EPI_ISL_2133560, EPI_ISL_2133568, EPI_ISL_2133569, EPI_ISL_2133571, EPI_ISL_2133573, EPI_ISL_2133583, EPI_ISL_2133587, EPI_ISL_2133588, EPI_ISL_2133589, EPI_ISL_2133593, EPI_ISL_2133601, EPI_ISL_2133603, EPI_ISL_2133604, EPI_ISL_2133605, EPI_ISL_2133643, EPI_ISL_2133676, EPI_ISL_2133679, EPI_ISL_2133716, EPI_ISL_2133725, EPI_ISL_2133743, EPI_ISL_2133753, EPI_ISL_2133772, EPI_ISL_2133776, EPI_ISL_2133778, EPI_ISL_2133799, EPI_ISL_2133818, EPI_ISL_2133836, EPI_ISL_2140760, EPI_ISL_2140761, EPI_ISL_2140762, EPI_ISL_2140763, EPI_ISL_2140764, EPI_ISL_2140767, EPI_ISL_2140769, EPI_ISL_2140770, EPI_ISL_2140771, EPI_ISL_2140772, EPI_ISL_2140773, EPI_ISL_2140776, EPI_ISL_2140792, EPI_ISL_2140793, EPI_ISL_2142880, EPI_ISL_2142881, EPI_ISL_2142884, EPI_ISL_2142886, EPI_ISL_2142890, EPI_ISL_2142892, EPI_ISL_2142893, EPI_ISL_2142896, EPI_ISL_2142898, EPI_ISL_2142902, EPI_ISL_2142914, EPI_ISL_2142920, EPI_ISL_2142921, EPI_ISL_2142929, EPI_ISL_2142931, EPI_ISL_2142932, EPI_ISL_2142940, EPI_ISL_2142941, EPI_ISL_2142942, EPI_ISL_2142945, EPI_ISL_2142946, EPI_ISL_2142949, EPI_ISL_2142950, EPI_ISL_2142951, EPI_ISL_2142954, EPI_ISL_2142955, EPI_ISL_2142956, EPI_ISL_2142957, EPI_ISL_2142964, EPI_ISL_2142967, EPI_ISL_2142968, EPI_ISL_2142969, EPI_ISL_2142970, EPI_ISL_2142971, EPI_ISL_2142974, EPI_ISL_2142975, EPI_ISL_2142981, EPI_ISL_2142988, EPI_ISL_2142989, EPI_ISL_2142991, EPI_ISL_2142998, EPI_ISL_2143000, EPI_ISL_2143002, EPI_ISL_2143005, EPI_ISL_2143007, EPI_ISL_2143009, EPI_ISL_2143010, EPI_ISL_2143011, EPI_ISL_2143012, EPI_ISL_2143016, EPI_ISL_2143021, EPI_ISL_2143022, EPI_ISL_2143036, EPI_ISL_2143057, EPI_ISL_2143059, EPI_ISL_2143081, EPI_ISL_2143111, EPI_ISL_2143115, EPI_ISL_2143124, EPI_ISL_2143128, EPI_ISL_2143173, EPI_ISL_2143174, EPI_ISL_2143175, EPI_ISL_2143177, EPI_ISL_2143178, EPI_ISL_2143179, EPI_ISL_2143181, EPI_ISL_2143182, EPI_ISL_2143184, EPI_ISL_2143186, EPI_ISL_2143187, EPI_ISL_2143189, EPI_ISL_2143193, EPI_ISL_2143195, EPI_ISL_2143197, EPI_ISL_2143198, EPI_ISL_2143201, EPI_ISL_2143203, EPI_ISL_2143204, EPI_ISL_2143205, EPI_ISL_2143206, EPI_ISL_2143207, EPI_ISL_2143208, EPI_ISL_2143214, EPI_ISL_2143217, EPI_ISL_2143218, EPI_ISL_2143220, EPI_ISL_2143223, EPI_ISL_2143226, EPI_ISL_2143227, EPI_ISL_2143229, EPI_ISL_2143231, EPI_ISL_2143233, EPI_ISL_2143235, EPI_ISL_2143236, EPI_ISL_2143238, EPI_ISL_2143240, EPI_ISL_2143242, EPI_ISL_2143244, EPI_ISL_2143245, EPI_ISL_2143246, EPI_ISL_2143249, EPI_ISL_2143255, EPI_ISL_2143258, EPI_ISL_2143261, EPI_ISL_2143262, EPI_ISL_2143264, EPI_ISL_2143266, EPI_ISL_2143268, EPI_ISL_2143270, EPI_ISL_2143271, EPI_ISL_2143273, EPI_ISL_2143274, EPI_ISL_2143275, EPI_ISL_2143276, EPI_ISL_2143279, EPI_ISL_2143283, EPI_ISL_2143286, EPI_ISL_2143287, EPI_ISL_2143288, EPI_ISL_2143289, EPI_ISL_2143290, EPI_ISL_2143291, EPI_ISL_2143295, EPI_ISL_2143296, EPI_ISL_2143298, EPI_ISL_2143301, EPI_ISL_2143315, EPI_ISL_2143326, EPI_ISL_2143339, EPI_ISL_2143348, EPI_ISL_2143362, EPI_ISL_2143369, EPI_ISL_2143371, EPI_ISL_2143379, EPI_ISL_2143389, EPI_ISL_2143412, EPI_ISL_2143414, EPI_ISL_2143436, EPI_ISL_2143455, EPI_ISL_2143461, EPI_ISL_2143481, EPI_ISL_2143509, EPI_ISL_2143522, EPI_ISL_2143699, EPI_ISL_2143760, EPI_ISL_2143789, EPI_ISL_2143848, EPI_ISL_2143899, EPI_ISL_2143908, EPI_ISL_2143936, EPI_ISL_2144009, EPI_ISL_2179931, EPI_ISL_2180479, EPI_ISL_2180501, EPI_ISL_2180503, EPI_ISL_2180508, EPI_ISL_2180513, EPI_ISL_2180520, EPI_ISL_2180527, EPI_ISL_2187230, EPI_ISL_2187231, EPI_ISL_2187232, EPI_ISL_2187234, EPI_ISL_2187236, EPI_ISL_2187237, EPI_ISL_2187238, EPI_ISL_2187239, EPI_ISL_2187240, EPI_ISL_2187241, EPI_ISL_2187242, EPI_ISL_2187252, EPI_ISL_2187258, EPI_ISL_2187259, EPI_ISL_2187262, EPI_ISL_2187264, EPI_ISL_2187266, EPI_ISL_2187267, EPI_ISL_2187268, EPI_ISL_2187269, EPI_ISL_2187270, EPI_ISL_2187272, EPI_ISL_2187273, EPI_ISL_2187274, EPI_ISL_2187275, EPI_ISL_2187276, EPI_ISL_2187278, EPI_ISL_2187279, EPI_ISL_2187281, EPI_ISL_2187282, EPI_ISL_2187283, EPI_ISL_2187284, EPI_ISL_2187285, EPI_ISL_2187286, EPI_ISL_2187287, EPI_ISL_2187288, EPI_ISL_2187300, EPI_ISL_2187302, EPI_ISL_2187307, EPI_ISL_2187314, EPI_ISL_2204125, EPI_ISL_2204208, EPI_ISL_2204249, EPI_ISL_2247071, EPI_ISL_2247092, EPI_ISL_2247174, EPI_ISL_2247179, EPI_ISL_2248518, EPI_ISL_2248559, EPI_ISL_2248666, EPI_ISL_2268159, EPI_ISL_2268590, EPI_ISL_2268594 | | | |
| see above | Quest Diagnostics Incorporated | Centers for Disease Control and Prevention Division of Viral Diseases, Pathogen Discovery | A. Gerasimova; A. Perez; Adrian Paskey; B. Anderson; Benjamin Rambo-Martin; Christopher Gulvick; Clinton R. Paden; Dakota Howard; Darlene Wagner; Dhwani Batra; Duncan MacCannell; F. Lacbawan; I. A. Shlyakhter; Jason Caravas; K.E. Livingston; Kara Moser; L.E. Bernstein; M. Hua; Matthew Schmerer; P. Tanpaiboon; Peter W. Cook; R. M. Kagan; R. Owen; R. V. Rolando; S. H. Rosenthal; Scott Sammons; Shatavia Morrison; Y. Liu; Yvette Unoarumhi |
| EPI_ISL_2628497, EPI_ISL_2628499, EPI_ISL_2628500, EPI_ISL_2628501, EPI_ISL_2628502, EPI_ISL_2628503, EPI_ISL_2628504 | | | |
| see above | Rady Children's Hospital - San Diego | Andersen lab at Scripps Research | Christina Clarke; David Dimmock; Denise Malicki; Kathryn Bouic; Linda Luo; SEARCH Alliance San Diego with Nanda Radamchar; Teresa Mueller |
| EPI_ISL_2319672 | Reditus Laboratories | Reditus Laboratories | Alexa Eichelberger; Cassy Philips; Joshua J. Geltz; M.S.; Ph.D.; Rex Dyer; Robert M. Sgambelluri |
| EPI_ISL_2253158, EPI_ISL_2253159, EPI_ISL_2253164, EPI_ISL_2253165, EPI_ISL_2253202, EPI_ISL_2254286, EPI_ISL_2254287, EPI_ISL_2254295, EPI_ISL_2254320, EPI_ISL_2254321, EPI_ISL_2254341, EPI_ISL_2254377, EPI_ISL_2254400, EPI_ISL_2254408, EPI_ISL_2254422, EPI_ISL_2254425, EPI_ISL_2254432, EPI_ISL_2254464 | | | |
| see above | Respiratory Viruses Branch, Centers for Disease Control and Prevention | Respiratory Viruses Branch, Centers for Disease Control and Prevention | Anderson, B.; Batra, D.; Bernstein, L.; C.R.; Campbell, P.; Caravas, J.; Case, R.; Clark, C.; Cook; Fierro, J.; Gao, H.; Gao, J.; Gerasimova, A.; Ghorpade, V.; Gulvick, C.; Hardison, M.; Houdeshell, H.; Howard, D.; Hua, M.; I.A.; Kagan; Kvalvaag, O.; Lacbawan, F.; Li, M.; Liu, Y.; Livingston, K.; MacCannell, D.; Meng, Y.; Morrison, S.; Moser, K.; Nall, D.; Ng, D.; Owen, R.; P.W.; Paden; Paskey, A.; Perez, A.; R.M.; R.V.; Rambo-Martin, B.; Rolando; Rosenthal; S.H.; Sammons, S.; Sanders, E.; Sapra, B.; Schmerer, M.; Shlyakhter; Tanpaiboon, P.; Tsai, B.; Unoarumhi, Y.; Vest, A.; Wagner, D.; Westlund, S.; Xie, J. |
| EPI_ISL_1405691, EPI_ISL_1405692, EPI_ISL_1405693, EPI_ISL_1405931, EPI_ISL_1405984, EPI_ISL_1406005, EPI_ISL_1406019, EPI_ISL_1406023, EPI_ISL_1794948, EPI_ISL_1794949, EPI_ISL_1794987, EPI_ISL_1794996, EPI_ISL_1795002, EPI_ISL_1795007, EPI_ISL_1795022, EPI_ISL_1795026, EPI_ISL_1795032, EPI_ISL_1795033, EPI_ISL_1795038, EPI_ISL_1969374, EPI_ISL_1969375, EPI_ISL_1969376, EPI_ISL_1969380, EPI_ISL_1969381, EPI_ISL_1969382, EPI_ISL_1969387, EPI_ISL_1969388, EPI_ISL_1969389, EPI_ISL_1969394, EPI_ISL_1969395, EPI_ISL_1969399, EPI_ISL_1969400, EPI_ISL_1969405, EPI_ISL_1969406, EPI_ISL_1969411, EPI_ISL_1969417, EPI_ISL_1969418, EPI_ISL_1969424, EPI_ISL_1969430, EPI_ISL_1969434, EPI_ISL_1969438, EPI_ISL_1969439, EPI_ISL_1969440, EPI_ISL_1969441, EPI_ISL_1969442, EPI_ISL_1969443, EPI_ISL_1969444, EPI_ISL_1969445, EPI_ISL_1969446, EPI_ISL_1969447, EPI_ISL_1969448, EPI_ISL_1969449, EPI_ISL_1969450, EPI_ISL_2231055, EPI_ISL_2231061, EPI_ISL_2231063, EPI_ISL_2231072, EPI_ISL_2231075, EPI_ISL_2231104, EPI_ISL_2231105, EPI_ISL_2231106, EPI_ISL_2231107, EPI_ISL_2231108, EPI_ISL_2231109, EPI_ISL_2231110, EPI_ISL_2231111, EPI_ISL_2231112, EPI_ISL_2231113, EPI_ISL_2231114, EPI_ISL_2231115, EPI_ISL_2231116, EPI_ISL_2231117, EPI_ISL_2231118, EPI_ISL_2231119, EPI_ISL_2231120, EPI_ISL_2231121, EPI_ISL_2231122, EPI_ISL_2231123, EPI_ISL_2231124, EPI_ISL_2231125, EPI_ISL_2231126, EPI_ISL_2231127, EPI_ISL_2231128, EPI_ISL_2231161, EPI_ISL_2231178, EPI_ISL_2231435, EPI_ISL_2231442, EPI_ISL_2329538, EPI_ISL_2329539, EPI_ISL_2329540, EPI_ISL_2329542, EPI_ISL_2329543, EPI_ISL_2329544, EPI_ISL_2329547, EPI_ISL_2329548, EPI_ISL_2329549, EPI_ISL_2329550, EPI_ISL_2329551, EPI_ISL_2329555, EPI_ISL_2329556, EPI_ISL_2329557, EPI_ISL_2329561, EPI_ISL_2329562, EPI_ISL_2329563, EPI_ISL_2329567, EPI_ISL_2329573, EPI_ISL_2329576, EPI_ISL_2329580, EPI_ISL_2329581, EPI_ISL_2329584, EPI_ISL_2329585, EPI_ISL_2329586, EPI_ISL_2329589, EPI_ISL_2329590, EPI_ISL_2329595, EPI_ISL_2329596, EPI_ISL_2329597, EPI_ISL_2329600, EPI_ISL_2329601, EPI_ISL_2329602, EPI_ISL_2329603, EPI_ISL_2329605, EPI_ISL_2329607, EPI_ISL_2329608, EPI_ISL_2422976, EPI_ISL_2422977, EPI_ISL_2422978, EPI_ISL_2422979, EPI_ISL_2422980, EPI_ISL_2422981, EPI_ISL_2422982, EPI_ISL_2422983, EPI_ISL_2422984, EPI_ISL_2422985, EPI_ISL_2422986, EPI_ISL_2422987, EPI_ISL_2422988, EPI_ISL_2422989, EPI_ISL_2422990, EPI_ISL_2422991, EPI_ISL_2422992, EPI_ISL_2422993, EPI_ISL_2422994, EPI_ISL_2422995, EPI_ISL_2422996, EPI_ISL_2422997, EPI_ISL_2422998, EPI_ISL_2422999, EPI_ISL_2423000, EPI_ISL_2423001, EPI_ISL_2423002, EPI_ISL_2423003, EPI_ISL_2423004, EPI_ISL_2423005, EPI_ISL_2423006, EPI_ISL_2423007, EPI_ISL_2423008, EPI_ISL_2423009, EPI_ISL_2423011, EPI_ISL_2423012, EPI_ISL_2423013, EPI_ISL_2423014, EPI_ISL_2423015, EPI_ISL_2423016, EPI_ISL_2423017, EPI_ISL_2423018, EPI_ISL_2423019, EPI_ISL_2423020, EPI_ISL_2423021, EPI_ISL_2423022, EPI_ISL_2423023, EPI_ISL_2423024, EPI_ISL_2423025, EPI_ISL_2423026, EPI_ISL_2423027, EPI_ISL_2423028, EPI_ISL_2423029, EPI_ISL_2423030, EPI_ISL_2423031, EPI_ISL_2423032, EPI_ISL_2628605, EPI_ISL_2628606, EPI_ISL_2628607, EPI_ISL_2628608, EPI_ISL_2628609, EPI_ISL_2628610, EPI_ISL_2628613, EPI_ISL_2628616, EPI_ISL_2628617, EPI_ISL_2628618, EPI_ISL_2628619, EPI_ISL_2628623, EPI_ISL_2628627, EPI_ISL_2628630, EPI_ISL_2628634, EPI_ISL_2628639, EPI_ISL_2628643, EPI_ISL_2628647, EPI_ISL_2628650, EPI_ISL_2628654, EPI_ISL_2628658, EPI_ISL_2628661, EPI_ISL_2628665, EPI_ISL_2628669, EPI_ISL_2628673, EPI_ISL_2628677, EPI_ISL_2628681 | | | |
| see above | San Diego County Public Health Laboratory | Andersen lab at Scripps Research | Brett Austin; Jovan Shephard; SEARCH Alliance San Diego with Ashleigh Murphy; SEARCH Alliance San Diego with Tracy Basler |
| EPI_ISL_2840925 | San Diego County Public Health Laboratory | San Diego County Public Health Laboratory | Ashleigh Murphy Schafer; Brett Austin |
| EPI_ISL_2103146, EPI_ISL_2103177, EPI_ISL_2341438, EPI_ISL_2341448, EPI_ISL_2341460, EPI_ISL_2341469, EPI_ISL_2341473, EPI_ISL_2341482, EPI_ISL_2341489, EPI_ISL_2341495, EPI_ISL_2341496, EPI_ISL_2341511, EPI_ISL_2341517, EPI_ISL_2341531, EPI_ISL_2341533, EPI_ISL_2341536, EPI_ISL_2341557, EPI_ISL_2341560, EPI_ISL_2341564, EPI_ISL_2341584, EPI_ISL_2341592, EPI_ISL_2341603, EPI_ISL_2341623, EPI_ISL_2341636, EPI_ISL_2341637, EPI_ISL_2341641, EPI_ISL_2341649, EPI_ISL_2496787, EPI_ISL_2496810, EPI_ISL_2496845, EPI_ISL_2496895, EPI_ISL_2496941, EPI_ISL_2500033, EPI_ISL_2500034, EPI_ISL_2500053, EPI_ISL_2500064, EPI_ISL_2500076, EPI_ISL_2500115, EPI_ISL_2500127, EPI_ISL_2500129, EPI_ISL_2500131, EPI_ISL_2500134, EPI_ISL_2500136, EPI_ISL_2500154, EPI_ISL_2500155, EPI_ISL_2500162, EPI_ISL_2500171, EPI_ISL_2500173, EPI_ISL_2500183, EPI_ISL_2500192, EPI_ISL_2500199, EPI_ISL_2500200, EPI_ISL_2500207, EPI_ISL_2658598, EPI_ISL_2658735, EPI_ISL_2659548 | | | |
| see above | Santa Clara County Public Health Laboratory | Chan-Zuckerberg Biohub | CZB Cliahub Consortium |
| EPI_ISL_2631189 | Santa Clara County Public Health Laboratory | Santa Clara County Public Health Laboratory | Santa Clara County Public Health Department |
| EPI_ISL_1664266, EPI_ISL_1664275, EPI_ISL_1913216, EPI_ISL_1913251 | Santa Clara County Public Health Laboratory | Santa Clara County public Health Laboratory | Santa Clara County Public Health Department |
| EPI_ISL_2631188, EPI_ISL_2658985, EPI_ISL_2658986, EPI_ISL_2658987, EPI_ISL_2658988, EPI_ISL_2658989, EPI_ISL_2658990, EPI_ISL_2658991, EPI_ISL_2658992, EPI_ISL_2658993, EPI_ISL_2658994, EPI_ISL_2658995, EPI_ISL_2658996, EPI_ISL_2658997, EPI_ISL_2658998, EPI_ISL_2658999, EPI_ISL_2659000, EPI_ISL_2659001, EPI_ISL_2659002, EPI_ISL_2659003, EPI_ISL_2659004, EPI_ISL_2659005, EPI_ISL_2659006, EPI_ISL_2659007, EPI_ISL_2659008, EPI_ISL_2659009, EPI_ISL_2659010, EPI_ISL_2659011, EPI_ISL_2659012, EPI_ISL_2659013, EPI_ISL_2659014, EPI_ISL_2659015, EPI_ISL_2659016, EPI_ISL_2659017, EPI_ISL_2659018, EPI_ISL_2659019, EPI_ISL_2659020, EPI_ISL_2659021, EPI_ISL_2659022, EPI_ISL_2659023, EPI_ISL_2659024, EPI_ISL_2659025, EPI_ISL_2659026, EPI_ISL_2659027, EPI_ISL_2659028, EPI_ISL_2659029, EPI_ISL_2659030, EPI_ISL_2659031, EPI_ISL_2659032, EPI_ISL_2659033, EPI_ISL_2659034, EPI_ISL_2659035, EPI_ISL_2659036, EPI_ISL_2659037, EPI_ISL_2659038, EPI_ISL_2659039, EPI_ISL_2659040, EPI_ISL_2659041, EPI_ISL_2659042, EPI_ISL_2659043, EPI_ISL_2659044, EPI_ISL_2659045, EPI_ISL_2659046, EPI_ISL_2659047, EPI_ISL_2659048, EPI_ISL_2659049, EPI_ISL_2659050, EPI_ISL_2659074, EPI_ISL_2659075, EPI_ISL_2659076, EPI_ISL_2659077, EPI_ISL_2659078, EPI_ISL_2659079, EPI_ISL_2659080, EPI_ISL_2659081, EPI_ISL_2659082, EPI_ISL_2659083, EPI_ISL_2659084, EPI_ISL_2659085, EPI_ISL_2659086, EPI_ISL_2659087, EPI_ISL_2659088, EPI_ISL_2659089, EPI_ISL_2659090, EPI_ISL_2659091, EPI_ISL_2659092, EPI_ISL_2659093, EPI_ISL_2659094, EPI_ISL_2659095, EPI_ISL_2659096, EPI_ISL_2659097, EPI_ISL_2659098, EPI_ISL_2659099, EPI_ISL_2659100, EPI_ISL_2659101, EPI_ISL_2659102, EPI_ISL_2659103, EPI_ISL_2659110, EPI_ISL_2659111, EPI_ISL_2659112, EPI_ISL_2659113, EPI_ISL_2659114, EPI_ISL_2659115, EPI_ISL_2691224, EPI_ISL_2691225, EPI_ISL_2691226, EPI_ISL_2691227, EPI_ISL_2691228, EPI_ISL_2691229, EPI_ISL_2691230, EPI_ISL_2691231, EPI_ISL_2691232, EPI_ISL_2691233, EPI_ISL_2691234, EPI_ISL_2691235, EPI_ISL_2691236, EPI_ISL_2691237, EPI_ISL_2691238, EPI_ISL_2691239, EPI_ISL_2691240, EPI_ISL_2691241, EPI_ISL_2691242, EPI_ISL_2691243, EPI_ISL_2691244, EPI_ISL_2691245, EPI_ISL_2691246, EPI_ISL_2691247, EPI_ISL_2691248, EPI_ISL_2691249, EPI_ISL_2691250, EPI_ISL_2691251, EPI_ISL_2691252, EPI_ISL_2691253, EPI_ISL_2691254, EPI_ISL_2691255, EPI_ISL_2691256, EPI_ISL_2691257, EPI_ISL_2691258, EPI_ISL_2691259, EPI_ISL_2691260, EPI_ISL_2691261, EPI_ISL_2691262, EPI_ISL_2691263, EPI_ISL_2691264, EPI_ISL_2691265, EPI_ISL_2691266, EPI_ISL_2691267, EPI_ISL_2691268, EPI_ISL_2691269, EPI_ISL_2691270, EPI_ISL_2691271, EPI_ISL_2691272, EPI_ISL_2691273, EPI_ISL_2691274, EPI_ISL_2691275, EPI_ISL_2691276, EPI_ISL_2691277, EPI_ISL_2691278, EPI_ISL_2691279, EPI_ISL_2691280, EPI_ISL_2691281, EPI_ISL_2691282, EPI_ISL_2691283, EPI_ISL_2691284, EPI_ISL_2691285, EPI_ISL_2691286, EPI_ISL_2691287, EPI_ISL_2691288, EPI_ISL_2691289, EPI_ISL_2691290, EPI_ISL_2691291, EPI_ISL_2691292, EPI_ISL_2691293, EPI_ISL_2691294, EPI_ISL_2774575, EPI_ISL_2774576, EPI_ISL_2774577, EPI_ISL_2774578, EPI_ISL_2774579, EPI_ISL_2933981, EPI_ISL_2933986, EPI_ISL_2933990, EPI_ISL_2934002, EPI_ISL_2934010, EPI_ISL_2934033, EPI_ISL_2934040, EPI_ISL_2934042, EPI_ISL_2934047, EPI_ISL_2934049, EPI_ISL_2934050, EPI_ISL_2934054, EPI_ISL_2934057, EPI_ISL_2934065, EPI_ISL_2934088, EPI_ISL_2934089, EPI_ISL_2934090, EPI_ISL_2934091, EPI_ISL_2934092 | | | |
| see above | Santa Clara Valley Medical Center | Santa Clara County Public Health Laboratory | Santa Clara County Public Health Department |
| EPI_ISL_1824084, EPI_ISL_1824085, EPI_ISL_1824086, EPI_ISL_1824087, EPI_ISL_1824088, EPI_ISL_1824089, EPI_ISL_1824090, EPI_ISL_1824091, EPI_ISL_1824092, EPI_ISL_1824093, EPI_ISL_1824094, EPI_ISL_1824095, EPI_ISL_1824096, EPI_ISL_1824097, EPI_ISL_1824098, EPI_ISL_1824099, EPI_ISL_1824100, EPI_ISL_1824101, EPI_ISL_1824102, EPI_ISL_1824103, EPI_ISL_1824104, EPI_ISL_1824105, EPI_ISL_1824106, EPI_ISL_1824107, EPI_ISL_1824108, EPI_ISL_1824109, EPI_ISL_1824110, EPI_ISL_1913218, EPI_ISL_1913226, EPI_ISL_1913227, EPI_ISL_1913228, EPI_ISL_1913229, EPI_ISL_1913230, EPI_ISL_1913231, EPI_ISL_1913232, EPI_ISL_1913233, EPI_ISL_1913234, EPI_ISL_1913235, EPI_ISL_1913236, EPI_ISL_1913237, EPI_ISL_1913238, EPI_ISL_1913239, EPI_ISL_1913240, EPI_ISL_1913241, EPI_ISL_1913242, EPI_ISL_1913243, EPI_ISL_1913244, EPI_ISL_1913245, EPI_ISL_1913246, EPI_ISL_1913248, EPI_ISL_1913249, EPI_ISL_1913250, EPI_ISL_1924631, EPI_ISL_1924632, EPI_ISL_2967928 | | | |
| see above | Santa Clara Valley Medical Center | Santa Clara County public Health Laboratory | Santa Clara County Public Health Department |
| EPI_ISL_1794840, EPI_ISL_1794841, EPI_ISL_1794843, EPI_ISL_1794844, EPI_ISL_1794849, EPI_ISL_1794935, EPI_ISL_1794936, EPI_ISL_1969507, EPI_ISL_1969508, EPI_ISL_1969509, EPI_ISL_1969510, EPI_ISL_1969511, EPI_ISL_1969512, EPI_ISL_1969513, EPI_ISL_1969514, EPI_ISL_1969515, EPI_ISL_1969516, EPI_ISL_1969517, EPI_ISL_1969518, EPI_ISL_1969519, EPI_ISL_1969520, EPI_ISL_1969521, EPI_ISL_1969522, EPI_ISL_1969523, EPI_ISL_1969524, EPI_ISL_1969525, EPI_ISL_1969526, EPI_ISL_1969527, EPI_ISL_1969528, EPI_ISL_1969529, EPI_ISL_1969530, EPI_ISL_1969531, EPI_ISL_1969532, EPI_ISL_1969533, EPI_ISL_1969534, EPI_ISL_1969535, EPI_ISL_1969536, EPI_ISL_1969537, EPI_ISL_1969538, EPI_ISL_1969539, EPI_ISL_1969540, EPI_ISL_1969541, EPI_ISL_1969542, EPI_ISL_1969543, EPI_ISL_1969544, EPI_ISL_1969545, EPI_ISL_1969546, EPI_ISL_1969547, EPI_ISL_1969548, EPI_ISL_1969549, EPI_ISL_1969550, EPI_ISL_1969551, EPI_ISL_1969552, EPI_ISL_1969553 | | | |
| see above | Scripps Medical Laboratory | Andersen lab at Scripps Research | Ellen Stefanski; Ian Mchardy; SEARCH Alliance San Diego with Michael Quigley |
| EPI_ISL_1794885, EPI_ISL_1794886, EPI_ISL_1794887, EPI_ISL_1794888, EPI_ISL_1794889, EPI_ISL_1794892, EPI_ISL_1794893, EPI_ISL_1794894, EPI_ISL_1794897, EPI_ISL_1794900, EPI_ISL_1794902, EPI_ISL_1794913, EPI_ISL_1794914, EPI_ISL_1794916, EPI_ISL_1794917, EPI_ISL_1794918, EPI_ISL_1969563, EPI_ISL_1969564, EPI_ISL_1969565, EPI_ISL_1969566, EPI_ISL_1969567, EPI_ISL_1969568, EPI_ISL_1969569, EPI_ISL_1969570, EPI_ISL_1969571, EPI_ISL_1969572, EPI_ISL_1969573, EPI_ISL_1969574, EPI_ISL_1969575, EPI_ISL_1969576, EPI_ISL_1969577, EPI_ISL_1969578, EPI_ISL_1969579, EPI_ISL_1969580, EPI_ISL_1969581, EPI_ISL_1969582, EPI_ISL_1969583, EPI_ISL_1969584, EPI_ISL_1969585, EPI_ISL_1969586, EPI_ISL_1969587, EPI_ISL_1969588, EPI_ISL_1969589, EPI_ISL_1969590, EPI_ISL_1969591, EPI_ISL_1969592, EPI_ISL_1969593, EPI_ISL_1969594, EPI_ISL_1969595, EPI_ISL_1969596, EPI_ISL_1969597, EPI_ISL_1969598, EPI_ISL_1969599, EPI_ISL_1969600, EPI_ISL_1969601, EPI_ISL_1969606, EPI_ISL_1969608, EPI_ISL_1969609, EPI_ISL_1969610, EPI_ISL_2231041, EPI_ISL_2231051, EPI_ISL_2231066, EPI_ISL_2231090, EPI_ISL_2231091, EPI_ISL_2231092, EPI_ISL_2231093, | | | |

| | | | |
|---|---|---|---|
| EPI_ISL_2231094, EPI_ISL_2231095, EPI_ISL_2231096, EPI_ISL_2231097, EPI_ISL_2231098, EPI_ISL_2231099, EPI_ISL_2231100, EPI_ISL_2231101, EPI_ISL_2231102, EPI_ISL_2231103, EPI_ISL_2231129, EPI_ISL_2231130, EPI_ISL_2231131, EPI_ISL_2231132, EPI_ISL_2231133, EPI_ISL_2231134, EPI_ISL_2231135, EPI_ISL_2231136, EPI_ISL_2231137, EPI_ISL_2231138, EPI_ISL_2231140, EPI_ISL_2231141, EPI_ISL_2231142, EPI_ISL_2231143, EPI_ISL_2231144, EPI_ISL_2231145, EPI_ISL_2231146, EPI_ISL_2231147, EPI_ISL_2231148, EPI_ISL_2231149, EPI_ISL_2231150, EPI_ISL_2231151, EPI_ISL_2231155, EPI_ISL_2231156, EPI_ISL_2231158, EPI_ISL_2231159, EPI_ISL_2231160, EPI_ISL_2231162, EPI_ISL_2231163, EPI_ISL_2231164, EPI_ISL_2231165, EPI_ISL_2231166, EPI_ISL_2231167, EPI_ISL_2231168, EPI_ISL_2231169, EPI_ISL_2231170, EPI_ISL_2231176, EPI_ISL_2231177, EPI_ISL_2231179, EPI_ISL_2231180, EPI_ISL_2231181, EPI_ISL_2231182, EPI_ISL_2231183, EPI_ISL_2231184, EPI_ISL_2231185, EPI_ISL_2231195, EPI_ISL_2231198, EPI_ISL_2231205, EPI_ISL_2231206, EPI_ISL_2231318, EPI_ISL_2231319, EPI_ISL_2231320, EPI_ISL_2231526, EPI_ISL_2231527, EPI_ISL_2231530, EPI_ISL_2329435, EPI_ISL_2329437, EPI_ISL_2329438, EPI_ISL_2329439, EPI_ISL_2329440 | | | |
| see above | Sharp HealthCare Laboratory | Andersen lab at Scripps Research | Art Mendoza; Cathy Woerle; Jacquelyn Berumen; Liam McGinnis; Omid Bakhtar; SEARCH Alliance San Diego with Aaron Harding |
| EPI_ISL_2659058, EPI_ISL_2659059, EPI_ISL_2659060, EPI_ISL_2659061, EPI_ISL_2659062, EPI_ISL_2659063 | Stanford Clinical Virology Laboratory | Santa Clara County Public Health Laboratory | Santa Clara County Public Health Department |
| EPI_ISL_1924633, EPI_ISL_1924634, EPI_ISL_1924636, EPI_ISL_1924642, EPI_ISL_1924643, EPI_ISL_1924644, EPI_ISL_1924645, EPI_ISL_1924646, EPI_ISL_1924647, EPI_ISL_1924648 | | | |
| see above | Stanford Clinical Virology Laboratory | Santa Clara County public Health Laboratory | Santa Clara County Public Health Department |
| EPI_ISL_1675227, EPI_ISL_2457058, EPI_ISL_2477504 | Stanford Health Care | Stanford University School of Medicine, Clinical Virology Laboratory | Becky Jiang; Bernadette Troung; ChunHong Huang; Daniel Solis; Fumiko Yamamoto; Hannah Wang; Jacob Miller; James Zehnder; Kenji O. Mfuh; Malaya K. Sahoo; Mamdouh Sibai; Marilyn Mar; Michelle Verghese; Naomi Iwai; and Benjamin A. Pinsky |
| EPI_ISL_2341054, EPI_ISL_2341058, EPI_ISL_2341061, EPI_ISL_2341062, EPI_ISL_2341063, EPI_ISL_2341064, EPI_ISL_2341065, EPI_ISL_2341082, EPI_ISL_2341083, EPI_ISL_2341084, EPI_ISL_2341245, EPI_ISL_2341246, EPI_ISL_2341259, EPI_ISL_2341264, EPI_ISL_2341265, EPI_ISL_2341266, EPI_ISL_2341267, EPI_ISL_3049207, EPI_ISL_3049208, EPI_ISL_3049209, EPI_ISL_3049210, EPI_ISL_3049211, EPI_ISL_3049212, EPI_ISL_3049213 | | | |
| see above | Tulare County Public Health Laboratory | Tulare County Public Health Laboratory | Denise L Lopez; Joshua Kootstra |
| EPI_ISL_2259436, EPI_ISL_2391760, EPI_ISL_2391762, EPI_ISL_2391764, EPI_ISL_2391808, EPI_ISL_2391809, EPI_ISL_2391811, EPI_ISL_2391813, EPI_ISL_2392171, EPI_ISL_2392173, EPI_ISL_2392175, EPI_ISL_2392176, EPI_ISL_2392178, EPI_ISL_2392192, EPI_ISL_2399613, EPI_ISL_2399614, EPI_ISL_2399615, EPI_ISL_2404848, EPI_ISL_2404849, EPI_ISL_2404850, EPI_ISL_2404851, EPI_ISL_2404852, EPI_ISL_2404853, EPI_ISL_2404854, EPI_ISL_2404856, EPI_ISL_2404858, EPI_ISL_2404859, EPI_ISL_2404862, EPI_ISL_2404865, EPI_ISL_2404868, EPI_ISL_2404869, EPI_ISL_2424166, EPI_ISL_2424168, EPI_ISL_2424169, EPI_ISL_2424171, EPI_ISL_2424173, EPI_ISL_2424177, EPI_ISL_2424178, EPI_ISL_2424179, EPI_ISL_2424180, EPI_ISL_2424181, EPI_ISL_2424182, EPI_ISL_2424188, EPI_ISL_2424192, EPI_ISL_2424194, EPI_ISL_2424195, EPI_ISL_2424196, EPI_ISL_2424197, EPI_ISL_2424200, EPI_ISL_2424201, EPI_ISL_2424202, EPI_ISL_2424207, EPI_ISL_2424208, EPI_ISL_2424209, EPI_ISL_2424210, EPI_ISL_2424211, EPI_ISL_2620746, EPI_ISL_2620747, EPI_ISL_2620748, EPI_ISL_2620749, EPI_ISL_2620751, EPI_ISL_2620752, EPI_ISL_2620753, EPI_ISL_2620754, EPI_ISL_2620755, EPI_ISL_2620756, EPI_ISL_2620757, EPI_ISL_2620758, EPI_ISL_2620759, EPI_ISL_2620760, EPI_ISL_2620762, EPI_ISL_2620763, EPI_ISL_2620764, EPI_ISL_2620765, EPI_ISL_2620766, EPI_ISL_2620767, EPI_ISL_2620768, EPI_ISL_2620769, EPI_ISL_3029869 | | | |
| see above | UCLA Clinical Micro Lab | Los Angeles County Public Health Laboratories | P. Hemarajata et al. |
| EPI_ISL_2031772, EPI_ISL_2031774, EPI_ISL_2031775, EPI_ISL_2031776, EPI_ISL_2031777, EPI_ISL_2031778, EPI_ISL_2031779, EPI_ISL_2031780, EPI_ISL_2031782, EPI_ISL_2031783, EPI_ISL_2031784, EPI_ISL_2031785, EPI_ISL_2031786, EPI_ISL_2081231, EPI_ISL_2081232, EPI_ISL_2081233, EPI_ISL_2081234, EPI_ISL_2081237, EPI_ISL_2081238, EPI_ISL_2081239, EPI_ISL_2081246, EPI_ISL_2081533, EPI_ISL_2081537, EPI_ISL_2081540, EPI_ISL_2081541, EPI_ISL_2081547, EPI_ISL_2081551, EPI_ISL_2081552, EPI_ISL_2082942, EPI_ISL_2083002, EPI_ISL_2179217 | | | |
| see above | UCSC Genomics Institute | UCSC Genomics Institute | A. Marm Kilpatrick; Angie Hinrichs; Beth Shapiro; Bryan Thornlow; Ciara Wanket; David Haussler; Eric Beraut; Hugh Olsen; Ikenna Anigbogu; Isabel Bjork; Jakob McBroome; Jeremy Sanford; Joshua Kapp; Mark Akeson; Maximilian Haeussler; Michael Stone; Miten Jain; Molly Cassatt-Johnstone; Namrita Dhillon; Russell Corbett-Detig; Terren Chang; Yatish Turakhia |
| EPI_ISL_2383378, EPI_ISL_2426731 | US Air Force School of Aerospace Medicine | US Air Force School of Aerospace Medicine | Amanda Javorina; Anthony Fries; Carol Garrett; Clarise Starr; Elizabeth Macias; Jennifer Meyer; Sarah Purves; William Gruner |
| EPI_ISL_1716881, EPI_ISL_1716882, EPI_ISL_1716883, EPI_ISL_1716884, EPI_ISL_1716885, EPI_ISL_1716886, EPI_ISL_1716887, EPI_ISL_1716888, EPI_ISL_1716889, EPI_ISL_1716890, EPI_ISL_1716891, EPI_ISL_1716892, EPI_ISL_2259249, EPI_ISL_2259432, EPI_ISL_2259433, EPI_ISL_2259434, EPI_ISL_2259435, EPI_ISL_2260370, EPI_ISL_2260371, EPI_ISL_2260373, EPI_ISL_2260375, EPI_ISL_2260377, EPI_ISL_2260379, EPI_ISL_2260381, EPI_ISL_2260383, EPI_ISL_2260385, EPI_ISL_2260386, EPI_ISL_2260388, EPI_ISL_2260390, EPI_ISL_2260392, EPI_ISL_2391781, EPI_ISL_2391782, EPI_ISL_2424203, EPI_ISL_2620770, EPI_ISL_2620771, EPI_ISL_2620772, EPI_ISL_2620773, EPI_ISL_2620774 | | | |
| see above | USC Clinical Lab | Los Angeles County Public Health Laboratories | P. Hemarajata et al. |
| EPI_ISL_2031858 | VA Medical Center | Los Angeles County Public Health Laboratories | P. Hemarajata et al. |
| EPI_ISL_2533751 | VA Palo Alto Health Care System | Genomics and Discovery, Respiratory Viruses Branch, Division of Viral Diseases, Centers for Disease Control and Prevention | Adam Retchless; Anna Kelleher; Anna Uehara; Brian Lynch; Clinton R. Paden; Dhwani Batra; Haibin Wang; Han Jia Justin Ng; Jasmine Padilla; Jing Zhang; Justin Lee; Krista Queen; Mark Burroughs; Mili Sheth; Morgan Davis; Peter Cook; Rachel Marine; Sarah Nobles; Suxiang Tong; Tara Coalter; Yan Li; Ying Tao |
| EPI_ISL_2404805, EPI_ISL_2404806, EPI_ISL_2404807, EPI_ISL_2404808, EPI_ISL_2404809, EPI_ISL_2404810, EPI_ISL_2404815 | | | |
| see above | VMC | Santa Clara County public Health Laboratory | Santa Clara County Public Health Department |
| EPI_ISL_2295379, EPI_ISL_2295380, EPI_ISL_2533758 | WLA VAMC | Genomics and Discovery, Respiratory Viruses Branch, Division of Viral Diseases, Centers for Disease Control and Prevention | Adam Retchless; Anna Kelleher; Anna Uehara; Brian Lynch; Clinton R. Paden; Dhwani Batra; Haibin Wang; Han Jia Justin Ng; Jasmine Padilla; Jing Zhang; Justin Lee; Krista Queen; Mark Burroughs; Mili Sheth; Morgan Davis; Peter Cook; Rachel Marine; Sarah Nobles; Suxiang Tong; Tara Coalter; Yan Li; Ying Tao |

We gratefully acknowledge the following Authors from the Originating laboratories responsible for obtaining the specimens, as well as the Submitting laboratories where the genome data were generated and shared via GISAID, on which this research is based.

All Submitters of data may be contacted directly via www.gisaid.org

Authors are sorted alphabetically.

| Accession ID | Originating Laboratory | Submitting Laboratory | Authors |
|---|---|---|---|
| EPI_ISL_2229147, EPI_ISL_2229157, EPI_ISL_2788100, EPI_ISL_2788103, EPI_ISL_2788106, EPI_ISL_2788109, EPI_ISL_2788112, EPI_ISL_2788115, EPI_ISL_2788121, EPI_ISL_2986990, EPI_ISL_2986991, EPI_ISL_2987103, EPI_ISL_2987104, EPI_ISL_2987105, EPI_ISL_2987106, EPI_ISL_2987107, EPI_ISL_2987108, EPI_ISL_2987109, EPI_ISL_2987110, EPI_ISL_2987111, EPI_ISL_2987112, EPI_ISL_2987113, EPI_ISL_2987114, EPI_ISL_2987115, EPI_ISL_2987116 | | | |
| see above | "CDPH, Viral and Rickettsial Disease Laboratory" | Centers for Disease Control and Prevention Division of Viral Diseases, Pathogen Discovery | Alison Laufer Halpin; Ben L. Rambo-Martin; Clinton R. Paden; Dakota Howard; Darlene Wagner; Dave Wentworth; Dhwani Batra; Jasmine Padilla; Justin Lee; Katie Dillon; Krista Queen; Kristen Knipe; Kristine Lacek; Mark Burroughs; Matthew Schmerer; Mili Sheth; Peter Cook; Sam Shepard; Sarah Nobles; Shoshona Le; Suxiang Tong; Vivien Dugan; Yvette Unoarumhi |
| EPI_ISL_2811941 | Adventist Health Glendale | Los Angeles County Public Health Laboratories | P. Hemarajata et al. |
| EPI_ISL_2146060, EPI_ISL_2146061, EPI_ISL_2146075, EPI_ISL_2146077, EPI_ISL_2146078, EPI_ISL_2146079, EPI_ISL_2146080, EPI_ISL_2146086, EPI_ISL_2146114, EPI_ISL_2146131, EPI_ISL_2146135, EPI_ISL_2146144, EPI_ISL_2146145, EPI_ISL_2146149, EPI_ISL_2146150, EPI_ISL_2146175, EPI_ISL_2146212, EPI_ISL_2146216, EPI_ISL_2146219, EPI_ISL_2146226, EPI_ISL_2146244, EPI_ISL_2146245, EPI_ISL_2146247, EPI_ISL_2146248, EPI_ISL_2146276, EPI_ISL_2146277, EPI_ISL_2146321, EPI_ISL_2146325, EPI_ISL_2146328, EPI_ISL_2146334, EPI_ISL_2146337, EPI_ISL_2146341, EPI_ISL_2146352, EPI_ISL_2146409, EPI_ISL_2146431, EPI_ISL_2146433, EPI_ISL_2146442, EPI_ISL_2146450, EPI_ISL_2146455, EPI_ISL_2146457, EPI_ISL_2146458, EPI_ISL_2146460, EPI_ISL_2146465, EPI_ISL_2146469, EPI_ISL_2146497, EPI_ISL_2146501, EPI_ISL_2146504, EPI_ISL_2146543, EPI_ISL_2146547, EPI_ISL_2146548, EPI_ISL_2146558, EPI_ISL_2146573, EPI_ISL_2146612, EPI_ISL_2146621, EPI_ISL_2146737, EPI_ISL_2146738, EPI_ISL_2146739, EPI_ISL_2146741, EPI_ISL_2146743, EPI_ISL_2146748, EPI_ISL_2146749, EPI_ISL_2146755, EPI_ISL_2146764, EPI_ISL_2146797, EPI_ISL_2146798, EPI_ISL_2146817, EPI_ISL_2146821, EPI_ISL_2146835, EPI_ISL_2146837, EPI_ISL_2146871, EPI_ISL_2146889, EPI_ISL_2146890, EPI_ISL_2146898, EPI_ISL_2146901, EPI_ISL_2146913, EPI_ISL_2146920, EPI_ISL_2146956, EPI_ISL_2146970, EPI_ISL_2146988, EPI_ISL_2146996, EPI_ISL_2147008, EPI_ISL_2147049, EPI_ISL_2147051, EPI_ISL_2147057, EPI_ISL_2147088, EPI_ISL_2147089, EPI_ISL_2147097, EPI_ISL_2147103, EPI_ISL_2147105, EPI_ISL_2147126, EPI_ISL_2147127, EPI_ISL_2147146, EPI_ISL_2147159, EPI_ISL_2147176, EPI_ISL_2147179, EPI_ISL_2147184, EPI_ISL_2147196, EPI_ISL_2147219, EPI_ISL_2147226, EPI_ISL_2147231, EPI_ISL_2147245, EPI_ISL_2147278, EPI_ISL_2147284, EPI_ISL_2147301, EPI_ISL_2147406, EPI_ISL_2147423, EPI_ISL_2147459, EPI_ISL_2147487, EPI_ISL_2147488, EPI_ISL_2147491, EPI_ISL_2147492, EPI_ISL_2147495, EPI_ISL_2147510, EPI_ISL_2147511, EPI_ISL_2147522, EPI_ISL_2147525, EPI_ISL_2147528, EPI_ISL_2147541, EPI_ISL_2147542, EPI_ISL_2147543, EPI_ISL_2147545, EPI_ISL_2147550, EPI_ISL_2147557, EPI_ISL_2147562, EPI_ISL_2147573, EPI_ISL_2147576, EPI_ISL_2147584, EPI_ISL_2147659, EPI_ISL_2147795, EPI_ISL_2147919, EPI_ISL_2147930, EPI_ISL_2147944, EPI_ISL_2147958, EPI_ISL_2147963, EPI_ISL_2148128, EPI_ISL_2148193, EPI_ISL_2148205, EPI_ISL_2148210, EPI_ISL_2148212, EPI_ISL_2148218, EPI_ISL_2148220, EPI_ISL_2148221, EPI_ISL_2148234, EPI_ISL_2148239, EPI_ISL_2148241, EPI_ISL_2148242, EPI_ISL_2148247, EPI_ISL_2148841, EPI_ISL_2148842, EPI_ISL_2148844, EPI_ISL_2148866, EPI_ISL_2148867, EPI_ISL_2148892, EPI_ISL_2148896, EPI_ISL_2149162, EPI_ISL_2158864, EPI_ISL_2158914, EPI_ISL_2158922, EPI_ISL_2158939, EPI_ISL_2158940, EPI_ISL_2158945, EPI_ISL_2158961, EPI_ISL_2158962, EPI_ISL_2158963, EPI_ISL_2158979, EPI_ISL_2158999, EPI_ISL_2159007, EPI_ISL_2159009, EPI_ISL_2159012, EPI_ISL_2159015, EPI_ISL_2159018, EPI_ISL_2159057, EPI_ISL_2159060, EPI_ISL_2159062, EPI_ISL_2159065, EPI_ISL_2159116, EPI_ISL_2159117, EPI_ISL_2159118, EPI_ISL_2159121, EPI_ISL_2159123, EPI_ISL_2159137, EPI_ISL_2159155, EPI_ISL_2159156, EPI_ISL_2159157, EPI_ISL_2159204, EPI_ISL_2159277, EPI_ISL_2159278, EPI_ISL_2159279, EPI_ISL_2159304, EPI_ISL_2159352, EPI_ISL_2159359, EPI_ISL_2159458, EPI_ISL_2159466, EPI_ISL_2159467, EPI_ISL_2180535, EPI_ISL_2180539, EPI_ISL_2180556, EPI_ISL_2180562, EPI_ISL_2180563, EPI_ISL_2180568, EPI_ISL_2180645, EPI_ISL_2180654, EPI_ISL_2180707, EPI_ISL_2180729, EPI_ISL_2180777, EPI_ISL_2180780, EPI_ISL_2180831, EPI_ISL_2180833, EPI_ISL_2180841, EPI_ISL_2180863, EPI_ISL_2180869, EPI_ISL_2180871, EPI_ISL_2180881, EPI_ISL_2180884, EPI_ISL_2180898, EPI_ISL_2180899, EPI_ISL_2180900, EPI_ISL_2180905, EPI_ISL_2180906, EPI_ISL_2180908, EPI_ISL_2180911, EPI_ISL_2180916, EPI_ISL_2180919, EPI_ISL_2180922, EPI_ISL_2180923, EPI_ISL_2180926, EPI_ISL_2180928, EPI_ISL_2180930, EPI_ISL_2180956, EPI_ISL_2180958, EPI_ISL_2180976, EPI_ISL_2181013, EPI_ISL_2181014, EPI_ISL_2181017, EPI_ISL_2181020, EPI_ISL_2181029, EPI_ISL_2181038, EPI_ISL_2181041, EPI_ISL_2181042, EPI_ISL_2181088, EPI_ISL_2181092, EPI_ISL_2181097, EPI_ISL_2181103, EPI_ISL_2181104, EPI_ISL_2181105, EPI_ISL_2181109, EPI_ISL_2181111, EPI_ISL_2181113, EPI_ISL_2181133, EPI_ISL_2181139, EPI_ISL_2181140, EPI_ISL_2181143, EPI_ISL_2181190, EPI_ISL_2181191, EPI_ISL_2181192, EPI_ISL_2181194, EPI_ISL_2181196, EPI_ISL_2181197, EPI_ISL_2181199, EPI_ISL_2181222, EPI_ISL_2181226, EPI_ISL_2181229, EPI_ISL_2181230, EPI_ISL_2181244, EPI_ISL_2181285, EPI_ISL_2181312, EPI_ISL_2181319, EPI_ISL_2181330, EPI_ISL_2181393, EPI_ISL_2181402, EPI_ISL_2181403, EPI_ISL_2181407, EPI_ISL_2181408, EPI_ISL_2181435, EPI_ISL_2181436, EPI_ISL_2181440, EPI_ISL_2181450, EPI_ISL_2181465, EPI_ISL_2181466, EPI_ISL_2181467, EPI_ISL_2181468, EPI_ISL_2181476, EPI_ISL_2181486, EPI_ISL_2181487, EPI_ISL_2181533, EPI_ISL_2181539, EPI_ISL_2181546, EPI_ISL_2181580, EPI_ISL_2181581, EPI_ISL_2181587, EPI_ISL_2181589, EPI_ISL_2181591, EPI_ISL_2181595, EPI_ISL_2181596, EPI_ISL_2181597, EPI_ISL_2187383, EPI_ISL_2187409, EPI_ISL_2187421, EPI_ISL_2187427, EPI_ISL_2187435, EPI_ISL_2187482, EPI_ISL_2187595, EPI_ISL_2187604, EPI_ISL_2187612, EPI_ISL_2204506, EPI_ISL_2204532, EPI_ISL_2204546, EPI_ISL_2204628, EPI_ISL_2204655, EPI_ISL_2204715, EPI_ISL_2204725, EPI_ISL_2204736, EPI_ISL_2204739, EPI_ISL_2204757, EPI_ISL_2204759, EPI_ISL_2204767, EPI_ISL_2204775, EPI_ISL_2204785, EPI_ISL_2204788, EPI_ISL_2204792, EPI_ISL_2204843, EPI_ISL_2204865, EPI_ISL_2204900, EPI_ISL_2204923, EPI_ISL_2204932, EPI_ISL_2204934, EPI_ISL_2204939, EPI_ISL_2204962, EPI_ISL_2204975, EPI_ISL_2204999, EPI_ISL_2241650, EPI_ISL_2268220, EPI_ISL_2268235, EPI_ISL_2268245, EPI_ISL_2280287, EPI_ISL_2280295, EPI_ISL_2280300, EPI_ISL_2280302, EPI_ISL_2280311, EPI_ISL_2280331, EPI_ISL_2280332, EPI_ISL_2280334, EPI_ISL_2280336, EPI_ISL_2280343, EPI_ISL_2280349, EPI_ISL_2280379, EPI_ISL_2280381, EPI_ISL_2280384, EPI_ISL_2280391, EPI_ISL_2280400, EPI_ISL_2280406, EPI_ISL_2280426, EPI_ISL_2280427, EPI_ISL_2280428, EPI_ISL_2280429, EPI_ISL_2280438, EPI_ISL_2280446, EPI_ISL_2280465, EPI_ISL_2280485, EPI_ISL_2280494, EPI_ISL_2280502, EPI_ISL_2280518, EPI_ISL_2280535, EPI_ISL_2280568, EPI_ISL_2280578, EPI_ISL_2280584, EPI_ISL_2280585, EPI_ISL_2280586, EPI_ISL_2280587, EPI_ISL_2280596, EPI_ISL_2280610, EPI_ISL_2280611, EPI_ISL_2280644, EPI_ISL_2280651, EPI_ISL_2280671, EPI_ISL_2280673, EPI_ISL_2280715, EPI_ISL_2280726, EPI_ISL_2280795, EPI_ISL_2280796, EPI_ISL_2280812, EPI_ISL_2280822, EPI_ISL_2280824, EPI_ISL_2280859, EPI_ISL_2280860, EPI_ISL_2280875, EPI_ISL_2280876, EPI_ISL_2280879, EPI_ISL_2280883, EPI_ISL_2280896, EPI_ISL_2280913, EPI_ISL_2280914, EPI_ISL_2280915, EPI_ISL_2280916, EPI_ISL_2280923, EPI_ISL_2280939, EPI_ISL_2280947, EPI_ISL_2280952, EPI_ISL_2280955, EPI_ISL_2280959, EPI_ISL_2280973, EPI_ISL_2280985, EPI_ISL_2280991, EPI_ISL_2280998, EPI_ISL_2281004, EPI_ISL_2281005, EPI_ISL_2281007, EPI_ISL_2281010, EPI_ISL_2281042, EPI_ISL_2281075, EPI_ISL_2281089, EPI_ISL_2281090, EPI_ISL_2281095, EPI_ISL_2281096, EPI_ISL_2281106, EPI_ISL_2281107, EPI_ISL_2281111, EPI_ISL_2281118, EPI_ISL_2281120, EPI_ISL_2281129, EPI_ISL_2281131, EPI_ISL_2281132, EPI_ISL_2281146, EPI_ISL_2281147, EPI_ISL_2281150, EPI_ISL_2281166, EPI_ISL_2281180, EPI_ISL_2281216, EPI_ISL_2281217, EPI_ISL_2281221, EPI_ISL_2283825, EPI_ISL_2283834, EPI_ISL_2283852, EPI_ISL_2283855, EPI_ISL_2283867, EPI_ISL_2283869, EPI_ISL_2283874, EPI_ISL_2321232, EPI_ISL_2321245, EPI_ISL_2321246, EPI_ISL_2321248, EPI_ISL_2321293, EPI_ISL_2321296, EPI_ISL_2321297, EPI_ISL_2321298, EPI_ISL_2321299, EPI_ISL_2321304, EPI_ISL_2321306, EPI_ISL_2321307, EPI_ISL_2321328, EPI_ISL_2321341, EPI_ISL_2321351, EPI_ISL_2321365, EPI_ISL_2321366, EPI_ISL_2321383, EPI_ISL_2321387, EPI_ISL_2367653, EPI_ISL_2367663, EPI_ISL_2367664, EPI_ISL_2367666, EPI_ISL_2367671, EPI_ISL_2367672, EPI_ISL_2367674, EPI_ISL_2367675, EPI_ISL_2368225, EPI_ISL_2368227, EPI_ISL_2368261, EPI_ISL_2368263, EPI_ISL_2368264, EPI_ISL_2368287, EPI_ISL_2368288, EPI_ISL_2368299, EPI_ISL_2368302, EPI_ISL_2368303, EPI_ISL_2368308, EPI_ISL_2368319, EPI_ISL_2368327, EPI_ISL_2368331, EPI_ISL_2368354, EPI_ISL_2368355, EPI_ISL_2368360, EPI_ISL_2368372, EPI_ISL_2368373, EPI_ISL_2368377, EPI_ISL_2368383, EPI_ISL_2368384, EPI_ISL_2368388, EPI_ISL_2368389, EPI_ISL_2368390, EPI_ISL_2368391, EPI_ISL_2368394, EPI_ISL_2368396, EPI_ISL_2368400, EPI_ISL_2368407, EPI_ISL_2368408, EPI_ISL_2368409, EPI_ISL_2368410, EPI_ISL_2368427, EPI_ISL_2368437, EPI_ISL_2368449, EPI_ISL_2368452, EPI_ISL_2368460, EPI_ISL_2368461, EPI_ISL_2368465, EPI_ISL_2368470, EPI_ISL_2368545, EPI_ISL_2368548, EPI_ISL_2368551, EPI_ISL_2368557, EPI_ISL_2368600, EPI_ISL_2368603, EPI_ISL_2368699, EPI_ISL_2368730, EPI_ISL_2368733, EPI_ISL_2368753, EPI_ISL_2368756, EPI_ISL_2368764, EPI_ISL_2368767, EPI_ISL_2368895, EPI_ISL_2368957, EPI_ISL_2368981, EPI_ISL_2369970, EPI_ISL_2370034, EPI_ISL_2370036, EPI_ISL_2370054, EPI_ISL_2370139, EPI_ISL_2370150, EPI_ISL_2370187, EPI_ISL_2370188, EPI_ISL_2370229, EPI_ISL_2370231, EPI_ISL_2370233, EPI_ISL_2370239, EPI_ISL_2370392, EPI_ISL_2370480, EPI_ISL_2370491, EPI_ISL_2370507, EPI_ISL_2370528, EPI_ISL_2370533, EPI_ISL_2370536, EPI_ISL_2370585, EPI_ISL_2370648, EPI_ISL_2370650, EPI_ISL_2370704, EPI_ISL_2372300, EPI_ISL_2372376, EPI_ISL_2372394, EPI_ISL_2372455, EPI_ISL_2372460, EPI_ISL_2372461, EPI_ISL_2372465, EPI_ISL_2422001, EPI_ISL_2422025, EPI_ISL_2422033, EPI_ISL_2422046, EPI_ISL_2422052, EPI_ISL_2422109, EPI_ISL_2422113, EPI_ISL_2422114, EPI_ISL_2422118, EPI_ISL_2422119, EPI_ISL_2442333, EPI_ISL_2489072, EPI_ISL_2489073, EPI_ISL_2489074, EPI_ISL_2489075, EPI_ISL_2489078, EPI_ISL_2489079, EPI_ISL_2489080, EPI_ISL_2489081, EPI_ISL_2489082, EPI_ISL_2489084, EPI_ISL_2489085, EPI_ISL_2489091, EPI_ISL_2489092, EPI_ISL_2489097, EPI_ISL_2489098, EPI_ISL_2489099, EPI_ISL_2489100, EPI_ISL_2489101, EPI_ISL_2489110, EPI_ISL_2489111, EPI_ISL_2489112, EPI_ISL_2489116, EPI_ISL_2489117, EPI_ISL_2489119, EPI_ISL_2489120, EPI_ISL_2489123, EPI_ISL_2489137, EPI_ISL_2489138, EPI_ISL_2489141, EPI_ISL_2489142, EPI_ISL_2489172, EPI_ISL_2489234, EPI_ISL_2489235, EPI_ISL_2489240, EPI_ISL_2489245, EPI_ISL_2489250, EPI_ISL_2489263, EPI_ISL_2489279, EPI_ISL_2489283, EPI_ISL_2489295, EPI_ISL_2489298, EPI_ISL_2489300, EPI_ISL_2489304, EPI_ISL_2489309, EPI_ISL_2489315, EPI_ISL_2489316, EPI_ISL_2489317, EPI_ISL_2489318, EPI_ISL_2489319, EPI_ISL_2489320, EPI_ISL_2489321, EPI_ISL_2489323, EPI_ISL_2489329, EPI_ISL_2489341, EPI_ISL_2489349, EPI_ISL_2489359, EPI_ISL_2489371, EPI_ISL_2489386, EPI_ISL_2489388, EPI_ISL_2489402, EPI_ISL_2489405, EPI_ISL_2489408, EPI_ISL_2489440, EPI_ISL_2489442, EPI_ISL_2489454, EPI_ISL_2489458, EPI_ISL_2489462, EPI_ISL_2489463, EPI_ISL_2489464, EPI_ISL_2489473, EPI_ISL_2489502, EPI_ISL_2489508, EPI_ISL_2489509, EPI_ISL_2489524, EPI_ISL_2489528, EPI_ISL_2489538, EPI_ISL_2489545, EPI_ISL_2489556, EPI_ISL_2489557, EPI_ISL_2489559, EPI_ISL_2489566, EPI_ISL_2489583, EPI_ISL_2489584, EPI_ISL_2489585, EPI_ISL_2489595, EPI_ISL_2489596, EPI_ISL_2489602, EPI_ISL_2489638, EPI_ISL_2489643, EPI_ISL_2489654, EPI_ISL_2489672, EPI_ISL_2489678, EPI_ISL_2489682, EPI_ISL_2489683, EPI_ISL_2489697, EPI_ISL_2489698, EPI_ISL_2489701, EPI_ISL_2489703, EPI_ISL_2489707, EPI_ISL_2489709, EPI_ISL_2489712, EPI_ISL_2489754, EPI_ISL_2489766, EPI_ISL_2489806, EPI_ISL_2489811, EPI_ISL_2527901, EPI_ISL_2527906, EPI_ISL_2527909, EPI_ISL_2527911, EPI_ISL_2527922, EPI_ISL_2527976, EPI_ISL_2527983, EPI_ISL_2528028, EPI_ISL_2528037, EPI_ISL_2528045, EPI_ISL_2528052, EPI_ISL_2528106, EPI_ISL_2528109, EPI_ISL_2528159, EPI_ISL_2528161, EPI_ISL_2528165, EPI_ISL_2528169, EPI_ISL_2528182, EPI_ISL_2528195, EPI_ISL_2528199, EPI_ISL_2528201, EPI_ISL_2528205, EPI_ISL_2528210, EPI_ISL_2528212, EPI_ISL_2528228, EPI_ISL_2528243, EPI_ISL_2528303, EPI_ISL_2528306, EPI_ISL_2528310, EPI_ISL_2528320, EPI_ISL_2528330, EPI_ISL_2528337, EPI_ISL_2528383, EPI_ISL_2528430, EPI_ISL_2528468, EPI_ISL_2528492, EPI_ISL_2528500, EPI_ISL_2528506, EPI_ISL_2528508, EPI_ISL_2528576, EPI_ISL_2528578, EPI_ISL_2528605, EPI_ISL_2528614, EPI_ISL_2528642, EPI_ISL_2528647, EPI_ISL_2528650, EPI_ISL_2528656, EPI_ISL_2528666, EPI_ISL_2528673, EPI_ISL_2528675, EPI_ISL_2528688, EPI_ISL_2528690, EPI_ISL_2528705, EPI_ISL_2528734, EPI_ISL_2528772, EPI_ISL_2528774, EPI_ISL_2528781, EPI_ISL_2528791, EPI_ISL_2528795, EPI_ISL_2528813, EPI_ISL_2528823, EPI_ISL_2528864, EPI_ISL_2528866, EPI_ISL_2528868, EPI_ISL_2528871, EPI_ISL_2528885, EPI_ISL_2528887, EPI_ISL_2528891, EPI_ISL_2528902, EPI_ISL_2528904, EPI_ISL_2528965, EPI_ISL_2528968, EPI_ISL_2528994, EPI_ISL_2528999, EPI_ISL_2530938, EPI_ISL_2530940, EPI_ISL_2530944, EPI_ISL_2530948, EPI_ISL_2530950, EPI_ISL_2530966, EPI_ISL_2531004, EPI_ISL_2531012, EPI_ISL_2531021, EPI_ISL_2531030, EPI_ISL_2531038, EPI_ISL_2531044, EPI_ISL_2531047, EPI_ISL_2531055, EPI_ISL_2531057, EPI_ISL_2534533, EPI_ISL_2534534, EPI_ISL_2686902, EPI_ISL_2686903, EPI_ISL_2686904, EPI_ISL_2686910, EPI_ISL_2686954, EPI_ISL_2686960, EPI_ISL_2686961, EPI_ISL_2686962, EPI_ISL_2686963, EPI_ISL_2686965, EPI_ISL_2686966, EPI_ISL_2686980, EPI_ISL_2686989, EPI_ISL_2686992, EPI_ISL_2686994, EPI_ISL_2686996, EPI_ISL_2687000, EPI_ISL_2687001, EPI_ISL_2687010, EPI_ISL_2687030, EPI_ISL_2687031, EPI_ISL_2687034, EPI_ISL_2687035, EPI_ISL_2687047, EPI_ISL_2687059, EPI_ISL_2687060, EPI_ISL_2687061, EPI_ISL_2687062, EPI_ISL_2687064, EPI_ISL_2687072, EPI_ISL_2687073, EPI_ISL_2687079, EPI_ISL_2687080, EPI_ISL_2687081, EPI_ISL_2687082, EPI_ISL_2687084, EPI_ISL_2687090, EPI_ISL_2687093, EPI_ISL_2687094, EPI_ISL_2687100, EPI_ISL_2687110, EPI_ISL_2687113, EPI_ISL_2687116, EPI_ISL_2687120, EPI_ISL_2687127, EPI_ISL_2687128, EPI_ISL_2687129, EPI_ISL_2687136, EPI_ISL_2687137, EPI_ISL_2687139, EPI_ISL_2687151, EPI_ISL_2687168, EPI_ISL_2687181, EPI_ISL_2687189, EPI_ISL_2687190, EPI_ISL_2687193, EPI_ISL_2687194, EPI_ISL_2687197, EPI_ISL_2687212, EPI_ISL_2687213, EPI_ISL_2687225, EPI_ISL_2687226, EPI_ISL_2687234, EPI_ISL_2687250, EPI_ISL_2687253, EPI_ISL_2687256, EPI_ISL_2687261, EPI_ISL_2687262, EPI_ISL_2687263, EPI_ISL_2687267, EPI_ISL_2687268, EPI_ISL_2687269, EPI_ISL_2687270, EPI_ISL_2687271, EPI_ISL_2687277, EPI_ISL_2687297, EPI_ISL_2687308, EPI_ISL_2687309, EPI_ISL_2687317, EPI_ISL_2687320, EPI_ISL_2687322, EPI_ISL_2687323, EPI_ISL_2687327, EPI_ISL_2687328, EPI_ISL_2687332, EPI_ISL_2687336, EPI_ISL_2687337, EPI_ISL_2687356, EPI_ISL_2687368, EPI_ISL_2687370, EPI_ISL_2687371, EPI_ISL_2687376, EPI_ISL_2687379, EPI_ISL_2687380, EPI_ISL_2687381, EPI_ISL_2687383, EPI_ISL_2687384, EPI_ISL_2687389, EPI_ISL_2687392, EPI_ISL_2687416, EPI_ISL_2687422, EPI_ISL_2687453, EPI_ISL_2687454, EPI_ISL_2687471, EPI_ISL_2687499, EPI_ISL_2687501, EPI_ISL_2687511, EPI_ISL_2687512, EPI_ISL_2687514, EPI_ISL_2687515, EPI_ISL_2687516, EPI_ISL_2687527, EPI_ISL_2687530, EPI_ISL_2687532, EPI_ISL_2687535, EPI_ISL_2687538, EPI_ISL_2687545, EPI_ISL_2687546, EPI_ISL_2687547, EPI_ISL_2687548, EPI_ISL_2687549, EPI_ISL_2687551, EPI_ISL_2687554, EPI_ISL_2687556, EPI_ISL_2687557, EPI_ISL_2687568, EPI_ISL_2687569, EPI_ISL_2687570, EPI_ISL_2687581, EPI_ISL_2687582, EPI_ISL_2709520, EPI_ISL_2709527, EPI_ISL_2709538, EPI_ISL_2709547, EPI_ISL_2709551, EPI_ISL_2784728, EPI_ISL_2784731, EPI_ISL_2784742, EPI_ISL_2784743, EPI_ISL_2784744, EPI_ISL_2784747, EPI_ISL_2784748, EPI_ISL_2784750, EPI_ISL_2784751, EPI_ISL_2784752, EPI_ISL_2784753, EPI_ISL_2784761, EPI_ISL_2784770, EPI_ISL_2784773, EPI_ISL_2784777, EPI_ISL_2784781, EPI_ISL_2784787, EPI_ISL_2784791, EPI_ISL_2784796, EPI_ISL_2784807, EPI_ISL_2784809, EPI_ISL_2784810, EPI_ISL_2784811, EPI_ISL_2784812, EPI_ISL_2784816, EPI_ISL_2784817, EPI_ISL_2784820, EPI_ISL_2784822, EPI_ISL_2784823, EPI_ISL_2784831, EPI_ISL_2784840, EPI_ISL_2784844, EPI_ISL_2784858, EPI_ISL_2784859, EPI_ISL_2784861, EPI_ISL_2784862, EPI_ISL_2784865, EPI_ISL_2784867, EPI_ISL_2784868, EPI_ISL_2784870, EPI_ISL_2784871, EPI_ISL_2784872, EPI_ISL_2784875, EPI_ISL_2784877, EPI_ISL_2784881, EPI_ISL_2784882, EPI_ISL_2784883, EPI_ISL_2784886, EPI_ISL_2784887, EPI_ISL_2784888, EPI_ISL_2784906, EPI_ISL_2784915, EPI_ISL_2784935, EPI_ISL_2784937, EPI_ISL_2784940, EPI_ISL_2784943, EPI_ISL_2784944, EPI_ISL_2784953, EPI_ISL_2784963, EPI_ISL_2784968, EPI_ISL_2784969, EPI_ISL_2784983, EPI_ISL_2784984, EPI_ISL_2784989, EPI_ISL_2784998, EPI_ISL_2785000, EPI_ISL_2785007, EPI_ISL_2785020, EPI_ISL_2785021, EPI_ISL_2785023, EPI_ISL_2785027, EPI_ISL_2785029, EPI_ISL_2785030, EPI_ISL_2785031, EPI_ISL_2785036, EPI_ISL_2785038, EPI_ISL_2785047, EPI_ISL_2785059, EPI_ISL_2785060, EPI_ISL_2785062, EPI_ISL_2785063, EPI_ISL_2785064, EPI_ISL_2785065, EPI_ISL_2785066, EPI_ISL_2785068, EPI_ISL_2785071, EPI_ISL_2785073, EPI_ISL_2785077, EPI_ISL_2785079, EPI_ISL_2785080, EPI_ISL_2785083, EPI_ISL_2785084, EPI_ISL_2785085, EPI_ISL_2785090, EPI_ISL_2785095, EPI_ISL_2785105, EPI_ISL_2785120, EPI_ISL_2785121, EPI_ISL_2785124, EPI_ISL_2785139, EPI_ISL_2785147, EPI_ISL_2785148, EPI_ISL_2785152, EPI_ISL_2785154, EPI_ISL_2785155, EPI_ISL_2785156, EPI_ISL_2785157, EPI_ISL_2785161, EPI_ISL_2785162, EPI_ISL_2785165, EPI_ISL_2785169, EPI_ISL_2785171, EPI_ISL_2785173, EPI_ISL_2785174, EPI_ISL_2785178, EPI_ISL_2785179, EPI_ISL_2785184, EPI_ISL_2785185, EPI_ISL_2785198, EPI_ISL_2785205, EPI_ISL_2785207, EPI_ISL_2785210, EPI_ISL_2785211, EPI_ISL_2785212, EPI_ISL_2785222, EPI_ISL_2785223, EPI_ISL_2785224, EPI_ISL_2785226, EPI_ISL_2785237, EPI_ISL_2785238, EPI_ISL_2785243, EPI_ISL_2785247, EPI_ISL_2785248, EPI_ISL_2785251, EPI_ISL_2785261, EPI_ISL_2785262, EPI_ISL_2785263, EPI_ISL_2785267, EPI_ISL_2785270, EPI_ISL_2785291, EPI_ISL_2785292, EPI_ISL_2785293, EPI_ISL_2785301, EPI_ISL_2785302, EPI_ISL_2785308, EPI_ISL_2785309, EPI_ISL_2785313, EPI_ISL_2785316, EPI_ISL_2785317, EPI_ISL_2785319, EPI_ISL_2785320, EPI_ISL_2785321, EPI_ISL_2785322, EPI_ISL_2785332, EPI_ISL_2785335, EPI_ISL_2785336, EPI_ISL_2785339, EPI_ISL_2785344, EPI_ISL_2785345, EPI_ISL_2785346, EPI_ISL_2785347, EPI_ISL_2785349, EPI_ISL_2785350, EPI_ISL_2785351, EPI_ISL_2785352, EPI_ISL_2785387, EPI_ISL_2785388, EPI_ISL_2785392, EPI_ISL_2785393, EPI_ISL_2785394, EPI_ISL_2785405, EPI_ISL_2785410, EPI_ISL_2785418, EPI_ISL_2785422, EPI_ISL_2785433, EPI_ISL_2785434, EPI_ISL_2785449, EPI_ISL_2785458, EPI_ISL_2785459, EPI_ISL_2785466, EPI_ISL_2785472, EPI_ISL_2785473, EPI_ISL_2785486, EPI_ISL_2785489, EPI_ISL_2785490, EPI_ISL_2785491, EPI_ISL_2785492, EPI_ISL_2785493, EPI_ISL_2785499, EPI_ISL_2785500, EPI_ISL_2785502, EPI_ISL_2832518, EPI_ISL_2832519, EPI_ISL_2832520, EPI_ISL_2832523, EPI_ISL_2832539, EPI_ISL_2832540, EPI_ISL_2832541, EPI_ISL_2832547, EPI_ISL_2832548, EPI_ISL_2832549, EPI_ISL_2832551, EPI_ISL_2832553, EPI_ISL_2832563, EPI_ISL_2832568, EPI_ISL_2832570, EPI_ISL_2832571, EPI_ISL_2832572, EPI_ISL_2832576, EPI_ISL_2832581, EPI_ISL_2875520, EPI_ISL_2875522, EPI_ISL_2875525, EPI_ISL_2875528, EPI_ISL_2875535, EPI_ISL_2875536, EPI_ISL_2875540, EPI_ISL_2875544, EPI_ISL_2875546, EPI_ISL_2875548, EPI_ISL_2875549, EPI_ISL_2875551, EPI_ISL_2875552, EPI_ISL_2875553, EPI_ISL_2875554, EPI_ISL_2875555, EPI_ISL_2875562, EPI_ISL_2875563, EPI_ISL_2875566, EPI_ISL_2875568, EPI_ISL_2875580, EPI_ISL_2875581, EPI_ISL_2875584, EPI_ISL_2875591, EPI_ISL_2875602, EPI_ISL_2875604, EPI_ISL_2875614, EPI_ISL_2875623, EPI_ISL_2875626, EPI_ISL_2875638, EPI_ISL_2875661, EPI_ISL_2875662, EPI_ISL_2875663, EPI_ISL_2875666, EPI_ISL_2875668, EPI_ISL_2875669, EPI_ISL_2875671, EPI_ISL_2875684, EPI_ISL_2875685, EPI_ISL_2875686, EPI_ISL_2875687, EPI_ISL_2875690, EPI_ISL_2875700, EPI_ISL_2875701, EPI_ISL_2875703, EPI_ISL_2875714, EPI_ISL_2875716, EPI_ISL_2875719, EPI_ISL_2875720, EPI_ISL_2875724, EPI_ISL_2875725, EPI_ISL_2875726, EPI_ISL_2875728, EPI_ISL_2875732, EPI_ISL_2875734, EPI_ISL_2875743, EPI_ISL_2875766, EPI_ISL_2875774, EPI_ISL_2875779, EPI_ISL_2875792, EPI_ISL_2875798, EPI_ISL_2875799, EPI_ISL_2875803, EPI_ISL_2875804, EPI_ISL_2875806, EPI_ISL_2875807, EPI_ISL_2875809, EPI_ISL_2875817, EPI_ISL_2875818, EPI_ISL_2875819, EPI_ISL_2875820, EPI_ISL_2875821, EPI_ISL_2875823, EPI_ISL_2875825, EPI_ISL_2875827, EPI_ISL_2875828, EPI_ISL_2875831, EPI_ISL_2875834, EPI_ISL_2875858, EPI_ISL_2875867, EPI_ISL_2875873, EPI_ISL_2875874, EPI_ISL_2875876, EPI_ISL_2875880, EPI_ISL_2875881, EPI_ISL_2875882, EPI_ISL_2875883, EPI_ISL_2875886, EPI_ISL_2875912, EPI_ISL_2875916, EPI_ISL_2875930, EPI_ISL_2875936, EPI_ISL_2875940, EPI_ISL_2875944, EPI_ISL_2875945, EPI_ISL_2875946, EPI_ISL_2875947, EPI_ISL_2875948, EPI_ISL_2875949, EPI_ISL_2875950, EPI_ISL_2875951, EPI_ISL_2875952, EPI_ISL_2875953, EPI_ISL_2875961, EPI_ISL_2875962, EPI_ISL_2875963, EPI_ISL_2875964, EPI_ISL_2875966, EPI_ISL_2875967, EPI_ISL_2875968, EPI_ISL_2875979, EPI_ISL_2875983, EPI_ISL_2875986, EPI_ISL_2875992, EPI_ISL_2875995, EPI_ISL_2875996, EPI_ISL_2875997, EPI_ISL_2876001, EPI_ISL_2876004, EPI_ISL_2876005, EPI_ISL_2876006, EPI_ISL_2876010, EPI_ISL_2876011, EPI_ISL_2876013, EPI_ISL_2876014, EPI_ISL_2877658, EPI_ISL_2877661, EPI_ISL_2877665, EPI_ISL_2877682, EPI_ISL_2877683, EPI_ISL_2877685, EPI_ISL_2877691, EPI_ISL_2877695 | | | |
| see above | Aegis Sciences Corporation | Centers for Disease Control and Prevention Division of Viral Diseases, Pathogen Discovery | Adrian Paskey; Alec Vest; Benjamin Rambo-Martin; Christopher Gulvick; Clinton R. Paden; Cyndi Clark; Dakota Howard; Darlene Wagner; Dhwani Batra; Dillon Nall; Duncan MacCannell; Ethan Sanders; Holly Houdeshell; Jason Caravas; Kara Moser; Matthew Hardison; Matthew Schmerer; Ola Kvalvaag; Patrick Campbell; Peter W. Cook; Rob Case; Scott Sammons; Shatavia Morrison; Shaun Westlund; Vikramsinha Ghorpade; Yvette Unoarumhi |
| EPI_ISL_2304324, EPI_ISL_2304327 | Alameda County Public Health Department | Alameda County Public Health Department | Kevin Libuit; Kristina Hsieh; Maria Elma |
| EPI_ISL_3047314, EPI_ISL_3047315 | Brentwood Biomedical Research | Genomics and Discovery, Respiratory Viruses Branch, Division of Viral Diseases, Centers for Disease Control and Prevention | Adam Retchless; Anna Kelleher; Anna Uehara; Brian Lynch; Clinton R. Paden; Dhwani Batra; Haibin Wang; Han Jia Justin Ng; Jasmine Padilla; Jing Zhang; Justin Lee; Mark Burroughs; Mili Sheth; Morgan Davis; Peter Cook; Rachel Marine; Sarah Nobles; Suxiang Tong; Tara Coalter; Yan Li; Ying Tao |
| EPI_ISL_2496770, EPI_ISL_2496771, EPI_ISL_2496772, EPI_ISL_2496773, EPI_ISL_2496774, EPI_ISL_2496775, EPI_ISL_2496778, EPI_ISL_2496779, EPI_ISL_2496780, EPI_ISL_2496781, EPI_ISL_2496782, EPI_ISL_2496783, EPI_ISL_2496785, EPI_ISL_2496786, EPI_ISL_2496789, EPI_ISL_2496790, EPI_ISL_2496791, EPI_ISL_2496792, EPI_ISL_2496794, EPI_ISL_2496796, EPI_ISL_2496797, EPI_ISL_2496798, EPI_ISL_2496799, EPI_ISL_2496801, EPI_ISL_2496802, EPI_ISL_2496803, EPI_ISL_2496804, EPI_ISL_2496806, EPI_ISL_2496807, EPI_ISL_2496808, EPI_ISL_2496809, EPI_ISL_2496811, EPI_ISL_2496813, EPI_ISL_2496815, EPI_ISL_2496816, EPI_ISL_2496817, EPI_ISL_2496818, EPI_ISL_2496819, EPI_ISL_2496820, EPI_ISL_2496822, EPI_ISL_2496823, EPI_ISL_2496824, EPI_ISL_2496825, EPI_ISL_2496826, EPI_ISL_2496830, EPI_ISL_2496831, EPI_ISL_2496833, EPI_ISL_2496835, EPI_ISL_2496836, EPI_ISL_2496837, EPI_ISL_2496839, EPI_ISL_2496840, EPI_ISL_2496841, EPI_ISL_2496842, EPI_ISL_2496844, EPI_ISL_2496846, EPI_ISL_2496848, EPI_ISL_2496849, EPI_ISL_2496850, EPI_ISL_2496851, EPI_ISL_2496854, EPI_ISL_2496856, EPI_ISL_2496857, EPI_ISL_2496859, EPI_ISL_2496860, EPI_ISL_2496861, EPI_ISL_2496862, EPI_ISL_2496863, EPI_ISL_2496865, EPI_ISL_2496868, EPI_ISL_2496869, EPI_ISL_2496870, EPI_ISL_2496871, EPI_ISL_2496873, EPI_ISL_2496874, EPI_ISL_2496875, EPI_ISL_2496876, EPI_ISL_2496880, EPI_ISL_2496884, EPI_ISL_2496889, EPI_ISL_2496890, EPI_ISL_2496891, EPI_ISL_2496893, EPI_ISL_2496894, EPI_ISL_2496896, EPI_ISL_2496897, EPI_ISL_2496898, EPI_ISL_2496899, EPI_ISL_2496900, EPI_ISL_2496901, EPI_ISL_2496902, EPI_ISL_2496903, EPI_ISL_2496905, EPI_ISL_2496906, EPI_ISL_2496907, EPI_ISL_2496908, EPI_ISL_2496909, EPI_ISL_2496911, EPI_ISL_2496913, EPI_ISL_2496915, EPI_ISL_2496918, EPI_ISL_2496922, EPI_ISL_2496926, EPI_ISL_2496927, EPI_ISL_2496928, EPI_ISL_2496929, EPI_ISL_2496930, EPI_ISL_2496934, EPI_ISL_2496937, EPI_ISL_2496940, EPI_ISL_2496944, EPI_ISL_2496946, EPI_ISL_2496948, EPI_ISL_2496949, EPI_ISL_2496950, EPI_ISL_2496951, EPI_ISL_2496952, EPI_ISL_2496953, EPI_ISL_2496954, EPI_ISL_2496955, EPI_ISL_2496957, EPI_ISL_2496958, EPI_ISL_2496959, EPI_ISL_2496961, EPI_ISL_2496962, EPI_ISL_2496963, EPI_ISL_2496964, EPI_ISL_2496966, EPI_ISL_2496967, EPI_ISL_2496970, EPI_ISL_2496974, EPI_ISL_2496978, EPI_ISL_2496982, EPI_ISL_2496983, EPI_ISL_2496984, EPI_ISL_2496985, EPI_ISL_2496986, EPI_ISL_2496988, EPI_ISL_2496991, EPI_ISL_2496993, EPI_ISL_2496994, EPI_ISL_2496997, EPI_ISL_2496998, EPI_ISL_2497544, EPI_ISL_2497546, EPI_ISL_2497548, EPI_ISL_2497556, EPI_ISL_2497557, EPI_ISL_2497560, EPI_ISL_2497561, EPI_ISL_2497563, EPI_ISL_2497564, EPI_ISL_2497568, EPI_ISL_2497570, EPI_ISL_2497575, EPI_ISL_2497580, EPI_ISL_2497582, EPI_ISL_2497583, EPI_ISL_2497585, EPI_ISL_2497588, EPI_ISL_2497594, EPI_ISL_2497597, EPI_ISL_2497598, EPI_ISL_2497600, EPI_ISL_2497601, EPI_ISL_2497606, EPI_ISL_2497608, EPI_ISL_2497609, EPI_ISL_2497610, EPI_ISL_2497613, EPI_ISL_2497615, EPI_ISL_2497616, EPI_ISL_2497617, EPI_ISL_2497620, EPI_ISL_2497623, EPI_ISL_2497624, EPI_ISL_2497626, EPI_ISL_2497627, EPI_ISL_2497630, EPI_ISL_2497631, EPI_ISL_2497632, EPI_ISL_2497634, EPI_ISL_2497636, EPI_ISL_2497637, EPI_ISL_2497640, EPI_ISL_2497645, EPI_ISL_2497646, EPI_ISL_2497647, EPI_ISL_2497648, EPI_ISL_2497649, EPI_ISL_2497650, EPI_ISL_2497651, EPI_ISL_2497652, EPI_ISL_2497654, EPI_ISL_2497655, EPI_ISL_2497656, EPI_ISL_2497658, EPI_ISL_2497665, EPI_ISL_2497666, EPI_ISL_2497668, EPI_ISL_2497670, EPI_ISL_2497671, EPI_ISL_2497672, EPI_ISL_2497673, EPI_ISL_2497674, EPI_ISL_2497685, EPI_ISL_2497689, EPI_ISL_2497694, EPI_ISL_2497710, EPI_ISL_2497737, EPI_ISL_2497768, EPI_ISL_2497845, EPI_ISL_2658681, EPI_ISL_2658683, EPI_ISL_2658685, EPI_ISL_2658705, EPI_ISL_2659200, EPI_ISL_2659300, EPI_ISL_2659349, EPI_ISL_2659392, EPI_ISL_2659400, EPI_ISL_2659457, EPI_ISL_2659503, EPI_ISL_2659514, EPI_ISL_2659522, EPI_ISL_2659551, EPI_ISL_2659579, EPI_ISL_2758534, EPI_ISL_2758535, EPI_ISL_2758536, EPI_ISL_2758537, EPI_ISL_2758538, EPI_ISL_2758539, EPI_ISL_2758540 | | | |
| see above | CA DPH Viral and Rickettsial Disease Laboratory | Chan-Zuckerberg Biohub | CZB Cliahub Consortium |
| EPI_ISL_2229189, EPI_ISL_2229190, EPI_ISL_2229191, EPI_ISL_2229192, EPI_ISL_2383986, EPI_ISL_2383987, EPI_ISL_2451881, EPI_ISL_2451883, EPI_ISL_2451884, EPI_ISL_2451886, EPI_ISL_2451888, EPI_ISL_2451890, EPI_ISL_2451891, EPI_ISL_2451893, EPI_ISL_2451895, EPI_ISL_2689932, EPI_ISL_2689933, EPI_ISL_2709557, EPI_ISL_2787321, EPI_ISL_2787323, EPI_ISL_2787325, EPI_ISL_2787328 | | | |
| see above | CA-Los Angeles County Public Health Laboratory | Centers for Disease Control and Prevention Division of Viral Diseases, Pathogen Discovery | Alison Laufer Halpin; Ben L. Rambo-Martin; Clinton R. Paden; Dakota Howard; Darlene Wagner; Dave Wentworth; Dhwani Batra; Jasmine Padilla; Justin Lee; Katie Dillon; Krista Queen; Kristen Knipe; Kristine Lacek; Mark Burroughs; Matthew Schmerer; Mili Sheth; Peter Cook; Sam Shepard; Sarah Nobles; Shoshona Le; Suxiang Tong; Vivien Dugan; Yvette Unoarumhi |
| EPI_ISL_2890369, EPI_ISL_2890370, EPI_ISL_2890544, EPI_ISL_2890545, EPI_ISL_2890943, EPI_ISL_2891608, EPI_ISL_2891708, EPI_ISL_2891709, EPI_ISL_2891710, EPI_ISL_2891711, EPI_ISL_2891712, EPI_ISL_2891713, EPI_ISL_2891714, EPI_ISL_2891715, EPI_ISL_2891716, EPI_ISL_2891717, EPI_ISL_2891722, EPI_ISL_2891723, EPI_ISL_2891724, EPI_ISL_2891725, EPI_ISL_2891726, EPI_ISL_2891727, EPI_ISL_2891730, EPI_ISL_2891731, EPI_ISL_2891732, EPI_ISL_2891733, EPI_ISL_2891734, EPI_ISL_2891735, EPI_ISL_2891736, EPI_ISL_2891737, EPI_ISL_2891738, EPI_ISL_2891739, EPI_ISL_2891740, EPI_ISL_2891743, EPI_ISL_2891745, EPI_ISL_2891746, EPI_ISL_2891747, EPI_ISL_2891748, EPI_ISL_2891749, EPI_ISL_2891753, EPI_ISL_2891754, EPI_ISL_2891757, EPI_ISL_2891760, EPI_ISL_2891761, EPI_ISL_2891762, EPI_ISL_2891763, EPI_ISL_2891764, EPI_ISL_2891767, EPI_ISL_2891769, EPI_ISL_2891770, EPI_ISL_2891771, EPI_ISL_2891772, EPI_ISL_2891773, EPI_ISL_2891774, EPI_ISL_2891775, EPI_ISL_2891776, EPI_ISL_2891777, EPI_ISL_2891778, EPI_ISL_2891779, EPI_ISL_2891780, EPI_ISL_2891782, EPI_ISL_2891783, EPI_ISL_2891784, EPI_ISL_2891785, EPI_ISL_2891786, EPI_ISL_2891787, EPI_ISL_2891788, EPI_ISL_2891789, EPI_ISL_2891790, EPI_ISL_2891791, EPI_ISL_2891792, EPI_ISL_2891793, EPI_ISL_2891794, EPI_ISL_2891795, EPI_ISL_2891796, EPI_ISL_2891797, EPI_ISL_2891798, EPI_ISL_2891799, EPI_ISL_2891800, EPI_ISL_2891801, EPI_ISL_2891803, EPI_ISL_2891804, EPI_ISL_2891805, EPI_ISL_2891806, EPI_ISL_2891807, EPI_ISL_2891808, EPI_ISL_2891809, EPI_ISL_2891810, | | | |

| | | | |
|---|---|---|---|
| EPI_ISL_2891811, EPI_ISL_2891812, EPI_ISL_2891813, EPI_ISL_2891814, EPI_ISL_2891815, EPI_ISL_2891816, EPI_ISL_2891817, EPI_ISL_2891818, EPI_ISL_2891819, EPI_ISL_2891820, EPI_ISL_2891821, EPI_ISL_2891822, EPI_ISL_2891823, EPI_ISL_2891824, EPI_ISL_2891825, EPI_ISL_2891826, EPI_ISL_2891827, EPI_ISL_2891828, EPI_ISL_2891829, EPI_ISL_2891830, EPI_ISL_2891831, EPI_ISL_2891832, EPI_ISL_2891833, EPI_ISL_2891834, EPI_ISL_2891835, EPI_ISL_2891836, EPI_ISL_2891837, EPI_ISL_2891838, EPI_ISL_2891839, EPI_ISL_2891840, EPI_ISL_2891841, EPI_ISL_2891842, EPI_ISL_2891843, EPI_ISL_2891844, EPI_ISL_2891845, EPI_ISL_2891846, EPI_ISL_2891847, EPI_ISL_2891848, EPI_ISL_2891849, EPI_ISL_2891850, EPI_ISL_2891851, EPI_ISL_2891852, EPI_ISL_2891853, EPI_ISL_2891854, EPI_ISL_2891855, EPI_ISL_2891856, EPI_ISL_2891859, EPI_ISL_2891861, EPI_ISL_2891862, EPI_ISL_2891863, EPI_ISL_2891864, EPI_ISL_2891865, EPI_ISL_2891866, EPI_ISL_2891867, EPI_ISL_2891868, EPI_ISL_2891869, EPI_ISL_2891870, EPI_ISL_2891876, EPI_ISL_2891881, EPI_ISL_2891882, EPI_ISL_2891883, EPI_ISL_2891884, EPI_ISL_2891887, EPI_ISL_2891888, EPI_ISL_2891889, EPI_ISL_2891890, EPI_ISL_2891891, EPI_ISL_2891892, EPI_ISL_2891893, EPI_ISL_2891894, EPI_ISL_2891895, EPI_ISL_2891896, EPI_ISL_2891897, EPI_ISL_2891898, EPI_ISL_2891899, EPI_ISL_2891900, EPI_ISL_2891901, EPI_ISL_2891902, EPI_ISL_2891903, EPI_ISL_2891904, EPI_ISL_2891905, EPI_ISL_2891906, EPI_ISL_2891907, EPI_ISL_2891908, EPI_ISL_2891909, EPI_ISL_2891910, EPI_ISL_2891911, EPI_ISL_2891912, EPI_ISL_2891914, EPI_ISL_2891915, EPI_ISL_2891916, EPI_ISL_2891917, EPI_ISL_2891918, EPI_ISL_2891919, EPI_ISL_2891920, EPI_ISL_2891921, EPI_ISL_2891922, EPI_ISL_2891923, EPI_ISL_2891924, EPI_ISL_2891925, EPI_ISL_2891926, EPI_ISL_2891927, EPI_ISL_2891928, EPI_ISL_2891929, EPI_ISL_2891930, EPI_ISL_2891931, EPI_ISL_2891932, EPI_ISL_2891933, EPI_ISL_2891934, EPI_ISL_2891935, EPI_ISL_2891936, EPI_ISL_2891937, EPI_ISL_2891938, EPI_ISL_2891939, EPI_ISL_2891940, EPI_ISL_2891941, EPI_ISL_2891942, EPI_ISL_2891943, EPI_ISL_2891944, EPI_ISL_2891945, EPI_ISL_2891946, EPI_ISL_2891947, EPI_ISL_2891948, EPI_ISL_2891949, EPI_ISL_2891950, EPI_ISL_2891951, EPI_ISL_2891952, EPI_ISL_2891953, EPI_ISL_2891956, EPI_ISL_2891958, EPI_ISL_2891959, EPI_ISL_2891960, EPI_ISL_2891961, EPI_ISL_2891962, EPI_ISL_2891963, EPI_ISL_2891964, EPI_ISL_2891965, EPI_ISL_2891966, EPI_ISL_2891967, EPI_ISL_2891968, EPI_ISL_2891969, EPI_ISL_2891974, EPI_ISL_2891975, EPI_ISL_2891976, EPI_ISL_2891977, EPI_ISL_2891978, EPI_ISL_2891983, EPI_ISL_2891984, EPI_ISL_2891985, EPI_ISL_2891989, EPI_ISL_2891991, EPI_ISL_2891992, EPI_ISL_2891994, EPI_ISL_2891995, EPI_ISL_2891996, EPI_ISL_2892002, EPI_ISL_2892003, EPI_ISL_2892004, EPI_ISL_2892005, EPI_ISL_2892006, EPI_ISL_2892007, EPI_ISL_2892008, EPI_ISL_2892009, EPI_ISL_2892010, EPI_ISL_2892011, EPI_ISL_2892012, EPI_ISL_2892013, EPI_ISL_2892014, EPI_ISL_2892016, EPI_ISL_2892018, EPI_ISL_2892021, EPI_ISL_2892022, EPI_ISL_2892023, EPI_ISL_2892024, EPI_ISL_2892025, EPI_ISL_2892026, EPI_ISL_2892027, EPI_ISL_2892028, EPI_ISL_2892029, EPI_ISL_2892030, EPI_ISL_2892031, EPI_ISL_2892032, EPI_ISL_2892033, EPI_ISL_2892034, EPI_ISL_2892035, EPI_ISL_2892036, EPI_ISL_2892037, EPI_ISL_2892038, EPI_ISL_2892039, EPI_ISL_2892042, EPI_ISL_2892043, EPI_ISL_2892044, EPI_ISL_2892045, EPI_ISL_2892046, EPI_ISL_2892047, EPI_ISL_2892048, EPI_ISL_2892049, EPI_ISL_2892050, EPI_ISL_2892051, EPI_ISL_2892052, EPI_ISL_2892053, EPI_ISL_2892055, EPI_ISL_2892056, EPI_ISL_2892059, EPI_ISL_2892060, EPI_ISL_2892062, EPI_ISL_2892063, EPI_ISL_2892064, EPI_ISL_2892065, EPI_ISL_2892066, EPI_ISL_2892067, EPI_ISL_2892068, EPI_ISL_2892070, EPI_ISL_2892073, EPI_ISL_2892074, EPI_ISL_2892075, EPI_ISL_2892076, EPI_ISL_2892077, EPI_ISL_2892078, EPI_ISL_2892079, EPI_ISL_2892080, EPI_ISL_2892081, EPI_ISL_2892082, EPI_ISL_2892083, EPI_ISL_2892084, EPI_ISL_2892085, EPI_ISL_2892086, EPI_ISL_2892087, EPI_ISL_2892088, EPI_ISL_2892089, EPI_ISL_2892090, EPI_ISL_2892093, EPI_ISL_2892094, EPI_ISL_2892095, EPI_ISL_2892096, EPI_ISL_2892097, EPI_ISL_2892098, EPI_ISL_2892099, EPI_ISL_2892100, EPI_ISL_2892101, EPI_ISL_2892102, EPI_ISL_2892103, EPI_ISL_2892105, EPI_ISL_2892107, EPI_ISL_2892108, EPI_ISL_2892110, EPI_ISL_2892111, EPI_ISL_2892112, EPI_ISL_2892113, EPI_ISL_2892114, EPI_ISL_2892115, EPI_ISL_2892116, EPI_ISL_2892117, EPI_ISL_2892119, EPI_ISL_2892120, EPI_ISL_2892121, EPI_ISL_2892123, EPI_ISL_2892124, EPI_ISL_2892127, EPI_ISL_2892128, EPI_ISL_2892130, EPI_ISL_2892131, EPI_ISL_2892133, EPI_ISL_2892134, EPI_ISL_2892140, EPI_ISL_2892141, EPI_ISL_2892142, EPI_ISL_2892143, EPI_ISL_2892144, EPI_ISL_2892145, EPI_ISL_2892146, EPI_ISL_2892147, EPI_ISL_2892148, EPI_ISL_2892149, EPI_ISL_2892150, EPI_ISL_2892152, EPI_ISL_2892153, EPI_ISL_2892154, EPI_ISL_2892155, EPI_ISL_2892156, EPI_ISL_2892159, EPI_ISL_2892160, EPI_ISL_2892161, EPI_ISL_2892162, EPI_ISL_2892163, EPI_ISL_2892164, EPI_ISL_2892165, EPI_ISL_2892166, EPI_ISL_2892170, EPI_ISL_2892171, EPI_ISL_2892174, EPI_ISL_2892175, EPI_ISL_2892176, EPI_ISL_2892177, EPI_ISL_2892178, EPI_ISL_2892180, EPI_ISL_2892181, EPI_ISL_2892182, EPI_ISL_2892183, EPI_ISL_2892184, EPI_ISL_2892185, EPI_ISL_2892186, EPI_ISL_2892187, EPI_ISL_2892189, EPI_ISL_2892190, EPI_ISL_2892191, EPI_ISL_2892192, EPI_ISL_2892193, EPI_ISL_2892195, EPI_ISL_2892196, EPI_ISL_2892197, EPI_ISL_2892198, EPI_ISL_2892199, EPI_ISL_2892200, EPI_ISL_2892201, EPI_ISL_2892202, EPI_ISL_2892204, EPI_ISL_2892205, EPI_ISL_2892206, EPI_ISL_2892207, EPI_ISL_2892208, EPI_ISL_2892209, EPI_ISL_2892211, EPI_ISL_2892213, EPI_ISL_2892215, EPI_ISL_2892216, EPI_ISL_2892217, EPI_ISL_2892221, EPI_ISL_2892224, EPI_ISL_2892225, EPI_ISL_2892231, EPI_ISL_2892232, EPI_ISL_2892241, EPI_ISL_2892242, EPI_ISL_2892243, EPI_ISL_2892244, EPI_ISL_2892248, EPI_ISL_2892249, EPI_ISL_2892251, EPI_ISL_2892253, EPI_ISL_2892254, EPI_ISL_2892255, EPI_ISL_2892256, EPI_ISL_2892257, EPI_ISL_2892259, EPI_ISL_2892262, EPI_ISL_2892263, EPI_ISL_2892265, EPI_ISL_2892266, EPI_ISL_2892267, EPI_ISL_2892268, EPI_ISL_2892269, EPI_ISL_2892270, EPI_ISL_2892271, EPI_ISL_2892274, EPI_ISL_2892275, EPI_ISL_2892276, EPI_ISL_2892277, EPI_ISL_2892283, EPI_ISL_2892284, EPI_ISL_2892285, EPI_ISL_2892290, EPI_ISL_2892291, EPI_ISL_2892293, EPI_ISL_2892294, EPI_ISL_2892295, EPI_ISL_2892296, EPI_ISL_2892297, EPI_ISL_2892298, EPI_ISL_2892299, EPI_ISL_2892300, EPI_ISL_2892301, EPI_ISL_2892302, EPI_ISL_2892303, EPI_ISL_2892304, EPI_ISL_2892305, EPI_ISL_2892306, EPI_ISL_2892307, EPI_ISL_2892311, EPI_ISL_2892312, EPI_ISL_2892313, EPI_ISL_2892314, EPI_ISL_2892315, EPI_ISL_2892316, EPI_ISL_2892317, EPI_ISL_2892318, EPI_ISL_2892319, EPI_ISL_2892320, EPI_ISL_2892323, EPI_ISL_2892326, EPI_ISL_2892327, EPI_ISL_2892331, EPI_ISL_2892333, EPI_ISL_2892334, EPI_ISL_2892335, EPI_ISL_2892336, EPI_ISL_2892344, EPI_ISL_2892345, EPI_ISL_2892346, EPI_ISL_2892347, EPI_ISL_2892348, EPI_ISL_2892349, EPI_ISL_2892350, EPI_ISL_2892351, EPI_ISL_2892353, EPI_ISL_2892354, EPI_ISL_2892355, EPI_ISL_2892356, EPI_ISL_2892361, EPI_ISL_2892362, EPI_ISL_2892363, EPI_ISL_2892364, EPI_ISL_2892365, EPI_ISL_2892366, EPI_ISL_2892367, EPI_ISL_2892368, EPI_ISL_2892371, EPI_ISL_2892399, EPI_ISL_2892400, EPI_ISL_2921852, EPI_ISL_2921861, EPI_ISL_2921864, EPI_ISL_2921865, EPI_ISL_2921866, EPI_ISL_2921867, EPI_ISL_2921869, EPI_ISL_2921871, EPI_ISL_2921872, EPI_ISL_2921885, EPI_ISL_2921887, EPI_ISL_2921888, EPI_ISL_2921894, EPI_ISL_2921895, EPI_ISL_2921896, EPI_ISL_2921909, EPI_ISL_2921910, EPI_ISL_2921921, EPI_ISL_2921927, EPI_ISL_2921935, EPI_ISL_2921936, EPI_ISL_2921937, EPI_ISL_2921939, EPI_ISL_2921941, EPI_ISL_2921945, EPI_ISL_2921946, EPI_ISL_2921947, EPI_ISL_2921948, EPI_ISL_2921965, EPI_ISL_2921966, EPI_ISL_2921969, EPI_ISL_2921971, EPI_ISL_2921972, EPI_ISL_2921973, EPI_ISL_2921975, EPI_ISL_2921980, EPI_ISL_2921981, EPI_ISL_2921986, EPI_ISL_2921989, EPI_ISL_2921990, EPI_ISL_2922003, EPI_ISL_2922005, EPI_ISL_2922021, EPI_ISL_2922034, EPI_ISL_2922043, EPI_ISL_2922044, EPI_ISL_2922046, EPI_ISL_2922047, EPI_ISL_2922048, EPI_ISL_2922049, EPI_ISL_2922051, EPI_ISL_2922053, EPI_ISL_2922054, EPI_ISL_2922055, EPI_ISL_2922056, EPI_ISL_2922079, EPI_ISL_2922081, EPI_ISL_2922086, EPI_ISL_2922102, EPI_ISL_2922103, EPI_ISL_2922104, EPI_ISL_2922113, EPI_ISL_2922115, EPI_ISL_2922118, EPI_ISL_2922119, EPI_ISL_2922127, EPI_ISL_2922128, EPI_ISL_2922135, EPI_ISL_2922142, EPI_ISL_2922163, EPI_ISL_2922166, EPI_ISL_2922171, EPI_ISL_2922182, EPI_ISL_2922183, EPI_ISL_2922184, EPI_ISL_2922189, EPI_ISL_2922190, EPI_ISL_2922205, EPI_ISL_2922206, EPI_ISL_2922207, EPI_ISL_2922208, EPI_ISL_2922209, EPI_ISL_2922210, EPI_ISL_2922211, EPI_ISL_2922221, EPI_ISL_2922225, EPI_ISL_2922226, EPI_ISL_2922227, EPI_ISL_2922228, EPI_ISL_2922249, EPI_ISL_2922250, EPI_ISL_2922251, EPI_ISL_2922262, EPI_ISL_2922263, EPI_ISL_2922264, EPI_ISL_2922269, EPI_ISL_2922273, EPI_ISL_2922285, EPI_ISL_2922289, EPI_ISL_2922293, EPI_ISL_2922294, EPI_ISL_2922295, EPI_ISL_2922296, EPI_ISL_2922297, EPI_ISL_2922298, EPI_ISL_2922299, EPI_ISL_2922300, EPI_ISL_2922301, EPI_ISL_2922302, EPI_ISL_2922303, EPI_ISL_2922304, EPI_ISL_2923285, EPI_ISL_2923292, EPI_ISL_2923293, EPI_ISL_2923294, EPI_ISL_2923295, EPI_ISL_2923301, EPI_ISL_2923302, EPI_ISL_2923303, EPI_ISL_2923304, EPI_ISL_2923307, EPI_ISL_2923318, EPI_ISL_2923319, EPI_ISL_2923320, EPI_ISL_2923321, EPI_ISL_2923323, EPI_ISL_2923326, EPI_ISL_2923327, EPI_ISL_2923328, EPI_ISL_2923329, EPI_ISL_2923332, EPI_ISL_2923333, EPI_ISL_2923334, EPI_ISL_2923335, EPI_ISL_2923336, EPI_ISL_2923337, EPI_ISL_2923340, EPI_ISL_2923341, EPI_ISL_2923342, EPI_ISL_2923343, EPI_ISL_2923355, EPI_ISL_2923356, EPI_ISL_2923357, EPI_ISL_2923358, EPI_ISL_2923359, EPI_ISL_2923360, EPI_ISL_2923361, EPI_ISL_2923363, EPI_ISL_2923364, EPI_ISL_2923365, EPI_ISL_2923367, EPI_ISL_2923368, EPI_ISL_2923371, EPI_ISL_2923372, EPI_ISL_2923373, EPI_ISL_2923379, EPI_ISL_2923380, EPI_ISL_2923384, EPI_ISL_2923385, EPI_ISL_2923387, EPI_ISL_2923389, EPI_ISL_2923390, EPI_ISL_2923401, EPI_ISL_2923405, EPI_ISL_2923406, EPI_ISL_2923407, EPI_ISL_2923408, EPI_ISL_2923409, EPI_ISL_2923410, EPI_ISL_2923411, EPI_ISL_2923433, EPI_ISL_2923434, EPI_ISL_2923435, EPI_ISL_2923436, EPI_ISL_2923455, EPI_ISL_2923456, EPI_ISL_2923458, EPI_ISL_2923459, EPI_ISL_2923460, EPI_ISL_2923461, EPI_ISL_2923472, EPI_ISL_2923473, EPI_ISL_2923474, EPI_ISL_2923475, EPI_ISL_2923476, EPI_ISL_2923477, EPI_ISL_2923479, EPI_ISL_2923486, EPI_ISL_2923488, EPI_ISL_2923492, EPI_ISL_2923493, EPI_ISL_2923494, EPI_ISL_2923502, EPI_ISL_2923503, EPI_ISL_2923504, EPI_ISL_2923505, EPI_ISL_2923506, EPI_ISL_2923507, EPI_ISL_2923511, EPI_ISL_2923515, EPI_ISL_2923523, EPI_ISL_2923525, EPI_ISL_2923541, EPI_ISL_2923555, EPI_ISL_2923558, EPI_ISL_2923560, EPI_ISL_2923561, EPI_ISL_2923562, EPI_ISL_2923564, EPI_ISL_2923565, EPI_ISL_2923566, EPI_ISL_2923567, EPI_ISL_2923568, EPI_ISL_2923569, EPI_ISL_2923570, EPI_ISL_2923571, EPI_ISL_2923572, EPI_ISL_2923573, EPI_ISL_2923574, EPI_ISL_2923575, EPI_ISL_2923576, EPI_ISL_2923577, EPI_ISL_2923578, EPI_ISL_2923579, EPI_ISL_2923614, EPI_ISL_2923615, EPI_ISL_2923620, EPI_ISL_2923623, EPI_ISL_2923633, EPI_ISL_2923634, EPI_ISL_2923635, EPI_ISL_2923636, EPI_ISL_2923637, EPI_ISL_2923638, EPI_ISL_2923639, EPI_ISL_2923643, EPI_ISL_2923644, EPI_ISL_2923652, EPI_ISL_2923653, EPI_ISL_2923659, EPI_ISL_2923660, EPI_ISL_2923661, EPI_ISL_2923662, EPI_ISL_2923663, EPI_ISL_2923664, EPI_ISL_2923665, EPI_ISL_2923666, EPI_ISL_2923667, EPI_ISL_2923668, EPI_ISL_2923671, EPI_ISL_2923672, EPI_ISL_2923673, EPI_ISL_2923679, EPI_ISL_2923682, EPI_ISL_2923683, EPI_ISL_2923684, EPI_ISL_2923688, EPI_ISL_2923689, EPI_ISL_2923690, EPI_ISL_2923691, EPI_ISL_2923693, EPI_ISL_2923710, EPI_ISL_2923711, EPI_ISL_2923713, EPI_ISL_2923722, EPI_ISL_2923730, EPI_ISL_2923731, EPI_ISL_2923732, EPI_ISL_2923738, EPI_ISL_2923739, EPI_ISL_2923741, EPI_ISL_2923742, EPI_ISL_2923744, EPI_ISL_2923745, EPI_ISL_2923746, EPI_ISL_2923747, EPI_ISL_2923756, EPI_ISL_2923759, EPI_ISL_2923764, EPI_ISL_2923765, EPI_ISL_2923769, EPI_ISL_2977096, EPI_ISL_2977097, EPI_ISL_2977098, EPI_ISL_2977103, EPI_ISL_2977104, EPI_ISL_2977105, EPI_ISL_2977111, EPI_ISL_2977112, EPI_ISL_2977113, EPI_ISL_2977118, EPI_ISL_2977119, EPI_ISL_2977120, EPI_ISL_2977125, EPI_ISL_2977126, EPI_ISL_2977132, EPI_ISL_2977133, EPI_ISL_2977134, EPI_ISL_2977140, EPI_ISL_2977141, EPI_ISL_2977146, EPI_ISL_2977147, EPI_ISL_2977151, EPI_ISL_2977152, EPI_ISL_2977153, EPI_ISL_2977154, EPI_ISL_2977159, EPI_ISL_2977160, EPI_ISL_2977161, EPI_ISL_2977162, EPI_ISL_2977167, EPI_ISL_2977168, EPI_ISL_2977169, EPI_ISL_2977174, EPI_ISL_2977175, EPI_ISL_2977176, EPI_ISL_2977177, EPI_ISL_2977178, EPI_ISL_2977179, EPI_ISL_2977180, EPI_ISL_2977181, EPI_ISL_2977182, EPI_ISL_2977183, EPI_ISL_2977184, EPI_ISL_2977185, EPI_ISL_2977186, EPI_ISL_2977187, EPI_ISL_2977188, EPI_ISL_2977189, EPI_ISL_2977190, EPI_ISL_2977191, EPI_ISL_2977192, EPI_ISL_2977193, EPI_ISL_2977194, EPI_ISL_2977195, EPI_ISL_2977196, EPI_ISL_2977197, EPI_ISL_2977198, EPI_ISL_2977199, EPI_ISL_2977200, EPI_ISL_2977201, EPI_ISL_2977202, EPI_ISL_2977203, EPI_ISL_2977204, EPI_ISL_2977205, EPI_ISL_2977206, EPI_ISL_2977207, EPI_ISL_2977208, EPI_ISL_2977209, EPI_ISL_2977210, EPI_ISL_2977211, EPI_ISL_2977212, EPI_ISL_2977213, EPI_ISL_2977214, EPI_ISL_2977215, EPI_ISL_2977216, EPI_ISL_2977217, EPI_ISL_2977218, EPI_ISL_2977219, EPI_ISL_2977220, EPI_ISL_2977221, EPI_ISL_2977222, EPI_ISL_2977223, EPI_ISL_2977224, EPI_ISL_2977225, EPI_ISL_2977226, EPI_ISL_2977227, EPI_ISL_2977228, EPI_ISL_2977229, EPI_ISL_2977230, EPI_ISL_2977231, EPI_ISL_2977232, EPI_ISL_2977233, EPI_ISL_2977234, EPI_ISL_2977235, EPI_ISL_2977236, EPI_ISL_2977237, EPI_ISL_2977238, EPI_ISL_2977239, EPI_ISL_2977240, EPI_ISL_2977241, EPI_ISL_2977242, EPI_ISL_2977243, EPI_ISL_2977244, EPI_ISL_2977245, EPI_ISL_2977246, EPI_ISL_2977247, EPI_ISL_2977248, EPI_ISL_2977249, EPI_ISL_2977250, EPI_ISL_2977251, EPI_ISL_2977252, EPI_ISL_2977253, EPI_ISL_2977254, EPI_ISL_2977255, EPI_ISL_2977256, EPI_ISL_2977257, EPI_ISL_2977258, EPI_ISL_2977259, EPI_ISL_2977260, EPI_ISL_2977261, EPI_ISL_2977262, EPI_ISL_2977263, EPI_ISL_2977264, EPI_ISL_2977265, EPI_ISL_2977266, EPI_ISL_2977267, EPI_ISL_2977268, EPI_ISL_2977269, EPI_ISL_2977270, EPI_ISL_2977271, EPI_ISL_2977272, EPI_ISL_2977273, EPI_ISL_2977274, EPI_ISL_2977275, EPI_ISL_2977276, EPI_ISL_2977277, EPI_ISL_2977278, EPI_ISL_2977279, EPI_ISL_2977280, EPI_ISL_2977281, EPI_ISL_2977282, EPI_ISL_2977283, EPI_ISL_2977284, EPI_ISL_2977285, EPI_ISL_2977286, EPI_ISL_2977287, EPI_ISL_2977288, EPI_ISL_2977289, EPI_ISL_2977290, EPI_ISL_2977291, EPI_ISL_2977292, EPI_ISL_2977293, EPI_ISL_2977294, EPI_ISL_2977295, EPI_ISL_2977296, EPI_ISL_2977297, EPI_ISL_2977298, EPI_ISL_2977299, EPI_ISL_2977300, EPI_ISL_2977301, EPI_ISL_2977302, EPI_ISL_2977303, EPI_ISL_2977304, EPI_ISL_2977305, EPI_ISL_2977306, EPI_ISL_2977307, EPI_ISL_2977308, EPI_ISL_2977309, EPI_ISL_2977310, EPI_ISL_2977311, EPI_ISL_2977312, EPI_ISL_2977313, EPI_ISL_2977314, EPI_ISL_2977315, EPI_ISL_2977316, EPI_ISL_2977317, EPI_ISL_2977318, EPI_ISL_2977319, EPI_ISL_2977320, EPI_ISL_2977321, EPI_ISL_2977322, EPI_ISL_2977323, EPI_ISL_2977324, EPI_ISL_2977325, EPI_ISL_2977326, EPI_ISL_2977327, EPI_ISL_2977328, EPI_ISL_2977329, EPI_ISL_2977330, EPI_ISL_2977331, EPI_ISL_2977332, EPI_ISL_2977333, EPI_ISL_2977334, EPI_ISL_2977335, EPI_ISL_2977336, EPI_ISL_2977337, EPI_ISL_2977338, EPI_ISL_2977339, EPI_ISL_2977340, EPI_ISL_2977341, EPI_ISL_2977342, EPI_ISL_2977343, EPI_ISL_2977344, EPI_ISL_2977345, EPI_ISL_2977346, EPI_ISL_2977347, EPI_ISL_2977348, EPI_ISL_2977349, EPI_ISL_2977350, EPI_ISL_2977351, EPI_ISL_2977352, EPI_ISL_2977353, EPI_ISL_2977354, EPI_ISL_2977355, EPI_ISL_2977356, EPI_ISL_2977357, EPI_ISL_2977358, EPI_ISL_2977359, EPI_ISL_2977360, EPI_ISL_2977361, EPI_ISL_2977362, EPI_ISL_2977363, EPI_ISL_2977364, EPI_ISL_2977365, EPI_ISL_2977366, EPI_ISL_2977367, EPI_ISL_2977368, EPI_ISL_2977369, EPI_ISL_2977370, EPI_ISL_2977371, EPI_ISL_2977372, EPI_ISL_2977373, EPI_ISL_2977374, EPI_ISL_2977375, EPI_ISL_2977376, EPI_ISL_2977377, EPI_ISL_2977378, EPI_ISL_2977379, EPI_ISL_2977380, EPI_ISL_2977381, EPI_ISL_2977382, EPI_ISL_2977383, EPI_ISL_2977384, EPI_ISL_2977385, EPI_ISL_2977386, EPI_ISL_2977387, EPI_ISL_2977388, EPI_ISL_2977389, EPI_ISL_2977390, EPI_ISL_2977391, EPI_ISL_2977392, EPI_ISL_2977393, EPI_ISL_2977394, EPI_ISL_2977395, EPI_ISL_2977396, EPI_ISL_2977397, EPI_ISL_2977398, EPI_ISL_2977399, EPI_ISL_2977400, EPI_ISL_2977401, EPI_ISL_2977402, EPI_ISL_2977403, EPI_ISL_2977404, EPI_ISL_2977405, EPI_ISL_2977406, EPI_ISL_2977407, EPI_ISL_2977408, EPI_ISL_2977409, EPI_ISL_2977410, EPI_ISL_2977411, EPI_ISL_2977412, EPI_ISL_2977413, EPI_ISL_2977414, EPI_ISL_2977415, EPI_ISL_2977416, EPI_ISL_2977417, EPI_ISL_2977418, EPI_ISL_2977419, EPI_ISL_2977420, EPI_ISL_2977421, EPI_ISL_2977422, EPI_ISL_2977423, EPI_ISL_2977424, EPI_ISL_2977425, EPI_ISL_2977426, EPI_ISL_2977427, EPI_ISL_2977428, EPI_ISL_2977429, EPI_ISL_2977430, EPI_ISL_2977431, EPI_ISL_2977432, EPI_ISL_2977433, EPI_ISL_2977434, EPI_ISL_2977435, EPI_ISL_2977436, EPI_ISL_2977437, EPI_ISL_2977438, EPI_ISL_2977439, EPI_ISL_2977440, EPI_ISL_2977441, EPI_ISL_2977442, EPI_ISL_2977443, EPI_ISL_2977444, EPI_ISL_2977445, EPI_ISL_2977446, EPI_ISL_2977447, EPI_ISL_2977448, EPI_ISL_2977449, EPI_ISL_2977450, EPI_ISL_2977451, EPI_ISL_2977452, EPI_ISL_2977453, EPI_ISL_2977454, EPI_ISL_2977455, EPI_ISL_2977456, EPI_ISL_2977457, EPI_ISL_2977458, EPI_ISL_2977459, EPI_ISL_2977460, EPI_ISL_2977461, EPI_ISL_2977462, EPI_ISL_2977463, EPI_ISL_2977464, EPI_ISL_2977465, EPI_ISL_2977466, EPI_ISL_2977467, EPI_ISL_2977468, EPI_ISL_2977469, EPI_ISL_2977470, EPI_ISL_2977471, EPI_ISL_2977472, EPI_ISL_2977473, EPI_ISL_2977474, EPI_ISL_2978157, EPI_ISL_2978160, EPI_ISL_2978162, EPI_ISL_2978164, EPI_ISL_2978165, EPI_ISL_2978166, EPI_ISL_2978167, EPI_ISL_2978168, EPI_ISL_2978169, EPI_ISL_2978170, EPI_ISL_2978171, EPI_ISL_2978172, EPI_ISL_2978173, EPI_ISL_2978174, EPI_ISL_2978175 | | | |
| see above | CDPH VBL | California Department of Public Health | CDPH-COVIDNet; UCSD EXCITE lab |
| EPI_ISL_2452028, EPI_ISL_2452030, EPI_ISL_2452031, EPI_ISL_2452035, EPI_ISL_2452037, EPI_ISL_2452038, EPI_ISL_2452040, EPI_ISL_2452042, EPI_ISL_2452043, EPI_ISL_2452045 | | | |
| see above | CDPH, Viral and Rickettsial Disease Laboratory | Centers for Disease Control and Prevention Division of Viral Diseases, Pathogen Discovery | Alison Laufer Halpin; Ben L. Rambo-Martin; Clinton R. Paden; Dakota Howard; Darlene Wagner; Dave Wentworth; Dhwani Batra; Jasmine Padilla; Justin Lee; Katie Dillon; Krista Queen; Kristen Knipe; Kristine Lacek; Mark Burroughs; Matthew Schmerer; Mili Sheth; Peter Cook; Sam Shepard; Sarah Nobles; Shoshona Le; Suxiang Tong; Vivien Dugan; Yvette Unoarumhi |
| EPI_ISL_2842811, EPI_ISL_2842812, EPI_ISL_2842813, EPI_ISL_2842814, EPI_ISL_2842815, EPI_ISL_2842816, EPI_ISL_2842818, EPI_ISL_2842832, EPI_ISL_2861257, EPI_ISL_2861258, EPI_ISL_2861259, EPI_ISL_2892405, EPI_ISL_2892406, EPI_ISL_2892407, EPI_ISL_2892409, EPI_ISL_2892410, EPI_ISL_2892411, EPI_ISL_2892413, EPI_ISL_2892415, EPI_ISL_2892416, EPI_ISL_2892417, EPI_ISL_2892418, EPI_ISL_2892419, EPI_ISL_2892436, EPI_ISL_2892439, EPI_ISL_2892440, EPI_ISL_2892441, EPI_ISL_2892442, EPI_ISL_2892443, EPI_ISL_2892444, EPI_ISL_2892445, EPI_ISL_2892446, EPI_ISL_2892447, EPI_ISL_2892448, EPI_ISL_2892449, EPI_ISL_2892450, EPI_ISL_2892451, EPI_ISL_2892452, EPI_ISL_2892453, EPI_ISL_2892458, EPI_ISL_2892459, EPI_ISL_2892460, EPI_ISL_2892461, EPI_ISL_2892462, EPI_ISL_2892463, EPI_ISL_2892464, EPI_ISL_2892465, EPI_ISL_2892466, EPI_ISL_2892467, EPI_ISL_2892468, EPI_ISL_2892469, EPI_ISL_2892470, EPI_ISL_2892471, EPI_ISL_2892472, EPI_ISL_2892473, EPI_ISL_2892474, EPI_ISL_2892475, EPI_ISL_2892476, EPI_ISL_2892477, EPI_ISL_2892478, EPI_ISL_2892479, EPI_ISL_2892480, EPI_ISL_2892481, EPI_ISL_2892483, EPI_ISL_2892484, EPI_ISL_2892485, EPI_ISL_2892486, EPI_ISL_2892487, EPI_ISL_2892488, EPI_ISL_2892489, EPI_ISL_2892490, EPI_ISL_2892503, EPI_ISL_2892504, EPI_ISL_2892507, EPI_ISL_2892508, EPI_ISL_2892509, EPI_ISL_2892510, EPI_ISL_2892512, EPI_ISL_2892513, EPI_ISL_2892514, EPI_ISL_2892515, EPI_ISL_2892516, EPI_ISL_2892517, EPI_ISL_2892518, EPI_ISL_2892519, EPI_ISL_2892520, EPI_ISL_2892521, EPI_ISL_2892522, EPI_ISL_2892523, EPI_ISL_2892525, EPI_ISL_2892526, EPI_ISL_2892527, EPI_ISL_2892528, EPI_ISL_2892529, EPI_ISL_2892531, EPI_ISL_2892532, EPI_ISL_2892533, EPI_ISL_2892534, EPI_ISL_2892535, EPI_ISL_2892536, EPI_ISL_2892540, EPI_ISL_2892550, EPI_ISL_2892552, EPI_ISL_2892553, EPI_ISL_2892554, EPI_ISL_2892555, EPI_ISL_2892556, EPI_ISL_2892557, EPI_ISL_2892558, EPI_ISL_2892559, EPI_ISL_2892560, EPI_ISL_2892561, EPI_ISL_2892562, EPI_ISL_2892563, EPI_ISL_2892564, EPI_ISL_2892565, EPI_ISL_2892576, EPI_ISL_2892578, EPI_ISL_2892579, EPI_ISL_2892580, EPI_ISL_2892581, EPI_ISL_2892582, EPI_ISL_2892583, EPI_ISL_2892584, EPI_ISL_2892585, EPI_ISL_2892586 | | | |
| see above | California Department of Public Health | California Department of Public Health | CDPH IDLB COVIDNet et al |
| EPI_ISL_2391753, EPI_ISL_2405179, EPI_ISL_2405205, EPI_ISL_2620776, EPI_ISL_2620777, EPI_ISL_2620778, EPI_ISL_2772502, EPI_ISL_2772503, EPI_ISL_2772504, EPI_ISL_2812001, EPI_ISL_2894571 | | | |
| see above | California Hospital Medical Center | Los Angeles County Public Health Laboratories | P. Hemarajata et al. |
| EPI_ISL_2907555, EPI_ISL_2907559, EPI_ISL_2907576 | Centinela Hospital Medical Center | Los Angeles County Public Health Laboratories | P. Hemarajata et al. |
| EPI_ISL_2391793 | City of Hope Comprehensive Cancer Center | Los Angeles County Public Health Laboratories | P. Hemarajata et al. |
| EPI_ISL_2341556, EPI_ISL_2497573, EPI_ISL_2497574, EPI_ISL_2497619, EPI_ISL_2497621, EPI_ISL_2500066, EPI_ISL_2500087, EPI_ISL_2500148 | | | |
| see above | County of San Luis Obispo Public Health Laboratory | Chan-Zuckerberg Biohub | CZB Cliahub Consortium |
| EPI_ISL_2614384, EPI_ISL_2652974, EPI_ISL_2658954, EPI_ISL_2658981, EPI_ISL_2658982, EPI_ISL_2658983, EPI_ISL_2659116, EPI_ISL_2921028, EPI_ISL_2921029, EPI_ISL_2921030, EPI_ISL_3021462 | | | |
| see above | County of San Luis Obispo Public Health Laboratory | County of San Luis Obispo Public Health Laboratory | Frances Sidhu; Katrina Erwin |
| EPI_ISL_2507257, EPI_ISL_2507258, EPI_ISL_2507260, EPI_ISL_2507266, EPI_ISL_2507272, EPI_ISL_2507281, EPI_ISL_2507287, EPI_ISL_2507295, EPI_ISL_2507296, EPI_ISL_2507299, EPI_ISL_2507300, EPI_ISL_2507304, EPI_ISL_2507305, EPI_ISL_2507307, EPI_ISL_2507312, EPI_ISL_2507315, EPI_ISL_2507316, EPI_ISL_2507317, EPI_ISL_2507318, EPI_ISL_2507325, EPI_ISL_2507328, EPI_ISL_2507329, EPI_ISL_2507330, EPI_ISL_2507332, EPI_ISL_2507336, EPI_ISL_2507338, EPI_ISL_2507342, EPI_ISL_2507343, EPI_ISL_2507344, EPI_ISL_2507345, EPI_ISL_2507346, EPI_ISL_2507347, EPI_ISL_2507348, EPI_ISL_2507350, EPI_ISL_2507351, EPI_ISL_2507355, EPI_ISL_2507362, EPI_ISL_2507363, EPI_ISL_2755141, EPI_ISL_2755142, EPI_ISL_2755143, EPI_ISL_2755144, EPI_ISL_2755152, EPI_ISL_2755153, EPI_ISL_2755154, EPI_ISL_2755155, EPI_ISL_2755156, EPI_ISL_2755157, EPI_ISL_2755158, EPI_ISL_2755159, EPI_ISL_2755160, EPI_ISL_2755161, EPI_ISL_2755162, EPI_ISL_2755163, EPI_ISL_2755164, EPI_ISL_2755165, EPI_ISL_2755166, EPI_ISL_2755180, EPI_ISL_2755181, EPI_ISL_2755182, EPI_ISL_2755183, EPI_ISL_2755184, EPI_ISL_2755186, EPI_ISL_2755187, EPI_ISL_2755188, EPI_ISL_2755189, EPI_ISL_2755190, EPI_ISL_2755191, EPI_ISL_2755192, EPI_ISL_2755193, EPI_ISL_2755194, EPI_ISL_2755195, EPI_ISL_2755196, EPI_ISL_2755197, EPI_ISL_2755198, EPI_ISL_2755199, EPI_ISL_2755216, EPI_ISL_2755217, EPI_ISL_2755218, EPI_ISL_2755219, EPI_ISL_2755220, EPI_ISL_2755221, EPI_ISL_2755222, EPI_ISL_2755223, EPI_ISL_2755224, EPI_ISL_2755225, EPI_ISL_2755226, EPI_ISL_2755227, EPI_ISL_2755228, EPI_ISL_2755229, EPI_ISL_2755230, EPI_ISL_2755240, EPI_ISL_2755241, EPI_ISL_2755242, EPI_ISL_2755243, EPI_ISL_2755244, EPI_ISL_2755245, EPI_ISL_2755246, EPI_ISL_2755247, EPI_ISL_2755248, EPI_ISL_2755249, EPI_ISL_2755250, EPI_ISL_2755251, EPI_ISL_2755252, EPI_ISL_2755253, EPI_ISL_2755254, EPI_ISL_2755255, EPI_ISL_2755256, EPI_ISL_2755257, EPI_ISL_2755258, EPI_ISL_2755259, EPI_ISL_2755260, EPI_ISL_2755261, EPI_ISL_2755262, EPI_ISL_2755263, EPI_ISL_2755268, EPI_ISL_2755269, EPI_ISL_2755270, EPI_ISL_2755271, EPI_ISL_2755272, EPI_ISL_2755273, EPI_ISL_2755274, EPI_ISL_2755275, EPI_ISL_2755276, EPI_ISL_2755277, EPI_ISL_2755278, EPI_ISL_2755279, EPI_ISL_2755280, EPI_ISL_2755281, EPI_ISL_2755282, EPI_ISL_2755283, EPI_ISL_2755284, EPI_ISL_2755285, EPI_ISL_2755286, EPI_ISL_2755287, EPI_ISL_2755288, EPI_ISL_2755289, EPI_ISL_2755290, EPI_ISL_2755291, EPI_ISL_2755292, EPI_ISL_2755293, EPI_ISL_2755294, EPI_ISL_2755295, EPI_ISL_2755296, EPI_ISL_2755297, EPI_ISL_2755298, EPI_ISL_2755299, EPI_ISL_2755300, EPI_ISL_2755304, EPI_ISL_2755306, EPI_ISL_2755307, EPI_ISL_2755308, EPI_ISL_2755309, EPI_ISL_2755310, EPI_ISL_2755311, EPI_ISL_2755312, EPI_ISL_2755313, EPI_ISL_2755314, EPI_ISL_2755315, EPI_ISL_2755316, EPI_ISL_2755317, EPI_ISL_2755318, EPI_ISL_2755319, EPI_ISL_2755320, EPI_ISL_2755321, EPI_ISL_2755322, EPI_ISL_2755323, EPI_ISL_2755324, EPI_ISL_2755325, EPI_ISL_2755326, EPI_ISL_2755328, EPI_ISL_2755329, EPI_ISL_2755330, EPI_ISL_2755331, EPI_ISL_2755332, EPI_ISL_2755333, EPI_ISL_2755334, EPI_ISL_2755335, EPI_ISL_2755336, EPI_ISL_2755337, EPI_ISL_2755341, EPI_ISL_2755342, EPI_ISL_2755343, EPI_ISL_2755344, EPI_ISL_2755345, EPI_ISL_2755346, EPI_ISL_2755347, EPI_ISL_2755348, EPI_ISL_2755349, EPI_ISL_2755350, EPI_ISL_2755351, EPI_ISL_2755352, EPI_ISL_2755353, EPI_ISL_2755354, EPI_ISL_2755355, EPI_ISL_2755356, EPI_ISL_2755357, EPI_ISL_2755358, EPI_ISL_2755359, EPI_ISL_2755360, EPI_ISL_2755361, EPI_ISL_2755362, EPI_ISL_2755363, EPI_ISL_2755364, EPI_ISL_2755365, EPI_ISL_2755366, EPI_ISL_2755367, EPI_ISL_2755368, EPI_ISL_2755369, EPI_ISL_2755370, EPI_ISL_2755371, EPI_ISL_2755372, EPI_ISL_2755373, EPI_ISL_2755378, EPI_ISL_2755379, EPI_ISL_2755380, EPI_ISL_2755381, EPI_ISL_2755382, EPI_ISL_2755383, EPI_ISL_2755384, EPI_ISL_2755385, EPI_ISL_2755386, EPI_ISL_2755387, EPI_ISL_2755388, EPI_ISL_2755389, EPI_ISL_2755390, EPI_ISL_2755391, EPI_ISL_2755392, EPI_ISL_2755393, EPI_ISL_2755394, EPI_ISL_2755395, EPI_ISL_2755396, EPI_ISL_2755397, EPI_ISL_2755398, EPI_ISL_2755399, EPI_ISL_2755400, EPI_ISL_2755401, EPI_ISL_2755402, EPI_ISL_2755403, EPI_ISL_2755404, EPI_ISL_2755405, EPI_ISL_2755406, EPI_ISL_2755407, EPI_ISL_2755408, EPI_ISL_2755409, EPI_ISL_2755410, EPI_ISL_2755411, EPI_ISL_2755412, EPI_ISL_2755413, EPI_ISL_2755414, EPI_ISL_2755415, EPI_ISL_2755422, EPI_ISL_2755423, EPI_ISL_2755424, EPI_ISL_2755425, EPI_ISL_2755426, EPI_ISL_2755427, EPI_ISL_2755428, EPI_ISL_2755429, EPI_ISL_2755430, EPI_ISL_2755431, EPI_ISL_2755432, EPI_ISL_2755433, EPI_ISL_2755434, EPI_ISL_2755435, EPI_ISL_2755436, EPI_ISL_2755437, EPI_ISL_2755438, EPI_ISL_2755439, EPI_ISL_2755440, EPI_ISL_2755441, EPI_ISL_2755442, EPI_ISL_2755443, EPI_ISL_2755445, EPI_ISL_2755446, EPI_ISL_2755447, EPI_ISL_2755448, EPI_ISL_2755449, EPI_ISL_2755450, EPI_ISL_2755451, EPI_ISL_2755452, EPI_ISL_2755453, EPI_ISL_2755454, EPI_ISL_2755455, EPI_ISL_2755456, EPI_ISL_2755457, EPI_ISL_2755458, EPI_ISL_2755459, EPI_ISL_2755460, EPI_ISL_2755461, EPI_ISL_2755462, EPI_ISL_2755463, EPI_ISL_2755464, EPI_ISL_2755465, EPI_ISL_2755466, EPI_ISL_2755467, EPI_ISL_2755468, EPI_ISL_2755469, EPI_ISL_2755470, EPI_ISL_2755471, EPI_ISL_2755472, EPI_ISL_2783647, EPI_ISL_2783648, EPI_ISL_2783649, EPI_ISL_2783652, EPI_ISL_2783653, EPI_ISL_2783654, EPI_ISL_2783655, EPI_ISL_2783656, EPI_ISL_2783657, EPI_ISL_2783658, EPI_ISL_2783659, EPI_ISL_2783660, EPI_ISL_2783661, EPI_ISL_2783662, EPI_ISL_2783663, EPI_ISL_2783664, EPI_ISL_2783795, EPI_ISL_2783976, EPI_ISL_2783977, EPI_ISL_2783978, EPI_ISL_2783979, EPI_ISL_2783980, EPI_ISL_2783981, EPI_ISL_2783982, EPI_ISL_2783983, EPI_ISL_2783984, EPI_ISL_2783985, EPI_ISL_2784169, EPI_ISL_2784180, EPI_ISL_2784193, EPI_ISL_2784201, EPI_ISL_2784216, EPI_ISL_2784217, EPI_ISL_2784218, EPI_ISL_2784219, EPI_ISL_2784220, EPI_ISL_2784221, EPI_ISL_2784225, EPI_ISL_2784226, EPI_ISL_2784227, EPI_ISL_2784228, EPI_ISL_2784229, EPI_ISL_2784230, EPI_ISL_2784231, EPI_ISL_2784233, EPI_ISL_2784237, EPI_ISL_2784238, EPI_ISL_2784239, EPI_ISL_2784240, EPI_ISL_2784241, EPI_ISL_2784243, EPI_ISL_2784244, EPI_ISL_2784245, EPI_ISL_2784246, EPI_ISL_2784247, EPI_ISL_2784248, EPI_ISL_2784249, EPI_ISL_2784250, EPI_ISL_2784251, EPI_ISL_2784252, EPI_ISL_2784253, EPI_ISL_2784254, EPI_ISL_2784255, EPI_ISL_2784256, EPI_ISL_2784264, EPI_ISL_2784265, EPI_ISL_2784266, EPI_ISL_2784267, EPI_ISL_2784268, EPI_ISL_2784269, EPI_ISL_2784270, EPI_ISL_2784271, EPI_ISL_2784272, EPI_ISL_2784273, EPI_ISL_2784277, EPI_ISL_2784278, EPI_ISL_2784279, EPI_ISL_2784280, EPI_ISL_2784281, EPI_ISL_2784282, EPI_ISL_2784283, EPI_ISL_2784287, EPI_ISL_2784288, EPI_ISL_2784289, EPI_ISL_2784290, EPI_ISL_2784291, EPI_ISL_2784292, EPI_ISL_2784293, EPI_ISL_2784294, EPI_ISL_2784297, EPI_ISL_2784301, EPI_ISL_2784302, EPI_ISL_2784303, EPI_ISL_2784309, EPI_ISL_2784310, EPI_ISL_2784312, EPI_ISL_2784313, EPI_ISL_2784314, EPI_ISL_2784315, EPI_ISL_2784316, EPI_ISL_2784317, EPI_ISL_2784318, EPI_ISL_2784319, EPI_ISL_2784320, EPI_ISL_2784321, EPI_ISL_2784322, EPI_ISL_2784323, EPI_ISL_2784324, EPI_ISL_2784325, EPI_ISL_2784326, EPI_ISL_2784327, EPI_ISL_2839678, EPI_ISL_2839685, EPI_ISL_2839686, EPI_ISL_2839693, EPI_ISL_2839699, EPI_ISL_2839700, EPI_ISL_2839717, EPI_ISL_2839720, EPI_ISL_2839724, EPI_ISL_2839730, EPI_ISL_2839739, EPI_ISL_2839747, EPI_ISL_2839763, EPI_ISL_2839774, EPI_ISL_2839782, EPI_ISL_2839788, EPI_ISL_2839790, EPI_ISL_2839791, EPI_ISL_2839792, EPI_ISL_2839800, EPI_ISL_2839804, EPI_ISL_2839807, EPI_ISL_2839808, EPI_ISL_2839811, EPI_ISL_2839812, EPI_ISL_2839813, EPI_ISL_2839826, EPI_ISL_2839829, EPI_ISL_2839831, EPI_ISL_2839833, EPI_ISL_2839835, EPI_ISL_2839836, EPI_ISL_2839839, EPI_ISL_2839842, EPI_ISL_2839843, EPI_ISL_2839852, EPI_ISL_2839857, EPI_ISL_2839862, EPI_ISL_2839864, EPI_ISL_2839878, EPI_ISL_2839881, EPI_ISL_2839882, EPI_ISL_2839884, EPI_ISL_2839887, EPI_ISL_2839888, EPI_ISL_2839897, EPI_ISL_2839898, EPI_ISL_2839899, EPI_ISL_2839901, EPI_ISL_2839902, EPI_ISL_2839903, EPI_ISL_2839904, EPI_ISL_2839909, EPI_ISL_2839910, EPI_ISL_2839912, EPI_ISL_2839915, EPI_ISL_2839916, EPI_ISL_2839924, EPI_ISL_2839927, EPI_ISL_2839937, EPI_ISL_2839946, EPI_ISL_2839948, EPI_ISL_2839954, EPI_ISL_2839961, EPI_ISL_2839962, EPI_ISL_2839963, EPI_ISL_2839964, EPI_ISL_2839965, EPI_ISL_2839967, EPI_ISL_2839968, EPI_ISL_2839970, EPI_ISL_2839971, EPI_ISL_2839972, EPI_ISL_2839973, EPI_ISL_2839974, EPI_ISL_2839975, EPI_ISL_2839977, EPI_ISL_2839978, EPI_ISL_2941840, EPI_ISL_2941853, EPI_ISL_2941854, EPI_ISL_2941855, EPI_ISL_2941856, EPI_ISL_2941857, EPI_ISL_2941858, EPI_ISL_2941859, EPI_ISL_2941882, EPI_ISL_2941883, EPI_ISL_2941884, EPI_ISL_2941894, EPI_ISL_2941895, EPI_ISL_2941896, EPI_ISL_2941897, EPI_ISL_2941898, EPI_ISL_2941901, EPI_ISL_2941902, EPI_ISL_2941903, EPI_ISL_2941904, EPI_ISL_2941916, EPI_ISL_2941917, EPI_ISL_2941918, EPI_ISL_2941919, EPI_ISL_2941920, EPI_ISL_2941921, EPI_ISL_2941922, EPI_ISL_2941923, EPI_ISL_2941924, EPI_ISL_2941925, EPI_ISL_2941926, EPI_ISL_2942016, EPI_ISL_2942029, EPI_ISL_2942031, EPI_ISL_2942037, EPI_ISL_2942040, EPI_ISL_2942047, EPI_ISL_2942064, EPI_ISL_2942092, EPI_ISL_2942094, EPI_ISL_2942097, EPI_ISL_2942098, EPI_ISL_2942099, EPI_ISL_2942100, EPI_ISL_3064416, EPI_ISL_3064417, EPI_ISL_3064418, EPI_ISL_3064419, EPI_ISL_3064420, EPI_ISL_3064421, EPI_ISL_3064422, EPI_ISL_3064423, EPI_ISL_3064424, EPI_ISL_3064425, EPI_ISL_3064426, EPI_ISL_3064469, EPI_ISL_3064470, EPI_ISL_3064471, EPI_ISL_3064472, EPI_ISL_3064473, EPI_ISL_3064474, EPI_ISL_3064478, EPI_ISL_3064479, EPI_ISL_3064480, EPI_ISL_3064481, EPI_ISL_3064482, EPI_ISL_3064483, EPI_ISL_3064484, EPI_ISL_3064485, EPI_ISL_3064486, EPI_ISL_3064487, EPI_ISL_3064488, EPI_ISL_3064489, EPI_ISL_3064490, EPI_ISL_3064491, EPI_ISL_3064492, EPI_ISL_3064493, EPI_ISL_3064494, EPI_ISL_3064495, EPI_ISL_3064496, EPI_ISL_3064498, EPI_ISL_3064499, EPI_ISL_3064500, EPI_ISL_3064505, EPI_ISL_3064506, EPI_ISL_3064507, EPI_ISL_3064508, EPI_ISL_3064509, EPI_ISL_3064510, EPI_ISL_3064511, EPI_ISL_3064512, EPI_ISL_3064513, EPI_ISL_3064514, EPI_ISL_3064515, EPI_ISL_3064516, | | | |

| | | | |
|---|---|---|---|
| EPI_ISL_3064656, EPI_ISL_3064660, EPI_ISL_3064662, EPI_ISL_3064663, EPI_ISL_3064664 | | | |
| see above | Curative Labs | Curative Labs | Elias L. Salfati; Eugenia Khorosheva; George Way; J.Cesar Ignacio-Espinoza; Janet Chen; Mikhail Hanewich-Hollatz; Nabjot Sandhu; Sophia Quasem; Vladimir Slepnev; Zhiyi Xie |
| EPI_ISL_2508359, EPI_ISL_2508360, EPI_ISL_2508361, EPI_ISL_2508362, EPI_ISL_2508363, EPI_ISL_2508364, EPI_ISL_2508365, EPI_ISL_2508366, EPI_ISL_2508367, EPI_ISL_2508368, EPI_ISL_2508369, EPI_ISL_2522093, EPI_ISL_2801556, EPI_ISL_2801557, EPI_ISL_2801558, EPI_ISL_2801559, EPI_ISL_2801560, EPI_ISL_2801561, EPI_ISL_2801562, EPI_ISL_2801563 | | | |
| see above | DeRisi Lab, University of California, San Francisco | DeRisi Lab, University of California, San Francisco | Amy Kistler; Anthea Mitchell; Carina Marquez; Diane Havlir; Diane Jones; Douglas Black; Eric Chow; Gabriel Chamie; Genay Pilarowski; IDseq Team; Jackie Martinez; James Peng; Jamin Liu; Jon Jacobo; Lucy Li; Luis Rubio; Manu Vanaerschot; Matthew Laurie; Matthias Hapte-Selassie; Maya Petersen; Patrick Ayscue; Sabrina Mann; Sara Sunshine; Susana Rojas; Susy Rojas; Valerie Tulier-Laiwa; and Joseph DeRisi |
| EPI_ISL_2650542, EPI_ISL_2650543, EPI_ISL_2753975, EPI_ISL_2774621, EPI_ISL_2934023, EPI_ISL_2934058 | El Camino Hospital | Santa Clara County Public Health Laboratory | Santa Clara County Public Health Department |
| EPI_ISL_2967968 | El Camino Hospital | Santa Clara County public Health Laboratory | Santa Clara County Public Health Department |
| EPI_ISL_2405184, EPI_ISL_2405185, EPI_ISL_2405207, EPI_ISL_2405208, EPI_ISL_2405209, EPI_ISL_2405210, EPI_ISL_2405213, EPI_ISL_2405214, EPI_ISL_2405215, EPI_ISL_2772522, EPI_ISL_2772523, EPI_ISL_2772524, EPI_ISL_2774568, EPI_ISL_2774569, EPI_ISL_2811981, EPI_ISL_2811982, EPI_ISL_2812002, EPI_ISL_2812003, EPI_ISL_3029853, EPI_ISL_3029868 | | | |
| see above | Flow Diagnostics Lab | Los Angeles County Public Health Laboratories | P. Hemarajata et al. |
| EPI_ISL_2362485, EPI_ISL_2362486, EPI_ISL_2362487 | Flow Health | Infectious Disease Program, Broad Institute of Harvard and MIT | Adams, G.; B.L.; B.W.; Bauer, M.; Birren; Carter, A.; Chaluvadi, S.; D.J.; DeRuff, K.; Gallagher, G.; Gladden-Young, A.; J.E.; K.J.; Lagerborg, K.; Lemieux; Loreth, C.; MacInnis; Normandin, E.; P.C.; Park; Reilly, S.; Rudy, M.; Siddle; Smole, S.; Tomkins-Tinch, C.; and Sabeti |
| EPI_ISL_1993216, EPI_ISL_1993217, EPI_ISL_1993218, EPI_ISL_1993219, EPI_ISL_1993220, EPI_ISL_1993221, EPI_ISL_1993224, EPI_ISL_1993227, EPI_ISL_1993229, EPI_ISL_1993230, EPI_ISL_1993233, EPI_ISL_1993237, EPI_ISL_1993238, EPI_ISL_1993242, EPI_ISL_1993244, EPI_ISL_1993245, EPI_ISL_1993246, EPI_ISL_1993247, EPI_ISL_1993250, EPI_ISL_1993251, EPI_ISL_1993253, EPI_ISL_1993260, EPI_ISL_1993262, EPI_ISL_1993263, EPI_ISL_1993264, EPI_ISL_1993267, EPI_ISL_1993268, EPI_ISL_1993270, EPI_ISL_1993272, EPI_ISL_1993273, EPI_ISL_1993274, EPI_ISL_1993275, EPI_ISL_1993276, EPI_ISL_1993277, EPI_ISL_1993278, EPI_ISL_2043709, EPI_ISL_2043710, EPI_ISL_2043711, EPI_ISL_2043714, EPI_ISL_2043715, EPI_ISL_2043716, EPI_ISL_2043717, EPI_ISL_2043718, EPI_ISL_2043719, EPI_ISL_2043720, EPI_ISL_2043721, EPI_ISL_2043722, EPI_ISL_2043723, EPI_ISL_2043724, EPI_ISL_2043725, EPI_ISL_2043726, EPI_ISL_2043727, EPI_ISL_2043728, EPI_ISL_2043729, EPI_ISL_2043730, EPI_ISL_2043731, EPI_ISL_2043732, EPI_ISL_2043733, EPI_ISL_2043734, EPI_ISL_2043735, EPI_ISL_2043736, EPI_ISL_2043738, EPI_ISL_2043739, EPI_ISL_2043740, EPI_ISL_2043741, EPI_ISL_2043742, EPI_ISL_2043743, EPI_ISL_2043744, EPI_ISL_2043745, EPI_ISL_2043746, EPI_ISL_2043747, EPI_ISL_2043748, EPI_ISL_2043749, EPI_ISL_2043750, EPI_ISL_2043751, EPI_ISL_2043752, EPI_ISL_2043753, EPI_ISL_2043754, EPI_ISL_2043755, EPI_ISL_2043756, EPI_ISL_2043757, EPI_ISL_2043758, EPI_ISL_2043759, EPI_ISL_2043760, EPI_ISL_2043761, EPI_ISL_2043762, EPI_ISL_2043763, EPI_ISL_2043764, EPI_ISL_2043765, EPI_ISL_2043766, EPI_ISL_2043767, EPI_ISL_2043768, EPI_ISL_2043769, EPI_ISL_2043770, EPI_ISL_2043771, EPI_ISL_2043772, EPI_ISL_2043773, EPI_ISL_2043938, EPI_ISL_2043942, EPI_ISL_2043943, EPI_ISL_2043944, EPI_ISL_2043945, EPI_ISL_2043946, EPI_ISL_2043947, EPI_ISL_2043948, EPI_ISL_2043949, EPI_ISL_2043950, EPI_ISL_2043951, EPI_ISL_2043952, EPI_ISL_2043953, EPI_ISL_2043954, EPI_ISL_2043956, EPI_ISL_2043957, EPI_ISL_2043958, EPI_ISL_2043959, EPI_ISL_2043960, EPI_ISL_2043961, EPI_ISL_2043962, EPI_ISL_2043963, EPI_ISL_2043964, EPI_ISL_2043965, EPI_ISL_2043966, EPI_ISL_2043967, EPI_ISL_2043968, EPI_ISL_2043969, EPI_ISL_2043970, EPI_ISL_2043971, EPI_ISL_2043972, EPI_ISL_2043973, EPI_ISL_2043974, EPI_ISL_2043975, EPI_ISL_2043976, EPI_ISL_2043977, EPI_ISL_2043978, EPI_ISL_2043979, EPI_ISL_2043980, EPI_ISL_2043981, EPI_ISL_2043983, EPI_ISL_2043984, EPI_ISL_2043985, EPI_ISL_2043986, EPI_ISL_2043987, EPI_ISL_2043988, EPI_ISL_2043989, EPI_ISL_2043990, EPI_ISL_2043991, EPI_ISL_2043992, EPI_ISL_2043993, EPI_ISL_2043994, EPI_ISL_2043995, EPI_ISL_2043996, EPI_ISL_2043997, EPI_ISL_2043998, EPI_ISL_2043999, EPI_ISL_2044000, EPI_ISL_2133929, EPI_ISL_2133930, EPI_ISL_2133931, EPI_ISL_2133932, EPI_ISL_2133933, EPI_ISL_2133934, EPI_ISL_2133935, EPI_ISL_2133936, EPI_ISL_2133937, EPI_ISL_2133938, EPI_ISL_2133939, EPI_ISL_2133940, EPI_ISL_2133941, EPI_ISL_2133942, EPI_ISL_2133943, EPI_ISL_2133944, EPI_ISL_2133945, EPI_ISL_2133946, EPI_ISL_2133947, EPI_ISL_2133948, EPI_ISL_2133949, EPI_ISL_2133950, EPI_ISL_2133951, EPI_ISL_2133952, EPI_ISL_2133953, EPI_ISL_2133954, EPI_ISL_2133955, EPI_ISL_2133956, EPI_ISL_2133957, EPI_ISL_2133958, EPI_ISL_2133960, EPI_ISL_2133961, EPI_ISL_2133962, EPI_ISL_2133963, EPI_ISL_2133964, EPI_ISL_2133965, EPI_ISL_2133966, EPI_ISL_2133967, EPI_ISL_2133968, EPI_ISL_2133969, EPI_ISL_2133970, EPI_ISL_2133971, EPI_ISL_2133972, EPI_ISL_2133973, EPI_ISL_2133974, EPI_ISL_2133975, EPI_ISL_2133976, EPI_ISL_2133977, EPI_ISL_2133978, EPI_ISL_2133979, EPI_ISL_2133980, EPI_ISL_2133981, EPI_ISL_2133982, EPI_ISL_2133983, EPI_ISL_2133984, EPI_ISL_2133985, EPI_ISL_2133986, EPI_ISL_2133987, EPI_ISL_2133988, EPI_ISL_2134173, EPI_ISL_2134176, EPI_ISL_2134177, EPI_ISL_2134178, EPI_ISL_2134179, EPI_ISL_2134180, EPI_ISL_2134181, EPI_ISL_2134182, EPI_ISL_2134183, EPI_ISL_2134185, EPI_ISL_2134186, EPI_ISL_2134187, EPI_ISL_2134188, EPI_ISL_2134189, EPI_ISL_2134190, EPI_ISL_2134191, EPI_ISL_2134192, EPI_ISL_2134193, EPI_ISL_2134194, EPI_ISL_2134195, EPI_ISL_2134196, EPI_ISL_2134197, EPI_ISL_2134198, EPI_ISL_2134199, EPI_ISL_2134200, EPI_ISL_2134201, EPI_ISL_2134202, EPI_ISL_2134203, EPI_ISL_2134204, EPI_ISL_2134205, EPI_ISL_2134206, EPI_ISL_2134207, EPI_ISL_2134208, EPI_ISL_2134209, EPI_ISL_2134210, EPI_ISL_2134211, EPI_ISL_2134212, EPI_ISL_2134213, EPI_ISL_2134214, EPI_ISL_2134215, EPI_ISL_2134216, EPI_ISL_2134217, EPI_ISL_2134218, EPI_ISL_2134219, EPI_ISL_2134220, EPI_ISL_2134221, EPI_ISL_2134222, EPI_ISL_2134223, EPI_ISL_2134224, EPI_ISL_2134225, EPI_ISL_2134226, EPI_ISL_2134227, EPI_ISL_2134228, EPI_ISL_2134229, EPI_ISL_2134230, EPI_ISL_2134231, EPI_ISL_2134232, EPI_ISL_2134233, EPI_ISL_2134234, EPI_ISL_2134235, EPI_ISL_2134339, EPI_ISL_2134340, EPI_ISL_2134341, EPI_ISL_2134342, EPI_ISL_2134343, EPI_ISL_2134344, EPI_ISL_2134345, EPI_ISL_2134346, EPI_ISL_2134347, EPI_ISL_2134348, EPI_ISL_2134349, EPI_ISL_2134350, EPI_ISL_2134351, EPI_ISL_2134352, EPI_ISL_2134353, EPI_ISL_2134354, EPI_ISL_2134355, EPI_ISL_2134356, EPI_ISL_2134357, EPI_ISL_2134358, EPI_ISL_2134359, EPI_ISL_2134360, EPI_ISL_2134361, EPI_ISL_2134362, EPI_ISL_2134363, EPI_ISL_2134364, EPI_ISL_2134365, EPI_ISL_2134366, EPI_ISL_2134367, EPI_ISL_2134368, EPI_ISL_2134369, EPI_ISL_2134370, EPI_ISL_2134371, EPI_ISL_2134372, EPI_ISL_2134373, EPI_ISL_2134374, EPI_ISL_2134375, EPI_ISL_2134376, EPI_ISL_2134377, EPI_ISL_2134378, EPI_ISL_2134379, EPI_ISL_2134380, EPI_ISL_2134381, EPI_ISL_2134382, EPI_ISL_2134383, EPI_ISL_2134384, EPI_ISL_2134385, EPI_ISL_2134386, EPI_ISL_2134387, EPI_ISL_2134388, EPI_ISL_2134389, EPI_ISL_2134390, EPI_ISL_2134391, EPI_ISL_2134392, EPI_ISL_2134393, EPI_ISL_2134394, EPI_ISL_2134395, EPI_ISL_2134396, EPI_ISL_2134397, EPI_ISL_2134398, EPI_ISL_2134399, EPI_ISL_2134400, EPI_ISL_2134401, EPI_ISL_2134402, EPI_ISL_2134403, EPI_ISL_2134404, EPI_ISL_2134405, EPI_ISL_2134406, EPI_ISL_2134407, EPI_ISL_2134408, EPI_ISL_2134409, EPI_ISL_2134410, EPI_ISL_2134411, EPI_ISL_2134412, EPI_ISL_2134413, EPI_ISL_2134414, EPI_ISL_2134415, EPI_ISL_2134416, EPI_ISL_2134417, EPI_ISL_2134418, EPI_ISL_2134419, EPI_ISL_2134420, EPI_ISL_2134421, EPI_ISL_2134422, EPI_ISL_2134423, EPI_ISL_2134424, EPI_ISL_2134425, EPI_ISL_2134426, EPI_ISL_2134427, EPI_ISL_2134428, EPI_ISL_2134429, EPI_ISL_2134430, EPI_ISL_2134431, EPI_ISL_2134432, EPI_ISL_2134433, EPI_ISL_2134434, EPI_ISL_2134435, EPI_ISL_2134436, EPI_ISL_2134437, EPI_ISL_2134438, EPI_ISL_2134439, EPI_ISL_2134440, EPI_ISL_2134441, EPI_ISL_2134442, EPI_ISL_2134443, EPI_ISL_2134444, EPI_ISL_2134445, EPI_ISL_2134446, EPI_ISL_2134447, EPI_ISL_2134448, EPI_ISL_2134449, EPI_ISL_2134450, EPI_ISL_2134451, EPI_ISL_2134452, EPI_ISL_2134453, EPI_ISL_2140799, EPI_ISL_2160793, EPI_ISL_2160794, EPI_ISL_2160795, EPI_ISL_2160796, EPI_ISL_2160797, EPI_ISL_2160798, EPI_ISL_2160799, EPI_ISL_2160800, EPI_ISL_2160801, EPI_ISL_2160802, EPI_ISL_2160803, EPI_ISL_2160804, EPI_ISL_2160805, EPI_ISL_2160806, EPI_ISL_2160807, EPI_ISL_2160808, EPI_ISL_2160809, EPI_ISL_2160811, EPI_ISL_2160812, EPI_ISL_2160813, EPI_ISL_2160814, EPI_ISL_2160815, EPI_ISL_2160816, EPI_ISL_2160817, EPI_ISL_2160818, EPI_ISL_2160819, EPI_ISL_2160820, EPI_ISL_2160821, EPI_ISL_2160822, EPI_ISL_2160823, EPI_ISL_2160824, EPI_ISL_2160825, EPI_ISL_2160826, EPI_ISL_2160827, EPI_ISL_2160828, EPI_ISL_2160829, EPI_ISL_2160830, EPI_ISL_2160831, EPI_ISL_2160832, EPI_ISL_2160833, EPI_ISL_2160834, EPI_ISL_2160835, EPI_ISL_2160836, EPI_ISL_2160837, EPI_ISL_2160838, EPI_ISL_2160839, EPI_ISL_2160840, EPI_ISL_2160841, EPI_ISL_2160842, EPI_ISL_2160843, EPI_ISL_2160844, EPI_ISL_2160845, EPI_ISL_2160846, EPI_ISL_2160847, EPI_ISL_2160848, EPI_ISL_2160849, EPI_ISL_2160850, EPI_ISL_2160851, EPI_ISL_2160852, EPI_ISL_2160853, EPI_ISL_2160854, EPI_ISL_2160855, EPI_ISL_2160856, EPI_ISL_2160857, EPI_ISL_2160858, EPI_ISL_2160859, EPI_ISL_2160860, EPI_ISL_2160961, EPI_ISL_2160962, EPI_ISL_2160963, EPI_ISL_2160964, EPI_ISL_2160965, EPI_ISL_2160966, EPI_ISL_2160967, EPI_ISL_2160968, EPI_ISL_2160969, EPI_ISL_2160970, EPI_ISL_2160971, EPI_ISL_2160972, EPI_ISL_2160973, EPI_ISL_2160974, EPI_ISL_2160975, EPI_ISL_2160976, EPI_ISL_2160977, EPI_ISL_2160978, EPI_ISL_2160979, EPI_ISL_2160980, EPI_ISL_2160981, EPI_ISL_2160982, EPI_ISL_2160983, EPI_ISL_2160984, EPI_ISL_2160985, EPI_ISL_2160986, EPI_ISL_2160987, EPI_ISL_2160988, EPI_ISL_2160989, EPI_ISL_2160990, EPI_ISL_2160991, EPI_ISL_2160992, EPI_ISL_2160993, EPI_ISL_2160994, EPI_ISL_2160995, EPI_ISL_2160996, EPI_ISL_2160997, EPI_ISL_2160998, EPI_ISL_2160999, EPI_ISL_2161000, EPI_ISL_2161001, EPI_ISL_2161002, EPI_ISL_2161003, EPI_ISL_2161004, EPI_ISL_2161005, EPI_ISL_2161006, EPI_ISL_2161007, EPI_ISL_2161008, EPI_ISL_2161009, EPI_ISL_2161010, EPI_ISL_2161011, EPI_ISL_2161012, EPI_ISL_2161013, EPI_ISL_2161014, EPI_ISL_2161015, EPI_ISL_2161016, EPI_ISL_2161017, EPI_ISL_2161018, EPI_ISL_2161019, EPI_ISL_2161020, EPI_ISL_2161021, EPI_ISL_2161022, EPI_ISL_2161023, EPI_ISL_2161024, EPI_ISL_2161025, EPI_ISL_2161026, EPI_ISL_2161027, EPI_ISL_2179850, EPI_ISL_2179851, EPI_ISL_2179852, EPI_ISL_2179853, EPI_ISL_2179854, EPI_ISL_2179855, EPI_ISL_2179856, EPI_ISL_2179857, EPI_ISL_2179858, EPI_ISL_2179859, EPI_ISL_2179860, EPI_ISL_2269264, EPI_ISL_2269266, EPI_ISL_2269268, EPI_ISL_2269269, EPI_ISL_2269270, EPI_ISL_2269271, EPI_ISL_2269272, EPI_ISL_2269273, EPI_ISL_2269274, EPI_ISL_2269275, EPI_ISL_2269276, EPI_ISL_2269277, EPI_ISL_2269278, EPI_ISL_2269279, EPI_ISL_2269280, EPI_ISL_2269281, EPI_ISL_2269282, EPI_ISL_2269283, EPI_ISL_2269284, EPI_ISL_2269285, EPI_ISL_2269286, EPI_ISL_2269287, EPI_ISL_2269288, EPI_ISL_2269289, EPI_ISL_2269290, EPI_ISL_2269291, EPI_ISL_2269292, EPI_ISL_2269293, EPI_ISL_2269294, EPI_ISL_2269295, EPI_ISL_2269296, EPI_ISL_2269297, EPI_ISL_2269298, EPI_ISL_2269299, EPI_ISL_2269300, EPI_ISL_2269301, EPI_ISL_2269302, EPI_ISL_2269303, EPI_ISL_2269304, EPI_ISL_2269305, EPI_ISL_2269306, EPI_ISL_2269307, EPI_ISL_2269308, EPI_ISL_2269309, EPI_ISL_2269310, EPI_ISL_2269311, EPI_ISL_2269312, EPI_ISL_2269314, EPI_ISL_2269315, EPI_ISL_2269316, EPI_ISL_2269317, EPI_ISL_2269318, EPI_ISL_2269319, EPI_ISL_2269320, EPI_ISL_2269321, EPI_ISL_2269322, EPI_ISL_2269323, EPI_ISL_2269324, EPI_ISL_2269325, EPI_ISL_2269326, EPI_ISL_2269327, EPI_ISL_2269328, EPI_ISL_2269329, EPI_ISL_2269330, EPI_ISL_2269331, EPI_ISL_2269332, EPI_ISL_2269333, EPI_ISL_2269334, EPI_ISL_2269335, EPI_ISL_2269336, EPI_ISL_2269337, EPI_ISL_2269338, EPI_ISL_2269339, EPI_ISL_2269340, EPI_ISL_2269341, EPI_ISL_2269342, EPI_ISL_2269343, EPI_ISL_2269344, EPI_ISL_2269345, EPI_ISL_2269346, EPI_ISL_2269347, EPI_ISL_2269348, EPI_ISL_2269349, EPI_ISL_2269350, EPI_ISL_2269351, EPI_ISL_2269352, EPI_ISL_2269353, EPI_ISL_2269354, EPI_ISL_2269355, EPI_ISL_2269356, EPI_ISL_2269357, EPI_ISL_2269358, EPI_ISL_2269359, EPI_ISL_2269360, EPI_ISL_2269361, EPI_ISL_2269362, EPI_ISL_2269363, EPI_ISL_2269364, EPI_ISL_2269365, EPI_ISL_2269366, EPI_ISL_2269367, EPI_ISL_2269368, EPI_ISL_2269369, EPI_ISL_2269370, EPI_ISL_2269371, EPI_ISL_2269372, EPI_ISL_2269373, EPI_ISL_2269374, EPI_ISL_2269375, EPI_ISL_2269429, EPI_ISL_2269450, EPI_ISL_2269471, EPI_ISL_2269472, EPI_ISL_2269473, EPI_ISL_2269474, EPI_ISL_2269475, EPI_ISL_2269476, EPI_ISL_2269477, EPI_ISL_2269478, EPI_ISL_2269479, EPI_ISL_2269480, EPI_ISL_2269481, EPI_ISL_2269482, EPI_ISL_2269483, EPI_ISL_2269484, EPI_ISL_2269485, EPI_ISL_2269486, EPI_ISL_2269487, EPI_ISL_2269488, EPI_ISL_2269489, EPI_ISL_2269490, EPI_ISL_2269492, EPI_ISL_2269493, EPI_ISL_2269494, EPI_ISL_2269495, EPI_ISL_2269496, EPI_ISL_2269497, EPI_ISL_2269498, EPI_ISL_2269499, EPI_ISL_2269500, EPI_ISL_2269501, EPI_ISL_2269502, EPI_ISL_2269503, EPI_ISL_2269504, EPI_ISL_2269505, EPI_ISL_2269506, EPI_ISL_2269507, EPI_ISL_2269508, EPI_ISL_2269509, EPI_ISL_2269510, EPI_ISL_2269511, EPI_ISL_2269512, EPI_ISL_2269513, EPI_ISL_2269514, EPI_ISL_2269515, EPI_ISL_2269516, EPI_ISL_2269517, EPI_ISL_2269518, EPI_ISL_2269519, EPI_ISL_2269520, EPI_ISL_2269521, EPI_ISL_2269522, EPI_ISL_2269523, EPI_ISL_2269524, EPI_ISL_2269525, EPI_ISL_2269526, EPI_ISL_2269527, EPI_ISL_2269528, EPI_ISL_2269529, EPI_ISL_2269530, EPI_ISL_2269531, EPI_ISL_2269532, EPI_ISL_2269533, EPI_ISL_2269534, EPI_ISL_2269535, EPI_ISL_2269536, EPI_ISL_2368134, EPI_ISL_2368135, EPI_ISL_2368136, EPI_ISL_2368137, EPI_ISL_2368138, EPI_ISL_2368139, EPI_ISL_2368140, EPI_ISL_2368141, EPI_ISL_2368142, EPI_ISL_2368143, EPI_ISL_2368144, EPI_ISL_2368145, EPI_ISL_2368146, EPI_ISL_2368147, EPI_ISL_2368148, EPI_ISL_2368149, EPI_ISL_2368150, EPI_ISL_2368151, EPI_ISL_2368152, EPI_ISL_2368153, EPI_ISL_2368154, EPI_ISL_2368155, EPI_ISL_2368156, EPI_ISL_2368157, EPI_ISL_2368158, EPI_ISL_2368159, EPI_ISL_2368162, EPI_ISL_2368163, EPI_ISL_2368164, EPI_ISL_2368165, EPI_ISL_2368166, EPI_ISL_2368167, EPI_ISL_2368168, EPI_ISL_2368169, EPI_ISL_2368170, EPI_ISL_2368171, EPI_ISL_2368172, EPI_ISL_2368173, EPI_ISL_2368174, EPI_ISL_2368175, EPI_ISL_2368176, EPI_ISL_2368177, EPI_ISL_2368178, EPI_ISL_2368179, EPI_ISL_2368180, EPI_ISL_2368181, EPI_ISL_2368182, EPI_ISL_2368183, EPI_ISL_2368184, EPI_ISL_2368185, EPI_ISL_2368186, EPI_ISL_2368187, EPI_ISL_2368188, EPI_ISL_2368189, EPI_ISL_2368190, EPI_ISL_2368191, EPI_ISL_2368192, EPI_ISL_2368193, EPI_ISL_2368195, EPI_ISL_2368196, EPI_ISL_2368197, EPI_ISL_2368198, EPI_ISL_2368199, EPI_ISL_2368200, EPI_ISL_2368201, EPI_ISL_2368202, EPI_ISL_2368203, EPI_ISL_2368204, EPI_ISL_2371514, EPI_ISL_2371517, EPI_ISL_2371518, EPI_ISL_2371519, EPI_ISL_2371520, EPI_ISL_2371521, EPI_ISL_2371522, EPI_ISL_2371523, EPI_ISL_2371524, EPI_ISL_2371525, EPI_ISL_2371526, EPI_ISL_2371527, EPI_ISL_2371528, EPI_ISL_2371529, EPI_ISL_2371530, EPI_ISL_2371531, EPI_ISL_2371532, EPI_ISL_2371533, EPI_ISL_2371534, EPI_ISL_2371535, EPI_ISL_2371536, EPI_ISL_2371537, EPI_ISL_2371538, EPI_ISL_2371539, EPI_ISL_2371540, EPI_ISL_2371541, EPI_ISL_2371542, EPI_ISL_2371543, EPI_ISL_2371544, EPI_ISL_2371545, EPI_ISL_2371546, EPI_ISL_2371547, EPI_ISL_2371548, EPI_ISL_2371549, EPI_ISL_2371550, EPI_ISL_2371551, EPI_ISL_2371552, EPI_ISL_2371553, EPI_ISL_2371554, EPI_ISL_2371555, EPI_ISL_2371556, EPI_ISL_2371557, EPI_ISL_2371558, EPI_ISL_2371559, EPI_ISL_2371560, EPI_ISL_2371561, EPI_ISL_2371562, EPI_ISL_2371563, EPI_ISL_2371564, EPI_ISL_2371565, EPI_ISL_2371566, EPI_ISL_2371567, EPI_ISL_2371568, EPI_ISL_2371569, EPI_ISL_2371570, EPI_ISL_2371571, EPI_ISL_2371572, EPI_ISL_2371573, EPI_ISL_2371627, EPI_ISL_2371628, EPI_ISL_2371629, EPI_ISL_2371630, EPI_ISL_2371631, EPI_ISL_2371632, EPI_ISL_2371633, EPI_ISL_2371634, EPI_ISL_2371635, EPI_ISL_2371636, EPI_ISL_2371637, EPI_ISL_2371638, EPI_ISL_2371639, EPI_ISL_2371640, EPI_ISL_2371641, EPI_ISL_2371642, EPI_ISL_2371643, EPI_ISL_2371644, EPI_ISL_2371645, EPI_ISL_2371646, EPI_ISL_2371647, EPI_ISL_2371648, EPI_ISL_2371649, EPI_ISL_2371650, EPI_ISL_2371651, EPI_ISL_2371652, EPI_ISL_2371653, EPI_ISL_2371654, EPI_ISL_2371655, EPI_ISL_2371656, EPI_ISL_2371657, EPI_ISL_2371658, EPI_ISL_2371659, EPI_ISL_2371660, EPI_ISL_2371661, EPI_ISL_2371662, EPI_ISL_2371663, EPI_ISL_2371664, EPI_ISL_2371665, EPI_ISL_2371666, EPI_ISL_2371667, EPI_ISL_2371668, EPI_ISL_2371669, EPI_ISL_2371670, EPI_ISL_2371671, EPI_ISL_2371672, EPI_ISL_2371673, EPI_ISL_2371674, EPI_ISL_2371675, EPI_ISL_2371676, EPI_ISL_2371677, EPI_ISL_2371678, EPI_ISL_2371679, EPI_ISL_2371680, EPI_ISL_2371681, EPI_ISL_2371682, EPI_ISL_2371684, EPI_ISL_2371685, EPI_ISL_2371686, EPI_ISL_2371687, EPI_ISL_2371688, EPI_ISL_2371689, EPI_ISL_2371690, EPI_ISL_2441902, EPI_ISL_2441903, EPI_ISL_2441904, EPI_ISL_2441905, EPI_ISL_2441906, EPI_ISL_2441907, EPI_ISL_2441908, EPI_ISL_2441909, EPI_ISL_2441910, EPI_ISL_2441911, EPI_ISL_2441912, EPI_ISL_2441913, EPI_ISL_2441914, EPI_ISL_2441915, EPI_ISL_2441916, EPI_ISL_2441917, EPI_ISL_2441918, EPI_ISL_2441919, EPI_ISL_2441920, EPI_ISL_2441921, EPI_ISL_2441922, EPI_ISL_2441923, EPI_ISL_2441924, EPI_ISL_2441925, EPI_ISL_2441926, EPI_ISL_2441927, EPI_ISL_2441928, EPI_ISL_2441929, EPI_ISL_2441930, EPI_ISL_2441931, EPI_ISL_2441932, EPI_ISL_2441933, EPI_ISL_2441934, EPI_ISL_2441935, EPI_ISL_2441936, EPI_ISL_2441937, EPI_ISL_2441938, EPI_ISL_2441939, EPI_ISL_2441940, EPI_ISL_2441941, EPI_ISL_2441942, EPI_ISL_2441943, EPI_ISL_2441944, EPI_ISL_2441945, EPI_ISL_2441946, EPI_ISL_2441947, EPI_ISL_2441948, EPI_ISL_2441949, EPI_ISL_2441950, EPI_ISL_2441951, EPI_ISL_2441952, EPI_ISL_2441953, EPI_ISL_2441954, EPI_ISL_2441955, EPI_ISL_2441956, EPI_ISL_2441957, EPI_ISL_2441958, EPI_ISL_2480088, EPI_ISL_2480089, EPI_ISL_2480090, EPI_ISL_2480091, EPI_ISL_2480092, EPI_ISL_2480093, EPI_ISL_2480094, EPI_ISL_2480095, EPI_ISL_2480096, EPI_ISL_2480097, EPI_ISL_2480098, EPI_ISL_2480099, EPI_ISL_2480100, EPI_ISL_2480101, EPI_ISL_2480102, EPI_ISL_2480103, EPI_ISL_2480104, EPI_ISL_2480105, EPI_ISL_2480106, EPI_ISL_2480107, EPI_ISL_2480108, EPI_ISL_2480109, EPI_ISL_2480110, EPI_ISL_2480111, EPI_ISL_2480112, EPI_ISL_2480113, EPI_ISL_2480114, EPI_ISL_2480115, EPI_ISL_2480116, EPI_ISL_2480117, EPI_ISL_2480118, EPI_ISL_2480119, EPI_ISL_2480120, EPI_ISL_2480121, EPI_ISL_2480122, EPI_ISL_2480123, EPI_ISL_2480124, EPI_ISL_2480125, EPI_ISL_2480126, EPI_ISL_2480127, EPI_ISL_2480128, EPI_ISL_2480129, EPI_ISL_2480130, EPI_ISL_2480131, EPI_ISL_2480132, EPI_ISL_2480133, EPI_ISL_2480134, EPI_ISL_2480135, EPI_ISL_2480136, EPI_ISL_2480137, EPI_ISL_2480138, EPI_ISL_2480139, EPI_ISL_2480140, EPI_ISL_2480141, EPI_ISL_2480142, EPI_ISL_2480143, EPI_ISL_2480144, EPI_ISL_2480145, EPI_ISL_2525648, EPI_ISL_2525652, EPI_ISL_2525658, EPI_ISL_2525660, EPI_ISL_2525662, EPI_ISL_2525665, EPI_ISL_2525667, EPI_ISL_2525669, EPI_ISL_2525671, EPI_ISL_2525674, EPI_ISL_2525676, EPI_ISL_2525678, EPI_ISL_2525679, EPI_ISL_2525680, EPI_ISL_2525681, EPI_ISL_2525683, EPI_ISL_2525685, EPI_ISL_2525687, EPI_ISL_2525689, EPI_ISL_2525691, EPI_ISL_2525693, EPI_ISL_2525695, EPI_ISL_2525698, EPI_ISL_2525700, EPI_ISL_2525702, EPI_ISL_2525704, EPI_ISL_2525706, EPI_ISL_2525708, EPI_ISL_2525710, EPI_ISL_2525712, EPI_ISL_2525714, EPI_ISL_2525716, EPI_ISL_2525718, EPI_ISL_2525720, EPI_ISL_2525722, EPI_ISL_2525725, EPI_ISL_2525727, EPI_ISL_2525729, EPI_ISL_2525731, EPI_ISL_2525733, EPI_ISL_2525735, EPI_ISL_2525737, EPI_ISL_2525739, EPI_ISL_2525741, EPI_ISL_2525744, EPI_ISL_2525746, EPI_ISL_2525748, EPI_ISL_2525750, EPI_ISL_2525752, EPI_ISL_2525754, EPI_ISL_2525756, EPI_ISL_2525759, EPI_ISL_2525761, EPI_ISL_2525763, EPI_ISL_2525765, EPI_ISL_2525767, EPI_ISL_2525769, EPI_ISL_2525771, EPI_ISL_2525773, EPI_ISL_2525776, EPI_ISL_2525778, EPI_ISL_2525787, EPI_ISL_2525973, EPI_ISL_2525975, EPI_ISL_2527563, EPI_ISL_2527565, EPI_ISL_2527567, EPI_ISL_2527570, EPI_ISL_2527573, EPI_ISL_2527575, EPI_ISL_2527577, EPI_ISL_2527579, EPI_ISL_2527581, EPI_ISL_2527584, EPI_ISL_2527586, EPI_ISL_2527588, EPI_ISL_2527590, EPI_ISL_2527592, EPI_ISL_2527594, EPI_ISL_2527597, EPI_ISL_2527599, EPI_ISL_2527601, EPI_ISL_2527603, EPI_ISL_2527605, EPI_ISL_2527608, EPI_ISL_2527610, EPI_ISL_2527612, EPI_ISL_2527614, EPI_ISL_2527616, EPI_ISL_2527618, EPI_ISL_2527620, EPI_ISL_2527622, EPI_ISL_2527624, EPI_ISL_2527627, EPI_ISL_2527629, EPI_ISL_2527632, EPI_ISL_2527634, EPI_ISL_2527636, EPI_ISL_2527638, EPI_ISL_2527641, EPI_ISL_2527643, EPI_ISL_2527645, EPI_ISL_2527648, EPI_ISL_2527650, EPI_ISL_2527652, EPI_ISL_2527654, EPI_ISL_2527656, EPI_ISL_2527658, EPI_ISL_2527660, EPI_ISL_2527663, EPI_ISL_2527665, EPI_ISL_2527667, EPI_ISL_2527669, EPI_ISL_2527672, EPI_ISL_2527712, EPI_ISL_2527714, EPI_ISL_2527716, EPI_ISL_2527719, EPI_ISL_2527721, EPI_ISL_2527723, EPI_ISL_2527725, EPI_ISL_2527727, EPI_ISL_2527729, EPI_ISL_2527731, EPI_ISL_2527733, EPI_ISL_2527735, EPI_ISL_2527737, EPI_ISL_2527739, EPI_ISL_2527741, EPI_ISL_2527743, EPI_ISL_2527745, EPI_ISL_2527747, EPI_ISL_2527749, EPI_ISL_2527751, EPI_ISL_2527753, EPI_ISL_2527755, EPI_ISL_2527757, EPI_ISL_2527760, EPI_ISL_2527762, EPI_ISL_2527764, EPI_ISL_2527766, EPI_ISL_2527768, EPI_ISL_2527770, EPI_ISL_2527772, EPI_ISL_2527774, EPI_ISL_2527775, EPI_ISL_2527777, EPI_ISL_2527779, EPI_ISL_2527781, EPI_ISL_2527784, EPI_ISL_2527786, EPI_ISL_2527788, EPI_ISL_2527790, EPI_ISL_2527792, EPI_ISL_2527794, EPI_ISL_2527796, EPI_ISL_2527798, EPI_ISL_2527801, EPI_ISL_2527803, EPI_ISL_2527805, EPI_ISL_2527807, EPI_ISL_2527809, EPI_ISL_2527811, EPI_ISL_2527813, EPI_ISL_2527816, EPI_ISL_2527818, EPI_ISL_2527820, EPI_ISL_2527823, EPI_ISL_2527825, EPI_ISL_2527827, EPI_ISL_2527829, EPI_ISL_2527831, EPI_ISL_2527833, EPI_ISL_2527835, EPI_ISL_2527837, EPI_ISL_2527839, EPI_ISL_2527841, EPI_ISL_2527843, EPI_ISL_2598710, EPI_ISL_2598711, EPI_ISL_2598713, EPI_ISL_2598714, EPI_ISL_2598715, EPI_ISL_2598716, EPI_ISL_2598717, EPI_ISL_2598718, EPI_ISL_2598719, EPI_ISL_2598720, EPI_ISL_2598721, EPI_ISL_2598722, EPI_ISL_2598723, EPI_ISL_2598724, EPI_ISL_2598725, EPI_ISL_2598726, EPI_ISL_2598727, EPI_ISL_2598728, EPI_ISL_2598729, EPI_ISL_2598731, EPI_ISL_2598732, EPI_ISL_2598733, EPI_ISL_2598734, EPI_ISL_2598735, EPI_ISL_2598736, EPI_ISL_2598737, EPI_ISL_2598738, EPI_ISL_2598739, EPI_ISL_2598740, EPI_ISL_2598741, EPI_ISL_2598742, EPI_ISL_2598743, EPI_ISL_2598744, EPI_ISL_2598745, EPI_ISL_2598746, EPI_ISL_2598747, EPI_ISL_2598748, EPI_ISL_2598749, EPI_ISL_2598750, EPI_ISL_2598751, EPI_ISL_2598752, EPI_ISL_2598753, EPI_ISL_2598754, EPI_ISL_2598755, EPI_ISL_2598756, EPI_ISL_2598757, EPI_ISL_2598758, EPI_ISL_2598759, EPI_ISL_2598760, EPI_ISL_2598761, EPI_ISL_2598762, EPI_ISL_2598763, EPI_ISL_2598764, EPI_ISL_2686394, EPI_ISL_2686395, EPI_ISL_2686396, EPI_ISL_2686397, EPI_ISL_2686398, EPI_ISL_2686399, EPI_ISL_2686400, EPI_ISL_2686401, EPI_ISL_2686402, EPI_ISL_2686403, EPI_ISL_2686404, EPI_ISL_2686405, EPI_ISL_2686406, EPI_ISL_2686407, EPI_ISL_2686408, EPI_ISL_2686409, EPI_ISL_2686410, EPI_ISL_2686411, EPI_ISL_2686412, EPI_ISL_2686413, EPI_ISL_2686414, EPI_ISL_2686415, EPI_ISL_2686416, EPI_ISL_2686417, EPI_ISL_2686418, EPI_ISL_2686419, EPI_ISL_2686420, EPI_ISL_2686421, EPI_ISL_2686422, EPI_ISL_2686423, EPI_ISL_2686424, EPI_ISL_2686425, EPI_ISL_2686426, EPI_ISL_2686427, EPI_ISL_2686428, EPI_ISL_2686429, EPI_ISL_2686430, EPI_ISL_2686431, EPI_ISL_2686432, EPI_ISL_2686433, EPI_ISL_2686434, EPI_ISL_2686435, EPI_ISL_2686436, EPI_ISL_2686437, EPI_ISL_2686438, EPI_ISL_2686439, EPI_ISL_2686440, EPI_ISL_2686441, EPI_ISL_2686442, EPI_ISL_2686443, EPI_ISL_2686550, EPI_ISL_2686551, EPI_ISL_2686552, EPI_ISL_2686553, EPI_ISL_2686554, EPI_ISL_2686555, EPI_ISL_2686556, EPI_ISL_2686557, EPI_ISL_2686558, EPI_ISL_2686559, EPI_ISL_2686560, EPI_ISL_2686561, EPI_ISL_2686562, EPI_ISL_2686563, EPI_ISL_2686564, EPI_ISL_2686565, EPI_ISL_2686566, EPI_ISL_2686567, EPI_ISL_2686568, EPI_ISL_2686569, EPI_ISL_2686570, EPI_ISL_2686571, EPI_ISL_2686572, EPI_ISL_2686573, EPI_ISL_2686574, EPI_ISL_2686575, EPI_ISL_2686576, EPI_ISL_2686577, EPI_ISL_2686578, EPI_ISL_2686579, EPI_ISL_2686580, EPI_ISL_2686581, EPI_ISL_2686582, EPI_ISL_2686583, EPI_ISL_2686584, EPI_ISL_2686585, EPI_ISL_2686586, EPI_ISL_2686587, EPI_ISL_2686588, EPI_ISL_2686589, EPI_ISL_2686590, EPI_ISL_2686591, EPI_ISL_2686592, EPI_ISL_2686593, EPI_ISL_2686594, EPI_ISL_2686595, EPI_ISL_2686596, EPI_ISL_2686597, EPI_ISL_2686598, EPI_ISL_2686599, EPI_ISL_2686600, EPI_ISL_2686601, EPI_ISL_2686602, EPI_ISL_2686603, EPI_ISL_2686604, EPI_ISL_2686605, EPI_ISL_2686606, EPI_ISL_2686607, EPI_ISL_2686608, EPI_ISL_2686609, EPI_ISL_2686610, EPI_ISL_2686611, EPI_ISL_2686612, EPI_ISL_2686613, EPI_ISL_2686614, EPI_ISL_2686615, EPI_ISL_2686616, EPI_ISL_2686617, EPI_ISL_2686623, EPI_ISL_2686624, EPI_ISL_2686625, EPI_ISL_2686626, EPI_ISL_2686627, EPI_ISL_2686628, EPI_ISL_2686630, EPI_ISL_2686631, EPI_ISL_2686632, EPI_ISL_2686633, EPI_ISL_2686634, EPI_ISL_2686635, EPI_ISL_2686636, EPI_ISL_2686637, EPI_ISL_2686638, EPI_ISL_2686639, EPI_ISL_2686640, EPI_ISL_2686641, EPI_ISL_2686642, EPI_ISL_2686643, EPI_ISL_2686644, EPI_ISL_2686645, EPI_ISL_2686646, EPI_ISL_2686647, EPI_ISL_2686648, EPI_ISL_2686649, EPI_ISL_2686650, EPI_ISL_2686651, EPI_ISL_2686652, EPI_ISL_2686653, EPI_ISL_2686654, EPI_ISL_2686655, EPI_ISL_2686656, EPI_ISL_2686657, EPI_ISL_2686658, EPI_ISL_2686659, EPI_ISL_2686660, EPI_ISL_2686661, EPI_ISL_2686662, EPI_ISL_2686663, EPI_ISL_2686664, EPI_ISL_2686665, EPI_ISL_2686666, EPI_ISL_2686667, EPI_ISL_2686668, EPI_ISL_2686669, EPI_ISL_2686670, EPI_ISL_2686671, EPI_ISL_2686672, EPI_ISL_2686673, EPI_ISL_2686674, EPI_ISL_2686675, EPI_ISL_2686676, EPI_ISL_2686677, EPI_ISL_2686678, EPI_ISL_2686679, EPI_ISL_2686680, EPI_ISL_2686681, EPI_ISL_2686682, EPI_ISL_2686683, EPI_ISL_2686684, EPI_ISL_2686685, EPI_ISL_2686686, EPI_ISL_2686687, EPI_ISL_2686688, EPI_ISL_2686689, EPI_ISL_2686690, EPI_ISL_2686691, EPI_ISL_2686692, EPI_ISL_2686693, EPI_ISL_2686694, EPI_ISL_2686695, EPI_ISL_2686729, EPI_ISL_2785535, EPI_ISL_2785545, EPI_ISL_2785546, EPI_ISL_2785547, EPI_ISL_2785549, EPI_ISL_2785550, EPI_ISL_2785551, EPI_ISL_2785552, EPI_ISL_2785553, EPI_ISL_2785554, EPI_ISL_2785555, EPI_ISL_2785556, EPI_ISL_2785557, EPI_ISL_2785558, EPI_ISL_2785559, EPI_ISL_2785560, EPI_ISL_2785561, EPI_ISL_2785562, EPI_ISL_2785563, EPI_ISL_2785564, EPI_ISL_2785565, EPI_ISL_2785566, EPI_ISL_2785567, EPI_ISL_2785568, EPI_ISL_2785569, EPI_ISL_2785570, EPI_ISL_2785571, EPI_ISL_2785572, EPI_ISL_2785573, EPI_ISL_2785574, EPI_ISL_2785575, EPI_ISL_2785576, EPI_ISL_2785577, EPI_ISL_2785578, EPI_ISL_2785579, EPI_ISL_2785580, EPI_ISL_2785581, EPI_ISL_2785582, EPI_ISL_2785583, EPI_ISL_2785584, EPI_ISL_2785585, EPI_ISL_2785586, EPI_ISL_2785587, EPI_ISL_2785588, EPI_ISL_2785589, EPI_ISL_2785590, EPI_ISL_2785591, EPI_ISL_2785592, EPI_ISL_2785593, EPI_ISL_2785594, EPI_ISL_2785595, EPI_ISL_2785596, EPI_ISL_2785597, EPI_ISL_2785598, EPI_ISL_2785599, EPI_ISL_2785600, EPI_ISL_2785601, EPI_ISL_2785602, EPI_ISL_2785603, EPI_ISL_2785604, EPI_ISL_2785605, EPI_ISL_2785606, EPI_ISL_2785607, EPI_ISL_2785608, EPI_ISL_2785609, EPI_ISL_2785610, EPI_ISL_2785611, EPI_ISL_2785612, EPI_ISL_2785613, EPI_ISL_2785614, EPI_ISL_2785615, EPI_ISL_2785616, EPI_ISL_2785617, EPI_ISL_2785618, EPI_ISL_2785619, EPI_ISL_2785620, EPI_ISL_2785621, EPI_ISL_2785622, EPI_ISL_2785623, EPI_ISL_2785624, EPI_ISL_2785625, EPI_ISL_2785626, EPI_ISL_2785627, EPI_ISL_2785628, EPI_ISL_2785629, EPI_ISL_2785630, EPI_ISL_2785631, EPI_ISL_2785632, EPI_ISL_2785633, EPI_ISL_2785634, EPI_ISL_2785635, EPI_ISL_2785636, EPI_ISL_2785637, EPI_ISL_2785638, EPI_ISL_2785639, EPI_ISL_2785640, EPI_ISL_2785641, EPI_ISL_2785642, EPI_ISL_2785643, EPI_ISL_2785644, EPI_ISL_2785645, EPI_ISL_2785646, EPI_ISL_2785647, EPI_ISL_2785648, EPI_ISL_2785649, EPI_ISL_2785650, EPI_ISL_2785651, EPI_ISL_2785652, EPI_ISL_2785653, EPI_ISL_2785654, EPI_ISL_2785655, EPI_ISL_2785656, EPI_ISL_2785657, EPI_ISL_2785658, EPI_ISL_2785659, EPI_ISL_2785660, EPI_ISL_2785661, EPI_ISL_2785662, EPI_ISL_2785663, EPI_ISL_2785664, EPI_ISL_2785665, EPI_ISL_2785666, EPI_ISL_2785667, EPI_ISL_2785668, EPI_ISL_2785669, EPI_ISL_2785670, EPI_ISL_2785671, EPI_ISL_2785672, EPI_ISL_2785673, EPI_ISL_2785674, EPI_ISL_2785675, EPI_ISL_2785676, EPI_ISL_2785677, EPI_ISL_2785678, EPI_ISL_2785679, EPI_ISL_2785680, EPI_ISL_2785704, EPI_ISL_2785729, EPI_ISL_2785730, EPI_ISL_2785731, EPI_ISL_2785732, EPI_ISL_2785733, EPI_ISL_2785734, EPI_ISL_2785735, EPI_ISL_2785736, EPI_ISL_2785737, EPI_ISL_2785738, EPI_ISL_2785739, EPI_ISL_2785740, EPI_ISL_2785741, EPI_ISL_2785742, EPI_ISL_2785743, EPI_ISL_2785744, EPI_ISL_2785745, EPI_ISL_2785746, EPI_ISL_2785747, EPI_ISL_2785748, EPI_ISL_2785749, EPI_ISL_2785750, EPI_ISL_2785751, EPI_ISL_2785752, EPI_ISL_2785753, EPI_ISL_2785754, EPI_ISL_2785755, EPI_ISL_2785756, EPI_ISL_2785757, EPI_ISL_2785758, EPI_ISL_2785759, EPI_ISL_2785760, EPI_ISL_2785761, EPI_ISL_2785762, EPI_ISL_2785763, EPI_ISL_2785764, EPI_ISL_2785765, EPI_ISL_2785766, EPI_ISL_2785767, EPI_ISL_2785768, EPI_ISL_2785769, EPI_ISL_2785770, EPI_ISL_2785771, EPI_ISL_2785772, EPI_ISL_2785773, EPI_ISL_2785774, EPI_ISL_2785775, EPI_ISL_2785776, EPI_ISL_2785777, EPI_ISL_2785778, EPI_ISL_2785779, EPI_ISL_2785780, EPI_ISL_2785781, EPI_ISL_2785782, EPI_ISL_2785783, EPI_ISL_2785784, EPI_ISL_2785785, EPI_ISL_2785786, EPI_ISL_2785787, EPI_ISL_2785788, EPI_ISL_2785789, EPI_ISL_2785790, EPI_ISL_2785791, EPI_ISL_2785792, EPI_ISL_2785794, EPI_ISL_2785795, EPI_ISL_2785797, EPI_ISL_2785798, EPI_ISL_2785799, EPI_ISL_2785800, EPI_ISL_2785801, EPI_ISL_2785802, EPI_ISL_2785803, EPI_ISL_2785804, EPI_ISL_2785805, EPI_ISL_2785806, EPI_ISL_2785807, EPI_ISL_2785808, EPI_ISL_2785809, EPI_ISL_2785810, EPI_ISL_2785811, EPI_ISL_2785812, EPI_ISL_2785813, EPI_ISL_2785814, EPI_ISL_2785815, EPI_ISL_2785816, EPI_ISL_2785817, EPI_ISL_2785818, EPI_ISL_2785819, EPI_ISL_2785821, EPI_ISL_2785822, EPI_ISL_2785823, EPI_ISL_2785824, EPI_ISL_2785825, EPI_ISL_2785826, EPI_ISL_2785827, EPI_ISL_2785828, EPI_ISL_2785829, EPI_ISL_2785830, EPI_ISL_2785831, EPI_ISL_2785832, EPI_ISL_2785833, EPI_ISL_2785834, EPI_ISL_2785835, EPI_ISL_2785836, EPI_ISL_2785837, EPI_ISL_2785838, EPI_ISL_2785839, EPI_ISL_2785840, EPI_ISL_2785841, EPI_ISL_2785842, EPI_ISL_2785843, EPI_ISL_2785844, EPI_ISL_2785845, EPI_ISL_2785846, EPI_ISL_2785847, EPI_ISL_2785848, EPI_ISL_2785849, EPI_ISL_2785850, EPI_ISL_2785851, EPI_ISL_2785852, EPI_ISL_2785853, EPI_ISL_2785854, EPI_ISL_2785855, EPI_ISL_2785856, EPI_ISL_2785857, EPI_ISL_2785858, EPI_ISL_2785859, EPI_ISL_2785860, EPI_ISL_2785886, EPI_ISL_2785887, EPI_ISL_2785888, EPI_ISL_2785889, EPI_ISL_2785890, EPI_ISL_2785891, EPI_ISL_2785892, EPI_ISL_2785893, EPI_ISL_2785894, EPI_ISL_2785895, EPI_ISL_2785896, EPI_ISL_2785897, EPI_ISL_2785898, EPI_ISL_2785899, EPI_ISL_2785900, EPI_ISL_2785901, EPI_ISL_2785902, EPI_ISL_2785903, EPI_ISL_2785904, EPI_ISL_2785905, EPI_ISL_2785906, EPI_ISL_2785907, EPI_ISL_2785908, EPI_ISL_2785909, EPI_ISL_2785910, EPI_ISL_2785911, EPI_ISL_2785912, EPI_ISL_2785913, EPI_ISL_2785914, EPI_ISL_2785915, EPI_ISL_2785916, EPI_ISL_2785917, EPI_ISL_2785918, EPI_ISL_2785919, EPI_ISL_2785921, EPI_ISL_2785922, EPI_ISL_2785923, EPI_ISL_2785925, EPI_ISL_2785926, EPI_ISL_2785927, EPI_ISL_2785928, EPI_ISL_2785929, EPI_ISL_2785930, EPI_ISL_2785931, EPI_ISL_2785932, EPI_ISL_2785933, EPI_ISL_2785934, EPI_ISL_2785935, EPI_ISL_2785936, EPI_ISL_2785937, EPI_ISL_2785938, EPI_ISL_2785939, EPI_ISL_2785940, EPI_ISL_2785941, EPI_ISL_2785942, EPI_ISL_2785943, EPI_ISL_2785944, EPI_ISL_2785945, EPI_ISL_2785946, EPI_ISL_2785947, EPI_ISL_2785948, EPI_ISL_2785949, EPI_ISL_2785950, EPI_ISL_2785951, EPI_ISL_2785952, EPI_ISL_2785953, EPI_ISL_2785954, EPI_ISL_2873346, EPI_ISL_2873358, EPI_ISL_2873359, EPI_ISL_2873360, EPI_ISL_2873361, EPI_ISL_2873362, EPI_ISL_2873363, EPI_ISL_2873364, EPI_ISL_2873365, EPI_ISL_2873366, EPI_ISL_2873367, EPI_ISL_2873368, EPI_ISL_2873369, EPI_ISL_2873370, EPI_ISL_2873371, EPI_ISL_2873372, EPI_ISL_2873373, EPI_ISL_2873374, EPI_ISL_2873375, EPI_ISL_2873376, EPI_ISL_2873377, EPI_ISL_2873378, EPI_ISL_2873379, EPI_ISL_2873380, EPI_ISL_2873381, EPI_ISL_2873382, EPI_ISL_2873383, EPI_ISL_2873384, EPI_ISL_2873385, EPI_ISL_2873386, EPI_ISL_2873387, EPI_ISL_2873388, EPI_ISL_2873389, EPI_ISL_2873390, EPI_ISL_2873391, EPI_ISL_2873392, EPI_ISL_2873393, EPI_ISL_2873394, EPI_ISL_2873395, EPI_ISL_2873396, EPI_ISL_2873397, EPI_ISL_2873398, EPI_ISL_2873399, EPI_ISL_2873400, EPI_ISL_2873401, EPI_ISL_2873402, EPI_ISL_2873403, EPI_ISL_2873404, EPI_ISL_2873405, EPI_ISL_2873406, EPI_ISL_2873407, EPI_ISL_2873408, EPI_ISL_2873409, EPI_ISL_2873410, EPI_ISL_2873411, EPI_ISL_2873412, EPI_ISL_2873413, EPI_ISL_2873414, EPI_ISL_2873415, EPI_ISL_2873416, EPI_ISL_2873417, EPI_ISL_2873418, EPI_ISL_2873419, EPI_ISL_2873420, EPI_ISL_2873421, EPI_ISL_2873422, EPI_ISL_2873423, EPI_ISL_2873424, EPI_ISL_2873425, EPI_ISL_2873426, EPI_ISL_2873427, EPI_ISL_2873428, EPI_ISL_2873429, EPI_ISL_2873430, EPI_ISL_2873431, EPI_ISL_2873432, EPI_ISL_2873433, EPI_ISL_2873434, EPI_ISL_2873435, EPI_ISL_2873436, EPI_ISL_2873437, EPI_ISL_2873438, EPI_ISL_2873439, EPI_ISL_2873440, EPI_ISL_2873441, EPI_ISL_2873442, EPI_ISL_2873443, EPI_ISL_2873444, EPI_ISL_2873445, EPI_ISL_2873446, EPI_ISL_2873447, EPI_ISL_2873448, EPI_ISL_2873449, EPI_ISL_2873450, EPI_ISL_2873451, EPI_ISL_2873452, EPI_ISL_2873453, EPI_ISL_2873454, EPI_ISL_2873455, EPI_ISL_2873456, EPI_ISL_2873457, EPI_ISL_2873458, EPI_ISL_2873459, EPI_ISL_2873460, EPI_ISL_2873461, EPI_ISL_2873462, EPI_ISL_2873463, EPI_ISL_2873464, EPI_ISL_2873465, EPI_ISL_2873466, EPI_ISL_2873467, EPI_ISL_2873468, EPI_ISL_2873469, EPI_ISL_2873470, EPI_ISL_2873471, EPI_ISL_2873472, EPI_ISL_2873473, EPI_ISL_2873474, EPI_ISL_2873475, EPI_ISL_2873476, EPI_ISL_2873477, EPI_ISL_2873478, EPI_ISL_2873479, EPI_ISL_2873480, EPI_ISL_2873481, EPI_ISL_2873482, EPI_ISL_2873483, EPI_ISL_2873484, EPI_ISL_2873485, EPI_ISL_2873486, EPI_ISL_2873487, EPI_ISL_2873488, EPI_ISL_2873489, EPI_ISL_2873490, EPI_ISL_2873491, EPI_ISL_2873492, EPI_ISL_2873493, EPI_ISL_2873494, EPI_ISL_2873495, EPI_ISL_2873496, EPI_ISL_2873497, EPI_ISL_2873498, EPI_ISL_2873499, EPI_ISL_2873500, | | | |

| | | | |
|---|---|---|---|
| EPI_ISL_2873508, EPI_ISL_2873509, EPI_ISL_2873510, EPI_ISL_2873511, EPI_ISL_2873512, EPI_ISL_2873513, EPI_ISL_2873514, EPI_ISL_2873515, EPI_ISL_2873516, EPI_ISL_2873517, EPI_ISL_2873518, EPI_ISL_2873519, EPI_ISL_2873520, EPI_ISL_2873521, EPI_ISL_2873522, EPI_ISL_2873523, EPI_ISL_2873524, EPI_ISL_2873525, EPI_ISL_2873526, EPI_ISL_2873527, EPI_ISL_2873528, EPI_ISL_2873529, EPI_ISL_2873530, EPI_ISL_2873531, EPI_ISL_2873532, EPI_ISL_2873533, EPI_ISL_2873534, EPI_ISL_2873535, EPI_ISL_2873536, EPI_ISL_2873537, EPI_ISL_2873538, EPI_ISL_2873539, EPI_ISL_2873540, EPI_ISL_2873541, EPI_ISL_2873542, EPI_ISL_2873543, EPI_ISL_2873544, EPI_ISL_2873545, EPI_ISL_2873546, EPI_ISL_2873547, EPI_ISL_2873548, EPI_ISL_2873549, EPI_ISL_2873550, EPI_ISL_2873551, EPI_ISL_2873552, EPI_ISL_2873553, EPI_ISL_2873554, EPI_ISL_2873555, EPI_ISL_2873556, EPI_ISL_2873557, EPI_ISL_2873558, EPI_ISL_2873559, EPI_ISL_2873560, EPI_ISL_2873561, EPI_ISL_2873562, EPI_ISL_2873563, EPI_ISL_2873564, EPI_ISL_2873565, EPI_ISL_2873566, EPI_ISL_2873567, EPI_ISL_2873568, EPI_ISL_2873569, EPI_ISL_2873570, EPI_ISL_2873571, EPI_ISL_2873572, EPI_ISL_2873573, EPI_ISL_2873574, EPI_ISL_2873575, EPI_ISL_2873576, EPI_ISL_2873577, EPI_ISL_2873578, EPI_ISL_2873579, EPI_ISL_2873580, EPI_ISL_2873581, EPI_ISL_2873582, EPI_ISL_2873583, EPI_ISL_2873584, EPI_ISL_2873585, EPI_ISL_2873586, EPI_ISL_2873587, EPI_ISL_2873588, EPI_ISL_2873589, EPI_ISL_2873590, EPI_ISL_2873591, EPI_ISL_2873592, EPI_ISL_2873593, EPI_ISL_2873594, EPI_ISL_2873595, EPI_ISL_2873596, EPI_ISL_2873597, EPI_ISL_2873598, EPI_ISL_2873599, EPI_ISL_2873600, EPI_ISL_2873601, EPI_ISL_2873602, EPI_ISL_2873603, EPI_ISL_2873604, EPI_ISL_2873605, EPI_ISL_2873606, EPI_ISL_2873607, EPI_ISL_2873608, EPI_ISL_2873609, EPI_ISL_2873610, EPI_ISL_2873611, EPI_ISL_2873612, EPI_ISL_2873613, EPI_ISL_2873614, EPI_ISL_2873615, EPI_ISL_2873616, EPI_ISL_2873617, EPI_ISL_2873618, EPI_ISL_2873619, EPI_ISL_2873620, EPI_ISL_2873621, EPI_ISL_2873622, EPI_ISL_2873623, EPI_ISL_2873624, EPI_ISL_2873625, EPI_ISL_2873626, EPI_ISL_2873627, EPI_ISL_2873628, EPI_ISL_2873629, EPI_ISL_2873630, EPI_ISL_2873631, EPI_ISL_2873632, EPI_ISL_2873633, EPI_ISL_2873634, EPI_ISL_2873635, EPI_ISL_2873636, EPI_ISL_2873637, EPI_ISL_2873638, EPI_ISL_2873639, EPI_ISL_2873640, EPI_ISL_2873641, EPI_ISL_2873643, EPI_ISL_2873644, EPI_ISL_2873645, EPI_ISL_2873646, EPI_ISL_2873647, EPI_ISL_2873648, EPI_ISL_2873649, EPI_ISL_2873650, EPI_ISL_2873651, EPI_ISL_2873652, EPI_ISL_2873653, EPI_ISL_2873654, EPI_ISL_2873655, EPI_ISL_2873656, EPI_ISL_2873657, EPI_ISL_2873658, EPI_ISL_2873659, EPI_ISL_2873660, EPI_ISL_2873661, EPI_ISL_2873662, EPI_ISL_2873663, EPI_ISL_2873664, EPI_ISL_2873665, EPI_ISL_2873666, EPI_ISL_2873667, EPI_ISL_2873668, EPI_ISL_2873669, EPI_ISL_2873670, EPI_ISL_2873671, EPI_ISL_2873672, EPI_ISL_2873673, EPI_ISL_2873674, EPI_ISL_2873675, EPI_ISL_2873676, EPI_ISL_2873677, EPI_ISL_2873678, EPI_ISL_2873679, EPI_ISL_2873680, EPI_ISL_2873681, EPI_ISL_2873682, EPI_ISL_2873683, EPI_ISL_2873684, EPI_ISL_2873685, EPI_ISL_2873686, EPI_ISL_2873687, EPI_ISL_2873688, EPI_ISL_2873689, EPI_ISL_2873690, EPI_ISL_2873691, EPI_ISL_2873692, EPI_ISL_2873693, EPI_ISL_2873694, EPI_ISL_2873695, EPI_ISL_2873696, EPI_ISL_2873697, EPI_ISL_2873698, EPI_ISL_2873699, EPI_ISL_2873700, EPI_ISL_2873701, EPI_ISL_2873702, EPI_ISL_2873703, EPI_ISL_2873704, EPI_ISL_2873705, EPI_ISL_2873706, EPI_ISL_2873715, EPI_ISL_2873716, EPI_ISL_2873717, EPI_ISL_2873718, EPI_ISL_2873719, EPI_ISL_2873720, EPI_ISL_2873721, EPI_ISL_2873722, EPI_ISL_2873723, EPI_ISL_2873724, EPI_ISL_2873725, EPI_ISL_2873726, EPI_ISL_2873727, EPI_ISL_2873728, EPI_ISL_2873729, EPI_ISL_2873730, EPI_ISL_2873731, EPI_ISL_2873732, EPI_ISL_2873733, EPI_ISL_2873734, EPI_ISL_2873735, EPI_ISL_2873736, EPI_ISL_2873737, EPI_ISL_2873738, EPI_ISL_2873739, EPI_ISL_2873740, EPI_ISL_2873741, EPI_ISL_2873742, EPI_ISL_2873743, EPI_ISL_2873744, EPI_ISL_2873745, EPI_ISL_2873746, EPI_ISL_2873747, EPI_ISL_2873748, EPI_ISL_2873749, EPI_ISL_2873750, EPI_ISL_2873751, EPI_ISL_2873752, EPI_ISL_2873753, EPI_ISL_2873754, EPI_ISL_2873755, EPI_ISL_2873756, EPI_ISL_2873757, EPI_ISL_2873758, EPI_ISL_2873759, EPI_ISL_2873760, EPI_ISL_2873761, EPI_ISL_2873762, EPI_ISL_2873763, EPI_ISL_2873764, EPI_ISL_2873765, EPI_ISL_2873766, EPI_ISL_2873767, EPI_ISL_2873768, EPI_ISL_2873769, EPI_ISL_2873770, EPI_ISL_2873771, EPI_ISL_2873772, EPI_ISL_2873773, EPI_ISL_2873774, EPI_ISL_2873775, EPI_ISL_2873776, EPI_ISL_2873777, EPI_ISL_2873778, EPI_ISL_2873779, EPI_ISL_2873780, EPI_ISL_2873781, EPI_ISL_2873782, EPI_ISL_2873783, EPI_ISL_2873784, EPI_ISL_2873785, EPI_ISL_2873786, EPI_ISL_2873787, EPI_ISL_2873788, EPI_ISL_2873789, EPI_ISL_2928580, EPI_ISL_2928581, EPI_ISL_2928582, EPI_ISL_2928590, EPI_ISL_2928591, EPI_ISL_2928592, EPI_ISL_2928593, EPI_ISL_2928594, EPI_ISL_2928595, EPI_ISL_2928597, EPI_ISL_2928598, EPI_ISL_2928600, EPI_ISL_2928601, EPI_ISL_2928603, EPI_ISL_2928611, EPI_ISL_2928612, EPI_ISL_2928613, EPI_ISL_2928614, EPI_ISL_2928616, EPI_ISL_2928621, EPI_ISL_2928627, EPI_ISL_2928628, EPI_ISL_2928629, EPI_ISL_2928634, EPI_ISL_2928655, EPI_ISL_2928673, EPI_ISL_2928699, EPI_ISL_2928701, EPI_ISL_2928925, EPI_ISL_2929130, EPI_ISL_2929131 | | | |
| see above | Fulgent Genetics | Centers for Disease Control and Prevention Division of Viral Diseases, Pathogen Discovery | Adrian Paskey; Becky Tsai; Benafsh Sapra; Benjamin Rambo-Martin; Christopher Gulvick; Clinton R. Paden; Dakota Howard; Darlene Wagner; Dhwani Batra; Doreen Ng; Duncan MacCannell; Harry Gao; James Xie; Jason Caravas; John Gao; Joseph Fierro; Kara Moser; Matthew Schmerer; Mickey Li; Peter W. Cook; Scott Sammons; Shatavia Morrison; Yan Meng; Yvette Unoarumhi |
| EPI_ISL_2404826 | GOOD SAMARITAN HOSPTIAL | Santa Clara County public Health Laboratory | Santa Clara County Public Health Department |
| EPI_ISL_2650566, EPI_ISL_2753977, EPI_ISL_2754022, EPI_ISL_2934001, EPI_ISL_2934053 | Good Samaritan Hospital | Santa Clara County Public Health Laboratory | Santa Clara County Public Health Department |
| EPI_ISL_2160397, EPI_ISL_2160400, EPI_ISL_2160407, EPI_ISL_2160414, EPI_ISL_2160429, EPI_ISL_2160443, EPI_ISL_2160446, EPI_ISL_2160454, EPI_ISL_2160463, EPI_ISL_2187635, EPI_ISL_2187636, EPI_ISL_2247574, EPI_ISL_2247613, EPI_ISL_2247620, EPI_ISL_2247660, EPI_ISL_2247776, EPI_ISL_2248020, EPI_ISL_2248042, EPI_ISL_2248059, EPI_ISL_2248074, EPI_ISL_2248085, EPI_ISL_2248097, EPI_ISL_2248117, EPI_ISL_2248118, EPI_ISL_2248146, EPI_ISL_2248433, EPI_ISL_2248451, EPI_ISL_2268355, EPI_ISL_2269641, EPI_ISL_2269679, EPI_ISL_2269685, EPI_ISL_2269704, EPI_ISL_2269713, EPI_ISL_2269722, EPI_ISL_2269731, EPI_ISL_2269752, EPI_ISL_2269773, EPI_ISL_2269830, EPI_ISL_2269847, EPI_ISL_2269863, EPI_ISL_2269873, EPI_ISL_2269885, EPI_ISL_2269889, EPI_ISL_2269896, EPI_ISL_2269913, EPI_ISL_2269937, EPI_ISL_2269938, EPI_ISL_2269950, EPI_ISL_2269999, EPI_ISL_2270028, EPI_ISL_2270034, EPI_ISL_2270036, EPI_ISL_2270055, EPI_ISL_2270083, EPI_ISL_2270135, EPI_ISL_2270156, EPI_ISL_2270296, EPI_ISL_2270315, EPI_ISL_2270318, EPI_ISL_2270319, EPI_ISL_2270328, EPI_ISL_2270330, EPI_ISL_2270346, EPI_ISL_2270406, EPI_ISL_2270412, EPI_ISL_2270422, EPI_ISL_2270429, EPI_ISL_2270539, EPI_ISL_2270630, EPI_ISL_2270632, EPI_ISL_2270638, EPI_ISL_2270685, EPI_ISL_2270742, EPI_ISL_2270751, EPI_ISL_2270763, EPI_ISL_2270768, EPI_ISL_2270795, EPI_ISL_2270806, EPI_ISL_2270807, EPI_ISL_2270817, EPI_ISL_2270834, EPI_ISL_2280186, EPI_ISL_2280188, EPI_ISL_2320000, EPI_ISL_2320001, EPI_ISL_2320011, EPI_ISL_2320014, EPI_ISL_2320022, EPI_ISL_2320023, EPI_ISL_2320025, EPI_ISL_2320060, EPI_ISL_2320096, EPI_ISL_2320100, EPI_ISL_2320103, EPI_ISL_2320119, EPI_ISL_2320123, EPI_ISL_2320163, EPI_ISL_2320226, EPI_ISL_2320255, EPI_ISL_2320272, EPI_ISL_2320280, EPI_ISL_2320285, EPI_ISL_2320293, EPI_ISL_2320295, EPI_ISL_2320297, EPI_ISL_2320314, EPI_ISL_2320345, EPI_ISL_2320356, EPI_ISL_2320378, EPI_ISL_2320443, EPI_ISL_2320446, EPI_ISL_2320455, EPI_ISL_2320487, EPI_ISL_2320524, EPI_ISL_2320526, EPI_ISL_2320528, EPI_ISL_2320530, EPI_ISL_2320590, EPI_ISL_2320646, EPI_ISL_2320676, EPI_ISL_2320774, EPI_ISL_2320826, EPI_ISL_2320838, EPI_ISL_2320847, EPI_ISL_2320854, EPI_ISL_2320862, EPI_ISL_2320863, EPI_ISL_2320866, EPI_ISL_2320873, EPI_ISL_2320901, EPI_ISL_2320921, EPI_ISL_2320922, EPI_ISL_2320928, EPI_ISL_2320930, EPI_ISL_2320943, EPI_ISL_2320945, EPI_ISL_2320955, EPI_ISL_2320961, EPI_ISL_2320991, EPI_ISL_2321048, EPI_ISL_2321081, EPI_ISL_2321091, EPI_ISL_2321101, EPI_ISL_2321135, EPI_ISL_2321146, EPI_ISL_2367608, EPI_ISL_2367620, EPI_ISL_2367641, EPI_ISL_2367644, EPI_ISL_2440055, EPI_ISL_2440066, EPI_ISL_2440079, EPI_ISL_2440089, EPI_ISL_2440195, EPI_ISL_2440206, EPI_ISL_2440223, EPI_ISL_2440366, EPI_ISL_2440508, EPI_ISL_2440520, EPI_ISL_2598841, EPI_ISL_2598852, EPI_ISL_2598873, EPI_ISL_2598889, EPI_ISL_2598946, EPI_ISL_2599112, EPI_ISL_2599139, EPI_ISL_2599158, EPI_ISL_2599162, EPI_ISL_2599173, EPI_ISL_2599212, EPI_ISL_2599214, EPI_ISL_2599223, EPI_ISL_2599239, EPI_ISL_2599241, EPI_ISL_2599252, EPI_ISL_2599273, EPI_ISL_2599308, EPI_ISL_2599309, EPI_ISL_2599400, EPI_ISL_2599408, EPI_ISL_2599416, EPI_ISL_2599420, EPI_ISL_2599447, EPI_ISL_2599472, EPI_ISL_2599475, EPI_ISL_2599540, EPI_ISL_2599575, EPI_ISL_2599606, EPI_ISL_2599608, EPI_ISL_2649941 | | | |
| see above | Helix/Illumina | Centers for Disease Control and Prevention Division of Viral Diseases, Pathogen Discovery | Adrian Paskey; Alexandre Bolze; Ary Ascencio; Benjamin Rambo-Martin; Brad Sickler; Charlotte Rivera-Garcia; Christine Tran; Christopher Gulvick; Clinton R. Paden; Dakota Howard; Darlene Wagner; David Becker; Dhwani Batra; Duncan MacCannell; Efren Sandoval; Eileen de Feo; Elizabeth Cirulli; Eric Allen; Geraint Levan; James Lu; Jan Antico; Jason Caravas; Jason Nguyen; Jimmy Ramirez; Jingtao Liu; Kara Moser; Kelly Schiabor Barrett; Kim Gietzen; Magnus Isaksson; Marc Laurent; Matthew Schmerer; Matthew Tolentino; Nicole L. Washington; Peter W. Cook; Phil Febbo; Ryan Cho; Scott Sammons; Shannon Wickline; Shatavia Morrison; Sherry Wang; Simon White; Tyler Cassens; William Lee; Yvette Unoarumhi |
| EPI_ISL_2907564, EPI_ISL_3029872 | Henry Mayo Newhall Hospital | Los Angeles County Public Health Laboratories | P. Hemarajata et al. |
| EPI_ISL_2811961 | Hollywood Presbyterian Medical Center | Los Angeles County Public Health Laboratories | P. Hemarajata et al. |
| EPI_ISL_2341415, EPI_ISL_2341421, EPI_ISL_2341436, EPI_ISL_2341449, EPI_ISL_2341458, EPI_ISL_2341463, EPI_ISL_2341471, EPI_ISL_2341472, EPI_ISL_2341480, EPI_ISL_2341487, EPI_ISL_2341498, EPI_ISL_2341501, EPI_ISL_2341543, EPI_ISL_2341563, EPI_ISL_2341565, EPI_ISL_2341572, EPI_ISL_2341595, EPI_ISL_2341613, EPI_ISL_2341616, EPI_ISL_2341621, EPI_ISL_2341625, EPI_ISL_2341627, EPI_ISL_2341629, EPI_ISL_2341650, EPI_ISL_2341655, EPI_ISL_2497549, EPI_ISL_2497567, EPI_ISL_2497622, EPI_ISL_2497625, EPI_ISL_2497628, EPI_ISL_2497633, EPI_ISL_2497653, EPI_ISL_2497657, EPI_ISL_2497660, EPI_ISL_2497662, EPI_ISL_2497663, EPI_ISL_2497664, EPI_ISL_2497669, EPI_ISL_2497686, EPI_ISL_2497690, EPI_ISL_2497691, EPI_ISL_2497693, EPI_ISL_2497697, EPI_ISL_2500025, EPI_ISL_2500048, EPI_ISL_2500054, EPI_ISL_2500058, EPI_ISL_2500060, EPI_ISL_2500061, EPI_ISL_2500063, EPI_ISL_2500069, EPI_ISL_2500074, EPI_ISL_2500078, EPI_ISL_2500100, EPI_ISL_2500108, EPI_ISL_2500116, EPI_ISL_2500125, EPI_ISL_2500133, EPI_ISL_2500142, EPI_ISL_2500150, EPI_ISL_2500151, EPI_ISL_2500157, EPI_ISL_2500159, EPI_ISL_2500182, EPI_ISL_2500185, EPI_ISL_2500189, EPI_ISL_2500196, EPI_ISL_2500201, EPI_ISL_2500202, EPI_ISL_2500213, EPI_ISL_2658738, EPI_ISL_2758541, EPI_ISL_2758544 | | | |
| see above | Humboldt County Public Health Laboratory | Chan-Zuckerberg Biohub | CZB Cliahub Consortium |
| EPI_ISL_2445617, EPI_ISL_2445618, EPI_ISL_2445619, EPI_ISL_2445620, EPI_ISL_2445621, EPI_ISL_2445622, EPI_ISL_2445623, EPI_ISL_2445624, EPI_ISL_2445625, EPI_ISL_2445626, EPI_ISL_2445627, EPI_ISL_2445628, EPI_ISL_2445629, EPI_ISL_2445630, EPI_ISL_2445631, EPI_ISL_2445632, EPI_ISL_2445633, EPI_ISL_2680469, EPI_ISL_2680471, EPI_ISL_2680473, EPI_ISL_2680474, EPI_ISL_2680476, EPI_ISL_2680478, EPI_ISL_2680480, EPI_ISL_2680482, EPI_ISL_2680483, EPI_ISL_2680485, EPI_ISL_2680487, EPI_ISL_2680488, EPI_ISL_2680490, EPI_ISL_2680492, EPI_ISL_2778969, EPI_ISL_2778970, EPI_ISL_2778971, EPI_ISL_2778972, EPI_ISL_2778973, EPI_ISL_2778974, EPI_ISL_2778975, EPI_ISL_2778976, EPI_ISL_2778977, EPI_ISL_2778978, EPI_ISL_2778979, EPI_ISL_2778980, EPI_ISL_2778981, EPI_ISL_2778982, EPI_ISL_2778983, EPI_ISL_2928419, EPI_ISL_2928420, EPI_ISL_2928421, EPI_ISL_2928422, EPI_ISL_2928423, EPI_ISL_2928424, EPI_ISL_2928425, EPI_ISL_2928426, EPI_ISL_2928427, EPI_ISL_2928428, EPI_ISL_2928429, EPI_ISL_2928430, EPI_ISL_2928431, EPI_ISL_2928432, EPI_ISL_2928433, EPI_ISL_2928434, EPI_ISL_2928435, EPI_ISL_2928436, EPI_ISL_2928437, EPI_ISL_2928438, EPI_ISL_3010281, EPI_ISL_3010282, EPI_ISL_3010283, EPI_ISL_3010284, EPI_ISL_3010285, EPI_ISL_3010286, EPI_ISL_3010287, EPI_ISL_3010288, EPI_ISL_3010289, EPI_ISL_3010290, EPI_ISL_3010291, EPI_ISL_3010292, EPI_ISL_3010293, EPI_ISL_3010294, EPI_ISL_3010295, EPI_ISL_3010296 | | | |
| see above | Humboldt County Public Health Laboratory | Humboldt County Public Health Laboratory | Jeremy Corrigan |
| EPI_ISL_2391788, EPI_ISL_2391807, EPI_ISL_2405182, EPI_ISL_2894586, EPI_ISL_3029863, EPI_ISL_3029864 | Huntington Memorial Hospital | Los Angeles County Public Health Laboratories | P. Hemarajata et al. |
| EPI_ISL_2801446 | IVY3 Central Lab, Vanderbilt University Medical Center | Lauring Lab, University of Michigan, Department of Microbiology and Immunology | Valesano |
| EPI_ISL_2604246 | Infectious Diseases, Chan-Zuckerberg Biohub Covidtracker | Infectious Diseases, Chan-Zuckerberg Biohub Covidtracker | CZB Cliahub Consortium |
| EPI_ISL_2088924, EPI_ISL_2088925, EPI_ISL_2186452, EPI_ISL_2186502, EPI_ISL_2282436, EPI_ISL_2282520, EPI_ISL_2282718, EPI_ISL_2282857, EPI_ISL_2324030, EPI_ISL_2324045, EPI_ISL_2324086, EPI_ISL_2324098, EPI_ISL_2324112, EPI_ISL_2324130, EPI_ISL_2324132, EPI_ISL_2324156, EPI_ISL_2375736, EPI_ISL_2557438, EPI_ISL_2557530, EPI_ISL_2557535, EPI_ISL_2557616, EPI_ISL_2652839, EPI_ISL_2652936, EPI_ISL_2652938, EPI_ISL_2686840, EPI_ISL_2786657, EPI_ISL_2876054 | | | |
| see above | Infinity Biologix | Centers for Disease Control and Prevention Division of Viral Diseases, Pathogen Discovery | Adrian Paskey; Benjamin Rambo-Martin; Chirayu Goswami; Christian Bixby; Christopher Gulvick; Clinton R. Paden; Dakota Howard; Darlene Wagner; Dhwani Batra; Duncan MacCannell; Jason Caravas; Jonathan Schultz; Kara Moser; Matthew Schmerer; Peter W. Cook; Robin Grimwood; Russ Hager; Scott Sammons; Shatavia Morrison; Yihe Wang; Yvette Unoarumhi |
| EPI_ISL_2753992, EPI_ISL_2753993, EPI_ISL_2753994, EPI_ISL_2753995, EPI_ISL_2753996, EPI_ISL_2753997, EPI_ISL_2753998, EPI_ISL_2753999, EPI_ISL_2754000, EPI_ISL_2754002, EPI_ISL_2754003, EPI_ISL_2754004, EPI_ISL_2754005, EPI_ISL_2754006, EPI_ISL_2754007, EPI_ISL_2754008, EPI_ISL_2754009, EPI_ISL_2754010, EPI_ISL_2754011, EPI_ISL_2754012, EPI_ISL_2754013, EPI_ISL_2754014, EPI_ISL_2754015, EPI_ISL_2754016, EPI_ISL_2754017, EPI_ISL_2754018, EPI_ISL_2754019, EPI_ISL_2754020, EPI_ISL_2754021, EPI_ISL_2933980, EPI_ISL_2933982, EPI_ISL_2933983, EPI_ISL_2933984, EPI_ISL_2933985, EPI_ISL_2933988, EPI_ISL_2933991, EPI_ISL_2933992, EPI_ISL_2933996, EPI_ISL_2933999, EPI_ISL_2934003, EPI_ISL_2934005, EPI_ISL_2934007, EPI_ISL_2934008, EPI_ISL_2934009, EPI_ISL_2934011, EPI_ISL_2934012, EPI_ISL_2934013, EPI_ISL_2934014, EPI_ISL_2934015, EPI_ISL_2934016, EPI_ISL_2934017, EPI_ISL_2934018, EPI_ISL_2934019, EPI_ISL_2934020, EPI_ISL_2934024, EPI_ISL_2934025, EPI_ISL_2934026, EPI_ISL_2934027, EPI_ISL_2934028, EPI_ISL_2934044, EPI_ISL_2934045, EPI_ISL_2934066, EPI_ISL_2934067, EPI_ISL_2934068, EPI_ISL_2934071, EPI_ISL_2934073, EPI_ISL_2934074, EPI_ISL_2934075, EPI_ISL_2934076, EPI_ISL_2934077, EPI_ISL_2934078, EPI_ISL_2934079, EPI_ISL_2934080, EPI_ISL_2934085, EPI_ISL_2934086, EPI_ISL_2934087, EPI_ISL_2934093 | | | |
| see above | Kaiser Regional | Santa Clara County Public Health Laboratory | Santa Clara County Public Health Department |
| EPI_ISL_2650539, EPI_ISL_2650540, EPI_ISL_2650581, EPI_ISL_2650584, EPI_ISL_2691295, EPI_ISL_2753981, EPI_ISL_2753982, EPI_ISL_2934004, EPI_ISL_2934061 | | | |
| see above | Kaiser San Jose | Santa Clara County Public Health Laboratory | Santa Clara County Public Health Department |
| EPI_ISL_2631210, EPI_ISL_2650541, EPI_ISL_2753978, EPI_ISL_2753979, EPI_ISL_2753980 | Kaiser Santa Clara | Santa Clara County Public Health Laboratory | Santa Clara County Public Health Department |
| EPI_ISL_2046772, EPI_ISL_2046773, EPI_ISL_2046774, EPI_ISL_2046775, EPI_ISL_2046777, EPI_ISL_2046791, EPI_ISL_2046820, EPI_ISL_2046822, EPI_ISL_2046823, EPI_ISL_2046824, EPI_ISL_2046825, EPI_ISL_2046826, EPI_ISL_2046827, EPI_ISL_2184941, EPI_ISL_2184942, EPI_ISL_2184943, EPI_ISL_2184944, EPI_ISL_2184945, EPI_ISL_2184946, EPI_ISL_2184996, EPI_ISL_2184997, EPI_ISL_2184998, EPI_ISL_2185011, EPI_ISL_2185012, EPI_ISL_2185013, EPI_ISL_2185014, EPI_ISL_2185015, EPI_ISL_2185016, EPI_ISL_2185017, EPI_ISL_2185027, EPI_ISL_2185028, EPI_ISL_2185029, EPI_ISL_2185030, EPI_ISL_2185031, EPI_ISL_2185032, EPI_ISL_2185033, EPI_ISL_2185034, EPI_ISL_2185035, EPI_ISL_2185170, EPI_ISL_2185175, EPI_ISL_2185176, EPI_ISL_2185177, EPI_ISL_2185203, EPI_ISL_2185204, EPI_ISL_2185215, EPI_ISL_2185232, EPI_ISL_2185233, EPI_ISL_2185242, EPI_ISL_2185243, EPI_ISL_2185244, EPI_ISL_2185245, EPI_ISL_2185246, EPI_ISL_2185247, EPI_ISL_2185248, EPI_ISL_2185249, EPI_ISL_2185250, EPI_ISL_2185251, EPI_ISL_2185252, EPI_ISL_2185274, EPI_ISL_2185275, EPI_ISL_2185276, EPI_ISL_2185277, EPI_ISL_2185278, EPI_ISL_2185279, EPI_ISL_2185280, EPI_ISL_2185281, EPI_ISL_2185282, EPI_ISL_2185283, EPI_ISL_2185284, EPI_ISL_2185285, EPI_ISL_2185286, EPI_ISL_2185418, EPI_ISL_2185419, EPI_ISL_2185446, EPI_ISL_2185447, EPI_ISL_2185448, EPI_ISL_2185499, EPI_ISL_2185511, EPI_ISL_2185512, EPI_ISL_2185528, EPI_ISL_2185529, EPI_ISL_2304865, EPI_ISL_2304866, EPI_ISL_2304867, EPI_ISL_2304978, EPI_ISL_2304979, EPI_ISL_2304980, EPI_ISL_2304981, EPI_ISL_2304982, EPI_ISL_2304983, EPI_ISL_2304984, EPI_ISL_2304985, EPI_ISL_2304986, EPI_ISL_2304987, EPI_ISL_2304988, EPI_ISL_2304989, EPI_ISL_2304990, EPI_ISL_2304991, EPI_ISL_2305018, EPI_ISL_2305019, EPI_ISL_2305088, EPI_ISL_2305089, EPI_ISL_2305090, EPI_ISL_2305091, EPI_ISL_2305092, EPI_ISL_2305093, EPI_ISL_2305103, EPI_ISL_2305104, EPI_ISL_2305105, EPI_ISL_2305106, EPI_ISL_2305107, EPI_ISL_2305108, EPI_ISL_2305109, EPI_ISL_2305110, EPI_ISL_2305111, EPI_ISL_2305112, EPI_ISL_2305113, EPI_ISL_2305114, EPI_ISL_2305115, EPI_ISL_2305116, EPI_ISL_2305117, EPI_ISL_2305118, EPI_ISL_2305119, EPI_ISL_2305120, EPI_ISL_2305139, EPI_ISL_2305140, EPI_ISL_2305141, EPI_ISL_2305142, EPI_ISL_2305143, EPI_ISL_2305144, EPI_ISL_2305145, EPI_ISL_2305146, EPI_ISL_2305147, EPI_ISL_2305148, EPI_ISL_2305202, EPI_ISL_2305204, EPI_ISL_2305205, EPI_ISL_2305284, EPI_ISL_2305285, EPI_ISL_2305286, EPI_ISL_2305287, EPI_ISL_2305315, EPI_ISL_2305316, EPI_ISL_2305317, EPI_ISL_2305318, EPI_ISL_2305319, EPI_ISL_2305320, EPI_ISL_2305321, EPI_ISL_2305322, EPI_ISL_2305323, EPI_ISL_2305364, EPI_ISL_2305365, EPI_ISL_2305366, EPI_ISL_2305367, EPI_ISL_2305368, EPI_ISL_2305369, EPI_ISL_2305372, EPI_ISL_2305373, EPI_ISL_2305375, EPI_ISL_2305376, EPI_ISL_2305383, EPI_ISL_2305384, EPI_ISL_2305405, EPI_ISL_2305406, EPI_ISL_2305443, EPI_ISL_2305444, EPI_ISL_2305445, EPI_ISL_2305494, EPI_ISL_2305495, EPI_ISL_2305497, EPI_ISL_2305498, EPI_ISL_2305499, EPI_ISL_2305539, EPI_ISL_2305540, EPI_ISL_2305541, EPI_ISL_2305617, EPI_ISL_2305618, EPI_ISL_2305619, EPI_ISL_2305620, EPI_ISL_2305622, EPI_ISL_2305623, EPI_ISL_2305624, EPI_ISL_2305625, EPI_ISL_2305626, EPI_ISL_2305628, EPI_ISL_2305656, EPI_ISL_2305657, EPI_ISL_2305658, EPI_ISL_2305659, EPI_ISL_2305660, EPI_ISL_2305661, EPI_ISL_2305662, EPI_ISL_2305663, EPI_ISL_2305664, EPI_ISL_2305665, EPI_ISL_2305666, EPI_ISL_2305950, EPI_ISL_2397822, EPI_ISL_2397823, EPI_ISL_2397824, EPI_ISL_2397825, EPI_ISL_2397879, EPI_ISL_2397881, EPI_ISL_2397882, EPI_ISL_2397883, EPI_ISL_2397884, EPI_ISL_2397885, EPI_ISL_2397886, EPI_ISL_2397887, EPI_ISL_2397888, EPI_ISL_2397890, EPI_ISL_2397891, EPI_ISL_2397892, EPI_ISL_2397926, EPI_ISL_2397927, EPI_ISL_2397928, EPI_ISL_2397972, EPI_ISL_2397973, EPI_ISL_2397995, EPI_ISL_2397996, EPI_ISL_2397997, EPI_ISL_2397998, EPI_ISL_2397999, EPI_ISL_2398000, EPI_ISL_2398001, EPI_ISL_2398002, EPI_ISL_2398003, EPI_ISL_2398004, EPI_ISL_2398005, EPI_ISL_2398026, EPI_ISL_2398027, EPI_ISL_2398028, EPI_ISL_2398029, EPI_ISL_2398039, EPI_ISL_2398040, EPI_ISL_2398041, EPI_ISL_2398042, EPI_ISL_2398043, EPI_ISL_2398044, EPI_ISL_2398045, EPI_ISL_2398046, EPI_ISL_2398067, EPI_ISL_2398068, EPI_ISL_2398069, EPI_ISL_2398070, EPI_ISL_2398071, EPI_ISL_2398072, EPI_ISL_2481478, EPI_ISL_2481550, EPI_ISL_2481590, EPI_ISL_2481591, EPI_ISL_2481592, EPI_ISL_2481593, EPI_ISL_2481594, EPI_ISL_2481595, EPI_ISL_2481646, EPI_ISL_2481647, EPI_ISL_2481648, EPI_ISL_2481649, EPI_ISL_2481650, EPI_ISL_2481651, EPI_ISL_2481652, EPI_ISL_2481654, EPI_ISL_2481655, EPI_ISL_2481677, EPI_ISL_2481678, EPI_ISL_2481736, EPI_ISL_2481756, EPI_ISL_2481758, EPI_ISL_2481759, EPI_ISL_2481760, EPI_ISL_2481761, EPI_ISL_2481762, EPI_ISL_2481763, EPI_ISL_2481764, EPI_ISL_2481775, EPI_ISL_2481776, EPI_ISL_2481777, EPI_ISL_2481778, EPI_ISL_2481779, EPI_ISL_2481780, EPI_ISL_2481781, EPI_ISL_2481782, EPI_ISL_2481783, EPI_ISL_2481804, EPI_ISL_2481812, EPI_ISL_2481813, EPI_ISL_2481814, EPI_ISL_2481815, EPI_ISL_2481816, EPI_ISL_2481817, EPI_ISL_2481818, EPI_ISL_2481993, EPI_ISL_2481994, EPI_ISL_2481995, EPI_ISL_2481996, EPI_ISL_2482017, EPI_ISL_2482018, EPI_ISL_2482019, EPI_ISL_2482048, EPI_ISL_2482066, EPI_ISL_2482067, EPI_ISL_2482068, EPI_ISL_2482069, EPI_ISL_2482070, EPI_ISL_2482092, EPI_ISL_2482093, EPI_ISL_2482094, EPI_ISL_2482095, EPI_ISL_2482096, EPI_ISL_2482097, EPI_ISL_2482098, EPI_ISL_2482099, EPI_ISL_2482164, EPI_ISL_2482165, EPI_ISL_2482166, EPI_ISL_2482167, EPI_ISL_2482168, EPI_ISL_2482169, EPI_ISL_2482170, EPI_ISL_2482171, EPI_ISL_2482172, EPI_ISL_2482173, EPI_ISL_2482174, EPI_ISL_2482175, EPI_ISL_2482176, EPI_ISL_2482177, EPI_ISL_2482178, EPI_ISL_2482190, EPI_ISL_2482191, EPI_ISL_2482192, EPI_ISL_2482207, EPI_ISL_2482231, EPI_ISL_2482232, EPI_ISL_2482233, EPI_ISL_2482235, EPI_ISL_2482236, EPI_ISL_2482237, EPI_ISL_2482262, EPI_ISL_2482263, EPI_ISL_2482264, EPI_ISL_2482265, EPI_ISL_2482266, EPI_ISL_2611298, EPI_ISL_2873146, EPI_ISL_2873147, EPI_ISL_2873148, EPI_ISL_2873149, EPI_ISL_2873150, EPI_ISL_2873155, EPI_ISL_2873156, EPI_ISL_2873157, EPI_ISL_2873158, EPI_ISL_2873159, EPI_ISL_2873160, EPI_ISL_2873161, EPI_ISL_2873219, EPI_ISL_2873220, EPI_ISL_2873224, EPI_ISL_2873225, EPI_ISL_2873226, EPI_ISL_2873227, EPI_ISL_2873229, EPI_ISL_2873235, EPI_ISL_2873254, EPI_ISL_2873268, EPI_ISL_2873269, EPI_ISL_2873270, EPI_ISL_2873271, EPI_ISL_2873272, EPI_ISL_2873273, EPI_ISL_2873282, EPI_ISL_2873283, EPI_ISL_2873303, EPI_ISL_2873304, EPI_ISL_2873305, EPI_ISL_2873306, EPI_ISL_2873307, EPI_ISL_2873314, EPI_ISL_2873315, EPI_ISL_2873316, EPI_ISL_2873317, EPI_ISL_2873322, EPI_ISL_2873323, EPI_ISL_2873324, EPI_ISL_2873325, EPI_ISL_2926342, EPI_ISL_2926684, EPI_ISL_2926685, EPI_ISL_2926686, EPI_ISL_2926687, EPI_ISL_2926688, EPI_ISL_2926689, EPI_ISL_2926690, EPI_ISL_2926697, EPI_ISL_2926702, | | | |

| | | | |
|---|---|---|---|
| EPI_ISL_2926731, EPI_ISL_2926732, EPI_ISL_2926733, EPI_ISL_2926734, EPI_ISL_2926735, EPI_ISL_2926736, EPI_ISL_2926737, EPI_ISL_2926738, EPI_ISL_2926739, EPI_ISL_2926740, EPI_ISL_2926741, EPI_ISL_2926742, EPI_ISL_2926743, EPI_ISL_2926744, EPI_ISL_2926753, EPI_ISL_2926754, EPI_ISL_2926755, EPI_ISL_2926756, EPI_ISL_2926764, EPI_ISL_2926765, EPI_ISL_2926766, EPI_ISL_2926767, EPI_ISL_2926768, EPI_ISL_2926773, EPI_ISL_2926782, EPI_ISL_2926820, EPI_ISL_2926821, EPI_ISL_2926822, EPI_ISL_2926823, EPI_ISL_2926824, EPI_ISL_2926825, EPI_ISL_2926826, EPI_ISL_2926827, EPI_ISL_2926828, EPI_ISL_2926829, EPI_ISL_2926830, EPI_ISL_2926840, EPI_ISL_2926845, EPI_ISL_2926846, EPI_ISL_2926847, EPI_ISL_2926848, EPI_ISL_2926849, EPI_ISL_2926850, EPI_ISL_2926851, EPI_ISL_2926852, EPI_ISL_2926853, EPI_ISL_2926854, EPI_ISL_2926855, EPI_ISL_2926856, EPI_ISL_2926857, EPI_ISL_2926858, EPI_ISL_2926895, EPI_ISL_2926896, EPI_ISL_2926897, EPI_ISL_2926898, EPI_ISL_2926899, EPI_ISL_2926900, EPI_ISL_2926901, EPI_ISL_2926902, EPI_ISL_2926905, EPI_ISL_2926913, EPI_ISL_2926914, EPI_ISL_2926915, EPI_ISL_2926916, EPI_ISL_2926917, EPI_ISL_2926918, EPI_ISL_2926919, EPI_ISL_2926920, EPI_ISL_2926921, EPI_ISL_2926922, EPI_ISL_2926927, EPI_ISL_2926928, EPI_ISL_2926929, EPI_ISL_2926931, EPI_ISL_2926932, EPI_ISL_2926958, EPI_ISL_2926972, EPI_ISL_2926973, EPI_ISL_2926987, EPI_ISL_2926988, EPI_ISL_2926991, EPI_ISL_2926992, EPI_ISL_2926993, EPI_ISL_2926994, EPI_ISL_2926997, EPI_ISL_2926998, EPI_ISL_2926999, EPI_ISL_2927018, EPI_ISL_2927019, EPI_ISL_2927034, EPI_ISL_2927055, EPI_ISL_2927056, EPI_ISL_2927057, EPI_ISL_2927058, EPI_ISL_2927059, EPI_ISL_2927109, EPI_ISL_2927125, EPI_ISL_2927132, EPI_ISL_2927133, EPI_ISL_2927158, EPI_ISL_2927159, EPI_ISL_2927160, EPI_ISL_2927161, EPI_ISL_2927162, EPI_ISL_2927163, EPI_ISL_2927164, EPI_ISL_2927170, EPI_ISL_2927171, EPI_ISL_2927172, EPI_ISL_2927173, EPI_ISL_2927174, EPI_ISL_2927175, EPI_ISL_2927176, EPI_ISL_2927178, EPI_ISL_2927179, EPI_ISL_2927193, EPI_ISL_2927194, EPI_ISL_2927195, EPI_ISL_2927196, EPI_ISL_2927197, EPI_ISL_2927198, EPI_ISL_2927199, EPI_ISL_2927215, EPI_ISL_2927216, EPI_ISL_2927217, EPI_ISL_2927218, EPI_ISL_2927219, EPI_ISL_2927255, EPI_ISL_2927258, EPI_ISL_2927259, EPI_ISL_2927260, EPI_ISL_2927261, EPI_ISL_2927262, EPI_ISL_2927263, EPI_ISL_2927264, EPI_ISL_2927265, EPI_ISL_2927266, EPI_ISL_2927267, EPI_ISL_2927285, EPI_ISL_2927293, EPI_ISL_2927294, EPI_ISL_2927296, EPI_ISL_2927297, EPI_ISL_2927298, EPI_ISL_2927299, EPI_ISL_2927300, EPI_ISL_2927301, EPI_ISL_2927302, EPI_ISL_2927303, EPI_ISL_2927360, EPI_ISL_2927361, EPI_ISL_2927362, EPI_ISL_2927363, EPI_ISL_2927364, EPI_ISL_2927365, EPI_ISL_2927366, EPI_ISL_2927367, EPI_ISL_2927368, EPI_ISL_2927369, EPI_ISL_2927376, EPI_ISL_2927377, EPI_ISL_2927378, EPI_ISL_2927379, EPI_ISL_2927386, EPI_ISL_2927433, EPI_ISL_2927434, EPI_ISL_2927435, EPI_ISL_2927436, EPI_ISL_2927437, EPI_ISL_2927438, EPI_ISL_2927439, EPI_ISL_2927440, EPI_ISL_2927441, EPI_ISL_2927442, EPI_ISL_2927443, EPI_ISL_2927444, EPI_ISL_2927445, EPI_ISL_3016782, EPI_ISL_3016783, EPI_ISL_3016787, EPI_ISL_3016788, EPI_ISL_3016794, EPI_ISL_3017610, EPI_ISL_3017611, EPI_ISL_3017612, EPI_ISL_3017614, EPI_ISL_3017615, EPI_ISL_3017616, EPI_ISL_3017617, EPI_ISL_3017618, EPI_ISL_3017619, EPI_ISL_3017620, EPI_ISL_3017621, EPI_ISL_3017645, EPI_ISL_3017653, EPI_ISL_3017654, EPI_ISL_3017655, EPI_ISL_3017656, EPI_ISL_3017657, EPI_ISL_3017658, EPI_ISL_3017659, EPI_ISL_3017661, EPI_ISL_3017662, EPI_ISL_3017663, EPI_ISL_3017664, EPI_ISL_3017671, EPI_ISL_3017672, EPI_ISL_3017673, EPI_ISL_3017674, EPI_ISL_3017675, EPI_ISL_3017676, EPI_ISL_3017677, EPI_ISL_3017678, EPI_ISL_3017679, EPI_ISL_3017680, EPI_ISL_3017681, EPI_ISL_3017682, EPI_ISL_3017683, EPI_ISL_3017685, EPI_ISL_3017686, EPI_ISL_3017687, EPI_ISL_3017688, EPI_ISL_3017689, EPI_ISL_3017690, EPI_ISL_3017692, EPI_ISL_3017720, EPI_ISL_3017764, EPI_ISL_3017765, EPI_ISL_3017766, EPI_ISL_3017767, EPI_ISL_3017768, EPI_ISL_3017769, EPI_ISL_3017770, EPI_ISL_3017771, EPI_ISL_3017772, EPI_ISL_3017773, EPI_ISL_3017774, EPI_ISL_3017780, EPI_ISL_3017790, EPI_ISL_3017791, EPI_ISL_3017792, EPI_ISL_3017793, EPI_ISL_3017794, EPI_ISL_3017795, EPI_ISL_3017796, EPI_ISL_3017797, EPI_ISL_3017798, EPI_ISL_3017799, EPI_ISL_3017800, EPI_ISL_3017801, EPI_ISL_3017802, EPI_ISL_3017816, EPI_ISL_3017817, EPI_ISL_3017819, EPI_ISL_3017820, EPI_ISL_3017821, EPI_ISL_3017822, EPI_ISL_3017823, EPI_ISL_3017824, EPI_ISL_3017825, EPI_ISL_3017826, EPI_ISL_3017827, EPI_ISL_3017828, EPI_ISL_3017829, EPI_ISL_3017830, EPI_ISL_3017831, EPI_ISL_3017832, EPI_ISL_3017833, EPI_ISL_3017834, EPI_ISL_3017835, EPI_ISL_3017836, EPI_ISL_3017923, EPI_ISL_3017924, EPI_ISL_3017926, EPI_ISL_3017927, EPI_ISL_3017928, EPI_ISL_3017929, EPI_ISL_3017930, EPI_ISL_3017931, EPI_ISL_3017932, EPI_ISL_3017938, EPI_ISL_3017939, EPI_ISL_3017940, EPI_ISL_3017941, EPI_ISL_3017942, EPI_ISL_3017943, EPI_ISL_3017986, EPI_ISL_3018052, EPI_ISL_3018058, EPI_ISL_3018059, EPI_ISL_3018060, EPI_ISL_3018061, EPI_ISL_3018062, EPI_ISL_3018063, EPI_ISL_3018065, EPI_ISL_3018066, EPI_ISL_3018067, EPI_ISL_3018068, EPI_ISL_3018069, EPI_ISL_3018128 | | | |
| see above | Laboratory Corporation of America | Centers for Disease Control and Prevention Division of Viral Diseases, Pathogen Discovery | Adrian Paskey; Amanda Douglas; Amanda Suchanek; Andrea Throop; Ayla Burns; Benjamin Rambo-Martin; Bobbi Croy; Brian Krueger; Brian Norvell; Christopher Gulvick; Christos Petropoulos; Clinton R. Paden; Craig Lukasik; Dakota Howard; Darlene Wagner; Debbie Boles; Dhwani Batra; Duncan MacCannell; Eyad Almasri; Goran Stevovic; Howard Engler; Hrushikesh Deshmukh; Jake Humphrey; Jana Schroth; Jason Caravas; Joe Voshell; John Pruitt; Jonathan Meltzer; Jonathan Williams; Kara Moser; Kimberly Wagner; Lax Iyer; Lisa Pfefferle; Lyndon Tilson; Manoj Jain; Marcia Eisenberg; Mary Ann Cristobal; Mary Williamson; Matthew Robinson; Matthew Schmerer; Michael Levandoski; Mike Sapeta; Mindy Nye; Minoo Agarwal; Mohan Kolli; Nuthawin Charoensri; Oren Cohen; Peter W. Cook; Prashant Gupta; Qian Zeng; Rama Ghatti; Scott Parker; Scott Ryan; Scott Sammons; Shatavia Morrison; Stanley Letovsky; Steven Ragan; Suresh Babu Selvaraju; Susan Countryman; Susan Hicks; Suzanne Dale; Thomas Urban; Tim Kuphal; Tricia Zwiefelhofer; Vincent Drouillon; Yvette Unoarumhi |
| EPI_ISL_2811922, EPI_ISL_2811939, EPI_ISL_2811940, EPI_ISL_2894574, EPI_ISL_2894575, EPI_ISL_2894576, EPI_ISL_2894577, EPI_ISL_2894578, EPI_ISL_2894584, EPI_ISL_2993431 | | | |
| see above | Los Angeles County Public Health Laboratories | Los Angeles County Public Health Laboratories | Hemarajata; P. Hemarajata et al.; P. and Green, N. |
| EPI_ISL_2525019, EPI_ISL_2786157, EPI_ISL_2875247 | Mako Medical | Centers for Disease Control and Prevention Division of Viral Diseases, Pathogen Discovery | Adrian Paskey; Benjamin Rambo-Martin; Christopher Gulvick; Clinton R. Paden; Dakota Howard; Darlene Wagner; Dhwani Batra; Duncan MacCannell; Jason Caravas; Kara Moser; Lauren Moon; Matthew Schmerer; Matthew Tugwell; Peter W. Cook; Scott Sammons; Shatavia Morrison; Yvette Unoarumhi |
| EPI_ISL_3021463 | Marian Medical Center Lab | County of San Luis Obispo Public Health Laboratory | Frances Sidhu; Katrina Erwin |
| EPI_ISL_2907577 | Martin Luther King Jr. Community Hospital | Los Angeles County Public Health Laboratories | P. Hemarajata et al. |
| EPI_ISL_2620826, EPI_ISL_2894590, EPI_ISL_2907562, EPI_ISL_2907563, EPI_ISL_3029854, EPI_ISL_3029870, EPI_ISL_3029871 | | | |
| see above | Martin Luther King, Jr. Outpatient Center | Los Angeles County Public Health Laboratories | P. Hemarajata et al. |
| EPI_ISL_2928302 | Moderna Inc. | Moderna Inc. | Groves Dixon; Rolando Pajon; Yamuna Paila |
| EPI_ISL_2716875, EPI_ISL_2759082, EPI_ISL_2759083, EPI_ISL_2759085, EPI_ISL_2759086, EPI_ISL_2759087, EPI_ISL_2759088, EPI_ISL_2774512, EPI_ISL_2790732, EPI_ISL_2790733, EPI_ISL_2955363, EPI_ISL_3037244 | | | |
| see above | Monterey County Public Health Laboratory | Monterey County Public Health Laboratory | Monterey County Public Health Laboratory |
| EPI_ISL_2663528 | NYU Langone Health | Departments of Pathology and Medicine, New York University School of Medicine | Adriana Heguy; Christian Marier; Dacia Dimartino; Emily Guzman; Gael Westby; Guiqing Wang; Paolo Cotzia; Paul Zappile; Peter Meyn; Sitharam Ramaswami; Yutong Zhang |
| EPI_ISL_2894579, EPI_ISL_2894580, EPI_ISL_2894581, EPI_ISL_2894582, EPI_ISL_2894583, EPI_ISL_3029850, EPI_ISL_3029851, EPI_ISL_3029855, EPI_ISL_3029856, EPI_ISL_3029857, EPI_ISL_3029858, EPI_ISL_3029859, EPI_ISL_3029860, EPI_ISL_3029861 | | | |
| see above | Olive View Medical Center | Los Angeles County Public Health Laboratories | P. Hemarajata et al. |
| EPI_ISL_2566346, EPI_ISL_2566347, EPI_ISL_2566348, EPI_ISL_2566349, EPI_ISL_2566350, EPI_ISL_2566351, EPI_ISL_2566352, EPI_ISL_2566353, EPI_ISL_2566359 | | | |
| see above | Operation Infectious Diseases, Naval Health Research Center | Operation Infectious Diseases, Naval Health Research Center | A.E.; Balansay-Ames; C.A.; Fierro, M.; Iniguez-Stevens, E.; Kriner, P.; M.S.; Moser, K.; Myers; Pan; Phippard; R.S.; R.W.; Underwood |
| EPI_ISL_2496882, EPI_ISL_2496883, EPI_ISL_2496888, EPI_ISL_2500029, EPI_ISL_2500059, EPI_ISL_2500104, EPI_ISL_2500130, EPI_ISL_2500211, EPI_ISL_2658724, EPI_ISL_2659357 | | | |
| see above | Orange County Public Health Lab | Chan-Zuckerberg Biohub | CZB Cliahub Consortium |
| EPI_ISL_2928149, EPI_ISL_2928150, EPI_ISL_2928152, EPI_ISL_2928153, EPI_ISL_2928154, EPI_ISL_2928155, EPI_ISL_2928156, EPI_ISL_2928157, EPI_ISL_2928158, EPI_ISL_2928159, EPI_ISL_2928160, EPI_ISL_2928161, EPI_ISL_2928162, EPI_ISL_2928163, EPI_ISL_2928164, EPI_ISL_2928165, EPI_ISL_2928166, EPI_ISL_2928167, EPI_ISL_2928168, EPI_ISL_2928169, EPI_ISL_2928170, EPI_ISL_2928171, EPI_ISL_2928172, EPI_ISL_2928173, EPI_ISL_2928174, EPI_ISL_2928175, EPI_ISL_2928176, EPI_ISL_2928177, EPI_ISL_2928178, EPI_ISL_2928179, EPI_ISL_2928180, EPI_ISL_2928181, EPI_ISL_2928182, EPI_ISL_2928183, EPI_ISL_2928184, EPI_ISL_2928185, EPI_ISL_2928186, EPI_ISL_2928187, EPI_ISL_2928188, EPI_ISL_2928189, EPI_ISL_2928190, EPI_ISL_2928191, EPI_ISL_2928192, EPI_ISL_2928193, EPI_ISL_2928194, EPI_ISL_2928195, EPI_ISL_2928196, EPI_ISL_2928197, EPI_ISL_2928198, EPI_ISL_2928199, EPI_ISL_2928200, EPI_ISL_2928201, EPI_ISL_2928202, EPI_ISL_2928203, EPI_ISL_2928204, EPI_ISL_2928205, EPI_ISL_2928206, EPI_ISL_2928207, EPI_ISL_2928208, EPI_ISL_2928209, EPI_ISL_2928210, EPI_ISL_2928211, EPI_ISL_2928212, EPI_ISL_2928213, EPI_ISL_2928214, EPI_ISL_2928215, EPI_ISL_2928216, EPI_ISL_2928217, EPI_ISL_2928218, EPI_ISL_2928219, EPI_ISL_3010297, EPI_ISL_3010298, EPI_ISL_3010299, EPI_ISL_3010300, EPI_ISL_3010301, EPI_ISL_3010302, EPI_ISL_3010303, EPI_ISL_3010304, EPI_ISL_3010305, EPI_ISL_3010306, EPI_ISL_3010307, EPI_ISL_3010308, EPI_ISL_3010309, EPI_ISL_3010310, EPI_ISL_3010311, EPI_ISL_3010312, EPI_ISL_3010313, EPI_ISL_3010314, EPI_ISL_3010315, EPI_ISL_3010316, EPI_ISL_3010317, EPI_ISL_3010318, EPI_ISL_3010319, EPI_ISL_3010320, EPI_ISL_3010321 | | | |
| see above | Orange County Public Health Laboratory | Orange County Public Health Laboratory | Angelica Torres; Julia Wolfe |
| EPI_ISL_3462057 | Orange County Public Health Laboratory, Orange County Public Health Laboratory submission group | Orange County Public Health Laboratory, Orange County Public Health Laboratory submission group | Elder; J.W. |
| EPI_ISL_2713066 | Oregon State Public Health Laboratory | Oregon State Public Health Laboratory | Eugene Yeboah; John Fontana and Shane Sevey; Laura Tsaknaridis; Rafia Razzaque; Vanda Makris |
| EPI_ISL_2692513 | Overlake Hospital | Seattle Flu Study | Amanda Adler; Barry R. Lutz; Benjamin Pelle; Brian Hiatt; Caitlin R. Wolf; Chris D. Frazar; Deborah A. Nickerson; Elisabeth Brandstetter; Erica Ryke; Geoff Melly; Helen Y. Chu; Janet A. Englund; Jay Shendure; Jover Lee; Kairsten Fay; Kirsten Lacombe; Lea M. Starita; Mark J. Rieder; Matthew Richardson; Matthew Thompson; Melissa Truong; Michael Boeckh; Michael Famulare; Misja Ilcisin; Peter D. Han; Philip Dykema; Romesh Gautom; Scott Lindquist; Thomas R. Sibley; Trevor Bedford |
| EPI_ISL_2404802, EPI_ISL_2404803, EPI_ISL_2404804, EPI_ISL_2404827 | PAMF PALO ALTO | Santa Clara County public Health Laboratory | Santa Clara County Public Health Department |
| EPI_ISL_2650544, EPI_ISL_2650545, EPI_ISL_2650562, EPI_ISL_2650585, EPI_ISL_2691296, EPI_ISL_2691297, EPI_ISL_2691298, EPI_ISL_2691301, EPI_ISL_2753983, EPI_ISL_2753984, EPI_ISL_2753985, EPI_ISL_2753986, EPI_ISL_2753987, EPI_ISL_2753988, EPI_ISL_2753989, EPI_ISL_2774622, EPI_ISL_2774623, EPI_ISL_2934038, EPI_ISL_2934048 | | | |
| see above | PAMF Palo Alto | Santa Clara County public Health Laboratory | Santa Clara County Public Health Department |
| EPI_ISL_2967972 | PAMF Palo Alto | Santa Clara County public Health Laboratory | Santa Clara County Public Health Department |
| EPI_ISL_2620827, EPI_ISL_2620828, EPI_ISL_2620829, EPI_ISL_2772525, EPI_ISL_2811967, EPI_ISL_2811968, EPI_ISL_2811969, EPI_ISL_2811970, EPI_ISL_2811971, EPI_ISL_2811972, EPI_ISL_2811973, EPI_ISL_2811974, EPI_ISL_2811975, EPI_ISL_2811976, EPI_ISL_2811977, EPI_ISL_2811978, EPI_ISL_2811979, EPI_ISL_2812006, EPI_ISL_2812007, EPI_ISL_2812008, EPI_ISL_2812009, EPI_ISL_2812010, EPI_ISL_2812011, EPI_ISL_2812012, EPI_ISL_2812013, EPI_ISL_2812014, EPI_ISL_2894591, EPI_ISL_2894592 | | | |
| see above | Palmdale Regional Medical Center | Los Angeles County Public Health Laboratories | P. Hemarajata et al. |
| EPI_ISL_2391751, EPI_ISL_2392168, EPI_ISL_2404847, EPI_ISL_2405178, EPI_ISL_2405202, EPI_ISL_2405203, EPI_ISL_2405204, EPI_ISL_2617942, EPI_ISL_2772505, EPI_ISL_2811999, EPI_ISL_2812000, EPI_ISL_2894572, EPI_ISL_2894573 | | | |
| see above | Pomona Valley Hospital Medical Center | Los Angeles County Public Health Laboratories | P. Hemarajata et al. |
| EPI_ISL_2907567, EPI_ISL_2907568 | Providence Saint John's Health Center | Los Angeles County Public Health Laboratories | P. Hemarajata et al. |
| EPI_ISL_2391792, EPI_ISL_2405206 | Providence Saint Joseph Medical Center | Los Angeles County Public Health Laboratories | P. Hemarajata et al. |
| EPI_ISL_2179946, EPI_ISL_2179950, EPI_ISL_2179965, EPI_ISL_2179981, EPI_ISL_2179983, EPI_ISL_2180495, EPI_ISL_2180519, EPI_ISL_2180530, EPI_ISL_2241634, EPI_ISL_2247062, EPI_ISL_2247065, EPI_ISL_2247067, EPI_ISL_2247074, EPI_ISL_2247075, EPI_ISL_2247079, EPI_ISL_2247080, EPI_ISL_2247084, EPI_ISL_2247085, EPI_ISL_2247087, EPI_ISL_2247090, EPI_ISL_2247094, EPI_ISL_2247106, EPI_ISL_2247112, EPI_ISL_2247138, EPI_ISL_2247139, EPI_ISL_2247143, EPI_ISL_2247144, EPI_ISL_2247157, EPI_ISL_2247161, EPI_ISL_2247162, EPI_ISL_2247172, EPI_ISL_2247183, EPI_ISL_2247186, EPI_ISL_2247187, EPI_ISL_2247190, EPI_ISL_2247191, EPI_ISL_2247198, EPI_ISL_2247205, EPI_ISL_2247337, EPI_ISL_2248473, EPI_ISL_2248560, EPI_ISL_2248580, EPI_ISL_2248585, EPI_ISL_2248589, EPI_ISL_2248602, EPI_ISL_2248603, EPI_ISL_2248606, EPI_ISL_2248607, EPI_ISL_2248612, EPI_ISL_2248613, EPI_ISL_2248614, EPI_ISL_2248621, EPI_ISL_2248622, EPI_ISL_2248624, EPI_ISL_2248628, EPI_ISL_2248637, EPI_ISL_2248641, EPI_ISL_2248642, EPI_ISL_2248655, EPI_ISL_2248667, EPI_ISL_2248669, EPI_ISL_2248687, EPI_ISL_2248689, EPI_ISL_2248690, EPI_ISL_2268287, EPI_ISL_2268291, EPI_ISL_2268295, EPI_ISL_2268379, EPI_ISL_2268380, EPI_ISL_2268389, EPI_ISL_2268598, EPI_ISL_2268672, EPI_ISL_2268690, EPI_ISL_2268746, EPI_ISL_2268807, EPI_ISL_2268809, EPI_ISL_2268829, EPI_ISL_2268874, EPI_ISL_2268882, EPI_ISL_2268883, EPI_ISL_2268899, EPI_ISL_2268916, EPI_ISL_2268937, EPI_ISL_2268942, EPI_ISL_2268945, EPI_ISL_2268946, EPI_ISL_2268950, EPI_ISL_2268951, EPI_ISL_2268952, EPI_ISL_2268958, EPI_ISL_2268960, EPI_ISL_2268961, EPI_ISL_2268962, EPI_ISL_2268963, EPI_ISL_2268964, EPI_ISL_2268969, EPI_ISL_2268970, EPI_ISL_2268972, EPI_ISL_2268974, EPI_ISL_2268975, EPI_ISL_2268977, EPI_ISL_2268979, EPI_ISL_2268980, EPI_ISL_2268981, EPI_ISL_2268983, EPI_ISL_2268984, EPI_ISL_2268985, EPI_ISL_2268987, EPI_ISL_2268989, EPI_ISL_2268990, EPI_ISL_2268992, EPI_ISL_2268993, EPI_ISL_2268995, EPI_ISL_2268996, EPI_ISL_2269000, EPI_ISL_2269001, EPI_ISL_2269004, EPI_ISL_2269005, EPI_ISL_2269007, EPI_ISL_2269008, EPI_ISL_2269014, EPI_ISL_2269017, EPI_ISL_2269019, EPI_ISL_2269020, EPI_ISL_2269022, EPI_ISL_2269023, EPI_ISL_2269024, EPI_ISL_2269025, EPI_ISL_2269029, EPI_ISL_2269031, EPI_ISL_2269034, EPI_ISL_2269044, EPI_ISL_2269045, EPI_ISL_2269046, EPI_ISL_2269048, EPI_ISL_2269051, EPI_ISL_2269053, EPI_ISL_2269054, EPI_ISL_2269055, EPI_ISL_2269058, EPI_ISL_2269059, EPI_ISL_2269060, EPI_ISL_2269061, EPI_ISL_2269062, EPI_ISL_2269063, EPI_ISL_2269064, EPI_ISL_2269065, EPI_ISL_2269066, EPI_ISL_2269067, EPI_ISL_2269068, EPI_ISL_2269069, EPI_ISL_2269070, EPI_ISL_2269071, EPI_ISL_2269072, EPI_ISL_2269073, EPI_ISL_2269093, EPI_ISL_2269125, EPI_ISL_2269210, EPI_ISL_2280114, EPI_ISL_2280123, EPI_ISL_2280126, EPI_ISL_2280136, EPI_ISL_2280138, EPI_ISL_2280139, EPI_ISL_2280140, EPI_ISL_2280143, EPI_ISL_2280144, EPI_ISL_2280145, EPI_ISL_2280148, EPI_ISL_2280149, EPI_ISL_2280150, EPI_ISL_2280152, EPI_ISL_2280154, EPI_ISL_2280155, EPI_ISL_2280156, EPI_ISL_2304503, EPI_ISL_2304506, EPI_ISL_2304510, EPI_ISL_2304511, EPI_ISL_2304518, EPI_ISL_2304523, EPI_ISL_2304526, EPI_ISL_2304527, EPI_ISL_2304529, EPI_ISL_2304531, EPI_ISL_2304533, EPI_ISL_2304534, EPI_ISL_2304543, EPI_ISL_2304554, EPI_ISL_2304559, EPI_ISL_2304561, EPI_ISL_2304563, EPI_ISL_2304568, EPI_ISL_2304571, EPI_ISL_2304574, EPI_ISL_2304576, EPI_ISL_2304579, EPI_ISL_2304580, EPI_ISL_2304588, EPI_ISL_2304591, EPI_ISL_2304593, EPI_ISL_2304600, EPI_ISL_2304612, EPI_ISL_2304614, EPI_ISL_2304618, EPI_ISL_2304619, EPI_ISL_2304623, EPI_ISL_2304629, EPI_ISL_2304633, EPI_ISL_2304634, EPI_ISL_2304635, EPI_ISL_2304639, EPI_ISL_2307140, EPI_ISL_2307141, EPI_ISL_2307142, EPI_ISL_2307145, EPI_ISL_2307146, EPI_ISL_2307151, EPI_ISL_2367680, EPI_ISL_2367688, EPI_ISL_2367689, EPI_ISL_2367694, EPI_ISL_2367695, EPI_ISL_2367697, EPI_ISL_2367702, EPI_ISL_2367711, EPI_ISL_2367714, EPI_ISL_2367718, EPI_ISL_2367722, EPI_ISL_2367726, EPI_ISL_2367728, EPI_ISL_2367732, EPI_ISL_2367736, EPI_ISL_2367738, EPI_ISL_2367741, EPI_ISL_2367745, EPI_ISL_2367748, EPI_ISL_2367750, EPI_ISL_2367751, EPI_ISL_2367806, EPI_ISL_2367807, EPI_ISL_2367811, | | | |

EPI_ISL_2367825, EPI_ISL_2367852, EPI_ISL_2367881, EPI_ISL_2367882, EPI_ISL_2367894, EPI_ISL_2367907, EPI_ISL_2367909, EPI_ISL_2367917, EPI_ISL_2367938, EPI_ISL_2367943, EPI_ISL_2367948, EPI_ISL_2367949, EPI_ISL_2367956, EPI_ISL_2367958, EPI_ISL_2367961, EPI_ISL_2367971, EPI_ISL_2367974, EPI_ISL_2367976, EPI_ISL_2367978, EPI_ISL_2367980, EPI_ISL_2367983, EPI_ISL_2367984,
EPI_ISL_2367985, EPI_ISL_2367987, EPI_ISL_2367996, EPI_ISL_2367999, EPI_ISL_2368001, EPI_ISL_2368002, EPI_ISL_2368003, EPI_ISL_2368006, EPI_ISL_2368010, EPI_ISL_2368012, EPI_ISL_2368016, EPI_ISL_2368021, EPI_ISL_2368023, EPI_ISL_2368024, EPI_ISL_2368025, EPI_ISL_2368026, EPI_ISL_2368031, EPI_ISL_2368033, EPI_ISL_2368043, EPI_ISL_2368045, EPI_ISL_2368046, EPI_ISL_2368048,
EPI_ISL_2368049, EPI_ISL_2368053, EPI_ISL_2368055, EPI_ISL_2368056, EPI_ISL_2368059, EPI_ISL_2368062, EPI_ISL_2368065, EPI_ISL_2369109, EPI_ISL_2369127, EPI_ISL_2369192, EPI_ISL_2369396, EPI_ISL_2371691, EPI_ISL_2371696, EPI_ISL_2371697, EPI_ISL_2371699, EPI_ISL_2371703, EPI_ISL_2371721, EPI_ISL_2371726, EPI_ISL_2371728, EPI_ISL_2371729, EPI_ISL_2371731, EPI_ISL_2371733,
EPI_ISL_2371736, EPI_ISL_2371748, EPI_ISL_2371774, EPI_ISL_2371869, EPI_ISL_2371872, EPI_ISL_2371873, EPI_ISL_2371875, EPI_ISL_2371876, EPI_ISL_2371877, EPI_ISL_2371878, EPI_ISL_2371886, EPI_ISL_2371892, EPI_ISL_2371909, EPI_ISL_2371910, EPI_ISL_2371923, EPI_ISL_2371927, EPI_ISL_2371934, EPI_ISL_2371938, EPI_ISL_2371942, EPI_ISL_2371946, EPI_ISL_2371948, EPI_ISL_2371950,
EPI_ISL_2371955, EPI_ISL_2371965, EPI_ISL_2371967, EPI_ISL_2371971, EPI_ISL_2371973, EPI_ISL_2371978, EPI_ISL_2372140, EPI_ISL_2372141, EPI_ISL_2372142, EPI_ISL_2372147, EPI_ISL_2372149, EPI_ISL_2372158, EPI_ISL_2372159, EPI_ISL_2372162, EPI_ISL_2372165, EPI_ISL_2372167, EPI_ISL_2372172, EPI_ISL_2372175, EPI_ISL_2372183, EPI_ISL_2372187, EPI_ISL_2372190, EPI_ISL_2372194,
EPI_ISL_2372206, EPI_ISL_2372207, EPI_ISL_2372211, EPI_ISL_2372219, EPI_ISL_2372220, EPI_ISL_2372224, EPI_ISL_2372227, EPI_ISL_2372228, EPI_ISL_2372229, EPI_ISL_2372234, EPI_ISL_2372240, EPI_ISL_2372248, EPI_ISL_2372250, EPI_ISL_2372251, EPI_ISL_2372253, EPI_ISL_2372255, EPI_ISL_2372258, EPI_ISL_2372922, EPI_ISL_2372927, EPI_ISL_2372944, EPI_ISL_2372951, EPI_ISL_2397413,
EPI_ISL_2397414, EPI_ISL_2397420, EPI_ISL_2397421, EPI_ISL_2397422, EPI_ISL_2397424, EPI_ISL_2397425, EPI_ISL_2397427, EPI_ISL_2397432, EPI_ISL_2397434, EPI_ISL_2397441, EPI_ISL_2397442, EPI_ISL_2397443, EPI_ISL_2397445, EPI_ISL_2397447, EPI_ISL_2397449, EPI_ISL_2397454, EPI_ISL_2397461, EPI_ISL_2397462, EPI_ISL_2397469, EPI_ISL_2397473, EPI_ISL_2397481, EPI_ISL_2397483,
EPI_ISL_2397488, EPI_ISL_2397492, EPI_ISL_2397497, EPI_ISL_2397502, EPI_ISL_2397505, EPI_ISL_2397510, EPI_ISL_2397514, EPI_ISL_2397520, EPI_ISL_2397523, EPI_ISL_2397524, EPI_ISL_2397526, EPI_ISL_2397529, EPI_ISL_2397532, EPI_ISL_2397537, EPI_ISL_2397541, EPI_ISL_2397548, EPI_ISL_2397550, EPI_ISL_2397638, EPI_ISL_2399476, EPI_ISL_2399478, EPI_ISL_2399480, EPI_ISL_2399484,
EPI_ISL_2439320, EPI_ISL_2439365, EPI_ISL_2439384, EPI_ISL_2439409, EPI_ISL_2439415, EPI_ISL_2439425, EPI_ISL_2439428, EPI_ISL_2439430, EPI_ISL_2439431, EPI_ISL_2439440, EPI_ISL_2439442, EPI_ISL_2439443, EPI_ISL_2439444, EPI_ISL_2439451, EPI_ISL_2439452, EPI_ISL_2439454, EPI_ISL_2439462, EPI_ISL_2439464, EPI_ISL_2439465, EPI_ISL_2439466, EPI_ISL_2439467, EPI_ISL_2439468,
EPI_ISL_2439469, EPI_ISL_2439471, EPI_ISL_2439474, EPI_ISL_2439475, EPI_ISL_2439481, EPI_ISL_2439486, EPI_ISL_2439488, EPI_ISL_2439490, EPI_ISL_2439495, EPI_ISL_2439497, EPI_ISL_2439501, EPI_ISL_2439502, EPI_ISL_2439503, EPI_ISL_2439514, EPI_ISL_2439516, EPI_ISL_2439521, EPI_ISL_2439522, EPI_ISL_2439523, EPI_ISL_2439524, EPI_ISL_2439525, EPI_ISL_2439528, EPI_ISL_2439529,
EPI_ISL_2439531, EPI_ISL_2439534, EPI_ISL_2439539, EPI_ISL_2439541, EPI_ISL_2439542, EPI_ISL_2439544, EPI_ISL_2439545, EPI_ISL_2439549, EPI_ISL_2439551, EPI_ISL_2439552, EPI_ISL_2439556, EPI_ISL_2439559, EPI_ISL_2439560, EPI_ISL_2439564, EPI_ISL_2439700, EPI_ISL_2439712, EPI_ISL_2439719, EPI_ISL_2439732, EPI_ISL_2439743, EPI_ISL_2439744, EPI_ISL_2439746, EPI_ISL_2439755,
EPI_ISL_2439757, EPI_ISL_2439762, EPI_ISL_2439770, EPI_ISL_2439773, EPI_ISL_2439777, EPI_ISL_2439778, EPI_ISL_2439787, EPI_ISL_2439801, EPI_ISL_2439813, EPI_ISL_2439814, EPI_ISL_2439818, EPI_ISL_2439827, EPI_ISL_2440687, EPI_ISL_2440777, EPI_ISL_2440781, EPI_ISL_2440794, EPI_ISL_2440815, EPI_ISL_2440820, EPI_ISL_2440862, EPI_ISL_2440876, EPI_ISL_2451278, EPI_ISL_2451279,
EPI_ISL_2451282, EPI_ISL_2451292, EPI_ISL_2451294, EPI_ISL_2451360, EPI_ISL_2451386, EPI_ISL_2451423, EPI_ISL_2451428, EPI_ISL_2451429, EPI_ISL_2451434, EPI_ISL_2451436, EPI_ISL_2451439, EPI_ISL_2463023, EPI_ISL_2463037, EPI_ISL_2463049, EPI_ISL_2463055, EPI_ISL_2463097, EPI_ISL_2463102, EPI_ISL_2463105, EPI_ISL_2463125, EPI_ISL_2463126, EPI_ISL_2463135, EPI_ISL_2463147,
EPI_ISL_2463149, EPI_ISL_2463152, EPI_ISL_2463154, EPI_ISL_2463157, EPI_ISL_2463160, EPI_ISL_2463161, EPI_ISL_2463168, EPI_ISL_2463170, EPI_ISL_2463171, EPI_ISL_2463172, EPI_ISL_2463174, EPI_ISL_2463175, EPI_ISL_2463176, EPI_ISL_2463180, EPI_ISL_2463181, EPI_ISL_2463184, EPI_ISL_2463186, EPI_ISL_2463187, EPI_ISL_2463188, EPI_ISL_2463190, EPI_ISL_2463192, EPI_ISL_2463197,
EPI_ISL_2463199, EPI_ISL_2463200, EPI_ISL_2463201, EPI_ISL_2463205, EPI_ISL_2463209, EPI_ISL_2463211, EPI_ISL_2463213, EPI_ISL_2463214, EPI_ISL_2463215, EPI_ISL_2463220, EPI_ISL_2463221, EPI_ISL_2463225, EPI_ISL_2463226, EPI_ISL_2463227, EPI_ISL_2463229, EPI_ISL_2463230, EPI_ISL_2463232, EPI_ISL_2463235, EPI_ISL_2463237, EPI_ISL_2463239, EPI_ISL_2463241, EPI_ISL_2463243,
EPI_ISL_2463244, EPI_ISL_2463246, EPI_ISL_2463248, EPI_ISL_2463251, EPI_ISL_2463253, EPI_ISL_2463254, EPI_ISL_2463256, EPI_ISL_2463258, EPI_ISL_2463262, EPI_ISL_2463265, EPI_ISL_2463266, EPI_ISL_2463268, EPI_ISL_2463269, EPI_ISL_2463270, EPI_ISL_2463281, EPI_ISL_2488824, EPI_ISL_2488833, EPI_ISL_2488835, EPI_ISL_2488845, EPI_ISL_2488846, EPI_ISL_2488847, EPI_ISL_2488849,
EPI_ISL_2488853, EPI_ISL_2488854, EPI_ISL_2488855, EPI_ISL_2526803, EPI_ISL_2526859, EPI_ISL_2526931, EPI_ISL_2527016, EPI_ISL_2529091, EPI_ISL_2529127, EPI_ISL_2529155, EPI_ISL_2529159, EPI_ISL_2529183, EPI_ISL_2529295, EPI_ISL_2529299, EPI_ISL_2529312, EPI_ISL_2529314, EPI_ISL_2529331, EPI_ISL_2529333, EPI_ISL_2534529, EPI_ISL_2534554, EPI_ISL_2534564, EPI_ISL_2599782,
EPI_ISL_2599858, EPI_ISL_2599886, EPI_ISL_2611825, EPI_ISL_2611835, EPI_ISL_2611836, EPI_ISL_2611839, EPI_ISL_2611853, EPI_ISL_2649976, EPI_ISL_2649978, EPI_ISL_2649985, EPI_ISL_2649990, EPI_ISL_2652368, EPI_ISL_2652376, EPI_ISL_2652377, EPI_ISL_2652382, EPI_ISL_2652401, EPI_ISL_2686378, EPI_ISL_2868689, EPI_ISL_2868691, EPI_ISL_2868695, EPI_ISL_2868702, EPI_ISL_2868709,
EPI_ISL_2868716, EPI_ISL_2868717, EPI_ISL_2868722, EPI_ISL_2868730, EPI_ISL_2868731, EPI_ISL_2868734, EPI_ISL_2868737, EPI_ISL_2868740, EPI_ISL_2868742, EPI_ISL_2868743, EPI_ISL_2868748, EPI_ISL_2868752, EPI_ISL_2868756, EPI_ISL_2868761, EPI_ISL_2868763, EPI_ISL_2868767, EPI_ISL_2868769, EPI_ISL_2868772, EPI_ISL_2868773, EPI_ISL_2868775, EPI_ISL_2868776, EPI_ISL_2868778,
EPI_ISL_2868780, EPI_ISL_2868782, EPI_ISL_2868787, EPI_ISL_2868789, EPI_ISL_2868795, EPI_ISL_2868797, EPI_ISL_2868802, EPI_ISL_2868804, EPI_ISL_2868806, EPI_ISL_2868810, EPI_ISL_2868812, EPI_ISL_2868814, EPI_ISL_2868823, EPI_ISL_2868824, EPI_ISL_2868826, EPI_ISL_2868830, EPI_ISL_2868832, EPI_ISL_2868834, EPI_ISL_2868836, EPI_ISL_2868839, EPI_ISL_2868841, EPI_ISL_2868842,
EPI_ISL_2868846, EPI_ISL_2868858, EPI_ISL_2868863, EPI_ISL_2868870, EPI_ISL_2868871, EPI_ISL_2868872, EPI_ISL_2868885, EPI_ISL_2868898, EPI_ISL_2868903, EPI_ISL_2868908, EPI_ISL_2868924, EPI_ISL_2868948, EPI_ISL_2868951, EPI_ISL_2868952, EPI_ISL_2868953, EPI_ISL_2868960, EPI_ISL_2868965, EPI_ISL_2868970, EPI_ISL_2868972, EPI_ISL_2868976, EPI_ISL_2868983, EPI_ISL_2868985,
EPI_ISL_2868988, EPI_ISL_2868994, EPI_ISL_2868999, EPI_ISL_2869004, EPI_ISL_2869005, EPI_ISL_2869011, EPI_ISL_2869012, EPI_ISL_2869013, EPI_ISL_2869017, EPI_ISL_2869025, EPI_ISL_2869033, EPI_ISL_2869039, EPI_ISL_2869042, EPI_ISL_2869044, EPI_ISL_2869048, EPI_ISL_2869058, EPI_ISL_2869061, EPI_ISL_2869070, EPI_ISL_2869074, EPI_ISL_2869081, EPI_ISL_2869085, EPI_ISL_2869086,
EPI_ISL_2869091, EPI_ISL_2869093, EPI_ISL_2869094, EPI_ISL_2869095, EPI_ISL_2869100, EPI_ISL_2869103, EPI_ISL_2869105, EPI_ISL_2869106, EPI_ISL_2869110, EPI_ISL_2869111, EPI_ISL_2869114, EPI_ISL_2869136, EPI_ISL_2869139, EPI_ISL_2869142, EPI_ISL_2869145, EPI_ISL_2869154, EPI_ISL_2869163, EPI_ISL_2869164, EPI_ISL_2869177, EPI_ISL_2869178, EPI_ISL_2869182, EPI_ISL_2869183,
EPI_ISL_2869192, EPI_ISL_2869194, EPI_ISL_2869195, EPI_ISL_2869207, EPI_ISL_2869210, EPI_ISL_2869212, EPI_ISL_2869221, EPI_ISL_2869231, EPI_ISL_2869243, EPI_ISL_2869245, EPI_ISL_2869250, EPI_ISL_2869258, EPI_ISL_2869265, EPI_ISL_2869276, EPI_ISL_2869280, EPI_ISL_2869284, EPI_ISL_2869289, EPI_ISL_2869313, EPI_ISL_2869322, EPI_ISL_2869331, EPI_ISL_2869337, EPI_ISL_2869357,
EPI_ISL_2869358, EPI_ISL_2869368, EPI_ISL_2869369, EPI_ISL_2869371, EPI_ISL_2869374, EPI_ISL_2869383, EPI_ISL_2869386, EPI_ISL_2869392, EPI_ISL_2869397, EPI_ISL_2869398, EPI_ISL_2869405, EPI_ISL_2869411, EPI_ISL_2869417, EPI_ISL_2869419, EPI_ISL_2869424, EPI_ISL_2869425, EPI_ISL_2869428, EPI_ISL_2869437, EPI_ISL_2869438, EPI_ISL_2869449, EPI_ISL_2869451, EPI_ISL_2869454,
EPI_ISL_2869465, EPI_ISL_2869476, EPI_ISL_2869487, EPI_ISL_2869495, EPI_ISL_2869496, EPI_ISL_2869502, EPI_ISL_2869503, EPI_ISL_2869509, EPI_ISL_2869519, EPI_ISL_2869528, EPI_ISL_2869529, EPI_ISL_2869536, EPI_ISL_2869540, EPI_ISL_2869569, EPI_ISL_2869573, EPI_ISL_2869578, EPI_ISL_2869585, EPI_ISL_2869587, EPI_ISL_2869589, EPI_ISL_2869594, EPI_ISL_2869600, EPI_ISL_2869602,
EPI_ISL_2869608, EPI_ISL_2869638, EPI_ISL_2869646, EPI_ISL_2869652, EPI_ISL_2869654, EPI_ISL_2869655, EPI_ISL_2869659, EPI_ISL_2869660, EPI_ISL_2869661, EPI_ISL_2869662, EPI_ISL_2869665, EPI_ISL_2869670, EPI_ISL_2869675, EPI_ISL_2869676, EPI_ISL_2869679, EPI_ISL_2869680, EPI_ISL_2869681, EPI_ISL_2869687, EPI_ISL_2869688, EPI_ISL_2869696, EPI_ISL_2869700, EPI_ISL_2869705,
EPI_ISL_2869707, EPI_ISL_2869712, EPI_ISL_2869714, EPI_ISL_2869716, EPI_ISL_2869719, EPI_ISL_2869725, EPI_ISL_2869727, EPI_ISL_2869729, EPI_ISL_2869732, EPI_ISL_2869733, EPI_ISL_2869739, EPI_ISL_2869748, EPI_ISL_2869760, EPI_ISL_2869763, EPI_ISL_2869773, EPI_ISL_2869789, EPI_ISL_2869792, EPI_ISL_2869795, EPI_ISL_2869810, EPI_ISL_2869817, EPI_ISL_2869833, EPI_ISL_2869839,
EPI_ISL_2869842, EPI_ISL_2869854, EPI_ISL_2869857, EPI_ISL_2869863, EPI_ISL_2869866, EPI_ISL_2869881, EPI_ISL_2869892, EPI_ISL_2869898, EPI_ISL_2869905, EPI_ISL_2869916, EPI_ISL_2869958, EPI_ISL_2869967, EPI_ISL_2869979, EPI_ISL_2869982, EPI_ISL_2869987, EPI_ISL_2869993, EPI_ISL_2870016, EPI_ISL_2870027, EPI_ISL_2870044, EPI_ISL_2870052, EPI_ISL_2870070, EPI_ISL_2870072,
EPI_ISL_2870086, EPI_ISL_2870089, EPI_ISL_2870092, EPI_ISL_2870098, EPI_ISL_2870106, EPI_ISL_2870120, EPI_ISL_2870127, EPI_ISL_2870138, EPI_ISL_2870164, EPI_ISL_2870173, EPI_ISL_2870176, EPI_ISL_2870199, EPI_ISL_2870205, EPI_ISL_2870208, EPI_ISL_2870217, EPI_ISL_2870240, EPI_ISL_2870246, EPI_ISL_2870254, EPI_ISL_2870263, EPI_ISL_2870266, EPI_ISL_2870269, EPI_ISL_2870274,
EPI_ISL_2870277, EPI_ISL_2870286, EPI_ISL_2870295, EPI_ISL_2870307, EPI_ISL_2870711, EPI_ISL_2870856, EPI_ISL_2870945, EPI_ISL_2870969, EPI_ISL_2871039, EPI_ISL_2871148, EPI_ISL_2871169, EPI_ISL_2871231, EPI_ISL_2871243, EPI_ISL_2871254, EPI_ISL_2871296, EPI_ISL_2871329, EPI_ISL_2871433, EPI_ISL_2871448, EPI_ISL_2871521, EPI_ISL_2871535, EPI_ISL_2871661, EPI_ISL_2871684,
EPI_ISL_2871693, EPI_ISL_2871710, EPI_ISL_2871890, EPI_ISL_2872001, EPI_ISL_2872115, EPI_ISL_2873888, EPI_ISL_2873901, EPI_ISL_2873903, EPI_ISL_2873911, EPI_ISL_2873987, EPI_ISL_2874006, EPI_ISL_2874034, EPI_ISL_2874069, EPI_ISL_2874076, EPI_ISL_2874096, EPI_ISL_2874112, EPI_ISL_2874117, EPI_ISL_2874147, EPI_ISL_2874154, EPI_ISL_2874162, EPI_ISL_2874163, EPI_ISL_2874165,
EPI_ISL_2874219, EPI_ISL_2874221, EPI_ISL_2874238, EPI_ISL_2874258, EPI_ISL_2874273, EPI_ISL_2874281, EPI_ISL_2874304, EPI_ISL_2874313, EPI_ISL_2874317, EPI_ISL_2874327, EPI_ISL_2874329, EPI_ISL_2874349, EPI_ISL_2874350, EPI_ISL_2874373, EPI_ISL_2874377, EPI_ISL_2874384, EPI_ISL_2874395, EPI_ISL_2874401, EPI_ISL_2874404, EPI_ISL_2874408, EPI_ISL_2874411, EPI_ISL_2874418,
EPI_ISL_2874437, EPI_ISL_2874445, EPI_ISL_2874450, EPI_ISL_2874455, EPI_ISL_2874463, EPI_ISL_2874466, EPI_ISL_2874469, EPI_ISL_2874484, EPI_ISL_2874500, EPI_ISL_2874501, EPI_ISL_2874514, EPI_ISL_2874515, EPI_ISL_2874518, EPI_ISL_2874528, EPI_ISL_2874529, EPI_ISL_2874531, EPI_ISL_2874533, EPI_ISL_2874538, EPI_ISL_2874549, EPI_ISL_2874554, EPI_ISL_2874562, EPI_ISL_2874564,
EPI_ISL_2874571, EPI_ISL_2874572, EPI_ISL_2874573, EPI_ISL_2874598, EPI_ISL_2874599, EPI_ISL_2874607, EPI_ISL_2874611, EPI_ISL_2874622, EPI_ISL_2874632, EPI_ISL_2874636, EPI_ISL_2874641, EPI_ISL_2874648, EPI_ISL_2874649, EPI_ISL_2874678, EPI_ISL_2874685, EPI_ISL_2874687, EPI_ISL_2874690, EPI_ISL_2874691, EPI_ISL_2874693, EPI_ISL_2874695, EPI_ISL_2874703, EPI_ISL_2874707,
EPI_ISL_2874708, EPI_ISL_2874711, EPI_ISL_2874718, EPI_ISL_2874743, EPI_ISL_2874756, EPI_ISL_2874758, EPI_ISL_2874760, EPI_ISL_2874767, EPI_ISL_2874768, EPI_ISL_2874769, EPI_ISL_2874776, EPI_ISL_2874779, EPI_ISL_2874784, EPI_ISL_2874786, EPI_ISL_2874787, EPI_ISL_2874792, EPI_ISL_2874796, EPI_ISL_2874802, EPI_ISL_2874806, EPI_ISL_2874809, EPI_ISL_2874810, EPI_ISL_2874811,
EPI_ISL_2874813, EPI_ISL_2874814, EPI_ISL_2874822, EPI_ISL_2874826, EPI_ISL_2874827, EPI_ISL_2874831, EPI_ISL_2874832, EPI_ISL_2874842, EPI_ISL_2874843, EPI_ISL_2874847, EPI_ISL_2874868, EPI_ISL_2874881, EPI_ISL_2874888, EPI_ISL_2874889, EPI_ISL_2874892, EPI_ISL_2874895, EPI_ISL_2874899, EPI_ISL_2874904, EPI_ISL_2874908, EPI_ISL_2874909, EPI_ISL_2874918, EPI_ISL_2874919,
EPI_ISL_2874925, EPI_ISL_2874927, EPI_ISL_2874929, EPI_ISL_2874930, EPI_ISL_2874939, EPI_ISL_2874941, EPI_ISL_2874943, EPI_ISL_2874946, EPI_ISL_2874948, EPI_ISL_2874956, EPI_ISL_2874957, EPI_ISL_2874962, EPI_ISL_2874964, EPI_ISL_2874972, EPI_ISL_2874973, EPI_ISL_2874974, EPI_ISL_2874975, EPI_ISL_2874982, EPI_ISL_2874983, EPI_ISL_2875000, EPI_ISL_2875006, EPI_ISL_2875012,
EPI_ISL_2875013, EPI_ISL_2875021, EPI_ISL_2875027, EPI_ISL_2875028, EPI_ISL_2875034, EPI_ISL_2875036, EPI_ISL_2875040, EPI_ISL_2875043, EPI_ISL_2875053, EPI_ISL_2875057, EPI_ISL_2875059, EPI_ISL_2875062, EPI_ISL_2875065, EPI_ISL_2875072, EPI_ISL_2875078, EPI_ISL_2875082, EPI_ISL_2875084, EPI_ISL_2875088, EPI_ISL_2875091, EPI_ISL_2875095, EPI_ISL_2875103, EPI_ISL_2875104,
EPI_ISL_2875105, EPI_ISL_2875106, EPI_ISL_2875119, EPI_ISL_2875123, EPI_ISL_2875124, EPI_ISL_2875129, EPI_ISL_2875130, EPI_ISL_2875135, EPI_ISL_2875144, EPI_ISL_2875151, EPI_ISL_2875152, EPI_ISL_2875155, EPI_ISL_2875165, EPI_ISL_2876105, EPI_ISL_2876117, EPI_ISL_2876124, EPI_ISL_2876126, EPI_ISL_2876136, EPI_ISL_2876138, EPI_ISL_2876142, EPI_ISL_2876146, EPI_ISL_2876149,
EPI_ISL_2876150, EPI_ISL_2876152, EPI_ISL_2876158, EPI_ISL_2876165, EPI_ISL_2876167, EPI_ISL_2876643, EPI_ISL_2876667, EPI_ISL_2876674, EPI_ISL_2876678, EPI_ISL_2876680, EPI_ISL_2876685, EPI_ISL_2876686, EPI_ISL_2876690, EPI_ISL_2876691, EPI_ISL_2876692, EPI_ISL_2876699, EPI_ISL_2876703, EPI_ISL_2876716, EPI_ISL_2876728, EPI_ISL_2876736, EPI_ISL_2876737, EPI_ISL_2876742,
EPI_ISL_2876746, EPI_ISL_2876748, EPI_ISL_2876754, EPI_ISL_2876759, EPI_ISL_2876763, EPI_ISL_2876766, EPI_ISL_2876767, EPI_ISL_2876777, EPI_ISL_2876785, EPI_ISL_2876787, EPI_ISL_2876794, EPI_ISL_2876796, EPI_ISL_2876800, EPI_ISL_2876802, EPI_ISL_2876809, EPI_ISL_2876812, EPI_ISL_2876820, EPI_ISL_2876821, EPI_ISL_2876836, EPI_ISL_2876840, EPI_ISL_2876850, EPI_ISL_2876854,
EPI_ISL_2876863, EPI_ISL_2876869, EPI_ISL_2876871, EPI_ISL_2876873, EPI_ISL_2876897, EPI_ISL_2876900, EPI_ISL_2876903, EPI_ISL_2876911, EPI_ISL_2876913, EPI_ISL_2876924, EPI_ISL_2876926, EPI_ISL_2876928, EPI_ISL_2876931, EPI_ISL_2929168, EPI_ISL_2929172, EPI_ISL_2929173, EPI_ISL_2929178, EPI_ISL_2929180, EPI_ISL_2929181, EPI_ISL_2929186, EPI_ISL_2929187, EPI_ISL_2929189,
EPI_ISL_2929192, EPI_ISL_2929212, EPI_ISL_2929217, EPI_ISL_2929222, EPI_ISL_2929223, EPI_ISL_2929232, EPI_ISL_2929239, EPI_ISL_2929242, EPI_ISL_2929243, EPI_ISL_2929246, EPI_ISL_2929247, EPI_ISL_2929248, EPI_ISL_2929256, EPI_ISL_2929268, EPI_ISL_2929269, EPI_ISL_2929270, EPI_ISL_2929272, EPI_ISL_2929273, EPI_ISL_2929274, EPI_ISL_2929284, EPI_ISL_2929289, EPI_ISL_2929292,
EPI_ISL_2929293, EPI_ISL_2929297, EPI_ISL_2929308, EPI_ISL_2929310, EPI_ISL_2929311, EPI_ISL_2929314, EPI_ISL_2929317, EPI_ISL_2929319, EPI_ISL_2929323, EPI_ISL_2929324, EPI_ISL_2929325, EPI_ISL_2929327, EPI_ISL_2929331, EPI_ISL_2929332, EPI_ISL_2929334, EPI_ISL_2929340, EPI_ISL_2929343, EPI_ISL_2929355, EPI_ISL_2929362, EPI_ISL_2929369, EPI_ISL_2929377, EPI_ISL_2929378,
EPI_ISL_2929381, EPI_ISL_2929382, EPI_ISL_2929385, EPI_ISL_2929388, EPI_ISL_2929394, EPI_ISL_2929395, EPI_ISL_2929396, EPI_ISL_2929402, EPI_ISL_2929414, EPI_ISL_2929415, EPI_ISL_2929417, EPI_ISL_2929419, EPI_ISL_2929422, EPI_ISL_2929423, EPI_ISL_2929425, EPI_ISL_2929433, EPI_ISL_2929435, EPI_ISL_2929440, EPI_ISL_2929442, EPI_ISL_2929445, EPI_ISL_2929448, EPI_ISL_2929456,
EPI_ISL_2929461, EPI_ISL_2929463, EPI_ISL_2929468, EPI_ISL_2929472, EPI_ISL_2929478, EPI_ISL_2929486, EPI_ISL_2929494, EPI_ISL_2929499, EPI_ISL_2929502, EPI_ISL_2929503, EPI_ISL_2929507, EPI_ISL_2929514, EPI_ISL_2929519, EPI_ISL_2929520, EPI_ISL_2929521, EPI_ISL_2929531, EPI_ISL_2929532, EPI_ISL_2929538, EPI_ISL_2929539, EPI_ISL_2929540, EPI_ISL_2929550, EPI_ISL_2929554,
EPI_ISL_2929555, EPI_ISL_2929557, EPI_ISL_2929559, EPI_ISL_2929564, EPI_ISL_2929565, EPI_ISL_2929566, EPI_ISL_2929570, EPI_ISL_2929572, EPI_ISL_2929574, EPI_ISL_2929577, EPI_ISL_2929578, EPI_ISL_2929579, EPI_ISL_2929580, EPI_ISL_2929585, EPI_ISL_2929586, EPI_ISL_2929589, EPI_ISL_2929591, EPI_ISL_2929593, EPI_ISL_2929604, EPI_ISL_2929605, EPI_ISL_2929609, EPI_ISL_2929611,
EPI_ISL_2929615, EPI_ISL_2929620, EPI_ISL_2929622, EPI_ISL_2929623, EPI_ISL_2929630, EPI_ISL_2929631, EPI_ISL_2929657, EPI_ISL_2929686, EPI_ISL_2929702, EPI_ISL_2929716, EPI_ISL_2929723, EPI_ISL_2929735, EPI_ISL_2929754, EPI_ISL_2929784, EPI_ISL_2929800, EPI_ISL_2929804, EPI_ISL_2929806, EPI_ISL_2929813, EPI_ISL_2929814, EPI_ISL_2929873, EPI_ISL_2929890, EPI_ISL_2929898,
EPI_ISL_2929902, EPI_ISL_2929903, EPI_ISL_2929904, EPI_ISL_2929928, EPI_ISL_2929953, EPI_ISL_2929976, EPI_ISL_2929981, EPI_ISL_2929982, EPI_ISL_2930000, EPI_ISL_2930002, EPI_ISL_2930006, EPI_ISL_2930016, EPI_ISL_2930034, EPI_ISL_2930090, EPI_ISL_2930099, EPI_ISL_2930130, EPI_ISL_2930151, EPI_ISL_2930173, EPI_ISL_2930203, EPI_ISL_2930206, EPI_ISL_2930217, EPI_ISL_2930250,
EPI_ISL_2930265, EPI_ISL_2930279, EPI_ISL_2930309, EPI_ISL_2930311, EPI_ISL_3018180, EPI_ISL_3018181, EPI_ISL_3018182, EPI_ISL_3018190, EPI_ISL_3018192, EPI_ISL_3018201, EPI_ISL_3018220, EPI_ISL_3018224, EPI_ISL_3018225, EPI_ISL_3018226, EPI_ISL_3018232, EPI_ISL_3018245, EPI_ISL_3018248, EPI_ISL_3018250, EPI_ISL_3018251, EPI_ISL_3018254, EPI_ISL_3018255, EPI_ISL_3018257,
EPI_ISL_3018259, EPI_ISL_3018260, EPI_ISL_3018264, EPI_ISL_3018266, EPI_ISL_3018270, EPI_ISL_3018272, EPI_ISL_3018273, EPI_ISL_3018277, EPI_ISL_3018286, EPI_ISL_3018287, EPI_ISL_3018305, EPI_ISL_3018306, EPI_ISL_3018313, EPI_ISL_3018320, EPI_ISL_3018326, EPI_ISL_3018338, EPI_ISL_3018345, EPI_ISL_3018352, EPI_ISL_3018360, EPI_ISL_3018361, EPI_ISL_3018371, EPI_ISL_3018372,
EPI_ISL_3018376, EPI_ISL_3018377, EPI_ISL_3018378, EPI_ISL_3018382, EPI_ISL_3018384, EPI_ISL_3018385, EPI_ISL_3018392, EPI_ISL_3018409, EPI_ISL_3018425, EPI_ISL_3018429, EPI_ISL_3018430, EPI_ISL_3018439, EPI_ISL_3018442, EPI_ISL_3018443, EPI_ISL_3018446, EPI_ISL_3018450, EPI_ISL_3018453, EPI_ISL_3018454, EPI_ISL_3018456, EPI_ISL_3018461, EPI_ISL_3018465, EPI_ISL_3018466,
EPI_ISL_3018468, EPI_ISL_3018469, EPI_ISL_3018472, EPI_ISL_3018476, EPI_ISL_3018480, EPI_ISL_3018482, EPI_ISL_3018484, EPI_ISL_3018485, EPI_ISL_3018488, EPI_ISL_3018489, EPI_ISL_3018492, EPI_ISL_3018494, EPI_ISL_3018498, EPI_ISL_3018505, EPI_ISL_3018521, EPI_ISL_3018523, EPI_ISL_3018527, EPI_ISL_3018529, EPI_ISL_3018533, EPI_ISL_3018535, EPI_ISL_3018540, EPI_ISL_3018546,
EPI_ISL_3018553, EPI_ISL_3018564, EPI_ISL_3018569, EPI_ISL_3018571, EPI_ISL_3018575, EPI_ISL_3018576, EPI_ISL_3018597, EPI_ISL_3018618, EPI_ISL_3018652, EPI_ISL_3018653, EPI_ISL_3018661, EPI_ISL_3018670, EPI_ISL_3018706, EPI_ISL_3018726, EPI_ISL_3018735, EPI_ISL_3018743, EPI_ISL_3018752, EPI_ISL_3018754, EPI_ISL_3018755, EPI_ISL_3018761, EPI_ISL_3018779, EPI_ISL_3018793,
EPI_ISL_3018795, EPI_ISL_3018798, EPI_ISL_3018820, EPI_ISL_3018825, EPI_ISL_3018828, EPI_ISL_3018836, EPI_ISL_3018841, EPI_ISL_3018850, EPI_ISL_3018860, EPI_ISL_3018883, EPI_ISL_3018885, EPI_ISL_3018900, EPI_ISL_3018906, EPI_ISL_3018915, EPI_ISL_3018920

| | | | |
|---|---|---|---|
| see above | Quest Diagnostics Incorporated | Centers for Disease Control and Prevention Division of Viral Diseases, Pathogen Discovery | A. Gerasimova; A. Perez; Adrian Paskey; B. Anderson; Benjamin Rambo-Martin; Christopher Gulvick; Clinton R. Paden; Dakota Howard; Darlene Wagner; Dhwani Batra; Duncan MacCannell; F. Lacbawan; I. A. Shlyakhter; Jason Caravas; K.E. Livingston; Kara Moser; L.E. Bernstein; M. Hua; Matthew Schmerer; P. Tanpaiboon; Peter W. Cook; R. M. Kagan; R. Owen; R. V. Rolando; S. H. Rosenthal; Scott Sammons; Shatavia Morrison; Y. Liu; Yvette Unoarumhi |
| EPI_ISL_2628498 | Rady Children's Hospital - San Diego | Andersen lab at Scripps Research | Christina Clarke; David Dimmock; Denise Malicki; Kathryn Bouic; Linda Luo; SEARCH Alliance San Diego with Nanda Radamchar; Teresa Mueller |
| EPI_ISL_2934052 | Regional Medical Center | Santa Clara County Public Health Laboratory | Santa Clara County Public Health Department |

EPI_ISL_2253203, EPI_ISL_3091564, EPI_ISL_3091570, EPI_ISL_3091571, EPI_ISL_3091577, EPI_ISL_3091588, EPI_ISL_3091589, EPI_ISL_3091597, EPI_ISL_3091598, EPI_ISL_3091603, EPI_ISL_3091604, EPI_ISL_3091610, EPI_ISL_3091611, EPI_ISL_3091615, EPI_ISL_3091620, EPI_ISL_3091640, EPI_ISL_3091646, EPI_ISL_3091651, EPI_ISL_3091653, EPI_ISL_3091655, EPI_ISL_3091665, EPI_ISL_3091668,
EPI_ISL_3091669, EPI_ISL_3091673, EPI_ISL_3091695, EPI_ISL_3091698, EPI_ISL_3091706, EPI_ISL_3091710, EPI_ISL_3091711, EPI_ISL_3091713, EPI_ISL_3091717, EPI_ISL_3091720, EPI_ISL_3091724, EPI_ISL_3091732, EPI_ISL_3091749, EPI_ISL_3091762, EPI_ISL_3091763, EPI_ISL_3091777, EPI_ISL_3091781, EPI_ISL_3091783, EPI_ISL_3091797, EPI_ISL_3091798, EPI_ISL_3091802, EPI_ISL_3091806,
EPI_ISL_3091813, EPI_ISL_3091819, EPI_ISL_3091823, EPI_ISL_3091830, EPI_ISL_3091837, EPI_ISL_3091852, EPI_ISL_3091854, EPI_ISL_3091867, EPI_ISL_3091872, EPI_ISL_3091875, EPI_ISL_3091878, EPI_ISL_3091879, EPI_ISL_3091887, EPI_ISL_3091891, EPI_ISL_3091903, EPI_ISL_3091908, EPI_ISL_3091913, EPI_ISL_3091917, EPI_ISL_3091931, EPI_ISL_3091935, EPI_ISL_3091979, EPI_ISL_3091980,
EPI_ISL_3091981, EPI_ISL_3091982, EPI_ISL_3091996, EPI_ISL_3092008, EPI_ISL_3092012, EPI_ISL_3092013, EPI_ISL_3092016, EPI_ISL_3092017, EPI_ISL_3092020, EPI_ISL_3092027, EPI_ISL_3092039, EPI_ISL_3092045

| | | | |
|---|---|---|---|
| see above | Respiratory Viruses Branch, Centers for Disease Control and Prevention | Respiratory Viruses Branch, Centers for Disease Control and Prevention | Anderson, B.; Batra, D.; Bernstein, L.; C.R.; Caravas, J.; Cook; Fierro, J.; Gao, H.; Gao, J.; Gerasimova, A.; Gulvick, C.; Howard, D.; Hua, M.; I.A.; Kagan; Lacbawan, F.; Li, M.; Liu, Y.; Livingston, K.; MacCannell, D.; Meng, Y.; Morrison, S.; Moser, K.; Ng, D.; Owen, R.; P.W.; Paden; Paskey, A.; Perez, A.; R.M.; R.V.; Rambo-Martin, B.; Rolando; Rosenthal; S.H.; Sammons, S.; Sapra, B.; Schmerer, M.; Shlyakhter; Tanpaiboon, P.; Tsai, B.; Unoarumhi, Y.; Wagner, D.; Xie, J. |
| EPI_ISL_2812005, EPI_ISL_2907565, EPI_ISL_2907566 | Saint Francis Medical Center | Los Angeles County Public Health Laboratories | P. Hemarajata et al. |
| EPI_ISL_2774497 | San Diego County Public Health Lab | San Diego County Public Health Lab | Ashleigh Murphy Schafer; Brett Austin |

EPI_ISL_2329357, EPI_ISL_2329364, EPI_ISL_2329370, EPI_ISL_2329381, EPI_ISL_2329470, EPI_ISL_2329479, EPI_ISL_2329492, EPI_ISL_2329503, EPI_ISL_2329509, EPI_ISL_2329522, EPI_ISL_2329523, EPI_ISL_2329529, EPI_ISL_2329530, EPI_ISL_2329536, EPI_ISL_2329537, EPI_ISL_2628329, EPI_ISL_2628334, EPI_ISL_2628346, EPI_ISL_2628350, EPI_ISL_2628439, EPI_ISL_2628440, EPI_ISL_2628443,
EPI_ISL_2628444, EPI_ISL_2628446, EPI_ISL_2628452, EPI_ISL_2628457, EPI_ISL_2628460, EPI_ISL_2628461, EPI_ISL_2628465, EPI_ISL_2628466, EPI_ISL_2628471, EPI_ISL_2628476, EPI_ISL_2628478, EPI_ISL_2628481, EPI_ISL_2628484, EPI_ISL_2628486, EPI_ISL_2628488, EPI_ISL_2628494, EPI_ISL_2628496, EPI_ISL_2628510, EPI_ISL_2628550, EPI_ISL_2628583, EPI_ISL_2628588, EPI_ISL_2628598,
EPI_ISL_2628611, EPI_ISL_2628612, EPI_ISL_2628614, EPI_ISL_2628723, EPI_ISL_2628739, EPI_ISL_2628742, EPI_ISL_2628745, EPI_ISL_2628749, EPI_ISL_2628753, EPI_ISL_2628757, EPI_ISL_2628760, EPI_ISL_2628764, EPI_ISL_2628768, EPI_ISL_2628771, EPI_ISL_2628775, EPI_ISL_2628779, EPI_ISL_2628782, EPI_ISL_2628786, EPI_ISL_2628790, EPI_ISL_2628793, EPI_ISL_2628797, EPI_ISL_2628801,
EPI_ISL_2628804, EPI_ISL_2628808, EPI_ISL_2628812, EPI_ISL_2628815, EPI_ISL_2628819, EPI_ISL_2628822, EPI_ISL_2628826, EPI_ISL_2628830, EPI_ISL_2628833, EPI_ISL_2628837, EPI_ISL_2628841, EPI_ISL_2628844, EPI_ISL_2628848, EPI_ISL_2628851, EPI_ISL_2628855, EPI_ISL_2628859, EPI_ISL_2628863, EPI_ISL_2628866, EPI_ISL_2628870, EPI_ISL_2628874, EPI_ISL_2628877, EPI_ISL_2628881,
EPI_ISL_2628884, EPI_ISL_2628887, EPI_ISL_2628891, EPI_ISL_2628895, EPI_ISL_2628897, EPI_ISL_2628901, EPI_ISL_2628904, EPI_ISL_2628908, EPI_ISL_2628911, EPI_ISL_2628915, EPI_ISL_2628918, EPI_ISL_2628922, EPI_ISL_2628926, EPI_ISL_2628929, EPI_ISL_2628933, EPI_ISL_2628937, EPI_ISL_2628940, EPI_ISL_2628944, EPI_ISL_2628948, EPI_ISL_2628951, EPI_ISL_2628955, EPI_ISL_2712831

| | | | |
|---|---|---|---|
| see above | San Diego County Public Health Laboratory | Andersen lab at Scripps Research | Brett Austin; Jovan Shephard; SEARCH Alliance San Diego with Ashleigh Murphy |

EPI_ISL_2834946, EPI_ISL_2834947, EPI_ISL_2834948, EPI_ISL_2834949, EPI_ISL_2834950, EPI_ISL_2834951, EPI_ISL_2834952, EPI_ISL_2834953, EPI_ISL_2834954, EPI_ISL_2834955, EPI_ISL_2834956, EPI_ISL_2834957, EPI_ISL_2840592, EPI_ISL_2840593, EPI_ISL_2840594, EPI_ISL_2840606, EPI_ISL_2840607, EPI_ISL_2840608, EPI_ISL_2840638, EPI_ISL_2840852, EPI_ISL_2840861, EPI_ISL_2840862,
EPI_ISL_2840863, EPI_ISL_2840864, EPI_ISL_2840867, EPI_ISL_2840895, EPI_ISL_2840896, EPI_ISL_2840897, EPI_ISL_2840898, EPI_ISL_2840899, EPI_ISL_2840900, EPI_ISL_2840901, EPI_ISL_2840902, EPI_ISL_2840903, EPI_ISL_2840917, EPI_ISL_2840918, EPI_ISL_2840920, EPI_ISL_2840921, EPI_ISL_2840922, EPI_ISL_2840926, EPI_ISL_2840927, EPI_ISL_2840928, EPI_ISL_2840930, EPI_ISL_2841488,
EPI_ISL_2841489, EPI_ISL_2841490, EPI_ISL_2841534, EPI_ISL_2841536, EPI_ISL_2841537, EPI_ISL_2841538, EPI_ISL_2841540, EPI_ISL_2841603, EPI_ISL_2841604, EPI_ISL_2841613, EPI_ISL_2841614, EPI_ISL_2841615, EPI_ISL_2842692, EPI_ISL_2842693, EPI_ISL_2842694, EPI_ISL_2842695, EPI_ISL_2842696, EPI_ISL_2842697, EPI_ISL_2842698, EPI_ISL_2842699, EPI_ISL_2842700, EPI_ISL_2842701,
EPI_ISL_2842702, EPI_ISL_2842703, EPI_ISL_2842704, EPI_ISL_2842705, EPI_ISL_2842706, EPI_ISL_2873837

| | | | |
|---|---|---|---|
| see above | San Diego County Public Health Laboratory | San Diego County Public Health Laboratory | Ashleigh Murphy Schafer; Brett Austin; San Diego County Public Health Laboratory |

EPI_ISL_2496776, EPI_ISL_2496777, EPI_ISL_2496788, EPI_ISL_2496892, EPI_ISL_2496923, EPI_ISL_2496931, EPI_ISL_2496976, EPI_ISL_2500085, EPI_ISL_2500156, EPI_ISL_2658589, EPI_ISL_2658592

| | | | |
|---|---|---|---|
| see above | Santa Clara County Public Health Laboratory | Chan-Zuckerberg Biohub | CZB Cliahub Consortium |

EPI_ISL_2631202, EPI_ISL_2631203, EPI_ISL_2631204, EPI_ISL_2631205, EPI_ISL_2631206, EPI_ISL_2631207, EPI_ISL_2650563, EPI_ISL_2659057, EPI_ISL_2659064

| | | | |
|---|---|---|---|
| see above | Santa Clara County Public Health Laboratory | Santa Clara County Public Health Laboratory | Santa Clara County Public Health Department |

EPI_ISL_2631208, EPI_ISL_2631209, EPI_ISL_2631211, EPI_ISL_2631212, EPI_ISL_2631213, EPI_ISL_2631214, EPI_ISL_2631215, EPI_ISL_2631216, EPI_ISL_2631217, EPI_ISL_2631218, EPI_ISL_2650546, EPI_ISL_2650547, EPI_ISL_2650548, EPI_ISL_2650549, EPI_ISL_2650550, EPI_ISL_2650551, EPI_ISL_2650552, EPI_ISL_2650553, EPI_ISL_2650554, EPI_ISL_2650555, EPI_ISL_2650556, EPI_ISL_2650557,
EPI_ISL_2650558, EPI_ISL_2650559, EPI_ISL_2650560, EPI_ISL_2650561, EPI_ISL_2650564, EPI_ISL_2650565, EPI_ISL_2650567, EPI_ISL_2650568, EPI_ISL_2650569, EPI_ISL_2650570, EPI_ISL_2650571, EPI_ISL_2650572, EPI_ISL_2650573, EPI_ISL_2650574, EPI_ISL_2650575, EPI_ISL_2650576, EPI_ISL_2650577, EPI_ISL_2650578, EPI_ISL_2650579, EPI_ISL_2650580, EPI_ISL_2650582, EPI_ISL_2650583,
EPI_ISL_2691299, EPI_ISL_2691300, EPI_ISL_2691302, EPI_ISL_2691303, EPI_ISL_2691304, EPI_ISL_2691305, EPI_ISL_2691306, EPI_ISL_2691307, EPI_ISL_2691308, EPI_ISL_2691309, EPI_ISL_2691310, EPI_ISL_2691311, EPI_ISL_2691312, EPI_ISL_2691313, EPI_ISL_2691314, EPI_ISL_2753976, EPI_ISL_2774591, EPI_ISL_2774592, EPI_ISL_2774593, EPI_ISL_2774594, EPI_ISL_2774595, EPI_ISL_2774596,
EPI_ISL_2774597, EPI_ISL_2774598, EPI_ISL_2774599, EPI_ISL_2774600, EPI_ISL_2774601, EPI_ISL_2774602, EPI_ISL_2774603, EPI_ISL_2774604, EPI_ISL_2774605, EPI_ISL_2774606, EPI_ISL_2774607, EPI_ISL_2774608, EPI_ISL_2774609, EPI_ISL_2774610, EPI_ISL_2774611, EPI_ISL_2774612, EPI_ISL_2774613, EPI_ISL_2774614, EPI_ISL_2774615, EPI_ISL_2774616, EPI_ISL_2774617, EPI_ISL_2774618,
EPI_ISL_2774619, EPI_ISL_2774620, EPI_ISL_2774624, EPI_ISL_2774625, EPI_ISL_2774626, EPI_ISL_2774627, EPI_ISL_2774628, EPI_ISL_2774629, EPI_ISL_2774630, EPI_ISL_2774631, EPI_ISL_2774632, EPI_ISL_2774633, EPI_ISL_2774634, EPI_ISL_2933987, EPI_ISL_2933989, EPI_ISL_2933993, EPI_ISL_2933994, EPI_ISL_2933995, EPI_ISL_2933997, EPI_ISL_2933998, EPI_ISL_2934000, EPI_ISL_2934006,
EPI_ISL_2934021, EPI_ISL_2934022, EPI_ISL_2934029, EPI_ISL_2934030, EPI_ISL_2934031, EPI_ISL_2934032, EPI_ISL_2934034, EPI_ISL_2934035, EPI_ISL_2934036, EPI_ISL_2934037, EPI_ISL_2934039, EPI_ISL_2934041, EPI_ISL_2934043, EPI_ISL_2934046, EPI_ISL_2934051, EPI_ISL_2934055, EPI_ISL_2934056, EPI_ISL_2934059, EPI_ISL_2934060, EPI_ISL_2934062, EPI_ISL_2934063, EPI_ISL_2934064,
EPI_ISL_2934069, EPI_ISL_2934070, EPI_ISL_2934072, EPI_ISL_2934081, EPI_ISL_2934082, EPI_ISL_2934083, EPI_ISL_2934084, EPI_ISL_3048871, EPI_ISL_3048872, EPI_ISL_3048873, EPI_ISL_3048874, EPI_ISL_3048877, EPI_ISL_3048878, EPI_ISL_3048879, EPI_ISL_3048880, EPI_ISL_3048881, EPI_ISL_3048882, EPI_ISL_3048883, EPI_ISL_3048884, EPI_ISL_3048885, EPI_ISL_3048886, EPI_ISL_3048887,
EPI_ISL_3048888, EPI_ISL_3048889, EPI_ISL_3048890, EPI_ISL_3048891, EPI_ISL_3048892, EPI_ISL_3048893, EPI_ISL_3048894, EPI_ISL_3048895, EPI_ISL_3048896, EPI_ISL_3048897, EPI_ISL_3048898, EPI_ISL_3048899, EPI_ISL_3048900, EPI_ISL_3048901, EPI_ISL_3048902, EPI_ISL_3048903, EPI_ISL_3048904, EPI_ISL_3048905, EPI_ISL_3048906, EPI_ISL_3048909, EPI_ISL_3048910, EPI_ISL_3048911,
EPI_ISL_3048912, EPI_ISL_3048913, EPI_ISL_3048914, EPI_ISL_3048915, EPI_ISL_3048916, EPI_ISL_3048917, EPI_ISL_3048918

| | | | |
|---|---|---|---|
| see above | Santa Clara Valley Medical Center | Santa Clara County Public Health Laboratory | Santa Clara County Public Health Department |
| EPI_ISL_2967929, EPI_ISL_2967930, EPI_ISL_2967932, EPI_ISL_2967933, EPI_ISL_2967934, EPI_ISL_2967935, EPI_ISL_2967936, EPI_ISL_2967937, EPI_ISL_2967938, EPI_ISL_2967939, EPI_ISL_2967940, EPI_ISL_2967941, EPI_ISL_2967942, EPI_ISL_2967943, EPI_ISL_2967944, EPI_ISL_2967945, EPI_ISL_2967946, EPI_ISL_2967947, EPI_ISL_2967948, EPI_ISL_2967949, EPI_ISL_2967950, EPI_ISL_2967951, EPI_ISL_2967952, EPI_ISL_2967953, EPI_ISL_2967954, EPI_ISL_2967955, EPI_ISL_2967956, EPI_ISL_2967957, EPI_ISL_2967958, EPI_ISL_2967959, EPI_ISL_2967960, EPI_ISL_2967961, EPI_ISL_2967962, EPI_ISL_2967963 | | | |
| see above | Santa Clara Valley Medical Center | Santa Clara County public Health Laboratory | Santa Clara County Public Health Department |
| EPI_ISL_2231139, EPI_ISL_2231157, EPI_ISL_2329427, EPI_ISL_2329428, EPI_ISL_2329429, EPI_ISL_2329430, EPI_ISL_2329431, EPI_ISL_2329432, EPI_ISL_2329433, EPI_ISL_2329434, EPI_ISL_2329436, EPI_ISL_2329441, EPI_ISL_2329442, EPI_ISL_2329443, EPI_ISL_2329444, EPI_ISL_2329445, EPI_ISL_2329446, EPI_ISL_2329450, EPI_ISL_2329451, EPI_ISL_2329452, EPI_ISL_2329453, EPI_ISL_2329454, EPI_ISL_2329455, EPI_ISL_2329456, EPI_ISL_2329457, EPI_ISL_2329459, EPI_ISL_2329460, EPI_ISL_2329461, EPI_ISL_2329462, EPI_ISL_2329463, EPI_ISL_2329464, EPI_ISL_2422815, EPI_ISL_2422816, EPI_ISL_2422817, EPI_ISL_2422819, EPI_ISL_2422820, EPI_ISL_2422821, EPI_ISL_2422822, EPI_ISL_2422823, EPI_ISL_2422824, EPI_ISL_2422825, EPI_ISL_2422826, EPI_ISL_2422827, EPI_ISL_2422831, EPI_ISL_2422833, EPI_ISL_2628380, EPI_ISL_2628381, EPI_ISL_2628382, EPI_ISL_2628383, EPI_ISL_2628384, EPI_ISL_2628385, EPI_ISL_2628386, EPI_ISL_2628387, EPI_ISL_2628388, EPI_ISL_2628389, EPI_ISL_2628390, EPI_ISL_2628391, EPI_ISL_2628392, EPI_ISL_2628393, EPI_ISL_2628394, EPI_ISL_2628395, EPI_ISL_2628536, EPI_ISL_2628537, EPI_ISL_2628538, EPI_ISL_2628539, EPI_ISL_2628541, EPI_ISL_2628542, EPI_ISL_2628543, EPI_ISL_2628544, EPI_ISL_2712535, EPI_ISL_2712536, EPI_ISL_2712537, EPI_ISL_2712538, EPI_ISL_2712539, EPI_ISL_2712540, EPI_ISL_2712541, EPI_ISL_2712542, EPI_ISL_2712543, EPI_ISL_2712544, EPI_ISL_2712545, EPI_ISL_2712546, EPI_ISL_2712547, EPI_ISL_2712548, EPI_ISL_2712549, EPI_ISL_2712550, EPI_ISL_2712551, EPI_ISL_2712552, EPI_ISL_2712553, EPI_ISL_2712554, EPI_ISL_2712555, EPI_ISL_2835552, EPI_ISL_2835553, EPI_ISL_2835554, EPI_ISL_2835555, EPI_ISL_2835556, EPI_ISL_2835557, EPI_ISL_2835558, EPI_ISL_2835559, EPI_ISL_2835560, EPI_ISL_2835561, EPI_ISL_2835562, EPI_ISL_2835563, EPI_ISL_2835564, EPI_ISL_2835565, EPI_ISL_2920432, EPI_ISL_2920433, EPI_ISL_2920434, EPI_ISL_2920435, EPI_ISL_2920436, EPI_ISL_2920437, EPI_ISL_2920438, EPI_ISL_2920439, EPI_ISL_2920440, EPI_ISL_2920441, EPI_ISL_2920442, EPI_ISL_2920443, EPI_ISL_2988836, EPI_ISL_2988840, EPI_ISL_2988841, EPI_ISL_2988842, EPI_ISL_2988843, EPI_ISL_2988844, EPI_ISL_2988845, EPI_ISL_2988846, EPI_ISL_2988847, EPI_ISL_2988848, EPI_ISL_2988849, EPI_ISL_2988850, EPI_ISL_2988851, EPI_ISL_2988852, EPI_ISL_2988853, EPI_ISL_2988854, EPI_ISL_2988855, EPI_ISL_2988856, EPI_ISL_2988857, EPI_ISL_2988858 | | | |
| see above | Sharp HealthCare Laboratory | Andersen lab at Scripps Research | Art Mendoza; Cathy Woerle; Jacquelyn Berumen; Liam McGinnis; Omid Bakhtar; SEARCH Alliance San Diego with Aaron Harding |
| EPI_ISL_3021461 | Sierra Vista Regional Medical Lab | County of San Luis Obispo Public Health Laboratory | Frances Sidhu; Katrina Erwin |
| EPI_ISL_2457061, EPI_ISL_2987140, EPI_ISL_2987141, EPI_ISL_2987142, EPI_ISL_3050180, EPI_ISL_3050181, EPI_ISL_3050182, EPI_ISL_3050183, EPI_ISL_3050184, EPI_ISL_3050185, EPI_ISL_3050186, EPI_ISL_3050187, EPI_ISL_3050188, EPI_ISL_3050189, EPI_ISL_3050190, EPI_ISL_3050191, EPI_ISL_3050192, EPI_ISL_3050193, EPI_ISL_3050194, EPI_ISL_3050195, EPI_ISL_3050196, EPI_ISL_3050197, EPI_ISL_3050198, EPI_ISL_3050199, EPI_ISL_3050200, EPI_ISL_3050201, EPI_ISL_3050202, EPI_ISL_3050203, EPI_ISL_3050204, EPI_ISL_3050205, EPI_ISL_3050206, EPI_ISL_3050207, EPI_ISL_3050208, EPI_ISL_3050209, EPI_ISL_3050210, EPI_ISL_3050211, EPI_ISL_3050212, EPI_ISL_3050213, EPI_ISL_3050214, EPI_ISL_3050215, EPI_ISL_3050216, EPI_ISL_3050219, EPI_ISL_3050223, EPI_ISL_3050257 | | | |
| see above | Stanford Health Care | Stanford University School of Medicine, Clinical Virology Laboratory | Becky Jiang; Bernadette Troung; ChunHong Huang; Daniel Solis; James Zehnder; Malaya K. Sahoo; Mamdouh Sibai; Michelle Verghese; Nathan Hammond; Selamawit Bihon; and Benjamin A. Pinsky |
| EPI_ISL_2391790, EPI_ISL_2391791 | Sun Clinical Laboratory | Los Angeles County Public Health Laboratories | P. Hemarajata et al. |
| EPI_ISL_2620779, EPI_ISL_2774572 | Torrance Memorial Medical Center | Los Angeles County Public Health Laboratories | P. Hemarajata et al. |
| EPI_ISL_3033638, EPI_ISL_3033639, EPI_ISL_3033640, EPI_ISL_3033641, EPI_ISL_3049228, EPI_ISL_3049229, EPI_ISL_3049230, EPI_ISL_3049231, EPI_ISL_3049232, EPI_ISL_3049233, EPI_ISL_3049256, EPI_ISL_3049257, EPI_ISL_3049258, EPI_ISL_3049259, EPI_ISL_3049260, EPI_ISL_3049261, EPI_ISL_3049262, EPI_ISL_3049263, EPI_ISL_3049264, EPI_ISL_3049265, EPI_ISL_3049266, EPI_ISL_3049267, EPI_ISL_3049268, EPI_ISL_3049269, EPI_ISL_3049270, EPI_ISL_3049271, EPI_ISL_3049272, EPI_ISL_3049273, EPI_ISL_3049274, EPI_ISL_3049275, EPI_ISL_3049276, EPI_ISL_3049277, EPI_ISL_3049278, EPI_ISL_3049279, EPI_ISL_3049280, EPI_ISL_3049281, EPI_ISL_3049282, EPI_ISL_3049283, EPI_ISL_3049284, EPI_ISL_3049285, EPI_ISL_3049286, EPI_ISL_3049287, EPI_ISL_3049288, EPI_ISL_3049289, EPI_ISL_3049290, EPI_ISL_3049291, EPI_ISL_3049292, EPI_ISL_3049293, EPI_ISL_3049294, EPI_ISL_3049295, EPI_ISL_3049296, EPI_ISL_3049297, EPI_ISL_3049298, EPI_ISL_3049299, EPI_ISL_3049300, EPI_ISL_3049301, EPI_ISL_3049302, EPI_ISL_3049303, EPI_ISL_3049304, EPI_ISL_3049305, EPI_ISL_3049306, EPI_ISL_3049307, EPI_ISL_3049308, EPI_ISL_3049309, EPI_ISL_3049310, EPI_ISL_3049311, EPI_ISL_3049312, EPI_ISL_3049313, EPI_ISL_3049314, EPI_ISL_3049315, EPI_ISL_3049316, EPI_ISL_3049317, EPI_ISL_3049318, EPI_ISL_3049319, EPI_ISL_3049320, EPI_ISL_3049321, EPI_ISL_3049322, EPI_ISL_3049323, EPI_ISL_3049324, EPI_ISL_3049325, EPI_ISL_3049326, EPI_ISL_3049327, EPI_ISL_3049328, EPI_ISL_3049329, EPI_ISL_3049330, EPI_ISL_3049331, EPI_ISL_3049332, EPI_ISL_3049333, EPI_ISL_3049334, EPI_ISL_3049335, EPI_ISL_3049336, EPI_ISL_3049337, EPI_ISL_3049338, EPI_ISL_3049339, EPI_ISL_3049340, EPI_ISL_3049341, EPI_ISL_3049342, EPI_ISL_3049343, EPI_ISL_3049344, EPI_ISL_3049345, EPI_ISL_3049346, EPI_ISL_3049347, EPI_ISL_3049348, EPI_ISL_3049349, EPI_ISL_3049350, EPI_ISL_3049351, EPI_ISL_3049352, EPI_ISL_3049353, EPI_ISL_3049354, EPI_ISL_3049355, EPI_ISL_3049356, EPI_ISL_3049357, EPI_ISL_3049358, EPI_ISL_3049359, EPI_ISL_3049360, EPI_ISL_3049361, EPI_ISL_3049362, EPI_ISL_3049363, EPI_ISL_3049364, EPI_ISL_3049365, EPI_ISL_3049366, EPI_ISL_3049367, EPI_ISL_3049368, EPI_ISL_3049369, EPI_ISL_3049370, EPI_ISL_3049371, EPI_ISL_3049372, EPI_ISL_3049373, EPI_ISL_3049374, EPI_ISL_3049375 | | | |
| see above | Tulare County Public Health Laboratory | Tulare County Public Health Laboratory | Denise L Lopez; Joshua Kootstra |
| EPI_ISL_3021460 | Twin Cities Community Hospital Lab | County of San Luis Obispo Public Health Laboratory | Frances Sidhu; Katrina Erwin |
| EPI_ISL_2426985, EPI_ISL_2426986, EPI_ISL_2426987, EPI_ISL_2426988, EPI_ISL_2426989, EPI_ISL_2426990, EPI_ISL_2426991, EPI_ISL_2426992, EPI_ISL_2426993, EPI_ISL_2426994 | | | |
| see above | UC Davis Genome Center and Healthy Davis Together | UC Davis Genome Center | UC Davis |
| EPI_ISL_2391761, EPI_ISL_2391763, EPI_ISL_2391810, EPI_ISL_2391812, EPI_ISL_2392172, EPI_ISL_2392174, EPI_ISL_2392177, EPI_ISL_2392179, EPI_ISL_2392180, EPI_ISL_2392191, EPI_ISL_2392193, EPI_ISL_2392194, EPI_ISL_2399612, EPI_ISL_2399616, EPI_ISL_2399619, EPI_ISL_2399620, EPI_ISL_2404855, EPI_ISL_2404857, EPI_ISL_2404860, EPI_ISL_2404861, EPI_ISL_2404863, EPI_ISL_2404864, EPI_ISL_2404866, EPI_ISL_2404867, EPI_ISL_2405167, EPI_ISL_2405183, EPI_ISL_2424170, EPI_ISL_2424172, EPI_ISL_2424175, EPI_ISL_2424183, EPI_ISL_2424189, EPI_ISL_2424190, EPI_ISL_2424193, EPI_ISL_2424198, EPI_ISL_2424199, EPI_ISL_2424204, EPI_ISL_2424205, EPI_ISL_2424206, EPI_ISL_2424234, EPI_ISL_2617943, EPI_ISL_2617944, EPI_ISL_2617945, EPI_ISL_2617946, EPI_ISL_2617947, EPI_ISL_2617948, EPI_ISL_2617949, EPI_ISL_2617950, EPI_ISL_2617951, EPI_ISL_2617952, EPI_ISL_2617953, EPI_ISL_2617954, EPI_ISL_2617955, EPI_ISL_2617956, EPI_ISL_2617957, EPI_ISL_2617958, EPI_ISL_2620750, EPI_ISL_2772506, EPI_ISL_2772507, EPI_ISL_2772508, EPI_ISL_2772509, EPI_ISL_2772510, EPI_ISL_2772511, EPI_ISL_2772512, EPI_ISL_2772513, EPI_ISL_2772514, EPI_ISL_2772515, EPI_ISL_2772516, EPI_ISL_2772517, EPI_ISL_2772518, EPI_ISL_2772519, EPI_ISL_2772520, EPI_ISL_2772521, EPI_ISL_2811920, EPI_ISL_2811921 | | | |
| see above | UCLA Clinical Micro Lab | Los Angeles County Public Health Laboratories | P. Hemarajata et al. |
| EPI_ISL_2179218, EPI_ISL_2179270, EPI_ISL_2179271, EPI_ISL_2179272, EPI_ISL_2295456, EPI_ISL_2295457, EPI_ISL_2378755, EPI_ISL_2378756, EPI_ISL_2494632, EPI_ISL_2494633, EPI_ISL_2494634, EPI_ISL_2494635, EPI_ISL_2982020, EPI_ISL_2982021, EPI_ISL_2982022, EPI_ISL_2982023, EPI_ISL_2982115, EPI_ISL_2982116, EPI_ISL_2982633 | | | |
| see above | UCSC Genomics Institute | UCSC Genomics Institute | A. Marm Kilpatrick; Angie Hinrichs; Beth Shapiro; Bryan Thornlow; Ciara Wanket; David Haussler; Eric Beraut; Hugh Olsen; Ikenna Anigbogu; Isabel Bjork; Jakob McBroome; Jeremy Sanford; Joshua Kapp; Mark Akeson; Maximilian Haeussler; Michael Stone; Miten Jain; Molly Cassatt-Johnstone; Namrita Dhillon; Russell Corbett-Detig; Terren Chang; Yatish Turakhia |
| EPI_ISL_2426732, EPI_ISL_2450384, EPI_ISL_2715313, EPI_ISL_3048147 | US Air Force School of Aerospace Medicine | US Air Force School of Aerospace Medicine | Amanda Javorina; Anthony Fries; Carol Garrett; Clarise Starr; Elizabeth Macias; Jennifer Meyer; Sarah Purves; William Gruner |
| EPI_ISL_2391783, EPI_ISL_2391784, EPI_ISL_2391785, EPI_ISL_2391786, EPI_ISL_2391787, EPI_ISL_2405180, EPI_ISL_2405181, EPI_ISL_2620783, EPI_ISL_2811942, EPI_ISL_2811943, EPI_ISL_2811966, EPI_ISL_2812004, EPI_ISL_2894587, EPI_ISL_2894588, EPI_ISL_2894589, EPI_ISL_2907573, EPI_ISL_2907574, EPI_ISL_2907575 | | | |
| see above | USC Clinical Lab | Los Angeles County Public Health Laboratories | P. Hemarajata et al. |
| EPI_ISL_3029862 | VA Medical Center | Los Angeles County Public Health Laboratories | P. Hemarajata et al. |
| EPI_ISL_2404811, EPI_ISL_2404812, EPI_ISL_2404813, EPI_ISL_2404814, EPI_ISL_2404816, EPI_ISL_2404817, EPI_ISL_2404818, EPI_ISL_2404819, EPI_ISL_2404820, EPI_ISL_2404821, EPI_ISL_2404822, EPI_ISL_2404823, EPI_ISL_2404824, EPI_ISL_2404825, EPI_ISL_2404828, EPI_ISL_2404829, EPI_ISL_2404830, EPI_ISL_2404831, EPI_ISL_2404832, EPI_ISL_2404833, EPI_ISL_2404834, EPI_ISL_2404835, EPI_ISL_2404836, EPI_ISL_2404837, EPI_ISL_2404838, EPI_ISL_2404839 | | | |
| see above | VMC | Santa Clara County public Health Laboratory | Santa Clara County Public Health Department |

We gratefully acknowledge the following Authors from the Originating laboratories responsible for obtaining the specimens, as well as the Submitting laboratories where the genome data were generated and shared via GISAID, on which this research is based.

All Submitters of data may be contacted directly via www.gisaid.org

Authors are sorted alphabetically.

| Accession ID | Originating Laboratory | Submitting Laboratory | Authors |
|---|---|---|---|
| EPI_ISL_2907554, EPI_ISL_2907556, EPI_ISL_2907557, EPI_ISL_2907558, EPI_ISL_2907560 | Centinela Hospital Medical Center | Los Angeles County Public Health Laboratories | P. Hemarajata et al. |
| EPI_ISL_3021464, EPI_ISL_3021465, EPI_ISL_3021466, EPI_ISL_3021468, EPI_ISL_3021469, EPI_ISL_3021470, EPI_ISL_3021471, EPI_ISL_3021472, EPI_ISL_3021473, EPI_ISL_3021475 | | | |
| see above | County of San Luis Obispo Public Health Laboratory | County of San Luis Obispo Public Health Laboratory | Frances Sidhu; Katrina Erwin |
| EPI_ISL_2942017, EPI_ISL_2942018, EPI_ISL_2942019, EPI_ISL_2942021, EPI_ISL_2942022, EPI_ISL_2942023, EPI_ISL_2942024, EPI_ISL_2942025, EPI_ISL_2942026, EPI_ISL_2942027, EPI_ISL_2942028, EPI_ISL_2942030, EPI_ISL_2942032, EPI_ISL_2942033, EPI_ISL_2942034, EPI_ISL_2942035, EPI_ISL_2942036, EPI_ISL_2942038, EPI_ISL_2942039, EPI_ISL_2942041, EPI_ISL_2942042, EPI_ISL_2942043, EPI_ISL_2942044, EPI_ISL_2942045, EPI_ISL_2942046, EPI_ISL_2942048, EPI_ISL_2942049, EPI_ISL_2942050, EPI_ISL_2942051, EPI_ISL_2942052, EPI_ISL_2942053, EPI_ISL_2942054, EPI_ISL_2942055, EPI_ISL_2942056, EPI_ISL_2942057, EPI_ISL_2942058, EPI_ISL_2942059, EPI_ISL_2942060, EPI_ISL_2942061, EPI_ISL_2942062, EPI_ISL_2942063, EPI_ISL_2942065, EPI_ISL_2942066, EPI_ISL_2942067, EPI_ISL_2942068, EPI_ISL_2942069, EPI_ISL_2942070, EPI_ISL_2942071, EPI_ISL_2942072, EPI_ISL_2942073, EPI_ISL_2942074, EPI_ISL_2942075, EPI_ISL_2942076, EPI_ISL_2942077, EPI_ISL_2942078, EPI_ISL_2942079, EPI_ISL_2942080, EPI_ISL_2942081, EPI_ISL_2942082, EPI_ISL_2942083, EPI_ISL_2942084, EPI_ISL_2942085, EPI_ISL_2942086, EPI_ISL_2942087, EPI_ISL_2942088, EPI_ISL_2942089, EPI_ISL_2942090, EPI_ISL_2942091, EPI_ISL_2942093, EPI_ISL_2942095, EPI_ISL_2942096, EPI_ISL_3064427, EPI_ISL_3064428, EPI_ISL_3064429, EPI_ISL_3064430, EPI_ISL_3064431, EPI_ISL_3064432, EPI_ISL_3064433, EPI_ISL_3064434, EPI_ISL_3064435, EPI_ISL_3064436, EPI_ISL_3064437, EPI_ISL_3064438, EPI_ISL_3064439, EPI_ISL_3064440, EPI_ISL_3064441, EPI_ISL_3064442, EPI_ISL_3064443, EPI_ISL_3064444, EPI_ISL_3064445, EPI_ISL_3064446, EPI_ISL_3064447, EPI_ISL_3064448, EPI_ISL_3064449, EPI_ISL_3064450, EPI_ISL_3064453, EPI_ISL_3064454, EPI_ISL_3064455, EPI_ISL_3064456, EPI_ISL_3064457, EPI_ISL_3064458, EPI_ISL_3064459, EPI_ISL_3064460, EPI_ISL_3064461, EPI_ISL_3064462, EPI_ISL_3064463, EPI_ISL_3064519, EPI_ISL_3064520, EPI_ISL_3064521, EPI_ISL_3064522, EPI_ISL_3064523, EPI_ISL_3064524, EPI_ISL_3064525, EPI_ISL_3064526, EPI_ISL_3064527, EPI_ISL_3064528, EPI_ISL_3064529, EPI_ISL_3064530, EPI_ISL_3064531, EPI_ISL_3064532, EPI_ISL_3064533, EPI_ISL_3064534, EPI_ISL_3064538, EPI_ISL_3064539, EPI_ISL_3064540, EPI_ISL_3064541, EPI_ISL_3064543, EPI_ISL_3064544, EPI_ISL_3064545, EPI_ISL_3064546, EPI_ISL_3064547, EPI_ISL_3064548, EPI_ISL_3064549, EPI_ISL_3064550, EPI_ISL_3064551, EPI_ISL_3064552, EPI_ISL_3064553, EPI_ISL_3064554, EPI_ISL_3064555, EPI_ISL_3064556, EPI_ISL_3064557, EPI_ISL_3064558, EPI_ISL_3064559, EPI_ISL_3064560, EPI_ISL_3064561, EPI_ISL_3064562, EPI_ISL_3064563, EPI_ISL_3064564, EPI_ISL_3064565, EPI_ISL_3064566, EPI_ISL_3064567, EPI_ISL_3064568, EPI_ISL_3064569, EPI_ISL_3064570, EPI_ISL_3064571, EPI_ISL_3064572, EPI_ISL_3064573, EPI_ISL_3064574, EPI_ISL_3064575, EPI_ISL_3064576, EPI_ISL_3064577, EPI_ISL_3064578, EPI_ISL_3064579, EPI_ISL_3064580, EPI_ISL_3064581, EPI_ISL_3064582, EPI_ISL_3064583, EPI_ISL_3064584, EPI_ISL_3064585, EPI_ISL_3064586, EPI_ISL_3064587, EPI_ISL_3064588, EPI_ISL_3064589, EPI_ISL_3064590, EPI_ISL_3064591, EPI_ISL_3064592, EPI_ISL_3064593, EPI_ISL_3064594, EPI_ISL_3064595, EPI_ISL_3064596, EPI_ISL_3064597, EPI_ISL_3064666, EPI_ISL_3064669, EPI_ISL_3064670, EPI_ISL_3064671, EPI_ISL_3064672, EPI_ISL_3064673, EPI_ISL_3064674, EPI_ISL_3064675, EPI_ISL_3064676, EPI_ISL_3064677, EPI_ISL_3064678, EPI_ISL_3064679, EPI_ISL_3064680, EPI_ISL_3064681, EPI_ISL_3064682, EPI_ISL_3064683, EPI_ISL_3064684, EPI_ISL_3064685, EPI_ISL_3064686, EPI_ISL_3064687, EPI_ISL_3064688, EPI_ISL_3064689, EPI_ISL_3064690, EPI_ISL_3064691, EPI_ISL_3064692, EPI_ISL_3064693, EPI_ISL_3064694, EPI_ISL_3064695, EPI_ISL_3064696, EPI_ISL_3064697, EPI_ISL_3064698, EPI_ISL_3064699, EPI_ISL_3064700 | | | |
| see above | Curative Labs | Curative Labs | Elias L. Salfati; Eugenia Khorosheva; George Way; J.Cesar Ignacio-Espinoza; Janet Chen; Mikhail Hanewich-Hollatz; Nabjot Sandhu; Sophia Quasem; Vladimir Slepnev; Zhiyi Xie |
| EPI_ISL_2967967 | El Camino Hospital | Santa Clara County public Health Laboratory | Santa Clara County Public Health Department |
| EPI_ISL_3021474 | French Hospital Laboratory | County of San Luis Obispo Public Health Laboratory | Frances Sidhu; Katrina Erwin |
| EPI_ISL_2928583, EPI_ISL_2928584, EPI_ISL_2928585, EPI_ISL_2928586, EPI_ISL_2928587, EPI_ISL_2928588, EPI_ISL_2928589, EPI_ISL_2928596, EPI_ISL_2928599, EPI_ISL_2928602, EPI_ISL_2928604, EPI_ISL_2928605, EPI_ISL_2928606, EPI_ISL_2928607, EPI_ISL_2928608, EPI_ISL_2928609, EPI_ISL_2928610, EPI_ISL_2928615, EPI_ISL_2928617, EPI_ISL_2928618, EPI_ISL_2928619, EPI_ISL_2928620, EPI_ISL_2928622, EPI_ISL_2928623, EPI_ISL_2928624, EPI_ISL_2928625, EPI_ISL_2928626, EPI_ISL_2928630, EPI_ISL_2928631, EPI_ISL_2928632, EPI_ISL_2928633, EPI_ISL_2928635, EPI_ISL_2928636, EPI_ISL_2928637, EPI_ISL_2928638, EPI_ISL_2928639, EPI_ISL_2928640, EPI_ISL_2928641, EPI_ISL_2928642, EPI_ISL_2928643, EPI_ISL_2928644, EPI_ISL_2928645, EPI_ISL_2928646, EPI_ISL_2928647, EPI_ISL_2928648, EPI_ISL_2928700, EPI_ISL_2928703, EPI_ISL_2928704, EPI_ISL_2928705, EPI_ISL_2928706, EPI_ISL_2928707, EPI_ISL_2928708, EPI_ISL_2928709, EPI_ISL_2928710, EPI_ISL_2928711, EPI_ISL_2928712, EPI_ISL_2928713, EPI_ISL_2928714, EPI_ISL_2928715, EPI_ISL_2928716, EPI_ISL_2928717, EPI_ISL_2928718, EPI_ISL_2928719, EPI_ISL_2928720, EPI_ISL_2928721, EPI_ISL_2928722, EPI_ISL_2928723, EPI_ISL_2928724, EPI_ISL_2928725, EPI_ISL_2928726, EPI_ISL_2928727, EPI_ISL_2928728, EPI_ISL_2928729, EPI_ISL_2928730, EPI_ISL_2928731, EPI_ISL_2928732, EPI_ISL_2928733, EPI_ISL_2928734, EPI_ISL_2928735, EPI_ISL_2928736, EPI_ISL_2928737, EPI_ISL_2928738, EPI_ISL_2928739, EPI_ISL_2928740, EPI_ISL_2928741, EPI_ISL_2928742, EPI_ISL_2928743, EPI_ISL_2928744, EPI_ISL_2928745, EPI_ISL_2928746, EPI_ISL_2928747, EPI_ISL_2928748, EPI_ISL_2928749, EPI_ISL_2928750, EPI_ISL_2928751, EPI_ISL_2928752, EPI_ISL_2928753, EPI_ISL_2928754, EPI_ISL_2928755, EPI_ISL_2928756, EPI_ISL_2928757, EPI_ISL_2928758, EPI_ISL_2928759, EPI_ISL_2928760, EPI_ISL_2928761, EPI_ISL_2928762, EPI_ISL_2928763, EPI_ISL_2928764, EPI_ISL_2928765, EPI_ISL_2928766, EPI_ISL_2928767, EPI_ISL_2928768, EPI_ISL_2928769, EPI_ISL_2928770, EPI_ISL_2928771, EPI_ISL_2928772, EPI_ISL_2928773, EPI_ISL_2928774, EPI_ISL_2928775, EPI_ISL_2928776, EPI_ISL_2928777, EPI_ISL_2928778, EPI_ISL_2928779, EPI_ISL_2928780, EPI_ISL_2928781, EPI_ISL_2928782, EPI_ISL_2928783, EPI_ISL_2928784, EPI_ISL_2928785, EPI_ISL_2928786, EPI_ISL_2928787, EPI_ISL_2928788, EPI_ISL_2928789, EPI_ISL_2928790, EPI_ISL_2928791, EPI_ISL_2928792, EPI_ISL_2928793, EPI_ISL_2928794, EPI_ISL_2928795, EPI_ISL_2928796, EPI_ISL_2928797, EPI_ISL_2928798, EPI_ISL_2928799, EPI_ISL_2928800, EPI_ISL_2928801, EPI_ISL_2928802, EPI_ISL_2928803, EPI_ISL_2928804, EPI_ISL_2928805, EPI_ISL_2928806, EPI_ISL_2928807, EPI_ISL_2928808, EPI_ISL_2928809, EPI_ISL_2928810, EPI_ISL_2928811, EPI_ISL_2928812, EPI_ISL_2928813, EPI_ISL_2928814, EPI_ISL_2928815, EPI_ISL_2928816, EPI_ISL_2928817, EPI_ISL_2928818, EPI_ISL_2928819, EPI_ISL_2928820, EPI_ISL_2928821, EPI_ISL_2928822, EPI_ISL_2928823, EPI_ISL_2928824, EPI_ISL_2928825, EPI_ISL_2928826, EPI_ISL_2928827, EPI_ISL_2928828, EPI_ISL_2928829, EPI_ISL_2928830, EPI_ISL_2928831, EPI_ISL_2928832, EPI_ISL_2928833, EPI_ISL_2928834, EPI_ISL_2928835, EPI_ISL_2928836, EPI_ISL_2928837, EPI_ISL_2928838, EPI_ISL_2928839, EPI_ISL_2928840, EPI_ISL_2928841, EPI_ISL_2928842, EPI_ISL_2928843, EPI_ISL_2928844, EPI_ISL_2928845, EPI_ISL_2928846, EPI_ISL_2928847, EPI_ISL_2928848, EPI_ISL_2928849, EPI_ISL_2928850, EPI_ISL_2928851, EPI_ISL_2928852, EPI_ISL_2928853, EPI_ISL_2928854, EPI_ISL_2928855, EPI_ISL_2928856, EPI_ISL_2928857, EPI_ISL_2928858, EPI_ISL_2928859, EPI_ISL_2928860, EPI_ISL_2928861, EPI_ISL_2928862, EPI_ISL_2928863, EPI_ISL_2928864, EPI_ISL_2928865, EPI_ISL_2928866, EPI_ISL_2928867, EPI_ISL_2928868, EPI_ISL_2928869, EPI_ISL_2928870, EPI_ISL_2928871, EPI_ISL_2928872, EPI_ISL_2928873, EPI_ISL_2928874, EPI_ISL_2928875, EPI_ISL_2928876, EPI_ISL_2928877, EPI_ISL_2928878, EPI_ISL_2928879, EPI_ISL_2928880, EPI_ISL_2928881, EPI_ISL_2928882, EPI_ISL_2928883, EPI_ISL_2928884, EPI_ISL_2928885, EPI_ISL_2928886, EPI_ISL_2928887, EPI_ISL_2928888, EPI_ISL_2928889, EPI_ISL_2928890, EPI_ISL_2928891, EPI_ISL_2928892, EPI_ISL_2928893, EPI_ISL_2928894, EPI_ISL_2928895, EPI_ISL_2928896, EPI_ISL_2928897, EPI_ISL_2928898, EPI_ISL_2928899, EPI_ISL_2928900, EPI_ISL_2928901, EPI_ISL_2928902, EPI_ISL_2928903, EPI_ISL_2928906, EPI_ISL_2928908, EPI_ISL_2928911, EPI_ISL_2928914, EPI_ISL_2928917, EPI_ISL_2928920, EPI_ISL_2928921, EPI_ISL_2928922, EPI_ISL_2928923, EPI_ISL_2928924, EPI_ISL_2928926, EPI_ISL_2928927, EPI_ISL_2928928, EPI_ISL_2928929, EPI_ISL_2928930, EPI_ISL_2928931, EPI_ISL_2928932, EPI_ISL_2928933, EPI_ISL_2928934, EPI_ISL_2928935, EPI_ISL_2928936, EPI_ISL_2928937, EPI_ISL_2928938, EPI_ISL_2928939, EPI_ISL_2928940, EPI_ISL_2928941, EPI_ISL_2928942, EPI_ISL_2928943, EPI_ISL_2928944, EPI_ISL_2928945, EPI_ISL_2928946, EPI_ISL_2928947, EPI_ISL_2928948, EPI_ISL_2928949, EPI_ISL_2928950, EPI_ISL_2928951, EPI_ISL_2928952, EPI_ISL_2928953, EPI_ISL_2928954, EPI_ISL_2928955, EPI_ISL_2928956, EPI_ISL_2928957, EPI_ISL_2928958, EPI_ISL_2928959, EPI_ISL_2928960, EPI_ISL_2928961, EPI_ISL_2928962, EPI_ISL_2928963, EPI_ISL_2928964, EPI_ISL_2928965, EPI_ISL_2928966, EPI_ISL_2928967, EPI_ISL_2928968, EPI_ISL_2928969, EPI_ISL_2928970, EPI_ISL_2928971, EPI_ISL_2928972, EPI_ISL_2928973, EPI_ISL_2928974, EPI_ISL_2928975, EPI_ISL_2928976, EPI_ISL_2928977, EPI_ISL_2928978, EPI_ISL_2928979, EPI_ISL_2928980, EPI_ISL_2928981, EPI_ISL_2928982, EPI_ISL_2928983, EPI_ISL_2928984, EPI_ISL_2928985, EPI_ISL_2928986, EPI_ISL_2928987, EPI_ISL_2928988, EPI_ISL_2928989, EPI_ISL_2928990, EPI_ISL_2928991, EPI_ISL_2928992, EPI_ISL_2928993, EPI_ISL_2928994, EPI_ISL_2928995, EPI_ISL_2928996, EPI_ISL_2928997, EPI_ISL_2928998, EPI_ISL_2928999, EPI_ISL_2929000, EPI_ISL_2929001, EPI_ISL_2929002, EPI_ISL_2929003, EPI_ISL_2929004, EPI_ISL_2929005, EPI_ISL_2929006, EPI_ISL_2929007, EPI_ISL_2929008, EPI_ISL_2929009, EPI_ISL_2929010, EPI_ISL_2929011, EPI_ISL_2929012, EPI_ISL_2929013, EPI_ISL_2929014, EPI_ISL_2929015, EPI_ISL_2929016, EPI_ISL_2929017, EPI_ISL_2929018, EPI_ISL_2929019, EPI_ISL_2929020, EPI_ISL_2929021, EPI_ISL_2929022, EPI_ISL_2929023, EPI_ISL_2929024, EPI_ISL_2929025, EPI_ISL_2929026, EPI_ISL_2929027, EPI_ISL_2929028, EPI_ISL_2929029, EPI_ISL_2929030, EPI_ISL_2929031, EPI_ISL_2929032, EPI_ISL_2929033, EPI_ISL_2929034, EPI_ISL_2929035, EPI_ISL_2929036, EPI_ISL_2929037, EPI_ISL_2929038, EPI_ISL_2929039, EPI_ISL_2929040, EPI_ISL_2929041, EPI_ISL_2929042, EPI_ISL_2929043, EPI_ISL_2929044, EPI_ISL_2929045, EPI_ISL_2929046, EPI_ISL_2929047, EPI_ISL_2929048, EPI_ISL_2929049, EPI_ISL_2929050, EPI_ISL_2929051, EPI_ISL_2929052, EPI_ISL_2929053, EPI_ISL_2929054, EPI_ISL_2929055, EPI_ISL_2929056, EPI_ISL_2929057, EPI_ISL_2929058, EPI_ISL_2929059, EPI_ISL_2929060, EPI_ISL_2929061, EPI_ISL_2929062, EPI_ISL_2929063, EPI_ISL_2929064, EPI_ISL_2929065, EPI_ISL_2929066, EPI_ISL_2929067, EPI_ISL_2929068, EPI_ISL_2929069, EPI_ISL_2929070, EPI_ISL_2929071, EPI_ISL_2929072, EPI_ISL_2929073, EPI_ISL_2929074, EPI_ISL_2929075, EPI_ISL_2929076, EPI_ISL_2929077, EPI_ISL_2929078, EPI_ISL_2929079, EPI_ISL_2929080, EPI_ISL_2929081, EPI_ISL_2929082, EPI_ISL_2929083, EPI_ISL_2929084, EPI_ISL_2929085, EPI_ISL_2929086, EPI_ISL_2929087, EPI_ISL_2929088, EPI_ISL_2929089, EPI_ISL_2929090, EPI_ISL_2929091, EPI_ISL_2929092, EPI_ISL_2929093, EPI_ISL_2929094, EPI_ISL_2929095, EPI_ISL_2929096, EPI_ISL_2929097, EPI_ISL_2929098, EPI_ISL_2929099, EPI_ISL_2929100, EPI_ISL_2929101, EPI_ISL_2929102, EPI_ISL_2929103, EPI_ISL_2929104, EPI_ISL_2929105, EPI_ISL_2929106, EPI_ISL_2929107, EPI_ISL_2929108, EPI_ISL_2929109, EPI_ISL_2929110, EPI_ISL_2929111, EPI_ISL_2929112, EPI_ISL_2929113, EPI_ISL_2929114, EPI_ISL_2929115, EPI_ISL_2929116, EPI_ISL_2929117, EPI_ISL_2929118, EPI_ISL_2929119, EPI_ISL_2929120, EPI_ISL_2929121, EPI_ISL_2929122, EPI_ISL_2929123, EPI_ISL_2929124, EPI_ISL_2929125, EPI_ISL_2929126, EPI_ISL_2929127, EPI_ISL_3016478, EPI_ISL_3016479, EPI_ISL_3016480, EPI_ISL_3016481, EPI_ISL_3016482, EPI_ISL_3016483, EPI_ISL_3016484, EPI_ISL_3016485, EPI_ISL_3016486, EPI_ISL_3016487, EPI_ISL_3016488, EPI_ISL_3016489, EPI_ISL_3016490, EPI_ISL_3016491, EPI_ISL_3016492, EPI_ISL_3016493, EPI_ISL_3016494, EPI_ISL_3016495, EPI_ISL_3016496, EPI_ISL_3016497, EPI_ISL_3016498, EPI_ISL_3016499, EPI_ISL_3016500, EPI_ISL_3016501, EPI_ISL_3016502, EPI_ISL_3016503, EPI_ISL_3016504, EPI_ISL_3016505, EPI_ISL_3016506, EPI_ISL_3016507, EPI_ISL_3016508, EPI_ISL_3016509, EPI_ISL_3016510, EPI_ISL_3016511, EPI_ISL_3016512, EPI_ISL_3016513, EPI_ISL_3016514, EPI_ISL_3016515, EPI_ISL_3016516, EPI_ISL_3016517, EPI_ISL_3016518, EPI_ISL_3016519, EPI_ISL_3016520, EPI_ISL_3016521, EPI_ISL_3016522, EPI_ISL_3016523, EPI_ISL_3016524, EPI_ISL_3016525, EPI_ISL_3016526, EPI_ISL_3016527, EPI_ISL_3016528, EPI_ISL_3016529, EPI_ISL_3016530, EPI_ISL_3016531, EPI_ISL_3016532, EPI_ISL_3016533, EPI_ISL_3016534, EPI_ISL_3016535, EPI_ISL_3016536, EPI_ISL_3016537, EPI_ISL_3016538, EPI_ISL_3016539, EPI_ISL_3016540, EPI_ISL_3016541, EPI_ISL_3016542, EPI_ISL_3016543, EPI_ISL_3016544, EPI_ISL_3016545, EPI_ISL_3016546, EPI_ISL_3016547, EPI_ISL_3016548, EPI_ISL_3016549, EPI_ISL_3016550, EPI_ISL_3016551, EPI_ISL_3016552, EPI_ISL_3016553, EPI_ISL_3016554, EPI_ISL_3016555, EPI_ISL_3016556, EPI_ISL_3016557, EPI_ISL_3016558, EPI_ISL_3016559, EPI_ISL_3016560, EPI_ISL_3016561, EPI_ISL_3016562, EPI_ISL_3016563, EPI_ISL_3016564, EPI_ISL_3016565, EPI_ISL_3016566, EPI_ISL_3016567, EPI_ISL_3016568, EPI_ISL_3016569, EPI_ISL_3016570, EPI_ISL_3016571, EPI_ISL_3016572, EPI_ISL_3016574, EPI_ISL_3016575, EPI_ISL_3016576, EPI_ISL_3016577, EPI_ISL_3016578, EPI_ISL_3016579, EPI_ISL_3016580, EPI_ISL_3016581, EPI_ISL_3016582, EPI_ISL_3016583, EPI_ISL_3016584, EPI_ISL_3016585, EPI_ISL_3016586, EPI_ISL_3016587, EPI_ISL_3016588, EPI_ISL_3016589, EPI_ISL_3016590, EPI_ISL_3016591, EPI_ISL_3016592, EPI_ISL_3016593, EPI_ISL_3016594, EPI_ISL_3016595, EPI_ISL_3016596, EPI_ISL_3016597, EPI_ISL_3016598, EPI_ISL_3016599, EPI_ISL_3016600, EPI_ISL_3016601, EPI_ISL_3016602, EPI_ISL_3016603, EPI_ISL_3016604, EPI_ISL_3016605, EPI_ISL_3016606, EPI_ISL_3016607, EPI_ISL_3016608, EPI_ISL_3016609, EPI_ISL_3016610, EPI_ISL_3016611, EPI_ISL_3016612, EPI_ISL_3016613, EPI_ISL_3016614, EPI_ISL_3016615, EPI_ISL_3016616, EPI_ISL_3016617, EPI_ISL_3016637, EPI_ISL_3016638, EPI_ISL_3016639, EPI_ISL_3016640, EPI_ISL_3016641, EPI_ISL_3016642, EPI_ISL_3016643, EPI_ISL_3016644, EPI_ISL_3016645, EPI_ISL_3016646, EPI_ISL_3016647, EPI_ISL_3016648, EPI_ISL_3016649, EPI_ISL_3016650, EPI_ISL_3016651, EPI_ISL_3016652, EPI_ISL_3016653, EPI_ISL_3016654, EPI_ISL_3016655, EPI_ISL_3016656, EPI_ISL_3016657, EPI_ISL_3016658, EPI_ISL_3016659, EPI_ISL_3016660, EPI_ISL_3016661, EPI_ISL_3016662, EPI_ISL_3016663, EPI_ISL_3016664, EPI_ISL_3016665, EPI_ISL_3016667, EPI_ISL_3016669, EPI_ISL_3016670, EPI_ISL_3016671, EPI_ISL_3016672, EPI_ISL_3016673, EPI_ISL_3016674, EPI_ISL_3016675, EPI_ISL_3016677, EPI_ISL_3016678, EPI_ISL_3016679, EPI_ISL_3016680, EPI_ISL_3016681, EPI_ISL_3016682, EPI_ISL_3016683, EPI_ISL_3016684, EPI_ISL_3016685, EPI_ISL_3016686, EPI_ISL_3016687, EPI_ISL_3016688, EPI_ISL_3016689, EPI_ISL_3016690, EPI_ISL_3016691, EPI_ISL_3016692, EPI_ISL_3016693, EPI_ISL_3016694, EPI_ISL_3016695, EPI_ISL_3016696, EPI_ISL_3016697, EPI_ISL_3016698, EPI_ISL_3016699, EPI_ISL_3016700, EPI_ISL_3016701, EPI_ISL_3016702, EPI_ISL_3016703, EPI_ISL_3016704, EPI_ISL_3016705, EPI_ISL_3016706, EPI_ISL_3064778, EPI_ISL_3064808, EPI_ISL_3064809, EPI_ISL_3064810, EPI_ISL_3064811, EPI_ISL_3064812, EPI_ISL_3064813, EPI_ISL_3064814, EPI_ISL_3064815, EPI_ISL_3064816, EPI_ISL_3064817, EPI_ISL_3064818, EPI_ISL_3064819, EPI_ISL_3064820, EPI_ISL_3064821, EPI_ISL_3064822, EPI_ISL_3064823, EPI_ISL_3064824, EPI_ISL_3064825, EPI_ISL_3064826, EPI_ISL_3064827, EPI_ISL_3064828, EPI_ISL_3064829, EPI_ISL_3064830, EPI_ISL_3064831, EPI_ISL_3064832, EPI_ISL_3064833, EPI_ISL_3064834, EPI_ISL_3064835, EPI_ISL_3064836, EPI_ISL_3064837, EPI_ISL_3064838, EPI_ISL_3064839, EPI_ISL_3064840, EPI_ISL_3064841, EPI_ISL_3064842, EPI_ISL_3064843, EPI_ISL_3064844, EPI_ISL_3064845, EPI_ISL_3064846, EPI_ISL_3064847, EPI_ISL_3064848, EPI_ISL_3064849, EPI_ISL_3064850, EPI_ISL_3064851, EPI_ISL_3064852, EPI_ISL_3064853, EPI_ISL_3064854, EPI_ISL_3064855, EPI_ISL_3064856, EPI_ISL_3064857, EPI_ISL_3064858, EPI_ISL_3064859, EPI_ISL_3064860, EPI_ISL_3064861, EPI_ISL_3064862, EPI_ISL_3064863, EPI_ISL_3064864, EPI_ISL_3064865, EPI_ISL_3064866, EPI_ISL_3064867, EPI_ISL_3064868, EPI_ISL_3064869, EPI_ISL_3064870, EPI_ISL_3064871, EPI_ISL_3064872, EPI_ISL_3064873, EPI_ISL_3064874, EPI_ISL_3064875, EPI_ISL_3064876, EPI_ISL_3064877, EPI_ISL_3064878, EPI_ISL_3064879, EPI_ISL_3064880, EPI_ISL_3064881, EPI_ISL_3064882, EPI_ISL_3064883, EPI_ISL_3064885, EPI_ISL_3064886, EPI_ISL_3064887, EPI_ISL_3064888, EPI_ISL_3064889, EPI_ISL_3064890, EPI_ISL_3064891, EPI_ISL_3064892, EPI_ISL_3064893, EPI_ISL_3064894, EPI_ISL_3064895, EPI_ISL_3064896, EPI_ISL_3064897, EPI_ISL_3064898, EPI_ISL_3064899, EPI_ISL_3064900, EPI_ISL_3064901, EPI_ISL_3064902, EPI_ISL_3064903, EPI_ISL_3064904, EPI_ISL_3064905, EPI_ISL_3064906, EPI_ISL_3064907, EPI_ISL_3064908, EPI_ISL_3064909, EPI_ISL_3064910, EPI_ISL_3064911, EPI_ISL_3064912, EPI_ISL_3064913, EPI_ISL_3064914, EPI_ISL_3064915, EPI_ISL_3064916, EPI_ISL_3064917, EPI_ISL_3064918, EPI_ISL_3064919, EPI_ISL_3064920, EPI_ISL_3064921, EPI_ISL_3064922, EPI_ISL_3064923, EPI_ISL_3064924, EPI_ISL_3064925, EPI_ISL_3064926, EPI_ISL_3064927, EPI_ISL_3064928, EPI_ISL_3064929, EPI_ISL_3064930, EPI_ISL_3064931, EPI_ISL_3064932, EPI_ISL_3064933, EPI_ISL_3064934, EPI_ISL_3064935, EPI_ISL_3064936, EPI_ISL_3064937, EPI_ISL_3064938, EPI_ISL_3064939, EPI_ISL_3064940, EPI_ISL_3064941, EPI_ISL_3064942, EPI_ISL_3064943, EPI_ISL_3064944, EPI_ISL_3064945, EPI_ISL_3064946, EPI_ISL_3064947, EPI_ISL_3064948, EPI_ISL_3064949, EPI_ISL_3064950, EPI_ISL_3064951, EPI_ISL_3064952, EPI_ISL_3064953, EPI_ISL_3064954, EPI_ISL_3064955, EPI_ISL_3064956, EPI_ISL_3064957, EPI_ISL_3064958, EPI_ISL_3064959, EPI_ISL_3064960, EPI_ISL_3064961, EPI_ISL_3064962, EPI_ISL_3064963, EPI_ISL_3064964, EPI_ISL_3064965, EPI_ISL_3064966, EPI_ISL_3064967, EPI_ISL_3064968, EPI_ISL_3064969, EPI_ISL_3064970, EPI_ISL_3064971, EPI_ISL_3064972, EPI_ISL_3064973, EPI_ISL_3064974, EPI_ISL_3064975, EPI_ISL_3064976, EPI_ISL_3064977, EPI_ISL_3064978, EPI_ISL_3064979, EPI_ISL_3064980, EPI_ISL_3064981, EPI_ISL_3064982, EPI_ISL_3064983, EPI_ISL_3064984, EPI_ISL_3064985, EPI_ISL_3064986, EPI_ISL_3064987, EPI_ISL_3064988, EPI_ISL_3064989, EPI_ISL_3064990, EPI_ISL_3064991, EPI_ISL_3064992, EPI_ISL_3064993, EPI_ISL_3064994, EPI_ISL_3064995, EPI_ISL_3064996, EPI_ISL_3064997, EPI_ISL_3064998, EPI_ISL_3064999, EPI_ISL_3065000, EPI_ISL_3065001, EPI_ISL_3065002, EPI_ISL_3065003, EPI_ISL_3065004, EPI_ISL_3065005, EPI_ISL_3065006, EPI_ISL_3065007, EPI_ISL_3065008, EPI_ISL_3065009, EPI_ISL_3065010, EPI_ISL_3065011, EPI_ISL_3065012, EPI_ISL_3065013, EPI_ISL_3065014, EPI_ISL_3065015, EPI_ISL_3065016, EPI_ISL_3065017, EPI_ISL_3065036 | | | |
| see above | Fulgent Genetics | Centers for Disease Control and Prevention Division of Viral Diseases, Pathogen Discovery | Adrian Paskey; Becky Tsai; Benafsh Sapra; Benjamin Rambo-Martin; Christopher Gulvick; Clinton R. Paden; Dakota Howard; Darlene Wagner; Dhwani Batra; Doreen Ng; Duncan MacCannell; Harry Gao; James Xie; Jason Caravas; John Gao; Joseph Fierro; Kara Moser; Matthew Schmerer; Mickey Li; Peter W. Cook; Scott Sammons; Shatavia Morrison; Yan Meng; Yvette Unoarumhi |
| EPI_ISL_2967965, EPI_ISL_2967966 | Good Samaritan Hospital | Santa Clara County public Health Laboratory | Santa Clara County Public Health Department |
| EPI_ISL_3020049, EPI_ISL_3020059 | Infinity Biologix | Centers for Disease Control and Prevention Division of Viral Diseases, Pathogen Discovery | Adrian Paskey; Benjamin Rambo-Martin; Chirayu Goswami; Christian Bixby; Christopher Gulvick; Clinton R. Paden; Dakota Howard; Darlene Wagner; Dhwani Batra; Duncan MacCannell; Jason Caravas; Jonathan Schultz; Kara Moser; Matthew Schmerer; Peter W. Cook; Robin Grimwood; Russ Hager; Scott Sammons; Shatavia Morrison; Yihe Wang; Yvette Unoarumhi |
| EPI_ISL_2967969, EPI_ISL_2967970, EPI_ISL_2967971 | Kaiser San Jose | Santa Clara County public Health Laboratory | Santa Clara County Public Health Department |
| EPI_ISL_2967964 | Kaiser Santa Clara | Santa Clara County public Health Laboratory | Santa Clara County Public Health Department |
| EPI_ISL_3016796, EPI_ISL_3018033, EPI_ISL_3018035, EPI_ISL_3018037, EPI_ISL_3018038, EPI_ISL_3018039, EPI_ISL_3018040, EPI_ISL_3018050, EPI_ISL_3018051, EPI_ISL_3018064, EPI_ISL_3018103, EPI_ISL_3018104, EPI_ISL_3018105, EPI_ISL_3018106, EPI_ISL_3018107, EPI_ISL_3018108, EPI_ISL_3018109, EPI_ISL_3018110, EPI_ISL_3018111, EPI_ISL_3018112, EPI_ISL_3018113, EPI_ISL_3018119, EPI_ISL_3018120, EPI_ISL_3018121, EPI_ISL_3018122, EPI_ISL_3018127, EPI_ISL_3018129, EPI_ISL_3018135, EPI_ISL_3018136, EPI_ISL_3018137, EPI_ISL_3018138, EPI_ISL_3018139, EPI_ISL_3018140, EPI_ISL_3018141, EPI_ISL_3018142, EPI_ISL_3018143 | | | |
| see above | Laboratory Corporation of America | Centers for Disease Control and Prevention Division of Viral Diseases, Pathogen Discovery | Adrian Paskey; Amanda Douglas; Amanda Suchanek; Andrea Throop; Ayla Burns; Benjamin Rambo-Martin; Bobbi Croy; Brian Krueger; Brian Norvell; Christopher Gulvick; Christos Petropoulos; Clinton R. Paden; Craig Lukasik; Dakota Howard; Darlene Wagner; Debbie Boles; Dhwani Batra; Duncan MacCannell; Eyad Almasri; Goran Stevovic; Howard Engler; Hrushikesh Deshmukh; Jake Humphrey; Jana Schroth; Jason Caravas; Joe Voshell; John Pruitt; Jonathan Meltzer; Jonathan Williams; Kara Moser; Kimberly Wagner; Lax Iyer; Lisa Pfefferle; Lyndon Tilson; Manoj Jain; Marcia Eisenberg; Mary Ann Cristobal; Mary Williamson; Matthew Robinson; Matthew Schmerer; Michael Levandoski; Mike Sapeta; Mindy Nye; Minoo Agarwal; Mohan Kolli; Nuthawin Charoensri; Oren Cohen; Peter W. Cook; Prashant Gupta; Qian Zeng; Rama Ghatti; Scott Parker; Scott Ryan; Scott Sammons; Shatavia Morrison; Stanley Letovsky; Steven Ragan; Suresh Babu Selvaraju; Susan Countryman; Susan Hicks; Suzanne Dale; Thomas Urban; Tim Kuphal; Tricia Zwiefelhofer; Vincent Drouillon; Yvette Unoarumhi |

| | | | |
|---|---|---|---|
| EPI_ISL_2907561 | Los Angeles County Public Health Laboratories | Los Angeles County Public Health Laboratories | P. Hemarajata et al. |
| EPI_ISL_3065488 | Mako Medical | Centers for Disease Control and Prevention Division of Viral Diseases, Pathogen Discovery | Adrian Paskey; Benjamin Rambo-Martin; Christopher Gulvick; Clinton R. Paden; Dakota Howard; Darlene Wagner; Dhwani Batra; Duncan MacCannell; Jason Caravas; Kara Moser; Lauren Moon; Matthew Schmerer; Matthew Tugwell; Peter W. Cook; Scott Sammons; Shatavia Morrison; Yvette Unoarumhi |
| EPI_ISL_2955364, EPI_ISL_2955365, EPI_ISL_2955366, EPI_ISL_3037245, EPI_ISL_3037247, EPI_ISL_3037248, EPI_ISL_3037249, EPI_ISL_3037251, EPI_ISL_3065507, EPI_ISL_3065508, EPI_ISL_3065509, EPI_ISL_3065510, EPI_ISL_3065511, EPI_ISL_3065512, EPI_ISL_3065513, EPI_ISL_3065514, EPI_ISL_3065515, EPI_ISL_3065516, EPI_ISL_3065517, EPI_ISL_3065518, EPI_ISL_3065519, EPI_ISL_3065520 | | | |
| see above | Monterey County Public Health Laboratory | Monterey County Public Health Laboratory | Monterey County Public Health Laboratory |
| EPI_ISL_2928220, EPI_ISL_3010322, EPI_ISL_3010323, EPI_ISL_3010324, EPI_ISL_3010325, EPI_ISL_3010326, EPI_ISL_3010327, EPI_ISL_3010328, EPI_ISL_3010329, EPI_ISL_3010330, EPI_ISL_3010331, EPI_ISL_3010332, EPI_ISL_3063042, EPI_ISL_3063043, EPI_ISL_3063044, EPI_ISL_3063045, EPI_ISL_3063046, EPI_ISL_3063047, EPI_ISL_3063048, EPI_ISL_3063049, EPI_ISL_3063050, EPI_ISL_3063051, EPI_ISL_3063052, EPI_ISL_3063053, EPI_ISL_3063054, EPI_ISL_3063055, EPI_ISL_3063056, EPI_ISL_3063057, EPI_ISL_3063058, EPI_ISL_3063059, EPI_ISL_3063060, EPI_ISL_3063061, EPI_ISL_3063062, EPI_ISL_3063063, EPI_ISL_3063064, EPI_ISL_3063065, EPI_ISL_3063066, EPI_ISL_3063067, EPI_ISL_3063068, EPI_ISL_3063069, EPI_ISL_3063070, EPI_ISL_3063071, EPI_ISL_3063072, EPI_ISL_3063073, EPI_ISL_3063074, EPI_ISL_3063075, EPI_ISL_3063076, EPI_ISL_3063077, EPI_ISL_3063078, EPI_ISL_3063079, EPI_ISL_3063080, EPI_ISL_3063081, EPI_ISL_3063082, EPI_ISL_3063083, EPI_ISL_3063084, EPI_ISL_3063085, EPI_ISL_3063086, EPI_ISL_3063087, EPI_ISL_3063088, EPI_ISL_3063089, EPI_ISL_3063090, EPI_ISL_3063091, EPI_ISL_3063092, EPI_ISL_3063093, EPI_ISL_3063094, EPI_ISL_3063095, EPI_ISL_3063096, EPI_ISL_3063097, EPI_ISL_3063098, EPI_ISL_3063099, EPI_ISL_3063100, EPI_ISL_3063101, EPI_ISL_3063102, EPI_ISL_3063103, EPI_ISL_3063104 | | | |
| see above | Orange County Public Health Laboratory | Orange County Public Health Laboratory | Angelica Torres; Julia Wolfe |
| EPI_ISL_2967973 | PAMF Palo Alto | Santa Clara County public Health Laboratory | Santa Clara County Public Health Department |
| EPI_ISL_2907569, EPI_ISL_2907570, EPI_ISL_2907571, EPI_ISL_2907572 | Providence Saint John's Health Center | Los Angeles County Public Health Laboratories | P. Hemarajata et al. |
| EPI_ISL_2930264, EPI_ISL_3018241, EPI_ISL_3018549, EPI_ISL_3018567, EPI_ISL_3018568, EPI_ISL_3018596, EPI_ISL_3018615, EPI_ISL_3018630, EPI_ISL_3018631, EPI_ISL_3018643, EPI_ISL_3018699, EPI_ISL_3018705, EPI_ISL_3018713, EPI_ISL_3018750, EPI_ISL_3018777, EPI_ISL_3018792, EPI_ISL_3018794, EPI_ISL_3018844, EPI_ISL_3018851, EPI_ISL_3018898, EPI_ISL_3018911, EPI_ISL_3018926 | | | |
| see above | Quest Diagnostics Incorporated | Centers for Disease Control and Prevention Division of Viral Diseases, Pathogen Discovery | A. Gerasimova; A. Perez; Adrian Paskey; B. Anderson; Benjamin Rambo-Martin; Christopher Gulvick; Clinton R. Paden; Dakota Howard; Darlene Wagner; Dhwani Batra; Duncan MacCannell; F. Lacbawan; I. A. Shlyakhter; Jason Caravas; K.E. Livingston; Kara Moser; L.E. Bernstein; M. Hua; Matthew Schmerer; P. Tanpaiboon; Peter W. Cook; R. M. Kagan; R. Owen; R. V. Rolando; S. H. Rosenthal; Scott Sammons; Shatavia Morrison; Y. Liu; Yvette Unoarumhi |
| EPI_ISL_3048870, EPI_ISL_3048908 | Santa Clara County Public Health Laboratory | Santa Clara County Public Health Laboratory | Santa Clara County Public Health Department |
| EPI_ISL_2967931 | Santa Clara County Public Health Laboratory | Santa Clara County public Health Laboratory | Santa Clara County Public Health Department |
| EPI_ISL_3048875, EPI_ISL_3048876, EPI_ISL_3048907, EPI_ISL_3048919, EPI_ISL_3048920, EPI_ISL_3048921, EPI_ISL_3048922, EPI_ISL_3048923, EPI_ISL_3048924, EPI_ISL_3048925, EPI_ISL_3048926, EPI_ISL_3048927, EPI_ISL_3048928, EPI_ISL_3048929, EPI_ISL_3048930, EPI_ISL_3048931, EPI_ISL_3048932, EPI_ISL_3048933, EPI_ISL_3048934, EPI_ISL_3048935, EPI_ISL_3048936, EPI_ISL_3048937, EPI_ISL_3048938, EPI_ISL_3048939, EPI_ISL_3048940, EPI_ISL_3048941, EPI_ISL_3048942, EPI_ISL_3048943, EPI_ISL_3048944, EPI_ISL_3048945, EPI_ISL_3048946, EPI_ISL_3067573, EPI_ISL_3067574, EPI_ISL_3067575, EPI_ISL_3067576, EPI_ISL_3067577, EPI_ISL_3067578, EPI_ISL_3067579, EPI_ISL_3067580, EPI_ISL_3067581, EPI_ISL_3067582, EPI_ISL_3067583, EPI_ISL_3067584, EPI_ISL_3067585, EPI_ISL_3067586, EPI_ISL_3067587, EPI_ISL_3067588, EPI_ISL_3067589, EPI_ISL_3067590, EPI_ISL_3067591, EPI_ISL_3067592, EPI_ISL_3067593, EPI_ISL_3067594, EPI_ISL_3067595, EPI_ISL_3067596, EPI_ISL_3067597, EPI_ISL_3067598, EPI_ISL_3067599, EPI_ISL_3067600, EPI_ISL_3067601, EPI_ISL_3067602, EPI_ISL_3067603, EPI_ISL_3067604, EPI_ISL_3067605, EPI_ISL_3067606, EPI_ISL_3067607, EPI_ISL_3067608, EPI_ISL_3067609, EPI_ISL_3067610, EPI_ISL_3067611, EPI_ISL_3067612, EPI_ISL_3067613, EPI_ISL_3067614, EPI_ISL_3067615, EPI_ISL_3067616, EPI_ISL_3067617, EPI_ISL_3067618, EPI_ISL_3067619, EPI_ISL_3067620, EPI_ISL_3067621, EPI_ISL_3067622, EPI_ISL_3067623, EPI_ISL_3067624, EPI_ISL_3067625, EPI_ISL_3067626, EPI_ISL_3067627, EPI_ISL_3067628, EPI_ISL_3067629, EPI_ISL_3067630, EPI_ISL_3067631, EPI_ISL_3067632, EPI_ISL_3067633, EPI_ISL_3067634, EPI_ISL_3067635, EPI_ISL_3067636, EPI_ISL_3067637, EPI_ISL_3067638, EPI_ISL_3067639, EPI_ISL_3067640, EPI_ISL_3067641, EPI_ISL_3067642, EPI_ISL_3067643, EPI_ISL_3067644, EPI_ISL_3067645, EPI_ISL_3067646, EPI_ISL_3067647, EPI_ISL_3067648, EPI_ISL_3067649, EPI_ISL_3067650, EPI_ISL_3067651 | | | |
| see above | Santa Clara Valley Medical Center | Santa Clara County Public Health Laboratory | Santa Clara County Public Health Department |
| EPI_ISL_3050217, EPI_ISL_3050218, EPI_ISL_3050220, EPI_ISL_3050221, EPI_ISL_3050222, EPI_ISL_3050224, EPI_ISL_3050225, EPI_ISL_3050226, EPI_ISL_3050227, EPI_ISL_3050228, EPI_ISL_3050229, EPI_ISL_3050230, EPI_ISL_3050231, EPI_ISL_3050232, EPI_ISL_3050233, EPI_ISL_3050234, EPI_ISL_3050235, EPI_ISL_3050236, EPI_ISL_3050237, EPI_ISL_3050238, EPI_ISL_3050239, EPI_ISL_3050240, EPI_ISL_3050241, EPI_ISL_3050242, EPI_ISL_3050243, EPI_ISL_3050244, EPI_ISL_3050245, EPI_ISL_3050246, EPI_ISL_3050247, EPI_ISL_3050248, EPI_ISL_3050249, EPI_ISL_3050250, EPI_ISL_3050251, EPI_ISL_3050252, EPI_ISL_3050253, EPI_ISL_3050254, EPI_ISL_3050255, EPI_ISL_3050256 | | | |
| see above | Stanford Health Care | Stanford University School of Medicine, Clinical Virology Laboratory | Becky Jiang; Bernadette Troung; Daniel Solis; James Zehnder; Malaya K. Sahoo; Mamdouh Sibai; Nathan Hammond; Selamawit Bihon; and Benjamin A. Pinsky |
| EPI_ISL_3021467 | Twin Cities Community Hospital Lab | County of San Luis Obispo Public Health Laboratory | Frances Sidhu; Katrina Erwin |
| EPI_ISL_3015204, EPI_ISL_3015205, EPI_ISL_3015206, EPI_ISL_3061931, EPI_ISL_3061932, EPI_ISL_3061933, EPI_ISL_3061934 | | | |
| see above | UCSC Genomics Institute | UCSC Genomics Institute | A. Marm Kilpatrick; Angie Hinrichs; Beth Shapiro; Bryan Thornlow; Ciara Wanket; David Haussler; Eric Beraut; Hugh Olsen; Ikenna Anigbogu; Isabel Bjork; Jakob McBroome; Jeremy Sanford; Joshua Kapp; Mark Akeson; Maximilian Haeussler; Michael Stone; Miten Jain; Molly Cassatt-Johnstone; Namrita Dhillon; Russell Corbett-Detig; Terren Chang; Yatish Turakhia |

We gratefully acknowledge the following Authors from the Originating laboratories responsible for obtaining the specimens, as well as the Submitting laboratories where the genome data were generated and shared via GISAID, on which this research is based.

All Submitters of data may be contacted directly via www.gisaid.org

Authors are sorted alphabetically.

| Accession ID | Originating Laboratory | Submitting Laboratory | Authors |
|---|---|---|---|
| EPI_ISL_1966393, EPI_ISL_1966394 | ADECIO POMPEU CARVALHO | Instituto Butantan / Mendelics | Antonio Jorge Martins; Bianca Cechetto Carlos. Mendelics: Bibiana Santos; Claudia Renata dos Santos Barros; Cíntia Bittar; David Schlesinger. Hemocentro Ribeirão Preto: Simone Kashima; Debora Botequio Moretti; Elaine Cristina Marqueze; Elaine Vieira dos Santos; Elisangela Chicaroni Mattos; Erika Freitas; Evandra Strazza Rodrigues; Felipe Allan da Silva da Costa; Flavia Aburjaile; Fábio Sossai Possebon; Guilherme Campos; Guilherme Targino Valente; Heidge Fukumasu. USP-Botucatu: Rejane Maria Tommasini Grotto; Helena Lage Ferreira; Instituto Butantan: Dimas Tadeu Covas; Jardelina de Souza Todao Bernardino; Jayme A. Souza-Neto; Jessika Cristina Chagas Lesbon; Jorge A. Petroli Marchesi; José Salvatore Leister Patané; João Paulo Kitajima; João Pessoa Araújo Jr.; Leila Sabrina Ullmann; Loyze Paola Oliveira de Lima; Luiz Aurelio de Campos Crispin. Centro de Genômica Funcional da ESALQ: Luiz Lehmann Coutinho; Luiz Carlos Junior de Alcantara; Lívia Sacchetto; Maisa C. Pereira Parra; Maria Carolina Elias; Marta Giovanetti; Marília Moraes; Maurício Lacerda Nogueira. Prefeitura de Sao Paulo: Melissa Palmieri.; Patricia Akemi Assato; Paula Rahal; Paulo Inacio da Costa; Rafael dos Santos Bezerra; Raquel de Lello Rocha Campos Cassano. NGS Soluções Genômicas: Pilar Drummond Sampaio Corrêa Mariani. FZEA-USP Pirassununga: Mirele Daiana Poleti; Raul Machado Neto; Ricardo Augusto Brassaloti; Ricardo Haddad; Rodrigo Tocantins Calado. FAMERP-SJRP: Cecília Artico Banho; Sandra Coccuzzo Sampaio; Svetoslav Nanev Slavov; Vagner Fonseca; Vincent Louis Viala |
| EPI_ISL_1966776 | AMB MEDICO DR FRANCISCO FELIX DE MENDONCA SAUDADE | Instituto Butantan / Mendelics | Antonio Jorge Martins; Bianca Cechetto Carlos. Mendelics: Bibiana Santos; Claudia Renata dos Santos Barros; Cíntia Bittar; David Schlesinger. Hemocentro Ribeirão Preto: Simone Kashima; Debora Botequio Moretti; Elaine Cristina Marqueze; Elaine Vieira dos Santos; Elisangela Chicaroni Mattos; Erika Freitas; Evandra Strazza Rodrigues; Felipe Allan da Silva da Costa; Flavia Aburjaile; Fábio Sossai Possebon; Guilherme Campos; Guilherme Targino Valente; Heidge Fukumasu. USP-Botucatu: Rejane Maria Tommasini Grotto; Helena Lage Ferreira; Instituto Butantan: Dimas Tadeu Covas; Jardelina de Souza Todao Bernardino; Jayme A. Souza-Neto; Jessika Cristina Chagas Lesbon; Jorge A. Petroli Marchesi; José Salvatore Leister Patané; João Paulo Kitajima; João Pessoa Araújo Jr.; Leila Sabrina Ullmann; Loyze Paola Oliveira de Lima; Luiz Aurelio de Campos Crispin. Centro de Genômica Funcional da ESALQ: Luiz Lehmann Coutinho; Luiz Carlos Junior de Alcantara; Lívia Sacchetto; Maisa C. Pereira Parra; Maria Carolina Elias; Marta Giovanetti; Marília Moraes; Maurício Lacerda Nogueira. Prefeitura de Sao Paulo: Melissa Palmieri.; Patricia Akemi Assato; Paula Rahal; Paulo Inacio da Costa; Rafael dos Santos Bezerra; Raquel de Lello Rocha Campos Cassano. NGS Soluções Genômicas: Pilar Drummond Sampaio Corrêa Mariani. FZEA-USP Pirassununga: Mirele Daiana Poleti; Raul Machado Neto; Ricardo Augusto Brassaloti; Ricardo Haddad; Rodrigo Tocantins Calado. FAMERP-SJRP: Cecília Artico Banho; Sandra Coccuzzo Sampaio; Svetoslav Nanev Slavov; Vagner Fonseca; Vincent Louis Viala |
| EPI_ISL_1966816 | AMBULATORIO COVID 19 | Instituto Butantan / Mendelics | Antonio Jorge Martins; Bianca Cechetto Carlos. Mendelics: Bibiana Santos; Claudia Renata dos Santos Barros; Cíntia Bittar; David Schlesinger. Hemocentro Ribeirão Preto: Simone Kashima; Debora Botequio Moretti; Elaine Cristina Marqueze; Elaine Vieira dos Santos; Elisangela Chicaroni Mattos; Erika Freitas; Evandra Strazza Rodrigues; Felipe Allan da Silva da Costa; Flavia Aburjaile; Fábio Sossai Possebon; Guilherme Campos; Guilherme Targino Valente; Heidge Fukumasu. USP-Botucatu: Rejane Maria Tommasini Grotto; Helena Lage Ferreira; Instituto Butantan: Dimas Tadeu Covas; Jardelina de Souza Todao Bernardino; Jayme A. Souza-Neto; Jessika Cristina Chagas Lesbon; Jorge A. Petroli Marchesi; José Salvatore Leister Patané; João Paulo Kitajima; João Pessoa Araújo Jr.; Leila Sabrina Ullmann; Loyze Paola Oliveira de Lima; Luiz Aurelio de Campos Crispin. Centro de Genômica Funcional da ESALQ: Luiz Lehmann Coutinho; Luiz Carlos Junior de Alcantara; Lívia Sacchetto; Maisa C. Pereira Parra; Maria Carolina Elias; Marta Giovanetti; Marília Moraes; Maurício Lacerda Nogueira. Prefeitura de Sao Paulo: Melissa Palmieri.; Patricia Akemi Assato; Paula Rahal; Paulo Inacio da Costa; Rafael dos Santos Bezerra; Raquel de Lello Rocha Campos Cassano. NGS Soluções Genômicas: Pilar Drummond Sampaio Corrêa Mariani. FZEA-USP Pirassununga: Mirele Daiana Poleti; Raul Machado Neto; Ricardo Augusto Brassaloti; Ricardo Haddad; Rodrigo Tocantins Calado. FAMERP-SJRP: Cecília Artico Banho; Sandra Coccuzzo Sampaio; Svetoslav Nanev Slavov; Vagner Fonseca; Vincent Louis Viala |
| EPI_ISL_1795113 | AMBULATORIO DE ATENDIMENTO DE DST DE GUARIBA | Instituto Butantan / ESALQ-Piracicaba | Antonio Jorge Martins; Bianca Cechetto Carlos. Mendelics: Bibiana Santos; Claudia Renata dos Santos Barros; David Schlesinger. Hemocentro Ribeirão Preto: Simone Kashima; Debora Botequio Moretti. Centro de Genômica Funcional da ESALQ: Luiz Lehmann Coutinho; Dimas Tadeu Covas; Elaine Cristina Marqueze; Elaine Vieira dos Santos; Elisangela Chicaroni Mattos; Erika Freitas; Evandra Strazza Rodrigues; Felipe Allan da Silva da Costa; Flavia Aburjaile; Guilherme Targino Valente; Heidge Fukumasu. USP-Botucatu: Rejane Maria Tommasini Grotto; Instituto Butantan: Alexander Roberto Precioso; Jayme A. Souza-Neto; Jessika Cristina Chagas Lesbon; José Salvatore Leister Patané; João Paulo Kitajima; Luiz Carlos Junior de Alcantara; Maria Carolina Elias; Marta Giovanetti; Patricia Akemi Assato; Rafael dos Santos Bezerra; Raquel de Lello Rocha Campos Cassano. NGS Soluções Genômicas: Pilar Drummond Sampaio Corrêa Mariani. FZEA-USP Pirassununga: Mirele Daiana Poleti; Raul Machado Neto; Ricardo Augusto Brassaloti; Ricardo Haddad; Rodrigo Tocantins Calado.; Sandra Coccuzzo Sampaio; Svetoslav Nanev Slavov; Vagner Fonseca; Vincent Louis Viala |
| EPI_ISL_1795062, EPI_ISL_1795194, EPI_ISL_1795195, EPI_ISL_1795196, EPI_ISL_1795197, EPI_ISL_1795198, EPI_ISL_1795199, EPI_ISL_1795200, EPI_ISL_1795201, EPI_ISL_1795202, EPI_ISL_1795203, EPI_ISL_1795204, EPI_ISL_1795205, EPI_ISL_1795206, EPI_ISL_1795207, EPI_ISL_1795210, EPI_ISL_1795291, EPI_ISL_1795292, EPI_ISL_1795417 | | | |
| see above | AMBULATORIO DE ESPECIALIDADE V E MOGI MIRIM | Instituto Butantan / ESALQ-Piracicaba | Antonio Jorge Martins; Bianca Cechetto Carlos. Mendelics: Bibiana Santos; Claudia Renata dos Santos Barros; David Schlesinger. Hemocentro Ribeirão Preto: Simone Kashima; Debora Botequio Moretti. Centro de Genômica Funcional da ESALQ: Luiz Lehmann Coutinho; Dimas Tadeu Covas; Elaine Cristina Marqueze; Elaine Vieira dos Santos; Elisangela Chicaroni Mattos; Erika Freitas; Evandra Strazza Rodrigues; Felipe Allan da Silva da Costa; Flavia Aburjaile; Guilherme Targino Valente; Heidge Fukumasu. USP-Botucatu: Rejane Maria Tommasini Grotto; Instituto Butantan: Alexander Roberto Precioso; Jayme A. Souza-Neto; Jessika Cristina Chagas Lesbon; José Salvatore Leister Patané; João Paulo Kitajima; Luiz Carlos Junior de Alcantara; Maria Carolina Elias; Marta Giovanetti; Patricia Akemi Assato; Rafael dos Santos Bezerra; Raquel de Lello Rocha Campos Cassano. NGS Soluções Genômicas: Pilar Drummond Sampaio Corrêa Mariani. FZEA-USP Pirassununga: Mirele Daiana Poleti; Raul Machado Neto; Ricardo Augusto Brassaloti; Ricardo Haddad; Rodrigo Tocantins Calado.; Sandra Coccuzzo Sampaio; Svetoslav Nanev Slavov; Vagner Fonseca; Vincent Louis Viala |
| EPI_ISL_1967296, EPI_ISL_1967297, EPI_ISL_2171018, EPI_ISL_2171019 | AMBULATORIO DE ESPECIALIDADE V E MOGI MIRIM | Instituto Butantan / Mendelics | Antonio Jorge Martins; Bianca Cechetto Carlos. Mendelics: Bibiana Santos; Claudia Renata dos Santos Barros; Cíntia Bittar; David Schlesinger. Hemocentro Ribeirão Preto: Simone Kashima; Debora Botequio Moretti; Elaine Cristina Marqueze; Elaine Vieira dos Santos; Elisangela Chicaroni Mattos; Erika Freitas; Evandra Strazza Rodrigues; Felipe Allan da Silva da Costa; Flavia Aburjaile; Fábio Sossai Possebon; Guilherme Campos; Guilherme Targino Valente; Heidge Fukumasu. USP-Botucatu: Rejane Maria Tommasini Grotto; Helena Lage Ferreira; Instituto Butantan: Dimas Tadeu Covas; Jardelina de Souza Todao Bernardino; Jayme A. Souza-Neto; Jessika Cristina Chagas Lesbon; Jorge A. Petroli Marchesi; José Salvatore Leister Patané; João Paulo Kitajima; João Pessoa Araújo Jr.; Leila Sabrina Ullmann; Loyze Paola Oliveira de Lima; Luiz Aurelio de Campos Crispin. Centro de Genômica Funcional da ESALQ: Luiz Lehmann Coutinho; Luiz Carlos Junior de Alcantara; Lívia Sacchetto; Maisa C. Pereira Parra; Maria Carolina Elias; Marta Giovanetti; Marília Moraes; Maurício Lacerda Nogueira. Prefeitura de Sao Paulo: Melissa Palmieri.; Patricia Akemi Assato; Paula Rahal; Paulo Inacio da Costa; Rafael dos Santos Bezerra; Raquel de Lello Rocha Campos Cassano. NGS Soluções Genômicas: Pilar Drummond Sampaio Corrêa Mariani. FZEA-USP Pirassununga: Mirele Daiana Poleti; Raul Machado Neto; Ricardo Augusto Brassaloti; Ricardo Haddad; Rodrigo Tocantins Calado. FAMERP-SJRP: Cecília Artico Banho; Sandra Coccuzzo Sampaio; Svetoslav Nanev Slavov; Vagner Fonseca; Vincent Louis Viala |
| EPI_ISL_1967028 | AMBULATORIO DE MOLESTIAS INFECCIOSAS DE SALTO | Instituto Butantan / ESALQ-USP (Piracicaba) | Antonio Jorge Martins; Bianca Cechetto Carlos. Mendelics: Bibiana Santos; Claudia Renata dos Santos Barros; Cíntia Bittar; David Schlesinger. Hemocentro Ribeirão Preto: Simone Kashima; Debora Botequio Moretti; Elaine Cristina Marqueze; Elaine Vieira dos Santos; Elisangela Chicaroni Mattos; Erika Freitas; Evandra Strazza Rodrigues; Felipe Allan da Silva da Costa; Flavia Aburjaile; Fábio Sossai Possebon; Guilherme Campos; Guilherme Targino Valente; Heidge Fukumasu. USP-Botucatu: Rejane Maria Tommasini Grotto; Helena Lage Ferreira; Instituto Butantan: Dimas Tadeu Covas; Jardelina de Souza Todao Bernardino; Jayme A. Souza-Neto; Jessika Cristina Chagas Lesbon; Jorge A. Petroli Marchesi; José Salvatore Leister Patané; João Paulo Kitajima; João Pessoa Araújo Jr.; Leila Sabrina Ullmann; Loyze Paola Oliveira de Lima; Luiz Aurelio de Campos Crispin. Centro de Genômica Funcional da ESALQ: Luiz Lehmann Coutinho; Luiz Carlos Junior de Alcantara; Lívia Sacchetto; Maisa C. Pereira Parra; Maria Carolina Elias; Marta Giovanetti; Marília Moraes; Maurício Lacerda Nogueira. Prefeitura de Sao Paulo: Melissa Palmieri.; Patricia Akemi Assato; Paula Rahal; Paulo Inacio da Costa; Rafael dos Santos Bezerra; Raquel de Lello Rocha Campos Cassano. NGS Soluções Genômicas: Pilar Drummond Sampaio Corrêa Mariani. FZEA-USP Pirassununga: Mirele Daiana Poleti; Raul Machado Neto; Ricardo Augusto Brassaloti; Ricardo Haddad; Rodrigo Tocantins Calado. FAMERP-SJRP: Cecília Artico Banho; Sandra Coccuzzo Sampaio; Svetoslav Nanev Slavov; Vagner Fonseca; Vincent Louis Viala |
| EPI_ISL_1966777, EPI_ISL_1966778, EPI_ISL_1966779, EPI_ISL_1966780, EPI_ISL_1966781, EPI_ISL_2170964, EPI_ISL_2170965 | | | |
| see above | AMBULATORIO DR JOAO LANIA CENTRO JOSE BONIFACIO | Instituto Butantan / Mendelics | Antonio Jorge Martins; Bianca Cechetto Carlos. Mendelics: Bibiana Santos; Claudia Renata dos Santos Barros; Cíntia Bittar; David Schlesinger. Hemocentro Ribeirão Preto: Simone Kashima; Debora Botequio Moretti; Elaine Cristina Marqueze; Elaine Vieira dos Santos; Elisangela Chicaroni Mattos; Erika Freitas; Evandra Strazza Rodrigues; Felipe Allan da Silva da Costa; Flavia Aburjaile; Fábio Sossai Possebon; Guilherme Campos; Guilherme Targino Valente; Heidge Fukumasu. USP-Botucatu: Rejane Maria Tommasini Grotto; Helena Lage Ferreira; Instituto Butantan: Dimas Tadeu Covas; Jardelina de Souza Todao Bernardino; Jayme A. Souza-Neto; Jessika Cristina Chagas Lesbon; Jorge A. Petroli Marchesi; José Salvatore Leister Patané; João Paulo Kitajima; João Pessoa Araújo Jr.; Leila Sabrina Ullmann; Loyze Paola Oliveira de Lima; Luiz Aurelio de Campos Crispin. Centro de Genômica Funcional da ESALQ: Luiz Lehmann Coutinho; Luiz Carlos Junior de Alcantara; Lívia Sacchetto; Maisa C. Pereira Parra; Maria Carolina Elias; Marta Giovanetti; Marília Moraes; Maurício Lacerda Nogueira. Prefeitura de Sao Paulo: Melissa Palmieri.; Patricia Akemi Assato; Paula Rahal; Paulo Inacio da Costa; Rafael dos Santos Bezerra; Raquel de Lello Rocha Campos Cassano. NGS Soluções Genômicas: Pilar Drummond Sampaio Corrêa Mariani. FZEA-USP Pirassununga: Mirele Daiana Poleti; Raul Machado Neto; Ricardo Augusto Brassaloti; Ricardo Haddad; Rodrigo Tocantins Calado. FAMERP-SJRP: Cecília Artico Banho; Sandra Coccuzzo Sampaio; Svetoslav Nanev Slavov; Vagner Fonseca; Vincent Louis Viala |
| EPI_ISL_1966697, EPI_ISL_1966698, EPI_ISL_1966699, EPI_ISL_1966700, EPI_ISL_1966701, EPI_ISL_1966702, EPI_ISL_1966703, EPI_ISL_2170956, EPI_ISL_2170957, EPI_ISL_2170958 | | | |
| see above | AMBULATORIO MEDICO DE ESPECIALIDADES DE PERUIBE | Instituto Butantan / Mendelics | Antonio Jorge Martins; Bianca Cechetto Carlos. Mendelics: Bibiana Santos; Claudia Renata dos Santos Barros; Cíntia Bittar; David Schlesinger. Hemocentro Ribeirão Preto: Simone Kashima; Debora Botequio Moretti; Elaine Cristina Marqueze; Elaine Vieira dos Santos; Elisangela Chicaroni Mattos; Erika Freitas; Evandra Strazza Rodrigues; Felipe Allan da Silva da Costa; Flavia Aburjaile; Fábio Sossai Possebon; Guilherme Campos; Guilherme Targino Valente; Heidge Fukumasu. USP-Botucatu: Rejane Maria Tommasini Grotto; Helena Lage Ferreira; Instituto Butantan: Dimas Tadeu Covas; Jardelina de Souza Todao Bernardino; Jayme A. Souza-Neto; Jessika Cristina Chagas Lesbon; Jorge A. Petroli Marchesi; José Salvatore Leister Patané; João Paulo Kitajima; João Pessoa Araújo Jr.; Leila Sabrina Ullmann; Loyze Paola Oliveira de Lima; Luiz Aurelio de Campos Crispin. Centro de Genômica Funcional da ESALQ: Luiz Lehmann Coutinho; Luiz Carlos Junior de Alcantara; Lívia Sacchetto; Maisa C. Pereira Parra; Maria Carolina Elias; Marta Giovanetti; Marília Moraes; Maurício Lacerda Nogueira. Prefeitura de Sao Paulo: Melissa Palmieri.; Patricia Akemi Assato; Paula Rahal; Paulo Inacio da Costa; Rafael dos Santos Bezerra; Raquel de Lello Rocha Campos Cassano. NGS Soluções Genômicas: Pilar Drummond Sampaio Corrêa Mariani. FZEA-USP Pirassununga: Mirele Daiana Poleti; Raul Machado Neto; Ricardo Augusto Brassaloti; Ricardo Haddad; Rodrigo Tocantins Calado. FAMERP-SJRP: Cecília Artico Banho; Sandra Coccuzzo Sampaio; Svetoslav Nanev Slavov; Vagner Fonseca; Vincent Louis Viala |
| EPI_ISL_1795367, EPI_ISL_1795369 | AMBULATORIO MEDICO MUNICIPAL DE AGUDOS | Instituto Butantan / ESALQ-Piracicaba | Antonio Jorge Martins; Bianca Cechetto Carlos. Mendelics: Bibiana Santos; Claudia Renata dos Santos Barros; David Schlesinger. Hemocentro Ribeirão Preto: Simone Kashima; Debora Botequio Moretti. Centro de Genômica Funcional da ESALQ: Luiz Lehmann Coutinho; Dimas Tadeu Covas; Elaine Cristina Marqueze; Elaine Vieira dos Santos; Elisangela Chicaroni Mattos; Erika Freitas; Evandra Strazza Rodrigues; Felipe Allan da Silva da Costa; Flavia Aburjaile; Guilherme Targino Valente; Heidge Fukumasu. USP-Botucatu: Rejane Maria Tommasini Grotto; Instituto Butantan: Alexander Roberto Precioso; Jayme A. Souza-Neto; Jessika Cristina Chagas Lesbon; José Salvatore Leister Patané; João Paulo Kitajima; Luiz Carlos Junior de Alcantara; Maria Carolina Elias; Marta Giovanetti; Patricia Akemi Assato; Rafael dos Santos Bezerra; Raquel de Lello Rocha Campos Cassano. NGS Soluções Genômicas: Pilar Drummond Sampaio Corrêa Mariani. FZEA-USP Pirassununga: Mirele Daiana Poleti; Raul Machado Neto; Ricardo Augusto Brassaloti; Ricardo Haddad; Rodrigo Tocantins Calado.; Sandra Coccuzzo Sampaio; Svetoslav Nanev Slavov; Vagner Fonseca; Vincent Louis Viala |
| EPI_ISL_1966706 | AMBULATORIO TUPI | Instituto Butantan / Mendelics | Antonio Jorge Martins; Bianca Cechetto Carlos. Mendelics: Bibiana Santos; Claudia Renata dos Santos Barros; Cíntia Bittar; David Schlesinger. Hemocentro Ribeirão Preto: Simone Kashima; Debora Botequio Moretti; Elaine Cristina Marqueze; Elaine Vieira dos Santos; Elisangela Chicaroni Mattos; Erika Freitas; Evandra Strazza Rodrigues; Felipe Allan da Silva da Costa; Flavia Aburjaile; Fábio Sossai Possebon; Guilherme Campos; Guilherme Targino Valente; Heidge Fukumasu. USP-Botucatu: Rejane Maria Tommasini Grotto; Helena Lage Ferreira; Instituto Butantan: Dimas Tadeu Covas; Jardelina de Souza Todao Bernardino; Jayme A. Souza-Neto; Jessika Cristina Chagas Lesbon; Jorge A. Petroli Marchesi; José Salvatore Leister Patané; João Paulo Kitajima; João Pessoa Araújo Jr.; Leila Sabrina Ullmann; Loyze Paola Oliveira de Lima; Luiz Aurelio de Campos Crispin. Centro de Genômica Funcional da ESALQ: Luiz Lehmann Coutinho; Luiz Carlos Junior de Alcantara; Lívia Sacchetto; Maisa C. Pereira Parra; Maria Carolina Elias; Marta Giovanetti; Marília Moraes; Maurício Lacerda Nogueira. Prefeitura de Sao Paulo: Melissa Palmieri.; Patricia Akemi Assato; Paula Rahal; Paulo Inacio da Costa; Rafael dos Santos Bezerra; Raquel de Lello Rocha Campos Cassano. NGS Soluções Genômicas: Pilar Drummond Sampaio Corrêa Mariani. FZEA-USP Pirassununga: Mirele Daiana Poleti; Raul Machado Neto; Ricardo Augusto Brassaloti; Ricardo Haddad; Rodrigo Tocantins Calado. FAMERP-SJRP: Cecília Artico Banho; Sandra Coccuzzo Sampaio; Svetoslav Nanev Slavov; Vagner Fonseca; Vincent Louis Viala |
| EPI_ISL_1625972 | Ama J Angela | Instituto Adolfo Lutz, Interdiciplinary Procedures Center, Strategic Laboratory | Caio Vinicius Dias Lopes; Claudia Regina Gonçalves; Claudio Tavares Sacchi; Erica Valessa Ramos Gomes; Karoline Rodrigues Campos; Katia Correa de Oliveira Santos; Leonardo Jose Tadeu de Araujo |
| EPI_ISL_1966323, EPI_ISL_1966324, EPI_ISL_1966325, EPI_ISL_1966326, EPI_ISL_1966327, EPI_ISL_1966328, EPI_ISL_1966329, EPI_ISL_1966330, EPI_ISL_1966331, EPI_ISL_1966332, EPI_ISL_1966333, EPI_ISL_1966334, EPI_ISL_1966335, EPI_ISL_1966336, EPI_ISL_1966337, EPI_ISL_1966338, EPI_ISL_1966339, EPI_ISL_1966340, EPI_ISL_1966341, EPI_ISL_1966497, EPI_ISL_1966498, EPI_ISL_1966499, EPI_ISL_1966500, EPI_ISL_1966501, EPI_ISL_1966502, EPI_ISL_1966503, EPI_ISL_1966504, EPI_ISL_1966505, EPI_ISL_1966506, EPI_ISL_2105677, EPI_ISL_2170929 | | | |
| see above | Ambulatório IB | Instituto Butantan / Mendelics | Antonio Jorge Martins; Bianca Cechetto Carlos. Mendelics: Bibiana Santos; Claudia Renata dos Santos Barros; Cíntia Bittar; David Schlesinger; David Schlesinger. Hemocentro Ribeirão Preto: Simone Kashima; Debora Botequio Moretti; Dimas Tadeu Covas; Elaine Cristina Marqueze; Elaine Vieira Santos; Elaine Vieira dos Santos; Elisangela Chicaroni Mattos; Erika Freitas; Evandra Strazza Rodrigues; Felipe Allan da Silva da Costa; Flavia Aburjaile; Fábio Sossai Possebon; Guilherme Campos; Guilherme Targino Valente; Heidge Fukumasu. USP-Botucatu: Rejane Maria Tommasini Grotto; Helena Lage Ferreira; Instituto Butantan: Dimas Tadeu Covas; Jardelina de Souza Todao Bernardino; Jayme A. Souza-Neto; Jayme Augusto de Souza-Neto; Jessika Cristina Chagas Lesbon; Jorge A. Petroli Marchesi; José Salvatore Leister Patané; João Paulo Kitajima; João Pessoa Araújo Jr.; Leila Sabrina Ullmann; Loyze Paola Oliveira de Lima; Luiz Alcantara; Luiz Aurelio de Campos Crispin. Centro de Genômica Funcional da ESALQ: Luiz Lehmann Coutinho; Luiz Carlos Junior de Alcantara; Luiz Lehmann Coutinho; Lívia Sacchetto; Maisa C. Pereira Parra; Maria Carolina Elias; Marta Giovanetti; Marília Moraes; Maurício Lacerda Nogueira; Maurício Lacerda Nogueira. Prefeitura de Sao Paulo: Melissa Palmieri.; Patricia Akemi Assato; Paula Rahal; Paulo Inacio da Costa; Rafael dos Santos Bezerra; Raquel de Lello Rocha Campos Cassano. NGS Soluções Genômicas: Pilar Drummond Sampaio Corrêa Mariani. FZEA-USP Pirassununga: Mirele Daiana Poleti; Raul Machado Neto; Rejane Maria Tommasini Grotto; Ricardo Augusto Brassaloti; Ricardo Haddad; Rodrigo Tocantins Calado. FAMERP-SJRP: Cecília Artico Banho; Sandra Coccuzzo Sampaio; Sandra Coccuzzo Sampaio Vessoni; Simone Kashima; Svetoslav Nanev Slavov; Vagner Fonseca; Vincent Louis Viala |
| EPI_ISL_1365747 | Associação Fundo de Incentivo a Pesquisa | Associação Fundo de Incentivo à Pesquisa (AFIP) | Debora Ribeiro Ramadan; Erika Rodrigues de Oliveira; Juliana Nogueira Martins Rodrigues; Priscila Farias Tempaku; Sergio Tufik; Soraya Sgambatti de Andrade |
| EPI_ISL_2106190 | Biology, UFLA, Universidade Federal de Lavras | Biology, UFLA, Universidade Federal de Lavras | Barcante, J.; Cherem, J.; Fernandes, G.; Luciano, P.; Melo, D.; Pylro, V. |
| EPI_ISL_1966895, EPI_ISL_1966896, EPI_ISL_1966897, EPI_ISL_1966898, EPI_ISL_1966899, EPI_ISL_1966900, EPI_ISL_1966901, EPI_ISL_1966902, EPI_ISL_1966903, EPI_ISL_1966904 | | | |
| see above | CENTRO DE APOIO EPIDEMIOLOGICO | Instituto Butantan / ESALQ-USP (Piracicaba) | Antonio Jorge Martins; Bianca Cechetto Carlos. Mendelics: Bibiana Santos; Claudia Renata dos Santos Barros; Cíntia Bittar; David Schlesinger. Hemocentro Ribeirão Preto: Simone Kashima; Debora Botequio Moretti; Elaine Cristina Marqueze; Elaine Vieira dos Santos; Elisangela Chicaroni Mattos; Erika Freitas; Evandra Strazza Rodrigues; Felipe Allan da Silva da Costa; Flavia Aburjaile; Fábio Sossai Possebon; Guilherme Campos; Guilherme Targino Valente; Heidge Fukumasu. USP-Botucatu: Rejane Maria Tommasini Grotto; Helena Lage Ferreira; Instituto Butantan: Dimas Tadeu Covas; Jardelina de Souza Todao Bernardino; Jayme A. Souza-Neto; Jessika Cristina Chagas Lesbon; Jorge A. Petroli Marchesi; José Salvatore Leister Patané; João Paulo Kitajima; João Pessoa Araújo Jr.; Leila Sabrina Ullmann; Loyze Paola Oliveira de Lima; Luiz Aurelio de Campos Crispin. Centro de Genômica Funcional da ESALQ: Luiz Lehmann Coutinho; Luiz Carlos Junior de Alcantara; Lívia Sacchetto; Maisa C. Pereira Parra; Maria Carolina Elias; Marta Giovanetti; Marília Moraes; Maurício Lacerda Nogueira. Prefeitura de Sao Paulo: Melissa Palmieri.; Patricia Akemi Assato; Paula Rahal; Paulo Inacio da Costa; Rafael dos Santos Bezerra; Raquel de Lello Rocha Campos Cassano. NGS Soluções Genômicas: Pilar Drummond Sampaio Corrêa Mariani. FZEA-USP Pirassununga: Mirele Daiana Poleti; Raul Machado Neto; Ricardo Augusto Brassaloti; Ricardo Haddad; Rodrigo Tocantins Calado. FAMERP-SJRP: Cecília Artico Banho; Sandra Coccuzzo Sampaio; Svetoslav Nanev Slavov; Vagner Fonseca; Vincent Louis Viala |
| EPI_ISL_1967251 | CENTRO DE DETENCAO PROVISORIA DE NOVA INDEPENDENCIA | Instituto Butantan / Mendelics | Antonio Jorge Martins; Bianca Cechetto Carlos. Mendelics: Bibiana Santos; Claudia Renata dos Santos Barros; Cíntia Bittar; David Schlesinger. Hemocentro Ribeirão Preto: Simone Kashima; Debora Botequio Moretti; Elaine Cristina Marqueze; Elaine Vieira dos Santos; Elisangela Chicaroni Mattos; Erika Freitas; Evandra Strazza Rodrigues; Felipe Allan da Silva da Costa; Flavia Aburjaile; Fábio Sossai Possebon; Guilherme Campos; Guilherme Targino Valente; Heidge Fukumasu. USP-Botucatu: Rejane Maria Tommasini Grotto; Helena Lage Ferreira; Instituto Butantan: Dimas Tadeu Covas; Jardelina de Souza Todao Bernardino; Jayme A. Souza-Neto; Jessika Cristina Chagas Lesbon; Jorge A. Petroli Marchesi; José Salvatore Leister Patané; João Paulo Kitajima; João Pessoa Araújo Jr.; Leila Sabrina Ullmann; Loyze Paola Oliveira de Lima; Luiz Aurelio de Campos Crispin. Centro de Genômica Funcional da ESALQ: Luiz Lehmann Coutinho; Luiz Carlos Junior de Alcantara; Lívia Sacchetto; Maisa C. Pereira Parra; Maria Carolina Elias; Marta Giovanetti; Marília Moraes; Maurício Lacerda Nogueira. Prefeitura de Sao Paulo: Melissa Palmieri.; Patricia Akemi Assato; Paula Rahal; Paulo Inacio da Costa; Rafael dos Santos Bezerra; Raquel de Lello Rocha Campos Cassano. NGS Soluções Genômicas: Pilar Drummond Sampaio Corrêa Mariani. FZEA-USP Pirassununga: Mirele Daiana Poleti; Raul Machado Neto; Ricardo Augusto Brassaloti; Ricardo Haddad; Rodrigo Tocantins Calado. FAMERP-SJRP: Cecília Artico Banho; Sandra Coccuzzo Sampaio; Svetoslav Nanev Slavov; Vagner Fonseca; Vincent Louis Viala |
| EPI_ISL_1795325, EPI_ISL_1795329, EPI_ISL_1795333 | CENTRO DE ESPECIALIDADES DE PRIMAVERA | Instituto Butantan / ESALQ-Piracicaba | Antonio Jorge Martins; Bianca Cechetto Carlos. Mendelics: Bibiana Santos; Claudia Renata dos Santos Barros; David Schlesinger. Hemocentro Ribeirão Preto: Simone Kashima; Debora Botequio Moretti. Centro de Genômica Funcional da ESALQ: Luiz Lehmann Coutinho; Dimas Tadeu Covas; Elaine Cristina Marqueze; Elaine Vieira dos Santos; Elisangela Chicaroni Mattos; Erika Freitas; Evandra Strazza Rodrigues; Felipe Allan da Silva da Costa; Flavia Aburjaile; Guilherme Targino Valente; Heidge Fukumasu. USP-Botucatu: Rejane Maria Tommasini Grotto; Instituto Butantan: Alexander Roberto Precioso; Jayme A. Souza-Neto; Jessika Cristina Chagas Lesbon; José Salvatore Leister Patané; João Paulo Kitajima; Luiz Carlos Junior de Alcantara; Maria Carolina Elias; Marta Giovanetti; Patricia Akemi Assato; Rafael dos Santos Bezerra; Raquel de Lello Rocha Campos Cassano. NGS Soluções Genômicas: Pilar Drummond Sampaio Corrêa Mariani. FZEA-USP Pirassununga: Mirele Daiana Poleti; Raul Machado Neto; Ricardo Augusto Brassaloti; Ricardo Haddad; Rodrigo Tocantins Calado.; Sandra Coccuzzo Sampaio; Svetoslav Nanev Slavov; Vagner Fonseca; Vincent Louis Viala |

| | | | |
|---|---|---|---|
| EPI_ISL_1795109 | CENTRO DE ESPECIALIDDS MEDICAS IRMA LEOPOLDINA PIRASSUNUNGA | Instituto Butantan / ESALQ-Piracicaba | Antonio Jorge Martins; Bianca Cechetto Carlos. Mendelics: Bibiana Santos; Claudia Renata dos Santos Barros; David Schlesinger. Hemocentro Ribeirão Preto: Simone Kashima; Debora Botequio Moretti. Centro de Genômica Funcional da ESALQ: Luiz Lehmann Coutinho; Dimas Tadeu Covas; Elaine Cristina Marqueze; Elaine Vieira dos Santos; Elisangela Chicaroni Mattos; Erika Freitas; Evandra Strazza Rodrigues; Felipe Allan da Silva da Costa; Flavia Aburjaile; Guilherme Targino Valente; Heidge Fukumasu. USP-Botucatu: Rejane Maria Tommasini Grotto; Instituto Butantan: Alexander Roberto Precioso; Jayme A. Souza-Neto; Jessika Cristina Chagas Lesbon; José Salvatore Leister Patané; João Paulo Kitajima; Luiz Carlos Junior de Alcantara; Maria Carolina Elias; Marta Giovanetti; Patricia Akemi Assato; Rafael dos Santos Bezerra; Raquel de Lello Rocha Campos Cassano. NGS Soluções Genômicas: Pilar Drummond Sampaio Corrêa Mariani. FZEA-USP Pirassununga: Mirele Daiana Poleti; Raul Machado Neto; Ricardo Augusto Brassaloti; Ricardo Haddad; Rodrigo Tocantins Calado.; Sandra Coccuzzo Sampaio; Svetoslav Nanev Slavov; Vagner Fonseca; Vincent Louis Viala |
| EPI_ISL_1795228, EPI_ISL_1795229 | CENTRO DE REFERENCIA DO IDOSO DR HUMBERTO MENDES DE CARVALHO | Instituto Butantan / ESALQ-Piracicaba | Antonio Jorge Martins; Bianca Cechetto Carlos. Mendelics: Bibiana Santos; Claudia Renata dos Santos Barros; David Schlesinger. Hemocentro Ribeirão Preto: Simone Kashima; Debora Botequio Moretti. Centro de Genômica Funcional da ESALQ: Luiz Lehmann Coutinho; Dimas Tadeu Covas; Elaine Cristina Marqueze; Elaine Vieira dos Santos; Elisangela Chicaroni Mattos; Erika Freitas; Evandra Strazza Rodrigues; Felipe Allan da Silva da Costa; Flavia Aburjaile; Guilherme Targino Valente; Heidge Fukumasu. USP-Botucatu: Rejane Maria Tommasini Grotto; Instituto Butantan: Alexander Roberto Precioso; Jayme A. Souza-Neto; Jessika Cristina Chagas Lesbon; José Salvatore Leister Patané; João Paulo Kitajima; Luiz Carlos Junior de Alcantara; Maria Carolina Elias; Marta Giovanetti; Patricia Akemi Assato; Rafael dos Santos Bezerra; Raquel de Lello Rocha Campos Cassano. NGS Soluções Genômicas: Pilar Drummond Sampaio Corrêa Mariani. FZEA-USP Pirassununga: Mirele Daiana Poleti; Raul Machado Neto; Ricardo Augusto Brassaloti; Ricardo Haddad; Rodrigo Tocantins Calado.; Sandra Coccuzzo Sampaio; Svetoslav Nanev Slavov; Vagner Fonseca; Vincent Louis Viala |
| EPI_ISL_1445147, EPI_ISL_1445148, EPI_ISL_1445149, EPI_ISL_1445150, EPI_ISL_1445151, EPI_ISL_1445152, EPI_ISL_1966118, EPI_ISL_1966119, EPI_ISL_1966120, EPI_ISL_1966121, EPI_ISL_1966122, EPI_ISL_1966123 | | | |
| see above | CENTRO DE REFERENCIA DO IDOSO DR HUMBERTO MENDES DE CARVALHO | Instituto Butantan / Mendelics | Antonio Jorge Martins; Bianca Cechetto Carlos. Mendelics: Bibiana Santos; Bibiana Santos; Claudia Renata dos Santos Barros; Cíntia Bittar; David Schlesinger; David Schlesinger. Hemocentro Ribeirão Preto: Simone Kashima; Debora Botequio Moretti; Dimas Tadeu Covas; Elaine Cristina Marqueze; Elaine Vieira dos Santos; Elisangela Chicaroni Mattos; Erika Freitas; Evandra Strazza Rodrigues; Felipe Allan da Silva da Costa; Flavia Aburjaile; Fábio Sossai Possebon; Guilherme Campos; Guilherme Targino Valente; Heidge Fukumasu. USP-Botucatu: Rejane Maria Tommasini Grotto; Helena Lage Ferreira; Instituto Butantan: Dimas Tadeu Covas; Jardelina de Souza Todao Bernardino; Jayme A. Souza-Neto; Jessika Cristina Chagas Lesbon; Jorge A. Petroli Marchesi; José Salvatore Leister Patané; João Paulo Kitajima; João Pessoa Araújo Jr.; Leila Sabrina Ullmann; Loyze Paola Oliveira de Lima; Luiz Aurelio de Campos Crispin. Centro de Genômica Funcional da ESALQ: Luiz Lehmann Coutinho; Luiz Carlos Junior de Alcantara; Lívia Sacchetto; Maisa C. Pereira Parra; Maria Carolina Elias; Marta Giovanetti; Marília Moraes; Maurício Lacerda Nogueira. Prefeitura de Sao Paulo: Melissa Palmieri.; Patricia Akemi Assato; Paula Rahal; Paulo Inacio da Costa; Rafael dos Santos Bezerra; Raquel de Lello Rocha Campos Cassano. NGS Soluções Genômicas: Pilar Drummond Sampaio Corrêa Mariani. FZEA-USP Pirassununga: Mirele Daiana Poleti; Raul Machado Neto; Ricardo Augusto Brassaloti; Ricardo Haddad; Rodrigo Tocantins Calado.; Rodrigo Tocantins Calado. FAMERP-SJRP: Cecília Artico Banho; Sandra Coccuzzo Sampaio; Simone Kashima; Svetoslav Nanev Slavov; Vagner Fonseca; Vincent Louis Viala |
| EPI_ISL_1966522 | CENTRO DE SAUDE DE ADAMANTINA | Instituto Butantan / FZEA-USP (Pirassununga) | Antonio Jorge Martins; Bianca Cechetto Carlos. Mendelics: Bibiana Santos; Claudia Renata dos Santos Barros; Cíntia Bittar; David Schlesinger. Hemocentro Ribeirão Preto: Simone Kashima; Debora Botequio Moretti; Elaine Cristina Marqueze; Elaine Vieira dos Santos; Elisangela Chicaroni Mattos; Erika Freitas; Evandra Strazza Rodrigues; Felipe Allan da Silva da Costa; Flavia Aburjaile; Fábio Sossai Possebon; Guilherme Campos; Guilherme Targino Valente; Heidge Fukumasu. USP-Botucatu: Rejane Maria Tommasini Grotto; Helena Lage Ferreira; Instituto Butantan: Dimas Tadeu Covas; Jardelina de Souza Todao Bernardino; Jayme A. Souza-Neto; Jessika Cristina Chagas Lesbon; Jorge A. Petroli Marchesi; José Salvatore Leister Patané; João Paulo Kitajima; João Pessoa Araújo Jr.; Leila Sabrina Ullmann; Loyze Paola Oliveira de Lima; Luiz Aurelio de Campos Crispin. Centro de Genômica Funcional da ESALQ: Luiz Lehmann Coutinho; Luiz Carlos Junior de Alcantara; Lívia Sacchetto; Maisa C. Pereira Parra; Maria Carolina Elias; Marta Giovanetti; Marília Moraes; Maurício Lacerda Nogueira. Prefeitura de Sao Paulo: Melissa Palmieri.; Patricia Akemi Assato; Paula Rahal; Paulo Inacio da Costa; Rafael dos Santos Bezerra; Raquel de Lello Rocha Campos Cassano. NGS Soluções Genômicas: Pilar Drummond Sampaio Corrêa Mariani. FZEA-USP Pirassununga: Mirele Daiana Poleti; Raul Machado Neto; Ricardo Augusto Brassaloti; Ricardo Haddad; Rodrigo Tocantins Calado. FAMERP-SJRP: Cecília Artico Banho; Sandra Coccuzzo Sampaio; Svetoslav Nanev Slavov; Vagner Fonseca; Vincent Louis Viala |
| EPI_ISL_1967212 | CENTRO DE SAUDE DE BARBOSA | Instituto Butantan / Mendelics | Antonio Jorge Martins; Bianca Cechetto Carlos. Mendelics: Bibiana Santos; Claudia Renata dos Santos Barros; Cíntia Bittar; David Schlesinger. Hemocentro Ribeirão Preto: Simone Kashima; Debora Botequio Moretti; Elaine Cristina Marqueze; Elaine Vieira dos Santos; Elisangela Chicaroni Mattos; Erika Freitas; Evandra Strazza Rodrigues; Felipe Allan da Silva da Costa; Flavia Aburjaile; Fábio Sossai Possebon; Guilherme Campos; Guilherme Targino Valente; Heidge Fukumasu. USP-Botucatu: Rejane Maria Tommasini Grotto; Helena Lage Ferreira; Instituto Butantan: Dimas Tadeu Covas; Jardelina de Souza Todao Bernardino; Jayme A. Souza-Neto; Jessika Cristina Chagas Lesbon; Jorge A. Petroli Marchesi; José Salvatore Leister Patané; João Paulo Kitajima; João Pessoa Araújo Jr.; Leila Sabrina Ullmann; Loyze Paola Oliveira de Lima; Luiz Aurelio de Campos Crispin. Centro de Genômica Funcional da ESALQ: Luiz Lehmann Coutinho; Luiz Carlos Junior de Alcantara; Lívia Sacchetto; Maisa C. Pereira Parra; Maria Carolina Elias; Marta Giovanetti; Marília Moraes; Maurício Lacerda Nogueira. Prefeitura de Sao Paulo: Melissa Palmieri.; Patricia Akemi Assato; Paula Rahal; Paulo Inacio da Costa; Rafael dos Santos Bezerra; Raquel de Lello Rocha Campos Cassano. NGS Soluções Genômicas: Pilar Drummond Sampaio Corrêa Mariani. FZEA-USP Pirassununga: Mirele Daiana Poleti; Raul Machado Neto; Ricardo Augusto Brassaloti; Ricardo Haddad; Rodrigo Tocantins Calado. FAMERP-SJRP: Cecília Artico Banho; Sandra Coccuzzo Sampaio; Svetoslav Nanev Slavov; Vagner Fonseca; Vincent Louis Viala |
| EPI_ISL_1445195, EPI_ISL_1966219 | CENTRO DE SAUDE DE BORA | Instituto Butantan / Mendelics | Antonio Jorge Martins; Bianca Cechetto Carlos. Mendelics: Bibiana Santos; Bibiana Santos; Claudia Renata dos Santos Barros; Cíntia Bittar; David Schlesinger; David Schlesinger. Hemocentro Ribeirão Preto: Simone Kashima; Debora Botequio Moretti; Dimas Tadeu Covas; Elaine Cristina Marqueze; Elaine Vieira dos Santos; Elisangela Chicaroni Mattos; Erika Freitas; Evandra Strazza Rodrigues; Felipe Allan da Silva da Costa; Flavia Aburjaile; Fábio Sossai Possebon; Guilherme Campos; Guilherme Targino Valente; Heidge Fukumasu. USP-Botucatu: Rejane Maria Tommasini Grotto; Helena Lage Ferreira; Instituto Butantan: Dimas Tadeu Covas; Jardelina de Souza Todao Bernardino; Jayme A. Souza-Neto; Jessika Cristina Chagas Lesbon; Jorge A. Petroli Marchesi; José Salvatore Leister Patané; João Paulo Kitajima; João Pessoa Araújo Jr.; Leila Sabrina Ullmann; Loyze Paola Oliveira de Lima; Luiz Aurelio de Campos Crispin. Centro de Genômica Funcional da ESALQ: Luiz Lehmann Coutinho; Luiz Carlos Junior de Alcantara; Lívia Sacchetto; Maisa C. Pereira Parra; Maria Carolina Elias; Marta Giovanetti; Marília Moraes; Maurício Lacerda Nogueira. Prefeitura de Sao Paulo: Melissa Palmieri.; Patricia Akemi Assato; Paula Rahal; Paulo Inacio da Costa; Rafael dos Santos Bezerra; Raquel de Lello Rocha Campos Cassano. NGS Soluções Genômicas: Pilar Drummond Sampaio Corrêa Mariani. FZEA-USP Pirassununga: Mirele Daiana Poleti; Raul Machado Neto; Ricardo Augusto Brassaloti; Ricardo Haddad; Rodrigo Tocantins Calado.; Rodrigo Tocantins Calado. FAMERP-SJRP: Cecília Artico Banho; Sandra Coccuzzo Sampaio; Simone Kashima; Svetoslav Nanev Slavov; Vagner Fonseca; Vincent Louis Viala |
| EPI_ISL_1795120, EPI_ISL_1795121, EPI_ISL_1795151 | CENTRO DE SAUDE DE CAJOBI | Instituto Butantan / ESALQ-Piracicaba | Antonio Jorge Martins; Bianca Cechetto Carlos. Mendelics: Bibiana Santos; Claudia Renata dos Santos Barros; David Schlesinger. Hemocentro Ribeirão Preto: Simone Kashima; Debora Botequio Moretti. Centro de Genômica Funcional da ESALQ: Luiz Lehmann Coutinho; Dimas Tadeu Covas; Elaine Cristina Marqueze; Elaine Vieira dos Santos; Elisangela Chicaroni Mattos; Erika Freitas; Evandra Strazza Rodrigues; Felipe Allan da Silva da Costa; Flavia Aburjaile; Guilherme Targino Valente; Heidge Fukumasu. USP-Botucatu: Rejane Maria Tommasini Grotto; Instituto Butantan: Alexander Roberto Precioso; Jayme A. Souza-Neto; Jessika Cristina Chagas Lesbon; José Salvatore Leister Patané; João Paulo Kitajima; Luiz Carlos Junior de Alcantara; Maria Carolina Elias; Marta Giovanetti; Patricia Akemi Assato; Rafael dos Santos Bezerra; Raquel de Lello Rocha Campos Cassano. NGS Soluções Genômicas: Pilar Drummond Sampaio Corrêa Mariani. FZEA-USP Pirassununga: Mirele Daiana Poleti; Raul Machado Neto; Ricardo Augusto Brassaloti; Ricardo Haddad; Rodrigo Tocantins Calado.; Sandra Coccuzzo Sampaio; Svetoslav Nanev Slavov; Vagner Fonseca; Vincent Louis Viala |
| EPI_ISL_1966365, EPI_ISL_1966366 | CENTRO DE SAUDE DE CANITAR | Instituto Butantan / Mendelics | Antonio Jorge Martins; Bianca Cechetto Carlos. Mendelics: Bibiana Santos; Claudia Renata dos Santos Barros; Cíntia Bittar; David Schlesinger. Hemocentro Ribeirão Preto: Simone Kashima; Debora Botequio Moretti; Elaine Cristina Marqueze; Elaine Vieira dos Santos; Elisangela Chicaroni Mattos; Erika Freitas; Evandra Strazza Rodrigues; Felipe Allan da Silva da Costa; Flavia Aburjaile; Fábio Sossai Possebon; Guilherme Campos; Guilherme Targino Valente; Heidge Fukumasu. USP-Botucatu: Rejane Maria Tommasini Grotto; Helena Lage Ferreira; Instituto Butantan: Dimas Tadeu Covas; Jardelina de Souza Todao Bernardino; Jayme A. Souza-Neto; Jessika Cristina Chagas Lesbon; Jorge A. Petroli Marchesi; José Salvatore Leister Patané; João Paulo Kitajima; João Pessoa Araújo Jr.; Leila Sabrina Ullmann; Loyze Paola Oliveira de Lima; Luiz Aurelio de Campos Crispin. Centro de Genômica Funcional da ESALQ: Luiz Lehmann Coutinho; Luiz Carlos Junior de Alcantara; Lívia Sacchetto; Maisa C. Pereira Parra; Maria Carolina Elias; Marta Giovanetti; Marília Moraes; Maurício Lacerda Nogueira. Prefeitura de Sao Paulo: Melissa Palmieri.; Patricia Akemi Assato; Paula Rahal; Paulo Inacio da Costa; Rafael dos Santos Bezerra; Raquel de Lello Rocha Campos Cassano. NGS Soluções Genômicas: Pilar Drummond Sampaio Corrêa Mariani. FZEA-USP Pirassununga: Mirele Daiana Poleti; Raul Machado Neto; Ricardo Augusto Brassaloti; Ricardo Haddad; Rodrigo Tocantins Calado. FAMERP-SJRP: Cecília Artico Banho; Sandra Coccuzzo Sampaio; Svetoslav Nanev Slavov; Vagner Fonseca; Vincent Louis Viala |
| EPI_ISL_1966367, EPI_ISL_1966368, EPI_ISL_2170930 | CENTRO DE SAUDE DE CHAVANTES | Instituto Butantan / Mendelics | Antonio Jorge Martins; Bianca Cechetto Carlos. Mendelics: Bibiana Santos; Claudia Renata dos Santos Barros; Cíntia Bittar; David Schlesinger. Hemocentro Ribeirão Preto: Simone Kashima; Debora Botequio Moretti; Elaine Cristina Marqueze; Elaine Vieira dos Santos; Elisangela Chicaroni Mattos; Erika Freitas; Evandra Strazza Rodrigues; Felipe Allan da Silva da Costa; Flavia Aburjaile; Fábio Sossai Possebon; Guilherme Campos; Guilherme Targino Valente; Heidge Fukumasu. USP-Botucatu: Rejane Maria Tommasini Grotto; Helena Lage Ferreira; Instituto Butantan: Dimas Tadeu Covas; Jardelina de Souza Todao Bernardino; Jayme A. Souza-Neto; Jessika Cristina Chagas Lesbon; Jorge A. Petroli Marchesi; José Salvatore Leister Patané; João Paulo Kitajima; João Pessoa Araújo Jr.; Leila Sabrina Ullmann; Loyze Paola Oliveira de Lima; Luiz Aurelio de Campos Crispin. Centro de Genômica Funcional da ESALQ: Luiz Lehmann Coutinho; Luiz Carlos Junior de Alcantara; Lívia Sacchetto; Maisa C. Pereira Parra; Maria Carolina Elias; Marta Giovanetti; Marília Moraes; Maurício Lacerda Nogueira. Prefeitura de Sao Paulo: Melissa Palmieri.; Patricia Akemi Assato; Paula Rahal; Paulo Inacio da Costa; Rafael dos Santos Bezerra; Raquel de Lello Rocha Campos Cassano. NGS Soluções Genômicas: Pilar Drummond Sampaio Corrêa Mariani. FZEA-USP Pirassununga: Mirele Daiana Poleti; Raul Machado Neto; Ricardo Augusto Brassaloti; Ricardo Haddad; Rodrigo Tocantins Calado. FAMERP-SJRP: Cecília Artico Banho; Sandra Coccuzzo Sampaio; Svetoslav Nanev Slavov; Vagner Fonseca; Vincent Louis Viala |
| EPI_ISL_1795167, EPI_ISL_1795168, EPI_ISL_1795169 | CENTRO DE SAUDE DE FLORINEA | Instituto Butantan / ESALQ-Piracicaba | Antonio Jorge Martins; Bianca Cechetto Carlos. Mendelics: Bibiana Santos; Claudia Renata dos Santos Barros; David Schlesinger. Hemocentro Ribeirão Preto: Simone Kashima; Debora Botequio Moretti. Centro de Genômica Funcional da ESALQ: Luiz Lehmann Coutinho; Dimas Tadeu Covas; Elaine Cristina Marqueze; Elaine Vieira dos Santos; Elisangela Chicaroni Mattos; Erika Freitas; Evandra Strazza Rodrigues; Felipe Allan da Silva da Costa; Flavia Aburjaile; Guilherme Targino Valente; Heidge Fukumasu. USP-Botucatu: Rejane Maria Tommasini Grotto; Instituto Butantan: Alexander Roberto Precioso; Jayme A. Souza-Neto; Jessika Cristina Chagas Lesbon; José Salvatore Leister Patané; João Paulo Kitajima; Luiz Carlos Junior de Alcantara; Maria Carolina Elias; Marta Giovanetti; Patricia Akemi Assato; Rafael dos Santos Bezerra; Raquel de Lello Rocha Campos Cassano. NGS Soluções Genômicas: Pilar Drummond Sampaio Corrêa Mariani. FZEA-USP Pirassununga: Mirele Daiana Poleti; Raul Machado Neto; Ricardo Augusto Brassaloti; Ricardo Haddad; Rodrigo Tocantins Calado.; Sandra Coccuzzo Sampaio; Svetoslav Nanev Slavov; Vagner Fonseca; Vincent Louis Viala |
| EPI_ISL_1966292 | CENTRO DE SAUDE DE FLORINEA | Instituto Butantan / Mendelics | Antonio Jorge Martins; Bianca Cechetto Carlos. Mendelics: Bibiana Santos; Claudia Renata dos Santos Barros; Cíntia Bittar; David Schlesinger. Hemocentro Ribeirão Preto: Simone Kashima; Debora Botequio Moretti; Elaine Cristina Marqueze; Elaine Vieira dos Santos; Elisangela Chicaroni Mattos; Erika Freitas; Evandra Strazza Rodrigues; Felipe Allan da Silva da Costa; Flavia Aburjaile; Fábio Sossai Possebon; Guilherme Campos; Guilherme Targino Valente; Heidge Fukumasu. USP-Botucatu: Rejane Maria Tommasini Grotto; Helena Lage Ferreira; Instituto Butantan: Dimas Tadeu Covas; Jardelina de Souza Todao Bernardino; Jayme A. Souza-Neto; Jessika Cristina Chagas Lesbon; Jorge A. Petroli Marchesi; José Salvatore Leister Patané; João Paulo Kitajima; João Pessoa Araújo Jr.; Leila Sabrina Ullmann; Loyze Paola Oliveira de Lima; Luiz Aurelio de Campos Crispin. Centro de Genômica Funcional da ESALQ: Luiz Lehmann Coutinho; Luiz Carlos Junior de Alcantara; Lívia Sacchetto; Maisa C. Pereira Parra; Maria Carolina Elias; Marta Giovanetti; Marília Moraes; Maurício Lacerda Nogueira. Prefeitura de Sao Paulo: Melissa Palmieri.; Patricia Akemi Assato; Paula Rahal; Paulo Inacio da Costa; Rafael dos Santos Bezerra; Raquel de Lello Rocha Campos Cassano. NGS Soluções Genômicas: Pilar Drummond Sampaio Corrêa Mariani. FZEA-USP Pirassununga: Mirele Daiana Poleti; Raul Machado Neto; Ricardo Augusto Brassaloti; Ricardo Haddad; Rodrigo Tocantins Calado. FAMERP-SJRP: Cecília Artico Banho; Sandra Coccuzzo Sampaio; Svetoslav Nanev Slavov; Vagner Fonseca; Vincent Louis Viala |
| EPI_ISL_1445197, EPI_ISL_1966228 | CENTRO DE SAUDE DE JULIO MESQUITA | Instituto Butantan / Mendelics | Antonio Jorge Martins; Bianca Cechetto Carlos. Mendelics: Bibiana Santos; Bibiana Santos; Claudia Renata dos Santos Barros; Cíntia Bittar; David Schlesinger; David Schlesinger. Hemocentro Ribeirão Preto: Simone Kashima; Debora Botequio Moretti; Dimas Tadeu Covas; Elaine Cristina Marqueze; Elaine Vieira dos Santos; Elisangela Chicaroni Mattos; Erika Freitas; Evandra Strazza Rodrigues; Felipe Allan da Silva da Costa; Flavia Aburjaile; Fábio Sossai Possebon; Guilherme Campos; Guilherme Targino Valente; Heidge Fukumasu. USP-Botucatu: Rejane Maria Tommasini Grotto; Helena Lage Ferreira; Instituto Butantan: Dimas Tadeu Covas; Jardelina de Souza Todao Bernardino; Jayme A. Souza-Neto; Jessika Cristina Chagas Lesbon; Jorge A. Petroli Marchesi; José Salvatore Leister Patané; João Paulo Kitajima; João Pessoa Araújo Jr.; Leila Sabrina Ullmann; Loyze Paola Oliveira de Lima; Luiz Aurelio de Campos Crispin. Centro de Genômica Funcional da ESALQ: Luiz Lehmann Coutinho; Luiz Carlos Junior de Alcantara; Lívia Sacchetto; Maisa C. Pereira Parra; Maria Carolina Elias; Marta Giovanetti; Marília Moraes; Maurício Lacerda Nogueira. Prefeitura de Sao Paulo: Melissa Palmieri.; Patricia Akemi Assato; Paula Rahal; Paulo Inacio da Costa; Rafael dos Santos Bezerra; Raquel de Lello Rocha Campos Cassano. NGS Soluções Genômicas: Pilar Drummond Sampaio Corrêa Mariani. FZEA-USP Pirassununga: Mirele Daiana Poleti; Raul Machado Neto; Ricardo Augusto Brassaloti; Ricardo Haddad; Rodrigo Tocantins Calado.; Rodrigo Tocantins Calado. FAMERP-SJRP: Cecília Artico Banho; Sandra Coccuzzo Sampaio; Simone Kashima; Svetoslav Nanev Slavov; Vagner Fonseca; Vincent Louis Viala |
| EPI_ISL_1795170, EPI_ISL_1795172, EPI_ISL_1795409, EPI_ISL_1795410 | CENTRO DE SAUDE DE MARACAI | Instituto Butantan / ESALQ-Piracicaba | Antonio Jorge Martins; Bianca Cechetto Carlos. Mendelics: Bibiana Santos; Claudia Renata dos Santos Barros; David Schlesinger. Hemocentro Ribeirão Preto: Simone Kashima; Debora Botequio Moretti. Centro de Genômica Funcional da ESALQ: Luiz Lehmann Coutinho; Dimas Tadeu Covas; Elaine Cristina Marqueze; Elaine Vieira dos Santos; Elisangela Chicaroni Mattos; Erika Freitas; Evandra Strazza Rodrigues; Felipe Allan da Silva da Costa; Flavia Aburjaile; Guilherme Targino Valente; Heidge Fukumasu. USP-Botucatu: Rejane Maria Tommasini Grotto; Instituto Butantan: Alexander Roberto Precioso; Jayme A. Souza-Neto; Jessika Cristina Chagas Lesbon; José Salvatore Leister Patané; João Paulo Kitajima; Luiz Carlos Junior de Alcantara; Maria Carolina Elias; Marta Giovanetti; Patricia Akemi Assato; Rafael dos Santos Bezerra; Raquel de Lello Rocha Campos Cassano. NGS Soluções Genômicas: Pilar Drummond Sampaio Corrêa Mariani. FZEA-USP Pirassununga: Mirele Daiana Poleti; Raul Machado Neto; Ricardo Augusto Brassaloti; Ricardo Haddad; Rodrigo Tocantins Calado.; Sandra Coccuzzo Sampaio; Svetoslav Nanev Slavov; Vagner Fonseca; Vincent Louis Viala |
| EPI_ISL_1966957, EPI_ISL_1966958, EPI_ISL_1966959, EPI_ISL_1966960 | CENTRO DE SAUDE DE MARACAI | Instituto Butantan / ESALQ-USP (Piracicaba) | Antonio Jorge Martins; Bianca Cechetto Carlos. Mendelics: Bibiana Santos; Claudia Renata dos Santos Barros; Cíntia Bittar; David Schlesinger. Hemocentro Ribeirão Preto: Simone Kashima; Debora Botequio Moretti; Elaine Cristina Marqueze; Elaine Vieira dos Santos; Elisangela Chicaroni Mattos; Erika Freitas; Evandra Strazza Rodrigues; Felipe Allan da Silva da Costa; Flavia Aburjaile; Fábio Sossai Possebon; Guilherme Campos; Guilherme Targino Valente; Heidge Fukumasu. USP-Botucatu: Rejane Maria Tommasini Grotto; Helena Lage Ferreira; Instituto Butantan: Dimas Tadeu Covas; Jardelina de Souza Todao Bernardino; Jayme A. Souza-Neto; Jessika Cristina Chagas Lesbon; Jorge A. Petroli Marchesi; José Salvatore Leister Patané; João Paulo Kitajima; João Pessoa Araújo Jr.; Leila Sabrina Ullmann; Loyze Paola Oliveira de Lima; Luiz Aurelio de Campos Crispin. Centro de Genômica Funcional da ESALQ: Luiz Lehmann Coutinho; Luiz Carlos Junior de Alcantara; Lívia Sacchetto; Maisa C. Pereira Parra; Maria Carolina Elias; Marta Giovanetti; Marília Moraes; Maurício Lacerda Nogueira. Prefeitura de Sao Paulo: Melissa Palmieri.; Patricia Akemi Assato; Paula Rahal; Paulo Inacio da Costa; Rafael dos Santos Bezerra; Raquel de Lello Rocha Campos Cassano. NGS Soluções Genômicas: Pilar Drummond Sampaio Corrêa Mariani. FZEA-USP Pirassununga: Mirele Daiana Poleti; Raul Machado Neto; Ricardo Augusto Brassaloti; Ricardo Haddad; Rodrigo Tocantins Calado. FAMERP-SJRP: Cecília Artico Banho; Sandra Coccuzzo Sampaio; Svetoslav Nanev Slavov; Vagner Fonseca; Vincent Louis Viala |
| EPI_ISL_1795427 | CENTRO DE SAUDE DE NATIVIDADE DA SERRA | Instituto Butantan / ESALQ-Piracicaba | Antonio Jorge Martins; Bianca Cechetto Carlos. Mendelics: Bibiana Santos; Claudia Renata dos Santos Barros; David Schlesinger. Hemocentro Ribeirão Preto: Simone Kashima; Debora Botequio Moretti. Centro de Genômica Funcional da ESALQ: Luiz Lehmann Coutinho; Dimas Tadeu Covas; Elaine Cristina Marqueze; Elaine Vieira dos Santos; Elisangela Chicaroni Mattos; Erika Freitas; Evandra Strazza Rodrigues; Felipe Allan da Silva da Costa; Flavia Aburjaile; Guilherme Targino Valente; Heidge Fukumasu. USP-Botucatu: Rejane Maria Tommasini Grotto; Instituto Butantan: Alexander Roberto Precioso; Jayme A. Souza-Neto; Jessika Cristina Chagas Lesbon; José Salvatore Leister Patané; João Paulo Kitajima; Luiz Carlos Junior de Alcantara; Maria Carolina Elias; Marta Giovanetti; Patricia Akemi Assato; Rafael dos Santos Bezerra; Raquel de Lello Rocha Campos Cassano. NGS Soluções Genômicas: Pilar Drummond Sampaio Corrêa Mariani. FZEA-USP Pirassununga: Mirele Daiana Poleti; Raul Machado Neto; Ricardo Augusto Brassaloti; Ricardo Haddad; Rodrigo Tocantins Calado.; Sandra Coccuzzo Sampaio; Svetoslav Nanev Slavov; Vagner Fonseca; Vincent Louis Viala |
| EPI_ISL_1966792, EPI_ISL_1966793, EPI_ISL_1966794 | CENTRO DE SAUDE DE OSVALDO CRUZ | Instituto Butantan / Mendelics | Antonio Jorge Martins; Bianca Cechetto Carlos. Mendelics: Bibiana Santos; Claudia Renata dos Santos Barros; Cíntia Bittar; David Schlesinger. Hemocentro Ribeirão Preto: Simone Kashima; Debora Botequio Moretti; Elaine Cristina Marqueze; Elaine Vieira dos Santos; Elisangela Chicaroni Mattos; Erika Freitas; Evandra Strazza Rodrigues; Felipe Allan da Silva da Costa; Flavia Aburjaile; Fábio Sossai Possebon; Guilherme Campos; Guilherme Targino Valente; Heidge Fukumasu. USP-Botucatu: Rejane Maria Tommasini Grotto; Helena Lage Ferreira; Instituto Butantan: Dimas Tadeu Covas; Jardelina de Souza Todao Bernardino; Jayme A. Souza-Neto; Jessika Cristina Chagas Lesbon; Jorge A. Petroli Marchesi; José Salvatore Leister Patané; João Paulo Kitajima; João Pessoa Araújo Jr.; Leila Sabrina Ullmann; Loyze Paola Oliveira de Lima; Luiz Aurelio de Campos Crispin. Centro de Genômica Funcional da ESALQ: Luiz Lehmann Coutinho; Luiz Carlos Junior de Alcantara; Lívia Sacchetto; Maisa C. Pereira Parra; Maria Carolina Elias; Marta Giovanetti; Marília Moraes; Maurício Lacerda Nogueira. Prefeitura de Sao Paulo: Melissa Palmieri.; Patricia Akemi Assato; Paula Rahal; Paulo Inacio da Costa; Rafael dos Santos Bezerra; Raquel de Lello Rocha Campos Cassano. NGS Soluções Genômicas: Pilar Drummond Sampaio Corrêa Mariani. FZEA-USP Pirassununga: Mirele Daiana Poleti; Raul Machado Neto; Ricardo Augusto Brassaloti; Ricardo Haddad; Rodrigo Tocantins Calado. FAMERP-SJRP: Cecília Artico Banho; Sandra Coccuzzo Sampaio; Svetoslav Nanev Slavov; Vagner Fonseca; Vincent Louis Viala |
| EPI_ISL_1795407 | CENTRO DE SAUDE DE RIBEIRAO DO SUL | Instituto Butantan / ESALQ-Piracicaba | Antonio Jorge Martins; Bianca Cechetto Carlos. Mendelics: Bibiana Santos; Claudia Renata dos Santos Barros; David Schlesinger. Hemocentro Ribeirão Preto: Simone Kashima; Debora Botequio Moretti. Centro de Genômica Funcional da ESALQ: Luiz Lehmann Coutinho; Dimas Tadeu Covas; Elaine Cristina Marqueze; Elaine Vieira dos Santos; Elisangela Chicaroni Mattos; Erika Freitas; Evandra Strazza Rodrigues; Felipe Allan da Silva da Costa; Flavia Aburjaile; Guilherme Targino Valente; Heidge Fukumasu. USP-Botucatu: Rejane Maria Tommasini Grotto; Instituto Butantan: Alexander Roberto Precioso; Jayme A. Souza-Neto; Jessika Cristina Chagas Lesbon; José Salvatore Leister Patané; João Paulo Kitajima; Luiz Carlos Junior de Alcantara; Maria Carolina Elias; Marta Giovanetti; Patricia Akemi Assato; Rafael dos Santos Bezerra; Raquel de Lello Rocha Campos Cassano. NGS Soluções Genômicas: Pilar Drummond Sampaio Corrêa Mariani. FZEA-USP Pirassununga: Mirele Daiana Poleti; Raul Machado Neto; Ricardo Augusto Brassaloti; Ricardo Haddad; Rodrigo Tocantins Calado.; Sandra Coccuzzo Sampaio; Svetoslav Nanev Slavov; Vagner Fonseca; Vincent Louis Viala |
| EPI_ISL_1966801, EPI_ISL_1966802, EPI_ISL_1966803, EPI_ISL_1966804, EPI_ISL_1966805, EPI_ISL_1966806 | CENTRO DE SAUDE DE RIBEIRAO DO SUL | Instituto Butantan / Mendelics | Antonio Jorge Martins; Bianca Cechetto Carlos. Mendelics: Bibiana Santos; Claudia Renata dos Santos Barros; Cíntia Bittar; David Schlesinger. Hemocentro Ribeirão Preto: Simone Kashima; Debora Botequio Moretti; Elaine Cristina Marqueze; Elaine Vieira dos Santos; Elisangela Chicaroni Mattos; Erika Freitas; Evandra Strazza Rodrigues; Felipe Allan da Silva da Costa; Flavia Aburjaile; Fábio Sossai Possebon; Guilherme Campos; Guilherme Targino Valente; Heidge Fukumasu. USP-Botucatu: Rejane Maria Tommasini Grotto; Helena Lage Ferreira; Instituto Butantan: Dimas Tadeu Covas; Jardelina de Souza Todao Bernardino; Jayme A. Souza-Neto; Jessika Cristina Chagas Lesbon; Jorge A. Petroli Marchesi; José Salvatore Leister Patané; João Paulo Kitajima; João Pessoa Araújo Jr.; Leila Sabrina Ullmann; Loyze Paola Oliveira de Lima; Luiz Aurelio de Campos Crispin. Centro de Genômica Funcional da ESALQ: Luiz Lehmann Coutinho; Luiz Carlos Junior de Alcantara; Lívia Sacchetto; Maisa C. Pereira Parra; Maria Carolina Elias; Marta Giovanetti; Marília Moraes; Maurício Lacerda Nogueira. Prefeitura de Sao Paulo: Melissa Palmieri.; Patricia Akemi Assato; Paula Rahal; Paulo Inacio da Costa; Rafael dos Santos Bezerra; Raquel de Lello Rocha Campos Cassano. NGS Soluções Genômicas: Pilar Drummond Sampaio Corrêa Mariani. FZEA-USP Pirassununga: Mirele Daiana Poleti; Raul Machado Neto; Ricardo Augusto Brassaloti; Ricardo Haddad; Rodrigo Tocantins Calado. FAMERP-SJRP: Cecília Artico Banho; Sandra Coccuzzo Sampaio; Svetoslav Nanev Slavov; Vagner Fonseca; Vincent Louis Viala |
| EPI_ISL_1966398 | CENTRO DE SAUDE DE SALMOURAO | Instituto Butantan / Mendelics | Antonio Jorge Martins; Bianca Cechetto Carlos. Mendelics: Bibiana Santos; Claudia Renata dos Santos Barros; Cíntia Bittar; David Schlesinger. Hemocentro Ribeirão Preto: Simone Kashima; Debora Botequio Moretti; Elaine Cristina Marqueze; Elaine Vieira dos Santos; Elisangela Chicaroni Mattos; Erika Freitas; Evandra Strazza Rodrigues; Felipe Allan da Silva da Costa; Flavia Aburjaile; Fábio Sossai Possebon; Guilherme Campos; Guilherme Targino Valente; Heidge Fukumasu. USP-Botucatu: Rejane Maria Tommasini Grotto; Helena Lage Ferreira; Instituto Butantan: Dimas Tadeu Covas; Jardelina de Souza Todao Bernardino; Jayme A. Souza-Neto; Jessika Cristina Chagas Lesbon; Jorge A. Petroli Marchesi; José Salvatore Leister Patané; João Paulo Kitajima; João Pessoa Araújo Jr.; Leila Sabrina Ullmann; Loyze Paola Oliveira de Lima; Luiz Aurelio de Campos Crispin. Centro de Genômica Funcional da ESALQ: Luiz Lehmann Coutinho; Luiz Carlos Junior de Alcantara; Lívia Sacchetto; Maisa C. Pereira Parra; Maria Carolina Elias; Marta Giovanetti; Marília Moraes; Maurício Lacerda Nogueira. Prefeitura de Sao Paulo: Melissa Palmieri.; Patricia Akemi Assato; Paula Rahal; Paulo Inacio da Costa; Rafael dos Santos Bezerra; Raquel de Lello Rocha Campos Cassano. NGS Soluções Genômicas: Pilar Drummond Sampaio Corrêa Mariani. FZEA-USP Pirassununga: Mirele Daiana Poleti; Raul Machado Neto; Ricardo Augusto Brassaloti; Ricardo Haddad; Rodrigo Tocantins Calado. FAMERP-SJRP: Cecília Artico Banho; Sandra Coccuzzo Sampaio; Svetoslav Nanev Slavov; Vagner Fonseca; Vincent Louis Viala |
| EPI_ISL_1966412 | CENTRO DE SAUDE DE SPEDRO DO TURVO | Instituto Butantan / Mendelics | Antonio Jorge Martins; Bianca Cechetto Carlos. Mendelics: Bibiana Santos; Claudia Renata dos Santos Barros; Cíntia Bittar; David Schlesinger. Hemocentro Ribeirão Preto: Simone Kashima; Debora Botequio Moretti; Elaine Cristina Marqueze; Elaine Vieira dos Santos; Elisangela Chicaroni Mattos; Erika Freitas; Evandra Strazza Rodrigues; Felipe Allan da Silva da Costa; Flavia Aburjaile; Fábio Sossai Possebon; Guilherme Campos; Guilherme Targino Valente; Heidge Fukumasu. USP-Botucatu: Rejane Maria Tommasini Grotto; Helena Lage Ferreira; Instituto Butantan: Dimas Tadeu Covas; Jardelina de Souza Todao Bernardino; Jayme A. Souza-Neto; Jessika Cristina Chagas Lesbon; Jorge A. Petroli Marchesi; José Salvatore Leister Patané; João Paulo Kitajima; João Pessoa Araújo Jr.; Leila Sabrina Ullmann; Loyze Paola Oliveira de Lima; Luiz Aurelio de Campos Crispin. Centro de Genômica Funcional da ESALQ: Luiz Lehmann Coutinho; Luiz Carlos Junior de Alcantara; Lívia Sacchetto; Maisa C. Pereira Parra; Maria Carolina Elias; Marta Giovanetti; Marília Moraes; Maurício Lacerda Nogueira. Prefeitura de Sao Paulo: Melissa Palmieri.; Patricia Akemi Assato; Paula Rahal; Paulo Inacio da Costa; Rafael dos Santos Bezerra; Raquel de Lello Rocha Campos Cassano. NGS Soluções Genômicas: Pilar Drummond Sampaio Corrêa Mariani. FZEA-USP Pirassununga: Mirele Daiana Poleti; Raul Machado Neto; Ricardo Augusto Brassaloti; Ricardo Haddad; Rodrigo Tocantins Calado. FAMERP-SJRP: Cecília Artico Banho; Sandra Coccuzzo Sampaio; Svetoslav Nanev Slavov; Vagner Fonseca; Vincent Louis Viala |
| EPI_ISL_1966239, | CENTRO DE SAUDE | Instituto Butantan / | Antonio Jorge Martins; Bianca Cechetto Carlos. Mendelics: Bibiana Santos; Claudia Renata dos Santos Barros; Cíntia Bittar; David Schlesinger. Hemocentro Ribeirão Preto: Simone Kashima; Debora Botequio Moretti; Elaine Cristina Marqueze; Elaine Vieira dos Santos; Elisangela Chicaroni Mattos; Erika Freitas; Evandra Strazza Rodrigues; |

| | | | |
|---|---|---|---|
| EPI_ISL_1966240, EPI_ISL_1966241 | DR MARIO DIAS DE AGUIAR CAPIVARI | Mendelics | Felipe Allan da Silva da Costa; Flavia Aburjaile; Fábio Sossai Possebon; Guilherme Campos; Guilherme Targino Valente; Heidge Fukumasu. USP-Botucatu: Rejane Maria Tommasini Grotto; Helena Lage Ferreira; Instituto Butantan: Dimas Tadeu Covas; Jardelina de Souza Todao Bernardino; Jayme A. Souza-Neto; Jessika Cristina Chagas Lesbon; Jorge A. Petroli Marchesi; José Salvatore Leister Patané; João Paulo Kitajima; João Pessoa Araújo Jr.; Leila Sabrina Ullmann; Loyze Paola Oliveira de Lima; Luiz Aurelio de Campos Crispin. Centro de Genômica Funcional da ESALQ: Luiz Lehmann Coutinho; Luiz Carlos Junior de Alcantara; Lívia Sacchetto; Maisa C. Pereira Parra; Maria Carolina Elias; Marta Giovanetti; Marília Moraes; Maurício Lacerda Nogueira. Prefeitura de Sao Paulo: Melissa Palmieri.; Patricia Akemi Assato; Paula Rahal; Paulo Inacio da Costa; Rafael dos Santos Bezerra; Raquel de Lello Rocha Campos Cassano. NGS Soluções Genômicas: Pilar Drummond Sampaio Corrêa Mariani. FZEA-USP Pirassununga: Mirele Daiana Poleti; Raul Machado Neto; Ricardo Augusto Brassaloti; Ricardo Haddad; Rodrigo Tocantins Calado. FAMERP-SJRP: Cecília Artico Banho; Sandra Coccuzzo Sampaio; Svetoslav Nanev Slavov; Vagner Fonseca; Vincent Louis Viala |
| EPI_ISL_1795162, EPI_ISL_1795163, EPI_ISL_1795164, EPI_ISL_1795166 | CENTRO DE SAUDE DR PLINIO ALBERS DE GALIA | Instituto Butantan / ESALQ-Piracicaba | Antonio Jorge Martins; Bianca Cechetto Carlos. Mendelics: Bibiana Santos; Claudia Renata dos Santos Barros; David Schlesinger. Hemocentro Ribeirão Preto: Simone Kashima; Debora Botequio Moretti. Centro de Genômica Funcional da ESALQ: Luiz Lehmann Coutinho; Dimas Tadeu Covas; Elaine Cristina Marqueze; Elaine Vieira dos Santos; Elisangela Chicaroni Mattos; Erika Freitas; Evandra Strazza Rodrigues; Felipe Allan da Silva da Costa; Flavia Aburjaile; Guilherme Targino Valente; Heidge Fukumasu. USP-Botucatu: Rejane Maria Tommasini Grotto; Instituto Butantan: Alexander Roberto Precioso; Jayme A. Souza-Neto; Jessika Cristina Chagas Lesbon; José Salvatore Leister Patané; João Paulo Kitajima; Luiz Carlos Junior de Alcantara; Maria Carolina Elias; Marta Giovanetti; Patricia Akemi Assato; Rafael dos Santos Bezerra; Raquel de Lello Rocha Campos Cassano. NGS Soluções Genômicas: Pilar Drummond Sampaio Corrêa Mariani. FZEA-USP Pirassununga: Mirele Daiana Poleti; Raul Machado Neto; Ricardo Augusto Brassaloti; Ricardo Haddad; Rodrigo Tocantins Calado.; Sandra Coccuzzo Sampaio; Svetoslav Nanev Slavov; Vagner Fonseca; Vincent Louis Viala |
| EPI_ISL_1966824, EPI_ISL_1966825, EPI_ISL_1966826, EPI_ISL_1966827, EPI_ISL_1966828, EPI_ISL_1966829 | CENTRO DE SAUDE DR RENATO DE CARVALHO RIBEIRO ANGATUBA | Instituto Butantan / ESALQ-USP (Piracicaba) | Antonio Jorge Martins; Bianca Cechetto Carlos. Mendelics: Bibiana Santos; Claudia Renata dos Santos Barros; Cíntia Bittar; David Schlesinger. Hemocentro Ribeirão Preto: Simone Kashima; Debora Botequio Moretti; Elaine Cristina Marqueze; Elaine Vieira dos Santos; Elisangela Chicaroni Mattos; Erika Freitas; Evandra Strazza Rodrigues; Felipe Allan da Silva da Costa; Flavia Aburjaile; Fábio Sossai Possebon; Guilherme Campos; Guilherme Targino Valente; Heidge Fukumasu. USP-Botucatu: Rejane Maria Tommasini Grotto; Helena Lage Ferreira; Instituto Butantan: Dimas Tadeu Covas; Jardelina de Souza Todao Bernardino; Jayme A. Souza-Neto; Jessika Cristina Chagas Lesbon; Jorge A. Petroli Marchesi; José Salvatore Leister Patané; João Paulo Kitajima; João Pessoa Araújo Jr.; Leila Sabrina Ullmann; Loyze Paola Oliveira de Lima; Luiz Aurelio de Campos Crispin. Centro de Genômica Funcional da ESALQ: Luiz Lehmann Coutinho; Luiz Carlos Junior de Alcantara; Lívia Sacchetto; Maisa C. Pereira Parra; Maria Carolina Elias; Marta Giovanetti; Marília Moraes; Maurício Lacerda Nogueira. Prefeitura de Sao Paulo: Melissa Palmieri.; Patricia Akemi Assato; Paula Rahal; Paulo Inacio da Costa; Rafael dos Santos Bezerra; Raquel de Lello Rocha Campos Cassano. NGS Soluções Genômicas: Pilar Drummond Sampaio Corrêa Mariani. FZEA-USP Pirassununga: Mirele Daiana Poleti; Raul Machado Neto; Ricardo Augusto Brassaloti; Ricardo Haddad; Rodrigo Tocantins Calado. FAMERP-SJRP: Cecília Artico Banho; Sandra Coccuzzo Sampaio; Svetoslav Nanev Slavov; Vagner Fonseca; Vincent Louis Viala |
| EPI_ISL_1966350, EPI_ISL_1966742, EPI_ISL_1966743, EPI_ISL_1966744 | CENTRO DE SAUDE DR RENATO DE CARVALHO RIBEIRO ANGATUBA | Instituto Butantan / Mendelics | Antonio Jorge Martins; Bianca Cechetto Carlos. Mendelics: Bibiana Santos; Claudia Renata dos Santos Barros; Cíntia Bittar; David Schlesinger. Hemocentro Ribeirão Preto: Simone Kashima; Debora Botequio Moretti; Elaine Cristina Marqueze; Elaine Vieira dos Santos; Elisangela Chicaroni Mattos; Erika Freitas; Evandra Strazza Rodrigues; Felipe Allan da Silva da Costa; Flavia Aburjaile; Fábio Sossai Possebon; Guilherme Campos; Guilherme Targino Valente; Heidge Fukumasu. USP-Botucatu: Rejane Maria Tommasini Grotto; Helena Lage Ferreira; Instituto Butantan: Dimas Tadeu Covas; Jardelina de Souza Todao Bernardino; Jayme A. Souza-Neto; Jessika Cristina Chagas Lesbon; Jorge A. Petroli Marchesi; José Salvatore Leister Patané; João Paulo Kitajima; João Pessoa Araújo Jr.; Leila Sabrina Ullmann; Loyze Paola Oliveira de Lima; Luiz Aurelio de Campos Crispin. Centro de Genômica Funcional da ESALQ: Luiz Lehmann Coutinho; Luiz Carlos Junior de Alcantara; Lívia Sacchetto; Maisa C. Pereira Parra; Maria Carolina Elias; Marta Giovanetti; Marília Moraes; Maurício Lacerda Nogueira. Prefeitura de Sao Paulo: Melissa Palmieri.; Patricia Akemi Assato; Paula Rahal; Paulo Inacio da Costa; Rafael dos Santos Bezerra; Raquel de Lello Rocha Campos Cassano. NGS Soluções Genômicas: Pilar Drummond Sampaio Corrêa Mariani. FZEA-USP Pirassununga: Mirele Daiana Poleti; Raul Machado Neto; Ricardo Augusto Brassaloti; Ricardo Haddad; Rodrigo Tocantins Calado. FAMERP-SJRP: Cecília Artico Banho; Sandra Coccuzzo Sampaio; Svetoslav Nanev Slavov; Vagner Fonseca; Vincent Louis Viala |
| EPI_ISL_1967058, EPI_ISL_1967059, EPI_ISL_1967060 | CENTRO DE SAUDE ESF IV ZONA RURAL DOMINGOS DE S SJRIOPARDO | Instituto Butantan / ESALQ-USP (Piracicaba) | Antonio Jorge Martins; Bianca Cechetto Carlos. Mendelics: Bibiana Santos; Claudia Renata dos Santos Barros; Cíntia Bittar; David Schlesinger. Hemocentro Ribeirão Preto: Simone Kashima; Debora Botequio Moretti; Elaine Cristina Marqueze; Elaine Vieira dos Santos; Elisangela Chicaroni Mattos; Erika Freitas; Evandra Strazza Rodrigues; Felipe Allan da Silva da Costa; Flavia Aburjaile; Fábio Sossai Possebon; Guilherme Campos; Guilherme Targino Valente; Heidge Fukumasu. USP-Botucatu: Rejane Maria Tommasini Grotto; Helena Lage Ferreira; Instituto Butantan: Dimas Tadeu Covas; Jardelina de Souza Todao Bernardino; Jayme A. Souza-Neto; Jessika Cristina Chagas Lesbon; Jorge A. Petroli Marchesi; José Salvatore Leister Patané; João Paulo Kitajima; João Pessoa Araújo Jr.; Leila Sabrina Ullmann; Loyze Paola Oliveira de Lima; Luiz Aurelio de Campos Crispin. Centro de Genômica Funcional da ESALQ: Luiz Lehmann Coutinho; Luiz Carlos Junior de Alcantara; Lívia Sacchetto; Maisa C. Pereira Parra; Maria Carolina Elias; Marta Giovanetti; Marília Moraes; Maurício Lacerda Nogueira. Prefeitura de Sao Paulo: Melissa Palmieri.; Patricia Akemi Assato; Paula Rahal; Paulo Inacio da Costa; Rafael dos Santos Bezerra; Raquel de Lello Rocha Campos Cassano. NGS Soluções Genômicas: Pilar Drummond Sampaio Corrêa Mariani. FZEA-USP Pirassununga: Mirele Daiana Poleti; Raul Machado Neto; Ricardo Augusto Brassaloti; Ricardo Haddad; Rodrigo Tocantins Calado. FAMERP-SJRP: Cecília Artico Banho; Sandra Coccuzzo Sampaio; Svetoslav Nanev Slavov; Vagner Fonseca; Vincent Louis Viala |
| EPI_ISL_1966833 | CENTRO DE SAUDE I AVARE | Instituto Butantan / ESALQ-USP (Piracicaba) | Antonio Jorge Martins; Bianca Cechetto Carlos. Mendelics: Bibiana Santos; Claudia Renata dos Santos Barros; Cíntia Bittar; David Schlesinger. Hemocentro Ribeirão Preto: Simone Kashima; Debora Botequio Moretti; Elaine Cristina Marqueze; Elaine Vieira dos Santos; Elisangela Chicaroni Mattos; Erika Freitas; Evandra Strazza Rodrigues; Felipe Allan da Silva da Costa; Flavia Aburjaile; Fábio Sossai Possebon; Guilherme Campos; Guilherme Targino Valente; Heidge Fukumasu. USP-Botucatu: Rejane Maria Tommasini Grotto; Helena Lage Ferreira; Instituto Butantan: Dimas Tadeu Covas; Jardelina de Souza Todao Bernardino; Jayme A. Souza-Neto; Jessika Cristina Chagas Lesbon; Jorge A. Petroli Marchesi; José Salvatore Leister Patané; João Paulo Kitajima; João Pessoa Araújo Jr.; Leila Sabrina Ullmann; Loyze Paola Oliveira de Lima; Luiz Aurelio de Campos Crispin. Centro de Genômica Funcional da ESALQ: Luiz Lehmann Coutinho; Luiz Carlos Junior de Alcantara; Lívia Sacchetto; Maisa C. Pereira Parra; Maria Carolina Elias; Marta Giovanetti; Marília Moraes; Maurício Lacerda Nogueira. Prefeitura de Sao Paulo: Melissa Palmieri.; Patricia Akemi Assato; Paula Rahal; Paulo Inacio da Costa; Rafael dos Santos Bezerra; Raquel de Lello Rocha Campos Cassano. NGS Soluções Genômicas: Pilar Drummond Sampaio Corrêa Mariani. FZEA-USP Pirassununga: Mirele Daiana Poleti; Raul Machado Neto; Ricardo Augusto Brassaloti; Ricardo Haddad; Rodrigo Tocantins Calado. FAMERP-SJRP: Cecília Artico Banho; Sandra Coccuzzo Sampaio; Svetoslav Nanev Slavov; Vagner Fonseca; Vincent Louis Viala |
| EPI_ISL_1966536, EPI_ISL_1966537, EPI_ISL_1966538, EPI_ISL_1966539, EPI_ISL_1966540 | CENTRO DE SAUDE I TACITO LEITE DE CARVALHO E SILVA | Instituto Butantan / FZEA-USP (Pirassununga) | Antonio Jorge Martins; Bianca Cechetto Carlos. Mendelics: Bibiana Santos; Claudia Renata dos Santos Barros; Cíntia Bittar; David Schlesinger. Hemocentro Ribeirão Preto: Simone Kashima; Debora Botequio Moretti; Elaine Cristina Marqueze; Elaine Vieira dos Santos; Elisangela Chicaroni Mattos; Erika Freitas; Evandra Strazza Rodrigues; Felipe Allan da Silva da Costa; Flavia Aburjaile; Fábio Sossai Possebon; Guilherme Campos; Guilherme Targino Valente; Heidge Fukumasu. USP-Botucatu: Rejane Maria Tommasini Grotto; Helena Lage Ferreira; Instituto Butantan: Dimas Tadeu Covas; Jardelina de Souza Todao Bernardino; Jayme A. Souza-Neto; Jessika Cristina Chagas Lesbon; Jorge A. Petroli Marchesi; José Salvatore Leister Patané; João Paulo Kitajima; João Pessoa Araújo Jr.; Leila Sabrina Ullmann; Loyze Paola Oliveira de Lima; Luiz Aurelio de Campos Crispin. Centro de Genômica Funcional da ESALQ: Luiz Lehmann Coutinho; Luiz Carlos Junior de Alcantara; Lívia Sacchetto; Maisa C. Pereira Parra; Maria Carolina Elias; Marta Giovanetti; Marília Moraes; Maurício Lacerda Nogueira. Prefeitura de Sao Paulo: Melissa Palmieri.; Patricia Akemi Assato; Paula Rahal; Paulo Inacio da Costa; Rafael dos Santos Bezerra; Raquel de Lello Rocha Campos Cassano. NGS Soluções Genômicas: Pilar Drummond Sampaio Corrêa Mariani. FZEA-USP Pirassununga: Mirele Daiana Poleti; Raul Machado Neto; Ricardo Augusto Brassaloti; Ricardo Haddad; Rodrigo Tocantins Calado. FAMERP-SJRP: Cecília Artico Banho; Sandra Coccuzzo Sampaio; Svetoslav Nanev Slavov; Vagner Fonseca; Vincent Louis Viala |
| EPI_ISL_1967027 | CENTRO DE SAUDE II | Instituto Butantan / ESALQ-USP (Piracicaba) | Antonio Jorge Martins; Bianca Cechetto Carlos. Mendelics: Bibiana Santos; Claudia Renata dos Santos Barros; Cíntia Bittar; David Schlesinger. Hemocentro Ribeirão Preto: Simone Kashima; Debora Botequio Moretti; Elaine Cristina Marqueze; Elaine Vieira dos Santos; Elisangela Chicaroni Mattos; Erika Freitas; Evandra Strazza Rodrigues; Felipe Allan da Silva da Costa; Flavia Aburjaile; Fábio Sossai Possebon; Guilherme Campos; Guilherme Targino Valente; Heidge Fukumasu. USP-Botucatu: Rejane Maria Tommasini Grotto; Helena Lage Ferreira; Instituto Butantan: Dimas Tadeu Covas; Jardelina de Souza Todao Bernardino; Jayme A. Souza-Neto; Jessika Cristina Chagas Lesbon; Jorge A. Petroli Marchesi; José Salvatore Leister Patané; João Paulo Kitajima; João Pessoa Araújo Jr.; Leila Sabrina Ullmann; Loyze Paola Oliveira de Lima; Luiz Aurelio de Campos Crispin. Centro de Genômica Funcional da ESALQ: Luiz Lehmann Coutinho; Luiz Carlos Junior de Alcantara; Lívia Sacchetto; Maisa C. Pereira Parra; Maria Carolina Elias; Marta Giovanetti; Marília Moraes; Maurício Lacerda Nogueira. Prefeitura de Sao Paulo: Melissa Palmieri.; Patricia Akemi Assato; Paula Rahal; Paulo Inacio da Costa; Rafael dos Santos Bezerra; Raquel de Lello Rocha Campos Cassano. NGS Soluções Genômicas: Pilar Drummond Sampaio Corrêa Mariani. FZEA-USP Pirassununga: Mirele Daiana Poleti; Raul Machado Neto; Ricardo Augusto Brassaloti; Ricardo Haddad; Rodrigo Tocantins Calado. FAMERP-SJRP: Cecília Artico Banho; Sandra Coccuzzo Sampaio; Svetoslav Nanev Slavov; Vagner Fonseca; Vincent Louis Viala |
| EPI_ISL_1966399 | CENTRO DE SAUDE II | Instituto Butantan / Mendelics | Antonio Jorge Martins; Bianca Cechetto Carlos. Mendelics: Bibiana Santos; Claudia Renata dos Santos Barros; Cíntia Bittar; David Schlesinger. Hemocentro Ribeirão Preto: Simone Kashima; Debora Botequio Moretti; Elaine Cristina Marqueze; Elaine Vieira dos Santos; Elisangela Chicaroni Mattos; Erika Freitas; Evandra Strazza Rodrigues; Felipe Allan da Silva da Costa; Flavia Aburjaile; Fábio Sossai Possebon; Guilherme Campos; Guilherme Targino Valente; Heidge Fukumasu. USP-Botucatu: Rejane Maria Tommasini Grotto; Helena Lage Ferreira; Instituto Butantan: Dimas Tadeu Covas; Jardelina de Souza Todao Bernardino; Jayme A. Souza-Neto; Jessika Cristina Chagas Lesbon; Jorge A. Petroli Marchesi; José Salvatore Leister Patané; João Paulo Kitajima; João Pessoa Araújo Jr.; Leila Sabrina Ullmann; Loyze Paola Oliveira de Lima; Luiz Aurelio de Campos Crispin. Centro de Genômica Funcional da ESALQ: Luiz Lehmann Coutinho; Luiz Carlos Junior de Alcantara; Lívia Sacchetto; Maisa C. Pereira Parra; Maria Carolina Elias; Marta Giovanetti; Marília Moraes; Maurício Lacerda Nogueira. Prefeitura de Sao Paulo: Melissa Palmieri.; Patricia Akemi Assato; Paula Rahal; Paulo Inacio da Costa; Rafael dos Santos Bezerra; Raquel de Lello Rocha Campos Cassano. NGS Soluções Genômicas: Pilar Drummond Sampaio Corrêa Mariani. FZEA-USP Pirassununga: Mirele Daiana Poleti; Raul Machado Neto; Ricardo Augusto Brassaloti; Ricardo Haddad; Rodrigo Tocantins Calado. FAMERP-SJRP: Cecília Artico Banho; Sandra Coccuzzo Sampaio; Svetoslav Nanev Slavov; Vagner Fonseca; Vincent Louis Viala |
| EPI_ISL_1795212 | CENTRO DE SAUDE II DR ALCIDES FACUNDO ARROYO | Instituto Butantan / ESALQ-Piracicaba | Antonio Jorge Martins; Bianca Cechetto Carlos. Mendelics: Bibiana Santos; Claudia Renata dos Santos Barros; David Schlesinger. Hemocentro Ribeirão Preto: Simone Kashima; Debora Botequio Moretti. Centro de Genômica Funcional da ESALQ: Luiz Lehmann Coutinho; Dimas Tadeu Covas; Elaine Cristina Marqueze; Elaine Vieira dos Santos; Elisangela Chicaroni Mattos; Erika Freitas; Evandra Strazza Rodrigues; Felipe Allan da Silva da Costa; Flavia Aburjaile; Guilherme Targino Valente; Heidge Fukumasu. USP-Botucatu: Rejane Maria Tommasini Grotto; Instituto Butantan: Alexander Roberto Precioso; Jayme A. Souza-Neto; Jessika Cristina Chagas Lesbon; José Salvatore Leister Patané; João Paulo Kitajima; Luiz Carlos Junior de Alcantara; Maria Carolina Elias; Marta Giovanetti; Patricia Akemi Assato; Rafael dos Santos Bezerra; Raquel de Lello Rocha Campos Cassano. NGS Soluções Genômicas: Pilar Drummond Sampaio Corrêa Mariani. FZEA-USP Pirassununga: Mirele Daiana Poleti; Raul Machado Neto; Ricardo Augusto Brassaloti; Ricardo Haddad; Rodrigo Tocantins Calado.; Sandra Coccuzzo Sampaio; Svetoslav Nanev Slavov; Vagner Fonseca; Vincent Louis Viala |
| EPI_ISL_1966472, EPI_ISL_1966473, EPI_ISL_2170936 | CENTRO DE SAUDE II DR ALCIDES FACUNDO ARROYO | Instituto Butantan / Mendelics | Antonio Jorge Martins; Bianca Cechetto Carlos. Mendelics: Bibiana Santos; Claudia Renata dos Santos Barros; Cíntia Bittar; David Schlesinger. Hemocentro Ribeirão Preto: Simone Kashima; Debora Botequio Moretti; Elaine Cristina Marqueze; Elaine Vieira dos Santos; Elisangela Chicaroni Mattos; Erika Freitas; Evandra Strazza Rodrigues; Felipe Allan da Silva da Costa; Flavia Aburjaile; Fábio Sossai Possebon; Guilherme Campos; Guilherme Targino Valente; Heidge Fukumasu. USP-Botucatu: Rejane Maria Tommasini Grotto; Helena Lage Ferreira; Instituto Butantan: Dimas Tadeu Covas; Jardelina de Souza Todao Bernardino; Jayme A. Souza-Neto; Jessika Cristina Chagas Lesbon; Jorge A. Petroli Marchesi; José Salvatore Leister Patané; João Paulo Kitajima; João Pessoa Araújo Jr.; Leila Sabrina Ullmann; Loyze Paola Oliveira de Lima; Luiz Aurelio de Campos Crispin. Centro de Genômica Funcional da ESALQ: Luiz Lehmann Coutinho; Luiz Carlos Junior de Alcantara; Lívia Sacchetto; Maisa C. Pereira Parra; Maria Carolina Elias; Marta Giovanetti; Marília Moraes; Maurício Lacerda Nogueira. Prefeitura de Sao Paulo: Melissa Palmieri.; Patricia Akemi Assato; Paula Rahal; Paulo Inacio da Costa; Rafael dos Santos Bezerra; Raquel de Lello Rocha Campos Cassano. NGS Soluções Genômicas: Pilar Drummond Sampaio Corrêa Mariani. FZEA-USP Pirassununga: Mirele Daiana Poleti; Raul Machado Neto; Ricardo Augusto Brassaloti; Ricardo Haddad; Rodrigo Tocantins Calado. FAMERP-SJRP: Cecília Artico Banho; Sandra Coccuzzo Sampaio; Svetoslav Nanev Slavov; Vagner Fonseca; Vincent Louis Viala |
| EPI_ISL_1966912 | CENTRO DE SAUDE II DR ARCY BANDEIRA IBIUNA SP | Instituto Butantan / ESALQ-USP (Piracicaba) | Antonio Jorge Martins; Bianca Cechetto Carlos. Mendelics: Bibiana Santos; Claudia Renata dos Santos Barros; Cíntia Bittar; David Schlesinger. Hemocentro Ribeirão Preto: Simone Kashima; Debora Botequio Moretti; Elaine Cristina Marqueze; Elaine Vieira dos Santos; Elisangela Chicaroni Mattos; Erika Freitas; Evandra Strazza Rodrigues; Felipe Allan da Silva da Costa; Flavia Aburjaile; Fábio Sossai Possebon; Guilherme Campos; Guilherme Targino Valente; Heidge Fukumasu. USP-Botucatu: Rejane Maria Tommasini Grotto; Helena Lage Ferreira; Instituto Butantan: Dimas Tadeu Covas; Jardelina de Souza Todao Bernardino; Jayme A. Souza-Neto; Jessika Cristina Chagas Lesbon; Jorge A. Petroli Marchesi; José Salvatore Leister Patané; João Paulo Kitajima; João Pessoa Araújo Jr.; Leila Sabrina Ullmann; Loyze Paola Oliveira de Lima; Luiz Aurelio de Campos Crispin. Centro de Genômica Funcional da ESALQ: Luiz Lehmann Coutinho; Luiz Carlos Junior de Alcantara; Lívia Sacchetto; Maisa C. Pereira Parra; Maria Carolina Elias; Marta Giovanetti; Marília Moraes; Maurício Lacerda Nogueira. Prefeitura de Sao Paulo: Melissa Palmieri.; Patricia Akemi Assato; Paula Rahal; Paulo Inacio da Costa; Rafael dos Santos Bezerra; Raquel de Lello Rocha Campos Cassano. NGS Soluções Genômicas: Pilar Drummond Sampaio Corrêa Mariani. FZEA-USP Pirassununga: Mirele Daiana Poleti; Raul Machado Neto; Ricardo Augusto Brassaloti; Ricardo Haddad; Rodrigo Tocantins Calado. FAMERP-SJRP: Cecília Artico Banho; Sandra Coccuzzo Sampaio; Svetoslav Nanev Slavov; Vagner Fonseca; Vincent Louis Viala |
| EPI_ISL_1445075, EPI_ISL_1445076, EPI_ISL_1445077, EPI_ISL_1445078, EPI_ISL_1445079, EPI_ISL_1966108, EPI_ISL_1966109, EPI_ISL_1966110, EPI_ISL_2170907 | | | |
| see above | CENTRO DE SAUDE II DR GABRIEL MESQUITA VARGEM GDE DO SUL | Instituto Butantan / Mendelics | Antonio Jorge Martins; Bianca Cechetto Carlos. Mendelics: Bibiana Santos; Bibiana Santos; Claudia Renata dos Santos Barros; Cíntia Bittar; David Schlesinger; David Schlesinger. Hemocentro Ribeirão Preto: Simone Kashima; Debora Botequio Moretti; Dimas Tadeu Covas; Elaine Cristina Marqueze; Elaine Vieira Santos; Elaine Vieira dos Santos; Elisangela Chicaroni Mattos; Erika Freitas; Evandra Strazza Rodrigues; Felipe Allan da Silva da Costa; Flavia Aburjaile; Fábio Sossai Possebon; Guilherme Campos; Guilherme Targino Valente; Heidge Fukumasu; Heidge Fukumasu. USP-Botucatu: Rejane Maria Tommasini Grotto; Helena Lage Ferreira; Instituto Butantan: Dimas Tadeu Covas; Jardelina de Souza Todao Bernardino; Jayme A. Souza-Neto; Jayme Augusto de Souza-Neto; Jessika Cristina Chagas Lesbon; Jorge A. Petroli Marchesi; José Salvatore Leister Patané; João Paulo Kitajima; João Pessoa Araújo Jr.; Leila Sabrina Ullmann; Loyze Paola Oliveira de Lima; Luiz Alcantara; Luiz Aurelio de Campos Crispin. Centro de Genômica Funcional da ESALQ: Luiz Lehmann Coutinho; Luiz Carlos Junior de Alcantara; Luiz Lehmann Coutinho; Lívia Sacchetto; Maisa C. Pereira Parra; Maria Carolina Elias; Marta Giovanetti; Marília Moraes; Maurício Lacerda Nogueira; Maurício Lacerda Nogueira. Prefeitura de Sao Paulo: Melissa Palmieri.; Patricia Akemi Assato; Paula Rahal; Paulo Inacio da Costa; Rafael dos Santos Bezerra; Raquel de Lello Rocha Campos Cassano. NGS Soluções Genômicas: Pilar Drummond Sampaio Corrêa Mariani. FZEA-USP Pirassununga: Mirele Daiana Poleti; Raul Machado Neto; Rejane Maria Tommasini Grotto; Ricardo Augusto Brassaloti; Ricardo Haddad; Rodrigo Tocantins Calado.; Rodrigo Tocantins Calado. FAMERP-SJRP: Cecília Artico Banho; Sandra Coccuzzo Sampaio; Sandra Coccuzzo Sampaio Vessoni; Simone Kashima; Svetoslav Nanev Slavov; Vagner Fonseca; Vincent Louis Viala |
| EPI_ISL_1966463 | CENTRO DE SAUDE II IBITINGA | Instituto Butantan / Mendelics | Antonio Jorge Martins; Bianca Cechetto Carlos. Mendelics: Bibiana Santos; Claudia Renata dos Santos Barros; Cíntia Bittar; David Schlesinger. Hemocentro Ribeirão Preto: Simone Kashima; Debora Botequio Moretti; Elaine Cristina Marqueze; Elaine Vieira dos Santos; Elisangela Chicaroni Mattos; Erika Freitas; Evandra Strazza Rodrigues; Felipe Allan da Silva da Costa; Flavia Aburjaile; Fábio Sossai Possebon; Guilherme Campos; Guilherme Targino Valente; Heidge Fukumasu. USP-Botucatu: Rejane Maria Tommasini Grotto; Helena Lage Ferreira; Instituto Butantan: Dimas Tadeu Covas; Jardelina de Souza Todao Bernardino; Jayme A. Souza-Neto; Jessika Cristina Chagas Lesbon; Jorge A. Petroli Marchesi; José Salvatore Leister Patané; João Paulo Kitajima; João Pessoa Araújo Jr.; Leila Sabrina Ullmann; Loyze Paola Oliveira de Lima; Luiz Aurelio de Campos Crispin. Centro de Genômica Funcional da ESALQ: Luiz Lehmann Coutinho; Luiz Carlos Junior de Alcantara; Lívia Sacchetto; Maisa C. Pereira Parra; Maria Carolina Elias; Marta Giovanetti; Marília Moraes; Maurício Lacerda Nogueira. Prefeitura de Sao Paulo: Melissa Palmieri.; Patricia Akemi Assato; Paula Rahal; Paulo Inacio da Costa; Rafael dos Santos Bezerra; Raquel de Lello Rocha Campos Cassano. NGS Soluções Genômicas: Pilar Drummond Sampaio Corrêa Mariani. FZEA-USP Pirassununga: Mirele Daiana Poleti; Raul Machado Neto; Ricardo Augusto Brassaloti; Ricardo Haddad; Rodrigo Tocantins Calado. FAMERP-SJRP: Cecília Artico Banho; Sandra Coccuzzo Sampaio; Svetoslav Nanev Slavov; Vagner Fonseca; Vincent Louis Viala |
| EPI_ISL_1795327, EPI_ISL_1795331 | CENTRO DE SAUDE II JUNQUEIROPOLIS | Instituto Butantan / ESALQ-Piracicaba | Antonio Jorge Martins; Bianca Cechetto Carlos. Mendelics: Bibiana Santos; Claudia Renata dos Santos Barros; David Schlesinger. Hemocentro Ribeirão Preto: Simone Kashima; Debora Botequio Moretti. Centro de Genômica Funcional da ESALQ: Luiz Lehmann Coutinho; Dimas Tadeu Covas; Elaine Cristina Marqueze; Elaine Vieira dos Santos; Elisangela Chicaroni Mattos; Erika Freitas; Evandra Strazza Rodrigues; Felipe Allan da Silva da Costa; Flavia Aburjaile; Guilherme Targino Valente; Heidge Fukumasu. USP-Botucatu: Rejane Maria Tommasini Grotto; Instituto Butantan: Alexander Roberto Precioso; Jayme A. Souza-Neto; Jessika Cristina Chagas Lesbon; José Salvatore Leister Patané; João Paulo Kitajima; Luiz Carlos Junior de Alcantara; Maria Carolina Elias; Marta Giovanetti; Patricia Akemi Assato; Rafael dos Santos Bezerra; Raquel de Lello Rocha Campos Cassano. NGS Soluções Genômicas: Pilar Drummond Sampaio Corrêa Mariani. FZEA-USP Pirassununga: Mirele Daiana Poleti; Raul Machado Neto; Ricardo Augusto Brassaloti; Ricardo Haddad; Rodrigo Tocantins Calado.; Sandra Coccuzzo Sampaio; Svetoslav Nanev Slavov; Vagner Fonseca; Vincent Louis Viala |
| EPI_ISL_1966955, EPI_ISL_1966956 | CENTRO DE SAUDE II MAIRINQUE MAIRINQUE | Instituto Butantan / ESALQ-USP (Piracicaba) | Antonio Jorge Martins; Bianca Cechetto Carlos. Mendelics: Bibiana Santos; Claudia Renata dos Santos Barros; Cíntia Bittar; David Schlesinger. Hemocentro Ribeirão Preto: Simone Kashima; Debora Botequio Moretti; Elaine Cristina Marqueze; Elaine Vieira dos Santos; Elisangela Chicaroni Mattos; Erika Freitas; Evandra Strazza Rodrigues; Felipe Allan da Silva da Costa; Flavia Aburjaile; Fábio Sossai Possebon; Guilherme Campos; Guilherme Targino Valente; Heidge Fukumasu. USP-Botucatu: Rejane Maria Tommasini Grotto; Helena Lage Ferreira; Instituto Butantan: Dimas Tadeu Covas; Jardelina de Souza Todao Bernardino; Jayme A. Souza-Neto; Jessika Cristina Chagas Lesbon; Jorge A. Petroli Marchesi; José Salvatore Leister Patané; João Paulo Kitajima; João Pessoa Araújo Jr.; Leila Sabrina Ullmann; Loyze Paola Oliveira de Lima; Luiz Aurelio de Campos Crispin. Centro de Genômica Funcional da ESALQ: Luiz Lehmann Coutinho; Luiz Carlos Junior de Alcantara; Lívia Sacchetto; Maisa C. Pereira Parra; Maria Carolina Elias; Marta Giovanetti; Marília Moraes; Maurício Lacerda Nogueira. Prefeitura de Sao Paulo: Melissa Palmieri.; Patricia Akemi Assato; Paula Rahal; Paulo Inacio da Costa; Rafael dos Santos Bezerra; Raquel de Lello Rocha Campos Cassano. NGS Soluções Genômicas: Pilar Drummond Sampaio Corrêa Mariani. FZEA-USP Pirassununga: Mirele Daiana Poleti; Raul Machado Neto; Ricardo Augusto Brassaloti; Ricardo Haddad; Rodrigo Tocantins Calado. FAMERP-SJRP: Cecília Artico Banho; Sandra Coccuzzo Sampaio; Svetoslav Nanev Slavov; Vagner Fonseca; Vincent Louis Viala |
| EPI_ISL_1445086, EPI_ISL_1445089, EPI_ISL_1966106, EPI_ISL_1966131, EPI_ISL_1966471 | CENTRO DE SAUDE II MAIRINQUE MAIRINQUE | Instituto Butantan / Mendelics | Antonio Jorge Martins; Bianca Cechetto Carlos. Mendelics: Bibiana Santos; Bibiana Santos; Claudia Renata dos Santos Barros; Cíntia Bittar; David Schlesinger; David Schlesinger. Hemocentro Ribeirão Preto: Simone Kashima; Debora Botequio Moretti; Dimas Tadeu Covas; Elaine Cristina Marqueze; Elaine Vieira dos Santos; Elisangela Chicaroni Mattos; Erika Freitas; Evandra Strazza Rodrigues; Felipe Allan da Silva da Costa; Flavia Aburjaile; Fábio Sossai Possebon; Guilherme Campos; Guilherme Targino Valente; Heidge Fukumasu. USP-Botucatu: Rejane Maria Tommasini Grotto; Helena Lage Ferreira; Instituto Butantan: Dimas Tadeu Covas; Jardelina de Souza Todao Bernardino; Jayme A. Souza-Neto; Jessika Cristina Chagas Lesbon; Jorge A. Petroli Marchesi; José Salvatore Leister Patané; João Paulo Kitajima; João Pessoa Araújo Jr.; Leila Sabrina Ullmann; Loyze Paola Oliveira de Lima; Luiz Aurelio de Campos Crispin. Centro de Genômica Funcional da ESALQ: Luiz Lehmann Coutinho; Luiz Carlos Junior de Alcantara; Lívia Sacchetto; Maisa C. Pereira Parra; Maria Carolina Elias; Marta Giovanetti; Marília Moraes; Maurício Lacerda Nogueira. Prefeitura de Sao Paulo: Melissa Palmieri.; Patricia Akemi Assato; Paula Rahal; Paulo Inacio da Costa; Rafael dos Santos Bezerra; Raquel de Lello Rocha Campos Cassano. NGS Soluções Genômicas: Pilar Drummond Sampaio Corrêa Mariani. FZEA-USP Pirassununga: Mirele Daiana Poleti; Raul Machado Neto; Ricardo Augusto Brassaloti; Ricardo Haddad; Rodrigo Tocantins Calado.; Rodrigo Tocantins Calado. FAMERP-SJRP: Cecília Artico Banho; Sandra Coccuzzo Sampaio; Simone Kashima; Svetoslav Nanev Slavov; Vagner Fonseca; Vincent Louis Viala |
| EPI_ISL_1967061, EPI_ISL_1967062, EPI_ISL_1967063, EPI_ISL_1967064 | CENTRO DE SAUDE II SAO MIGUEL ARCANJO | Instituto Butantan / ESALQ-USP (Piracicaba) | Antonio Jorge Martins; Bianca Cechetto Carlos. Mendelics: Bibiana Santos; Claudia Renata dos Santos Barros; Cíntia Bittar; David Schlesinger. Hemocentro Ribeirão Preto: Simone Kashima; Debora Botequio Moretti; Elaine Cristina Marqueze; Elaine Vieira dos Santos; Elisangela Chicaroni Mattos; Erika Freitas; Evandra Strazza Rodrigues; Felipe Allan da Silva da Costa; Flavia Aburjaile; Fábio Sossai Possebon; Guilherme Campos; Guilherme Targino Valente; Heidge Fukumasu. USP-Botucatu: Rejane Maria Tommasini Grotto; Helena Lage Ferreira; Instituto Butantan: Dimas Tadeu Covas; Jardelina de Souza Todao Bernardino; Jayme A. Souza-Neto; Jessika Cristina Chagas Lesbon; Jorge A. Petroli Marchesi; José Salvatore Leister Patané; João Paulo Kitajima; João Pessoa Araújo Jr.; Leila Sabrina Ullmann; Loyze Paola Oliveira de Lima; Luiz Aurelio de Campos Crispin. Centro de Genômica Funcional da ESALQ: Luiz Lehmann Coutinho; Luiz Carlos Junior de Alcantara; Lívia Sacchetto; Maisa C. Pereira Parra; Maria Carolina Elias; Marta Giovanetti; Marília Moraes; Maurício Lacerda Nogueira. Prefeitura de Sao Paulo: Melissa Palmieri.; Patricia Akemi Assato; Paula Rahal; Paulo Inacio da Costa; Rafael dos Santos Bezerra; Raquel de Lello Rocha Campos Cassano. NGS Soluções Genômicas: Pilar Drummond Sampaio Corrêa Mariani. FZEA-USP Pirassununga: Mirele Daiana Poleti; Raul Machado Neto; Ricardo Augusto Brassaloti; Ricardo Haddad; Rodrigo Tocantins Calado. FAMERP-SJRP: Cecília Artico Banho; Sandra Coccuzzo Sampaio; Svetoslav Nanev Slavov; Vagner Fonseca; Vincent Louis Viala |
| EPI_ISL_1966814, EPI_ISL_1966815 | CENTRO DE SAUDE II SAO MIGUEL ARCANJO | Instituto Butantan / Mendelics | Antonio Jorge Martins; Bianca Cechetto Carlos. Mendelics: Bibiana Santos; Claudia Renata dos Santos Barros; Cíntia Bittar; David Schlesinger. Hemocentro Ribeirão Preto: Simone Kashima; Debora Botequio Moretti; Elaine Cristina Marqueze; Elaine Vieira dos Santos; Elisangela Chicaroni Mattos; Erika Freitas; Evandra Strazza Rodrigues; Felipe Allan da Silva da Costa; Flavia Aburjaile; Fábio Sossai Possebon; Guilherme Campos; Guilherme Targino Valente; Heidge Fukumasu. USP-Botucatu: Rejane Maria Tommasini Grotto; Helena Lage Ferreira; Instituto Butantan: Dimas Tadeu Covas; Jardelina de Souza Todao Bernardino; Jayme A. Souza-Neto; Jessika Cristina Chagas Lesbon; Jorge A. Petroli Marchesi; José Salvatore Leister Patané; João Paulo Kitajima; João Pessoa Araújo Jr.; Leila Sabrina Ullmann; Loyze Paola Oliveira de Lima; Luiz Aurelio de Campos Crispin. Centro de Genômica Funcional da ESALQ: Luiz Lehmann Coutinho; Luiz Carlos Junior de Alcantara; Lívia Sacchetto; Maisa C. Pereira Parra; Maria Carolina Elias; Marta Giovanetti; Marília Moraes; Maurício Lacerda Nogueira. Prefeitura de Sao Paulo: Melissa Palmieri.; Patricia Akemi Assato; Paula Rahal; Paulo Inacio da Costa; Rafael dos Santos Bezerra; Raquel de Lello Rocha Campos Cassano. NGS Soluções Genômicas: Pilar Drummond Sampaio Corrêa Mariani. FZEA-USP Pirassununga: Mirele Daiana Poleti; Raul Machado Neto; Ricardo Augusto Brassaloti; Ricardo Haddad; Rodrigo Tocantins Calado. FAMERP-SJRP: Cecília Artico Banho; Sandra Coccuzzo Sampaio; Svetoslav Nanev Slavov; Vagner Fonseca; Vincent Louis Viala |
| EPI_ISL_1966554 | CENTRO DE SAUDE II SIZENANDO NABUCO TAPIRATIBA | Instituto Butantan / FZEA-USP (Pirassununga) | Antonio Jorge Martins; Bianca Cechetto Carlos. Mendelics: Bibiana Santos; Claudia Renata dos Santos Barros; Cíntia Bittar; David Schlesinger. Hemocentro Ribeirão Preto: Simone Kashima; Debora Botequio Moretti; Elaine Cristina Marqueze; Elaine Vieira dos Santos; Elisangela Chicaroni Mattos; Erika Freitas; Evandra Strazza Rodrigues; Felipe Allan da Silva da Costa; Flavia Aburjaile; Fábio Sossai Possebon; Guilherme Campos; Guilherme Targino Valente; Heidge Fukumasu. USP-Botucatu: Rejane Maria Tommasini Grotto; Helena Lage Ferreira; Instituto Butantan: Dimas Tadeu Covas; Jardelina de Souza Todao Bernardino; Jayme A. Souza-Neto; Jessika Cristina Chagas Lesbon; Jorge A. Petroli Marchesi; José Salvatore Leister Patané; João Paulo Kitajima; João Pessoa Araújo Jr.; Leila Sabrina Ullmann; Loyze Paola Oliveira de Lima; Luiz Aurelio de Campos Crispin. Centro de Genômica Funcional da ESALQ: Luiz Lehmann Coutinho; Luiz Carlos Junior de Alcantara; Lívia Sacchetto; Maisa C. Pereira Parra; Maria Carolina Elias; Marta Giovanetti; Marília Moraes; Maurício Lacerda Nogueira. Prefeitura de Sao Paulo: Melissa Palmieri.; Patricia Akemi Assato; Paula Rahal; Paulo Inacio da Costa; Rafael dos Santos Bezerra; Raquel de Lello Rocha Campos Cassano. NGS Soluções Genômicas: Pilar Drummond Sampaio Corrêa Mariani. FZEA-USP Pirassununga: Mirele Daiana Poleti; Raul Machado Neto; Ricardo Augusto Brassaloti; Ricardo Haddad; Rodrigo Tocantins Calado. FAMERP-SJRP: Cecília Artico Banho; Sandra Coccuzzo Sampaio; Svetoslav Nanev Slavov; Vagner Fonseca; Vincent Louis Viala |
| EPI_ISL_1966818, | CENTRO DE SAUDE | Instituto Butantan / | Antonio Jorge Martins; Bianca Cechetto Carlos. Mendelics: Bibiana Santos; Claudia Renata dos Santos Barros; Cíntia Bittar; David Schlesinger. Hemocentro Ribeirão Preto: Simone Kashima; Debora Botequio Moretti; Elaine Cristina Marqueze; Elaine Vieira dos Santos; Elisangela Chicaroni Mattos; Erika Freitas; Evandra Strazza Rodrigues; |

| | | | |
|---|---|---|---|
| EPI_ISL_1966819, EPI_ISL_1966820, EPI_ISL_1966821 | II TIETE DR RUI SILVEIRA MELO | Mendelics | Felipe Allan da Silva da Costa; Flavia Aburjaile; Fábio Sossai Possebon; Guilherme Campos; Guilherme Targino Valente; Heidge Fukumasu. USP-Botucatu: Rejane Maria Tommasini Grotto; Helena Lage Ferreira; Instituto Butantan: Dimas Tadeu Covas; Jardelina de Souza Todao Bernardino; Jayme A. Souza-Neto; Jessika Cristina Chagas Lesbon; Jorge A. Petroli Marchesi; José Salvatore Leister Patané; João Paulo Kitajima; João Pessoa Araújo Jr.; Leila Sabrina Ullmann; Loyze Paola Oliveira de Lima; Luiz Aurelio de Campos Crispin. Centro de Genômica Funcional da ESALQ: Luiz Lehmann Coutinho; Luiz Carlos Junior de Alcantara; Lívia Sacchetto; Maisa C. Pereira Parra; Maria Carolina Elias; Marta Giovanetti; Marília Moraes; Maurício Lacerda Nogueira. Prefeitura de Sao Paulo: Melissa Palmieri.; Patricia Akemi Assato; Paula Rahal; Paulo Inacio da Costa; Rafael dos Santos Bezerra; Raquel de Lello Rocha Campos Cassano. NGS Soluções Genômicas: Pilar Drummond Sampaio Corrêa Mariani. FZEA-USP Pirassununga: Mirele Daiana Poleti; Raul Machado Neto; Ricardo Augusto Brassaloti; Ricardo Haddad; Rodrigo Tocantins Calado. FAMERP-SJRP: Cecília Artico Banho; Sandra Coccuzzo Sampaio; Svetoslav Nanev Slavov; Vagner Fonseca; Vincent Louis Viala |
| EPI_ISL_1795081, EPI_ISL_1795087, EPI_ISL_1795088, EPI_ISL_1795089, EPI_ISL_1795090, EPI_ISL_1795296, EPI_ISL_1795297, EPI_ISL_1795299 | | | |
| see above | CENTRO DE SAUDE III AFFONSO LUZZI SANTA CRUZ DAS PALMEIRAS | Instituto Butantan / ESALQ-Piracicaba | Antonio Jorge Martins; Bianca Cechetto Carlos. Mendelics: Bibiana Santos; Claudia Renata dos Santos Barros; David Schlesinger. Hemocentro Ribeirão Preto: Simone Kashima; Debora Botequio Moretti. Centro de Genômica Funcional da ESALQ: Luiz Lehmann Coutinho; Dimas Tadeu Covas; Elaine Cristina Marqueze; Elaine Vieira dos Santos; Elisangela Chicaroni Mattos; Erika Freitas; Evandra Strazza Rodrigues; Felipe Allan da Silva da Costa; Flavia Aburjaile; Guilherme Targino Valente; Heidge Fukumasu. USP-Botucatu: Rejane Maria Tommasini Grotto; Instituto Butantan: Alexander Roberto Precioso; Jayme A. Souza-Neto; Jessika Cristina Chagas Lesbon; José Salvatore Leister Patané; João Paulo Kitajima; Luiz Carlos Junior de Alcantara; Maria Carolina Elias; Marta Giovanetti; Patricia Akemi Assato; Rafael dos Santos Bezerra; Raquel de Lello Rocha Campos Cassano. NGS Soluções Genômicas: Pilar Drummond Sampaio Corrêa Mariani. FZEA-USP Pirassununga: Mirele Daiana Poleti; Raul Machado Neto; Ricardo Augusto Brassaloti; Ricardo Haddad; Rodrigo Tocantins Calado.; Sandra Coccuzzo Sampaio; Svetoslav Nanev Slavov; Vagner Fonseca; Vincent Louis Viala |
| EPI_ISL_1966541 | CENTRO DE SAUDE III AFFONSO LUZZI SANTA CRUZ DAS PALMEIRAS | Instituto Butantan / FZEA-USP (Pirassununga) | Antonio Jorge Martins; Bianca Cechetto Carlos. Mendelics: Bibiana Santos; Claudia Renata dos Santos Barros; Cíntia Bittar; David Schlesinger. Hemocentro Ribeirão Preto: Simone Kashima; Debora Botequio Moretti; Elaine Cristina Marqueze; Elaine Vieira dos Santos; Elisangela Chicaroni Mattos; Erika Freitas; Evandra Strazza Rodrigues; Felipe Allan da Silva da Costa; Flavia Aburjaile; Fábio Sossai Possebon; Guilherme Campos; Guilherme Targino Valente; Heidge Fukumasu. USP-Botucatu: Rejane Maria Tommasini Grotto; Helena Lage Ferreira; Instituto Butantan: Dimas Tadeu Covas; Jardelina de Souza Todao Bernardino; Jayme A. Souza-Neto; Jessika Cristina Chagas Lesbon; Jorge A. Petroli Marchesi; José Salvatore Leister Patané; João Paulo Kitajima; João Pessoa Araújo Jr.; Leila Sabrina Ullmann; Loyze Paola Oliveira de Lima; Luiz Aurelio de Campos Crispin. Centro de Genômica Funcional da ESALQ: Luiz Lehmann Coutinho; Luiz Carlos Junior de Alcantara; Lívia Sacchetto; Maisa C. Pereira Parra; Maria Carolina Elias; Marta Giovanetti; Marília Moraes; Maurício Lacerda Nogueira. Prefeitura de Sao Paulo: Melissa Palmieri.; Patricia Akemi Assato; Paula Rahal; Paulo Inacio da Costa; Rafael dos Santos Bezerra; Raquel de Lello Rocha Campos Cassano. NGS Soluções Genômicas: Pilar Drummond Sampaio Corrêa Mariani. FZEA-USP Pirassununga: Mirele Daiana Poleti; Raul Machado Neto; Ricardo Augusto Brassaloti; Ricardo Haddad; Rodrigo Tocantins Calado. FAMERP-SJRP: Cecília Artico Banho; Sandra Coccuzzo Sampaio; Svetoslav Nanev Slavov; Vagner Fonseca; Vincent Louis Viala |
| EPI_ISL_1795114, EPI_ISL_1795115 | CENTRO DE SAUDE III BORBOREMA | Instituto Butantan / ESALQ-Piracicaba | Antonio Jorge Martins; Bianca Cechetto Carlos. Mendelics: Bibiana Santos; Claudia Renata dos Santos Barros; David Schlesinger. Hemocentro Ribeirão Preto: Simone Kashima; Debora Botequio Moretti. Centro de Genômica Funcional da ESALQ: Luiz Lehmann Coutinho; Dimas Tadeu Covas; Elaine Cristina Marqueze; Elaine Vieira dos Santos; Elisangela Chicaroni Mattos; Erika Freitas; Evandra Strazza Rodrigues; Felipe Allan da Silva da Costa; Flavia Aburjaile; Guilherme Targino Valente; Heidge Fukumasu. USP-Botucatu: Rejane Maria Tommasini Grotto; Instituto Butantan: Alexander Roberto Precioso; Jayme A. Souza-Neto; Jessika Cristina Chagas Lesbon; José Salvatore Leister Patané; João Paulo Kitajima; Luiz Carlos Junior de Alcantara; Maria Carolina Elias; Marta Giovanetti; Patricia Akemi Assato; Rafael dos Santos Bezerra; Raquel de Lello Rocha Campos Cassano. NGS Soluções Genômicas: Pilar Drummond Sampaio Corrêa Mariani. FZEA-USP Pirassununga: Mirele Daiana Poleti; Raul Machado Neto; Ricardo Augusto Brassaloti; Ricardo Haddad; Rodrigo Tocantins Calado.; Sandra Coccuzzo Sampaio; Svetoslav Nanev Slavov; Vagner Fonseca; Vincent Louis Viala |
| EPI_ISL_1966749, EPI_ISL_1966750 | CENTRO DE SAUDE III BORBOREMA | Instituto Butantan / Mendelics | Antonio Jorge Martins; Bianca Cechetto Carlos. Mendelics: Bibiana Santos; Claudia Renata dos Santos Barros; Cíntia Bittar; David Schlesinger. Hemocentro Ribeirão Preto: Simone Kashima; Debora Botequio Moretti; Elaine Cristina Marqueze; Elaine Vieira dos Santos; Elisangela Chicaroni Mattos; Erika Freitas; Evandra Strazza Rodrigues; Felipe Allan da Silva da Costa; Flavia Aburjaile; Fábio Sossai Possebon; Guilherme Campos; Guilherme Targino Valente; Heidge Fukumasu. USP-Botucatu: Rejane Maria Tommasini Grotto; Helena Lage Ferreira; Instituto Butantan: Dimas Tadeu Covas; Jardelina de Souza Todao Bernardino; Jayme A. Souza-Neto; Jessika Cristina Chagas Lesbon; Jorge A. Petroli Marchesi; José Salvatore Leister Patané; João Paulo Kitajima; João Pessoa Araújo Jr.; Leila Sabrina Ullmann; Loyze Paola Oliveira de Lima; Luiz Aurelio de Campos Crispin. Centro de Genômica Funcional da ESALQ: Luiz Lehmann Coutinho; Luiz Carlos Junior de Alcantara; Lívia Sacchetto; Maisa C. Pereira Parra; Maria Carolina Elias; Marta Giovanetti; Marília Moraes; Maurício Lacerda Nogueira. Prefeitura de Sao Paulo: Melissa Palmieri.; Patricia Akemi Assato; Paula Rahal; Paulo Inacio da Costa; Rafael dos Santos Bezerra; Raquel de Lello Rocha Campos Cassano. NGS Soluções Genômicas: Pilar Drummond Sampaio Corrêa Mariani. FZEA-USP Pirassununga: Mirele Daiana Poleti; Raul Machado Neto; Ricardo Augusto Brassaloti; Ricardo Haddad; Rodrigo Tocantins Calado. FAMERP-SJRP: Cecília Artico Banho; Sandra Coccuzzo Sampaio; Svetoslav Nanev Slavov; Vagner Fonseca; Vincent Louis Viala |
| EPI_ISL_1795125, EPI_ISL_1795127 | CENTRO DE SAUDE III CANDIDO RODRIGUES | Instituto Butantan / ESALQ-Piracicaba | Antonio Jorge Martins; Bianca Cechetto Carlos. Mendelics: Bibiana Santos; Claudia Renata dos Santos Barros; David Schlesinger. Hemocentro Ribeirão Preto: Simone Kashima; Debora Botequio Moretti. Centro de Genômica Funcional da ESALQ: Luiz Lehmann Coutinho; Dimas Tadeu Covas; Elaine Cristina Marqueze; Elaine Vieira dos Santos; Elisangela Chicaroni Mattos; Erika Freitas; Evandra Strazza Rodrigues; Felipe Allan da Silva da Costa; Flavia Aburjaile; Guilherme Targino Valente; Heidge Fukumasu. USP-Botucatu: Rejane Maria Tommasini Grotto; Instituto Butantan: Alexander Roberto Precioso; Jayme A. Souza-Neto; Jessika Cristina Chagas Lesbon; José Salvatore Leister Patané; João Paulo Kitajima; Luiz Carlos Junior de Alcantara; Maria Carolina Elias; Marta Giovanetti; Patricia Akemi Assato; Rafael dos Santos Bezerra; Raquel de Lello Rocha Campos Cassano. NGS Soluções Genômicas: Pilar Drummond Sampaio Corrêa Mariani. FZEA-USP Pirassununga: Mirele Daiana Poleti; Raul Machado Neto; Ricardo Augusto Brassaloti; Ricardo Haddad; Rodrigo Tocantins Calado.; Sandra Coccuzzo Sampaio; Svetoslav Nanev Slavov; Vagner Fonseca; Vincent Louis Viala |
| EPI_ISL_2170976 | CENTRO DE SAUDE III DR NICOLA JORGE CARNEIRO GUARACAI | Instituto Butantan / Mendelics | Antonio Jorge Martins; Bianca Cechetto Carlos. Mendelics: Bibiana Santos; Claudia Renata dos Santos Barros; Cíntia Bittar; David Schlesinger. Hemocentro Ribeirão Preto: Simone Kashima; Debora Botequio Moretti; Elaine Cristina Marqueze; Elaine Vieira dos Santos; Elisangela Chicaroni Mattos; Erika Freitas; Evandra Strazza Rodrigues; Felipe Allan da Silva da Costa; Flavia Aburjaile; Fábio Sossai Possebon; Guilherme Campos; Guilherme Targino Valente; Heidge Fukumasu. USP-Botucatu: Rejane Maria Tommasini Grotto; Helena Lage Ferreira; Instituto Butantan: Dimas Tadeu Covas; Jardelina de Souza Todao Bernardino; Jayme A. Souza-Neto; Jessika Cristina Chagas Lesbon; Jorge A. Petroli Marchesi; José Salvatore Leister Patané; João Paulo Kitajima; João Pessoa Araújo Jr.; Leila Sabrina Ullmann; Loyze Paola Oliveira de Lima; Luiz Aurelio de Campos Crispin. Centro de Genômica Funcional da ESALQ: Luiz Lehmann Coutinho; Luiz Carlos Junior de Alcantara; Lívia Sacchetto; Maisa C. Pereira Parra; Maria Carolina Elias; Marta Giovanetti; Marília Moraes; Maurício Lacerda Nogueira. Prefeitura de Sao Paulo: Melissa Palmieri.; Patricia Akemi Assato; Paula Rahal; Paulo Inacio da Costa; Rafael dos Santos Bezerra; Raquel de Lello Rocha Campos Cassano. NGS Soluções Genômicas: Pilar Drummond Sampaio Corrêa Mariani. FZEA-USP Pirassununga: Mirele Daiana Poleti; Raul Machado Neto; Ricardo Augusto Brassaloti; Ricardo Haddad; Rodrigo Tocantins Calado. FAMERP-SJRP: Cecília Artico Banho; Sandra Coccuzzo Sampaio; Svetoslav Nanev Slavov; Vagner Fonseca; Vincent Louis Viala |
| EPI_ISL_1966293 | CENTRO DE SAUDE III GLICERIO GIMAIEL ELIAS GIMAIEL | Instituto Butantan / Mendelics | Antonio Jorge Martins; Bianca Cechetto Carlos. Mendelics: Bibiana Santos; Claudia Renata dos Santos Barros; Cíntia Bittar; David Schlesinger. Hemocentro Ribeirão Preto: Simone Kashima; Debora Botequio Moretti; Elaine Cristina Marqueze; Elaine Vieira dos Santos; Elisangela Chicaroni Mattos; Erika Freitas; Evandra Strazza Rodrigues; Felipe Allan da Silva da Costa; Flavia Aburjaile; Fábio Sossai Possebon; Guilherme Campos; Guilherme Targino Valente; Heidge Fukumasu. USP-Botucatu: Rejane Maria Tommasini Grotto; Helena Lage Ferreira; Instituto Butantan: Dimas Tadeu Covas; Jardelina de Souza Todao Bernardino; Jayme A. Souza-Neto; Jessika Cristina Chagas Lesbon; Jorge A. Petroli Marchesi; José Salvatore Leister Patané; João Paulo Kitajima; João Pessoa Araújo Jr.; Leila Sabrina Ullmann; Loyze Paola Oliveira de Lima; Luiz Aurelio de Campos Crispin. Centro de Genômica Funcional da ESALQ: Luiz Lehmann Coutinho; Luiz Carlos Junior de Alcantara; Lívia Sacchetto; Maisa C. Pereira Parra; Maria Carolina Elias; Marta Giovanetti; Marília Moraes; Maurício Lacerda Nogueira. Prefeitura de Sao Paulo: Melissa Palmieri.; Patricia Akemi Assato; Paula Rahal; Paulo Inacio da Costa; Rafael dos Santos Bezerra; Raquel de Lello Rocha Campos Cassano. NGS Soluções Genômicas: Pilar Drummond Sampaio Corrêa Mariani. FZEA-USP Pirassununga: Mirele Daiana Poleti; Raul Machado Neto; Ricardo Augusto Brassaloti; Ricardo Haddad; Rodrigo Tocantins Calado. FAMERP-SJRP: Cecília Artico Banho; Sandra Coccuzzo Sampaio; Svetoslav Nanev Slavov; Vagner Fonseca; Vincent Louis Viala |
| EPI_ISL_2170925 | CENTRO DE SAUDE III NELCIDIO DA SILVEIRA BASTOS | Instituto Butantan / Mendelics | Antonio Jorge Martins; Claudia Renata dos Santos Barros; David Schlesinger; Debora Botequio Moretti; Dimas Tadeu Covas; Elaine Cristina Marqueze; Elaine Vieira Santos; Evandra Strazza Rodrigues; Heidge Fukumasu; Jayme Augusto de Souza-Neto; José Salvatore Leister Patané; Luiz Alcantara; Luiz Lehmann Coutinho; Maria Carolina Elias; Maurício Lacerda Nogueira; Rafael dos Santos Bezerra; Raul Machado Neto; Rejane Maria Tommasini Grotto; Ricardo Haddad; Sandra Coccuzzo Sampaio Vessoni; Simone Kashima; Svetoslav Nanev Slavov; Vincent Louis Viala |
| EPI_ISL_1966477, EPI_ISL_1966478, EPI_ISL_1966479 | CENTRO DE SAUDE III RINCAO | Instituto Butantan / Mendelics | Antonio Jorge Martins; Bianca Cechetto Carlos. Mendelics: Bibiana Santos; Claudia Renata dos Santos Barros; Cíntia Bittar; David Schlesinger. Hemocentro Ribeirão Preto: Simone Kashima; Debora Botequio Moretti; Elaine Cristina Marqueze; Elaine Vieira dos Santos; Elisangela Chicaroni Mattos; Erika Freitas; Evandra Strazza Rodrigues; Felipe Allan da Silva da Costa; Flavia Aburjaile; Fábio Sossai Possebon; Guilherme Campos; Guilherme Targino Valente; Heidge Fukumasu. USP-Botucatu: Rejane Maria Tommasini Grotto; Helena Lage Ferreira; Instituto Butantan: Dimas Tadeu Covas; Jardelina de Souza Todao Bernardino; Jayme A. Souza-Neto; Jessika Cristina Chagas Lesbon; Jorge A. Petroli Marchesi; José Salvatore Leister Patané; João Paulo Kitajima; João Pessoa Araújo Jr.; Leila Sabrina Ullmann; Loyze Paola Oliveira de Lima; Luiz Aurelio de Campos Crispin. Centro de Genômica Funcional da ESALQ: Luiz Lehmann Coutinho; Luiz Carlos Junior de Alcantara; Lívia Sacchetto; Maisa C. Pereira Parra; Maria Carolina Elias; Marta Giovanetti; Marília Moraes; Maurício Lacerda Nogueira. Prefeitura de Sao Paulo: Melissa Palmieri.; Patricia Akemi Assato; Paula Rahal; Paulo Inacio da Costa; Rafael dos Santos Bezerra; Raquel de Lello Rocha Campos Cassano. NGS Soluções Genômicas: Pilar Drummond Sampaio Corrêa Mariani. FZEA-USP Pirassununga: Mirele Daiana Poleti; Raul Machado Neto; Ricardo Augusto Brassaloti; Ricardo Haddad; Rodrigo Tocantins Calado. FAMERP-SJRP: Cecília Artico Banho; Sandra Coccuzzo Sampaio; Svetoslav Nanev Slavov; Vagner Fonseca; Vincent Louis Viala |
| EPI_ISL_1795126 | CENTRO DE SAUDE III SALES OLIVEIRA | Instituto Butantan / ESALQ-Piracicaba | Antonio Jorge Martins; Bianca Cechetto Carlos. Mendelics: Bibiana Santos; Claudia Renata dos Santos Barros; David Schlesinger. Hemocentro Ribeirão Preto: Simone Kashima; Debora Botequio Moretti. Centro de Genômica Funcional da ESALQ: Luiz Lehmann Coutinho; Dimas Tadeu Covas; Elaine Cristina Marqueze; Elaine Vieira dos Santos; Elisangela Chicaroni Mattos; Erika Freitas; Evandra Strazza Rodrigues; Felipe Allan da Silva da Costa; Flavia Aburjaile; Guilherme Targino Valente; Heidge Fukumasu. USP-Botucatu: Rejane Maria Tommasini Grotto; Instituto Butantan: Alexander Roberto Precioso; Jayme A. Souza-Neto; Jessika Cristina Chagas Lesbon; José Salvatore Leister Patané; João Paulo Kitajima; Luiz Carlos Junior de Alcantara; Maria Carolina Elias; Marta Giovanetti; Patricia Akemi Assato; Rafael dos Santos Bezerra; Raquel de Lello Rocha Campos Cassano. NGS Soluções Genômicas: Pilar Drummond Sampaio Corrêa Mariani. FZEA-USP Pirassununga: Mirele Daiana Poleti; Raul Machado Neto; Ricardo Augusto Brassaloti; Ricardo Haddad; Rodrigo Tocantins Calado.; Sandra Coccuzzo Sampaio; Svetoslav Nanev Slavov; Vagner Fonseca; Vincent Louis Viala |
| EPI_ISL_1445199, EPI_ISL_1966235 | CENTRO DE SAUDE III SALES OLIVEIRA | Instituto Butantan / Mendelics | Antonio Jorge Martins; Bianca Cechetto Carlos. Mendelics: Bibiana Santos; Bibiana Santos; Claudia Renata dos Santos Barros; Cíntia Bittar; David Schlesinger; David Schlesinger. Hemocentro Ribeirão Preto: Simone Kashima; Debora Botequio Moretti; Dimas Tadeu Covas; Elaine Cristina Marqueze; Elaine Vieira dos Santos; Elisangela Chicaroni Mattos; Erika Freitas; Evandra Strazza Rodrigues; Felipe Allan da Silva da Costa; Flavia Aburjaile; Fábio Sossai Possebon; Guilherme Campos; Guilherme Targino Valente; Heidge Fukumasu. USP-Botucatu: Rejane Maria Tommasini Grotto; Helena Lage Ferreira; Instituto Butantan: Dimas Tadeu Covas; Jardelina de Souza Todao Bernardino; Jayme A. Souza-Neto; Jessika Cristina Chagas Lesbon; Jorge A. Petroli Marchesi; José Salvatore Leister Patané; João Paulo Kitajima; João Pessoa Araújo Jr.; Leila Sabrina Ullmann; Loyze Paola Oliveira de Lima; Luiz Aurelio de Campos Crispin. Centro de Genômica Funcional da ESALQ: Luiz Lehmann Coutinho; Luiz Carlos Junior de Alcantara; Lívia Sacchetto; Maisa C. Pereira Parra; Maria Carolina Elias; Marta Giovanetti; Marília Moraes; Maurício Lacerda Nogueira. Prefeitura de Sao Paulo: Melissa Palmieri.; Patricia Akemi Assato; Paula Rahal; Paulo Inacio da Costa; Rafael dos Santos Bezerra; Raquel de Lello Rocha Campos Cassano. NGS Soluções Genômicas: Pilar Drummond Sampaio Corrêa Mariani. FZEA-USP Pirassununga: Mirele Daiana Poleti; Raul Machado Neto; Ricardo Augusto Brassaloti; Ricardo Haddad; Rodrigo Tocantins Calado.; Rodrigo Tocantins Calado. FAMERP-SJRP: Cecília Artico Banho; Sandra Coccuzzo Sampaio; Simone Kashima; Svetoslav Nanev Slavov; Vagner Fonseca; Vincent Louis Viala |
| EPI_ISL_1966343, EPI_ISL_1966344, EPI_ISL_1966345, EPI_ISL_1966634, EPI_ISL_1966635, EPI_ISL_1966636, EPI_ISL_1966637, EPI_ISL_1966638, EPI_ISL_1966639, EPI_ISL_1967277, EPI_ISL_1967278, EPI_ISL_1967279 | | | |
| see above | CENTRO DE SAUDE III TABATINGA | Instituto Butantan / Mendelics | Antonio Jorge Martins; Bianca Cechetto Carlos. Mendelics: Bibiana Santos; Claudia Renata dos Santos Barros; Cíntia Bittar; David Schlesinger. Hemocentro Ribeirão Preto: Simone Kashima; Debora Botequio Moretti; Elaine Cristina Marqueze; Elaine Vieira dos Santos; Elisangela Chicaroni Mattos; Erika Freitas; Evandra Strazza Rodrigues; Felipe Allan da Silva da Costa; Flavia Aburjaile; Fábio Sossai Possebon; Guilherme Campos; Guilherme Targino Valente; Heidge Fukumasu. USP-Botucatu: Rejane Maria Tommasini Grotto; Helena Lage Ferreira; Instituto Butantan: Dimas Tadeu Covas; Jardelina de Souza Todao Bernardino; Jayme A. Souza-Neto; Jessika Cristina Chagas Lesbon; Jorge A. Petroli Marchesi; José Salvatore Leister Patané; João Paulo Kitajima; João Pessoa Araújo Jr.; Leila Sabrina Ullmann; Loyze Paola Oliveira de Lima; Luiz Aurelio de Campos Crispin. Centro de Genômica Funcional da ESALQ: Luiz Lehmann Coutinho; Luiz Carlos Junior de Alcantara; Lívia Sacchetto; Maisa C. Pereira Parra; Maria Carolina Elias; Marta Giovanetti; Marília Moraes; Maurício Lacerda Nogueira. Prefeitura de Sao Paulo: Melissa Palmieri.; Patricia Akemi Assato; Paula Rahal; Paulo Inacio da Costa; Rafael dos Santos Bezerra; Raquel de Lello Rocha Campos Cassano. NGS Soluções Genômicas: Pilar Drummond Sampaio Corrêa Mariani. FZEA-USP Pirassununga: Mirele Daiana Poleti; Raul Machado Neto; Ricardo Augusto Brassaloti; Ricardo Haddad; Rodrigo Tocantins Calado. FAMERP-SJRP: Cecília Artico Banho; Sandra Coccuzzo Sampaio; Svetoslav Nanev Slavov; Vagner Fonseca; Vincent Louis Viala |
| EPI_ISL_1967096, EPI_ISL_1967097 | CENTRO DE SAUDE SAO ROQUE DR JOSE CARVALHO BRITO | Instituto Butantan / ESALQ-USP (Piracicaba) | Antonio Jorge Martins; Bianca Cechetto Carlos. Mendelics: Bibiana Santos; Claudia Renata dos Santos Barros; Cíntia Bittar; David Schlesinger. Hemocentro Ribeirão Preto: Simone Kashima; Debora Botequio Moretti; Elaine Cristina Marqueze; Elaine Vieira dos Santos; Elisangela Chicaroni Mattos; Erika Freitas; Evandra Strazza Rodrigues; Felipe Allan da Silva da Costa; Flavia Aburjaile; Fábio Sossai Possebon; Guilherme Campos; Guilherme Targino Valente; Heidge Fukumasu. USP-Botucatu: Rejane Maria Tommasini Grotto; Helena Lage Ferreira; Instituto Butantan: Dimas Tadeu Covas; Jardelina de Souza Todao Bernardino; Jayme A. Souza-Neto; Jessika Cristina Chagas Lesbon; Jorge A. Petroli Marchesi; José Salvatore Leister Patané; João Paulo Kitajima; João Pessoa Araújo Jr.; Leila Sabrina Ullmann; Loyze Paola Oliveira de Lima; Luiz Aurelio de Campos Crispin. Centro de Genômica Funcional da ESALQ: Luiz Lehmann Coutinho; Luiz Carlos Junior de Alcantara; Lívia Sacchetto; Maisa C. Pereira Parra; Maria Carolina Elias; Marta Giovanetti; Marília Moraes; Maurício Lacerda Nogueira. Prefeitura de Sao Paulo: Melissa Palmieri.; Patricia Akemi Assato; Paula Rahal; Paulo Inacio da Costa; Rafael dos Santos Bezerra; Raquel de Lello Rocha Campos Cassano. NGS Soluções Genômicas: Pilar Drummond Sampaio Corrêa Mariani. FZEA-USP Pirassununga: Mirele Daiana Poleti; Raul Machado Neto; Ricardo Augusto Brassaloti; Ricardo Haddad; Rodrigo Tocantins Calado. FAMERP-SJRP: Cecília Artico Banho; Sandra Coccuzzo Sampaio; Svetoslav Nanev Slavov; Vagner Fonseca; Vincent Louis Viala |
| EPI_ISL_1445081, EPI_ISL_1966507, EPI_ISL_2170904 | CENTRO DE SAUDE SAO ROQUE DR JOSE CARVALHO BRITO | Instituto Butantan / Mendelics | Antonio Jorge Martins; Bianca Cechetto Carlos. Mendelics: Bibiana Santos; Bibiana Santos; Claudia Renata dos Santos Barros; Cíntia Bittar; David Schlesinger; David Schlesinger. Hemocentro Ribeirão Preto: Simone Kashima; Debora Botequio Moretti; Dimas Tadeu Covas; Elaine Cristina Marqueze; Elaine Vieira dos Santos; Elisangela Chicaroni Mattos; Erika Freitas; Evandra Strazza Rodrigues; Felipe Allan da Silva da Costa; Flavia Aburjaile; Fábio Sossai Possebon; Guilherme Campos; Guilherme Targino Valente; Heidge Fukumasu. USP-Botucatu: Rejane Maria Tommasini Grotto; Helena Lage Ferreira; Instituto Butantan: Dimas Tadeu Covas; Jardelina de Souza Todao Bernardino; Jayme A. Souza-Neto; Jessika Cristina Chagas Lesbon; Jorge A. Petroli Marchesi; José Salvatore Leister Patané; João Paulo Kitajima; João Pessoa Araújo Jr.; Leila Sabrina Ullmann; Loyze Paola Oliveira de Lima; Luiz Aurelio de Campos Crispin. Centro de Genômica Funcional da ESALQ: Luiz Lehmann Coutinho; Luiz Carlos Junior de Alcantara; Lívia Sacchetto; Maisa C. Pereira Parra; Maria Carolina Elias; Marta Giovanetti; Marília Moraes; Maurício Lacerda Nogueira. Prefeitura de Sao Paulo: Melissa Palmieri.; Patricia Akemi Assato; Paula Rahal; Paulo Inacio da Costa; Rafael dos Santos Bezerra; Raquel de Lello Rocha Campos Cassano. NGS Soluções Genômicas: Pilar Drummond Sampaio Corrêa Mariani. FZEA-USP Pirassununga: Mirele Daiana Poleti; Raul Machado Neto; Ricardo Augusto Brassaloti; Ricardo Haddad; Rodrigo Tocantins Calado.; Rodrigo Tocantins Calado. FAMERP-SJRP: Cecília Artico Banho; Sandra Coccuzzo Sampaio; Simone Kashima; Svetoslav Nanev Slavov; Vagner Fonseca; Vincent Louis Viala |
| EPI_ISL_1966796 | CENTRO DE SAUDE TEREZINHA DE MORAES ARSILLA | Instituto Butantan / Mendelics | Antonio Jorge Martins; Bianca Cechetto Carlos. Mendelics: Bibiana Santos; Claudia Renata dos Santos Barros; Cíntia Bittar; David Schlesinger. Hemocentro Ribeirão Preto: Simone Kashima; Debora Botequio Moretti; Elaine Cristina Marqueze; Elaine Vieira dos Santos; Elisangela Chicaroni Mattos; Erika Freitas; Evandra Strazza Rodrigues; Felipe Allan da Silva da Costa; Flavia Aburjaile; Fábio Sossai Possebon; Guilherme Campos; Guilherme Targino Valente; Heidge Fukumasu. USP-Botucatu: Rejane Maria Tommasini Grotto; Helena Lage Ferreira; Instituto Butantan: Dimas Tadeu Covas; Jardelina de Souza Todao Bernardino; Jayme A. Souza-Neto; Jessika Cristina Chagas Lesbon; Jorge A. Petroli Marchesi; José Salvatore Leister Patané; João Paulo Kitajima; João Pessoa Araújo Jr.; Leila Sabrina Ullmann; Loyze Paola Oliveira de Lima; Luiz Aurelio de Campos Crispin. Centro de Genômica Funcional da ESALQ: Luiz Lehmann Coutinho; Luiz Carlos Junior de Alcantara; Lívia Sacchetto; Maisa C. Pereira Parra; Maria Carolina Elias; Marta Giovanetti; Marília Moraes; Maurício Lacerda Nogueira. Prefeitura de Sao Paulo: Melissa Palmieri.; Patricia Akemi Assato; Paula Rahal; Paulo Inacio da Costa; Rafael dos Santos Bezerra; Raquel de Lello Rocha Campos Cassano. NGS Soluções Genômicas: Pilar Drummond Sampaio Corrêa Mariani. FZEA-USP Pirassununga: Mirele Daiana Poleti; Raul Machado Neto; Ricardo Augusto Brassaloti; Ricardo Haddad; Rodrigo Tocantins Calado. FAMERP-SJRP: Cecília Artico Banho; Sandra Coccuzzo Sampaio; Svetoslav Nanev Slavov; Vagner Fonseca; Vincent Louis Viala |
| EPI_ISL_1795093, EPI_ISL_1795368, EPI_ISL_1795370, EPI_ISL_1795429 | CENTRO INTEGRADO DE SAUDE | Instituto Butantan / ESALQ-Piracicaba | Antonio Jorge Martins; Bianca Cechetto Carlos. Mendelics: Bibiana Santos; Claudia Renata dos Santos Barros; David Schlesinger. Hemocentro Ribeirão Preto: Simone Kashima; Debora Botequio Moretti. Centro de Genômica Funcional da ESALQ: Luiz Lehmann Coutinho; Dimas Tadeu Covas; Elaine Cristina Marqueze; Elaine Vieira dos Santos; Elisangela Chicaroni Mattos; Erika Freitas; Evandra Strazza Rodrigues; Felipe Allan da Silva da Costa; Flavia Aburjaile; Guilherme Targino Valente; Heidge Fukumasu. USP-Botucatu: Rejane Maria Tommasini Grotto; Instituto Butantan: Alexander Roberto Precioso; Jayme A. Souza-Neto; Jessika Cristina Chagas Lesbon; José Salvatore Leister Patané; João Paulo Kitajima; Luiz Carlos Junior de Alcantara; Maria Carolina Elias; Marta Giovanetti; Patricia Akemi Assato; Rafael dos Santos Bezerra; Raquel de Lello Rocha Campos Cassano. NGS Soluções Genômicas: Pilar Drummond Sampaio Corrêa Mariani. FZEA-USP Pirassununga: Mirele Daiana Poleti; Raul Machado Neto; Ricardo Augusto Brassaloti; Ricardo Haddad; Rodrigo Tocantins Calado.; Sandra Coccuzzo Sampaio; Svetoslav Nanev Slavov; Vagner Fonseca; Vincent Louis Viala |
| EPI_ISL_1966256 | CENTRO INTEGRADO DE SAUDE | Instituto Butantan / Mendelics | Antonio Jorge Martins; Bianca Cechetto Carlos. Mendelics: Bibiana Santos; Claudia Renata dos Santos Barros; Cíntia Bittar; David Schlesinger. Hemocentro Ribeirão Preto: Simone Kashima; Debora Botequio Moretti; Elaine Cristina Marqueze; Elaine Vieira dos Santos; Elisangela Chicaroni Mattos; Erika Freitas; Evandra Strazza Rodrigues; Felipe Allan da Silva da Costa; Flavia Aburjaile; Fábio Sossai Possebon; Guilherme Campos; Guilherme Targino Valente; Heidge Fukumasu. USP-Botucatu: Rejane Maria Tommasini Grotto; Helena Lage Ferreira; Instituto Butantan: Dimas Tadeu Covas; Jardelina de Souza Todao Bernardino; Jayme A. Souza-Neto; Jessika Cristina Chagas Lesbon; Jorge A. Petroli Marchesi; José Salvatore Leister Patané; João Paulo Kitajima; João Pessoa Araújo Jr.; Leila Sabrina Ullmann; Loyze Paola Oliveira de Lima; Luiz Aurelio de Campos Crispin. Centro de Genômica Funcional da ESALQ: Luiz Lehmann Coutinho; Luiz Carlos Junior de Alcantara; Lívia Sacchetto; Maisa C. Pereira Parra; Maria Carolina Elias; Marta Giovanetti; Marília Moraes; Maurício Lacerda Nogueira. Prefeitura de Sao Paulo: Melissa Palmieri.; Patricia Akemi Assato; Paula Rahal; Paulo Inacio da Costa; Rafael dos Santos Bezerra; Raquel de Lello Rocha Campos Cassano. NGS Soluções Genômicas: Pilar Drummond Sampaio Corrêa Mariani. FZEA-USP Pirassununga: Mirele Daiana Poleti; Raul Machado Neto; Ricardo Augusto Brassaloti; Ricardo Haddad; Rodrigo Tocantins Calado. FAMERP-SJRP: Cecília Artico Banho; Sandra Coccuzzo Sampaio; Svetoslav Nanev Slavov; Vagner Fonseca; Vincent Louis Viala |
| EPI_ISL_1795080, EPI_ISL_1795274, EPI_ISL_1795275 | CENTRO MEDICO PMESP | Instituto Butantan / ESALQ-Piracicaba | Antonio Jorge Martins; Bianca Cechetto Carlos. Mendelics: Bibiana Santos; Claudia Renata dos Santos Barros; David Schlesinger. Hemocentro Ribeirão Preto: Simone Kashima; Debora Botequio Moretti. Centro de Genômica Funcional da ESALQ: Luiz Lehmann Coutinho; Dimas Tadeu Covas; Elaine Cristina Marqueze; Elaine Vieira dos Santos; Elisangela Chicaroni Mattos; Erika Freitas; Evandra Strazza Rodrigues; Felipe Allan da Silva da Costa; Flavia Aburjaile; Guilherme Targino Valente; Heidge Fukumasu. USP-Botucatu: Rejane Maria Tommasini Grotto; Instituto Butantan: Alexander Roberto Precioso; Jayme A. Souza-Neto; Jessika Cristina Chagas Lesbon; José Salvatore Leister Patané; João Paulo Kitajima; Luiz Carlos Junior de Alcantara; Maria Carolina Elias; Marta Giovanetti; Patricia Akemi Assato; Rafael dos Santos Bezerra; Raquel de Lello Rocha Campos Cassano. NGS Soluções Genômicas: Pilar Drummond Sampaio Corrêa Mariani. FZEA-USP Pirassununga: Mirele Daiana Poleti; Raul Machado Neto; Ricardo Augusto Brassaloti; Ricardo Haddad; Rodrigo Tocantins Calado.; Sandra Coccuzzo Sampaio; Svetoslav Nanev Slavov; Vagner Fonseca; Vincent Louis Viala |
| EPI_ISL_1716879 | CENTRO MEDICO PMESP | Instituto Butantan / ESALQ-USP | Antonio Jorge Martins; Bianca Cechetto Carlos. Mendelics: Bibiana Santos; Claudia Renata dos Santos Barros; David Schlesinger. Hemocentro Ribeirão Preto: Simone Kashima; Debora Botequio Moretti. Centro de Genômica Funcional da ESALQ: Luiz Lehmann Coutinho; Dimas Tadeu Covas; Elaine Cristina Marqueze; Elaine Vieira dos Santos; Elisangela Chicaroni Mattos; Erika Freitas; Evandra Strazza Rodrigues; Felipe Allan da Silva da Costa; Flavia Aburjaile; Guilherme Targino Valente; Heidge Fukumasu. USP-Botucatu: Rejane Maria Tommasini Grotto; Instituto Butantan: Alexander Roberto Precioso; Jayme A. Souza-Neto; Jessika Cristina Chagas Lesbon; José Salvatore Leister Patané; João Paulo Kitajima; Luiz Carlos Junior de Alcantara; Maria Carolina Elias; Marta Giovanetti; Patricia Akemi Assato; Rafael dos Santos Bezerra; Raquel de Lello Rocha Campos Cassano. NGS Soluções Genômicas: Pilar Drummond Sampaio Corrêa Mariani. FZEA-USP Pirassununga: Mirele Daiana Poleti; Raul Machado Neto; Ricardo Augusto Brassaloti; Ricardo Haddad; Rodrigo Tocantins Calado.; Sandra Coccuzzo Sampaio; Svetoslav Nanev Slavov; Vagner Fonseca; Vincent Louis Viala |
| EPI_ISL_1966407, EPI_ISL_1966411 | CENTRO MEDICO PMESP | Instituto Butantan / Mendelics | Antonio Jorge Martins; Bianca Cechetto Carlos. Mendelics: Bibiana Santos; Claudia Renata dos Santos Barros; Cíntia Bittar; David Schlesinger. Hemocentro Ribeirão Preto: Simone Kashima; Debora Botequio Moretti; Elaine Cristina Marqueze; Elaine Vieira dos Santos; Elisangela Chicaroni Mattos; Erika Freitas; Evandra Strazza Rodrigues; Felipe Allan da Silva da Costa; Flavia Aburjaile; Fábio Sossai Possebon; Guilherme Campos; Guilherme Targino Valente; Heidge Fukumasu. USP-Botucatu: Rejane Maria Tommasini Grotto; Helena Lage Ferreira; Instituto Butantan: Dimas Tadeu Covas; Jardelina de Souza Todao Bernardino; Jayme A. Souza-Neto; Jessika Cristina Chagas Lesbon; Jorge A. Petroli Marchesi; José Salvatore Leister Patané; João Paulo Kitajima; João Pessoa Araújo Jr.; Leila Sabrina Ullmann; Loyze Paola Oliveira de Lima; Luiz Aurelio de Campos Crispin. Centro de Genômica Funcional da ESALQ: Luiz Lehmann Coutinho; Luiz Carlos Junior de Alcantara; Lívia Sacchetto; Maisa C. Pereira Parra; Maria Carolina Elias; Marta Giovanetti; Marília Moraes; Maurício Lacerda Nogueira. Prefeitura de Sao Paulo: Melissa Palmieri.; Patricia Akemi Assato; Paula Rahal; Paulo Inacio da Costa; Rafael dos Santos Bezerra; Raquel de Lello Rocha Campos Cassano. NGS Soluções Genômicas: Pilar Drummond Sampaio Corrêa Mariani. FZEA-USP Pirassununga: Mirele Daiana Poleti; Raul Machado Neto; Ricardo Augusto Brassaloti; Ricardo Haddad; Rodrigo Tocantins Calado. FAMERP-SJRP: Cecília Artico Banho; Sandra Coccuzzo Sampaio; Svetoslav Nanev Slavov; Vagner Fonseca; Vincent Louis Viala |
| EPI_ISL_1966400 | CLINICA SALTO | Instituto Butantan / | Antonio Jorge Martins; Bianca Cechetto Carlos. Mendelics: Bibiana Santos; Claudia Renata dos Santos Barros; Cíntia Bittar; David Schlesinger. Hemocentro Ribeirão Preto: Simone Kashima; Debora Botequio Moretti; Elaine Cristina Marqueze; Elaine Vieira dos Santos; Elisangela Chicaroni Mattos; Erika Freitas; Evandra Strazza Rodrigues; |

| | | | |
|---|---|---|---|
| | SAUDE SANTA CRUZ | Mendelics | Felipe Allan da Silva da Costa; Flavia Aburjaile; Fábio Sossai Possebon; Guilherme Campos; Guilherme Targino Valente; Heidge Fukumasu. USP-Botucatu: Rejane Maria Tommasini Grotto; Helena Lage Ferreira; Instituto Butantan: Dimas Tadeu Covas; Jardelina de Souza Todao Bernardino; Jayme A. Souza-Neto; Jessika Cristina Chagas Lesbon; Jorge A. Petroli Marchesi; José Salvatore Leister Patané; João Paulo Kitajima; João Pessoa Araújo Jr.; Leila Sabrina Ullmann; Loyze Paola Oliveira de Lima; Luiz Aurelio de Campos Crispin. Centro de Genômica Funcional da ESALQ: Luiz Lehmann Coutinho; Luiz Carlos Junior de Alcantara; Lívia Sacchetto; Maisa C. Pereira Parra; Maria Carolina Elias; Marta Giovanetti; Marília Moraes; Maurício Lacerda Nogueira. Prefeitura de Sao Paulo: Melissa Palmieri.; Patricia Akemi Assato; Paula Rahal; Paulo Inacio da Costa; Rafael dos Santos Bezerra; Raquel de Lello Rocha Campos Cassano. NGS Soluções Genômicas: Pilar Drummond Sampaio Corrêa Mariani. FZEA-USP Pirassununga: Mirele Daiana Poleti; Raul Machado Neto; Ricardo Augusto Brassaloti; Ricardo Haddad; Rodrigo Tocantins Calado. FAMERP-SJRP: Cecília Artico Banho; Sandra Coccuzzo Sampaio; Svetoslav Nanev Slavov; Vagner Fonseca; Vincent Louis Viala |
| EPI_ISL_1445109, EPI_ISL_1445110, EPI_ISL_1445114, EPI_ISL_1445115, EPI_ISL_1445116, EPI_ISL_1445117, EPI_ISL_1445118, EPI_ISL_1966145, EPI_ISL_1966146, EPI_ISL_1966157, EPI_ISL_1966158, EPI_ISL_1966159, EPI_ISL_1966160, EPI_ISL_1966161 | | | |
| see above | COMPLEXO HOSPITALAR OURO VERDE DE CAMPINAS | Instituto Butantan / Mendelics | Antonio Jorge Martins; Bianca Cechetto Carlos. Mendelics: Bibiana Santos; Bibiana Santos; Claudia Renata dos Santos Barros; Cíntia Bittar; David Schlesinger; David Schlesinger. Hemocentro Ribeirão Preto: Simone Kashima; Debora Botequio Moretti; Dimas Tadeu Covas; Elaine Cristina Marqueze; Elaine Vieira dos Santos; Elisangela Chicaroni Mattos; Erika Freitas; Evandra Strazza Rodrigues; Felipe Allan da Silva da Costa; Flavia Aburjaile; Fábio Sossai Possebon; Guilherme Campos; Guilherme Targino Valente; Heidge Fukumasu. USP-Botucatu: Rejane Maria Tommasini Grotto; Helena Lage Ferreira; Instituto Butantan: Dimas Tadeu Covas; Jardelina de Souza Todao Bernardino; Jayme A. Souza-Neto; Jessika Cristina Chagas Lesbon; Jorge A. Petroli Marchesi; José Salvatore Leister Patané; João Paulo Kitajima; João Pessoa Araújo Jr.; Leila Sabrina Ullmann; Loyze Paola Oliveira de Lima; Luiz Aurelio de Campos Crispin. Centro de Genômica Funcional da ESALQ: Luiz Lehmann Coutinho; Luiz Carlos Junior de Alcantara; Lívia Sacchetto; Maisa C. Pereira Parra; Maria Carolina Elias; Marta Giovanetti; Marília Moraes; Maurício Lacerda Nogueira. Prefeitura de Sao Paulo: Melissa Palmieri.; Patricia Akemi Assato; Paula Rahal; Paulo Inacio da Costa; Rafael dos Santos Bezerra; Raquel de Lello Rocha Campos Cassano. NGS Soluções Genômicas: Pilar Drummond Sampaio Corrêa Mariani. FZEA-USP Pirassununga: Mirele Daiana Poleti; Raul Machado Neto; Ricardo Augusto Brassaloti; Ricardo Haddad; Rodrigo Tocantins Calado.; Rodrigo Tocantins Calado. FAMERP-SJRP: Cecília Artico Banho; Sandra Coccuzzo Sampaio; Simone Kashima; Svetoslav Nanev Slavov; Vagner Fonseca; Vincent Louis Viala |
| EPI_ISL_1795107 | COORDENADORIA MUNICIPAL DE SAUDE DE IRACEMAPOLIS | Instituto Butantan / ESALQ-Piracicaba | Antonio Jorge Martins; Bianca Cechetto Carlos. Mendelics: Bibiana Santos; Claudia Renata dos Santos Barros; David Schlesinger. Hemocentro Ribeirão Preto: Simone Kashima; Debora Botequio Moretti. Centro de Genômica Funcional da ESALQ: Luiz Lehmann Coutinho; Dimas Tadeu Covas; Elaine Cristina Marqueze; Elaine Vieira dos Santos; Elisangela Chicaroni Mattos; Erika Freitas; Evandra Strazza Rodrigues; Felipe Allan da Silva da Costa; Flavia Aburjaile; Guilherme Targino Valente; Heidge Fukumasu. USP-Botucatu: Rejane Maria Tommasini Grotto; Instituto Butantan: Alexander Roberto Precioso; Jayme A. Souza-Neto; Jessika Cristina Chagas Lesbon; José Salvatore Leister Patané; João Paulo Kitajima; Luiz Carlos Junior de Alcantara; Maria Carolina Elias; Marta Giovanetti; Patricia Akemi Assato; Rafael dos Santos Bezerra; Raquel de Lello Rocha Campos Cassano. NGS Soluções Genômicas: Pilar Drummond Sampaio Corrêa Mariani. FZEA-USP Pirassununga: Mirele Daiana Poleti; Raul Machado Neto; Ricardo Augusto Brassaloti; Ricardo Haddad; Rodrigo Tocantins Calado.; Sandra Coccuzzo Sampaio; Svetoslav Nanev Slavov; Vagner Fonseca; Vincent Louis Viala |
| EPI_ISL_1795165 | CS DE ALVARO CARVALHO | Instituto Butantan / ESALQ-Piracicaba | Antonio Jorge Martins; Bianca Cechetto Carlos. Mendelics: Bibiana Santos; Claudia Renata dos Santos Barros; David Schlesinger. Hemocentro Ribeirão Preto: Simone Kashima; Debora Botequio Moretti. Centro de Genômica Funcional da ESALQ: Luiz Lehmann Coutinho; Dimas Tadeu Covas; Elaine Cristina Marqueze; Elaine Vieira dos Santos; Elisangela Chicaroni Mattos; Erika Freitas; Evandra Strazza Rodrigues; Felipe Allan da Silva da Costa; Flavia Aburjaile; Guilherme Targino Valente; Heidge Fukumasu. USP-Botucatu: Rejane Maria Tommasini Grotto; Instituto Butantan: Alexander Roberto Precioso; Jayme A. Souza-Neto; Jessika Cristina Chagas Lesbon; José Salvatore Leister Patané; João Paulo Kitajima; Luiz Carlos Junior de Alcantara; Maria Carolina Elias; Marta Giovanetti; Patricia Akemi Assato; Rafael dos Santos Bezerra; Raquel de Lello Rocha Campos Cassano. NGS Soluções Genômicas: Pilar Drummond Sampaio Corrêa Mariani. FZEA-USP Pirassununga: Mirele Daiana Poleti; Raul Machado Neto; Ricardo Augusto Brassaloti; Ricardo Haddad; Rodrigo Tocantins Calado.; Sandra Coccuzzo Sampaio; Svetoslav Nanev Slavov; Vagner Fonseca; Vincent Louis Viala |
| EPI_ISL_1966739, EPI_ISL_1966740, EPI_ISL_1966741 | CS DE AMERICO DE CAMPOS | Instituto Butantan / Mendelics | Antonio Jorge Martins; Bianca Cechetto Carlos. Mendelics: Bibiana Santos; Claudia Renata dos Santos Barros; Cíntia Bittar; David Schlesinger. Hemocentro Ribeirão Preto: Simone Kashima; Debora Botequio Moretti; Elaine Cristina Marqueze; Elaine Vieira dos Santos; Elisangela Chicaroni Mattos; Erika Freitas; Evandra Strazza Rodrigues; Felipe Allan da Silva da Costa; Flavia Aburjaile; Fábio Sossai Possebon; Guilherme Campos; Guilherme Targino Valente; Heidge Fukumasu. USP-Botucatu: Rejane Maria Tommasini Grotto; Helena Lage Ferreira; Instituto Butantan: Dimas Tadeu Covas; Jardelina de Souza Todao Bernardino; Jayme A. Souza-Neto; Jessika Cristina Chagas Lesbon; Jorge A. Petroli Marchesi; José Salvatore Leister Patané; João Paulo Kitajima; João Pessoa Araújo Jr.; Leila Sabrina Ullmann; Loyze Paola Oliveira de Lima; Luiz Aurelio de Campos Crispin. Centro de Genômica Funcional da ESALQ: Luiz Lehmann Coutinho; Luiz Carlos Junior de Alcantara; Lívia Sacchetto; Maisa C. Pereira Parra; Maria Carolina Elias; Marta Giovanetti; Marília Moraes; Maurício Lacerda Nogueira. Prefeitura de Sao Paulo: Melissa Palmieri.; Patricia Akemi Assato; Paula Rahal; Paulo Inacio da Costa; Rafael dos Santos Bezerra; Raquel de Lello Rocha Campos Cassano. NGS Soluções Genômicas: Pilar Drummond Sampaio Corrêa Mariani. FZEA-USP Pirassununga: Mirele Daiana Poleti; Raul Machado Neto; Ricardo Augusto Brassaloti; Ricardo Haddad; Rodrigo Tocantins Calado. FAMERP-SJRP: Cecília Artico Banho; Sandra Coccuzzo Sampaio; Svetoslav Nanev Slavov; Vagner Fonseca; Vincent Louis Viala |
| EPI_ISL_1795234, EPI_ISL_1795235, EPI_ISL_1795236, EPI_ISL_1795237, EPI_ISL_1795238, EPI_ISL_1795241, EPI_ISL_1795389 | | | |
| see above | CS DE BALSAMO | Instituto Butantan / ESALQ-Piracicaba | Antonio Jorge Martins; Bianca Cechetto Carlos. Mendelics: Bibiana Santos; Claudia Renata dos Santos Barros; David Schlesinger. Hemocentro Ribeirão Preto: Simone Kashima; Debora Botequio Moretti. Centro de Genômica Funcional da ESALQ: Luiz Lehmann Coutinho; Dimas Tadeu Covas; Elaine Cristina Marqueze; Elaine Vieira dos Santos; Elisangela Chicaroni Mattos; Erika Freitas; Evandra Strazza Rodrigues; Felipe Allan da Silva da Costa; Flavia Aburjaile; Guilherme Targino Valente; Heidge Fukumasu. USP-Botucatu: Rejane Maria Tommasini Grotto; Instituto Butantan: Alexander Roberto Precioso; Jayme A. Souza-Neto; Jessika Cristina Chagas Lesbon; José Salvatore Leister Patané; João Paulo Kitajima; Luiz Carlos Junior de Alcantara; Maria Carolina Elias; Marta Giovanetti; Patricia Akemi Assato; Rafael dos Santos Bezerra; Raquel de Lello Rocha Campos Cassano. NGS Soluções Genômicas: Pilar Drummond Sampaio Corrêa Mariani. FZEA-USP Pirassununga: Mirele Daiana Poleti; Raul Machado Neto; Ricardo Augusto Brassaloti; Ricardo Haddad; Rodrigo Tocantins Calado.; Sandra Coccuzzo Sampaio; Svetoslav Nanev Slavov; Vagner Fonseca; Vincent Louis Viala |
| EPI_ISL_1966834 | CS DE BALSAMO | Instituto Butantan / ESALQ-USP (Piracicaba) | Antonio Jorge Martins; Bianca Cechetto Carlos. Mendelics: Bibiana Santos; Claudia Renata dos Santos Barros; Cíntia Bittar; David Schlesinger. Hemocentro Ribeirão Preto: Simone Kashima; Debora Botequio Moretti; Elaine Cristina Marqueze; Elaine Vieira dos Santos; Elisangela Chicaroni Mattos; Erika Freitas; Evandra Strazza Rodrigues; Felipe Allan da Silva da Costa; Flavia Aburjaile; Fábio Sossai Possebon; Guilherme Campos; Guilherme Targino Valente; Heidge Fukumasu. USP-Botucatu: Rejane Maria Tommasini Grotto; Helena Lage Ferreira; Instituto Butantan: Dimas Tadeu Covas; Jardelina de Souza Todao Bernardino; Jayme A. Souza-Neto; Jessika Cristina Chagas Lesbon; Jorge A. Petroli Marchesi; José Salvatore Leister Patané; João Paulo Kitajima; João Pessoa Araújo Jr.; Leila Sabrina Ullmann; Loyze Paola Oliveira de Lima; Luiz Aurelio de Campos Crispin. Centro de Genômica Funcional da ESALQ: Luiz Lehmann Coutinho; Luiz Carlos Junior de Alcantara; Lívia Sacchetto; Maisa C. Pereira Parra; Maria Carolina Elias; Marta Giovanetti; Marília Moraes; Maurício Lacerda Nogueira. Prefeitura de Sao Paulo: Melissa Palmieri.; Patricia Akemi Assato; Paula Rahal; Paulo Inacio da Costa; Rafael dos Santos Bezerra; Raquel de Lello Rocha Campos Cassano. NGS Soluções Genômicas: Pilar Drummond Sampaio Corrêa Mariani. FZEA-USP Pirassununga: Mirele Daiana Poleti; Raul Machado Neto; Ricardo Augusto Brassaloti; Ricardo Haddad; Rodrigo Tocantins Calado. FAMERP-SJRP: Cecília Artico Banho; Sandra Coccuzzo Sampaio; Svetoslav Nanev Slavov; Vagner Fonseca; Vincent Louis Viala |
| EPI_ISL_1966523, EPI_ISL_1966524 | CS DE BALSAMO | Instituto Butantan / FZEA-USP (Pirassunungа) | Antonio Jorge Martins; Bianca Cechetto Carlos. Mendelics: Bibiana Santos; Claudia Renata dos Santos Barros; Cíntia Bittar; David Schlesinger. Hemocentro Ribeirão Preto: Simone Kashima; Debora Botequio Moretti; Elaine Cristina Marqueze; Elaine Vieira dos Santos; Elisangela Chicaroni Mattos; Erika Freitas; Evandra Strazza Rodrigues; Felipe Allan da Silva da Costa; Flavia Aburjaile; Fábio Sossai Possebon; Guilherme Campos; Guilherme Targino Valente; Heidge Fukumasu. USP-Botucatu: Rejane Maria Tommasini Grotto; Helena Lage Ferreira; Instituto Butantan: Dimas Tadeu Covas; Jardelina de Souza Todao Bernardino; Jayme A. Souza-Neto; Jessika Cristina Chagas Lesbon; Jorge A. Petroli Marchesi; José Salvatore Leister Patané; João Paulo Kitajima; João Pessoa Araújo Jr.; Leila Sabrina Ullmann; Loyze Paola Oliveira de Lima; Luiz Aurelio de Campos Crispin. Centro de Genômica Funcional da ESALQ: Luiz Lehmann Coutinho; Luiz Carlos Junior de Alcantara; Lívia Sacchetto; Maisa C. Pereira Parra; Maria Carolina Elias; Marta Giovanetti; Marília Moraes; Maurício Lacerda Nogueira. Prefeitura de Sao Paulo: Melissa Palmieri.; Patricia Akemi Assato; Paula Rahal; Paulo Inacio da Costa; Rafael dos Santos Bezerra; Raquel de Lello Rocha Campos Cassano. NGS Soluções Genômicas: Pilar Drummond Sampaio Corrêa Mariani. FZEA-USP Pirassununga: Mirele Daiana Poleti; Raul Machado Neto; Ricardo Augusto Brassaloti; Ricardo Haddad; Rodrigo Tocantins Calado. FAMERP-SJRP: Cecília Artico Banho; Sandra Coccuzzo Sampaio; Svetoslav Nanev Slavov; Vagner Fonseca; Vincent Louis Viala |
| EPI_ISL_1966868, EPI_ISL_1966869 | CS DE CATIGUA | Instituto Butantan / ESALQ-USP (Piracicaba) | Antonio Jorge Martins; Bianca Cechetto Carlos. Mendelics: Bibiana Santos; Claudia Renata dos Santos Barros; Cíntia Bittar; David Schlesinger. Hemocentro Ribeirão Preto: Simone Kashima; Debora Botequio Moretti; Elaine Cristina Marqueze; Elaine Vieira dos Santos; Elisangela Chicaroni Mattos; Erika Freitas; Evandra Strazza Rodrigues; Felipe Allan da Silva da Costa; Flavia Aburjaile; Fábio Sossai Possebon; Guilherme Campos; Guilherme Targino Valente; Heidge Fukumasu. USP-Botucatu: Rejane Maria Tommasini Grotto; Helena Lage Ferreira; Instituto Butantan: Dimas Tadeu Covas; Jardelina de Souza Todao Bernardino; Jayme A. Souza-Neto; Jessika Cristina Chagas Lesbon; Jorge A. Petroli Marchesi; José Salvatore Leister Patané; João Paulo Kitajima; João Pessoa Araújo Jr.; Leila Sabrina Ullmann; Loyze Paola Oliveira de Lima; Luiz Aurelio de Campos Crispin. Centro de Genômica Funcional da ESALQ: Luiz Lehmann Coutinho; Luiz Carlos Junior de Alcantara; Lívia Sacchetto; Maisa C. Pereira Parra; Maria Carolina Elias; Marta Giovanetti; Marília Moraes; Maurício Lacerda Nogueira. Prefeitura de Sao Paulo: Melissa Palmieri.; Patricia Akemi Assato; Paula Rahal; Paulo Inacio da Costa; Rafael dos Santos Bezerra; Raquel de Lello Rocha Campos Cassano. NGS Soluções Genômicas: Pilar Drummond Sampaio Corrêa Mariani. FZEA-USP Pirassununga: Mirele Daiana Poleti; Raul Machado Neto; Ricardo Augusto Brassaloti; Ricardo Haddad; Rodrigo Tocantins Calado. FAMERP-SJRP: Cecília Artico Banho; Sandra Coccuzzo Sampaio; Svetoslav Nanev Slavov; Vagner Fonseca; Vincent Louis Viala |
| EPI_ISL_1795259, EPI_ISL_1795260, EPI_ISL_1795261, EPI_ISL_1795262, EPI_ISL_1795263, EPI_ISL_1795265 | CS DE ICEM | Instituto Butantan / ESALQ-Piracicaba | Antonio Jorge Martins; Bianca Cechetto Carlos. Mendelics: Bibiana Santos; Claudia Renata dos Santos Barros; David Schlesinger. Hemocentro Ribeirão Preto: Simone Kashima; Debora Botequio Moretti. Centro de Genômica Funcional da ESALQ: Luiz Lehmann Coutinho; Dimas Tadeu Covas; Elaine Cristina Marqueze; Elaine Vieira dos Santos; Elisangela Chicaroni Mattos; Erika Freitas; Evandra Strazza Rodrigues; Felipe Allan da Silva da Costa; Flavia Aburjaile; Guilherme Targino Valente; Heidge Fukumasu. USP-Botucatu: Rejane Maria Tommasini Grotto; Instituto Butantan: Alexander Roberto Precioso; Jayme A. Souza-Neto; Jessika Cristina Chagas Lesbon; José Salvatore Leister Patané; João Paulo Kitajima; Luiz Carlos Junior de Alcantara; Maria Carolina Elias; Marta Giovanetti; Patricia Akemi Assato; Rafael dos Santos Bezerra; Raquel de Lello Rocha Campos Cassano. NGS Soluções Genômicas: Pilar Drummond Sampaio Corrêa Mariani. FZEA-USP Pirassununga: Mirele Daiana Poleti; Raul Machado Neto; Ricardo Augusto Brassaloti; Ricardo Haddad; Rodrigo Tocantins Calado.; Sandra Coccuzzo Sampaio; Svetoslav Nanev Slavov; Vagner Fonseca; Vincent Louis Viala |
| EPI_ISL_1966764, EPI_ISL_1966765 | CS DE ICEM | Instituto Butantan / Mendelics | Antonio Jorge Martins; Bianca Cechetto Carlos. Mendelics: Bibiana Santos; Claudia Renata dos Santos Barros; Cíntia Bittar; David Schlesinger. Hemocentro Ribeirão Preto: Simone Kashima; Debora Botequio Moretti; Elaine Cristina Marqueze; Elaine Vieira dos Santos; Elisangela Chicaroni Mattos; Erika Freitas; Evandra Strazza Rodrigues; Felipe Allan da Silva da Costa; Flavia Aburjaile; Fábio Sossai Possebon; Guilherme Campos; Guilherme Targino Valente; Heidge Fukumasu. USP-Botucatu: Rejane Maria Tommasini Grotto; Helena Lage Ferreira; Instituto Butantan: Dimas Tadeu Covas; Jardelina de Souza Todao Bernardino; Jayme A. Souza-Neto; Jessika Cristina Chagas Lesbon; Jorge A. Petroli Marchesi; José Salvatore Leister Patané; João Paulo Kitajima; João Pessoa Araújo Jr.; Leila Sabrina Ullmann; Loyze Paola Oliveira de Lima; Luiz Aurelio de Campos Crispin. Centro de Genômica Funcional da ESALQ: Luiz Lehmann Coutinho; Luiz Carlos Junior de Alcantara; Lívia Sacchetto; Maisa C. Pereira Parra; Maria Carolina Elias; Marta Giovanetti; Marília Moraes; Maurício Lacerda Nogueira. Prefeitura de Sao Paulo: Melissa Palmieri.; Patricia Akemi Assato; Paula Rahal; Paulo Inacio da Costa; Rafael dos Santos Bezerra; Raquel de Lello Rocha Campos Cassano. NGS Soluções Genômicas: Pilar Drummond Sampaio Corrêa Mariani. FZEA-USP Pirassununga: Mirele Daiana Poleti; Raul Machado Neto; Ricardo Augusto Brassaloti; Ricardo Haddad; Rodrigo Tocantins Calado. FAMERP-SJRP: Cecília Artico Banho; Sandra Coccuzzo Sampaio; Svetoslav Nanev Slavov; Vagner Fonseca; Vincent Louis Viala |
| EPI_ISL_1795416 | CS DE MARAPOAMA | Instituto Butantan / ESALQ-Piracicaba | Antonio Jorge Martins; Bianca Cechetto Carlos. Mendelics: Bibiana Santos; Claudia Renata dos Santos Barros; David Schlesinger. Hemocentro Ribeirão Preto: Simone Kashima; Debora Botequio Moretti. Centro de Genômica Funcional da ESALQ: Luiz Lehmann Coutinho; Dimas Tadeu Covas; Elaine Cristina Marqueze; Elaine Vieira dos Santos; Elisangela Chicaroni Mattos; Erika Freitas; Evandra Strazza Rodrigues; Felipe Allan da Silva da Costa; Flavia Aburjaile; Guilherme Targino Valente; Heidge Fukumasu. USP-Botucatu: Rejane Maria Tommasini Grotto; Instituto Butantan: Alexander Roberto Precioso; Jayme A. Souza-Neto; Jessika Cristina Chagas Lesbon; José Salvatore Leister Patané; João Paulo Kitajima; Luiz Carlos Junior de Alcantara; Maria Carolina Elias; Marta Giovanetti; Patricia Akemi Assato; Rafael dos Santos Bezerra; Raquel de Lello Rocha Campos Cassano. NGS Soluções Genômicas: Pilar Drummond Sampaio Corrêa Mariani. FZEA-USP Pirassununga: Mirele Daiana Poleti; Raul Machado Neto; Ricardo Augusto Brassaloti; Ricardo Haddad; Rodrigo Tocantins Calado.; Sandra Coccuzzo Sampaio; Svetoslav Nanev Slavov; Vagner Fonseca; Vincent Louis Viala |
| EPI_ISL_1966784, EPI_ISL_1966785, EPI_ISL_2170966 | CS DE MIRASSOLANDIA | Instituto Butantan / Mendelics | Antonio Jorge Martins; Bianca Cechetto Carlos. Mendelics: Bibiana Santos; Claudia Renata dos Santos Barros; Cíntia Bittar; David Schlesinger. Hemocentro Ribeirão Preto: Simone Kashima; Debora Botequio Moretti; Elaine Cristina Marqueze; Elaine Vieira dos Santos; Elisangela Chicaroni Mattos; Erika Freitas; Evandra Strazza Rodrigues; Felipe Allan da Silva da Costa; Flavia Aburjaile; Fábio Sossai Possebon; Guilherme Campos; Guilherme Targino Valente; Heidge Fukumasu. USP-Botucatu: Rejane Maria Tommasini Grotto; Helena Lage Ferreira; Instituto Butantan: Dimas Tadeu Covas; Jardelina de Souza Todao Bernardino; Jayme A. Souza-Neto; Jessika Cristina Chagas Lesbon; Jorge A. Petroli Marchesi; José Salvatore Leister Patané; João Paulo Kitajima; João Pessoa Araújo Jr.; Leila Sabrina Ullmann; Loyze Paola Oliveira de Lima; Luiz Aurelio de Campos Crispin. Centro de Genômica Funcional da ESALQ: Luiz Lehmann Coutinho; Luiz Carlos Junior de Alcantara; Lívia Sacchetto; Maisa C. Pereira Parra; Maria Carolina Elias; Marta Giovanetti; Marília Moraes; Maurício Lacerda Nogueira. Prefeitura de Sao Paulo: Melissa Palmieri.; Patricia Akemi Assato; Paula Rahal; Paulo Inacio da Costa; Rafael dos Santos Bezerra; Raquel de Lello Rocha Campos Cassano. NGS Soluções Genômicas: Pilar Drummond Sampaio Corrêa Mariani. FZEA-USP Pirassununga: Mirele Daiana Poleti; Raul Machado Neto; Ricardo Augusto Brassaloti; Ricardo Haddad; Rodrigo Tocantins Calado. FAMERP-SJRP: Cecília Artico Banho; Sandra Coccuzzo Sampaio; Svetoslav Nanev Slavov; Vagner Fonseca; Vincent Louis Viala |
| EPI_ISL_2170895, EPI_ISL_2170896 | CS DE MONTE APRAZIVEL | Instituto Butantan / ESALQ-Piracicaba | Antonio Jorge Martins; Bianca Cechetto Carlos. Mendelics: Bibiana Santos; Claudia Renata dos Santos Barros; Cíntia Bittar; David Schlesinger. Hemocentro Ribeirão Preto: Simone Kashima; Debora Botequio Moretti; Elaine Cristina Marqueze; Elaine Vieira dos Santos; Elisangela Chicaroni Mattos; Erika Freitas; Evandra Strazza Rodrigues; Felipe Allan da Silva da Costa; Flavia Aburjaile; Fábio Sossai Possebon; Guilherme Campos; Guilherme Targino Valente; Heidge Fukumasu. USP-Botucatu: Rejane Maria Tommasini Grotto; Helena Lage Ferreira; Instituto Butantan: Dimas Tadeu Covas; Jardelina de Souza Todao Bernardino; Jayme A. Souza-Neto; Jessika Cristina Chagas Lesbon; Jorge A. Petroli Marchesi; José Salvatore Leister Patané; João Paulo Kitajima; João Pessoa Araújo Jr.; Leila Sabrina Ullmann; Loyze Paola Oliveira de Lima; Luiz Aurelio de Campos Crispin. Centro de Genômica Funcional da ESALQ: Luiz Lehmann Coutinho; Luiz Carlos Junior de Alcantara; Lívia Sacchetto; Maisa C. Pereira Parra; Maria Carolina Elias; Marta Giovanetti; Marília Moraes; Maurício Lacerda Nogueira. Prefeitura de Sao Paulo: Melissa Palmieri.; Patricia Akemi Assato; Paula Rahal; Paulo Inacio da Costa; Rafael dos Santos Bezerra; Raquel de Lello Rocha Campos Cassano. NGS Soluções Genômicas: Pilar Drummond Sampaio Corrêa Mariani. FZEA-USP Pirassununga: Mirele Daiana Poleti; Raul Machado Neto; Ricardo Augusto Brassaloti; Ricardo Haddad; Rodrigo Tocantins Calado. FAMERP-SJRP: Cecília Artico Banho; Sandra Coccuzzo Sampaio; Svetoslav Nanev Slavov; Vagner Fonseca; Vincent Louis Viala |
| EPI_ISL_1966961, EPI_ISL_1966962, EPI_ISL_1966963, EPI_ISL_1966964 | CS DE MONTE APRAZIVEL | Instituto Butantan / ESALQ-USP (Piracicaba) | Antonio Jorge Martins; Bianca Cechetto Carlos. Mendelics: Bibiana Santos; Claudia Renata dos Santos Barros; Cíntia Bittar; David Schlesinger. Hemocentro Ribeirão Preto: Simone Kashima; Debora Botequio Moretti; Elaine Cristina Marqueze; Elaine Vieira dos Santos; Elisangela Chicaroni Mattos; Erika Freitas; Evandra Strazza Rodrigues; Felipe Allan da Silva da Costa; Flavia Aburjaile; Fábio Sossai Possebon; Guilherme Campos; Guilherme Targino Valente; Heidge Fukumasu. USP-Botucatu: Rejane Maria Tommasini Grotto; Helena Lage Ferreira; Instituto Butantan: Dimas Tadeu Covas; Jardelina de Souza Todao Bernardino; Jayme A. Souza-Neto; Jessika Cristina Chagas Lesbon; Jorge A. Petroli Marchesi; José Salvatore Leister Patané; João Paulo Kitajima; João Pessoa Araújo Jr.; Leila Sabrina Ullmann; Loyze Paola Oliveira de Lima; Luiz Aurelio de Campos Crispin. Centro de Genômica Funcional da ESALQ: Luiz Lehmann Coutinho; Luiz Carlos Junior de Alcantara; Lívia Sacchetto; Maisa C. Pereira Parra; Maria Carolina Elias; Marta Giovanetti; Marília Moraes; Maurício Lacerda Nogueira. Prefeitura de Sao Paulo: Melissa Palmieri.; Patricia Akemi Assato; Paula Rahal; Paulo Inacio da Costa; Rafael dos Santos Bezerra; Raquel de Lello Rocha Campos Cassano. NGS Soluções Genômicas: Pilar Drummond Sampaio Corrêa Mariani. FZEA-USP Pirassununga: Mirele Daiana Poleti; Raul Machado Neto; Ricardo Augusto Brassaloti; Ricardo Haddad; Rodrigo Tocantins Calado. FAMERP-SJRP: Cecília Artico Banho; Sandra Coccuzzo Sampaio; Svetoslav Nanev Slavov; Vagner Fonseca; Vincent Louis Viala |
| EPI_ISL_1966984, EPI_ISL_1966985 | CS DE NEVES PAULISTA | Instituto Butantan / ESALQ-USP (Piracicaba) | Antonio Jorge Martins; Bianca Cechetto Carlos. Mendelics: Bibiana Santos; Claudia Renata dos Santos Barros; Cíntia Bittar; David Schlesinger. Hemocentro Ribeirão Preto: Simone Kashima; Debora Botequio Moretti; Elaine Cristina Marqueze; Elaine Vieira dos Santos; Elisangela Chicaroni Mattos; Erika Freitas; Evandra Strazza Rodrigues; Felipe Allan da Silva da Costa; Flavia Aburjaile; Fábio Sossai Possebon; Guilherme Campos; Guilherme Targino Valente; Heidge Fukumasu. USP-Botucatu: Rejane Maria Tommasini Grotto; Helena Lage Ferreira; Instituto Butantan: Dimas Tadeu Covas; Jardelina de Souza Todao Bernardino; Jayme A. Souza-Neto; Jessika Cristina Chagas Lesbon; Jorge A. Petroli Marchesi; José Salvatore Leister Patané; João Paulo Kitajima; João Pessoa Araújo Jr.; Leila Sabrina Ullmann; Loyze Paola Oliveira de Lima; Luiz Aurelio de Campos Crispin. Centro de Genômica Funcional da ESALQ: Luiz Lehmann Coutinho; Luiz Carlos Junior de Alcantara; Lívia Sacchetto; Maisa C. Pereira Parra; Maria Carolina Elias; Marta Giovanetti; Marília Moraes; Maurício Lacerda Nogueira. Prefeitura de Sao Paulo: Melissa Palmieri.; Patricia Akemi Assato; Paula Rahal; Paulo Inacio da Costa; Rafael dos Santos Bezerra; Raquel de Lello Rocha Campos Cassano. NGS Soluções Genômicas: Pilar Drummond Sampaio Corrêa Mariani. FZEA-USP Pirassununga: Mirele Daiana Poleti; Raul Machado Neto; Ricardo Augusto Brassaloti; Ricardo Haddad; Rodrigo Tocantins Calado. FAMERP-SJRP: Cecília Artico Banho; Sandra Coccuzzo Sampaio; Svetoslav Nanev Slavov; Vagner Fonseca; Vincent Louis Viala |
| EPI_ISL_1795288 | CS DE NIPOA | Instituto Butantan / ESALQ-Piracicaba | Antonio Jorge Martins; Bianca Cechetto Carlos. Mendelics: Bibiana Santos; Claudia Renata dos Santos Barros; David Schlesinger. Hemocentro Ribeirão Preto: Simone Kashima; Debora Botequio Moretti. Centro de Genômica Funcional da ESALQ: Luiz Lehmann Coutinho; Dimas Tadeu Covas; Elaine Cristina Marqueze; Elaine Vieira dos Santos; Elisangela Chicaroni Mattos; Erika Freitas; Evandra Strazza Rodrigues; Felipe Allan da Silva da Costa; Flavia Aburjaile; Guilherme Targino Valente; Heidge Fukumasu. USP-Botucatu: Rejane Maria Tommasini Grotto; Instituto Butantan: Alexander Roberto Precioso; Jayme A. Souza-Neto; Jessika Cristina Chagas Lesbon; José Salvatore Leister Patané; João Paulo Kitajima; Luiz Carlos Junior de Alcantara; Maria Carolina Elias; Marta Giovanetti; Patricia Akemi Assato; Rafael dos Santos Bezerra; Raquel de Lello Rocha Campos Cassano. NGS Soluções Genômicas: Pilar Drummond Sampaio Corrêa Mariani. FZEA-USP Pirassununga: Mirele Daiana Poleti; Raul Machado Neto; Ricardo Augusto Brassaloti; Ricardo Haddad; Rodrigo Tocantins Calado.; Sandra Coccuzzo Sampaio; Svetoslav Nanev Slavov; Vagner Fonseca; Vincent Louis Viala |
| EPI_ISL_1966787, EPI_ISL_1966788, EPI_ISL_1966789, EPI_ISL_1966790, EPI_ISL_1966791, EPI_ISL_2170967 | CS DE NIPOA | Instituto Butantan / Mendelics | Antonio Jorge Martins; Bianca Cechetto Carlos. Mendelics: Bibiana Santos; Claudia Renata dos Santos Barros; Cíntia Bittar; David Schlesinger. Hemocentro Ribeirão Preto: Simone Kashima; Debora Botequio Moretti; Elaine Cristina Marqueze; Elaine Vieira dos Santos; Elisangela Chicaroni Mattos; Erika Freitas; Evandra Strazza Rodrigues; Felipe Allan da Silva da Costa; Flavia Aburjaile; Fábio Sossai Possebon; Guilherme Campos; Guilherme Targino Valente; Heidge Fukumasu. USP-Botucatu: Rejane Maria Tommasini Grotto; Helena Lage Ferreira; Instituto Butantan: Dimas Tadeu Covas; Jardelina de Souza Todao Bernardino; Jayme A. Souza-Neto; Jessika Cristina Chagas Lesbon; Jorge A. Petroli Marchesi; José Salvatore Leister Patané; João Paulo Kitajima; João Pessoa Araújo Jr.; Leila Sabrina Ullmann; Loyze Paola Oliveira de Lima; Luiz Aurelio de Campos Crispin. Centro de Genômica Funcional da ESALQ: Luiz Lehmann Coutinho; Luiz Carlos Junior de Alcantara; Lívia Sacchetto; Maisa C. Pereira Parra; Maria Carolina Elias; Marta Giovanetti; Marília Moraes; Maurício Lacerda Nogueira. Prefeitura de Sao Paulo: Melissa Palmieri.; Patricia Akemi Assato; Paula Rahal; Paulo Inacio da Costa; Rafael dos Santos Bezerra; Raquel de Lello Rocha Campos Cassano. NGS Soluções Genômicas: Pilar Drummond Sampaio Corrêa Mariani. FZEA-USP Pirassununga: Mirele Daiana Poleti; Raul Machado Neto; Ricardo Augusto Brassaloti; Ricardo Haddad; Rodrigo Tocantins Calado. FAMERP-SJRP: Cecília Artico Banho; Sandra Coccuzzo Sampaio; Svetoslav Nanev Slavov; Vagner Fonseca; Vincent Louis Viala |
| EPI_ISL_1966988 | CS DE ORINDIUVA | Instituto Butantan / ESALQ-USP (Piracicaba) | Antonio Jorge Martins; Bianca Cechetto Carlos. Mendelics: Bibiana Santos; Claudia Renata dos Santos Barros; Cíntia Bittar; David Schlesinger. Hemocentro Ribeirão Preto: Simone Kashima; Debora Botequio Moretti; Elaine Cristina Marqueze; Elaine Vieira dos Santos; Elisangela Chicaroni Mattos; Erika Freitas; Evandra Strazza Rodrigues; Felipe Allan da Silva da Costa; Flavia Aburjaile; Fábio Sossai Possebon; Guilherme Campos; Guilherme Targino Valente; Heidge Fukumasu. USP-Botucatu: Rejane Maria Tommasini Grotto; Helena Lage Ferreira; Instituto Butantan: Dimas Tadeu Covas; Jardelina de Souza Todao Bernardino; Jayme A. Souza-Neto; Jessika Cristina Chagas Lesbon; Jorge A. Petroli Marchesi; José Salvatore Leister Patané; João Paulo Kitajima; João Pessoa Araújo Jr.; Leila Sabrina Ullmann; Loyze Paola Oliveira de Lima; Luiz Aurelio de Campos Crispin. Centro de Genômica Funcional da ESALQ: Luiz Lehmann Coutinho; Luiz Carlos Junior de Alcantara; Lívia Sacchetto; Maisa C. Pereira Parra; Maria Carolina Elias; Marta Giovanetti; Marília Moraes; Maurício Lacerda Nogueira. Prefeitura de Sao Paulo: Melissa Palmieri.; Patricia Akemi Assato; Paula Rahal; Paulo Inacio da Costa; Rafael dos Santos Bezerra; Raquel de Lello Rocha Campos Cassano. NGS Soluções Genômicas: Pilar Drummond Sampaio Corrêa Mariani. FZEA-USP Pirassununga: Mirele Daiana Poleti; Raul Machado Neto; Ricardo Augusto Brassaloti; Ricardo Haddad; Rodrigo Tocantins Calado. FAMERP-SJRP: Cecília Artico Banho; Sandra Coccuzzo Sampaio; Svetoslav Nanev Slavov; Vagner Fonseca; Vincent Louis Viala |
| EPI_ISL_1795171, EPI_ISL_1795173, EPI_ISL_1795390, EPI_ISL_1795406, EPI_ISL_1795408 | CS DE OSCAR BRESSANE PSF | Instituto Butantan / ESALQ-Piracicaba | Antonio Jorge Martins; Bianca Cechetto Carlos. Mendelics: Bibiana Santos; Claudia Renata dos Santos Barros; David Schlesinger. Hemocentro Ribeirão Preto: Simone Kashima; Debora Botequio Moretti. Centro de Genômica Funcional da ESALQ: Luiz Lehmann Coutinho; Dimas Tadeu Covas; Elaine Cristina Marqueze; Elaine Vieira dos Santos; Elisangela Chicaroni Mattos; Erika Freitas; Evandra Strazza Rodrigues; Felipe Allan da Silva da Costa; Flavia Aburjaile; Guilherme Targino Valente; Heidge Fukumasu. USP-Botucatu: Rejane Maria Tommasini Grotto; Instituto Butantan: Alexander Roberto Precioso; Jayme A. Souza-Neto; Jessika Cristina Chagas Lesbon; José Salvatore Leister Patané; João Paulo Kitajima; Luiz Carlos Junior de Alcantara; Maria Carolina Elias; Marta Giovanetti; Patricia Akemi Assato; Rafael dos Santos Bezerra; Raquel de Lello Rocha Campos Cassano. NGS Soluções Genômicas: Pilar Drummond Sampaio Corrêa Mariani. FZEA-USP Pirassununga: Mirele Daiana Poleti; Raul Machado Neto; Ricardo Augusto Brassaloti; Ricardo Haddad; Rodrigo Tocantins Calado.; Sandra Coccuzzo Sampaio; Svetoslav Nanev Slavov; Vagner Fonseca; Vincent Louis Viala |
| EPI_ISL_1795276, EPI_ISL_1795277, EPI_ISL_1795278, EPI_ISL_1795279, EPI_ISL_1795280, EPI_ISL_1795281, EPI_ISL_1795282, EPI_ISL_1795283, EPI_ISL_1795284, EPI_ISL_1795285, EPI_ISL_1795286 | | | |

| | | | |
|---|---|---|---|
| see above | CS DE PALESTINA | Instituto Butantan / ESALQ-Piracicaba | Antonio Jorge Martins; Bianca Cechetto Carlos. Mendelics: Bibiana Santos; Claudia Renata dos Santos Barros; David Schlesinger. Hemocentro Ribeirão Preto: Simone Kashima; Debora Botequio Moretti. Centro de Genômica Funcional da ESALQ: Luiz Lehmann Coutinho; Dimas Tadeu Covas; Elaine Cristina Marqueze; Elaine Vieira dos Santos; Elisangela Chicaroni Mattos; Erika Freitas; Evandra Strazza Rodrigues; Felipe Allan da Silva da Costa; Flavia Aburjaile; Guilherme Targino Valente; Heidge Fukumasu. USP-Botucatu: Rejane Maria Tommasini Grotto; Instituto Butantan: Alexander Roberto Precioso; Jayme A. Souza-Neto; Jessika Cristina Chagas Lesbon; José Salvatore Leister Patané; João Paulo Kitajima; Luiz Carlos Junior de Alcantara; Maria Carolina Elias; Marta Giovanetti; Patricia Akemi Assato; Rafael dos Santos Bezerra; Raquel de Lello Rocha Campos Cassano. NGS Soluções Genômicas: Pilar Drummond Sampaio Corrêa Mariani. FZEA-USP Pirassununga: Mirele Daiana Poleti; Raul Machado Neto; Ricardo Augusto Brassaloti; Ricardo Haddad; Rodrigo Tocantins Calado.; Sandra Coccuzzo Sampaio; Svetoslav Nanev Slavov; Vagner Fonseca; Vincent Louis Viala |
| EPI_ISL_1966795 | CS DE PALESTINA | Instituto Butantan / Mendelics | Antonio Jorge Martins; Bianca Cechetto Carlos. Mendelics: Bibiana Santos; Claudia Renata dos Santos Barros; Cíntia Bittar; David Schlesinger. Hemocentro Ribeirão Preto: Simone Kashima; Debora Botequio Moretti; Elaine Cristina Marqueze; Elaine Vieira dos Santos; Elisangela Chicaroni Mattos; Erika Freitas; Evandra Strazza Rodrigues; Felipe Allan da Silva da Costa; Flavia Aburjaile; Fábio Sossai Possebon; Guilherme Campos; Guilherme Targino Valente; Heidge Fukumasu. USP-Botucatu: Rejane Maria Tommasini Grotto; Helena Lage Ferreira; Instituto Butantan: Dimas Tadeu Covas; Jardelina de Souza Todao Bernardino; Jayme A. Souza-Neto; Jessika Cristina Chagas Lesbon; Jorge A. Petroli Marchesi; José Salvatore Leister Patané; João Paulo Kitajima; João Pessoa Araújo Jr.; Leila Sabrina Ullmann; Loyze Paola Oliveira de Lima; Luiz Aurelio de Campos Crispin. Centro de Genômica Funcional da ESALQ: Luiz Lehmann Coutinho; Luiz Carlos Junior de Alcantara; Lívia Sacchetto; Maisa C. Pereira Parra; Maria Carolina Elias; Marta Giovanetti; Marília Moraes; Maurício Lacerda Nogueira. Prefeitura de Sao Paulo: Melissa Palmieri.; Patricia Akemi Assato; Paula Rahal; Paulo Inacio da Costa; Rafael dos Santos Bezerra; Raquel de Lello Rocha Campos Cassano. NGS Soluções Genômicas: Pilar Drummond Sampaio Corrêa Mariani. FZEA-USP Pirassununga: Mirele Daiana Poleti; Raul Machado Neto; Ricardo Augusto Brassaloti; Ricardo Haddad; Rodrigo Tocantins Calado. FAMERP-SJRP: Cecília Artico Banho; Sandra Coccuzzo Sampaio; Svetoslav Nanev Slavov; Vagner Fonseca; Vincent Louis Viala |
| EPI_ISL_1795079, EPI_ISL_1795264, EPI_ISL_1795271 | CS DE PARANAPUA | Instituto Butantan / ESALQ-Piracicaba | Antonio Jorge Martins; Bianca Cechetto Carlos. Mendelics: Bibiana Santos; Claudia Renata dos Santos Barros; David Schlesinger. Hemocentro Ribeirão Preto: Simone Kashima; Debora Botequio Moretti. Centro de Genômica Funcional da ESALQ: Luiz Lehmann Coutinho; Dimas Tadeu Covas; Elaine Cristina Marqueze; Elaine Vieira dos Santos; Elisangela Chicaroni Mattos; Erika Freitas; Evandra Strazza Rodrigues; Felipe Allan da Silva da Costa; Flavia Aburjaile; Guilherme Targino Valente; Heidge Fukumasu. USP-Botucatu: Rejane Maria Tommasini Grotto; Instituto Butantan: Alexander Roberto Precioso; Jayme A. Souza-Neto; Jessika Cristina Chagas Lesbon; José Salvatore Leister Patané; João Paulo Kitajima; Luiz Carlos Junior de Alcantara; Maria Carolina Elias; Marta Giovanetti; Patricia Akemi Assato; Rafael dos Santos Bezerra; Raquel de Lello Rocha Campos Cassano. NGS Soluções Genômicas: Pilar Drummond Sampaio Corrêa Mariani. FZEA-USP Pirassununga: Mirele Daiana Poleti; Raul Machado Neto; Ricardo Augusto Brassaloti; Ricardo Haddad; Rodrigo Tocantins Calado.; Sandra Coccuzzo Sampaio; Svetoslav Nanev Slavov; Vagner Fonseca; Vincent Louis Viala |
| EPI_ISL_1795258 | CS DE PLANALTO | Instituto Butantan / ESALQ-Piracicaba | Antonio Jorge Martins; Bianca Cechetto Carlos. Mendelics: Bibiana Santos; Claudia Renata dos Santos Barros; David Schlesinger. Hemocentro Ribeirão Preto: Simone Kashima; Debora Botequio Moretti. Centro de Genômica Funcional da ESALQ: Luiz Lehmann Coutinho; Dimas Tadeu Covas; Elaine Cristina Marqueze; Elaine Vieira dos Santos; Elisangela Chicaroni Mattos; Erika Freitas; Evandra Strazza Rodrigues; Felipe Allan da Silva da Costa; Flavia Aburjaile; Guilherme Targino Valente; Heidge Fukumasu. USP-Botucatu: Rejane Maria Tommasini Grotto; Instituto Butantan: Alexander Roberto Precioso; Jayme A. Souza-Neto; Jessika Cristina Chagas Lesbon; José Salvatore Leister Patané; João Paulo Kitajima; Luiz Carlos Junior de Alcantara; Maria Carolina Elias; Marta Giovanetti; Patricia Akemi Assato; Rafael dos Santos Bezerra; Raquel de Lello Rocha Campos Cassano. NGS Soluções Genômicas: Pilar Drummond Sampaio Corrêa Mariani. FZEA-USP Pirassununga: Mirele Daiana Poleti; Raul Machado Neto; Ricardo Augusto Brassaloti; Ricardo Haddad; Rodrigo Tocantins Calado.; Sandra Coccuzzo Sampaio; Svetoslav Nanev Slavov; Vagner Fonseca; Vincent Louis Viala |
| EPI_ISL_1966257, EPI_ISL_1966258 | CS DE RUBINEIA | Instituto Butantan / Mendelics | Antonio Jorge Martins; Bianca Cechetto Carlos. Mendelics: Bibiana Santos; Claudia Renata dos Santos Barros; Cíntia Bittar; David Schlesinger. Hemocentro Ribeirão Preto: Simone Kashima; Debora Botequio Moretti; Elaine Cristina Marqueze; Elaine Vieira dos Santos; Elisangela Chicaroni Mattos; Erika Freitas; Evandra Strazza Rodrigues; Felipe Allan da Silva da Costa; Flavia Aburjaile; Fábio Sossai Possebon; Guilherme Campos; Guilherme Targino Valente; Heidge Fukumasu. USP-Botucatu: Rejane Maria Tommasini Grotto; Helena Lage Ferreira; Instituto Butantan: Dimas Tadeu Covas; Jardelina de Souza Todao Bernardino; Jayme A. Souza-Neto; Jessika Cristina Chagas Lesbon; Jorge A. Petroli Marchesi; José Salvatore Leister Patané; João Paulo Kitajima; João Pessoa Araújo Jr.; Leila Sabrina Ullmann; Loyze Paola Oliveira de Lima; Luiz Aurelio de Campos Crispin. Centro de Genômica Funcional da ESALQ: Luiz Lehmann Coutinho; Luiz Carlos Junior de Alcantara; Lívia Sacchetto; Maisa C. Pereira Parra; Maria Carolina Elias; Marta Giovanetti; Marília Moraes; Maurício Lacerda Nogueira. Prefeitura de Sao Paulo: Melissa Palmieri.; Patricia Akemi Assato; Paula Rahal; Paulo Inacio da Costa; Rafael dos Santos Bezerra; Raquel de Lello Rocha Campos Cassano. NGS Soluções Genômicas: Pilar Drummond Sampaio Corrêa Mariani. FZEA-USP Pirassununga: Mirele Daiana Poleti; Raul Machado Neto; Ricardo Augusto Brassaloti; Ricardo Haddad; Rodrigo Tocantins Calado. FAMERP-SJRP: Cecília Artico Banho; Sandra Coccuzzo Sampaio; Svetoslav Nanev Slavov; Vagner Fonseca; Vincent Louis Viala |
| EPI_ISL_1967050 | CS DE SANTA RITA DOESTE | Instituto Butantan / ESALQ-USP (Piracicaba) | Antonio Jorge Martins; Bianca Cechetto Carlos. Mendelics: Bibiana Santos; Claudia Renata dos Santos Barros; Cíntia Bittar; David Schlesinger. Hemocentro Ribeirão Preto: Simone Kashima; Debora Botequio Moretti; Elaine Cristina Marqueze; Elaine Vieira dos Santos; Elisangela Chicaroni Mattos; Erika Freitas; Evandra Strazza Rodrigues; Felipe Allan da Silva da Costa; Flavia Aburjaile; Fábio Sossai Possebon; Guilherme Campos; Guilherme Targino Valente; Heidge Fukumasu. USP-Botucatu: Rejane Maria Tommasini Grotto; Helena Lage Ferreira; Instituto Butantan: Dimas Tadeu Covas; Jardelina de Souza Todao Bernardino; Jayme A. Souza-Neto; Jessika Cristina Chagas Lesbon; Jorge A. Petroli Marchesi; José Salvatore Leister Patané; João Paulo Kitajima; João Pessoa Araújo Jr.; Leila Sabrina Ullmann; Loyze Paola Oliveira de Lima; Luiz Aurelio de Campos Crispin. Centro de Genômica Funcional da ESALQ: Luiz Lehmann Coutinho; Luiz Carlos Junior de Alcantara; Lívia Sacchetto; Maisa C. Pereira Parra; Maria Carolina Elias; Marta Giovanetti; Marília Moraes; Maurício Lacerda Nogueira. Prefeitura de Sao Paulo: Melissa Palmieri.; Patricia Akemi Assato; Paula Rahal; Paulo Inacio da Costa; Rafael dos Santos Bezerra; Raquel de Lello Rocha Campos Cassano. NGS Soluções Genômicas: Pilar Drummond Sampaio Corrêa Mariani. FZEA-USP Pirassununga: Mirele Daiana Poleti; Raul Machado Neto; Ricardo Augusto Brassaloti; Ricardo Haddad; Rodrigo Tocantins Calado. FAMERP-SJRP: Cecília Artico Banho; Sandra Coccuzzo Sampaio; Svetoslav Nanev Slavov; Vagner Fonseca; Vincent Louis Viala |
| EPI_ISL_1966547 | CS DE SANTA RITA DOESTE | Instituto Butantan / FZEA-USP (Pirassununga) | Antonio Jorge Martins; Bianca Cechetto Carlos. Mendelics: Bibiana Santos; Claudia Renata dos Santos Barros; Cíntia Bittar; David Schlesinger. Hemocentro Ribeirão Preto: Simone Kashima; Debora Botequio Moretti; Elaine Cristina Marqueze; Elaine Vieira dos Santos; Elisangela Chicaroni Mattos; Erika Freitas; Evandra Strazza Rodrigues; Felipe Allan da Silva da Costa; Flavia Aburjaile; Fábio Sossai Possebon; Guilherme Campos; Guilherme Targino Valente; Heidge Fukumasu. USP-Botucatu: Rejane Maria Tommasini Grotto; Helena Lage Ferreira; Instituto Butantan: Dimas Tadeu Covas; Jardelina de Souza Todao Bernardino; Jayme A. Souza-Neto; Jessika Cristina Chagas Lesbon; Jorge A. Petroli Marchesi; José Salvatore Leister Patané; João Paulo Kitajima; João Pessoa Araújo Jr.; Leila Sabrina Ullmann; Loyze Paola Oliveira de Lima; Luiz Aurelio de Campos Crispin. Centro de Genômica Funcional da ESALQ: Luiz Lehmann Coutinho; Luiz Carlos Junior de Alcantara; Lívia Sacchetto; Maisa C. Pereira Parra; Maria Carolina Elias; Marta Giovanetti; Marília Moraes; Maurício Lacerda Nogueira. Prefeitura de Sao Paulo: Melissa Palmieri.; Patricia Akemi Assato; Paula Rahal; Paulo Inacio da Costa; Rafael dos Santos Bezerra; Raquel de Lello Rocha Campos Cassano. NGS Soluções Genômicas: Pilar Drummond Sampaio Corrêa Mariani. FZEA-USP Pirassununga: Mirele Daiana Poleti; Raul Machado Neto; Ricardo Augusto Brassaloti; Ricardo Haddad; Rodrigo Tocantins Calado. FAMERP-SJRP: Cecília Artico Banho; Sandra Coccuzzo Sampaio; Svetoslav Nanev Slavov; Vagner Fonseca; Vincent Louis Viala |
| EPI_ISL_1795240 | CS DE SEBASTIANOPOLIS DO SUL | Instituto Butantan / ESALQ-Piracicaba | Antonio Jorge Martins; Bianca Cechetto Carlos. Mendelics: Bibiana Santos; Claudia Renata dos Santos Barros; David Schlesinger. Hemocentro Ribeirão Preto: Simone Kashima; Debora Botequio Moretti. Centro de Genômica Funcional da ESALQ: Luiz Lehmann Coutinho; Dimas Tadeu Covas; Elaine Cristina Marqueze; Elaine Vieira dos Santos; Elisangela Chicaroni Mattos; Erika Freitas; Evandra Strazza Rodrigues; Felipe Allan da Silva da Costa; Flavia Aburjaile; Guilherme Targino Valente; Heidge Fukumasu. USP-Botucatu: Rejane Maria Tommasini Grotto; Instituto Butantan: Alexander Roberto Precioso; Jayme A. Souza-Neto; Jessika Cristina Chagas Lesbon; José Salvatore Leister Patané; João Paulo Kitajima; Luiz Carlos Junior de Alcantara; Maria Carolina Elias; Marta Giovanetti; Patricia Akemi Assato; Rafael dos Santos Bezerra; Raquel de Lello Rocha Campos Cassano. NGS Soluções Genômicas: Pilar Drummond Sampaio Corrêa Mariani. FZEA-USP Pirassununga: Mirele Daiana Poleti; Raul Machado Neto; Ricardo Augusto Brassaloti; Ricardo Haddad; Rodrigo Tocantins Calado.; Sandra Coccuzzo Sampaio; Svetoslav Nanev Slavov; Vagner Fonseca; Vincent Louis Viala |
| EPI_ISL_1795232, EPI_ISL_1795233, EPI_ISL_1795239, EPI_ISL_1795242, EPI_ISL_1795243, EPI_ISL_1795244 | CS DE URUPES | Instituto Butantan / ESALQ-Piracicaba | Antonio Jorge Martins; Bianca Cechetto Carlos. Mendelics: Bibiana Santos; Claudia Renata dos Santos Barros; David Schlesinger. Hemocentro Ribeirão Preto: Simone Kashima; Debora Botequio Moretti. Centro de Genômica Funcional da ESALQ: Luiz Lehmann Coutinho; Dimas Tadeu Covas; Elaine Cristina Marqueze; Elaine Vieira dos Santos; Elisangela Chicaroni Mattos; Erika Freitas; Evandra Strazza Rodrigues; Felipe Allan da Silva da Costa; Flavia Aburjaile; Guilherme Targino Valente; Heidge Fukumasu. USP-Botucatu: Rejane Maria Tommasini Grotto; Instituto Butantan: Alexander Roberto Precioso; Jayme A. Souza-Neto; Jessika Cristina Chagas Lesbon; José Salvatore Leister Patané; João Paulo Kitajima; Luiz Carlos Junior de Alcantara; Maria Carolina Elias; Marta Giovanetti; Patricia Akemi Assato; Rafael dos Santos Bezerra; Raquel de Lello Rocha Campos Cassano. NGS Soluções Genômicas: Pilar Drummond Sampaio Corrêa Mariani. FZEA-USP Pirassununga: Mirele Daiana Poleti; Raul Machado Neto; Ricardo Augusto Brassaloti; Ricardo Haddad; Rodrigo Tocantins Calado.; Sandra Coccuzzo Sampaio; Svetoslav Nanev Slavov; Vagner Fonseca; Vincent Louis Viala |
| EPI_ISL_1966992, EPI_ISL_1966993 | CS I DE OURINHOS | Instituto Butantan / ESALQ-USP (Piracicaba) | Antonio Jorge Martins; Bianca Cechetto Carlos. Mendelics: Bibiana Santos; Claudia Renata dos Santos Barros; Cíntia Bittar; David Schlesinger. Hemocentro Ribeirão Preto: Simone Kashima; Debora Botequio Moretti; Elaine Cristina Marqueze; Elaine Vieira dos Santos; Elisangela Chicaroni Mattos; Erika Freitas; Evandra Strazza Rodrigues; Felipe Allan da Silva da Costa; Flavia Aburjaile; Fábio Sossai Possebon; Guilherme Campos; Guilherme Targino Valente; Heidge Fukumasu. USP-Botucatu: Rejane Maria Tommasini Grotto; Helena Lage Ferreira; Instituto Butantan: Dimas Tadeu Covas; Jardelina de Souza Todao Bernardino; Jayme A. Souza-Neto; Jessika Cristina Chagas Lesbon; Jorge A. Petroli Marchesi; José Salvatore Leister Patané; João Paulo Kitajima; João Pessoa Araújo Jr.; Leila Sabrina Ullmann; Loyze Paola Oliveira de Lima; Luiz Aurelio de Campos Crispin. Centro de Genômica Funcional da ESALQ: Luiz Lehmann Coutinho; Luiz Carlos Junior de Alcantara; Lívia Sacchetto; Maisa C. Pereira Parra; Maria Carolina Elias; Marta Giovanetti; Marília Moraes; Maurício Lacerda Nogueira. Prefeitura de Sao Paulo: Melissa Palmieri.; Patricia Akemi Assato; Paula Rahal; Paulo Inacio da Costa; Rafael dos Santos Bezerra; Raquel de Lello Rocha Campos Cassano. NGS Soluções Genômicas: Pilar Drummond Sampaio Corrêa Mariani. FZEA-USP Pirassununga: Mirele Daiana Poleti; Raul Machado Neto; Ricardo Augusto Brassaloti; Ricardo Haddad; Rodrigo Tocantins Calado. FAMERP-SJRP: Cecília Artico Banho; Sandra Coccuzzo Sampaio; Svetoslav Nanev Slavov; Vagner Fonseca; Vincent Louis Viala |
| EPI_ISL_1966909 | CS II DR JAHYR DE PAULA RIBEIRO GUARA | Instituto Butantan / ESALQ-USP (Piracicaba) | Antonio Jorge Martins; Bianca Cechetto Carlos. Mendelics: Bibiana Santos; Claudia Renata dos Santos Barros; Cíntia Bittar; David Schlesinger. Hemocentro Ribeirão Preto: Simone Kashima; Debora Botequio Moretti; Elaine Cristina Marqueze; Elaine Vieira dos Santos; Elisangela Chicaroni Mattos; Erika Freitas; Evandra Strazza Rodrigues; Felipe Allan da Silva da Costa; Flavia Aburjaile; Fábio Sossai Possebon; Guilherme Campos; Guilherme Targino Valente; Heidge Fukumasu. USP-Botucatu: Rejane Maria Tommasini Grotto; Helena Lage Ferreira; Instituto Butantan: Dimas Tadeu Covas; Jardelina de Souza Todao Bernardino; Jayme A. Souza-Neto; Jessika Cristina Chagas Lesbon; Jorge A. Petroli Marchesi; José Salvatore Leister Patané; João Paulo Kitajima; João Pessoa Araújo Jr.; Leila Sabrina Ullmann; Loyze Paola Oliveira de Lima; Luiz Aurelio de Campos Crispin. Centro de Genômica Funcional da ESALQ: Luiz Lehmann Coutinho; Luiz Carlos Junior de Alcantara; Lívia Sacchetto; Maisa C. Pereira Parra; Maria Carolina Elias; Marta Giovanetti; Marília Moraes; Maurício Lacerda Nogueira. Prefeitura de Sao Paulo: Melissa Palmieri.; Patricia Akemi Assato; Paula Rahal; Paulo Inacio da Costa; Rafael dos Santos Bezerra; Raquel de Lello Rocha Campos Cassano. NGS Soluções Genômicas: Pilar Drummond Sampaio Corrêa Mariani. FZEA-USP Pirassununga: Mirele Daiana Poleti; Raul Machado Neto; Ricardo Augusto Brassaloti; Ricardo Haddad; Rodrigo Tocantins Calado. FAMERP-SJRP: Cecília Artico Banho; Sandra Coccuzzo Sampaio; Svetoslav Nanev Slavov; Vagner Fonseca; Vincent Louis Viala |
| EPI_ISL_1966808 | CS II DR JOSE CARQUEIJO SCRPARDO | Instituto Butantan / Mendelics | Antonio Jorge Martins; Bianca Cechetto Carlos. Mendelics: Bibiana Santos; Claudia Renata dos Santos Barros; Cíntia Bittar; David Schlesinger. Hemocentro Ribeirão Preto: Simone Kashima; Debora Botequio Moretti; Elaine Cristina Marqueze; Elaine Vieira dos Santos; Elisangela Chicaroni Mattos; Erika Freitas; Evandra Strazza Rodrigues; Felipe Allan da Silva da Costa; Flavia Aburjaile; Fábio Sossai Possebon; Guilherme Campos; Guilherme Targino Valente; Heidge Fukumasu. USP-Botucatu: Rejane Maria Tommasini Grotto; Helena Lage Ferreira; Instituto Butantan: Dimas Tadeu Covas; Jardelina de Souza Todao Bernardino; Jayme A. Souza-Neto; Jessika Cristina Chagas Lesbon; Jorge A. Petroli Marchesi; José Salvatore Leister Patané; João Paulo Kitajima; João Pessoa Araújo Jr.; Leila Sabrina Ullmann; Loyze Paola Oliveira de Lima; Luiz Aurelio de Campos Crispin. Centro de Genômica Funcional da ESALQ: Luiz Lehmann Coutinho; Luiz Carlos Junior de Alcantara; Lívia Sacchetto; Maisa C. Pereira Parra; Maria Carolina Elias; Marta Giovanetti; Marília Moraes; Maurício Lacerda Nogueira. Prefeitura de Sao Paulo: Melissa Palmieri.; Patricia Akemi Assato; Paula Rahal; Paulo Inacio da Costa; Rafael dos Santos Bezerra; Raquel de Lello Rocha Campos Cassano. NGS Soluções Genômicas: Pilar Drummond Sampaio Corrêa Mariani. FZEA-USP Pirassununga: Mirele Daiana Poleti; Raul Machado Neto; Ricardo Augusto Brassaloti; Ricardo Haddad; Rodrigo Tocantins Calado. FAMERP-SJRP: Cecília Artico Banho; Sandra Coccuzzo Sampaio; Svetoslav Nanev Slavov; Vagner Fonseca; Vincent Louis Viala |
| EPI_ISL_1966989 | CS II DR MIGUEL VITALIANO ORLANDIA | Instituto Butantan / ESALQ-USP (Piracicaba) | Antonio Jorge Martins; Bianca Cechetto Carlos. Mendelics: Bibiana Santos; Claudia Renata dos Santos Barros; Cíntia Bittar; David Schlesinger. Hemocentro Ribeirão Preto: Simone Kashima; Debora Botequio Moretti; Elaine Cristina Marqueze; Elaine Vieira dos Santos; Elisangela Chicaroni Mattos; Erika Freitas; Evandra Strazza Rodrigues; Felipe Allan da Silva da Costa; Flavia Aburjaile; Fábio Sossai Possebon; Guilherme Campos; Guilherme Targino Valente; Heidge Fukumasu. USP-Botucatu: Rejane Maria Tommasini Grotto; Helena Lage Ferreira; Instituto Butantan: Dimas Tadeu Covas; Jardelina de Souza Todao Bernardino; Jayme A. Souza-Neto; Jessika Cristina Chagas Lesbon; Jorge A. Petroli Marchesi; José Salvatore Leister Patané; João Paulo Kitajima; João Pessoa Araújo Jr.; Leila Sabrina Ullmann; Loyze Paola Oliveira de Lima; Luiz Aurelio de Campos Crispin. Centro de Genômica Funcional da ESALQ: Luiz Lehmann Coutinho; Luiz Carlos Junior de Alcantara; Lívia Sacchetto; Maisa C. Pereira Parra; Maria Carolina Elias; Marta Giovanetti; Marília Moraes; Maurício Lacerda Nogueira. Prefeitura de Sao Paulo: Melissa Palmieri.; Patricia Akemi Assato; Paula Rahal; Paulo Inacio da Costa; Rafael dos Santos Bezerra; Raquel de Lello Rocha Campos Cassano. NGS Soluções Genômicas: Pilar Drummond Sampaio Corrêa Mariani. FZEA-USP Pirassununga: Mirele Daiana Poleti; Raul Machado Neto; Ricardo Augusto Brassaloti; Ricardo Haddad; Rodrigo Tocantins Calado. FAMERP-SJRP: Cecília Artico Banho; Sandra Coccuzzo Sampaio; Svetoslav Nanev Slavov; Vagner Fonseca; Vincent Louis Viala |
| EPI_ISL_1966238 | CS II DR SEBASTIAO RIBEIRO DO VALLE | Instituto Butantan / Mendelics | Antonio Jorge Martins; Bianca Cechetto Carlos. Mendelics: Bibiana Santos; Claudia Renata dos Santos Barros; Cíntia Bittar; David Schlesinger. Hemocentro Ribeirão Preto: Simone Kashima; Debora Botequio Moretti; Elaine Cristina Marqueze; Elaine Vieira dos Santos; Elisangela Chicaroni Mattos; Erika Freitas; Evandra Strazza Rodrigues; Felipe Allan da Silva da Costa; Flavia Aburjaile; Fábio Sossai Possebon; Guilherme Campos; Guilherme Targino Valente; Heidge Fukumasu. USP-Botucatu: Rejane Maria Tommasini Grotto; Helena Lage Ferreira; Instituto Butantan: Dimas Tadeu Covas; Jardelina de Souza Todao Bernardino; Jayme A. Souza-Neto; Jessika Cristina Chagas Lesbon; Jorge A. Petroli Marchesi; José Salvatore Leister Patané; João Paulo Kitajima; João Pessoa Araújo Jr.; Leila Sabrina Ullmann; Loyze Paola Oliveira de Lima; Luiz Aurelio de Campos Crispin. Centro de Genômica Funcional da ESALQ: Luiz Lehmann Coutinho; Luiz Carlos Junior de Alcantara; Lívia Sacchetto; Maisa C. Pereira Parra; Maria Carolina Elias; Marta Giovanetti; Marília Moraes; Maurício Lacerda Nogueira. Prefeitura de Sao Paulo: Melissa Palmieri.; Patricia Akemi Assato; Paula Rahal; Paulo Inacio da Costa; Rafael dos Santos Bezerra; Raquel de Lello Rocha Campos Cassano. NGS Soluções Genômicas: Pilar Drummond Sampaio Corrêa Mariani. FZEA-USP Pirassununga: Mirele Daiana Poleti; Raul Machado Neto; Ricardo Augusto Brassaloti; Ricardo Haddad; Rodrigo Tocantins Calado. FAMERP-SJRP: Cecília Artico Banho; Sandra Coccuzzo Sampaio; Svetoslav Nanev Slavov; Vagner Fonseca; Vincent Louis Viala |
| EPI_ISL_1625975 | CS II Dr Antonio Vicoso Moreira De Rezende Sumare | Instituto Adolfo Lutz, Interdiciplinary Procedures Center, Strategic Laboratory | Caio Vinicius Dias Lopes; Claudia Regina Gonçalves; Claudio Tavares Sacchi; Erica Valessa Ramos Gomes; Karoline Rodrigues Campos; Katia Correa de Oliveira Santos; Leonardo Jose Tadeu de Araujo |
| EPI_ISL_1715135, EPI_ISL_1715136 | CS II Dr Jose Ferreira Telles | Instituto Adolfo Lutz, Interdiciplinary Procedures Center, Strategic Laboratory | Caio Vinicius Dias Lopes; Claudia Regina Gonçalves; Claudio Tavares Sacchi; Erica Valessa Ramos Gomes; Karoline Rodrigues Campos; Katia Correa de Oliveira Santos; Leonardo Jose Tadeu de Araujo |
| EPI_ISL_2170897 | CS II EGIDIO BRUNHARA MORRO AGUDO | Instituto Butantan / ESALQ-Piracicaba | Antonio Jorge Martins; Bianca Cechetto Carlos. Mendelics: Bibiana Santos; Claudia Renata dos Santos Barros; Cíntia Bittar; David Schlesinger. Hemocentro Ribeirão Preto: Simone Kashima; Debora Botequio Moretti; Elaine Cristina Marqueze; Elaine Vieira dos Santos; Elisangela Chicaroni Mattos; Erika Freitas; Evandra Strazza Rodrigues; Felipe Allan da Silva da Costa; Flavia Aburjaile; Fábio Sossai Possebon; Guilherme Campos; Guilherme Targino Valente; Heidge Fukumasu. USP-Botucatu: Rejane Maria Tommasini Grotto; Helena Lage Ferreira; Instituto Butantan: Dimas Tadeu Covas; Jardelina de Souza Todao Bernardino; Jayme A. Souza-Neto; Jessika Cristina Chagas Lesbon; Jorge A. Petroli Marchesi; José Salvatore Leister Patané; João Paulo Kitajima; João Pessoa Araújo Jr.; Leila Sabrina Ullmann; Loyze Paola Oliveira de Lima; Luiz Aurelio de Campos Crispin. Centro de Genômica Funcional da ESALQ: Luiz Lehmann Coutinho; Luiz Carlos Junior de Alcantara; Lívia Sacchetto; Maisa C. Pereira Parra; Maria Carolina Elias; Marta Giovanetti; Marília Moraes; Maurício Lacerda Nogueira. Prefeitura de Sao Paulo: Melissa Palmieri.; Patricia Akemi Assato; Paula Rahal; Paulo Inacio da Costa; Rafael dos Santos Bezerra; Raquel de Lello Rocha Campos Cassano. NGS Soluções Genômicas: Pilar Drummond Sampaio Corrêa Mariani. FZEA-USP Pirassununga: Mirele Daiana Poleti; Raul Machado Neto; Ricardo Augusto Brassaloti; Ricardo Haddad; Rodrigo Tocantins Calado. FAMERP-SJRP: Cecília Artico Banho; Sandra Coccuzzo Sampaio; Svetoslav Nanev Slavov; Vagner Fonseca; Vincent Louis Viala |
| EPI_ISL_1966965, EPI_ISL_1966966, EPI_ISL_1966967, EPI_ISL_1966968, EPI_ISL_1966969, EPI_ISL_1966970, EPI_ISL_1966971, EPI_ISL_1966972, EPI_ISL_1966973, EPI_ISL_1966974, EPI_ISL_1966975, EPI_ISL_1966976, EPI_ISL_1966977, EPI_ISL_1966978, EPI_ISL_1966979, EPI_ISL_1966980, EPI_ISL_1966981, EPI_ISL_1966982, EPI_ISL_1966983 | | | |
| see above | CS II EGIDIO BRUNHARA MORRO AGUDO | Instituto Butantan / ESALQ-USP (Piracicaba) | Antonio Jorge Martins; Bianca Cechetto Carlos. Mendelics: Bibiana Santos; Claudia Renata dos Santos Barros; Cíntia Bittar; David Schlesinger. Hemocentro Ribeirão Preto: Simone Kashima; Debora Botequio Moretti; Elaine Cristina Marqueze; Elaine Vieira dos Santos; Elisangela Chicaroni Mattos; Erika Freitas; Evandra Strazza Rodrigues; Felipe Allan da Silva da Costa; Flavia Aburjaile; Fábio Sossai Possebon; Guilherme Campos; Guilherme Targino Valente; Heidge Fukumasu. USP-Botucatu: Rejane Maria Tommasini Grotto; Helena Lage Ferreira; Instituto Butantan: Dimas Tadeu Covas; Jardelina de Souza Todao Bernardino; Jayme A. Souza-Neto; Jessika Cristina Chagas Lesbon; Jorge A. Petroli Marchesi; José Salvatore Leister Patané; João Paulo Kitajima; João Pessoa Araújo Jr.; Leila Sabrina Ullmann; Loyze Paola Oliveira de Lima; Luiz Aurelio de Campos Crispin. Centro de Genômica Funcional da ESALQ: Luiz Lehmann Coutinho; Luiz Carlos Junior de Alcantara; Lívia Sacchetto; Maisa C. Pereira Parra; Maria Carolina Elias; Marta Giovanetti; Marília Moraes; Maurício Lacerda Nogueira. Prefeitura de Sao Paulo: Melissa Palmieri.; Patricia Akemi Assato; Paula Rahal; Paulo Inacio da Costa; Rafael dos Santos Bezerra; Raquel de Lello Rocha Campos Cassano. NGS Soluções Genômicas: Pilar Drummond Sampaio Corrêa Mariani. FZEA-USP Pirassununga: Mirele Daiana Poleti; Raul Machado Neto; Ricardo Augusto Brassaloti; Ricardo Haddad; Rodrigo Tocantins Calado. FAMERP-SJRP: Cecília Artico Banho; Sandra Coccuzzo Sampaio; Svetoslav Nanev Slavov; Vagner Fonseca; Vincent Louis Viala |
| EPI_ISL_1445142, EPI_ISL_1445143, EPI_ISL_1445144, EPI_ISL_1445145, EPI_ISL_1445146, EPI_ISL_1966113, EPI_ISL_1966114, EPI_ISL_1966115, EPI_ISL_1966116, EPI_ISL_1966117 | | | |
| see above | CS II EGIDIO BRUNHARA MORRO AGUDO | Instituto Butantan / Mendelics | Antonio Jorge Martins; Bianca Cechetto Carlos. Mendelics: Bibiana Santos; Bibiana Santos; Claudia Renata dos Santos Barros; Cíntia Bittar; David Schlesinger; David Schlesinger. Hemocentro Ribeirão Preto: Simone Kashima; Debora Botequio Moretti; Dimas Tadeu Covas; Elaine Cristina Marqueze; Elaine Vieira dos Santos; Elisangela Chicaroni Mattos; Erika Freitas; Evandra Strazza Rodrigues; Felipe Allan da Silva da Costa; Flavia Aburjaile; Fábio Sossai Possebon; Guilherme Campos; Guilherme Targino Valente; Heidge Fukumasu. USP-Botucatu: Rejane Maria Tommasini Grotto; Helena Lage Ferreira; Instituto Butantan: Dimas Tadeu Covas; Jardelina de Souza Todao Bernardino; Jayme A. Souza-Neto; Jessika Cristina Chagas Lesbon; Jorge A. Petroli Marchesi; José Salvatore Leister Patané; João Paulo Kitajima; João Pessoa Araújo Jr.; Leila Sabrina Ullmann; Loyze Paola Oliveira de Lima; Luiz Aurelio de Campos Crispin. Centro de Genômica Funcional da ESALQ: Luiz Lehmann Coutinho; Luiz Carlos Junior de Alcantara; Lívia Sacchetto; Maisa C. Pereira Parra; Maria Carolina Elias; Marta Giovanetti; Marília Moraes; Maurício Lacerda Nogueira. Prefeitura de Sao Paulo: Melissa Palmieri.; Patricia Akemi Assato; Paula Rahal; Paulo Inacio da Costa; Rafael dos Santos Bezerra; Raquel de Lello Rocha Campos Cassano. NGS Soluções Genômicas: Pilar Drummond Sampaio Corrêa Mariani. FZEA-USP Pirassununga: Mirele Daiana Poleti; Raul Machado Neto; Ricardo Augusto Brassaloti; Ricardo Haddad; Rodrigo Tocantins Calado.; Rodrigo Tocantins Calado. FAMERP-SJRP: Cecília Artico Banho; Sandra Coccuzzo Sampaio; Simone Kashima; Svetoslav Nanev Slavov; Vagner Fonseca; Vincent Louis Viala |
| EPI_ISL_1966480 | CS III SANTOPOLIS DO AGUAPEI | Instituto Butantan / Mendelics | Antonio Jorge Martins; Bianca Cechetto Carlos. Mendelics: Bibiana Santos; Claudia Renata dos Santos Barros; Cíntia Bittar; David Schlesinger. Hemocentro Ribeirão Preto: Simone Kashima; Debora Botequio Moretti; Elaine Cristina Marqueze; Elaine Vieira dos Santos; Elisangela Chicaroni Mattos; Erika Freitas; Evandra Strazza Rodrigues; Felipe Allan da Silva da Costa; Flavia Aburjaile; Fábio Sossai Possebon; Guilherme Campos; Guilherme Targino Valente; Heidge Fukumasu. USP-Botucatu: Rejane Maria Tommasini Grotto; Helena Lage Ferreira; Instituto Butantan: Dimas Tadeu Covas; Jardelina de Souza Todao Bernardino; Jayme A. Souza-Neto; Jessika Cristina Chagas Lesbon; Jorge A. Petroli Marchesi; José Salvatore Leister Patané; João Paulo Kitajima; João Pessoa Araújo Jr.; Leila Sabrina Ullmann; Loyze Paola Oliveira de Lima; Luiz Aurelio de Campos Crispin. Centro de Genômica Funcional da ESALQ: Luiz Lehmann Coutinho; Luiz Carlos Junior de Alcantara; Lívia Sacchetto; Maisa C. Pereira Parra; Maria Carolina Elias; Marta Giovanetti; Marília Moraes; Maurício Lacerda Nogueira. Prefeitura de Sao Paulo: Melissa Palmieri.; Patricia Akemi Assato; Paula Rahal; Paulo Inacio da Costa; Rafael dos Santos Bezerra; Raquel de Lello Rocha Campos Cassano. NGS Soluções Genômicas: Pilar Drummond Sampaio Corrêa Mariani. FZEA-USP Pirassununga: Mirele Daiana Poleti; Raul Machado Neto; Ricardo Augusto Brassaloti; Ricardo Haddad; Rodrigo Tocantins Calado. FAMERP-SJRP: Cecília Artico Banho; Sandra Coccuzzo Sampaio; Svetoslav Nanev Slavov; Vagner Fonseca; Vincent Louis Viala |
| EPI_ISL_1795161 | CS III VILA ODILON | Instituto Butantan / ESALQ-Piracicaba | Antonio Jorge Martins; Bianca Cechetto Carlos. Mendelics: Bibiana Santos; Claudia Renata dos Santos Barros; David Schlesinger. Hemocentro Ribeirão Preto: Simone Kashima; Debora Botequio Moretti. Centro de Genômica Funcional da ESALQ: Luiz Lehmann Coutinho; Dimas Tadeu Covas; Elaine Cristina Marqueze; Elaine Vieira dos Santos; Elisangela Chicaroni Mattos; Erika Freitas; Evandra Strazza Rodrigues; Felipe Allan da Silva da Costa; Flavia Aburjaile; Guilherme Targino Valente; Heidge Fukumasu. USP-Botucatu: Rejane Maria Tommasini Grotto; Instituto Butantan: Alexander Roberto Precioso; Jayme A. Souza-Neto; Jessika Cristina Chagas Lesbon; José Salvatore Leister Patané; João Paulo Kitajima; Luiz Carlos Junior de Alcantara; Maria Carolina Elias; Marta Giovanetti; Patricia Akemi Assato; Rafael dos Santos Bezerra; Raquel de Lello Rocha Campos Cassano. NGS Soluções Genômicas: Pilar Drummond Sampaio Corrêa Mariani. FZEA-USP Pirassununga: Mirele Daiana Poleti; Raul Machado Neto; Ricardo |

| | | | |
|---|---|---|---|
| | | | Augusto Brassaloti; Ricardo Haddad; Rodrigo Tocantins Calado.; Sandra Coccuzzo Sampaio; Svetoslav Nanev Slavov; Vagner Fonseca; Vincent Louis Viala |
| EPI_ISL_1628366, EPI_ISL_1715142 | CS III de Patrocinio Paulista | Instituto Adolfo Lutz, Interdiciplinary Procedures Center, Strategic Laboratory | Caio Vinicius Dias Lopes; Claudia Regina Gonçalves; Claudio Tavares Sacchi; Erica Valessa Ramos Gomes; Karoline Rodrigues Campos; Katia Correa de Oliveira Santos; Leonardo Jose Tadeu de Araujo |
| EPI_ISL_1966695 | CS IOLANDA FLEURY DE CAMPOS GUEDES NUPORANGA | Instituto Butantan / Mendelics | Antonio Jorge Martins; Bianca Cechetto Carlos. Mendelics: Bibiana Santos; Claudia Renata dos Santos Barros; Cíntia Bittar; David Schlesinger. Hemocentro Ribeirão Preto: Simone Kashima; Debora Botequio Moretti; Elaine Cristina Marqueze; Elaine Vieira dos Santos; Elisangela Chicaroni Mattos; Erika Freitas; Evandra Strazza Rodrigues; Felipe Allan da Silva da Costa; Flavia Aburjaile; Fábio Sossai Possebon; Guilherme Campos; Guilherme Targino Valente; Heidge Fukumasu. USP-Botucatu: Rejane Maria Tommasini Grotto; Helena Lage Ferreira; Instituto Butantan: Dimas Tadeu Covas; Jardelina de Souza Todao Bernardino; Jayme A. Souza-Neto; Jessika Cristina Chagas Lesbon; Jorge A. Petroli Marchesi; José Salvatore Leister Patané; João Paulo Kitajima; João Pessoa Araújo Jr.; Leila Sabrina Ullmann; Loyze Paola Oliveira de Lima; Luiz Aurelio de Campos Crispin. Centro de Genômica Funcional da ESALQ: Luiz Lehmann Coutinho; Luiz Carlos Junior de Alcantara; Lívia Sacchetto; Maisa C. Pereira Parra; Maria Carolina Elias; Marta Giovanetti; Marília Moraes; Maurício Lacerda Nogueira. Prefeitura de Sao Paulo: Melissa Palmieri.; Patricia Akemi Assato; Paula Rahal; Paulo Inacio da Costa; Rafael dos Santos Bezerra; Raquel de Lello Rocha Campos Cassano. NGS Soluções Genômicas: Pilar Drummond Sampaio Corrêa Mariani. FZEA-USP Pirassununga: Mirele Daiana Poleti; Raul Machado Neto; Ricardo Augusto Brassaloti; Ricardo Haddad; Rodrigo Tocantins Calado. FAMERP-SJRP: Cecília Artico Banho; Sandra Coccuzzo Sampaio; Svetoslav Nanev Slavov; Vagner Fonseca; Vincent Louis Viala |
| EPI_ISL_1821209, EPI_ISL_2003112 | CS de Paulo de Faria | Instituto Adolfo Lutz, Interdiciplinary Procedures Center, Strategic Laboratory | Caio Vinicius Dias Lopes; Claudia Regina Gonçalves; Claudio Tavares Sacchi; Erica Valessa Ramos Gomes; Karoline Rodrigues Campos; Leonardo Jose Tadeu de Araujo |
| EPI_ISL_1967004, EPI_ISL_1967006, EPI_ISL_1967007, EPI_ISL_1967008, EPI_ISL_1967009, EPI_ISL_1967010, EPI_ISL_1967011, EPI_ISL_1967012, EPI_ISL_1967013 | | | |
| see above | CSII DR WASHINGTON LUIS M RODRIGUES DA SILVA PITANGUEIRAS | Instituto Butantan / ESALQ-USP (Piracicaba) | Antonio Jorge Martins; Bianca Cechetto Carlos. Mendelics: Bibiana Santos; Claudia Renata dos Santos Barros; Cíntia Bittar; David Schlesinger. Hemocentro Ribeirão Preto: Simone Kashima; Debora Botequio Moretti; Elaine Cristina Marqueze; Elaine Vieira dos Santos; Elisangela Chicaroni Mattos; Erika Freitas; Evandra Strazza Rodrigues; Felipe Allan da Silva da Costa; Flavia Aburjaile; Fábio Sossai Possebon; Guilherme Campos; Guilherme Targino Valente; Heidge Fukumasu. USP-Botucatu: Rejane Maria Tommasini Grotto; Helena Lage Ferreira; Instituto Butantan: Dimas Tadeu Covas; Jardelina de Souza Todao Bernardino; Jayme A. Souza-Neto; Jessika Cristina Chagas Lesbon; Jorge A. Petroli Marchesi; José Salvatore Leister Patané; João Paulo Kitajima; João Pessoa Araújo Jr.; Leila Sabrina Ullmann; Loyze Paola Oliveira de Lima; Luiz Aurelio de Campos Crispin. Centro de Genômica Funcional da ESALQ: Luiz Lehmann Coutinho; Luiz Carlos Junior de Alcantara; Lívia Sacchetto; Maisa C. Pereira Parra; Maria Carolina Elias; Marta Giovanetti; Marília Moraes; Maurício Lacerda Nogueira. Prefeitura de Sao Paulo: Melissa Palmieri.; Patricia Akemi Assato; Paula Rahal; Paulo Inacio da Costa; Rafael dos Santos Bezerra; Raquel de Lello Rocha Campos Cassano. NGS Soluções Genômicas: Pilar Drummond Sampaio Corrêa Mariani. FZEA-USP Pirassununga: Mirele Daiana Poleti; Raul Machado Neto; Ricardo Augusto Brassaloti; Ricardo Haddad; Rodrigo Tocantins Calado. FAMERP-SJRP: Cecília Artico Banho; Sandra Coccuzzo Sampaio; Svetoslav Nanev Slavov; Vagner Fonseca; Vincent Louis Viala |
| EPI_ISL_1731575, EPI_ISL_1752638 | Casa de Caridade Sao Vicente de Paulo Cajuru | Instituto Adolfo Lutz, Interdiciplinary Procedures Center, Strategic Laboratory | Caio Vinicius Dias Lopes; Claudia Regina Gonçalves; Claudio Tavares Sacchi; Erica Valessa Ramos Gomes; Karoline Rodrigues Campos; Katia Correa de Oliveira Santos; Leonardo Jose Tadeu de Araujo |
| EPI_ISL_1583672, EPI_ISL_1583673, EPI_ISL_1583676, EPI_ISL_1583679, EPI_ISL_1583681, EPI_ISL_1583689, EPI_ISL_1583694, EPI_ISL_1583705, EPI_ISL_1583714, EPI_ISL_1583716, EPI_ISL_1583725, EPI_ISL_1583727, EPI_ISL_1583730 | | | |
| see above | Central Public Health Laboratory - LACEN -Bahia, Salvador, Brazil | Central Public Health Laboratory - LACEN - Bahia, Salvador, Brazil | Arabela Leal; Breno Dominguez; Felicidade Pereira; Jaqueline Gomes; Luciana Oliveira; Luiz Alcantara; Marcela Gómez; Marta Giovanetti; Patrícia Cajado; Stephane Tosta; Vagner Fonseca; Vanessa Nardy |
| EPI_ISL_1628371, EPI_ISL_1628377, EPI_ISL_1715141 | Centro De Saude II Ibitinga | Instituto Adolfo Lutz, Interdiciplinary Procedures Center, Strategic Laboratory | Caio Vinicius Dias Lopes; Claudia Regina Gonçalves; Claudio Tavares Sacchi; Erica Valessa Ramos Gomes; Karoline Rodrigues Campos; Katia Correa de Oliveira Santos; Leonardo Jose Tadeu de Araujo |
| EPI_ISL_1628368 | Centro Medico Social Comunitario Januario Teodoro de Souza | Instituto Adolfo Lutz, Interdiciplinary Procedures Center, Strategic Laboratory | Caio Vinicius Dias Lopes; Claudia Regina Gonçalves; Claudio Tavares Sacchi; Erica Valessa Ramos Gomes; Karoline Rodrigues Campos; Katia Correa de Oliveira Santos; Leonardo Jose Tadeu de Araujo |
| EPI_ISL_1715134 | Centro de Saude II Ibitinga | Instituto Adolfo Lutz, Interdiciplinary Procedures Center, Strategic Laboratory | Caio Vinicius Dias Lopes; Claudia Regina Gonçalves; Claudio Tavares Sacchi; Erica Valessa Ramos Gomes; Karoline Rodrigues Campos; Katia Correa de Oliveira Santos; Leonardo Jose Tadeu de Araujo |
| EPI_ISL_1625976 | Centro de Saude III de Divinolandia | Instituto Adolfo Lutz, Interdiciplinary Procedures Center, Strategic Laboratory | Caio Vinicius Dias Lopes; Claudia Regina Gonçalves; Claudio Tavares Sacchi; Erica Valessa Ramos Gomes; Karoline Rodrigues Campos; Katia Correa de Oliveira Santos; Leonardo Jose Tadeu de Araujo |
| EPI_ISL_1381068 | Conjunto Hospitalar do Mandaqui de Sao Paulo | Instituto Adolfo Lutz, Interdiciplinary Procedures Center, Strategic Laboratory | Caio Vinicius Dias Lopes; Claudia Regina Gonçalves; Claudio Tavares Sacchi; Erica Valessa Ramos Gomes; Karoline Rodrigues Campos |
| EPI_ISL_1967196, EPI_ISL_1967197 | DEPARTAMENTO DE SAUDE COLETIVA | Instituto Butantan / ESALQ-USP (Piracicaba) | Antonio Jorge Martins; Bianca Cechetto Carlos. Mendelics: Bibiana Santos; Claudia Renata dos Santos Barros; Cíntia Bittar; David Schlesinger. Hemocentro Ribeirão Preto: Simone Kashima; Debora Botequio Moretti; Elaine Cristina Marqueze; Elaine Vieira dos Santos; Elisangela Chicaroni Mattos; Erika Freitas; Evandra Strazza Rodrigues; Felipe Allan da Silva da Costa; Flavia Aburjaile; Fábio Sossai Possebon; Guilherme Campos; Guilherme Targino Valente; Heidge Fukumasu. USP-Botucatu: Rejane Maria Tommasini Grotto; Helena Lage Ferreira; Instituto Butantan: Dimas Tadeu Covas; Jardelina de Souza Todao Bernardino; Jayme A. Souza-Neto; Jessika Cristina Chagas Lesbon; Jorge A. Petroli Marchesi; José Salvatore Leister Patané; João Paulo Kitajima; João Pessoa Araújo Jr.; Leila Sabrina Ullmann; Loyze Paola Oliveira de Lima; Luiz Aurelio de Campos Crispin. Centro de Genômica Funcional da ESALQ: Luiz Lehmann Coutinho; Luiz Carlos Junior de Alcantara; Lívia Sacchetto; Maisa C. Pereira Parra; Maria Carolina Elias; Marta Giovanetti; Marília Moraes; Maurício Lacerda Nogueira. Prefeitura de Sao Paulo: Melissa Palmieri.; Patricia Akemi Assato; Paula Rahal; Paulo Inacio da Costa; Rafael dos Santos Bezerra; Raquel de Lello Rocha Campos Cassano. NGS Soluções Genômicas: Pilar Drummond Sampaio Corrêa Mariani. FZEA-USP Pirassununga: Mirele Daiana Poleti; Raul Machado Neto; Ricardo Augusto Brassaloti; Ricardo Haddad; Rodrigo Tocantins Calado. FAMERP-SJRP: Cecília Artico Banho; Sandra Coccuzzo Sampaio; Svetoslav Nanev Slavov; Vagner Fonseca; Vincent Louis Viala |
| EPI_ISL_1445175, EPI_ISL_1966130, EPI_ISL_1966439 | DEPARTAMENTO DE SAUDE COLETIVA | Instituto Butantan / Mendelics | Antonio Jorge Martins; Bianca Cechetto Carlos. Mendelics: Bibiana Santos; Bibiana Santos; Claudia Renata dos Santos Barros; Cíntia Bittar; David Schlesinger; David Schlesinger. Hemocentro Ribeirão Preto: Simone Kashima; Debora Botequio Moretti; Dimas Tadeu Covas; Elaine Cristina Marqueze; Elaine Vieira dos Santos; Elisangela Chicaroni Mattos; Erika Freitas; Evandra Strazza Rodrigues; Felipe Allan da Silva da Costa; Flavia Aburjaile; Fábio Sossai Possebon; Guilherme Campos; Guilherme Targino Valente; Heidge Fukumasu. USP-Botucatu: Rejane Maria Tommasini Grotto; Helena Lage Ferreira; Instituto Butantan: Dimas Tadeu Covas; Jardelina de Souza Todao Bernardino; Jayme A. Souza-Neto; Jessika Cristina Chagas Lesbon; Jorge A. Petroli Marchesi; José Salvatore Leister Patané; João Paulo Kitajima; João Pessoa Araújo Jr.; Leila Sabrina Ullmann; Loyze Paola Oliveira de Lima; Luiz Aurelio de Campos Crispin. Centro de Genômica Funcional da ESALQ: Luiz Lehmann Coutinho; Luiz Carlos Junior de Alcantara; Lívia Sacchetto; Maisa C. Pereira Parra; Maria Carolina Elias; Marta Giovanetti; Marília Moraes; Maurício Lacerda Nogueira. Prefeitura de Sao Paulo: Melissa Palmieri.; Patricia Akemi Assato; Paula Rahal; Paulo Inacio da Costa; Rafael dos Santos Bezerra; Raquel de Lello Rocha Campos Cassano. NGS Soluções Genômicas: Pilar Drummond Sampaio Corrêa Mariani. FZEA-USP Pirassununga: Mirele Daiana Poleti; Raul Machado Neto; Ricardo Augusto Brassaloti; Ricardo Haddad; Rodrigo Tocantins Calado.; Rodrigo Tocantins Calado. FAMERP-SJRP: Cecília Artico Banho; Sandra Coccuzzo Sampaio; Simone Kashima; Svetoslav Nanev Slavov; Vagner Fonseca; Vincent Louis Viala |
| EPI_ISL_1795182 | DEPARTAMENTO DE SAUDE MUNICIPAL SOCORRO SP | Instituto Butantan / ESALQ-Piracicaba | Antonio Jorge Martins; Bianca Cechetto Carlos. Mendelics: Bibiana Santos; Claudia Renata dos Santos Barros; David Schlesinger. Hemocentro Ribeirão Preto: Simone Kashima; Debora Botequio Moretti. Centro de Genômica Funcional da ESALQ: Luiz Lehmann Coutinho; Dimas Tadeu Covas; Elaine Cristina Marqueze; Elaine Vieira dos Santos; Elisangela Chicaroni Mattos; Erika Freitas; Evandra Strazza Rodrigues; Felipe Allan da Silva da Costa; Flavia Aburjaile; Guilherme Targino Valente; Heidge Fukumasu. USP-Botucatu: Rejane Maria Tommasini Grotto; Instituto Butantan: Alexander Roberto Precioso; Jayme A. Souza-Neto; Jessika Cristina Chagas Lesbon; José Salvatore Leister Patané; João Paulo Kitajima; Luiz Carlos Junior de Alcantara; Maria Carolina Elias; Marta Giovanetti; Patricia Akemi Assato; Rafael dos Santos Bezerra; Raquel de Lello Rocha Campos Cassano. NGS Soluções Genômicas: Pilar Drummond Sampaio Corrêa Mariani. FZEA-USP Pirassununga: Mirele Daiana Poleti; Raul Machado Neto; Ricardo Augusto Brassaloti; Ricardo Haddad; Rodrigo Tocantins Calado.; Sandra Coccuzzo Sampaio; Svetoslav Nanev Slavov; Vagner Fonseca; Vincent Louis Viala |
| EPI_ISL_1583675 | DNA Laboratory | Central Public Health Laboratory - LACEN - Bahia, Salvador, Brazil | Arabela Leal; Breno Dominguez; Felicidade Pereira; Jaqueline Gomes; Luciana Oliveira; Luiz Alcantara; Marcela Gómez; Marta Giovanetti; Patrícia Cajado; Stephane Tosta; Vagner Fonseca; Vanessa Nardy |
| EPI_ISL_1821211, EPI_ISL_1821212, EPI_ISL_1821213 | Diagnóstico da America S/A | Instituto Adolfo Lutz, Interdiciplinary Procedures Center, Strategic Laboratory | Caio Vinicius Dias Lopes; Claudia Regina Gonçalves; Claudio Tavares Sacchi; Erica Valessa Ramos Gomes; Karoline Rodrigues Campos; Leonardo Jose Tadeu de Araujo |
| EPI_ISL_1795245, EPI_ISL_1795246, EPI_ISL_1795247 | EMERGENCIA RESPIRATORIA DE NOVA GRANADA | Instituto Butantan / ESALQ-Piracicaba | Antonio Jorge Martins; Bianca Cechetto Carlos. Mendelics: Bibiana Santos; Claudia Renata dos Santos Barros; David Schlesinger. Hemocentro Ribeirão Preto: Simone Kashima; Debora Botequio Moretti. Centro de Genômica Funcional da ESALQ: Luiz Lehmann Coutinho; Dimas Tadeu Covas; Elaine Cristina Marqueze; Elaine Vieira dos Santos; Elisangela Chicaroni Mattos; Erika Freitas; Evandra Strazza Rodrigues; Felipe Allan da Silva da Costa; Flavia Aburjaile; Guilherme Targino Valente; Heidge Fukumasu. USP-Botucatu: Rejane Maria Tommasini Grotto; Instituto Butantan: Alexander Roberto Precioso; Jayme A. Souza-Neto; Jessika Cristina Chagas Lesbon; José Salvatore Leister Patané; João Paulo Kitajima; Luiz Carlos Junior de Alcantara; Maria Carolina Elias; Marta Giovanetti; Patricia Akemi Assato; Rafael dos Santos Bezerra; Raquel de Lello Rocha Campos Cassano. NGS Soluções Genômicas: Pilar Drummond Sampaio Corrêa Mariani. FZEA-USP Pirassununga: Mirele Daiana Poleti; Raul Machado Neto; Ricardo Augusto Brassaloti; Ricardo Haddad; Rodrigo Tocantins Calado.; Sandra Coccuzzo Sampaio; Svetoslav Nanev Slavov; Vagner Fonseca; Vincent Louis Viala |
| EPI_ISL_1966531, EPI_ISL_1966532, EPI_ISL_1966533 | EMERGENCIA RESPIRATORIA DE NOVA GRANADA | Instituto Butantan / FZEA-USP (Pirassununga) | Antonio Jorge Martins; Bianca Cechetto Carlos. Mendelics: Bibiana Santos; Claudia Renata dos Santos Barros; Cíntia Bittar; David Schlesinger. Hemocentro Ribeirão Preto: Simone Kashima; Debora Botequio Moretti; Elaine Cristina Marqueze; Elaine Vieira dos Santos; Elisangela Chicaroni Mattos; Erika Freitas; Evandra Strazza Rodrigues; Felipe Allan da Silva da Costa; Flavia Aburjaile; Fábio Sossai Possebon; Guilherme Campos; Guilherme Targino Valente; Heidge Fukumasu. USP-Botucatu: Rejane Maria Tommasini Grotto; Helena Lage Ferreira; Instituto Butantan: Dimas Tadeu Covas; Jardelina de Souza Todao Bernardino; Jayme A. Souza-Neto; Jessika Cristina Chagas Lesbon; Jorge A. Petroli Marchesi; José Salvatore Leister Patané; João Paulo Kitajima; João Pessoa Araújo Jr.; Leila Sabrina Ullmann; Loyze Paola Oliveira de Lima; Luiz Aurelio de Campos Crispin. Centro de Genômica Funcional da ESALQ: Luiz Lehmann Coutinho; Luiz Carlos Junior de Alcantara; Lívia Sacchetto; Maisa C. Pereira Parra; Maria Carolina Elias; Marta Giovanetti; Marília Moraes; Maurício Lacerda Nogueira. Prefeitura de Sao Paulo: Melissa Palmieri.; Patricia Akemi Assato; Paula Rahal; Paulo Inacio da Costa; Rafael dos Santos Bezerra; Raquel de Lello Rocha Campos Cassano. NGS Soluções Genômicas: Pilar Drummond Sampaio Corrêa Mariani. FZEA-USP Pirassununga: Mirele Daiana Poleti; Raul Machado Neto; Ricardo Augusto Brassaloti; Ricardo Haddad; Rodrigo Tocantins Calado. FAMERP-SJRP: Cecília Artico Banho; Sandra Coccuzzo Sampaio; Svetoslav Nanev Slavov; Vagner Fonseca; Vincent Louis Viala |
| EPI_ISL_1795098, EPI_ISL_1795099 | ESALQ | Instituto Butantan / ESALQ-Piracicaba | Antonio Jorge Martins; Bianca Cechetto Carlos. Mendelics: Bibiana Santos; Claudia Renata dos Santos Barros; David Schlesinger. Hemocentro Ribeirão Preto: Simone Kashima; Debora Botequio Moretti. Centro de Genômica Funcional da ESALQ: Luiz Lehmann Coutinho; Dimas Tadeu Covas; Elaine Cristina Marqueze; Elaine Vieira dos Santos; Elisangela Chicaroni Mattos; Erika Freitas; Evandra Strazza Rodrigues; Felipe Allan da Silva da Costa; Flavia Aburjaile; Guilherme Targino Valente; Heidge Fukumasu. USP-Botucatu: Rejane Maria Tommasini Grotto; Instituto Butantan: Alexander Roberto Precioso; Jayme A. Souza-Neto; Jessika Cristina Chagas Lesbon; José Salvatore Leister Patané; João Paulo Kitajima; Luiz Carlos Junior de Alcantara; Maria Carolina Elias; Marta Giovanetti; Patricia Akemi Assato; Rafael dos Santos Bezerra; Raquel de Lello Rocha Campos Cassano. NGS Soluções Genômicas: Pilar Drummond Sampaio Corrêa Mariani. FZEA-USP Pirassununga: Mirele Daiana Poleti; Raul Machado Neto; Ricardo Augusto Brassaloti; Ricardo Haddad; Rodrigo Tocantins Calado.; Sandra Coccuzzo Sampaio; Svetoslav Nanev Slavov; Vagner Fonseca; Vincent Louis Viala |
| EPI_ISL_1966251 | ESF DR LUIS ERNESTO SANDI MORI JALES | Instituto Butantan / Mendelics | Antonio Jorge Martins; Bianca Cechetto Carlos. Mendelics: Bibiana Santos; Claudia Renata dos Santos Barros; Cíntia Bittar; David Schlesinger. Hemocentro Ribeirão Preto: Simone Kashima; Debora Botequio Moretti; Elaine Cristina Marqueze; Elaine Vieira dos Santos; Elisangela Chicaroni Mattos; Erika Freitas; Evandra Strazza Rodrigues; Felipe Allan da Silva da Costa; Flavia Aburjaile; Fábio Sossai Possebon; Guilherme Campos; Guilherme Targino Valente; Heidge Fukumasu. USP-Botucatu: Rejane Maria Tommasini Grotto; Helena Lage Ferreira; Instituto Butantan: Dimas Tadeu Covas; Jardelina de Souza Todao Bernardino; Jayme A. Souza-Neto; Jessika Cristina Chagas Lesbon; Jorge A. Petroli Marchesi; José Salvatore Leister Patané; João Paulo Kitajima; João Pessoa Araújo Jr.; Leila Sabrina Ullmann; Loyze Paola Oliveira de Lima; Luiz Aurelio de Campos Crispin. Centro de Genômica Funcional da ESALQ: Luiz Lehmann Coutinho; Luiz Carlos Junior de Alcantara; Lívia Sacchetto; Maisa C. Pereira Parra; Maria Carolina Elias; Marta Giovanetti; Marília Moraes; Maurício Lacerda Nogueira. Prefeitura de Sao Paulo: Melissa Palmieri.; Patricia Akemi Assato; Paula Rahal; Paulo Inacio da Costa; Rafael dos Santos Bezerra; Raquel de Lello Rocha Campos Cassano. NGS Soluções Genômicas: Pilar Drummond Sampaio Corrêa Mariani. FZEA-USP Pirassununga: Mirele Daiana Poleti; Raul Machado Neto; Ricardo Augusto Brassaloti; Ricardo Haddad; Rodrigo Tocantins Calado. FAMERP-SJRP: Cecília Artico Banho; Sandra Coccuzzo Sampaio; Svetoslav Nanev Slavov; Vagner Fonseca; Vincent Louis Viala |
| EPI_ISL_1795376, EPI_ISL_1795377, EPI_ISL_1795378 | ESF MINEIROS DO TIETE | Instituto Butantan / ESALQ-Piracicaba | Antonio Jorge Martins; Bianca Cechetto Carlos. Mendelics: Bibiana Santos; Claudia Renata dos Santos Barros; David Schlesinger. Hemocentro Ribeirão Preto: Simone Kashima; Debora Botequio Moretti. Centro de Genômica Funcional da ESALQ: Luiz Lehmann Coutinho; Dimas Tadeu Covas; Elaine Cristina Marqueze; Elaine Vieira dos Santos; Elisangela Chicaroni Mattos; Erika Freitas; Evandra Strazza Rodrigues; Felipe Allan da Silva da Costa; Flavia Aburjaile; Guilherme Targino Valente; Heidge Fukumasu. USP-Botucatu: Rejane Maria Tommasini Grotto; Instituto Butantan: Alexander Roberto Precioso; Jayme A. Souza-Neto; Jessika Cristina Chagas Lesbon; José Salvatore Leister Patané; João Paulo Kitajima; Luiz Carlos Junior de Alcantara; Maria Carolina Elias; Marta Giovanetti; Patricia Akemi Assato; Rafael dos Santos Bezerra; Raquel de Lello Rocha Campos Cassano. NGS Soluções Genômicas: Pilar Drummond Sampaio Corrêa Mariani. FZEA-USP Pirassununga: Mirele Daiana Poleti; Raul Machado Neto; Ricardo Augusto Brassaloti; Ricardo Haddad; Rodrigo Tocantins Calado.; Sandra Coccuzzo Sampaio; Svetoslav Nanev Slavov; Vagner Fonseca; Vincent Louis Viala |
| EPI_ISL_1966674 | ESF NOVA ALEXANDRIA | Instituto Butantan / Mendelics | Antonio Jorge Martins; Bianca Cechetto Carlos. Mendelics: Bibiana Santos; Claudia Renata dos Santos Barros; Cíntia Bittar; David Schlesinger. Hemocentro Ribeirão Preto: Simone Kashima; Debora Botequio Moretti; Elaine Cristina Marqueze; Elaine Vieira dos Santos; Elisangela Chicaroni Mattos; Erika Freitas; Evandra Strazza Rodrigues; Felipe Allan da Silva da Costa; Flavia Aburjaile; Fábio Sossai Possebon; Guilherme Campos; Guilherme Targino Valente; Heidge Fukumasu. USP-Botucatu: Rejane Maria Tommasini Grotto; Helena Lage Ferreira; Instituto Butantan: Dimas Tadeu Covas; Jardelina de Souza Todao Bernardino; Jayme A. Souza-Neto; Jessika Cristina Chagas Lesbon; Jorge A. Petroli Marchesi; José Salvatore Leister Patané; João Paulo Kitajima; João Pessoa Araújo Jr.; Leila Sabrina Ullmann; Loyze Paola Oliveira de Lima; Luiz Aurelio de Campos Crispin. Centro de Genômica Funcional da ESALQ: Luiz Lehmann Coutinho; Luiz Carlos Junior de Alcantara; Lívia Sacchetto; Maisa C. Pereira Parra; Maria Carolina Elias; Marta Giovanetti; Marília Moraes; Maurício Lacerda Nogueira. Prefeitura de Sao Paulo: Melissa Palmieri.; Patricia Akemi Assato; Paula Rahal; Paulo Inacio da Costa; Rafael dos Santos Bezerra; Raquel de Lello Rocha Campos Cassano. NGS Soluções Genômicas: Pilar Drummond Sampaio Corrêa Mariani. FZEA-USP Pirassununga: Mirele Daiana Poleti; Raul Machado Neto; Ricardo Augusto Brassaloti; Ricardo Haddad; Rodrigo Tocantins Calado. FAMERP-SJRP: Cecília Artico Banho; Sandra Coccuzzo Sampaio; Svetoslav Nanev Slavov; Vagner Fonseca; Vincent Louis Viala |
| EPI_ISL_1795248, | ESF NOVA TANABI II | Instituto Butantan / | Antonio Jorge Martins; Bianca Cechetto Carlos. Mendelics: Bibiana Santos; Claudia Renata dos Santos Barros; David Schlesinger. Hemocentro Ribeirão Preto: Simone Kashima; Debora Botequio Moretti. Centro de Genômica Funcional da ESALQ: Luiz Lehmann Coutinho; Dimas Tadeu Covas; Elaine Cristina Marqueze; Elaine Vieira dos Santos; |

| | | | |
|---|---|---|---|
| EPI_ISL_1795249, EPI_ISL_1795250 | | ESALQ-Piracicaba | Elisangela Chicaroni Mattos; Erika Freitas; Evandra Strazza Rodrigues; Felipe Allan da Silva da Costa; Flavia Aburjaile; Guilherme Targino Valente; Heidge Fukumasu. USP-Botucatu: Rejane Maria Tommasini Grotto; Instituto Butantan: Alexander Roberto Precioso; Jayme A. Souza-Neto; Jessika Cristina Chagas Lesbon; José Salvatore Leister Patané; João Paulo Kitajima; Luiz Carlos Junior de Alcantara; Maria Carolina Elias; Marta Giovanetti; Patricia Akemi Assato; Rafael dos Santos Bezerra; Raquel de Lello Rocha Campos Cassano. NGS Soluções Genômicas: Pilar Drummond Sampaio Corrêa Mariani. FZEA-USP Pirassununga: Mirele Daiana Poleti; Raul Machado Neto; Ricardo Augusto Brassaloti; Ricardo Haddad; Rodrigo Tocantins Calado.; Sandra Coccuzzo Sampaio; Svetoslav Nanev Slavov; Vagner Fonseca; Vincent Louis Viala |
| EPI_ISL_1966853 | ESF ROSA ROSSI MAIA | Instituto Butantan / ESALQ-USP (Piracicaba) | Antonio Jorge Martins; Bianca Cechetto Carlos. Mendelics: Bibiana Santos; Claudia Renata dos Santos Barros; Cíntia Bittar; David Schlesinger. Hemocentro Ribeirão Preto: Simone Kashima; Debora Botequio Moretti; Elaine Cristina Marqueze; Elaine Vieira dos Santos; Elisangela Chicaroni Mattos; Erika Freitas; Evandra Strazza Rodrigues; Felipe Allan da Silva da Costa; Flavia Aburjaile; Fábio Sossai Possebon; Guilherme Campos; Guilherme Targino Valente; Heidge Fukumasu. USP-Botucatu: Rejane Maria Tommasini Grotto; Helena Lage Ferreira; Instituto Butantan: Dimas Tadeu Covas; Jardelina de Souza Todao Bernardino; Jayme A. Souza-Neto; Jessika Cristina Chagas Lesbon; Jorge A. Petroli Marchesi; José Salvatore Leister Patané; João Paulo Kitajima; João Pessoa Araújo Jr.; Leila Sabrina Ullmann; Loyze Paola Oliveira de Lima; Luiz Aurelio de Campos Crispin. Centro de Genômica Funcional da ESALQ: Luiz Lehmann Coutinho; Luiz Carlos Junior de Alcantara; Lívia Sacchetto; Maisa C. Pereira Parra; Maria Carolina Elias; Marta Giovanetti; Marília Moraes; Maurício Lacerda Nogueira. Prefeitura de Sao Paulo: Melissa Palmieri.; Patricia Akemi Assato; Paula Rahal; Paulo Inacio da Costa; Rafael dos Santos Bezerra; Raquel de Lello Rocha Campos Cassano. NGS Soluções Genômicas: Pilar Drummond Sampaio Corrêa Mariani. FZEA-USP Pirassununga: Mirele Daiana Poleti; Raul Machado Neto; Ricardo Augusto Brassaloti; Ricardo Haddad; Rodrigo Tocantins Calado. FAMERP-SJRP: Cecília Artico Banho; Sandra Coccuzzo Sampaio; Svetoslav Nanev Slavov; Vagner Fonseca; Vincent Louis Viala |
| EPI_ISL_1966351 | ESF VILA GLORIA I | Instituto Butantan / Mendelics | Antonio Jorge Martins; Bianca Cechetto Carlos. Mendelics: Bibiana Santos; Claudia Renata dos Santos Barros; Cíntia Bittar; David Schlesinger. Hemocentro Ribeirão Preto: Simone Kashima; Debora Botequio Moretti; Elaine Cristina Marqueze; Elaine Vieira dos Santos; Elisangela Chicaroni Mattos; Erika Freitas; Evandra Strazza Rodrigues; Felipe Allan da Silva da Costa; Flavia Aburjaile; Fábio Sossai Possebon; Guilherme Campos; Guilherme Targino Valente; Heidge Fukumasu. USP-Botucatu: Rejane Maria Tommasini Grotto; Helena Lage Ferreira; Instituto Butantan: Dimas Tadeu Covas; Jardelina de Souza Todao Bernardino; Jayme A. Souza-Neto; Jessika Cristina Chagas Lesbon; Jorge A. Petroli Marchesi; José Salvatore Leister Patané; João Paulo Kitajima; João Pessoa Araújo Jr.; Leila Sabrina Ullmann; Loyze Paola Oliveira de Lima; Luiz Aurelio de Campos Crispin. Centro de Genômica Funcional da ESALQ: Luiz Lehmann Coutinho; Luiz Carlos Junior de Alcantara; Lívia Sacchetto; Maisa C. Pereira Parra; Maria Carolina Elias; Marta Giovanetti; Marília Moraes; Maurício Lacerda Nogueira. Prefeitura de Sao Paulo: Melissa Palmieri.; Patricia Akemi Assato; Paula Rahal; Paulo Inacio da Costa; Rafael dos Santos Bezerra; Raquel de Lello Rocha Campos Cassano. NGS Soluções Genômicas: Pilar Drummond Sampaio Corrêa Mariani. FZEA-USP Pirassununga: Mirele Daiana Poleti; Raul Machado Neto; Ricardo Augusto Brassaloti; Ricardo Haddad; Rodrigo Tocantins Calado. FAMERP-SJRP: Cecília Artico Banho; Sandra Coccuzzo Sampaio; Svetoslav Nanev Slavov; Vagner Fonseca; Vincent Louis Viala |
| EPI_ISL_1966832 | ESTRATEGIA DE SAUDE DA FAMILIA DE ARCO IRIS | Instituto Butantan / ESALQ-USP (Piracicaba) | Antonio Jorge Martins; Bianca Cechetto Carlos. Mendelics: Bibiana Santos; Claudia Renata dos Santos Barros; Cíntia Bittar; David Schlesinger. Hemocentro Ribeirão Preto: Simone Kashima; Debora Botequio Moretti; Elaine Cristina Marqueze; Elaine Vieira dos Santos; Elisangela Chicaroni Mattos; Erika Freitas; Evandra Strazza Rodrigues; Felipe Allan da Silva da Costa; Flavia Aburjaile; Fábio Sossai Possebon; Guilherme Campos; Guilherme Targino Valente; Heidge Fukumasu. USP-Botucatu: Rejane Maria Tommasini Grotto; Helena Lage Ferreira; Instituto Butantan: Dimas Tadeu Covas; Jardelina de Souza Todao Bernardino; Jayme A. Souza-Neto; Jessika Cristina Chagas Lesbon; Jorge A. Petroli Marchesi; José Salvatore Leister Patané; João Paulo Kitajima; João Pessoa Araújo Jr.; Leila Sabrina Ullmann; Loyze Paola Oliveira de Lima; Luiz Aurelio de Campos Crispin. Centro de Genômica Funcional da ESALQ: Luiz Lehmann Coutinho; Luiz Carlos Junior de Alcantara; Lívia Sacchetto; Maisa C. Pereira Parra; Maria Carolina Elias; Marta Giovanetti; Marília Moraes; Maurício Lacerda Nogueira. Prefeitura de Sao Paulo: Melissa Palmieri.; Patricia Akemi Assato; Paula Rahal; Paulo Inacio da Costa; Rafael dos Santos Bezerra; Raquel de Lello Rocha Campos Cassano. NGS Soluções Genômicas: Pilar Drummond Sampaio Corrêa Mariani. FZEA-USP Pirassununga: Mirele Daiana Poleti; Raul Machado Neto; Ricardo Augusto Brassaloti; Ricardo Haddad; Rodrigo Tocantins Calado. FAMERP-SJRP: Cecília Artico Banho; Sandra Coccuzzo Sampaio; Svetoslav Nanev Slavov; Vagner Fonseca; Vincent Louis Viala |
| EPI_ISL_1620639, EPI_ISL_1652237 | Fleury | Instituto Butantan / Mendelics | Alexander Roberto Precioso; Antonio Jorge Martins; Bibiana Santos; Claudia Renata dos Santos Barros; David Schlesinger; Debora Botequio Moretti; Dimas Tadeu Covas; Elaine Cristina Marqueze; Elaine Vieira dos Santos; Erika Freitas; Evandra Strazza Rodrigues; Flavia Aburjaile; José Salvatore Leister Patané; João Paulo Kitajima; Luiz Carlos Junior de Alcantara; Maria Carolina Elias; Marta Giovanetti; Rafael dos Santos Bezerra; Raul Machado Neto; Ricardo Haddad; Rodrigo Tocantins Calado.; Sandra Coccuzzo Sampaio; Simone Kashima; Svetoslav Nanev Slavov; Vagner Fonseca; Vincent Louis Viala |
| EPI_ISL_1239137, EPI_ISL_1239138, EPI_ISL_1240642 | Fundação Ezequiel Dias | Coordenação Geral de Laboratórios de Saúde Pública (CGLAB) | ; Vagner Fonseca et al; Vagner Fonseca et al. |
| EPI_ISL_1966724, EPI_ISL_1966725, EPI_ISL_1966726 | GRUPO TECNICO DE VIGILANCIA SANITARIA E EPIDEMIOLOGICA | Instituto Butantan | Antonio Jorge Martins; Bianca Cechetto Carlos. Mendelics: Bibiana Santos; Claudia Renata dos Santos Barros; Cíntia Bittar; David Schlesinger. Hemocentro Ribeirão Preto: Simone Kashima; Debora Botequio Moretti; Elaine Cristina Marqueze; Elaine Vieira dos Santos; Elisangela Chicaroni Mattos; Erika Freitas; Evandra Strazza Rodrigues; Felipe Allan da Silva da Costa; Flavia Aburjaile; Fábio Sossai Possebon; Guilherme Campos; Guilherme Targino Valente; Heidge Fukumasu. USP-Botucatu: Rejane Maria Tommasini Grotto; Helena Lage Ferreira; Instituto Butantan: Dimas Tadeu Covas; Jardelina de Souza Todao Bernardino; Jayme A. Souza-Neto; Jessika Cristina Chagas Lesbon; Jorge A. Petroli Marchesi; José Salvatore Leister Patané; João Paulo Kitajima; João Pessoa Araújo Jr.; Leila Sabrina Ullmann; Loyze Paola Oliveira de Lima; Luiz Aurelio de Campos Crispin. Centro de Genômica Funcional da ESALQ: Luiz Lehmann Coutinho; Luiz Carlos Junior de Alcantara; Lívia Sacchetto; Maisa C. Pereira Parra; Maria Carolina Elias; Marta Giovanetti; Marília Moraes; Maurício Lacerda Nogueira. Prefeitura de Sao Paulo: Melissa Palmieri.; Patricia Akemi Assato; Paula Rahal; Paulo Inacio da Costa; Rafael dos Santos Bezerra; Raquel de Lello Rocha Campos Cassano. NGS Soluções Genômicas: Pilar Drummond Sampaio Corrêa Mariani. FZEA-USP Pirassununga: Mirele Daiana Poleti; Raul Machado Neto; Ricardo Augusto Brassaloti; Ricardo Haddad; Rodrigo Tocantins Calado. FAMERP-SJRP: Cecília Artico Banho; Sandra Coccuzzo Sampaio; Svetoslav Nanev Slavov; Vagner Fonseca; Vincent Louis Viala |
| EPI_ISL_1966835, EPI_ISL_1966836, EPI_ISL_1966837, EPI_ISL_1966838, EPI_ISL_1966839 | GRUPO TECNICO DE VIGILANCIA SANITARIA E EPIDEMIOLOGICA | Instituto Butantan / ESALQ-USP (Piracicaba) | Antonio Jorge Martins; Bianca Cechetto Carlos. Mendelics: Bibiana Santos; Claudia Renata dos Santos Barros; Cíntia Bittar; David Schlesinger. Hemocentro Ribeirão Preto: Simone Kashima; Debora Botequio Moretti; Elaine Cristina Marqueze; Elaine Vieira dos Santos; Elisangela Chicaroni Mattos; Erika Freitas; Evandra Strazza Rodrigues; Felipe Allan da Silva da Costa; Flavia Aburjaile; Fábio Sossai Possebon; Guilherme Campos; Guilherme Targino Valente; Heidge Fukumasu. USP-Botucatu: Rejane Maria Tommasini Grotto; Helena Lage Ferreira; Instituto Butantan: Dimas Tadeu Covas; Jardelina de Souza Todao Bernardino; Jayme A. Souza-Neto; Jessika Cristina Chagas Lesbon; Jorge A. Petroli Marchesi; José Salvatore Leister Patané; João Paulo Kitajima; João Pessoa Araújo Jr.; Leila Sabrina Ullmann; Loyze Paola Oliveira de Lima; Luiz Aurelio de Campos Crispin. Centro de Genômica Funcional da ESALQ: Luiz Lehmann Coutinho; Luiz Carlos Junior de Alcantara; Lívia Sacchetto; Maisa C. Pereira Parra; Maria Carolina Elias; Marta Giovanetti; Marília Moraes; Maurício Lacerda Nogueira. Prefeitura de Sao Paulo: Melissa Palmieri.; Patricia Akemi Assato; Paula Rahal; Paulo Inacio da Costa; Rafael dos Santos Bezerra; Raquel de Lello Rocha Campos Cassano. NGS Soluções Genômicas: Pilar Drummond Sampaio Corrêa Mariani. FZEA-USP Pirassununga: Mirele Daiana Poleti; Raul Machado Neto; Ricardo Augusto Brassaloti; Ricardo Haddad; Rodrigo Tocantins Calado. FAMERP-SJRP: Cecília Artico Banho; Sandra Coccuzzo Sampaio; Svetoslav Nanev Slavov; Vagner Fonseca; Vincent Louis Viala |
| EPI_ISL_2017282, EPI_ISL_2017283, EPI_ISL_2017286, EPI_ISL_2017294, EPI_ISL_2017324, EPI_ISL_2017325, EPI_ISL_2017326, EPI_ISL_2017327, EPI_ISL_2017328, EPI_ISL_2017329, EPI_ISL_2017330, EPI_ISL_2017331, EPI_ISL_2017332, EPI_ISL_2017333, EPI_ISL_2017334, EPI_ISL_2017335, EPI_ISL_2017336, EPI_ISL_2017337, EPI_ISL_2017338, EPI_ISL_2017339, EPI_ISL_2017340, EPI_ISL_2017341, EPI_ISL_2017342, EPI_ISL_2017343, EPI_ISL_2017344, EPI_ISL_2017345, EPI_ISL_2017346, EPI_ISL_2017347, EPI_ISL_2017348, EPI_ISL_2017349, EPI_ISL_2017350, EPI_ISL_2017351, EPI_ISL_2017352, EPI_ISL_2017353, EPI_ISL_2017354, EPI_ISL_2017355, EPI_ISL_2017356, EPI_ISL_2017357, EPI_ISL_2017358, EPI_ISL_2017359, EPI_ISL_2017360, EPI_ISL_2017361, EPI_ISL_2017362, EPI_ISL_2017363, EPI_ISL_2017364, EPI_ISL_2017365, EPI_ISL_2017366, EPI_ISL_2017367, EPI_ISL_2017368, EPI_ISL_2017369, EPI_ISL_2017370, EPI_ISL_2017371, EPI_ISL_2017372, EPI_ISL_2017373, EPI_ISL_2017374, EPI_ISL_2017375, EPI_ISL_2017376, EPI_ISL_2017377, EPI_ISL_2017378, EPI_ISL_2017379, EPI_ISL_2017380, EPI_ISL_2017381, EPI_ISL_2017382, EPI_ISL_2017383, EPI_ISL_2017384, EPI_ISL_2017385, EPI_ISL_2017386, EPI_ISL_2017387, EPI_ISL_2017388, EPI_ISL_2017389, EPI_ISL_2017390, EPI_ISL_2017391, EPI_ISL_2017392, EPI_ISL_2017393, EPI_ISL_2017394, EPI_ISL_2017395, EPI_ISL_2017396, EPI_ISL_2017397, EPI_ISL_2017398, EPI_ISL_2017399, EPI_ISL_2017400, EPI_ISL_2017401, EPI_ISL_2017402, EPI_ISL_2017403, EPI_ISL_2017404, EPI_ISL_2017405, EPI_ISL_2017406, EPI_ISL_2017407, EPI_ISL_2017408, EPI_ISL_2017409, EPI_ISL_2017410, EPI_ISL_2017411, EPI_ISL_2017412, EPI_ISL_2017413, EPI_ISL_2017414, EPI_ISL_2017415, EPI_ISL_2017416, EPI_ISL_2017417, EPI_ISL_2017418, EPI_ISL_2017419, EPI_ISL_2017420, EPI_ISL_2017421, EPI_ISL_2017422, EPI_ISL_2017423, EPI_ISL_2017424, EPI_ISL_2017425, EPI_ISL_2017426, EPI_ISL_2017427, EPI_ISL_2017428, EPI_ISL_2017429, EPI_ISL_2017430, EPI_ISL_2017431, EPI_ISL_2017432, EPI_ISL_2017433, EPI_ISL_2017434, EPI_ISL_2017435, EPI_ISL_2017436, EPI_ISL_2017437, EPI_ISL_2017438, EPI_ISL_2017439, EPI_ISL_2017440, EPI_ISL_2017441, EPI_ISL_2017442, EPI_ISL_2017443, EPI_ISL_2017444, EPI_ISL_2017445, EPI_ISL_2017446, EPI_ISL_2017448, EPI_ISL_2017451, EPI_ISL_2017452, EPI_ISL_2017453, EPI_ISL_2017454, EPI_ISL_2017455, EPI_ISL_2017456, EPI_ISL_2017462, EPI_ISL_2017463, EPI_ISL_2017464, EPI_ISL_2017471, EPI_ISL_2017472, EPI_ISL_2017473, EPI_ISL_2017474, EPI_ISL_2017475, EPI_ISL_2017476, EPI_ISL_2017477, EPI_ISL_2017480, EPI_ISL_2017482, EPI_ISL_2017483, EPI_ISL_2017484, EPI_ISL_2102518, EPI_ISL_2102519, EPI_ISL_2102520, EPI_ISL_2102521, EPI_ISL_2187826, EPI_ISL_2187827, EPI_ISL_2187828, EPI_ISL_2187829, EPI_ISL_2187830, EPI_ISL_2187831, EPI_ISL_2187832, EPI_ISL_2187833, EPI_ISL_2187834, EPI_ISL_2187835, EPI_ISL_2187836, EPI_ISL_2187837, EPI_ISL_2187838, EPI_ISL_2187839, EPI_ISL_2187840, EPI_ISL_2187841, EPI_ISL_2187842, EPI_ISL_2187843, EPI_ISL_2187844, EPI_ISL_2187845, EPI_ISL_2187846, EPI_ISL_2187847, EPI_ISL_2187848, EPI_ISL_2187849, EPI_ISL_2187850, EPI_ISL_2187851, EPI_ISL_2187852, EPI_ISL_2187853, EPI_ISL_2187854, EPI_ISL_2187855, EPI_ISL_2187856, EPI_ISL_2187857, EPI_ISL_2187858, EPI_ISL_2187859, EPI_ISL_2187860, EPI_ISL_2187861, EPI_ISL_2187862, EPI_ISL_2187863, EPI_ISL_2187864, EPI_ISL_2187865, EPI_ISL_2187866, EPI_ISL_2187867, EPI_ISL_2187868, EPI_ISL_2187869, EPI_ISL_2187870, EPI_ISL_2187871, EPI_ISL_2187872, EPI_ISL_2187873, EPI_ISL_2187874, EPI_ISL_2187875, EPI_ISL_2187876, EPI_ISL_2187878, EPI_ISL_2187879, EPI_ISL_2187880, EPI_ISL_2187881, EPI_ISL_2187882, EPI_ISL_2187883, EPI_ISL_2187884, EPI_ISL_2187885, EPI_ISL_2187886, EPI_ISL_2187887, EPI_ISL_2187888, EPI_ISL_2187889, EPI_ISL_2187890, EPI_ISL_2187891, EPI_ISL_2187892, EPI_ISL_2187893, EPI_ISL_2187894, EPI_ISL_2187895, EPI_ISL_2187896, EPI_ISL_2187897, EPI_ISL_2187898, EPI_ISL_2187899, EPI_ISL_2187900, EPI_ISL_2187901, EPI_ISL_2187902, EPI_ISL_2187903, EPI_ISL_2187904, EPI_ISL_2187905, EPI_ISL_2187906, EPI_ISL_2187907, EPI_ISL_2187908, EPI_ISL_2187909, EPI_ISL_2187910, EPI_ISL_2187911, EPI_ISL_2187912, EPI_ISL_2187913, EPI_ISL_2187914, EPI_ISL_2187915, EPI_ISL_2187916, EPI_ISL_2187917, EPI_ISL_2187918, EPI_ISL_2187919, EPI_ISL_2187920, EPI_ISL_2187921, EPI_ISL_2187922, EPI_ISL_2187923, EPI_ISL_2187924, EPI_ISL_2187925, EPI_ISL_2187926, EPI_ISL_2187927, EPI_ISL_2187928, EPI_ISL_2187929, EPI_ISL_2187930, EPI_ISL_2187931, EPI_ISL_2187932, EPI_ISL_2187933, EPI_ISL_2187934, EPI_ISL_2187935, EPI_ISL_2187936, EPI_ISL_2187937, EPI_ISL_2187938, EPI_ISL_2187939, EPI_ISL_2187940, EPI_ISL_2187941, EPI_ISL_2187942, EPI_ISL_2187943, EPI_ISL_2187944, EPI_ISL_2187945, EPI_ISL_2187946, EPI_ISL_2187947, EPI_ISL_2187948, EPI_ISL_2187949, EPI_ISL_2187950, EPI_ISL_2187951, EPI_ISL_2187952, EPI_ISL_2187953, EPI_ISL_2187954, EPI_ISL_2187955, EPI_ISL_2187956, EPI_ISL_2187957, EPI_ISL_2187958, EPI_ISL_2187959, EPI_ISL_2187960, EPI_ISL_2187961, EPI_ISL_2187962, EPI_ISL_2187963, EPI_ISL_2187964, EPI_ISL_2187965, EPI_ISL_2187966, EPI_ISL_2187967, EPI_ISL_2187968, EPI_ISL_2187969, EPI_ISL_2187970, EPI_ISL_2187971, EPI_ISL_2187972, EPI_ISL_2187973, EPI_ISL_2187974, EPI_ISL_2187975, EPI_ISL_2187976, EPI_ISL_2187977, EPI_ISL_2187978, EPI_ISL_2187979, EPI_ISL_2187980, EPI_ISL_2187981, EPI_ISL_2187982, EPI_ISL_2187983, EPI_ISL_2187984, EPI_ISL_2187985, EPI_ISL_2187986, EPI_ISL_2187987, EPI_ISL_2187988, EPI_ISL_2187990 | | | |
| see above | HLAGYN - Laboratorio de Imunologia de Transplantes de Goias | HLAGYN - Laboratorio de Imunologia de Transplantes de Goias | Alessandro Leonardo Alvares Magalhaes; Daniel Ferreira de Sousa; Danielle de Paiva Rezende; Erika Lopes Rocha Batista; Fernando Antonio Vinhal dos Santos; Frederico Rodrigues Vinhal; Lucas Carlos Gomes Pereira; Paola Cristina Resende Silva; Raphael Bessa Parmigiane; Sabrina Sara Moreira Duarte |
| EPI_ISL_1967092, EPI_ISL_1967093, EPI_ISL_1967094, EPI_ISL_1967095 | HOSP DO SERV PUB ESTFCO MORATO DE OLIVEIRA SAO PAULO | Instituto Butantan / ESALQ-USP (Piracicaba) | Antonio Jorge Martins; Bianca Cechetto Carlos. Mendelics: Bibiana Santos; Claudia Renata dos Santos Barros; Cíntia Bittar; David Schlesinger. Hemocentro Ribeirão Preto: Simone Kashima; Debora Botequio Moretti; Elaine Cristina Marqueze; Elaine Vieira dos Santos; Elisangela Chicaroni Mattos; Erika Freitas; Evandra Strazza Rodrigues; Felipe Allan da Silva da Costa; Flavia Aburjaile; Fábio Sossai Possebon; Guilherme Campos; Guilherme Targino Valente; Heidge Fukumasu. USP-Botucatu: Rejane Maria Tommasini Grotto; Helena Lage Ferreira; Instituto Butantan: Dimas Tadeu Covas; Jardelina de Souza Todao Bernardino; Jayme A. Souza-Neto; Jessika Cristina Chagas Lesbon; Jorge A. Petroli Marchesi; José Salvatore Leister Patané; João Paulo Kitajima; João Pessoa Araújo Jr.; Leila Sabrina Ullmann; Loyze Paola Oliveira de Lima; Luiz Aurelio de Campos Crispin. Centro de Genômica Funcional da ESALQ: Luiz Lehmann Coutinho; Luiz Carlos Junior de Alcantara; Lívia Sacchetto; Maisa C. Pereira Parra; Maria Carolina Elias; Marta Giovanetti; Marília Moraes; Maurício Lacerda Nogueira. Prefeitura de Sao Paulo: Melissa Palmieri.; Patricia Akemi Assato; Paula Rahal; Paulo Inacio da Costa; Rafael dos Santos Bezerra; Raquel de Lello Rocha Campos Cassano. NGS Soluções Genômicas: Pilar Drummond Sampaio Corrêa Mariani. FZEA-USP Pirassununga: Mirele Daiana Poleti; Raul Machado Neto; Ricardo Augusto Brassaloti; Ricardo Haddad; Rodrigo Tocantins Calado. FAMERP-SJRP: Cecília Artico Banho; Sandra Coccuzzo Sampaio; Svetoslav Nanev Slavov; Vagner Fonseca; Vincent Louis Viala |
| EPI_ISL_1967029, EPI_ISL_1967030, EPI_ISL_1967031, EPI_ISL_1967032 | HOSP E MATERNIDADE MUNICIPAL N SRA MONTE SERRAT | Instituto Butantan / ESALQ-USP (Piracicaba) | Antonio Jorge Martins; Bianca Cechetto Carlos. Mendelics: Bibiana Santos; Claudia Renata dos Santos Barros; Cíntia Bittar; David Schlesinger. Hemocentro Ribeirão Preto: Simone Kashima; Debora Botequio Moretti; Elaine Cristina Marqueze; Elaine Vieira dos Santos; Elisangela Chicaroni Mattos; Erika Freitas; Evandra Strazza Rodrigues; Felipe Allan da Silva da Costa; Flavia Aburjaile; Fábio Sossai Possebon; Guilherme Campos; Guilherme Targino Valente; Heidge Fukumasu. USP-Botucatu: Rejane Maria Tommasini Grotto; Helena Lage Ferreira; Instituto Butantan: Dimas Tadeu Covas; Jardelina de Souza Todao Bernardino; Jayme A. Souza-Neto; Jessika Cristina Chagas Lesbon; Jorge A. Petroli Marchesi; José Salvatore Leister Patané; João Paulo Kitajima; João Pessoa Araújo Jr.; Leila Sabrina Ullmann; Loyze Paola Oliveira de Lima; Luiz Aurelio de Campos Crispin. Centro de Genômica Funcional da ESALQ: Luiz Lehmann Coutinho; Luiz Carlos Junior de Alcantara; Lívia Sacchetto; Maisa C. Pereira Parra; Maria Carolina Elias; Marta Giovanetti; Marília Moraes; Maurício Lacerda Nogueira. Prefeitura de Sao Paulo: Melissa Palmieri.; Patricia Akemi Assato; Paula Rahal; Paulo Inacio da Costa; Rafael dos Santos Bezerra; Raquel de Lello Rocha Campos Cassano. NGS Soluções Genômicas: Pilar Drummond Sampaio Corrêa Mariani. FZEA-USP Pirassununga: Mirele Daiana Poleti; Raul Machado Neto; Ricardo Augusto Brassaloti; Ricardo Haddad; Rodrigo Tocantins Calado. FAMERP-SJRP: Cecília Artico Banho; Sandra Coccuzzo Sampaio; Svetoslav Nanev Slavov; Vagner Fonseca; Vincent Louis Viala |
| EPI_ISL_1966401, EPI_ISL_2170934, EPI_ISL_2170937 | HOSP E MATERNIDADE MUNICIPAL N SRA MONTE SERRAT | Instituto Butantan / Mendelics | Antonio Jorge Martins; Bianca Cechetto Carlos. Mendelics: Bibiana Santos; Claudia Renata dos Santos Barros; Cíntia Bittar; David Schlesinger. Hemocentro Ribeirão Preto: Simone Kashima; Debora Botequio Moretti; Elaine Cristina Marqueze; Elaine Vieira dos Santos; Elisangela Chicaroni Mattos; Erika Freitas; Evandra Strazza Rodrigues; Felipe Allan da Silva da Costa; Flavia Aburjaile; Fábio Sossai Possebon; Guilherme Campos; Guilherme Targino Valente; Heidge Fukumasu. USP-Botucatu: Rejane Maria Tommasini Grotto; Helena Lage Ferreira; Instituto Butantan: Dimas Tadeu Covas; Jardelina de Souza Todao Bernardino; Jayme A. Souza-Neto; Jessika Cristina Chagas Lesbon; Jorge A. Petroli Marchesi; José Salvatore Leister Patané; João Paulo Kitajima; João Pessoa Araújo Jr.; Leila Sabrina Ullmann; Loyze Paola Oliveira de Lima; Luiz Aurelio de Campos Crispin. Centro de Genômica Funcional da ESALQ: Luiz Lehmann Coutinho; Luiz Carlos Junior de Alcantara; Lívia Sacchetto; Maisa C. Pereira Parra; Maria Carolina Elias; Marta Giovanetti; Marília Moraes; Maurício Lacerda Nogueira. Prefeitura de Sao Paulo: Melissa Palmieri.; Patricia Akemi Assato; Paula Rahal; Paulo Inacio da Costa; Rafael dos Santos Bezerra; Raquel de Lello Rocha Campos Cassano. NGS Soluções Genômicas: Pilar Drummond Sampaio Corrêa Mariani. FZEA-USP Pirassununga: Mirele Daiana Poleti; Raul Machado Neto; Ricardo Augusto Brassaloti; Ricardo Haddad; Rodrigo Tocantins Calado. FAMERP-SJRP: Cecília Artico Banho; Sandra Coccuzzo Sampaio; Svetoslav Nanev Slavov; Vagner Fonseca; Vincent Louis Viala |
| EPI_ISL_1967068, EPI_ISL_1967069, EPI_ISL_1967070, EPI_ISL_1967071, EPI_ISL_1967072, EPI_ISL_1967073, EPI_ISL_1967074, EPI_ISL_1967091 | | | |
| see above | HOSP MUN CIDADE TIRADENTES CARMEN PRUDENTE | Instituto Butantan / ESALQ-USP (Piracicaba) | Antonio Jorge Martins; Bianca Cechetto Carlos. Mendelics: Bibiana Santos; Claudia Renata dos Santos Barros; Cíntia Bittar; David Schlesinger. Hemocentro Ribeirão Preto: Simone Kashima; Debora Botequio Moretti; Elaine Cristina Marqueze; Elaine Vieira dos Santos; Elisangela Chicaroni Mattos; Erika Freitas; Evandra Strazza Rodrigues; Felipe Allan da Silva da Costa; Flavia Aburjaile; Fábio Sossai Possebon; Guilherme Campos; Guilherme Targino Valente; Heidge Fukumasu. USP-Botucatu: Rejane Maria Tommasini Grotto; Helena Lage Ferreira; Instituto Butantan: Dimas Tadeu Covas; Jardelina de Souza Todao Bernardino; Jayme A. Souza-Neto; Jessika Cristina Chagas Lesbon; Jorge A. Petroli Marchesi; José Salvatore Leister Patané; João Paulo Kitajima; João Pessoa Araújo Jr.; Leila Sabrina Ullmann; Loyze Paola Oliveira de Lima; Luiz Aurelio de Campos Crispin. Centro de Genômica Funcional da ESALQ: Luiz Lehmann Coutinho; Luiz Carlos Junior de Alcantara; Lívia Sacchetto; Maisa C. Pereira Parra; Maria Carolina Elias; Marta Giovanetti; Marília Moraes; Maurício Lacerda Nogueira. Prefeitura de Sao Paulo: Melissa Palmieri.; Patricia Akemi Assato; Paula Rahal; Paulo Inacio da Costa; Rafael dos Santos Bezerra; Raquel de Lello Rocha Campos Cassano. NGS Soluções Genômicas: Pilar Drummond Sampaio Corrêa Mariani. FZEA-USP Pirassununga: Mirele Daiana Poleti; Raul Machado Neto; Ricardo Augusto Brassaloti; Ricardo Haddad; Rodrigo Tocantins Calado. FAMERP-SJRP: Cecília Artico Banho; Sandra Coccuzzo Sampaio; Svetoslav Nanev Slavov; Vagner Fonseca; Vincent Louis Viala |
| EPI_ISL_1966737 | HOSP MUN JOSANIAS CASTANHA BRAGA | Instituto Butantan | Antonio Jorge Martins; Bianca Cechetto Carlos. Mendelics: Bibiana Santos; Claudia Renata dos Santos Barros; Cíntia Bittar; David Schlesinger. Hemocentro Ribeirão Preto: Simone Kashima; Debora Botequio Moretti; Elaine Cristina Marqueze; Elaine Vieira dos Santos; Elisangela Chicaroni Mattos; Erika Freitas; Evandra Strazza Rodrigues; Felipe Allan da Silva da Costa; Flavia Aburjaile; Fábio Sossai Possebon; Guilherme Campos; Guilherme Targino Valente; Heidge Fukumasu. USP-Botucatu: Rejane Maria Tommasini Grotto; Helena Lage Ferreira; Instituto Butantan: Dimas Tadeu Covas; Jardelina de Souza Todao Bernardino; Jayme A. Souza-Neto; Jessika Cristina Chagas Lesbon; Jorge A. Petroli Marchesi; José Salvatore Leister Patané; João Paulo Kitajima; João Pessoa Araújo Jr.; Leila Sabrina Ullmann; Loyze Paola Oliveira de Lima; Luiz Aurelio de Campos Crispin. Centro de Genômica Funcional da ESALQ: Luiz Lehmann Coutinho; Luiz Carlos Junior de Alcantara; Lívia Sacchetto; Maisa C. Pereira Parra; Maria Carolina Elias; Marta Giovanetti; Marília Moraes; Maurício Lacerda Nogueira. Prefeitura de Sao Paulo: Melissa Palmieri.; Patricia Akemi Assato; Paula Rahal; Paulo Inacio da Costa; Rafael dos Santos Bezerra; Raquel de Lello Rocha Campos Cassano. NGS Soluções Genômicas: Pilar Drummond Sampaio Corrêa Mariani. FZEA-USP Pirassununga: Mirele Daiana Poleti; Raul Machado Neto; Ricardo Augusto Brassaloti; Ricardo Haddad; Rodrigo Tocantins Calado. FAMERP-SJRP: Cecília Artico Banho; Sandra Coccuzzo Sampaio; Svetoslav Nanev Slavov; Vagner Fonseca; Vincent Louis Viala |
| EPI_ISL_1967075 | HOSP MUN JOSANIAS CASTANHA BRAGA | Instituto Butantan / ESALQ-USP (Piracicaba) | Antonio Jorge Martins; Bianca Cechetto Carlos. Mendelics: Bibiana Santos; Claudia Renata dos Santos Barros; Cíntia Bittar; David Schlesinger. Hemocentro Ribeirão Preto: Simone Kashima; Debora Botequio Moretti; Elaine Cristina Marqueze; Elaine Vieira dos Santos; Elisangela Chicaroni Mattos; Erika Freitas; Evandra Strazza Rodrigues; Felipe Allan da Silva da Costa; Flavia Aburjaile; Fábio Sossai Possebon; Guilherme Campos; Guilherme Targino Valente; Heidge Fukumasu. USP-Botucatu: Rejane Maria Tommasini Grotto; Helena Lage Ferreira; Instituto Butantan: Dimas Tadeu Covas; Jardelina de Souza Todao Bernardino; Jayme A. Souza-Neto; Jessika Cristina Chagas Lesbon; Jorge A. Petroli Marchesi; José Salvatore Leister Patané; João Paulo Kitajima; João Pessoa Araújo Jr.; Leila Sabrina Ullmann; Loyze Paola Oliveira de Lima; Luiz Aurelio de Campos Crispin. Centro de Genômica Funcional da ESALQ: Luiz Lehmann Coutinho; Luiz Carlos Junior de Alcantara; Lívia Sacchetto; Maisa C. Pereira Parra; Maria Carolina Elias; Marta Giovanetti; Marília Moraes; Maurício Lacerda Nogueira. Prefeitura de Sao Paulo: Melissa Palmieri.; Patricia Akemi Assato; Paula Rahal; Paulo Inacio da Costa; Rafael dos Santos Bezerra; Raquel de Lello Rocha Campos Cassano. NGS Soluções Genômicas: Pilar Drummond Sampaio Corrêa Mariani. FZEA-USP Pirassununga: Mirele Daiana Poleti; Raul Machado Neto; Ricardo Augusto Brassaloti; Ricardo Haddad; Rodrigo Tocantins Calado. FAMERP-SJRP: Cecília Artico Banho; Sandra Coccuzzo Sampaio; Svetoslav Nanev Slavov; Vagner Fonseca; Vincent Louis Viala |
| EPI_ISL_1967067 | HOSP PRONTO SOCORRO ITAQUERA | Instituto Butantan / ESALQ-USP (Piracicaba) | Antonio Jorge Martins; Bianca Cechetto Carlos. Mendelics: Bibiana Santos; Claudia Renata dos Santos Barros; Cíntia Bittar; David Schlesinger. Hemocentro Ribeirão Preto: Simone Kashima; Debora Botequio Moretti; Elaine Cristina Marqueze; Elaine Vieira dos Santos; Elisangela Chicaroni Mattos; Erika Freitas; Evandra Strazza Rodrigues; Felipe Allan da Silva da Costa; Flavia Aburjaile; Fábio Sossai Possebon; Guilherme Campos; Guilherme Targino Valente; Heidge Fukumasu. USP-Botucatu: Rejane Maria Tommasini Grotto; Helena Lage Ferreira; Instituto Butantan: Dimas Tadeu Covas; Jardelina de Souza Todao Bernardino; Jayme A. Souza-Neto; Jessika Cristina Chagas Lesbon; Jorge A. Petroli Marchesi; José Salvatore Leister Patané; João Paulo Kitajima; João Pessoa Araújo Jr.; Leila Sabrina Ullmann; Loyze Paola Oliveira de Lima; Luiz Aurelio de Campos Crispin. Centro de Genômica Funcional da ESALQ: Luiz Lehmann Coutinho; Luiz Carlos Junior de Alcantara; Lívia Sacchetto; Maisa C. Pereira Parra; Maria Carolina Elias; Marta Giovanetti; Marília Moraes; Maurício Lacerda Nogueira. Prefeitura de Sao Paulo: Melissa Palmieri.; Patricia Akemi Assato; Paula Rahal; Paulo Inacio da Costa; Rafael dos Santos Bezerra; Raquel de Lello Rocha Campos Cassano. NGS Soluções Genômicas: Pilar Drummond Sampaio Corrêa Mariani. FZEA-USP Pirassununga: Mirele Daiana Poleti; Raul Machado Neto; Ricardo Augusto Brassaloti; Ricardo Haddad; Rodrigo Tocantins Calado. FAMERP-SJRP: Cecília Artico Banho; Sandra Coccuzzo Sampaio; Svetoslav Nanev Slavov; Vagner Fonseca; Vincent Louis Viala |
| EPI_ISL_1966408 | HOSP STA VIRGINIA | Instituto Butantan / Mendelics | Antonio Jorge Martins; Bianca Cechetto Carlos. Mendelics: Bibiana Santos; Claudia Renata dos Santos Barros; Cíntia Bittar; David Schlesinger. Hemocentro Ribeirão Preto: Simone Kashima; Debora Botequio Moretti; Elaine Cristina Marqueze; Elaine Vieira dos Santos; Elisangela Chicaroni Mattos; Erika Freitas; Evandra Strazza Rodrigues; Felipe Allan da Silva da Costa; Flavia Aburjaile; Fábio Sossai Possebon; Guilherme Campos; Guilherme Targino Valente; Heidge Fukumasu. USP-Botucatu: Rejane Maria Tommasini Grotto; Helena Lage Ferreira; Instituto Butantan: Dimas Tadeu Covas; Jardelina de Souza Todao Bernardino; Jayme A. Souza-Neto; Jessika Cristina Chagas Lesbon; Jorge A. Petroli Marchesi; José Salvatore Leister Patané; João Paulo Kitajima; João Pessoa Araújo Jr.; Leila Sabrina Ullmann; Loyze Paola Oliveira de Lima; Luiz Aurelio de Campos Crispin. Centro de Genômica Funcional da ESALQ: Luiz Lehmann Coutinho; Luiz Carlos Junior de Alcantara; Lívia Sacchetto; Maisa C. Pereira Parra; Maria Carolina Elias; Marta Giovanetti; Marília Moraes; Maurício Lacerda Nogueira. Prefeitura de Sao Paulo: Melissa Palmieri.; Patricia Akemi Assato; Paula Rahal; Paulo Inacio da Costa; Rafael dos Santos Bezerra; Raquel de Lello Rocha Campos Cassano. NGS Soluções Genômicas: Pilar Drummond Sampaio Corrêa Mariani. FZEA-USP Pirassununga: Mirele Daiana Poleti; Raul Machado Neto; Ricardo Augusto Brassaloti; Ricardo Haddad; Rodrigo Tocantins Calado. FAMERP-SJRP: Cecília Artico Banho; Sandra Coccuzzo Sampaio; Svetoslav Nanev Slavov; Vagner Fonseca; Vincent Louis Viala |
| EPI_ISL_1795345 | HOSPITAL DE CAMPANHA COVID 19 MUNICIPIO DE TAUBATE | Instituto Butantan / ESALQ-Piracicaba | Antonio Jorge Martins; Bianca Cechetto Carlos. Mendelics: Bibiana Santos; Claudia Renata dos Santos Barros; David Schlesinger. Hemocentro Ribeirão Preto: Simone Kashima; Debora Botequio Moretti. Centro de Genômica Funcional da ESALQ: Luiz Lehmann Coutinho; Dimas Tadeu Covas; Elaine Cristina Marqueze; Elaine Vieira dos Santos; Elisangela Chicaroni Mattos; Erika Freitas; Evandra Strazza Rodrigues; Felipe Allan da Silva da Costa; Flavia Aburjaile; Guilherme Targino Valente; Heidge Fukumasu. USP-Botucatu: Rejane Maria Tommasini Grotto; Instituto Butantan: Alexander Roberto Precioso; Jayme A. Souza-Neto; Jessika Cristina Chagas Lesbon; José Salvatore Leister Patané; João Paulo Kitajima; Luiz Carlos Junior de Alcantara; Maria Carolina Elias; Marta Giovanetti; Patricia Akemi Assato; Rafael dos Santos Bezerra; Raquel de Lello Rocha Campos Cassano. NGS Soluções Genômicas: Pilar Drummond Sampaio Corrêa Mariani. FZEA-USP Pirassununga: Mirele Daiana Poleti; Raul Machado Neto; Ricardo Augusto Brassaloti; Ricardo Haddad; Rodrigo Tocantins Calado.; Sandra Coccuzzo Sampaio; Svetoslav Nanev Slavov; Vagner Fonseca; Vincent Louis Viala |
| EPI_ISL_1967165, EPI_ISL_1967166, EPI_ISL_1967167, EPI_ISL_1967168, EPI_ISL_1967169, EPI_ISL_1967170, EPI_ISL_1967171, EPI_ISL_1967172, EPI_ISL_1967173, EPI_ISL_1967174, EPI_ISL_1967175, EPI_ISL_1967176, EPI_ISL_1967177, EPI_ISL_1967178, EPI_ISL_1967179, EPI_ISL_1967180, EPI_ISL_1967181, EPI_ISL_1967182, EPI_ISL_1967183, EPI_ISL_1967184, EPI_ISL_1967185, EPI_ISL_1967186, EPI_ISL_1967187, EPI_ISL_1967188, EPI_ISL_1967189, EPI_ISL_1967190, EPI_ISL_1967191 | | | |

| | | | |
|---|---|---|---|
| see above | HOSPITAL DE CAMPANHA COVID 19 MUNICIPIO DE TAUBATE | Instituto Butantan / ESALQ-USP (Piracicaba) | Antonio Jorge Martins; Bianca Cechetto Carlos. Mendelics: Bibiana Santos; Claudia Renata dos Santos Barros; Cíntia Bittar; David Schlesinger. Hemocentro Ribeirão Preto: Simone Kashima; Debora Botequio Moretti; Elaine Cristina Marqueze; Elaine Vieira dos Santos; Elisangela Chicaroni Mattos; Erika Freitas; Evandra Strazza Rodrigues; Felipe Allan da Silva da Costa; Flavia Aburjaile; Fábio Sossai Possebon; Guilherme Campos; Guilherme Targino Valente; Heidge Fukumasu. USP-Botucatu: Rejane Maria Tommasini Grotto; Helena Lage Ferreira; Instituto Butantan: Dimas Tadeu Covas; Jardelina de Souza Todao Bernardino; Jayme A. Souza-Neto; Jessika Cristina Chagas Lesbon; Jorge A. Petroli Marchesi; José Salvatore Leister Patané; João Paulo Kitajima; João Pessoa Araújo Jr.; Leila Sabrina Ullmann; Loyze Paola Oliveira de Lima; Luiz Aurelio de Campos Crispin. Centro de Genômica Funcional da ESALQ: Luiz Lehmann Coutinho; Luiz Carlos Junior de Alcantara; Lívia Sacchetto; Maisa C. Pereira Parra; Maria Carolina Elias; Marta Giovanetti; Marília Moraes; Maurício Lacerda Nogueira. Prefeitura de Sao Paulo: Melissa Palmieri.; Patricia Akemi Assato; Paula Rahal; Paulo Inacio da Costa; Rafael dos Santos Bezerra; Raquel de Lello Rocha Campos Cassano. NGS Soluções Genômicas: Pilar Drummond Sampaio Corrêa Mariani. FZEA-USP Pirassununga: Mirele Daiana Poleti; Raul Machado Neto; Ricardo Augusto Brassaloti; Ricardo Haddad; Rodrigo Tocantins Calado. FAMERP-SJRP: Cecília Artico Banho; Sandra Coccuzzo Sampaio; Svetoslav Nanev Slavov; Vagner Fonseca; Vincent Louis Viala |
| EPI_ISL_1795095, EPI_ISL_1795101, EPI_ISL_1795103, EPI_ISL_1795106 | HOSPITAL DOS FORNCEDORES | Instituto Butantan / ESALQ-Piracicaba | Antonio Jorge Martins; Bianca Cechetto Carlos. Mendelics: Bibiana Santos; Claudia Renata dos Santos Barros; David Schlesinger. Hemocentro Ribeirão Preto: Simone Kashima; Debora Botequio Moretti. Centro de Genômica Funcional da ESALQ: Luiz Lehmann Coutinho; Dimas Tadeu Covas; Elaine Cristina Marqueze; Elaine Vieira dos Santos; Elisangela Chicaroni Mattos; Erika Freitas; Evandra Strazza Rodrigues; Felipe Allan da Silva da Costa; Flavia Aburjaile; Guilherme Targino Valente; Heidge Fukumasu. USP-Botucatu: Rejane Maria Tommasini Grotto; Instituto Butantan: Alexander Roberto Precioso; Jayme A. Souza-Neto; Jessika Cristina Chagas Lesbon; José Salvatore Leister Patané; João Paulo Kitajima; Luiz Carlos Junior de Alcantara; Maria Carolina Elias; Marta Giovanetti; Patricia Akemi Assato; Rafael dos Santos Bezerra; Raquel de Lello Rocha Campos Cassano. NGS Soluções Genômicas: Pilar Drummond Sampaio Corrêa Mariani. FZEA-USP Pirassununga: Mirele Daiana Poleti; Raul Machado Neto; Ricardo Augusto Brassaloti; Ricardo Haddad; Rodrigo Tocantins Calado.; Sandra Coccuzzo Sampaio; Svetoslav Nanev Slavov; Vagner Fonseca; Vincent Louis Viala |
| EPI_ISL_1795344 | HOSPITAL E MATERNIDADE NOSSA SENHORA DA AJUDA | Instituto Butantan / ESALQ-Piracicaba | Antonio Jorge Martins; Bianca Cechetto Carlos. Mendelics: Bibiana Santos; Claudia Renata dos Santos Barros; David Schlesinger. Hemocentro Ribeirão Preto: Simone Kashima; Debora Botequio Moretti. Centro de Genômica Funcional da ESALQ: Luiz Lehmann Coutinho; Dimas Tadeu Covas; Elaine Cristina Marqueze; Elaine Vieira dos Santos; Elisangela Chicaroni Mattos; Erika Freitas; Evandra Strazza Rodrigues; Felipe Allan da Silva da Costa; Flavia Aburjaile; Guilherme Targino Valente; Heidge Fukumasu. USP-Botucatu: Rejane Maria Tommasini Grotto; Instituto Butantan: Alexander Roberto Precioso; Jayme A. Souza-Neto; Jessika Cristina Chagas Lesbon; José Salvatore Leister Patané; João Paulo Kitajima; Luiz Carlos Junior de Alcantara; Maria Carolina Elias; Marta Giovanetti; Patricia Akemi Assato; Rafael dos Santos Bezerra; Raquel de Lello Rocha Campos Cassano. NGS Soluções Genômicas: Pilar Drummond Sampaio Corrêa Mariani. FZEA-USP Pirassununga: Mirele Daiana Poleti; Raul Machado Neto; Ricardo Augusto Brassaloti; Ricardo Haddad; Rodrigo Tocantins Calado.; Sandra Coccuzzo Sampaio; Svetoslav Nanev Slavov; Vagner Fonseca; Vincent Louis Viala |
| EPI_ISL_1445200, EPI_ISL_2170916 | HOSPITAL E MATERNIDADE NOSSA SENHORA DA AJUDA | Instituto Butantan / Mendelics | Antonio Jorge Martins; Bianca Cechetto Carlos. Mendelics: Bibiana Santos; Bibiana Santos; Claudia Renata dos Santos Barros; Cíntia Bittar; David Schlesinger; David Schlesinger. Hemocentro Ribeirão Preto: Simone Kashima; Debora Botequio Moretti; Dimas Tadeu Covas; Elaine Cristina Marqueze; Elaine Vieira dos Santos; Elisangela Chicaroni Mattos; Erika Freitas; Evandra Strazza Rodrigues; Felipe Allan da Silva da Costa; Flavia Aburjaile; Fábio Sossai Possebon; Guilherme Campos; Guilherme Targino Valente; Heidge Fukumasu. USP-Botucatu: Rejane Maria Tommasini Grotto; Helena Lage Ferreira; Instituto Butantan: Dimas Tadeu Covas; Jardelina de Souza Todao Bernardino; Jayme A. Souza-Neto; Jessika Cristina Chagas Lesbon; Jorge A. Petroli Marchesi; José Salvatore Leister Patané; João Paulo Kitajima; João Pessoa Araújo Jr.; Leila Sabrina Ullmann; Loyze Paola Oliveira de Lima; Luiz Aurelio de Campos Crispin. Centro de Genômica Funcional da ESALQ: Luiz Lehmann Coutinho; Luiz Carlos Junior de Alcantara; Lívia Sacchetto; Maisa C. Pereira Parra; Maria Carolina Elias; Marta Giovanetti; Marília Moraes; Maurício Lacerda Nogueira. Prefeitura de Sao Paulo: Melissa Palmieri.; Patricia Akemi Assato; Paula Rahal; Paulo Inacio da Costa; Rafael dos Santos Bezerra; Raquel de Lello Rocha Campos Cassano. NGS Soluções Genômicas: Pilar Drummond Sampaio Corrêa Mariani. FZEA-USP Pirassununga: Mirele Daiana Poleti; Raul Machado Neto; Ricardo Augusto Brassaloti; Ricardo Haddad; Rodrigo Tocantins Calado.; Rodrigo Tocantins Calado. FAMERP-SJRP: Cecília Artico Banho; Sandra Coccuzzo Sampaio; Simone Kashima; Svetoslav Nanev Slavov; Vagner Fonseca; Vincent Louis Viala |
| EPI_ISL_1967299 | HOSPITAL E PRONTO SOCORRO PORTINARI | Instituto Butantan / Mendelics | Antonio Jorge Martins; Bianca Cechetto Carlos. Mendelics: Bibiana Santos; Claudia Renata dos Santos Barros; Cíntia Bittar; David Schlesinger. Hemocentro Ribeirão Preto: Simone Kashima; Debora Botequio Moretti; Elaine Cristina Marqueze; Elaine Vieira dos Santos; Elisangela Chicaroni Mattos; Erika Freitas; Evandra Strazza Rodrigues; Felipe Allan da Silva da Costa; Flavia Aburjaile; Fábio Sossai Possebon; Guilherme Campos; Guilherme Targino Valente; Heidge Fukumasu. USP-Botucatu: Rejane Maria Tommasini Grotto; Helena Lage Ferreira; Instituto Butantan: Dimas Tadeu Covas; Jardelina de Souza Todao Bernardino; Jayme A. Souza-Neto; Jessika Cristina Chagas Lesbon; Jorge A. Petroli Marchesi; José Salvatore Leister Patané; João Paulo Kitajima; João Pessoa Araújo Jr.; Leila Sabrina Ullmann; Loyze Paola Oliveira de Lima; Luiz Aurelio de Campos Crispin. Centro de Genômica Funcional da ESALQ: Luiz Lehmann Coutinho; Luiz Carlos Junior de Alcantara; Lívia Sacchetto; Maisa C. Pereira Parra; Maria Carolina Elias; Marta Giovanetti; Marília Moraes; Maurício Lacerda Nogueira. Prefeitura de Sao Paulo: Melissa Palmieri.; Patricia Akemi Assato; Paula Rahal; Paulo Inacio da Costa; Rafael dos Santos Bezerra; Raquel de Lello Rocha Campos Cassano. NGS Soluções Genômicas: Pilar Drummond Sampaio Corrêa Mariani. FZEA-USP Pirassununga: Mirele Daiana Poleti; Raul Machado Neto; Ricardo Augusto Brassaloti; Ricardo Haddad; Rodrigo Tocantins Calado. FAMERP-SJRP: Cecília Artico Banho; Sandra Coccuzzo Sampaio; Svetoslav Nanev Slavov; Vagner Fonseca; Vincent Louis Viala |
| EPI_ISL_1966738 | HOSPITAL GERAL DE VILA NOVA CACHOEIRINHA SAO PAULO | Instituto Butantan | Antonio Jorge Martins; Bianca Cechetto Carlos. Mendelics: Bibiana Santos; Claudia Renata dos Santos Barros; Cíntia Bittar; David Schlesinger. Hemocentro Ribeirão Preto: Simone Kashima; Debora Botequio Moretti; Elaine Cristina Marqueze; Elaine Vieira dos Santos; Elisangela Chicaroni Mattos; Erika Freitas; Evandra Strazza Rodrigues; Felipe Allan da Silva da Costa; Flavia Aburjaile; Fábio Sossai Possebon; Guilherme Campos; Guilherme Targino Valente; Heidge Fukumasu. USP-Botucatu: Rejane Maria Tommasini Grotto; Helena Lage Ferreira; Instituto Butantan: Dimas Tadeu Covas; Jardelina de Souza Todao Bernardino; Jayme A. Souza-Neto; Jessika Cristina Chagas Lesbon; Jorge A. Petroli Marchesi; José Salvatore Leister Patané; João Paulo Kitajima; João Pessoa Araújo Jr.; Leila Sabrina Ullmann; Loyze Paola Oliveira de Lima; Luiz Aurelio de Campos Crispin. Centro de Genômica Funcional da ESALQ: Luiz Lehmann Coutinho; Luiz Carlos Junior de Alcantara; Lívia Sacchetto; Maisa C. Pereira Parra; Maria Carolina Elias; Marta Giovanetti; Marília Moraes; Maurício Lacerda Nogueira. Prefeitura de Sao Paulo: Melissa Palmieri.; Patricia Akemi Assato; Paula Rahal; Paulo Inacio da Costa; Rafael dos Santos Bezerra; Raquel de Lello Rocha Campos Cassano. NGS Soluções Genômicas: Pilar Drummond Sampaio Corrêa Mariani. FZEA-USP Pirassununga: Mirele Daiana Poleti; Raul Machado Neto; Ricardo Augusto Brassaloti; Ricardo Haddad; Rodrigo Tocantins Calado. FAMERP-SJRP: Cecília Artico Banho; Sandra Coccuzzo Sampaio; Svetoslav Nanev Slavov; Vagner Fonseca; Vincent Louis Viala |
| EPI_ISL_1966406, EPI_ISL_1966409, EPI_ISL_1966410 | HOSPITAL GERAL DE VILA NOVA CACHOEIRINHA SAO PAULO | Instituto Butantan / Mendelics | Antonio Jorge Martins; Bianca Cechetto Carlos. Mendelics: Bibiana Santos; Claudia Renata dos Santos Barros; Cíntia Bittar; David Schlesinger. Hemocentro Ribeirão Preto: Simone Kashima; Debora Botequio Moretti; Elaine Cristina Marqueze; Elaine Vieira dos Santos; Elisangela Chicaroni Mattos; Erika Freitas; Evandra Strazza Rodrigues; Felipe Allan da Silva da Costa; Flavia Aburjaile; Fábio Sossai Possebon; Guilherme Campos; Guilherme Targino Valente; Heidge Fukumasu. USP-Botucatu: Rejane Maria Tommasini Grotto; Helena Lage Ferreira; Instituto Butantan: Dimas Tadeu Covas; Jardelina de Souza Todao Bernardino; Jayme A. Souza-Neto; Jessika Cristina Chagas Lesbon; Jorge A. Petroli Marchesi; José Salvatore Leister Patané; João Paulo Kitajima; João Pessoa Araújo Jr.; Leila Sabrina Ullmann; Loyze Paola Oliveira de Lima; Luiz Aurelio de Campos Crispin. Centro de Genômica Funcional da ESALQ: Luiz Lehmann Coutinho; Luiz Carlos Junior de Alcantara; Lívia Sacchetto; Maisa C. Pereira Parra; Maria Carolina Elias; Marta Giovanetti; Marília Moraes; Maurício Lacerda Nogueira. Prefeitura de Sao Paulo: Melissa Palmieri.; Patricia Akemi Assato; Paula Rahal; Paulo Inacio da Costa; Rafael dos Santos Bezerra; Raquel de Lello Rocha Campos Cassano. NGS Soluções Genômicas: Pilar Drummond Sampaio Corrêa Mariani. FZEA-USP Pirassununga: Mirele Daiana Poleti; Raul Machado Neto; Ricardo Augusto Brassaloti; Ricardo Haddad; Rodrigo Tocantins Calado. FAMERP-SJRP: Cecília Artico Banho; Sandra Coccuzzo Sampaio; Svetoslav Nanev Slavov; Vagner Fonseca; Vincent Louis Viala |
| EPI_ISL_2105675 | HOSPITAL GERAL JESUS TEIXEIRA DA COSTA GUAIANASES SAO PAULO | Instituto Butantan / ESALQ-Piracicaba | Antonio Jorge Martins; Bianca Cechetto Carlos. Mendelics: Bibiana Santos; Claudia Renata dos Santos Barros; Cíntia Bittar; David Schlesinger. Hemocentro Ribeirão Preto: Simone Kashima; Debora Botequio Moretti; Elaine Cristina Marqueze; Elaine Vieira dos Santos; Elisangela Chicaroni Mattos; Erika Freitas; Evandra Strazza Rodrigues; Felipe Allan da Silva da Costa; Flavia Aburjaile; Fábio Sossai Possebon; Guilherme Campos; Guilherme Targino Valente; Heidge Fukumasu. USP-Botucatu: Rejane Maria Tommasini Grotto; Helena Lage Ferreira; Instituto Butantan: Dimas Tadeu Covas; Jardelina de Souza Todao Bernardino; Jayme A. Souza-Neto; Jessika Cristina Chagas Lesbon; Jorge A. Petroli Marchesi; José Salvatore Leister Patané; João Paulo Kitajima; João Pessoa Araújo Jr.; Leila Sabrina Ullmann; Loyze Paola Oliveira de Lima; Luiz Aurelio de Campos Crispin. Centro de Genômica Funcional da ESALQ: Luiz Lehmann Coutinho; Luiz Carlos Junior de Alcantara; Lívia Sacchetto; Maisa C. Pereira Parra; Maria Carolina Elias; Marta Giovanetti; Marília Moraes; Maurício Lacerda Nogueira. Prefeitura de Sao Paulo: Melissa Palmieri.; Patricia Akemi Assato; Paula Rahal; Paulo Inacio da Costa; Rafael dos Santos Bezerra; Raquel de Lello Rocha Campos Cassano. NGS Soluções Genômicas: Pilar Drummond Sampaio Corrêa Mariani. FZEA-USP Pirassununga: Mirele Daiana Poleti; Raul Machado Neto; Ricardo Augusto Brassaloti; Ricardo Haddad; Rodrigo Tocantins Calado. FAMERP-SJRP: Cecília Artico Banho; Sandra Coccuzzo Sampaio; Svetoslav Nanev Slavov; Vagner Fonseca; Vincent Louis Viala |
| EPI_ISL_1967077, EPI_ISL_1967078, EPI_ISL_1967079, EPI_ISL_1967080, EPI_ISL_1967081, EPI_ISL_1967082, EPI_ISL_1967083, EPI_ISL_1967084, EPI_ISL_1967085, EPI_ISL_1967086, EPI_ISL_1967087, EPI_ISL_1967088, EPI_ISL_1967089, EPI_ISL_1967090 | | | |
| see above | HOSPITAL GERAL JESUS TEIXEIRA DA COSTA GUAIANASES SAO PAULO | Instituto Butantan / ESALQ-USP (Piracicaba) | Antonio Jorge Martins; Bianca Cechetto Carlos. Mendelics: Bibiana Santos; Claudia Renata dos Santos Barros; Cíntia Bittar; David Schlesinger. Hemocentro Ribeirão Preto: Simone Kashima; Debora Botequio Moretti; Elaine Cristina Marqueze; Elaine Vieira dos Santos; Elisangela Chicaroni Mattos; Erika Freitas; Evandra Strazza Rodrigues; Felipe Allan da Silva da Costa; Flavia Aburjaile; Fábio Sossai Possebon; Guilherme Campos; Guilherme Targino Valente; Heidge Fukumasu. USP-Botucatu: Rejane Maria Tommasini Grotto; Helena Lage Ferreira; Instituto Butantan: Dimas Tadeu Covas; Jardelina de Souza Todao Bernardino; Jayme A. Souza-Neto; Jessika Cristina Chagas Lesbon; Jorge A. Petroli Marchesi; José Salvatore Leister Patané; João Paulo Kitajima; João Pessoa Araújo Jr.; Leila Sabrina Ullmann; Loyze Paola Oliveira de Lima; Luiz Aurelio de Campos Crispin. Centro de Genômica Funcional da ESALQ: Luiz Lehmann Coutinho; Luiz Carlos Junior de Alcantara; Lívia Sacchetto; Maisa C. Pereira Parra; Maria Carolina Elias; Marta Giovanetti; Marília Moraes; Maurício Lacerda Nogueira. Prefeitura de Sao Paulo: Melissa Palmieri.; Patricia Akemi Assato; Paula Rahal; Paulo Inacio da Costa; Rafael dos Santos Bezerra; Raquel de Lello Rocha Campos Cassano. NGS Soluções Genômicas: Pilar Drummond Sampaio Corrêa Mariani. FZEA-USP Pirassununga: Mirele Daiana Poleti; Raul Machado Neto; Ricardo Augusto Brassaloti; Ricardo Haddad; Rodrigo Tocantins Calado. FAMERP-SJRP: Cecília Artico Banho; Sandra Coccuzzo Sampaio; Svetoslav Nanev Slavov; Vagner Fonseca; Vincent Louis Viala |
| EPI_ISL_1795380 | HOSPITAL MATERNIDADE SAO JOSE ITAPUI | Instituto Butantan / ESALQ-Piracicaba | Antonio Jorge Martins; Bianca Cechetto Carlos. Mendelics: Bibiana Santos; Claudia Renata dos Santos Barros; David Schlesinger. Hemocentro Ribeirão Preto: Simone Kashima; Debora Botequio Moretti. Centro de Genômica Funcional da ESALQ: Luiz Lehmann Coutinho; Dimas Tadeu Covas; Elaine Cristina Marqueze; Elaine Vieira dos Santos; Elisangela Chicaroni Mattos; Erika Freitas; Evandra Strazza Rodrigues; Felipe Allan da Silva da Costa; Flavia Aburjaile; Guilherme Targino Valente; Heidge Fukumasu. USP-Botucatu: Rejane Maria Tommasini Grotto; Instituto Butantan: Alexander Roberto Precioso; Jayme A. Souza-Neto; Jessika Cristina Chagas Lesbon; José Salvatore Leister Patané; João Paulo Kitajima; Luiz Carlos Junior de Alcantara; Maria Carolina Elias; Marta Giovanetti; Patricia Akemi Assato; Rafael dos Santos Bezerra; Raquel de Lello Rocha Campos Cassano. NGS Soluções Genômicas: Pilar Drummond Sampaio Corrêa Mariani. FZEA-USP Pirassununga: Mirele Daiana Poleti; Raul Machado Neto; Ricardo Augusto Brassaloti; Ricardo Haddad; Rodrigo Tocantins Calado.; Sandra Coccuzzo Sampaio; Svetoslav Nanev Slavov; Vagner Fonseca; Vincent Louis Viala |
| EPI_ISL_1795414 | HOSPITAL MUNICIPAL DE IBIUNA IBIUNA SP | Instituto Butantan / ESALQ-Piracicaba | Antonio Jorge Martins; Bianca Cechetto Carlos. Mendelics: Bibiana Santos; Claudia Renata dos Santos Barros; David Schlesinger. Hemocentro Ribeirão Preto: Simone Kashima; Debora Botequio Moretti. Centro de Genômica Funcional da ESALQ: Luiz Lehmann Coutinho; Dimas Tadeu Covas; Elaine Cristina Marqueze; Elaine Vieira dos Santos; Elisangela Chicaroni Mattos; Erika Freitas; Evandra Strazza Rodrigues; Felipe Allan da Silva da Costa; Flavia Aburjaile; Guilherme Targino Valente; Heidge Fukumasu. USP-Botucatu: Rejane Maria Tommasini Grotto; Instituto Butantan: Alexander Roberto Precioso; Jayme A. Souza-Neto; Jessika Cristina Chagas Lesbon; José Salvatore Leister Patané; João Paulo Kitajima; Luiz Carlos Junior de Alcantara; Maria Carolina Elias; Marta Giovanetti; Patricia Akemi Assato; Rafael dos Santos Bezerra; Raquel de Lello Rocha Campos Cassano. NGS Soluções Genômicas: Pilar Drummond Sampaio Corrêa Mariani. FZEA-USP Pirassununga: Mirele Daiana Poleti; Raul Machado Neto; Ricardo Augusto Brassaloti; Ricardo Haddad; Rodrigo Tocantins Calado.; Sandra Coccuzzo Sampaio; Svetoslav Nanev Slavov; Vagner Fonseca; Vincent Louis Viala |
| EPI_ISL_1966913, EPI_ISL_1966914 | HOSPITAL MUNICIPAL DE IBIUNA IBIUNA SP | Instituto Butantan / ESALQ-USP (Piracicaba) | Antonio Jorge Martins; Bianca Cechetto Carlos. Mendelics: Bibiana Santos; Claudia Renata dos Santos Barros; Cíntia Bittar; David Schlesinger. Hemocentro Ribeirão Preto: Simone Kashima; Debora Botequio Moretti; Elaine Cristina Marqueze; Elaine Vieira dos Santos; Elisangela Chicaroni Mattos; Erika Freitas; Evandra Strazza Rodrigues; Felipe Allan da Silva da Costa; Flavia Aburjaile; Fábio Sossai Possebon; Guilherme Campos; Guilherme Targino Valente; Heidge Fukumasu. USP-Botucatu: Rejane Maria Tommasini Grotto; Helena Lage Ferreira; Instituto Butantan: Dimas Tadeu Covas; Jardelina de Souza Todao Bernardino; Jayme A. Souza-Neto; Jessika Cristina Chagas Lesbon; Jorge A. Petroli Marchesi; José Salvatore Leister Patané; João Paulo Kitajima; João Pessoa Araújo Jr.; Leila Sabrina Ullmann; Loyze Paola Oliveira de Lima; Luiz Aurelio de Campos Crispin. Centro de Genômica Funcional da ESALQ: Luiz Lehmann Coutinho; Luiz Carlos Junior de Alcantara; Lívia Sacchetto; Maisa C. Pereira Parra; Maria Carolina Elias; Marta Giovanetti; Marília Moraes; Maurício Lacerda Nogueira. Prefeitura de Sao Paulo: Melissa Palmieri.; Patricia Akemi Assato; Paula Rahal; Paulo Inacio da Costa; Rafael dos Santos Bezerra; Raquel de Lello Rocha Campos Cassano. NGS Soluções Genômicas: Pilar Drummond Sampaio Corrêa Mariani. FZEA-USP Pirassununga: Mirele Daiana Poleti; Raul Machado Neto; Ricardo Augusto Brassaloti; Ricardo Haddad; Rodrigo Tocantins Calado. FAMERP-SJRP: Cecília Artico Banho; Sandra Coccuzzo Sampaio; Svetoslav Nanev Slavov; Vagner Fonseca; Vincent Louis Viala |
| EPI_ISL_1445173, EPI_ISL_1966763 | HOSPITAL MUNICIPAL DE IBIUNA IBIUNA SP | Instituto Butantan / Mendelics | Antonio Jorge Martins; Bianca Cechetto Carlos. Mendelics: Bibiana Santos; Bibiana Santos; Claudia Renata dos Santos Barros; Cíntia Bittar; David Schlesinger; David Schlesinger. Hemocentro Ribeirão Preto: Simone Kashima; Debora Botequio Moretti; Dimas Tadeu Covas; Elaine Cristina Marqueze; Elaine Vieira dos Santos; Elisangela Chicaroni Mattos; Erika Freitas; Evandra Strazza Rodrigues; Felipe Allan da Silva da Costa; Flavia Aburjaile; Fábio Sossai Possebon; Guilherme Campos; Guilherme Targino Valente; Heidge Fukumasu. USP-Botucatu: Rejane Maria Tommasini Grotto; Helena Lage Ferreira; Instituto Butantan: Dimas Tadeu Covas; Jardelina de Souza Todao Bernardino; Jayme A. Souza-Neto; Jessika Cristina Chagas Lesbon; Jorge A. Petroli Marchesi; José Salvatore Leister Patané; João Paulo Kitajima; João Pessoa Araújo Jr.; Leila Sabrina Ullmann; Loyze Paola Oliveira de Lima; Luiz Aurelio de Campos Crispin. Centro de Genômica Funcional da ESALQ: Luiz Lehmann Coutinho; Luiz Carlos Junior de Alcantara; Lívia Sacchetto; Maisa C. Pereira Parra; Maria Carolina Elias; Marta Giovanetti; Marília Moraes; Maurício Lacerda Nogueira. Prefeitura de Sao Paulo: Melissa Palmieri.; Patricia Akemi Assato; Paula Rahal; Paulo Inacio da Costa; Rafael dos Santos Bezerra; Raquel de Lello Rocha Campos Cassano. NGS Soluções Genômicas: Pilar Drummond Sampaio Corrêa Mariani. FZEA-USP Pirassununga: Mirele Daiana Poleti; Raul Machado Neto; Ricardo Augusto Brassaloti; Ricardo Haddad; Rodrigo Tocantins Calado.; Rodrigo Tocantins Calado. FAMERP-SJRP: Cecília Artico Banho; Sandra Coccuzzo Sampaio; Simone Kashima; Svetoslav Nanev Slavov; Vagner Fonseca; Vincent Louis Viala |
| EPI_ISL_1966919, EPI_ISL_1966920, EPI_ISL_1966921, EPI_ISL_1966922 | HOSPITAL MUNICIPAL DE ITABERA | Instituto Butantan / ESALQ-USP (Piracicaba) | Antonio Jorge Martins; Bianca Cechetto Carlos. Mendelics: Bibiana Santos; Claudia Renata dos Santos Barros; Cíntia Bittar; David Schlesinger. Hemocentro Ribeirão Preto: Simone Kashima; Debora Botequio Moretti; Elaine Cristina Marqueze; Elaine Vieira dos Santos; Elisangela Chicaroni Mattos; Erika Freitas; Evandra Strazza Rodrigues; Felipe Allan da Silva da Costa; Flavia Aburjaile; Fábio Sossai Possebon; Guilherme Campos; Guilherme Targino Valente; Heidge Fukumasu. USP-Botucatu: Rejane Maria Tommasini Grotto; Helena Lage Ferreira; Instituto Butantan: Dimas Tadeu Covas; Jardelina de Souza Todao Bernardino; Jayme A. Souza-Neto; Jessika Cristina Chagas Lesbon; Jorge A. Petroli Marchesi; José Salvatore Leister Patané; João Paulo Kitajima; João Pessoa Araújo Jr.; Leila Sabrina Ullmann; Loyze Paola Oliveira de Lima; Luiz Aurelio de Campos Crispin. Centro de Genômica Funcional da ESALQ: Luiz Lehmann Coutinho; Luiz Carlos Junior de Alcantara; Lívia Sacchetto; Maisa C. Pereira Parra; Maria Carolina Elias; Marta Giovanetti; Marília Moraes; Maurício Lacerda Nogueira. Prefeitura de Sao Paulo: Melissa Palmieri.; Patricia Akemi Assato; Paula Rahal; Paulo Inacio da Costa; Rafael dos Santos Bezerra; Raquel de Lello Rocha Campos Cassano. NGS Soluções Genômicas: Pilar Drummond Sampaio Corrêa Mariani. FZEA-USP Pirassununga: Mirele Daiana Poleti; Raul Machado Neto; Ricardo Augusto Brassaloti; Ricardo Haddad; Rodrigo Tocantins Calado. FAMERP-SJRP: Cecília Artico Banho; Sandra Coccuzzo Sampaio; Svetoslav Nanev Slavov; Vagner Fonseca; Vincent Louis Viala |
| EPI_ISL_1966942, EPI_ISL_1966943, EPI_ISL_1966944, EPI_ISL_1966945, EPI_ISL_1966946, EPI_ISL_1966947, EPI_ISL_1966948, EPI_ISL_1966949 | | | |
| see above | HOSPITAL MUNICIPAL DE ITAPIRA | Instituto Butantan / ESALQ-USP (Piracicaba) | Antonio Jorge Martins; Bianca Cechetto Carlos. Mendelics: Bibiana Santos; Claudia Renata dos Santos Barros; Cíntia Bittar; David Schlesinger. Hemocentro Ribeirão Preto: Simone Kashima; Debora Botequio Moretti; Elaine Cristina Marqueze; Elaine Vieira dos Santos; Elisangela Chicaroni Mattos; Erika Freitas; Evandra Strazza Rodrigues; Felipe Allan da Silva da Costa; Flavia Aburjaile; Fábio Sossai Possebon; Guilherme Campos; Guilherme Targino Valente; Heidge Fukumasu. USP-Botucatu: Rejane Maria Tommasini Grotto; Helena Lage Ferreira; Instituto Butantan: Dimas Tadeu Covas; Jardelina de Souza Todao Bernardino; Jayme A. Souza-Neto; Jessika Cristina Chagas Lesbon; Jorge A. Petroli Marchesi; José Salvatore Leister Patané; João Paulo Kitajima; João Pessoa Araújo Jr.; Leila Sabrina Ullmann; Loyze Paola Oliveira de Lima; Luiz Aurelio de Campos Crispin. Centro de Genômica Funcional da ESALQ: Luiz Lehmann Coutinho; Luiz Carlos Junior de Alcantara; Lívia Sacchetto; Maisa C. Pereira Parra; Maria Carolina Elias; Marta Giovanetti; Marília Moraes; Maurício Lacerda Nogueira. Prefeitura de Sao Paulo: Melissa Palmieri.; Patricia Akemi Assato; Paula Rahal; Paulo Inacio da Costa; Rafael dos Santos Bezerra; Raquel de Lello Rocha Campos Cassano. NGS Soluções Genômicas: Pilar Drummond Sampaio Corrêa Mariani. FZEA-USP Pirassununga: Mirele Daiana Poleti; Raul Machado Neto; Ricardo Augusto Brassaloti; Ricardo Haddad; Rodrigo Tocantins Calado. FAMERP-SJRP: Cecília Artico Banho; Sandra Coccuzzo Sampaio; Svetoslav Nanev Slavov; Vagner Fonseca; Vincent Louis Viala |
| EPI_ISL_1795211 | HOSPITAL MUNICIPAL DR MARIO GATTI CAMPINAS | Instituto Butantan / ESALQ-Piracicaba | Antonio Jorge Martins; Bianca Cechetto Carlos. Mendelics: Bibiana Santos; Claudia Renata dos Santos Barros; David Schlesinger. Hemocentro Ribeirão Preto: Simone Kashima; Debora Botequio Moretti. Centro de Genômica Funcional da ESALQ: Luiz Lehmann Coutinho; Dimas Tadeu Covas; Elaine Cristina Marqueze; Elaine Vieira dos Santos; Elisangela Chicaroni Mattos; Erika Freitas; Evandra Strazza Rodrigues; Felipe Allan da Silva da Costa; Flavia Aburjaile; Guilherme Targino Valente; Heidge Fukumasu. USP-Botucatu: Rejane Maria Tommasini Grotto; Instituto Butantan: Alexander Roberto Precioso; Jayme A. Souza-Neto; Jessika Cristina Chagas Lesbon; José Salvatore Leister Patané; João Paulo Kitajima; Luiz Carlos Junior de Alcantara; Maria Carolina Elias; Marta Giovanetti; Patricia Akemi Assato; Rafael dos Santos Bezerra; Raquel de Lello Rocha Campos Cassano. NGS Soluções Genômicas: Pilar Drummond Sampaio Corrêa Mariani. FZEA-USP Pirassununga: Mirele Daiana Poleti; Raul Machado Neto; Ricardo Augusto Brassaloti; Ricardo Haddad; Rodrigo Tocantins Calado.; Sandra Coccuzzo Sampaio; Svetoslav Nanev Slavov; Vagner Fonseca; Vincent Louis Viala |
| EPI_ISL_1966364 | HOSPITAL MUNICIPAL DR MARIO GATTI CAMPINAS | Instituto Butantan / Mendelics | Antonio Jorge Martins; Bianca Cechetto Carlos. Mendelics: Bibiana Santos; Claudia Renata dos Santos Barros; Cíntia Bittar; David Schlesinger. Hemocentro Ribeirão Preto: Simone Kashima; Debora Botequio Moretti; Elaine Cristina Marqueze; Elaine Vieira dos Santos; Elisangela Chicaroni Mattos; Erika Freitas; Evandra Strazza Rodrigues; Felipe Allan da Silva da Costa; Flavia Aburjaile; Fábio Sossai Possebon; Guilherme Campos; Guilherme Targino Valente; Heidge Fukumasu. USP-Botucatu: Rejane Maria Tommasini Grotto; Helena Lage Ferreira; Instituto Butantan: Dimas Tadeu Covas; Jardelina de Souza Todao Bernardino; Jayme A. Souza-Neto; Jessika Cristina Chagas Lesbon; Jorge A. Petroli Marchesi; José Salvatore Leister Patané; João Paulo Kitajima; João Pessoa Araújo Jr.; Leila Sabrina Ullmann; Loyze Paola Oliveira de Lima; Luiz Aurelio de Campos Crispin. Centro de Genômica Funcional da ESALQ: Luiz Lehmann Coutinho; Luiz Carlos Junior de Alcantara; Lívia Sacchetto; Maisa C. Pereira Parra; Maria Carolina Elias; Marta Giovanetti; Marília Moraes; Maurício Lacerda Nogueira. Prefeitura de Sao Paulo: Melissa Palmieri.; Patricia Akemi Assato; Paula Rahal; Paulo Inacio da Costa; Rafael dos Santos Bezerra; Raquel de Lello Rocha Campos Cassano. NGS Soluções Genômicas: Pilar Drummond Sampaio Corrêa Mariani. FZEA-USP Pirassununga: Mirele Daiana Poleti; Raul Machado Neto; Ricardo Augusto Brassaloti; Ricardo Haddad; Rodrigo Tocantins Calado. FAMERP-SJRP: Cecília Artico Banho; Sandra Coccuzzo Sampaio; Svetoslav Nanev Slavov; Vagner Fonseca; Vincent Louis Viala |
| EPI_ISL_1966530 | HOSPITAL MUNICIPAL DR TABAJARA RAMOS | Instituto Butantan / FZEA-USP (Pirassununga) | Antonio Jorge Martins; Bianca Cechetto Carlos. Mendelics: Bibiana Santos; Claudia Renata dos Santos Barros; Cíntia Bittar; David Schlesinger. Hemocentro Ribeirão Preto: Simone Kashima; Debora Botequio Moretti; Elaine Cristina Marqueze; Elaine Vieira dos Santos; Elisangela Chicaroni Mattos; Erika Freitas; Evandra Strazza Rodrigues; Felipe Allan da Silva da Costa; Flavia Aburjaile; Fábio Sossai Possebon; Guilherme Campos; Guilherme Targino Valente; Heidge Fukumasu. USP-Botucatu: Rejane Maria Tommasini Grotto; Helena Lage Ferreira; Instituto Butantan: Dimas Tadeu Covas; Jardelina de Souza Todao Bernardino; Jayme A. Souza-Neto; Jessika Cristina Chagas Lesbon; Jorge A. Petroli Marchesi; José Salvatore Leister Patané; João Paulo Kitajima; João Pessoa Araújo Jr.; Leila Sabrina Ullmann; Loyze Paola Oliveira de Lima; Luiz Aurelio de Campos Crispin. Centro de Genômica Funcional da ESALQ: Luiz Lehmann Coutinho; Luiz Carlos Junior de Alcantara; Lívia Sacchetto; Maisa C. Pereira Parra; Maria Carolina Elias; Marta Giovanetti; Marília Moraes; Maurício Lacerda Nogueira. Prefeitura de Sao Paulo: Melissa Palmieri.; Patricia Akemi Assato; Paula Rahal; Paulo Inacio da Costa; Rafael dos Santos Bezerra; Raquel de Lello Rocha Campos Cassano. NGS Soluções Genômicas: Pilar Drummond Sampaio Corrêa Mariani. FZEA-USP Pirassununga: Mirele Daiana Poleti; Raul Machado Neto; Ricardo Augusto Brassaloti; Ricardo Haddad; Rodrigo Tocantins Calado. FAMERP-SJRP: Cecília Artico Banho; Sandra Coccuzzo Sampaio; Svetoslav Nanev Slavov; Vagner Fonseca; Vincent Louis Viala |
| EPI_ISL_2171017 | HOSPITAL MUNICIPAL DR TABAJARA RAMOS | Instituto Butantan / Mendelics | Antonio Jorge Martins; Bianca Cechetto Carlos. Mendelics: Bibiana Santos; Claudia Renata dos Santos Barros; Cíntia Bittar; David Schlesinger. Hemocentro Ribeirão Preto: Simone Kashima; Debora Botequio Moretti; Elaine Cristina Marqueze; Elaine Vieira dos Santos; Elisangela Chicaroni Mattos; Erika Freitas; Evandra Strazza Rodrigues; Felipe Allan da Silva da Costa; Flavia Aburjaile; Fábio Sossai Possebon; Guilherme Campos; Guilherme Targino Valente; Heidge Fukumasu. USP-Botucatu: Rejane Maria Tommasini Grotto; Helena Lage Ferreira; Instituto Butantan: Dimas Tadeu Covas; Jardelina de Souza Todao Bernardino; Jayme A. Souza-Neto; Jessika Cristina Chagas Lesbon; Jorge A. Petroli Marchesi; José Salvatore Leister Patané; João Paulo Kitajima; João Pessoa Araújo Jr.; Leila Sabrina Ullmann; Loyze Paola Oliveira de Lima; Luiz Aurelio de Campos Crispin. Centro de Genômica Funcional da ESALQ: Luiz Lehmann Coutinho; Luiz Carlos Junior de Alcantara; Lívia Sacchetto; Maisa C. Pereira Parra; Maria Carolina Elias; Marta Giovanetti; Marília Moraes; Maurício Lacerda Nogueira. Prefeitura de Sao Paulo: Melissa Palmieri.; Patricia Akemi Assato; Paula Rahal; Paulo Inacio da Costa; Rafael dos Santos Bezerra; Raquel de Lello Rocha Campos Cassano. NGS Soluções Genômicas: Pilar Drummond Sampaio Corrêa Mariani. FZEA-USP Pirassununga: Mirele Daiana Poleti; Raul Machado Neto; Ricardo Augusto Brassaloti; Ricardo Haddad; Rodrigo Tocantins Calado. FAMERP-SJRP: Cecília Artico Banho; Sandra Coccuzzo Sampaio; Svetoslav Nanev Slavov; Vagner Fonseca; Vincent Louis Viala |
| EPI_ISL_1966644, EPI_ISL_1966645, EPI_ISL_1966646 | HOSPITAL MUNICIPAL DR WALDEMAR TEBALDI | Instituto Butantan / Mendelics | Antonio Jorge Martins; Bianca Cechetto Carlos. Mendelics: Bibiana Santos; Claudia Renata dos Santos Barros; Cíntia Bittar; David Schlesinger; David Schlesinger. Hemocentro Ribeirão Preto: Simone Kashima; Debora Botequio Moretti; Dimas Tadeu Covas; Elaine Cristina Marqueze; Elaine Vieira Santos; Elaine Vieira dos Santos; Elisangela Chicaroni Mattos; Erika Freitas; Evandra Strazza Rodrigues; Felipe Allan da Silva da Costa; Flavia Aburjaile; Fábio Sossai Possebon; Guilherme Campos; Guilherme Targino Valente; Heidge Fukumasu; Heidge Fukumasu. USP-Botucatu: Rejane Maria Tommasini Grotto; Helena Lage Ferreira; Instituto Butantan: Dimas Tadeu Covas; Jardelina de Souza Todao Bernardino; Jayme A. Souza-Neto; Jayme Augusto de Souza-Neto; Jessika Cristina Chagas Lesbon; Jorge A. Petroli Marchesi; José Salvatore Leister Patané; João Paulo Kitajima; João Pessoa Araújo Jr.; Leila Sabrina Ullmann; Loyze Paola Oliveira de Lima; Luiz Alcantara; Luiz Aurelio de Campos Crispin. Centro de Genômica Funcional da ESALQ: Luiz Lehmann Coutinho; Luiz Carlos Junior de Alcantara; Luiz Lehmann Coutinho; Lívia Sacchetto; Maisa C. Pereira Parra; Maria Carolina Elias; Marta Giovanetti; Marília Moraes; Maurício Lacerda Nogueira; Maurício Lacerda Nogueira. Prefeitura de Sao Paulo: Melissa Palmieri.; Patricia Akemi Assato; Paula Rahal; Paulo Inacio da Costa; Rafael dos Santos Bezerra; Raquel de Lello Rocha Campos Cassano. NGS Soluções Genômicas: Pilar Drummond Sampaio Corrêa Mariani. FZEA-USP Pirassununga: Mirele Daiana Poleti; Raul Machado Neto; Rejane Maria Tommasini Grotto; Ricardo Augusto Brassaloti; Ricardo Haddad; Rodrigo Tocantins Calado. FAMERP-SJRP: Cecília Artico Banho; Sandra Coccuzzo Sampaio; Sandra Coccuzzo Sampaio Vessoni; Simone Kashima; Svetoslav Nanev Slavov; Vagner Fonseca; Vincent Louis Viala |

| | | | |
|---|---|---|---|
| EPI_ISL_1967025, EPI_ISL_1967026 | HOSPITAL MUNICIPAL MARIA ROSA CARDOSO | Instituto Butantan / ESALQ-USP (Piracicaba) | Antonio Jorge Martins; Bianca Cechetto Carlos. Mendelics: Bibiana Santos; Claudia Renata dos Santos Barros; Cíntia Bittar; David Schlesinger. Hemocentro Ribeirão Preto: Simone Kashima; Debora Botequio Moretti; Elaine Cristina Marqueze; Elaine Vieira dos Santos; Elisangela Chicaroni Mattos; Erika Freitas; Evandra Strazza Rodrigues; Felipe Allan da Silva da Costa; Flavia Aburjaile; Fábio Sossai Possebon; Guilherme Campos; Guilherme Targino Valente; Heidge Fukumasu. USP-Botucatu: Rejane Maria Tommasini Grotto; Helena Lage Ferreira; Instituto Butantan: Dimas Tadeu Covas; Jardelina de Souza Todao Bernardino; Jayme A. Souza-Neto; Jessika Cristina Chagas Lesbon; Jorge A. Petroli Marchesi; José Salvatore Leister Patané; João Paulo Kitajima; João Pessoa Araújo Jr.; Leila Sabrina Ullmann; Loyze Paola Oliveira de Lima; Luiz Aurelio de Campos Crispin. Centro de Genômica Funcional da ESALQ: Luiz Lehmann Coutinho; Luiz Carlos Junior de Alcantara; Lívia Sacchetto; Maisa C. Pereira Parra; Maria Carolina Elias; Marta Giovanetti; Marília Moraes; Maurício Lacerda Nogueira. Prefeitura de Sao Paulo: Melissa Palmieri.; Patricia Akemi Assato; Paula Rahal; Paulo Inacio da Costa; Rafael dos Santos Bezerra; Raquel de Lello Rocha Campos Cassano. NGS Soluções Genômicas: Pilar Drummond Sampaio Corrêa Mariani. FZEA-USP Pirassununga: Mirele Daiana Poleti; Raul Machado Neto; Ricardo Augusto Brassaloti; Ricardo Haddad; Rodrigo Tocantins Calado. FAMERP-SJRP: Cecília Artico Banho; Sandra Coccuzzo Sampaio; Svetoslav Nanev Slavov; Vagner Fonseca; Vincent Louis Viala |
| EPI_ISL_1795337, EPI_ISL_1795338, EPI_ISL_1795341 | HOSPITAL MUNICIPAL REYNALDO GUERRA CAJATI | Instituto Butantan / ESALQ-Piracicaba | Antonio Jorge Martins; Bianca Cechetto Carlos. Mendelics: Bibiana Santos; Claudia Renata dos Santos Barros; David Schlesinger. Hemocentro Ribeirão Preto: Simone Kashima; Debora Botequio Moretti. Centro de Genômica Funcional da ESALQ: Luiz Lehmann Coutinho; Dimas Tadeu Covas; Elaine Cristina Marqueze; Elaine Vieira dos Santos; Elisangela Chicaroni Mattos; Erika Freitas; Evandra Strazza Rodrigues; Felipe Allan da Silva da Costa; Flavia Aburjaile; Guilherme Targino Valente; Heidge Fukumasu. USP-Botucatu: Rejane Maria Tommasini Grotto; Instituto Butantan: Alexander Roberto Precioso; Jayme A. Souza-Neto; Jessika Cristina Chagas Lesbon; José Salvatore Leister Patané; João Paulo Kitajima; Luiz Carlos Junior de Alcantara; Maria Carolina Elias; Marta Giovanetti; Patricia Akemi Assato; Rafael dos Santos Bezerra; Raquel de Lello Rocha Campos Cassano. NGS Soluções Genômicas: Pilar Drummond Sampaio Corrêa Mariani. FZEA-USP Pirassununga: Mirele Daiana Poleti; Raul Machado Neto; Ricardo Augusto Brassaloti; Ricardo Haddad; Rodrigo Tocantins Calado.; Sandra Coccuzzo Sampaio; Svetoslav Nanev Slavov; Vagner Fonseca; Vincent Louis Viala |
| EPI_ISL_1966846 | HOSPITAL MUNICIPAL REYNALDO GUERRA CAJATI | Instituto Butantan / ESALQ-USP (Piracicaba) | Antonio Jorge Martins; Bianca Cechetto Carlos. Mendelics: Bibiana Santos; Claudia Renata dos Santos Barros; Cíntia Bittar; David Schlesinger. Hemocentro Ribeirão Preto: Simone Kashima; Debora Botequio Moretti; Elaine Cristina Marqueze; Elaine Vieira dos Santos; Elisangela Chicaroni Mattos; Erika Freitas; Evandra Strazza Rodrigues; Felipe Allan da Silva da Costa; Flavia Aburjaile; Fábio Sossai Possebon; Guilherme Campos; Guilherme Targino Valente; Heidge Fukumasu. USP-Botucatu: Rejane Maria Tommasini Grotto; Helena Lage Ferreira; Instituto Butantan: Dimas Tadeu Covas; Jardelina de Souza Todao Bernardino; Jayme A. Souza-Neto; Jessika Cristina Chagas Lesbon; Jorge A. Petroli Marchesi; José Salvatore Leister Patané; João Paulo Kitajima; João Pessoa Araújo Jr.; Leila Sabrina Ullmann; Loyze Paola Oliveira de Lima; Luiz Aurelio de Campos Crispin. Centro de Genômica Funcional da ESALQ: Luiz Lehmann Coutinho; Luiz Carlos Junior de Alcantara; Lívia Sacchetto; Maisa C. Pereira Parra; Maria Carolina Elias; Marta Giovanetti; Marília Moraes; Maurício Lacerda Nogueira. Prefeitura de Sao Paulo: Melissa Palmieri.; Patricia Akemi Assato; Paula Rahal; Paulo Inacio da Costa; Rafael dos Santos Bezerra; Raquel de Lello Rocha Campos Cassano. NGS Soluções Genômicas: Pilar Drummond Sampaio Corrêa Mariani. FZEA-USP Pirassununga: Mirele Daiana Poleti; Raul Machado Neto; Ricardo Augusto Brassaloti; Ricardo Haddad; Rodrigo Tocantins Calado. FAMERP-SJRP: Cecília Artico Banho; Sandra Coccuzzo Sampaio; Svetoslav Nanev Slavov; Vagner Fonseca; Vincent Louis Viala |
| EPI_ISL_1966670, EPI_ISL_1966671, EPI_ISL_1966672, EPI_ISL_1966673 | HOSPITAL MUNICIPAL REYNALDO GUERRA CAJATI | Instituto Butantan / Mendelics | Antonio Jorge Martins; Bianca Cechetto Carlos. Mendelics: Bibiana Santos; Claudia Renata dos Santos Barros; Cíntia Bittar; David Schlesinger. Hemocentro Ribeirão Preto: Simone Kashima; Debora Botequio Moretti; Elaine Cristina Marqueze; Elaine Vieira dos Santos; Elisangela Chicaroni Mattos; Erika Freitas; Evandra Strazza Rodrigues; Felipe Allan da Silva da Costa; Flavia Aburjaile; Fábio Sossai Possebon; Guilherme Campos; Guilherme Targino Valente; Heidge Fukumasu. USP-Botucatu: Rejane Maria Tommasini Grotto; Helena Lage Ferreira; Instituto Butantan: Dimas Tadeu Covas; Jardelina de Souza Todao Bernardino; Jayme A. Souza-Neto; Jessika Cristina Chagas Lesbon; Jorge A. Petroli Marchesi; José Salvatore Leister Patané; João Paulo Kitajima; João Pessoa Araújo Jr.; Leila Sabrina Ullmann; Loyze Paola Oliveira de Lima; Luiz Aurelio de Campos Crispin. Centro de Genômica Funcional da ESALQ: Luiz Lehmann Coutinho; Luiz Carlos Junior de Alcantara; Lívia Sacchetto; Maisa C. Pereira Parra; Maria Carolina Elias; Marta Giovanetti; Marília Moraes; Maurício Lacerda Nogueira. Prefeitura de Sao Paulo: Melissa Palmieri.; Patricia Akemi Assato; Paula Rahal; Paulo Inacio da Costa; Rafael dos Santos Bezerra; Raquel de Lello Rocha Campos Cassano. NGS Soluções Genômicas: Pilar Drummond Sampaio Corrêa Mariani. FZEA-USP Pirassununga: Mirele Daiana Poleti; Raul Machado Neto; Ricardo Augusto Brassaloti; Ricardo Haddad; Rodrigo Tocantins Calado. FAMERP-SJRP: Cecília Artico Banho; Sandra Coccuzzo Sampaio; Svetoslav Nanev Slavov; Vagner Fonseca; Vincent Louis Viala |
| EPI_ISL_1966640, EPI_ISL_1966641, EPI_ISL_1966642, EPI_ISL_1966643 | HOSPITAL MUNICIPAL SANTO ANTONIO | Instituto Butantan / Mendelics | Antonio Jorge Martins; Bianca Cechetto Carlos. Mendelics: Bibiana Santos; Claudia Renata dos Santos Barros; Cíntia Bittar; David Schlesinger. Hemocentro Ribeirão Preto: Simone Kashima; Debora Botequio Moretti; Elaine Cristina Marqueze; Elaine Vieira dos Santos; Elisangela Chicaroni Mattos; Erika Freitas; Evandra Strazza Rodrigues; Felipe Allan da Silva da Costa; Flavia Aburjaile; Fábio Sossai Possebon; Guilherme Campos; Guilherme Targino Valente; Heidge Fukumasu. USP-Botucatu: Rejane Maria Tommasini Grotto; Helena Lage Ferreira; Instituto Butantan: Dimas Tadeu Covas; Jardelina de Souza Todao Bernardino; Jayme A. Souza-Neto; Jessika Cristina Chagas Lesbon; Jorge A. Petroli Marchesi; José Salvatore Leister Patané; João Paulo Kitajima; João Pessoa Araújo Jr.; Leila Sabrina Ullmann; Loyze Paola Oliveira de Lima; Luiz Aurelio de Campos Crispin. Centro de Genômica Funcional da ESALQ: Luiz Lehmann Coutinho; Luiz Carlos Junior de Alcantara; Lívia Sacchetto; Maisa C. Pereira Parra; Maria Carolina Elias; Marta Giovanetti; Marília Moraes; Maurício Lacerda Nogueira. Prefeitura de Sao Paulo: Melissa Palmieri.; Patricia Akemi Assato; Paula Rahal; Paulo Inacio da Costa; Rafael dos Santos Bezerra; Raquel de Lello Rocha Campos Cassano. NGS Soluções Genômicas: Pilar Drummond Sampaio Corrêa Mariani. FZEA-USP Pirassununga: Mirele Daiana Poleti; Raul Machado Neto; Ricardo Augusto Brassaloti; Ricardo Haddad; Rodrigo Tocantins Calado. FAMERP-SJRP: Cecília Artico Banho; Sandra Coccuzzo Sampaio; Svetoslav Nanev Slavov; Vagner Fonseca; Vincent Louis Viala |
| EPI_ISL_1795221, EPI_ISL_1795223 | HOSPITAL REGIONAL DE ITAPETININGA | Instituto Butantan / ESALQ-Piracicaba | Antonio Jorge Martins; Bianca Cechetto Carlos. Mendelics: Bibiana Santos; Claudia Renata dos Santos Barros; David Schlesinger. Hemocentro Ribeirão Preto: Simone Kashima; Debora Botequio Moretti. Centro de Genômica Funcional da ESALQ: Luiz Lehmann Coutinho; Dimas Tadeu Covas; Elaine Cristina Marqueze; Elaine Vieira dos Santos; Elisangela Chicaroni Mattos; Erika Freitas; Evandra Strazza Rodrigues; Felipe Allan da Silva da Costa; Flavia Aburjaile; Guilherme Targino Valente; Heidge Fukumasu. USP-Botucatu: Rejane Maria Tommasini Grotto; Instituto Butantan: Alexander Roberto Precioso; Jayme A. Souza-Neto; Jessika Cristina Chagas Lesbon; José Salvatore Leister Patané; João Paulo Kitajima; Luiz Carlos Junior de Alcantara; Maria Carolina Elias; Marta Giovanetti; Patricia Akemi Assato; Rafael dos Santos Bezerra; Raquel de Lello Rocha Campos Cassano. NGS Soluções Genômicas: Pilar Drummond Sampaio Corrêa Mariani. FZEA-USP Pirassununga: Mirele Daiana Poleti; Raul Machado Neto; Ricardo Augusto Brassaloti; Ricardo Haddad; Rodrigo Tocantins Calado.; Sandra Coccuzzo Sampaio; Svetoslav Nanev Slavov; Vagner Fonseca; Vincent Louis Viala |
| EPI_ISL_1966923, EPI_ISL_1966924, EPI_ISL_1966925, EPI_ISL_1966926 | HOSPITAL REGIONAL DE ITAPETININGA | Instituto Butantan / ESALQ-USP (Piracicaba) | Antonio Jorge Martins; Bianca Cechetto Carlos. Mendelics: Bibiana Santos; Claudia Renata dos Santos Barros; Cíntia Bittar; David Schlesinger. Hemocentro Ribeirão Preto: Simone Kashima; Debora Botequio Moretti; Elaine Cristina Marqueze; Elaine Vieira dos Santos; Elisangela Chicaroni Mattos; Erika Freitas; Evandra Strazza Rodrigues; Felipe Allan da Silva da Costa; Flavia Aburjaile; Fábio Sossai Possebon; Guilherme Campos; Guilherme Targino Valente; Heidge Fukumasu. USP-Botucatu: Rejane Maria Tommasini Grotto; Helena Lage Ferreira; Instituto Butantan: Dimas Tadeu Covas; Jardelina de Souza Todao Bernardino; Jayme A. Souza-Neto; Jessika Cristina Chagas Lesbon; Jorge A. Petroli Marchesi; José Salvatore Leister Patané; João Paulo Kitajima; João Pessoa Araújo Jr.; Leila Sabrina Ullmann; Loyze Paola Oliveira de Lima; Luiz Aurelio de Campos Crispin. Centro de Genômica Funcional da ESALQ: Luiz Lehmann Coutinho; Luiz Carlos Junior de Alcantara; Lívia Sacchetto; Maisa C. Pereira Parra; Maria Carolina Elias; Marta Giovanetti; Marília Moraes; Maurício Lacerda Nogueira. Prefeitura de Sao Paulo: Melissa Palmieri.; Patricia Akemi Assato; Paula Rahal; Paulo Inacio da Costa; Rafael dos Santos Bezerra; Raquel de Lello Rocha Campos Cassano. NGS Soluções Genômicas: Pilar Drummond Sampaio Corrêa Mariani. FZEA-USP Pirassununga: Mirele Daiana Poleti; Raul Machado Neto; Ricardo Augusto Brassaloti; Ricardo Haddad; Rodrigo Tocantins Calado. FAMERP-SJRP: Cecília Artico Banho; Sandra Coccuzzo Sampaio; Svetoslav Nanev Slavov; Vagner Fonseca; Vincent Louis Viala |
| EPI_ISL_1445083, EPI_ISL_1445181, EPI_ISL_1966105, EPI_ISL_1966147, EPI_ISL_1966380, EPI_ISL_1966466, EPI_ISL_1966769 | | | |
| see above | HOSPITAL REGIONAL DE ITAPETININGA | Instituto Butantan / Mendelics | Antonio Jorge Martins; Bianca Cechetto Carlos. Mendelics: Bibiana Santos; Bibiana Santos; Claudia Renata dos Santos Barros; Cíntia Bittar; David Schlesinger; David Schlesinger. Hemocentro Ribeirão Preto: Simone Kashima; Debora Botequio Moretti; Dimas Tadeu Covas; Elaine Cristina Marqueze; Elaine Vieira dos Santos; Elisangela Chicaroni Mattos; Erika Freitas; Evandra Strazza Rodrigues; Felipe Allan da Silva da Costa; Flavia Aburjaile; Fábio Sossai Possebon; Guilherme Campos; Guilherme Targino Valente; Heidge Fukumasu. USP-Botucatu: Rejane Maria Tommasini Grotto; Helena Lage Ferreira; Instituto Butantan: Dimas Tadeu Covas; Jardelina de Souza Todao Bernardino; Jayme A. Souza-Neto; Jessika Cristina Chagas Lesbon; Jorge A. Petroli Marchesi; José Salvatore Leister Patané; João Paulo Kitajima; João Pessoa Araújo Jr.; Leila Sabrina Ullmann; Loyze Paola Oliveira de Lima; Luiz Aurelio de Campos Crispin. Centro de Genômica Funcional da ESALQ: Luiz Lehmann Coutinho; Luiz Carlos Junior de Alcantara; Lívia Sacchetto; Maisa C. Pereira Parra; Maria Carolina Elias; Marta Giovanetti; Marília Moraes; Maurício Lacerda Nogueira. Prefeitura de Sao Paulo: Melissa Palmieri.; Patricia Akemi Assato; Paula Rahal; Paulo Inacio da Costa; Rafael dos Santos Bezerra; Raquel de Lello Rocha Campos Cassano. NGS Soluções Genômicas: Pilar Drummond Sampaio Corrêa Mariani. FZEA-USP Pirassununga: Mirele Daiana Poleti; Raul Machado Neto; Ricardo Augusto Brassaloti; Ricardo Haddad; Rodrigo Tocantins Calado.; Rodrigo Tocantins Calado. FAMERP-SJRP: Cecília Artico Banho; Sandra Coccuzzo Sampaio; Simone Kashima; Svetoslav Nanev Slavov; Vagner Fonseca; Vincent Louis Viala |
| EPI_ISL_1795371, EPI_ISL_1795372 | HOSPITAL SANTA THEREZINHA BROTAS | Instituto Butantan / ESALQ-Piracicaba | Antonio Jorge Martins; Bianca Cechetto Carlos. Mendelics: Bibiana Santos; Claudia Renata dos Santos Barros; David Schlesinger. Hemocentro Ribeirão Preto: Simone Kashima; Debora Botequio Moretti. Centro de Genômica Funcional da ESALQ: Luiz Lehmann Coutinho; Dimas Tadeu Covas; Elaine Cristina Marqueze; Elaine Vieira dos Santos; Elisangela Chicaroni Mattos; Erika Freitas; Evandra Strazza Rodrigues; Felipe Allan da Silva da Costa; Flavia Aburjaile; Guilherme Targino Valente; Heidge Fukumasu. USP-Botucatu: Rejane Maria Tommasini Grotto; Instituto Butantan: Alexander Roberto Precioso; Jayme A. Souza-Neto; Jessika Cristina Chagas Lesbon; José Salvatore Leister Patané; João Paulo Kitajima; Luiz Carlos Junior de Alcantara; Maria Carolina Elias; Marta Giovanetti; Patricia Akemi Assato; Rafael dos Santos Bezerra; Raquel de Lello Rocha Campos Cassano. NGS Soluções Genômicas: Pilar Drummond Sampaio Corrêa Mariani. FZEA-USP Pirassununga: Mirele Daiana Poleti; Raul Machado Neto; Ricardo Augusto Brassaloti; Ricardo Haddad; Rodrigo Tocantins Calado.; Sandra Coccuzzo Sampaio; Svetoslav Nanev Slavov; Vagner Fonseca; Vincent Louis Viala |
| EPI_ISL_1966954 | HOSPITAL SANTO ANTONIO DE JUQUIA JUQUIA | Instituto Butantan / ESALQ-USP (Piracicaba) | Antonio Jorge Martins; Bianca Cechetto Carlos. Mendelics: Bibiana Santos; Claudia Renata dos Santos Barros; Cíntia Bittar; David Schlesinger. Hemocentro Ribeirão Preto: Simone Kashima; Debora Botequio Moretti; Elaine Cristina Marqueze; Elaine Vieira dos Santos; Elisangela Chicaroni Mattos; Erika Freitas; Evandra Strazza Rodrigues; Felipe Allan da Silva da Costa; Flavia Aburjaile; Fábio Sossai Possebon; Guilherme Campos; Guilherme Targino Valente; Heidge Fukumasu. USP-Botucatu: Rejane Maria Tommasini Grotto; Helena Lage Ferreira; Instituto Butantan: Dimas Tadeu Covas; Jardelina de Souza Todao Bernardino; Jayme A. Souza-Neto; Jessika Cristina Chagas Lesbon; Jorge A. Petroli Marchesi; José Salvatore Leister Patané; João Paulo Kitajima; João Pessoa Araújo Jr.; Leila Sabrina Ullmann; Loyze Paola Oliveira de Lima; Luiz Aurelio de Campos Crispin. Centro de Genômica Funcional da ESALQ: Luiz Lehmann Coutinho; Luiz Carlos Junior de Alcantara; Lívia Sacchetto; Maisa C. Pereira Parra; Maria Carolina Elias; Marta Giovanetti; Marília Moraes; Maurício Lacerda Nogueira. Prefeitura de Sao Paulo: Melissa Palmieri.; Patricia Akemi Assato; Paula Rahal; Paulo Inacio da Costa; Rafael dos Santos Bezerra; Raquel de Lello Rocha Campos Cassano. NGS Soluções Genômicas: Pilar Drummond Sampaio Corrêa Mariani. FZEA-USP Pirassununga: Mirele Daiana Poleti; Raul Machado Neto; Ricardo Augusto Brassaloti; Ricardo Haddad; Rodrigo Tocantins Calado. FAMERP-SJRP: Cecília Artico Banho; Sandra Coccuzzo Sampaio; Svetoslav Nanev Slavov; Vagner Fonseca; Vincent Louis Viala |
| EPI_ISL_1966694, EPI_ISL_2170955 | HOSPITAL SANTO ANTONIO DE JUQUIA JUQUIA | Instituto Butantan / Mendelics | Antonio Jorge Martins; Bianca Cechetto Carlos. Mendelics: Bibiana Santos; Claudia Renata dos Santos Barros; Cíntia Bittar; David Schlesinger. Hemocentro Ribeirão Preto: Simone Kashima; Debora Botequio Moretti; Elaine Cristina Marqueze; Elaine Vieira dos Santos; Elisangela Chicaroni Mattos; Erika Freitas; Evandra Strazza Rodrigues; Felipe Allan da Silva da Costa; Flavia Aburjaile; Fábio Sossai Possebon; Guilherme Campos; Guilherme Targino Valente; Heidge Fukumasu. USP-Botucatu: Rejane Maria Tommasini Grotto; Helena Lage Ferreira; Instituto Butantan: Dimas Tadeu Covas; Jardelina de Souza Todao Bernardino; Jayme A. Souza-Neto; Jessika Cristina Chagas Lesbon; Jorge A. Petroli Marchesi; José Salvatore Leister Patané; João Paulo Kitajima; João Pessoa Araújo Jr.; Leila Sabrina Ullmann; Loyze Paola Oliveira de Lima; Luiz Aurelio de Campos Crispin. Centro de Genômica Funcional da ESALQ: Luiz Lehmann Coutinho; Luiz Carlos Junior de Alcantara; Lívia Sacchetto; Maisa C. Pereira Parra; Maria Carolina Elias; Marta Giovanetti; Marília Moraes; Maurício Lacerda Nogueira. Prefeitura de Sao Paulo: Melissa Palmieri.; Patricia Akemi Assato; Paula Rahal; Paulo Inacio da Costa; Rafael dos Santos Bezerra; Raquel de Lello Rocha Campos Cassano. NGS Soluções Genômicas: Pilar Drummond Sampaio Corrêa Mariani. FZEA-USP Pirassununga: Mirele Daiana Poleti; Raul Machado Neto; Ricardo Augusto Brassaloti; Ricardo Haddad; Rodrigo Tocantins Calado. FAMERP-SJRP: Cecília Artico Banho; Sandra Coccuzzo Sampaio; Svetoslav Nanev Slavov; Vagner Fonseca; Vincent Louis Viala |
| EPI_ISL_1967065, EPI_ISL_1967066 | HOSPITAL SAO MIGUEL ARCANJO | Instituto Butantan / ESALQ-USP (Piracicaba) | Antonio Jorge Martins; Bianca Cechetto Carlos. Mendelics: Bibiana Santos; Claudia Renata dos Santos Barros; Cíntia Bittar; David Schlesinger. Hemocentro Ribeirão Preto: Simone Kashima; Debora Botequio Moretti; Elaine Cristina Marqueze; Elaine Vieira dos Santos; Elisangela Chicaroni Mattos; Erika Freitas; Evandra Strazza Rodrigues; Felipe Allan da Silva da Costa; Flavia Aburjaile; Fábio Sossai Possebon; Guilherme Campos; Guilherme Targino Valente; Heidge Fukumasu. USP-Botucatu: Rejane Maria Tommasini Grotto; Helena Lage Ferreira; Instituto Butantan: Dimas Tadeu Covas; Jardelina de Souza Todao Bernardino; Jayme A. Souza-Neto; Jessika Cristina Chagas Lesbon; Jorge A. Petroli Marchesi; José Salvatore Leister Patané; João Paulo Kitajima; João Pessoa Araújo Jr.; Leila Sabrina Ullmann; Loyze Paola Oliveira de Lima; Luiz Aurelio de Campos Crispin. Centro de Genômica Funcional da ESALQ: Luiz Lehmann Coutinho; Luiz Carlos Junior de Alcantara; Lívia Sacchetto; Maisa C. Pereira Parra; Maria Carolina Elias; Marta Giovanetti; Marília Moraes; Maurício Lacerda Nogueira. Prefeitura de Sao Paulo: Melissa Palmieri.; Patricia Akemi Assato; Paula Rahal; Paulo Inacio da Costa; Rafael dos Santos Bezerra; Raquel de Lello Rocha Campos Cassano. NGS Soluções Genômicas: Pilar Drummond Sampaio Corrêa Mariani. FZEA-USP Pirassununga: Mirele Daiana Poleti; Raul Machado Neto; Ricardo Augusto Brassaloti; Ricardo Haddad; Rodrigo Tocantins Calado. FAMERP-SJRP: Cecília Artico Banho; Sandra Coccuzzo Sampaio; Svetoslav Nanev Slavov; Vagner Fonseca; Vincent Louis Viala |
| EPI_ISL_2170983, EPI_ISL_2170984 | HOSPITAL UNIVERSITARIO SAO FRANCISCO BRAGANCA PAULISTA | Instituto Butantan / Mendelics | Antonio Jorge Martins; Bianca Cechetto Carlos. Mendelics: Bibiana Santos; Claudia Renata dos Santos Barros; Cíntia Bittar; David Schlesinger. Hemocentro Ribeirão Preto: Simone Kashima; Debora Botequio Moretti; Elaine Cristina Marqueze; Elaine Vieira dos Santos; Elisangela Chicaroni Mattos; Erika Freitas; Evandra Strazza Rodrigues; Felipe Allan da Silva da Costa; Flavia Aburjaile; Fábio Sossai Possebon; Guilherme Campos; Guilherme Targino Valente; Heidge Fukumasu. USP-Botucatu: Rejane Maria Tommasini Grotto; Helena Lage Ferreira; Instituto Butantan: Dimas Tadeu Covas; Jardelina de Souza Todao Bernardino; Jayme A. Souza-Neto; Jessika Cristina Chagas Lesbon; Jorge A. Petroli Marchesi; José Salvatore Leister Patané; João Paulo Kitajima; João Pessoa Araújo Jr.; Leila Sabrina Ullmann; Loyze Paola Oliveira de Lima; Luiz Aurelio de Campos Crispin. Centro de Genômica Funcional da ESALQ: Luiz Lehmann Coutinho; Luiz Carlos Junior de Alcantara; Lívia Sacchetto; Maisa C. Pereira Parra; Maria Carolina Elias; Marta Giovanetti; Marília Moraes; Maurício Lacerda Nogueira. Prefeitura de Sao Paulo: Melissa Palmieri.; Patricia Akemi Assato; Paula Rahal; Paulo Inacio da Costa; Rafael dos Santos Bezerra; Raquel de Lello Rocha Campos Cassano. NGS Soluções Genômicas: Pilar Drummond Sampaio Corrêa Mariani. FZEA-USP Pirassununga: Mirele Daiana Poleti; Raul Machado Neto; Ricardo Augusto Brassaloti; Ricardo Haddad; Rodrigo Tocantins Calado. FAMERP-SJRP: Cecília Artico Banho; Sandra Coccuzzo Sampaio; Svetoslav Nanev Slavov; Vagner Fonseca; Vincent Louis Viala |
| EPI_ISL_1533726 | Hospital Estadual de Campanha Covid 19 Barradas | Instituto Adolfo Lutz, Interdiciplinary Procedures Center, Strategic Laboratory | Caio Vinicius Dias Lopes; Claudia Regina Gonçalves; Claudio Tavares Sacchi; Erica Valessa Ramos Gomes; Karoline Rodrigues Campos; Leonardo Jose Tadeu de Araujo |
| EPI_ISL_1533709, EPI_ISL_1533710 | Hospital Estadual de Vila Alpina Org Social Seconci Sao Paulo | Instituto Adolfo Lutz, Interdiciplinary Procedures Center, Strategic Laboratory | Caio Vinicius Dias Lopes; Claudia Regina Gonçalves; Claudio Tavares Sacchi; Erica Valessa Ramos Gomes; Karoline Rodrigues Campos; Leonardo Jose Tadeu de Araujo |
| EPI_ISL_1533725 | Hospital Geral de Guarulhos | Instituto Adolfo Lutz, Interdiciplinary Procedures Center, Strategic Laboratory | Caio Vinicius Dias Lopes; Claudia Regina Gonçalves; Claudio Tavares Sacchi; Erica Valessa Ramos Gomes; Karoline Rodrigues Campos; Leonardo Jose Tadeu de Araujo |
| EPI_ISL_1533723 | Hospital Geral de Itaquaquecetuba | Instituto Adolfo Lutz, Interdiciplinary Procedures Center, Strategic Laboratory | Caio Vinicius Dias Lopes; Claudia Regina Gonçalves; Claudio Tavares Sacchi; Erica Valessa Ramos Gomes; Karoline Rodrigues Campos; Leonardo Jose Tadeu de Araujo |
| EPI_ISL_1628344 | Hospital Geral de Pedreira | Instituto Adolfo Lutz, Interdiciplinary Procedures Center, Strategic Laboratory | Caio Vinicius Dias Lopes; Claudia Regina Gonçalves; Claudio Tavares Sacchi; Erica Valessa Ramos Gomes; Karoline Rodrigues Campos; Katia Correa de Oliveira Santos; Leonardo Jose Tadeu de Araujo |
| EPI_ISL_1381069 | Hospital Heliopolis | Instituto Adolfo Lutz, Interdiciplinary Procedures Center, Strategic Laboratory | Caio Vinicius Dias Lopes; Claudia Regina Gonçalves; Claudio Tavares Sacchi; Erica Valessa Ramos Gomes; Karoline Rodrigues Campos |
| EPI_ISL_1381070, EPI_ISL_1381071 | Hospital Municipal Cidade Tiradentes Carmem Prudente | Instituto Adolfo Lutz, Interdiciplinary Procedures Center, Strategic Laboratory | Caio Vinicius Dias Lopes; Claudia Regina Gonçalves; Claudio Tavares Sacchi; Erica Valessa Ramos Gomes; Karoline Rodrigues Campos |
| EPI_ISL_1731577, EPI_ISL_1752640 | Hospital Municipal Dr Mario Gatti Campinas | Instituto Adolfo Lutz, Interdiciplinary Procedures Center, Strategic Laboratory | Caio Vinicius Dias Lopes; Claudia Regina Gonçalves; Claudio Tavares Sacchi; Erica Valessa Ramos Gomes; Karoline Rodrigues Campos; Katia Correa de Oliveira Santos; Leonardo Jose Tadeu de Araujo |
| EPI_ISL_1620638 | Hospital Municipal Dr. Ignacio Proença de Gouvea | Instituto Butantan / Mendelics | Alexander Roberto Precioso; Antonio Jorge Martins; Bibiana Santos; Claudia Renata dos Santos Barros; David Schlesinger; Debora Botequio Moretti; Dimas Tadeu Covas; Elaine Cristina Marqueze; Elaine Vieira dos Santos; Erika Freitas; Evandra Strazza Rodrigues; Flavia Aburjaile; José Salvatore Leister Patané; João Paulo Kitajima; Luiz Carlos Junior de Alcantara; Maria Carolina Elias; Marta Giovanetti; Rafael dos Santos Bezerra; Raul Machado Neto; Ricardo Haddad; Rodrigo Tocantins Calado.; Sandra Coccuzzo Sampaio; Simone Kashima; Svetoslav Nanev Slavov; Vagner Fonseca; Vincent Louis Viala |
| EPI_ISL_1358287 | Hospital Nipo | Instituto Adolfo Lutz, | Caio Vinicius Dias Lopes; Claudia Regina Gonçalves; Claudio Tavares Sacchi; Erica Valessa Ramos Gomes; Karoline Rodrigues Campos |

| | | | |
|---|---|---|---|
| | Brasileiro | Interdiciplinary Procedures Center, Strategic Laboratory | |
| EPI_ISL_1628372 | Hospital Sao Marcos da Sama Morro Agudo | Instituto Adolfo Lutz, Interdiciplinary Procedures Center, Strategic Laboratory | Caio Vinicius Dias Lopes; Claudia Regina Gonçalves; Claudio Tavares Sacchi; Erica Valessa Ramos Gomes; Karoline Rodrigues Campos; Katia Correa de Oliveira Santos; Leonardo Jose Tadeu de Araujo |
| EPI_ISL_1533712, EPI_ISL_1533727 | Hospital Universitario da USP Sao Paulo | Instituto Adolfo Lutz, Interdiciplinary Procedures Center, Strategic Laboratory | Caio Vinicius Dias Lopes; Claudia Regina Gonçalves; Claudio Tavares Sacchi; Erica Valessa Ramos Gomes; Karoline Rodrigues Campos; Leonardo Jose Tadeu de Araujo |
| EPI_ISL_1821210 | Hospital Universitario de Marilia | Instituto Adolfo Lutz, Interdiciplinary Procedures Center, Strategic Laboratory | Caio Vinicius Dias Lopes; Claudia Regina Gonçalves; Claudio Tavares Sacchi; Erica Valessa Ramos Gomes; Karoline Rodrigues Campos; Leonardo Jose Tadeu de Araujo |
| EPI_ISL_1358288, EPI_ISL_1358289, EPI_ISL_1358290 | IAL Regional de Aracatuba | Instituto Adolfo Lutz, Interdiciplinary Procedures Center, Strategic Laboratory | Caio Vinicius Dias Lopes; Claudia Regina Gonçalves; Claudio Tavares Sacchi; Erica Valessa Ramos Gomes; Karoline Rodrigues Campos |
| EPI_ISL_1715146, EPI_ISL_1715147, EPI_ISL_1731578, EPI_ISL_1731579, EPI_ISL_1731580, EPI_ISL_1752641, EPI_ISL_1752642, EPI_ISL_1752643, EPI_ISL_1821214, EPI_ISL_1821215, EPI_ISL_1821216, EPI_ISL_1821217, EPI_ISL_1821218, EPI_ISL_1821219, EPI_ISL_1821220, EPI_ISL_1821221, EPI_ISL_1821222, EPI_ISL_1821223, EPI_ISL_1821224 | | | |
| see above | Instituto Adolfo Lutz - Regional de Aracatuba | Instituto Adolfo Lutz, Interdiciplinary Procedures Center, Strategic Laboratory | Caio Vinicius Dias Lopes; Claudia Regina Gonçalves; Claudio Tavares Sacchi; Erica Valessa Ramos Gomes; Karoline Rodrigues Campos; Katia Correa de Oliveira Santos; Leonardo Jose Tadeu de Araujo |
| EPI_ISL_2003150, EPI_ISL_2003151 | Instituto Adolfo Lutz - Regional de Bauru | Instituto Adolfo Lutz, Interdiciplinary Procedures Center, Strategic Laboratory | Caio Vinicius Dias Lopes; Claudia Regina Gonçalves; Claudio Tavares Sacchi; Erica Valessa Ramos Gomes; Karoline Rodrigues Campos; Leonardo Jose Tadeu de Araujo |
| EPI_ISL_2003162, EPI_ISL_2003163, EPI_ISL_2003164, EPI_ISL_2003165, EPI_ISL_2003166, EPI_ISL_2003167, EPI_ISL_2003168, EPI_ISL_2003169, EPI_ISL_2003170 | | | |
| see above | Instituto Adolfo Lutz - Regional de Campinas | Instituto Adolfo Lutz, Interdiciplinary Procedures Center, Strategic Laboratory | Caio Vinicius Dias Lopes; Claudia Regina Gonçalves; Claudio Tavares Sacchi; Erica Valessa Ramos Gomes; Karoline Rodrigues Campos; Leonardo Jose Tadeu de Araujo |
| EPI_ISL_1821237, EPI_ISL_1821238, EPI_ISL_1821239, EPI_ISL_1821240, EPI_ISL_1821241, EPI_ISL_1821242, EPI_ISL_1821243, EPI_ISL_1821244, EPI_ISL_1821245, EPI_ISL_2003152 | | | |
| see above | Instituto Adolfo Lutz - Regional de Marilia | Instituto Adolfo Lutz, Interdiciplinary Procedures Center, Strategic Laboratory | Caio Vinicius Dias Lopes; Claudia Regina Gonçalves; Claudio Tavares Sacchi; Erica Valessa Ramos Gomes; Karoline Rodrigues Campos; Leonardo Jose Tadeu de Araujo |
| EPI_ISL_1715148, EPI_ISL_1731581, EPI_ISL_1731582, EPI_ISL_1752644, EPI_ISL_1752645, EPI_ISL_1821246, EPI_ISL_1821247, EPI_ISL_1821248, EPI_ISL_1821249, EPI_ISL_1821250, EPI_ISL_1821251, EPI_ISL_1821252, EPI_ISL_1821253, EPI_ISL_1821254, EPI_ISL_1821255, EPI_ISL_1821256, EPI_ISL_1821257, EPI_ISL_1821258, EPI_ISL_1821259, EPI_ISL_1821260, EPI_ISL_1821261, EPI_ISL_1821262, EPI_ISL_1821263, EPI_ISL_1821264, EPI_ISL_1821265 | | | |
| see above | Instituto Adolfo Lutz - Regional de Ribeirao Preto | Instituto Adolfo Lutz, Interdiciplinary Procedures Center, Strategic Laboratory | Caio Vinicius Dias Lopes; Claudia Regina Gonçalves; Claudio Tavares Sacchi; Erica Valessa Ramos Gomes; Karoline Rodrigues Campos; Katia Correa de Oliveira Santos; Leonardo Jose Tadeu de Araujo |
| EPI_ISL_2003134, EPI_ISL_2003135, EPI_ISL_2003136, EPI_ISL_2003137, EPI_ISL_2003138, EPI_ISL_2003139, EPI_ISL_2003140, EPI_ISL_2003141, EPI_ISL_2003142, EPI_ISL_2003143, EPI_ISL_2003144, EPI_ISL_2003145, EPI_ISL_2003146, EPI_ISL_2003147, EPI_ISL_2003148, EPI_ISL_2003149 | | | |
| see above | Instituto Adolfo Lutz - Regional de Santos | Instituto Adolfo Lutz, Interdiciplinary Procedures Center, Strategic Laboratory | Caio Vinicius Dias Lopes; Claudia Regina Gonçalves; Claudio Tavares Sacchi; Erica Valessa Ramos Gomes; Karoline Rodrigues Campos; Leonardo Jose Tadeu de Araujo |
| EPI_ISL_1707689, EPI_ISL_1707690, EPI_ISL_1707691, EPI_ISL_1707692 | Instituto Adolfo Lutz - Regional de Taubate | Instituto Adolfo Lutz, Interdiciplinary Procedures Center, Strategic Laboratory | Caio Vinicius Dias Lopes; Claudia Regina Gonçalves; Claudio Tavares Sacchi; Erica Valessa Ramos Gomes; Karoline Rodrigues Campos; Katia Correa de Oliveira Santos; Leonardo Jose Tadeu de Araujo |
| EPI_ISL_1628347, EPI_ISL_1628348, EPI_ISL_1628349, EPI_ISL_1628350, EPI_ISL_1628351, EPI_ISL_1628352, EPI_ISL_1628353, EPI_ISL_1628354, EPI_ISL_1628355, EPI_ISL_1628356, EPI_ISL_1628357, EPI_ISL_1628358, EPI_ISL_1628359, EPI_ISL_1628360, EPI_ISL_1628361, EPI_ISL_1628362, EPI_ISL_1715149, EPI_ISL_1715150, EPI_ISL_1715151, EPI_ISL_1715152, EPI_ISL_1715153, EPI_ISL_1715154, EPI_ISL_1715155, EPI_ISL_1715156, EPI_ISL_1715157, EPI_ISL_1715158, EPI_ISL_1715159, EPI_ISL_1731583, EPI_ISL_1731584, EPI_ISL_1731585, EPI_ISL_1731586, EPI_ISL_1731587, EPI_ISL_1731588, EPI_ISL_1731589, EPI_ISL_1731590, EPI_ISL_1731591, EPI_ISL_1731592, EPI_ISL_1731593, EPI_ISL_1731594, EPI_ISL_1731595, EPI_ISL_1731596, EPI_ISL_1731597, EPI_ISL_1731598, EPI_ISL_1731599, EPI_ISL_1731600, EPI_ISL_1731601, EPI_ISL_1731602, EPI_ISL_1731603, EPI_ISL_1731604, EPI_ISL_1731605, EPI_ISL_1731606, EPI_ISL_1731607, EPI_ISL_1731608, EPI_ISL_1731609, EPI_ISL_1731610, EPI_ISL_1731611, EPI_ISL_1731612, EPI_ISL_1752646, EPI_ISL_1752647, EPI_ISL_1752648, EPI_ISL_1752649, EPI_ISL_1752650, EPI_ISL_1752651, EPI_ISL_1752652, EPI_ISL_1752653, EPI_ISL_1752654, EPI_ISL_1752655, EPI_ISL_1752656, EPI_ISL_1752657, EPI_ISL_1752658, EPI_ISL_1752659, EPI_ISL_1752660, EPI_ISL_1752661, EPI_ISL_1752662, EPI_ISL_1752663, EPI_ISL_1752664, EPI_ISL_1752665, EPI_ISL_1752666, EPI_ISL_1752667, EPI_ISL_1752668, EPI_ISL_1821266, EPI_ISL_1821267, EPI_ISL_1821268, EPI_ISL_1821269, EPI_ISL_1821270, EPI_ISL_1821271, EPI_ISL_1821272, EPI_ISL_1821273, EPI_ISL_2003114, EPI_ISL_2003115, EPI_ISL_2003116, EPI_ISL_2003117, EPI_ISL_2003118, EPI_ISL_2003119, EPI_ISL_2003120, EPI_ISL_2003121, EPI_ISL_2003122, EPI_ISL_2003123, EPI_ISL_2003124, EPI_ISL_2003125, EPI_ISL_2003126, EPI_ISL_2003127, EPI_ISL_2003128, EPI_ISL_2003129, EPI_ISL_2003130, EPI_ISL_2003131, EPI_ISL_2003132, EPI_ISL_2003133 | | | |
| see above | Instituto Adolfo Lutz Central | Instituto Adolfo Lutz, Interdiciplinary Procedures Center, Strategic Laboratory | Caio Vinicius Dias Lopes; Claudia Regina Gonçalves; Claudio Tavares Sacchi; Erica Valessa Ramos Gomes; Karoline Rodrigues Campos; Katia Correa de Oliveira Santos; Leonardo Jose Tadeu de Araujo |
| EPI_ISL_1734841, EPI_ISL_1734843, EPI_ISL_1734844, EPI_ISL_1734846, EPI_ISL_1734847, EPI_ISL_1734849, EPI_ISL_1734850, EPI_ISL_1734852, EPI_ISL_1734853, EPI_ISL_1734855, EPI_ISL_1734857, EPI_ISL_1734858, EPI_ISL_1734860, EPI_ISL_1734861, EPI_ISL_1734863, EPI_ISL_1734864, EPI_ISL_1734866, EPI_ISL_1734868, EPI_ISL_1734869, EPI_ISL_1734871, EPI_ISL_1734872, EPI_ISL_1734874, EPI_ISL_1734875, EPI_ISL_1734877, EPI_ISL_1734878, EPI_ISL_1734880, EPI_ISL_1734881, EPI_ISL_1734883, EPI_ISL_1734884, EPI_ISL_1911958, EPI_ISL_1911959, EPI_ISL_1911960, EPI_ISL_1911961, EPI_ISL_1911962, EPI_ISL_1911963, EPI_ISL_1911964 | | | |
| see above | Instituto de Biotecnologia - UNESP-Botucatu-SP | Instituto de Biotecnologia - UNESP-Botucatu-SP | Cecília Artico Banho; Cíntia Bittar; Fábio Sossai Possebon; Guilherme Campos; Helena Lage Ferreira; Jorge A. Petroli Marchesi; João Pessoa Araújo Jr.; Leila Sabrina Ullmann; Lívia Sacchetto; Maisa C. Pereira Parra; Marília Moraes; Maurício L. Nogueira; Paula Rahal; Paulo Inacio da Costa |
| EPI_ISL_2086610, EPI_ISL_2086611, EPI_ISL_2086612, EPI_ISL_2086613, EPI_ISL_2086614, EPI_ISL_2086615, EPI_ISL_2086616, EPI_ISL_2086617, EPI_ISL_2086618, EPI_ISL_2086619, EPI_ISL_2086620, EPI_ISL_2086621, EPI_ISL_2086622, EPI_ISL_2086765, EPI_ISL_2086766, EPI_ISL_2086767 | | | |
| see above | LABCOVID_HCPA | LABRESIS_HCPA | Barth AL; Martins AF; Monteiro F; Volpato F; Wink PL; Zavascki AP |
| EPI_ISL_1795066, EPI_ISL_1795067, EPI_ISL_1795068, EPI_ISL_1795069, EPI_ISL_1795070, EPI_ISL_1795071, EPI_ISL_1795078, EPI_ISL_1795303, EPI_ISL_1795306, EPI_ISL_1795307, EPI_ISL_1795308, EPI_ISL_1795309, EPI_ISL_1795310, EPI_ISL_1795311, EPI_ISL_1795312, EPI_ISL_1795313, EPI_ISL_1795314, EPI_ISL_1795315, EPI_ISL_1795316, EPI_ISL_1795317, EPI_ISL_1795318, EPI_ISL_1795319, EPI_ISL_1795320, EPI_ISL_1795321, EPI_ISL_1795322, EPI_ISL_1795323, EPI_ISL_1795393, EPI_ISL_1795394, EPI_ISL_1795418, EPI_ISL_1795419, EPI_ISL_1795420, EPI_ISL_1795421, EPI_ISL_1795422, EPI_ISL_1795423 | | | |
| see above | LABORATORIO DE FRANCA | Instituto Butantan / ESALQ-Piracicaba | Antonio Jorge Martins; Bianca Cechetto Carlos. Mendelics: Bibiana Santos; Claudia Renata dos Santos Barros; David Schlesinger. Hemocentro Ribeirão Preto: Simone Kashima; Debora Botequio Moretti. Centro de Genômica Funcional da ESALQ: Luiz Lehmann Coutinho; Dimas Tadeu Covas; Elaine Cristina Marqueze; Elaine Vieira dos Santos; Elisangela Chicaroni Mattos; Erika Freitas; Evandra Strazza Rodrigues; Felipe Allan da Silva da Costa; Flavia Aburjaile; Guilherme Targino Valente; Heidge Fukumasu. USP-Botucatu: Rejane Maria Tommasini Grotto; Instituto Butantan: Alexander Roberto Precioso; Jayme A. Souza-Neto; Jessika Cristina Chagas Lesbon; José Salvatore Leister Patané; João Paulo Kitajima; Luiz Carlos Junior de Alcantara; Maria Carolina Elias; Marta Giovanetti; Patricia Akemi Assato; Rafael dos Santos Bezerra; Raquel de Lello Rocha Campos Cassano. NGS Soluções Genômicas: Pilar Drummond Sampaio Corrêa Mariani. FZEA-USP Pirassununga: Mirele Daiana Poleti; Raul Machado Neto; Ricardo Augusto Brassaloti; Ricardo Haddad; Rodrigo Tocantins Calado.; Sandra Coccuzzo Sampaio; Svetoslav Nanev Slavov; Vagner Fonseca; Vincent Louis Viala |
| EPI_ISL_1966905, EPI_ISL_1966906 | LABORATORIO DE FRANCA | Instituto Butantan / ESALQ-USP (Piracicaba) | Antonio Jorge Martins; Bianca Cechetto Carlos. Mendelics: Bibiana Santos; Claudia Renata dos Santos Barros; Cíntia Bittar; David Schlesinger. Hemocentro Ribeirão Preto: Simone Kashima; Debora Botequio Moretti; Elaine Cristina Marqueze; Elaine Vieira dos Santos; Elisangela Chicaroni Mattos; Erika Freitas; Evandra Strazza Rodrigues; Felipe Allan da Silva da Costa; Flavia Aburjaile; Fábio Sossai Possebon; Guilherme Campos; Guilherme Targino Valente; Heidge Fukumasu. USP-Botucatu: Rejane Maria Tommasini Grotto; Helena Lage Ferreira; Instituto Butantan: Dimas Tadeu Covas; Jardelina de Souza Todao Bernardino; Jayme A. Souza-Neto; Jessika Cristina Chagas Lesbon; Jorge A. Petroli Marchesi; José Salvatore Leister Patané; João Paulo Kitajima; João Pessoa Araújo Jr.; Leila Sabrina Ullmann; Loyze Paola Oliveira de Lima; Luiz Aurelio de Campos Crispin. Centro de Genômica Funcional da ESALQ: Luiz Lehmann Coutinho; Luiz Carlos Junior de Alcantara; Lívia Sacchetto; Maisa C. Pereira Parra; Maria Carolina Elias; Marta Giovanetti; Marília Moraes; Maurício Lacerda Nogueira. Prefeitura de Sao Paulo: Melissa Palmieri.; Patricia Akemi Assato; Paula Rahal; Paulo Inacio da Costa; Rafael dos Santos Bezerra; Raquel de Lello Rocha Campos Cassano. NGS Soluções Genômicas: Pilar Drummond Sampaio Corrêa Mariani. FZEA-USP Pirassununga: Mirele Daiana Poleti; Raul Machado Neto; Ricardo Augusto Brassaloti; Ricardo Haddad; Rodrigo Tocantins Calado. FAMERP-SJRP: Cecília Artico Banho; Sandra Coccuzzo Sampaio; Svetoslav Nanev Slavov; Vagner Fonseca; Vincent Louis Viala |
| EPI_ISL_1445155, EPI_ISL_1966675, EPI_ISL_1966676, EPI_ISL_1966677, EPI_ISL_1966678, EPI_ISL_1966679, EPI_ISL_1966680, EPI_ISL_1966756, EPI_ISL_1966757, EPI_ISL_1966758, EPI_ISL_1966759, EPI_ISL_1966760, EPI_ISL_2170908, EPI_ISL_2170954 | | | |
| see above | LABORATORIO DE FRANCA | Instituto Butantan / Mendelics | Antonio Jorge Martins; Bianca Cechetto Carlos. Mendelics: Bibiana Santos; Bibiana Santos; Claudia Renata dos Santos Barros; Cíntia Bittar; David Schlesinger; David Schlesinger. Hemocentro Ribeirão Preto: Simone Kashima; Debora Botequio Moretti; Dimas Tadeu Covas; Elaine Cristina Marqueze; Elaine Vieira dos Santos; Elisangela Chicaroni Mattos; Erika Freitas; Evandra Strazza Rodrigues; Felipe Allan da Silva da Costa; Flavia Aburjaile; Fábio Sossai Possebon; Guilherme Campos; Guilherme Targino Valente; Heidge Fukumasu. USP-Botucatu: Rejane Maria Tommasini Grotto; Helena Lage Ferreira; Instituto Butantan: Dimas Tadeu Covas; Jardelina de Souza Todao Bernardino; Jayme A. Souza-Neto; Jessika Cristina Chagas Lesbon; Jorge A. Petroli Marchesi; José Salvatore Leister Patané; João Paulo Kitajima; João Pessoa Araújo Jr.; Leila Sabrina Ullmann; Loyze Paola Oliveira de Lima; Luiz Aurelio de Campos Crispin. Centro de Genômica Funcional da ESALQ: Luiz Lehmann Coutinho; Luiz Carlos Junior de Alcantara; Lívia Sacchetto; Maisa C. Pereira Parra; Maria Carolina Elias; Marta Giovanetti; Marília Moraes; Maurício Lacerda Nogueira. Prefeitura de Sao Paulo: Melissa Palmieri.; Patricia Akemi Assato; Paula Rahal; Paulo Inacio da Costa; Rafael dos Santos Bezerra; Raquel de Lello Rocha Campos Cassano. NGS Soluções Genômicas: Pilar Drummond Sampaio Corrêa Mariani. FZEA-USP Pirassununga: Mirele Daiana Poleti; Raul Machado Neto; Ricardo Augusto Brassaloti; Ricardo Haddad; Rodrigo Tocantins Calado.; Rodrigo Tocantins Calado. FAMERP-SJRP: Cecília Artico Banho; Sandra Coccuzzo Sampaio; Simone Kashima; Svetoslav Nanev Slavov; Vagner Fonseca; Vincent Louis Viala |
| EPI_ISL_1795072, EPI_ISL_1795073, EPI_ISL_1795074, EPI_ISL_1795097, EPI_ISL_1795346, EPI_ISL_1795347, EPI_ISL_1795348, EPI_ISL_1795349, EPI_ISL_1795350, EPI_ISL_1795351, EPI_ISL_1795352, EPI_ISL_1795353, EPI_ISL_1795354, EPI_ISL_1795355, EPI_ISL_1795356, EPI_ISL_1795357, EPI_ISL_1795358, EPI_ISL_1795359, EPI_ISL_1795360, EPI_ISL_1795365, EPI_ISL_1795366, EPI_ISL_1795382, EPI_ISL_1795383, EPI_ISL_1795428 | | | |
| see above | LABORATORIO DR PAULO EMILIO DALESSANDRO PINDAMONHANGABA | Instituto Butantan / ESALQ-Piracicaba | Antonio Jorge Martins; Bianca Cechetto Carlos. Mendelics: Bibiana Santos; Claudia Renata dos Santos Barros; David Schlesinger. Hemocentro Ribeirão Preto: Simone Kashima; Debora Botequio Moretti. Centro de Genômica Funcional da ESALQ: Luiz Lehmann Coutinho; Dimas Tadeu Covas; Elaine Cristina Marqueze; Elaine Vieira dos Santos; Elisangela Chicaroni Mattos; Erika Freitas; Evandra Strazza Rodrigues; Felipe Allan da Silva da Costa; Flavia Aburjaile; Guilherme Targino Valente; Heidge Fukumasu. USP-Botucatu: Rejane Maria Tommasini Grotto; Instituto Butantan: Alexander Roberto Precioso; Jayme A. Souza-Neto; Jessika Cristina Chagas Lesbon; José Salvatore Leister Patané; João Paulo Kitajima; Luiz Carlos Junior de Alcantara; Maria Carolina Elias; Marta Giovanetti; Patricia Akemi Assato; Rafael dos Santos Bezerra; Raquel de Lello Rocha Campos Cassano. NGS Soluções Genômicas: Pilar Drummond Sampaio Corrêa Mariani. FZEA-USP Pirassununga: Mirele Daiana Poleti; Raul Machado Neto; Ricardo Augusto Brassaloti; Ricardo Haddad; Rodrigo Tocantins Calado.; Sandra Coccuzzo Sampaio; Svetoslav Nanev Slavov; Vagner Fonseca; Vincent Louis Viala |
| EPI_ISL_1966608, EPI_ISL_1966609, EPI_ISL_1966610, EPI_ISL_1966611, EPI_ISL_1966683, EPI_ISL_1966684, EPI_ISL_1966685, EPI_ISL_1966686, EPI_ISL_1966687, EPI_ISL_1966688, EPI_ISL_1966689, EPI_ISL_1966690, EPI_ISL_1967244, EPI_ISL_1967245, EPI_ISL_1967246, EPI_ISL_1967247, EPI_ISL_1967248, EPI_ISL_1967249, EPI_ISL_1967250, EPI_ISL_2170942, EPI_ISL_2170943 | | | |
| see above | LABORATORIO MUNICIPAL DE ANALISES CLINICAS | Instituto Butantan / Mendelics | Antonio Jorge Martins; Bianca Cechetto Carlos. Mendelics: Bibiana Santos; Claudia Renata dos Santos Barros; Cíntia Bittar; David Schlesinger. Hemocentro Ribeirão Preto: Simone Kashima; Debora Botequio Moretti; Elaine Cristina Marqueze; Elaine Vieira dos Santos; Elisangela Chicaroni Mattos; Erika Freitas; Evandra Strazza Rodrigues; Felipe Allan da Silva da Costa; Flavia Aburjaile; Fábio Sossai Possebon; Guilherme Campos; Guilherme Targino Valente; Heidge Fukumasu. USP-Botucatu: Rejane Maria Tommasini Grotto; Helena Lage Ferreira; Instituto Butantan: Dimas Tadeu Covas; Jardelina de Souza Todao Bernardino; Jayme A. Souza-Neto; Jessika Cristina Chagas Lesbon; Jorge A. Petroli Marchesi; José Salvatore Leister Patané; João Paulo Kitajima; João Pessoa Araújo Jr.; Leila Sabrina Ullmann; Loyze Paola Oliveira de Lima; Luiz Aurelio de Campos Crispin. Centro de Genômica Funcional da ESALQ: Luiz Lehmann Coutinho; Luiz Carlos Junior de Alcantara; Lívia Sacchetto; Maisa C. Pereira Parra; Maria |

| | | | |
|---|---|---|---|
| | DE ITANHAEM | | Carolina Elias; Marta Giovanetti; Marília Moraes; Maurício Lacerda Nogueira. Prefeitura de Sao Paulo: Melissa Palmieri.; Patricia Akemi Assato; Paula Rahal; Paulo Inacio da Costa; Rafael dos Santos Bezerra; Raquel de Lello Rocha Campos Cassano. NGS Soluções Genômicas: Pilar Drummond Sampaio Corrêa Mariani. FZEA-USP Pirassununga: Mirele Daiana Poleti; Raul Machado Neto; Ricardo Augusto Brassaloti; Ricardo Haddad; Rodrigo Tocantins Calado. FAMERP-SJRP: Cecília Artico Banho; Sandra Coccuzzo Sampaio; Svetoslav Nanev Slavov; Vagner Fonseca; Vincent Louis Viala |
| EPI_ISL_1795400, EPI_ISL_1795401 | LABORATORIO MUNICIPAL DE ANALISES CLINICAS DE RIO CLARO | Instituto Butantan / ESALQ-Piracicaba | Antonio Jorge Martins; Bianca Cechetto Carlos. Mendelics: Bibiana Santos; Claudia Renata dos Santos Barros; David Schlesinger. Hemocentro Ribeirão Preto: Simone Kashima; Debora Botequio Moretti. Centro de Genômica Funcional da ESALQ: Luiz Lehmann Coutinho; Dimas Tadeu Covas; Elaine Cristina Marqueze; Elaine Vieira dos Santos; Elisangela Chicaroni Mattos; Erika Freitas; Evandra Strazza Rodrigues; Felipe Allan da Silva da Costa; Flavia Aburjaile; Guilherme Targino Valente; Heidge Fukumasu. USP-Botucatu: Rejane Maria Tommasini Grotto; Instituto Butantan: Alexander Roberto Precioso; Jayme A. Souza-Neto; Jessika Cristina Chagas Lesbon; José Salvatore Leister Patané; João Paulo Kitajima; Luiz Carlos Junior de Alcantara; Maria Carolina Elias; Marta Giovanetti; Patricia Akemi Assato; Rafael dos Santos Bezerra; Raquel de Lello Rocha Campos Cassano. NGS Soluções Genômicas: Pilar Drummond Sampaio Corrêa Mariani. FZEA-USP Pirassununga: Mirele Daiana Poleti; Raul Machado Neto; Ricardo Augusto Brassaloti; Ricardo Haddad; Rodrigo Tocantins Calado.; Sandra Coccuzzo Sampaio; Svetoslav Nanev Slavov; Vagner Fonseca; Vincent Louis Viala |
| EPI_ISL_1795100, EPI_ISL_1795102, EPI_ISL_1795104, EPI_ISL_1795105 | LABORATORIO MUNICIPAL DE PIRACICABA | Instituto Butantan / ESALQ-Piracicaba | Antonio Jorge Martins; Bianca Cechetto Carlos. Mendelics: Bibiana Santos; Claudia Renata dos Santos Barros; David Schlesinger. Hemocentro Ribeirão Preto: Simone Kashima; Debora Botequio Moretti. Centro de Genômica Funcional da ESALQ: Luiz Lehmann Coutinho; Dimas Tadeu Covas; Elaine Cristina Marqueze; Elaine Vieira dos Santos; Elisangela Chicaroni Mattos; Erika Freitas; Evandra Strazza Rodrigues; Felipe Allan da Silva da Costa; Flavia Aburjaile; Guilherme Targino Valente; Heidge Fukumasu. USP-Botucatu: Rejane Maria Tommasini Grotto; Instituto Butantan: Alexander Roberto Precioso; Jayme A. Souza-Neto; Jessika Cristina Chagas Lesbon; José Salvatore Leister Patané; João Paulo Kitajima; Luiz Carlos Junior de Alcantara; Maria Carolina Elias; Marta Giovanetti; Patricia Akemi Assato; Rafael dos Santos Bezerra; Raquel de Lello Rocha Campos Cassano. NGS Soluções Genômicas: Pilar Drummond Sampaio Corrêa Mariani. FZEA-USP Pirassununga: Mirele Daiana Poleti; Raul Machado Neto; Ricardo Augusto Brassaloti; Ricardo Haddad; Rodrigo Tocantins Calado.; Sandra Coccuzzo Sampaio; Svetoslav Nanev Slavov; Vagner Fonseca; Vincent Louis Viala |
| EPI_ISL_1966996, EPI_ISL_1966997, EPI_ISL_1966998, EPI_ISL_1966999, EPI_ISL_1967000, EPI_ISL_1967001, EPI_ISL_1967002, EPI_ISL_1967003 | | | |
| see above | LABORATORIO MUNICIPAL DE PIRACICABA | Instituto Butantan / ESALQ-USP (Piracicaba) | Antonio Jorge Martins; Bianca Cechetto Carlos. Mendelics: Bibiana Santos; Claudia Renata dos Santos Barros; Cíntia Bittar; David Schlesinger. Hemocentro Ribeirão Preto: Simone Kashima; Debora Botequio Moretti; Elaine Cristina Marqueze; Elaine Vieira dos Santos; Elisangela Chicaroni Mattos; Erika Freitas; Evandra Strazza Rodrigues; Felipe Allan da Silva da Costa; Flavia Aburjaile; Fábio Sossai Possebon; Guilherme Campos; Guilherme Targino Valente; Heidge Fukumasu. USP-Botucatu: Rejane Maria Tommasini Grotto; Helena Lage Ferreira; Instituto Butantan: Dimas Tadeu Covas; Jardelina de Souza Todao Bernardino; Jayme A. Souza-Neto; Jessika Cristina Chagas Lesbon; Jorge A. Petroli Marchesi; José Salvatore Leister Patané; João Paulo Kitajima; João Pessoa Araújo Jr.; Leila Sabrina Ullmann; Loyze Paola Oliveira de Lima; Luiz Aurelio de Campos Crispin. Centro de Genômica Funcional da ESALQ: Luiz Lehmann Coutinho; Luiz Carlos Junior de Alcantara; Lívia Sacchetto; Maisa C. Pereira Parra; Maria Carolina Elias; Marta Giovanetti; Marília Moraes; Maurício Lacerda Nogueira. Prefeitura de Sao Paulo: Melissa Palmieri.; Patricia Akemi Assato; Paula Rahal; Paulo Inacio da Costa; Rafael dos Santos Bezerra; Raquel de Lello Rocha Campos Cassano. NGS Soluções Genômicas: Pilar Drummond Sampaio Corrêa Mariani. FZEA-USP Pirassununga: Mirele Daiana Poleti; Raul Machado Neto; Ricardo Augusto Brassaloti; Ricardo Haddad; Rodrigo Tocantins Calado. FAMERP-SJRP: Cecília Artico Banho; Sandra Coccuzzo Sampaio; Svetoslav Nanev Slavov; Vagner Fonseca; Vincent Louis Viala |
| EPI_ISL_1967148, EPI_ISL_1967149, EPI_ISL_1967150, EPI_ISL_1967151, EPI_ISL_1967152, EPI_ISL_1967153, EPI_ISL_1967154, EPI_ISL_1967155, EPI_ISL_1967156, EPI_ISL_1967157, EPI_ISL_1967158 | | | |
| see above | LABORATORIO MUNICIPAL DE SUZANO | Instituto Butantan / ESALQ-USP (Piracicaba) | Antonio Jorge Martins; Bianca Cechetto Carlos. Mendelics: Bibiana Santos; Claudia Renata dos Santos Barros; Cíntia Bittar; David Schlesinger. Hemocentro Ribeirão Preto: Simone Kashima; Debora Botequio Moretti; Elaine Cristina Marqueze; Elaine Vieira dos Santos; Elisangela Chicaroni Mattos; Erika Freitas; Evandra Strazza Rodrigues; Felipe Allan da Silva da Costa; Flavia Aburjaile; Fábio Sossai Possebon; Guilherme Campos; Guilherme Targino Valente; Heidge Fukumasu. USP-Botucatu: Rejane Maria Tommasini Grotto; Helena Lage Ferreira; Instituto Butantan: Dimas Tadeu Covas; Jardelina de Souza Todao Bernardino; Jayme A. Souza-Neto; Jessika Cristina Chagas Lesbon; Jorge A. Petroli Marchesi; José Salvatore Leister Patané; João Paulo Kitajima; João Pessoa Araújo Jr.; Leila Sabrina Ullmann; Loyze Paola Oliveira de Lima; Luiz Aurelio de Campos Crispin. Centro de Genômica Funcional da ESALQ: Luiz Lehmann Coutinho; Luiz Carlos Junior de Alcantara; Lívia Sacchetto; Maisa C. Pereira Parra; Maria Carolina Elias; Marta Giovanetti; Marília Moraes; Maurício Lacerda Nogueira. Prefeitura de Sao Paulo: Melissa Palmieri.; Patricia Akemi Assato; Paula Rahal; Paulo Inacio da Costa; Rafael dos Santos Bezerra; Raquel de Lello Rocha Campos Cassano. NGS Soluções Genômicas: Pilar Drummond Sampaio Corrêa Mariani. FZEA-USP Pirassununga: Mirele Daiana Poleti; Raul Machado Neto; Ricardo Augusto Brassaloti; Ricardo Haddad; Rodrigo Tocantins Calado. FAMERP-SJRP: Cecília Artico Banho; Sandra Coccuzzo Sampaio; Svetoslav Nanev Slavov; Vagner Fonseca; Vincent Louis Viala |
| EPI_ISL_1716458 | LACEN (Laboratorio de Saude Publica Dr. Giovanni Cysneiros) | LGBio (Laboratorio de Genetica & Biodiversidade) | Amanda Alves de Melo; Cintia Pelegrineti Targueta de Azevedo Brito; Daniela de Melo e Silva; Elisangela de Paula Silveira Lacerda; Francyelli Mello Andrade; Mariana Pires de Campos Telles; Ramilla dos Santos Braga; Renata de Oliveira Dias; Rhewter Nunes; Thais Guimarães Castro; Thays Millena Alves Pedroso |
| EPI_ISL_1468413, EPI_ISL_1468414, EPI_ISL_1468415, EPI_ISL_1468431, EPI_ISL_1493573, EPI_ISL_1493574, EPI_ISL_1628363, EPI_ISL_1628364, EPI_ISL_1628365 | | | |
| see above | LACEN do Estado de Goias | Instituto Adolfo Lutz, Interdiciplinary Procedures Center, Strategic Laboratory | Caio Vinicius Dias Lopes; Claudia Regina Gonçalves; Claudio Tavares Sacchi; Erica Valessa Ramos Gomes; Karoline Rodrigues Campos; Katia Correa de Oliveira Santos; Leonardo Jose Tadeu de Araujo |
| EPI_ISL_1520108, EPI_ISL_1520109 | LACEN do Estado de Rondonia | Instituto Adolfo Lutz, Interdiciplinary Procedures Center, Strategic Laboratory | Caio Vinicius Dias Lopes; Claudia Regina Gonçalves; Claudio Tavares Sacchi; Erica Valessa Ramos Gomes; Karoline Rodrigues Campos |
| EPI_ISL_1358305 | LACEN do Mato Grosso do Sul | Instituto Adolfo Lutz, Interdiciplinary Procedures Center, Strategic Laboratory | Caio Vinicius Dias Lopes; Claudia Regina Gonçalves; Claudio Tavares Sacchi; Erica Valessa Ramos Gomes; Karoline Rodrigues Campos |
| EPI_ISL_1446194 | LATE - Laboratório de Técnicas Especiais - Hospital Israelita Albert Einstein | LATE - Laboratório de Técnicas Especiais - Hospital Israelita Albert Einstein | Ana Paula Moreira Salles; Deyvid Amgarten; Fernanda de Mello Malta; João Renato Rebello Pinho; Pedro Henrique Sebe Rodrigues; Raquel Riyuzo |
| EPI_ISL_2157480, EPI_ISL_2157481, EPI_ISL_2157491 | Laboratorio Central de Saude Publica do Estado de Minas Gerais (LACEN/MG) | Laboratory of Respiratory Viruses and Measles, Oswaldo Cruz Institute, FIOCRUZ | Alice Sampaio Rocha; Ana Carolina Mendonca; Andre Felipe Leal Bernardes; Anna Carolina Paixao; Elisa Cavalcante Pereira; Fernando Motta; Luciana Appolinario; Marilda Siqueira on behalf of the Fiocruz COVID-19 Genomic Surveillance Network; Paola Resende; Renata Serrano Lopes; Taina Venas |
| EPI_ISL_1664113, EPI_ISL_1664114, EPI_ISL_1664115, EPI_ISL_1664116, EPI_ISL_1664117, EPI_ISL_1664118, EPI_ISL_1664119, EPI_ISL_1664120, EPI_ISL_1664121, EPI_ISL_1664122, EPI_ISL_1664123, EPI_ISL_1664124, EPI_ISL_1664125, EPI_ISL_1664126, EPI_ISL_1664127, EPI_ISL_1664128, EPI_ISL_1664129, EPI_ISL_1664130, EPI_ISL_1664131, EPI_ISL_1664132, EPI_ISL_1664133, EPI_ISL_1664134, EPI_ISL_1664135, EPI_ISL_1664136, EPI_ISL_1664137, EPI_ISL_1664138, EPI_ISL_1664139, EPI_ISL_1664140, EPI_ISL_1664141, EPI_ISL_1664142, EPI_ISL_1664143, EPI_ISL_1664144, EPI_ISL_1664145, EPI_ISL_1664146, EPI_ISL_1664147, EPI_ISL_1664148, EPI_ISL_1664149, EPI_ISL_1664150, EPI_ISL_1664151, EPI_ISL_1664152, EPI_ISL_1664153, EPI_ISL_1664154, EPI_ISL_1664155, EPI_ISL_1664156, EPI_ISL_1664157, EPI_ISL_1664158, EPI_ISL_1664159, EPI_ISL_1664160, EPI_ISL_1664161, EPI_ISL_1664162, EPI_ISL_1664163, EPI_ISL_1664164, EPI_ISL_1664165, EPI_ISL_1664166, EPI_ISL_1664167, EPI_ISL_1664168, EPI_ISL_1664169, EPI_ISL_1664170, EPI_ISL_1664171, EPI_ISL_1664172, EPI_ISL_1664173, EPI_ISL_1664174, EPI_ISL_1664175, EPI_ISL_1664176, EPI_ISL_1664177, EPI_ISL_1664178, EPI_ISL_1664179, EPI_ISL_1664180, EPI_ISL_1664181, EPI_ISL_1664182, EPI_ISL_1664183, EPI_ISL_1664184, EPI_ISL_1664185, EPI_ISL_1664186, EPI_ISL_1664187, EPI_ISL_1664188, EPI_ISL_1664189, EPI_ISL_1664190, EPI_ISL_1664191, EPI_ISL_1664192, EPI_ISL_1664193, EPI_ISL_1664194, EPI_ISL_1664195, EPI_ISL_1664196, EPI_ISL_1664197, EPI_ISL_1664198, EPI_ISL_1664199, EPI_ISL_1664200, EPI_ISL_1664201, EPI_ISL_1664202, EPI_ISL_1858267, EPI_ISL_1858269, EPI_ISL_1858271, EPI_ISL_1858273, EPI_ISL_1858275, EPI_ISL_1858277, EPI_ISL_1858279, EPI_ISL_1858281, EPI_ISL_1858282, EPI_ISL_1858284, EPI_ISL_1858286, EPI_ISL_1858288, EPI_ISL_1858290, EPI_ISL_1858292, EPI_ISL_1858294, EPI_ISL_1858295, EPI_ISL_1858297, EPI_ISL_1858299, EPI_ISL_1858301, EPI_ISL_1858303, EPI_ISL_1858304, EPI_ISL_1858306, EPI_ISL_1858308, EPI_ISL_1858310, EPI_ISL_1858312, EPI_ISL_1858314, EPI_ISL_1858315, EPI_ISL_1858317, EPI_ISL_1858319, EPI_ISL_1858321, EPI_ISL_1858323, EPI_ISL_1858325, EPI_ISL_1858327, EPI_ISL_1858329, EPI_ISL_1858331, EPI_ISL_1858333, EPI_ISL_1858334, EPI_ISL_1858336, EPI_ISL_1858338, EPI_ISL_1858340, EPI_ISL_1858342, EPI_ISL_1858344, EPI_ISL_1858346, EPI_ISL_1858348, EPI_ISL_1858350, EPI_ISL_1858352, EPI_ISL_1858354, EPI_ISL_1858356, EPI_ISL_1858357, EPI_ISL_1858359, EPI_ISL_1858361, EPI_ISL_1858363, EPI_ISL_1858365, EPI_ISL_1858367, EPI_ISL_1858369, EPI_ISL_1858371, EPI_ISL_1858373, EPI_ISL_1858375, EPI_ISL_1858377, EPI_ISL_1858379, EPI_ISL_1858380, EPI_ISL_1858383, EPI_ISL_1858384, EPI_ISL_1858387, EPI_ISL_1858389, EPI_ISL_1858391, EPI_ISL_1858392, EPI_ISL_1858395, EPI_ISL_1858397, EPI_ISL_1858399, EPI_ISL_1858401, EPI_ISL_1858403, EPI_ISL_1858405, EPI_ISL_1858407, EPI_ISL_1858409, EPI_ISL_1858411, EPI_ISL_1858414, EPI_ISL_1858416, EPI_ISL_1858418, EPI_ISL_1858420, EPI_ISL_1858422, EPI_ISL_1858423, EPI_ISL_1858426, EPI_ISL_1858428, EPI_ISL_1858430, EPI_ISL_1858432, EPI_ISL_1858434, EPI_ISL_1858436, EPI_ISL_1858438, EPI_ISL_1858440, EPI_ISL_1858442, EPI_ISL_1858444, EPI_ISL_1858446, EPI_ISL_1858449, EPI_ISL_1858451, EPI_ISL_1858453, EPI_ISL_1858454, EPI_ISL_1858456, EPI_ISL_1858458, EPI_ISL_1858460, EPI_ISL_1858462, EPI_ISL_1858464, EPI_ISL_1858466, EPI_ISL_1858468, EPI_ISL_1858470, EPI_ISL_1858473, EPI_ISL_1858475, EPI_ISL_1858477, EPI_ISL_1858479, EPI_ISL_1858481, EPI_ISL_1858483, EPI_ISL_1858486, EPI_ISL_1858488, EPI_ISL_1858490, EPI_ISL_1858492, EPI_ISL_1858494, EPI_ISL_1858496, EPI_ISL_1858498, EPI_ISL_1858500, EPI_ISL_1858502, EPI_ISL_1858504, EPI_ISL_1858506, EPI_ISL_1858508, EPI_ISL_1858510, EPI_ISL_1858512, EPI_ISL_1858514, EPI_ISL_1858516, EPI_ISL_1858518, EPI_ISL_1858520, EPI_ISL_1858522, EPI_ISL_1858524, EPI_ISL_1858526, EPI_ISL_1858528, EPI_ISL_1858530, EPI_ISL_1858532, EPI_ISL_1858534, EPI_ISL_1858536, EPI_ISL_1858538, EPI_ISL_1858540, EPI_ISL_1858542, EPI_ISL_1858544, EPI_ISL_1858546, EPI_ISL_1858548, EPI_ISL_1858549, EPI_ISL_1858551, EPI_ISL_1858554, EPI_ISL_1858555, EPI_ISL_1858557, EPI_ISL_1858559, EPI_ISL_1858561, EPI_ISL_1858563, EPI_ISL_1858565, EPI_ISL_1858567, EPI_ISL_1858569, EPI_ISL_1858571, EPI_ISL_1858572, EPI_ISL_1858575, EPI_ISL_1858577, EPI_ISL_1858578, EPI_ISL_1858580, EPI_ISL_1858582, EPI_ISL_1858584, EPI_ISL_1858586, EPI_ISL_1858588, EPI_ISL_1858590, EPI_ISL_1858592, EPI_ISL_1858594, EPI_ISL_1858596, EPI_ISL_1858598, EPI_ISL_1858599, EPI_ISL_1858602, EPI_ISL_1858603, EPI_ISL_1858606, EPI_ISL_1858607, EPI_ISL_1858610, EPI_ISL_1858611, EPI_ISL_1858613, EPI_ISL_1858615, EPI_ISL_1858617, EPI_ISL_1858619, EPI_ISL_1858621, EPI_ISL_1858623, EPI_ISL_1858625, EPI_ISL_1858627, EPI_ISL_1858629, EPI_ISL_1858631, EPI_ISL_1858633, EPI_ISL_1858636, EPI_ISL_1858637, EPI_ISL_1858639, EPI_ISL_1858641, EPI_ISL_1858643, EPI_ISL_1858645, EPI_ISL_1858648, EPI_ISL_1858650, EPI_ISL_1858651, EPI_ISL_1858653, EPI_ISL_1858655, EPI_ISL_1858656, EPI_ISL_1858658, EPI_ISL_1858660, EPI_ISL_1858662, EPI_ISL_1858664, EPI_ISL_1858666, EPI_ISL_1858668, EPI_ISL_1858670, EPI_ISL_1858672, EPI_ISL_1858674, EPI_ISL_1858675, EPI_ISL_1858677, EPI_ISL_1858680, EPI_ISL_1858682, EPI_ISL_1858684, EPI_ISL_1858686, EPI_ISL_1858688, EPI_ISL_1858691, EPI_ISL_1858693, EPI_ISL_1858694, EPI_ISL_1858697, EPI_ISL_1858699, EPI_ISL_1858701, EPI_ISL_1858703, EPI_ISL_1858705, EPI_ISL_1858707, EPI_ISL_1858709, EPI_ISL_1858711, EPI_ISL_1858713, EPI_ISL_1858715, EPI_ISL_1858717, EPI_ISL_1858719, EPI_ISL_1858721, EPI_ISL_1858722, EPI_ISL_1858724, EPI_ISL_1858726, EPI_ISL_1858728, EPI_ISL_1858730, EPI_ISL_1858732, EPI_ISL_1858734, EPI_ISL_1858736, EPI_ISL_1858738, EPI_ISL_1858740, EPI_ISL_1858741, EPI_ISL_1858743, EPI_ISL_1858745, EPI_ISL_1858747, EPI_ISL_1858749, EPI_ISL_1858751, EPI_ISL_1858752, EPI_ISL_1858754, EPI_ISL_1858756, EPI_ISL_2101388, EPI_ISL_2101389, EPI_ISL_2101393, EPI_ISL_2101395, EPI_ISL_2101396, EPI_ISL_2101397, EPI_ISL_2101398, EPI_ISL_2101399, EPI_ISL_2101400, EPI_ISL_2101401, EPI_ISL_2101402, EPI_ISL_2101403, EPI_ISL_2101404, EPI_ISL_2101405, EPI_ISL_2101406, EPI_ISL_2101408, EPI_ISL_2101409, EPI_ISL_2101410, EPI_ISL_2101411, EPI_ISL_2101412, EPI_ISL_2101413, EPI_ISL_2101414, EPI_ISL_2101415, EPI_ISL_2101416, EPI_ISL_2101417, EPI_ISL_2101418, EPI_ISL_2101419, EPI_ISL_2101420, EPI_ISL_2101421, EPI_ISL_2101422, EPI_ISL_2101423, EPI_ISL_2101424, EPI_ISL_2101425, EPI_ISL_2101426, EPI_ISL_2101427, EPI_ISL_2101428, EPI_ISL_2101434, EPI_ISL_2101435, EPI_ISL_2101436, EPI_ISL_2101437, EPI_ISL_2101438, EPI_ISL_2101439, EPI_ISL_2101440, EPI_ISL_2101441, EPI_ISL_2101442, EPI_ISL_2101444, EPI_ISL_2101445, EPI_ISL_2101446, EPI_ISL_2101447, EPI_ISL_2101448, EPI_ISL_2101449, EPI_ISL_2101450, EPI_ISL_2101451, EPI_ISL_2101452, EPI_ISL_2101453, EPI_ISL_2101454, EPI_ISL_2101464, EPI_ISL_2101465, EPI_ISL_2101466, EPI_ISL_2101467, EPI_ISL_2101468, EPI_ISL_2101469, EPI_ISL_2101474, EPI_ISL_2101476, EPI_ISL_2101477, EPI_ISL_2101478, EPI_ISL_2101479, EPI_ISL_2101480, EPI_ISL_2101481, EPI_ISL_2101482, EPI_ISL_2101485, EPI_ISL_2101486, EPI_ISL_2101487, EPI_ISL_2101488, EPI_ISL_2101489, EPI_ISL_2101490, EPI_ISL_2101491, EPI_ISL_2101492, EPI_ISL_2101493, EPI_ISL_2101496, EPI_ISL_2101497, EPI_ISL_2101498, EPI_ISL_2101499, EPI_ISL_2101500, EPI_ISL_2101501, EPI_ISL_2101502, EPI_ISL_2101503, EPI_ISL_2101504, EPI_ISL_2101507, EPI_ISL_2101514, EPI_ISL_2101515, EPI_ISL_2101516, EPI_ISL_2101517, EPI_ISL_2101518, EPI_ISL_2101519, EPI_ISL_2101528, EPI_ISL_2101529, EPI_ISL_2101530, EPI_ISL_2101531, EPI_ISL_2101532, EPI_ISL_2101533, EPI_ISL_2101534, EPI_ISL_2101535, EPI_ISL_2101536, EPI_ISL_2101537, EPI_ISL_2101538, EPI_ISL_2101539, EPI_ISL_2101540, EPI_ISL_2101541, EPI_ISL_2101542, EPI_ISL_2101543, EPI_ISL_2101544, EPI_ISL_2101545, EPI_ISL_2101546, EPI_ISL_2101547, EPI_ISL_2101548, EPI_ISL_2101549, EPI_ISL_2101550, EPI_ISL_2101551, EPI_ISL_2101552, EPI_ISL_2101553, EPI_ISL_2101554, EPI_ISL_2101555, EPI_ISL_2101556, EPI_ISL_2101557, EPI_ISL_2101558, EPI_ISL_2101559, EPI_ISL_2101560, EPI_ISL_2101561, EPI_ISL_2101562, EPI_ISL_2101563, EPI_ISL_2101564, EPI_ISL_2101565, EPI_ISL_2101573, EPI_ISL_2101574, EPI_ISL_2101575, EPI_ISL_2101576, EPI_ISL_2101577, EPI_ISL_2101578, EPI_ISL_2101579, EPI_ISL_2101580, EPI_ISL_2101581, EPI_ISL_2101582, EPI_ISL_2101583, EPI_ISL_2101584, EPI_ISL_2101585, EPI_ISL_2101586, EPI_ISL_2101587, EPI_ISL_2101588, EPI_ISL_2101589, EPI_ISL_2101590, EPI_ISL_2101591, EPI_ISL_2101592, EPI_ISL_2101593, EPI_ISL_2101594, EPI_ISL_2101595, EPI_ISL_2101596, EPI_ISL_2101597, EPI_ISL_2101598, EPI_ISL_2101599, EPI_ISL_2101600, EPI_ISL_2101601, EPI_ISL_2101602, EPI_ISL_2101603, EPI_ISL_2101604, EPI_ISL_2101605, EPI_ISL_2101606, EPI_ISL_2101607, EPI_ISL_2101608, EPI_ISL_2101609, EPI_ISL_2101610, EPI_ISL_2101611, EPI_ISL_2101613, EPI_ISL_2101615, EPI_ISL_2101616, EPI_ISL_2101617, EPI_ISL_2101618, EPI_ISL_2101619, EPI_ISL_2101620, EPI_ISL_2101621, EPI_ISL_2101622, EPI_ISL_2101623, EPI_ISL_2101624, EPI_ISL_2101625, EPI_ISL_2101626, EPI_ISL_2101627, EPI_ISL_2101628, EPI_ISL_2101629, EPI_ISL_2101630, EPI_ISL_2101631, EPI_ISL_2101632, EPI_ISL_2101633, EPI_ISL_2101634, EPI_ISL_2101635, EPI_ISL_2101636, EPI_ISL_2101637, EPI_ISL_2101672, EPI_ISL_2101673, EPI_ISL_2101674, EPI_ISL_2101675, EPI_ISL_2101689, EPI_ISL_2101690, EPI_ISL_2101691, EPI_ISL_2101697, EPI_ISL_2101698, EPI_ISL_2101699, EPI_ISL_2101700, EPI_ISL_2101701, EPI_ISL_2101702, EPI_ISL_2101703, EPI_ISL_2101704, EPI_ISL_2101705, EPI_ISL_2101706, EPI_ISL_2101707, EPI_ISL_2101708, EPI_ISL_2101709, EPI_ISL_2101710, EPI_ISL_2101711, EPI_ISL_2101712, EPI_ISL_2101713, EPI_ISL_2101716, EPI_ISL_2101717, EPI_ISL_2101718, EPI_ISL_2101719, EPI_ISL_2101720, EPI_ISL_2101721, EPI_ISL_2101723, EPI_ISL_2101725, EPI_ISL_2101726, EPI_ISL_2101728, EPI_ISL_2101729, EPI_ISL_2101730, EPI_ISL_2101732, EPI_ISL_2101733, EPI_ISL_2101735, EPI_ISL_2101736, EPI_ISL_2101737, EPI_ISL_2101738, EPI_ISL_2101739, EPI_ISL_2101742, EPI_ISL_2101743, EPI_ISL_2101745, EPI_ISL_2101746, EPI_ISL_2101747 | | | |
| see above | Laboratorio Central Noel Nutels | Bioinformatics Laboratory / LNCC | Alessandra P Lamarca; Alexandra L Gerber; Amilcar Tanuri; Ana Paula de C Guimaraes; Ana Paula de C Guimarães; Ana Tereza R Vasconcelos; Andrea Cony Cavalcanti; Andréa Cony Cavalcanti; Caio Luiz Pereira Ribeiro; Cassia Alves; Cintia Policarpo; Claudia Maria Braga de Mello; Cristiane Gomes da Silva; Diana Mariani; Douglas Terra Machado; Flavio Dias da Silva; Flávio Dias da Silva; Gleidson da Silva de Oliveira; Leandro Magalhaes de Souza; Leandro Magalhães de Souza; Liliane Cavalcante; Luiz G P de Almeida; Marcio Henrique de Oliveira Garcia; Mario Sergio Ribeiro; Ronaldo da Silva F Jr; Silvia Carvalho; Thais Felix Cruz |
| EPI_ISL_2157412, EPI_ISL_2157414, EPI_ISL_2157415, EPI_ISL_2157422, EPI_ISL_2157451, EPI_ISL_2157461, EPI_ISL_2157489, EPI_ISL_2157490, EPI_ISL_2157507, EPI_ISL_2157508, EPI_ISL_2157509, EPI_ISL_2157512, EPI_ISL_2157514, EPI_ISL_2157519, EPI_ISL_2157520, EPI_ISL_2157521, EPI_ISL_2157522, EPI_ISL_2157523, EPI_ISL_2157541 | | | |
| see above | Laboratorio Central de Saude Publica do Esatado de Alagoas (LACEN/AL) | Laboratory of Respiratory Viruses and Measles, Oswaldo Cruz Institute, FIOCRUZ | Alice Sampaio Rocha; Ana Carolina Mendonca; Anderson Brandao Leite; Anna Carolina Paixao; Elisa Cavalcante Pereira; Fernando Motta; Luciana Appolinario; Marilda Siqueira on behalf of the Fiocruz COVID-19 Genomic Surveillance Network; Paola Resende; Renata Serrano Lopes; Taina Venas |
| EPI_ISL_2157531 | Laboratorio Central de Saude Publica do Estado da Bahia (LACEN/BA) | Laboratory of Respiratory Viruses and Measles, Oswaldo Cruz Institute, FIOCRUZ | Alice Sampaio Rocha; Ana Carolina Mendonca; Anna Carolina Paixao; Elisa Cavalcante Pereira; Felicidade Pereira; Fernando Motta; Luciana Appolinario; Marilda Siqueira on behalf of the Fiocruz COVID-19 Genomic Surveillance Network; Paola Resende; Renata Serrano Lopes; Taina Venas |
| EPI_ISL_2157394, EPI_ISL_2157395, EPI_ISL_2157401, EPI_ISL_2157425, EPI_ISL_2157589, EPI_ISL_2157591 | Laboratorio Central de Saude Publica do Estado da Paraiba (LACEN-PB) | Laboratory of Respiratory Viruses and Measles, Oswaldo Cruz Institute, FIOCRUZ | Alice Sampaio Rocha; Ana Carolina Mendonca; Anna Carolina Paixao; Dalane Loudal Florentino Teixeira; Elisa Cavalcante Pereira; Fernando Motta; Joao Felipe Bezerra; Luciana Appolinario; Marilda Siqueira on behalf of the Fiocruz COVID-19 Genomic Surveillance Network; Paola Resende; Renata Serrano Lopes; Taina Venas |
| EPI_ISL_2157374, EPI_ISL_2157427, EPI_ISL_2157428, EPI_ISL_2157429, EPI_ISL_2157430, EPI_ISL_2157432, EPI_ISL_2157433, EPI_ISL_2157434, EPI_ISL_2157435, EPI_ISL_2157436, EPI_ISL_2157437, EPI_ISL_2157438, EPI_ISL_2157439, EPI_ISL_2157440, EPI_ISL_2157441, EPI_ISL_2157442, EPI_ISL_2157443, EPI_ISL_2157444, EPI_ISL_2157445, EPI_ISL_2157446, EPI_ISL_2157449, EPI_ISL_2157453, EPI_ISL_2157454, EPI_ISL_2157455, EPI_ISL_2157456, EPI_ISL_2157457, EPI_ISL_2157458, EPI_ISL_2157459, EPI_ISL_2157460, EPI_ISL_2157462, EPI_ISL_2157463, EPI_ISL_2157464, EPI_ISL_2157465, EPI_ISL_2157466, EPI_ISL_2157467, EPI_ISL_2157468, EPI_ISL_2157469, EPI_ISL_2157470, EPI_ISL_2157471, EPI_ISL_2157472, EPI_ISL_2157473, EPI_ISL_2157474, EPI_ISL_2157475, EPI_ISL_2157476, EPI_ISL_2157477, EPI_ISL_2157478, EPI_ISL_2157546, EPI_ISL_2157594, EPI_ISL_2157596 | | | |
| see above | Laboratorio Central de Saude Publica do Estado de Sergipe (LACEN/SE) | Laboratory of Respiratory Viruses and Measles, Oswaldo Cruz Institute, | Alice Sampaio Rocha; Ana Carolina Mendonca; Anna Carolina Paixao; Cliomar Alves dos Santos; Elisa Cavalcante Pereira; Fernando Motta; Luciana Appolinario; Marilda Siqueira on behalf of the Fiocruz COVID-19 Genomic Surveillance Network; Paola Resende; Renata Serrano Lopes; Tainá Moreira Martins Venas |

| | | | |
|---|---|---|---|
| | | FIOCRUZ | |
| EPI_ISL_2157375, EPI_ISL_2157376 | Laboratorio Central de Saude Publica do Estado do Espirito Santo (LACEN/ES) | Laboratory of Respiratory Viruses and Measles, Oswaldo Cruz Institute, FIOCRUZ | Alice Sampaio Rocha; Ana Carolina Mendonca; Anna Carolina Paixao; Eilisa Cavalcante Pereira; Fernando Motta; Luciana Appolinario; Marilda Siqueira on behalf of the Fiocruz COVID-19 Genomic Surveillance Network; Paola Resende; Renata Serrano Lopes; Rodrigo Ribeiro Rodrigues; Taina Venas |
| EPI_ISL_2038968 | Laboratorio Central de Saude Publica do Estado do Espito Santo (LACEN-ES) | Laboratory of Respiratory Viruses and Measles, Oswaldo Cruz Institute, FIOCRUZ | Alice Sampaio Rocha; Ana Carolina Mendonca; Anna Carolina Paixao; Fernando Motta; Luciana Appolinario; Marilda Siqueira on behalf of the Fiocruz COVID-19 Genomic Surveillance Network; Paola Resende; Renata Serrano Lopes; Rodrigo Ribeiro Rodrigues |
| EPI_ISL_2157482 | Laboratorio Central de Saude Publica do Estado do Parana (LACEN/PR) | Laboratory of Respiratory Viruses and Measles, Oswaldo Cruz Institute, FIOCRUZ | Alice Sampaio Rocha; Ana Carolina Mendonca; Anna Carolina Paixao; Elisa Cavalcante Pereira; Fernando Motta; Irina Riediger; Luciana Appolinario; Marilda Siqueira on behalf of the Fiocruz COVID-19 Genomic Surveillance Network; Paola Resende; Renata Serrano Lopes; Taina Venas |
| EPI_ISL_1534013, EPI_ISL_2038959, EPI_ISL_2038960 | Laboratorio Central de Saude Publica do Estado do Rio Grande do Sul (LACEN-RS) | Laboratory of Respiratory Viruses and Measles, Oswaldo Cruz Institute, FIOCRUZ | Alice Sampaio Rocha; Ana Carolina Mendonca; Anna Carolina Paixao; Elisa Cavalcante Pereira; Fernando Motta; Luciana Appolinario; Marilda Siqueira on behalf of the Fiocruz COVID-19 Genomic Surveillance Network; Paola Resende; Renata Serrano Lopes; Richard Salvato; Taina Venas; Tatiana Schaffer Gregianini |
| EPI_ISL_1534012, EPI_ISL_2038956 | Laboratorio Central de Saude Publica do Estado do Rio de Janeiro (LACEN-RJ) | Laboratory of Respiratory Viruses and Measles, Oswaldo Cruz Institute, FIOCRUZ | Alice Sampaio Rocha; Ana Carolina Mendonca; Andrea Cony Cavalcanti; Anna Carolina Paixao; Elisa Cavalcante Pereira; Fernando Motta; Luciana Appolinario; Marilda Siqueira on behalf of the Fiocruz COVID-19 Genomic Surveillance Network; Paola Resende; Renata Serrano Lopes; Taina Venas |
| EPI_ISL_2139494, EPI_ISL_2139495, EPI_ISL_2139496, EPI_ISL_2139497, EPI_ISL_2139498, EPI_ISL_2139499, EPI_ISL_2139500, EPI_ISL_2139501, EPI_ISL_2139502, EPI_ISL_2139503, EPI_ISL_2139504, EPI_ISL_2139505, EPI_ISL_2139506, EPI_ISL_2139507, EPI_ISL_2139508, EPI_ISL_2139509, EPI_ISL_2139510, EPI_ISL_2139511, EPI_ISL_2139512, EPI_ISL_2139513, EPI_ISL_2139514, EPI_ISL_2139515, EPI_ISL_2139516, EPI_ISL_2139517, EPI_ISL_2139518, EPI_ISL_2139519, EPI_ISL_2139520, EPI_ISL_2139521, EPI_ISL_2139522, EPI_ISL_2139523, EPI_ISL_2139524, EPI_ISL_2139525, EPI_ISL_2139526, EPI_ISL_2139527, EPI_ISL_2139528, EPI_ISL_2139529, EPI_ISL_2139530, EPI_ISL_2139531, EPI_ISL_2139532, EPI_ISL_2139533, EPI_ISL_2139534, EPI_ISL_2139535, EPI_ISL_2139536, EPI_ISL_2139537, EPI_ISL_2139538, EPI_ISL_2139539, EPI_ISL_2139540, EPI_ISL_2139541, EPI_ISL_2139542, EPI_ISL_2139543, EPI_ISL_2139544, EPI_ISL_2139545, EPI_ISL_2139546, EPI_ISL_2139547, EPI_ISL_2139548, EPI_ISL_2139549 | | | |
| see above | Laboratorio Exame | Universidade Federal de Ciencias da Saude de Porto Alegre | Gabriel Dickin Caldana et al.; Vinícius Bonetti Franceschi |
| EPI_ISL_1700737 | Laboratorio Local de Itapecerica da Serra | Instituto Butantan / USP-Pirassununga | Antonio Jorge Martins; Bianca Cechetto Carlos. Mendelics: Bibiana Santos; Claudia Renata dos Santos Barros; David Schlesinger. Hemocentro Ribeirão Preto: Simone Kashima; Debora Botequio Moretti. Centro de Genômica Funcional da ESALQ: Luiz Lehmann Coutinho; Elaine Cristina Marqueze; Elaine Vieira dos Santos; Elisangela Chicaroni Mattos; Erika Freitas; Evandra Strazza Rodrigues; Felipe Allan da Silva da Costa; Guilherme Targino Valente; Heidge Fukumasu. USP-Botucatu: Rejane Maria Tommasini Grotto; Instituto Butantan: Dimas Tadeu Covas; Jayme A. Souza-Neto; Jessika Cristina Chagas Lesbon; José Salvatore Leister Patané; João Paulo Kitajima; Luiz Carlos Junior de Alcant; Maria Carolina Elias; Patricia Akemi Assato; Rafael dos Santos Bezerra; Raquel de Lello Rocha Campos Cassano. NGS Soluções Genômicas: Pilar Drummond Sampaio Corrêa Mariani. FZEA-USP Pirassununga: Mirele Daiana Poleti; Raul Machado Neto; Ricardo Augusto Brassaloti; Ricardo Haddad; Sandra Coccuzzo Sampaio; Svetoslav Nanev Slavov; Vincent Louis Viala |
| EPI_ISL_1716397, EPI_ISL_1716398, EPI_ISL_1716399, EPI_ISL_1716400, EPI_ISL_1716401, EPI_ISL_1716402, EPI_ISL_1716403, EPI_ISL_1716404, EPI_ISL_1716405, EPI_ISL_1716406, EPI_ISL_1716407, EPI_ISL_1716408, EPI_ISL_1716409, EPI_ISL_1716410, EPI_ISL_1716411, EPI_ISL_1716412, EPI_ISL_1716413, EPI_ISL_1716414, EPI_ISL_1716415, EPI_ISL_1716416, EPI_ISL_1716417, EPI_ISL_1716418, EPI_ISL_1716419, EPI_ISL_1716420, EPI_ISL_1716421, EPI_ISL_1716422, EPI_ISL_1716423, EPI_ISL_1716424, EPI_ISL_1716425, EPI_ISL_1716426, EPI_ISL_1716427, EPI_ISL_1716428, EPI_ISL_1716429, EPI_ISL_1716430, EPI_ISL_1716431, EPI_ISL_1716432, EPI_ISL_1716433, EPI_ISL_1716434, EPI_ISL_1716435, EPI_ISL_1716436, EPI_ISL_1716437, EPI_ISL_1716438, EPI_ISL_1716439, EPI_ISL_1716440, EPI_ISL_1716441, EPI_ISL_1716442, EPI_ISL_1716443, EPI_ISL_1716444, EPI_ISL_1716445, EPI_ISL_1716446, EPI_ISL_1716447, EPI_ISL_1716448, EPI_ISL_1716449, EPI_ISL_1716450, EPI_ISL_1716451, EPI_ISL_1716452, EPI_ISL_1716453, EPI_ISL_1716454, EPI_ISL_1716455, EPI_ISL_1716456 | | | |
| see above | Laboratorio Saude | LGBio (Laboratorio de Genetica & Biodiversidade) | Amanda Alves de Melo; Cintia Pelegrineti Targueta de Azevedo Brito; Daniela de Melo e Silva; Elisangela de Paula Silveira Lacerda; Francyelli Mello Andrade; Mariana Pires de Campos Telles; Ramilla dos Santos Braga; Renata de Oliveira Dias; Rhewter Nunes; Thais Guimarães Castro; Thays Millena Alves Pedroso |
| EPI_ISL_2008906, EPI_ISL_2008907, EPI_ISL_2008908, EPI_ISL_2008909, EPI_ISL_2008910, EPI_ISL_2008911, EPI_ISL_2008912, EPI_ISL_2008913, EPI_ISL_2008914, EPI_ISL_2008915, EPI_ISL_2008916, EPI_ISL_2008917, EPI_ISL_2008918, EPI_ISL_2008919, EPI_ISL_2008920, EPI_ISL_2008921, EPI_ISL_2008922, EPI_ISL_2008923, EPI_ISL_2008924, EPI_ISL_2008925, EPI_ISL_2008926, EPI_ISL_2008927, EPI_ISL_2008928, EPI_ISL_2008929, EPI_ISL_2008930, EPI_ISL_2008931, EPI_ISL_2008932, EPI_ISL_2008933, EPI_ISL_2008934, EPI_ISL_2008935, EPI_ISL_2008940, EPI_ISL_2008941, EPI_ISL_2008944, EPI_ISL_2008945, EPI_ISL_2008946, EPI_ISL_2008947, EPI_ISL_2008948, EPI_ISL_2008949, EPI_ISL_2008950, EPI_ISL_2008951, EPI_ISL_2008952, EPI_ISL_2008953, EPI_ISL_2008954, EPI_ISL_2008955, EPI_ISL_2008956, EPI_ISL_2008957, EPI_ISL_2008958, EPI_ISL_2008959, EPI_ISL_2008960, EPI_ISL_2008961, EPI_ISL_2008962, EPI_ISL_2008963, EPI_ISL_2008964, EPI_ISL_2008965, EPI_ISL_2008966, EPI_ISL_2008967, EPI_ISL_2107292, EPI_ISL_2107293, EPI_ISL_2107294, EPI_ISL_2107295, EPI_ISL_2107296, EPI_ISL_2107297, EPI_ISL_2107298, EPI_ISL_2107301, EPI_ISL_2107302, EPI_ISL_2107303, EPI_ISL_2107305, EPI_ISL_2154703, EPI_ISL_2154956 | | | |
| see above | Laboratorio de Pesquisa em Virologia, FAMERP, SJRP | Laboratorio de Pesquisa em Virologia, FAMERP, SJRP | Cecília Artico Banho; Cíntia Bittar; Fábio Sossai Possebon; Guilherme Campos; Helena Lage Ferreira; Jorge A. Petroli Marchesi; João Pessoa Araújo Jr.; Leila Sabrina Ullmann; Lívia Sacchetto; Maisa C. Pereira Parra; Marília Moraes; Maurício L. Nogueira.; Paula Rahal; Paulo Inacio da Costa |
| EPI_ISL_1786560, EPI_ISL_1786561, EPI_ISL_1786562, EPI_ISL_1786563, EPI_ISL_1786564, EPI_ISL_1786565, EPI_ISL_1786566, EPI_ISL_1786567, EPI_ISL_1786568, EPI_ISL_1786569, EPI_ISL_1786570 | | | |
| see above | Laboratorio de Virologia Clinica do HCFMRP-USP | Hocentro de Ribeirao Preto - Laboratório de Biologia Molecular | Aparecida Yulie Yamamoto; Diego Villa Clé; Dimas Tadeu Covas; Elaine Vieira Santos; Evandra Strazza Rodrigues; Glauco de Carvalho Pereira; Joilson Xavier; Josiane Serrano Borges; Luiz Carlos Junior Alcantara; Mariane Evaristo; Marta Giovanetti; Rafael dos Santos Bezerra; Rodrigo Tocantins Calado; Simone Kashima; Svetoslav Nanev Slavov; Talita Adelino; Vagner Fonseca |
| EPI_ISL_1534011, EPI_ISL_1534014, EPI_ISL_1534015, EPI_ISL_1534016, EPI_ISL_2157540, EPI_ISL_2157545 | Laboratory of Respiratory Viruses and Measles, Oswaldo Cruz Institute, FIOCRUZ | Laboratory of Respiratory Viruses and Measles, Oswaldo Cruz Institute, FIOCRUZ | Alice Sampaio Rocha; Ana Carolina Mendonca; Andrea Cony Cavalcanti; Anna Carolina Paixao; Elisa Cavalcante Pereira; Fernando Motta; Luciana Appolinario; Marilda Siqueira on behalf of the Fiocruz COVID-19 Genomic Surveillance Network; Paola Resende; Renata Serrano Lopes; Taina Venas |
| EPI_ISL_2157532, EPI_ISL_2157533, EPI_ISL_2157534, EPI_ISL_2157535, EPI_ISL_2157542, EPI_ISL_2157543, EPI_ISL_2157544, EPI_ISL_2157598 | | | |
| see above | Laboratório Central de Saude Publica do Estado da Bahia (LACEN/BA) | Laboratory of Respiratory Viruses and Measles, Oswaldo Cruz Institute, FIOCRUZ | Alice Sampaio Rocha; Ana Carolina Mendonca; Anna Carolina Paixao; Elisa Cavalcante Pereira; Felicidade Pereira; Fernando Motta; Luciana Appolinario; Marilda Siqueira on behalf of the Fiocruz COVID-19 Genomic Surveillance Network; Paola Resende; Renata Serrano Lopes; Taina Venas |
| EPI_ISL_2157377, EPI_ISL_2157391, EPI_ISL_2157392, EPI_ISL_2157393, EPI_ISL_2157396, EPI_ISL_2157398, EPI_ISL_2157399, EPI_ISL_2157400, EPI_ISL_2157405, EPI_ISL_2157406, EPI_ISL_2157407, EPI_ISL_2157409, EPI_ISL_2157410, EPI_ISL_2157411, EPI_ISL_2157418, EPI_ISL_2157426, EPI_ISL_2157447, EPI_ISL_2157448, EPI_ISL_2157450, EPI_ISL_2157484, EPI_ISL_2157493, EPI_ISL_2157494, EPI_ISL_2157495, EPI_ISL_2157497, EPI_ISL_2157498, EPI_ISL_2157499, EPI_ISL_2157500, EPI_ISL_2157501, EPI_ISL_2157502, EPI_ISL_2157503, EPI_ISL_2157510, EPI_ISL_2157511, EPI_ISL_2157513, EPI_ISL_2157515, EPI_ISL_2157518, EPI_ISL_2157547, EPI_ISL_2157548, EPI_ISL_2157590, EPI_ISL_2157592 | | | |
| see above | Laboratório Central de Saude Publica do Estado de Santa Catarina (LACEN/SC) | Laboratory of Respiratory Viruses and Measles, Oswaldo Cruz Institute, FIOCRUZ | Alice Sampaio Rocha; Ana Carolina Mendonca; Anna Carolina Paixao; Darcita Buerger Rovaris; Elisa Cavalcante Pereira; Fernando Motta; Luciana Appolinario; Marilda Siqueira on behalf of the Fiocruz COVID-19 Genomic Surveillance Network; Paola Resende; Renata Serrano Lopes; Sandra Bianchini Fernandes; Taina Venas |
| EPI_ISL_2157506, EPI_ISL_2157516, EPI_ISL_2157517 | Laboratório Central de Saude Publica do Estado do Paraná (LACEN/PR) | Laboratory of Respiratory Viruses and Measles, Oswaldo Cruz Institute, FIOCRUZ | Alice Sampaio Rocha; Ana Carolina Mendonca; Anna Carolina Paixao; Elisa Cavalcante Pereira; Fernando Motta; Irina Nastassja Riediger; Luciana Appolinario; Maria do Carmo Debur; Marilda Siqueira on behalf of the Fiocruz COVID-19 Genomic Surveillance Network; Paola Resende; Renata Serrano Lopes; Taina Venas |
| EPI_ISL_2157524, EPI_ISL_2157525, EPI_ISL_2157526, EPI_ISL_2157528, EPI_ISL_2157529, EPI_ISL_2157530, EPI_ISL_2157536, EPI_ISL_2157537, EPI_ISL_2157538, EPI_ISL_2157539, EPI_ISL_2157549, EPI_ISL_2157599 | | | |
| see above | Laboratório Central de Saude Publica do Estado do Rio de Janeiro (LACEN/RJ) | Laboratory of Respiratory Viruses and Measles, Oswaldo Cruz Institute, FIOCRUZ | Alice Sampaio Rocha; Ana Carolina Mendonca; Andrea Cony Cavalcanti; Anna Carolina Paixao; Elisa Cavalcante Pereira; Fernando Motta; Luciana Appolinario; Marilda Siqueira on behalf of the Fiocruz COVID-19 Genomic Surveillance Network; Paola Resende; Renata Serrano Lopes; Taina Venas |
| EPI_ISL_2102018, EPI_ISL_2102063 | Laboratório Central de Saúde Pública do Estado do Amazonas (LACEN-AM) | Laboratorio de Ecologia de Doencas Transmissiveis na Amazonia, Instituto Leonidas e Maria Deane - Fiocruz Amazonia | André Corado; Debora Duarte; Felipe Naveca; Fernanda Nascimento; George Silva; Karina Pessoa; Luciana Gonçalves; Maria Júlia Brandão; Matilde Mejía; Valdinete Nascimento; Victor Souza; Ágatha Costa |
| EPI_ISL_1592227, EPI_ISL_1592228, EPI_ISL_1592229 | Laboratório Hermes Pardini | Laboratório de Biologia Integrativa, Instituto de Ciências Biológicas, Universidade Federal de Minas Gerais | Alessandro Clayton de Souza Ferreira; Aline Brito de Lima; Carolina Moreira Voloch; Daniel Costa Queiroz; Danielle Alves Gomes Zauli; Diego Menezes Bonfim; Filipe Romero Rebello Moreira; Frederico Scott Varella Malta; Joice do Prado Silva; Rafael Marques de Souza; Renan Pedra de Souza; Renato Santana Aguiar.; Rennan Garcias Moreira; Victor Cavalcanti Pardini; Victor Emmanuel Viana Geddes; Wagner Carlos Santos Magalhaes; Walyson Coelho Costa |
| EPI_ISL_2181398 | Laboratório de Agentes Infecciosos | Laboratório Baculovírus | Aline Belmok; Bergmann M Ribeiro; Fernando L Melo; Jaime Amorim; Josilene Pinheiro |

| | | | |
|---|---|---|---|
| | e Vetores -LAIVE- Bahia, Barreiras, Brazil | | |
| EPI_ISL_1495009, EPI_ISL_1495010, EPI_ISL_1495011, EPI_ISL_1495012, EPI_ISL_1495013, EPI_ISL_1495014, EPI_ISL_1495015, EPI_ISL_1495016, EPI_ISL_1495017, EPI_ISL_1495018, EPI_ISL_1495019, EPI_ISL_1495020, EPI_ISL_1495021, EPI_ISL_1495022, EPI_ISL_1495023, EPI_ISL_1495024, EPI_ISL_1495025, EPI_ISL_1495026, EPI_ISL_1495027, EPI_ISL_1495028, EPI_ISL_1495029, EPI_ISL_1495030, EPI_ISL_1495031, EPI_ISL_1495035, EPI_ISL_1495036, EPI_ISL_1495037, EPI_ISL_1495038 | | | |
| see above | Laboratório de Biologia Integrativa | Laboratório de Biologia Integrativa | Alessandro Clayton de Souza Ferreira; Aline Brito de Lima; Carolina Moreira Voloch; Daniel Costa Queiroz; Danielle Alves Gomes Zauli; Diego Menezes Bonfim; Filipe Romero Rebello Moreira; Frederico Scott Varella Malta; Joice do Prado Silva; Lucyene Miguita Luiz; Nuno Rodrigues Faria; Paula Luize Camargos Fonseca; Rafael Marques de Souza; Renan Pedra de Souza; Renato Santana Aguiar; Rennan Garcias Moreira; Victor Cavalcanti Pardini; Victor Emmanuel Viana Geddes |
| EPI_ISL_1578453 | Laboratório de Biologia Molecular do Hospital das Clínicas da Faculdade de Medicina de Botucatu/SP | Laboratórios Genômica Funcional (FCA/UNESP) e Biologia Molecular (FMB-HC/UNESP) - Rede de Vigilância Genômica (Vigenômica)/UNESP | Bianca Cechetto Carlos; Felipe Allan da Silva da Costa; Flavia Hebner Barbosa Trovão; Guilherme Targino Valente; Jayme A. Souza-Neto.; Patrícia Akemi Assato; Rejane Maria Tommasini Grotto |
| EPI_ISL_1578740, EPI_ISL_1579258, EPI_ISL_1580269, EPI_ISL_1580502 | Laboratório de Biologia Molecular do Hospital das Clínicas da Faculdade de Medicina de Botucatu/SP | Laboratórios de Genômica Funcional (FCA/UNESP) e Biologia Molecular (FMB-HC/UNESP) - Rede de Vigilância Genômica (Vigenômica)/UNESP | Bianca Cechetto Carlos; Felipe Allan da Silva da Costa; Flavia Hebner Barbosa Trovão; Guilherme Targino Valente; Jayme A. Souza-Neto.; Patrícia Akemi Assato; Rejane Maria Tommasini Grotto |
| EPI_ISL_1629809 | Laboratório de Microbiologia Molecular - Universidade FEEVALE | Molecular Microbiology Laboratory | Alana Witt Hansen; Fernando Rosado Spilki; Flávio Silveira; Fágner Henrique Heldt; Juliana Schons Gulart; Juliane Deise Fleck; Mariana Soares da Silva; Matheus Nunes Weber; Meriane Demoliner; Michele Filippi.; Paula Rodrigues de Almeida |
| EPI_ISL_1754186, EPI_ISL_1785612 | Laboratório de Pesquisa em Virologia, FAMERP, SJRP | Laboratório de Pesquisa em Virologia, FAMERP, SJRP | Cecília Artico Banho; Cíntia Bittar; Fábio Sossai Possebon; Guilherme Campos; Helena Lage Ferreira; Jorge A. Petroli Marchesi; João Pessoa Araújo Jr.; Leila Sabrina Ullmann; Lívia Sacchetto; Maisa C. Pereira Parra; Marília Moraes; Maurício L. Nogueira; Paula Rahal; Paulo Inacio da Costa |
| EPI_ISL_1464626, EPI_ISL_1464627, EPI_ISL_1464628, EPI_ISL_1464629, EPI_ISL_1464630, EPI_ISL_1464631, EPI_ISL_1464632, EPI_ISL_1464633, EPI_ISL_1464634, EPI_ISL_1464635, EPI_ISL_1464636, EPI_ISL_1464637, EPI_ISL_1464638, EPI_ISL_1464639, EPI_ISL_1464640, EPI_ISL_1464641, EPI_ISL_1464642, EPI_ISL_1464643, EPI_ISL_1464644, EPI_ISL_1464645, EPI_ISL_1464646, EPI_ISL_1464647, EPI_ISL_1464648, EPI_ISL_1464649, EPI_ISL_1464650, EPI_ISL_1464651, EPI_ISL_1464652, EPI_ISL_1464653, EPI_ISL_1464654, EPI_ISL_1464655, EPI_ISL_1464656, EPI_ISL_1464657, EPI_ISL_1464658, EPI_ISL_1464659, EPI_ISL_1464660, EPI_ISL_1464661, EPI_ISL_1464662, EPI_ISL_1464663, EPI_ISL_1464664, EPI_ISL_1464665, EPI_ISL_1464666, EPI_ISL_1464667, EPI_ISL_1464668, EPI_ISL_1464669, EPI_ISL_1464670, EPI_ISL_1464671, EPI_ISL_1464672, EPI_ISL_1464673, EPI_ISL_1464674, EPI_ISL_1464675, EPI_ISL_1464676, EPI_ISL_1464677 | | | |
| see above | Laboratório de Virologia - UNIFESP | Laboratory of Respiratory Viruses and Measles, Oswaldo Cruz Institute, FIOCRUZ | Alice Sampaio Rocha; Ana Carolina Mendonca; Anna Carolina Paixao; Fernando Motta; Luciana Appolinario; Marilda Siqueira on behalf of the Fiocruz COVID-19 Genomic Surveillance Network; Nancy Belei; Paola Resende; Renata Serrano Lopes |
| EPI_ISL_2157343, EPI_ISL_2157397, EPI_ISL_2157402, EPI_ISL_2157403, EPI_ISL_2157404, EPI_ISL_2157408, EPI_ISL_2157413, EPI_ISL_2157416, EPI_ISL_2157417, EPI_ISL_2157419, EPI_ISL_2157420, EPI_ISL_2157421, EPI_ISL_2157424, EPI_ISL_2157431, EPI_ISL_2157452, EPI_ISL_2157479, EPI_ISL_2157483, EPI_ISL_2157485, EPI_ISL_2157486, EPI_ISL_2157487, EPI_ISL_2157488, EPI_ISL_2157492, EPI_ISL_2157496, EPI_ISL_2157504, EPI_ISL_2157527, EPI_ISL_2157593, EPI_ISL_2157595, EPI_ISL_2157597 | | | |
| see above | Lboratorio Central de Saude Publica do Estado do Parana (LACEN/PR) | Laboratory of Respiratory Viruses and Measles, Oswaldo Cruz Institute, FIOCRUZ | Alice Sampaio Rocha; Ana Carolina Mendonca; Anna Carolina Paixao; Elisa Cavalcante Pereira; Fernando Motta; Irina Riediger; Luciana Appolinario; Marilda Siqueira on behalf of the Fiocruz COVID-19 Genomic Surveillance Network; Paola Resende; Renata Serrano Lopes; Taina Venas |
| EPI_ISL_1795077 | MARIA TEREZINHA BOGNAR CHECHETO | Instituto Butantan / ESALQ-Piracicaba | Antonio Jorge Martins; Bianca Cechetto Carlos. Mendelics: Bibiana Santos; Claudia Renata dos Santos Barros; David Schlesinger. Hemocentro Ribeirão Preto: Simone Kashima; Debora Botequio Moretti. Centro de Genômica Funcional da ESALQ: Luiz Lehmann Coutinho; Dimas Tadeu Covas; Elaine Cristina Marqueze; Elaine Vieira dos Santos; Elisangela Chicaroni Mattos; Erika Freitas; Evandra Strazza Rodrigues; Felipe Allan da Silva da Costa; Flavia Aburjaile; Guilherme Targino Valente; Heidge Fukumasu. USP-Botucatu: Rejane Maria Tommasini Grotto; Instituto Butantan: Alexander Roberto Precioso; Jayme A. Souza-Neto; Jessika Cristina Chagas Lesbon; José Salvatore Leister Patané; João Paulo Kitajima; Luiz Carlos Junior de Alcantara; Maria Carolina Elias; Marta Giovanetti; Patricia Akemi Assato; Rafael dos Santos Bezerra; Raquel de Lello Rocha Campos Cassano. NGS Soluções Genômicas: Pilar Drummond Sampaio Corrêa Mariani. FZEA-USP Pirassununga: Mirele Daiana Poleti; Raul Machado Neto; Ricardo Augusto Brassaloti; Ricardo Haddad; Rodrigo Tocantins Calado.; Sandra Coccuzzo Sampaio; Svetoslav Nanev Slavov; Vagner Fonseca; Vincent Louis Viala |
| EPI_ISL_2170926 | NUCLEO CENTRAL DE SAUDE JALES | Instituto Butantan / Mendelics | Antonio Jorge Martins; Bianca Cechetto Carlos. Mendelics: Bibiana Santos; Claudia Renata dos Santos Barros; Cíntia Bittar; David Schlesinger. Hemocentro Ribeirão Preto: Simone Kashima; Debora Botequio Moretti; Elaine Cristina Marqueze; Elaine Vieira dos Santos; Elisangela Chicaroni Mattos; Erika Freitas; Evandra Strazza Rodrigues; Felipe Allan da Silva da Costa; Flavia Aburjaile; Fábio Sossai Possebon; Guilherme Campos; Guilherme Targino Valente; Heidge Fukumasu. USP-Botucatu: Rejane Maria Tommasini Grotto; Helena Lage Ferreira; Instituto Butantan: Dimas Tadeu Covas; Jardelina de Souza Todao Bernardino; Jayme A. Souza-Neto; Jessika Cristina Chagas Lesbon; Jorge A. Petroli Marchesi; José Salvatore Leister Patané; João Paulo Kitajima; João Pessoa Araújo Jr.; Leila Sabrina Ullmann; Loyze Paola Oliveira de Lima; Luiz Aurelio de Campos Crispin. Centro de Genômica Funcional da ESALQ: Luiz Lehmann Coutinho; Luiz Carlos Junior de Alcantara; Lívia Sacchetto; Maisa C. Pereira Parra; Maria Carolina Elias; Marta Giovanetti; Marília Moraes; Maurício Lacerda Nogueira. Prefeitura de Sao Paulo: Melissa Palmieri.; Patricia Akemi Assato; Paula Rahal; Paulo Inacio da Costa; Rafael dos Santos Bezerra; Raquel de Lello Rocha Campos Cassano. NGS Soluções Genômicas: Pilar Drummond Sampaio Corrêa Mariani. FZEA-USP Pirassununga: Mirele Daiana Poleti; Raul Machado Neto; Ricardo Augusto Brassaloti; Ricardo Haddad; Rodrigo Tocantins Calado. FAMERP-SJRP: Cecília Artico Banho; Sandra Coccuzzo Sampaio; Svetoslav Nanev Slavov; Vagner Fonseca; Vincent Louis Viala |
| EPI_ISL_1967213, EPI_ISL_1967214, EPI_ISL_1967215, EPI_ISL_1967217 | NUCLEO DE SAUDE MARY DOTA DE BAURU | Instituto Butantan / Mendelics | Antonio Jorge Martins; Bianca Cechetto Carlos. Mendelics: Bibiana Santos; Claudia Renata dos Santos Barros; Cíntia Bittar; David Schlesinger. Hemocentro Ribeirão Preto: Simone Kashima; Debora Botequio Moretti; Elaine Cristina Marqueze; Elaine Vieira dos Santos; Elisangela Chicaroni Mattos; Erika Freitas; Evandra Strazza Rodrigues; Felipe Allan da Silva da Costa; Flavia Aburjaile; Fábio Sossai Possebon; Guilherme Campos; Guilherme Targino Valente; Heidge Fukumasu. USP-Botucatu: Rejane Maria Tommasini Grotto; Helena Lage Ferreira; Instituto Butantan: Dimas Tadeu Covas; Jardelina de Souza Todao Bernardino; Jayme A. Souza-Neto; Jessika Cristina Chagas Lesbon; Jorge A. Petroli Marchesi; José Salvatore Leister Patané; João Paulo Kitajima; João Pessoa Araújo Jr.; Leila Sabrina Ullmann; Loyze Paola Oliveira de Lima; Luiz Aurelio de Campos Crispin. Centro de Genômica Funcional da ESALQ: Luiz Lehmann Coutinho; Luiz Carlos Junior de Alcantara; Lívia Sacchetto; Maisa C. Pereira Parra; Maria Carolina Elias; Marta Giovanetti; Marília Moraes; Maurício Lacerda Nogueira. Prefeitura de Sao Paulo: Melissa Palmieri.; Patricia Akemi Assato; Paula Rahal; Paulo Inacio da Costa; Rafael dos Santos Bezerra; Raquel de Lello Rocha Campos Cassano. NGS Soluções Genômicas: Pilar Drummond Sampaio Corrêa Mariani. FZEA-USP Pirassununga: Mirele Daiana Poleti; Raul Machado Neto; Ricardo Augusto Brassaloti; Ricardo Haddad; Rodrigo Tocantins Calado. FAMERP-SJRP: Cecília Artico Banho; Sandra Coccuzzo Sampaio; Svetoslav Nanev Slavov; Vagner Fonseca; Vincent Louis Viala |
| EPI_ISL_1795379 | NUCLEO DE SAUDE VILA FALCAO DE BAURU | Instituto Butantan / ESALQ-Piracicaba | Antonio Jorge Martins; Bianca Cechetto Carlos. Mendelics: Bibiana Santos; Claudia Renata dos Santos Barros; David Schlesinger. Hemocentro Ribeirão Preto: Simone Kashima; Debora Botequio Moretti. Centro de Genômica Funcional da ESALQ: Luiz Lehmann Coutinho; Dimas Tadeu Covas; Elaine Cristina Marqueze; Elaine Vieira dos Santos; Elisangela Chicaroni Mattos; Erika Freitas; Evandra Strazza Rodrigues; Felipe Allan da Silva da Costa; Flavia Aburjaile; Guilherme Targino Valente; Heidge Fukumasu. USP-Botucatu: Rejane Maria Tommasini Grotto; Instituto Butantan: Alexander Roberto Precioso; Jayme A. Souza-Neto; Jessika Cristina Chagas Lesbon; José Salvatore Leister Patané; João Paulo Kitajima; Luiz Carlos Junior de Alcantara; Maria Carolina Elias; Marta Giovanetti; Patricia Akemi Assato; Rafael dos Santos Bezerra; Raquel de Lello Rocha Campos Cassano. NGS Soluções Genômicas: Pilar Drummond Sampaio Corrêa Mariani. FZEA-USP Pirassununga: Mirele Daiana Poleti; Raul Machado Neto; Ricardo Augusto Brassaloti; Ricardo Haddad; Rodrigo Tocantins Calado.; Sandra Coccuzzo Sampaio; Svetoslav Nanev Slavov; Vagner Fonseca; Vincent Louis Viala |
| EPI_ISL_1966446, EPI_ISL_1966447, EPI_ISL_1966448, EPI_ISL_1966449, EPI_ISL_1966450, EPI_ISL_1966451, EPI_ISL_1966452, EPI_ISL_1966453, EPI_ISL_1966454, EPI_ISL_1966455, EPI_ISL_1966456, EPI_ISL_1966457, EPI_ISL_1966458, EPI_ISL_1966459, EPI_ISL_1966562, EPI_ISL_1966563, EPI_ISL_1966564, EPI_ISL_1966566, EPI_ISL_1966567, EPI_ISL_1966568, EPI_ISL_1966570, EPI_ISL_1966571, EPI_ISL_1966572, EPI_ISL_1966574, EPI_ISL_1966575, EPI_ISL_1966576, EPI_ISL_1966577, EPI_ISL_1966578, EPI_ISL_1966579, EPI_ISL_1966580, EPI_ISL_1966581, EPI_ISL_1966582, EPI_ISL_1966583, EPI_ISL_1966584, EPI_ISL_1966585, EPI_ISL_1966586, EPI_ISL_1966587, EPI_ISL_1966588, EPI_ISL_1966589, EPI_ISL_1966590, EPI_ISL_1966591, EPI_ISL_1966592, EPI_ISL_2170935, EPI_ISL_2170940 | | | |
| see above | NUCLEO DE SAUDE VILA FALCAO DE BAURU | Instituto Butantan / Mendelics | Antonio Jorge Martins; Bianca Cechetto Carlos. Mendelics: Bibiana Santos; Claudia Renata dos Santos Barros; Cíntia Bittar; David Schlesinger. Hemocentro Ribeirão Preto: Simone Kashima; Debora Botequio Moretti; Elaine Cristina Marqueze; Elaine Vieira dos Santos; Elisangela Chicaroni Mattos; Erika Freitas; Evandra Strazza Rodrigues; Felipe Allan da Silva da Costa; Flavia Aburjaile; Fábio Sossai Possebon; Guilherme Campos; Guilherme Targino Valente; Heidge Fukumasu. USP-Botucatu: Rejane Maria Tommasini Grotto; Helena Lage Ferreira; Instituto Butantan: Dimas Tadeu Covas; Jardelina de Souza Todao Bernardino; Jayme A. Souza-Neto; Jessika Cristina Chagas Lesbon; Jorge A. Petroli Marchesi; José Salvatore Leister Patané; João Paulo Kitajima; João Pessoa Araújo Jr.; Leila Sabrina Ullmann; Loyze Paola Oliveira de Lima; Luiz Aurelio de Campos Crispin. Centro de Genômica Funcional da ESALQ: Luiz Lehmann Coutinho; Luiz Carlos Junior de Alcantara; Lívia Sacchetto; Maisa C. Pereira Parra; Maria Carolina Elias; Marta Giovanetti; Marília Moraes; Maurício Lacerda Nogueira. Prefeitura de Sao Paulo: Melissa Palmieri.; Patricia Akemi Assato; Paula Rahal; Paulo Inacio da Costa; Rafael dos Santos Bezerra; Raquel de Lello Rocha Campos Cassano. NGS Soluções Genômicas: Pilar Drummond Sampaio Corrêa Mariani. FZEA-USP Pirassununga: Mirele Daiana Poleti; Raul Machado Neto; Ricardo Augusto Brassaloti; Ricardo Haddad; Rodrigo Tocantins Calado. FAMERP-SJRP: Cecília Artico Banho; Sandra Coccuzzo Sampaio; Svetoslav Nanev Slavov; Vagner Fonseca; Vincent Louis Viala |
| EPI_ISL_1795075, EPI_ISL_1795076 | NUCLEO SAUDE III N HABITACIONAL PRESIDENTE GEISEL BAURU | Instituto Butantan / ESALQ-Piracicaba | Antonio Jorge Martins; Bianca Cechetto Carlos. Mendelics: Bibiana Santos; Claudia Renata dos Santos Barros; David Schlesinger. Hemocentro Ribeirão Preto: Simone Kashima; Debora Botequio Moretti. Centro de Genômica Funcional da ESALQ: Luiz Lehmann Coutinho; Dimas Tadeu Covas; Elaine Cristina Marqueze; Elaine Vieira dos Santos; Elisangela Chicaroni Mattos; Erika Freitas; Evandra Strazza Rodrigues; Felipe Allan da Silva da Costa; Flavia Aburjaile; Guilherme Targino Valente; Heidge Fukumasu. USP-Botucatu: Rejane Maria Tommasini Grotto; Instituto Butantan: Alexander Roberto Precioso; Jayme A. Souza-Neto; Jessika Cristina Chagas Lesbon; José Salvatore Leister Patané; João Paulo Kitajima; Luiz Carlos Junior de Alcantara; Maria Carolina Elias; Marta Giovanetti; Patricia Akemi Assato; Rafael dos Santos Bezerra; Raquel de Lello Rocha Campos Cassano. NGS Soluções Genômicas: Pilar Drummond Sampaio Corrêa Mariani. FZEA-USP Pirassununga: Mirele Daiana Poleti; Raul Machado Neto; Ricardo Augusto Brassaloti; Ricardo Haddad; Rodrigo Tocantins Calado.; Sandra Coccuzzo Sampaio; Svetoslav Nanev Slavov; Vagner Fonseca; Vincent Louis Viala |
| EPI_ISL_1967005 | PA DE IBITIUVA DR OTAVIO BENETTI PITANGUEIRAS | Instituto Butantan / ESALQ-USP (Piracicaba) | Antonio Jorge Martins; Bianca Cechetto Carlos. Mendelics: Bibiana Santos; Claudia Renata dos Santos Barros; Cíntia Bittar; David Schlesinger. Hemocentro Ribeirão Preto: Simone Kashima; Debora Botequio Moretti; Elaine Cristina Marqueze; Elaine Vieira dos Santos; Elisangela Chicaroni Mattos; Erika Freitas; Evandra Strazza Rodrigues; Felipe Allan da Silva da Costa; Flavia Aburjaile; Fábio Sossai Possebon; Guilherme Campos; Guilherme Targino Valente; Heidge Fukumasu. USP-Botucatu: Rejane Maria Tommasini Grotto; Helena Lage Ferreira; Instituto Butantan: Dimas Tadeu Covas; Jardelina de Souza Todao Bernardino; Jayme A. Souza-Neto; Jessika Cristina Chagas Lesbon; Jorge A. Petroli Marchesi; José Salvatore Leister Patané; João Paulo Kitajima; João Pessoa Araújo Jr.; Leila Sabrina Ullmann; Loyze Paola Oliveira de Lima; Luiz Aurelio de Campos Crispin. Centro de Genômica Funcional da ESALQ: Luiz Lehmann Coutinho; Luiz Carlos Junior de Alcantara; Lívia Sacchetto; Maisa C. Pereira Parra; Maria Carolina Elias; Marta Giovanetti; Marília Moraes; Maurício Lacerda Nogueira. Prefeitura de Sao Paulo: Melissa Palmieri.; Patricia Akemi Assato; Paula Rahal; Paulo Inacio da Costa; Rafael dos Santos Bezerra; Raquel de Lello Rocha Campos Cassano. NGS Soluções Genômicas: Pilar Drummond Sampaio Corrêa Mariani. FZEA-USP Pirassununga: Mirele Daiana Poleti; Raul Machado Neto; Ricardo Augusto Brassaloti; Ricardo Haddad; Rodrigo Tocantins Calado. FAMERP-SJRP: Cecília Artico Banho; Sandra Coccuzzo Sampaio; Svetoslav Nanev Slavov; Vagner Fonseca; Vincent Louis Viala |
| EPI_ISL_1966475 | PAS CIDADE JARDIM | Instituto Butantan / Mendelics | Antonio Jorge Martins; Bianca Cechetto Carlos. Mendelics: Bibiana Santos; Claudia Renata dos Santos Barros; Cíntia Bittar; David Schlesinger. Hemocentro Ribeirão Preto: Simone Kashima; Debora Botequio Moretti; Elaine Cristina Marqueze; Elaine Vieira dos Santos; Elisangela Chicaroni Mattos; Erika Freitas; Evandra Strazza Rodrigues; Felipe Allan da Silva da Costa; Flavia Aburjaile; Fábio Sossai Possebon; Guilherme Campos; Guilherme Targino Valente; Heidge Fukumasu. USP-Botucatu: Rejane Maria Tommasini Grotto; Helena Lage Ferreira; Instituto Butantan: Dimas Tadeu Covas; Jardelina de Souza Todao Bernardino; Jayme A. Souza-Neto; Jessika Cristina Chagas Lesbon; Jorge A. Petroli Marchesi; José Salvatore Leister Patané; João Paulo Kitajima; João Pessoa Araújo Jr.; Leila Sabrina Ullmann; Loyze Paola Oliveira de Lima; Luiz Aurelio de Campos Crispin. Centro de Genômica Funcional da ESALQ: Luiz Lehmann Coutinho; Luiz Carlos Junior de Alcantara; Lívia Sacchetto; Maisa C. Pereira Parra; Maria Carolina Elias; Marta Giovanetti; Marília Moraes; Maurício Lacerda Nogueira. Prefeitura de Sao Paulo: Melissa Palmieri.; Patricia Akemi Assato; Paula Rahal; Paulo Inacio da Costa; Rafael dos Santos Bezerra; Raquel de Lello Rocha Campos Cassano. NGS Soluções Genômicas: Pilar Drummond Sampaio Corrêa Mariani. FZEA-USP Pirassununga: Mirele Daiana Poleti; Raul Machado Neto; Ricardo Augusto Brassaloti; Ricardo Haddad; Rodrigo Tocantins Calado. FAMERP-SJRP: Cecília Artico Banho; Sandra Coccuzzo Sampaio; Svetoslav Nanev Slavov; Vagner Fonseca; Vincent Louis Viala |
| EPI_ISL_1795336 | PAS JOAO ANTONIO DO NASCIMENTO | Instituto Butantan / ESALQ-Piracicaba | Antonio Jorge Martins; Bianca Cechetto Carlos. Mendelics: Bibiana Santos; Claudia Renata dos Santos Barros; David Schlesinger. Hemocentro Ribeirão Preto: Simone Kashima; Debora Botequio Moretti. Centro de Genômica Funcional da ESALQ: Luiz Lehmann Coutinho; Dimas Tadeu Covas; Elaine Cristina Marqueze; Elaine Vieira dos Santos; Elisangela Chicaroni Mattos; Erika Freitas; Evandra Strazza Rodrigues; Felipe Allan da Silva da Costa; Flavia Aburjaile; Guilherme Targino Valente; Heidge Fukumasu. USP-Botucatu: Rejane Maria Tommasini Grotto; Instituto Butantan: Alexander Roberto Precioso; Jayme A. Souza-Neto; Jessika Cristina Chagas Lesbon; José Salvatore Leister Patané; João Paulo Kitajima; Luiz Carlos Junior de Alcantara; Maria Carolina Elias; Marta Giovanetti; Patricia Akemi Assato; Rafael dos Santos Bezerra; Raquel de Lello Rocha Campos Cassano. NGS Soluções Genômicas: Pilar Drummond Sampaio Corrêa Mariani. FZEA-USP Pirassununga: Mirele Daiana Poleti; Raul Machado Neto; Ricardo Augusto Brassaloti; Ricardo Haddad; Rodrigo Tocantins Calado.; Sandra Coccuzzo Sampaio; Svetoslav Nanev Slavov; Vagner Fonseca; Vincent Louis Viala |
| EPI_ISL_1966474 | PAS SILVIA COVAS | Instituto Butantan / Mendelics | Antonio Jorge Martins; Bianca Cechetto Carlos. Mendelics: Bibiana Santos; Claudia Renata dos Santos Barros; Cíntia Bittar; David Schlesinger. Hemocentro Ribeirão Preto: Simone Kashima; Debora Botequio Moretti; Elaine Cristina Marqueze; Elaine Vieira dos Santos; Elisangela Chicaroni Mattos; Erika Freitas; Evandra Strazza Rodrigues; Felipe Allan da Silva da Costa; Flavia Aburjaile; Fábio Sossai Possebon; Guilherme Campos; Guilherme Targino Valente; Heidge Fukumasu. USP-Botucatu: Rejane Maria Tommasini Grotto; Helena Lage Ferreira; Instituto Butantan: Dimas Tadeu Covas; Jardelina de Souza Todao Bernardino; Jayme A. Souza-Neto; Jessika Cristina Chagas Lesbon; Jorge A. Petroli Marchesi; José Salvatore Leister Patané; João Paulo Kitajima; João Pessoa Araújo Jr.; Leila Sabrina Ullmann; Loyze Paola Oliveira de Lima; Luiz Aurelio de Campos Crispin. Centro de Genômica Funcional da ESALQ: Luiz Lehmann Coutinho; Luiz Carlos Junior de Alcantara; Lívia Sacchetto; Maisa C. Pereira Parra; Maria Carolina Elias; Marta Giovanetti; Marília Moraes; Maurício Lacerda Nogueira. Prefeitura de Sao Paulo: Melissa Palmieri.; Patricia Akemi Assato; Paula Rahal; Paulo Inacio da Costa; Rafael dos Santos Bezerra; Raquel de Lello Rocha Campos Cassano. NGS Soluções Genômicas: Pilar Drummond Sampaio Corrêa Mariani. FZEA-USP Pirassununga: Mirele Daiana Poleti; Raul Machado Neto; Ricardo Augusto Brassaloti; Ricardo Haddad; Rodrigo Tocantins Calado. FAMERP-SJRP: Cecília Artico Banho; Sandra Coccuzzo Sampaio; Svetoslav Nanev Slavov; Vagner Fonseca; Vincent Louis Viala |
| EPI_ISL_1966916, EPI_ISL_1966917 | PENITENCIARIA ODON RAMOS MARANHAO IPERO | Instituto Butantan / ESALQ-USP (Piracicaba) | Antonio Jorge Martins; Bianca Cechetto Carlos. Mendelics: Bibiana Santos; Claudia Renata dos Santos Barros; Cíntia Bittar; David Schlesinger. Hemocentro Ribeirão Preto: Simone Kashima; Debora Botequio Moretti; Elaine Cristina Marqueze; Elaine Vieira dos Santos; Elisangela Chicaroni Mattos; Erika Freitas; Evandra Strazza Rodrigues; Felipe Allan da Silva da Costa; Flavia Aburjaile; Fábio Sossai Possebon; Guilherme Campos; Guilherme Targino Valente; Heidge Fukumasu. USP-Botucatu: Rejane Maria Tommasini Grotto; Helena Lage Ferreira; Instituto Butantan: Dimas Tadeu Covas; Jardelina de Souza Todao Bernardino; Jayme A. Souza-Neto; Jessika Cristina Chagas Lesbon; Jorge A. Petroli Marchesi; José Salvatore Leister Patané; João Paulo Kitajima; João Pessoa Araújo Jr.; Leila Sabrina Ullmann; Loyze Paola Oliveira de Lima; Luiz Aurelio de Campos Crispin. Centro de Genômica Funcional da ESALQ: Luiz Lehmann Coutinho; Luiz Carlos Junior de Alcantara; Lívia Sacchetto; Maisa C. Pereira Parra; Maria Carolina Elias; Marta Giovanetti; Marília Moraes; Maurício Lacerda Nogueira. Prefeitura de Sao Paulo: Melissa Palmieri.; Patricia Akemi Assato; Paula Rahal; Paulo Inacio da Costa; Rafael dos Santos Bezerra; Raquel de Lello Rocha Campos Cassano. NGS Soluções Genômicas: Pilar Drummond Sampaio Corrêa Mariani. FZEA-USP Pirassununga: Mirele Daiana Poleti; Raul Machado Neto; Ricardo Augusto Brassaloti; Ricardo Haddad; Rodrigo Tocantins Calado. FAMERP-SJRP: Cecília Artico Banho; Sandra Coccuzzo Sampaio; Svetoslav Nanev Slavov; Vagner Fonseca; Vincent Louis Viala |

| | | | |
|---|---|---|---|
| EPI_ISL_1966927, EPI_ISL_1966928, EPI_ISL_1966929, EPI_ISL_1966930, EPI_ISL_1966931, EPI_ISL_1966932, EPI_ISL_1966933, EPI_ISL_1966934 | | | |
| see above | POLICLINICA COVID 19 ITAPETININGA | Instituto Butantan / ESALQ-USP (Piracicaba) | Antonio Jorge Martins; Bianca Cechetto Carlos. Mendelics: Bibiana Santos; Claudia Renata dos Santos Barros; Cíntia Bittar; David Schlesinger. Hemocentro Ribeirão Preto: Simone Kashima; Debora Botequio Moretti; Elaine Cristina Marqueze; Elaine Vieira dos Santos; Elisangela Chicaroni Mattos; Erika Freitas; Evandra Strazza Rodrigues; Felipe Allan da Silva da Costa; Flavia Aburjaile; Fábio Sossai Possebon; Guilherme Campos; Guilherme Targino Valente; Heidge Fukumasu. USP-Botucatu: Rejane Maria Tommasini Grotto; Helena Lage Ferreira; Instituto Butantan: Dimas Tadeu Covas; Jardelina de Souza Todao Bernardino; Jayme A. Souza-Neto; Jessika Cristina Chagas Lesbon; Jorge A. Petroli Marchesi; José Salvatore Leister Patané; João Paulo Kitajima; João Pessoa Araújo Jr.; Leila Sabrina Ullmann; Loyze Paola Oliveira de Lima; Luiz Aurelio de Campos Crispin. Centro de Genômica Funcional da ESALQ: Luiz Lehmann Coutinho; Luiz Carlos Junior de Alcantara; Lívia Sacchetto; Maisa C. Pereira Parra; Maria Carolina Elias; Marta Giovanetti; Marília Moraes; Maurício Lacerda Nogueira. Prefeitura de Sao Paulo: Melissa Palmieri.; Patricia Akemi Assato; Paula Rahal; Paulo Inacio da Costa; Rafael dos Santos Bezerra; Raquel de Lello Rocha Campos Cassano. NGS Soluções Genômicas: Pilar Drummond Sampaio Corrêa Mariani. FZEA-USP Pirassununga: Mirele Daiana Poleti; Raul Machado Neto; Ricardo Augusto Brassaloti; Ricardo Haddad; Rodrigo Tocantins Calado. FAMERP-SJRP: Cecília Artico Banho; Sandra Coccuzzo Sampaio; Svetoslav Nanev Slavov; Vagner Fonseca; Vincent Louis Viala |
| EPI_ISL_1445090, EPI_ISL_1966104, EPI_ISL_1966379, EPI_ISL_1966381, EPI_ISL_1966382, EPI_ISL_1966383, EPI_ISL_1966384, EPI_ISL_1966464, EPI_ISL_1966465, EPI_ISL_1966467, EPI_ISL_1966766, EPI_ISL_1966767, EPI_ISL_1966768, EPI_ISL_1966770 | | | |
| see above | POLICLINICA COVID 19 ITAPETININGA | Instituto Butantan / Mendelics | Antonio Jorge Martins; Bianca Cechetto Carlos. Mendelics: Bibiana Santos; Bibiana Santos; Claudia Renata dos Santos Barros; Cíntia Bittar; David Schlesinger; David Schlesinger. Hemocentro Ribeirão Preto: Simone Kashima; Debora Botequio Moretti; Dimas Tadeu Covas; Elaine Cristina Marqueze; Elaine Vieira dos Santos; Elisangela Chicaroni Mattos; Erika Freitas; Evandra Strazza Rodrigues; Felipe Allan da Silva da Costa; Flavia Aburjaile; Fábio Sossai Possebon; Guilherme Campos; Guilherme Targino Valente; Heidge Fukumasu. USP-Botucatu: Rejane Maria Tommasini Grotto; Helena Lage Ferreira; Instituto Butantan: Dimas Tadeu Covas; Jardelina de Souza Todao Bernardino; Jayme A. Souza-Neto; Jessika Cristina Chagas Lesbon; Jorge A. Petroli Marchesi; José Salvatore Leister Patané; João Paulo Kitajima; João Pessoa Araújo Jr.; Leila Sabrina Ullmann; Loyze Paola Oliveira de Lima; Luiz Aurelio de Campos Crispin. Centro de Genômica Funcional da ESALQ: Luiz Lehmann Coutinho; Luiz Carlos Junior de Alcantara; Lívia Sacchetto; Maisa C. Pereira Parra; Maria Carolina Elias; Marta Giovanetti; Marília Moraes; Maurício Lacerda Nogueira. Prefeitura de Sao Paulo: Melissa Palmieri.; Patricia Akemi Assato; Paula Rahal; Paulo Inacio da Costa; Rafael dos Santos Bezerra; Raquel de Lello Rocha Campos Cassano. NGS Soluções Genômicas: Pilar Drummond Sampaio Corrêa Mariani. FZEA-USP Pirassununga: Mirele Daiana Poleti; Raul Machado Neto; Ricardo Augusto Brassaloti; Ricardo Haddad; Rodrigo Tocantins Calado.; Rodrigo Tocantins Calado. FAMERP-SJRP: Cecília Artico Banho; Sandra Coccuzzo Sampaio; Simone Kashima; Svetoslav Nanev Slavov; Vagner Fonseca; Vincent Louis Viala |
| EPI_ISL_1795061, EPI_ISL_1795082, EPI_ISL_1795083, EPI_ISL_1795141, EPI_ISL_1795142, EPI_ISL_1795143, EPI_ISL_1795144, EPI_ISL_1795145, EPI_ISL_1795146, EPI_ISL_1795147, EPI_ISL_1795148, EPI_ISL_1795152, EPI_ISL_1795153, EPI_ISL_1795154, EPI_ISL_1795155, EPI_ISL_1795156, EPI_ISL_1795157, EPI_ISL_1795158, EPI_ISL_1795159, EPI_ISL_1795177, EPI_ISL_1795178, EPI_ISL_1795179, EPI_ISL_1795180, EPI_ISL_1795181, EPI_ISL_1795183, EPI_ISL_1795184, EPI_ISL_1795185, EPI_ISL_1795186, EPI_ISL_1795188, EPI_ISL_1795190, EPI_ISL_1795191, EPI_ISL_1795387, EPI_ISL_1795388, EPI_ISL_1795392, EPI_ISL_1795399, EPI_ISL_1795404, EPI_ISL_1795411 | | | |
| see above | POLICLINICA HORTOLANDIA | Instituto Butantan / ESALQ-Piracicaba | Antonio Jorge Martins; Bianca Cechetto Carlos. Mendelics: Bibiana Santos; Claudia Renata dos Santos Barros; David Schlesinger. Hemocentro Ribeirão Preto: Simone Kashima; Debora Botequio Moretti. Centro de Genômica Funcional da ESALQ: Luiz Lehmann Coutinho; Dimas Tadeu Covas; Elaine Cristina Marqueze; Elaine Vieira dos Santos; Elisangela Chicaroni Mattos; Erika Freitas; Evandra Strazza Rodrigues; Felipe Allan da Silva da Costa; Flavia Aburjaile; Guilherme Targino Valente; Heidge Fukumasu. USP-Botucatu: Rejane Maria Tommasini Grotto; Instituto Butantan: Alexander Roberto Precioso; Jayme A. Souza-Neto; Jessika Cristina Chagas Lesbon; José Salvatore Leister Patané; João Paulo Kitajima; Luiz Carlos Junior de Alcantara; Maria Carolina Elias; Marta Giovanetti; Patricia Akemi Assato; Rafael dos Santos Bezerra; Raquel de Lello Rocha Campos Cassano. NGS Soluções Genômicas: Pilar Drummond Sampaio Corrêa Mariani. FZEA-USP Pirassununga: Mirele Daiana Poleti; Raul Machado Neto; Ricardo Augusto Brassaloti; Ricardo Haddad; Rodrigo Tocantins Calado.; Sandra Coccuzzo Sampaio; Svetoslav Nanev Slavov; Vagner Fonseca; Vincent Louis Viala |
| EPI_ISL_1966616, EPI_ISL_1966617, EPI_ISL_1966618, EPI_ISL_1966619, EPI_ISL_1966620, EPI_ISL_1966621, EPI_ISL_1966622, EPI_ISL_1966623, EPI_ISL_1966624, EPI_ISL_1966625, EPI_ISL_2170945, EPI_ISL_2171007, EPI_ISL_2171008, EPI_ISL_2171009, EPI_ISL_2171010, EPI_ISL_2171012, EPI_ISL_2171013, EPI_ISL_2171014, EPI_ISL_2171015, EPI_ISL_2171016 | | | |
| see above | POLICLINICA HORTOLANDIA | Instituto Butantan / Mendelics | Antonio Jorge Martins; Bianca Cechetto Carlos. Mendelics: Bibiana Santos; Claudia Renata dos Santos Barros; Cíntia Bittar; David Schlesinger. Hemocentro Ribeirão Preto: Simone Kashima; Debora Botequio Moretti; Elaine Cristina Marqueze; Elaine Vieira dos Santos; Elisangela Chicaroni Mattos; Erika Freitas; Evandra Strazza Rodrigues; Felipe Allan da Silva da Costa; Flavia Aburjaile; Fábio Sossai Possebon; Guilherme Campos; Guilherme Targino Valente; Heidge Fukumasu. USP-Botucatu: Rejane Maria Tommasini Grotto; Helena Lage Ferreira; Instituto Butantan: Dimas Tadeu Covas; Jardelina de Souza Todao Bernardino; Jayme A. Souza-Neto; Jessika Cristina Chagas Lesbon; Jorge A. Petroli Marchesi; José Salvatore Leister Patané; João Paulo Kitajima; João Pessoa Araújo Jr.; Leila Sabrina Ullmann; Loyze Paola Oliveira de Lima; Luiz Aurelio de Campos Crispin. Centro de Genômica Funcional da ESALQ: Luiz Lehmann Coutinho; Luiz Carlos Junior de Alcantara; Lívia Sacchetto; Maisa C. Pereira Parra; Maria Carolina Elias; Marta Giovanetti; Marília Moraes; Maurício Lacerda Nogueira. Prefeitura de Sao Paulo: Melissa Palmieri.; Patricia Akemi Assato; Paula Rahal; Paulo Inacio da Costa; Rafael dos Santos Bezerra; Raquel de Lello Rocha Campos Cassano. NGS Soluções Genômicas: Pilar Drummond Sampaio Corrêa Mariani. FZEA-USP Pirassununga: Mirele Daiana Poleti; Raul Machado Neto; Ricardo Augusto Brassaloti; Ricardo Haddad; Rodrigo Tocantins Calado. FAMERP-SJRP: Cecília Artico Banho; Sandra Coccuzzo Sampaio; Svetoslav Nanev Slavov; Vagner Fonseca; Vincent Louis Viala |
| EPI_ISL_1966244 | POSTO MEDICO NATAL DIEGUES DE ESTIVA GERBI | Instituto Butantan / Mendelics | Antonio Jorge Martins; Bianca Cechetto Carlos. Mendelics: Bibiana Santos; Claudia Renata dos Santos Barros; Cíntia Bittar; David Schlesinger. Hemocentro Ribeirão Preto: Simone Kashima; Debora Botequio Moretti; Elaine Cristina Marqueze; Elaine Vieira dos Santos; Elisangela Chicaroni Mattos; Erika Freitas; Evandra Strazza Rodrigues; Felipe Allan da Silva da Costa; Flavia Aburjaile; Fábio Sossai Possebon; Guilherme Campos; Guilherme Targino Valente; Heidge Fukumasu. USP-Botucatu: Rejane Maria Tommasini Grotto; Helena Lage Ferreira; Instituto Butantan: Dimas Tadeu Covas; Jardelina de Souza Todao Bernardino; Jayme A. Souza-Neto; Jessika Cristina Chagas Lesbon; Jorge A. Petroli Marchesi; José Salvatore Leister Patané; João Paulo Kitajima; João Pessoa Araújo Jr.; Leila Sabrina Ullmann; Loyze Paola Oliveira de Lima; Luiz Aurelio de Campos Crispin. Centro de Genômica Funcional da ESALQ: Luiz Lehmann Coutinho; Luiz Carlos Junior de Alcantara; Lívia Sacchetto; Maisa C. Pereira Parra; Maria Carolina Elias; Marta Giovanetti; Marília Moraes; Maurício Lacerda Nogueira. Prefeitura de Sao Paulo: Melissa Palmieri.; Patricia Akemi Assato; Paula Rahal; Paulo Inacio da Costa; Rafael dos Santos Bezerra; Raquel de Lello Rocha Campos Cassano. NGS Soluções Genômicas: Pilar Drummond Sampaio Corrêa Mariani. FZEA-USP Pirassununga: Mirele Daiana Poleti; Raul Machado Neto; Ricardo Augusto Brassaloti; Ricardo Haddad; Rodrigo Tocantins Calado. FAMERP-SJRP: Cecília Artico Banho; Sandra Coccuzzo Sampaio; Svetoslav Nanev Slavov; Vagner Fonseca; Vincent Louis Viala |
| EPI_ISL_1445224, EPI_ISL_1966218 | PPA FENELON GUEDES PEREIRA | Instituto Butantan / Mendelics | Antonio Jorge Martins; Bianca Cechetto Carlos. Mendelics: Bibiana Santos; Bibiana Santos; Claudia Renata dos Santos Barros; Cíntia Bittar; David Schlesinger; David Schlesinger. Hemocentro Ribeirão Preto: Simone Kashima; Debora Botequio Moretti; Dimas Tadeu Covas; Elaine Cristina Marqueze; Elaine Vieira dos Santos; Elisangela Chicaroni Mattos; Erika Freitas; Evandra Strazza Rodrigues; Felipe Allan da Silva da Costa; Flavia Aburjaile; Fábio Sossai Possebon; Guilherme Campos; Guilherme Targino Valente; Heidge Fukumasu. USP-Botucatu: Rejane Maria Tommasini Grotto; Helena Lage Ferreira; Instituto Butantan: Dimas Tadeu Covas; Jardelina de Souza Todao Bernardino; Jayme A. Souza-Neto; Jessika Cristina Chagas Lesbon; Jorge A. Petroli Marchesi; José Salvatore Leister Patané; João Paulo Kitajima; João Pessoa Araújo Jr.; Leila Sabrina Ullmann; Loyze Paola Oliveira de Lima; Luiz Aurelio de Campos Crispin. Centro de Genômica Funcional da ESALQ: Luiz Lehmann Coutinho; Luiz Carlos Junior de Alcantara; Lívia Sacchetto; Maisa C. Pereira Parra; Maria Carolina Elias; Marta Giovanetti; Marília Moraes; Maurício Lacerda Nogueira. Prefeitura de Sao Paulo: Melissa Palmieri.; Patricia Akemi Assato; Paula Rahal; Paulo Inacio da Costa; Rafael dos Santos Bezerra; Raquel de Lello Rocha Campos Cassano. NGS Soluções Genômicas: Pilar Drummond Sampaio Corrêa Mariani. FZEA-USP Pirassununga: Mirele Daiana Poleti; Raul Machado Neto; Ricardo Augusto Brassaloti; Ricardo Haddad; Rodrigo Tocantins Calado.; Rodrigo Tocantins Calado. FAMERP-SJRP: Cecília Artico Banho; Sandra Coccuzzo Sampaio; Simone Kashima; Svetoslav Nanev Slavov; Vagner Fonseca; Vincent Louis Viala |
| EPI_ISL_1966727 | PPA MARCIO VALDEVINO BATISTA | Instituto Butantan | Antonio Jorge Martins; Bianca Cechetto Carlos. Mendelics: Bibiana Santos; Claudia Renata dos Santos Barros; Cíntia Bittar; David Schlesinger. Hemocentro Ribeirão Preto: Simone Kashima; Debora Botequio Moretti; Elaine Cristina Marqueze; Elaine Vieira dos Santos; Elisangela Chicaroni Mattos; Erika Freitas; Evandra Strazza Rodrigues; Felipe Allan da Silva da Costa; Flavia Aburjaile; Fábio Sossai Possebon; Guilherme Campos; Guilherme Targino Valente; Heidge Fukumasu. USP-Botucatu: Rejane Maria Tommasini Grotto; Helena Lage Ferreira; Instituto Butantan: Dimas Tadeu Covas; Jardelina de Souza Todao Bernardino; Jayme A. Souza-Neto; Jessika Cristina Chagas Lesbon; Jorge A. Petroli Marchesi; José Salvatore Leister Patané; João Paulo Kitajima; João Pessoa Araújo Jr.; Leila Sabrina Ullmann; Loyze Paola Oliveira de Lima; Luiz Aurelio de Campos Crispin. Centro de Genômica Funcional da ESALQ: Luiz Lehmann Coutinho; Luiz Carlos Junior de Alcantara; Lívia Sacchetto; Maisa C. Pereira Parra; Maria Carolina Elias; Marta Giovanetti; Marília Moraes; Maurício Lacerda Nogueira. Prefeitura de Sao Paulo: Melissa Palmieri.; Patricia Akemi Assato; Paula Rahal; Paulo Inacio da Costa; Rafael dos Santos Bezerra; Raquel de Lello Rocha Campos Cassano. NGS Soluções Genômicas: Pilar Drummond Sampaio Corrêa Mariani. FZEA-USP Pirassununga: Mirele Daiana Poleti; Raul Machado Neto; Ricardo Augusto Brassaloti; Ricardo Haddad; Rodrigo Tocantins Calado. FAMERP-SJRP: Cecília Artico Banho; Sandra Coccuzzo Sampaio; Svetoslav Nanev Slavov; Vagner Fonseca; Vincent Louis Viala |
| EPI_ISL_1445207, EPI_ISL_2170913 | PPA MARCIO VALDEVINO BATISTA | Instituto Butantan / Mendelics | Antonio Jorge Martins; Bianca Cechetto Carlos. Mendelics: Bibiana Santos; Bibiana Santos; Claudia Renata dos Santos Barros; Cíntia Bittar; David Schlesinger; David Schlesinger. Hemocentro Ribeirão Preto: Simone Kashima; Debora Botequio Moretti; Dimas Tadeu Covas; Elaine Cristina Marqueze; Elaine Vieira dos Santos; Elisangela Chicaroni Mattos; Erika Freitas; Evandra Strazza Rodrigues; Felipe Allan da Silva da Costa; Flavia Aburjaile; Fábio Sossai Possebon; Guilherme Campos; Guilherme Targino Valente; Heidge Fukumasu. USP-Botucatu: Rejane Maria Tommasini Grotto; Helena Lage Ferreira; Instituto Butantan: Dimas Tadeu Covas; Jardelina de Souza Todao Bernardino; Jayme A. Souza-Neto; Jessika Cristina Chagas Lesbon; Jorge A. Petroli Marchesi; José Salvatore Leister Patané; João Paulo Kitajima; João Pessoa Araújo Jr.; Leila Sabrina Ullmann; Loyze Paola Oliveira de Lima; Luiz Aurelio de Campos Crispin. Centro de Genômica Funcional da ESALQ: Luiz Lehmann Coutinho; Luiz Carlos Junior de Alcantara; Lívia Sacchetto; Maisa C. Pereira Parra; Maria Carolina Elias; Marta Giovanetti; Marília Moraes; Maurício Lacerda Nogueira. Prefeitura de Sao Paulo: Melissa Palmieri.; Patricia Akemi Assato; Paula Rahal; Paulo Inacio da Costa; Rafael dos Santos Bezerra; Raquel de Lello Rocha Campos Cassano. NGS Soluções Genômicas: Pilar Drummond Sampaio Corrêa Mariani. FZEA-USP Pirassununga: Mirele Daiana Poleti; Raul Machado Neto; Ricardo Augusto Brassaloti; Ricardo Haddad; Rodrigo Tocantins Calado.; Rodrigo Tocantins Calado. FAMERP-SJRP: Cecília Artico Banho; Sandra Coccuzzo Sampaio; Simone Kashima; Svetoslav Nanev Slavov; Vagner Fonseca; Vincent Louis Viala |
| EPI_ISL_1966910, EPI_ISL_1966911 | PR S DA FAMILIA UNIDADE DE SAUDE ADALBERTO ROCHA GUAREI | Instituto Butantan / ESALQ-USP (Piracicaba) | Antonio Jorge Martins; Bianca Cechetto Carlos. Mendelics: Bibiana Santos; Claudia Renata dos Santos Barros; Cíntia Bittar; David Schlesinger. Hemocentro Ribeirão Preto: Simone Kashima; Debora Botequio Moretti; Elaine Cristina Marqueze; Elaine Vieira dos Santos; Elisangela Chicaroni Mattos; Erika Freitas; Evandra Strazza Rodrigues; Felipe Allan da Silva da Costa; Flavia Aburjaile; Fábio Sossai Possebon; Guilherme Campos; Guilherme Targino Valente; Heidge Fukumasu. USP-Botucatu: Rejane Maria Tommasini Grotto; Helena Lage Ferreira; Instituto Butantan: Dimas Tadeu Covas; Jardelina de Souza Todao Bernardino; Jayme A. Souza-Neto; Jessika Cristina Chagas Lesbon; Jorge A. Petroli Marchesi; José Salvatore Leister Patané; João Paulo Kitajima; João Pessoa Araújo Jr.; Leila Sabrina Ullmann; Loyze Paola Oliveira de Lima; Luiz Aurelio de Campos Crispin. Centro de Genômica Funcional da ESALQ: Luiz Lehmann Coutinho; Luiz Carlos Junior de Alcantara; Lívia Sacchetto; Maisa C. Pereira Parra; Maria Carolina Elias; Marta Giovanetti; Marília Moraes; Maurício Lacerda Nogueira. Prefeitura de Sao Paulo: Melissa Palmieri.; Patricia Akemi Assato; Paula Rahal; Paulo Inacio da Costa; Rafael dos Santos Bezerra; Raquel de Lello Rocha Campos Cassano. NGS Soluções Genômicas: Pilar Drummond Sampaio Corrêa Mariani. FZEA-USP Pirassununga: Mirele Daiana Poleti; Raul Machado Neto; Ricardo Augusto Brassaloti; Ricardo Haddad; Rodrigo Tocantins Calado. FAMERP-SJRP: Cecília Artico Banho; Sandra Coccuzzo Sampaio; Svetoslav Nanev Slavov; Vagner Fonseca; Vincent Louis Viala |
| EPI_ISL_1445130, EPI_ISL_1445131, EPI_ISL_1445132, EPI_ISL_1966174, EPI_ISL_1966175, EPI_ISL_1966176 | PRONTO ATENDIMENTO DA PONTE DE SAO JOAO | Instituto Butantan / Mendelics | Antonio Jorge Martins; Bianca Cechetto Carlos. Mendelics: Bibiana Santos; Bibiana Santos; Claudia Renata dos Santos Barros; Cíntia Bittar; David Schlesinger; David Schlesinger. Hemocentro Ribeirão Preto: Simone Kashima; Debora Botequio Moretti; Dimas Tadeu Covas; Elaine Cristina Marqueze; Elaine Vieira dos Santos; Elisangela Chicaroni Mattos; Erika Freitas; Evandra Strazza Rodrigues; Felipe Allan da Silva da Costa; Flavia Aburjaile; Fábio Sossai Possebon; Guilherme Campos; Guilherme Targino Valente; Heidge Fukumasu. USP-Botucatu: Rejane Maria Tommasini Grotto; Helena Lage Ferreira; Instituto Butantan: Dimas Tadeu Covas; Jardelina de Souza Todao Bernardino; Jayme A. Souza-Neto; Jessika Cristina Chagas Lesbon; Jorge A. Petroli Marchesi; José Salvatore Leister Patané; João Paulo Kitajima; João Pessoa Araújo Jr.; Leila Sabrina Ullmann; Loyze Paola Oliveira de Lima; Luiz Aurelio de Campos Crispin. Centro de Genômica Funcional da ESALQ: Luiz Lehmann Coutinho; Luiz Carlos Junior de Alcantara; Lívia Sacchetto; Maisa C. Pereira Parra; Maria Carolina Elias; Marta Giovanetti; Marília Moraes; Maurício Lacerda Nogueira. Prefeitura de Sao Paulo: Melissa Palmieri.; Patricia Akemi Assato; Paula Rahal; Paulo Inacio da Costa; Rafael dos Santos Bezerra; Raquel de Lello Rocha Campos Cassano. NGS Soluções Genômicas: Pilar Drummond Sampaio Corrêa Mariani. FZEA-USP Pirassununga: Mirele Daiana Poleti; Raul Machado Neto; Ricardo Augusto Brassaloti; Ricardo Haddad; Rodrigo Tocantins Calado.; Rodrigo Tocantins Calado. FAMERP-SJRP: Cecília Artico Banho; Sandra Coccuzzo Sampaio; Simone Kashima; Svetoslav Nanev Slavov; Vagner Fonseca; Vincent Louis Viala |
| EPI_ISL_1967281 | PRONTO ATENDIMENTO MUNICIPAL | Instituto Butantan / Mendelics | Antonio Jorge Martins; Bianca Cechetto Carlos. Mendelics: Bibiana Santos; Claudia Renata dos Santos Barros; Cíntia Bittar; David Schlesinger. Hemocentro Ribeirão Preto: Simone Kashima; Debora Botequio Moretti; Elaine Cristina Marqueze; Elaine Vieira dos Santos; Elisangela Chicaroni Mattos; Erika Freitas; Evandra Strazza Rodrigues; Felipe Allan da Silva da Costa; Flavia Aburjaile; Fábio Sossai Possebon; Guilherme Campos; Guilherme Targino Valente; Heidge Fukumasu. USP-Botucatu: Rejane Maria Tommasini Grotto; Helena Lage Ferreira; Instituto Butantan: Dimas Tadeu Covas; Jardelina de Souza Todao Bernardino; Jayme A. Souza-Neto; Jessika Cristina Chagas Lesbon; Jorge A. Petroli Marchesi; José Salvatore Leister Patané; João Paulo Kitajima; João Pessoa Araújo Jr.; Leila Sabrina Ullmann; Loyze Paola Oliveira de Lima; Luiz Aurelio de Campos Crispin. Centro de Genômica Funcional da ESALQ: Luiz Lehmann Coutinho; Luiz Carlos Junior de Alcantara; Lívia Sacchetto; Maisa C. Pereira Parra; Maria Carolina Elias; Marta Giovanetti; Marília Moraes; Maurício Lacerda Nogueira. Prefeitura de Sao Paulo: Melissa Palmieri.; Patricia Akemi Assato; Paula Rahal; Paulo Inacio da Costa; Rafael dos Santos Bezerra; Raquel de Lello Rocha Campos Cassano. NGS Soluções Genômicas: Pilar Drummond Sampaio Corrêa Mariani. FZEA-USP Pirassununga: Mirele Daiana Poleti; Raul Machado Neto; Ricardo Augusto Brassaloti; Ricardo Haddad; Rodrigo Tocantins Calado. FAMERP-SJRP: Cecília Artico Banho; Sandra Coccuzzo Sampaio; Svetoslav Nanev Slavov; Vagner Fonseca; Vincent Louis Viala |
| EPI_ISL_1966823 | PRONTO ATENDIMENTO MUNICIPAL ALUMINIO | Instituto Butantan / ESALQ-USP (Piracicaba) | Antonio Jorge Martins; Bianca Cechetto Carlos. Mendelics: Bibiana Santos; Claudia Renata dos Santos Barros; Cíntia Bittar; David Schlesinger. Hemocentro Ribeirão Preto: Simone Kashima; Debora Botequio Moretti; Elaine Cristina Marqueze; Elaine Vieira dos Santos; Elisangela Chicaroni Mattos; Erika Freitas; Evandra Strazza Rodrigues; Felipe Allan da Silva da Costa; Flavia Aburjaile; Fábio Sossai Possebon; Guilherme Campos; Guilherme Targino Valente; Heidge Fukumasu. USP-Botucatu: Rejane Maria Tommasini Grotto; Helena Lage Ferreira; Instituto Butantan: Dimas Tadeu Covas; Jardelina de Souza Todao Bernardino; Jayme A. Souza-Neto; Jessika Cristina Chagas Lesbon; Jorge A. Petroli Marchesi; José Salvatore Leister Patané; João Paulo Kitajima; João Pessoa Araújo Jr.; Leila Sabrina Ullmann; Loyze Paola Oliveira de Lima; Luiz Aurelio de Campos Crispin. Centro de Genômica Funcional da ESALQ: Luiz Lehmann Coutinho; Luiz Carlos Junior de Alcantara; Lívia Sacchetto; Maisa C. Pereira Parra; Maria Carolina Elias; Marta Giovanetti; Marília Moraes; Maurício Lacerda Nogueira. Prefeitura de Sao Paulo: Melissa Palmieri.; Patricia Akemi Assato; Paula Rahal; Paulo Inacio da Costa; Rafael dos Santos Bezerra; Raquel de Lello Rocha Campos Cassano. NGS Soluções Genômicas: Pilar Drummond Sampaio Corrêa Mariani. FZEA-USP Pirassununga: Mirele Daiana Poleti; Raul Machado Neto; Ricardo Augusto Brassaloti; Ricardo Haddad; Rodrigo Tocantins Calado. FAMERP-SJRP: Cecília Artico Banho; Sandra Coccuzzo Sampaio; Svetoslav Nanev Slavov; Vagner Fonseca; Vincent Louis Viala |
| EPI_ISL_1966952, EPI_ISL_1966953 | PRONTO ATENDIMENTO MUNICIPAL DE JACUPIRANGA | Instituto Butantan / ESALQ-USP (Piracicaba) | Antonio Jorge Martins; Bianca Cechetto Carlos. Mendelics: Bibiana Santos; Claudia Renata dos Santos Barros; Cíntia Bittar; David Schlesinger. Hemocentro Ribeirão Preto: Simone Kashima; Debora Botequio Moretti; Elaine Cristina Marqueze; Elaine Vieira dos Santos; Elisangela Chicaroni Mattos; Erika Freitas; Evandra Strazza Rodrigues; Felipe Allan da Silva da Costa; Flavia Aburjaile; Fábio Sossai Possebon; Guilherme Campos; Guilherme Targino Valente; Heidge Fukumasu. USP-Botucatu: Rejane Maria Tommasini Grotto; Helena Lage Ferreira; Instituto Butantan: Dimas Tadeu Covas; Jardelina de Souza Todao Bernardino; Jayme A. Souza-Neto; Jessika Cristina Chagas Lesbon; Jorge A. Petroli Marchesi; José Salvatore Leister Patané; João Paulo Kitajima; João Pessoa Araújo Jr.; Leila Sabrina Ullmann; Loyze Paola Oliveira de Lima; Luiz Aurelio de Campos Crispin. Centro de Genômica Funcional da ESALQ: Luiz Lehmann Coutinho; Luiz Carlos Junior de Alcantara; Lívia Sacchetto; Maisa C. Pereira Parra; Maria Carolina Elias; Marta Giovanetti; Marília Moraes; Maurício Lacerda Nogueira. Prefeitura de Sao Paulo: Melissa Palmieri.; Patricia Akemi Assato; Paula Rahal; Paulo Inacio da Costa; Rafael dos Santos Bezerra; Raquel de Lello Rocha Campos Cassano. NGS Soluções Genômicas: Pilar Drummond Sampaio Corrêa Mariani. FZEA-USP Pirassununga: Mirele Daiana Poleti; Raul Machado Neto; Ricardo Augusto Brassaloti; Ricardo Haddad; Rodrigo Tocantins Calado. FAMERP-SJRP: Cecília Artico Banho; Sandra Coccuzzo Sampaio; Svetoslav Nanev Slavov; Vagner Fonseca; Vincent Louis Viala |
| EPI_ISL_1966691, EPI_ISL_1966692 | PRONTO ATENDIMENTO MUNICIPAL DE JACUPIRANGA | Instituto Butantan / Mendelics | Antonio Jorge Martins; Bianca Cechetto Carlos. Mendelics: Bibiana Santos; Claudia Renata dos Santos Barros; Cíntia Bittar; David Schlesinger. Hemocentro Ribeirão Preto: Simone Kashima; Debora Botequio Moretti; Elaine Cristina Marqueze; Elaine Vieira dos Santos; Elisangela Chicaroni Mattos; Erika Freitas; Evandra Strazza Rodrigues; Felipe Allan da Silva da Costa; Flavia Aburjaile; Fábio Sossai Possebon; Guilherme Campos; Guilherme Targino Valente; Heidge Fukumasu. USP-Botucatu: Rejane Maria Tommasini Grotto; Helena Lage Ferreira; Instituto Butantan: Dimas Tadeu Covas; Jardelina de Souza Todao Bernardino; Jayme A. Souza-Neto; Jessika Cristina Chagas Lesbon; Jorge A. Petroli Marchesi; José Salvatore Leister Patané; João Paulo Kitajima; João Pessoa Araújo Jr.; Leila Sabrina Ullmann; Loyze Paola Oliveira de Lima; Luiz Aurelio de Campos Crispin. Centro de Genômica Funcional da ESALQ: Luiz Lehmann Coutinho; Luiz Carlos Junior de Alcantara; Lívia Sacchetto; Maisa C. Pereira Parra; Maria Carolina Elias; Marta Giovanetti; Marília Moraes; Maurício Lacerda Nogueira. Prefeitura de Sao Paulo: Melissa Palmieri.; Patricia Akemi Assato; Paula Rahal; Paulo Inacio da Costa; Rafael dos Santos Bezerra; Raquel de Lello Rocha Campos Cassano. NGS Soluções Genômicas: Pilar Drummond Sampaio Corrêa Mariani. FZEA-USP Pirassununga: Mirele Daiana Poleti; Raul Machado Neto; Ricardo Augusto Brassaloti; Ricardo Haddad; Rodrigo Tocantins Calado. FAMERP-SJRP: Cecília Artico Banho; Sandra Coccuzzo Sampaio; Svetoslav Nanev Slavov; Vagner Fonseca; Vincent Louis Viala |
| EPI_ISL_1966845 | PRONTO ATENDIMENTO MUNICIPAL ITALO SANTUCCI | Instituto Butantan / ESALQ-USP (Piracicaba) | Antonio Jorge Martins; Bianca Cechetto Carlos. Mendelics: Bibiana Santos; Claudia Renata dos Santos Barros; Cíntia Bittar; David Schlesinger. Hemocentro Ribeirão Preto: Simone Kashima; Debora Botequio Moretti; Elaine Cristina Marqueze; Elaine Vieira dos Santos; Elisangela Chicaroni Mattos; Erika Freitas; Evandra Strazza Rodrigues; Felipe Allan da Silva da Costa; Flavia Aburjaile; Fábio Sossai Possebon; Guilherme Campos; Guilherme Targino Valente; Heidge Fukumasu. USP-Botucatu: Rejane Maria Tommasini Grotto; Helena Lage Ferreira; Instituto Butantan: Dimas Tadeu Covas; Jardelina de Souza Todao Bernardino; Jayme A. Souza-Neto; Jessika Cristina Chagas Lesbon; Jorge A. Petroli Marchesi; José Salvatore Leister Patané; João Paulo Kitajima; João Pessoa Araújo Jr.; Leila Sabrina Ullmann; Loyze Paola Oliveira de Lima; Luiz Aurelio de Campos Crispin. Centro de Genômica Funcional da ESALQ: Luiz Lehmann Coutinho; Luiz Carlos Junior de Alcantara; Lívia Sacchetto; Maisa C. Pereira Parra; Maria Carolina Elias; Marta Giovanetti; Marília Moraes; Maurício Lacerda Nogueira. Prefeitura de Sao Paulo: Melissa Palmieri.; Patricia Akemi Assato; Paula Rahal; Paulo Inacio da Costa; Rafael dos Santos Bezerra; Raquel de Lello Rocha Campos Cassano. NGS Soluções Genômicas: Pilar Drummond Sampaio Corrêa Mariani. FZEA-USP Pirassununga: Mirele Daiana Poleti; Raul Machado Neto; Ricardo Augusto Brassaloti; Ricardo Haddad; Rodrigo Tocantins Calado. FAMERP-SJRP: Cecília Artico Banho; Sandra Coccuzzo Sampaio; Svetoslav Nanev Slavov; Vagner Fonseca; Vincent Louis Viala |
| EPI_ISL_1966751 | PRONTO ATENDIMENTO MUNICIPAL ITALO SANTUCCI | Instituto Butantan / Mendelics | Antonio Jorge Martins; Bianca Cechetto Carlos. Mendelics: Bibiana Santos; Claudia Renata dos Santos Barros; Cíntia Bittar; David Schlesinger. Hemocentro Ribeirão Preto: Simone Kashima; Debora Botequio Moretti; Elaine Cristina Marqueze; Elaine Vieira dos Santos; Elisangela Chicaroni Mattos; Erika Freitas; Evandra Strazza Rodrigues; Felipe Allan da Silva da Costa; Flavia Aburjaile; Fábio Sossai Possebon; Guilherme Campos; Guilherme Targino Valente; Heidge Fukumasu. USP-Botucatu: Rejane Maria Tommasini Grotto; Helena Lage Ferreira; Instituto Butantan: Dimas Tadeu Covas; Jardelina de Souza Todao Bernardino; Jayme A. Souza-Neto; Jessika Cristina Chagas Lesbon; Jorge A. Petroli Marchesi; José Salvatore Leister Patané; João Paulo Kitajima; João Pessoa Araújo Jr.; Leila Sabrina Ullmann; Loyze Paola Oliveira de Lima; Luiz Aurelio de Campos Crispin. Centro de Genômica Funcional da ESALQ: Luiz Lehmann Coutinho; Luiz Carlos Junior de Alcantara; Lívia Sacchetto; Maisa C. Pereira Parra; Maria Carolina Elias; Marta Giovanetti; Marília Moraes; Maurício Lacerda Nogueira. Prefeitura de Sao Paulo: Melissa Palmieri.; Patricia Akemi Assato; Paula Rahal; Paulo Inacio da Costa; Rafael dos Santos Bezerra; Raquel de Lello Rocha Campos Cassano. NGS Soluções Genômicas: Pilar Drummond Sampaio Corrêa Mariani. FZEA-USP Pirassununga: Mirele Daiana Poleti; Raul Machado Neto; Ricardo Augusto Brassaloti; Ricardo Haddad; Rodrigo Tocantins Calado. FAMERP-SJRP: Cecília Artico Banho; Sandra Coccuzzo Sampaio; Svetoslav Nanev Slavov; Vagner Fonseca; Vincent Louis Viala |
| EPI_ISL_1445185, EPI_ISL_1445187, EPI_ISL_1445193, EPI_ISL_1445194, EPI_ISL_1966220, EPI_ISL_1966222, EPI_ISL_1966226, EPI_ISL_1966361, EPI_ISL_1966362, EPI_ISL_1966363, EPI_ISL_2170919 | | | |
| see above | PRONTO ATENDIMENTO SAO JOSE | Instituto Butantan / Mendelics | Antonio Jorge Martins; Bianca Cechetto Carlos. Mendelics: Bibiana Santos; Bibiana Santos; Claudia Renata dos Santos Barros; Cíntia Bittar; David Schlesinger; David Schlesinger. Hemocentro Ribeirão Preto: Simone Kashima; Debora Botequio Moretti; Dimas Tadeu Covas; Elaine Cristina Marqueze; Elaine Vieira dos Santos; Elisangela Chicaroni Mattos; Erika Freitas; Evandra Strazza Rodrigues; Felipe Allan da Silva da Costa; Flavia Aburjaile; Fábio Sossai Possebon; Guilherme Campos; Guilherme Targino Valente; Heidge Fukumasu. USP-Botucatu: Rejane Maria Tommasini Grotto; Helena Lage Ferreira; Instituto Butantan: Dimas Tadeu Covas; Jardelina de Souza Todao Bernardino; Jayme A. Souza-Neto; Jessika Cristina Chagas Lesbon; Jorge A. Petroli Marchesi; José Salvatore Leister Patané; João Paulo Kitajima; João Pessoa Araújo Jr.; Leila Sabrina Ullmann; Loyze Paola Oliveira de Lima; Luiz Aurelio de Campos Crispin. Centro de Genômica Funcional da ESALQ: Luiz Lehmann Coutinho; Luiz Carlos Junior de Alcantara; Lívia Sacchetto; Maisa C. Pereira Parra; Maria Carolina Elias; Marta Giovanetti; Marília Moraes; Maurício Lacerda Nogueira. Prefeitura de Sao Paulo: Melissa Palmieri.; Patricia Akemi Assato; Paula Rahal; Paulo Inacio da Costa; Rafael dos Santos Bezerra; Raquel de Lello Rocha Campos Cassano. NGS Soluções Genômicas: Pilar Drummond Sampaio Corrêa Mariani. FZEA-USP Pirassununga: Mirele Daiana Poleti; Raul Machado Neto; Ricardo Augusto Brassaloti; Ricardo Haddad; Rodrigo Tocantins Calado.; Rodrigo Tocantins Calado. FAMERP-SJRP: Cecília Artico Banho; Sandra Coccuzzo Sampaio; Simone Kashima; Svetoslav Nanev Slavov; Vagner Fonseca; Vincent Louis Viala |
| EPI_ISL_1795219 | PRONTO ATENDIMENTO UNIDADE SAUDE ADALBERTO ROCHA GUAREI | Instituto Butantan / ESALQ-Piracicaba | Antonio Jorge Martins; Bianca Cechetto Carlos. Mendelics: Bibiana Santos; Claudia Renata dos Santos Barros; David Schlesinger. Hemocentro Ribeirão Preto: Simone Kashima; Debora Botequio Moretti. Centro de Genômica Funcional da ESALQ: Luiz Lehmann Coutinho; Dimas Tadeu Covas; Elaine Cristina Marqueze; Elaine Vieira dos Santos; Elisangela Chicaroni Mattos; Erika Freitas; Evandra Strazza Rodrigues; Felipe Allan da Silva da Costa; Flavia Aburjaile; Guilherme Targino Valente; Heidge Fukumasu. USP-Botucatu: Rejane Maria Tommasini Grotto; Instituto Butantan: Alexander Roberto Precioso; Jayme A. Souza-Neto; Jessika Cristina Chagas Lesbon; José Salvatore Leister Patané; João Paulo Kitajima; Luiz Carlos Junior de Alcantara; Maria Carolina Elias; Marta Giovanetti; Patricia Akemi Assato; Rafael dos Santos Bezerra; Raquel de Lello Rocha Campos Cassano. NGS Soluções Genômicas: Pilar Drummond Sampaio Corrêa Mariani. FZEA-USP Pirassununga: Mirele Daiana Poleti; Raul Machado Neto; Ricardo Augusto Brassaloti; Ricardo Haddad; Rodrigo Tocantins Calado.; Sandra Coccuzzo Sampaio; Svetoslav Nanev Slavov; Vagner Fonseca; Vincent Louis Viala |

| | | | |
|---|---|---|---|
| EPI_ISL_1445096, EPI_ISL_1445097, EPI_ISL_1445098, EPI_ISL_1445099, EPI_ISL_1445100, EPI_ISL_1445102, EPI_ISL_1445103, EPI_ISL_1445104, EPI_ISL_1445105, EPI_ISL_1445106, EPI_ISL_1445107, EPI_ISL_1445108, EPI_ISL_1445111, EPI_ISL_1445112, EPI_ISL_1445186, EPI_ISL_1445188, EPI_ISL_1445189, EPI_ISL_1445190, EPI_ISL_1445191, EPI_ISL_1445192, EPI_ISL_1966133, EPI_ISL_1966134, EPI_ISL_1966135, EPI_ISL_1966136, EPI_ISL_1966137, EPI_ISL_1966138, EPI_ISL_1966139, EPI_ISL_1966140, EPI_ISL_1966141, EPI_ISL_1966142, EPI_ISL_1966143, EPI_ISL_1966144, EPI_ISL_1966156, EPI_ISL_1966179, EPI_ISL_1966221, EPI_ISL_1966223, EPI_ISL_1966224, EPI_ISL_1966225, EPI_ISL_2170917, EPI_ISL_2170918 | | | |
| see above | PRONTO ATENDIMENTO VILA PADRE ANCHIETA | Instituto Butantan / Mendelics | Antonio Jorge Martins; Bianca Cechetto Carlos. Mendelics: Bibiana Santos; Bibiana Santos; Claudia Renata dos Santos Barros; Cíntia Bittar; David Schlesinger; David Schlesinger. Hemocentro Ribeirão Preto: Simone Kashima; Debora Botequio Moretti; Dimas Tadeu Covas; Elaine Cristina Marqueze; Elaine Vieira dos Santos; Elisangela Chicaroni Mattos; Erika Freitas; Evandra Strazza Rodrigues; Felipe Allan da Silva da Costa; Flavia Aburjaile; Fábio Sossai Possebon; Guilherme Campos; Guilherme Targino Valente; Heidge Fukumasu. USP-Botucatu: Rejane Maria Tommasini Grotto; Helena Lage Ferreira; Instituto Butantan: Dimas Tadeu Covas; Jardelina de Souza Todao Bernardino; Jayme A. Souza-Neto; Jessika Cristina Chagas Lesbon; Jorge A. Petroli Marchesi; José Salvatore Leister Patané; João Paulo Kitajima; João Pessoa Araújo Jr.; Leila Sabrina Ullmann; Loyze Paola Oliveira de Lima; Luiz Aurelio de Campos Crispin. Centro de Genômica Funcional da ESALQ: Luiz Lehmann Coutinho; Luiz Carlos Junior de Alcantara; Lívia Sacchetto; Maisa C. Pereira Parra; Maria Carolina Elias; Marta Giovanetti; Marília Moraes; Maurício Lacerda Nogueira. Prefeitura de Sao Paulo: Melissa Palmieri.; Patricia Akemi Assato; Paula Rahal; Paulo Inacio da Costa; Rafael dos Santos Bezerra; Raquel de Lello Rocha Campos Cassano. NGS Soluções Genômicas: Pilar Drummond Sampaio Corrêa Mariani. FZEA-USP Pirassununga: Mirele Daiana Poleti; Raul Machado Neto; Ricardo Augusto Brassaloti; Ricardo Haddad; Rodrigo Tocantins Calado.; Rodrigo Tocantins Calado. FAMERP-SJRP: Cecília Artico Banho; Sandra Coccuzzo Sampaio; Simone Kashima; Svetoslav Nanev Slavov; Vagner Fonseca; Vincent Louis Viala |
| EPI_ISL_1966876 | PRONTO SOCORRO CAUCAIA DO ALTO | Instituto Butantan / ESALQ-USP (Piracicaba) | Antonio Jorge Martins; Bianca Cechetto Carlos. Mendelics: Bibiana Santos; Claudia Renata dos Santos Barros; Cíntia Bittar; David Schlesinger. Hemocentro Ribeirão Preto: Simone Kashima; Debora Botequio Moretti; Elaine Cristina Marqueze; Elaine Vieira dos Santos; Elisangela Chicaroni Mattos; Erika Freitas; Evandra Strazza Rodrigues; Felipe Allan da Silva da Costa; Flavia Aburjaile; Fábio Sossai Possebon; Guilherme Campos; Guilherme Targino Valente; Heidge Fukumasu. USP-Botucatu: Rejane Maria Tommasini Grotto; Helena Lage Ferreira; Instituto Butantan: Dimas Tadeu Covas; Jardelina de Souza Todao Bernardino; Jayme A. Souza-Neto; Jessika Cristina Chagas Lesbon; Jorge A. Petroli Marchesi; José Salvatore Leister Patané; João Paulo Kitajima; João Pessoa Araújo Jr.; Leila Sabrina Ullmann; Loyze Paola Oliveira de Lima; Luiz Aurelio de Campos Crispin. Centro de Genômica Funcional da ESALQ: Luiz Lehmann Coutinho; Luiz Carlos Junior de Alcantara; Lívia Sacchetto; Maisa C. Pereira Parra; Maria Carolina Elias; Marta Giovanetti; Marília Moraes; Maurício Lacerda Nogueira. Prefeitura de Sao Paulo: Melissa Palmieri.; Patricia Akemi Assato; Paula Rahal; Paulo Inacio da Costa; Rafael dos Santos Bezerra; Raquel de Lello Rocha Campos Cassano. NGS Soluções Genômicas: Pilar Drummond Sampaio Corrêa Mariani. FZEA-USP Pirassununga: Mirele Daiana Poleti; Raul Machado Neto; Ricardo Augusto Brassaloti; Ricardo Haddad; Rodrigo Tocantins Calado. FAMERP-SJRP: Cecília Artico Banho; Sandra Coccuzzo Sampaio; Svetoslav Nanev Slavov; Vagner Fonseca; Vincent Louis Viala |
| EPI_ISL_1967236, EPI_ISL_1967237, EPI_ISL_1967238, EPI_ISL_1967239, EPI_ISL_1967240, EPI_ISL_1967241, EPI_ISL_2170974, EPI_ISL_2170975 | | | |
| see above | PRONTO SOCORRO CENTRAL GUIOMAR FERREIRA ROEBBELEN | Instituto Butantan / Mendelics | Antonio Jorge Martins; Bianca Cechetto Carlos. Mendelics: Bibiana Santos; Claudia Renata dos Santos Barros; Cíntia Bittar; David Schlesinger. Hemocentro Ribeirão Preto: Simone Kashima; Debora Botequio Moretti; Elaine Cristina Marqueze; Elaine Vieira dos Santos; Elisangela Chicaroni Mattos; Erika Freitas; Evandra Strazza Rodrigues; Felipe Allan da Silva da Costa; Flavia Aburjaile; Fábio Sossai Possebon; Guilherme Campos; Guilherme Targino Valente; Heidge Fukumasu. USP-Botucatu: Rejane Maria Tommasini Grotto; Helena Lage Ferreira; Instituto Butantan: Dimas Tadeu Covas; Jardelina de Souza Todao Bernardino; Jayme A. Souza-Neto; Jessika Cristina Chagas Lesbon; Jorge A. Petroli Marchesi; José Salvatore Leister Patané; João Paulo Kitajima; João Pessoa Araújo Jr.; Leila Sabrina Ullmann; Loyze Paola Oliveira de Lima; Luiz Aurelio de Campos Crispin. Centro de Genômica Funcional da ESALQ: Luiz Lehmann Coutinho; Luiz Carlos Junior de Alcantara; Lívia Sacchetto; Maisa C. Pereira Parra; Maria Carolina Elias; Marta Giovanetti; Marília Moraes; Maurício Lacerda Nogueira. Prefeitura de Sao Paulo: Melissa Palmieri.; Patricia Akemi Assato; Paula Rahal; Paulo Inacio da Costa; Rafael dos Santos Bezerra; Raquel de Lello Rocha Campos Cassano. NGS Soluções Genômicas: Pilar Drummond Sampaio Corrêa Mariani. FZEA-USP Pirassununga: Mirele Daiana Poleti; Raul Machado Neto; Ricardo Augusto Brassaloti; Ricardo Haddad; Rodrigo Tocantins Calado. FAMERP-SJRP: Cecília Artico Banho; Sandra Coccuzzo Sampaio; Svetoslav Nanev Slavov; Vagner Fonseca; Vincent Louis Viala |
| EPI_ISL_1966302, EPI_ISL_1966303, EPI_ISL_1966304, EPI_ISL_1966305, EPI_ISL_1966308, EPI_ISL_1966311, EPI_ISL_1966312, EPI_ISL_1966313, EPI_ISL_1966314, EPI_ISL_1966315, EPI_ISL_1966317, EPI_ISL_1966318, EPI_ISL_1966320, EPI_ISL_2170927, EPI_ISL_2170928 | | | |
| see above | PRONTO SOCORRO DE AGENOR DE CAMPOS MONGAGUA | Instituto Butantan / Mendelics | Antonio Jorge Martins; Bianca Cechetto Carlos. Mendelics: Bibiana Santos; Claudia Renata dos Santos Barros; Cíntia Bittar; David Schlesinger. Hemocentro Ribeirão Preto: Simone Kashima; Debora Botequio Moretti; Elaine Cristina Marqueze; Elaine Vieira dos Santos; Elisangela Chicaroni Mattos; Erika Freitas; Evandra Strazza Rodrigues; Felipe Allan da Silva da Costa; Flavia Aburjaile; Fábio Sossai Possebon; Guilherme Campos; Guilherme Targino Valente; Heidge Fukumasu. USP-Botucatu: Rejane Maria Tommasini Grotto; Helena Lage Ferreira; Instituto Butantan: Dimas Tadeu Covas; Jardelina de Souza Todao Bernardino; Jayme A. Souza-Neto; Jessika Cristina Chagas Lesbon; Jorge A. Petroli Marchesi; José Salvatore Leister Patané; João Paulo Kitajima; João Pessoa Araújo Jr.; Leila Sabrina Ullmann; Loyze Paola Oliveira de Lima; Luiz Aurelio de Campos Crispin. Centro de Genômica Funcional da ESALQ: Luiz Lehmann Coutinho; Luiz Carlos Junior de Alcantara; Lívia Sacchetto; Maisa C. Pereira Parra; Maria Carolina Elias; Marta Giovanetti; Marília Moraes; Maurício Lacerda Nogueira. Prefeitura de Sao Paulo: Melissa Palmieri.; Patricia Akemi Assato; Paula Rahal; Paulo Inacio da Costa; Rafael dos Santos Bezerra; Raquel de Lello Rocha Campos Cassano. NGS Soluções Genômicas: Pilar Drummond Sampaio Corrêa Mariani. FZEA-USP Pirassununga: Mirele Daiana Poleti; Raul Machado Neto; Ricardo Augusto Brassaloti; Ricardo Haddad; Rodrigo Tocantins Calado. FAMERP-SJRP: Cecília Artico Banho; Sandra Coccuzzo Sampaio; Svetoslav Nanev Slavov; Vagner Fonseca; Vincent Louis Viala |
| EPI_ISL_1445225, EPI_ISL_2170915 | PRONTO SOCORRO DR ANTONIO FLAVIO FRANCA | Instituto Butantan / Mendelics | Antonio Jorge Martins; Bianca Cechetto Carlos. Mendelics: Bibiana Santos; Bibiana Santos; Claudia Renata dos Santos Barros; Cíntia Bittar; David Schlesinger; David Schlesinger. Hemocentro Ribeirão Preto: Simone Kashima; Debora Botequio Moretti; Dimas Tadeu Covas; Elaine Cristina Marqueze; Elaine Vieira dos Santos; Elisangela Chicaroni Mattos; Erika Freitas; Evandra Strazza Rodrigues; Felipe Allan da Silva da Costa; Flavia Aburjaile; Fábio Sossai Possebon; Guilherme Campos; Guilherme Targino Valente; Heidge Fukumasu. USP-Botucatu: Rejane Maria Tommasini Grotto; Helena Lage Ferreira; Instituto Butantan: Dimas Tadeu Covas; Jardelina de Souza Todao Bernardino; Jayme A. Souza-Neto; Jessika Cristina Chagas Lesbon; Jorge A. Petroli Marchesi; José Salvatore Leister Patané; João Paulo Kitajima; João Pessoa Araújo Jr.; Leila Sabrina Ullmann; Loyze Paola Oliveira de Lima; Luiz Aurelio de Campos Crispin. Centro de Genômica Funcional da ESALQ: Luiz Lehmann Coutinho; Luiz Carlos Junior de Alcantara; Lívia Sacchetto; Maisa C. Pereira Parra; Maria Carolina Elias; Marta Giovanetti; Marília Moraes; Maurício Lacerda Nogueira. Prefeitura de Sao Paulo: Melissa Palmieri.; Patricia Akemi Assato; Paula Rahal; Paulo Inacio da Costa; Rafael dos Santos Bezerra; Raquel de Lello Rocha Campos Cassano. NGS Soluções Genômicas: Pilar Drummond Sampaio Corrêa Mariani. FZEA-USP Pirassununga: Mirele Daiana Poleti; Raul Machado Neto; Ricardo Augusto Brassaloti; Ricardo Haddad; Rodrigo Tocantins Calado.; Rodrigo Tocantins Calado. FAMERP-SJRP: Cecília Artico Banho; Sandra Coccuzzo Sampaio; Simone Kashima; Svetoslav Nanev Slavov; Vagner Fonseca; Vincent Louis Viala |
| EPI_ISL_1795426 | PRONTO SOCORRO MUNICIPAL DE PARAIBUNA | Instituto Butantan / ESALQ-Piracicaba | Antonio Jorge Martins; Bianca Cechetto Carlos. Mendelics: Bibiana Santos; Claudia Renata dos Santos Barros; David Schlesinger. Hemocentro Ribeirão Preto: Simone Kashima; Debora Botequio Moretti. Centro de Genômica Funcional da ESALQ: Luiz Lehmann Coutinho; Dimas Tadeu Covas; Elaine Cristina Marqueze; Elaine Vieira dos Santos; Elisangela Chicaroni Mattos; Erika Freitas; Evandra Strazza Rodrigues; Felipe Allan da Silva da Costa; Flavia Aburjaile; Guilherme Targino Valente; Heidge Fukumasu. USP-Botucatu: Rejane Maria Tommasini Grotto; Instituto Butantan: Alexander Roberto Precioso; Jayme A. Souza-Neto; Jessika Cristina Chagas Lesbon; José Salvatore Leister Patané; João Paulo Kitajima; Luiz Carlos Junior de Alcantara; Maria Carolina Elias; Marta Giovanetti; Patricia Akemi Assato; Rafael dos Santos Bezerra; Raquel de Lello Rocha Campos Cassano. NGS Soluções Genômicas: Pilar Drummond Sampaio Corrêa Mariani. FZEA-USP Pirassununga: Mirele Daiana Poleti; Raul Machado Neto; Ricardo Augusto Brassaloti; Ricardo Haddad; Rodrigo Tocantins Calado.; Sandra Coccuzzo Sampaio; Svetoslav Nanev Slavov; Vagner Fonseca; Vincent Louis Viala |
| EPI_ISL_1795213, EPI_ISL_1795214, EPI_ISL_1795215, EPI_ISL_1795216 | PRONTO SOCORRO MUNICIPAL DE SEVERINIA | Instituto Butantan / ESALQ-Piracicaba | Antonio Jorge Martins; Bianca Cechetto Carlos. Mendelics: Bibiana Santos; Claudia Renata dos Santos Barros; David Schlesinger. Hemocentro Ribeirão Preto: Simone Kashima; Debora Botequio Moretti. Centro de Genômica Funcional da ESALQ: Luiz Lehmann Coutinho; Dimas Tadeu Covas; Elaine Cristina Marqueze; Elaine Vieira dos Santos; Elisangela Chicaroni Mattos; Erika Freitas; Evandra Strazza Rodrigues; Felipe Allan da Silva da Costa; Flavia Aburjaile; Guilherme Targino Valente; Heidge Fukumasu. USP-Botucatu: Rejane Maria Tommasini Grotto; Instituto Butantan: Alexander Roberto Precioso; Jayme A. Souza-Neto; Jessika Cristina Chagas Lesbon; José Salvatore Leister Patané; João Paulo Kitajima; Luiz Carlos Junior de Alcantara; Maria Carolina Elias; Marta Giovanetti; Patricia Akemi Assato; Rafael dos Santos Bezerra; Raquel de Lello Rocha Campos Cassano. NGS Soluções Genômicas: Pilar Drummond Sampaio Corrêa Mariani. FZEA-USP Pirassununga: Mirele Daiana Poleti; Raul Machado Neto; Ricardo Augusto Brassaloti; Ricardo Haddad; Rodrigo Tocantins Calado.; Sandra Coccuzzo Sampaio; Svetoslav Nanev Slavov; Vagner Fonseca; Vincent Louis Viala |
| EPI_ISL_1966508, EPI_ISL_1966509 | PRONTO SOCORRO MUNICIPAL DE SEVERINIA | Instituto Butantan / Mendelics | Antonio Jorge Martins; Bianca Cechetto Carlos. Mendelics: Bibiana Santos; Claudia Renata dos Santos Barros; Cíntia Bittar; David Schlesinger. Hemocentro Ribeirão Preto: Simone Kashima; Debora Botequio Moretti; Elaine Cristina Marqueze; Elaine Vieira dos Santos; Elisangela Chicaroni Mattos; Erika Freitas; Evandra Strazza Rodrigues; Felipe Allan da Silva da Costa; Flavia Aburjaile; Fábio Sossai Possebon; Guilherme Campos; Guilherme Targino Valente; Heidge Fukumasu. USP-Botucatu: Rejane Maria Tommasini Grotto; Helena Lage Ferreira; Instituto Butantan: Dimas Tadeu Covas; Jardelina de Souza Todao Bernardino; Jayme A. Souza-Neto; Jessika Cristina Chagas Lesbon; Jorge A. Petroli Marchesi; José Salvatore Leister Patané; João Paulo Kitajima; João Pessoa Araújo Jr.; Leila Sabrina Ullmann; Loyze Paola Oliveira de Lima; Luiz Aurelio de Campos Crispin. Centro de Genômica Funcional da ESALQ: Luiz Lehmann Coutinho; Luiz Carlos Junior de Alcantara; Lívia Sacchetto; Maisa C. Pereira Parra; Maria Carolina Elias; Marta Giovanetti; Marília Moraes; Maurício Lacerda Nogueira. Prefeitura de Sao Paulo: Melissa Palmieri.; Patricia Akemi Assato; Paula Rahal; Paulo Inacio da Costa; Rafael dos Santos Bezerra; Raquel de Lello Rocha Campos Cassano. NGS Soluções Genômicas: Pilar Drummond Sampaio Corrêa Mariani. FZEA-USP Pirassununga: Mirele Daiana Poleti; Raul Machado Neto; Ricardo Augusto Brassaloti; Ricardo Haddad; Rodrigo Tocantins Calado. FAMERP-SJRP: Cecília Artico Banho; Sandra Coccuzzo Sampaio; Svetoslav Nanev Slavov; Vagner Fonseca; Vincent Louis Viala |
| EPI_ISL_1445174, EPI_ISL_1966129 | PRONTO SOCORRO MUNICIPAL DE TATUI | Instituto Butantan / Mendelics | Antonio Jorge Martins; Bianca Cechetto Carlos. Mendelics: Bibiana Santos; Bibiana Santos; Claudia Renata dos Santos Barros; Cíntia Bittar; David Schlesinger; David Schlesinger. Hemocentro Ribeirão Preto: Simone Kashima; Debora Botequio Moretti; Dimas Tadeu Covas; Elaine Cristina Marqueze; Elaine Vieira dos Santos; Elisangela Chicaroni Mattos; Erika Freitas; Evandra Strazza Rodrigues; Felipe Allan da Silva da Costa; Flavia Aburjaile; Fábio Sossai Possebon; Guilherme Campos; Guilherme Targino Valente; Heidge Fukumasu. USP-Botucatu: Rejane Maria Tommasini Grotto; Helena Lage Ferreira; Instituto Butantan: Dimas Tadeu Covas; Jardelina de Souza Todao Bernardino; Jayme A. Souza-Neto; Jessika Cristina Chagas Lesbon; Jorge A. Petroli Marchesi; José Salvatore Leister Patané; João Paulo Kitajima; João Pessoa Araújo Jr.; Leila Sabrina Ullmann; Loyze Paola Oliveira de Lima; Luiz Aurelio de Campos Crispin. Centro de Genômica Funcional da ESALQ: Luiz Lehmann Coutinho; Luiz Carlos Junior de Alcantara; Lívia Sacchetto; Maisa C. Pereira Parra; Maria Carolina Elias; Marta Giovanetti; Marília Moraes; Maurício Lacerda Nogueira. Prefeitura de Sao Paulo: Melissa Palmieri.; Patricia Akemi Assato; Paula Rahal; Paulo Inacio da Costa; Rafael dos Santos Bezerra; Raquel de Lello Rocha Campos Cassano. NGS Soluções Genômicas: Pilar Drummond Sampaio Corrêa Mariani. FZEA-USP Pirassununga: Mirele Daiana Poleti; Raul Machado Neto; Ricardo Augusto Brassaloti; Ricardo Haddad; Rodrigo Tocantins Calado.; Rodrigo Tocantins Calado. FAMERP-SJRP: Cecília Artico Banho; Sandra Coccuzzo Sampaio; Simone Kashima; Svetoslav Nanev Slavov; Vagner Fonseca; Vincent Louis Viala |
| EPI_ISL_1795084, EPI_ISL_1795085, EPI_ISL_1795086, EPI_ISL_1795091, EPI_ISL_1795092, EPI_ISL_1795208, EPI_ISL_1795209, EPI_ISL_1795300, EPI_ISL_1795301, EPI_ISL_1795302, EPI_ISL_1795304, EPI_ISL_1795413 | | | |
| see above | PRONTO SOCORRO MUNICIPAL TAMBAU | Instituto Butantan / ESALQ-Piracicaba | Antonio Jorge Martins; Bianca Cechetto Carlos. Mendelics: Bibiana Santos; Claudia Renata dos Santos Barros; David Schlesinger. Hemocentro Ribeirão Preto: Simone Kashima; Debora Botequio Moretti. Centro de Genômica Funcional da ESALQ: Luiz Lehmann Coutinho; Dimas Tadeu Covas; Elaine Cristina Marqueze; Elaine Vieira dos Santos; Elisangela Chicaroni Mattos; Erika Freitas; Evandra Strazza Rodrigues; Felipe Allan da Silva da Costa; Flavia Aburjaile; Guilherme Targino Valente; Heidge Fukumasu. USP-Botucatu: Rejane Maria Tommasini Grotto; Instituto Butantan: Alexander Roberto Precioso; Jayme A. Souza-Neto; Jessika Cristina Chagas Lesbon; José Salvatore Leister Patané; João Paulo Kitajima; Luiz Carlos Junior de Alcantara; Maria Carolina Elias; Marta Giovanetti; Patricia Akemi Assato; Rafael dos Santos Bezerra; Raquel de Lello Rocha Campos Cassano. NGS Soluções Genômicas: Pilar Drummond Sampaio Corrêa Mariani. FZEA-USP Pirassununga: Mirele Daiana Poleti; Raul Machado Neto; Ricardo Augusto Brassaloti; Ricardo Haddad; Rodrigo Tocantins Calado.; Sandra Coccuzzo Sampaio; Svetoslav Nanev Slavov; Vagner Fonseca; Vincent Louis Viala |
| EPI_ISL_1967310, EPI_ISL_1967311, EPI_ISL_2171030, EPI_ISL_2171031 | PRONTO SOCORRO MUNICIPAL TAMBAU | Instituto Butantan / Mendelics | Antonio Jorge Martins; Bianca Cechetto Carlos. Mendelics: Bibiana Santos; Claudia Renata dos Santos Barros; Cíntia Bittar; David Schlesinger. Hemocentro Ribeirão Preto: Simone Kashima; Debora Botequio Moretti; Elaine Cristina Marqueze; Elaine Vieira dos Santos; Elisangela Chicaroni Mattos; Erika Freitas; Evandra Strazza Rodrigues; Felipe Allan da Silva da Costa; Flavia Aburjaile; Fábio Sossai Possebon; Guilherme Campos; Guilherme Targino Valente; Heidge Fukumasu. USP-Botucatu: Rejane Maria Tommasini Grotto; Helena Lage Ferreira; Instituto Butantan: Dimas Tadeu Covas; Jardelina de Souza Todao Bernardino; Jayme A. Souza-Neto; Jessika Cristina Chagas Lesbon; Jorge A. Petroli Marchesi; José Salvatore Leister Patané; João Paulo Kitajima; João Pessoa Araújo Jr.; Leila Sabrina Ullmann; Loyze Paola Oliveira de Lima; Luiz Aurelio de Campos Crispin. Centro de Genômica Funcional da ESALQ: Luiz Lehmann Coutinho; Luiz Carlos Junior de Alcantara; Lívia Sacchetto; Maisa C. Pereira Parra; Maria Carolina Elias; Marta Giovanetti; Marília Moraes; Maurício Lacerda Nogueira. Prefeitura de Sao Paulo: Melissa Palmieri.; Patricia Akemi Assato; Paula Rahal; Paulo Inacio da Costa; Rafael dos Santos Bezerra; Raquel de Lello Rocha Campos Cassano. NGS Soluções Genômicas: Pilar Drummond Sampaio Corrêa Mariani. FZEA-USP Pirassununga: Mirele Daiana Poleti; Raul Machado Neto; Ricardo Augusto Brassaloti; Ricardo Haddad; Rodrigo Tocantins Calado. FAMERP-SJRP: Cecília Artico Banho; Sandra Coccuzzo Sampaio; Svetoslav Nanev Slavov; Vagner Fonseca; Vincent Louis Viala |
| EPI_ISL_1966735, EPI_ISL_1966736 | PRONTO SOCORRO PESTANA P S ANDRE SACCO | Instituto Butantan | Antonio Jorge Martins; Bianca Cechetto Carlos. Mendelics: Bibiana Santos; Claudia Renata dos Santos Barros; Cíntia Bittar; David Schlesinger. Hemocentro Ribeirão Preto: Simone Kashima; Debora Botequio Moretti; Elaine Cristina Marqueze; Elaine Vieira dos Santos; Elisangela Chicaroni Mattos; Erika Freitas; Evandra Strazza Rodrigues; Felipe Allan da Silva da Costa; Flavia Aburjaile; Fábio Sossai Possebon; Guilherme Campos; Guilherme Targino Valente; Heidge Fukumasu. USP-Botucatu: Rejane Maria Tommasini Grotto; Helena Lage Ferreira; Instituto Butantan: Dimas Tadeu Covas; Jardelina de Souza Todao Bernardino; Jayme A. Souza-Neto; Jessika Cristina Chagas Lesbon; Jorge A. Petroli Marchesi; José Salvatore Leister Patané; João Paulo Kitajima; João Pessoa Araújo Jr.; Leila Sabrina Ullmann; Loyze Paola Oliveira de Lima; Luiz Aurelio de Campos Crispin. Centro de Genômica Funcional da ESALQ: Luiz Lehmann Coutinho; Luiz Carlos Junior de Alcantara; Lívia Sacchetto; Maisa C. Pereira Parra; Maria Carolina Elias; Marta Giovanetti; Marília Moraes; Maurício Lacerda Nogueira. Prefeitura de Sao Paulo: Melissa Palmieri.; Patricia Akemi Assato; Paula Rahal; Paulo Inacio da Costa; Rafael dos Santos Bezerra; Raquel de Lello Rocha Campos Cassano. NGS Soluções Genômicas: Pilar Drummond Sampaio Corrêa Mariani. FZEA-USP Pirassununga: Mirele Daiana Poleti; Raul Machado Neto; Ricardo Augusto Brassaloti; Ricardo Haddad; Rodrigo Tocantins Calado. FAMERP-SJRP: Cecília Artico Banho; Sandra Coccuzzo Sampaio; Svetoslav Nanev Slavov; Vagner Fonseca; Vincent Louis Viala |
| EPI_ISL_1967020, EPI_ISL_1967021, EPI_ISL_1967022, EPI_ISL_1967023, EPI_ISL_1967024 | PSF DR ANTONIO PIRES DE ALMEIDA PORTO FELIZ | Instituto Butantan / ESALQ-USP (Piracicaba) | Antonio Jorge Martins; Bianca Cechetto Carlos. Mendelics: Bibiana Santos; Claudia Renata dos Santos Barros; Cíntia Bittar; David Schlesinger. Hemocentro Ribeirão Preto: Simone Kashima; Debora Botequio Moretti; Elaine Cristina Marqueze; Elaine Vieira dos Santos; Elisangela Chicaroni Mattos; Erika Freitas; Evandra Strazza Rodrigues; Felipe Allan da Silva da Costa; Flavia Aburjaile; Fábio Sossai Possebon; Guilherme Campos; Guilherme Targino Valente; Heidge Fukumasu. USP-Botucatu: Rejane Maria Tommasini Grotto; Helena Lage Ferreira; Instituto Butantan: Dimas Tadeu Covas; Jardelina de Souza Todao Bernardino; Jayme A. Souza-Neto; Jessika Cristina Chagas Lesbon; Jorge A. Petroli Marchesi; José Salvatore Leister Patané; João Paulo Kitajima; João Pessoa Araújo Jr.; Leila Sabrina Ullmann; Loyze Paola Oliveira de Lima; Luiz Aurelio de Campos Crispin. Centro de Genômica Funcional da ESALQ: Luiz Lehmann Coutinho; Luiz Carlos Junior de Alcantara; Lívia Sacchetto; Maisa C. Pereira Parra; Maria Carolina Elias; Marta Giovanetti; Marília Moraes; Maurício Lacerda Nogueira. Prefeitura de Sao Paulo: Melissa Palmieri.; Patricia Akemi Assato; Paula Rahal; Paulo Inacio da Costa; Rafael dos Santos Bezerra; Raquel de Lello Rocha Campos Cassano. NGS Soluções Genômicas: Pilar Drummond Sampaio Corrêa Mariani. FZEA-USP Pirassununga: Mirele Daiana Poleti; Raul Machado Neto; Ricardo Augusto Brassaloti; Ricardo Haddad; Rodrigo Tocantins Calado. FAMERP-SJRP: Cecília Artico Banho; Sandra Coccuzzo Sampaio; Svetoslav Nanev Slavov; Vagner Fonseca; Vincent Louis Viala |
| EPI_ISL_1966396, EPI_ISL_1966397, EPI_ISL_1966797, EPI_ISL_1966798, EPI_ISL_1966799 | PSF DR ANTONIO PIRES DE ALMEIDA PORTO FELIZ | Instituto Butantan / Mendelics | Antonio Jorge Martins; Bianca Cechetto Carlos. Mendelics: Bibiana Santos; Claudia Renata dos Santos Barros; Cíntia Bittar; David Schlesinger. Hemocentro Ribeirão Preto: Simone Kashima; Debora Botequio Moretti; Elaine Cristina Marqueze; Elaine Vieira dos Santos; Elisangela Chicaroni Mattos; Erika Freitas; Evandra Strazza Rodrigues; Felipe Allan da Silva da Costa; Flavia Aburjaile; Fábio Sossai Possebon; Guilherme Campos; Guilherme Targino Valente; Heidge Fukumasu. USP-Botucatu: Rejane Maria Tommasini Grotto; Helena Lage Ferreira; Instituto Butantan: Dimas Tadeu Covas; Jardelina de Souza Todao Bernardino; Jayme A. Souza-Neto; Jessika Cristina Chagas Lesbon; Jorge A. Petroli Marchesi; José Salvatore Leister Patané; João Paulo Kitajima; João Pessoa Araújo Jr.; Leila Sabrina Ullmann; Loyze Paola Oliveira de Lima; Luiz Aurelio de Campos Crispin. Centro de Genômica Funcional da ESALQ: Luiz Lehmann Coutinho; Luiz Carlos Junior de Alcantara; Lívia Sacchetto; Maisa C. Pereira Parra; Maria Carolina Elias; Marta Giovanetti; Marília Moraes; Maurício Lacerda Nogueira. Prefeitura de Sao Paulo: Melissa Palmieri.; Patricia Akemi Assato; Paula Rahal; Paulo Inacio da Costa; Rafael dos Santos Bezerra; Raquel de Lello Rocha Campos Cassano. NGS Soluções Genômicas: Pilar Drummond Sampaio Corrêa Mariani. FZEA-USP Pirassununga: Mirele Daiana Poleti; Raul Machado Neto; Ricardo Augusto Brassaloti; Ricardo Haddad; Rodrigo Tocantins Calado. FAMERP-SJRP: Cecília Artico Banho; Sandra Coccuzzo Sampaio; Svetoslav Nanev Slavov; Vagner Fonseca; Vincent Louis Viala |
| EPI_ISL_1967033, EPI_ISL_1967034, EPI_ISL_1967035, EPI_ISL_1967036, EPI_ISL_1967037, EPI_ISL_1967038, EPI_ISL_1967039, EPI_ISL_1967040, EPI_ISL_1967041 | | | |
| see above | PSF MARIA JOSE SALTO DE PIRAPORA | Instituto Butantan / ESALQ-USP (Piracicaba) | Antonio Jorge Martins; Bianca Cechetto Carlos. Mendelics: Bibiana Santos; Claudia Renata dos Santos Barros; Cíntia Bittar; David Schlesinger. Hemocentro Ribeirão Preto: Simone Kashima; Debora Botequio Moretti; Elaine Cristina Marqueze; Elaine Vieira dos Santos; Elisangela Chicaroni Mattos; Erika Freitas; Evandra Strazza Rodrigues; Felipe Allan da Silva da Costa; Flavia Aburjaile; Fábio Sossai Possebon; Guilherme Campos; Guilherme Targino Valente; Heidge Fukumasu. USP-Botucatu: Rejane Maria Tommasini Grotto; Helena Lage Ferreira; Instituto Butantan: Dimas Tadeu Covas; Jardelina de Souza Todao Bernardino; Jayme A. Souza-Neto; Jessika Cristina Chagas Lesbon; Jorge A. Petroli Marchesi; José Salvatore Leister Patané; João Paulo Kitajima; João Pessoa Araújo Jr.; Leila Sabrina Ullmann; Loyze Paola Oliveira de Lima; Luiz Aurelio de Campos Crispin. Centro de Genômica Funcional da ESALQ: Luiz Lehmann Coutinho; Luiz Carlos Junior de Alcantara; Lívia Sacchetto; Maisa C. Pereira Parra; Maria Carolina Elias; Marta Giovanetti; Marília Moraes; Maurício Lacerda Nogueira. Prefeitura de Sao Paulo: Melissa Palmieri.; Patricia Akemi Assato; Paula Rahal; Paulo Inacio da Costa; Rafael dos Santos Bezerra; Raquel de Lello Rocha Campos Cassano. NGS Soluções Genômicas: Pilar Drummond Sampaio Corrêa Mariani. FZEA-USP Pirassununga: Mirele Daiana Poleti; Raul Machado Neto; Ricardo Augusto Brassaloti; Ricardo Haddad; Rodrigo Tocantins Calado. FAMERP-SJRP: Cecília Artico Banho; Sandra Coccuzzo Sampaio; Svetoslav Nanev Slavov; Vagner Fonseca; Vincent Louis Viala |
| EPI_ISL_1966413, EPI_ISL_1966414, EPI_ISL_1966415, EPI_ISL_1966416, EPI_ISL_1966417, EPI_ISL_1966418, EPI_ISL_1966419, EPI_ISL_1966420, EPI_ISL_1966421, EPI_ISL_1966422, EPI_ISL_1966423, EPI_ISL_1966424, EPI_ISL_1966425, EPI_ISL_1966426, EPI_ISL_1966427, EPI_ISL_1966428, EPI_ISL_1966429, EPI_ISL_1966430, EPI_ISL_1966431, EPI_ISL_1966432, EPI_ISL_1966433, EPI_ISL_1967275, EPI_ISL_1967276, EPI_ISL_1967304, EPI_ISL_1967305 | | | |
| see above | PSF PARQUE EMILIA | Instituto Butantan / Mendelics | Antonio Jorge Martins; Bianca Cechetto Carlos. Mendelics: Bibiana Santos; Claudia Renata dos Santos Barros; Cíntia Bittar; David Schlesinger. Hemocentro Ribeirão Preto: Simone Kashima; Debora Botequio Moretti; Elaine Cristina Marqueze; Elaine Vieira dos Santos; Elisangela Chicaroni Mattos; Erika Freitas; Evandra Strazza Rodrigues; Felipe Allan da Silva da Costa; Flavia Aburjaile; Fábio Sossai Possebon; Guilherme Campos; Guilherme Targino Valente; Heidge Fukumasu. USP-Botucatu: Rejane Maria Tommasini Grotto; Helena Lage Ferreira; Instituto Butantan: Dimas Tadeu Covas; Jardelina de Souza Todao Bernardino; Jayme A. Souza-Neto; Jessika Cristina Chagas Lesbon; Jorge A. Petroli Marchesi; José Salvatore Leister Patané; João Paulo Kitajima; João Pessoa Araújo Jr.; Leila Sabrina Ullmann; Loyze Paola Oliveira de Lima; Luiz Aurelio de Campos Crispin. Centro de Genômica Funcional da ESALQ: Luiz Lehmann Coutinho; Luiz Carlos Junior de Alcantara; Lívia Sacchetto; Maisa C. Pereira Parra; Maria Carolina Elias; Marta Giovanetti; Marília Moraes; Maurício Lacerda Nogueira. Prefeitura de Sao Paulo: Melissa Palmieri.; Patricia Akemi Assato; Paula Rahal; Paulo Inacio da Costa; Rafael dos Santos Bezerra; Raquel de Lello Rocha Campos Cassano. NGS Soluções Genômicas: Pilar Drummond Sampaio Corrêa Mariani. FZEA-USP Pirassununga: Mirele Daiana Poleti; Raul Machado Neto; Ricardo Augusto Brassaloti; Ricardo Haddad; Rodrigo Tocantins Calado. FAMERP-SJRP: Cecília Artico Banho; Sandra Coccuzzo Sampaio; Svetoslav Nanev Slavov; Vagner Fonseca; Vincent Louis Viala |
| EPI_ISL_1966786 | PSF SETOR LESTE NHANDEARA | Instituto Butantan / Mendelics | Antonio Jorge Martins; Bianca Cechetto Carlos. Mendelics: Bibiana Santos; Claudia Renata dos Santos Barros; Cíntia Bittar; David Schlesinger. Hemocentro Ribeirão Preto: Simone Kashima; Debora Botequio Moretti; Elaine Cristina Marqueze; Elaine Vieira dos Santos; Elisangela Chicaroni Mattos; Erika Freitas; Evandra Strazza Rodrigues; Felipe Allan da Silva da Costa; Flavia Aburjaile; Fábio Sossai Possebon; Guilherme Campos; Guilherme Targino Valente; Heidge Fukumasu. USP-Botucatu: Rejane Maria Tommasini Grotto; Helena Lage Ferreira; Instituto Butantan: Dimas Tadeu Covas; Jardelina de Souza Todao Bernardino; Jayme A. Souza-Neto; Jessika Cristina Chagas Lesbon; Jorge A. Petroli Marchesi; José Salvatore Leister Patané; João Paulo Kitajima; João Pessoa Araújo Jr.; Leila Sabrina Ullmann; Loyze Paola Oliveira de Lima; Luiz Aurelio de Campos Crispin. Centro de Genômica Funcional da ESALQ: Luiz Lehmann Coutinho; Luiz Carlos Junior de Alcantara; Lívia Sacchetto; Maisa C. Pereira Parra; Maria Carolina Elias; Marta Giovanetti; Marília Moraes; Maurício Lacerda Nogueira. Prefeitura de Sao Paulo: Melissa Palmieri.; Patricia Akemi Assato; Paula Rahal; Paulo Inacio da Costa; Rafael dos Santos Bezerra; Raquel de Lello Rocha Campos Cassano. NGS Soluções Genômicas: Pilar Drummond Sampaio Corrêa Mariani. FZEA-USP Pirassununga: Mirele Daiana Poleti; Raul Machado Neto; Ricardo Augusto Brassaloti; Ricardo Haddad; Rodrigo Tocantins Calado. FAMERP-SJRP: Cecília Artico Banho; Sandra Coccuzzo Sampaio; Svetoslav Nanev Slavov; Vagner Fonseca; Vincent Louis Viala |
| EPI_ISL_1533719 | Policlinica Munic da Est Turistica de Holambra | Instituto Adolfo Lutz, Interdiciplinary Procedures Center, Strategic Laboratory | Caio Vinicius Dias Lopes; Claudia Regina Gonçalves; Claudio Tavares Sacchi; Erica Valessa Ramos Gomes; Karoline Rodrigues Campos; Leonardo Jose Tadeu de Araujo |
| EPI_ISL_2003111 | Pronto Socorro Central de Bauru | Instituto Adolfo Lutz, Interdiciplinary Procedures Center, | Caio Vinicius Dias Lopes; Claudia Regina Gonçalves; Claudio Tavares Sacchi; Erica Valessa Ramos Gomes; Karoline Rodrigues Campos; Leonardo Jose Tadeu de Araujo |

| | | | |
|---|---|---|---|
| | | Strategic Laboratory | |
| EPI_ISL_1533714 | Pronto Socorro Dr Osmar Mesquita | Instituto Adolfo Lutz, Interdiciplinary Procedures Center, Strategic Laboratory | Caio Vinicius Dias Lopes; Claudia Regina Gonçalves; Claudio Tavares Sacchi; Erica Valessa Ramos Gomes; Karoline Rodrigues Campos; Leonardo Jose Tadeu de Araujo |
| EPI_ISL_1966395 | SAE CTA SERV ATEND ESPEC TESTAGEM E ACONSELHAMENTO | Instituto Butantan / Mendelics | Antonio Jorge Martins; Bianca Cechetto Carlos. Mendelics: Bibiana Santos; Claudia Renata dos Santos Barros; Cíntia Bittar; David Schlesinger. Hemocentro Ribeirão Preto: Simone Kashima; Debora Botequio Moretti; Elaine Cristina Marqueze; Elaine Vieira dos Santos; Elisangela Chicaroni Mattos; Erika Freitas; Evandra Strazza Rodrigues; Felipe Allan da Silva da Costa; Flavia Aburjaile; Fábio Sossai Possebon; Guilherme Campos; Guilherme Targino Valente; Heidge Fukumasu. USP-Botucatu: Rejane Maria Tommasini Grotto; Helena Lage Ferreira; Instituto Butantan: Dimas Tadeu Covas; Jardelina de Souza Todao Bernardino; Jayme A. Souza-Neto; Jessika Cristina Chagas Lesbon; Jorge A. Petroli Marchesi; José Salvatore Leister Patané; João Paulo Kitajima; João Pessoa Araújo Jr.; Leila Sabrina Ullmann; Loyze Paola Oliveira de Lima; Luiz Aurelio de Campos Crispin. Centro de Genômica Funcional da ESALQ: Luiz Lehmann Coutinho; Luiz Carlos Junior de Alcantara; Lívia Sacchetto; Maisa C. Pereira Parra; Maria Carolina Elias; Marta Giovanetti; Marília Moraes; Maurício Lacerda Nogueira. Prefeitura de Sao Paulo: Melissa Palmieri.; Patricia Akemi Assato; Paula Rahal; Paulo Inacio da Costa; Rafael dos Santos Bezerra; Raquel de Lello Rocha Campos Cassano. NGS Soluções Genômicas: Pilar Drummond Sampaio Corrêa Mariani. FZEA-USP Pirassununga: Mirele Daiana Poleti; Raul Machado Neto; Ricardo Augusto Brassaloti; Ricardo Haddad; Rodrigo Tocantins Calado. FAMERP-SJRP: Cecília Artico Banho; Sandra Coccuzzo Sampaio; Svetoslav Nanev Slavov; Vagner Fonseca; Vincent Louis Viala |
| EPI_ISL_1445153, EPI_ISL_1445154, EPI_ISL_1445156, EPI_ISL_1966126, EPI_ISL_1966127, EPI_ISL_1966128, EPI_ISL_1966517, EPI_ISL_1966518, EPI_ISL_1966519, EPI_ISL_1966520, EPI_ISL_2170939 | | | |
| see above | SAE SERVICO DE ATENDIMENTO ESPECIALIZADO | Instituto Butantan / Mendelics | Antonio Jorge Martins; Bianca Cechetto Carlos. Mendelics: Bibiana Santos; Bibiana Santos; Claudia Renata dos Santos Barros; Cíntia Bittar; David Schlesinger; David Schlesinger. Hemocentro Ribeirão Preto: Simone Kashima; Debora Botequio Moretti; Dimas Tadeu Covas; Elaine Cristina Marqueze; Elaine Vieira dos Santos; Elisangela Chicaroni Mattos; Erika Freitas; Evandra Strazza Rodrigues; Felipe Allan da Silva da Costa; Flavia Aburjaile; Fábio Sossai Possebon; Guilherme Campos; Guilherme Targino Valente; Heidge Fukumasu. USP-Botucatu: Rejane Maria Tommasini Grotto; Helena Lage Ferreira; Instituto Butantan: Dimas Tadeu Covas; Jardelina de Souza Todao Bernardino; Jayme A. Souza-Neto; Jessika Cristina Chagas Lesbon; Jorge A. Petroli Marchesi; José Salvatore Leister Patané; João Paulo Kitajima; João Pessoa Araújo Jr.; Leila Sabrina Ullmann; Loyze Paola Oliveira de Lima; Luiz Aurelio de Campos Crispin. Centro de Genômica Funcional da ESALQ: Luiz Lehmann Coutinho; Luiz Carlos Junior de Alcantara; Lívia Sacchetto; Maisa C. Pereira Parra; Maria Carolina Elias; Marta Giovanetti; Marília Moraes; Maurício Lacerda Nogueira. Prefeitura de Sao Paulo: Melissa Palmieri.; Patricia Akemi Assato; Paula Rahal; Paulo Inacio da Costa; Rafael dos Santos Bezerra; Raquel de Lello Rocha Campos Cassano. NGS Soluções Genômicas: Pilar Drummond Sampaio Corrêa Mariani. FZEA-USP Pirassununga: Mirele Daiana Poleti; Raul Machado Neto; Ricardo Augusto Brassaloti; Ricardo Haddad; Rodrigo Tocantins Calado.; Rodrigo Tocantins Calado. FAMERP-SJRP: Cecília Artico Banho; Sandra Coccuzzo Sampaio; Simone Kashima; Svetoslav Nanev Slavov; Vagner Fonseca; Vincent Louis Viala |
| EPI_ISL_1967282, EPI_ISL_2170982 | SANTA CASA DE ATIBAIA PRO SAUDE | Instituto Butantan / Mendelics | Antonio Jorge Martins; Bianca Cechetto Carlos. Mendelics: Bibiana Santos; Claudia Renata dos Santos Barros; Cíntia Bittar; David Schlesinger. Hemocentro Ribeirão Preto: Simone Kashima; Debora Botequio Moretti; Elaine Cristina Marqueze; Elaine Vieira dos Santos; Elisangela Chicaroni Mattos; Erika Freitas; Evandra Strazza Rodrigues; Felipe Allan da Silva da Costa; Flavia Aburjaile; Fábio Sossai Possebon; Guilherme Campos; Guilherme Targino Valente; Heidge Fukumasu. USP-Botucatu: Rejane Maria Tommasini Grotto; Helena Lage Ferreira; Instituto Butantan: Dimas Tadeu Covas; Jardelina de Souza Todao Bernardino; Jayme A. Souza-Neto; Jessika Cristina Chagas Lesbon; Jorge A. Petroli Marchesi; José Salvatore Leister Patané; João Paulo Kitajima; João Pessoa Araújo Jr.; Leila Sabrina Ullmann; Loyze Paola Oliveira de Lima; Luiz Aurelio de Campos Crispin. Centro de Genômica Funcional da ESALQ: Luiz Lehmann Coutinho; Luiz Carlos Junior de Alcantara; Lívia Sacchetto; Maisa C. Pereira Parra; Maria Carolina Elias; Marta Giovanetti; Marília Moraes; Maurício Lacerda Nogueira. Prefeitura de Sao Paulo: Melissa Palmieri.; Patricia Akemi Assato; Paula Rahal; Paulo Inacio da Costa; Rafael dos Santos Bezerra; Raquel de Lello Rocha Campos Cassano. NGS Soluções Genômicas: Pilar Drummond Sampaio Corrêa Mariani. FZEA-USP Pirassununga: Mirele Daiana Poleti; Raul Machado Neto; Ricardo Augusto Brassaloti; Ricardo Haddad; Rodrigo Tocantins Calado. FAMERP-SJRP: Cecília Artico Banho; Sandra Coccuzzo Sampaio; Svetoslav Nanev Slavov; Vagner Fonseca; Vincent Louis Viala |
| EPI_ISL_1795374, EPI_ISL_1795375 | SANTA CASA DE DOIS CORREGOS | Instituto Butantan / ESALQ-Piracicaba | Antonio Jorge Martins; Bianca Cechetto Carlos. Mendelics: Bibiana Santos; Claudia Renata dos Santos Barros; David Schlesinger. Hemocentro Ribeirão Preto: Simone Kashima; Debora Botequio Moretti. Centro de Genômica Funcional da ESALQ: Luiz Lehmann Coutinho; Dimas Tadeu Covas; Elaine Cristina Marqueze; Elaine Vieira dos Santos; Elisangela Chicaroni Mattos; Erika Freitas; Evandra Strazza Rodrigues; Felipe Allan da Silva da Costa; Flavia Aburjaile; Guilherme Targino Valente; Heidge Fukumasu. USP-Botucatu: Rejane Maria Tommasini Grotto; Instituto Butantan: Alexander Roberto Precioso; Jayme A. Souza-Neto; Jessika Cristina Chagas Lesbon; José Salvatore Leister Patané; João Paulo Kitajima; Luiz Carlos Junior de Alcantara; Maria Carolina Elias; Marta Giovanetti; Patricia Akemi Assato; Rafael dos Santos Bezerra; Raquel de Lello Rocha Campos Cassano. NGS Soluções Genômicas: Pilar Drummond Sampaio Corrêa Mariani. FZEA-USP Pirassununga: Mirele Daiana Poleti; Raul Machado Neto; Ricardo Augusto Brassaloti; Ricardo Haddad; Rodrigo Tocantins Calado.; Sandra Coccuzzo Sampaio; Svetoslav Nanev Slavov; Vagner Fonseca; Vincent Louis Viala |
| EPI_ISL_2170997 | SANTA CASA DE GUARAREMA | Instituto Butantan / Mendelics | Antonio Jorge Martins; Bianca Cechetto Carlos. Mendelics: Bibiana Santos; Claudia Renata dos Santos Barros; Cíntia Bittar; David Schlesinger. Hemocentro Ribeirão Preto: Simone Kashima; Debora Botequio Moretti; Elaine Cristina Marqueze; Elaine Vieira dos Santos; Elisangela Chicaroni Mattos; Erika Freitas; Evandra Strazza Rodrigues; Felipe Allan da Silva da Costa; Flavia Aburjaile; Fábio Sossai Possebon; Guilherme Campos; Guilherme Targino Valente; Heidge Fukumasu. USP-Botucatu: Rejane Maria Tommasini Grotto; Helena Lage Ferreira; Instituto Butantan: Dimas Tadeu Covas; Jardelina de Souza Todao Bernardino; Jayme A. Souza-Neto; Jessika Cristina Chagas Lesbon; Jorge A. Petroli Marchesi; José Salvatore Leister Patané; João Paulo Kitajima; João Pessoa Araújo Jr.; Leila Sabrina Ullmann; Loyze Paola Oliveira de Lima; Luiz Aurelio de Campos Crispin. Centro de Genômica Funcional da ESALQ: Luiz Lehmann Coutinho; Luiz Carlos Junior de Alcantara; Lívia Sacchetto; Maisa C. Pereira Parra; Maria Carolina Elias; Marta Giovanetti; Marília Moraes; Maurício Lacerda Nogueira. Prefeitura de Sao Paulo: Melissa Palmieri.; Patricia Akemi Assato; Paula Rahal; Paulo Inacio da Costa; Rafael dos Santos Bezerra; Raquel de Lello Rocha Campos Cassano. NGS Soluções Genômicas: Pilar Drummond Sampaio Corrêa Mariani. FZEA-USP Pirassununga: Mirele Daiana Poleti; Raul Machado Neto; Ricardo Augusto Brassaloti; Ricardo Haddad; Rodrigo Tocantins Calado. FAMERP-SJRP: Cecília Artico Banho; Sandra Coccuzzo Sampaio; Svetoslav Nanev Slavov; Vagner Fonseca; Vincent Louis Viala |
| EPI_ISL_1966242 | SANTA CASA DE MIS E MATERNIDADE N SRA DA CONCEICAO CUNHA | Instituto Butantan / Mendelics | Antonio Jorge Martins; Bianca Cechetto Carlos. Mendelics: Bibiana Santos; Claudia Renata dos Santos Barros; Cíntia Bittar; David Schlesinger. Hemocentro Ribeirão Preto: Simone Kashima; Debora Botequio Moretti; Elaine Cristina Marqueze; Elaine Vieira dos Santos; Elisangela Chicaroni Mattos; Erika Freitas; Evandra Strazza Rodrigues; Felipe Allan da Silva da Costa; Flavia Aburjaile; Fábio Sossai Possebon; Guilherme Campos; Guilherme Targino Valente; Heidge Fukumasu. USP-Botucatu: Rejane Maria Tommasini Grotto; Helena Lage Ferreira; Instituto Butantan: Dimas Tadeu Covas; Jardelina de Souza Todao Bernardino; Jayme A. Souza-Neto; Jessika Cristina Chagas Lesbon; Jorge A. Petroli Marchesi; José Salvatore Leister Patané; João Paulo Kitajima; João Pessoa Araújo Jr.; Leila Sabrina Ullmann; Loyze Paola Oliveira de Lima; Luiz Aurelio de Campos Crispin. Centro de Genômica Funcional da ESALQ: Luiz Lehmann Coutinho; Luiz Carlos Junior de Alcantara; Lívia Sacchetto; Maisa C. Pereira Parra; Maria Carolina Elias; Marta Giovanetti; Marília Moraes; Maurício Lacerda Nogueira. Prefeitura de Sao Paulo: Melissa Palmieri.; Patricia Akemi Assato; Paula Rahal; Paulo Inacio da Costa; Rafael dos Santos Bezerra; Raquel de Lello Rocha Campos Cassano. NGS Soluções Genômicas: Pilar Drummond Sampaio Corrêa Mariani. FZEA-USP Pirassununga: Mirele Daiana Poleti; Raul Machado Neto; Ricardo Augusto Brassaloti; Ricardo Haddad; Rodrigo Tocantins Calado. FAMERP-SJRP: Cecília Artico Banho; Sandra Coccuzzo Sampaio; Svetoslav Nanev Slavov; Vagner Fonseca; Vincent Louis Viala |
| EPI_ISL_1967192, EPI_ISL_1967193, EPI_ISL_1967194, EPI_ISL_1967195 | SANTA CASA DE MISERICORDIA DE TIETE | Instituto Butantan / ESALQ-USP (Piracicaba) | Antonio Jorge Martins; Bianca Cechetto Carlos. Mendelics: Bibiana Santos; Claudia Renata dos Santos Barros; Cíntia Bittar; David Schlesinger. Hemocentro Ribeirão Preto: Simone Kashima; Debora Botequio Moretti; Elaine Cristina Marqueze; Elaine Vieira dos Santos; Elisangela Chicaroni Mattos; Erika Freitas; Evandra Strazza Rodrigues; Felipe Allan da Silva da Costa; Flavia Aburjaile; Fábio Sossai Possebon; Guilherme Campos; Guilherme Targino Valente; Heidge Fukumasu. USP-Botucatu: Rejane Maria Tommasini Grotto; Helena Lage Ferreira; Instituto Butantan: Dimas Tadeu Covas; Jardelina de Souza Todao Bernardino; Jayme A. Souza-Neto; Jessika Cristina Chagas Lesbon; Jorge A. Petroli Marchesi; José Salvatore Leister Patané; João Paulo Kitajima; João Pessoa Araújo Jr.; Leila Sabrina Ullmann; Loyze Paola Oliveira de Lima; Luiz Aurelio de Campos Crispin. Centro de Genômica Funcional da ESALQ: Luiz Lehmann Coutinho; Luiz Carlos Junior de Alcantara; Lívia Sacchetto; Maisa C. Pereira Parra; Maria Carolina Elias; Marta Giovanetti; Marília Moraes; Maurício Lacerda Nogueira. Prefeitura de Sao Paulo: Melissa Palmieri.; Patricia Akemi Assato; Paula Rahal; Paulo Inacio da Costa; Rafael dos Santos Bezerra; Raquel de Lello Rocha Campos Cassano. NGS Soluções Genômicas: Pilar Drummond Sampaio Corrêa Mariani. FZEA-USP Pirassununga: Mirele Daiana Poleti; Raul Machado Neto; Ricardo Augusto Brassaloti; Ricardo Haddad; Rodrigo Tocantins Calado. FAMERP-SJRP: Cecília Artico Banho; Sandra Coccuzzo Sampaio; Svetoslav Nanev Slavov; Vagner Fonseca; Vincent Louis Viala |
| EPI_ISL_1795361 | SANTA CASA DE MISERICORDIA DE UBATUBA | Instituto Butantan / ESALQ-Piracicaba | Antonio Jorge Martins; Bianca Cechetto Carlos. Mendelics: Bibiana Santos; Claudia Renata dos Santos Barros; David Schlesinger. Hemocentro Ribeirão Preto: Simone Kashima; Debora Botequio Moretti. Centro de Genômica Funcional da ESALQ: Luiz Lehmann Coutinho; Dimas Tadeu Covas; Elaine Cristina Marqueze; Elaine Vieira dos Santos; Elisangela Chicaroni Mattos; Erika Freitas; Evandra Strazza Rodrigues; Felipe Allan da Silva da Costa; Flavia Aburjaile; Guilherme Targino Valente; Heidge Fukumasu. USP-Botucatu: Rejane Maria Tommasini Grotto; Instituto Butantan: Alexander Roberto Precioso; Jayme A. Souza-Neto; Jessika Cristina Chagas Lesbon; José Salvatore Leister Patané; João Paulo Kitajima; Luiz Carlos Junior de Alcantara; Maria Carolina Elias; Marta Giovanetti; Patricia Akemi Assato; Rafael dos Santos Bezerra; Raquel de Lello Rocha Campos Cassano. NGS Soluções Genômicas: Pilar Drummond Sampaio Corrêa Mariani. FZEA-USP Pirassununga: Mirele Daiana Poleti; Raul Machado Neto; Ricardo Augusto Brassaloti; Ricardo Haddad; Rodrigo Tocantins Calado.; Sandra Coccuzzo Sampaio; Svetoslav Nanev Slavov; Vagner Fonseca; Vincent Louis Viala |
| EPI_ISL_1966259 | SANTA CASA DE MISERICORDIA DE UBATUBA | Instituto Butantan / Mendelics | Antonio Jorge Martins; Bianca Cechetto Carlos. Mendelics: Bibiana Santos; Claudia Renata dos Santos Barros; Cíntia Bittar; David Schlesinger. Hemocentro Ribeirão Preto: Simone Kashima; Debora Botequio Moretti; Elaine Cristina Marqueze; Elaine Vieira dos Santos; Elisangela Chicaroni Mattos; Erika Freitas; Evandra Strazza Rodrigues; Felipe Allan da Silva da Costa; Flavia Aburjaile; Fábio Sossai Possebon; Guilherme Campos; Guilherme Targino Valente; Heidge Fukumasu. USP-Botucatu: Rejane Maria Tommasini Grotto; Helena Lage Ferreira; Instituto Butantan: Dimas Tadeu Covas; Jardelina de Souza Todao Bernardino; Jayme A. Souza-Neto; Jessika Cristina Chagas Lesbon; Jorge A. Petroli Marchesi; José Salvatore Leister Patané; João Paulo Kitajima; João Pessoa Araújo Jr.; Leila Sabrina Ullmann; Loyze Paola Oliveira de Lima; Luiz Aurelio de Campos Crispin. Centro de Genômica Funcional da ESALQ: Luiz Lehmann Coutinho; Luiz Carlos Junior de Alcantara; Lívia Sacchetto; Maisa C. Pereira Parra; Maria Carolina Elias; Marta Giovanetti; Marília Moraes; Maurício Lacerda Nogueira. Prefeitura de Sao Paulo: Melissa Palmieri.; Patricia Akemi Assato; Paula Rahal; Paulo Inacio da Costa; Rafael dos Santos Bezerra; Raquel de Lello Rocha Campos Cassano. NGS Soluções Genômicas: Pilar Drummond Sampaio Corrêa Mariani. FZEA-USP Pirassununga: Mirele Daiana Poleti; Raul Machado Neto; Ricardo Augusto Brassaloti; Ricardo Haddad; Rodrigo Tocantins Calado. FAMERP-SJRP: Cecília Artico Banho; Sandra Coccuzzo Sampaio; Svetoslav Nanev Slavov; Vagner Fonseca; Vincent Louis Viala |
| EPI_ISL_2170924 | SANTA CASA DE MISERICORDIA SAO JOSE | Instituto Butantan / Mendelics | Antonio Jorge Martins; Bianca Cechetto Carlos. Mendelics: Bibiana Santos; Claudia Renata dos Santos Barros; Cíntia Bittar; David Schlesinger. Hemocentro Ribeirão Preto: Simone Kashima; Debora Botequio Moretti; Elaine Cristina Marqueze; Elaine Vieira dos Santos; Elisangela Chicaroni Mattos; Erika Freitas; Evandra Strazza Rodrigues; Felipe Allan da Silva da Costa; Flavia Aburjaile; Fábio Sossai Possebon; Guilherme Campos; Guilherme Targino Valente; Heidge Fukumasu. USP-Botucatu: Rejane Maria Tommasini Grotto; Helena Lage Ferreira; Instituto Butantan: Dimas Tadeu Covas; Jardelina de Souza Todao Bernardino; Jayme A. Souza-Neto; Jessika Cristina Chagas Lesbon; Jorge A. Petroli Marchesi; José Salvatore Leister Patané; João Paulo Kitajima; João Pessoa Araújo Jr.; Leila Sabrina Ullmann; Loyze Paola Oliveira de Lima; Luiz Aurelio de Campos Crispin. Centro de Genômica Funcional da ESALQ: Luiz Lehmann Coutinho; Luiz Carlos Junior de Alcantara; Lívia Sacchetto; Maisa C. Pereira Parra; Maria Carolina Elias; Marta Giovanetti; Marília Moraes; Maurício Lacerda Nogueira. Prefeitura de Sao Paulo: Melissa Palmieri.; Patricia Akemi Assato; Paula Rahal; Paulo Inacio da Costa; Rafael dos Santos Bezerra; Raquel de Lello Rocha Campos Cassano. NGS Soluções Genômicas: Pilar Drummond Sampaio Corrêa Mariani. FZEA-USP Pirassununga: Mirele Daiana Poleti; Raul Machado Neto; Ricardo Augusto Brassaloti; Ricardo Haddad; Rodrigo Tocantins Calado. FAMERP-SJRP: Cecília Artico Banho; Sandra Coccuzzo Sampaio; Svetoslav Nanev Slavov; Vagner Fonseca; Vincent Louis Viala |
| EPI_ISL_1967014, EPI_ISL_1967015, EPI_ISL_1967016, EPI_ISL_1967017, EPI_ISL_1967018, EPI_ISL_1967019 | SANTA CASA DE PORTO FELIZ | Instituto Butantan / ESALQ-USP (Piracicaba) | Antonio Jorge Martins; Bianca Cechetto Carlos. Mendelics: Bibiana Santos; Claudia Renata dos Santos Barros; Cíntia Bittar; David Schlesinger. Hemocentro Ribeirão Preto: Simone Kashima; Debora Botequio Moretti; Elaine Cristina Marqueze; Elaine Vieira dos Santos; Elisangela Chicaroni Mattos; Erika Freitas; Evandra Strazza Rodrigues; Felipe Allan da Silva da Costa; Flavia Aburjaile; Fábio Sossai Possebon; Guilherme Campos; Guilherme Targino Valente; Heidge Fukumasu. USP-Botucatu: Rejane Maria Tommasini Grotto; Helena Lage Ferreira; Instituto Butantan: Dimas Tadeu Covas; Jardelina de Souza Todao Bernardino; Jayme A. Souza-Neto; Jessika Cristina Chagas Lesbon; Jorge A. Petroli Marchesi; José Salvatore Leister Patané; João Paulo Kitajima; João Pessoa Araújo Jr.; Leila Sabrina Ullmann; Loyze Paola Oliveira de Lima; Luiz Aurelio de Campos Crispin. Centro de Genômica Funcional da ESALQ: Luiz Lehmann Coutinho; Luiz Carlos Junior de Alcantara; Lívia Sacchetto; Maisa C. Pereira Parra; Maria Carolina Elias; Marta Giovanetti; Marília Moraes; Maurício Lacerda Nogueira. Prefeitura de Sao Paulo: Melissa Palmieri.; Patricia Akemi Assato; Paula Rahal; Paulo Inacio da Costa; Rafael dos Santos Bezerra; Raquel de Lello Rocha Campos Cassano. NGS Soluções Genômicas: Pilar Drummond Sampaio Corrêa Mariani. FZEA-USP Pirassununga: Mirele Daiana Poleti; Raul Machado Neto; Ricardo Augusto Brassaloti; Ricardo Haddad; Rodrigo Tocantins Calado. FAMERP-SJRP: Cecília Artico Banho; Sandra Coccuzzo Sampaio; Svetoslav Nanev Slavov; Vagner Fonseca; Vincent Louis Viala |
| EPI_ISL_1445080, EPI_ISL_1445085, EPI_ISL_1445177, EPI_ISL_1966112, EPI_ISL_2170903 | SAUDE COLETIVA CAPAO BONITO | Instituto Butantan / Mendelics | Antonio Jorge Martins; Bianca Cechetto Carlos. Mendelics: Bibiana Santos; Bibiana Santos; Claudia Renata dos Santos Barros; Cíntia Bittar; David Schlesinger; David Schlesinger. Hemocentro Ribeirão Preto: Simone Kashima; Debora Botequio Moretti; Dimas Tadeu Covas; Elaine Cristina Marqueze; Elaine Vieira Santos; Elaine Vieira dos Santos; Elisangela Chicaroni Mattos; Erika Freitas; Evandra Strazza Rodrigues; Felipe Allan da Silva da Costa; Flavia Aburjaile; Fábio Sossai Possebon; Guilherme Campos; Guilherme Targino Valente; Heidge Fukumasu; Heidge Fukumasu. USP-Botucatu: Rejane Maria Tommasini Grotto; Helena Lage Ferreira; Instituto Butantan: Dimas Tadeu Covas; Jardelina de Souza Todao Bernardino; Jayme A. Souza-Neto; Jayme Augusto de Souza-Neto; Jessika Cristina Chagas Lesbon; Jorge A. Petroli Marchesi; José Salvatore Leister Patané; João Paulo Kitajima; João Pessoa Araújo Jr.; Leila Sabrina Ullmann; Loyze Paola Oliveira de Lima; Luiz Alcantara; Luiz Aurelio de Campos Crispin. Centro de Genômica Funcional da ESALQ: Luiz Lehmann Coutinho; Luiz Carlos Junior de Alcantara; Luiz Lehmann Coutinho; Lívia Sacchetto; Maisa C. Pereira Parra; Maria Carolina Elias; Marta Giovanetti; Marília Moraes; Maurício Lacerda Nogueira; Maurício Lacerda Nogueira. Prefeitura de Sao Paulo: Melissa Palmieri.; Patricia Akemi Assato; Paula Rahal; Paulo Inacio da Costa; Rafael dos Santos Bezerra; Raquel de Lello Rocha Campos Cassano. NGS Soluções Genômicas: Pilar Drummond Sampaio Corrêa Mariani. FZEA-USP Pirassununga: Mirele Daiana Poleti; Raul Machado Neto; Rejane Maria Tommasini Grotto; Ricardo Augusto Brassaloti; Ricardo Haddad; Rodrigo Tocantins Calado.; Rodrigo Tocantins Calado. FAMERP-SJRP: Cecília Artico Banho; Sandra Coccuzzo Sampaio; Sandra Coccuzzo Sampaio Vessoni; Simone Kashima; Svetoslav Nanev Slavov; Vagner Fonseca; Vincent Louis Viala |
| EPI_ISL_1445157, EPI_ISL_1445158, EPI_ISL_1445159, EPI_ISL_1445160, EPI_ISL_1445161, EPI_ISL_1445168, EPI_ISL_1445169, EPI_ISL_1445170, EPI_ISL_1732275, EPI_ISL_1966189, EPI_ISL_1966190, EPI_ISL_1966191, EPI_ISL_1966192, EPI_ISL_1966193, EPI_ISL_1966194, EPI_ISL_1966195, EPI_ISL_1966196, EPI_ISL_1966481, EPI_ISL_1966482, EPI_ISL_1966483, EPI_ISL_1966484, EPI_ISL_1966485, EPI_ISL_1966486, EPI_ISL_1966487, EPI_ISL_1966488, EPI_ISL_1966489, EPI_ISL_1966490, EPI_ISL_1966491, EPI_ISL_1966492, EPI_ISL_1966493, EPI_ISL_1966494, EPI_ISL_1966495, EPI_ISL_1966496, EPI_ISL_1966627, EPI_ISL_1966628, EPI_ISL_1966629, EPI_ISL_1966630, EPI_ISL_1966631, EPI_ISL_1966632, EPI_ISL_1966633, EPI_ISL_1966716, EPI_ISL_1966717, EPI_ISL_1966718, EPI_ISL_1966719, EPI_ISL_1967258, EPI_ISL_1967259, EPI_ISL_1967260, EPI_ISL_1967261, EPI_ISL_1967262, EPI_ISL_1967263, EPI_ISL_1967264, EPI_ISL_1967265, EPI_ISL_1967266, EPI_ISL_1967267, EPI_ISL_1967268, EPI_ISL_1967269, EPI_ISL_1967270, EPI_ISL_1967271, EPI_ISL_1967272, EPI_ISL_1967273, EPI_ISL_1967274, EPI_ISL_2170979, EPI_ISL_2170980, EPI_ISL_2170981 | | | |
| see above | SECAO CENTRO DE DIAGNOSTICO SECEDI | Instituto Butantan / Mendelics | Antonio Jorge Martins; Bianca Cechetto Carlos. Mendelics: Bibiana Santos; Bibiana Santos; Claudia Renata dos Santos Barros; Cíntia Bittar; David Schlesinger; David Schlesinger. Hemocentro Ribeirão Preto: Simone Kashima; Debora Botequio Moretti; Debora Botequio Moretti. Centro de Genômica Funcional da ESALQ: Luiz Lehmann Coutinho; Dimas Tadeu Covas; Elaine Cristina Marqueze; Elaine Vieira dos Santos; Elisangela Chicaroni Mattos; Erika Freitas; Evandra Strazza Rodrigues; Felipe Allan da Silva da Costa; Flavia Aburjaile; Fábio Sossai Possebon; Guilherme Campos; Guilherme Targino Valente; Heidge Fukumasu. USP-Botucatu: Rejane Maria Tommasini Grotto; Helena Lage Ferreira; Instituto Butantan: Alexander Roberto Precioso; Instituto Butantan: Dimas Tadeu Covas; Jardelina de Souza Todao Bernardino; Jayme A. Souza-Neto; Jessika Cristina Chagas Lesbon; Jorge A. Petroli Marchesi; José Salvatore Leister Patané; João Paulo Kitajima; João Pessoa Araújo Jr.; Leila Sabrina Ullmann; Loyze Paola Oliveira de Lima; Luiz Aurelio de Campos Crispin. Centro de Genômica Funcional da ESALQ: Luiz Lehmann Coutinho; Luiz Carlos Junior de Alcantara; Lívia Sacchetto; Maisa C. Pereira Parra; Maria Carolina Elias; Marta Giovanetti; Marília Moraes; Maurício Lacerda Nogueira. Prefeitura de Sao Paulo: Melissa Palmieri.; Patricia Akemi Assato; Paula Rahal; Paulo Inacio da Costa; Rafael dos Santos Bezerra; Raquel de Lello Rocha Campos Cassano. NGS Soluções Genômicas: Pilar Drummond Sampaio Corrêa Mariani. FZEA-USP Pirassununga: Mirele Daiana Poleti; Raul Machado Neto; Ricardo Augusto Brassaloti; Ricardo Haddad; Rodrigo Tocantins Calado.; Rodrigo Tocantins Calado. FAMERP-SJRP: Cecília Artico Banho; Sandra Coccuzzo Sampaio; Simone Kashima; Svetoslav Nanev Slavov; Vagner Fonseca; Vincent Louis Viala |
| EPI_ISL_1795364 | SECRETARIA DE SAUDE | Instituto Butantan / ESALQ-Piracicaba | Antonio Jorge Martins; Bianca Cechetto Carlos. Mendelics: Bibiana Santos; Claudia Renata dos Santos Barros; David Schlesinger. Hemocentro Ribeirão Preto: Simone Kashima; Debora Botequio Moretti. Centro de Genômica Funcional da ESALQ: Luiz Lehmann Coutinho; Dimas Tadeu Covas; Elaine Cristina Marqueze; Elaine Vieira dos Santos; Elisangela Chicaroni Mattos; Erika Freitas; Evandra Strazza Rodrigues; Felipe Allan da Silva da Costa; Flavia Aburjaile; Guilherme Targino Valente; Heidge Fukumasu. USP-Botucatu: Rejane Maria Tommasini Grotto; Instituto Butantan: Alexander Roberto Precioso; Jayme A. Souza-Neto; Jessika Cristina Chagas Lesbon; José Salvatore Leister Patané; João Paulo Kitajima; Luiz Carlos Junior de Alcantara; Maria Carolina Elias; Marta Giovanetti; Patricia Akemi Assato; Rafael dos Santos Bezerra; Raquel de Lello Rocha Campos Cassano. NGS Soluções Genômicas: Pilar Drummond Sampaio Corrêa Mariani. FZEA-USP Pirassununga: Mirele Daiana Poleti; Raul Machado Neto; Ricardo Augusto Brassaloti; Ricardo Haddad; Rodrigo Tocantins Calado.; Sandra Coccuzzo Sampaio; Svetoslav Nanev Slavov; Vagner Fonseca; Vincent Louis Viala |
| EPI_ISL_1966360 | SECRETARIA DE SAUDE CABREUVA | Instituto Butantan / Mendelics | Antonio Jorge Martins; Bianca Cechetto Carlos. Mendelics: Bibiana Santos; Claudia Renata dos Santos Barros; Cíntia Bittar; David Schlesinger. Hemocentro Ribeirão Preto: Simone Kashima; Debora Botequio Moretti; Elaine Cristina Marqueze; Elaine Vieira dos Santos; Elisangela Chicaroni Mattos; Erika Freitas; Evandra Strazza Rodrigues; Felipe Allan da Silva da Costa; Flavia Aburjaile; Fábio Sossai Possebon; Guilherme Campos; Guilherme Targino Valente; Heidge Fukumasu. USP-Botucatu: Rejane Maria Tommasini Grotto; Helena Lage Ferreira; Instituto Butantan: Dimas Tadeu Covas; Jardelina de Souza Todao Bernardino; Jayme A. Souza-Neto; Jessika Cristina Chagas Lesbon; Jorge A. Petroli Marchesi; José Salvatore Leister Patané; João Paulo Kitajima; João Pessoa Araújo Jr.; Leila Sabrina Ullmann; Loyze Paola Oliveira de Lima; Luiz Aurelio de Campos Crispin. Centro de Genômica Funcional da ESALQ: Luiz Lehmann Coutinho; Luiz Carlos Junior de Alcantara; Lívia Sacchetto; Maisa C. Pereira Parra; Maria Carolina Elias; Marta Giovanetti; Marília Moraes; Maurício Lacerda Nogueira. Prefeitura de Sao Paulo: Melissa Palmieri.; Patricia Akemi Assato; Paula Rahal; Paulo Inacio da Costa; Rafael dos Santos Bezerra; Raquel de Lello Rocha Campos Cassano. NGS Soluções Genômicas: Pilar Drummond Sampaio Corrêa Mariani. FZEA-USP Pirassununga: Mirele Daiana Poleti; Raul Machado Neto; Ricardo Augusto Brassaloti; Ricardo Haddad; Rodrigo Tocantins Calado. FAMERP-SJRP: Cecília Artico Banho; Sandra Coccuzzo Sampaio; Svetoslav Nanev Slavov; Vagner Fonseca; Vincent Louis Viala |
| EPI_ISL_1795108 | SECRETARIA DE SAUDE DE SAO PEDRO | Instituto Butantan / ESALQ-Piracicaba | Antonio Jorge Martins; Bianca Cechetto Carlos. Mendelics: Bibiana Santos; Claudia Renata dos Santos Barros; David Schlesinger. Hemocentro Ribeirão Preto: Simone Kashima; Debora Botequio Moretti. Centro de Genômica Funcional da ESALQ: Luiz Lehmann Coutinho; Dimas Tadeu Covas; Elaine Cristina Marqueze; Elaine Vieira dos Santos; Elisangela Chicaroni Mattos; Erika Freitas; Evandra Strazza Rodrigues; Felipe Allan da Silva da Costa; Flavia Aburjaile; Guilherme Targino Valente; Heidge Fukumasu. USP-Botucatu: Rejane Maria Tommasini Grotto; Instituto Butantan: Alexander Roberto Precioso; Jayme A. Souza-Neto; Jessika Cristina Chagas Lesbon; José Salvatore Leister Patané; João Paulo Kitajima; Luiz Carlos Junior de Alcantara; Maria Carolina Elias; Marta Giovanetti; Patricia Akemi Assato; Rafael dos Santos Bezerra; Raquel de Lello Rocha Campos Cassano. NGS Soluções Genômicas: Pilar Drummond Sampaio Corrêa Mariani. FZEA-USP Pirassununga: Mirele Daiana Poleti; Raul Machado Neto; Ricardo Augusto Brassaloti; Ricardo Haddad; Rodrigo Tocantins Calado.; Sandra Coccuzzo Sampaio; Svetoslav Nanev Slavov; Vagner Fonseca; Vincent Louis Viala |
| EPI_ISL_1966247 | SECRETARIA DE SAUDE DO MUNICIPIO DE IPUA | Instituto Butantan / Mendelics | Antonio Jorge Martins; Bianca Cechetto Carlos. Mendelics: Bibiana Santos; Claudia Renata dos Santos Barros; Cíntia Bittar; David Schlesinger. Hemocentro Ribeirão Preto: Simone Kashima; Debora Botequio Moretti; Elaine Cristina Marqueze; Elaine Vieira dos Santos; Elisangela Chicaroni Mattos; Erika Freitas; Evandra Strazza Rodrigues; Felipe Allan da Silva da Costa; Flavia Aburjaile; Fábio Sossai Possebon; Guilherme Campos; Guilherme Targino Valente; Heidge Fukumasu. USP-Botucatu: Rejane Maria Tommasini Grotto; Helena Lage Ferreira; Instituto Butantan: Dimas Tadeu Covas; Jardelina de Souza Todao Bernardino; Jayme A. Souza-Neto; Jessika Cristina Chagas Lesbon; Jorge A. Petroli Marchesi; José Salvatore Leister Patané; João Paulo Kitajima; João Pessoa Araújo Jr.; Leila Sabrina Ullmann; Loyze Paola Oliveira de Lima; Luiz Aurelio de Campos Crispin. Centro de Genômica Funcional da ESALQ: Luiz Lehmann Coutinho; Luiz Carlos Junior de Alcantara; Lívia Sacchetto; Maisa C. Pereira Parra; Maria Carolina Elias; Marta Giovanetti; Marília Moraes; Maurício Lacerda Nogueira. Prefeitura de Sao Paulo: Melissa Palmieri.; Patricia Akemi Assato; Paula Rahal; Paulo Inacio da Costa; Rafael dos Santos Bezerra; Raquel de Lello Rocha Campos Cassano. NGS Soluções Genômicas: Pilar Drummond Sampaio Corrêa Mariani. FZEA-USP Pirassununga: Mirele Daiana Poleti; Raul Machado Neto; Ricardo Augusto Brassaloti; Ricardo Haddad; Rodrigo Tocantins Calado. FAMERP-SJRP: Cecília Artico Banho; Sandra Coccuzzo Sampaio; Svetoslav Nanev Slavov; Vagner Fonseca; Vincent Louis Viala |
| EPI_ISL_1966476 | SECRETARIA DE SAUDE PUBLICA DE PRAIA GRANDE | Instituto Butantan / Mendelics | Antonio Jorge Martins; Bianca Cechetto Carlos. Mendelics: Bibiana Santos; Claudia Renata dos Santos Barros; Cíntia Bittar; David Schlesinger. Hemocentro Ribeirão Preto: Simone Kashima; Debora Botequio Moretti; Elaine Cristina Marqueze; Elaine Vieira dos Santos; Elisangela Chicaroni Mattos; Erika Freitas; Evandra Strazza Rodrigues; Felipe Allan da Silva da Costa; Flavia Aburjaile; Fábio Sossai Possebon; Guilherme Campos; Guilherme Targino Valente; Heidge Fukumasu. USP-Botucatu: Rejane Maria Tommasini Grotto; Helena Lage Ferreira; Instituto Butantan: Dimas Tadeu Covas; Jardelina de Souza Todao Bernardino; Jayme A. Souza-Neto; Jessika Cristina Chagas Lesbon; Jorge A. Petroli Marchesi; José Salvatore Leister Patané; João Paulo Kitajima; João Pessoa Araújo Jr.; Leila Sabrina Ullmann; Loyze Paola Oliveira de Lima; Luiz Aurelio de Campos Crispin. Centro de Genômica Funcional da ESALQ: Luiz Lehmann Coutinho; Luiz Carlos Junior de Alcantara; Lívia Sacchetto; Maisa C. Pereira Parra; Maria Carolina Elias; Marta Giovanetti; Marília Moraes; Maurício Lacerda Nogueira. Prefeitura de Sao Paulo: Melissa Palmieri.; Patricia Akemi Assato; Paula Rahal; Paulo Inacio da Costa; Rafael dos Santos Bezerra; Raquel de Lello Rocha Campos Cassano. NGS Soluções Genômicas: Pilar Drummond Sampaio Corrêa Mariani. FZEA-USP Pirassununga: Mirele Daiana Poleti; Raul Machado Neto; Ricardo Augusto Brassaloti; Ricardo Haddad; Rodrigo Tocantins Calado. FAMERP-SJRP: Cecília Artico Banho; Sandra Coccuzzo Sampaio; Svetoslav Nanev Slavov; Vagner Fonseca; Vincent Louis Viala |
| EPI_ISL_1966709, EPI_ISL_1966710, EPI_ISL_1966711, EPI_ISL_1966712, EPI_ISL_1966713, EPI_ISL_1966714, EPI_ISL_1966715, EPI_ISL_2170960 | | | |

| | | | |
|---|---|---|---|
| see above | SECRETARIA MUNICIPAL DE SAUDE DE SANTA BARBARA D OESTE | Instituto Butantan / Mendelics | Antonio Jorge Martins; Bianca Cechetto Carlos. Mendelics: Bibiana Santos; Claudia Renata dos Santos Barros; Cíntia Bittar; David Schlesinger. Hemocentro Ribeirão Preto: Simone Kashima; Debora Botequio Moretti; Elaine Cristina Marqueze; Elaine Vieira dos Santos; Elisangela Chicaroni Mattos; Erika Freitas; Evandra Strazza Rodrigues; Felipe Allan da Silva da Costa; Flavia Aburjaile; Fábio Sossai Possebon; Guilherme Campos; Guilherme Targino Valente; Heidge Fukumasu. USP-Botucatu: Rejane Maria Tommasini Grotto; Helena Lage Ferreira; Instituto Butantan: Dimas Tadeu Covas; Jardelina de Souza Todao Bernardino; Jayme A. Souza-Neto; Jessika Cristina Chagas Lesbon; Jorge A. Petroli Marchesi; José Salvatore Leister Patané; João Paulo Kitajima; João Pessoa Araújo Jr.; Leila Sabrina Ullmann; Loyze Paola Oliveira de Lima; Luiz Aurelio de Campos Crispin. Centro de Genômica Funcional da ESALQ: Luiz Lehmann Coutinho; Luiz Carlos Junior de Alcantara; Lívia Sacchetto; Maisa C. Pereira Parra; Maria Carolina Elias; Marta Giovanetti; Marília Moraes; Maurício Lacerda Nogueira. Prefeitura de Sao Paulo: Melissa Palmieri.; Patricia Akemi Assato; Paula Rahal; Paulo Inacio da Costa; Rafael dos Santos Bezerra; Raquel de Lello Rocha Campos Cassano. NGS Soluções Genômicas: Pilar Drummond Sampaio Corrêa Mariani. FZEA-USP Pirassununga: Mirele Daiana Poleti; Raul Machado Neto; Ricardo Augusto Brassaloti; Ricardo Haddad; Rodrigo Tocantins Calado. FAMERP-SJRP: Cecília Artico Banho; Sandra Coccuzzo Sampaio; Svetoslav Nanev Slavov; Vagner Fonseca; Vincent Louis Viala |
| EPI_ISL_1795111, EPI_ISL_1795112 | SECRETARIA MUNICIPAL DA SAUDE DE GUARIBA | Instituto Butantan / ESALQ-Piracicaba | Antonio Jorge Martins; Bianca Cechetto Carlos. Mendelics: Bibiana Santos; Claudia Renata dos Santos Barros; David Schlesinger. Hemocentro Ribeirão Preto: Simone Kashima; Debora Botequio Moretti. Centro de Genômica Funcional da ESALQ: Luiz Lehmann Coutinho; Dimas Tadeu Covas; Elaine Cristina Marqueze; Elaine Vieira dos Santos; Elisangela Chicaroni Mattos; Erika Freitas; Evandra Strazza Rodrigues; Felipe Allan da Silva da Costa; Flavia Aburjaile; Guilherme Targino Valente; Heidge Fukumasu. USP-Botucatu: Rejane Maria Tommasini Grotto; Instituto Butantan: Alexander Roberto Precioso; Jayme A. Souza-Neto; Jessika Cristina Chagas Lesbon; José Salvatore Leister Patané; João Paulo Kitajima; Luiz Carlos Junior de Alcantara; Maria Carolina Elias; Marta Giovanetti; Patricia Akemi Assato; Rafael dos Santos Bezerra; Raquel de Lello Rocha Campos Cassano. NGS Soluções Genômicas: Pilar Drummond Sampaio Corrêa Mariani. FZEA-USP Pirassununga: Mirele Daiana Poleti; Raul Machado Neto; Ricardo Augusto Brassaloti; Ricardo Haddad; Rodrigo Tocantins Calado.; Sandra Coccuzzo Sampaio; Svetoslav Nanev Slavov; Vagner Fonseca; Vincent Louis Viala |
| EPI_ISL_1445066, EPI_ISL_2170910 | SECRETARIA MUNICIPAL DA SAUDE DE GUARIBA | Instituto Butantan / Mendelics | Antonio Jorge Martins; Bianca Cechetto Carlos. Mendelics: Bibiana Santos; Bibiana Santos; Claudia Renata dos Santos Barros; Cíntia Bittar; David Schlesinger; David Schlesinger. Hemocentro Ribeirão Preto: Simone Kashima; Debora Botequio Moretti; Dimas Tadeu Covas; Elaine Cristina Marqueze; Elaine Vieira dos Santos; Elisangela Chicaroni Mattos; Erika Freitas; Evandra Strazza Rodrigues; Felipe Allan da Silva da Costa; Flavia Aburjaile; Fábio Sossai Possebon; Guilherme Campos; Guilherme Targino Valente; Heidge Fukumasu. USP-Botucatu: Rejane Maria Tommasini Grotto; Helena Lage Ferreira; Instituto Butantan: Dimas Tadeu Covas; Jardelina de Souza Todao Bernardino; Jayme A. Souza-Neto; Jessika Cristina Chagas Lesbon; Jorge A. Petroli Marchesi; José Salvatore Leister Patané; João Paulo Kitajima; João Pessoa Araújo Jr.; Leila Sabrina Ullmann; Loyze Paola Oliveira de Lima; Luiz Aurelio de Campos Crispin. Centro de Genômica Funcional da ESALQ: Luiz Lehmann Coutinho; Luiz Carlos Junior de Alcantara; Lívia Sacchetto; Maisa C. Pereira Parra; Maria Carolina Elias; Marta Giovanetti; Marília Moraes; Maurício Lacerda Nogueira. Prefeitura de Sao Paulo: Melissa Palmieri.; Patricia Akemi Assato; Paula Rahal; Paulo Inacio da Costa; Rafael dos Santos Bezerra; Raquel de Lello Rocha Campos Cassano. NGS Soluções Genômicas: Pilar Drummond Sampaio Corrêa Mariani. FZEA-USP Pirassununga: Mirele Daiana Poleti; Raul Machado Neto; Ricardo Augusto Brassaloti; Ricardo Haddad; Rodrigo Tocantins Calado.; Rodrigo Tocantins Calado. FAMERP-SJRP: Cecília Artico Banho; Sandra Coccuzzo Sampaio; Simone Kashima; Svetoslav Nanev Slavov; Vagner Fonseca; Vincent Louis Viala |
| EPI_ISL_1966771, EPI_ISL_2170962 | SECRETARIA MUNICIPAL DA SAUDE DE ITAPEVA SP | Instituto Butantan / Mendelics | Antonio Jorge Martins; Bianca Cechetto Carlos. Mendelics: Bibiana Santos; Claudia Renata dos Santos Barros; Cíntia Bittar; David Schlesinger. Hemocentro Ribeirão Preto: Simone Kashima; Debora Botequio Moretti; Elaine Cristina Marqueze; Elaine Vieira dos Santos; Elisangela Chicaroni Mattos; Erika Freitas; Evandra Strazza Rodrigues; Felipe Allan da Silva da Costa; Flavia Aburjaile; Fábio Sossai Possebon; Guilherme Campos; Guilherme Targino Valente; Heidge Fukumasu. USP-Botucatu: Rejane Maria Tommasini Grotto; Helena Lage Ferreira; Instituto Butantan: Dimas Tadeu Covas; Jardelina de Souza Todao Bernardino; Jayme A. Souza-Neto; Jessika Cristina Chagas Lesbon; Jorge A. Petroli Marchesi; José Salvatore Leister Patané; João Paulo Kitajima; João Pessoa Araújo Jr.; Leila Sabrina Ullmann; Loyze Paola Oliveira de Lima; Luiz Aurelio de Campos Crispin. Centro de Genômica Funcional da ESALQ: Luiz Lehmann Coutinho; Luiz Carlos Junior de Alcantara; Lívia Sacchetto; Maisa C. Pereira Parra; Maria Carolina Elias; Marta Giovanetti; Marília Moraes; Maurício Lacerda Nogueira. Prefeitura de Sao Paulo: Melissa Palmieri.; Patricia Akemi Assato; Paula Rahal; Paulo Inacio da Costa; Rafael dos Santos Bezerra; Raquel de Lello Rocha Campos Cassano. NGS Soluções Genômicas: Pilar Drummond Sampaio Corrêa Mariani. FZEA-USP Pirassununga: Mirele Daiana Poleti; Raul Machado Neto; Ricardo Augusto Brassaloti; Ricardo Haddad; Rodrigo Tocantins Calado. FAMERP-SJRP: Cecília Artico Banho; Sandra Coccuzzo Sampaio; Svetoslav Nanev Slavov; Vagner Fonseca; Vincent Louis Viala |
| EPI_ISL_1966950, EPI_ISL_1966951 | SECRETARIA MUNICIPAL DA SAUDE DE ITARARE | Instituto Butantan / ESALQ-USP (Piracicaba) | Antonio Jorge Martins; Bianca Cechetto Carlos. Mendelics: Bibiana Santos; Claudia Renata dos Santos Barros; Cíntia Bittar; David Schlesinger. Hemocentro Ribeirão Preto: Simone Kashima; Debora Botequio Moretti; Elaine Cristina Marqueze; Elaine Vieira dos Santos; Elisangela Chicaroni Mattos; Erika Freitas; Evandra Strazza Rodrigues; Felipe Allan da Silva da Costa; Flavia Aburjaile; Fábio Sossai Possebon; Guilherme Campos; Guilherme Targino Valente; Heidge Fukumasu. USP-Botucatu: Rejane Maria Tommasini Grotto; Helena Lage Ferreira; Instituto Butantan: Dimas Tadeu Covas; Jardelina de Souza Todao Bernardino; Jayme A. Souza-Neto; Jessika Cristina Chagas Lesbon; Jorge A. Petroli Marchesi; José Salvatore Leister Patané; João Paulo Kitajima; João Pessoa Araújo Jr.; Leila Sabrina Ullmann; Loyze Paola Oliveira de Lima; Luiz Aurelio de Campos Crispin. Centro de Genômica Funcional da ESALQ: Luiz Lehmann Coutinho; Luiz Carlos Junior de Alcantara; Lívia Sacchetto; Maisa C. Pereira Parra; Maria Carolina Elias; Marta Giovanetti; Marília Moraes; Maurício Lacerda Nogueira. Prefeitura de Sao Paulo: Melissa Palmieri.; Patricia Akemi Assato; Paula Rahal; Paulo Inacio da Costa; Rafael dos Santos Bezerra; Raquel de Lello Rocha Campos Cassano. NGS Soluções Genômicas: Pilar Drummond Sampaio Corrêa Mariani. FZEA-USP Pirassununga: Mirele Daiana Poleti; Raul Machado Neto; Ricardo Augusto Brassaloti; Ricardo Haddad; Rodrigo Tocantins Calado. FAMERP-SJRP: Cecília Artico Banho; Sandra Coccuzzo Sampaio; Svetoslav Nanev Slavov; Vagner Fonseca; Vincent Louis Viala |
| EPI_ISL_1966772 | SECRETARIA MUNICIPAL DA SAUDE DE ITARARE | Instituto Butantan / Mendelics | Antonio Jorge Martins; Bianca Cechetto Carlos. Mendelics: Bibiana Santos; Claudia Renata dos Santos Barros; Cíntia Bittar; David Schlesinger. Hemocentro Ribeirão Preto: Simone Kashima; Debora Botequio Moretti; Elaine Cristina Marqueze; Elaine Vieira dos Santos; Elisangela Chicaroni Mattos; Erika Freitas; Evandra Strazza Rodrigues; Felipe Allan da Silva da Costa; Flavia Aburjaile; Fábio Sossai Possebon; Guilherme Campos; Guilherme Targino Valente; Heidge Fukumasu. USP-Botucatu: Rejane Maria Tommasini Grotto; Helena Lage Ferreira; Instituto Butantan: Dimas Tadeu Covas; Jardelina de Souza Todao Bernardino; Jayme A. Souza-Neto; Jessika Cristina Chagas Lesbon; Jorge A. Petroli Marchesi; José Salvatore Leister Patané; João Paulo Kitajima; João Pessoa Araújo Jr.; Leila Sabrina Ullmann; Loyze Paola Oliveira de Lima; Luiz Aurelio de Campos Crispin. Centro de Genômica Funcional da ESALQ: Luiz Lehmann Coutinho; Luiz Carlos Junior de Alcantara; Lívia Sacchetto; Maisa C. Pereira Parra; Maria Carolina Elias; Marta Giovanetti; Marília Moraes; Maurício Lacerda Nogueira. Prefeitura de Sao Paulo: Melissa Palmieri.; Patricia Akemi Assato; Paula Rahal; Paulo Inacio da Costa; Rafael dos Santos Bezerra; Raquel de Lello Rocha Campos Cassano. NGS Soluções Genômicas: Pilar Drummond Sampaio Corrêa Mariani. FZEA-USP Pirassununga: Mirele Daiana Poleti; Raul Machado Neto; Ricardo Augusto Brassaloti; Ricardo Haddad; Rodrigo Tocantins Calado. FAMERP-SJRP: Cecília Artico Banho; Sandra Coccuzzo Sampaio; Svetoslav Nanev Slavov; Vagner Fonseca; Vincent Louis Viala |
| EPI_ISL_1966389, EPI_ISL_1966390, EPI_ISL_1966391, EPI_ISL_1966392 | SECRETARIA MUNICIPAL DA SAUDE DE JOANOPOLIS | Instituto Butantan / Mendelics | Antonio Jorge Martins; Bianca Cechetto Carlos. Mendelics: Bibiana Santos; Claudia Renata dos Santos Barros; Cíntia Bittar; David Schlesinger. Hemocentro Ribeirão Preto: Simone Kashima; Debora Botequio Moretti; Elaine Cristina Marqueze; Elaine Vieira dos Santos; Elisangela Chicaroni Mattos; Erika Freitas; Evandra Strazza Rodrigues; Felipe Allan da Silva da Costa; Flavia Aburjaile; Fábio Sossai Possebon; Guilherme Campos; Guilherme Targino Valente; Heidge Fukumasu. USP-Botucatu: Rejane Maria Tommasini Grotto; Helena Lage Ferreira; Instituto Butantan: Dimas Tadeu Covas; Jardelina de Souza Todao Bernardino; Jayme A. Souza-Neto; Jessika Cristina Chagas Lesbon; Jorge A. Petroli Marchesi; José Salvatore Leister Patané; João Paulo Kitajima; João Pessoa Araújo Jr.; Leila Sabrina Ullmann; Loyze Paola Oliveira de Lima; Luiz Aurelio de Campos Crispin. Centro de Genômica Funcional da ESALQ: Luiz Lehmann Coutinho; Luiz Carlos Junior de Alcantara; Lívia Sacchetto; Maisa C. Pereira Parra; Maria Carolina Elias; Marta Giovanetti; Marília Moraes; Maurício Lacerda Nogueira. Prefeitura de Sao Paulo: Melissa Palmieri.; Patricia Akemi Assato; Paula Rahal; Paulo Inacio da Costa; Rafael dos Santos Bezerra; Raquel de Lello Rocha Campos Cassano. NGS Soluções Genômicas: Pilar Drummond Sampaio Corrêa Mariani. FZEA-USP Pirassununga: Mirele Daiana Poleti; Raul Machado Neto; Ricardo Augusto Brassaloti; Ricardo Haddad; Rodrigo Tocantins Calado. FAMERP-SJRP: Cecília Artico Banho; Sandra Coccuzzo Sampaio; Svetoslav Nanev Slavov; Vagner Fonseca; Vincent Louis Viala |
| EPI_ISL_1966551 | SECRETARIA MUNICIPAL DE SAUDE | Instituto Butantan / FZEA-USP (Pirassununga) | Antonio Jorge Martins; Bianca Cechetto Carlos. Mendelics: Bibiana Santos; Claudia Renata dos Santos Barros; Cíntia Bittar; David Schlesinger. Hemocentro Ribeirão Preto: Simone Kashima; Debora Botequio Moretti; Elaine Cristina Marqueze; Elaine Vieira dos Santos; Elisangela Chicaroni Mattos; Erika Freitas; Evandra Strazza Rodrigues; Felipe Allan da Silva da Costa; Flavia Aburjaile; Fábio Sossai Possebon; Guilherme Campos; Guilherme Targino Valente; Heidge Fukumasu. USP-Botucatu: Rejane Maria Tommasini Grotto; Helena Lage Ferreira; Instituto Butantan: Dimas Tadeu Covas; Jardelina de Souza Todao Bernardino; Jayme A. Souza-Neto; Jessika Cristina Chagas Lesbon; Jorge A. Petroli Marchesi; José Salvatore Leister Patané; João Paulo Kitajima; João Pessoa Araújo Jr.; Leila Sabrina Ullmann; Loyze Paola Oliveira de Lima; Luiz Aurelio de Campos Crispin. Centro de Genômica Funcional da ESALQ: Luiz Lehmann Coutinho; Luiz Carlos Junior de Alcantara; Lívia Sacchetto; Maisa C. Pereira Parra; Maria Carolina Elias; Marta Giovanetti; Marília Moraes; Maurício Lacerda Nogueira. Prefeitura de Sao Paulo: Melissa Palmieri.; Patricia Akemi Assato; Paula Rahal; Paulo Inacio da Costa; Rafael dos Santos Bezerra; Raquel de Lello Rocha Campos Cassano. NGS Soluções Genômicas: Pilar Drummond Sampaio Corrêa Mariani. FZEA-USP Pirassununga: Mirele Daiana Poleti; Raul Machado Neto; Ricardo Augusto Brassaloti; Ricardo Haddad; Rodrigo Tocantins Calado. FAMERP-SJRP: Cecília Artico Banho; Sandra Coccuzzo Sampaio; Svetoslav Nanev Slavov; Vagner Fonseca; Vincent Louis Viala |
| EPI_ISL_1966468, EPI_ISL_1967300, EPI_ISL_1967301, EPI_ISL_1967302, EPI_ISL_1967303, EPI_ISL_2171023, EPI_ISL_2171024 | | | |
| see above | SECRETARIA MUNICIPAL DE SAUDE | Instituto Butantan / Mendelics | Antonio Jorge Martins; Bianca Cechetto Carlos. Mendelics: Bibiana Santos; Claudia Renata dos Santos Barros; Cíntia Bittar; David Schlesinger; David Schlesinger. Hemocentro Ribeirão Preto: Simone Kashima; Debora Botequio Moretti; Dimas Tadeu Covas; Elaine Cristina Marqueze; Elaine Vieira Santos; Elaine Vieira dos Santos; Elisangela Chicaroni Mattos; Erika Freitas; Evandra Strazza Rodrigues; Felipe Allan da Silva da Costa; Flavia Aburjaile; Fábio Sossai Possebon; Guilherme Campos; Guilherme Targino Valente; Heidge Fukumasu. USP-Botucatu: Rejane Maria Tommasini Grotto; Helena Lage Ferreira; Instituto Butantan: Dimas Tadeu Covas; Jardelina de Souza Todao Bernardino; Jayme A. Souza-Neto; Jayme Augusto de Souza-Neto; Jessika Cristina Chagas Lesbon; Jorge A. Petroli Marchesi; José Salvatore Leister Patané; João Paulo Kitajima; João Pessoa Araújo Jr.; Leila Sabrina Ullmann; Loyze Paola Oliveira de Lima; Luiz Alcantara; Luiz Aurelio de Campos Crispin. Centro de Genômica Funcional da ESALQ: Luiz Lehmann Coutinho; Luiz Carlos Junior de Alcantara; Luiz Lehmann Coutinho; Lívia Sacchetto; Maisa C. Pereira Parra; Maria Carolina Elias; Marta Giovanetti; Marília Moraes; Maurício Lacerda Nogueira; Maurício Lacerda Nogueira. Prefeitura de Sao Paulo: Melissa Palmieri.; Patricia Akemi Assato; Paula Rahal; Paulo Inacio da Costa; Rafael dos Santos Bezerra; Raquel de Lello Rocha Campos Cassano. NGS Soluções Genômicas: Pilar Drummond Sampaio Corrêa Mariani. FZEA-USP Pirassununga: Mirele Daiana Poleti; Raul Machado Neto; Rejane Maria Tommasini Grotto; Ricardo Augusto Brassaloti; Ricardo Haddad; Rodrigo Tocantins Calado. FAMERP-SJRP: Cecília Artico Banho; Sandra Coccuzzo Sampaio; Sandra Coccuzzo Sampaio Vessoni; Simone Kashima; Svetoslav Nanev Slavov; Vagner Fonseca; Vincent Louis Viala |
| EPI_ISL_1966270, EPI_ISL_1966271, EPI_ISL_1966272, EPI_ISL_1966273, EPI_ISL_1966274, EPI_ISL_1966275, EPI_ISL_1966276, EPI_ISL_1966277 | | | |
| see above | SECRETARIA MUNICIPAL DA SAUDE DE ANDRADINA | Instituto Butantan / Mendelics | Antonio Jorge Martins; Bianca Cechetto Carlos. Mendelics: Bibiana Santos; Claudia Renata dos Santos Barros; Cíntia Bittar; David Schlesinger. Hemocentro Ribeirão Preto: Simone Kashima; Debora Botequio Moretti; Elaine Cristina Marqueze; Elaine Vieira dos Santos; Elisangela Chicaroni Mattos; Erika Freitas; Evandra Strazza Rodrigues; Felipe Allan da Silva da Costa; Flavia Aburjaile; Fábio Sossai Possebon; Guilherme Campos; Guilherme Targino Valente; Heidge Fukumasu. USP-Botucatu: Rejane Maria Tommasini Grotto; Helena Lage Ferreira; Instituto Butantan: Dimas Tadeu Covas; Jardelina de Souza Todao Bernardino; Jayme A. Souza-Neto; Jessika Cristina Chagas Lesbon; Jorge A. Petroli Marchesi; José Salvatore Leister Patané; João Paulo Kitajima; João Pessoa Araújo Jr.; Leila Sabrina Ullmann; Loyze Paola Oliveira de Lima; Luiz Aurelio de Campos Crispin. Centro de Genômica Funcional da ESALQ: Luiz Lehmann Coutinho; Luiz Carlos Junior de Alcantara; Lívia Sacchetto; Maisa C. Pereira Parra; Maria Carolina Elias; Marta Giovanetti; Marília Moraes; Maurício Lacerda Nogueira. Prefeitura de Sao Paulo: Melissa Palmieri.; Patricia Akemi Assato; Paula Rahal; Paulo Inacio da Costa; Rafael dos Santos Bezerra; Raquel de Lello Rocha Campos Cassano. NGS Soluções Genômicas: Pilar Drummond Sampaio Corrêa Mariani. FZEA-USP Pirassununga: Mirele Daiana Poleti; Raul Machado Neto; Ricardo Augusto Brassaloti; Ricardo Haddad; Rodrigo Tocantins Calado. FAMERP-SJRP: Cecília Artico Banho; Sandra Coccuzzo Sampaio; Svetoslav Nanev Slavov; Vagner Fonseca; Vincent Louis Viala |
| EPI_ISL_1795402 | SECRETARIA MUNICIPAL DA SAUDE DE CORDEIROPOLIS | Instituto Butantan / ESALQ-Piracicaba | Antonio Jorge Martins; Bianca Cechetto Carlos. Mendelics: Bibiana Santos; Claudia Renata dos Santos Barros; David Schlesinger. Hemocentro Ribeirão Preto: Simone Kashima; Debora Botequio Moretti. Centro de Genômica Funcional da ESALQ: Luiz Lehmann Coutinho; Dimas Tadeu Covas; Elaine Cristina Marqueze; Elaine Vieira dos Santos; Elisangela Chicaroni Mattos; Erika Freitas; Evandra Strazza Rodrigues; Felipe Allan da Silva da Costa; Flavia Aburjaile; Guilherme Targino Valente; Heidge Fukumasu. USP-Botucatu: Rejane Maria Tommasini Grotto; Instituto Butantan: Alexander Roberto Precioso; Jayme A. Souza-Neto; Jessika Cristina Chagas Lesbon; José Salvatore Leister Patané; João Paulo Kitajima; Luiz Carlos Junior de Alcantara; Maria Carolina Elias; Marta Giovanetti; Patricia Akemi Assato; Rafael dos Santos Bezerra; Raquel de Lello Rocha Campos Cassano. NGS Soluções Genômicas: Pilar Drummond Sampaio Corrêa Mariani. FZEA-USP Pirassununga: Mirele Daiana Poleti; Raul Machado Neto; Ricardo Augusto Brassaloti; Ricardo Haddad; Rodrigo Tocantins Calado.; Sandra Coccuzzo Sampaio; Svetoslav Nanev Slavov; Vagner Fonseca; Vincent Louis Viala |
| EPI_ISL_1966693 | SECRETARIA MUNICIPAL DA SAUDE DE JARINU | Instituto Butantan / Mendelics | Antonio Jorge Martins; Bianca Cechetto Carlos. Mendelics: Bibiana Santos; Claudia Renata dos Santos Barros; Cíntia Bittar; David Schlesinger. Hemocentro Ribeirão Preto: Simone Kashima; Debora Botequio Moretti; Elaine Cristina Marqueze; Elaine Vieira dos Santos; Elisangela Chicaroni Mattos; Erika Freitas; Evandra Strazza Rodrigues; Felipe Allan da Silva da Costa; Flavia Aburjaile; Fábio Sossai Possebon; Guilherme Campos; Guilherme Targino Valente; Heidge Fukumasu. USP-Botucatu: Rejane Maria Tommasini Grotto; Helena Lage Ferreira; Instituto Butantan: Dimas Tadeu Covas; Jardelina de Souza Todao Bernardino; Jayme A. Souza-Neto; Jessika Cristina Chagas Lesbon; Jorge A. Petroli Marchesi; José Salvatore Leister Patané; João Paulo Kitajima; João Pessoa Araújo Jr.; Leila Sabrina Ullmann; Loyze Paola Oliveira de Lima; Luiz Aurelio de Campos Crispin. Centro de Genômica Funcional da ESALQ: Luiz Lehmann Coutinho; Luiz Carlos Junior de Alcantara; Lívia Sacchetto; Maisa C. Pereira Parra; Maria Carolina Elias; Marta Giovanetti; Marília Moraes; Maurício Lacerda Nogueira. Prefeitura de Sao Paulo: Melissa Palmieri.; Patricia Akemi Assato; Paula Rahal; Paulo Inacio da Costa; Rafael dos Santos Bezerra; Raquel de Lello Rocha Campos Cassano. NGS Soluções Genômicas: Pilar Drummond Sampaio Corrêa Mariani. FZEA-USP Pirassununga: Mirele Daiana Poleti; Raul Machado Neto; Ricardo Augusto Brassaloti; Ricardo Haddad; Rodrigo Tocantins Calado. FAMERP-SJRP: Cecília Artico Banho; Sandra Coccuzzo Sampaio; Svetoslav Nanev Slavov; Vagner Fonseca; Vincent Louis Viala |
| EPI_ISL_1795381 | SECRETARIA MUNICIPAL DA SAUDE DE MACATUBA | Instituto Butantan / ESALQ-Piracicaba | Antonio Jorge Martins; Bianca Cechetto Carlos. Mendelics: Bibiana Santos; Claudia Renata dos Santos Barros; David Schlesinger. Hemocentro Ribeirão Preto: Simone Kashima; Debora Botequio Moretti. Centro de Genômica Funcional da ESALQ: Luiz Lehmann Coutinho; Dimas Tadeu Covas; Elaine Cristina Marqueze; Elaine Vieira dos Santos; Elisangela Chicaroni Mattos; Erika Freitas; Evandra Strazza Rodrigues; Felipe Allan da Silva da Costa; Flavia Aburjaile; Guilherme Targino Valente; Heidge Fukumasu. USP-Botucatu: Rejane Maria Tommasini Grotto; Instituto Butantan: Alexander Roberto Precioso; Jayme A. Souza-Neto; Jessika Cristina Chagas Lesbon; José Salvatore Leister Patané; João Paulo Kitajima; Luiz Carlos Junior de Alcantara; Maria Carolina Elias; Marta Giovanetti; Patricia Akemi Assato; Rafael dos Santos Bezerra; Raquel de Lello Rocha Campos Cassano. NGS Soluções Genômicas: Pilar Drummond Sampaio Corrêa Mariani. FZEA-USP Pirassununga: Mirele Daiana Poleti; Raul Machado Neto; Ricardo Augusto Brassaloti; Ricardo Haddad; Rodrigo Tocantins Calado.; Sandra Coccuzzo Sampaio; Svetoslav Nanev Slavov; Vagner Fonseca; Vincent Louis Viala |
| EPI_ISL_1795373 | SECRETARIA MUNICIPAL DA SAUDE DE PEDERNEIRAS | Instituto Butantan / ESALQ-Piracicaba | Antonio Jorge Martins; Bianca Cechetto Carlos. Mendelics: Bibiana Santos; Claudia Renata dos Santos Barros; David Schlesinger. Hemocentro Ribeirão Preto: Simone Kashima; Debora Botequio Moretti. Centro de Genômica Funcional da ESALQ: Luiz Lehmann Coutinho; Dimas Tadeu Covas; Elaine Cristina Marqueze; Elaine Vieira dos Santos; Elisangela Chicaroni Mattos; Erika Freitas; Evandra Strazza Rodrigues; Felipe Allan da Silva da Costa; Flavia Aburjaile; Guilherme Targino Valente; Heidge Fukumasu. USP-Botucatu: Rejane Maria Tommasini Grotto; Instituto Butantan: Alexander Roberto Precioso; Jayme A. Souza-Neto; Jessika Cristina Chagas Lesbon; José Salvatore Leister Patané; João Paulo Kitajima; Luiz Carlos Junior de Alcantara; Maria Carolina Elias; Marta Giovanetti; Patricia Akemi Assato; Rafael dos Santos Bezerra; Raquel de Lello Rocha Campos Cassano. NGS Soluções Genômicas: Pilar Drummond Sampaio Corrêa Mariani. FZEA-USP Pirassununga: Mirele Daiana Poleti; Raul Machado Neto; Ricardo Augusto Brassaloti; Ricardo Haddad; Rodrigo Tocantins Calado.; Sandra Coccuzzo Sampaio; Svetoslav Nanev Slavov; Vagner Fonseca; Vincent Louis Viala |
| EPI_ISL_1445091, EPI_ISL_1445092, EPI_ISL_1445093, EPI_ISL_1445094, EPI_ISL_1445095, EPI_ISL_1445101, EPI_ISL_1966148, EPI_ISL_1966149, EPI_ISL_1966150, EPI_ISL_1966151, EPI_ISL_1966152, EPI_ISL_1966153 | | | |
| see above | SECRETARIA MUNICIPAL DE SAUDE DE PIRACAIA | Instituto Butantan / Mendelics | Antonio Jorge Martins; Bianca Cechetto Carlos. Mendelics: Bibiana Santos; Bibiana Santos; Claudia Renata dos Santos Barros; Cíntia Bittar; David Schlesinger; David Schlesinger. Hemocentro Ribeirão Preto: Simone Kashima; Debora Botequio Moretti; Dimas Tadeu Covas; Elaine Cristina Marqueze; Elaine Vieira dos Santos; Elisangela Chicaroni Mattos; Erika Freitas; Evandra Strazza Rodrigues; Felipe Allan da Silva da Costa; Flavia Aburjaile; Fábio Sossai Possebon; Guilherme Campos; Guilherme Targino Valente; Heidge Fukumasu. USP-Botucatu: Rejane Maria Tommasini Grotto; Helena Lage Ferreira; Instituto Butantan: Dimas Tadeu Covas; Jardelina de Souza Todao Bernardino; Jayme A. Souza-Neto; Jessika Cristina Chagas Lesbon; Jorge A. Petroli Marchesi; José Salvatore Leister Patané; João Paulo Kitajima; João Pessoa Araújo Jr.; Leila Sabrina Ullmann; Loyze Paola Oliveira de Lima; Luiz Aurelio de Campos Crispin. Centro de Genômica Funcional da ESALQ: Luiz Lehmann Coutinho; Luiz Carlos Junior de Alcantara; Lívia Sacchetto; Maisa C. Pereira Parra; Maria Carolina Elias; Marta Giovanetti; Marília Moraes; Maurício Lacerda Nogueira. Prefeitura de Sao Paulo: Melissa Palmieri.; Patricia Akemi Assato; Paula Rahal; Paulo Inacio da Costa; Rafael dos Santos Bezerra; Raquel de Lello Rocha Campos Cassano. NGS Soluções Genômicas: Pilar Drummond Sampaio Corrêa Mariani. FZEA-USP Pirassununga: Mirele Daiana Poleti; Raul Machado Neto; Ricardo Augusto Brassaloti; Ricardo Haddad; Rodrigo Tocantins Calado.; Rodrigo Tocantins Calado. FAMERP-SJRP: Cecília Artico Banho; Sandra Coccuzzo Sampaio; Simone Kashima; Svetoslav Nanev Slavov; Vagner Fonseca; Vincent Louis Viala |
| EPI_ISL_1966346, EPI_ISL_1966347, EPI_ISL_1966348, EPI_ISL_1966349 | SECRETARIA MUNICIPAL DE SAUDE DE VALPARAISO SP | Instituto Butantan / Mendelics | Antonio Jorge Martins; Bianca Cechetto Carlos. Mendelics: Bibiana Santos; Claudia Renata dos Santos Barros; Cíntia Bittar; David Schlesinger. Hemocentro Ribeirão Preto: Simone Kashima; Debora Botequio Moretti; Elaine Cristina Marqueze; Elaine Vieira dos Santos; Elisangela Chicaroni Mattos; Erika Freitas; Evandra Strazza Rodrigues; Felipe Allan da Silva da Costa; Flavia Aburjaile; Fábio Sossai Possebon; Guilherme Campos; Guilherme Targino Valente; Heidge Fukumasu. USP-Botucatu: Rejane Maria Tommasini Grotto; Helena Lage Ferreira; Instituto Butantan: Dimas Tadeu Covas; Jardelina de Souza Todao Bernardino; Jayme A. Souza-Neto; Jessika Cristina Chagas Lesbon; Jorge A. Petroli Marchesi; José Salvatore Leister Patané; João Paulo Kitajima; João Pessoa Araújo Jr.; Leila Sabrina Ullmann; Loyze Paola Oliveira de Lima; Luiz Aurelio de Campos Crispin. Centro de Genômica Funcional da ESALQ: Luiz Lehmann Coutinho; Luiz Carlos Junior de Alcantara; Lívia Sacchetto; Maisa C. Pereira Parra; Maria Carolina Elias; Marta Giovanetti; Marília Moraes; Maurício Lacerda Nogueira. Prefeitura de Sao Paulo: Melissa Palmieri.; Patricia Akemi Assato; Paula Rahal; Paulo Inacio da Costa; Rafael dos Santos Bezerra; Raquel de Lello Rocha Campos Cassano. NGS Soluções Genômicas: Pilar Drummond Sampaio Corrêa Mariani. FZEA-USP Pirassununga: Mirele Daiana Poleti; Raul Machado Neto; Ricardo Augusto Brassaloti; Ricardo Haddad; Rodrigo Tocantins Calado. FAMERP-SJRP: Cecília Artico Banho; Sandra Coccuzzo Sampaio; Svetoslav Nanev Slavov; Vagner Fonseca; Vincent Louis Viala |
| EPI_ISL_1967252, EPI_ISL_1967253, EPI_ISL_1967254, EPI_ISL_1967255, EPI_ISL_1967256, EPI_ISL_1967257, EPI_ISL_2170978 | | | |
| see above | SECRETARIA MUNICIPAL DE SAUDE PORTO FERREIRA | Instituto Butantan / Mendelics | Antonio Jorge Martins; Bianca Cechetto Carlos. Mendelics: Bibiana Santos; Claudia Renata dos Santos Barros; Cíntia Bittar; David Schlesinger. Hemocentro Ribeirão Preto: Simone Kashima; Debora Botequio Moretti; Elaine Cristina Marqueze; Elaine Vieira dos Santos; Elisangela Chicaroni Mattos; Erika Freitas; Evandra Strazza Rodrigues; Felipe Allan da Silva da Costa; Flavia Aburjaile; Fábio Sossai Possebon; Guilherme Campos; Guilherme Targino Valente; Heidge Fukumasu. USP-Botucatu: Rejane Maria Tommasini Grotto; Helena Lage Ferreira; Instituto Butantan: Dimas Tadeu Covas; Jardelina de Souza Todao Bernardino; Jayme A. Souza-Neto; Jessika Cristina Chagas Lesbon; Jorge A. Petroli Marchesi; José Salvatore Leister Patané; João Paulo Kitajima; João Pessoa Araújo Jr.; Leila Sabrina Ullmann; Loyze Paola Oliveira de Lima; Luiz Aurelio de Campos Crispin. Centro de Genômica Funcional da ESALQ: Luiz Lehmann Coutinho; Luiz Carlos Junior de Alcantara; Lívia Sacchetto; Maisa C. Pereira Parra; Maria Carolina Elias; Marta Giovanetti; Marília Moraes; Maurício Lacerda Nogueira. Prefeitura de Sao Paulo: Melissa Palmieri.; Patricia Akemi Assato; Paula Rahal; Paulo Inacio da Costa; Rafael dos Santos Bezerra; Raquel de Lello Rocha Campos Cassano. NGS Soluções Genômicas: Pilar Drummond Sampaio Corrêa Mariani. FZEA-USP Pirassununga: Mirele Daiana Poleti; Raul Machado Neto; Ricardo Augusto Brassaloti; Ricardo Haddad; Rodrigo Tocantins Calado. FAMERP-SJRP: Cecília Artico Banho; Sandra Coccuzzo Sampaio; Svetoslav Nanev Slavov; Vagner Fonseca; Vincent Louis Viala |
| EPI_ISL_1967098, EPI_ISL_1967099, EPI_ISL_1967100, EPI_ISL_1967101, EPI_ISL_1967102, EPI_ISL_1967103, EPI_ISL_1967104, EPI_ISL_1967105, EPI_ISL_1967106, EPI_ISL_1967107, EPI_ISL_1967108, EPI_ISL_1967109, EPI_ISL_1967110, EPI_ISL_1967111, EPI_ISL_1967112, EPI_ISL_1967113, EPI_ISL_1967114, EPI_ISL_1967115, EPI_ISL_1967116, EPI_ISL_1967117, EPI_ISL_1967118, EPI_ISL_1967119, EPI_ISL_1967120, EPI_ISL_1967121, EPI_ISL_1967122, EPI_ISL_1967123, EPI_ISL_1967124, EPI_ISL_1967125, EPI_ISL_1967126, EPI_ISL_1967127, EPI_ISL_1967128, EPI_ISL_1967129, EPI_ISL_1967130, EPI_ISL_1967131, EPI_ISL_1967132, EPI_ISL_1967133, EPI_ISL_1967134, EPI_ISL_1967135, EPI_ISL_1967136, EPI_ISL_1967137, EPI_ISL_1967138, EPI_ISL_1967139, EPI_ISL_1967140, EPI_ISL_1967141, EPI_ISL_1967142, EPI_ISL_1967143, EPI_ISL_1967144, EPI_ISL_1967145, EPI_ISL_1967146, EPI_ISL_1967147 | | | |
| see above | SECRETARIA MUNICIPAL DE SAUDE SOROCABA | Instituto Butantan / ESALQ-USP (Piracicaba) | Antonio Jorge Martins; Bianca Cechetto Carlos. Mendelics: Bibiana Santos; Claudia Renata dos Santos Barros; Cíntia Bittar; David Schlesinger. Hemocentro Ribeirão Preto: Simone Kashima; Debora Botequio Moretti; Elaine Cristina Marqueze; Elaine Vieira dos Santos; Elisangela Chicaroni Mattos; Erika Freitas; Evandra Strazza Rodrigues; Felipe Allan da Silva da Costa; Flavia Aburjaile; Fábio Sossai Possebon; Guilherme Campos; Guilherme Targino Valente; Heidge Fukumasu. USP-Botucatu: Rejane Maria Tommasini Grotto; Helena Lage Ferreira; Instituto Butantan: Dimas Tadeu Covas; Jardelina de Souza Todao Bernardino; Jayme A. Souza-Neto; Jessika Cristina Chagas Lesbon; Jorge A. Petroli Marchesi; José Salvatore Leister Patané; João Paulo Kitajima; João Pessoa Araújo Jr.; Leila Sabrina Ullmann; Loyze Paola Oliveira de Lima; Luiz Aurelio de Campos Crispin. Centro de Genômica Funcional da ESALQ: Luiz Lehmann Coutinho; Luiz Carlos Junior de Alcantara; Lívia Sacchetto; Maisa C. Pereira Parra; Maria Carolina Elias; Marta Giovanetti; Marília Moraes; Maurício Lacerda Nogueira. Prefeitura de Sao Paulo: Melissa Palmieri.; Patricia Akemi Assato; Paula Rahal; Paulo Inacio da Costa; Rafael dos Santos Bezerra; Raquel de Lello Rocha Campos Cassano. NGS Soluções Genômicas: Pilar Drummond Sampaio Corrêa Mariani. FZEA-USP Pirassununga: Mirele Daiana Poleti; Raul Machado Neto; Ricardo Augusto Brassaloti; Ricardo Haddad; Rodrigo Tocantins Calado. FAMERP-SJRP: Cecília Artico Banho; Sandra Coccuzzo Sampaio; Svetoslav Nanev Slavov; Vagner Fonseca; Vincent Louis Viala |
| EPI_ISL_1445133, EPI_ISL_1445134, EPI_ISL_1445135, EPI_ISL_1445137, EPI_ISL_1445138, EPI_ISL_1445139, EPI_ISL_1445171, EPI_ISL_1445176, EPI_ISL_1445178, EPI_ISL_1445179, EPI_ISL_1445180, EPI_ISL_1445182, EPI_ISL_1716877, EPI_ISL_1966107, EPI_ISL_1966124, EPI_ISL_1966125, EPI_ISL_1966154, EPI_ISL_1966155, EPI_ISL_1966177, EPI_ISL_1966178, EPI_ISL_1966197, EPI_ISL_1966198, EPI_ISL_1966199, EPI_ISL_1966200, EPI_ISL_2170905, EPI_ISL_2170906, EPI_ISL_2170909 | | | |
| see above | SECRETARIA | Instituto Butantan / | Antonio Jorge Martins; Bianca Cechetto Carlos. Mendelics: Bibiana Santos; Bibiana Santos; Claudia Renata dos Santos Barros; Cíntia Bittar; David Schlesinger; David Schlesinger. Hemocentro Ribeirão Preto: Simone Kashima; Debora Botequio Moretti; Debora Botequio Moretti. Centro de Genômica Funcional da ESALQ: Luiz Lehmann |

| | | | |
|---|---|---|---|
| | MUNICIPAL DE SAUDE SOROCABA | Mendelics | Coutinho; Dimas Tadeu Covas; Elaine Cristina Marqueze; Elaine Vieira Santos; Elaine Vieira dos Santos; Elisangela Chicaroni Mattos; Erika Freitas; Evandra Strazza Rodrigues; Felipe Allan da Silva da Costa; Flavia Aburjaile; Fábio Sossai Possebon; Guilherme Campos; Guilherme Targino Valente; Heidge Fukumasu; Heidge Fukumasu. USP-Botucatu: Rejane Maria Tommasini Grotto; Helena Lage Ferreira; Instituto Butantan: Alexander Roberto Precioso; Instituto Butantan: Dimas Tadeu Covas; Jardelina de Souza Todao Bernardino; Jayme A. Souza-Neto; Jayme Augusto de Souza-Neto; Jessika Cristina Chagas Lesbon; Jorge A. Petroli Marchesi; José Salvatore Leister Patané; João Paulo Kitajima; João Pessoa Araújo Jr.; Leila Sabrina Ullmann; Loyze Paola Oliveira de Lima; Luiz Alcantara; Luiz Aurelio de Campos Crispin. Centro de Genômica Funcional da ESALQ: Luiz Lehmann Coutinho; Luiz Carlos Junior de Alcantara; Luiz Lehmann Coutinho; Lívia Sacchetto; Maisa C. Pereira Parra; Maria Carolina Elias; Marta Giovanetti; Marília Moraes; Maurício Lacerda Nogueira; Maurício Lacerda Nogueira. Prefeitura de Sao Paulo: Melissa Palmieri.; Patricia Akemi Assato; Paula Rahal; Paulo Inacio da Costa; Rafael dos Santos Bezerra; Raquel de Lello Rocha Campos Cassano. NGS Soluções Genômicas: Pilar Drummond Sampaio Corrêa Mariani. FZEA-USP Pirassununga: Mirele Daiana Poleti; Raul Machado Neto; Rejane Maria Tommasini Grotto; Ricardo Augusto Brassaloti; Ricardo Haddad; Rodrigo Tocantins Calado.; Rodrigo Tocantins Calado. FAMERP-SJRP: Cecília Artico Banho; Sandra Coccuzzo Sampaio; Sandra Coccuzzo Sampaio Vessoni; Simone Kashima; Svetoslav Nanev Slavov; Vagner Fonseca; Vincent Louis Viala |
| EPI_ISL_1445162, EPI_ISL_1445163, EPI_ISL_1445164, EPI_ISL_1445165, EPI_ISL_1445166, EPI_ISL_1445167, EPI_ISL_1966181, EPI_ISL_1966182, EPI_ISL_1966183, EPI_ISL_1966184, EPI_ISL_1966185, EPI_ISL_1966186 | | | |
| see above | SERV DE VIG SANITARIA EPIDEMIO E CTRL DE ZOONOZES GUARUJA | Instituto Butantan / Mendelics | Antonio Jorge Martins; Bianca Cechetto Carlos. Mendelics: Bibiana Santos; Bibiana Santos; Claudia Renata dos Santos Barros; Cíntia Bittar; David Schlesinger; David Schlesinger. Hemocentro Ribeirão Preto: Simone Kashima; Debora Botequio Moretti; Dimas Tadeu Covas; Elaine Cristina Marqueze; Elaine Vieira dos Santos; Elisangela Chicaroni Mattos; Erika Freitas; Evandra Strazza Rodrigues; Felipe Allan da Silva da Costa; Flavia Aburjaile; Fábio Sossai Possebon; Guilherme Campos; Guilherme Targino Valente; Heidge Fukumasu. USP-Botucatu: Rejane Maria Tommasini Grotto; Helena Lage Ferreira; Instituto Butantan: Dimas Tadeu Covas; Jardelina de Souza Todao Bernardino; Jayme A. Souza-Neto; Jessika Cristina Chagas Lesbon; Jorge A. Petroli Marchesi; José Salvatore Leister Patané; João Paulo Kitajima; João Pessoa Araújo Jr.; Leila Sabrina Ullmann; Loyze Paola Oliveira de Lima; Luiz Aurelio de Campos Crispin. Centro de Genômica Funcional da ESALQ: Luiz Lehmann Coutinho; Luiz Carlos Junior de Alcantara; Lívia Sacchetto; Maisa C. Pereira Parra; Maria Carolina Elias; Marta Giovanetti; Marília Moraes; Maurício Lacerda Nogueira. Prefeitura de Sao Paulo: Melissa Palmieri.; Patricia Akemi Assato; Paula Rahal; Paulo Inacio da Costa; Rafael dos Santos Bezerra; Raquel de Lello Rocha Campos Cassano. NGS Soluções Genômicas: Pilar Drummond Sampaio Corrêa Mariani. FZEA-USP Pirassununga: Mirele Daiana Poleti; Raul Machado Neto; Ricardo Augusto Brassaloti; Ricardo Haddad; Rodrigo Tocantins Calado.; Rodrigo Tocantins Calado. FAMERP-SJRP: Cecília Artico Banho; Sandra Coccuzzo Sampaio; Simone Kashima; Svetoslav Nanev Slavov; Vagner Fonseca; Vincent Louis Viala |
| EPI_ISL_1445136, EPI_ISL_1966187, EPI_ISL_1966377, EPI_ISL_1966378 | SMS IPERO | Instituto Butantan / Mendelics | Antonio Jorge Martins; Bianca Cechetto Carlos. Mendelics: Bibiana Santos; Bibiana Santos; Claudia Renata dos Santos Barros; Cíntia Bittar; David Schlesinger; David Schlesinger. Hemocentro Ribeirão Preto: Simone Kashima; Debora Botequio Moretti; Dimas Tadeu Covas; Elaine Cristina Marqueze; Elaine Vieira dos Santos; Elisangela Chicaroni Mattos; Erika Freitas; Evandra Strazza Rodrigues; Felipe Allan da Silva da Costa; Flavia Aburjaile; Fábio Sossai Possebon; Guilherme Campos; Guilherme Targino Valente; Heidge Fukumasu. USP-Botucatu: Rejane Maria Tommasini Grotto; Helena Lage Ferreira; Instituto Butantan: Dimas Tadeu Covas; Jardelina de Souza Todao Bernardino; Jayme A. Souza-Neto; Jessika Cristina Chagas Lesbon; Jorge A. Petroli Marchesi; José Salvatore Leister Patané; João Paulo Kitajima; João Pessoa Araújo Jr.; Leila Sabrina Ullmann; Loyze Paola Oliveira de Lima; Luiz Aurelio de Campos Crispin. Centro de Genômica Funcional da ESALQ: Luiz Lehmann Coutinho; Luiz Carlos Junior de Alcantara; Lívia Sacchetto; Maisa C. Pereira Parra; Maria Carolina Elias; Marta Giovanetti; Marília Moraes; Maurício Lacerda Nogueira. Prefeitura de Sao Paulo: Melissa Palmieri.; Patricia Akemi Assato; Paula Rahal; Paulo Inacio da Costa; Rafael dos Santos Bezerra; Raquel de Lello Rocha Campos Cassano. NGS Soluções Genômicas: Pilar Drummond Sampaio Corrêa Mariani. FZEA-USP Pirassununga: Mirele Daiana Poleti; Raul Machado Neto; Ricardo Augusto Brassaloti; Ricardo Haddad; Rodrigo Tocantins Calado.; Rodrigo Tocantins Calado. FAMERP-SJRP: Cecília Artico Banho; Sandra Coccuzzo Sampaio; Simone Kashima; Svetoslav Nanev Slavov; Vagner Fonseca; Vincent Louis Viala |
| EPI_ISL_1795398 | SMS SECRETARIA MUNICIPAL DE SAUDE DE BOITUVA | Instituto Butantan / ESALQ-Piracicaba | Antonio Jorge Martins; Bianca Cechetto Carlos. Mendelics: Bibiana Santos; Claudia Renata dos Santos Barros; David Schlesinger. Hemocentro Ribeirão Preto: Simone Kashima; Debora Botequio Moretti. Centro de Genômica Funcional da ESALQ: Luiz Lehmann Coutinho; Dimas Tadeu Covas; Elaine Cristina Marqueze; Elaine Vieira dos Santos; Elisangela Chicaroni Mattos; Erika Freitas; Evandra Strazza Rodrigues; Felipe Allan da Silva da Costa; Flavia Aburjaile; Guilherme Targino Valente; Heidge Fukumasu. USP-Botucatu: Rejane Maria Tommasini Grotto; Instituto Butantan: Alexander Roberto Precioso; Jayme A. Souza-Neto; Jessika Cristina Chagas Lesbon; José Salvatore Leister Patané; João Paulo Kitajima; Luiz Carlos Junior de Alcantara; Maria Carolina Elias; Marta Giovanetti; Patricia Akemi Assato; Rafael dos Santos Bezerra; Raquel de Lello Rocha Campos Cassano. NGS Soluções Genômicas: Pilar Drummond Sampaio Corrêa Mariani. FZEA-USP Pirassununga: Mirele Daiana Poleti; Raul Machado Neto; Ricardo Augusto Brassaloti; Ricardo Haddad; Rodrigo Tocantins Calado.; Sandra Coccuzzo Sampaio; Svetoslav Nanev Slavov; Vagner Fonseca; Vincent Louis Viala |
| EPI_ISL_1966840, EPI_ISL_1966841, EPI_ISL_1966842, EPI_ISL_1966843, EPI_ISL_1966844 | SMS SECRETARIA MUNICIPAL DE SAUDE DE BOITUVA | Instituto Butantan / ESALQ-USP (Piracicaba) | Antonio Jorge Martins; Bianca Cechetto Carlos. Mendelics: Bibiana Santos; Claudia Renata dos Santos Barros; Cíntia Bittar; David Schlesinger. Hemocentro Ribeirão Preto: Simone Kashima; Debora Botequio Moretti; Elaine Cristina Marqueze; Elaine Vieira dos Santos; Elisangela Chicaroni Mattos; Erika Freitas; Evandra Strazza Rodrigues; Felipe Allan da Silva da Costa; Flavia Aburjaile; Fábio Sossai Possebon; Guilherme Campos; Guilherme Targino Valente; Heidge Fukumasu. USP-Botucatu: Rejane Maria Tommasini Grotto; Helena Lage Ferreira; Instituto Butantan: Dimas Tadeu Covas; Jardelina de Souza Todao Bernardino; Jayme A. Souza-Neto; Jessika Cristina Chagas Lesbon; Jorge A. Petroli Marchesi; José Salvatore Leister Patané; João Paulo Kitajima; João Pessoa Araújo Jr.; Leila Sabrina Ullmann; Loyze Paola Oliveira de Lima; Luiz Aurelio de Campos Crispin. Centro de Genômica Funcional da ESALQ: Luiz Lehmann Coutinho; Luiz Carlos Junior de Alcantara; Lívia Sacchetto; Maisa C. Pereira Parra; Maria Carolina Elias; Marta Giovanetti; Marília Moraes; Maurício Lacerda Nogueira. Prefeitura de Sao Paulo: Melissa Palmieri.; Patricia Akemi Assato; Paula Rahal; Paulo Inacio da Costa; Rafael dos Santos Bezerra; Raquel de Lello Rocha Campos Cassano. NGS Soluções Genômicas: Pilar Drummond Sampaio Corrêa Mariani. FZEA-USP Pirassununga: Mirele Daiana Poleti; Raul Machado Neto; Ricardo Augusto Brassaloti; Ricardo Haddad; Rodrigo Tocantins Calado. FAMERP-SJRP: Cecília Artico Banho; Sandra Coccuzzo Sampaio; Svetoslav Nanev Slavov; Vagner Fonseca; Vincent Louis Viala |
| EPI_ISL_1445084, EPI_ISL_1966103, EPI_ISL_1966260, EPI_ISL_1966261, EPI_ISL_1966262, EPI_ISL_1966263, EPI_ISL_1966264, EPI_ISL_1966357, EPI_ISL_1966358, EPI_ISL_1966359, EPI_ISL_1966746, EPI_ISL_1966747, EPI_ISL_1966748, EPI_ISL_2170921, EPI_ISL_2170922, EPI_ISL_2170923 | | | |
| see above | SMS SECRETARIA MUNICIPAL DE SAUDE DE BOITUVA | Instituto Butantan / Mendelics | Antonio Jorge Martins; Bianca Cechetto Carlos. Mendelics: Bibiana Santos; Bibiana Santos; Claudia Renata dos Santos Barros; Cíntia Bittar; David Schlesinger; David Schlesinger. Hemocentro Ribeirão Preto: Simone Kashima; Debora Botequio Moretti; Dimas Tadeu Covas; Elaine Cristina Marqueze; Elaine Vieira dos Santos; Elisangela Chicaroni Mattos; Erika Freitas; Evandra Strazza Rodrigues; Felipe Allan da Silva da Costa; Flavia Aburjaile; Fábio Sossai Possebon; Guilherme Campos; Guilherme Targino Valente; Heidge Fukumasu. USP-Botucatu: Rejane Maria Tommasini Grotto; Helena Lage Ferreira; Instituto Butantan: Dimas Tadeu Covas; Jardelina de Souza Todao Bernardino; Jayme A. Souza-Neto; Jessika Cristina Chagas Lesbon; Jorge A. Petroli Marchesi; José Salvatore Leister Patané; João Paulo Kitajima; João Pessoa Araújo Jr.; Leila Sabrina Ullmann; Loyze Paola Oliveira de Lima; Luiz Aurelio de Campos Crispin. Centro de Genômica Funcional da ESALQ: Luiz Lehmann Coutinho; Luiz Carlos Junior de Alcantara; Lívia Sacchetto; Maisa C. Pereira Parra; Maria Carolina Elias; Marta Giovanetti; Marília Moraes; Maurício Lacerda Nogueira. Prefeitura de Sao Paulo: Melissa Palmieri.; Patricia Akemi Assato; Paula Rahal; Paulo Inacio da Costa; Rafael dos Santos Bezerra; Raquel de Lello Rocha Campos Cassano. NGS Soluções Genômicas: Pilar Drummond Sampaio Corrêa Mariani. FZEA-USP Pirassununga: Mirele Daiana Poleti; Raul Machado Neto; Ricardo Augusto Brassaloti; Ricardo Haddad; Rodrigo Tocantins Calado.; Rodrigo Tocantins Calado. FAMERP-SJRP: Cecília Artico Banho; Sandra Coccuzzo Sampaio; Simone Kashima; Svetoslav Nanev Slavov; Vagner Fonseca; Vincent Louis Viala |
| EPI_ISL_1628370, EPI_ISL_1715139 | Sae Servico De Atendimento Especializado | Instituto Adolfo Lutz, Interdiciplinary Procedures Center, Strategic Laboratory | Caio Vinicius Dias Lopes; Claudia Regina Gonçalves; Claudio Tavares Sacchi; Erica Valessa Ramos Gomes; Karoline Rodrigues Campos; Katia Correa de Oliveira Santos; Leonardo Jose Tadeu de Araujo |
| EPI_ISL_1821206 | Sae Servico de Atendimento Especializado | Instituto Adolfo Lutz, Interdiciplinary Procedures Center, Strategic Laboratory | Caio Vinicius Dias Lopes; Claudia Regina Gonçalves; Claudio Tavares Sacchi; Erica Valessa Ramos Gomes; Karoline Rodrigues Campos; Leonardo Jose Tadeu de Araujo |
| EPI_ISL_1628374, EPI_ISL_1715138 | Santa Casa De Cravinhos | Instituto Adolfo Lutz, Interdiciplinary Procedures Center, Strategic Laboratory | Caio Vinicius Dias Lopes; Claudia Regina Gonçalves; Claudio Tavares Sacchi; Erica Valessa Ramos Gomes; Karoline Rodrigues Campos; Katia Correa de Oliveira Santos; Leonardo Jose Tadeu de Araujo |
| EPI_ISL_1628380 | Santa Casa De Pitangueiras | Instituto Adolfo Lutz, Interdiciplinary Procedures Center, Strategic Laboratory | Caio Vinicius Dias Lopes; Claudia Regina Gonçalves; Claudio Tavares Sacchi; Erica Valessa Ramos Gomes; Karoline Rodrigues Campos; Katia Correa de Oliveira Santos; Leonardo Jose Tadeu de Araujo |
| EPI_ISL_1533721 | Santa Casa de Aracatuba Hospital Sagrado Coracao De Jesus | Instituto Adolfo Lutz, Interdiciplinary Procedures Center, Strategic Laboratory | Caio Vinicius Dias Lopes; Claudia Regina Gonçalves; Claudio Tavares Sacchi; Erica Valessa Ramos Gomes; Karoline Rodrigues Campos; Leonardo Jose Tadeu de Araujo |
| EPI_ISL_1533715, EPI_ISL_1625973, EPI_ISL_1625974 | Santa Casa de Aracatuba Hospital Sagrado Coracao de Jesus | Instituto Adolfo Lutz, Interdiciplinary Procedures Center, Strategic Laboratory | Caio Vinicius Dias Lopes; Claudia Regina Gonçalves; Claudio Tavares Sacchi; Erica Valessa Ramos Gomes; Karoline Rodrigues Campos; Katia Correa de Oliveira Santos; Leonardo Jose Tadeu de Araujo |
| EPI_ISL_1821204 | Santa Casa de Cravinhos | Instituto Adolfo Lutz, Interdiciplinary Procedures Center, Strategic Laboratory | Caio Vinicius Dias Lopes; Claudia Regina Gonçalves; Claudio Tavares Sacchi; Erica Valessa Ramos Gomes; Karoline Rodrigues Campos; Leonardo Jose Tadeu de Araujo |
| EPI_ISL_1628367 | Santa Casa de Guaira | Instituto Adolfo Lutz, Interdiciplinary Procedures Center, Strategic Laboratory | Caio Vinicius Dias Lopes; Claudia Regina Gonçalves; Claudio Tavares Sacchi; Erica Valessa Ramos Gomes; Karoline Rodrigues Campos; Katia Correa de Oliveira Santos; Leonardo Jose Tadeu de Araujo |
| EPI_ISL_1533720 | Santa Casa de Penapolis | Instituto Adolfo Lutz, Interdiciplinary Procedures Center, Strategic Laboratory | Caio Vinicius Dias Lopes; Claudia Regina Gonçalves; Claudio Tavares Sacchi; Erica Valessa Ramos Gomes; Karoline Rodrigues Campos; Leonardo Jose Tadeu de Araujo |
| EPI_ISL_1715137 | Secretaria Municipal De Saude | Instituto Adolfo Lutz, Interdiciplinary Procedures Center, Strategic Laboratory | Caio Vinicius Dias Lopes; Claudia Regina Gonçalves; Claudio Tavares Sacchi; Erica Valessa Ramos Gomes; Karoline Rodrigues Campos; Katia Correa de Oliveira Santos; Leonardo Jose Tadeu de Araujo |
| EPI_ISL_1715143 | Secretaria Municipal de Saude De Guariba | Instituto Adolfo Lutz, Interdiciplinary Procedures Center, Strategic Laboratory | Caio Vinicius Dias Lopes; Claudia Regina Gonçalves; Claudio Tavares Sacchi; Erica Valessa Ramos Gomes; Karoline Rodrigues Campos; Katia Correa de Oliveira Santos; Leonardo Jose Tadeu de Araujo |
| EPI_ISL_1533722 | Secretaria Municipal de Saude De Piracaia | Instituto Adolfo Lutz, Interdiciplinary Procedures Center, Strategic Laboratory | Caio Vinicius Dias Lopes; Claudia Regina Gonçalves; Claudio Tavares Sacchi; Erica Valessa Ramos Gomes; Karoline Rodrigues Campos; Leonardo Jose Tadeu de Araujo |
| EPI_ISL_1628375, EPI_ISL_1628376 | Secretaria Municipal de Saude de Guariba | Instituto Adolfo Lutz, Interdiciplinary Procedures Center, Strategic Laboratory | Caio Vinicius Dias Lopes; Claudia Regina Gonçalves; Claudio Tavares Sacchi; Erica Valessa Ramos Gomes; Karoline Rodrigues Campos; Katia Correa de Oliveira Santos; Leonardo Jose Tadeu de Araujo |
| EPI_ISL_1821207 | Secretaria Municipal de Saúde de Lins | Instituto Adolfo Lutz, Interdiciplinary Procedures Center, Strategic Laboratory | Caio Vinicius Dias Lopes; Claudia Regina Gonçalves; Claudio Tavares Sacchi; Erica Valessa Ramos Gomes; Karoline Rodrigues Campos; Leonardo Jose Tadeu de Araujo |
| EPI_ISL_1533718 | Servico de Verificacao de Obitos Svo Guarulhos | Instituto Adolfo Lutz, Interdiciplinary Procedures Center, Strategic Laboratory | Caio Vinicius Dias Lopes; Claudia Regina Gonçalves; Claudio Tavares Sacchi; Erica Valessa Ramos Gomes; Karoline Rodrigues Campos; Leonardo Jose Tadeu de Araujo |
| EPI_ISL_1715144 | UBDS Dr Joao Baptista Quartim Central | Instituto Adolfo Lutz, Interdiciplinary Procedures Center, Strategic Laboratory | Caio Vinicius Dias Lopes; Claudia Regina Gonçalves; Claudio Tavares Sacchi; Erica Valessa Ramos Gomes; Karoline Rodrigues Campos; Katia Correa de Oliveira Santos; Leonardo Jose Tadeu de Araujo |

| | | | |
|---|---|---|---|
| EPI_ISL_1707688, EPI_ISL_1731574, EPI_ISL_1752637 | UBDS Dr Marco Antonio Sahao Vila Virginia | Instituto Adolfo Lutz, Interdiciplinary Procedures Center, Strategic Laboratory | Caio Vinicius Dias Lopes; Claudia Regina Gonçalves; Claudio Tavares Sacchi; Erica Valessa Ramos Gomes; Karoline Rodrigues Campos; Katia Correa de Oliveira Santos; Leonardo Jose Tadeu de Araujo |
| EPI_ISL_1967229, EPI_ISL_1967233 | UBS 01 VILA BANDEIRANTES BIRIGUI | Instituto Butantan / Mendelics | Antonio Jorge Martins; Bianca Cechetto Carlos. Mendelics: Bibiana Santos; Claudia Renata dos Santos Barros; Cíntia Bittar; David Schlesinger. Hemocentro Ribeirão Preto: Simone Kashima; Debora Botequio Moretti; Elaine Cristina Marqueze; Elaine Vieira dos Santos; Elisangela Chicaroni Mattos; Erika Freitas; Evandra Strazza Rodrigues; Felipe Allan da Silva da Costa; Flavia Aburjaile; Fábio Sossai Possebon; Guilherme Campos; Guilherme Targino Valente; Heidge Fukumasu. USP-Botucatu: Rejane Maria Tommasini Grotto; Helena Lage Ferreira; Instituto Butantan: Dimas Tadeu Covas; Jardelina de Souza Todao Bernardino; Jayme A. Souza-Neto; Jessika Cristina Chagas Lesbon; Jorge A. Petroli Marchesi; José Salvatore Leister Patané; João Paulo Kitajima; João Pessoa Araújo Jr.; Leila Sabrina Ullmann; Loyze Paola Oliveira de Lima; Luiz Aurelio de Campos Crispin. Centro de Genômica Funcional da ESALQ: Luiz Lehmann Coutinho; Luiz Carlos Junior de Alcantara; Lívia Sacchetto; Maisa C. Pereira Parra; Maria Carolina Elias; Marta Giovanetti; Marília Moraes; Maurício Lacerda Nogueira. Prefeitura de Sao Paulo: Melissa Palmieri.; Patricia Akemi Assato; Paula Rahal; Paulo Inacio da Costa; Rafael dos Santos Bezerra; Raquel de Lello Rocha Campos Cassano. NGS Soluções Genômicas: Pilar Drummond Sampaio Corrêa Mariani. FZEA-USP Pirassununga: Mirele Daiana Poleti; Raul Machado Neto; Ricardo Augusto Brassaloti; Ricardo Haddad; Rodrigo Tocantins Calado. FAMERP-SJRP: Cecília Artico Banho; Sandra Coccuzzo Sampaio; Svetoslav Nanev Slavov; Vagner Fonseca; Vincent Louis Viala |
| EPI_ISL_1967234 | UBS 02 JARDIM TOSELAR BIRIGUI | Instituto Butantan / Mendelics | Antonio Jorge Martins; Bianca Cechetto Carlos. Mendelics: Bibiana Santos; Claudia Renata dos Santos Barros; Cíntia Bittar; David Schlesinger. Hemocentro Ribeirão Preto: Simone Kashima; Debora Botequio Moretti; Elaine Cristina Marqueze; Elaine Vieira dos Santos; Elisangela Chicaroni Mattos; Erika Freitas; Evandra Strazza Rodrigues; Felipe Allan da Silva da Costa; Flavia Aburjaile; Fábio Sossai Possebon; Guilherme Campos; Guilherme Targino Valente; Heidge Fukumasu. USP-Botucatu: Rejane Maria Tommasini Grotto; Helena Lage Ferreira; Instituto Butantan: Dimas Tadeu Covas; Jardelina de Souza Todao Bernardino; Jayme A. Souza-Neto; Jessika Cristina Chagas Lesbon; Jorge A. Petroli Marchesi; José Salvatore Leister Patané; João Paulo Kitajima; João Pessoa Araújo Jr.; Leila Sabrina Ullmann; Loyze Paola Oliveira de Lima; Luiz Aurelio de Campos Crispin. Centro de Genômica Funcional da ESALQ: Luiz Lehmann Coutinho; Luiz Carlos Junior de Alcantara; Lívia Sacchetto; Maisa C. Pereira Parra; Maria Carolina Elias; Marta Giovanetti; Marília Moraes; Maurício Lacerda Nogueira. Prefeitura de Sao Paulo: Melissa Palmieri.; Patricia Akemi Assato; Paula Rahal; Paulo Inacio da Costa; Rafael dos Santos Bezerra; Raquel de Lello Rocha Campos Cassano. NGS Soluções Genômicas: Pilar Drummond Sampaio Corrêa Mariani. FZEA-USP Pirassununga: Mirele Daiana Poleti; Raul Machado Neto; Ricardo Augusto Brassaloti; Ricardo Haddad; Rodrigo Tocantins Calado. FAMERP-SJRP: Cecília Artico Banho; Sandra Coccuzzo Sampaio; Svetoslav Nanev Slavov; Vagner Fonseca; Vincent Louis Viala |
| EPI_ISL_1967231 | UBS 04 JARDIM COSTA RICA BIRIGUI | Instituto Butantan / Mendelics | Antonio Jorge Martins; Bianca Cechetto Carlos. Mendelics: Bibiana Santos; Claudia Renata dos Santos Barros; Cíntia Bittar; David Schlesinger. Hemocentro Ribeirão Preto: Simone Kashima; Debora Botequio Moretti; Elaine Cristina Marqueze; Elaine Vieira dos Santos; Elisangela Chicaroni Mattos; Erika Freitas; Evandra Strazza Rodrigues; Felipe Allan da Silva da Costa; Flavia Aburjaile; Fábio Sossai Possebon; Guilherme Campos; Guilherme Targino Valente; Heidge Fukumasu. USP-Botucatu: Rejane Maria Tommasini Grotto; Helena Lage Ferreira; Instituto Butantan: Dimas Tadeu Covas; Jardelina de Souza Todao Bernardino; Jayme A. Souza-Neto; Jessika Cristina Chagas Lesbon; Jorge A. Petroli Marchesi; José Salvatore Leister Patané; João Paulo Kitajima; João Pessoa Araújo Jr.; Leila Sabrina Ullmann; Loyze Paola Oliveira de Lima; Luiz Aurelio de Campos Crispin. Centro de Genômica Funcional da ESALQ: Luiz Lehmann Coutinho; Luiz Carlos Junior de Alcantara; Lívia Sacchetto; Maisa C. Pereira Parra; Maria Carolina Elias; Marta Giovanetti; Marília Moraes; Maurício Lacerda Nogueira. Prefeitura de Sao Paulo: Melissa Palmieri.; Patricia Akemi Assato; Paula Rahal; Paulo Inacio da Costa; Rafael dos Santos Bezerra; Raquel de Lello Rocha Campos Cassano. NGS Soluções Genômicas: Pilar Drummond Sampaio Corrêa Mariani. FZEA-USP Pirassununga: Mirele Daiana Poleti; Raul Machado Neto; Ricardo Augusto Brassaloti; Ricardo Haddad; Rodrigo Tocantins Calado. FAMERP-SJRP: Cecília Artico Banho; Sandra Coccuzzo Sampaio; Svetoslav Nanev Slavov; Vagner Fonseca; Vincent Louis Viala |
| EPI_ISL_1967228, EPI_ISL_1967232 | UBS 09 JOAO CREVELARO BIRIGUI | Instituto Butantan / Mendelics | Antonio Jorge Martins; Bianca Cechetto Carlos. Mendelics: Bibiana Santos; Claudia Renata dos Santos Barros; Cíntia Bittar; David Schlesinger. Hemocentro Ribeirão Preto: Simone Kashima; Debora Botequio Moretti; Elaine Cristina Marqueze; Elaine Vieira dos Santos; Elisangela Chicaroni Mattos; Erika Freitas; Evandra Strazza Rodrigues; Felipe Allan da Silva da Costa; Flavia Aburjaile; Fábio Sossai Possebon; Guilherme Campos; Guilherme Targino Valente; Heidge Fukumasu. USP-Botucatu: Rejane Maria Tommasini Grotto; Helena Lage Ferreira; Instituto Butantan: Dimas Tadeu Covas; Jardelina de Souza Todao Bernardino; Jayme A. Souza-Neto; Jessika Cristina Chagas Lesbon; Jorge A. Petroli Marchesi; José Salvatore Leister Patané; João Paulo Kitajima; João Pessoa Araújo Jr.; Leila Sabrina Ullmann; Loyze Paola Oliveira de Lima; Luiz Aurelio de Campos Crispin. Centro de Genômica Funcional da ESALQ: Luiz Lehmann Coutinho; Luiz Carlos Junior de Alcantara; Lívia Sacchetto; Maisa C. Pereira Parra; Maria Carolina Elias; Marta Giovanetti; Marília Moraes; Maurício Lacerda Nogueira. Prefeitura de Sao Paulo: Melissa Palmieri.; Patricia Akemi Assato; Paula Rahal; Paulo Inacio da Costa; Rafael dos Santos Bezerra; Raquel de Lello Rocha Campos Cassano. NGS Soluções Genômicas: Pilar Drummond Sampaio Corrêa Mariani. FZEA-USP Pirassununga: Mirele Daiana Poleti; Raul Machado Neto; Ricardo Augusto Brassaloti; Ricardo Haddad; Rodrigo Tocantins Calado. FAMERP-SJRP: Cecília Artico Banho; Sandra Coccuzzo Sampaio; Svetoslav Nanev Slavov; Vagner Fonseca; Vincent Louis Viala |
| EPI_ISL_1967230 | UBS 10 MANOEL SANCHES GRANEIRO COLINAS BIRIGUI | Instituto Butantan / Mendelics | Antonio Jorge Martins; Bianca Cechetto Carlos. Mendelics: Bibiana Santos; Claudia Renata dos Santos Barros; Cíntia Bittar; David Schlesinger. Hemocentro Ribeirão Preto: Simone Kashima; Debora Botequio Moretti; Elaine Cristina Marqueze; Elaine Vieira dos Santos; Elisangela Chicaroni Mattos; Erika Freitas; Evandra Strazza Rodrigues; Felipe Allan da Silva da Costa; Flavia Aburjaile; Fábio Sossai Possebon; Guilherme Campos; Guilherme Targino Valente; Heidge Fukumasu. USP-Botucatu: Rejane Maria Tommasini Grotto; Helena Lage Ferreira; Instituto Butantan: Dimas Tadeu Covas; Jardelina de Souza Todao Bernardino; Jayme A. Souza-Neto; Jessika Cristina Chagas Lesbon; Jorge A. Petroli Marchesi; José Salvatore Leister Patané; João Paulo Kitajima; João Pessoa Araújo Jr.; Leila Sabrina Ullmann; Loyze Paola Oliveira de Lima; Luiz Aurelio de Campos Crispin. Centro de Genômica Funcional da ESALQ: Luiz Lehmann Coutinho; Luiz Carlos Junior de Alcantara; Lívia Sacchetto; Maisa C. Pereira Parra; Maria Carolina Elias; Marta Giovanetti; Marília Moraes; Maurício Lacerda Nogueira. Prefeitura de Sao Paulo: Melissa Palmieri.; Patricia Akemi Assato; Paula Rahal; Paulo Inacio da Costa; Rafael dos Santos Bezerra; Raquel de Lello Rocha Campos Cassano. NGS Soluções Genômicas: Pilar Drummond Sampaio Corrêa Mariani. FZEA-USP Pirassununga: Mirele Daiana Poleti; Raul Machado Neto; Ricardo Augusto Brassaloti; Ricardo Haddad; Rodrigo Tocantins Calado. FAMERP-SJRP: Cecília Artico Banho; Sandra Coccuzzo Sampaio; Svetoslav Nanev Slavov; Vagner Fonseca; Vincent Louis Viala |
| EPI_ISL_1967235 | UBS 11 DR SERGIO AUGUSTO CLARK XAVIER SOARES PORTAL II | Instituto Butantan / Mendelics | Antonio Jorge Martins; Bianca Cechetto Carlos. Mendelics: Bibiana Santos; Claudia Renata dos Santos Barros; Cíntia Bittar; David Schlesinger. Hemocentro Ribeirão Preto: Simone Kashima; Debora Botequio Moretti; Elaine Cristina Marqueze; Elaine Vieira dos Santos; Elisangela Chicaroni Mattos; Erika Freitas; Evandra Strazza Rodrigues; Felipe Allan da Silva da Costa; Flavia Aburjaile; Fábio Sossai Possebon; Guilherme Campos; Guilherme Targino Valente; Heidge Fukumasu. USP-Botucatu: Rejane Maria Tommasini Grotto; Helena Lage Ferreira; Instituto Butantan: Dimas Tadeu Covas; Jardelina de Souza Todao Bernardino; Jayme A. Souza-Neto; Jessika Cristina Chagas Lesbon; Jorge A. Petroli Marchesi; José Salvatore Leister Patané; João Paulo Kitajima; João Pessoa Araújo Jr.; Leila Sabrina Ullmann; Loyze Paola Oliveira de Lima; Luiz Aurelio de Campos Crispin. Centro de Genômica Funcional da ESALQ: Luiz Lehmann Coutinho; Luiz Carlos Junior de Alcantara; Lívia Sacchetto; Maisa C. Pereira Parra; Maria Carolina Elias; Marta Giovanetti; Marília Moraes; Maurício Lacerda Nogueira. Prefeitura de Sao Paulo: Melissa Palmieri.; Patricia Akemi Assato; Paula Rahal; Paulo Inacio da Costa; Rafael dos Santos Bezerra; Raquel de Lello Rocha Campos Cassano. NGS Soluções Genômicas: Pilar Drummond Sampaio Corrêa Mariani. FZEA-USP Pirassununga: Mirele Daiana Poleti; Raul Machado Neto; Ricardo Augusto Brassaloti; Ricardo Haddad; Rodrigo Tocantins Calado. FAMERP-SJRP: Cecília Artico Banho; Sandra Coccuzzo Sampaio; Svetoslav Nanev Slavov; Vagner Fonseca; Vincent Louis Viala |
| EPI_ISL_1966511 | UBS 2 DE SUZANAPOLIS | Instituto Butantan / Mendelics | Antonio Jorge Martins; Bianca Cechetto Carlos. Mendelics: Bibiana Santos; Claudia Renata dos Santos Barros; Cíntia Bittar; David Schlesinger. Hemocentro Ribeirão Preto: Simone Kashima; Debora Botequio Moretti; Elaine Cristina Marqueze; Elaine Vieira dos Santos; Elisangela Chicaroni Mattos; Erika Freitas; Evandra Strazza Rodrigues; Felipe Allan da Silva da Costa; Flavia Aburjaile; Fábio Sossai Possebon; Guilherme Campos; Guilherme Targino Valente; Heidge Fukumasu. USP-Botucatu: Rejane Maria Tommasini Grotto; Helena Lage Ferreira; Instituto Butantan: Dimas Tadeu Covas; Jardelina de Souza Todao Bernardino; Jayme A. Souza-Neto; Jessika Cristina Chagas Lesbon; Jorge A. Petroli Marchesi; José Salvatore Leister Patané; João Paulo Kitajima; João Pessoa Araújo Jr.; Leila Sabrina Ullmann; Loyze Paola Oliveira de Lima; Luiz Aurelio de Campos Crispin. Centro de Genômica Funcional da ESALQ: Luiz Lehmann Coutinho; Luiz Carlos Junior de Alcantara; Lívia Sacchetto; Maisa C. Pereira Parra; Maria Carolina Elias; Marta Giovanetti; Marília Moraes; Maurício Lacerda Nogueira. Prefeitura de Sao Paulo: Melissa Palmieri.; Patricia Akemi Assato; Paula Rahal; Paulo Inacio da Costa; Rafael dos Santos Bezerra; Raquel de Lello Rocha Campos Cassano. NGS Soluções Genômicas: Pilar Drummond Sampaio Corrêa Mariani. FZEA-USP Pirassununga: Mirele Daiana Poleti; Raul Machado Neto; Ricardo Augusto Brassaloti; Ricardo Haddad; Rodrigo Tocantins Calado. FAMERP-SJRP: Cecília Artico Banho; Sandra Coccuzzo Sampaio; Svetoslav Nanev Slavov; Vagner Fonseca; Vincent Louis Viala |
| EPI_ISL_1795287, EPI_ISL_1795289 | UBS ALCIMINIO DE ASSIS LOURENCO BADY BASSITT | Instituto Butantan / ESALQ-Piracicaba | Antonio Jorge Martins; Bianca Cechetto Carlos. Mendelics: Bibiana Santos; Claudia Renata dos Santos Barros; David Schlesinger. Hemocentro Ribeirão Preto: Simone Kashima; Debora Botequio Moretti. Centro de Genômica Funcional da ESALQ: Luiz Lehmann Coutinho; Dimas Tadeu Covas; Elaine Cristina Marqueze; Elaine Vieira dos Santos; Elisangela Chicaroni Mattos; Erika Freitas; Evandra Strazza Rodrigues; Felipe Allan da Silva da Costa; Flavia Aburjaile; Guilherme Targino Valente; Heidge Fukumasu. USP-Botucatu: Rejane Maria Tommasini Grotto; Instituto Butantan: Alexander Roberto Precioso; Jayme A. Souza-Neto; Jessika Cristina Chagas Lesbon; José Salvatore Leister Patané; João Paulo Kitajima; Luiz Carlos Junior de Alcantara; Maria Carolina Elias; Marta Giovanetti; Patricia Akemi Assato; Rafael dos Santos Bezerra; Raquel de Lello Rocha Campos Cassano. NGS Soluções Genômicas: Pilar Drummond Sampaio Corrêa Mariani. FZEA-USP Pirassununga: Mirele Daiana Poleti; Raul Machado Neto; Ricardo Augusto Brassaloti; Ricardo Haddad; Rodrigo Tocantins Calado.; Sandra Coccuzzo Sampaio; Svetoslav Nanev Slavov; Vagner Fonseca; Vincent Louis Viala |
| EPI_ISL_1966870, EPI_ISL_1966871, EPI_ISL_1966872, EPI_ISL_1966873, EPI_ISL_1966874, EPI_ISL_1966875, EPI_ISL_1966877, EPI_ISL_1966878, EPI_ISL_1966879, EPI_ISL_1966880, EPI_ISL_1966881, EPI_ISL_1966882, EPI_ISL_1966883, EPI_ISL_1966884, EPI_ISL_1966885, EPI_ISL_1966886, EPI_ISL_1966887, EPI_ISL_1966888, EPI_ISL_1966890 | | | |
| see above | UBS ATALAIA DR MARIO AGUIAR FILHO | Instituto Butantan / ESALQ-USP (Piracicaba) | Antonio Jorge Martins; Bianca Cechetto Carlos. Mendelics: Bibiana Santos; Claudia Renata dos Santos Barros; Cíntia Bittar; David Schlesinger. Hemocentro Ribeirão Preto: Simone Kashima; Debora Botequio Moretti; Elaine Cristina Marqueze; Elaine Vieira dos Santos; Elisangela Chicaroni Mattos; Erika Freitas; Evandra Strazza Rodrigues; Felipe Allan da Silva da Costa; Flavia Aburjaile; Fábio Sossai Possebon; Guilherme Campos; Guilherme Targino Valente; Heidge Fukumasu. USP-Botucatu: Rejane Maria Tommasini Grotto; Helena Lage Ferreira; Instituto Butantan: Dimas Tadeu Covas; Jardelina de Souza Todao Bernardino; Jayme A. Souza-Neto; Jessika Cristina Chagas Lesbon; Jorge A. Petroli Marchesi; José Salvatore Leister Patané; João Paulo Kitajima; João Pessoa Araújo Jr.; Leila Sabrina Ullmann; Loyze Paola Oliveira de Lima; Luiz Aurelio de Campos Crispin. Centro de Genômica Funcional da ESALQ: Luiz Lehmann Coutinho; Luiz Carlos Junior de Alcantara; Lívia Sacchetto; Maisa C. Pereira Parra; Maria Carolina Elias; Marta Giovanetti; Marília Moraes; Maurício Lacerda Nogueira. Prefeitura de Sao Paulo: Melissa Palmieri.; Patricia Akemi Assato; Paula Rahal; Paulo Inacio da Costa; Rafael dos Santos Bezerra; Raquel de Lello Rocha Campos Cassano. NGS Soluções Genômicas: Pilar Drummond Sampaio Corrêa Mariani. FZEA-USP Pirassununga: Mirele Daiana Poleti; Raul Machado Neto; Ricardo Augusto Brassaloti; Ricardo Haddad; Rodrigo Tocantins Calado. FAMERP-SJRP: Cecília Artico Banho; Sandra Coccuzzo Sampaio; Svetoslav Nanev Slavov; Vagner Fonseca; Vincent Louis Viala |
| EPI_ISL_1795405 | UBS CAIC OURINHOS | Instituto Butantan / ESALQ-Piracicaba | Antonio Jorge Martins; Bianca Cechetto Carlos. Mendelics: Bibiana Santos; Claudia Renata dos Santos Barros; David Schlesinger. Hemocentro Ribeirão Preto: Simone Kashima; Debora Botequio Moretti. Centro de Genômica Funcional da ESALQ: Luiz Lehmann Coutinho; Dimas Tadeu Covas; Elaine Cristina Marqueze; Elaine Vieira dos Santos; Elisangela Chicaroni Mattos; Erika Freitas; Evandra Strazza Rodrigues; Felipe Allan da Silva da Costa; Flavia Aburjaile; Guilherme Targino Valente; Heidge Fukumasu. USP-Botucatu: Rejane Maria Tommasini Grotto; Instituto Butantan: Alexander Roberto Precioso; Jayme A. Souza-Neto; Jessika Cristina Chagas Lesbon; José Salvatore Leister Patané; João Paulo Kitajima; Luiz Carlos Junior de Alcantara; Maria Carolina Elias; Marta Giovanetti; Patricia Akemi Assato; Rafael dos Santos Bezerra; Raquel de Lello Rocha Campos Cassano. NGS Soluções Genômicas: Pilar Drummond Sampaio Corrêa Mariani. FZEA-USP Pirassununga: Mirele Daiana Poleti; Raul Machado Neto; Ricardo Augusto Brassaloti; Ricardo Haddad; Rodrigo Tocantins Calado.; Sandra Coccuzzo Sampaio; Svetoslav Nanev Slavov; Vagner Fonseca; Vincent Louis Viala |
| EPI_ISL_1967288 | UBS CAVADAS | Instituto Butantan / Mendelics | Antonio Jorge Martins; Bianca Cechetto Carlos. Mendelics: Bibiana Santos; Claudia Renata dos Santos Barros; Cíntia Bittar; David Schlesinger. Hemocentro Ribeirão Preto: Simone Kashima; Debora Botequio Moretti; Elaine Cristina Marqueze; Elaine Vieira dos Santos; Elisangela Chicaroni Mattos; Erika Freitas; Evandra Strazza Rodrigues; Felipe Allan da Silva da Costa; Flavia Aburjaile; Fábio Sossai Possebon; Guilherme Campos; Guilherme Targino Valente; Heidge Fukumasu. USP-Botucatu: Rejane Maria Tommasini Grotto; Helena Lage Ferreira; Instituto Butantan: Dimas Tadeu Covas; Jardelina de Souza Todao Bernardino; Jayme A. Souza-Neto; Jessika Cristina Chagas Lesbon; Jorge A. Petroli Marchesi; José Salvatore Leister Patané; João Paulo Kitajima; João Pessoa Araújo Jr.; Leila Sabrina Ullmann; Loyze Paola Oliveira de Lima; Luiz Aurelio de Campos Crispin. Centro de Genômica Funcional da ESALQ: Luiz Lehmann Coutinho; Luiz Carlos Junior de Alcantara; Lívia Sacchetto; Maisa C. Pereira Parra; Maria Carolina Elias; Marta Giovanetti; Marília Moraes; Maurício Lacerda Nogueira. Prefeitura de Sao Paulo: Melissa Palmieri.; Patricia Akemi Assato; Paula Rahal; Paulo Inacio da Costa; Rafael dos Santos Bezerra; Raquel de Lello Rocha Campos Cassano. NGS Soluções Genômicas: Pilar Drummond Sampaio Corrêa Mariani. FZEA-USP Pirassununga: Mirele Daiana Poleti; Raul Machado Neto; Ricardo Augusto Brassaloti; Ricardo Haddad; Rodrigo Tocantins Calado. FAMERP-SJRP: Cecília Artico Banho; Sandra Coccuzzo Sampaio; Svetoslav Nanev Slavov; Vagner Fonseca; Vincent Louis Viala |
| EPI_ISL_2170977 | UBS CIRCA FERREIRA SOARES MATTOS | Instituto Butantan / Mendelics | Antonio Jorge Martins; Bianca Cechetto Carlos. Mendelics: Bibiana Santos; Claudia Renata dos Santos Barros; Cíntia Bittar; David Schlesinger. Hemocentro Ribeirão Preto: Simone Kashima; Debora Botequio Moretti; Elaine Cristina Marqueze; Elaine Vieira dos Santos; Elisangela Chicaroni Mattos; Erika Freitas; Evandra Strazza Rodrigues; Felipe Allan da Silva da Costa; Flavia Aburjaile; Fábio Sossai Possebon; Guilherme Campos; Guilherme Targino Valente; Heidge Fukumasu. USP-Botucatu: Rejane Maria Tommasini Grotto; Helena Lage Ferreira; Instituto Butantan: Dimas Tadeu Covas; Jardelina de Souza Todao Bernardino; Jayme A. Souza-Neto; Jessika Cristina Chagas Lesbon; Jorge A. Petroli Marchesi; José Salvatore Leister Patané; João Paulo Kitajima; João Pessoa Araújo Jr.; Leila Sabrina Ullmann; Loyze Paola Oliveira de Lima; Luiz Aurelio de Campos Crispin. Centro de Genômica Funcional da ESALQ: Luiz Lehmann Coutinho; Luiz Carlos Junior de Alcantara; Lívia Sacchetto; Maisa C. Pereira Parra; Maria Carolina Elias; Marta Giovanetti; Marília Moraes; Maurício Lacerda Nogueira. Prefeitura de Sao Paulo: Melissa Palmieri.; Patricia Akemi Assato; Paula Rahal; Paulo Inacio da Costa; Rafael dos Santos Bezerra; Raquel de Lello Rocha Campos Cassano. NGS Soluções Genômicas: Pilar Drummond Sampaio Corrêa Mariani. FZEA-USP Pirassununga: Mirele Daiana Poleti; Raul Machado Neto; Ricardo Augusto Brassaloti; Ricardo Haddad; Rodrigo Tocantins Calado. FAMERP-SJRP: Cecília Artico Banho; Sandra Coccuzzo Sampaio; Svetoslav Nanev Slavov; Vagner Fonseca; Vincent Louis Viala |
| EPI_ISL_1445206, EPI_ISL_1445208, EPI_ISL_1966205, EPI_ISL_2170912 | UBS DARCY ALVES E ROBALINHO | Instituto Butantan / Mendelics | Antonio Jorge Martins; Bianca Cechetto Carlos. Mendelics: Bibiana Santos; Bibiana Santos; Claudia Renata dos Santos Barros; Cíntia Bittar; David Schlesinger; David Schlesinger. Hemocentro Ribeirão Preto: Simone Kashima; Debora Botequio Moretti; Dimas Tadeu Covas; Elaine Cristina Marqueze; Elaine Vieira dos Santos; Elisangela Chicaroni Mattos; Erika Freitas; Evandra Strazza Rodrigues; Felipe Allan da Silva da Costa; Flavia Aburjaile; Fábio Sossai Possebon; Guilherme Campos; Guilherme Targino Valente; Heidge Fukumasu. USP-Botucatu: Rejane Maria Tommasini Grotto; Helena Lage Ferreira; Instituto Butantan: Dimas Tadeu Covas; Jardelina de Souza Todao Bernardino; Jayme A. Souza-Neto; Jessika Cristina Chagas Lesbon; Jorge A. Petroli Marchesi; José Salvatore Leister Patané; João Paulo Kitajima; João Pessoa Araújo Jr.; Leila Sabrina Ullmann; Loyze Paola Oliveira de Lima; Luiz Aurelio de Campos Crispin. Centro de Genômica Funcional da ESALQ: Luiz Lehmann Coutinho; Luiz Carlos Junior de Alcantara; Lívia Sacchetto; Maisa C. Pereira Parra; Maria Carolina Elias; Marta Giovanetti; Marília Moraes; Maurício Lacerda Nogueira. Prefeitura de Sao Paulo: Melissa Palmieri.; Patricia Akemi Assato; Paula Rahal; Paulo Inacio da Costa; Rafael dos Santos Bezerra; Raquel de Lello Rocha Campos Cassano. NGS Soluções Genômicas: Pilar Drummond Sampaio Corrêa Mariani. FZEA-USP Pirassununga: Mirele Daiana Poleti; Raul Machado Neto; Ricardo Augusto Brassaloti; Ricardo Haddad; Rodrigo Tocantins Calado.; Rodrigo Tocantins Calado. FAMERP-SJRP: Cecília Artico Banho; Sandra Coccuzzo Sampaio; Simone Kashima; Svetoslav Nanev Slavov; Vagner Fonseca; Vincent Louis Viala |
| EPI_ISL_1966287, EPI_ISL_1966444 | UBS DE AURIFLAMA | Instituto Butantan / Mendelics | Antonio Jorge Martins; Bianca Cechetto Carlos. Mendelics: Bibiana Santos; Claudia Renata dos Santos Barros; Cíntia Bittar; David Schlesinger. Hemocentro Ribeirão Preto: Simone Kashima; Debora Botequio Moretti; Elaine Cristina Marqueze; Elaine Vieira dos Santos; Elisangela Chicaroni Mattos; Erika Freitas; Evandra Strazza Rodrigues; Felipe Allan da Silva da Costa; Flavia Aburjaile; Fábio Sossai Possebon; Guilherme Campos; Guilherme Targino Valente; Heidge Fukumasu. USP-Botucatu: Rejane Maria Tommasini Grotto; Helena Lage Ferreira; Instituto Butantan: Dimas Tadeu Covas; Jardelina de Souza Todao Bernardino; Jayme A. Souza-Neto; Jessika Cristina Chagas Lesbon; Jorge A. Petroli Marchesi; José Salvatore Leister Patané; João Paulo Kitajima; João Pessoa Araújo Jr.; Leila Sabrina Ullmann; Loyze Paola Oliveira de Lima; Luiz Aurelio de Campos Crispin. Centro de Genômica Funcional da ESALQ: Luiz Lehmann Coutinho; Luiz Carlos Junior de Alcantara; Lívia Sacchetto; Maisa C. Pereira Parra; Maria Carolina Elias; Marta Giovanetti; Marília Moraes; Maurício Lacerda Nogueira. Prefeitura de Sao Paulo: Melissa Palmieri.; Patricia Akemi Assato; Paula Rahal; Paulo Inacio da Costa; Rafael dos Santos Bezerra; Raquel de Lello Rocha Campos Cassano. NGS Soluções Genômicas: Pilar Drummond Sampaio Corrêa Mariani. FZEA-USP Pirassununga: Mirele Daiana Poleti; Raul Machado Neto; Ricardo Augusto Brassaloti; Ricardo Haddad; Rodrigo Tocantins Calado. FAMERP-SJRP: Cecília Artico Banho; Sandra Coccuzzo Sampaio; Svetoslav Nanev Slavov; Vagner Fonseca; Vincent Louis Viala |
| EPI_ISL_1966782, EPI_ISL_1966783 | UBS DE MENDONCA | Instituto Butantan / Mendelics | Antonio Jorge Martins; Bianca Cechetto Carlos. Mendelics: Bibiana Santos; Claudia Renata dos Santos Barros; Cíntia Bittar; David Schlesinger. Hemocentro Ribeirão Preto: Simone Kashima; Debora Botequio Moretti; Elaine Cristina Marqueze; Elaine Vieira dos Santos; Elisangela Chicaroni Mattos; Erika Freitas; Evandra Strazza Rodrigues; Felipe Allan da Silva da Costa; Flavia Aburjaile; Fábio Sossai Possebon; Guilherme Campos; Guilherme Targino Valente; Heidge Fukumasu. USP-Botucatu: Rejane Maria Tommasini Grotto; Helena Lage Ferreira; Instituto Butantan: Dimas Tadeu Covas; Jardelina de Souza Todao Bernardino; Jayme A. Souza-Neto; Jessika Cristina Chagas Lesbon; Jorge A. Petroli Marchesi; José Salvatore Leister Patané; João Paulo Kitajima; João Pessoa Araújo Jr.; Leila Sabrina Ullmann; Loyze Paola Oliveira de Lima; Luiz Aurelio de Campos Crispin. Centro de Genômica Funcional da ESALQ: Luiz Lehmann Coutinho; Luiz Carlos Junior de Alcantara; Lívia Sacchetto; Maisa C. Pereira Parra; Maria Carolina Elias; Marta Giovanetti; Marília Moraes; Maurício Lacerda Nogueira. Prefeitura de Sao Paulo: Melissa Palmieri.; Patricia Akemi Assato; Paula Rahal; Paulo Inacio da Costa; Rafael dos Santos Bezerra; Raquel de Lello Rocha Campos Cassano. NGS Soluções Genômicas: Pilar Drummond Sampaio Corrêa Mariani. FZEA-USP Pirassununga: Mirele Daiana Poleti; Raul Machado Neto; Ricardo Augusto Brassaloti; Ricardo Haddad; Rodrigo Tocantins Calado. FAMERP-SJRP: Cecília Artico Banho; Sandra Coccuzzo Sampaio; Svetoslav Nanev Slavov; Vagner Fonseca; Vincent Louis Viala |
| EPI_ISL_1795415 | UBS DE SANTA SALETE | Instituto Butantan / ESALQ-Piracicaba | Antonio Jorge Martins; Bianca Cechetto Carlos. Mendelics: Bibiana Santos; Claudia Renata dos Santos Barros; David Schlesinger. Hemocentro Ribeirão Preto: Simone Kashima; Debora Botequio Moretti. Centro de Genômica Funcional da ESALQ: Luiz Lehmann Coutinho; Dimas Tadeu Covas; Elaine Cristina Marqueze; Elaine Vieira dos Santos; Elisangela Chicaroni Mattos; Erika Freitas; Evandra Strazza Rodrigues; Felipe Allan da Silva da Costa; Flavia Aburjaile; Guilherme Targino Valente; Heidge Fukumasu. USP-Botucatu: Rejane Maria Tommasini Grotto; Instituto Butantan: Alexander Roberto Precioso; Jayme A. Souza-Neto; Jessika Cristina Chagas Lesbon; José Salvatore Leister Patané; João Paulo Kitajima; Luiz Carlos Junior de Alcantara; Maria Carolina Elias; Marta Giovanetti; Patricia Akemi Assato; Rafael dos Santos Bezerra; Raquel de Lello Rocha Campos Cassano. NGS Soluções Genômicas: Pilar Drummond Sampaio Corrêa Mariani. FZEA-USP Pirassununga: Mirele Daiana Poleti; Raul Machado Neto; Ricardo Augusto Brassaloti; Ricardo Haddad; Rodrigo Tocantins Calado.; Sandra Coccuzzo Sampaio; Svetoslav Nanev Slavov; Vagner Fonseca; Vincent Louis Viala |
| EPI_ISL_1967051 | UBS DE SANTA SALETE | Instituto Butantan / ESALQ-USP (Piracicaba) | Antonio Jorge Martins; Bianca Cechetto Carlos. Mendelics: Bibiana Santos; Claudia Renata dos Santos Barros; Cíntia Bittar; David Schlesinger. Hemocentro Ribeirão Preto: Simone Kashima; Debora Botequio Moretti; Elaine Cristina Marqueze; Elaine Vieira dos Santos; Elisangela Chicaroni Mattos; Erika Freitas; Evandra Strazza Rodrigues; Felipe Allan da Silva da Costa; Flavia Aburjaile; Fábio Sossai Possebon; Guilherme Campos; Guilherme Targino Valente; Heidge Fukumasu. USP-Botucatu: Rejane Maria Tommasini Grotto; Helena Lage Ferreira; Instituto Butantan: Dimas Tadeu Covas; Jardelina de Souza Todao Bernardino; Jayme A. Souza-Neto; Jessika Cristina Chagas Lesbon; Jorge A. Petroli Marchesi; José Salvatore Leister Patané; João Paulo Kitajima; João Pessoa Araújo Jr.; Leila Sabrina Ullmann; Loyze Paola Oliveira de Lima; Luiz Aurelio de Campos Crispin. Centro de Genômica Funcional da ESALQ: Luiz Lehmann Coutinho; Luiz Carlos Junior de Alcantara; Lívia Sacchetto; Maisa C. Pereira Parra; Maria Carolina Elias; Marta Giovanetti; Marília Moraes; Maurício Lacerda Nogueira. Prefeitura de Sao Paulo: Melissa Palmieri.; Patricia Akemi Assato; Paula Rahal; Paulo Inacio da Costa; Rafael dos Santos Bezerra; Raquel de Lello Rocha Campos Cassano. NGS Soluções Genômicas: Pilar Drummond Sampaio Corrêa Mariani. FZEA-USP Pirassununga: Mirele Daiana Poleti; Raul Machado Neto; Ricardo Augusto Brassaloti; Ricardo Haddad; Rodrigo Tocantins Calado. FAMERP-SJRP: Cecília Artico Banho; Sandra Coccuzzo Sampaio; Svetoslav Nanev Slavov; Vagner Fonseca; Vincent Louis Viala |
| EPI_ISL_1966510, EPI_ISL_2170938 | UBS DE SUD MENNUCCI | Instituto Butantan / Mendelics | Antonio Jorge Martins; Bianca Cechetto Carlos. Mendelics: Bibiana Santos; Claudia Renata dos Santos Barros; Cíntia Bittar; David Schlesinger. Hemocentro Ribeirão Preto: Simone Kashima; Debora Botequio Moretti; Elaine Cristina Marqueze; Elaine Vieira dos Santos; Elisangela Chicaroni Mattos; Erika Freitas; Evandra Strazza Rodrigues; Felipe Allan da Silva da Costa; Flavia Aburjaile; Fábio Sossai Possebon; Guilherme Campos; Guilherme Targino Valente; Heidge Fukumasu. USP-Botucatu: Rejane Maria Tommasini Grotto; Helena Lage Ferreira; Instituto Butantan: Dimas Tadeu Covas; Jardelina de Souza Todao Bernardino; Jayme A. Souza-Neto; Jessika Cristina Chagas Lesbon; Jorge A. Petroli Marchesi; José Salvatore Leister Patané; João Paulo Kitajima; João Pessoa Araújo Jr.; Leila Sabrina Ullmann; Loyze Paola Oliveira de Lima; Luiz Aurelio de Campos Crispin. Centro de Genômica Funcional da ESALQ: Luiz Lehmann Coutinho; Luiz Carlos Junior de Alcantara; Lívia Sacchetto; Maisa C. Pereira Parra; Maria Carolina Elias; Marta Giovanetti; Marília Moraes; Maurício Lacerda Nogueira. Prefeitura de Sao Paulo: Melissa Palmieri.; Patricia Akemi Assato; Paula Rahal; Paulo Inacio da Costa; Rafael dos Santos Bezerra; Raquel de Lello Rocha Campos Cassano. NGS Soluções Genômicas: Pilar Drummond Sampaio Corrêa Mariani. FZEA-USP Pirassununga: Mirele Daiana Poleti; Raul Machado Neto; Ricardo Augusto Brassaloti; Ricardo Haddad; Rodrigo Tocantins Calado. FAMERP-SJRP: Cecília Artico Banho; Sandra Coccuzzo Sampaio; Svetoslav Nanev Slavov; Vagner Fonseca; Vincent Louis Viala |
| EPI_ISL_1967159, EPI_ISL_1967160, EPI_ISL_1967161, EPI_ISL_1967162 | UBS DE TABAPUA | Instituto Butantan / ESALQ-USP (Piracicaba) | Antonio Jorge Martins; Bianca Cechetto Carlos. Mendelics: Bibiana Santos; Claudia Renata dos Santos Barros; Cíntia Bittar; David Schlesinger. Hemocentro Ribeirão Preto: Simone Kashima; Debora Botequio Moretti; Elaine Cristina Marqueze; Elaine Vieira dos Santos; Elisangela Chicaroni Mattos; Erika Freitas; Evandra Strazza Rodrigues; Felipe Allan da Silva da Costa; Flavia Aburjaile; Fábio Sossai Possebon; Guilherme Campos; Guilherme Targino Valente; Heidge Fukumasu. USP-Botucatu: Rejane Maria Tommasini Grotto; Helena Lage Ferreira; Instituto Butantan: Dimas Tadeu Covas; Jardelina de Souza Todao Bernardino; Jayme A. Souza-Neto; Jessika Cristina Chagas Lesbon; Jorge A. Petroli Marchesi; José Salvatore Leister Patané; João Paulo Kitajima; João Pessoa Araújo Jr.; Leila Sabrina Ullmann; Loyze Paola Oliveira de Lima; Luiz Aurelio de Campos Crispin. Centro de Genômica Funcional da ESALQ: Luiz Lehmann Coutinho; Luiz Carlos Junior de Alcantara; Lívia Sacchetto; Maisa C. Pereira Parra; Maria Carolina Elias; Marta Giovanetti; Marília Moraes; Maurício Lacerda Nogueira. Prefeitura de Sao Paulo: Melissa Palmieri.; Patricia Akemi Assato; Paula Rahal; Paulo Inacio da Costa; Rafael dos Santos Bezerra; Raquel de Lello Rocha Campos Cassano. NGS Soluções Genômicas: Pilar Drummond Sampaio Corrêa Mariani. FZEA-USP Pirassununga: Mirele Daiana Poleti; Raul Machado Neto; Ricardo Augusto Brassaloti; Ricardo Haddad; Rodrigo Tocantins Calado. FAMERP-SJRP: Cecília Artico Banho; Sandra Coccuzzo Sampaio; Svetoslav Nanev Slavov; Vagner Fonseca; Vincent Louis Viala |
| EPI_ISL_1966543, EPI_ISL_1966544 | UBS DR ABELARDO PINHEIRO GUIMARAES | Instituto Butantan / FZEA-USP (Pirassununga) | Antonio Jorge Martins; Bianca Cechetto Carlos. Mendelics: Bibiana Santos; Claudia Renata dos Santos Barros; Cíntia Bittar; David Schlesinger. Hemocentro Ribeirão Preto: Simone Kashima; Debora Botequio Moretti; Elaine Cristina Marqueze; Elaine Vieira dos Santos; Elisangela Chicaroni Mattos; Erika Freitas; Evandra Strazza Rodrigues; Felipe Allan da Silva da Costa; Flavia Aburjaile; Fábio Sossai Possebon; Guilherme Campos; Guilherme Targino Valente; Heidge Fukumasu. USP-Botucatu: Rejane Maria Tommasini Grotto; Helena Lage Ferreira; Instituto Butantan: Dimas Tadeu Covas; Jardelina de Souza Todao Bernardino; Jayme A. Souza-Neto; Jessika Cristina Chagas Lesbon; Jorge A. Petroli Marchesi; José Salvatore Leister Patané; João Paulo Kitajima; João Pessoa Araújo Jr.; Leila Sabrina Ullmann; Loyze Paola Oliveira de Lima; Luiz Aurelio de Campos Crispin. Centro de Genômica Funcional da ESALQ: Luiz Lehmann Coutinho; Luiz Carlos Junior de Alcantara; Lívia Sacchetto; Maisa C. Pereira Parra; Maria |

| | | | |
|---|---|---|---|
| | SCRPARDO | | Carolina Elias; Marta Giovanetti; Marília Moraes; Maurício Lacerda Nogueira. Prefeitura de Sao Paulo: Melissa Palmieri.; Patricia Akemi Assato; Paula Rahal; Paulo Inacio da Costa; Rafael dos Santos Bezerra; Raquel de Lello Rocha Campos Cassano. NGS Soluções Genômicas: Pilar Drummond Sampaio Corrêa Mariani. FZEA-USP Pirassununga: Mirele Daiana Poletti; Raul Machado Neto; Ricardo Augusto Brassaloti; Ricardo Haddad; Rodrigo Tocantins Calado. FAMERP-SJRP: Cecília Artico Banho; Sandra Coccuzzo Sampaio; Svetoslav Nanev Slavov; Vagner Fonseca; Vincent Louis Viala |
| EPI_ISL_1966809, EPI_ISL_1966811 | UBS DR ABELARDO PINHEIRO GUIMARAES SCRPARDO | Instituto Butantan / Mendelics | Antonio Jorge Martins; Bianca Cechetto Carlos. Mendelics: Bibiana Santos; Claudia Renata dos Santos Barros; Cíntia Bittar; David Schlesinger. Hemocentro Ribeirão Preto: Simone Kashima; Debora Botequio Moretti; Elaine Cristina Marqueze; Elaine Vieira dos Santos; Elisangela Chicaroni Mattos; Erika Freitas; Evandra Strazza Rodrigues; Felipe Allan da Silva da Costa; Flavia Aburjaile; Fábio Sossai Possebon; Guilherme Campos; Guilherme Targino Valente; Heidge Fukumasu. USP-Botucatu: Rejane Maria Tommasini Grotto; Helena Lage Ferreira; Instituto Butantan: Dimas Tadeu Covas; Jardelina de Souza Todao Bernardino; Jayme A. Souza-Neto; Jessika Cristina Chagas Lesbon; Jorge A. Petroli Marchesi; José Salvatore Leister Patané; João Paulo Kitajima; João Pessoa Araújo Jr.; Leila Sabrina Ullmann; Loyze Paola Oliveira de Lima; Luiz Aurelio de Campos Crispin. Centro de Genômica Funcional da ESALQ: Luiz Lehmann Coutinho; Luiz Carlos Junior de Alcantara; Lívia Sacchetto; Maisa C. Pereira Parra; Maria Carolina Elias; Marta Giovanetti; Marília Moraes; Maurício Lacerda Nogueira. Prefeitura de Sao Paulo: Melissa Palmieri.; Patricia Akemi Assato; Paula Rahal; Paulo Inacio da Costa; Rafael dos Santos Bezerra; Raquel de Lello Rocha Campos Cassano. NGS Soluções Genômicas: Pilar Drummond Sampaio Corrêa Mariani. FZEA-USP Pirassununga: Mirele Daiana Poletti; Raul Machado Neto; Ricardo Augusto Brassaloti; Ricardo Haddad; Rodrigo Tocantins Calado. FAMERP-SJRP: Cecília Artico Banho; Sandra Coccuzzo Sampaio; Svetoslav Nanev Slavov; Vagner Fonseca; Vincent Louis Viala |
| EPI_ISL_1966278, EPI_ISL_1966279, EPI_ISL_1966280, EPI_ISL_1966281, EPI_ISL_1966282, EPI_ISL_1966283, EPI_ISL_1966284, EPI_ISL_1966285, EPI_ISL_1966286, EPI_ISL_1966440, EPI_ISL_1966441, EPI_ISL_1966442, EPI_ISL_1966443, EPI_ISL_1967204, EPI_ISL_1967205, EPI_ISL_1967206, EPI_ISL_1967207, EPI_ISL_1967208, EPI_ISL_1967209, EPI_ISL_1967210, EPI_ISL_1967211 | | | |
| see above | UBS DR ALFREDO DANTAS DE SOUZA UMUARAMA | Instituto Butantan / Mendelics | Antonio Jorge Martins; Bianca Cechetto Carlos. Mendelics: Bibiana Santos; Claudia Renata dos Santos Barros; Cíntia Bittar; David Schlesinger. Hemocentro Ribeirão Preto: Simone Kashima; Debora Botequio Moretti; Elaine Cristina Marqueze; Elaine Vieira dos Santos; Elisangela Chicaroni Mattos; Erika Freitas; Evandra Strazza Rodrigues; Felipe Allan da Silva da Costa; Flavia Aburjaile; Fábio Sossai Possebon; Guilherme Campos; Guilherme Targino Valente; Heidge Fukumasu. USP-Botucatu: Rejane Maria Tommasini Grotto; Helena Lage Ferreira; Instituto Butantan: Dimas Tadeu Covas; Jardelina de Souza Todao Bernardino; Jayme A. Souza-Neto; Jessika Cristina Chagas Lesbon; Jorge A. Petroli Marchesi; José Salvatore Leister Patané; João Paulo Kitajima; João Pessoa Araújo Jr.; Leila Sabrina Ullmann; Loyze Paola Oliveira de Lima; Luiz Aurelio de Campos Crispin. Centro de Genômica Funcional da ESALQ: Luiz Lehmann Coutinho; Luiz Carlos Junior de Alcantara; Lívia Sacchetto; Maisa C. Pereira Parra; Maria Carolina Elias; Marta Giovanetti; Marília Moraes; Maurício Lacerda Nogueira. Prefeitura de Sao Paulo: Melissa Palmieri.; Patricia Akemi Assato; Paula Rahal; Paulo Inacio da Costa; Rafael dos Santos Bezerra; Raquel de Lello Rocha Campos Cassano. NGS Soluções Genômicas: Pilar Drummond Sampaio Corrêa Mariani. FZEA-USP Pirassununga: Mirele Daiana Poletti; Raul Machado Neto; Ricardo Augusto Brassaloti; Ricardo Haddad; Rodrigo Tocantins Calado. FAMERP-SJRP: Cecília Artico Banho; Sandra Coccuzzo Sampaio; Svetoslav Nanev Slavov; Vagner Fonseca; Vincent Louis Viala |
| EPI_ISL_1966289, EPI_ISL_1966445 | UBS DR ALLY ALAHMAR DE BARRETOS | Instituto Butantan / Mendelics | Antonio Jorge Martins; Bianca Cechetto Carlos. Mendelics: Bibiana Santos; Claudia Renata dos Santos Barros; Cíntia Bittar; David Schlesinger. Hemocentro Ribeirão Preto: Simone Kashima; Debora Botequio Moretti; Elaine Cristina Marqueze; Elaine Vieira dos Santos; Elisangela Chicaroni Mattos; Erika Freitas; Evandra Strazza Rodrigues; Felipe Allan da Silva da Costa; Flavia Aburjaile; Fábio Sossai Possebon; Guilherme Campos; Guilherme Targino Valente; Heidge Fukumasu. USP-Botucatu: Rejane Maria Tommasini Grotto; Helena Lage Ferreira; Instituto Butantan: Dimas Tadeu Covas; Jardelina de Souza Todao Bernardino; Jayme A. Souza-Neto; Jessika Cristina Chagas Lesbon; Jorge A. Petroli Marchesi; José Salvatore Leister Patané; João Paulo Kitajima; João Pessoa Araújo Jr.; Leila Sabrina Ullmann; Loyze Paola Oliveira de Lima; Luiz Aurelio de Campos Crispin. Centro de Genômica Funcional da ESALQ: Luiz Lehmann Coutinho; Luiz Carlos Junior de Alcantara; Lívia Sacchetto; Maisa C. Pereira Parra; Maria Carolina Elias; Marta Giovanetti; Marília Moraes; Maurício Lacerda Nogueira. Prefeitura de Sao Paulo: Melissa Palmieri.; Patricia Akemi Assato; Paula Rahal; Paulo Inacio da Costa; Rafael dos Santos Bezerra; Raquel de Lello Rocha Campos Cassano. NGS Soluções Genômicas: Pilar Drummond Sampaio Corrêa Mariani. FZEA-USP Pirassununga: Mirele Daiana Poletti; Raul Machado Neto; Ricardo Augusto Brassaloti; Ricardo Haddad; Rodrigo Tocantins Calado. FAMERP-SJRP: Cecília Artico Banho; Sandra Coccuzzo Sampaio; Svetoslav Nanev Slavov; Vagner Fonseca; Vincent Louis Viala |
| EPI_ISL_1966990, EPI_ISL_1966991, EPI_ISL_1966994 | UBS DR HELIO MIGLIARI | Instituto Butantan / ESALQ-USP (Piracicaba) | Antonio Jorge Martins; Bianca Cechetto Carlos. Mendelics: Bibiana Santos; Claudia Renata dos Santos Barros; Cíntia Bittar; David Schlesinger. Hemocentro Ribeirão Preto: Simone Kashima; Debora Botequio Moretti; Elaine Cristina Marqueze; Elaine Vieira dos Santos; Elisangela Chicaroni Mattos; Erika Freitas; Evandra Strazza Rodrigues; Felipe Allan da Silva da Costa; Flavia Aburjaile; Fábio Sossai Possebon; Guilherme Campos; Guilherme Targino Valente; Heidge Fukumasu. USP-Botucatu: Rejane Maria Tommasini Grotto; Helena Lage Ferreira; Instituto Butantan: Dimas Tadeu Covas; Jardelina de Souza Todao Bernardino; Jayme A. Souza-Neto; Jessika Cristina Chagas Lesbon; Jorge A. Petroli Marchesi; José Salvatore Leister Patané; João Paulo Kitajima; João Pessoa Araújo Jr.; Leila Sabrina Ullmann; Loyze Paola Oliveira de Lima; Luiz Aurelio de Campos Crispin. Centro de Genômica Funcional da ESALQ: Luiz Lehmann Coutinho; Luiz Carlos Junior de Alcantara; Lívia Sacchetto; Maisa C. Pereira Parra; Maria Carolina Elias; Marta Giovanetti; Marília Moraes; Maurício Lacerda Nogueira. Prefeitura de Sao Paulo: Melissa Palmieri.; Patricia Akemi Assato; Paula Rahal; Paulo Inacio da Costa; Rafael dos Santos Bezerra; Raquel de Lello Rocha Campos Cassano. NGS Soluções Genômicas: Pilar Drummond Sampaio Corrêa Mariani. FZEA-USP Pirassununga: Mirele Daiana Poletti; Raul Machado Neto; Ricardo Augusto Brassaloti; Ricardo Haddad; Rodrigo Tocantins Calado. FAMERP-SJRP: Cecília Artico Banho; Sandra Coccuzzo Sampaio; Svetoslav Nanev Slavov; Vagner Fonseca; Vincent Louis Viala |
| EPI_ISL_1966534 | UBS DR HELIO MIGLIARI | Instituto Butantan / FZEA-USP (Pirassununga) | Antonio Jorge Martins; Bianca Cechetto Carlos. Mendelics: Bibiana Santos; Claudia Renata dos Santos Barros; Cíntia Bittar; David Schlesinger. Hemocentro Ribeirão Preto: Simone Kashima; Debora Botequio Moretti; Elaine Cristina Marqueze; Elaine Vieira dos Santos; Elisangela Chicaroni Mattos; Erika Freitas; Evandra Strazza Rodrigues; Felipe Allan da Silva da Costa; Flavia Aburjaile; Fábio Sossai Possebon; Guilherme Campos; Guilherme Targino Valente; Heidge Fukumasu. USP-Botucatu: Rejane Maria Tommasini Grotto; Helena Lage Ferreira; Instituto Butantan: Dimas Tadeu Covas; Jardelina de Souza Todao Bernardino; Jayme A. Souza-Neto; Jessika Cristina Chagas Lesbon; Jorge A. Petroli Marchesi; José Salvatore Leister Patané; João Paulo Kitajima; João Pessoa Araújo Jr.; Leila Sabrina Ullmann; Loyze Paola Oliveira de Lima; Luiz Aurelio de Campos Crispin. Centro de Genômica Funcional da ESALQ: Luiz Lehmann Coutinho; Luiz Carlos Junior de Alcantara; Lívia Sacchetto; Maisa C. Pereira Parra; Maria Carolina Elias; Marta Giovanetti; Marília Moraes; Maurício Lacerda Nogueira. Prefeitura de Sao Paulo: Melissa Palmieri.; Patricia Akemi Assato; Paula Rahal; Paulo Inacio da Costa; Rafael dos Santos Bezerra; Raquel de Lello Rocha Campos Cassano. NGS Soluções Genômicas: Pilar Drummond Sampaio Corrêa Mariani. FZEA-USP Pirassununga: Mirele Daiana Poletti; Raul Machado Neto; Ricardo Augusto Brassaloti; Ricardo Haddad; Rodrigo Tocantins Calado. FAMERP-SJRP: Cecília Artico Banho; Sandra Coccuzzo Sampaio; Svetoslav Nanev Slavov; Vagner Fonseca; Vincent Louis Viala |
| EPI_ISL_1966252, EPI_ISL_1966253, EPI_ISL_1966254, EPI_ISL_1966255 | UBS DR MOHANNA ADAS | Instituto Butantan / Mendelics | Antonio Jorge Martins; Bianca Cechetto Carlos. Mendelics: Bibiana Santos; Claudia Renata dos Santos Barros; Cíntia Bittar; David Schlesinger. Hemocentro Ribeirão Preto: Simone Kashima; Debora Botequio Moretti; Elaine Cristina Marqueze; Elaine Vieira dos Santos; Elisangela Chicaroni Mattos; Erika Freitas; Evandra Strazza Rodrigues; Felipe Allan da Silva da Costa; Flavia Aburjaile; Fábio Sossai Possebon; Guilherme Campos; Guilherme Targino Valente; Heidge Fukumasu. USP-Botucatu: Rejane Maria Tommasini Grotto; Helena Lage Ferreira; Instituto Butantan: Dimas Tadeu Covas; Jardelina de Souza Todao Bernardino; Jayme A. Souza-Neto; Jessika Cristina Chagas Lesbon; Jorge A. Petroli Marchesi; José Salvatore Leister Patané; João Paulo Kitajima; João Pessoa Araújo Jr.; Leila Sabrina Ullmann; Loyze Paola Oliveira de Lima; Luiz Aurelio de Campos Crispin. Centro de Genômica Funcional da ESALQ: Luiz Lehmann Coutinho; Luiz Carlos Junior de Alcantara; Lívia Sacchetto; Maisa C. Pereira Parra; Maria Carolina Elias; Marta Giovanetti; Marília Moraes; Maurício Lacerda Nogueira. Prefeitura de Sao Paulo: Melissa Palmieri.; Patricia Akemi Assato; Paula Rahal; Paulo Inacio da Costa; Rafael dos Santos Bezerra; Raquel de Lello Rocha Campos Cassano. NGS Soluções Genômicas: Pilar Drummond Sampaio Corrêa Mariani. FZEA-USP Pirassununga: Mirele Daiana Poletti; Raul Machado Neto; Ricardo Augusto Brassaloti; Ricardo Haddad; Rodrigo Tocantins Calado. FAMERP-SJRP: Cecília Artico Banho; Sandra Coccuzzo Sampaio; Svetoslav Nanev Slavov; Vagner Fonseca; Vincent Louis Viala |
| EPI_ISL_1966545, EPI_ISL_1966546 | UBS DR WALDOMIRO FERREIRA NEVES SCRPARDO | Instituto Butantan / FZEA-USP (Pirassununga) | Antonio Jorge Martins; Bianca Cechetto Carlos. Mendelics: Bibiana Santos; Claudia Renata dos Santos Barros; Cíntia Bittar; David Schlesinger. Hemocentro Ribeirão Preto: Simone Kashima; Debora Botequio Moretti; Elaine Cristina Marqueze; Elaine Vieira dos Santos; Elisangela Chicaroni Mattos; Erika Freitas; Evandra Strazza Rodrigues; Felipe Allan da Silva da Costa; Flavia Aburjaile; Fábio Sossai Possebon; Guilherme Campos; Guilherme Targino Valente; Heidge Fukumasu. USP-Botucatu: Rejane Maria Tommasini Grotto; Helena Lage Ferreira; Instituto Butantan: Dimas Tadeu Covas; Jardelina de Souza Todao Bernardino; Jayme A. Souza-Neto; Jessika Cristina Chagas Lesbon; Jorge A. Petroli Marchesi; José Salvatore Leister Patané; João Paulo Kitajima; João Pessoa Araújo Jr.; Leila Sabrina Ullmann; Loyze Paola Oliveira de Lima; Luiz Aurelio de Campos Crispin. Centro de Genômica Funcional da ESALQ: Luiz Lehmann Coutinho; Luiz Carlos Junior de Alcantara; Lívia Sacchetto; Maisa C. Pereira Parra; Maria Carolina Elias; Marta Giovanetti; Marília Moraes; Maurício Lacerda Nogueira. Prefeitura de Sao Paulo: Melissa Palmieri.; Patricia Akemi Assato; Paula Rahal; Paulo Inacio da Costa; Rafael dos Santos Bezerra; Raquel de Lello Rocha Campos Cassano. NGS Soluções Genômicas: Pilar Drummond Sampaio Corrêa Mariani. FZEA-USP Pirassununga: Mirele Daiana Poletti; Raul Machado Neto; Ricardo Augusto Brassaloti; Ricardo Haddad; Rodrigo Tocantins Calado. FAMERP-SJRP: Cecília Artico Banho; Sandra Coccuzzo Sampaio; Svetoslav Nanev Slavov; Vagner Fonseca; Vincent Louis Viala |
| EPI_ISL_1966807, EPI_ISL_1966813 | UBS DR WALDOMIRO FERREIRA NEVES SCRPARDO | Instituto Butantan / Mendelics | Antonio Jorge Martins; Bianca Cechetto Carlos. Mendelics: Bibiana Santos; Claudia Renata dos Santos Barros; Cíntia Bittar; David Schlesinger. Hemocentro Ribeirão Preto: Simone Kashima; Debora Botequio Moretti; Elaine Cristina Marqueze; Elaine Vieira dos Santos; Elisangela Chicaroni Mattos; Erika Freitas; Evandra Strazza Rodrigues; Felipe Allan da Silva da Costa; Flavia Aburjaile; Fábio Sossai Possebon; Guilherme Campos; Guilherme Targino Valente; Heidge Fukumasu. USP-Botucatu: Rejane Maria Tommasini Grotto; Helena Lage Ferreira; Instituto Butantan: Dimas Tadeu Covas; Jardelina de Souza Todao Bernardino; Jayme A. Souza-Neto; Jessika Cristina Chagas Lesbon; Jorge A. Petroli Marchesi; José Salvatore Leister Patané; João Paulo Kitajima; João Pessoa Araújo Jr.; Leila Sabrina Ullmann; Loyze Paola Oliveira de Lima; Luiz Aurelio de Campos Crispin. Centro de Genômica Funcional da ESALQ: Luiz Lehmann Coutinho; Luiz Carlos Junior de Alcantara; Lívia Sacchetto; Maisa C. Pereira Parra; Maria Carolina Elias; Marta Giovanetti; Marília Moraes; Maurício Lacerda Nogueira. Prefeitura de Sao Paulo: Melissa Palmieri.; Patricia Akemi Assato; Paula Rahal; Paulo Inacio da Costa; Rafael dos Santos Bezerra; Raquel de Lello Rocha Campos Cassano. NGS Soluções Genômicas: Pilar Drummond Sampaio Corrêa Mariani. FZEA-USP Pirassununga: Mirele Daiana Poletti; Raul Machado Neto; Ricardo Augusto Brassaloti; Ricardo Haddad; Rodrigo Tocantins Calado. FAMERP-SJRP: Cecília Artico Banho; Sandra Coccuzzo Sampaio; Svetoslav Nanev Slavov; Vagner Fonseca; Vincent Louis Viala |
| EPI_ISL_1445217, EPI_ISL_1445219, EPI_ISL_1445220, EPI_ISL_1445227, EPI_ISL_1966211, EPI_ISL_1966213, EPI_ISL_1966214, EPI_ISL_1966230 | | | |
| see above | UBS EMILIA COSME CERQUEIRA | Instituto Butantan / Mendelics | Antonio Jorge Martins; Bianca Cechetto Carlos. Mendelics: Bibiana Santos; Bibiana Santos; Claudia Renata dos Santos Barros; Cíntia Bittar; David Schlesinger; David Schlesinger. Hemocentro Ribeirão Preto: Simone Kashima; Debora Botequio Moretti; Dimas Tadeu Covas; Elaine Cristina Marqueze; Elaine Vieira dos Santos; Elisangela Chicaroni Mattos; Erika Freitas; Evandra Strazza Rodrigues; Felipe Allan da Silva da Costa; Flavia Aburjaile; Fábio Sossai Possebon; Guilherme Campos; Guilherme Targino Valente; Heidge Fukumasu. USP-Botucatu: Rejane Maria Tommasini Grotto; Helena Lage Ferreira; Instituto Butantan: Dimas Tadeu Covas; Jardelina de Souza Todao Bernardino; Jayme A. Souza-Neto; Jessika Cristina Chagas Lesbon; Jorge A. Petroli Marchesi; José Salvatore Leister Patané; João Paulo Kitajima; João Pessoa Araújo Jr.; Leila Sabrina Ullmann; Loyze Paola Oliveira de Lima; Luiz Aurelio de Campos Crispin. Centro de Genômica Funcional da ESALQ: Luiz Lehmann Coutinho; Luiz Carlos Junior de Alcantara; Lívia Sacchetto; Maisa C. Pereira Parra; Maria Carolina Elias; Marta Giovanetti; Marília Moraes; Maurício Lacerda Nogueira. Prefeitura de Sao Paulo: Melissa Palmieri.; Patricia Akemi Assato; Paula Rahal; Paulo Inacio da Costa; Rafael dos Santos Bezerra; Raquel de Lello Rocha Campos Cassano. NGS Soluções Genômicas: Pilar Drummond Sampaio Corrêa Mariani. FZEA-USP Pirassununga: Mirele Daiana Poletti; Raul Machado Neto; Ricardo Augusto Brassaloti; Ricardo Haddad; Rodrigo Tocantins Calado.; Rodrigo Tocantins Calado. FAMERP-SJRP: Cecília Artico Banho; Sandra Coccuzzo Sampaio; Simone Kashima; Svetoslav Nanev Slavov; Vagner Fonseca; Vincent Louis Viala |
| EPI_ISL_1967163, EPI_ISL_1967164 | UBS ENRIQUE GUILHERME ERNESTO MARZEUSKI | Instituto Butantan / ESALQ-USP (Piracicaba) | Antonio Jorge Martins; Bianca Cechetto Carlos. Mendelics: Bibiana Santos; Claudia Renata dos Santos Barros; Cíntia Bittar; David Schlesinger. Hemocentro Ribeirão Preto: Simone Kashima; Debora Botequio Moretti; Elaine Cristina Marqueze; Elaine Vieira dos Santos; Elisangela Chicaroni Mattos; Erika Freitas; Evandra Strazza Rodrigues; Felipe Allan da Silva da Costa; Flavia Aburjaile; Fábio Sossai Possebon; Guilherme Campos; Guilherme Targino Valente; Heidge Fukumasu. USP-Botucatu: Rejane Maria Tommasini Grotto; Helena Lage Ferreira; Instituto Butantan: Dimas Tadeu Covas; Jardelina de Souza Todao Bernardino; Jayme A. Souza-Neto; Jessika Cristina Chagas Lesbon; Jorge A. Petroli Marchesi; José Salvatore Leister Patané; João Paulo Kitajima; João Pessoa Araújo Jr.; Leila Sabrina Ullmann; Loyze Paola Oliveira de Lima; Luiz Aurelio de Campos Crispin. Centro de Genômica Funcional da ESALQ: Luiz Lehmann Coutinho; Luiz Carlos Junior de Alcantara; Lívia Sacchetto; Maisa C. Pereira Parra; Maria Carolina Elias; Marta Giovanetti; Marília Moraes; Maurício Lacerda Nogueira. Prefeitura de Sao Paulo: Melissa Palmieri.; Patricia Akemi Assato; Paula Rahal; Paulo Inacio da Costa; Rafael dos Santos Bezerra; Raquel de Lello Rocha Campos Cassano. NGS Soluções Genômicas: Pilar Drummond Sampaio Corrêa Mariani. FZEA-USP Pirassununga: Mirele Daiana Poletti; Raul Machado Neto; Ricardo Augusto Brassaloti; Ricardo Haddad; Rodrigo Tocantins Calado. FAMERP-SJRP: Cecília Artico Banho; Sandra Coccuzzo Sampaio; Svetoslav Nanev Slavov; Vagner Fonseca; Vincent Louis Viala |
| EPI_ISL_1445205, EPI_ISL_1966207 | UBS FRANCISCA LIMA DE LIRA | Instituto Butantan / Mendelics | Antonio Jorge Martins; Bianca Cechetto Carlos. Mendelics: Bibiana Santos; Bibiana Santos; Claudia Renata dos Santos Barros; Cíntia Bittar; David Schlesinger; David Schlesinger. Hemocentro Ribeirão Preto: Simone Kashima; Debora Botequio Moretti; Dimas Tadeu Covas; Elaine Cristina Marqueze; Elaine Vieira dos Santos; Elisangela Chicaroni Mattos; Erika Freitas; Evandra Strazza Rodrigues; Felipe Allan da Silva da Costa; Flavia Aburjaile; Fábio Sossai Possebon; Guilherme Campos; Guilherme Targino Valente; Heidge Fukumasu. USP-Botucatu: Rejane Maria Tommasini Grotto; Helena Lage Ferreira; Instituto Butantan: Dimas Tadeu Covas; Jardelina de Souza Todao Bernardino; Jayme A. Souza-Neto; Jessika Cristina Chagas Lesbon; Jorge A. Petroli Marchesi; José Salvatore Leister Patané; João Paulo Kitajima; João Pessoa Araújo Jr.; Leila Sabrina Ullmann; Loyze Paola Oliveira de Lima; Luiz Aurelio de Campos Crispin. Centro de Genômica Funcional da ESALQ: Luiz Lehmann Coutinho; Luiz Carlos Junior de Alcantara; Lívia Sacchetto; Maisa C. Pereira Parra; Maria Carolina Elias; Marta Giovanetti; Marília Moraes; Maurício Lacerda Nogueira. Prefeitura de Sao Paulo: Melissa Palmieri.; Patricia Akemi Assato; Paula Rahal; Paulo Inacio da Costa; Rafael dos Santos Bezerra; Raquel de Lello Rocha Campos Cassano. NGS Soluções Genômicas: Pilar Drummond Sampaio Corrêa Mariani. FZEA-USP Pirassununga: Mirele Daiana Poletti; Raul Machado Neto; Ricardo Augusto Brassaloti; Ricardo Haddad; Rodrigo Tocantins Calado.; Rodrigo Tocantins Calado. FAMERP-SJRP: Cecília Artico Banho; Sandra Coccuzzo Sampaio; Simone Kashima; Svetoslav Nanev Slavov; Vagner Fonseca; Vincent Louis Viala |
| EPI_ISL_2133093 | UBS GETULINO JOSE DIAS | Instituto Butantan / Mendelics | Antonio Jorge Martins; Bianca Cechetto Carlos. Mendelics: Bibiana Santos; Claudia Renata dos Santos Barros; Cíntia Bittar; David Schlesinger. Hemocentro Ribeirão Preto: Simone Kashima; Debora Botequio Moretti; Elaine Cristina Marqueze; Elaine Vieira dos Santos; Elisangela Chicaroni Mattos; Erika Freitas; Evandra Strazza Rodrigues; Felipe Allan da Silva da Costa; Flavia Aburjaile; Fábio Sossai Possebon; Guilherme Campos; Guilherme Targino Valente; Heidge Fukumasu. USP-Botucatu: Rejane Maria Tommasini Grotto; Helena Lage Ferreira; Instituto Butantan: Dimas Tadeu Covas; Jardelina de Souza Todao Bernardino; Jayme A. Souza-Neto; Jessika Cristina Chagas Lesbon; Jorge A. Petroli Marchesi; José Salvatore Leister Patané; João Paulo Kitajima; João Pessoa Araújo Jr.; Leila Sabrina Ullmann; Loyze Paola Oliveira de Lima; Luiz Aurelio de Campos Crispin. Centro de Genômica Funcional da ESALQ: Luiz Lehmann Coutinho; Luiz Carlos Junior de Alcantara; Lívia Sacchetto; Maisa C. Pereira Parra; Maria Carolina Elias; Marta Giovanetti; Marília Moraes; Maurício Lacerda Nogueira. Prefeitura de Sao Paulo: Melissa Palmieri.; Patricia Akemi Assato; Paula Rahal; Paulo Inacio da Costa; Rafael dos Santos Bezerra; Raquel de Lello Rocha Campos Cassano. NGS Soluções Genômicas: Pilar Drummond Sampaio Corrêa Mariani. FZEA-USP Pirassununga: Mirele Daiana Poletti; Raul Machado Neto; Ricardo Augusto Brassaloti; Ricardo Haddad; Rodrigo Tocantins Calado. FAMERP-SJRP: Cecília Artico Banho; Sandra Coccuzzo Sampaio; Svetoslav Nanev Slavov; Vagner Fonseca; Vincent Louis Viala |
| EPI_ISL_1966761, EPI_ISL_1966762, EPI_ISL_2170961 | UBS GLENDA RODELLA DUMAS | Instituto Butantan / Mendelics | Antonio Jorge Martins; Bianca Cechetto Carlos. Mendelics: Bibiana Santos; Claudia Renata dos Santos Barros; Cíntia Bittar; David Schlesinger. Hemocentro Ribeirão Preto: Simone Kashima; Debora Botequio Moretti; Elaine Cristina Marqueze; Elaine Vieira dos Santos; Elisangela Chicaroni Mattos; Erika Freitas; Evandra Strazza Rodrigues; Felipe Allan da Silva da Costa; Flavia Aburjaile; Fábio Sossai Possebon; Guilherme Campos; Guilherme Targino Valente; Heidge Fukumasu. USP-Botucatu: Rejane Maria Tommasini Grotto; Helena Lage Ferreira; Instituto Butantan: Dimas Tadeu Covas; Jardelina de Souza Todao Bernardino; Jayme A. Souza-Neto; Jessika Cristina Chagas Lesbon; Jorge A. Petroli Marchesi; José Salvatore Leister Patané; João Paulo Kitajima; João Pessoa Araújo Jr.; Leila Sabrina Ullmann; Loyze Paola Oliveira de Lima; Luiz Aurelio de Campos Crispin. Centro de Genômica Funcional da ESALQ: Luiz Lehmann Coutinho; Luiz Carlos Junior de Alcantara; Lívia Sacchetto; Maisa C. Pereira Parra; Maria Carolina Elias; Marta Giovanetti; Marília Moraes; Maurício Lacerda Nogueira. Prefeitura de Sao Paulo: Melissa Palmieri.; Patricia Akemi Assato; Paula Rahal; Paulo Inacio da Costa; Rafael dos Santos Bezerra; Raquel de Lello Rocha Campos Cassano. NGS Soluções Genômicas: Pilar Drummond Sampaio Corrêa Mariani. FZEA-USP Pirassununga: Mirele Daiana Poletti; Raul Machado Neto; Ricardo Augusto Brassaloti; Ricardo Haddad; Rodrigo Tocantins Calado. FAMERP-SJRP: Cecília Artico Banho; Sandra Coccuzzo Sampaio; Svetoslav Nanev Slavov; Vagner Fonseca; Vincent Louis Viala |
| EPI_ISL_1445215, EPI_ISL_1445216, EPI_ISL_1445221, EPI_ISL_1445222, EPI_ISL_1445223, EPI_ISL_1966209, EPI_ISL_1966210, EPI_ISL_1966215, EPI_ISL_1966216, EPI_ISL_1966217 | | | |
| see above | UBS HELENA MARREY | Instituto Butantan / Mendelics | Antonio Jorge Martins; Bianca Cechetto Carlos. Mendelics: Bibiana Santos; Bibiana Santos; Claudia Renata dos Santos Barros; Cíntia Bittar; David Schlesinger. Hemocentro Ribeirão Preto: Simone Kashima; Debora Botequio Moretti; Dimas Tadeu Covas; Elaine Cristina Marqueze; Elaine Vieira dos Santos; Elisangela Chicaroni Mattos; Erika Freitas; Evandra Strazza Rodrigues; Felipe Allan da Silva da Costa; Flavia Aburjaile; Fábio Sossai Possebon; Guilherme Campos; Guilherme Targino Valente; Heidge Fukumasu. USP-Botucatu: Rejane Maria Tommasini Grotto; Helena Lage Ferreira; Instituto Butantan: Dimas Tadeu Covas; Jardelina de Souza Todao Bernardino; Jayme A. Souza-Neto; Jessika Cristina Chagas Lesbon; Jorge A. Petroli Marchesi; José Salvatore Leister Patané; João Paulo Kitajima; João Pessoa Araújo Jr.; Leila Sabrina Ullmann; Loyze Paola Oliveira de Lima; Luiz Aurelio de Campos Crispin. Centro de Genômica Funcional da ESALQ: Luiz Lehmann Coutinho; Luiz Carlos Junior de Alcantara; Lívia Sacchetto; Maisa C. Pereira Parra; Maria Carolina Elias; Marta Giovanetti; Marília Moraes; Maurício Lacerda Nogueira. Prefeitura de Sao Paulo: Melissa Palmieri.; Patricia Akemi Assato; Paula Rahal; Paulo Inacio da Costa; Rafael dos Santos Bezerra; Raquel de Lello Rocha Campos Cassano. NGS Soluções Genômicas: Pilar Drummond Sampaio Corrêa Mariani. FZEA-USP Pirassununga: Mirele Daiana Poletti; Raul Machado Neto; Ricardo Augusto Brassaloti; Ricardo Haddad; Rodrigo Tocantins Calado.; Rodrigo Tocantins Calado. FAMERP-SJRP: Cecília Artico Banho; Sandra Coccuzzo Sampaio; Simone Kashima; Svetoslav Nanev Slavov; Vagner Fonseca; Vincent Louis Viala |
| EPI_ISL_1966469, EPI_ISL_1966470 | UBS II DA FAMILIA SEGISFREDO MAMEDE PINTO LOURDES | Instituto Butantan / Mendelics | Antonio Jorge Martins; Bianca Cechetto Carlos. Mendelics: Bibiana Santos; Claudia Renata dos Santos Barros; Cíntia Bittar; David Schlesinger. Hemocentro Ribeirão Preto: Simone Kashima; Debora Botequio Moretti; Elaine Cristina Marqueze; Elaine Vieira dos Santos; Elisangela Chicaroni Mattos; Erika Freitas; Evandra Strazza Rodrigues; Felipe Allan da Silva da Costa; Flavia Aburjaile; Fábio Sossai Possebon; Guilherme Campos; Guilherme Targino Valente; Heidge Fukumasu. USP-Botucatu: Rejane Maria Tommasini Grotto; Helena Lage Ferreira; Instituto Butantan: Dimas Tadeu Covas; Jardelina de Souza Todao Bernardino; Jayme A. Souza-Neto; Jessika Cristina Chagas Lesbon; Jorge A. Petroli Marchesi; José Salvatore Leister Patané; João Paulo Kitajima; João Pessoa Araújo Jr.; Leila Sabrina Ullmann; Loyze Paola Oliveira de Lima; Luiz Aurelio de Campos Crispin. Centro de Genômica Funcional da ESALQ: Luiz Lehmann Coutinho; Luiz Carlos Junior de Alcantara; Lívia Sacchetto; Maisa C. Pereira Parra; Maria Carolina Elias; Marta Giovanetti; Marília Moraes; Maurício Lacerda Nogueira. Prefeitura de Sao Paulo: Melissa Palmieri.; Patricia Akemi Assato; Paula Rahal; Paulo Inacio da Costa; Rafael dos Santos Bezerra; Raquel de Lello Rocha Campos Cassano. NGS Soluções Genômicas: Pilar Drummond Sampaio Corrêa Mariani. FZEA-USP Pirassununga: Mirele Daiana Poletti; Raul Machado Neto; Ricardo Augusto Brassaloti; Ricardo Haddad; Rodrigo Tocantins Calado. FAMERP-SJRP: Cecília Artico Banho; Sandra Coccuzzo Sampaio; Svetoslav Nanev Slavov; Vagner Fonseca; Vincent Louis Viala |
| EPI_ISL_1795324 | UBS II DE NARANDIBA | Instituto Butantan / ESALQ-Piracicaba | Antonio Jorge Martins; Bianca Cechetto Carlos. Mendelics: Bibiana Santos; Claudia Renata dos Santos Barros; David Schlesinger. Hemocentro Ribeirão Preto: Simone Kashima; Debora Botequio Moretti. Centro de Genômica Funcional da ESALQ: Luiz Lehmann Coutinho; Dimas Tadeu Covas; Elaine Cristina Marqueze; Elaine Vieira dos Santos; Elisangela Chicaroni Mattos; Erika Freitas; Evandra Strazza Rodrigues; Felipe Allan da Silva da Costa; Flavia Aburjaile; Guilherme Targino Valente; Heidge Fukumasu. USP-Botucatu: Rejane Maria Tommasini Grotto; Instituto Butantan: Alexander Roberto Precioso; Jayme A. Souza-Neto; Jessika Cristina Chagas Lesbon; José Salvatore Leister Patané; João Paulo Kitajima; Luiz Carlos Junior de Alcantara; Maria Carolina Elias; Marta Giovanetti; Patricia Akemi Assato; Rafael dos Santos Bezerra; Raquel de Lello Rocha Campos Cassano. NGS Soluções Genômicas: Pilar Drummond Sampaio Corrêa Mariani. FZEA-USP Pirassununga: Mirele Daiana Poletti; Raul Machado Neto; Ricardo Augusto Brassaloti; Ricardo Haddad; Rodrigo Tocantins Calado.; Sandra Coccuzzo Sampaio; Svetoslav Nanev Slavov; Vagner Fonseca; Vincent Louis Viala |
| EPI_ISL_1966265 | UBS II DE NARANDIBA | Instituto Butantan / Mendelics | Antonio Jorge Martins; Bianca Cechetto Carlos. Mendelics: Bibiana Santos; Claudia Renata dos Santos Barros; Cíntia Bittar; David Schlesinger. Hemocentro Ribeirão Preto: Simone Kashima; Debora Botequio Moretti; Elaine Cristina Marqueze; Elaine Vieira dos Santos; Elisangela Chicaroni Mattos; Erika Freitas; Evandra Strazza Rodrigues; Felipe Allan da Silva da Costa; Flavia Aburjaile; Fábio Sossai Possebon; Guilherme Campos; Guilherme Targino Valente; Heidge Fukumasu. USP-Botucatu: Rejane Maria Tommasini Grotto; Helena Lage Ferreira; Instituto Butantan: Dimas Tadeu Covas; Jardelina de Souza Todao Bernardino; Jayme A. Souza-Neto; Jessika Cristina Chagas Lesbon; Jorge A. Petroli Marchesi; José Salvatore Leister Patané; João Paulo Kitajima; João Pessoa Araújo Jr.; Leila Sabrina Ullmann; Loyze Paola Oliveira de Lima; Luiz Aurelio de Campos Crispin. Centro de Genômica Funcional da ESALQ: Luiz Lehmann Coutinho; Luiz Carlos Junior de Alcantara; Lívia Sacchetto; Maisa C. Pereira Parra; Maria Carolina Elias; Marta Giovanetti; Marília Moraes; Maurício Lacerda Nogueira. Prefeitura de Sao Paulo: Melissa Palmieri.; Patricia Akemi Assato; Paula Rahal; Paulo Inacio da Costa; Rafael dos Santos Bezerra; Raquel de Lello Rocha Campos Cassano. NGS Soluções Genômicas: Pilar Drummond Sampaio Corrêa Mariani. FZEA-USP Pirassununga: Mirele Daiana Poletti; Raul Machado Neto; Ricardo Augusto Brassaloti; Ricardo Haddad; Rodrigo Tocantins Calado. FAMERP-SJRP: Cecília Artico Banho; Sandra Coccuzzo Sampaio; Svetoslav Nanev Slavov; Vagner Fonseca; Vincent Louis Viala |
| EPI_ISL_1795326, EPI_ISL_1795424 | UBS II DE REGENTE FEIJO | Instituto Butantan / ESALQ-Piracicaba | Antonio Jorge Martins; Bianca Cechetto Carlos. Mendelics: Bibiana Santos; Claudia Renata dos Santos Barros; David Schlesinger. Hemocentro Ribeirão Preto: Simone Kashima; Debora Botequio Moretti. Centro de Genômica Funcional da ESALQ: Luiz Lehmann Coutinho; Dimas Tadeu Covas; Elaine Cristina Marqueze; Elaine Vieira dos Santos; Elisangela Chicaroni Mattos; Erika Freitas; Evandra Strazza Rodrigues; Felipe Allan da Silva da Costa; Flavia Aburjaile; Guilherme Targino Valente; Heidge Fukumasu. USP-Botucatu: Rejane Maria Tommasini Grotto; Instituto Butantan: Alexander Roberto Precioso; Jayme A. Souza-Neto; Jessika Cristina Chagas Lesbon; José Salvatore Leister Patané; João Paulo Kitajima; Luiz Carlos Junior de Alcantara; Maria Carolina Elias; Marta Giovanetti; Patricia Akemi Assato; Rafael dos Santos Bezerra; Raquel de Lello Rocha Campos Cassano. NGS Soluções Genômicas: Pilar Drummond Sampaio Corrêa Mariani. FZEA-USP Pirassununga: Mirele Daiana Poletti; Raul Machado Neto; Ricardo Augusto Brassaloti; Ricardo Haddad; Rodrigo Tocantins Calado.; Sandra Coccuzzo Sampaio; Svetoslav Nanev Slavov; Vagner Fonseca; Vincent Louis Viala |
| EPI_ISL_1795251, EPI_ISL_1795252, EPI_ISL_1795253, EPI_ISL_1795254, EPI_ISL_1795255, EPI_ISL_1795256, EPI_ISL_1795257 | | | |
| see above | UBS II DE TANABI MILTON MARTINS | Instituto Butantan / ESALQ-Piracicaba | Antonio Jorge Martins; Bianca Cechetto Carlos. Mendelics: Bibiana Santos; Claudia Renata dos Santos Barros; David Schlesinger. Hemocentro Ribeirão Preto: Simone Kashima; Debora Botequio Moretti. Centro de Genômica Funcional da ESALQ: Luiz Lehmann Coutinho; Dimas Tadeu Covas; Elaine Cristina Marqueze; Elaine Vieira dos Santos; Elisangela Chicaroni Mattos; Erika Freitas; Evandra Strazza Rodrigues; Felipe Allan da Silva da Costa; Flavia Aburjaile; Guilherme Targino Valente; Heidge Fukumasu. USP-Botucatu: Rejane Maria Tommasini Grotto; Instituto Butantan: Alexander Roberto Precioso; Jayme A. Souza-Neto; Jessika Cristina Chagas Lesbon; José Salvatore Leister |

| | | | |
|---|---|---|---|
| | PERCHES | | Patané; João Paulo Kitajima; Luiz Carlos Junior de Alcantara; Maria Carolina Elias; Marta Giovanetti; Patricia Akemi Assato; Rafael dos Santos Bezerra; Raquel de Lello Rocha Campos Cassano. NGS Soluções Genômicas: Pilar Drummond Sampaio Corrêa Mariani. FZEA-USP Pirassununga: Mirele Daiana Poleti; Raul Machado Neto; Ricardo Augusto Brassaloti; Ricardo Haddad; Rodrigo Tocantins Calado.; Sandra Coccuzzo Sampaio; Svetoslav Nanev Slavov; Vagner Fonseca; Vincent Louis Viala |
| EPI_ISL_1966552, EPI_ISL_1966553 | UBS II DE TANABI MILTON MARTINS PERCHES | Instituto Butantan / FZEA-USP (Pirassununga) | Antonio Jorge Martins; Bianca Cechetto Carlos. Mendelics: Bibiana Santos; Claudia Renata dos Santos Barros; Cíntia Bittar; David Schlesinger. Hemocentro Ribeirão Preto: Simone Kashima; Debora Botequio Moretti; Elaine Cristina Marqueze; Elaine Vieira dos Santos; Elisangela Chicaroni Mattos; Erika Freitas; Evandra Strazza Rodrigues; Felipe Allan da Silva da Costa; Flavia Aburjaile; Fábio Sossai Possebon; Guilherme Campos; Guilherme Targino Valente; Heidge Fukumasu. USP-Botucatu: Rejane Maria Tommasini Grotto; Helena Lage Ferreira; Instituto Butantan: Dimas Tadeu Covas; Jardelina de Souza Todao Bernardino; Jayme A. Souza-Neto; Jessika Cristina Chagas Lesbon; Jorge A. Petroli Marchesi; José Salvatore Leister Patané; João Paulo Kitajima; João Pessoa Araújo Jr.; Leila Sabrina Ullmann; Loyze Paola Oliveira de Lima; Luiz Aurelio de Campos Crispin. Centro de Genômica Funcional da ESALQ: Luiz Lehmann Coutinho; Luiz Carlos Junior de Alcantara; Lívia Sacchetto; Maisa C. Pereira Parra; Maria Carolina Elias; Marta Giovanetti; Marília Moraes; Maurício Lacerda Nogueira. Prefeitura de Sao Paulo: Melissa Palmieri.; Patricia Akemi Assato; Paula Rahal; Paulo Inacio da Costa; Rafael dos Santos Bezerra; Raquel de Lello Rocha Campos Cassano. NGS Soluções Genômicas: Pilar Drummond Sampaio Corrêa Mariani. FZEA-USP Pirassununga: Mirele Daiana Poleti; Raul Machado Neto; Ricardo Augusto Brassaloti; Ricardo Haddad; Rodrigo Tocantins Calado. FAMERP-SJRP: Cecília Artico Banho; Sandra Coccuzzo Sampaio; Svetoslav Nanev Slavov; Vagner Fonseca; Vincent Louis Viala |
| EPI_ISL_1966548 | UBS II DOUTOR ORLANDO BERTOLLI | Instituto Butantan / FZEA-USP (Pirassununga) | Antonio Jorge Martins; Bianca Cechetto Carlos. Mendelics: Bibiana Santos; Claudia Renata dos Santos Barros; Cíntia Bittar; David Schlesinger. Hemocentro Ribeirão Preto: Simone Kashima; Debora Botequio Moretti; Elaine Cristina Marqueze; Elaine Vieira dos Santos; Elisangela Chicaroni Mattos; Erika Freitas; Evandra Strazza Rodrigues; Felipe Allan da Silva da Costa; Flavia Aburjaile; Fábio Sossai Possebon; Guilherme Campos; Guilherme Targino Valente; Heidge Fukumasu. USP-Botucatu: Rejane Maria Tommasini Grotto; Helena Lage Ferreira; Instituto Butantan: Dimas Tadeu Covas; Jardelina de Souza Todao Bernardino; Jayme A. Souza-Neto; Jessika Cristina Chagas Lesbon; Jorge A. Petroli Marchesi; José Salvatore Leister Patané; João Paulo Kitajima; João Pessoa Araújo Jr.; Leila Sabrina Ullmann; Loyze Paola Oliveira de Lima; Luiz Aurelio de Campos Crispin. Centro de Genômica Funcional da ESALQ: Luiz Lehmann Coutinho; Luiz Carlos Junior de Alcantara; Lívia Sacchetto; Maisa C. Pereira Parra; Maria Carolina Elias; Marta Giovanetti; Marília Moraes; Maurício Lacerda Nogueira. Prefeitura de Sao Paulo: Melissa Palmieri.; Patricia Akemi Assato; Paula Rahal; Paulo Inacio da Costa; Rafael dos Santos Bezerra; Raquel de Lello Rocha Campos Cassano. NGS Soluções Genômicas: Pilar Drummond Sampaio Corrêa Mariani. FZEA-USP Pirassununga: Mirele Daiana Poleti; Raul Machado Neto; Ricardo Augusto Brassaloti; Ricardo Haddad; Rodrigo Tocantins Calado. FAMERP-SJRP: Cecília Artico Banho; Sandra Coccuzzo Sampaio; Svetoslav Nanev Slavov; Vagner Fonseca; Vincent Louis Viala |
| EPI_ISL_1795328, EPI_ISL_1795330 | UBS II DR EXPEDITO SHIZUO KUROCE | Instituto Butantan / ESALQ-Piracicaba | Antonio Jorge Martins; Bianca Cechetto Carlos. Mendelics: Bibiana Santos; Claudia Renata dos Santos Barros; David Schlesinger. Hemocentro Ribeirão Preto: Simone Kashima; Debora Botequio Moretti. Centro de Genômica Funcional da ESALQ: Luiz Lehmann Coutinho; Dimas Tadeu Covas; Elaine Cristina Marqueze; Elaine Vieira dos Santos; Elisangela Chicaroni Mattos; Erika Freitas; Evandra Strazza Rodrigues; Felipe Allan da Silva da Costa; Flavia Aburjaile; Guilherme Targino Valente; Heidge Fukumasu. USP-Botucatu: Rejane Maria Tommasini Grotto; Instituto Butantan: Alexander Roberto Precioso; Jayme A. Souza-Neto; Jessika Cristina Chagas Lesbon; José Salvatore Leister Patané; João Paulo Kitajima; Luiz Carlos Junior de Alcantara; Maria Carolina Elias; Marta Giovanetti; Patricia Akemi Assato; Rafael dos Santos Bezerra; Raquel de Lello Rocha Campos Cassano. NGS Soluções Genômicas: Pilar Drummond Sampaio Corrêa Mariani. FZEA-USP Pirassununga: Mirele Daiana Poleti; Raul Machado Neto; Ricardo Augusto Brassaloti; Ricardo Haddad; Rodrigo Tocantins Calado.; Sandra Coccuzzo Sampaio; Svetoslav Nanev Slavov; Vagner Fonseca; Vincent Louis Viala |
| EPI_ISL_1966995 | UBS III DE PARIQUERA ACU PARIQUERA ACU | Instituto Butantan / ESALQ-USP (Piracicaba) | Antonio Jorge Martins; Bianca Cechetto Carlos. Mendelics: Bibiana Santos; Claudia Renata dos Santos Barros; Cíntia Bittar; David Schlesinger. Hemocentro Ribeirão Preto: Simone Kashima; Debora Botequio Moretti; Elaine Cristina Marqueze; Elaine Vieira dos Santos; Elisangela Chicaroni Mattos; Erika Freitas; Evandra Strazza Rodrigues; Felipe Allan da Silva da Costa; Flavia Aburjaile; Fábio Sossai Possebon; Guilherme Campos; Guilherme Targino Valente; Heidge Fukumasu. USP-Botucatu: Rejane Maria Tommasini Grotto; Helena Lage Ferreira; Instituto Butantan: Dimas Tadeu Covas; Jardelina de Souza Todao Bernardino; Jayme A. Souza-Neto; Jessika Cristina Chagas Lesbon; Jorge A. Petroli Marchesi; José Salvatore Leister Patané; João Paulo Kitajima; João Pessoa Araújo Jr.; Leila Sabrina Ullmann; Loyze Paola Oliveira de Lima; Luiz Aurelio de Campos Crispin. Centro de Genômica Funcional da ESALQ: Luiz Lehmann Coutinho; Luiz Carlos Junior de Alcantara; Lívia Sacchetto; Maisa C. Pereira Parra; Maria Carolina Elias; Marta Giovanetti; Marília Moraes; Maurício Lacerda Nogueira. Prefeitura de Sao Paulo: Melissa Palmieri.; Patricia Akemi Assato; Paula Rahal; Paulo Inacio da Costa; Rafael dos Santos Bezerra; Raquel de Lello Rocha Campos Cassano. NGS Soluções Genômicas: Pilar Drummond Sampaio Corrêa Mariani. FZEA-USP Pirassununga: Mirele Daiana Poleti; Raul Machado Neto; Ricardo Augusto Brassaloti; Ricardo Haddad; Rodrigo Tocantins Calado. FAMERP-SJRP: Cecília Artico Banho; Sandra Coccuzzo Sampaio; Svetoslav Nanev Slavov; Vagner Fonseca; Vincent Louis Viala |
| EPI_ISL_1966696 | UBS III DE PARIQUERA ACU PARIQUERA ACU | Instituto Butantan / Mendelics | Antonio Jorge Martins; Bianca Cechetto Carlos. Mendelics: Bibiana Santos; Claudia Renata dos Santos Barros; Cíntia Bittar; David Schlesinger. Hemocentro Ribeirão Preto: Simone Kashima; Debora Botequio Moretti; Elaine Cristina Marqueze; Elaine Vieira dos Santos; Elisangela Chicaroni Mattos; Erika Freitas; Evandra Strazza Rodrigues; Felipe Allan da Silva da Costa; Flavia Aburjaile; Fábio Sossai Possebon; Guilherme Campos; Guilherme Targino Valente; Heidge Fukumasu. USP-Botucatu: Rejane Maria Tommasini Grotto; Helena Lage Ferreira; Instituto Butantan: Dimas Tadeu Covas; Jardelina de Souza Todao Bernardino; Jayme A. Souza-Neto; Jessika Cristina Chagas Lesbon; Jorge A. Petroli Marchesi; José Salvatore Leister Patané; João Paulo Kitajima; João Pessoa Araújo Jr.; Leila Sabrina Ullmann; Loyze Paola Oliveira de Lima; Luiz Aurelio de Campos Crispin. Centro de Genômica Funcional da ESALQ: Luiz Lehmann Coutinho; Luiz Carlos Junior de Alcantara; Lívia Sacchetto; Maisa C. Pereira Parra; Maria Carolina Elias; Marta Giovanetti; Marília Moraes; Maurício Lacerda Nogueira. Prefeitura de Sao Paulo: Melissa Palmieri.; Patricia Akemi Assato; Paula Rahal; Paulo Inacio da Costa; Rafael dos Santos Bezerra; Raquel de Lello Rocha Campos Cassano. NGS Soluções Genômicas: Pilar Drummond Sampaio Corrêa Mariani. FZEA-USP Pirassununga: Mirele Daiana Poleti; Raul Machado Neto; Ricardo Augusto Brassaloti; Ricardo Haddad; Rodrigo Tocantins Calado. FAMERP-SJRP: Cecília Artico Banho; Sandra Coccuzzo Sampaio; Svetoslav Nanev Slavov; Vagner Fonseca; Vincent Louis Viala |
| EPI_ISL_1795332, EPI_ISL_1795335, EPI_ISL_1795395, EPI_ISL_1795396, EPI_ISL_1795397, EPI_ISL_1795425 | UBS III DE RANCHARIA | Instituto Butantan / ESALQ-Piracicaba | Antonio Jorge Martins; Bianca Cechetto Carlos. Mendelics: Bibiana Santos; Claudia Renata dos Santos Barros; David Schlesinger. Hemocentro Ribeirão Preto: Simone Kashima; Debora Botequio Moretti. Centro de Genômica Funcional da ESALQ: Luiz Lehmann Coutinho; Dimas Tadeu Covas; Elaine Cristina Marqueze; Elaine Vieira dos Santos; Elisangela Chicaroni Mattos; Erika Freitas; Evandra Strazza Rodrigues; Felipe Allan da Silva da Costa; Flavia Aburjaile; Guilherme Targino Valente; Heidge Fukumasu. USP-Botucatu: Rejane Maria Tommasini Grotto; Instituto Butantan: Alexander Roberto Precioso; Jayme A. Souza-Neto; Jessika Cristina Chagas Lesbon; José Salvatore Leister Patané; João Paulo Kitajima; Luiz Carlos Junior de Alcantara; Maria Carolina Elias; Marta Giovanetti; Patricia Akemi Assato; Rafael dos Santos Bezerra; Raquel de Lello Rocha Campos Cassano. NGS Soluções Genômicas: Pilar Drummond Sampaio Corrêa Mariani. FZEA-USP Pirassununga: Mirele Daiana Poleti; Raul Machado Neto; Ricardo Augusto Brassaloti; Ricardo Haddad; Rodrigo Tocantins Calado.; Sandra Coccuzzo Sampaio; Svetoslav Nanev Slavov; Vagner Fonseca; Vincent Louis Viala |
| EPI_ISL_1445212, EPI_ISL_1445228, EPI_ISL_1966203, EPI_ISL_1966231, EPI_ISL_1966266 | UBS IRMA AGUEDA MARIA JAIME | Instituto Butantan / Mendelics | Antonio Jorge Martins; Bianca Cechetto Carlos. Mendelics: Bibiana Santos; Bibiana Santos; Claudia Renata dos Santos Barros; Cíntia Bittar; David Schlesinger; David Schlesinger. Hemocentro Ribeirão Preto: Simone Kashima; Debora Botequio Moretti; Dimas Tadeu Covas; Elaine Cristina Marqueze; Elaine Vieira dos Santos; Elisangela Chicaroni Mattos; Erika Freitas; Evandra Strazza Rodrigues; Felipe Allan da Silva da Costa; Flavia Aburjaile; Fábio Sossai Possebon; Guilherme Campos; Guilherme Targino Valente; Heidge Fukumasu. USP-Botucatu: Rejane Maria Tommasini Grotto; Helena Lage Ferreira; Instituto Butantan: Dimas Tadeu Covas; Jardelina de Souza Todao Bernardino; Jayme A. Souza-Neto; Jessika Cristina Chagas Lesbon; Jorge A. Petroli Marchesi; José Salvatore Leister Patané; João Paulo Kitajima; João Pessoa Araújo Jr.; Leila Sabrina Ullmann; Loyze Paola Oliveira de Lima; Luiz Aurelio de Campos Crispin. Centro de Genômica Funcional da ESALQ: Luiz Lehmann Coutinho; Luiz Carlos Junior de Alcantara; Lívia Sacchetto; Maisa C. Pereira Parra; Maria Carolina Elias; Marta Giovanetti; Marília Moraes; Maurício Lacerda Nogueira. Prefeitura de Sao Paulo: Melissa Palmieri.; Patricia Akemi Assato; Paula Rahal; Paulo Inacio da Costa; Rafael dos Santos Bezerra; Raquel de Lello Rocha Campos Cassano. NGS Soluções Genômicas: Pilar Drummond Sampaio Corrêa Mariani. FZEA-USP Pirassununga: Mirele Daiana Poleti; Raul Machado Neto; Ricardo Augusto Brassaloti; Ricardo Haddad; Rodrigo Tocantins Calado.; Rodrigo Tocantins Calado. FAMERP-SJRP: Cecília Artico Banho; Sandra Coccuzzo Sampaio; Simone Kashima; Svetoslav Nanev Slavov; Vagner Fonseca; Vincent Louis Viala |
| EPI_ISL_1966460, EPI_ISL_1966461, EPI_ISL_1966462 | UBS IV GUARARAPES | Instituto Butantan / Mendelics | Antonio Jorge Martins; Bianca Cechetto Carlos. Mendelics: Bibiana Santos; Claudia Renata dos Santos Barros; Cíntia Bittar; David Schlesinger. Hemocentro Ribeirão Preto: Simone Kashima; Debora Botequio Moretti; Elaine Cristina Marqueze; Elaine Vieira dos Santos; Elisangela Chicaroni Mattos; Erika Freitas; Evandra Strazza Rodrigues; Felipe Allan da Silva da Costa; Flavia Aburjaile; Fábio Sossai Possebon; Guilherme Campos; Guilherme Targino Valente; Heidge Fukumasu. USP-Botucatu: Rejane Maria Tommasini Grotto; Helena Lage Ferreira; Instituto Butantan: Dimas Tadeu Covas; Jardelina de Souza Todao Bernardino; Jayme A. Souza-Neto; Jessika Cristina Chagas Lesbon; Jorge A. Petroli Marchesi; José Salvatore Leister Patané; João Paulo Kitajima; João Pessoa Araújo Jr.; Leila Sabrina Ullmann; Loyze Paola Oliveira de Lima; Luiz Aurelio de Campos Crispin. Centro de Genômica Funcional da ESALQ: Luiz Lehmann Coutinho; Luiz Carlos Junior de Alcantara; Lívia Sacchetto; Maisa C. Pereira Parra; Maria Carolina Elias; Marta Giovanetti; Marília Moraes; Maurício Lacerda Nogueira. Prefeitura de Sao Paulo: Melissa Palmieri.; Patricia Akemi Assato; Paula Rahal; Paulo Inacio da Costa; Rafael dos Santos Bezerra; Raquel de Lello Rocha Campos Cassano. NGS Soluções Genômicas: Pilar Drummond Sampaio Corrêa Mariani. FZEA-USP Pirassununga: Mirele Daiana Poleti; Raul Machado Neto; Ricardo Augusto Brassaloti; Ricardo Haddad; Rodrigo Tocantins Calado. FAMERP-SJRP: Cecília Artico Banho; Sandra Coccuzzo Sampaio; Svetoslav Nanev Slavov; Vagner Fonseca; Vincent Louis Viala |
| EPI_ISL_1967285, EPI_ISL_1967286 | UBS JARDIM CABUCU | Instituto Butantan / Mendelics | Antonio Jorge Martins; Bianca Cechetto Carlos. Mendelics: Bibiana Santos; Claudia Renata dos Santos Barros; Cíntia Bittar; David Schlesinger. Hemocentro Ribeirão Preto: Simone Kashima; Debora Botequio Moretti; Elaine Cristina Marqueze; Elaine Vieira dos Santos; Elisangela Chicaroni Mattos; Erika Freitas; Evandra Strazza Rodrigues; Felipe Allan da Silva da Costa; Flavia Aburjaile; Fábio Sossai Possebon; Guilherme Campos; Guilherme Targino Valente; Heidge Fukumasu. USP-Botucatu: Rejane Maria Tommasini Grotto; Helena Lage Ferreira; Instituto Butantan: Dimas Tadeu Covas; Jardelina de Souza Todao Bernardino; Jayme A. Souza-Neto; Jessika Cristina Chagas Lesbon; Jorge A. Petroli Marchesi; José Salvatore Leister Patané; João Paulo Kitajima; João Pessoa Araújo Jr.; Leila Sabrina Ullmann; Loyze Paola Oliveira de Lima; Luiz Aurelio de Campos Crispin. Centro de Genômica Funcional da ESALQ: Luiz Lehmann Coutinho; Luiz Carlos Junior de Alcantara; Lívia Sacchetto; Maisa C. Pereira Parra; Maria Carolina Elias; Marta Giovanetti; Marília Moraes; Maurício Lacerda Nogueira. Prefeitura de Sao Paulo: Melissa Palmieri.; Patricia Akemi Assato; Paula Rahal; Paulo Inacio da Costa; Rafael dos Santos Bezerra; Raquel de Lello Rocha Campos Cassano. NGS Soluções Genômicas: Pilar Drummond Sampaio Corrêa Mariani. FZEA-USP Pirassununga: Mirele Daiana Poleti; Raul Machado Neto; Ricardo Augusto Brassaloti; Ricardo Haddad; Rodrigo Tocantins Calado. FAMERP-SJRP: Cecília Artico Banho; Sandra Coccuzzo Sampaio; Svetoslav Nanev Slavov; Vagner Fonseca; Vincent Louis Viala |
| EPI_ISL_1795160 | UBS JARDIM ITAMARATY | Instituto Butantan / ESALQ-Piracicaba | Antonio Jorge Martins; Bianca Cechetto Carlos. Mendelics: Bibiana Santos; Claudia Renata dos Santos Barros; David Schlesinger. Hemocentro Ribeirão Preto: Simone Kashima; Debora Botequio Moretti. Centro de Genômica Funcional da ESALQ: Luiz Lehmann Coutinho; Dimas Tadeu Covas; Elaine Cristina Marqueze; Elaine Vieira dos Santos; Elisangela Chicaroni Mattos; Erika Freitas; Evandra Strazza Rodrigues; Felipe Allan da Silva da Costa; Flavia Aburjaile; Guilherme Targino Valente; Heidge Fukumasu. USP-Botucatu: Rejane Maria Tommasini Grotto; Instituto Butantan: Alexander Roberto Precioso; Jayme A. Souza-Neto; Jessika Cristina Chagas Lesbon; José Salvatore Leister Patané; João Paulo Kitajima; Luiz Carlos Junior de Alcantara; Maria Carolina Elias; Marta Giovanetti; Patricia Akemi Assato; Rafael dos Santos Bezerra; Raquel de Lello Rocha Campos Cassano. NGS Soluções Genômicas: Pilar Drummond Sampaio Corrêa Mariani. FZEA-USP Pirassununga: Mirele Daiana Poleti; Raul Machado Neto; Ricardo Augusto Brassaloti; Ricardo Haddad; Rodrigo Tocantins Calado.; Sandra Coccuzzo Sampaio; Svetoslav Nanev Slavov; Vagner Fonseca; Vincent Louis Viala |
| EPI_ISL_2171000 | UBS JARDIM MUNHOZ | Instituto Butantan / Mendelics | Antonio Jorge Martins; Bianca Cechetto Carlos. Mendelics: Bibiana Santos; Claudia Renata dos Santos Barros; Cíntia Bittar; David Schlesinger. Hemocentro Ribeirão Preto: Simone Kashima; Debora Botequio Moretti; Elaine Cristina Marqueze; Elaine Vieira dos Santos; Elisangela Chicaroni Mattos; Erika Freitas; Evandra Strazza Rodrigues; Felipe Allan da Silva da Costa; Flavia Aburjaile; Fábio Sossai Possebon; Guilherme Campos; Guilherme Targino Valente; Heidge Fukumasu. USP-Botucatu: Rejane Maria Tommasini Grotto; Helena Lage Ferreira; Instituto Butantan: Dimas Tadeu Covas; Jardelina de Souza Todao Bernardino; Jayme A. Souza-Neto; Jessika Cristina Chagas Lesbon; Jorge A. Petroli Marchesi; José Salvatore Leister Patané; João Paulo Kitajima; João Pessoa Araújo Jr.; Leila Sabrina Ullmann; Loyze Paola Oliveira de Lima; Luiz Aurelio de Campos Crispin. Centro de Genômica Funcional da ESALQ: Luiz Lehmann Coutinho; Luiz Carlos Junior de Alcantara; Lívia Sacchetto; Maisa C. Pereira Parra; Maria Carolina Elias; Marta Giovanetti; Marília Moraes; Maurício Lacerda Nogueira. Prefeitura de Sao Paulo: Melissa Palmieri.; Patricia Akemi Assato; Paula Rahal; Paulo Inacio da Costa; Rafael dos Santos Bezerra; Raquel de Lello Rocha Campos Cassano. NGS Soluções Genômicas: Pilar Drummond Sampaio Corrêa Mariani. FZEA-USP Pirassununga: Mirele Daiana Poleti; Raul Machado Neto; Ricardo Augusto Brassaloti; Ricardo Haddad; Rodrigo Tocantins Calado. FAMERP-SJRP: Cecília Artico Banho; Sandra Coccuzzo Sampaio; Svetoslav Nanev Slavov; Vagner Fonseca; Vincent Louis Viala |
| EPI_ISL_1966918 | UBS JARDIM VITORINO IPERO | Instituto Butantan / ESALQ-USP (Piracicaba) | Antonio Jorge Martins; Bianca Cechetto Carlos. Mendelics: Bibiana Santos; Claudia Renata dos Santos Barros; Cíntia Bittar; David Schlesinger. Hemocentro Ribeirão Preto: Simone Kashima; Debora Botequio Moretti; Elaine Cristina Marqueze; Elaine Vieira dos Santos; Elisangela Chicaroni Mattos; Erika Freitas; Evandra Strazza Rodrigues; Felipe Allan da Silva da Costa; Flavia Aburjaile; Fábio Sossai Possebon; Guilherme Campos; Guilherme Targino Valente; Heidge Fukumasu. USP-Botucatu: Rejane Maria Tommasini Grotto; Helena Lage Ferreira; Instituto Butantan: Dimas Tadeu Covas; Jardelina de Souza Todao Bernardino; Jayme A. Souza-Neto; Jessika Cristina Chagas Lesbon; Jorge A. Petroli Marchesi; José Salvatore Leister Patané; João Paulo Kitajima; João Pessoa Araújo Jr.; Leila Sabrina Ullmann; Loyze Paola Oliveira de Lima; Luiz Aurelio de Campos Crispin. Centro de Genômica Funcional da ESALQ: Luiz Lehmann Coutinho; Luiz Carlos Junior de Alcantara; Lívia Sacchetto; Maisa C. Pereira Parra; Maria Carolina Elias; Marta Giovanetti; Marília Moraes; Maurício Lacerda Nogueira. Prefeitura de Sao Paulo: Melissa Palmieri.; Patricia Akemi Assato; Paula Rahal; Paulo Inacio da Costa; Rafael dos Santos Bezerra; Raquel de Lello Rocha Campos Cassano. NGS Soluções Genômicas: Pilar Drummond Sampaio Corrêa Mariani. FZEA-USP Pirassununga: Mirele Daiana Poleti; Raul Machado Neto; Ricardo Augusto Brassaloti; Ricardo Haddad; Rodrigo Tocantins Calado. FAMERP-SJRP: Cecília Artico Banho; Sandra Coccuzzo Sampaio; Svetoslav Nanev Slavov; Vagner Fonseca; Vincent Louis Viala |
| EPI_ISL_1445214, EPI_ISL_1966204 | UBS JOSE HILARIO DOS SANTOS | Instituto Butantan / Mendelics | Antonio Jorge Martins; Bianca Cechetto Carlos. Mendelics: Bibiana Santos; Bibiana Santos; Claudia Renata dos Santos Barros; Cíntia Bittar; David Schlesinger; David Schlesinger. Hemocentro Ribeirão Preto: Simone Kashima; Debora Botequio Moretti; Dimas Tadeu Covas; Elaine Cristina Marqueze; Elaine Vieira dos Santos; Elisangela Chicaroni Mattos; Erika Freitas; Evandra Strazza Rodrigues; Felipe Allan da Silva da Costa; Flavia Aburjaile; Fábio Sossai Possebon; Guilherme Campos; Guilherme Targino Valente; Heidge Fukumasu. USP-Botucatu: Rejane Maria Tommasini Grotto; Helena Lage Ferreira; Instituto Butantan: Dimas Tadeu Covas; Jardelina de Souza Todao Bernardino; Jayme A. Souza-Neto; Jessika Cristina Chagas Lesbon; Jorge A. Petroli Marchesi; José Salvatore Leister Patané; João Paulo Kitajima; João Pessoa Araújo Jr.; Leila Sabrina Ullmann; Loyze Paola Oliveira de Lima; Luiz Aurelio de Campos Crispin. Centro de Genômica Funcional da ESALQ: Luiz Lehmann Coutinho; Luiz Carlos Junior de Alcantara; Lívia Sacchetto; Maisa C. Pereira Parra; Maria Carolina Elias; Marta Giovanetti; Marília Moraes; Maurício Lacerda Nogueira. Prefeitura de Sao Paulo: Melissa Palmieri.; Patricia Akemi Assato; Paula Rahal; Paulo Inacio da Costa; Rafael dos Santos Bezerra; Raquel de Lello Rocha Campos Cassano. NGS Soluções Genômicas: Pilar Drummond Sampaio Corrêa Mariani. FZEA-USP Pirassununga: Mirele Daiana Poleti; Raul Machado Neto; Ricardo Augusto Brassaloti; Ricardo Haddad; Rodrigo Tocantins Calado.; Rodrigo Tocantins Calado. FAMERP-SJRP: Cecília Artico Banho; Sandra Coccuzzo Sampaio; Simone Kashima; Svetoslav Nanev Slavov; Vagner Fonseca; Vincent Louis Viala |
| EPI_ISL_1966728, EPI_ISL_1966730 | UBS JOSE SABINO FERREIRA | Instituto Butantan | Antonio Jorge Martins; Bianca Cechetto Carlos. Mendelics: Bibiana Santos; Claudia Renata dos Santos Barros; Cíntia Bittar; David Schlesinger. Hemocentro Ribeirão Preto: Simone Kashima; Debora Botequio Moretti; Elaine Cristina Marqueze; Elaine Vieira dos Santos; Elisangela Chicaroni Mattos; Erika Freitas; Evandra Strazza Rodrigues; Felipe Allan da Silva da Costa; Flavia Aburjaile; Fábio Sossai Possebon; Guilherme Campos; Guilherme Targino Valente; Heidge Fukumasu. USP-Botucatu: Rejane Maria Tommasini Grotto; Helena Lage Ferreira; Instituto Butantan: Dimas Tadeu Covas; Jardelina de Souza Todao Bernardino; Jayme A. Souza-Neto; Jessika Cristina Chagas Lesbon; Jorge A. Petroli Marchesi; José Salvatore Leister Patané; João Paulo Kitajima; João Pessoa Araújo Jr.; Leila Sabrina Ullmann; Loyze Paola Oliveira de Lima; Luiz Aurelio de Campos Crispin. Centro de Genômica Funcional da ESALQ: Luiz Lehmann Coutinho; Luiz Carlos Junior de Alcantara; Lívia Sacchetto; Maisa C. Pereira Parra; Maria Carolina Elias; Marta Giovanetti; Marília Moraes; Maurício Lacerda Nogueira. Prefeitura de Sao Paulo: Melissa Palmieri.; Patricia Akemi Assato; Paula Rahal; Paulo Inacio da Costa; Rafael dos Santos Bezerra; Raquel de Lello Rocha Campos Cassano. NGS Soluções Genômicas: Pilar Drummond Sampaio Corrêa Mariani. FZEA-USP Pirassununga: Mirele Daiana Poleti; Raul Machado Neto; Ricardo Augusto Brassaloti; Ricardo Haddad; Rodrigo Tocantins Calado. FAMERP-SJRP: Cecília Artico Banho; Sandra Coccuzzo Sampaio; Svetoslav Nanev Slavov; Vagner Fonseca; Vincent Louis Viala |
| EPI_ISL_1445210, EPI_ISL_1445211, EPI_ISL_1966202, EPI_ISL_2170914 | UBS JOSE SABINO FERREIRA | Instituto Butantan / Mendelics | Antonio Jorge Martins; Bianca Cechetto Carlos. Mendelics: Bibiana Santos; Bibiana Santos; Claudia Renata dos Santos Barros; Cíntia Bittar; David Schlesinger; David Schlesinger. Hemocentro Ribeirão Preto: Simone Kashima; Debora Botequio Moretti; Dimas Tadeu Covas; Elaine Cristina Marqueze; Elaine Vieira dos Santos; Elisangela Chicaroni Mattos; Erika Freitas; Evandra Strazza Rodrigues; Felipe Allan da Silva da Costa; Flavia Aburjaile; Fábio Sossai Possebon; Guilherme Campos; Guilherme Targino Valente; Heidge Fukumasu. USP-Botucatu: Rejane Maria Tommasini Grotto; Helena Lage Ferreira; Instituto Butantan: Dimas Tadeu Covas; Jardelina de Souza Todao Bernardino; Jayme A. Souza-Neto; Jessika Cristina Chagas Lesbon; Jorge A. Petroli Marchesi; José Salvatore Leister Patané; João Paulo Kitajima; João Pessoa Araújo Jr.; Leila Sabrina Ullmann; Loyze Paola Oliveira de Lima; Luiz Aurelio de Campos Crispin. Centro de Genômica Funcional da ESALQ: Luiz Lehmann Coutinho; Luiz Carlos Junior de Alcantara; Lívia Sacchetto; Maisa C. Pereira Parra; Maria Carolina Elias; Marta Giovanetti; Marília Moraes; Maurício Lacerda Nogueira. Prefeitura de Sao Paulo: Melissa Palmieri.; Patricia Akemi Assato; Paula Rahal; Paulo Inacio da Costa; Rafael dos Santos Bezerra; Raquel de Lello Rocha Campos Cassano. NGS Soluções Genômicas: Pilar Drummond Sampaio Corrêa Mariani. FZEA-USP Pirassununga: Mirele Daiana Poleti; Raul Machado Neto; Ricardo Augusto Brassaloti; Ricardo Haddad; Rodrigo Tocantins Calado.; Rodrigo Tocantins Calado. FAMERP-SJRP: Cecília Artico Banho; Sandra Coccuzzo Sampaio; Simone Kashima; Svetoslav Nanev Slavov; Vagner Fonseca; Vincent Louis Viala |
| EPI_ISL_2170999 | UBS JOVAIA | Instituto Butantan / Mendelics | Antonio Jorge Martins; Bianca Cechetto Carlos. Mendelics: Bibiana Santos; Claudia Renata dos Santos Barros; Cíntia Bittar; David Schlesinger. Hemocentro Ribeirão Preto: Simone Kashima; Debora Botequio Moretti; Elaine Cristina Marqueze; Elaine Vieira dos Santos; Elisangela Chicaroni Mattos; Erika Freitas; Evandra Strazza Rodrigues; Felipe Allan da Silva da Costa; Flavia Aburjaile; Fábio Sossai Possebon; Guilherme Campos; Guilherme Targino Valente; Heidge Fukumasu. USP-Botucatu: Rejane Maria Tommasini Grotto; Helena Lage Ferreira; Instituto Butantan: Dimas Tadeu Covas; Jardelina de Souza Todao Bernardino; Jayme A. Souza-Neto; Jessika Cristina Chagas Lesbon; Jorge A. Petroli Marchesi; José Salvatore Leister Patané; João Paulo Kitajima; João Pessoa Araújo Jr.; Leila Sabrina Ullmann; Loyze Paola Oliveira de Lima; Luiz Aurelio de Campos Crispin. Centro de Genômica Funcional da ESALQ: Luiz Lehmann Coutinho; Luiz Carlos Junior de Alcantara; Lívia Sacchetto; Maisa C. Pereira Parra; Maria Carolina Elias; Marta Giovanetti; Marília Moraes; Maurício Lacerda Nogueira. Prefeitura de Sao Paulo: Melissa Palmieri.; Patricia Akemi Assato; Paula Rahal; Paulo Inacio da Costa; Rafael dos Santos Bezerra; Raquel de Lello Rocha Campos Cassano. NGS Soluções Genômicas: Pilar Drummond Sampaio Corrêa Mariani. FZEA-USP Pirassununga: Mirele Daiana Poleti; Raul Machado Neto; Ricardo Augusto Brassaloti; Ricardo Haddad; Rodrigo Tocantins Calado. FAMERP-SJRP: Cecília Artico Banho; Sandra Coccuzzo Sampaio; Svetoslav Nanev Slavov; Vagner Fonseca; Vincent Louis Viala |
| EPI_ISL_1795266, EPI_ISL_1795267, EPI_ISL_1795268 | UBS LUIS FACHIN IPIGUA | Instituto Butantan / ESALQ-Piracicaba | Antonio Jorge Martins; Bianca Cechetto Carlos. Mendelics: Bibiana Santos; Claudia Renata dos Santos Barros; David Schlesinger. Hemocentro Ribeirão Preto: Simone Kashima; Debora Botequio Moretti. Centro de Genômica Funcional da ESALQ: Luiz Lehmann Coutinho; Dimas Tadeu Covas; Elaine Cristina Marqueze; Elaine Vieira dos Santos; Elisangela Chicaroni Mattos; Erika Freitas; Evandra Strazza Rodrigues; Felipe Allan da Silva da Costa; Flavia Aburjaile; Guilherme Targino Valente; Heidge Fukumasu. USP-Botucatu: Rejane Maria Tommasini Grotto; Instituto Butantan: Alexander Roberto Precioso; Jayme A. Souza-Neto; Jessika Cristina Chagas Lesbon; José Salvatore Leister Patané; João Paulo Kitajima; Luiz Carlos Junior de Alcantara; Maria Carolina Elias; Marta Giovanetti; Patricia Akemi Assato; Rafael dos Santos Bezerra; Raquel de Lello Rocha Campos Cassano. NGS Soluções Genômicas: Pilar Drummond Sampaio Corrêa Mariani. FZEA-USP Pirassununga: Mirele Daiana Poleti; Raul Machado Neto; Ricardo Augusto Brassaloti; Ricardo Haddad; Rodrigo Tocantins Calado.; Sandra Coccuzzo Sampaio; Svetoslav Nanev Slavov; Vagner Fonseca; Vincent Louis Viala |
| EPI_ISL_1795334 | UBS MARCIA CRISTIANE DA SILVA DE OURO VERDE | Instituto Butantan / ESALQ-Piracicaba | Antonio Jorge Martins; Bianca Cechetto Carlos. Mendelics: Bibiana Santos; Claudia Renata dos Santos Barros; David Schlesinger. Hemocentro Ribeirão Preto: Simone Kashima; Debora Botequio Moretti. Centro de Genômica Funcional da ESALQ: Luiz Lehmann Coutinho; Dimas Tadeu Covas; Elaine Cristina Marqueze; Elaine Vieira dos Santos; Elisangela Chicaroni Mattos; Erika Freitas; Evandra Strazza Rodrigues; Felipe Allan da Silva da Costa; Flavia Aburjaile; Guilherme Targino Valente; Heidge Fukumasu. USP-Botucatu: Rejane Maria Tommasini Grotto; Instituto Butantan: Alexander Roberto Precioso; Jayme A. Souza-Neto; Jessika Cristina Chagas Lesbon; José Salvatore Leister Patané; João Paulo Kitajima; Luiz Carlos Junior de Alcantara; Maria Carolina Elias; Marta Giovanetti; Patricia Akemi Assato; Rafael dos Santos Bezerra; Raquel de Lello Rocha Campos Cassano. NGS Soluções Genômicas: Pilar Drummond Sampaio Corrêa Mariani. FZEA-USP Pirassununga: Mirele Daiana Poleti; Raul Machado Neto; Ricardo Augusto Brassaloti; Ricardo Haddad; Rodrigo Tocantins Calado.; Sandra Coccuzzo Sampaio; Svetoslav Nanev Slavov; Vagner Fonseca; Vincent Louis Viala |
| EPI_ISL_1445209, EPI_ISL_1445213, EPI_ISL_1966206, EPI_ISL_1966208 | UBS OTACILIO FIRMINO LOPES | Instituto Butantan / Mendelics | Antonio Jorge Martins; Bianca Cechetto Carlos. Mendelics: Bibiana Santos; Bibiana Santos; Claudia Renata dos Santos Barros; Cíntia Bittar; David Schlesinger; David Schlesinger. Hemocentro Ribeirão Preto: Simone Kashima; Debora Botequio Moretti; Dimas Tadeu Covas; Elaine Cristina Marqueze; Elaine Vieira dos Santos; Elisangela Chicaroni Mattos; Erika Freitas; Evandra Strazza Rodrigues; Felipe Allan da Silva da Costa; Flavia Aburjaile; Fábio Sossai Possebon; Guilherme Campos; Guilherme Targino Valente; Heidge Fukumasu. USP-Botucatu: Rejane Maria Tommasini Grotto; Helena Lage Ferreira; Instituto Butantan: Dimas Tadeu Covas; Jardelina de Souza Todao Bernardino; Jayme A. Souza-Neto; Jessika Cristina Chagas Lesbon; Jorge A. Petroli Marchesi; José Salvatore Leister Patané; João Paulo Kitajima; João Pessoa Araújo Jr.; Leila Sabrina Ullmann; Loyze Paola Oliveira de Lima; Luiz Aurelio de Campos Crispin. Centro de Genômica Funcional da ESALQ: Luiz Lehmann Coutinho; Luiz Carlos Junior de Alcantara; Lívia Sacchetto; Maisa C. Pereira Parra; Maria Carolina Elias; Marta Giovanetti; Marília Moraes; Maurício Lacerda Nogueira. Prefeitura de Sao Paulo: Melissa Palmieri.; Patricia Akemi Assato; Paula Rahal; Paulo Inacio da Costa; Rafael dos Santos Bezerra; Raquel de Lello Rocha Campos Cassano. NGS Soluções Genômicas: Pilar Drummond Sampaio Corrêa Mariani. FZEA-USP Pirassununga: Mirele Daiana Poleti; Raul Machado Neto; Ricardo Augusto Brassaloti; Ricardo Haddad; Rodrigo Tocantins Calado.; Rodrigo Tocantins Calado. FAMERP-SJRP: Cecília Artico Banho; Sandra Coccuzzo Sampaio; Simone Kashima; Svetoslav Nanev Slavov; Vagner Fonseca; Vincent Louis Viala |
| EPI_ISL_1966731, EPI_ISL_1966732, EPI_ISL_1966733, | UBS RAIMUNDA CAVALCANTE DE SOUZA | Instituto Butantan | Antonio Jorge Martins; Bianca Cechetto Carlos. Mendelics: Bibiana Santos; Claudia Renata dos Santos Barros; Cíntia Bittar; David Schlesinger. Hemocentro Ribeirão Preto: Simone Kashima; Debora Botequio Moretti; Elaine Cristina Marqueze; Elaine Vieira dos Santos; Elisangela Chicaroni Mattos; Erika Freitas; Evandra Strazza Rodrigues; Felipe Allan da Silva da Costa; Flavia Aburjaile; Fábio Sossai Possebon; Guilherme Campos; Guilherme Targino Valente; Heidge Fukumasu. USP-Botucatu: Rejane Maria Tommasini Grotto; Helena Lage Ferreira; Instituto Butantan: Dimas Tadeu Covas; Jardelina de Souza Todao Bernardino; Jayme A. Souza-Neto; Jessika Cristina Chagas Lesbon; Jorge A. Petroli Marchesi; José Salvatore Leister Patané; João Paulo Kitajima; João Pessoa Araújo Jr.; Leila Sabrina Ullmann; Loyze Paola Oliveira de Lima; Luiz Aurelio de Campos Crispin. Centro de Genômica Funcional da ESALQ: Luiz Lehmann Coutinho; Luiz Carlos Junior de Alcantara; Lívia Sacchetto; Maisa C. Pereira Parra; Maria |

| | | | |
|---|---|---|---|
| EPI_ISL_1966734 | | | Carolina Elias; Marta Giovanetti; Marília Moraes; Maurício Lacerda Nogueira. Prefeitura de Sao Paulo: Melissa Palmieri.; Patricia Akemi Assato; Paula Rahal; Paulo Inacio da Costa; Rafael dos Santos Bezerra; Raquel de Lello Rocha Campos Cassano. NGS Soluções Genômicas: Pilar Drummond Sampaio Corrêa Mariani. FZEA-USP Pirassununga: Mirele Daiana Poleti; Raul Machado Neto; Ricardo Augusto Brassaloti; Ricardo Haddad; Rodrigo Tocantins Calado. FAMERP-SJRP: Cecília Artico Banho; Sandra Coccuzzo Sampaio; Svetoslav Nanev Slavov; Vagner Fonseca; Vincent Louis Viala |
| EPI_ISL_1967284, EPI_ISL_2170998 | UBS RECREIO SAO JORGE | Instituto Butantan / Mendelics | Antonio Jorge Martins; Bianca Cechetto Carlos. Mendelics: Bibiana Santos; Claudia Renata dos Santos Barros; Cíntia Bittar; David Schlesinger. Hemocentro Ribeirão Preto: Simone Kashima; Debora Botequio Moretti; Elaine Cristina Marqueze; Elaine Vieira dos Santos; Elisangela Chicaroni Mattos; Erika Freitas; Evandra Strazza Rodrigues; Felipe Allan da Silva da Costa; Flavia Aburjaile; Fábio Sossai Possebon; Guilherme Campos; Guilherme Targino Valente; Heidge Fukumasu. USP-Botucatu: Rejane Maria Tommasini Grotto; Helena Lage Ferreira; Instituto Butantan: Dimas Tadeu Covas; Jardelina de Souza Todao Bernardino; Jayme A. Souza-Neto; Jessika Cristina Chagas Lesbon; Jorge A. Petroli Marchesi; José Salvatore Leister Patané; João Paulo Kitajima; João Pessoa Araújo Jr.; Leila Sabrina Ullmann; Loyze Paola Oliveira de Lima; Luiz Aurelio de Campos Crispin. Centro de Genômica Funcional da ESALQ: Luiz Lehmann Coutinho; Luiz Carlos Junior de Alcantara; Lívia Sacchetto; Maisa C. Pereira Parra; Maria Carolina Elias; Marta Giovanetti; Marília Moraes; Maurício Lacerda Nogueira. Prefeitura de Sao Paulo: Melissa Palmieri.; Patricia Akemi Assato; Paula Rahal; Paulo Inacio da Costa; Rafael dos Santos Bezerra; Raquel de Lello Rocha Campos Cassano. NGS Soluções Genômicas: Pilar Drummond Sampaio Corrêa Mariani. FZEA-USP Pirassununga: Mirele Daiana Poleti; Raul Machado Neto; Ricardo Augusto Brassaloti; Ricardo Haddad; Rodrigo Tocantins Calado. FAMERP-SJRP: Cecília Artico Banho; Sandra Coccuzzo Sampaio; Svetoslav Nanev Slavov; Vagner Fonseca; Vincent Louis Viala |
| EPI_ISL_1966535 | UBS REGIAO OESTE DE OURINHOS | Instituto Butantan / FZEA-USP (Pirassununga) | Antonio Jorge Martins; Bianca Cechetto Carlos. Mendelics: Bibiana Santos; Claudia Renata dos Santos Barros; Cíntia Bittar; David Schlesinger. Hemocentro Ribeirão Preto: Simone Kashima; Debora Botequio Moretti; Elaine Cristina Marqueze; Elaine Vieira dos Santos; Elisangela Chicaroni Mattos; Erika Freitas; Evandra Strazza Rodrigues; Felipe Allan da Silva da Costa; Flavia Aburjaile; Fábio Sossai Possebon; Guilherme Campos; Guilherme Targino Valente; Heidge Fukumasu. USP-Botucatu: Rejane Maria Tommasini Grotto; Helena Lage Ferreira; Instituto Butantan: Dimas Tadeu Covas; Jardelina de Souza Todao Bernardino; Jayme A. Souza-Neto; Jessika Cristina Chagas Lesbon; Jorge A. Petroli Marchesi; José Salvatore Leister Patané; João Paulo Kitajima; João Pessoa Araújo Jr.; Leila Sabrina Ullmann; Loyze Paola Oliveira de Lima; Luiz Aurelio de Campos Crispin. Centro de Genômica Funcional da ESALQ: Luiz Lehmann Coutinho; Luiz Carlos Junior de Alcantara; Lívia Sacchetto; Maisa C. Pereira Parra; Maria Carolina Elias; Marta Giovanetti; Marília Moraes; Maurício Lacerda Nogueira. Prefeitura de Sao Paulo: Melissa Palmieri.; Patricia Akemi Assato; Paula Rahal; Paulo Inacio da Costa; Rafael dos Santos Bezerra; Raquel de Lello Rocha Campos Cassano. NGS Soluções Genômicas: Pilar Drummond Sampaio Corrêa Mariani. FZEA-USP Pirassununga: Mirele Daiana Poleti; Raul Machado Neto; Ricardo Augusto Brassaloti; Ricardo Haddad; Rodrigo Tocantins Calado. FAMERP-SJRP: Cecília Artico Banho; Sandra Coccuzzo Sampaio; Svetoslav Nanev Slavov; Vagner Fonseca; Vincent Louis Viala |
| EPI_ISL_1445141, EPI_ISL_1966188 | UBS SALTO DE SAO JOSE | Instituto Butantan / Mendelics | Antonio Jorge Martins; Bianca Cechetto Carlos. Mendelics: Bibiana Santos; Bibiana Santos; Claudia Renata dos Santos Barros; Cíntia Bittar; David Schlesinger; David Schlesinger. Hemocentro Ribeirão Preto: Simone Kashima; Debora Botequio Moretti; Dimas Tadeu Covas; Elaine Cristina Marqueze; Elaine Vieira dos Santos; Elisangela Chicaroni Mattos; Erika Freitas; Evandra Strazza Rodrigues; Felipe Allan da Silva da Costa; Flavia Aburjaile; Fábio Sossai Possebon; Guilherme Campos; Guilherme Targino Valente; Heidge Fukumasu. USP-Botucatu: Rejane Maria Tommasini Grotto; Helena Lage Ferreira; Instituto Butantan: Dimas Tadeu Covas; Jardelina de Souza Todao Bernardino; Jayme A. Souza-Neto; Jessika Cristina Chagas Lesbon; Jorge A. Petroli Marchesi; José Salvatore Leister Patané; João Paulo Kitajima; João Pessoa Araújo Jr.; Leila Sabrina Ullmann; Loyze Paola Oliveira de Lima; Luiz Aurelio de Campos Crispin. Centro de Genômica Funcional da ESALQ: Luiz Lehmann Coutinho; Luiz Carlos Junior de Alcantara; Lívia Sacchetto; Maisa C. Pereira Parra; Maria Carolina Elias; Marta Giovanetti; Marília Moraes; Maurício Lacerda Nogueira. Prefeitura de Sao Paulo: Melissa Palmieri.; Patricia Akemi Assato; Paula Rahal; Paulo Inacio da Costa; Rafael dos Santos Bezerra; Raquel de Lello Rocha Campos Cassano. NGS Soluções Genômicas: Pilar Drummond Sampaio Corrêa Mariani. FZEA-USP Pirassununga: Mirele Daiana Poleti; Raul Machado Neto; Ricardo Augusto Brassaloti; Ricardo Haddad; Rodrigo Tocantins Calado.; Rodrigo Tocantins Calado. FAMERP-SJRP: Cecília Artico Banho; Sandra Coccuzzo Sampaio; Simone Kashima; Svetoslav Nanev Slavov; Vagner Fonseca; Vincent Louis Viala |
| EPI_ISL_1966847, EPI_ISL_1966848, EPI_ISL_1966849, EPI_ISL_1966850, EPI_ISL_1966851, EPI_ISL_1966852 | UBS SAO JUDAS | Instituto Butantan / ESALQ-USP (Piracicaba) | Antonio Jorge Martins; Bianca Cechetto Carlos. Mendelics: Bibiana Santos; Claudia Renata dos Santos Barros; Cíntia Bittar; David Schlesinger. Hemocentro Ribeirão Preto: Simone Kashima; Debora Botequio Moretti; Elaine Cristina Marqueze; Elaine Vieira dos Santos; Elisangela Chicaroni Mattos; Erika Freitas; Evandra Strazza Rodrigues; Felipe Allan da Silva da Costa; Flavia Aburjaile; Fábio Sossai Possebon; Guilherme Campos; Guilherme Targino Valente; Heidge Fukumasu. USP-Botucatu: Rejane Maria Tommasini Grotto; Helena Lage Ferreira; Instituto Butantan: Dimas Tadeu Covas; Jardelina de Souza Todao Bernardino; Jayme A. Souza-Neto; Jessika Cristina Chagas Lesbon; Jorge A. Petroli Marchesi; José Salvatore Leister Patané; João Paulo Kitajima; João Pessoa Araújo Jr.; Leila Sabrina Ullmann; Loyze Paola Oliveira de Lima; Luiz Aurelio de Campos Crispin. Centro de Genômica Funcional da ESALQ: Luiz Lehmann Coutinho; Luiz Carlos Junior de Alcantara; Lívia Sacchetto; Maisa C. Pereira Parra; Maria Carolina Elias; Marta Giovanetti; Marília Moraes; Maurício Lacerda Nogueira. Prefeitura de Sao Paulo: Melissa Palmieri.; Patricia Akemi Assato; Paula Rahal; Paulo Inacio da Costa; Rafael dos Santos Bezerra; Raquel de Lello Rocha Campos Cassano. NGS Soluções Genômicas: Pilar Drummond Sampaio Corrêa Mariani. FZEA-USP Pirassununga: Mirele Daiana Poleti; Raul Machado Neto; Ricardo Augusto Brassaloti; Ricardo Haddad; Rodrigo Tocantins Calado. FAMERP-SJRP: Cecília Artico Banho; Sandra Coccuzzo Sampaio; Svetoslav Nanev Slavov; Vagner Fonseca; Vincent Louis Viala |
| EPI_ISL_1445196, EPI_ISL_1966227 | UBS SAO JUDAS | Instituto Butantan / Mendelics | Antonio Jorge Martins; Bianca Cechetto Carlos. Mendelics: Bibiana Santos; Bibiana Santos; Claudia Renata dos Santos Barros; Cíntia Bittar; David Schlesinger; David Schlesinger. Hemocentro Ribeirão Preto: Simone Kashima; Debora Botequio Moretti; Dimas Tadeu Covas; Elaine Cristina Marqueze; Elaine Vieira dos Santos; Elisangela Chicaroni Mattos; Erika Freitas; Evandra Strazza Rodrigues; Felipe Allan da Silva da Costa; Flavia Aburjaile; Fábio Sossai Possebon; Guilherme Campos; Guilherme Targino Valente; Heidge Fukumasu. USP-Botucatu: Rejane Maria Tommasini Grotto; Helena Lage Ferreira; Instituto Butantan: Dimas Tadeu Covas; Jardelina de Souza Todao Bernardino; Jayme A. Souza-Neto; Jessika Cristina Chagas Lesbon; Jorge A. Petroli Marchesi; José Salvatore Leister Patané; João Paulo Kitajima; João Pessoa Araújo Jr.; Leila Sabrina Ullmann; Loyze Paola Oliveira de Lima; Luiz Aurelio de Campos Crispin. Centro de Genômica Funcional da ESALQ: Luiz Lehmann Coutinho; Luiz Carlos Junior de Alcantara; Lívia Sacchetto; Maisa C. Pereira Parra; Maria Carolina Elias; Marta Giovanetti; Marília Moraes; Maurício Lacerda Nogueira. Prefeitura de Sao Paulo: Melissa Palmieri.; Patricia Akemi Assato; Paula Rahal; Paulo Inacio da Costa; Rafael dos Santos Bezerra; Raquel de Lello Rocha Campos Cassano. NGS Soluções Genômicas: Pilar Drummond Sampaio Corrêa Mariani. FZEA-USP Pirassununga: Mirele Daiana Poleti; Raul Machado Neto; Ricardo Augusto Brassaloti; Ricardo Haddad; Rodrigo Tocantins Calado.; Rodrigo Tocantins Calado. FAMERP-SJRP: Cecília Artico Banho; Sandra Coccuzzo Sampaio; Simone Kashima; Svetoslav Nanev Slavov; Vagner Fonseca; Vincent Louis Viala |
| EPI_ISL_1445218, EPI_ISL_1445226, EPI_ISL_1966212, EPI_ISL_1966229 | UBS SYLVIO JOAO L DE LUCIA | Instituto Butantan / Mendelics | Antonio Jorge Martins; Bianca Cechetto Carlos. Mendelics: Bibiana Santos; Bibiana Santos; Claudia Renata dos Santos Barros; Cíntia Bittar; David Schlesinger; David Schlesinger. Hemocentro Ribeirão Preto: Simone Kashima; Debora Botequio Moretti; Dimas Tadeu Covas; Elaine Cristina Marqueze; Elaine Vieira dos Santos; Elisangela Chicaroni Mattos; Erika Freitas; Evandra Strazza Rodrigues; Felipe Allan da Silva da Costa; Flavia Aburjaile; Fábio Sossai Possebon; Guilherme Campos; Guilherme Targino Valente; Heidge Fukumasu. USP-Botucatu: Rejane Maria Tommasini Grotto; Helena Lage Ferreira; Instituto Butantan: Dimas Tadeu Covas; Jardelina de Souza Todao Bernardino; Jayme A. Souza-Neto; Jessika Cristina Chagas Lesbon; Jorge A. Petroli Marchesi; José Salvatore Leister Patané; João Paulo Kitajima; João Pessoa Araújo Jr.; Leila Sabrina Ullmann; Loyze Paola Oliveira de Lima; Luiz Aurelio de Campos Crispin. Centro de Genômica Funcional da ESALQ: Luiz Lehmann Coutinho; Luiz Carlos Junior de Alcantara; Lívia Sacchetto; Maisa C. Pereira Parra; Maria Carolina Elias; Marta Giovanetti; Marília Moraes; Maurício Lacerda Nogueira. Prefeitura de Sao Paulo: Melissa Palmieri.; Patricia Akemi Assato; Paula Rahal; Paulo Inacio da Costa; Rafael dos Santos Bezerra; Raquel de Lello Rocha Campos Cassano. NGS Soluções Genômicas: Pilar Drummond Sampaio Corrêa Mariani. FZEA-USP Pirassununga: Mirele Daiana Poleti; Raul Machado Neto; Ricardo Augusto Brassaloti; Ricardo Haddad; Rodrigo Tocantins Calado.; Rodrigo Tocantins Calado. FAMERP-SJRP: Cecília Artico Banho; Sandra Coccuzzo Sampaio; Simone Kashima; Svetoslav Nanev Slavov; Vagner Fonseca; Vincent Louis Viala |
| EPI_ISL_1795269, EPI_ISL_1795270, EPI_ISL_1795272, EPI_ISL_1795273 | UBS TEREZA GALLO UCHOA | Instituto Butantan / ESALQ-Piracicaba | Antonio Jorge Martins; Bianca Cechetto Carlos. Mendelics: Bibiana Santos; Claudia Renata dos Santos Barros; David Schlesinger. Hemocentro Ribeirão Preto: Simone Kashima; Debora Botequio Moretti. Centro de Genômica Funcional da ESALQ: Luiz Lehmann Coutinho; Dimas Tadeu Covas; Elaine Cristina Marqueze; Elaine Vieira dos Santos; Elisangela Chicaroni Mattos; Erika Freitas; Evandra Strazza Rodrigues; Felipe Allan da Silva da Costa; Flavia Aburjaile; Guilherme Targino Valente; Heidge Fukumasu. USP-Botucatu: Rejane Maria Tommasini Grotto; Instituto Butantan: Alexander Roberto Precioso; Jayme A. Souza-Neto; Jessika Cristina Chagas Lesbon; José Salvatore Leister Patané; João Paulo Kitajima; Luiz Carlos Junior de Alcantara; Maria Carolina Elias; Marta Giovanetti; Patricia Akemi Assato; Rafael dos Santos Bezerra; Raquel de Lello Rocha Campos Cassano. NGS Soluções Genômicas: Pilar Drummond Sampaio Corrêa Mariani. FZEA-USP Pirassununga: Mirele Daiana Poleti; Raul Machado Neto; Ricardo Augusto Brassaloti; Ricardo Haddad; Rodrigo Tocantins Calado.; Sandra Coccuzzo Sampaio; Svetoslav Nanev Slavov; Vagner Fonseca; Vincent Louis Viala |
| EPI_ISL_1967287 | UBS VILA GALVAO | Instituto Butantan / Mendelics | Antonio Jorge Martins; Bianca Cechetto Carlos. Mendelics: Bibiana Santos; Claudia Renata dos Santos Barros; Cíntia Bittar; David Schlesinger. Hemocentro Ribeirão Preto: Simone Kashima; Debora Botequio Moretti; Elaine Cristina Marqueze; Elaine Vieira dos Santos; Elisangela Chicaroni Mattos; Erika Freitas; Evandra Strazza Rodrigues; Felipe Allan da Silva da Costa; Flavia Aburjaile; Fábio Sossai Possebon; Guilherme Campos; Guilherme Targino Valente; Heidge Fukumasu. USP-Botucatu: Rejane Maria Tommasini Grotto; Helena Lage Ferreira; Instituto Butantan: Dimas Tadeu Covas; Jardelina de Souza Todao Bernardino; Jayme A. Souza-Neto; Jessika Cristina Chagas Lesbon; Jorge A. Petroli Marchesi; José Salvatore Leister Patané; João Paulo Kitajima; João Pessoa Araújo Jr.; Leila Sabrina Ullmann; Loyze Paola Oliveira de Lima; Luiz Aurelio de Campos Crispin. Centro de Genômica Funcional da ESALQ: Luiz Lehmann Coutinho; Luiz Carlos Junior de Alcantara; Lívia Sacchetto; Maisa C. Pereira Parra; Maria Carolina Elias; Marta Giovanetti; Marília Moraes; Maurício Lacerda Nogueira. Prefeitura de Sao Paulo: Melissa Palmieri.; Patricia Akemi Assato; Paula Rahal; Paulo Inacio da Costa; Rafael dos Santos Bezerra; Raquel de Lello Rocha Campos Cassano. NGS Soluções Genômicas: Pilar Drummond Sampaio Corrêa Mariani. FZEA-USP Pirassununga: Mirele Daiana Poleti; Raul Machado Neto; Ricardo Augusto Brassaloti; Ricardo Haddad; Rodrigo Tocantins Calado. FAMERP-SJRP: Cecília Artico Banho; Sandra Coccuzzo Sampaio; Svetoslav Nanev Slavov; Vagner Fonseca; Vincent Louis Viala |
| EPI_ISL_1445183, EPI_ISL_1445184, EPI_ISL_1966201, EPI_ISL_1966290, EPI_ISL_1966291, EPI_ISL_2170911 | UNIDADE BASICA DE SAUDE DA FAMILIA BILAC | Instituto Butantan / Mendelics | Antonio Jorge Martins; Bianca Cechetto Carlos. Mendelics: Bibiana Santos; Bibiana Santos; Claudia Renata dos Santos Barros; Cíntia Bittar; David Schlesinger; David Schlesinger. Hemocentro Ribeirão Preto: Simone Kashima; Debora Botequio Moretti; Dimas Tadeu Covas; Elaine Cristina Marqueze; Elaine Vieira dos Santos; Elisangela Chicaroni Mattos; Erika Freitas; Evandra Strazza Rodrigues; Felipe Allan da Silva da Costa; Flavia Aburjaile; Fábio Sossai Possebon; Guilherme Campos; Guilherme Targino Valente; Heidge Fukumasu. USP-Botucatu: Rejane Maria Tommasini Grotto; Helena Lage Ferreira; Instituto Butantan: Dimas Tadeu Covas; Jardelina de Souza Todao Bernardino; Jayme A. Souza-Neto; Jessika Cristina Chagas Lesbon; Jorge A. Petroli Marchesi; José Salvatore Leister Patané; João Paulo Kitajima; João Pessoa Araújo Jr.; Leila Sabrina Ullmann; Loyze Paola Oliveira de Lima; Luiz Aurelio de Campos Crispin. Centro de Genômica Funcional da ESALQ: Luiz Lehmann Coutinho; Luiz Carlos Junior de Alcantara; Lívia Sacchetto; Maisa C. Pereira Parra; Maria Carolina Elias; Marta Giovanetti; Marília Moraes; Maurício Lacerda Nogueira. Prefeitura de Sao Paulo: Melissa Palmieri.; Patricia Akemi Assato; Paula Rahal; Paulo Inacio da Costa; Rafael dos Santos Bezerra; Raquel de Lello Rocha Campos Cassano. NGS Soluções Genômicas: Pilar Drummond Sampaio Corrêa Mariani. FZEA-USP Pirassununga: Mirele Daiana Poleti; Raul Machado Neto; Ricardo Augusto Brassaloti; Ricardo Haddad; Rodrigo Tocantins Calado.; Rodrigo Tocantins Calado. FAMERP-SJRP: Cecília Artico Banho; Sandra Coccuzzo Sampaio; Simone Kashima; Svetoslav Nanev Slavov; Vagner Fonseca; Vincent Louis Viala |
| EPI_ISL_1795362, EPI_ISL_1795363 | UNIDADE BASICA DE SAUDE DE PIQUETE | Instituto Butantan / ESALQ-Piracicaba | Antonio Jorge Martins; Bianca Cechetto Carlos. Mendelics: Bibiana Santos; Claudia Renata dos Santos Barros; David Schlesinger. Hemocentro Ribeirão Preto: Simone Kashima; Debora Botequio Moretti. Centro de Genômica Funcional da ESALQ: Luiz Lehmann Coutinho; Dimas Tadeu Covas; Elaine Cristina Marqueze; Elaine Vieira dos Santos; Elisangela Chicaroni Mattos; Erika Freitas; Evandra Strazza Rodrigues; Felipe Allan da Silva da Costa; Flavia Aburjaile; Guilherme Targino Valente; Heidge Fukumasu. USP-Botucatu: Rejane Maria Tommasini Grotto; Instituto Butantan: Alexander Roberto Precioso; Jayme A. Souza-Neto; Jessika Cristina Chagas Lesbon; José Salvatore Leister Patané; João Paulo Kitajima; Luiz Carlos Junior de Alcantara; Maria Carolina Elias; Marta Giovanetti; Patricia Akemi Assato; Rafael dos Santos Bezerra; Raquel de Lello Rocha Campos Cassano. NGS Soluções Genômicas: Pilar Drummond Sampaio Corrêa Mariani. FZEA-USP Pirassununga: Mirele Daiana Poleti; Raul Machado Neto; Ricardo Augusto Brassaloti; Ricardo Haddad; Rodrigo Tocantins Calado.; Sandra Coccuzzo Sampaio; Svetoslav Nanev Slavov; Vagner Fonseca; Vincent Louis Viala |
| EPI_ISL_1966513, EPI_ISL_1966514, EPI_ISL_1966515, EPI_ISL_1966516, EPI_ISL_1967280 | UNIDADE BASICA DE SAUDE DE TAQUARAL CAETANO PITELLI | Instituto Butantan / Mendelics | Antonio Jorge Martins; Bianca Cechetto Carlos. Mendelics: Bibiana Santos; Claudia Renata dos Santos Barros; Cíntia Bittar; David Schlesinger. Hemocentro Ribeirão Preto: Simone Kashima; Debora Botequio Moretti; Elaine Cristina Marqueze; Elaine Vieira dos Santos; Elisangela Chicaroni Mattos; Erika Freitas; Evandra Strazza Rodrigues; Felipe Allan da Silva da Costa; Flavia Aburjaile; Fábio Sossai Possebon; Guilherme Campos; Guilherme Targino Valente; Heidge Fukumasu. USP-Botucatu: Rejane Maria Tommasini Grotto; Helena Lage Ferreira; Instituto Butantan: Dimas Tadeu Covas; Jardelina de Souza Todao Bernardino; Jayme A. Souza-Neto; Jessika Cristina Chagas Lesbon; Jorge A. Petroli Marchesi; José Salvatore Leister Patané; João Paulo Kitajima; João Pessoa Araújo Jr.; Leila Sabrina Ullmann; Loyze Paola Oliveira de Lima; Luiz Aurelio de Campos Crispin. Centro de Genômica Funcional da ESALQ: Luiz Lehmann Coutinho; Luiz Carlos Junior de Alcantara; Lívia Sacchetto; Maisa C. Pereira Parra; Maria Carolina Elias; Marta Giovanetti; Marília Moraes; Maurício Lacerda Nogueira. Prefeitura de Sao Paulo: Melissa Palmieri.; Patricia Akemi Assato; Paula Rahal; Paulo Inacio da Costa; Rafael dos Santos Bezerra; Raquel de Lello Rocha Campos Cassano. NGS Soluções Genômicas: Pilar Drummond Sampaio Corrêa Mariani. FZEA-USP Pirassununga: Mirele Daiana Poleti; Raul Machado Neto; Ricardo Augusto Brassaloti; Ricardo Haddad; Rodrigo Tocantins Calado. FAMERP-SJRP: Cecília Artico Banho; Sandra Coccuzzo Sampaio; Svetoslav Nanev Slavov; Vagner Fonseca; Vincent Louis Viala |
| EPI_ISL_1966817 | UNIDADE BASICA DE SAUDE DE TARUMA | Instituto Butantan / Mendelics | Antonio Jorge Martins; Bianca Cechetto Carlos. Mendelics: Bibiana Santos; Claudia Renata dos Santos Barros; Cíntia Bittar; David Schlesinger. Hemocentro Ribeirão Preto: Simone Kashima; Debora Botequio Moretti; Elaine Cristina Marqueze; Elaine Vieira dos Santos; Elisangela Chicaroni Mattos; Erika Freitas; Evandra Strazza Rodrigues; Felipe Allan da Silva da Costa; Flavia Aburjaile; Fábio Sossai Possebon; Guilherme Campos; Guilherme Targino Valente; Heidge Fukumasu. USP-Botucatu: Rejane Maria Tommasini Grotto; Helena Lage Ferreira; Instituto Butantan: Dimas Tadeu Covas; Jardelina de Souza Todao Bernardino; Jayme A. Souza-Neto; Jessika Cristina Chagas Lesbon; Jorge A. Petroli Marchesi; José Salvatore Leister Patané; João Paulo Kitajima; João Pessoa Araújo Jr.; Leila Sabrina Ullmann; Loyze Paola Oliveira de Lima; Luiz Aurelio de Campos Crispin. Centro de Genômica Funcional da ESALQ: Luiz Lehmann Coutinho; Luiz Carlos Junior de Alcantara; Lívia Sacchetto; Maisa C. Pereira Parra; Maria Carolina Elias; Marta Giovanetti; Marília Moraes; Maurício Lacerda Nogueira. Prefeitura de Sao Paulo: Melissa Palmieri.; Patricia Akemi Assato; Paula Rahal; Paulo Inacio da Costa; Rafael dos Santos Bezerra; Raquel de Lello Rocha Campos Cassano. NGS Soluções Genômicas: Pilar Drummond Sampaio Corrêa Mariani. FZEA-USP Pirassununga: Mirele Daiana Poleti; Raul Machado Neto; Ricardo Augusto Brassaloti; Ricardo Haddad; Rodrigo Tocantins Calado. FAMERP-SJRP: Cecília Artico Banho; Sandra Coccuzzo Sampaio; Svetoslav Nanev Slavov; Vagner Fonseca; Vincent Louis Viala |
| EPI_ISL_1795224 | UNIDADE BASICA DE SAUDE DR APOLONIO MORAES E SOUZA | Instituto Butantan / ESALQ-Piracicaba | Antonio Jorge Martins; Bianca Cechetto Carlos. Mendelics: Bibiana Santos; Claudia Renata dos Santos Barros; David Schlesinger. Hemocentro Ribeirão Preto: Simone Kashima; Debora Botequio Moretti. Centro de Genômica Funcional da ESALQ: Luiz Lehmann Coutinho; Dimas Tadeu Covas; Elaine Cristina Marqueze; Elaine Vieira dos Santos; Elisangela Chicaroni Mattos; Erika Freitas; Evandra Strazza Rodrigues; Felipe Allan da Silva da Costa; Flavia Aburjaile; Guilherme Targino Valente; Heidge Fukumasu. USP-Botucatu: Rejane Maria Tommasini Grotto; Instituto Butantan: Alexander Roberto Precioso; Jayme A. Souza-Neto; Jessika Cristina Chagas Lesbon; José Salvatore Leister Patané; João Paulo Kitajima; Luiz Carlos Junior de Alcantara; Maria Carolina Elias; Marta Giovanetti; Patricia Akemi Assato; Rafael dos Santos Bezerra; Raquel de Lello Rocha Campos Cassano. NGS Soluções Genômicas: Pilar Drummond Sampaio Corrêa Mariani. FZEA-USP Pirassununga: Mirele Daiana Poleti; Raul Machado Neto; Ricardo Augusto Brassaloti; Ricardo Haddad; Rodrigo Tocantins Calado.; Sandra Coccuzzo Sampaio; Svetoslav Nanev Slavov; Vagner Fonseca; Vincent Louis Viala |
| EPI_ISL_1966288 | UNIDADE BASICA DE SAUDE DR BARTOLOMEU MARAGLIANO VENERE | Instituto Butantan / Mendelics | Antonio Jorge Martins; Bianca Cechetto Carlos. Mendelics: Bibiana Santos; Claudia Renata dos Santos Barros; Cíntia Bittar; David Schlesinger. Hemocentro Ribeirão Preto: Simone Kashima; Debora Botequio Moretti; Elaine Cristina Marqueze; Elaine Vieira dos Santos; Elisangela Chicaroni Mattos; Erika Freitas; Evandra Strazza Rodrigues; Felipe Allan da Silva da Costa; Flavia Aburjaile; Fábio Sossai Possebon; Guilherme Campos; Guilherme Targino Valente; Heidge Fukumasu. USP-Botucatu: Rejane Maria Tommasini Grotto; Helena Lage Ferreira; Instituto Butantan: Dimas Tadeu Covas; Jardelina de Souza Todao Bernardino; Jayme A. Souza-Neto; Jessika Cristina Chagas Lesbon; Jorge A. Petroli Marchesi; José Salvatore Leister Patané; João Paulo Kitajima; João Pessoa Araújo Jr.; Leila Sabrina Ullmann; Loyze Paola Oliveira de Lima; Luiz Aurelio de Campos Crispin. Centro de Genômica Funcional da ESALQ: Luiz Lehmann Coutinho; Luiz Carlos Junior de Alcantara; Lívia Sacchetto; Maisa C. Pereira Parra; Maria Carolina Elias; Marta Giovanetti; Marília Moraes; Maurício Lacerda Nogueira. Prefeitura de Sao Paulo: Melissa Palmieri.; Patricia Akemi Assato; Paula Rahal; Paulo Inacio da Costa; Rafael dos Santos Bezerra; Raquel de Lello Rocha Campos Cassano. NGS Soluções Genômicas: Pilar Drummond Sampaio Corrêa Mariani. FZEA-USP Pirassununga: Mirele Daiana Poleti; Raul Machado Neto; Ricardo Augusto Brassaloti; Ricardo Haddad; Rodrigo Tocantins Calado. FAMERP-SJRP: Cecília Artico Banho; Sandra Coccuzzo Sampaio; Svetoslav Nanev Slavov; Vagner Fonseca; Vincent Louis Viala |
| EPI_ISL_1795227 | UNIDADE BASICA DE SAUDE DR LOTFALLAH MIZIARA | Instituto Butantan / ESALQ-Piracicaba | Antonio Jorge Martins; Bianca Cechetto Carlos. Mendelics: Bibiana Santos; Claudia Renata dos Santos Barros; David Schlesinger. Hemocentro Ribeirão Preto: Simone Kashima; Debora Botequio Moretti. Centro de Genômica Funcional da ESALQ: Luiz Lehmann Coutinho; Dimas Tadeu Covas; Elaine Cristina Marqueze; Elaine Vieira dos Santos; Elisangela Chicaroni Mattos; Erika Freitas; Evandra Strazza Rodrigues; Felipe Allan da Silva da Costa; Flavia Aburjaile; Guilherme Targino Valente; Heidge Fukumasu. USP-Botucatu: Rejane Maria Tommasini Grotto; Instituto Butantan: Alexander Roberto Precioso; Jayme A. Souza-Neto; Jessika Cristina Chagas Lesbon; José Salvatore Leister Patané; João Paulo Kitajima; Luiz Carlos Junior de Alcantara; Maria Carolina Elias; Marta Giovanetti; Patricia Akemi Assato; Rafael dos Santos Bezerra; Raquel de Lello Rocha Campos Cassano. NGS Soluções Genômicas: Pilar Drummond Sampaio Corrêa Mariani. FZEA-USP Pirassununga: Mirele Daiana Poleti; Raul Machado Neto; Ricardo Augusto Brassaloti; Ricardo Haddad; Rodrigo Tocantins Calado.; Sandra Coccuzzo Sampaio; Svetoslav Nanev Slavov; Vagner Fonseca; Vincent Louis Viala |
| EPI_ISL_1966245 | UNIDADE BASICA DE SAUDE DR ORESTES DE MOURA PINTO | Instituto Butantan / Mendelics | Antonio Jorge Martins; Bianca Cechetto Carlos. Mendelics: Bibiana Santos; Claudia Renata dos Santos Barros; Cíntia Bittar; David Schlesinger. Hemocentro Ribeirão Preto: Simone Kashima; Debora Botequio Moretti; Elaine Cristina Marqueze; Elaine Vieira dos Santos; Elisangela Chicaroni Mattos; Erika Freitas; Evandra Strazza Rodrigues; Felipe Allan da Silva da Costa; Flavia Aburjaile; Fábio Sossai Possebon; Guilherme Campos; Guilherme Targino Valente; Heidge Fukumasu. USP-Botucatu: Rejane Maria Tommasini Grotto; Helena Lage Ferreira; Instituto Butantan: Dimas Tadeu Covas; Jardelina de Souza Todao Bernardino; Jayme A. Souza-Neto; Jessika Cristina Chagas Lesbon; Jorge A. Petroli Marchesi; José Salvatore Leister Patané; João Paulo Kitajima; João Pessoa Araújo Jr.; Leila Sabrina Ullmann; Loyze Paola Oliveira de Lima; Luiz Aurelio de Campos Crispin. Centro de Genômica Funcional da ESALQ: Luiz Lehmann Coutinho; Luiz Carlos Junior de Alcantara; Lívia Sacchetto; Maisa C. Pereira Parra; Maria Carolina Elias; Marta Giovanetti; Marília Moraes; Maurício Lacerda Nogueira. Prefeitura de Sao Paulo: Melissa Palmieri.; Patricia Akemi Assato; Paula Rahal; Paulo Inacio da Costa; Rafael dos Santos Bezerra; Raquel de Lello Rocha Campos Cassano. NGS Soluções Genômicas: Pilar Drummond Sampaio Corrêa Mariani. FZEA-USP Pirassununga: Mirele Daiana Poleti; Raul Machado Neto; Ricardo Augusto Brassaloti; Ricardo Haddad; Rodrigo Tocantins Calado. FAMERP-SJRP: Cecília Artico Banho; Sandra Coccuzzo Sampaio; Svetoslav Nanev Slavov; Vagner Fonseca; Vincent Louis Viala |
| EPI_ISL_1795225 | UNIDADE BASICA DE SAUDE DR WILSON HAYEK SAIGH | Instituto Butantan / ESALQ-Piracicaba | Antonio Jorge Martins; Bianca Cechetto Carlos. Mendelics: Bibiana Santos; Claudia Renata dos Santos Barros; David Schlesinger. Hemocentro Ribeirão Preto: Simone Kashima; Debora Botequio Moretti. Centro de Genômica Funcional da ESALQ: Luiz Lehmann Coutinho; Dimas Tadeu Covas; Elaine Cristina Marqueze; Elaine Vieira dos Santos; Elisangela Chicaroni Mattos; Erika Freitas; Evandra Strazza Rodrigues; Felipe Allan da Silva da Costa; Flavia Aburjaile; Guilherme Targino Valente; Heidge Fukumasu. USP-Botucatu: Rejane Maria Tommasini Grotto; Instituto Butantan: Alexander Roberto Precioso; Jayme A. Souza-Neto; Jessika Cristina Chagas Lesbon; José Salvatore Leister Patané; João Paulo Kitajima; Luiz Carlos Junior de Alcantara; Maria Carolina Elias; Marta Giovanetti; Patricia Akemi Assato; Rafael dos Santos Bezerra; Raquel de Lello Rocha Campos Cassano. NGS Soluções Genômicas: Pilar Drummond Sampaio Corrêa Mariani. FZEA-USP Pirassununga: Mirele Daiana Poleti; Raul Machado Neto; Ricardo Augusto Brassaloti; Ricardo Haddad; Rodrigo Tocantins Calado.; Sandra Coccuzzo Sampaio; Svetoslav Nanev Slavov; Vagner Fonseca; Vincent Louis Viala |
| EPI_ISL_1966555 | UNIDADE BASICA DE SAUDE II DE TARABAI | Instituto Butantan / FZEA-USP (Pirassununga) | Antonio Jorge Martins; Bianca Cechetto Carlos. Mendelics: Bibiana Santos; Claudia Renata dos Santos Barros; Cíntia Bittar; David Schlesinger. Hemocentro Ribeirão Preto: Simone Kashima; Debora Botequio Moretti; Elaine Cristina Marqueze; Elaine Vieira dos Santos; Elisangela Chicaroni Mattos; Erika Freitas; Evandra Strazza Rodrigues; Felipe Allan da Silva da Costa; Flavia Aburjaile; Fábio Sossai Possebon; Guilherme Campos; Guilherme Targino Valente; Heidge Fukumasu. USP-Botucatu: Rejane Maria Tommasini Grotto; Helena Lage Ferreira; Instituto Butantan: Dimas Tadeu Covas; Jardelina de Souza Todao Bernardino; Jayme A. Souza-Neto; Jessika Cristina Chagas Lesbon; Jorge A. Petroli Marchesi; José Salvatore Leister Patané; João Paulo Kitajima; João Pessoa Araújo Jr.; Leila Sabrina Ullmann; Loyze Paola Oliveira de Lima; Luiz Aurelio de Campos Crispin. Centro de Genômica Funcional da ESALQ: Luiz Lehmann Coutinho; Luiz Carlos Junior de Alcantara; Lívia Sacchetto; Maisa C. Pereira Parra; Maria Carolina Elias; Marta Giovanetti; Marília Moraes; Maurício Lacerda Nogueira. Prefeitura de Sao Paulo: Melissa Palmieri.; Patricia Akemi Assato; Paula Rahal; Paulo Inacio da Costa; Rafael dos Santos Bezerra; Raquel de Lello Rocha Campos Cassano. NGS Soluções Genômicas: Pilar Drummond Sampaio Corrêa Mariani. FZEA-USP Pirassununga: Mirele Daiana Poleti; Raul Machado Neto; Ricardo Augusto Brassaloti; Ricardo Haddad; Rodrigo Tocantins Calado. FAMERP-SJRP: Cecília Artico Banho; Sandra Coccuzzo Sampaio; Svetoslav Nanev Slavov; Vagner Fonseca; Vincent Louis Viala |
| EPI_ISL_1795226 | UNIDADE BASICA DR JOSE PARASSU CARVALHO | Instituto Butantan / ESALQ-Piracicaba | Antonio Jorge Martins; Bianca Cechetto Carlos. Mendelics: Bibiana Santos; Claudia Renata dos Santos Barros; David Schlesinger. Hemocentro Ribeirão Preto: Simone Kashima; Debora Botequio Moretti. Centro de Genômica Funcional da ESALQ: Luiz Lehmann Coutinho; Dimas Tadeu Covas; Elaine Cristina Marqueze; Elaine Vieira dos Santos; Elisangela Chicaroni Mattos; Erika Freitas; Evandra Strazza Rodrigues; Felipe Allan da Silva da Costa; Flavia Aburjaile; Guilherme Targino Valente; Heidge Fukumasu. USP-Botucatu: Rejane Maria Tommasini Grotto; Instituto Butantan: Alexander Roberto Precioso; Jayme A. Souza-Neto; Jessika Cristina Chagas Lesbon; José Salvatore Leister Patané; João Paulo Kitajima; Luiz Carlos Junior de Alcantara; Maria Carolina Elias; Marta Giovanetti; Patricia Akemi Assato; Rafael dos Santos Bezerra; Raquel de Lello Rocha Campos Cassano. NGS Soluções Genômicas: Pilar Drummond Sampaio Corrêa Mariani. FZEA-USP Pirassununga: Mirele Daiana Poleti; Raul Machado Neto; Ricardo Augusto Brassaloti; Ricardo Haddad; Rodrigo Tocantins Calado.; Sandra Coccuzzo Sampaio; Svetoslav Nanev Slavov; Vagner Fonseca; Vincent Louis Viala |
| EPI_ISL_1795305 | UNIDADE DA SAUDE DO ADULTO CASA BRANCA PREFEITURA | Instituto Butantan / ESALQ-Piracicaba | Antonio Jorge Martins; Bianca Cechetto Carlos. Mendelics: Bibiana Santos; Claudia Renata dos Santos Barros; David Schlesinger. Hemocentro Ribeirão Preto: Simone Kashima; Debora Botequio Moretti. Centro de Genômica Funcional da ESALQ: Luiz Lehmann Coutinho; Dimas Tadeu Covas; Elaine Cristina Marqueze; Elaine Vieira dos Santos; Elisangela Chicaroni Mattos; Erika Freitas; Evandra Strazza Rodrigues; Felipe Allan da Silva da Costa; Flavia Aburjaile; Guilherme Targino Valente; Heidge Fukumasu. USP-Botucatu: Rejane Maria Tommasini Grotto; Instituto Butantan: Alexander Roberto Precioso; Jayme A. Souza-Neto; Jessika Cristina Chagas Lesbon; José Salvatore Leister Patané; João Paulo Kitajima; Luiz Carlos Junior de Alcantara; Maria Carolina Elias; Marta Giovanetti; Patricia Akemi Assato; Rafael dos Santos Bezerra; Raquel de Lello Rocha Campos Cassano. NGS Soluções Genômicas: Pilar Drummond Sampaio Corrêa Mariani. FZEA-USP Pirassununga: Mirele Daiana Poleti; Raul Machado Neto; Ricardo Augusto Brassaloti; Ricardo Haddad; Rodrigo Tocantins Calado.; Sandra Coccuzzo Sampaio; Svetoslav Nanev Slavov; Vagner Fonseca; Vincent Louis Viala |
| EPI_ISL_1966306, | UNIDADE DE | Instituto Butantan / | Antonio Jorge Martins; Bianca Cechetto Carlos. Mendelics: Bibiana Santos; Claudia Renata dos Santos Barros; Cíntia Bittar; David Schlesinger. Hemocentro Ribeirão Preto: Simone Kashima; Debora Botequio Moretti; Elaine Cristina Marqueze; Elaine Vieira dos Santos; Elisangela Chicaroni Mattos; Erika Freitas; Evandra Strazza Rodrigues; |

| | | | |
|---|---|---|---|
| EPI_ISL_1966307, EPI_ISL_1966309, EPI_ISL_1966310, EPI_ISL_1966316, EPI_ISL_1966319 | PRONTO ATENDIMENTO DE AGENOR DE CAMPOS | Mendelics | Felipe Allan da Silva da Costa; Flavia Aburjaile; Fábio Sossai Possebon; Guilherme Campos; Guilherme Targino Valente; Heidge Fukumasu. USP-Botucatu: Rejane Maria Tommasini Grotto; Helena Lage Ferreira; Instituto Butantan: Dimas Tadeu Covas; Jardelina de Souza Todao Bernardino; Jayme A. Souza-Neto; Jessika Cristina Chagas Lesbon; Jorge A. Petroli Marchesi; José Salvatore Leister Patané; João Paulo Kitajima; João Pessoa Araújo Jr.; Leila Sabrina Ullmann; Loyze Paola Oliveira de Lima; Luiz Aurelio de Campos Crispin. Centro de Genômica Funcional da ESALQ: Luiz Lehmann Coutinho; Luiz Carlos Junior de Alcantara; Lívia Sacchetto; Maisa C. Pereira Parra; Maria Carolina Elias; Marta Giovanetti; Marília Moraes; Maurício Lacerda Nogueira. Prefeitura de Sao Paulo: Melissa Palmieri.; Patricia Akemi Assato; Paula Rahal; Paulo Inacio da Costa; Rafael dos Santos Bezerra; Raquel de Lello Rocha Campos Cassano. NGS Soluções Genômicas: Pilar Drummond Sampaio Corrêa Mariani. FZEA-USP Pirassununga: Mirele Daiana Poleti; Raul Machado Neto; Ricardo Augusto Brassaloti; Ricardo Haddad; Rodrigo Tocantins Calado. FAMERP-SJRP: Cecília Artico Banho; Sandra Coccuzzo Sampaio; Svetoslav Nanev Slavov; Vagner Fonseca; Vincent Louis Viala |
| EPI_ISL_1966248, EPI_ISL_1966249 | UNIDADE DE PRONTO ATENDIMENTO DE JALES | Instituto Butantan / Mendelics | Antonio Jorge Martins; Bianca Cechetto Carlos. Mendelics: Bibiana Santos; Claudia Renata dos Santos Barros; Cíntia Bittar; David Schlesinger. Hemocentro Ribeirão Preto: Simone Kashima; Debora Botequio Moretti; Elaine Cristina Marqueze; Elaine Vieira dos Santos; Elisangela Chicaroni Mattos; Erika Freitas; Evandra Strazza Rodrigues; Felipe Allan da Silva da Costa; Flavia Aburjaile; Fábio Sossai Possebon; Guilherme Campos; Guilherme Targino Valente; Heidge Fukumasu. USP-Botucatu: Rejane Maria Tommasini Grotto; Helena Lage Ferreira; Instituto Butantan: Dimas Tadeu Covas; Jardelina de Souza Todao Bernardino; Jayme A. Souza-Neto; Jessika Cristina Chagas Lesbon; Jorge A. Petroli Marchesi; José Salvatore Leister Patané; João Paulo Kitajima; João Pessoa Araújo Jr.; Leila Sabrina Ullmann; Loyze Paola Oliveira de Lima; Luiz Aurelio de Campos Crispin. Centro de Genômica Funcional da ESALQ: Luiz Lehmann Coutinho; Luiz Carlos Junior de Alcantara; Lívia Sacchetto; Maisa C. Pereira Parra; Maria Carolina Elias; Marta Giovanetti; Marília Moraes; Maurício Lacerda Nogueira. Prefeitura de Sao Paulo: Melissa Palmieri.; Patricia Akemi Assato; Paula Rahal; Paulo Inacio da Costa; Rafael dos Santos Bezerra; Raquel de Lello Rocha Campos Cassano. NGS Soluções Genômicas: Pilar Drummond Sampaio Corrêa Mariani. FZEA-USP Pirassununga: Mirele Daiana Poleti; Raul Machado Neto; Ricardo Augusto Brassaloti; Ricardo Haddad; Rodrigo Tocantins Calado. FAMERP-SJRP: Cecília Artico Banho; Sandra Coccuzzo Sampaio; Svetoslav Nanev Slavov; Vagner Fonseca; Vincent Louis Viala |
| EPI_ISL_1966721, EPI_ISL_1966722, EPI_ISL_1966723, EPI_ISL_1967312, EPI_ISL_1967313, EPI_ISL_1967314, EPI_ISL_2171032, EPI_ISL_2171033, EPI_ISL_2171034, EPI_ISL_2171035, EPI_ISL_2171036, EPI_ISL_2171037, EPI_ISL_2171038, EPI_ISL_2171039 | | | |
| see above | UNIDADE DE PRONTO ATENDIMENTO DE VARZEA PAULISTA UPA II | Instituto Butantan / Mendelics | Antonio Jorge Martins; Bianca Cechetto Carlos. Mendelics: Bibiana Santos; Claudia Renata dos Santos Barros; Cíntia Bittar; David Schlesinger. Hemocentro Ribeirão Preto: Simone Kashima; Debora Botequio Moretti; Elaine Cristina Marqueze; Elaine Vieira dos Santos; Elisangela Chicaroni Mattos; Erika Freitas; Evandra Strazza Rodrigues; Felipe Allan da Silva da Costa; Flavia Aburjaile; Fábio Sossai Possebon; Guilherme Campos; Guilherme Targino Valente; Heidge Fukumasu. USP-Botucatu: Rejane Maria Tommasini Grotto; Helena Lage Ferreira; Instituto Butantan: Dimas Tadeu Covas; Jardelina de Souza Todao Bernardino; Jayme A. Souza-Neto; Jessika Cristina Chagas Lesbon; Jorge A. Petroli Marchesi; José Salvatore Leister Patané; João Paulo Kitajima; João Pessoa Araújo Jr.; Leila Sabrina Ullmann; Loyze Paola Oliveira de Lima; Luiz Aurelio de Campos Crispin. Centro de Genômica Funcional da ESALQ: Luiz Lehmann Coutinho; Luiz Carlos Junior de Alcantara; Lívia Sacchetto; Maisa C. Pereira Parra; Maria Carolina Elias; Marta Giovanetti; Marília Moraes; Maurício Lacerda Nogueira. Prefeitura de Sao Paulo: Melissa Palmieri.; Patricia Akemi Assato; Paula Rahal; Paulo Inacio da Costa; Rafael dos Santos Bezerra; Raquel de Lello Rocha Campos Cassano. NGS Soluções Genômicas: Pilar Drummond Sampaio Corrêa Mariani. FZEA-USP Pirassununga: Mirele Daiana Poleti; Raul Machado Neto; Ricardo Augusto Brassaloti; Ricardo Haddad; Rodrigo Tocantins Calado. FAMERP-SJRP: Cecília Artico Banho; Sandra Coccuzzo Sampaio; Svetoslav Nanev Slavov; Vagner Fonseca; Vincent Louis Viala |
| EPI_ISL_1966708, EPI_ISL_1966800, EPI_ISL_2170970 | UNIDADE DE PRONTO ATENDIMENTO UPA | Instituto Butantan / Mendelics | Antonio Jorge Martins; Bianca Cechetto Carlos. Mendelics: Bibiana Santos; Claudia Renata dos Santos Barros; Cíntia Bittar; David Schlesinger. Hemocentro Ribeirão Preto: Simone Kashima; Debora Botequio Moretti; Elaine Cristina Marqueze; Elaine Vieira dos Santos; Elisangela Chicaroni Mattos; Erika Freitas; Evandra Strazza Rodrigues; Felipe Allan da Silva da Costa; Flavia Aburjaile; Fábio Sossai Possebon; Guilherme Campos; Guilherme Targino Valente; Heidge Fukumasu. USP-Botucatu: Rejane Maria Tommasini Grotto; Helena Lage Ferreira; Instituto Butantan: Dimas Tadeu Covas; Jardelina de Souza Todao Bernardino; Jayme A. Souza-Neto; Jessika Cristina Chagas Lesbon; Jorge A. Petroli Marchesi; José Salvatore Leister Patané; João Paulo Kitajima; João Pessoa Araújo Jr.; Leila Sabrina Ullmann; Loyze Paola Oliveira de Lima; Luiz Aurelio de Campos Crispin. Centro de Genômica Funcional da ESALQ: Luiz Lehmann Coutinho; Luiz Carlos Junior de Alcantara; Lívia Sacchetto; Maisa C. Pereira Parra; Maria Carolina Elias; Marta Giovanetti; Marília Moraes; Maurício Lacerda Nogueira. Prefeitura de Sao Paulo: Melissa Palmieri.; Patricia Akemi Assato; Paula Rahal; Paulo Inacio da Costa; Rafael dos Santos Bezerra; Raquel de Lello Rocha Campos Cassano. NGS Soluções Genômicas: Pilar Drummond Sampaio Corrêa Mariani. FZEA-USP Pirassununga: Mirele Daiana Poleti; Raul Machado Neto; Ricardo Augusto Brassaloti; Ricardo Haddad; Rodrigo Tocantins Calado. FAMERP-SJRP: Cecília Artico Banho; Sandra Coccuzzo Sampaio; Svetoslav Nanev Slavov; Vagner Fonseca; Vincent Louis Viala |
| EPI_ISL_2172524 | UNIDADE DE PRONTO ATENDIMENTO UPA DRA ANA OLIVIA BENTIVOGLIO | Instituto Butantan / ESALQ-Piracicaba | Antonio Jorge Martins; Bianca Cechetto Carlos. Mendelics: Bibiana Santos; Claudia Renata dos Santos Barros; Cíntia Bittar; David Schlesinger. Hemocentro Ribeirão Preto: Simone Kashima; Debora Botequio Moretti; Elaine Cristina Marqueze; Elaine Vieira dos Santos; Elisangela Chicaroni Mattos; Erika Freitas; Evandra Strazza Rodrigues; Felipe Allan da Silva da Costa; Flavia Aburjaile; Fábio Sossai Possebon; Guilherme Campos; Guilherme Targino Valente; Heidge Fukumasu. USP-Botucatu: Rejane Maria Tommasini Grotto; Helena Lage Ferreira; Instituto Butantan: Dimas Tadeu Covas; Jardelina de Souza Todao Bernardino; Jayme A. Souza-Neto; Jessika Cristina Chagas Lesbon; Jorge A. Petroli Marchesi; José Salvatore Leister Patané; João Paulo Kitajima; João Pessoa Araújo Jr.; Leila Sabrina Ullmann; Loyze Paola Oliveira de Lima; Luiz Aurelio de Campos Crispin. Centro de Genômica Funcional da ESALQ: Luiz Lehmann Coutinho; Luiz Carlos Junior de Alcantara; Lívia Sacchetto; Maisa C. Pereira Parra; Maria Carolina Elias; Marta Giovanetti; Marília Moraes; Maurício Lacerda Nogueira. Prefeitura de Sao Paulo: Melissa Palmieri.; Patricia Akemi Assato; Paula Rahal; Paulo Inacio da Costa; Rafael dos Santos Bezerra; Raquel de Lello Rocha Campos Cassano. NGS Soluções Genômicas: Pilar Drummond Sampaio Corrêa Mariani. FZEA-USP Pirassununga: Mirele Daiana Poleti; Raul Machado Neto; Ricardo Augusto Brassaloti; Ricardo Haddad; Rodrigo Tocantins Calado. FAMERP-SJRP: Cecília Artico Banho; Sandra Coccuzzo Sampaio; Svetoslav Nanev Slavov; Vagner Fonseca; Vincent Louis Viala |
| EPI_ISL_1445120, EPI_ISL_1445121, EPI_ISL_1445122, EPI_ISL_1445123, EPI_ISL_1445124, EPI_ISL_1445125, EPI_ISL_1445126, EPI_ISL_1966167, EPI_ISL_1966168, EPI_ISL_1966169, EPI_ISL_1966170, EPI_ISL_1966171, EPI_ISL_1966172, EPI_ISL_1966173, EPI_ISL_1966385, EPI_ISL_1966386, EPI_ISL_1966387, EPI_ISL_1966388, EPI_ISL_1966612, EPI_ISL_1966613, EPI_ISL_1966614, EPI_ISL_1966615, EPI_ISL_2170932, EPI_ISL_2170933, EPI_ISL_2170944 | | | |
| see above | UNIDADE DE PRONTO ATENDIMENTO UPA DRA ANA OLIVIA BENTIVOGLIO | Instituto Butantan / Mendelics | Antonio Jorge Martins; Bianca Cechetto Carlos. Mendelics: Bibiana Santos; Bibiana Santos; Claudia Renata dos Santos Barros; Cíntia Bittar; David Schlesinger; David Schlesinger. Hemocentro Ribeirão Preto: Simone Kashima; Debora Botequio Moretti; Dimas Tadeu Covas; Elaine Cristina Marqueze; Elaine Vieira dos Santos; Elisangela Chicaroni Mattos; Erika Freitas; Evandra Strazza Rodrigues; Felipe Allan da Silva da Costa; Flavia Aburjaile; Fábio Sossai Possebon; Guilherme Campos; Guilherme Targino Valente; Heidge Fukumasu. USP-Botucatu: Rejane Maria Tommasini Grotto; Helena Lage Ferreira; Instituto Butantan: Dimas Tadeu Covas; Jardelina de Souza Todao Bernardino; Jayme A. Souza-Neto; Jessika Cristina Chagas Lesbon; Jorge A. Petroli Marchesi; José Salvatore Leister Patané; João Paulo Kitajima; João Pessoa Araújo Jr.; Leila Sabrina Ullmann; Loyze Paola Oliveira de Lima; Luiz Aurelio de Campos Crispin. Centro de Genômica Funcional da ESALQ: Luiz Lehmann Coutinho; Luiz Carlos Junior de Alcantara; Lívia Sacchetto; Maisa C. Pereira Parra; Maria Carolina Elias; Marta Giovanetti; Marília Moraes; Maurício Lacerda Nogueira. Prefeitura de Sao Paulo: Melissa Palmieri.; Patricia Akemi Assato; Paula Rahal; Paulo Inacio da Costa; Rafael dos Santos Bezerra; Raquel de Lello Rocha Campos Cassano. NGS Soluções Genômicas: Pilar Drummond Sampaio Corrêa Mariani. FZEA-USP Pirassununga: Mirele Daiana Poleti; Raul Machado Neto; Ricardo Augusto Brassaloti; Ricardo Haddad; Rodrigo Tocantins Calado.; Rodrigo Tocantins Calado. FAMERP-SJRP: Cecília Artico Banho; Sandra Coccuzzo Sampaio; Simone Kashima; Svetoslav Nanev Slavov; Vagner Fonseca; Vincent Louis Viala |
| EPI_ISL_1967042, EPI_ISL_1967043, EPI_ISL_1967044, EPI_ISL_1967045, EPI_ISL_1967046, EPI_ISL_1967047, EPI_ISL_1967048, EPI_ISL_1967049 | | | |
| see above | UNIDADE DE PRONTO ATENDIMENTO UPA I SANTA ISABEL | Instituto Butantan / ESALQ-USP (Piracicaba) | Antonio Jorge Martins; Bianca Cechetto Carlos. Mendelics: Bibiana Santos; Claudia Renata dos Santos Barros; Cíntia Bittar; David Schlesinger. Hemocentro Ribeirão Preto: Simone Kashima; Debora Botequio Moretti; Elaine Cristina Marqueze; Elaine Vieira dos Santos; Elisangela Chicaroni Mattos; Erika Freitas; Evandra Strazza Rodrigues; Felipe Allan da Silva da Costa; Flavia Aburjaile; Fábio Sossai Possebon; Guilherme Campos; Guilherme Targino Valente; Heidge Fukumasu. USP-Botucatu: Rejane Maria Tommasini Grotto; Helena Lage Ferreira; Instituto Butantan: Dimas Tadeu Covas; Jardelina de Souza Todao Bernardino; Jayme A. Souza-Neto; Jessika Cristina Chagas Lesbon; Jorge A. Petroli Marchesi; José Salvatore Leister Patané; João Paulo Kitajima; João Pessoa Araújo Jr.; Leila Sabrina Ullmann; Loyze Paola Oliveira de Lima; Luiz Aurelio de Campos Crispin. Centro de Genômica Funcional da ESALQ: Luiz Lehmann Coutinho; Luiz Carlos Junior de Alcantara; Lívia Sacchetto; Maisa C. Pereira Parra; Maria Carolina Elias; Marta Giovanetti; Marília Moraes; Maurício Lacerda Nogueira. Prefeitura de Sao Paulo: Melissa Palmieri.; Patricia Akemi Assato; Paula Rahal; Paulo Inacio da Costa; Rafael dos Santos Bezerra; Raquel de Lello Rocha Campos Cassano. NGS Soluções Genômicas: Pilar Drummond Sampaio Corrêa Mariani. FZEA-USP Pirassununga: Mirele Daiana Poleti; Raul Machado Neto; Ricardo Augusto Brassaloti; Ricardo Haddad; Rodrigo Tocantins Calado. FAMERP-SJRP: Cecília Artico Banho; Sandra Coccuzzo Sampaio; Svetoslav Nanev Slavov; Vagner Fonseca; Vincent Louis Viala |
| EPI_ISL_1966402, EPI_ISL_1966403, EPI_ISL_1966404, EPI_ISL_1966405 | UNIDADE DE PRONTO ATENDIMENTO UPA I SANTA ISABEL | Instituto Butantan / Mendelics | Antonio Jorge Martins; Bianca Cechetto Carlos. Mendelics: Bibiana Santos; Claudia Renata dos Santos Barros; Cíntia Bittar; David Schlesinger. Hemocentro Ribeirão Preto: Simone Kashima; Debora Botequio Moretti; Elaine Cristina Marqueze; Elaine Vieira dos Santos; Elisangela Chicaroni Mattos; Erika Freitas; Evandra Strazza Rodrigues; Felipe Allan da Silva da Costa; Flavia Aburjaile; Fábio Sossai Possebon; Guilherme Campos; Guilherme Targino Valente; Heidge Fukumasu. USP-Botucatu: Rejane Maria Tommasini Grotto; Helena Lage Ferreira; Instituto Butantan: Dimas Tadeu Covas; Jardelina de Souza Todao Bernardino; Jayme A. Souza-Neto; Jessika Cristina Chagas Lesbon; Jorge A. Petroli Marchesi; José Salvatore Leister Patané; João Paulo Kitajima; João Pessoa Araújo Jr.; Leila Sabrina Ullmann; Loyze Paola Oliveira de Lima; Luiz Aurelio de Campos Crispin. Centro de Genômica Funcional da ESALQ: Luiz Lehmann Coutinho; Luiz Carlos Junior de Alcantara; Lívia Sacchetto; Maisa C. Pereira Parra; Maria Carolina Elias; Marta Giovanetti; Marília Moraes; Maurício Lacerda Nogueira. Prefeitura de Sao Paulo: Melissa Palmieri.; Patricia Akemi Assato; Paula Rahal; Paulo Inacio da Costa; Rafael dos Santos Bezerra; Raquel de Lello Rocha Campos Cassano. NGS Soluções Genômicas: Pilar Drummond Sampaio Corrêa Mariani. FZEA-USP Pirassununga: Mirele Daiana Poleti; Raul Machado Neto; Ricardo Augusto Brassaloti; Ricardo Haddad; Rodrigo Tocantins Calado. FAMERP-SJRP: Cecília Artico Banho; Sandra Coccuzzo Sampaio; Svetoslav Nanev Slavov; Vagner Fonseca; Vincent Louis Viala |
| EPI_ISL_1966822 | UNIDADE DE SAUDE DA FAMILIA DE UBIRAJARA | Instituto Butantan / Mendelics | Antonio Jorge Martins; Bianca Cechetto Carlos. Mendelics: Bibiana Santos; Claudia Renata dos Santos Barros; Cíntia Bittar; David Schlesinger. Hemocentro Ribeirão Preto: Simone Kashima; Debora Botequio Moretti; Elaine Cristina Marqueze; Elaine Vieira dos Santos; Elisangela Chicaroni Mattos; Erika Freitas; Evandra Strazza Rodrigues; Felipe Allan da Silva da Costa; Flavia Aburjaile; Fábio Sossai Possebon; Guilherme Campos; Guilherme Targino Valente; Heidge Fukumasu. USP-Botucatu: Rejane Maria Tommasini Grotto; Helena Lage Ferreira; Instituto Butantan: Dimas Tadeu Covas; Jardelina de Souza Todao Bernardino; Jayme A. Souza-Neto; Jessika Cristina Chagas Lesbon; Jorge A. Petroli Marchesi; José Salvatore Leister Patané; João Paulo Kitajima; João Pessoa Araújo Jr.; Leila Sabrina Ullmann; Loyze Paola Oliveira de Lima; Luiz Aurelio de Campos Crispin. Centro de Genômica Funcional da ESALQ: Luiz Lehmann Coutinho; Luiz Carlos Junior de Alcantara; Lívia Sacchetto; Maisa C. Pereira Parra; Maria Carolina Elias; Marta Giovanetti; Marília Moraes; Maurício Lacerda Nogueira. Prefeitura de Sao Paulo: Melissa Palmieri.; Patricia Akemi Assato; Paula Rahal; Paulo Inacio da Costa; Rafael dos Santos Bezerra; Raquel de Lello Rocha Campos Cassano. NGS Soluções Genômicas: Pilar Drummond Sampaio Corrêa Mariani. FZEA-USP Pirassununga: Mirele Daiana Poleti; Raul Machado Neto; Ricardo Augusto Brassaloti; Ricardo Haddad; Rodrigo Tocantins Calado. FAMERP-SJRP: Cecília Artico Banho; Sandra Coccuzzo Sampaio; Svetoslav Nanev Slavov; Vagner Fonseca; Vincent Louis Viala |
| EPI_ISL_1966755 | UNIDADE DE SAUDE DA FAMILIA I COLOMBIA | Instituto Butantan / Mendelics | Antonio Jorge Martins; Bianca Cechetto Carlos. Mendelics: Bibiana Santos; Claudia Renata dos Santos Barros; Cíntia Bittar; David Schlesinger. Hemocentro Ribeirão Preto: Simone Kashima; Debora Botequio Moretti; Elaine Cristina Marqueze; Elaine Vieira dos Santos; Elisangela Chicaroni Mattos; Erika Freitas; Evandra Strazza Rodrigues; Felipe Allan da Silva da Costa; Flavia Aburjaile; Fábio Sossai Possebon; Guilherme Campos; Guilherme Targino Valente; Heidge Fukumasu. USP-Botucatu: Rejane Maria Tommasini Grotto; Helena Lage Ferreira; Instituto Butantan: Dimas Tadeu Covas; Jardelina de Souza Todao Bernardino; Jayme A. Souza-Neto; Jessika Cristina Chagas Lesbon; Jorge A. Petroli Marchesi; José Salvatore Leister Patané; João Paulo Kitajima; João Pessoa Araújo Jr.; Leila Sabrina Ullmann; Loyze Paola Oliveira de Lima; Luiz Aurelio de Campos Crispin. Centro de Genômica Funcional da ESALQ: Luiz Lehmann Coutinho; Luiz Carlos Junior de Alcantara; Lívia Sacchetto; Maisa C. Pereira Parra; Maria Carolina Elias; Marta Giovanetti; Marília Moraes; Maurício Lacerda Nogueira. Prefeitura de Sao Paulo: Melissa Palmieri.; Patricia Akemi Assato; Paula Rahal; Paulo Inacio da Costa; Rafael dos Santos Bezerra; Raquel de Lello Rocha Campos Cassano. NGS Soluções Genômicas: Pilar Drummond Sampaio Corrêa Mariani. FZEA-USP Pirassununga: Mirele Daiana Poleti; Raul Machado Neto; Ricardo Augusto Brassaloti; Ricardo Haddad; Rodrigo Tocantins Calado. FAMERP-SJRP: Cecília Artico Banho; Sandra Coccuzzo Sampaio; Svetoslav Nanev Slavov; Vagner Fonseca; Vincent Louis Viala |
| EPI_ISL_1795065, EPI_ISL_1795230, EPI_ISL_1795231 | UNIDADE DE SAUDE DA FAMILIA JOSE ADALBERTO LELLIS GARCIA | Instituto Butantan / ESALQ-Piracicaba | Antonio Jorge Martins; Bianca Cechetto Carlos. Mendelics: Bibiana Santos; Claudia Renata dos Santos Barros; David Schlesinger. Hemocentro Ribeirão Preto: Simone Kashima; Debora Botequio Moretti. Centro de Genômica Funcional da ESALQ: Luiz Lehmann Coutinho; Dimas Tadeu Covas; Elaine Cristina Marqueze; Elaine Vieira dos Santos; Elisangela Chicaroni Mattos; Erika Freitas; Evandra Strazza Rodrigues; Felipe Allan da Silva da Costa; Flavia Aburjaile; Guilherme Targino Valente; Heidge Fukumasu. USP-Botucatu: Rejane Maria Tommasini Grotto; Instituto Butantan: Alexander Roberto Precioso; Jayme A. Souza-Neto; Jessika Cristina Chagas Lesbon; José Salvatore Leister Patané; João Paulo Kitajima; Luiz Carlos Junior de Alcantara; Maria Carolina Elias; Marta Giovanetti; Patricia Akemi Assato; Rafael dos Santos Bezerra; Raquel de Lello Rocha Campos Cassano. NGS Soluções Genômicas: Pilar Drummond Sampaio Corrêa Mariani. FZEA-USP Pirassununga: Mirele Daiana Poleti; Raul Machado Neto; Ricardo Augusto Brassaloti; Ricardo Haddad; Rodrigo Tocantins Calado.; Sandra Coccuzzo Sampaio; Svetoslav Nanev Slavov; Vagner Fonseca; Vincent Louis Viala |
| EPI_ISL_1795293, EPI_ISL_1795294, EPI_ISL_1795298 | UNIDADE DE SAUDE DE ITOBI ALCIBIADES PIRES | Instituto Butantan / ESALQ-Piracicaba | Antonio Jorge Martins; Bianca Cechetto Carlos. Mendelics: Bibiana Santos; Claudia Renata dos Santos Barros; David Schlesinger. Hemocentro Ribeirão Preto: Simone Kashima; Debora Botequio Moretti. Centro de Genômica Funcional da ESALQ: Luiz Lehmann Coutinho; Dimas Tadeu Covas; Elaine Cristina Marqueze; Elaine Vieira dos Santos; Elisangela Chicaroni Mattos; Erika Freitas; Evandra Strazza Rodrigues; Felipe Allan da Silva da Costa; Flavia Aburjaile; Guilherme Targino Valente; Heidge Fukumasu. USP-Botucatu: Rejane Maria Tommasini Grotto; Instituto Butantan: Alexander Roberto Precioso; Jayme A. Souza-Neto; Jessika Cristina Chagas Lesbon; José Salvatore Leister Patané; João Paulo Kitajima; Luiz Carlos Junior de Alcantara; Maria Carolina Elias; Marta Giovanetti; Patricia Akemi Assato; Rafael dos Santos Bezerra; Raquel de Lello Rocha Campos Cassano. NGS Soluções Genômicas: Pilar Drummond Sampaio Corrêa Mariani. FZEA-USP Pirassununga: Mirele Daiana Poleti; Raul Machado Neto; Ricardo Augusto Brassaloti; Ricardo Haddad; Rodrigo Tocantins Calado.; Sandra Coccuzzo Sampaio; Svetoslav Nanev Slavov; Vagner Fonseca; Vincent Louis Viala |
| EPI_ISL_1795063, EPI_ISL_1795122, EPI_ISL_1795123, EPI_ISL_1795124, EPI_ISL_1795128, EPI_ISL_1795129, EPI_ISL_1795130, EPI_ISL_1795131, EPI_ISL_1795132, EPI_ISL_1795133, EPI_ISL_1795134, EPI_ISL_1795135, EPI_ISL_1795385, EPI_ISL_1795386 | | | |
| see above | UNIDADE DE VIGILANCIA EM SAUDE | Instituto Butantan / ESALQ-Piracicaba | Antonio Jorge Martins; Bianca Cechetto Carlos. Mendelics: Bibiana Santos; Claudia Renata dos Santos Barros; David Schlesinger. Hemocentro Ribeirão Preto: Simone Kashima; Debora Botequio Moretti. Centro de Genômica Funcional da ESALQ: Luiz Lehmann Coutinho; Dimas Tadeu Covas; Elaine Cristina Marqueze; Elaine Vieira dos Santos; Elisangela Chicaroni Mattos; Erika Freitas; Evandra Strazza Rodrigues; Felipe Allan da Silva da Costa; Flavia Aburjaile; Guilherme Targino Valente; Heidge Fukumasu. USP-Botucatu: Rejane Maria Tommasini Grotto; Instituto Butantan: Alexander Roberto Precioso; Jayme A. Souza-Neto; Jessika Cristina Chagas Lesbon; José Salvatore Leister Patané; João Paulo Kitajima; Luiz Carlos Junior de Alcantara; Maria Carolina Elias; Marta Giovanetti; Patricia Akemi Assato; Rafael dos Santos Bezerra; Raquel de Lello Rocha Campos Cassano. NGS Soluções Genômicas: Pilar Drummond Sampaio Corrêa Mariani. FZEA-USP Pirassununga: Mirele Daiana Poleti; Raul Machado Neto; Ricardo Augusto Brassaloti; Ricardo Haddad; Rodrigo Tocantins Calado.; Sandra Coccuzzo Sampaio; Svetoslav Nanev Slavov; Vagner Fonseca; Vincent Louis Viala |
| EPI_ISL_1967052, EPI_ISL_1967053, EPI_ISL_1967054, EPI_ISL_1967055, EPI_ISL_1967056, EPI_ISL_1967057 | UNIDADE DE VIGILANCIA EM SAUDE | Instituto Butantan / ESALQ-USP (Piracicaba) | Antonio Jorge Martins; Bianca Cechetto Carlos. Mendelics: Bibiana Santos; Claudia Renata dos Santos Barros; Cíntia Bittar; David Schlesinger. Hemocentro Ribeirão Preto: Simone Kashima; Debora Botequio Moretti; Elaine Cristina Marqueze; Elaine Vieira dos Santos; Elisangela Chicaroni Mattos; Erika Freitas; Evandra Strazza Rodrigues; Felipe Allan da Silva da Costa; Flavia Aburjaile; Fábio Sossai Possebon; Guilherme Campos; Guilherme Targino Valente; Heidge Fukumasu. USP-Botucatu: Rejane Maria Tommasini Grotto; Helena Lage Ferreira; Instituto Butantan: Dimas Tadeu Covas; Jardelina de Souza Todao Bernardino; Jayme A. Souza-Neto; Jessika Cristina Chagas Lesbon; Jorge A. Petroli Marchesi; José Salvatore Leister Patané; João Paulo Kitajima; João Pessoa Araújo Jr.; Leila Sabrina Ullmann; Loyze Paola Oliveira de Lima; Luiz Aurelio de Campos Crispin. Centro de Genômica Funcional da ESALQ: Luiz Lehmann Coutinho; Luiz Carlos Junior de Alcantara; Lívia Sacchetto; Maisa C. Pereira Parra; Maria Carolina Elias; Marta Giovanetti; Marília Moraes; Maurício Lacerda Nogueira. Prefeitura de Sao Paulo: Melissa Palmieri.; Patricia Akemi Assato; Paula Rahal; Paulo Inacio da Costa; Rafael dos Santos Bezerra; Raquel de Lello Rocha Campos Cassano. NGS Soluções Genômicas: Pilar Drummond Sampaio Corrêa Mariani. FZEA-USP Pirassununga: Mirele Daiana Poleti; Raul Machado Neto; Ricardo Augusto Brassaloti; Ricardo Haddad; Rodrigo Tocantins Calado. FAMERP-SJRP: Cecília Artico Banho; Sandra Coccuzzo Sampaio; Svetoslav Nanev Slavov; Vagner Fonseca; Vincent Louis Viala |
| EPI_ISL_1966549, EPI_ISL_1966550 | UNIDADE DE VIGILANCIA EM SAUDE | Instituto Butantan / FZEA-USP (Pirassununga) | Antonio Jorge Martins; Bianca Cechetto Carlos. Mendelics: Bibiana Santos; Claudia Renata dos Santos Barros; Cíntia Bittar; David Schlesinger. Hemocentro Ribeirão Preto: Simone Kashima; Debora Botequio Moretti; Elaine Cristina Marqueze; Elaine Vieira dos Santos; Elisangela Chicaroni Mattos; Erika Freitas; Evandra Strazza Rodrigues; Felipe Allan da Silva da Costa; Flavia Aburjaile; Fábio Sossai Possebon; Guilherme Campos; Guilherme Targino Valente; Heidge Fukumasu. USP-Botucatu: Rejane Maria Tommasini Grotto; Helena Lage Ferreira; Instituto Butantan: Dimas Tadeu Covas; Jardelina de Souza Todao Bernardino; Jayme A. Souza-Neto; Jessika Cristina Chagas Lesbon; Jorge A. Petroli Marchesi; José Salvatore Leister Patané; João Paulo Kitajima; João Pessoa Araújo Jr.; Leila Sabrina Ullmann; Loyze Paola Oliveira de Lima; Luiz Aurelio de Campos Crispin. Centro de Genômica Funcional da ESALQ: Luiz Lehmann Coutinho; Luiz Carlos Junior de Alcantara; Lívia Sacchetto; Maisa C. Pereira Parra; Maria Carolina Elias; Marta Giovanetti; Marília Moraes; Maurício Lacerda Nogueira. Prefeitura de Sao Paulo: Melissa Palmieri.; Patricia Akemi Assato; Paula Rahal; Paulo Inacio da Costa; Rafael dos Santos Bezerra; Raquel de Lello Rocha Campos Cassano. NGS Soluções Genômicas: Pilar Drummond Sampaio Corrêa Mariani. FZEA-USP Pirassununga: Mirele Daiana Poleti; Raul Machado Neto; Ricardo Augusto Brassaloti; Ricardo Haddad; Rodrigo Tocantins Calado. FAMERP-SJRP: Cecília Artico Banho; Sandra Coccuzzo Sampaio; Svetoslav Nanev Slavov; Vagner Fonseca; Vincent Louis Viala |
| EPI_ISL_1966720 | UNIDADE DE VIGILANCIA EM SAUDE | Instituto Butantan / Mendelics | Antonio Jorge Martins; Bianca Cechetto Carlos. Mendelics: Bibiana Santos; Claudia Renata dos Santos Barros; Cíntia Bittar; David Schlesinger. Hemocentro Ribeirão Preto: Simone Kashima; Debora Botequio Moretti; Elaine Cristina Marqueze; Elaine Vieira dos Santos; Elisangela Chicaroni Mattos; Erika Freitas; Evandra Strazza Rodrigues; Felipe Allan da Silva da Costa; Flavia Aburjaile; Fábio Sossai Possebon; Guilherme Campos; Guilherme Targino Valente; Heidge Fukumasu. USP-Botucatu: Rejane Maria Tommasini Grotto; Helena Lage Ferreira; Instituto Butantan: Dimas Tadeu Covas; Jardelina de Souza Todao Bernardino; Jayme A. Souza-Neto; Jessika Cristina Chagas Lesbon; Jorge A. Petroli Marchesi; José Salvatore Leister Patané; João Paulo Kitajima; João Pessoa Araújo Jr.; Leila Sabrina Ullmann; Loyze Paola Oliveira de Lima; Luiz Aurelio de Campos Crispin. Centro de Genômica Funcional da ESALQ: Luiz Lehmann Coutinho; Luiz Carlos Junior de Alcantara; Lívia Sacchetto; Maisa C. Pereira Parra; Maria Carolina Elias; Marta Giovanetti; Marília Moraes; Maurício Lacerda Nogueira. Prefeitura de Sao Paulo: Melissa Palmieri.; Patricia Akemi Assato; Paula Rahal; Paulo Inacio da Costa; Rafael dos Santos Bezerra; Raquel de Lello Rocha Campos Cassano. NGS Soluções Genômicas: Pilar Drummond Sampaio Corrêa Mariani. FZEA-USP Pirassununga: Mirele Daiana Poleti; Raul Machado Neto; Ricardo Augusto Brassaloti; Ricardo Haddad; Rodrigo Tocantins Calado. FAMERP-SJRP: Cecília Artico Banho; Sandra Coccuzzo Sampaio; Svetoslav Nanev Slavov; Vagner Fonseca; Vincent Louis Viala |
| EPI_ISL_1795110 | UNIDADE ESF DR LUIZ SPINA | Instituto Butantan / ESALQ-Piracicaba | Antonio Jorge Martins; Bianca Cechetto Carlos. Mendelics: Bibiana Santos; Claudia Renata dos Santos Barros; David Schlesinger. Hemocentro Ribeirão Preto: Simone Kashima; Debora Botequio Moretti. Centro de Genômica Funcional da ESALQ: Luiz Lehmann Coutinho; Dimas Tadeu Covas; Elaine Cristina Marqueze; Elaine Vieira dos Santos; Elisangela Chicaroni Mattos; Erika Freitas; Evandra Strazza Rodrigues; Felipe Allan da Silva da Costa; Flavia Aburjaile; Guilherme Targino Valente; Heidge Fukumasu. USP-Botucatu: Rejane Maria Tommasini Grotto; Instituto Butantan: Alexander Roberto Precioso; Jayme A. Souza-Neto; Jessika Cristina Chagas Lesbon; José Salvatore Leister Patané; João Paulo Kitajima; Luiz Carlos Junior de Alcantara; Maria Carolina Elias; Marta Giovanetti; Patricia Akemi Assato; Rafael dos Santos Bezerra; Raquel de Lello Rocha Campos Cassano. NGS Soluções Genômicas: Pilar Drummond Sampaio Corrêa Mariani. FZEA-USP Pirassununga: Mirele Daiana Poleti; Raul Machado Neto; Ricardo Augusto Brassaloti; Ricardo Haddad; Rodrigo Tocantins Calado.; Sandra Coccuzzo Sampaio; Svetoslav Nanev Slavov; Vagner Fonseca; Vincent Louis Viala |
| EPI_ISL_1795222 | UNIDADE MISTA ARACOIABA DA SERRA | Instituto Butantan / ESALQ-Piracicaba | Antonio Jorge Martins; Bianca Cechetto Carlos. Mendelics: Bibiana Santos; Claudia Renata dos Santos Barros; David Schlesinger. Hemocentro Ribeirão Preto: Simone Kashima; Debora Botequio Moretti. Centro de Genômica Funcional da ESALQ: Luiz Lehmann Coutinho; Dimas Tadeu Covas; Elaine Cristina Marqueze; Elaine Vieira dos Santos; Elisangela Chicaroni Mattos; Erika Freitas; Evandra Strazza Rodrigues; Felipe Allan da Silva da Costa; Flavia Aburjaile; Guilherme Targino Valente; Heidge Fukumasu. USP-Botucatu: Rejane Maria Tommasini Grotto; Instituto Butantan: Alexander Roberto Precioso; Jayme A. Souza-Neto; Jessika Cristina Chagas Lesbon; José Salvatore Leister Patané; João Paulo Kitajima; Luiz Carlos Junior de Alcantara; Maria Carolina Elias; Marta Giovanetti; Patricia Akemi Assato; Rafael dos Santos Bezerra; Raquel de Lello Rocha Campos Cassano. NGS Soluções Genômicas: Pilar Drummond Sampaio Corrêa Mariani. FZEA-USP Pirassununga: Mirele Daiana Poleti; Raul Machado Neto; Ricardo Augusto Brassaloti; Ricardo Haddad; Rodrigo Tocantins Calado.; Sandra Coccuzzo Sampaio; Svetoslav Nanev Slavov; Vagner Fonseca; Vincent Louis Viala |
| EPI_ISL_1445172, EPI_ISL_1966111 | UNIDADE MISTA ARACOIABA DA SERRA | Instituto Butantan / Mendelics | Antonio Jorge Martins; Bibiana Santos; Claudia Renata dos Santos Barros; David Schlesinger; Debora Botequio Moretti; Dimas Tadeu Covas; Elaine Cristina Marqueze; Elaine Vieira Santos; Elaine Vieira dos Santos; Erika Freitas; Evandra Strazza Rodrigues; Flavia Aburjaile; Heidge Fukumasu; Jayme Augusto de Souza-Neto; José Salvatore Leister Patané; João Paulo Kitajima; Luiz Alcantara; Luiz Carlos Junior de Alcantara; Luiz Lehmann Coutinho; Maria Carolina Elias; Marta Giovanetti; Maurício Lacerda Nogueira; Rafael dos Santos Bezerra; Raul Machado Neto; Rejane Maria Tommasini Grotto; Ricardo Haddad; Rodrigo Tocantins Calado.; Sandra Coccuzzo Sampaio; Sandra Coccuzzo Sampaio Vessoni; Simone Kashima; Svetoslav Nanev Slavov; Vagner Fonseca; Vincent Louis Viala |
| EPI_ISL_1966647, EPI_ISL_1966648, EPI_ISL_1966649 | UNIDADE MISTA DE BOM JESUS DOS PERDOES | Instituto Butantan / Mendelics | Antonio Jorge Martins; Bianca Cechetto Carlos. Mendelics: Bibiana Santos; Claudia Renata dos Santos Barros; Cíntia Bittar; David Schlesinger. Hemocentro Ribeirão Preto: Simone Kashima; Debora Botequio Moretti; Elaine Cristina Marqueze; Elaine Vieira dos Santos; Elisangela Chicaroni Mattos; Erika Freitas; Evandra Strazza Rodrigues; Felipe Allan da Silva da Costa; Flavia Aburjaile; Fábio Sossai Possebon; Guilherme Campos; Guilherme Targino Valente; Heidge Fukumasu. USP-Botucatu: Rejane Maria Tommasini Grotto; Helena Lage Ferreira; Instituto Butantan: Dimas Tadeu Covas; Jardelina de Souza Todao Bernardino; Jayme A. Souza-Neto; Jessika Cristina Chagas Lesbon; Jorge A. Petroli Marchesi; José Salvatore Leister Patané; João Paulo Kitajima; João Pessoa Araújo Jr.; Leila Sabrina Ullmann; Loyze Paola Oliveira de Lima; Luiz Aurelio de Campos Crispin. Centro de Genômica Funcional da ESALQ: Luiz Lehmann Coutinho; Luiz Carlos Junior de Alcantara; Lívia Sacchetto; Maisa C. Pereira Parra; Maria Carolina Elias; Marta Giovanetti; Marília Moraes; Maurício Lacerda Nogueira. Prefeitura de Sao Paulo: Melissa Palmieri.; Patricia Akemi Assato; Paula Rahal; Paulo Inacio da Costa; Rafael dos Santos Bezerra; Raquel de Lello Rocha Campos Cassano. NGS Soluções Genômicas: Pilar Drummond Sampaio Corrêa Mariani. FZEA-USP Pirassununga: |

| | | | |
|---|---|---|---|
| | | | Mirele Daiana Poleti; Raul Machado Neto; Ricardo Augusto Brassaloti; Ricardo Haddad; Rodrigo Tocantins Calado. FAMERP-SJRP: Cecília Artico Banho; Sandra Coccuzzo Sampaio; Svetoslav Nanev Slavov; Vagner Fonseca; Vincent Louis Viala |
| EPI_ISL_1966915 | UNIDADE MISTA DE IGUAPE | Instituto Butantan / ESALQ-USP (Piracicaba) | Antonio Jorge Martins; Bianca Cechetto Carlos. Mendelics: Bibiana Santos; Claudia Renata dos Santos Barros; Cíntia Bittar; David Schlesinger. Hemocentro Ribeirão Preto: Simone Kashima; Debora Botequio Moretti; Elaine Cristina Marqueze; Elaine Vieira dos Santos; Elisangela Chicaroni Mattos; Erika Freitas; Evandra Strazza Rodrigues; Felipe Allan da Silva da Costa; Flavia Aburjaile; Fábio Sossai Possebon; Guilherme Campos; Guilherme Targino Valente; Heidge Fukumasu. USP-Botucatu: Rejane Maria Tommasini Grotto; Helena Lage Ferreira; Instituto Butantan: Dimas Tadeu Covas; Jardelina de Souza Todao Bernardino; Jayme A. Souza-Neto; Jessika Cristina Chagas Lesbon; Jorge A. Petroli Marchesi; José Salvatore Leister Patané; João Paulo Kitajima; João Pessoa Araújo Jr.; Leila Sabrina Ullmann; Loyze Paola Oliveira de Lima; Luiz Aurelio de Campos Crispin. Centro de Genômica Funcional da ESALQ: Luiz Lehmann Coutinho; Luiz Carlos Junior de Alcantara; Lívia Sacchetto; Maisa C. Pereira Parra; Maria Carolina Elias; Marta Giovanetti; Marília Moraes; Maurício Lacerda Nogueira. Prefeitura de Sao Paulo: Melissa Palmieri.; Patricia Akemi Assato; Paula Rahal; Paulo Inacio da Costa; Rafael dos Santos Bezerra; Raquel de Lello Rocha Campos Cassano. NGS Soluções Genômicas: Pilar Drummond Sampaio Corrêa Mariani. FZEA-USP Pirassununga: Mirele Daiana Poleti; Raul Machado Neto; Ricardo Augusto Brassaloti; Ricardo Haddad; Rodrigo Tocantins Calado. FAMERP-SJRP: Cecília Artico Banho; Sandra Coccuzzo Sampaio; Svetoslav Nanev Slavov; Vagner Fonseca; Vincent Louis Viala |
| EPI_ISL_1966246, EPI_ISL_1966681, EPI_ISL_1966682 | UNIDADE MISTA DE IGUAPE | Instituto Butantan / Mendelics | Antonio Jorge Martins; Bianca Cechetto Carlos. Mendelics: Bibiana Santos; Claudia Renata dos Santos Barros; Cíntia Bittar; David Schlesinger. Hemocentro Ribeirão Preto: Simone Kashima; Debora Botequio Moretti; Elaine Cristina Marqueze; Elaine Vieira dos Santos; Elisangela Chicaroni Mattos; Erika Freitas; Evandra Strazza Rodrigues; Felipe Allan da Silva da Costa; Flavia Aburjaile; Fábio Sossai Possebon; Guilherme Campos; Guilherme Targino Valente; Heidge Fukumasu. USP-Botucatu: Rejane Maria Tommasini Grotto; Helena Lage Ferreira; Instituto Butantan: Dimas Tadeu Covas; Jardelina de Souza Todao Bernardino; Jayme A. Souza-Neto; Jessika Cristina Chagas Lesbon; Jorge A. Petroli Marchesi; José Salvatore Leister Patané; João Paulo Kitajima; João Pessoa Araújo Jr.; Leila Sabrina Ullmann; Loyze Paola Oliveira de Lima; Luiz Aurelio de Campos Crispin. Centro de Genômica Funcional da ESALQ: Luiz Lehmann Coutinho; Luiz Carlos Junior de Alcantara; Lívia Sacchetto; Maisa C. Pereira Parra; Maria Carolina Elias; Marta Giovanetti; Marília Moraes; Maurício Lacerda Nogueira. Prefeitura de Sao Paulo: Melissa Palmieri.; Patricia Akemi Assato; Paula Rahal; Paulo Inacio da Costa; Rafael dos Santos Bezerra; Raquel de Lello Rocha Campos Cassano. NGS Soluções Genômicas: Pilar Drummond Sampaio Corrêa Mariani. FZEA-USP Pirassununga: Mirele Daiana Poleti; Raul Machado Neto; Ricardo Augusto Brassaloti; Ricardo Haddad; Rodrigo Tocantins Calado. FAMERP-SJRP: Cecília Artico Banho; Sandra Coccuzzo Sampaio; Svetoslav Nanev Slavov; Vagner Fonseca; Vincent Louis Viala |
| EPI_ISL_1795116, EPI_ISL_1795117, EPI_ISL_1795118, EPI_ISL_1795119, EPI_ISL_1795403 | UNIDADE MISTA DE LUIZ ANTONIO | Instituto Butantan / ESALQ-Piracicaba | Antonio Jorge Martins; Bianca Cechetto Carlos. Mendelics: Bibiana Santos; Claudia Renata dos Santos Barros; David Schlesinger. Hemocentro Ribeirão Preto: Simone Kashima; Debora Botequio Moretti. Centro de Genômica Funcional da ESALQ: Luiz Lehmann Coutinho; Dimas Tadeu Covas; Elaine Cristina Marqueze; Elaine Vieira dos Santos; Elisangela Chicaroni Mattos; Erika Freitas; Evandra Strazza Rodrigues; Felipe Allan da Silva da Costa; Flavia Aburjaile; Guilherme Targino Valente; Heidge Fukumasu. USP-Botucatu: Rejane Maria Tommasini Grotto; Instituto Butantan: Alexander Roberto Precioso; Jayme A. Souza-Neto; Jessika Cristina Chagas Lesbon; José Salvatore Leister Patané; João Paulo Kitajima; Luiz Carlos Junior de Alcantara; Maria Carolina Elias; Marta Giovanetti; Patricia Akemi Assato; Rafael dos Santos Bezerra; Raquel de Lello Rocha Campos Cassano. NGS Soluções Genômicas: Pilar Drummond Sampaio Corrêa Mariani. FZEA-USP Pirassununga: Mirele Daiana Poleti; Raul Machado Neto; Ricardo Augusto Brassaloti; Ricardo Haddad; Rodrigo Tocantins Calado.; Sandra Coccuzzo Sampaio; Svetoslav Nanev Slavov; Vagner Fonseca; Vincent Louis Viala |
| EPI_ISL_1966863, EPI_ISL_1966864, EPI_ISL_1966865, EPI_ISL_1966866, EPI_ISL_1966867 | UNIDADE MISTA DE SAUDE CAPELA DO ALTO | Instituto Butantan / ESALQ-USP (Piracicaba) | Antonio Jorge Martins; Bianca Cechetto Carlos. Mendelics: Bibiana Santos; Claudia Renata dos Santos Barros; Cíntia Bittar; David Schlesinger. Hemocentro Ribeirão Preto: Simone Kashima; Debora Botequio Moretti; Elaine Cristina Marqueze; Elaine Vieira dos Santos; Elisangela Chicaroni Mattos; Erika Freitas; Evandra Strazza Rodrigues; Felipe Allan da Silva da Costa; Flavia Aburjaile; Fábio Sossai Possebon; Guilherme Campos; Guilherme Targino Valente; Heidge Fukumasu. USP-Botucatu: Rejane Maria Tommasini Grotto; Helena Lage Ferreira; Instituto Butantan: Dimas Tadeu Covas; Jardelina de Souza Todao Bernardino; Jayme A. Souza-Neto; Jessika Cristina Chagas Lesbon; Jorge A. Petroli Marchesi; José Salvatore Leister Patané; João Paulo Kitajima; João Pessoa Araújo Jr.; Leila Sabrina Ullmann; Loyze Paola Oliveira de Lima; Luiz Aurelio de Campos Crispin. Centro de Genômica Funcional da ESALQ: Luiz Lehmann Coutinho; Luiz Carlos Junior de Alcantara; Lívia Sacchetto; Maisa C. Pereira Parra; Maria Carolina Elias; Marta Giovanetti; Marília Moraes; Maurício Lacerda Nogueira. Prefeitura de Sao Paulo: Melissa Palmieri.; Patricia Akemi Assato; Paula Rahal; Paulo Inacio da Costa; Rafael dos Santos Bezerra; Raquel de Lello Rocha Campos Cassano. NGS Soluções Genômicas: Pilar Drummond Sampaio Corrêa Mariani. FZEA-USP Pirassununga: Mirele Daiana Poleti; Raul Machado Neto; Ricardo Augusto Brassaloti; Ricardo Haddad; Rodrigo Tocantins Calado. FAMERP-SJRP: Cecília Artico Banho; Sandra Coccuzzo Sampaio; Svetoslav Nanev Slavov; Vagner Fonseca; Vincent Louis Viala |
| EPI_ISL_1445140, EPI_ISL_1966180 | UNIDADE MISTA DE SAUDE CAPELA DO ALTO | Instituto Butantan / Mendelics | Antonio Jorge Martins; Bianca Cechetto Carlos. Mendelics: Bibiana Santos; Bibiana Santos; Claudia Renata dos Santos Barros; Cíntia Bittar; David Schlesinger; David Schlesinger. Hemocentro Ribeirão Preto: Simone Kashima; Debora Botequio Moretti; Dimas Tadeu Covas; Elaine Cristina Marqueze; Elaine Vieira dos Santos; Elisangela Chicaroni Mattos; Erika Freitas; Evandra Strazza Rodrigues; Felipe Allan da Silva da Costa; Flavia Aburjaile; Fábio Sossai Possebon; Guilherme Campos; Guilherme Targino Valente; Heidge Fukumasu. USP-Botucatu: Rejane Maria Tommasini Grotto; Helena Lage Ferreira; Instituto Butantan: Dimas Tadeu Covas; Jardelina de Souza Todao Bernardino; Jayme A. Souza-Neto; Jessika Cristina Chagas Lesbon; Jorge A. Petroli Marchesi; José Salvatore Leister Patané; João Paulo Kitajima; João Pessoa Araújo Jr.; Leila Sabrina Ullmann; Loyze Paola Oliveira de Lima; Luiz Aurelio de Campos Crispin. Centro de Genômica Funcional da ESALQ: Luiz Lehmann Coutinho; Luiz Carlos Junior de Alcantara; Lívia Sacchetto; Maisa C. Pereira Parra; Maria Carolina Elias; Marta Giovanetti; Marília Moraes; Maurício Lacerda Nogueira. Prefeitura de Sao Paulo: Melissa Palmieri.; Patricia Akemi Assato; Paula Rahal; Paulo Inacio da Costa; Rafael dos Santos Bezerra; Raquel de Lello Rocha Campos Cassano. NGS Soluções Genômicas: Pilar Drummond Sampaio Corrêa Mariani. FZEA-USP Pirassununga: Mirele Daiana Poleti; Raul Machado Neto; Ricardo Augusto Brassaloti; Ricardo Haddad; Rodrigo Tocantins Calado.; Rodrigo Tocantins Calado. FAMERP-SJRP: Cecília Artico Banho; Sandra Coccuzzo Sampaio; Simone Kashima; Svetoslav Nanev Slavov; Vagner Fonseca; Vincent Louis Viala |
| EPI_ISL_1966243 | UNIDADE MISTA DE SAUDE DE DUMONT | Instituto Butantan / Mendelics | Antonio Jorge Martins; Bianca Cechetto Carlos. Mendelics: Bibiana Santos; Claudia Renata dos Santos Barros; Cíntia Bittar; David Schlesinger. Hemocentro Ribeirão Preto: Simone Kashima; Debora Botequio Moretti; Elaine Cristina Marqueze; Elaine Vieira dos Santos; Elisangela Chicaroni Mattos; Erika Freitas; Evandra Strazza Rodrigues; Felipe Allan da Silva da Costa; Flavia Aburjaile; Fábio Sossai Possebon; Guilherme Campos; Guilherme Targino Valente; Heidge Fukumasu. USP-Botucatu: Rejane Maria Tommasini Grotto; Helena Lage Ferreira; Instituto Butantan: Dimas Tadeu Covas; Jardelina de Souza Todao Bernardino; Jayme A. Souza-Neto; Jessika Cristina Chagas Lesbon; Jorge A. Petroli Marchesi; José Salvatore Leister Patané; João Paulo Kitajima; João Pessoa Araújo Jr.; Leila Sabrina Ullmann; Loyze Paola Oliveira de Lima; Luiz Aurelio de Campos Crispin. Centro de Genômica Funcional da ESALQ: Luiz Lehmann Coutinho; Luiz Carlos Junior de Alcantara; Lívia Sacchetto; Maisa C. Pereira Parra; Maria Carolina Elias; Marta Giovanetti; Marília Moraes; Maurício Lacerda Nogueira. Prefeitura de Sao Paulo: Melissa Palmieri.; Patricia Akemi Assato; Paula Rahal; Paulo Inacio da Costa; Rafael dos Santos Bezerra; Raquel de Lello Rocha Campos Cassano. NGS Soluções Genômicas: Pilar Drummond Sampaio Corrêa Mariani. FZEA-USP Pirassununga: Mirele Daiana Poleti; Raul Machado Neto; Ricardo Augusto Brassaloti; Ricardo Haddad; Rodrigo Tocantins Calado. FAMERP-SJRP: Cecília Artico Banho; Sandra Coccuzzo Sampaio; Svetoslav Nanev Slavov; Vagner Fonseca; Vincent Louis Viala |
| EPI_ISL_1966986, EPI_ISL_1966987 | UNIDADE MISTA DE SAUDE DE ORIENTE | Instituto Butantan / ESALQ-USP (Piracicaba) | Antonio Jorge Martins; Bianca Cechetto Carlos. Mendelics: Bibiana Santos; Claudia Renata dos Santos Barros; Cíntia Bittar; David Schlesinger. Hemocentro Ribeirão Preto: Simone Kashima; Debora Botequio Moretti; Elaine Cristina Marqueze; Elaine Vieira dos Santos; Elisangela Chicaroni Mattos; Erika Freitas; Evandra Strazza Rodrigues; Felipe Allan da Silva da Costa; Flavia Aburjaile; Fábio Sossai Possebon; Guilherme Campos; Guilherme Targino Valente; Heidge Fukumasu. USP-Botucatu: Rejane Maria Tommasini Grotto; Helena Lage Ferreira; Instituto Butantan: Dimas Tadeu Covas; Jardelina de Souza Todao Bernardino; Jayme A. Souza-Neto; Jessika Cristina Chagas Lesbon; Jorge A. Petroli Marchesi; José Salvatore Leister Patané; João Paulo Kitajima; João Pessoa Araújo Jr.; Leila Sabrina Ullmann; Loyze Paola Oliveira de Lima; Luiz Aurelio de Campos Crispin. Centro de Genômica Funcional da ESALQ: Luiz Lehmann Coutinho; Luiz Carlos Junior de Alcantara; Lívia Sacchetto; Maisa C. Pereira Parra; Maria Carolina Elias; Marta Giovanetti; Marília Moraes; Maurício Lacerda Nogueira. Prefeitura de Sao Paulo: Melissa Palmieri.; Patricia Akemi Assato; Paula Rahal; Paulo Inacio da Costa; Rafael dos Santos Bezerra; Raquel de Lello Rocha Campos Cassano. NGS Soluções Genômicas: Pilar Drummond Sampaio Corrêa Mariani. FZEA-USP Pirassununga: Mirele Daiana Poleti; Raul Machado Neto; Ricardo Augusto Brassaloti; Ricardo Haddad; Rodrigo Tocantins Calado. FAMERP-SJRP: Cecília Artico Banho; Sandra Coccuzzo Sampaio; Svetoslav Nanev Slavov; Vagner Fonseca; Vincent Louis Viala |
| EPI_ISL_1445065, EPI_ISL_1445198, EPI_ISL_1966132, EPI_ISL_1966236, EPI_ISL_1966512 | UNIDADE MISTA DE TAIACU | Instituto Butantan / Mendelics | Antonio Jorge Martins; Bianca Cechetto Carlos. Mendelics: Bibiana Santos; Bibiana Santos; Claudia Renata dos Santos Barros; Cíntia Bittar; David Schlesinger; David Schlesinger. Hemocentro Ribeirão Preto: Simone Kashima; Debora Botequio Moretti; Dimas Tadeu Covas; Elaine Cristina Marqueze; Elaine Vieira Santos; Elaine Vieira dos Santos; Elisangela Chicaroni Mattos; Erika Freitas; Evandra Strazza Rodrigues; Felipe Allan da Silva da Costa; Flavia Aburjaile; Fábio Sossai Possebon; Guilherme Campos; Guilherme Targino Valente; Heidge Fukumasu; Heidge Fukumasu. USP-Botucatu: Rejane Maria Tommasini Grotto; Helena Lage Ferreira; Instituto Butantan: Dimas Tadeu Covas; Jardelina de Souza Todao Bernardino; Jayme A. Souza-Neto; Jayme Augusto de Souza-Neto; Jessika Cristina Chagas Lesbon; Jorge A. Petroli Marchesi; José Salvatore Leister Patané; João Paulo Kitajima; João Pessoa Araújo Jr.; Leila Sabrina Ullmann; Loyze Paola Oliveira de Lima; Luiz Alcantara; Luiz Aurelio de Campos Crispin. Centro de Genômica Funcional da ESALQ: Luiz Lehmann Coutinho; Luiz Carlos Junior de Alcantara; Luiz Lehmann Coutinho; Lívia Sacchetto; Maisa C. Pereira Parra; Maria Carolina Elias; Marta Giovanetti; Marília Moraes; Maurício Lacerda Nogueira; Maurício Lacerda Nogueira. Prefeitura de Sao Paulo: Melissa Palmieri.; Patricia Akemi Assato; Paula Rahal; Paulo Inacio da Costa; Rafael dos Santos Bezerra; Raquel de Lello Rocha Campos Cassano. NGS Soluções Genômicas: Pilar Drummond Sampaio Corrêa Mariani. FZEA-USP Pirassununga: Mirele Daiana Poleti; Raul Machado Neto; Rejane Maria Tommasini Grotto; Ricardo Augusto Brassaloti; Ricardo Haddad; Rodrigo Tocantins Calado.; Rodrigo Tocantins Calado. FAMERP-SJRP: Cecília Artico Banho; Sandra Coccuzzo Sampaio; Sandra Coccuzzo Sampaio Vessoni; Simone Kashima; Svetoslav Nanev Slavov; Vagner Fonseca; Vincent Louis Viala |
| EPI_ISL_1795136, EPI_ISL_1795137, EPI_ISL_1795138, EPI_ISL_1795139, EPI_ISL_1795140, EPI_ISL_1795149, EPI_ISL_1795150 | | | |
| see above | UNIDADE MISTA DE VISTA ALEGRE DO ALTO VISTA ALEGRE DO ALTO | Instituto Butantan / ESALQ-Piracicaba | Antonio Jorge Martins; Bianca Cechetto Carlos. Mendelics: Bibiana Santos; Claudia Renata dos Santos Barros; David Schlesinger. Hemocentro Ribeirão Preto: Simone Kashima; Debora Botequio Moretti. Centro de Genômica Funcional da ESALQ: Luiz Lehmann Coutinho; Dimas Tadeu Covas; Elaine Cristina Marqueze; Elaine Vieira dos Santos; Elisangela Chicaroni Mattos; Erika Freitas; Evandra Strazza Rodrigues; Felipe Allan da Silva da Costa; Flavia Aburjaile; Guilherme Targino Valente; Heidge Fukumasu. USP-Botucatu: Rejane Maria Tommasini Grotto; Instituto Butantan: Alexander Roberto Precioso; Jayme A. Souza-Neto; Jessika Cristina Chagas Lesbon; José Salvatore Leister Patané; João Paulo Kitajima; Luiz Carlos Junior de Alcantara; Maria Carolina Elias; Marta Giovanetti; Patricia Akemi Assato; Rafael dos Santos Bezerra; Raquel de Lello Rocha Campos Cassano. NGS Soluções Genômicas: Pilar Drummond Sampaio Corrêa Mariani. FZEA-USP Pirassununga: Mirele Daiana Poleti; Raul Machado Neto; Ricardo Augusto Brassaloti; Ricardo Haddad; Rodrigo Tocantins Calado.; Sandra Coccuzzo Sampaio; Svetoslav Nanev Slavov; Vagner Fonseca; Vincent Louis Viala |
| EPI_ISL_1966521 | UNIDADE MISTA DE VISTA ALEGRE DO ALTO VISTA ALEGRE DO ALTO | Instituto Butantan / Mendelics | Antonio Jorge Martins; Bianca Cechetto Carlos. Mendelics: Bibiana Santos; Claudia Renata dos Santos Barros; Cíntia Bittar; David Schlesinger. Hemocentro Ribeirão Preto: Simone Kashima; Debora Botequio Moretti; Elaine Cristina Marqueze; Elaine Vieira dos Santos; Elisangela Chicaroni Mattos; Erika Freitas; Evandra Strazza Rodrigues; Felipe Allan da Silva da Costa; Flavia Aburjaile; Fábio Sossai Possebon; Guilherme Campos; Guilherme Targino Valente; Heidge Fukumasu. USP-Botucatu: Rejane Maria Tommasini Grotto; Helena Lage Ferreira; Instituto Butantan: Dimas Tadeu Covas; Jardelina de Souza Todao Bernardino; Jayme A. Souza-Neto; Jessika Cristina Chagas Lesbon; Jorge A. Petroli Marchesi; José Salvatore Leister Patané; João Paulo Kitajima; João Pessoa Araújo Jr.; Leila Sabrina Ullmann; Loyze Paola Oliveira de Lima; Luiz Aurelio de Campos Crispin. Centro de Genômica Funcional da ESALQ: Luiz Lehmann Coutinho; Luiz Carlos Junior de Alcantara; Lívia Sacchetto; Maisa C. Pereira Parra; Maria Carolina Elias; Marta Giovanetti; Marília Moraes; Maurício Lacerda Nogueira. Prefeitura de Sao Paulo: Melissa Palmieri.; Patricia Akemi Assato; Paula Rahal; Paulo Inacio da Costa; Rafael dos Santos Bezerra; Raquel de Lello Rocha Campos Cassano. NGS Soluções Genômicas: Pilar Drummond Sampaio Corrêa Mariani. FZEA-USP Pirassununga: Mirele Daiana Poleti; Raul Machado Neto; Ricardo Augusto Brassaloti; Ricardo Haddad; Rodrigo Tocantins Calado. FAMERP-SJRP: Cecília Artico Banho; Sandra Coccuzzo Sampaio; Svetoslav Nanev Slavov; Vagner Fonseca; Vincent Louis Viala |
| EPI_ISL_1966352, EPI_ISL_1966353, EPI_ISL_1966354, EPI_ISL_1966355, EPI_ISL_1966356 | UNIDADE REFERENCIAL SUDOESTE | Instituto Butantan / Mendelics | Antonio Jorge Martins; Bianca Cechetto Carlos. Mendelics: Bibiana Santos; Claudia Renata dos Santos Barros; Cíntia Bittar; David Schlesinger. Hemocentro Ribeirão Preto: Simone Kashima; Debora Botequio Moretti; Elaine Cristina Marqueze; Elaine Vieira dos Santos; Elisangela Chicaroni Mattos; Erika Freitas; Evandra Strazza Rodrigues; Felipe Allan da Silva da Costa; Flavia Aburjaile; Fábio Sossai Possebon; Guilherme Campos; Guilherme Targino Valente; Heidge Fukumasu. USP-Botucatu: Rejane Maria Tommasini Grotto; Helena Lage Ferreira; Instituto Butantan: Dimas Tadeu Covas; Jardelina de Souza Todao Bernardino; Jayme A. Souza-Neto; Jessika Cristina Chagas Lesbon; Jorge A. Petroli Marchesi; José Salvatore Leister Patané; João Paulo Kitajima; João Pessoa Araújo Jr.; Leila Sabrina Ullmann; Loyze Paola Oliveira de Lima; Luiz Aurelio de Campos Crispin. Centro de Genômica Funcional da ESALQ: Luiz Lehmann Coutinho; Luiz Carlos Junior de Alcantara; Lívia Sacchetto; Maisa C. Pereira Parra; Maria Carolina Elias; Marta Giovanetti; Marília Moraes; Maurício Lacerda Nogueira. Prefeitura de Sao Paulo: Melissa Palmieri.; Patricia Akemi Assato; Paula Rahal; Paulo Inacio da Costa; Rafael dos Santos Bezerra; Raquel de Lello Rocha Campos Cassano. NGS Soluções Genômicas: Pilar Drummond Sampaio Corrêa Mariani. FZEA-USP Pirassununga: Mirele Daiana Poleti; Raul Machado Neto; Ricardo Augusto Brassaloti; Ricardo Haddad; Rodrigo Tocantins Calado. FAMERP-SJRP: Cecília Artico Banho; Sandra Coccuzzo Sampaio; Svetoslav Nanev Slavov; Vagner Fonseca; Vincent Louis Viala |
| EPI_ISL_1445113, EPI_ISL_1445119, EPI_ISL_1445127, EPI_ISL_1445128, EPI_ISL_1445129, EPI_ISL_1966162, EPI_ISL_1966163, EPI_ISL_1966164, EPI_ISL_1966165, EPI_ISL_1966166, EPI_ISL_1966375, EPI_ISL_1966376, EPI_ISL_1966602, EPI_ISL_1966603, EPI_ISL_1966604, EPI_ISL_1966605, EPI_ISL_1966606, EPI_ISL_1967289, EPI_ISL_1967290, EPI_ISL_1967291, EPI_ISL_1967292, EPI_ISL_1967293, EPI_ISL_2170931, EPI_ISL_2171001, EPI_ISL_2171002, EPI_ISL_2171003, EPI_ISL_2171004, EPI_ISL_2171005 | | | |
| see above | UNIDADE RESPIRATORIA NOVA HORTOLANDIA | Instituto Butantan / Mendelics | Antonio Jorge Martins; Bianca Cechetto Carlos. Mendelics: Bibiana Santos; Bibiana Santos; Claudia Renata dos Santos Barros; Cíntia Bittar; David Schlesinger; David Schlesinger. Hemocentro Ribeirão Preto: Simone Kashima; Debora Botequio Moretti; Dimas Tadeu Covas; Elaine Cristina Marqueze; Elaine Vieira Santos; Elaine Vieira dos Santos; Elisangela Chicaroni Mattos; Erika Freitas; Evandra Strazza Rodrigues; Felipe Allan da Silva da Costa; Flavia Aburjaile; Fábio Sossai Possebon; Guilherme Campos; Guilherme Targino Valente; Heidge Fukumasu; Heidge Fukumasu. USP-Botucatu: Rejane Maria Tommasini Grotto; Helena Lage Ferreira; Instituto Butantan: Dimas Tadeu Covas; Jardelina de Souza Todao Bernardino; Jayme A. Souza-Neto; Jayme Augusto de Souza-Neto; Jessika Cristina Chagas Lesbon; Jorge A. Petroli Marchesi; José Salvatore Leister Patané; João Paulo Kitajima; João Pessoa Araújo Jr.; Leila Sabrina Ullmann; Loyze Paola Oliveira de Lima; Luiz Alcantara; Luiz Aurelio de Campos Crispin. Centro de Genômica Funcional da ESALQ: Luiz Lehmann Coutinho; Luiz Carlos Junior de Alcantara; Luiz Lehmann Coutinho; Lívia Sacchetto; Maisa C. Pereira Parra; Maria Carolina Elias; Marta Giovanetti; Marília Moraes; Maurício Lacerda Nogueira; Maurício Lacerda Nogueira. Prefeitura de Sao Paulo: Melissa Palmieri.; Patricia Akemi Assato; Paula Rahal; Paulo Inacio da Costa; Rafael dos Santos Bezerra; Raquel de Lello Rocha Campos Cassano. NGS Soluções Genômicas: Pilar Drummond Sampaio Corrêa Mariani. FZEA-USP Pirassununga: Mirele Daiana Poleti; Raul Machado Neto; Rejane Maria Tommasini Grotto; Ricardo Augusto Brassaloti; Ricardo Haddad; Rodrigo Tocantins Calado.; Rodrigo Tocantins Calado. FAMERP-SJRP: Cecília Artico Banho; Sandra Coccuzzo Sampaio; Sandra Coccuzzo Sampaio Vessoni; Simone Kashima; Svetoslav Nanev Slavov; Vagner Fonseca; Vincent Louis Viala |
| EPI_ISL_1966250 | UNIDADE SAUDE DA FAMILIA DRA ZILDA ARNS NEUMANN JALES | Instituto Butantan / Mendelics | Antonio Jorge Martins; Bianca Cechetto Carlos. Mendelics: Bibiana Santos; Claudia Renata dos Santos Barros; Cíntia Bittar; David Schlesinger. Hemocentro Ribeirão Preto: Simone Kashima; Debora Botequio Moretti; Elaine Cristina Marqueze; Elaine Vieira dos Santos; Elisangela Chicaroni Mattos; Erika Freitas; Evandra Strazza Rodrigues; Felipe Allan da Silva da Costa; Flavia Aburjaile; Fábio Sossai Possebon; Guilherme Campos; Guilherme Targino Valente; Heidge Fukumasu. USP-Botucatu: Rejane Maria Tommasini Grotto; Helena Lage Ferreira; Instituto Butantan: Dimas Tadeu Covas; Jardelina de Souza Todao Bernardino; Jayme A. Souza-Neto; Jessika Cristina Chagas Lesbon; Jorge A. Petroli Marchesi; José Salvatore Leister Patané; João Paulo Kitajima; João Pessoa Araújo Jr.; Leila Sabrina Ullmann; Loyze Paola Oliveira de Lima; Luiz Aurelio de Campos Crispin. Centro de Genômica Funcional da ESALQ: Luiz Lehmann Coutinho; Luiz Carlos Junior de Alcantara; Lívia Sacchetto; Maisa C. Pereira Parra; Maria Carolina Elias; Marta Giovanetti; Marília Moraes; Maurício Lacerda Nogueira. Prefeitura de Sao Paulo: Melissa Palmieri.; Patricia Akemi Assato; Paula Rahal; Paulo Inacio da Costa; Rafael dos Santos Bezerra; Raquel de Lello Rocha Campos Cassano. NGS Soluções Genômicas: Pilar Drummond Sampaio Corrêa Mariani. FZEA-USP Pirassununga: Mirele Daiana Poleti; Raul Machado Neto; Ricardo Augusto Brassaloti; Ricardo Haddad; Rodrigo Tocantins Calado. FAMERP-SJRP: Cecília Artico Banho; Sandra Coccuzzo Sampaio; Svetoslav Nanev Slavov; Vagner Fonseca; Vincent Louis Viala |
| EPI_ISL_1795220 | UNIDADE SENTINELA COVID19 | Instituto Butantan / ESALQ-Piracicaba | Antonio Jorge Martins; Bianca Cechetto Carlos. Mendelics: Bibiana Santos; Claudia Renata dos Santos Barros; David Schlesinger. Hemocentro Ribeirão Preto: Simone Kashima; Debora Botequio Moretti. Centro de Genômica Funcional da ESALQ: Luiz Lehmann Coutinho; Dimas Tadeu Covas; Elaine Cristina Marqueze; Elaine Vieira dos Santos; Elisangela Chicaroni Mattos; Erika Freitas; Evandra Strazza Rodrigues; Felipe Allan da Silva da Costa; Flavia Aburjaile; Guilherme Targino Valente; Heidge Fukumasu. USP-Botucatu: Rejane Maria Tommasini Grotto; Instituto Butantan: Alexander Roberto Precioso; Jayme A. Souza-Neto; Jessika Cristina Chagas Lesbon; José Salvatore Leister Patané; João Paulo Kitajima; Luiz Carlos Junior de Alcantara; Maria Carolina Elias; Marta Giovanetti; Patricia Akemi Assato; Rafael dos Santos Bezerra; Raquel de Lello Rocha Campos Cassano. NGS Soluções Genômicas: Pilar Drummond Sampaio Corrêa Mariani. FZEA-USP Pirassununga: Mirele Daiana Poleti; Raul Machado Neto; Ricardo Augusto Brassaloti; Ricardo Haddad; Rodrigo Tocantins Calado.; Sandra Coccuzzo Sampaio; Svetoslav Nanev Slavov; Vagner Fonseca; Vincent Louis Viala |
| EPI_ISL_1966854, EPI_ISL_1966855, EPI_ISL_1966856, EPI_ISL_1966857, EPI_ISL_1966861, EPI_ISL_1966862 | UNIDADE SENTINELA COVID19 | Instituto Butantan / ESALQ-USP (Piracicaba) | Antonio Jorge Martins; Bianca Cechetto Carlos. Mendelics: Bibiana Santos; Claudia Renata dos Santos Barros; Cíntia Bittar; David Schlesinger. Hemocentro Ribeirão Preto: Simone Kashima; Debora Botequio Moretti; Elaine Cristina Marqueze; Elaine Vieira dos Santos; Elisangela Chicaroni Mattos; Erika Freitas; Evandra Strazza Rodrigues; Felipe Allan da Silva da Costa; Flavia Aburjaile; Fábio Sossai Possebon; Guilherme Campos; Guilherme Targino Valente; Heidge Fukumasu. USP-Botucatu: Rejane Maria Tommasini Grotto; Helena Lage Ferreira; Instituto Butantan: Dimas Tadeu Covas; Jardelina de Souza Todao Bernardino; Jayme A. Souza-Neto; Jessika Cristina Chagas Lesbon; Jorge A. Petroli Marchesi; José Salvatore Leister Patané; João Paulo Kitajima; João Pessoa Araújo Jr.; Leila Sabrina Ullmann; Loyze Paola Oliveira de Lima; Luiz Aurelio de Campos Crispin. Centro de Genômica Funcional da ESALQ: Luiz Lehmann Coutinho; Luiz Carlos Junior de Alcantara; Lívia Sacchetto; Maisa C. Pereira Parra; Maria Carolina Elias; Marta Giovanetti; Marília Moraes; Maurício Lacerda Nogueira. Prefeitura de Sao Paulo: Melissa Palmieri.; Patricia Akemi Assato; Paula Rahal; Paulo Inacio da Costa; Rafael dos Santos Bezerra; Raquel de Lello Rocha Campos Cassano. NGS Soluções Genômicas: Pilar Drummond Sampaio Corrêa Mariani. FZEA-USP Pirassununga: Mirele Daiana Poleti; Raul Machado Neto; Ricardo Augusto Brassaloti; Ricardo Haddad; Rodrigo Tocantins Calado. FAMERP-SJRP: Cecília Artico Banho; Sandra Coccuzzo Sampaio; Svetoslav Nanev Slavov; Vagner Fonseca; Vincent Louis Viala |
| EPI_ISL_1966752, EPI_ISL_1966753, EPI_ISL_1966754 | UNIDADE SENTINELA COVID19 | Instituto Butantan / Mendelics | Antonio Jorge Martins; Bianca Cechetto Carlos. Mendelics: Bibiana Santos; Claudia Renata dos Santos Barros; Cíntia Bittar; David Schlesinger. Hemocentro Ribeirão Preto: Simone Kashima; Debora Botequio Moretti; Elaine Cristina Marqueze; Elaine Vieira dos Santos; Elisangela Chicaroni Mattos; Erika Freitas; Evandra Strazza Rodrigues; Felipe Allan da Silva da Costa; Flavia Aburjaile; Fábio Sossai Possebon; Guilherme Campos; Guilherme Targino Valente; Heidge Fukumasu. USP-Botucatu: Rejane Maria Tommasini Grotto; Helena Lage Ferreira; Instituto Butantan: Dimas Tadeu Covas; Jardelina de Souza Todao Bernardino; Jayme A. Souza-Neto; Jessika Cristina Chagas Lesbon; Jorge A. Petroli Marchesi; José Salvatore Leister Patané; João Paulo Kitajima; João Pessoa Araújo Jr.; Leila Sabrina Ullmann; Loyze Paola Oliveira de Lima; Luiz Aurelio de Campos Crispin. Centro de Genômica Funcional da ESALQ: Luiz Lehmann Coutinho; Luiz Carlos Junior de Alcantara; Lívia Sacchetto; Maisa C. Pereira Parra; Maria Carolina Elias; Marta Giovanetti; Marília Moraes; Maurício Lacerda Nogueira. Prefeitura de Sao Paulo: Melissa Palmieri.; Patricia Akemi Assato; Paula Rahal; Paulo Inacio da Costa; Rafael dos Santos Bezerra; Raquel de Lello Rocha Campos Cassano. NGS Soluções Genômicas: Pilar Drummond Sampaio Corrêa Mariani. FZEA-USP Pirassununga: Mirele Daiana Poleti; Raul Machado Neto; Ricardo Augusto Brassaloti; Ricardo Haddad; Rodrigo Tocantins Calado. FAMERP-SJRP: Cecília Artico Banho; Sandra Coccuzzo Sampaio; Svetoslav Nanev Slavov; Vagner Fonseca; Vincent Louis Viala |
| EPI_ISL_1966858, EPI_ISL_1966859, EPI_ISL_1966860 | UNIDADE SENTINELA VILA APARECIDA | Instituto Butantan / ESALQ-USP (Piracicaba) | Antonio Jorge Martins; Bianca Cechetto Carlos. Mendelics: Bibiana Santos; Claudia Renata dos Santos Barros; Cíntia Bittar; David Schlesinger. Hemocentro Ribeirão Preto: Simone Kashima; Debora Botequio Moretti; Elaine Cristina Marqueze; Elaine Vieira dos Santos; Elisangela Chicaroni Mattos; Erika Freitas; Evandra Strazza Rodrigues; Felipe Allan da Silva da Costa; Flavia Aburjaile; Fábio Sossai Possebon; Guilherme Campos; Guilherme Targino Valente; Heidge Fukumasu. USP-Botucatu: Rejane Maria Tommasini Grotto; Helena Lage Ferreira; Instituto Butantan: Dimas Tadeu Covas; Jardelina de Souza Todao Bernardino; Jayme A. Souza-Neto; Jessika Cristina Chagas Lesbon; Jorge A. Petroli Marchesi; José Salvatore Leister Patané; João Paulo Kitajima; João Pessoa Araújo Jr.; Leila Sabrina Ullmann; Loyze Paola Oliveira de Lima; Luiz Aurelio de Campos Crispin. Centro de Genômica Funcional da ESALQ: Luiz Lehmann Coutinho; Luiz Carlos Junior de Alcantara; Lívia Sacchetto; Maisa C. Pereira Parra; Maria Carolina Elias; Marta Giovanetti; Marília Moraes; Maurício Lacerda Nogueira. Prefeitura de Sao Paulo: Melissa Palmieri.; Patricia Akemi Assato; Paula Rahal; Paulo Inacio da Costa; Rafael dos Santos Bezerra; Raquel de Lello Rocha Campos Cassano. NGS Soluções Genômicas: Pilar Drummond Sampaio Corrêa Mariani. FZEA-USP Pirassununga: Mirele Daiana Poleti; Raul Machado Neto; Ricardo Augusto Brassaloti; Ricardo Haddad; Rodrigo Tocantins Calado. FAMERP-SJRP: Cecília Artico Banho; Sandra Coccuzzo Sampaio; Svetoslav Nanev Slavov; Vagner Fonseca; Vincent Louis Viala |
| EPI_ISL_2157423 | UNIVERSIDADE FEDERAL DE VIÇOSA | Laboratory of Respiratory Viruses and Measles, Oswaldo Cruz Institute, FIOCRUZ | Alice Sampaio Rocha; Ana Carolina Mendonca; Anna Carolina Paixao; Elisa Cavalcante Pereira; Fernando Motta; Luciana Appolinario; Marilda Siqueira on behalf of the Fiocruz COVID-19 Genomic Surveillance Network; Paola Resende; Renata Serrano Lopes; Rubens Pasa; Taina Venas |
| EPI_ISL_1966342 | UPA 24 HORAS CENTRO | Instituto Butantan / Mendelics | Antonio Jorge Martins; Bianca Cechetto Carlos. Mendelics: Bibiana Santos; Claudia Renata dos Santos Barros; Cíntia Bittar; David Schlesinger. Hemocentro Ribeirão Preto: Simone Kashima; Debora Botequio Moretti; Elaine Cristina Marqueze; Elaine Vieira dos Santos; Elisangela Chicaroni Mattos; Erika Freitas; Evandra Strazza Rodrigues; Felipe Allan da Silva da Costa; Flavia Aburjaile; Fábio Sossai Possebon; Guilherme Campos; Guilherme Targino Valente; Heidge Fukumasu. USP-Botucatu: Rejane Maria Tommasini Grotto; Helena Lage Ferreira; Instituto Butantan: Dimas Tadeu Covas; Jardelina de Souza Todao Bernardino; Jayme A. Souza-Neto; Jessika Cristina Chagas Lesbon; Jorge A. Petroli Marchesi; José Salvatore Leister Patané; João Paulo Kitajima; João Pessoa Araújo Jr.; Leila Sabrina Ullmann; Loyze Paola Oliveira de Lima; Luiz Aurelio de Campos Crispin. Centro de Genômica Funcional da ESALQ: Luiz Lehmann Coutinho; Luiz Carlos Junior de Alcantara; Lívia Sacchetto; Maisa C. Pereira Parra; Maria Carolina Elias; Marta Giovanetti; Marília Moraes; Maurício Lacerda Nogueira. Prefeitura de Sao Paulo: Melissa Palmieri.; Patricia Akemi Assato; Paula Rahal; Paulo Inacio da Costa; Rafael dos Santos Bezerra; Raquel de Lello Rocha Campos Cassano. NGS Soluções Genômicas: Pilar Drummond Sampaio Corrêa Mariani. FZEA-USP Pirassununga: |

| | | | |
|---|---|---|---|
| | | | Mirele Daiana Poleti; Raul Machado Neto; Ricardo Augusto Brassaloti; Ricardo Haddad; Rodrigo Tocantins Calado. FAMERP-SJRP: Cecília Artico Banho; Sandra Coccuzzo Sampaio; Svetoslav Nanev Slavov; Vagner Fonseca; Vincent Louis Viala |
| EPI_ISL_1966889, EPI_ISL_1966891, EPI_ISL_1966892, EPI_ISL_1966893, EPI_ISL_1966894 | UPA ATALAIA DRA RITA DE CASSIA SORIO | Instituto Butantan / ESALQ-USP (Piracicaba) | Antonio Jorge Martins; Bianca Cechetto Carlos. Mendelics: Bibiana Santos; Claudia Renata dos Santos Barros; Cíntia Bittar; David Schlesinger. Hemocentro Ribeirão Preto: Simone Kashima; Debora Botequio Moretti; Elaine Cristina Marqueze; Elaine Vieira dos Santos; Elisangela Chicaroni Mattos; Erika Freitas; Evandra Strazza Rodrigues; Felipe Allan da Silva da Costa; Flavia Aburjaile; Fábio Sossai Possebon; Guilherme Campos; Guilherme Targino Valente; Heidge Fukumasu. USP-Botucatu: Rejane Maria Tommasini Grotto; Helena Lage Ferreira; Instituto Butantan: Dimas Tadeu Covas; Jardelina de Souza Todao Bernardino; Jayme A. Souza-Neto; Jessika Cristina Chagas Lesbon; Jorge A. Petroli Marchesi; José Salvatore Leister Patané; João Paulo Kitajima; João Pessoa Araújo Jr.; Leila Sabrina Ullmann; Loyze Paola Oliveira de Lima; Luiz Aurelio de Campos Crispin. Centro de Genômica Funcional da ESALQ: Luiz Lehmann Coutinho; Luiz Carlos Junior de Alcantara; Lívia Sacchetto; Maisa C. Pereira Parra; Maria Carolina Elias; Marta Giovanetti; Marília Moraes; Maurício Lacerda Nogueira. Prefeitura de Sao Paulo: Melissa Palmieri.; Patricia Akemi Assato; Paula Rahal; Paulo Inacio da Costa; Rafael dos Santos Bezerra; Raquel de Lello Rocha Campos Cassano. NGS Soluções Genômicas: Pilar Drummond Sampaio Corrêa Mariani. FZEA-USP Pirassununga: Mirele Daiana Poleti; Raul Machado Neto; Ricardo Augusto Brassaloti; Ricardo Haddad; Rodrigo Tocantins Calado. FAMERP-SJRP: Cecília Artico Banho; Sandra Coccuzzo Sampaio; Svetoslav Nanev Slavov; Vagner Fonseca; Vincent Louis Viala |
| EPI_ISL_1966565, EPI_ISL_1966569, EPI_ISL_1966573 | UPA BELA VISTA | Instituto Butantan / Mendelics | Antonio Jorge Martins; Bianca Cechetto Carlos. Mendelics: Bibiana Santos; Claudia Renata dos Santos Barros; Cíntia Bittar; David Schlesinger. Hemocentro Ribeirão Preto: Simone Kashima; Debora Botequio Moretti; Elaine Cristina Marqueze; Elaine Vieira dos Santos; Elisangela Chicaroni Mattos; Erika Freitas; Evandra Strazza Rodrigues; Felipe Allan da Silva da Costa; Flavia Aburjaile; Fábio Sossai Possebon; Guilherme Campos; Guilherme Targino Valente; Heidge Fukumasu. USP-Botucatu: Rejane Maria Tommasini Grotto; Helena Lage Ferreira; Instituto Butantan: Dimas Tadeu Covas; Jardelina de Souza Todao Bernardino; Jayme A. Souza-Neto; Jessika Cristina Chagas Lesbon; Jorge A. Petroli Marchesi; José Salvatore Leister Patané; João Paulo Kitajima; João Pessoa Araújo Jr.; Leila Sabrina Ullmann; Loyze Paola Oliveira de Lima; Luiz Aurelio de Campos Crispin. Centro de Genômica Funcional da ESALQ: Luiz Lehmann Coutinho; Luiz Carlos Junior de Alcantara; Lívia Sacchetto; Maisa C. Pereira Parra; Maria Carolina Elias; Marta Giovanetti; Marília Moraes; Maurício Lacerda Nogueira. Prefeitura de Sao Paulo: Melissa Palmieri.; Patricia Akemi Assato; Paula Rahal; Paulo Inacio da Costa; Rafael dos Santos Bezerra; Raquel de Lello Rocha Campos Cassano. NGS Soluções Genômicas: Pilar Drummond Sampaio Corrêa Mariani. FZEA-USP Pirassununga: Mirele Daiana Poleti; Raul Machado Neto; Ricardo Augusto Brassaloti; Ricardo Haddad; Rodrigo Tocantins Calado. FAMERP-SJRP: Cecília Artico Banho; Sandra Coccuzzo Sampaio; Svetoslav Nanev Slavov; Vagner Fonseca; Vincent Louis Viala |
| EPI_ISL_1966237, EPI_ISL_1966593, EPI_ISL_1966594, EPI_ISL_1966595, EPI_ISL_1966596, EPI_ISL_1966597, EPI_ISL_1966598, EPI_ISL_1966599, EPI_ISL_1966600, EPI_ISL_1966601, EPI_ISL_1967218, EPI_ISL_1967219, EPI_ISL_1967220, EPI_ISL_1967221, EPI_ISL_1967222, EPI_ISL_1967223, EPI_ISL_1967224, EPI_ISL_1967225, EPI_ISL_1967226, EPI_ISL_1967227, EPI_ISL_2170941, EPI_ISL_2170972, EPI_ISL_2170973 | | | |
| see above | UPA DE BEBEDOURO | Instituto Butantan / Mendelics | Antonio Jorge Martins; Bianca Cechetto Carlos. Mendelics: Bibiana Santos; Claudia Renata dos Santos Barros; Cíntia Bittar; David Schlesinger. Hemocentro Ribeirão Preto: Simone Kashima; Debora Botequio Moretti; Elaine Cristina Marqueze; Elaine Vieira dos Santos; Elisangela Chicaroni Mattos; Erika Freitas; Evandra Strazza Rodrigues; Felipe Allan da Silva da Costa; Flavia Aburjaile; Fábio Sossai Possebon; Guilherme Campos; Guilherme Targino Valente; Heidge Fukumasu. USP-Botucatu: Rejane Maria Tommasini Grotto; Helena Lage Ferreira; Instituto Butantan: Dimas Tadeu Covas; Jardelina de Souza Todao Bernardino; Jayme A. Souza-Neto; Jessika Cristina Chagas Lesbon; Jorge A. Petroli Marchesi; José Salvatore Leister Patané; João Paulo Kitajima; João Pessoa Araújo Jr.; Leila Sabrina Ullmann; Loyze Paola Oliveira de Lima; Luiz Aurelio de Campos Crispin. Centro de Genômica Funcional da ESALQ: Luiz Lehmann Coutinho; Luiz Carlos Junior de Alcantara; Lívia Sacchetto; Maisa C. Pereira Parra; Maria Carolina Elias; Marta Giovanetti; Marília Moraes; Maurício Lacerda Nogueira. Prefeitura de Sao Paulo: Melissa Palmieri.; Patricia Akemi Assato; Paula Rahal; Paulo Inacio da Costa; Rafael dos Santos Bezerra; Raquel de Lello Rocha Campos Cassano. NGS Soluções Genômicas: Pilar Drummond Sampaio Corrêa Mariani. FZEA-USP Pirassununga: Mirele Daiana Poleti; Raul Machado Neto; Ricardo Augusto Brassaloti; Ricardo Haddad; Rodrigo Tocantins Calado. FAMERP-SJRP: Cecília Artico Banho; Sandra Coccuzzo Sampaio; Svetoslav Nanev Slavov; Vagner Fonseca; Vincent Louis Viala |
| EPI_ISL_1966434, EPI_ISL_1966435, EPI_ISL_1966436, EPI_ISL_1966437, EPI_ISL_1966438, EPI_ISL_1967306, EPI_ISL_1967307, EPI_ISL_1967308, EPI_ISL_1967309, EPI_ISL_2171025, EPI_ISL_2171026, EPI_ISL_2171027, EPI_ISL_2171028, EPI_ISL_2171029 | | | |
| see above | UPA DR AKIRA TADA | Instituto Butantan / Mendelics | Antonio Jorge Martins; Bianca Cechetto Carlos. Mendelics: Bibiana Santos; Claudia Renata dos Santos Barros; Cíntia Bittar; David Schlesinger. Hemocentro Ribeirão Preto: Simone Kashima; Debora Botequio Moretti; Elaine Cristina Marqueze; Elaine Vieira dos Santos; Elisangela Chicaroni Mattos; Erika Freitas; Evandra Strazza Rodrigues; Felipe Allan da Silva da Costa; Flavia Aburjaile; Fábio Sossai Possebon; Guilherme Campos; Guilherme Targino Valente; Heidge Fukumasu. USP-Botucatu: Rejane Maria Tommasini Grotto; Helena Lage Ferreira; Instituto Butantan: Dimas Tadeu Covas; Jardelina de Souza Todao Bernardino; Jayme A. Souza-Neto; Jessika Cristina Chagas Lesbon; Jorge A. Petroli Marchesi; José Salvatore Leister Patané; João Paulo Kitajima; João Pessoa Araújo Jr.; Leila Sabrina Ullmann; Loyze Paola Oliveira de Lima; Luiz Aurelio de Campos Crispin. Centro de Genômica Funcional da ESALQ: Luiz Lehmann Coutinho; Luiz Carlos Junior de Alcantara; Lívia Sacchetto; Maisa C. Pereira Parra; Maria Carolina Elias; Marta Giovanetti; Marília Moraes; Maurício Lacerda Nogueira. Prefeitura de Sao Paulo: Melissa Palmieri.; Patricia Akemi Assato; Paula Rahal; Paulo Inacio da Costa; Rafael dos Santos Bezerra; Raquel de Lello Rocha Campos Cassano. NGS Soluções Genômicas: Pilar Drummond Sampaio Corrêa Mariani. FZEA-USP Pirassununga: Mirele Daiana Poleti; Raul Machado Neto; Ricardo Augusto Brassaloti; Ricardo Haddad; Rodrigo Tocantins Calado. FAMERP-SJRP: Cecília Artico Banho; Sandra Coccuzzo Sampaio; Svetoslav Nanev Slavov; Vagner Fonseca; Vincent Louis Viala |
| EPI_ISL_1966369, EPI_ISL_1966370, EPI_ISL_1966371, EPI_ISL_1966372, EPI_ISL_1966373, EPI_ISL_2170996 | UPA DR FRANCO DA ROCHA | Instituto Butantan / Mendelics | Antonio Jorge Martins; Bianca Cechetto Carlos. Mendelics: Bibiana Santos; Claudia Renata dos Santos Barros; Cíntia Bittar; David Schlesinger. Hemocentro Ribeirão Preto: Simone Kashima; Debora Botequio Moretti; Elaine Cristina Marqueze; Elaine Vieira dos Santos; Elisangela Chicaroni Mattos; Erika Freitas; Evandra Strazza Rodrigues; Felipe Allan da Silva da Costa; Flavia Aburjaile; Fábio Sossai Possebon; Guilherme Campos; Guilherme Targino Valente; Heidge Fukumasu. USP-Botucatu: Rejane Maria Tommasini Grotto; Helena Lage Ferreira; Instituto Butantan: Dimas Tadeu Covas; Jardelina de Souza Todao Bernardino; Jayme A. Souza-Neto; Jessika Cristina Chagas Lesbon; Jorge A. Petroli Marchesi; José Salvatore Leister Patané; João Paulo Kitajima; João Pessoa Araújo Jr.; Leila Sabrina Ullmann; Loyze Paola Oliveira de Lima; Luiz Aurelio de Campos Crispin. Centro de Genômica Funcional da ESALQ: Luiz Lehmann Coutinho; Luiz Carlos Junior de Alcantara; Lívia Sacchetto; Maisa C. Pereira Parra; Maria Carolina Elias; Marta Giovanetti; Marília Moraes; Maurício Lacerda Nogueira. Prefeitura de Sao Paulo: Melissa Palmieri.; Patricia Akemi Assato; Paula Rahal; Paulo Inacio da Costa; Rafael dos Santos Bezerra; Raquel de Lello Rocha Campos Cassano. NGS Soluções Genômicas: Pilar Drummond Sampaio Corrêa Mariani. FZEA-USP Pirassununga: Mirele Daiana Poleti; Raul Machado Neto; Ricardo Augusto Brassaloti; Ricardo Haddad; Rodrigo Tocantins Calado. FAMERP-SJRP: Cecília Artico Banho; Sandra Coccuzzo Sampaio; Svetoslav Nanev Slavov; Vagner Fonseca; Vincent Louis Viala |
| EPI_ISL_1628373 | UPA De Bebedouro | Instituto Adolfo Lutz, Interdiciplinary Procedures Center, Strategic Laboratory | Caio Vinicius Dias Lopes; Claudia Regina Gonçalves; Claudio Tavares Sacchi; Erica Valessa Ramos Gomes; Karoline Rodrigues Campos; Katia Correa de Oliveira Santos; Leonardo Jose Tadeu de Araujo |
| EPI_ISL_1628369, EPI_ISL_1628379, EPI_ISL_1715140, EPI_ISL_1731576, EPI_ISL_1752639, EPI_ISL_1821205 | UPA Dr Luis Atilio Losi Viana Ribeirao Preto | Instituto Adolfo Lutz, Interdiciplinary Procedures Center, Strategic Laboratory | Caio Vinicius Dias Lopes; Claudia Regina Gonçalves; Claudio Tavares Sacchi; Erica Valessa Ramos Gomes; Karoline Rodrigues Campos; Katia Correa de Oliveira Santos; Leonardo Jose Tadeu de Araujo |
| EPI_ISL_1967294, EPI_ISL_1967295, EPI_ISL_2171006, EPI_ISL_2171011 | UPA II JUNDIAI | Instituto Butantan / Mendelics | Antonio Jorge Martins; Bianca Cechetto Carlos. Mendelics: Bibiana Santos; Claudia Renata dos Santos Barros; Cíntia Bittar; David Schlesinger. Hemocentro Ribeirão Preto: Simone Kashima; Debora Botequio Moretti; Elaine Cristina Marqueze; Elaine Vieira dos Santos; Elisangela Chicaroni Mattos; Erika Freitas; Evandra Strazza Rodrigues; Felipe Allan da Silva da Costa; Flavia Aburjaile; Fábio Sossai Possebon; Guilherme Campos; Guilherme Targino Valente; Heidge Fukumasu. USP-Botucatu: Rejane Maria Tommasini Grotto; Helena Lage Ferreira; Instituto Butantan: Dimas Tadeu Covas; Jardelina de Souza Todao Bernardino; Jayme A. Souza-Neto; Jessika Cristina Chagas Lesbon; Jorge A. Petroli Marchesi; José Salvatore Leister Patané; João Paulo Kitajima; João Pessoa Araújo Jr.; Leila Sabrina Ullmann; Loyze Paola Oliveira de Lima; Luiz Aurelio de Campos Crispin. Centro de Genômica Funcional da ESALQ: Luiz Lehmann Coutinho; Luiz Carlos Junior de Alcantara; Lívia Sacchetto; Maisa C. Pereira Parra; Maria Carolina Elias; Marta Giovanetti; Marília Moraes; Maurício Lacerda Nogueira. Prefeitura de Sao Paulo: Melissa Palmieri.; Patricia Akemi Assato; Paula Rahal; Paulo Inacio da Costa; Rafael dos Santos Bezerra; Raquel de Lello Rocha Campos Cassano. NGS Soluções Genômicas: Pilar Drummond Sampaio Corrêa Mariani. FZEA-USP Pirassununga: Mirele Daiana Poleti; Raul Machado Neto; Ricardo Augusto Brassaloti; Ricardo Haddad; Rodrigo Tocantins Calado. FAMERP-SJRP: Cecília Artico Banho; Sandra Coccuzzo Sampaio; Svetoslav Nanev Slavov; Vagner Fonseca; Vincent Louis Viala |
| EPI_ISL_1966729 | UPA JOSE CAMPOS BARRETO ZEQUINHA BARRETO | Instituto Butantan | Antonio Jorge Martins; Bianca Cechetto Carlos. Mendelics: Bibiana Santos; Claudia Renata dos Santos Barros; Cíntia Bittar; David Schlesinger. Hemocentro Ribeirão Preto: Simone Kashima; Debora Botequio Moretti; Elaine Cristina Marqueze; Elaine Vieira dos Santos; Elisangela Chicaroni Mattos; Erika Freitas; Evandra Strazza Rodrigues; Felipe Allan da Silva da Costa; Flavia Aburjaile; Fábio Sossai Possebon; Guilherme Campos; Guilherme Targino Valente; Heidge Fukumasu. USP-Botucatu: Rejane Maria Tommasini Grotto; Helena Lage Ferreira; Instituto Butantan: Dimas Tadeu Covas; Jardelina de Souza Todao Bernardino; Jayme A. Souza-Neto; Jessika Cristina Chagas Lesbon; Jorge A. Petroli Marchesi; José Salvatore Leister Patané; João Paulo Kitajima; João Pessoa Araújo Jr.; Leila Sabrina Ullmann; Loyze Paola Oliveira de Lima; Luiz Aurelio de Campos Crispin. Centro de Genômica Funcional da ESALQ: Luiz Lehmann Coutinho; Luiz Carlos Junior de Alcantara; Lívia Sacchetto; Maisa C. Pereira Parra; Maria Carolina Elias; Marta Giovanetti; Marília Moraes; Maurício Lacerda Nogueira. Prefeitura de Sao Paulo: Melissa Palmieri.; Patricia Akemi Assato; Paula Rahal; Paulo Inacio da Costa; Rafael dos Santos Bezerra; Raquel de Lello Rocha Campos Cassano. NGS Soluções Genômicas: Pilar Drummond Sampaio Corrêa Mariani. FZEA-USP Pirassununga: Mirele Daiana Poleti; Raul Machado Neto; Ricardo Augusto Brassaloti; Ricardo Haddad; Rodrigo Tocantins Calado. FAMERP-SJRP: Cecília Artico Banho; Sandra Coccuzzo Sampaio; Svetoslav Nanev Slavov; Vagner Fonseca; Vincent Louis Viala |
| EPI_ISL_1967216, EPI_ISL_2170971 | UPA MARY DOTA | Instituto Butantan / Mendelics | Antonio Jorge Martins; Bianca Cechetto Carlos. Mendelics: Bibiana Santos; Claudia Renata dos Santos Barros; Cíntia Bittar; David Schlesinger. Hemocentro Ribeirão Preto: Simone Kashima; Debora Botequio Moretti; Elaine Cristina Marqueze; Elaine Vieira dos Santos; Elisangela Chicaroni Mattos; Erika Freitas; Evandra Strazza Rodrigues; Felipe Allan da Silva da Costa; Flavia Aburjaile; Fábio Sossai Possebon; Guilherme Campos; Guilherme Targino Valente; Heidge Fukumasu. USP-Botucatu: Rejane Maria Tommasini Grotto; Helena Lage Ferreira; Instituto Butantan: Dimas Tadeu Covas; Jardelina de Souza Todao Bernardino; Jayme A. Souza-Neto; Jessika Cristina Chagas Lesbon; Jorge A. Petroli Marchesi; José Salvatore Leister Patané; João Paulo Kitajima; João Pessoa Araújo Jr.; Leila Sabrina Ullmann; Loyze Paola Oliveira de Lima; Luiz Aurelio de Campos Crispin. Centro de Genômica Funcional da ESALQ: Luiz Lehmann Coutinho; Luiz Carlos Junior de Alcantara; Lívia Sacchetto; Maisa C. Pereira Parra; Maria Carolina Elias; Marta Giovanetti; Marília Moraes; Maurício Lacerda Nogueira. Prefeitura de Sao Paulo: Melissa Palmieri.; Patricia Akemi Assato; Paula Rahal; Paulo Inacio da Costa; Rafael dos Santos Bezerra; Raquel de Lello Rocha Campos Cassano. NGS Soluções Genômicas: Pilar Drummond Sampaio Corrêa Mariani. FZEA-USP Pirassununga: Mirele Daiana Poleti; Raul Machado Neto; Ricardo Augusto Brassaloti; Ricardo Haddad; Rodrigo Tocantins Calado. FAMERP-SJRP: Cecília Artico Banho; Sandra Coccuzzo Sampaio; Svetoslav Nanev Slavov; Vagner Fonseca; Vincent Louis Viala |
| EPI_ISL_1966321, EPI_ISL_1966322 | UPA SILVINA | Instituto Butantan / Mendelics | Antonio Jorge Martins; Bianca Cechetto Carlos. Mendelics: Bibiana Santos; Claudia Renata dos Santos Barros; Cíntia Bittar; David Schlesinger. Hemocentro Ribeirão Preto: Simone Kashima; Debora Botequio Moretti; Elaine Cristina Marqueze; Elaine Vieira dos Santos; Elisangela Chicaroni Mattos; Erika Freitas; Evandra Strazza Rodrigues; Felipe Allan da Silva da Costa; Flavia Aburjaile; Fábio Sossai Possebon; Guilherme Campos; Guilherme Targino Valente; Heidge Fukumasu. USP-Botucatu: Rejane Maria Tommasini Grotto; Helena Lage Ferreira; Instituto Butantan: Dimas Tadeu Covas; Jardelina de Souza Todao Bernardino; Jayme A. Souza-Neto; Jessika Cristina Chagas Lesbon; Jorge A. Petroli Marchesi; José Salvatore Leister Patané; João Paulo Kitajima; João Pessoa Araújo Jr.; Leila Sabrina Ullmann; Loyze Paola Oliveira de Lima; Luiz Aurelio de Campos Crispin. Centro de Genômica Funcional da ESALQ: Luiz Lehmann Coutinho; Luiz Carlos Junior de Alcantara; Lívia Sacchetto; Maisa C. Pereira Parra; Maria Carolina Elias; Marta Giovanetti; Marília Moraes; Maurício Lacerda Nogueira. Prefeitura de Sao Paulo: Melissa Palmieri.; Patricia Akemi Assato; Paula Rahal; Paulo Inacio da Costa; Rafael dos Santos Bezerra; Raquel de Lello Rocha Campos Cassano. NGS Soluções Genômicas: Pilar Drummond Sampaio Corrêa Mariani. FZEA-USP Pirassununga: Mirele Daiana Poleti; Raul Machado Neto; Ricardo Augusto Brassaloti; Ricardo Haddad; Rodrigo Tocantins Calado. FAMERP-SJRP: Cecília Artico Banho; Sandra Coccuzzo Sampaio; Svetoslav Nanev Slavov; Vagner Fonseca; Vincent Louis Viala |
| EPI_ISL_1966650, EPI_ISL_1966651, EPI_ISL_1966652, EPI_ISL_1966653, EPI_ISL_1966654, EPI_ISL_1966655, EPI_ISL_1966656, EPI_ISL_1966657, EPI_ISL_1966658, EPI_ISL_1966659, EPI_ISL_1966660, EPI_ISL_1966661, EPI_ISL_1966662, EPI_ISL_1966663, EPI_ISL_1966664, EPI_ISL_1966665, EPI_ISL_1966666, EPI_ISL_1966667, EPI_ISL_1966668, EPI_ISL_1966669, EPI_ISL_2170946, EPI_ISL_2170947, EPI_ISL_2170948, EPI_ISL_2170949, EPI_ISL_2170950, EPI_ISL_2170951, EPI_ISL_2170952, EPI_ISL_2170953 | | | |
| see above | UPA UNIDADE DE PRONTO ATENDIMENTO 24 HORAS BOM JESUS | Instituto Butantan / Mendelics | Antonio Jorge Martins; Bianca Cechetto Carlos. Mendelics: Bibiana Santos; Claudia Renata dos Santos Barros; Cíntia Bittar; David Schlesinger; David Schlesinger. Hemocentro Ribeirão Preto: Simone Kashima; Debora Botequio Moretti; Dimas Tadeu Covas; Elaine Cristina Marqueze; Elaine Vieira Santos; Elaine Vieira dos Santos; Elisangela Chicaroni Mattos; Erika Freitas; Evandra Strazza Rodrigues; Felipe Allan da Silva da Costa; Flavia Aburjaile; Fábio Sossai Possebon; Guilherme Campos; Guilherme Targino Valente; Heidge Fukumasu; Heidge Fukumasu. USP-Botucatu: Rejane Maria Tommasini Grotto; Helena Lage Ferreira; Instituto Butantan: Dimas Tadeu Covas; Jardelina de Souza Todao Bernardino; Jayme A. Souza-Neto; Jayme Augusto de Souza-Neto; Jessika Cristina Chagas Lesbon; Jorge A. Petroli Marchesi; José Salvatore Leister Patané; João Paulo Kitajima; João Pessoa Araújo Jr.; Leila Sabrina Ullmann; Loyze Paola Oliveira de Lima; Luiz Alcantara; Luiz Aurelio de Campos Crispin. Centro de Genômica Funcional da ESALQ: Luiz Lehmann Coutinho; Luiz Carlos Junior de Alcantara; Luiz Lehmann Coutinho; Lívia Sacchetto; Maisa C. Pereira Parra; Maria Carolina Elias; Marta Giovanetti; Marília Moraes; Maurício Lacerda Nogueira; Maurício Lacerda Nogueira. Prefeitura de Sao Paulo: Melissa Palmieri.; Patricia Akemi Assato; Paula Rahal; Paulo Inacio da Costa; Rafael dos Santos Bezerra; Raquel de Lello Rocha Campos Cassano. NGS Soluções Genômicas: Pilar Drummond Sampaio Corrêa Mariani. FZEA-USP Pirassununga: Mirele Daiana Poleti; Raul Machado Neto; Rejane Maria Tommasini Grotto; Ricardo Augusto Brassaloti; Ricardo Haddad; Rodrigo Tocantins Calado. FAMERP-SJRP: Cecília Artico Banho; Sandra Coccuzzo Sampaio; Sandra Coccuzzo Sampaio Vessoni; Simone Kashima; Svetoslav Nanev Slavov; Vagner Fonseca; Vincent Louis Viala |
| EPI_ISL_1967076 | UPA VILA SANTA CATARINA | Instituto Butantan / ESALQ-USP (Piracicaba) | Antonio Jorge Martins; Bianca Cechetto Carlos. Mendelics: Bibiana Santos; Claudia Renata dos Santos Barros; Cíntia Bittar; David Schlesinger. Hemocentro Ribeirão Preto: Simone Kashima; Debora Botequio Moretti; Elaine Cristina Marqueze; Elaine Vieira dos Santos; Elisangela Chicaroni Mattos; Erika Freitas; Evandra Strazza Rodrigues; Felipe Allan da Silva da Costa; Flavia Aburjaile; Fábio Sossai Possebon; Guilherme Campos; Guilherme Targino Valente; Heidge Fukumasu. USP-Botucatu: Rejane Maria Tommasini Grotto; Helena Lage Ferreira; Instituto Butantan: Dimas Tadeu Covas; Jardelina de Souza Todao Bernardino; Jayme A. Souza-Neto; Jessika Cristina Chagas Lesbon; Jorge A. Petroli Marchesi; José Salvatore Leister Patané; João Paulo Kitajima; João Pessoa Araújo Jr.; Leila Sabrina Ullmann; Loyze Paola Oliveira de Lima; Luiz Aurelio de Campos Crispin. Centro de Genômica Funcional da ESALQ: Luiz Lehmann Coutinho; Luiz Carlos Junior de Alcantara; Lívia Sacchetto; Maisa C. Pereira Parra; Maria Carolina Elias; Marta Giovanetti; Marília Moraes; Maurício Lacerda Nogueira. Prefeitura de Sao Paulo: Melissa Palmieri.; Patricia Akemi Assato; Paula Rahal; Paulo Inacio da Costa; Rafael dos Santos Bezerra; Raquel de Lello Rocha Campos Cassano. NGS Soluções Genômicas: Pilar Drummond Sampaio Corrêa Mariani. FZEA-USP Pirassununga: Mirele Daiana Poleti; Raul Machado Neto; Ricardo Augusto Brassaloti; Ricardo Haddad; Rodrigo Tocantins Calado. FAMERP-SJRP: Cecília Artico Banho; Sandra Coccuzzo Sampaio; Svetoslav Nanev Slavov; Vagner Fonseca; Vincent Louis Viala |
| EPI_ISL_1967298, EPI_ISL_2171020, EPI_ISL_2171021, EPI_ISL_2171022 | UPA VILA SANTA CATARINA | Instituto Butantan / Mendelics | Antonio Jorge Martins; Bianca Cechetto Carlos. Mendelics: Bibiana Santos; Claudia Renata dos Santos Barros; Cíntia Bittar; David Schlesinger. Hemocentro Ribeirão Preto: Simone Kashima; Debora Botequio Moretti; Elaine Cristina Marqueze; Elaine Vieira dos Santos; Elisangela Chicaroni Mattos; Erika Freitas; Evandra Strazza Rodrigues; Felipe Allan da Silva da Costa; Flavia Aburjaile; Fábio Sossai Possebon; Guilherme Campos; Guilherme Targino Valente; Heidge Fukumasu. USP-Botucatu: Rejane Maria Tommasini Grotto; Helena Lage Ferreira; Instituto Butantan: Dimas Tadeu Covas; Jardelina de Souza Todao Bernardino; Jayme A. Souza-Neto; Jessika Cristina Chagas Lesbon; Jorge A. Petroli Marchesi; José Salvatore Leister Patané; João Paulo Kitajima; João Pessoa Araújo Jr.; Leila Sabrina Ullmann; Loyze Paola Oliveira de Lima; Luiz Aurelio de Campos Crispin. Centro de Genômica Funcional da ESALQ: Luiz Lehmann Coutinho; Luiz Carlos Junior de Alcantara; Lívia Sacchetto; Maisa C. Pereira Parra; Maria Carolina Elias; Marta Giovanetti; Marília Moraes; Maurício Lacerda Nogueira. Prefeitura de Sao Paulo: Melissa Palmieri.; Patricia Akemi Assato; Paula Rahal; Paulo Inacio da Costa; Rafael dos Santos Bezerra; Raquel de Lello Rocha Campos Cassano. NGS Soluções Genômicas: Pilar Drummond Sampaio Corrêa Mariani. FZEA-USP Pirassununga: Mirele Daiana Poleti; Raul Machado Neto; Ricardo Augusto Brassaloti; Ricardo Haddad; Rodrigo Tocantins Calado. FAMERP-SJRP: Cecília Artico Banho; Sandra Coccuzzo Sampaio; Svetoslav Nanev Slavov; Vagner Fonseca; Vincent Louis Viala |
| EPI_ISL_1715145 | UPA de Bebedouro | Instituto Adolfo Lutz, Interdiciplinary Procedures Center, Strategic Laboratory | Caio Vinicius Dias Lopes; Claudia Regina Gonçalves; Claudio Tavares Sacchi; Erica Valessa Ramos Gomes; Karoline Rodrigues Campos; Katia Correa de Oliveira Santos; Leonardo Jose Tadeu de Araujo |
| EPI_ISL_1966705 | USAFA ALOHA | Instituto Butantan / Mendelics | Antonio Jorge Martins; Bianca Cechetto Carlos. Mendelics: Bibiana Santos; Claudia Renata dos Santos Barros; Cíntia Bittar; David Schlesinger. Hemocentro Ribeirão Preto: Simone Kashima; Debora Botequio Moretti; Elaine Cristina Marqueze; Elaine Vieira dos Santos; Elisangela Chicaroni Mattos; Erika Freitas; Evandra Strazza Rodrigues; Felipe Allan da Silva da Costa; Flavia Aburjaile; Fábio Sossai Possebon; Guilherme Campos; Guilherme Targino Valente; Heidge Fukumasu. USP-Botucatu: Rejane Maria Tommasini Grotto; Helena Lage Ferreira; Instituto Butantan: Dimas Tadeu Covas; Jardelina de Souza Todao Bernardino; Jayme A. Souza-Neto; Jessika Cristina Chagas Lesbon; Jorge A. Petroli Marchesi; José Salvatore Leister Patané; João Paulo Kitajima; João Pessoa Araújo Jr.; Leila Sabrina Ullmann; Loyze Paola Oliveira de Lima; Luiz Aurelio de Campos Crispin. Centro de Genômica Funcional da ESALQ: Luiz Lehmann Coutinho; Luiz Carlos Junior de Alcantara; Lívia Sacchetto; Maisa C. Pereira Parra; Maria Carolina Elias; Marta Giovanetti; Marília Moraes; Maurício Lacerda Nogueira. Prefeitura de Sao Paulo: Melissa Palmieri.; Patricia Akemi Assato; Paula Rahal; Paulo Inacio da Costa; Rafael dos Santos Bezerra; Raquel de Lello Rocha Campos Cassano. NGS Soluções Genômicas: Pilar Drummond Sampaio Corrêa Mariani. FZEA-USP Pirassununga: Mirele Daiana Poleti; Raul Machado Neto; Ricardo Augusto Brassaloti; Ricardo Haddad; Rodrigo Tocantins Calado. FAMERP-SJRP: Cecília Artico Banho; Sandra Coccuzzo Sampaio; Svetoslav Nanev Slavov; Vagner Fonseca; Vincent Louis Viala |
| EPI_ISL_1966707, EPI_ISL_2170959 | USAFA GUARAMAR | Instituto Butantan / Mendelics | Antonio Jorge Martins; Bianca Cechetto Carlos. Mendelics: Bibiana Santos; Claudia Renata dos Santos Barros; Cíntia Bittar; David Schlesinger. Hemocentro Ribeirão Preto: Simone Kashima; Debora Botequio Moretti; Elaine Cristina Marqueze; Elaine Vieira dos Santos; Elisangela Chicaroni Mattos; Erika Freitas; Evandra Strazza Rodrigues; Felipe Allan da Silva da Costa; Flavia Aburjaile; Fábio Sossai Possebon; Guilherme Campos; Guilherme Targino Valente; Heidge Fukumasu. USP-Botucatu: Rejane Maria Tommasini Grotto; Helena Lage Ferreira; Instituto Butantan: Dimas Tadeu Covas; Jardelina de Souza Todao Bernardino; Jayme A. Souza-Neto; Jessika Cristina Chagas Lesbon; Jorge A. Petroli Marchesi; José Salvatore Leister Patané; João Paulo Kitajima; João Pessoa Araújo Jr.; Leila Sabrina Ullmann; Loyze Paola Oliveira de Lima; Luiz Aurelio de Campos Crispin. Centro de Genômica Funcional da ESALQ: Luiz Lehmann Coutinho; Luiz Carlos Junior de Alcantara; Lívia Sacchetto; Maisa C. Pereira Parra; Maria Carolina Elias; Marta Giovanetti; Marília Moraes; Maurício Lacerda Nogueira. Prefeitura de Sao Paulo: Melissa Palmieri.; Patricia Akemi Assato; Paula Rahal; Paulo Inacio da Costa; Rafael dos Santos Bezerra; Raquel de Lello Rocha Campos Cassano. NGS Soluções Genômicas: Pilar Drummond Sampaio Corrêa Mariani. FZEA-USP Pirassununga: Mirele Daiana Poleti; Raul Machado Neto; Ricardo Augusto Brassaloti; Ricardo Haddad; Rodrigo Tocantins Calado. FAMERP-SJRP: Cecília Artico Banho; Sandra Coccuzzo Sampaio; Svetoslav Nanev Slavov; Vagner Fonseca; Vincent Louis Viala |
| EPI_ISL_1445203, EPI_ISL_1966232 | USAFA GUILHERMINA | Instituto Butantan / Mendelics | Antonio Jorge Martins; Bianca Cechetto Carlos. Mendelics: Bibiana Santos; Bibiana Santos; Claudia Renata dos Santos Barros; Cíntia Bittar; David Schlesinger; David Schlesinger. Hemocentro Ribeirão Preto: Simone Kashima; Debora Botequio Moretti; Dimas Tadeu Covas; Elaine Cristina Marqueze; Elaine Vieira dos Santos; Elisangela Chicaroni Mattos; Erika Freitas; Evandra Strazza Rodrigues; Felipe Allan da Silva da Costa; Flavia Aburjaile; Fábio Sossai Possebon; Guilherme Campos; Guilherme Targino Valente; Heidge Fukumasu. USP-Botucatu: Rejane Maria Tommasini Grotto; Helena Lage Ferreira; Instituto Butantan: Dimas Tadeu Covas; Jardelina de Souza Todao Bernardino; Jayme A. Souza-Neto; Jessika Cristina Chagas Lesbon; Jorge A. Petroli Marchesi; José Salvatore Leister Patané; João Paulo Kitajima; João Pessoa Araújo Jr.; Leila Sabrina Ullmann; Loyze Paola Oliveira de Lima; Luiz Aurelio de Campos Crispin. Centro de Genômica Funcional da ESALQ: Luiz Lehmann Coutinho; Luiz Carlos Junior de Alcantara; Lívia Sacchetto; Maisa C. Pereira Parra; Maria Carolina Elias; Marta Giovanetti; Marília Moraes; Maurício Lacerda Nogueira. Prefeitura de Sao Paulo: Melissa Palmieri.; Patricia Akemi Assato; Paula Rahal; Paulo Inacio da Costa; Rafael dos Santos Bezerra; Raquel de Lello Rocha Campos Cassano. NGS Soluções Genômicas: Pilar Drummond Sampaio Corrêa Mariani. FZEA-USP Pirassununga: Mirele Daiana Poleti; Raul Machado Neto; Ricardo Augusto Brassaloti; Ricardo Haddad; Rodrigo Tocantins Calado.; Rodrigo Tocantins Calado. FAMERP-SJRP: Cecília Artico Banho; Sandra Coccuzzo Sampaio; Simone Kashima; Svetoslav Nanev Slavov; Vagner Fonseca; Vincent Louis Viala |
| EPI_ISL_1445202, EPI_ISL_1445204, EPI_ISL_1966233, | USAFA MARACANA | Instituto Butantan / Mendelics | Antonio Jorge Martins; Bianca Cechetto Carlos. Mendelics: Bibiana Santos; Bibiana Santos; Claudia Renata dos Santos Barros; Cíntia Bittar; David Schlesinger; David Schlesinger. Hemocentro Ribeirão Preto: Simone Kashima; Debora Botequio Moretti; Dimas Tadeu Covas; Elaine Cristina Marqueze; Elaine Vieira dos Santos; Elisangela Chicaroni Mattos; Erika Freitas; Evandra Strazza Rodrigues; Felipe Allan da Silva da Costa; Flavia Aburjaile; Fábio Sossai Possebon; Guilherme Campos; Guilherme Targino Valente; Heidge Fukumasu. USP-Botucatu: Rejane Maria Tommasini Grotto; Helena Lage Ferreira; Instituto Butantan: Dimas Tadeu Covas; Jardelina de Souza Todao Bernardino; Jayme A. Souza-Neto; Jessika Cristina Chagas Lesbon; Jorge A. Petroli Marchesi; José Salvatore Leister Patané; João Paulo Kitajima; João Pessoa Araújo Jr.; Leila Sabrina Ullmann; Loyze Paola Oliveira de Lima; Luiz Aurelio de Campos Crispin. Centro de Genômica Funcional da ESALQ: Luiz Lehmann Coutinho; Luiz Carlos Junior de |

| | | | |
|---|---|---|---|
| EPI_ISL_2170920 | | | Alcantara; Lívia Sacchetto; Maisa C. Pereira Parra; Maria Carolina Elias; Marta Giovanetti; Marília Moraes; Maurício Lacerda Nogueira. Prefeitura de Sao Paulo: Melissa Palmieri.; Patricia Akemi Assato; Paula Rahal; Paulo Inacio da Costa; Rafael dos Santos Bezerra; Raquel de Lello Rocha Campos Cassano. NGS Soluções Genômicas: Pilar Drummond Sampaio Corrêa Mariani. FZEA-USP Pirassununga: Mirele Daiana Poleti; Raul Machado Neto; Ricardo Augusto Brassaloti; Ricardo Haddad; Rodrigo Tocantins Calado.; Rodrigo Tocantins Calado. FAMERP-SJRP: Cecília Artico Banho; Sandra Coccuzzo Sampaio; Simone Kashima; Svetoslav Nanev Slavov; Vagner Fonseca; Vincent Louis Viala |
| EPI_ISL_1966704 | USAFA REAL | Instituto Butantan / Mendelics | Antonio Jorge Martins; Bianca Cechetto Carlos. Mendelics: Bibiana Santos; Claudia Renata dos Santos Barros; Cíntia Bittar; David Schlesinger. Hemocentro Ribeirão Preto: Simone Kashima; Debora Botequio Moretti; Elaine Cristina Marqueze; Elaine Vieira dos Santos; Elisangela Chicaroni Mattos; Erika Freitas; Evandra Strazza Rodrigues; Felipe Allan da Silva da Costa; Flavia Aburjaile; Fábio Sossai Possebon; Guilherme Campos; Guilherme Targino Valente; Heidge Fukumasu. USP-Botucatu: Rejane Maria Tommasini Grotto; Helena Lage Ferreira; Instituto Butantan: Dimas Tadeu Covas; Jardelina de Souza Todao Bernardino; Jayme A. Souza-Neto; Jessika Cristina Chagas Lesbon; Jorge A. Petroli Marchesi; José Salvatore Leister Patané; João Paulo Kitajima; João Pessoa Araújo Jr.; Leila Sabrina Ullmann; Loyze Paola Oliveira de Lima; Luiz Aurelio de Campos Crispin. Centro de Genômica Funcional da ESALQ: Luiz Lehmann Coutinho; Luiz Carlos Junior de Alcantara; Lívia Sacchetto; Maisa C. Pereira Parra; Maria Carolina Elias; Marta Giovanetti; Marília Moraes; Maurício Lacerda Nogueira. Prefeitura de Sao Paulo: Melissa Palmieri.; Patricia Akemi Assato; Paula Rahal; Paulo Inacio da Costa; Rafael dos Santos Bezerra; Raquel de Lello Rocha Campos Cassano. NGS Soluções Genômicas: Pilar Drummond Sampaio Corrêa Mariani. FZEA-USP Pirassununga: Mirele Daiana Poleti; Raul Machado Neto; Ricardo Augusto Brassaloti; Ricardo Haddad; Rodrigo Tocantins Calado. FAMERP-SJRP: Cecília Artico Banho; Sandra Coccuzzo Sampaio; Svetoslav Nanev Slavov; Vagner Fonseca; Vincent Louis Viala |
| EPI_ISL_1445201, EPI_ISL_1966234 | USAFA TUDE BASTOS | Instituto Butantan / Mendelics | Antonio Jorge Martins; Bianca Cechetto Carlos. Mendelics: Bibiana Santos; Bibiana Santos; Claudia Renata dos Santos Barros; Cíntia Bittar; David Schlesinger; David Schlesinger. Hemocentro Ribeirão Preto: Simone Kashima; Debora Botequio Moretti; Dimas Tadeu Covas; Elaine Cristina Marqueze; Elaine Vieira dos Santos; Elisangela Chicaroni Mattos; Erika Freitas; Evandra Strazza Rodrigues; Felipe Allan da Silva da Costa; Flavia Aburjaile; Fábio Sossai Possebon; Guilherme Campos; Guilherme Targino Valente; Heidge Fukumasu. USP-Botucatu: Rejane Maria Tommasini Grotto; Helena Lage Ferreira; Instituto Butantan: Dimas Tadeu Covas; Jardelina de Souza Todao Bernardino; Jayme A. Souza-Neto; Jessika Cristina Chagas Lesbon; Jorge A. Petroli Marchesi; José Salvatore Leister Patané; João Paulo Kitajima; João Pessoa Araújo Jr.; Leila Sabrina Ullmann; Loyze Paola Oliveira de Lima; Luiz Aurelio de Campos Crispin. Centro de Genômica Funcional da ESALQ: Luiz Lehmann Coutinho; Luiz Carlos Junior de Alcantara; Lívia Sacchetto; Maisa C. Pereira Parra; Maria Carolina Elias; Marta Giovanetti; Marília Moraes; Maurício Lacerda Nogueira. Prefeitura de Sao Paulo: Melissa Palmieri.; Patricia Akemi Assato; Paula Rahal; Paulo Inacio da Costa; Rafael dos Santos Bezerra; Raquel de Lello Rocha Campos Cassano. NGS Soluções Genômicas: Pilar Drummond Sampaio Corrêa Mariani. FZEA-USP Pirassununga: Mirele Daiana Poleti; Raul Machado Neto; Ricardo Augusto Brassaloti; Ricardo Haddad; Rodrigo Tocantins Calado.; Rodrigo Tocantins Calado. FAMERP-SJRP: Cecília Artico Banho; Sandra Coccuzzo Sampaio; Simone Kashima; Svetoslav Nanev Slavov; Vagner Fonseca; Vincent Louis Viala |
| EPI_ISL_1445070, EPI_ISL_2170901 | USF ARCADAS | Instituto Butantan / Mendelics | Antonio Jorge Martins; Bianca Cechetto Carlos. Mendelics: Bibiana Santos; Bibiana Santos; Claudia Renata dos Santos Barros; Cíntia Bittar; David Schlesinger; David Schlesinger. Hemocentro Ribeirão Preto: Simone Kashima; Debora Botequio Moretti; Dimas Tadeu Covas; Elaine Cristina Marqueze; Elaine Vieira dos Santos; Elisangela Chicaroni Mattos; Erika Freitas; Evandra Strazza Rodrigues; Felipe Allan da Silva da Costa; Flavia Aburjaile; Fábio Sossai Possebon; Guilherme Campos; Guilherme Targino Valente; Heidge Fukumasu. USP-Botucatu: Rejane Maria Tommasini Grotto; Helena Lage Ferreira; Instituto Butantan: Dimas Tadeu Covas; Jardelina de Souza Todao Bernardino; Jayme A. Souza-Neto; Jessika Cristina Chagas Lesbon; Jorge A. Petroli Marchesi; José Salvatore Leister Patané; João Paulo Kitajima; João Pessoa Araújo Jr.; Leila Sabrina Ullmann; Loyze Paola Oliveira de Lima; Luiz Aurelio de Campos Crispin. Centro de Genômica Funcional da ESALQ: Luiz Lehmann Coutinho; Luiz Carlos Junior de Alcantara; Lívia Sacchetto; Maisa C. Pereira Parra; Maria Carolina Elias; Marta Giovanetti; Marília Moraes; Maurício Lacerda Nogueira. Prefeitura de Sao Paulo: Melissa Palmieri.; Patricia Akemi Assato; Paula Rahal; Paulo Inacio da Costa; Rafael dos Santos Bezerra; Raquel de Lello Rocha Campos Cassano. NGS Soluções Genômicas: Pilar Drummond Sampaio Corrêa Mariani. FZEA-USP Pirassununga: Mirele Daiana Poleti; Raul Machado Neto; Ricardo Augusto Brassaloti; Ricardo Haddad; Rodrigo Tocantins Calado.; Rodrigo Tocantins Calado. FAMERP-SJRP: Cecília Artico Banho; Sandra Coccuzzo Sampaio; Simone Kashima; Svetoslav Nanev Slavov; Vagner Fonseca; Vincent Louis Viala |
| EPI_ISL_1966542 | USF DR APARECIDO RODRIGUES MOUCO SCRPARDO | Instituto Butantan / FZEA-USP (Pirassununga) | Antonio Jorge Martins; Bianca Cechetto Carlos. Mendelics: Bibiana Santos; Claudia Renata dos Santos Barros; Cíntia Bittar; David Schlesinger. Hemocentro Ribeirão Preto: Simone Kashima; Debora Botequio Moretti; Elaine Cristina Marqueze; Elaine Vieira dos Santos; Elisangela Chicaroni Mattos; Erika Freitas; Evandra Strazza Rodrigues; Felipe Allan da Silva da Costa; Flavia Aburjaile; Fábio Sossai Possebon; Guilherme Campos; Guilherme Targino Valente; Heidge Fukumasu. USP-Botucatu: Rejane Maria Tommasini Grotto; Helena Lage Ferreira; Instituto Butantan: Dimas Tadeu Covas; Jardelina de Souza Todao Bernardino; Jayme A. Souza-Neto; Jessika Cristina Chagas Lesbon; Jorge A. Petroli Marchesi; José Salvatore Leister Patané; João Paulo Kitajima; João Pessoa Araújo Jr.; Leila Sabrina Ullmann; Loyze Paola Oliveira de Lima; Luiz Aurelio de Campos Crispin. Centro de Genômica Funcional da ESALQ: Luiz Lehmann Coutinho; Luiz Carlos Junior de Alcantara; Lívia Sacchetto; Maisa C. Pereira Parra; Maria Carolina Elias; Marta Giovanetti; Marília Moraes; Maurício Lacerda Nogueira. Prefeitura de Sao Paulo: Melissa Palmieri.; Patricia Akemi Assato; Paula Rahal; Paulo Inacio da Costa; Rafael dos Santos Bezerra; Raquel de Lello Rocha Campos Cassano. NGS Soluções Genômicas: Pilar Drummond Sampaio Corrêa Mariani. FZEA-USP Pirassununga: Mirele Daiana Poleti; Raul Machado Neto; Ricardo Augusto Brassaloti; Ricardo Haddad; Rodrigo Tocantins Calado. FAMERP-SJRP: Cecília Artico Banho; Sandra Coccuzzo Sampaio; Svetoslav Nanev Slavov; Vagner Fonseca; Vincent Louis Viala |
| EPI_ISL_1795290 | USF EUCALIPTOS | Instituto Butantan / ESALQ-Piracicaba | Antonio Jorge Martins; Bianca Cechetto Carlos. Mendelics: Bibiana Santos; Claudia Renata dos Santos Barros; David Schlesinger. Hemocentro Ribeirão Preto: Simone Kashima; Debora Botequio Moretti. Centro de Genômica Funcional da ESALQ: Luiz Lehmann Coutinho; Dimas Tadeu Covas; Elaine Cristina Marqueze; Elaine Vieira dos Santos; Elisangela Chicaroni Mattos; Erika Freitas; Evandra Strazza Rodrigues; Felipe Allan da Silva da Costa; Flavia Aburjaile; Guilherme Targino Valente; Heidge Fukumasu. USP-Botucatu: Rejane Maria Tommasini Grotto; Instituto Butantan: Alexander Roberto Precioso; Jayme A. Souza-Neto; Jessika Cristina Chagas Lesbon; José Salvatore Leister Patané; João Paulo Kitajima; Luiz Carlos Junior de Alcantara; Maria Carolina Elias; Marta Giovanetti; Patricia Akemi Assato; Rafael dos Santos Bezerra; Raquel de Lello Rocha Campos Cassano. NGS Soluções Genômicas: Pilar Drummond Sampaio Corrêa Mariani. FZEA-USP Pirassununga: Mirele Daiana Poleti; Raul Machado Neto; Ricardo Augusto Brassaloti; Ricardo Haddad; Rodrigo Tocantins Calado.; Sandra Coccuzzo Sampaio; Svetoslav Nanev Slavov; Vagner Fonseca; Vincent Louis Viala |
| EPI_ISL_1795295 | USF GUACUANO | Instituto Butantan / ESALQ-Piracicaba | Antonio Jorge Martins; Bianca Cechetto Carlos. Mendelics: Bibiana Santos; Claudia Renata dos Santos Barros; David Schlesinger. Hemocentro Ribeirão Preto: Simone Kashima; Debora Botequio Moretti. Centro de Genômica Funcional da ESALQ: Luiz Lehmann Coutinho; Dimas Tadeu Covas; Elaine Cristina Marqueze; Elaine Vieira dos Santos; Elisangela Chicaroni Mattos; Erika Freitas; Evandra Strazza Rodrigues; Felipe Allan da Silva da Costa; Flavia Aburjaile; Guilherme Targino Valente; Heidge Fukumasu. USP-Botucatu: Rejane Maria Tommasini Grotto; Instituto Butantan: Alexander Roberto Precioso; Jayme A. Souza-Neto; Jessika Cristina Chagas Lesbon; José Salvatore Leister Patané; João Paulo Kitajima; Luiz Carlos Junior de Alcantara; Maria Carolina Elias; Marta Giovanetti; Patricia Akemi Assato; Rafael dos Santos Bezerra; Raquel de Lello Rocha Campos Cassano. NGS Soluções Genômicas: Pilar Drummond Sampaio Corrêa Mariani. FZEA-USP Pirassununga: Mirele Daiana Poleti; Raul Machado Neto; Ricardo Augusto Brassaloti; Ricardo Haddad; Rodrigo Tocantins Calado.; Sandra Coccuzzo Sampaio; Svetoslav Nanev Slavov; Vagner Fonseca; Vincent Louis Viala |
| EPI_ISL_1966907, EPI_ISL_1966908 | USF I DE GUAIMBE | Instituto Butantan / ESALQ-USP (Piracicaba) | Antonio Jorge Martins; Bianca Cechetto Carlos. Mendelics: Bibiana Santos; Claudia Renata dos Santos Barros; Cíntia Bittar; David Schlesinger. Hemocentro Ribeirão Preto: Simone Kashima; Debora Botequio Moretti; Elaine Cristina Marqueze; Elaine Vieira dos Santos; Elisangela Chicaroni Mattos; Erika Freitas; Evandra Strazza Rodrigues; Felipe Allan da Silva da Costa; Flavia Aburjaile; Fábio Sossai Possebon; Guilherme Campos; Guilherme Targino Valente; Heidge Fukumasu. USP-Botucatu: Rejane Maria Tommasini Grotto; Helena Lage Ferreira; Instituto Butantan: Dimas Tadeu Covas; Jardelina de Souza Todao Bernardino; Jayme A. Souza-Neto; Jessika Cristina Chagas Lesbon; Jorge A. Petroli Marchesi; José Salvatore Leister Patané; João Paulo Kitajima; João Pessoa Araújo Jr.; Leila Sabrina Ullmann; Loyze Paola Oliveira de Lima; Luiz Aurelio de Campos Crispin. Centro de Genômica Funcional da ESALQ: Luiz Lehmann Coutinho; Luiz Carlos Junior de Alcantara; Lívia Sacchetto; Maisa C. Pereira Parra; Maria Carolina Elias; Marta Giovanetti; Marília Moraes; Maurício Lacerda Nogueira. Prefeitura de Sao Paulo: Melissa Palmieri.; Patricia Akemi Assato; Paula Rahal; Paulo Inacio da Costa; Rafael dos Santos Bezerra; Raquel de Lello Rocha Campos Cassano. NGS Soluções Genômicas: Pilar Drummond Sampaio Corrêa Mariani. FZEA-USP Pirassununga: Mirele Daiana Poleti; Raul Machado Neto; Ricardo Augusto Brassaloti; Ricardo Haddad; Rodrigo Tocantins Calado. FAMERP-SJRP: Cecília Artico Banho; Sandra Coccuzzo Sampaio; Svetoslav Nanev Slavov; Vagner Fonseca; Vincent Louis Viala |
| EPI_ISL_1966374 | USF II DE HERCULANDIA | Instituto Butantan / Mendelics | Antonio Jorge Martins; Bianca Cechetto Carlos. Mendelics: Bibiana Santos; Claudia Renata dos Santos Barros; Cíntia Bittar; David Schlesinger. Hemocentro Ribeirão Preto: Simone Kashima; Debora Botequio Moretti; Elaine Cristina Marqueze; Elaine Vieira dos Santos; Elisangela Chicaroni Mattos; Erika Freitas; Evandra Strazza Rodrigues; Felipe Allan da Silva da Costa; Flavia Aburjaile; Fábio Sossai Possebon; Guilherme Campos; Guilherme Targino Valente; Heidge Fukumasu. USP-Botucatu: Rejane Maria Tommasini Grotto; Helena Lage Ferreira; Instituto Butantan: Dimas Tadeu Covas; Jardelina de Souza Todao Bernardino; Jayme A. Souza-Neto; Jessika Cristina Chagas Lesbon; Jorge A. Petroli Marchesi; José Salvatore Leister Patané; João Paulo Kitajima; João Pessoa Araújo Jr.; Leila Sabrina Ullmann; Loyze Paola Oliveira de Lima; Luiz Aurelio de Campos Crispin. Centro de Genômica Funcional da ESALQ: Luiz Lehmann Coutinho; Luiz Carlos Junior de Alcantara; Lívia Sacchetto; Maisa C. Pereira Parra; Maria Carolina Elias; Marta Giovanetti; Marília Moraes; Maurício Lacerda Nogueira. Prefeitura de Sao Paulo: Melissa Palmieri.; Patricia Akemi Assato; Paula Rahal; Paulo Inacio da Costa; Rafael dos Santos Bezerra; Raquel de Lello Rocha Campos Cassano. NGS Soluções Genômicas: Pilar Drummond Sampaio Corrêa Mariani. FZEA-USP Pirassununga: Mirele Daiana Poleti; Raul Machado Neto; Ricardo Augusto Brassaloti; Ricardo Haddad; Rodrigo Tocantins Calado. FAMERP-SJRP: Cecília Artico Banho; Sandra Coccuzzo Sampaio; Svetoslav Nanev Slavov; Vagner Fonseca; Vincent Louis Viala |
| EPI_ISL_1445068 | USF JARDIM DAS AVES MOREIRINHA | Instituto Butantan / Mendelics | Antonio Jorge Martins; Bibiana Santos; Claudia Renata dos Santos Barros; David Schlesinger; Debora Botequio Moretti; Dimas Tadeu Covas; Elaine Cristina Marqueze; Elaine Vieira dos Santos; Erika Freitas; Evandra Strazza Rodrigues; Flavia Aburjaile; José Salvatore Leister Patané; João Paulo Kitajima; Luiz Carlos Junior de Alcantara; Maria Carolina Elias; Marta Giovanetti; Rafael dos Santos Bezerra; Raul Machado Neto; Ricardo Haddad; Rodrigo Tocantins Calado.; Sandra Coccuzzo Sampaio; Simone Kashima; Svetoslav Nanev Slavov; Vagner Fonseca; Vincent Louis Viala |
| EPI_ISL_1445069, EPI_ISL_1445072, EPI_ISL_2170902 | USF JARDIM SAO DIMAS | Instituto Butantan / Mendelics | Antonio Jorge Martins; Bianca Cechetto Carlos. Mendelics: Bibiana Santos; Bibiana Santos; Claudia Renata dos Santos Barros; Cíntia Bittar; David Schlesinger; David Schlesinger. Hemocentro Ribeirão Preto: Simone Kashima; Debora Botequio Moretti; Dimas Tadeu Covas; Elaine Cristina Marqueze; Elaine Vieira dos Santos; Elisangela Chicaroni Mattos; Erika Freitas; Evandra Strazza Rodrigues; Felipe Allan da Silva da Costa; Flavia Aburjaile; Fábio Sossai Possebon; Guilherme Campos; Guilherme Targino Valente; Heidge Fukumasu. USP-Botucatu: Rejane Maria Tommasini Grotto; Helena Lage Ferreira; Instituto Butantan: Dimas Tadeu Covas; Jardelina de Souza Todao Bernardino; Jayme A. Souza-Neto; Jessika Cristina Chagas Lesbon; Jorge A. Petroli Marchesi; José Salvatore Leister Patané; João Paulo Kitajima; João Pessoa Araújo Jr.; Leila Sabrina Ullmann; Loyze Paola Oliveira de Lima; Luiz Aurelio de Campos Crispin. Centro de Genômica Funcional da ESALQ: Luiz Lehmann Coutinho; Luiz Carlos Junior de Alcantara; Lívia Sacchetto; Maisa C. Pereira Parra; Maria Carolina Elias; Marta Giovanetti; Marília Moraes; Maurício Lacerda Nogueira. Prefeitura de Sao Paulo: Melissa Palmieri.; Patricia Akemi Assato; Paula Rahal; Paulo Inacio da Costa; Rafael dos Santos Bezerra; Raquel de Lello Rocha Campos Cassano. NGS Soluções Genômicas: Pilar Drummond Sampaio Corrêa Mariani. FZEA-USP Pirassununga: Mirele Daiana Poleti; Raul Machado Neto; Ricardo Augusto Brassaloti; Ricardo Haddad; Rodrigo Tocantins Calado.; Rodrigo Tocantins Calado. FAMERP-SJRP: Cecília Artico Banho; Sandra Coccuzzo Sampaio; Simone Kashima; Svetoslav Nanev Slavov; Vagner Fonseca; Vincent Louis Viala |
| EPI_ISL_1966812 | USF JARDIM SAO JOAO SCRPARDO | Instituto Butantan / Mendelics | Antonio Jorge Martins; Bianca Cechetto Carlos. Mendelics: Bibiana Santos; Claudia Renata dos Santos Barros; Cíntia Bittar; David Schlesinger. Hemocentro Ribeirão Preto: Simone Kashima; Debora Botequio Moretti; Elaine Cristina Marqueze; Elaine Vieira dos Santos; Elisangela Chicaroni Mattos; Erika Freitas; Evandra Strazza Rodrigues; Felipe Allan da Silva da Costa; Flavia Aburjaile; Fábio Sossai Possebon; Guilherme Campos; Guilherme Targino Valente; Heidge Fukumasu. USP-Botucatu: Rejane Maria Tommasini Grotto; Helena Lage Ferreira; Instituto Butantan: Dimas Tadeu Covas; Jardelina de Souza Todao Bernardino; Jayme A. Souza-Neto; Jessika Cristina Chagas Lesbon; Jorge A. Petroli Marchesi; José Salvatore Leister Patané; João Paulo Kitajima; João Pessoa Araújo Jr.; Leila Sabrina Ullmann; Loyze Paola Oliveira de Lima; Luiz Aurelio de Campos Crispin. Centro de Genômica Funcional da ESALQ: Luiz Lehmann Coutinho; Luiz Carlos Junior de Alcantara; Lívia Sacchetto; Maisa C. Pereira Parra; Maria Carolina Elias; Marta Giovanetti; Marília Moraes; Maurício Lacerda Nogueira. Prefeitura de Sao Paulo: Melissa Palmieri.; Patricia Akemi Assato; Paula Rahal; Paulo Inacio da Costa; Rafael dos Santos Bezerra; Raquel de Lello Rocha Campos Cassano. NGS Soluções Genômicas: Pilar Drummond Sampaio Corrêa Mariani. FZEA-USP Pirassununga: Mirele Daiana Poleti; Raul Machado Neto; Ricardo Augusto Brassaloti; Ricardo Haddad; Rodrigo Tocantins Calado. FAMERP-SJRP: Cecília Artico Banho; Sandra Coccuzzo Sampaio; Svetoslav Nanev Slavov; Vagner Fonseca; Vincent Louis Viala |
| EPI_ISL_2170968 | USF JOAO NASCIMENTO TELLES | Instituto Butantan / Mendelics | Antonio Jorge Martins; Bianca Cechetto Carlos. Mendelics: Bibiana Santos; Claudia Renata dos Santos Barros; Cíntia Bittar; David Schlesinger. Hemocentro Ribeirão Preto: Simone Kashima; Debora Botequio Moretti; Elaine Cristina Marqueze; Elaine Vieira dos Santos; Elisangela Chicaroni Mattos; Erika Freitas; Evandra Strazza Rodrigues; Felipe Allan da Silva da Costa; Flavia Aburjaile; Fábio Sossai Possebon; Guilherme Campos; Guilherme Targino Valente; Heidge Fukumasu. USP-Botucatu: Rejane Maria Tommasini Grotto; Helena Lage Ferreira; Instituto Butantan: Dimas Tadeu Covas; Jardelina de Souza Todao Bernardino; Jayme A. Souza-Neto; Jessika Cristina Chagas Lesbon; Jorge A. Petroli Marchesi; José Salvatore Leister Patané; João Paulo Kitajima; João Pessoa Araújo Jr.; Leila Sabrina Ullmann; Loyze Paola Oliveira de Lima; Luiz Aurelio de Campos Crispin. Centro de Genômica Funcional da ESALQ: Luiz Lehmann Coutinho; Luiz Carlos Junior de Alcantara; Lívia Sacchetto; Maisa C. Pereira Parra; Maria Carolina Elias; Marta Giovanetti; Marília Moraes; Maurício Lacerda Nogueira. Prefeitura de Sao Paulo: Melissa Palmieri.; Patricia Akemi Assato; Paula Rahal; Paulo Inacio da Costa; Rafael dos Santos Bezerra; Raquel de Lello Rocha Campos Cassano. NGS Soluções Genômicas: Pilar Drummond Sampaio Corrêa Mariani. FZEA-USP Pirassununga: Mirele Daiana Poleti; Raul Machado Neto; Ricardo Augusto Brassaloti; Ricardo Haddad; Rodrigo Tocantins Calado. FAMERP-SJRP: Cecília Artico Banho; Sandra Coccuzzo Sampaio; Svetoslav Nanev Slavov; Vagner Fonseca; Vincent Louis Viala |
| EPI_ISL_1966626 | USF MARGARIDA JANETE FERRARI GANZAROLI | Instituto Butantan / Mendelics | Antonio Jorge Martins; Bianca Cechetto Carlos. Mendelics: Bibiana Santos; Claudia Renata dos Santos Barros; Cíntia Bittar; David Schlesinger. Hemocentro Ribeirão Preto: Simone Kashima; Debora Botequio Moretti; Elaine Cristina Marqueze; Elaine Vieira dos Santos; Elisangela Chicaroni Mattos; Erika Freitas; Evandra Strazza Rodrigues; Felipe Allan da Silva da Costa; Flavia Aburjaile; Fábio Sossai Possebon; Guilherme Campos; Guilherme Targino Valente; Heidge Fukumasu. USP-Botucatu: Rejane Maria Tommasini Grotto; Helena Lage Ferreira; Instituto Butantan: Dimas Tadeu Covas; Jardelina de Souza Todao Bernardino; Jayme A. Souza-Neto; Jessika Cristina Chagas Lesbon; Jorge A. Petroli Marchesi; José Salvatore Leister Patané; João Paulo Kitajima; João Pessoa Araújo Jr.; Leila Sabrina Ullmann; Loyze Paola Oliveira de Lima; Luiz Aurelio de Campos Crispin. Centro de Genômica Funcional da ESALQ: Luiz Lehmann Coutinho; Luiz Carlos Junior de Alcantara; Lívia Sacchetto; Maisa C. Pereira Parra; Maria Carolina Elias; Marta Giovanetti; Marília Moraes; Maurício Lacerda Nogueira. Prefeitura de Sao Paulo: Melissa Palmieri.; Patricia Akemi Assato; Paula Rahal; Paulo Inacio da Costa; Rafael dos Santos Bezerra; Raquel de Lello Rocha Campos Cassano. NGS Soluções Genômicas: Pilar Drummond Sampaio Corrêa Mariani. FZEA-USP Pirassununga: Mirele Daiana Poleti; Raul Machado Neto; Ricardo Augusto Brassaloti; Ricardo Haddad; Rodrigo Tocantins Calado. FAMERP-SJRP: Cecília Artico Banho; Sandra Coccuzzo Sampaio; Svetoslav Nanev Slavov; Vagner Fonseca; Vincent Louis Viala |
| EPI_ISL_2170969 | USF ODAIR APARECIDO ROQUE BOTTER | Instituto Butantan / Mendelics | Antonio Jorge Martins; Bianca Cechetto Carlos. Mendelics: Bibiana Santos; Claudia Renata dos Santos Barros; Cíntia Bittar; David Schlesinger. Hemocentro Ribeirão Preto: Simone Kashima; Debora Botequio Moretti; Elaine Cristina Marqueze; Elaine Vieira dos Santos; Elisangela Chicaroni Mattos; Erika Freitas; Evandra Strazza Rodrigues; Felipe Allan da Silva da Costa; Flavia Aburjaile; Fábio Sossai Possebon; Guilherme Campos; Guilherme Targino Valente; Heidge Fukumasu. USP-Botucatu: Rejane Maria Tommasini Grotto; Helena Lage Ferreira; Instituto Butantan: Dimas Tadeu Covas; Jardelina de Souza Todao Bernardino; Jayme A. Souza-Neto; Jessika Cristina Chagas Lesbon; Jorge A. Petroli Marchesi; José Salvatore Leister Patané; João Paulo Kitajima; João Pessoa Araújo Jr.; Leila Sabrina Ullmann; Loyze Paola Oliveira de Lima; Luiz Aurelio de Campos Crispin. Centro de Genômica Funcional da ESALQ: Luiz Lehmann Coutinho; Luiz Carlos Junior de Alcantara; Lívia Sacchetto; Maisa C. Pereira Parra; Maria Carolina Elias; Marta Giovanetti; Marília Moraes; Maurício Lacerda Nogueira. Prefeitura de Sao Paulo: Melissa Palmieri.; Patricia Akemi Assato; Paula Rahal; Paulo Inacio da Costa; Rafael dos Santos Bezerra; Raquel de Lello Rocha Campos Cassano. NGS Soluções Genômicas: Pilar Drummond Sampaio Corrêa Mariani. FZEA-USP Pirassununga: Mirele Daiana Poleti; Raul Machado Neto; Ricardo Augusto Brassaloti; Ricardo Haddad; Rodrigo Tocantins Calado. FAMERP-SJRP: Cecília Artico Banho; Sandra Coccuzzo Sampaio; Svetoslav Nanev Slavov; Vagner Fonseca; Vincent Louis Viala |
| EPI_ISL_1966810 | USF PARQUE DAS NACOES SCRPARDO | Instituto Butantan / Mendelics | Antonio Jorge Martins; Bianca Cechetto Carlos. Mendelics: Bibiana Santos; Claudia Renata dos Santos Barros; Cíntia Bittar; David Schlesinger. Hemocentro Ribeirão Preto: Simone Kashima; Debora Botequio Moretti; Elaine Cristina Marqueze; Elaine Vieira dos Santos; Elisangela Chicaroni Mattos; Erika Freitas; Evandra Strazza Rodrigues; Felipe Allan da Silva da Costa; Flavia Aburjaile; Fábio Sossai Possebon; Guilherme Campos; Guilherme Targino Valente; Heidge Fukumasu. USP-Botucatu: Rejane Maria Tommasini Grotto; Helena Lage Ferreira; Instituto Butantan: Dimas Tadeu Covas; Jardelina de Souza Todao Bernardino; Jayme A. Souza-Neto; Jessika Cristina Chagas Lesbon; Jorge A. Petroli Marchesi; José Salvatore Leister Patané; João Paulo Kitajima; João Pessoa Araújo Jr.; Leila Sabrina Ullmann; Loyze Paola Oliveira de Lima; Luiz Aurelio de Campos Crispin. Centro de Genômica Funcional da ESALQ: Luiz Lehmann Coutinho; Luiz Carlos Junior de Alcantara; Lívia Sacchetto; Maisa C. Pereira Parra; Maria Carolina Elias; Marta Giovanetti; Marília Moraes; Maurício Lacerda Nogueira. Prefeitura de Sao Paulo: Melissa Palmieri.; Patricia Akemi Assato; Paula Rahal; Paulo Inacio da Costa; Rafael dos Santos Bezerra; Raquel de Lello Rocha Campos Cassano. NGS Soluções Genômicas: Pilar Drummond Sampaio Corrêa Mariani. FZEA-USP Pirassununga: Mirele Daiana Poleti; Raul Machado Neto; Ricardo Augusto Brassaloti; Ricardo Haddad; Rodrigo Tocantins Calado. FAMERP-SJRP: Cecília Artico Banho; Sandra Coccuzzo Sampaio; Svetoslav Nanev Slavov; Vagner Fonseca; Vincent Louis Viala |
| EPI_ISL_1966526, EPI_ISL_1966527, EPI_ISL_1966528, EPI_ISL_1966529 | USF PAULISTA FERNANDOPOLIS ANTONIO PIVATO | Instituto Butantan / FZEA-USP (Pirassununga) | Antonio Jorge Martins; Bianca Cechetto Carlos. Mendelics: Bibiana Santos; Claudia Renata dos Santos Barros; Cíntia Bittar; David Schlesinger. Hemocentro Ribeirão Preto: Simone Kashima; Debora Botequio Moretti; Elaine Cristina Marqueze; Elaine Vieira dos Santos; Elisangela Chicaroni Mattos; Erika Freitas; Evandra Strazza Rodrigues; Felipe Allan da Silva da Costa; Flavia Aburjaile; Fábio Sossai Possebon; Guilherme Campos; Guilherme Targino Valente; Heidge Fukumasu. USP-Botucatu: Rejane Maria Tommasini Grotto; Helena Lage Ferreira; Instituto Butantan: Dimas Tadeu Covas; Jardelina de Souza Todao Bernardino; Jayme A. Souza-Neto; Jessika Cristina Chagas Lesbon; Jorge A. Petroli Marchesi; José Salvatore Leister Patané; João Paulo Kitajima; João Pessoa Araújo Jr.; Leila Sabrina Ullmann; Loyze Paola Oliveira de Lima; Luiz Aurelio de Campos Crispin. Centro de Genômica Funcional da ESALQ: Luiz Lehmann Coutinho; Luiz Carlos Junior de Alcantara; Lívia Sacchetto; Maisa C. Pereira Parra; Maria Carolina Elias; Marta Giovanetti; Marília Moraes; Maurício Lacerda Nogueira. Prefeitura de Sao Paulo: Melissa Palmieri.; Patricia Akemi Assato; Paula Rahal; Paulo Inacio da Costa; Rafael dos Santos Bezerra; Raquel de Lello Rocha Campos Cassano. NGS Soluções Genômicas: Pilar Drummond Sampaio Corrêa Mariani. FZEA-USP Pirassununga: Mirele Daiana Poleti; Raul Machado Neto; Ricardo Augusto Brassaloti; Ricardo Haddad; Rodrigo Tocantins Calado. FAMERP-SJRP: Cecília Artico Banho; Sandra Coccuzzo Sampaio; Svetoslav Nanev Slavov; Vagner Fonseca; Vincent Louis Viala |
| EPI_ISL_1795391 | USF ROSA CRUZ | Instituto Butantan / ESALQ-Piracicaba | Antonio Jorge Martins; Bianca Cechetto Carlos. Mendelics: Bibiana Santos; Claudia Renata dos Santos Barros; David Schlesinger. Hemocentro Ribeirão Preto: Simone Kashima; Debora Botequio Moretti. Centro de Genômica Funcional da ESALQ: Luiz Lehmann Coutinho; Dimas Tadeu Covas; Elaine Cristina Marqueze; Elaine Vieira dos Santos; Elisangela Chicaroni Mattos; Erika Freitas; Evandra Strazza Rodrigues; Felipe Allan da Silva da Costa; Flavia Aburjaile; Guilherme Targino Valente; Heidge Fukumasu. USP-Botucatu: Rejane Maria Tommasini Grotto; Instituto Butantan: Alexander Roberto Precioso; Jayme A. Souza-Neto; Jessika Cristina Chagas Lesbon; José Salvatore Leister Patané; João Paulo Kitajima; Luiz Carlos Junior de Alcantara; Maria Carolina Elias; Marta Giovanetti; Patricia Akemi Assato; Rafael dos Santos Bezerra; Raquel de Lello Rocha Campos Cassano. NGS Soluções Genômicas: Pilar Drummond Sampaio Corrêa Mariani. FZEA-USP Pirassununga: Mirele Daiana Poleti; Raul Machado Neto; Ricardo Augusto Brassaloti; Ricardo Haddad; Rodrigo Tocantins Calado.; Sandra Coccuzzo Sampaio; Svetoslav Nanev Slavov; Vagner Fonseca; Vincent Louis Viala |
| EPI_ISL_1795064, EPI_ISL_1795096, EPI_ISL_1795174, EPI_ISL_1795175, EPI_ISL_1795176, EPI_ISL_1795187, EPI_ISL_1795189, EPI_ISL_1795192, EPI_ISL_1795193, EPI_ISL_1795384, EPI_ISL_1795412 | | | |
| see above | USF SALERNO | Instituto Butantan / ESALQ-Piracicaba | Antonio Jorge Martins; Bianca Cechetto Carlos. Mendelics: Bibiana Santos; Claudia Renata dos Santos Barros; David Schlesinger. Hemocentro Ribeirão Preto: Simone Kashima; Debora Botequio Moretti. Centro de Genômica Funcional da ESALQ: Luiz Lehmann Coutinho; Dimas Tadeu Covas; Elaine Cristina Marqueze; Elaine Vieira dos Santos; Elisangela Chicaroni Mattos; Erika Freitas; Evandra Strazza Rodrigues; Felipe Allan da Silva da Costa; Flavia Aburjaile; Guilherme Targino Valente; Heidge Fukumasu. USP-Botucatu: Rejane Maria Tommasini Grotto; Instituto Butantan: Alexander Roberto Precioso; Jayme A. Souza-Neto; Jessika Cristina Chagas Lesbon; José Salvatore Leister Patané; João Paulo Kitajima; Luiz Carlos Junior de Alcantara; Maria Carolina Elias; Marta Giovanetti; Patricia Akemi Assato; Rafael dos Santos Bezerra; Raquel de Lello Rocha Campos Cassano. NGS Soluções Genômicas: Pilar Drummond Sampaio Corrêa Mariani. FZEA-USP Pirassununga: Mirele Daiana Poleti; Raul Machado Neto; Ricardo Augusto Brassaloti; Ricardo Haddad; Rodrigo Tocantins Calado.; Sandra Coccuzzo Sampaio; Svetoslav Nanev Slavov; Vagner Fonseca; Vincent Louis Viala |
| EPI_ISL_1445073 | USF SANTA MARIA DO AMPARO | Instituto Butantan / Mendelics | Antonio Jorge Martins; Bibiana Santos; Claudia Renata dos Santos Barros; David Schlesinger; Debora Botequio Moretti; Dimas Tadeu Covas; Elaine Cristina Marqueze; Elaine Vieira dos Santos; Erika Freitas; Evandra Strazza Rodrigues; Flavia Aburjaile; José Salvatore Leister Patané; João Paulo Kitajima; Luiz Carlos Junior de Alcantara; Maria Carolina Elias; Marta Giovanetti; Rafael dos Santos Bezerra; Raul Machado Neto; Ricardo Haddad; Rodrigo Tocantins Calado.; Sandra Coccuzzo Sampaio; Simone Kashima; Svetoslav Nanev Slavov; Vagner Fonseca; Vincent Louis Viala |
| EPI_ISL_1445067, EPI_ISL_1445071, EPI_ISL_1445074, EPI_ISL_1966102 | USF TRES PONTES | Instituto Butantan / Mendelics | Antonio Jorge Martins; Bianca Cechetto Carlos. Mendelics: Bibiana Santos; Bibiana Santos; Claudia Renata dos Santos Barros; Cíntia Bittar; David Schlesinger; David Schlesinger. Hemocentro Ribeirão Preto: Simone Kashima; Debora Botequio Moretti; Dimas Tadeu Covas; Elaine Cristina Marqueze; Elaine Vieira dos Santos; Elisangela Chicaroni Mattos; Erika Freitas; Evandra Strazza Rodrigues; Felipe Allan da Silva da Costa; Flavia Aburjaile; Fábio Sossai Possebon; Guilherme Campos; Guilherme Targino Valente; Heidge Fukumasu. USP-Botucatu: Rejane Maria Tommasini Grotto; Helena Lage Ferreira; Instituto Butantan: Dimas Tadeu Covas; Jardelina de Souza Todao Bernardino; Jayme A. Souza-Neto; Jessika Cristina Chagas Lesbon; Jorge A. Petroli Marchesi; José Salvatore Leister Patané; João Paulo Kitajima; João Pessoa Araújo Jr.; Leila Sabrina Ullmann; Loyze Paola Oliveira de Lima; Luiz Aurelio de Campos Crispin. Centro de Genômica Funcional da ESALQ: Luiz Lehmann Coutinho; Luiz Carlos Junior de Alcantara; Lívia Sacchetto; Maisa C. Pereira Parra; Maria Carolina Elias; Marta Giovanetti; Marília Moraes; Maurício Lacerda Nogueira. Prefeitura de Sao Paulo: Melissa Palmieri.; Patricia Akemi Assato; Paula Rahal; Paulo Inacio da Costa; Rafael dos Santos Bezerra; Raquel de Lello Rocha Campos Cassano. NGS Soluções Genômicas: Pilar Drummond Sampaio Corrêa Mariani. FZEA-USP Pirassununga: Mirele Daiana Poleti; Raul Machado Neto; Ricardo Augusto Brassaloti; Ricardo Haddad; Rodrigo Tocantins Calado.; Rodrigo Tocantins Calado. FAMERP-SJRP: Cecília Artico Banho; Sandra Coccuzzo Sampaio; Simone Kashima; Svetoslav Nanev Slavov; Vagner Fonseca; Vincent Louis Viala |
| EPI_ISL_1533713, EPI_ISL_1533717 | Unidade de Pronto Atendimento Jd | Instituto Adolfo Lutz, Interdiciplinary | Caio Vinicius Dias Lopes; Claudia Regina Gonçalves; Claudio Tavares Sacchi; Erica Valessa Ramos Gomes; Karoline Rodrigues Campos; Leonardo Jose Tadeu de Araujo |

| | | | |
|---|---|---|---|
| | Amanda | Procedures Center, Strategic Laboratory | |
| EPI_ISL_1858758, EPI_ISL_1858761, EPI_ISL_1858763, EPI_ISL_1858765, EPI_ISL_1858767, EPI_ISL_1858769, EPI_ISL_1858771, EPI_ISL_1858773, EPI_ISL_1858775, EPI_ISL_1858777, EPI_ISL_1858779, EPI_ISL_1858781, EPI_ISL_1858783, EPI_ISL_1858786, EPI_ISL_1858788, EPI_ISL_1858790, EPI_ISL_1858792, EPI_ISL_1858794, EPI_ISL_1858796, EPI_ISL_1858798, EPI_ISL_1858800, EPI_ISL_1858802, EPI_ISL_1858803, EPI_ISL_1858805, EPI_ISL_1858807, EPI_ISL_1858809, EPI_ISL_1858811, EPI_ISL_1858814, EPI_ISL_1858816, EPI_ISL_1858818, EPI_ISL_1858819, EPI_ISL_1858822, EPI_ISL_1858824, EPI_ISL_1858826, EPI_ISL_1858828, EPI_ISL_1858830, EPI_ISL_1858832, EPI_ISL_1858834, EPI_ISL_1858836, EPI_ISL_1858838, EPI_ISL_1858840, EPI_ISL_1858842, EPI_ISL_1858843, EPI_ISL_1858846, EPI_ISL_1858847, EPI_ISL_1858850, EPI_ISL_1858852, EPI_ISL_1858853, EPI_ISL_1858856, EPI_ISL_1858858, EPI_ISL_1858860, EPI_ISL_1858862, EPI_ISL_1858864, EPI_ISL_1858866, EPI_ISL_1858868, EPI_ISL_1858870, EPI_ISL_1858872, EPI_ISL_1858874, EPI_ISL_1858876, EPI_ISL_1858878, EPI_ISL_1858880, EPI_ISL_1858882, EPI_ISL_1858884, EPI_ISL_1858886, EPI_ISL_1858888, EPI_ISL_1858890, EPI_ISL_1858892, EPI_ISL_1858894, EPI_ISL_1858896, EPI_ISL_1858898, EPI_ISL_1858900, EPI_ISL_1858902, EPI_ISL_1858904, EPI_ISL_1858906, EPI_ISL_1858908, EPI_ISL_1858910, EPI_ISL_1858912, EPI_ISL_1858914, EPI_ISL_1858916, EPI_ISL_1858918, EPI_ISL_1858920, EPI_ISL_1858922, EPI_ISL_1858924, EPI_ISL_1858926, EPI_ISL_1858928, EPI_ISL_1858930, EPI_ISL_1858933, EPI_ISL_1858934, EPI_ISL_1858936, EPI_ISL_1858938, EPI_ISL_1858940, EPI_ISL_1858942, EPI_ISL_1858944, EPI_ISL_1858946, EPI_ISL_1858948, EPI_ISL_1858950, EPI_ISL_1858952, EPI_ISL_1858954, EPI_ISL_1858957, EPI_ISL_1858958, EPI_ISL_1858960, EPI_ISL_1858963, EPI_ISL_1858964, EPI_ISL_1858966, EPI_ISL_1858968, EPI_ISL_1858969, EPI_ISL_1858970, EPI_ISL_1858971, EPI_ISL_1858973, EPI_ISL_1858975, EPI_ISL_1858977, EPI_ISL_1858979, EPI_ISL_1858981, EPI_ISL_1858984, EPI_ISL_1858986, EPI_ISL_1858988, EPI_ISL_1858990, EPI_ISL_1858992, EPI_ISL_1858994, EPI_ISL_1858996, EPI_ISL_1858998, EPI_ISL_1859000, EPI_ISL_1859002, EPI_ISL_1859005, EPI_ISL_1859007, EPI_ISL_1859008, EPI_ISL_2101385, EPI_ISL_2101386, EPI_ISL_2101387, EPI_ISL_2101390, EPI_ISL_2101391, EPI_ISL_2101392, EPI_ISL_2101394, EPI_ISL_2101407, EPI_ISL_2101429, EPI_ISL_2101430, EPI_ISL_2101431, EPI_ISL_2101432, EPI_ISL_2101433, EPI_ISL_2101443, EPI_ISL_2101455, EPI_ISL_2101456, EPI_ISL_2101457, EPI_ISL_2101458, EPI_ISL_2101459, EPI_ISL_2101460, EPI_ISL_2101461, EPI_ISL_2101462, EPI_ISL_2101463, EPI_ISL_2101470, EPI_ISL_2101471, EPI_ISL_2101472, EPI_ISL_2101473, EPI_ISL_2101475, EPI_ISL_2101483, EPI_ISL_2101484, EPI_ISL_2101494, EPI_ISL_2101495, EPI_ISL_2101505, EPI_ISL_2101506, EPI_ISL_2101508, EPI_ISL_2101509, EPI_ISL_2101510, EPI_ISL_2101511, EPI_ISL_2101512, EPI_ISL_2101513, EPI_ISL_2101520, EPI_ISL_2101521, EPI_ISL_2101522, EPI_ISL_2101523, EPI_ISL_2101524, EPI_ISL_2101525, EPI_ISL_2101526, EPI_ISL_2101527, EPI_ISL_2101566, EPI_ISL_2101567, EPI_ISL_2101568, EPI_ISL_2101569, EPI_ISL_2101570, EPI_ISL_2101571, EPI_ISL_2101572, EPI_ISL_2101612, EPI_ISL_2101638, EPI_ISL_2101639, EPI_ISL_2101640, EPI_ISL_2101641, EPI_ISL_2101642, EPI_ISL_2101643, EPI_ISL_2101644, EPI_ISL_2101645, EPI_ISL_2101646, EPI_ISL_2101647, EPI_ISL_2101648, EPI_ISL_2101649, EPI_ISL_2101650, EPI_ISL_2101651, EPI_ISL_2101652, EPI_ISL_2101653, EPI_ISL_2101654, EPI_ISL_2101655, EPI_ISL_2101656, EPI_ISL_2101657, EPI_ISL_2101658, EPI_ISL_2101659, EPI_ISL_2101660, EPI_ISL_2101661, EPI_ISL_2101662, EPI_ISL_2101663, EPI_ISL_2101664, EPI_ISL_2101665, EPI_ISL_2101666, EPI_ISL_2101667, EPI_ISL_2101668, EPI_ISL_2101669, EPI_ISL_2101670, EPI_ISL_2101671, EPI_ISL_2101676, EPI_ISL_2101677, EPI_ISL_2101678, EPI_ISL_2101679, EPI_ISL_2101680, EPI_ISL_2101681, EPI_ISL_2101682, EPI_ISL_2101683, EPI_ISL_2101684, EPI_ISL_2101685, EPI_ISL_2101686, EPI_ISL_2101687, EPI_ISL_2101688, EPI_ISL_2101692, EPI_ISL_2101693, EPI_ISL_2101694, EPI_ISL_2101695, EPI_ISL_2101696, EPI_ISL_2101714, EPI_ISL_2101715, EPI_ISL_2101722, EPI_ISL_2101724, EPI_ISL_2101727, EPI_ISL_2101731, EPI_ISL_2101734, EPI_ISL_2101740, EPI_ISL_2101741, EPI_ISL_2101744 | | | |
| see above | Unidade de apoio ao diagnóstico da COVID – UNADIG | Bioinformatics Laboratory / LNCC | Alessandra P Lamarca; Alexandra L Gerber; Amilcar Tanuri; Ana Paula de C Guimaraes; Ana Paula de C Guimarães; Ana Tereza R Vasconcelos; Andrea Cony Cavalcanti; Andréa Cony Cavalcanti; Caio Luiz Pereira Ribeiro; Cassia Alves; Cintia Policarpo; Claudia Maria Braga de Mello; Cristiane Gomes da Silva; Diana Mariani; Douglas Terra Machado; Flavio Dias da Silva; Flávio Dias da Silva; Gleidson da Silva de Oliveira; Leandro Magalhaes de Souza; Leandro Magalhães de Souza; Liliane Cavalcante; Luiz G P de Almeida; Marcio Henrique de Oliveira Garcia; Mario Sergio Ribeiro; Ronaldo da Silva F Jr; Silvia Carvalho; Thais Felix Cruz |
| EPI_ISL_1795339, EPI_ISL_1795340, EPI_ISL_1795342, EPI_ISL_1795343 | VIGILANCIA EM SAUDE | Instituto Butantan / ESALQ-Piracicaba | Antonio Jorge Martins; Bianca Cechetto Carlos. Mendelics: Bibiana Santos; Claudia Renata dos Santos Barros; David Schlesinger. Hemocentro Ribeirão Preto: Simone Kashima; Debora Botequio Moretti. Centro de Genômica Funcional da ESALQ: Luiz Lehmann Coutinho; Dimas Tadeu Covas; Elaine Cristina Marqueze; Elaine Vieira dos Santos; Elisangela Chicaroni Mattos; Erika Freitas; Evandra Strazza Rodrigues; Felipe Allan da Silva da Costa; Flavia Aburjaile; Guilherme Targino Valente; Heidge Fukumasu. USP-Botucatu: Rejane Maria Tommasini Grotto; Instituto Butantan: Alexander Roberto Precioso; Jayme A. Souza-Neto; Jessika Cristina Chagas Lesbon; José Salvatore Leister Patané; João Paulo Kitajima; Luiz Carlos Junior de Alcantara; Maria Carolina Elias; Marta Giovanetti; Patricia Akemi Assato; Rafael dos Santos Bezerra; Raquel de Lello Rocha Campos Cassano. NGS Soluções Genômicas: Pilar Drummond Sampaio Corrêa Mariani. FZEA-USP Pirassununga: Mirele Daiana Poleti; Raul Machado Neto; Ricardo Augusto Brassaloti; Ricardo Haddad; Rodrigo Tocantins Calado.; Sandra Coccuzzo Sampaio; Svetoslav Nanev Slavov; Vagner Fonseca; Vincent Louis Viala |
| EPI_ISL_1966525 | VIGILANCIA EM SAUDE | Instituto Butantan / FZEA-USP (Pirassununga) | Antonio Jorge Martins; Bianca Cechetto Carlos. Mendelics: Bibiana Santos; Claudia Renata dos Santos Barros; Cíntia Bittar; David Schlesinger. Hemocentro Ribeirão Preto: Simone Kashima; Debora Botequio Moretti; Elaine Cristina Marqueze; Elaine Vieira dos Santos; Elisangela Chicaroni Mattos; Erika Freitas; Evandra Strazza Rodrigues; Felipe Allan da Silva da Costa; Flavia Aburjaile; Fábio Sossai Possebon; Guilherme Campos; Guilherme Targino Valente; Heidge Fukumasu. USP-Botucatu: Rejane Maria Tommasini Grotto; Helena Lage Ferreira; Instituto Butantan: Dimas Tadeu Covas; Jardelina de Souza Todao Bernardino; Jayme A. Souza-Neto; Jessika Cristina Chagas Lesbon; Jorge A. Petroli Marchesi; José Salvatore Leister Patané; João Paulo Kitajima; João Pessoa Araújo Jr.; Leila Sabrina Ullmann; Loyze Paola Oliveira de Lima; Luiz Aurelio de Campos Crispin. Centro de Genômica Funcional da ESALQ: Luiz Lehmann Coutinho; Luiz Carlos Junior de Alcantara; Lívia Sacchetto; Maisa C. Pereira Parra; Maria Carolina Elias; Marta Giovanetti; Marília Moraes; Maurício Lacerda Nogueira. Prefeitura de Sao Paulo: Melissa Palmieri.; Patricia Akemi Assato; Paula Rahal; Paulo Inacio da Costa; Rafael dos Santos Bezerra; Raquel de Lello Rocha Campos Cassano. NGS Soluções Genômicas: Pilar Drummond Sampaio Corrêa Mariani. FZEA-USP Pirassununga: Mirele Daiana Poleti; Raul Machado Neto; Ricardo Augusto Brassaloti; Ricardo Haddad; Rodrigo Tocantins Calado. FAMERP-SJRP: Cecília Artico Banho; Sandra Coccuzzo Sampaio; Svetoslav Nanev Slavov; Vagner Fonseca; Vincent Louis Viala |
| EPI_ISL_1967283, EPI_ISL_2170985, EPI_ISL_2170986, EPI_ISL_2170987, EPI_ISL_2170988, EPI_ISL_2170989, EPI_ISL_2170990, EPI_ISL_2170991, EPI_ISL_2170992, EPI_ISL_2170993, EPI_ISL_2170994, EPI_ISL_2170995 | | | |
| see above | VIGILANCIA EM SAUDE DE FCO MORATO | Instituto Butantan / Mendelics | Antonio Jorge Martins; Bianca Cechetto Carlos. Mendelics: Bibiana Santos; Claudia Renata dos Santos Barros; Cíntia Bittar; David Schlesinger; David Schlesinger. Hemocentro Ribeirão Preto: Simone Kashima; Debora Botequio Moretti; Dimas Tadeu Covas; Elaine Cristina Marqueze; Elaine Vieira Santos; Elaine Vieira dos Santos; Elisangela Chicaroni Mattos; Erika Freitas; Evandra Strazza Rodrigues; Felipe Allan da Silva da Costa; Flavia Aburjaile; Fábio Sossai Possebon; Guilherme Campos; Guilherme Targino Valente; Heidge Fukumasu; Heidge Fukumasu. USP-Botucatu: Rejane Maria Tommasini Grotto; Helena Lage Ferreira; Instituto Butantan: Dimas Tadeu Covas; Jardelina de Souza Todao Bernardino; Jayme A. Souza-Neto; Jayme Augusto de Souza-Neto; Jessika Cristina Chagas Lesbon; Jorge A. Petroli Marchesi; José Salvatore Leister Patané; João Paulo Kitajima; João Pessoa Araújo Jr.; Leila Sabrina Ullmann; Loyze Paola Oliveira de Lima; Luiz Alcantara; Luiz Aurelio de Campos Crispin. Centro de Genômica Funcional da ESALQ: Luiz Lehmann Coutinho; Luiz Carlos Junior de Alcantara; Luiz Lehmann Coutinho; Lívia Sacchetto; Maisa C. Pereira Parra; Maria Carolina Elias; Marta Giovanetti; Marília Moraes; Maurício Lacerda Nogueira; Maurício Lacerda Nogueira. Prefeitura de Sao Paulo: Melissa Palmieri.; Patricia Akemi Assato; Paula Rahal; Paulo Inacio da Costa; Rafael dos Santos Bezerra; Raquel de Lello Rocha Campos Cassano. NGS Soluções Genômicas: Pilar Drummond Sampaio Corrêa Mariani. FZEA-USP Pirassununga: Mirele Daiana Poleti; Raul Machado Neto; Rejane Maria Tommasini Grotto; Ricardo Augusto Brassaloti; Ricardo Haddad; Rodrigo Tocantins Calado. FAMERP-SJRP: Cecília Artico Banho; Sandra Coccuzzo Sampaio; Sandra Coccuzzo Sampaio Vessoni; Simone Kashima; Svetoslav Nanev Slavov; Vagner Fonseca; Vincent Louis Viala |
| EPI_ISL_1966267, EPI_ISL_1966268, EPI_ISL_1966269, EPI_ISL_1966556, EPI_ISL_1966557, EPI_ISL_1966558, EPI_ISL_1966559, EPI_ISL_1966560, EPI_ISL_1966561, EPI_ISL_1967198, EPI_ISL_1967199, EPI_ISL_1967200, EPI_ISL_1967201, EPI_ISL_1967202, EPI_ISL_1967203 | | | |
| see above | VIGILANCIA EM SAUDE SAE VIGILANCIA EPIDEMIOLOGICA | Instituto Butantan / Mendelics | Antonio Jorge Martins; Bianca Cechetto Carlos. Mendelics: Bibiana Santos; Claudia Renata dos Santos Barros; Cíntia Bittar; David Schlesinger; David Schlesinger. Hemocentro Ribeirão Preto: Simone Kashima; Debora Botequio Moretti; Dimas Tadeu Covas; Elaine Cristina Marqueze; Elaine Vieira Santos; Elaine Vieira dos Santos; Elisangela Chicaroni Mattos; Erika Freitas; Evandra Strazza Rodrigues; Felipe Allan da Silva da Costa; Flavia Aburjaile; Fábio Sossai Possebon; Guilherme Campos; Guilherme Targino Valente; Heidge Fukumasu; Heidge Fukumasu. USP-Botucatu: Rejane Maria Tommasini Grotto; Helena Lage Ferreira; Instituto Butantan: Dimas Tadeu Covas; Jardelina de Souza Todao Bernardino; Jayme A. Souza-Neto; Jayme Augusto de Souza-Neto; Jessika Cristina Chagas Lesbon; Jorge A. Petroli Marchesi; José Salvatore Leister Patané; João Paulo Kitajima; João Pessoa Araújo Jr.; Leila Sabrina Ullmann; Loyze Paola Oliveira de Lima; Luiz Alcantara; Luiz Aurelio de Campos Crispin. Centro de Genômica Funcional da ESALQ: Luiz Lehmann Coutinho; Luiz Carlos Junior de Alcantara; Luiz Lehmann Coutinho; Lívia Sacchetto; Maisa C. Pereira Parra; Maria Carolina Elias; Marta Giovanetti; Marília Moraes; Maurício Lacerda Nogueira; Maurício Lacerda Nogueira. Prefeitura de Sao Paulo: Melissa Palmieri.; Patricia Akemi Assato; Paula Rahal; Paulo Inacio da Costa; Rafael dos Santos Bezerra; Raquel de Lello Rocha Campos Cassano. NGS Soluções Genômicas: Pilar Drummond Sampaio Corrêa Mariani. FZEA-USP Pirassununga: Mirele Daiana Poleti; Raul Machado Neto; Rejane Maria Tommasini Grotto; Ricardo Augusto Brassaloti; Ricardo Haddad; Rodrigo Tocantins Calado. FAMERP-SJRP: Cecília Artico Banho; Sandra Coccuzzo Sampaio; Sandra Coccuzzo Sampaio Vessoni; Simone Kashima; Svetoslav Nanev Slavov; Vagner Fonseca; Vincent Louis Viala |
| EPI_ISL_1966082, EPI_ISL_1966086, EPI_ISL_1966087, EPI_ISL_1966090, EPI_ISL_1966092 | VIGILANCIA EPIDEMIOLOGICA | Instituto Butantan / Mendelics | Antonio Jorge Martins; Bianca Cechetto Carlos. Mendelics: Bibiana Santos; Claudia Renata dos Santos Barros; Cíntia Bittar; David Schlesinger. Hemocentro Ribeirão Preto: Simone Kashima; Debora Botequio Moretti; Elaine Cristina Marqueze; Elaine Vieira dos Santos; Elisangela Chicaroni Mattos; Erika Freitas; Evandra Strazza Rodrigues; Felipe Allan da Silva da Costa; Flavia Aburjaile; Fábio Sossai Possebon; Guilherme Campos; Guilherme Targino Valente; Heidge Fukumasu. USP-Botucatu: Rejane Maria Tommasini Grotto; Helena Lage Ferreira; Instituto Butantan: Dimas Tadeu Covas; Jardelina de Souza Todao Bernardino; Jayme A. Souza-Neto; Jessika Cristina Chagas Lesbon; Jorge A. Petroli Marchesi; José Salvatore Leister Patané; João Paulo Kitajima; João Pessoa Araújo Jr.; Leila Sabrina Ullmann; Loyze Paola Oliveira de Lima; Luiz Aurelio de Campos Crispin. Centro de Genômica Funcional da ESALQ: Luiz Lehmann Coutinho; Luiz Carlos Junior de Alcantara; Lívia Sacchetto; Maisa C. Pereira Parra; Maria Carolina Elias; Marta Giovanetti; Marília Moraes; Maurício Lacerda Nogueira. Prefeitura de Sao Paulo: Melissa Palmieri.; Patricia Akemi Assato; Paula Rahal; Paulo Inacio da Costa; Rafael dos Santos Bezerra; Raquel de Lello Rocha Campos Cassano. NGS Soluções Genômicas: Pilar Drummond Sampaio Corrêa Mariani. FZEA-USP Pirassununga: Mirele Daiana Poleti; Raul Machado Neto; Ricardo Augusto Brassaloti; Ricardo Haddad; Rodrigo Tocantins Calado. FAMERP-SJRP: Cecília Artico Banho; Sandra Coccuzzo Sampaio; Svetoslav Nanev Slavov; Vagner Fonseca; Vincent Louis Viala |
| EPI_ISL_1966294, EPI_ISL_1966295, EPI_ISL_1966296, EPI_ISL_1966297, EPI_ISL_1966298, EPI_ISL_1966299, EPI_ISL_1966300, EPI_ISL_1966301, EPI_ISL_1966607, EPI_ISL_1967242, EPI_ISL_1967243 | | | |
| see above | VIGILANCIA EPIDEMIOLOGICA DE IBATE | Instituto Butantan / Mendelics | Antonio Jorge Martins; Bianca Cechetto Carlos. Mendelics: Bibiana Santos; Claudia Renata dos Santos Barros; Cíntia Bittar; David Schlesinger. Hemocentro Ribeirão Preto: Simone Kashima; Debora Botequio Moretti; Elaine Cristina Marqueze; Elaine Vieira dos Santos; Elisangela Chicaroni Mattos; Erika Freitas; Evandra Strazza Rodrigues; Felipe Allan da Silva da Costa; Flavia Aburjaile; Fábio Sossai Possebon; Guilherme Campos; Guilherme Targino Valente; Heidge Fukumasu. USP-Botucatu: Rejane Maria Tommasini Grotto; Helena Lage Ferreira; Instituto Butantan: Dimas Tadeu Covas; Jardelina de Souza Todao Bernardino; Jayme A. Souza-Neto; Jessika Cristina Chagas Lesbon; Jorge A. Petroli Marchesi; José Salvatore Leister Patané; João Paulo Kitajima; João Pessoa Araújo Jr.; Leila Sabrina Ullmann; Loyze Paola Oliveira de Lima; Luiz Aurelio de Campos Crispin. Centro de Genômica Funcional da ESALQ: Luiz Lehmann Coutinho; Luiz Carlos Junior de Alcantara; Lívia Sacchetto; Maisa C. Pereira Parra; Maria Carolina Elias; Marta Giovanetti; Marília Moraes; Maurício Lacerda Nogueira. Prefeitura de Sao Paulo: Melissa Palmieri.; Patricia Akemi Assato; Paula Rahal; Paulo Inacio da Costa; Rafael dos Santos Bezerra; Raquel de Lello Rocha Campos Cassano. NGS Soluções Genômicas: Pilar Drummond Sampaio Corrêa Mariani. FZEA-USP Pirassununga: Mirele Daiana Poleti; Raul Machado Neto; Ricardo Augusto Brassaloti; Ricardo Haddad; Rodrigo Tocantins Calado. FAMERP-SJRP: Cecília Artico Banho; Sandra Coccuzzo Sampaio; Svetoslav Nanev Slavov; Vagner Fonseca; Vincent Louis Viala |
| EPI_ISL_1795094 | VIGILANCIA EPIDEMIOLOGICA DE LEME | Instituto Butantan / ESALQ-Piracicaba | Antonio Jorge Martins; Bianca Cechetto Carlos. Mendelics: Bibiana Santos; Claudia Renata dos Santos Barros; David Schlesinger. Hemocentro Ribeirão Preto: Simone Kashima; Debora Botequio Moretti. Centro de Genômica Funcional da ESALQ: Luiz Lehmann Coutinho; Dimas Tadeu Covas; Elaine Cristina Marqueze; Elaine Vieira dos Santos; Elisangela Chicaroni Mattos; Erika Freitas; Evandra Strazza Rodrigues; Felipe Allan da Silva da Costa; Flavia Aburjaile; Guilherme Targino Valente; Heidge Fukumasu. USP-Botucatu: Rejane Maria Tommasini Grotto; Instituto Butantan: Alexander Roberto Precioso; Jayme A. Souza-Neto; Jessika Cristina Chagas Lesbon; José Salvatore Leister Patané; João Paulo Kitajima; Luiz Carlos Junior de Alcantara; Maria Carolina Elias; Marta Giovanetti; Patricia Akemi Assato; Rafael dos Santos Bezerra; Raquel de Lello Rocha Campos Cassano. NGS Soluções Genômicas: Pilar Drummond Sampaio Corrêa Mariani. FZEA-USP Pirassununga: Mirele Daiana Poleti; Raul Machado Neto; Ricardo Augusto Brassaloti; Ricardo Haddad; Rodrigo Tocantins Calado.; Sandra Coccuzzo Sampaio; Svetoslav Nanev Slavov; Vagner Fonseca; Vincent Louis Viala |
| EPI_ISL_1795217 | VIGILANCIA EPIDEMIOLOGICA JARDINOPOLIS SP | Instituto Butantan / ESALQ-Piracicaba | Antonio Jorge Martins; Bianca Cechetto Carlos. Mendelics: Bibiana Santos; Claudia Renata dos Santos Barros; David Schlesinger. Hemocentro Ribeirão Preto: Simone Kashima; Debora Botequio Moretti. Centro de Genômica Funcional da ESALQ: Luiz Lehmann Coutinho; Dimas Tadeu Covas; Elaine Cristina Marqueze; Elaine Vieira dos Santos; Elisangela Chicaroni Mattos; Erika Freitas; Evandra Strazza Rodrigues; Felipe Allan da Silva da Costa; Flavia Aburjaile; Guilherme Targino Valente; Heidge Fukumasu. USP-Botucatu: Rejane Maria Tommasini Grotto; Instituto Butantan: Alexander Roberto Precioso; Jayme A. Souza-Neto; Jessika Cristina Chagas Lesbon; José Salvatore Leister Patané; João Paulo Kitajima; Luiz Carlos Junior de Alcantara; Maria Carolina Elias; Marta Giovanetti; Patricia Akemi Assato; Rafael dos Santos Bezerra; Raquel de Lello Rocha Campos Cassano. NGS Soluções Genômicas: Pilar Drummond Sampaio Corrêa Mariani. FZEA-USP Pirassununga: Mirele Daiana Poleti; Raul Machado Neto; Ricardo Augusto Brassaloti; Ricardo Haddad; Rodrigo Tocantins Calado.; Sandra Coccuzzo Sampaio; Svetoslav Nanev Slavov; Vagner Fonseca; Vincent Louis Viala |
| EPI_ISL_1716878 | VIGILANCIA EPIDEMIOLOGICA JARDINOPOLIS SP | Instituto Butantan / ESALQ-USP | Antonio Jorge Martins; Bianca Cechetto Carlos. Mendelics: Bibiana Santos; Claudia Renata dos Santos Barros; David Schlesinger. Hemocentro Ribeirão Preto: Simone Kashima; Debora Botequio Moretti. Centro de Genômica Funcional da ESALQ: Luiz Lehmann Coutinho; Dimas Tadeu Covas; Elaine Cristina Marqueze; Elaine Vieira dos Santos; Elisangela Chicaroni Mattos; Erika Freitas; Evandra Strazza Rodrigues; Felipe Allan da Silva da Costa; Flavia Aburjaile; Guilherme Targino Valente; Heidge Fukumasu. USP-Botucatu: Rejane Maria Tommasini Grotto; Instituto Butantan: Alexander Roberto Precioso; Jayme A. Souza-Neto; Jessika Cristina Chagas Lesbon; José Salvatore Leister Patané; João Paulo Kitajima; Luiz Carlos Junior de Alcantara; Maria Carolina Elias; Marta Giovanetti; Patricia Akemi Assato; Rafael dos Santos Bezerra; Raquel de Lello Rocha Campos Cassano. NGS Soluções Genômicas: Pilar Drummond Sampaio Corrêa Mariani. FZEA-USP Pirassununga: Mirele Daiana Poleti; Raul Machado Neto; Ricardo Augusto Brassaloti; Ricardo Haddad; Rodrigo Tocantins Calado.; Sandra Coccuzzo Sampaio; Svetoslav Nanev Slavov; Vagner Fonseca; Vincent Louis Viala |
| EPI_ISL_1966773, EPI_ISL_1966774, EPI_ISL_1966775, EPI_ISL_2170963 | VIGILANCIA EPIDEMIOLOGICA JARDINOPOLIS SP | Instituto Butantan / Mendelics | Antonio Jorge Martins; Bianca Cechetto Carlos. Mendelics: Bibiana Santos; Claudia Renata dos Santos Barros; Cíntia Bittar; David Schlesinger. Hemocentro Ribeirão Preto: Simone Kashima; Debora Botequio Moretti; Elaine Cristina Marqueze; Elaine Vieira dos Santos; Elisangela Chicaroni Mattos; Erika Freitas; Evandra Strazza Rodrigues; Felipe Allan da Silva da Costa; Flavia Aburjaile; Fábio Sossai Possebon; Guilherme Campos; Guilherme Targino Valente; Heidge Fukumasu. USP-Botucatu: Rejane Maria Tommasini Grotto; Helena Lage Ferreira; Instituto Butantan: Dimas Tadeu Covas; Jardelina de Souza Todao Bernardino; Jayme A. Souza-Neto; Jessika Cristina Chagas Lesbon; Jorge A. Petroli Marchesi; José Salvatore Leister Patané; João Paulo Kitajima; João Pessoa Araújo Jr.; Leila Sabrina Ullmann; Loyze Paola Oliveira de Lima; Luiz Aurelio de Campos Crispin. Centro de Genômica Funcional da ESALQ: Luiz Lehmann Coutinho; Luiz Carlos Junior de Alcantara; Lívia Sacchetto; Maisa C. Pereira Parra; Maria Carolina Elias; Marta Giovanetti; Marília Moraes; Maurício Lacerda Nogueira. Prefeitura de Sao Paulo: Melissa Palmieri.; Patricia Akemi Assato; Paula Rahal; Paulo Inacio da Costa; Rafael dos Santos Bezerra; Raquel de Lello Rocha Campos Cassano. NGS Soluções Genômicas: Pilar Drummond Sampaio Corrêa Mariani. FZEA-USP Pirassununga: Mirele Daiana Poleti; Raul Machado Neto; Ricardo Augusto Brassaloti; Ricardo Haddad; Rodrigo Tocantins Calado. FAMERP-SJRP: Cecília Artico Banho; Sandra Coccuzzo Sampaio; Svetoslav Nanev Slavov; Vagner Fonseca; Vincent Louis Viala |
| EPI_ISL_1795218 | VIGILANCIA EPIDIMIOLOGICA DE ARACARIGUAMA | Instituto Butantan / ESALQ-Piracicaba | Antonio Jorge Martins; Bianca Cechetto Carlos. Mendelics: Bibiana Santos; Claudia Renata dos Santos Barros; David Schlesinger. Hemocentro Ribeirão Preto: Simone Kashima; Debora Botequio Moretti. Centro de Genômica Funcional da ESALQ: Luiz Lehmann Coutinho; Dimas Tadeu Covas; Elaine Cristina Marqueze; Elaine Vieira dos Santos; Elisangela Chicaroni Mattos; Erika Freitas; Evandra Strazza Rodrigues; Felipe Allan da Silva da Costa; Flavia Aburjaile; Guilherme Targino Valente; Heidge Fukumasu. USP-Botucatu: Rejane Maria Tommasini Grotto; Instituto Butantan: Alexander Roberto Precioso; Jayme A. Souza-Neto; Jessika Cristina Chagas Lesbon; José Salvatore Leister Patané; João Paulo Kitajima; Luiz Carlos Junior de Alcantara; Maria Carolina Elias; Marta Giovanetti; Patricia Akemi Assato; Rafael dos Santos Bezerra; Raquel de Lello Rocha Campos Cassano. NGS Soluções Genômicas: Pilar Drummond Sampaio Corrêa Mariani. FZEA-USP Pirassununga: Mirele Daiana Poleti; Raul Machado Neto; Ricardo Augusto Brassaloti; Ricardo Haddad; Rodrigo Tocantins Calado.; Sandra Coccuzzo Sampaio; Svetoslav Nanev Slavov; Vagner Fonseca; Vincent Louis Viala |
| EPI_ISL_1966830, EPI_ISL_1966831 | VIGILANCIA EPIDIMIOLOGICA DE ARACARIGUAMA | Instituto Butantan / ESALQ-USP (Piracicaba) | Antonio Jorge Martins; Bianca Cechetto Carlos. Mendelics: Bibiana Santos; Claudia Renata dos Santos Barros; Cíntia Bittar; David Schlesinger. Hemocentro Ribeirão Preto: Simone Kashima; Debora Botequio Moretti; Elaine Cristina Marqueze; Elaine Vieira dos Santos; Elisangela Chicaroni Mattos; Erika Freitas; Evandra Strazza Rodrigues; Felipe Allan da Silva da Costa; Flavia Aburjaile; Fábio Sossai Possebon; Guilherme Campos; Guilherme Targino Valente; Heidge Fukumasu. USP-Botucatu: Rejane Maria Tommasini Grotto; Helena Lage Ferreira; Instituto Butantan: Dimas Tadeu Covas; Jardelina de Souza Todao Bernardino; Jayme A. Souza-Neto; Jessika Cristina Chagas Lesbon; Jorge A. Petroli Marchesi; José Salvatore Leister Patané; João Paulo Kitajima; João Pessoa Araújo Jr.; Leila Sabrina Ullmann; Loyze Paola Oliveira de Lima; Luiz Aurelio de Campos Crispin. Centro de Genômica Funcional da ESALQ: Luiz Lehmann Coutinho; Luiz Carlos Junior de Alcantara; Lívia Sacchetto; Maisa C. Pereira Parra; Maria Carolina Elias; Marta Giovanetti; Marília Moraes; Maurício Lacerda Nogueira. Prefeitura de Sao Paulo: Melissa Palmieri.; Patricia Akemi Assato; Paula Rahal; Paulo Inacio da Costa; Rafael dos Santos Bezerra; Raquel de Lello Rocha Campos Cassano. NGS Soluções Genômicas: Pilar Drummond Sampaio Corrêa Mariani. FZEA-USP Pirassununga: Mirele Daiana Poleti; Raul Machado Neto; Ricardo Augusto Brassaloti; Ricardo Haddad; Rodrigo Tocantins Calado. FAMERP-SJRP: Cecília Artico Banho; Sandra Coccuzzo Sampaio; Svetoslav Nanev Slavov; Vagner Fonseca; Vincent Louis Viala |
| EPI_ISL_1966745 | VIGILANCIA EPIDIMIOLOGICA DE ARACARIGUAMA | Instituto Butantan / Mendelics | Antonio Jorge Martins; Bianca Cechetto Carlos. Mendelics: Bibiana Santos; Claudia Renata dos Santos Barros; Cíntia Bittar; David Schlesinger. Hemocentro Ribeirão Preto: Simone Kashima; Debora Botequio Moretti; Elaine Cristina Marqueze; Elaine Vieira dos Santos; Elisangela Chicaroni Mattos; Erika Freitas; Evandra Strazza Rodrigues; Felipe Allan da Silva da Costa; Flavia Aburjaile; Fábio Sossai Possebon; Guilherme Campos; Guilherme Targino Valente; Heidge Fukumasu. USP-Botucatu: Rejane Maria Tommasini Grotto; Helena Lage Ferreira; Instituto Butantan: Dimas Tadeu Covas; Jardelina de Souza Todao Bernardino; Jayme A. Souza-Neto; Jessika Cristina Chagas Lesbon; Jorge A. Petroli Marchesi; José Salvatore Leister Patané; João Paulo Kitajima; João Pessoa Araújo Jr.; Leila Sabrina Ullmann; Loyze Paola Oliveira de Lima; Luiz Aurelio de Campos Crispin. Centro de Genômica Funcional da ESALQ: Luiz Lehmann Coutinho; Luiz Carlos Junior de Alcantara; Lívia Sacchetto; Maisa C. Pereira Parra; Maria Carolina Elias; Marta Giovanetti; Marília Moraes; Maurício Lacerda Nogueira. Prefeitura de Sao Paulo: Melissa Palmieri.; Patricia Akemi Assato; Paula Rahal; Paulo Inacio da Costa; Rafael dos Santos Bezerra; Raquel de Lello Rocha Campos Cassano. NGS Soluções Genômicas: Pilar Drummond Sampaio Corrêa Mariani. FZEA-USP Pirassununga: Mirele Daiana Poleti; Raul Machado Neto; Ricardo Augusto Brassaloti; Ricardo Haddad; Rodrigo Tocantins Calado. FAMERP-SJRP: Cecília Artico Banho; Sandra Coccuzzo Sampaio; Svetoslav Nanev Slavov; Vagner Fonseca; Vincent Louis Viala |
| EPI_ISL_1966935, EPI_ISL_1966936, EPI_ISL_1966937, EPI_ISL_1966938, EPI_ISL_1966939, EPI_ISL_1966940, EPI_ISL_1966941 | | | |
| see above | VIGILANCIA SANITARIA E VIG EPIDEMIOLOGICA DE ITAPEVI | Instituto Butantan / ESALQ-USP (Piracicaba) | Antonio Jorge Martins; Bianca Cechetto Carlos. Mendelics: Bibiana Santos; Claudia Renata dos Santos Barros; Cíntia Bittar; David Schlesinger. Hemocentro Ribeirão Preto: Simone Kashima; Debora Botequio Moretti; Elaine Cristina Marqueze; Elaine Vieira dos Santos; Elisangela Chicaroni Mattos; Erika Freitas; Evandra Strazza Rodrigues; Felipe Allan da Silva da Costa; Flavia Aburjaile; Fábio Sossai Possebon; Guilherme Campos; Guilherme Targino Valente; Heidge Fukumasu. USP-Botucatu: Rejane Maria Tommasini Grotto; Helena Lage Ferreira; Instituto Butantan: Dimas Tadeu Covas; Jardelina de Souza Todao Bernardino; Jayme A. Souza-Neto; Jessika Cristina Chagas Lesbon; Jorge A. Petroli Marchesi; José Salvatore Leister Patané; João Paulo Kitajima; João Pessoa Araújo Jr.; Leila Sabrina Ullmann; Loyze Paola Oliveira de Lima; Luiz Aurelio de Campos Crispin. Centro de Genômica Funcional da ESALQ: Luiz Lehmann Coutinho; Luiz Carlos Junior de Alcantara; Lívia Sacchetto; Maisa C. Pereira Parra; Maria Carolina Elias; Marta Giovanetti; Marília Moraes; Maurício Lacerda Nogueira. Prefeitura de Sao Paulo: Melissa Palmieri.; Patricia Akemi Assato; Paula Rahal; Paulo Inacio da Costa; Rafael dos Santos Bezerra; Raquel de Lello Rocha Campos Cassano. NGS Soluções Genômicas: Pilar Drummond Sampaio Corrêa Mariani. FZEA-USP Pirassununga: Mirele Daiana Poleti; Raul Machado Neto; Ricardo Augusto Brassaloti; Ricardo Haddad; Rodrigo Tocantins Calado. FAMERP-SJRP: Cecília Artico Banho; Sandra Coccuzzo Sampaio; Svetoslav Nanev Slavov; Vagner Fonseca; Vincent Louis Viala |
| EPI_ISL_1533716 | Vigilancia Em Saude | Instituto Adolfo Lutz, Interdiciplinary Procedures Center, Strategic Laboratory | Caio Vinicius Dias Lopes; Claudia Regina Gonçalves; Claudio Tavares Sacchi; Erica Valessa Ramos Gomes; Karoline Rodrigues Campos; Leonardo Jose Tadeu de Araujo |
| EPI_ISL_1628378 | Vigilancia Epidemiologica Jardinopolis | Instituto Adolfo Lutz, Interdiciplinary Procedures Center, Strategic Laboratory | Caio Vinicius Dias Lopes; Claudia Regina Gonçalves; Claudio Tavares Sacchi; Erica Valessa Ramos Gomes; Karoline Rodrigues Campos; Katia Correa de Oliveira Santos; Leonardo Jose Tadeu de Araujo |
| EPI_ISL_1857098 | Virology Lab- IMTSP | Virology Laboratory, Institute of Tropical Medicine of Sao Paulo-USP | Ester C Sabino; Flavia Salles; Ingra Morales Claro; Maria Cassia J M Correa |

We gratefully acknowledge the following Authors from the Originating laboratories responsible for obtaining the specimens, as well as the Submitting laboratories where the genome data were generated and shared via GISAID, on which this research is based.

All Submitters of data may be contacted directly via www.gisaid.org

Authors are sorted alphabetically.

| Accession ID | Originating Laboratory | Submitting Laboratory | Authors |
|---|---|---|---|
| EPI_ISL_2493256, EPI_ISL_2493285, EPI_ISL_2493739, EPI_ISL_2493740, EPI_ISL_2493741, EPI_ISL_2493742, EPI_ISL_2493743, EPI_ISL_2493744, EPI_ISL_2493745, EPI_ISL_2493746, EPI_ISL_2493747, EPI_ISL_2493748, EPI_ISL_2493749, EPI_ISL_2493750, EPI_ISL_2493751, EPI_ISL_2493752, EPI_ISL_2493753, EPI_ISL_2493754, EPI_ISL_2493755, EPI_ISL_2493756, EPI_ISL_2493757, EPI_ISL_2493758, EPI_ISL_2493759, EPI_ISL_2493760, EPI_ISL_2493761, EPI_ISL_2493762, EPI_ISL_2494293 | | | |
| see above | AFIP | Instituto Butantan | Antonio Jorge Martins; Claudia Renata dos Santos Barros; David Schlesinger; Debora Botequio Moretti; Dimas Tadeu Covas; Elaine Cristina Marqueze; Elaine Vieira Santos; Evandra Strazza Rodrigues; Heidge Fukumasu; Jayme Augusto de Souza-Neto; José Salvatore Leister Patané; Luiz Alcantara; Luiz Lehmann Coutinho; Maria Carolina Elias; Maurício Lacerda Nogueira; Rafael dos Santos Bezerra; Raul Machado Neto; Rejane Maria Tommasini Grotto; Ricardo Haddad; Sandra Coccuzzo Sampaio Vessoni; Simone Kashima; Svetoslav Nanev Slavov; Vincent Louis Viala |
| EPI_ISL_2494052, EPI_ISL_2494053, EPI_ISL_2494054, EPI_ISL_2494055, EPI_ISL_2494056, EPI_ISL_2494057 | AFIP LESTE | Instituto Butantan | Antonio Jorge Martins; Claudia Renata dos Santos Barros; David Schlesinger; Debora Botequio Moretti; Dimas Tadeu Covas; Elaine Cristina Marqueze; Elaine Vieira Santos; Evandra Strazza Rodrigues; Heidge Fukumasu; Jayme Augusto de Souza-Neto; José Salvatore Leister Patané; Luiz Alcantara; Luiz Lehmann Coutinho; Maria Carolina Elias; Maurício Lacerda Nogueira; Rafael dos Santos Bezerra; Raul Machado Neto; Rejane Maria Tommasini Grotto; Ricardo Haddad; Sandra Coccuzzo Sampaio Vessoni; Simone Kashima; Svetoslav Nanev Slavov; Vincent Louis Viala |
| EPI_ISL_2493183, EPI_ISL_2493266, EPI_ISL_2493269, EPI_ISL_2494058, EPI_ISL_2494059, EPI_ISL_2494060, EPI_ISL_2494061, EPI_ISL_2494062, EPI_ISL_2494063, EPI_ISL_2494064, EPI_ISL_2494065, EPI_ISL_2494066 | | | |
| see above | AFIP SUDESTE | Instituto Butantan | Antonio Jorge Martins; Claudia Renata dos Santos Barros; David Schlesinger; Debora Botequio Moretti; Dimas Tadeu Covas; Elaine Cristina Marqueze; Elaine Vieira Santos; Evandra Strazza Rodrigues; Heidge Fukumasu; Jayme Augusto de Souza-Neto; José Salvatore Leister Patané; Luiz Alcantara; Luiz Lehmann Coutinho; Maria Carolina Elias; Maurício Lacerda Nogueira; Rafael dos Santos Bezerra; Raul Machado Neto; Rejane Maria Tommasini Grotto; Ricardo Haddad; Sandra Coccuzzo Sampaio Vessoni; Simone Kashima; Svetoslav Nanev Slavov; Vincent Louis Viala |
| EPI_ISL_2493169, EPI_ISL_2493279, EPI_ISL_2493287, EPI_ISL_2494067, EPI_ISL_2494068, EPI_ISL_2494069, EPI_ISL_2494070, EPI_ISL_2494071, EPI_ISL_2494072, EPI_ISL_2494073, EPI_ISL_2494074, EPI_ISL_2494075, EPI_ISL_2494076, EPI_ISL_2494077, EPI_ISL_2494078, EPI_ISL_2494079, EPI_ISL_2494080, EPI_ISL_2494081, EPI_ISL_2494082, EPI_ISL_2494083, EPI_ISL_2494084, EPI_ISL_2494085, EPI_ISL_2494086, EPI_ISL_2494087, EPI_ISL_2494088, EPI_ISL_2494089, EPI_ISL_2494090, EPI_ISL_2494091, EPI_ISL_2494092, EPI_ISL_2494093, EPI_ISL_2494094, EPI_ISL_2494095, EPI_ISL_2494096, EPI_ISL_2494097, EPI_ISL_2494098, EPI_ISL_2494311, EPI_ISL_2494312, EPI_ISL_2494330 | | | |
| see above | AFIP SUL | Instituto Butantan | Antonio Jorge Martins; Claudia Renata dos Santos Barros; David Schlesinger; Debora Botequio Moretti; Dimas Tadeu Covas; Elaine Cristina Marqueze; Elaine Vieira Santos; Evandra Strazza Rodrigues; Heidge Fukumasu; Jayme Augusto de Souza-Neto; José Salvatore Leister Patané; Luiz Alcantara; Luiz Lehmann Coutinho; Maria Carolina Elias; Maurício Lacerda Nogueira; Rafael dos Santos Bezerra; Raul Machado Neto; Rejane Maria Tommasini Grotto; Ricardo Haddad; Sandra Coccuzzo Sampaio Vessoni; Simone Kashima; Svetoslav Nanev Slavov; Vincent Louis Viala |
| EPI_ISL_2378739, EPI_ISL_2493553 | AMBULATORIO AVIACAO | Instituto Butantan | Antonio Jorge Martins; Claudia Renata dos Santos Barros; David Schlesinger; Debora Botequio Moretti; Dimas Tadeu Covas; Elaine Cristina Marqueze; Elaine Vieira Santos; Evandra Strazza Rodrigues; Heidge Fukumasu; Jayme Augusto de Souza-Neto; José Salvatore Leister Patané; Luiz Alcantara; Luiz Lehmann Coutinho; Maria Carolina Elias; Maurício Lacerda Nogueira; Rafael dos Santos Bezerra; Raul Machado Neto; Rejane Maria Tommasini Grotto; Ricardo Haddad; Sandra Coccuzzo Sampaio Vessoni; Simone Kashima; Svetoslav Nanev Slavov; Vincent Louis Viala |
| EPI_ISL_2493549, EPI_ISL_2493554 | AMBULATORIO BOQUEIRAO | Instituto Butantan | Antonio Jorge Martins; Claudia Renata dos Santos Barros; David Schlesinger; Debora Botequio Moretti; Dimas Tadeu Covas; Elaine Cristina Marqueze; Elaine Vieira Santos; Evandra Strazza Rodrigues; Heidge Fukumasu; Jayme Augusto de Souza-Neto; José Salvatore Leister Patané; Luiz Alcantara; Luiz Lehmann Coutinho; Maria Carolina Elias; Maurício Lacerda Nogueira; Rafael dos Santos Bezerra; Raul Machado Neto; Rejane Maria Tommasini Grotto; Ricardo Haddad; Sandra Coccuzzo Sampaio Vessoni; Simone Kashima; Svetoslav Nanev Slavov; Vincent Louis Viala |
| EPI_ISL_2345686, EPI_ISL_2345736, EPI_ISL_2345780 | AMBULATORIO CENTRAL DE ESPECIALIDADES ITATIBA | Instituto Butantan / Mendelics | Antonio Jorge Martins; Claudia Renata dos Santos Barros; David Schlesinger; Debora Botequio Moretti; Dimas Tadeu Covas; Elaine Cristina Marqueze; Elaine Vieira Santos; Evandra Strazza Rodrigues; Heidge Fukumasu; Jayme Augusto de Souza-Neto; José Salvatore Leister Patané; Luiz Alcantara; Luiz Lehmann Coutinho; Maria Carolina Elias; Maurício Lacerda Nogueira; Rafael dos Santos Bezerra; Raul Machado Neto; Rejane Maria Tommasini Grotto; Ricardo Haddad; Sandra Coccuzzo Sampaio Vessoni; Simone Kashima; Svetoslav Nanev Slavov; Vincent Louis Viala |
| EPI_ISL_2493608, EPI_ISL_2493615 | AMBULATORIO COVID 19 | Instituto Butantan | Antonio Jorge Martins; Claudia Renata dos Santos Barros; David Schlesinger; Debora Botequio Moretti; Dimas Tadeu Covas; Elaine Cristina Marqueze; Elaine Vieira Santos; Evandra Strazza Rodrigues; Heidge Fukumasu; Jayme Augusto de Souza-Neto; José Salvatore Leister Patané; Luiz Alcantara; Luiz Lehmann Coutinho; Maria Carolina Elias; Maurício Lacerda Nogueira; Rafael dos Santos Bezerra; Raul Machado Neto; Rejane Maria Tommasini Grotto; Ricardo Haddad; Sandra Coccuzzo Sampaio Vessoni; Simone Kashima; Svetoslav Nanev Slavov; Vincent Louis Viala |
| EPI_ISL_2445415, EPI_ISL_2445419, EPI_ISL_2445420, EPI_ISL_2445421, EPI_ISL_2445422, EPI_ISL_2445423, EPI_ISL_2445424, EPI_ISL_2462327, EPI_ISL_2493170, EPI_ISL_2493255, EPI_ISL_2493528, EPI_ISL_2493529, EPI_ISL_2493530, EPI_ISL_2493532, EPI_ISL_2493533, EPI_ISL_2493534, EPI_ISL_2493538, EPI_ISL_2494133, EPI_ISL_2494134, EPI_ISL_2494137, EPI_ISL_2494138, EPI_ISL_2494331, EPI_ISL_2500980 | | | |
| see above | AMBULATORIO DE ESPECIALIDADE V E MOGI MIRIM | Instituto Butantan | Antonio Jorge Martins; Claudia Renata dos Santos Barros; David Schlesinger; Debora Botequio Moretti; Dimas Tadeu Covas; Elaine Cristina Marqueze; Elaine Vieira Santos; Evandra Strazza Rodrigues; Heidge Fukumasu; Jayme Augusto de Souza-Neto; José Salvatore Leister Patané; Luiz Alcantara; Luiz Lehmann Coutinho; Maria Carolina Elias; Maurício Lacerda Nogueira; Rafael dos Santos Bezerra; Raul Machado Neto; Rejane Maria Tommasini Grotto; Ricardo Haddad; Sandra Coccuzzo Sampaio Vessoni; Simone Kashima; Svetoslav Nanev Slavov; Vincent Louis Viala |
| EPI_ISL_2493701 | AMBULATORIO MEDICO E ODONTOLOGICO MUNICIPAL DE PIEDADE | Instituto Butantan | Antonio Jorge Martins; Claudia Renata dos Santos Barros; David Schlesinger; Debora Botequio Moretti; Dimas Tadeu Covas; Elaine Cristina Marqueze; Elaine Vieira Santos; Evandra Strazza Rodrigues; Heidge Fukumasu; Jayme Augusto de Souza-Neto; José Salvatore Leister Patané; Luiz Alcantara; Luiz Lehmann Coutinho; Maria Carolina Elias; Maurício Lacerda Nogueira; Rafael dos Santos Bezerra; Raul Machado Neto; Rejane Maria Tommasini Grotto; Ricardo Haddad; Sandra Coccuzzo Sampaio Vessoni; Simone Kashima; Svetoslav Nanev Slavov; Vincent Louis Viala |
| EPI_ISL_2445148, EPI_ISL_2445149, EPI_ISL_2445151, EPI_ISL_2445152, EPI_ISL_2473705, EPI_ISL_2473762, EPI_ISL_2473763, EPI_ISL_2473764, EPI_ISL_2473766, EPI_ISL_2473767, EPI_ISL_2473768, EPI_ISL_2473769, EPI_ISL_2473770, EPI_ISL_2473771, EPI_ISL_2473772, EPI_ISL_2473773, EPI_ISL_2473774, EPI_ISL_2473775, EPI_ISL_2473776, EPI_ISL_2493364 | | | |
| see above | AMBULATÓRIO INSTITUTO BUTANTAN | Instituto Butantan | Antonio Jorge Martins; Claudia Renata dos Santos Barros; David Schlesinger; Debora Botequio Moretti; Dimas Tadeu Covas; Elaine Cristina Marqueze; Elaine Vieira Santos; Evandra Strazza Rodrigues; Heidge Fukumasu; Jayme Augusto de Souza-Neto; José Salvatore Leister Patané; Luiz Alcantara; Luiz Lehmann Coutinho; Maria Carolina Elias; Maurício Lacerda Nogueira; Rafael dos Santos Bezerra; Raul Machado Neto; Rejane Maria Tommasini Grotto; Ricardo Haddad; Sandra Coccuzzo Sampaio Vessoni; Simone Kashima; Svetoslav Nanev Slavov; Vincent Louis Viala |
| EPI_ISL_2274979 | Aeroporto Internacional de Guarulhos | Instituto Adolfo Lutz, Interdiciplinary Procedures Center, Strategic Laboratory | Caio Vinicius Dias Lopes; Claudia Regina Gonçalves; Claudio Tavares Sacchi; Erica Valessa Ramos Gomes; Karoline Rodrigues Campos; Leonardo Jose Tadeu de Araujo |
| EPI_ISL_2493193, EPI_ISL_2493203, EPI_ISL_2493286, EPI_ISL_2494039, EPI_ISL_2494040, EPI_ISL_2494041, EPI_ISL_2494042, EPI_ISL_2494043, EPI_ISL_2494044, EPI_ISL_2494045, EPI_ISL_2494046, EPI_ISL_2494047, EPI_ISL_2494048, EPI_ISL_2494049, EPI_ISL_2494050, EPI_ISL_2494051, EPI_ISL_2494329, EPI_ISL_2500985 | | | |
| see above | Ambulatório IB | Instituto Butantan | Antonio Jorge Martins; Claudia Renata dos Santos Barros; David Schlesinger; Debora Botequio Moretti; Dimas Tadeu Covas; Elaine Cristina Marqueze; Elaine Vieira Santos; Evandra Strazza Rodrigues; Heidge Fukumasu; Jayme Augusto de Souza-Neto; José Salvatore Leister Patané; Luiz Alcantara; Luiz Lehmann Coutinho; Maria Carolina Elias; Maurício Lacerda Nogueira; Rafael dos Santos Bezerra; Raul Machado Neto; Rejane Maria Tommasini Grotto; Ricardo Haddad; Sandra Coccuzzo Sampaio Vessoni; Simone Kashima; Svetoslav Nanev Slavov; Vincent Louis Viala |
| EPI_ISL_2444202 | BADY BASSITT | Instituto Butantan / FAMERP | Antonio Jorge Martins; Claudia Renata dos Santos Barros; David Schlesinger; Debora Botequio Moretti; Dimas Tadeu Covas; Elaine Cristina Marqueze; Elaine Vieira Santos; Evandra Strazza Rodrigues; Heidge Fukumasu; Jayme Augusto de Souza-Neto; José Salvatore Leister Patané; Luiz Alcantara; Luiz Lehmann Coutinho; Maria Carolina Elias; Maurício Lacerda Nogueira; Rafael dos Santos Bezerra; Raul Machado Neto; Rejane Maria Tommasini Grotto; Ricardo Haddad; Sandra Coccuzzo Sampaio Vessoni; Simone Kashima; Svetoslav Nanev Slavov; Vincent Louis Viala |
| EPI_ISL_2444212 | BARRETOS | Instituto Butantan / FAMERP | Antonio Jorge Martins; Claudia Renata dos Santos Barros; David Schlesinger; Debora Botequio Moretti; Dimas Tadeu Covas; Elaine Cristina Marqueze; Elaine Vieira Santos; Evandra Strazza Rodrigues; Heidge Fukumasu; Jayme Augusto de Souza-Neto; José Salvatore Leister Patané; Luiz Alcantara; Luiz Lehmann Coutinho; Maria Carolina Elias; Maurício Lacerda Nogueira; Rafael dos Santos Bezerra; Raul Machado Neto; Rejane Maria Tommasini Grotto; Ricardo Haddad; Sandra Coccuzzo Sampaio Vessoni; Simone Kashima; Svetoslav Nanev Slavov; Vincent Louis Viala |
| EPI_ISL_2493187, EPI_ISL_2493199, EPI_ISL_2493202, EPI_ISL_2493763, EPI_ISL_2493764, EPI_ISL_2493765, EPI_ISL_2493766, EPI_ISL_2493767, EPI_ISL_2493768, EPI_ISL_2493769, EPI_ISL_2493770, EPI_ISL_2493771, EPI_ISL_2493772, EPI_ISL_2493773, EPI_ISL_2493774, EPI_ISL_2493775, EPI_ISL_2493776, EPI_ISL_2493777, EPI_ISL_2493778, EPI_ISL_2493779, EPI_ISL_2493780, EPI_ISL_2493781, EPI_ISL_2493782, EPI_ISL_2493783, EPI_ISL_2493784, EPI_ISL_2493785, EPI_ISL_2493786, EPI_ISL_2493787, EPI_ISL_2493788, EPI_ISL_2494314, EPI_ISL_2494315, EPI_ISL_2494319, EPI_ISL_2500974 | | | |
| see above | BIOFAST | Instituto Butantan | Antonio Jorge Martins; Claudia Renata dos Santos Barros; David Schlesinger; Debora Botequio Moretti; Dimas Tadeu Covas; Elaine Cristina Marqueze; Elaine Vieira Santos; Evandra Strazza Rodrigues; Heidge Fukumasu; Jayme Augusto de Souza-Neto; José Salvatore Leister Patané; Luiz Alcantara; Luiz Lehmann Coutinho; Maria Carolina Elias; Maurício Lacerda Nogueira; Rafael dos Santos Bezerra; Raul Machado Neto; Rejane Maria Tommasini Grotto; Ricardo Haddad; Sandra Coccuzzo Sampaio Vessoni; Simone Kashima; Svetoslav Nanev Slavov; Vincent Louis Viala |
| EPI_ISL_2493261, EPI_ISL_2493262, EPI_ISL_2493268, EPI_ISL_2494220, EPI_ISL_2494221, EPI_ISL_2494222, EPI_ISL_2494223, EPI_ISL_2494224 | | | |
| see above | BIOFAST CENTRO | Instituto Butantan | Antonio Jorge Martins; Claudia Renata dos Santos Barros; David Schlesinger; Debora Botequio Moretti; Dimas Tadeu Covas; Elaine Cristina Marqueze; Elaine Vieira Santos; Evandra Strazza Rodrigues; Heidge Fukumasu; Jayme Augusto de Souza-Neto; José Salvatore Leister Patané; Luiz Alcantara; Luiz Lehmann Coutinho; Maria Carolina Elias; Maurício Lacerda Nogueira; Rafael dos Santos Bezerra; Raul Machado Neto; Rejane Maria Tommasini Grotto; Ricardo Haddad; Sandra Coccuzzo Sampaio Vessoni; Simone Kashima; Svetoslav Nanev Slavov; Vincent Louis Viala |
| EPI_ISL_2445085 | BIOFAST CENTRO SANTA CECÍLIA | Instituto Butantan | Antonio Jorge Martins; Claudia Renata dos Santos Barros; David Schlesinger; Debora Botequio Moretti; Dimas Tadeu Covas; Elaine Cristina Marqueze; Elaine Vieira Santos; Evandra Strazza Rodrigues; Heidge Fukumasu; Jayme Augusto de Souza-Neto; José Salvatore Leister Patané; Luiz Alcantara; Luiz Lehmann Coutinho; Maria Carolina Elias; Maurício Lacerda Nogueira; Rafael dos Santos Bezerra; Raul Machado Neto; Rejane Maria Tommasini Grotto; Ricardo Haddad; Sandra Coccuzzo Sampaio Vessoni; Simone Kashima; Svetoslav Nanev Slavov; Vincent Louis Viala |
| EPI_ISL_2445086 | BIOFAST CENTRO SÉ | Instituto Butantan | Antonio Jorge Martins; Claudia Renata dos Santos Barros; David Schlesinger; Debora Botequio Moretti; Dimas Tadeu Covas; Elaine Cristina Marqueze; Elaine Vieira Santos; Evandra Strazza Rodrigues; Heidge Fukumasu; Jayme Augusto de Souza-Neto; José Salvatore Leister Patané; Luiz Alcantara; Luiz Lehmann Coutinho; Maria Carolina Elias; Maurício Lacerda Nogueira; Rafael dos Santos Bezerra; Raul Machado Neto; Rejane Maria Tommasini Grotto; Ricardo Haddad; Sandra Coccuzzo Sampaio Vessoni; Simone Kashima; Svetoslav Nanev Slavov; Vincent Louis Viala |
| EPI_ISL_2493191, EPI_ISL_2493245, EPI_ISL_2493248, EPI_ISL_2493291, EPI_ISL_2494225, EPI_ISL_2494226, EPI_ISL_2494227, EPI_ISL_2494228, EPI_ISL_2494229, EPI_ISL_2494230, EPI_ISL_2494231, EPI_ISL_2494232, EPI_ISL_2494233, EPI_ISL_2494234, EPI_ISL_2494235, EPI_ISL_2494236, EPI_ISL_2494237, EPI_ISL_2494238, EPI_ISL_2494239, EPI_ISL_2494240, EPI_ISL_2494241, EPI_ISL_2494242, EPI_ISL_2494243, EPI_ISL_2494244, EPI_ISL_2494245, EPI_ISL_2494246, EPI_ISL_2494247, EPI_ISL_2494248, EPI_ISL_2494249, EPI_ISL_2494250, EPI_ISL_2494251, EPI_ISL_2494252, EPI_ISL_2494253, EPI_ISL_2494254, EPI_ISL_2494255, EPI_ISL_2494256, EPI_ISL_2494257, EPI_ISL_2494258, EPI_ISL_2494259, EPI_ISL_2494260, EPI_ISL_2494261, EPI_ISL_2494262, EPI_ISL_2494263, EPI_ISL_2494264, EPI_ISL_2494265, EPI_ISL_2494266, EPI_ISL_2494301, EPI_ISL_2494353 | | | |
| see above | BIOFAST LESTE | Instituto Butantan | Antonio Jorge Martins; Claudia Renata dos Santos Barros; David Schlesinger; Debora Botequio Moretti; Dimas Tadeu Covas; Elaine Cristina Marqueze; Elaine Vieira Santos; Evandra Strazza Rodrigues; Heidge Fukumasu; Jayme Augusto de Souza-Neto; José Salvatore Leister Patané; Luiz Alcantara; Luiz Lehmann Coutinho; Maria Carolina Elias; Maurício Lacerda Nogueira; Rafael dos Santos Bezerra; Raul Machado Neto; Rejane Maria Tommasini Grotto; Ricardo Haddad; Sandra Coccuzzo Sampaio Vessoni; Simone Kashima; Svetoslav Nanev Slavov; Vincent Louis Viala |
| EPI_ISL_2445093, EPI_ISL_2445094, EPI_ISL_2445095 | BIOFAST LESTE GUAIANASES | Instituto Butantan | Antonio Jorge Martins; Claudia Renata dos Santos Barros; David Schlesinger; Debora Botequio Moretti; Dimas Tadeu Covas; Elaine Cristina Marqueze; Elaine Vieira Santos; Evandra Strazza Rodrigues; Heidge Fukumasu; Jayme Augusto de Souza-Neto; José Salvatore Leister Patané; Luiz Alcantara; Luiz Lehmann Coutinho; Maria Carolina Elias; Maurício Lacerda Nogueira; Rafael dos Santos Bezerra; Raul Machado Neto; Rejane Maria Tommasini Grotto; Ricardo Haddad; Sandra Coccuzzo Sampaio Vessoni; Simone Kashima; Svetoslav Nanev Slavov; Vincent Louis Viala |
| EPI_ISL_2445099, EPI_ISL_2445100, EPI_ISL_2445101, EPI_ISL_2445102 | BIOFAST LESTE ITAIM PAULISTA | Instituto Butantan | Antonio Jorge Martins; Claudia Renata dos Santos Barros; David Schlesinger; Debora Botequio Moretti; Dimas Tadeu Covas; Elaine Cristina Marqueze; Elaine Vieira Santos; Evandra Strazza Rodrigues; Heidge Fukumasu; Jayme Augusto de Souza-Neto; José Salvatore Leister Patané; Luiz Alcantara; Luiz Lehmann Coutinho; Maria Carolina Elias; Maurício Lacerda Nogueira; Rafael dos Santos Bezerra; Raul Machado Neto; Rejane Maria Tommasini Grotto; Ricardo Haddad; Sandra Coccuzzo Sampaio Vessoni; Simone Kashima; Svetoslav Nanev Slavov; Vincent Louis Viala |
| EPI_ISL_2445087, EPI_ISL_2445088, EPI_ISL_2445089, EPI_ISL_2445090, EPI_ISL_2445091, EPI_ISL_2445092 | BIOFAST LESTE ITAQUERA | Instituto Butantan | Antonio Jorge Martins; Claudia Renata dos Santos Barros; David Schlesinger; Debora Botequio Moretti; Dimas Tadeu Covas; Elaine Cristina Marqueze; Elaine Vieira Santos; Evandra Strazza Rodrigues; Heidge Fukumasu; Jayme Augusto de Souza-Neto; José Salvatore Leister Patané; Luiz Alcantara; Luiz Lehmann Coutinho; Maria Carolina Elias; Maurício Lacerda Nogueira; Rafael dos Santos Bezerra; Raul Machado Neto; Rejane Maria Tommasini Grotto; Ricardo Haddad; Sandra Coccuzzo Sampaio Vessoni; Simone Kashima; Svetoslav Nanev Slavov; Vincent Louis Viala |
| EPI_ISL_2445096, EPI_ISL_2445097, EPI_ISL_2445098 | BIOFAST LESTE SÃO MIGUEL PAULISTA | Instituto Butantan | Antonio Jorge Martins; Claudia Renata dos Santos Barros; David Schlesinger; Debora Botequio Moretti; Dimas Tadeu Covas; Elaine Cristina Marqueze; Elaine Vieira Santos; Evandra Strazza Rodrigues; Heidge Fukumasu; Jayme Augusto de Souza-Neto; José Salvatore Leister Patané; Luiz Alcantara; Luiz Lehmann Coutinho; Maria Carolina Elias; Maurício Lacerda Nogueira; Rafael dos Santos Bezerra; Raul Machado Neto; Rejane Maria Tommasini Grotto; Ricardo Haddad; Sandra Coccuzzo Sampaio Vessoni; Simone Kashima; Svetoslav Nanev Slavov; Vincent Louis Viala |
| EPI_ISL_2445103, EPI_ISL_2445104 | BIOFAST LESTE TIRADENTES | Instituto Butantan | Antonio Jorge Martins; Claudia Renata dos Santos Barros; David Schlesinger; Debora Botequio Moretti; Dimas Tadeu Covas; Elaine Cristina Marqueze; Elaine Vieira Santos; Evandra Strazza Rodrigues; Heidge Fukumasu; Jayme Augusto de Souza-Neto; José Salvatore Leister Patané; Luiz Alcantara; Luiz Lehmann Coutinho; Maria Carolina Elias; Maurício Lacerda Nogueira; Rafael dos Santos Bezerra; Raul Machado Neto; Rejane Maria Tommasini Grotto; Ricardo Haddad; Sandra Coccuzzo Sampaio Vessoni; Simone Kashima; Svetoslav Nanev Slavov; Vincent Louis Viala |
| EPI_ISL_2493188, EPI_ISL_2493270, EPI_ISL_2494213, EPI_ISL_2494214, EPI_ISL_2494215, EPI_ISL_2494216, EPI_ISL_2494217, EPI_ISL_2494218, EPI_ISL_2494219, EPI_ISL_2494333 | | | |
| see above | BIOFAST NORTE | Instituto Butantan | Antonio Jorge Martins; Claudia Renata dos Santos Barros; David Schlesinger; Debora Botequio Moretti; Dimas Tadeu Covas; Elaine Cristina Marqueze; Elaine Vieira Santos; Evandra Strazza Rodrigues; Heidge Fukumasu; Jayme Augusto de Souza-Neto; José Salvatore Leister Patané; Luiz Alcantara; Luiz Lehmann |

| | | | |
|---|---|---|---|
| | | | Coutinho; Maria Carolina Elias; Maurício Lacerda Nogueira; Rafael dos Santos Bezerra; Raul Machado Neto; Rejane Maria Tommasini Grotto; Ricardo Haddad; Sandra Coccuzzo Sampaio Vessoni; Simone Kashima; Svetoslav Nanev Slavov; Vincent Louis Viala |
| EPI_ISL_2445105, EPI_ISL_2445106, EPI_ISL_2445107, EPI_ISL_2473811 | BIOFAST NORTE SANTANA/JAÇANA/TUCURUVI | Instituto Butantan | Antonio Jorge Martins; Claudia Renata dos Santos Barros; David Schlesinger; Debora Botequio Moretti; Dimas Tadeu Covas; Elaine Cristina Marqueze; Elaine Vieira Santos; Evandra Strazza Rodrigues; Heidge Fukumasu; Jayme Augusto de Souza-Neto; José Salvatore Leister Patané; Luiz Alcantara; Luiz Lehmann Coutinho; Maria Carolina Elias; Maurício Lacerda Nogueira; Rafael dos Santos Bezerra; Raul Machado Neto; Rejane Maria Tommasini Grotto; Ricardo Haddad; Sandra Coccuzzo Sampaio Vessoni; Simone Kashima; Svetoslav Nanev Slavov; Vincent Louis Viala |
| EPI_ISL_2493260, EPI_ISL_2493267, EPI_ISL_2494201, EPI_ISL_2494202, EPI_ISL_2494203, EPI_ISL_2494204, EPI_ISL_2494205, EPI_ISL_2494206, EPI_ISL_2494207, EPI_ISL_2494208, EPI_ISL_2494209, EPI_ISL_2494210, EPI_ISL_2494211, EPI_ISL_2494212, EPI_ISL_2494275, EPI_ISL_2494313 | | | |
| see above | BIOFAST SUL | Instituto Butantan | Antonio Jorge Martins; Claudia Renata dos Santos Barros; David Schlesinger; Debora Botequio Moretti; Dimas Tadeu Covas; Elaine Cristina Marqueze; Elaine Vieira Santos; Evandra Strazza Rodrigues; Heidge Fukumasu; Jayme Augusto de Souza-Neto; José Salvatore Leister Patané; Luiz Alcantara; Luiz Lehmann Coutinho; Maria Carolina Elias; Maurício Lacerda Nogueira; Rafael dos Santos Bezerra; Raul Machado Neto; Rejane Maria Tommasini Grotto; Ricardo Haddad; Sandra Coccuzzo Sampaio Vessoni; Simone Kashima; Svetoslav Nanev Slavov; Vincent Louis Viala |
| EPI_ISL_2445108, EPI_ISL_2445109, EPI_ISL_2445110, EPI_ISL_2445111, EPI_ISL_2445112, EPI_ISL_2445113 | BIOFAST SUL SANTO AMARO/CIDADE ADEMAR | Instituto Butantan | Antonio Jorge Martins; Claudia Renata dos Santos Barros; David Schlesinger; Debora Botequio Moretti; Dimas Tadeu Covas; Elaine Cristina Marqueze; Elaine Vieira Santos; Evandra Strazza Rodrigues; Heidge Fukumasu; Jayme Augusto de Souza-Neto; José Salvatore Leister Patané; Luiz Alcantara; Luiz Lehmann Coutinho; Maria Carolina Elias; Maurício Lacerda Nogueira; Rafael dos Santos Bezerra; Raul Machado Neto; Rejane Maria Tommasini Grotto; Ricardo Haddad; Sandra Coccuzzo Sampaio Vessoni; Simone Kashima; Svetoslav Nanev Slavov; Vincent Louis Viala |
| EPI_ISL_2698105, EPI_ISL_2698106 | Biology, UFLA, Universidade Federal de Lavras | Biology, UFLA | Barcante, J.; Cherem, J.; Fernandes, G.; Luciano, P.; Melo, D.; Pylro, V. |
| EPI_ISL_2445508, EPI_ISL_2445512, EPI_ISL_2445513, EPI_ISL_2445514, EPI_ISL_2473723, EPI_ISL_2473786, EPI_ISL_2473788, EPI_ISL_2473818 | | | |
| see above | CADIP VILA REGINA FERNANDOPOLIS | Instituto Butantan | Antonio Jorge Martins; Claudia Renata dos Santos Barros; David Schlesinger; Debora Botequio Moretti; Dimas Tadeu Covas; Elaine Cristina Marqueze; Elaine Vieira Santos; Evandra Strazza Rodrigues; Heidge Fukumasu; Jayme Augusto de Souza-Neto; José Salvatore Leister Patané; Luiz Alcantara; Luiz Lehmann Coutinho; Maria Carolina Elias; Maurício Lacerda Nogueira; Rafael dos Santos Bezerra; Raul Machado Neto; Rejane Maria Tommasini Grotto; Ricardo Haddad; Sandra Coccuzzo Sampaio Vessoni; Simone Kashima; Svetoslav Nanev Slavov; Vincent Louis Viala |
| EPI_ISL_2801337, EPI_ISL_2801341, EPI_ISL_2801343, EPI_ISL_2801344, EPI_ISL_2801346, EPI_ISL_2801363, EPI_ISL_2801364 | | | |
| see above | CENTRO DE ATENDIMENTO PARA ENFRENTAMENTO AO COVID 19 | Oswaldo Cruz Institute, FIOCRUZ/CE | Cleber Furtado Aksenen e Suzana Porto Almeida; Fabio Miyajima; Fernando Braga Stehling; Francisco Eder de Moura Lopes; Jamille Maria Mendes Bezerra; Joaquim César do Nascimento Sousa Junior; Pedro Miguel Carneiro Jeronimo; Thais Ferreira de Oliveira; Thais de Oliveira Costa; Ticiane Cavalcante de Souza; Veridiana Pessoa Miyajima |
| EPI_ISL_2493548, EPI_ISL_2494347 | CENTRO DE ESPECIALIDADES DE AGUAI AGUAI | Instituto Butantan | Antonio Jorge Martins; Claudia Renata dos Santos Barros; David Schlesinger; Debora Botequio Moretti; Dimas Tadeu Covas; Elaine Cristina Marqueze; Elaine Vieira Santos; Evandra Strazza Rodrigues; Heidge Fukumasu; Jayme Augusto de Souza-Neto; José Salvatore Leister Patané; Luiz Alcantara; Luiz Lehmann Coutinho; Maria Carolina Elias; Maurício Lacerda Nogueira; Rafael dos Santos Bezerra; Raul Machado Neto; Rejane Maria Tommasini Grotto; Ricardo Haddad; Sandra Coccuzzo Sampaio Vessoni; Simone Kashima; Svetoslav Nanev Slavov; Vincent Louis Viala |
| EPI_ISL_2445551, EPI_ISL_2445553 | CENTRO DE ESPECIALIDADES DE PRIMAVERA | Instituto Butantan | Antonio Jorge Martins; Claudia Renata dos Santos Barros; David Schlesinger; Debora Botequio Moretti; Dimas Tadeu Covas; Elaine Cristina Marqueze; Elaine Vieira Santos; Evandra Strazza Rodrigues; Heidge Fukumasu; Jayme Augusto de Souza-Neto; José Salvatore Leister Patané; Luiz Alcantara; Luiz Lehmann Coutinho; Maria Carolina Elias; Maurício Lacerda Nogueira; Rafael dos Santos Bezerra; Raul Machado Neto; Rejane Maria Tommasini Grotto; Ricardo Haddad; Sandra Coccuzzo Sampaio Vessoni; Simone Kashima; Svetoslav Nanev Slavov; Vincent Louis Viala |
| EPI_ISL_2345715, EPI_ISL_2345811 | CENTRO DE ESPECIALIDADES DE PRIMAVERA | Instituto Butantan / Mendelics | Antonio Jorge Martins; Claudia Renata dos Santos Barros; David Schlesinger; Debora Botequio Moretti; Dimas Tadeu Covas; Elaine Cristina Marqueze; Elaine Vieira Santos; Evandra Strazza Rodrigues; Heidge Fukumasu; Jayme Augusto de Souza-Neto; José Salvatore Leister Patané; Luiz Alcantara; Luiz Lehmann Coutinho; Maria Carolina Elias; Maurício Lacerda Nogueira; Rafael dos Santos Bezerra; Raul Machado Neto; Rejane Maria Tommasini Grotto; Ricardo Haddad; Sandra Coccuzzo Sampaio Vessoni; Simone Kashima; Svetoslav Nanev Slavov; Vincent Louis Viala |
| EPI_ISL_2445539 | CENTRO DE ESPECIALIDADES MARIAS DA GLORIA | Instituto Butantan | Antonio Jorge Martins; Claudia Renata dos Santos Barros; David Schlesinger; Debora Botequio Moretti; Dimas Tadeu Covas; Elaine Cristina Marqueze; Elaine Vieira Santos; Evandra Strazza Rodrigues; Heidge Fukumasu; Jayme Augusto de Souza-Neto; José Salvatore Leister Patané; Luiz Alcantara; Luiz Lehmann Coutinho; Maria Carolina Elias; Maurício Lacerda Nogueira; Rafael dos Santos Bezerra; Raul Machado Neto; Rejane Maria Tommasini Grotto; Ricardo Haddad; Sandra Coccuzzo Sampaio Vessoni; Simone Kashima; Svetoslav Nanev Slavov; Vincent Louis Viala |
| EPI_ISL_2445340, EPI_ISL_2445341 | CENTRO DE ESPECIALIDADES MUNICIPAL CESAR TORREZAN R BONITO | Instituto Butantan | Antonio Jorge Martins; Claudia Renata dos Santos Barros; David Schlesinger; Debora Botequio Moretti; Dimas Tadeu Covas; Elaine Cristina Marqueze; Elaine Vieira Santos; Evandra Strazza Rodrigues; Heidge Fukumasu; Jayme Augusto de Souza-Neto; José Salvatore Leister Patané; Luiz Alcantara; Luiz Lehmann Coutinho; Maria Carolina Elias; Maurício Lacerda Nogueira; Rafael dos Santos Bezerra; Raul Machado Neto; Rejane Maria Tommasini Grotto; Ricardo Haddad; Sandra Coccuzzo Sampaio Vessoni; Simone Kashima; Svetoslav Nanev Slavov; Vincent Louis Viala |
| EPI_ISL_2345656, EPI_ISL_2345845 | CENTRO DE ESPECIALIDADES MUNICIPAL CESAR TORREZAN R BONITO | Instituto Butantan / Mendelics | Antonio Jorge Martins; Claudia Renata dos Santos Barros; David Schlesinger; Debora Botequio Moretti; Dimas Tadeu Covas; Elaine Cristina Marqueze; Elaine Vieira Santos; Evandra Strazza Rodrigues; Heidge Fukumasu; Jayme Augusto de Souza-Neto; José Salvatore Leister Patané; Luiz Alcantara; Luiz Lehmann Coutinho; Maria Carolina Elias; Maurício Lacerda Nogueira; Rafael dos Santos Bezerra; Raul Machado Neto; Rejane Maria Tommasini Grotto; Ricardo Haddad; Sandra Coccuzzo Sampaio Vessoni; Simone Kashima; Svetoslav Nanev Slavov; Vincent Louis Viala |
| EPI_ISL_2493375 | CENTRO DE ESPECIALIDDS MEDICAS IRMA LEOPOLDINA PIRASSUNUNGA | Instituto Butantan | Antonio Jorge Martins; Claudia Renata dos Santos Barros; David Schlesinger; Debora Botequio Moretti; Dimas Tadeu Covas; Elaine Cristina Marqueze; Elaine Vieira Santos; Evandra Strazza Rodrigues; Heidge Fukumasu; Jayme Augusto de Souza-Neto; José Salvatore Leister Patané; Luiz Alcantara; Luiz Lehmann Coutinho; Maria Carolina Elias; Maurício Lacerda Nogueira; Rafael dos Santos Bezerra; Raul Machado Neto; Rejane Maria Tommasini Grotto; Ricardo Haddad; Sandra Coccuzzo Sampaio Vessoni; Simone Kashima; Svetoslav Nanev Slavov; Vincent Louis Viala |
| EPI_ISL_2445615, EPI_ISL_2445616, EPI_ISL_2473809, EPI_ISL_2473810, EPI_ISL_2493280, EPI_ISL_2493789, EPI_ISL_2493790, EPI_ISL_2493791 | | | |
| see above | CENTRO DE GENOMAS | Instituto Butantan | Antonio Jorge Martins; Claudia Renata dos Santos Barros; David Schlesinger; Debora Botequio Moretti; Dimas Tadeu Covas; Elaine Cristina Marqueze; Elaine Vieira Santos; Evandra Strazza Rodrigues; Heidge Fukumasu; Jayme Augusto de Souza-Neto; José Salvatore Leister Patané; Luiz Alcantara; Luiz Lehmann Coutinho; Maria Carolina Elias; Maurício Lacerda Nogueira; Rafael dos Santos Bezerra; Raul Machado Neto; Rejane Maria Tommasini Grotto; Ricardo Haddad; Sandra Coccuzzo Sampaio Vessoni; Simone Kashima; Svetoslav Nanev Slavov; Vincent Louis Viala |
| EPI_ISL_2493155, EPI_ISL_2493435, EPI_ISL_2493436, EPI_ISL_2493437, EPI_ISL_2493438 | CENTRO DE GENOMAS SUL | Instituto Butantan | Antonio Jorge Martins; Claudia Renata dos Santos Barros; David Schlesinger; Debora Botequio Moretti; Dimas Tadeu Covas; Elaine Cristina Marqueze; Elaine Vieira Santos; Evandra Strazza Rodrigues; Heidge Fukumasu; Jayme Augusto de Souza-Neto; José Salvatore Leister Patané; Luiz Alcantara; Luiz Lehmann Coutinho; Maria Carolina Elias; Maurício Lacerda Nogueira; Rafael dos Santos Bezerra; Raul Machado Neto; Rejane Maria Tommasini Grotto; Ricardo Haddad; Sandra Coccuzzo Sampaio Vessoni; Simone Kashima; Svetoslav Nanev Slavov; Vincent Louis Viala |
| EPI_ISL_2445338, EPI_ISL_2445339, EPI_ISL_2493153, EPI_ISL_2493824, EPI_ISL_2493826, EPI_ISL_2494127, EPI_ISL_2494128 | | | |
| see above | CENTRO DE REFERENCIA DO IDOSO DR HUMBERTO MENDES DE CARVALHO | Instituto Butantan | Antonio Jorge Martins; Claudia Renata dos Santos Barros; David Schlesinger; Debora Botequio Moretti; Dimas Tadeu Covas; Elaine Cristina Marqueze; Elaine Vieira Santos; Evandra Strazza Rodrigues; Heidge Fukumasu; Jayme Augusto de Souza-Neto; José Salvatore Leister Patané; Luiz Alcantara; Luiz Lehmann Coutinho; Maria Carolina Elias; Maurício Lacerda Nogueira; Rafael dos Santos Bezerra; Raul Machado Neto; Rejane Maria Tommasini Grotto; Ricardo Haddad; Sandra Coccuzzo Sampaio Vessoni; Simone Kashima; Svetoslav Nanev Slavov; Vincent Louis Viala |
| EPI_ISL_2345669, EPI_ISL_2345815, EPI_ISL_2345860, EPI_ISL_2345865 | CENTRO DE REFERENCIA DO IDOSO DR HUMBERTO MENDES DE CARVALHO | Instituto Butantan / Mendelics | Antonio Jorge Martins; Claudia Renata dos Santos Barros; David Schlesinger; Debora Botequio Moretti; Dimas Tadeu Covas; Elaine Cristina Marqueze; Elaine Vieira Santos; Evandra Strazza Rodrigues; Heidge Fukumasu; Jayme Augusto de Souza-Neto; José Salvatore Leister Patané; Luiz Alcantara; Luiz Lehmann Coutinho; Maria Carolina Elias; Maurício Lacerda Nogueira; Rafael dos Santos Bezerra; Raul Machado Neto; Rejane Maria Tommasini Grotto; Ricardo Haddad; Sandra Coccuzzo Sampaio Vessoni; Simone Kashima; Svetoslav Nanev Slavov; Vincent Louis Viala |
| EPI_ISL_2493917, EPI_ISL_2494309 | CENTRO DE REFERENCIA E TREINAMENTO DSTAIDS SAO PAULO | Instituto Butantan | Antonio Jorge Martins; Claudia Renata dos Santos Barros; David Schlesinger; Debora Botequio Moretti; Dimas Tadeu Covas; Elaine Cristina Marqueze; Elaine Vieira Santos; Evandra Strazza Rodrigues; Heidge Fukumasu; Jayme Augusto de Souza-Neto; José Salvatore Leister Patané; Luiz Alcantara; Luiz Lehmann Coutinho; Maria Carolina Elias; Maurício Lacerda Nogueira; Rafael dos Santos Bezerra; Raul Machado Neto; Rejane Maria Tommasini Grotto; Ricardo Haddad; Sandra Coccuzzo Sampaio Vessoni; Simone Kashima; Svetoslav Nanev Slavov; Vincent Louis Viala |
| EPI_ISL_2493394 | CENTRO DE SAUDE | Instituto Butantan | Antonio Jorge Martins; Claudia Renata dos Santos Barros; David Schlesinger; Debora Botequio Moretti; Dimas Tadeu Covas; Elaine Cristina Marqueze; Elaine Vieira Santos; Evandra Strazza Rodrigues; Heidge Fukumasu; Jayme Augusto de Souza-Neto; José Salvatore Leister Patané; Luiz Alcantara; Luiz Lehmann Coutinho; Maria Carolina Elias; Maurício Lacerda Nogueira; Rafael dos Santos Bezerra; Raul Machado Neto; Rejane Maria Tommasini Grotto; Ricardo Haddad; Sandra Coccuzzo Sampaio Vessoni; Simone Kashima; Svetoslav Nanev Slavov; Vincent Louis Viala |
| EPI_ISL_2493282, EPI_ISL_2494179 | CENTRO DE SAUDE DE ANALANDIA | Instituto Butantan | Antonio Jorge Martins; Claudia Renata dos Santos Barros; David Schlesinger; Debora Botequio Moretti; Dimas Tadeu Covas; Elaine Cristina Marqueze; Elaine Vieira Santos; Evandra Strazza Rodrigues; Heidge Fukumasu; Jayme Augusto de Souza-Neto; José Salvatore Leister Patané; Luiz Alcantara; Luiz Lehmann Coutinho; Maria Carolina Elias; Maurício Lacerda Nogueira; Rafael dos Santos Bezerra; Raul Machado Neto; Rejane Maria Tommasini Grotto; Ricardo Haddad; Sandra Coccuzzo Sampaio Vessoni; Simone Kashima; Svetoslav Nanev Slavov; Vincent Louis Viala |
| EPI_ISL_2445165, EPI_ISL_2473820 | CENTRO DE SAUDE DE CANITAR | Instituto Butantan | Antonio Jorge Martins; Claudia Renata dos Santos Barros; David Schlesinger; Debora Botequio Moretti; Dimas Tadeu Covas; Elaine Cristina Marqueze; Elaine Vieira Santos; Evandra Strazza Rodrigues; Heidge Fukumasu; Jayme Augusto de Souza-Neto; José Salvatore Leister Patané; Luiz Alcantara; Luiz Lehmann Coutinho; Maria Carolina Elias; Maurício Lacerda Nogueira; Rafael dos Santos Bezerra; Raul Machado Neto; Rejane Maria Tommasini Grotto; Ricardo Haddad; Sandra Coccuzzo Sampaio Vessoni; Simone Kashima; Svetoslav Nanev Slavov; Vincent Louis Viala |
| EPI_ISL_2445197 | CENTRO DE SAUDE DE CRUZALIA | Instituto Butantan | Antonio Jorge Martins; Claudia Renata dos Santos Barros; David Schlesinger; Debora Botequio Moretti; Dimas Tadeu Covas; Elaine Cristina Marqueze; Elaine Vieira Santos; Evandra Strazza Rodrigues; Heidge Fukumasu; Jayme Augusto de Souza-Neto; José Salvatore Leister Patané; Luiz Alcantara; Luiz Lehmann Coutinho; Maria Carolina Elias; Maurício Lacerda Nogueira; Rafael dos Santos Bezerra; Raul Machado Neto; Rejane Maria Tommasini Grotto; Ricardo Haddad; Sandra Coccuzzo Sampaio Vessoni; Simone Kashima; Svetoslav Nanev Slavov; Vincent Louis Viala |
| EPI_ISL_2445153 | CENTRO DE SAUDE DE FLORIDA PAULISTA | Instituto Butantan | Antonio Jorge Martins; Claudia Renata dos Santos Barros; David Schlesinger; Debora Botequio Moretti; Dimas Tadeu Covas; Elaine Cristina Marqueze; Elaine Vieira Santos; Evandra Strazza Rodrigues; Heidge Fukumasu; Jayme Augusto de Souza-Neto; José Salvatore Leister Patané; Luiz Alcantara; Luiz Lehmann Coutinho; Maria Carolina Elias; Maurício Lacerda Nogueira; Rafael dos Santos Bezerra; Raul Machado Neto; Rejane Maria Tommasini Grotto; Ricardo Haddad; Sandra Coccuzzo Sampaio Vessoni; Simone Kashima; Svetoslav Nanev Slavov; Vincent Louis Viala |
| EPI_ISL_2445444, EPI_ISL_2473713 | CENTRO DE SAUDE DE IBIRAREMA | Instituto Butantan | Antonio Jorge Martins; Claudia Renata dos Santos Barros; David Schlesinger; Debora Botequio Moretti; Dimas Tadeu Covas; Elaine Cristina Marqueze; Elaine Vieira Santos; Evandra Strazza Rodrigues; Heidge Fukumasu; Jayme Augusto de Souza-Neto; José Salvatore Leister Patané; Luiz Alcantara; Luiz Lehmann Coutinho; Maria Carolina Elias; Maurício Lacerda Nogueira; Rafael dos Santos Bezerra; Raul Machado Neto; Rejane Maria Tommasini Grotto; Ricardo Haddad; Sandra Coccuzzo Sampaio Vessoni; Simone Kashima; Svetoslav Nanev Slavov; Vincent Louis Viala |
| EPI_ISL_2445189, EPI_ISL_2445428, EPI_ISL_2445429, EPI_ISL_2445430, EPI_ISL_2445431, EPI_ISL_2445432, EPI_ISL_2445433, EPI_ISL_2445434, EPI_ISL_2493586 | | | |
| see above | CENTRO DE SAUDE DE LUCELIA | Instituto Butantan | Antonio Jorge Martins; Claudia Renata dos Santos Barros; David Schlesinger; Debora Botequio Moretti; Dimas Tadeu Covas; Elaine Cristina Marqueze; Elaine Vieira Santos; Evandra Strazza Rodrigues; Heidge Fukumasu; Jayme Augusto de Souza-Neto; José Salvatore Leister Patané; Luiz Alcantara; Luiz Lehmann Coutinho; Maria Carolina Elias; Maurício Lacerda Nogueira; Rafael dos Santos Bezerra; Raul Machado Neto; Rejane Maria Tommasini Grotto; Ricardo Haddad; Sandra Coccuzzo Sampaio Vessoni; Simone Kashima; Svetoslav Nanev Slavov; Vincent Louis Viala |
| EPI_ISL_2209754, EPI_ISL_2210292 | CENTRO DE SAUDE DE LUPERCIO | Instituto Butantan / Mendelics | Antonio Jorge Martins; Claudia Renata dos Santos Barros; David Schlesinger; Debora Botequio Moretti; Dimas Tadeu Covas; Elaine Cristina Marqueze; Elaine Vieira Santos; Evandra Strazza Rodrigues; Heidge Fukumasu; Jayme Augusto de Souza-Neto; José Salvatore Leister Patané; Luiz Alcantara; Luiz Lehmann Coutinho; Maria Carolina Elias; Maurício Lacerda Nogueira; Rafael dos Santos Bezerra; Raul Machado Neto; Rejane Maria Tommasini Grotto; Ricardo Haddad; Sandra Coccuzzo Sampaio Vessoni; Simone Kashima; Svetoslav Nanev Slavov; Vincent Louis Viala |
| EPI_ISL_2209464, EPI_ISL_2209593, EPI_ISL_2345657, EPI_ISL_2345702, EPI_ISL_2345753, EPI_ISL_2345846 | CENTRO DE SAUDE DE MARACAI | Instituto Butantan / Mendelics | Antonio Jorge Martins; Claudia Renata dos Santos Barros; David Schlesinger; Debora Botequio Moretti; Dimas Tadeu Covas; Elaine Cristina Marqueze; Elaine Vieira Santos; Evandra Strazza Rodrigues; Heidge Fukumasu; Jayme Augusto de Souza-Neto; José Salvatore Leister Patané; Luiz Alcantara; Luiz Lehmann Coutinho; Maria Carolina Elias; Maurício Lacerda Nogueira; Rafael dos Santos Bezerra; Raul Machado Neto; Rejane Maria Tommasini Grotto; Ricardo Haddad; Sandra Coccuzzo Sampaio Vessoni; Simone Kashima; Svetoslav Nanev Slavov; Vincent Louis Viala |
| EPI_ISL_2473793 | CENTRO DE SAUDE DE NOVA GUATAPORANGA | Instituto Butantan | Antonio Jorge Martins; Claudia Renata dos Santos Barros; David Schlesinger; Debora Botequio Moretti; Dimas Tadeu Covas; Elaine Cristina Marqueze; Elaine Vieira Santos; Evandra Strazza Rodrigues; Heidge Fukumasu; Jayme Augusto de Souza-Neto; José Salvatore Leister Patané; Luiz Alcantara; Luiz Lehmann Coutinho; Maria Carolina Elias; Maurício Lacerda Nogueira; Rafael dos Santos Bezerra; Raul Machado Neto; Rejane Maria Tommasini Grotto; Ricardo Haddad; Sandra Coccuzzo Sampaio Vessoni; Simone Kashima; Svetoslav Nanev Slavov; Vincent Louis Viala |
| EPI_ISL_2345857 | CENTRO DE SAUDE DE NOVA GUATAPORANGA | Instituto Butantan / Mendelics | Antonio Jorge Martins; Claudia Renata dos Santos Barros; David Schlesinger; Debora Botequio Moretti; Dimas Tadeu Covas; Elaine Cristina Marqueze; Elaine Vieira Santos; Evandra Strazza Rodrigues; Heidge Fukumasu; Jayme Augusto de Souza-Neto; José Salvatore Leister Patané; Luiz Alcantara; Luiz Lehmann Coutinho; Maria Carolina Elias; Maurício Lacerda Nogueira; Rafael dos Santos Bezerra; Raul Machado Neto; Rejane Maria Tommasini Grotto; Ricardo Haddad; Sandra Coccuzzo Sampaio Vessoni; Simone Kashima; Svetoslav Nanev Slavov; Vincent Louis Viala |
| EPI_ISL_2445129, EPI_ISL_2462314, EPI_ISL_2473744, EPI_ISL_2473745, EPI_ISL_2473746, EPI_ISL_2493792, EPI_ISL_2493793, EPI_ISL_2493794, EPI_ISL_2493795, EPI_ISL_2493796, EPI_ISL_2493797 | | | |
| see above | CENTRO DE SAUDE DE PIRAJUI | Instituto Butantan | Antonio Jorge Martins; Claudia Renata dos Santos Barros; David Schlesinger; Debora Botequio Moretti; Dimas Tadeu Covas; Elaine Cristina Marqueze; Elaine Vieira Santos; Evandra Strazza Rodrigues; Heidge Fukumasu; Jayme Augusto de Souza-Neto; José Salvatore Leister Patané; Luiz Alcantara; Luiz Lehmann Coutinho; Maria Carolina Elias; Maurício Lacerda Nogueira; Rafael dos Santos Bezerra; Raul Machado Neto; Rejane Maria Tommasini Grotto; Ricardo Haddad; Sandra Coccuzzo Sampaio Vessoni; Simone Kashima; Svetoslav Nanev Slavov; Vincent Louis Viala |
| EPI_ISL_2445193, EPI_ISL_2445194 | CENTRO DE SAUDE DE RIBEIRAO DO SUL | Instituto Butantan | Antonio Jorge Martins; Claudia Renata dos Santos Barros; David Schlesinger; Debora Botequio Moretti; Dimas Tadeu Covas; Elaine Cristina Marqueze; Elaine Vieira Santos; Evandra Strazza Rodrigues; Heidge Fukumasu; Jayme Augusto de Souza-Neto; José Salvatore Leister Patané; Luiz Alcantara; Luiz Lehmann Coutinho; Maria Carolina Elias; Maurício Lacerda Nogueira; Rafael dos Santos Bezerra; Raul Machado Neto; Rejane Maria Tommasini Grotto; Ricardo Haddad; Sandra Coccuzzo Sampaio Vessoni; Simone Kashima; Svetoslav Nanev Slavov; Vincent Louis Viala |
| EPI_ISL_2493354 | CENTRO DE SAUDE DR JOSE AGOSTINHO NOGUEIRA PAULICEIA | Instituto Butantan | Antonio Jorge Martins; Claudia Renata dos Santos Barros; David Schlesinger; Debora Botequio Moretti; Dimas Tadeu Covas; Elaine Cristina Marqueze; Elaine Vieira Santos; Evandra Strazza Rodrigues; Heidge Fukumasu; Jayme Augusto de Souza-Neto; José Salvatore Leister Patané; Luiz Alcantara; Luiz Lehmann Coutinho; Maria Carolina Elias; Maurício Lacerda Nogueira; Rafael dos Santos Bezerra; Raul Machado Neto; Rejane Maria Tommasini Grotto; Ricardo Haddad; Sandra Coccuzzo Sampaio Vessoni; Simone Kashima; Svetoslav Nanev Slavov; Vincent Louis Viala |
| EPI_ISL_2493377, EPI_ISL_2493378, EPI_ISL_2494150 | CENTRO DE SAUDE DR MARIO DIAS DE AGUIAR CAPIVARI | Instituto Butantan | Antonio Jorge Martins; Claudia Renata dos Santos Barros; David Schlesinger; Debora Botequio Moretti; Dimas Tadeu Covas; Elaine Cristina Marqueze; Elaine Vieira Santos; Evandra Strazza Rodrigues; Heidge Fukumasu; Jayme Augusto de Souza-Neto; José Salvatore Leister Patané; Luiz Alcantara; Luiz Lehmann Coutinho; Maria Carolina Elias; Maurício Lacerda Nogueira; Rafael dos Santos Bezerra; Raul Machado Neto; Rejane Maria Tommasini Grotto; Ricardo Haddad; Sandra Coccuzzo Sampaio Vessoni; Simone Kashima; Svetoslav Nanev Slavov; Vincent Louis Viala |
| EPI_ISL_2445441, EPI_ISL_2445442, | CENTRO DE SAUDE DR NELSON DA CUNHA BASTOS | Instituto Butantan | Antonio Jorge Martins; Claudia Renata dos Santos Barros; David Schlesinger; Debora Botequio Moretti; Dimas Tadeu Covas; Elaine Cristina Marqueze; Elaine Vieira Santos; Evandra Strazza Rodrigues; Heidge Fukumasu; Jayme Augusto de Souza-Neto; José Salvatore Leister Patané; Luiz Alcantara; Luiz Lehmann Coutinho; Maria Carolina Elias; Maurício Lacerda Nogueira; Rafael dos Santos Bezerra; Raul Machado Neto; Rejane Maria Tommasini Grotto; Ricardo Haddad; Sandra Coccuzzo Sampaio Vessoni; Simone Kashima; Svetoslav Nanev Slavov; Vincent Louis Viala |

| | | | |
|---|---|---|---|
| EPI_ISL_2445445, EPI_ISL_2445450, EPI_ISL_2445453 | | | |
| EPI_ISL_2445440 | CENTRO DE SAUDE ECHAPORA | Instituto Butantan | Antonio Jorge Martins; Claudia Renata dos Santos Barros; David Schlesinger; Debora Botequio Moretti; Dimas Tadeu Covas; Elaine Cristina Marqueze; Elaine Vieira Santos; Evandra Strazza Rodrigues; Heidge Fukumasu; Jayme Augusto de Souza-Neto; José Salvatore Leister Patané; Luiz Alcantara; Luiz Lehmann Coutinho; Maria Carolina Elias; Maurício Lacerda Nogueira; Rafael dos Santos Bezerra; Raul Machado Neto; Rejane Maria Tommasini Grotto; Ricardo Haddad; Sandra Coccuzzo Sampaio Vessoni; Simone Kashima; Svetoslav Nanev Slavov; Vincent Louis Viala |
| EPI_ISL_2445517, EPI_ISL_2445556 | CENTRO DE SAUDE I TACITO LEITE DE CARVALHO E SILVA | Instituto Butantan | Antonio Jorge Martins; Claudia Renata dos Santos Barros; David Schlesinger; Debora Botequio Moretti; Dimas Tadeu Covas; Elaine Cristina Marqueze; Elaine Vieira Santos; Evandra Strazza Rodrigues; Heidge Fukumasu; Jayme Augusto de Souza-Neto; José Salvatore Leister Patané; Luiz Alcantara; Luiz Lehmann Coutinho; Maria Carolina Elias; Maurício Lacerda Nogueira; Rafael dos Santos Bezerra; Raul Machado Neto; Rejane Maria Tommasini Grotto; Ricardo Haddad; Sandra Coccuzzo Sampaio Vessoni; Simone Kashima; Svetoslav Nanev Slavov; Vincent Louis Viala |
| EPI_ISL_2473787 | CENTRO DE SAUDE II CARDOSO | Instituto Butantan | Antonio Jorge Martins; Claudia Renata dos Santos Barros; David Schlesinger; Debora Botequio Moretti; Dimas Tadeu Covas; Elaine Cristina Marqueze; Elaine Vieira Santos; Evandra Strazza Rodrigues; Heidge Fukumasu; Jayme Augusto de Souza-Neto; José Salvatore Leister Patané; Luiz Alcantara; Luiz Lehmann Coutinho; Maria Carolina Elias; Maurício Lacerda Nogueira; Rafael dos Santos Bezerra; Raul Machado Neto; Rejane Maria Tommasini Grotto; Ricardo Haddad; Sandra Coccuzzo Sampaio Vessoni; Simone Kashima; Svetoslav Nanev Slavov; Vincent Louis Viala |
| EPI_ISL_2445529, EPI_ISL_2445530, EPI_ISL_2445531, EPI_ISL_2445532, EPI_ISL_2445533 | CENTRO DE SAUDE II DE POTIRENDABA CESARINO BENFATTI | Instituto Butantan | Antonio Jorge Martins; Claudia Renata dos Santos Barros; David Schlesinger; Debora Botequio Moretti; Dimas Tadeu Covas; Elaine Cristina Marqueze; Elaine Vieira Santos; Evandra Strazza Rodrigues; Heidge Fukumasu; Jayme Augusto de Souza-Neto; José Salvatore Leister Patané; Luiz Alcantara; Luiz Lehmann Coutinho; Maria Carolina Elias; Maurício Lacerda Nogueira; Rafael dos Santos Bezerra; Raul Machado Neto; Rejane Maria Tommasini Grotto; Ricardo Haddad; Sandra Coccuzzo Sampaio Vessoni; Simone Kashima; Svetoslav Nanev Slavov; Vincent Louis Viala |
| EPI_ISL_2493643 | CENTRO DE SAUDE II DR GABRIEL MESQUITA VARGEM GDE DO SUL | Instituto Butantan | Antonio Jorge Martins; Claudia Renata dos Santos Barros; David Schlesinger; Debora Botequio Moretti; Dimas Tadeu Covas; Elaine Cristina Marqueze; Elaine Vieira Santos; Evandra Strazza Rodrigues; Heidge Fukumasu; Jayme Augusto de Souza-Neto; José Salvatore Leister Patané; Luiz Alcantara; Luiz Lehmann Coutinho; Maria Carolina Elias; Maurício Lacerda Nogueira; Rafael dos Santos Bezerra; Raul Machado Neto; Rejane Maria Tommasini Grotto; Ricardo Haddad; Sandra Coccuzzo Sampaio Vessoni; Simone Kashima; Svetoslav Nanev Slavov; Vincent Louis Viala |
| EPI_ISL_2494160, EPI_ISL_2494195 | CENTRO DE SAUDE II DR JOSE DE FELIPE E S PINHAL SP | Instituto Butantan | Antonio Jorge Martins; Claudia Renata dos Santos Barros; David Schlesinger; Debora Botequio Moretti; Dimas Tadeu Covas; Elaine Cristina Marqueze; Elaine Vieira Santos; Evandra Strazza Rodrigues; Heidge Fukumasu; Jayme Augusto de Souza-Neto; José Salvatore Leister Patané; Luiz Alcantara; Luiz Lehmann Coutinho; Maria Carolina Elias; Maurício Lacerda Nogueira; Rafael dos Santos Bezerra; Raul Machado Neto; Rejane Maria Tommasini Grotto; Ricardo Haddad; Sandra Coccuzzo Sampaio Vessoni; Simone Kashima; Svetoslav Nanev Slavov; Vincent Louis Viala |
| EPI_ISL_2494192 | CENTRO DE SAUDE II DR JOSE PAIONE MOCOCA | Instituto Butantan | Antonio Jorge Martins; Claudia Renata dos Santos Barros; David Schlesinger; Debora Botequio Moretti; Dimas Tadeu Covas; Elaine Cristina Marqueze; Elaine Vieira Santos; Evandra Strazza Rodrigues; Heidge Fukumasu; Jayme Augusto de Souza-Neto; José Salvatore Leister Patané; Luiz Alcantara; Luiz Lehmann Coutinho; Maria Carolina Elias; Maurício Lacerda Nogueira; Rafael dos Santos Bezerra; Raul Machado Neto; Rejane Maria Tommasini Grotto; Ricardo Haddad; Sandra Coccuzzo Sampaio Vessoni; Simone Kashima; Svetoslav Nanev Slavov; Vincent Louis Viala |
| EPI_ISL_2445312, EPI_ISL_2445313, EPI_ISL_2445314, EPI_ISL_2445315, EPI_ISL_2445316, EPI_ISL_2445317, EPI_ISL_2445318, EPI_ISL_2445319, EPI_ISL_2445320, EPI_ISL_2473699, EPI_ISL_2473823, EPI_ISL_2493537, EPI_ISL_2493540, EPI_ISL_2493542, EPI_ISL_2493571, EPI_ISL_2493572, EPI_ISL_2493574, EPI_ISL_2493575, EPI_ISL_2493576, EPI_ISL_2493578, EPI_ISL_2493580, EPI_ISL_2493582, EPI_ISL_2493583, EPI_ISL_2493675, EPI_ISL_2493676, EPI_ISL_2493680, EPI_ISL_2493682, EPI_ISL_2493683, EPI_ISL_2494289, EPI_ISL_2494290 | | | |
| see above | CENTRO DE SAUDE II IBITINGA | Instituto Butantan | Antonio Jorge Martins; Claudia Renata dos Santos Barros; David Schlesinger; Debora Botequio Moretti; Dimas Tadeu Covas; Elaine Cristina Marqueze; Elaine Vieira Santos; Evandra Strazza Rodrigues; Heidge Fukumasu; Jayme Augusto de Souza-Neto; José Salvatore Leister Patané; Luiz Alcantara; Luiz Lehmann Coutinho; Maria Carolina Elias; Maurício Lacerda Nogueira; Rafael dos Santos Bezerra; Raul Machado Neto; Rejane Maria Tommasini Grotto; Ricardo Haddad; Sandra Coccuzzo Sampaio Vessoni; Simone Kashima; Svetoslav Nanev Slavov; Vincent Louis Viala |
| EPI_ISL_2208974, EPI_ISL_2209353, EPI_ISL_2209368, EPI_ISL_2209590, EPI_ISL_2209707, EPI_ISL_2345674, EPI_ISL_2345720, EPI_ISL_2345724, EPI_ISL_2345766, EPI_ISL_2345768, EPI_ISL_2345816, EPI_ISL_2345821, EPI_ISL_2345861 | | | |
| see above | CENTRO DE SAUDE II IBITINGA | Instituto Butantan / Mendelics | Antonio Jorge Martins; Claudia Renata dos Santos Barros; David Schlesinger; Debora Botequio Moretti; Dimas Tadeu Covas; Elaine Cristina Marqueze; Elaine Vieira Santos; Evandra Strazza Rodrigues; Heidge Fukumasu; Jayme Augusto de Souza-Neto; José Salvatore Leister Patané; Luiz Alcantara; Luiz Lehmann Coutinho; Maria Carolina Elias; Maurício Lacerda Nogueira; Rafael dos Santos Bezerra; Raul Machado Neto; Rejane Maria Tommasini Grotto; Ricardo Haddad; Sandra Coccuzzo Sampaio Vessoni; Simone Kashima; Svetoslav Nanev Slavov; Vincent Louis Viala |
| EPI_ISL_2493336, EPI_ISL_2493337, EPI_ISL_2493338, EPI_ISL_2493339, EPI_ISL_2493342, EPI_ISL_2493344, EPI_ISL_2493347 | | | |
| see above | CENTRO DE SAUDE II JUNQUEIROPOLIS | Instituto Butantan | Antonio Jorge Martins; Claudia Renata dos Santos Barros; David Schlesinger; Debora Botequio Moretti; Dimas Tadeu Covas; Elaine Cristina Marqueze; Elaine Vieira Santos; Evandra Strazza Rodrigues; Heidge Fukumasu; Jayme Augusto de Souza-Neto; José Salvatore Leister Patané; Luiz Alcantara; Luiz Lehmann Coutinho; Maria Carolina Elias; Maurício Lacerda Nogueira; Rafael dos Santos Bezerra; Raul Machado Neto; Rejane Maria Tommasini Grotto; Ricardo Haddad; Sandra Coccuzzo Sampaio Vessoni; Simone Kashima; Svetoslav Nanev Slavov; Vincent Louis Viala |
| EPI_ISL_2473807 | CENTRO DE SAUDE II MAIRINQUE MAIRINQUE | Instituto Butantan | Antonio Jorge Martins; Claudia Renata dos Santos Barros; David Schlesinger; Debora Botequio Moretti; Dimas Tadeu Covas; Elaine Cristina Marqueze; Elaine Vieira Santos; Evandra Strazza Rodrigues; Heidge Fukumasu; Jayme Augusto de Souza-Neto; José Salvatore Leister Patané; Luiz Alcantara; Luiz Lehmann Coutinho; Maria Carolina Elias; Maurício Lacerda Nogueira; Rafael dos Santos Bezerra; Raul Machado Neto; Rejane Maria Tommasini Grotto; Ricardo Haddad; Sandra Coccuzzo Sampaio Vessoni; Simone Kashima; Svetoslav Nanev Slavov; Vincent Louis Viala |
| EPI_ISL_2445603 | CENTRO DE SAUDE II SAO MIGUEL ARCANJO | Instituto Butantan | Antonio Jorge Martins; Claudia Renata dos Santos Barros; David Schlesinger; Debora Botequio Moretti; Dimas Tadeu Covas; Elaine Cristina Marqueze; Elaine Vieira Santos; Evandra Strazza Rodrigues; Heidge Fukumasu; Jayme Augusto de Souza-Neto; José Salvatore Leister Patané; Luiz Alcantara; Luiz Lehmann Coutinho; Maria Carolina Elias; Maurício Lacerda Nogueira; Rafael dos Santos Bezerra; Raul Machado Neto; Rejane Maria Tommasini Grotto; Ricardo Haddad; Sandra Coccuzzo Sampaio Vessoni; Simone Kashima; Svetoslav Nanev Slavov; Vincent Louis Viala |
| EPI_ISL_2445594 | CENTRO DE SAUDE II TIETE DR RUI SILVEIRA MELO | Instituto Butantan | Antonio Jorge Martins; Claudia Renata dos Santos Barros; David Schlesinger; Debora Botequio Moretti; Dimas Tadeu Covas; Elaine Cristina Marqueze; Elaine Vieira Santos; Evandra Strazza Rodrigues; Heidge Fukumasu; Jayme Augusto de Souza-Neto; José Salvatore Leister Patané; Luiz Alcantara; Luiz Lehmann Coutinho; Maria Carolina Elias; Maurício Lacerda Nogueira; Rafael dos Santos Bezerra; Raul Machado Neto; Rejane Maria Tommasini Grotto; Ricardo Haddad; Sandra Coccuzzo Sampaio Vessoni; Simone Kashima; Svetoslav Nanev Slavov; Vincent Louis Viala |
| EPI_ISL_2493263 | CENTRO DE SAUDE III AFFONSO LUZZI SANTA CRUZ DAS PALMEIRAS | Instituto Butantan | Antonio Jorge Martins; Claudia Renata dos Santos Barros; David Schlesinger; Debora Botequio Moretti; Dimas Tadeu Covas; Elaine Cristina Marqueze; Elaine Vieira Santos; Evandra Strazza Rodrigues; Heidge Fukumasu; Jayme Augusto de Souza-Neto; José Salvatore Leister Patané; Luiz Alcantara; Luiz Lehmann Coutinho; Maria Carolina Elias; Maurício Lacerda Nogueira; Rafael dos Santos Bezerra; Raul Machado Neto; Rejane Maria Tommasini Grotto; Ricardo Haddad; Sandra Coccuzzo Sampaio Vessoni; Simone Kashima; Svetoslav Nanev Slavov; Vincent Louis Viala |
| EPI_ISL_2493539, EPI_ISL_2493544, EPI_ISL_2493547, EPI_ISL_2493570 | CENTRO DE SAUDE III BORBOREMA | Instituto Butantan | Antonio Jorge Martins; Claudia Renata dos Santos Barros; David Schlesinger; Debora Botequio Moretti; Dimas Tadeu Covas; Elaine Cristina Marqueze; Elaine Vieira Santos; Evandra Strazza Rodrigues; Heidge Fukumasu; Jayme Augusto de Souza-Neto; José Salvatore Leister Patané; Luiz Alcantara; Luiz Lehmann Coutinho; Maria Carolina Elias; Maurício Lacerda Nogueira; Rafael dos Santos Bezerra; Raul Machado Neto; Rejane Maria Tommasini Grotto; Ricardo Haddad; Sandra Coccuzzo Sampaio Vessoni; Simone Kashima; Svetoslav Nanev Slavov; Vincent Louis Viala |
| EPI_ISL_2494281, EPI_ISL_2494282, EPI_ISL_2494284, EPI_ISL_2494285, EPI_ISL_2494317, EPI_ISL_2494332 | CENTRO DE SAUDE III DE AGUAS DE SAO PEDRO | Instituto Butantan | Antonio Jorge Martins; Claudia Renata dos Santos Barros; David Schlesinger; Debora Botequio Moretti; Dimas Tadeu Covas; Elaine Cristina Marqueze; Elaine Vieira Santos; Evandra Strazza Rodrigues; Heidge Fukumasu; Jayme Augusto de Souza-Neto; José Salvatore Leister Patané; Luiz Alcantara; Luiz Lehmann Coutinho; Maria Carolina Elias; Maurício Lacerda Nogueira; Rafael dos Santos Bezerra; Raul Machado Neto; Rejane Maria Tommasini Grotto; Ricardo Haddad; Sandra Coccuzzo Sampaio Vessoni; Simone Kashima; Svetoslav Nanev Slavov; Vincent Louis Viala |
| EPI_ISL_2494165, EPI_ISL_2494186 | CENTRO DE SAUDE III DE DIVINOLANDIA | Instituto Butantan | Antonio Jorge Martins; Claudia Renata dos Santos Barros; David Schlesinger; Debora Botequio Moretti; Dimas Tadeu Covas; Elaine Cristina Marqueze; Elaine Vieira Santos; Evandra Strazza Rodrigues; Heidge Fukumasu; Jayme Augusto de Souza-Neto; José Salvatore Leister Patané; Luiz Alcantara; Luiz Lehmann Coutinho; Maria Carolina Elias; Maurício Lacerda Nogueira; Rafael dos Santos Bezerra; Raul Machado Neto; Rejane Maria Tommasini Grotto; Ricardo Haddad; Sandra Coccuzzo Sampaio Vessoni; Simone Kashima; Svetoslav Nanev Slavov; Vincent Louis Viala |
| EPI_ISL_2493236, EPI_ISL_2493669, EPI_ISL_2494108 | CENTRO DE SAUDE III DOBRADA | Instituto Butantan | Antonio Jorge Martins; Claudia Renata dos Santos Barros; David Schlesinger; Debora Botequio Moretti; Dimas Tadeu Covas; Elaine Cristina Marqueze; Elaine Vieira Santos; Evandra Strazza Rodrigues; Heidge Fukumasu; Jayme Augusto de Souza-Neto; José Salvatore Leister Patané; Luiz Alcantara; Luiz Lehmann Coutinho; Maria Carolina Elias; Maurício Lacerda Nogueira; Rafael dos Santos Bezerra; Raul Machado Neto; Rejane Maria Tommasini Grotto; Ricardo Haddad; Sandra Coccuzzo Sampaio Vessoni; Simone Kashima; Svetoslav Nanev Slavov; Vincent Louis Viala |
| EPI_ISL_2445548, EPI_ISL_2445549, EPI_ISL_2445557 | CENTRO DE SAUDE III NELCIDIO DA SILVEIRA BASTOS | Instituto Butantan | Antonio Jorge Martins; Claudia Renata dos Santos Barros; David Schlesinger; Debora Botequio Moretti; Dimas Tadeu Covas; Elaine Cristina Marqueze; Elaine Vieira Santos; Evandra Strazza Rodrigues; Heidge Fukumasu; Jayme Augusto de Souza-Neto; José Salvatore Leister Patané; Luiz Alcantara; Luiz Lehmann Coutinho; Maria Carolina Elias; Maurício Lacerda Nogueira; Rafael dos Santos Bezerra; Raul Machado Neto; Rejane Maria Tommasini Grotto; Ricardo Haddad; Sandra Coccuzzo Sampaio Vessoni; Simone Kashima; Svetoslav Nanev Slavov; Vincent Louis Viala |
| EPI_ISL_2209702 | CENTRO DE SAUDE III NELCIDIO DA SILVEIRA BASTOS | Instituto Butantan / Mendelics | Antonio Jorge Martins; Claudia Renata dos Santos Barros; David Schlesinger; Debora Botequio Moretti; Dimas Tadeu Covas; Elaine Cristina Marqueze; Elaine Vieira Santos; Evandra Strazza Rodrigues; Heidge Fukumasu; Jayme Augusto de Souza-Neto; José Salvatore Leister Patané; Luiz Alcantara; Luiz Lehmann Coutinho; Maria Carolina Elias; Maurício Lacerda Nogueira; Rafael dos Santos Bezerra; Raul Machado Neto; Rejane Maria Tommasini Grotto; Ricardo Haddad; Sandra Coccuzzo Sampaio Vessoni; Simone Kashima; Svetoslav Nanev Slavov; Vincent Louis Viala |
| EPI_ISL_2208938, EPI_ISL_2209968, EPI_ISL_2210119, EPI_ISL_2345652, EPI_ISL_2345697, EPI_ISL_2345842 | CENTRO DE SAUDE III SALES OLIVEIRA | Instituto Butantan / Mendelics | Antonio Jorge Martins; Claudia Renata dos Santos Barros; David Schlesinger; Debora Botequio Moretti; Dimas Tadeu Covas; Elaine Cristina Marqueze; Elaine Vieira Santos; Evandra Strazza Rodrigues; Heidge Fukumasu; Jayme Augusto de Souza-Neto; José Salvatore Leister Patané; Luiz Alcantara; Luiz Lehmann Coutinho; Maria Carolina Elias; Maurício Lacerda Nogueira; Rafael dos Santos Bezerra; Raul Machado Neto; Rejane Maria Tommasini Grotto; Ricardo Haddad; Sandra Coccuzzo Sampaio Vessoni; Simone Kashima; Svetoslav Nanev Slavov; Vincent Louis Viala |
| EPI_ISL_2345696, EPI_ISL_2345748 | CENTRO DE SAUDE III SANTA ERNESTINA | Instituto Butantan / Mendelics | Antonio Jorge Martins; Claudia Renata dos Santos Barros; David Schlesinger; Debora Botequio Moretti; Dimas Tadeu Covas; Elaine Cristina Marqueze; Elaine Vieira Santos; Evandra Strazza Rodrigues; Heidge Fukumasu; Jayme Augusto de Souza-Neto; José Salvatore Leister Patané; Luiz Alcantara; Luiz Lehmann Coutinho; Maria Carolina Elias; Maurício Lacerda Nogueira; Rafael dos Santos Bezerra; Raul Machado Neto; Rejane Maria Tommasini Grotto; Ricardo Haddad; Sandra Coccuzzo Sampaio Vessoni; Simone Kashima; Svetoslav Nanev Slavov; Vincent Louis Viala |
| EPI_ISL_2493822 | CENTRO DE SAUDE III SANTA LUCIA | Instituto Butantan | Antonio Jorge Martins; Claudia Renata dos Santos Barros; David Schlesinger; Debora Botequio Moretti; Dimas Tadeu Covas; Elaine Cristina Marqueze; Elaine Vieira Santos; Evandra Strazza Rodrigues; Heidge Fukumasu; Jayme Augusto de Souza-Neto; José Salvatore Leister Patané; Luiz Alcantara; Luiz Lehmann Coutinho; Maria Carolina Elias; Maurício Lacerda Nogueira; Rafael dos Santos Bezerra; Raul Machado Neto; Rejane Maria Tommasini Grotto; Ricardo Haddad; Sandra Coccuzzo Sampaio Vessoni; Simone Kashima; Svetoslav Nanev Slavov; Vincent Louis Viala |
| EPI_ISL_2445328, EPI_ISL_2493154, EPI_ISL_2493677, EPI_ISL_2493678, EPI_ISL_2494109, EPI_ISL_2494115, EPI_ISL_2494117, EPI_ISL_2494123 | | | |
| see above | CENTRO DE SAUDE III TABATINGA | Instituto Butantan | Antonio Jorge Martins; Claudia Renata dos Santos Barros; David Schlesinger; Debora Botequio Moretti; Dimas Tadeu Covas; Elaine Cristina Marqueze; Elaine Vieira Santos; Evandra Strazza Rodrigues; Heidge Fukumasu; Jayme Augusto de Souza-Neto; José Salvatore Leister Patané; Luiz Alcantara; Luiz Lehmann Coutinho; Maria Carolina Elias; Maurício Lacerda Nogueira; Rafael dos Santos Bezerra; Raul Machado Neto; Rejane Maria Tommasini Grotto; Ricardo Haddad; Sandra Coccuzzo Sampaio Vessoni; Simone Kashima; Svetoslav Nanev Slavov; Vincent Louis Viala |
| EPI_ISL_2209891, EPI_ISL_2209927, EPI_ISL_2345793 | CENTRO DE SAUDE III TABATINGA | Instituto Butantan / Mendelics | Antonio Jorge Martins; Claudia Renata dos Santos Barros; David Schlesinger; Debora Botequio Moretti; Dimas Tadeu Covas; Elaine Cristina Marqueze; Elaine Vieira Santos; Evandra Strazza Rodrigues; Heidge Fukumasu; Jayme Augusto de Souza-Neto; José Salvatore Leister Patané; Luiz Alcantara; Luiz Lehmann Coutinho; Maria Carolina Elias; Maurício Lacerda Nogueira; Rafael dos Santos Bezerra; Raul Machado Neto; Rejane Maria Tommasini Grotto; Ricardo Haddad; Sandra Coccuzzo Sampaio Vessoni; Simone Kashima; Svetoslav Nanev Slavov; Vincent Louis Viala |
| EPI_ISL_2493950, EPI_ISL_2493951 | CENTRO DE SAUDE SAO ROQUE DR JOSE CARVALHO BRITO | Instituto Butantan | Antonio Jorge Martins; Claudia Renata dos Santos Barros; David Schlesinger; Debora Botequio Moretti; Dimas Tadeu Covas; Elaine Cristina Marqueze; Elaine Vieira Santos; Evandra Strazza Rodrigues; Heidge Fukumasu; Jayme Augusto de Souza-Neto; José Salvatore Leister Patané; Luiz Alcantara; Luiz Lehmann Coutinho; Maria Carolina Elias; Maurício Lacerda Nogueira; Rafael dos Santos Bezerra; Raul Machado Neto; Rejane Maria Tommasini Grotto; Ricardo Haddad; Sandra Coccuzzo Sampaio Vessoni; Simone Kashima; Svetoslav Nanev Slavov; Vincent Louis Viala |
| EPI_ISL_2340842, EPI_ISL_2445345, EPI_ISL_2445348, EPI_ISL_2445351, EPI_ISL_2445354, EPI_ISL_2445356, EPI_ISL_2445358, EPI_ISL_2445359, EPI_ISL_2445360, EPI_ISL_2445361, EPI_ISL_2445363, EPI_ISL_2445365, EPI_ISL_2445366, EPI_ISL_2445367, EPI_ISL_2445446, EPI_ISL_2445447, EPI_ISL_2445448, EPI_ISL_2445449, EPI_ISL_2445451, EPI_ISL_2445452, EPI_ISL_2473697, EPI_ISL_2473780 | | | |
| see above | CENTRO INTEGRADO DE SAUDE | Instituto Butantan | Antonio Jorge Martins; Claudia Renata dos Santos Barros; David Schlesinger; Debora Botequio Moretti; Dimas Tadeu Covas; Elaine Cristina Marqueze; Elaine Vieira Santos; Evandra Strazza Rodrigues; Heidge Fukumasu; Jayme Augusto de Souza-Neto; José Salvatore Leister Patané; Luiz Alcantara; Luiz Lehmann Coutinho; Maria Carolina Elias; Maurício Lacerda Nogueira; Rafael dos Santos Bezerra; Raul Machado Neto; Rejane Maria Tommasini Grotto; Ricardo Haddad; Sandra Coccuzzo Sampaio Vessoni; Simone Kashima; Svetoslav Nanev Slavov; Vincent Louis Viala |
| EPI_ISL_2494276, EPI_ISL_2494291, EPI_ISL_2494342, EPI_ISL_2494343 | CENTRO MEDICO DR NELSON SALOME DE CONCHAL | Instituto Butantan | Antonio Jorge Martins; Claudia Renata dos Santos Barros; David Schlesinger; Debora Botequio Moretti; Dimas Tadeu Covas; Elaine Cristina Marqueze; Elaine Vieira Santos; Evandra Strazza Rodrigues; Heidge Fukumasu; Jayme Augusto de Souza-Neto; José Salvatore Leister Patané; Luiz Alcantara; Luiz Lehmann Coutinho; Maria Carolina Elias; Maurício Lacerda Nogueira; Rafael dos Santos Bezerra; Raul Machado Neto; Rejane Maria Tommasini Grotto; Ricardo Haddad; Sandra Coccuzzo Sampaio Vessoni; Simone Kashima; Svetoslav Nanev Slavov; Vincent Louis Viala |
| EPI_ISL_2493164, EPI_ISL_2493182, EPI_ISL_2493292, EPI_ISL_2493300 | CENTRO MEDICO PMESP | Instituto Butantan | Antonio Jorge Martins; Claudia Renata dos Santos Barros; David Schlesinger; Debora Botequio Moretti; Dimas Tadeu Covas; Elaine Cristina Marqueze; Elaine Vieira Santos; Evandra Strazza Rodrigues; Heidge Fukumasu; Jayme Augusto de Souza-Neto; José Salvatore Leister Patané; Luiz Alcantara; Luiz Lehmann Coutinho; Maria Carolina Elias; Maurício Lacerda Nogueira; Rafael dos Santos Bezerra; Raul Machado Neto; Rejane Maria Tommasini Grotto; Ricardo Haddad; Sandra Coccuzzo Sampaio Vessoni; Simone Kashima; Svetoslav Nanev Slavov; Vincent Louis Viala |
| EPI_ISL_2493591, EPI_ISL_2493596, EPI_ISL_2493597, EPI_ISL_2493600, EPI_ISL_2493606, EPI_ISL_2493625 | CENTRO MEDICO SOCIAL COMUNITARIO JANUARIO TEODORO DE SOUZA | Instituto Butantan | Antonio Jorge Martins; Claudia Renata dos Santos Barros; David Schlesinger; Debora Botequio Moretti; Dimas Tadeu Covas; Elaine Cristina Marqueze; Elaine Vieira Santos; Evandra Strazza Rodrigues; Heidge Fukumasu; Jayme Augusto de Souza-Neto; José Salvatore Leister Patané; Luiz Alcantara; Luiz Lehmann Coutinho; Maria Carolina Elias; Maurício Lacerda Nogueira; Rafael dos Santos Bezerra; Raul Machado Neto; Rejane Maria Tommasini Grotto; Ricardo Haddad; Sandra Coccuzzo Sampaio Vessoni; Simone Kashima; Svetoslav Nanev Slavov; Vincent Louis Viala |
| EPI_ISL_2462325, | CENTRO MUNICIPAL DE SAUDE RUY DE | Instituto Butantan | Antonio Jorge Martins; Claudia Renata dos Santos Barros; David Schlesinger; Debora Botequio Moretti; Dimas Tadeu Covas; Elaine Cristina Marqueze; Elaine Vieira Santos; Evandra Strazza Rodrigues; Heidge Fukumasu; Jayme Augusto de Souza-Neto; José Salvatore Leister Patané; Luiz Alcantara; Luiz Lehmann |

| | | | |
|---|---|---|---|
| EPI_ISL_2494130 | CAMARGO | | Coutinho; Maria Carolina Elias; Maurício Lacerda Nogueira; Rafael dos Santos Bezerra; Raul Machado Neto; Rejane Maria Tommasini Grotto; Ricardo Haddad; Sandra Coccuzzo Sampaio Vessoni; Simone Kashima; Svetoslav Nanev Slavov; Vincent Louis Viala |
| EPI_ISL_2493433, EPI_ISL_2493445, EPI_ISL_2494159, EPI_ISL_2494188 | CENTRO SAUDE III DR JOSE TADEU MOURAO | Instituto Butantan | Antonio Jorge Martins; Claudia Renata dos Santos Barros; David Schlesinger; Debora Botequio Moretti; Dimas Tadeu Covas; Elaine Cristina Marqueze; Elaine Vieira Santos; Evandra Strazza Rodrigues; Heidge Fukumasu; Jayme Augusto de Souza-Neto; José Salvatore Leister Patané; Luiz Alcantara; Luiz Lehmann Coutinho; Maria Carolina Elias; Maurício Lacerda Nogueira; Rafael dos Santos Bezerra; Raul Machado Neto; Rejane Maria Tommasini Grotto; Ricardo Haddad; Sandra Coccuzzo Sampaio Vessoni; Simone Kashima; Svetoslav Nanev Slavov; Vincent Louis Viala |
| EPI_ISL_2493898 | CLINICA SALTO SAUDE BELA VISTA | Instituto Butantan | Antonio Jorge Martins; Claudia Renata dos Santos Barros; David Schlesinger; Debora Botequio Moretti; Dimas Tadeu Covas; Elaine Cristina Marqueze; Elaine Vieira Santos; Evandra Strazza Rodrigues; Heidge Fukumasu; Jayme Augusto de Souza-Neto; José Salvatore Leister Patané; Luiz Alcantara; Luiz Lehmann Coutinho; Maria Carolina Elias; Maurício Lacerda Nogueira; Rafael dos Santos Bezerra; Raul Machado Neto; Rejane Maria Tommasini Grotto; Ricardo Haddad; Sandra Coccuzzo Sampaio Vessoni; Simone Kashima; Svetoslav Nanev Slavov; Vincent Louis Viala |
| EPI_ISL_2493384, EPI_ISL_2493385, EPI_ISL_2493446, EPI_ISL_2494153 | COORDENADORIA MUNICIPAL DE SAUDE DE IRACEMAPOLIS | Instituto Butantan | Antonio Jorge Martins; Claudia Renata dos Santos Barros; David Schlesinger; Debora Botequio Moretti; Dimas Tadeu Covas; Elaine Cristina Marqueze; Elaine Vieira Santos; Evandra Strazza Rodrigues; Heidge Fukumasu; Jayme Augusto de Souza-Neto; José Salvatore Leister Patané; Luiz Alcantara; Luiz Lehmann Coutinho; Maria Carolina Elias; Maurício Lacerda Nogueira; Rafael dos Santos Bezerra; Raul Machado Neto; Rejane Maria Tommasini Grotto; Ricardo Haddad; Sandra Coccuzzo Sampaio Vessoni; Simone Kashima; Svetoslav Nanev Slavov; Vincent Louis Viala |
| EPI_ISL_2209929 | CS ALVINLANDIA | Instituto Butantan / Mendelics | Antonio Jorge Martins; Claudia Renata dos Santos Barros; David Schlesinger; Debora Botequio Moretti; Dimas Tadeu Covas; Elaine Cristina Marqueze; Elaine Vieira Santos; Evandra Strazza Rodrigues; Heidge Fukumasu; Jayme Augusto de Souza-Neto; José Salvatore Leister Patané; Luiz Alcantara; Luiz Lehmann Coutinho; Maria Carolina Elias; Maurício Lacerda Nogueira; Rafael dos Santos Bezerra; Raul Machado Neto; Rejane Maria Tommasini Grotto; Ricardo Haddad; Sandra Coccuzzo Sampaio Vessoni; Simone Kashima; Svetoslav Nanev Slavov; Vincent Louis Viala |
| EPI_ISL_2378744, EPI_ISL_2378753, EPI_ISL_2378754 | CS DE APARECIDA DOESTE | Instituto Butantan | Antonio Jorge Martins; Claudia Renata dos Santos Barros; David Schlesinger; Debora Botequio Moretti; Dimas Tadeu Covas; Elaine Cristina Marqueze; Elaine Vieira Santos; Evandra Strazza Rodrigues; Heidge Fukumasu; Jayme Augusto de Souza-Neto; José Salvatore Leister Patané; Luiz Alcantara; Luiz Lehmann Coutinho; Maria Carolina Elias; Maurício Lacerda Nogueira; Rafael dos Santos Bezerra; Raul Machado Neto; Rejane Maria Tommasini Grotto; Ricardo Haddad; Sandra Coccuzzo Sampaio Vessoni; Simone Kashima; Svetoslav Nanev Slavov; Vincent Louis Viala |
| EPI_ISL_2445209, EPI_ISL_2445210, EPI_ISL_2473720 | CS DE BALSAMO | Instituto Butantan | Antonio Jorge Martins; Claudia Renata dos Santos Barros; David Schlesinger; Debora Botequio Moretti; Dimas Tadeu Covas; Elaine Cristina Marqueze; Elaine Vieira Santos; Evandra Strazza Rodrigues; Heidge Fukumasu; Jayme Augusto de Souza-Neto; José Salvatore Leister Patané; Luiz Alcantara; Luiz Lehmann Coutinho; Maria Carolina Elias; Maurício Lacerda Nogueira; Rafael dos Santos Bezerra; Raul Machado Neto; Rejane Maria Tommasini Grotto; Ricardo Haddad; Sandra Coccuzzo Sampaio Vessoni; Simone Kashima; Svetoslav Nanev Slavov; Vincent Louis Viala |
| EPI_ISL_2493161 | CS DE CATIGUA | Instituto Butantan | Antonio Jorge Martins; Claudia Renata dos Santos Barros; David Schlesinger; Debora Botequio Moretti; Dimas Tadeu Covas; Elaine Cristina Marqueze; Elaine Vieira Santos; Evandra Strazza Rodrigues; Heidge Fukumasu; Jayme Augusto de Souza-Neto; José Salvatore Leister Patané; Luiz Alcantara; Luiz Lehmann Coutinho; Maria Carolina Elias; Maurício Lacerda Nogueira; Rafael dos Santos Bezerra; Raul Machado Neto; Rejane Maria Tommasini Grotto; Ricardo Haddad; Sandra Coccuzzo Sampaio Vessoni; Simone Kashima; Svetoslav Nanev Slavov; Vincent Louis Viala |
| EPI_ISL_2445212, EPI_ISL_2494297 | CS DE COSMORAMA | Instituto Butantan | Antonio Jorge Martins; Claudia Renata dos Santos Barros; David Schlesinger; Debora Botequio Moretti; Dimas Tadeu Covas; Elaine Cristina Marqueze; Elaine Vieira Santos; Evandra Strazza Rodrigues; Heidge Fukumasu; Jayme Augusto de Souza-Neto; José Salvatore Leister Patané; Luiz Alcantara; Luiz Lehmann Coutinho; Maria Carolina Elias; Maurício Lacerda Nogueira; Rafael dos Santos Bezerra; Raul Machado Neto; Rejane Maria Tommasini Grotto; Ricardo Haddad; Sandra Coccuzzo Sampaio Vessoni; Simone Kashima; Svetoslav Nanev Slavov; Vincent Louis Viala |
| EPI_ISL_2493424 | CS DE MONTE APRAZIVEL | Instituto Butantan | Antonio Jorge Martins; Claudia Renata dos Santos Barros; David Schlesinger; Debora Botequio Moretti; Dimas Tadeu Covas; Elaine Cristina Marqueze; Elaine Vieira Santos; Evandra Strazza Rodrigues; Heidge Fukumasu; Jayme Augusto de Souza-Neto; José Salvatore Leister Patané; Luiz Alcantara; Luiz Lehmann Coutinho; Maria Carolina Elias; Maurício Lacerda Nogueira; Rafael dos Santos Bezerra; Raul Machado Neto; Rejane Maria Tommasini Grotto; Ricardo Haddad; Sandra Coccuzzo Sampaio Vessoni; Simone Kashima; Svetoslav Nanev Slavov; Vincent Louis Viala |
| EPI_ISL_2493180, EPI_ISL_2493405, EPI_ISL_2493406, EPI_ISL_2493407, EPI_ISL_2493412, EPI_ISL_2493414, EPI_ISL_2493416 | | | |
| see above | CS DE NEVES PAULISTA | Instituto Butantan | Antonio Jorge Martins; Claudia Renata dos Santos Barros; David Schlesinger; Debora Botequio Moretti; Dimas Tadeu Covas; Elaine Cristina Marqueze; Elaine Vieira Santos; Evandra Strazza Rodrigues; Heidge Fukumasu; Jayme Augusto de Souza-Neto; José Salvatore Leister Patané; Luiz Alcantara; Luiz Lehmann Coutinho; Maria Carolina Elias; Maurício Lacerda Nogueira; Rafael dos Santos Bezerra; Raul Machado Neto; Rejane Maria Tommasini Grotto; Ricardo Haddad; Sandra Coccuzzo Sampaio Vessoni; Simone Kashima; Svetoslav Nanev Slavov; Vincent Louis Viala |
| EPI_ISL_2445213, EPI_ISL_2445214, EPI_ISL_2445215, EPI_ISL_2445217, EPI_ISL_2445218, EPI_ISL_2473721, EPI_ISL_2473722, EPI_ISL_2493661, EPI_ISL_2493662 | | | |
| see above | CS DE NOVO HORIZONTE | Instituto Butantan | Antonio Jorge Martins; Claudia Renata dos Santos Barros; David Schlesinger; Debora Botequio Moretti; Dimas Tadeu Covas; Elaine Cristina Marqueze; Elaine Vieira Santos; Evandra Strazza Rodrigues; Heidge Fukumasu; Jayme Augusto de Souza-Neto; José Salvatore Leister Patané; Luiz Alcantara; Luiz Lehmann Coutinho; Maria Carolina Elias; Maurício Lacerda Nogueira; Rafael dos Santos Bezerra; Raul Machado Neto; Rejane Maria Tommasini Grotto; Ricardo Haddad; Sandra Coccuzzo Sampaio Vessoni; Simone Kashima; Svetoslav Nanev Slavov; Vincent Louis Viala |
| EPI_ISL_2493411, EPI_ISL_2493419, EPI_ISL_2493429 | CS DE ORINDIUVA | Instituto Butantan | Antonio Jorge Martins; Claudia Renata dos Santos Barros; David Schlesinger; Debora Botequio Moretti; Dimas Tadeu Covas; Elaine Cristina Marqueze; Elaine Vieira Santos; Evandra Strazza Rodrigues; Heidge Fukumasu; Jayme Augusto de Souza-Neto; José Salvatore Leister Patané; Luiz Alcantara; Luiz Lehmann Coutinho; Maria Carolina Elias; Maurício Lacerda Nogueira; Rafael dos Santos Bezerra; Raul Machado Neto; Rejane Maria Tommasini Grotto; Ricardo Haddad; Sandra Coccuzzo Sampaio Vessoni; Simone Kashima; Svetoslav Nanev Slavov; Vincent Louis Viala |
| EPI_ISL_2493410, EPI_ISL_2493413, EPI_ISL_2493415, EPI_ISL_2493417, EPI_ISL_2493420 | CS DE PALMARES PAULISTA | Instituto Butantan | Antonio Jorge Martins; Claudia Renata dos Santos Barros; David Schlesinger; Debora Botequio Moretti; Dimas Tadeu Covas; Elaine Cristina Marqueze; Elaine Vieira Santos; Evandra Strazza Rodrigues; Heidge Fukumasu; Jayme Augusto de Souza-Neto; José Salvatore Leister Patané; Luiz Alcantara; Luiz Lehmann Coutinho; Maria Carolina Elias; Maurício Lacerda Nogueira; Rafael dos Santos Bezerra; Raul Machado Neto; Rejane Maria Tommasini Grotto; Ricardo Haddad; Sandra Coccuzzo Sampaio Vessoni; Simone Kashima; Svetoslav Nanev Slavov; Vincent Louis Viala |
| EPI_ISL_2445211, EPI_ISL_2445538 | CS DE PLANALTO | Instituto Butantan | Antonio Jorge Martins; Claudia Renata dos Santos Barros; David Schlesinger; Debora Botequio Moretti; Dimas Tadeu Covas; Elaine Cristina Marqueze; Elaine Vieira Santos; Evandra Strazza Rodrigues; Heidge Fukumasu; Jayme Augusto de Souza-Neto; José Salvatore Leister Patané; Luiz Alcantara; Luiz Lehmann Coutinho; Maria Carolina Elias; Maurício Lacerda Nogueira; Rafael dos Santos Bezerra; Raul Machado Neto; Rejane Maria Tommasini Grotto; Ricardo Haddad; Sandra Coccuzzo Sampaio Vessoni; Simone Kashima; Svetoslav Nanev Slavov; Vincent Louis Viala |
| EPI_ISL_2445436, EPI_ISL_2445437, EPI_ISL_2445438, EPI_ISL_2445443, EPI_ISL_2493587 | CS DE QUINTANA | Instituto Butantan | Antonio Jorge Martins; Claudia Renata dos Santos Barros; David Schlesinger; Debora Botequio Moretti; Dimas Tadeu Covas; Elaine Cristina Marqueze; Elaine Vieira Santos; Evandra Strazza Rodrigues; Heidge Fukumasu; Jayme Augusto de Souza-Neto; José Salvatore Leister Patané; Luiz Alcantara; Luiz Lehmann Coutinho; Maria Carolina Elias; Maurício Lacerda Nogueira; Rafael dos Santos Bezerra; Raul Machado Neto; Rejane Maria Tommasini Grotto; Ricardo Haddad; Sandra Coccuzzo Sampaio Vessoni; Simone Kashima; Svetoslav Nanev Slavov; Vincent Louis Viala |
| EPI_ISL_2473795, EPI_ISL_2493362, EPI_ISL_2493363 | CS DE ROSANA | Instituto Butantan | Antonio Jorge Martins; Claudia Renata dos Santos Barros; David Schlesinger; Debora Botequio Moretti; Dimas Tadeu Covas; Elaine Cristina Marqueze; Elaine Vieira Santos; Evandra Strazza Rodrigues; Heidge Fukumasu; Jayme Augusto de Souza-Neto; José Salvatore Leister Patané; Luiz Alcantara; Luiz Lehmann Coutinho; Maria Carolina Elias; Maurício Lacerda Nogueira; Rafael dos Santos Bezerra; Raul Machado Neto; Rejane Maria Tommasini Grotto; Ricardo Haddad; Sandra Coccuzzo Sampaio Vessoni; Simone Kashima; Svetoslav Nanev Slavov; Vincent Louis Viala |
| EPI_ISL_2493400 | CS DE SANTA ALBERTINA | Instituto Butantan | Antonio Jorge Martins; Claudia Renata dos Santos Barros; David Schlesinger; Debora Botequio Moretti; Dimas Tadeu Covas; Elaine Cristina Marqueze; Elaine Vieira Santos; Evandra Strazza Rodrigues; Heidge Fukumasu; Jayme Augusto de Souza-Neto; José Salvatore Leister Patané; Luiz Alcantara; Luiz Lehmann Coutinho; Maria Carolina Elias; Maurício Lacerda Nogueira; Rafael dos Santos Bezerra; Raul Machado Neto; Rejane Maria Tommasini Grotto; Ricardo Haddad; Sandra Coccuzzo Sampaio Vessoni; Simone Kashima; Svetoslav Nanev Slavov; Vincent Louis Viala |
| EPI_ISL_2493421, EPI_ISL_2493422 | CS DE SANTA RITA DOESTE | Instituto Butantan | Antonio Jorge Martins; Claudia Renata dos Santos Barros; David Schlesinger; Debora Botequio Moretti; Dimas Tadeu Covas; Elaine Cristina Marqueze; Elaine Vieira Santos; Evandra Strazza Rodrigues; Heidge Fukumasu; Jayme Augusto de Souza-Neto; José Salvatore Leister Patané; Luiz Alcantara; Luiz Lehmann Coutinho; Maria Carolina Elias; Maurício Lacerda Nogueira; Rafael dos Santos Bezerra; Raul Machado Neto; Rejane Maria Tommasini Grotto; Ricardo Haddad; Sandra Coccuzzo Sampaio Vessoni; Simone Kashima; Svetoslav Nanev Slavov; Vincent Louis Viala |
| EPI_ISL_2493430, EPI_ISL_2493431 | CS DE VALENTIM GENTIL | Instituto Butantan | Antonio Jorge Martins; Claudia Renata dos Santos Barros; David Schlesinger; Debora Botequio Moretti; Dimas Tadeu Covas; Elaine Cristina Marqueze; Elaine Vieira Santos; Evandra Strazza Rodrigues; Heidge Fukumasu; Jayme Augusto de Souza-Neto; José Salvatore Leister Patané; Luiz Alcantara; Luiz Lehmann Coutinho; Maria Carolina Elias; Maurício Lacerda Nogueira; Rafael dos Santos Bezerra; Raul Machado Neto; Rejane Maria Tommasini Grotto; Ricardo Haddad; Sandra Coccuzzo Sampaio Vessoni; Simone Kashima; Svetoslav Nanev Slavov; Vincent Louis Viala |
| EPI_ISL_2493299 | CS II DE IGARAPAVA | Instituto Butantan | Antonio Jorge Martins; Claudia Renata dos Santos Barros; David Schlesinger; Debora Botequio Moretti; Dimas Tadeu Covas; Elaine Cristina Marqueze; Elaine Vieira Santos; Evandra Strazza Rodrigues; Heidge Fukumasu; Jayme Augusto de Souza-Neto; José Salvatore Leister Patané; Luiz Alcantara; Luiz Lehmann Coutinho; Maria Carolina Elias; Maurício Lacerda Nogueira; Rafael dos Santos Bezerra; Raul Machado Neto; Rejane Maria Tommasini Grotto; Ricardo Haddad; Sandra Coccuzzo Sampaio Vessoni; Simone Kashima; Svetoslav Nanev Slavov; Vincent Louis Viala |
| EPI_ISL_2445398, EPI_ISL_2445399, EPI_ISL_2445401 | CS II DR ANTONIO VICOSO MOREIRA DE REZENDE SUMARE | Instituto Butantan | Antonio Jorge Martins; Claudia Renata dos Santos Barros; David Schlesinger; Debora Botequio Moretti; Dimas Tadeu Covas; Elaine Cristina Marqueze; Elaine Vieira Santos; Evandra Strazza Rodrigues; Heidge Fukumasu; Jayme Augusto de Souza-Neto; José Salvatore Leister Patané; Luiz Alcantara; Luiz Lehmann Coutinho; Maria Carolina Elias; Maurício Lacerda Nogueira; Rafael dos Santos Bezerra; Raul Machado Neto; Rejane Maria Tommasini Grotto; Ricardo Haddad; Sandra Coccuzzo Sampaio Vessoni; Simone Kashima; Svetoslav Nanev Slavov; Vincent Louis Viala |
| EPI_ISL_2445455, EPI_ISL_2445458, EPI_ISL_2445459, EPI_ISL_2445460, EPI_ISL_2445461, EPI_ISL_2445462 | CS II DR JAHYR DE PAULA RIBEIRO GUARA | Instituto Butantan | Antonio Jorge Martins; Claudia Renata dos Santos Barros; David Schlesinger; Debora Botequio Moretti; Dimas Tadeu Covas; Elaine Cristina Marqueze; Elaine Vieira Santos; Evandra Strazza Rodrigues; Heidge Fukumasu; Jayme Augusto de Souza-Neto; José Salvatore Leister Patané; Luiz Alcantara; Luiz Lehmann Coutinho; Maria Carolina Elias; Maurício Lacerda Nogueira; Rafael dos Santos Bezerra; Raul Machado Neto; Rejane Maria Tommasini Grotto; Ricardo Haddad; Sandra Coccuzzo Sampaio Vessoni; Simone Kashima; Svetoslav Nanev Slavov; Vincent Louis Viala |
| EPI_ISL_2445406 | CS II DR MIGUEL VITALIANO ORLANDIA | Instituto Butantan | Antonio Jorge Martins; Claudia Renata dos Santos Barros; David Schlesinger; Debora Botequio Moretti; Dimas Tadeu Covas; Elaine Cristina Marqueze; Elaine Vieira Santos; Evandra Strazza Rodrigues; Heidge Fukumasu; Jayme Augusto de Souza-Neto; José Salvatore Leister Patané; Luiz Alcantara; Luiz Lehmann Coutinho; Maria Carolina Elias; Maurício Lacerda Nogueira; Rafael dos Santos Bezerra; Raul Machado Neto; Rejane Maria Tommasini Grotto; Ricardo Haddad; Sandra Coccuzzo Sampaio Vessoni; Simone Kashima; Svetoslav Nanev Slavov; Vincent Louis Viala |
| EPI_ISL_2345794 | CS II DR MIGUEL VITALIANO ORLANDIA | Instituto Butantan / Mendelics | Antonio Jorge Martins; Claudia Renata dos Santos Barros; David Schlesinger; Debora Botequio Moretti; Dimas Tadeu Covas; Elaine Cristina Marqueze; Elaine Vieira Santos; Evandra Strazza Rodrigues; Heidge Fukumasu; Jayme Augusto de Souza-Neto; José Salvatore Leister Patané; Luiz Alcantara; Luiz Lehmann Coutinho; Maria Carolina Elias; Maurício Lacerda Nogueira; Rafael dos Santos Bezerra; Raul Machado Neto; Rejane Maria Tommasini Grotto; Ricardo Haddad; Sandra Coccuzzo Sampaio Vessoni; Simone Kashima; Svetoslav Nanev Slavov; Vincent Louis Viala |
| EPI_ISL_2493543 | CS II DR SEBASTIAO RIBEIRO DO VALLE | Instituto Butantan | Antonio Jorge Martins; Claudia Renata dos Santos Barros; David Schlesinger; Debora Botequio Moretti; Dimas Tadeu Covas; Elaine Cristina Marqueze; Elaine Vieira Santos; Evandra Strazza Rodrigues; Heidge Fukumasu; Jayme Augusto de Souza-Neto; José Salvatore Leister Patané; Luiz Alcantara; Luiz Lehmann Coutinho; Maria Carolina Elias; Maurício Lacerda Nogueira; Rafael dos Santos Bezerra; Raul Machado Neto; Rejane Maria Tommasini Grotto; Ricardo Haddad; Sandra Coccuzzo Sampaio Vessoni; Simone Kashima; Svetoslav Nanev Slavov; Vincent Louis Viala |
| EPI_ISL_2445454 | CS II EGIDIO BRUNHARA MORRO AGUDO | Instituto Butantan | Antonio Jorge Martins; Claudia Renata dos Santos Barros; David Schlesinger; Debora Botequio Moretti; Dimas Tadeu Covas; Elaine Cristina Marqueze; Elaine Vieira Santos; Evandra Strazza Rodrigues; Heidge Fukumasu; Jayme Augusto de Souza-Neto; José Salvatore Leister Patané; Luiz Alcantara; Luiz Lehmann Coutinho; Maria Carolina Elias; Maurício Lacerda Nogueira; Rafael dos Santos Bezerra; Raul Machado Neto; Rejane Maria Tommasini Grotto; Ricardo Haddad; Sandra Coccuzzo Sampaio Vessoni; Simone Kashima; Svetoslav Nanev Slavov; Vincent Louis Viala |
| EPI_ISL_2493573, EPI_ISL_2494183 | CS III DR ANTONIO ANADAO SSGRAMA | Instituto Butantan | Antonio Jorge Martins; Claudia Renata dos Santos Barros; David Schlesinger; Debora Botequio Moretti; Dimas Tadeu Covas; Elaine Cristina Marqueze; Elaine Vieira Santos; Evandra Strazza Rodrigues; Heidge Fukumasu; Jayme Augusto de Souza-Neto; José Salvatore Leister Patané; Luiz Alcantara; Luiz Lehmann Coutinho; Maria Carolina Elias; Maurício Lacerda Nogueira; Rafael dos Santos Bezerra; Raul Machado Neto; Rejane Maria Tommasini Grotto; Ricardo Haddad; Sandra Coccuzzo Sampaio Vessoni; Simone Kashima; Svetoslav Nanev Slavov; Vincent Louis Viala |
| EPI_ISL_2493288 | CS LUIZ FIOD DE BURITIZAL | Instituto Butantan | Antonio Jorge Martins; Claudia Renata dos Santos Barros; David Schlesinger; Debora Botequio Moretti; Dimas Tadeu Covas; Elaine Cristina Marqueze; Elaine Vieira Santos; Evandra Strazza Rodrigues; Heidge Fukumasu; Jayme Augusto de Souza-Neto; José Salvatore Leister Patané; Luiz Alcantara; Luiz Lehmann Coutinho; Maria Carolina Elias; Maurício Lacerda Nogueira; Rafael dos Santos Bezerra; Raul Machado Neto; Rejane Maria Tommasini Grotto; Ricardo Haddad; Sandra Coccuzzo Sampaio Vessoni; Simone Kashima; Svetoslav Nanev Slavov; Vincent Louis Viala |
| EPI_ISL_2493301 | CSII DR JOSE FERREIRA TELLES | Instituto Butantan | Antonio Jorge Martins; Claudia Renata dos Santos Barros; David Schlesinger; Debora Botequio Moretti; Dimas Tadeu Covas; Elaine Cristina Marqueze; Elaine Vieira Santos; Evandra Strazza Rodrigues; Heidge Fukumasu; Jayme Augusto de Souza-Neto; José Salvatore Leister Patané; Luiz Alcantara; Luiz Lehmann Coutinho; Maria Carolina Elias; Maurício Lacerda Nogueira; Rafael dos Santos Bezerra; Raul Machado Neto; Rejane Maria Tommasini Grotto; Ricardo Haddad; Sandra Coccuzzo Sampaio Vessoni; Simone Kashima; Svetoslav Nanev Slavov; Vincent Louis Viala |
| EPI_ISL_2378743, EPI_ISL_2493622, EPI_ISL_2493829, EPI_ISL_2493830, EPI_ISL_2494321 | CSII DR WASHINGTON LUIS M RODRIGUES DA SILVA PITANGUEIRAS | Instituto Butantan | Antonio Jorge Martins; Claudia Renata dos Santos Barros; David Schlesinger; Debora Botequio Moretti; Dimas Tadeu Covas; Elaine Cristina Marqueze; Elaine Vieira Santos; Evandra Strazza Rodrigues; Heidge Fukumasu; Jayme Augusto de Souza-Neto; José Salvatore Leister Patané; Luiz Alcantara; Luiz Lehmann Coutinho; Maria Carolina Elias; Maurício Lacerda Nogueira; Rafael dos Santos Bezerra; Raul Machado Neto; Rejane Maria Tommasini Grotto; Ricardo Haddad; Sandra Coccuzzo Sampaio Vessoni; Simone Kashima; Svetoslav Nanev Slavov; Vincent Louis Viala |
| EPI_ISL_2208992, EPI_ISL_2210279, EPI_ISL_2345680, EPI_ISL_2345771, EPI_ISL_2345824, EPI_ISL_2345868, EPI_ISL_2362268 | | | |
| see above | CSII DR WASHINGTON LUIS M RODRIGUES DA SILVA PITANGUEIRAS | Instituto Butantan / Mendelics | Antonio Jorge Martins; Claudia Renata dos Santos Barros; David Schlesinger; Debora Botequio Moretti; Dimas Tadeu Covas; Elaine Cristina Marqueze; Elaine Vieira Santos; Evandra Strazza Rodrigues; Heidge Fukumasu; Jayme Augusto de Souza-Neto; José Salvatore Leister Patané; Luiz Alcantara; Luiz Lehmann Coutinho; Maria Carolina Elias; Maurício Lacerda Nogueira; Rafael dos Santos Bezerra; Raul Machado Neto; Rejane Maria Tommasini Grotto; Ricardo Haddad; Sandra Coccuzzo Sampaio Vessoni; Simone Kashima; Svetoslav Nanev Slavov; Vincent Louis Viala |
| EPI_ISL_2493631, EPI_ISL_2493632, EPI_ISL_2493634, EPI_ISL_2493636, EPI_ISL_2493638, EPI_ISL_2493639, EPI_ISL_2493641, EPI_ISL_2493642 | | | |
| see above | DEPARTAMENTO DE SAUDE MUNICIPAL SOCORRO SP | Instituto Butantan | Antonio Jorge Martins; Claudia Renata dos Santos Barros; David Schlesinger; Debora Botequio Moretti; Dimas Tadeu Covas; Elaine Cristina Marqueze; Elaine Vieira Santos; Evandra Strazza Rodrigues; Heidge Fukumasu; Jayme Augusto de Souza-Neto; José Salvatore Leister Patané; Luiz Alcantara; Luiz Lehmann Coutinho; Maria Carolina Elias; Maurício Lacerda Nogueira; Rafael dos Santos Bezerra; Raul Machado Neto; Rejane Maria Tommasini Grotto; Ricardo Haddad; Sandra Coccuzzo Sampaio Vessoni; Simone Kashima; Svetoslav Nanev Slavov; Vincent Louis Viala |
| EPI_ISL_2209045, EPI_ISL_2209370, EPI_ISL_2209739, EPI_ISL_2210228, EPI_ISL_2345679, EPI_ISL_2345683, EPI_ISL_2345728, EPI_ISL_2345732, EPI_ISL_2345773, EPI_ISL_2345776, EPI_ISL_2345799, EPI_ISL_2345827, EPI_ISL_2345871 | | | |
| see above | DEPARTAMENTO DE SAUDE MUNICIPAL | Instituto Butantan / | Antonio Jorge Martins; Claudia Renata dos Santos Barros; David Schlesinger; Debora Botequio Moretti; Dimas Tadeu Covas; Elaine Cristina Marqueze; Elaine Vieira Santos; Evandra Strazza Rodrigues; Heidge Fukumasu; Jayme Augusto de Souza-Neto; José Salvatore Leister Patané; Luiz Alcantara; Luiz Lehmann |

| | | | |
|---|---|---|---|
| | SOCORRO SP | Mendelics | Coutinho; Maria Carolina Elias; Maurício Lacerda Nogueira; Rafael dos Santos Bezerra; Raul Machado Neto; Rejane Maria Tommasini Grotto; Ricardo Haddad; Sandra Coccuzzo Sampaio Vessoni; Simone Kashima; Svetoslav Nanev Slavov; Vincent Louis Viala |
| EPI_ISL_2445117, EPI_ISL_2445177, EPI_ISL_2445178, EPI_ISL_2445180, EPI_ISL_2445182, EPI_ISL_2445183, EPI_ISL_2445184, EPI_ISL_2445186, EPI_ISL_2473733 | | | |
| see above | DIRETORIA MUNICIPAL DE SAUDE | Instituto Butantan | Antonio Jorge Martins; Claudia Renata dos Santos Barros; David Schlesinger; Debora Botequio Moretti; Dimas Tadeu Covas; Elaine Cristina Marqueze; Elaine Vieira Santos; Evandra Strazza Rodrigues; Heidge Fukumasu; Jayme Augusto de Souza-Neto; José Salvatore Leister Patané; Luiz Alcantara; Luiz Lehmann Coutinho; Maria Carolina Elias; Maurício Lacerda Nogueira; Rafael dos Santos Bezerra; Raul Machado Neto; Rejane Maria Tommasini Grotto; Ricardo Haddad; Sandra Coccuzzo Sampaio Vessoni; Simone Kashima; Svetoslav Nanev Slavov; Vincent Louis Viala |
| EPI_ISL_2493374 | DIRETORIA MUNICIPAL DE SAUDE DE ENGENHEIRO COELHO | Instituto Butantan | Antonio Jorge Martins; Claudia Renata dos Santos Barros; David Schlesinger; Debora Botequio Moretti; Dimas Tadeu Covas; Elaine Cristina Marqueze; Elaine Vieira Santos; Evandra Strazza Rodrigues; Heidge Fukumasu; Jayme Augusto de Souza-Neto; José Salvatore Leister Patané; Luiz Alcantara; Luiz Lehmann Coutinho; Maria Carolina Elias; Maurício Lacerda Nogueira; Rafael dos Santos Bezerra; Raul Machado Neto; Rejane Maria Tommasini Grotto; Ricardo Haddad; Sandra Coccuzzo Sampaio Vessoni; Simone Kashima; Svetoslav Nanev Slavov; Vincent Louis Viala |
| EPI_ISL_2493401, EPI_ISL_2493408 | EMERGENCIA RESPIRATORIA DE NOVA GRANADA | Instituto Butantan | Antonio Jorge Martins; Claudia Renata dos Santos Barros; David Schlesinger; Debora Botequio Moretti; Dimas Tadeu Covas; Elaine Cristina Marqueze; Elaine Vieira Santos; Evandra Strazza Rodrigues; Heidge Fukumasu; Jayme Augusto de Souza-Neto; José Salvatore Leister Patané; Luiz Alcantara; Luiz Lehmann Coutinho; Maria Carolina Elias; Maurício Lacerda Nogueira; Rafael dos Santos Bezerra; Raul Machado Neto; Rejane Maria Tommasini Grotto; Ricardo Haddad; Sandra Coccuzzo Sampaio Vessoni; Simone Kashima; Svetoslav Nanev Slavov; Vincent Louis Viala |
| EPI_ISL_2445535, EPI_ISL_2473790, EPI_ISL_2473792 | ESF I E II | Instituto Butantan | Antonio Jorge Martins; Claudia Renata dos Santos Barros; David Schlesinger; Debora Botequio Moretti; Dimas Tadeu Covas; Elaine Cristina Marqueze; Elaine Vieira Santos; Evandra Strazza Rodrigues; Heidge Fukumasu; Jayme Augusto de Souza-Neto; José Salvatore Leister Patané; Luiz Alcantara; Luiz Lehmann Coutinho; Maria Carolina Elias; Maurício Lacerda Nogueira; Rafael dos Santos Bezerra; Raul Machado Neto; Rejane Maria Tommasini Grotto; Ricardo Haddad; Sandra Coccuzzo Sampaio Vessoni; Simone Kashima; Svetoslav Nanev Slavov; Vincent Louis Viala |
| EPI_ISL_2445515, EPI_ISL_2473691 | ESF NOVA TANABI II | Instituto Butantan | Antonio Jorge Martins; Claudia Renata dos Santos Barros; David Schlesinger; Debora Botequio Moretti; Dimas Tadeu Covas; Elaine Cristina Marqueze; Elaine Vieira Santos; Evandra Strazza Rodrigues; Heidge Fukumasu; Jayme Augusto de Souza-Neto; José Salvatore Leister Patané; Luiz Alcantara; Luiz Lehmann Coutinho; Maria Carolina Elias; Maurício Lacerda Nogueira; Rafael dos Santos Bezerra; Raul Machado Neto; Rejane Maria Tommasini Grotto; Ricardo Haddad; Sandra Coccuzzo Sampaio Vessoni; Simone Kashima; Svetoslav Nanev Slavov; Vincent Louis Viala |
| EPI_ISL_2445157, EPI_ISL_2473706 | ESF PARQUE COLINAS | Instituto Butantan | Antonio Jorge Martins; Claudia Renata dos Santos Barros; David Schlesinger; Debora Botequio Moretti; Dimas Tadeu Covas; Elaine Cristina Marqueze; Elaine Vieira Santos; Evandra Strazza Rodrigues; Heidge Fukumasu; Jayme Augusto de Souza-Neto; José Salvatore Leister Patané; Luiz Alcantara; Luiz Lehmann Coutinho; Maria Carolina Elias; Maurício Lacerda Nogueira; Rafael dos Santos Bezerra; Raul Machado Neto; Rejane Maria Tommasini Grotto; Ricardo Haddad; Sandra Coccuzzo Sampaio Vessoni; Simone Kashima; Svetoslav Nanev Slavov; Vincent Louis Viala |
| EPI_ISL_2445190, EPI_ISL_2445191, EPI_ISL_2445192, EPI_ISL_2473712 | ESF PQ UNIVERSITARIO DE ASSIS | Instituto Butantan | Antonio Jorge Martins; Claudia Renata dos Santos Barros; David Schlesinger; Debora Botequio Moretti; Dimas Tadeu Covas; Elaine Cristina Marqueze; Elaine Vieira Santos; Evandra Strazza Rodrigues; Heidge Fukumasu; Jayme Augusto de Souza-Neto; José Salvatore Leister Patané; Luiz Alcantara; Luiz Lehmann Coutinho; Maria Carolina Elias; Maurício Lacerda Nogueira; Rafael dos Santos Bezerra; Raul Machado Neto; Rejane Maria Tommasini Grotto; Ricardo Haddad; Sandra Coccuzzo Sampaio Vessoni; Simone Kashima; Svetoslav Nanev Slavov; Vincent Louis Viala |
| EPI_ISL_2445169, EPI_ISL_2445176 | ESF VILA GLORIA I | Instituto Butantan | Antonio Jorge Martins; Claudia Renata dos Santos Barros; David Schlesinger; Debora Botequio Moretti; Dimas Tadeu Covas; Elaine Cristina Marqueze; Elaine Vieira Santos; Evandra Strazza Rodrigues; Heidge Fukumasu; Jayme Augusto de Souza-Neto; José Salvatore Leister Patané; Luiz Alcantara; Luiz Lehmann Coutinho; Maria Carolina Elias; Maurício Lacerda Nogueira; Rafael dos Santos Bezerra; Raul Machado Neto; Rejane Maria Tommasini Grotto; Ricardo Haddad; Sandra Coccuzzo Sampaio Vessoni; Simone Kashima; Svetoslav Nanev Slavov; Vincent Louis Viala |
| EPI_ISL_2445171, EPI_ISL_2445187, EPI_ISL_2473816 | ESF VILA PROGRESSO | Instituto Butantan | Antonio Jorge Martins; Claudia Renata dos Santos Barros; David Schlesinger; Debora Botequio Moretti; Dimas Tadeu Covas; Elaine Cristina Marqueze; Elaine Vieira Santos; Evandra Strazza Rodrigues; Heidge Fukumasu; Jayme Augusto de Souza-Neto; José Salvatore Leister Patané; Luiz Alcantara; Luiz Lehmann Coutinho; Maria Carolina Elias; Maurício Lacerda Nogueira; Rafael dos Santos Bezerra; Raul Machado Neto; Rejane Maria Tommasini Grotto; Ricardo Haddad; Sandra Coccuzzo Sampaio Vessoni; Simone Kashima; Svetoslav Nanev Slavov; Vincent Louis Viala |
| EPI_ISL_2445158, EPI_ISL_2445160 | ESF VITORIA | Instituto Butantan | Antonio Jorge Martins; Claudia Renata dos Santos Barros; David Schlesinger; Debora Botequio Moretti; Dimas Tadeu Covas; Elaine Cristina Marqueze; Elaine Vieira Santos; Evandra Strazza Rodrigues; Heidge Fukumasu; Jayme Augusto de Souza-Neto; José Salvatore Leister Patané; Luiz Alcantara; Luiz Lehmann Coutinho; Maria Carolina Elias; Maurício Lacerda Nogueira; Rafael dos Santos Bezerra; Raul Machado Neto; Rejane Maria Tommasini Grotto; Ricardo Haddad; Sandra Coccuzzo Sampaio Vessoni; Simone Kashima; Svetoslav Nanev Slavov; Vincent Louis Viala |
| EPI_ISL_2445114, EPI_ISL_2445115, EPI_ISL_2445116, EPI_ISL_2493708, EPI_ISL_2493709, EPI_ISL_2493710, EPI_ISL_2493711, EPI_ISL_2493712, EPI_ISL_2494267, EPI_ISL_2494268, EPI_ISL_2494269, EPI_ISL_2494270, EPI_ISL_2494271, EPI_ISL_2494272, EPI_ISL_2494273, EPI_ISL_2494296, EPI_ISL_2494302, EPI_ISL_2494341 | | | |
| see above | FACULDADE DE MEDICINA ABC | Instituto Butantan | Antonio Jorge Martins; Claudia Renata dos Santos Barros; David Schlesinger; Debora Botequio Moretti; Dimas Tadeu Covas; Elaine Cristina Marqueze; Elaine Vieira Santos; Evandra Strazza Rodrigues; Heidge Fukumasu; Jayme Augusto de Souza-Neto; José Salvatore Leister Patané; Luiz Alcantara; Luiz Lehmann Coutinho; Maria Carolina Elias; Maurício Lacerda Nogueira; Rafael dos Santos Bezerra; Raul Machado Neto; Rejane Maria Tommasini Grotto; Ricardo Haddad; Sandra Coccuzzo Sampaio Vessoni; Simone Kashima; Svetoslav Nanev Slavov; Vincent Louis Viala |
| EPI_ISL_2473694 | FUNDAÇÃO ABC | Instituto Butantan | Antonio Jorge Martins; Claudia Renata dos Santos Barros; David Schlesinger; Debora Botequio Moretti; Dimas Tadeu Covas; Elaine Cristina Marqueze; Elaine Vieira Santos; Evandra Strazza Rodrigues; Heidge Fukumasu; Jayme Augusto de Souza-Neto; José Salvatore Leister Patané; Luiz Alcantara; Luiz Lehmann Coutinho; Maria Carolina Elias; Maurício Lacerda Nogueira; Rafael dos Santos Bezerra; Raul Machado Neto; Rejane Maria Tommasini Grotto; Ricardo Haddad; Sandra Coccuzzo Sampaio Vessoni; Simone Kashima; Svetoslav Nanev Slavov; Vincent Louis Viala |
| EPI_ISL_2828668 | FoxEs Enterprise Solutions | Instituto Adolfo Lutz, Interdiciplinary Procedures Center, Strategic Laboratory | Caio Vinicius Dias Lopes; Claudia Regina Gonçalves; Claudio Tavares Sacchi; Erica Valessa Ramos Gomes; Karoline Rodrigues Campos |
| EPI_ISL_2311861 | Fundação Ezequiel Dias | Coordenação Geral de Laboratórios de Saúde Pública (CGLAB/DAEVS/SVS/MS) | Vagner Fonseca; et al. |
| EPI_ISL_2209586, EPI_ISL_2345701, EPI_ISL_2345709, EPI_ISL_2345756, EPI_ISL_2345797, EPI_ISL_2345806 | GRIPARIO FALCAO | Instituto Butantan / Mendelics | Antonio Jorge Martins; Claudia Renata dos Santos Barros; David Schlesinger; Debora Botequio Moretti; Dimas Tadeu Covas; Elaine Cristina Marqueze; Elaine Vieira Santos; Evandra Strazza Rodrigues; Heidge Fukumasu; Jayme Augusto de Souza-Neto; José Salvatore Leister Patané; Luiz Alcantara; Luiz Lehmann Coutinho; Maria Carolina Elias; Maurício Lacerda Nogueira; Rafael dos Santos Bezerra; Raul Machado Neto; Rejane Maria Tommasini Grotto; Ricardo Haddad; Sandra Coccuzzo Sampaio Vessoni; Simone Kashima; Svetoslav Nanev Slavov; Vincent Louis Viala |
| EPI_ISL_2493943 | GRUPO TECNICO DE VIGILANCIA SANITARIA E EPIDEMIOLOGICA | Instituto Butantan | Antonio Jorge Martins; Claudia Renata dos Santos Barros; David Schlesinger; Debora Botequio Moretti; Dimas Tadeu Covas; Elaine Cristina Marqueze; Elaine Vieira Santos; Evandra Strazza Rodrigues; Heidge Fukumasu; Jayme Augusto de Souza-Neto; José Salvatore Leister Patané; Luiz Alcantara; Luiz Lehmann Coutinho; Maria Carolina Elias; Maurício Lacerda Nogueira; Rafael dos Santos Bezerra; Raul Machado Neto; Rejane Maria Tommasini Grotto; Ricardo Haddad; Sandra Coccuzzo Sampaio Vessoni; Simone Kashima; Svetoslav Nanev Slavov; Vincent Louis Viala |
| EPI_ISL_2187991, EPI_ISL_2187992, EPI_ISL_2187993, EPI_ISL_2187994, EPI_ISL_2187995, EPI_ISL_2187996, EPI_ISL_2187997, EPI_ISL_2187998, EPI_ISL_2187999, EPI_ISL_2188001, EPI_ISL_2188002, EPI_ISL_2188003, EPI_ISL_2188004, EPI_ISL_2188005, EPI_ISL_2188006, EPI_ISL_2188007, EPI_ISL_2188008, EPI_ISL_2188009, EPI_ISL_2188010, EPI_ISL_2188011, EPI_ISL_2188012, EPI_ISL_2188013, EPI_ISL_2188014, EPI_ISL_2188015, EPI_ISL_2188016, EPI_ISL_2188017, EPI_ISL_2188018, EPI_ISL_2188019, EPI_ISL_2188020, EPI_ISL_2188021, EPI_ISL_2188022, EPI_ISL_2188023, EPI_ISL_2188024, EPI_ISL_2188025, EPI_ISL_2188026, EPI_ISL_2188027, EPI_ISL_2222900, EPI_ISL_2222902, EPI_ISL_2222905, EPI_ISL_2222908, EPI_ISL_2222911, EPI_ISL_2222913, EPI_ISL_2222916, EPI_ISL_2222917, EPI_ISL_2222918, EPI_ISL_2227564, EPI_ISL_2227565, EPI_ISL_2348604, EPI_ISL_2348605, EPI_ISL_2348606, EPI_ISL_2348607, EPI_ISL_2348608, EPI_ISL_2348609, EPI_ISL_2348610, EPI_ISL_2348611, EPI_ISL_2348612, EPI_ISL_2348613, EPI_ISL_2348614, EPI_ISL_2348615, EPI_ISL_2466446, EPI_ISL_2466447, EPI_ISL_2466448, EPI_ISL_2466449, EPI_ISL_2466450, EPI_ISL_2466451, EPI_ISL_2466452, EPI_ISL_2466453, EPI_ISL_2466454, EPI_ISL_2466455, EPI_ISL_2466456, EPI_ISL_2466457, EPI_ISL_2466458, EPI_ISL_2466459, EPI_ISL_2466460, EPI_ISL_2466461, EPI_ISL_2466462, EPI_ISL_2466463, EPI_ISL_2466464, EPI_ISL_2466465, EPI_ISL_2466466, EPI_ISL_2497440, EPI_ISL_2497441, EPI_ISL_2497442, EPI_ISL_2497443, EPI_ISL_2497444, EPI_ISL_2497445, EPI_ISL_2497446, EPI_ISL_2497447, EPI_ISL_2497448, EPI_ISL_2497449, EPI_ISL_2497450, EPI_ISL_2497451, EPI_ISL_2497452, EPI_ISL_2497453, EPI_ISL_2497454, EPI_ISL_2497455, EPI_ISL_2497456, EPI_ISL_2497457, EPI_ISL_2497458, EPI_ISL_2497459, EPI_ISL_2497460, EPI_ISL_2497461, EPI_ISL_2497462, EPI_ISL_2497463, EPI_ISL_2497464, EPI_ISL_2497465, EPI_ISL_2497466, EPI_ISL_2497467, EPI_ISL_2497468, EPI_ISL_2497469, EPI_ISL_2497470, EPI_ISL_2497471, EPI_ISL_2497472, EPI_ISL_2497473, EPI_ISL_2497474, EPI_ISL_2497475, EPI_ISL_2497476, EPI_ISL_2617584, EPI_ISL_2617585, EPI_ISL_2617586, EPI_ISL_2617587, EPI_ISL_2617588, EPI_ISL_2617589, EPI_ISL_2617590, EPI_ISL_2617591, EPI_ISL_2617592, EPI_ISL_2617593, EPI_ISL_2617594, EPI_ISL_2617595, EPI_ISL_2617596, EPI_ISL_2617597, EPI_ISL_2617598, EPI_ISL_2617599, EPI_ISL_2617600, EPI_ISL_2617601, EPI_ISL_2617602, EPI_ISL_2617603, EPI_ISL_2617604, EPI_ISL_2617605, EPI_ISL_2617606, EPI_ISL_2617607, EPI_ISL_2617608, EPI_ISL_2617609, EPI_ISL_2617610, EPI_ISL_2617611, EPI_ISL_2617612, EPI_ISL_2617613, EPI_ISL_2617614, EPI_ISL_2617615, EPI_ISL_2617616, EPI_ISL_2617617, EPI_ISL_2617618, EPI_ISL_2617619, EPI_ISL_2617620, EPI_ISL_2617621, EPI_ISL_2617622, EPI_ISL_2617623, EPI_ISL_2617624, EPI_ISL_2617625, EPI_ISL_2617627, EPI_ISL_2680895, EPI_ISL_2680896, EPI_ISL_2680897, EPI_ISL_2680898, EPI_ISL_2680899, EPI_ISL_2680900, EPI_ISL_2680901, EPI_ISL_2680902, EPI_ISL_2680903, EPI_ISL_2680904, EPI_ISL_2680905, EPI_ISL_2680906, EPI_ISL_2680907, EPI_ISL_2680908, EPI_ISL_2680909, EPI_ISL_2680910, EPI_ISL_2680911, EPI_ISL_2680912, EPI_ISL_2680913, EPI_ISL_2680914, EPI_ISL_2680915, EPI_ISL_2921533, EPI_ISL_2921534, EPI_ISL_2921535, EPI_ISL_2921536, EPI_ISL_2921537, EPI_ISL_2921538, EPI_ISL_2921539, EPI_ISL_2921540, EPI_ISL_2921541, EPI_ISL_2921542, EPI_ISL_2921543, EPI_ISL_2921544, EPI_ISL_2921545, EPI_ISL_2921546, EPI_ISL_2921547, EPI_ISL_2921548, EPI_ISL_2921549, EPI_ISL_2921550, EPI_ISL_2921551, EPI_ISL_2921552, EPI_ISL_2921553, EPI_ISL_2921554, EPI_ISL_2921555, EPI_ISL_2921556, EPI_ISL_2921557, EPI_ISL_2921558, EPI_ISL_2921559, EPI_ISL_2921560, EPI_ISL_2921561, EPI_ISL_2921562, EPI_ISL_2921563, EPI_ISL_2921564, EPI_ISL_2921565, EPI_ISL_2921566, EPI_ISL_2921567, EPI_ISL_2921568, EPI_ISL_2921569, EPI_ISL_2921570, EPI_ISL_2921571, EPI_ISL_2921572, EPI_ISL_2921573, EPI_ISL_2921574, EPI_ISL_2921575, EPI_ISL_2921576, EPI_ISL_2921577, EPI_ISL_2921578, EPI_ISL_2921579, EPI_ISL_2921580, EPI_ISL_2921581, EPI_ISL_2921582, EPI_ISL_2921583, EPI_ISL_2921584, EPI_ISL_2921585, EPI_ISL_2921586, EPI_ISL_2921587, EPI_ISL_2921588, EPI_ISL_2921589, EPI_ISL_2921590, EPI_ISL_2921591, EPI_ISL_2921592, EPI_ISL_2921593, EPI_ISL_2921594, EPI_ISL_2921595, EPI_ISL_2921596, EPI_ISL_2921597, EPI_ISL_2921598, EPI_ISL_2921599, EPI_ISL_2921600, EPI_ISL_2921601, EPI_ISL_2921602, EPI_ISL_2921604, EPI_ISL_2921606, EPI_ISL_2921607, EPI_ISL_2921609, EPI_ISL_2928511 | | | |
| see above | HLAGYN - Laboratorio de Imunologia de Transplantes de Goias | HLAGYN - Laboratorio de Imunologia de Transplantes de Goias | Alessandro Leonardo Alvares Magalhaes; Daniel Ferreira de Sousa; Danielle de Paiva Rezende; Erika Lopes Rocha Batista; Fernando Antonio Vinhal dos Santos; Frederico Rodrigues Vinhal; Lucas Carlos Gomes Pereira; Sabrina Sara Moreira Duarte |
| EPI_ISL_2445547, EPI_ISL_2473805 | HOSP E MATERNIDADE MUNICIPAL N SRA MONTE SERRAT | Instituto Butantan | Antonio Jorge Martins; Claudia Renata dos Santos Barros; David Schlesinger; Debora Botequio Moretti; Dimas Tadeu Covas; Elaine Cristina Marqueze; Elaine Vieira Santos; Evandra Strazza Rodrigues; Heidge Fukumasu; Jayme Augusto de Souza-Neto; José Salvatore Leister Patané; Luiz Alcantara; Luiz Lehmann Coutinho; Maria Carolina Elias; Maurício Lacerda Nogueira; Rafael dos Santos Bezerra; Raul Machado Neto; Rejane Maria Tommasini Grotto; Ricardo Haddad; Sandra Coccuzzo Sampaio Vessoni; Simone Kashima; Svetoslav Nanev Slavov; Vincent Louis Viala |
| EPI_ISL_2445144, EPI_ISL_2445145, EPI_ISL_2445146, EPI_ISL_2473702, EPI_ISL_2473703, EPI_ISL_2473726, EPI_ISL_2473727, EPI_ISL_2473728, EPI_ISL_2473729, EPI_ISL_2473730, EPI_ISL_2473734, EPI_ISL_2473736, EPI_ISL_2473749, EPI_ISL_2473757, EPI_ISL_2473758, EPI_ISL_2473759, EPI_ISL_2473760, EPI_ISL_2473761, EPI_ISL_2473821 | | | |
| see above | HOSP GERAL DE SAO PAULO | Instituto Butantan | Antonio Jorge Martins; Claudia Renata dos Santos Barros; David Schlesinger; Debora Botequio Moretti; Dimas Tadeu Covas; Elaine Cristina Marqueze; Elaine Vieira Santos; Evandra Strazza Rodrigues; Heidge Fukumasu; Jayme Augusto de Souza-Neto; José Salvatore Leister Patané; Luiz Alcantara; Luiz Lehmann Coutinho; Maria Carolina Elias; Maurício Lacerda Nogueira; Rafael dos Santos Bezerra; Raul Machado Neto; Rejane Maria Tommasini Grotto; Ricardo Haddad; Sandra Coccuzzo Sampaio Vessoni; Simone Kashima; Svetoslav Nanev Slavov; Vincent Louis Viala |
| EPI_ISL_2493243, EPI_ISL_2493845, EPI_ISL_2493850, EPI_ISL_2493851, EPI_ISL_2493853, EPI_ISL_2493854, EPI_ISL_2493855, EPI_ISL_2493856, EPI_ISL_2493857, EPI_ISL_2493859, EPI_ISL_2493860, EPI_ISL_2493905, EPI_ISL_2493918, EPI_ISL_2493919, EPI_ISL_2493920, EPI_ISL_2493921, EPI_ISL_2493922, EPI_ISL_2493923, EPI_ISL_2493924, EPI_ISL_2493925, EPI_ISL_2493926, EPI_ISL_2493927, EPI_ISL_2493928, EPI_ISL_2493929, EPI_ISL_2493930, EPI_ISL_2493937, EPI_ISL_2493938, EPI_ISL_2493939, EPI_ISL_2493954, EPI_ISL_2494300 | | | |
| see above | HOSP MUN DE MOGI DAS CRUZES PREF WALDEMAR COSTA FILHO | Instituto Butantan | Antonio Jorge Martins; Claudia Renata dos Santos Barros; David Schlesinger; Debora Botequio Moretti; Dimas Tadeu Covas; Elaine Cristina Marqueze; Elaine Vieira Santos; Evandra Strazza Rodrigues; Heidge Fukumasu; Jayme Augusto de Souza-Neto; José Salvatore Leister Patané; Luiz Alcantara; Luiz Lehmann Coutinho; Maria Carolina Elias; Maurício Lacerda Nogueira; Rafael dos Santos Bezerra; Raul Machado Neto; Rejane Maria Tommasini Grotto; Ricardo Haddad; Sandra Coccuzzo Sampaio Vessoni; Simone Kashima; Svetoslav Nanev Slavov; Vincent Louis Viala |
| EPI_ISL_2105676, EPI_ISL_2209295, EPI_ISL_2345839 | HOSP MUN DE MOGI DAS CRUZES PREF WALDEMAR COSTA FILHO | Instituto Butantan / Mendelics | Antonio Jorge Martins; Bianca Cechetto Carlos. Mendelics: Bibiana Santos; Claudia Renata dos Santos Barros; Cíntia Bittar; David Schlesinger; David Schlesinger. Hemocentro Ribeirão Preto: Simone Kashima; Debora Botequio Moretti; Dimas Tadeu Covas; Elaine Cristina Marqueze; Elaine Vieira Santos; Elaine Vieira dos Santos; Elisangela Chicaroni Mattos; Erika Freitas; Evandra Strazza Rodrigues; Felipe Allan da Silva da Costa; Flavia Aburjaile; Fábio Sossai Possebon; Guilherme Campos; Guilherme Targino Valente; Heidge Fukumasu; Heidge Fukumasu. USP-Botucatu: Rejane Maria Tommasini Grotto; Helena Lage Ferreira; Instituto Butantan: Dimas Tadeu Covas; Jardelina de Souza Todao Bernardino; Jayme A. Souza-Neto; Jayme Augusto de Souza-Neto; Jessika Cristina Chagas Lesbon; Jorge A. Petroli Marchesi; José Salvatore Leister Patané; João Paulo Kitajima; João Pessoa Araújo Jr.; Leila Sabrina Ullmann; Loyze Paola Oliveira de Lima; Luiz Alcantara; Luiz Aurelio de Campos Crispin. Centro de Genômica Funcional da ESALQ: Luiz Lehmann Coutinho; Luiz Carlos Junior de Alcantara; Luiz Lehmann Coutinho; Lívia Sacchetto; Maisa C. Pereira Parra; Maria Carolina Elias; Marta Giovanetti; Marília Moraes; Maurício Lacerda Nogueira; Maurício Lacerda Nogueira. Prefeitura de Sao Paulo: Melissa Palmieri.; Patricia Akemi Assato; Paula Rahal; Paulo Inacio da Costa; Rafael dos Santos Bezerra; Raquel de Lello Rocha Campos Cassano. NGS Soluções Genômicas: Pilar Drummond Sampaio Corrêa Mariani. FZEA-USP Pirassununga: Mirele Daiana Poleti; Raul Machado Neto; Rejane Maria Tommasini Grotto; Ricardo Augusto Brassaloti; Ricardo Haddad; Rodrigo Tocantins Calado. FAMERP-SJRP: Cecília Artico Banho; Sandra Coccuzzo Sampaio; Sandra Coccuzzo Sampaio Vessoni; Simone Kashima; Svetoslav Nanev Slavov; Vagner Fonseca; Vincent Louis Viala |
| EPI_ISL_2493941 | HOSP MUN JOSANIAS CASTANHA BRAGA | Instituto Butantan | Antonio Jorge Martins; Claudia Renata dos Santos Barros; David Schlesinger; Debora Botequio Moretti; Dimas Tadeu Covas; Elaine Cristina Marqueze; Elaine Vieira Santos; Evandra Strazza Rodrigues; Heidge Fukumasu; Jayme Augusto de Souza-Neto; José Salvatore Leister Patané; Luiz Alcantara; Luiz Lehmann Coutinho; Maria Carolina Elias; Maurício Lacerda Nogueira; Rafael dos Santos Bezerra; Raul Machado Neto; Rejane Maria Tommasini Grotto; Ricardo Haddad; Sandra Coccuzzo Sampaio Vessoni; Simone Kashima; Svetoslav Nanev Slavov; Vincent Louis Viala |
| EPI_ISL_2445305 | HOSP PRESIDENTE | Instituto Butantan | Antonio Jorge Martins; Claudia Renata dos Santos Barros; David Schlesinger; Debora Botequio Moretti; Dimas Tadeu Covas; Elaine Cristina Marqueze; Elaine Vieira Santos; Evandra Strazza Rodrigues; Heidge Fukumasu; Jayme Augusto de Souza-Neto; José Salvatore Leister Patané; Luiz Alcantara; Luiz Lehmann Coutinho; Maria Carolina Elias; Maurício Lacerda Nogueira; Rafael dos Santos Bezerra; Raul Machado Neto; Rejane Maria Tommasini Grotto; Ricardo Haddad; Sandra Coccuzzo Sampaio Vessoni; Simone Kashima; Svetoslav Nanev Slavov; Vincent Louis Viala |
| EPI_ISL_2493956, EPI_ISL_2493959 | HOSPITAL DE CAMPANHA COVID 19 DE MAIRIPORA | Instituto Butantan | Antonio Jorge Martins; Claudia Renata dos Santos Barros; David Schlesinger; Debora Botequio Moretti; Dimas Tadeu Covas; Elaine Cristina Marqueze; Elaine Vieira Santos; Evandra Strazza Rodrigues; Heidge Fukumasu; Jayme Augusto de Souza-Neto; José Salvatore Leister Patané; Luiz Alcantara; Luiz Lehmann Coutinho; Maria Carolina Elias; Maurício Lacerda Nogueira; Rafael dos Santos Bezerra; Raul Machado Neto; Rejane Maria Tommasini Grotto; Ricardo Haddad; Sandra Coccuzzo Sampaio Vessoni; Simone Kashima; Svetoslav Nanev Slavov; Vincent Louis Viala |
| EPI_ISL_2445571, EPI_ISL_2445572, EPI_ISL_2445577, EPI_ISL_2445578, EPI_ISL_2445579, EPI_ISL_2445581, EPI_ISL_2445583, EPI_ISL_2445584, EPI_ISL_2445588, EPI_ISL_2445591, EPI_ISL_2473799, EPI_ISL_2473800, EPI_ISL_2473802, EPI_ISL_2473803, EPI_ISL_2493240, EPI_ISL_2493873, EPI_ISL_2493876, EPI_ISL_2493877 | | | |
| see above | HOSPITAL DE CAMPANHA COVID 19 MUNICIPIO DE TAUBATE | Instituto Butantan | Antonio Jorge Martins; Claudia Renata dos Santos Barros; David Schlesinger; Debora Botequio Moretti; Dimas Tadeu Covas; Elaine Cristina Marqueze; Elaine Vieira Santos; Evandra Strazza Rodrigues; Heidge Fukumasu; Jayme Augusto de Souza-Neto; José Salvatore Leister Patané; Luiz Alcantara; Luiz Lehmann Coutinho; Maria Carolina Elias; Maurício Lacerda Nogueira; Rafael dos Santos Bezerra; Raul Machado Neto; Rejane Maria Tommasini Grotto; Ricardo Haddad; Sandra Coccuzzo Sampaio Vessoni; Simone Kashima; Svetoslav Nanev Slavov; Vincent Louis Viala |
| EPI_ISL_2378745, EPI_ISL_2493192, EPI_ISL_2493303, EPI_ISL_2493304, EPI_ISL_2493305, EPI_ISL_2493306, EPI_ISL_2493307, EPI_ISL_2493308, EPI_ISL_2493309, EPI_ISL_2493310, EPI_ISL_2493312, EPI_ISL_2493316, EPI_ISL_2493318, EPI_ISL_2493320, EPI_ISL_2493321, EPI_ISL_2493322, EPI_ISL_2493323, EPI_ISL_2493324, EPI_ISL_2493325, EPI_ISL_2493330, EPI_ISL_2493331, EPI_ISL_2493332, EPI_ISL_2493333, EPI_ISL_2493334 | | | |
| see above | HOSPITAL DO RIM E HIPERTENSAO | Instituto Butantan | Antonio Jorge Martins; Claudia Renata dos Santos Barros; David Schlesinger; Debora Botequio Moretti; Dimas Tadeu Covas; Elaine Cristina Marqueze; Elaine Vieira Santos; Evandra Strazza Rodrigues; Heidge Fukumasu; Jayme Augusto de Souza-Neto; José Salvatore Leister Patané; Luiz Alcantara; Luiz Lehmann Coutinho; Maria Carolina Elias; Maurício Lacerda Nogueira; Rafael dos Santos Bezerra; Raul Machado Neto; Rejane Maria Tommasini Grotto; Ricardo Haddad; Sandra Coccuzzo Sampaio Vessoni; Simone Kashima; Svetoslav Nanev Slavov; Vincent Louis Viala |
| EPI_ISL_2445274 | HOSPITAL DR OSIRIS FLORINDO COELHO FERRAZ DE VASCONCELOS | Instituto Butantan | Antonio Jorge Martins; Claudia Renata dos Santos Barros; David Schlesinger; Debora Botequio Moretti; Dimas Tadeu Covas; Elaine Cristina Marqueze; Elaine Vieira Santos; Evandra Strazza Rodrigues; Heidge Fukumasu; Jayme Augusto de Souza-Neto; José Salvatore Leister Patané; Luiz Alcantara; Luiz Lehmann Coutinho; Maria Carolina Elias; Maurício Lacerda Nogueira; Rafael dos Santos Bezerra; Raul Machado Neto; Rejane Maria Tommasini Grotto; Ricardo Haddad; Sandra Coccuzzo Sampaio Vessoni; Simone Kashima; Svetoslav Nanev Slavov; Vincent Louis Viala |
| EPI_ISL_2493370 | HOSPITAL E MATERNIDADE MADRE VANNINI | Instituto Butantan | Antonio Jorge Martins; Claudia Renata dos Santos Barros; David Schlesinger; Debora Botequio Moretti; Dimas Tadeu Covas; Elaine Cristina Marqueze; Elaine Vieira Santos; Evandra Strazza Rodrigues; Heidge Fukumasu; Jayme Augusto de Souza-Neto; José Salvatore Leister Patané; Luiz Alcantara; Luiz Lehmann Coutinho; Maria Carolina Elias; Maurício Lacerda Nogueira; Rafael dos Santos Bezerra; Raul Machado Neto; Rejane Maria Tommasini Grotto; Ricardo Haddad; Sandra Coccuzzo Sampaio Vessoni; Simone Kashima; Svetoslav Nanev Slavov; Vincent Louis Viala |

| | | | |
|---|---|---|---|
| EPI_ISL_2493257, EPI_ISL_2493258, EPI_ISL_2493863, EPI_ISL_2493864, EPI_ISL_2493865, EPI_ISL_2493870, EPI_ISL_2493871, EPI_ISL_2494340, EPI_ISL_2500986 | | | |
| see above | HOSPITAL E MATERNIDADE NOSSA SENHORA DA AJUDA | Instituto Butantan | Antonio Jorge Martins; Claudia Renata dos Santos Barros; David Schlesinger; Debora Botequio Moretti; Dimas Tadeu Covas; Elaine Cristina Marqueze; Elaine Vieira Santos; Evandra Strazza Rodrigues; Heidge Fukumasu; Jayme Augusto de Souza-Neto; José Salvatore Leister Patané; Luiz Alcantara; Luiz Lehmann Coutinho; Maria Carolina Elias; Maurício Lacerda Nogueira; Rafael dos Santos Bezerra; Raul Machado Neto; Rejane Maria Tommasini Grotto; Ricardo Haddad; Sandra Coccuzzo Sampaio Vessoni; Simone Kashima; Svetoslav Nanev Slavov; Vincent Louis Viala |
| EPI_ISL_2445384 | HOSPITAL ESTADUAL DE MIRANDOPOLIS | Instituto Butantan | Antonio Jorge Martins; Claudia Renata dos Santos Barros; David Schlesinger; Debora Botequio Moretti; Dimas Tadeu Covas; Elaine Cristina Marqueze; Elaine Vieira Santos; Evandra Strazza Rodrigues; Heidge Fukumasu; Jayme Augusto de Souza-Neto; José Salvatore Leister Patané; Luiz Alcantara; Luiz Lehmann Coutinho; Maria Carolina Elias; Maurício Lacerda Nogueira; Rafael dos Santos Bezerra; Raul Machado Neto; Rejane Maria Tommasini Grotto; Ricardo Haddad; Sandra Coccuzzo Sampaio Vessoni; Simone Kashima; Svetoslav Nanev Slavov; Vincent Louis Viala |
| EPI_ISL_2445554 | HOSPITAL ESTADUAL PORTO PRIMAVERA ROSANA | Instituto Butantan | Antonio Jorge Martins; Claudia Renata dos Santos Barros; David Schlesinger; Debora Botequio Moretti; Dimas Tadeu Covas; Elaine Cristina Marqueze; Elaine Vieira Santos; Evandra Strazza Rodrigues; Heidge Fukumasu; Jayme Augusto de Souza-Neto; José Salvatore Leister Patané; Luiz Alcantara; Luiz Lehmann Coutinho; Maria Carolina Elias; Maurício Lacerda Nogueira; Rafael dos Santos Bezerra; Raul Machado Neto; Rejane Maria Tommasini Grotto; Ricardo Haddad; Sandra Coccuzzo Sampaio Vessoni; Simone Kashima; Svetoslav Nanev Slavov; Vincent Louis Viala |
| EPI_ISL_2493940 | HOSPITAL GERAL JESUS TEIXEIRA DA COSTA GUAIANASES SAO PAULO | Instituto Butantan | Antonio Jorge Martins; Claudia Renata dos Santos Barros; David Schlesinger; Debora Botequio Moretti; Dimas Tadeu Covas; Elaine Cristina Marqueze; Elaine Vieira Santos; Evandra Strazza Rodrigues; Heidge Fukumasu; Jayme Augusto de Souza-Neto; José Salvatore Leister Patané; Luiz Alcantara; Luiz Lehmann Coutinho; Maria Carolina Elias; Maurício Lacerda Nogueira; Rafael dos Santos Bezerra; Raul Machado Neto; Rejane Maria Tommasini Grotto; Ricardo Haddad; Sandra Coccuzzo Sampaio Vessoni; Simone Kashima; Svetoslav Nanev Slavov; Vincent Louis Viala |
| EPI_ISL_2493861, EPI_ISL_2493957 | HOSPITAL HELIOPOLIS UNIDADE DE GESTAO ASSISTENCIAL I SP | Instituto Butantan | Antonio Jorge Martins; Claudia Renata dos Santos Barros; David Schlesinger; Debora Botequio Moretti; Dimas Tadeu Covas; Elaine Cristina Marqueze; Elaine Vieira Santos; Evandra Strazza Rodrigues; Heidge Fukumasu; Jayme Augusto de Souza-Neto; José Salvatore Leister Patané; Luiz Alcantara; Luiz Lehmann Coutinho; Maria Carolina Elias; Maurício Lacerda Nogueira; Rafael dos Santos Bezerra; Raul Machado Neto; Rejane Maria Tommasini Grotto; Ricardo Haddad; Sandra Coccuzzo Sampaio Vessoni; Simone Kashima; Svetoslav Nanev Slavov; Vincent Louis Viala |
| EPI_ISL_2493882 | HOSPITAL MUN DR GUIDO GUIDA | Instituto Butantan | Antonio Jorge Martins; Claudia Renata dos Santos Barros; David Schlesinger; Debora Botequio Moretti; Dimas Tadeu Covas; Elaine Cristina Marqueze; Elaine Vieira Santos; Evandra Strazza Rodrigues; Heidge Fukumasu; Jayme Augusto de Souza-Neto; José Salvatore Leister Patané; Luiz Alcantara; Luiz Lehmann Coutinho; Maria Carolina Elias; Maurício Lacerda Nogueira; Rafael dos Santos Bezerra; Raul Machado Neto; Rejane Maria Tommasini Grotto; Ricardo Haddad; Sandra Coccuzzo Sampaio Vessoni; Simone Kashima; Svetoslav Nanev Slavov; Vincent Louis Viala |
| EPI_ISL_2445605, EPI_ISL_2445609, EPI_ISL_2445610, EPI_ISL_2445612, EPI_ISL_2445613, EPI_ISL_2473698, EPI_ISL_2473808, EPI_ISL_2493685, EPI_ISL_2493687, EPI_ISL_2493689, EPI_ISL_2493955 | | | |
| see above | HOSPITAL MUNICIPAL DE IBIUNA IBIUNA SP | Instituto Butantan | Antonio Jorge Martins; Claudia Renata dos Santos Barros; David Schlesinger; Debora Botequio Moretti; Dimas Tadeu Covas; Elaine Cristina Marqueze; Elaine Vieira Santos; Evandra Strazza Rodrigues; Heidge Fukumasu; Jayme Augusto de Souza-Neto; José Salvatore Leister Patané; Luiz Alcantara; Luiz Lehmann Coutinho; Maria Carolina Elias; Maurício Lacerda Nogueira; Rafael dos Santos Bezerra; Raul Machado Neto; Rejane Maria Tommasini Grotto; Ricardo Haddad; Sandra Coccuzzo Sampaio Vessoni; Simone Kashima; Svetoslav Nanev Slavov; Vincent Louis Viala |
| EPI_ISL_2493209, EPI_ISL_2493210, EPI_ISL_2493211, EPI_ISL_2493212, EPI_ISL_2493213, EPI_ISL_2493214, EPI_ISL_2493215, EPI_ISL_2493216, EPI_ISL_2493217, EPI_ISL_2493218, EPI_ISL_2493219, EPI_ISL_2493220, EPI_ISL_2493221, EPI_ISL_2493222, EPI_ISL_2493223, EPI_ISL_2493224, EPI_ISL_2493225, EPI_ISL_2493226, EPI_ISL_2493227, EPI_ISL_2493228, EPI_ISL_2493229, EPI_ISL_2493230, EPI_ISL_2493231, EPI_ISL_2493232, EPI_ISL_2493233, EPI_ISL_2493234, EPI_ISL_2493235, EPI_ISL_2493253, EPI_ISL_2493254, EPI_ISL_2493592, EPI_ISL_2493593, EPI_ISL_2493609, EPI_ISL_2493650, EPI_ISL_2493651, EPI_ISL_2493652, EPI_ISL_2500978 | | | |
| see above | HOSPITAL MUNICIPAL DE ILHABELA GOV MARIO COVAS JR | Instituto Butantan | Antonio Jorge Martins; Claudia Renata dos Santos Barros; David Schlesinger; Debora Botequio Moretti; Dimas Tadeu Covas; Elaine Cristina Marqueze; Elaine Vieira Santos; Evandra Strazza Rodrigues; Heidge Fukumasu; Jayme Augusto de Souza-Neto; José Salvatore Leister Patané; Luiz Alcantara; Luiz Lehmann Coutinho; Maria Carolina Elias; Maurício Lacerda Nogueira; Rafael dos Santos Bezerra; Raul Machado Neto; Rejane Maria Tommasini Grotto; Ricardo Haddad; Sandra Coccuzzo Sampaio Vessoni; Simone Kashima; Svetoslav Nanev Slavov; Vincent Louis Viala |
| EPI_ISL_2723770 | HOSPITAL MUNICIPAL DE ILHABELA GOV MARIO COVAS JR, ILHABELA, SP | Instituto Butantan | ; Antonio Jorge Martins; Bianca Cechetto Carlos. Mendelics: Bibiana Santos; Claudia Renata dos Santos Barros; David Schlesinger. Hemocentro Ribeirão Preto: Simone Kashima; Debora Botequio Moretti. Centro de Genômica Funcional da ESALQ: Luiz Lehmann Coutinho; Dimas Tadeu Covas; Elaine Cristina Marqueze; Elaine Vieira dos Santos; Elisangela Chicaroni Mattos; Erika Freitas; Evandra Strazza Rodrigues; Felipe Allan da Silva da Costa; Guilherme Targino Valente; Heidge Fukumasu. USP-Botucatu: Rejane Maria Tommasini Grotto; Instituto Butantan: Alexander Roberto Precioso; Jayme A. Souza-Neto; Jessika Cristina Chagas Lesbon; José Salvatore Leister Patané; João Paulo Kitajima; Maria Carolina Elias; Patricia Akemi Assato; Rafael dos Santos Bezerra; Raquel de Lello Rocha Campos Cassano. NGS Soluções Genômicas: Pilar Drummond Sampaio Corrêa Mariani. FZEA-USP Pirassununga: Mirele Daiana Poleti; Raul Machado Neto; Ricardo Augusto Brassaloti; Ricardo Haddad; Sandra Coccuzzo Sampaio; Svetoslav Nanev Slavov; Vincent Louis Viala |
| EPI_ISL_2462326, EPI_ISL_2493149, EPI_ISL_2493158, EPI_ISL_2493172, EPI_ISL_2493174, EPI_ISL_2493628, EPI_ISL_2494131, EPI_ISL_2494132, EPI_ISL_2494135, EPI_ISL_2494136, EPI_ISL_2494139, EPI_ISL_2494140 | | | |
| see above | HOSPITAL MUNICIPAL DE ITAPIRA | Instituto Butantan | Antonio Jorge Martins; Claudia Renata dos Santos Barros; David Schlesinger; Debora Botequio Moretti; Dimas Tadeu Covas; Elaine Cristina Marqueze; Elaine Vieira Santos; Evandra Strazza Rodrigues; Heidge Fukumasu; Jayme Augusto de Souza-Neto; José Salvatore Leister Patané; Luiz Alcantara; Luiz Lehmann Coutinho; Maria Carolina Elias; Maurício Lacerda Nogueira; Rafael dos Santos Bezerra; Raul Machado Neto; Rejane Maria Tommasini Grotto; Ricardo Haddad; Sandra Coccuzzo Sampaio Vessoni; Simone Kashima; Svetoslav Nanev Slavov; Vincent Louis Viala |
| EPI_ISL_2208870, EPI_ISL_2209499, EPI_ISL_2209603, EPI_ISL_2345690, EPI_ISL_2345740, EPI_ISL_2345774, EPI_ISL_2345784, EPI_ISL_2345833, EPI_ISL_2345879, EPI_ISL_2362248 | | | |
| see above | HOSPITAL MUNICIPAL DE ITAPIRA | Instituto Butantan / Mendelics | Antonio Jorge Martins; Claudia Renata dos Santos Barros; David Schlesinger; Debora Botequio Moretti; Dimas Tadeu Covas; Elaine Cristina Marqueze; Elaine Vieira Santos; Evandra Strazza Rodrigues; Heidge Fukumasu; Jayme Augusto de Souza-Neto; José Salvatore Leister Patané; Luiz Alcantara; Luiz Lehmann Coutinho; Maria Carolina Elias; Maurício Lacerda Nogueira; Rafael dos Santos Bezerra; Raul Machado Neto; Rejane Maria Tommasini Grotto; Ricardo Haddad; Sandra Coccuzzo Sampaio Vessoni; Simone Kashima; Svetoslav Nanev Slavov; Vincent Louis Viala |
| EPI_ISL_2801347 | HOSPITAL MUNICIPAL DR EDUARDO DIAS | Oswaldo Cruz Institute, FIOCRUZ/CE | Cleber Furtado Aksenen e Suzana Porto Almeida; Fabio Miyajima; Fernando Braga Stehling; Francisco Eder de Moura Lopes; Jamille Maria Mendes Bezerra; Joaquim César do Nascimento Sousa Junior; Pedro Miguel Carneiro Jeronimo; Thais Ferreira de Oliveira; Thais de Oliveira Costa; Ticiane Cavalcante de Souza; Veridiana Pessoa Miyajima |
| EPI_ISL_2445393, EPI_ISL_2445394, EPI_ISL_2445395, EPI_ISL_2445396, EPI_ISL_2445397, EPI_ISL_2445400, EPI_ISL_2445403, EPI_ISL_2445404, EPI_ISL_2445408, EPI_ISL_2445410, EPI_ISL_2445412, EPI_ISL_2445414, EPI_ISL_2462315, EPI_ISL_2493519, EPI_ISL_2493520, EPI_ISL_2493521, EPI_ISL_2493522, EPI_ISL_2493821, EPI_ISL_2493827 | | | |
| see above | HOSPITAL MUNICIPAL DR MARIO GATTI CAMPINAS | Instituto Butantan | Antonio Jorge Martins; Claudia Renata dos Santos Barros; David Schlesinger; Debora Botequio Moretti; Dimas Tadeu Covas; Elaine Cristina Marqueze; Elaine Vieira Santos; Evandra Strazza Rodrigues; Heidge Fukumasu; Jayme Augusto de Souza-Neto; José Salvatore Leister Patané; Luiz Alcantara; Luiz Lehmann Coutinho; Maria Carolina Elias; Maurício Lacerda Nogueira; Rafael dos Santos Bezerra; Raul Machado Neto; Rejane Maria Tommasini Grotto; Ricardo Haddad; Sandra Coccuzzo Sampaio Vessoni; Simone Kashima; Svetoslav Nanev Slavov; Vincent Louis Viala |
| EPI_ISL_2445500, EPI_ISL_2445501, EPI_ISL_2445502, EPI_ISL_2445503, EPI_ISL_2445504, EPI_ISL_2445505, EPI_ISL_2445506, EPI_ISL_2445507, EPI_ISL_2445509, EPI_ISL_2445510, EPI_ISL_2445511, EPI_ISL_2473785, EPI_ISL_2493392, EPI_ISL_2493523, EPI_ISL_2493524, EPI_ISL_2500976 | | | |
| see above | HOSPITAL MUNICIPAL DR TABAJARA RAMOS | Instituto Butantan | Antonio Jorge Martins; Claudia Renata dos Santos Barros; David Schlesinger; Debora Botequio Moretti; Dimas Tadeu Covas; Elaine Cristina Marqueze; Elaine Vieira Santos; Evandra Strazza Rodrigues; Heidge Fukumasu; Jayme Augusto de Souza-Neto; José Salvatore Leister Patané; Luiz Alcantara; Luiz Lehmann Coutinho; Maria Carolina Elias; Maurício Lacerda Nogueira; Rafael dos Santos Bezerra; Raul Machado Neto; Rejane Maria Tommasini Grotto; Ricardo Haddad; Sandra Coccuzzo Sampaio Vessoni; Simone Kashima; Svetoslav Nanev Slavov; Vincent Louis Viala |
| EPI_ISL_2345647, EPI_ISL_2345693, EPI_ISL_2345838, EPI_ISL_2362259, EPI_ISL_2362272 | HOSPITAL MUNICIPAL DR WALDEMAR TEBALDI | Instituto Butantan / Mendelics | Antonio Jorge Martins; Claudia Renata dos Santos Barros; David Schlesinger; Debora Botequio Moretti; Dimas Tadeu Covas; Elaine Cristina Marqueze; Elaine Vieira Santos; Evandra Strazza Rodrigues; Heidge Fukumasu; Jayme Augusto de Souza-Neto; José Salvatore Leister Patané; Luiz Alcantara; Luiz Lehmann Coutinho; Maria Carolina Elias; Maurício Lacerda Nogueira; Rafael dos Santos Bezerra; Raul Machado Neto; Rejane Maria Tommasini Grotto; Ricardo Haddad; Sandra Coccuzzo Sampaio Vessoni; Simone Kashima; Svetoslav Nanev Slavov; Vincent Louis Viala |
| EPI_ISL_2493441, EPI_ISL_2494164 | HOSPITAL MUNICIPAL JORGE CURY DE SANTA MARIA DA SERRA | Instituto Butantan | Antonio Jorge Martins; Claudia Renata dos Santos Barros; David Schlesinger; Debora Botequio Moretti; Dimas Tadeu Covas; Elaine Cristina Marqueze; Elaine Vieira Santos; Evandra Strazza Rodrigues; Heidge Fukumasu; Jayme Augusto de Souza-Neto; José Salvatore Leister Patané; Luiz Alcantara; Luiz Lehmann Coutinho; Maria Carolina Elias; Maurício Lacerda Nogueira; Rafael dos Santos Bezerra; Raul Machado Neto; Rejane Maria Tommasini Grotto; Ricardo Haddad; Sandra Coccuzzo Sampaio Vessoni; Simone Kashima; Svetoslav Nanev Slavov; Vincent Louis Viala |
| EPI_ISL_2445592, EPI_ISL_2473804, EPI_ISL_2493688 | HOSPITAL MUNICIPAL LEONARDUS VAN MELLIS | Instituto Butantan | Antonio Jorge Martins; Claudia Renata dos Santos Barros; David Schlesinger; Debora Botequio Moretti; Dimas Tadeu Covas; Elaine Cristina Marqueze; Elaine Vieira Santos; Evandra Strazza Rodrigues; Heidge Fukumasu; Jayme Augusto de Souza-Neto; José Salvatore Leister Patané; Luiz Alcantara; Luiz Lehmann Coutinho; Maria Carolina Elias; Maurício Lacerda Nogueira; Rafael dos Santos Bezerra; Raul Machado Neto; Rejane Maria Tommasini Grotto; Ricardo Haddad; Sandra Coccuzzo Sampaio Vessoni; Simone Kashima; Svetoslav Nanev Slavov; Vincent Louis Viala |
| EPI_ISL_2801305, EPI_ISL_2801335, EPI_ISL_2801336, EPI_ISL_2801345, EPI_ISL_2801348, EPI_ISL_2801349, EPI_ISL_2801352, EPI_ISL_2801355, EPI_ISL_2801358, EPI_ISL_2801362, EPI_ISL_2801370 | | | |
| see above | HOSPITAL MUNICIPAL NOSSA SENHORA DOS MILAGRES | Oswaldo Cruz Institute, FIOCRUZ/CE | Cleber Furtado Aksenen e Suzana Porto Almeida; Fabio Miyajima; Fernando Braga Stehling; Francisco Eder de Moura Lopes; Jamille Maria Mendes Bezerra; Joaquim César do Nascimento Sousa Junior; Pedro Miguel Carneiro Jeronimo; Thais Ferreira de Oliveira; Thais de Oliveira Costa; Ticiane Cavalcante de Souza; Veridiana Pessoa Miyajima |
| EPI_ISL_2445223, EPI_ISL_2445224, EPI_ISL_2445225, EPI_ISL_2445226, EPI_ISL_2445227, EPI_ISL_2445228, EPI_ISL_2445229, EPI_ISL_2445230, EPI_ISL_2445231, EPI_ISL_2445232, EPI_ISL_2445233, EPI_ISL_2445235, EPI_ISL_2445238, EPI_ISL_2445239, EPI_ISL_2445240 | | | |
| see above | HOSPITAL MUNICIPAL REYNALDO GUERRA CAJATI | Instituto Butantan | Antonio Jorge Martins; Claudia Renata dos Santos Barros; David Schlesinger; Debora Botequio Moretti; Dimas Tadeu Covas; Elaine Cristina Marqueze; Elaine Vieira Santos; Evandra Strazza Rodrigues; Heidge Fukumasu; Jayme Augusto de Souza-Neto; José Salvatore Leister Patané; Luiz Alcantara; Luiz Lehmann Coutinho; Maria Carolina Elias; Maurício Lacerda Nogueira; Rafael dos Santos Bezerra; Raul Machado Neto; Rejane Maria Tommasini Grotto; Ricardo Haddad; Sandra Coccuzzo Sampaio Vessoni; Simone Kashima; Svetoslav Nanev Slavov; Vincent Louis Viala |
| EPI_ISL_2493296 | HOSPITAL MUNICIPAL SANTO ANTONIO | Instituto Butantan | Antonio Jorge Martins; Claudia Renata dos Santos Barros; David Schlesinger; Debora Botequio Moretti; Dimas Tadeu Covas; Elaine Cristina Marqueze; Elaine Vieira Santos; Evandra Strazza Rodrigues; Heidge Fukumasu; Jayme Augusto de Souza-Neto; José Salvatore Leister Patané; Luiz Alcantara; Luiz Lehmann Coutinho; Maria Carolina Elias; Maurício Lacerda Nogueira; Rafael dos Santos Bezerra; Raul Machado Neto; Rejane Maria Tommasini Grotto; Ricardo Haddad; Sandra Coccuzzo Sampaio Vessoni; Simone Kashima; Svetoslav Nanev Slavov; Vincent Louis Viala |
| EPI_ISL_2801367 | HOSPITAL OTOCLINICA | Oswaldo Cruz Institute, FIOCRUZ/CE | Cleber Furtado Aksenen e Suzana Porto Almeida; Fabio Miyajima; Fernando Braga Stehling; Francisco Eder de Moura Lopes; Jamille Maria Mendes Bezerra; Joaquim César do Nascimento Sousa Junior; Pedro Miguel Carneiro Jeronimo; Thais Ferreira de Oliveira; Thais de Oliveira Costa; Ticiane Cavalcante de Souza; Veridiana Pessoa Miyajima |
| EPI_ISL_2445550, EPI_ISL_2597993 | HOSPITAL REGIONAL DE ITAPETININGA | Instituto Butantan | Antonio Jorge Martins; Claudia Renata dos Santos Barros; David Schlesinger; Debora Botequio Moretti; Dimas Tadeu Covas; Elaine Cristina Marqueze; Elaine Vieira Santos; Evandra Strazza Rodrigues; Heidge Fukumasu; Jayme Augusto de Souza-Neto; José Salvatore Leister Patané; Luiz Alcantara; Luiz Lehmann Coutinho; Maria Carolina Elias; Maurício Lacerda Nogueira; Rafael dos Santos Bezerra; Raul Machado Neto; Rejane Maria Tommasini Grotto; Ricardo Haddad; Sandra Coccuzzo Sampaio Vessoni; Simone Kashima; Svetoslav Nanev Slavov; Vincent Louis Viala |
| EPI_ISL_2445540, EPI_ISL_2445545, EPI_ISL_2473690 | HOSPITAL REGIONAL DE REGISTRO REGISTRO | Instituto Butantan | Antonio Jorge Martins; Claudia Renata dos Santos Barros; David Schlesinger; Debora Botequio Moretti; Dimas Tadeu Covas; Elaine Cristina Marqueze; Elaine Vieira Santos; Evandra Strazza Rodrigues; Heidge Fukumasu; Jayme Augusto de Souza-Neto; José Salvatore Leister Patané; Luiz Alcantara; Luiz Lehmann Coutinho; Maria Carolina Elias; Maurício Lacerda Nogueira; Rafael dos Santos Bezerra; Raul Machado Neto; Rejane Maria Tommasini Grotto; Ricardo Haddad; Sandra Coccuzzo Sampaio Vessoni; Simone Kashima; Svetoslav Nanev Slavov; Vincent Louis Viala |
| EPI_ISL_2801334 | HOSPITAL REGIONAL DO CARIRI | Oswaldo Cruz Institute, FIOCRUZ/CE | Cleber Furtado Aksenen e Suzana Porto Almeida; Fabio Miyajima; Fernando Braga Stehling; Francisco Eder de Moura Lopes; Jamille Maria Mendes Bezerra; Joaquim César do Nascimento Sousa Junior; Pedro Miguel Carneiro Jeronimo; Thais Ferreira de Oliveira; Thais de Oliveira Costa; Ticiane Cavalcante de Souza; Veridiana Pessoa Miyajima |
| EPI_ISL_2445219, EPI_ISL_2445220 | HOSPITAL SANTO ANTONIO DE JUQUIA JUQUIA | Instituto Butantan | Antonio Jorge Martins; Claudia Renata dos Santos Barros; David Schlesinger; Debora Botequio Moretti; Dimas Tadeu Covas; Elaine Cristina Marqueze; Elaine Vieira Santos; Evandra Strazza Rodrigues; Heidge Fukumasu; Jayme Augusto de Souza-Neto; José Salvatore Leister Patané; Luiz Alcantara; Luiz Lehmann Coutinho; Maria Carolina Elias; Maurício Lacerda Nogueira; Rafael dos Santos Bezerra; Raul Machado Neto; Rejane Maria Tommasini Grotto; Ricardo Haddad; Sandra Coccuzzo Sampaio Vessoni; Simone Kashima; Svetoslav Nanev Slavov; Vincent Louis Viala |
| EPI_ISL_2445242 | HOSPITAL SAO FRANCISCO SC LTDA | Instituto Butantan | Antonio Jorge Martins; Claudia Renata dos Santos Barros; David Schlesinger; Debora Botequio Moretti; Dimas Tadeu Covas; Elaine Cristina Marqueze; Elaine Vieira Santos; Evandra Strazza Rodrigues; Heidge Fukumasu; Jayme Augusto de Souza-Neto; José Salvatore Leister Patané; Luiz Alcantara; Luiz Lehmann Coutinho; Maria Carolina Elias; Maurício Lacerda Nogueira; Rafael dos Santos Bezerra; Raul Machado Neto; Rejane Maria Tommasini Grotto; Ricardo Haddad; Sandra Coccuzzo Sampaio Vessoni; Simone Kashima; Svetoslav Nanev Slavov; Vincent Louis Viala |
| EPI_ISL_2494168, EPI_ISL_2494172 | HOSPITAL SAO JOSE ITIRAPINA | Instituto Butantan | Antonio Jorge Martins; Claudia Renata dos Santos Barros; David Schlesinger; Debora Botequio Moretti; Dimas Tadeu Covas; Elaine Cristina Marqueze; Elaine Vieira Santos; Evandra Strazza Rodrigues; Heidge Fukumasu; Jayme Augusto de Souza-Neto; José Salvatore Leister Patané; Luiz Alcantara; Luiz Lehmann Coutinho; Maria Carolina Elias; Maurício Lacerda Nogueira; Rafael dos Santos Bezerra; Raul Machado Neto; Rejane Maria Tommasini Grotto; Ricardo Haddad; Sandra Coccuzzo Sampaio Vessoni; Simone Kashima; Svetoslav Nanev Slavov; Vincent Louis Viala |
| EPI_ISL_2493987, EPI_ISL_2493994, EPI_ISL_2494001, EPI_ISL_2494022, EPI_ISL_2494327 | INSIDE CRSN CACHOEIRINHA/CASA VERDE | Instituto Butantan | Antonio Jorge Martins; Claudia Renata dos Santos Barros; David Schlesinger; Debora Botequio Moretti; Dimas Tadeu Covas; Elaine Cristina Marqueze; Elaine Vieira Santos; Evandra Strazza Rodrigues; Heidge Fukumasu; Jayme Augusto de Souza-Neto; José Salvatore Leister Patané; Luiz Alcantara; Luiz Lehmann Coutinho; Maria Carolina Elias; Maurício Lacerda Nogueira; Rafael dos Santos Bezerra; Raul Machado Neto; Rejane Maria Tommasini Grotto; Ricardo Haddad; Sandra Coccuzzo Sampaio Vessoni; Simone Kashima; Svetoslav Nanev Slavov; Vincent Louis Viala |
| EPI_ISL_2493969, EPI_ISL_2493973, EPI_ISL_2493982, EPI_ISL_2493988, EPI_ISL_2493992, EPI_ISL_2493998, EPI_ISL_2494024, EPI_ISL_2494035, EPI_ISL_2494037 | | | |
| see above | INSIDE CRSN FREGUESIA/BRASILÂNDIA | Instituto Butantan | Antonio Jorge Martins; Claudia Renata dos Santos Barros; David Schlesinger; Debora Botequio Moretti; Dimas Tadeu Covas; Elaine Cristina Marqueze; Elaine Vieira Santos; Evandra Strazza Rodrigues; Heidge Fukumasu; Jayme Augusto de Souza-Neto; José Salvatore Leister Patané; Luiz Alcantara; Luiz Lehmann Coutinho; Maria Carolina Elias; Maurício Lacerda Nogueira; Rafael dos Santos Bezerra; Raul Machado Neto; Rejane Maria Tommasini Grotto; Ricardo Haddad; Sandra Coccuzzo Sampaio Vessoni; Simone Kashima; Svetoslav Nanev Slavov; Vincent Louis Viala |
| EPI_ISL_2493974, EPI_ISL_2493979, EPI_ISL_2493986, EPI_ISL_2494026, EPI_ISL_2494274 | INSIDE CRSN PERUS | Instituto Butantan | Antonio Jorge Martins; Claudia Renata dos Santos Barros; David Schlesinger; Debora Botequio Moretti; Dimas Tadeu Covas; Elaine Cristina Marqueze; Elaine Vieira Santos; Evandra Strazza Rodrigues; Heidge Fukumasu; Jayme Augusto de Souza-Neto; José Salvatore Leister Patané; Luiz Alcantara; Luiz Lehmann Coutinho; Maria Carolina Elias; Maurício Lacerda Nogueira; Rafael dos Santos Bezerra; Raul Machado Neto; Rejane Maria Tommasini Grotto; Ricardo Haddad; Sandra Coccuzzo Sampaio Vessoni; Simone Kashima; Svetoslav Nanev Slavov; Vincent Louis Viala |
| EPI_ISL_2493201, EPI_ISL_2493967, EPI_ISL_2493971, EPI_ISL_2493993, EPI_ISL_2494003, EPI_ISL_2494008, EPI_ISL_2494011, EPI_ISL_2494027 | | | |
| see above | INSIDE CRSN PIRITUBA | Instituto Butantan | Antonio Jorge Martins; Claudia Renata dos Santos Barros; David Schlesinger; Debora Botequio Moretti; Dimas Tadeu Covas; Elaine Cristina Marqueze; Elaine Vieira Santos; Evandra Strazza Rodrigues; Heidge Fukumasu; Jayme Augusto de Souza-Neto; José Salvatore Leister Patané; Luiz Alcantara; Luiz Lehmann Coutinho; Maria Carolina Elias; Maurício Lacerda Nogueira; Rafael dos Santos Bezerra; Raul Machado Neto; Rejane Maria Tommasini Grotto; Ricardo Haddad; Sandra Coccuzzo Sampaio Vessoni; Simone Kashima; Svetoslav Nanev Slavov; Vincent Louis Viala |
| EPI_ISL_2493259, EPI_ISL_2493966, EPI_ISL_2493984, EPI_ISL_2494006, | INSIDE CRSN VILA MARIA/VILA GUILHERME | Instituto Butantan | Antonio Jorge Martins; Claudia Renata dos Santos Barros; David Schlesinger; Debora Botequio Moretti; Dimas Tadeu Covas; Elaine Cristina Marqueze; Elaine Vieira Santos; Evandra Strazza Rodrigues; Heidge Fukumasu; Jayme Augusto de Souza-Neto; José Salvatore Leister Patané; Luiz Alcantara; Luiz Lehmann Coutinho; Maria Carolina Elias; Maurício Lacerda Nogueira; Rafael dos Santos Bezerra; Raul Machado Neto; Rejane Maria Tommasini Grotto; Ricardo Haddad; Sandra Coccuzzo Sampaio Vessoni; Simone Kashima; Svetoslav Nanev Slavov; Vincent Louis Viala |

| | | | |
|---|---|---|---|
| EPI_ISL_2494007 | | | |
| EPI_ISL_2493265, EPI_ISL_2493968, EPI_ISL_2494000, EPI_ISL_2494002, EPI_ISL_2494010, EPI_ISL_2494023, EPI_ISL_2494030, EPI_ISL_2494033 | | | |
| see above | INSIDE CRSO BUTANTÃ | Instituto Butantan | Antonio Jorge Martins; Claudia Renata dos Santos Barros; David Schlesinger; Debora Botequio Moretti; Dimas Tadeu Covas; Elaine Cristina Marqueze; Elaine Vieira Santos; Evandra Strazza Rodrigues; Heidge Fukumasu; Jayme Augusto de Souza-Neto; José Salvatore Leister Patané; Luiz Alcantara; Luiz Lehmann Coutinho; Maria Carolina Elias; Maurício Lacerda Nogueira; Rafael dos Santos Bezerra; Raul Machado Neto; Rejane Maria Tommasini Grotto; Ricardo Haddad; Sandra Coccuzzo Sampaio Vessoni; Simone Kashima; Svetoslav Nanev Slavov; Vincent Louis Viala |
| EPI_ISL_2493200, EPI_ISL_2493981, EPI_ISL_2494014, EPI_ISL_2494015 | INSIDE CRSO LAPA/PINHEIROS | Instituto Butantan | Antonio Jorge Martins; Claudia Renata dos Santos Barros; David Schlesinger; Debora Botequio Moretti; Dimas Tadeu Covas; Elaine Cristina Marqueze; Elaine Vieira Santos; Evandra Strazza Rodrigues; Heidge Fukumasu; Jayme Augusto de Souza-Neto; José Salvatore Leister Patané; Luiz Alcantara; Luiz Lehmann Coutinho; Maria Carolina Elias; Maurício Lacerda Nogueira; Rafael dos Santos Bezerra; Raul Machado Neto; Rejane Maria Tommasini Grotto; Ricardo Haddad; Sandra Coccuzzo Sampaio Vessoni; Simone Kashima; Svetoslav Nanev Slavov; Vincent Louis Viala |
| EPI_ISL_2493964, EPI_ISL_2493972, EPI_ISL_2493976, EPI_ISL_2493978, EPI_ISL_2493980, EPI_ISL_2493995, EPI_ISL_2494012, EPI_ISL_2494016, EPI_ISL_2494017, EPI_ISL_2494018, EPI_ISL_2494029, EPI_ISL_2494034, EPI_ISL_2494038 | | | |
| see above | INSIDE CRSS CAPELA DO SOCORRO | Instituto Butantan | Antonio Jorge Martins; Claudia Renata dos Santos Barros; David Schlesinger; Debora Botequio Moretti; Dimas Tadeu Covas; Elaine Cristina Marqueze; Elaine Vieira Santos; Evandra Strazza Rodrigues; Heidge Fukumasu; Jayme Augusto de Souza-Neto; José Salvatore Leister Patané; Luiz Alcantara; Luiz Lehmann Coutinho; Maria Carolina Elias; Maurício Lacerda Nogueira; Rafael dos Santos Bezerra; Raul Machado Neto; Rejane Maria Tommasini Grotto; Ricardo Haddad; Sandra Coccuzzo Sampaio Vessoni; Simone Kashima; Svetoslav Nanev Slavov; Vincent Louis Viala |
| EPI_ISL_2462331, EPI_ISL_2493960, EPI_ISL_2493961, EPI_ISL_2493977, EPI_ISL_2493989, EPI_ISL_2493990, EPI_ISL_2493997, EPI_ISL_2494005, EPI_ISL_2494036 | | | |
| see above | INSIDE CRSS PARELHEIROS | Instituto Butantan | Antonio Jorge Martins; Claudia Renata dos Santos Barros; David Schlesinger; Debora Botequio Moretti; Dimas Tadeu Covas; Elaine Cristina Marqueze; Elaine Vieira Santos; Evandra Strazza Rodrigues; Heidge Fukumasu; Jayme Augusto de Souza-Neto; José Salvatore Leister Patané; Luiz Alcantara; Luiz Lehmann Coutinho; Maria Carolina Elias; Maurício Lacerda Nogueira; Rafael dos Santos Bezerra; Raul Machado Neto; Rejane Maria Tommasini Grotto; Ricardo Haddad; Sandra Coccuzzo Sampaio Vessoni; Simone Kashima; Svetoslav Nanev Slavov; Vincent Louis Viala |
| EPI_ISL_2493274, EPI_ISL_2493975, EPI_ISL_2493996, EPI_ISL_2494025, EPI_ISL_2494279, EPI_ISL_2494316 | INSIDE CRSSU IPIRANGA | Instituto Butantan | Antonio Jorge Martins; Claudia Renata dos Santos Barros; David Schlesinger; Debora Botequio Moretti; Dimas Tadeu Covas; Elaine Cristina Marqueze; Elaine Vieira Santos; Evandra Strazza Rodrigues; Heidge Fukumasu; Jayme Augusto de Souza-Neto; José Salvatore Leister Patané; Luiz Alcantara; Luiz Lehmann Coutinho; Maria Carolina Elias; Maurício Lacerda Nogueira; Rafael dos Santos Bezerra; Raul Machado Neto; Rejane Maria Tommasini Grotto; Ricardo Haddad; Sandra Coccuzzo Sampaio Vessoni; Simone Kashima; Svetoslav Nanev Slavov; Vincent Louis Viala |
| EPI_ISL_2493963, EPI_ISL_2493970, EPI_ISL_2494328 | INSIDE CRSSU JABAQUARA/VILA MARIANA | Instituto Butantan | Antonio Jorge Martins; Claudia Renata dos Santos Barros; David Schlesinger; Debora Botequio Moretti; Dimas Tadeu Covas; Elaine Cristina Marqueze; Elaine Vieira Santos; Evandra Strazza Rodrigues; Heidge Fukumasu; Jayme Augusto de Souza-Neto; José Salvatore Leister Patané; Luiz Alcantara; Luiz Lehmann Coutinho; Maria Carolina Elias; Maurício Lacerda Nogueira; Rafael dos Santos Bezerra; Raul Machado Neto; Rejane Maria Tommasini Grotto; Ricardo Haddad; Sandra Coccuzzo Sampaio Vessoni; Simone Kashima; Svetoslav Nanev Slavov; Vincent Louis Viala |
| EPI_ISL_2493991, EPI_ISL_2494004, EPI_ISL_2494028, EPI_ISL_2494031, EPI_ISL_2494032, EPI_ISL_2494310 | INSIDE CRSSU MOOCA/ARICANDUVA | Instituto Butantan | Antonio Jorge Martins; Claudia Renata dos Santos Barros; David Schlesinger; Debora Botequio Moretti; Dimas Tadeu Covas; Elaine Cristina Marqueze; Elaine Vieira Santos; Evandra Strazza Rodrigues; Heidge Fukumasu; Jayme Augusto de Souza-Neto; José Salvatore Leister Patané; Luiz Alcantara; Luiz Lehmann Coutinho; Maria Carolina Elias; Maurício Lacerda Nogueira; Rafael dos Santos Bezerra; Raul Machado Neto; Rejane Maria Tommasini Grotto; Ricardo Haddad; Sandra Coccuzzo Sampaio Vessoni; Simone Kashima; Svetoslav Nanev Slavov; Vincent Louis Viala |
| EPI_ISL_2493962, EPI_ISL_2493965, EPI_ISL_2493983, EPI_ISL_2493999, EPI_ISL_2494009, EPI_ISL_2494013, EPI_ISL_2494019, EPI_ISL_2494020, EPI_ISL_2494021 | | | |
| see above | INSIDE CRSSU SAPOPEMBA/VILA PRUDENTE | Instituto Butantan | Antonio Jorge Martins; Claudia Renata dos Santos Barros; David Schlesinger; Debora Botequio Moretti; Dimas Tadeu Covas; Elaine Cristina Marqueze; Elaine Vieira Santos; Evandra Strazza Rodrigues; Heidge Fukumasu; Jayme Augusto de Souza-Neto; José Salvatore Leister Patané; Luiz Alcantara; Luiz Lehmann Coutinho; Maria Carolina Elias; Maurício Lacerda Nogueira; Rafael dos Santos Bezerra; Raul Machado Neto; Rejane Maria Tommasini Grotto; Ricardo Haddad; Sandra Coccuzzo Sampaio Vessoni; Simone Kashima; Svetoslav Nanev Slavov; Vincent Louis Viala |
| EPI_ISL_2493162, EPI_ISL_2493178, EPI_ISL_2493239, EPI_ISL_2493247, EPI_ISL_2493713, EPI_ISL_2493714, EPI_ISL_2493715, EPI_ISL_2493716, EPI_ISL_2493717, EPI_ISL_2493718, EPI_ISL_2493719, EPI_ISL_2493720, EPI_ISL_2493721, EPI_ISL_2493722, EPI_ISL_2493723, EPI_ISL_2493724, EPI_ISL_2493725, EPI_ISL_2493726, EPI_ISL_2493727, EPI_ISL_2493728, EPI_ISL_2493729, EPI_ISL_2493730, EPI_ISL_2493731, EPI_ISL_2493732, EPI_ISL_2493733, EPI_ISL_2493734, EPI_ISL_2493735, EPI_ISL_2493736, EPI_ISL_2493737, EPI_ISL_2493738, EPI_ISL_2494318, EPI_ISL_2494352, EPI_ISL_2500972, EPI_ISL_2500973, EPI_ISL_2820952 | | | |
| see above | INSIDE DIAGNÓSTICOS | Instituto Butantan | Antonio Jorge Martins; Claudia Renata dos Santos Barros; David Schlesinger; Debora Botequio Moretti; Dimas Tadeu Covas; Elaine Cristina Marqueze; Elaine Vieira Santos; Evandra Strazza Rodrigues; Heidge Fukumasu; Jayme Augusto de Souza-Neto; José Salvatore Leister Patané; Luiz Alcantara; Luiz Lehmann Coutinho; Maria Carolina Elias; Maurício Lacerda Nogueira; Rafael dos Santos Bezerra; Raul Machado Neto; Rejane Maria Tommasini Grotto; Ricardo Haddad; Sandra Coccuzzo Sampaio Vessoni; Simone Kashima; Svetoslav Nanev Slavov; Vincent Louis Viala |
| EPI_ISL_2493289, EPI_ISL_2493704, EPI_ISL_2493705, EPI_ISL_2493707 | INSIDE DIAGNÓSTICOS NORTE BRASILÂNDIA/FREGUESIA | Instituto Butantan | Antonio Jorge Martins; Claudia Renata dos Santos Barros; David Schlesinger; Debora Botequio Moretti; Dimas Tadeu Covas; Elaine Cristina Marqueze; Elaine Vieira Santos; Evandra Strazza Rodrigues; Heidge Fukumasu; Jayme Augusto de Souza-Neto; José Salvatore Leister Patané; Luiz Alcantara; Luiz Lehmann Coutinho; Maria Carolina Elias; Maurício Lacerda Nogueira; Rafael dos Santos Bezerra; Raul Machado Neto; Rejane Maria Tommasini Grotto; Ricardo Haddad; Sandra Coccuzzo Sampaio Vessoni; Simone Kashima; Svetoslav Nanev Slavov; Vincent Louis Viala |
| EPI_ISL_2445050, EPI_ISL_2445061, EPI_ISL_2445066, EPI_ISL_2445081 | INSIDE DIAGNÓSTICOS NORTE BRASILÂNDIA/FREGUESIA DO Ó | Instituto Butantan | Antonio Jorge Martins; Claudia Renata dos Santos Barros; David Schlesinger; Debora Botequio Moretti; Dimas Tadeu Covas; Elaine Cristina Marqueze; Elaine Vieira Santos; Evandra Strazza Rodrigues; Heidge Fukumasu; Jayme Augusto de Souza-Neto; José Salvatore Leister Patané; Luiz Alcantara; Luiz Lehmann Coutinho; Maria Carolina Elias; Maurício Lacerda Nogueira; Rafael dos Santos Bezerra; Raul Machado Neto; Rejane Maria Tommasini Grotto; Ricardo Haddad; Sandra Coccuzzo Sampaio Vessoni; Simone Kashima; Svetoslav Nanev Slavov; Vincent Louis Viala |
| EPI_ISL_2445059, EPI_ISL_2445065, EPI_ISL_2445070, EPI_ISL_2493706 | INSIDE DIAGNÓSTICOS NORTE CACHOEIRINHA/CASA VERDE | Instituto Butantan | Antonio Jorge Martins; Claudia Renata dos Santos Barros; David Schlesinger; Debora Botequio Moretti; Dimas Tadeu Covas; Elaine Cristina Marqueze; Elaine Vieira Santos; Evandra Strazza Rodrigues; Heidge Fukumasu; Jayme Augusto de Souza-Neto; José Salvatore Leister Patané; Luiz Alcantara; Luiz Lehmann Coutinho; Maria Carolina Elias; Maurício Lacerda Nogueira; Rafael dos Santos Bezerra; Raul Machado Neto; Rejane Maria Tommasini Grotto; Ricardo Haddad; Sandra Coccuzzo Sampaio Vessoni; Simone Kashima; Svetoslav Nanev Slavov; Vincent Louis Viala |
| EPI_ISL_2445062, EPI_ISL_2445071 | INSIDE DIAGNÓSTICOS NORTE PERUS | Instituto Butantan | Antonio Jorge Martins; Claudia Renata dos Santos Barros; David Schlesinger; Debora Botequio Moretti; Dimas Tadeu Covas; Elaine Cristina Marqueze; Elaine Vieira Santos; Evandra Strazza Rodrigues; Heidge Fukumasu; Jayme Augusto de Souza-Neto; José Salvatore Leister Patané; Luiz Alcantara; Luiz Lehmann Coutinho; Maria Carolina Elias; Maurício Lacerda Nogueira; Rafael dos Santos Bezerra; Raul Machado Neto; Rejane Maria Tommasini Grotto; Ricardo Haddad; Sandra Coccuzzo Sampaio Vessoni; Simone Kashima; Svetoslav Nanev Slavov; Vincent Louis Viala |
| EPI_ISL_2445058, EPI_ISL_2445078 | INSIDE DIAGNÓSTICOS NORTE PIRITUBA | Instituto Butantan | Antonio Jorge Martins; Claudia Renata dos Santos Barros; David Schlesinger; Debora Botequio Moretti; Dimas Tadeu Covas; Elaine Cristina Marqueze; Elaine Vieira Santos; Evandra Strazza Rodrigues; Heidge Fukumasu; Jayme Augusto de Souza-Neto; José Salvatore Leister Patané; Luiz Alcantara; Luiz Lehmann Coutinho; Maria Carolina Elias; Maurício Lacerda Nogueira; Rafael dos Santos Bezerra; Raul Machado Neto; Rejane Maria Tommasini Grotto; Ricardo Haddad; Sandra Coccuzzo Sampaio Vessoni; Simone Kashima; Svetoslav Nanev Slavov; Vincent Louis Viala |
| EPI_ISL_2445047, EPI_ISL_2445060, EPI_ISL_2445064 | INSIDE DIAGNÓSTICOS NORTE VILA MARIA/VILA GUILHERME | Instituto Butantan | Antonio Jorge Martins; Claudia Renata dos Santos Barros; David Schlesinger; Debora Botequio Moretti; Dimas Tadeu Covas; Elaine Cristina Marqueze; Elaine Vieira Santos; Evandra Strazza Rodrigues; Heidge Fukumasu; Jayme Augusto de Souza-Neto; José Salvatore Leister Patané; Luiz Alcantara; Luiz Lehmann Coutinho; Maria Carolina Elias; Maurício Lacerda Nogueira; Rafael dos Santos Bezerra; Raul Machado Neto; Rejane Maria Tommasini Grotto; Ricardo Haddad; Sandra Coccuzzo Sampaio Vessoni; Simone Kashima; Svetoslav Nanev Slavov; Vincent Louis Viala |
| EPI_ISL_2445053, EPI_ISL_2445074, EPI_ISL_2445077 | INSIDE DIAGNÓSTICOS OESTE BUTANTÃ | Instituto Butantan | Antonio Jorge Martins; Claudia Renata dos Santos Barros; David Schlesinger; Debora Botequio Moretti; Dimas Tadeu Covas; Elaine Cristina Marqueze; Elaine Vieira Santos; Evandra Strazza Rodrigues; Heidge Fukumasu; Jayme Augusto de Souza-Neto; José Salvatore Leister Patané; Luiz Alcantara; Luiz Lehmann Coutinho; Maria Carolina Elias; Maurício Lacerda Nogueira; Rafael dos Santos Bezerra; Raul Machado Neto; Rejane Maria Tommasini Grotto; Ricardo Haddad; Sandra Coccuzzo Sampaio Vessoni; Simone Kashima; Svetoslav Nanev Slavov; Vincent Louis Viala |
| EPI_ISL_2445048, EPI_ISL_2445073 | INSIDE DIAGNÓSTICOS OESTE LAPA/PINHEIROS | Instituto Butantan | Antonio Jorge Martins; Claudia Renata dos Santos Barros; David Schlesinger; Debora Botequio Moretti; Dimas Tadeu Covas; Elaine Cristina Marqueze; Elaine Vieira Santos; Evandra Strazza Rodrigues; Heidge Fukumasu; Jayme Augusto de Souza-Neto; José Salvatore Leister Patané; Luiz Alcantara; Luiz Lehmann Coutinho; Maria Carolina Elias; Maurício Lacerda Nogueira; Rafael dos Santos Bezerra; Raul Machado Neto; Rejane Maria Tommasini Grotto; Ricardo Haddad; Sandra Coccuzzo Sampaio Vessoni; Simone Kashima; Svetoslav Nanev Slavov; Vincent Louis Viala |
| EPI_ISL_2445067, EPI_ISL_2445072, EPI_ISL_2445076, EPI_ISL_2445084 | INSIDE DIAGNÓSTICOS SUDESTE IPIRANGA | Instituto Butantan | Antonio Jorge Martins; Claudia Renata dos Santos Barros; David Schlesinger; Debora Botequio Moretti; Dimas Tadeu Covas; Elaine Cristina Marqueze; Elaine Vieira Santos; Evandra Strazza Rodrigues; Heidge Fukumasu; Jayme Augusto de Souza-Neto; José Salvatore Leister Patané; Luiz Alcantara; Luiz Lehmann Coutinho; Maria Carolina Elias; Maurício Lacerda Nogueira; Rafael dos Santos Bezerra; Raul Machado Neto; Rejane Maria Tommasini Grotto; Ricardo Haddad; Sandra Coccuzzo Sampaio Vessoni; Simone Kashima; Svetoslav Nanev Slavov; Vincent Louis Viala |
| EPI_ISL_2445056 | INSIDE DIAGNÓSTICOS SUDESTE MOOCA/ARICANDUVA | Instituto Butantan | Antonio Jorge Martins; Claudia Renata dos Santos Barros; David Schlesinger; Debora Botequio Moretti; Dimas Tadeu Covas; Elaine Cristina Marqueze; Elaine Vieira Santos; Evandra Strazza Rodrigues; Heidge Fukumasu; Jayme Augusto de Souza-Neto; José Salvatore Leister Patané; Luiz Alcantara; Luiz Lehmann Coutinho; Maria Carolina Elias; Maurício Lacerda Nogueira; Rafael dos Santos Bezerra; Raul Machado Neto; Rejane Maria Tommasini Grotto; Ricardo Haddad; Sandra Coccuzzo Sampaio Vessoni; Simone Kashima; Svetoslav Nanev Slavov; Vincent Louis Viala |
| EPI_ISL_2445052, EPI_ISL_2473815 | INSIDE DIAGNÓSTICOS SUDESTE VILA MARIANA/JABAQUARA | Instituto Butantan | Antonio Jorge Martins; Claudia Renata dos Santos Barros; David Schlesinger; Debora Botequio Moretti; Dimas Tadeu Covas; Elaine Cristina Marqueze; Elaine Vieira Santos; Evandra Strazza Rodrigues; Heidge Fukumasu; Jayme Augusto de Souza-Neto; José Salvatore Leister Patané; Luiz Alcantara; Luiz Lehmann Coutinho; Maria Carolina Elias; Maurício Lacerda Nogueira; Rafael dos Santos Bezerra; Raul Machado Neto; Rejane Maria Tommasini Grotto; Ricardo Haddad; Sandra Coccuzzo Sampaio Vessoni; Simone Kashima; Svetoslav Nanev Slavov; Vincent Louis Viala |
| EPI_ISL_2445063, EPI_ISL_2445079, EPI_ISL_2445082, EPI_ISL_2445083 | INSIDE DIAGNÓSTICOS SUDESTE VILA PRUDENTE/SAPOPEMBA | Instituto Butantan | Antonio Jorge Martins; Claudia Renata dos Santos Barros; David Schlesinger; Debora Botequio Moretti; Dimas Tadeu Covas; Elaine Cristina Marqueze; Elaine Vieira Santos; Evandra Strazza Rodrigues; Heidge Fukumasu; Jayme Augusto de Souza-Neto; José Salvatore Leister Patané; Luiz Alcantara; Luiz Lehmann Coutinho; Maria Carolina Elias; Maurício Lacerda Nogueira; Rafael dos Santos Bezerra; Raul Machado Neto; Rejane Maria Tommasini Grotto; Ricardo Haddad; Sandra Coccuzzo Sampaio Vessoni; Simone Kashima; Svetoslav Nanev Slavov; Vincent Louis Viala |
| EPI_ISL_2445054, EPI_ISL_2445055, EPI_ISL_2445057, EPI_ISL_2445069, EPI_ISL_2445080 | INSIDE DIAGNÓSTICOS SUL CAPELA DO SOCORRO | Instituto Butantan | Antonio Jorge Martins; Claudia Renata dos Santos Barros; David Schlesinger; Debora Botequio Moretti; Dimas Tadeu Covas; Elaine Cristina Marqueze; Elaine Vieira Santos; Evandra Strazza Rodrigues; Heidge Fukumasu; Jayme Augusto de Souza-Neto; José Salvatore Leister Patané; Luiz Alcantara; Luiz Lehmann Coutinho; Maria Carolina Elias; Maurício Lacerda Nogueira; Rafael dos Santos Bezerra; Raul Machado Neto; Rejane Maria Tommasini Grotto; Ricardo Haddad; Sandra Coccuzzo Sampaio Vessoni; Simone Kashima; Svetoslav Nanev Slavov; Vincent Louis Viala |
| EPI_ISL_2445049, EPI_ISL_2445051, EPI_ISL_2445068, EPI_ISL_2445075 | INSIDE DIAGNÓSTICOS SUL PARELHEIROS | Instituto Butantan | Antonio Jorge Martins; Claudia Renata dos Santos Barros; David Schlesinger; Debora Botequio Moretti; Dimas Tadeu Covas; Elaine Cristina Marqueze; Elaine Vieira Santos; Evandra Strazza Rodrigues; Heidge Fukumasu; Jayme Augusto de Souza-Neto; José Salvatore Leister Patané; Luiz Alcantara; Luiz Lehmann Coutinho; Maria Carolina Elias; Maurício Lacerda Nogueira; Rafael dos Santos Bezerra; Raul Machado Neto; Rejane Maria Tommasini Grotto; Ricardo Haddad; Sandra Coccuzzo Sampaio Vessoni; Simone Kashima; Svetoslav Nanev Slavov; Vincent Louis Viala |
| EPI_ISL_2445046 | INSIDE LABORATÓRIOS NORTE PIRITUBA | Instituto Butantan | Antonio Jorge Martins; Claudia Renata dos Santos Barros; David Schlesinger; Debora Botequio Moretti; Dimas Tadeu Covas; Elaine Cristina Marqueze; Elaine Vieira Santos; Evandra Strazza Rodrigues; Heidge Fukumasu; Jayme Augusto de Souza-Neto; José Salvatore Leister Patané; Luiz Alcantara; Luiz Lehmann Coutinho; Maria Carolina Elias; Maurício Lacerda Nogueira; Rafael dos Santos Bezerra; Raul Machado Neto; Rejane Maria Tommasini Grotto; Ricardo Haddad; Sandra Coccuzzo Sampaio Vessoni; Simone Kashima; Svetoslav Nanev Slavov; Vincent Louis Viala |
| EPI_ISL_2445147, EPI_ISL_2473731, EPI_ISL_2473756 | INST DO CANCER ARNALDO VIEIRA DE CARVALHO | Instituto Butantan | Antonio Jorge Martins; Claudia Renata dos Santos Barros; David Schlesinger; Debora Botequio Moretti; Dimas Tadeu Covas; Elaine Cristina Marqueze; Elaine Vieira Santos; Evandra Strazza Rodrigues; Heidge Fukumasu; Jayme Augusto de Souza-Neto; José Salvatore Leister Patané; Luiz Alcantara; Luiz Lehmann Coutinho; Maria Carolina Elias; Maurício Lacerda Nogueira; Rafael dos Santos Bezerra; Raul Machado Neto; Rejane Maria Tommasini Grotto; Ricardo Haddad; Sandra Coccuzzo Sampaio Vessoni; Simone Kashima; Svetoslav Nanev Slavov; Vincent Louis Viala |
| EPI_ISL_2445564, EPI_ISL_2445565, EPI_ISL_2445567, EPI_ISL_2445568, EPI_ISL_2445569, EPI_ISL_2445570 | IRMANDADE DA SANTA CASA DE MISERICORDIA LORENA | Instituto Butantan | Antonio Jorge Martins; Claudia Renata dos Santos Barros; David Schlesinger; Debora Botequio Moretti; Dimas Tadeu Covas; Elaine Cristina Marqueze; Elaine Vieira Santos; Evandra Strazza Rodrigues; Heidge Fukumasu; Jayme Augusto de Souza-Neto; José Salvatore Leister Patané; Luiz Alcantara; Luiz Lehmann Coutinho; Maria Carolina Elias; Maurício Lacerda Nogueira; Rafael dos Santos Bezerra; Raul Machado Neto; Rejane Maria Tommasini Grotto; Ricardo Haddad; Sandra Coccuzzo Sampaio Vessoni; Simone Kashima; Svetoslav Nanev Slavov; Vincent Louis Viala |
| EPI_ISL_2828661, EPI_ISL_2828662, EPI_ISL_2828663, EPI_ISL_2828664, | Instituo Adolfo Lutz - Reginal de Ribeirao Preto | Instituto Adolfo Lutz, Interdiciplinary Procedures Center, Strategic Laboratory | Caio Vinicius Dias Lopes; Claudia Regina Gonçalves; Claudio Tavares Sacchi; Erica Valessa Ramos Gomes; Karoline Rodrigues Campos |

| | | | |
|---|---|---|---|
| EPI_ISL_2828665 | | | |
| EPI_ISL_2691151, EPI_ISL_2691152, EPI_ISL_2691153, EPI_ISL_2691154, EPI_ISL_2691155, EPI_ISL_2691156, EPI_ISL_2691159, EPI_ISL_2691160, EPI_ISL_2691161, EPI_ISL_2691162, EPI_ISL_2691163, EPI_ISL_2691164, EPI_ISL_2691165, EPI_ISL_2691166, EPI_ISL_2691167, EPI_ISL_2691168, EPI_ISL_2691169, EPI_ISL_2691170, EPI_ISL_2828654, EPI_ISL_2828655, EPI_ISL_2828656, EPI_ISL_2828657, EPI_ISL_2828658, EPI_ISL_2828659, EPI_ISL_2828660 | | | |
| see above | Instituo Adolfo Lutz Central | Instituto Adolfo Lutz, Interdiciplinary Procedures Center, Strategic Laboratory | Caio Vinicius Dias Lopes; Claudia Regina Gonçalves; Claudio Tavares Sacchi; Erica Valessa Ramos Gomes; Karoline Rodrigues Campos; Leonardo Jose Tadeu de Araujo |
| EPI_ISL_2691171, EPI_ISL_2691172, EPI_ISL_2691173, EPI_ISL_2691174, EPI_ISL_2691175, EPI_ISL_2691176, EPI_ISL_2691177, EPI_ISL_2691178, EPI_ISL_2691179, EPI_ISL_2691180, EPI_ISL_2691181, EPI_ISL_2691182, EPI_ISL_2691183, EPI_ISL_2691184, EPI_ISL_2691185, EPI_ISL_2691186, EPI_ISL_2691187, EPI_ISL_2691188, EPI_ISL_2691189, EPI_ISL_2691190, EPI_ISL_2691191, EPI_ISL_2691192, EPI_ISL_2691193, EPI_ISL_2691194, EPI_ISL_2691195, EPI_ISL_2691196, EPI_ISL_2691197, EPI_ISL_2691198, EPI_ISL_2691199, EPI_ISL_2691200, EPI_ISL_2691201, EPI_ISL_2691202, EPI_ISL_2691203, EPI_ISL_2828647, EPI_ISL_2828648, EPI_ISL_2828649, EPI_ISL_2828650, EPI_ISL_2828651, EPI_ISL_2828652, EPI_ISL_2828653 | | | |
| see above | Instituto Adolfo Lutz - Reginal de Sao José do Rio Preto | Instituto Adolfo Lutz, Interdiciplinary Procedures Center, Strategic Laboratory | Caio Vinicius Dias Lopes; Claudia Regina Gonçalves; Claudio Tavares Sacchi; Erica Valessa Ramos Gomes; Karoline Rodrigues Campos; Leonardo Jose Tadeu de Araujo |
| EPI_ISL_2691110, EPI_ISL_2691111, EPI_ISL_2691112, EPI_ISL_2691113, EPI_ISL_2691120, EPI_ISL_2919241, EPI_ISL_2919242, EPI_ISL_2919243, EPI_ISL_2919244, EPI_ISL_2919245, EPI_ISL_2919246, EPI_ISL_2919250, EPI_ISL_2919251, EPI_ISL_2919252, EPI_ISL_2919253, EPI_ISL_2919254, EPI_ISL_2919255, EPI_ISL_2919256, EPI_ISL_2919257, EPI_ISL_2919259, EPI_ISL_2919260 | | | |
| see above | Instituto Adolfo Lutz - Regional de Bauru | Instituto Adolfo Lutz, Interdiciplinary Procedures Center, Strategic Laboratory | Caio Vinicius Dias Lopes; Claudia Regina Gonçalves; Claudio Tavares Sacchi; Erica Valessa Ramos Gomes; Karoline Rodrigues Campos; Leonardo Jose Tadeu de Araujo |
| EPI_ISL_2828684, EPI_ISL_2828685, EPI_ISL_2919263 | Instituto Adolfo Lutz - Regional de Campinas | Instituto Adolfo Lutz, Interdiciplinary Procedures Center, Strategic Laboratory | Caio Vinicius Dias Lopes; Claudia Regina Gonçalves; Claudio Tavares Sacchi; Erica Valessa Ramos Gomes; Karoline Rodrigues Campos |
| EPI_ISL_2614617, EPI_ISL_2614619, EPI_ISL_2614620, EPI_ISL_2614621, EPI_ISL_2614623, EPI_ISL_2614624, EPI_ISL_2614625 | | | |
| see above | Instituto Adolfo Lutz - Regional de Presidente Prudente | Instituto Adolfo Lutz, Interdiciplinary Procedures Center, Strategic Laboratory | Caio Vinicius Dias Lopes; Claudia Regina Gonçalves; Claudio Tavares Sacchi; Erica Valessa Ramos Gomes; Karoline Rodrigues Campos; Leonardo Jose Tadeu de Araujo |
| EPI_ISL_2691124, EPI_ISL_2691125 | Instituto Adolfo Lutz - Regional de Ribeirao Preto | Instituto Adolfo Lutz, Interdiciplinary Procedures Center, Strategic Laboratory | Caio Vinicius Dias Lopes; Claudia Regina Gonçalves; Claudio Tavares Sacchi; Erica Valessa Ramos Gomes; Karoline Rodrigues Campos; Leonardo Jose Tadeu de Araujo |
| EPI_ISL_2828669, EPI_ISL_2828670, EPI_ISL_2828671, EPI_ISL_2828672, EPI_ISL_2828673, EPI_ISL_2828674, EPI_ISL_2828675, EPI_ISL_2828676, EPI_ISL_2828677, EPI_ISL_2828678, EPI_ISL_2828679, EPI_ISL_2828680, EPI_ISL_2828681, EPI_ISL_2828682, EPI_ISL_2828683 | | | |
| see above | Instituto Adolfo Lutz - Regional de Santo Andre | Instituto Adolfo Lutz, Interdiciplinary Procedures Center, Strategic Laboratory | Caio Vinicius Dias Lopes; Claudia Regina Gonçalves; Claudio Tavares Sacchi; Erica Valessa Ramos Gomes; Karoline Rodrigues Campos |
| EPI_ISL_2828698, EPI_ISL_2828699, EPI_ISL_2828701, EPI_ISL_2828702 | Instituto Adolfo Lutz - Regional de Santos | Instituto Adolfo Lutz, Interdiciplinary Procedures Center, Strategic Laboratory | Caio Vinicius Dias Lopes; Claudia Regina Gonçalves; Claudio Tavares Sacchi; Erica Valessa Ramos Gomes; Karoline Rodrigues Campos |
| EPI_ISL_2691145, EPI_ISL_2919265, EPI_ISL_2919266, EPI_ISL_2919267 | Instituto Adolfo Lutz - Regional de Sao Jose do Rio Preto | Instituto Adolfo Lutz, Interdiciplinary Procedures Center, Strategic Laboratory | Caio Vinicius Dias Lopes; Claudia Regina Gonçalves; Claudio Tavares Sacchi; Erica Valessa Ramos Gomes; Karoline Rodrigues Campos; Leonardo Jose Tadeu de Araujo |
| EPI_ISL_2691126, EPI_ISL_2691127, EPI_ISL_2691128, EPI_ISL_2691129, EPI_ISL_2691131, EPI_ISL_2691132, EPI_ISL_2691133, EPI_ISL_2691134, EPI_ISL_2691135, EPI_ISL_2691136, EPI_ISL_2691137, EPI_ISL_2691139, EPI_ISL_2691142, EPI_ISL_2691143, EPI_ISL_2691144, EPI_ISL_2756426, EPI_ISL_2756428, EPI_ISL_2756431, EPI_ISL_2919233, EPI_ISL_2919234, EPI_ISL_2919268, EPI_ISL_2919269, EPI_ISL_2919270, EPI_ISL_2919271, EPI_ISL_2919272, EPI_ISL_2919273, EPI_ISL_2919274, EPI_ISL_2919275, EPI_ISL_2919276, EPI_ISL_2919277, EPI_ISL_2919278, EPI_ISL_2919279, EPI_ISL_2919280, EPI_ISL_2919281, EPI_ISL_2919282, EPI_ISL_2919283, EPI_ISL_2919284, EPI_ISL_2919285, EPI_ISL_2919286, EPI_ISL_2919287, EPI_ISL_2919288, EPI_ISL_2919289, EPI_ISL_2919290, EPI_ISL_2919291, EPI_ISL_2919292, EPI_ISL_2919293, EPI_ISL_2919294 | | | |
| see above | Instituto Adolfo Lutz Central | Instituto Adolfo Lutz, Interdiciplinary Procedures Center, Strategic Laboratory | Caio Vinicius Dias Lopes; Claudia Regina Gonçalves; Claudio Tavares Sacchi; Erica Valessa Ramos Gomes; Karoline Rodrigues Campos; Leonardo Jose Tadeu de Araujo |
| EPI_ISL_2924058 | Instituto Adolfo Lutz Central | Instituto Adolfo Lutz, Rapid Response Center, Strategic Laboratory | Caio Vinicius Dias Lopes; Claudia Regina Gonçalves; Claudio Tavares Sacchi; Erica Valessa Ramos Gomes; Karoline Rodrigues Campos; Leonardo Jose Tadeu de Araujo |
| EPI_ISL_2241317, EPI_ISL_2241318, EPI_ISL_2241319, EPI_ISL_2241320, EPI_ISL_2241321, EPI_ISL_2241322, EPI_ISL_2241323, EPI_ISL_2241324, EPI_ISL_2241325, EPI_ISL_2241326 | | | |
| see above | Instituto Butantan | Instituto Butantan | Antonio Jorge Martins; Claudia Renata dos Santos Barros; David Schlesinger; Debora Botequio Moretti; Dimas Tadeu Covas; Elaine Cristina Marqueze; Elaine Vieira Santos; Evandra Strazza Rodrigues; Heidge Fukumasu; Jayme Augusto de Souza-Neto; José Salvatore Leister Patané; Luiz Alcantara; Luiz Lehmann Coutinho; Maria Carolina Elias; Maurício Lacerda Nogueira; Rafael dos Santos Bezerra; Raul Machado Neto; Rejane Maria Tommasini Grotto; Ricardo Haddad; Sandra Coccuzzo Sampaio Vessoni; Simone Kashima; Svetoslav Nanev Slavov; Vincent Louis Viala |
| EPI_ISL_2295428, EPI_ISL_2295429, EPI_ISL_2295430, EPI_ISL_2295431, EPI_ISL_2295432, EPI_ISL_2295433, EPI_ISL_2295434, EPI_ISL_2295435, EPI_ISL_2295436, EPI_ISL_2295437, EPI_ISL_2295438, EPI_ISL_2295439, EPI_ISL_2295440, EPI_ISL_2295441, EPI_ISL_2295442, EPI_ISL_2295443, EPI_ISL_2295444, EPI_ISL_2295445, EPI_ISL_2295446, EPI_ISL_2295447, EPI_ISL_2295448, EPI_ISL_2295449, EPI_ISL_2295450, EPI_ISL_2295451, EPI_ISL_2295452, EPI_ISL_2308610, EPI_ISL_2323128, EPI_ISL_2323129, EPI_ISL_2323130, EPI_ISL_2323131, EPI_ISL_2323132, EPI_ISL_2323133, EPI_ISL_2323134, EPI_ISL_2323135, EPI_ISL_2323136, EPI_ISL_2323137, EPI_ISL_2323138, EPI_ISL_2323139, EPI_ISL_2336150, EPI_ISL_2378644, EPI_ISL_2378645, EPI_ISL_2378646, EPI_ISL_2378647, EPI_ISL_2378648, EPI_ISL_2378649, EPI_ISL_2378650, EPI_ISL_2378651, EPI_ISL_2378652, EPI_ISL_2378653, EPI_ISL_2378654, EPI_ISL_2378655, EPI_ISL_2378656, EPI_ISL_2378657, EPI_ISL_2378658, EPI_ISL_2378659, EPI_ISL_2378660, EPI_ISL_2378661, EPI_ISL_2378662, EPI_ISL_2378663, EPI_ISL_2378664, EPI_ISL_2378665, EPI_ISL_2378666, EPI_ISL_2378667, EPI_ISL_2378668, EPI_ISL_2378669, EPI_ISL_2378670, EPI_ISL_2378671, EPI_ISL_2378672, EPI_ISL_2378673, EPI_ISL_2378674, EPI_ISL_2378675, EPI_ISL_2378676, EPI_ISL_2378677, EPI_ISL_2378678, EPI_ISL_2378679, EPI_ISL_2378680, EPI_ISL_2378681, EPI_ISL_2378682, EPI_ISL_2378683, EPI_ISL_2378684, EPI_ISL_2378685, EPI_ISL_2378686, EPI_ISL_2378687, EPI_ISL_2378688, EPI_ISL_2378689, EPI_ISL_2378690, EPI_ISL_2378691, EPI_ISL_2378692, EPI_ISL_2378693, EPI_ISL_2378694, EPI_ISL_2378695, EPI_ISL_2378696, EPI_ISL_2378697, EPI_ISL_2378698, EPI_ISL_2382534, EPI_ISL_2843903, EPI_ISL_2843904, EPI_ISL_2843905, EPI_ISL_2843906, EPI_ISL_2843907, EPI_ISL_2843908, EPI_ISL_2843909, EPI_ISL_2843910, EPI_ISL_2843911, EPI_ISL_2843912, EPI_ISL_2843913, EPI_ISL_2843914, EPI_ISL_2843915, EPI_ISL_2843916, EPI_ISL_2843917, EPI_ISL_2843918, EPI_ISL_2843919, EPI_ISL_2843920, EPI_ISL_2843921, EPI_ISL_2843922, EPI_ISL_2843923, EPI_ISL_2843924, EPI_ISL_2843925, EPI_ISL_2843926, EPI_ISL_2843927, EPI_ISL_2843928, EPI_ISL_2843929, EPI_ISL_2843930, EPI_ISL_2843931, EPI_ISL_2843932, EPI_ISL_2843933, EPI_ISL_2843934, EPI_ISL_2843935, EPI_ISL_2843936, EPI_ISL_2843937, EPI_ISL_2843938, EPI_ISL_2843939, EPI_ISL_2843940, EPI_ISL_2843941, EPI_ISL_2843942, EPI_ISL_2843943, EPI_ISL_2843944, EPI_ISL_2843945, EPI_ISL_2843946, EPI_ISL_2843947, EPI_ISL_2843948, EPI_ISL_2843949, EPI_ISL_2843950, EPI_ISL_2843951, EPI_ISL_2843952, EPI_ISL_2843953, EPI_ISL_2843954, EPI_ISL_2843955, EPI_ISL_2843956, EPI_ISL_2843957, EPI_ISL_2843958, EPI_ISL_2843959, EPI_ISL_2843960, EPI_ISL_2843961, EPI_ISL_2843962, EPI_ISL_2843963, EPI_ISL_2843964, EPI_ISL_2843965, EPI_ISL_2843966, EPI_ISL_2843967, EPI_ISL_2843968, EPI_ISL_2843969, EPI_ISL_2843970, EPI_ISL_2843971, EPI_ISL_2843972, EPI_ISL_2843973, EPI_ISL_2843974, EPI_ISL_2843975, EPI_ISL_2843976, EPI_ISL_2843977, EPI_ISL_2843978, EPI_ISL_2843979, EPI_ISL_2843980, EPI_ISL_2843981, EPI_ISL_2843982, EPI_ISL_2843983, EPI_ISL_2843984, EPI_ISL_2843985, EPI_ISL_2843986, EPI_ISL_2843987, EPI_ISL_2843988, EPI_ISL_2843989, EPI_ISL_2843990, EPI_ISL_2843991, EPI_ISL_2843992, EPI_ISL_2843993, EPI_ISL_2843994, EPI_ISL_2843995, EPI_ISL_2843996, EPI_ISL_2843997, EPI_ISL_2843998, EPI_ISL_2843999, EPI_ISL_2844000, EPI_ISL_2844001, EPI_ISL_2844002, EPI_ISL_2844003, EPI_ISL_2844004, EPI_ISL_2844005, EPI_ISL_2844006, EPI_ISL_2844007, EPI_ISL_2844008, EPI_ISL_2844009, EPI_ISL_2844010, EPI_ISL_2844011, EPI_ISL_2844012, EPI_ISL_2844013, EPI_ISL_2844014, EPI_ISL_2844015, EPI_ISL_2844016, EPI_ISL_2844017, EPI_ISL_2844018, EPI_ISL_2844019, EPI_ISL_2844020, EPI_ISL_2844021, EPI_ISL_2844022, EPI_ISL_2844023, EPI_ISL_2844024, EPI_ISL_2844025, EPI_ISL_2844026, EPI_ISL_2844027, EPI_ISL_2844028, EPI_ISL_2844029, EPI_ISL_2844030, EPI_ISL_2844031, EPI_ISL_2844032, EPI_ISL_2844033, EPI_ISL_2844034, EPI_ISL_2844035, EPI_ISL_2844036, EPI_ISL_2844037, EPI_ISL_2844038, EPI_ISL_2844039, EPI_ISL_2844040, EPI_ISL_2844041, EPI_ISL_2844042, EPI_ISL_2844043, EPI_ISL_2844044, EPI_ISL_2844045, EPI_ISL_2844046, EPI_ISL_2844047, EPI_ISL_2844048, EPI_ISL_2844049, EPI_ISL_2844050, EPI_ISL_2844051, EPI_ISL_2844052, EPI_ISL_2844053, EPI_ISL_2844054, EPI_ISL_2844055, EPI_ISL_2844056, EPI_ISL_2844057, EPI_ISL_2844058, EPI_ISL_2844059, EPI_ISL_2844060, EPI_ISL_2844061, EPI_ISL_2844062, EPI_ISL_2844063, EPI_ISL_2844064, EPI_ISL_2844065, EPI_ISL_2844066, EPI_ISL_2844067, EPI_ISL_2844068, EPI_ISL_2844069, EPI_ISL_2844070, EPI_ISL_2844071, EPI_ISL_2844072, EPI_ISL_2844073, EPI_ISL_2844074, EPI_ISL_2844075, EPI_ISL_2844076, EPI_ISL_2844077, EPI_ISL_2844078, EPI_ISL_2844079, EPI_ISL_2844080, EPI_ISL_2844081, EPI_ISL_2844082, EPI_ISL_2844083, EPI_ISL_2844084, EPI_ISL_2844085, EPI_ISL_2844086, EPI_ISL_2844087, EPI_ISL_2844088, EPI_ISL_2844089, EPI_ISL_2844090, EPI_ISL_2844091, EPI_ISL_2844092, EPI_ISL_2844093, EPI_ISL_2844094, EPI_ISL_2844095, EPI_ISL_2844096, EPI_ISL_2844097, EPI_ISL_2844098, EPI_ISL_2844099, EPI_ISL_2844100, EPI_ISL_2844101, EPI_ISL_2844102, EPI_ISL_2844103, EPI_ISL_2844104, EPI_ISL_2844105, EPI_ISL_2844106, EPI_ISL_2844107, EPI_ISL_2844108, EPI_ISL_2844109, EPI_ISL_2844110, EPI_ISL_2844111, EPI_ISL_2844112, EPI_ISL_2844113, EPI_ISL_2844114, EPI_ISL_2844115, EPI_ISL_2844116, EPI_ISL_2844117, EPI_ISL_2844118, EPI_ISL_2844119, EPI_ISL_2844120, EPI_ISL_2844121, EPI_ISL_2844122, EPI_ISL_2844123, EPI_ISL_2844124, EPI_ISL_2844125, EPI_ISL_2844126, EPI_ISL_2844127, EPI_ISL_2844128, EPI_ISL_2844129, EPI_ISL_2844130, EPI_ISL_2844131, EPI_ISL_2844132, EPI_ISL_2844133, EPI_ISL_2844134, EPI_ISL_2844135, EPI_ISL_2844136, EPI_ISL_2844137, EPI_ISL_2844138, EPI_ISL_2844139, EPI_ISL_2844140, EPI_ISL_2844141, EPI_ISL_2844142, EPI_ISL_2844143, EPI_ISL_2844144, EPI_ISL_2844145, EPI_ISL_2844146, EPI_ISL_2844147, EPI_ISL_2844148, EPI_ISL_2844149, EPI_ISL_2844150, EPI_ISL_2844151, EPI_ISL_2844152, EPI_ISL_2844153, EPI_ISL_2844154, EPI_ISL_2844155, EPI_ISL_2844156, EPI_ISL_2844157, EPI_ISL_2844158, EPI_ISL_2844159, EPI_ISL_2844160, EPI_ISL_2844161, EPI_ISL_2844162, EPI_ISL_2844163, EPI_ISL_2844164, EPI_ISL_2844165, EPI_ISL_2844166, EPI_ISL_2844167, EPI_ISL_2844168, EPI_ISL_2844169, EPI_ISL_2844170, EPI_ISL_2844171, EPI_ISL_2844172, EPI_ISL_2844173, EPI_ISL_2844174, EPI_ISL_2844175, EPI_ISL_2844176, EPI_ISL_2844177, EPI_ISL_2844178, EPI_ISL_2844179, EPI_ISL_2844180, EPI_ISL_2844181, EPI_ISL_2844182, EPI_ISL_2844183, EPI_ISL_2844184, EPI_ISL_2844185, EPI_ISL_2844186, EPI_ISL_2844187, EPI_ISL_2844188, EPI_ISL_2844189, EPI_ISL_2844190, EPI_ISL_2844191, EPI_ISL_2844192, EPI_ISL_2844193, EPI_ISL_2844194, EPI_ISL_2844195 | | | |
| see above | Instituto de Biotecnologia - UNESP-Botucatu-SP | Instituto de Biotecnologia - UNESP-Botucatu-SP | Cecília Artico Banho; Cíntia Bittar; Fábio Sossai Possebon; Guilherme Campos; Helena Lage Ferreira; Jorge A. Petroli Marchesi; João Pessoa Araújo Jr.; Leila Sabrina Ullmann; Lívia Sacchetto; Maisa C. Pereira Parra; Marília Moraes; Maurício L. Nogueira; Paula Rahal; Paulo Inacio da Costa |
| EPI_ISL_2445492, EPI_ISL_2445493, EPI_ISL_2445494, EPI_ISL_2445495, EPI_ISL_2445496, EPI_ISL_2445497, EPI_ISL_2445498, EPI_ISL_2462323, EPI_ISL_2473783, EPI_ISL_2493151, EPI_ISL_2493165, EPI_ISL_2493185, EPI_ISL_2493302, EPI_ISL_2493646, EPI_ISL_2493647, EPI_ISL_2493648, EPI_ISL_2494141, EPI_ISL_2494142, EPI_ISL_2494143 | | | |
| see above | LABORATORIO DE FRANCA | Instituto Butantan | Antonio Jorge Martins; Claudia Renata dos Santos Barros; David Schlesinger; Debora Botequio Moretti; Dimas Tadeu Covas; Elaine Cristina Marqueze; Elaine Vieira Santos; Evandra Strazza Rodrigues; Heidge Fukumasu; Jayme Augusto de Souza-Neto; José Salvatore Leister Patané; Luiz Alcantara; Luiz Lehmann Coutinho; Maria Carolina Elias; Maurício Lacerda Nogueira; Rafael dos Santos Bezerra; Raul Machado Neto; Rejane Maria Tommasini Grotto; Ricardo Haddad; Sandra Coccuzzo Sampaio Vessoni; Simone Kashima; Svetoslav Nanev Slavov; Vincent Louis Viala |
| EPI_ISL_2445560, EPI_ISL_2445561, EPI_ISL_2445562, EPI_ISL_2445563, EPI_ISL_2445566, EPI_ISL_2473797, EPI_ISL_2473798, EPI_ISL_2938095 | | | |
| see above | LABORATORIO DR PAULO EMILIO DALESSANDRO PINDAMONHANGABA | Instituto Butantan | Antonio Jorge Martins; Claudia Renata dos Santos Barros; David Schlesinger; Debora Botequio Moretti; Dimas Tadeu Covas; Elaine Cristina Marqueze; Elaine Vieira Santos; Evandra Strazza Rodrigues; Heidge Fukumasu; Jayme Augusto de Souza-Neto; José Salvatore Leister Patané; Luiz Alcantara; Luiz Lehmann Coutinho; Maria Carolina Elias; Maurício Lacerda Nogueira; Rafael dos Santos Bezerra; Raul Machado Neto; Rejane Maria Tommasini Grotto; Ricardo Haddad; Sandra Coccuzzo Sampaio Vessoni; Simone Kashima; Svetoslav Nanev Slavov; Vincent Louis Viala |
| EPI_ISL_2445298, EPI_ISL_2445299, EPI_ISL_2445301, EPI_ISL_2445304 | LABORATORIO LOCAL DE ITAPECERICA DA SERRA | Instituto Butantan | Antonio Jorge Martins; Claudia Renata dos Santos Barros; David Schlesinger; Debora Botequio Moretti; Dimas Tadeu Covas; Elaine Cristina Marqueze; Elaine Vieira Santos; Evandra Strazza Rodrigues; Heidge Fukumasu; Jayme Augusto de Souza-Neto; José Salvatore Leister Patané; Luiz Alcantara; Luiz Lehmann Coutinho; Maria Carolina Elias; Maurício Lacerda Nogueira; Rafael dos Santos Bezerra; Raul Machado Neto; Rejane Maria Tommasini Grotto; Ricardo Haddad; Sandra Coccuzzo Sampaio Vessoni; Simone Kashima; Svetoslav Nanev Slavov; Vincent Louis Viala |
| EPI_ISL_2445331, EPI_ISL_2445335, EPI_ISL_2445337, EPI_ISL_2445343, EPI_ISL_2445344, EPI_ISL_2445347, EPI_ISL_2445349, EPI_ISL_2445350, EPI_ISL_2445352, EPI_ISL_2445353, EPI_ISL_2445355, EPI_ISL_2445357, EPI_ISL_2445362, EPI_ISL_2473692, EPI_ISL_2473700, EPI_ISL_2473779, EPI_ISL_2473781, EPI_ISL_2493684, EPI_ISL_2493702, EPI_ISL_2493703 | | | |
| see above | LABORATORIO MUNICIPAL DE ANALISES CLINICAS DE ITANHAEM | Instituto Butantan | Antonio Jorge Martins; Claudia Renata dos Santos Barros; David Schlesinger; Debora Botequio Moretti; Dimas Tadeu Covas; Elaine Cristina Marqueze; Elaine Vieira Santos; Evandra Strazza Rodrigues; Heidge Fukumasu; Jayme Augusto de Souza-Neto; José Salvatore Leister Patané; Luiz Alcantara; Luiz Lehmann Coutinho; Maria Carolina Elias; Maurício Lacerda Nogueira; Rafael dos Santos Bezerra; Raul Machado Neto; Rejane Maria Tommasini Grotto; Ricardo Haddad; Sandra Coccuzzo Sampaio Vessoni; Simone Kashima; Svetoslav Nanev Slavov; Vincent Louis Viala |
| EPI_ISL_2493173, EPI_ISL_2493368, EPI_ISL_2493369, EPI_ISL_2493379, EPI_ISL_2493380, EPI_ISL_2493381, EPI_ISL_2493397, EPI_ISL_2493398, EPI_ISL_2493403, EPI_ISL_2493440, EPI_ISL_2494151, EPI_ISL_2494166, EPI_ISL_2494176, EPI_ISL_2494184, EPI_ISL_2494299 | | | |
| see above | LABORATORIO MUNICIPAL DE ANALISES CLINICAS DE RIO CLARO | Instituto Butantan | Antonio Jorge Martins; Claudia Renata dos Santos Barros; David Schlesinger; Debora Botequio Moretti; Dimas Tadeu Covas; Elaine Cristina Marqueze; Elaine Vieira Santos; Evandra Strazza Rodrigues; Heidge Fukumasu; Jayme Augusto de Souza-Neto; José Salvatore Leister Patané; Luiz Alcantara; Luiz Lehmann Coutinho; Maria Carolina Elias; Maurício Lacerda Nogueira; Rafael dos Santos Bezerra; Raul Machado Neto; Rejane Maria Tommasini Grotto; Ricardo Haddad; Sandra Coccuzzo Sampaio Vessoni; Simone Kashima; Svetoslav Nanev Slavov; Vincent Louis Viala |
| EPI_ISL_2383033, EPI_ISL_2493150, EPI_ISL_2493186, EPI_ISL_2493190, EPI_ISL_2493449, EPI_ISL_2493451, EPI_ISL_2493453, EPI_ISL_2493454, EPI_ISL_2493455, EPI_ISL_2493649, EPI_ISL_2493653, EPI_ISL_2493696, EPI_ISL_2494154, EPI_ISL_2494157, EPI_ISL_2494163, EPI_ISL_2494180, EPI_ISL_2494182, EPI_ISL_2494193, EPI_ISL_2494200, EPI_ISL_2494338 | | | |
| see above | LABORATORIO MUNICIPAL DE PIRACICABA | Instituto Butantan | Antonio Jorge Martins; Claudia Renata dos Santos Barros; David Schlesinger; Debora Botequio Moretti; Dimas Tadeu Covas; Elaine Cristina Marqueze; Elaine Vieira Santos; Evandra Strazza Rodrigues; Heidge Fukumasu; Jayme Augusto de Souza-Neto; José Salvatore Leister Patané; Luiz Alcantara; Luiz Lehmann |

| | | | |
|---|---|---|---|
| | | | Coutinho; Maria Carolina Elias; Maurício Lacerda Nogueira; Rafael dos Santos Bezerra; Raul Machado Neto; Rejane Maria Tommasini Grotto; Ricardo Haddad; Sandra Coccuzzo Sampaio Vessoni; Simone Kashima; Svetoslav Nanev Slavov; Vincent Louis Viala |
| EPI_ISL_2493241, EPI_ISL_2493242, EPI_ISL_2493881, EPI_ISL_2493884, EPI_ISL_2493885, EPI_ISL_2493886, EPI_ISL_2493887, EPI_ISL_2493888, EPI_ISL_2493906, EPI_ISL_2493908, EPI_ISL_2493948, EPI_ISL_2494323, EPI_ISL_2500987 | | | |
| see above | LABORATORIO MUNICIPAL DE SUZANO | Instituto Butantan | Antonio Jorge Martins; Claudia Renata dos Santos Barros; David Schlesinger; Debora Botequio Moretti; Dimas Tadeu Covas; Elaine Cristina Marqueze; Elaine Vieira Santos; Evandra Strazza Rodrigues; Heidge Fukumasu; Jayme Augusto de Souza-Neto; José Salvatore Leister Patané; Luiz Alcantara; Luiz Lehmann Coutinho; Maria Carolina Elias; Maurício Lacerda Nogueira; Rafael dos Santos Bezerra; Raul Machado Neto; Rejane Maria Tommasini Grotto; Ricardo Haddad; Sandra Coccuzzo Sampaio Vessoni; Simone Kashima; Svetoslav Nanev Slavov; Vincent Louis Viala |
| EPI_ISL_2209355, EPI_ISL_2210259, EPI_ISL_2345691, EPI_ISL_2345741, EPI_ISL_2345785 | LABORATORIO MUNICIPAL DE SUZANO | Instituto Butantan / Mendelics | Antonio Jorge Martins; Claudia Renata dos Santos Barros; David Schlesinger; Debora Botequio Moretti; Dimas Tadeu Covas; Elaine Cristina Marqueze; Elaine Vieira Santos; Evandra Strazza Rodrigues; Heidge Fukumasu; Jayme Augusto de Souza-Neto; José Salvatore Leister Patané; Luiz Alcantara; Luiz Lehmann Coutinho; Maria Carolina Elias; Maurício Lacerda Nogueira; Rafael dos Santos Bezerra; Raul Machado Neto; Rejane Maria Tommasini Grotto; Ricardo Haddad; Sandra Coccuzzo Sampaio Vessoni; Simone Kashima; Svetoslav Nanev Slavov; Vincent Louis Viala |
| EPI_ISL_2488814, EPI_ISL_2488815, EPI_ISL_2488816, EPI_ISL_2488817, EPI_ISL_2488818, EPI_ISL_2488819 | LACEN - Laboratório Central de Saúde Pública do Para | Evandro Chagas Institute | A.M.; Barbagelata; E.C.; E.M.A.; Ferreira; J.A.; Junior; K.C.; L.C.; L.S.; M.C.; P.S.; Pinheiro; Santos; Silva; Sousa; Sousa Junior; W.D.C.; da Silva |
| EPI_ISL_2691206, EPI_ISL_2691207, EPI_ISL_2691209, EPI_ISL_2691210, EPI_ISL_2691211 | LACEN do Estado de Goias | Instituto Adolfo Lutz, Interdiciplinary Procedures Center, Strategic Laboratory | Caio Vinicius Dias Lopes; Claudia Regina Gonçalves; Claudio Tavares Sacchi; Erica Valessa Ramos Gomes; Karoline Rodrigues Campos; Leonardo Jose Tadeu de Araujo |
| EPI_ISL_2894547, EPI_ISL_2894548, EPI_ISL_2896232, EPI_ISL_2896233, EPI_ISL_2896234, EPI_ISL_2896235, EPI_ISL_2896236, EPI_ISL_2896237, EPI_ISL_2896238, EPI_ISL_2896239, EPI_ISL_2896240, EPI_ISL_2896241 | | | |
| see above | LACEN do Estado de Mato Grosso | Instituto Adolfo Lutz, Interdiciplinary Procedures Center, Strategic Laboratory | Caio Vinicius Dias Lopes; Claudia Regina Gonçalves; Claudio Tavares Sacchi; Karoline Rodrigues Campos |
| EPI_ISL_2919205, EPI_ISL_2919206, EPI_ISL_2919207, EPI_ISL_2919208, EPI_ISL_2919209, EPI_ISL_2919211, EPI_ISL_2919212, EPI_ISL_2919213, EPI_ISL_2919214, EPI_ISL_2919215 | | | |
| see above | LACEN do Estado de Tocantins | Instituto Adolfo Lutz, Interdiciplinary Procedures Center, Strategic Laboratory | Caio Vinicius Dias Lopes; Claudia Regina Gonçalves; Claudio Tavares Sacchi; Erica Valessa Ramos Gomes; Karoline Rodrigues Campos |
| EPI_ISL_2821274 | LACEN/PE | WallauLab on behalf of Fiocruz COVID-19 Genomic Surveillance Network | Alexandre Freitas da Silva; Antonio Mauro Rezende; Constância Flávia Junqueira Ayres; Cássia Docena; Duschinka Ribeiro Duarte Guedes; Elisama Helvecio; Filipe Zimmer Dezordi; Gabriel Luz Wallau; Larissa Krokovsky; Laís Ceschini Machado; Marcelo Henrique dos Santos Paiva; Matheus Filgueira Bezerra; Sinval Pinto Brandão Filho |
| EPI_ISL_2674368 | LATE - Laboratório de Técnicas Especiais - Hospital Israelita Albert Einstein | LATE - Laboratório de Técnicas Especiais - Hospital Israelita Albert Einstein | Alexandre Hideaki Takara; Ana Paula Moreira Salles; Anelisie da Silva Santos; Deyvid Amgarten; Fernanda de Mello Malta; João Renato Rebello Pinho; Marcio Anunciacao Menezes; Pedro Henrique Sebe Rodrigues; Raquel Riyuzo |
| EPI_ISL_2645421, EPI_ISL_2645423, EPI_ISL_2645424, EPI_ISL_2645425, EPI_ISL_2645426, EPI_ISL_2645427, EPI_ISL_2645428, EPI_ISL_2645429, EPI_ISL_2645430, EPI_ISL_2645431, EPI_ISL_2645432, EPI_ISL_2645433, EPI_ISL_2645434, EPI_ISL_2645435, EPI_ISL_2645436, EPI_ISL_2645437, EPI_ISL_2645438, EPI_ISL_2645439, EPI_ISL_2731453, EPI_ISL_2731454, EPI_ISL_2731455, EPI_ISL_2731456, EPI_ISL_2863578, EPI_ISL_2863580, EPI_ISL_2863582, EPI_ISL_2863706, EPI_ISL_2863707, EPI_ISL_2863709, EPI_ISL_2863711, EPI_ISL_2863801, EPI_ISL_2863802, EPI_ISL_2863803, EPI_ISL_2863804, EPI_ISL_2863805, EPI_ISL_2863806, EPI_ISL_2863807, EPI_ISL_2863808, EPI_ISL_2863809, EPI_ISL_2863810, EPI_ISL_2863811, EPI_ISL_2863812, EPI_ISL_2863813, EPI_ISL_2863814, EPI_ISL_2863815, EPI_ISL_2863816, EPI_ISL_2863817, EPI_ISL_2863818, EPI_ISL_2863819, EPI_ISL_2863820, EPI_ISL_2863821, EPI_ISL_2863822, EPI_ISL_2863823, EPI_ISL_2863824, EPI_ISL_2863825, EPI_ISL_2863826, EPI_ISL_2863827, EPI_ISL_2863828, EPI_ISL_2863829, EPI_ISL_2863830, EPI_ISL_2863831, EPI_ISL_2863832, EPI_ISL_2863833, EPI_ISL_2863834, EPI_ISL_2863835, EPI_ISL_2863836, EPI_ISL_2863837, EPI_ISL_2863838, EPI_ISL_2863839, EPI_ISL_2863840, EPI_ISL_2863841, EPI_ISL_2863842, EPI_ISL_2863843, EPI_ISL_2863844, EPI_ISL_2863845, EPI_ISL_2863846, EPI_ISL_2863847, EPI_ISL_2863848, EPI_ISL_2863849, EPI_ISL_2863850, EPI_ISL_2863851, EPI_ISL_2863852, EPI_ISL_2863853, EPI_ISL_2863854, EPI_ISL_2863892, EPI_ISL_2863894 | | | |
| see above | Laboratorio Central de Saude Publica do Estado do Para (LACEN/PA) | Laboratory of Respiratory Viruses and Measles, Oswaldo Cruz Institute, FIOCRUZ | Agatha Cristinne Prudencio; Agatha Cristinne Prudencio Soares; Alice Sampaio Rocha; Ana Carolina Mendonca; Anna Carolina Paixao; Elisa Cavalcante Pereira; Fernando Motta; Ighor Leonardo Arantes Gomes; Luciana Appolinario; Marilda Siqueira on behalf of the Fiocruz COVID-19 Genomic Surveillance Network; Paola Resende; Renata Serrano Lopes; Taina Venas; Valnete Andrade |
| EPI_ISL_2385464, EPI_ISL_2385468, EPI_ISL_2385469, EPI_ISL_2385470, EPI_ISL_2385471, EPI_ISL_2385472, EPI_ISL_2385473, EPI_ISL_2385474, EPI_ISL_2385475, EPI_ISL_2385476, EPI_ISL_2385477, EPI_ISL_2385488, EPI_ISL_2385489, EPI_ISL_2385490, EPI_ISL_2385497, EPI_ISL_2385498, EPI_ISL_2385499, EPI_ISL_2385500, EPI_ISL_2385501, EPI_ISL_2385502, EPI_ISL_2385503, EPI_ISL_2385504, EPI_ISL_2385505, EPI_ISL_2385508, EPI_ISL_2385509, EPI_ISL_2385512, EPI_ISL_2385514, EPI_ISL_2385515, EPI_ISL_2385516, EPI_ISL_2385518, EPI_ISL_2385519, EPI_ISL_2385520, EPI_ISL_2385521, EPI_ISL_2385522, EPI_ISL_2385523, EPI_ISL_2385524, EPI_ISL_2385525, EPI_ISL_2385526, EPI_ISL_2385527, EPI_ISL_2385528, EPI_ISL_2385529, EPI_ISL_2385530, EPI_ISL_2385531, EPI_ISL_2385532, EPI_ISL_2385533, EPI_ISL_2385534, EPI_ISL_2385535, EPI_ISL_2385536, EPI_ISL_2385538, EPI_ISL_2385539, EPI_ISL_2385540, EPI_ISL_2385541, EPI_ISL_2385542, EPI_ISL_2385543, EPI_ISL_2385544, EPI_ISL_2385545, EPI_ISL_2385546, EPI_ISL_2385547, EPI_ISL_2385548, EPI_ISL_2385549, EPI_ISL_2385550, EPI_ISL_2385551, EPI_ISL_2385552, EPI_ISL_2385553, EPI_ISL_2385554, EPI_ISL_2385555, EPI_ISL_2385556, EPI_ISL_2385557, EPI_ISL_2385558, EPI_ISL_2385559, EPI_ISL_2385560, EPI_ISL_2385561, EPI_ISL_2385562, EPI_ISL_2385563, EPI_ISL_2385564, EPI_ISL_2385565, EPI_ISL_2385566, EPI_ISL_2385567, EPI_ISL_2385568, EPI_ISL_2385569, EPI_ISL_2385570, EPI_ISL_2385571, EPI_ISL_2385572, EPI_ISL_2385573, EPI_ISL_2385574, EPI_ISL_2385575, EPI_ISL_2385576, EPI_ISL_2385577, EPI_ISL_2385578, EPI_ISL_2385579, EPI_ISL_2385580, EPI_ISL_2385581, EPI_ISL_2385582, EPI_ISL_2385583, EPI_ISL_2385584, EPI_ISL_2385585, EPI_ISL_2385586, EPI_ISL_2385587, EPI_ISL_2385588, EPI_ISL_2385589, EPI_ISL_2385590, EPI_ISL_2385591, EPI_ISL_2385592, EPI_ISL_2385593, EPI_ISL_2385594, EPI_ISL_2385595, EPI_ISL_2385596, EPI_ISL_2385597, EPI_ISL_2385598, EPI_ISL_2385599, EPI_ISL_2385600, EPI_ISL_2385601, EPI_ISL_2385602, EPI_ISL_2385603, EPI_ISL_2385604, EPI_ISL_2385605, EPI_ISL_2385606, EPI_ISL_2385607, EPI_ISL_2385608, EPI_ISL_2385609, EPI_ISL_2385610, EPI_ISL_2385611, EPI_ISL_2385612, EPI_ISL_2385613, EPI_ISL_2385614, EPI_ISL_2385615, EPI_ISL_2385616, EPI_ISL_2385617, EPI_ISL_2385618, EPI_ISL_2385619, EPI_ISL_2385620, EPI_ISL_2385621, EPI_ISL_2385622, EPI_ISL_2385623, EPI_ISL_2385624, EPI_ISL_2385625, EPI_ISL_2385626, EPI_ISL_2385627, EPI_ISL_2385628, EPI_ISL_2385629, EPI_ISL_2385630, EPI_ISL_2385631, EPI_ISL_2385632, EPI_ISL_2385633, EPI_ISL_2385634, EPI_ISL_2385635, EPI_ISL_2385636, EPI_ISL_2385637, EPI_ISL_2385638, EPI_ISL_2385639, EPI_ISL_2385640, EPI_ISL_2385641, EPI_ISL_2385642, EPI_ISL_2385643, EPI_ISL_2385644, EPI_ISL_2385645, EPI_ISL_2385646, EPI_ISL_2385647, EPI_ISL_2385648, EPI_ISL_2385649, EPI_ISL_2385650, EPI_ISL_2385651, EPI_ISL_2385652, EPI_ISL_2385653, EPI_ISL_2385654, EPI_ISL_2385655, EPI_ISL_2385656, EPI_ISL_2385657, EPI_ISL_2385658, EPI_ISL_2385659, EPI_ISL_2385660, EPI_ISL_2385661, EPI_ISL_2385662, EPI_ISL_2385663, EPI_ISL_2385664, EPI_ISL_2385665, EPI_ISL_2385666, EPI_ISL_2385667, EPI_ISL_2385668, EPI_ISL_2534862, EPI_ISL_2534863, EPI_ISL_2534864, EPI_ISL_2534865, EPI_ISL_2534866, EPI_ISL_2534867, EPI_ISL_2534868, EPI_ISL_2534869, EPI_ISL_2534870, EPI_ISL_2534871, EPI_ISL_2534872, EPI_ISL_2534873, EPI_ISL_2534874, EPI_ISL_2534875, EPI_ISL_2534876, EPI_ISL_2534877, EPI_ISL_2534878, EPI_ISL_2534879, EPI_ISL_2534880, EPI_ISL_2534881, EPI_ISL_2534882, EPI_ISL_2534883, EPI_ISL_2534884, EPI_ISL_2534885, EPI_ISL_2534886, EPI_ISL_2534887, EPI_ISL_2534888, EPI_ISL_2534889, EPI_ISL_2534890, EPI_ISL_2534891, EPI_ISL_2534892, EPI_ISL_2534893, EPI_ISL_2534894, EPI_ISL_2534895, EPI_ISL_2534896, EPI_ISL_2534897, EPI_ISL_2534898, EPI_ISL_2534899, EPI_ISL_2534900, EPI_ISL_2534901, EPI_ISL_2534902, EPI_ISL_2534903, EPI_ISL_2534904, EPI_ISL_2534905, EPI_ISL_2534906, EPI_ISL_2534907, EPI_ISL_2534908, EPI_ISL_2534909, EPI_ISL_2534910, EPI_ISL_2534911, EPI_ISL_2534912, EPI_ISL_2534913, EPI_ISL_2534914, EPI_ISL_2534915, EPI_ISL_2534916, EPI_ISL_2534917, EPI_ISL_2534918, EPI_ISL_2534919, EPI_ISL_2534920, EPI_ISL_2534921, EPI_ISL_2534922, EPI_ISL_2534923, EPI_ISL_2534924, EPI_ISL_2534925, EPI_ISL_2534926, EPI_ISL_2534927, EPI_ISL_2534928, EPI_ISL_2534929, EPI_ISL_2534930, EPI_ISL_2534931, EPI_ISL_2534932, EPI_ISL_2534933, EPI_ISL_2534934, EPI_ISL_2534935, EPI_ISL_2534936, EPI_ISL_2534937, EPI_ISL_2534938, EPI_ISL_2534939, EPI_ISL_2534940, EPI_ISL_2534941, EPI_ISL_2534942, EPI_ISL_2534943, EPI_ISL_2534944, EPI_ISL_2534945, EPI_ISL_2534946, EPI_ISL_2534947, EPI_ISL_2534948, EPI_ISL_2534949, EPI_ISL_2534950, EPI_ISL_2534951, EPI_ISL_2534952, EPI_ISL_2534953, EPI_ISL_2534954, EPI_ISL_2534955, EPI_ISL_2534956, EPI_ISL_2534957, EPI_ISL_2534958, EPI_ISL_2534959, EPI_ISL_2534960, EPI_ISL_2534961, EPI_ISL_2534962, EPI_ISL_2534963, EPI_ISL_2534964, EPI_ISL_2534965, EPI_ISL_2534966, EPI_ISL_2534967, EPI_ISL_2534968, EPI_ISL_2534969, EPI_ISL_2534970, EPI_ISL_2534971, EPI_ISL_2534972, EPI_ISL_2534973, EPI_ISL_2534974, EPI_ISL_2534975, EPI_ISL_2534976, EPI_ISL_2534977, EPI_ISL_2534978, EPI_ISL_2534979, EPI_ISL_2534980, EPI_ISL_2534981, EPI_ISL_2534982, EPI_ISL_2534983, EPI_ISL_2534984, EPI_ISL_2534985, EPI_ISL_2534986, EPI_ISL_2534987, EPI_ISL_2534988, EPI_ISL_2534989, EPI_ISL_2534990, EPI_ISL_2534991, EPI_ISL_2534992, EPI_ISL_2534993, EPI_ISL_2534994, EPI_ISL_2534995, EPI_ISL_2534996, EPI_ISL_2534997, EPI_ISL_2534998, EPI_ISL_2534999, EPI_ISL_2535000, EPI_ISL_2535001, EPI_ISL_2535002, EPI_ISL_2535003, EPI_ISL_2535004, EPI_ISL_2535005, EPI_ISL_2535006, EPI_ISL_2535007, EPI_ISL_2535008, EPI_ISL_2535009, EPI_ISL_2535010, EPI_ISL_2535011, EPI_ISL_2535012, EPI_ISL_2535013, EPI_ISL_2535014, EPI_ISL_2535015, EPI_ISL_2535016, EPI_ISL_2535017, EPI_ISL_2535018, EPI_ISL_2535019, EPI_ISL_2535020, EPI_ISL_2535021, EPI_ISL_2535022, EPI_ISL_2535023, EPI_ISL_2535024, EPI_ISL_2535025, EPI_ISL_2535026, EPI_ISL_2535027, EPI_ISL_2535028, EPI_ISL_2535029, EPI_ISL_2535030, EPI_ISL_2535031, EPI_ISL_2535032, EPI_ISL_2535033, EPI_ISL_2535034, EPI_ISL_2535035, EPI_ISL_2535036, EPI_ISL_2535037, EPI_ISL_2535038, EPI_ISL_2535039, EPI_ISL_2535040, EPI_ISL_2535041, EPI_ISL_2535042, EPI_ISL_2535043, EPI_ISL_2535044, EPI_ISL_2535045, EPI_ISL_2535046, EPI_ISL_2535047, EPI_ISL_2535048, EPI_ISL_2535049, EPI_ISL_2535050, EPI_ISL_2535051, EPI_ISL_2535052, EPI_ISL_2535053, EPI_ISL_2535054, EPI_ISL_2535055, EPI_ISL_2535056, EPI_ISL_2535057, EPI_ISL_2535058, EPI_ISL_2535059, EPI_ISL_2535060, EPI_ISL_2535061, EPI_ISL_2535062, EPI_ISL_2535063, EPI_ISL_2535064, EPI_ISL_2535065, EPI_ISL_2535066, EPI_ISL_2535067, EPI_ISL_2535068, EPI_ISL_2535069, EPI_ISL_2535070, EPI_ISL_2535071, EPI_ISL_2535072, EPI_ISL_2535073, EPI_ISL_2535074, EPI_ISL_2535075, EPI_ISL_2535076, EPI_ISL_2535077, EPI_ISL_2535078, EPI_ISL_2535079, EPI_ISL_2535080, EPI_ISL_2535081, EPI_ISL_2535082, EPI_ISL_2535083, EPI_ISL_2535084, EPI_ISL_2535085, EPI_ISL_2535086, EPI_ISL_2535087, EPI_ISL_2535088, EPI_ISL_2535089, EPI_ISL_2535090, EPI_ISL_2535091, EPI_ISL_2535092, EPI_ISL_2535093, EPI_ISL_2535094, EPI_ISL_2535095, EPI_ISL_2535096, EPI_ISL_2535097, EPI_ISL_2535098, EPI_ISL_2535099, EPI_ISL_2535100, EPI_ISL_2535101, EPI_ISL_2535102, EPI_ISL_2535103, EPI_ISL_2535104, EPI_ISL_2535105, EPI_ISL_2535106, EPI_ISL_2535107, EPI_ISL_2535108, EPI_ISL_2535109, EPI_ISL_2535110, EPI_ISL_2691315, EPI_ISL_2691316, EPI_ISL_2691317, EPI_ISL_2691318, EPI_ISL_2691319, EPI_ISL_2691320, EPI_ISL_2691321, EPI_ISL_2691322, EPI_ISL_2691323, EPI_ISL_2691324, EPI_ISL_2691325, EPI_ISL_2691326, EPI_ISL_2691327, EPI_ISL_2691328, EPI_ISL_2691329, EPI_ISL_2691330, EPI_ISL_2691331, EPI_ISL_2691332, EPI_ISL_2691333, EPI_ISL_2691334, EPI_ISL_2691335, EPI_ISL_2691336, EPI_ISL_2691337, EPI_ISL_2691338, EPI_ISL_2691339, EPI_ISL_2691340, EPI_ISL_2691341, EPI_ISL_2691342, EPI_ISL_2691343, EPI_ISL_2691344, EPI_ISL_2691345, EPI_ISL_2691346, EPI_ISL_2691347, EPI_ISL_2691348, EPI_ISL_2691349, EPI_ISL_2691350, EPI_ISL_2691351, EPI_ISL_2691352, EPI_ISL_2691353, EPI_ISL_2691354, EPI_ISL_2691355, EPI_ISL_2691356, EPI_ISL_2691357, EPI_ISL_2691358, EPI_ISL_2691359, EPI_ISL_2691360, EPI_ISL_2691361, EPI_ISL_2691362, EPI_ISL_2691363, EPI_ISL_2691364, EPI_ISL_2691365, EPI_ISL_2691366, EPI_ISL_2691367, EPI_ISL_2691368, EPI_ISL_2691369, EPI_ISL_2691370, EPI_ISL_2691371, EPI_ISL_2691372, EPI_ISL_2691373, EPI_ISL_2691374, EPI_ISL_2691375, EPI_ISL_2691376, EPI_ISL_2691377, EPI_ISL_2691378, EPI_ISL_2691379, EPI_ISL_2691380, EPI_ISL_2691381, EPI_ISL_2691382, EPI_ISL_2691383, EPI_ISL_2691384, EPI_ISL_2691385, EPI_ISL_2691386, EPI_ISL_2691387, EPI_ISL_2691388, EPI_ISL_2691389, EPI_ISL_2691390, EPI_ISL_2691391, EPI_ISL_2691392, EPI_ISL_2691393, EPI_ISL_2691394, EPI_ISL_2691395, EPI_ISL_2691396, EPI_ISL_2691397, EPI_ISL_2691398, EPI_ISL_2691399, EPI_ISL_2691400, EPI_ISL_2691401, EPI_ISL_2691402, EPI_ISL_2691403, EPI_ISL_2691404, EPI_ISL_2691405, EPI_ISL_2691406, EPI_ISL_2691407, EPI_ISL_2691408, EPI_ISL_2691409, EPI_ISL_2691410, EPI_ISL_2691411, EPI_ISL_2691412, EPI_ISL_2691413, EPI_ISL_2691414, EPI_ISL_2691415, EPI_ISL_2691416, EPI_ISL_2691417, EPI_ISL_2691418, EPI_ISL_2691419, EPI_ISL_2691420, EPI_ISL_2691421, EPI_ISL_2691422, EPI_ISL_2691423, EPI_ISL_2691424, EPI_ISL_2691425, EPI_ISL_2691426, EPI_ISL_2691427, EPI_ISL_2691428, EPI_ISL_2691429, EPI_ISL_2691430, EPI_ISL_2691431, EPI_ISL_2691432, EPI_ISL_2691433, EPI_ISL_2691434, EPI_ISL_2691435, EPI_ISL_2691436, EPI_ISL_2691437, EPI_ISL_2691438, EPI_ISL_2691439, EPI_ISL_2691440, EPI_ISL_2691441, EPI_ISL_2691442, EPI_ISL_2691443, EPI_ISL_2691444, EPI_ISL_2691445, EPI_ISL_2691446, EPI_ISL_2691447, EPI_ISL_2691448, EPI_ISL_2691449, EPI_ISL_2691450, EPI_ISL_2691451, EPI_ISL_2691452, EPI_ISL_2691453, EPI_ISL_2691454, EPI_ISL_2691455, EPI_ISL_2691456, EPI_ISL_2691457, EPI_ISL_2691458, EPI_ISL_2691459, EPI_ISL_2691460, EPI_ISL_2691461, EPI_ISL_2691462, EPI_ISL_2691463, EPI_ISL_2691464, EPI_ISL_2691465, EPI_ISL_2691466, EPI_ISL_2691467, EPI_ISL_2691468, EPI_ISL_2691469, EPI_ISL_2691470, EPI_ISL_2691471, EPI_ISL_2691472, EPI_ISL_2691473, EPI_ISL_2691474, EPI_ISL_2691475, EPI_ISL_2691476, EPI_ISL_2691477, EPI_ISL_2691478, EPI_ISL_2691479, EPI_ISL_2691480, EPI_ISL_2691481, EPI_ISL_2691482, EPI_ISL_2691483, EPI_ISL_2691484, EPI_ISL_2691485, EPI_ISL_2691486, EPI_ISL_2691487, EPI_ISL_2691488, EPI_ISL_2691489, EPI_ISL_2691490, EPI_ISL_2691491, EPI_ISL_2691492, EPI_ISL_2691493, EPI_ISL_2691494, EPI_ISL_2691495, EPI_ISL_2691496, EPI_ISL_2691497, EPI_ISL_2691498, EPI_ISL_2691499, EPI_ISL_2691500, EPI_ISL_2691501, EPI_ISL_2691502, EPI_ISL_2691503, EPI_ISL_2691504, EPI_ISL_2691505, EPI_ISL_2691506, EPI_ISL_2691507, EPI_ISL_2691508, EPI_ISL_2691509, EPI_ISL_2691510, EPI_ISL_2691511, EPI_ISL_2691512, EPI_ISL_2691513, EPI_ISL_2691514, EPI_ISL_2691515, EPI_ISL_2691516, EPI_ISL_2691517, EPI_ISL_2691518, EPI_ISL_2691519, EPI_ISL_2691520, EPI_ISL_2691521, EPI_ISL_2691522, EPI_ISL_2691523, EPI_ISL_2691524, EPI_ISL_2691525, EPI_ISL_2691526, EPI_ISL_2691527, EPI_ISL_2691528, EPI_ISL_2691529, EPI_ISL_2691530, EPI_ISL_2691531, EPI_ISL_2691532, EPI_ISL_2691533, EPI_ISL_2691534, EPI_ISL_2691535, EPI_ISL_2691536, EPI_ISL_2691537, EPI_ISL_2691538, EPI_ISL_2691539, EPI_ISL_2691540, EPI_ISL_2691541, EPI_ISL_2691542, EPI_ISL_2691543, EPI_ISL_2691544, EPI_ISL_2691545, EPI_ISL_2691546, EPI_ISL_2691547, EPI_ISL_2691548, EPI_ISL_2691549, EPI_ISL_2691550, EPI_ISL_2691551, EPI_ISL_2691552, EPI_ISL_2691553, EPI_ISL_2691554, EPI_ISL_2691555, EPI_ISL_2691556, EPI_ISL_2691557, EPI_ISL_2691558, EPI_ISL_2691559, EPI_ISL_2691560, EPI_ISL_2691561, EPI_ISL_2691562, EPI_ISL_2691563, EPI_ISL_2691564, EPI_ISL_2691565, EPI_ISL_2691566, EPI_ISL_2691567, EPI_ISL_2691568, EPI_ISL_2836972, EPI_ISL_2836973, EPI_ISL_2836974, EPI_ISL_2836975, EPI_ISL_2836976, EPI_ISL_2836977, EPI_ISL_2836978, EPI_ISL_2836979, EPI_ISL_2836980, EPI_ISL_2836981, EPI_ISL_2836982, EPI_ISL_2836983, EPI_ISL_2836984, EPI_ISL_2836985, EPI_ISL_2836986, EPI_ISL_2836987, EPI_ISL_2836988, EPI_ISL_2836989, EPI_ISL_2836990, EPI_ISL_2836991, EPI_ISL_2836992, EPI_ISL_2836993, EPI_ISL_2836994, EPI_ISL_2836995, EPI_ISL_2836996, EPI_ISL_2836997, EPI_ISL_2836998, EPI_ISL_2836999, EPI_ISL_2837000, EPI_ISL_2837001, EPI_ISL_2837002, EPI_ISL_2837003, EPI_ISL_2837004, EPI_ISL_2837005, EPI_ISL_2837006, EPI_ISL_2837007, EPI_ISL_2837008, EPI_ISL_2837009, EPI_ISL_2837010, EPI_ISL_2837011, EPI_ISL_2837012, EPI_ISL_2837013, EPI_ISL_2837014, EPI_ISL_2837015, EPI_ISL_2837016, EPI_ISL_2837017, EPI_ISL_2837018, EPI_ISL_2837019, EPI_ISL_2837020, EPI_ISL_2837021, EPI_ISL_2837022, EPI_ISL_2837023, EPI_ISL_2837024, EPI_ISL_2837025, EPI_ISL_2837026, EPI_ISL_2837027, EPI_ISL_2837028, EPI_ISL_2837029, EPI_ISL_2837030, EPI_ISL_2837031, EPI_ISL_2837032, EPI_ISL_2837033, EPI_ISL_2837034, EPI_ISL_2837035, EPI_ISL_2837036, EPI_ISL_2837037, EPI_ISL_2837038, EPI_ISL_2837039, EPI_ISL_2837040, EPI_ISL_2837042, EPI_ISL_2837043, EPI_ISL_2837044, EPI_ISL_2837045, EPI_ISL_2837046, EPI_ISL_2837047, EPI_ISL_2837048, EPI_ISL_2837049, EPI_ISL_2837050, EPI_ISL_2837051, EPI_ISL_2837052, EPI_ISL_2837053, EPI_ISL_2837054, EPI_ISL_2837055, EPI_ISL_2837056, EPI_ISL_2837057, EPI_ISL_2837058, EPI_ISL_2837059, EPI_ISL_2837060, EPI_ISL_2837061, EPI_ISL_2837062, EPI_ISL_2837063, EPI_ISL_2837064, EPI_ISL_2837065, EPI_ISL_2837066, EPI_ISL_2837067, EPI_ISL_2837068, EPI_ISL_2837069, EPI_ISL_2837070, EPI_ISL_2837071, EPI_ISL_2837072, EPI_ISL_2837073, EPI_ISL_2837074, EPI_ISL_2837075, EPI_ISL_2837076, EPI_ISL_2837077, EPI_ISL_2837078, EPI_ISL_2837079, EPI_ISL_2837080, EPI_ISL_2837081, EPI_ISL_2837082, EPI_ISL_2837083, EPI_ISL_2837084, EPI_ISL_2837085, EPI_ISL_2837086, EPI_ISL_2837087, EPI_ISL_2837088, EPI_ISL_2837089, EPI_ISL_2837090, EPI_ISL_2837091, EPI_ISL_2837092, EPI_ISL_2837093, EPI_ISL_2837094, EPI_ISL_2837095, EPI_ISL_2837096, EPI_ISL_2837097, EPI_ISL_2837098, EPI_ISL_2837099, EPI_ISL_2837100, EPI_ISL_2837101, EPI_ISL_2837102, EPI_ISL_2837103, EPI_ISL_2837104, EPI_ISL_2837105, EPI_ISL_2837106, EPI_ISL_2837107, EPI_ISL_2837108, EPI_ISL_2837109, EPI_ISL_2837110, EPI_ISL_2837111, EPI_ISL_2837112, EPI_ISL_2837113, EPI_ISL_2837114, EPI_ISL_2837115, EPI_ISL_2837116, EPI_ISL_2837117, EPI_ISL_2837118, EPI_ISL_2837119, EPI_ISL_2837120, EPI_ISL_2837121, EPI_ISL_2837122, EPI_ISL_2837123, EPI_ISL_2837124, EPI_ISL_2837125, EPI_ISL_2837126, EPI_ISL_2837127, EPI_ISL_2837128, EPI_ISL_2837129, EPI_ISL_2837130, EPI_ISL_2837131, EPI_ISL_2837132, EPI_ISL_2837133, EPI_ISL_2837134, EPI_ISL_2837135, EPI_ISL_2837136, EPI_ISL_2837137, EPI_ISL_2837138, EPI_ISL_2837139, EPI_ISL_2837140, EPI_ISL_2837141, EPI_ISL_2837142, EPI_ISL_2837143, EPI_ISL_2837144, EPI_ISL_2837145, EPI_ISL_2837146, EPI_ISL_2837147, EPI_ISL_2837148, EPI_ISL_2837149, EPI_ISL_2837150, EPI_ISL_2837151, EPI_ISL_2837152, EPI_ISL_2837153, EPI_ISL_2837154, EPI_ISL_2837155, EPI_ISL_2837156, EPI_ISL_2837157, EPI_ISL_2837158, EPI_ISL_2837159, EPI_ISL_2837160, EPI_ISL_2837161, EPI_ISL_2837162, EPI_ISL_2837163, EPI_ISL_2837164, EPI_ISL_2837165, EPI_ISL_2837166, EPI_ISL_2837167, EPI_ISL_2837168, EPI_ISL_2837169, EPI_ISL_2837170, EPI_ISL_2837171, EPI_ISL_2837172, EPI_ISL_2837173, EPI_ISL_2837174, EPI_ISL_2837175, EPI_ISL_2837176, EPI_ISL_2837177, EPI_ISL_2837179, EPI_ISL_2837180, EPI_ISL_2837181, EPI_ISL_2837182, EPI_ISL_2837183, EPI_ISL_2837184, EPI_ISL_2837185, EPI_ISL_2837186, EPI_ISL_2837187, EPI_ISL_2837188, EPI_ISL_2837189, EPI_ISL_2837190, EPI_ISL_2837191, EPI_ISL_2837192, EPI_ISL_2837193, EPI_ISL_2837194, EPI_ISL_2837195, EPI_ISL_2837196, EPI_ISL_2837197, EPI_ISL_2837198, EPI_ISL_2837199, EPI_ISL_2837200, EPI_ISL_2837201, EPI_ISL_2837202, EPI_ISL_2837203, EPI_ISL_2837204, EPI_ISL_2837205, EPI_ISL_2837206, EPI_ISL_2837207 | | | |
| see above | Laboratorio Central Noel Nutels | Bioinformatics Laboratory / LNCC | Alessandra P Lamarca; Alexandra L Gerber; Amilcar Tanuri; Ana Paula de C Guimaraes; Ana Tereza R Vasconcelos; Andrea Cony Cavalcanti; Caio Luiz Pereira Ribeiro; Cassia Alves; Cintia Policarpo; Claudia Maria Braga de Mello; Cristiane Gomes da Silva; Diana Mariani; Douglas Terra Machado; Flavio Dias da Silva; Gleidson da Silva de Oliveira; Leandro Magalhaes de Souza; Liliane Cavalcante; Luiz G P de Almeida; Marcio Henrique de Oliveira Garcia; Mario Sergio Ribeiro; Ronaldo da Silva F Jr; Silvia Carvalho |
| EPI_ISL_2645411, EPI_ISL_2645412, EPI_ISL_2645413, EPI_ISL_2645414, EPI_ISL_2645415, EPI_ISL_2645416 | Laboratorio Central de Saude Publica do Estado Maranhao (LACEN-MA) | Laboratory of Respiratory Viruses and Measles, Oswaldo Cruz Institute, FIOCRUZ | Alex Pauvolid-Corrêa; Alice Sampaio Rocha; Ana Beatriz Machado Lima; Ana Carolina Mendonca; Anna Carolina Paixao; Elisa Cavalcante Pereira; Fernando Motta; Lidio Gonçalves Lima Neto; Luciana Appolinario; Marilda Siqueira on behalf of the Fiocruz COVID-19 Genomic Surveillance Network; Mia Ferreira de Araujo; Paola Resende; Renata Serrano Lopes; Taina Venas |
| EPI_ISL_2274072, EPI_ISL_2274073, EPI_ISL_2274074, EPI_ISL_2274075, EPI_ISL_2466262, EPI_ISL_2466263, EPI_ISL_2466264, EPI_ISL_2557337, EPI_ISL_2557338, EPI_ISL_2557339, EPI_ISL_2645408, EPI_ISL_2645409, EPI_ISL_2645410, EPI_ISL_2863589, EPI_ISL_2863591, EPI_ISL_2863593, EPI_ISL_2863695, EPI_ISL_2863714, EPI_ISL_2863716 | | | |
| see above | Laboratorio Central de Saude Publica do Estado da Bahia (LACEN/BA) | Laboratory of Respiratory Viruses and Measles, Oswaldo Cruz Institute, FIOCRUZ | Alice Sampaio Rocha; Ana Carolina Mendonca; Anderson Brandao Leite; Anna Carolina Paixao; Elisa Cavalcante Pereira; Felicidade Pereira; Fernando Motta; Luciana Appolinario; Marilda Siqueira on behalf of the Fiocruz COVID-19 Genomic Surveillance Network; Paola Resende; Renata Serrano Lopes; Taina Venas |
| EPI_ISL_2536323 | Laboratorio Central de Saude Publica do | Laboratory of | Alice Sampaio Rocha; Ana Carolina Mendonca; Anna Carolina Paixao; Dalane Loudal Florentino Teixeira; Elisa Cavalcante Pereira; Fernando Motta; Joao Felipe Bezerra; Luciana Appolinario; Marilda Siqueira on behalf of the Fiocruz COVID-19 Genomic Surveillance Network; Paola Resende; Renata Serrano Lopes; Taina |

| | | | |
|---|---|---|---|
| | Estado da Paraiba (LACEN-PB) | Respiratory Viruses and Measles, Oswaldo Cruz Institute, FIOCRUZ | Venas |
| EPI_ISL_2466231, EPI_ISL_2557310, EPI_ISL_2557311, EPI_ISL_2557312, EPI_ISL_2557313, EPI_ISL_2557314, EPI_ISL_2557315, EPI_ISL_2557317, EPI_ISL_2557318, EPI_ISL_2557319, EPI_ISL_2557320, EPI_ISL_2557321, EPI_ISL_2557322, EPI_ISL_2557323, EPI_ISL_2557324, EPI_ISL_2557325, EPI_ISL_2557326, EPI_ISL_2557327, EPI_ISL_2557328, EPI_ISL_2557329, EPI_ISL_2557330, EPI_ISL_2557331, EPI_ISL_2557333, EPI_ISL_2557334, EPI_ISL_2645384, EPI_ISL_2645385, EPI_ISL_2645386, EPI_ISL_2645387, EPI_ISL_2645388, EPI_ISL_2645389, EPI_ISL_2645390, EPI_ISL_2645391, EPI_ISL_2645392, EPI_ISL_2645393, EPI_ISL_2645394, EPI_ISL_2645395, EPI_ISL_2645396, EPI_ISL_2645397, EPI_ISL_2645398, EPI_ISL_2645399, EPI_ISL_2645400, EPI_ISL_2645401, EPI_ISL_2645402, EPI_ISL_2645403, EPI_ISL_2645404, EPI_ISL_2645405, EPI_ISL_2645406, EPI_ISL_2645407, EPI_ISL_2645444 | | | |
| see above | Laboratorio Central de Saude Publica do Estado de Alagoas (LACEN/AL) | Laboratory of Respiratory Viruses and Measles, Oswaldo Cruz Institute, FIOCRUZ | Alice Sampaio Rocha; Ana Carolina Mendonca; Anderson Brandao Leite; Anna Carolina Paixao; Elisa Cavalcante Pereira; Fernando Motta; Luciana Appolinario; Marilda Siqueira on behalf of the Fiocruz COVID-19 Genomic Surveillance Network; Paola Resende; Renata Serrano Lopes; Taina Venas |
| EPI_ISL_2863647, EPI_ISL_2863648, EPI_ISL_2863651, EPI_ISL_2863653, EPI_ISL_2863655, EPI_ISL_2863656, EPI_ISL_2863658, EPI_ISL_2863660, EPI_ISL_2863662, EPI_ISL_2863664, EPI_ISL_2863666, EPI_ISL_2863712 | | | |
| see above | Laboratorio Central de Saude Publica do Estado de Santa Catarina (LACEN-SC) | Laboratory of Respiratory Viruses and Measles, Oswaldo Cruz Institute, FIOCRUZ | Alice Sampaio Rocha; Ana Carolina Mendonca; Anna Carolina Paixao; Darcita Buerger Rovaris; Elisa Cavalcante Pereira; Fernando Motta; Luciana Appolinario; Marilda Siqueira on behalf of the Fiocruz COVID-19 Genomic Surveillance Network; Paola Resende; Renata Serrano Lopes; Sandra Bianchini Fernandes; Taina Venas |
| EPI_ISL_2863573, EPI_ISL_2863575, EPI_ISL_2863618, EPI_ISL_2863619, EPI_ISL_2863621, EPI_ISL_2863623, EPI_ISL_2863625, EPI_ISL_2863626, EPI_ISL_2863628, EPI_ISL_2863630, EPI_ISL_2863632, EPI_ISL_2863634, EPI_ISL_2863635, EPI_ISL_2863637, EPI_ISL_2863639, EPI_ISL_2863641, EPI_ISL_2863643, EPI_ISL_2863645, EPI_ISL_2863719, EPI_ISL_2863729 | | | |
| see above | Laboratorio Central de Saude Publica do Estado de Sergipe (LACEN/SE) | Laboratory of Respiratory Viruses and Measles, Oswaldo Cruz Institute, FIOCRUZ | Alice Sampaio Rocha; Ana Carolina Mendonca; Anna Carolina Paixao; Cliomar Alves dos Santos; Elisa Cavalcante Pereira; Fernando Motta; Luciana Appolinario; Marilda Siqueira on behalf of the Fiocruz COVID-19 Genomic Surveillance Network; Paola Resende; Renata Serrano Lopes; Tainá Moreira Martins Venas |
| EPI_ISL_2375476, EPI_ISL_2375477 | Laboratorio Central de Saude Publica do Estado do Espirito Santo (LACEN/ES) | Laboratory of Respiratory Viruses and Measles, Oswaldo Cruz Institute, FIOCRUZ | Alice Sampaio Rocha; Ana Carolina Mendonca; Anna Carolina Paixao; Eilisa Cavalcante Pereira; Fernando Motta; Luciana Appolinario; Marilda Siqueira on behalf of the Fiocruz COVID-19 Genomic Surveillance Network; Paola Resende; Renata Serrano Lopes; Rodrigo Ribeiro Rodrigues; Taina Venas |
| EPI_ISL_2645417, EPI_ISL_2645418 | Laboratorio Central de Saude Publica do Estado do Maranhao (LACEN-MA) | Laboratory of Respiratory Viruses and Measles, Oswaldo Cruz Institute, FIOCRUZ | Alex Pauvolid-Corrêa; Alice Sampaio Rocha; Ana Beatriz Machado Lima; Ana Carolina Mendonca; Anna Carolina Paixao; Elisa Cavalcante Pereira; Fernando Motta; Lidio Gonçalves Lima Neto; Luciana Appolinario; Marilda Siqueira on behalf of the Fiocruz COVID-19 Genomic Surveillance Network; Mia Ferreira de Araujo; Paola Resende; Renata Serrano Lopes; Taina Venas |
| EPI_ISL_2645445, EPI_ISL_2645446, EPI_ISL_2645447, EPI_ISL_2645448, EPI_ISL_2645449, EPI_ISL_2645450, EPI_ISL_2645451, EPI_ISL_2645452, EPI_ISL_2645453, EPI_ISL_2645454, EPI_ISL_2645455, EPI_ISL_2645456, EPI_ISL_2645457, EPI_ISL_2645458, EPI_ISL_2645459, EPI_ISL_2645460, EPI_ISL_2645461, EPI_ISL_2645462, EPI_ISL_2645463, EPI_ISL_2645464, EPI_ISL_2645465, EPI_ISL_2645466, EPI_ISL_2645467, EPI_ISL_2645468, EPI_ISL_2645469, EPI_ISL_2645470, EPI_ISL_2645471, EPI_ISL_2645472, EPI_ISL_2645473, EPI_ISL_2645474, EPI_ISL_2645475, EPI_ISL_2645476, EPI_ISL_2645477, EPI_ISL_2645478, EPI_ISL_2645479, EPI_ISL_2645480, EPI_ISL_2645481, EPI_ISL_2645482, EPI_ISL_2645483, EPI_ISL_2645484, EPI_ISL_2645485, EPI_ISL_2645486, EPI_ISL_2645487, EPI_ISL_2645491, EPI_ISL_2645492, EPI_ISL_2645493, EPI_ISL_2645494, EPI_ISL_2645495, EPI_ISL_2645496, EPI_ISL_2645497, EPI_ISL_2645498, EPI_ISL_2645499, EPI_ISL_2645500, EPI_ISL_2645501, EPI_ISL_2645502, EPI_ISL_2645503, EPI_ISL_2645504 | | | |
| see above | Laboratorio Central de Saude Publica do Estado do Parana (LACEN/PR) | Laboratory of Respiratory Viruses and Measles, Oswaldo Cruz Institute, FIOCRUZ | Alice Sampaio Rocha; Ana Carolina Mendonca; Anna Carolina Paixao; Elisa Cavalcante Pereira; Fernando Motta; Irina Riediger; Luciana Appolinario; Marilda Siqueira on behalf of the Fiocruz COVID-19 Genomic Surveillance Network; Paola Resende; Renata Serrano Lopes; Taina Venas |
| EPI_ISL_2443673, EPI_ISL_2603432, EPI_ISL_2603433, EPI_ISL_2603434, EPI_ISL_2603441, EPI_ISL_2603442, EPI_ISL_2603443, EPI_ISL_2603444, EPI_ISL_2603445, EPI_ISL_2603446, EPI_ISL_2603447, EPI_ISL_2603448, EPI_ISL_2603529, EPI_ISL_2603530, EPI_ISL_2603531, EPI_ISL_2603532, EPI_ISL_2603533, EPI_ISL_2603534, EPI_ISL_2603535, EPI_ISL_2603536, EPI_ISL_2603537, EPI_ISL_2603538, EPI_ISL_2603539, EPI_ISL_2603540, EPI_ISL_2603541, EPI_ISL_2603542, EPI_ISL_2603547, EPI_ISL_2603548, EPI_ISL_2603549, EPI_ISL_2603550, EPI_ISL_2603551, EPI_ISL_2603552, EPI_ISL_2603553, EPI_ISL_2603554, EPI_ISL_2603555, EPI_ISL_2603556, EPI_ISL_2603557, EPI_ISL_2603558, EPI_ISL_2603559, EPI_ISL_2603560, EPI_ISL_2603561, EPI_ISL_2603562, EPI_ISL_2603563, EPI_ISL_2603564, EPI_ISL_2603565, EPI_ISL_2603566, EPI_ISL_2603567, EPI_ISL_2603568, EPI_ISL_2603569, EPI_ISL_2603570, EPI_ISL_2603571, EPI_ISL_2603572, EPI_ISL_2603573, EPI_ISL_2603574, EPI_ISL_2603575, EPI_ISL_2603576 | | | |
| see above | Laboratorio Central de Saude Publica do Estado do Rio Grande do Sul (LACEN-RS) | Laboratory of Respiratory Viruses and Measles, Oswaldo Cruz Institute, FIOCRUZ | Alice Sampaio Rocha; Ana Carolina Mendonca; Anderson Brandao Leite; Anna Carolina Paixao; Elisa Cavalcante Pereira; Fernando Motta; Luciana Appolinario; Marilda Siqueira on behalf of the Fiocruz COVID-19 Genomic Surveillance Network; Paola Resende; Renata Serrano Lopes; Richard Salvato; Taina Venas; Tatiana Schaffer Gregianini |
| EPI_ISL_2466268, EPI_ISL_2863584, EPI_ISL_2863585, EPI_ISL_2863586, EPI_ISL_2863587, EPI_ISL_2863588, EPI_ISL_2863602, EPI_ISL_2863604, EPI_ISL_2863667, EPI_ISL_2863669, EPI_ISL_2863671, EPI_ISL_2863673, EPI_ISL_2863675, EPI_ISL_2863677, EPI_ISL_2863679, EPI_ISL_2863680, EPI_ISL_2863696, EPI_ISL_2863698, EPI_ISL_2863700, EPI_ISL_2863702, EPI_ISL_2863704, EPI_ISL_2863717, EPI_ISL_2863721, EPI_ISL_2863723, EPI_ISL_2863727 | | | |
| see above | Laboratorio Central de Saude Publica do Estado do Rio de Janeiro (LACEN-RJ) | Laboratory of Respiratory Viruses and Measles, Oswaldo Cruz Institute, FIOCRUZ | Agatha Cristinne Prudencio; Alice Sampaio Rocha; Ana Carolina Mendonca; Andrea Cony Cavalcanti; Anna Carolina Paixao; Elisa Cavalcante Pereira; Fernando Motta; Ighor Leonardo Arantes Gomes; Luciana Appolinario; Marilda Siqueira on behalf of the Fiocruz COVID-19 Genomic Surveillance Network; Paola Resende; Renata Serrano Lopes; Taina Venas |
| EPI_ISL_2777433, EPI_ISL_2777434, EPI_ISL_2777435, EPI_ISL_2777436, EPI_ISL_2777437, EPI_ISL_2777438, EPI_ISL_2777439, EPI_ISL_2777440, EPI_ISL_2777441, EPI_ISL_2777442, EPI_ISL_2777443, EPI_ISL_2777444, EPI_ISL_2777445, EPI_ISL_2777446, EPI_ISL_2777447, EPI_ISL_2777448, EPI_ISL_2777449, EPI_ISL_2777450, EPI_ISL_2777451, EPI_ISL_2777452, EPI_ISL_2777453 | | | |
| see above | Laboratorio de Ecologia de Doencas Transmissiveis na Amazonia, Instituto Leonidas e Maria Deane - Fiocruz Amazonia | Laboratorio de Ecologia de Doencas Transmissiveis na Amazonia, Instituto Leonidas e Maria Deane - Fiocruz Amazonia | André Corado; Debora Duarte; Felipe Naveca; Fernanda Nascimento; George Silva; Karina Pessoa; Luciana Gonçalves; Maria Júlia Brandão; Matilde Mejía; Michele Jesus; Valdinete Nascimento; Victor Souza; Ágatha Costa |
| EPI_ISL_2443610, EPI_ISL_2443611, EPI_ISL_2443613, EPI_ISL_2443634, EPI_ISL_2443659, EPI_ISL_2443660, EPI_ISL_2443661, EPI_ISL_2443662, EPI_ISL_2557405, EPI_ISL_2557407, EPI_ISL_2557409, EPI_ISL_2557410, EPI_ISL_2645506, EPI_ISL_2731468, EPI_ISL_2863569, EPI_ISL_2863605, EPI_ISL_2863607, EPI_ISL_2863609, EPI_ISL_2863611, EPI_ISL_2863613, EPI_ISL_2863614, EPI_ISL_2863616, EPI_ISL_2863689, EPI_ISL_2863691, EPI_ISL_2863693, EPI_ISL_2863797, EPI_ISL_2863798, EPI_ISL_2863799, EPI_ISL_2863800, EPI_ISL_2863897 | | | |
| see above | Laboratory of Respiratory Viruses and Measles, Oswaldo Cruz Institute, FIOCRUZ | Laboratory of Respiratory Viruses and Measles, Oswaldo Cruz Institute, FIOCRUZ | Agatha Cristinne Prudencio; Agatha Cristinne Prudencio Soares; Alex Pauvolid-Corrêa; Alice Sampaio Rocha; Ana Beatriz Machado Lima; Ana Carolina Mendonca; Anna Carolina Paixao; Elisa Cavalcante Pereira; Fernando Motta; Ighor Leonardo Arantes Gomes; Luciana Appolinario; Marilda Siqueira on behalf of the Fiocruz COVID-19 Genomic Surveillance Network; Mia Ferreira de Araujo; Paola Resende; Renata Serrano Lopes; Taina Venas |
| EPI_ISL_2777242, EPI_ISL_2777243, EPI_ISL_2777244, EPI_ISL_2777245, EPI_ISL_2777246, EPI_ISL_2777283, EPI_ISL_2777284, EPI_ISL_2777285, EPI_ISL_2777287, EPI_ISL_2777288, EPI_ISL_2777293, EPI_ISL_2777295, EPI_ISL_2777297, EPI_ISL_2777299, EPI_ISL_2777316, EPI_ISL_2777318, EPI_ISL_2777335, EPI_ISL_2777336, EPI_ISL_2777337, EPI_ISL_2777338, EPI_ISL_2777339, EPI_ISL_2777341, EPI_ISL_2777342, EPI_ISL_2777343, EPI_ISL_2777344, EPI_ISL_2777345, EPI_ISL_2777348, EPI_ISL_2777349, EPI_ISL_2777350, EPI_ISL_2777351, EPI_ISL_2777352, EPI_ISL_2777353, EPI_ISL_2777354, EPI_ISL_2777355, EPI_ISL_2777359, EPI_ISL_2777511, EPI_ISL_2777512, EPI_ISL_2777523, EPI_ISL_2777524, EPI_ISL_2777527, EPI_ISL_2777531, EPI_ISL_2777532, EPI_ISL_2777546, EPI_ISL_2777553, EPI_ISL_2777555, EPI_ISL_2777556, EPI_ISL_2777559, EPI_ISL_2777561, EPI_ISL_2777562, EPI_ISL_2777564, EPI_ISL_2777570, EPI_ISL_2777578, EPI_ISL_2777580, EPI_ISL_2777584, EPI_ISL_2777586, EPI_ISL_2777587, EPI_ISL_2777590, EPI_ISL_2777592, EPI_ISL_2777593, EPI_ISL_2777598, EPI_ISL_2777602, EPI_ISL_2777606, EPI_ISL_2777607, EPI_ISL_2777608, EPI_ISL_2777611, EPI_ISL_2777612, EPI_ISL_2777613, EPI_ISL_2777616, EPI_ISL_2777617, EPI_ISL_2777619, EPI_ISL_2777624, EPI_ISL_2777625, EPI_ISL_2777626, EPI_ISL_2777627, EPI_ISL_2777629, EPI_ISL_2777631, EPI_ISL_2777632, EPI_ISL_2777635, EPI_ISL_2777636, EPI_ISL_2777637, EPI_ISL_2777639, EPI_ISL_2777640, EPI_ISL_2777641, EPI_ISL_2777642, EPI_ISL_2777643, EPI_ISL_2777644, EPI_ISL_2777645, EPI_ISL_2777648, EPI_ISL_2777649, EPI_ISL_2777650, EPI_ISL_2777651, EPI_ISL_2777652, EPI_ISL_2777653, EPI_ISL_2777654, EPI_ISL_2777656, EPI_ISL_2777658, EPI_ISL_2777659, EPI_ISL_2777661, EPI_ISL_2777663, EPI_ISL_2777664, EPI_ISL_2777665, EPI_ISL_2777667, EPI_ISL_2777669, EPI_ISL_2777671, EPI_ISL_2777672, EPI_ISL_2777673, EPI_ISL_2777674, EPI_ISL_2777675, EPI_ISL_2777677, EPI_ISL_2777678, EPI_ISL_2777679, EPI_ISL_2777680, EPI_ISL_2777681, EPI_ISL_2777682, EPI_ISL_2777684, EPI_ISL_2777685, EPI_ISL_2777687, EPI_ISL_2777688, EPI_ISL_2777690, EPI_ISL_2777692, EPI_ISL_2777693, EPI_ISL_2777694, EPI_ISL_2777695, EPI_ISL_2777702, EPI_ISL_2777703, EPI_ISL_2777711, EPI_ISL_2777712, EPI_ISL_2777773, EPI_ISL_2777774, EPI_ISL_2777777, EPI_ISL_2777778, EPI_ISL_2777806, EPI_ISL_2777810, EPI_ISL_2777814, EPI_ISL_2777826, EPI_ISL_2777831, EPI_ISL_2777844, EPI_ISL_2777845, EPI_ISL_2777846, EPI_ISL_2777849, EPI_ISL_2777850, EPI_ISL_2777852, EPI_ISL_2777853, EPI_ISL_2777856, EPI_ISL_2777863, EPI_ISL_2777865, EPI_ISL_2777866, EPI_ISL_2777884, EPI_ISL_2777885, EPI_ISL_2777886, EPI_ISL_2777887, EPI_ISL_2777888, EPI_ISL_2777889, EPI_ISL_2777890, EPI_ISL_2777891, EPI_ISL_2777892, EPI_ISL_2777893, EPI_ISL_2777895, EPI_ISL_2777896, EPI_ISL_2777897, EPI_ISL_2777898, EPI_ISL_2777900, EPI_ISL_2777901, EPI_ISL_2777902, EPI_ISL_2777903, EPI_ISL_2777904, EPI_ISL_2777905, EPI_ISL_2777906, EPI_ISL_2777907, EPI_ISL_2777908, EPI_ISL_2777909, EPI_ISL_2777910, EPI_ISL_2777911, EPI_ISL_2777912, EPI_ISL_2777913, EPI_ISL_2777914, EPI_ISL_2777915, EPI_ISL_2777916, EPI_ISL_2777917, EPI_ISL_2777918, EPI_ISL_2777919, EPI_ISL_2777920, EPI_ISL_2777921, EPI_ISL_2777922, EPI_ISL_2777923, EPI_ISL_2777924, EPI_ISL_2777925, EPI_ISL_2777926, EPI_ISL_2777927, EPI_ISL_2777928, EPI_ISL_2777929, EPI_ISL_2777930, EPI_ISL_2777932, EPI_ISL_2777933, EPI_ISL_2777934, EPI_ISL_2777935, EPI_ISL_2777937, EPI_ISL_2777938, EPI_ISL_2777939, EPI_ISL_2777940, EPI_ISL_2777941, EPI_ISL_2777942, EPI_ISL_2777943, EPI_ISL_2777944, EPI_ISL_2777945, EPI_ISL_2777946, EPI_ISL_2777947, EPI_ISL_2777948, EPI_ISL_2777949, EPI_ISL_2777950, EPI_ISL_2777951, EPI_ISL_2777952, EPI_ISL_2777953, EPI_ISL_2777954, EPI_ISL_2777955, EPI_ISL_2777956, EPI_ISL_2777957, EPI_ISL_2777958, EPI_ISL_2777959, EPI_ISL_2777960, EPI_ISL_2777961, EPI_ISL_2777962, EPI_ISL_2777963, EPI_ISL_2777964, EPI_ISL_2777965, EPI_ISL_2777966, EPI_ISL_2777967, EPI_ISL_2777968, EPI_ISL_2777983, EPI_ISL_2777984, EPI_ISL_2777985, EPI_ISL_2777991, EPI_ISL_2778006, EPI_ISL_2778007, EPI_ISL_2778009 | | | |
| see above | Laboratório Central de Saúde Pública do Amazonas - LACEN-AM | Laboratorio de Ecologia de Doencas Transmissiveis na Amazonia, Instituto Leonidas e Maria Deane - Fiocruz Amazonia | André Corado; Debora Duarte; Felipe Naveca; Fernanda Nascimento; George Silva; Karina Pessoa; Luciana Gonçalves; Maria Júlia Brandão; Matilde Mejía; Michele Jesus; Valdinete Nascimento; Victor Souza; Ágatha Costa |
| EPI_ISL_2466190, EPI_ISL_2466191, EPI_ISL_2466192, EPI_ISL_2466193, EPI_ISL_2466194, EPI_ISL_2466195, EPI_ISL_2466196, EPI_ISL_2466197, EPI_ISL_2466198, EPI_ISL_2466199 | | | |
| see above | Laboratório de Biologia Molecular de Doenças Infecciosas e do Câncer (LADIC - UFRN) | Laboratory of Respiratory Viruses and Measles, Oswaldo Cruz Institute, FIOCRUZ | Alice Sampaio Rocha; Ana Carolina Mendonca; Anna Carolina Paixao; Elisa Cavalcante Pereira; Fernando Motta; Josélio Araújo; Luciana Appolinario; Marilda Siqueira on behalf of the Fiocruz COVID-19 Genomic Surveillance Network; Paola Resende; Renata Serrano Lopes; Taina Venas |
| EPI_ISL_2731508, EPI_ISL_2731509, EPI_ISL_2731510, EPI_ISL_2731511, EPI_ISL_2731512, EPI_ISL_2731513, EPI_ISL_2731514, EPI_ISL_2731515, EPI_ISL_2731516, EPI_ISL_2731517, EPI_ISL_2731518, EPI_ISL_2731519, EPI_ISL_2731520, EPI_ISL_2731521, EPI_ISL_2731522, EPI_ISL_2731523, EPI_ISL_2731524, EPI_ISL_2731525, EPI_ISL_2731526, EPI_ISL_2731527, EPI_ISL_2731528, EPI_ISL_2731529, EPI_ISL_2731530, EPI_ISL_2731531, EPI_ISL_2731532, EPI_ISL_2731533, EPI_ISL_2731534, EPI_ISL_2731535, EPI_ISL_2731536, EPI_ISL_2731537, EPI_ISL_2731538, EPI_ISL_2731539, EPI_ISL_2731540, EPI_ISL_2731541, EPI_ISL_2731542, EPI_ISL_2731543, EPI_ISL_2731544, EPI_ISL_2731545, EPI_ISL_2731546, EPI_ISL_2731547, EPI_ISL_2731548, EPI_ISL_2731549, EPI_ISL_2731550, EPI_ISL_2731551, EPI_ISL_2731552, EPI_ISL_2731553, EPI_ISL_2731554, EPI_ISL_2731555, EPI_ISL_2731556, EPI_ISL_2731557, EPI_ISL_2731558, EPI_ISL_2731559, EPI_ISL_2731560, EPI_ISL_2731561, EPI_ISL_2731562, EPI_ISL_2731563, EPI_ISL_2731564, EPI_ISL_2731565, EPI_ISL_2731566, EPI_ISL_2731567, EPI_ISL_2731568, EPI_ISL_2731569, EPI_ISL_2731570, EPI_ISL_2731571, EPI_ISL_2731572, EPI_ISL_2731573, EPI_ISL_2731574, EPI_ISL_2731575, EPI_ISL_2731576, EPI_ISL_2731577, EPI_ISL_2731578, EPI_ISL_2731579, EPI_ISL_2731580, EPI_ISL_2731581, EPI_ISL_2731582, EPI_ISL_2731583, EPI_ISL_2731584, EPI_ISL_2731585, EPI_ISL_2731586, EPI_ISL_2731587, EPI_ISL_2731588, EPI_ISL_2731589, EPI_ISL_2731590, EPI_ISL_2731591, EPI_ISL_2731592, EPI_ISL_2731593, EPI_ISL_2731594, EPI_ISL_2731595, EPI_ISL_2731596, EPI_ISL_2731597, EPI_ISL_2731598, EPI_ISL_2731599, EPI_ISL_2731600, EPI_ISL_2731601, EPI_ISL_2731602, EPI_ISL_2731603, EPI_ISL_2731604, EPI_ISL_2731605, EPI_ISL_2731606, EPI_ISL_2731607, EPI_ISL_2731608, EPI_ISL_2731609, EPI_ISL_2731610, EPI_ISL_2731611, EPI_ISL_2731612, EPI_ISL_2731613, EPI_ISL_2731614, EPI_ISL_2731615, EPI_ISL_2731616, EPI_ISL_2731617, EPI_ISL_2731618, EPI_ISL_2731619, EPI_ISL_2731620, EPI_ISL_2731621, EPI_ISL_2731622, EPI_ISL_2731623, EPI_ISL_2731624, EPI_ISL_2731625, EPI_ISL_2731626, EPI_ISL_2731627, EPI_ISL_2731628, EPI_ISL_2731629, EPI_ISL_2731630, EPI_ISL_2731631, EPI_ISL_2731632, EPI_ISL_2731633, EPI_ISL_2731634, EPI_ISL_2731635, EPI_ISL_2731636, EPI_ISL_2731637, EPI_ISL_2731638, EPI_ISL_2731639, EPI_ISL_2731640, EPI_ISL_2731641, EPI_ISL_2731642, EPI_ISL_2731643, EPI_ISL_2731644, EPI_ISL_2731645, EPI_ISL_2731646, EPI_ISL_2731647, EPI_ISL_2731648, EPI_ISL_2731649, EPI_ISL_2731650, EPI_ISL_2731651, EPI_ISL_2731652, EPI_ISL_2731653, EPI_ISL_2731654, EPI_ISL_2731655, EPI_ISL_2731656, EPI_ISL_2731657, EPI_ISL_2731658, EPI_ISL_2731659, EPI_ISL_2731660, EPI_ISL_2731661, EPI_ISL_2731662, EPI_ISL_2731663, EPI_ISL_2731664, EPI_ISL_2731665, EPI_ISL_2731666, EPI_ISL_2731667, EPI_ISL_2731668, EPI_ISL_2731669, EPI_ISL_2731670, EPI_ISL_2731671, EPI_ISL_2731672, EPI_ISL_2731673, EPI_ISL_2731674, EPI_ISL_2731675, EPI_ISL_2731676, EPI_ISL_2731677, EPI_ISL_2790137, EPI_ISL_2790138, EPI_ISL_2790139, EPI_ISL_2790140, EPI_ISL_2790141, EPI_ISL_2790142, EPI_ISL_2790143, EPI_ISL_2790144, EPI_ISL_2790145, EPI_ISL_2790146, EPI_ISL_2790147, EPI_ISL_2790148, EPI_ISL_2790149, EPI_ISL_2790150, EPI_ISL_2790151, EPI_ISL_2790152, EPI_ISL_2790153, EPI_ISL_2790154, EPI_ISL_2790155, EPI_ISL_2790156, EPI_ISL_2790157, EPI_ISL_2790158, EPI_ISL_2790159, EPI_ISL_2790160, EPI_ISL_2790161, EPI_ISL_2790162, EPI_ISL_2790163, EPI_ISL_2790164, EPI_ISL_2790165, EPI_ISL_2790166, EPI_ISL_2790167, EPI_ISL_2790168, EPI_ISL_2790169, EPI_ISL_2790170, EPI_ISL_2790171, EPI_ISL_2790172, EPI_ISL_2790173, EPI_ISL_2790174, EPI_ISL_2835107, EPI_ISL_2835108, EPI_ISL_2835109, EPI_ISL_2835110, EPI_ISL_2835111, EPI_ISL_2835112, EPI_ISL_2835113, EPI_ISL_2835114, EPI_ISL_2835115, EPI_ISL_2835116, EPI_ISL_2835117, EPI_ISL_2835118, EPI_ISL_2835119, EPI_ISL_2835120, EPI_ISL_2835121, EPI_ISL_2835122, EPI_ISL_2835123, EPI_ISL_2835124, EPI_ISL_2835125, EPI_ISL_2835126, EPI_ISL_2835127, EPI_ISL_2835128, EPI_ISL_2835129, EPI_ISL_2835130, EPI_ISL_2835131, EPI_ISL_2835132, EPI_ISL_2835133, EPI_ISL_2835134, EPI_ISL_2835135, EPI_ISL_2835136, EPI_ISL_2835137, EPI_ISL_2835138, EPI_ISL_2835139, EPI_ISL_2835140, EPI_ISL_2835141, EPI_ISL_2835142, EPI_ISL_2835143, EPI_ISL_2835144, EPI_ISL_2835145, EPI_ISL_2835146, EPI_ISL_2835147, EPI_ISL_2835148, EPI_ISL_2835149, EPI_ISL_2835150, EPI_ISL_2835151, EPI_ISL_2835152, EPI_ISL_2835153, EPI_ISL_2835154, EPI_ISL_2835155, EPI_ISL_2835156, EPI_ISL_2835157, EPI_ISL_2835158, EPI_ISL_2835159, EPI_ISL_2835160, EPI_ISL_2835161, EPI_ISL_2835162, EPI_ISL_2835163, EPI_ISL_2835164, EPI_ISL_2835165, EPI_ISL_2835166, EPI_ISL_2835167, EPI_ISL_2835168, EPI_ISL_2835169, EPI_ISL_2835170, EPI_ISL_2835171, EPI_ISL_2835172, EPI_ISL_2835173, EPI_ISL_2835174, EPI_ISL_2835175, EPI_ISL_2835176, EPI_ISL_2835177, EPI_ISL_2835178, EPI_ISL_2835179, EPI_ISL_2835180, EPI_ISL_2835181, EPI_ISL_2835182, EPI_ISL_2835183, EPI_ISL_2835184, EPI_ISL_2835185, EPI_ISL_2835186, EPI_ISL_2835187, EPI_ISL_2835188, EPI_ISL_2835189, EPI_ISL_2835190, EPI_ISL_2835191, EPI_ISL_2835192, EPI_ISL_2835193, EPI_ISL_2835194, EPI_ISL_2835195, EPI_ISL_2835196, EPI_ISL_2835197, EPI_ISL_2835198, EPI_ISL_2835199, EPI_ISL_2835200, EPI_ISL_2835201, EPI_ISL_2835202, EPI_ISL_2835203, EPI_ISL_2835204, EPI_ISL_2835205, EPI_ISL_2835206, EPI_ISL_2835207, EPI_ISL_2835208, EPI_ISL_2835209, EPI_ISL_2835210, EPI_ISL_2835211, EPI_ISL_2835212, EPI_ISL_2835213, EPI_ISL_2835214, EPI_ISL_2835215, EPI_ISL_2835216, EPI_ISL_2835217, EPI_ISL_2835218, EPI_ISL_2835219, EPI_ISL_2835220, EPI_ISL_2835221, EPI_ISL_2835222, EPI_ISL_2835223 | | | |

| | | | |
|---|---|---|---|
| see above | Laboratório de Biotecnologia Aplicada (LBA) - Laboratório de Biologia Molecular - Hospital das Clínicas, Faculdade de Medicina de Botucatu, Departamento de Bioprocessos e Biotecnologia - Faculdade de Ciências Agronômicas, UNESP – Botucatu/SP | Laboratory of Respiratory Viruses and Measles, Oswaldo Cruz Institute, FIOCRUZ | Alice Sampaio Rocha; Ana Carolina Mendonca; Anna Carolina Paixao; Elisa Cavalcante Pereira; Felipe Allan da Silva Costa; Fernando Motta; Jayme Augusto de Souza Neto; Leonardo Nazario de Moraes; Luciana Appolinario; Marilda Siqueira on behalf of the Fiocruz COVID-19 Genomic Surveillance Network; Paola Resende; Patricia Akemi Assato; Rejane Maria Tommasini; Renata Serrano Lopes; Taina Venas |
| EPI_ISL_2918996, EPI_ISL_2918997, EPI_ISL_2918998, EPI_ISL_2918999, EPI_ISL_2919000, EPI_ISL_2919001, EPI_ISL_2919002, EPI_ISL_2919003, EPI_ISL_2919004, EPI_ISL_2919005, EPI_ISL_2919006, EPI_ISL_2919007, EPI_ISL_2919008, EPI_ISL_2919010, EPI_ISL_2919011, EPI_ISL_2919012, EPI_ISL_2919015, EPI_ISL_2919016, EPI_ISL_2925736, EPI_ISL_2928507, EPI_ISL_2928508 | | | |
| see above | Laboratório de Microbiologia Molecular - Universidade FEEVALE | Laboratório de Microbiologia Molecular - Universidade FEEVALE | Alana Witt Hansen; Fernando Rosado Spilki; Flávio Silveira; Fágner Henrique Heldt; Juliana Schons Gulart; Juliana Schons Gularte; Juliane Deise Fleck; Mariana Soares da Silva; Matheus Nunes Weber; Meriane Demoliner; Michele Filippi.; Micheli Filippi.; Paula Rodrigues de Almeida |
| EPI_ISL_2629836, EPI_ISL_2629837, EPI_ISL_2629838 | Laboratório de Virologia Molecular - Universidade Federal do Rio de Janeiro | Laboratório de Virologia Molecular - Universidade Federal do Rio de Janeiro | ; Alice Laschuk Herlinger; Amilcar Tanuri; André Felipe Andrade dos Santos; Carolina Moreira Voloch; Cássia Cristina Alves Gonçalves; Diana Mariani; Débora Souza Faffe; Filipe Romero Rebello Moreira; Francine Bittencourt Schiffler; Isabela de Carvalho Leitão; Marcelo Calado de Paula Tôrres; Matheus Augusto Calvano Cosentino; Mirela D'arc; Orlando da Costa Ferreira Junior; Rafael Mello Galliez; Raíssa Mirella dos Santos Cunha da Costa; Renato Santana de Aguiar; Terezinha Marta Pereira Pinto Castineiras; Thamiris dos Santos Miranda; Átila Duque Rossi |
| EPI_ISL_2536227, EPI_ISL_2536229, EPI_ISL_2536232, EPI_ISL_2536233, EPI_ISL_2536235, EPI_ISL_2536236, EPI_ISL_2536238, EPI_ISL_2536240, EPI_ISL_2536243, EPI_ISL_2536245, EPI_ISL_2536246, EPI_ISL_2536248, EPI_ISL_2536249, EPI_ISL_2536250, EPI_ISL_2536253, EPI_ISL_2536258, EPI_ISL_2536259, EPI_ISL_2536260, EPI_ISL_2557411, EPI_ISL_2557412, EPI_ISL_2557413, EPI_ISL_2557414, EPI_ISL_2557416, EPI_ISL_2557417, EPI_ISL_2557418, EPI_ISL_2557419, EPI_ISL_2557420, EPI_ISL_2557421, EPI_ISL_2731506 | | | |
| see above | Labortorio Central de Saude Publica do Estado de Santa Catarina (LACEN/SC) | Laboratory of Respiratory Viruses and Measles, Oswaldo Cruz Institute, FIOCRUZ | Alice Sampaio Rocha; Ana Carolina Mendonca; Anna Carolina Paixao; Darcita Buerger Rovaris; Elisa Cavalcante Pereira; Fernando Motta; Luciana Appolinario; Marilda Siqueira on behalf of the Fiocruz COVID-19 Genomic Surveillance Network; Paola Resende; Renata Serrano Lopes; Sandra Bianchini Fernandes; Taina Venas |
| EPI_ISL_2466230, EPI_ISL_2536275, EPI_ISL_2536276, EPI_ISL_2536277, EPI_ISL_2536278, EPI_ISL_2536279, EPI_ISL_2536280, EPI_ISL_2536281, EPI_ISL_2536282, EPI_ISL_2536283, EPI_ISL_2536284, EPI_ISL_2536285, EPI_ISL_2536286, EPI_ISL_2536287, EPI_ISL_2536288, EPI_ISL_2536289, EPI_ISL_2536290, EPI_ISL_2536291, EPI_ISL_2536292, EPI_ISL_2536293, EPI_ISL_2536294, EPI_ISL_2536295, EPI_ISL_2536296, EPI_ISL_2536297, EPI_ISL_2536298, EPI_ISL_2536299, EPI_ISL_2536300, EPI_ISL_2536301, EPI_ISL_2536302, EPI_ISL_2557380, EPI_ISL_2557381, EPI_ISL_2557382, EPI_ISL_2557383, EPI_ISL_2557406, EPI_ISL_2645505, EPI_ISL_2677318, EPI_ISL_2863891 | | | |
| see above | Labortorio Central de Saude Publica do Estado do Rio de Janeiro (LACEN/RJ) | Laboratory of Respiratory Viruses and Measles, Oswaldo Cruz Institute, FIOCRUZ | Agatha Cristinne Prudencio Soares; Alex Pauvolid-Corrêa; Alice Sampaio Rocha; Ana Beatriz Machado Lima; Ana Carolina Mendonca; Andrea Cony Cavalcanti; Anna Carolina Paixao; Elisa Cavalcante Pereira; Fernando Motta; Ighor Leonardo Arantes Gomes; Luciana Appolinario; Marilda Siqueira on behalf of the Fiocruz COVID-19 Genomic Surveillance Network; Mia Ferreira de Araujo; Paola Resende; Renata Serrano Lopes; Taina Venas |
| EPI_ISL_2645507, EPI_ISL_2645508, EPI_ISL_2645509, EPI_ISL_2645510, EPI_ISL_2645511, EPI_ISL_2645512, EPI_ISL_2645513, EPI_ISL_2863595, EPI_ISL_2863596, EPI_ISL_2863598, EPI_ISL_2863600 | | | |
| see above | Labortorio Central de Saude Publica do Estado do Tocantins (LACEN/TO) | Laboratory of Respiratory Viruses and Measles, Oswaldo Cruz Institute, FIOCRUZ | Alice Sampaio Rocha; Ana Carolina Mendonca; Anna Carolina Paixao; Elisa Cavalcante Pereira; Fernando Motta; Jucimaria Dantas Galvao; Luciana Appolinario; Marilda Siqueira on behalf of the Fiocruz COVID-19 Genomic Surveillance Network; Paola Resende; Renata Serrano Lopes; Taina Venas |
| EPI_ISL_2557359, EPI_ISL_2557360, EPI_ISL_2557361, EPI_ISL_2557362, EPI_ISL_2557363, EPI_ISL_2557364, EPI_ISL_2557365, EPI_ISL_2557366, EPI_ISL_2557367, EPI_ISL_2557368, EPI_ISL_2557369, EPI_ISL_2557370, EPI_ISL_2557371, EPI_ISL_2557372, EPI_ISL_2557373, EPI_ISL_2557374, EPI_ISL_2557375, EPI_ISL_2603430, EPI_ISL_2603451, EPI_ISL_2603454, EPI_ISL_2603455, EPI_ISL_2613809, EPI_ISL_2613810, EPI_ISL_2613811, EPI_ISL_2613812, EPI_ISL_2613813, EPI_ISL_2613814, EPI_ISL_2613815, EPI_ISL_2645440, EPI_ISL_2645441, EPI_ISL_2645442, EPI_ISL_2645443, EPI_ISL_2863571, EPI_ISL_2863576, EPI_ISL_2863682, EPI_ISL_2863684, EPI_ISL_2863685, EPI_ISL_2863687, EPI_ISL_2863725, EPI_ISL_2863795, EPI_ISL_2863883, EPI_ISL_2863884, EPI_ISL_2863887, EPI_ISL_2863888, EPI_ISL_2863889, EPI_ISL_2863893 | | | |
| see above | Lboratorio Central de Saude Publica do Estado do Parana (LACEN/PR) | Laboratory of Respiratory Viruses and Measles, Oswaldo Cruz Institute, FIOCRUZ | Agatha Cristinne Prudencio; Agatha Cristinne Prudencio Soares; Alice Sampaio Rocha; Ana Carolina Mendonca; Anna Carolina Paixao; Elisa Cavalcante Pereira; Fernando Motta; Ighor Leonardo Arantes Gomes; Irina Riediger; Luciana Appolinario; Marilda Siqueira on behalf of the Fiocruz COVID-19 Genomic Surveillance Network; Paola Resende; Renata Serrano Lopes; Taina Venas |
| EPI_ISL_2801332, EPI_ISL_2801333 | MATERNIDADE ESCOLA ASSIS CHATEAUBRIAND | Oswaldo Cruz Institute, FIOCRUZ/CE | Cleber Furtado Aksenen e Suzana Porto Almeida; Fabio Miyajima; Fernando Braga Stehling; Francisco Eder de Moura Lopes; Jamille Maria Mendes Bezerra; Joaquim César do Nascimento Sousa Junior; Pedro Miguel Carneiro Jeronimo; Thais Ferreira de Oliveira; Thais de Oliveira Costa; Ticiane Cavalcante de Souza; Veridiana Pessoa Miyajima |
| EPI_ISL_2444199 | MIRASSOL | Instituto Butantan / FAMERP | Antonio Jorge Martins; Claudia Renata dos Santos Barros; David Schlesinger; Debora Botequio Moretti; Dimas Tadeu Covas; Elaine Cristina Marqueze; Elaine Vieira Santos; Evandra Strazza Rodrigues; Heidge Fukumasu; Jayme Augusto de Souza-Neto; José Salvatore Leister Patané; Luiz Alcantara; Luiz Lehmann Coutinho; Maria Carolina Elias; Maurício Lacerda Nogueira; Rafael dos Santos Bezerra; Raul Machado Neto; Rejane Maria Tommasini Grotto; Ricardo Haddad; Sandra Coccuzzo Sampaio Vessoni; Simone Kashima; Svetoslav Nanev Slavov; Vincent Louis Viala |
| EPI_ISL_2444203, EPI_ISL_2444210 | MONTE APRAZIVEL | Instituto Butantan / FAMERP | Antonio Jorge Martins; Claudia Renata dos Santos Barros; David Schlesinger; Debora Botequio Moretti; Dimas Tadeu Covas; Elaine Cristina Marqueze; Elaine Vieira Santos; Evandra Strazza Rodrigues; Heidge Fukumasu; Jayme Augusto de Souza-Neto; José Salvatore Leister Patané; Luiz Alcantara; Luiz Lehmann Coutinho; Maria Carolina Elias; Maurício Lacerda Nogueira; Rafael dos Santos Bezerra; Raul Machado Neto; Rejane Maria Tommasini Grotto; Ricardo Haddad; Sandra Coccuzzo Sampaio Vessoni; Simone Kashima; Svetoslav Nanev Slavov; Vincent Louis Viala |
| EPI_ISL_2445118, EPI_ISL_2445119, EPI_ISL_2445120, EPI_ISL_2445121, EPI_ISL_2445122, EPI_ISL_2445123, EPI_ISL_2445124, EPI_ISL_2473732, EPI_ISL_2473735 | | | |
| see above | NUCLEO DE ESPECIALIDADES EM SAUDE | Instituto Butantan | Antonio Jorge Martins; Claudia Renata dos Santos Barros; David Schlesinger; Debora Botequio Moretti; Dimas Tadeu Covas; Elaine Cristina Marqueze; Elaine Vieira Santos; Evandra Strazza Rodrigues; Heidge Fukumasu; Jayme Augusto de Souza-Neto; José Salvatore Leister Patané; Luiz Alcantara; Luiz Lehmann Coutinho; Maria Carolina Elias; Maurício Lacerda Nogueira; Rafael dos Santos Bezerra; Raul Machado Neto; Rejane Maria Tommasini Grotto; Ricardo Haddad; Sandra Coccuzzo Sampaio Vessoni; Simone Kashima; Svetoslav Nanev Slavov; Vincent Louis Viala |
| EPI_ISL_2209052, EPI_ISL_2209565, EPI_ISL_2209571, EPI_ISL_2209607, EPI_ISL_2210238, EPI_ISL_2345687, EPI_ISL_2345733, EPI_ISL_2345737, EPI_ISL_2345777, EPI_ISL_2345781, EPI_ISL_2345800, EPI_ISL_2345876 | | | |
| see above | NUCLEO DE SAUDE MARY DOTA DE BAURU | Instituto Butantan / Mendelics | Antonio Jorge Martins; Claudia Renata dos Santos Barros; David Schlesinger; Debora Botequio Moretti; Dimas Tadeu Covas; Elaine Cristina Marqueze; Elaine Vieira Santos; Evandra Strazza Rodrigues; Heidge Fukumasu; Jayme Augusto de Souza-Neto; José Salvatore Leister Patané; Luiz Alcantara; Luiz Lehmann Coutinho; Maria Carolina Elias; Maurício Lacerda Nogueira; Rafael dos Santos Bezerra; Raul Machado Neto; Rejane Maria Tommasini Grotto; Ricardo Haddad; Sandra Coccuzzo Sampaio Vessoni; Simone Kashima; Svetoslav Nanev Slavov; Vincent Louis Viala |
| EPI_ISL_2445125, EPI_ISL_2445126, EPI_ISL_2445127, EPI_ISL_2445128, EPI_ISL_2445130, EPI_ISL_2445131, EPI_ISL_2445132, EPI_ISL_2445134, EPI_ISL_2445135, EPI_ISL_2445136, EPI_ISL_2445138, EPI_ISL_2473704, EPI_ISL_2473737, EPI_ISL_2473738, EPI_ISL_2473739, EPI_ISL_2473740, EPI_ISL_2473741, EPI_ISL_2473742, EPI_ISL_2473743, EPI_ISL_2473747, EPI_ISL_2473748, EPI_ISL_2473750, EPI_ISL_2473751, EPI_ISL_2473753, EPI_ISL_2473754, EPI_ISL_2473813, EPI_ISL_2473814 | | | |
| see above | NUCLEO DE SAUDE VILA FALCAO DE BAURU | Instituto Butantan | Antonio Jorge Martins; Claudia Renata dos Santos Barros; David Schlesinger; Debora Botequio Moretti; Dimas Tadeu Covas; Elaine Cristina Marqueze; Elaine Vieira Santos; Evandra Strazza Rodrigues; Heidge Fukumasu; Jayme Augusto de Souza-Neto; José Salvatore Leister Patané; Luiz Alcantara; Luiz Lehmann Coutinho; Maria Carolina Elias; Maurício Lacerda Nogueira; Rafael dos Santos Bezerra; Raul Machado Neto; Rejane Maria Tommasini Grotto; Ricardo Haddad; Sandra Coccuzzo Sampaio Vessoni; Simone Kashima; Svetoslav Nanev Slavov; Vincent Louis Viala |
| EPI_ISL_2821280, EPI_ISL_2821281, EPI_ISL_2821282, EPI_ISL_2821283, EPI_ISL_2821284 | NUPIT/UFPE | WallauLab on behalf of Fiocruz COVID-19 Genomic Surveillance Network | Alexandre Freitas da Silva; Antonio Mauro Rezende; Constância Flávia Junqueira Ayres; Cássia Docena; Duschinka Ribeiro Duarte Guedes; Elisama Helvecio; Filipe Zimmer Dezordi; Gabriel Luz Wallau; Larissa Krokovsky; Laís Ceschini Machado; Maira Galdino da Rocha Pitta; Marcelo Henrique dos Santos Paiva; Matheus Filgueira Bezerra; Michelly Cristiny Pereira; Rômulo Pessoa e Silva; Sinval Pinto Brandão Filho |
| EPI_ISL_2445407, EPI_ISL_2445427, EPI_ISL_2445435, EPI_ISL_2445439 | PA CAPELA | Instituto Butantan | Antonio Jorge Martins; Claudia Renata dos Santos Barros; David Schlesinger; Debora Botequio Moretti; Dimas Tadeu Covas; Elaine Cristina Marqueze; Elaine Vieira Santos; Evandra Strazza Rodrigues; Heidge Fukumasu; Jayme Augusto de Souza-Neto; José Salvatore Leister Patané; Luiz Alcantara; Luiz Lehmann Coutinho; Maria Carolina Elias; Maurício Lacerda Nogueira; Rafael dos Santos Bezerra; Raul Machado Neto; Rejane Maria Tommasini Grotto; Ricardo Haddad; Sandra Coccuzzo Sampaio Vessoni; Simone Kashima; Svetoslav Nanev Slavov; Vincent Louis Viala |
| EPI_ISL_2493828 | PA DE IBITIUVA DR OTAVIO BENETTI PITANGUEIRAS | Instituto Butantan | Antonio Jorge Martins; Claudia Renata dos Santos Barros; David Schlesinger; Debora Botequio Moretti; Dimas Tadeu Covas; Elaine Cristina Marqueze; Elaine Vieira Santos; Evandra Strazza Rodrigues; Heidge Fukumasu; Jayme Augusto de Souza-Neto; José Salvatore Leister Patané; Luiz Alcantara; Luiz Lehmann Coutinho; Maria Carolina Elias; Maurício Lacerda Nogueira; Rafael dos Santos Bezerra; Raul Machado Neto; Rejane Maria Tommasini Grotto; Ricardo Haddad; Sandra Coccuzzo Sampaio Vessoni; Simone Kashima; Svetoslav Nanev Slavov; Vincent Louis Viala |
| EPI_ISL_2493472, EPI_ISL_2493485, EPI_ISL_2493489, EPI_ISL_2493490, EPI_ISL_2493497, EPI_ISL_2493498, EPI_ISL_2493501, EPI_ISL_2493504 | | | |
| see above | PA MATAO | Instituto Butantan | Antonio Jorge Martins; Claudia Renata dos Santos Barros; David Schlesinger; Debora Botequio Moretti; Dimas Tadeu Covas; Elaine Cristina Marqueze; Elaine Vieira Santos; Evandra Strazza Rodrigues; Heidge Fukumasu; Jayme Augusto de Souza-Neto; José Salvatore Leister Patané; Luiz Alcantara; Luiz Lehmann Coutinho; Maria Carolina Elias; Maurício Lacerda Nogueira; Rafael dos Santos Bezerra; Raul Machado Neto; Rejane Maria Tommasini Grotto; Ricardo Haddad; Sandra Coccuzzo Sampaio Vessoni; Simone Kashima; Svetoslav Nanev Slavov; Vincent Louis Viala |
| EPI_ISL_2378748, EPI_ISL_2378749, EPI_ISL_2493207, EPI_ISL_2493356, EPI_ISL_2493357, EPI_ISL_2493358, EPI_ISL_2493359 | | | |
| see above | PAS JOAO ANTONIO DO NASCIMENTO | Instituto Butantan | Antonio Jorge Martins; Claudia Renata dos Santos Barros; David Schlesinger; Debora Botequio Moretti; Dimas Tadeu Covas; Elaine Cristina Marqueze; Elaine Vieira Santos; Evandra Strazza Rodrigues; Heidge Fukumasu; Jayme Augusto de Souza-Neto; José Salvatore Leister Patané; Luiz Alcantara; Luiz Lehmann Coutinho; Maria Carolina Elias; Maurício Lacerda Nogueira; Rafael dos Santos Bezerra; Raul Machado Neto; Rejane Maria Tommasini Grotto; Ricardo Haddad; Sandra Coccuzzo Sampaio Vessoni; Simone Kashima; Svetoslav Nanev Slavov; Vincent Louis Viala |
| EPI_ISL_2500981 | PAS JULIANIA DE HERCULANDIA | Instituto Butantan | Antonio Jorge Martins; Claudia Renata dos Santos Barros; David Schlesinger; Debora Botequio Moretti; Dimas Tadeu Covas; Elaine Cristina Marqueze; Elaine Vieira Santos; Evandra Strazza Rodrigues; Heidge Fukumasu; Jayme Augusto de Souza-Neto; José Salvatore Leister Patané; Luiz Alcantara; Luiz Lehmann Coutinho; Maria Carolina Elias; Maurício Lacerda Nogueira; Rafael dos Santos Bezerra; Raul Machado Neto; Rejane Maria Tommasini Grotto; Ricardo Haddad; Sandra Coccuzzo Sampaio Vessoni; Simone Kashima; Svetoslav Nanev Slavov; Vincent Louis Viala |
| EPI_ISL_2345670 | POLICLINICA ALCIA LINEIA VITTI REJANI | Instituto Butantan / Mendelics | Antonio Jorge Martins; Claudia Renata dos Santos Barros; David Schlesinger; Debora Botequio Moretti; Dimas Tadeu Covas; Elaine Cristina Marqueze; Elaine Vieira Santos; Evandra Strazza Rodrigues; Heidge Fukumasu; Jayme Augusto de Souza-Neto; José Salvatore Leister Patané; Luiz Alcantara; Luiz Lehmann Coutinho; Maria Carolina Elias; Maurício Lacerda Nogueira; Rafael dos Santos Bezerra; Raul Machado Neto; Rejane Maria Tommasini Grotto; Ricardo Haddad; Sandra Coccuzzo Sampaio Vessoni; Simone Kashima; Svetoslav Nanev Slavov; Vincent Louis Viala |
| EPI_ISL_2445595, EPI_ISL_2445596, EPI_ISL_2445601, EPI_ISL_2493667, EPI_ISL_2493673, EPI_ISL_2493913 | POLICLINICA COVID 19 ITAPETININGA | Instituto Butantan | Antonio Jorge Martins; Claudia Renata dos Santos Barros; David Schlesinger; Debora Botequio Moretti; Dimas Tadeu Covas; Elaine Cristina Marqueze; Elaine Vieira Santos; Evandra Strazza Rodrigues; Heidge Fukumasu; Jayme Augusto de Souza-Neto; José Salvatore Leister Patané; Luiz Alcantara; Luiz Lehmann Coutinho; Maria Carolina Elias; Maurício Lacerda Nogueira; Rafael dos Santos Bezerra; Raul Machado Neto; Rejane Maria Tommasini Grotto; Ricardo Haddad; Sandra Coccuzzo Sampaio Vessoni; Simone Kashima; Svetoslav Nanev Slavov; Vincent Louis Viala |
| EPI_ISL_2493195, EPI_ISL_2493198, EPI_ISL_2493246, EPI_ISL_2493456, EPI_ISL_2493460, EPI_ISL_2493464, EPI_ISL_2493467, EPI_ISL_2493468, EPI_ISL_2493471, EPI_ISL_2493478, EPI_ISL_2493481, EPI_ISL_2493491, EPI_ISL_2493493, EPI_ISL_2493495, EPI_ISL_2493499, EPI_ISL_2493500, EPI_ISL_2493502, EPI_ISL_2493503 | | | |
| see above | POLICLINICA HORTOLANDIA | Instituto Butantan | Antonio Jorge Martins; Claudia Renata dos Santos Barros; David Schlesinger; Debora Botequio Moretti; Dimas Tadeu Covas; Elaine Cristina Marqueze; Elaine Vieira Santos; Evandra Strazza Rodrigues; Heidge Fukumasu; Jayme Augusto de Souza-Neto; José Salvatore Leister Patané; Luiz Alcantara; Luiz Lehmann Coutinho; Maria Carolina Elias; Maurício Lacerda Nogueira; Rafael dos Santos Bezerra; Raul Machado Neto; Rejane Maria Tommasini Grotto; Ricardo Haddad; Sandra Coccuzzo Sampaio Vessoni; Simone Kashima; Svetoslav Nanev Slavov; Vincent Louis Viala |
| EPI_ISL_2445558, EPI_ISL_2445559, EPI_ISL_2445574, EPI_ISL_2445575, EPI_ISL_2445576 | POSTO DE SAUDE BENVIRA E S F 4 | Instituto Butantan | Antonio Jorge Martins; Claudia Renata dos Santos Barros; David Schlesinger; Debora Botequio Moretti; Dimas Tadeu Covas; Elaine Cristina Marqueze; Elaine Vieira Santos; Evandra Strazza Rodrigues; Heidge Fukumasu; Jayme Augusto de Souza-Neto; José Salvatore Leister Patané; Luiz Alcantara; Luiz Lehmann Coutinho; Maria Carolina Elias; Maurício Lacerda Nogueira; Rafael dos Santos Bezerra; Raul Machado Neto; Rejane Maria Tommasini Grotto; Ricardo Haddad; Sandra Coccuzzo Sampaio Vessoni; Simone Kashima; Svetoslav Nanev Slavov; Vincent Louis Viala |
| EPI_ISL_2445426 | POSTO DE SAUDE DO MARACANA ATIBAIA | Instituto Butantan | Antonio Jorge Martins; Claudia Renata dos Santos Barros; David Schlesinger; Debora Botequio Moretti; Dimas Tadeu Covas; Elaine Cristina Marqueze; Elaine Vieira Santos; Evandra Strazza Rodrigues; Heidge Fukumasu; Jayme Augusto de Souza-Neto; José Salvatore Leister Patané; Luiz Alcantara; Luiz Lehmann Coutinho; Maria Carolina Elias; Maurício Lacerda Nogueira; Rafael dos Santos Bezerra; Raul Machado Neto; Rejane Maria Tommasini Grotto; Ricardo Haddad; Sandra Coccuzzo Sampaio Vessoni; Simone Kashima; Svetoslav Nanev Slavov; Vincent Louis Viala |
| EPI_ISL_2445402 | POSTO DE SAUDE DO TANQUE ATIBAIA | Instituto Butantan | Antonio Jorge Martins; Claudia Renata dos Santos Barros; David Schlesinger; Debora Botequio Moretti; Dimas Tadeu Covas; Elaine Cristina Marqueze; Elaine Vieira Santos; Evandra Strazza Rodrigues; Heidge Fukumasu; Jayme Augusto de Souza-Neto; José Salvatore Leister Patané; Luiz Alcantara; Luiz Lehmann Coutinho; Maria Carolina Elias; Maurício Lacerda Nogueira; Rafael dos Santos Bezerra; Raul Machado Neto; Rejane Maria Tommasini Grotto; Ricardo Haddad; Sandra Coccuzzo Sampaio Vessoni; Simone Kashima; Svetoslav Nanev Slavov; Vincent Louis Viala |

| | | | |
|---|---|---|---|
| EPI_ISL_2493393 | POSTO MEDICO CIDADE NOVA | Instituto Butantan | Antonio Jorge Martins; Claudia Renata dos Santos Barros; David Schlesinger; Debora Botequio Moretti; Dimas Tadeu Covas; Elaine Cristina Marqueze; Elaine Vieira Santos; Evandra Strazza Rodrigues; Heidge Fukumasu; Jayme Augusto de Souza-Neto; José Salvatore Leister Patané; Luiz Alcantara; Luiz Lehmann Coutinho; Maria Carolina Elias; Maurício Lacerda Nogueira; Rafael dos Santos Bezerra; Raul Machado Neto; Rejane Maria Tommasini Grotto; Ricardo Haddad; Sandra Coccuzzo Sampaio Vessoni; Simone Kashima; Svetoslav Nanev Slavov; Vincent Louis Viala |
| EPI_ISL_2493589, EPI_ISL_2493590, EPI_ISL_2493629 | POSTO MEDICO NATAL DIEGUES DE ESTIVA GERBI | Instituto Butantan | Antonio Jorge Martins; Claudia Renata dos Santos Barros; David Schlesinger; Debora Botequio Moretti; Dimas Tadeu Covas; Elaine Cristina Marqueze; Elaine Vieira Santos; Evandra Strazza Rodrigues; Heidge Fukumasu; Jayme Augusto de Souza-Neto; José Salvatore Leister Patané; Luiz Alcantara; Luiz Lehmann Coutinho; Maria Carolina Elias; Maurício Lacerda Nogueira; Rafael dos Santos Bezerra; Raul Machado Neto; Rejane Maria Tommasini Grotto; Ricardo Haddad; Sandra Coccuzzo Sampaio Vessoni; Simone Kashima; Svetoslav Nanev Slavov; Vincent Louis Viala |
| EPI_ISL_2378747, EPI_ISL_2493277, EPI_ISL_2493355, EPI_ISL_2493694, EPI_ISL_2493695 | PROGRAMA SAUDE DA FAMILIA I IEPE | Instituto Butantan | Antonio Jorge Martins; Claudia Renata dos Santos Barros; David Schlesinger; Debora Botequio Moretti; Dimas Tadeu Covas; Elaine Cristina Marqueze; Elaine Vieira Santos; Evandra Strazza Rodrigues; Heidge Fukumasu; Jayme Augusto de Souza-Neto; José Salvatore Leister Patané; Luiz Alcantara; Luiz Lehmann Coutinho; Maria Carolina Elias; Maurício Lacerda Nogueira; Rafael dos Santos Bezerra; Raul Machado Neto; Rejane Maria Tommasini Grotto; Ricardo Haddad; Sandra Coccuzzo Sampaio Vessoni; Simone Kashima; Svetoslav Nanev Slavov; Vincent Louis Viala |
| EPI_ISL_2445241, EPI_ISL_2445244, EPI_ISL_2445252, EPI_ISL_2445368, EPI_ISL_2445369, EPI_ISL_2445370, EPI_ISL_2493252, EPI_ISL_2493875, EPI_ISL_2493878, EPI_ISL_2493883, EPI_ISL_2493891, EPI_ISL_2493896, EPI_ISL_2493899, EPI_ISL_2493901, EPI_ISL_2493902, EPI_ISL_2493904, EPI_ISL_2493910, EPI_ISL_2493911, EPI_ISL_2493915, EPI_ISL_2493931, EPI_ISL_2493932, EPI_ISL_2493933, EPI_ISL_2493934, EPI_ISL_2493935, EPI_ISL_2493936, EPI_ISL_2494295, EPI_ISL_2494325, EPI_ISL_2494326 | | | |
| see above | PRONTO ATENDIMENTO DE VARGEM GRANDE PAULISTA | Instituto Butantan | Antonio Jorge Martins; Claudia Renata dos Santos Barros; David Schlesinger; Debora Botequio Moretti; Dimas Tadeu Covas; Elaine Cristina Marqueze; Elaine Vieira Santos; Evandra Strazza Rodrigues; Heidge Fukumasu; Jayme Augusto de Souza-Neto; José Salvatore Leister Patané; Luiz Alcantara; Luiz Lehmann Coutinho; Maria Carolina Elias; Maurício Lacerda Nogueira; Rafael dos Santos Bezerra; Raul Machado Neto; Rejane Maria Tommasini Grotto; Ricardo Haddad; Sandra Coccuzzo Sampaio Vessoni; Simone Kashima; Svetoslav Nanev Slavov; Vincent Louis Viala |
| EPI_ISL_2345714 | PRONTO ATENDIMENTO MUNICIPAL | Instituto Butantan / Mendelics | Antonio Jorge Martins; Claudia Renata dos Santos Barros; David Schlesinger; Debora Botequio Moretti; Dimas Tadeu Covas; Elaine Cristina Marqueze; Elaine Vieira Santos; Evandra Strazza Rodrigues; Heidge Fukumasu; Jayme Augusto de Souza-Neto; José Salvatore Leister Patané; Luiz Alcantara; Luiz Lehmann Coutinho; Maria Carolina Elias; Maurício Lacerda Nogueira; Rafael dos Santos Bezerra; Raul Machado Neto; Rejane Maria Tommasini Grotto; Ricardo Haddad; Sandra Coccuzzo Sampaio Vessoni; Simone Kashima; Svetoslav Nanev Slavov; Vincent Louis Viala |
| EPI_ISL_2445606 | PRONTO ATENDIMENTO MUNICIPAL ALUMINIO | Instituto Butantan | Antonio Jorge Martins; Claudia Renata dos Santos Barros; David Schlesinger; Debora Botequio Moretti; Dimas Tadeu Covas; Elaine Cristina Marqueze; Elaine Vieira Santos; Evandra Strazza Rodrigues; Heidge Fukumasu; Jayme Augusto de Souza-Neto; José Salvatore Leister Patané; Luiz Alcantara; Luiz Lehmann Coutinho; Maria Carolina Elias; Maurício Lacerda Nogueira; Rafael dos Santos Bezerra; Raul Machado Neto; Rejane Maria Tommasini Grotto; Ricardo Haddad; Sandra Coccuzzo Sampaio Vessoni; Simone Kashima; Svetoslav Nanev Slavov; Vincent Louis Viala |
| EPI_ISL_2493470 | PRONTO ATENDIMENTO SAO JOSE | Instituto Butantan | Antonio Jorge Martins; Claudia Renata dos Santos Barros; David Schlesinger; Debora Botequio Moretti; Dimas Tadeu Covas; Elaine Cristina Marqueze; Elaine Vieira Santos; Evandra Strazza Rodrigues; Heidge Fukumasu; Jayme Augusto de Souza-Neto; José Salvatore Leister Patané; Luiz Alcantara; Luiz Lehmann Coutinho; Maria Carolina Elias; Maurício Lacerda Nogueira; Rafael dos Santos Bezerra; Raul Machado Neto; Rejane Maria Tommasini Grotto; Ricardo Haddad; Sandra Coccuzzo Sampaio Vessoni; Simone Kashima; Svetoslav Nanev Slavov; Vincent Louis Viala |
| EPI_ISL_2493552, EPI_ISL_2493556, EPI_ISL_2493564 | PRONTO SOCORRO DE AGENOR DE CAMPOS MONGAGUA | Instituto Butantan | Antonio Jorge Martins; Claudia Renata dos Santos Barros; David Schlesinger; Debora Botequio Moretti; Dimas Tadeu Covas; Elaine Cristina Marqueze; Elaine Vieira Santos; Evandra Strazza Rodrigues; Heidge Fukumasu; Jayme Augusto de Souza-Neto; José Salvatore Leister Patané; Luiz Alcantara; Luiz Lehmann Coutinho; Maria Carolina Elias; Maurício Lacerda Nogueira; Rafael dos Santos Bezerra; Raul Machado Neto; Rejane Maria Tommasini Grotto; Ricardo Haddad; Sandra Coccuzzo Sampaio Vessoni; Simone Kashima; Svetoslav Nanev Slavov; Vincent Louis Viala |
| EPI_ISL_2493594, EPI_ISL_2493595, EPI_ISL_2493613, EPI_ISL_2493618, EPI_ISL_2493624, EPI_ISL_2493626 | PRONTO SOCORRO MUNICIPAL DE CRAVINHOS | Instituto Butantan | Antonio Jorge Martins; Claudia Renata dos Santos Barros; David Schlesinger; Debora Botequio Moretti; Dimas Tadeu Covas; Elaine Cristina Marqueze; Elaine Vieira Santos; Evandra Strazza Rodrigues; Heidge Fukumasu; Jayme Augusto de Souza-Neto; José Salvatore Leister Patané; Luiz Alcantara; Luiz Lehmann Coutinho; Maria Carolina Elias; Maurício Lacerda Nogueira; Rafael dos Santos Bezerra; Raul Machado Neto; Rejane Maria Tommasini Grotto; Ricardo Haddad; Sandra Coccuzzo Sampaio Vessoni; Simone Kashima; Svetoslav Nanev Slavov; Vincent Louis Viala |
| EPI_ISL_2445374, EPI_ISL_2445375, EPI_ISL_2445376, EPI_ISL_2445377, EPI_ISL_2445378, EPI_ISL_2445379, EPI_ISL_2445380, EPI_ISL_2473695, EPI_ISL_2473782 | | | |
| see above | PRONTO SOCORRO MUNICIPAL DE SEVERINIA | Instituto Butantan | Antonio Jorge Martins; Claudia Renata dos Santos Barros; David Schlesinger; Debora Botequio Moretti; Dimas Tadeu Covas; Elaine Cristina Marqueze; Elaine Vieira Santos; Evandra Strazza Rodrigues; Heidge Fukumasu; Jayme Augusto de Souza-Neto; José Salvatore Leister Patané; Luiz Alcantara; Luiz Lehmann Coutinho; Maria Carolina Elias; Maurício Lacerda Nogueira; Rafael dos Santos Bezerra; Raul Machado Neto; Rejane Maria Tommasini Grotto; Ricardo Haddad; Sandra Coccuzzo Sampaio Vessoni; Simone Kashima; Svetoslav Nanev Slavov; Vincent Louis Viala |
| EPI_ISL_2493525, EPI_ISL_2493527 | PRONTO SOCORRO MUNICIPAL TAMBAU | Instituto Butantan | Antonio Jorge Martins; Claudia Renata dos Santos Barros; David Schlesinger; Debora Botequio Moretti; Dimas Tadeu Covas; Elaine Cristina Marqueze; Elaine Vieira Santos; Evandra Strazza Rodrigues; Heidge Fukumasu; Jayme Augusto de Souza-Neto; José Salvatore Leister Patané; Luiz Alcantara; Luiz Lehmann Coutinho; Maria Carolina Elias; Maurício Lacerda Nogueira; Rafael dos Santos Bezerra; Raul Machado Neto; Rejane Maria Tommasini Grotto; Ricardo Haddad; Sandra Coccuzzo Sampaio Vessoni; Simone Kashima; Svetoslav Nanev Slavov; Vincent Louis Viala |
| EPI_ISL_2345729 | PRONTO SOCORRO MUNICIPAL TAMBAU | Instituto Butantan / Mendelics | Antonio Jorge Martins; Claudia Renata dos Santos Barros; David Schlesinger; Debora Botequio Moretti; Dimas Tadeu Covas; Elaine Cristina Marqueze; Elaine Vieira Santos; Evandra Strazza Rodrigues; Heidge Fukumasu; Jayme Augusto de Souza-Neto; José Salvatore Leister Patané; Luiz Alcantara; Luiz Lehmann Coutinho; Maria Carolina Elias; Maurício Lacerda Nogueira; Rafael dos Santos Bezerra; Raul Machado Neto; Rejane Maria Tommasini Grotto; Ricardo Haddad; Sandra Coccuzzo Sampaio Vessoni; Simone Kashima; Svetoslav Nanev Slavov; Vincent Louis Viala |
| EPI_ISL_2493846, EPI_ISL_2493847 | PS E AMBULATORIO ENGENHEIRO CARDOSO | Instituto Butantan | Antonio Jorge Martins; Claudia Renata dos Santos Barros; David Schlesinger; Debora Botequio Moretti; Dimas Tadeu Covas; Elaine Cristina Marqueze; Elaine Vieira Santos; Evandra Strazza Rodrigues; Heidge Fukumasu; Jayme Augusto de Souza-Neto; José Salvatore Leister Patané; Luiz Alcantara; Luiz Lehmann Coutinho; Maria Carolina Elias; Maurício Lacerda Nogueira; Rafael dos Santos Bezerra; Raul Machado Neto; Rejane Maria Tommasini Grotto; Ricardo Haddad; Sandra Coccuzzo Sampaio Vessoni; Simone Kashima; Svetoslav Nanev Slavov; Vincent Louis Viala |
| EPI_ISL_2445546, EPI_ISL_2445589, EPI_ISL_2445590, EPI_ISL_2493900 | PSF DR ANTONIO PIRES DE ALMEIDA PORTO FELIZ | Instituto Butantan | Antonio Jorge Martins; Claudia Renata dos Santos Barros; David Schlesinger; Debora Botequio Moretti; Dimas Tadeu Covas; Elaine Cristina Marqueze; Elaine Vieira Santos; Evandra Strazza Rodrigues; Heidge Fukumasu; Jayme Augusto de Souza-Neto; José Salvatore Leister Patané; Luiz Alcantara; Luiz Lehmann Coutinho; Maria Carolina Elias; Maurício Lacerda Nogueira; Rafael dos Santos Bezerra; Raul Machado Neto; Rejane Maria Tommasini Grotto; Ricardo Haddad; Sandra Coccuzzo Sampaio Vessoni; Simone Kashima; Svetoslav Nanev Slavov; Vincent Louis Viala |
| EPI_ISL_2445582 | PSF MARIA JOSE SALTO DE PIRAPORA | Instituto Butantan | Antonio Jorge Martins; Claudia Renata dos Santos Barros; David Schlesinger; Debora Botequio Moretti; Dimas Tadeu Covas; Elaine Cristina Marqueze; Elaine Vieira Santos; Evandra Strazza Rodrigues; Heidge Fukumasu; Jayme Augusto de Souza-Neto; José Salvatore Leister Patané; Luiz Alcantara; Luiz Lehmann Coutinho; Maria Carolina Elias; Maurício Lacerda Nogueira; Rafael dos Santos Bezerra; Raul Machado Neto; Rejane Maria Tommasini Grotto; Ricardo Haddad; Sandra Coccuzzo Sampaio Vessoni; Simone Kashima; Svetoslav Nanev Slavov; Vincent Louis Viala |
| EPI_ISL_2493179, EPI_ISL_2493189, EPI_ISL_2493640, EPI_ISL_2493644, EPI_ISL_2493645 | PSF VIRGINIO BASSO | Instituto Butantan | Antonio Jorge Martins; Claudia Renata dos Santos Barros; David Schlesinger; Debora Botequio Moretti; Dimas Tadeu Covas; Elaine Cristina Marqueze; Elaine Vieira Santos; Evandra Strazza Rodrigues; Heidge Fukumasu; Jayme Augusto de Souza-Neto; José Salvatore Leister Patané; Luiz Alcantara; Luiz Lehmann Coutinho; Maria Carolina Elias; Maurício Lacerda Nogueira; Rafael dos Santos Bezerra; Raul Machado Neto; Rejane Maria Tommasini Grotto; Ricardo Haddad; Sandra Coccuzzo Sampaio Vessoni; Simone Kashima; Svetoslav Nanev Slavov; Vincent Louis Viala |
| EPI_ISL_2663311 | Plataforma de Vigilancia Molecular (PVM) - FIOCRUZ/BA | Plataforma de Vigilancia Molecular (PVM) - FIOCRUZ/BA | Bruno Bezerril Andrade; Camila I. de Oliveira on behalf of the Fiocruz COVID-19 Genomic Surveillance Network.; Clarissa Araújo Gurgel; Leonardo Paiva Farias; Marina Cucco; Ricardo Khouri; Tiago Graf |
| EPI_ISL_2445155, EPI_ISL_2445156, EPI_ISL_2445159, EPI_ISL_2445161, EPI_ISL_2445162, EPI_ISL_2445163, EPI_ISL_2445166, EPI_ISL_2445167, EPI_ISL_2445170, EPI_ISL_2445172, EPI_ISL_2445174, EPI_ISL_2445179, EPI_ISL_2445405, EPI_ISL_2445409, EPI_ISL_2445411, EPI_ISL_2445413, EPI_ISL_2445418, EPI_ISL_2473708, EPI_ISL_2473709 | | | |
| see above | SAE CTA SERV ATEND ESPEC TESTAGEM E ACONSELHAMENTO | Instituto Butantan | Antonio Jorge Martins; Claudia Renata dos Santos Barros; David Schlesinger; Debora Botequio Moretti; Dimas Tadeu Covas; Elaine Cristina Marqueze; Elaine Vieira Santos; Evandra Strazza Rodrigues; Heidge Fukumasu; Jayme Augusto de Souza-Neto; José Salvatore Leister Patané; Luiz Alcantara; Luiz Lehmann Coutinho; Maria Carolina Elias; Maurício Lacerda Nogueira; Rafael dos Santos Bezerra; Raul Machado Neto; Rejane Maria Tommasini Grotto; Ricardo Haddad; Sandra Coccuzzo Sampaio Vessoni; Simone Kashima; Svetoslav Nanev Slavov; Vincent Louis Viala |
| EPI_ISL_2445425 | SANTA CASA DE ATIBAIA PRO SAUDE | Instituto Butantan | Antonio Jorge Martins; Claudia Renata dos Santos Barros; David Schlesinger; Debora Botequio Moretti; Dimas Tadeu Covas; Elaine Cristina Marqueze; Elaine Vieira Santos; Evandra Strazza Rodrigues; Heidge Fukumasu; Jayme Augusto de Souza-Neto; José Salvatore Leister Patané; Luiz Alcantara; Luiz Lehmann Coutinho; Maria Carolina Elias; Maurício Lacerda Nogueira; Rafael dos Santos Bezerra; Raul Machado Neto; Rejane Maria Tommasini Grotto; Ricardo Haddad; Sandra Coccuzzo Sampaio Vessoni; Simone Kashima; Svetoslav Nanev Slavov; Vincent Louis Viala |
| EPI_ISL_2445133, EPI_ISL_2445137, EPI_ISL_2445139, EPI_ISL_2445140, EPI_ISL_2445141, EPI_ISL_2445142, EPI_ISL_2445143, EPI_ISL_2473752, EPI_ISL_2473755 | | | |
| see above | SANTA CASA DE DOIS CORREGOS | Instituto Butantan | Antonio Jorge Martins; Claudia Renata dos Santos Barros; David Schlesinger; Debora Botequio Moretti; Dimas Tadeu Covas; Elaine Cristina Marqueze; Elaine Vieira Santos; Evandra Strazza Rodrigues; Heidge Fukumasu; Jayme Augusto de Souza-Neto; José Salvatore Leister Patané; Luiz Alcantara; Luiz Lehmann Coutinho; Maria Carolina Elias; Maurício Lacerda Nogueira; Rafael dos Santos Bezerra; Raul Machado Neto; Rejane Maria Tommasini Grotto; Ricardo Haddad; Sandra Coccuzzo Sampaio Vessoni; Simone Kashima; Svetoslav Nanev Slavov; Vincent Louis Viala |
| EPI_ISL_2473801 | SANTA CASA DE MISERICORDIA DE SAO LUIZ DO PARAITINGA | Instituto Butantan | Antonio Jorge Martins; Claudia Renata dos Santos Barros; David Schlesinger; Debora Botequio Moretti; Dimas Tadeu Covas; Elaine Cristina Marqueze; Elaine Vieira Santos; Evandra Strazza Rodrigues; Heidge Fukumasu; Jayme Augusto de Souza-Neto; José Salvatore Leister Patané; Luiz Alcantara; Luiz Lehmann Coutinho; Maria Carolina Elias; Maurício Lacerda Nogueira; Rafael dos Santos Bezerra; Raul Machado Neto; Rejane Maria Tommasini Grotto; Ricardo Haddad; Sandra Coccuzzo Sampaio Vessoni; Simone Kashima; Svetoslav Nanev Slavov; Vincent Louis Viala |
| EPI_ISL_2445604, EPI_ISL_2493681, EPI_ISL_2493867 | SANTA CASA DE MISERICORDIA DE TIETE | Instituto Butantan | Antonio Jorge Martins; Claudia Renata dos Santos Barros; David Schlesinger; Debora Botequio Moretti; Dimas Tadeu Covas; Elaine Cristina Marqueze; Elaine Vieira Santos; Evandra Strazza Rodrigues; Heidge Fukumasu; Jayme Augusto de Souza-Neto; José Salvatore Leister Patané; Luiz Alcantara; Luiz Lehmann Coutinho; Maria Carolina Elias; Maurício Lacerda Nogueira; Rafael dos Santos Bezerra; Raul Machado Neto; Rejane Maria Tommasini Grotto; Ricardo Haddad; Sandra Coccuzzo Sampaio Vessoni; Simone Kashima; Svetoslav Nanev Slavov; Vincent Louis Viala |
| EPI_ISL_2445586, EPI_ISL_2445587, EPI_ISL_2493880 | SANTA CASA DE PORTO FELIZ | Instituto Butantan | Antonio Jorge Martins; Claudia Renata dos Santos Barros; David Schlesinger; Debora Botequio Moretti; Dimas Tadeu Covas; Elaine Cristina Marqueze; Elaine Vieira Santos; Evandra Strazza Rodrigues; Heidge Fukumasu; Jayme Augusto de Souza-Neto; José Salvatore Leister Patané; Luiz Alcantara; Luiz Lehmann Coutinho; Maria Carolina Elias; Maurício Lacerda Nogueira; Rafael dos Santos Bezerra; Raul Machado Neto; Rejane Maria Tommasini Grotto; Ricardo Haddad; Sandra Coccuzzo Sampaio Vessoni; Simone Kashima; Svetoslav Nanev Slavov; Vincent Louis Viala |
| EPI_ISL_2493909 | SANTA CASA SALTO DE PIRAPORA | Instituto Butantan | Antonio Jorge Martins; Claudia Renata dos Santos Barros; David Schlesinger; Debora Botequio Moretti; Dimas Tadeu Covas; Elaine Cristina Marqueze; Elaine Vieira Santos; Evandra Strazza Rodrigues; Heidge Fukumasu; Jayme Augusto de Souza-Neto; José Salvatore Leister Patané; Luiz Alcantara; Luiz Lehmann Coutinho; Maria Carolina Elias; Maurício Lacerda Nogueira; Rafael dos Santos Bezerra; Raul Machado Neto; Rejane Maria Tommasini Grotto; Ricardo Haddad; Sandra Coccuzzo Sampaio Vessoni; Simone Kashima; Svetoslav Nanev Slavov; Vincent Louis Viala |
| EPI_ISL_2494169, EPI_ISL_2494177, EPI_ISL_2494189, EPI_ISL_2494196 | SANTA CASA SANTA RITA DO PASSA QUATRO | Instituto Butantan | Antonio Jorge Martins; Claudia Renata dos Santos Barros; David Schlesinger; Debora Botequio Moretti; Dimas Tadeu Covas; Elaine Cristina Marqueze; Elaine Vieira Santos; Evandra Strazza Rodrigues; Heidge Fukumasu; Jayme Augusto de Souza-Neto; José Salvatore Leister Patané; Luiz Alcantara; Luiz Lehmann Coutinho; Maria Carolina Elias; Maurício Lacerda Nogueira; Rafael dos Santos Bezerra; Raul Machado Neto; Rejane Maria Tommasini Grotto; Ricardo Haddad; Sandra Coccuzzo Sampaio Vessoni; Simone Kashima; Svetoslav Nanev Slavov; Vincent Louis Viala |
| EPI_ISL_2444193, EPI_ISL_2444194, EPI_ISL_2444196, EPI_ISL_2444197, EPI_ISL_2444198, EPI_ISL_2444200, EPI_ISL_2444201, EPI_ISL_2444204, EPI_ISL_2444205, EPI_ISL_2444206, EPI_ISL_2444207, EPI_ISL_2444208, EPI_ISL_2444209, EPI_ISL_2444211, EPI_ISL_2444213, EPI_ISL_2444214, EPI_ISL_2444215, EPI_ISL_2444216, EPI_ISL_2444217, EPI_ISL_2444218, EPI_ISL_2444219, EPI_ISL_2444220, EPI_ISL_2444221, EPI_ISL_2444222, EPI_ISL_2455397 | | | |
| see above | SAO JOSE DO RIO PRETO | Instituto Butantan / FAMERP | Antonio Jorge Martins; Claudia Renata dos Santos Barros; David Schlesinger; Debora Botequio Moretti; Dimas Tadeu Covas; Elaine Cristina Marqueze; Elaine Vieira Santos; Evandra Strazza Rodrigues; Heidge Fukumasu; Jayme Augusto de Souza-Neto; José Salvatore Leister Patané; Luiz Alcantara; Luiz Lehmann Coutinho; Maria Carolina Elias; Maurício Lacerda Nogueira; Rafael dos Santos Bezerra; Raul Machado Neto; Rejane Maria Tommasini Grotto; Ricardo Haddad; Sandra Coccuzzo Sampaio Vessoni; Simone Kashima; Svetoslav Nanev Slavov; Vincent Louis Viala |
| EPI_ISL_2445323, EPI_ISL_2445325, EPI_ISL_2493157, EPI_ISL_2494100 | SECAO CENTRO DE DIAGNOSTICO SECEDI | Instituto Butantan | Antonio Jorge Martins; Claudia Renata dos Santos Barros; David Schlesinger; Debora Botequio Moretti; Dimas Tadeu Covas; Elaine Cristina Marqueze; Elaine Vieira Santos; Evandra Strazza Rodrigues; Heidge Fukumasu; Jayme Augusto de Souza-Neto; José Salvatore Leister Patané; Luiz Alcantara; Luiz Lehmann Coutinho; Maria Carolina Elias; Maurício Lacerda Nogueira; Rafael dos Santos Bezerra; Raul Machado Neto; Rejane Maria Tommasini Grotto; Ricardo Haddad; Sandra Coccuzzo Sampaio Vessoni; Simone Kashima; Svetoslav Nanev Slavov; Vincent Louis Viala |
| EPI_ISL_2493619, EPI_ISL_2493621, EPI_ISL_2493627 | SECRETARIA DE SAUDE | Instituto Butantan | Antonio Jorge Martins; Claudia Renata dos Santos Barros; David Schlesinger; Debora Botequio Moretti; Dimas Tadeu Covas; Elaine Cristina Marqueze; Elaine Vieira Santos; Evandra Strazza Rodrigues; Heidge Fukumasu; Jayme Augusto de Souza-Neto; José Salvatore Leister Patané; Luiz Alcantara; Luiz Lehmann Coutinho; Maria Carolina Elias; Maurício Lacerda Nogueira; Rafael dos Santos Bezerra; Raul Machado Neto; Rejane Maria Tommasini Grotto; Ricardo Haddad; Sandra Coccuzzo Sampaio Vessoni; Simone Kashima; Svetoslav Nanev Slavov; Vincent Louis Viala |
| EPI_ISL_2445464, EPI_ISL_2445465, EPI_ISL_2445466, EPI_ISL_2445467, EPI_ISL_2445468, EPI_ISL_2445469, EPI_ISL_2445470, EPI_ISL_2445471, EPI_ISL_2445472, EPI_ISL_2445473, EPI_ISL_2445474, EPI_ISL_2445475, EPI_ISL_2445476, EPI_ISL_2445477, EPI_ISL_2445478 | | | |
| see above | SECRETARIA DE SAUDE DE LOUVEIRA | Instituto Butantan | Antonio Jorge Martins; Claudia Renata dos Santos Barros; David Schlesinger; Debora Botequio Moretti; Dimas Tadeu Covas; Elaine Cristina Marqueze; Elaine Vieira Santos; Evandra Strazza Rodrigues; Heidge Fukumasu; Jayme Augusto de Souza-Neto; José Salvatore Leister Patané; Luiz Alcantara; Luiz Lehmann Coutinho; Maria Carolina Elias; Maurício Lacerda Nogueira; Rafael dos Santos Bezerra; Raul Machado Neto; Rejane Maria Tommasini Grotto; Ricardo Haddad; Sandra Coccuzzo Sampaio Vessoni; Simone Kashima; Svetoslav Nanev Slavov; Vincent Louis Viala |
| EPI_ISL_2493439, EPI_ISL_2494147, EPI_ISL_2494148 | SECRETARIA DE SAUDE DE RAFARD | Instituto Butantan | Antonio Jorge Martins; Claudia Renata dos Santos Barros; David Schlesinger; Debora Botequio Moretti; Dimas Tadeu Covas; Elaine Cristina Marqueze; Elaine Vieira Santos; Evandra Strazza Rodrigues; Heidge Fukumasu; Jayme Augusto de Souza-Neto; José Salvatore Leister Patané; Luiz Alcantara; Luiz Lehmann Coutinho; Maria Carolina Elias; Maurício Lacerda Nogueira; Rafael dos Santos Bezerra; Raul Machado Neto; Rejane Maria Tommasini Grotto; Ricardo Haddad; Sandra Coccuzzo Sampaio Vessoni; Simone Kashima; Svetoslav Nanev Slavov; Vincent Louis Viala |
| EPI_ISL_2493373, EPI_ISL_2493389, EPI_ISL_2493390, EPI_ISL_2493447, EPI_ISL_2494149, EPI_ISL_2494156, EPI_ISL_2494161, EPI_ISL_2494170, EPI_ISL_2494178 | | | |
| see above | SECRETARIA DE SAUDE DE SANTA GERTRUDES | Instituto Butantan | Antonio Jorge Martins; Claudia Renata dos Santos Barros; David Schlesinger; Debora Botequio Moretti; Dimas Tadeu Covas; Elaine Cristina Marqueze; Elaine Vieira Santos; Evandra Strazza Rodrigues; Heidge Fukumasu; Jayme Augusto de Souza-Neto; José Salvatore Leister Patané; Luiz Alcantara; Luiz Lehmann Coutinho; Maria Carolina Elias; Maurício Lacerda Nogueira; Rafael dos Santos Bezerra; Raul Machado Neto; Rejane Maria Tommasini Grotto; Ricardo Haddad; Sandra Coccuzzo Sampaio Vessoni; Simone Kashima; Svetoslav Nanev Slavov; Vincent Louis Viala |

| | | | |
|---|---|---|---|
| EPI_ISL_2493281, EPI_ISL_2493371, EPI_ISL_2493388, EPI_ISL_2493444, EPI_ISL_2494155, EPI_ISL_2494167, EPI_ISL_2494190, EPI_ISL_2494280, EPI_ISL_2494286, EPI_ISL_2494287 | | | |
| see above | SECRETARIA DE SAUDE DE SAO PEDRO | Instituto Butantan | Antonio Jorge Martins; Claudia Renata dos Santos Barros; David Schlesinger; Debora Botequio Moretti; Dimas Tadeu Covas; Elaine Cristina Marqueze; Elaine Vieira Santos; Evandra Strazza Rodrigues; Heidge Fukumasu; Jayme Augusto de Souza-Neto; José Salvatore Leister Patané; Luiz Alcantara; Luiz Lehmann Coutinho; Maria Carolina Elias; Maurício Lacerda Nogueira; Rafael dos Santos Bezerra; Raul Machado Neto; Rejane Maria Tommasini Grotto; Ricardo Haddad; Sandra Coccuzzo Sampaio Vessoni; Simone Kashima; Svetoslav Nanev Slavov; Vincent Louis Viala |
| EPI_ISL_2209516, EPI_ISL_2209668, EPI_ISL_2209905, EPI_ISL_2210305, EPI_ISL_2345661, EPI_ISL_2345665, EPI_ISL_2345704, EPI_ISL_2345752, EPI_ISL_2345760, EPI_ISL_2345802, EPI_ISL_2345849 | | | |
| see above | SECRETARIA DE SAUDE PUBLICA DE PRAIA GRANDE | Instituto Butantan / Mendelics | Antonio Jorge Martins; Claudia Renata dos Santos Barros; David Schlesinger; Debora Botequio Moretti; Dimas Tadeu Covas; Elaine Cristina Marqueze; Elaine Vieira Santos; Evandra Strazza Rodrigues; Heidge Fukumasu; Jayme Augusto de Souza-Neto; José Salvatore Leister Patané; Luiz Alcantara; Luiz Lehmann Coutinho; Maria Carolina Elias; Maurício Lacerda Nogueira; Rafael dos Santos Bezerra; Raul Machado Neto; Rejane Maria Tommasini Grotto; Ricardo Haddad; Sandra Coccuzzo Sampaio Vessoni; Simone Kashima; Svetoslav Nanev Slavov; Vincent Louis Viala |
| EPI_ISL_2445573 | SECRETARIA MUNICIPAL DA SAUDE DE GUARATINGUETA | Instituto Butantan | Antonio Jorge Martins; Claudia Renata dos Santos Barros; David Schlesinger; Debora Botequio Moretti; Dimas Tadeu Covas; Elaine Cristina Marqueze; Elaine Vieira Santos; Evandra Strazza Rodrigues; Heidge Fukumasu; Jayme Augusto de Souza-Neto; José Salvatore Leister Patané; Luiz Alcantara; Luiz Lehmann Coutinho; Maria Carolina Elias; Maurício Lacerda Nogueira; Rafael dos Santos Bezerra; Raul Machado Neto; Rejane Maria Tommasini Grotto; Ricardo Haddad; Sandra Coccuzzo Sampaio Vessoni; Simone Kashima; Svetoslav Nanev Slavov; Vincent Louis Viala |
| EPI_ISL_2938096 | SECRETARIA MUNICIPAL DA SAUDE DE GUARATINGUETA LABORATÓRIO 01 | Instituto Butantan | Antonio Jorge Martins; Claudia Renata dos Santos Barros; David Schlesinger; Debora Botequio Moretti; Dimas Tadeu Covas; Elaine Cristina Marqueze; Elaine Vieira Santos; Evandra Strazza Rodrigues; Heidge Fukumasu; Jayme Augusto de Souza-Neto; José Salvatore Leister Patané; Luiz Alcantara; Luiz Lehmann Coutinho; Maria Carolina Elias; Maurício Lacerda Nogueira; Rafael dos Santos Bezerra; Raul Machado Neto; Rejane Maria Tommasini Grotto; Ricardo Haddad; Sandra Coccuzzo Sampaio Vessoni; Simone Kashima; Svetoslav Nanev Slavov; Vincent Louis Viala |
| EPI_ISL_2493598, EPI_ISL_2493601, EPI_ISL_2493603, EPI_ISL_2493612 | SECRETARIA MUNICIPAL DA SAUDE DE GUARIBA | Instituto Butantan | Antonio Jorge Martins; Claudia Renata dos Santos Barros; David Schlesinger; Debora Botequio Moretti; Dimas Tadeu Covas; Elaine Cristina Marqueze; Elaine Vieira Santos; Evandra Strazza Rodrigues; Heidge Fukumasu; Jayme Augusto de Souza-Neto; José Salvatore Leister Patané; Luiz Alcantara; Luiz Lehmann Coutinho; Maria Carolina Elias; Maurício Lacerda Nogueira; Rafael dos Santos Bezerra; Raul Machado Neto; Rejane Maria Tommasini Grotto; Ricardo Haddad; Sandra Coccuzzo Sampaio Vessoni; Simone Kashima; Svetoslav Nanev Slavov; Vincent Louis Viala |
| EPI_ISL_2493290 | SECRETARIA MUNICIPAL DA SAUDE DE ITAPEVA SP | Instituto Butantan | Antonio Jorge Martins; Claudia Renata dos Santos Barros; David Schlesinger; Debora Botequio Moretti; Dimas Tadeu Covas; Elaine Cristina Marqueze; Elaine Vieira Santos; Evandra Strazza Rodrigues; Heidge Fukumasu; Jayme Augusto de Souza-Neto; José Salvatore Leister Patané; Luiz Alcantara; Luiz Lehmann Coutinho; Maria Carolina Elias; Maurício Lacerda Nogueira; Rafael dos Santos Bezerra; Raul Machado Neto; Rejane Maria Tommasini Grotto; Ricardo Haddad; Sandra Coccuzzo Sampaio Vessoni; Simone Kashima; Svetoslav Nanev Slavov; Vincent Louis Viala |
| EPI_ISL_2445598, EPI_ISL_2445599, EPI_ISL_2473806, EPI_ISL_2493903 | SECRETARIA MUNICIPAL DA SAUDE DE ITARARE | Instituto Butantan | Antonio Jorge Martins; Claudia Renata dos Santos Barros; David Schlesinger; Debora Botequio Moretti; Dimas Tadeu Covas; Elaine Cristina Marqueze; Elaine Vieira Santos; Evandra Strazza Rodrigues; Heidge Fukumasu; Jayme Augusto de Souza-Neto; José Salvatore Leister Patané; Luiz Alcantara; Luiz Lehmann Coutinho; Maria Carolina Elias; Maurício Lacerda Nogueira; Rafael dos Santos Bezerra; Raul Machado Neto; Rejane Maria Tommasini Grotto; Ricardo Haddad; Sandra Coccuzzo Sampaio Vessoni; Simone Kashima; Svetoslav Nanev Slavov; Vincent Louis Viala |
| EPI_ISL_2445416, EPI_ISL_2445417 | SECRETARIA MUNICIPAL DA SAUDE DE JOANOPOLIS | Instituto Butantan | Antonio Jorge Martins; Claudia Renata dos Santos Barros; David Schlesinger; Debora Botequio Moretti; Dimas Tadeu Covas; Elaine Cristina Marqueze; Elaine Vieira Santos; Evandra Strazza Rodrigues; Heidge Fukumasu; Jayme Augusto de Souza-Neto; José Salvatore Leister Patané; Luiz Alcantara; Luiz Lehmann Coutinho; Maria Carolina Elias; Maurício Lacerda Nogueira; Rafael dos Santos Bezerra; Raul Machado Neto; Rejane Maria Tommasini Grotto; Ricardo Haddad; Sandra Coccuzzo Sampaio Vessoni; Simone Kashima; Svetoslav Nanev Slavov; Vincent Louis Viala |
| EPI_ISL_2208862, EPI_ISL_2345831, EPI_ISL_2345875 | SECRETARIA MUNICIPAL DA SAUDE DE JOANOPOLIS | Instituto Butantan / Mendelics | Antonio Jorge Martins; Claudia Renata dos Santos Barros; David Schlesinger; Debora Botequio Moretti; Dimas Tadeu Covas; Elaine Cristina Marqueze; Elaine Vieira Santos; Evandra Strazza Rodrigues; Heidge Fukumasu; Jayme Augusto de Souza-Neto; José Salvatore Leister Patané; Luiz Alcantara; Luiz Lehmann Coutinho; Maria Carolina Elias; Maurício Lacerda Nogueira; Rafael dos Santos Bezerra; Raul Machado Neto; Rejane Maria Tommasini Grotto; Ricardo Haddad; Sandra Coccuzzo Sampaio Vessoni; Simone Kashima; Svetoslav Nanev Slavov; Vincent Louis Viala |
| EPI_ISL_2462324, EPI_ISL_2493176, EPI_ISL_2493275, EPI_ISL_2493276, EPI_ISL_2493513, EPI_ISL_2493577, EPI_ISL_2493579, EPI_ISL_2493581, EPI_ISL_2493630, EPI_ISL_2493633, EPI_ISL_2493635, EPI_ISL_2493637, EPI_ISL_2494110, EPI_ISL_2494277, EPI_ISL_2494278 | | | |
| see above | SECRETARIA MUNICIPAL DE SAUDE | Instituto Butantan | Antonio Jorge Martins; Claudia Renata dos Santos Barros; David Schlesinger; Debora Botequio Moretti; Dimas Tadeu Covas; Elaine Cristina Marqueze; Elaine Vieira Santos; Evandra Strazza Rodrigues; Heidge Fukumasu; Jayme Augusto de Souza-Neto; José Salvatore Leister Patané; Luiz Alcantara; Luiz Lehmann Coutinho; Maria Carolina Elias; Maurício Lacerda Nogueira; Rafael dos Santos Bezerra; Raul Machado Neto; Rejane Maria Tommasini Grotto; Ricardo Haddad; Sandra Coccuzzo Sampaio Vessoni; Simone Kashima; Svetoslav Nanev Slavov; Vincent Louis Viala |
| EPI_ISL_2493156, EPI_ISL_2493184, EPI_ISL_2493283, EPI_ISL_2493367, EPI_ISL_2493382, EPI_ISL_2493396, EPI_ISL_2493427, EPI_ISL_2493428, EPI_ISL_2493452, EPI_ISL_2494158, EPI_ISL_2494185, EPI_ISL_2494194 | | | |
| see above | SECRETARIA MUNICIPAL DE SAUDE DE CORDEIROPOLIS | Instituto Butantan | Antonio Jorge Martins; Claudia Renata dos Santos Barros; David Schlesinger; Debora Botequio Moretti; Dimas Tadeu Covas; Elaine Cristina Marqueze; Elaine Vieira Santos; Evandra Strazza Rodrigues; Heidge Fukumasu; Jayme Augusto de Souza-Neto; José Salvatore Leister Patané; Luiz Alcantara; Luiz Lehmann Coutinho; Maria Carolina Elias; Maurício Lacerda Nogueira; Rafael dos Santos Bezerra; Raul Machado Neto; Rejane Maria Tommasini Grotto; Ricardo Haddad; Sandra Coccuzzo Sampaio Vessoni; Simone Kashima; Svetoslav Nanev Slavov; Vincent Louis Viala |
| EPI_ISL_2493825 | SECRETARIA MUNICIPAL DE SAUDE DE MORUNGABA | Instituto Butantan | Antonio Jorge Martins; Claudia Renata dos Santos Barros; David Schlesinger; Debora Botequio Moretti; Dimas Tadeu Covas; Elaine Cristina Marqueze; Elaine Vieira Santos; Evandra Strazza Rodrigues; Heidge Fukumasu; Jayme Augusto de Souza-Neto; José Salvatore Leister Patané; Luiz Alcantara; Luiz Lehmann Coutinho; Maria Carolina Elias; Maurício Lacerda Nogueira; Rafael dos Santos Bezerra; Raul Machado Neto; Rejane Maria Tommasini Grotto; Ricardo Haddad; Sandra Coccuzzo Sampaio Vessoni; Simone Kashima; Svetoslav Nanev Slavov; Vincent Louis Viala |
| EPI_ISL_2445342, EPI_ISL_2493802, EPI_ISL_2493823, EPI_ISL_2494306, EPI_ISL_2494307, EPI_ISL_2494308 | SECRETARIA MUNICIPAL DE SAUDE DE PEDERNEIRAS | Instituto Butantan | Antonio Jorge Martins; Claudia Renata dos Santos Barros; David Schlesinger; Debora Botequio Moretti; Dimas Tadeu Covas; Elaine Cristina Marqueze; Elaine Vieira Santos; Evandra Strazza Rodrigues; Heidge Fukumasu; Jayme Augusto de Souza-Neto; José Salvatore Leister Patané; Luiz Alcantara; Luiz Lehmann Coutinho; Maria Carolina Elias; Maurício Lacerda Nogueira; Rafael dos Santos Bezerra; Raul Machado Neto; Rejane Maria Tommasini Grotto; Ricardo Haddad; Sandra Coccuzzo Sampaio Vessoni; Simone Kashima; Svetoslav Nanev Slavov; Vincent Louis Viala |
| EPI_ISL_2209325, EPI_ISL_2345823, EPI_ISL_2345867 | SECRETARIA MUNICIPAL DE SAUDE DE PIRACAIA | Instituto Butantan / Mendelics | Antonio Jorge Martins; Claudia Renata dos Santos Barros; David Schlesinger; Debora Botequio Moretti; Dimas Tadeu Covas; Elaine Cristina Marqueze; Elaine Vieira Santos; Evandra Strazza Rodrigues; Heidge Fukumasu; Jayme Augusto de Souza-Neto; José Salvatore Leister Patané; Luiz Alcantara; Luiz Lehmann Coutinho; Maria Carolina Elias; Maurício Lacerda Nogueira; Rafael dos Santos Bezerra; Raul Machado Neto; Rejane Maria Tommasini Grotto; Ricardo Haddad; Sandra Coccuzzo Sampaio Vessoni; Simone Kashima; Svetoslav Nanev Slavov; Vincent Louis Viala |
| EPI_ISL_2462329, EPI_ISL_2494146 | SECRETARIA MUNICIPAL DE SAUDE DE PRES BERNARDES SP | Instituto Butantan | Antonio Jorge Martins; Claudia Renata dos Santos Barros; David Schlesinger; Debora Botequio Moretti; Dimas Tadeu Covas; Elaine Cristina Marqueze; Elaine Vieira Santos; Evandra Strazza Rodrigues; Heidge Fukumasu; Jayme Augusto de Souza-Neto; José Salvatore Leister Patané; Luiz Alcantara; Luiz Lehmann Coutinho; Maria Carolina Elias; Maurício Lacerda Nogueira; Rafael dos Santos Bezerra; Raul Machado Neto; Rejane Maria Tommasini Grotto; Ricardo Haddad; Sandra Coccuzzo Sampaio Vessoni; Simone Kashima; Svetoslav Nanev Slavov; Vincent Louis Viala |
| EPI_ISL_2345798 | SECRETARIA MUNICIPAL DE SAUDE DE PRES BERNARDES SP | Instituto Butantan / Mendelics | Antonio Jorge Martins; Claudia Renata dos Santos Barros; David Schlesinger; Debora Botequio Moretti; Dimas Tadeu Covas; Elaine Cristina Marqueze; Elaine Vieira Santos; Evandra Strazza Rodrigues; Heidge Fukumasu; Jayme Augusto de Souza-Neto; José Salvatore Leister Patané; Luiz Alcantara; Luiz Lehmann Coutinho; Maria Carolina Elias; Maurício Lacerda Nogueira; Rafael dos Santos Bezerra; Raul Machado Neto; Rejane Maria Tommasini Grotto; Ricardo Haddad; Sandra Coccuzzo Sampaio Vessoni; Simone Kashima; Svetoslav Nanev Slavov; Vincent Louis Viala |
| EPI_ISL_2801338, EPI_ISL_2801339, EPI_ISL_2801340, EPI_ISL_2801342, EPI_ISL_2801350, EPI_ISL_2801351, EPI_ISL_2801353, EPI_ISL_2801354, EPI_ISL_2801359, EPI_ISL_2801366 | | | |
| see above | SECRETARIA MUNICIPAL DE SAUDE DE TIANGUA | Oswaldo Cruz Institute, FIOCRUZ/CE | Cleber Furtado Aksenen e Suzana Porto Almeida; Fabio Miyajima; Fernando Braga Stehling; Francisco Eder de Moura Lopes; Jamille Maria Mendes Bezerra; Joaquim César do Nascimento Sousa Junior; Pedro Miguel Carneiro Jeronimo; Thais Ferreira de Oliveira; Thais de Oliveira Costa; Ticiane Cavalcante de Souza; Veridiana Pessoa Miyajima |
| EPI_ISL_2210198 | SECRETARIA MUNICIPAL DE SAUDE DE TUPI PAULISTA | Instituto Butantan / Mendelics | Antonio Jorge Martins; Claudia Renata dos Santos Barros; David Schlesinger; Debora Botequio Moretti; Dimas Tadeu Covas; Elaine Cristina Marqueze; Elaine Vieira Santos; Evandra Strazza Rodrigues; Heidge Fukumasu; Jayme Augusto de Souza-Neto; José Salvatore Leister Patané; Luiz Alcantara; Luiz Lehmann Coutinho; Maria Carolina Elias; Maurício Lacerda Nogueira; Rafael dos Santos Bezerra; Raul Machado Neto; Rejane Maria Tommasini Grotto; Ricardo Haddad; Sandra Coccuzzo Sampaio Vessoni; Simone Kashima; Svetoslav Nanev Slavov; Vincent Louis Viala |
| EPI_ISL_2445385, EPI_ISL_2445386, EPI_ISL_2445388, EPI_ISL_2445389, EPI_ISL_2445390, EPI_ISL_2445391 | SECRETARIA MUNICIPAL DE SAUDE DE VALPARAISO SP | Instituto Butantan | Antonio Jorge Martins; Claudia Renata dos Santos Barros; David Schlesinger; Debora Botequio Moretti; Dimas Tadeu Covas; Elaine Cristina Marqueze; Elaine Vieira Santos; Evandra Strazza Rodrigues; Heidge Fukumasu; Jayme Augusto de Souza-Neto; José Salvatore Leister Patané; Luiz Alcantara; Luiz Lehmann Coutinho; Maria Carolina Elias; Maurício Lacerda Nogueira; Rafael dos Santos Bezerra; Raul Machado Neto; Rejane Maria Tommasini Grotto; Ricardo Haddad; Sandra Coccuzzo Sampaio Vessoni; Simone Kashima; Svetoslav Nanev Slavov; Vincent Louis Viala |
| EPI_ISL_2493264, EPI_ISL_2493679, EPI_ISL_2494348, EPI_ISL_2494349, EPI_ISL_2494350, EPI_ISL_2494351 | SECRETARIA MUNICIPAL DE SAUDE PORTO FERREIRA | Instituto Butantan | Antonio Jorge Martins; Claudia Renata dos Santos Barros; David Schlesinger; Debora Botequio Moretti; Dimas Tadeu Covas; Elaine Cristina Marqueze; Elaine Vieira Santos; Evandra Strazza Rodrigues; Heidge Fukumasu; Jayme Augusto de Souza-Neto; José Salvatore Leister Patané; Luiz Alcantara; Luiz Lehmann Coutinho; Maria Carolina Elias; Maurício Lacerda Nogueira; Rafael dos Santos Bezerra; Raul Machado Neto; Rejane Maria Tommasini Grotto; Ricardo Haddad; Sandra Coccuzzo Sampaio Vessoni; Simone Kashima; Svetoslav Nanev Slavov; Vincent Louis Viala |
| EPI_ISL_2445597, EPI_ISL_2445602, EPI_ISL_2473701, EPI_ISL_2493654, EPI_ISL_2493657, EPI_ISL_2493660, EPI_ISL_2493668, EPI_ISL_2493674, EPI_ISL_2493690, EPI_ISL_2493691, EPI_ISL_2493692, EPI_ISL_2493693, EPI_ISL_2493697, EPI_ISL_2493698, EPI_ISL_2493700, EPI_ISL_2500977, EPI_ISL_2500979, EPI_ISL_2500983 | | | |
| see above | SECRETARIA MUNICIPAL DE SAUDE SOROCABA | Instituto Butantan | Antonio Jorge Martins; Claudia Renata dos Santos Barros; David Schlesinger; Debora Botequio Moretti; Dimas Tadeu Covas; Elaine Cristina Marqueze; Elaine Vieira Santos; Evandra Strazza Rodrigues; Heidge Fukumasu; Jayme Augusto de Souza-Neto; José Salvatore Leister Patané; Luiz Alcantara; Luiz Lehmann Coutinho; Maria Carolina Elias; Maurício Lacerda Nogueira; Rafael dos Santos Bezerra; Raul Machado Neto; Rejane Maria Tommasini Grotto; Ricardo Haddad; Sandra Coccuzzo Sampaio Vessoni; Simone Kashima; Svetoslav Nanev Slavov; Vincent Louis Viala |
| EPI_ISL_2462330, EPI_ISL_2493562, EPI_ISL_2493565, EPI_ISL_2493567, EPI_ISL_2493568, EPI_ISL_2493569, EPI_ISL_2493832, EPI_ISL_2493833, EPI_ISL_2493834, EPI_ISL_2493836, EPI_ISL_2493837, EPI_ISL_2493838, EPI_ISL_2493839, EPI_ISL_2493840, EPI_ISL_2493842, EPI_ISL_2493889, EPI_ISL_2493890, EPI_ISL_2493892, EPI_ISL_2493893, EPI_ISL_2493894, EPI_ISL_2494294, EPI_ISL_2494322 | | | |
| see above | SERV DE VIG SANITARIA EPIDEMIO E CTRL DE ZOONOZES GUARUJA | Instituto Butantan | Antonio Jorge Martins; Claudia Renata dos Santos Barros; David Schlesinger; Debora Botequio Moretti; Dimas Tadeu Covas; Elaine Cristina Marqueze; Elaine Vieira Santos; Evandra Strazza Rodrigues; Heidge Fukumasu; Jayme Augusto de Souza-Neto; José Salvatore Leister Patané; Luiz Alcantara; Luiz Lehmann Coutinho; Maria Carolina Elias; Maurício Lacerda Nogueira; Rafael dos Santos Bezerra; Raul Machado Neto; Rejane Maria Tommasini Grotto; Ricardo Haddad; Sandra Coccuzzo Sampaio Vessoni; Simone Kashima; Svetoslav Nanev Slavov; Vincent Louis Viala |
| EPI_ISL_2208897, EPI_ISL_2209341, EPI_ISL_2209617, EPI_ISL_2345810, EPI_ISL_2345856 | SERV DE VIG SANITARIA EPIDEMIO E CTRL DE ZOONOZES GUARUJA | Instituto Butantan / Mendelics | Antonio Jorge Martins; Claudia Renata dos Santos Barros; David Schlesinger; Debora Botequio Moretti; Dimas Tadeu Covas; Elaine Cristina Marqueze; Elaine Vieira Santos; Evandra Strazza Rodrigues; Heidge Fukumasu; Jayme Augusto de Souza-Neto; José Salvatore Leister Patané; Luiz Alcantara; Luiz Lehmann Coutinho; Maria Carolina Elias; Maurício Lacerda Nogueira; Rafael dos Santos Bezerra; Raul Machado Neto; Rejane Maria Tommasini Grotto; Ricardo Haddad; Sandra Coccuzzo Sampaio Vessoni; Simone Kashima; Svetoslav Nanev Slavov; Vincent Louis Viala |
| EPI_ISL_2462317, EPI_ISL_2493862, EPI_ISL_2493872, EPI_ISL_2493874, EPI_ISL_2493942 | SMS ARUJA | Instituto Butantan | Antonio Jorge Martins; Claudia Renata dos Santos Barros; David Schlesinger; Debora Botequio Moretti; Dimas Tadeu Covas; Elaine Cristina Marqueze; Elaine Vieira Santos; Evandra Strazza Rodrigues; Heidge Fukumasu; Jayme Augusto de Souza-Neto; José Salvatore Leister Patané; Luiz Alcantara; Luiz Lehmann Coutinho; Maria Carolina Elias; Maurício Lacerda Nogueira; Rafael dos Santos Bezerra; Raul Machado Neto; Rejane Maria Tommasini Grotto; Ricardo Haddad; Sandra Coccuzzo Sampaio Vessoni; Simone Kashima; Svetoslav Nanev Slavov; Vincent Louis Viala |
| EPI_ISL_2445541, EPI_ISL_2445580, EPI_ISL_2445593, EPI_ISL_2445600, EPI_ISL_2493671 | SMS SECRETARIA MUNICIPAL DE SAUDE DE BOITUVA | Instituto Butantan | Antonio Jorge Martins; Claudia Renata dos Santos Barros; David Schlesinger; Debora Botequio Moretti; Dimas Tadeu Covas; Elaine Cristina Marqueze; Elaine Vieira Santos; Evandra Strazza Rodrigues; Heidge Fukumasu; Jayme Augusto de Souza-Neto; José Salvatore Leister Patané; Luiz Alcantara; Luiz Lehmann Coutinho; Maria Carolina Elias; Maurício Lacerda Nogueira; Rafael dos Santos Bezerra; Raul Machado Neto; Rejane Maria Tommasini Grotto; Ricardo Haddad; Sandra Coccuzzo Sampaio Vessoni; Simone Kashima; Svetoslav Nanev Slavov; Vincent Louis Viala |
| EPI_ISL_2919013, EPI_ISL_2919014 | Secretaria Municipal da Saúde- Porto Alegre | Laboratório de Microbiologia Molecular - Universidade FEEVALE | Alana Witt Hansen; Fernando Rosado Spilki; Flávio Silveira; Fágner Henrique Heldt; Juliana Schons Gulart; Juliana Schons Gularte; Juliane Deise Fleck; Mariana Soares da Silva; Matheus Nunes Weber; Meriane Demoliner; Michele Filippi.; Micheli Filippi.; Paula Rodrigues de Almeida |
| EPI_ISL_2445614 | UBS ALAMBARI | Instituto Butantan | Antonio Jorge Martins; Claudia Renata dos Santos Barros; David Schlesinger; Debora Botequio Moretti; Dimas Tadeu Covas; Elaine Cristina Marqueze; Elaine Vieira Santos; Evandra Strazza Rodrigues; Heidge Fukumasu; Jayme Augusto de Souza-Neto; José Salvatore Leister Patané; Luiz Alcantara; Luiz Lehmann Coutinho; Maria Carolina Elias; Maurício Lacerda Nogueira; Rafael dos Santos Bezerra; Raul Machado Neto; Rejane Maria Tommasini Grotto; Ricardo Haddad; Sandra Coccuzzo Sampaio Vessoni; Simone Kashima; Svetoslav Nanev Slavov; Vincent Louis Viala |
| EPI_ISL_2445544, EPI_ISL_2473693 | UBS ALCIMINIO DE ASSIS LOURENCO BADY BASSITT | Instituto Butantan | Antonio Jorge Martins; Claudia Renata dos Santos Barros; David Schlesinger; Debora Botequio Moretti; Dimas Tadeu Covas; Elaine Cristina Marqueze; Elaine Vieira Santos; Evandra Strazza Rodrigues; Heidge Fukumasu; Jayme Augusto de Souza-Neto; José Salvatore Leister Patané; Luiz Alcantara; Luiz Lehmann Coutinho; Maria Carolina Elias; Maurício Lacerda Nogueira; Rafael dos Santos Bezerra; Raul Machado Neto; Rejane Maria Tommasini Grotto; Ricardo Haddad; Sandra Coccuzzo Sampaio Vessoni; Simone Kashima; Svetoslav Nanev Slavov; Vincent Louis Viala |
| EPI_ISL_2445150, EPI_ISL_2473765 | UBS ATALAIA DR MARIO AGUIAR FILHO | Instituto Butantan | Antonio Jorge Martins; Claudia Renata dos Santos Barros; David Schlesinger; Debora Botequio Moretti; Dimas Tadeu Covas; Elaine Cristina Marqueze; Elaine Vieira Santos; Evandra Strazza Rodrigues; Heidge Fukumasu; Jayme Augusto de Souza-Neto; José Salvatore Leister Patané; Luiz Alcantara; Luiz Lehmann Coutinho; Maria Carolina Elias; Maurício Lacerda Nogueira; Rafael dos Santos Bezerra; Raul Machado Neto; Rejane Maria Tommasini Grotto; Ricardo Haddad; Sandra Coccuzzo Sampaio Vessoni; Simone Kashima; Svetoslav Nanev Slavov; Vincent Louis Viala |
| EPI_ISL_2445173 | UBS CAIC OURINHOS | Instituto Butantan | Antonio Jorge Martins; Claudia Renata dos Santos Barros; David Schlesinger; Debora Botequio Moretti; Dimas Tadeu Covas; Elaine Cristina Marqueze; Elaine Vieira Santos; Evandra Strazza Rodrigues; Heidge Fukumasu; Jayme Augusto de Souza-Neto; José Salvatore Leister Patané; Luiz Alcantara; Luiz Lehmann |

| | | | |
|---|---|---|---|
| | | | Coutinho; Maria Carolina Elias; Maurício Lacerda Nogueira; Rafael dos Santos Bezerra; Raul Machado Neto; Rejane Maria Tommasini Grotto; Ricardo Haddad; Sandra Coccuzzo Sampaio Vessoni; Simone Kashima; Svetoslav Nanev Slavov; Vincent Louis Viala |
| EPI_ISL_2445207, EPI_ISL_2462328, EPI_ISL_2473718, EPI_ISL_2494144 | UBS CELESTINO CARNIELO | Instituto Butantan | Antonio Jorge Martins; Claudia Renata dos Santos Barros; David Schlesinger; Debora Botequio Moretti; Dimas Tadeu Covas; Elaine Cristina Marqueze; Elaine Vieira Santos; Evandra Strazza Rodrigues; Heidge Fukumasu; Jayme Augusto de Souza-Neto; José Salvatore Leister Patané; Luiz Alcantara; Luiz Lehmann Coutinho; Maria Carolina Elias; Maurício Lacerda Nogueira; Rafael dos Santos Bezerra; Raul Machado Neto; Rejane Maria Tommasini Grotto; Ricardo Haddad; Sandra Coccuzzo Sampaio Vessoni; Simone Kashima; Svetoslav Nanev Slavov; Vincent Louis Viala |
| EPI_ISL_2493866, EPI_ISL_2493868, EPI_ISL_2493869 | UBS DE JAMBEIRO JOCELINA MEIRELES | Instituto Butantan | Antonio Jorge Martins; Claudia Renata dos Santos Barros; David Schlesinger; Debora Botequio Moretti; Dimas Tadeu Covas; Elaine Cristina Marqueze; Elaine Vieira Santos; Evandra Strazza Rodrigues; Heidge Fukumasu; Jayme Augusto de Souza-Neto; José Salvatore Leister Patané; Luiz Alcantara; Luiz Lehmann Coutinho; Maria Carolina Elias; Maurício Lacerda Nogueira; Rafael dos Santos Bezerra; Raul Machado Neto; Rejane Maria Tommasini Grotto; Ricardo Haddad; Sandra Coccuzzo Sampaio Vessoni; Simone Kashima; Svetoslav Nanev Slavov; Vincent Louis Viala |
| EPI_ISL_2445208, EPI_ISL_2445216, EPI_ISL_2473719, EPI_ISL_2493655, EPI_ISL_2493656, EPI_ISL_2493658 | UBS DE PARAISO | Instituto Butantan | Antonio Jorge Martins; Claudia Renata dos Santos Barros; David Schlesinger; Debora Botequio Moretti; Dimas Tadeu Covas; Elaine Cristina Marqueze; Elaine Vieira Santos; Evandra Strazza Rodrigues; Heidge Fukumasu; Jayme Augusto de Souza-Neto; José Salvatore Leister Patané; Luiz Alcantara; Luiz Lehmann Coutinho; Maria Carolina Elias; Maurício Lacerda Nogueira; Rafael dos Santos Bezerra; Raul Machado Neto; Rejane Maria Tommasini Grotto; Ricardo Haddad; Sandra Coccuzzo Sampaio Vessoni; Simone Kashima; Svetoslav Nanev Slavov; Vincent Louis Viala |
| EPI_ISL_2445528, EPI_ISL_2493418 | UBS DE PEDRANOPOLIS | Instituto Butantan | Antonio Jorge Martins; Claudia Renata dos Santos Barros; David Schlesinger; Debora Botequio Moretti; Dimas Tadeu Covas; Elaine Cristina Marqueze; Elaine Vieira Santos; Evandra Strazza Rodrigues; Heidge Fukumasu; Jayme Augusto de Souza-Neto; José Salvatore Leister Patané; Luiz Alcantara; Luiz Lehmann Coutinho; Maria Carolina Elias; Maurício Lacerda Nogueira; Rafael dos Santos Bezerra; Raul Machado Neto; Rejane Maria Tommasini Grotto; Ricardo Haddad; Sandra Coccuzzo Sampaio Vessoni; Simone Kashima; Svetoslav Nanev Slavov; Vincent Louis Viala |
| EPI_ISL_2445195 | UBS DE SUD MENNUCCI | Instituto Butantan | Antonio Jorge Martins; Claudia Renata dos Santos Barros; David Schlesinger; Debora Botequio Moretti; Dimas Tadeu Covas; Elaine Cristina Marqueze; Elaine Vieira Santos; Evandra Strazza Rodrigues; Heidge Fukumasu; Jayme Augusto de Souza-Neto; José Salvatore Leister Patané; Luiz Alcantara; Luiz Lehmann Coutinho; Maria Carolina Elias; Maurício Lacerda Nogueira; Rafael dos Santos Bezerra; Raul Machado Neto; Rejane Maria Tommasini Grotto; Ricardo Haddad; Sandra Coccuzzo Sampaio Vessoni; Simone Kashima; Svetoslav Nanev Slavov; Vincent Louis Viala |
| EPI_ISL_2445206, EPI_ISL_2493177, EPI_ISL_2493402, EPI_ISL_2493404, EPI_ISL_2493409, EPI_ISL_2493423, EPI_ISL_2493425, EPI_ISL_2493426, EPI_ISL_2494337, EPI_ISL_2500975 | | | |
| see above | UBS DE TABAPUA | Instituto Butantan | Antonio Jorge Martins; Claudia Renata dos Santos Barros; David Schlesinger; Debora Botequio Moretti; Dimas Tadeu Covas; Elaine Cristina Marqueze; Elaine Vieira Santos; Evandra Strazza Rodrigues; Heidge Fukumasu; Jayme Augusto de Souza-Neto; José Salvatore Leister Patané; Luiz Alcantara; Luiz Lehmann Coutinho; Maria Carolina Elias; Maurício Lacerda Nogueira; Rafael dos Santos Bezerra; Raul Machado Neto; Rejane Maria Tommasini Grotto; Ricardo Haddad; Sandra Coccuzzo Sampaio Vessoni; Simone Kashima; Svetoslav Nanev Slavov; Vincent Louis Viala |
| EPI_ISL_2445536, EPI_ISL_2445537, EPI_ISL_2493659, EPI_ISL_2493665, EPI_ISL_2493666 | UBS DE UBARANA | Instituto Butantan | Antonio Jorge Martins; Claudia Renata dos Santos Barros; David Schlesinger; Debora Botequio Moretti; Dimas Tadeu Covas; Elaine Cristina Marqueze; Elaine Vieira Santos; Evandra Strazza Rodrigues; Heidge Fukumasu; Jayme Augusto de Souza-Neto; José Salvatore Leister Patané; Luiz Alcantara; Luiz Lehmann Coutinho; Maria Carolina Elias; Maurício Lacerda Nogueira; Rafael dos Santos Bezerra; Raul Machado Neto; Rejane Maria Tommasini Grotto; Ricardo Haddad; Sandra Coccuzzo Sampaio Vessoni; Simone Kashima; Svetoslav Nanev Slavov; Vincent Louis Viala |
| EPI_ISL_2462320, EPI_ISL_2462321, EPI_ISL_2493163, EPI_ISL_2493244, EPI_ISL_2494114, EPI_ISL_2494118, EPI_ISL_2494119, EPI_ISL_2494120, EPI_ISL_2494121, EPI_ISL_2494122, EPI_ISL_2494126, EPI_ISL_2494129 | | | |
| see above | UBS DR ALFREDO DANTAS DE SOUZA UMUARAMA | Instituto Butantan | Antonio Jorge Martins; Claudia Renata dos Santos Barros; David Schlesinger; Debora Botequio Moretti; Dimas Tadeu Covas; Elaine Cristina Marqueze; Elaine Vieira Santos; Evandra Strazza Rodrigues; Heidge Fukumasu; Jayme Augusto de Souza-Neto; José Salvatore Leister Patané; Luiz Alcantara; Luiz Lehmann Coutinho; Maria Carolina Elias; Maurício Lacerda Nogueira; Rafael dos Santos Bezerra; Raul Machado Neto; Rejane Maria Tommasini Grotto; Ricardo Haddad; Sandra Coccuzzo Sampaio Vessoni; Simone Kashima; Svetoslav Nanev Slavov; Vincent Louis Viala |
| EPI_ISL_2345646, EPI_ISL_2345692, EPI_ISL_2345745, EPI_ISL_2345788 | UBS DR ALFREDO DANTAS DE SOUZA UMUARAMA | Instituto Butantan / Mendelics | Antonio Jorge Martins; Claudia Renata dos Santos Barros; David Schlesinger; Debora Botequio Moretti; Dimas Tadeu Covas; Elaine Cristina Marqueze; Elaine Vieira Santos; Evandra Strazza Rodrigues; Heidge Fukumasu; Jayme Augusto de Souza-Neto; José Salvatore Leister Patané; Luiz Alcantara; Luiz Lehmann Coutinho; Maria Carolina Elias; Maurício Lacerda Nogueira; Rafael dos Santos Bezerra; Raul Machado Neto; Rejane Maria Tommasini Grotto; Ricardo Haddad; Sandra Coccuzzo Sampaio Vessoni; Simone Kashima; Svetoslav Nanev Slavov; Vincent Louis Viala |
| EPI_ISL_2345803 | UBS DR ERNESTO GAION | Instituto Butantan / Mendelics | Antonio Jorge Martins; Claudia Renata dos Santos Barros; David Schlesinger; Debora Botequio Moretti; Dimas Tadeu Covas; Elaine Cristina Marqueze; Elaine Vieira Santos; Evandra Strazza Rodrigues; Heidge Fukumasu; Jayme Augusto de Souza-Neto; José Salvatore Leister Patané; Luiz Alcantara; Luiz Lehmann Coutinho; Maria Carolina Elias; Maurício Lacerda Nogueira; Rafael dos Santos Bezerra; Raul Machado Neto; Rejane Maria Tommasini Grotto; Ricardo Haddad; Sandra Coccuzzo Sampaio Vessoni; Simone Kashima; Svetoslav Nanev Slavov; Vincent Louis Viala |
| EPI_ISL_2209532, EPI_ISL_2210186, EPI_ISL_2345757 | UBS DR WALDOMIRO FERREIRA NEVES SCRPARDO | Instituto Butantan / Mendelics | Antonio Jorge Martins; Claudia Renata dos Santos Barros; David Schlesinger; Debora Botequio Moretti; Dimas Tadeu Covas; Elaine Cristina Marqueze; Elaine Vieira Santos; Evandra Strazza Rodrigues; Heidge Fukumasu; Jayme Augusto de Souza-Neto; José Salvatore Leister Patané; Luiz Alcantara; Luiz Lehmann Coutinho; Maria Carolina Elias; Maurício Lacerda Nogueira; Rafael dos Santos Bezerra; Raul Machado Neto; Rejane Maria Tommasini Grotto; Ricardo Haddad; Sandra Coccuzzo Sampaio Vessoni; Simone Kashima; Svetoslav Nanev Slavov; Vincent Louis Viala |
| EPI_ISL_2445608 | UBS ENRIQUE GUILHERME ERNESTO MARZEUSKI | Instituto Butantan | Antonio Jorge Martins; Claudia Renata dos Santos Barros; David Schlesinger; Debora Botequio Moretti; Dimas Tadeu Covas; Elaine Cristina Marqueze; Elaine Vieira Santos; Evandra Strazza Rodrigues; Heidge Fukumasu; Jayme Augusto de Souza-Neto; José Salvatore Leister Patané; Luiz Alcantara; Luiz Lehmann Coutinho; Maria Carolina Elias; Maurício Lacerda Nogueira; Rafael dos Santos Bezerra; Raul Machado Neto; Rejane Maria Tommasini Grotto; Ricardo Haddad; Sandra Coccuzzo Sampaio Vessoni; Simone Kashima; Svetoslav Nanev Slavov; Vincent Louis Viala |
| EPI_ISL_2345662, EPI_ISL_2345705, EPI_ISL_2345850 | UBS GLENDA RODELLA DUMAS | Instituto Butantan / Mendelics | Antonio Jorge Martins; Claudia Renata dos Santos Barros; David Schlesinger; Debora Botequio Moretti; Dimas Tadeu Covas; Elaine Cristina Marqueze; Elaine Vieira Santos; Evandra Strazza Rodrigues; Heidge Fukumasu; Jayme Augusto de Souza-Neto; José Salvatore Leister Patané; Luiz Alcantara; Luiz Lehmann Coutinho; Maria Carolina Elias; Maurício Lacerda Nogueira; Rafael dos Santos Bezerra; Raul Machado Neto; Rejane Maria Tommasini Grotto; Ricardo Haddad; Sandra Coccuzzo Sampaio Vessoni; Simone Kashima; Svetoslav Nanev Slavov; Vincent Louis Viala |
| EPI_ISL_2378746, EPI_ISL_2493168, EPI_ISL_2493206, EPI_ISL_2493278, EPI_ISL_2493326, EPI_ISL_2493327, EPI_ISL_2493328, EPI_ISL_2493329, EPI_ISL_2493335, EPI_ISL_2493340, EPI_ISL_2493341, EPI_ISL_2493343, EPI_ISL_2493345, EPI_ISL_2493346, EPI_ISL_2493348, EPI_ISL_2493349, EPI_ISL_2493350, EPI_ISL_2493351, EPI_ISL_2493352, EPI_ISL_2493353, EPI_ISL_2500984 | | | |
| see above | UBS II DE PIRAPOZINHO C SERVICO DE EACS E ESF | Instituto Butantan | Antonio Jorge Martins; Claudia Renata dos Santos Barros; David Schlesinger; Debora Botequio Moretti; Dimas Tadeu Covas; Elaine Cristina Marqueze; Elaine Vieira Santos; Evandra Strazza Rodrigues; Heidge Fukumasu; Jayme Augusto de Souza-Neto; José Salvatore Leister Patané; Luiz Alcantara; Luiz Lehmann Coutinho; Maria Carolina Elias; Maurício Lacerda Nogueira; Rafael dos Santos Bezerra; Raul Machado Neto; Rejane Maria Tommasini Grotto; Ricardo Haddad; Sandra Coccuzzo Sampaio Vessoni; Simone Kashima; Svetoslav Nanev Slavov; Vincent Louis Viala |
| EPI_ISL_2445481, EPI_ISL_2445482, EPI_ISL_2445483, EPI_ISL_2445484, EPI_ISL_2445485 | UBS II DE REGENTE FEIJO | Instituto Butantan | Antonio Jorge Martins; Claudia Renata dos Santos Barros; David Schlesinger; Debora Botequio Moretti; Dimas Tadeu Covas; Elaine Cristina Marqueze; Elaine Vieira Santos; Evandra Strazza Rodrigues; Heidge Fukumasu; Jayme Augusto de Souza-Neto; José Salvatore Leister Patané; Luiz Alcantara; Luiz Lehmann Coutinho; Maria Carolina Elias; Maurício Lacerda Nogueira; Rafael dos Santos Bezerra; Raul Machado Neto; Rejane Maria Tommasini Grotto; Ricardo Haddad; Sandra Coccuzzo Sampaio Vessoni; Simone Kashima; Svetoslav Nanev Slavov; Vincent Louis Viala |
| EPI_ISL_2209618 | UBS II DE REGENTE FEIJO | Instituto Butantan / Mendelics | Antonio Jorge Martins; Claudia Renata dos Santos Barros; David Schlesinger; Debora Botequio Moretti; Dimas Tadeu Covas; Elaine Cristina Marqueze; Elaine Vieira Santos; Evandra Strazza Rodrigues; Heidge Fukumasu; Jayme Augusto de Souza-Neto; José Salvatore Leister Patané; Luiz Alcantara; Luiz Lehmann Coutinho; Maria Carolina Elias; Maurício Lacerda Nogueira; Rafael dos Santos Bezerra; Raul Machado Neto; Rejane Maria Tommasini Grotto; Ricardo Haddad; Sandra Coccuzzo Sampaio Vessoni; Simone Kashima; Svetoslav Nanev Slavov; Vincent Louis Viala |
| EPI_ISL_2445202, EPI_ISL_2445516, EPI_ISL_2445518, EPI_ISL_2445519, EPI_ISL_2445520, EPI_ISL_2445534, EPI_ISL_2445542, EPI_ISL_2473696, EPI_ISL_2473791 | | | |
| see above | UBS II DE TANABI MILTON MARTINS PERCHES | Instituto Butantan | Antonio Jorge Martins; Claudia Renata dos Santos Barros; David Schlesinger; Debora Botequio Moretti; Dimas Tadeu Covas; Elaine Cristina Marqueze; Elaine Vieira Santos; Evandra Strazza Rodrigues; Heidge Fukumasu; Jayme Augusto de Souza-Neto; José Salvatore Leister Patané; Luiz Alcantara; Luiz Lehmann Coutinho; Maria Carolina Elias; Maurício Lacerda Nogueira; Rafael dos Santos Bezerra; Raul Machado Neto; Rejane Maria Tommasini Grotto; Ricardo Haddad; Sandra Coccuzzo Sampaio Vessoni; Simone Kashima; Svetoslav Nanev Slavov; Vincent Louis Viala |
| EPI_ISL_2445552, EPI_ISL_2445555, EPI_ISL_2473796, EPI_ISL_2493360 | UBS II DR JOSE CARLOS MACUCO JANINI | Instituto Butantan | Antonio Jorge Martins; Claudia Renata dos Santos Barros; David Schlesinger; Debora Botequio Moretti; Dimas Tadeu Covas; Elaine Cristina Marqueze; Elaine Vieira Santos; Evandra Strazza Rodrigues; Heidge Fukumasu; Jayme Augusto de Souza-Neto; José Salvatore Leister Patané; Luiz Alcantara; Luiz Lehmann Coutinho; Maria Carolina Elias; Maurício Lacerda Nogueira; Rafael dos Santos Bezerra; Raul Machado Neto; Rejane Maria Tommasini Grotto; Ricardo Haddad; Sandra Coccuzzo Sampaio Vessoni; Simone Kashima; Svetoslav Nanev Slavov; Vincent Louis Viala |
| EPI_ISL_2445387 | UBS III DE BENTO DE ABREU | Instituto Butantan | Antonio Jorge Martins; Claudia Renata dos Santos Barros; David Schlesinger; Debora Botequio Moretti; Dimas Tadeu Covas; Elaine Cristina Marqueze; Elaine Vieira Santos; Evandra Strazza Rodrigues; Heidge Fukumasu; Jayme Augusto de Souza-Neto; José Salvatore Leister Patané; Luiz Alcantara; Luiz Lehmann Coutinho; Maria Carolina Elias; Maurício Lacerda Nogueira; Rafael dos Santos Bezerra; Raul Machado Neto; Rejane Maria Tommasini Grotto; Ricardo Haddad; Sandra Coccuzzo Sampaio Vessoni; Simone Kashima; Svetoslav Nanev Slavov; Vincent Louis Viala |
| EPI_ISL_2445383 | UBS III DE COROADOS | Instituto Butantan | Antonio Jorge Martins; Claudia Renata dos Santos Barros; David Schlesinger; Debora Botequio Moretti; Dimas Tadeu Covas; Elaine Cristina Marqueze; Elaine Vieira Santos; Evandra Strazza Rodrigues; Heidge Fukumasu; Jayme Augusto de Souza-Neto; José Salvatore Leister Patané; Luiz Alcantara; Luiz Lehmann Coutinho; Maria Carolina Elias; Maurício Lacerda Nogueira; Rafael dos Santos Bezerra; Raul Machado Neto; Rejane Maria Tommasini Grotto; Ricardo Haddad; Sandra Coccuzzo Sampaio Vessoni; Simone Kashima; Svetoslav Nanev Slavov; Vincent Louis Viala |
| EPI_ISL_2462332 | UBS III DE ELDORADO ELDORADO S P | Instituto Butantan | Antonio Jorge Martins; Claudia Renata dos Santos Barros; David Schlesinger; Debora Botequio Moretti; Dimas Tadeu Covas; Elaine Cristina Marqueze; Elaine Vieira Santos; Evandra Strazza Rodrigues; Heidge Fukumasu; Jayme Augusto de Souza-Neto; José Salvatore Leister Patané; Luiz Alcantara; Luiz Lehmann Coutinho; Maria Carolina Elias; Maurício Lacerda Nogueira; Rafael dos Santos Bezerra; Raul Machado Neto; Rejane Maria Tommasini Grotto; Ricardo Haddad; Sandra Coccuzzo Sampaio Vessoni; Simone Kashima; Svetoslav Nanev Slavov; Vincent Louis Viala |
| EPI_ISL_2493531, EPI_ISL_2493545, EPI_ISL_2493550 | UBS III DE PARIQUERA ACU PARIQUERA ACU | Instituto Butantan | Antonio Jorge Martins; Claudia Renata dos Santos Barros; David Schlesinger; Debora Botequio Moretti; Dimas Tadeu Covas; Elaine Cristina Marqueze; Elaine Vieira Santos; Evandra Strazza Rodrigues; Heidge Fukumasu; Jayme Augusto de Souza-Neto; José Salvatore Leister Patané; Luiz Alcantara; Luiz Lehmann Coutinho; Maria Carolina Elias; Maurício Lacerda Nogueira; Rafael dos Santos Bezerra; Raul Machado Neto; Rejane Maria Tommasini Grotto; Ricardo Haddad; Sandra Coccuzzo Sampaio Vessoni; Simone Kashima; Svetoslav Nanev Slavov; Vincent Louis Viala |
| EPI_ISL_2466733, EPI_ISL_2493686, EPI_ISL_2493699 | UBS III DE RANCHARIA | Instituto Butantan | Antonio Jorge Martins; Claudia Renata dos Santos Barros; David Schlesinger; Debora Botequio Moretti; Dimas Tadeu Covas; Elaine Cristina Marqueze; Elaine Vieira Santos; Evandra Strazza Rodrigues; Heidge Fukumasu; Jayme Augusto de Souza-Neto; José Salvatore Leister Patané; Luiz Alcantara; Luiz Lehmann Coutinho; Maria Carolina Elias; Maurício Lacerda Nogueira; Rafael dos Santos Bezerra; Raul Machado Neto; Rejane Maria Tommasini Grotto; Ricardo Haddad; Sandra Coccuzzo Sampaio Vessoni; Simone Kashima; Svetoslav Nanev Slavov; Vincent Louis Viala |
| EPI_ISL_2345666, EPI_ISL_2345710, EPI_ISL_2345853, EPI_ISL_2362265 | UBS III DE RANCHARIA | Instituto Butantan / Mendelics | Antonio Jorge Martins; Claudia Renata dos Santos Barros; David Schlesinger; Debora Botequio Moretti; Dimas Tadeu Covas; Elaine Cristina Marqueze; Elaine Vieira Santos; Evandra Strazza Rodrigues; Heidge Fukumasu; Jayme Augusto de Souza-Neto; José Salvatore Leister Patané; Luiz Alcantara; Luiz Lehmann Coutinho; Maria Carolina Elias; Maurício Lacerda Nogueira; Rafael dos Santos Bezerra; Raul Machado Neto; Rejane Maria Tommasini Grotto; Ricardo Haddad; Sandra Coccuzzo Sampaio Vessoni; Simone Kashima; Svetoslav Nanev Slavov; Vincent Louis Viala |
| EPI_ISL_2445543 | UBS III DR BENITO MALZONE | Instituto Butantan | Antonio Jorge Martins; Claudia Renata dos Santos Barros; David Schlesinger; Debora Botequio Moretti; Dimas Tadeu Covas; Elaine Cristina Marqueze; Elaine Vieira Santos; Evandra Strazza Rodrigues; Heidge Fukumasu; Jayme Augusto de Souza-Neto; José Salvatore Leister Patané; Luiz Alcantara; Luiz Lehmann Coutinho; Maria Carolina Elias; Maurício Lacerda Nogueira; Rafael dos Santos Bezerra; Raul Machado Neto; Rejane Maria Tommasini Grotto; Ricardo Haddad; Sandra Coccuzzo Sampaio Vessoni; Simone Kashima; Svetoslav Nanev Slavov; Vincent Louis Viala |
| EPI_ISL_2493820, EPI_ISL_2494320 | UBS JARDIM CAIUBY | Instituto Butantan | Antonio Jorge Martins; Claudia Renata dos Santos Barros; David Schlesinger; Debora Botequio Moretti; Dimas Tadeu Covas; Elaine Cristina Marqueze; Elaine Vieira Santos; Evandra Strazza Rodrigues; Heidge Fukumasu; Jayme Augusto de Souza-Neto; José Salvatore Leister Patané; Luiz Alcantara; Luiz Lehmann Coutinho; Maria Carolina Elias; Maurício Lacerda Nogueira; Rafael dos Santos Bezerra; Raul Machado Neto; Rejane Maria Tommasini Grotto; Ricardo Haddad; Sandra Coccuzzo Sampaio Vessoni; Simone Kashima; Svetoslav Nanev Slavov; Vincent Louis Viala |
| EPI_ISL_2493849, EPI_ISL_2494324 | UBS JARDIM ODETE | Instituto Butantan | Antonio Jorge Martins; Claudia Renata dos Santos Barros; David Schlesinger; Debora Botequio Moretti; Dimas Tadeu Covas; Elaine Cristina Marqueze; Elaine Vieira Santos; Evandra Strazza Rodrigues; Heidge Fukumasu; Jayme Augusto de Souza-Neto; José Salvatore Leister Patané; Luiz Alcantara; Luiz Lehmann Coutinho; Maria Carolina Elias; Maurício Lacerda Nogueira; Rafael dos Santos Bezerra; Raul Machado Neto; Rejane Maria Tommasini Grotto; Ricardo Haddad; Sandra Coccuzzo Sampaio Vessoni; Simone Kashima; Svetoslav Nanev Slavov; Vincent Louis Viala |
| EPI_ISL_2493663, EPI_ISL_2493664 | UBS LUIS FACHIN IPIGUA | Instituto Butantan | Antonio Jorge Martins; Claudia Renata dos Santos Barros; David Schlesinger; Debora Botequio Moretti; Dimas Tadeu Covas; Elaine Cristina Marqueze; Elaine Vieira Santos; Evandra Strazza Rodrigues; Heidge Fukumasu; Jayme Augusto de Souza-Neto; José Salvatore Leister Patané; Luiz Alcantara; Luiz Lehmann Coutinho; Maria Carolina Elias; Maurício Lacerda Nogueira; Rafael dos Santos Bezerra; Raul Machado Neto; Rejane Maria Tommasini Grotto; Ricardo Haddad; Sandra Coccuzzo Sampaio Vessoni; Simone Kashima; Svetoslav Nanev Slavov; Vincent Louis Viala |
| EPI_ISL_2493584 | UBS MARIA IZABEL | Instituto Butantan | Antonio Jorge Martins; Claudia Renata dos Santos Barros; David Schlesinger; Debora Botequio Moretti; Dimas Tadeu Covas; Elaine Cristina Marqueze; Elaine Vieira Santos; Evandra Strazza Rodrigues; Heidge Fukumasu; Jayme Augusto de Souza-Neto; José Salvatore Leister Patané; Luiz Alcantara; Luiz Lehmann Coutinho; Maria Carolina Elias; Maurício Lacerda Nogueira; Rafael dos Santos Bezerra; Raul Machado Neto; Rejane Maria Tommasini Grotto; Ricardo Haddad; Sandra Coccuzzo Sampaio Vessoni; Simone Kashima; Svetoslav Nanev Slavov; Vincent Louis Viala |
| EPI_ISL_2445198, EPI_ISL_2445199, EPI_ISL_2445200, EPI_ISL_2445201, EPI_ISL_2445521, EPI_ISL_2445522, EPI_ISL_2445523, EPI_ISL_2445524, EPI_ISL_2445525, EPI_ISL_2473714, EPI_ISL_2473715, EPI_ISL_2473789 | | | |
| see above | UBS PASTOR MARCELINO DEUNGARO RIO GRANDE | Instituto Butantan | Antonio Jorge Martins; Claudia Renata dos Santos Barros; David Schlesinger; Debora Botequio Moretti; Dimas Tadeu Covas; Elaine Cristina Marqueze; Elaine Vieira Santos; Evandra Strazza Rodrigues; Heidge Fukumasu; Jayme Augusto de Souza-Neto; José Salvatore Leister Patané; Luiz Alcantara; Luiz Lehmann Coutinho; Maria Carolina Elias; Maurício Lacerda Nogueira; Rafael dos Santos Bezerra; Raul Machado Neto; Rejane Maria Tommasini Grotto; Ricardo Haddad; Sandra Coccuzzo Sampaio Vessoni; Simone Kashima; Svetoslav Nanev Slavov; Vincent Louis Viala |
| EPI_ISL_2493293 | UBS PAULO HUMBERTO DE FIGUEIREDO | Instituto Butantan | Antonio Jorge Martins; Claudia Renata dos Santos Barros; David Schlesinger; Debora Botequio Moretti; Dimas Tadeu Covas; Elaine Cristina Marqueze; Elaine Vieira Santos; Evandra Strazza Rodrigues; Heidge Fukumasu; Jayme Augusto de Souza-Neto; José Salvatore Leister Patané; Luiz Alcantara; Luiz Lehmann Coutinho; Maria Carolina Elias; Maurício Lacerda Nogueira; Rafael dos Santos Bezerra; Raul Machado Neto; Rejane Maria Tommasini Grotto; Ricardo Haddad; Sandra Coccuzzo Sampaio Vessoni; Simone Kashima; Svetoslav Nanev Slavov; Vincent Louis Viala |
| EPI_ISL_2445254, EPI_ISL_2445276 | UBS SAO LOURENCO DA SERRA | Instituto Butantan | Antonio Jorge Martins; Claudia Renata dos Santos Barros; David Schlesinger; Debora Botequio Moretti; Dimas Tadeu Covas; Elaine Cristina Marqueze; Elaine Vieira Santos; Evandra Strazza Rodrigues; Heidge Fukumasu; Jayme Augusto de Souza-Neto; José Salvatore Leister Patané; Luiz Alcantara; Luiz Lehmann Coutinho; Maria Carolina Elias; Maurício Lacerda Nogueira; Rafael dos Santos Bezerra; Raul Machado Neto; Rejane Maria Tommasini Grotto; Ricardo Haddad; Sandra Coccuzzo Sampaio Vessoni; Simone Kashima; Svetoslav Nanev Slavov; Vincent Louis Viala |

| | | | |
|---|---|---|---|
| EPI_ISL_2445188, EPI_ISL_2473710 | UBS VILA RIBEIRO | Instituto Butantan | Antonio Jorge Martins; Claudia Renata dos Santos Barros; David Schlesinger; Debora Botequio Moretti; Dimas Tadeu Covas; Elaine Cristina Marqueze; Elaine Vieira Santos; Evandra Strazza Rodrigues; Heidge Fukumasu; Jayme Augusto de Souza-Neto; José Salvatore Leister Patané; Luiz Alcantara; Luiz Lehmann Coutinho; Maria Carolina Elias; Maurício Lacerda Nogueira; Rafael dos Santos Bezerra; Raul Machado Neto; Rejane Maria Tommasini Grotto; Ricardo Haddad; Sandra Coccuzzo Sampaio Vessoni; Simone Kashima; Svetoslav Nanev Slavov; Vincent Louis Viala |
| EPI_ISL_2444195 | UCHOA | Instituto Butantan / FAMERP | Antonio Jorge Martins; Claudia Renata dos Santos Barros; David Schlesinger; Debora Botequio Moretti; Dimas Tadeu Covas; Elaine Cristina Marqueze; Elaine Vieira Santos; Evandra Strazza Rodrigues; Heidge Fukumasu; Jayme Augusto de Souza-Neto; José Salvatore Leister Patané; Luiz Alcantara; Luiz Lehmann Coutinho; Maria Carolina Elias; Maurício Lacerda Nogueira; Rafael dos Santos Bezerra; Raul Machado Neto; Rejane Maria Tommasini Grotto; Ricardo Haddad; Sandra Coccuzzo Sampaio Vessoni; Simone Kashima; Svetoslav Nanev Slavov; Vincent Louis Viala |
| EPI_ISL_2493585, EPI_ISL_2493588 | UNIDADE BASICA DE SAUDE DE TARUMA | Instituto Butantan | Antonio Jorge Martins; Claudia Renata dos Santos Barros; David Schlesinger; Debora Botequio Moretti; Dimas Tadeu Covas; Elaine Cristina Marqueze; Elaine Vieira Santos; Evandra Strazza Rodrigues; Heidge Fukumasu; Jayme Augusto de Souza-Neto; José Salvatore Leister Patané; Luiz Alcantara; Luiz Lehmann Coutinho; Maria Carolina Elias; Maurício Lacerda Nogueira; Rafael dos Santos Bezerra; Raul Machado Neto; Rejane Maria Tommasini Grotto; Ricardo Haddad; Sandra Coccuzzo Sampaio Vessoni; Simone Kashima; Svetoslav Nanev Slavov; Vincent Louis Viala |
| EPI_ISL_2383035, EPI_ISL_2493383, EPI_ISL_2494152, EPI_ISL_2494175, EPI_ISL_2494187 | UNIDADE BASICA DE SAUDE DR MATHEUS GABRIEL BONASSA | Instituto Butantan | Antonio Jorge Martins; Claudia Renata dos Santos Barros; David Schlesinger; Debora Botequio Moretti; Dimas Tadeu Covas; Elaine Cristina Marqueze; Elaine Vieira Santos; Evandra Strazza Rodrigues; Heidge Fukumasu; Jayme Augusto de Souza-Neto; José Salvatore Leister Patané; Luiz Alcantara; Luiz Lehmann Coutinho; Maria Carolina Elias; Maurício Lacerda Nogueira; Rafael dos Santos Bezerra; Raul Machado Neto; Rejane Maria Tommasini Grotto; Ricardo Haddad; Sandra Coccuzzo Sampaio Vessoni; Simone Kashima; Svetoslav Nanev Slavov; Vincent Louis Viala |
| EPI_ISL_2493196, EPI_ISL_2493197, EPI_ISL_2493311, EPI_ISL_2493313, EPI_ISL_2493314, EPI_ISL_2493315, EPI_ISL_2493317, EPI_ISL_2493319, EPI_ISL_2493457, EPI_ISL_2493458, EPI_ISL_2493459, EPI_ISL_2493462, EPI_ISL_2493465, EPI_ISL_2493466 | | | |
| see above | UNIDADE BASICA DE SAUDE HORTOLANDIA PACS | Instituto Butantan | Antonio Jorge Martins; Claudia Renata dos Santos Barros; David Schlesinger; Debora Botequio Moretti; Dimas Tadeu Covas; Elaine Cristina Marqueze; Elaine Vieira Santos; Evandra Strazza Rodrigues; Heidge Fukumasu; Jayme Augusto de Souza-Neto; José Salvatore Leister Patané; Luiz Alcantara; Luiz Lehmann Coutinho; Maria Carolina Elias; Maurício Lacerda Nogueira; Rafael dos Santos Bezerra; Raul Machado Neto; Rejane Maria Tommasini Grotto; Ricardo Haddad; Sandra Coccuzzo Sampaio Vessoni; Simone Kashima; Svetoslav Nanev Slavov; Vincent Louis Viala |
| EPI_ISL_2473794, EPI_ISL_2493361 | UNIDADE BASICA DE SAUDE II ESTRELA DO NORTE | Instituto Butantan | Antonio Jorge Martins; Claudia Renata dos Santos Barros; David Schlesinger; Debora Botequio Moretti; Dimas Tadeu Covas; Elaine Cristina Marqueze; Elaine Vieira Santos; Evandra Strazza Rodrigues; Heidge Fukumasu; Jayme Augusto de Souza-Neto; José Salvatore Leister Patané; Luiz Alcantara; Luiz Lehmann Coutinho; Maria Carolina Elias; Maurício Lacerda Nogueira; Rafael dos Santos Bezerra; Raul Machado Neto; Rejane Maria Tommasini Grotto; Ricardo Haddad; Sandra Coccuzzo Sampaio Vessoni; Simone Kashima; Svetoslav Nanev Slavov; Vincent Louis Viala |
| EPI_ISL_2209351, EPI_ISL_2209844, EPI_ISL_2210141, EPI_ISL_2345673, EPI_ISL_2345719, EPI_ISL_2345723 | UNIDADE BASICA DE SAUDE JARDIM DO LAGO | Instituto Butantan / Mendelics | Antonio Jorge Martins; Claudia Renata dos Santos Barros; David Schlesinger; Debora Botequio Moretti; Dimas Tadeu Covas; Elaine Cristina Marqueze; Elaine Vieira Santos; Evandra Strazza Rodrigues; Heidge Fukumasu; Jayme Augusto de Souza-Neto; José Salvatore Leister Patané; Luiz Alcantara; Luiz Lehmann Coutinho; Maria Carolina Elias; Maurício Lacerda Nogueira; Rafael dos Santos Bezerra; Raul Machado Neto; Rejane Maria Tommasini Grotto; Ricardo Haddad; Sandra Coccuzzo Sampaio Vessoni; Simone Kashima; Svetoslav Nanev Slavov; Vincent Louis Viala |
| EPI_ISL_2445463, EPI_ISL_2493294, EPI_ISL_2493295 | UNIDADE BASICA SAUDE NATAL ABADIO LACERDA | Instituto Butantan | Antonio Jorge Martins; Claudia Renata dos Santos Barros; David Schlesinger; Debora Botequio Moretti; Dimas Tadeu Covas; Elaine Cristina Marqueze; Elaine Vieira Santos; Evandra Strazza Rodrigues; Heidge Fukumasu; Jayme Augusto de Souza-Neto; José Salvatore Leister Patané; Luiz Alcantara; Luiz Lehmann Coutinho; Maria Carolina Elias; Maurício Lacerda Nogueira; Rafael dos Santos Bezerra; Raul Machado Neto; Rejane Maria Tommasini Grotto; Ricardo Haddad; Sandra Coccuzzo Sampaio Vessoni; Simone Kashima; Svetoslav Nanev Slavov; Vincent Louis Viala |
| EPI_ISL_2345749, EPI_ISL_2345789 | UNIDADE BASICA SAUDE NATAL ABADIO LACERDA | Instituto Butantan / Mendelics | Antonio Jorge Martins; Claudia Renata dos Santos Barros; David Schlesinger; Debora Botequio Moretti; Dimas Tadeu Covas; Elaine Cristina Marqueze; Elaine Vieira Santos; Evandra Strazza Rodrigues; Heidge Fukumasu; Jayme Augusto de Souza-Neto; José Salvatore Leister Patané; Luiz Alcantara; Luiz Lehmann Coutinho; Maria Carolina Elias; Maurício Lacerda Nogueira; Rafael dos Santos Bezerra; Raul Machado Neto; Rejane Maria Tommasini Grotto; Ricardo Haddad; Sandra Coccuzzo Sampaio Vessoni; Simone Kashima; Svetoslav Nanev Slavov; Vincent Louis Viala |
| EPI_ISL_2493473, EPI_ISL_2493480, EPI_ISL_2493482, EPI_ISL_2493484, EPI_ISL_2493486, EPI_ISL_2493487, EPI_ISL_2493488, EPI_ISL_2493492, EPI_ISL_2493494, EPI_ISL_2493496, EPI_ISL_2493505, EPI_ISL_2493512 | | | |
| see above | UNIDADE DE PRONTO ATENDIMENTO | Instituto Butantan | Antonio Jorge Martins; Claudia Renata dos Santos Barros; David Schlesinger; Debora Botequio Moretti; Dimas Tadeu Covas; Elaine Cristina Marqueze; Elaine Vieira Santos; Evandra Strazza Rodrigues; Heidge Fukumasu; Jayme Augusto de Souza-Neto; José Salvatore Leister Patané; Luiz Alcantara; Luiz Lehmann Coutinho; Maria Carolina Elias; Maurício Lacerda Nogueira; Rafael dos Santos Bezerra; Raul Machado Neto; Rejane Maria Tommasini Grotto; Ricardo Haddad; Sandra Coccuzzo Sampaio Vessoni; Simone Kashima; Svetoslav Nanev Slavov; Vincent Louis Viala |
| EPI_ISL_2462318, EPI_ISL_2462319, EPI_ISL_2493160, EPI_ISL_2493175, EPI_ISL_2494099, EPI_ISL_2494101, EPI_ISL_2494102, EPI_ISL_2494103, EPI_ISL_2494104, EPI_ISL_2494105, EPI_ISL_2494106, EPI_ISL_2494107, EPI_ISL_2494334 | | | |
| see above | UNIDADE DE PRONTO ATENDIMENTO DE AGENOR DE CAMPOS | Instituto Butantan | Antonio Jorge Martins; Claudia Renata dos Santos Barros; David Schlesinger; Debora Botequio Moretti; Dimas Tadeu Covas; Elaine Cristina Marqueze; Elaine Vieira Santos; Evandra Strazza Rodrigues; Heidge Fukumasu; Jayme Augusto de Souza-Neto; José Salvatore Leister Patané; Luiz Alcantara; Luiz Lehmann Coutinho; Maria Carolina Elias; Maurício Lacerda Nogueira; Rafael dos Santos Bezerra; Raul Machado Neto; Rejane Maria Tommasini Grotto; Ricardo Haddad; Sandra Coccuzzo Sampaio Vessoni; Simone Kashima; Svetoslav Nanev Slavov; Vincent Louis Viala |
| EPI_ISL_2493835 | UNIDADE DE PRONTO ATENDIMENTO DE PITANGUEIRAS | Instituto Butantan | Antonio Jorge Martins; Claudia Renata dos Santos Barros; David Schlesinger; Debora Botequio Moretti; Dimas Tadeu Covas; Elaine Cristina Marqueze; Elaine Vieira Santos; Evandra Strazza Rodrigues; Heidge Fukumasu; Jayme Augusto de Souza-Neto; José Salvatore Leister Patané; Luiz Alcantara; Luiz Lehmann Coutinho; Maria Carolina Elias; Maurício Lacerda Nogueira; Rafael dos Santos Bezerra; Raul Machado Neto; Rejane Maria Tommasini Grotto; Ricardo Haddad; Sandra Coccuzzo Sampaio Vessoni; Simone Kashima; Svetoslav Nanev Slavov; Vincent Louis Viala |
| EPI_ISL_2473819 | UNIDADE DE PRONTO ATENDIMENTO UPA | Instituto Butantan | Antonio Jorge Martins; Claudia Renata dos Santos Barros; David Schlesinger; Debora Botequio Moretti; Dimas Tadeu Covas; Elaine Cristina Marqueze; Elaine Vieira Santos; Evandra Strazza Rodrigues; Heidge Fukumasu; Jayme Augusto de Souza-Neto; José Salvatore Leister Patané; Luiz Alcantara; Luiz Lehmann Coutinho; Maria Carolina Elias; Maurício Lacerda Nogueira; Rafael dos Santos Bezerra; Raul Machado Neto; Rejane Maria Tommasini Grotto; Ricardo Haddad; Sandra Coccuzzo Sampaio Vessoni; Simone Kashima; Svetoslav Nanev Slavov; Vincent Louis Viala |
| EPI_ISL_2378750 | UNIDADE DE SAUDE DA FAMILIA DE FLORIDA PAULISTA II | Instituto Butantan | Antonio Jorge Martins; Claudia Renata dos Santos Barros; David Schlesinger; Debora Botequio Moretti; Dimas Tadeu Covas; Elaine Cristina Marqueze; Elaine Vieira Santos; Evandra Strazza Rodrigues; Heidge Fukumasu; Jayme Augusto de Souza-Neto; José Salvatore Leister Patané; Luiz Alcantara; Luiz Lehmann Coutinho; Maria Carolina Elias; Maurício Lacerda Nogueira; Rafael dos Santos Bezerra; Raul Machado Neto; Rejane Maria Tommasini Grotto; Ricardo Haddad; Sandra Coccuzzo Sampaio Vessoni; Simone Kashima; Svetoslav Nanev Slavov; Vincent Louis Viala |
| EPI_ISL_2493803, EPI_ISL_2493804, EPI_ISL_2493805, EPI_ISL_2493806 | UNIDADE DE SAUDE DA FAMILIA DR AMELIO TANGANELLI | Instituto Butantan | Antonio Jorge Martins; Claudia Renata dos Santos Barros; David Schlesinger; Debora Botequio Moretti; Dimas Tadeu Covas; Elaine Cristina Marqueze; Elaine Vieira Santos; Evandra Strazza Rodrigues; Heidge Fukumasu; Jayme Augusto de Souza-Neto; José Salvatore Leister Patané; Luiz Alcantara; Luiz Lehmann Coutinho; Maria Carolina Elias; Maurício Lacerda Nogueira; Rafael dos Santos Bezerra; Raul Machado Neto; Rejane Maria Tommasini Grotto; Ricardo Haddad; Sandra Coccuzzo Sampaio Vessoni; Simone Kashima; Svetoslav Nanev Slavov; Vincent Louis Viala |
| EPI_ISL_2493844 | UNIDADE DE SAUDE DA FAMILIA JARDIM AMERICA ITAQUAQUECETUBA | Instituto Butantan | Antonio Jorge Martins; Claudia Renata dos Santos Barros; David Schlesinger; Debora Botequio Moretti; Dimas Tadeu Covas; Elaine Cristina Marqueze; Elaine Vieira Santos; Evandra Strazza Rodrigues; Heidge Fukumasu; Jayme Augusto de Souza-Neto; José Salvatore Leister Patané; Luiz Alcantara; Luiz Lehmann Coutinho; Maria Carolina Elias; Maurício Lacerda Nogueira; Rafael dos Santos Bezerra; Raul Machado Neto; Rejane Maria Tommasini Grotto; Ricardo Haddad; Sandra Coccuzzo Sampaio Vessoni; Simone Kashima; Svetoslav Nanev Slavov; Vincent Louis Viala |
| EPI_ISL_2494305 | UNIDADE DE SAUDE DA FAMILIA NOVO HORIZONTE | Instituto Butantan | Antonio Jorge Martins; Claudia Renata dos Santos Barros; David Schlesinger; Debora Botequio Moretti; Dimas Tadeu Covas; Elaine Cristina Marqueze; Elaine Vieira Santos; Evandra Strazza Rodrigues; Heidge Fukumasu; Jayme Augusto de Souza-Neto; José Salvatore Leister Patané; Luiz Alcantara; Luiz Lehmann Coutinho; Maria Carolina Elias; Maurício Lacerda Nogueira; Rafael dos Santos Bezerra; Raul Machado Neto; Rejane Maria Tommasini Grotto; Ricardo Haddad; Sandra Coccuzzo Sampaio Vessoni; Simone Kashima; Svetoslav Nanev Slavov; Vincent Louis Viala |
| EPI_ISL_2493526 | UNIDADE DE SAUDE DE ITOBI ALCIBIADES PIRES | Instituto Butantan | Antonio Jorge Martins; Claudia Renata dos Santos Barros; David Schlesinger; Debora Botequio Moretti; Dimas Tadeu Covas; Elaine Cristina Marqueze; Elaine Vieira Santos; Evandra Strazza Rodrigues; Heidge Fukumasu; Jayme Augusto de Souza-Neto; José Salvatore Leister Patané; Luiz Alcantara; Luiz Lehmann Coutinho; Maria Carolina Elias; Maurício Lacerda Nogueira; Rafael dos Santos Bezerra; Raul Machado Neto; Rejane Maria Tommasini Grotto; Ricardo Haddad; Sandra Coccuzzo Sampaio Vessoni; Simone Kashima; Svetoslav Nanev Slavov; Vincent Louis Viala |
| EPI_ISL_2378742, EPI_ISL_2445486, EPI_ISL_2445487, EPI_ISL_2445488, EPI_ISL_2445489, EPI_ISL_2445490, EPI_ISL_2445491, EPI_ISL_2445499, EPI_ISL_2473784, EPI_ISL_2493297, EPI_ISL_2493298 | | | |
| see above | UNIDADE DE SAUDE DR PHEBO DE OLIVEIRA ROGE FERREIRA | Instituto Butantan | Antonio Jorge Martins; Claudia Renata dos Santos Barros; David Schlesinger; Debora Botequio Moretti; Dimas Tadeu Covas; Elaine Cristina Marqueze; Elaine Vieira Santos; Evandra Strazza Rodrigues; Heidge Fukumasu; Jayme Augusto de Souza-Neto; José Salvatore Leister Patané; Luiz Alcantara; Luiz Lehmann Coutinho; Maria Carolina Elias; Maurício Lacerda Nogueira; Rafael dos Santos Bezerra; Raul Machado Neto; Rejane Maria Tommasini Grotto; Ricardo Haddad; Sandra Coccuzzo Sampaio Vessoni; Simone Kashima; Svetoslav Nanev Slavov; Vincent Louis Viala |
| EPI_ISL_2445479, EPI_ISL_2445480 | UNIDADE DE VIGILANCIA EM SAUDE | Instituto Butantan | Antonio Jorge Martins; Claudia Renata dos Santos Barros; David Schlesinger; Debora Botequio Moretti; Dimas Tadeu Covas; Elaine Cristina Marqueze; Elaine Vieira Santos; Evandra Strazza Rodrigues; Heidge Fukumasu; Jayme Augusto de Souza-Neto; José Salvatore Leister Patané; Luiz Alcantara; Luiz Lehmann Coutinho; Maria Carolina Elias; Maurício Lacerda Nogueira; Rafael dos Santos Bezerra; Raul Machado Neto; Rejane Maria Tommasini Grotto; Ricardo Haddad; Sandra Coccuzzo Sampaio Vessoni; Simone Kashima; Svetoslav Nanev Slavov; Vincent Louis Viala |
| EPI_ISL_2493167, EPI_ISL_2493376, EPI_ISL_2494181 | UNIDADE DE VIGILANCIA EPIDEMIOLOGICA | Instituto Butantan | Antonio Jorge Martins; Claudia Renata dos Santos Barros; David Schlesinger; Debora Botequio Moretti; Dimas Tadeu Covas; Elaine Cristina Marqueze; Elaine Vieira Santos; Evandra Strazza Rodrigues; Heidge Fukumasu; Jayme Augusto de Souza-Neto; José Salvatore Leister Patané; Luiz Alcantara; Luiz Lehmann Coutinho; Maria Carolina Elias; Maurício Lacerda Nogueira; Rafael dos Santos Bezerra; Raul Machado Neto; Rejane Maria Tommasini Grotto; Ricardo Haddad; Sandra Coccuzzo Sampaio Vessoni; Simone Kashima; Svetoslav Nanev Slavov; Vincent Louis Viala |
| EPI_ISL_2493947 | UNIDADE MISTA ARACOIABA DA SERRA | Instituto Butantan | Antonio Jorge Martins; Claudia Renata dos Santos Barros; David Schlesinger; Debora Botequio Moretti; Dimas Tadeu Covas; Elaine Cristina Marqueze; Elaine Vieira Santos; Evandra Strazza Rodrigues; Heidge Fukumasu; Jayme Augusto de Souza-Neto; José Salvatore Leister Patané; Luiz Alcantara; Luiz Lehmann Coutinho; Maria Carolina Elias; Maurício Lacerda Nogueira; Rafael dos Santos Bezerra; Raul Machado Neto; Rejane Maria Tommasini Grotto; Ricardo Haddad; Sandra Coccuzzo Sampaio Vessoni; Simone Kashima; Svetoslav Nanev Slavov; Vincent Louis Viala |
| EPI_ISL_2445221, EPI_ISL_2445222, EPI_ISL_2445234, EPI_ISL_2445236, EPI_ISL_2445237 | UNIDADE MISTA DE IGUAPE | Instituto Butantan | Antonio Jorge Martins; Claudia Renata dos Santos Barros; David Schlesinger; Debora Botequio Moretti; Dimas Tadeu Covas; Elaine Cristina Marqueze; Elaine Vieira Santos; Evandra Strazza Rodrigues; Heidge Fukumasu; Jayme Augusto de Souza-Neto; José Salvatore Leister Patané; Luiz Alcantara; Luiz Lehmann Coutinho; Maria Carolina Elias; Maurício Lacerda Nogueira; Rafael dos Santos Bezerra; Raul Machado Neto; Rejane Maria Tommasini Grotto; Ricardo Haddad; Sandra Coccuzzo Sampaio Vessoni; Simone Kashima; Svetoslav Nanev Slavov; Vincent Louis Viala |
| EPI_ISL_2493181, EPI_ISL_2493602, EPI_ISL_2493604, EPI_ISL_2493605, EPI_ISL_2493607, EPI_ISL_2493611, EPI_ISL_2493614, EPI_ISL_2493616, EPI_ISL_2493617, EPI_ISL_2493620, EPI_ISL_2493623 | | | |
| see above | UNIDADE MISTA DE LUIZ ANTONIO | Instituto Butantan | Antonio Jorge Martins; Claudia Renata dos Santos Barros; David Schlesinger; Debora Botequio Moretti; Dimas Tadeu Covas; Elaine Cristina Marqueze; Elaine Vieira Santos; Evandra Strazza Rodrigues; Heidge Fukumasu; Jayme Augusto de Souza-Neto; José Salvatore Leister Patané; Luiz Alcantara; Luiz Lehmann Coutinho; Maria Carolina Elias; Maurício Lacerda Nogueira; Rafael dos Santos Bezerra; Raul Machado Neto; Rejane Maria Tommasini Grotto; Ricardo Haddad; Sandra Coccuzzo Sampaio Vessoni; Simone Kashima; Svetoslav Nanev Slavov; Vincent Louis Viala |
| EPI_ISL_2445611 | UNIDADE MISTA DE SAUDE CAPELA DO ALTO | Instituto Butantan | Antonio Jorge Martins; Claudia Renata dos Santos Barros; David Schlesinger; Debora Botequio Moretti; Dimas Tadeu Covas; Elaine Cristina Marqueze; Elaine Vieira Santos; Evandra Strazza Rodrigues; Heidge Fukumasu; Jayme Augusto de Souza-Neto; José Salvatore Leister Patané; Luiz Alcantara; Luiz Lehmann Coutinho; Maria Carolina Elias; Maurício Lacerda Nogueira; Rafael dos Santos Bezerra; Raul Machado Neto; Rejane Maria Tommasini Grotto; Ricardo Haddad; Sandra Coccuzzo Sampaio Vessoni; Simone Kashima; Svetoslav Nanev Slavov; Vincent Louis Viala |
| EPI_ISL_2445196 | UNIDADE MISTA DE SAUDE DE ORIENTE | Instituto Butantan | Antonio Jorge Martins; Claudia Renata dos Santos Barros; David Schlesinger; Debora Botequio Moretti; Dimas Tadeu Covas; Elaine Cristina Marqueze; Elaine Vieira Santos; Evandra Strazza Rodrigues; Heidge Fukumasu; Jayme Augusto de Souza-Neto; José Salvatore Leister Patané; Luiz Alcantara; Luiz Lehmann Coutinho; Maria Carolina Elias; Maurício Lacerda Nogueira; Rafael dos Santos Bezerra; Raul Machado Neto; Rejane Maria Tommasini Grotto; Ricardo Haddad; Sandra Coccuzzo Sampaio Vessoni; Simone Kashima; Svetoslav Nanev Slavov; Vincent Louis Viala |
| EPI_ISL_2493386, EPI_ISL_2493387, EPI_ISL_2493442, EPI_ISL_2493443 | UNIDADE MISTA DE SAUDE MOMBUCA | Instituto Butantan | Antonio Jorge Martins; Claudia Renata dos Santos Barros; David Schlesinger; Debora Botequio Moretti; Dimas Tadeu Covas; Elaine Cristina Marqueze; Elaine Vieira Santos; Evandra Strazza Rodrigues; Heidge Fukumasu; Jayme Augusto de Souza-Neto; José Salvatore Leister Patané; Luiz Alcantara; Luiz Lehmann Coutinho; Maria Carolina Elias; Maurício Lacerda Nogueira; Rafael dos Santos Bezerra; Raul Machado Neto; Rejane Maria Tommasini Grotto; Ricardo Haddad; Sandra Coccuzzo Sampaio Vessoni; Simone Kashima; Svetoslav Nanev Slavov; Vincent Louis Viala |
| EPI_ISL_2494174 | UNIDADE MISTA DE SAUDE RAFARD | Instituto Butantan | Antonio Jorge Martins; Claudia Renata dos Santos Barros; David Schlesinger; Debora Botequio Moretti; Dimas Tadeu Covas; Elaine Cristina Marqueze; Elaine Vieira Santos; Evandra Strazza Rodrigues; Heidge Fukumasu; Jayme Augusto de Souza-Neto; José Salvatore Leister Patané; Luiz Alcantara; Luiz Lehmann Coutinho; Maria Carolina Elias; Maurício Lacerda Nogueira; Rafael dos Santos Bezerra; Raul Machado Neto; Rejane Maria Tommasini Grotto; Ricardo Haddad; Sandra Coccuzzo Sampaio Vessoni; Simone Kashima; Svetoslav Nanev Slavov; Vincent Louis Viala |
| EPI_ISL_2445585 | UNIDADE MISTA DE SAUDE ROSEIRA DR PAULO GUIMARAES CASTRO | Instituto Butantan | Antonio Jorge Martins; Claudia Renata dos Santos Barros; David Schlesinger; Debora Botequio Moretti; Dimas Tadeu Covas; Elaine Cristina Marqueze; Elaine Vieira Santos; Evandra Strazza Rodrigues; Heidge Fukumasu; Jayme Augusto de Souza-Neto; José Salvatore Leister Patané; Luiz Alcantara; Luiz Lehmann Coutinho; Maria Carolina Elias; Maurício Lacerda Nogueira; Rafael dos Santos Bezerra; Raul Machado Neto; Rejane Maria Tommasini Grotto; Ricardo Haddad; Sandra Coccuzzo Sampaio Vessoni; Simone Kashima; Svetoslav Nanev Slavov; Vincent Louis Viala |
| EPI_ISL_2493372 | UNIDADE MISTA OSVALDO P CAMARGO ELIAS FAUSTO | Instituto Butantan | Antonio Jorge Martins; Claudia Renata dos Santos Barros; David Schlesinger; Debora Botequio Moretti; Dimas Tadeu Covas; Elaine Cristina Marqueze; Elaine Vieira Santos; Evandra Strazza Rodrigues; Heidge Fukumasu; Jayme Augusto de Souza-Neto; José Salvatore Leister Patané; Luiz Alcantara; Luiz Lehmann Coutinho; Maria Carolina Elias; Maurício Lacerda Nogueira; Rafael dos Santos Bezerra; Raul Machado Neto; Rejane Maria Tommasini Grotto; Ricardo Haddad; Sandra Coccuzzo Sampaio Vessoni; Simone Kashima; Svetoslav Nanev Slavov; Vincent Louis Viala |
| EPI_ISL_2801360 | UNIDADE PRONTO ATENDIMENTO CONJUNTO CEARA | Oswaldo Cruz Institute, FIOCRUZ/CE | Cleber Furtado Aksenen e Suzana Porto Almeida; Fabio Miyajima; Fernando Braga Stehling; Francisco Eder de Moura Lopes; Jamille Maria Mendes Bezerra; Joaquim César do Nascimento Sousa Junior; Pedro Miguel Carneiro Jeronimo; Thais Ferreira de Oliveira; Thais de Oliveira Costa; Ticiane Cavalcante de Souza; Veridiana Pessoa Miyajima |
| EPI_ISL_2801357 | UNIDADE PRONTO ATENDIMENTO ITAPERI | Oswaldo Cruz Institute, FIOCRUZ/CE | Cleber Furtado Aksenen e Suzana Porto Almeida; Fabio Miyajima; Fernando Braga Stehling; Francisco Eder de Moura Lopes; Jamille Maria Mendes Bezerra; Joaquim César do Nascimento Sousa Junior; Pedro Miguel Carneiro Jeronimo; Thais Ferreira de Oliveira; Thais de Oliveira Costa; Ticiane Cavalcante de Souza; Veridiana Pessoa Miyajima |
| EPI_ISL_2378752, EPI_ISL_2445154, EPI_ISL_2445164, EPI_ISL_2445168, EPI_ISL_2445175, EPI_ISL_2445181, EPI_ISL_2445185, EPI_ISL_2473707, EPI_ISL_2473724, EPI_ISL_2473812 | | | |
| see above | UNIDADE REFERENCIAL SUDOESTE | Instituto Butantan | Antonio Jorge Martins; Claudia Renata dos Santos Barros; David Schlesinger; Debora Botequio Moretti; Dimas Tadeu Covas; Elaine Cristina Marqueze; Elaine Vieira Santos; Evandra Strazza Rodrigues; Heidge Fukumasu; Jayme Augusto de Souza-Neto; José Salvatore Leister Patané; Luiz Alcantara; Luiz Lehmann Coutinho; Maria Carolina Elias; Maurício Lacerda Nogueira; Rafael dos Santos Bezerra; Raul Machado Neto; Rejane Maria Tommasini Grotto; Ricardo Haddad; Sandra Coccuzzo Sampaio Vessoni; Simone Kashima; Svetoslav Nanev Slavov; Vincent Louis Viala |
| EPI_ISL_2493249, EPI_ISL_2493250, EPI_ISL_2493271, EPI_ISL_2493461, EPI_ISL_2493474, EPI_ISL_2493476, EPI_ISL_2493477, EPI_ISL_2493483, EPI_ISL_2493507, EPI_ISL_2493508, EPI_ISL_2493509, EPI_ISL_2493511, EPI_ISL_2493515, EPI_ISL_2493516, EPI_ISL_2493517, EPI_ISL_2493518, EPI_ISL_2494292, EPI_ISL_2494344, EPI_ISL_2494345, EPI_ISL_2494346 | | | |
| see above | UNIDADE RESPIRATORIA NOVA HORTOLANDIA | Instituto Butantan | Antonio Jorge Martins; Claudia Renata dos Santos Barros; David Schlesinger; Debora Botequio Moretti; Dimas Tadeu Covas; Elaine Cristina Marqueze; Elaine Vieira Santos; Evandra Strazza Rodrigues; Heidge Fukumasu; Jayme Augusto de Souza-Neto; José Salvatore Leister Patané; Luiz Alcantara; Luiz Lehmann Coutinho; Maria Carolina Elias; Maurício Lacerda Nogueira; Rafael dos Santos Bezerra; Raul Machado Neto; Rejane Maria Tommasini Grotto; Ricardo Haddad; Sandra Coccuzzo Sampaio Vessoni; Simone Kashima; Svetoslav Nanev Slavov; Vincent Louis Viala |
| EPI_ISL_2493807, | UNIDADE SAUDE FAMILIA DR PIERRE | Instituto Butantan | Antonio Jorge Martins; Claudia Renata dos Santos Barros; David Schlesinger; Debora Botequio Moretti; Dimas Tadeu Covas; Elaine Cristina Marqueze; Elaine Vieira Santos; Evandra Strazza Rodrigues; Heidge Fukumasu; Jayme Augusto de Souza-Neto; José Salvatore Leister Patané; Luiz Alcantara; Luiz Lehmann |

| | | | |
|---|---|---|---|
| EPI_ISL_2493808 | ANTONIO COPPIETERS AREALVA | | Coutinho; Maria Carolina Elias; Maurício Lacerda Nogueira; Rafael dos Santos Bezerra; Raul Machado Neto; Rejane Maria Tommasini Grotto; Ricardo Haddad; Sandra Coccuzzo Sampaio Vessoni; Simone Kashima; Svetoslav Nanev Slavov; Vincent Louis Viala |
| EPI_ISL_2493897, EPI_ISL_2493912 | UNIDADE SENTINELA COVID19 | Instituto Butantan | Antonio Jorge Martins; Claudia Renata dos Santos Barros; David Schlesinger; Debora Botequio Moretti; Dimas Tadeu Covas; Elaine Cristina Marqueze; Elaine Vieira Santos; Evandra Strazza Rodrigues; Heidge Fukumasu; Jayme Augusto de Souza-Neto; José Salvatore Leister Patané; Luiz Alcantara; Luiz Lehmann Coutinho; Maria Carolina Elias; Maurício Lacerda Nogueira; Rafael dos Santos Bezerra; Raul Machado Neto; Rejane Maria Tommasini Grotto; Ricardo Haddad; Sandra Coccuzzo Sampaio Vessoni; Simone Kashima; Svetoslav Nanev Slavov; Vincent Louis Viala |
| EPI_ISL_2493907 | UNIDADE SENTINELA VILA APARECIDA | Instituto Butantan | Antonio Jorge Martins; Claudia Renata dos Santos Barros; David Schlesinger; Debora Botequio Moretti; Dimas Tadeu Covas; Elaine Cristina Marqueze; Elaine Vieira Santos; Evandra Strazza Rodrigues; Heidge Fukumasu; Jayme Augusto de Souza-Neto; José Salvatore Leister Patané; Luiz Alcantara; Luiz Lehmann Coutinho; Maria Carolina Elias; Maurício Lacerda Nogueira; Rafael dos Santos Bezerra; Raul Machado Neto; Rejane Maria Tommasini Grotto; Ricardo Haddad; Sandra Coccuzzo Sampaio Vessoni; Simone Kashima; Svetoslav Nanev Slavov; Vincent Louis Viala |
| EPI_ISL_2928137 | UPA - LUIS ATÍLIO LOSI VIANA | Hemocentro de Ribeirao Preto | Brazil; Rede de Alerta das Variantes da COVID-19 coordinated by Butantan Institute; São Paulo |
| EPI_ISL_2378737, EPI_ISL_2378740, EPI_ISL_2378741, EPI_ISL_2445329, EPI_ISL_2462322, EPI_ISL_2493272, EPI_ISL_2493273, EPI_ISL_2493566, EPI_ISL_2494116, EPI_ISL_2494124, EPI_ISL_2494125, EPI_ISL_2494298 | | | |
| see above | UPA DE BEBEDOURO | Instituto Butantan | Antonio Jorge Martins; Claudia Renata dos Santos Barros; David Schlesinger; Debora Botequio Moretti; Dimas Tadeu Covas; Elaine Cristina Marqueze; Elaine Vieira Santos; Evandra Strazza Rodrigues; Heidge Fukumasu; Jayme Augusto de Souza-Neto; José Salvatore Leister Patané; Luiz Alcantara; Luiz Lehmann Coutinho; Maria Carolina Elias; Maurício Lacerda Nogueira; Rafael dos Santos Bezerra; Raul Machado Neto; Rejane Maria Tommasini Grotto; Ricardo Haddad; Sandra Coccuzzo Sampaio Vessoni; Simone Kashima; Svetoslav Nanev Slavov; Vincent Louis Viala |
| EPI_ISL_2208944, EPI_ISL_2209579, EPI_ISL_2209671, EPI_ISL_2345765 | UPA DE BEBEDOURO | Instituto Butantan / Mendelics | Antonio Jorge Martins; Claudia Renata dos Santos Barros; David Schlesinger; Debora Botequio Moretti; Dimas Tadeu Covas; Elaine Cristina Marqueze; Elaine Vieira Santos; Evandra Strazza Rodrigues; Heidge Fukumasu; Jayme Augusto de Souza-Neto; José Salvatore Leister Patané; Luiz Alcantara; Luiz Lehmann Coutinho; Maria Carolina Elias; Maurício Lacerda Nogueira; Rafael dos Santos Bezerra; Raul Machado Neto; Rejane Maria Tommasini Grotto; Ricardo Haddad; Sandra Coccuzzo Sampaio Vessoni; Simone Kashima; Svetoslav Nanev Slavov; Vincent Louis Viala |
| EPI_ISL_2445243, EPI_ISL_2445245, EPI_ISL_2445246, EPI_ISL_2445247, EPI_ISL_2445248, EPI_ISL_2445249, EPI_ISL_2445250, EPI_ISL_2445251, EPI_ISL_2445253, EPI_ISL_2445255, EPI_ISL_2445256, EPI_ISL_2445257, EPI_ISL_2445258, EPI_ISL_2445259, EPI_ISL_2445260, EPI_ISL_2445261, EPI_ISL_2445262, EPI_ISL_2445263, EPI_ISL_2445264, EPI_ISL_2445265, EPI_ISL_2445266, EPI_ISL_2445267, EPI_ISL_2445268, EPI_ISL_2445269, EPI_ISL_2445270, EPI_ISL_2445271 | | | |
| see above | UPA DR AKIRA TADA | Instituto Butantan | Antonio Jorge Martins; Claudia Renata dos Santos Barros; David Schlesinger; Debora Botequio Moretti; Dimas Tadeu Covas; Elaine Cristina Marqueze; Elaine Vieira Santos; Evandra Strazza Rodrigues; Heidge Fukumasu; Jayme Augusto de Souza-Neto; José Salvatore Leister Patané; Luiz Alcantara; Luiz Lehmann Coutinho; Maria Carolina Elias; Maurício Lacerda Nogueira; Rafael dos Santos Bezerra; Raul Machado Neto; Rejane Maria Tommasini Grotto; Ricardo Haddad; Sandra Coccuzzo Sampaio Vessoni; Simone Kashima; Svetoslav Nanev Slavov; Vincent Louis Viala |
| EPI_ISL_2445272, EPI_ISL_2445273, EPI_ISL_2445288, EPI_ISL_2445371, EPI_ISL_2445382 | UPA DR FRANCO DA ROCHA | Instituto Butantan | Antonio Jorge Martins; Claudia Renata dos Santos Barros; David Schlesinger; Debora Botequio Moretti; Dimas Tadeu Covas; Elaine Cristina Marqueze; Elaine Vieira Santos; Evandra Strazza Rodrigues; Heidge Fukumasu; Jayme Augusto de Souza-Neto; José Salvatore Leister Patané; Luiz Alcantara; Luiz Lehmann Coutinho; Maria Carolina Elias; Maurício Lacerda Nogueira; Rafael dos Santos Bezerra; Raul Machado Neto; Rejane Maria Tommasini Grotto; Ricardo Haddad; Sandra Coccuzzo Sampaio Vessoni; Simone Kashima; Svetoslav Nanev Slavov; Vincent Louis Viala |
| EPI_ISL_2445324, EPI_ISL_2445326, EPI_ISL_2445336, EPI_ISL_2473777 | UPA GEISEL | Instituto Butantan | Antonio Jorge Martins; Claudia Renata dos Santos Barros; David Schlesinger; Debora Botequio Moretti; Dimas Tadeu Covas; Elaine Cristina Marqueze; Elaine Vieira Santos; Evandra Strazza Rodrigues; Heidge Fukumasu; Jayme Augusto de Souza-Neto; José Salvatore Leister Patané; Luiz Alcantara; Luiz Lehmann Coutinho; Maria Carolina Elias; Maurício Lacerda Nogueira; Rafael dos Santos Bezerra; Raul Machado Neto; Rejane Maria Tommasini Grotto; Ricardo Haddad; Sandra Coccuzzo Sampaio Vessoni; Simone Kashima; Svetoslav Nanev Slavov; Vincent Louis Viala |
| EPI_ISL_2378736, EPI_ISL_2493194, EPI_ISL_2493506, EPI_ISL_2493510, EPI_ISL_2493514, EPI_ISL_2494288, EPI_ISL_2494303 | | | |
| see above | UPA II JUNDIAI | Instituto Butantan | Antonio Jorge Martins; Claudia Renata dos Santos Barros; David Schlesinger; Debora Botequio Moretti; Dimas Tadeu Covas; Elaine Cristina Marqueze; Elaine Vieira Santos; Evandra Strazza Rodrigues; Heidge Fukumasu; Jayme Augusto de Souza-Neto; José Salvatore Leister Patané; Luiz Alcantara; Luiz Lehmann Coutinho; Maria Carolina Elias; Maurício Lacerda Nogueira; Rafael dos Santos Bezerra; Raul Machado Neto; Rejane Maria Tommasini Grotto; Ricardo Haddad; Sandra Coccuzzo Sampaio Vessoni; Simone Kashima; Svetoslav Nanev Slavov; Vincent Louis Viala |
| EPI_ISL_2345880 | UPA II OROPO | Instituto Butantan / Mendelics | Antonio Jorge Martins; Claudia Renata dos Santos Barros; David Schlesinger; Debora Botequio Moretti; Dimas Tadeu Covas; Elaine Cristina Marqueze; Elaine Vieira Santos; Evandra Strazza Rodrigues; Heidge Fukumasu; Jayme Augusto de Souza-Neto; José Salvatore Leister Patané; Luiz Alcantara; Luiz Lehmann Coutinho; Maria Carolina Elias; Maurício Lacerda Nogueira; Rafael dos Santos Bezerra; Raul Machado Neto; Rejane Maria Tommasini Grotto; Ricardo Haddad; Sandra Coccuzzo Sampaio Vessoni; Simone Kashima; Svetoslav Nanev Slavov; Vincent Louis Viala |
| EPI_ISL_2445330, EPI_ISL_2445332, EPI_ISL_2445333, EPI_ISL_2445334, EPI_ISL_2445346, EPI_ISL_2445364, EPI_ISL_2473778 | | | |
| see above | UPA MARY DOTA | Instituto Butantan | Antonio Jorge Martins; Claudia Renata dos Santos Barros; David Schlesinger; Debora Botequio Moretti; Dimas Tadeu Covas; Elaine Cristina Marqueze; Elaine Vieira Santos; Evandra Strazza Rodrigues; Heidge Fukumasu; Jayme Augusto de Souza-Neto; José Salvatore Leister Patané; Luiz Alcantara; Luiz Lehmann Coutinho; Maria Carolina Elias; Maurício Lacerda Nogueira; Rafael dos Santos Bezerra; Raul Machado Neto; Rejane Maria Tommasini Grotto; Ricardo Haddad; Sandra Coccuzzo Sampaio Vessoni; Simone Kashima; Svetoslav Nanev Slavov; Vincent Louis Viala |
| EPI_ISL_2209332, EPI_ISL_2209374, EPI_ISL_2209746, EPI_ISL_2345684, EPI_ISL_2345828, EPI_ISL_2345872 | UPA MARY DOTA | Instituto Butantan / Mendelics | Antonio Jorge Martins; Claudia Renata dos Santos Barros; David Schlesinger; Debora Botequio Moretti; Dimas Tadeu Covas; Elaine Cristina Marqueze; Elaine Vieira Santos; Evandra Strazza Rodrigues; Heidge Fukumasu; Jayme Augusto de Souza-Neto; José Salvatore Leister Patané; Luiz Alcantara; Luiz Lehmann Coutinho; Maria Carolina Elias; Maurício Lacerda Nogueira; Rafael dos Santos Bezerra; Raul Machado Neto; Rejane Maria Tommasini Grotto; Ricardo Haddad; Sandra Coccuzzo Sampaio Vessoni; Simone Kashima; Svetoslav Nanev Slavov; Vincent Louis Viala |
| EPI_ISL_2493816, EPI_ISL_2493848, EPI_ISL_2493852, EPI_ISL_2493858 | UPA SADAKO SEDOGUTI | Instituto Butantan | Antonio Jorge Martins; Claudia Renata dos Santos Barros; David Schlesinger; Debora Botequio Moretti; Dimas Tadeu Covas; Elaine Cristina Marqueze; Elaine Vieira Santos; Evandra Strazza Rodrigues; Heidge Fukumasu; Jayme Augusto de Souza-Neto; José Salvatore Leister Patané; Luiz Alcantara; Luiz Lehmann Coutinho; Maria Carolina Elias; Maurício Lacerda Nogueira; Rafael dos Santos Bezerra; Raul Machado Neto; Rejane Maria Tommasini Grotto; Ricardo Haddad; Sandra Coccuzzo Sampaio Vessoni; Simone Kashima; Svetoslav Nanev Slavov; Vincent Louis Viala |
| EPI_ISL_2479923 | UPA UNIDADE DE PRONTO ATENDIMENTO 24 HORAS BOM JESUS | Instituto Butantan | Antonio Jorge Martins; Claudia Renata dos Santos Barros; David Schlesinger; Debora Botequio Moretti; Dimas Tadeu Covas; Elaine Cristina Marqueze; Elaine Vieira Santos; Evandra Strazza Rodrigues; Heidge Fukumasu; Jayme Augusto de Souza-Neto; José Salvatore Leister Patané; Luiz Alcantara; Luiz Lehmann Coutinho; Maria Carolina Elias; Maurício Lacerda Nogueira; Rafael dos Santos Bezerra; Raul Machado Neto; Rejane Maria Tommasini Grotto; Ricardo Haddad; Sandra Coccuzzo Sampaio Vessoni; Simone Kashima; Svetoslav Nanev Slavov; Vincent Louis Viala |
| EPI_ISL_2210275, EPI_ISL_2345677, EPI_ISL_2345820, EPI_ISL_2345864, EPI_ISL_2362243 | UPA UNIDADE DE PRONTO ATENDIMENTO 24 HORAS BOM JESUS | Instituto Butantan / Mendelics | Antonio Jorge Martins; Claudia Renata dos Santos Barros; David Schlesinger; Debora Botequio Moretti; Dimas Tadeu Covas; Elaine Cristina Marqueze; Elaine Vieira Santos; Evandra Strazza Rodrigues; Heidge Fukumasu; Jayme Augusto de Souza-Neto; José Salvatore Leister Patané; Luiz Alcantara; Luiz Lehmann Coutinho; Maria Carolina Elias; Maurício Lacerda Nogueira; Rafael dos Santos Bezerra; Raul Machado Neto; Rejane Maria Tommasini Grotto; Ricardo Haddad; Sandra Coccuzzo Sampaio Vessoni; Simone Kashima; Svetoslav Nanev Slavov; Vincent Louis Viala |
| EPI_ISL_2445275, EPI_ISL_2445277, EPI_ISL_2445278, EPI_ISL_2445279, EPI_ISL_2445297, EPI_ISL_2445300, EPI_ISL_2445302, EPI_ISL_2445310, EPI_ISL_2445311, EPI_ISL_2445372, EPI_ISL_2445373, EPI_ISL_2445381 | | | |
| see above | UPA VILA SANTA CATARINA | Instituto Butantan | Antonio Jorge Martins; Claudia Renata dos Santos Barros; David Schlesinger; Debora Botequio Moretti; Dimas Tadeu Covas; Elaine Cristina Marqueze; Elaine Vieira Santos; Evandra Strazza Rodrigues; Heidge Fukumasu; Jayme Augusto de Souza-Neto; José Salvatore Leister Patané; Luiz Alcantara; Luiz Lehmann Coutinho; Maria Carolina Elias; Maurício Lacerda Nogueira; Rafael dos Santos Bezerra; Raul Machado Neto; Rejane Maria Tommasini Grotto; Ricardo Haddad; Sandra Coccuzzo Sampaio Vessoni; Simone Kashima; Svetoslav Nanev Slavov; Vincent Louis Viala |
| EPI_ISL_2378738, EPI_ISL_2493546 | USAFA FORTE | Instituto Butantan | Antonio Jorge Martins; Claudia Renata dos Santos Barros; David Schlesinger; Debora Botequio Moretti; Dimas Tadeu Covas; Elaine Cristina Marqueze; Elaine Vieira Santos; Evandra Strazza Rodrigues; Heidge Fukumasu; Jayme Augusto de Souza-Neto; José Salvatore Leister Patané; Luiz Alcantara; Luiz Lehmann Coutinho; Maria Carolina Elias; Maurício Lacerda Nogueira; Rafael dos Santos Bezerra; Raul Machado Neto; Rejane Maria Tommasini Grotto; Ricardo Haddad; Sandra Coccuzzo Sampaio Vessoni; Simone Kashima; Svetoslav Nanev Slavov; Vincent Louis Viala |
| EPI_ISL_2493536, EPI_ISL_2494304 | USAFA GUILHERMINA | Instituto Butantan | Antonio Jorge Martins; Claudia Renata dos Santos Barros; David Schlesinger; Debora Botequio Moretti; Dimas Tadeu Covas; Elaine Cristina Marqueze; Elaine Vieira Santos; Evandra Strazza Rodrigues; Heidge Fukumasu; Jayme Augusto de Souza-Neto; José Salvatore Leister Patané; Luiz Alcantara; Luiz Lehmann Coutinho; Maria Carolina Elias; Maurício Lacerda Nogueira; Rafael dos Santos Bezerra; Raul Machado Neto; Rejane Maria Tommasini Grotto; Ricardo Haddad; Sandra Coccuzzo Sampaio Vessoni; Simone Kashima; Svetoslav Nanev Slavov; Vincent Louis Viala |
| EPI_ISL_2493879 | USAFA MARACANA | Instituto Butantan | Antonio Jorge Martins; Claudia Renata dos Santos Barros; David Schlesinger; Debora Botequio Moretti; Dimas Tadeu Covas; Elaine Cristina Marqueze; Elaine Vieira Santos; Evandra Strazza Rodrigues; Heidge Fukumasu; Jayme Augusto de Souza-Neto; José Salvatore Leister Patané; Luiz Alcantara; Luiz Lehmann Coutinho; Maria Carolina Elias; Maurício Lacerda Nogueira; Rafael dos Santos Bezerra; Raul Machado Neto; Rejane Maria Tommasini Grotto; Ricardo Haddad; Sandra Coccuzzo Sampaio Vessoni; Simone Kashima; Svetoslav Nanev Slavov; Vincent Louis Viala |
| EPI_ISL_2493555 | USAFA MELVI | Instituto Butantan | Antonio Jorge Martins; Claudia Renata dos Santos Barros; David Schlesinger; Debora Botequio Moretti; Dimas Tadeu Covas; Elaine Cristina Marqueze; Elaine Vieira Santos; Evandra Strazza Rodrigues; Heidge Fukumasu; Jayme Augusto de Souza-Neto; José Salvatore Leister Patané; Luiz Alcantara; Luiz Lehmann Coutinho; Maria Carolina Elias; Maurício Lacerda Nogueira; Rafael dos Santos Bezerra; Raul Machado Neto; Rejane Maria Tommasini Grotto; Ricardo Haddad; Sandra Coccuzzo Sampaio Vessoni; Simone Kashima; Svetoslav Nanev Slavov; Vincent Louis Viala |
| EPI_ISL_2493558, EPI_ISL_2493895 | USAFA MIRIM | Instituto Butantan | Antonio Jorge Martins; Claudia Renata dos Santos Barros; David Schlesinger; Debora Botequio Moretti; Dimas Tadeu Covas; Elaine Cristina Marqueze; Elaine Vieira Santos; Evandra Strazza Rodrigues; Heidge Fukumasu; Jayme Augusto de Souza-Neto; José Salvatore Leister Patané; Luiz Alcantara; Luiz Lehmann Coutinho; Maria Carolina Elias; Maurício Lacerda Nogueira; Rafael dos Santos Bezerra; Raul Machado Neto; Rejane Maria Tommasini Grotto; Ricardo Haddad; Sandra Coccuzzo Sampaio Vessoni; Simone Kashima; Svetoslav Nanev Slavov; Vincent Louis Viala |
| EPI_ISL_2493557 | USAFA OCIAN | Instituto Butantan | Antonio Jorge Martins; Claudia Renata dos Santos Barros; David Schlesinger; Debora Botequio Moretti; Dimas Tadeu Covas; Elaine Cristina Marqueze; Elaine Vieira Santos; Evandra Strazza Rodrigues; Heidge Fukumasu; Jayme Augusto de Souza-Neto; José Salvatore Leister Patané; Luiz Alcantara; Luiz Lehmann Coutinho; Maria Carolina Elias; Maurício Lacerda Nogueira; Rafael dos Santos Bezerra; Raul Machado Neto; Rejane Maria Tommasini Grotto; Ricardo Haddad; Sandra Coccuzzo Sampaio Vessoni; Simone Kashima; Svetoslav Nanev Slavov; Vincent Louis Viala |
| EPI_ISL_2493559, EPI_ISL_2493561 | USAFA PRINCESA | Instituto Butantan | Antonio Jorge Martins; Claudia Renata dos Santos Barros; David Schlesinger; Debora Botequio Moretti; Dimas Tadeu Covas; Elaine Cristina Marqueze; Elaine Vieira Santos; Evandra Strazza Rodrigues; Heidge Fukumasu; Jayme Augusto de Souza-Neto; José Salvatore Leister Patané; Luiz Alcantara; Luiz Lehmann Coutinho; Maria Carolina Elias; Maurício Lacerda Nogueira; Rafael dos Santos Bezerra; Raul Machado Neto; Rejane Maria Tommasini Grotto; Ricardo Haddad; Sandra Coccuzzo Sampaio Vessoni; Simone Kashima; Svetoslav Nanev Slavov; Vincent Louis Viala |
| EPI_ISL_2493535, EPI_ISL_2493541, EPI_ISL_2494339 | USAFA SAMAMBAIA | Instituto Butantan | Antonio Jorge Martins; Claudia Renata dos Santos Barros; David Schlesinger; Debora Botequio Moretti; Dimas Tadeu Covas; Elaine Cristina Marqueze; Elaine Vieira Santos; Evandra Strazza Rodrigues; Heidge Fukumasu; Jayme Augusto de Souza-Neto; José Salvatore Leister Patané; Luiz Alcantara; Luiz Lehmann Coutinho; Maria Carolina Elias; Maurício Lacerda Nogueira; Rafael dos Santos Bezerra; Raul Machado Neto; Rejane Maria Tommasini Grotto; Ricardo Haddad; Sandra Coccuzzo Sampaio Vessoni; Simone Kashima; Svetoslav Nanev Slavov; Vincent Louis Viala |
| EPI_ISL_2493560, EPI_ISL_2493563, EPI_ISL_2493831 | USAFA SAO JORGE | Instituto Butantan | Antonio Jorge Martins; Claudia Renata dos Santos Barros; David Schlesinger; Debora Botequio Moretti; Dimas Tadeu Covas; Elaine Cristina Marqueze; Elaine Vieira Santos; Evandra Strazza Rodrigues; Heidge Fukumasu; Jayme Augusto de Souza-Neto; José Salvatore Leister Patané; Luiz Alcantara; Luiz Lehmann Coutinho; Maria Carolina Elias; Maurício Lacerda Nogueira; Rafael dos Santos Bezerra; Raul Machado Neto; Rejane Maria Tommasini Grotto; Ricardo Haddad; Sandra Coccuzzo Sampaio Vessoni; Simone Kashima; Svetoslav Nanev Slavov; Vincent Louis Viala |
| EPI_ISL_2493551 | USAFA TUDE BASTOS | Instituto Butantan | Antonio Jorge Martins; Claudia Renata dos Santos Barros; David Schlesinger; Debora Botequio Moretti; Dimas Tadeu Covas; Elaine Cristina Marqueze; Elaine Vieira Santos; Evandra Strazza Rodrigues; Heidge Fukumasu; Jayme Augusto de Souza-Neto; José Salvatore Leister Patané; Luiz Alcantara; Luiz Lehmann Coutinho; Maria Carolina Elias; Maurício Lacerda Nogueira; Rafael dos Santos Bezerra; Raul Machado Neto; Rejane Maria Tommasini Grotto; Ricardo Haddad; Sandra Coccuzzo Sampaio Vessoni; Simone Kashima; Svetoslav Nanev Slavov; Vincent Louis Viala |
| EPI_ISL_2345746 | USF ARCADAS | Instituto Butantan / Mendelics | Antonio Jorge Martins; Claudia Renata dos Santos Barros; David Schlesinger; Debora Botequio Moretti; Dimas Tadeu Covas; Elaine Cristina Marqueze; Elaine Vieira Santos; Evandra Strazza Rodrigues; Heidge Fukumasu; Jayme Augusto de Souza-Neto; José Salvatore Leister Patané; Luiz Alcantara; Luiz Lehmann Coutinho; Maria Carolina Elias; Maurício Lacerda Nogueira; Rafael dos Santos Bezerra; Raul Machado Neto; Rejane Maria Tommasini Grotto; Ricardo Haddad; Sandra Coccuzzo Sampaio Vessoni; Simone Kashima; Svetoslav Nanev Slavov; Vincent Louis Viala |
| EPI_ISL_2345761 | USF II ALADIA ARAN RODRIGUES | Instituto Butantan / Mendelics | Antonio Jorge Martins; Claudia Renata dos Santos Barros; David Schlesinger; Debora Botequio Moretti; Dimas Tadeu Covas; Elaine Cristina Marqueze; Elaine Vieira Santos; Evandra Strazza Rodrigues; Heidge Fukumasu; Jayme Augusto de Souza-Neto; José Salvatore Leister Patané; Luiz Alcantara; Luiz Lehmann Coutinho; Maria Carolina Elias; Maurício Lacerda Nogueira; Rafael dos Santos Bezerra; Raul Machado Neto; Rejane Maria Tommasini Grotto; Ricardo Haddad; Sandra Coccuzzo Sampaio Vessoni; Simone Kashima; Svetoslav Nanev Slavov; Vincent Louis Viala |
| EPI_ISL_2445456, EPI_ISL_2445457 | USF II DR JOSE RODOLFO ZIMMERMANN DIAS | Instituto Butantan | Antonio Jorge Martins; Claudia Renata dos Santos Barros; David Schlesinger; Debora Botequio Moretti; Dimas Tadeu Covas; Elaine Cristina Marqueze; Elaine Vieira Santos; Evandra Strazza Rodrigues; Heidge Fukumasu; Jayme Augusto de Souza-Neto; José Salvatore Leister Patané; Luiz Alcantara; Luiz Lehmann Coutinho; Maria Carolina Elias; Maurício Lacerda Nogueira; Rafael dos Santos Bezerra; Raul Machado Neto; Rejane Maria Tommasini Grotto; Ricardo Haddad; Sandra Coccuzzo Sampaio Vessoni; Simone Kashima; Svetoslav Nanev Slavov; Vincent Louis Viala |
| EPI_ISL_2473711 | USF ITAJUBY | Instituto Butantan | Antonio Jorge Martins; Claudia Renata dos Santos Barros; David Schlesinger; Debora Botequio Moretti; Dimas Tadeu Covas; Elaine Cristina Marqueze; Elaine Vieira Santos; Evandra Strazza Rodrigues; Heidge Fukumasu; Jayme Augusto de Souza-Neto; José Salvatore Leister Patané; Luiz Alcantara; Luiz Lehmann Coutinho; Maria Carolina Elias; Maurício Lacerda Nogueira; Rafael dos Santos Bezerra; Raul Machado Neto; Rejane Maria Tommasini Grotto; Ricardo Haddad; Sandra Coccuzzo Sampaio Vessoni; Simone Kashima; Svetoslav Nanev Slavov; Vincent Louis Viala |
| EPI_ISL_2493801 | USF JARDIM AMERICA FIGUEIRA | Instituto Butantan | Antonio Jorge Martins; Claudia Renata dos Santos Barros; David Schlesinger; Debora Botequio Moretti; Dimas Tadeu Covas; Elaine Cristina Marqueze; Elaine Vieira Santos; Evandra Strazza Rodrigues; Heidge Fukumasu; Jayme Augusto de Souza-Neto; José Salvatore Leister Patané; Luiz Alcantara; Luiz Lehmann Coutinho; Maria Carolina Elias; Maurício Lacerda Nogueira; Rafael dos Santos Bezerra; Raul Machado Neto; Rejane Maria Tommasini Grotto; Ricardo Haddad; Sandra Coccuzzo Sampaio Vessoni; Simone Kashima; Svetoslav Nanev Slavov; Vincent Louis Viala |
| EPI_ISL_2345834 | USF JARDIM BRASIL | Instituto Butantan / Mendelics | Antonio Jorge Martins; Claudia Renata dos Santos Barros; David Schlesinger; Debora Botequio Moretti; Dimas Tadeu Covas; Elaine Cristina Marqueze; Elaine Vieira Santos; Evandra Strazza Rodrigues; Heidge Fukumasu; Jayme Augusto de Souza-Neto; José Salvatore Leister Patané; Luiz Alcantara; Luiz Lehmann Coutinho; Maria Carolina Elias; Maurício Lacerda Nogueira; Rafael dos Santos Bezerra; Raul Machado Neto; Rejane Maria Tommasini Grotto; Ricardo Haddad; Sandra Coccuzzo Sampaio Vessoni; Simone Kashima; Svetoslav Nanev Slavov; Vincent Louis Viala |
| EPI_ISL_2493811, EPI_ISL_2493814, EPI_ISL_2493817 | USF JARDIM CAMANDUCAIA | Instituto Butantan | Antonio Jorge Martins; Claudia Renata dos Santos Barros; David Schlesinger; Debora Botequio Moretti; Dimas Tadeu Covas; Elaine Cristina Marqueze; Elaine Vieira Santos; Evandra Strazza Rodrigues; Heidge Fukumasu; Jayme Augusto de Souza-Neto; José Salvatore Leister Patané; Luiz Alcantara; Luiz Lehmann Coutinho; Maria Carolina Elias; Maurício Lacerda Nogueira; Rafael dos Santos Bezerra; Raul Machado Neto; Rejane Maria Tommasini Grotto; Ricardo Haddad; Sandra Coccuzzo Sampaio Vessoni; Simone Kashima; Svetoslav Nanev Slavov; Vincent Louis Viala |
| EPI_ISL_2493799, | USF JARDIM DAS AVES MOREIRINHA | Instituto Butantan | Antonio Jorge Martins; Claudia Renata dos Santos Barros; David Schlesinger; Debora Botequio Moretti; Dimas Tadeu Covas; Elaine Cristina Marqueze; Elaine Vieira Santos; Evandra Strazza Rodrigues; Heidge Fukumasu; Jayme Augusto de Souza-Neto; José Salvatore Leister Patané; Luiz Alcantara; Luiz Lehmann |

| | | | |
|---|---|---|---|
| EPI_ISL_2493810, EPI_ISL_2493812, EPI_ISL_2493813 | | | Coutinho; Maria Carolina Elias; Maurício Lacerda Nogueira; Rafael dos Santos Bezerra; Raul Machado Neto; Rejane Maria Tommasini Grotto; Ricardo Haddad; Sandra Coccuzzo Sampaio Vessoni; Simone Kashima; Svetoslav Nanev Slavov; Vincent Louis Viala |
| EPI_ISL_2493819 | USF JARDIM JOSELY | Instituto Butantan | Antonio Jorge Martins; Claudia Renata dos Santos Barros; David Schlesinger; Debora Botequio Moretti; Dimas Tadeu Covas; Elaine Cristina Marqueze; Elaine Vieira Santos; Evandra Strazza Rodrigues; Heidge Fukumasu; Jayme Augusto de Souza-Neto; José Salvatore Leister Patané; Luiz Alcantara; Luiz Lehmann Coutinho; Maria Carolina Elias; Maurício Lacerda Nogueira; Rafael dos Santos Bezerra; Raul Machado Neto; Rejane Maria Tommasini Grotto; Ricardo Haddad; Sandra Coccuzzo Sampaio Vessoni; Simone Kashima; Svetoslav Nanev Slavov; Vincent Louis Viala |
| EPI_ISL_2493798, EPI_ISL_2493809 | USF JARDIM SAO DIMAS | Instituto Butantan | Antonio Jorge Martins; Claudia Renata dos Santos Barros; David Schlesinger; Debora Botequio Moretti; Dimas Tadeu Covas; Elaine Cristina Marqueze; Elaine Vieira Santos; Evandra Strazza Rodrigues; Heidge Fukumasu; Jayme Augusto de Souza-Neto; José Salvatore Leister Patané; Luiz Alcantara; Luiz Lehmann Coutinho; Maria Carolina Elias; Maurício Lacerda Nogueira; Rafael dos Santos Bezerra; Raul Machado Neto; Rejane Maria Tommasini Grotto; Ricardo Haddad; Sandra Coccuzzo Sampaio Vessoni; Simone Kashima; Svetoslav Nanev Slavov; Vincent Louis Viala |
| EPI_ISL_2493815 | USF JARDIM SILVESTRE II | Instituto Butantan | Antonio Jorge Martins; Claudia Renata dos Santos Barros; David Schlesinger; Debora Botequio Moretti; Dimas Tadeu Covas; Elaine Cristina Marqueze; Elaine Vieira Santos; Evandra Strazza Rodrigues; Heidge Fukumasu; Jayme Augusto de Souza-Neto; José Salvatore Leister Patané; Luiz Alcantara; Luiz Lehmann Coutinho; Maria Carolina Elias; Maurício Lacerda Nogueira; Rafael dos Santos Bezerra; Raul Machado Neto; Rejane Maria Tommasini Grotto; Ricardo Haddad; Sandra Coccuzzo Sampaio Vessoni; Simone Kashima; Svetoslav Nanev Slavov; Vincent Louis Viala |
| EPI_ISL_2378751, EPI_ISL_2445203, EPI_ISL_2445204, EPI_ISL_2445205, EPI_ISL_2445526, EPI_ISL_2445527, EPI_ISL_2473716, EPI_ISL_2473717, EPI_ISL_2473817 | | | |
| see above | USF PAULISTA FERNANDOPOLIS ANTONIO PIVATO | Instituto Butantan | Antonio Jorge Martins; Claudia Renata dos Santos Barros; David Schlesinger; Debora Botequio Moretti; Dimas Tadeu Covas; Elaine Cristina Marqueze; Elaine Vieira Santos; Evandra Strazza Rodrigues; Heidge Fukumasu; Jayme Augusto de Souza-Neto; José Salvatore Leister Patané; Luiz Alcantara; Luiz Lehmann Coutinho; Maria Carolina Elias; Maurício Lacerda Nogueira; Rafael dos Santos Bezerra; Raul Machado Neto; Rejane Maria Tommasini Grotto; Ricardo Haddad; Sandra Coccuzzo Sampaio Vessoni; Simone Kashima; Svetoslav Nanev Slavov; Vincent Louis Viala |
| EPI_ISL_2445392, EPI_ISL_2493463, EPI_ISL_2493469, EPI_ISL_2493475, EPI_ISL_2493479 | USF SALERNO | Instituto Butantan | Antonio Jorge Martins; Claudia Renata dos Santos Barros; David Schlesinger; Debora Botequio Moretti; Dimas Tadeu Covas; Elaine Cristina Marqueze; Elaine Vieira Santos; Evandra Strazza Rodrigues; Heidge Fukumasu; Jayme Augusto de Souza-Neto; José Salvatore Leister Patané; Luiz Alcantara; Luiz Lehmann Coutinho; Maria Carolina Elias; Maurício Lacerda Nogueira; Rafael dos Santos Bezerra; Raul Machado Neto; Rejane Maria Tommasini Grotto; Ricardo Haddad; Sandra Coccuzzo Sampaio Vessoni; Simone Kashima; Svetoslav Nanev Slavov; Vincent Louis Viala |
| EPI_ISL_2493818 | USF SANTA MARIA DO AMPARO | Instituto Butantan | Antonio Jorge Martins; Claudia Renata dos Santos Barros; David Schlesinger; Debora Botequio Moretti; Dimas Tadeu Covas; Elaine Cristina Marqueze; Elaine Vieira Santos; Evandra Strazza Rodrigues; Heidge Fukumasu; Jayme Augusto de Souza-Neto; José Salvatore Leister Patané; Luiz Alcantara; Luiz Lehmann Coutinho; Maria Carolina Elias; Maurício Lacerda Nogueira; Rafael dos Santos Bezerra; Raul Machado Neto; Rejane Maria Tommasini Grotto; Ricardo Haddad; Sandra Coccuzzo Sampaio Vessoni; Simone Kashima; Svetoslav Nanev Slavov; Vincent Louis Viala |
| EPI_ISL_2494145 | USF SERROTE REGISTRO | Instituto Butantan | Antonio Jorge Martins; Claudia Renata dos Santos Barros; David Schlesinger; Debora Botequio Moretti; Dimas Tadeu Covas; Elaine Cristina Marqueze; Elaine Vieira Santos; Evandra Strazza Rodrigues; Heidge Fukumasu; Jayme Augusto de Souza-Neto; José Salvatore Leister Patané; Luiz Alcantara; Luiz Lehmann Coutinho; Maria Carolina Elias; Maurício Lacerda Nogueira; Rafael dos Santos Bezerra; Raul Machado Neto; Rejane Maria Tommasini Grotto; Ricardo Haddad; Sandra Coccuzzo Sampaio Vessoni; Simone Kashima; Svetoslav Nanev Slavov; Vincent Louis Viala |
| EPI_ISL_2493800 | USF TRES PONTES | Instituto Butantan | Antonio Jorge Martins; Claudia Renata dos Santos Barros; David Schlesinger; Debora Botequio Moretti; Dimas Tadeu Covas; Elaine Cristina Marqueze; Elaine Vieira Santos; Evandra Strazza Rodrigues; Heidge Fukumasu; Jayme Augusto de Souza-Neto; José Salvatore Leister Patané; Luiz Alcantara; Luiz Lehmann Coutinho; Maria Carolina Elias; Maurício Lacerda Nogueira; Rafael dos Santos Bezerra; Raul Machado Neto; Rejane Maria Tommasini Grotto; Ricardo Haddad; Sandra Coccuzzo Sampaio Vessoni; Simone Kashima; Svetoslav Nanev Slavov; Vincent Louis Viala |
| EPI_ISL_2493914 | USF YEN WANN SHENG | Instituto Butantan | Antonio Jorge Martins; Claudia Renata dos Santos Barros; David Schlesinger; Debora Botequio Moretti; Dimas Tadeu Covas; Elaine Cristina Marqueze; Elaine Vieira Santos; Evandra Strazza Rodrigues; Heidge Fukumasu; Jayme Augusto de Souza-Neto; José Salvatore Leister Patané; Luiz Alcantara; Luiz Lehmann Coutinho; Maria Carolina Elias; Maurício Lacerda Nogueira; Rafael dos Santos Bezerra; Raul Machado Neto; Rejane Maria Tommasini Grotto; Ricardo Haddad; Sandra Coccuzzo Sampaio Vessoni; Simone Kashima; Svetoslav Nanev Slavov; Vincent Louis Viala |
| EPI_ISL_2535111, EPI_ISL_2535112, EPI_ISL_2535113, EPI_ISL_2535114, EPI_ISL_2535115, EPI_ISL_2535116, EPI_ISL_2535117, EPI_ISL_2535118, EPI_ISL_2535119, EPI_ISL_2535120, EPI_ISL_2535121, EPI_ISL_2535122, EPI_ISL_2535123, EPI_ISL_2535124, EPI_ISL_2535125, EPI_ISL_2535126, EPI_ISL_2535127, EPI_ISL_2535128, EPI_ISL_2535129, EPI_ISL_2535130, EPI_ISL_2535131, EPI_ISL_2535132, EPI_ISL_2535133, EPI_ISL_2535134, EPI_ISL_2535135, EPI_ISL_2535136, EPI_ISL_2535137, EPI_ISL_2535138, EPI_ISL_2535139, EPI_ISL_2535140, EPI_ISL_2535141, EPI_ISL_2535142, EPI_ISL_2535143, EPI_ISL_2535144, EPI_ISL_2535145, EPI_ISL_2535146, EPI_ISL_2535147, EPI_ISL_2535148, EPI_ISL_2535149, EPI_ISL_2535150, EPI_ISL_2535151, EPI_ISL_2535152, EPI_ISL_2535153, EPI_ISL_2535154, EPI_ISL_2535155, EPI_ISL_2535156, EPI_ISL_2535157, EPI_ISL_2535158, EPI_ISL_2535159, EPI_ISL_2535160, EPI_ISL_2535161, EPI_ISL_2535162, EPI_ISL_2535163, EPI_ISL_2535164, EPI_ISL_2535165, EPI_ISL_2535166, EPI_ISL_2535167, EPI_ISL_2535168, EPI_ISL_2535169, EPI_ISL_2535170, EPI_ISL_2535171, EPI_ISL_2535172, EPI_ISL_2535173, EPI_ISL_2535174, EPI_ISL_2535175, EPI_ISL_2535176, EPI_ISL_2535177, EPI_ISL_2535178, EPI_ISL_2535179, EPI_ISL_2535180, EPI_ISL_2535181, EPI_ISL_2535182, EPI_ISL_2535183, EPI_ISL_2535184, EPI_ISL_2535185, EPI_ISL_2535186, EPI_ISL_2535187, EPI_ISL_2535188, EPI_ISL_2535189, EPI_ISL_2535190, EPI_ISL_2535191, EPI_ISL_2535192, EPI_ISL_2535193, EPI_ISL_2535194, EPI_ISL_2535195, EPI_ISL_2535196, EPI_ISL_2535197, EPI_ISL_2535198, EPI_ISL_2535199, EPI_ISL_2535200, EPI_ISL_2535201, EPI_ISL_2535202, EPI_ISL_2535203, EPI_ISL_2535204, EPI_ISL_2535205, EPI_ISL_2535206, EPI_ISL_2535207, EPI_ISL_2535208, EPI_ISL_2535209, EPI_ISL_2535210, EPI_ISL_2535211, EPI_ISL_2535212, EPI_ISL_2535213, EPI_ISL_2535214, EPI_ISL_2535215, EPI_ISL_2535216, EPI_ISL_2535217, EPI_ISL_2535218, EPI_ISL_2535219, EPI_ISL_2535220, EPI_ISL_2535221, EPI_ISL_2535222, EPI_ISL_2535223, EPI_ISL_2535224, EPI_ISL_2535225, EPI_ISL_2535226, EPI_ISL_2535227, EPI_ISL_2535228, EPI_ISL_2535229, EPI_ISL_2535230, EPI_ISL_2535231, EPI_ISL_2535232, EPI_ISL_2535233, EPI_ISL_2535234, EPI_ISL_2535235, EPI_ISL_2691569, EPI_ISL_2691570, EPI_ISL_2691571, EPI_ISL_2691572, EPI_ISL_2691573, EPI_ISL_2691574, EPI_ISL_2691575, EPI_ISL_2691576, EPI_ISL_2691577, EPI_ISL_2691578, EPI_ISL_2691579, EPI_ISL_2691580, EPI_ISL_2691581, EPI_ISL_2691582, EPI_ISL_2691583, EPI_ISL_2691584, EPI_ISL_2691585, EPI_ISL_2691586, EPI_ISL_2691587, EPI_ISL_2691588, EPI_ISL_2691589, EPI_ISL_2691590, EPI_ISL_2691591, EPI_ISL_2691592, EPI_ISL_2691593, EPI_ISL_2691594, EPI_ISL_2691595, EPI_ISL_2691596, EPI_ISL_2691597, EPI_ISL_2691598, EPI_ISL_2691599, EPI_ISL_2691600, EPI_ISL_2691601, EPI_ISL_2691602, EPI_ISL_2691603, EPI_ISL_2691604, EPI_ISL_2691605, EPI_ISL_2691606, EPI_ISL_2691607, EPI_ISL_2691608, EPI_ISL_2691609, EPI_ISL_2691610, EPI_ISL_2691611, EPI_ISL_2691612, EPI_ISL_2691613, EPI_ISL_2691614, EPI_ISL_2691615, EPI_ISL_2691616, EPI_ISL_2691617, EPI_ISL_2691618, EPI_ISL_2691619, EPI_ISL_2691620, EPI_ISL_2691621, EPI_ISL_2691622, EPI_ISL_2691623, EPI_ISL_2691624, EPI_ISL_2691625, EPI_ISL_2691626, EPI_ISL_2691627, EPI_ISL_2691628, EPI_ISL_2691629, EPI_ISL_2691630, EPI_ISL_2691631, EPI_ISL_2691632, EPI_ISL_2691633, EPI_ISL_2691634, EPI_ISL_2691635, EPI_ISL_2691636, EPI_ISL_2691637, EPI_ISL_2691638, EPI_ISL_2691639, EPI_ISL_2691640, EPI_ISL_2691642, EPI_ISL_2691643, EPI_ISL_2691644, EPI_ISL_2691645, EPI_ISL_2691646, EPI_ISL_2691647, EPI_ISL_2691648, EPI_ISL_2691649, EPI_ISL_2691650, EPI_ISL_2691651, EPI_ISL_2691652, EPI_ISL_2691653, EPI_ISL_2691654, EPI_ISL_2691655, EPI_ISL_2691656, EPI_ISL_2691657, EPI_ISL_2691658, EPI_ISL_2691659, EPI_ISL_2691660, EPI_ISL_2691661, EPI_ISL_2691662, EPI_ISL_2691663, EPI_ISL_2691664, EPI_ISL_2691665, EPI_ISL_2691666, EPI_ISL_2691667, EPI_ISL_2691668, EPI_ISL_2691669, EPI_ISL_2691670, EPI_ISL_2691671, EPI_ISL_2691672, EPI_ISL_2691673, EPI_ISL_2691674, EPI_ISL_2691675, EPI_ISL_2691676, EPI_ISL_2691677, EPI_ISL_2691678, EPI_ISL_2691679, EPI_ISL_2691680, EPI_ISL_2691681, EPI_ISL_2691682, EPI_ISL_2691683, EPI_ISL_2691684, EPI_ISL_2837208, EPI_ISL_2837209, EPI_ISL_2837210, EPI_ISL_2837211, EPI_ISL_2837212, EPI_ISL_2837213, EPI_ISL_2837214, EPI_ISL_2837215, EPI_ISL_2837216, EPI_ISL_2837217, EPI_ISL_2837218, EPI_ISL_2837219, EPI_ISL_2837220, EPI_ISL_2837221, EPI_ISL_2837222, EPI_ISL_2837223, EPI_ISL_2837224, EPI_ISL_2837225, EPI_ISL_2837226, EPI_ISL_2837227, EPI_ISL_2837228, EPI_ISL_2837229, EPI_ISL_2837230, EPI_ISL_2837231, EPI_ISL_2837232, EPI_ISL_2837233, EPI_ISL_2837234, EPI_ISL_2837235, EPI_ISL_2837236, EPI_ISL_2837237, EPI_ISL_2837238, EPI_ISL_2837239, EPI_ISL_2837240, EPI_ISL_2837241, EPI_ISL_2837242, EPI_ISL_2837243, EPI_ISL_2837244, EPI_ISL_2837245, EPI_ISL_2837246, EPI_ISL_2837248, EPI_ISL_2837249, EPI_ISL_2837250, EPI_ISL_2837251, EPI_ISL_2837252, EPI_ISL_2837253, EPI_ISL_2837254, EPI_ISL_2837255, EPI_ISL_2837256, EPI_ISL_2837257, EPI_ISL_2837258, EPI_ISL_2837259, EPI_ISL_2837260, EPI_ISL_2837261, EPI_ISL_2837262, EPI_ISL_2837263, EPI_ISL_2837264, EPI_ISL_2837265, EPI_ISL_2837266, EPI_ISL_2837267, EPI_ISL_2837268, EPI_ISL_2837269, EPI_ISL_2837270, EPI_ISL_2837271, EPI_ISL_2837272, EPI_ISL_2837273, EPI_ISL_2837274, EPI_ISL_2837275, EPI_ISL_2837276, EPI_ISL_2837277, EPI_ISL_2837278, EPI_ISL_2837279, EPI_ISL_2837280, EPI_ISL_2837281, EPI_ISL_2837282, EPI_ISL_2837283, EPI_ISL_2837284, EPI_ISL_2837285, EPI_ISL_2837286, EPI_ISL_2837287, EPI_ISL_2837288, EPI_ISL_2837289, EPI_ISL_2837290, EPI_ISL_2837291, EPI_ISL_2837292, EPI_ISL_2837293, EPI_ISL_2837294, EPI_ISL_2837295, EPI_ISL_2837296, EPI_ISL_2837297, EPI_ISL_2837298, EPI_ISL_2837299, EPI_ISL_2837300, EPI_ISL_2837301, EPI_ISL_2837302, EPI_ISL_2837303, EPI_ISL_2837304, EPI_ISL_2837305, EPI_ISL_2837306, EPI_ISL_2837307, EPI_ISL_2837308, EPI_ISL_2837309, EPI_ISL_2837310, EPI_ISL_2837311, EPI_ISL_2837312, EPI_ISL_2837313, EPI_ISL_2837314 | | | |
| see above | Unidade de apoio ao diagnostico da COVID - UNADIG | Bioinformatics Laboratory / LNCC | Alessandra P Lamarca; Alexandra L Gerber; Amilcar Tanuri; Ana Paula de C Guimaraes; Ana Tereza R Vasconcelos; Andrea Cony Cavalcanti; Caio Luiz Pereira Ribeiro; Cassia Alves; Cintia Policarpo; Claudia Maria Braga de Mello; Cristiane Gomes da Silva; Diana Mariani; Douglas Terra Machado; Flavio Dias da Silva; Gleidson da Silva de Oliveira; Leandro Magalhaes de Souza; Liliane Cavalcante; Luiz G P de Almeida; Marcio Henrique de Oliveira Garcia; Mario Sergio Ribeiro; Ronaldo da Silva F Jr; Silvia Carvalho |
| EPI_ISL_2385697, EPI_ISL_2385698, EPI_ISL_2385699, EPI_ISL_2385712, EPI_ISL_2385714, EPI_ISL_2385715, EPI_ISL_2385716, EPI_ISL_2385722, EPI_ISL_2385723 | | | |
| see above | Unidade de apoio ao diagnóstico da COVID – UNADIG | Bioinformatics Laboratory / LNCC | Alessandra P Lamarca; Alexandra L Gerber; Amilcar Tanuri; Ana Paula de C Guimaraes; Ana Tereza R Vasconcelos; Andrea Cony Cavalcanti; Caio Luiz Pereira Ribeiro; Cassia Alves; Cintia Policarpo; Claudia Maria Braga de Mello; Cristiane Gomes da Silva; Diana Mariani; Douglas Terra Machado; Flavio Dias da Silva; Gleidson da Silva de Oliveira; Leandro Magalhaes de Souza; Liliane Cavalcante; Luiz G P de Almeida; Marcio Henrique de Oliveira Garcia; Mario Sergio Ribeiro; Ronaldo da Silva F Jr; Silvia Carvalho |
| EPI_ISL_2462333 | VIGILANCIA EM SAUDE | Instituto Butantan | Antonio Jorge Martins; Claudia Renata dos Santos Barros; David Schlesinger; Debora Botequio Moretti; Dimas Tadeu Covas; Elaine Cristina Marqueze; Elaine Vieira Santos; Evandra Strazza Rodrigues; Heidge Fukumasu; Jayme Augusto de Souza-Neto; José Salvatore Leister Patané; Luiz Alcantara; Luiz Lehmann Coutinho; Maria Carolina Elias; Maurício Lacerda Nogueira; Rafael dos Santos Bezerra; Raul Machado Neto; Rejane Maria Tommasini Grotto; Ricardo Haddad; Sandra Coccuzzo Sampaio Vessoni; Simone Kashima; Svetoslav Nanev Slavov; Vincent Louis Viala |
| EPI_ISL_2462316, EPI_ISL_2493841, EPI_ISL_2493843 | VIGILANCIA EM SAUDE DE FCO MORATO | Instituto Butantan | Antonio Jorge Martins; Claudia Renata dos Santos Barros; David Schlesinger; Debora Botequio Moretti; Dimas Tadeu Covas; Elaine Cristina Marqueze; Elaine Vieira Santos; Evandra Strazza Rodrigues; Heidge Fukumasu; Jayme Augusto de Souza-Neto; José Salvatore Leister Patané; Luiz Alcantara; Luiz Lehmann Coutinho; Maria Carolina Elias; Maurício Lacerda Nogueira; Rafael dos Santos Bezerra; Raul Machado Neto; Rejane Maria Tommasini Grotto; Ricardo Haddad; Sandra Coccuzzo Sampaio Vessoni; Simone Kashima; Svetoslav Nanev Slavov; Vincent Louis Viala |
| EPI_ISL_2479921, EPI_ISL_2493159, EPI_ISL_2493166, EPI_ISL_2493204, EPI_ISL_2494111, EPI_ISL_2494112, EPI_ISL_2494113 | | | |
| see above | VIGILANCIA EM SAUDE SAE VIGILANCIA EPIDEMIOLOGICA | Instituto Butantan | Antonio Jorge Martins; Claudia Renata dos Santos Barros; David Schlesinger; Debora Botequio Moretti; Dimas Tadeu Covas; Elaine Cristina Marqueze; Elaine Vieira Santos; Evandra Strazza Rodrigues; Heidge Fukumasu; Jayme Augusto de Souza-Neto; José Salvatore Leister Patané; Luiz Alcantara; Luiz Lehmann Coutinho; Maria Carolina Elias; Maurício Lacerda Nogueira; Rafael dos Santos Bezerra; Raul Machado Neto; Rejane Maria Tommasini Grotto; Ricardo Haddad; Sandra Coccuzzo Sampaio Vessoni; Simone Kashima; Svetoslav Nanev Slavov; Vincent Louis Viala |
| EPI_ISL_2209961, EPI_ISL_2210110, EPI_ISL_2345651, EPI_ISL_2345841, EPI_ISL_2362250 | VIGILANCIA EM SAUDE SAE VIGILANCIA EPIDEMIOLOGICA | Instituto Butantan / Mendelics | Antonio Jorge Martins; Claudia Renata dos Santos Barros; David Schlesinger; Debora Botequio Moretti; Dimas Tadeu Covas; Elaine Cristina Marqueze; Elaine Vieira Santos; Evandra Strazza Rodrigues; Heidge Fukumasu; Jayme Augusto de Souza-Neto; José Salvatore Leister Patané; Luiz Alcantara; Luiz Lehmann Coutinho; Maria Carolina Elias; Maurício Lacerda Nogueira; Rafael dos Santos Bezerra; Raul Machado Neto; Rejane Maria Tommasini Grotto; Ricardo Haddad; Sandra Coccuzzo Sampaio Vessoni; Simone Kashima; Svetoslav Nanev Slavov; Vincent Louis Viala |
| EPI_ISL_2493916 | VIGILANCIA EPIDEMIOLOGICA DE FERRAZ DE VASCONCELOS | Instituto Butantan | Antonio Jorge Martins; Claudia Renata dos Santos Barros; David Schlesinger; Debora Botequio Moretti; Dimas Tadeu Covas; Elaine Cristina Marqueze; Elaine Vieira Santos; Evandra Strazza Rodrigues; Heidge Fukumasu; Jayme Augusto de Souza-Neto; José Salvatore Leister Patané; Luiz Alcantara; Luiz Lehmann Coutinho; Maria Carolina Elias; Maurício Lacerda Nogueira; Rafael dos Santos Bezerra; Raul Machado Neto; Rejane Maria Tommasini Grotto; Ricardo Haddad; Sandra Coccuzzo Sampaio Vessoni; Simone Kashima; Svetoslav Nanev Slavov; Vincent Louis Viala |
| EPI_ISL_2445321, EPI_ISL_2445322, EPI_ISL_2445327, EPI_ISL_2493152, EPI_ISL_2493171, EPI_ISL_2493205, EPI_ISL_2493237, EPI_ISL_2493238, EPI_ISL_2493670, EPI_ISL_2493672 | | | |
| see above | VIGILANCIA EPIDEMIOLOGICA DE IBATE | Instituto Butantan | Antonio Jorge Martins; Claudia Renata dos Santos Barros; David Schlesinger; Debora Botequio Moretti; Dimas Tadeu Covas; Elaine Cristina Marqueze; Elaine Vieira Santos; Evandra Strazza Rodrigues; Heidge Fukumasu; Jayme Augusto de Souza-Neto; José Salvatore Leister Patané; Luiz Alcantara; Luiz Lehmann Coutinho; Maria Carolina Elias; Maurício Lacerda Nogueira; Rafael dos Santos Bezerra; Raul Machado Neto; Rejane Maria Tommasini Grotto; Ricardo Haddad; Sandra Coccuzzo Sampaio Vessoni; Simone Kashima; Svetoslav Nanev Slavov; Vincent Louis Viala |
| EPI_ISL_2210288 | VIGILANCIA EPIDEMIOLOGICA DE IBATE | Instituto Butantan / Mendelics | Antonio Jorge Martins; Claudia Renata dos Santos Barros; David Schlesinger; Debora Botequio Moretti; Dimas Tadeu Covas; Elaine Cristina Marqueze; Elaine Vieira Santos; Evandra Strazza Rodrigues; Heidge Fukumasu; Jayme Augusto de Souza-Neto; José Salvatore Leister Patané; Luiz Alcantara; Luiz Lehmann Coutinho; Maria Carolina Elias; Maurício Lacerda Nogueira; Rafael dos Santos Bezerra; Raul Machado Neto; Rejane Maria Tommasini Grotto; Ricardo Haddad; Sandra Coccuzzo Sampaio Vessoni; Simone Kashima; Svetoslav Nanev Slavov; Vincent Louis Viala |
| EPI_ISL_2493366, EPI_ISL_2493395, EPI_ISL_2493399, EPI_ISL_2493450, EPI_ISL_2494162, EPI_ISL_2494173, EPI_ISL_2494335, EPI_ISL_2494336 | | | |
| see above | VIGILANCIA EPIDEMIOLOGICA DE LEME | Instituto Butantan | Antonio Jorge Martins; Claudia Renata dos Santos Barros; David Schlesinger; Debora Botequio Moretti; Dimas Tadeu Covas; Elaine Cristina Marqueze; Elaine Vieira Santos; Evandra Strazza Rodrigues; Heidge Fukumasu; Jayme Augusto de Souza-Neto; José Salvatore Leister Patané; Luiz Alcantara; Luiz Lehmann Coutinho; Maria Carolina Elias; Maurício Lacerda Nogueira; Rafael dos Santos Bezerra; Raul Machado Neto; Rejane Maria Tommasini Grotto; Ricardo Haddad; Sandra Coccuzzo Sampaio Vessoni; Simone Kashima; Svetoslav Nanev Slavov; Vincent Louis Viala |
| EPI_ISL_2383034, EPI_ISL_2493208, EPI_ISL_2493251, EPI_ISL_2493284, EPI_ISL_2493365, EPI_ISL_2493391, EPI_ISL_2493432, EPI_ISL_2493434, EPI_ISL_2493448, EPI_ISL_2494171, EPI_ISL_2494191, EPI_ISL_2494197, EPI_ISL_2494198, EPI_ISL_2494199, EPI_ISL_2494283, EPI_ISL_2500982 | | | |
| see above | VIGILANCIA EPIDEMIOLOGICA E CONTROLE DE VETORES PIRASSUNUN | Instituto Butantan | Antonio Jorge Martins; Claudia Renata dos Santos Barros; David Schlesinger; Debora Botequio Moretti; Dimas Tadeu Covas; Elaine Cristina Marqueze; Elaine Vieira Santos; Evandra Strazza Rodrigues; Heidge Fukumasu; Jayme Augusto de Souza-Neto; José Salvatore Leister Patané; Luiz Alcantara; Luiz Lehmann Coutinho; Maria Carolina Elias; Maurício Lacerda Nogueira; Rafael dos Santos Bezerra; Raul Machado Neto; Rejane Maria Tommasini Grotto; Ricardo Haddad; Sandra Coccuzzo Sampaio Vessoni; Simone Kashima; Svetoslav Nanev Slavov; Vincent Louis Viala |
| EPI_ISL_2493599, EPI_ISL_2493610 | VIGILANCIA EPIDEMIOLOGICA JARDINOPOLIS SP | Instituto Butantan | Antonio Jorge Martins; Claudia Renata dos Santos Barros; David Schlesinger; Debora Botequio Moretti; Dimas Tadeu Covas; Elaine Cristina Marqueze; Elaine Vieira Santos; Evandra Strazza Rodrigues; Heidge Fukumasu; Jayme Augusto de Souza-Neto; José Salvatore Leister Patané; Luiz Alcantara; Luiz Lehmann Coutinho; Maria Carolina Elias; Maurício Lacerda Nogueira; Rafael dos Santos Bezerra; Raul Machado Neto; Rejane Maria Tommasini Grotto; Ricardo Haddad; Sandra Coccuzzo Sampaio Vessoni; Simone Kashima; Svetoslav Nanev Slavov; Vincent Louis Viala |
| EPI_ISL_2445607 | VIGILANCIA EPIDIMIOLOGICA DE ARACARIGUAMA | Instituto Butantan | Antonio Jorge Martins; Claudia Renata dos Santos Barros; David Schlesinger; Debora Botequio Moretti; Dimas Tadeu Covas; Elaine Cristina Marqueze; Elaine Vieira Santos; Evandra Strazza Rodrigues; Heidge Fukumasu; Jayme Augusto de Souza-Neto; José Salvatore Leister Patané; Luiz Alcantara; Luiz Lehmann Coutinho; Maria Carolina Elias; Maurício Lacerda Nogueira; Rafael dos Santos Bezerra; Raul Machado Neto; Rejane Maria Tommasini Grotto; Ricardo Haddad; Sandra Coccuzzo Sampaio Vessoni; Simone Kashima; Svetoslav Nanev Slavov; Vincent Louis Viala |
| EPI_ISL_2445280, EPI_ISL_2445281, EPI_ISL_2445282, EPI_ISL_2445283, EPI_ISL_2445284, EPI_ISL_2445285, EPI_ISL_2445286, EPI_ISL_2445287, EPI_ISL_2445289, EPI_ISL_2445290, EPI_ISL_2445291, EPI_ISL_2445292, EPI_ISL_2445293, EPI_ISL_2445294, EPI_ISL_2445295, EPI_ISL_2445296, EPI_ISL_2445303, EPI_ISL_2445306, EPI_ISL_2445307, EPI_ISL_2445308, EPI_ISL_2445309, EPI_ISL_2473725, EPI_ISL_2473822, EPI_ISL_2493944, EPI_ISL_2493945, EPI_ISL_2493946, EPI_ISL_2493949, EPI_ISL_2493952, EPI_ISL_2493953, EPI_ISL_2493958 | | | |
| see above | VIGILANCIA SANITARIA E VIG EPIDEMIOLOGICA DE ITAPEVI | Instituto Butantan | Antonio Jorge Martins; Claudia Renata dos Santos Barros; David Schlesinger; Debora Botequio Moretti; Dimas Tadeu Covas; Elaine Cristina Marqueze; Elaine Vieira Santos; Evandra Strazza Rodrigues; Heidge Fukumasu; Jayme Augusto de Souza-Neto; José Salvatore Leister Patané; Luiz Alcantara; Luiz Lehmann Coutinho; Maria Carolina Elias; Maurício Lacerda Nogueira; Rafael dos Santos Bezerra; Raul Machado Neto; Rejane Maria Tommasini Grotto; Ricardo Haddad; Sandra Coccuzzo Sampaio Vessoni; Simone Kashima; Svetoslav Nanev Slavov; Vincent Louis Viala |
| EPI_ISL_2444192 | VOTUPORANGA | Instituto Butantan / FAMERP | Antonio Jorge Martins; Claudia Renata dos Santos Barros; David Schlesinger; Debora Botequio Moretti; Dimas Tadeu Covas; Elaine Cristina Marqueze; Elaine Vieira Santos; Evandra Strazza Rodrigues; Heidge Fukumasu; Jayme Augusto de Souza-Neto; José Salvatore Leister Patané; Luiz Alcantara; Luiz Lehmann Coutinho; Maria Carolina Elias; Maurício Lacerda Nogueira; Rafael dos Santos Bezerra; Raul Machado Neto; Rejane Maria Tommasini Grotto; Ricardo Haddad; Sandra Coccuzzo Sampaio Vessoni; Simone Kashima; Svetoslav Nanev Slavov; Vincent Louis Viala |
| EPI_ISL_2488813 | Evandro Chagas Institute | Evandro Chagas Institute | A.M.; Barbagelata; E.C.; E.M.A.; Ferreira; J.A.; Junior; K.C.; L.C.; L.S.; M.C.; P.S.; Pinheiro; Santos; Silva; Sousa; Sousa Junior; W.D.C.; da Silva |

We gratefully acknowledge the following Authors from the Originating laboratories responsible for obtaining the specimens, as well as the Submitting laboratories where the genome data were generated and shared via GISAID, on which this research is based.

All Submitters of data may be contacted directly via www.gisaid.org

Authors are sorted alphabetically.

| Accession ID | Originating Laboratory | Submitting Laboratory | Authors |
|---|---|---|---|
| EPI_ISL_1446773, EPI_ISL_1446774, EPI_ISL_1446775, EPI_ISL_1446776, EPI_ISL_1446777, EPI_ISL_1446778, EPI_ISL_1446779, EPI_ISL_1446780, EPI_ISL_1446782, EPI_ISL_1446783, EPI_ISL_1446786, EPI_ISL_1446788, EPI_ISL_1446789, EPI_ISL_1446793, EPI_ISL_1446795, EPI_ISL_1446796, EPI_ISL_1446797, EPI_ISL_1446799, EPI_ISL_1446800, EPI_ISL_1446802, EPI_ISL_1446840, EPI_ISL_1446841, EPI_ISL_1446842, EPI_ISL_1446843, EPI_ISL_1446844, EPI_ISL_1446845, EPI_ISL_1678143, EPI_ISL_1678144, EPI_ISL_1678145, EPI_ISL_1678146, EPI_ISL_1678148, EPI_ISL_1678149, EPI_ISL_1678150, EPI_ISL_1678151, EPI_ISL_1678152, EPI_ISL_1678154, EPI_ISL_1678156, EPI_ISL_1678157, EPI_ISL_1678158, EPI_ISL_1678159, EPI_ISL_1678160, EPI_ISL_1678161, EPI_ISL_1678162, EPI_ISL_1678313, EPI_ISL_1678314, EPI_ISL_1678315 | | | |
| see above | "CDPH, Viral and Rickettsial Disease Laboratory" | Centers for Disease Control and Prevention Division of Viral Diseases, Pathogen Discovery | Alison Laufer Halpin; Ben L. Rambo-Martin; Clinton R. Paden; Dakota Howard; Darlene Wagner; Dave Wentworth; Dhwani Batra; Jasmine Padilla; Justin Lee; Katie Dillon; Krista Queen; Kristen Knipe; Kristine Lacek; Mark Burroughs; Matthew Schmerer; Mili Sheth; Peter Cook; Sam Shepard; Sarah Nobles; Shoshona Le; Suxiang Tong; Vivien Dugan; Yvette Unoarumhi |
| EPI_ISL_1445279, EPI_ISL_1445280, EPI_ISL_1445289, EPI_ISL_1445292, EPI_ISL_1445293, EPI_ISL_1445294, EPI_ISL_1445384, EPI_ISL_1445425, EPI_ISL_1445426, EPI_ISL_1445427, EPI_ISL_1445429, EPI_ISL_1445431, EPI_ISL_1445434, EPI_ISL_1445454, EPI_ISL_1445455, EPI_ISL_1445461, EPI_ISL_1445462, EPI_ISL_1445468, EPI_ISL_1445472, EPI_ISL_1445491, EPI_ISL_1445506, EPI_ISL_1445508, EPI_ISL_1445509, EPI_ISL_1445511, EPI_ISL_1445537, EPI_ISL_1445541, EPI_ISL_1445545, EPI_ISL_1445546, EPI_ISL_1445547, EPI_ISL_1445550, EPI_ISL_1445551, EPI_ISL_1445554, EPI_ISL_1445555, EPI_ISL_1445556, EPI_ISL_1445557, EPI_ISL_1445558, EPI_ISL_1445559, EPI_ISL_1445560, EPI_ISL_1445561, EPI_ISL_1445562, EPI_ISL_1445565, EPI_ISL_1445566, EPI_ISL_1445568, EPI_ISL_1445569, EPI_ISL_1445573, EPI_ISL_1445584, EPI_ISL_1445593, EPI_ISL_1445595, EPI_ISL_1445598, EPI_ISL_1445605, EPI_ISL_1445608, EPI_ISL_1445609, EPI_ISL_1445610, EPI_ISL_1445613, EPI_ISL_1445615, EPI_ISL_1445616, EPI_ISL_1445620, EPI_ISL_1445622, EPI_ISL_1445627, EPI_ISL_1445652, EPI_ISL_1445653, EPI_ISL_1445655, EPI_ISL_1445656, EPI_ISL_1445657, EPI_ISL_1445658, EPI_ISL_1445659, EPI_ISL_1445671, EPI_ISL_1445673, EPI_ISL_1445675, EPI_ISL_1445685, EPI_ISL_1445690, EPI_ISL_1445691, EPI_ISL_1445694, EPI_ISL_1445695, EPI_ISL_1445703, EPI_ISL_1445709, EPI_ISL_1445720, EPI_ISL_1445725, EPI_ISL_1445731, EPI_ISL_1445735, EPI_ISL_1445754, EPI_ISL_1445764, EPI_ISL_1445769, EPI_ISL_1445774, EPI_ISL_1445775, EPI_ISL_1445776, EPI_ISL_1445789, EPI_ISL_1445790, EPI_ISL_1445791, EPI_ISL_1445796, EPI_ISL_1445797, EPI_ISL_1445798, EPI_ISL_1445799, EPI_ISL_1445800, EPI_ISL_1445801, EPI_ISL_1445802, EPI_ISL_1445803, EPI_ISL_1445805, EPI_ISL_1445806, EPI_ISL_1445809, EPI_ISL_1445815, EPI_ISL_1445816, EPI_ISL_1445817, EPI_ISL_1445818, EPI_ISL_1445819, EPI_ISL_1445820, EPI_ISL_1445821, EPI_ISL_1445835, EPI_ISL_1445843, EPI_ISL_1445845, EPI_ISL_1445858, EPI_ISL_1445871, EPI_ISL_1445904, EPI_ISL_1445913, EPI_ISL_1445931, EPI_ISL_1445932, EPI_ISL_1445940, EPI_ISL_1445941, EPI_ISL_1445942, EPI_ISL_1445950, EPI_ISL_1445959, EPI_ISL_1446032, EPI_ISL_1446035, EPI_ISL_1446083, EPI_ISL_1446084, EPI_ISL_1446090, EPI_ISL_1446091, EPI_ISL_1446092, EPI_ISL_1446096, EPI_ISL_1446097, EPI_ISL_1446110, EPI_ISL_1446116, EPI_ISL_1446118, EPI_ISL_1446122, EPI_ISL_1446123, EPI_ISL_1446132, EPI_ISL_1446147, EPI_ISL_1446162, EPI_ISL_1446179, EPI_ISL_1446181, EPI_ISL_1446192, EPI_ISL_1479498, EPI_ISL_1479512, EPI_ISL_1479518, EPI_ISL_1479587, EPI_ISL_1479590, EPI_ISL_1479591, EPI_ISL_1479608, EPI_ISL_1479613, EPI_ISL_1479656, EPI_ISL_1479805, EPI_ISL_1479825, EPI_ISL_1479828, EPI_ISL_1479830, EPI_ISL_1479834, EPI_ISL_1479842, EPI_ISL_1491734, EPI_ISL_1491744, EPI_ISL_1491786, EPI_ISL_1491787, EPI_ISL_1491788, EPI_ISL_1491814, EPI_ISL_1491817, EPI_ISL_1491829, EPI_ISL_1491842, EPI_ISL_1491878, EPI_ISL_1491884, EPI_ISL_1491885, EPI_ISL_1491888, EPI_ISL_1491898, EPI_ISL_1491899, EPI_ISL_1491917, EPI_ISL_1491918, EPI_ISL_1491929, EPI_ISL_1491939, EPI_ISL_1491940, EPI_ISL_1493502, EPI_ISL_1493503, EPI_ISL_1493535, EPI_ISL_1493550, EPI_ISL_1493571, EPI_ISL_1513139, EPI_ISL_1513152, EPI_ISL_1513153, EPI_ISL_1513160, EPI_ISL_1513172, EPI_ISL_1513183, EPI_ISL_1513185, EPI_ISL_1513186, EPI_ISL_1513190, EPI_ISL_1513199, EPI_ISL_1513200, EPI_ISL_1513220, EPI_ISL_1513222, EPI_ISL_1513236, EPI_ISL_1513239, EPI_ISL_1513240, EPI_ISL_1513242, EPI_ISL_1513262, EPI_ISL_1513263, EPI_ISL_1513294, EPI_ISL_1513296, EPI_ISL_1513297, EPI_ISL_1513301, EPI_ISL_1513303, EPI_ISL_1513306, EPI_ISL_1513307, EPI_ISL_1513309, EPI_ISL_1513323, EPI_ISL_1513324, EPI_ISL_1513325, EPI_ISL_1513352, EPI_ISL_1513359, EPI_ISL_1513360, EPI_ISL_1513361, EPI_ISL_1513362, EPI_ISL_1513386, EPI_ISL_1513387, EPI_ISL_1513410, EPI_ISL_1513487, EPI_ISL_1513488, EPI_ISL_1513489, EPI_ISL_1513513, EPI_ISL_1513518, EPI_ISL_1513524, EPI_ISL_1513541, EPI_ISL_1513542, EPI_ISL_1513548, EPI_ISL_1513570, EPI_ISL_1513583, EPI_ISL_1513622, EPI_ISL_1513625, EPI_ISL_1513627, EPI_ISL_1525033, EPI_ISL_1525034, EPI_ISL_1525035, EPI_ISL_1525040, EPI_ISL_1525041, EPI_ISL_1525042, EPI_ISL_1525048, EPI_ISL_1525049, EPI_ISL_1525066, EPI_ISL_1525075, EPI_ISL_1525123, EPI_ISL_1525146, EPI_ISL_1525147, EPI_ISL_1525152, EPI_ISL_1525153, EPI_ISL_1525155, EPI_ISL_1525161, EPI_ISL_1525162, EPI_ISL_1525163, EPI_ISL_1525176, EPI_ISL_1525186, EPI_ISL_1525187, EPI_ISL_1525220, EPI_ISL_1550455, EPI_ISL_1550465, EPI_ISL_1550466, EPI_ISL_1550492, EPI_ISL_1550516, EPI_ISL_1550519, EPI_ISL_1550521, EPI_ISL_1550522, EPI_ISL_1550550, EPI_ISL_1550565, EPI_ISL_1550669, EPI_ISL_1550670, EPI_ISL_1550690, EPI_ISL_1550739, EPI_ISL_1550742, EPI_ISL_1550743, EPI_ISL_1550794, EPI_ISL_1550845, EPI_ISL_1550972, EPI_ISL_1550975, EPI_ISL_1550976, EPI_ISL_1550982, EPI_ISL_1550984, EPI_ISL_1551127, EPI_ISL_1551128, EPI_ISL_1551220, EPI_ISL_1551266, EPI_ISL_1551393, EPI_ISL_1551422, EPI_ISL_1551543, EPI_ISL_1551556, EPI_ISL_1551585, EPI_ISL_1551636, EPI_ISL_1559656, EPI_ISL_1559657, EPI_ISL_1559658, EPI_ISL_1559659, EPI_ISL_1559660, EPI_ISL_1559661, EPI_ISL_1559662, EPI_ISL_1559663, EPI_ISL_1559664, EPI_ISL_1559665, EPI_ISL_1559667, EPI_ISL_1559668, EPI_ISL_1559669, EPI_ISL_1559670, EPI_ISL_1559672, EPI_ISL_1559740, EPI_ISL_1559741, EPI_ISL_1559758, EPI_ISL_1559759, EPI_ISL_1559760, EPI_ISL_1559761, EPI_ISL_1559806, EPI_ISL_1559807, EPI_ISL_1559808, EPI_ISL_1559809, EPI_ISL_1559828, EPI_ISL_1559829, EPI_ISL_1559847, EPI_ISL_1559877, EPI_ISL_1559880, EPI_ISL_1559881, EPI_ISL_1559883, EPI_ISL_1559887, EPI_ISL_1559903, EPI_ISL_1560009, EPI_ISL_1560010, EPI_ISL_1560011, EPI_ISL_1560018, EPI_ISL_1560037, EPI_ISL_1560106, EPI_ISL_1560123, EPI_ISL_1560137, EPI_ISL_1560142, EPI_ISL_1560150, EPI_ISL_1560177, EPI_ISL_1560179, EPI_ISL_1560255, EPI_ISL_1560350, EPI_ISL_1560382, EPI_ISL_1560383, EPI_ISL_1560491, EPI_ISL_1560496, EPI_ISL_1560519, EPI_ISL_1560527, EPI_ISL_1560603, EPI_ISL_1560606, EPI_ISL_1560644, EPI_ISL_1560645, EPI_ISL_1560672, EPI_ISL_1560679, EPI_ISL_1560681, EPI_ISL_1560702, EPI_ISL_1560707, EPI_ISL_1560717, EPI_ISL_1560720, EPI_ISL_1560726, EPI_ISL_1560740, EPI_ISL_1560787, EPI_ISL_1560788, EPI_ISL_1560942, EPI_ISL_1560949, EPI_ISL_1561002, EPI_ISL_1561020, EPI_ISL_1561022, EPI_ISL_1561030, EPI_ISL_1561043, EPI_ISL_1561202, EPI_ISL_1561222, EPI_ISL_1561223, EPI_ISL_1561224, EPI_ISL_1561225, EPI_ISL_1561226, EPI_ISL_1561237, EPI_ISL_1561248, EPI_ISL_1561249, EPI_ISL_1561260, EPI_ISL_1561289, EPI_ISL_1561298, EPI_ISL_1561328, EPI_ISL_1561330, EPI_ISL_1561335, EPI_ISL_1561342, EPI_ISL_1561345, EPI_ISL_1561366, EPI_ISL_1561374, EPI_ISL_1561400, EPI_ISL_1561404, EPI_ISL_1561449, EPI_ISL_1561496, EPI_ISL_1561525, EPI_ISL_1561541, EPI_ISL_1561551, EPI_ISL_1561553, EPI_ISL_1561563, EPI_ISL_1561580, EPI_ISL_1561588, EPI_ISL_1561595, EPI_ISL_1561596, EPI_ISL_1561614, EPI_ISL_1561615, EPI_ISL_1561622, EPI_ISL_1561637, EPI_ISL_1561638, EPI_ISL_1561647, EPI_ISL_1561682, EPI_ISL_1561693, EPI_ISL_1561717, EPI_ISL_1561719, EPI_ISL_1561788, EPI_ISL_1561794, EPI_ISL_1561802, EPI_ISL_1561803, EPI_ISL_1561804, EPI_ISL_1561805, EPI_ISL_1561806, EPI_ISL_1561809, EPI_ISL_1561810, EPI_ISL_1561811, EPI_ISL_1561817, EPI_ISL_1561818, EPI_ISL_1561819, EPI_ISL_1561828, EPI_ISL_1561866, EPI_ISL_1561870, EPI_ISL_1561891, EPI_ISL_1561893, EPI_ISL_1561902, EPI_ISL_1561904, EPI_ISL_1561907, EPI_ISL_1561918, EPI_ISL_1561926, EPI_ISL_1561927, EPI_ISL_1561975, EPI_ISL_1561976, EPI_ISL_1561983, EPI_ISL_1561987, EPI_ISL_1561998, EPI_ISL_1562015, EPI_ISL_1562017, EPI_ISL_1562018, EPI_ISL_1562025, EPI_ISL_1562043, EPI_ISL_1562046, EPI_ISL_1562054, EPI_ISL_1562073, EPI_ISL_1562096, EPI_ISL_1562102, EPI_ISL_1562103, EPI_ISL_1562104, EPI_ISL_1562145, EPI_ISL_1562146, EPI_ISL_1562152, EPI_ISL_1562156, EPI_ISL_1562181, EPI_ISL_1562182, EPI_ISL_1562183, EPI_ISL_1562184, EPI_ISL_1562198, EPI_ISL_1562219, EPI_ISL_1562225, EPI_ISL_1562254, EPI_ISL_1562255, EPI_ISL_1562267, EPI_ISL_1562268, EPI_ISL_1562343, EPI_ISL_1562352, EPI_ISL_1562353, EPI_ISL_1562364, EPI_ISL_1562377, EPI_ISL_1562378, EPI_ISL_1562380, EPI_ISL_1562381, EPI_ISL_1562384, EPI_ISL_1562387, EPI_ISL_1562391, EPI_ISL_1562392, EPI_ISL_1562400, EPI_ISL_1562411, EPI_ISL_1562412, EPI_ISL_1562416, EPI_ISL_1562418, EPI_ISL_1562426, EPI_ISL_1562450, EPI_ISL_1562451, EPI_ISL_1562454, EPI_ISL_1562456, EPI_ISL_1562503, EPI_ISL_1562506, EPI_ISL_1562554, EPI_ISL_1562567, EPI_ISL_1562601, EPI_ISL_1562631, EPI_ISL_1562791, EPI_ISL_1562812, EPI_ISL_1562820, EPI_ISL_1562823, EPI_ISL_1562827, EPI_ISL_1562831, EPI_ISL_1562836, EPI_ISL_1562848, EPI_ISL_1562945, EPI_ISL_1562950, EPI_ISL_1562987, EPI_ISL_1562990, EPI_ISL_1563006, EPI_ISL_1563044, EPI_ISL_1563048, EPI_ISL_1563055, EPI_ISL_1563071, EPI_ISL_1563072, EPI_ISL_1563128, EPI_ISL_1563132, EPI_ISL_1563154, EPI_ISL_1563224, EPI_ISL_1563225, EPI_ISL_1563226, EPI_ISL_1563229, EPI_ISL_1563239, EPI_ISL_1563281, EPI_ISL_1563329, EPI_ISL_1563350, EPI_ISL_1563353, EPI_ISL_1563398, EPI_ISL_1563409, EPI_ISL_1563410, EPI_ISL_1563411, EPI_ISL_1563417, EPI_ISL_1563418, EPI_ISL_1563454, EPI_ISL_1563549, EPI_ISL_1563550, EPI_ISL_1563558, EPI_ISL_1563573, EPI_ISL_1563607, EPI_ISL_1563608, EPI_ISL_1648968, EPI_ISL_1649028, EPI_ISL_1649029, EPI_ISL_1649030, EPI_ISL_1649031, EPI_ISL_1649048, EPI_ISL_1649126, EPI_ISL_1649150, EPI_ISL_1649159, EPI_ISL_1649210, EPI_ISL_1649211, EPI_ISL_1649263, EPI_ISL_1649264, EPI_ISL_1649336, EPI_ISL_1649381, EPI_ISL_1649382, EPI_ISL_1649414, EPI_ISL_1649447, EPI_ISL_1649448, EPI_ISL_1649471, EPI_ISL_1649525, EPI_ISL_1649546, EPI_ISL_1649547, EPI_ISL_1649548, EPI_ISL_1649550, EPI_ISL_1649556, EPI_ISL_1649566, EPI_ISL_1649598, EPI_ISL_1649599, EPI_ISL_1649622, EPI_ISL_1649652, EPI_ISL_1649694, EPI_ISL_1649732, EPI_ISL_1649734, EPI_ISL_1649737, EPI_ISL_1649738, EPI_ISL_1649757, EPI_ISL_1649764, EPI_ISL_1649766, EPI_ISL_1649767, EPI_ISL_1649769, EPI_ISL_1649777, EPI_ISL_1649787, EPI_ISL_1649796, EPI_ISL_1649804, EPI_ISL_1649858, EPI_ISL_1649894, EPI_ISL_1649897, EPI_ISL_1649907, EPI_ISL_1649944, EPI_ISL_1650002, EPI_ISL_1650041, EPI_ISL_1650088, EPI_ISL_1650205, EPI_ISL_1650271, EPI_ISL_1650278, EPI_ISL_1650292, EPI_ISL_1650409, EPI_ISL_1650412, EPI_ISL_1650472, EPI_ISL_1650534, EPI_ISL_1650538, EPI_ISL_1650632, EPI_ISL_1650638, EPI_ISL_1650659, EPI_ISL_1650672, EPI_ISL_1650693, EPI_ISL_1650695, EPI_ISL_1650703, EPI_ISL_1650729, EPI_ISL_1650730, EPI_ISL_1650944, EPI_ISL_1650954, EPI_ISL_1650966, EPI_ISL_1650967, EPI_ISL_1651036, EPI_ISL_1651045, EPI_ISL_1651061, EPI_ISL_1651075, EPI_ISL_1651094, EPI_ISL_1651099, EPI_ISL_1651133, EPI_ISL_1651152, EPI_ISL_1651169, EPI_ISL_1651180, EPI_ISL_1651182, EPI_ISL_1685472, EPI_ISL_1685852, EPI_ISL_1686708, EPI_ISL_1687815, EPI_ISL_1695152, EPI_ISL_1695225, EPI_ISL_1695275, EPI_ISL_1695302, EPI_ISL_1695422, EPI_ISL_1695448, EPI_ISL_1925479, EPI_ISL_2185633, EPI_ISL_2185669, EPI_ISL_2185680, EPI_ISL_2185681, EPI_ISL_2185682, EPI_ISL_2185683, EPI_ISL_2185704, EPI_ISL_2185708, EPI_ISL_2185714, EPI_ISL_2185716, EPI_ISL_2185736, EPI_ISL_2185752, EPI_ISL_2185754, EPI_ISL_2185755, EPI_ISL_2185756, EPI_ISL_2185757, EPI_ISL_2185760, EPI_ISL_2185762, EPI_ISL_2185764, EPI_ISL_2185780, EPI_ISL_2185782, EPI_ISL_2185783, EPI_ISL_2185784, EPI_ISL_2185785, EPI_ISL_2185786, EPI_ISL_2185804, EPI_ISL_2185847, EPI_ISL_2185848, EPI_ISL_2185853, EPI_ISL_2185873, EPI_ISL_2185875, EPI_ISL_2185877, EPI_ISL_2185878, EPI_ISL_2185879, EPI_ISL_2185891, EPI_ISL_2185901, EPI_ISL_2185905, EPI_ISL_2185910, EPI_ISL_2185924, EPI_ISL_2185942, EPI_ISL_2185949, EPI_ISL_2185960, EPI_ISL_2185963, EPI_ISL_2185965, EPI_ISL_2185976, EPI_ISL_2185983, EPI_ISL_2185998, EPI_ISL_2186002, EPI_ISL_2186008, EPI_ISL_2186035, EPI_ISL_2186043, EPI_ISL_2186162, EPI_ISL_2186166 | | | |
| see above | Aegis Sciences Corporation | Centers for Disease Control and Prevention Division of Viral Diseases, Pathogen Discovery | Adrian Paskey; Alec Vest; Benjamin Rambo-Martin; Christopher Gulvick; Clinton R. Paden; Cyndi Clark; Dakota Howard; Darlene Wagner; Dhwani Batra; Dillon Nall; Duncan MacCannell; Ethan Sanders; Holly Houdeshell; Jason Caravas; Kara Moser; Matthew Hardison; Matthew Schmerer; Ola Kvalvaag; Patrick Campbell; Peter W. Cook; Rob Case; Scott Sammons; Shatavia Morrison; Shaun Westlund; Vikramsinha Ghorpade; Yvette Unoarumhi |
| EPI_ISL_1575114, EPI_ISL_1575115, EPI_ISL_1575116, EPI_ISL_1575117, EPI_ISL_1575118, EPI_ISL_1575119, EPI_ISL_1575121, EPI_ISL_1575122, EPI_ISL_1575123, EPI_ISL_1575124, EPI_ISL_1575125, EPI_ISL_1575126, EPI_ISL_1576991, EPI_ISL_1577000, EPI_ISL_1577001, EPI_ISL_1577002 | | | |
| see above | Alameda County Public Health Department | Alameda County Public Health Department | Kevin Libuit; Kristna Hsieh; Maria Elma |
| EPI_ISL_1853453, EPI_ISL_1853470, EPI_ISL_1853565 | Avellino Labs | Avellino Labs | Milos Pjanic |
| EPI_ISL_1701042, EPI_ISL_1701043, EPI_ISL_1701044, EPI_ISL_1701045, EPI_ISL_1701047, EPI_ISL_1701048, EPI_ISL_1701049, EPI_ISL_1701051, EPI_ISL_1701053, EPI_ISL_1701054, EPI_ISL_1701055, EPI_ISL_1701056, EPI_ISL_1701058, EPI_ISL_1701059, EPI_ISL_1701060, EPI_ISL_1701062, EPI_ISL_1701063, EPI_ISL_1701064, EPI_ISL_1701067, EPI_ISL_1701068, EPI_ISL_1701070, EPI_ISL_1701075, EPI_ISL_1701076, EPI_ISL_1701077, EPI_ISL_1701085, EPI_ISL_1701089, EPI_ISL_1701094, EPI_ISL_1701095, EPI_ISL_1701097, EPI_ISL_1701098, EPI_ISL_1701100, EPI_ISL_1701101, EPI_ISL_1701102, EPI_ISL_1701105, EPI_ISL_1701107, EPI_ISL_1701108, EPI_ISL_1701109, EPI_ISL_1701110, EPI_ISL_1701111, EPI_ISL_1701112, EPI_ISL_1701113, EPI_ISL_1701114, EPI_ISL_1701116, EPI_ISL_1701118, EPI_ISL_1701121, EPI_ISL_1701122, EPI_ISL_1701125, EPI_ISL_1701126, EPI_ISL_1701128, EPI_ISL_1701129, EPI_ISL_1701130, EPI_ISL_1701131, EPI_ISL_1701133, EPI_ISL_1701134, EPI_ISL_1701138, EPI_ISL_1701139, EPI_ISL_1701140, EPI_ISL_1701141, EPI_ISL_1701143, EPI_ISL_1701144, EPI_ISL_1701145, EPI_ISL_1701146, EPI_ISL_1701149, EPI_ISL_1701151, EPI_ISL_1701152, EPI_ISL_1701153, EPI_ISL_1701154, EPI_ISL_1701158, EPI_ISL_1701159, EPI_ISL_1701160, EPI_ISL_1701163, EPI_ISL_1701164, EPI_ISL_1701165, EPI_ISL_1701166, EPI_ISL_1701169, EPI_ISL_1701170, EPI_ISL_1701173, EPI_ISL_1701181, EPI_ISL_1701182, EPI_ISL_1701183, EPI_ISL_1701184, EPI_ISL_1701185, EPI_ISL_1701186, EPI_ISL_1701187, EPI_ISL_1701190, EPI_ISL_1701191, EPI_ISL_1701192, EPI_ISL_1701193, EPI_ISL_1701195, EPI_ISL_1701196, EPI_ISL_1701197, EPI_ISL_1701198, EPI_ISL_1701199, EPI_ISL_1701200, EPI_ISL_1701202, EPI_ISL_1701203, EPI_ISL_1701206, EPI_ISL_1701207, EPI_ISL_1701209, EPI_ISL_1701215, EPI_ISL_1701216, EPI_ISL_1701220, EPI_ISL_1701221, EPI_ISL_1701223, EPI_ISL_1701225, EPI_ISL_1701226, EPI_ISL_1701227, EPI_ISL_1701229, EPI_ISL_1701231, EPI_ISL_1701232, EPI_ISL_1701237, EPI_ISL_1701238, EPI_ISL_1701239, EPI_ISL_1701240, EPI_ISL_1701241, EPI_ISL_1701242, EPI_ISL_1701243, EPI_ISL_1701244, EPI_ISL_1701246, EPI_ISL_1701248, EPI_ISL_1701252, EPI_ISL_1701253, EPI_ISL_1701254, EPI_ISL_1701255, EPI_ISL_1701256, EPI_ISL_1701257, EPI_ISL_1701258, EPI_ISL_1701259, EPI_ISL_1701260, EPI_ISL_1701261, EPI_ISL_1701265, EPI_ISL_1701269, EPI_ISL_1701270, EPI_ISL_1701271, EPI_ISL_1701272, EPI_ISL_1701275, EPI_ISL_1701276, EPI_ISL_1701278, EPI_ISL_1701279, EPI_ISL_1701282, EPI_ISL_1701283, EPI_ISL_1701284, EPI_ISL_1701286, EPI_ISL_1701287, EPI_ISL_1701288, EPI_ISL_1701292, EPI_ISL_1701294, EPI_ISL_1701296, EPI_ISL_1701298, EPI_ISL_1701300, EPI_ISL_1701304, EPI_ISL_1701305, EPI_ISL_1701308, EPI_ISL_1701309, EPI_ISL_1701310, EPI_ISL_1821315, EPI_ISL_1821340, EPI_ISL_1821345, EPI_ISL_1821356, EPI_ISL_1821357, EPI_ISL_1821360, EPI_ISL_1821361, EPI_ISL_1821364, EPI_ISL_1821369, EPI_ISL_1821379, EPI_ISL_1821382, EPI_ISL_1821403, EPI_ISL_1821410, EPI_ISL_1821412, EPI_ISL_1821424, EPI_ISL_1821440, EPI_ISL_1821442, EPI_ISL_1821466, EPI_ISL_2102525, EPI_ISL_2102527, EPI_ISL_2102539, EPI_ISL_2102540, EPI_ISL_2102542, EPI_ISL_2102546, EPI_ISL_2102551, EPI_ISL_2102552, EPI_ISL_2102562, EPI_ISL_2102564, EPI_ISL_2102566, EPI_ISL_2102568, EPI_ISL_2102570, EPI_ISL_2102572, EPI_ISL_2102573, EPI_ISL_2102575, EPI_ISL_2102580, EPI_ISL_2102584, EPI_ISL_2102586, EPI_ISL_2102587, EPI_ISL_2102592, EPI_ISL_2102595, EPI_ISL_2102597, EPI_ISL_2102741, EPI_ISL_2102743, EPI_ISL_2102750, EPI_ISL_2102754, EPI_ISL_2102756, EPI_ISL_2102765, EPI_ISL_2102771, EPI_ISL_2102773, EPI_ISL_2102775, EPI_ISL_2102777, EPI_ISL_2102778, EPI_ISL_2102780, EPI_ISL_2102783, EPI_ISL_2102786, EPI_ISL_2102787, EPI_ISL_2102789, EPI_ISL_2102793, EPI_ISL_2102794, EPI_ISL_2102799, EPI_ISL_2102807, EPI_ISL_2102816, EPI_ISL_2102818, EPI_ISL_2102819, EPI_ISL_2102829, EPI_ISL_2102834, EPI_ISL_2102835, EPI_ISL_2102839, EPI_ISL_2102846, EPI_ISL_2102854, EPI_ISL_2102857, EPI_ISL_2102860, EPI_ISL_2102864, EPI_ISL_2102867, EPI_ISL_2102873, EPI_ISL_2102874, EPI_ISL_2102877, EPI_ISL_2102879, EPI_ISL_2102881, EPI_ISL_2102884, EPI_ISL_2102897, EPI_ISL_2102900, EPI_ISL_2102910, EPI_ISL_2102915, EPI_ISL_2102921, EPI_ISL_2102924, EPI_ISL_2102928, EPI_ISL_2102929, EPI_ISL_2102934, EPI_ISL_2102940, EPI_ISL_2102942, EPI_ISL_2102944, EPI_ISL_2102957, EPI_ISL_2102962, EPI_ISL_2102965, EPI_ISL_2102968, EPI_ISL_2102974, EPI_ISL_2102977, EPI_ISL_2102980, EPI_ISL_2102983, EPI_ISL_2102984, EPI_ISL_2102987, EPI_ISL_2102989, EPI_ISL_2102991, EPI_ISL_2102993, EPI_ISL_2103000, EPI_ISL_2103004, EPI_ISL_2103015, EPI_ISL_2103027, EPI_ISL_2103047, EPI_ISL_2103050, EPI_ISL_2103053, EPI_ISL_2103066, EPI_ISL_2103075, EPI_ISL_2103095, EPI_ISL_2103098, EPI_ISL_2103103, EPI_ISL_2103116, EPI_ISL_2103118, EPI_ISL_2103120, EPI_ISL_2103124, EPI_ISL_2103135, EPI_ISL_2103139, EPI_ISL_2103142, EPI_ISL_2103155, EPI_ISL_2103160, EPI_ISL_2103163, EPI_ISL_2103166, EPI_ISL_2103180, EPI_ISL_2103181, EPI_ISL_2103193 | | | |
| see above | CA DPH Viral and Rickettsial Disease Laboratory | Chan-Zuckerberg Biohub | CZB Cliahub Consortium |
| EPI_ISL_1318378, EPI_ISL_1318379, EPI_ISL_1446714, EPI_ISL_1446715, EPI_ISL_1516361, EPI_ISL_1678085, EPI_ISL_1678086, EPI_ISL_1678088, EPI_ISL_1678089, EPI_ISL_1678090, EPI_ISL_1678091, EPI_ISL_1678092, EPI_ISL_1678276, EPI_ISL_1678277, EPI_ISL_1678278, EPI_ISL_1678279, EPI_ISL_1678280, EPI_ISL_1678281, EPI_ISL_1678283, EPI_ISL_1678284, EPI_ISL_1678285, EPI_ISL_1678286, EPI_ISL_1678287, EPI_ISL_1798830, EPI_ISL_1798832, EPI_ISL_1798836, EPI_ISL_1798839, EPI_ISL_1798843, EPI_ISL_1798844, EPI_ISL_1798846, EPI_ISL_1798848, EPI_ISL_1798850, EPI_ISL_1798858, EPI_ISL_1798862, EPI_ISL_1798864, EPI_ISL_1798866, EPI_ISL_1823714, EPI_ISL_1823721, EPI_ISL_1823723, EPI_ISL_1823731, EPI_ISL_1823735, EPI_ISL_1823740, EPI_ISL_1823742, EPI_ISL_1823746 | | | |
| see above | CA-Los Angeles County Public Health Laboratory | Centers for Disease Control and Prevention Division of Viral Diseases, Pathogen Discovery | Alison Laufer Halpin; Anna Montmayeur; Anna Uehara; Ben L. Rambo-Martin; Clinton R. Paden; Dakota Howard; Darlene Wagner; Dave Wentworth; Dhwani Batra; Haibin Wang; Jasmine Padilla; Jing Zhang; Justin Lee; Katie Dillon; Krista Queen; Kristen Knipe; Kristine Lacek; Lori Rowe; Mark Burroughs; Matthew Schmerer; Mili Sheth; Peter Cook; Peter W. Cook; Rachel Marine; Sam Shepard; Sarah Nobles; Shoshona Le; Suxiang Tong; Vivien Dugan; Yan Li; Ying Tao; Yvette Unoarumhi |
| EPI_ISL_1798789, EPI_ISL_1798797, EPI_ISL_1798799, EPI_ISL_1798801 | CDPH, Viral and Rickettsial Disease Laboratory | Centers for Disease Control and Prevention Division of Viral Diseases, Pathogen Discovery | Alison Laufer Halpin; Ben L. Rambo-Martin; Clinton R. Paden; Dakota Howard; Darlene Wagner; Dave Wentworth; Dhwani Batra; Jasmine Padilla; Justin Lee; Katie Dillon; Krista Queen; Kristen Knipe; Kristine Lacek; Mark Burroughs; Matthew Schmerer; Mili Sheth; Peter Cook; Sam Shepard; Sarah Nobles; Shoshona Le; Suxiang Tong; Vivien Dugan; Yvette Unoarumhi |
| EPI_ISL_1373737, EPI_ISL_1373786 | CDPH, Viral and Rickettsial Disease Laboratory | Genomics and Discovery, Respiratory Viruses Branch, Division of Viral Diseases, Centers for Disease Control and Prevention | Anna Kelleher; Anna Uehara; Brian Lynch; Clinton R. Paden; Haibin Wang; Han Jia Justin Ng; Jing Zhang; Krista Queen; Peter Cook; Suxiang Tong; Yan Li; Ying Tao |
| EPI_ISL_1591240, EPI_ISL_1591241, EPI_ISL_1591242, EPI_ISL_1623836, EPI_ISL_1911349, EPI_ISL_1911350, EPI_ISL_1911351, EPI_ISL_1911352, EPI_ISL_1911353, EPI_ISL_1911354, EPI_ISL_1911355, EPI_ISL_1911356, EPI_ISL_1911357, EPI_ISL_1911358, EPI_ISL_1911359, EPI_ISL_1911360, EPI_ISL_1911361, EPI_ISL_1911362, EPI_ISL_1911363, EPI_ISL_1911364, EPI_ISL_1911365, EPI_ISL_1911366, EPI_ISL_1911367, EPI_ISL_1911368 | | | |
| see above | CDPH, Viral and Rickettsial Disease Laboratory | Pathogen Discovery, Respiratory Viruses Branch, Division of Viral Diseases, Centers for Disease Control and Prevention | Adam Retchless; Anna Kelleher; Anna Montmayeur; Anna Uehara; Brian Lynch; Clinton R. Paden; Dhwani Batra; Haibin Wang; Han Jia Justin Ng; Jasmine Padilla; Jing Zhang; Justin Lee; Krista Queen; Mark Burroughs; Mili Sheth; Morgan Davis; Peter Cook; Rachel Marine; Sarah Nobles; Suxiang Tong; Tara Coalter; Yan Li; Ying Tao |
| EPI_ISL_1701494, EPI_ISL_1701495 | California Institute of Technology | Chan-Zuckerberg Biohub | CZB Cliahub Consortium |
| EPI_ISL_1661979 | Centro de Investigacion Biomedica de Occidente (CIBO) | Unidad de Genomica Avanzada | Alejandro Sanchez-Flores; Alfredo Herrera-Estrella; Alicia Ocana-Mondragon; Angel Gustavo Salas-Lais; Bernardo Martinez-Miguel; Blanca Taboada; Brenda Irasema Maldonado-Meza; Carla Ivon Herrera-Najera; Carlos F. Arias; Celia Boukadida; Celida Duque Molina; Clara Esperanza Santacruz-Tinoco; Concepcion Grajales-Muniz; Consorcio Mexicano de Vigilancia Genomica (CoViGen-Mex). Authors (in alphabetical order): Julio Elias Alvarado-Yaah; Fernando Fontove-Herrera; Francisco Pulido; Gloria Elena Espinosa-Ayala; Gloria Maria Molina-Salinas; Gloria Vazquez; Hector Esteban Paz-Juarez; Hector Montoya-Fuentes; Helen Haydee Fernanda Ramirez-Plascencia; Jose Antonio Enciso-Moreno; Jose Esteban Munoz-Medina; Jose de Jesus Nunez-Contreras; Juan Bautista Chale-Dzul; Luis Alberto Ochoa-Carrera; Margarita Matias-Florentino; Maria Guadalupe Santiago-Mauricio; Maria Guadalupe de Jesus Mireles-Rivera; Nelly Selem-Mojica; Pavel Isa; Ricardo Grande; Santiago Ávila-Rios; Victor Eduardo Garcia-Arias; Victor Hugo Borja-Aburto |
| EPI_ISL_1824350, EPI_ISL_1824352, EPI_ISL_1824359, EPI_ISL_1824360, EPI_ISL_1824361, EPI_ISL_1824364, EPI_ISL_1824367, EPI_ISL_1824368, EPI_ISL_1824369, EPI_ISL_1824370, EPI_ISL_1824371, EPI_ISL_1824372, EPI_ISL_1824373, EPI_ISL_1824374, EPI_ISL_1824375, EPI_ISL_1824376, EPI_ISL_1824377, EPI_ISL_1824378, EPI_ISL_1824379, EPI_ISL_1824380, EPI_ISL_1824381, EPI_ISL_1824382, | | | |

| | | | |
|---|---|---|---|
| EPI_ISL_1824383, EPI_ISL_1824384, EPI_ISL_1824385, EPI_ISL_1824386, EPI_ISL_1824387, EPI_ISL_1824388, EPI_ISL_1824389, EPI_ISL_1824390, EPI_ISL_1824391, EPI_ISL_1824392, EPI_ISL_1824393, EPI_ISL_1824394, EPI_ISL_1824395, EPI_ISL_1824396, EPI_ISL_1824397, EPI_ISL_1824398, EPI_ISL_1824399, EPI_ISL_1824400, EPI_ISL_1824401, EPI_ISL_1824402, EPI_ISL_1824403, EPI_ISL_1824404, EPI_ISL_1824405, EPI_ISL_1824406, EPI_ISL_1824407, EPI_ISL_1824408, EPI_ISL_1824413, EPI_ISL_1824416, EPI_ISL_1824417 | | | |
| see above | Chiu Laboratory, University of California, San Francisco | Chiu Laboratory, University of California, San Francisco | Alicia Sotomayor-Gonzalez; Candace Wang; Charles Chiu; Daniel Wong; Venice Servellita; Yueyuan Zhang |
| EPI_ISL_1825028, EPI_ISL_1825029, EPI_ISL_1825030, EPI_ISL_1825031, EPI_ISL_1825032, EPI_ISL_1825033, EPI_ISL_1825034, EPI_ISL_1825035, EPI_ISL_1825036, EPI_ISL_1825037, EPI_ISL_1825038, EPI_ISL_1825039, EPI_ISL_1825040, EPI_ISL_1825041, EPI_ISL_1825042, EPI_ISL_1825043, EPI_ISL_1825044, EPI_ISL_1825045, EPI_ISL_1825046, EPI_ISL_1825047, EPI_ISL_1825048, EPI_ISL_1825049, EPI_ISL_1825050, EPI_ISL_1825051, EPI_ISL_1825052, EPI_ISL_1825053, EPI_ISL_1825054, EPI_ISL_1825055, EPI_ISL_1825056, EPI_ISL_1825057, EPI_ISL_1825058, EPI_ISL_1825059, EPI_ISL_1825060, EPI_ISL_1825061, EPI_ISL_1825062, EPI_ISL_1825063, EPI_ISL_1825064, EPI_ISL_1825065, EPI_ISL_1825066, EPI_ISL_1825067, EPI_ISL_1825068, EPI_ISL_1825069, EPI_ISL_1825070, EPI_ISL_1825071, EPI_ISL_1825072, EPI_ISL_1825073, EPI_ISL_1825074, EPI_ISL_1825075, EPI_ISL_1825076, EPI_ISL_1825078 | | | |
| see above | Color Genomics | Chiu Laboratory, University of California, San Francisco | Alicia Sotomayor-Gonzalez; Alicia Zhou; Candace Wang; Charles Chiu; Daniel Wong; Erika Torres; Madyson Wanner; Scott Topper; Venice Servellita; Yueyuan Zhang |
| EPI_ISL_1529122, EPI_ISL_1529123, EPI_ISL_1529124, EPI_ISL_1529131, EPI_ISL_1529133, EPI_ISL_1529134, EPI_ISL_1529135, EPI_ISL_1529155, EPI_ISL_1529176, EPI_ISL_1529179, EPI_ISL_1529185, EPI_ISL_1529190, EPI_ISL_1529202, EPI_ISL_1529209, EPI_ISL_1529235, EPI_ISL_1529247, EPI_ISL_1529251, EPI_ISL_1529300, EPI_ISL_1529308, EPI_ISL_1529318, EPI_ISL_1529322, EPI_ISL_1529323, EPI_ISL_1529327, EPI_ISL_1529345, EPI_ISL_1529358, EPI_ISL_1529364, EPI_ISL_1529367, EPI_ISL_1529368, EPI_ISL_1664020, EPI_ISL_1664070, EPI_ISL_1664086, EPI_ISL_1664091, EPI_ISL_1664093, EPI_ISL_2102751, EPI_ISL_2102824, EPI_ISL_2102843, EPI_ISL_2102866, EPI_ISL_2102945, EPI_ISL_2102946, EPI_ISL_2102982 | | | |
| see above | Contra Costa County Public Health Lab | Chan-Zuckerberg Biohub | CZB Cliahub Consortium |
| EPI_ISL_1805719, EPI_ISL_1805720, EPI_ISL_1805721, EPI_ISL_1805722, EPI_ISL_1805723, EPI_ISL_1805727, EPI_ISL_1805728, EPI_ISL_1805729, EPI_ISL_1910451, EPI_ISL_1910452, EPI_ISL_1910453, EPI_ISL_1910454, EPI_ISL_1910455, EPI_ISL_1910456, EPI_ISL_1910457, EPI_ISL_1910458, EPI_ISL_1910459, EPI_ISL_1910460, EPI_ISL_1910461, EPI_ISL_1910463 | | | |
| see above | County of San Diego Health and Human Services Agency | Pathogen Discovery, Respiratory Viruses Branch, Division of Viral Diseases, Centers for Disease Control and Prevention | Adam Retchless; Anna Kelleher; Anna Montmayeur; Anna Uehara; Brian Lynch; Clinton R. Paden; Dhwani Batra; Haibin Wang; Han Jia Justin Ng; Jasmine Padilla; Jing Zhang; Justin Lee; Krista Queen; Mark Burroughs; Mili Sheth; Morgan Davis; Peter Cook; Rachel Marine; Sarah Nobles; Suxiang Tong; Tara Coalter; Yan Li; Ying Tao |
| EPI_ISL_1477550, EPI_ISL_1477992, EPI_ISL_1478174, EPI_ISL_1529127, EPI_ISL_1529128, EPI_ISL_1529129, EPI_ISL_1529161, EPI_ISL_1529208, EPI_ISL_1529213, EPI_ISL_1529329, EPI_ISL_1529341, EPI_ISL_1529342, EPI_ISL_1701136, EPI_ISL_1701266, EPI_ISL_1701297, EPI_ISL_1821359, EPI_ISL_1821363, EPI_ISL_1821367, EPI_ISL_1821378, EPI_ISL_1821404, EPI_ISL_1821408, EPI_ISL_1821447, EPI_ISL_1821468, EPI_ISL_2102998, EPI_ISL_2103073, EPI_ISL_2103172 | | | |
| see above | County of San Luis Obispo Public Health Laboratory | Chan-Zuckerberg Biohub | CZB Cliahub Consortium |
| EPI_ISL_1503991 | Curative Labs | Los Angeles County PHL | P. Hemarajata et al. |
| EPI_ISL_1318697, EPI_ISL_1318711, EPI_ISL_1318727, EPI_ISL_1318748, EPI_ISL_1318775, EPI_ISL_1318777, EPI_ISL_1318802, EPI_ISL_1318803, EPI_ISL_1318820, EPI_ISL_1318823, EPI_ISL_1318832, EPI_ISL_1318845, EPI_ISL_1318871, EPI_ISL_1318886, EPI_ISL_1318896, EPI_ISL_1318898, EPI_ISL_1318900, EPI_ISL_1318906, EPI_ISL_1318908, EPI_ISL_1318917, EPI_ISL_1318937, EPI_ISL_1318945, EPI_ISL_1318952, EPI_ISL_1349078, EPI_ISL_1349093, EPI_ISL_2158080, EPI_ISL_2158081, EPI_ISL_2158082, EPI_ISL_2158083, EPI_ISL_2158084, EPI_ISL_2158085, EPI_ISL_2158086, EPI_ISL_2158087, EPI_ISL_2158088, EPI_ISL_2158089, EPI_ISL_2158090, EPI_ISL_2158091, EPI_ISL_2158092, EPI_ISL_2158093, EPI_ISL_2158094, EPI_ISL_2158095, EPI_ISL_2158096, EPI_ISL_2158097, EPI_ISL_2158098, EPI_ISL_2158099, EPI_ISL_2158100, EPI_ISL_2158101, EPI_ISL_2158102, EPI_ISL_2158103, EPI_ISL_2158104, EPI_ISL_2158106, EPI_ISL_2158107 | | | |
| see above | DeRisi Lab, University of California, San Francisco | DeRisi Lab, University of California, San Francisco | Amy Kistler; Anthea Mitchell; Carina Marquez; Diane Havlir; Diane Jones; Douglas Black; Eric Chow; Gabriel Chamie; Genay Pilarowski; IDseq Team; Jackie Martinez; James Peng; Jamin Liu; Jon Jacobo; Lucy Li; Luis Rubio; Manu Vanaerschot; Matthew Laurie; Maya Petersen; Patrick Ayscue; Sabrina Mann; Sara Sunshine; Susana Rojas; Susy Rojas; Valerie Tulier-Laiwa; and Joseph DeRisi |
| EPI_ISL_1811653, EPI_ISL_1811662, EPI_ISL_1811669, EPI_ISL_1811693, EPI_ISL_1811702, EPI_ISL_1811706, EPI_ISL_1811732, EPI_ISL_1811763, EPI_ISL_1811807, EPI_ISL_1811817, EPI_ISL_1811828, EPI_ISL_1811831, EPI_ISL_1811836, EPI_ISL_1811847, EPI_ISL_1811853, EPI_ISL_1811914, EPI_ISL_1811924, EPI_ISL_1811980, EPI_ISL_1811994, EPI_ISL_1812171, EPI_ISL_1812365, EPI_ISL_1812406, EPI_ISL_1812497, EPI_ISL_1812540, EPI_ISL_1812564, EPI_ISL_1812571, EPI_ISL_1812587, EPI_ISL_1812592, EPI_ISL_1812675, EPI_ISL_1812695, EPI_ISL_1812756, EPI_ISL_1812760, EPI_ISL_1812763, EPI_ISL_1812792, EPI_ISL_1812841, EPI_ISL_1812852, EPI_ISL_1812858, EPI_ISL_1812874, EPI_ISL_1812882, EPI_ISL_1812897, EPI_ISL_1812970, EPI_ISL_1812978, EPI_ISL_1812990, EPI_ISL_1813008, EPI_ISL_1813009, EPI_ISL_1813038, EPI_ISL_1813051, EPI_ISL_1813075, EPI_ISL_1813081, EPI_ISL_1813104, EPI_ISL_1813119, EPI_ISL_1813126, EPI_ISL_1813144, EPI_ISL_1813149, EPI_ISL_1813166, EPI_ISL_1813198, EPI_ISL_1813217, EPI_ISL_1813237, EPI_ISL_1813238, EPI_ISL_1813257, EPI_ISL_1813261, EPI_ISL_1813296, EPI_ISL_1813364, EPI_ISL_1813367, EPI_ISL_1813371, EPI_ISL_1813385, EPI_ISL_1813387, EPI_ISL_1813391, EPI_ISL_1813428, EPI_ISL_1813440, EPI_ISL_1813476, EPI_ISL_1813480, EPI_ISL_1813485, EPI_ISL_1813498, EPI_ISL_1813528, EPI_ISL_1813542, EPI_ISL_1813561, EPI_ISL_1813579, EPI_ISL_1813581, EPI_ISL_1813582, EPI_ISL_1813594, EPI_ISL_1813601, EPI_ISL_1813606, EPI_ISL_1813628, EPI_ISL_1813634, EPI_ISL_1813648, EPI_ISL_1813666, EPI_ISL_1813673, EPI_ISL_1813678, EPI_ISL_1813701, EPI_ISL_1813705, EPI_ISL_1813707, EPI_ISL_1813714, EPI_ISL_1813726, EPI_ISL_1813734, EPI_ISL_1813756, EPI_ISL_1813760, EPI_ISL_1813761, EPI_ISL_1813787, EPI_ISL_1813792, EPI_ISL_1813795, EPI_ISL_1813821, EPI_ISL_1813827, EPI_ISL_1813837, EPI_ISL_1813840, EPI_ISL_1813849, EPI_ISL_1813855, EPI_ISL_1813863, EPI_ISL_1813865, EPI_ISL_1813877, EPI_ISL_1813878, EPI_ISL_1813884, EPI_ISL_1813891, EPI_ISL_1813893, EPI_ISL_1813899, EPI_ISL_1813901, EPI_ISL_1813906, EPI_ISL_1813943, EPI_ISL_1813957, EPI_ISL_1813974, EPI_ISL_1813976, EPI_ISL_1813986, EPI_ISL_1813987, EPI_ISL_1813994, EPI_ISL_1814009, EPI_ISL_1814014, EPI_ISL_1814043, EPI_ISL_1814046, EPI_ISL_1814060, EPI_ISL_1814065, EPI_ISL_1814066, EPI_ISL_1814082, EPI_ISL_1814089, EPI_ISL_1814098, EPI_ISL_1814101, EPI_ISL_1814117, EPI_ISL_1814122, EPI_ISL_1814130, EPI_ISL_1814131, EPI_ISL_1814135, EPI_ISL_1814140, EPI_ISL_1814145, EPI_ISL_1814147, EPI_ISL_1814153, EPI_ISL_1814170, EPI_ISL_1814197, EPI_ISL_1814212, EPI_ISL_1814214, EPI_ISL_1814226, EPI_ISL_1814227, EPI_ISL_1814234, EPI_ISL_1814252, EPI_ISL_1814259, EPI_ISL_1814260, EPI_ISL_1814261, EPI_ISL_1814269, EPI_ISL_1814276, EPI_ISL_1814279, EPI_ISL_1814287, EPI_ISL_1814297, EPI_ISL_1814298, EPI_ISL_1814301, EPI_ISL_1814306, EPI_ISL_1814310, EPI_ISL_1814313, EPI_ISL_1814314, EPI_ISL_1814320, EPI_ISL_1814324, EPI_ISL_1814325, EPI_ISL_1814329, EPI_ISL_1814330, EPI_ISL_1814333, EPI_ISL_1814334, EPI_ISL_1814338, EPI_ISL_1814341, EPI_ISL_1814346, EPI_ISL_1814353, EPI_ISL_1814359, EPI_ISL_1814361, EPI_ISL_1814364, EPI_ISL_1814378, EPI_ISL_1814381, EPI_ISL_1814386, EPI_ISL_1814392, EPI_ISL_1814397, EPI_ISL_1814401, EPI_ISL_1814402, EPI_ISL_1814414, EPI_ISL_1814427, EPI_ISL_1814443, EPI_ISL_1814446, EPI_ISL_1814449, EPI_ISL_1814457, EPI_ISL_1814462, EPI_ISL_1814465, EPI_ISL_1814483, EPI_ISL_1814493, EPI_ISL_1814494, EPI_ISL_1814504, EPI_ISL_1814519, EPI_ISL_1814522, EPI_ISL_1814524, EPI_ISL_1814527, EPI_ISL_1814537, EPI_ISL_1814540, EPI_ISL_1814545, EPI_ISL_1814552, EPI_ISL_1814553, EPI_ISL_1814556, EPI_ISL_1814558, EPI_ISL_1814561, EPI_ISL_1814569, EPI_ISL_1814600, EPI_ISL_1814612, EPI_ISL_1814628, EPI_ISL_1814630, EPI_ISL_1814634, EPI_ISL_1814635, EPI_ISL_1814642, EPI_ISL_1814652, EPI_ISL_1814655, EPI_ISL_1814656, EPI_ISL_1814658, EPI_ISL_1814662, EPI_ISL_1814669, EPI_ISL_1814670, EPI_ISL_1814673, EPI_ISL_1814674, EPI_ISL_1814682, EPI_ISL_1814686, EPI_ISL_1814687, EPI_ISL_1814697, EPI_ISL_1814704, EPI_ISL_1814710, EPI_ISL_1814718, EPI_ISL_1814720, EPI_ISL_1814721, EPI_ISL_1814736, EPI_ISL_1814739, EPI_ISL_1814742, EPI_ISL_1814746, EPI_ISL_1814747, EPI_ISL_1814757, EPI_ISL_1814762, EPI_ISL_1814763, EPI_ISL_1814771, EPI_ISL_1814775, EPI_ISL_1814781, EPI_ISL_1814783, EPI_ISL_1814788, EPI_ISL_1814792, EPI_ISL_1814795, EPI_ISL_1814798, EPI_ISL_1814806, EPI_ISL_1814812, EPI_ISL_1814814, EPI_ISL_1814816, EPI_ISL_1814825, EPI_ISL_1814826, EPI_ISL_1814833, EPI_ISL_1814836, EPI_ISL_1814845, EPI_ISL_1814848, EPI_ISL_1814850, EPI_ISL_1814851, EPI_ISL_1814856, EPI_ISL_1814861, EPI_ISL_1814872, EPI_ISL_1814877, EPI_ISL_1814878, EPI_ISL_1814879, EPI_ISL_1814881, EPI_ISL_1814883, EPI_ISL_1814892, EPI_ISL_1814893, EPI_ISL_1814894, EPI_ISL_1814895, EPI_ISL_1814896, EPI_ISL_1814905, EPI_ISL_1814906, EPI_ISL_1814914, EPI_ISL_1814918, EPI_ISL_1814924, EPI_ISL_1814930, EPI_ISL_1814939, EPI_ISL_1814950, EPI_ISL_1814957, EPI_ISL_1814962, EPI_ISL_1814971, EPI_ISL_1814984, EPI_ISL_1814989, EPI_ISL_1814998, EPI_ISL_1815000, EPI_ISL_1815002, EPI_ISL_1815008, EPI_ISL_1815017, EPI_ISL_1815018, EPI_ISL_1815026, EPI_ISL_1815031, EPI_ISL_1815040, EPI_ISL_1815048, EPI_ISL_1815060, EPI_ISL_1815063, EPI_ISL_1815070, EPI_ISL_1815075, EPI_ISL_1815084, EPI_ISL_1815088, EPI_ISL_1815092, EPI_ISL_1815108, EPI_ISL_1815109, EPI_ISL_1815110, EPI_ISL_1815112, EPI_ISL_1815114, EPI_ISL_1815115, EPI_ISL_1815121, EPI_ISL_1815126, EPI_ISL_1815142, EPI_ISL_1815147, EPI_ISL_1815151, EPI_ISL_1815155, EPI_ISL_1815158, EPI_ISL_1815160, EPI_ISL_1815162, EPI_ISL_1815164, EPI_ISL_1815166, EPI_ISL_1815169, EPI_ISL_1815178, EPI_ISL_1815183, EPI_ISL_1815197, EPI_ISL_1815198, EPI_ISL_1815200, EPI_ISL_1815201, EPI_ISL_1815203, EPI_ISL_1815207, EPI_ISL_1815212, EPI_ISL_1815213, EPI_ISL_1815220, EPI_ISL_1815230, EPI_ISL_1815235, EPI_ISL_1815237, EPI_ISL_1815251, EPI_ISL_1815253, EPI_ISL_1815255, EPI_ISL_1815257, EPI_ISL_1815264, EPI_ISL_1815270, EPI_ISL_1815274, EPI_ISL_1815278, EPI_ISL_1815283, EPI_ISL_1815287, EPI_ISL_1815288, EPI_ISL_1815290, EPI_ISL_1815293, EPI_ISL_1815296, EPI_ISL_1815298, EPI_ISL_1815301, EPI_ISL_1815318, EPI_ISL_1815320, EPI_ISL_1815326, EPI_ISL_1815329, EPI_ISL_1815335, EPI_ISL_1815336, EPI_ISL_1815340, EPI_ISL_1815341, EPI_ISL_1815346, EPI_ISL_1815350, EPI_ISL_1815354, EPI_ISL_1815357, EPI_ISL_1815363, EPI_ISL_1815368, EPI_ISL_1815371, EPI_ISL_1815378, EPI_ISL_1815385, EPI_ISL_1815386, EPI_ISL_1815387, EPI_ISL_1815388, EPI_ISL_1815390, EPI_ISL_1815393, EPI_ISL_1815396, EPI_ISL_1815402, EPI_ISL_1815405, EPI_ISL_1815415, EPI_ISL_1815420, EPI_ISL_1815422, EPI_ISL_1815429, EPI_ISL_1815435, EPI_ISL_1815441, EPI_ISL_1815446, EPI_ISL_1815450, EPI_ISL_1815452, EPI_ISL_1815453, EPI_ISL_1815454, EPI_ISL_1815455, EPI_ISL_1815457, EPI_ISL_1815463, EPI_ISL_1815464, EPI_ISL_1815491, EPI_ISL_1815493, EPI_ISL_1815496, EPI_ISL_1815502, EPI_ISL_1815509, EPI_ISL_1815513, EPI_ISL_1815514, EPI_ISL_1815526, EPI_ISL_1815534, EPI_ISL_1815535, EPI_ISL_1815544, EPI_ISL_1815546, EPI_ISL_1815548, EPI_ISL_1815550, EPI_ISL_1815554, EPI_ISL_1815564, EPI_ISL_1815573, EPI_ISL_1815574, EPI_ISL_1815575, EPI_ISL_1815576, EPI_ISL_1815577, EPI_ISL_1815579, EPI_ISL_1815584, EPI_ISL_1815605, EPI_ISL_1815606, EPI_ISL_1815607, EPI_ISL_1815616, EPI_ISL_1815617, EPI_ISL_1815618, EPI_ISL_1815619, EPI_ISL_1815622, EPI_ISL_1815623, EPI_ISL_1815631, EPI_ISL_1815634, EPI_ISL_1815635, EPI_ISL_1815638, EPI_ISL_1815641, EPI_ISL_1815647, EPI_ISL_1815653, EPI_ISL_1815654, EPI_ISL_1815656, EPI_ISL_1815660, EPI_ISL_1815663, EPI_ISL_1815665, EPI_ISL_1815667, EPI_ISL_1815670, EPI_ISL_1815672, EPI_ISL_1815673, EPI_ISL_1815681, EPI_ISL_1815683, EPI_ISL_1815691, EPI_ISL_1815698, EPI_ISL_1815702, EPI_ISL_1815705, EPI_ISL_1815706, EPI_ISL_1815708, EPI_ISL_1815712, EPI_ISL_1815713, EPI_ISL_1815714, EPI_ISL_1815720, EPI_ISL_1815725, EPI_ISL_1815727, EPI_ISL_1815735, EPI_ISL_1815741, EPI_ISL_1815744, EPI_ISL_1815747, EPI_ISL_1815749, EPI_ISL_1815751, EPI_ISL_1815753, EPI_ISL_1815754, EPI_ISL_1815759, EPI_ISL_1815760, EPI_ISL_1815761, EPI_ISL_1815765, EPI_ISL_1815768, EPI_ISL_1815770, EPI_ISL_1815774, EPI_ISL_1815775, EPI_ISL_1815776, EPI_ISL_1815778, EPI_ISL_1815781, EPI_ISL_1815784, EPI_ISL_1815786, EPI_ISL_1815788, EPI_ISL_1815794, EPI_ISL_1815809, EPI_ISL_1815812, EPI_ISL_1815813, EPI_ISL_1815818, EPI_ISL_1815821, EPI_ISL_1815823, EPI_ISL_1815825, EPI_ISL_1815832, EPI_ISL_1815833, EPI_ISL_1815838, EPI_ISL_1815847, EPI_ISL_1815848, EPI_ISL_1815849, EPI_ISL_1815856, EPI_ISL_1815863, EPI_ISL_1815868, EPI_ISL_1815870, EPI_ISL_1815878, EPI_ISL_1815879, EPI_ISL_1815887, EPI_ISL_1815890, EPI_ISL_1815893, EPI_ISL_1815897, EPI_ISL_1815901, EPI_ISL_1815902, EPI_ISL_1815904, EPI_ISL_1815905, EPI_ISL_1815910, EPI_ISL_1815914, EPI_ISL_1815915, EPI_ISL_1815921, EPI_ISL_1815924, EPI_ISL_1815932, EPI_ISL_1815937, EPI_ISL_1815942, EPI_ISL_1815946, EPI_ISL_1815948, EPI_ISL_1815957, EPI_ISL_1815963, EPI_ISL_1815970, EPI_ISL_1815972, EPI_ISL_1815973, EPI_ISL_1815976, EPI_ISL_1815980, EPI_ISL_1815985, EPI_ISL_1815988, EPI_ISL_1815991, EPI_ISL_1816001, EPI_ISL_1816002, EPI_ISL_1816003, EPI_ISL_1816010, EPI_ISL_1816016, EPI_ISL_1816019, EPI_ISL_1816020, EPI_ISL_1816043, EPI_ISL_1816047, EPI_ISL_1816053, EPI_ISL_1816055, EPI_ISL_1816058, EPI_ISL_1816060, EPI_ISL_1816062, EPI_ISL_1816064, EPI_ISL_1816065, EPI_ISL_1816071, EPI_ISL_1816085, EPI_ISL_1816087, EPI_ISL_1816090, EPI_ISL_1816091, EPI_ISL_1816095, EPI_ISL_1816096, EPI_ISL_1816098, EPI_ISL_1816099, EPI_ISL_1816100, EPI_ISL_1816101, EPI_ISL_1816104, EPI_ISL_1816105, EPI_ISL_1816112, EPI_ISL_1816114, EPI_ISL_1816116, EPI_ISL_1816117, EPI_ISL_1816126, EPI_ISL_1816133, EPI_ISL_1816136, EPI_ISL_1816141, EPI_ISL_1816144, EPI_ISL_1816146, EPI_ISL_1816150, EPI_ISL_1816162, EPI_ISL_1816166, EPI_ISL_1816174, EPI_ISL_1816176, EPI_ISL_1816180, EPI_ISL_1816183, EPI_ISL_1816186, EPI_ISL_1816192, EPI_ISL_1816196, EPI_ISL_1816202, EPI_ISL_1816203, EPI_ISL_1816204, EPI_ISL_1816205, EPI_ISL_1816206, EPI_ISL_1816209, EPI_ISL_1816210, EPI_ISL_1816211, EPI_ISL_1816212, EPI_ISL_1816214, EPI_ISL_1816226, EPI_ISL_1816234, EPI_ISL_1816239, EPI_ISL_1816241, EPI_ISL_1816245, EPI_ISL_1816246, EPI_ISL_1816253, EPI_ISL_1816256, EPI_ISL_1816260, EPI_ISL_1816261, EPI_ISL_1816266, EPI_ISL_1816269, EPI_ISL_1816271, EPI_ISL_1816274, EPI_ISL_1816275, EPI_ISL_1816278, EPI_ISL_1816280, EPI_ISL_1816284, EPI_ISL_1816299, EPI_ISL_1816302, EPI_ISL_1816308, EPI_ISL_1816309, EPI_ISL_1816311, EPI_ISL_1816312, EPI_ISL_1816314, EPI_ISL_1816316, EPI_ISL_1816319, EPI_ISL_1816320, EPI_ISL_1816324, EPI_ISL_1816326, EPI_ISL_1816331, EPI_ISL_1816335, EPI_ISL_1816339, EPI_ISL_1816346, EPI_ISL_1816348, EPI_ISL_1816349, EPI_ISL_1816351, EPI_ISL_1816355, EPI_ISL_1816356, EPI_ISL_1816366, EPI_ISL_1816369, EPI_ISL_1816373, EPI_ISL_1816375, EPI_ISL_1816377, EPI_ISL_1816384, EPI_ISL_1816392, EPI_ISL_1816398, EPI_ISL_1816399, EPI_ISL_1816400, EPI_ISL_1816401, EPI_ISL_1816411, EPI_ISL_1816414, EPI_ISL_1816415, EPI_ISL_1816420, EPI_ISL_1816428, EPI_ISL_1816433, EPI_ISL_1816444, EPI_ISL_1816450, EPI_ISL_1816454, EPI_ISL_1816458, EPI_ISL_1816459, EPI_ISL_1816461, EPI_ISL_1816465, EPI_ISL_1816470, EPI_ISL_1816473, EPI_ISL_1816498, EPI_ISL_1816500, EPI_ISL_1816503, EPI_ISL_1816511, EPI_ISL_1816512, EPI_ISL_1816516, EPI_ISL_1816519, EPI_ISL_1816522, EPI_ISL_1816523, EPI_ISL_1816525, EPI_ISL_1816530, EPI_ISL_1816537, EPI_ISL_1816540, EPI_ISL_1816541, EPI_ISL_1816542, EPI_ISL_1816551, EPI_ISL_1816552, EPI_ISL_1816553, EPI_ISL_1816559, EPI_ISL_1816567, EPI_ISL_1816569, EPI_ISL_1816584, EPI_ISL_1816587, EPI_ISL_1816594, EPI_ISL_1816598, EPI_ISL_1816603, EPI_ISL_1816610, EPI_ISL_1816611, EPI_ISL_1816613, EPI_ISL_1816615, EPI_ISL_1816634, EPI_ISL_1816636, EPI_ISL_1816637, EPI_ISL_1816639, EPI_ISL_1816640, EPI_ISL_1816650, EPI_ISL_1816652, EPI_ISL_1816666, EPI_ISL_1816669, EPI_ISL_1816671, EPI_ISL_1816675, EPI_ISL_1816678, EPI_ISL_1816680, EPI_ISL_1816681, EPI_ISL_1816699, EPI_ISL_1816700, EPI_ISL_1816708, EPI_ISL_1816716, EPI_ISL_1816718, EPI_ISL_1816719, EPI_ISL_1816722, EPI_ISL_1816723, EPI_ISL_1816725, EPI_ISL_1816727, EPI_ISL_1816733, EPI_ISL_1816737, EPI_ISL_1816741, EPI_ISL_1816748, EPI_ISL_1816752, EPI_ISL_1816754, EPI_ISL_1816759, EPI_ISL_1816772, EPI_ISL_1816773, EPI_ISL_1816774, EPI_ISL_1816780, EPI_ISL_1816785, EPI_ISL_1816786, EPI_ISL_1816793, EPI_ISL_1816795, EPI_ISL_1816796, EPI_ISL_1816797, EPI_ISL_1816802, EPI_ISL_1816805, EPI_ISL_1816813, EPI_ISL_1816818, EPI_ISL_1816819, EPI_ISL_1816824, EPI_ISL_1816827, EPI_ISL_1816829, EPI_ISL_1816835, EPI_ISL_1816839, EPI_ISL_1816840, EPI_ISL_1816841, EPI_ISL_1816842, EPI_ISL_1816843, EPI_ISL_1816848, EPI_ISL_1816849, EPI_ISL_1816855, EPI_ISL_1816861, EPI_ISL_1816864, EPI_ISL_1816867, EPI_ISL_1816875, EPI_ISL_1816879, EPI_ISL_1816881, EPI_ISL_1816884, EPI_ISL_1816887, EPI_ISL_1816891, EPI_ISL_1816892, EPI_ISL_1816893, EPI_ISL_1816894, EPI_ISL_1816903, EPI_ISL_1816904, EPI_ISL_1816905, EPI_ISL_1816907, EPI_ISL_1816909, EPI_ISL_1816911, EPI_ISL_1816913, EPI_ISL_1816915 | | | |
| see above | EXCITE Lab | Andersen lab at Scripps Research | Abigail Schnapper; Alexandre Bolze; Alice Summerfield; Angela Scioscia; Araz Majnoonian; Ashkan Hassani; Celena Andrade; Charlotte Rivera-Garcia; Cheryl Anderson; Chip Schooley; Christine Mitchell + SEARCH; David Becker; David Pride; Efren Sandoval; Elizabeth Cirulli; Fitri Carrissa Wijaya; Francisco Tanudjaja; Fred Wu; Geraint Levan; Greg Humphrey; Helena Tubb; James Lu + SEARCH; Jason Nguyen; Jimmy Ramirez; Kelly Schiabor Barrett; Laura Kohn; Magnus Isaksson; Marc Laurent; Marlene Flores; Megan Nguyen; Natasha Martin; Natasha Martin Cheryl Anderson; Nicole L Washington; Patricia Gonzalez-Zuniga; Rebecca Fielding-Miller; Rodolfo Salido; Ryan Cho; Sawyer Farmer; Sharon Reed; Sherry Wang; Simon White; Smruthi Karthikeyan; Tommy Valles + SEARCH; Tyler Cassens; Victor Cantu; Vinton Omaleki; William Lee |
| EPI_ISL_1483789, EPI_ISL_1483790, EPI_ISL_1483791, EPI_ISL_1483792, EPI_ISL_1483793, EPI_ISL_1483794, EPI_ISL_1483800, EPI_ISL_1483801 | | | |
| see above | El Camino Hospital | Santa Clara County Public Health Laboratory | Santa Clara County Public Health Department |
| EPI_ISL_1303474, EPI_ISL_1303475, EPI_ISL_1303476, EPI_ISL_1303477, EPI_ISL_1303478, EPI_ISL_1303479, EPI_ISL_1303484, EPI_ISL_1303485, EPI_ISL_1303486, EPI_ISL_1303487, EPI_ISL_1303488, EPI_ISL_1307636, EPI_ISL_1307637, EPI_ISL_1379978, EPI_ISL_1497542, EPI_ISL_1669341, EPI_ISL_1669342, EPI_ISL_1669343, EPI_ISL_1669344, EPI_ISL_1669345, EPI_ISL_1669346, EPI_ISL_1669347, EPI_ISL_1697372, EPI_ISL_1706605, EPI_ISL_1706606, EPI_ISL_1706607, EPI_ISL_1707675, EPI_ISL_1707676 | | | |
| see above | Flow Diagnostics Lab | Los Angeles County PHL | P. Hemarajata et al. |
| EPI_ISL_1516746, EPI_ISL_1578215, EPI_ISL_1578327, EPI_ISL_1578328, EPI_ISL_1578329, EPI_ISL_1578331, EPI_ISL_1578339, EPI_ISL_1578451 | | | |
| see above | Flow Health | Infectious Disease Program, Broad Institute of Harvard and MIT | Adams, G.; B.L.; B.W.; Bauer, M.; Birren; Carter, A.; Chaluvadi, S.; D.J.; DeRuff, K.; Gallagher, G.; Gladden-Young, A.; J.E.; K.J.; Lagerborg, K.; Lemieux; Loreth, C.; MacInnis; Normandin, E.; P.C.; Park; Reilly, S.; Rudy, M.; Siddle; Smole, S.; Tomkins-Tinch, C.; and Sabeti |
| EPI_ISL_1457792, EPI_ISL_1457793, EPI_ISL_1457794, EPI_ISL_1457795, EPI_ISL_1457796, EPI_ISL_1457797, EPI_ISL_1457798, EPI_ISL_1457799, EPI_ISL_1457800, EPI_ISL_1457801, EPI_ISL_1457802, EPI_ISL_1457803, EPI_ISL_1457804, EPI_ISL_1457805, EPI_ISL_1457806, EPI_ISL_1457807, EPI_ISL_1457808, EPI_ISL_1457809, EPI_ISL_1457810, EPI_ISL_1457811, EPI_ISL_1457812, EPI_ISL_1457813, EPI_ISL_1457814, EPI_ISL_1457815, EPI_ISL_1457816, EPI_ISL_1457817, EPI_ISL_1457818, EPI_ISL_1457819, EPI_ISL_1457820, EPI_ISL_1457821, EPI_ISL_1457822, EPI_ISL_1457823, EPI_ISL_1457824, EPI_ISL_1457825, EPI_ISL_1457826, EPI_ISL_1457827, EPI_ISL_1457828, EPI_ISL_1457829, EPI_ISL_1457830, EPI_ISL_1457831, EPI_ISL_1457832, EPI_ISL_1457833, EPI_ISL_1457834, EPI_ISL_1457835, EPI_ISL_1457836, EPI_ISL_1457837, EPI_ISL_1457838, EPI_ISL_1457839, EPI_ISL_1457840, EPI_ISL_1457841, EPI_ISL_1457842, EPI_ISL_1457843, EPI_ISL_1457844, EPI_ISL_1457845, EPI_ISL_1457846, EPI_ISL_1457847, EPI_ISL_1457848, EPI_ISL_1457849, EPI_ISL_1457850, EPI_ISL_1457851, EPI_ISL_1457852, EPI_ISL_1457853, EPI_ISL_1457854, EPI_ISL_1457855, EPI_ISL_1457856, EPI_ISL_1457857, EPI_ISL_1457858, EPI_ISL_1457859, EPI_ISL_1457861, EPI_ISL_1457862, EPI_ISL_1457863, EPI_ISL_1457864, EPI_ISL_1457865, EPI_ISL_1457866, EPI_ISL_1457867, EPI_ISL_1457868, EPI_ISL_1457869, EPI_ISL_1457870, EPI_ISL_1457871, EPI_ISL_1457872, EPI_ISL_1457873, EPI_ISL_1457874, EPI_ISL_1457875, EPI_ISL_1457876, EPI_ISL_1457877, EPI_ISL_1457878, EPI_ISL_1457879, EPI_ISL_1457880, EPI_ISL_1457881, EPI_ISL_1457882, EPI_ISL_1457883, EPI_ISL_1457884, EPI_ISL_1457885, EPI_ISL_1457886, EPI_ISL_1457887, EPI_ISL_1457888, EPI_ISL_1457889, EPI_ISL_1457890, EPI_ISL_1457891, EPI_ISL_1457892, EPI_ISL_1457893, EPI_ISL_1457894, EPI_ISL_1457895, EPI_ISL_1457896, EPI_ISL_1457897, EPI_ISL_1457898, EPI_ISL_1457899, EPI_ISL_1457900, EPI_ISL_1457901, EPI_ISL_1457902, EPI_ISL_1457903, EPI_ISL_1457904, EPI_ISL_1457905, EPI_ISL_1457906, EPI_ISL_1457907, EPI_ISL_1457908, EPI_ISL_1457909, EPI_ISL_1457910, EPI_ISL_1457911, EPI_ISL_1457912, EPI_ISL_1457913, EPI_ISL_1457914, EPI_ISL_1457915, EPI_ISL_1457916, EPI_ISL_1457917, EPI_ISL_1457918, EPI_ISL_1457919, EPI_ISL_1457920, EPI_ISL_1457921, EPI_ISL_1457922, EPI_ISL_1457923, EPI_ISL_1457924, EPI_ISL_1457925, EPI_ISL_1457926, EPI_ISL_1457927, EPI_ISL_1457928, EPI_ISL_1457929, EPI_ISL_1457930, EPI_ISL_1457931, EPI_ISL_1457932, EPI_ISL_1457933, EPI_ISL_1457934, EPI_ISL_1457935, EPI_ISL_1457936, EPI_ISL_1457937, EPI_ISL_1457938, EPI_ISL_1457939, EPI_ISL_1457940, EPI_ISL_1457941, EPI_ISL_1457942, EPI_ISL_1457943, EPI_ISL_1457944, EPI_ISL_1457945, EPI_ISL_1457946, EPI_ISL_1457947, EPI_ISL_1457948, EPI_ISL_1457949, EPI_ISL_1457950, EPI_ISL_1457951, EPI_ISL_1457952, EPI_ISL_1457953, EPI_ISL_1457954, EPI_ISL_1457955, EPI_ISL_1457956, EPI_ISL_1457957, EPI_ISL_1457958, EPI_ISL_1457959, EPI_ISL_1457960, EPI_ISL_1457961, EPI_ISL_1457962, EPI_ISL_1457963, EPI_ISL_1457964, EPI_ISL_1457965, EPI_ISL_1457966, EPI_ISL_1457968, EPI_ISL_1457969, EPI_ISL_1457970, EPI_ISL_1457971, EPI_ISL_1457972, EPI_ISL_1457973, EPI_ISL_1457974, EPI_ISL_1457975, EPI_ISL_1457976, EPI_ISL_1457977, EPI_ISL_1457978, EPI_ISL_1457980, EPI_ISL_1457981, EPI_ISL_1457982, EPI_ISL_1457983, EPI_ISL_1457984, EPI_ISL_1457985, EPI_ISL_1457986, EPI_ISL_1457987, EPI_ISL_1457988, EPI_ISL_1457989, EPI_ISL_1457990, EPI_ISL_1457991, EPI_ISL_1457992, EPI_ISL_1457993, EPI_ISL_1457994, EPI_ISL_1457995, EPI_ISL_1457996, EPI_ISL_1457997, EPI_ISL_1457998, EPI_ISL_1457999, EPI_ISL_1458000, EPI_ISL_1458001, EPI_ISL_1458002, EPI_ISL_1458003, EPI_ISL_1458004, EPI_ISL_1458005, EPI_ISL_1458006, EPI_ISL_1458007, EPI_ISL_1458008, EPI_ISL_1458009, EPI_ISL_1458010, EPI_ISL_1458011, EPI_ISL_1458012, EPI_ISL_1458013, EPI_ISL_1458014, EPI_ISL_1458015, EPI_ISL_1458016, EPI_ISL_1458017, EPI_ISL_1458018, EPI_ISL_1458019, EPI_ISL_1458020, EPI_ISL_1458021, EPI_ISL_1458022, EPI_ISL_1458023, EPI_ISL_1458024, EPI_ISL_1458025, EPI_ISL_1458026, EPI_ISL_1458027, EPI_ISL_1458028, EPI_ISL_1458029, EPI_ISL_1458030, EPI_ISL_1458031, EPI_ISL_1458032, EPI_ISL_1458033, EPI_ISL_1458034, EPI_ISL_1458035, EPI_ISL_1458036, EPI_ISL_1458037, EPI_ISL_1458038, EPI_ISL_1458039, EPI_ISL_1458040, EPI_ISL_1458041, EPI_ISL_1458042, EPI_ISL_1458043, EPI_ISL_1458044, EPI_ISL_1458045, EPI_ISL_1458046, EPI_ISL_1458047, EPI_ISL_1458048, EPI_ISL_1458049, EPI_ISL_1458050, EPI_ISL_1458051, EPI_ISL_1458052, EPI_ISL_1458053, EPI_ISL_1458054, EPI_ISL_1458055, EPI_ISL_1458056, EPI_ISL_1458057, EPI_ISL_1458058, EPI_ISL_1458059, EPI_ISL_1458060, EPI_ISL_1458061, EPI_ISL_1458062, EPI_ISL_1458063, EPI_ISL_1458064, EPI_ISL_1458065, EPI_ISL_1458066, EPI_ISL_1458067, EPI_ISL_1458068, EPI_ISL_1458069, EPI_ISL_1458070, EPI_ISL_1458071, EPI_ISL_1458072, EPI_ISL_1458073, EPI_ISL_1458074, EPI_ISL_1458075, EPI_ISL_1458076, EPI_ISL_1458077, EPI_ISL_1458078, EPI_ISL_1458079, EPI_ISL_1458080, EPI_ISL_1458081, EPI_ISL_1458082, EPI_ISL_1458083, EPI_ISL_1458084, EPI_ISL_1458086, EPI_ISL_1458087, EPI_ISL_1458088, EPI_ISL_1458089, EPI_ISL_1458090, EPI_ISL_1458091, EPI_ISL_1458092, EPI_ISL_1458093, EPI_ISL_1458094, EPI_ISL_1458095, EPI_ISL_1458096, EPI_ISL_1458097, EPI_ISL_1458098, EPI_ISL_1458099, EPI_ISL_1458100, EPI_ISL_1458101, EPI_ISL_1458102, EPI_ISL_1458103, EPI_ISL_1458104, EPI_ISL_1458105, EPI_ISL_1458106, EPI_ISL_1458107, EPI_ISL_1458108, EPI_ISL_1458109, EPI_ISL_1458110, EPI_ISL_1458111, EPI_ISL_1458112, EPI_ISL_1458113, EPI_ISL_1458114, EPI_ISL_1458115, EPI_ISL_1458116, EPI_ISL_1458117, EPI_ISL_1458118, EPI_ISL_1458119, EPI_ISL_1458120, EPI_ISL_1458121, EPI_ISL_1458122, EPI_ISL_1458123, EPI_ISL_1458124, EPI_ISL_1458125, EPI_ISL_1458126, EPI_ISL_1458127, EPI_ISL_1458128, EPI_ISL_1458129, EPI_ISL_1458130, EPI_ISL_1458131, EPI_ISL_1458132, EPI_ISL_1458133, EPI_ISL_1458134, EPI_ISL_1458135, EPI_ISL_1458136, EPI_ISL_1458137, EPI_ISL_1458138, EPI_ISL_1458139, EPI_ISL_1458140, EPI_ISL_1458141, EPI_ISL_1458142, EPI_ISL_1458143, EPI_ISL_1458144, EPI_ISL_1458145, EPI_ISL_1458146, EPI_ISL_1458147, EPI_ISL_1458148, EPI_ISL_1458149, EPI_ISL_1458150, EPI_ISL_1458151, EPI_ISL_1458152, EPI_ISL_1458153, EPI_ISL_1458154, EPI_ISL_1458155, EPI_ISL_1458156, EPI_ISL_1458157, EPI_ISL_1458158, EPI_ISL_1458159, EPI_ISL_1458160, EPI_ISL_1458161, EPI_ISL_1458162, EPI_ISL_1458163, EPI_ISL_1458164, EPI_ISL_1458165, EPI_ISL_1458166, EPI_ISL_1458167, EPI_ISL_1458168, EPI_ISL_1458169, EPI_ISL_1458170, EPI_ISL_1458171, EPI_ISL_1458172, EPI_ISL_1458173, EPI_ISL_1458174, EPI_ISL_1458175, EPI_ISL_1458176, EPI_ISL_1458177, EPI_ISL_1458178, EPI_ISL_1458179, EPI_ISL_1458180, EPI_ISL_1458181, EPI_ISL_1458182, EPI_ISL_1458183, EPI_ISL_1458184, EPI_ISL_1458185, EPI_ISL_1458186, EPI_ISL_1458187, EPI_ISL_1458188, EPI_ISL_1458189, EPI_ISL_1458190, EPI_ISL_1458191, | | | |

EPI_ISL_1458192, EPI_ISL_1458193, EPI_ISL_1458194, EPI_ISL_1458195, EPI_ISL_1458196, EPI_ISL_1458197, EPI_ISL_1458198, EPI_ISL_1458199, EPI_ISL_1458200, EPI_ISL_1458201, EPI_ISL_1458202, EPI_ISL_1458203, EPI_ISL_1458204, EPI_ISL_1458205, EPI_ISL_1458206, EPI_ISL_1458207, EPI_ISL_1458208, EPI_ISL_1458209, EPI_ISL_1458210, EPI_ISL_1458211, EPI_ISL_1458212, EPI_ISL_1458213,
EPI_ISL_1458214, EPI_ISL_1458215, EPI_ISL_1458216, EPI_ISL_1458217, EPI_ISL_1458218, EPI_ISL_1458219, EPI_ISL_1458220, EPI_ISL_1458221, EPI_ISL_1458222, EPI_ISL_1458223, EPI_ISL_1458224, EPI_ISL_1458225, EPI_ISL_1458226, EPI_ISL_1458227, EPI_ISL_1458228, EPI_ISL_1458229, EPI_ISL_1458230, EPI_ISL_1458231, EPI_ISL_1458232, EPI_ISL_1458233, EPI_ISL_1458234, EPI_ISL_1458235,
EPI_ISL_1458236, EPI_ISL_1458237, EPI_ISL_1458238, EPI_ISL_1458239, EPI_ISL_1458240, EPI_ISL_1458241, EPI_ISL_1458242, EPI_ISL_1458243, EPI_ISL_1458244, EPI_ISL_1458245, EPI_ISL_1458246, EPI_ISL_1458247, EPI_ISL_1458248, EPI_ISL_1458249, EPI_ISL_1458250, EPI_ISL_1458251, EPI_ISL_1458252, EPI_ISL_1458253, EPI_ISL_1458254, EPI_ISL_1458255, EPI_ISL_1458256, EPI_ISL_1458257,
EPI_ISL_1458258, EPI_ISL_1458259, EPI_ISL_1458260, EPI_ISL_1458261, EPI_ISL_1458262, EPI_ISL_1458263, EPI_ISL_1458264, EPI_ISL_1458265, EPI_ISL_1458266, EPI_ISL_1458267, EPI_ISL_1458268, EPI_ISL_1458269, EPI_ISL_1458270, EPI_ISL_1458271, EPI_ISL_1458272, EPI_ISL_1458273, EPI_ISL_1458274, EPI_ISL_1458275, EPI_ISL_1458276, EPI_ISL_1458277, EPI_ISL_1458278, EPI_ISL_1458279,
EPI_ISL_1458280, EPI_ISL_1458281, EPI_ISL_1458282, EPI_ISL_1458283, EPI_ISL_1458285, EPI_ISL_1458286, EPI_ISL_1458289, EPI_ISL_1458290, EPI_ISL_1458291, EPI_ISL_1458292, EPI_ISL_1458293, EPI_ISL_1458294, EPI_ISL_1458295, EPI_ISL_1458296, EPI_ISL_1458297, EPI_ISL_1458298, EPI_ISL_1458299, EPI_ISL_1458300, EPI_ISL_1458301, EPI_ISL_1458302, EPI_ISL_1458303, EPI_ISL_1458304,
EPI_ISL_1458305, EPI_ISL_1458306, EPI_ISL_1458307, EPI_ISL_1458308, EPI_ISL_1458309, EPI_ISL_1458310, EPI_ISL_1458311, EPI_ISL_1458312, EPI_ISL_1458313, EPI_ISL_1458314, EPI_ISL_1458315, EPI_ISL_1458316, EPI_ISL_1458317, EPI_ISL_1458318, EPI_ISL_1458319, EPI_ISL_1458320, EPI_ISL_1458321, EPI_ISL_1458322, EPI_ISL_1458323, EPI_ISL_1458324, EPI_ISL_1458325, EPI_ISL_1458326,
EPI_ISL_1458327, EPI_ISL_1458328, EPI_ISL_1458329, EPI_ISL_1458330, EPI_ISL_1458331, EPI_ISL_1458332, EPI_ISL_1458333, EPI_ISL_1458334, EPI_ISL_1458335, EPI_ISL_1458336, EPI_ISL_1458337, EPI_ISL_1458338, EPI_ISL_1458339, EPI_ISL_1458340, EPI_ISL_1458341, EPI_ISL_1458342, EPI_ISL_1458343, EPI_ISL_1458344, EPI_ISL_1458345, EPI_ISL_1458346, EPI_ISL_1458347, EPI_ISL_1458348,
EPI_ISL_1458349, EPI_ISL_1458350, EPI_ISL_1458351, EPI_ISL_1458352, EPI_ISL_1458353, EPI_ISL_1458354, EPI_ISL_1458355, EPI_ISL_1458356, EPI_ISL_1458357, EPI_ISL_1458358, EPI_ISL_1458359, EPI_ISL_1458360, EPI_ISL_1458361, EPI_ISL_1458362, EPI_ISL_1458363, EPI_ISL_1458364, EPI_ISL_1458365, EPI_ISL_1458366, EPI_ISL_1458367, EPI_ISL_1458368, EPI_ISL_1458369, EPI_ISL_1458370,
EPI_ISL_1458371, EPI_ISL_1458372, EPI_ISL_1458373, EPI_ISL_1458374, EPI_ISL_1458375, EPI_ISL_1458376, EPI_ISL_1458377, EPI_ISL_1458378, EPI_ISL_1458379, EPI_ISL_1458380, EPI_ISL_1458381, EPI_ISL_1458382, EPI_ISL_1458383, EPI_ISL_1458384, EPI_ISL_1458385, EPI_ISL_1458386, EPI_ISL_1458387, EPI_ISL_1458388, EPI_ISL_1458389, EPI_ISL_1458390, EPI_ISL_1458391, EPI_ISL_1458392,
EPI_ISL_1458393, EPI_ISL_1458394, EPI_ISL_1458395, EPI_ISL_1458396, EPI_ISL_1458397, EPI_ISL_1458398, EPI_ISL_1458399, EPI_ISL_1458400, EPI_ISL_1458401, EPI_ISL_1458402, EPI_ISL_1458403, EPI_ISL_1458404, EPI_ISL_1458405, EPI_ISL_1458406, EPI_ISL_1458407, EPI_ISL_1458408, EPI_ISL_1458409, EPI_ISL_1458410, EPI_ISL_1458411, EPI_ISL_1458412, EPI_ISL_1458413, EPI_ISL_1458414,
EPI_ISL_1458415, EPI_ISL_1458416, EPI_ISL_1458417, EPI_ISL_1458418, EPI_ISL_1458481, EPI_ISL_1458482, EPI_ISL_1458483, EPI_ISL_1458484, EPI_ISL_1458485, EPI_ISL_1458486, EPI_ISL_1458487, EPI_ISL_1458488, EPI_ISL_1458489, EPI_ISL_1458490, EPI_ISL_1458491, EPI_ISL_1458492, EPI_ISL_1458493, EPI_ISL_1458494, EPI_ISL_1458495, EPI_ISL_1458496, EPI_ISL_1458497, EPI_ISL_1458498,
EPI_ISL_1458499, EPI_ISL_1458500, EPI_ISL_1458501, EPI_ISL_1458502, EPI_ISL_1458503, EPI_ISL_1458504, EPI_ISL_1458505, EPI_ISL_1458506, EPI_ISL_1458507, EPI_ISL_1458508, EPI_ISL_1458509, EPI_ISL_1458510, EPI_ISL_1458511, EPI_ISL_1458512, EPI_ISL_1458513, EPI_ISL_1458514, EPI_ISL_1458515, EPI_ISL_1458516, EPI_ISL_1458517, EPI_ISL_1458518, EPI_ISL_1458519, EPI_ISL_1458520,
EPI_ISL_1458521, EPI_ISL_1458522, EPI_ISL_1458523, EPI_ISL_1458524, EPI_ISL_1458525, EPI_ISL_1458526, EPI_ISL_1458527, EPI_ISL_1458528, EPI_ISL_1458529, EPI_ISL_1458530, EPI_ISL_1458531, EPI_ISL_1458532, EPI_ISL_1458533, EPI_ISL_1458534, EPI_ISL_1458535, EPI_ISL_1458536, EPI_ISL_1458537, EPI_ISL_1458538, EPI_ISL_1458539, EPI_ISL_1458540, EPI_ISL_1458541, EPI_ISL_1458542,
EPI_ISL_1458543, EPI_ISL_1458544, EPI_ISL_1458545, EPI_ISL_1458546, EPI_ISL_1458547, EPI_ISL_1458548, EPI_ISL_1458549, EPI_ISL_1458550, EPI_ISL_1458551, EPI_ISL_1458552, EPI_ISL_1458553, EPI_ISL_1458554, EPI_ISL_1458555, EPI_ISL_1458556, EPI_ISL_1458557, EPI_ISL_1458558, EPI_ISL_1458559, EPI_ISL_1458560, EPI_ISL_1458561, EPI_ISL_1458562, EPI_ISL_1458563, EPI_ISL_1458564,
EPI_ISL_1458565, EPI_ISL_1458566, EPI_ISL_1458567, EPI_ISL_1458568, EPI_ISL_1458569, EPI_ISL_1458570, EPI_ISL_1458571, EPI_ISL_1458572, EPI_ISL_1458573, EPI_ISL_1458574, EPI_ISL_1458575, EPI_ISL_1458576, EPI_ISL_1458577, EPI_ISL_1458578, EPI_ISL_1458579, EPI_ISL_1458580, EPI_ISL_1458581, EPI_ISL_1458582, EPI_ISL_1458583, EPI_ISL_1458584, EPI_ISL_1458585, EPI_ISL_1458586,
EPI_ISL_1458587, EPI_ISL_1458588, EPI_ISL_1458589, EPI_ISL_1458590, EPI_ISL_1458591, EPI_ISL_1458592, EPI_ISL_1458593, EPI_ISL_1458594, EPI_ISL_1458595, EPI_ISL_1458596, EPI_ISL_1458597, EPI_ISL_1458598, EPI_ISL_1458599, EPI_ISL_1458600, EPI_ISL_1458601, EPI_ISL_1458602, EPI_ISL_1458603, EPI_ISL_1458604, EPI_ISL_1458605, EPI_ISL_1458606, EPI_ISL_1458607, EPI_ISL_1458608,
EPI_ISL_1458609, EPI_ISL_1458610, EPI_ISL_1458611, EPI_ISL_1458612, EPI_ISL_1458614, EPI_ISL_1458615, EPI_ISL_1458616, EPI_ISL_1458617, EPI_ISL_1458618, EPI_ISL_1458619, EPI_ISL_1458620, EPI_ISL_1458621, EPI_ISL_1458622, EPI_ISL_1458623, EPI_ISL_1458624, EPI_ISL_1458625, EPI_ISL_1458626, EPI_ISL_1458627, EPI_ISL_1458628, EPI_ISL_1458629, EPI_ISL_1458630, EPI_ISL_1458631,
EPI_ISL_1458632, EPI_ISL_1458633, EPI_ISL_1458634, EPI_ISL_1458635, EPI_ISL_1458636, EPI_ISL_1458637, EPI_ISL_1458638, EPI_ISL_1458639, EPI_ISL_1458640, EPI_ISL_1458641, EPI_ISL_1458642, EPI_ISL_1458643, EPI_ISL_1458644, EPI_ISL_1458645, EPI_ISL_1458646, EPI_ISL_1458647, EPI_ISL_1458648, EPI_ISL_1458649, EPI_ISL_1458650, EPI_ISL_1458651, EPI_ISL_1458652, EPI_ISL_1458653,
EPI_ISL_1458654, EPI_ISL_1458655, EPI_ISL_1458656, EPI_ISL_1458657, EPI_ISL_1458658, EPI_ISL_1458659, EPI_ISL_1458660, EPI_ISL_1458661, EPI_ISL_1458662, EPI_ISL_1458663, EPI_ISL_1458664, EPI_ISL_1458665, EPI_ISL_1458666, EPI_ISL_1458667, EPI_ISL_1458668, EPI_ISL_1458669, EPI_ISL_1458670, EPI_ISL_1458671, EPI_ISL_1458672, EPI_ISL_1458673, EPI_ISL_1458674, EPI_ISL_1458675,
EPI_ISL_1458676, EPI_ISL_1458677, EPI_ISL_1458678, EPI_ISL_1458679, EPI_ISL_1458680, EPI_ISL_1458681, EPI_ISL_1458682, EPI_ISL_1458683, EPI_ISL_1458684, EPI_ISL_1458685, EPI_ISL_1458686, EPI_ISL_1458687, EPI_ISL_1458688, EPI_ISL_1458689, EPI_ISL_1458690, EPI_ISL_1458691, EPI_ISL_1458692, EPI_ISL_1458693, EPI_ISL_1458694, EPI_ISL_1458695, EPI_ISL_1458696, EPI_ISL_1458697,
EPI_ISL_1458698, EPI_ISL_1458699, EPI_ISL_1458700, EPI_ISL_1458701, EPI_ISL_1458702, EPI_ISL_1458703, EPI_ISL_1458704, EPI_ISL_1458705, EPI_ISL_1458706, EPI_ISL_1458707, EPI_ISL_1458708, EPI_ISL_1458709, EPI_ISL_1458710, EPI_ISL_1458711, EPI_ISL_1458714, EPI_ISL_1458715, EPI_ISL_1458716, EPI_ISL_1458717, EPI_ISL_1458718, EPI_ISL_1458719, EPI_ISL_1458720, EPI_ISL_1458721,
EPI_ISL_1458722, EPI_ISL_1458723, EPI_ISL_1458724, EPI_ISL_1458725, EPI_ISL_1458726, EPI_ISL_1458727, EPI_ISL_1458728, EPI_ISL_1458729, EPI_ISL_1458730, EPI_ISL_1458731, EPI_ISL_1458732, EPI_ISL_1458733, EPI_ISL_1458734, EPI_ISL_1458735, EPI_ISL_1458736, EPI_ISL_1458737, EPI_ISL_1458738, EPI_ISL_1458739, EPI_ISL_1458740, EPI_ISL_1458741, EPI_ISL_1458742, EPI_ISL_1458743,
EPI_ISL_1458744, EPI_ISL_1458745, EPI_ISL_1458746, EPI_ISL_1458747, EPI_ISL_1458748, EPI_ISL_1458749, EPI_ISL_1458750, EPI_ISL_1458751, EPI_ISL_1458752, EPI_ISL_1458753, EPI_ISL_1458754, EPI_ISL_1458755, EPI_ISL_1458756, EPI_ISL_1458757, EPI_ISL_1458758, EPI_ISL_1458759, EPI_ISL_1458760, EPI_ISL_1458761, EPI_ISL_1458762, EPI_ISL_1458763, EPI_ISL_1458764, EPI_ISL_1458765,
EPI_ISL_1458766, EPI_ISL_1458767, EPI_ISL_1458768, EPI_ISL_1458769, EPI_ISL_1458770, EPI_ISL_1458771, EPI_ISL_1458772, EPI_ISL_1458773, EPI_ISL_1458774, EPI_ISL_1458775, EPI_ISL_1458776, EPI_ISL_1458777, EPI_ISL_1458778, EPI_ISL_1458779, EPI_ISL_1458780, EPI_ISL_1458781, EPI_ISL_1458782, EPI_ISL_1458783, EPI_ISL_1458784, EPI_ISL_1458785, EPI_ISL_1458786, EPI_ISL_1458787,
EPI_ISL_1458788, EPI_ISL_1458789, EPI_ISL_1458790, EPI_ISL_1458791, EPI_ISL_1458792, EPI_ISL_1458793, EPI_ISL_1458794, EPI_ISL_1458795, EPI_ISL_1458796, EPI_ISL_1458797, EPI_ISL_1458798, EPI_ISL_1458799, EPI_ISL_1458800, EPI_ISL_1458801, EPI_ISL_1458802, EPI_ISL_1458803, EPI_ISL_1458804, EPI_ISL_1458805, EPI_ISL_1458806, EPI_ISL_1458807, EPI_ISL_1458808, EPI_ISL_1458809,
EPI_ISL_1458810, EPI_ISL_1458811, EPI_ISL_1458812, EPI_ISL_1458813, EPI_ISL_1458814, EPI_ISL_1458815, EPI_ISL_1458816, EPI_ISL_1458817, EPI_ISL_1458818, EPI_ISL_1458819, EPI_ISL_1458844, EPI_ISL_1458845, EPI_ISL_1458847, EPI_ISL_1458850, EPI_ISL_1458851, EPI_ISL_1458852, EPI_ISL_1458853, EPI_ISL_1458854, EPI_ISL_1458855, EPI_ISL_1458856, EPI_ISL_1458857, EPI_ISL_1458858,
EPI_ISL_1458859, EPI_ISL_1458900, EPI_ISL_1458901, EPI_ISL_1458902, EPI_ISL_1458903, EPI_ISL_1458904, EPI_ISL_1458905, EPI_ISL_1458906, EPI_ISL_1458907, EPI_ISL_1458908, EPI_ISL_1458909, EPI_ISL_1458910, EPI_ISL_1458911, EPI_ISL_1458912, EPI_ISL_1458913, EPI_ISL_1458914, EPI_ISL_1458915, EPI_ISL_1458916, EPI_ISL_1458917, EPI_ISL_1458918, EPI_ISL_1458919, EPI_ISL_1458920,
EPI_ISL_1458921, EPI_ISL_1458922, EPI_ISL_1458923, EPI_ISL_1458924, EPI_ISL_1458925, EPI_ISL_1458926, EPI_ISL_1458927, EPI_ISL_1458928, EPI_ISL_1458929, EPI_ISL_1458930, EPI_ISL_1458931, EPI_ISL_1458932, EPI_ISL_1458933, EPI_ISL_1458934, EPI_ISL_1458935, EPI_ISL_1458936, EPI_ISL_1458937, EPI_ISL_1458938, EPI_ISL_1458939, EPI_ISL_1458965, EPI_ISL_1458968, EPI_ISL_1458970,
EPI_ISL_1458971, EPI_ISL_1458972, EPI_ISL_1458973, EPI_ISL_1458974, EPI_ISL_1458975, EPI_ISL_1458976, EPI_ISL_1458977, EPI_ISL_1458978, EPI_ISL_1458979, EPI_ISL_1459207, EPI_ISL_1459208, EPI_ISL_1459209, EPI_ISL_1459256, EPI_ISL_1459257, EPI_ISL_1459258, EPI_ISL_1459259, EPI_ISL_1459260, EPI_ISL_1459261, EPI_ISL_1459262, EPI_ISL_1459263, EPI_ISL_1459264, EPI_ISL_1459265,
EPI_ISL_1459266, EPI_ISL_1459267, EPI_ISL_1459268, EPI_ISL_1459269, EPI_ISL_1459292, EPI_ISL_1459293, EPI_ISL_1459294, EPI_ISL_1459295, EPI_ISL_1459296, EPI_ISL_1459297, EPI_ISL_1459298, EPI_ISL_1459299, EPI_ISL_1459300, EPI_ISL_1459301, EPI_ISL_1459302, EPI_ISL_1459303, EPI_ISL_1459304, EPI_ISL_1459305, EPI_ISL_1459306, EPI_ISL_1459307, EPI_ISL_1459308, EPI_ISL_1459309,
EPI_ISL_1459310, EPI_ISL_1459311, EPI_ISL_1459312, EPI_ISL_1459313, EPI_ISL_1459314, EPI_ISL_1459315, EPI_ISL_1459316, EPI_ISL_1459317, EPI_ISL_1459318, EPI_ISL_1459319, EPI_ISL_1459320, EPI_ISL_1459321, EPI_ISL_1459322, EPI_ISL_1459323, EPI_ISL_1459324, EPI_ISL_1459325, EPI_ISL_1459326, EPI_ISL_1459327, EPI_ISL_1459328, EPI_ISL_1459329, EPI_ISL_1459330, EPI_ISL_1459331,
EPI_ISL_1459332, EPI_ISL_1459333, EPI_ISL_1459334, EPI_ISL_1459335, EPI_ISL_1459336, EPI_ISL_1459337, EPI_ISL_1459338, EPI_ISL_1459339, EPI_ISL_1459340, EPI_ISL_1459341, EPI_ISL_1459342, EPI_ISL_1459343, EPI_ISL_1459344, EPI_ISL_1459345, EPI_ISL_1459346, EPI_ISL_1459347, EPI_ISL_1459348, EPI_ISL_1459349, EPI_ISL_1459350, EPI_ISL_1459351, EPI_ISL_1459352, EPI_ISL_1459353,
EPI_ISL_1459354, EPI_ISL_1459355, EPI_ISL_1459356, EPI_ISL_1459357, EPI_ISL_1459358, EPI_ISL_1459359, EPI_ISL_1459360, EPI_ISL_1459361, EPI_ISL_1459362, EPI_ISL_1459363, EPI_ISL_1459364, EPI_ISL_1459365, EPI_ISL_1459366, EPI_ISL_1459367, EPI_ISL_1459368, EPI_ISL_1459369, EPI_ISL_1459370, EPI_ISL_1459371, EPI_ISL_1459372, EPI_ISL_1459373, EPI_ISL_1459374, EPI_ISL_1459375,
EPI_ISL_1459376, EPI_ISL_1459377, EPI_ISL_1459378, EPI_ISL_1459379, EPI_ISL_1459381, EPI_ISL_1459382, EPI_ISL_1459383, EPI_ISL_1459384, EPI_ISL_1459385, EPI_ISL_1459386, EPI_ISL_1459387, EPI_ISL_1459388, EPI_ISL_1459389, EPI_ISL_1459390, EPI_ISL_1459391, EPI_ISL_1459392, EPI_ISL_1459393, EPI_ISL_1459394, EPI_ISL_1459395, EPI_ISL_1459396, EPI_ISL_1459397, EPI_ISL_1459398,
EPI_ISL_1459399, EPI_ISL_1459400, EPI_ISL_1459401, EPI_ISL_1459402, EPI_ISL_1459403, EPI_ISL_1459404, EPI_ISL_1459405, EPI_ISL_1459406, EPI_ISL_1459407, EPI_ISL_1459408, EPI_ISL_1459409, EPI_ISL_1459410, EPI_ISL_1459411, EPI_ISL_1459412, EPI_ISL_1459413, EPI_ISL_1459414, EPI_ISL_1459415, EPI_ISL_1459416, EPI_ISL_1459417, EPI_ISL_1459418, EPI_ISL_1459419, EPI_ISL_1459420,
EPI_ISL_1459421, EPI_ISL_1459422, EPI_ISL_1459423, EPI_ISL_1459424, EPI_ISL_1459425, EPI_ISL_1459426, EPI_ISL_1459427, EPI_ISL_1459428, EPI_ISL_1459429, EPI_ISL_1459430, EPI_ISL_1459431, EPI_ISL_1459432, EPI_ISL_1459433, EPI_ISL_1459434, EPI_ISL_1459435, EPI_ISL_1459436, EPI_ISL_1459437, EPI_ISL_1459438, EPI_ISL_1459439, EPI_ISL_1459440, EPI_ISL_1459441, EPI_ISL_1459442,
EPI_ISL_1459443, EPI_ISL_1459444, EPI_ISL_1459445, EPI_ISL_1459446, EPI_ISL_1459447, EPI_ISL_1459448, EPI_ISL_1459449, EPI_ISL_1459450, EPI_ISL_1459451, EPI_ISL_1459452, EPI_ISL_1459453, EPI_ISL_1459454, EPI_ISL_1459455, EPI_ISL_1459456, EPI_ISL_1459457, EPI_ISL_1459458, EPI_ISL_1459459, EPI_ISL_1459460, EPI_ISL_1459461, EPI_ISL_1459462, EPI_ISL_1459463, EPI_ISL_1459464,
EPI_ISL_1459465, EPI_ISL_1459466, EPI_ISL_1459467, EPI_ISL_1459468, EPI_ISL_1459469, EPI_ISL_1459470, EPI_ISL_1459471, EPI_ISL_1459472, EPI_ISL_1459473, EPI_ISL_1459474, EPI_ISL_1459475, EPI_ISL_1459476, EPI_ISL_1459477, EPI_ISL_1459478, EPI_ISL_1459479, EPI_ISL_1459480, EPI_ISL_1459481, EPI_ISL_1459482, EPI_ISL_1459483, EPI_ISL_1459484, EPI_ISL_1459485, EPI_ISL_1459486,
EPI_ISL_1459487, EPI_ISL_1459488, EPI_ISL_1459489, EPI_ISL_1459490, EPI_ISL_1459491, EPI_ISL_1459492, EPI_ISL_1459493, EPI_ISL_1459494, EPI_ISL_1459495, EPI_ISL_1459496, EPI_ISL_1459497, EPI_ISL_1459498, EPI_ISL_1459499, EPI_ISL_1459500, EPI_ISL_1459501, EPI_ISL_1459502, EPI_ISL_1459503, EPI_ISL_1459504, EPI_ISL_1459505, EPI_ISL_1459506, EPI_ISL_1459507, EPI_ISL_1459508,
EPI_ISL_1459529, EPI_ISL_1459530, EPI_ISL_1459531, EPI_ISL_1459532, EPI_ISL_1459533, EPI_ISL_1459534, EPI_ISL_1459535, EPI_ISL_1459536, EPI_ISL_1459537, EPI_ISL_1459538, EPI_ISL_1459539, EPI_ISL_1459540, EPI_ISL_1459541, EPI_ISL_1459542, EPI_ISL_1459543, EPI_ISL_1459544, EPI_ISL_1459545, EPI_ISL_1459546, EPI_ISL_1459547, EPI_ISL_1459548, EPI_ISL_1459549, EPI_ISL_1459550,
EPI_ISL_1459551, EPI_ISL_1459552, EPI_ISL_1459553, EPI_ISL_1459554, EPI_ISL_1459555, EPI_ISL_1459556, EPI_ISL_1459557, EPI_ISL_1459558, EPI_ISL_1459559, EPI_ISL_1459560, EPI_ISL_1459561, EPI_ISL_1459562, EPI_ISL_1459563, EPI_ISL_1459564, EPI_ISL_1459565, EPI_ISL_1459566, EPI_ISL_1459567, EPI_ISL_1459568, EPI_ISL_1459569, EPI_ISL_1459570, EPI_ISL_1459571, EPI_ISL_1459572,
EPI_ISL_1459573, EPI_ISL_1459574, EPI_ISL_1459575, EPI_ISL_1459576, EPI_ISL_1459577, EPI_ISL_1459578, EPI_ISL_1459579, EPI_ISL_1459580, EPI_ISL_1459581, EPI_ISL_1459582, EPI_ISL_1459583, EPI_ISL_1459584, EPI_ISL_1459585, EPI_ISL_1459586, EPI_ISL_1459587, EPI_ISL_1459588, EPI_ISL_1459589, EPI_ISL_1459590, EPI_ISL_1459591, EPI_ISL_1459592, EPI_ISL_1459593, EPI_ISL_1459594,
EPI_ISL_1459595, EPI_ISL_1459596, EPI_ISL_1459597, EPI_ISL_1459598, EPI_ISL_1459599, EPI_ISL_1459600, EPI_ISL_1459601, EPI_ISL_1459602, EPI_ISL_1459603, EPI_ISL_1459604, EPI_ISL_1459605, EPI_ISL_1459606, EPI_ISL_1459607, EPI_ISL_1459608, EPI_ISL_1459609, EPI_ISL_1459610, EPI_ISL_1459611, EPI_ISL_1459612, EPI_ISL_1459613, EPI_ISL_1459614, EPI_ISL_1459615, EPI_ISL_1459616,
EPI_ISL_1459617, EPI_ISL_1459618, EPI_ISL_1459619, EPI_ISL_1459620, EPI_ISL_1459621, EPI_ISL_1459622, EPI_ISL_1459623, EPI_ISL_1459624, EPI_ISL_1459625, EPI_ISL_1459626, EPI_ISL_1459627, EPI_ISL_1459628, EPI_ISL_1459629, EPI_ISL_1459630, EPI_ISL_1459631, EPI_ISL_1459632, EPI_ISL_1459633, EPI_ISL_1459634, EPI_ISL_1459635, EPI_ISL_1459636, EPI_ISL_1459637, EPI_ISL_1459638,
EPI_ISL_1459639, EPI_ISL_1459640, EPI_ISL_1459641, EPI_ISL_1459642, EPI_ISL_1459643, EPI_ISL_1459644, EPI_ISL_1459645, EPI_ISL_1459646, EPI_ISL_1459647, EPI_ISL_1459648, EPI_ISL_1459649, EPI_ISL_1459650, EPI_ISL_1459651, EPI_ISL_1459652, EPI_ISL_1459653, EPI_ISL_1459654, EPI_ISL_1459655, EPI_ISL_1459656, EPI_ISL_1459657, EPI_ISL_1459658, EPI_ISL_1459659, EPI_ISL_1459660,
EPI_ISL_1459661, EPI_ISL_1459662, EPI_ISL_1459663, EPI_ISL_1459664, EPI_ISL_1459665, EPI_ISL_1459666, EPI_ISL_1459667, EPI_ISL_1459668, EPI_ISL_1459669, EPI_ISL_1459670, EPI_ISL_1459671, EPI_ISL_1459672, EPI_ISL_1459673, EPI_ISL_1459674, EPI_ISL_1459675, EPI_ISL_1459676, EPI_ISL_1459677, EPI_ISL_1459678, EPI_ISL_1459679, EPI_ISL_1459680, EPI_ISL_1459681, EPI_ISL_1459682,
EPI_ISL_1459683, EPI_ISL_1459684, EPI_ISL_1459685, EPI_ISL_1459686, EPI_ISL_1459687, EPI_ISL_1459688, EPI_ISL_1459689, EPI_ISL_1459690, EPI_ISL_1459691, EPI_ISL_1459692, EPI_ISL_1459693, EPI_ISL_1459694, EPI_ISL_1459695, EPI_ISL_1459696, EPI_ISL_1459697, EPI_ISL_1459698, EPI_ISL_1459699, EPI_ISL_1459700, EPI_ISL_1459701, EPI_ISL_1459702, EPI_ISL_1459703, EPI_ISL_1459704,
EPI_ISL_1459705, EPI_ISL_1459706, EPI_ISL_1459707, EPI_ISL_1459708, EPI_ISL_1459709, EPI_ISL_1459710, EPI_ISL_1459711, EPI_ISL_1459712, EPI_ISL_1459713, EPI_ISL_1459714, EPI_ISL_1459715, EPI_ISL_1459716, EPI_ISL_1459717, EPI_ISL_1459718, EPI_ISL_1459719, EPI_ISL_1459720, EPI_ISL_1459721, EPI_ISL_1459722, EPI_ISL_1459723, EPI_ISL_1459724, EPI_ISL_1459725, EPI_ISL_1459726,
EPI_ISL_1459727, EPI_ISL_1459728, EPI_ISL_1459729, EPI_ISL_1459730, EPI_ISL_1459731, EPI_ISL_1459732, EPI_ISL_1459733, EPI_ISL_1459734, EPI_ISL_1459735, EPI_ISL_1459736, EPI_ISL_1459737, EPI_ISL_1459738, EPI_ISL_1459739, EPI_ISL_1459740, EPI_ISL_1459741, EPI_ISL_1459742, EPI_ISL_1459743, EPI_ISL_1459744, EPI_ISL_1459745, EPI_ISL_1459746, EPI_ISL_1459747, EPI_ISL_1459748,
EPI_ISL_1459749, EPI_ISL_1459750, EPI_ISL_1459751, EPI_ISL_1459752, EPI_ISL_1459753, EPI_ISL_1459754, EPI_ISL_1459755, EPI_ISL_1459756, EPI_ISL_1459757, EPI_ISL_1459758, EPI_ISL_1459759, EPI_ISL_1459760, EPI_ISL_1459761, EPI_ISL_1459762, EPI_ISL_1459763, EPI_ISL_1459764, EPI_ISL_1459765, EPI_ISL_1459766, EPI_ISL_1459767, EPI_ISL_1459768, EPI_ISL_1459769, EPI_ISL_1459771,
EPI_ISL_1459772, EPI_ISL_1459773, EPI_ISL_1459774, EPI_ISL_1459775, EPI_ISL_1459776, EPI_ISL_1459777, EPI_ISL_1459778, EPI_ISL_1459779, EPI_ISL_1459780, EPI_ISL_1459781, EPI_ISL_1459782, EPI_ISL_1459783, EPI_ISL_1459784, EPI_ISL_1459785, EPI_ISL_1459786, EPI_ISL_1459787, EPI_ISL_1459788, EPI_ISL_1459789, EPI_ISL_1459790, EPI_ISL_1459791, EPI_ISL_1459792, EPI_ISL_1459793,
EPI_ISL_1459794, EPI_ISL_1459795, EPI_ISL_1459796, EPI_ISL_1459797, EPI_ISL_1459798, EPI_ISL_1459799, EPI_ISL_1459800, EPI_ISL_1459801, EPI_ISL_1459802, EPI_ISL_1459803, EPI_ISL_1459804, EPI_ISL_1459805, EPI_ISL_1459806, EPI_ISL_1459807, EPI_ISL_1459808, EPI_ISL_1459809, EPI_ISL_1459810, EPI_ISL_1459811, EPI_ISL_1459812, EPI_ISL_1459813, EPI_ISL_1459814, EPI_ISL_1459815,
EPI_ISL_1459816, EPI_ISL_1459817, EPI_ISL_1459818, EPI_ISL_1459819, EPI_ISL_1459820, EPI_ISL_1459821, EPI_ISL_1459822, EPI_ISL_1459823, EPI_ISL_1459824, EPI_ISL_1459825, EPI_ISL_1459826, EPI_ISL_1459827, EPI_ISL_1459828, EPI_ISL_1459829, EPI_ISL_1459830, EPI_ISL_1459831, EPI_ISL_1459832, EPI_ISL_1459833, EPI_ISL_1459834, EPI_ISL_1459835, EPI_ISL_1459836, EPI_ISL_1459837,
EPI_ISL_1459838, EPI_ISL_1459839, EPI_ISL_1459840, EPI_ISL_1459842, EPI_ISL_1459843, EPI_ISL_1459844, EPI_ISL_1459845, EPI_ISL_1459846, EPI_ISL_1459848, EPI_ISL_1459849, EPI_ISL_1459850, EPI_ISL_1459851, EPI_ISL_1459852, EPI_ISL_1459853, EPI_ISL_1459854, EPI_ISL_1459855, EPI_ISL_1459856, EPI_ISL_1459857, EPI_ISL_1459858, EPI_ISL_1459859, EPI_ISL_1459860, EPI_ISL_1459861,
EPI_ISL_1459862, EPI_ISL_1459863, EPI_ISL_1459864, EPI_ISL_1459865, EPI_ISL_1459866, EPI_ISL_1459867, EPI_ISL_1459868, EPI_ISL_1459869, EPI_ISL_1459870, EPI_ISL_1459871, EPI_ISL_1459872, EPI_ISL_1459873, EPI_ISL_1459874, EPI_ISL_1459875, EPI_ISL_1459876, EPI_ISL_1459877, EPI_ISL_1459878, EPI_ISL_1459879, EPI_ISL_1459880, EPI_ISL_1459881, EPI_ISL_1459882, EPI_ISL_1459883,
EPI_ISL_1459884, EPI_ISL_1459885, EPI_ISL_1459886, EPI_ISL_1459887, EPI_ISL_1459888, EPI_ISL_1459889, EPI_ISL_1459890, EPI_ISL_1459891, EPI_ISL_1459892, EPI_ISL_1459893, EPI_ISL_1459894, EPI_ISL_1459895, EPI_ISL_1459896, EPI_ISL_1459897, EPI_ISL_1459898, EPI_ISL_1459899, EPI_ISL_1459900, EPI_ISL_1459901, EPI_ISL_1459902, EPI_ISL_1459903, EPI_ISL_1459904, EPI_ISL_1459905,
EPI_ISL_1459906, EPI_ISL_1459907, EPI_ISL_1459908, EPI_ISL_1459909, EPI_ISL_1459910, EPI_ISL_1459911, EPI_ISL_1459912, EPI_ISL_1459913, EPI_ISL_1459914, EPI_ISL_1459915, EPI_ISL_1459916, EPI_ISL_1459917, EPI_ISL_1459918, EPI_ISL_1459919, EPI_ISL_1459920, EPI_ISL_1459921, EPI_ISL_1459922, EPI_ISL_1459923, EPI_ISL_1459924, EPI_ISL_1459925, EPI_ISL_1459926, EPI_ISL_1459927,
EPI_ISL_1459928, EPI_ISL_1459929, EPI_ISL_1459930, EPI_ISL_1459931, EPI_ISL_1459932, EPI_ISL_1459933, EPI_ISL_1459934, EPI_ISL_1459935, EPI_ISL_1459936, EPI_ISL_1459937, EPI_ISL_1459938, EPI_ISL_1459939, EPI_ISL_1459940, EPI_ISL_1459941, EPI_ISL_1459942, EPI_ISL_1459943, EPI_ISL_1459944, EPI_ISL_1459945, EPI_ISL_1459946, EPI_ISL_1459947, EPI_ISL_1459948, EPI_ISL_1459949,
EPI_ISL_1459950, EPI_ISL_1459951, EPI_ISL_1459952, EPI_ISL_1459953, EPI_ISL_1459954, EPI_ISL_1459955, EPI_ISL_1459956, EPI_ISL_1459957, EPI_ISL_1459958, EPI_ISL_1459959, EPI_ISL_1459960, EPI_ISL_1459961, EPI_ISL_1459962, EPI_ISL_1459963, EPI_ISL_1459964, EPI_ISL_1459965, EPI_ISL_1459966, EPI_ISL_1459967, EPI_ISL_1459968, EPI_ISL_1459969, EPI_ISL_1459970, EPI_ISL_1459971,
EPI_ISL_1459972, EPI_ISL_1459973, EPI_ISL_1459974, EPI_ISL_1459975, EPI_ISL_1459976, EPI_ISL_1459977, EPI_ISL_1459978, EPI_ISL_1459979, EPI_ISL_1459980, EPI_ISL_1459981, EPI_ISL_1459982, EPI_ISL_1459983, EPI_ISL_1459984, EPI_ISL_1459985, EPI_ISL_1459986, EPI_ISL_1459987, EPI_ISL_1459988, EPI_ISL_1459989, EPI_ISL_1459990, EPI_ISL_1459991, EPI_ISL_1459992, EPI_ISL_1459993,
EPI_ISL_1459994, EPI_ISL_1459995, EPI_ISL_1459996, EPI_ISL_1459997, EPI_ISL_1459998, EPI_ISL_1459999, EPI_ISL_1460000, EPI_ISL_1460001, EPI_ISL_1460002, EPI_ISL_1460003, EPI_ISL_1460004, EPI_ISL_1460005, EPI_ISL_1460006, EPI_ISL_1460007, EPI_ISL_1460008, EPI_ISL_1460009, EPI_ISL_1460010, EPI_ISL_1460011, EPI_ISL_1460012, EPI_ISL_1460013, EPI_ISL_1460014, EPI_ISL_1460015,
EPI_ISL_1460016, EPI_ISL_1460017, EPI_ISL_1460018, EPI_ISL_1460019, EPI_ISL_1460020, EPI_ISL_1460021, EPI_ISL_1460022, EPI_ISL_1460023, EPI_ISL_1460024, EPI_ISL_1460025, EPI_ISL_1460026, EPI_ISL_1460027, EPI_ISL_1460028, EPI_ISL_1460029, EPI_ISL_1460030, EPI_ISL_1460031, EPI_ISL_1460032, EPI_ISL_1460033, EPI_ISL_1460034, EPI_ISL_1460035, EPI_ISL_1460036, EPI_ISL_1460037,
EPI_ISL_1460038, EPI_ISL_1460039, EPI_ISL_1460040, EPI_ISL_1460041, EPI_ISL_1460042, EPI_ISL_1460043, EPI_ISL_1460044, EPI_ISL_1460045, EPI_ISL_1460046, EPI_ISL_1460047, EPI_ISL_1460048, EPI_ISL_1460049, EPI_ISL_1460050, EPI_ISL_1460051, EPI_ISL_1460052, EPI_ISL_1460053, EPI_ISL_1460054, EPI_ISL_1460055, EPI_ISL_1460056, EPI_ISL_1460057, EPI_ISL_1460058, EPI_ISL_1460059,
EPI_ISL_1460060, EPI_ISL_1460061, EPI_ISL_1460062, EPI_ISL_1460063, EPI_ISL_1460064, EPI_ISL_1460065, EPI_ISL_1460066, EPI_ISL_1460067, EPI_ISL_1460068, EPI_ISL_1460069, EPI_ISL_1460070, EPI_ISL_1460071, EPI_ISL_1460072, EPI_ISL_1460073, EPI_ISL_1460074, EPI_ISL_1460075, EPI_ISL_1460076, EPI_ISL_1460077, EPI_ISL_1460078, EPI_ISL_1460079, EPI_ISL_1460080, EPI_ISL_1460081,
EPI_ISL_1460082, EPI_ISL_1460083, EPI_ISL_1460084, EPI_ISL_1460085, EPI_ISL_1460086, EPI_ISL_1460087, EPI_ISL_1460088, EPI_ISL_1460089, EPI_ISL_1460090, EPI_ISL_1460091, EPI_ISL_1460092, EPI_ISL_1460093, EPI_ISL_1460094, EPI_ISL_1460095, EPI_ISL_1460096, EPI_ISL_1460097, EPI_ISL_1460098, EPI_ISL_1460099, EPI_ISL_1460100, EPI_ISL_1460101, EPI_ISL_1460102, EPI_ISL_1460103,
EPI_ISL_1460104, EPI_ISL_1460105, EPI_ISL_1460106, EPI_ISL_1460107, EPI_ISL_1460108, EPI_ISL_1460109, EPI_ISL_1460110, EPI_ISL_1460111, EPI_ISL_1460112, EPI_ISL_1460113, EPI_ISL_1460114, EPI_ISL_1460115, EPI_ISL_1460116, EPI_ISL_1460117, EPI_ISL_1460118, EPI_ISL_1460119, EPI_ISL_1460120, EPI_ISL_1460121, EPI_ISL_1460123, EPI_ISL_1460124, EPI_ISL_1460125, EPI_ISL_1460126,
EPI_ISL_1460127, EPI_ISL_1460128, EPI_ISL_1460129, EPI_ISL_1460130, EPI_ISL_1460131, EPI_ISL_1460132, EPI_ISL_1460133, EPI_ISL_1460134, EPI_ISL_1460135, EPI_ISL_1460136, EPI_ISL_1460137, EPI_ISL_1460138, EPI_ISL_1460139, EPI_ISL_1460140, EPI_ISL_1460141, EPI_ISL_1460142, EPI_ISL_1460143, EPI_ISL_1460144, EPI_ISL_1460145, EPI_ISL_1460146, EPI_ISL_1460147, EPI_ISL_1460148,
EPI_ISL_1460149, EPI_ISL_1460150, EPI_ISL_1460151, EPI_ISL_1460152, EPI_ISL_1460153, EPI_ISL_1460154, EPI_ISL_1460155, EPI_ISL_1460156, EPI_ISL_1460157, EPI_ISL_1460158, EPI_ISL_1460159, EPI_ISL_1460160, EPI_ISL_1460161, EPI_ISL_1460162, EPI_ISL_1460163, EPI_ISL_1460164, EPI_ISL_1460165, EPI_ISL_1460166, EPI_ISL_1460167, EPI_ISL_1460168, EPI_ISL_1460169, EPI_ISL_1460170,
EPI_ISL_1460171, EPI_ISL_1460172, EPI_ISL_1460173, EPI_ISL_1460174, EPI_ISL_1460175, EPI_ISL_1460176, EPI_ISL_1460177, EPI_ISL_1460178, EPI_ISL_1460179, EPI_ISL_1460180, EPI_ISL_1460181, EPI_ISL_1460182, EPI_ISL_1460183, EPI_ISL_1460184, EPI_ISL_1460185, EPI_ISL_1460186, EPI_ISL_1460187, EPI_ISL_1460188, EPI_ISL_1460189, EPI_ISL_1460190, EPI_ISL_1460191, EPI_ISL_1460192,
EPI_ISL_1460193, EPI_ISL_1460194, EPI_ISL_1460195, EPI_ISL_1460196, EPI_ISL_1460197, EPI_ISL_1460198, EPI_ISL_1460199, EPI_ISL_1460200, EPI_ISL_1460201, EPI_ISL_1460202, EPI_ISL_1460203, EPI_ISL_1460204, EPI_ISL_1460205, EPI_ISL_1460206, EPI_ISL_1460207, EPI_ISL_1460208, EPI_ISL_1460209, EPI_ISL_1460210, EPI_ISL_1460211, EPI_ISL_1460212, EPI_ISL_1460213, EPI_ISL_1460214,
EPI_ISL_1460263, EPI_ISL_1460295, EPI_ISL_1460296, EPI_ISL_1460297, EPI_ISL_1460298, EPI_ISL_1460299, EPI_ISL_1460300, EPI_ISL_1460301, EPI_ISL_1460302, EPI_ISL_1460303, EPI_ISL_1460304, EPI_ISL_1460305, EPI_ISL_1460306, EPI_ISL_1460307, EPI_ISL_1460308, EPI_ISL_1460309, EPI_ISL_1460310, EPI_ISL_1460311, EPI_ISL_1460312, EPI_ISL_1460313, EPI_ISL_1460314, EPI_ISL_1460315,
EPI_ISL_1460316, EPI_ISL_1460317, EPI_ISL_1460318, EPI_ISL_1460319, EPI_ISL_1460320, EPI_ISL_1460321, EPI_ISL_1460322, EPI_ISL_1460323, EPI_ISL_1460324, EPI_ISL_1460325, EPI_ISL_1460326, EPI_ISL_1460327, EPI_ISL_1460328, EPI_ISL_1460329, EPI_ISL_1460330, EPI_ISL_1460331, EPI_ISL_1460332, EPI_ISL_1460333, EPI_ISL_1460334, EPI_ISL_1460335, EPI_ISL_1460336, EPI_ISL_1460337,
EPI_ISL_1460338, EPI_ISL_1460339, EPI_ISL_1460340, EPI_ISL_1460341, EPI_ISL_1460342, EPI_ISL_1460343, EPI_ISL_1460344, EPI_ISL_1460345, EPI_ISL_1460346, EPI_ISL_1460347, EPI_ISL_1460348, EPI_ISL_1460349, EPI_ISL_1460350, EPI_ISL_1460351, EPI_ISL_1460352, EPI_ISL_1460353, EPI_ISL_1460356, EPI_ISL_1460357, EPI_ISL_1460358, EPI_ISL_1460359, EPI_ISL_1460360, EPI_ISL_1460361,
EPI_ISL_1460362, EPI_ISL_1460363, EPI_ISL_1460364, EPI_ISL_1460365, EPI_ISL_1460366, EPI_ISL_1460367, EPI_ISL_1460368, EPI_ISL_1460369, EPI_ISL_1460370, EPI_ISL_1460371, EPI_ISL_1460372, EPI_ISL_1460373, EPI_ISL_1460374, EPI_ISL_1460375, EPI_ISL_1460376, EPI_ISL_1460377, EPI_ISL_1460378, EPI_ISL_1460379, EPI_ISL_1460380, EPI_ISL_1460381, EPI_ISL_1460382, EPI_ISL_1460383,
EPI_ISL_1460384, EPI_ISL_1460385, EPI_ISL_1460386, EPI_ISL_1460387, EPI_ISL_1460388, EPI_ISL_1460389, EPI_ISL_1460390, EPI_ISL_1460391, EPI_ISL_1460392, EPI_ISL_1460393, EPI_ISL_1460394, EPI_ISL_1460395, EPI_ISL_1460396, EPI_ISL_1460397, EPI_ISL_1460398, EPI_ISL_1460399, EPI_ISL_1460400, EPI_ISL_1460401, EPI_ISL_1460402, EPI_ISL_1460403, EPI_ISL_1460404, EPI_ISL_1460405,
EPI_ISL_1460406, EPI_ISL_1460407, EPI_ISL_1460408, EPI_ISL_1460409, EPI_ISL_1460410, EPI_ISL_1460411, EPI_ISL_1460412, EPI_ISL_1460413, EPI_ISL_1460414, EPI_ISL_1460415, EPI_ISL_1460416, EPI_ISL_1460417, EPI_ISL_1460418, EPI_ISL_1460419, EPI_ISL_1460420, EPI_ISL_1460421, EPI_ISL_1460422, EPI_ISL_1460423, EPI_ISL_1460424, EPI_ISL_1460425, EPI_ISL_1460426, EPI_ISL_1460427,
EPI_ISL_1460428, EPI_ISL_1460429, EPI_ISL_1460430, EPI_ISL_1460431, EPI_ISL_1460432, EPI_ISL_1460433, EPI_ISL_1460434, EPI_ISL_1460435, EPI_ISL_1460436, EPI_ISL_1460437, EPI_ISL_1460438, EPI_ISL_1460439, EPI_ISL_1460440, EPI_ISL_1460442, EPI_ISL_1460443, EPI_ISL_1460444, EPI_ISL_1460445, EPI_ISL_1460446, EPI_ISL_1460447, EPI_ISL_1460448, EPI_ISL_1460449, EPI_ISL_1460450,
EPI_ISL_1460451, EPI_ISL_1460452, EPI_ISL_1460453, EPI_ISL_1460454, EPI_ISL_1460455, EPI_ISL_1460456, EPI_ISL_1460457, EPI_ISL_1460458, EPI_ISL_1460459, EPI_ISL_1460460, EPI_ISL_1460461, EPI_ISL_1460462, EPI_ISL_1460463, EPI_ISL_1460464, EPI_ISL_1460465, EPI_ISL_1460466, EPI_ISL_1460467, EPI_ISL_1460468, EPI_ISL_1460469, EPI_ISL_1460470, EPI_ISL_1460471, EPI_ISL_1460472,
EPI_ISL_1460473, EPI_ISL_1460474, EPI_ISL_1460475, EPI_ISL_1460476, EPI_ISL_1460477, EPI_ISL_1460478, EPI_ISL_1460479, EPI_ISL_1460480, EPI_ISL_1460481, EPI_ISL_1460482, EPI_ISL_1460483, EPI_ISL_1460484, EPI_ISL_1460485, EPI_ISL_1460486, EPI_ISL_1460487, EPI_ISL_1460488, EPI_ISL_1460489, EPI_ISL_1460490, EPI_ISL_1460491, EPI_ISL_1460492, EPI_ISL_1460493, EPI_ISL_1460494,
EPI_ISL_1460495, EPI_ISL_1460496, EPI_ISL_1460497, EPI_ISL_1460498, EPI_ISL_1460499, EPI_ISL_1460500, EPI_ISL_1460501, EPI_ISL_1460502, EPI_ISL_1460503, EPI_ISL_1460504, EPI_ISL_1460505, EPI_ISL_1460506, EPI_ISL_1460507, EPI_ISL_1460508, EPI_ISL_1460509, EPI_ISL_1460510, EPI_ISL_1460511, EPI_ISL_1460512, EPI_ISL_1460513, EPI_ISL_1460514, EPI_ISL_1460515, EPI_ISL_1460516,
EPI_ISL_1460517, EPI_ISL_1460518, EPI_ISL_1460519, EPI_ISL_1460520, EPI_ISL_1460521, EPI_ISL_1460522, EPI_ISL_1460523, EPI_ISL_1460524, EPI_ISL_1460525, EPI_ISL_1460526, EPI_ISL_1460527, EPI_ISL_1460528, EPI_ISL_1460529, EPI_ISL_1460530, EPI_ISL_1460531, EPI_ISL_1460532, EPI_ISL_1460533, EPI_ISL_1460534, EPI_ISL_1460535, EPI_ISL_1460536, EPI_ISL_1460537, EPI_ISL_1460538,
EPI_ISL_1460539, EPI_ISL_1460540, EPI_ISL_1460541, EPI_ISL_1460542, EPI_ISL_1460543, EPI_ISL_1460544, EPI_ISL_1460545, EPI_ISL_1460546, EPI_ISL_1460547, EPI_ISL_1460548, EPI_ISL_1460549, EPI_ISL_1460550, EPI_ISL_1460551, EPI_ISL_1460552, EPI_ISL_1460553, EPI_ISL_1460554, EPI_ISL_1460555, EPI_ISL_1460556, EPI_ISL_1460557, EPI_ISL_1460558, EPI_ISL_1460559, EPI_ISL_1460560,
EPI_ISL_1460561, EPI_ISL_1460562, EPI_ISL_1460563, EPI_ISL_1460564, EPI_ISL_1460565, EPI_ISL_1460566, EPI_ISL_1460567, EPI_ISL_1460568, EPI_ISL_1460569, EPI_ISL_1460570, EPI_ISL_1460571, EPI_ISL_1460572, EPI_ISL_1460573, EPI_ISL_1460574, EPI_ISL_1460575, EPI_ISL_1460576, EPI_ISL_1460577, EPI_ISL_1460578, EPI_ISL_1460580, EPI_ISL_1460581, EPI_ISL_1460582, EPI_ISL_1460583,
EPI_ISL_1460584, EPI_ISL_1460585, EPI_ISL_1460586, EPI_ISL_1460587, EPI_ISL_1460588, EPI_ISL_1460589, EPI_ISL_1460590, EPI_ISL_1460591, EPI_ISL_1460592, EPI_ISL_1460593, EPI_ISL_1460594, EPI_ISL_1460595, EPI_ISL_1460596, EPI_ISL_1460597, EPI_ISL_1460598, EPI_ISL_1460599, EPI_ISL_1460600, EPI_ISL_1460601, EPI_ISL_1460602, EPI_ISL_1460603, EPI_ISL_1460604, EPI_ISL_1460605,
EPI_ISL_1460606, EPI_ISL_1460607, EPI_ISL_1460608, EPI_ISL_1460609, EPI_ISL_1460610, EPI_ISL_1460611, EPI_ISL_1460612, EPI_ISL_1460613, EPI_ISL_1460614, EPI_ISL_1460615, EPI_ISL_1460616, EPI_ISL_1460617, EPI_ISL_1460618, EPI_ISL_1460619, EPI_ISL_1460620, EPI_ISL_1460621, EPI_ISL_1460622, EPI_ISL_1460623, EPI_ISL_1460624, EPI_ISL_1460625, EPI_ISL_1460626, EPI_ISL_1460627,
EPI_ISL_1460628, EPI_ISL_1460629, EPI_ISL_1460630, EPI_ISL_1460631, EPI_ISL_1460632, EPI_ISL_1460633, EPI_ISL_1460634, EPI_ISL_1460635, EPI_ISL_1460636, EPI_ISL_1460637, EPI_ISL_1460638, EPI_ISL_1460639, EPI_ISL_1460640, EPI_ISL_1460641, EPI_ISL_1460642, EPI_ISL_1460643, EPI_ISL_1460644, EPI_ISL_1460645, EPI_ISL_1460646, EPI_ISL_1460647, EPI_ISL_1460648, EPI_ISL_1460649,
EPI_ISL_1460650, EPI_ISL_1460651, EPI_ISL_1460652, EPI_ISL_1460653, EPI_ISL_1460654, EPI_ISL_1460655, EPI_ISL_1460656, EPI_ISL_1460657, EPI_ISL_1460658, EPI_ISL_1460659, EPI_ISL_1460660, EPI_ISL_1460661, EPI_ISL_1460662, EPI_ISL_1460663, EPI_ISL_1460664, EPI_ISL_1460665, EPI_ISL_1460666, EPI_ISL_1460667, EPI_ISL_1460668, EPI_ISL_1460669, EPI_ISL_1460670, EPI_ISL_1460671,
EPI_ISL_1460672, EPI_ISL_1460673, EPI_ISL_1460674, EPI_ISL_1460675, EPI_ISL_1460676, EPI_ISL_1460677, EPI_ISL_1460678, EPI_ISL_1460679, EPI_ISL_1460680, EPI_ISL_1460681, EPI_ISL_1460682, EPI_ISL_1460683, EPI_ISL_1460684, EPI_ISL_1460685, EPI_ISL_1460686, EPI_ISL_1460687, EPI_ISL_1460688, EPI_ISL_1460689, EPI_ISL_1460690, EPI_ISL_1460691, EPI_ISL_1460692, EPI_ISL_1460693,
EPI_ISL_1460694, EPI_ISL_1460695, EPI_ISL_1460696, EPI_ISL_1460697, EPI_ISL_1460698, EPI_ISL_1460699, EPI_ISL_1460700, EPI_ISL_1460701, EPI_ISL_1460702, EPI_ISL_1460703, EPI_ISL_1460704, EPI_ISL_1460705, EPI_ISL_1460706, EPI_ISL_1460707, EPI_ISL_1460708, EPI_ISL_1460709, EPI_ISL_1460710, EPI_ISL_1460711, EPI_ISL_1460712, EPI_ISL_1460713, EPI_ISL_1460714, EPI_ISL_1460715,
EPI_ISL_1460716, EPI_ISL_1460717, EPI_ISL_1460718, EPI_ISL_1460719, EPI_ISL_1460720, EPI_ISL_1460721, EPI_ISL_1460722, EPI_ISL_1460723, EPI_ISL_1460724, EPI_ISL_1460725, EPI_ISL_1460726, EPI_ISL_1460727, EPI_ISL_1460728, EPI_ISL_1460729, EPI_ISL_1481550, EPI_ISL_1481551, EPI_ISL_1481552, EPI_ISL_1481553, EPI_ISL_1481555, EPI_ISL_1481556, EPI_ISL_1481557, EPI_ISL_1481558,
EPI_ISL_1481559, EPI_ISL_1481560, EPI_ISL_1481561, EPI_ISL_1481562, EPI_ISL_1481563, EPI_ISL_1481564, EPI_ISL_1481565, EPI_ISL_1481566, EPI_ISL_1481567, EPI_ISL_1481568, EPI_ISL_1481569, EPI_ISL_1481570, EPI_ISL_1481571, EPI_ISL_1481572, EPI_ISL_1481573, EPI_ISL_1481574, EPI_ISL_1481575, EPI_ISL_1481576, EPI_ISL_1481577, EPI_ISL_1481578, EPI_ISL_1481579, EPI_ISL_1481580,
EPI_ISL_1481581, EPI_ISL_1481582, EPI_ISL_1481583, EPI_ISL_1481584, EPI_ISL_1481585, EPI_ISL_1481586, EPI_ISL_1481587, EPI_ISL_1481588, EPI_ISL_1481589, EPI_ISL_1481590, EPI_ISL_1481591, EPI_ISL_1481592, EPI_ISL_1481593, EPI_ISL_1481594, EPI_ISL_1481595, EPI_ISL_1481596, EPI_ISL_1481597, EPI_ISL_1481679, EPI_ISL_1481716, EPI_ISL_1481717, EPI_ISL_1481718, EPI_ISL_1481719,
EPI_ISL_1481720, EPI_ISL_1481721, EPI_ISL_1481722, EPI_ISL_1481723, EPI_ISL_1481724, EPI_ISL_1481725, EPI_ISL_1481726, EPI_ISL_1481727, EPI_ISL_1481728, EPI_ISL_1481729, EPI_ISL_1481730, EPI_ISL_1481731, EPI_ISL_1481732, EPI_ISL_1481733, EPI_ISL_1481734, EPI_ISL_1481735, EPI_ISL_1481736, EPI_ISL_1481737, EPI_ISL_1481738, EPI_ISL_1481739, EPI_ISL_1481740, EPI_ISL_1481741,
EPI_ISL_1481742, EPI_ISL_1481743, EPI_ISL_1481744, EPI_ISL_1481745, EPI_ISL_1481746, EPI_ISL_1481747, EPI_ISL_1481748, EPI_ISL_1481749, EPI_ISL_1481750, EPI_ISL_1481751, EPI_ISL_1481752, EPI_ISL_1481753, EPI_ISL_1481754, EPI_ISL_1481755, EPI_ISL_1481756, EPI_ISL_1481757, EPI_ISL_1481758, EPI_ISL_1481759, EPI_ISL_1481760, EPI_ISL_1481761, EPI_ISL_1481762, EPI_ISL_1481763,
EPI_ISL_1481764, EPI_ISL_1481765, EPI_ISL_1481766, EPI_ISL_1481767, EPI_ISL_1481768, EPI_ISL_1481769, EPI_ISL_1481770, EPI_ISL_1481771, EPI_ISL_1481772, EPI_ISL_1481773, EPI_ISL_1481774, EPI_ISL_1481775, EPI_ISL_1481776, EPI_ISL_1481777, EPI_ISL_1481778, EPI_ISL_1481779, EPI_ISL_1481780, EPI_ISL_1481781, EPI_ISL_1481782, EPI_ISL_1481783, EPI_ISL_1481784, EPI_ISL_1481785,
EPI_ISL_1481786, EPI_ISL_1481787, EPI_ISL_1481788, EPI_ISL_1481789, EPI_ISL_1481790, EPI_ISL_1481791, EPI_ISL_1481792, EPI_ISL_1481793, EPI_ISL_1481794, EPI_ISL_1481795, EPI_ISL_1481796, EPI_ISL_1481797, EPI_ISL_1481798, EPI_ISL_1481799, EPI_ISL_1481800, EPI_ISL_1481801, EPI_ISL_1481802, EPI_ISL_1481803, EPI_ISL_1481804, EPI_ISL_1481805, EPI_ISL_1481806, EPI_ISL_1481807,
EPI_ISL_1481808, EPI_ISL_1481809, EPI_ISL_1481810, EPI_ISL_1481811, EPI_ISL_1481812, EPI_ISL_1481813, EPI_ISL_1481814, EPI_ISL_1481815, EPI_ISL_1481816, EPI_ISL_1481817, EPI_ISL_1481818, EPI_ISL_1481819, EPI_ISL_1481837, EPI_ISL_1481901, EPI_ISL_1481904, EPI_ISL_1481905, EPI_ISL_1481906, EPI_ISL_1481907, EPI_ISL_1481908, EPI_ISL_1481909, EPI_ISL_1481910, EPI_ISL_1481911,
EPI_ISL_1481912, EPI_ISL_1481913, EPI_ISL_1481914, EPI_ISL_1481915, EPI_ISL_1481916, EPI_ISL_1481917, EPI_ISL_1481918, EPI_ISL_1481919, EPI_ISL_1481920, EPI_ISL_1481921, EPI_ISL_1481922, EPI_ISL_1481923, EPI_ISL_1481924, EPI_ISL_1481925, EPI_ISL_1481926, EPI_ISL_1481927, EPI_ISL_1481928, EPI_ISL_1481929, EPI_ISL_1481930, EPI_ISL_1481931, EPI_ISL_1481932, EPI_ISL_1481933,
EPI_ISL_1481934, EPI_ISL_1481935, EPI_ISL_1481936, EPI_ISL_1481937, EPI_ISL_1481938, EPI_ISL_1481939, EPI_ISL_1481940, EPI_ISL_1481941, EPI_ISL_1481942, EPI_ISL_1481943, EPI_ISL_1481944, EPI_ISL_1481945, EPI_ISL_1481946, EPI_ISL_1481947, EPI_ISL_1481948, EPI_ISL_1481949, EPI_ISL_1481950, EPI_ISL_1481951, EPI_ISL_1481952, EPI_ISL_1481953, EPI_ISL_1481954, EPI_ISL_1481955,
EPI_ISL_1481956, EPI_ISL_1481957, EPI_ISL_1481958, EPI_ISL_1481959, EPI_ISL_1481960, EPI_ISL_1481961, EPI_ISL_1481962, EPI_ISL_1481963, EPI_ISL_1481964, EPI_ISL_1481965, EPI_ISL_1481966, EPI_ISL_1481967, EPI_ISL_1481968, EPI_ISL_1481969, EPI_ISL_1481970, EPI_ISL_1481971, EPI_ISL_1481972, EPI_ISL_1481973, EPI_ISL_1481974, EPI_ISL_1481975, EPI_ISL_1481976, EPI_ISL_1481977,
EPI_ISL_1481978, EPI_ISL_1481979, EPI_ISL_1481980, EPI_ISL_1481981, EPI_ISL_1481982, EPI_ISL_1481983, EPI_ISL_1481984, EPI_ISL_1481985, EPI_ISL_1481986, EPI_ISL_1481987, EPI_ISL_1481988, EPI_ISL_1481989, EPI_ISL_1481990, EPI_ISL_1481991, EPI_ISL_1481992, EPI_ISL_1481993, EPI_ISL_1481994, EPI_ISL_1481995, EPI_ISL_1481996, EPI_ISL_1481997, EPI_ISL_1482000, EPI_ISL_1482001,
EPI_ISL_1482002, EPI_ISL_1482003, EPI_ISL_1482004, EPI_ISL_1482005, EPI_ISL_1482006, EPI_ISL_1482007, EPI_ISL_1482008, EPI_ISL_1482009, EPI_ISL_1482010, EPI_ISL_1482011, EPI_ISL_1482012, EPI_ISL_1482013, EPI_ISL_1482014, EPI_ISL_1482015, EPI_ISL_1482016, EPI_ISL_1482017, EPI_ISL_1482018, EPI_ISL_1482019, EPI_ISL_1482020, EPI_ISL_1482021, EPI_ISL_1482022, EPI_ISL_1482023,
EPI_ISL_1482024, EPI_ISL_1482025, EPI_ISL_1482026, EPI_ISL_1482027, EPI_ISL_1482028, EPI_ISL_1482029, EPI_ISL_1482030, EPI_ISL_1482031, EPI_ISL_1482032, EPI_ISL_1482033, EPI_ISL_1482034, EPI_ISL_1482035, EPI_ISL_1482042, EPI_ISL_1482043, EPI_ISL_1482044, EPI_ISL_1482045, EPI_ISL_1482046, EPI_ISL_1482047, EPI_ISL_1482048, EPI_ISL_1482050, EPI_ISL_1482051, EPI_ISL_1482052,
EPI_ISL_1482053, EPI_ISL_1482054, EPI_ISL_1482055, EPI_ISL_1482056, EPI_ISL_1482057, EPI_ISL_1482058, EPI_ISL_1482059, EPI_ISL_1482060, EPI_ISL_1482061, EPI_ISL_1482062, EPI_ISL_1482063, EPI_ISL_1482064, EPI_ISL_1482065, EPI_ISL_1482066, EPI_ISL_1482067, EPI_ISL_1482068, EPI_ISL_1482069, EPI_ISL_1482070, EPI_ISL_1482071, EPI_ISL_1482072, EPI_ISL_1482073, EPI_ISL_1482074,
EPI_ISL_1482075, EPI_ISL_1482076, EPI_ISL_1482077, EPI_ISL_1482078, EPI_ISL_1482079, EPI_ISL_1482080, EPI_ISL_1482081, EPI_ISL_1482082, EPI_ISL_1482083, EPI_ISL_1482084, EPI_ISL_1482085, EPI_ISL_1482086, EPI_ISL_1482087, EPI_ISL_1482088, EPI_ISL_1482089, EPI_ISL_1482090, EPI_ISL_1482091, EPI_ISL_1482092, EPI_ISL_1482093, EPI_ISL_1482094, EPI_ISL_1482095, EPI_ISL_1482096,
EPI_ISL_1482097, EPI_ISL_1482098, EPI_ISL_1482099, EPI_ISL_1482101, EPI_ISL_1482102, EPI_ISL_1482103, EPI_ISL_1482104, EPI_ISL_1482105, EPI_ISL_1482106, EPI_ISL_1482107, EPI_ISL_1482108, EPI_ISL_1482109, EPI_ISL_1482110, EPI_ISL_1482111, EPI_ISL_1482112, EPI_ISL_1482113, EPI_ISL_1482114, EPI_ISL_1482115, EPI_ISL_1482116, EPI_ISL_1482117, EPI_ISL_1482118, EPI_ISL_1482119,
EPI_ISL_1482120, EPI_ISL_1482121, EPI_ISL_1482122, EPI_ISL_1482123, EPI_ISL_1482124, EPI_ISL_1482125, EPI_ISL_1482126, EPI_ISL_1482127, EPI_ISL_1482128, EPI_ISL_1482129, EPI_ISL_1482130, EPI_ISL_1482131, EPI_ISL_1482132, EPI_ISL_1482133, EPI_ISL_1482134, EPI_ISL_1482135, EPI_ISL_1482136, EPI_ISL_1482137, EPI_ISL_1482138, EPI_ISL_1482139, EPI_ISL_1482140, EPI_ISL_1482141,
EPI_ISL_1482142, EPI_ISL_1482143, EPI_ISL_1482144, EPI_ISL_1482145, EPI_ISL_1482146, EPI_ISL_1482147, EPI_ISL_1482148, EPI_ISL_1482149, EPI_ISL_1482150, EPI_ISL_1482151, EPI_ISL_1482152, EPI_ISL_1482153, EPI_ISL_1482154, EPI_ISL_1482155, EPI_ISL_1482156, EPI_ISL_1482157, EPI_ISL_1482158, EPI_ISL_1482159, EPI_ISL_1482160, EPI_ISL_1482161, EPI_ISL_1482162, EPI_ISL_1482163,
EPI_ISL_1482164, EPI_ISL_1482165, EPI_ISL_1482166, EPI_ISL_1482167, EPI_ISL_1482168, EPI_ISL_1482169, EPI_ISL_1482170, EPI_ISL_1482171, EPI_ISL_1482172, EPI_ISL_1482173, EPI_ISL_1482174, EPI_ISL_1482175, EPI_ISL_1482176, EPI_ISL_1482194, EPI_ISL_1525545, EPI_ISL_1525546, EPI_ISL_1525547, EPI_ISL_1525548, EPI_ISL_1525549, EPI_ISL_1525550, EPI_ISL_1525551, EPI_ISL_1525552,
EPI_ISL_1525553, EPI_ISL_1525554, EPI_ISL_1525555, EPI_ISL_1525556, EPI_ISL_1525557, EPI_ISL_1525558, EPI_ISL_1525559, EPI_ISL_1525560, EPI_ISL_1525561, EPI_ISL_1525562, EPI_ISL_1525563, EPI_ISL_1525564, EPI_ISL_1525565, EPI_ISL_1525566, EPI_ISL_1525567, EPI_ISL_1525568, EPI_ISL_1525569, EPI_ISL_1525570, EPI_ISL_1525571, EPI_ISL_1525572, EPI_ISL_1525573, EPI_ISL_1525574,

| | | | |
|---|---|---|---|
| EPI_ISL_1525575, EPI_ISL_1525576, EPI_ISL_1525577, EPI_ISL_1525578, EPI_ISL_1525579, EPI_ISL_1525580, EPI_ISL_1525581, EPI_ISL_1525582, EPI_ISL_1525583, EPI_ISL_1525584, EPI_ISL_1525585, EPI_ISL_1525586, EPI_ISL_1525587, EPI_ISL_1525588, EPI_ISL_1525589, EPI_ISL_1525590, EPI_ISL_1525591, EPI_ISL_1525592, EPI_ISL_1525593, EPI_ISL_1525594, EPI_ISL_1525595, EPI_ISL_1525596, EPI_ISL_1525597, EPI_ISL_1525598, EPI_ISL_1525599, EPI_ISL_1525600, EPI_ISL_1525601, EPI_ISL_1525602, EPI_ISL_1525603, EPI_ISL_1525604, EPI_ISL_1525605, EPI_ISL_1525606, EPI_ISL_1525607, EPI_ISL_1525608, EPI_ISL_1525609, EPI_ISL_1525610, EPI_ISL_1525611, EPI_ISL_1525612, EPI_ISL_1525613, EPI_ISL_1525614, EPI_ISL_1525615, EPI_ISL_1525616, EPI_ISL_1525617, EPI_ISL_1525618, EPI_ISL_1525619, EPI_ISL_1525620, EPI_ISL_1525621, EPI_ISL_1525622, EPI_ISL_1525623, EPI_ISL_1525624, EPI_ISL_1525625, EPI_ISL_1525626, EPI_ISL_1525627, EPI_ISL_1525635, EPI_ISL_1525641, EPI_ISL_1525654, EPI_ISL_1525657, EPI_ISL_1525664, EPI_ISL_1525665, EPI_ISL_1525693, EPI_ISL_1525715, EPI_ISL_1554950, EPI_ISL_1554951, EPI_ISL_1554953, EPI_ISL_1554954, EPI_ISL_1554956, EPI_ISL_1554957, EPI_ISL_1554959, EPI_ISL_1554962, EPI_ISL_1554963, EPI_ISL_1554964, EPI_ISL_1554965, EPI_ISL_1554966, EPI_ISL_1554967, EPI_ISL_1554968, EPI_ISL_1554969, EPI_ISL_1554970, EPI_ISL_1554971, EPI_ISL_1554972, EPI_ISL_1554973, EPI_ISL_1554974, EPI_ISL_1554975, EPI_ISL_1554976, EPI_ISL_1554977, EPI_ISL_1554978, EPI_ISL_1554979, EPI_ISL_1554980, EPI_ISL_1554982, EPI_ISL_1554983, EPI_ISL_1554984, EPI_ISL_1554985, EPI_ISL_1554986, EPI_ISL_1554987, EPI_ISL_1554988, EPI_ISL_1554989, EPI_ISL_1554990, EPI_ISL_1554991, EPI_ISL_1554992, EPI_ISL_1554993, EPI_ISL_1554994, EPI_ISL_1554995, EPI_ISL_1554996, EPI_ISL_1554997, EPI_ISL_1554998, EPI_ISL_1554999, EPI_ISL_1555000, EPI_ISL_1555001, EPI_ISL_1555002, EPI_ISL_1555003, EPI_ISL_1555004, EPI_ISL_1555005, EPI_ISL_1555006, EPI_ISL_1555007, EPI_ISL_1555010, EPI_ISL_1555011, EPI_ISL_1555012, EPI_ISL_1555013, EPI_ISL_1555014, EPI_ISL_1555015, EPI_ISL_1555016, EPI_ISL_1555017, EPI_ISL_1555018, EPI_ISL_1555019, EPI_ISL_1555020, EPI_ISL_1555021, EPI_ISL_1555022, EPI_ISL_1555023, EPI_ISL_1555024, EPI_ISL_1555025, EPI_ISL_1555026, EPI_ISL_1555027, EPI_ISL_1555028, EPI_ISL_1555029, EPI_ISL_1555030, EPI_ISL_1555031, EPI_ISL_1555032, EPI_ISL_1555033, EPI_ISL_1555034, EPI_ISL_1555035, EPI_ISL_1555036, EPI_ISL_1555037, EPI_ISL_1555042, EPI_ISL_1555043, EPI_ISL_1555044, EPI_ISL_1555045, EPI_ISL_1555046, EPI_ISL_1555047, EPI_ISL_1555048, EPI_ISL_1555049, EPI_ISL_1555050, EPI_ISL_1555052, EPI_ISL_1555053, EPI_ISL_1555054, EPI_ISL_1555055, EPI_ISL_1555056, EPI_ISL_1555057, EPI_ISL_1555058, EPI_ISL_1555059, EPI_ISL_1555060, EPI_ISL_1555061, EPI_ISL_1555062, EPI_ISL_1555063, EPI_ISL_1555064, EPI_ISL_1555065, EPI_ISL_1555066, EPI_ISL_1555067, EPI_ISL_1555068, EPI_ISL_1555069, EPI_ISL_1555070, EPI_ISL_1555071, EPI_ISL_1555072, EPI_ISL_1555073, EPI_ISL_1555074, EPI_ISL_1555075, EPI_ISL_1555076, EPI_ISL_1555077, EPI_ISL_1555078, EPI_ISL_1555079, EPI_ISL_1555080, EPI_ISL_1555081, EPI_ISL_1555082, EPI_ISL_1555083, EPI_ISL_1555084, EPI_ISL_1555085, EPI_ISL_1555086, EPI_ISL_1555087, EPI_ISL_1555088, EPI_ISL_1555089, EPI_ISL_1555090, EPI_ISL_1555091, EPI_ISL_1555092, EPI_ISL_1555093, EPI_ISL_1555094, EPI_ISL_1555095, EPI_ISL_1555096, EPI_ISL_1555097, EPI_ISL_1555098, EPI_ISL_1555099, EPI_ISL_1555100, EPI_ISL_1555101, EPI_ISL_1555102, EPI_ISL_1555103, EPI_ISL_1555104, EPI_ISL_1555105, EPI_ISL_1555106, EPI_ISL_1555107, EPI_ISL_1555108, EPI_ISL_1555109, EPI_ISL_1555110, EPI_ISL_1555112, EPI_ISL_1555113, EPI_ISL_1555114, EPI_ISL_1555117, EPI_ISL_1555118, EPI_ISL_1555119, EPI_ISL_1555120, EPI_ISL_1555121, EPI_ISL_1555122, EPI_ISL_1555123, EPI_ISL_1555124, EPI_ISL_1555125, EPI_ISL_1555126, EPI_ISL_1555127, EPI_ISL_1555128, EPI_ISL_1555129, EPI_ISL_1555130, EPI_ISL_1555131, EPI_ISL_1555132, EPI_ISL_1555133, EPI_ISL_1555134, EPI_ISL_1555135, EPI_ISL_1555136, EPI_ISL_1555137, EPI_ISL_1555138, EPI_ISL_1555139, EPI_ISL_1555140, EPI_ISL_1555141, EPI_ISL_1555142, EPI_ISL_1555143, EPI_ISL_1555144, EPI_ISL_1555145, EPI_ISL_1555146, EPI_ISL_1555147, EPI_ISL_1555148, EPI_ISL_1555149, EPI_ISL_1555150, EPI_ISL_1555151, EPI_ISL_1555152, EPI_ISL_1555153, EPI_ISL_1555154, EPI_ISL_1555155, EPI_ISL_1555156, EPI_ISL_1555157, EPI_ISL_1555158, EPI_ISL_1555159, EPI_ISL_1555264, EPI_ISL_1555265, EPI_ISL_1555266, EPI_ISL_1555267, EPI_ISL_1555268, EPI_ISL_1555269, EPI_ISL_1555270, EPI_ISL_1555271, EPI_ISL_1555272, EPI_ISL_1555273, EPI_ISL_1555274, EPI_ISL_1555275, EPI_ISL_1555276, EPI_ISL_1555277, EPI_ISL_1555278, EPI_ISL_1555279, EPI_ISL_1555280, EPI_ISL_1555281, EPI_ISL_1555282, EPI_ISL_1555283, EPI_ISL_1555284, EPI_ISL_1555285, EPI_ISL_1555286, EPI_ISL_1555287, EPI_ISL_1555289, EPI_ISL_1555290, EPI_ISL_1555291, EPI_ISL_1555292, EPI_ISL_1555293, EPI_ISL_1555294, EPI_ISL_1555295, EPI_ISL_1555296, EPI_ISL_1555297, EPI_ISL_1555298, EPI_ISL_1555299, EPI_ISL_1555300, EPI_ISL_1555301, EPI_ISL_1555302, EPI_ISL_1555303, EPI_ISL_1555306, EPI_ISL_1555307, EPI_ISL_1555308, EPI_ISL_1555309, EPI_ISL_1555310, EPI_ISL_1555311, EPI_ISL_1555312, EPI_ISL_1555313, EPI_ISL_1555314, EPI_ISL_1555315, EPI_ISL_1555316, EPI_ISL_1555317, EPI_ISL_1555318, EPI_ISL_1555319, EPI_ISL_1555320, EPI_ISL_1555321, EPI_ISL_1555322, EPI_ISL_1555323, EPI_ISL_1555324, EPI_ISL_1555325, EPI_ISL_1555326, EPI_ISL_1555327, EPI_ISL_1555328, EPI_ISL_1555329, EPI_ISL_1555330, EPI_ISL_1555331, EPI_ISL_1555332, EPI_ISL_1555333, EPI_ISL_1555334, EPI_ISL_1555335, EPI_ISL_1555336, EPI_ISL_1555337, EPI_ISL_1555338, EPI_ISL_1555339, EPI_ISL_1555340, EPI_ISL_1555539, EPI_ISL_1555540, EPI_ISL_1555563, EPI_ISL_1555566, EPI_ISL_1555567, EPI_ISL_1555571, EPI_ISL_1555572, EPI_ISL_1555573, EPI_ISL_1555574, EPI_ISL_1555575, EPI_ISL_1555577, EPI_ISL_1555579, EPI_ISL_1555580, EPI_ISL_1555581, EPI_ISL_1555582, EPI_ISL_1555583, EPI_ISL_1555584, EPI_ISL_1555585, EPI_ISL_1555586, EPI_ISL_1555587, EPI_ISL_1555588, EPI_ISL_1555589, EPI_ISL_1555590, EPI_ISL_1555591, EPI_ISL_1555592, EPI_ISL_1555593, EPI_ISL_1555594, EPI_ISL_1555595, EPI_ISL_1555596, EPI_ISL_1555597, EPI_ISL_1555598, EPI_ISL_1555599, EPI_ISL_1555600, EPI_ISL_1555601, EPI_ISL_1555602, EPI_ISL_1555603, EPI_ISL_1555604, EPI_ISL_1555605, EPI_ISL_1555606, EPI_ISL_1555607, EPI_ISL_1555608, EPI_ISL_1555609, EPI_ISL_1555610, EPI_ISL_1555611, EPI_ISL_1555612, EPI_ISL_1555613, EPI_ISL_1555614, EPI_ISL_1555615, EPI_ISL_1555616, EPI_ISL_1555617, EPI_ISL_1555618, EPI_ISL_1555619, EPI_ISL_1555620, EPI_ISL_1555621, EPI_ISL_1555622, EPI_ISL_1555623, EPI_ISL_1555624, EPI_ISL_1555625, EPI_ISL_1555626, EPI_ISL_1555627, EPI_ISL_1555628, EPI_ISL_1555629, EPI_ISL_1555630, EPI_ISL_1555631, EPI_ISL_1555632, EPI_ISL_1555633, EPI_ISL_1555635, EPI_ISL_1555636, EPI_ISL_1555637, EPI_ISL_1555638, EPI_ISL_1555639, EPI_ISL_1555640, EPI_ISL_1555641, EPI_ISL_1555642, EPI_ISL_1555643, EPI_ISL_1555644, EPI_ISL_1555645, EPI_ISL_1555646, EPI_ISL_1555647, EPI_ISL_1555648, EPI_ISL_1555649, EPI_ISL_1555650, EPI_ISL_1555651, EPI_ISL_1555652, EPI_ISL_1555653, EPI_ISL_1555654, EPI_ISL_1555655, EPI_ISL_1555656, EPI_ISL_1555657, EPI_ISL_1555658, EPI_ISL_1555660, EPI_ISL_1555661, EPI_ISL_1555662, EPI_ISL_1555663, EPI_ISL_1555664, EPI_ISL_1555665, EPI_ISL_1555666, EPI_ISL_1555667, EPI_ISL_1555668, EPI_ISL_1555669, EPI_ISL_1555670, EPI_ISL_1555671, EPI_ISL_1555672, EPI_ISL_1555673, EPI_ISL_1555674, EPI_ISL_1555675, EPI_ISL_1555676, EPI_ISL_1555677, EPI_ISL_1555678, EPI_ISL_1555679, EPI_ISL_1555680, EPI_ISL_1555681, EPI_ISL_1555682, EPI_ISL_1555683, EPI_ISL_1555684, EPI_ISL_1555685, EPI_ISL_1555686, EPI_ISL_1555687, EPI_ISL_1555688, EPI_ISL_1555689, EPI_ISL_1555690, EPI_ISL_1555691, EPI_ISL_1555692, EPI_ISL_1555693, EPI_ISL_1555694, EPI_ISL_1555699, EPI_ISL_1555700, EPI_ISL_1555701, EPI_ISL_1555702, EPI_ISL_1555703, EPI_ISL_1555704, EPI_ISL_1555705, EPI_ISL_1555706, EPI_ISL_1555707, EPI_ISL_1555708, EPI_ISL_1555709, EPI_ISL_1555710, EPI_ISL_1555711, EPI_ISL_1555712, EPI_ISL_1555713, EPI_ISL_1555714, EPI_ISL_1555715, EPI_ISL_1555716, EPI_ISL_1555717, EPI_ISL_1555718, EPI_ISL_1555719, EPI_ISL_1555720, EPI_ISL_1555721, EPI_ISL_1555722, EPI_ISL_1555723, EPI_ISL_1555724, EPI_ISL_1555725, EPI_ISL_1555726, EPI_ISL_1555727, EPI_ISL_1555728, EPI_ISL_1555729, EPI_ISL_1555730, EPI_ISL_1555731, EPI_ISL_1555732, EPI_ISL_1555733, EPI_ISL_1555734, EPI_ISL_1555735, EPI_ISL_1555736, EPI_ISL_1555737, EPI_ISL_1555738, EPI_ISL_1555739, EPI_ISL_1555740, EPI_ISL_1555741, EPI_ISL_1555742, EPI_ISL_1555743, EPI_ISL_1555744, EPI_ISL_1555745, EPI_ISL_1555746, EPI_ISL_1555747, EPI_ISL_1555748, EPI_ISL_1555749, EPI_ISL_1555877, EPI_ISL_1555878, EPI_ISL_1555879, EPI_ISL_1555880, EPI_ISL_1555881, EPI_ISL_1555882, EPI_ISL_1555883, EPI_ISL_1555884, EPI_ISL_1555886, EPI_ISL_1555887, EPI_ISL_1555888, EPI_ISL_1555889, EPI_ISL_1555890, EPI_ISL_1555891, EPI_ISL_1555892, EPI_ISL_1555894, EPI_ISL_1555895, EPI_ISL_1555896, EPI_ISL_1555897, EPI_ISL_1555898, EPI_ISL_1555899, EPI_ISL_1555900, EPI_ISL_1555901, EPI_ISL_1555902, EPI_ISL_1555903, EPI_ISL_1555904, EPI_ISL_1555905, EPI_ISL_1555906, EPI_ISL_1555907, EPI_ISL_1555908, EPI_ISL_1555909, EPI_ISL_1555910, EPI_ISL_1555911, EPI_ISL_1555912, EPI_ISL_1555913, EPI_ISL_1555914, EPI_ISL_1555915, EPI_ISL_1555916, EPI_ISL_1555917, EPI_ISL_1555918, EPI_ISL_1555919, EPI_ISL_1555920, EPI_ISL_1555921, EPI_ISL_1555922, EPI_ISL_1555923, EPI_ISL_1555924, EPI_ISL_1555925, EPI_ISL_1555926, EPI_ISL_1555927, EPI_ISL_1555928, EPI_ISL_1555929, EPI_ISL_1555930, EPI_ISL_1555931, EPI_ISL_1555932, EPI_ISL_1555933, EPI_ISL_1555934, EPI_ISL_1555935, EPI_ISL_1555936, EPI_ISL_1555937, EPI_ISL_1555938, EPI_ISL_1555939, EPI_ISL_1555940, EPI_ISL_1555941, EPI_ISL_1555942, EPI_ISL_1555943, EPI_ISL_1555944, EPI_ISL_1555945, EPI_ISL_1555946, EPI_ISL_1555947, EPI_ISL_1555948, EPI_ISL_1555949, EPI_ISL_1555950, EPI_ISL_1555951, EPI_ISL_1555952, EPI_ISL_1555953, EPI_ISL_1555954, EPI_ISL_1555955, EPI_ISL_1555956, EPI_ISL_1555957, EPI_ISL_1555958, EPI_ISL_1555959, EPI_ISL_1555960, EPI_ISL_1555961, EPI_ISL_1555962, EPI_ISL_1555963, EPI_ISL_1555964, EPI_ISL_1555965, EPI_ISL_1555966, EPI_ISL_1555967, EPI_ISL_1555968, EPI_ISL_1556078, EPI_ISL_1556079, EPI_ISL_1556080, EPI_ISL_1556081, EPI_ISL_1556082, EPI_ISL_1556083, EPI_ISL_1556084, EPI_ISL_1556085, EPI_ISL_1556086, EPI_ISL_1556087, EPI_ISL_1556088, EPI_ISL_1556089, EPI_ISL_1556090, EPI_ISL_1556091, EPI_ISL_1556092, EPI_ISL_1556093, EPI_ISL_1556094, EPI_ISL_1556095, EPI_ISL_1556096, EPI_ISL_1556097, EPI_ISL_1556098, EPI_ISL_1556099, EPI_ISL_1556100, EPI_ISL_1556101, EPI_ISL_1556102, EPI_ISL_1556103, EPI_ISL_1556104, EPI_ISL_1556105, EPI_ISL_1556106, EPI_ISL_1556107, EPI_ISL_1556108, EPI_ISL_1556109, EPI_ISL_1556110, EPI_ISL_1556111, EPI_ISL_1556112, EPI_ISL_1556113, EPI_ISL_1556114, EPI_ISL_1556115, EPI_ISL_1556116, EPI_ISL_1556119, EPI_ISL_1556197, EPI_ISL_1556198, EPI_ISL_1556199, EPI_ISL_1556200, EPI_ISL_1556202, EPI_ISL_1556203, EPI_ISL_1556204, EPI_ISL_1556205, EPI_ISL_1556206, EPI_ISL_1556207, EPI_ISL_1556208, EPI_ISL_1556209, EPI_ISL_1556210, EPI_ISL_1556211, EPI_ISL_1556212, EPI_ISL_1556213, EPI_ISL_1556214, EPI_ISL_1556215, EPI_ISL_1556216, EPI_ISL_1556217, EPI_ISL_1556218, EPI_ISL_1556219, EPI_ISL_1556220, EPI_ISL_1556221, EPI_ISL_1556222, EPI_ISL_1556223, EPI_ISL_1556224, EPI_ISL_1556225, EPI_ISL_1556230, EPI_ISL_1556231, EPI_ISL_1556232, EPI_ISL_1556233, EPI_ISL_1556243, EPI_ISL_1556244, EPI_ISL_1556245, EPI_ISL_1556246, EPI_ISL_1556247, EPI_ISL_1556248, EPI_ISL_1556249, EPI_ISL_1556250, EPI_ISL_1556251, EPI_ISL_1556254, EPI_ISL_1556255, EPI_ISL_1556256, EPI_ISL_1556257, EPI_ISL_1556281, EPI_ISL_1556282, EPI_ISL_1556875, EPI_ISL_1556876, EPI_ISL_1556877, EPI_ISL_1556878, EPI_ISL_1556879, EPI_ISL_1556880, EPI_ISL_1556881, EPI_ISL_1556882, EPI_ISL_1556883, EPI_ISL_1556884, EPI_ISL_1556885, EPI_ISL_1556886, EPI_ISL_1556887, EPI_ISL_1556888, EPI_ISL_1556889, EPI_ISL_1556890, EPI_ISL_1556891, EPI_ISL_1556892, EPI_ISL_1556893, EPI_ISL_1556894, EPI_ISL_1556895, EPI_ISL_1556896, EPI_ISL_1556897, EPI_ISL_1556898, EPI_ISL_1556899, EPI_ISL_1556900, EPI_ISL_1556901, EPI_ISL_1556902, EPI_ISL_1556903, EPI_ISL_1556904, EPI_ISL_1556905, EPI_ISL_1556906, EPI_ISL_1556907, EPI_ISL_1556908, EPI_ISL_1556909, EPI_ISL_1556910, EPI_ISL_1556911, EPI_ISL_1556912, EPI_ISL_1556913, EPI_ISL_1556914, EPI_ISL_1556915, EPI_ISL_1556916, EPI_ISL_1556917, EPI_ISL_1556918, EPI_ISL_1556919, EPI_ISL_1556920, EPI_ISL_1556921, EPI_ISL_1556939, EPI_ISL_1557186, EPI_ISL_1667103, EPI_ISL_1667105, EPI_ISL_1667106, EPI_ISL_1667107, EPI_ISL_1694957, EPI_ISL_1694960, EPI_ISL_1694961, EPI_ISL_1694962, EPI_ISL_1694964, EPI_ISL_1694965, EPI_ISL_1694966, EPI_ISL_1694967, EPI_ISL_1694968, EPI_ISL_1694973 | | | |
| see above | Fulgent Genetics | Centers for Disease Control and Prevention Division of Viral Diseases, Pathogen Discovery | Adrian Paskey; Becky Tsai; Benafsh Sapra; Benjamin Rambo-Martin; Christopher Gulvick; Clinton R. Paden; Dakota Howard; Darlene Wagner; Dhwani Batra; Doreen Ng; Duncan MacCannell; Harry Gao; James Xie; Jason Caravas; John Gao; Joseph Fierro; Kara Moser; Matthew Schmerer; Mickey Li; Peter W. Cook; Scott Sammons; Shatavia Morrison; Yan Meng; Yvette Unoarumhi |
| EPI_ISL_1231437, EPI_ISL_1231438, EPI_ISL_1231439, EPI_ISL_1231440, EPI_ISL_1231441, EPI_ISL_1231442, EPI_ISL_1231443, EPI_ISL_1231444, EPI_ISL_1231445, EPI_ISL_1231446, EPI_ISL_1231447, EPI_ISL_1231448, EPI_ISL_1231449, EPI_ISL_1231450, EPI_ISL_1231451, EPI_ISL_1231452, EPI_ISL_1231453, EPI_ISL_1231454, EPI_ISL_1231455, EPI_ISL_1231456, EPI_ISL_1231457, EPI_ISL_1231458, EPI_ISL_1231459, EPI_ISL_1231460, EPI_ISL_1231461, EPI_ISL_1231462, EPI_ISL_1231463, EPI_ISL_1231464, EPI_ISL_1231465, EPI_ISL_1231466, EPI_ISL_1231467, EPI_ISL_1231468, EPI_ISL_1231469, EPI_ISL_1231470, EPI_ISL_1231471, EPI_ISL_1231472, EPI_ISL_1231473, EPI_ISL_1231474, EPI_ISL_1231475, EPI_ISL_1231476, EPI_ISL_1231477, EPI_ISL_1231478, EPI_ISL_1231479, EPI_ISL_1231480, EPI_ISL_1231481, EPI_ISL_1231482, EPI_ISL_1231483, EPI_ISL_1231484, EPI_ISL_1231485, EPI_ISL_1231486, EPI_ISL_1231487, EPI_ISL_1231488, EPI_ISL_1231489, EPI_ISL_1231490, EPI_ISL_1231491, EPI_ISL_1231492, EPI_ISL_1231493, EPI_ISL_1231494, EPI_ISL_1231495, EPI_ISL_1231496, EPI_ISL_1231497, EPI_ISL_1231498, EPI_ISL_1231499, EPI_ISL_1231500, EPI_ISL_1231501, EPI_ISL_1231502, EPI_ISL_1231503, EPI_ISL_1231504, EPI_ISL_1231505, EPI_ISL_1231506, EPI_ISL_1231507, EPI_ISL_1231508, EPI_ISL_1231509, EPI_ISL_1231510, EPI_ISL_1231512, EPI_ISL_1231513, EPI_ISL_1231514, EPI_ISL_1231515, EPI_ISL_1231516, EPI_ISL_1231517, EPI_ISL_1231518, EPI_ISL_1231519, EPI_ISL_1231520, EPI_ISL_1231521, EPI_ISL_1231522, EPI_ISL_1231523, EPI_ISL_1231524, EPI_ISL_1231525, EPI_ISL_1231526, EPI_ISL_1231527, EPI_ISL_1231528, EPI_ISL_1231529, EPI_ISL_1231530, EPI_ISL_1231533, EPI_ISL_1231537, EPI_ISL_1231541, EPI_ISL_1231545, EPI_ISL_1231549, EPI_ISL_1231553, EPI_ISL_1231557, EPI_ISL_1231561, EPI_ISL_1231565, EPI_ISL_1231569, EPI_ISL_1231573, EPI_ISL_1231577, EPI_ISL_1231581, EPI_ISL_1231585, EPI_ISL_1231590, EPI_ISL_1231594, EPI_ISL_1231598, EPI_ISL_1231602, EPI_ISL_1231606, EPI_ISL_1231610, EPI_ISL_1231615, EPI_ISL_1231618, EPI_ISL_1231623, EPI_ISL_1231626, EPI_ISL_1231630, EPI_ISL_1231634, EPI_ISL_1231638, EPI_ISL_1231642, EPI_ISL_1231646, EPI_ISL_1231651, EPI_ISL_1231655, EPI_ISL_1231659, EPI_ISL_1231663, EPI_ISL_1231666, EPI_ISL_1231670, EPI_ISL_1231675, EPI_ISL_1231678, EPI_ISL_1231682, EPI_ISL_1231686, EPI_ISL_1231690, EPI_ISL_1231694, EPI_ISL_1231698, EPI_ISL_1231702, EPI_ISL_1231706, EPI_ISL_1231710, EPI_ISL_1231714, EPI_ISL_1231719, EPI_ISL_1231723, EPI_ISL_1231727, EPI_ISL_1231730, EPI_ISL_1231735, EPI_ISL_1231739, EPI_ISL_1231743, EPI_ISL_1231747, EPI_ISL_1231751, EPI_ISL_1231755, EPI_ISL_1231758, EPI_ISL_1231762, EPI_ISL_1231767, EPI_ISL_1231771, EPI_ISL_1231774, EPI_ISL_1231775, EPI_ISL_1231776, EPI_ISL_1231777, EPI_ISL_1231778, EPI_ISL_1231779, EPI_ISL_1231780, EPI_ISL_1231781, EPI_ISL_1231782, EPI_ISL_1231783, EPI_ISL_1231784, EPI_ISL_1231785, EPI_ISL_1231786, EPI_ISL_1231787, EPI_ISL_1231788, EPI_ISL_1231789, EPI_ISL_1231790, EPI_ISL_1231791, EPI_ISL_1231793, EPI_ISL_1231794, EPI_ISL_1231795, EPI_ISL_1231796, EPI_ISL_1231797, EPI_ISL_1231798, EPI_ISL_1231799, EPI_ISL_1231800, EPI_ISL_1231802, EPI_ISL_1231803, EPI_ISL_1231804, EPI_ISL_1231805, EPI_ISL_1231806, EPI_ISL_1231807, EPI_ISL_1231808, EPI_ISL_1231809, EPI_ISL_1231810, EPI_ISL_1231811, EPI_ISL_1231812, EPI_ISL_1231813, EPI_ISL_1231814, EPI_ISL_1231815, EPI_ISL_1231816, EPI_ISL_1231817, EPI_ISL_1231818, EPI_ISL_1231819, EPI_ISL_1231820, EPI_ISL_1231821, EPI_ISL_1231822, EPI_ISL_1231823, EPI_ISL_1231824, EPI_ISL_1231825, EPI_ISL_1231826, EPI_ISL_1231827, EPI_ISL_1231828, EPI_ISL_1231829, EPI_ISL_1231830, EPI_ISL_1231831, EPI_ISL_1231832, EPI_ISL_1231833, EPI_ISL_1231834, EPI_ISL_1231835, EPI_ISL_1231836, EPI_ISL_1231837, EPI_ISL_1231838, EPI_ISL_1231839, EPI_ISL_1231840, EPI_ISL_1231841, EPI_ISL_1231842, EPI_ISL_1231843, EPI_ISL_1231844, EPI_ISL_1231845, EPI_ISL_1231846, EPI_ISL_1231847, EPI_ISL_1231848, EPI_ISL_1231849, EPI_ISL_1231850, EPI_ISL_1231851, EPI_ISL_1231853, EPI_ISL_1231854, EPI_ISL_1231855, EPI_ISL_1231856, EPI_ISL_1231857, EPI_ISL_1231858, EPI_ISL_1231859, EPI_ISL_1231860, EPI_ISL_1231861, EPI_ISL_1231862, EPI_ISL_1231863, EPI_ISL_1231864, EPI_ISL_1231865, EPI_ISL_1231866, EPI_ISL_1231867, EPI_ISL_1231868, EPI_ISL_1231869, EPI_ISL_1231870, EPI_ISL_1231871, EPI_ISL_1231872, EPI_ISL_1231873, EPI_ISL_1231874, EPI_ISL_1231875, EPI_ISL_1231876, EPI_ISL_1231877, EPI_ISL_1231878, EPI_ISL_1231879, EPI_ISL_1231880, EPI_ISL_1231881, EPI_ISL_1231882, EPI_ISL_1231883, EPI_ISL_1231884, EPI_ISL_1231885, EPI_ISL_1231886, EPI_ISL_1231887, EPI_ISL_1231888, EPI_ISL_1231889, EPI_ISL_1231890, EPI_ISL_1231891, EPI_ISL_1231892, EPI_ISL_1231893, EPI_ISL_1231894, EPI_ISL_1231895, EPI_ISL_1231896, EPI_ISL_1231897, EPI_ISL_1231898, EPI_ISL_1231899, EPI_ISL_1231900, EPI_ISL_1231901, EPI_ISL_1231902, EPI_ISL_1231903, EPI_ISL_1231904, EPI_ISL_1231905, EPI_ISL_1231906, EPI_ISL_1231907, EPI_ISL_1231908, EPI_ISL_1231909, EPI_ISL_1231910, EPI_ISL_1231911, EPI_ISL_1231912, EPI_ISL_1231913, EPI_ISL_1251273 | | | |
| see above | Fulgent Genetics | Fulgent Genetics | Becky Tsai; Benafsh Sapra; Doreen Ng; Harry Gao; James Xie; John Gao; Joseph Fierro; Mickey Li; Yan Meng |
| EPI_ISL_1532123 | Good Samaritan Hospital | Santa Clara County Public Health Laboratory | Santa Clara County Public Health Department |
| EPI_ISL_1283377, EPI_ISL_1283443, EPI_ISL_1283456, EPI_ISL_1283639, EPI_ISL_1283692, EPI_ISL_1283707, EPI_ISL_1283742, EPI_ISL_1297941, EPI_ISL_1298016, EPI_ISL_1298067, EPI_ISL_1298069, EPI_ISL_1298074, EPI_ISL_1298077, EPI_ISL_1298080, EPI_ISL_1298084, EPI_ISL_1298098, EPI_ISL_1298111, EPI_ISL_1298114, EPI_ISL_1298122, EPI_ISL_1298168, EPI_ISL_1298171, EPI_ISL_1298172, EPI_ISL_1298182, EPI_ISL_1338057, EPI_ISL_1338070, EPI_ISL_1338074, EPI_ISL_1338085, EPI_ISL_1338087, EPI_ISL_1338091, EPI_ISL_1338097, EPI_ISL_1338101, EPI_ISL_1338122, EPI_ISL_1338124, EPI_ISL_1338130, EPI_ISL_1338133, EPI_ISL_1338140, EPI_ISL_1338156, EPI_ISL_1338166, EPI_ISL_1338170, EPI_ISL_1338194, EPI_ISL_1338204, EPI_ISL_1338210, EPI_ISL_1338212, EPI_ISL_1338226, EPI_ISL_1338230, EPI_ISL_1338233, EPI_ISL_1338245, EPI_ISL_1338289, EPI_ISL_1338307, EPI_ISL_1338333, EPI_ISL_1338394, EPI_ISL_1338397, EPI_ISL_1340060, EPI_ISL_1340061, EPI_ISL_1340068, EPI_ISL_1340083, EPI_ISL_1340084, EPI_ISL_1340088, EPI_ISL_1340090, EPI_ISL_1340096, EPI_ISL_1340124, EPI_ISL_1340129, EPI_ISL_1340225, EPI_ISL_1340249, EPI_ISL_1340251, EPI_ISL_1340254, EPI_ISL_1340258, EPI_ISL_1340260, EPI_ISL_1340264, EPI_ISL_1340265, EPI_ISL_1340275, EPI_ISL_1340301, EPI_ISL_1340305, EPI_ISL_1340308, EPI_ISL_1340337, EPI_ISL_1340339, EPI_ISL_1340368, EPI_ISL_1340373, EPI_ISL_1340377, EPI_ISL_1340384, EPI_ISL_1340447, EPI_ISL_1391729, EPI_ISL_1391750, EPI_ISL_1391839, EPI_ISL_1391843, EPI_ISL_1391848, EPI_ISL_1391853, EPI_ISL_1391864, EPI_ISL_1391865, EPI_ISL_1391888, EPI_ISL_1391895, EPI_ISL_1391905, EPI_ISL_1391919, EPI_ISL_1391929, EPI_ISL_1391934, EPI_ISL_1391939, EPI_ISL_1391943, EPI_ISL_1391949, EPI_ISL_1391962, EPI_ISL_1421655, EPI_ISL_1443819, EPI_ISL_1443827, EPI_ISL_1443831, EPI_ISL_1443835, EPI_ISL_1443842, EPI_ISL_1443846, EPI_ISL_1443865, EPI_ISL_1443918, EPI_ISL_1443932, EPI_ISL_1443938, EPI_ISL_1443950, EPI_ISL_1444022, EPI_ISL_1444024, EPI_ISL_1444030, EPI_ISL_1444041, EPI_ISL_1444046, EPI_ISL_1444064, EPI_ISL_1444074, EPI_ISL_1444092, EPI_ISL_1444098, EPI_ISL_1444188, EPI_ISL_1444190, EPI_ISL_1444192, EPI_ISL_1444200, EPI_ISL_1444227, EPI_ISL_1444242, EPI_ISL_1460798, EPI_ISL_1460801, EPI_ISL_1460802, EPI_ISL_1460831, EPI_ISL_1460835, EPI_ISL_1460853, EPI_ISL_1460917, EPI_ISL_1460959, EPI_ISL_1460975, EPI_ISL_1460981, EPI_ISL_1460985, EPI_ISL_1460997, EPI_ISL_1460998, EPI_ISL_1479226, EPI_ISL_1479271, EPI_ISL_1479275, EPI_ISL_1479283, EPI_ISL_1479333, EPI_ISL_1479353, EPI_ISL_1479425, EPI_ISL_1480009, EPI_ISL_1480066, EPI_ISL_1480190, EPI_ISL_1480238, EPI_ISL_1480260, EPI_ISL_1480271, EPI_ISL_1480341, EPI_ISL_1480451, EPI_ISL_1480513, EPI_ISL_1493758, EPI_ISL_1493886, EPI_ISL_1493898, EPI_ISL_1493921, EPI_ISL_1511861, EPI_ISL_1511862, EPI_ISL_1511867, EPI_ISL_1511871, EPI_ISL_1511872, EPI_ISL_1511875, EPI_ISL_1511879, EPI_ISL_1511883, EPI_ISL_1511888, EPI_ISL_1511889, EPI_ISL_1511897, EPI_ISL_1511903, EPI_ISL_1511905, EPI_ISL_1511986, EPI_ISL_1511989, EPI_ISL_1512001, EPI_ISL_1512003, EPI_ISL_1512005, EPI_ISL_1512011, EPI_ISL_1512027, EPI_ISL_1512028, EPI_ISL_1512034, EPI_ISL_1512044, EPI_ISL_1512048, EPI_ISL_1512049, EPI_ISL_1512083, EPI_ISL_1512089, EPI_ISL_1512090, EPI_ISL_1512093, EPI_ISL_1512095, EPI_ISL_1512096, EPI_ISL_1512098, EPI_ISL_1512100, EPI_ISL_1512101, EPI_ISL_1512102, EPI_ISL_1512105, EPI_ISL_1512107, EPI_ISL_1512109, EPI_ISL_1512111, EPI_ISL_1512112, EPI_ISL_1512113, EPI_ISL_1512115, EPI_ISL_1512116, EPI_ISL_1512119, EPI_ISL_1512122, EPI_ISL_1512125, EPI_ISL_1512148, EPI_ISL_1512203, EPI_ISL_1512233, EPI_ISL_1512247, EPI_ISL_1512248, EPI_ISL_1512249, EPI_ISL_1512252, EPI_ISL_1512253, EPI_ISL_1512255, EPI_ISL_1512259, EPI_ISL_1512266, EPI_ISL_1512267, EPI_ISL_1512268, EPI_ISL_1512314, EPI_ISL_1512317, EPI_ISL_1512319, EPI_ISL_1512321, EPI_ISL_1512322, EPI_ISL_1512324, EPI_ISL_1512326, EPI_ISL_1512329, EPI_ISL_1512331, EPI_ISL_1512335, EPI_ISL_1512349, EPI_ISL_1512351, EPI_ISL_1512352, EPI_ISL_1512358, EPI_ISL_1512366, EPI_ISL_1512376, EPI_ISL_1512377, EPI_ISL_1512378, EPI_ISL_1512383, EPI_ISL_1512386, EPI_ISL_1512395, EPI_ISL_1512402, EPI_ISL_1512404, EPI_ISL_1512419, EPI_ISL_1512420, EPI_ISL_1512423, EPI_ISL_1512424, EPI_ISL_1512429, EPI_ISL_1512431, EPI_ISL_1512432, EPI_ISL_1512436, EPI_ISL_1512438, EPI_ISL_1512442, EPI_ISL_1512443, EPI_ISL_1512451, EPI_ISL_1512459, EPI_ISL_1512463, EPI_ISL_1512464, EPI_ISL_1512466, EPI_ISL_1512469, EPI_ISL_1512471, EPI_ISL_1512475, EPI_ISL_1512478, EPI_ISL_1512479, EPI_ISL_1512484, EPI_ISL_1512489, EPI_ISL_1512492, EPI_ISL_1512519, EPI_ISL_1512520, EPI_ISL_1512554, EPI_ISL_1512562, EPI_ISL_1512584, EPI_ISL_1512589, EPI_ISL_1512593, EPI_ISL_1512634, EPI_ISL_1512642, EPI_ISL_1512652, EPI_ISL_1512664, EPI_ISL_1512667, EPI_ISL_1512672, EPI_ISL_1512686, EPI_ISL_1512691, EPI_ISL_1512707, EPI_ISL_1512718, EPI_ISL_1512734, EPI_ISL_1512735, EPI_ISL_1512736, EPI_ISL_1512742, EPI_ISL_1512744, EPI_ISL_1553291, EPI_ISL_1553301, EPI_ISL_1553418, EPI_ISL_1553432, EPI_ISL_1553447, EPI_ISL_1553474, EPI_ISL_1553500, EPI_ISL_1553518, EPI_ISL_1553595, EPI_ISL_1553931, EPI_ISL_1553965, EPI_ISL_1554030, EPI_ISL_1554035, EPI_ISL_1554197, EPI_ISL_1554200, EPI_ISL_1554204, EPI_ISL_1554216, EPI_ISL_1554217, EPI_ISL_1554224, EPI_ISL_1554304, EPI_ISL_1554343, EPI_ISL_1554361, EPI_ISL_1554367, EPI_ISL_1554375, EPI_ISL_1554401, EPI_ISL_1554406, EPI_ISL_1554419, EPI_ISL_1554422, EPI_ISL_1554425, EPI_ISL_1554465, EPI_ISL_1554466, EPI_ISL_1554467, EPI_ISL_1554468, EPI_ISL_1554469, EPI_ISL_1554470, EPI_ISL_1554471, EPI_ISL_1554472, EPI_ISL_1554475, EPI_ISL_1554476, EPI_ISL_1554477, EPI_ISL_1554479, EPI_ISL_1554480, EPI_ISL_1554481, EPI_ISL_1554486, EPI_ISL_1554487, EPI_ISL_1554488, EPI_ISL_1554490, EPI_ISL_1554493, EPI_ISL_1554494, EPI_ISL_1554495, EPI_ISL_1554499, EPI_ISL_1554500, EPI_ISL_1554501, EPI_ISL_1554502, EPI_ISL_1554506, EPI_ISL_1554507, EPI_ISL_1554508, EPI_ISL_1554523, EPI_ISL_1554525, EPI_ISL_1554526, EPI_ISL_1554528, EPI_ISL_1554537, EPI_ISL_1554545, EPI_ISL_1554570, EPI_ISL_1554621, EPI_ISL_1554645, EPI_ISL_1554649, EPI_ISL_1554653, EPI_ISL_1554659, EPI_ISL_1554662, EPI_ISL_1554663, EPI_ISL_1554664, EPI_ISL_1554685, EPI_ISL_1554686, EPI_ISL_1554687, EPI_ISL_1554688, EPI_ISL_1554689, EPI_ISL_1554701, EPI_ISL_1554715, EPI_ISL_1554720, EPI_ISL_1554725, EPI_ISL_1554728, EPI_ISL_1554744, EPI_ISL_1554747, EPI_ISL_1554769, EPI_ISL_1554770, EPI_ISL_1554771, EPI_ISL_1554776, EPI_ISL_1554778, EPI_ISL_1554780, EPI_ISL_1554784, EPI_ISL_1554790, EPI_ISL_1554791, EPI_ISL_1554794, EPI_ISL_1554801, EPI_ISL_1554806, EPI_ISL_1554818, EPI_ISL_1554819, EPI_ISL_1554820, EPI_ISL_1554824, EPI_ISL_1554827, EPI_ISL_1554829, EPI_ISL_1554831, EPI_ISL_1554834, EPI_ISL_1554839, EPI_ISL_1554844, EPI_ISL_1554845, EPI_ISL_1554849, EPI_ISL_1554850, EPI_ISL_1554857, EPI_ISL_1554859, EPI_ISL_1554860, EPI_ISL_1554865, EPI_ISL_1554866, EPI_ISL_1554872, EPI_ISL_1575494, EPI_ISL_1575499, EPI_ISL_1575506, EPI_ISL_1575512, EPI_ISL_1575521, EPI_ISL_1575524, EPI_ISL_1575527, EPI_ISL_1575534, EPI_ISL_1575546, EPI_ISL_1575549, EPI_ISL_1575551, EPI_ISL_1575552, EPI_ISL_1575573, EPI_ISL_1575581, EPI_ISL_1575582, EPI_ISL_1575595, EPI_ISL_1575606, EPI_ISL_1575617, EPI_ISL_1575623, EPI_ISL_1575628, EPI_ISL_1575631, EPI_ISL_1575640, EPI_ISL_1575653, EPI_ISL_1575665, EPI_ISL_1575836, EPI_ISL_1575837, EPI_ISL_1575847, EPI_ISL_1575848, EPI_ISL_1575851, EPI_ISL_1575863, EPI_ISL_1575882, EPI_ISL_1575895, EPI_ISL_1575900, EPI_ISL_1575909, EPI_ISL_1575910, EPI_ISL_1575919, EPI_ISL_1575933, EPI_ISL_1575954, EPI_ISL_1575975, EPI_ISL_1575977, EPI_ISL_1575992, EPI_ISL_1575997, EPI_ISL_1576016, EPI_ISL_1576032, EPI_ISL_1576041, EPI_ISL_1576042, EPI_ISL_1576046, EPI_ISL_1576047, EPI_ISL_1576048, EPI_ISL_1576049, EPI_ISL_1576051, EPI_ISL_1576053, EPI_ISL_1576058, EPI_ISL_1576059, EPI_ISL_1576060, EPI_ISL_1576062, EPI_ISL_1576066, EPI_ISL_1576070, EPI_ISL_1576072, EPI_ISL_1576076, EPI_ISL_1576078, EPI_ISL_1576082, EPI_ISL_1576084, EPI_ISL_1576086, EPI_ISL_1576087, EPI_ISL_1576094, EPI_ISL_1576102, EPI_ISL_1576104, EPI_ISL_1576115, EPI_ISL_1576122, EPI_ISL_1576132, EPI_ISL_1576168, EPI_ISL_1576171, EPI_ISL_1576183, EPI_ISL_1576278, EPI_ISL_1576279, EPI_ISL_1576302, EPI_ISL_1576309, EPI_ISL_1576320, EPI_ISL_1576328, EPI_ISL_1576330, EPI_ISL_1576340, EPI_ISL_1576351, EPI_ISL_1576352, EPI_ISL_1576354, EPI_ISL_1576357, EPI_ISL_1576366, EPI_ISL_1576374, EPI_ISL_1576376, EPI_ISL_1576378, EPI_ISL_1576391, EPI_ISL_1576395, EPI_ISL_1576587, EPI_ISL_1576615, EPI_ISL_1576637, EPI_ISL_1576658, EPI_ISL_1576666, EPI_ISL_1581154, EPI_ISL_1581158, EPI_ISL_1581169, EPI_ISL_1581175, EPI_ISL_1581190, EPI_ISL_1581193, EPI_ISL_1581213, EPI_ISL_1581297, EPI_ISL_1581309, EPI_ISL_1581704, EPI_ISL_1581740, EPI_ISL_1581751, EPI_ISL_1694635, EPI_ISL_1694637, EPI_ISL_1694638, EPI_ISL_1694641, EPI_ISL_1694664 | | | |
| see above | Helix/Illumina | Centers for Disease Control and Prevention Division of Viral Diseases, Pathogen Discovery | Adrian Paskey; Alexandre Bolze; Ary Ascencio; Ben L. Rambo-Martin; Benjamin Rambo-Martin; Brad Sickler; Charlotte Rivera-Garcia; Christine Tran; Christopher Gulvick; Clinton R. Paden; Dakota Howard; Darlene Wagner; David Becker; Dhwani Batra; Duncan MacCannell; Efren Sandoval; Eileen de Feo; Elizabeth Cirulli; Eric Allen; Geraint Levan; James Lu; Jan Antico; Jason Caravas; Jason Nguyen; Jimmy Ramirez; Jingtao Liu; Kara Moser; Kelly Schiabor Barrett; Kim Gietzen; Magnus Isaksson; Marc Laurent; Matthew Schmerer; Matthew Tolentino; Nicole L. Washington; Peter W. Cook; Phil Febbo; Ryan Cho; Scott Sammons; Shannon Wickline; Shatavia Morrison; Sherry Wang; Simon White; Summer Galloway; Suxiang Tong; Tyler Cassens; William Lee; Yvette Unoarumhi |
| EPI_ISL_1303471 | Henry Mayo Newhall Hospital | Los Angeles County PHL | P. Hemarajata et al. |
| EPI_ISL_1477951, EPI_ISL_1477954, EPI_ISL_1477955, EPI_ISL_1477970, EPI_ISL_1477973, EPI_ISL_1477975, EPI_ISL_1477976, EPI_ISL_1477981, EPI_ISL_1477987, EPI_ISL_1477991, EPI_ISL_1477997, EPI_ISL_1478009, EPI_ISL_1478010, EPI_ISL_1478026, EPI_ISL_1478041, EPI_ISL_1478062, EPI_ISL_1478065, EPI_ISL_1478066, EPI_ISL_1478075, EPI_ISL_1478077, EPI_ISL_1478082, EPI_ISL_1478083, EPI_ISL_1478093, EPI_ISL_1478094, EPI_ISL_1478096, EPI_ISL_1478105, EPI_ISL_1478107, EPI_ISL_1478121, EPI_ISL_1478134, EPI_ISL_1478135, EPI_ISL_1478138, EPI_ISL_1478140, EPI_ISL_1478143, EPI_ISL_1478150, EPI_ISL_1478156, EPI_ISL_1478163, EPI_ISL_1478164, EPI_ISL_1478165, EPI_ISL_1478169, EPI_ISL_1478172, EPI_ISL_1478176, EPI_ISL_1478177, EPI_ISL_1478186, EPI_ISL_1478190, EPI_ISL_1701396, EPI_ISL_1701398, EPI_ISL_1701402, EPI_ISL_1701404, EPI_ISL_1701412, EPI_ISL_1701420, EPI_ISL_1701422, EPI_ISL_1701428, EPI_ISL_1701429, EPI_ISL_1701433, EPI_ISL_1701435, EPI_ISL_1701441, EPI_ISL_1701446, EPI_ISL_1701448, EPI_ISL_1701451, EPI_ISL_1701454, EPI_ISL_1701458, EPI_ISL_1701472, EPI_ISL_1701474, EPI_ISL_1701484, EPI_ISL_1701500, EPI_ISL_1701507, EPI_ISL_1701509, EPI_ISL_1701527, EPI_ISL_1701534, EPI_ISL_1701538, EPI_ISL_1701549, EPI_ISL_1701558, EPI_ISL_1701568, EPI_ISL_1701577, EPI_ISL_1701584, EPI_ISL_1701588, EPI_ISL_1701590, EPI_ISL_1701595, EPI_ISL_1701597, EPI_ISL_1701600, EPI_ISL_1701603, EPI_ISL_1701608, EPI_ISL_1701617, EPI_ISL_1701618, EPI_ISL_1701624, EPI_ISL_1701627, EPI_ISL_1701637, EPI_ISL_1701643, EPI_ISL_1701658 | | | |
| see above | Humboldt County Public Health Laboratory | Chan-Zuckerberg Biohub | CZB Cliahub Consortium |
| EPI_ISL_1497544, EPI_ISL_1497545, EPI_ISL_1497546, EPI_ISL_1506116 | Huntington Memorial Hospital | Los Angeles County PHL | P. Hemarajata et al. |
| EPI_ISL_2253167, EPI_ISL_2253169, EPI_ISL_2253170, EPI_ISL_2253172, EPI_ISL_2253174, EPI_ISL_2253175 | Infectious Diseases, Chan-Zuckerberg Biohub Covidtracker | Infectious Diseases, Chan-Zuckerberg Biohub Covidtracker | CZB Cliahub Consortium |
| EPI_ISL_1512824, EPI_ISL_1512945, EPI_ISL_1691713, EPI_ISL_1692004, EPI_ISL_1692184, EPI_ISL_1692231, EPI_ISL_1692270, EPI_ISL_1692487, EPI_ISL_1702257 | | | |
| see above | Infinity Biologix | Centers for Disease Control and Prevention Division of Viral | Adrian Paskey; Benjamin Rambo-Martin; Chirayu Goswami; Christian Bixby; Christopher Gulvick; Clinton R. Paden; Dakota Howard; Darlene Wagner; Dhwani Batra; Duncan MacCannell; Jason Caravas; Jonathan Schultz; Kara Moser; Matthew Schmerer; Peter W. Cook; Robin Grimwood; Russ Hager; Scott Sammons; Shatavia Morrison; Yihe Wang; Yvette Unoarumhi |

| | | | |
|---|---|---|---|
| | | Diseases, Pathogen Discovery | |
| EPI_ISL_1712422, EPI_ISL_1712423, EPI_ISL_1712424, EPI_ISL_1712425, EPI_ISL_1712426, EPI_ISL_1712427, EPI_ISL_1712429, EPI_ISL_1712430, EPI_ISL_1712431, EPI_ISL_1712462, EPI_ISL_1712463, EPI_ISL_1712464, EPI_ISL_1712465, EPI_ISL_1712466, EPI_ISL_1712467, EPI_ISL_1712468, EPI_ISL_1712469, EPI_ISL_1712470, EPI_ISL_1712471, EPI_ISL_1712472, EPI_ISL_1712474, EPI_ISL_1712475, EPI_ISL_1712476, EPI_ISL_1712477, EPI_ISL_1712478, EPI_ISL_1712479, EPI_ISL_1712542, EPI_ISL_1712543, EPI_ISL_1712544, EPI_ISL_1712545, EPI_ISL_1712547, EPI_ISL_1712548, EPI_ISL_1712549 | | | |
| see above | Innovative Genomics Institute, UC Berkeley | Innovative Genomics Institute, UC Berkeley | Holly Gildea; John Boyle; Phil Frankino; Stacia Wyman |
| EPI_ISL_1508918 | Kaiser Permanente Downey Medical Center | Los Angeles County PHL | P. Hemarajata et al. |
| EPI_ISL_1508902, EPI_ISL_1508917 | Kaiser Permanente West Los Angeles Medical Center | Los Angeles County PHL | P. Hemarajata et al. |
| EPI_ISL_1508966, EPI_ISL_1508967, EPI_ISL_1508970, EPI_ISL_1508972, EPI_ISL_1508975 | Kaiser Santa Clara | Santa Clara County Public Health Laboratory | Santa Clara County Public Health Department |
| EPI_ISL_1337390, EPI_ISL_1399280, EPI_ISL_1504108, EPI_ISL_1532264, EPI_ISL_1558798, EPI_ISL_1661550, EPI_ISL_1700837, EPI_ISL_1710971, EPI_ISL_1710972 | | | |
| see above | LESP Baja California | Instituto de Diagnostico y Referencia Epidemiologicos (INDRE) | Abril Rodriguez-Maldonado; Ariadna Medina-Benitez; Claudia Wong-Arambula; Ernesto Ramirez-Gonzalez.; Gisela Barrera-Badillo; Irma Lopez-Martinez; Joaquin Quiroz-Mercado; Lucia Hernandez-Rivas; Natividad Cruz-Ortiz; Sergio Rangel-Guerrero; Tatiana Nunez-Garcia; Vanessa Rivero-Arredondo |
| EPI_ISL_1504105, EPI_ISL_1532243, EPI_ISL_1532244, EPI_ISL_1532245, EPI_ISL_1558801, EPI_ISL_1626811, EPI_ISL_1626812, EPI_ISL_1626813, EPI_ISL_1626814, EPI_ISL_1626815, EPI_ISL_1651890, EPI_ISL_1651891, EPI_ISL_1651892, EPI_ISL_1651893, EPI_ISL_1651894, EPI_ISL_1651895, EPI_ISL_1651900, EPI_ISL_1700819, EPI_ISL_1700822 | | | |
| see above | LESP Baja California Sur | Instituto de Diagnostico y Referencia Epidemiologicos (INDRE) | Abril Rodriguez-Maldonado; Ariadna Medina-Benitez; Claudia Wong-Arambula; Ernesto Ramirez-Gonzalez.; Gisela Barrera-Badillo; Irma Lopez-Martinez; Joaquin Quiroz-Mercado; Lucia Hernandez-Rivas; Natividad Cruz-Ortiz; Sergio Rangel-Guerrero; Tatiana Nunez-Garcia; Vanessa Rivero-Arredondo |
| EPI_ISL_1661980, EPI_ISL_1661981 | Laboratorio Central de Epidemiologia (LCE) | Unidad de Genomica Avanzada | Alejandro Sanchez-Flores; Alfredo Herrera-Estrella; Alicia Ocana-Mondragon; Angel Gustavo Salas-Lais; Bernardo Martinez-Miguel; Blanca Taboada; Brenda Irasema Maldonado-Meza; Carla Ivon Herrera-Najera; Carlos F. Arias; Celia Boukadida; Celida Duque Molina; Clara Esperanza Santacruz-Tinoco; Concepcion Grajales-Muniz; Consorcio Mexicano de Vigilancia Genomica (CoViGen-Mex). Authors (in alphabetical order): Julio Elias Alvarado-Yaah; Fernando Fontove-Herrera; Francisco Pulido; Gloria Elena Espinosa-Ayala; Gloria Maria Molina-Salinas; Gloria Vazquez; Hector Esteban Paz-Juarez; Hector Montoya-Fuentes; Helen Haydee Fernanda Ramirez-Plascencia; Jose Antonio Enciso-Moreno; Jose Esteban Munoz-Medina; Jose de Jesus Nunez-Contreras; Juan Bautista Chale-Dzul; Luis Alberto Ochoa-Carrera; Margarita Matias-Florentino; Maria Guadalupe Santiago-Mauricio; Maria Guadalupe de Jesus Mireles-Rivera; Nelly Selem-Mojica; Pavel Isa; Ricardo Grande; Santiago Ávila-Rios; Victor Eduardo Garcia-Arias; Victor Hugo Borja-Aburto |
| EPI_ISL_1351981, EPI_ISL_1351984, EPI_ISL_1351987, EPI_ISL_1351990, EPI_ISL_1352048 | Laboratorio Central de EpidemiologÃa (LCE) | Unidad de Genomica Avanzada | Alejandro Sanchez-Flores; Alfredo Herrera-Estrella; Alicia Ocana-Mondragon; Angel Gustavo Salas-Lais; Bernardo Martinez-Miguel; Blanca Taboada; Brenda Irasema Maldonado-Meza; Carla Ivon Herrera-Najera; Carlos F. Arias; Celia Boukadida; Clara Esperanza Santacruz-Tinoco; Concepcion Grajales-Muniz; Consorcio Mexicano de Vigilancia Genomica (CoViGen-Mex). Authors (in alphabetical order): Julio Elias Alvarado-Yaah; Fernando Fontove-Herrera; Francisco Pulido; Gloria Elena Espinoza-Ayala; Gloria Maria Molina-Salinas; Gloria Vazquez; Hector Esteban Paz-Juarez; Hector Montoya-Fuentes; Helen Haydee Fernanda Ramirez-Plascencia; Jorge Ivan Salinal-Nevarez; Jose Antonio Enciso-Moreno; Jose Esteban Munoz-Medina; Jose de Jesus Nunez-Contreras; Juan Bautista Chale-Dzul; Luis Alberto Ochoa-Carrera; Margarita Matias-Florentino; Maria Guadalupe Santiago-Mauricio; Maria Guadalupe de Jesus Mireles-Rivera; Nelly Selem-Mojica; Pavel Isa; Ricardo Grande; Santiago Avila-Rios; Victor Hugo Borja-Aburto |
| EPI_ISL_1585658 | Laboratorio Central de Epidemiología (LCE) | Instituto Nacional de Enfermedades Respiratorias (INER): Centro de Investigación en Enfermedades Infecciosas (CIENI) | Alejandro Sanchez-Flores; Alfredo Herrera-Estrella; Alicia Ocaña-Mondragón; Angel Gustavo Salas-Lais; Bernardo Martínez-Miguel; Blanca Taboada; Brenda Irasema Maldonado-Meza; Carla Ivón Herrera-Najera; Carlos F. Arias; Celia Boukadida; Clara Esperanza Santacruz-Tinoco; Concepción Grajales-Muñiz; Consorcio Mexicano de Vigilancia Genómica (CoViGen-Mex). Authors (in alphabetical order): Julio Elias Alvarado-Yaah; Célida Duque Molina; Fernando Fontove-Herrera; Francisco Pulido; Gloria Elena Espinoza-Ayala; Gloria María Molina-Salinas; Gloria Vazquez; Hector Esteban Paz-Juárez; Hector Montoya-Fuentes; Helen Haydee Fernanda Ramírez-Plascencia; José Antonio Enciso-Moreno; José Esteban Muñoz-Medina; José de Jesús Nuñez-Contreras; Juan Bautista Chale-Dzul; Luis Alberto Ochoa-Carrera; Margarita Matías-Florentino; María Guadalupe Santiago-Mauricio; María Guadalupe de Jesús Mireles-Rivera; Nelly Sélem-Mojica; Pavel Isa; Ricardo Grande; Santiago Ávila-Ríos; Victor Eduardo Garcia-Arias; Víctor Hugo Borja-Aburto |
| EPI_ISL_1416468, EPI_ISL_1416469, EPI_ISL_1416470, EPI_ISL_1416471, EPI_ISL_1416472, EPI_ISL_1416473, EPI_ISL_1416474, EPI_ISL_1416547, EPI_ISL_1416548, EPI_ISL_1416549, EPI_ISL_1416594, EPI_ISL_1416599 | | | |
| see above | Laboratorio Central de Epidemiología (LCE) | Instituto de Biotecnología de la UNAM | Alejandro Sanchez-Flores; Alfredo Herrera-Estrella; Alicia Ocaña-Mondragón; Angel Gustavo Salas-Lais; Bernardo Martínez-Miguel; Blanca Taboada; Brenda Irasema Maldonado-Meza; Carla Ivón Herrera-Najera; Carlos F. Arias; Celia Boukadida; Clara Esperanza Santacruz-Tinoco; Concepción Grajales-Muñiz; Consorcio Mexicano de Vigilancia Genómica (CoViGen-Mex). Authors (in alphabetical order): Julio Elias Alvarado-Yaah; Fernando Fontove-Herrera; Francisco Pulido; Gloria Elena Espinoza-Ayala; Gloria María Molina-Salinas; Gloria Vazquez; Hector Esteban Paz-Juárez; Hector Montoya-Fuentes; Helen Haydee Fernanda Ramírez-Plascencia; Jorge Ivan Salinal-Nevarez; José Antonio Enciso-Moreno; José Esteban Muñoz-Medina; José de Jesús Nuñez-Contreras; Juan Bautista Chale-Dzul; Luis Alberto Ochoa-Carrera; Margarita Matías-Florentino; María Guadalupe Santiago-Mauricio; María Guadalupe de Jesús Mireles-Rivera; Nelly Sélem-Mojica; Pavel Isa; Ricardo Grande; Santiago Ávila-Ríos; Víctor Hugo Borja-Aburto |
| EPI_ISL_1320008, EPI_ISL_1320274, EPI_ISL_1320275, EPI_ISL_1320277, EPI_ISL_1320278, EPI_ISL_1320279, EPI_ISL_1320280, EPI_ISL_1320281, EPI_ISL_1320283, EPI_ISL_1320284, EPI_ISL_1320285, EPI_ISL_1320286, EPI_ISL_1320288, EPI_ISL_1320289, EPI_ISL_1320290, EPI_ISL_1320291, EPI_ISL_1320292, EPI_ISL_1320293, EPI_ISL_1320294, EPI_ISL_1320295, EPI_ISL_1320296, EPI_ISL_1320297, EPI_ISL_1320298, EPI_ISL_1320302, EPI_ISL_1320303, EPI_ISL_1320304, EPI_ISL_1320305, EPI_ISL_1320307, EPI_ISL_1320308, EPI_ISL_1320310, EPI_ISL_1320311, EPI_ISL_1320312, EPI_ISL_1320315, EPI_ISL_1320316, EPI_ISL_1320317, EPI_ISL_1320318, EPI_ISL_1320319, EPI_ISL_1320320, EPI_ISL_1320321, EPI_ISL_1320322, EPI_ISL_1320323, EPI_ISL_1320324, EPI_ISL_1320325, EPI_ISL_1320326, EPI_ISL_1320327, EPI_ISL_1320328, EPI_ISL_1320329, EPI_ISL_1320585, EPI_ISL_1321166, EPI_ISL_1321167, EPI_ISL_1321168, EPI_ISL_1321176, EPI_ISL_1321177, EPI_ISL_1321178, EPI_ISL_1321179, EPI_ISL_1321180, EPI_ISL_1321190, EPI_ISL_1321191, EPI_ISL_1321192, EPI_ISL_1321193, EPI_ISL_1321194, EPI_ISL_1321195, EPI_ISL_1321197, EPI_ISL_1321198, EPI_ISL_1321199, EPI_ISL_1321200, EPI_ISL_1321201, EPI_ISL_1321202, EPI_ISL_1321203, EPI_ISL_1321204, EPI_ISL_1321205, EPI_ISL_1321206, EPI_ISL_1321207, EPI_ISL_1321208, EPI_ISL_1321209, EPI_ISL_1321210, EPI_ISL_1321211, EPI_ISL_1321212, EPI_ISL_1321213, EPI_ISL_1321214, EPI_ISL_1321215, EPI_ISL_1321216, EPI_ISL_1321217, EPI_ISL_1321218, EPI_ISL_1321228, EPI_ISL_1321229, EPI_ISL_1321230, EPI_ISL_1321231, EPI_ISL_1321232, EPI_ISL_1321233, EPI_ISL_1321234, EPI_ISL_1321235, EPI_ISL_1321236, EPI_ISL_1321237, EPI_ISL_1321238, EPI_ISL_1321239, EPI_ISL_1321240, EPI_ISL_1321241, EPI_ISL_1321242, EPI_ISL_1321243, EPI_ISL_1321244, EPI_ISL_1321245, EPI_ISL_1321246, EPI_ISL_1321247, EPI_ISL_1321248, EPI_ISL_1321249, EPI_ISL_1321250, EPI_ISL_1321251, EPI_ISL_1321252, EPI_ISL_1321253, EPI_ISL_1321254, EPI_ISL_1321255, EPI_ISL_1321256, EPI_ISL_1321259, EPI_ISL_1321260, EPI_ISL_1321261, EPI_ISL_1321262, EPI_ISL_1321263, EPI_ISL_1321264, EPI_ISL_1321265, EPI_ISL_1321266, EPI_ISL_1321267, EPI_ISL_1321268, EPI_ISL_1321269, EPI_ISL_1321274, EPI_ISL_1321275, EPI_ISL_1321276, EPI_ISL_1321277, EPI_ISL_1321278, EPI_ISL_1321279, EPI_ISL_1321280, EPI_ISL_1321281, EPI_ISL_1321282, EPI_ISL_1321283, EPI_ISL_1321284, EPI_ISL_1321285, EPI_ISL_1321286, EPI_ISL_1321287, EPI_ISL_1321288, EPI_ISL_1321289, EPI_ISL_1321290, EPI_ISL_1321291, EPI_ISL_1321292, EPI_ISL_1321293, EPI_ISL_1321294, EPI_ISL_1321295, EPI_ISL_1321296, EPI_ISL_1321297, EPI_ISL_1321299, EPI_ISL_1321300, EPI_ISL_1321301, EPI_ISL_1321302, EPI_ISL_1321303, EPI_ISL_1321304, EPI_ISL_1321305, EPI_ISL_1321306, EPI_ISL_1321307, EPI_ISL_1321316, EPI_ISL_1321317, EPI_ISL_1321318, EPI_ISL_1321319, EPI_ISL_1321320, EPI_ISL_1321321, EPI_ISL_1321322, EPI_ISL_1321323, EPI_ISL_1321324, EPI_ISL_1321325, EPI_ISL_1321326, EPI_ISL_1321327, EPI_ISL_1321328, EPI_ISL_1321329, EPI_ISL_1321330, EPI_ISL_1321331, EPI_ISL_1321332, EPI_ISL_1321333, EPI_ISL_1321334, EPI_ISL_1321335, EPI_ISL_1321336, EPI_ISL_1321337, EPI_ISL_1321345, EPI_ISL_1321347, EPI_ISL_1321354, EPI_ISL_1338888, EPI_ISL_1338896, EPI_ISL_1338897, EPI_ISL_1338898, EPI_ISL_1338899, EPI_ISL_1338900, EPI_ISL_1338901, EPI_ISL_1338902, EPI_ISL_1338903, EPI_ISL_1338904, EPI_ISL_1338906, EPI_ISL_1338907, EPI_ISL_1338908, EPI_ISL_1338909, EPI_ISL_1338910, EPI_ISL_1338912, EPI_ISL_1338921, EPI_ISL_1338923, EPI_ISL_1338924, EPI_ISL_1338925, EPI_ISL_1338926, EPI_ISL_1338927, EPI_ISL_1338928, EPI_ISL_1338930, EPI_ISL_1338931, EPI_ISL_1338932, EPI_ISL_1338933, EPI_ISL_1338934, EPI_ISL_1338935, EPI_ISL_1338937, EPI_ISL_1338938, EPI_ISL_1338939, EPI_ISL_1338948, EPI_ISL_1338949, EPI_ISL_1338950, EPI_ISL_1338951, EPI_ISL_1338952, EPI_ISL_1338953, EPI_ISL_1338954, EPI_ISL_1338955, EPI_ISL_1338956, EPI_ISL_1338957, EPI_ISL_1338958, EPI_ISL_1338959, EPI_ISL_1338991, EPI_ISL_1338992, EPI_ISL_1339004, EPI_ISL_1339005, EPI_ISL_1339006, EPI_ISL_1339038, EPI_ISL_1339039, EPI_ISL_1339051, EPI_ISL_1339052, EPI_ISL_1339053, EPI_ISL_1339054, EPI_ISL_1339055, EPI_ISL_1339056, EPI_ISL_1339057, EPI_ISL_1339090, EPI_ISL_1339091, EPI_ISL_1339092, EPI_ISL_1339093, EPI_ISL_1339094, EPI_ISL_1339095, EPI_ISL_1339149, EPI_ISL_1339158, EPI_ISL_1339159, EPI_ISL_1339160, EPI_ISL_1339161, EPI_ISL_1339162, EPI_ISL_1339182, EPI_ISL_1339183, EPI_ISL_1339184, EPI_ISL_1339185, EPI_ISL_1339186, EPI_ISL_1339187, EPI_ISL_1339188, EPI_ISL_1339207, EPI_ISL_1339234, EPI_ISL_1339251, EPI_ISL_1339253, EPI_ISL_1421988, EPI_ISL_1421989, EPI_ISL_1421990, EPI_ISL_1421991, EPI_ISL_1421992, EPI_ISL_1422037, EPI_ISL_1422183, EPI_ISL_1422184, EPI_ISL_1422190, EPI_ISL_1422191, EPI_ISL_1422194, EPI_ISL_1422195, EPI_ISL_1422196, EPI_ISL_1422199, EPI_ISL_1422200, EPI_ISL_1422201, EPI_ISL_1422203, EPI_ISL_1422204, EPI_ISL_1422205, EPI_ISL_1422206, EPI_ISL_1461710, EPI_ISL_1461711, EPI_ISL_1461712, EPI_ISL_1461713, EPI_ISL_1461719, EPI_ISL_1461720, EPI_ISL_1461721, EPI_ISL_1461722, EPI_ISL_1461723, EPI_ISL_1461724, EPI_ISL_1461725, EPI_ISL_1461726, EPI_ISL_1461727, EPI_ISL_1461728, EPI_ISL_1461729, EPI_ISL_1461730, EPI_ISL_1461731, EPI_ISL_1461741, EPI_ISL_1461742, EPI_ISL_1461743, EPI_ISL_1461744, EPI_ISL_1461745, EPI_ISL_1461746, EPI_ISL_1461750, EPI_ISL_1461751, EPI_ISL_1461753, EPI_ISL_1461754, EPI_ISL_1461755, EPI_ISL_1461756, EPI_ISL_1461757, EPI_ISL_1461759, EPI_ISL_1461760, EPI_ISL_1461761, EPI_ISL_1461786, EPI_ISL_1461787, EPI_ISL_1461788, EPI_ISL_1461789, EPI_ISL_1461790, EPI_ISL_1461791, EPI_ISL_1461792, EPI_ISL_1461793, EPI_ISL_1461794, EPI_ISL_1461795, EPI_ISL_1461796, EPI_ISL_1461797, EPI_ISL_1461798, EPI_ISL_1461799, EPI_ISL_1461800, EPI_ISL_1461801, EPI_ISL_1461802, EPI_ISL_1461803, EPI_ISL_1461804, EPI_ISL_1461805, EPI_ISL_1461806, EPI_ISL_1461807, EPI_ISL_1461808, EPI_ISL_1461809, EPI_ISL_1461810, EPI_ISL_1461811, EPI_ISL_1461827, EPI_ISL_1461859, EPI_ISL_1461877, EPI_ISL_1461878, EPI_ISL_1461879, EPI_ISL_1481185, EPI_ISL_1481186, EPI_ISL_1481189, EPI_ISL_1481190, EPI_ISL_1481191, EPI_ISL_1481192, EPI_ISL_1481194, EPI_ISL_1481202, EPI_ISL_1481214, EPI_ISL_1481265, EPI_ISL_1481270, EPI_ISL_1513757, EPI_ISL_1513762, EPI_ISL_1513768, EPI_ISL_1513769, EPI_ISL_1513801, EPI_ISL_1513802, EPI_ISL_1513803, EPI_ISL_1513804, EPI_ISL_1513805, EPI_ISL_1513818, EPI_ISL_1513819, EPI_ISL_1513820, EPI_ISL_1513821, EPI_ISL_1513822, EPI_ISL_1513823, EPI_ISL_1513824, EPI_ISL_1513825, EPI_ISL_1513826, EPI_ISL_1513827, EPI_ISL_1513833, EPI_ISL_1513859, EPI_ISL_1513860, EPI_ISL_1513862, EPI_ISL_1513863, EPI_ISL_1513864, EPI_ISL_1513865, EPI_ISL_1513866, EPI_ISL_1513867, EPI_ISL_1513868, EPI_ISL_1513869, EPI_ISL_1513870, EPI_ISL_1513871, EPI_ISL_1513872, EPI_ISL_1513873, EPI_ISL_1513874, EPI_ISL_1513875, EPI_ISL_1513876, EPI_ISL_1513877, EPI_ISL_1513878, EPI_ISL_1513879, EPI_ISL_1513880, EPI_ISL_1513881, EPI_ISL_1513882, EPI_ISL_1513883, EPI_ISL_1513884, EPI_ISL_1513885, EPI_ISL_1513886, EPI_ISL_1513887, EPI_ISL_1513888, EPI_ISL_1513889, EPI_ISL_1513890, EPI_ISL_1513891, EPI_ISL_1513892, EPI_ISL_1513893, EPI_ISL_1513894, EPI_ISL_1513895, EPI_ISL_1513896, EPI_ISL_1513897, EPI_ISL_1513898, EPI_ISL_1513899, EPI_ISL_1513900, EPI_ISL_1513901, EPI_ISL_1513902, EPI_ISL_1513903, EPI_ISL_1513904, EPI_ISL_1513905, EPI_ISL_1513906, EPI_ISL_1513907, EPI_ISL_1513908, EPI_ISL_1513909, EPI_ISL_1514723, EPI_ISL_1514724, EPI_ISL_1514725, EPI_ISL_1514726, EPI_ISL_1514727, EPI_ISL_1514731, EPI_ISL_1514752, EPI_ISL_1514753, EPI_ISL_1514754, EPI_ISL_1514755, EPI_ISL_1514756, EPI_ISL_1514758, EPI_ISL_1514759, EPI_ISL_1514760, EPI_ISL_1514771, EPI_ISL_1514772, EPI_ISL_1514773, EPI_ISL_1514774, EPI_ISL_1514813, EPI_ISL_1514825, EPI_ISL_1514826, EPI_ISL_1514827, EPI_ISL_1514828, EPI_ISL_1514832, EPI_ISL_1514838, EPI_ISL_1514839, EPI_ISL_1514840, EPI_ISL_1514842, EPI_ISL_1514843, EPI_ISL_1514844, EPI_ISL_1514845, EPI_ISL_1514846, EPI_ISL_1514847, EPI_ISL_1514848, EPI_ISL_1514849, EPI_ISL_1514850, EPI_ISL_1514851, EPI_ISL_1514852, EPI_ISL_1514853, EPI_ISL_1514854, EPI_ISL_1514855, EPI_ISL_1514856, EPI_ISL_1514857, EPI_ISL_1514858, EPI_ISL_1514859, EPI_ISL_1514860, EPI_ISL_1514861, EPI_ISL_1514862, EPI_ISL_1514863, EPI_ISL_1514864, EPI_ISL_1514865, EPI_ISL_1514866, EPI_ISL_1514867, EPI_ISL_1514868, EPI_ISL_1514871, EPI_ISL_1514884, EPI_ISL_1514895, EPI_ISL_1514896, EPI_ISL_1514897, EPI_ISL_1514898, EPI_ISL_1514899, EPI_ISL_1514927, EPI_ISL_1514928, EPI_ISL_1514929, EPI_ISL_1514930, EPI_ISL_1514931, EPI_ISL_1514939, EPI_ISL_1514940, EPI_ISL_1514941, EPI_ISL_1514942, EPI_ISL_1514943, EPI_ISL_1514961, EPI_ISL_1514974, EPI_ISL_1514975, EPI_ISL_1514976, EPI_ISL_1514977, EPI_ISL_1514978, EPI_ISL_1514979, EPI_ISL_1514980, EPI_ISL_1514982, EPI_ISL_1514997, EPI_ISL_1514998, EPI_ISL_1514999, EPI_ISL_1515000, EPI_ISL_1515001, EPI_ISL_1515002, EPI_ISL_1515003, EPI_ISL_1515004, EPI_ISL_1515005, EPI_ISL_1515006, EPI_ISL_1515007, EPI_ISL_1515008, EPI_ISL_1515009, EPI_ISL_1515010, EPI_ISL_1515011, EPI_ISL_1515012, EPI_ISL_1515013, EPI_ISL_1515014, EPI_ISL_1515015, EPI_ISL_1515016, EPI_ISL_1515017, EPI_ISL_1515018, EPI_ISL_1515019, EPI_ISL_1515020, EPI_ISL_1515021, EPI_ISL_1515022, EPI_ISL_1515051, EPI_ISL_1515052, EPI_ISL_1515053, EPI_ISL_1515054, EPI_ISL_1515055, EPI_ISL_1515091, EPI_ISL_1515117, EPI_ISL_1515118, EPI_ISL_1515119, EPI_ISL_1515120, EPI_ISL_1515121, EPI_ISL_1515139, EPI_ISL_1515140, EPI_ISL_1515141, EPI_ISL_1515142, EPI_ISL_1515143, EPI_ISL_1515144, EPI_ISL_1515145, EPI_ISL_1515146, EPI_ISL_1515147, EPI_ISL_1515148, EPI_ISL_1515149, EPI_ISL_1515157, EPI_ISL_1515158, EPI_ISL_1515159, EPI_ISL_1515160, EPI_ISL_1515161, EPI_ISL_1515162, EPI_ISL_1515163, EPI_ISL_1515164, EPI_ISL_1515166, EPI_ISL_1515167, EPI_ISL_1515168, EPI_ISL_1515169, EPI_ISL_1515170, EPI_ISL_1515171, EPI_ISL_1515350, EPI_ISL_1515365, EPI_ISL_1515374, EPI_ISL_1515375, EPI_ISL_1515389, EPI_ISL_1515390, EPI_ISL_1515391, EPI_ISL_1515392, EPI_ISL_1515393, EPI_ISL_1515394, EPI_ISL_1515395, EPI_ISL_1515405, EPI_ISL_1515406, EPI_ISL_1515407, EPI_ISL_1515408, EPI_ISL_1515409, EPI_ISL_1515410, EPI_ISL_1515411, EPI_ISL_1515412, EPI_ISL_1515413, EPI_ISL_1515414, EPI_ISL_1515415, EPI_ISL_1515416, EPI_ISL_1515417, EPI_ISL_1515418, EPI_ISL_1515419, EPI_ISL_1515420, EPI_ISL_1515421, EPI_ISL_1515422, EPI_ISL_1515423, EPI_ISL_1515424, EPI_ISL_1515425, EPI_ISL_1515426, EPI_ISL_1515427, EPI_ISL_1515428, EPI_ISL_1515429, EPI_ISL_1515430, EPI_ISL_1515431, EPI_ISL_1515432, EPI_ISL_1515433, EPI_ISL_1515586, EPI_ISL_1515615, EPI_ISL_1515618, EPI_ISL_1515619, EPI_ISL_1515620, EPI_ISL_1515621, EPI_ISL_1515622, EPI_ISL_1515623, EPI_ISL_1515632, EPI_ISL_1515633, EPI_ISL_1515634, EPI_ISL_1515635, EPI_ISL_1515682, EPI_ISL_1515733, EPI_ISL_1548117, EPI_ISL_1548118, EPI_ISL_1548125, EPI_ISL_1548126, EPI_ISL_1548139, EPI_ISL_1548140, EPI_ISL_1549203, EPI_ISL_1549205, EPI_ISL_1549245, EPI_ISL_1549246, EPI_ISL_1549247, EPI_ISL_1549248, EPI_ISL_1549809, EPI_ISL_1549811, EPI_ISL_1549825, EPI_ISL_1549826, EPI_ISL_1549827, EPI_ISL_1549828, EPI_ISL_1549829, EPI_ISL_1549830, EPI_ISL_1549831, EPI_ISL_1549849, EPI_ISL_1549858, EPI_ISL_1549859, EPI_ISL_1549877, EPI_ISL_1549883, EPI_ISL_1549884, EPI_ISL_1549885, EPI_ISL_1549886, EPI_ISL_1549887, EPI_ISL_1549888, EPI_ISL_1549889, EPI_ISL_1549890, EPI_ISL_1549903, EPI_ISL_1549904, EPI_ISL_1549905, EPI_ISL_1549934, EPI_ISL_1549935, EPI_ISL_1549944, EPI_ISL_1549945, EPI_ISL_1549952, EPI_ISL_1550057, EPI_ISL_1550058, EPI_ISL_1550068, EPI_ISL_1550093, EPI_ISL_1550094, EPI_ISL_1550095, EPI_ISL_1550096, EPI_ISL_1550097, EPI_ISL_1550098, EPI_ISL_1550099, EPI_ISL_1550101, EPI_ISL_1550102, EPI_ISL_1550103, EPI_ISL_1550104, EPI_ISL_1550105, EPI_ISL_1550184, EPI_ISL_1550204, EPI_ISL_1550205, EPI_ISL_1550224, EPI_ISL_1550232, EPI_ISL_1550246, EPI_ISL_1550247, EPI_ISL_1695016, EPI_ISL_1695018 | | | |
| see above | Laboratory Corporation of America | Centers for Disease Control and Prevention Division of Viral Diseases, Pathogen Discovery | Adrian Paskey; Amanda Douglas; Amanda Suchanek; Andrea Throop; Ayla Burns; Ben L. Rambo-Martin; Benjamin Rambo-Martin; Bobbi Croy; Brian Krueger; Brian Norvell; Christopher Gulvick; Christos Petropoulos; Clinton R. Paden; Craig Lukasik; Dakota Howard; Darlene Wagner; Debbie Boles; Dhwani Batra; Duncan MacCannell; Eyad Almasri; Goran Stevovic; Howard Engler; Hrushikesh Deshmukh; Jake Humphrey; Jana Schroth; Jason Caravas; Joe Voshell; John Pruitt; Jonathan Meltzer; Jonathan Williams; Kara Moser; Kimberly Wagner; Lax Iyer; Lyndon Tilson; Manoj Jain; Marcia Eisenberg; Mary Ann Cristobal; Mary Williamson; Matthew Schmerer; Michael Levandoski; Mike Sapeta; Mindy Nye; Minoo Agarwal; Mohan Kolli; Nuthawin Charoensri; Oren Cohen; Peter W. Cook; Prashant Gupta; Qian Zeng; Rama Ghatti; Scott Parker; Scott Ryan; Scott Sammons; Shatavia Morrison; Stanley Letovsky; Steven Ragan; Suresh Babu Selvaraju; Susan Countryman; Susan Hicks; Suxiang Tong; Suzanne Dale; Thomas Urban; Tim Kuphal; Tricia Zwiefelhofer; Vincent Drouillon; Yvette Unoarumhi |
| EPI_ISL_1504027, EPI_ISL_1508909 | Los Angeles County PHL | Los Angeles County PHL | P. Hemarajata et al. |
| EPI_ISL_1502804, EPI_ISL_1502805, EPI_ISL_1502806, EPI_ISL_1502807, EPI_ISL_1502808, EPI_ISL_1502809, EPI_ISL_1506117, EPI_ISL_1506118, EPI_ISL_1506119 | | | |
| see above | MD Laboratories | Los Angeles County PHL | P. Hemarajata et al. |
| EPI_ISL_1664594, EPI_ISL_1664595, EPI_ISL_1664596, EPI_ISL_1664597, EPI_ISL_1664598, EPI_ISL_1664618, EPI_ISL_1664619, EPI_ISL_1669862, EPI_ISL_1921891 | | | |
| see above | Monterey County Public Health Laboratory | Monterey County Public Health Laboratory | Monterey County Public Health Laboratory |
| EPI_ISL_1477495, EPI_ISL_1477498, EPI_ISL_1477986, EPI_ISL_1478024, EPI_ISL_1478030, EPI_ISL_1478106, EPI_ISL_1478110, EPI_ISL_1478175, EPI_ISL_1529158, EPI_ISL_1529336, EPI_ISL_1701251 | | | |
| see above | Orange County Public Health Lab | Chan-Zuckerberg Biohub | CZB Cliahub Consortium |
| EPI_ISL_1303404, EPI_ISL_1303405, EPI_ISL_1303472, EPI_ISL_1303473, EPI_ISL_1497525, EPI_ISL_1506115, EPI_ISL_1508898, EPI_ISL_1508899, EPI_ISL_1508900, EPI_ISL_1508901 | | | |
| see above | Pomona Valley Hospital Medical Center | Los Angeles County PHL | P. Hemarajata et al. |
| EPI_ISL_1265930, EPI_ISL_1265954, EPI_ISL_1265979, EPI_ISL_1266205, EPI_ISL_1266216, EPI_ISL_1267283, EPI_ISL_1267391, EPI_ISL_1267412, EPI_ISL_1267422, EPI_ISL_1267430, EPI_ISL_1267432, EPI_ISL_1267434, EPI_ISL_1267445, EPI_ISL_1267479, EPI_ISL_1267481, EPI_ISL_1291002, EPI_ISL_1291005, EPI_ISL_1291015, EPI_ISL_1291028, EPI_ISL_1291032, EPI_ISL_1291042, EPI_ISL_1291091, EPI_ISL_1291092, EPI_ISL_1291094, EPI_ISL_1291096, EPI_ISL_1291099, EPI_ISL_1291100, EPI_ISL_1291105, EPI_ISL_1291106, EPI_ISL_1291109, EPI_ISL_1291110, EPI_ISL_1291111, EPI_ISL_1291112, EPI_ISL_1291114, EPI_ISL_1291115, EPI_ISL_1291116, EPI_ISL_1291117, EPI_ISL_1291118, EPI_ISL_1291121, EPI_ISL_1291123, EPI_ISL_1291124, EPI_ISL_1291125, EPI_ISL_1291127, EPI_ISL_1291128, EPI_ISL_1291129, EPI_ISL_1291130, EPI_ISL_1291131, EPI_ISL_1291132, EPI_ISL_1291134, EPI_ISL_1291135, EPI_ISL_1291137, EPI_ISL_1291138, EPI_ISL_1291140, EPI_ISL_1291141, EPI_ISL_1291142, EPI_ISL_1291143, EPI_ISL_1291144, EPI_ISL_1291145, EPI_ISL_1291146, EPI_ISL_1291147, EPI_ISL_1291149, EPI_ISL_1291150, EPI_ISL_1291151, EPI_ISL_1291152, EPI_ISL_1291153, EPI_ISL_1291154, EPI_ISL_1291155, EPI_ISL_1291156, EPI_ISL_1291157, EPI_ISL_1291159, EPI_ISL_1291160, EPI_ISL_1291161, EPI_ISL_1291162, EPI_ISL_1291163, EPI_ISL_1291164, EPI_ISL_1291165, EPI_ISL_1291166, EPI_ISL_1291167, EPI_ISL_1291170, EPI_ISL_1291171, EPI_ISL_1291172, EPI_ISL_1291173, EPI_ISL_1291179, EPI_ISL_1291180, EPI_ISL_1291182, EPI_ISL_1291186, EPI_ISL_1291187, EPI_ISL_1291188, EPI_ISL_1297683, EPI_ISL_1297688, EPI_ISL_1297689, EPI_ISL_1297691, EPI_ISL_1297701, EPI_ISL_1297705, EPI_ISL_1297708, EPI_ISL_1297719, EPI_ISL_1297735, EPI_ISL_1297737, EPI_ISL_1297738, EPI_ISL_1297741, EPI_ISL_1297744, EPI_ISL_1297745, EPI_ISL_1297747, EPI_ISL_1297751, EPI_ISL_1297754, EPI_ISL_1297757, EPI_ISL_1297758, EPI_ISL_1297760, EPI_ISL_1314735, EPI_ISL_1339830, | | | |

EPI_ISL_1339837, EPI_ISL_1339839, EPI_ISL_1339850, EPI_ISL_1339866, EPI_ISL_1339876, EPI_ISL_1339877, EPI_ISL_1339881, EPI_ISL_1339892, EPI_ISL_1339893, EPI_ISL_1339896, EPI_ISL_1339910, EPI_ISL_1339913, EPI_ISL_1339916, EPI_ISL_1339917, EPI_ISL_1339928, EPI_ISL_1339935, EPI_ISL_1339949, EPI_ISL_1339964, EPI_ISL_1340679, EPI_ISL_1367239, EPI_ISL_1367241, EPI_ISL_1367242, EPI_ISL_1367249, EPI_ISL_1367252, EPI_ISL_1367253, EPI_ISL_1367257, EPI_ISL_1367261, EPI_ISL_1367262, EPI_ISL_1367263, EPI_ISL_1367267, EPI_ISL_1367273, EPI_ISL_1367277, EPI_ISL_1367278, EPI_ISL_1367281, EPI_ISL_1367282, EPI_ISL_1367283, EPI_ISL_1367286, EPI_ISL_1367288, EPI_ISL_1367290, EPI_ISL_1367293, EPI_ISL_1367298, EPI_ISL_1367300, EPI_ISL_1367302, EPI_ISL_1367307, EPI_ISL_1367311, EPI_ISL_1367312, EPI_ISL_1367314, EPI_ISL_1367315, EPI_ISL_1367319, EPI_ISL_1367321, EPI_ISL_1367324, EPI_ISL_1367326, EPI_ISL_1367328, EPI_ISL_1391459, EPI_ISL_1391464, EPI_ISL_1391608, EPI_ISL_1391672, EPI_ISL_1391692, EPI_ISL_1422623, EPI_ISL_1422635, EPI_ISL_1422642, EPI_ISL_1422653, EPI_ISL_1422655, EPI_ISL_1422663, EPI_ISL_1422669, EPI_ISL_1422671, EPI_ISL_1422674, EPI_ISL_1422675, EPI_ISL_1422679, EPI_ISL_1422680, EPI_ISL_1422682, EPI_ISL_1422683, EPI_ISL_1422684, EPI_ISL_1422685, EPI_ISL_1422687, EPI_ISL_1422688, EPI_ISL_1422692, EPI_ISL_1422693, EPI_ISL_1422694, EPI_ISL_1422697, EPI_ISL_1422699, EPI_ISL_1422702, EPI_ISL_1422705, EPI_ISL_1422706, EPI_ISL_1422707, EPI_ISL_1422708, EPI_ISL_1422710, EPI_ISL_1422787, EPI_ISL_1422795, EPI_ISL_1422798, EPI_ISL_1444408, EPI_ISL_1444409, EPI_ISL_1444410, EPI_ISL_1444411, EPI_ISL_1444412, EPI_ISL_1444414, EPI_ISL_1444417, EPI_ISL_1444419, EPI_ISL_1444421, EPI_ISL_1444422, EPI_ISL_1444426, EPI_ISL_1444427, EPI_ISL_1444428, EPI_ISL_1444429, EPI_ISL_1444430, EPI_ISL_1444431, EPI_ISL_1444433, EPI_ISL_1444434, EPI_ISL_1444435, EPI_ISL_1444436, EPI_ISL_1444437, EPI_ISL_1444438, EPI_ISL_1444439, EPI_ISL_1444440, EPI_ISL_1444441, EPI_ISL_1444442, EPI_ISL_1444443, EPI_ISL_1444444, EPI_ISL_1444445, EPI_ISL_1444446, EPI_ISL_1444447, EPI_ISL_1444448, EPI_ISL_1444449, EPI_ISL_1444452, EPI_ISL_1444453, EPI_ISL_1444454, EPI_ISL_1444455, EPI_ISL_1444456, EPI_ISL_1444457, EPI_ISL_1444459, EPI_ISL_1444461, EPI_ISL_1444462, EPI_ISL_1444463, EPI_ISL_1444465, EPI_ISL_1444466, EPI_ISL_1444468, EPI_ISL_1444469, EPI_ISL_1444470, EPI_ISL_1444473, EPI_ISL_1444474, EPI_ISL_1444475, EPI_ISL_1444476, EPI_ISL_1444477, EPI_ISL_1444478, EPI_ISL_1444479, EPI_ISL_1444481, EPI_ISL_1444482, EPI_ISL_1444483, EPI_ISL_1444484, EPI_ISL_1444485, EPI_ISL_1444486, EPI_ISL_1444487, EPI_ISL_1444489, EPI_ISL_1444490, EPI_ISL_1444491, EPI_ISL_1444493, EPI_ISL_1444494, EPI_ISL_1444495, EPI_ISL_1444496, EPI_ISL_1444497, EPI_ISL_1444499, EPI_ISL_1444523, EPI_ISL_1444527, EPI_ISL_1444566, EPI_ISL_1444568, EPI_ISL_1444569, EPI_ISL_1444571, EPI_ISL_1444573, EPI_ISL_1444574, EPI_ISL_1444576, EPI_ISL_1444578, EPI_ISL_1444579, EPI_ISL_1444580, EPI_ISL_1444581, EPI_ISL_1444583, EPI_ISL_1444585, EPI_ISL_1444588, EPI_ISL_1444589, EPI_ISL_1444590, EPI_ISL_1444592, EPI_ISL_1444593, EPI_ISL_1444594, EPI_ISL_1444595, EPI_ISL_1444596, EPI_ISL_1444597, EPI_ISL_1444601, EPI_ISL_1444602, EPI_ISL_1444604, EPI_ISL_1444605, EPI_ISL_1444606, EPI_ISL_1444608, EPI_ISL_1444609, EPI_ISL_1444611, EPI_ISL_1444612, EPI_ISL_1444613, EPI_ISL_1444614, EPI_ISL_1444615, EPI_ISL_1444617, EPI_ISL_1444618, EPI_ISL_1444619, EPI_ISL_1444620, EPI_ISL_1444621, EPI_ISL_1444622, EPI_ISL_1444623, EPI_ISL_1444624, EPI_ISL_1444625, EPI_ISL_1444626, EPI_ISL_1444627, EPI_ISL_1444628, EPI_ISL_1444631, EPI_ISL_1444632, EPI_ISL_1444633, EPI_ISL_1444638, EPI_ISL_1444639, EPI_ISL_1444640, EPI_ISL_1444642, EPI_ISL_1444643, EPI_ISL_1444644, EPI_ISL_1444645, EPI_ISL_1444646, EPI_ISL_1444647, EPI_ISL_1444648, EPI_ISL_1444649, EPI_ISL_1444650, EPI_ISL_1444651, EPI_ISL_1444652, EPI_ISL_1444653, EPI_ISL_1444654, EPI_ISL_1444655, EPI_ISL_1444656, EPI_ISL_1444657, EPI_ISL_1444658, EPI_ISL_1444659, EPI_ISL_1444660, EPI_ISL_1444661, EPI_ISL_1444662, EPI_ISL_1444663, EPI_ISL_1444664, EPI_ISL_1444665, EPI_ISL_1444666, EPI_ISL_1444667, EPI_ISL_1444669, EPI_ISL_1444714, EPI_ISL_1460730, EPI_ISL_1460731, EPI_ISL_1460733, EPI_ISL_1460735, EPI_ISL_1460737, EPI_ISL_1460739, EPI_ISL_1460740, EPI_ISL_1460741, EPI_ISL_1460742, EPI_ISL_1460748, EPI_ISL_1460751, EPI_ISL_1460752, EPI_ISL_1460753, EPI_ISL_1460754, EPI_ISL_1460755, EPI_ISL_1460756, EPI_ISL_1460757, EPI_ISL_1460760, EPI_ISL_1460761, EPI_ISL_1460762, EPI_ISL_1460763, EPI_ISL_1460764, EPI_ISL_1460765, EPI_ISL_1460766, EPI_ISL_1460767, EPI_ISL_1460768, EPI_ISL_1460769, EPI_ISL_1460770, EPI_ISL_1460771, EPI_ISL_1460772, EPI_ISL_1460774, EPI_ISL_1460776, EPI_ISL_1460777, EPI_ISL_1460778, EPI_ISL_1460779, EPI_ISL_1460780, EPI_ISL_1460782, EPI_ISL_1460783, EPI_ISL_1460784, EPI_ISL_1479739, EPI_ISL_1479757, EPI_ISL_1479758, EPI_ISL_1479759, EPI_ISL_1479767, EPI_ISL_1479768, EPI_ISL_1479769, EPI_ISL_1479770, EPI_ISL_1479774, EPI_ISL_1479776, EPI_ISL_1479779, EPI_ISL_1479782, EPI_ISL_1479783, EPI_ISL_1479784, EPI_ISL_1479789, EPI_ISL_1479793, EPI_ISL_1479798, EPI_ISL_1479801, EPI_ISL_1479950, EPI_ISL_1480817, EPI_ISL_1480838, EPI_ISL_1480839, EPI_ISL_1480842, EPI_ISL_1480848, EPI_ISL_1480858, EPI_ISL_1480862, EPI_ISL_1491950, EPI_ISL_1493939, EPI_ISL_1493941, EPI_ISL_1493949, EPI_ISL_1493951, EPI_ISL_1493960, EPI_ISL_1493962, EPI_ISL_1493963, EPI_ISL_1493966, EPI_ISL_1493976, EPI_ISL_1493978, EPI_ISL_1493983, EPI_ISL_1494011, EPI_ISL_1494021, EPI_ISL_1511667, EPI_ISL_1511706, EPI_ISL_1511711, EPI_ISL_1511721, EPI_ISL_1511735, EPI_ISL_1525910, EPI_ISL_1525912, EPI_ISL_1525939, EPI_ISL_1525955, EPI_ISL_1525969, EPI_ISL_1525971, EPI_ISL_1525977, EPI_ISL_1525978, EPI_ISL_1525979, EPI_ISL_1525980, EPI_ISL_1525981, EPI_ISL_1525983, EPI_ISL_1525984, EPI_ISL_1525986, EPI_ISL_1525987, EPI_ISL_1525988, EPI_ISL_1525989, EPI_ISL_1525992, EPI_ISL_1525993, EPI_ISL_1525994, EPI_ISL_1525996, EPI_ISL_1525998, EPI_ISL_1525999, EPI_ISL_1526000, EPI_ISL_1526001, EPI_ISL_1526002, EPI_ISL_1526003, EPI_ISL_1526004, EPI_ISL_1526005, EPI_ISL_1526008, EPI_ISL_1526009, EPI_ISL_1526010, EPI_ISL_1526013, EPI_ISL_1526016, EPI_ISL_1526017, EPI_ISL_1526018, EPI_ISL_1526019, EPI_ISL_1526020, EPI_ISL_1526021, EPI_ISL_1526022, EPI_ISL_1526024, EPI_ISL_1526025, EPI_ISL_1526026, EPI_ISL_1526027, EPI_ISL_1526028, EPI_ISL_1526032, EPI_ISL_1526033, EPI_ISL_1526034, EPI_ISL_1526035, EPI_ISL_1526037, EPI_ISL_1526038, EPI_ISL_1526039, EPI_ISL_1526040, EPI_ISL_1526041, EPI_ISL_1526042, EPI_ISL_1526043, EPI_ISL_1526044, EPI_ISL_1526045, EPI_ISL_1526046, EPI_ISL_1526049, EPI_ISL_1526050, EPI_ISL_1526051, EPI_ISL_1526052, EPI_ISL_1526054, EPI_ISL_1526056, EPI_ISL_1526057, EPI_ISL_1526058, EPI_ISL_1526061, EPI_ISL_1526062, EPI_ISL_1526063, EPI_ISL_1526064, EPI_ISL_1526065, EPI_ISL_1526068, EPI_ISL_1526070, EPI_ISL_1551776, EPI_ISL_1551778, EPI_ISL_1551780, EPI_ISL_1551781, EPI_ISL_1551782, EPI_ISL_1551783, EPI_ISL_1551784, EPI_ISL_1551785, EPI_ISL_1551788, EPI_ISL_1551789, EPI_ISL_1551791, EPI_ISL_1551792, EPI_ISL_1551793, EPI_ISL_1551794, EPI_ISL_1551795, EPI_ISL_1551799, EPI_ISL_1551801, EPI_ISL_1551802, EPI_ISL_1551803, EPI_ISL_1551805, EPI_ISL_1551806, EPI_ISL_1551807, EPI_ISL_1551808, EPI_ISL_1551809, EPI_ISL_1551810, EPI_ISL_1551813, EPI_ISL_1551814, EPI_ISL_1551816, EPI_ISL_1551817, EPI_ISL_1551820, EPI_ISL_1551881, EPI_ISL_1551883, EPI_ISL_1551885, EPI_ISL_1551886, EPI_ISL_1551887, EPI_ISL_1551890, EPI_ISL_1551891, EPI_ISL_1551892, EPI_ISL_1551895, EPI_ISL_1551899, EPI_ISL_1551900, EPI_ISL_1551901, EPI_ISL_1551903, EPI_ISL_1551905, EPI_ISL_1551906, EPI_ISL_1551908, EPI_ISL_1551911, EPI_ISL_1551912, EPI_ISL_1551913, EPI_ISL_1551916, EPI_ISL_1551917, EPI_ISL_1551919, EPI_ISL_1551920, EPI_ISL_1551921, EPI_ISL_1551922, EPI_ISL_1551923, EPI_ISL_1552010, EPI_ISL_1552012, EPI_ISL_1552013, EPI_ISL_1552020, EPI_ISL_1552021, EPI_ISL_1552025, EPI_ISL_1552028, EPI_ISL_1552029, EPI_ISL_1552031, EPI_ISL_1552032, EPI_ISL_1552033, EPI_ISL_1552037, EPI_ISL_1552038, EPI_ISL_1552040, EPI_ISL_1552047, EPI_ISL_1552051, EPI_ISL_1552055, EPI_ISL_1552057, EPI_ISL_1552062, EPI_ISL_1552063, EPI_ISL_1552064, EPI_ISL_1552070, EPI_ISL_1552074, EPI_ISL_1552174, EPI_ISL_1552202, EPI_ISL_1552214, EPI_ISL_1552721, EPI_ISL_1552738, EPI_ISL_1552762, EPI_ISL_1552763, EPI_ISL_1552765, EPI_ISL_1552767, EPI_ISL_1552768, EPI_ISL_1552769, EPI_ISL_1552774, EPI_ISL_1552781, EPI_ISL_1552785, EPI_ISL_1552787, EPI_ISL_1552788, EPI_ISL_1552789, EPI_ISL_1552791, EPI_ISL_1552798, EPI_ISL_1552799, EPI_ISL_1552800, EPI_ISL_1552801, EPI_ISL_1552802, EPI_ISL_1552803, EPI_ISL_1552804, EPI_ISL_1552806, EPI_ISL_1552809, EPI_ISL_1552813, EPI_ISL_1552815, EPI_ISL_1552816, EPI_ISL_1559413, EPI_ISL_1559416, EPI_ISL_1559418, EPI_ISL_1559425, EPI_ISL_1559431, EPI_ISL_1559432, EPI_ISL_1559437, EPI_ISL_1559443, EPI_ISL_1559450, EPI_ISL_1559454, EPI_ISL_1559455, EPI_ISL_1559457, EPI_ISL_1559461, EPI_ISL_1559463, EPI_ISL_1559464, EPI_ISL_1559480, EPI_ISL_1559485, EPI_ISL_1559486, EPI_ISL_1559488, EPI_ISL_1559491, EPI_ISL_1559495, EPI_ISL_1559496, EPI_ISL_1559498, EPI_ISL_1559506, EPI_ISL_1559510, EPI_ISL_1559512, EPI_ISL_1559515, EPI_ISL_1559517, EPI_ISL_1559518, EPI_ISL_1559534, EPI_ISL_1559535, EPI_ISL_1559536, EPI_ISL_1559537, EPI_ISL_1559538, EPI_ISL_1559540, EPI_ISL_1559541, EPI_ISL_1559542, EPI_ISL_1559543, EPI_ISL_1559545, EPI_ISL_1559549, EPI_ISL_1559550, EPI_ISL_1559551, EPI_ISL_1559553, EPI_ISL_1559555, EPI_ISL_1559559, EPI_ISL_1559561, EPI_ISL_1559562, EPI_ISL_1559563, EPI_ISL_1559565, EPI_ISL_1559569, EPI_ISL_1559577, EPI_ISL_1559579, EPI_ISL_1559582, EPI_ISL_1559583, EPI_ISL_1559586, EPI_ISL_1582093, EPI_ISL_1582122, EPI_ISL_1582123, EPI_ISL_1582135, EPI_ISL_1582136, EPI_ISL_1582139, EPI_ISL_1582140, EPI_ISL_1582146, EPI_ISL_1582163, EPI_ISL_1694666, EPI_ISL_1694667, EPI_ISL_1694668, EPI_ISL_1694692, EPI_ISL_1694694

| | | | |
|---|---|---|---|
| see above | Quest Diagnostics Incorporated | Centers for Disease Control and Prevention Division of Viral Diseases, Pathogen Discovery | A. Gerasimova; A. Perez; Adrian Paskey; B. Anderson; Ben L. Rambo-Martin; Benjamin Rambo-Martin; Christopher Gulvick; Clinton R. Paden; Dakota Howard; Darlene Wagner; Dhwani Batra; Duncan MacCannell; F. Lacbawan; I. A. Shlyakhter; Jason Caravas; K.E. Livingston; Kara Moser; L.E. Bernstein; M. Hua; Matthew Schmerer; P. Tanpaiboon; Peter W. Cook; R. M. Kagan; R. Owen; R. V. Rolando; S. H. Rosenthal; Scott Sammons; Shatavia Morrison; Suxiang Tong; Y. Liu; Yvette Unoarumhi |

EPI_ISL_1794769, EPI_ISL_1794770, EPI_ISL_1794772, EPI_ISL_1794777, EPI_ISL_1794778, EPI_ISL_1794779, EPI_ISL_1794781

| | | | |
|---|---|---|---|
| see above | Rady Children's Hospital - San Diego | Andersen lab at Scripps Research | Christina Clarke; David Dimmock; Denise Malicki; Kathryn Bouic; Linda Luo; SEARCH Alliance San Diego with Nanda Radamchar; Teresa Mueller |
| EPI_ISL_2313501, EPI_ISL_2313691, EPI_ISL_2313799, EPI_ISL_2313878 | Respiratory Viruses Branch, Centers for Disease Control and Prevention | Respiratory Viruses Branch, Centers for Disease Control and Prevention | Batra, D.; Bixby, C.; C.R.; Caravas, J.; Cook; Goswami, C.; Grimwood, R.; Gulvick, C.; Hager, R.; Howard, D.; MacCannell, D.; Morrison, S.; Moser, K.; P.W.; Paden; Paskey, A.; Rambo-Martin, B.; Sammons, S.; Schmerer, M.; Schultz, J.; Unoarumhi, Y.; Wagner, D.; Wang, Y. |

EPI_ISL_1315073, EPI_ISL_1366749, EPI_ISL_1366750, EPI_ISL_1366751, EPI_ISL_1468832, EPI_ISL_1468844, EPI_ISL_1468877, EPI_ISL_1468878, EPI_ISL_1468879, EPI_ISL_1468880, EPI_ISL_1468881, EPI_ISL_1468914, EPI_ISL_1468915, EPI_ISL_1468916, EPI_ISL_1468922, EPI_ISL_1468923, EPI_ISL_1468924, EPI_ISL_1468925, EPI_ISL_1468926, EPI_ISL_1468927, EPI_ISL_1468928, EPI_ISL_1468929, EPI_ISL_1468930, EPI_ISL_1468931, EPI_ISL_1468932, EPI_ISL_1468933, EPI_ISL_1468934, EPI_ISL_1468935, EPI_ISL_1468936, EPI_ISL_1468937, EPI_ISL_1468938, EPI_ISL_1468939, EPI_ISL_1468940, EPI_ISL_1468941, EPI_ISL_1468942, EPI_ISL_1468943, EPI_ISL_1468944, EPI_ISL_1468945, EPI_ISL_1468946, EPI_ISL_1468948, EPI_ISL_1468949, EPI_ISL_1468951, EPI_ISL_1468982, EPI_ISL_1468985, EPI_ISL_1468988, EPI_ISL_1468992, EPI_ISL_1468993, EPI_ISL_1468998, EPI_ISL_1469003, EPI_ISL_1469007, EPI_ISL_1469011, EPI_ISL_1469018, EPI_ISL_1469023, EPI_ISL_1469024, EPI_ISL_1469025, EPI_ISL_1469026, EPI_ISL_1469027, EPI_ISL_1469028, EPI_ISL_1469029, EPI_ISL_1469030, EPI_ISL_1469031, EPI_ISL_1469032, EPI_ISL_1469033, EPI_ISL_1469034, EPI_ISL_1469035, EPI_ISL_1469036, EPI_ISL_1469037, EPI_ISL_1469038, EPI_ISL_1469039, EPI_ISL_1469040, EPI_ISL_1469041, EPI_ISL_1469042, EPI_ISL_1791403, EPI_ISL_1791404, EPI_ISL_1791405, EPI_ISL_1791406, EPI_ISL_1791407, EPI_ISL_1791408, EPI_ISL_1791409, EPI_ISL_1791410, EPI_ISL_1791411, EPI_ISL_1791412, EPI_ISL_1791413, EPI_ISL_1791414, EPI_ISL_1791415, EPI_ISL_1791416, EPI_ISL_1791417, EPI_ISL_1791418, EPI_ISL_1791419, EPI_ISL_1791420, EPI_ISL_1791421, EPI_ISL_1791422, EPI_ISL_1791423, EPI_ISL_1791424, EPI_ISL_1791425, EPI_ISL_1791426, EPI_ISL_1791427, EPI_ISL_1791428, EPI_ISL_1794619, EPI_ISL_1794649, EPI_ISL_1794654, EPI_ISL_1794997, EPI_ISL_1795012, EPI_ISL_1795023, EPI_ISL_1795034, EPI_ISL_1969251, EPI_ISL_1969256, EPI_ISL_1969261, EPI_ISL_1969264, EPI_ISL_1969267, EPI_ISL_1969269

| | | | |
|---|---|---|---|
| see above | San Diego County Public Health Laboratory | Andersen lab at Scripps Research | Brett Austin; Jovan Shephard; SEARCH Alliance San Diego with Ashleigh Murphy; SEARCH Alliance San Diego with Tracy Basler |

EPI_ISL_1529148, EPI_ISL_1529193, EPI_ISL_1529205, EPI_ISL_1529234, EPI_ISL_1529240, EPI_ISL_1529245, EPI_ISL_1529259, EPI_ISL_1529263, EPI_ISL_1529267, EPI_ISL_1529284, EPI_ISL_1529317, EPI_ISL_1663931, EPI_ISL_1663933, EPI_ISL_1663939, EPI_ISL_1663963, EPI_ISL_1663979, EPI_ISL_1663982, EPI_ISL_1663993, EPI_ISL_1664004, EPI_ISL_1664016, EPI_ISL_1664025, EPI_ISL_1664036, EPI_ISL_1664043, EPI_ISL_1664062, EPI_ISL_1664073, EPI_ISL_1664079, EPI_ISL_1664080, EPI_ISL_1664083, EPI_ISL_1664089, EPI_ISL_1664094, EPI_ISL_1664095, EPI_ISL_1664107, EPI_ISL_1701401, EPI_ISL_1701413, EPI_ISL_1701437, EPI_ISL_1701439, EPI_ISL_1701440, EPI_ISL_1701445, EPI_ISL_1701457, EPI_ISL_1701462, EPI_ISL_1701477, EPI_ISL_1701480, EPI_ISL_1701481, EPI_ISL_1701486, EPI_ISL_1701496, EPI_ISL_1701498, EPI_ISL_1701501, EPI_ISL_1701510, EPI_ISL_1701516, EPI_ISL_1701518, EPI_ISL_1701523, EPI_ISL_1701526, EPI_ISL_1701528, EPI_ISL_1701532, EPI_ISL_1701544, EPI_ISL_1701550, EPI_ISL_1701554, EPI_ISL_1701564, EPI_ISL_1701567, EPI_ISL_1701573, EPI_ISL_1701575, EPI_ISL_1701583, EPI_ISL_1701593, EPI_ISL_1701596, EPI_ISL_1701598, EPI_ISL_1701609, EPI_ISL_1701612, EPI_ISL_1701625, EPI_ISL_1701630, EPI_ISL_1701631, EPI_ISL_1701640, EPI_ISL_1701645, EPI_ISL_1701653, EPI_ISL_1821319, EPI_ISL_1821320, EPI_ISL_1821324, EPI_ISL_1821327, EPI_ISL_1821329, EPI_ISL_1821330, EPI_ISL_1821331, EPI_ISL_1821333, EPI_ISL_1821337, EPI_ISL_1821338, EPI_ISL_1821339, EPI_ISL_1821341, EPI_ISL_1821342, EPI_ISL_1821350, EPI_ISL_1821355, EPI_ISL_1821392, EPI_ISL_1821396, EPI_ISL_1821397, EPI_ISL_1821398, EPI_ISL_1821402, EPI_ISL_1821405, EPI_ISL_1821413, EPI_ISL_1821414, EPI_ISL_1821419, EPI_ISL_1821426, EPI_ISL_1821427, EPI_ISL_1821437, EPI_ISL_1821443, EPI_ISL_1821445, EPI_ISL_1821455, EPI_ISL_1821458, EPI_ISL_1821460, EPI_ISL_1821461, EPI_ISL_1821470, EPI_ISL_2102744, EPI_ISL_2102766, EPI_ISL_2102774, EPI_ISL_2102781, EPI_ISL_2102784, EPI_ISL_2102785, EPI_ISL_2102803, EPI_ISL_2102810, EPI_ISL_2102823, EPI_ISL_2102827, EPI_ISL_2102831, EPI_ISL_2102851, EPI_ISL_2102862, EPI_ISL_2102865, EPI_ISL_2102871, EPI_ISL_2102875, EPI_ISL_2102876, EPI_ISL_2102882, EPI_ISL_2102911, EPI_ISL_2102920, EPI_ISL_2102941, EPI_ISL_2102950, EPI_ISL_2102956, EPI_ISL_2102959, EPI_ISL_2102969, EPI_ISL_2103014, EPI_ISL_2103021, EPI_ISL_2103042, EPI_ISL_2103059, EPI_ISL_2103083, EPI_ISL_2103089, EPI_ISL_2103123, EPI_ISL_2103144, EPI_ISL_2103148, EPI_ISL_2103190

| | | | |
|---|---|---|---|
| see above | Santa Clara County Public Health Laboratory | Chan-Zuckerberg Biohub | CZB Cliahub Consortium |

EPI_ISL_1203817, EPI_ISL_1222757, EPI_ISL_1335868, EPI_ISL_1335869, EPI_ISL_1335870, EPI_ISL_1372415, EPI_ISL_1406155, EPI_ISL_1406168, EPI_ISL_1406169, EPI_ISL_1483802, EPI_ISL_1920832

| | | | |
|---|---|---|---|
| see above | Santa Clara County Public Health Laboratory | Santa Clara County Public Health Laboratory | Santa Clara County Public Health Department |

EPI_ISL_1483799, EPI_ISL_1532131, EPI_ISL_1532133, EPI_ISL_1532134, EPI_ISL_1532135, EPI_ISL_1532136, EPI_ISL_1532137, EPI_ISL_1532138

| | | | |
|---|---|---|---|
| see above | Santa Clara Valley Medical Center | Santa Clara County Public Health Laboratory | Santa Clara County Public Health Department |
| EPI_ISL_1664288 | Santa Clara Valley Medical Center | Santa Clara County public Health Laboratory | Santa Clara County Public Health Department |

EPI_ISL_1468829, EPI_ISL_1468830, EPI_ISL_1468831, EPI_ISL_1468833, EPI_ISL_1468834, EPI_ISL_1468835, EPI_ISL_1468836, EPI_ISL_1468838, EPI_ISL_1468840, EPI_ISL_1468845, EPI_ISL_1468846, EPI_ISL_1468848, EPI_ISL_1468850, EPI_ISL_1468851, EPI_ISL_1468852, EPI_ISL_1468853, EPI_ISL_1468855, EPI_ISL_1468856, EPI_ISL_1468857, EPI_ISL_1468858, EPI_ISL_1468859, EPI_ISL_1468860, EPI_ISL_1468861, EPI_ISL_1468862, EPI_ISL_1468863, EPI_ISL_1468864, EPI_ISL_1468865, EPI_ISL_1468866, EPI_ISL_1468867, EPI_ISL_1468868, EPI_ISL_1468869, EPI_ISL_1468870, EPI_ISL_1468871, EPI_ISL_1468872, EPI_ISL_1468873, EPI_ISL_1468889, EPI_ISL_1468891, EPI_ISL_1468892, EPI_ISL_1468893, EPI_ISL_1468894, EPI_ISL_1468895, EPI_ISL_1468896, EPI_ISL_1468897, EPI_ISL_1468898, EPI_ISL_1468899, EPI_ISL_1468900, EPI_ISL_1468901, EPI_ISL_1468902, EPI_ISL_1468903, EPI_ISL_1468904, EPI_ISL_1468905, EPI_ISL_1468910, EPI_ISL_1468911, EPI_ISL_1468912, EPI_ISL_1468913, EPI_ISL_1468917, EPI_ISL_1468918, EPI_ISL_1468919, EPI_ISL_1468920, EPI_ISL_1468921, EPI_ISL_1468952, EPI_ISL_1468954, EPI_ISL_1468957, EPI_ISL_1468958, EPI_ISL_1468960, EPI_ISL_1468961, EPI_ISL_1468962, EPI_ISL_1468963, EPI_ISL_1468964, EPI_ISL_1468965, EPI_ISL_1468966, EPI_ISL_1468967, EPI_ISL_1468968, EPI_ISL_1468969, EPI_ISL_1468970, EPI_ISL_1468971, EPI_ISL_1468972, EPI_ISL_1468973, EPI_ISL_1468974, EPI_ISL_1468975, EPI_ISL_1468976, EPI_ISL_1468977, EPI_ISL_1468978, EPI_ISL_1469061, EPI_ISL_1469062, EPI_ISL_1469063, EPI_ISL_1469064, EPI_ISL_1469065, EPI_ISL_1469066, EPI_ISL_1469067, EPI_ISL_1469068, EPI_ISL_1469069, EPI_ISL_1469070, EPI_ISL_1469071, EPI_ISL_1469072, EPI_ISL_1469073, EPI_ISL_1469074, EPI_ISL_1469075, EPI_ISL_1469076, EPI_ISL_1469077, EPI_ISL_1469078, EPI_ISL_1469079, EPI_ISL_1469080, EPI_ISL_1469081, EPI_ISL_1469082, EPI_ISL_1469083, EPI_ISL_1469084, EPI_ISL_1469085, EPI_ISL_1469086, EPI_ISL_1469087, EPI_ISL_1531777, EPI_ISL_1531778, EPI_ISL_1531779, EPI_ISL_1531780, EPI_ISL_1531781, EPI_ISL_1531782, EPI_ISL_1531783, EPI_ISL_1531784, EPI_ISL_1531785, EPI_ISL_1531786, EPI_ISL_1531787, EPI_ISL_1531788, EPI_ISL_1531789, EPI_ISL_1531790, EPI_ISL_1531791, EPI_ISL_1531792, EPI_ISL_1531793, EPI_ISL_1531794, EPI_ISL_1531795, EPI_ISL_1531796, EPI_ISL_1531797, EPI_ISL_1531798, EPI_ISL_1531799, EPI_ISL_1531800, EPI_ISL_1531801, EPI_ISL_1531802, EPI_ISL_1531803, EPI_ISL_1531804, EPI_ISL_1531805, EPI_ISL_1531806, EPI_ISL_1794685, EPI_ISL_1794686, EPI_ISL_1794687, EPI_ISL_1794688, EPI_ISL_1794689, EPI_ISL_1794690, EPI_ISL_1794691, EPI_ISL_1794692, EPI_ISL_1794693, EPI_ISL_1794694, EPI_ISL_1794695, EPI_ISL_1794696, EPI_ISL_1794697, EPI_ISL_1794698, EPI_ISL_1794699, EPI_ISL_1794703, EPI_ISL_1794704, EPI_ISL_1794713, EPI_ISL_1794714, EPI_ISL_1794715, EPI_ISL_1794716, EPI_ISL_1794717, EPI_ISL_1794718, EPI_ISL_1794719, EPI_ISL_1794720, EPI_ISL_1969312, EPI_ISL_1969313

| | | | |
|---|---|---|---|
| see above | Sharp HealthCare Laboratory | Andersen lab at Scripps Research | Art Mendoza; Cathy Woerle; Jacquelyn Berumen; Liam McGinnis; Omid Bakhtar; SEARCH Alliance San Diego with Aaron Harding |

EPI_ISL_1379889, EPI_ISL_1675223, EPI_ISL_1675224, EPI_ISL_1675225, EPI_ISL_1701679, EPI_ISL_2027136, EPI_ISL_2027138

| | | | |
|---|---|---|---|
| see above | Stanford Health Care | Stanford University School of Medicine, Clinical Virology Laboratory | Becky Jiang; Bernadette Troung; Bernadette Truong; Daniel Solis; Fumiko Yamamoto; Hannah Wang; Jacob Miller; James Zehnder; Kenji O. Mfuh; Malaya K. Sahoo; Mamdouh Sibai; Marilyn Mar; Michelle Verghese; Naomi Iwai; and Benjamin A. Pinsky |

EPI_ISL_1303459, EPI_ISL_1303460, EPI_ISL_1303461, EPI_ISL_1379958, EPI_ISL_1379959, EPI_ISL_1379960, EPI_ISL_1379961, EPI_ISL_1379962, EPI_ISL_1379963, EPI_ISL_1379964, EPI_ISL_1379965, EPI_ISL_1379966, EPI_ISL_1379967, EPI_ISL_1379968, EPI_ISL_1379969, EPI_ISL_1379970, EPI_ISL_1379971, EPI_ISL_1379972, EPI_ISL_1379973, EPI_ISL_1379974, EPI_ISL_1379975, EPI_ISL_1497541, EPI_ISL_1506114, EPI_ISL_1707043, EPI_ISL_1707049

| | | | |
|---|---|---|---|
| see above | UCLA Clinical Micro Lab | Los Angeles County PHL | P. Hemarajata et al. |

EPI_ISL_1402625, EPI_ISL_1402626, EPI_ISL_1402627, EPI_ISL_1402628, EPI_ISL_1402629, EPI_ISL_1402630, EPI_ISL_1402631, EPI_ISL_1402632, EPI_ISL_1402633, EPI_ISL_1402634, EPI_ISL_1402635, EPI_ISL_1402636, EPI_ISL_1402637, EPI_ISL_1402638, EPI_ISL_1402639, EPI_ISL_1402640, EPI_ISL_1402641, EPI_ISL_1402642, EPI_ISL_1402643, EPI_ISL_1402644, EPI_ISL_1402645, EPI_ISL_1402646, EPI_ISL_1402647, EPI_ISL_1402648, EPI_ISL_1402649, EPI_ISL_1402650, EPI_ISL_1402651, EPI_ISL_1402652, EPI_ISL_1402653, EPI_ISL_1402654, EPI_ISL_1405052, EPI_ISL_1405053, EPI_ISL_1405054, EPI_ISL_1405055, EPI_ISL_1405056, EPI_ISL_1405057, EPI_ISL_1405058, EPI_ISL_1405059, EPI_ISL_1405060, EPI_ISL_1469853, EPI_ISL_1469877, EPI_ISL_1469882

| | | | |
|---|---|---|---|
| see above | UCSC Genomics Institute | UCSC Genomics Institute | A. Marm Kilpatrick; Angie Hinrichs; Beth Shapiro; Bryan Thornlow; David Haussler; Eric Beraut; Hugh Olsen; Ikenna Anigbogu; Isabel Bjork; Jakob McBroome; Jeremy Sanford; Joshua Kapp; Mark Akeson; Maximilian Haeussler; Michael Stone; Miten Jain; Molly Cassatt-Johnstone; Namrita Dhillon; Russell Corbett-Detig; Terren Chang; Yatish Turakhia |

EPI_ISL_1303409, EPI_ISL_1303410, EPI_ISL_1303411, EPI_ISL_1303412, EPI_ISL_1303413, EPI_ISL_1303425, EPI_ISL_1303441, EPI_ISL_1303442, EPI_ISL_1303443, EPI_ISL_1303444, EPI_ISL_1303445, EPI_ISL_1303446, EPI_ISL_1303464, EPI_ISL_1303465, EPI_ISL_1303466, EPI_ISL_1303467, EPI_ISL_1303468, EPI_ISL_1303469, EPI_ISL_1303470, EPI_ISL_1303489, EPI_ISL_1303490, EPI_ISL_1303491, EPI_ISL_1303492, EPI_ISL_1303493, EPI_ISL_1303494, EPI_ISL_1303495, EPI_ISL_1303496, EPI_ISL_1303497, EPI_ISL_1303498, EPI_ISL_1379977, EPI_ISL_1497526, EPI_ISL_1497527, EPI_ISL_1497528, EPI_ISL_1497529, EPI_ISL_1497530, EPI_ISL_1497531, EPI_ISL_1497532, EPI_ISL_1497533, EPI_ISL_1497534, EPI_ISL_1497535, EPI_ISL_1497536, EPI_ISL_1497537, EPI_ISL_1497538, EPI_ISL_1497539, EPI_ISL_1497540, EPI_ISL_1502810, EPI_ISL_1502811, EPI_ISL_1502812, EPI_ISL_1503989, EPI_ISL_1503990, EPI_ISL_1503994, EPI_ISL_1503996, EPI_ISL_1503997, EPI_ISL_1504025, EPI_ISL_1504028, EPI_ISL_1504029, EPI_ISL_1504030, EPI_ISL_1504031, EPI_ISL_1504032, EPI_ISL_1504033, EPI_ISL_1504034, EPI_ISL_1504035, EPI_ISL_1504037, EPI_ISL_1504038, EPI_ISL_1504039, EPI_ISL_1504040, EPI_ISL_1504041, EPI_ISL_1506120, EPI_ISL_1506121, EPI_ISL_1506122, EPI_ISL_1506123, EPI_ISL_1506124, EPI_ISL_1506125, EPI_ISL_1506126, EPI_ISL_1508903, EPI_ISL_1508904, EPI_ISL_1508905, EPI_ISL_1508906, EPI_ISL_1508907, EPI_ISL_1508908, EPI_ISL_1508910, EPI_ISL_1508911, EPI_ISL_1508912, EPI_ISL_1508913, EPI_ISL_1508914, EPI_ISL_1508919, EPI_ISL_1508920, EPI_ISL_1508929, EPI_ISL_1508931, EPI_ISL_1508932, EPI_ISL_1508933, EPI_ISL_1508934, EPI_ISL_1508935, EPI_ISL_1508936, EPI_ISL_1508937, EPI_ISL_1508938, EPI_ISL_1508939, EPI_ISL_1509274, EPI_ISL_1509275, EPI_ISL_1509276, EPI_ISL_1509277, EPI_ISL_1509278, EPI_ISL_1509279, EPI_ISL_1509280, EPI_ISL_1509281, EPI_ISL_1509282, EPI_ISL_1509283, EPI_ISL_1509284, EPI_ISL_1509285, EPI_ISL_1509286, EPI_ISL_1509287, EPI_ISL_1509288, EPI_ISL_1509289, EPI_ISL_1509290, EPI_ISL_1509291, EPI_ISL_1509292, EPI_ISL_1509293, EPI_ISL_1509294, EPI_ISL_1509295, EPI_ISL_1509296, EPI_ISL_1509297, EPI_ISL_1515800, EPI_ISL_1706608

| | | | |
|---|---|---|---|
| see above | USC Clinical Lab | Los Angeles County PHL | P. Hemarajata et al. |
| EPI_ISL_1504026, EPI_ISL_1506127 | VA Medical Center | Los Angeles County PHL | P. Hemarajata et al. |

We gratefully acknowledge the following Authors from the Originating laboratories responsible for obtaining the specimens, as well as the Submitting laboratories where the genome data were generated and shared via GISAID, on which this research is based.

All Submitters of data may be contacted directly via www.gisaid.org

Authors are sorted alphabetically.

| Accession ID | Originating Laboratory | Submitting Laboratory | Authors |
|---|---|---|---|
| EPI_ISL_1650051, EPI_ISL_1650058, EPI_ISL_1650388, EPI_ISL_1650403, EPI_ISL_1650417, EPI_ISL_1650449, EPI_ISL_1650453, EPI_ISL_1650454, EPI_ISL_1650455, EPI_ISL_1650457, EPI_ISL_1650459, EPI_ISL_1650460, EPI_ISL_1650471, EPI_ISL_1667437, EPI_ISL_1667497, EPI_ISL_1667498, EPI_ISL_1667525, EPI_ISL_1667542, EPI_ISL_1667573, EPI_ISL_1667576, EPI_ISL_1667579, EPI_ISL_1667580, EPI_ISL_1667586, EPI_ISL_1667604, EPI_ISL_1667605, EPI_ISL_1667623, EPI_ISL_1667647, EPI_ISL_1667677, EPI_ISL_1667764, EPI_ISL_1667768, EPI_ISL_1667770, EPI_ISL_1667779, EPI_ISL_1667788, EPI_ISL_1667861, EPI_ISL_1667862, EPI_ISL_1667863, EPI_ISL_1667866, EPI_ISL_1667919, EPI_ISL_1667965, EPI_ISL_1667975, EPI_ISL_1667976, EPI_ISL_1667977, EPI_ISL_1667986, EPI_ISL_1667987, EPI_ISL_1668046, EPI_ISL_1668058, EPI_ISL_1668073, EPI_ISL_1668075, EPI_ISL_1668080, EPI_ISL_1668082, EPI_ISL_1668084, EPI_ISL_1668087, EPI_ISL_1668090, EPI_ISL_1668096, EPI_ISL_1668099, EPI_ISL_1668136, EPI_ISL_1668139, EPI_ISL_1668150, EPI_ISL_1668155, EPI_ISL_1668158, EPI_ISL_1668159, EPI_ISL_1668163, EPI_ISL_1668165, EPI_ISL_1668175, EPI_ISL_1668177, EPI_ISL_1668179, EPI_ISL_1668225, EPI_ISL_1668257, EPI_ISL_1668280, EPI_ISL_1668320, EPI_ISL_1668324, EPI_ISL_1668335, EPI_ISL_1685474, EPI_ISL_1685487, EPI_ISL_1685548, EPI_ISL_1685552, EPI_ISL_1685553, EPI_ISL_1685555, EPI_ISL_1685556, EPI_ISL_1685566, EPI_ISL_1685619, EPI_ISL_1685621, EPI_ISL_1685625, EPI_ISL_1685634, EPI_ISL_1685680, EPI_ISL_1685681, EPI_ISL_1685762, EPI_ISL_1685764, EPI_ISL_1685770, EPI_ISL_1685786, EPI_ISL_1685843, EPI_ISL_1685860, EPI_ISL_1685942, EPI_ISL_1685943, EPI_ISL_1686023, EPI_ISL_1686079, EPI_ISL_1686204, EPI_ISL_1686233, EPI_ISL_1686290, EPI_ISL_1686462, EPI_ISL_1686508, EPI_ISL_1686542, EPI_ISL_1686543, EPI_ISL_1686573, EPI_ISL_1686576, EPI_ISL_1686587, EPI_ISL_1686589, EPI_ISL_1686627, EPI_ISL_1686635, EPI_ISL_1686699, EPI_ISL_1686700, EPI_ISL_1686768, EPI_ISL_1686901, EPI_ISL_1686907, EPI_ISL_1686926, EPI_ISL_1686938, EPI_ISL_1686939, EPI_ISL_1686940, EPI_ISL_1686941, EPI_ISL_1686991, EPI_ISL_1686998, EPI_ISL_1687070, EPI_ISL_1687113, EPI_ISL_1687157, EPI_ISL_1687159, EPI_ISL_1687160, EPI_ISL_1687161, EPI_ISL_1687162, EPI_ISL_1687164, EPI_ISL_1687172, EPI_ISL_1687180, EPI_ISL_1687181, EPI_ISL_1687190, EPI_ISL_1687241, EPI_ISL_1687242, EPI_ISL_1687269, EPI_ISL_1687309, EPI_ISL_1687332, EPI_ISL_1687346, EPI_ISL_1687416, EPI_ISL_1687435, EPI_ISL_1687446, EPI_ISL_1687454, EPI_ISL_1687460, EPI_ISL_1687470, EPI_ISL_1687474, EPI_ISL_1687481, EPI_ISL_1687484, EPI_ISL_1687485, EPI_ISL_1687486, EPI_ISL_1687510, EPI_ISL_1687512, EPI_ISL_1687517, EPI_ISL_1687633, EPI_ISL_1687701, EPI_ISL_1687702, EPI_ISL_1687731, EPI_ISL_1687734, EPI_ISL_1687758, EPI_ISL_1687764, EPI_ISL_1687770, EPI_ISL_1687827, EPI_ISL_1687837, EPI_ISL_1687898, EPI_ISL_1687942, EPI_ISL_1687964, EPI_ISL_1687971, EPI_ISL_1687980, EPI_ISL_1687981, EPI_ISL_1687983, EPI_ISL_1687988, EPI_ISL_1687994, EPI_ISL_1688003, EPI_ISL_1688017, EPI_ISL_1688028, EPI_ISL_1688045, EPI_ISL_1688074, EPI_ISL_1688077, EPI_ISL_1688078, EPI_ISL_1688097, EPI_ISL_1688104, EPI_ISL_1688111, EPI_ISL_1688117, EPI_ISL_1688118, EPI_ISL_1688203, EPI_ISL_1688274, EPI_ISL_1688361, EPI_ISL_1688370, EPI_ISL_1688460, EPI_ISL_1688512, EPI_ISL_1688587, EPI_ISL_1688590, EPI_ISL_1688593, EPI_ISL_1688705, EPI_ISL_1688709, EPI_ISL_1688768, EPI_ISL_1688859, EPI_ISL_1688862, EPI_ISL_1688866, EPI_ISL_1689033, EPI_ISL_1689061, EPI_ISL_1689075, EPI_ISL_1689194, EPI_ISL_1689333, EPI_ISL_1689494, EPI_ISL_1689560, EPI_ISL_1689590, EPI_ISL_1689592, EPI_ISL_1689633, EPI_ISL_1689634, EPI_ISL_1689636, EPI_ISL_1689637, EPI_ISL_1689638, EPI_ISL_1689640, EPI_ISL_1689649, EPI_ISL_1689655, EPI_ISL_1689684, EPI_ISL_1689738, EPI_ISL_1689754, EPI_ISL_1689757, EPI_ISL_1689760, EPI_ISL_1689813, EPI_ISL_1689827, EPI_ISL_1689829, EPI_ISL_1689832, EPI_ISL_1689858, EPI_ISL_1689992, EPI_ISL_1690028, EPI_ISL_1690106, EPI_ISL_1690127, EPI_ISL_1690178, EPI_ISL_1690179, EPI_ISL_1690180, EPI_ISL_1690190, EPI_ISL_1690229, EPI_ISL_1690235, EPI_ISL_1690238, EPI_ISL_1690241, EPI_ISL_1695389, EPI_ISL_1695506, EPI_ISL_1695560, EPI_ISL_1701908, EPI_ISL_1701914, EPI_ISL_1701921, EPI_ISL_1701924, EPI_ISL_1701979, EPI_ISL_1701993, EPI_ISL_1702008, EPI_ISL_1702047, EPI_ISL_1702143, EPI_ISL_1702725, EPI_ISL_1702726, EPI_ISL_1702738, EPI_ISL_1702745, EPI_ISL_1702759, EPI_ISL_1702769, EPI_ISL_1702792, EPI_ISL_1702797, EPI_ISL_1702798, EPI_ISL_1702799, EPI_ISL_1702800, EPI_ISL_1702803, EPI_ISL_1702812, EPI_ISL_1702813, EPI_ISL_1702952, EPI_ISL_1735687, EPI_ISL_1735689, EPI_ISL_1735765, EPI_ISL_1735767, EPI_ISL_1735789, EPI_ISL_1735804, EPI_ISL_1735805, EPI_ISL_1735822, EPI_ISL_1735846, EPI_ISL_1735873, EPI_ISL_1735874, EPI_ISL_1735877, EPI_ISL_1735908, EPI_ISL_1735956, EPI_ISL_1735958, EPI_ISL_1735967, EPI_ISL_1735977, EPI_ISL_1736013, EPI_ISL_1736085, EPI_ISL_1736088, EPI_ISL_1736101, EPI_ISL_1736102, EPI_ISL_1736103, EPI_ISL_1736137, EPI_ISL_1736141, EPI_ISL_1736150, EPI_ISL_1736185, EPI_ISL_1736302, EPI_ISL_1736355, EPI_ISL_1736404, EPI_ISL_1736427, EPI_ISL_1736428, EPI_ISL_1736440, EPI_ISL_1736451, EPI_ISL_1736458, EPI_ISL_1736461, EPI_ISL_1736482, EPI_ISL_1736489, EPI_ISL_1736498, EPI_ISL_1736499, EPI_ISL_1736504, EPI_ISL_1736505, EPI_ISL_1736543, EPI_ISL_1736544, EPI_ISL_1736561, EPI_ISL_1736566, EPI_ISL_1736567, EPI_ISL_1736572, EPI_ISL_1736576, EPI_ISL_1736645, EPI_ISL_1736648, EPI_ISL_1736681, EPI_ISL_1736753, EPI_ISL_1736787, EPI_ISL_1736790, EPI_ISL_1736813, EPI_ISL_1736823, EPI_ISL_1736870, EPI_ISL_1736873, EPI_ISL_1736875, EPI_ISL_1736877, EPI_ISL_1736878, EPI_ISL_1737032, EPI_ISL_1737064, EPI_ISL_1737071, EPI_ISL_1737072, EPI_ISL_1737073, EPI_ISL_1737075, EPI_ISL_1737086, EPI_ISL_1737087, EPI_ISL_1737154, EPI_ISL_1737182, EPI_ISL_1737199, EPI_ISL_1737217, EPI_ISL_1737218, EPI_ISL_1737224, EPI_ISL_1737241, EPI_ISL_1737252, EPI_ISL_1737256, EPI_ISL_1737258, EPI_ISL_1737264, EPI_ISL_1737265, EPI_ISL_1737266, EPI_ISL_1737267, EPI_ISL_1737268, EPI_ISL_1737322, EPI_ISL_1737323, EPI_ISL_1737434, EPI_ISL_1737438, EPI_ISL_1737446, EPI_ISL_1737454, EPI_ISL_1737474, EPI_ISL_1737479, EPI_ISL_1737490, EPI_ISL_1737500, EPI_ISL_1737511, EPI_ISL_1737514, EPI_ISL_1737553, EPI_ISL_1737592, EPI_ISL_1737609, EPI_ISL_1737653, EPI_ISL_1737654, EPI_ISL_1737655, EPI_ISL_1737679, EPI_ISL_1737693, EPI_ISL_1737694, EPI_ISL_1737696, EPI_ISL_1737715, EPI_ISL_1737723, EPI_ISL_1737739, EPI_ISL_1737740, EPI_ISL_1737742, EPI_ISL_1737750, EPI_ISL_1737755, EPI_ISL_1737777, EPI_ISL_1737778, EPI_ISL_1737782, EPI_ISL_1737783, EPI_ISL_1737787, EPI_ISL_1737788, EPI_ISL_1737798, EPI_ISL_1737805, EPI_ISL_1737831, EPI_ISL_1737833, EPI_ISL_1737834, EPI_ISL_1737838, EPI_ISL_1737840, EPI_ISL_1738763, EPI_ISL_1753554, EPI_ISL_1753593, EPI_ISL_1753597, EPI_ISL_1753605, EPI_ISL_1753609, EPI_ISL_1834453, EPI_ISL_1834455, EPI_ISL_1834494, EPI_ISL_1834499, EPI_ISL_1834515, EPI_ISL_1834516, EPI_ISL_1834522, EPI_ISL_1834581, EPI_ISL_1834602, EPI_ISL_1834616, EPI_ISL_1834650, EPI_ISL_1834733, EPI_ISL_1834743, EPI_ISL_1834762, EPI_ISL_1834765, EPI_ISL_1834778, EPI_ISL_1834809, EPI_ISL_1834824, EPI_ISL_1834831, EPI_ISL_1834880, EPI_ISL_1834918, EPI_ISL_1834945, EPI_ISL_1834971, EPI_ISL_1835057, EPI_ISL_1835058, EPI_ISL_1835059, EPI_ISL_1835084, EPI_ISL_1835085, EPI_ISL_1835156, EPI_ISL_1835183, EPI_ISL_1835223, EPI_ISL_1835232, EPI_ISL_1835273, EPI_ISL_1835332, EPI_ISL_1835350, EPI_ISL_1835372, EPI_ISL_1835392, EPI_ISL_1835404, EPI_ISL_1835451, EPI_ISL_1835455, EPI_ISL_1835478, EPI_ISL_1835495, EPI_ISL_1835513, EPI_ISL_1835517, EPI_ISL_1835545, EPI_ISL_1835546, EPI_ISL_1835556, EPI_ISL_1835561, EPI_ISL_1835605, EPI_ISL_1835643, EPI_ISL_1835655, EPI_ISL_1835676, EPI_ISL_1835694, EPI_ISL_1835702, EPI_ISL_1835733, EPI_ISL_1835734, EPI_ISL_1835791, EPI_ISL_1835792, EPI_ISL_1835793, EPI_ISL_1835796, EPI_ISL_1835807, EPI_ISL_1835809, EPI_ISL_1835829, EPI_ISL_1835830, EPI_ISL_1835850, EPI_ISL_1835878, EPI_ISL_1835896, EPI_ISL_1835945, EPI_ISL_1835946, EPI_ISL_1835964, EPI_ISL_1835966, EPI_ISL_1836010, EPI_ISL_1836032, EPI_ISL_1836085, EPI_ISL_1836087, EPI_ISL_1836095, EPI_ISL_1836098, EPI_ISL_1836100, EPI_ISL_1836105, EPI_ISL_1836107, EPI_ISL_1836119, EPI_ISL_1836335, EPI_ISL_1836338, EPI_ISL_1836339, EPI_ISL_1836354, EPI_ISL_1836426, EPI_ISL_1836428, EPI_ISL_1836429, EPI_ISL_1836430, EPI_ISL_1836464, EPI_ISL_1836465, EPI_ISL_1836477, EPI_ISL_1836483, EPI_ISL_1836490, EPI_ISL_1836493, EPI_ISL_1836499, EPI_ISL_1836518, EPI_ISL_1836521, EPI_ISL_1836536, EPI_ISL_1836580, EPI_ISL_1836610, EPI_ISL_1836611, EPI_ISL_1836612, EPI_ISL_1836639, EPI_ISL_1836640, EPI_ISL_1836644, EPI_ISL_1836668, EPI_ISL_1836687, EPI_ISL_1836826, EPI_ISL_1836840, EPI_ISL_1836853, EPI_ISL_1836886, EPI_ISL_1836909, EPI_ISL_1836913, EPI_ISL_1836914, EPI_ISL_1836916, EPI_ISL_1836918, EPI_ISL_1836952, EPI_ISL_1836958, EPI_ISL_1836986, EPI_ISL_1837008, EPI_ISL_1837039, EPI_ISL_1837068, EPI_ISL_1837097, EPI_ISL_1837098, EPI_ISL_1837112, EPI_ISL_1837113, EPI_ISL_1837115, EPI_ISL_1837116, EPI_ISL_1837118, EPI_ISL_1837119, EPI_ISL_1837120, EPI_ISL_1837176, EPI_ISL_1837314, EPI_ISL_1837385, EPI_ISL_1837417, EPI_ISL_1837432, EPI_ISL_1837447, EPI_ISL_1837448, EPI_ISL_1837449, EPI_ISL_1837452, EPI_ISL_1837454, EPI_ISL_1837455, EPI_ISL_1837456, EPI_ISL_1837477, EPI_ISL_1837479, EPI_ISL_1837480, EPI_ISL_1837499, EPI_ISL_1837514, EPI_ISL_1837565, EPI_ISL_1837617, EPI_ISL_1837619, EPI_ISL_1837639, EPI_ISL_1837644, EPI_ISL_1837664, EPI_ISL_1837670, EPI_ISL_1837701, EPI_ISL_1837702, EPI_ISL_1837703, EPI_ISL_1837717, EPI_ISL_1837758, EPI_ISL_1837957, EPI_ISL_1837959, EPI_ISL_1837970, EPI_ISL_1837971, EPI_ISL_1838211, EPI_ISL_1838407, EPI_ISL_1900357, EPI_ISL_1900378, EPI_ISL_1900408, EPI_ISL_1922910, EPI_ISL_1922912, EPI_ISL_1922913, EPI_ISL_1922917, EPI_ISL_1922934, EPI_ISL_1922968, EPI_ISL_1922970, EPI_ISL_1923038, EPI_ISL_1923154, EPI_ISL_1923155, EPI_ISL_1925142, EPI_ISL_1925194, EPI_ISL_1925208, EPI_ISL_1925211, EPI_ISL_1925229, EPI_ISL_1925233, EPI_ISL_1925286, EPI_ISL_1925375, EPI_ISL_1925386, EPI_ISL_1925434, EPI_ISL_1925443, EPI_ISL_1925444, EPI_ISL_1925449, EPI_ISL_1925469, EPI_ISL_1925472, EPI_ISL_1925587, EPI_ISL_1925588, EPI_ISL_1995006, EPI_ISL_1995061, EPI_ISL_1995067, EPI_ISL_1995107, EPI_ISL_1995113, EPI_ISL_1995116, EPI_ISL_1995136, EPI_ISL_1995177, EPI_ISL_1995188, EPI_ISL_1995226, EPI_ISL_1995230, EPI_ISL_1995235, EPI_ISL_1995244, EPI_ISL_1995259, EPI_ISL_1995260, EPI_ISL_1995261, EPI_ISL_1995264, EPI_ISL_1995322, EPI_ISL_1995323, EPI_ISL_1995341, EPI_ISL_1995348, EPI_ISL_1995410, EPI_ISL_1995455, EPI_ISL_1995476, EPI_ISL_1995483, EPI_ISL_1995490, EPI_ISL_1995491, EPI_ISL_1995507, EPI_ISL_1995541, EPI_ISL_1995552, EPI_ISL_1995570, EPI_ISL_1995582, EPI_ISL_1995599, EPI_ISL_1995722, EPI_ISL_1995831, EPI_ISL_1995863, EPI_ISL_1995872, EPI_ISL_1995942, EPI_ISL_1995964, EPI_ISL_1995990, EPI_ISL_1995991, EPI_ISL_1996059, EPI_ISL_1996083, EPI_ISL_1996085, EPI_ISL_1996086, EPI_ISL_1996088, EPI_ISL_1996089, EPI_ISL_1996090, EPI_ISL_1996092, EPI_ISL_1996093, EPI_ISL_1996188, EPI_ISL_1996203, EPI_ISL_1996264, EPI_ISL_1996267, EPI_ISL_1996311, EPI_ISL_1996440, EPI_ISL_1996486, EPI_ISL_1996523, EPI_ISL_1996594, EPI_ISL_1996596, EPI_ISL_1996598, EPI_ISL_1996600, EPI_ISL_1996626, EPI_ISL_1996657, EPI_ISL_1996661, EPI_ISL_1996664, EPI_ISL_1996665, EPI_ISL_1996667, EPI_ISL_1996671, EPI_ISL_1996672, EPI_ISL_1996673, EPI_ISL_1996681, EPI_ISL_1996707, EPI_ISL_1996719, EPI_ISL_1996730, EPI_ISL_1996731, EPI_ISL_1996732, EPI_ISL_1996745, EPI_ISL_1996746, EPI_ISL_1996855, EPI_ISL_1996857, EPI_ISL_1996861, EPI_ISL_1996872, EPI_ISL_1996874, EPI_ISL_1996882, EPI_ISL_1996892, EPI_ISL_1996944, EPI_ISL_1996949, EPI_ISL_1996973, EPI_ISL_1996987, EPI_ISL_1996990, EPI_ISL_1997023, EPI_ISL_1997063, EPI_ISL_1997079, EPI_ISL_1997088, EPI_ISL_1997101, EPI_ISL_1997107, EPI_ISL_1997108, EPI_ISL_1997122, EPI_ISL_1997123, EPI_ISL_1997124, EPI_ISL_1997126, EPI_ISL_1997130, EPI_ISL_1997132, EPI_ISL_1997133, EPI_ISL_1997134, EPI_ISL_1997135, EPI_ISL_1997136, EPI_ISL_1997138, EPI_ISL_1997156, EPI_ISL_1997159, EPI_ISL_1997168, EPI_ISL_1997247, EPI_ISL_1997262, EPI_ISL_1997314, EPI_ISL_1997362, EPI_ISL_1997363, EPI_ISL_1997399, EPI_ISL_1997400, EPI_ISL_1997404, EPI_ISL_1997416, EPI_ISL_1997426, EPI_ISL_1997427, EPI_ISL_1997428, EPI_ISL_1997431, EPI_ISL_1997432, EPI_ISL_1997436, EPI_ISL_1997439, EPI_ISL_1997440, EPI_ISL_1997496, EPI_ISL_1997497, EPI_ISL_1997498, EPI_ISL_1997528, EPI_ISL_1997613, EPI_ISL_1997614, EPI_ISL_1997615, EPI_ISL_1997616, EPI_ISL_1997617, EPI_ISL_1997618, EPI_ISL_1997620, EPI_ISL_1997621, EPI_ISL_1997622, EPI_ISL_1997628, EPI_ISL_1997629, EPI_ISL_1997630, EPI_ISL_1997641, EPI_ISL_1997722, EPI_ISL_1997739, EPI_ISL_1997740, EPI_ISL_1997745, EPI_ISL_1997747, EPI_ISL_1997751, EPI_ISL_1997754, EPI_ISL_1997756, EPI_ISL_1997762, EPI_ISL_1997763, EPI_ISL_1997769, EPI_ISL_1997805, EPI_ISL_1997806, EPI_ISL_1997824, EPI_ISL_1997851, EPI_ISL_1997869, EPI_ISL_1997922, EPI_ISL_1997935, EPI_ISL_1997949, EPI_ISL_1997950, EPI_ISL_1997986, EPI_ISL_1998013, EPI_ISL_1998025, EPI_ISL_1998054, EPI_ISL_1998061, EPI_ISL_1998088, EPI_ISL_1998111, EPI_ISL_1998112, EPI_ISL_1998120, EPI_ISL_1998122, EPI_ISL_1998123, EPI_ISL_1998187, EPI_ISL_1998212, EPI_ISL_1998217, EPI_ISL_1998237, EPI_ISL_1998238, EPI_ISL_1998240, EPI_ISL_1998268, EPI_ISL_1998269, EPI_ISL_1998283, EPI_ISL_1998285, EPI_ISL_1998307, EPI_ISL_1998375, EPI_ISL_1998377, EPI_ISL_1998381, EPI_ISL_1998407, EPI_ISL_1998408, EPI_ISL_1998410, EPI_ISL_1998411, EPI_ISL_1998412, EPI_ISL_1998419, EPI_ISL_1998432, EPI_ISL_1998439, EPI_ISL_1998467, EPI_ISL_1998513, EPI_ISL_1998514, EPI_ISL_1998518, EPI_ISL_1998537, EPI_ISL_1998538, EPI_ISL_1998620, EPI_ISL_1998668, EPI_ISL_1998672, EPI_ISL_1998722, EPI_ISL_1998755, EPI_ISL_1998774, EPI_ISL_1998789, EPI_ISL_1998799, EPI_ISL_1998831, EPI_ISL_1998848, EPI_ISL_1998859, EPI_ISL_1998860, EPI_ISL_1998862, EPI_ISL_1998863, EPI_ISL_1998882, EPI_ISL_1998896, EPI_ISL_1998965, EPI_ISL_1998975, EPI_ISL_1999023, EPI_ISL_1999024, EPI_ISL_1999034, EPI_ISL_1999040, EPI_ISL_1999098, EPI_ISL_1999132, EPI_ISL_1999141, EPI_ISL_1999162, EPI_ISL_1999189, EPI_ISL_1999195, EPI_ISL_1999196, EPI_ISL_1999200, EPI_ISL_1999211, EPI_ISL_1999212, EPI_ISL_1999227, EPI_ISL_1999231, EPI_ISL_1999248, EPI_ISL_1999293, EPI_ISL_1999299, EPI_ISL_1999302, EPI_ISL_1999309, EPI_ISL_1999311, EPI_ISL_1999366, EPI_ISL_1999368, EPI_ISL_1999375, EPI_ISL_1999446, EPI_ISL_1999491, EPI_ISL_1999569, EPI_ISL_1999600, EPI_ISL_1999716, EPI_ISL_1999717, EPI_ISL_1999780, EPI_ISL_1999878, EPI_ISL_1999914, EPI_ISL_2000128, EPI_ISL_2000130, EPI_ISL_2000131, EPI_ISL_2000175, EPI_ISL_2000180, EPI_ISL_2000209, EPI_ISL_2000210, EPI_ISL_2000215, EPI_ISL_2000220, EPI_ISL_2000222, EPI_ISL_2000226, EPI_ISL_2000233, EPI_ISL_2000234, EPI_ISL_2000237, EPI_ISL_2000262, EPI_ISL_2000270, EPI_ISL_2000271, EPI_ISL_2000275, EPI_ISL_2000296, EPI_ISL_2000333, EPI_ISL_2000343, EPI_ISL_2000393, EPI_ISL_2000407, EPI_ISL_2000410, EPI_ISL_2000412, EPI_ISL_2000417, EPI_ISL_2000483, EPI_ISL_2000484, EPI_ISL_2000509, EPI_ISL_2000512, EPI_ISL_2000520, EPI_ISL_2035046, EPI_ISL_2035069, EPI_ISL_2035110, EPI_ISL_2035138, EPI_ISL_2035143, EPI_ISL_2035184, EPI_ISL_2035189, EPI_ISL_2035223, EPI_ISL_2035225, EPI_ISL_2039034, EPI_ISL_2039064, EPI_ISL_2039065, EPI_ISL_2039067, EPI_ISL_2039088, EPI_ISL_2039089, EPI_ISL_2039095, EPI_ISL_2039101, EPI_ISL_2039125, EPI_ISL_2039126, EPI_ISL_2039131, EPI_ISL_2039135, EPI_ISL_2039141, EPI_ISL_2039143, EPI_ISL_2039152, EPI_ISL_2039154, EPI_ISL_2039155, EPI_ISL_2039393, EPI_ISL_2039395, EPI_ISL_2039397, EPI_ISL_2039445, EPI_ISL_2039481, EPI_ISL_2039484, EPI_ISL_2039487, EPI_ISL_2039503, EPI_ISL_2039507, EPI_ISL_2039521, EPI_ISL_2039522, EPI_ISL_2039523, EPI_ISL_2039564, EPI_ISL_2039577, EPI_ISL_2039579, EPI_ISL_2039580, EPI_ISL_2039592, EPI_ISL_2039632, EPI_ISL_2039700, EPI_ISL_2039710, EPI_ISL_2039711, EPI_ISL_2039715, EPI_ISL_2039719, EPI_ISL_2039734, EPI_ISL_2039736, EPI_ISL_2039737, EPI_ISL_2039765, EPI_ISL_2039818, EPI_ISL_2039824, EPI_ISL_2039827, EPI_ISL_2039880, EPI_ISL_2039893, EPI_ISL_2039954, EPI_ISL_2039988, EPI_ISL_2039989, EPI_ISL_2040082, EPI_ISL_2040090, EPI_ISL_2040187, EPI_ISL_2040189, EPI_ISL_2040190, EPI_ISL_2040191, EPI_ISL_2040254, EPI_ISL_2040265, EPI_ISL_2040317, EPI_ISL_2040325, EPI_ISL_2040326, EPI_ISL_2040328, EPI_ISL_2040331, EPI_ISL_2040332, EPI_ISL_2040333, EPI_ISL_2040352, EPI_ISL_2040357, EPI_ISL_2040361, EPI_ISL_2040410, EPI_ISL_2040424, EPI_ISL_2040429, EPI_ISL_2040477, EPI_ISL_2040478, EPI_ISL_2040555, EPI_ISL_2040573, EPI_ISL_2040581, EPI_ISL_2040592, EPI_ISL_2040607, EPI_ISL_2040614, EPI_ISL_2040615, EPI_ISL_2040616, EPI_ISL_2040617, EPI_ISL_2040634, EPI_ISL_2040636, EPI_ISL_2040640, EPI_ISL_2040657, EPI_ISL_2040746, EPI_ISL_2040747, EPI_ISL_2040778, EPI_ISL_2040812, EPI_ISL_2040829, EPI_ISL_2040905, EPI_ISL_2040906, EPI_ISL_2040942, EPI_ISL_2040943, EPI_ISL_2040949, EPI_ISL_2040950, EPI_ISL_2040955, EPI_ISL_2040996, EPI_ISL_2041016, EPI_ISL_2041038, EPI_ISL_2041070, EPI_ISL_2041075, EPI_ISL_2041096, EPI_ISL_2041104, EPI_ISL_2041125, EPI_ISL_2041209, EPI_ISL_2041215, EPI_ISL_2041216, EPI_ISL_2041217, EPI_ISL_2041218, EPI_ISL_2041234, EPI_ISL_2041235, EPI_ISL_2041239, EPI_ISL_2041240, EPI_ISL_2041247, EPI_ISL_2041285, EPI_ISL_2041286, EPI_ISL_2041309, EPI_ISL_2041327, EPI_ISL_2041347, EPI_ISL_2041403, EPI_ISL_2041416, EPI_ISL_2041418, EPI_ISL_2041484, EPI_ISL_2041500, EPI_ISL_2041544, EPI_ISL_2041554, EPI_ISL_2041556, EPI_ISL_2041557, EPI_ISL_2041558, EPI_ISL_2041560, EPI_ISL_2041614, EPI_ISL_2041621, EPI_ISL_2041674, EPI_ISL_2041675, EPI_ISL_2041676, EPI_ISL_2041678, EPI_ISL_2041704, EPI_ISL_2041751, EPI_ISL_2041821, EPI_ISL_2041838, EPI_ISL_2041841, EPI_ISL_2041873, EPI_ISL_2041874, EPI_ISL_2041895, EPI_ISL_2041955, EPI_ISL_2041959, EPI_ISL_2041960, EPI_ISL_2041961, EPI_ISL_2041962, EPI_ISL_2041964, EPI_ISL_2042042, EPI_ISL_2042053, EPI_ISL_2042057, EPI_ISL_2042065, EPI_ISL_2042074, EPI_ISL_2042199, EPI_ISL_2042258, EPI_ISL_2042292, EPI_ISL_2042293, EPI_ISL_2042298, EPI_ISL_2042299, EPI_ISL_2042315, EPI_ISL_2042346, EPI_ISL_2042351, EPI_ISL_2042352, EPI_ISL_2042353, EPI_ISL_2042370, EPI_ISL_2042459, EPI_ISL_2042494, EPI_ISL_2042502, EPI_ISL_2042505, EPI_ISL_2042517, EPI_ISL_2042530, EPI_ISL_2042541, EPI_ISL_2042603, EPI_ISL_2042626, EPI_ISL_2042650, EPI_ISL_2042661, EPI_ISL_2042662, EPI_ISL_2042663, EPI_ISL_2042664, EPI_ISL_2042665, EPI_ISL_2042669, EPI_ISL_2042677, EPI_ISL_2042685, EPI_ISL_2042709, EPI_ISL_2042710, EPI_ISL_2042712, EPI_ISL_2042717, EPI_ISL_2042718, EPI_ISL_2042751, EPI_ISL_2042756, EPI_ISL_2042771, EPI_ISL_2042790, EPI_ISL_2042795, EPI_ISL_2042830, EPI_ISL_2042832, EPI_ISL_2042845, EPI_ISL_2042860, EPI_ISL_2042868, EPI_ISL_2042892, EPI_ISL_2042907, EPI_ISL_2042909, EPI_ISL_2042934, EPI_ISL_2042950, EPI_ISL_2043014, EPI_ISL_2043020, EPI_ISL_2043032, EPI_ISL_2043036, EPI_ISL_2043037, EPI_ISL_2043043, EPI_ISL_2043044, EPI_ISL_2043045, EPI_ISL_2043054, EPI_ISL_2043073, EPI_ISL_2043130, EPI_ISL_2043133, EPI_ISL_2043134, EPI_ISL_2043146, EPI_ISL_2043156, EPI_ISL_2043167, EPI_ISL_2043170, EPI_ISL_2043181, EPI_ISL_2043190, EPI_ISL_2043196, EPI_ISL_2043228, EPI_ISL_2043235, EPI_ISL_2043238, EPI_ISL_2043293, EPI_ISL_2043296, EPI_ISL_2043304, EPI_ISL_2043328, EPI_ISL_2043329, EPI_ISL_2043330, EPI_ISL_2043390, EPI_ISL_2043399, EPI_ISL_2043400, EPI_ISL_2043403, EPI_ISL_2043416, EPI_ISL_2043426, EPI_ISL_2043427, EPI_ISL_2043435, EPI_ISL_2043443, EPI_ISL_2043445, EPI_ISL_2043448, EPI_ISL_2043460, EPI_ISL_2043463, EPI_ISL_2043488, EPI_ISL_2043493, EPI_ISL_2043504, EPI_ISL_2043528, EPI_ISL_2043534, EPI_ISL_2043549, EPI_ISL_2089613, EPI_ISL_2089660, EPI_ISL_2089680, EPI_ISL_2089686, EPI_ISL_2089693, EPI_ISL_2089694, EPI_ISL_2089707, EPI_ISL_2089709, EPI_ISL_2089710, EPI_ISL_2089785, EPI_ISL_2089818, EPI_ISL_2089829, EPI_ISL_2089842, EPI_ISL_2089896, EPI_ISL_2089911, EPI_ISL_2089913, EPI_ISL_2089924, EPI_ISL_2089930, EPI_ISL_2089934, EPI_ISL_2089951, EPI_ISL_2089953, EPI_ISL_2089956, EPI_ISL_2089959, EPI_ISL_2089972, EPI_ISL_2089994, EPI_ISL_2089995, EPI_ISL_2089997, EPI_ISL_2090045, EPI_ISL_2090103, EPI_ISL_2090105, EPI_ISL_2090141, EPI_ISL_2090142, EPI_ISL_2090143, EPI_ISL_2090154, EPI_ISL_2090157, EPI_ISL_2090158, EPI_ISL_2090160, EPI_ISL_2090169, EPI_ISL_2090183, EPI_ISL_2090225, EPI_ISL_2090268, EPI_ISL_2090269, EPI_ISL_2090272, EPI_ISL_2090273, EPI_ISL_2090274, EPI_ISL_2090275, EPI_ISL_2090311, EPI_ISL_2090323, EPI_ISL_2090421, EPI_ISL_2090428, EPI_ISL_2090430, EPI_ISL_2090431, EPI_ISL_2090434, EPI_ISL_2090436, EPI_ISL_2090437, EPI_ISL_2090438, EPI_ISL_2090443, EPI_ISL_2132537, EPI_ISL_2132553, EPI_ISL_2132567, EPI_ISL_2145804, EPI_ISL_2145814, EPI_ISL_2145825, EPI_ISL_2145832, EPI_ISL_2145836, EPI_ISL_2145839, EPI_ISL_2145864, EPI_ISL_2145882, EPI_ISL_2145888, EPI_ISL_2145889, EPI_ISL_2145893, EPI_ISL_2145928, EPI_ISL_2145942, EPI_ISL_2145986, EPI_ISL_2146001, EPI_ISL_2146011, EPI_ISL_2148007, EPI_ISL_2148024, EPI_ISL_2148028, EPI_ISL_2148029, EPI_ISL_2148045, EPI_ISL_2148048, EPI_ISL_2148058, EPI_ISL_2148413, EPI_ISL_2148551, EPI_ISL_2148572, EPI_ISL_2148579, EPI_ISL_2148600, EPI_ISL_2148604, EPI_ISL_2148610, EPI_ISL_2148617, EPI_ISL_2148638, EPI_ISL_2148646, EPI_ISL_2148700, EPI_ISL_2148701, EPI_ISL_2148717, EPI_ISL_2148721, EPI_ISL_2148732, EPI_ISL_2148735, EPI_ISL_2148736, EPI_ISL_2148925, EPI_ISL_2149221, EPI_ISL_2149222, EPI_ISL_2149223, EPI_ISL_2149224, EPI_ISL_2149225, EPI_ISL_2149226, EPI_ISL_2149227, EPI_ISL_2149228, EPI_ISL_2149231, EPI_ISL_2149256, EPI_ISL_2149257, EPI_ISL_2149258, EPI_ISL_2149262, EPI_ISL_2149296, EPI_ISL_2149302, EPI_ISL_2149306, EPI_ISL_2149327, EPI_ISL_2149328, EPI_ISL_2149329, EPI_ISL_2149330, EPI_ISL_2149334, EPI_ISL_2149379, EPI_ISL_2149395, EPI_ISL_2149399, EPI_ISL_2149420, EPI_ISL_2149423, EPI_ISL_2149427, EPI_ISL_2149428, EPI_ISL_2149441, EPI_ISL_2149448, EPI_ISL_2149474, EPI_ISL_2149475, EPI_ISL_2149531, EPI_ISL_2149549, EPI_ISL_2149558, EPI_ISL_2149559, EPI_ISL_2149567, EPI_ISL_2149568, EPI_ISL_2149569, EPI_ISL_2149605, EPI_ISL_2149611, EPI_ISL_2149628, EPI_ISL_2149629, EPI_ISL_2149634, EPI_ISL_2149654, EPI_ISL_2149658, EPI_ISL_2149659, EPI_ISL_2149666, EPI_ISL_2149668, EPI_ISL_2149672, EPI_ISL_2149677, EPI_ISL_2149691, EPI_ISL_2149733, EPI_ISL_2149745, EPI_ISL_2149772, EPI_ISL_2149774, EPI_ISL_2149829, EPI_ISL_2149856, EPI_ISL_2149858, EPI_ISL_2149886, EPI_ISL_2149897, EPI_ISL_2150038, EPI_ISL_2150071, EPI_ISL_2150121, EPI_ISL_2150221, EPI_ISL_2150222, EPI_ISL_2150235, EPI_ISL_2150251, EPI_ISL_2150296, EPI_ISL_2150309, EPI_ISL_2150326, EPI_ISL_2150328, EPI_ISL_2150342, EPI_ISL_2150343, EPI_ISL_2150344, EPI_ISL_2150354, EPI_ISL_2150357, EPI_ISL_2150377, EPI_ISL_2150384, EPI_ISL_2150385, EPI_ISL_2150386, EPI_ISL_2150405, EPI_ISL_2150415, EPI_ISL_2150432, EPI_ISL_2150446, EPI_ISL_2150452, EPI_ISL_2150462, EPI_ISL_2150463, EPI_ISL_2150480, EPI_ISL_2150513, EPI_ISL_2150514, EPI_ISL_2150547, EPI_ISL_2150548, EPI_ISL_2150556, EPI_ISL_2150663, EPI_ISL_2150707, EPI_ISL_2150718, EPI_ISL_2150730, EPI_ISL_2150744, EPI_ISL_2150819, EPI_ISL_2150937, EPI_ISL_2150962, EPI_ISL_2150965, EPI_ISL_2150966, EPI_ISL_2159214, EPI_ISL_2159223, EPI_ISL_2159225, EPI_ISL_2159226, EPI_ISL_2159248, EPI_ISL_2159261, EPI_ISL_2159272, EPI_ISL_2159273, EPI_ISL_2159275, EPI_ISL_2159427, EPI_ISL_2187362, EPI_ISL_2187531, EPI_ISL_2187539, EPI_ISL_2187579, EPI_ISL_2187589 | | | |
| see above | Aegis Sciences Corporation | Centers for Disease Control and Prevention Division of Viral Diseases, Pathogen Discovery | Adrian Paskey; Alec Vest; Benjamin Rambo-Martin; Christopher Gulvick; Clinton R. Paden; Cyndi Clark; Dakota Howard; Darlene Wagner; Dhwani Batra; Dillon Nall; Duncan MacCannell; Ethan Sanders; Holly Houdeshell; Jason Caravas; Kara Moser; Matthew Hardison; Matthew Schmerer; Ola Kvalvaag; Patrick Campbell; Peter W. Cook; Rob Case; Scott Sammons; Shatavia Morrison; Shaun Westlund; Vikramsinha Ghorpade; Yvette Unoarumhi |
| EPI_ISL_1585085 | Arizona State University | Arizona State University | Efrem S. Lim; Joshua LaBaer; Joy M. Blain; LaRinda A. Holland; Nicholas J. Mellor; Peter T. Skidmore; Rabia Maqsood; Valerie Harris; Vel Murugan |
| EPI_ISL_2102999, EPI_ISL_2103001, EPI_ISL_2103002, EPI_ISL_2103018, EPI_ISL_2103019, EPI_ISL_2103020, EPI_ISL_2103032, EPI_ISL_2103034, EPI_ISL_2103038, EPI_ISL_2103046, EPI_ISL_2103051, EPI_ISL_2103064, EPI_ISL_2103071, EPI_ISL_2103074, EPI_ISL_2103088, EPI_ISL_2103093, EPI_ISL_2103107, EPI_ISL_2103119, EPI_ISL_2103128, EPI_ISL_2103129, EPI_ISL_2103132, EPI_ISL_2103137, EPI_ISL_2103140, EPI_ISL_2103147, EPI_ISL_2103151, EPI_ISL_2103162, EPI_ISL_2103165, EPI_ISL_2103167, EPI_ISL_2103192, EPI_ISL_2103194 | | | |
| see above | CA DPH Viral and Rickettsial Disease Laboratory | Chan-Zuckerberg Biohub | CZB Cliahub Consortium |
| EPI_ISL_1678087, EPI_ISL_1678093, EPI_ISL_1678094, EPI_ISL_1678095, EPI_ISL_1678282, EPI_ISL_1798834, EPI_ISL_1798841, EPI_ISL_1798853, EPI_ISL_1798855, EPI_ISL_1798857, EPI_ISL_1798860, EPI_ISL_1798868, EPI_ISL_1823713, EPI_ISL_1823715, EPI_ISL_1823716, EPI_ISL_1823717, EPI_ISL_1823718, EPI_ISL_1823719, EPI_ISL_1823720, EPI_ISL_1823722, EPI_ISL_1823724, EPI_ISL_1823725, EPI_ISL_1823726, EPI_ISL_1823727, EPI_ISL_1823728, EPI_ISL_1823729, EPI_ISL_1823730, EPI_ISL_1823732, EPI_ISL_1823733, EPI_ISL_1823734, EPI_ISL_1823736, EPI_ISL_1823737, EPI_ISL_1823738, EPI_ISL_1823739, EPI_ISL_1823741, EPI_ISL_1823743, EPI_ISL_1823744, EPI_ISL_1823745, EPI_ISL_1823747, EPI_ISL_1937812, EPI_ISL_1937813, EPI_ISL_1937814, EPI_ISL_1937815, EPI_ISL_1937816, EPI_ISL_1937817, EPI_ISL_1937818, EPI_ISL_1937819, EPI_ISL_1937820, EPI_ISL_1937821, EPI_ISL_1937822, EPI_ISL_1937823, EPI_ISL_1937824, EPI_ISL_1937825, EPI_ISL_1937826, EPI_ISL_1937827, EPI_ISL_1937828, EPI_ISL_1937829, EPI_ISL_2105107, EPI_ISL_2105108, EPI_ISL_2105109, EPI_ISL_2105128, EPI_ISL_2105134, EPI_ISL_2105135, EPI_ISL_2105158 | | | |
| see above | CA-Los Angeles County Public Health Laboratory | Centers for Disease Control and Prevention Division of Viral Diseases, Pathogen Discovery | Alison Laufer Halpin; Ben L. Rambo-Martin; Clinton R. Paden; Dakota Howard; Darlene Wagner; Dave Wentworth; Dhwani Batra; Jasmine Padilla; Justin Lee; Katie Dillon; Krista Queen; Kristen Knipe; Kristine Lacek; Mark Burroughs; Matthew Schmerer; Mili Sheth; Peter Cook; Sam Shepard; Sarah Nobles; Shoshona Le; Suxiang Tong; Vivien Dugan; Yvette Unoarumhi |
| EPI_ISL_1798766, EPI_ISL_1798768, EPI_ISL_1798770, EPI_ISL_1798772, EPI_ISL_1798777, EPI_ISL_1798779, EPI_ISL_1798781, EPI_ISL_1798783, EPI_ISL_1798787, EPI_ISL_1798791, EPI_ISL_1798793, EPI_ISL_2018310, EPI_ISL_2018311, EPI_ISL_2018312, EPI_ISL_2018313, EPI_ISL_2018314, EPI_ISL_2018315, EPI_ISL_2018316, EPI_ISL_2018317, EPI_ISL_2018318, EPI_ISL_2018319, EPI_ISL_2018320, EPI_ISL_2018321, EPI_ISL_2018322, EPI_ISL_2018323, EPI_ISL_2018324, EPI_ISL_2018325 | | | |
| see above | CDPH, Viral and Rickettsial Disease Laboratory | Centers for Disease Control and Prevention Division of Viral Diseases, Pathogen Discovery | Alison Laufer Halpin; Ben L. Rambo-Martin; Clinton R. Paden; Dakota Howard; Darlene Wagner; Dave Wentworth; Dhwani Batra; Jasmine Padilla; Justin Lee; Katie Dillon; Krista Queen; Kristen Knipe; Kristine Lacek; Mark Burroughs; Matthew Schmerer; Mili Sheth; Peter Cook; Sam Shepard; Sarah Nobles; Shoshona Le; Suxiang Tong; Vivien Dugan; Yvette Unoarumhi |
| EPI_ISL_1924630 | CHILDRENS HOSPITAL OF NORCAL | Santa Clara County public Health Laboratory | Santa Clara County Public Health Department |
| EPI_ISL_1824409, EPI_ISL_1824410, EPI_ISL_1824411, EPI_ISL_1824412, EPI_ISL_1824414, EPI_ISL_1824415 | Chiu Laboratory, University of California, San Francisco | Chiu Laboratory, University of California, San Francisco | Alicia Sotomayor-Gonzalez; Candace Wang; Charles Chiu; Daniel Wong; Venice Servellita; Yueyuan Zhang |
| EPI_ISL_1825079, EPI_ISL_1825080, EPI_ISL_1825081, | Color Genomics | Chiu Laboratory, University of California, San Francisco | Alicia Sotomayor-Gonzalez; Alicia Zhou; Candace Wang; Charles Chiu; Daniel Wong; Erika Torres; Madyson Wanner; Scott Topper; Venice Servellita; Yueyuan Zhang |

| | | | |
|---|---|---|---|
| EPI_ISL_1825082, EPI_ISL_1825083 | | | |
| EPI_ISL_2102749, EPI_ISL_2102763, EPI_ISL_2102772, EPI_ISL_2102798, EPI_ISL_2102826, EPI_ISL_2102861, EPI_ISL_2102887, EPI_ISL_2102891, EPI_ISL_2102932, EPI_ISL_2102954, EPI_ISL_2102960, EPI_ISL_2102972, EPI_ISL_2102988, EPI_ISL_2102997, EPI_ISL_2103009, EPI_ISL_2103031, EPI_ISL_2103062, EPI_ISL_2103097, EPI_ISL_2103106, EPI_ISL_2103138, EPI_ISL_2103169, EPI_ISL_2103173 | | | |
| see above | Contra Costa County Public Health Lab | Chan-Zuckerberg Biohub | CZB Cliahub Consortium |
| EPI_ISL_1805724, EPI_ISL_1805725, EPI_ISL_1805726, EPI_ISL_1805730, EPI_ISL_1805731, EPI_ISL_1805732, EPI_ISL_1805733, EPI_ISL_1805734, EPI_ISL_1805735, EPI_ISL_1805736, EPI_ISL_1805737, EPI_ISL_1805738, EPI_ISL_1805739, EPI_ISL_1841392, EPI_ISL_1910462, EPI_ISL_1910464, EPI_ISL_1910465, EPI_ISL_1910466, EPI_ISL_1968661 | | | |
| see above | County of San Diego Health and Human Services Agency | Pathogen Discovery, Respiratory Viruses Branch, Division of Viral Diseases, Centers for Disease Control and Prevention | Adam Retchless; Anna Kelleher; Anna Montmayeur; Anna Uehara; Brian Lynch; Clinton R. Paden; Dhwani Batra; Haibin Wang; Han Jia Justin Ng; Jasmine Padilla; Jing Zhang; Justin Lee; Krista Queen; Mark Burroughs; Mili Sheth; Morgan Davis; Peter Cook; Rachel Marine; Sarah Nobles; Suxiang Tong; Tara Coalter; Yan Li; Ying Tao |
| EPI_ISL_1701423, EPI_ISL_1701591, EPI_ISL_1701607, EPI_ISL_1821323, EPI_ISL_1821332, EPI_ISL_1821354, EPI_ISL_1821358, EPI_ISL_1821362, EPI_ISL_1821365, EPI_ISL_1821366, EPI_ISL_1821368, EPI_ISL_1821371, EPI_ISL_1821372, EPI_ISL_1821373, EPI_ISL_1821374, EPI_ISL_1821375, EPI_ISL_1821376, EPI_ISL_1821377, EPI_ISL_1821380, EPI_ISL_1821381, EPI_ISL_1821383, EPI_ISL_1821384, EPI_ISL_1821385, EPI_ISL_1821386, EPI_ISL_1821387, EPI_ISL_1821388, EPI_ISL_1821389, EPI_ISL_1821390, EPI_ISL_1821391, EPI_ISL_1821394, EPI_ISL_1821399, EPI_ISL_1821434, EPI_ISL_1821454, EPI_ISL_1821463, EPI_ISL_2102747, EPI_ISL_2102748, EPI_ISL_2102755, EPI_ISL_2102776, EPI_ISL_2102790, EPI_ISL_2102809, EPI_ISL_2102811, EPI_ISL_2102820, EPI_ISL_2102830, EPI_ISL_2102837, EPI_ISL_2102842, EPI_ISL_2102844, EPI_ISL_2102845, EPI_ISL_2102848, EPI_ISL_2102849, EPI_ISL_2102850, EPI_ISL_2102869, EPI_ISL_2102890, EPI_ISL_2102893, EPI_ISL_2102902, EPI_ISL_2102905, EPI_ISL_2102912, EPI_ISL_2102933, EPI_ISL_2102947, EPI_ISL_2102955, EPI_ISL_2102958, EPI_ISL_2102976, EPI_ISL_2102978, EPI_ISL_2102994, EPI_ISL_2103008, EPI_ISL_2103026, EPI_ISL_2103045, EPI_ISL_2103090, EPI_ISL_2103092, EPI_ISL_2103112, EPI_ISL_2103126, EPI_ISL_2103130, EPI_ISL_2103133, EPI_ISL_2103153, EPI_ISL_2103161, EPI_ISL_2103176, EPI_ISL_2103189 | | | |
| see above | County of San Luis Obispo Public Health Laboratory | Chan-Zuckerberg Biohub | CZB Cliahub Consortium |
| EPI_ISL_2158079, EPI_ISL_2178765, EPI_ISL_2178766, EPI_ISL_2178767, EPI_ISL_2178768, EPI_ISL_2178769, EPI_ISL_2178770, EPI_ISL_2178771, EPI_ISL_2178772, EPI_ISL_2178773, EPI_ISL_2178774, EPI_ISL_2178775, EPI_ISL_2178776, EPI_ISL_2178777, EPI_ISL_2178778 | | | |
| see above | DeRisi Lab, University of California, San Francisco | DeRisi Lab, University of California, San Francisco | Amy Kistler; Anthea Mitchell; Carina Marquez; Diane Havlir; Diane Jones; Douglas Black; Eric Chow; Gabriel Chamie; Genay Pilarowski; IDseq Team; Jackie Martinez; James Peng; Jamin Liu; Jon Jacobo; Lucy Li; Luis Rubio; Manu Vanaerschot; Matthew Laurie; Maya Petersen; Patrick Ayscue; Sabrina Mann; Sara Sunshine; Susana Rojas; Susy Rojas; Valerie Tulier-Laiwa; and Joseph DeRisi |
| EPI_ISL_1913219, EPI_ISL_1913220, EPI_ISL_1913221, EPI_ISL_1913222, EPI_ISL_1913223, EPI_ISL_1913224 | EL CAMINO HOSPITAL | Santa Clara County public Health Laboratory | Santa Clara County Public Health Department |
| EPI_ISL_1812119, EPI_ISL_1813447, EPI_ISL_1813669, EPI_ISL_1814286, EPI_ISL_1814396, EPI_ISL_1816811 | EXCITE Lab | Andersen lab at Scripps Research | Abigail Schnapper; Angela Scioscia; Cheryl Anderson; Chip Schooley; Greg Humphrey; Helena Tubb; Natasha Martin; Sawyer Farmer; Smruthi Karthikeyan; Tommy Valles + SEARCH |
| EPI_ISL_1532124, EPI_ISL_1532125, EPI_ISL_1532126 | El Camino Hospital | Santa Clara County Public Health Laboratory | Santa Clara County Public Health Department |
| EPI_ISL_1664270, EPI_ISL_1664271, EPI_ISL_1664272, EPI_ISL_1664273, EPI_ISL_1664274, EPI_ISL_1824117, EPI_ISL_1824118, EPI_ISL_1824119 | | | |
| see above | El Camino Hospital | Santa Clara County public Health Laboratory | Santa Clara County Public Health Department |
| EPI_ISL_2031859, EPI_ISL_2031860 | Flow Diagnostics Lab | Los Angeles County PHL | P. Hemarajata et al. |
| EPI_ISL_1578136, EPI_ISL_1578139, EPI_ISL_1578144, EPI_ISL_1578153, EPI_ISL_1578157, EPI_ISL_1578162, EPI_ISL_1578176, EPI_ISL_1578180, EPI_ISL_1578299, EPI_ISL_1578330, EPI_ISL_1578333, EPI_ISL_1578336, EPI_ISL_1578338, EPI_ISL_1918991, EPI_ISL_1918992 | | | |
| see above | Flow Health | Infectious Disease Program, Broad Institute of Harvard and MIT | Adams, G.; B.L.; B.W.; Bauer, M.; Birren; Carter, A.; Chaluvadi, S.; D.J.; DeRuff, K.; Gallagher, G.; Gladden-Young, A.; J.E.; K.J.; Lagerborg, K.; Lemieux; Loreth, C.; MacInnis; Normandin, E.; P.C.; Park; Reilly, S.; Rudy, M.; Siddle; Smole, S.; Tomkins-Tinch, C.; and Sabeti |

EPI_ISL_1525267, EPI_ISL_1525268, EPI_ISL_1525269, EPI_ISL_1525270, EPI_ISL_1525271, EPI_ISL_1525272, EPI_ISL_1525273, EPI_ISL_1525274, EPI_ISL_1525275, EPI_ISL_1525276, EPI_ISL_1525277, EPI_ISL_1525278, EPI_ISL_1525279, EPI_ISL_1525280, EPI_ISL_1525281, EPI_ISL_1525282, EPI_ISL_1525283, EPI_ISL_1525284, EPI_ISL_1525285, EPI_ISL_1525286, EPI_ISL_1525287, EPI_ISL_1525288, EPI_ISL_1525289, EPI_ISL_1525290, EPI_ISL_1525291, EPI_ISL_1525292, EPI_ISL_1525293, EPI_ISL_1525294, EPI_ISL_1525295, EPI_ISL_1525296, EPI_ISL_1525297, EPI_ISL_1525298, EPI_ISL_1525299, EPI_ISL_1525300, EPI_ISL_1525301, EPI_ISL_1525302, EPI_ISL_1525303, EPI_ISL_1525304, EPI_ISL_1525305, EPI_ISL_1525306, EPI_ISL_1525307, EPI_ISL_1525308, EPI_ISL_1525309, EPI_ISL_1525310, EPI_ISL_1525311, EPI_ISL_1525312, EPI_ISL_1525313, EPI_ISL_1525314, EPI_ISL_1525628, EPI_ISL_1525629, EPI_ISL_1525630, EPI_ISL_1525631, EPI_ISL_1525632, EPI_ISL_1525633, EPI_ISL_1525634, EPI_ISL_1525636, EPI_ISL_1525637, EPI_ISL_1525638, EPI_ISL_1525639, EPI_ISL_1525640, EPI_ISL_1525642, EPI_ISL_1525643, EPI_ISL_1525644, EPI_ISL_1525645, EPI_ISL_1525646, EPI_ISL_1525647, EPI_ISL_1525648, EPI_ISL_1525649, EPI_ISL_1525650, EPI_ISL_1525651, EPI_ISL_1525652, EPI_ISL_1525653, EPI_ISL_1525655, EPI_ISL_1525656, EPI_ISL_1525658, EPI_ISL_1525659, EPI_ISL_1525660, EPI_ISL_1525661, EPI_ISL_1525662, EPI_ISL_1525663, EPI_ISL_1525666, EPI_ISL_1525667, EPI_ISL_1525668, EPI_ISL_1525669, EPI_ISL_1525670, EPI_ISL_1525671, EPI_ISL_1525672, EPI_ISL_1525673, EPI_ISL_1525674, EPI_ISL_1525675, EPI_ISL_1525676, EPI_ISL_1525677, EPI_ISL_1525678, EPI_ISL_1525679, EPI_ISL_1525680, EPI_ISL_1525681, EPI_ISL_1525682, EPI_ISL_1525683, EPI_ISL_1525684, EPI_ISL_1525685, EPI_ISL_1525686, EPI_ISL_1525687, EPI_ISL_1525688, EPI_ISL_1525689, EPI_ISL_1525690, EPI_ISL_1525691, EPI_ISL_1525692, EPI_ISL_1525694, EPI_ISL_1525695, EPI_ISL_1525696, EPI_ISL_1525697, EPI_ISL_1525698, EPI_ISL_1525699, EPI_ISL_1525700, EPI_ISL_1525701, EPI_ISL_1525702, EPI_ISL_1525703, EPI_ISL_1525704, EPI_ISL_1525705, EPI_ISL_1525706, EPI_ISL_1525707, EPI_ISL_1525708, EPI_ISL_1525709, EPI_ISL_1525710, EPI_ISL_1525711, EPI_ISL_1525712, EPI_ISL_1525713, EPI_ISL_1525714, EPI_ISL_1525716, EPI_ISL_1525717, EPI_ISL_1525718, EPI_ISL_1525719, EPI_ISL_1525720, EPI_ISL_1525721, EPI_ISL_1525722, EPI_ISL_1525723, EPI_ISL_1525724, EPI_ISL_1525725, EPI_ISL_1525726, EPI_ISL_1525727, EPI_ISL_1525728, EPI_ISL_1525729, EPI_ISL_1525730, EPI_ISL_1525731, EPI_ISL_1525732, EPI_ISL_1525733, EPI_ISL_1525734, EPI_ISL_1525735, EPI_ISL_1525736, EPI_ISL_1525737, EPI_ISL_1525738, EPI_ISL_1525739, EPI_ISL_1525740, EPI_ISL_1525741, EPI_ISL_1525742, EPI_ISL_1525743, EPI_ISL_1525744, EPI_ISL_1525745, EPI_ISL_1525746, EPI_ISL_1525747, EPI_ISL_1525748, EPI_ISL_1525749, EPI_ISL_1525750, EPI_ISL_1525751, EPI_ISL_1525752, EPI_ISL_1525753, EPI_ISL_1525754, EPI_ISL_1525755, EPI_ISL_1525756, EPI_ISL_1525757, EPI_ISL_1525758, EPI_ISL_1525759, EPI_ISL_1525760, EPI_ISL_1525761, EPI_ISL_1525762, EPI_ISL_1525763, EPI_ISL_1525764, EPI_ISL_1525765, EPI_ISL_1525766, EPI_ISL_1525767, EPI_ISL_1525768, EPI_ISL_1525769, EPI_ISL_1525770, EPI_ISL_1525771, EPI_ISL_1525772, EPI_ISL_1525773, EPI_ISL_1525774, EPI_ISL_1525775, EPI_ISL_1525776, EPI_ISL_1525777, EPI_ISL_1525778, EPI_ISL_1525779, EPI_ISL_1525780, EPI_ISL_1525781, EPI_ISL_1525782, EPI_ISL_1525783, EPI_ISL_1525784, EPI_ISL_1525785, EPI_ISL_1525786, EPI_ISL_1525787, EPI_ISL_1525788, EPI_ISL_1525789, EPI_ISL_1525790, EPI_ISL_1525791, EPI_ISL_1525792, EPI_ISL_1525793, EPI_ISL_1525794, EPI_ISL_1525795, EPI_ISL_1525796, EPI_ISL_1525797, EPI_ISL_1525798, EPI_ISL_1525799, EPI_ISL_1556394, EPI_ISL_1556395, EPI_ISL_1556396, EPI_ISL_1556397, EPI_ISL_1556398, EPI_ISL_1556399, EPI_ISL_1556400, EPI_ISL_1556401, EPI_ISL_1556402, EPI_ISL_1556403, EPI_ISL_1556404, EPI_ISL_1556405, EPI_ISL_1556406, EPI_ISL_1556407, EPI_ISL_1556408, EPI_ISL_1556409, EPI_ISL_1556410, EPI_ISL_1556411, EPI_ISL_1556412, EPI_ISL_1556413, EPI_ISL_1556414, EPI_ISL_1556415, EPI_ISL_1556416, EPI_ISL_1556417, EPI_ISL_1556418, EPI_ISL_1556419, EPI_ISL_1556922, EPI_ISL_1556923, EPI_ISL_1556924, EPI_ISL_1556925, EPI_ISL_1556926, EPI_ISL_1556927, EPI_ISL_1556928, EPI_ISL_1556929, EPI_ISL_1556930, EPI_ISL_1556931, EPI_ISL_1556932, EPI_ISL_1556933, EPI_ISL_1556934, EPI_ISL_1556935, EPI_ISL_1556936, EPI_ISL_1556937, EPI_ISL_1556938, EPI_ISL_1556940, EPI_ISL_1556941, EPI_ISL_1556942, EPI_ISL_1556943, EPI_ISL_1556944, EPI_ISL_1556945, EPI_ISL_1556946, EPI_ISL_1556947, EPI_ISL_1556948, EPI_ISL_1556949, EPI_ISL_1556950, EPI_ISL_1556951, EPI_ISL_1556952, EPI_ISL_1556953, EPI_ISL_1556954, EPI_ISL_1556955, EPI_ISL_1556956, EPI_ISL_1556957, EPI_ISL_1556958, EPI_ISL_1556959, EPI_ISL_1556960, EPI_ISL_1556961, EPI_ISL_1556962, EPI_ISL_1556963, EPI_ISL_1556964, EPI_ISL_1556965, EPI_ISL_1556966, EPI_ISL_1556967, EPI_ISL_1556968, EPI_ISL_1556969, EPI_ISL_1556970, EPI_ISL_1556971, EPI_ISL_1556972, EPI_ISL_1556973, EPI_ISL_1556974, EPI_ISL_1556976, EPI_ISL_1556977, EPI_ISL_1556978, EPI_ISL_1556979, EPI_ISL_1556980, EPI_ISL_1556981, EPI_ISL_1556982, EPI_ISL_1556983, EPI_ISL_1556984, EPI_ISL_1556985, EPI_ISL_1556986, EPI_ISL_1556987, EPI_ISL_1556988, EPI_ISL_1556989, EPI_ISL_1556990, EPI_ISL_1556991, EPI_ISL_1556992, EPI_ISL_1556993, EPI_ISL_1556994, EPI_ISL_1556995, EPI_ISL_1556996, EPI_ISL_1556997, EPI_ISL_1556998, EPI_ISL_1556999, EPI_ISL_1557000, EPI_ISL_1557001, EPI_ISL_1557002, EPI_ISL_1557003, EPI_ISL_1557004, EPI_ISL_1557005, EPI_ISL_1557006, EPI_ISL_1557007, EPI_ISL_1557008, EPI_ISL_1557009, EPI_ISL_1557010, EPI_ISL_1557011, EPI_ISL_1557012, EPI_ISL_1557013, EPI_ISL_1557014, EPI_ISL_1557015, EPI_ISL_1557016, EPI_ISL_1557017, EPI_ISL_1557018, EPI_ISL_1557019, EPI_ISL_1557020, EPI_ISL_1557021, EPI_ISL_1557022, EPI_ISL_1557023, EPI_ISL_1557024, EPI_ISL_1557025, EPI_ISL_1557026, EPI_ISL_1557027, EPI_ISL_1557028, EPI_ISL_1557029, EPI_ISL_1557030, EPI_ISL_1557031, EPI_ISL_1557032, EPI_ISL_1612850, EPI_ISL_1612953, EPI_ISL_1612954, EPI_ISL_1612955, EPI_ISL_1612956, EPI_ISL_1612957, EPI_ISL_1612958, EPI_ISL_1612959, EPI_ISL_1612960, EPI_ISL_1612961, EPI_ISL_1612962, EPI_ISL_1612963, EPI_ISL_1612964, EPI_ISL_1612965, EPI_ISL_1612966, EPI_ISL_1612967, EPI_ISL_1612968, EPI_ISL_1612969, EPI_ISL_1612970, EPI_ISL_1612971, EPI_ISL_1612972, EPI_ISL_1612973, EPI_ISL_1612974, EPI_ISL_1612975, EPI_ISL_1612976, EPI_ISL_1612977, EPI_ISL_1612978, EPI_ISL_1612979, EPI_ISL_1612980, EPI_ISL_1612981, EPI_ISL_1612982, EPI_ISL_1612983, EPI_ISL_1612984, EPI_ISL_1612985, EPI_ISL_1612986, EPI_ISL_1612987, EPI_ISL_1612988, EPI_ISL_1612989, EPI_ISL_1612990, EPI_ISL_1612991, EPI_ISL_1612992, EPI_ISL_1612993, EPI_ISL_1612994, EPI_ISL_1612995, EPI_ISL_1612996, EPI_ISL_1612997, EPI_ISL_1612998, EPI_ISL_1612999, EPI_ISL_1613000, EPI_ISL_1613001, EPI_ISL_1613002, EPI_ISL_1613003, EPI_ISL_1613004, EPI_ISL_1613005, EPI_ISL_1613006, EPI_ISL_1613007, EPI_ISL_1613008, EPI_ISL_1613009, EPI_ISL_1613010, EPI_ISL_1613011, EPI_ISL_1613012, EPI_ISL_1613013, EPI_ISL_1613014, EPI_ISL_1613015, EPI_ISL_1613016, EPI_ISL_1613017, EPI_ISL_1613018, EPI_ISL_1613019, EPI_ISL_1613020, EPI_ISL_1613021, EPI_ISL_1613022, EPI_ISL_1613023, EPI_ISL_1613024, EPI_ISL_1613025, EPI_ISL_1613026, EPI_ISL_1613027, EPI_ISL_1613028, EPI_ISL_1613029, EPI_ISL_1613030, EPI_ISL_1613031, EPI_ISL_1613032, EPI_ISL_1613033, EPI_ISL_1613034, EPI_ISL_1613035, EPI_ISL_1613036, EPI_ISL_1613037, EPI_ISL_1613038, EPI_ISL_1613039, EPI_ISL_1613040, EPI_ISL_1613041, EPI_ISL_1613042, EPI_ISL_1613043, EPI_ISL_1613044, EPI_ISL_1613045, EPI_ISL_1613046, EPI_ISL_1613047, EPI_ISL_1613048, EPI_ISL_1613049, EPI_ISL_1613050, EPI_ISL_1613051, EPI_ISL_1613052, EPI_ISL_1613053, EPI_ISL_1613054, EPI_ISL_1613055, EPI_ISL_1613056, EPI_ISL_1613057, EPI_ISL_1613058, EPI_ISL_1613059, EPI_ISL_1613060, EPI_ISL_1613061, EPI_ISL_1613062, EPI_ISL_1613063, EPI_ISL_1613064, EPI_ISL_1613065, EPI_ISL_1613066, EPI_ISL_1613067, EPI_ISL_1613068, EPI_ISL_1613069, EPI_ISL_1613070, EPI_ISL_1613071, EPI_ISL_1613072, EPI_ISL_1613073, EPI_ISL_1613074, EPI_ISL_1613075, EPI_ISL_1613076, EPI_ISL_1613077, EPI_ISL_1613078, EPI_ISL_1613079, EPI_ISL_1613182, EPI_ISL_1613185, EPI_ISL_1613188, EPI_ISL_1613191, EPI_ISL_1613194, EPI_ISL_1613197, EPI_ISL_1613200, EPI_ISL_1613203, EPI_ISL_1613206, EPI_ISL_1613209, EPI_ISL_1613212, EPI_ISL_1613215, EPI_ISL_1613218, EPI_ISL_1613221, EPI_ISL_1613224, EPI_ISL_1613226, EPI_ISL_1613229, EPI_ISL_1613235, EPI_ISL_1613238, EPI_ISL_1613241, EPI_ISL_1613245, EPI_ISL_1613248, EPI_ISL_1613251, EPI_ISL_1613254, EPI_ISL_1613257, EPI_ISL_1613260, EPI_ISL_1613263, EPI_ISL_1613266, EPI_ISL_1613269, EPI_ISL_1613272, EPI_ISL_1613275, EPI_ISL_1613277, EPI_ISL_1613280, EPI_ISL_1613284, EPI_ISL_1613287, EPI_ISL_1613290, EPI_ISL_1613292, EPI_ISL_1613295, EPI_ISL_1613298, EPI_ISL_1613301, EPI_ISL_1613304, EPI_ISL_1613307, EPI_ISL_1613310, EPI_ISL_1613313, EPI_ISL_1613316, EPI_ISL_1613319, EPI_ISL_1613321, EPI_ISL_1613324, EPI_ISL_1613327, EPI_ISL_1613331, EPI_ISL_1613334, EPI_ISL_1613339, EPI_ISL_1613342, EPI_ISL_1613345, EPI_ISL_1613348, EPI_ISL_1613351, EPI_ISL_1613354, EPI_ISL_1613357, EPI_ISL_1613360, EPI_ISL_1613363, EPI_ISL_1613366, EPI_ISL_1613369, EPI_ISL_1613372, EPI_ISL_1613375, EPI_ISL_1613378, EPI_ISL_1613381, EPI_ISL_1613384, EPI_ISL_1613386, EPI_ISL_1613389, EPI_ISL_1613392, EPI_ISL_1613401, EPI_ISL_1613404, EPI_ISL_1613407, EPI_ISL_1613410, EPI_ISL_1613413, EPI_ISL_1613416, EPI_ISL_1613419, EPI_ISL_1613422, EPI_ISL_1613425, EPI_ISL_1613428, EPI_ISL_1613431, EPI_ISL_1613433, EPI_ISL_1613436, EPI_ISL_1613439, EPI_ISL_1613442, EPI_ISL_1613445, EPI_ISL_1613448, EPI_ISL_1613451, EPI_ISL_1613454, EPI_ISL_1613458, EPI_ISL_1613461, EPI_ISL_1613464, EPI_ISL_1613467, EPI_ISL_1613469, EPI_ISL_1613472, EPI_ISL_1613475, EPI_ISL_1613478, EPI_ISL_1613481, EPI_ISL_1613485, EPI_ISL_1613487, EPI_ISL_1613490, EPI_ISL_1613493, EPI_ISL_1613496, EPI_ISL_1613499, EPI_ISL_1613502, EPI_ISL_1613505, EPI_ISL_1613508, EPI_ISL_1613511, EPI_ISL_1613514, EPI_ISL_1613517, EPI_ISL_1613520, EPI_ISL_1613522, EPI_ISL_1613529, EPI_ISL_1613532, EPI_ISL_1613534, EPI_ISL_1613538, EPI_ISL_1613540, EPI_ISL_1613543, EPI_ISL_1613546, EPI_ISL_1613549, EPI_ISL_1613552, EPI_ISL_1613555, EPI_ISL_1613558, EPI_ISL_1613562, EPI_ISL_1613565, EPI_ISL_1613567, EPI_ISL_1613570, EPI_ISL_1613573, EPI_ISL_1613576, EPI_ISL_1613579, EPI_ISL_1613582, EPI_ISL_1613585, EPI_ISL_1613588, EPI_ISL_1613591, EPI_ISL_1613593, EPI_ISL_1613596, EPI_ISL_1613599, EPI_ISL_1613602, EPI_ISL_1613605, EPI_ISL_1613608, EPI_ISL_1613805, EPI_ISL_1613808, EPI_ISL_1613810, EPI_ISL_1613813, EPI_ISL_1613816, EPI_ISL_1613819, EPI_ISL_1613822, EPI_ISL_1613825, EPI_ISL_1613828, EPI_ISL_1613831, EPI_ISL_1613834, EPI_ISL_1613836, EPI_ISL_1613839, EPI_ISL_1613843, EPI_ISL_1613846, EPI_ISL_1613849, EPI_ISL_1613852, EPI_ISL_1613854, EPI_ISL_1613857, EPI_ISL_1613859, EPI_ISL_1613862, EPI_ISL_1613865, EPI_ISL_1613868, EPI_ISL_1613871, EPI_ISL_1613874, EPI_ISL_1613877, EPI_ISL_1613880, EPI_ISL_1613883, EPI_ISL_1613886, EPI_ISL_1613889, EPI_ISL_1613891, EPI_ISL_1613894, EPI_ISL_1613897, EPI_ISL_1613900, EPI_ISL_1613902, EPI_ISL_1613905, EPI_ISL_1613908, EPI_ISL_1613911, EPI_ISL_1613914, EPI_ISL_1613916, EPI_ISL_1613919, EPI_ISL_1613922, EPI_ISL_1613925, EPI_ISL_1613928, EPI_ISL_1613931, EPI_ISL_1613934, EPI_ISL_1613937, EPI_ISL_1613938, EPI_ISL_1613939, EPI_ISL_1613940, EPI_ISL_1613941, EPI_ISL_1613942, EPI_ISL_1613943, EPI_ISL_1613944, EPI_ISL_1613945, EPI_ISL_1613946, EPI_ISL_1613947, EPI_ISL_1613948, EPI_ISL_1613949, EPI_ISL_1613950, EPI_ISL_1613951, EPI_ISL_1613952, EPI_ISL_1613953, EPI_ISL_1613954, EPI_ISL_1613955, EPI_ISL_1613956, EPI_ISL_1613957, EPI_ISL_1613958, EPI_ISL_1613959, EPI_ISL_1613960, EPI_ISL_1613961, EPI_ISL_1613962, EPI_ISL_1613963, EPI_ISL_1613964, EPI_ISL_1613965, EPI_ISL_1613966, EPI_ISL_1613967, EPI_ISL_1613968, EPI_ISL_1613969, EPI_ISL_1613970, EPI_ISL_1613973, EPI_ISL_1613974, EPI_ISL_1613975, EPI_ISL_1613976, EPI_ISL_1613977, EPI_ISL_1613978, EPI_ISL_1613979, EPI_ISL_1613980, EPI_ISL_1613981, EPI_ISL_1613982, EPI_ISL_1613983, EPI_ISL_1613984, EPI_ISL_1613985, EPI_ISL_1613986, EPI_ISL_1613987, EPI_ISL_1613988, EPI_ISL_1613989, EPI_ISL_1613990, EPI_ISL_1613991, EPI_ISL_1613992, EPI_ISL_1613993, EPI_ISL_1613994, EPI_ISL_1613995, EPI_ISL_1613996, EPI_ISL_1613997, EPI_ISL_1613998, EPI_ISL_1613999, EPI_ISL_1614000, EPI_ISL_1614001, EPI_ISL_1614002, EPI_ISL_1614003, EPI_ISL_1614004, EPI_ISL_1614005, EPI_ISL_1614006, EPI_ISL_1614007, EPI_ISL_1614008, EPI_ISL_1614009, EPI_ISL_1614010, EPI_ISL_1614011, EPI_ISL_1614012, EPI_ISL_1614013, EPI_ISL_1614014, EPI_ISL_1614015, EPI_ISL_1614016, EPI_ISL_1614017, EPI_ISL_1614018, EPI_ISL_1614019, EPI_ISL_1614020, EPI_ISL_1614021, EPI_ISL_1614022, EPI_ISL_1614023, EPI_ISL_1614024, EPI_ISL_1614025, EPI_ISL_1614026, EPI_ISL_1614027, EPI_ISL_1614028, EPI_ISL_1614029, EPI_ISL_1614030, EPI_ISL_1614031, EPI_ISL_1614032, EPI_ISL_1614033, EPI_ISL_1614034, EPI_ISL_1614035, EPI_ISL_1614036, EPI_ISL_1614037, EPI_ISL_1614038, EPI_ISL_1614039, EPI_ISL_1614040, EPI_ISL_1614041, EPI_ISL_1614042, EPI_ISL_1614043, EPI_ISL_1614044, EPI_ISL_1614045, EPI_ISL_1614046, EPI_ISL_1614047, EPI_ISL_1614048, EPI_ISL_1614049, EPI_ISL_1614050, EPI_ISL_1614051, EPI_ISL_1614052, EPI_ISL_1614053, EPI_ISL_1614054, EPI_ISL_1614055, EPI_ISL_1614056, EPI_ISL_1614057, EPI_ISL_1614058, EPI_ISL_1614059, EPI_ISL_1614060, EPI_ISL_1614061, EPI_ISL_1614062, EPI_ISL_1614063, EPI_ISL_1614064, EPI_ISL_1614065, EPI_ISL_1614066, EPI_ISL_1614067, EPI_ISL_1614068, EPI_ISL_1614069, EPI_ISL_1614070, EPI_ISL_1614071, EPI_ISL_1614072, EPI_ISL_1614073, EPI_ISL_1614074, EPI_ISL_1614075, EPI_ISL_1614077, EPI_ISL_1614078, EPI_ISL_1614080, EPI_ISL_1614081, EPI_ISL_1614082, EPI_ISL_1614083, EPI_ISL_1614084, EPI_ISL_1614085, EPI_ISL_1614086, EPI_ISL_1614087, EPI_ISL_1614088, EPI_ISL_1614089, EPI_ISL_1614090, EPI_ISL_1614091, EPI_ISL_1614092, EPI_ISL_1614093, EPI_ISL_1614094, EPI_ISL_1614095, EPI_ISL_1614096, EPI_ISL_1614097, EPI_ISL_1614098, EPI_ISL_1614099, EPI_ISL_1614100, EPI_ISL_1614101, EPI_ISL_1614102, EPI_ISL_1614103, EPI_ISL_1614104, EPI_ISL_1614105, EPI_ISL_1614106, EPI_ISL_1614107, EPI_ISL_1614108, EPI_ISL_1614109, EPI_ISL_1614110, EPI_ISL_1614111, EPI_ISL_1614112, EPI_ISL_1614113, EPI_ISL_1614114, EPI_ISL_1614115, EPI_ISL_1614116, EPI_ISL_1614117, EPI_ISL_1614118, EPI_ISL_1614119, EPI_ISL_1614120, EPI_ISL_1614121, EPI_ISL_1614122, EPI_ISL_1614123, EPI_ISL_1614124, EPI_ISL_1614125, EPI_ISL_1614126, EPI_ISL_1614127, EPI_ISL_1614128, EPI_ISL_1614129, EPI_ISL_1614130, EPI_ISL_1614131, EPI_ISL_1614132, EPI_ISL_1614133, EPI_ISL_1614134, EPI_ISL_1614135, EPI_ISL_1614136, EPI_ISL_1614137, EPI_ISL_1614138, EPI_ISL_1614139, EPI_ISL_1614140, EPI_ISL_1614141, EPI_ISL_1614142, EPI_ISL_1614143, EPI_ISL_1614144, EPI_ISL_1614145, EPI_ISL_1614146, EPI_ISL_1614147, EPI_ISL_1614148, EPI_ISL_1614149, EPI_ISL_1614150, EPI_ISL_1614151, EPI_ISL_1614152, EPI_ISL_1614153, EPI_ISL_1614154, EPI_ISL_1614155, EPI_ISL_1614156, EPI_ISL_1614157, EPI_ISL_1614158, EPI_ISL_1614159, EPI_ISL_1614160, EPI_ISL_1614161, EPI_ISL_1614162, EPI_ISL_1614163, EPI_ISL_1614164, EPI_ISL_1614165, EPI_ISL_1614166, EPI_ISL_1614167, EPI_ISL_1614168, EPI_ISL_1614169, EPI_ISL_1614170, EPI_ISL_1614226, EPI_ISL_1614348, EPI_ISL_1614460, EPI_ISL_1614462, EPI_ISL_1614463, EPI_ISL_1614464, EPI_ISL_1614465, EPI_ISL_1614466, EPI_ISL_1614467, EPI_ISL_1614468, EPI_ISL_1614469, EPI_ISL_1614470, EPI_ISL_1614471, EPI_ISL_1614472, EPI_ISL_1614473, EPI_ISL_1614474, EPI_ISL_1614475, EPI_ISL_1614476, EPI_ISL_1614477, EPI_ISL_1614478, EPI_ISL_1614479, EPI_ISL_1614480, EPI_ISL_1614481, EPI_ISL_1614482, EPI_ISL_1614483, EPI_ISL_1614484, EPI_ISL_1614485, EPI_ISL_1614486, EPI_ISL_1614487, EPI_ISL_1614488, EPI_ISL_1614489, EPI_ISL_1614490, EPI_ISL_1614491, EPI_ISL_1614492, EPI_ISL_1614493, EPI_ISL_1614494, EPI_ISL_1614495, EPI_ISL_1614496, EPI_ISL_1614497, EPI_ISL_1614498, EPI_ISL_1614499, EPI_ISL_1614500, EPI_ISL_1614501, EPI_ISL_1614502, EPI_ISL_1614503, EPI_ISL_1614504, EPI_ISL_1614505, EPI_ISL_1614506, EPI_ISL_1614507, EPI_ISL_1614508, EPI_ISL_1614509, EPI_ISL_1614510, EPI_ISL_1614511, EPI_ISL_1614512, EPI_ISL_1614513, EPI_ISL_1614514, EPI_ISL_1614515, EPI_ISL_1614516, EPI_ISL_1614517, EPI_ISL_1614518, EPI_ISL_1614519, EPI_ISL_1614520, EPI_ISL_1614521, EPI_ISL_1614522, EPI_ISL_1614523, EPI_ISL_1614524, EPI_ISL_1614525, EPI_ISL_1614526, EPI_ISL_1614527, EPI_ISL_1614528, EPI_ISL_1614529, EPI_ISL_1614530, EPI_ISL_1614531, EPI_ISL_1614532, EPI_ISL_1614533, EPI_ISL_1614534, EPI_ISL_1614536, EPI_ISL_1614537, EPI_ISL_1614538, EPI_ISL_1614539, EPI_ISL_1614540, EPI_ISL_1614541, EPI_ISL_1614542, EPI_ISL_1614543, EPI_ISL_1614544, EPI_ISL_1614545, EPI_ISL_1614546, EPI_ISL_1614547, EPI_ISL_1614548, EPI_ISL_1614549, EPI_ISL_1614550, EPI_ISL_1614551, EPI_ISL_1614552, EPI_ISL_1614553, EPI_ISL_1614554, EPI_ISL_1614555, EPI_ISL_1614556, EPI_ISL_1614557, EPI_ISL_1614558, EPI_ISL_1614559, EPI_ISL_1614560, EPI_ISL_1614561, EPI_ISL_1614562, EPI_ISL_1614563, EPI_ISL_1614564, EPI_ISL_1614565, EPI_ISL_1614566, EPI_ISL_1614567, EPI_ISL_1614568, EPI_ISL_1614569, EPI_ISL_1614570, EPI_ISL_1614571, EPI_ISL_1614572, EPI_ISL_1614573, EPI_ISL_1614574, EPI_ISL_1614575, EPI_ISL_1614576, EPI_ISL_1614577, EPI_ISL_1614578, EPI_ISL_1614579, EPI_ISL_1614580, EPI_ISL_1614581, EPI_ISL_1614582, EPI_ISL_1614583, EPI_ISL_1614584, EPI_ISL_1614585, EPI_ISL_1614586, EPI_ISL_1614587, EPI_ISL_1614588, EPI_ISL_1614589, EPI_ISL_1614590, EPI_ISL_1614591, EPI_ISL_1614592, EPI_ISL_1614593, EPI_ISL_1614594, EPI_ISL_1614595, EPI_ISL_1614596, EPI_ISL_1614597, EPI_ISL_1614598, EPI_ISL_1614599, EPI_ISL_1614600, EPI_ISL_1614601, EPI_ISL_1614602, EPI_ISL_1614603, EPI_ISL_1614604, EPI_ISL_1614605, EPI_ISL_1614606, EPI_ISL_1614607, EPI_ISL_1614608, EPI_ISL_1614609, EPI_ISL_1614610, EPI_ISL_1614611, EPI_ISL_1614612, EPI_ISL_1614613, EPI_ISL_1614614, EPI_ISL_1614615, EPI_ISL_1614616, EPI_ISL_1614617, EPI_ISL_1614618, EPI_ISL_1614619, EPI_ISL_1614620, EPI_ISL_1614621, EPI_ISL_1614622, EPI_ISL_1614623, EPI_ISL_1614624, EPI_ISL_1614625, EPI_ISL_1614626, EPI_ISL_1614627, EPI_ISL_1614628, EPI_ISL_1614629, EPI_ISL_1614630, EPI_ISL_1614631, EPI_ISL_1614632, EPI_ISL_1614633, EPI_ISL_1614634, EPI_ISL_1614635, EPI_ISL_1614636, EPI_ISL_1614637, EPI_ISL_1614638, EPI_ISL_1614639, EPI_ISL_1614640, EPI_ISL_1614641, EPI_ISL_1614642, EPI_ISL_1614643, EPI_ISL_1614644, EPI_ISL_1614645, EPI_ISL_1614646, EPI_ISL_1614647, EPI_ISL_1614648, EPI_ISL_1614649, EPI_ISL_1614650, EPI_ISL_1614651, EPI_ISL_1614652, EPI_ISL_1614653, EPI_ISL_1614654, EPI_ISL_1614655, EPI_ISL_1614656, EPI_ISL_1614657, EPI_ISL_1614658, EPI_ISL_1614659, EPI_ISL_1614660, EPI_ISL_1614661, EPI_ISL_1614662, EPI_ISL_1614663, EPI_ISL_1614664, EPI_ISL_1614665, EPI_ISL_1614666, EPI_ISL_1614667, EPI_ISL_1614668, EPI_ISL_1614669, EPI_ISL_1614670, EPI_ISL_1614671, EPI_ISL_1614672, EPI_ISL_1614673, EPI_ISL_1614674, EPI_ISL_1614675, EPI_ISL_1614676, EPI_ISL_1614677, EPI_ISL_1614678, EPI_ISL_1614679, EPI_ISL_1614680, EPI_ISL_1614681, EPI_ISL_1614682, EPI_ISL_1614683, EPI_ISL_1614684, EPI_ISL_1614685, EPI_ISL_1614686, EPI_ISL_1614687, EPI_ISL_1614688, EPI_ISL_1614689, EPI_ISL_1614690, EPI_ISL_1614691, EPI_ISL_1614692, EPI_ISL_1614693, EPI_ISL_1614694, EPI_ISL_1614695, EPI_ISL_1614696, EPI_ISL_1614697, EPI_ISL_1614698, EPI_ISL_1614699, EPI_ISL_1614700, EPI_ISL_1614702, EPI_ISL_1614703, EPI_ISL_1614704, EPI_ISL_1614705, EPI_ISL_1614706, EPI_ISL_1614707, EPI_ISL_1614708, EPI_ISL_1614709, EPI_ISL_1614710, EPI_ISL_1614711, EPI_ISL_1614712, EPI_ISL_1614713, EPI_ISL_1614714, EPI_ISL_1614715, EPI_ISL_1614716, EPI_ISL_1614717, EPI_ISL_1614718, EPI_ISL_1614719, EPI_ISL_1614720, EPI_ISL_1614721, EPI_ISL_1614722, EPI_ISL_1614723, EPI_ISL_1614724, EPI_ISL_1614725, EPI_ISL_1614726, EPI_ISL_1614727, EPI_ISL_1614728, EPI_ISL_1614729, EPI_ISL_1614730, EPI_ISL_1614731, EPI_ISL_1614732, EPI_ISL_1614733, EPI_ISL_1614734, EPI_ISL_1614735, EPI_ISL_1614736, EPI_ISL_1614737, EPI_ISL_1614738, EPI_ISL_1614739, EPI_ISL_1614740, EPI_ISL_1614741, EPI_ISL_1614742, EPI_ISL_1614743, EPI_ISL_1614745, EPI_ISL_1614746, EPI_ISL_1614747, EPI_ISL_1614748, EPI_ISL_1614749, EPI_ISL_1614750, EPI_ISL_1614751, EPI_ISL_1614752, EPI_ISL_1614753, EPI_ISL_1614754, EPI_ISL_1614755, EPI_ISL_1614756, EPI_ISL_1614757, EPI_ISL_1614760, EPI_ISL_1614761, EPI_ISL_1614762, EPI_ISL_1614763, EPI_ISL_1614764, EPI_ISL_1614765, EPI_ISL_1614766, EPI_ISL_1614767, EPI_ISL_1614768, EPI_ISL_1614769, EPI_ISL_1614770, EPI_ISL_1614771, EPI_ISL_1614772, EPI_ISL_1614773, EPI_ISL_1614774, EPI_ISL_1614776, EPI_ISL_1614777, EPI_ISL_1614778, EPI_ISL_1614779, EPI_ISL_1614780, EPI_ISL_1614781, EPI_ISL_1614782, EPI_ISL_1614783, EPI_ISL_1614784, EPI_ISL_1614785, EPI_ISL_1614786, EPI_ISL_1614787, EPI_ISL_1614788, EPI_ISL_1614789, EPI_ISL_1614790, EPI_ISL_1614791, EPI_ISL_1614792, EPI_ISL_1614793, EPI_ISL_1614794, EPI_ISL_1614795, EPI_ISL_1614796, EPI_ISL_1614797, EPI_ISL_1614798, EPI_ISL_1614799, EPI_ISL_1614800, EPI_ISL_1614801, EPI_ISL_1614802, EPI_ISL_1614803, EPI_ISL_1614804, EPI_ISL_1614805, EPI_ISL_1614806, EPI_ISL_1614807, EPI_ISL_1614808, EPI_ISL_1614809, EPI_ISL_1614810, EPI_ISL_1614811, EPI_ISL_1614812, EPI_ISL_1614813, EPI_ISL_1614814, EPI_ISL_1614815, EPI_ISL_1614816, EPI_ISL_1614817, EPI_ISL_1614818, EPI_ISL_1614819, EPI_ISL_1614820, EPI_ISL_1614821, EPI_ISL_1614822, EPI_ISL_1614823, EPI_ISL_1614824, EPI_ISL_1614825, EPI_ISL_1614826, EPI_ISL_1614827, EPI_ISL_1614828, EPI_ISL_1614829, EPI_ISL_1614830, EPI_ISL_1614831, EPI_ISL_1614832, EPI_ISL_1614833, EPI_ISL_1614834, EPI_ISL_1614835, EPI_ISL_1614836, EPI_ISL_1614837, EPI_ISL_1614838, EPI_ISL_1614839, EPI_ISL_1614840, EPI_ISL_1614841, EPI_ISL_1614842, EPI_ISL_1614843, EPI_ISL_1614844, EPI_ISL_1614845, EPI_ISL_1614846, EPI_ISL_1614847, EPI_ISL_1614848, EPI_ISL_1614849, EPI_ISL_1614850, EPI_ISL_1614851, EPI_ISL_1614852, EPI_ISL_1614853, EPI_ISL_1614854, EPI_ISL_1614855, EPI_ISL_1614856, EPI_ISL_1614857, EPI_ISL_1614858, EPI_ISL_1614859, EPI_ISL_1614860, EPI_ISL_1614861, EPI_ISL_1614862, EPI_ISL_1614863, EPI_ISL_1614864, EPI_ISL_1614865, EPI_ISL_1614866, EPI_ISL_1614867, EPI_ISL_1614868, EPI_ISL_1614869, EPI_ISL_1614870, EPI_ISL_1614871, EPI_ISL_1614872, EPI_ISL_1614873, EPI_ISL_1614874, EPI_ISL_1614875, EPI_ISL_1614876, EPI_ISL_1614877, EPI_ISL_1614878, EPI_ISL_1614879, EPI_ISL_1614880, EPI_ISL_1614881, EPI_ISL_1614882, EPI_ISL_1614883, EPI_ISL_1614884, EPI_ISL_1614885, EPI_ISL_1614886, EPI_ISL_1614887, EPI_ISL_1614888, EPI_ISL_1614889, EPI_ISL_1614890, EPI_ISL_1614891, EPI_ISL_1614893, EPI_ISL_1614895, EPI_ISL_1614898, EPI_ISL_1614900, EPI_ISL_1614901, EPI_ISL_1614902, EPI_ISL_1614903, EPI_ISL_1614904, EPI_ISL_1665940, EPI_ISL_1666000, EPI_ISL_1666001, EPI_ISL_1666002, EPI_ISL_1666003, EPI_ISL_1666004, EPI_ISL_1666005, EPI_ISL_1666006, EPI_ISL_1666007, EPI_ISL_1666008, EPI_ISL_1666009, EPI_ISL_1666010, EPI_ISL_1666011, EPI_ISL_1666012, EPI_ISL_1666013, EPI_ISL_1666014,

EPI_ISL_1666015, EPI_ISL_1666016, EPI_ISL_1666017, EPI_ISL_1666018, EPI_ISL_1666019, EPI_ISL_1666020, EPI_ISL_1666021, EPI_ISL_1666022, EPI_ISL_1666023, EPI_ISL_1666024, EPI_ISL_1666025, EPI_ISL_1666026, EPI_ISL_1666027, EPI_ISL_1666028, EPI_ISL_1666029, EPI_ISL_1666030, EPI_ISL_1666031, EPI_ISL_1666032, EPI_ISL_1666033, EPI_ISL_1666034, EPI_ISL_1666035, EPI_ISL_1666036,
EPI_ISL_1666037, EPI_ISL_1666038, EPI_ISL_1666039, EPI_ISL_1666040, EPI_ISL_1666041, EPI_ISL_1666042, EPI_ISL_1666043, EPI_ISL_1666044, EPI_ISL_1666045, EPI_ISL_1666046, EPI_ISL_1666047, EPI_ISL_1666048, EPI_ISL_1666049, EPI_ISL_1666050, EPI_ISL_1666051, EPI_ISL_1666052, EPI_ISL_1666053, EPI_ISL_1666054, EPI_ISL_1666055, EPI_ISL_1666056, EPI_ISL_1666060, EPI_ISL_1666061,
EPI_ISL_1666062, EPI_ISL_1666063, EPI_ISL_1666064, EPI_ISL_1666065, EPI_ISL_1666066, EPI_ISL_1666067, EPI_ISL_1666068, EPI_ISL_1666069, EPI_ISL_1666070, EPI_ISL_1666071, EPI_ISL_1666072, EPI_ISL_1666073, EPI_ISL_1666074, EPI_ISL_1666075, EPI_ISL_1666076, EPI_ISL_1666077, EPI_ISL_1666078, EPI_ISL_1666079, EPI_ISL_1666080, EPI_ISL_1666081, EPI_ISL_1666082, EPI_ISL_1666083,
EPI_ISL_1666084, EPI_ISL_1666085, EPI_ISL_1666086, EPI_ISL_1666087, EPI_ISL_1666088, EPI_ISL_1666089, EPI_ISL_1666090, EPI_ISL_1666091, EPI_ISL_1666092, EPI_ISL_1666093, EPI_ISL_1666094, EPI_ISL_1666095, EPI_ISL_1666096, EPI_ISL_1666097, EPI_ISL_1666098, EPI_ISL_1666099, EPI_ISL_1666100, EPI_ISL_1666101, EPI_ISL_1666102, EPI_ISL_1666103, EPI_ISL_1666104, EPI_ISL_1666105,
EPI_ISL_1666106, EPI_ISL_1666107, EPI_ISL_1666108, EPI_ISL_1666109, EPI_ISL_1666110, EPI_ISL_1666111, EPI_ISL_1666112, EPI_ISL_1666113, EPI_ISL_1666114, EPI_ISL_1666115, EPI_ISL_1666116, EPI_ISL_1666117, EPI_ISL_1666118, EPI_ISL_1666119, EPI_ISL_1666120, EPI_ISL_1666121, EPI_ISL_1666122, EPI_ISL_1666123, EPI_ISL_1666124, EPI_ISL_1666125, EPI_ISL_1666126, EPI_ISL_1666127,
EPI_ISL_1666128, EPI_ISL_1666129, EPI_ISL_1666130, EPI_ISL_1666131, EPI_ISL_1666132, EPI_ISL_1666133, EPI_ISL_1666134, EPI_ISL_1666135, EPI_ISL_1666136, EPI_ISL_1666137, EPI_ISL_1666138, EPI_ISL_1666139, EPI_ISL_1666140, EPI_ISL_1666141, EPI_ISL_1666142, EPI_ISL_1666143, EPI_ISL_1666144, EPI_ISL_1666145, EPI_ISL_1666146, EPI_ISL_1666147, EPI_ISL_1666148, EPI_ISL_1666149,
EPI_ISL_1666150, EPI_ISL_1666151, EPI_ISL_1666152, EPI_ISL_1666153, EPI_ISL_1666154, EPI_ISL_1666155, EPI_ISL_1666156, EPI_ISL_1666157, EPI_ISL_1666158, EPI_ISL_1666159, EPI_ISL_1666160, EPI_ISL_1666161, EPI_ISL_1666162, EPI_ISL_1666163, EPI_ISL_1666164, EPI_ISL_1666165, EPI_ISL_1666166, EPI_ISL_1666167, EPI_ISL_1666168, EPI_ISL_1666169, EPI_ISL_1666170, EPI_ISL_1666171,
EPI_ISL_1666172, EPI_ISL_1666173, EPI_ISL_1666174, EPI_ISL_1666176, EPI_ISL_1666177, EPI_ISL_1666178, EPI_ISL_1666179, EPI_ISL_1666180, EPI_ISL_1666181, EPI_ISL_1666182, EPI_ISL_1666183, EPI_ISL_1666184, EPI_ISL_1666185, EPI_ISL_1666187, EPI_ISL_1666188, EPI_ISL_1666189, EPI_ISL_1666190, EPI_ISL_1666191, EPI_ISL_1666192, EPI_ISL_1666193, EPI_ISL_1666194, EPI_ISL_1666195,
EPI_ISL_1666196, EPI_ISL_1666197, EPI_ISL_1666198, EPI_ISL_1666199, EPI_ISL_1666200, EPI_ISL_1666201, EPI_ISL_1666202, EPI_ISL_1666203, EPI_ISL_1666204, EPI_ISL_1666205, EPI_ISL_1666206, EPI_ISL_1666207, EPI_ISL_1666208, EPI_ISL_1666209, EPI_ISL_1666210, EPI_ISL_1666211, EPI_ISL_1666212, EPI_ISL_1666213, EPI_ISL_1666214, EPI_ISL_1666215, EPI_ISL_1666216, EPI_ISL_1666217,
EPI_ISL_1666218, EPI_ISL_1666219, EPI_ISL_1666221, EPI_ISL_1666222, EPI_ISL_1666223, EPI_ISL_1666224, EPI_ISL_1666225, EPI_ISL_1666226, EPI_ISL_1666227, EPI_ISL_1666228, EPI_ISL_1666229, EPI_ISL_1666230, EPI_ISL_1666231, EPI_ISL_1666232, EPI_ISL_1666233, EPI_ISL_1666234, EPI_ISL_1666235, EPI_ISL_1666236, EPI_ISL_1666237, EPI_ISL_1666238, EPI_ISL_1666239, EPI_ISL_1666240,
EPI_ISL_1666241, EPI_ISL_1666242, EPI_ISL_1666243, EPI_ISL_1666244, EPI_ISL_1666245, EPI_ISL_1666246, EPI_ISL_1666247, EPI_ISL_1666248, EPI_ISL_1666249, EPI_ISL_1666250, EPI_ISL_1666251, EPI_ISL_1666252, EPI_ISL_1666253, EPI_ISL_1666254, EPI_ISL_1666255, EPI_ISL_1666256, EPI_ISL_1666257, EPI_ISL_1666258, EPI_ISL_1666259, EPI_ISL_1666260, EPI_ISL_1666261, EPI_ISL_1666262,
EPI_ISL_1666263, EPI_ISL_1666264, EPI_ISL_1666265, EPI_ISL_1666266, EPI_ISL_1666267, EPI_ISL_1666268, EPI_ISL_1666269, EPI_ISL_1666270, EPI_ISL_1666271, EPI_ISL_1666272, EPI_ISL_1666273, EPI_ISL_1666274, EPI_ISL_1666275, EPI_ISL_1666276, EPI_ISL_1666277, EPI_ISL_1666278, EPI_ISL_1666279, EPI_ISL_1666280, EPI_ISL_1666281, EPI_ISL_1666282, EPI_ISL_1666283, EPI_ISL_1666284,
EPI_ISL_1666285, EPI_ISL_1666286, EPI_ISL_1666287, EPI_ISL_1666288, EPI_ISL_1666289, EPI_ISL_1666290, EPI_ISL_1666291, EPI_ISL_1666292, EPI_ISL_1666293, EPI_ISL_1666294, EPI_ISL_1666295, EPI_ISL_1666296, EPI_ISL_1666297, EPI_ISL_1666298, EPI_ISL_1666299, EPI_ISL_1666300, EPI_ISL_1666301, EPI_ISL_1666302, EPI_ISL_1666303, EPI_ISL_1666304, EPI_ISL_1666305, EPI_ISL_1666306,
EPI_ISL_1666307, EPI_ISL_1666308, EPI_ISL_1666309, EPI_ISL_1666310, EPI_ISL_1666311, EPI_ISL_1666312, EPI_ISL_1666313, EPI_ISL_1666314, EPI_ISL_1666315, EPI_ISL_1666316, EPI_ISL_1666317, EPI_ISL_1666318, EPI_ISL_1666319, EPI_ISL_1666320, EPI_ISL_1666321, EPI_ISL_1666322, EPI_ISL_1666323, EPI_ISL_1666324, EPI_ISL_1666325, EPI_ISL_1666326, EPI_ISL_1666327, EPI_ISL_1666329,
EPI_ISL_1666330, EPI_ISL_1666331, EPI_ISL_1666332, EPI_ISL_1666333, EPI_ISL_1666334, EPI_ISL_1666335, EPI_ISL_1666336, EPI_ISL_1666337, EPI_ISL_1666338, EPI_ISL_1666339, EPI_ISL_1666340, EPI_ISL_1666341, EPI_ISL_1666342, EPI_ISL_1666343, EPI_ISL_1666344, EPI_ISL_1666345, EPI_ISL_1666346, EPI_ISL_1666347, EPI_ISL_1666348, EPI_ISL_1666349, EPI_ISL_1666350, EPI_ISL_1666351,
EPI_ISL_1666352, EPI_ISL_1666353, EPI_ISL_1666354, EPI_ISL_1666355, EPI_ISL_1666356, EPI_ISL_1666357, EPI_ISL_1666358, EPI_ISL_1666359, EPI_ISL_1666360, EPI_ISL_1666361, EPI_ISL_1666362, EPI_ISL_1666363, EPI_ISL_1666365, EPI_ISL_1666366, EPI_ISL_1666367, EPI_ISL_1666368, EPI_ISL_1666369, EPI_ISL_1666370, EPI_ISL_1666371, EPI_ISL_1666372, EPI_ISL_1666373, EPI_ISL_1666374,
EPI_ISL_1666375, EPI_ISL_1666376, EPI_ISL_1666377, EPI_ISL_1666378, EPI_ISL_1666379, EPI_ISL_1666380, EPI_ISL_1666381, EPI_ISL_1666382, EPI_ISL_1666383, EPI_ISL_1666384, EPI_ISL_1666385, EPI_ISL_1666386, EPI_ISL_1666387, EPI_ISL_1666388, EPI_ISL_1666389, EPI_ISL_1666390, EPI_ISL_1666391, EPI_ISL_1666392, EPI_ISL_1666393, EPI_ISL_1666394, EPI_ISL_1666395, EPI_ISL_1666396,
EPI_ISL_1666397, EPI_ISL_1666398, EPI_ISL_1666399, EPI_ISL_1666400, EPI_ISL_1666401, EPI_ISL_1666402, EPI_ISL_1666404, EPI_ISL_1666405, EPI_ISL_1666406, EPI_ISL_1666407, EPI_ISL_1666408, EPI_ISL_1666409, EPI_ISL_1666410, EPI_ISL_1666411, EPI_ISL_1666412, EPI_ISL_1666415, EPI_ISL_1666416, EPI_ISL_1666417, EPI_ISL_1666418, EPI_ISL_1666419, EPI_ISL_1666420, EPI_ISL_1666421,
EPI_ISL_1666422, EPI_ISL_1666423, EPI_ISL_1666424, EPI_ISL_1666425, EPI_ISL_1666426, EPI_ISL_1666427, EPI_ISL_1666428, EPI_ISL_1666429, EPI_ISL_1666430, EPI_ISL_1666431, EPI_ISL_1666432, EPI_ISL_1666433, EPI_ISL_1666434, EPI_ISL_1666435, EPI_ISL_1666436, EPI_ISL_1666437, EPI_ISL_1666438, EPI_ISL_1666439, EPI_ISL_1666440, EPI_ISL_1666441, EPI_ISL_1666442, EPI_ISL_1666443,
EPI_ISL_1666444, EPI_ISL_1666445, EPI_ISL_1666446, EPI_ISL_1666447, EPI_ISL_1666448, EPI_ISL_1666449, EPI_ISL_1666450, EPI_ISL_1666451, EPI_ISL_1666452, EPI_ISL_1666453, EPI_ISL_1666454, EPI_ISL_1666455, EPI_ISL_1666456, EPI_ISL_1666457, EPI_ISL_1666458, EPI_ISL_1666459, EPI_ISL_1666460, EPI_ISL_1666461, EPI_ISL_1666462, EPI_ISL_1666463, EPI_ISL_1666464, EPI_ISL_1666465,
EPI_ISL_1666466, EPI_ISL_1666467, EPI_ISL_1666468, EPI_ISL_1666469, EPI_ISL_1666470, EPI_ISL_1666471, EPI_ISL_1666472, EPI_ISL_1666474, EPI_ISL_1666475, EPI_ISL_1666476, EPI_ISL_1666477, EPI_ISL_1666478, EPI_ISL_1666479, EPI_ISL_1666480, EPI_ISL_1666481, EPI_ISL_1666482, EPI_ISL_1666483, EPI_ISL_1666484, EPI_ISL_1666485, EPI_ISL_1666486, EPI_ISL_1666487, EPI_ISL_1666488,
EPI_ISL_1666489, EPI_ISL_1666490, EPI_ISL_1666491, EPI_ISL_1666492, EPI_ISL_1666493, EPI_ISL_1666494, EPI_ISL_1666495, EPI_ISL_1666496, EPI_ISL_1666497, EPI_ISL_1666498, EPI_ISL_1666499, EPI_ISL_1666500, EPI_ISL_1666501, EPI_ISL_1666502, EPI_ISL_1666503, EPI_ISL_1666504, EPI_ISL_1666505, EPI_ISL_1666507, EPI_ISL_1666508, EPI_ISL_1666509, EPI_ISL_1666510, EPI_ISL_1666511,
EPI_ISL_1666512, EPI_ISL_1666513, EPI_ISL_1666514, EPI_ISL_1666515, EPI_ISL_1666516, EPI_ISL_1666517, EPI_ISL_1666518, EPI_ISL_1666519, EPI_ISL_1666520, EPI_ISL_1666521, EPI_ISL_1666522, EPI_ISL_1666523, EPI_ISL_1666524, EPI_ISL_1666525, EPI_ISL_1666526, EPI_ISL_1666527, EPI_ISL_1666528, EPI_ISL_1666529, EPI_ISL_1666530, EPI_ISL_1666531, EPI_ISL_1666532, EPI_ISL_1666533,
EPI_ISL_1666534, EPI_ISL_1666535, EPI_ISL_1666536, EPI_ISL_1666537, EPI_ISL_1666538, EPI_ISL_1666539, EPI_ISL_1666540, EPI_ISL_1666541, EPI_ISL_1666542, EPI_ISL_1666655, EPI_ISL_1666658, EPI_ISL_1666702, EPI_ISL_1666724, EPI_ISL_1666934, EPI_ISL_1667018, EPI_ISL_1667077, EPI_ISL_1667078, EPI_ISL_1667083, EPI_ISL_1667084, EPI_ISL_1667085, EPI_ISL_1667086, EPI_ISL_1667087,
EPI_ISL_1667088, EPI_ISL_1667089, EPI_ISL_1667090, EPI_ISL_1667091, EPI_ISL_1667092, EPI_ISL_1667093, EPI_ISL_1667094, EPI_ISL_1667095, EPI_ISL_1667096, EPI_ISL_1667097, EPI_ISL_1667098, EPI_ISL_1667099, EPI_ISL_1667100, EPI_ISL_1667101, EPI_ISL_1667102, EPI_ISL_1667104, EPI_ISL_1667108, EPI_ISL_1667109, EPI_ISL_1667110, EPI_ISL_1667111, EPI_ISL_1667112, EPI_ISL_1667113,
EPI_ISL_1667114, EPI_ISL_1667115, EPI_ISL_1667116, EPI_ISL_1667117, EPI_ISL_1667118, EPI_ISL_1667119, EPI_ISL_1667120, EPI_ISL_1667121, EPI_ISL_1667122, EPI_ISL_1667123, EPI_ISL_1667124, EPI_ISL_1667125, EPI_ISL_1667126, EPI_ISL_1667127, EPI_ISL_1667128, EPI_ISL_1667129, EPI_ISL_1667130, EPI_ISL_1667131, EPI_ISL_1667132, EPI_ISL_1667133, EPI_ISL_1667134, EPI_ISL_1667135,
EPI_ISL_1667136, EPI_ISL_1667137, EPI_ISL_1667138, EPI_ISL_1667139, EPI_ISL_1667140, EPI_ISL_1667141, EPI_ISL_1667142, EPI_ISL_1667143, EPI_ISL_1667144, EPI_ISL_1667145, EPI_ISL_1667146, EPI_ISL_1667147, EPI_ISL_1667148, EPI_ISL_1667149, EPI_ISL_1667150, EPI_ISL_1667151, EPI_ISL_1667152, EPI_ISL_1667153, EPI_ISL_1667154, EPI_ISL_1667155, EPI_ISL_1667156, EPI_ISL_1667157,
EPI_ISL_1667158, EPI_ISL_1667159, EPI_ISL_1667160, EPI_ISL_1667161, EPI_ISL_1667162, EPI_ISL_1667163, EPI_ISL_1667164, EPI_ISL_1667165, EPI_ISL_1667166, EPI_ISL_1667167, EPI_ISL_1667168, EPI_ISL_1667169, EPI_ISL_1667170, EPI_ISL_1667171, EPI_ISL_1667172, EPI_ISL_1667174, EPI_ISL_1667175, EPI_ISL_1667176, EPI_ISL_1667177, EPI_ISL_1667178, EPI_ISL_1667179, EPI_ISL_1667180,
EPI_ISL_1667181, EPI_ISL_1667182, EPI_ISL_1667183, EPI_ISL_1667184, EPI_ISL_1667185, EPI_ISL_1667186, EPI_ISL_1667187, EPI_ISL_1667188, EPI_ISL_1667189, EPI_ISL_1667190, EPI_ISL_1667191, EPI_ISL_1667192, EPI_ISL_1667193, EPI_ISL_1667194, EPI_ISL_1667195, EPI_ISL_1667196, EPI_ISL_1667197, EPI_ISL_1667198, EPI_ISL_1667199, EPI_ISL_1667200, EPI_ISL_1667202, EPI_ISL_1667203,
EPI_ISL_1667204, EPI_ISL_1667205, EPI_ISL_1667206, EPI_ISL_1667207, EPI_ISL_1667208, EPI_ISL_1667209, EPI_ISL_1667210, EPI_ISL_1667211, EPI_ISL_1667212, EPI_ISL_1667213, EPI_ISL_1667214, EPI_ISL_1667215, EPI_ISL_1667216, EPI_ISL_1667217, EPI_ISL_1667218, EPI_ISL_1667219, EPI_ISL_1667220, EPI_ISL_1667221, EPI_ISL_1667222, EPI_ISL_1667223, EPI_ISL_1667224, EPI_ISL_1667225,
EPI_ISL_1667226, EPI_ISL_1667227, EPI_ISL_1667228, EPI_ISL_1667229, EPI_ISL_1667230, EPI_ISL_1667231, EPI_ISL_1667232, EPI_ISL_1667233, EPI_ISL_1667234, EPI_ISL_1667235, EPI_ISL_1667236, EPI_ISL_1667237, EPI_ISL_1667238, EPI_ISL_1667239, EPI_ISL_1667240, EPI_ISL_1667241, EPI_ISL_1667242, EPI_ISL_1667243, EPI_ISL_1667244, EPI_ISL_1667245, EPI_ISL_1667246, EPI_ISL_1667247,
EPI_ISL_1667248, EPI_ISL_1667249, EPI_ISL_1667250, EPI_ISL_1667251, EPI_ISL_1667252, EPI_ISL_1667253, EPI_ISL_1667254, EPI_ISL_1667255, EPI_ISL_1667256, EPI_ISL_1667257, EPI_ISL_1667258, EPI_ISL_1667259, EPI_ISL_1667260, EPI_ISL_1667261, EPI_ISL_1667262, EPI_ISL_1667263, EPI_ISL_1667264, EPI_ISL_1667265, EPI_ISL_1667266, EPI_ISL_1667267, EPI_ISL_1667268, EPI_ISL_1667269,
EPI_ISL_1667270, EPI_ISL_1667271, EPI_ISL_1667272, EPI_ISL_1667273, EPI_ISL_1667274, EPI_ISL_1667275, EPI_ISL_1667276, EPI_ISL_1667277, EPI_ISL_1667278, EPI_ISL_1667279, EPI_ISL_1667280, EPI_ISL_1667281, EPI_ISL_1667282, EPI_ISL_1667283, EPI_ISL_1667284, EPI_ISL_1667285, EPI_ISL_1667286, EPI_ISL_1667287, EPI_ISL_1667288, EPI_ISL_1667289, EPI_ISL_1667290, EPI_ISL_1667291,
EPI_ISL_1667292, EPI_ISL_1667293, EPI_ISL_1667294, EPI_ISL_1667295, EPI_ISL_1667296, EPI_ISL_1667297, EPI_ISL_1667298, EPI_ISL_1667299, EPI_ISL_1667300, EPI_ISL_1667302, EPI_ISL_1667303, EPI_ISL_1667304, EPI_ISL_1667305, EPI_ISL_1667306, EPI_ISL_1667307, EPI_ISL_1667308, EPI_ISL_1667309, EPI_ISL_1667310, EPI_ISL_1667311, EPI_ISL_1667312, EPI_ISL_1667313, EPI_ISL_1667314,
EPI_ISL_1667315, EPI_ISL_1667316, EPI_ISL_1667317, EPI_ISL_1667318, EPI_ISL_1667319, EPI_ISL_1667320, EPI_ISL_1667321, EPI_ISL_1667322, EPI_ISL_1667323, EPI_ISL_1667324, EPI_ISL_1667325, EPI_ISL_1667326, EPI_ISL_1667327, EPI_ISL_1667328, EPI_ISL_1667329, EPI_ISL_1667330, EPI_ISL_1667331, EPI_ISL_1667332, EPI_ISL_1667333, EPI_ISL_1667334, EPI_ISL_1667335, EPI_ISL_1667336,
EPI_ISL_1667337, EPI_ISL_1667338, EPI_ISL_1667339, EPI_ISL_1667340, EPI_ISL_1667341, EPI_ISL_1667342, EPI_ISL_1667343, EPI_ISL_1667344, EPI_ISL_1667345, EPI_ISL_1667346, EPI_ISL_1667347, EPI_ISL_1667348, EPI_ISL_1667349, EPI_ISL_1667350, EPI_ISL_1667351, EPI_ISL_1667352, EPI_ISL_1667353, EPI_ISL_1667354, EPI_ISL_1667355, EPI_ISL_1667356, EPI_ISL_1667357, EPI_ISL_1667358,
EPI_ISL_1667359, EPI_ISL_1667360, EPI_ISL_1667361, EPI_ISL_1667362, EPI_ISL_1667363, EPI_ISL_1667364, EPI_ISL_1667365, EPI_ISL_1667366, EPI_ISL_1667367, EPI_ISL_1667368, EPI_ISL_1667369, EPI_ISL_1667370, EPI_ISL_1667371, EPI_ISL_1667372, EPI_ISL_1667373, EPI_ISL_1667374, EPI_ISL_1667375, EPI_ISL_1667376, EPI_ISL_1667377, EPI_ISL_1667378, EPI_ISL_1667379, EPI_ISL_1667380,
EPI_ISL_1667381, EPI_ISL_1694974, EPI_ISL_1695033, EPI_ISL_1695036, EPI_ISL_1733806, EPI_ISL_1733838, EPI_ISL_1733839, EPI_ISL_1733840, EPI_ISL_1733841, EPI_ISL_1733842, EPI_ISL_1733843, EPI_ISL_1733844, EPI_ISL_1733845, EPI_ISL_1733846, EPI_ISL_1733847, EPI_ISL_1733848, EPI_ISL_1733849, EPI_ISL_1733850, EPI_ISL_1733851, EPI_ISL_1733852, EPI_ISL_1733853, EPI_ISL_1733854,
EPI_ISL_1733855, EPI_ISL_1733856, EPI_ISL_1733857, EPI_ISL_1733858, EPI_ISL_1733859, EPI_ISL_1733860, EPI_ISL_1733861, EPI_ISL_1733862, EPI_ISL_1733863, EPI_ISL_1733864, EPI_ISL_1733865, EPI_ISL_1733866, EPI_ISL_1733867, EPI_ISL_1733868, EPI_ISL_1733869, EPI_ISL_1733870, EPI_ISL_1733871, EPI_ISL_1733872, EPI_ISL_1733873, EPI_ISL_1733874, EPI_ISL_1733875, EPI_ISL_1733876,
EPI_ISL_1733877, EPI_ISL_1733878, EPI_ISL_1733879, EPI_ISL_1733880, EPI_ISL_1733881, EPI_ISL_1733882, EPI_ISL_1733883, EPI_ISL_1733884, EPI_ISL_1733885, EPI_ISL_1733886, EPI_ISL_1733887, EPI_ISL_1733888, EPI_ISL_1733889, EPI_ISL_1733890, EPI_ISL_1733891, EPI_ISL_1733892, EPI_ISL_1733893, EPI_ISL_1733894, EPI_ISL_1733895, EPI_ISL_1733896, EPI_ISL_1733897, EPI_ISL_1733898,
EPI_ISL_1733899, EPI_ISL_1733900, EPI_ISL_1733901, EPI_ISL_1733902, EPI_ISL_1733903, EPI_ISL_1733904, EPI_ISL_1733905, EPI_ISL_1733906, EPI_ISL_1733907, EPI_ISL_1733908, EPI_ISL_1733909, EPI_ISL_1733910, EPI_ISL_1733911, EPI_ISL_1733912, EPI_ISL_1733913, EPI_ISL_1733914, EPI_ISL_1733915, EPI_ISL_1733916, EPI_ISL_1733919, EPI_ISL_1733920, EPI_ISL_1733921, EPI_ISL_1733922,
EPI_ISL_1733923, EPI_ISL_1733924, EPI_ISL_1733925, EPI_ISL_1733926, EPI_ISL_1733927, EPI_ISL_1733928, EPI_ISL_1733929, EPI_ISL_1733931, EPI_ISL_1733932, EPI_ISL_1733933, EPI_ISL_1733934, EPI_ISL_1733935, EPI_ISL_1733936, EPI_ISL_1733937, EPI_ISL_1733938, EPI_ISL_1733939, EPI_ISL_1733940, EPI_ISL_1733941, EPI_ISL_1733942, EPI_ISL_1733943, EPI_ISL_1733944, EPI_ISL_1733945,
EPI_ISL_1733946, EPI_ISL_1733947, EPI_ISL_1733948, EPI_ISL_1733949, EPI_ISL_1733950, EPI_ISL_1733951, EPI_ISL_1733952, EPI_ISL_1733953, EPI_ISL_1733954, EPI_ISL_1733955, EPI_ISL_1733956, EPI_ISL_1733957, EPI_ISL_1733958, EPI_ISL_1733959, EPI_ISL_1733960, EPI_ISL_1733961, EPI_ISL_1733962, EPI_ISL_1733963, EPI_ISL_1733964, EPI_ISL_1733965, EPI_ISL_1733966, EPI_ISL_1733967,
EPI_ISL_1733968, EPI_ISL_1733969, EPI_ISL_1733970, EPI_ISL_1733971, EPI_ISL_1733972, EPI_ISL_1733973, EPI_ISL_1733974, EPI_ISL_1733975, EPI_ISL_1733976, EPI_ISL_1734413, EPI_ISL_1839210, EPI_ISL_1839211, EPI_ISL_1839212, EPI_ISL_1839215, EPI_ISL_1839216, EPI_ISL_1839217, EPI_ISL_1839218, EPI_ISL_1839219, EPI_ISL_1839220, EPI_ISL_1839221, EPI_ISL_1839222, EPI_ISL_1839223,
EPI_ISL_1839224, EPI_ISL_1839225, EPI_ISL_1839226, EPI_ISL_1839227, EPI_ISL_1839228, EPI_ISL_1839229, EPI_ISL_1839230, EPI_ISL_1839231, EPI_ISL_1839232, EPI_ISL_1839233, EPI_ISL_1839234, EPI_ISL_1839235, EPI_ISL_1839236, EPI_ISL_1839237, EPI_ISL_1839238, EPI_ISL_1839239, EPI_ISL_1839240, EPI_ISL_1839241, EPI_ISL_1839242, EPI_ISL_1839243, EPI_ISL_1839244, EPI_ISL_1839245,
EPI_ISL_1839246, EPI_ISL_1839247, EPI_ISL_1839248, EPI_ISL_1839249, EPI_ISL_1839250, EPI_ISL_1839251, EPI_ISL_1839253, EPI_ISL_1839254, EPI_ISL_1839255, EPI_ISL_1839256, EPI_ISL_1839257, EPI_ISL_1839258, EPI_ISL_1839259, EPI_ISL_1839260, EPI_ISL_1839261, EPI_ISL_1839262, EPI_ISL_1839263, EPI_ISL_1839264, EPI_ISL_1839265, EPI_ISL_1839266, EPI_ISL_1839267, EPI_ISL_1839268,
EPI_ISL_1839269, EPI_ISL_1839270, EPI_ISL_1839271, EPI_ISL_1839272, EPI_ISL_1839273, EPI_ISL_1839274, EPI_ISL_1839275, EPI_ISL_1839276, EPI_ISL_1839277, EPI_ISL_1839402, EPI_ISL_1839546, EPI_ISL_1839547, EPI_ISL_1839548, EPI_ISL_1839549, EPI_ISL_1839550, EPI_ISL_1839551, EPI_ISL_1839552, EPI_ISL_1839553, EPI_ISL_1839554, EPI_ISL_1839555, EPI_ISL_1839556, EPI_ISL_1839557,
EPI_ISL_1839558, EPI_ISL_1839559, EPI_ISL_1839560, EPI_ISL_1839561, EPI_ISL_1839562, EPI_ISL_1839563, EPI_ISL_1839564, EPI_ISL_1839565, EPI_ISL_1839566, EPI_ISL_1839567, EPI_ISL_1839568, EPI_ISL_1839569, EPI_ISL_1839570, EPI_ISL_1839571, EPI_ISL_1839572, EPI_ISL_1839573, EPI_ISL_1839574, EPI_ISL_1839576, EPI_ISL_1839577, EPI_ISL_1839578, EPI_ISL_1839579, EPI_ISL_1839580,
EPI_ISL_1839581, EPI_ISL_1839582, EPI_ISL_1839584, EPI_ISL_1839585, EPI_ISL_1839586, EPI_ISL_1839587, EPI_ISL_1839588, EPI_ISL_1839589, EPI_ISL_1839590, EPI_ISL_1839591, EPI_ISL_1839592, EPI_ISL_1839593, EPI_ISL_1839594, EPI_ISL_1839595, EPI_ISL_1839596, EPI_ISL_1839597, EPI_ISL_1839598, EPI_ISL_1839599, EPI_ISL_1839600, EPI_ISL_1839601, EPI_ISL_1839602, EPI_ISL_1839603,
EPI_ISL_1839604, EPI_ISL_1839605, EPI_ISL_1839606, EPI_ISL_1839607, EPI_ISL_1839608, EPI_ISL_1839609, EPI_ISL_1839610, EPI_ISL_1839611, EPI_ISL_1839612, EPI_ISL_1839613, EPI_ISL_1839614, EPI_ISL_1839615, EPI_ISL_1839843, EPI_ISL_1839847, EPI_ISL_1839848, EPI_ISL_1839849, EPI_ISL_1839850, EPI_ISL_1839851, EPI_ISL_1839852, EPI_ISL_1839853, EPI_ISL_1839854, EPI_ISL_1839855,
EPI_ISL_1839856, EPI_ISL_1839857, EPI_ISL_1839858, EPI_ISL_1839859, EPI_ISL_1839860, EPI_ISL_1839861, EPI_ISL_1839862, EPI_ISL_1839863, EPI_ISL_1839864, EPI_ISL_1839865, EPI_ISL_1839866, EPI_ISL_1839867, EPI_ISL_1839868, EPI_ISL_1839869, EPI_ISL_1839870, EPI_ISL_1839871, EPI_ISL_1839872, EPI_ISL_1839873, EPI_ISL_1839874, EPI_ISL_1839875, EPI_ISL_1839876, EPI_ISL_1839877,
EPI_ISL_1839878, EPI_ISL_1839879, EPI_ISL_1839880, EPI_ISL_1839881, EPI_ISL_1839882, EPI_ISL_1839884, EPI_ISL_1839885, EPI_ISL_1839886, EPI_ISL_1839888, EPI_ISL_1839889, EPI_ISL_1839890, EPI_ISL_1839891, EPI_ISL_1839892, EPI_ISL_1839893, EPI_ISL_1839894, EPI_ISL_1839895, EPI_ISL_1839896, EPI_ISL_1839897, EPI_ISL_1839898, EPI_ISL_1839899, EPI_ISL_1839900, EPI_ISL_1839942,
EPI_ISL_1839943, EPI_ISL_1839944, EPI_ISL_1839945, EPI_ISL_1839946, EPI_ISL_1839947, EPI_ISL_1839948, EPI_ISL_1839949, EPI_ISL_1839950, EPI_ISL_1839951, EPI_ISL_1839952, EPI_ISL_1839953, EPI_ISL_1839954, EPI_ISL_1839955, EPI_ISL_1839956, EPI_ISL_1839957, EPI_ISL_1839958, EPI_ISL_1839959, EPI_ISL_1839960, EPI_ISL_1839961, EPI_ISL_1839962, EPI_ISL_1839964, EPI_ISL_1839965,
EPI_ISL_1839966, EPI_ISL_1839967, EPI_ISL_1839968, EPI_ISL_1839969, EPI_ISL_1839970, EPI_ISL_1839972, EPI_ISL_1839973, EPI_ISL_1839974, EPI_ISL_1839975, EPI_ISL_1839976, EPI_ISL_1839977, EPI_ISL_1839978, EPI_ISL_1839979, EPI_ISL_1839980, EPI_ISL_1839981, EPI_ISL_1839982, EPI_ISL_1839983, EPI_ISL_1839984, EPI_ISL_1839985, EPI_ISL_1839986, EPI_ISL_1839987, EPI_ISL_1839988,
EPI_ISL_1839989, EPI_ISL_1839990, EPI_ISL_1839991, EPI_ISL_1839992, EPI_ISL_1839993, EPI_ISL_1839994, EPI_ISL_1839995, EPI_ISL_1839996, EPI_ISL_1839997, EPI_ISL_1839998, EPI_ISL_1839999, EPI_ISL_1840000, EPI_ISL_1840001, EPI_ISL_1840002, EPI_ISL_1840003, EPI_ISL_1840005, EPI_ISL_1840006, EPI_ISL_1840007, EPI_ISL_1840008, EPI_ISL_1840009, EPI_ISL_1840010, EPI_ISL_1840011,
EPI_ISL_1840012, EPI_ISL_1840013, EPI_ISL_1840014, EPI_ISL_1840015, EPI_ISL_1840016, EPI_ISL_1840017, EPI_ISL_1840018, EPI_ISL_1840019, EPI_ISL_1840020, EPI_ISL_1840021, EPI_ISL_1840022, EPI_ISL_1840023, EPI_ISL_1840024, EPI_ISL_1840025, EPI_ISL_1840026, EPI_ISL_1840027, EPI_ISL_1840028, EPI_ISL_1840029, EPI_ISL_1840030, EPI_ISL_1840031, EPI_ISL_1840032, EPI_ISL_1840033,
EPI_ISL_1840034, EPI_ISL_1840035, EPI_ISL_1840036, EPI_ISL_1840037, EPI_ISL_1840038, EPI_ISL_1840039, EPI_ISL_1840040, EPI_ISL_1840041, EPI_ISL_1840042, EPI_ISL_1840043, EPI_ISL_1840044, EPI_ISL_1840045, EPI_ISL_1840046, EPI_ISL_1840047, EPI_ISL_1840048, EPI_ISL_1840049, EPI_ISL_1840050, EPI_ISL_1840051, EPI_ISL_1840052, EPI_ISL_1840053, EPI_ISL_1840054, EPI_ISL_1840055,
EPI_ISL_1840056, EPI_ISL_1840057, EPI_ISL_1840058, EPI_ISL_1840059, EPI_ISL_1840060, EPI_ISL_1840061, EPI_ISL_1840062, EPI_ISL_1840063, EPI_ISL_1840064, EPI_ISL_1840065, EPI_ISL_1840066, EPI_ISL_1840067, EPI_ISL_1840068, EPI_ISL_1840069, EPI_ISL_1840070, EPI_ISL_1840071, EPI_ISL_1840072, EPI_ISL_1840073, EPI_ISL_1840074, EPI_ISL_1840075, EPI_ISL_1840076, EPI_ISL_1840077,
EPI_ISL_1840078, EPI_ISL_1840079, EPI_ISL_1840080, EPI_ISL_1840081, EPI_ISL_1840082, EPI_ISL_1840083, EPI_ISL_1840084, EPI_ISL_1840085, EPI_ISL_1840086, EPI_ISL_1840087, EPI_ISL_1840088, EPI_ISL_1840089, EPI_ISL_1840091, EPI_ISL_1840092, EPI_ISL_1840093, EPI_ISL_1840094, EPI_ISL_1840095, EPI_ISL_1840096, EPI_ISL_1840097, EPI_ISL_1840098, EPI_ISL_1840099, EPI_ISL_1840100,
EPI_ISL_1840101, EPI_ISL_1840102, EPI_ISL_1840103, EPI_ISL_1840104, EPI_ISL_1840105, EPI_ISL_1840106, EPI_ISL_1840107, EPI_ISL_1840108, EPI_ISL_1840109, EPI_ISL_1840110, EPI_ISL_1840111, EPI_ISL_1840112, EPI_ISL_1840113, EPI_ISL_1840114, EPI_ISL_1840115, EPI_ISL_1840116, EPI_ISL_1840117, EPI_ISL_1840118, EPI_ISL_1840119, EPI_ISL_1840120, EPI_ISL_1840121, EPI_ISL_1840122,
EPI_ISL_1840123, EPI_ISL_1840124, EPI_ISL_1840125, EPI_ISL_1992018, EPI_ISL_1992019, EPI_ISL_1992022, EPI_ISL_1992023, EPI_ISL_1992024, EPI_ISL_1992025, EPI_ISL_1992026, EPI_ISL_1992027, EPI_ISL_1992028, EPI_ISL_1992029, EPI_ISL_1992030, EPI_ISL_1992031, EPI_ISL_1992032, EPI_ISL_1992033, EPI_ISL_1992034, EPI_ISL_1992035, EPI_ISL_1992036, EPI_ISL_1992037, EPI_ISL_1992038,
EPI_ISL_1992039, EPI_ISL_1992040, EPI_ISL_1992041, EPI_ISL_1992042, EPI_ISL_1992043, EPI_ISL_1992044, EPI_ISL_1992045, EPI_ISL_1992046, EPI_ISL_1992047, EPI_ISL_1992048, EPI_ISL_1992049, EPI_ISL_1992050, EPI_ISL_1992051, EPI_ISL_1992052, EPI_ISL_1992053, EPI_ISL_1992054, EPI_ISL_1992055, EPI_ISL_1992056, EPI_ISL_1992057, EPI_ISL_1992058, EPI_ISL_1992059, EPI_ISL_1992060,
EPI_ISL_1992061, EPI_ISL_1992062, EPI_ISL_1992063, EPI_ISL_1992064, EPI_ISL_1992065, EPI_ISL_1992066, EPI_ISL_1992067, EPI_ISL_1992068, EPI_ISL_1992069, EPI_ISL_1992070, EPI_ISL_1992071, EPI_ISL_1992072, EPI_ISL_1992073, EPI_ISL_1992074, EPI_ISL_1992075, EPI_ISL_1992076, EPI_ISL_1992077, EPI_ISL_1992078, EPI_ISL_1992079, EPI_ISL_1992080, EPI_ISL_1992081, EPI_ISL_1992082,
EPI_ISL_1992083, EPI_ISL_1992084, EPI_ISL_1992085, EPI_ISL_1992086, EPI_ISL_1992087, EPI_ISL_1992088, EPI_ISL_1992092, EPI_ISL_1992093, EPI_ISL_1992096, EPI_ISL_1992097, EPI_ISL_1992098, EPI_ISL_1992099, EPI_ISL_1992101, EPI_ISL_1992102, EPI_ISL_1992103, EPI_ISL_1992104, EPI_ISL_1992105, EPI_ISL_1992106, EPI_ISL_1992107, EPI_ISL_1992108, EPI_ISL_1992109, EPI_ISL_1992110,
EPI_ISL_1992111, EPI_ISL_1992112, EPI_ISL_1992113, EPI_ISL_1992114, EPI_ISL_1992115, EPI_ISL_1992116, EPI_ISL_1992118, EPI_ISL_1992119, EPI_ISL_1992120, EPI_ISL_1992121, EPI_ISL_1992122, EPI_ISL_1992123, EPI_ISL_1992124, EPI_ISL_1992125, EPI_ISL_1992127, EPI_ISL_1992128, EPI_ISL_1992129, EPI_ISL_1992130, EPI_ISL_1992131, EPI_ISL_1992132, EPI_ISL_1992133, EPI_ISL_1992134,
EPI_ISL_1992135, EPI_ISL_1992136, EPI_ISL_1992137, EPI_ISL_1992138, EPI_ISL_1992139, EPI_ISL_1992140, EPI_ISL_1992141, EPI_ISL_1992142, EPI_ISL_1992143, EPI_ISL_1992144, EPI_ISL_1992145, EPI_ISL_1992146, EPI_ISL_1992147, EPI_ISL_1992148, EPI_ISL_1992149, EPI_ISL_1992150, EPI_ISL_1992151, EPI_ISL_1992152, EPI_ISL_1992153, EPI_ISL_1992154, EPI_ISL_1992265, EPI_ISL_1992266,
EPI_ISL_1992267, EPI_ISL_1992268, EPI_ISL_1992269, EPI_ISL_1992270, EPI_ISL_1992271, EPI_ISL_1992272, EPI_ISL_1992273, EPI_ISL_1992274, EPI_ISL_1992275, EPI_ISL_1992276, EPI_ISL_1992277, EPI_ISL_1992278, EPI_ISL_1992279, EPI_ISL_1992280, EPI_ISL_1992281, EPI_ISL_1992282, EPI_ISL_1992283, EPI_ISL_1992284, EPI_ISL_1992285, EPI_ISL_1992286, EPI_ISL_1992287, EPI_ISL_1992288,
EPI_ISL_1992289, EPI_ISL_1992290, EPI_ISL_1992291, EPI_ISL_1992292, EPI_ISL_1992293, EPI_ISL_1992294, EPI_ISL_1992295, EPI_ISL_1992296, EPI_ISL_1992297, EPI_ISL_1992298, EPI_ISL_1992299, EPI_ISL_1992300, EPI_ISL_1992301, EPI_ISL_1992302, EPI_ISL_1992303, EPI_ISL_1992304, EPI_ISL_1992305, EPI_ISL_1992306, EPI_ISL_1992307, EPI_ISL_1992308, EPI_ISL_1992309, EPI_ISL_1992310,
EPI_ISL_1992311, EPI_ISL_1992312, EPI_ISL_1992313, EPI_ISL_1992314, EPI_ISL_1992315, EPI_ISL_1992316, EPI_ISL_1992317, EPI_ISL_1992318, EPI_ISL_1992319, EPI_ISL_1992320, EPI_ISL_1992321, EPI_ISL_1992322, EPI_ISL_1992323, EPI_ISL_1992324, EPI_ISL_1992329, EPI_ISL_1992330, EPI_ISL_1992331, EPI_ISL_1992332, EPI_ISL_1992333, EPI_ISL_1992334, EPI_ISL_1992335, EPI_ISL_1992336,
EPI_ISL_1992337, EPI_ISL_1992338, EPI_ISL_1992339, EPI_ISL_1992340, EPI_ISL_1992341, EPI_ISL_1992342, EPI_ISL_1992343, EPI_ISL_1992344, EPI_ISL_1992345, EPI_ISL_1992346, EPI_ISL_1992347, EPI_ISL_1992348, EPI_ISL_1992349, EPI_ISL_1992350, EPI_ISL_1992351, EPI_ISL_1992352, EPI_ISL_1992354, EPI_ISL_1992355, EPI_ISL_1992356, EPI_ISL_1992357, EPI_ISL_1992358,
EPI_ISL_1992359, EPI_ISL_1992360, EPI_ISL_1992361, EPI_ISL_1992362, EPI_ISL_1992363, EPI_ISL_1992364, EPI_ISL_1992365, EPI_ISL_1992366, EPI_ISL_1992367, EPI_ISL_1992368, EPI_ISL_1992369, EPI_ISL_1992370, EPI_ISL_1992371, EPI_ISL_1992372, EPI_ISL_1992373, EPI_ISL_1992374, EPI_ISL_1992375, EPI_ISL_1992376, EPI_ISL_1992377, EPI_ISL_1992378, EPI_ISL_1992379, EPI_ISL_1992380,
EPI_ISL_1992381, EPI_ISL_1992382, EPI_ISL_1992383, EPI_ISL_1992384, EPI_ISL_1992385, EPI_ISL_1992386, EPI_ISL_1992387, EPI_ISL_1992388, EPI_ISL_1992389, EPI_ISL_1992390, EPI_ISL_1992391, EPI_ISL_1992392, EPI_ISL_1992393, EPI_ISL_1992407, EPI_ISL_1992665, EPI_ISL_1992777, EPI_ISL_1992980, EPI_ISL_1992984, EPI_ISL_1992985, EPI_ISL_1992986, EPI_ISL_1992987, EPI_ISL_1992988,
EPI_ISL_1992989, EPI_ISL_1992990, EPI_ISL_1992991, EPI_ISL_1992992, EPI_ISL_1992993, EPI_ISL_1992994, EPI_ISL_1992995, EPI_ISL_1992996, EPI_ISL_1992997, EPI_ISL_1992998, EPI_ISL_1992999, EPI_ISL_1993000, EPI_ISL_1993001, EPI_ISL_1993002, EPI_ISL_1993003, EPI_ISL_1993004, EPI_ISL_1993005, EPI_ISL_1993006, EPI_ISL_1993007, EPI_ISL_1993008, EPI_ISL_1993009, EPI_ISL_1993010,
EPI_ISL_1993011, EPI_ISL_1993012, EPI_ISL_1993013, EPI_ISL_1993014, EPI_ISL_1993015, EPI_ISL_1993016, EPI_ISL_1993017, EPI_ISL_1993018, EPI_ISL_1993019, EPI_ISL_1993020, EPI_ISL_1993021, EPI_ISL_1993022, EPI_ISL_1993023, EPI_ISL_1993024, EPI_ISL_1993025, EPI_ISL_1993026, EPI_ISL_1993027, EPI_ISL_1993028, EPI_ISL_1993029, EPI_ISL_1993030, EPI_ISL_1993031, EPI_ISL_1993032,
EPI_ISL_1993033, EPI_ISL_1993034, EPI_ISL_1993035, EPI_ISL_1993036, EPI_ISL_1993037, EPI_ISL_1993040, EPI_ISL_1993042, EPI_ISL_1993043, EPI_ISL_1993044, EPI_ISL_1993045, EPI_ISL_1993046, EPI_ISL_1993047, EPI_ISL_1993048, EPI_ISL_1993049, EPI_ISL_1993050, EPI_ISL_1993051, EPI_ISL_1993052, EPI_ISL_1993053, EPI_ISL_1993054, EPI_ISL_1993055, EPI_ISL_1993056, EPI_ISL_1993057,
EPI_ISL_1993058, EPI_ISL_1993059, EPI_ISL_1993060, EPI_ISL_1993061, EPI_ISL_1993062, EPI_ISL_1993063, EPI_ISL_1993064, EPI_ISL_1993065, EPI_ISL_1993066, EPI_ISL_1993067, EPI_ISL_1993068, EPI_ISL_1993069, EPI_ISL_1993070, EPI_ISL_1993071, EPI_ISL_1993072, EPI_ISL_1993073, EPI_ISL_1993074, EPI_ISL_1993075, EPI_ISL_1993076, EPI_ISL_1993077, EPI_ISL_1993078, EPI_ISL_1993079,
EPI_ISL_1993080, EPI_ISL_1993081, EPI_ISL_1993082, EPI_ISL_1993083, EPI_ISL_1993084, EPI_ISL_1993085, EPI_ISL_1993086, EPI_ISL_1993087, EPI_ISL_1993088, EPI_ISL_1993089, EPI_ISL_1993090, EPI_ISL_1993091, EPI_ISL_1993092, EPI_ISL_1993093, EPI_ISL_1993094, EPI_ISL_1993097, EPI_ISL_1993103, EPI_ISL_1993104, EPI_ISL_1993105, EPI_ISL_1993111, EPI_ISL_1993112, EPI_ISL_1993113,
EPI_ISL_1993114, EPI_ISL_1993115, EPI_ISL_1993118, EPI_ISL_1993119, EPI_ISL_1993121, EPI_ISL_1993124, EPI_ISL_1993125, EPI_ISL_1993126, EPI_ISL_1993127, EPI_ISL_1993128, EPI_ISL_1993129, EPI_ISL_1993130, EPI_ISL_1993132, EPI_ISL_1993135, EPI_ISL_1993136, EPI_ISL_1993141, EPI_ISL_1993142, EPI_ISL_1993145, EPI_ISL_1993146, EPI_ISL_1993148, EPI_ISL_1993149, EPI_ISL_1993150,
EPI_ISL_1993151, EPI_ISL_1993152, EPI_ISL_1993153, EPI_ISL_1993210, EPI_ISL_1993287, EPI_ISL_2034996, EPI_ISL_2035000, EPI_ISL_2037981, EPI_ISL_2038260, EPI_ISL_2038261, EPI_ISL_2038262, EPI_ISL_2038263, EPI_ISL_2038266, EPI_ISL_2038267, EPI_ISL_2038268, EPI_ISL_2038269, EPI_ISL_2038270, EPI_ISL_2038271, EPI_ISL_2038272, EPI_ISL_2038273, EPI_ISL_2038274, EPI_ISL_2038275,
EPI_ISL_2038276, EPI_ISL_2038277, EPI_ISL_2038278, EPI_ISL_2038279, EPI_ISL_2038280, EPI_ISL_2038281, EPI_ISL_2038282, EPI_ISL_2038283, EPI_ISL_2038284, EPI_ISL_2038285, EPI_ISL_2038286, EPI_ISL_2038287, EPI_ISL_2038288, EPI_ISL_2038289, EPI_ISL_2038290, EPI_ISL_2038291, EPI_ISL_2038292, EPI_ISL_2038293, EPI_ISL_2038294, EPI_ISL_2038295, EPI_ISL_2038296, EPI_ISL_2038297,
EPI_ISL_2038298, EPI_ISL_2038299, EPI_ISL_2038300, EPI_ISL_2038301, EPI_ISL_2038303, EPI_ISL_2038304, EPI_ISL_2038305, EPI_ISL_2038306, EPI_ISL_2038307, EPI_ISL_2038309, EPI_ISL_2038310, EPI_ISL_2038311, EPI_ISL_2038312, EPI_ISL_2038313, EPI_ISL_2038314, EPI_ISL_2038315, EPI_ISL_2038316, EPI_ISL_2038317, EPI_ISL_2038318, EPI_ISL_2038319, EPI_ISL_2038320, EPI_ISL_2038321,
EPI_ISL_2038322, EPI_ISL_2038323, EPI_ISL_2038324, EPI_ISL_2038325, EPI_ISL_2038326, EPI_ISL_2038327, EPI_ISL_2038328, EPI_ISL_2038329, EPI_ISL_2038459, EPI_ISL_2038460, EPI_ISL_2038461, EPI_ISL_2038462, EPI_ISL_2038463, EPI_ISL_2038464, EPI_ISL_2038465, EPI_ISL_2038466, EPI_ISL_2038467, EPI_ISL_2038468, EPI_ISL_2038469, EPI_ISL_2038470, EPI_ISL_2038471, EPI_ISL_2038472,
EPI_ISL_2038473, EPI_ISL_2038474, EPI_ISL_2038475, EPI_ISL_2038476, EPI_ISL_2038477, EPI_ISL_2038478, EPI_ISL_2038479, EPI_ISL_2038480, EPI_ISL_2038481, EPI_ISL_2038482, EPI_ISL_2038483, EPI_ISL_2038484, EPI_ISL_2038485, EPI_ISL_2038486, EPI_ISL_2038487, EPI_ISL_2038488, EPI_ISL_2038489, EPI_ISL_2038490, EPI_ISL_2038491, EPI_ISL_2038492, EPI_ISL_2038493, EPI_ISL_2038494,
EPI_ISL_2038495, EPI_ISL_2038496, EPI_ISL_2038497, EPI_ISL_2038498, EPI_ISL_2038499, EPI_ISL_2038500, EPI_ISL_2038501, EPI_ISL_2038502, EPI_ISL_2038503, EPI_ISL_2038504, EPI_ISL_2038505, EPI_ISL_2038506, EPI_ISL_2038507, EPI_ISL_2038508, EPI_ISL_2038509, EPI_ISL_2038510, EPI_ISL_2038511, EPI_ISL_2038512, EPI_ISL_2038513, EPI_ISL_2038514, EPI_ISL_2038515, EPI_ISL_2038516,
EPI_ISL_2038517, EPI_ISL_2038518, EPI_ISL_2038519, EPI_ISL_2038520, EPI_ISL_2038521, EPI_ISL_2038522, EPI_ISL_2038524, EPI_ISL_2038525, EPI_ISL_2038526, EPI_ISL_2038527, EPI_ISL_2038528, EPI_ISL_2038529, EPI_ISL_2038530, EPI_ISL_2038531, EPI_ISL_2038532, EPI_ISL_2038533, EPI_ISL_2038534, EPI_ISL_2038535, EPI_ISL_2038536, EPI_ISL_2038537, EPI_ISL_2038538, EPI_ISL_2038539,
EPI_ISL_2038540, EPI_ISL_2038541, EPI_ISL_2038542, EPI_ISL_2038543, EPI_ISL_2038544, EPI_ISL_2038545, EPI_ISL_2038546, EPI_ISL_2038547, EPI_ISL_2038548, EPI_ISL_2038549, EPI_ISL_2038550, EPI_ISL_2038551, EPI_ISL_2038552, EPI_ISL_2038553, EPI_ISL_2038554, EPI_ISL_2038555, EPI_ISL_2038556, EPI_ISL_2038557, EPI_ISL_2038558, EPI_ISL_2038559, EPI_ISL_2038560, EPI_ISL_2038561,
EPI_ISL_2038562, EPI_ISL_2038563, EPI_ISL_2038565, EPI_ISL_2038566, EPI_ISL_2038567, EPI_ISL_2038568, EPI_ISL_2038569, EPI_ISL_2038570, EPI_ISL_2038571, EPI_ISL_2038572, EPI_ISL_2038573, EPI_ISL_2038574, EPI_ISL_2038575, EPI_ISL_2038576, EPI_ISL_2038577, EPI_ISL_2038578, EPI_ISL_2038579, EPI_ISL_2038580, EPI_ISL_2038581, EPI_ISL_2038582, EPI_ISL_2038583, EPI_ISL_2038584,
EPI_ISL_2038585, EPI_ISL_2038586, EPI_ISL_2038587, EPI_ISL_2038588, EPI_ISL_2038591, EPI_ISL_2038592, EPI_ISL_2038593, EPI_ISL_2038594, EPI_ISL_2038595, EPI_ISL_2038596, EPI_ISL_2038597, EPI_ISL_2038598, EPI_ISL_2038599, EPI_ISL_2038600, EPI_ISL_2038601, EPI_ISL_2038602, EPI_ISL_2038603, EPI_ISL_2038604, EPI_ISL_2038605, EPI_ISL_2038606, EPI_ISL_2038607, EPI_ISL_2038608,
EPI_ISL_2038609, EPI_ISL_2038610, EPI_ISL_2038611, EPI_ISL_2038612, EPI_ISL_2038613, EPI_ISL_2038614, EPI_ISL_2038615, EPI_ISL_2038616, EPI_ISL_2038617, EPI_ISL_2038618, EPI_ISL_2038619, EPI_ISL_2038620, EPI_ISL_2038621, EPI_ISL_2038622, EPI_ISL_2038624, EPI_ISL_2038625, EPI_ISL_2038626, EPI_ISL_2038627, EPI_ISL_2038628, EPI_ISL_2038629, EPI_ISL_2038630, EPI_ISL_2038631,
EPI_ISL_2038632, EPI_ISL_2038633, EPI_ISL_2038634, EPI_ISL_2038635, EPI_ISL_2038636, EPI_ISL_2038637, EPI_ISL_2038638, EPI_ISL_2038639, EPI_ISL_2038640, EPI_ISL_2038641, EPI_ISL_2038642, EPI_ISL_2038643, EPI_ISL_2038644, EPI_ISL_2038645, EPI_ISL_2038646, EPI_ISL_2038647, EPI_ISL_2038648, EPI_ISL_2038649, EPI_ISL_2038650, EPI_ISL_2038651, EPI_ISL_2038652, EPI_ISL_2038653,
EPI_ISL_2038654, EPI_ISL_2038655, EPI_ISL_2038656, EPI_ISL_2038657, EPI_ISL_2038659, EPI_ISL_2038660, EPI_ISL_2038661, EPI_ISL_2038662, EPI_ISL_2038663, EPI_ISL_2038664, EPI_ISL_2038665, EPI_ISL_2038666, EPI_ISL_2038667, EPI_ISL_2038668, EPI_ISL_2038669, EPI_ISL_2038670, EPI_ISL_2038671, EPI_ISL_2038672, EPI_ISL_2038673, EPI_ISL_2038674, EPI_ISL_2038675, EPI_ISL_2038676,
EPI_ISL_2038677, EPI_ISL_2038678, EPI_ISL_2038679, EPI_ISL_2038680, EPI_ISL_2038681, EPI_ISL_2038682, EPI_ISL_2038683, EPI_ISL_2038684, EPI_ISL_2038685, EPI_ISL_2038686, EPI_ISL_2038687, EPI_ISL_2038688, EPI_ISL_2038689, EPI_ISL_2038690, EPI_ISL_2038691, EPI_ISL_2038692, EPI_ISL_2038693, EPI_ISL_2038694, EPI_ISL_2038695, EPI_ISL_2038696, EPI_ISL_2038697, EPI_ISL_2038698,
EPI_ISL_2038699, EPI_ISL_2038700, EPI_ISL_2038701, EPI_ISL_2038702, EPI_ISL_2038703, EPI_ISL_2038704, EPI_ISL_2038705, EPI_ISL_2038706, EPI_ISL_2038707, EPI_ISL_2038708, EPI_ISL_2038709, EPI_ISL_2038710, EPI_ISL_2038711, EPI_ISL_2038712, EPI_ISL_2038713, EPI_ISL_2038714, EPI_ISL_2038715, EPI_ISL_2038716, EPI_ISL_2038717, EPI_ISL_2038718, EPI_ISL_2038719, EPI_ISL_2038720,
EPI_ISL_2038721, EPI_ISL_2038722, EPI_ISL_2038723, EPI_ISL_2038724, EPI_ISL_2038725, EPI_ISL_2038726, EPI_ISL_2038727, EPI_ISL_2038728, EPI_ISL_2038729, EPI_ISL_2038730, EPI_ISL_2038731, EPI_ISL_2038732, EPI_ISL_2038733, EPI_ISL_2038734, EPI_ISL_2038735, EPI_ISL_2038736, EPI_ISL_2038737, EPI_ISL_2038738, EPI_ISL_2038739, EPI_ISL_2038740, EPI_ISL_2038741, EPI_ISL_2038742,
EPI_ISL_2038743, EPI_ISL_2038744, EPI_ISL_2038745, EPI_ISL_2038746, EPI_ISL_2038747, EPI_ISL_2038748, EPI_ISL_2038749, EPI_ISL_2038750, EPI_ISL_2038752, EPI_ISL_2038753, EPI_ISL_2038754, EPI_ISL_2038755, EPI_ISL_2038756, EPI_ISL_2038757, EPI_ISL_2038758, EPI_ISL_2038759, EPI_ISL_2038760, EPI_ISL_2038761, EPI_ISL_2038762, EPI_ISL_2038763, EPI_ISL_2038764, EPI_ISL_2038765,
EPI_ISL_2038766, EPI_ISL_2038767, EPI_ISL_2038768, EPI_ISL_2038769, EPI_ISL_2038770, EPI_ISL_2038771, EPI_ISL_2038772, EPI_ISL_2038773, EPI_ISL_2038774, EPI_ISL_2038775, EPI_ISL_2038776, EPI_ISL_2038777, EPI_ISL_2038778, EPI_ISL_2038779, EPI_ISL_2038780, EPI_ISL_2038781, EPI_ISL_2038782, EPI_ISL_2038783, EPI_ISL_2038784, EPI_ISL_2038785, EPI_ISL_2038786, EPI_ISL_2038787,
EPI_ISL_2038788, EPI_ISL_2038789, EPI_ISL_2038790, EPI_ISL_2038791, EPI_ISL_2038792, EPI_ISL_2038793, EPI_ISL_2038794, EPI_ISL_2038795, EPI_ISL_2038796, EPI_ISL_2038797, EPI_ISL_2038798, EPI_ISL_2043633

| | | | |
|---|---|---|---|
| see above | Fulgent Genetics | Centers for Disease Control and Prevention Division of Viral Diseases, Pathogen Discovery | Adrian Paskey; Becky Tsai; Benafsh Sapra; Benjamin Rambo-Martin; Christopher Gulvick; Clinton R. Paden; Dakota Howard; Darlene Wagner; Dhwani Batra; Doreen Ng; Duncan MacCannell; Harry Gao; James Xie; Jason Caravas; John Gao; Joseph Fierro; Kara Moser; Matthew Schmerer; Mickey Li; Peter W. Cook; Scott Sammons; Shatavia Morrison; Yan Meng; Yvette Unoarumhi |
| EPI_ISL_1707680, EPI_ISL_1707681, EPI_ISL_1707682, EPI_ISL_1707683, EPI_ISL_1707684, EPI_ISL_1707685 | Fulgent Genetics | Los Angeles County PHL | P. Hemarajata et al. |
| EPI_ISL_1664267, EPI_ISL_1824116 | Good Samaritan Hospital | Santa Clara County public Health Laboratory | Santa Clara County Public Health Department |

EPI_ISL_1575872, EPI_ISL_1576607, EPI_ISL_1576614, EPI_ISL_1576619, EPI_ISL_1576629, EPI_ISL_1576636, EPI_ISL_1576641, EPI_ISL_1576657, EPI_ISL_1576669, EPI_ISL_1576678, EPI_ISL_1576681, EPI_ISL_1576683, EPI_ISL_1576689, EPI_ISL_1576692, EPI_ISL_1576701, EPI_ISL_1576704, EPI_ISL_1576713, EPI_ISL_1576721, EPI_ISL_1576723, EPI_ISL_1576734, EPI_ISL_1576735, EPI_ISL_1576744,

| | | | |
|---|---|---|---|
| EPI_ISL_1576746, EPI_ISL_1576747, EPI_ISL_1576754, EPI_ISL_1576756, EPI_ISL_1576757, EPI_ISL_1580871, EPI_ISL_1580873, EPI_ISL_1580880, EPI_ISL_1580885, EPI_ISL_1580887, EPI_ISL_1580895, EPI_ISL_1580904, EPI_ISL_1580907, EPI_ISL_1580908, EPI_ISL_1580919, EPI_ISL_1580925, EPI_ISL_1580935, EPI_ISL_1580949, EPI_ISL_1581240, EPI_ISL_1581321, EPI_ISL_1581334, EPI_ISL_1581357, EPI_ISL_1581366, EPI_ISL_1581368, EPI_ISL_1581369, EPI_ISL_1581378, EPI_ISL_1581379, EPI_ISL_1581402, EPI_ISL_1581561, EPI_ISL_1581817, EPI_ISL_1592405, EPI_ISL_1592408, EPI_ISL_1592409, EPI_ISL_1592413, EPI_ISL_1592415, EPI_ISL_1592422, EPI_ISL_1592423, EPI_ISL_1592424, EPI_ISL_1592432, EPI_ISL_1592435, EPI_ISL_1592436, EPI_ISL_1592440, EPI_ISL_1592441, EPI_ISL_1592448, EPI_ISL_1592461, EPI_ISL_1592463, EPI_ISL_1592471, EPI_ISL_1592477, EPI_ISL_1592479, EPI_ISL_1592483, EPI_ISL_1592484, EPI_ISL_1592487, EPI_ISL_1592489, EPI_ISL_1592496, EPI_ISL_1592497, EPI_ISL_1592573, EPI_ISL_1592593, EPI_ISL_1592594, EPI_ISL_1592595, EPI_ISL_1592596, EPI_ISL_1592597, EPI_ISL_1592604, EPI_ISL_1592605, EPI_ISL_1592606, EPI_ISL_1592608, EPI_ISL_1592609, EPI_ISL_1592614, EPI_ISL_1592615, EPI_ISL_1592616, EPI_ISL_1592617, EPI_ISL_1592618, EPI_ISL_1592619, EPI_ISL_1592620, EPI_ISL_1592626, EPI_ISL_1592627, EPI_ISL_1592628, EPI_ISL_1592629, EPI_ISL_1592630, EPI_ISL_1592631, EPI_ISL_1592638, EPI_ISL_1592639, EPI_ISL_1592642, EPI_ISL_1592643, EPI_ISL_1592644, EPI_ISL_1592650, EPI_ISL_1592652, EPI_ISL_1592653, EPI_ISL_1592654, EPI_ISL_1592662, EPI_ISL_1592664, EPI_ISL_1592665, EPI_ISL_1592667, EPI_ISL_1592674, EPI_ISL_1592677, EPI_ISL_1592678, EPI_ISL_1592896, EPI_ISL_1592940, EPI_ISL_1593021, EPI_ISL_1593054, EPI_ISL_1593063, EPI_ISL_1593074, EPI_ISL_1593162, EPI_ISL_1593166, EPI_ISL_1593182, EPI_ISL_1593197, EPI_ISL_1615101, EPI_ISL_1615136, EPI_ISL_1615233, EPI_ISL_1615234, EPI_ISL_1615240, EPI_ISL_1615241, EPI_ISL_1615295, EPI_ISL_1615322, EPI_ISL_1615323, EPI_ISL_1615370, EPI_ISL_1615372, EPI_ISL_1615393, EPI_ISL_1615409, EPI_ISL_1615430, EPI_ISL_1615436, EPI_ISL_1615441, EPI_ISL_1615450, EPI_ISL_1615478, EPI_ISL_1615531, EPI_ISL_1615537, EPI_ISL_1615555, EPI_ISL_1678894, EPI_ISL_1678901, EPI_ISL_1678928, EPI_ISL_1678936, EPI_ISL_1678974, EPI_ISL_1678983, EPI_ISL_1678984, EPI_ISL_1679020, EPI_ISL_1679021, EPI_ISL_1679159, EPI_ISL_1679214, EPI_ISL_1679218, EPI_ISL_1679220, EPI_ISL_1679236, EPI_ISL_1679239, EPI_ISL_1679247, EPI_ISL_1679249, EPI_ISL_1679258, EPI_ISL_1679267, EPI_ISL_1679288, EPI_ISL_1679301, EPI_ISL_1679303, EPI_ISL_1679309, EPI_ISL_1679363, EPI_ISL_1679370, EPI_ISL_1679384, EPI_ISL_1679399, EPI_ISL_1679450, EPI_ISL_1679451, EPI_ISL_1679472, EPI_ISL_1679523, EPI_ISL_1679529, EPI_ISL_1679613, EPI_ISL_1679616, EPI_ISL_1679677, EPI_ISL_1679702, EPI_ISL_1679713, EPI_ISL_1679720, EPI_ISL_1679738, EPI_ISL_1679771, EPI_ISL_1679775, EPI_ISL_1679776, EPI_ISL_1679781, EPI_ISL_1679791, EPI_ISL_1679792, EPI_ISL_1679841, EPI_ISL_1679862, EPI_ISL_1679878, EPI_ISL_1679900, EPI_ISL_1679908, EPI_ISL_1679911, EPI_ISL_1679915, EPI_ISL_1679922, EPI_ISL_1680073, EPI_ISL_1680144, EPI_ISL_1680178, EPI_ISL_1690283, EPI_ISL_1690290, EPI_ISL_1690295, EPI_ISL_1690301, EPI_ISL_1690311, EPI_ISL_1690312, EPI_ISL_1690330, EPI_ISL_1690342, EPI_ISL_1690435, EPI_ISL_1690452, EPI_ISL_1690457, EPI_ISL_1690459, EPI_ISL_1690498, EPI_ISL_1690584, EPI_ISL_1690600, EPI_ISL_1690617, EPI_ISL_1690649, EPI_ISL_1690708, EPI_ISL_1690772, EPI_ISL_1690817, EPI_ISL_1690833, EPI_ISL_1690899, EPI_ISL_1690984, EPI_ISL_1691022, EPI_ISL_1691032, EPI_ISL_1691062, EPI_ISL_1691063, EPI_ISL_1691082, EPI_ISL_1691133, EPI_ISL_1691155, EPI_ISL_1691162, EPI_ISL_1691163, EPI_ISL_1691190, EPI_ISL_1691192, EPI_ISL_1691199, EPI_ISL_1691201, EPI_ISL_1691216, EPI_ISL_1691218, EPI_ISL_1691222, EPI_ISL_1691310, EPI_ISL_1691312, EPI_ISL_1691340, EPI_ISL_1691343, EPI_ISL_1695048, EPI_ISL_1701848, EPI_ISL_1701852, EPI_ISL_1701854, EPI_ISL_1702158, EPI_ISL_1734681, EPI_ISL_1734703, EPI_ISL_1734784, EPI_ISL_1734798, EPI_ISL_1734832, EPI_ISL_1734848, EPI_ISL_1734876, EPI_ISL_1734894, EPI_ISL_1734905, EPI_ISL_1734906, EPI_ISL_1734911, EPI_ISL_1734915, EPI_ISL_1734935, EPI_ISL_1734962, EPI_ISL_1734980, EPI_ISL_1734992, EPI_ISL_1735013, EPI_ISL_1735015, EPI_ISL_1735018, EPI_ISL_1735174, EPI_ISL_1735217, EPI_ISL_1735227, EPI_ISL_1735228, EPI_ISL_1735238, EPI_ISL_1735291, EPI_ISL_1735300, EPI_ISL_1735323, EPI_ISL_1735416, EPI_ISL_1735427, EPI_ISL_1735429, EPI_ISL_1735438, EPI_ISL_1735604, EPI_ISL_1735621, EPI_ISL_1735631, EPI_ISL_1735674, EPI_ISL_1753503, EPI_ISL_1753510, EPI_ISL_1796139, EPI_ISL_1796218, EPI_ISL_1796229, EPI_ISL_1796231, EPI_ISL_1796237, EPI_ISL_1796241, EPI_ISL_1796250, EPI_ISL_1796251, EPI_ISL_1796259, EPI_ISL_1796275, EPI_ISL_1796285, EPI_ISL_1796341, EPI_ISL_1796428, EPI_ISL_1796435, EPI_ISL_1796454, EPI_ISL_1796474, EPI_ISL_1796487, EPI_ISL_1796500, EPI_ISL_1796595, EPI_ISL_1796613, EPI_ISL_1796621, EPI_ISL_1796630, EPI_ISL_1796639, EPI_ISL_1796641, EPI_ISL_1796708, EPI_ISL_1796718, EPI_ISL_1796853, EPI_ISL_1796869, EPI_ISL_1796882, EPI_ISL_1796894, EPI_ISL_1796897, EPI_ISL_1796911, EPI_ISL_1796922, EPI_ISL_1796977, EPI_ISL_1796992, EPI_ISL_1796993, EPI_ISL_1797102, EPI_ISL_1797124, EPI_ISL_1797134, EPI_ISL_1797265, EPI_ISL_1797276, EPI_ISL_1797277, EPI_ISL_1797299, EPI_ISL_1797335, EPI_ISL_1797371, EPI_ISL_1797385, EPI_ISL_1797392, EPI_ISL_1797395, EPI_ISL_1797416, EPI_ISL_1797425, EPI_ISL_1803526, EPI_ISL_1803535, EPI_ISL_1803554, EPI_ISL_1803592, EPI_ISL_1803593, EPI_ISL_1803614, EPI_ISL_1803627, EPI_ISL_1803663, EPI_ISL_1803669, EPI_ISL_1803673, EPI_ISL_1803674, EPI_ISL_1803676, EPI_ISL_1803694, EPI_ISL_1803697, EPI_ISL_1803704, EPI_ISL_1803709, EPI_ISL_1803722, EPI_ISL_1803729, EPI_ISL_1803815, EPI_ISL_1803820, EPI_ISL_1803964, EPI_ISL_1803987, EPI_ISL_1803990, EPI_ISL_1803997, EPI_ISL_1804000, EPI_ISL_1804083, EPI_ISL_1804094, EPI_ISL_1804095, EPI_ISL_1804103, EPI_ISL_1804108, EPI_ISL_1804122, EPI_ISL_1804135, EPI_ISL_1804143, EPI_ISL_1804169, EPI_ISL_1804179, EPI_ISL_1804191, EPI_ISL_1804202, EPI_ISL_1804213, EPI_ISL_1804256, EPI_ISL_1804518, EPI_ISL_1804524, EPI_ISL_1804534, EPI_ISL_1804535, EPI_ISL_1804554, EPI_ISL_1804564, EPI_ISL_1804568, EPI_ISL_1804583, EPI_ISL_1804589, EPI_ISL_1804601, EPI_ISL_1804611, EPI_ISL_1804625, EPI_ISL_1804643, EPI_ISL_1804676, EPI_ISL_1804681, EPI_ISL_1804702, EPI_ISL_1804713, EPI_ISL_1804722, EPI_ISL_1804729, EPI_ISL_1818008, EPI_ISL_1818009, EPI_ISL_1818028, EPI_ISL_1906970, EPI_ISL_1907014, EPI_ISL_1907023, EPI_ISL_1907031, EPI_ISL_1907068, EPI_ISL_1907098, EPI_ISL_1907130, EPI_ISL_1907133, EPI_ISL_1907221, EPI_ISL_1907252, EPI_ISL_1907255, EPI_ISL_1907262, EPI_ISL_1907283, EPI_ISL_1907384, EPI_ISL_1907395, EPI_ISL_1907424, EPI_ISL_1907455, EPI_ISL_1907456, EPI_ISL_1907464, EPI_ISL_1907469, EPI_ISL_1907475, EPI_ISL_1907480, EPI_ISL_1907492, EPI_ISL_1907493, EPI_ISL_1907513, EPI_ISL_1907520, EPI_ISL_1907521, EPI_ISL_1907531, EPI_ISL_1907544, EPI_ISL_1907546, EPI_ISL_1907679, EPI_ISL_1907692, EPI_ISL_1907702, EPI_ISL_1907745, EPI_ISL_1907785, EPI_ISL_1907797, EPI_ISL_1907831, EPI_ISL_1907851, EPI_ISL_1907860, EPI_ISL_1907873, EPI_ISL_1907875, EPI_ISL_1907879, EPI_ISL_1908068, EPI_ISL_1908103, EPI_ISL_1908114, EPI_ISL_1908138, EPI_ISL_1923183, EPI_ISL_1923209, EPI_ISL_1923235, EPI_ISL_1923287, EPI_ISL_1923355, EPI_ISL_1923393, EPI_ISL_1923399, EPI_ISL_1923415, EPI_ISL_1923426, EPI_ISL_1923446, EPI_ISL_1923453, EPI_ISL_1923459, EPI_ISL_1923638, EPI_ISL_1923639, EPI_ISL_1923650, EPI_ISL_1923711, EPI_ISL_1923743, EPI_ISL_1923763, EPI_ISL_1923768, EPI_ISL_1923784, EPI_ISL_1923785, EPI_ISL_1923793, EPI_ISL_1923832, EPI_ISL_1923833, EPI_ISL_1923847, EPI_ISL_1923848, EPI_ISL_1923861, EPI_ISL_1923926, EPI_ISL_1923936, EPI_ISL_1923979, EPI_ISL_1923990, EPI_ISL_1923999, EPI_ISL_1924035, EPI_ISL_1924060, EPI_ISL_1924080, EPI_ISL_1924110, EPI_ISL_1924143, EPI_ISL_1924231, EPI_ISL_1990551, EPI_ISL_1990555, EPI_ISL_1990560, EPI_ISL_1990590, EPI_ISL_1990594, EPI_ISL_1990613, EPI_ISL_1990620, EPI_ISL_1990678, EPI_ISL_1990723, EPI_ISL_1990836, EPI_ISL_1990848, EPI_ISL_1990859, EPI_ISL_1990889, EPI_ISL_1990901, EPI_ISL_1990902, EPI_ISL_1990913, EPI_ISL_1990918, EPI_ISL_1990924, EPI_ISL_1990932, EPI_ISL_1990943, EPI_ISL_1990954, EPI_ISL_1990957, EPI_ISL_1990975, EPI_ISL_1990986, EPI_ISL_1991010, EPI_ISL_1991022, EPI_ISL_1991061, EPI_ISL_1991073, EPI_ISL_1991087, EPI_ISL_1991100, EPI_ISL_1991113, EPI_ISL_1991134, EPI_ISL_1991160, EPI_ISL_1991162, EPI_ISL_1991269, EPI_ISL_1991300, EPI_ISL_1991301, EPI_ISL_1991308, EPI_ISL_1991487, EPI_ISL_1991501, EPI_ISL_1991513, EPI_ISL_1991524, EPI_ISL_1991545, EPI_ISL_1991725, EPI_ISL_1991734, EPI_ISL_1991740, EPI_ISL_1991745, EPI_ISL_1991748, EPI_ISL_1991758, EPI_ISL_1991784, EPI_ISL_1991785, EPI_ISL_1991805, EPI_ISL_1991824, EPI_ISL_1991826, EPI_ISL_2009725, EPI_ISL_2009845, EPI_ISL_2009856, EPI_ISL_2009887, EPI_ISL_2009896, EPI_ISL_2009907, EPI_ISL_2009917, EPI_ISL_2010005, EPI_ISL_2010020, EPI_ISL_2010031, EPI_ISL_2010062, EPI_ISL_2010175, EPI_ISL_2010179, EPI_ISL_2010181, EPI_ISL_2010186, EPI_ISL_2010192, EPI_ISL_2010201, EPI_ISL_2010209, EPI_ISL_2010210, EPI_ISL_2010215, EPI_ISL_2010217, EPI_ISL_2010229, EPI_ISL_2010237, EPI_ISL_2010273, EPI_ISL_2010282, EPI_ISL_2010316, EPI_ISL_2010413, EPI_ISL_2010427, EPI_ISL_2010497, EPI_ISL_2010552, EPI_ISL_2010553, EPI_ISL_2010560, EPI_ISL_2010561, EPI_ISL_2010589, EPI_ISL_2010714, EPI_ISL_2010747, EPI_ISL_2010758, EPI_ISL_2010778, EPI_ISL_2010785, EPI_ISL_2010796, EPI_ISL_2010797, EPI_ISL_2035278, EPI_ISL_2035285, EPI_ISL_2035287, EPI_ISL_2097374, EPI_ISL_2097388, EPI_ISL_2097391, EPI_ISL_2097395, EPI_ISL_2097422, EPI_ISL_2097430, EPI_ISL_2097469, EPI_ISL_2097496, EPI_ISL_2097624, EPI_ISL_2097638, EPI_ISL_2097693, EPI_ISL_2097702, EPI_ISL_2097845, EPI_ISL_2097898, EPI_ISL_2097899, EPI_ISL_2097966, EPI_ISL_2098005, EPI_ISL_2098011, EPI_ISL_2098029, EPI_ISL_2098049, EPI_ISL_2098077, EPI_ISL_2098095, EPI_ISL_2098106, EPI_ISL_2132607, EPI_ISL_2132613, EPI_ISL_2144118, EPI_ISL_2144141, EPI_ISL_2144340, EPI_ISL_2144371, EPI_ISL_2144401, EPI_ISL_2144411, EPI_ISL_2144413, EPI_ISL_2144520, EPI_ISL_2144583, EPI_ISL_2144601, EPI_ISL_2144619, EPI_ISL_2144655, EPI_ISL_2144747, EPI_ISL_2144793, EPI_ISL_2144803, EPI_ISL_2144843, EPI_ISL_2144929, EPI_ISL_2145000, EPI_ISL_2145463, EPI_ISL_2145501, EPI_ISL_2145576, EPI_ISL_2145685, EPI_ISL_2159490, EPI_ISL_2159518, EPI_ISL_2159520, EPI_ISL_2159539, EPI_ISL_2159686, EPI_ISL_2159811, EPI_ISL_2159812, EPI_ISL_2159823, EPI_ISL_2159830, EPI_ISL_2159839, EPI_ISL_2159853, EPI_ISL_2159872, EPI_ISL_2159891, EPI_ISL_2159899, EPI_ISL_2159910, EPI_ISL_2160089, EPI_ISL_2160105, EPI_ISL_2160115, EPI_ISL_2160116, EPI_ISL_2160125, EPI_ISL_2160131, EPI_ISL_2160156, EPI_ISL_2160572, EPI_ISL_2160576 | | | |
| see above | Helix/Illumina | Centers for Disease Control and Prevention Division of Viral Diseases, Pathogen Discovery | Adrian Paskey; Alexandre Bolze; Ary Ascencio; Benjamin Rambo-Martin; Brad Sickler; Charlotte Rivera-Garcia; Christine Tran; Christopher Gulvick; Clinton R. Paden; Dakota Howard; Darlene Wagner; David Becker; Dhwani Batra; Duncan MacCannell; Efren Sandoval; Eileen de Feo; Elizabeth Cirulli; Eric Allen; Geraint Levan; James Lu; Jan Antico; Jason Caravas; Jason Nguyen; Jimmy Ramirez; Jingtao Liu; Kara Moser; Kelly Schiabor Barrett; Kim Gietzen; Magnus Isaksson; Marc Laurent; Matthew Schmerer; Matthew Tolentino; Nicole L. Washington; Peter W. Cook; Phil Febbo; Ryan Cho; Scott Sammons; Shannon Wickline; Shatavia Morrison; Sherry Wang; Simon White; Tyler Cassens; William Lee; Yvette Unoarumhi |
| EPI_ISL_1701411, EPI_ISL_1701444, EPI_ISL_1701447, EPI_ISL_1701479, EPI_ISL_1701493, EPI_ISL_1701497, EPI_ISL_1701555, EPI_ISL_1701560, EPI_ISL_1701587, EPI_ISL_1701594, EPI_ISL_1701601, EPI_ISL_1701605, EPI_ISL_1701614, EPI_ISL_2102528, EPI_ISL_2102533, EPI_ISL_2102537, EPI_ISL_2102550, EPI_ISL_2102561, EPI_ISL_2102585, EPI_ISL_2102752, EPI_ISL_2102753, EPI_ISL_2102791, EPI_ISL_2102797, EPI_ISL_2102806, EPI_ISL_2102808, EPI_ISL_2102822, EPI_ISL_2102832, EPI_ISL_2102838, EPI_ISL_2102847, EPI_ISL_2102852, EPI_ISL_2102853, EPI_ISL_2102872, EPI_ISL_2102888, EPI_ISL_2102903, EPI_ISL_2102906, EPI_ISL_2102935, EPI_ISL_2102948, EPI_ISL_2102964, EPI_ISL_2102979, EPI_ISL_2102986, EPI_ISL_2102992, EPI_ISL_2103203, EPI_ISL_2103204, EPI_ISL_2103205, EPI_ISL_2103206, EPI_ISL_2103207, EPI_ISL_2103208, EPI_ISL_2103209, EPI_ISL_2103210, EPI_ISL_2103211, EPI_ISL_2103212, EPI_ISL_2103213, EPI_ISL_2103214, EPI_ISL_2103215, EPI_ISL_2103216, EPI_ISL_2103217, EPI_ISL_2103218, EPI_ISL_2103219, EPI_ISL_2103220, EPI_ISL_2103221, EPI_ISL_2103222, EPI_ISL_2103223, EPI_ISL_2103224, EPI_ISL_2103225, EPI_ISL_2103226, EPI_ISL_2103227, EPI_ISL_2103228 | | | |
| see above | Humboldt County Public Health Laboratory | Chan-Zuckerberg Biohub | CZB Cliahub Consortium |
| EPI_ISL_1512777, EPI_ISL_1512805, EPI_ISL_1512823, EPI_ISL_1512827, EPI_ISL_1512844, EPI_ISL_1512880, EPI_ISL_1512898, EPI_ISL_1513116, EPI_ISL_1513119, EPI_ISL_1513120, EPI_ISL_1513127, EPI_ISL_1513134, EPI_ISL_1513137, EPI_ISL_1513138, EPI_ISL_1587592, EPI_ISL_1587898, EPI_ISL_1587916, EPI_ISL_1587932, EPI_ISL_1587948, EPI_ISL_1588416, EPI_ISL_1692123, EPI_ISL_1693196, EPI_ISL_1693463, EPI_ISL_1693826, EPI_ISL_1694297, EPI_ISL_1694349, EPI_ISL_1694535, EPI_ISL_1795476, EPI_ISL_1795480, EPI_ISL_1795494, EPI_ISL_1795606, EPI_ISL_1802878, EPI_ISL_1818093, EPI_ISL_2013193, EPI_ISL_2013277, EPI_ISL_2013332, EPI_ISL_2014190, EPI_ISL_2087709, EPI_ISL_2087772, EPI_ISL_2087845, EPI_ISL_2087859, EPI_ISL_2088291, EPI_ISL_2088565, EPI_ISL_2089006, EPI_ISL_2089217, EPI_ISL_2089299, EPI_ISL_2089312, EPI_ISL_2089408, EPI_ISL_2132366 | | | |
| see above | Infinity Biologix | Centers for Disease Control and Prevention Division of Viral Diseases, Pathogen Discovery | Adrian Paskey; Benjamin Rambo-Martin; Chirayu Goswami; Christian Bixby; Christopher Gulvick; Clinton R. Paden; Dakota Howard; Darlene Wagner; Dhwani Batra; Duncan MacCannell; Jason Caravas; Jonathan Schultz; Kara Moser; Matthew Schmerer; Peter W. Cook; Robin Grimwood; Russ Hager; Scott Sammons; Shatavia Morrison; Yihe Wang; Yvette Unoarumhi |
| EPI_ISL_1712523, EPI_ISL_1712524, EPI_ISL_1712525, EPI_ISL_1712526, EPI_ISL_1712527, EPI_ISL_1712528, EPI_ISL_1712529, EPI_ISL_1712530, EPI_ISL_1712531, EPI_ISL_1712532, EPI_ISL_1712533, EPI_ISL_1712534, EPI_ISL_1712535, EPI_ISL_1712536, EPI_ISL_1712537, EPI_ISL_1712538, EPI_ISL_1712539, EPI_ISL_1712540, EPI_ISL_1712541, EPI_ISL_1712550, EPI_ISL_1712551, EPI_ISL_1712552, EPI_ISL_1712553, EPI_ISL_1712554, EPI_ISL_1712555, EPI_ISL_1712556, EPI_ISL_1712557, EPI_ISL_1712558, EPI_ISL_1712559, EPI_ISL_1712560, EPI_ISL_1712561, EPI_ISL_1712562 | | | |
| see above | Innovative Genomics Institute, UC Berkeley | Innovative Genomics Institute, UC Berkeley | Holly Gildea; John Boyle; Phil Frankino; Stacia Wyman |
| EPI_ISL_1913217, EPI_ISL_1913225 | KAISER SAN JOSE | Santa Clara County public Health Laboratory | Santa Clara County Public Health Department |
| EPI_ISL_1824111, EPI_ISL_1824112, EPI_ISL_1824113, EPI_ISL_1824114, EPI_ISL_1824115 | Kaiser San Jose | Santa Clara County public Health Laboratory | Santa Clara County Public Health Department |
| EPI_ISL_1508968, EPI_ISL_1508974 | Kaiser Santa Clara | Santa Clara County Public Health Laboratory | Santa Clara County Public Health Department |
| EPI_ISL_1664268, EPI_ISL_1664269, EPI_ISL_1824120 | Kaiser Santa Clara | Santa Clara County public Health Laboratory | Santa Clara County Public Health Department |
| EPI_ISL_1700838, EPI_ISL_1754141, EPI_ISL_1805493, EPI_ISL_1821201, EPI_ISL_1821202, EPI_ISL_1821203, EPI_ISL_1857271, EPI_ISL_2101880, EPI_ISL_2105688, EPI_ISL_2139908, EPI_ISL_2139909, EPI_ISL_2139914, EPI_ISL_2157328, EPI_ISL_2158108 | | | |
| see above | LESP Baja California | Instituto de Diagnostico y Referencia Epidemiologicos (INDRE) | Abril Rodriguez-Maldonado; Ariadna Medina-Benitez; Claudia Wong-Arambula; Ernesto Ramirez-Gonzalez.; Gisela Barrera-Badillo; Irma Lopez-Martinez; Joaquin Quiroz-Mercado; Lucia Hernandez-Rivas; Natividad Cruz-Ortiz; Sergio Rangel-Guerrero; Tatiana Nunez-Garcia; Vanessa Rivero-Arredondo |
| EPI_ISL_1805487, EPI_ISL_1821104, EPI_ISL_1821105, EPI_ISL_1821180, EPI_ISL_1821181, EPI_ISL_1821182, EPI_ISL_1821183, EPI_ISL_1821184, EPI_ISL_1857224, EPI_ISL_1857225, EPI_ISL_1857226, EPI_ISL_1857227, EPI_ISL_1857235, EPI_ISL_1857266, EPI_ISL_1857267, EPI_ISL_2139913, EPI_ISL_2158231 | | | |
| see above | LESP Baja California Sur | Instituto de Diagnostico y Referencia Epidemiologicos (INDRE) | Abril Rodriguez-Maldonado; Ariadna Medina-Benitez; Claudia Wong-Arambula; Ernesto Ramirez-Gonzalez.; Gisela Barrera-Badillo; Irma Lopez-Martinez; Joaquin Quiroz-Mercado; Lucia Hernandez-Rivas; Natividad Cruz-Ortiz; Sergio Rangel-Guerrero; Tatiana Nunez-Garcia; Vanessa Rivero-Arredondo |
| EPI_ISL_1811524, EPI_ISL_1811531 | Laboratorio Central de Epidemiología (LCE) | Instituto de Biotecnología de la UNAM | Alejandro Sanchez-Flores; Alfredo Herrera-Estrella; Alicia Ocaña-Mondragón; Angel Gustavo Salas-Lais; Bernardo Martínez-Miguel; Blanca Taboada; Brenda Irasema Maldonado-Meza; Carla Ivón Herrera-Najera; Carlos F. Arias; Celia Boukadida; Clara Esperanza Santacruz-Tinoco; Concepción Grajales-Muñiz; Célida Duque-Molina; Fernando Fontove-Herrera; Francisco Pulido; Gloria Elena Espinosa-Ayala; Gloria María Molina-Salinas; Gloria Vazquez; Hector Esteban Paz-Juárez; Hector Montoya-Fuentes; Helen Haydee Fernanda Ramírez-Plascencia; José Antonio Enciso-Moreno; José Esteban Muñoz-Medina; José de Jesús Nuñez-Contreras; Juan Bautista Chale-Dzul; Julio Elias Alvarado-Yaah; Luis Alberto Ochoa-Carrera; Margarita Matías-Florentino; María Guadalupe Santiago-Mauricio; María Guadalupe de Jesús Mireles-Rivera; Nelly Sélem-Mojica; Pavel Isa; Ricardo Grande; Santiago Ávila-Ríos; Victor Eduardo Garcia-Arias; Víctor Hugo Borja-Aburto |
| EPI_ISL_1515636, EPI_ISL_1515637, EPI_ISL_1515638, EPI_ISL_1515640, EPI_ISL_1515744, EPI_ISL_1550248, EPI_ISL_1550361, EPI_ISL_1612132, EPI_ISL_1612231, EPI_ISL_1612243, EPI_ISL_1612244, EPI_ISL_1612245, EPI_ISL_1612259, EPI_ISL_1612263, EPI_ISL_1612394, EPI_ISL_1612395, EPI_ISL_1612396, EPI_ISL_1612397, EPI_ISL_1612398, EPI_ISL_1612399, EPI_ISL_1612401, EPI_ISL_1612406, EPI_ISL_1612441, EPI_ISL_1612442, EPI_ISL_1612443, EPI_ISL_1612444, EPI_ISL_1612445, EPI_ISL_1612446, EPI_ISL_1612447, EPI_ISL_1612448, EPI_ISL_1612449, EPI_ISL_1612450, EPI_ISL_1612451, EPI_ISL_1612452, EPI_ISL_1612453, EPI_ISL_1612454, EPI_ISL_1612455, EPI_ISL_1612456, EPI_ISL_1612457, EPI_ISL_1612458, EPI_ISL_1612459, EPI_ISL_1612510, EPI_ISL_1612616, EPI_ISL_1612617, EPI_ISL_1612618, EPI_ISL_1612619, EPI_ISL_1612620, EPI_ISL_1612621, EPI_ISL_1612622, EPI_ISL_1612623, EPI_ISL_1612624, EPI_ISL_1612625, EPI_ISL_1612626, EPI_ISL_1612627, EPI_ISL_1612628, EPI_ISL_1612629, EPI_ISL_1612671, EPI_ISL_1612672, EPI_ISL_1612673, EPI_ISL_1612677, EPI_ISL_1612678, EPI_ISL_1612679, EPI_ISL_1612680, EPI_ISL_1612681, EPI_ISL_1612682, EPI_ISL_1612683, EPI_ISL_1612684, EPI_ISL_1612685, EPI_ISL_1612686, EPI_ISL_1612687, EPI_ISL_1612688, EPI_ISL_1612689, EPI_ISL_1612690, EPI_ISL_1612691, EPI_ISL_1612692, EPI_ISL_1612694, EPI_ISL_1612695, EPI_ISL_1612696, EPI_ISL_1612697, EPI_ISL_1612698, EPI_ISL_1612699, EPI_ISL_1612700, EPI_ISL_1612701, EPI_ISL_1612702, EPI_ISL_1612703, EPI_ISL_1612704, EPI_ISL_1612705, EPI_ISL_1612706, EPI_ISL_1612707, EPI_ISL_1612708, EPI_ISL_1612709, EPI_ISL_1612710, EPI_ISL_1612711, EPI_ISL_1612712, EPI_ISL_1612713, EPI_ISL_1612714, EPI_ISL_1612715, EPI_ISL_1612716, EPI_ISL_1612717, EPI_ISL_1612718, EPI_ISL_1612719, EPI_ISL_1612720, EPI_ISL_1612722, EPI_ISL_1612723, EPI_ISL_1612724, EPI_ISL_1612728, EPI_ISL_1612729, EPI_ISL_1612731, EPI_ISL_1612732, EPI_ISL_1612733, EPI_ISL_1612734, EPI_ISL_1612735, EPI_ISL_1612736, EPI_ISL_1612737, EPI_ISL_1612738, EPI_ISL_1612739, EPI_ISL_1612740, EPI_ISL_1612741, EPI_ISL_1612742, EPI_ISL_1612743, EPI_ISL_1612744, EPI_ISL_1612745, EPI_ISL_1612746, EPI_ISL_1612747, EPI_ISL_1612748, EPI_ISL_1612749, EPI_ISL_1612750, EPI_ISL_1612751, EPI_ISL_1612752, EPI_ISL_1612753, EPI_ISL_1612759, EPI_ISL_1612760, EPI_ISL_1612769, EPI_ISL_1612770, EPI_ISL_1612771, EPI_ISL_1612772, EPI_ISL_1612773, EPI_ISL_1612781, EPI_ISL_1612782, EPI_ISL_1612783, EPI_ISL_1612784, EPI_ISL_1612785, EPI_ISL_1612786, EPI_ISL_1612794, EPI_ISL_1612795, EPI_ISL_1612796, EPI_ISL_1612797, EPI_ISL_1612798, EPI_ISL_1612799, EPI_ISL_1612800, EPI_ISL_1612801, EPI_ISL_1612802, EPI_ISL_1612803, EPI_ISL_1612804, EPI_ISL_1612805, EPI_ISL_1681623, EPI_ISL_1684343, EPI_ISL_1684344, EPI_ISL_1684345, EPI_ISL_1684346, EPI_ISL_1684347, EPI_ISL_1684348, EPI_ISL_1684354, EPI_ISL_1684359, EPI_ISL_1684360, EPI_ISL_1684364, EPI_ISL_1684365, EPI_ISL_1684366, EPI_ISL_1684367, EPI_ISL_1684368, EPI_ISL_1684369, EPI_ISL_1684370, EPI_ISL_1684371, EPI_ISL_1684372, EPI_ISL_1684373, EPI_ISL_1684374, EPI_ISL_1684375, EPI_ISL_1684376, EPI_ISL_1684377, EPI_ISL_1684378, EPI_ISL_1684379, EPI_ISL_1684380, EPI_ISL_1684383, EPI_ISL_1684384, EPI_ISL_1684401, EPI_ISL_1684402, EPI_ISL_1684403, EPI_ISL_1684404, EPI_ISL_1684405, EPI_ISL_1684406, EPI_ISL_1684407, EPI_ISL_1684409, EPI_ISL_1684410, EPI_ISL_1684411, EPI_ISL_1684412, EPI_ISL_1684413, EPI_ISL_1684414, EPI_ISL_1684415, EPI_ISL_1684454, EPI_ISL_1684455, EPI_ISL_1684456, EPI_ISL_1684457, EPI_ISL_1684458, EPI_ISL_1684459, EPI_ISL_1684460, EPI_ISL_1684461, EPI_ISL_1684462, EPI_ISL_1684500, EPI_ISL_1684632, EPI_ISL_1684635, EPI_ISL_1684758, EPI_ISL_1684786, EPI_ISL_1684833, EPI_ISL_1684834, EPI_ISL_1684836, EPI_ISL_1684848, EPI_ISL_1684853, EPI_ISL_1684854, EPI_ISL_1684855, EPI_ISL_1684883, EPI_ISL_1684894, EPI_ISL_1684895, EPI_ISL_1684896, EPI_ISL_1684897, EPI_ISL_1684898, EPI_ISL_1684971, EPI_ISL_1684997, EPI_ISL_1695023, EPI_ISL_1798209, EPI_ISL_1798210, EPI_ISL_1798211, EPI_ISL_1798258, EPI_ISL_1798259, EPI_ISL_1798260, EPI_ISL_1798292, EPI_ISL_1798297, EPI_ISL_1798298, EPI_ISL_1798299, EPI_ISL_1798300, EPI_ISL_1798301, EPI_ISL_1798302, EPI_ISL_1798308, EPI_ISL_1798309, EPI_ISL_1798310, EPI_ISL_1798313, EPI_ISL_1798317, EPI_ISL_1798318, EPI_ISL_1798321, EPI_ISL_1798322, EPI_ISL_1798326, EPI_ISL_1798328, EPI_ISL_1798330, EPI_ISL_1798435, EPI_ISL_1798436, EPI_ISL_1798438, EPI_ISL_1798440, EPI_ISL_1798471, EPI_ISL_1798474, EPI_ISL_1798475, EPI_ISL_1798477, EPI_ISL_1798479, EPI_ISL_1798493, EPI_ISL_1798495, EPI_ISL_1798529, EPI_ISL_1798531, EPI_ISL_1798537, EPI_ISL_1798539, EPI_ISL_1798541, EPI_ISL_1798544, EPI_ISL_1798546, EPI_ISL_1798548, EPI_ISL_1798550, EPI_ISL_1798552, EPI_ISL_1798556, EPI_ISL_1798558, EPI_ISL_1798560, EPI_ISL_1798562, EPI_ISL_1798564, EPI_ISL_1798567, EPI_ISL_1798569, EPI_ISL_1798570, EPI_ISL_1798572, EPI_ISL_1798574, EPI_ISL_1798587, EPI_ISL_1798588, EPI_ISL_1798590, EPI_ISL_1798592, EPI_ISL_1798594, EPI_ISL_1798596, EPI_ISL_1798598, EPI_ISL_1798601, EPI_ISL_1798602, EPI_ISL_1798629, EPI_ISL_1798897, EPI_ISL_1798949, EPI_ISL_1798950, EPI_ISL_1798951, EPI_ISL_1798952, EPI_ISL_1798953, EPI_ISL_1798954, EPI_ISL_1798959, EPI_ISL_1798960, EPI_ISL_1798976, EPI_ISL_1798978, EPI_ISL_1798979, EPI_ISL_1798980, EPI_ISL_1798981, EPI_ISL_1798982, EPI_ISL_1798983, EPI_ISL_1798984, EPI_ISL_1798985, EPI_ISL_1798986, EPI_ISL_1798987, EPI_ISL_1798988, EPI_ISL_1798989, EPI_ISL_1798991, EPI_ISL_1798992, EPI_ISL_1798993, EPI_ISL_1798994, EPI_ISL_1798995, EPI_ISL_1798996, EPI_ISL_1798997, EPI_ISL_1798998, EPI_ISL_1798999, EPI_ISL_1799000, EPI_ISL_1799001, EPI_ISL_1799002, EPI_ISL_1799003, EPI_ISL_1799004, EPI_ISL_1799006, EPI_ISL_1799007, EPI_ISL_1799008, EPI_ISL_1799009, EPI_ISL_1799079, EPI_ISL_1799142, EPI_ISL_1799143, EPI_ISL_1799153, EPI_ISL_1799167, EPI_ISL_1799168, EPI_ISL_1799169, EPI_ISL_1799259, EPI_ISL_1799277, EPI_ISL_1799290, EPI_ISL_1799291, EPI_ISL_1799292, EPI_ISL_1799335, EPI_ISL_1799336, EPI_ISL_1799337, EPI_ISL_1799399, EPI_ISL_1799400, EPI_ISL_1799401, EPI_ISL_1799425, EPI_ISL_1799426, EPI_ISL_1799427, EPI_ISL_1799428, EPI_ISL_1799447, EPI_ISL_1799448, EPI_ISL_1799449, EPI_ISL_1799454, EPI_ISL_1799455, EPI_ISL_1799456, EPI_ISL_1799457, EPI_ISL_1799471, EPI_ISL_1799472, EPI_ISL_1799473, EPI_ISL_1799474, EPI_ISL_1799484, EPI_ISL_1799485, EPI_ISL_1799535, EPI_ISL_1799551, EPI_ISL_1799552, EPI_ISL_1799564, EPI_ISL_1799605, EPI_ISL_1799620, EPI_ISL_1799621, EPI_ISL_1799640, EPI_ISL_1925654, EPI_ISL_1926318, EPI_ISL_1926668, EPI_ISL_1930643, EPI_ISL_1930646, EPI_ISL_1930650, EPI_ISL_1930653, EPI_ISL_1930656, EPI_ISL_1930659, EPI_ISL_1930668, EPI_ISL_1930669, EPI_ISL_1930674, EPI_ISL_1930675, EPI_ISL_1930676, EPI_ISL_1930679, EPI_ISL_1930681, EPI_ISL_1930682, EPI_ISL_1930687, EPI_ISL_1930688, EPI_ISL_1930689, EPI_ISL_1930701, EPI_ISL_1930702, EPI_ISL_1930703, EPI_ISL_1930704, EPI_ISL_1930705, EPI_ISL_1930706, EPI_ISL_1930707, EPI_ISL_1930708, EPI_ISL_1930709, EPI_ISL_1930710, EPI_ISL_1930727, EPI_ISL_1930745, EPI_ISL_1930786, EPI_ISL_1930787, EPI_ISL_1930788, EPI_ISL_1930790, EPI_ISL_1930791, EPI_ISL_1930792, EPI_ISL_1930829, EPI_ISL_1930830, EPI_ISL_1930831, EPI_ISL_1930832, EPI_ISL_1930833, EPI_ISL_1930834, EPI_ISL_1930835, EPI_ISL_1930836, EPI_ISL_1930848, EPI_ISL_1930849, EPI_ISL_1930850, EPI_ISL_1930851, EPI_ISL_1930852, EPI_ISL_1930853, EPI_ISL_1930854, EPI_ISL_1930855, EPI_ISL_1930857, EPI_ISL_1930858, EPI_ISL_1930859, EPI_ISL_1930860, EPI_ISL_1930861, EPI_ISL_1930970, EPI_ISL_1930989, EPI_ISL_1930992, EPI_ISL_1930995, EPI_ISL_1930998, EPI_ISL_1931001, EPI_ISL_1931003, EPI_ISL_1931034, EPI_ISL_1931037, EPI_ISL_1931040, EPI_ISL_1931042, EPI_ISL_1931046, EPI_ISL_1931049, EPI_ISL_1931052, EPI_ISL_1931122, EPI_ISL_1931123, EPI_ISL_1931124, EPI_ISL_1931126, EPI_ISL_1931127, EPI_ISL_1931129, EPI_ISL_1931130, EPI_ISL_1931131, EPI_ISL_1931133, EPI_ISL_1931134, EPI_ISL_1931136, EPI_ISL_1931418, EPI_ISL_1931484, EPI_ISL_1931487, EPI_ISL_1931490, EPI_ISL_1931492, EPI_ISL_1931495, EPI_ISL_1931606, EPI_ISL_1931609, EPI_ISL_1931612, EPI_ISL_1931615, EPI_ISL_1931616, EPI_ISL_1931623, EPI_ISL_1931672, EPI_ISL_1931673, EPI_ISL_1931675, EPI_ISL_1931676, EPI_ISL_1931678, EPI_ISL_1931679, EPI_ISL_1931680, EPI_ISL_1931683, EPI_ISL_1931685, EPI_ISL_1931708, EPI_ISL_1931710, EPI_ISL_1931711, EPI_ISL_1931713, EPI_ISL_1931714, EPI_ISL_1931716, EPI_ISL_1931717, EPI_ISL_1931718, EPI_ISL_1931742, EPI_ISL_1931745, EPI_ISL_1931748, EPI_ISL_1931751, EPI_ISL_1931754, EPI_ISL_1931755, EPI_ISL_1931756, EPI_ISL_1931758, EPI_ISL_1931759, EPI_ISL_1931760, EPI_ISL_1931762, EPI_ISL_1931994, EPI_ISL_1931996, EPI_ISL_1932045, EPI_ISL_1932048, EPI_ISL_1932050, EPI_ISL_1932051, EPI_ISL_1932053, EPI_ISL_1932054, EPI_ISL_2044694, EPI_ISL_2044742, EPI_ISL_2045197, EPI_ISL_2046011, EPI_ISL_2046027, EPI_ISL_2046028, EPI_ISL_2046029, EPI_ISL_2046064, EPI_ISL_2046071, EPI_ISL_2046072, EPI_ISL_2046190, EPI_ISL_2046216, EPI_ISL_2046217, EPI_ISL_2046218, EPI_ISL_2046219, EPI_ISL_2046220, EPI_ISL_2046221, EPI_ISL_2046252, EPI_ISL_2046253, EPI_ISL_2046254, EPI_ISL_2046312, EPI_ISL_2046356, EPI_ISL_2046357, EPI_ISL_2046358, EPI_ISL_2046359, EPI_ISL_2046360, EPI_ISL_2046361, EPI_ISL_2046362, EPI_ISL_2046363, EPI_ISL_2046364, EPI_ISL_2046409, EPI_ISL_2046410, EPI_ISL_2046411, EPI_ISL_2046412, EPI_ISL_2046413, EPI_ISL_2046414, EPI_ISL_2046415, EPI_ISL_2046416, EPI_ISL_2046417, EPI_ISL_2046418, EPI_ISL_2046419, EPI_ISL_2046420, EPI_ISL_2046421, EPI_ISL_2046422, EPI_ISL_2046423, EPI_ISL_2046424, EPI_ISL_2046425, EPI_ISL_2046426, EPI_ISL_2046427, EPI_ISL_2046428, EPI_ISL_2046429, EPI_ISL_2046430, EPI_ISL_2046431, EPI_ISL_2046488, EPI_ISL_2046490, EPI_ISL_2046491, EPI_ISL_2046492, EPI_ISL_2046493, EPI_ISL_2046564, EPI_ISL_2046565, EPI_ISL_2046566, EPI_ISL_2046567, EPI_ISL_2046568, EPI_ISL_2046569, EPI_ISL_2046570, EPI_ISL_2046571, EPI_ISL_2046572, EPI_ISL_2046573, EPI_ISL_2046574, EPI_ISL_2046614, EPI_ISL_2046615 | | | |
| see above | Laboratory Corporation of America | Centers for Disease Control and Prevention Division of Viral Diseases, Pathogen Discovery | Adrian Paskey; Amanda Douglas; Amanda Suchanek; Andrea Throop; Ayla Burns; Benjamin Rambo-Martin; Bobbi Croy; Brian Krueger; Brian Norvell; Christopher Gulvick; Christos Petropoulos; Clinton R. Paden; Craig Lukasik; Dakota Howard; Darlene Wagner; Debbie Boles; Dhwani Batra; Duncan MacCannell; Eyad Almasri; Goran Stevovic; Howard Engler; Hrushikesh Deshmukh; Jake Humphrey; Jana Schroth; Jason Caravas; Joe Voshell; John Pruitt; Jonathan Meltzer; Jonathan Williams; Kara Moser; Kimberly Wagner; Lax Iyer; Lyndon Tilson; Manoj Jain; Marcia Eisenberg; Mary Ann Cristobal; Mary Williamson; Matthew Schmerer; Michael Levandoski; Mike Sapeta; Mindy Nye; Minoo Agarwal; Mohan Kolli; Nuthawin Charoensri; Oren Cohen; Peter W. Cook; Prashant Gupta; Qian Zeng; Rama Ghatti; Scott Parker; Scott Ryan; Scott Sammons; Shatavia Morrison; Stanley Letovsky; Steven Ragan; |

| | | | |
|---|---|---|---|
| | | | Suresh Babu Selvaraju; Susan Countryman; Susan Hicks; Suzanne Dale; Thomas Urban; Tim Kuphal; Tricia Zwiefelhofer; Vincent Drouillon; Yvette Unoarumhi |
| EPI_ISL_1707686, EPI_ISL_1707687 | Los Angeles County PHL | Los Angeles County PHL | P. Hemarajata et al. |
| EPI_ISL_2106181 | Los Angeles County Public Health Laboratories, Los Angeles County Public Health Lab microbial pathogen submission group | Los Angeles County Public Health Laboratories, Los Angeles County Public Health Lab microbial pathogen submission group | Hemarajata; P. and Green, N. |
| EPI_ISL_2009392 | Lurie Children's Hospital of Chicago | Northwestern University - Ozer Lab | Egon A. Ozer; Judd F. Hultquist; Lacy M. Simons; Larry K. Kociolek; Michael G. Ison; Ramon Lorenzo-Redondo; Taylor J. Dean; William J. Muller; Xiaotian; Zheng |
| EPI_ISL_1716896, EPI_ISL_1716899 | Martin Luther King Jr. Community Hospital | Los Angeles County PHL | P. Hemarajata et al. |
| EPI_ISL_1677791, EPI_ISL_1700688, EPI_ISL_1700689, EPI_ISL_1700690, EPI_ISL_1700691, EPI_ISL_1700692, EPI_ISL_1700693, EPI_ISL_1700694, EPI_ISL_2001029, EPI_ISL_2001032, EPI_ISL_2001034, EPI_ISL_2001036, EPI_ISL_2001039 | | | |
| see above | Monterey County Public Health Laboratory | Monterey County Public Health Laboratory | Monterey County Public Health Laboratory |
| EPI_ISL_2009353 | Northwestern Memorial Hospital | Northwestern University - Ozer Lab | Chad J. Achenbach; Chao Qi; Egon A. Ozer; Judd F. Hultquist; Lacy M. Simons; Lawrence J. Jennings; Michael G. Ison; Ramon Lorenzo-Redondo; Taylor J. Dean |
| EPI_ISL_2080756 | OHSU Lab Services Molecular Microbiology Lab | Oregon SARS-CoV-2 Genome Sequencing Center | Alec J. Hirsch; Andrew C. Adey; Benjamin N. Bimber; Brendan L. O'Connell; Brian J. O'Roak; Cierra LaBlanc; Daniel N. Streblow; Donna Hansel; Guang Fan; Kayla Carter; Ruth V. Nichols; Sally Grindstaff; Sonia Acharya; William B. Messer; Xuan Qin |
| EPI_ISL_2103197 | Orange County Public Health Lab | Chan-Zuckerberg Biohub | CZB Cliahub Consortium |
| EPI_ISL_1824248 | Pathology and Laboratory Medicine Institute, Cleveland Clinic, Ohio, USA | Pathology and Laboratory Medicine Institute, Cleveland Clinic, Ohio, USA | Daniel H. Farkas; Daniel Rhoads; David Bosler; David Plunkett; Gary Procop; Jay Brock; Jessica Spildener; Joy Nakitandwe; Kristen McDonnell; Yu-Wei Cheng; Zheng Jin Tu |
| EPI_ISL_1669340 | Providence Little Company of Mary Medical Center | Los Angeles County PHL | P. Hemarajata et al. |
| EPI_ISL_1525956, EPI_ISL_1526012, EPI_ISL_1526029, EPI_ISL_1526059, EPI_ISL_1552703, EPI_ISL_1552752, EPI_ISL_1552761, EPI_ISL_1552792, EPI_ISL_1552793, EPI_ISL_1552794, EPI_ISL_1552855, EPI_ISL_1559458, EPI_ISL_1559476, EPI_ISL_1559481, EPI_ISL_1559514, EPI_ISL_1559560, EPI_ISL_1581859, EPI_ISL_1581889, EPI_ISL_1581943, EPI_ISL_1581945, EPI_ISL_1581947, EPI_ISL_1581958, EPI_ISL_1581978, EPI_ISL_1582001, EPI_ISL_1582032, EPI_ISL_1582080, EPI_ISL_1622104, EPI_ISL_1648141, EPI_ISL_1648142, EPI_ISL_1648143, EPI_ISL_1648154, EPI_ISL_1648155, EPI_ISL_1648156, EPI_ISL_1648157, EPI_ISL_1648160, EPI_ISL_1648166, EPI_ISL_1648172, EPI_ISL_1648175, EPI_ISL_1648180, EPI_ISL_1648182, EPI_ISL_1648185, EPI_ISL_1648186, EPI_ISL_1648188, EPI_ISL_1648190, EPI_ISL_1648191, EPI_ISL_1648192, EPI_ISL_1648196, EPI_ISL_1648197, EPI_ISL_1648198, EPI_ISL_1648199, EPI_ISL_1648201, EPI_ISL_1648203, EPI_ISL_1648205, EPI_ISL_1648210, EPI_ISL_1648214, EPI_ISL_1648217, EPI_ISL_1648221, EPI_ISL_1648222, EPI_ISL_1648224, EPI_ISL_1648228, EPI_ISL_1648234, EPI_ISL_1648235, EPI_ISL_1648239, EPI_ISL_1648242, EPI_ISL_1648243, EPI_ISL_1648247, EPI_ISL_1648254, EPI_ISL_1648258, EPI_ISL_1648261, EPI_ISL_1648262, EPI_ISL_1648266, EPI_ISL_1648268, EPI_ISL_1648269, EPI_ISL_1648270, EPI_ISL_1648271, EPI_ISL_1648273, EPI_ISL_1648275, EPI_ISL_1648277, EPI_ISL_1648278, EPI_ISL_1648281, EPI_ISL_1648282, EPI_ISL_1648283, EPI_ISL_1648290, EPI_ISL_1648291, EPI_ISL_1648292, EPI_ISL_1648298, EPI_ISL_1648299, EPI_ISL_1648306, EPI_ISL_1648311, EPI_ISL_1648312, EPI_ISL_1648319, EPI_ISL_1648321, EPI_ISL_1648323, EPI_ISL_1648324, EPI_ISL_1648326, EPI_ISL_1648330, EPI_ISL_1648331, EPI_ISL_1648333, EPI_ISL_1648334, EPI_ISL_1648335, EPI_ISL_1648337, EPI_ISL_1648343, EPI_ISL_1648345, EPI_ISL_1648346, EPI_ISL_1648347, EPI_ISL_1648349, EPI_ISL_1648351, EPI_ISL_1648352, EPI_ISL_1648355, EPI_ISL_1648364, EPI_ISL_1648366, EPI_ISL_1648369, EPI_ISL_1648376, EPI_ISL_1648380, EPI_ISL_1648381, EPI_ISL_1648385, EPI_ISL_1648391, EPI_ISL_1648393, EPI_ISL_1648394, EPI_ISL_1648395, EPI_ISL_1648398, EPI_ISL_1648400, EPI_ISL_1648403, EPI_ISL_1648406, EPI_ISL_1648407, EPI_ISL_1648416, EPI_ISL_1648419, EPI_ISL_1648420, EPI_ISL_1648421, EPI_ISL_1648422, EPI_ISL_1648423, EPI_ISL_1648424, EPI_ISL_1648427, EPI_ISL_1648431, EPI_ISL_1648432, EPI_ISL_1648435, EPI_ISL_1648436, EPI_ISL_1648438, EPI_ISL_1648441, EPI_ISL_1648444, EPI_ISL_1648447, EPI_ISL_1648448, EPI_ISL_1648449, EPI_ISL_1648452, EPI_ISL_1648453, EPI_ISL_1648454, EPI_ISL_1648618, EPI_ISL_1648629, EPI_ISL_1648631, EPI_ISL_1648639, EPI_ISL_1648643, EPI_ISL_1648645, EPI_ISL_1648654, EPI_ISL_1648666, EPI_ISL_1648667, EPI_ISL_1648670, EPI_ISL_1648679, EPI_ISL_1648690, EPI_ISL_1648693, EPI_ISL_1648699, EPI_ISL_1648702, EPI_ISL_1648714, EPI_ISL_1648723, EPI_ISL_1648725, EPI_ISL_1648728, EPI_ISL_1648742, EPI_ISL_1648754, EPI_ISL_1648766, EPI_ISL_1648770, EPI_ISL_1648833, EPI_ISL_1648893, EPI_ISL_1668548, EPI_ISL_1668614, EPI_ISL_1668634, EPI_ISL_1668635, EPI_ISL_1668641, EPI_ISL_1668657, EPI_ISL_1668659, EPI_ISL_1668661, EPI_ISL_1668667, EPI_ISL_1668668, EPI_ISL_1668669, EPI_ISL_1668681, EPI_ISL_1668687, EPI_ISL_1668691, EPI_ISL_1668694, EPI_ISL_1668713, EPI_ISL_1668728, EPI_ISL_1668729, EPI_ISL_1668733, EPI_ISL_1668737, EPI_ISL_1668753, EPI_ISL_1668760, EPI_ISL_1668776, EPI_ISL_1668777, EPI_ISL_1668781, EPI_ISL_1668789, EPI_ISL_1668790, EPI_ISL_1668798, EPI_ISL_1668803, EPI_ISL_1668843, EPI_ISL_1668845, EPI_ISL_1668871, EPI_ISL_1668889, EPI_ISL_1668944, EPI_ISL_1668991, EPI_ISL_1669000, EPI_ISL_1669004, EPI_ISL_1669009, EPI_ISL_1669021, EPI_ISL_1669033, EPI_ISL_1669046, EPI_ISL_1669061, EPI_ISL_1669064, EPI_ISL_1669074, EPI_ISL_1669080, EPI_ISL_1669092, EPI_ISL_1669102, EPI_ISL_1669111, EPI_ISL_1669129, EPI_ISL_1669148, EPI_ISL_1669190, EPI_ISL_1669204, EPI_ISL_1669224, EPI_ISL_1669260, EPI_ISL_1669264, EPI_ISL_1669304, EPI_ISL_1691398, EPI_ISL_1691472, EPI_ISL_1691476, EPI_ISL_1691482, EPI_ISL_1691488, EPI_ISL_1694695, EPI_ISL_1695040, EPI_ISL_1695057, EPI_ISL_1695073, EPI_ISL_1695076, EPI_ISL_1695077, EPI_ISL_1695078, EPI_ISL_1695080, EPI_ISL_1695082, EPI_ISL_1695084, EPI_ISL_1695085, EPI_ISL_1695087, EPI_ISL_1695091, EPI_ISL_1695674, EPI_ISL_1695736, EPI_ISL_1702691, EPI_ISL_1738064, EPI_ISL_1738069, EPI_ISL_1738087, EPI_ISL_1738101, EPI_ISL_1738123, EPI_ISL_1738178, EPI_ISL_1738195, EPI_ISL_1738204, EPI_ISL_1738207, EPI_ISL_1738210, EPI_ISL_1738222, EPI_ISL_1738231, EPI_ISL_1738238, EPI_ISL_1738246, EPI_ISL_1738250, EPI_ISL_1738263, EPI_ISL_1738280, EPI_ISL_1738292, EPI_ISL_1738296, EPI_ISL_1738329, EPI_ISL_1738383, EPI_ISL_1738401, EPI_ISL_1738467, EPI_ISL_1738474, EPI_ISL_1738477, EPI_ISL_1738481, EPI_ISL_1738482, EPI_ISL_1738487, EPI_ISL_1738491, EPI_ISL_1738492, EPI_ISL_1738494, EPI_ISL_1738497, EPI_ISL_1738506, EPI_ISL_1738507, EPI_ISL_1738508, EPI_ISL_1738511, EPI_ISL_1738515, EPI_ISL_1738518, EPI_ISL_1738523, EPI_ISL_1738524, EPI_ISL_1738526, EPI_ISL_1738531, EPI_ISL_1738532, EPI_ISL_1738536, EPI_ISL_1738544, EPI_ISL_1738547, EPI_ISL_1738550, EPI_ISL_1738562, EPI_ISL_1738565, EPI_ISL_1738583, EPI_ISL_1738597, EPI_ISL_1738598, EPI_ISL_1738599, EPI_ISL_1738600, EPI_ISL_1738606, EPI_ISL_1738620, EPI_ISL_1738622, EPI_ISL_1738630, EPI_ISL_1738641, EPI_ISL_1738652, EPI_ISL_1738662, EPI_ISL_1738663, EPI_ISL_1738684, EPI_ISL_1738688, EPI_ISL_1738690, EPI_ISL_1738713, EPI_ISL_1738721, EPI_ISL_1738736, EPI_ISL_1738739, EPI_ISL_1738747, EPI_ISL_1753033, EPI_ISL_1753034, EPI_ISL_1753035, EPI_ISL_1753038, EPI_ISL_1753041, EPI_ISL_1753045, EPI_ISL_1753062, EPI_ISL_1753063, EPI_ISL_1753068, EPI_ISL_1753070, EPI_ISL_1753084, EPI_ISL_1753086, EPI_ISL_1753091, EPI_ISL_1753105, EPI_ISL_1753108, EPI_ISL_1753111, EPI_ISL_1753113, EPI_ISL_1753126, EPI_ISL_1753139, EPI_ISL_1753143, EPI_ISL_1753145, EPI_ISL_1753149, EPI_ISL_1753154, EPI_ISL_1753156, EPI_ISL_1753161, EPI_ISL_1753163, EPI_ISL_1753165, EPI_ISL_1753170, EPI_ISL_1753172, EPI_ISL_1753175, EPI_ISL_1753177, EPI_ISL_1753178, EPI_ISL_1753179, EPI_ISL_1753182, EPI_ISL_1753183, EPI_ISL_1753184, EPI_ISL_1753185, EPI_ISL_1753186, EPI_ISL_1753187, EPI_ISL_1753188, EPI_ISL_1753190, EPI_ISL_1753193, EPI_ISL_1753196, EPI_ISL_1753197, EPI_ISL_1753200, EPI_ISL_1753203, EPI_ISL_1753205, EPI_ISL_1753207, EPI_ISL_1753211, EPI_ISL_1753212, EPI_ISL_1753213, EPI_ISL_1753219, EPI_ISL_1753220, EPI_ISL_1753221, EPI_ISL_1753222, EPI_ISL_1753223, EPI_ISL_1753226, EPI_ISL_1753228, EPI_ISL_1753233, EPI_ISL_1753238, EPI_ISL_1753240, EPI_ISL_1753242, EPI_ISL_1753243, EPI_ISL_1753248, EPI_ISL_1753250, EPI_ISL_1753252, EPI_ISL_1753255, EPI_ISL_1753257, EPI_ISL_1753260, EPI_ISL_1753261, EPI_ISL_1753262, EPI_ISL_1753264, EPI_ISL_1753265, EPI_ISL_1753266, EPI_ISL_1753267, EPI_ISL_1753269, EPI_ISL_1753274, EPI_ISL_1753276, EPI_ISL_1753279, EPI_ISL_1753280, EPI_ISL_1753284, EPI_ISL_1753287, EPI_ISL_1753300, EPI_ISL_1753306, EPI_ISL_1753307, EPI_ISL_1753316, EPI_ISL_1753326, EPI_ISL_1753332, EPI_ISL_1753344, EPI_ISL_1753347, EPI_ISL_1753349, EPI_ISL_1753352, EPI_ISL_1753356, EPI_ISL_1753361, EPI_ISL_1753365, EPI_ISL_1753372, EPI_ISL_1753375, EPI_ISL_1753379, EPI_ISL_1753380, EPI_ISL_1753382, EPI_ISL_1753383, EPI_ISL_1753389, EPI_ISL_1753393, EPI_ISL_1753396, EPI_ISL_1753404, EPI_ISL_1753416, EPI_ISL_1753621, EPI_ISL_1753645, EPI_ISL_1753646, EPI_ISL_1753647, EPI_ISL_1753653, EPI_ISL_1753655, EPI_ISL_1753656, EPI_ISL_1753657, EPI_ISL_1761871, EPI_ISL_1761872, EPI_ISL_1761873, EPI_ISL_1761877, EPI_ISL_1761879, EPI_ISL_1761880, EPI_ISL_1761881, EPI_ISL_1761885, EPI_ISL_1761891, EPI_ISL_1761902, EPI_ISL_1761904, EPI_ISL_1761909, EPI_ISL_1761911, EPI_ISL_1761912, EPI_ISL_1761916, EPI_ISL_1761917, EPI_ISL_1761930, EPI_ISL_1761934, EPI_ISL_1761935, EPI_ISL_1761938, EPI_ISL_1761943, EPI_ISL_1761945, EPI_ISL_1761948, EPI_ISL_1761950, EPI_ISL_1761956, EPI_ISL_1761958, EPI_ISL_1761959, EPI_ISL_1761960, EPI_ISL_1761964, EPI_ISL_1761966, EPI_ISL_1761969, EPI_ISL_1761973, EPI_ISL_1761979, EPI_ISL_1761982, EPI_ISL_1761987, EPI_ISL_1761991, EPI_ISL_1761998, EPI_ISL_1762007, EPI_ISL_1762009, EPI_ISL_1762011, EPI_ISL_1762022, EPI_ISL_1762025, EPI_ISL_1762026, EPI_ISL_1797926, EPI_ISL_1798036, EPI_ISL_1798071, EPI_ISL_1798086, EPI_ISL_1798096, EPI_ISL_1798104, EPI_ISL_1798118, EPI_ISL_1798129, EPI_ISL_1798151, EPI_ISL_1798161, EPI_ISL_1798183, EPI_ISL_1804812, EPI_ISL_1804815, EPI_ISL_1804816, EPI_ISL_1804819, EPI_ISL_1818064, EPI_ISL_1818069, EPI_ISL_1818309, EPI_ISL_1818410, EPI_ISL_1834201, EPI_ISL_1834227, EPI_ISL_1834293, EPI_ISL_1834301, EPI_ISL_1834310, EPI_ISL_1834311, EPI_ISL_1834402, EPI_ISL_1840260, EPI_ISL_1840275, EPI_ISL_1840279, EPI_ISL_1840284, EPI_ISL_1840286, EPI_ISL_1840294, EPI_ISL_1840304, EPI_ISL_1840315, EPI_ISL_1840336, EPI_ISL_1840339, EPI_ISL_1840346, EPI_ISL_1840375, EPI_ISL_1840380, EPI_ISL_1840385, EPI_ISL_1840398, EPI_ISL_1840406, EPI_ISL_1900132, EPI_ISL_1900147, EPI_ISL_1900442, EPI_ISL_1900445, EPI_ISL_1900449, EPI_ISL_1900451, EPI_ISL_1905848, EPI_ISL_1905852, EPI_ISL_1905859, EPI_ISL_1905866, EPI_ISL_1905869, EPI_ISL_1905901, EPI_ISL_1905905, EPI_ISL_1905925, EPI_ISL_1905927, EPI_ISL_1906009, EPI_ISL_1906030, EPI_ISL_1906036, EPI_ISL_1906074, EPI_ISL_1906102, EPI_ISL_1906126, EPI_ISL_1906143, EPI_ISL_1906145, EPI_ISL_1906168, EPI_ISL_1906301, EPI_ISL_1906332, EPI_ISL_1906450, EPI_ISL_1906571, EPI_ISL_1906584, EPI_ISL_1906586, EPI_ISL_1906589, EPI_ISL_1906591, EPI_ISL_1906613, EPI_ISL_1906617, EPI_ISL_1906621, EPI_ISL_1906625, EPI_ISL_1906641, EPI_ISL_1906650, EPI_ISL_1906654, EPI_ISL_1906663, EPI_ISL_1906672, EPI_ISL_1906684, EPI_ISL_1906736, EPI_ISL_1906742, EPI_ISL_1906744, EPI_ISL_1906749, EPI_ISL_1906751, EPI_ISL_1906753, EPI_ISL_1906757, EPI_ISL_1906763, EPI_ISL_1906768, EPI_ISL_1906772, EPI_ISL_1906778, EPI_ISL_1906790, EPI_ISL_1906804, EPI_ISL_1906808, EPI_ISL_1906819, EPI_ISL_1906825, EPI_ISL_1906830, EPI_ISL_1906846, EPI_ISL_1906847, EPI_ISL_1906848, EPI_ISL_1906856, EPI_ISL_1906863, EPI_ISL_1906864, EPI_ISL_1906867, EPI_ISL_1906871, EPI_ISL_1906874, EPI_ISL_1906882, EPI_ISL_1924317, EPI_ISL_1924347, EPI_ISL_1924356, EPI_ISL_1924651, EPI_ISL_1924652, EPI_ISL_1924653, EPI_ISL_1924660, EPI_ISL_1924661, EPI_ISL_1924663, EPI_ISL_1924669, EPI_ISL_1924671, EPI_ISL_1924672, EPI_ISL_1924677, EPI_ISL_1924680, EPI_ISL_1924686, EPI_ISL_1924690, EPI_ISL_1924694, EPI_ISL_1924696, EPI_ISL_1924699, EPI_ISL_1924704, EPI_ISL_1924709, EPI_ISL_1924714, EPI_ISL_1924716, EPI_ISL_1924719, EPI_ISL_1924724, EPI_ISL_1924733, EPI_ISL_1924741, EPI_ISL_1924742, EPI_ISL_1924744, EPI_ISL_1924746, EPI_ISL_1924749, EPI_ISL_1924751, EPI_ISL_1924753, EPI_ISL_1924757, EPI_ISL_1924760, EPI_ISL_1924761, EPI_ISL_1924762, EPI_ISL_1924767, EPI_ISL_1924769, EPI_ISL_1924778, EPI_ISL_1924785, EPI_ISL_1924786, EPI_ISL_1924787, EPI_ISL_1924788, EPI_ISL_1924792, EPI_ISL_1924795, EPI_ISL_1924810, EPI_ISL_1924819, EPI_ISL_1924821, EPI_ISL_1924835, EPI_ISL_1924847, EPI_ISL_1924866, EPI_ISL_1924867, EPI_ISL_1924877, EPI_ISL_1924881, EPI_ISL_1924887, EPI_ISL_1924888, EPI_ISL_1924900, EPI_ISL_1924912, EPI_ISL_1935558, EPI_ISL_1935563, EPI_ISL_1935571, EPI_ISL_1935576, EPI_ISL_1935582, EPI_ISL_1942184, EPI_ISL_1942187, EPI_ISL_1942189, EPI_ISL_1942190, EPI_ISL_1942199, EPI_ISL_1942201, EPI_ISL_1942203, EPI_ISL_1942205, EPI_ISL_1942206, EPI_ISL_1942208, EPI_ISL_1942209, EPI_ISL_1942210, EPI_ISL_1942214, EPI_ISL_1989696, EPI_ISL_1989710, EPI_ISL_1989720, EPI_ISL_1989721, EPI_ISL_1989726, EPI_ISL_1989727, EPI_ISL_1989729, EPI_ISL_1989738, EPI_ISL_1989744, EPI_ISL_1989758, EPI_ISL_1989773, EPI_ISL_1989776, EPI_ISL_1989782, EPI_ISL_1989787, EPI_ISL_1989790, EPI_ISL_1989793, EPI_ISL_1989796, EPI_ISL_1989805, EPI_ISL_1989806, EPI_ISL_1989807, EPI_ISL_1989814, EPI_ISL_1989847, EPI_ISL_1989853, EPI_ISL_1989868, EPI_ISL_1989881, EPI_ISL_1989975, EPI_ISL_1989985, EPI_ISL_1989991, EPI_ISL_1990052, EPI_ISL_1990160, EPI_ISL_1990166, EPI_ISL_1990180, EPI_ISL_1990181, EPI_ISL_1990216, EPI_ISL_1990258, EPI_ISL_1990280, EPI_ISL_1990281, EPI_ISL_1990290, EPI_ISL_1990291, EPI_ISL_1990294, EPI_ISL_1990295, EPI_ISL_1990298, EPI_ISL_1990311, EPI_ISL_1990322, EPI_ISL_1990326, EPI_ISL_1990328, EPI_ISL_1990330, EPI_ISL_1990333, EPI_ISL_1990334, EPI_ISL_1990350, EPI_ISL_1990368, EPI_ISL_1990376, EPI_ISL_1990394, EPI_ISL_1990401, EPI_ISL_1990406, EPI_ISL_2010890, EPI_ISL_2010895, EPI_ISL_2010903, EPI_ISL_2010906, EPI_ISL_2010909, EPI_ISL_2010912, EPI_ISL_2010915, EPI_ISL_2010916, EPI_ISL_2010918, EPI_ISL_2010919, EPI_ISL_2010920, EPI_ISL_2010922, EPI_ISL_2010924, EPI_ISL_2010925, EPI_ISL_2010926, EPI_ISL_2010927, EPI_ISL_2010928, EPI_ISL_2010933, EPI_ISL_2010937, EPI_ISL_2010938, EPI_ISL_2010940, EPI_ISL_2010942, EPI_ISL_2010943, EPI_ISL_2010944, EPI_ISL_2010945, EPI_ISL_2010946, EPI_ISL_2010948, EPI_ISL_2010949, EPI_ISL_2010950, EPI_ISL_2010953, EPI_ISL_2010955, EPI_ISL_2010958, EPI_ISL_2010960, EPI_ISL_2010964, EPI_ISL_2010965, EPI_ISL_2010967, EPI_ISL_2010968, EPI_ISL_2010974, EPI_ISL_2010976, EPI_ISL_2010978, EPI_ISL_2010979, EPI_ISL_2010980, EPI_ISL_2010981, EPI_ISL_2010982, EPI_ISL_2010985, EPI_ISL_2010986, EPI_ISL_2010992, EPI_ISL_2010994, EPI_ISL_2010996, EPI_ISL_2010997, EPI_ISL_2010998, EPI_ISL_2011001, EPI_ISL_2011002, EPI_ISL_2011005, EPI_ISL_2011007, EPI_ISL_2011008, EPI_ISL_2011009, EPI_ISL_2011011, EPI_ISL_2011012, EPI_ISL_2011014, EPI_ISL_2011019, EPI_ISL_2011020, EPI_ISL_2011023, EPI_ISL_2011171, EPI_ISL_2011221, EPI_ISL_2034722, EPI_ISL_2034729, EPI_ISL_2034736, EPI_ISL_2034762, EPI_ISL_2034766, EPI_ISL_2034769, EPI_ISL_2034776, EPI_ISL_2034779, EPI_ISL_2034782, EPI_ISL_2035302, EPI_ISL_2035303, EPI_ISL_2035304, EPI_ISL_2035306, EPI_ISL_2035307, EPI_ISL_2035309, EPI_ISL_2035310, EPI_ISL_2035311, EPI_ISL_2035312, EPI_ISL_2035313, EPI_ISL_2035314, EPI_ISL_2035315, EPI_ISL_2035316, EPI_ISL_2035317, EPI_ISL_2035318, EPI_ISL_2035319, EPI_ISL_2035320, EPI_ISL_2035322, EPI_ISL_2046840, EPI_ISL_2046844, EPI_ISL_2046884, EPI_ISL_2046891, EPI_ISL_2046976, EPI_ISL_2047006, EPI_ISL_2047022, EPI_ISL_2047026, EPI_ISL_2047028, EPI_ISL_2047100, EPI_ISL_2047151, EPI_ISL_2047154, EPI_ISL_2047180, EPI_ISL_2047199, EPI_ISL_2047200, EPI_ISL_2047249, EPI_ISL_2090474, EPI_ISL_2090477, EPI_ISL_2090478, EPI_ISL_2090482, EPI_ISL_2090485, EPI_ISL_2090516, EPI_ISL_2090529, EPI_ISL_2090531, EPI_ISL_2090532, EPI_ISL_2090539, EPI_ISL_2090552, EPI_ISL_2090560, EPI_ISL_2090566, EPI_ISL_2090599, EPI_ISL_2098355, EPI_ISL_2098401, EPI_ISL_2132575, EPI_ISL_2132581, EPI_ISL_2132582, EPI_ISL_2132584, EPI_ISL_2132585, EPI_ISL_2132619, EPI_ISL_2133176, EPI_ISL_2133281, EPI_ISL_2133318, EPI_ISL_2133319, EPI_ISL_2133326, EPI_ISL_2133392, EPI_ISL_2133393, EPI_ISL_2133404, EPI_ISL_2133414, EPI_ISL_2133416, EPI_ISL_2133417, EPI_ISL_2133421, EPI_ISL_2133423, EPI_ISL_2133426, EPI_ISL_2133428, EPI_ISL_2133429, EPI_ISL_2133430, EPI_ISL_2133431, EPI_ISL_2133433, EPI_ISL_2133437, EPI_ISL_2133445, EPI_ISL_2133447, EPI_ISL_2133449, EPI_ISL_2133453, EPI_ISL_2133461, EPI_ISL_2133463, EPI_ISL_2133464, EPI_ISL_2133467, EPI_ISL_2133471, EPI_ISL_2133472, EPI_ISL_2133475, EPI_ISL_2133476, EPI_ISL_2133477, EPI_ISL_2133489, EPI_ISL_2133490, EPI_ISL_2133496, EPI_ISL_2133497, EPI_ISL_2133498, EPI_ISL_2133504, EPI_ISL_2133509, EPI_ISL_2133511, EPI_ISL_2133515, EPI_ISL_2133527, EPI_ISL_2133528, EPI_ISL_2133530, EPI_ISL_2133533, EPI_ISL_2133535, EPI_ISL_2133540, EPI_ISL_2133543, EPI_ISL_2133544, EPI_ISL_2133547, EPI_ISL_2133549, EPI_ISL_2133551, EPI_ISL_2133553, EPI_ISL_2133555, EPI_ISL_2133556, EPI_ISL_2133557, EPI_ISL_2133560, EPI_ISL_2133568, EPI_ISL_2133569, EPI_ISL_2133571, EPI_ISL_2133573, EPI_ISL_2133583, EPI_ISL_2133587, EPI_ISL_2133588, EPI_ISL_2133589, EPI_ISL_2133593, EPI_ISL_2133601, EPI_ISL_2133603, EPI_ISL_2133604, EPI_ISL_2133605, EPI_ISL_2133643, EPI_ISL_2133676, EPI_ISL_2133679, EPI_ISL_2133716, EPI_ISL_2133725, EPI_ISL_2133743, EPI_ISL_2133753, EPI_ISL_2133772, EPI_ISL_2133776, EPI_ISL_2133778, EPI_ISL_2133799, EPI_ISL_2133818, EPI_ISL_2133836, EPI_ISL_2140760, EPI_ISL_2140761, EPI_ISL_2140762, EPI_ISL_2140763, EPI_ISL_2140764, EPI_ISL_2140767, EPI_ISL_2140769, EPI_ISL_2140770, EPI_ISL_2140771, EPI_ISL_2140772, EPI_ISL_2140773, EPI_ISL_2140776, EPI_ISL_2140792, EPI_ISL_2140793, EPI_ISL_2142880, EPI_ISL_2142881, EPI_ISL_2142884, EPI_ISL_2142886, EPI_ISL_2142890, EPI_ISL_2142893, EPI_ISL_2142896, EPI_ISL_2142902, EPI_ISL_2142914, EPI_ISL_2142920, EPI_ISL_2142921, EPI_ISL_2142929, EPI_ISL_2142940, EPI_ISL_2142942, EPI_ISL_2142946, EPI_ISL_2142949, EPI_ISL_2142956, EPI_ISL_2142957, EPI_ISL_2142964, EPI_ISL_2142967, EPI_ISL_2142968, EPI_ISL_2142969, EPI_ISL_2142970, EPI_ISL_2142974, EPI_ISL_2142975, EPI_ISL_2142981, EPI_ISL_2142989, EPI_ISL_2143000, EPI_ISL_2143002, EPI_ISL_2143005, EPI_ISL_2143007, EPI_ISL_2143009, EPI_ISL_2143010, EPI_ISL_2143021, EPI_ISL_2143022, EPI_ISL_2143115, EPI_ISL_2143124, EPI_ISL_2143128, EPI_ISL_2143173, EPI_ISL_2143174, EPI_ISL_2143175, EPI_ISL_2143177, EPI_ISL_2143178, EPI_ISL_2143179, EPI_ISL_2143181, EPI_ISL_2143182, EPI_ISL_2143184, EPI_ISL_2143186, EPI_ISL_2143187, EPI_ISL_2143189, EPI_ISL_2143193, EPI_ISL_2143195, EPI_ISL_2143197, EPI_ISL_2143198, EPI_ISL_2143201, EPI_ISL_2143203, EPI_ISL_2143204, EPI_ISL_2143205, EPI_ISL_2143206, EPI_ISL_2143207, EPI_ISL_2143208, EPI_ISL_2143214, EPI_ISL_2143217, EPI_ISL_2143218, EPI_ISL_2143220, EPI_ISL_2143223, EPI_ISL_2143226, EPI_ISL_2143227, EPI_ISL_2143229, EPI_ISL_2143231, EPI_ISL_2143233, EPI_ISL_2143235, EPI_ISL_2143236, EPI_ISL_2143238, EPI_ISL_2143240, EPI_ISL_2143242, EPI_ISL_2143244, EPI_ISL_2143245, EPI_ISL_2143246, EPI_ISL_2143249, EPI_ISL_2143255, EPI_ISL_2143258, EPI_ISL_2143261, EPI_ISL_2143262, EPI_ISL_2143264, EPI_ISL_2143266, EPI_ISL_2143268, EPI_ISL_2143270, EPI_ISL_2143271, EPI_ISL_2143273, EPI_ISL_2143274, EPI_ISL_2143275, EPI_ISL_2143276, EPI_ISL_2143279, EPI_ISL_2143283, EPI_ISL_2143286, EPI_ISL_2143287, EPI_ISL_2143288, EPI_ISL_2143289, EPI_ISL_2143290, EPI_ISL_2143291, EPI_ISL_2143295, EPI_ISL_2143296, EPI_ISL_2143298, EPI_ISL_2143301, EPI_ISL_2143315, EPI_ISL_2143326, EPI_ISL_2143339, EPI_ISL_2143348, EPI_ISL_2143362, EPI_ISL_2143369, EPI_ISL_2143371, EPI_ISL_2143379, EPI_ISL_2143389, EPI_ISL_2143412, EPI_ISL_2143414, EPI_ISL_2143436, EPI_ISL_2143455, EPI_ISL_2143461, EPI_ISL_2143481, EPI_ISL_2143509, EPI_ISL_2143522, EPI_ISL_2143699, EPI_ISL_2143760, EPI_ISL_2143789, EPI_ISL_2143848, EPI_ISL_2143899, EPI_ISL_2143908, EPI_ISL_2143936, EPI_ISL_2144009, EPI_ISL_2179931, EPI_ISL_2180479, EPI_ISL_2180501, EPI_ISL_2180503, EPI_ISL_2180508, EPI_ISL_2180513, EPI_ISL_2180520, EPI_ISL_2180527, EPI_ISL_2187232, EPI_ISL_2187236, EPI_ISL_2187237, EPI_ISL_2187239, EPI_ISL_2187240, EPI_ISL_2187241, EPI_ISL_2187258, EPI_ISL_2187259, EPI_ISL_2187262, EPI_ISL_2187264, EPI_ISL_2187266, EPI_ISL_2187267, EPI_ISL_2187268, EPI_ISL_2187269, EPI_ISL_2187270, EPI_ISL_2187272, EPI_ISL_2187273, EPI_ISL_2187274, EPI_ISL_2187275, EPI_ISL_2187278, EPI_ISL_2187281, EPI_ISL_2187282, EPI_ISL_2187283, EPI_ISL_2187284, EPI_ISL_2187285, EPI_ISL_2187286, EPI_ISL_2187287, EPI_ISL_2187288, EPI_ISL_2187300, EPI_ISL_2187302, EPI_ISL_2187307, EPI_ISL_2187314 | | | |
| see above | Quest Diagnostics Incorporated | Centers for Disease Control and Prevention Division of Viral Diseases, Pathogen Discovery | A. Gerasimova; A. Perez; Adrian Paskey; B. Anderson; Benjamin Rambo-Martin; Christopher Gulvick; Clinton R. Paden; Dakota Howard; Darlene Wagner; Dhwani Batra; Duncan MacCannell; F. Lacbawan; I. A. Shlyakhter; Jason Caravas; K.E. Livingston; Kara Moser; L.E. Bernstein; M. Hua; Matthew Schmerer; P. Tanpaiboon; Peter W. Cook; R. M. Kagan; R. Owen; R. V. Rolando; S. H. Rosenthal; Scott Sammons; Shatavia Morrison; Y. Liu; Yvette Unoarumhi |
| EPI_ISL_1794768 | Rady Children's Hospital - San Diego | Andersen lab at Scripps Research | Christina Clarke; David Dimmock; Denise Malicki; Kathryn Bouic; Linda Luo; SEARCH Alliance San Diego with Nanda Radamchar; Teresa Mueller |
| EPI_ISL_2253158, EPI_ISL_2253159, EPI_ISL_2253164, EPI_ISL_2253165, EPI_ISL_2253202, EPI_ISL_2254286, EPI_ISL_2254287, EPI_ISL_2254295, EPI_ISL_2254320, EPI_ISL_2254321, EPI_ISL_2254341, EPI_ISL_2254377, EPI_ISL_2254400, EPI_ISL_2254408, EPI_ISL_2254422, EPI_ISL_2254425, EPI_ISL_2254432, EPI_ISL_2254464, EPI_ISL_2313477, EPI_ISL_2313488, EPI_ISL_2313523, EPI_ISL_2313525, EPI_ISL_2313527, EPI_ISL_2313528, EPI_ISL_2313564, EPI_ISL_2313657 | | | |
| see above | Respiratory Viruses Branch, Centers for Disease Control and Prevention | Respiratory Viruses Branch, Centers for Disease Control and Prevention | Anderson, B.; Batra, D.; Bernstein, L.; Bixby, C.; C.R.; Campbell, P.; Caravas, J.; Case, R.; Clark, C.; Cook; Fierro, J.; Gao, H.; Gao, J.; Gerasimova, A.; Ghorpade, V.; Goswami, C.; Grimwood, R.; Gulvick, C.; Hager, R.; Hardison, M.; Houdeshell, H.; Howard, D.; Hua, M.; I.A.; Kagan; Kvalvaag, O.; Lacbawan, F.; Li, M.; Liu, Y.; Livingston, K.; MacCannell, D.; Meng, Y.; Morrison, S.; Moser, K.; Nall, D.; Ng, D.; Owen, R.; P.W.; Paden; Paskey, A.; Perez, A.; R.M.; R.V.; Rambo-Martin, B.; Rolando; Rosenthal; S.H.; Sammons, S.; Sanders, E.; Sapra, B.; Schmerer, M.; Schultz, J.; Shlyakhter; Tanpaiboon, P.; Tsai, B.; Unoarumhi, Y.; Vest, A.; Wagner, D.; Wang, Y.; Westlund, S.; Xie, J. |
| EPI_ISL_1791429, EPI_ISL_1791430, EPI_ISL_1791431, EPI_ISL_1791432, EPI_ISL_1791433, EPI_ISL_1791434, EPI_ISL_1791435, EPI_ISL_1791436, EPI_ISL_1791437, EPI_ISL_1791438, EPI_ISL_1791439, EPI_ISL_1791440, EPI_ISL_1791441, EPI_ISL_1791442, EPI_ISL_1791443, EPI_ISL_1791444, EPI_ISL_1791445, EPI_ISL_1791446, EPI_ISL_1791447, EPI_ISL_1791448, EPI_ISL_1791449, EPI_ISL_1791450, EPI_ISL_1791451, EPI_ISL_1791452, EPI_ISL_1791453, EPI_ISL_1791454, EPI_ISL_1791455, EPI_ISL_1791456, EPI_ISL_1794616, EPI_ISL_1794617, EPI_ISL_1794622, EPI_ISL_1794623, EPI_ISL_1794624, EPI_ISL_1794625, EPI_ISL_1794626, EPI_ISL_1794630, EPI_ISL_1794631, EPI_ISL_1794632, EPI_ISL_1794633, EPI_ISL_1794637, EPI_ISL_1794640, EPI_ISL_1794642, EPI_ISL_1794644, EPI_ISL_1794646, EPI_ISL_1794647, EPI_ISL_1794651, EPI_ISL_1794652, EPI_ISL_1794937, EPI_ISL_1794938, EPI_ISL_1794942, EPI_ISL_1794944, EPI_ISL_1794948, EPI_ISL_1794949, EPI_ISL_1794950, EPI_ISL_1794952, EPI_ISL_1794953, EPI_ISL_1794954, EPI_ISL_1794957, EPI_ISL_1794958, EPI_ISL_1794959, EPI_ISL_1794961, EPI_ISL_1794965, EPI_ISL_1794966, EPI_ISL_1794969, EPI_ISL_1794970, EPI_ISL_1794973, EPI_ISL_1794974, EPI_ISL_1794975, EPI_ISL_1794979, EPI_ISL_1794987, EPI_ISL_1794996, EPI_ISL_1795001, EPI_ISL_1795002, EPI_ISL_1795006, EPI_ISL_1795007, EPI_ISL_1795011, EPI_ISL_1795017, EPI_ISL_1795022, EPI_ISL_1795025, EPI_ISL_1795026, EPI_ISL_1795032, EPI_ISL_1795033, EPI_ISL_1795038, EPI_ISL_1969283, EPI_ISL_1969373, EPI_ISL_1969374, EPI_ISL_1969375, EPI_ISL_1969376, EPI_ISL_1969380, EPI_ISL_1969381, EPI_ISL_1969382, EPI_ISL_1969386, EPI_ISL_1969387, EPI_ISL_1969388, EPI_ISL_1969389, EPI_ISL_1969393, EPI_ISL_1969394, EPI_ISL_1969395, EPI_ISL_1969399, EPI_ISL_1969400, EPI_ISL_1969405, EPI_ISL_1969406, EPI_ISL_1969410, EPI_ISL_1969411, EPI_ISL_1969416, EPI_ISL_1969417, EPI_ISL_1969418, EPI_ISL_1969423, EPI_ISL_1969424, EPI_ISL_1969429, EPI_ISL_1969430, EPI_ISL_1969434, EPI_ISL_1969437, EPI_ISL_1969438, EPI_ISL_1969439, EPI_ISL_1969440, EPI_ISL_1969441, EPI_ISL_1969442, EPI_ISL_1969443, EPI_ISL_1969444, EPI_ISL_1969445, EPI_ISL_1969446, EPI_ISL_1969447, EPI_ISL_1969448, EPI_ISL_1969449, EPI_ISL_1969450, EPI_ISL_1969499, EPI_ISL_1969500 | | | |
| see above | San Diego County Public Health Laboratory | Andersen lab at Scripps Research | Brett Austin; Jovan Shephard; SEARCH Alliance San Diego with Ashleigh Murphy; SEARCH Alliance San Diego with Tracy Basler |
| EPI_ISL_2102995, EPI_ISL_2103023, EPI_ISL_2103033, EPI_ISL_2103058, EPI_ISL_2103109, EPI_ISL_2103121, EPI_ISL_2103146, EPI_ISL_2103177 | | | |
| see above | Santa Clara County Public Health Laboratory | Chan-Zuckerberg Biohub | CZB Cliahub Consortium |
| EPI_ISL_1508964, EPI_ISL_1508965, EPI_ISL_1532139, EPI_ISL_1532140, EPI_ISL_1532144, | Santa Clara County Public Health Laboratory | Santa Clara County Public Health Laboratory | Santa Clara County Public Health Department |

| | | | |
|---|---|---|---|
| EPI_ISL_1620461 | | | |
| EPI_ISL_1664263, EPI_ISL_1664264, EPI_ISL_1664265, EPI_ISL_1664266, EPI_ISL_1664275, EPI_ISL_1913216, EPI_ISL_1913251 | | | |
| see above | Santa Clara County Public Health Laboratory | Santa Clara County public Health Laboratory | Santa Clara County Public Health Department |
| EPI_ISL_1532127, EPI_ISL_1532128, EPI_ISL_1532129, EPI_ISL_1532130, EPI_ISL_1532132, EPI_ISL_1620456 | Santa Clara Valley Medical Center | Santa Clara County Public Health Laboratory | Santa Clara County Public Health Department |
| EPI_ISL_1664276, EPI_ISL_1664277, EPI_ISL_1664278, EPI_ISL_1664279, EPI_ISL_1664280, EPI_ISL_1664281, EPI_ISL_1664282, EPI_ISL_1664283, EPI_ISL_1664284, EPI_ISL_1664285, EPI_ISL_1664286, EPI_ISL_1664287, EPI_ISL_1664289, EPI_ISL_1664290, EPI_ISL_1664291, EPI_ISL_1664292, EPI_ISL_1664293, EPI_ISL_1664294, EPI_ISL_1664295, EPI_ISL_1664296, EPI_ISL_1664297, EPI_ISL_1664298, EPI_ISL_1664299, EPI_ISL_1824084, EPI_ISL_1824085, EPI_ISL_1824086, EPI_ISL_1824087, EPI_ISL_1824088, EPI_ISL_1824089, EPI_ISL_1824090, EPI_ISL_1824091, EPI_ISL_1824092, EPI_ISL_1824093, EPI_ISL_1824094, EPI_ISL_1824095, EPI_ISL_1824096, EPI_ISL_1824097, EPI_ISL_1824098, EPI_ISL_1824099, EPI_ISL_1824100, EPI_ISL_1824101, EPI_ISL_1824102, EPI_ISL_1824103, EPI_ISL_1824104, EPI_ISL_1824105, EPI_ISL_1824106, EPI_ISL_1824107, EPI_ISL_1824108, EPI_ISL_1824109, EPI_ISL_1824110, EPI_ISL_1913215, EPI_ISL_1913218, EPI_ISL_1913226, EPI_ISL_1913227, EPI_ISL_1913228, EPI_ISL_1913229, EPI_ISL_1913230, EPI_ISL_1913231, EPI_ISL_1913232, EPI_ISL_1913233, EPI_ISL_1913234, EPI_ISL_1913235, EPI_ISL_1913236, EPI_ISL_1913237, EPI_ISL_1913238, EPI_ISL_1913239, EPI_ISL_1913240, EPI_ISL_1913241, EPI_ISL_1913242, EPI_ISL_1913243, EPI_ISL_1913244, EPI_ISL_1913245, EPI_ISL_1913246, EPI_ISL_1913247, EPI_ISL_1913248, EPI_ISL_1913249, EPI_ISL_1913250, EPI_ISL_1924631, EPI_ISL_1924632 | | | |
| see above | Santa Clara Valley Medical Center | Santa Clara County public Health Laboratory | Santa Clara County Public Health Department |
| EPI_ISL_1794721, EPI_ISL_1794722, EPI_ISL_1794723, EPI_ISL_1794724, EPI_ISL_1794725, EPI_ISL_1794726, EPI_ISL_1794727, EPI_ISL_1794728, EPI_ISL_1794729, EPI_ISL_1794730, EPI_ISL_1794731, EPI_ISL_1794732, EPI_ISL_1794733, EPI_ISL_1794734, EPI_ISL_1794735, EPI_ISL_1794736, EPI_ISL_1794737, EPI_ISL_1794738, EPI_ISL_1794739, EPI_ISL_1794740, EPI_ISL_1794741, EPI_ISL_1794839, EPI_ISL_1794840, EPI_ISL_1794841, EPI_ISL_1794842, EPI_ISL_1794843, EPI_ISL_1794844, EPI_ISL_1794845, EPI_ISL_1794846, EPI_ISL_1794847, EPI_ISL_1794848, EPI_ISL_1794849, EPI_ISL_1794850, EPI_ISL_1794851, EPI_ISL_1794852, EPI_ISL_1794853, EPI_ISL_1794934, EPI_ISL_1794935, EPI_ISL_1794936, EPI_ISL_1969507, EPI_ISL_1969508, EPI_ISL_1969509, EPI_ISL_1969510, EPI_ISL_1969511, EPI_ISL_1969512, EPI_ISL_1969513, EPI_ISL_1969514, EPI_ISL_1969515, EPI_ISL_1969516, EPI_ISL_1969517, EPI_ISL_1969518, EPI_ISL_1969519, EPI_ISL_1969520, EPI_ISL_1969521, EPI_ISL_1969522, EPI_ISL_1969523, EPI_ISL_1969524, EPI_ISL_1969525, EPI_ISL_1969526, EPI_ISL_1969527, EPI_ISL_1969528, EPI_ISL_1969529, EPI_ISL_1969530, EPI_ISL_1969531, EPI_ISL_1969532, EPI_ISL_1969533, EPI_ISL_1969534, EPI_ISL_1969535, EPI_ISL_1969536, EPI_ISL_1969537, EPI_ISL_1969538, EPI_ISL_1969539, EPI_ISL_1969540, EPI_ISL_1969541, EPI_ISL_1969542, EPI_ISL_1969543, EPI_ISL_1969544, EPI_ISL_1969545, EPI_ISL_1969546, EPI_ISL_1969547, EPI_ISL_1969548, EPI_ISL_1969549, EPI_ISL_1969550, EPI_ISL_1969551, EPI_ISL_1969552, EPI_ISL_1969553 | | | |
| see above | Scripps Medical Laboratory | Andersen lab at Scripps Research | Ellen Stefanski; Ian Mchardy; SEARCH Alliance San Diego with Michael Quigley |
| EPI_ISL_1794700, EPI_ISL_1794701, EPI_ISL_1794702, EPI_ISL_1794705, EPI_ISL_1794706, EPI_ISL_1794707, EPI_ISL_1794708, EPI_ISL_1794709, EPI_ISL_1794710, EPI_ISL_1794711, EPI_ISL_1794712, EPI_ISL_1794742, EPI_ISL_1794743, EPI_ISL_1794744, EPI_ISL_1794745, EPI_ISL_1794746, EPI_ISL_1794747, EPI_ISL_1794748, EPI_ISL_1794749, EPI_ISL_1794750, EPI_ISL_1794751, EPI_ISL_1794752, EPI_ISL_1794753, EPI_ISL_1794754, EPI_ISL_1794755, EPI_ISL_1794756, EPI_ISL_1794757, EPI_ISL_1794758, EPI_ISL_1794759, EPI_ISL_1794760, EPI_ISL_1794761, EPI_ISL_1794762, EPI_ISL_1794763, EPI_ISL_1794764, EPI_ISL_1794765, EPI_ISL_1794766, EPI_ISL_1794767, EPI_ISL_1794782, EPI_ISL_1794783, EPI_ISL_1794784, EPI_ISL_1794785, EPI_ISL_1794786, EPI_ISL_1794885, EPI_ISL_1794886, EPI_ISL_1794887, EPI_ISL_1794888, EPI_ISL_1794889, EPI_ISL_1794890, EPI_ISL_1794891, EPI_ISL_1794892, EPI_ISL_1794893, EPI_ISL_1794894, EPI_ISL_1794895, EPI_ISL_1794896, EPI_ISL_1794897, EPI_ISL_1794898, EPI_ISL_1794899, EPI_ISL_1794900, EPI_ISL_1794901, EPI_ISL_1794902, EPI_ISL_1794903, EPI_ISL_1794904, EPI_ISL_1794905, EPI_ISL_1794906, EPI_ISL_1794907, EPI_ISL_1794908, EPI_ISL_1794909, EPI_ISL_1794910, EPI_ISL_1794911, EPI_ISL_1794912, EPI_ISL_1794913, EPI_ISL_1794914, EPI_ISL_1794915, EPI_ISL_1794916, EPI_ISL_1794917, EPI_ISL_1794918, EPI_ISL_1969563, EPI_ISL_1969564, EPI_ISL_1969565, EPI_ISL_1969566, EPI_ISL_1969567, EPI_ISL_1969568, EPI_ISL_1969569, EPI_ISL_1969570, EPI_ISL_1969571, EPI_ISL_1969572, EPI_ISL_1969573, EPI_ISL_1969574, EPI_ISL_1969575, EPI_ISL_1969576, EPI_ISL_1969577, EPI_ISL_1969578, EPI_ISL_1969579, EPI_ISL_1969580, EPI_ISL_1969581, EPI_ISL_1969582, EPI_ISL_1969583, EPI_ISL_1969584, EPI_ISL_1969585, EPI_ISL_1969586, EPI_ISL_1969587, EPI_ISL_1969588, EPI_ISL_1969589, EPI_ISL_1969590, EPI_ISL_1969591, EPI_ISL_1969592, EPI_ISL_1969593, EPI_ISL_1969594, EPI_ISL_1969595, EPI_ISL_1969596, EPI_ISL_1969597, EPI_ISL_1969598, EPI_ISL_1969599, EPI_ISL_1969600, EPI_ISL_1969601, EPI_ISL_1969606, EPI_ISL_1969608, EPI_ISL_1969609, EPI_ISL_1969610 | | | |
| see above | Sharp HealthCare Laboratory | Andersen lab at Scripps Research | Art Mendoza; Cathy Woerle; Jacquelyn Berumen; Liam McGinnis; Omid Bakhtar; SEARCH Alliance San Diego with Aaron Harding |
| EPI_ISL_1620455, EPI_ISL_1620457, EPI_ISL_1620458, EPI_ISL_1620459, EPI_ISL_1620460, EPI_ISL_1620462, EPI_ISL_1620463, EPI_ISL_1620464, EPI_ISL_1620465 | | | |
| see above | Stanford Clinical Virology Laboratory | Santa Clara County Public Health Laboratory | Santa Clara County Public Health Department |
| EPI_ISL_1924633, EPI_ISL_1924634, EPI_ISL_1924635, EPI_ISL_1924636, EPI_ISL_1924637, EPI_ISL_1924638, EPI_ISL_1924639, EPI_ISL_1924640, EPI_ISL_1924641, EPI_ISL_1924642, EPI_ISL_1924643, EPI_ISL_1924644, EPI_ISL_1924645, EPI_ISL_1924646, EPI_ISL_1924647, EPI_ISL_1924648 | | | |
| see above | Stanford Clinical Virology Laboratory | Santa Clara County public Health Laboratory | Santa Clara County Public Health Department |
| EPI_ISL_1675226, EPI_ISL_1675227, EPI_ISL_1701680, EPI_ISL_1701681 | Stanford Health Care | Stanford University School of Medicine, Clinical Virology Laboratory | Becky Jiang; Bernadette Truong; Daniel Solis; Fumiko Yamamoto; Hannah Wang; Jacob Miller; James Zehnder; Kenji O. Mfuh; Malaya K. Sahoo; Mamdouh Sibai; Marilyn Mar; Michelle Verghese; Naomi Iwai; and Benjamin A. Pinsky |
| EPI_ISL_1540751 | The Jackson Laboratory | The Jackson Laboratory | Adams M; Bergeron D; Kelly K; Li L; Long J; Omerza G; Renzette N |
| EPI_ISL_1707042, EPI_ISL_1707044, EPI_ISL_1707045, EPI_ISL_1707046, EPI_ISL_1707047, EPI_ISL_1707048, EPI_ISL_1707050, EPI_ISL_1707051, EPI_ISL_1707052, EPI_ISL_1707053, EPI_ISL_1707054, EPI_ISL_1707055, EPI_ISL_1707056, EPI_ISL_1707057, EPI_ISL_1707058, EPI_ISL_1707059, EPI_ISL_1707060, EPI_ISL_1707061, EPI_ISL_1716873, EPI_ISL_1716874, EPI_ISL_1716875, EPI_ISL_1716876 | | | |
| see above | UCLA Clinical Micro Lab | Los Angeles County PHL | P. Hemarajata et al. |
| EPI_ISL_1469852, EPI_ISL_1469875, EPI_ISL_1469876, EPI_ISL_1469878, EPI_ISL_1469879, EPI_ISL_1469880, EPI_ISL_1469881, EPI_ISL_1469883, EPI_ISL_1921080, EPI_ISL_1921081, EPI_ISL_1921082, EPI_ISL_1921083, EPI_ISL_1921084, EPI_ISL_1921112, EPI_ISL_1921113, EPI_ISL_1921114, EPI_ISL_1921115, EPI_ISL_1921128, EPI_ISL_1921146, EPI_ISL_2031772, EPI_ISL_2031773, EPI_ISL_2031774, EPI_ISL_2031775, EPI_ISL_2031776, EPI_ISL_2031777, EPI_ISL_2031778, EPI_ISL_2031779, EPI_ISL_2031780, EPI_ISL_2031781, EPI_ISL_2031782, EPI_ISL_2031783, EPI_ISL_2031784, EPI_ISL_2031785, EPI_ISL_2031786, EPI_ISL_2081231, EPI_ISL_2081232, EPI_ISL_2081233, EPI_ISL_2081234, EPI_ISL_2081237, EPI_ISL_2081238, EPI_ISL_2081239, EPI_ISL_2081246, EPI_ISL_2081533, EPI_ISL_2081537, EPI_ISL_2081540, EPI_ISL_2081541, EPI_ISL_2081547, EPI_ISL_2081551, EPI_ISL_2081552, EPI_ISL_2082942, EPI_ISL_2083002 | | | |
| see above | UCSC Genomics Institute | UCSC Genomics Institute | A. Marm Kilpatrick; Angie Hinrichs; Beth Shapiro; Bryan Thornlow; Ciara Wanket; David Haussler; Eric Beraut; Hugh Olsen; Ikenna Anigbogu; Isabel Bjork; Jakob McBroome; Jeremy Sanford; Joshua Kapp; Mark Akeson; Maximilian Haeussler; Michael Stone; Miten Jain; Molly Cassatt-Johnstone; Namrita Dhillon; Russell Corbett-Detig; Terren Chang; Yatish Turakhia |
| EPI_ISL_1508921, EPI_ISL_1508922, EPI_ISL_1508923, EPI_ISL_1508924, EPI_ISL_1508925, EPI_ISL_1508926, EPI_ISL_1508927, EPI_ISL_1508928, EPI_ISL_1669348, EPI_ISL_1706609, EPI_ISL_1707078, EPI_ISL_1707079, EPI_ISL_1707080, EPI_ISL_1707081, EPI_ISL_1707082, EPI_ISL_1707083, EPI_ISL_1707084, EPI_ISL_1707085, EPI_ISL_1707086, EPI_ISL_1707087, EPI_ISL_1707088, EPI_ISL_1707089, EPI_ISL_1707090, EPI_ISL_1707091, EPI_ISL_1707092, EPI_ISL_1707093, EPI_ISL_1707094, EPI_ISL_1707095, EPI_ISL_1707096, EPI_ISL_1707097, EPI_ISL_1707098, EPI_ISL_1716881, EPI_ISL_1716882, EPI_ISL_1716883, EPI_ISL_1716884, EPI_ISL_1716885, EPI_ISL_1716886, EPI_ISL_1716887, EPI_ISL_1716888, EPI_ISL_1716889, EPI_ISL_1716890, EPI_ISL_1716891, EPI_ISL_1716892, EPI_ISL_1716893, EPI_ISL_1716894, EPI_ISL_1716895, EPI_ISL_1716897, EPI_ISL_1716898 | | | |
| see above | USC Clinical Lab | Los Angeles County PHL | P. Hemarajata et al. |
| EPI_ISL_2031858 | VA Medical Center | Los Angeles County PHL | P. Hemarajata et al. |

We gratefully acknowledge the following Authors from the Originating laboratories responsible for obtaining the specimens, as well as the Submitting laboratories where the genome data were generated and shared via GISAID, on which this research is based.

All Submitters of data may be contacted directly via www.gisaid.org

Authors are sorted alphabetically.

| Accession ID | Originating Laboratory | Submitting Laboratory | Authors |
|---|---|---|---|
| EPI_ISL_2229145, EPI_ISL_2229146, EPI_ISL_2229147, EPI_ISL_2229149, EPI_ISL_2229156, EPI_ISL_2229157, EPI_ISL_2788100, EPI_ISL_2788103, EPI_ISL_2788106, EPI_ISL_2788109, EPI_ISL_2788112, EPI_ISL_2788115, EPI_ISL_2788121 | | | |
| see above | "CDPH, Viral and Rickettsial Disease Laboratory" | Centers for Disease Control and Prevention Division of Viral Diseases, Pathogen Discovery | Alison Laufer Halpin; Ben L. Rambo-Martin; Clinton R. Paden; Dakota Howard; Darlene Wagner; Dave Wentworth; Dhwani Batra; Jasmine Padilla; Justin Lee; Katie Dillon; Krista Queen; Kristen Knipe; Kristine Lacek; Mark Burroughs; Matthew Schmerer; Mili Sheth; Peter Cook; Sam Shepard; Sarah Nobles; Shoshona Le; Suxiang Tong; Vivien Dugan; Yvette Unoarumhi |
| EPI_ISL_2811941 | Adventist Health Glendale | Los Angeles County PHL | P. Hemarajata et al. |
| EPI_ISL_2146060, EPI_ISL_2146061, EPI_ISL_2146075, EPI_ISL_2146077, EPI_ISL_2146078, EPI_ISL_2146079, EPI_ISL_2146080, EPI_ISL_2146086, EPI_ISL_2146091, EPI_ISL_2146094, EPI_ISL_2146095, EPI_ISL_2146100, EPI_ISL_2146114, EPI_ISL_2146131, EPI_ISL_2146135, EPI_ISL_2146144, EPI_ISL_2146145, EPI_ISL_2146149, EPI_ISL_2146150, EPI_ISL_2146169, EPI_ISL_2146175, EPI_ISL_2146212, EPI_ISL_2146216, EPI_ISL_2146219, EPI_ISL_2146226, EPI_ISL_2146244, EPI_ISL_2146245, EPI_ISL_2146247, EPI_ISL_2146248, EPI_ISL_2146276, EPI_ISL_2146277, EPI_ISL_2146321, EPI_ISL_2146325, EPI_ISL_2146328, EPI_ISL_2146334, EPI_ISL_2146337, EPI_ISL_2146341, EPI_ISL_2146352, EPI_ISL_2146409, EPI_ISL_2146431, EPI_ISL_2146433, EPI_ISL_2146442, EPI_ISL_2146450, EPI_ISL_2146455, EPI_ISL_2146457, EPI_ISL_2146458, EPI_ISL_2146460, EPI_ISL_2146465, EPI_ISL_2146469, EPI_ISL_2146497, EPI_ISL_2146501, EPI_ISL_2146504, EPI_ISL_2146543, EPI_ISL_2146547, EPI_ISL_2146548, EPI_ISL_2146558, EPI_ISL_2146573, EPI_ISL_2146612, EPI_ISL_2146621, EPI_ISL_2146737, EPI_ISL_2146738, EPI_ISL_2146739, EPI_ISL_2146741, EPI_ISL_2146743, EPI_ISL_2146748, EPI_ISL_2146749, EPI_ISL_2146755, EPI_ISL_2146764, EPI_ISL_2146797, EPI_ISL_2146798, EPI_ISL_2146817, EPI_ISL_2146821, EPI_ISL_2146835, EPI_ISL_2146837, EPI_ISL_2146871, EPI_ISL_2146889, EPI_ISL_2146890, EPI_ISL_2146898, EPI_ISL_2146901, EPI_ISL_2146913, EPI_ISL_2146920, EPI_ISL_2146956, EPI_ISL_2146970, EPI_ISL_2146988, EPI_ISL_2146996, EPI_ISL_2147008, EPI_ISL_2147049, EPI_ISL_2147051, EPI_ISL_2147057, EPI_ISL_2147080, EPI_ISL_2147081, EPI_ISL_2147088, EPI_ISL_2147089, EPI_ISL_2147097, EPI_ISL_2147103, EPI_ISL_2147105, EPI_ISL_2147126, EPI_ISL_2147127, EPI_ISL_2147146, EPI_ISL_2147159, EPI_ISL_2147176, EPI_ISL_2147179, EPI_ISL_2147184, EPI_ISL_2147196, EPI_ISL_2147219, EPI_ISL_2147226, EPI_ISL_2147231, EPI_ISL_2147245, EPI_ISL_2147278, EPI_ISL_2147284, EPI_ISL_2147301, EPI_ISL_2147406, EPI_ISL_2147423, EPI_ISL_2147459, EPI_ISL_2147487, EPI_ISL_2147488, EPI_ISL_2147491, EPI_ISL_2147492, EPI_ISL_2147495, EPI_ISL_2147510, EPI_ISL_2147511, EPI_ISL_2147522, EPI_ISL_2147525, EPI_ISL_2147528, EPI_ISL_2147541, EPI_ISL_2147542, EPI_ISL_2147543, EPI_ISL_2147545, EPI_ISL_2147550, EPI_ISL_2147557, EPI_ISL_2147562, EPI_ISL_2147573, EPI_ISL_2147576, EPI_ISL_2147584, EPI_ISL_2147659, EPI_ISL_2147795, EPI_ISL_2147797, EPI_ISL_2147919, EPI_ISL_2147930, EPI_ISL_2147944, EPI_ISL_2147958, EPI_ISL_2147963, EPI_ISL_2148120, EPI_ISL_2148128, EPI_ISL_2148144, EPI_ISL_2148145, EPI_ISL_2148148, EPI_ISL_2148149, EPI_ISL_2148151, EPI_ISL_2148152, EPI_ISL_2148153, EPI_ISL_2148164, EPI_ISL_2148165, EPI_ISL_2148166, EPI_ISL_2148167, EPI_ISL_2148168, EPI_ISL_2148169, EPI_ISL_2148193, EPI_ISL_2148205, EPI_ISL_2148210, EPI_ISL_2148212, EPI_ISL_2148218, EPI_ISL_2148220, EPI_ISL_2148221, EPI_ISL_2148234, EPI_ISL_2148239, EPI_ISL_2148241, EPI_ISL_2148242, EPI_ISL_2148247, EPI_ISL_2148275, EPI_ISL_2148283, EPI_ISL_2148287, EPI_ISL_2148297, EPI_ISL_2148298, EPI_ISL_2148304, EPI_ISL_2148403, EPI_ISL_2148404, EPI_ISL_2148406, EPI_ISL_2148409, EPI_ISL_2148410, EPI_ISL_2148412, EPI_ISL_2148415, EPI_ISL_2148457, EPI_ISL_2148465, EPI_ISL_2148470, EPI_ISL_2148840, EPI_ISL_2148841, EPI_ISL_2148842, EPI_ISL_2148844, EPI_ISL_2148866, EPI_ISL_2148867, EPI_ISL_2148877, EPI_ISL_2148889, EPI_ISL_2148892, EPI_ISL_2148894, EPI_ISL_2148896, EPI_ISL_2148909, EPI_ISL_2148910, EPI_ISL_2149147, EPI_ISL_2149151, EPI_ISL_2149153, EPI_ISL_2149155, EPI_ISL_2149162, EPI_ISL_2149165, EPI_ISL_2149166, EPI_ISL_2149167, EPI_ISL_2149172, EPI_ISL_2149173, EPI_ISL_2149174, EPI_ISL_2149183, EPI_ISL_2149185, EPI_ISL_2158864, EPI_ISL_2158914, EPI_ISL_2158922, EPI_ISL_2158939, EPI_ISL_2158940, EPI_ISL_2158945, EPI_ISL_2158961, EPI_ISL_2158962, EPI_ISL_2158963, EPI_ISL_2158979, EPI_ISL_2158999, EPI_ISL_2159007, EPI_ISL_2159009, EPI_ISL_2159012, EPI_ISL_2159015, EPI_ISL_2159018, EPI_ISL_2159021, EPI_ISL_2159057, EPI_ISL_2159060, EPI_ISL_2159062, EPI_ISL_2159065, EPI_ISL_2159116, EPI_ISL_2159117, EPI_ISL_2159118, EPI_ISL_2159121, EPI_ISL_2159123, EPI_ISL_2159137, EPI_ISL_2159155, EPI_ISL_2159156, EPI_ISL_2159157, EPI_ISL_2159204, EPI_ISL_2159277, EPI_ISL_2159278, EPI_ISL_2159279, EPI_ISL_2159304, EPI_ISL_2159327, EPI_ISL_2159328, EPI_ISL_2159340, EPI_ISL_2159352, EPI_ISL_2159359, EPI_ISL_2159364, EPI_ISL_2159377, EPI_ISL_2159378, EPI_ISL_2159458, EPI_ISL_2159466, EPI_ISL_2159467, EPI_ISL_2180535, EPI_ISL_2180539, EPI_ISL_2180556, EPI_ISL_2180562, EPI_ISL_2180563, EPI_ISL_2180568, EPI_ISL_2180645, EPI_ISL_2180654, EPI_ISL_2180707, EPI_ISL_2180729, EPI_ISL_2180777, EPI_ISL_2180780, EPI_ISL_2180831, EPI_ISL_2180833, EPI_ISL_2180841, EPI_ISL_2180863, EPI_ISL_2180869, EPI_ISL_2180871, EPI_ISL_2180881, EPI_ISL_2180884, EPI_ISL_2180898, EPI_ISL_2180899, EPI_ISL_2180900, EPI_ISL_2180905, EPI_ISL_2180906, EPI_ISL_2180908, EPI_ISL_2180911, EPI_ISL_2180916, EPI_ISL_2180919, EPI_ISL_2180922, EPI_ISL_2180923, EPI_ISL_2180926, EPI_ISL_2180928, EPI_ISL_2180930, EPI_ISL_2180956, EPI_ISL_2180958, EPI_ISL_2180976, EPI_ISL_2181013, EPI_ISL_2181014, EPI_ISL_2181017, EPI_ISL_2181020, EPI_ISL_2181029, EPI_ISL_2181038, EPI_ISL_2181041, EPI_ISL_2181042, EPI_ISL_2181088, EPI_ISL_2181092, EPI_ISL_2181097, EPI_ISL_2181103, EPI_ISL_2181104, EPI_ISL_2181105, EPI_ISL_2181109, EPI_ISL_2181111, EPI_ISL_2181113, EPI_ISL_2181133, EPI_ISL_2181139, EPI_ISL_2181140, EPI_ISL_2181143, EPI_ISL_2181190, EPI_ISL_2181191, EPI_ISL_2181192, EPI_ISL_2181194, EPI_ISL_2181196, EPI_ISL_2181197, EPI_ISL_2181199, EPI_ISL_2181222, EPI_ISL_2181226, EPI_ISL_2181229, EPI_ISL_2181230, EPI_ISL_2181244, EPI_ISL_2181285, EPI_ISL_2181312, EPI_ISL_2181319, EPI_ISL_2181330, EPI_ISL_2181393, EPI_ISL_2181402, EPI_ISL_2181403, EPI_ISL_2181407, EPI_ISL_2181408, EPI_ISL_2181435, EPI_ISL_2181436, EPI_ISL_2181440, EPI_ISL_2181450, EPI_ISL_2181465, EPI_ISL_2181466, EPI_ISL_2181467, EPI_ISL_2181468, EPI_ISL_2181476, EPI_ISL_2181486, EPI_ISL_2181487, EPI_ISL_2181533, EPI_ISL_2181539, EPI_ISL_2181546, EPI_ISL_2181580, EPI_ISL_2181581, EPI_ISL_2181587, EPI_ISL_2181589, EPI_ISL_2181591, EPI_ISL_2181595, EPI_ISL_2181596, EPI_ISL_2181597, EPI_ISL_2187383, EPI_ISL_2187409, EPI_ISL_2187419, EPI_ISL_2187421, EPI_ISL_2187427, EPI_ISL_2187435, EPI_ISL_2187448, EPI_ISL_2187481, EPI_ISL_2187482, EPI_ISL_2187500, EPI_ISL_2187526, EPI_ISL_2187595, EPI_ISL_2187604, EPI_ISL_2187612, EPI_ISL_2204506, EPI_ISL_2204532, EPI_ISL_2204546, EPI_ISL_2204628, EPI_ISL_2204655, EPI_ISL_2204715, EPI_ISL_2204725, EPI_ISL_2204736, EPI_ISL_2204739, EPI_ISL_2204757, EPI_ISL_2204759, EPI_ISL_2204767, EPI_ISL_2204775, EPI_ISL_2204785, EPI_ISL_2204788, EPI_ISL_2204792, EPI_ISL_2204843, EPI_ISL_2204865, EPI_ISL_2204900, EPI_ISL_2204923, EPI_ISL_2204932, EPI_ISL_2204934, EPI_ISL_2204939, EPI_ISL_2204962, EPI_ISL_2204975, EPI_ISL_2204999, EPI_ISL_2241650, EPI_ISL_2268220, EPI_ISL_2268235, EPI_ISL_2268245, EPI_ISL_2280287, EPI_ISL_2280295, EPI_ISL_2280300, EPI_ISL_2280302, EPI_ISL_2280311, EPI_ISL_2280331, EPI_ISL_2280332, EPI_ISL_2280334, EPI_ISL_2280336, EPI_ISL_2280343, EPI_ISL_2280349, EPI_ISL_2280379, EPI_ISL_2280381, EPI_ISL_2280384, EPI_ISL_2280391, EPI_ISL_2280400, EPI_ISL_2280406, EPI_ISL_2280426, EPI_ISL_2280427, EPI_ISL_2280428, EPI_ISL_2280429, EPI_ISL_2280438, EPI_ISL_2280446, EPI_ISL_2280465, EPI_ISL_2280485, EPI_ISL_2280494, EPI_ISL_2280502, EPI_ISL_2280518, EPI_ISL_2280535, EPI_ISL_2280568, EPI_ISL_2280578, EPI_ISL_2280584, EPI_ISL_2280585, EPI_ISL_2280586, EPI_ISL_2280587, EPI_ISL_2280596, EPI_ISL_2280610, EPI_ISL_2280611, EPI_ISL_2280644, EPI_ISL_2280651, EPI_ISL_2280671, EPI_ISL_2280673, EPI_ISL_2280715, EPI_ISL_2280726, EPI_ISL_2280795, EPI_ISL_2280796, EPI_ISL_2280812, EPI_ISL_2280822, EPI_ISL_2280824, EPI_ISL_2280859, EPI_ISL_2280860, EPI_ISL_2280875, EPI_ISL_2280876, EPI_ISL_2280879, EPI_ISL_2280883, EPI_ISL_2280896, EPI_ISL_2280913, EPI_ISL_2280914, EPI_ISL_2280915, EPI_ISL_2280916, EPI_ISL_2280923, EPI_ISL_2280939, EPI_ISL_2280947, EPI_ISL_2280952, EPI_ISL_2280955, EPI_ISL_2280959, EPI_ISL_2280973, EPI_ISL_2280985, EPI_ISL_2280991, EPI_ISL_2280998, EPI_ISL_2281004, EPI_ISL_2281005, EPI_ISL_2281007, EPI_ISL_2281010, EPI_ISL_2281042, EPI_ISL_2281075, EPI_ISL_2281089, EPI_ISL_2281090, EPI_ISL_2281095, EPI_ISL_2281096, EPI_ISL_2281106, EPI_ISL_2281107, EPI_ISL_2281111, EPI_ISL_2281118, EPI_ISL_2281120, EPI_ISL_2281129, EPI_ISL_2281131, EPI_ISL_2281132, EPI_ISL_2281146, EPI_ISL_2281147, EPI_ISL_2281150, EPI_ISL_2281166, EPI_ISL_2281180, EPI_ISL_2281216, EPI_ISL_2281217, EPI_ISL_2281221, EPI_ISL_2283825, EPI_ISL_2283834, EPI_ISL_2283852, EPI_ISL_2283855, EPI_ISL_2283867, EPI_ISL_2283869, EPI_ISL_2283874, EPI_ISL_2321232, EPI_ISL_2321245, EPI_ISL_2321246, EPI_ISL_2321248, EPI_ISL_2321293, EPI_ISL_2321296, EPI_ISL_2321297, EPI_ISL_2321298, EPI_ISL_2321299, EPI_ISL_2321304, EPI_ISL_2321306, EPI_ISL_2321307, EPI_ISL_2321328, EPI_ISL_2321341, EPI_ISL_2321351, EPI_ISL_2321365, EPI_ISL_2321366, EPI_ISL_2321383, EPI_ISL_2321387, EPI_ISL_2367653, EPI_ISL_2367663, EPI_ISL_2367664, EPI_ISL_2367666, EPI_ISL_2367671, EPI_ISL_2367672, EPI_ISL_2367674, EPI_ISL_2367675, EPI_ISL_2368225, EPI_ISL_2368227, EPI_ISL_2368261, EPI_ISL_2368263, EPI_ISL_2368264, EPI_ISL_2368287, EPI_ISL_2368288, EPI_ISL_2368299, EPI_ISL_2368302, EPI_ISL_2368303, EPI_ISL_2368308, EPI_ISL_2368319, EPI_ISL_2368327, EPI_ISL_2368331, EPI_ISL_2368354, EPI_ISL_2368355, EPI_ISL_2368360, EPI_ISL_2368372, EPI_ISL_2368373, EPI_ISL_2368377, EPI_ISL_2368383, EPI_ISL_2368384, EPI_ISL_2368388, EPI_ISL_2368389, EPI_ISL_2368390, EPI_ISL_2368391, EPI_ISL_2368394, EPI_ISL_2368396, EPI_ISL_2368400, EPI_ISL_2368407, EPI_ISL_2368408, EPI_ISL_2368409, EPI_ISL_2368410, EPI_ISL_2368427, EPI_ISL_2368437, EPI_ISL_2368449, EPI_ISL_2368452, EPI_ISL_2368460, EPI_ISL_2368461, EPI_ISL_2368465, EPI_ISL_2368470, EPI_ISL_2368545, EPI_ISL_2368548, EPI_ISL_2368551, EPI_ISL_2368557, EPI_ISL_2368600, EPI_ISL_2368603, EPI_ISL_2368699, EPI_ISL_2368730, EPI_ISL_2368733, EPI_ISL_2368753, EPI_ISL_2368756, EPI_ISL_2368764, EPI_ISL_2368767, EPI_ISL_2368895, EPI_ISL_2368957, EPI_ISL_2368981, EPI_ISL_2369970, EPI_ISL_2370034, EPI_ISL_2370036, EPI_ISL_2370054, EPI_ISL_2370139, EPI_ISL_2370150, EPI_ISL_2370187, EPI_ISL_2370188, EPI_ISL_2370229, EPI_ISL_2370231, EPI_ISL_2370233, EPI_ISL_2370239, EPI_ISL_2370392, EPI_ISL_2370480, EPI_ISL_2370491, EPI_ISL_2370507, EPI_ISL_2370528, EPI_ISL_2370533, EPI_ISL_2370536, EPI_ISL_2370585, EPI_ISL_2370648, EPI_ISL_2370650, EPI_ISL_2370704, EPI_ISL_2372300, EPI_ISL_2372376, EPI_ISL_2372394, EPI_ISL_2372455, EPI_ISL_2372460, EPI_ISL_2372461, EPI_ISL_2372465, EPI_ISL_2422001, EPI_ISL_2422025, EPI_ISL_2422033, EPI_ISL_2422046, EPI_ISL_2422052, EPI_ISL_2422109, EPI_ISL_2422113, EPI_ISL_2422114, EPI_ISL_2422118, EPI_ISL_2422119, EPI_ISL_2442333, EPI_ISL_2489072, EPI_ISL_2489073, EPI_ISL_2489074, EPI_ISL_2489075, EPI_ISL_2489078, EPI_ISL_2489079, EPI_ISL_2489080, EPI_ISL_2489081, EPI_ISL_2489082, EPI_ISL_2489084, EPI_ISL_2489085, EPI_ISL_2489091, EPI_ISL_2489092, EPI_ISL_2489097, EPI_ISL_2489098, EPI_ISL_2489099, EPI_ISL_2489100, EPI_ISL_2489101, EPI_ISL_2489110, EPI_ISL_2489111, EPI_ISL_2489112, EPI_ISL_2489116, EPI_ISL_2489117, EPI_ISL_2489119, EPI_ISL_2489120, EPI_ISL_2489123, EPI_ISL_2489137, EPI_ISL_2489138, EPI_ISL_2489141, EPI_ISL_2489142, EPI_ISL_2489172, EPI_ISL_2489234, EPI_ISL_2489235, EPI_ISL_2489240, EPI_ISL_2489245, EPI_ISL_2489250, EPI_ISL_2489263, EPI_ISL_2489279, EPI_ISL_2489283, EPI_ISL_2489295, EPI_ISL_2489298, EPI_ISL_2489300, EPI_ISL_2489304, EPI_ISL_2489309, EPI_ISL_2489315, EPI_ISL_2489316, EPI_ISL_2489317, EPI_ISL_2489318, EPI_ISL_2489319, EPI_ISL_2489320, EPI_ISL_2489321, EPI_ISL_2489323, EPI_ISL_2489329, EPI_ISL_2489341, EPI_ISL_2489349, EPI_ISL_2489359, EPI_ISL_2489371, EPI_ISL_2489386, EPI_ISL_2489388, EPI_ISL_2489402, EPI_ISL_2489405, EPI_ISL_2489408, EPI_ISL_2489440, EPI_ISL_2489442, EPI_ISL_2489454, EPI_ISL_2489458, EPI_ISL_2489462, EPI_ISL_2489463, EPI_ISL_2489464, EPI_ISL_2489473, EPI_ISL_2489502, EPI_ISL_2489508, EPI_ISL_2489509, EPI_ISL_2489524, EPI_ISL_2489528, EPI_ISL_2489538, EPI_ISL_2489545, EPI_ISL_2489556, EPI_ISL_2489557, EPI_ISL_2489559, EPI_ISL_2489566, EPI_ISL_2489583, EPI_ISL_2489584, EPI_ISL_2489585, EPI_ISL_2489595, EPI_ISL_2489596, EPI_ISL_2489602, EPI_ISL_2489638, EPI_ISL_2489643, EPI_ISL_2489654, EPI_ISL_2489672, EPI_ISL_2489678, EPI_ISL_2489682, EPI_ISL_2489683, EPI_ISL_2489697, EPI_ISL_2489698, EPI_ISL_2489701, EPI_ISL_2489703, EPI_ISL_2489707, EPI_ISL_2489709, EPI_ISL_2489712, EPI_ISL_2489754, EPI_ISL_2489766, EPI_ISL_2489806, EPI_ISL_2489811, EPI_ISL_2527901, EPI_ISL_2527906, EPI_ISL_2527909, EPI_ISL_2527911, EPI_ISL_2527922, EPI_ISL_2527976, EPI_ISL_2527983, EPI_ISL_2528028, EPI_ISL_2528037, EPI_ISL_2528045, EPI_ISL_2528052, EPI_ISL_2528106, EPI_ISL_2528109, EPI_ISL_2528159, EPI_ISL_2528161, EPI_ISL_2528165, EPI_ISL_2528169, EPI_ISL_2528182, EPI_ISL_2528195, EPI_ISL_2528199, EPI_ISL_2528201, EPI_ISL_2528205, EPI_ISL_2528210, EPI_ISL_2528212, EPI_ISL_2528228, EPI_ISL_2528243, EPI_ISL_2528303, EPI_ISL_2528306, EPI_ISL_2528310, EPI_ISL_2528320, EPI_ISL_2528330, EPI_ISL_2528337, EPI_ISL_2528383, EPI_ISL_2528430, EPI_ISL_2528468, EPI_ISL_2528492, EPI_ISL_2528500, EPI_ISL_2528506, EPI_ISL_2528508, EPI_ISL_2528576, EPI_ISL_2528578, EPI_ISL_2528605, EPI_ISL_2528614, EPI_ISL_2528642, EPI_ISL_2528647, EPI_ISL_2528650, EPI_ISL_2528656, EPI_ISL_2528666, EPI_ISL_2528673, EPI_ISL_2528675, EPI_ISL_2528688, EPI_ISL_2528690, EPI_ISL_2528705, EPI_ISL_2528734, EPI_ISL_2528772, EPI_ISL_2528774, EPI_ISL_2528781, EPI_ISL_2528791, EPI_ISL_2528795, EPI_ISL_2528813, EPI_ISL_2528823, EPI_ISL_2528864, EPI_ISL_2528866, EPI_ISL_2528868, EPI_ISL_2528871, EPI_ISL_2528885, EPI_ISL_2528887, EPI_ISL_2528891, EPI_ISL_2528902, EPI_ISL_2528904, EPI_ISL_2528965, EPI_ISL_2528968, EPI_ISL_2528994, EPI_ISL_2528999, EPI_ISL_2530938, EPI_ISL_2530940, EPI_ISL_2530944, EPI_ISL_2530948, EPI_ISL_2530950, EPI_ISL_2530966, EPI_ISL_2531004, EPI_ISL_2531012, EPI_ISL_2531021, EPI_ISL_2531030, EPI_ISL_2531038, EPI_ISL_2531044, EPI_ISL_2531047, EPI_ISL_2531055, EPI_ISL_2531057, EPI_ISL_2534533, EPI_ISL_2534534, EPI_ISL_2686902, EPI_ISL_2686903, EPI_ISL_2686904, EPI_ISL_2686910, EPI_ISL_2686954, EPI_ISL_2686960, EPI_ISL_2686961, EPI_ISL_2686962, EPI_ISL_2686963, EPI_ISL_2686965, EPI_ISL_2686966, EPI_ISL_2686980, EPI_ISL_2686989, EPI_ISL_2686992, EPI_ISL_2686994, EPI_ISL_2686996, EPI_ISL_2687000, EPI_ISL_2687001, EPI_ISL_2687010, EPI_ISL_2687030, EPI_ISL_2687031, EPI_ISL_2687034, EPI_ISL_2687035, EPI_ISL_2687047, EPI_ISL_2687059, EPI_ISL_2687060, EPI_ISL_2687061, EPI_ISL_2687062, EPI_ISL_2687064, EPI_ISL_2687072, EPI_ISL_2687073, EPI_ISL_2687079, EPI_ISL_2687080, EPI_ISL_2687081, EPI_ISL_2687082, EPI_ISL_2687084, EPI_ISL_2687090, EPI_ISL_2687093, EPI_ISL_2687094, EPI_ISL_2687100, EPI_ISL_2687110, EPI_ISL_2687113, EPI_ISL_2687116, EPI_ISL_2687120, EPI_ISL_2687127, EPI_ISL_2687128, EPI_ISL_2687129, EPI_ISL_2687136, EPI_ISL_2687137, EPI_ISL_2687139, EPI_ISL_2687151, EPI_ISL_2687168, EPI_ISL_2687181, EPI_ISL_2687189, EPI_ISL_2687190, EPI_ISL_2687193, EPI_ISL_2687194, EPI_ISL_2687197, EPI_ISL_2687212, EPI_ISL_2687213, EPI_ISL_2687225, EPI_ISL_2687226, EPI_ISL_2687234, EPI_ISL_2687250, EPI_ISL_2687253, EPI_ISL_2687256, EPI_ISL_2687261, EPI_ISL_2687262, EPI_ISL_2687263, EPI_ISL_2687267, EPI_ISL_2687268, EPI_ISL_2687269, EPI_ISL_2687270, EPI_ISL_2687271, EPI_ISL_2687277, EPI_ISL_2687297, EPI_ISL_2687308, EPI_ISL_2687309, EPI_ISL_2687317, EPI_ISL_2687320, EPI_ISL_2687322, EPI_ISL_2687323, EPI_ISL_2687327, EPI_ISL_2687328, EPI_ISL_2687332, EPI_ISL_2687336, EPI_ISL_2687337, EPI_ISL_2687356, EPI_ISL_2687368, EPI_ISL_2687370, EPI_ISL_2687371, EPI_ISL_2687376, EPI_ISL_2687379, EPI_ISL_2687380, EPI_ISL_2687381, EPI_ISL_2687383, EPI_ISL_2687384, EPI_ISL_2687389, EPI_ISL_2687392, EPI_ISL_2687416, EPI_ISL_2687422, EPI_ISL_2687453, EPI_ISL_2687454, EPI_ISL_2687471, EPI_ISL_2687499, EPI_ISL_2687501, EPI_ISL_2687511, EPI_ISL_2687512, EPI_ISL_2687514, EPI_ISL_2687515, EPI_ISL_2687516, EPI_ISL_2687527, EPI_ISL_2687530, EPI_ISL_2687532, EPI_ISL_2687535, EPI_ISL_2687538, EPI_ISL_2687545, EPI_ISL_2687546, EPI_ISL_2687547, EPI_ISL_2687548, EPI_ISL_2687549, EPI_ISL_2687551, EPI_ISL_2687554, EPI_ISL_2687556, EPI_ISL_2687557, EPI_ISL_2687568, EPI_ISL_2687569, EPI_ISL_2687570, EPI_ISL_2687581, EPI_ISL_2687582, EPI_ISL_2709520, EPI_ISL_2709527, EPI_ISL_2709538, EPI_ISL_2709547, EPI_ISL_2709551, EPI_ISL_2784728, EPI_ISL_2784731, EPI_ISL_2784742, EPI_ISL_2784743, EPI_ISL_2784744, EPI_ISL_2784747, EPI_ISL_2784748, EPI_ISL_2784750, EPI_ISL_2784751, EPI_ISL_2784752, EPI_ISL_2784753, EPI_ISL_2784761, EPI_ISL_2784770, EPI_ISL_2784773, EPI_ISL_2784777, EPI_ISL_2784781, EPI_ISL_2784787, EPI_ISL_2784791, EPI_ISL_2784796, EPI_ISL_2784807, EPI_ISL_2784809, EPI_ISL_2784810, EPI_ISL_2784811, EPI_ISL_2784812, EPI_ISL_2784816, EPI_ISL_2784817, EPI_ISL_2784820, EPI_ISL_2784822, EPI_ISL_2784823, EPI_ISL_2784831, EPI_ISL_2784840, EPI_ISL_2784844, EPI_ISL_2784858, EPI_ISL_2784859, EPI_ISL_2784861, EPI_ISL_2784862, EPI_ISL_2784865, EPI_ISL_2784867, EPI_ISL_2784868, EPI_ISL_2784870, EPI_ISL_2784871, EPI_ISL_2784872, EPI_ISL_2784875, EPI_ISL_2784877, EPI_ISL_2784881, EPI_ISL_2784882, EPI_ISL_2784883, EPI_ISL_2784886, EPI_ISL_2784887, EPI_ISL_2784888, EPI_ISL_2784906, EPI_ISL_2784915, EPI_ISL_2784935, EPI_ISL_2784937, EPI_ISL_2784940, EPI_ISL_2784943, EPI_ISL_2784944, EPI_ISL_2784953, EPI_ISL_2784963, EPI_ISL_2784968, EPI_ISL_2784969, EPI_ISL_2784983, EPI_ISL_2784984, EPI_ISL_2784989, EPI_ISL_2784998, EPI_ISL_2785000, EPI_ISL_2785007, EPI_ISL_2785020, EPI_ISL_2785021, EPI_ISL_2785023, EPI_ISL_2785027, EPI_ISL_2785029, EPI_ISL_2785030, EPI_ISL_2785031, EPI_ISL_2785036, EPI_ISL_2785038, EPI_ISL_2785047, EPI_ISL_2785059, EPI_ISL_2785060, EPI_ISL_2785062, EPI_ISL_2785063, EPI_ISL_2785064, EPI_ISL_2785065, EPI_ISL_2785066, EPI_ISL_2785068, EPI_ISL_2785071, EPI_ISL_2785073, EPI_ISL_2785077, EPI_ISL_2785079, EPI_ISL_2785080, EPI_ISL_2785083, EPI_ISL_2785084, EPI_ISL_2785085, EPI_ISL_2785090, EPI_ISL_2785095, EPI_ISL_2785105, EPI_ISL_2785120, EPI_ISL_2785121, EPI_ISL_2785124, EPI_ISL_2785139, EPI_ISL_2785147, EPI_ISL_2785148, EPI_ISL_2785152, EPI_ISL_2785154, EPI_ISL_2785155, EPI_ISL_2785156, EPI_ISL_2785157, EPI_ISL_2785161, EPI_ISL_2785162, EPI_ISL_2785165, EPI_ISL_2785169, EPI_ISL_2785171, EPI_ISL_2785173, EPI_ISL_2785174, EPI_ISL_2785178, EPI_ISL_2785179, EPI_ISL_2785184, EPI_ISL_2785185, EPI_ISL_2785198, EPI_ISL_2785205, EPI_ISL_2785207, EPI_ISL_2785210, EPI_ISL_2785211, EPI_ISL_2785212, EPI_ISL_2785222, EPI_ISL_2785223, EPI_ISL_2785224, EPI_ISL_2785226, EPI_ISL_2785237, EPI_ISL_2785238, EPI_ISL_2785243, EPI_ISL_2785247, EPI_ISL_2785248, EPI_ISL_2785251, EPI_ISL_2785261, EPI_ISL_2785262, EPI_ISL_2785263, EPI_ISL_2785267, EPI_ISL_2785270, EPI_ISL_2785291, EPI_ISL_2785292, EPI_ISL_2785293, EPI_ISL_2785301, EPI_ISL_2785302, EPI_ISL_2785308, EPI_ISL_2785309, EPI_ISL_2785313, EPI_ISL_2785316, EPI_ISL_2785317, EPI_ISL_2785319, EPI_ISL_2785320, EPI_ISL_2785321, EPI_ISL_2785322, EPI_ISL_2785332, EPI_ISL_2785335, EPI_ISL_2785336, EPI_ISL_2785339, EPI_ISL_2785344, EPI_ISL_2785345, EPI_ISL_2785346, EPI_ISL_2785347, EPI_ISL_2785349, EPI_ISL_2785350, EPI_ISL_2785351, EPI_ISL_2785352, EPI_ISL_2785387, EPI_ISL_2785388, EPI_ISL_2785392, EPI_ISL_2785393, EPI_ISL_2785394, EPI_ISL_2785405, EPI_ISL_2785410, EPI_ISL_2785418, EPI_ISL_2785422, EPI_ISL_2785433, EPI_ISL_2785434, EPI_ISL_2785449, EPI_ISL_2785458, EPI_ISL_2785459, EPI_ISL_2785466, EPI_ISL_2785472, EPI_ISL_2785473, EPI_ISL_2785486, EPI_ISL_2785489, EPI_ISL_2785490, EPI_ISL_2785491, EPI_ISL_2785492, EPI_ISL_2785493, EPI_ISL_2785499, EPI_ISL_2785500, EPI_ISL_2785502, EPI_ISL_2832518, EPI_ISL_2832519, EPI_ISL_2832520, EPI_ISL_2832523, EPI_ISL_2832539, EPI_ISL_2832540, EPI_ISL_2832541, EPI_ISL_2832547, EPI_ISL_2832548, EPI_ISL_2832549, EPI_ISL_2832551, EPI_ISL_2832553, EPI_ISL_2832563, EPI_ISL_2832568, EPI_ISL_2832570, EPI_ISL_2832571, EPI_ISL_2832572, EPI_ISL_2832576, EPI_ISL_2832581, EPI_ISL_2875520, EPI_ISL_2875522, EPI_ISL_2875525, EPI_ISL_2875528, EPI_ISL_2875535, EPI_ISL_2875536, EPI_ISL_2875540, EPI_ISL_2875544, EPI_ISL_2875546, EPI_ISL_2875548, EPI_ISL_2875549, EPI_ISL_2875551, EPI_ISL_2875552, EPI_ISL_2875553, EPI_ISL_2875554, EPI_ISL_2875555, EPI_ISL_2875562, EPI_ISL_2875563, EPI_ISL_2875566, EPI_ISL_2875568, EPI_ISL_2875580, EPI_ISL_2875581, EPI_ISL_2875584, EPI_ISL_2875591, EPI_ISL_2875602, EPI_ISL_2875604, EPI_ISL_2875614, EPI_ISL_2875623, EPI_ISL_2875626, EPI_ISL_2875638, EPI_ISL_2875661, EPI_ISL_2875662, EPI_ISL_2875663, EPI_ISL_2875666, EPI_ISL_2875668, EPI_ISL_2875669, EPI_ISL_2875671, EPI_ISL_2875684, EPI_ISL_2875685, EPI_ISL_2875686, EPI_ISL_2875687, EPI_ISL_2875690, EPI_ISL_2875700, EPI_ISL_2875701, EPI_ISL_2875703, EPI_ISL_2875714, EPI_ISL_2875716, EPI_ISL_2875719, EPI_ISL_2875720, EPI_ISL_2875724, EPI_ISL_2875725, EPI_ISL_2875726, EPI_ISL_2875728, EPI_ISL_2875732, EPI_ISL_2875734, EPI_ISL_2875743, EPI_ISL_2875766, EPI_ISL_2875774, EPI_ISL_2875779, EPI_ISL_2875792, EPI_ISL_2875798, EPI_ISL_2875799, EPI_ISL_2875803, EPI_ISL_2875804, EPI_ISL_2875806, EPI_ISL_2875807, EPI_ISL_2875809, EPI_ISL_2875817, EPI_ISL_2875818, EPI_ISL_2875819, EPI_ISL_2875820, EPI_ISL_2875821, EPI_ISL_2875823, EPI_ISL_2875825, EPI_ISL_2875827, EPI_ISL_2875828, EPI_ISL_2875831, EPI_ISL_2875834, EPI_ISL_2875858, EPI_ISL_2875867, EPI_ISL_2875873, EPI_ISL_2875874, EPI_ISL_2875876, EPI_ISL_2875880, EPI_ISL_2875881, EPI_ISL_2875882, EPI_ISL_2875883, EPI_ISL_2875886, EPI_ISL_2875912, EPI_ISL_2875916, EPI_ISL_2875930, EPI_ISL_2875936, EPI_ISL_2875940, EPI_ISL_2875944, EPI_ISL_2875945, EPI_ISL_2875946, EPI_ISL_2875947, EPI_ISL_2875948, EPI_ISL_2875949, EPI_ISL_2875950, EPI_ISL_2875951, EPI_ISL_2875952, EPI_ISL_2875953, EPI_ISL_2875961, EPI_ISL_2875962, EPI_ISL_2875963, EPI_ISL_2875964, EPI_ISL_2875966, EPI_ISL_2875967, EPI_ISL_2875968, EPI_ISL_2875979, EPI_ISL_2875983, EPI_ISL_2875986, EPI_ISL_2875992, EPI_ISL_2875995, EPI_ISL_2875996, EPI_ISL_2875997, EPI_ISL_2876001, EPI_ISL_2876004, EPI_ISL_2876005, EPI_ISL_2876006, EPI_ISL_2876010, EPI_ISL_2876011, EPI_ISL_2876013, EPI_ISL_2876014, EPI_ISL_2877658, EPI_ISL_2877661, EPI_ISL_2877665, EPI_ISL_2877682, EPI_ISL_2877683, EPI_ISL_2877685, EPI_ISL_2877691, EPI_ISL_2877695 | | | |
| see above | Aegis Sciences Corporation | Centers for Disease Control and Prevention Division of Viral Diseases, Pathogen Discovery | Adrian Paskey; Alec Vest; Benjamin Rambo-Martin; Christopher Gulvick; Clinton R. Paden; Cyndi Clark; Dakota Howard; Darlene Wagner; Dhwani Batra; Dillon Nall; Duncan MacCannell; Ethan Sanders; Holly Houdeshell; Jason Caravas; Kara Moser; Matthew Hardison; Matthew Schmerer; Ola Kvalvaag; Patrick Campbell; Peter W. Cook; Rob Case; Scott Sammons; Shatavia Morrison; Shaun Westlund; Vikramsinha Ghorpade; Yvette Unoarumhi |
| EPI_ISL_2304324, EPI_ISL_2304327 | Alameda County Public Health Department | Alameda County Public Health Department | Kevin Libuit; Kristina Hsieh; Maria Elma |
| EPI_ISL_2496770, EPI_ISL_2496771, EPI_ISL_2496772, EPI_ISL_2496773, EPI_ISL_2496774, EPI_ISL_2496775, EPI_ISL_2496778, EPI_ISL_2496779, EPI_ISL_2496780, EPI_ISL_2496781, EPI_ISL_2496782, EPI_ISL_2496783, EPI_ISL_2496785, EPI_ISL_2496786, EPI_ISL_2496789, EPI_ISL_2496790, EPI_ISL_2496791, EPI_ISL_2496792, EPI_ISL_2496794, EPI_ISL_2496796, EPI_ISL_2496797, EPI_ISL_2496798, EPI_ISL_2496799, EPI_ISL_2496801, EPI_ISL_2496802, EPI_ISL_2496803, EPI_ISL_2496804, EPI_ISL_2496806, EPI_ISL_2496807, EPI_ISL_2496808, EPI_ISL_2496809, EPI_ISL_2496811, EPI_ISL_2496813, EPI_ISL_2496815, EPI_ISL_2496816, EPI_ISL_2496817, EPI_ISL_2496818, EPI_ISL_2496819, EPI_ISL_2496820, EPI_ISL_2496822, EPI_ISL_2496823, EPI_ISL_2496824, EPI_ISL_2496825, EPI_ISL_2496826, EPI_ISL_2496830, EPI_ISL_2496831, EPI_ISL_2496833, EPI_ISL_2496835, EPI_ISL_2496836, EPI_ISL_2496837, EPI_ISL_2496839, EPI_ISL_2496840, EPI_ISL_2496841, EPI_ISL_2496842, EPI_ISL_2496844, EPI_ISL_2496846, EPI_ISL_2496848, EPI_ISL_2496849, EPI_ISL_2496850, EPI_ISL_2496851, EPI_ISL_2496854, EPI_ISL_2496856, EPI_ISL_2496857, EPI_ISL_2496859, EPI_ISL_2496860, EPI_ISL_2496861, EPI_ISL_2496862, EPI_ISL_2496863, EPI_ISL_2496865, EPI_ISL_2496868, EPI_ISL_2496869, EPI_ISL_2496870, EPI_ISL_2496871, EPI_ISL_2496873, EPI_ISL_2496874, EPI_ISL_2496875, EPI_ISL_2496876, EPI_ISL_2496880, EPI_ISL_2496884, EPI_ISL_2496889, EPI_ISL_2496890, EPI_ISL_2496891, EPI_ISL_2496893, EPI_ISL_2496894, EPI_ISL_2496896, EPI_ISL_2496897, EPI_ISL_2496898, EPI_ISL_2496899, EPI_ISL_2496900, EPI_ISL_2496901, EPI_ISL_2496902, EPI_ISL_2496903, EPI_ISL_2496905, EPI_ISL_2496906, EPI_ISL_2496907, EPI_ISL_2496908, EPI_ISL_2496909, EPI_ISL_2496911, EPI_ISL_2496913, EPI_ISL_2496915, EPI_ISL_2496918, EPI_ISL_2496922, EPI_ISL_2496926, EPI_ISL_2496927, EPI_ISL_2496928, EPI_ISL_2496929, EPI_ISL_2496930, EPI_ISL_2496934, EPI_ISL_2496937, EPI_ISL_2496940, EPI_ISL_2496944, EPI_ISL_2496946, EPI_ISL_2496948, EPI_ISL_2496949, EPI_ISL_2496950, EPI_ISL_2496951, EPI_ISL_2496952, EPI_ISL_2496953, EPI_ISL_2496954, EPI_ISL_2496955, EPI_ISL_2496957, EPI_ISL_2496958, EPI_ISL_2496959, EPI_ISL_2496961, EPI_ISL_2496962, EPI_ISL_2496963, EPI_ISL_2496964, EPI_ISL_2496966, EPI_ISL_2496967, EPI_ISL_2496970, EPI_ISL_2496974, EPI_ISL_2496978, EPI_ISL_2496982, EPI_ISL_2496983, EPI_ISL_2496984, EPI_ISL_2496985, EPI_ISL_2496986, EPI_ISL_2496988, EPI_ISL_2496991, EPI_ISL_2496993, EPI_ISL_2496994, EPI_ISL_2496997, EPI_ISL_2496998, EPI_ISL_2497544, EPI_ISL_2497546, EPI_ISL_2497548, EPI_ISL_2497556, EPI_ISL_2497557, EPI_ISL_2497560, EPI_ISL_2497561, EPI_ISL_2497563, EPI_ISL_2497564, EPI_ISL_2497568, EPI_ISL_2497570, EPI_ISL_2497575, EPI_ISL_2497580, EPI_ISL_2497582, EPI_ISL_2497583, EPI_ISL_2497585, EPI_ISL_2497588, EPI_ISL_2497594, EPI_ISL_2497597, EPI_ISL_2497598, EPI_ISL_2497600, EPI_ISL_2497601, EPI_ISL_2497606, EPI_ISL_2497608, EPI_ISL_2497609, EPI_ISL_2497610, EPI_ISL_2497613, EPI_ISL_2497615, EPI_ISL_2497616, EPI_ISL_2497617, EPI_ISL_2497620, EPI_ISL_2497623, EPI_ISL_2497624, EPI_ISL_2497626, EPI_ISL_2497627, EPI_ISL_2497630, EPI_ISL_2497631, EPI_ISL_2497632, EPI_ISL_2497634, EPI_ISL_2497636, EPI_ISL_2497637, EPI_ISL_2497640, EPI_ISL_2497645, EPI_ISL_2497646, EPI_ISL_2497647, EPI_ISL_2497648, EPI_ISL_2497649, EPI_ISL_2497650, EPI_ISL_2497651, EPI_ISL_2497652, EPI_ISL_2497654, EPI_ISL_2497655, EPI_ISL_2497656, EPI_ISL_2497658, EPI_ISL_2497665, EPI_ISL_2497666, EPI_ISL_2497668, EPI_ISL_2497670, EPI_ISL_2497671, EPI_ISL_2497672, EPI_ISL_2497673, EPI_ISL_2497674, EPI_ISL_2497685, EPI_ISL_2497689, EPI_ISL_2497694, EPI_ISL_2497710, EPI_ISL_2497737, EPI_ISL_2497768, EPI_ISL_2497845, EPI_ISL_2658681, EPI_ISL_2658683, EPI_ISL_2658685, EPI_ISL_2658705, EPI_ISL_2659200, EPI_ISL_2659300, EPI_ISL_2659349, EPI_ISL_2659392, EPI_ISL_2659400, EPI_ISL_2659457, EPI_ISL_2659503, EPI_ISL_2659514, EPI_ISL_2659522, EPI_ISL_2659551, EPI_ISL_2659579, EPI_ISL_2758534, EPI_ISL_2758535, EPI_ISL_2758536, EPI_ISL_2758537, EPI_ISL_2758538, EPI_ISL_2758539, EPI_ISL_2758540 | | | |
| see above | CA DPH Viral and Rickettsial Disease Laboratory | Chan-Zuckerberg Biohub | CZB Cliahub Consortium |
| EPI_ISL_2229189, EPI_ISL_2229190, EPI_ISL_2229191, EPI_ISL_2229192, EPI_ISL_2383986, EPI_ISL_2383987, EPI_ISL_2451881, EPI_ISL_2451883, EPI_ISL_2451884, EPI_ISL_2451886, EPI_ISL_2451888, EPI_ISL_2451890, EPI_ISL_2451891, EPI_ISL_2451893, EPI_ISL_2451895, EPI_ISL_2689932, EPI_ISL_2689933, EPI_ISL_2709557, EPI_ISL_2787321, EPI_ISL_2787323, EPI_ISL_2787325, EPI_ISL_2787328 | | | |
| see above | CA-Los Angeles County Public Health Laboratory | Centers for Disease Control and Prevention Division of Viral | Alison Laufer Halpin; Ben L. Rambo-Martin; Clinton R. Paden; Dakota Howard; Darlene Wagner; Dave Wentworth; Dhwani Batra; Jasmine Padilla; Justin Lee; Katie Dillon; Krista Queen; Kristen Knipe; Kristine Lacek; Mark Burroughs; Matthew Schmerer; Mili Sheth; Peter Cook; Sam Shepard; Sarah Nobles; Shoshona Le; Suxiang Tong; Vivien Dugan; Yvette Unoarumhi |

Diseases, Pathogen Discovery

EPI_ISL_2890369, EPI_ISL_2890370, EPI_ISL_2890544, EPI_ISL_2890545, EPI_ISL_2890943, EPI_ISL_2891608, EPI_ISL_2891708, EPI_ISL_2891709, EPI_ISL_2891710, EPI_ISL_2891711, EPI_ISL_2891712, EPI_ISL_2891713, EPI_ISL_2891714, EPI_ISL_2891715, EPI_ISL_2891716, EPI_ISL_2891717, EPI_ISL_2891722, EPI_ISL_2891723, EPI_ISL_2891724, EPI_ISL_2891725, EPI_ISL_2891726, EPI_ISL_2891727, EPI_ISL_2891730, EPI_ISL_2891731, EPI_ISL_2891732, EPI_ISL_2891733, EPI_ISL_2891734, EPI_ISL_2891735, EPI_ISL_2891736, EPI_ISL_2891737, EPI_ISL_2891738, EPI_ISL_2891739, EPI_ISL_2891740, EPI_ISL_2891743, EPI_ISL_2891745, EPI_ISL_2891746, EPI_ISL_2891747, EPI_ISL_2891748, EPI_ISL_2891749, EPI_ISL_2891753, EPI_ISL_2891754, EPI_ISL_2891757, EPI_ISL_2891760, EPI_ISL_2891761, EPI_ISL_2891762, EPI_ISL_2891763, EPI_ISL_2891764, EPI_ISL_2891767, EPI_ISL_2891769, EPI_ISL_2891770, EPI_ISL_2891771, EPI_ISL_2891772, EPI_ISL_2891773, EPI_ISL_2891774, EPI_ISL_2891775, EPI_ISL_2891776, EPI_ISL_2891777, EPI_ISL_2891778, EPI_ISL_2891779, EPI_ISL_2891780, EPI_ISL_2891782, EPI_ISL_2891783, EPI_ISL_2891784, EPI_ISL_2891785, EPI_ISL_2891786, EPI_ISL_2891787, EPI_ISL_2891788, EPI_ISL_2891789, EPI_ISL_2891790, EPI_ISL_2891791, EPI_ISL_2891792, EPI_ISL_2891793, EPI_ISL_2891794, EPI_ISL_2891795, EPI_ISL_2891796, EPI_ISL_2891797, EPI_ISL_2891798, EPI_ISL_2891799, EPI_ISL_2891800, EPI_ISL_2891801, EPI_ISL_2891803, EPI_ISL_2891804, EPI_ISL_2891805, EPI_ISL_2891806, EPI_ISL_2891807, EPI_ISL_2891808, EPI_ISL_2891809, EPI_ISL_2891810, EPI_ISL_2891811, EPI_ISL_2891812, EPI_ISL_2891813, EPI_ISL_2891814, EPI_ISL_2891815, EPI_ISL_2891816, EPI_ISL_2891817, EPI_ISL_2891818, EPI_ISL_2891819, EPI_ISL_2891820, EPI_ISL_2891821, EPI_ISL_2891822, EPI_ISL_2891823, EPI_ISL_2891824, EPI_ISL_2891825, EPI_ISL_2891826, EPI_ISL_2891827, EPI_ISL_2891828, EPI_ISL_2891829, EPI_ISL_2891830, EPI_ISL_2891831, EPI_ISL_2891832, EPI_ISL_2891833, EPI_ISL_2891834, EPI_ISL_2891835, EPI_ISL_2891836, EPI_ISL_2891837, EPI_ISL_2891838, EPI_ISL_2891839, EPI_ISL_2891840, EPI_ISL_2891841, EPI_ISL_2891842, EPI_ISL_2891843, EPI_ISL_2891844, EPI_ISL_2891845, EPI_ISL_2891846, EPI_ISL_2891847, EPI_ISL_2891848, EPI_ISL_2891849, EPI_ISL_2891850, EPI_ISL_2891851, EPI_ISL_2891852, EPI_ISL_2891853, EPI_ISL_2891854, EPI_ISL_2891855, EPI_ISL_2891856, EPI_ISL_2891859, EPI_ISL_2891861, EPI_ISL_2891862, EPI_ISL_2891863, EPI_ISL_2891864, EPI_ISL_2891865, EPI_ISL_2891866, EPI_ISL_2891867, EPI_ISL_2891868, EPI_ISL_2891869, EPI_ISL_2891870, EPI_ISL_2891876, EPI_ISL_2891881, EPI_ISL_2891882, EPI_ISL_2891883, EPI_ISL_2891884, EPI_ISL_2891887, EPI_ISL_2891888, EPI_ISL_2891889, EPI_ISL_2891890, EPI_ISL_2891891, EPI_ISL_2891892, EPI_ISL_2891893, EPI_ISL_2891894, EPI_ISL_2891895, EPI_ISL_2891896, EPI_ISL_2891897, EPI_ISL_2891898, EPI_ISL_2891899, EPI_ISL_2891900, EPI_ISL_2891901, EPI_ISL_2891902, EPI_ISL_2891903, EPI_ISL_2891904, EPI_ISL_2891905, EPI_ISL_2891906, EPI_ISL_2891907, EPI_ISL_2891908, EPI_ISL_2891909, EPI_ISL_2891910, EPI_ISL_2891911, EPI_ISL_2891912, EPI_ISL_2891914, EPI_ISL_2891915, EPI_ISL_2891916, EPI_ISL_2891917, EPI_ISL_2891918, EPI_ISL_2891919, EPI_ISL_2891920, EPI_ISL_2891921, EPI_ISL_2891922, EPI_ISL_2891923, EPI_ISL_2891924, EPI_ISL_2891925, EPI_ISL_2891926, EPI_ISL_2891927, EPI_ISL_2891928, EPI_ISL_2891929, EPI_ISL_2891930, EPI_ISL_2891931, EPI_ISL_2891932, EPI_ISL_2891933, EPI_ISL_2891934, EPI_ISL_2891935, EPI_ISL_2891936, EPI_ISL_2891937, EPI_ISL_2891938, EPI_ISL_2891939, EPI_ISL_2891940, EPI_ISL_2891941, EPI_ISL_2891942, EPI_ISL_2891943, EPI_ISL_2891944, EPI_ISL_2891945, EPI_ISL_2891946, EPI_ISL_2891947, EPI_ISL_2891948, EPI_ISL_2891949, EPI_ISL_2891950, EPI_ISL_2891951, EPI_ISL_2891952, EPI_ISL_2891953, EPI_ISL_2891956, EPI_ISL_2891958, EPI_ISL_2891959, EPI_ISL_2891960, EPI_ISL_2891961, EPI_ISL_2891962, EPI_ISL_2891963, EPI_ISL_2891964, EPI_ISL_2891965, EPI_ISL_2891966, EPI_ISL_2891967, EPI_ISL_2891968, EPI_ISL_2891969, EPI_ISL_2891974, EPI_ISL_2891975, EPI_ISL_2891976, EPI_ISL_2891977, EPI_ISL_2891978, EPI_ISL_2891983, EPI_ISL_2891984, EPI_ISL_2891985, EPI_ISL_2891988, EPI_ISL_2891989, EPI_ISL_2891991, EPI_ISL_2891992, EPI_ISL_2891994, EPI_ISL_2891995, EPI_ISL_2891996, EPI_ISL_2892002, EPI_ISL_2892003, EPI_ISL_2892004, EPI_ISL_2892005, EPI_ISL_2892006, EPI_ISL_2892007, EPI_ISL_2892008, EPI_ISL_2892009, EPI_ISL_2892010, EPI_ISL_2892011, EPI_ISL_2892012, EPI_ISL_2892013, EPI_ISL_2892014, EPI_ISL_2892016, EPI_ISL_2892018, EPI_ISL_2892021, EPI_ISL_2892022, EPI_ISL_2892023, EPI_ISL_2892024, EPI_ISL_2892025, EPI_ISL_2892026, EPI_ISL_2892027, EPI_ISL_2892028, EPI_ISL_2892029, EPI_ISL_2892030, EPI_ISL_2892031, EPI_ISL_2892032, EPI_ISL_2892033, EPI_ISL_2892034, EPI_ISL_2892035, EPI_ISL_2892036, EPI_ISL_2892037, EPI_ISL_2892038, EPI_ISL_2892039, EPI_ISL_2892042, EPI_ISL_2892043, EPI_ISL_2892044, EPI_ISL_2892045, EPI_ISL_2892046, EPI_ISL_2892047, EPI_ISL_2892048, EPI_ISL_2892049, EPI_ISL_2892050, EPI_ISL_2892051, EPI_ISL_2892052, EPI_ISL_2892053, EPI_ISL_2892055, EPI_ISL_2892056, EPI_ISL_2892059, EPI_ISL_2892060, EPI_ISL_2892062, EPI_ISL_2892063, EPI_ISL_2892064, EPI_ISL_2892065, EPI_ISL_2892066, EPI_ISL_2892067, EPI_ISL_2892068, EPI_ISL_2892070, EPI_ISL_2892073, EPI_ISL_2892074, EPI_ISL_2892075, EPI_ISL_2892076, EPI_ISL_2892077, EPI_ISL_2892078, EPI_ISL_2892079, EPI_ISL_2892080, EPI_ISL_2892081, EPI_ISL_2892083, EPI_ISL_2892084, EPI_ISL_2892085, EPI_ISL_2892086, EPI_ISL_2892087, EPI_ISL_2892088, EPI_ISL_2892089, EPI_ISL_2892090, EPI_ISL_2892093, EPI_ISL_2892094, EPI_ISL_2892095, EPI_ISL_2892096, EPI_ISL_2892097, EPI_ISL_2892098, EPI_ISL_2892099, EPI_ISL_2892100, EPI_ISL_2892101, EPI_ISL_2892102, EPI_ISL_2892103, EPI_ISL_2892105, EPI_ISL_2892107, EPI_ISL_2892108, EPI_ISL_2892110, EPI_ISL_2892111, EPI_ISL_2892112, EPI_ISL_2892113, EPI_ISL_2892114, EPI_ISL_2892115, EPI_ISL_2892116, EPI_ISL_2892117, EPI_ISL_2892119, EPI_ISL_2892120, EPI_ISL_2892121, EPI_ISL_2892123, EPI_ISL_2892124, EPI_ISL_2892127, EPI_ISL_2892128, EPI_ISL_2892130, EPI_ISL_2892131, EPI_ISL_2892133, EPI_ISL_2892134, EPI_ISL_2892140, EPI_ISL_2892141, EPI_ISL_2892142, EPI_ISL_2892143, EPI_ISL_2892144, EPI_ISL_2892145, EPI_ISL_2892146, EPI_ISL_2892147, EPI_ISL_2892148, EPI_ISL_2892149, EPI_ISL_2892150, EPI_ISL_2892152, EPI_ISL_2892153, EPI_ISL_2892154, EPI_ISL_2892155, EPI_ISL_2892156, EPI_ISL_2892159, EPI_ISL_2892160, EPI_ISL_2892161, EPI_ISL_2892162, EPI_ISL_2892163, EPI_ISL_2892164, EPI_ISL_2892165, EPI_ISL_2892166, EPI_ISL_2892170, EPI_ISL_2892171, EPI_ISL_2892174, EPI_ISL_2892175, EPI_ISL_2892176, EPI_ISL_2892177, EPI_ISL_2892178, EPI_ISL_2892180, EPI_ISL_2892181, EPI_ISL_2892182, EPI_ISL_2892183, EPI_ISL_2892184, EPI_ISL_2892185, EPI_ISL_2892186, EPI_ISL_2892187, EPI_ISL_2892188, EPI_ISL_2892189, EPI_ISL_2892190, EPI_ISL_2892191, EPI_ISL_2892192, EPI_ISL_2892193, EPI_ISL_2892195, EPI_ISL_2892196, EPI_ISL_2892197, EPI_ISL_2892198, EPI_ISL_2892199, EPI_ISL_2892200, EPI_ISL_2892201, EPI_ISL_2892202, EPI_ISL_2892204, EPI_ISL_2892205, EPI_ISL_2892206, EPI_ISL_2892207, EPI_ISL_2892208, EPI_ISL_2892209, EPI_ISL_2892211, EPI_ISL_2892213, EPI_ISL_2892215, EPI_ISL_2892216, EPI_ISL_2892217, EPI_ISL_2892221, EPI_ISL_2892224, EPI_ISL_2892225, EPI_ISL_2892231, EPI_ISL_2892232, EPI_ISL_2892241, EPI_ISL_2892242, EPI_ISL_2892243, EPI_ISL_2892244, EPI_ISL_2892248, EPI_ISL_2892249, EPI_ISL_2892251, EPI_ISL_2892253, EPI_ISL_2892254, EPI_ISL_2892255, EPI_ISL_2892256, EPI_ISL_2892257, EPI_ISL_2892259, EPI_ISL_2892262, EPI_ISL_2892263, EPI_ISL_2892265, EPI_ISL_2892266, EPI_ISL_2892267, EPI_ISL_2892268, EPI_ISL_2892269, EPI_ISL_2892270, EPI_ISL_2892271, EPI_ISL_2892274, EPI_ISL_2892275, EPI_ISL_2892276, EPI_ISL_2892277, EPI_ISL_2892283, EPI_ISL_2892284, EPI_ISL_2892285, EPI_ISL_2892290, EPI_ISL_2892291, EPI_ISL_2892293, EPI_ISL_2892294, EPI_ISL_2892295, EPI_ISL_2892296, EPI_ISL_2892297, EPI_ISL_2892298, EPI_ISL_2892299, EPI_ISL_2892300, EPI_ISL_2892301, EPI_ISL_2892302, EPI_ISL_2892303, EPI_ISL_2892304, EPI_ISL_2892305, EPI_ISL_2892306, EPI_ISL_2892307, EPI_ISL_2892311, EPI_ISL_2892312, EPI_ISL_2892313, EPI_ISL_2892314, EPI_ISL_2892315, EPI_ISL_2892316, EPI_ISL_2892317, EPI_ISL_2892318, EPI_ISL_2892319, EPI_ISL_2892320, EPI_ISL_2892321, EPI_ISL_2892323, EPI_ISL_2892326, EPI_ISL_2892327, EPI_ISL_2892331, EPI_ISL_2892333, EPI_ISL_2892334, EPI_ISL_2892335, EPI_ISL_2892336, EPI_ISL_2892344, EPI_ISL_2892345, EPI_ISL_2892346, EPI_ISL_2892347, EPI_ISL_2892348, EPI_ISL_2892349, EPI_ISL_2892350, EPI_ISL_2892351, EPI_ISL_2892353, EPI_ISL_2892354, EPI_ISL_2892355, EPI_ISL_2892356, EPI_ISL_2892361, EPI_ISL_2892362, EPI_ISL_2892363, EPI_ISL_2892364, EPI_ISL_2892365, EPI_ISL_2892366, EPI_ISL_2892367, EPI_ISL_2892368, EPI_ISL_2892371, EPI_ISL_2892399, EPI_ISL_2892400, EPI_ISL_2921850, EPI_ISL_2921851, EPI_ISL_2921852, EPI_ISL_2921857, EPI_ISL_2921858, EPI_ISL_2921859, EPI_ISL_2921860, EPI_ISL_2921861, EPI_ISL_2921864, EPI_ISL_2921865, EPI_ISL_2921866, EPI_ISL_2921867, EPI_ISL_2921869, EPI_ISL_2921871, EPI_ISL_2921872, EPI_ISL_2921875, EPI_ISL_2921885, EPI_ISL_2921887, EPI_ISL_2921888, EPI_ISL_2921894, EPI_ISL_2921895, EPI_ISL_2921896, EPI_ISL_2921909, EPI_ISL_2921910, EPI_ISL_2921913, EPI_ISL_2921915, EPI_ISL_2921916, EPI_ISL_2921917, EPI_ISL_2921921, EPI_ISL_2921927, EPI_ISL_2921935, EPI_ISL_2921936, EPI_ISL_2921937, EPI_ISL_2921939, EPI_ISL_2921940, EPI_ISL_2921941, EPI_ISL_2921945, EPI_ISL_2921946, EPI_ISL_2921947, EPI_ISL_2921948, EPI_ISL_2921955, EPI_ISL_2921956, EPI_ISL_2921964, EPI_ISL_2921965, EPI_ISL_2921966, EPI_ISL_2921967, EPI_ISL_2921968, EPI_ISL_2921969, EPI_ISL_2921970, EPI_ISL_2921971, EPI_ISL_2921972, EPI_ISL_2921973, EPI_ISL_2921975, EPI_ISL_2921976, EPI_ISL_2921977, EPI_ISL_2921978, EPI_ISL_2921979, EPI_ISL_2921980, EPI_ISL_2921981, EPI_ISL_2921986, EPI_ISL_2921987, EPI_ISL_2921989, EPI_ISL_2921990, EPI_ISL_2921991, EPI_ISL_2921992, EPI_ISL_2921993, EPI_ISL_2922002, EPI_ISL_2922003, EPI_ISL_2922005, EPI_ISL_2922013, EPI_ISL_2922020, EPI_ISL_2922021, EPI_ISL_2922034, EPI_ISL_2922039, EPI_ISL_2922040, EPI_ISL_2922041, EPI_ISL_2922042, EPI_ISL_2922043, EPI_ISL_2922044, EPI_ISL_2922046, EPI_ISL_2922047, EPI_ISL_2922048, EPI_ISL_2922049, EPI_ISL_2922051, EPI_ISL_2922053, EPI_ISL_2922054, EPI_ISL_2922055, EPI_ISL_2922056, EPI_ISL_2922063, EPI_ISL_2922079, EPI_ISL_2922081, EPI_ISL_2922086, EPI_ISL_2922090, EPI_ISL_2922102, EPI_ISL_2922103, EPI_ISL_2922104, EPI_ISL_2922106, EPI_ISL_2922113, EPI_ISL_2922115, EPI_ISL_2922117, EPI_ISL_2922118, EPI_ISL_2922119, EPI_ISL_2922120, EPI_ISL_2922124, EPI_ISL_2922127, EPI_ISL_2922128, EPI_ISL_2922132, EPI_ISL_2922135, EPI_ISL_2922142, EPI_ISL_2922148, EPI_ISL_2922163, EPI_ISL_2922165, EPI_ISL_2922166, EPI_ISL_2922171, EPI_ISL_2922178, EPI_ISL_2922180, EPI_ISL_2922181, EPI_ISL_2922182, EPI_ISL_2922183, EPI_ISL_2922184, EPI_ISL_2922189, EPI_ISL_2922190, EPI_ISL_2922194, EPI_ISL_2922195, EPI_ISL_2922205, EPI_ISL_2922206, EPI_ISL_2922207, EPI_ISL_2922208, EPI_ISL_2922209, EPI_ISL_2922210, EPI_ISL_2922211, EPI_ISL_2922221, EPI_ISL_2922222, EPI_ISL_2922223, EPI_ISL_2922225, EPI_ISL_2922226, EPI_ISL_2922227, EPI_ISL_2922228, EPI_ISL_2922246, EPI_ISL_2922248, EPI_ISL_2922249, EPI_ISL_2922250, EPI_ISL_2922251, EPI_ISL_2922255, EPI_ISL_2922256, EPI_ISL_2922257, EPI_ISL_2922262, EPI_ISL_2922263, EPI_ISL_2922264, EPI_ISL_2922269, EPI_ISL_2922273, EPI_ISL_2922274, EPI_ISL_2922275, EPI_ISL_2922276, EPI_ISL_2922277, EPI_ISL_2922278, EPI_ISL_2922279, EPI_ISL_2922280, EPI_ISL_2922285, EPI_ISL_2922289, EPI_ISL_2922290, EPI_ISL_2922293, EPI_ISL_2922294, EPI_ISL_2922295, EPI_ISL_2922296, EPI_ISL_2922297, EPI_ISL_2922298, EPI_ISL_2922299, EPI_ISL_2922300, EPI_ISL_2922301, EPI_ISL_2922302, EPI_ISL_2922303, EPI_ISL_2922304, EPI_ISL_2923285, EPI_ISL_2923292, EPI_ISL_2923293, EPI_ISL_2923294, EPI_ISL_2923295, EPI_ISL_2923301, EPI_ISL_2923302, EPI_ISL_2923303, EPI_ISL_2923304, EPI_ISL_2923307, EPI_ISL_2923318, EPI_ISL_2923319, EPI_ISL_2923320, EPI_ISL_2923321, EPI_ISL_2923323, EPI_ISL_2923326, EPI_ISL_2923327, EPI_ISL_2923328, EPI_ISL_2923329, EPI_ISL_2923332, EPI_ISL_2923333, EPI_ISL_2923334, EPI_ISL_2923335, EPI_ISL_2923336, EPI_ISL_2923337, EPI_ISL_2923340, EPI_ISL_2923341, EPI_ISL_2923342, EPI_ISL_2923343, EPI_ISL_2923355, EPI_ISL_2923356, EPI_ISL_2923357, EPI_ISL_2923358, EPI_ISL_2923359, EPI_ISL_2923360, EPI_ISL_2923361, EPI_ISL_2923363, EPI_ISL_2923364, EPI_ISL_2923365, EPI_ISL_2923367, EPI_ISL_2923368, EPI_ISL_2923371, EPI_ISL_2923372, EPI_ISL_2923373, EPI_ISL_2923379, EPI_ISL_2923380, EPI_ISL_2923384, EPI_ISL_2923385, EPI_ISL_2923386, EPI_ISL_2923387, EPI_ISL_2923388, EPI_ISL_2923389, EPI_ISL_2923390, EPI_ISL_2923401, EPI_ISL_2923405, EPI_ISL_2923406, EPI_ISL_2923407, EPI_ISL_2923408, EPI_ISL_2923409, EPI_ISL_2923410, EPI_ISL_2923411, EPI_ISL_2923433, EPI_ISL_2923434, EPI_ISL_2923435, EPI_ISL_2923436, EPI_ISL_2923455, EPI_ISL_2923456, EPI_ISL_2923458, EPI_ISL_2923459, EPI_ISL_2923460, EPI_ISL_2923461, EPI_ISL_2923472, EPI_ISL_2923473, EPI_ISL_2923474, EPI_ISL_2923475, EPI_ISL_2923476, EPI_ISL_2923477, EPI_ISL_2923479, EPI_ISL_2923486, EPI_ISL_2923488, EPI_ISL_2923492, EPI_ISL_2923493, EPI_ISL_2923494, EPI_ISL_2923502, EPI_ISL_2923503, EPI_ISL_2923504, EPI_ISL_2923505, EPI_ISL_2923506, EPI_ISL_2923507, EPI_ISL_2923511, EPI_ISL_2923515, EPI_ISL_2923523, EPI_ISL_2923525, EPI_ISL_2923541, EPI_ISL_2923555, EPI_ISL_2923558, EPI_ISL_2923560, EPI_ISL_2923561, EPI_ISL_2923562, EPI_ISL_2923563, EPI_ISL_2923564, EPI_ISL_2923565, EPI_ISL_2923566, EPI_ISL_2923567, EPI_ISL_2923568, EPI_ISL_2923569, EPI_ISL_2923570, EPI_ISL_2923571, EPI_ISL_2923572, EPI_ISL_2923573, EPI_ISL_2923574, EPI_ISL_2923575, EPI_ISL_2923576, EPI_ISL_2923577, EPI_ISL_2923578, EPI_ISL_2923579, EPI_ISL_2923614, EPI_ISL_2923615, EPI_ISL_2923620, EPI_ISL_2923623, EPI_ISL_2923633, EPI_ISL_2923634, EPI_ISL_2923635, EPI_ISL_2923636, EPI_ISL_2923637, EPI_ISL_2923638, EPI_ISL_2923639, EPI_ISL_2923643, EPI_ISL_2923644, EPI_ISL_2923652, EPI_ISL_2923653, EPI_ISL_2923659, EPI_ISL_2923660, EPI_ISL_2923661, EPI_ISL_2923662, EPI_ISL_2923663, EPI_ISL_2923664, EPI_ISL_2923665, EPI_ISL_2923666, EPI_ISL_2923667, EPI_ISL_2923668, EPI_ISL_2923671, EPI_ISL_2923672, EPI_ISL_2923673, EPI_ISL_2923679, EPI_ISL_2923682, EPI_ISL_2923683, EPI_ISL_2923684, EPI_ISL_2923688, EPI_ISL_2923689, EPI_ISL_2923690, EPI_ISL_2923691, EPI_ISL_2923693, EPI_ISL_2923710, EPI_ISL_2923711, EPI_ISL_2923713, EPI_ISL_2923722, EPI_ISL_2923730, EPI_ISL_2923731, EPI_ISL_2923732, EPI_ISL_2923738, EPI_ISL_2923739, EPI_ISL_2923741, EPI_ISL_2923742, EPI_ISL_2923744, EPI_ISL_2923745, EPI_ISL_2923746, EPI_ISL_2923747, EPI_ISL_2923756, EPI_ISL_2923759, EPI_ISL_2923764, EPI_ISL_2923765, EPI_ISL_2923769

| | | | |
|---|---|---|---|
| see above | CDPH VBL | California Department of Public Health | CDPH-COVIDNet |

EPI_ISL_2452028, EPI_ISL_2452030, EPI_ISL_2452031, EPI_ISL_2452035, EPI_ISL_2452037, EPI_ISL_2452038, EPI_ISL_2452040, EPI_ISL_2452042, EPI_ISL_2452043, EPI_ISL_2452045

| | | | |
|---|---|---|---|
| see above | CDPH, Viral and Rickettsial Disease Laboratory | Centers for Disease Control and Prevention Division of Viral Diseases, Pathogen Discovery | Alison Laufer Halpin; Ben L. Rambo-Martin; Clinton R. Paden; Dakota Howard; Darlene Wagner; Dave Wentworth; Dhwani Batra; Jasmine Padilla; Justin Lee; Katie Dillon; Krista Queen; Kristen Knipe; Kristine Lacek; Mark Burroughs; Matthew Schmerer; Mili Sheth; Peter Cook; Sam Shepard; Sarah Nobles; Shoshona Le; Suxiang Tong; Vivien Dugan; Yvette Unoarumhi |

EPI_ISL_2842807, EPI_ISL_2842811, EPI_ISL_2842812, EPI_ISL_2842813, EPI_ISL_2842814, EPI_ISL_2842815, EPI_ISL_2842816, EPI_ISL_2842818, EPI_ISL_2842830, EPI_ISL_2842832, EPI_ISL_2892405, EPI_ISL_2892406, EPI_ISL_2892407, EPI_ISL_2892409, EPI_ISL_2892410, EPI_ISL_2892411, EPI_ISL_2892412, EPI_ISL_2892413, EPI_ISL_2892414, EPI_ISL_2892415, EPI_ISL_2892416, EPI_ISL_2892417, EPI_ISL_2892418, EPI_ISL_2892419, EPI_ISL_2892436, EPI_ISL_2892437, EPI_ISL_2892438, EPI_ISL_2892439, EPI_ISL_2892440, EPI_ISL_2892441, EPI_ISL_2892442, EPI_ISL_2892443, EPI_ISL_2892444, EPI_ISL_2892445, EPI_ISL_2892446, EPI_ISL_2892447, EPI_ISL_2892448, EPI_ISL_2892449, EPI_ISL_2892450, EPI_ISL_2892451, EPI_ISL_2892452, EPI_ISL_2892453, EPI_ISL_2892458, EPI_ISL_2892459, EPI_ISL_2892460, EPI_ISL_2892461, EPI_ISL_2892462, EPI_ISL_2892463, EPI_ISL_2892464, EPI_ISL_2892465, EPI_ISL_2892466, EPI_ISL_2892467, EPI_ISL_2892468, EPI_ISL_2892469, EPI_ISL_2892470, EPI_ISL_2892471, EPI_ISL_2892472, EPI_ISL_2892473, EPI_ISL_2892474, EPI_ISL_2892475, EPI_ISL_2892476, EPI_ISL_2892477, EPI_ISL_2892478, EPI_ISL_2892479, EPI_ISL_2892480, EPI_ISL_2892481, EPI_ISL_2892482, EPI_ISL_2892483, EPI_ISL_2892484, EPI_ISL_2892485, EPI_ISL_2892486, EPI_ISL_2892487, EPI_ISL_2892488, EPI_ISL_2892489, EPI_ISL_2892490, EPI_ISL_2892501, EPI_ISL_2892502, EPI_ISL_2892503, EPI_ISL_2892504, EPI_ISL_2892507, EPI_ISL_2892508, EPI_ISL_2892509, EPI_ISL_2892510, EPI_ISL_2892512, EPI_ISL_2892513, EPI_ISL_2892514, EPI_ISL_2892515, EPI_ISL_2892516, EPI_ISL_2892517, EPI_ISL_2892518, EPI_ISL_2892519, EPI_ISL_2892520, EPI_ISL_2892521, EPI_ISL_2892522, EPI_ISL_2892523, EPI_ISL_2892524, EPI_ISL_2892525, EPI_ISL_2892526, EPI_ISL_2892527, EPI_ISL_2892528, EPI_ISL_2892529, EPI_ISL_2892530, EPI_ISL_2892531, EPI_ISL_2892532, EPI_ISL_2892533, EPI_ISL_2892534, EPI_ISL_2892536, EPI_ISL_2892540, EPI_ISL_2892550, EPI_ISL_2892552, EPI_ISL_2892553, EPI_ISL_2892554, EPI_ISL_2892555, EPI_ISL_2892556, EPI_ISL_2892557, EPI_ISL_2892558, EPI_ISL_2892559, EPI_ISL_2892560, EPI_ISL_2892561, EPI_ISL_2892562, EPI_ISL_2892563, EPI_ISL_2892564, EPI_ISL_2892565, EPI_ISL_2892576, EPI_ISL_2892577, EPI_ISL_2892578, EPI_ISL_2892579, EPI_ISL_2892580, EPI_ISL_2892581, EPI_ISL_2892582, EPI_ISL_2892583, EPI_ISL_2892584, EPI_ISL_2892585, EPI_ISL_2892586, EPI_ISL_2892587, EPI_ISL_2892588, EPI_ISL_2892589, EPI_ISL_2892590, EPI_ISL_2892591, EPI_ISL_2892592

| | | | |
|---|---|---|---|
| see above | California Department of Public Health | California Department of Public Health | CDPH IDLB COVIDNet et al |

EPI_ISL_2391753, EPI_ISL_2405179, EPI_ISL_2405205, EPI_ISL_2620776, EPI_ISL_2620777, EPI_ISL_2620778, EPI_ISL_2772502, EPI_ISL_2772503, EPI_ISL_2772504, EPI_ISL_2812001, EPI_ISL_2894571

| | | | |
|---|---|---|---|
| see above | California Hospital Medical Center | Los Angeles County PHL | P. Hemarajata et al. |
| EPI_ISL_2907555, EPI_ISL_2907559, EPI_ISL_2907576 | Centinela Hospital Medical Center | Los Angeles County Public Health Laboratories | P. Hemarajata et al. |
| EPI_ISL_2671484, EPI_ISL_2942729 | Centro de Investigacion Biomedica de Occidente (CIBO) | Unidad de Genomica Avanzada | ; Alejandra Garcia-Gasca; Alejandra Hernandez-Teran; Alejandro Sanchez-Flores; Alfredo Herrera-Estrella; Alicia Ocaña-Mondragon; Andreu Comas-Garcia; Angel Gustavo Salas-Lais; Antonio Loza Roman; Bernardo Martinez-Miguel; Blanca Taboada; Brenda Irasema Maldonado-Meza; Bruno Gomez-Gil; Carla Ivon Herrera-Najera; Carlos F. Arias; Celia Boukadida; Celida Duque Molina; Celida Martinez- Rodriguez; Clara Esperanza Santacruz-Tinoco; Concepcion Grajales-Muñiz; Consorcio Mexicano de Vigilancia Genomica (CoViGen-Mex). Authors (in alphabetical order): Julio Elias Alvarado-Yaah; Cristobal Chaidez-Quiroz; Daniel Fregoso-Rueda; Daniel Lira Morales; Eduardo Becerril-Vargas; Fernando Fontove-Herrera; Fidencio Mejia-Nepomuceno; Francisco Pulido; Gloria Elena Espinosa-Ayala; Gloria Maria Molina-Salinas; Gloria Vazquez; Hector Esteban Paz-Juarez; Hector Montoya-Fuentes; Helen Haydee Fernanda Ramirez-Plascencia; Irvin Gonzalez-Lopez; Jean Pierre Gonzalez; Jesus Hernandez; Joel Armando Vazquez-Perez.; Jorge Salas-Hernandez; Jose Antonio Enciso-Moreno; Jose Arturo Martinez-Orozco; Jose Esteban Muñoz-Medina; Jose de Jesus Nuñez-Contreras; Juan Bautista Chale-Dzul; Julissa Enciso-Ibarra; Luis Alberto Ochoa-Carrera; Margarita Matias-Florentino; Maria Guadalupe Santiago-Mauricio; Maria Guadalupe de Jesus Mireles-Rivera; Mario Mujica-Sanchez; Marissa Perez-Garcia; Nelly Selem-Mojica; Pavel Isa; Ricardo Ciria Merce; Ricardo Grande; Rosa Maria Gutierrez Rios; Santiago avila-Rios; Selene Zarate; Susana Lopez; Veronica Mata-Haro; Victor Eduardo Garcia-Arias; Victor Hugo Borja-Aburto |
| EPI_ISL_2391793 | City of Hope Comprehensive Cancer Center | Los Angeles County PHL | P. Hemarajata et al. |

EPI_ISL_2341556, EPI_ISL_2497573, EPI_ISL_2497574, EPI_ISL_2497619, EPI_ISL_2497621, EPI_ISL_2500066, EPI_ISL_2500087, EPI_ISL_2500148

| | | | |
|---|---|---|---|
| see above | County of San Luis Obispo Public Health Laboratory | Chan-Zuckerberg Biohub | CZB Cliahub Consortium |

EPI_ISL_2614384, EPI_ISL_2652974, EPI_ISL_2658954, EPI_ISL_2658981, EPI_ISL_2658982, EPI_ISL_2658983, EPI_ISL_2659116, EPI_ISL_2921028, EPI_ISL_2921029, EPI_ISL_2921030

| | | | |
|---|---|---|---|
| see above | County of San Luis Obispo Public Health Laboratory | County of San Luis Obispo Public Health Laboratory | Frances Sidhu; Katrina Erwin |

EPI_ISL_2507257, EPI_ISL_2507258, EPI_ISL_2507260, EPI_ISL_2507265, EPI_ISL_2507266, EPI_ISL_2507272, EPI_ISL_2507281, EPI_ISL_2507287, EPI_ISL_2507295, EPI_ISL_2507296, EPI_ISL_2507299, EPI_ISL_2507300, EPI_ISL_2507303, EPI_ISL_2507304, EPI_ISL_2507305, EPI_ISL_2507307, EPI_ISL_2507312, EPI_ISL_2507315, EPI_ISL_2507316, EPI_ISL_2507317, EPI_ISL_2507318, EPI_ISL_2507323, EPI_ISL_2507324, EPI_ISL_2507325, EPI_ISL_2507328, EPI_ISL_2507329, EPI_ISL_2507330, EPI_ISL_2507332, EPI_ISL_2507335, EPI_ISL_2507336, EPI_ISL_2507338, EPI_ISL_2507342, EPI_ISL_2507343, EPI_ISL_2507344, EPI_ISL_2507345, EPI_ISL_2507346, EPI_ISL_2507347, EPI_ISL_2507348, EPI_ISL_2507350, EPI_ISL_2507351, EPI_ISL_2507355, EPI_ISL_2507362, EPI_ISL_2507363, EPI_ISL_2755141, EPI_ISL_2755142, EPI_ISL_2755143, EPI_ISL_2755144, EPI_ISL_2755148, EPI_ISL_2755149, EPI_ISL_2755150, EPI_ISL_2755151, EPI_ISL_2755152, EPI_ISL_2755153, EPI_ISL_2755154, EPI_ISL_2755155, EPI_ISL_2755156, EPI_ISL_2755157, EPI_ISL_2755158, EPI_ISL_2755159, EPI_ISL_2755160, EPI_ISL_2755161, EPI_ISL_2755162, EPI_ISL_2755163, EPI_ISL_2755164, EPI_ISL_2755165, EPI_ISL_2755166, EPI_ISL_2755177, EPI_ISL_2755178, EPI_ISL_2755179, EPI_ISL_2755180, EPI_ISL_2755181, EPI_ISL_2755182, EPI_ISL_2755183, EPI_ISL_2755184, EPI_ISL_2755185, EPI_ISL_2755186, EPI_ISL_2755187, EPI_ISL_2755188, EPI_ISL_2755189, EPI_ISL_2755190, EPI_ISL_2755191, EPI_ISL_2755192, EPI_ISL_2755193, EPI_ISL_2755194, EPI_ISL_2755195, EPI_ISL_2755196, EPI_ISL_2755197, EPI_ISL_2755198, EPI_ISL_2755199, EPI_ISL_2755209, EPI_ISL_2755210, EPI_ISL_2755211, EPI_ISL_2755212, EPI_ISL_2755213, EPI_ISL_2755214, EPI_ISL_2755215, EPI_ISL_2755216, EPI_ISL_2755217, EPI_ISL_2755218, EPI_ISL_2755219, EPI_ISL_2755220, EPI_ISL_2755221, EPI_ISL_2755222, EPI_ISL_2755223, EPI_ISL_2755224, EPI_ISL_2755225, EPI_ISL_2755226, EPI_ISL_2755227, EPI_ISL_2755228, EPI_ISL_2755229, EPI_ISL_2755230, EPI_ISL_2755237, EPI_ISL_2755238, EPI_ISL_2755239, EPI_ISL_2755240, EPI_ISL_2755241, EPI_ISL_2755242, EPI_ISL_2755243, EPI_ISL_2755244, EPI_ISL_2755245, EPI_ISL_2755246, EPI_ISL_2755247, EPI_ISL_2755248, EPI_ISL_2755249, EPI_ISL_2755250, EPI_ISL_2755251, EPI_ISL_2755252, EPI_ISL_2755253, EPI_ISL_2755254, EPI_ISL_2755255, EPI_ISL_2755256, EPI_ISL_2755257, EPI_ISL_2755258, EPI_ISL_2755259, EPI_ISL_2755260, EPI_ISL_2755261, EPI_ISL_2755262, EPI_ISL_2755263, EPI_ISL_2755266, EPI_ISL_2755267, EPI_ISL_2755268, EPI_ISL_2755269, EPI_ISL_2755270, EPI_ISL_2755271, EPI_ISL_2755273, EPI_ISL_2755274, EPI_ISL_2755275, EPI_ISL_2755276, EPI_ISL_2755277, EPI_ISL_2755278, EPI_ISL_2755279, EPI_ISL_2755280, EPI_ISL_2755281, EPI_ISL_2755282, EPI_ISL_2755283, EPI_ISL_2755284, EPI_ISL_2755285, EPI_ISL_2755286, EPI_ISL_2755287, EPI_ISL_2755288, EPI_ISL_2755289, EPI_ISL_2755290, EPI_ISL_2755291, EPI_ISL_2755292, EPI_ISL_2755293, EPI_ISL_2755294, EPI_ISL_2755295, EPI_ISL_2755296, EPI_ISL_2755297, EPI_ISL_2755298, EPI_ISL_2755299, EPI_ISL_2755300, EPI_ISL_2755302, EPI_ISL_2755303, EPI_ISL_2755304, EPI_ISL_2755305, EPI_ISL_2755306, EPI_ISL_2755307, EPI_ISL_2755308, EPI_ISL_2755309, EPI_ISL_2755310, EPI_ISL_2755311, EPI_ISL_2755312, EPI_ISL_2755313, EPI_ISL_2755314, EPI_ISL_2755315, EPI_ISL_2755316, EPI_ISL_2755317, EPI_ISL_2755318, EPI_ISL_2755319, EPI_ISL_2755320, EPI_ISL_2755321, EPI_ISL_2755322, EPI_ISL_2755323, EPI_ISL_2755324, EPI_ISL_2755325, EPI_ISL_2755326, EPI_ISL_2755328, EPI_ISL_2755329, EPI_ISL_2755330, EPI_ISL_2755331, EPI_ISL_2755332, EPI_ISL_2755333, EPI_ISL_2755334, EPI_ISL_2755335, EPI_ISL_2755336, EPI_ISL_2755337, EPI_ISL_2755341, EPI_ISL_2755342, EPI_ISL_2755343, EPI_ISL_2755344, EPI_ISL_2755345, EPI_ISL_2755346, EPI_ISL_2755347, EPI_ISL_2755348, EPI_ISL_2755349, EPI_ISL_2755350, EPI_ISL_2755351, EPI_ISL_2755352, EPI_ISL_2755353, EPI_ISL_2755354, EPI_ISL_2755355, EPI_ISL_2755356, EPI_ISL_2755357, EPI_ISL_2755358, EPI_ISL_2755359, EPI_ISL_2755360, EPI_ISL_2755361, EPI_ISL_2755362, EPI_ISL_2755363, EPI_ISL_2755364, EPI_ISL_2755365, EPI_ISL_2755366, EPI_ISL_2755367, EPI_ISL_2755368, EPI_ISL_2755369, EPI_ISL_2755370, EPI_ISL_2755371, EPI_ISL_2755372, EPI_ISL_2755373, EPI_ISL_2755377, EPI_ISL_2755378, EPI_ISL_2755379, EPI_ISL_2755380, EPI_ISL_2755381, EPI_ISL_2755382, EPI_ISL_2755383, EPI_ISL_2755384, EPI_ISL_2755385, EPI_ISL_2755386, EPI_ISL_2755387, EPI_ISL_2755388, EPI_ISL_2755389, EPI_ISL_2755390, EPI_ISL_2755391, EPI_ISL_2755392, EPI_ISL_2755393, EPI_ISL_2755394, EPI_ISL_2755395, EPI_ISL_2755396, EPI_ISL_2755397, EPI_ISL_2755398, EPI_ISL_2755399, EPI_ISL_2755400, EPI_ISL_2755401, EPI_ISL_2755402, EPI_ISL_2755403, EPI_ISL_2755404, EPI_ISL_2755405, EPI_ISL_2755406, EPI_ISL_2755407, EPI_ISL_2755408, EPI_ISL_2755409, EPI_ISL_2755410, EPI_ISL_2755411, EPI_ISL_2755412, EPI_ISL_2755413, EPI_ISL_2755414, EPI_ISL_2755415, EPI_ISL_2755422, EPI_ISL_2755423, EPI_ISL_2755424, EPI_ISL_2755425, EPI_ISL_2755426, EPI_ISL_2755427, EPI_ISL_2755428, EPI_ISL_2755429, EPI_ISL_2755430, EPI_ISL_2755431, EPI_ISL_2755432, EPI_ISL_2755433, EPI_ISL_2755434, EPI_ISL_2755435, EPI_ISL_2755436, EPI_ISL_2755437, EPI_ISL_2755438, EPI_ISL_2755439, EPI_ISL_2755440, EPI_ISL_2755441, EPI_ISL_2755442, EPI_ISL_2755443, EPI_ISL_2755445, EPI_ISL_2755446, EPI_ISL_2755447, EPI_ISL_2755448, EPI_ISL_2755449, EPI_ISL_2755450, EPI_ISL_2755451, EPI_ISL_2755452, EPI_ISL_2755453, EPI_ISL_2755454, EPI_ISL_2755455, EPI_ISL_2755456, EPI_ISL_2755457, EPI_ISL_2755458, EPI_ISL_2755459, EPI_ISL_2755460, EPI_ISL_2755461, EPI_ISL_2755462, EPI_ISL_2755463, EPI_ISL_2755464, EPI_ISL_2755465, EPI_ISL_2755466, EPI_ISL_2755467, EPI_ISL_2755468, EPI_ISL_2755469, EPI_ISL_2755470, EPI_ISL_2755471, EPI_ISL_2755472, EPI_ISL_2783647, EPI_ISL_2783648, EPI_ISL_2783649, EPI_ISL_2783652, EPI_ISL_2783653, EPI_ISL_2783654, EPI_ISL_2783655, EPI_ISL_2783656, EPI_ISL_2783657, EPI_ISL_2783658, EPI_ISL_2783659, EPI_ISL_2783660, EPI_ISL_2783661, EPI_ISL_2783662, EPI_ISL_2783663, EPI_ISL_2783664, EPI_ISL_2783795, EPI_ISL_2783975, EPI_ISL_2783976, EPI_ISL_2783978, EPI_ISL_2783979, EPI_ISL_2783980, EPI_ISL_2783981, EPI_ISL_2783982, EPI_ISL_2783983, EPI_ISL_2783985, EPI_ISL_2784169, EPI_ISL_2784180, EPI_ISL_2784193, EPI_ISL_2784201, EPI_ISL_2784216, EPI_ISL_2784217, EPI_ISL_2784218, EPI_ISL_2784219, EPI_ISL_2784220, EPI_ISL_2784221, EPI_ISL_2784225, EPI_ISL_2784226, EPI_ISL_2784227, EPI_ISL_2784228, EPI_ISL_2784229, EPI_ISL_2784230, EPI_ISL_2784231, EPI_ISL_2784233, EPI_ISL_2784237, EPI_ISL_2784238, EPI_ISL_2784239, EPI_ISL_2784240, EPI_ISL_2784241, EPI_ISL_2784242, EPI_ISL_2784243, EPI_ISL_2784244, EPI_ISL_2784245, EPI_ISL_2784246, EPI_ISL_2784247, EPI_ISL_2784248, EPI_ISL_2784249, EPI_ISL_2784250, EPI_ISL_2784251, EPI_ISL_2784252, EPI_ISL_2784253, EPI_ISL_2784254, EPI_ISL_2784255, EPI_ISL_2784256, EPI_ISL_2784264, EPI_ISL_2784265, EPI_ISL_2784266, EPI_ISL_2784267, EPI_ISL_2784268, EPI_ISL_2784269, EPI_ISL_2784270, EPI_ISL_2784271, EPI_ISL_2784272, EPI_ISL_2784273, EPI_ISL_2784277, EPI_ISL_2784278, EPI_ISL_2784279, EPI_ISL_2784280, EPI_ISL_2784281, EPI_ISL_2784282, EPI_ISL_2784283, EPI_ISL_2784287, EPI_ISL_2784288, EPI_ISL_2784289, EPI_ISL_2784290, EPI_ISL_2784291, EPI_ISL_2784292, EPI_ISL_2784293, EPI_ISL_2784294, EPI_ISL_2784297, EPI_ISL_2784301, EPI_ISL_2784302, EPI_ISL_2784303, EPI_ISL_2784309, EPI_ISL_2784310, EPI_ISL_2784312, EPI_ISL_2784313, EPI_ISL_2784314, EPI_ISL_2784315, EPI_ISL_2784316, EPI_ISL_2784317, EPI_ISL_2784318, EPI_ISL_2784319, EPI_ISL_2784320, EPI_ISL_2784321, EPI_ISL_2784322, EPI_ISL_2784323, EPI_ISL_2784324, EPI_ISL_2784325, EPI_ISL_2784326, EPI_ISL_2784327, EPI_ISL_2839678, EPI_ISL_2839685, EPI_ISL_2839686, EPI_ISL_2839693, EPI_ISL_2839699, EPI_ISL_2839700, EPI_ISL_2839717, EPI_ISL_2839720, EPI_ISL_2839724, EPI_ISL_2839730, EPI_ISL_2839739, EPI_ISL_2839747, EPI_ISL_2839763, EPI_ISL_2839774, EPI_ISL_2839782, EPI_ISL_2839788, EPI_ISL_2839790, EPI_ISL_2839791, EPI_ISL_2839792, EPI_ISL_2839800, EPI_ISL_2839804, EPI_ISL_2839807, EPI_ISL_2839808, EPI_ISL_2839811, EPI_ISL_2839812, EPI_ISL_2839813, EPI_ISL_2839823, EPI_ISL_2839826, EPI_ISL_2839829, EPI_ISL_2839831, EPI_ISL_2839833, EPI_ISL_2839835, EPI_ISL_2839836, EPI_ISL_2839839, EPI_ISL_2839842, EPI_ISL_2839843, EPI_ISL_2839844, EPI_ISL_2839852, EPI_ISL_2839857, EPI_ISL_2839862, EPI_ISL_2839864, EPI_ISL_2839878, EPI_ISL_2839881, EPI_ISL_2839882, EPI_ISL_2839884, EPI_ISL_2839887, EPI_ISL_2839888, EPI_ISL_2839897, EPI_ISL_2839898, EPI_ISL_2839899, EPI_ISL_2839901, EPI_ISL_2839902, EPI_ISL_2839903, EPI_ISL_2839904, EPI_ISL_2839909, EPI_ISL_2839910, EPI_ISL_2839912, EPI_ISL_2839915, EPI_ISL_2839916, EPI_ISL_2839924, EPI_ISL_2839927, EPI_ISL_2839937, EPI_ISL_2839946, EPI_ISL_2839948, EPI_ISL_2839954,

| | | | |
|---|---|---|---|
| EPI_ISL_2839961, EPI_ISL_2839962, EPI_ISL_2839963, EPI_ISL_2839964, EPI_ISL_2839965, EPI_ISL_2839966, EPI_ISL_2839967, EPI_ISL_2839968, EPI_ISL_2839969, EPI_ISL_2839970, EPI_ISL_2839971, EPI_ISL_2839972, EPI_ISL_2839973, EPI_ISL_2839974, EPI_ISL_2839975, EPI_ISL_2839976, EPI_ISL_2839977, EPI_ISL_2839978, EPI_ISL_2941840, EPI_ISL_2941853, EPI_ISL_2941854, EPI_ISL_2941855, EPI_ISL_2941856, EPI_ISL_2941857, EPI_ISL_2941858, EPI_ISL_2941859, EPI_ISL_2941882, EPI_ISL_2941883, EPI_ISL_2941884, EPI_ISL_2941894, EPI_ISL_2941895, EPI_ISL_2941896, EPI_ISL_2941897, EPI_ISL_2941898, EPI_ISL_2941901, EPI_ISL_2941902, EPI_ISL_2941903, EPI_ISL_2941904, EPI_ISL_2941916, EPI_ISL_2941917, EPI_ISL_2941918, EPI_ISL_2941919, EPI_ISL_2941920, EPI_ISL_2941921, EPI_ISL_2941922, EPI_ISL_2941923, EPI_ISL_2941924, EPI_ISL_2941925, EPI_ISL_2941926, EPI_ISL_2942016, EPI_ISL_2942029, EPI_ISL_2942031, EPI_ISL_2942037, EPI_ISL_2942040, EPI_ISL_2942047, EPI_ISL_2942064, EPI_ISL_2942092, EPI_ISL_2942094, EPI_ISL_2942097, EPI_ISL_2942098, EPI_ISL_2942099, EPI_ISL_2942100 | | | |
| see above | Curative Labs | Curative Labs | Elias L. Salfati; Eugenia Khorosheva; George Way; J.Cesar Ignacio-Espinoza; Janet Chen; Mikhail Hanewich-Hollatz; Nabjot Sandhu; Sophia Quasem; Vladimir Slepnev; Zhiyi Xie |
| EPI_ISL_2508357, EPI_ISL_2508358, EPI_ISL_2508359, EPI_ISL_2508360, EPI_ISL_2508361, EPI_ISL_2508362, EPI_ISL_2508363, EPI_ISL_2508364, EPI_ISL_2508365, EPI_ISL_2508366, EPI_ISL_2508367, EPI_ISL_2508368, EPI_ISL_2508369, EPI_ISL_2522093, EPI_ISL_2801556, EPI_ISL_2801557, EPI_ISL_2801558, EPI_ISL_2801559, EPI_ISL_2801560, EPI_ISL_2801561, EPI_ISL_2801562, EPI_ISL_2801563 | | | |
| see above | DeRisi Lab, University of California, San Francisco | DeRisi Lab, University of California, San Francisco | Amy Kistler; Anthea Mitchell; Carina Marquez; Diane Havlir; Diane Jones; Douglas Black; Eric Chow; Gabriel Chamie; Genay Pilarowski; IDseq Team; Jackie Martinez; James Peng; Jamin Liu; Jon Jacobo; Lucy Li; Luis Rubio; Manu Vanaerschot; Matthew Laurie; Matthias Hapte-Selassie; Maya Petersen; Patrick Ayscue; Sabrina Mann; Sara Sunshine; Susana Rojas; Susy Rojas; Valerie Tulier-Laiwa; and Joseph DeRisi |
| EPI_ISL_2650542, EPI_ISL_2650543, EPI_ISL_2753975, EPI_ISL_2774621, EPI_ISL_2934023, EPI_ISL_2934058 | El Camino Hospital | Santa Clara County Public Health Laboratory | Santa Clara County Public Health Department |
| EPI_ISL_2405184, EPI_ISL_2405185, EPI_ISL_2405207, EPI_ISL_2405208, EPI_ISL_2405209, EPI_ISL_2405210, EPI_ISL_2405213, EPI_ISL_2405214, EPI_ISL_2405215, EPI_ISL_2772522, EPI_ISL_2772523, EPI_ISL_2772524, EPI_ISL_2774568, EPI_ISL_2774569, EPI_ISL_2811981, EPI_ISL_2811982, EPI_ISL_2812002, EPI_ISL_2812003 | | | |
| see above | Flow Diagnostics Lab | Los Angeles County PHL | P. Hemarajata et al. |
| EPI_ISL_2362485, EPI_ISL_2362486, EPI_ISL_2362487 | Flow Health | Infectious Disease Program, Broad Institute of Harvard and MIT | Adams, G.; B.L.; B.W.; Bauer, M.; Birren; Carter, A.; Chaluvadi, S.; D.J.; DeRuff, K.; Gallagher, G.; Gladden-Young, A.; J.E.; K.J.; Lagerborg, K.; Lemieux; Loreth, C.; MacInnis; Normandin, E.; P.C.; Park; Reilly, S.; Rudy, M.; Siddle; Smole, S.; Tomkins-Tinch, C.; and Sabeti |
| EPI_ISL_1993098, EPI_ISL_1993099, EPI_ISL_1993100, EPI_ISL_1993101, EPI_ISL_1993102, EPI_ISL_1993106, EPI_ISL_1993107, EPI_ISL_1993108, EPI_ISL_1993109, EPI_ISL_1993110, EPI_ISL_1993116, EPI_ISL_1993117, EPI_ISL_1993120, EPI_ISL_1993122, EPI_ISL_1993123, EPI_ISL_1993131, EPI_ISL_1993133, EPI_ISL_1993134, EPI_ISL_1993137, EPI_ISL_1993138, EPI_ISL_1993139, EPI_ISL_1993140, EPI_ISL_1993143, EPI_ISL_1993144, EPI_ISL_1993147, EPI_ISL_1993216, EPI_ISL_1993217, EPI_ISL_1993218, EPI_ISL_1993219, EPI_ISL_1993220, EPI_ISL_1993221, EPI_ISL_1993222, EPI_ISL_1993223, EPI_ISL_1993224, EPI_ISL_1993225, EPI_ISL_1993226, EPI_ISL_1993227, EPI_ISL_1993228, EPI_ISL_1993229, EPI_ISL_1993230, EPI_ISL_1993231, EPI_ISL_1993232, EPI_ISL_1993233, EPI_ISL_1993234, EPI_ISL_1993235, EPI_ISL_1993236, EPI_ISL_1993237, EPI_ISL_1993238, EPI_ISL_1993239, EPI_ISL_1993240, EPI_ISL_1993241, EPI_ISL_1993242, EPI_ISL_1993243, EPI_ISL_1993244, EPI_ISL_1993245, EPI_ISL_1993246, EPI_ISL_1993247, EPI_ISL_1993248, EPI_ISL_1993249, EPI_ISL_1993250, EPI_ISL_1993251, EPI_ISL_1993252, EPI_ISL_1993253, EPI_ISL_1993254, EPI_ISL_1993255, EPI_ISL_1993256, EPI_ISL_1993257, EPI_ISL_1993258, EPI_ISL_1993259, EPI_ISL_1993260, EPI_ISL_1993261, EPI_ISL_1993262, EPI_ISL_1993263, EPI_ISL_1993264, EPI_ISL_1993265, EPI_ISL_1993266, EPI_ISL_1993267, EPI_ISL_1993268, EPI_ISL_1993269, EPI_ISL_1993270, EPI_ISL_1993271, EPI_ISL_1993272, EPI_ISL_1993273, EPI_ISL_1993274, EPI_ISL_1993275, EPI_ISL_1993276, EPI_ISL_1993277, EPI_ISL_1993278, EPI_ISL_2043709, EPI_ISL_2043710, EPI_ISL_2043711, EPI_ISL_2043714, EPI_ISL_2043715, EPI_ISL_2043716, EPI_ISL_2043717, EPI_ISL_2043718, EPI_ISL_2043719, EPI_ISL_2043720, EPI_ISL_2043721, EPI_ISL_2043722, EPI_ISL_2043723, EPI_ISL_2043724, EPI_ISL_2043725, EPI_ISL_2043726, EPI_ISL_2043727, EPI_ISL_2043728, EPI_ISL_2043729, EPI_ISL_2043730, EPI_ISL_2043731, EPI_ISL_2043732, EPI_ISL_2043733, EPI_ISL_2043734, EPI_ISL_2043735, EPI_ISL_2043736, EPI_ISL_2043738, EPI_ISL_2043739, EPI_ISL_2043740, EPI_ISL_2043741, EPI_ISL_2043742, EPI_ISL_2043743, EPI_ISL_2043744, EPI_ISL_2043745, EPI_ISL_2043746, EPI_ISL_2043747, EPI_ISL_2043748, EPI_ISL_2043749, EPI_ISL_2043750, EPI_ISL_2043751, EPI_ISL_2043752, EPI_ISL_2043753, EPI_ISL_2043754, EPI_ISL_2043755, EPI_ISL_2043756, EPI_ISL_2043757, EPI_ISL_2043758, EPI_ISL_2043759, EPI_ISL_2043760, EPI_ISL_2043761, EPI_ISL_2043762, EPI_ISL_2043763, EPI_ISL_2043764, EPI_ISL_2043765, EPI_ISL_2043766, EPI_ISL_2043767, EPI_ISL_2043768, EPI_ISL_2043769, EPI_ISL_2043770, EPI_ISL_2043771, EPI_ISL_2043772, EPI_ISL_2043773, EPI_ISL_2043938, EPI_ISL_2043942, EPI_ISL_2043943, EPI_ISL_2043944, EPI_ISL_2043945, EPI_ISL_2043946, EPI_ISL_2043947, EPI_ISL_2043948, EPI_ISL_2043949, EPI_ISL_2043950, EPI_ISL_2043951, EPI_ISL_2043952, EPI_ISL_2043953, EPI_ISL_2043954, EPI_ISL_2043955, EPI_ISL_2043956, EPI_ISL_2043957, EPI_ISL_2043958, EPI_ISL_2043959, EPI_ISL_2043960, EPI_ISL_2043961, EPI_ISL_2043962, EPI_ISL_2043963, EPI_ISL_2043964, EPI_ISL_2043965, EPI_ISL_2043966, EPI_ISL_2043967, EPI_ISL_2043968, EPI_ISL_2043969, EPI_ISL_2043970, EPI_ISL_2043971, EPI_ISL_2043972, EPI_ISL_2043973, EPI_ISL_2043974, EPI_ISL_2043975, EPI_ISL_2043976, EPI_ISL_2043977, EPI_ISL_2043978, EPI_ISL_2043979, EPI_ISL_2043980, EPI_ISL_2043981, EPI_ISL_2043982, EPI_ISL_2043983, EPI_ISL_2043984, EPI_ISL_2043985, EPI_ISL_2043986, EPI_ISL_2043987, EPI_ISL_2043988, EPI_ISL_2043989, EPI_ISL_2043990, EPI_ISL_2043991, EPI_ISL_2043992, EPI_ISL_2043993, EPI_ISL_2043994, EPI_ISL_2043995, EPI_ISL_2043996, EPI_ISL_2043997, EPI_ISL_2043998, EPI_ISL_2043999, EPI_ISL_2044000, EPI_ISL_2133929, EPI_ISL_2133930, EPI_ISL_2133931, EPI_ISL_2133932, EPI_ISL_2133933, EPI_ISL_2133934, EPI_ISL_2133935, EPI_ISL_2133936, EPI_ISL_2133937, EPI_ISL_2133938, EPI_ISL_2133939, EPI_ISL_2133940, EPI_ISL_2133941, EPI_ISL_2133942, EPI_ISL_2133943, EPI_ISL_2133944, EPI_ISL_2133945, EPI_ISL_2133946, EPI_ISL_2133947, EPI_ISL_2133948, EPI_ISL_2133949, EPI_ISL_2133950, EPI_ISL_2133951, EPI_ISL_2133952, EPI_ISL_2133953, EPI_ISL_2133954, EPI_ISL_2133955, EPI_ISL_2133956, EPI_ISL_2133957, EPI_ISL_2133958, EPI_ISL_2133960, EPI_ISL_2133961, EPI_ISL_2133962, EPI_ISL_2133963, EPI_ISL_2133964, EPI_ISL_2133965, EPI_ISL_2133966, EPI_ISL_2133967, EPI_ISL_2133968, EPI_ISL_2133969, EPI_ISL_2133970, EPI_ISL_2133971, EPI_ISL_2133972, EPI_ISL_2133973, EPI_ISL_2133974, EPI_ISL_2133975, EPI_ISL_2133976, EPI_ISL_2133977, EPI_ISL_2133978, EPI_ISL_2133979, EPI_ISL_2133980, EPI_ISL_2133981, EPI_ISL_2133982, EPI_ISL_2133983, EPI_ISL_2133984, EPI_ISL_2133985, EPI_ISL_2133986, EPI_ISL_2133987, EPI_ISL_2133988, EPI_ISL_2134173, EPI_ISL_2134176, EPI_ISL_2134177, EPI_ISL_2134178, EPI_ISL_2134179, EPI_ISL_2134180, EPI_ISL_2134181, EPI_ISL_2134182, EPI_ISL_2134183, EPI_ISL_2134185, EPI_ISL_2134186, EPI_ISL_2134187, EPI_ISL_2134188, EPI_ISL_2134189, EPI_ISL_2134190, EPI_ISL_2134191, EPI_ISL_2134192, EPI_ISL_2134193, EPI_ISL_2134194, EPI_ISL_2134195, EPI_ISL_2134196, EPI_ISL_2134197, EPI_ISL_2134198, EPI_ISL_2134199, EPI_ISL_2134200, EPI_ISL_2134201, EPI_ISL_2134202, EPI_ISL_2134203, EPI_ISL_2134204, EPI_ISL_2134205, EPI_ISL_2134206, EPI_ISL_2134207, EPI_ISL_2134208, EPI_ISL_2134209, EPI_ISL_2134210, EPI_ISL_2134211, EPI_ISL_2134212, EPI_ISL_2134213, EPI_ISL_2134214, EPI_ISL_2134215, EPI_ISL_2134216, EPI_ISL_2134217, EPI_ISL_2134218, EPI_ISL_2134219, EPI_ISL_2134220, EPI_ISL_2134221, EPI_ISL_2134222, EPI_ISL_2134223, EPI_ISL_2134224, EPI_ISL_2134225, EPI_ISL_2134226, EPI_ISL_2134227, EPI_ISL_2134228, EPI_ISL_2134229, EPI_ISL_2134230, EPI_ISL_2134231, EPI_ISL_2134232, EPI_ISL_2134233, EPI_ISL_2134234, EPI_ISL_2134235, EPI_ISL_2134339, EPI_ISL_2134340, EPI_ISL_2134341, EPI_ISL_2134342, EPI_ISL_2134343, EPI_ISL_2134344, EPI_ISL_2134345, EPI_ISL_2134346, EPI_ISL_2134347, EPI_ISL_2134348, EPI_ISL_2134349, EPI_ISL_2134350, EPI_ISL_2134351, EPI_ISL_2134352, EPI_ISL_2134353, EPI_ISL_2134354, EPI_ISL_2134355, EPI_ISL_2134356, EPI_ISL_2134357, EPI_ISL_2134358, EPI_ISL_2134359, EPI_ISL_2134360, EPI_ISL_2134361, EPI_ISL_2134362, EPI_ISL_2134363, EPI_ISL_2134364, EPI_ISL_2134365, EPI_ISL_2134366, EPI_ISL_2134367, EPI_ISL_2134368, EPI_ISL_2134369, EPI_ISL_2134370, EPI_ISL_2134371, EPI_ISL_2134372, EPI_ISL_2134373, EPI_ISL_2134374, EPI_ISL_2134375, EPI_ISL_2134376, EPI_ISL_2134377, EPI_ISL_2134378, EPI_ISL_2134379, EPI_ISL_2134380, EPI_ISL_2134381, EPI_ISL_2134382, EPI_ISL_2134383, EPI_ISL_2134384, EPI_ISL_2134385, EPI_ISL_2134386, EPI_ISL_2134387, EPI_ISL_2134388, EPI_ISL_2134389, EPI_ISL_2134390, EPI_ISL_2134391, EPI_ISL_2134392, EPI_ISL_2134393, EPI_ISL_2134394, EPI_ISL_2134395, EPI_ISL_2134396, EPI_ISL_2134397, EPI_ISL_2134398, EPI_ISL_2134399, EPI_ISL_2134400, EPI_ISL_2134401, EPI_ISL_2134402, EPI_ISL_2134403, EPI_ISL_2134404, EPI_ISL_2134405, EPI_ISL_2134406, EPI_ISL_2134407, EPI_ISL_2134408, EPI_ISL_2134409, EPI_ISL_2134410, EPI_ISL_2134411, EPI_ISL_2134412, EPI_ISL_2134413, EPI_ISL_2134414, EPI_ISL_2134415, EPI_ISL_2134416, EPI_ISL_2134417, EPI_ISL_2134418, EPI_ISL_2134419, EPI_ISL_2134420, EPI_ISL_2134421, EPI_ISL_2134422, EPI_ISL_2134423, EPI_ISL_2134424, EPI_ISL_2134425, EPI_ISL_2134426, EPI_ISL_2134427, EPI_ISL_2134428, EPI_ISL_2134429, EPI_ISL_2134430, EPI_ISL_2134431, EPI_ISL_2134432, EPI_ISL_2134433, EPI_ISL_2134434, EPI_ISL_2134435, EPI_ISL_2134436, EPI_ISL_2134437, EPI_ISL_2134438, EPI_ISL_2134439, EPI_ISL_2134440, EPI_ISL_2134441, EPI_ISL_2134442, EPI_ISL_2134443, EPI_ISL_2134444, EPI_ISL_2134445, EPI_ISL_2134446, EPI_ISL_2134447, EPI_ISL_2134448, EPI_ISL_2134449, EPI_ISL_2134450, EPI_ISL_2134451, EPI_ISL_2134452, EPI_ISL_2134453, EPI_ISL_2140799, EPI_ISL_2160793, EPI_ISL_2160794, EPI_ISL_2160795, EPI_ISL_2160796, EPI_ISL_2160797, EPI_ISL_2160798, EPI_ISL_2160799, EPI_ISL_2160800, EPI_ISL_2160801, EPI_ISL_2160802, EPI_ISL_2160803, EPI_ISL_2160804, EPI_ISL_2160805, EPI_ISL_2160806, EPI_ISL_2160807, EPI_ISL_2160808, EPI_ISL_2160809, EPI_ISL_2160811, EPI_ISL_2160812, EPI_ISL_2160813, EPI_ISL_2160814, EPI_ISL_2160815, EPI_ISL_2160816, EPI_ISL_2160817, EPI_ISL_2160818, EPI_ISL_2160819, EPI_ISL_2160820, EPI_ISL_2160821, EPI_ISL_2160822, EPI_ISL_2160823, EPI_ISL_2160824, EPI_ISL_2160825, EPI_ISL_2160826, EPI_ISL_2160827, EPI_ISL_2160828, EPI_ISL_2160829, EPI_ISL_2160830, EPI_ISL_2160831, EPI_ISL_2160832, EPI_ISL_2160833, EPI_ISL_2160834, EPI_ISL_2160835, EPI_ISL_2160836, EPI_ISL_2160837, EPI_ISL_2160838, EPI_ISL_2160839, EPI_ISL_2160840, EPI_ISL_2160841, EPI_ISL_2160842, EPI_ISL_2160843, EPI_ISL_2160844, EPI_ISL_2160845, EPI_ISL_2160846, EPI_ISL_2160847, EPI_ISL_2160848, EPI_ISL_2160849, EPI_ISL_2160850, EPI_ISL_2160851, EPI_ISL_2160852, EPI_ISL_2160853, EPI_ISL_2160854, EPI_ISL_2160855, EPI_ISL_2160856, EPI_ISL_2160857, EPI_ISL_2160858, EPI_ISL_2160859, EPI_ISL_2160860, EPI_ISL_2160961, EPI_ISL_2160962, EPI_ISL_2160963, EPI_ISL_2160964, EPI_ISL_2160965, EPI_ISL_2160966, EPI_ISL_2160967, EPI_ISL_2160968, EPI_ISL_2160969, EPI_ISL_2160970, EPI_ISL_2160971, EPI_ISL_2160972, EPI_ISL_2160973, EPI_ISL_2160974, EPI_ISL_2160975, EPI_ISL_2160976, EPI_ISL_2160977, EPI_ISL_2160978, EPI_ISL_2160979, EPI_ISL_2160980, EPI_ISL_2160981, EPI_ISL_2160982, EPI_ISL_2160983, EPI_ISL_2160984, EPI_ISL_2160985, EPI_ISL_2160986, EPI_ISL_2160987, EPI_ISL_2160988, EPI_ISL_2160989, EPI_ISL_2160990, EPI_ISL_2160991, EPI_ISL_2160992, EPI_ISL_2160993, EPI_ISL_2160994, EPI_ISL_2160995, EPI_ISL_2160996, EPI_ISL_2160997, EPI_ISL_2160998, EPI_ISL_2160999, EPI_ISL_2161000, EPI_ISL_2161001, EPI_ISL_2161002, EPI_ISL_2161003, EPI_ISL_2161004, EPI_ISL_2161005, EPI_ISL_2161006, EPI_ISL_2161007, EPI_ISL_2161008, EPI_ISL_2161009, EPI_ISL_2161010, EPI_ISL_2161011, EPI_ISL_2161012, EPI_ISL_2161013, EPI_ISL_2161014, EPI_ISL_2161015, EPI_ISL_2161016, EPI_ISL_2161017, EPI_ISL_2161018, EPI_ISL_2161019, EPI_ISL_2161020, EPI_ISL_2161021, EPI_ISL_2161022, EPI_ISL_2161023, EPI_ISL_2161024, EPI_ISL_2161025, EPI_ISL_2161026, EPI_ISL_2161027, EPI_ISL_2179850, EPI_ISL_2179851, EPI_ISL_2179852, EPI_ISL_2179853, EPI_ISL_2179854, EPI_ISL_2179855, EPI_ISL_2179856, EPI_ISL_2179857, EPI_ISL_2179858, EPI_ISL_2179859, EPI_ISL_2179860, EPI_ISL_2269264, EPI_ISL_2269266, EPI_ISL_2269268, EPI_ISL_2269269, EPI_ISL_2269270, EPI_ISL_2269271, EPI_ISL_2269272, EPI_ISL_2269273, EPI_ISL_2269274, EPI_ISL_2269275, EPI_ISL_2269276, EPI_ISL_2269277, EPI_ISL_2269278, EPI_ISL_2269279, EPI_ISL_2269280, EPI_ISL_2269281, EPI_ISL_2269282, EPI_ISL_2269283, EPI_ISL_2269284, EPI_ISL_2269285, EPI_ISL_2269286, EPI_ISL_2269287, EPI_ISL_2269288, EPI_ISL_2269289, EPI_ISL_2269290, EPI_ISL_2269291, EPI_ISL_2269292, EPI_ISL_2269293, EPI_ISL_2269294, EPI_ISL_2269295, EPI_ISL_2269296, EPI_ISL_2269297, EPI_ISL_2269298, EPI_ISL_2269299, EPI_ISL_2269300, EPI_ISL_2269301, EPI_ISL_2269302, EPI_ISL_2269303, EPI_ISL_2269304, EPI_ISL_2269305, EPI_ISL_2269306, EPI_ISL_2269307, EPI_ISL_2269308, EPI_ISL_2269309, EPI_ISL_2269310, EPI_ISL_2269311, EPI_ISL_2269312, EPI_ISL_2269314, EPI_ISL_2269315, EPI_ISL_2269316, EPI_ISL_2269317, EPI_ISL_2269318, EPI_ISL_2269319, EPI_ISL_2269320, EPI_ISL_2269321, EPI_ISL_2269322, EPI_ISL_2269323, EPI_ISL_2269324, EPI_ISL_2269325, EPI_ISL_2269326, EPI_ISL_2269327, EPI_ISL_2269328, EPI_ISL_2269329, EPI_ISL_2269330, EPI_ISL_2269331, EPI_ISL_2269332, EPI_ISL_2269333, EPI_ISL_2269334, EPI_ISL_2269335, EPI_ISL_2269336, EPI_ISL_2269337, EPI_ISL_2269338, EPI_ISL_2269339, EPI_ISL_2269340, EPI_ISL_2269341, EPI_ISL_2269342, EPI_ISL_2269343, EPI_ISL_2269344, EPI_ISL_2269345, EPI_ISL_2269346, EPI_ISL_2269347, EPI_ISL_2269348, EPI_ISL_2269349, EPI_ISL_2269350, EPI_ISL_2269351, EPI_ISL_2269352, EPI_ISL_2269353, EPI_ISL_2269354, EPI_ISL_2269355, EPI_ISL_2269356, EPI_ISL_2269357, EPI_ISL_2269358, EPI_ISL_2269359, EPI_ISL_2269360, EPI_ISL_2269361, EPI_ISL_2269362, EPI_ISL_2269363, EPI_ISL_2269364, EPI_ISL_2269365, EPI_ISL_2269366, EPI_ISL_2269367, EPI_ISL_2269368, EPI_ISL_2269369, EPI_ISL_2269370, EPI_ISL_2269371, EPI_ISL_2269372, EPI_ISL_2269373, EPI_ISL_2269374, EPI_ISL_2269375, EPI_ISL_2269429, EPI_ISL_2269450, EPI_ISL_2269471, EPI_ISL_2269472, EPI_ISL_2269473, EPI_ISL_2269474, EPI_ISL_2269475, EPI_ISL_2269476, EPI_ISL_2269477, EPI_ISL_2269478, EPI_ISL_2269479, EPI_ISL_2269480, EPI_ISL_2269481, EPI_ISL_2269482, EPI_ISL_2269483, EPI_ISL_2269484, EPI_ISL_2269485, EPI_ISL_2269486, EPI_ISL_2269487, EPI_ISL_2269488, EPI_ISL_2269489, EPI_ISL_2269490, EPI_ISL_2269492, EPI_ISL_2269493, EPI_ISL_2269494, EPI_ISL_2269495, EPI_ISL_2269496, EPI_ISL_2269497, EPI_ISL_2269498, EPI_ISL_2269499, EPI_ISL_2269500, EPI_ISL_2269501, EPI_ISL_2269502, EPI_ISL_2269503, EPI_ISL_2269504, EPI_ISL_2269505, EPI_ISL_2269506, EPI_ISL_2269507, EPI_ISL_2269508, EPI_ISL_2269509, EPI_ISL_2269510, EPI_ISL_2269511, EPI_ISL_2269512, EPI_ISL_2269513, EPI_ISL_2269514, EPI_ISL_2269515, EPI_ISL_2269516, EPI_ISL_2269517, EPI_ISL_2269518, EPI_ISL_2269519, EPI_ISL_2269520, EPI_ISL_2269521, EPI_ISL_2269522, EPI_ISL_2269523, EPI_ISL_2269524, EPI_ISL_2269525, EPI_ISL_2269526, EPI_ISL_2269527, EPI_ISL_2269528, EPI_ISL_2269529, EPI_ISL_2269530, EPI_ISL_2269531, EPI_ISL_2269532, EPI_ISL_2269533, EPI_ISL_2269534, EPI_ISL_2269535, EPI_ISL_2269536, EPI_ISL_2368134, EPI_ISL_2368135, EPI_ISL_2368136, EPI_ISL_2368137, EPI_ISL_2368138, EPI_ISL_2368139, EPI_ISL_2368140, EPI_ISL_2368141, EPI_ISL_2368142, EPI_ISL_2368143, EPI_ISL_2368144, EPI_ISL_2368145, EPI_ISL_2368146, EPI_ISL_2368147, EPI_ISL_2368148, EPI_ISL_2368149, EPI_ISL_2368150, EPI_ISL_2368151, EPI_ISL_2368152, EPI_ISL_2368153, EPI_ISL_2368154, EPI_ISL_2368155, EPI_ISL_2368156, EPI_ISL_2368157, EPI_ISL_2368158, EPI_ISL_2368159, EPI_ISL_2368162, EPI_ISL_2368163, EPI_ISL_2368164, EPI_ISL_2368165, EPI_ISL_2368166, EPI_ISL_2368167, EPI_ISL_2368168, EPI_ISL_2368169, EPI_ISL_2368170, EPI_ISL_2368171, EPI_ISL_2368172, EPI_ISL_2368173, EPI_ISL_2368174, EPI_ISL_2368175, EPI_ISL_2368176, EPI_ISL_2368177, EPI_ISL_2368178, EPI_ISL_2368179, EPI_ISL_2368180, EPI_ISL_2368181, EPI_ISL_2368182, EPI_ISL_2368183, EPI_ISL_2368184, EPI_ISL_2368185, EPI_ISL_2368186, EPI_ISL_2368187, EPI_ISL_2368188, EPI_ISL_2368189, EPI_ISL_2368190, EPI_ISL_2368191, EPI_ISL_2368192, EPI_ISL_2368193, EPI_ISL_2368195, EPI_ISL_2368196, EPI_ISL_2368197, EPI_ISL_2368198, EPI_ISL_2368199, EPI_ISL_2368200, EPI_ISL_2368201, EPI_ISL_2368202, EPI_ISL_2368203, EPI_ISL_2368204, EPI_ISL_2371514, EPI_ISL_2371517, EPI_ISL_2371518, EPI_ISL_2371519, EPI_ISL_2371520, EPI_ISL_2371521, EPI_ISL_2371522, EPI_ISL_2371523, EPI_ISL_2371524, EPI_ISL_2371525, EPI_ISL_2371526, EPI_ISL_2371527, EPI_ISL_2371528, EPI_ISL_2371529, EPI_ISL_2371530, EPI_ISL_2371531, EPI_ISL_2371532, EPI_ISL_2371533, EPI_ISL_2371534, EPI_ISL_2371535, EPI_ISL_2371536, EPI_ISL_2371537, EPI_ISL_2371538, EPI_ISL_2371539, EPI_ISL_2371540, EPI_ISL_2371541, EPI_ISL_2371542, EPI_ISL_2371543, EPI_ISL_2371544, EPI_ISL_2371545, EPI_ISL_2371546, EPI_ISL_2371547, EPI_ISL_2371548, EPI_ISL_2371549, EPI_ISL_2371550, EPI_ISL_2371551, EPI_ISL_2371552, EPI_ISL_2371553, EPI_ISL_2371554, EPI_ISL_2371555, EPI_ISL_2371556, EPI_ISL_2371557, EPI_ISL_2371558, EPI_ISL_2371559, EPI_ISL_2371560, EPI_ISL_2371561, EPI_ISL_2371562, EPI_ISL_2371563, EPI_ISL_2371564, EPI_ISL_2371565, EPI_ISL_2371566, EPI_ISL_2371567, EPI_ISL_2371568, EPI_ISL_2371569, EPI_ISL_2371570, EPI_ISL_2371571, EPI_ISL_2371572, EPI_ISL_2371573, EPI_ISL_2371627, EPI_ISL_2371628, EPI_ISL_2371629, EPI_ISL_2371630, EPI_ISL_2371631, EPI_ISL_2371632, EPI_ISL_2371633, EPI_ISL_2371634, EPI_ISL_2371635, EPI_ISL_2371636, EPI_ISL_2371637, EPI_ISL_2371638, EPI_ISL_2371639, EPI_ISL_2371640, EPI_ISL_2371641, EPI_ISL_2371642, EPI_ISL_2371643, EPI_ISL_2371644, EPI_ISL_2371645, EPI_ISL_2371646, EPI_ISL_2371647, EPI_ISL_2371648, EPI_ISL_2371649, EPI_ISL_2371650, EPI_ISL_2371651, EPI_ISL_2371652, EPI_ISL_2371653, EPI_ISL_2371654, EPI_ISL_2371655, EPI_ISL_2371656, EPI_ISL_2371657, EPI_ISL_2371658, EPI_ISL_2371659, EPI_ISL_2371660, EPI_ISL_2371661, EPI_ISL_2371662, EPI_ISL_2371663, EPI_ISL_2371664, EPI_ISL_2371665, EPI_ISL_2371666, EPI_ISL_2371667, EPI_ISL_2371668, EPI_ISL_2371669, EPI_ISL_2371670, EPI_ISL_2371671, EPI_ISL_2371672, EPI_ISL_2371673, EPI_ISL_2371674, EPI_ISL_2371675, EPI_ISL_2371676, EPI_ISL_2371677, EPI_ISL_2371678, EPI_ISL_2371679, EPI_ISL_2371680, EPI_ISL_2371681, EPI_ISL_2371682, EPI_ISL_2371684, EPI_ISL_2371685, EPI_ISL_2371686, EPI_ISL_2371687, EPI_ISL_2371688, EPI_ISL_2371689, EPI_ISL_2371690, EPI_ISL_2441902, EPI_ISL_2441903, EPI_ISL_2441904, EPI_ISL_2441905, EPI_ISL_2441906, EPI_ISL_2441907, EPI_ISL_2441908, EPI_ISL_2441909, EPI_ISL_2441910, EPI_ISL_2441911, EPI_ISL_2441912, EPI_ISL_2441913, EPI_ISL_2441914, EPI_ISL_2441915, EPI_ISL_2441916, EPI_ISL_2441917, EPI_ISL_2441918, EPI_ISL_2441919, EPI_ISL_2441920, EPI_ISL_2441921, EPI_ISL_2441922, EPI_ISL_2441923, EPI_ISL_2441924, EPI_ISL_2441925, EPI_ISL_2441926, EPI_ISL_2441927, EPI_ISL_2441928, EPI_ISL_2441929, EPI_ISL_2441930, EPI_ISL_2441931, EPI_ISL_2441932, EPI_ISL_2441933, EPI_ISL_2441934, EPI_ISL_2441935, EPI_ISL_2441936, EPI_ISL_2441937, EPI_ISL_2441938, EPI_ISL_2441939, EPI_ISL_2441940, EPI_ISL_2441941, EPI_ISL_2441942, EPI_ISL_2441943, EPI_ISL_2441944, EPI_ISL_2441945, EPI_ISL_2441946, EPI_ISL_2441947, EPI_ISL_2441948, EPI_ISL_2441949, EPI_ISL_2441950, EPI_ISL_2441951, EPI_ISL_2441952, EPI_ISL_2441953, EPI_ISL_2441954, EPI_ISL_2441955, EPI_ISL_2441956, EPI_ISL_2441957, EPI_ISL_2441958, EPI_ISL_2480088, EPI_ISL_2480089, EPI_ISL_2480090, EPI_ISL_2480091, EPI_ISL_2480092, EPI_ISL_2480093, EPI_ISL_2480094, EPI_ISL_2480095, EPI_ISL_2480096, EPI_ISL_2480097, EPI_ISL_2480098, EPI_ISL_2480099, EPI_ISL_2480100, EPI_ISL_2480101, EPI_ISL_2480102, EPI_ISL_2480103, EPI_ISL_2480104, EPI_ISL_2480105, EPI_ISL_2480106, EPI_ISL_2480107, EPI_ISL_2480108, EPI_ISL_2480109, EPI_ISL_2480110, EPI_ISL_2480111, EPI_ISL_2480112, EPI_ISL_2480113, EPI_ISL_2480114, EPI_ISL_2480115, EPI_ISL_2480116, EPI_ISL_2480117, EPI_ISL_2480118, EPI_ISL_2480119, EPI_ISL_2480120, EPI_ISL_2480121, EPI_ISL_2480122, EPI_ISL_2480123, EPI_ISL_2480124, EPI_ISL_2480125, EPI_ISL_2480126, EPI_ISL_2480127, EPI_ISL_2480128, EPI_ISL_2480129, EPI_ISL_2480130, EPI_ISL_2480131, EPI_ISL_2480132, EPI_ISL_2480133, EPI_ISL_2480134, EPI_ISL_2480135, EPI_ISL_2480136, EPI_ISL_2480137, EPI_ISL_2480138, EPI_ISL_2480139, EPI_ISL_2480140, EPI_ISL_2480141, EPI_ISL_2480142, EPI_ISL_2480143, EPI_ISL_2480144, EPI_ISL_2480145, EPI_ISL_2525648, EPI_ISL_2525652, EPI_ISL_2525658, EPI_ISL_2525660, EPI_ISL_2525662, EPI_ISL_2525665, EPI_ISL_2525667, EPI_ISL_2525669, EPI_ISL_2525671, EPI_ISL_2525674, EPI_ISL_2525676, EPI_ISL_2525678, EPI_ISL_2525679, EPI_ISL_2525680, EPI_ISL_2525681, EPI_ISL_2525683, EPI_ISL_2525685, EPI_ISL_2525687, EPI_ISL_2525689, EPI_ISL_2525691, EPI_ISL_2525693, EPI_ISL_2525695, EPI_ISL_2525698, EPI_ISL_2525700, EPI_ISL_2525702, EPI_ISL_2525704, EPI_ISL_2525706, EPI_ISL_2525708, EPI_ISL_2525710, EPI_ISL_2525712, EPI_ISL_2525714, EPI_ISL_2525716, EPI_ISL_2525718, EPI_ISL_2525720, EPI_ISL_2525722, EPI_ISL_2525725, EPI_ISL_2525727, EPI_ISL_2525729, EPI_ISL_2525731, EPI_ISL_2525733, EPI_ISL_2525735, EPI_ISL_2525737, EPI_ISL_2525739, EPI_ISL_2525741, EPI_ISL_2525744, EPI_ISL_2525746, EPI_ISL_2525748, EPI_ISL_2525750, EPI_ISL_2525752, EPI_ISL_2525754, EPI_ISL_2525756, EPI_ISL_2525759, EPI_ISL_2525761, EPI_ISL_2525763, EPI_ISL_2525765, EPI_ISL_2525767, EPI_ISL_2525769, EPI_ISL_2525771, EPI_ISL_2525773, EPI_ISL_2525776, EPI_ISL_2525778, EPI_ISL_2525787, EPI_ISL_2525973, EPI_ISL_2525975, EPI_ISL_2527563, EPI_ISL_2527565, EPI_ISL_2527567, EPI_ISL_2527570, EPI_ISL_2527573, EPI_ISL_2527575, EPI_ISL_2527577, EPI_ISL_2527579, EPI_ISL_2527581, EPI_ISL_2527584, EPI_ISL_2527586, EPI_ISL_2527588, EPI_ISL_2527590, EPI_ISL_2527592, EPI_ISL_2527594, EPI_ISL_2527597, EPI_ISL_2527599, EPI_ISL_2527601, EPI_ISL_2527603, EPI_ISL_2527605, EPI_ISL_2527608, EPI_ISL_2527610, EPI_ISL_2527612, EPI_ISL_2527614, EPI_ISL_2527616, EPI_ISL_2527618, EPI_ISL_2527620, EPI_ISL_2527622, EPI_ISL_2527624, EPI_ISL_2527627, EPI_ISL_2527629, EPI_ISL_2527632, EPI_ISL_2527634, EPI_ISL_2527636, EPI_ISL_2527638, EPI_ISL_2527641, EPI_ISL_2527643, EPI_ISL_2527645, EPI_ISL_2527648, EPI_ISL_2527650, EPI_ISL_2527652, EPI_ISL_2527654, EPI_ISL_2527656, EPI_ISL_2527658, EPI_ISL_2527660, EPI_ISL_2527663, EPI_ISL_2527665, EPI_ISL_2527667, EPI_ISL_2527669, EPI_ISL_2527672, EPI_ISL_2527712, EPI_ISL_2527714, EPI_ISL_2527716, EPI_ISL_2527719, EPI_ISL_2527721, EPI_ISL_2527723, EPI_ISL_2527725, EPI_ISL_2527727, EPI_ISL_2527729, EPI_ISL_2527731, EPI_ISL_2527733, EPI_ISL_2527735, EPI_ISL_2527737, EPI_ISL_2527739, EPI_ISL_2527741, EPI_ISL_2527743, EPI_ISL_2527745, EPI_ISL_2527747, EPI_ISL_2527749, EPI_ISL_2527751, EPI_ISL_2527753, EPI_ISL_2527755, EPI_ISL_2527757, EPI_ISL_2527760, EPI_ISL_2527762, EPI_ISL_2527764, EPI_ISL_2527766, EPI_ISL_2527768, EPI_ISL_2527770, EPI_ISL_2527772, EPI_ISL_2527774, EPI_ISL_2527775, EPI_ISL_2527777, EPI_ISL_2527779, EPI_ISL_2527781, EPI_ISL_2527784, EPI_ISL_2527786, EPI_ISL_2527788, EPI_ISL_2527790, EPI_ISL_2527792, EPI_ISL_2527794, EPI_ISL_2527796, EPI_ISL_2527798, EPI_ISL_2527801, EPI_ISL_2527803, EPI_ISL_2527805, EPI_ISL_2527807, EPI_ISL_2527809, EPI_ISL_2527811, EPI_ISL_2527813, EPI_ISL_2527816, EPI_ISL_2527818, EPI_ISL_2527820, EPI_ISL_2527823, EPI_ISL_2527825, EPI_ISL_2527827, EPI_ISL_2527829, EPI_ISL_2527831, EPI_ISL_2527833, EPI_ISL_2527835, EPI_ISL_2527837, EPI_ISL_2527839, EPI_ISL_2527841, EPI_ISL_2527843, EPI_ISL_2598710, EPI_ISL_2598711, EPI_ISL_2598713, EPI_ISL_2598714, EPI_ISL_2598715, EPI_ISL_2598716, EPI_ISL_2598717, EPI_ISL_2598718, EPI_ISL_2598719, EPI_ISL_2598720, EPI_ISL_2598721, EPI_ISL_2598722, EPI_ISL_2598723, EPI_ISL_2598724, EPI_ISL_2598725, EPI_ISL_2598726, EPI_ISL_2598727, EPI_ISL_2598728, EPI_ISL_2598729, EPI_ISL_2598731, EPI_ISL_2598732, EPI_ISL_2598733, EPI_ISL_2598734, EPI_ISL_2598735, EPI_ISL_2598736, EPI_ISL_2598737, EPI_ISL_2598738, EPI_ISL_2598739, EPI_ISL_2598740, EPI_ISL_2598741, EPI_ISL_2598742, EPI_ISL_2598743, EPI_ISL_2598744, EPI_ISL_2598745, EPI_ISL_2598746, EPI_ISL_2598747, EPI_ISL_2598748, EPI_ISL_2598749, EPI_ISL_2598750, EPI_ISL_2598751, EPI_ISL_2598752, EPI_ISL_2598753, EPI_ISL_2598754, EPI_ISL_2598755, EPI_ISL_2598756, EPI_ISL_2598757, EPI_ISL_2598758, EPI_ISL_2598759, EPI_ISL_2598760, EPI_ISL_2598761, EPI_ISL_2598762, EPI_ISL_2598763, EPI_ISL_2598764, EPI_ISL_2686394, EPI_ISL_2686395, EPI_ISL_2686396, EPI_ISL_2686397, EPI_ISL_2686398, EPI_ISL_2686399, EPI_ISL_2686400, EPI_ISL_2686401, EPI_ISL_2686402, EPI_ISL_2686403, EPI_ISL_2686404, EPI_ISL_2686405, EPI_ISL_2686406, EPI_ISL_2686407, EPI_ISL_2686408, EPI_ISL_2686409, EPI_ISL_2686410, EPI_ISL_2686411, EPI_ISL_2686412, EPI_ISL_2686413, EPI_ISL_2686414, EPI_ISL_2686415, EPI_ISL_2686416, EPI_ISL_2686417, EPI_ISL_2686418, EPI_ISL_2686419, EPI_ISL_2686420, EPI_ISL_2686421, EPI_ISL_2686422, EPI_ISL_2686423, EPI_ISL_2686424, EPI_ISL_2686425, EPI_ISL_2686426, EPI_ISL_2686427, EPI_ISL_2686428, EPI_ISL_2686429, EPI_ISL_2686430, EPI_ISL_2686431, EPI_ISL_2686432, EPI_ISL_2686433, EPI_ISL_2686434, EPI_ISL_2686435, EPI_ISL_2686436, EPI_ISL_2686437, EPI_ISL_2686438, EPI_ISL_2686439, EPI_ISL_2686440, EPI_ISL_2686441, EPI_ISL_2686442, EPI_ISL_2686443, EPI_ISL_2686550, EPI_ISL_2686551, EPI_ISL_2686552, EPI_ISL_2686553, EPI_ISL_2686554, EPI_ISL_2686555, EPI_ISL_2686556, EPI_ISL_2686557, EPI_ISL_2686558, EPI_ISL_2686559, EPI_ISL_2686560, EPI_ISL_2686561, EPI_ISL_2686562, EPI_ISL_2686563, EPI_ISL_2686564, EPI_ISL_2686565, EPI_ISL_2686566, EPI_ISL_2686567, EPI_ISL_2686568, EPI_ISL_2686569, EPI_ISL_2686570, EPI_ISL_2686571, EPI_ISL_2686572, EPI_ISL_2686573, EPI_ISL_2686574, EPI_ISL_2686575, EPI_ISL_2686576, EPI_ISL_2686577, EPI_ISL_2686578, EPI_ISL_2686579, EPI_ISL_2686580, EPI_ISL_2686581, EPI_ISL_2686582, EPI_ISL_2686583, EPI_ISL_2686584, EPI_ISL_2686585, EPI_ISL_2686586, EPI_ISL_2686587, EPI_ISL_2686588, EPI_ISL_2686589, EPI_ISL_2686590, EPI_ISL_2686591, EPI_ISL_2686592, EPI_ISL_2686593, EPI_ISL_2686594, EPI_ISL_2686595, EPI_ISL_2686596, EPI_ISL_2686597, EPI_ISL_2686598, EPI_ISL_2686599, EPI_ISL_2686600, EPI_ISL_2686601, EPI_ISL_2686602, EPI_ISL_2686603, EPI_ISL_2686604, EPI_ISL_2686605, EPI_ISL_2686606, EPI_ISL_2686607, EPI_ISL_2686608, EPI_ISL_2686609, EPI_ISL_2686610, EPI_ISL_2686611, EPI_ISL_2686612, EPI_ISL_2686613, EPI_ISL_2686614, EPI_ISL_2686615, EPI_ISL_2686616, EPI_ISL_2686617, EPI_ISL_2686623, EPI_ISL_2686624, EPI_ISL_2686625, EPI_ISL_2686626, EPI_ISL_2686627, EPI_ISL_2686628, EPI_ISL_2686630, EPI_ISL_2686631, EPI_ISL_2686632, EPI_ISL_2686633, EPI_ISL_2686634, EPI_ISL_2686635, EPI_ISL_2686636, EPI_ISL_2686637, EPI_ISL_2686638, EPI_ISL_2686639, EPI_ISL_2686640, EPI_ISL_2686641, EPI_ISL_2686642, EPI_ISL_2686643, EPI_ISL_2686644, EPI_ISL_2686645, EPI_ISL_2686646, EPI_ISL_2686647, EPI_ISL_2686648, EPI_ISL_2686649, EPI_ISL_2686650, EPI_ISL_2686651, EPI_ISL_2686652, EPI_ISL_2686653, EPI_ISL_2686654, EPI_ISL_2686655, EPI_ISL_2686656, EPI_ISL_2686657, EPI_ISL_2686658, EPI_ISL_2686659, EPI_ISL_2686660, EPI_ISL_2686661, EPI_ISL_2686662, EPI_ISL_2686663, EPI_ISL_2686664, EPI_ISL_2686665, EPI_ISL_2686666, EPI_ISL_2686667, EPI_ISL_2686668, EPI_ISL_2686669, EPI_ISL_2686670, EPI_ISL_2686671, EPI_ISL_2686672, EPI_ISL_2686673, EPI_ISL_2686674, EPI_ISL_2686675, EPI_ISL_2686676, EPI_ISL_2686677, EPI_ISL_2686678, EPI_ISL_2686679, EPI_ISL_2686680, EPI_ISL_2686681, EPI_ISL_2686682, EPI_ISL_2686683, EPI_ISL_2686684, EPI_ISL_2686685, EPI_ISL_2686686, EPI_ISL_2686687, EPI_ISL_2686688, EPI_ISL_2686689, EPI_ISL_2686690, EPI_ISL_2686691, EPI_ISL_2686692, EPI_ISL_2686693, EPI_ISL_2686694, EPI_ISL_2686695, EPI_ISL_2686729, EPI_ISL_2785535, EPI_ISL_2785545, EPI_ISL_2785546, EPI_ISL_2785547, EPI_ISL_2785549, EPI_ISL_2785550, EPI_ISL_2785551, EPI_ISL_2785552, EPI_ISL_2785553, EPI_ISL_2785554, EPI_ISL_2785555, EPI_ISL_2785556, EPI_ISL_2785557, EPI_ISL_2785558, EPI_ISL_2785559, EPI_ISL_2785560, EPI_ISL_2785561, EPI_ISL_2785562, EPI_ISL_2785563, EPI_ISL_2785564, EPI_ISL_2785565, EPI_ISL_2785566, EPI_ISL_2785567, EPI_ISL_2785568, EPI_ISL_2785569, EPI_ISL_2785570, EPI_ISL_2785571, EPI_ISL_2785572, EPI_ISL_2785573, EPI_ISL_2785574, EPI_ISL_2785575, EPI_ISL_2785576, EPI_ISL_2785577, EPI_ISL_2785578, EPI_ISL_2785579, EPI_ISL_2785580, EPI_ISL_2785581, EPI_ISL_2785582, EPI_ISL_2785583, EPI_ISL_2785584, EPI_ISL_2785585, EPI_ISL_2785586, EPI_ISL_2785587, EPI_ISL_2785588, EPI_ISL_2785589, EPI_ISL_2785590, EPI_ISL_2785591, EPI_ISL_2785592, EPI_ISL_2785593, EPI_ISL_2785594, EPI_ISL_2785595, EPI_ISL_2785596, EPI_ISL_2785597, EPI_ISL_2785598, EPI_ISL_2785599, EPI_ISL_2785600, EPI_ISL_2785601, EPI_ISL_2785602, EPI_ISL_2785603, EPI_ISL_2785604, EPI_ISL_2785605, EPI_ISL_2785606, EPI_ISL_2785607, EPI_ISL_2785608, EPI_ISL_2785609, EPI_ISL_2785610, EPI_ISL_2785611, EPI_ISL_2785612, EPI_ISL_2785613, EPI_ISL_2785614, EPI_ISL_2785615, EPI_ISL_2785616, EPI_ISL_2785617, EPI_ISL_2785618, EPI_ISL_2785619, EPI_ISL_2785620, EPI_ISL_2785621, EPI_ISL_2785622, EPI_ISL_2785623, EPI_ISL_2785624, EPI_ISL_2785625, EPI_ISL_2785626, EPI_ISL_2785627, EPI_ISL_2785628, EPI_ISL_2785629, EPI_ISL_2785630, EPI_ISL_2785631, EPI_ISL_2785632, EPI_ISL_2785633, EPI_ISL_2785634, EPI_ISL_2785635, EPI_ISL_2785636, EPI_ISL_2785637, EPI_ISL_2785638, EPI_ISL_2785639, EPI_ISL_2785640, EPI_ISL_2785641, EPI_ISL_2785642, EPI_ISL_2785643, EPI_ISL_2785644, EPI_ISL_2785645, EPI_ISL_2785646, EPI_ISL_2785647, EPI_ISL_2785648, EPI_ISL_2785649, EPI_ISL_2785650, EPI_ISL_2785651, EPI_ISL_2785652, EPI_ISL_2785653, EPI_ISL_2785654, EPI_ISL_2785655, EPI_ISL_2785656, EPI_ISL_2785657, EPI_ISL_2785658, EPI_ISL_2785659, EPI_ISL_2785660, EPI_ISL_2785661, EPI_ISL_2785662, EPI_ISL_2785663, EPI_ISL_2785664, EPI_ISL_2785665, EPI_ISL_2785666, EPI_ISL_2785667, EPI_ISL_2785668, EPI_ISL_2785669, EPI_ISL_2785670, EPI_ISL_2785671, EPI_ISL_2785672, EPI_ISL_2785673, EPI_ISL_2785674, EPI_ISL_2785675, EPI_ISL_2785676, EPI_ISL_2785677, EPI_ISL_2785678, EPI_ISL_2785679, EPI_ISL_2785680, EPI_ISL_2785704, EPI_ISL_2785729, EPI_ISL_2785730, EPI_ISL_2785731, EPI_ISL_2785732, EPI_ISL_2785733, EPI_ISL_2785734, EPI_ISL_2785735, EPI_ISL_2785736, EPI_ISL_2785737, EPI_ISL_2785738, EPI_ISL_2785739, EPI_ISL_2785740, EPI_ISL_2785741, EPI_ISL_2785742, EPI_ISL_2785743, EPI_ISL_2785744, EPI_ISL_2785745, EPI_ISL_2785746, EPI_ISL_2785747, EPI_ISL_2785748, EPI_ISL_2785749, EPI_ISL_2785750, EPI_ISL_2785751, EPI_ISL_2785752, EPI_ISL_2785753, EPI_ISL_2785754, EPI_ISL_2785755, EPI_ISL_2785756, EPI_ISL_2785757, EPI_ISL_2785758, EPI_ISL_2785759, EPI_ISL_2785760, EPI_ISL_2785761, EPI_ISL_2785762, EPI_ISL_2785763, EPI_ISL_2785764, EPI_ISL_2785765, EPI_ISL_2785766, EPI_ISL_2785767, EPI_ISL_2785768, EPI_ISL_2785769, EPI_ISL_2785770, EPI_ISL_2785771, EPI_ISL_2785772, EPI_ISL_2785773, EPI_ISL_2785774, EPI_ISL_2785775, EPI_ISL_2785776, EPI_ISL_2785777, EPI_ISL_2785778, EPI_ISL_2785779, EPI_ISL_2785780, EPI_ISL_2785781, EPI_ISL_2785782, EPI_ISL_2785783, EPI_ISL_2785784, EPI_ISL_2785785, EPI_ISL_2785786, EPI_ISL_2785787, EPI_ISL_2785788, EPI_ISL_2785789, EPI_ISL_2785790, EPI_ISL_2785791, EPI_ISL_2785792, EPI_ISL_2785794, EPI_ISL_2785795, EPI_ISL_2785797, EPI_ISL_2785798, EPI_ISL_2785799, EPI_ISL_2785800, EPI_ISL_2785801, EPI_ISL_2785802, EPI_ISL_2785803, EPI_ISL_2785804, EPI_ISL_2785805, EPI_ISL_2785806, EPI_ISL_2785807, EPI_ISL_2785808, EPI_ISL_2785809, EPI_ISL_2785810, EPI_ISL_2785811, EPI_ISL_2785812, EPI_ISL_2785813, EPI_ISL_2785814, EPI_ISL_2785815, EPI_ISL_2785816, EPI_ISL_2785817, EPI_ISL_2785818, EPI_ISL_2785819, EPI_ISL_2785821, EPI_ISL_2785822, EPI_ISL_2785823, EPI_ISL_2785824, EPI_ISL_2785825, EPI_ISL_2785826, EPI_ISL_2785827, EPI_ISL_2785828, EPI_ISL_2785829, EPI_ISL_2785830, EPI_ISL_2785831, EPI_ISL_2785832, EPI_ISL_2785833, EPI_ISL_2785834, EPI_ISL_2785835, EPI_ISL_2785836, EPI_ISL_2785837, EPI_ISL_2785838, EPI_ISL_2785839, EPI_ISL_2785840, EPI_ISL_2785841, EPI_ISL_2785842, EPI_ISL_2785843, EPI_ISL_2785844, EPI_ISL_2785845, EPI_ISL_2785846, EPI_ISL_2785847, EPI_ISL_2785848, EPI_ISL_2785849, EPI_ISL_2785850, EPI_ISL_2785851, EPI_ISL_2785852, EPI_ISL_2785853, EPI_ISL_2785854, EPI_ISL_2785855, EPI_ISL_2785856, EPI_ISL_2785857, EPI_ISL_2785858, EPI_ISL_2785859, EPI_ISL_2785860, EPI_ISL_2785886, EPI_ISL_2785887, EPI_ISL_2785888, EPI_ISL_2785889, EPI_ISL_2785890, EPI_ISL_2785891, EPI_ISL_2785892, EPI_ISL_2785893, EPI_ISL_2785894, EPI_ISL_2785895, EPI_ISL_2785896, EPI_ISL_2785897, EPI_ISL_2785898, EPI_ISL_2785899, EPI_ISL_2785900, EPI_ISL_2785901, EPI_ISL_2785902, EPI_ISL_2785903, EPI_ISL_2785904, EPI_ISL_2785905, EPI_ISL_2785906, EPI_ISL_2785907, EPI_ISL_2785908, EPI_ISL_2785909, EPI_ISL_2785910, EPI_ISL_2785911, EPI_ISL_2785912, EPI_ISL_2785913, EPI_ISL_2785914, EPI_ISL_2785915, EPI_ISL_2785916, EPI_ISL_2785917, EPI_ISL_2785918, EPI_ISL_2785919, EPI_ISL_2785921, EPI_ISL_2785922, EPI_ISL_2785923, EPI_ISL_2785925, EPI_ISL_2785926, EPI_ISL_2785927, EPI_ISL_2785928, EPI_ISL_2785929, EPI_ISL_2785930, EPI_ISL_2785931, EPI_ISL_2785932, EPI_ISL_2785933, EPI_ISL_2785934, EPI_ISL_2785935, EPI_ISL_2785936, EPI_ISL_2785937, EPI_ISL_2785938, EPI_ISL_2785939, EPI_ISL_2785940, EPI_ISL_2785941, EPI_ISL_2785942, EPI_ISL_2785943, EPI_ISL_2785944, EPI_ISL_2785945, EPI_ISL_2785946, EPI_ISL_2785947, EPI_ISL_2785948, EPI_ISL_2785949, EPI_ISL_2785950, EPI_ISL_2785951, EPI_ISL_2785952, EPI_ISL_2785953, EPI_ISL_2785954, EPI_ISL_2873346, EPI_ISL_2873358, EPI_ISL_2873359, EPI_ISL_2873360, EPI_ISL_2873361, EPI_ISL_2873362, EPI_ISL_2873363, EPI_ISL_2873364, EPI_ISL_2873365, EPI_ISL_2873366, EPI_ISL_2873367, EPI_ISL_2873368, EPI_ISL_2873369, EPI_ISL_2873370, EPI_ISL_2873371, EPI_ISL_2873372, EPI_ISL_2873373, EPI_ISL_2873374, EPI_ISL_2873375, EPI_ISL_2873376, EPI_ISL_2873377, EPI_ISL_2873378, EPI_ISL_2873379, EPI_ISL_2873380, EPI_ISL_2873381, EPI_ISL_2873382, EPI_ISL_2873383, EPI_ISL_2873384, EPI_ISL_2873385, EPI_ISL_2873386, EPI_ISL_2873387, EPI_ISL_2873388, EPI_ISL_2873389, EPI_ISL_2873390, EPI_ISL_2873391, EPI_ISL_2873392, EPI_ISL_2873393, EPI_ISL_2873394, EPI_ISL_2873395, EPI_ISL_2873396, EPI_ISL_2873397, EPI_ISL_2873398, EPI_ISL_2873399, EPI_ISL_2873400, EPI_ISL_2873401, EPI_ISL_2873402, EPI_ISL_2873403, EPI_ISL_2873404, EPI_ISL_2873405, EPI_ISL_2873406, EPI_ISL_2873407, EPI_ISL_2873408, EPI_ISL_2873409, EPI_ISL_2873410, EPI_ISL_2873411, EPI_ISL_2873412, EPI_ISL_2873413, EPI_ISL_2873414, EPI_ISL_2873415, EPI_ISL_2873416, EPI_ISL_2873417, EPI_ISL_2873418, EPI_ISL_2873419, EPI_ISL_2873420, EPI_ISL_2873421, EPI_ISL_2873422, EPI_ISL_2873423, | | | |

| | | | |
|---|---|---|---|
| EPI_ISL_2873424, EPI_ISL_2873425, EPI_ISL_2873426, EPI_ISL_2873427, EPI_ISL_2873428, EPI_ISL_2873429, EPI_ISL_2873430, EPI_ISL_2873431, EPI_ISL_2873432, EPI_ISL_2873433, EPI_ISL_2873434, EPI_ISL_2873435, EPI_ISL_2873436, EPI_ISL_2873437, EPI_ISL_2873438, EPI_ISL_2873439, EPI_ISL_2873440, EPI_ISL_2873441, EPI_ISL_2873442, EPI_ISL_2873443, EPI_ISL_2873444, EPI_ISL_2873445, EPI_ISL_2873446, EPI_ISL_2873447, EPI_ISL_2873448, EPI_ISL_2873449, EPI_ISL_2873450, EPI_ISL_2873451, EPI_ISL_2873452, EPI_ISL_2873453, EPI_ISL_2873454, EPI_ISL_2873455, EPI_ISL_2873456, EPI_ISL_2873457, EPI_ISL_2873458, EPI_ISL_2873459, EPI_ISL_2873460, EPI_ISL_2873461, EPI_ISL_2873462, EPI_ISL_2873463, EPI_ISL_2873464, EPI_ISL_2873465, EPI_ISL_2873466, EPI_ISL_2873467, EPI_ISL_2873468, EPI_ISL_2873469, EPI_ISL_2873470, EPI_ISL_2873471, EPI_ISL_2873472, EPI_ISL_2873473, EPI_ISL_2873474, EPI_ISL_2873475, EPI_ISL_2873476, EPI_ISL_2873477, EPI_ISL_2873478, EPI_ISL_2873479, EPI_ISL_2873480, EPI_ISL_2873481, EPI_ISL_2873482, EPI_ISL_2873483, EPI_ISL_2873484, EPI_ISL_2873485, EPI_ISL_2873486, EPI_ISL_2873487, EPI_ISL_2873488, EPI_ISL_2873489, EPI_ISL_2873490, EPI_ISL_2873491, EPI_ISL_2873492, EPI_ISL_2873493, EPI_ISL_2873494, EPI_ISL_2873495, EPI_ISL_2873496, EPI_ISL_2873497, EPI_ISL_2873498, EPI_ISL_2873499, EPI_ISL_2873500, EPI_ISL_2873508, EPI_ISL_2873509, EPI_ISL_2873510, EPI_ISL_2873511, EPI_ISL_2873512, EPI_ISL_2873513, EPI_ISL_2873514, EPI_ISL_2873515, EPI_ISL_2873516, EPI_ISL_2873517, EPI_ISL_2873518, EPI_ISL_2873519, EPI_ISL_2873520, EPI_ISL_2873521, EPI_ISL_2873522, EPI_ISL_2873523, EPI_ISL_2873524, EPI_ISL_2873525, EPI_ISL_2873526, EPI_ISL_2873527, EPI_ISL_2873528, EPI_ISL_2873529, EPI_ISL_2873530, EPI_ISL_2873531, EPI_ISL_2873532, EPI_ISL_2873533, EPI_ISL_2873534, EPI_ISL_2873535, EPI_ISL_2873536, EPI_ISL_2873537, EPI_ISL_2873538, EPI_ISL_2873539, EPI_ISL_2873540, EPI_ISL_2873541, EPI_ISL_2873542, EPI_ISL_2873543, EPI_ISL_2873544, EPI_ISL_2873545, EPI_ISL_2873546, EPI_ISL_2873547, EPI_ISL_2873548, EPI_ISL_2873549, EPI_ISL_2873550, EPI_ISL_2873551, EPI_ISL_2873552, EPI_ISL_2873553, EPI_ISL_2873554, EPI_ISL_2873555, EPI_ISL_2873556, EPI_ISL_2873557, EPI_ISL_2873558, EPI_ISL_2873559, EPI_ISL_2873560, EPI_ISL_2873561, EPI_ISL_2873562, EPI_ISL_2873563, EPI_ISL_2873564, EPI_ISL_2873565, EPI_ISL_2873566, EPI_ISL_2873567, EPI_ISL_2873568, EPI_ISL_2873569, EPI_ISL_2873570, EPI_ISL_2873571, EPI_ISL_2873572, EPI_ISL_2873573, EPI_ISL_2873574, EPI_ISL_2873575, EPI_ISL_2873576, EPI_ISL_2873577, EPI_ISL_2873578, EPI_ISL_2873579, EPI_ISL_2873580, EPI_ISL_2873581, EPI_ISL_2873582, EPI_ISL_2873583, EPI_ISL_2873584, EPI_ISL_2873585, EPI_ISL_2873586, EPI_ISL_2873587, EPI_ISL_2873588, EPI_ISL_2873589, EPI_ISL_2873590, EPI_ISL_2873591, EPI_ISL_2873592, EPI_ISL_2873593, EPI_ISL_2873594, EPI_ISL_2873595, EPI_ISL_2873596, EPI_ISL_2873597, EPI_ISL_2873598, EPI_ISL_2873599, EPI_ISL_2873600, EPI_ISL_2873601, EPI_ISL_2873602, EPI_ISL_2873603, EPI_ISL_2873604, EPI_ISL_2873605, EPI_ISL_2873606, EPI_ISL_2873607, EPI_ISL_2873608, EPI_ISL_2873609, EPI_ISL_2873610, EPI_ISL_2873611, EPI_ISL_2873612, EPI_ISL_2873613, EPI_ISL_2873614, EPI_ISL_2873615, EPI_ISL_2873616, EPI_ISL_2873617, EPI_ISL_2873618, EPI_ISL_2873619, EPI_ISL_2873620, EPI_ISL_2873621, EPI_ISL_2873622, EPI_ISL_2873623, EPI_ISL_2873624, EPI_ISL_2873625, EPI_ISL_2873626, EPI_ISL_2873627, EPI_ISL_2873628, EPI_ISL_2873629, EPI_ISL_2873630, EPI_ISL_2873631, EPI_ISL_2873632, EPI_ISL_2873633, EPI_ISL_2873634, EPI_ISL_2873635, EPI_ISL_2873636, EPI_ISL_2873637, EPI_ISL_2873638, EPI_ISL_2873639, EPI_ISL_2873640, EPI_ISL_2873641, EPI_ISL_2873643, EPI_ISL_2873644, EPI_ISL_2873645, EPI_ISL_2873646, EPI_ISL_2873647, EPI_ISL_2873648, EPI_ISL_2873649, EPI_ISL_2873650, EPI_ISL_2873651, EPI_ISL_2873652, EPI_ISL_2873653, EPI_ISL_2873654, EPI_ISL_2873655, EPI_ISL_2873656, EPI_ISL_2873657, EPI_ISL_2873658, EPI_ISL_2873659, EPI_ISL_2873660, EPI_ISL_2873661, EPI_ISL_2873662, EPI_ISL_2873663, EPI_ISL_2873664, EPI_ISL_2873665, EPI_ISL_2873666, EPI_ISL_2873667, EPI_ISL_2873668, EPI_ISL_2873669, EPI_ISL_2873670, EPI_ISL_2873671, EPI_ISL_2873672, EPI_ISL_2873673, EPI_ISL_2873674, EPI_ISL_2873675, EPI_ISL_2873676, EPI_ISL_2873677, EPI_ISL_2873678, EPI_ISL_2873679, EPI_ISL_2873680, EPI_ISL_2873681, EPI_ISL_2873682, EPI_ISL_2873683, EPI_ISL_2873684, EPI_ISL_2873685, EPI_ISL_2873686, EPI_ISL_2873687, EPI_ISL_2873688, EPI_ISL_2873689, EPI_ISL_2873690, EPI_ISL_2873691, EPI_ISL_2873692, EPI_ISL_2873693, EPI_ISL_2873694, EPI_ISL_2873695, EPI_ISL_2873696, EPI_ISL_2873697, EPI_ISL_2873698, EPI_ISL_2873699, EPI_ISL_2873700, EPI_ISL_2873701, EPI_ISL_2873702, EPI_ISL_2873703, EPI_ISL_2873704, EPI_ISL_2873705, EPI_ISL_2873706, EPI_ISL_2873715, EPI_ISL_2873716, EPI_ISL_2873717, EPI_ISL_2873718, EPI_ISL_2873719, EPI_ISL_2873720, EPI_ISL_2873721, EPI_ISL_2873722, EPI_ISL_2873723, EPI_ISL_2873724, EPI_ISL_2873725, EPI_ISL_2873726, EPI_ISL_2873727, EPI_ISL_2873728, EPI_ISL_2873729, EPI_ISL_2873730, EPI_ISL_2873731, EPI_ISL_2873732, EPI_ISL_2873733, EPI_ISL_2873734, EPI_ISL_2873735, EPI_ISL_2873736, EPI_ISL_2873737, EPI_ISL_2873738, EPI_ISL_2873739, EPI_ISL_2873740, EPI_ISL_2873741, EPI_ISL_2873742, EPI_ISL_2873743, EPI_ISL_2873744, EPI_ISL_2873745, EPI_ISL_2873746, EPI_ISL_2873747, EPI_ISL_2873748, EPI_ISL_2873749, EPI_ISL_2873750, EPI_ISL_2873751, EPI_ISL_2873752, EPI_ISL_2873753, EPI_ISL_2873754, EPI_ISL_2873755, EPI_ISL_2873756, EPI_ISL_2873757, EPI_ISL_2873758, EPI_ISL_2873759, EPI_ISL_2873760, EPI_ISL_2873761, EPI_ISL_2873762, EPI_ISL_2873763, EPI_ISL_2873764, EPI_ISL_2873765, EPI_ISL_2873766, EPI_ISL_2873767, EPI_ISL_2873768, EPI_ISL_2873769, EPI_ISL_2873770, EPI_ISL_2873771, EPI_ISL_2873772, EPI_ISL_2873773, EPI_ISL_2873774, EPI_ISL_2873775, EPI_ISL_2873776, EPI_ISL_2873777, EPI_ISL_2873778, EPI_ISL_2873779, EPI_ISL_2873780, EPI_ISL_2873781, EPI_ISL_2873782, EPI_ISL_2873783, EPI_ISL_2873784, EPI_ISL_2873785, EPI_ISL_2873786, EPI_ISL_2873787, EPI_ISL_2873788, EPI_ISL_2873789, EPI_ISL_2928580, EPI_ISL_2928581, EPI_ISL_2928582, EPI_ISL_2928590, EPI_ISL_2928591, EPI_ISL_2928592, EPI_ISL_2928593, EPI_ISL_2928594, EPI_ISL_2928595, EPI_ISL_2928597, EPI_ISL_2928598, EPI_ISL_2928600, EPI_ISL_2928601, EPI_ISL_2928603, EPI_ISL_2928611, EPI_ISL_2928612, EPI_ISL_2928613, EPI_ISL_2928614, EPI_ISL_2928616, EPI_ISL_2928621, EPI_ISL_2928627, EPI_ISL_2928628, EPI_ISL_2928629, EPI_ISL_2928634, EPI_ISL_2928655, EPI_ISL_2928673, EPI_ISL_2928699, EPI_ISL_2928701, EPI_ISL_2928925, EPI_ISL_2929130, EPI_ISL_2929131 | | | |
| see above | Fulgent Genetics | Centers for Disease Control and Prevention Division of Viral Diseases, Pathogen Discovery | Adrian Paskey; Becky Tsai; Benafsh Sapra; Benjamin Rambo-Martin; Christopher Gulvick; Clinton R. Paden; Dakota Howard; Darlene Wagner; Dhwani Batra; Doreen Ng; Duncan MacCannell; Harry Gao; James Xie; Jason Caravas; John Gao; Joseph Fierro; Kara Moser; Matthew Schmerer; Mickey Li; Peter W. Cook; Scott Sammons; Shatavia Morrison; Yan Meng; Yvette Unoarumhi |
| EPI_ISL_2404801, EPI_ISL_2404826 | GOOD SAMARITAN HOSPTIAL | Santa Clara County public Health Laboratory | Santa Clara County Public Health Department |
| EPI_ISL_2650566, EPI_ISL_2753977, EPI_ISL_2754022, EPI_ISL_2934001, EPI_ISL_2934053 | Good Samaritan Hospital | Santa Clara County Public Health Laboratory | Santa Clara County Public Health Department |
| EPI_ISL_2159541, EPI_ISL_2159774, EPI_ISL_2159787, EPI_ISL_2159794, EPI_ISL_2159795, EPI_ISL_2159806, EPI_ISL_2159807, EPI_ISL_2159815, EPI_ISL_2159817, EPI_ISL_2159842, EPI_ISL_2160094, EPI_ISL_2160224, EPI_ISL_2160231, EPI_ISL_2160327, EPI_ISL_2160335, EPI_ISL_2160348, EPI_ISL_2160360, EPI_ISL_2160375, EPI_ISL_2160397, EPI_ISL_2160400, EPI_ISL_2160407, EPI_ISL_2160414, EPI_ISL_2160429, EPI_ISL_2160443, EPI_ISL_2160446, EPI_ISL_2160454, EPI_ISL_2160459, EPI_ISL_2160463, EPI_ISL_2160481, EPI_ISL_2160490, EPI_ISL_2160514, EPI_ISL_2187635, EPI_ISL_2187636, EPI_ISL_2247574, EPI_ISL_2247613, EPI_ISL_2247620, EPI_ISL_2247660, EPI_ISL_2247687, EPI_ISL_2247690, EPI_ISL_2247776, EPI_ISL_2247842, EPI_ISL_2248020, EPI_ISL_2248042, EPI_ISL_2248059, EPI_ISL_2248074, EPI_ISL_2248085, EPI_ISL_2248097, EPI_ISL_2248117, EPI_ISL_2248118, EPI_ISL_2248146, EPI_ISL_2248330, EPI_ISL_2248433, EPI_ISL_2248451, EPI_ISL_2268355, EPI_ISL_2269641, EPI_ISL_2269679, EPI_ISL_2269685, EPI_ISL_2269704, EPI_ISL_2269713, EPI_ISL_2269722, EPI_ISL_2269731, EPI_ISL_2269752, EPI_ISL_2269773, EPI_ISL_2269830, EPI_ISL_2269847, EPI_ISL_2269863, EPI_ISL_2269873, EPI_ISL_2269885, EPI_ISL_2269889, EPI_ISL_2269896, EPI_ISL_2269913, EPI_ISL_2269937, EPI_ISL_2269938, EPI_ISL_2269950, EPI_ISL_2269999, EPI_ISL_2270028, EPI_ISL_2270034, EPI_ISL_2270036, EPI_ISL_2270055, EPI_ISL_2270083, EPI_ISL_2270135, EPI_ISL_2270156, EPI_ISL_2270296, EPI_ISL_2270315, EPI_ISL_2270318, EPI_ISL_2270319, EPI_ISL_2270328, EPI_ISL_2270330, EPI_ISL_2270346, EPI_ISL_2270406, EPI_ISL_2270412, EPI_ISL_2270422, EPI_ISL_2270429, EPI_ISL_2270539, EPI_ISL_2270630, EPI_ISL_2270632, EPI_ISL_2270638, EPI_ISL_2270685, EPI_ISL_2270742, EPI_ISL_2270751, EPI_ISL_2270763, EPI_ISL_2270768, EPI_ISL_2270795, EPI_ISL_2270806, EPI_ISL_2270807, EPI_ISL_2270817, EPI_ISL_2270834, EPI_ISL_2280186, EPI_ISL_2280188, EPI_ISL_2320000, EPI_ISL_2320001, EPI_ISL_2320011, EPI_ISL_2320014, EPI_ISL_2320022, EPI_ISL_2320023, EPI_ISL_2320025, EPI_ISL_2320060, EPI_ISL_2320096, EPI_ISL_2320100, EPI_ISL_2320103, EPI_ISL_2320119, EPI_ISL_2320123, EPI_ISL_2320163, EPI_ISL_2320226, EPI_ISL_2320255, EPI_ISL_2320272, EPI_ISL_2320280, EPI_ISL_2320285, EPI_ISL_2320293, EPI_ISL_2320295, EPI_ISL_2320297, EPI_ISL_2320314, EPI_ISL_2320345, EPI_ISL_2320356, EPI_ISL_2320378, EPI_ISL_2320443, EPI_ISL_2320446, EPI_ISL_2320455, EPI_ISL_2320487, EPI_ISL_2320524, EPI_ISL_2320526, EPI_ISL_2320528, EPI_ISL_2320530, EPI_ISL_2320590, EPI_ISL_2320646, EPI_ISL_2320676, EPI_ISL_2320774, EPI_ISL_2320826, EPI_ISL_2320838, EPI_ISL_2320847, EPI_ISL_2320854, EPI_ISL_2320862, EPI_ISL_2320863, EPI_ISL_2320866, EPI_ISL_2320873, EPI_ISL_2320901, EPI_ISL_2320921, EPI_ISL_2320922, EPI_ISL_2320928, EPI_ISL_2320930, EPI_ISL_2320943, EPI_ISL_2320945, EPI_ISL_2320955, EPI_ISL_2320961, EPI_ISL_2320991, EPI_ISL_2321048, EPI_ISL_2321081, EPI_ISL_2321091, EPI_ISL_2321101, EPI_ISL_2321135, EPI_ISL_2321146, EPI_ISL_2367608, EPI_ISL_2367620, EPI_ISL_2367641, EPI_ISL_2367644, EPI_ISL_2440055, EPI_ISL_2440066, EPI_ISL_2440079, EPI_ISL_2440089, EPI_ISL_2440195, EPI_ISL_2440206, EPI_ISL_2440223, EPI_ISL_2440366, EPI_ISL_2440508, EPI_ISL_2440520, EPI_ISL_2598841, EPI_ISL_2598852, EPI_ISL_2598873, EPI_ISL_2598889, EPI_ISL_2598946, EPI_ISL_2599112, EPI_ISL_2599139, EPI_ISL_2599158, EPI_ISL_2599162, EPI_ISL_2599173, EPI_ISL_2599212, EPI_ISL_2599214, EPI_ISL_2599223, EPI_ISL_2599239, EPI_ISL_2599241, EPI_ISL_2599252, EPI_ISL_2599273, EPI_ISL_2599308, EPI_ISL_2599309, EPI_ISL_2599400, EPI_ISL_2599408, EPI_ISL_2599416, EPI_ISL_2599420, EPI_ISL_2599447, EPI_ISL_2599472, EPI_ISL_2599475, EPI_ISL_2599540, EPI_ISL_2599575, EPI_ISL_2599606, EPI_ISL_2599608, EPI_ISL_2649941 | | | |
| see above | Helix/Illumina | Centers for Disease Control and Prevention Division of Viral Diseases, Pathogen Discovery | Adrian Paskey; Alexandre Bolze; Ary Ascencio; Benjamin Rambo-Martin; Brad Sickler; Charlotte Rivera-Garcia; Christine Tran; Christopher Gulvick; Clinton R. Paden; Dakota Howard; Darlene Wagner; David Becker; Dhwani Batra; Duncan MacCannell; Efren Sandoval; Eileen de Feo; Elizabeth Cirulli; Eric Allen; Geraint Levan; James Lu; Jan Antico; Jason Caravas; Jason Nguyen; Jimmy Ramirez; Jingtao Liu; Kara Moser; Kelly Schiabor Barrett; Kim Gietzen; Magnus Isaksson; Marc Laurent; Matthew Schmerer; Matthew Tolentino; Nicole L. Washington; Peter W. Cook; Phil Febbo; Ryan Cho; Scott Sammons; Shannon Wickline; Shatavia Morrison; Sherry Wang; Simon White; Tyler Cassens; William Lee; Yvette Unoarumhi |
| EPI_ISL_2907564 | Henry Mayo Newhall Hospital | Los Angeles County Public Health Laboratories | P. Hemarajata et al. |
| EPI_ISL_2811961 | Hollywood Presbyterian Medical Center | Los Angeles County PHL | P. Hemarajata et al. |
| EPI_ISL_2341415, EPI_ISL_2341421, EPI_ISL_2341436, EPI_ISL_2341449, EPI_ISL_2341458, EPI_ISL_2341463, EPI_ISL_2341471, EPI_ISL_2341472, EPI_ISL_2341480, EPI_ISL_2341487, EPI_ISL_2341498, EPI_ISL_2341501, EPI_ISL_2341543, EPI_ISL_2341563, EPI_ISL_2341565, EPI_ISL_2341572, EPI_ISL_2341595, EPI_ISL_2341613, EPI_ISL_2341616, EPI_ISL_2341621, EPI_ISL_2341625, EPI_ISL_2341627, EPI_ISL_2341629, EPI_ISL_2341650, EPI_ISL_2341655, EPI_ISL_2497549, EPI_ISL_2497567, EPI_ISL_2497622, EPI_ISL_2497625, EPI_ISL_2497628, EPI_ISL_2497633, EPI_ISL_2497653, EPI_ISL_2497657, EPI_ISL_2497660, EPI_ISL_2497662, EPI_ISL_2497663, EPI_ISL_2497664, EPI_ISL_2497669, EPI_ISL_2497686, EPI_ISL_2497690, EPI_ISL_2497691, EPI_ISL_2497693, EPI_ISL_2497697, EPI_ISL_2500025, EPI_ISL_2500048, EPI_ISL_2500054, EPI_ISL_2500058, EPI_ISL_2500060, EPI_ISL_2500061, EPI_ISL_2500063, EPI_ISL_2500069, EPI_ISL_2500074, EPI_ISL_2500078, EPI_ISL_2500100, EPI_ISL_2500108, EPI_ISL_2500116, EPI_ISL_2500125, EPI_ISL_2500133, EPI_ISL_2500142, EPI_ISL_2500150, EPI_ISL_2500151, EPI_ISL_2500157, EPI_ISL_2500159, EPI_ISL_2500182, EPI_ISL_2500185, EPI_ISL_2500189, EPI_ISL_2500196, EPI_ISL_2500201, EPI_ISL_2500202, EPI_ISL_2500213, EPI_ISL_2658738, EPI_ISL_2758541, EPI_ISL_2758544 | | | |
| see above | Humboldt County Public Health Laboratory | Chan-Zuckerberg Biohub | CZB Cliahub Consortium |
| EPI_ISL_2445617, EPI_ISL_2445618, EPI_ISL_2445619, EPI_ISL_2445620, EPI_ISL_2445621, EPI_ISL_2445622, EPI_ISL_2445623, EPI_ISL_2445624, EPI_ISL_2445625, EPI_ISL_2445626, EPI_ISL_2445627, EPI_ISL_2445628, EPI_ISL_2445629, EPI_ISL_2445630, EPI_ISL_2445631, EPI_ISL_2445632, EPI_ISL_2445633, EPI_ISL_2680469, EPI_ISL_2680471, EPI_ISL_2680473, EPI_ISL_2680474, EPI_ISL_2680476, EPI_ISL_2680478, EPI_ISL_2680480, EPI_ISL_2680482, EPI_ISL_2680483, EPI_ISL_2680485, EPI_ISL_2680487, EPI_ISL_2680488, EPI_ISL_2680490, EPI_ISL_2680492, EPI_ISL_2778969, EPI_ISL_2778970, EPI_ISL_2778971, EPI_ISL_2778972, EPI_ISL_2778973, EPI_ISL_2778974, EPI_ISL_2778975, EPI_ISL_2778976, EPI_ISL_2778977, EPI_ISL_2778978, EPI_ISL_2778979, EPI_ISL_2778980, EPI_ISL_2778981, EPI_ISL_2778982, EPI_ISL_2778983, EPI_ISL_2928419, EPI_ISL_2928420, EPI_ISL_2928421, EPI_ISL_2928422, EPI_ISL_2928423, EPI_ISL_2928424, EPI_ISL_2928425, EPI_ISL_2928426, EPI_ISL_2928427, EPI_ISL_2928428, EPI_ISL_2928429, EPI_ISL_2928430, EPI_ISL_2928431, EPI_ISL_2928432, EPI_ISL_2928433, EPI_ISL_2928434, EPI_ISL_2928435, EPI_ISL_2928436, EPI_ISL_2928437, EPI_ISL_2928438 | | | |
| see above | Humboldt County Public Health Laboratory | Humboldt County Public Health Laboratory | Jeremy Corrigan |
| EPI_ISL_2391788, EPI_ISL_2391807, EPI_ISL_2405182, EPI_ISL_2894586 | Huntington Memorial Hospital | Los Angeles County PHL | P. Hemarajata et al. |
| EPI_ISL_2801446 | IVY3 Central Lab, Vanderbilt University Medical Center | Lauring Lab, University of Michigan, Department of Microbiology and Immunology | Valesano |
| EPI_ISL_2604246 | Infectious Diseases, Chan-Zuckerberg Biohub Covidtracker | Infectious Diseases, Chan-Zuckerberg Biohub Covidtracker | CZB Cliahub Consortium |
| EPI_ISL_2088924, EPI_ISL_2088925, EPI_ISL_2186452, EPI_ISL_2186502, EPI_ISL_2282436, EPI_ISL_2282520, EPI_ISL_2282718, EPI_ISL_2282857, EPI_ISL_2324030, EPI_ISL_2324045, EPI_ISL_2324086, EPI_ISL_2324098, EPI_ISL_2324112, EPI_ISL_2324130, EPI_ISL_2324132, EPI_ISL_2324156, EPI_ISL_2375736, EPI_ISL_2557438, EPI_ISL_2557530, EPI_ISL_2557535, EPI_ISL_2557616, EPI_ISL_2652839, EPI_ISL_2652936, EPI_ISL_2652938, EPI_ISL_2686840, EPI_ISL_2786657, EPI_ISL_2876054 | | | |
| see above | Infinity Biologix | Centers for Disease Control and Prevention Division of Viral Diseases, Pathogen Discovery | Adrian Paskey; Benjamin Rambo-Martin; Chirayu Goswami; Christian Bixby; Christopher Gulvick; Clinton R. Paden; Dakota Howard; Darlene Wagner; Dhwani Batra; Duncan MacCannell; Jason Caravas; Jonathan Schultz; Kara Moser; Matthew Schmerer; Peter W. Cook; Robin Grimwood; Russ Hager; Scott Sammons; Shatavia Morrison; Yihe Wang; Yvette Unoarumhi |
| EPI_ISL_2404800 | KAISER SANTA CLARA | Santa Clara County public Health Laboratory | Santa Clara County Public Health Department |
| EPI_ISL_2753992, EPI_ISL_2753993, EPI_ISL_2753994, EPI_ISL_2753995, EPI_ISL_2753996, EPI_ISL_2753997, EPI_ISL_2753998, EPI_ISL_2753999, EPI_ISL_2754000, EPI_ISL_2754002, EPI_ISL_2754003, EPI_ISL_2754004, EPI_ISL_2754005, EPI_ISL_2754006, EPI_ISL_2754007, EPI_ISL_2754008, EPI_ISL_2754009, EPI_ISL_2754010, EPI_ISL_2754011, EPI_ISL_2754012, EPI_ISL_2754013, EPI_ISL_2754014, EPI_ISL_2754015, EPI_ISL_2754016, EPI_ISL_2754017, EPI_ISL_2754018, EPI_ISL_2754019, EPI_ISL_2754020, EPI_ISL_2754021, EPI_ISL_2933980, EPI_ISL_2933982, EPI_ISL_2933983, EPI_ISL_2933984, EPI_ISL_2933985, EPI_ISL_2933988, EPI_ISL_2933991, EPI_ISL_2933992, EPI_ISL_2933996, EPI_ISL_2933999, EPI_ISL_2934003, EPI_ISL_2934005, EPI_ISL_2934007, EPI_ISL_2934008, EPI_ISL_2934009, EPI_ISL_2934011, EPI_ISL_2934012, EPI_ISL_2934013, EPI_ISL_2934014, EPI_ISL_2934015, EPI_ISL_2934016, EPI_ISL_2934017, EPI_ISL_2934018, EPI_ISL_2934019, EPI_ISL_2934020, EPI_ISL_2934024, EPI_ISL_2934025, EPI_ISL_2934026, EPI_ISL_2934027, EPI_ISL_2934028, EPI_ISL_2934044, EPI_ISL_2934045, EPI_ISL_2934066, EPI_ISL_2934067, EPI_ISL_2934068, EPI_ISL_2934071, EPI_ISL_2934073, EPI_ISL_2934074, EPI_ISL_2934075, EPI_ISL_2934076, EPI_ISL_2934077, EPI_ISL_2934078, EPI_ISL_2934079, EPI_ISL_2934080, EPI_ISL_2934085, EPI_ISL_2934086, EPI_ISL_2934087, EPI_ISL_2934093 | | | |
| see above | Kaiser Regional | Santa Clara County Public Health Laboratory | Santa Clara County Public Health Department |
| EPI_ISL_2650539, EPI_ISL_2650540, EPI_ISL_2650581, EPI_ISL_2650584, EPI_ISL_2691295, EPI_ISL_2753981, EPI_ISL_2753982, EPI_ISL_2934004, EPI_ISL_2934061 | | | |
| see above | Kaiser San Jose | Santa Clara County Public Health Laboratory | Santa Clara County Public Health Department |
| EPI_ISL_2631210, EPI_ISL_2650541, | Kaiser Santa Clara | Santa Clara County Public Health | Santa Clara County Public Health Department |

| | | | |
|---|---|---|---|
| EPI_ISL_2753978, EPI_ISL_2753979, EPI_ISL_2753980 | | Laboratory | |
| EPI_ISL_2296064, EPI_ISL_2296104, EPI_ISL_2455998, EPI_ISL_2455999, EPI_ISL_2456000, EPI_ISL_2476439, EPI_ISL_2476443, EPI_ISL_2479884, EPI_ISL_2479888, EPI_ISL_2545705, EPI_ISL_2545807, EPI_ISL_2709687, EPI_ISL_2736832, EPI_ISL_2835023, EPI_ISL_2858980, EPI_ISL_2920685 | | | |
| see above | LESP Baja California | Instituto de Diagnostico y Referencia Epidemiologicos (INDRE) | Abril Rodriguez-Maldonado; Ariadna Medina-Benitez; Claudia Wong-Arambula; Ernesto Ramirez-Gonzalez.; Gisela Barrera-Badillo; Irma Lopez-Martinez; Joaquin Quiroz-Mercado; Lucia Hernandez-Rivas; Maribel Gonzalez-Villa; Natividad Cruz-Ortiz; Sergio Rangel-Guerrero; Tatiana Nunez-Garcia; Vanessa Rivero-Arredondo |
| EPI_ISL_2341013, EPI_ISL_2533818, EPI_ISL_2545729, EPI_ISL_2545730, EPI_ISL_2663353, EPI_ISL_2779197, EPI_ISL_2937836, EPI_ISL_2937847, EPI_ISL_2937848 | | | |
| see above | LESP Baja California Sur | Instituto de Diagnostico y Referencia Epidemiologicos (INDRE) | Abril Rodriguez-Maldonado; Ariadna Medina-Benitez; Claudia Wong-Arambula; Ernesto Ramirez-Gonzalez.; Gisela Barrera-Badillo; Irma Lopez-Martinez; Joaquin Quiroz-Mercado; Lucia Hernandez-Rivas; Maribel Gonzalez-Villa; Natividad Cruz-Ortiz; Sergio Rangel-Guerrero; Tatiana Nunez-Garcia; Vanessa Rivero-Arredondo |
| EPI_ISL_2942560, EPI_ISL_2942561, EPI_ISL_2942579, EPI_ISL_2942598, EPI_ISL_2942599, EPI_ISL_2942600, EPI_ISL_2942601, EPI_ISL_2942602, EPI_ISL_2942603, EPI_ISL_2942610, EPI_ISL_2942611, EPI_ISL_2942612, EPI_ISL_2942613, EPI_ISL_2942614, EPI_ISL_2942615, EPI_ISL_2942616, EPI_ISL_2942617, EPI_ISL_2942618, EPI_ISL_2942619, EPI_ISL_2942620, EPI_ISL_2942621, EPI_ISL_2942622, EPI_ISL_2942623, EPI_ISL_2942624, EPI_ISL_2942799, EPI_ISL_2942800, EPI_ISL_2942801, EPI_ISL_2942802, EPI_ISL_2942803, EPI_ISL_2942809, EPI_ISL_2942810, EPI_ISL_2942811, EPI_ISL_2942852, EPI_ISL_2942853, EPI_ISL_2942854, EPI_ISL_2942855, EPI_ISL_2942856, EPI_ISL_2942857, EPI_ISL_2942858, EPI_ISL_2942859, EPI_ISL_2942860, EPI_ISL_2942877 | | | |
| see above | Laboratorio Central de Epidemiologia (LCE) | Unidad de Genomica Avanzada | ; Alejandra Garcia-Gasca; Alejandra Hernandez-Teran; Alejandro Sanchez-Flores; Alfredo Herrera-Estrella; Alicia Ocaña-Mondragon; Andreu Comas-Garcia; Angel Gustavo Salas-Lais; Antonio Loza Roman; Bernardo Martinez-Miguel; Blanca Taboada; Brenda Irasema Maldonado-Meza; Bruno Gomez-Gil; Carla Ivon Herrera-Najera; Carlos F. Arias; Celia Boukadida; Celida Duque Molina; Celida Martinez- Rodriguez; Clara Esperanza Santacruz-Tinoco; Concepcion Grajales-Muñiz; Consorcio Mexicano de Vigilancia Genomica (CoViGen-Mex). Authors (in alphabetical order): Julio Elias Alvarado-Yaah; Cristobal Chaidez-Quiroz; Daniel Fregoso-Rueda; Daniel Lira Morales; Eduardo Becerril-Vargas; Fernando Fontove-Herrera; Fidencio Mejia-Nepomuceno; Francisco Pulido; Gloria Elena Espinosa-Ayala; Gloria Maria Molina-Salinas; Gloria Vazquez; Hector Esteban Paz-Juarez; Hector Montoya-Fuentes; Helen Haydee Fernanda Ramirez-Plascencia; Irvin Gonzalez-Lopez; Jean Pierre Gonzalez; Jesus Hernandez; Joel Armando Vazquez-Perez.; Jorge Salas-Hernandez; Jose Antonio Enciso-Moreno; Jose Arturo Martinez-Orozco; Jose Esteban Muñoz-Medina; Jose de Jesus Nuñez-Contreras; Juan Bautista Chale-Dzul; Julissa Enciso-Ibarra; Luis Alberto Ochoa-Carrera; Margarita Matias-Florentino; Maria Guadalupe Santiago-Mauricio; Maria Guadalupe de Jesus Mireles-Rivera; Mario Mujica-Sanchez; Marissa Perez-Garcia; Nelly Selem-Mojica; Pavel Isa; Ricardo Ciria Merce; Ricardo Grande; Rosa Maria Gutierrez Rios; Santiago avila-Rios; Selene Zarate; Susana Lopez; Veronica Mata-Haro; Victor Eduardo Garcia-Arias; Victor Hugo Borja-Aburto |
| EPI_ISL_2671485, EPI_ISL_2671486, EPI_ISL_2671487, EPI_ISL_2671488, EPI_ISL_2671489, EPI_ISL_2671490, EPI_ISL_2671491, EPI_ISL_2671492 | | | |
| see above | Laboratorio Central de Epidemiologia (LCE) | Unidad de Genomica Avanzada | ; Alejandra Garcia-Gasca; Alejandra Hernandez-Teran; Alejandro Sanchez-Flores; Alfredo Herrera-Estrella; Alicia Ocaña-Mondragon; Andreu Comas-Garcia; Angel Gustavo Salas-Lais; Antonio Loza Roman; Bernardo Martinez-Miguel; Blanca Taboada; Brenda Irasema Maldonado-Meza; Bruno Gomez-Gil; Carla Ivon Herrera-Najera; Carlos F. Arias; Celia Boukadida; Celida Duque Molina; Celida Martinez- Rodriguez; Clara Esperanza Santacruz-Tinoco; Concepcion Grajales-Muñiz; Consorcio Mexicano de Vigilancia Genomica (CoViGen-Mex). Authors (in alphabetical order): Julio Elias Alvarado-Yaah; Cristobal Chaidez-Quiroz; Daniel Fregoso-Rueda; Daniel Lira Morales; Eduardo Becerril-Vargas; Fernando Fontove-Herrera; Fidencio Mejia-Nepomuceno; Francisco Pulido; Gloria Elena Espinosa-Ayala; Gloria Maria Molina-Salinas; Gloria Vazquez; Hector Esteban Paz-Juarez; Hector Montoya-Fuentes; Helen Haydee Fernanda Ramirez-Plascencia; Irvin Gonzalez-Lopez; Jean Pierre Gonzalez; Jesus Hernandez; Joel Armando Vazquez-Perez.; Jorge Salas-Hernandez; Jose Antonio Enciso-Moreno; Jose Arturo Martinez-Orozco; Jose Esteban Muñoz-Medina; Jose de Jesus Nuñez-Contreras; Juan Bautista Chale-Dzul; Julissa Enciso-Ibarra; Luis Alberto Ochoa-Carrera; Margarita Matias-Florentino; Maria Guadalupe Santiago-Mauricio; Maria Guadalupe de Jesus Mireles-Rivera; Mario Mujica-Sanchez; Marissa Perez-Garcia; Nelly Selem-Mojica; Pavel Isa; Ricardo Ciria Merce; Ricardo Grande; Rosa Maria Gutierrez Rios; Santiago avila-Rios; Selene Zarate; Susana Lopez; Veronica Mata-Haro; Victor Eduardo Garcia-Arias; Victor Hugo Borja-Aburto |
| EPI_ISL_2490392 | Laboratorio Central de Epidemiología (LCE) | Centro de Investigación en Enfermedades Infecciosas (CIENI), Instituto Nacional de Enfermedades Respiratorias (INER) | ; Alejandra Garcia-Gasca; Alejandra Hernández-Terán; Alejandro Sanchez-Flores; Alfredo Herrera-Estrella; Alicia Ocaña-Mondragón; Andreu Comas-García; Angel Gustavo Salas-Lais; Antonio Loza Román; Bernardo Martínez-Miguel; Blanca Taboada; Brenda Irasema Maldonado-Meza; Bruno Gomez-Gil; Carla Ivón Herrera-Najera; Carlos F. Arias; Celia Boukadida; Clara Esperanza Santacruz-Tinoco; Concepción Grajales-Muñiz; Consorcio Mexicano de Vigilancia Genómica (CoViGen-Mex). Authors (in alphabetical order): Julio Elias Alvarado-Yaah; Cristóbal Cháidez-Quiróz; Célida Duque Molina; Célida Martínez- Rodríguez; Daniel Fregoso-Rueda; Daniel Lira Morales; Eduardo Becerril-Vargas; Fernando Fontove-Herrera; Fidencio Mejía-Nepomuceno; Francisco Pulido; Gloria Elena Espinosa-Ayala; Gloria María Molina-Salinas; Gloria Vazquez; Hector Esteban Paz-Juárez; Hector Montoya-Fuentes; Helen Haydee Fernanda Ramírez-Plascencia; Irvin González-López; Jean Pierre González; Joel Armando Vázquez-Pérez.; Jorge Salas-Hernández; José Antonio Enciso-Moreno; José Arturo Martínez-Orozco; José Esteban Muñoz-Medina; José de Jesús Nuñez-Contreras; Juan Bautista Chale-Dzul; Julissa Enciso-Ibarra; Luis Alberto Ochoa-Carrera; Margarita Matías-Florentino; Mario Mújica-Sánchez; Marissa Perez-Garcia; María Guadalupe Santiago-Mauricio; María Guadalupe de Jesús Mireles-Rivera; Nelly Sélem-Mojica; Pavel Isa; Ricardo Ciria Merce; Ricardo Grande; Rosa María Gutierrez Rios; Santiago Ávila-Ríos; Selene Zárate; Susana Lopez; Victor Eduardo Garcia-Arias; Víctor Hugo Borja-Aburto |
| EPI_ISL_2391587, EPI_ISL_2681046, EPI_ISL_2681059, EPI_ISL_2681064, EPI_ISL_2681065, EPI_ISL_2681071, EPI_ISL_2681113, EPI_ISL_2681129, EPI_ISL_2681131, EPI_ISL_2681140, EPI_ISL_2681146, EPI_ISL_2681151, EPI_ISL_2681152, EPI_ISL_2681155, EPI_ISL_2681156, EPI_ISL_2681157, EPI_ISL_2681172, EPI_ISL_2681175, EPI_ISL_2681181, EPI_ISL_2681187, EPI_ISL_2681188, EPI_ISL_2681193, EPI_ISL_2681196, EPI_ISL_2681205, EPI_ISL_2681213, EPI_ISL_2681214, EPI_ISL_2681229, EPI_ISL_2681230, EPI_ISL_2681231, EPI_ISL_2681237, EPI_ISL_2681251, EPI_ISL_2681266, EPI_ISL_2681267, EPI_ISL_2681284, EPI_ISL_2681297, EPI_ISL_2681306, EPI_ISL_2681314, EPI_ISL_2681337, EPI_ISL_2681371, EPI_ISL_2681386, EPI_ISL_2801757, EPI_ISL_2801773, EPI_ISL_2801774, EPI_ISL_2801775, EPI_ISL_2801801, EPI_ISL_2801802, EPI_ISL_2801803, EPI_ISL_2801804, EPI_ISL_2801805, EPI_ISL_2801809, EPI_ISL_2801810, EPI_ISL_2801811, EPI_ISL_2801812, EPI_ISL_2801813, EPI_ISL_2801814, EPI_ISL_2801815, EPI_ISL_2801816, EPI_ISL_2801817, EPI_ISL_2801818, EPI_ISL_2801819, EPI_ISL_2801820, EPI_ISL_2801821, EPI_ISL_2801822, EPI_ISL_2801823, EPI_ISL_2801824, EPI_ISL_2801825, EPI_ISL_2801826, EPI_ISL_2801827, EPI_ISL_2801828, EPI_ISL_2801829, EPI_ISL_2801830, EPI_ISL_2801846, EPI_ISL_2801856, EPI_ISL_2801878 | | | |
| see above | Laboratorio Central de Epidemiología (LCE) | Instituto de Biotecnología de la UNAM | ; Alejandra Garcia-Gasca; Alejandra García-Gasca; Alejandra Hernández-Terán; Alejandro Sanchez-Flores; Alejandro Sánchez-Flores; Alfredo Herrera-Estrella; Alicia Ocaña-Mondragón; Andreu Comas-García; Angel Gustavo Salas-Lais; Antonio Loza Román; Bernardo Martínez-Miguel; Blanca Taboada; Brenda Irasema Maldonado-Meza; Bruno Gomez-Gil; Bruno Gómez-Gil; Carla Ivón Herrera-Najera; Carlos F. Arias; Celia Boukadida; Clara Esperanza Santacruz-Tinoco; Concepción Grajales-Muñiz; Consorcio Mexicano de Vigilancia Genómica (CoViGen-Mex). Authors (in alphabetical order): Julio Elias Alvarado-Yaah; Cristóbal Cháidez-Quiróz; Célida Duque Molina; Célida Martínez- Rodríguez; Daniel Fregoso-Rueda; Daniel Lira Morales; Eduardo Becerril-Vargas; Fernando Fontove-Herrera; Fidencio Mejía-Nepomuceno; Francisco Pulido; Gloria Elena Espinosa-Ayala; Gloria María Molina-Salinas; Gloria Vazquez; Hector Esteban Paz-Juárez; Hector Montoya-Fuentes; Helen Haydee Fernanda Ramírez-Plascencia; Irvin González-López; Jean Pierre González; Jesús Hernández; Joel Armando Vázquez-Pérez.; Jorge Salas-Hernández; José Antonio Enciso-Moreno; José Arturo Martínez-Orozco; José Esteban Muñoz-Medina; José de Jesús Nuñez-Contreras; Juan Bautista Chale-Dzul; Julissa Enciso-Ibarra; Luis Alberto Ochoa-Carrera; Margarita Matías-Florentino; Mario Mújica-Sánchez; Marissa Perez-Garcia; María Guadalupe Santiago-Mauricio; María Guadalupe de Jesús Mireles-Rivera; Nelly Sélem-Mojica; Pavel Isa; Ricardo Ciria Merce; Ricardo Grande; Rosa María Gutierrez Rios; Rosa María Gutiérrez Rios; Santiago Ávila-Ríos; Selene Zárate; Susana Lopez; Verónica Mata-Haro; Victor Eduardo Garcia-Arias; Victor Eduardo García-Arias; Víctor Hugo Borja-Aburto |
| EPI_ISL_2046611, EPI_ISL_2046612, EPI_ISL_2046613, EPI_ISL_2046680, EPI_ISL_2046716, EPI_ISL_2046717, EPI_ISL_2046772, EPI_ISL_2046773, EPI_ISL_2046774, EPI_ISL_2046775, EPI_ISL_2046776, EPI_ISL_2046777, EPI_ISL_2046778, EPI_ISL_2046790, EPI_ISL_2046791, EPI_ISL_2046792, EPI_ISL_2046793, EPI_ISL_2046820, EPI_ISL_2046821, EPI_ISL_2046822, EPI_ISL_2046823, EPI_ISL_2046824, EPI_ISL_2046825, EPI_ISL_2046826, EPI_ISL_2046827, EPI_ISL_2184941, EPI_ISL_2184942, EPI_ISL_2184943, EPI_ISL_2184944, EPI_ISL_2184945, EPI_ISL_2184946, EPI_ISL_2184996, EPI_ISL_2184997, EPI_ISL_2184998, EPI_ISL_2185011, EPI_ISL_2185012, EPI_ISL_2185013, EPI_ISL_2185014, EPI_ISL_2185015, EPI_ISL_2185016, EPI_ISL_2185017, EPI_ISL_2185027, EPI_ISL_2185028, EPI_ISL_2185029, EPI_ISL_2185030, EPI_ISL_2185031, EPI_ISL_2185032, EPI_ISL_2185033, EPI_ISL_2185034, EPI_ISL_2185035, EPI_ISL_2185170, EPI_ISL_2185175, EPI_ISL_2185176, EPI_ISL_2185177, EPI_ISL_2185203, EPI_ISL_2185204, EPI_ISL_2185215, EPI_ISL_2185232, EPI_ISL_2185233, EPI_ISL_2185242, EPI_ISL_2185243, EPI_ISL_2185244, EPI_ISL_2185245, EPI_ISL_2185246, EPI_ISL_2185247, EPI_ISL_2185248, EPI_ISL_2185249, EPI_ISL_2185250, EPI_ISL_2185251, EPI_ISL_2185252, EPI_ISL_2185274, EPI_ISL_2185275, EPI_ISL_2185276, EPI_ISL_2185277, EPI_ISL_2185278, EPI_ISL_2185279, EPI_ISL_2185280, EPI_ISL_2185281, EPI_ISL_2185282, EPI_ISL_2185283, EPI_ISL_2185284, EPI_ISL_2185285, EPI_ISL_2185286, EPI_ISL_2185418, EPI_ISL_2185419, EPI_ISL_2185446, EPI_ISL_2185447, EPI_ISL_2185448, EPI_ISL_2185499, EPI_ISL_2185511, EPI_ISL_2185512, EPI_ISL_2185528, EPI_ISL_2185529, EPI_ISL_2304865, EPI_ISL_2304866, EPI_ISL_2304867, EPI_ISL_2304978, EPI_ISL_2304979, EPI_ISL_2304980, EPI_ISL_2304981, EPI_ISL_2304982, EPI_ISL_2304983, EPI_ISL_2304984, EPI_ISL_2304985, EPI_ISL_2304986, EPI_ISL_2304987, EPI_ISL_2304988, EPI_ISL_2304989, EPI_ISL_2304990, EPI_ISL_2304991, EPI_ISL_2305018, EPI_ISL_2305019, EPI_ISL_2305088, EPI_ISL_2305089, EPI_ISL_2305090, EPI_ISL_2305091, EPI_ISL_2305092, EPI_ISL_2305093, EPI_ISL_2305103, EPI_ISL_2305104, EPI_ISL_2305105, EPI_ISL_2305106, EPI_ISL_2305107, EPI_ISL_2305108, EPI_ISL_2305109, EPI_ISL_2305110, EPI_ISL_2305111, EPI_ISL_2305112, EPI_ISL_2305113, EPI_ISL_2305114, EPI_ISL_2305115, EPI_ISL_2305116, EPI_ISL_2305117, EPI_ISL_2305118, EPI_ISL_2305119, EPI_ISL_2305120, EPI_ISL_2305139, EPI_ISL_2305140, EPI_ISL_2305141, EPI_ISL_2305142, EPI_ISL_2305143, EPI_ISL_2305144, EPI_ISL_2305145, EPI_ISL_2305146, EPI_ISL_2305147, EPI_ISL_2305148, EPI_ISL_2305202, EPI_ISL_2305204, EPI_ISL_2305205, EPI_ISL_2305284, EPI_ISL_2305285, EPI_ISL_2305286, EPI_ISL_2305287, EPI_ISL_2305315, EPI_ISL_2305316, EPI_ISL_2305317, EPI_ISL_2305318, EPI_ISL_2305319, EPI_ISL_2305320, EPI_ISL_2305321, EPI_ISL_2305322, EPI_ISL_2305323, EPI_ISL_2305364, EPI_ISL_2305365, EPI_ISL_2305366, EPI_ISL_2305367, EPI_ISL_2305368, EPI_ISL_2305369, EPI_ISL_2305372, EPI_ISL_2305373, EPI_ISL_2305375, EPI_ISL_2305376, EPI_ISL_2305383, EPI_ISL_2305384, EPI_ISL_2305405, EPI_ISL_2305406, EPI_ISL_2305443, EPI_ISL_2305444, EPI_ISL_2305445, EPI_ISL_2305494, EPI_ISL_2305495, EPI_ISL_2305497, EPI_ISL_2305498, EPI_ISL_2305499, EPI_ISL_2305539, EPI_ISL_2305540, EPI_ISL_2305541, EPI_ISL_2305617, EPI_ISL_2305618, EPI_ISL_2305619, EPI_ISL_2305620, EPI_ISL_2305622, EPI_ISL_2305623, EPI_ISL_2305624, EPI_ISL_2305625, EPI_ISL_2305626, EPI_ISL_2305628, EPI_ISL_2305656, EPI_ISL_2305657, EPI_ISL_2305658, EPI_ISL_2305659, EPI_ISL_2305660, EPI_ISL_2305661, EPI_ISL_2305662, EPI_ISL_2305663, EPI_ISL_2305664, EPI_ISL_2305665, EPI_ISL_2305666, EPI_ISL_2305950, EPI_ISL_2397822, EPI_ISL_2397823, EPI_ISL_2397824, EPI_ISL_2397825, EPI_ISL_2397879, EPI_ISL_2397881, EPI_ISL_2397882, EPI_ISL_2397883, EPI_ISL_2397884, EPI_ISL_2397885, EPI_ISL_2397886, EPI_ISL_2397887, EPI_ISL_2397888, EPI_ISL_2397890, EPI_ISL_2397891, EPI_ISL_2397892, EPI_ISL_2397926, EPI_ISL_2397927, EPI_ISL_2397928, EPI_ISL_2397972, EPI_ISL_2397973, EPI_ISL_2397995, EPI_ISL_2397996, EPI_ISL_2397997, EPI_ISL_2397998, EPI_ISL_2397999, EPI_ISL_2398000, EPI_ISL_2398001, EPI_ISL_2398002, EPI_ISL_2398003, EPI_ISL_2398004, EPI_ISL_2398005, EPI_ISL_2398026, EPI_ISL_2398027, EPI_ISL_2398028, EPI_ISL_2398029, EPI_ISL_2398039, EPI_ISL_2398040, EPI_ISL_2398041, EPI_ISL_2398042, EPI_ISL_2398043, EPI_ISL_2398044, EPI_ISL_2398045, EPI_ISL_2398046, EPI_ISL_2398067, EPI_ISL_2398068, EPI_ISL_2398069, EPI_ISL_2398070, EPI_ISL_2398071, EPI_ISL_2398072, EPI_ISL_2481478, EPI_ISL_2481550, EPI_ISL_2481590, EPI_ISL_2481591, EPI_ISL_2481592, EPI_ISL_2481593, EPI_ISL_2481594, EPI_ISL_2481595, EPI_ISL_2481646, EPI_ISL_2481647, EPI_ISL_2481648, EPI_ISL_2481649, EPI_ISL_2481650, EPI_ISL_2481651, EPI_ISL_2481652, EPI_ISL_2481654, EPI_ISL_2481655, EPI_ISL_2481677, EPI_ISL_2481678, EPI_ISL_2481736, EPI_ISL_2481756, EPI_ISL_2481758, EPI_ISL_2481759, EPI_ISL_2481760, EPI_ISL_2481761, EPI_ISL_2481762, EPI_ISL_2481763, EPI_ISL_2481764, EPI_ISL_2481775, EPI_ISL_2481776, EPI_ISL_2481777, EPI_ISL_2481778, EPI_ISL_2481779, EPI_ISL_2481780, EPI_ISL_2481781, EPI_ISL_2481782, EPI_ISL_2481783, EPI_ISL_2481804, EPI_ISL_2481812, EPI_ISL_2481813, EPI_ISL_2481814, EPI_ISL_2481815, EPI_ISL_2481816, EPI_ISL_2481817, EPI_ISL_2481818, EPI_ISL_2481993, EPI_ISL_2481994, EPI_ISL_2481995, EPI_ISL_2481996, EPI_ISL_2482017, EPI_ISL_2482018, EPI_ISL_2482019, EPI_ISL_2482048, EPI_ISL_2482066, EPI_ISL_2482067, EPI_ISL_2482068, EPI_ISL_2482069, EPI_ISL_2482070, EPI_ISL_2482092, EPI_ISL_2482093, EPI_ISL_2482094, EPI_ISL_2482095, EPI_ISL_2482096, EPI_ISL_2482097, EPI_ISL_2482098, EPI_ISL_2482099, EPI_ISL_2482164, EPI_ISL_2482165, EPI_ISL_2482166, EPI_ISL_2482167, EPI_ISL_2482168, EPI_ISL_2482169, EPI_ISL_2482170, EPI_ISL_2482171, EPI_ISL_2482172, EPI_ISL_2482173, EPI_ISL_2482174, EPI_ISL_2482175, EPI_ISL_2482176, EPI_ISL_2482177, EPI_ISL_2482178, EPI_ISL_2482190, EPI_ISL_2482191, EPI_ISL_2482192, EPI_ISL_2482207, EPI_ISL_2482231, EPI_ISL_2482232, EPI_ISL_2482233, EPI_ISL_2482235, EPI_ISL_2482236, EPI_ISL_2482237, EPI_ISL_2482262, EPI_ISL_2482263, EPI_ISL_2482264, EPI_ISL_2482265, EPI_ISL_2482266, EPI_ISL_2611298, EPI_ISL_2873146, EPI_ISL_2873147, EPI_ISL_2873148, EPI_ISL_2873149, EPI_ISL_2873150, EPI_ISL_2873155, EPI_ISL_2873156, EPI_ISL_2873157, EPI_ISL_2873158, EPI_ISL_2873159, EPI_ISL_2873160, EPI_ISL_2873161, EPI_ISL_2873219, EPI_ISL_2873220, EPI_ISL_2873224, EPI_ISL_2873225, EPI_ISL_2873226, EPI_ISL_2873227, EPI_ISL_2873229, EPI_ISL_2873235, EPI_ISL_2873254, EPI_ISL_2873268, EPI_ISL_2873269, EPI_ISL_2873270, EPI_ISL_2873271, EPI_ISL_2873272, EPI_ISL_2873273, EPI_ISL_2873282, EPI_ISL_2873283, EPI_ISL_2873303, EPI_ISL_2873304, EPI_ISL_2873305, EPI_ISL_2873306, EPI_ISL_2873307, EPI_ISL_2873314, EPI_ISL_2873315, EPI_ISL_2873316, EPI_ISL_2873317, EPI_ISL_2873322, EPI_ISL_2873323, EPI_ISL_2873324, EPI_ISL_2873325, EPI_ISL_2926342, EPI_ISL_2926684, EPI_ISL_2926685, EPI_ISL_2926686, EPI_ISL_2926687, EPI_ISL_2926688, EPI_ISL_2926689, EPI_ISL_2926690, EPI_ISL_2926697, EPI_ISL_2926702, EPI_ISL_2926731, EPI_ISL_2926732, EPI_ISL_2926733, EPI_ISL_2926734, EPI_ISL_2926735, EPI_ISL_2926736, EPI_ISL_2926737, EPI_ISL_2926738, EPI_ISL_2926739, EPI_ISL_2926740, EPI_ISL_2926741, EPI_ISL_2926742, EPI_ISL_2926743, EPI_ISL_2926744, EPI_ISL_2926753, EPI_ISL_2926754, EPI_ISL_2926755, EPI_ISL_2926756, EPI_ISL_2926764, EPI_ISL_2926765, EPI_ISL_2926766, EPI_ISL_2926767, EPI_ISL_2926768, EPI_ISL_2926773, EPI_ISL_2926782, EPI_ISL_2926820, EPI_ISL_2926821, EPI_ISL_2926822, EPI_ISL_2926823, EPI_ISL_2926824, EPI_ISL_2926825, EPI_ISL_2926826, EPI_ISL_2926827, EPI_ISL_2926828, EPI_ISL_2926829, EPI_ISL_2926830, EPI_ISL_2926840, EPI_ISL_2926845, EPI_ISL_2926846, EPI_ISL_2926847, EPI_ISL_2926848, EPI_ISL_2926849, EPI_ISL_2926850, EPI_ISL_2926851, EPI_ISL_2926852, EPI_ISL_2926853, EPI_ISL_2926854, EPI_ISL_2926855, EPI_ISL_2926856, EPI_ISL_2926857, EPI_ISL_2926858, EPI_ISL_2926895, EPI_ISL_2926896, EPI_ISL_2926897, EPI_ISL_2926898, EPI_ISL_2926899, EPI_ISL_2926900, EPI_ISL_2926901, EPI_ISL_2926902, EPI_ISL_2926905, EPI_ISL_2926913, EPI_ISL_2926914, EPI_ISL_2926915, EPI_ISL_2926916, EPI_ISL_2926917, EPI_ISL_2926918, EPI_ISL_2926919, EPI_ISL_2926920, EPI_ISL_2926921, EPI_ISL_2926922, EPI_ISL_2926927, EPI_ISL_2926928, EPI_ISL_2926929, EPI_ISL_2926931, EPI_ISL_2926932, EPI_ISL_2926958, EPI_ISL_2926972, EPI_ISL_2926973, EPI_ISL_2926987, EPI_ISL_2926988, EPI_ISL_2926991, EPI_ISL_2926992, EPI_ISL_2926993, EPI_ISL_2926994, EPI_ISL_2926997, EPI_ISL_2926998, EPI_ISL_2926999, EPI_ISL_2927018, EPI_ISL_2927019, EPI_ISL_2927034, EPI_ISL_2927055, EPI_ISL_2927056, EPI_ISL_2927057, EPI_ISL_2927058, EPI_ISL_2927059, EPI_ISL_2927109, EPI_ISL_2927125, EPI_ISL_2927132, EPI_ISL_2927133, EPI_ISL_2927158, EPI_ISL_2927159, EPI_ISL_2927160, EPI_ISL_2927161, EPI_ISL_2927162, EPI_ISL_2927163, EPI_ISL_2927164, EPI_ISL_2927170, EPI_ISL_2927171, EPI_ISL_2927172, EPI_ISL_2927173, EPI_ISL_2927174, EPI_ISL_2927175, EPI_ISL_2927176, EPI_ISL_2927178, EPI_ISL_2927179, EPI_ISL_2927193, EPI_ISL_2927194, EPI_ISL_2927195, EPI_ISL_2927196, EPI_ISL_2927197, EPI_ISL_2927198, EPI_ISL_2927199, EPI_ISL_2927215, EPI_ISL_2927216, EPI_ISL_2927217, EPI_ISL_2927218, EPI_ISL_2927219, EPI_ISL_2927255, EPI_ISL_2927258, EPI_ISL_2927259, EPI_ISL_2927260, EPI_ISL_2927261, EPI_ISL_2927262, EPI_ISL_2927263, EPI_ISL_2927264, EPI_ISL_2927265, EPI_ISL_2927266, EPI_ISL_2927267, EPI_ISL_2927285, EPI_ISL_2927293, EPI_ISL_2927294, EPI_ISL_2927296, EPI_ISL_2927297, EPI_ISL_2927298, EPI_ISL_2927299, EPI_ISL_2927300, EPI_ISL_2927301, EPI_ISL_2927302, EPI_ISL_2927303, EPI_ISL_2927360, EPI_ISL_2927361, EPI_ISL_2927362, EPI_ISL_2927363, EPI_ISL_2927364, EPI_ISL_2927365, EPI_ISL_2927366, EPI_ISL_2927367, EPI_ISL_2927368, EPI_ISL_2927369, EPI_ISL_2927376, EPI_ISL_2927377, EPI_ISL_2927378, EPI_ISL_2927379, EPI_ISL_2927386, EPI_ISL_2927433, EPI_ISL_2927434, EPI_ISL_2927435, EPI_ISL_2927436, EPI_ISL_2927437, EPI_ISL_2927438, EPI_ISL_2927439, EPI_ISL_2927440, EPI_ISL_2927441, EPI_ISL_2927442, EPI_ISL_2927443, EPI_ISL_2927444, EPI_ISL_2927445 | | | |
| see above | Laboratory Corporation of America | Centers for Disease Control and Prevention Division of Viral Diseases, Pathogen Discovery | Adrian Paskey; Amanda Douglas; Amanda Suchanek; Andrea Throop; Ayla Burns; Benjamin Rambo-Martin; Bobbi Croy; Brian Krueger; Brian Norvell; Christopher Gulvick; Christos Petropoulos; Clinton R. Paden; Craig Lukasik; Dakota Howard; Darlene Wagner; Debbie Boles; Dhwani Batra; Duncan MacCannell; Eyad Almasri; Goran Stevovic; Howard Engler; Hrushikesh Deshmukh; Jake Humphrey; Jana Schroth; Jason Caravas; Joe Voshell; John Pruitt; Jonathan Meltzer; Jonathan Williams; Kara Moser; Kimberly Wagner; Lax Iyer; Lisa Pfefferle; Lyndon Tilson; Manoj Jain; Marcia Eisenberg; Mary Ann Cristobal; Mary Williamson; Matthew Robinson; Matthew Schmerer; Michael Levandoski; Mike Sapeta; Mindy Nye; Minoo Agarwal; Mohan Kolli; Nuthawin Charoensri; Oren Cohen; Peter W. Cook; Prashant Gupta; Qian Zeng; Rama Ghatti; Scott Parker; Scott Ryan; Scott Sammons; Shatavia Morrison; Stanley Letovsky; Steven Ragan; Suresh Babu Selvaraju; Susan Countryman; Susan Hicks; Suzanne Dale; Thomas Urban; Tim Kuphal; Tricia Zwiefelhofer; Vincent Drouillon; Yvette Unoarumhi |
| EPI_ISL_2811922, EPI_ISL_2811939, EPI_ISL_2811940, EPI_ISL_2894574, EPI_ISL_2894575, EPI_ISL_2894576, EPI_ISL_2894577, EPI_ISL_2894578, EPI_ISL_2894584 | | | |
| see above | Los Angeles County PHL | Los Angeles County PHL | P. Hemarajata et al. |
| EPI_ISL_2993431 | Los Angeles County Public Health Laboratories, Los Angeles County Public Health Lab microbial pathogen submission group | Los Angeles County Public Health Laboratories, Los Angeles County Public Health Lab microbial pathogen submission group | Hemarajata; P. and Green, N. |
| EPI_ISL_2525019, EPI_ISL_2786157, EPI_ISL_2875247 | Mako Medical | Centers for Disease Control and Prevention Division of Viral Diseases, Pathogen Discovery | Adrian Paskey; Benjamin Rambo-Martin; Christopher Gulvick; Clinton R. Paden; Dakota Howard; Darlene Wagner; Dhwani Batra; Duncan MacCannell; Jason Caravas; Kara Moser; Lauren Moon; Matthew Schmerer; Matthew Tugwell; Peter W. Cook; Scott Sammons; Shatavia Morrison; Yvette Unoarumhi |
| EPI_ISL_2424212 | Martin Luther King Jr. Community Hospital | Los Angeles County PHL | P. Hemarajata et al. |
| EPI_ISL_2907577 | Martin Luther King Jr. Community Hospital | Los Angeles County Public Health Laboratories | P. Hemarajata et al. |
| EPI_ISL_2620826, EPI_ISL_2894590 | Martin Luther King, Jr. Outpatient Center | Los Angeles County PHL | P. Hemarajata et al. |
| EPI_ISL_2907562, EPI_ISL_2907563 | Martin Luther King, Jr. Outpatient Center | Los Angeles County Public Health | P. Hemarajata et al. |

| | | | |
|---|---|---|---|
| | | Laboratories | |
| EPI_ISL_2928302 | Moderna Inc. | Moderna Inc. | Groves Dixon; Rolando Pajon; Yamuna Paila |
| EPI_ISL_2716875, EPI_ISL_2759082, EPI_ISL_2759083, EPI_ISL_2759085, EPI_ISL_2759086, EPI_ISL_2759087, EPI_ISL_2759088, EPI_ISL_2774512, EPI_ISL_2790732, EPI_ISL_2790733, EPI_ISL_2955363 | | | |
| see above | Monterey County Public Health Laboratory | Monterey County Public Health Laboratory | Monterey County Public Health Laboratory |
| EPI_ISL_2663528 | NYU Langone Health | Departments of Pathology and Medicine, New York University School of Medicine | Adriana Heguy; Christian Marier; Dacia Dimartino; Emily Guzman; Gael Westby; Guiqing Wang; Paolo Cotzia; Paul Zappile; Peter Meyn; Sitharam Ramaswami; Yutong Zhang |
| EPI_ISL_2894579, EPI_ISL_2894580, EPI_ISL_2894581, EPI_ISL_2894582, EPI_ISL_2894583 | Olive View Medical Center | Los Angeles County PHL | P. Hemarajata et al. |
| EPI_ISL_2566346, EPI_ISL_2566347, EPI_ISL_2566348, EPI_ISL_2566349, EPI_ISL_2566350, EPI_ISL_2566351, EPI_ISL_2566352, EPI_ISL_2566353, EPI_ISL_2566354, EPI_ISL_2566355, EPI_ISL_2566356, EPI_ISL_2566357, EPI_ISL_2566358, EPI_ISL_2566359, EPI_ISL_2566360 | | | |
| see above | Operation Infectious Diseases, Naval Health Research Center | Operation Infectious Diseases, Naval Health Research Center | A.E.; Balansay-Ames; C.A.; Fierro, M.; Iniguez-Stevens, E.; Kriner, P.; M.S.; Moser, K.; Myers; Pan; Phippard; R.S.; R.W.; Underwood |
| EPI_ISL_2496882, EPI_ISL_2496883, EPI_ISL_2496888, EPI_ISL_2500029, EPI_ISL_2500059, EPI_ISL_2500104, EPI_ISL_2500130, EPI_ISL_2500211, EPI_ISL_2658724, EPI_ISL_2659357 | | | |
| see above | Orange County Public Health Lab | Chan-Zuckerberg Biohub | CZB Cliahub Consortium |
| EPI_ISL_2928149, EPI_ISL_2928150, EPI_ISL_2928152, EPI_ISL_2928153, EPI_ISL_2928154, EPI_ISL_2928155, EPI_ISL_2928156, EPI_ISL_2928157, EPI_ISL_2928158, EPI_ISL_2928159, EPI_ISL_2928160, EPI_ISL_2928161, EPI_ISL_2928162, EPI_ISL_2928163, EPI_ISL_2928164, EPI_ISL_2928165, EPI_ISL_2928166, EPI_ISL_2928167, EPI_ISL_2928168, EPI_ISL_2928169, EPI_ISL_2928170, EPI_ISL_2928171, EPI_ISL_2928172, EPI_ISL_2928173, EPI_ISL_2928174, EPI_ISL_2928175, EPI_ISL_2928176, EPI_ISL_2928177, EPI_ISL_2928178, EPI_ISL_2928179, EPI_ISL_2928180, EPI_ISL_2928181, EPI_ISL_2928182, EPI_ISL_2928183, EPI_ISL_2928184, EPI_ISL_2928185, EPI_ISL_2928186, EPI_ISL_2928187, EPI_ISL_2928188, EPI_ISL_2928189, EPI_ISL_2928190, EPI_ISL_2928191, EPI_ISL_2928192, EPI_ISL_2928193, EPI_ISL_2928194, EPI_ISL_2928195, EPI_ISL_2928196, EPI_ISL_2928197, EPI_ISL_2928198, EPI_ISL_2928199, EPI_ISL_2928200, EPI_ISL_2928201, EPI_ISL_2928202, EPI_ISL_2928203, EPI_ISL_2928204, EPI_ISL_2928205, EPI_ISL_2928206, EPI_ISL_2928207, EPI_ISL_2928208, EPI_ISL_2928209, EPI_ISL_2928210, EPI_ISL_2928211, EPI_ISL_2928212, EPI_ISL_2928213, EPI_ISL_2928214, EPI_ISL_2928215, EPI_ISL_2928216, EPI_ISL_2928217, EPI_ISL_2928218, EPI_ISL_2928219 | | | |
| see above | Orange County Public Health Laboratory | Orange County Public Health Laboratory | Angelica Torres; Julia Wolfe |
| EPI_ISL_2713066 | Oregon State Public Health Laboratory | Oregon State Public Health Laboratory | Eugene Yeboah; John Fontana and Shane Sevey; Laura Tsaknaridis; Rafia Razzaque; Vanda Makris |
| EPI_ISL_2692513 | Overlake Hospital | Seattle Flu Study | Amanda Adler; Barry R. Lutz; Benjamin Pelle; Brian Hiatt; Caitlin R. Wolf; Chris D. Frazar; Deborah A. Nickerson; Elisabeth Brandstetter; Erica Ryke; Geoff Melly; Helen Y. Chu; Janet A. Englund; Jay Shendure; Jover Lee; Kairsten Fay; Kirsten Lacombe; Lea M. Starita; Mark J. Rieder; Matthew Richardson; Matthew Thompson; Melissa Truong; Michael Boeckh; Michael Famulare; Misja Ilcisin; Peter D. Han; Philip Dykema; Romesh Gautom; Scott Lindquist; Thomas R. Sibley; Trevor Bedford |
| EPI_ISL_2404802, EPI_ISL_2404803, EPI_ISL_2404804, EPI_ISL_2404827 | PAMF PALO ALTO | Santa Clara County public Health Laboratory | Santa Clara County Public Health Department |
| EPI_ISL_2650544, EPI_ISL_2650545, EPI_ISL_2650562, EPI_ISL_2650585, EPI_ISL_2691296, EPI_ISL_2691297, EPI_ISL_2691298, EPI_ISL_2691301, EPI_ISL_2753983, EPI_ISL_2753984, EPI_ISL_2753985, EPI_ISL_2753986, EPI_ISL_2753987, EPI_ISL_2753988, EPI_ISL_2753989, EPI_ISL_2774622, EPI_ISL_2774623, EPI_ISL_2934038, EPI_ISL_2934048 | | | |
| see above | PAMF Palo Alto | Santa Clara County Public Health Laboratory | Santa Clara County Public Health Department |
| EPI_ISL_2620827, EPI_ISL_2620828, EPI_ISL_2620829, EPI_ISL_2772525, EPI_ISL_2811967, EPI_ISL_2811968, EPI_ISL_2811969, EPI_ISL_2811970, EPI_ISL_2811971, EPI_ISL_2811972, EPI_ISL_2811973, EPI_ISL_2811974, EPI_ISL_2811975, EPI_ISL_2811976, EPI_ISL_2811977, EPI_ISL_2811978, EPI_ISL_2811979, EPI_ISL_2812006, EPI_ISL_2812007, EPI_ISL_2812008, EPI_ISL_2812009, EPI_ISL_2812010, EPI_ISL_2812011, EPI_ISL_2812012, EPI_ISL_2812013, EPI_ISL_2812014, EPI_ISL_2894591, EPI_ISL_2894592 | | | |
| see above | Palmdale Regional Medical Center | Los Angeles County PHL | P. Hemarajata et al. |
| EPI_ISL_2391750, EPI_ISL_2391751, EPI_ISL_2391752, EPI_ISL_2392168, EPI_ISL_2404847, EPI_ISL_2405178, EPI_ISL_2405202, EPI_ISL_2405203, EPI_ISL_2405204, EPI_ISL_2617942, EPI_ISL_2772505, EPI_ISL_2811999, EPI_ISL_2812000, EPI_ISL_2894572, EPI_ISL_2894573 | | | |
| see above | Pomona Valley Hospital Medical Center | Los Angeles County PHL | P. Hemarajata et al. |
| EPI_ISL_2907567, EPI_ISL_2907568 | Providence Saint John's Health Center | Los Angeles County Public Health Laboratories | P. Hemarajata et al. |
| EPI_ISL_2391792, EPI_ISL_2405206 | Providence Saint Joseph Medical Center | Los Angeles County PHL | P. Hemarajata et al. |
| EPI_ISL_2142892, EPI_ISL_2142898, EPI_ISL_2142931, EPI_ISL_2142932, EPI_ISL_2142941, EPI_ISL_2142945, EPI_ISL_2142950, EPI_ISL_2142951, EPI_ISL_2142954, EPI_ISL_2142955, EPI_ISL_2142971, EPI_ISL_2142988, EPI_ISL_2142991, EPI_ISL_2142998, EPI_ISL_2143011, EPI_ISL_2143012, EPI_ISL_2143016, EPI_ISL_2143036, EPI_ISL_2143057, EPI_ISL_2143059, EPI_ISL_2143081, EPI_ISL_2143111, EPI_ISL_2179946, EPI_ISL_2179950, EPI_ISL_2179965, EPI_ISL_2179981, EPI_ISL_2179983, EPI_ISL_2180495, EPI_ISL_2180519, EPI_ISL_2180530, EPI_ISL_2187230, EPI_ISL_2187231, EPI_ISL_2187234, EPI_ISL_2187238, EPI_ISL_2187242, EPI_ISL_2187252, EPI_ISL_2187276, EPI_ISL_2241634, EPI_ISL_2247062, EPI_ISL_2247065, EPI_ISL_2247067, EPI_ISL_2247071, EPI_ISL_2247074, EPI_ISL_2247075, EPI_ISL_2247079, EPI_ISL_2247080, EPI_ISL_2247084, EPI_ISL_2247085, EPI_ISL_2247087, EPI_ISL_2247090, EPI_ISL_2247092, EPI_ISL_2247094, EPI_ISL_2247106, EPI_ISL_2247112, EPI_ISL_2247138, EPI_ISL_2247139, EPI_ISL_2247143, EPI_ISL_2247144, EPI_ISL_2247157, EPI_ISL_2247161, EPI_ISL_2247162, EPI_ISL_2247172, EPI_ISL_2247174, EPI_ISL_2247179, EPI_ISL_2247183, EPI_ISL_2247186, EPI_ISL_2247187, EPI_ISL_2247190, EPI_ISL_2247191, EPI_ISL_2247198, EPI_ISL_2247205, EPI_ISL_2247337, EPI_ISL_2248473, EPI_ISL_2248559, EPI_ISL_2248560, EPI_ISL_2248580, EPI_ISL_2248585, EPI_ISL_2248589, EPI_ISL_2248602, EPI_ISL_2248603, EPI_ISL_2248606, EPI_ISL_2248607, EPI_ISL_2248612, EPI_ISL_2248613, EPI_ISL_2248614, EPI_ISL_2248621, EPI_ISL_2248622, EPI_ISL_2248624, EPI_ISL_2248628, EPI_ISL_2248637, EPI_ISL_2248641, EPI_ISL_2248642, EPI_ISL_2248655, EPI_ISL_2248666, EPI_ISL_2248667, EPI_ISL_2248669, EPI_ISL_2248687, EPI_ISL_2248689, EPI_ISL_2248690, EPI_ISL_2268287, EPI_ISL_2268291, EPI_ISL_2268295, EPI_ISL_2268379, EPI_ISL_2268380, EPI_ISL_2268389, EPI_ISL_2268590, EPI_ISL_2268594, EPI_ISL_2268598, EPI_ISL_2268672, EPI_ISL_2268690, EPI_ISL_2268746, EPI_ISL_2268807, EPI_ISL_2268809, EPI_ISL_2268829, EPI_ISL_2268874, EPI_ISL_2268882, EPI_ISL_2268883, EPI_ISL_2268899, EPI_ISL_2268916, EPI_ISL_2268937, EPI_ISL_2268942, EPI_ISL_2268945, EPI_ISL_2268946, EPI_ISL_2268950, EPI_ISL_2268951, EPI_ISL_2268952, EPI_ISL_2268958, EPI_ISL_2268960, EPI_ISL_2268961, EPI_ISL_2268962, EPI_ISL_2268963, EPI_ISL_2268964, EPI_ISL_2268969, EPI_ISL_2268970, EPI_ISL_2268972, EPI_ISL_2268974, EPI_ISL_2268975, EPI_ISL_2268977, EPI_ISL_2268979, EPI_ISL_2268980, EPI_ISL_2268981, EPI_ISL_2268983, EPI_ISL_2268984, EPI_ISL_2268985, EPI_ISL_2268987, EPI_ISL_2268989, EPI_ISL_2268990, EPI_ISL_2268992, EPI_ISL_2268993, EPI_ISL_2268995, EPI_ISL_2268996, EPI_ISL_2269000, EPI_ISL_2269001, EPI_ISL_2269004, EPI_ISL_2269005, EPI_ISL_2269007, EPI_ISL_2269008, EPI_ISL_2269014, EPI_ISL_2269017, EPI_ISL_2269019, EPI_ISL_2269020, EPI_ISL_2269022, EPI_ISL_2269023, EPI_ISL_2269024, EPI_ISL_2269025, EPI_ISL_2269029, EPI_ISL_2269031, EPI_ISL_2269034, EPI_ISL_2269044, EPI_ISL_2269045, EPI_ISL_2269046, EPI_ISL_2269048, EPI_ISL_2269051, EPI_ISL_2269053, EPI_ISL_2269054, EPI_ISL_2269055, EPI_ISL_2269058, EPI_ISL_2269059, EPI_ISL_2269060, EPI_ISL_2269061, EPI_ISL_2269062, EPI_ISL_2269063, EPI_ISL_2269064, EPI_ISL_2269065, EPI_ISL_2269066, EPI_ISL_2269067, EPI_ISL_2269068, EPI_ISL_2269069, EPI_ISL_2269070, EPI_ISL_2269071, EPI_ISL_2269072, EPI_ISL_2269073, EPI_ISL_2269093, EPI_ISL_2269125, EPI_ISL_2269210, EPI_ISL_2280114, EPI_ISL_2280123, EPI_ISL_2280126, EPI_ISL_2280136, EPI_ISL_2280138, EPI_ISL_2280139, EPI_ISL_2280140, EPI_ISL_2280143, EPI_ISL_2280144, EPI_ISL_2280145, EPI_ISL_2280148, EPI_ISL_2280149, EPI_ISL_2280150, EPI_ISL_2280152, EPI_ISL_2280154, EPI_ISL_2280155, EPI_ISL_2280156, EPI_ISL_2304503, EPI_ISL_2304506, EPI_ISL_2304510, EPI_ISL_2304511, EPI_ISL_2304518, EPI_ISL_2304523, EPI_ISL_2304526, EPI_ISL_2304527, EPI_ISL_2304529, EPI_ISL_2304531, EPI_ISL_2304533, EPI_ISL_2304534, EPI_ISL_2304543, EPI_ISL_2304554, EPI_ISL_2304559, EPI_ISL_2304561, EPI_ISL_2304563, EPI_ISL_2304568, EPI_ISL_2304571, EPI_ISL_2304574, EPI_ISL_2304576, EPI_ISL_2304579, EPI_ISL_2304580, EPI_ISL_2304588, EPI_ISL_2304591, EPI_ISL_2304593, EPI_ISL_2304600, EPI_ISL_2304612, EPI_ISL_2304614, EPI_ISL_2304618, EPI_ISL_2304619, EPI_ISL_2304623, EPI_ISL_2304629, EPI_ISL_2304633, EPI_ISL_2304634, EPI_ISL_2304635, EPI_ISL_2304639, EPI_ISL_2307140, EPI_ISL_2307141, EPI_ISL_2307142, EPI_ISL_2307145, EPI_ISL_2307146, EPI_ISL_2307151, EPI_ISL_2367680, EPI_ISL_2367688, EPI_ISL_2367689, EPI_ISL_2367694, EPI_ISL_2367695, EPI_ISL_2367697, EPI_ISL_2367702, EPI_ISL_2367711, EPI_ISL_2367714, EPI_ISL_2367718, EPI_ISL_2367722, EPI_ISL_2367726, EPI_ISL_2367728, EPI_ISL_2367732, EPI_ISL_2367736, EPI_ISL_2367738, EPI_ISL_2367741, EPI_ISL_2367745, EPI_ISL_2367748, EPI_ISL_2367750, EPI_ISL_2367751, EPI_ISL_2367806, EPI_ISL_2367807, EPI_ISL_2367811, EPI_ISL_2367825, EPI_ISL_2367852, EPI_ISL_2367881, EPI_ISL_2367882, EPI_ISL_2367894, EPI_ISL_2367907, EPI_ISL_2367909, EPI_ISL_2367917, EPI_ISL_2367938, EPI_ISL_2367943, EPI_ISL_2367948, EPI_ISL_2367949, EPI_ISL_2367956, EPI_ISL_2367958, EPI_ISL_2367961, EPI_ISL_2367971, EPI_ISL_2367974, EPI_ISL_2367976, EPI_ISL_2367978, EPI_ISL_2367980, EPI_ISL_2367983, EPI_ISL_2367984, EPI_ISL_2367985, EPI_ISL_2367987, EPI_ISL_2367996, EPI_ISL_2367999, EPI_ISL_2368001, EPI_ISL_2368002, EPI_ISL_2368003, EPI_ISL_2368006, EPI_ISL_2368010, EPI_ISL_2368012, EPI_ISL_2368016, EPI_ISL_2368021, EPI_ISL_2368023, EPI_ISL_2368024, EPI_ISL_2368025, EPI_ISL_2368026, EPI_ISL_2368031, EPI_ISL_2368033, EPI_ISL_2368043, EPI_ISL_2368045, EPI_ISL_2368046, EPI_ISL_2368048, EPI_ISL_2368049, EPI_ISL_2368053, EPI_ISL_2368055, EPI_ISL_2368056, EPI_ISL_2368059, EPI_ISL_2368062, EPI_ISL_2368065, EPI_ISL_2369109, EPI_ISL_2369127, EPI_ISL_2369192, EPI_ISL_2369396, EPI_ISL_2371691, EPI_ISL_2371696, EPI_ISL_2371697, EPI_ISL_2371699, EPI_ISL_2371703, EPI_ISL_2371721, EPI_ISL_2371726, EPI_ISL_2371728, EPI_ISL_2371729, EPI_ISL_2371731, EPI_ISL_2371733, EPI_ISL_2371736, EPI_ISL_2371748, EPI_ISL_2371774, EPI_ISL_2371869, EPI_ISL_2371872, EPI_ISL_2371873, EPI_ISL_2371875, EPI_ISL_2371876, EPI_ISL_2371877, EPI_ISL_2371878, EPI_ISL_2371886, EPI_ISL_2371892, EPI_ISL_2371909, EPI_ISL_2371910, EPI_ISL_2371923, EPI_ISL_2371927, EPI_ISL_2371934, EPI_ISL_2371938, EPI_ISL_2371942, EPI_ISL_2371946, EPI_ISL_2371948, EPI_ISL_2371950, EPI_ISL_2371955, EPI_ISL_2371965, EPI_ISL_2371967, EPI_ISL_2371971, EPI_ISL_2371973, EPI_ISL_2371978, EPI_ISL_2372140, EPI_ISL_2372141, EPI_ISL_2372142, EPI_ISL_2372147, EPI_ISL_2372149, EPI_ISL_2372158, EPI_ISL_2372159, EPI_ISL_2372162, EPI_ISL_2372165, EPI_ISL_2372167, EPI_ISL_2372172, EPI_ISL_2372175, EPI_ISL_2372183, EPI_ISL_2372187, EPI_ISL_2372190, EPI_ISL_2372194, EPI_ISL_2372206, EPI_ISL_2372207, EPI_ISL_2372211, EPI_ISL_2372219, EPI_ISL_2372220, EPI_ISL_2372224, EPI_ISL_2372227, EPI_ISL_2372228, EPI_ISL_2372229, EPI_ISL_2372234, EPI_ISL_2372240, EPI_ISL_2372248, EPI_ISL_2372250, EPI_ISL_2372251, EPI_ISL_2372253, EPI_ISL_2372255, EPI_ISL_2372258, EPI_ISL_2372922, EPI_ISL_2372927, EPI_ISL_2372944, EPI_ISL_2372951, EPI_ISL_2397413, EPI_ISL_2397414, EPI_ISL_2397420, EPI_ISL_2397421, EPI_ISL_2397422, EPI_ISL_2397424, EPI_ISL_2397425, EPI_ISL_2397427, EPI_ISL_2397432, EPI_ISL_2397434, EPI_ISL_2397441, EPI_ISL_2397442, EPI_ISL_2397443, EPI_ISL_2397445, EPI_ISL_2397447, EPI_ISL_2397449, EPI_ISL_2397454, EPI_ISL_2397461, EPI_ISL_2397462, EPI_ISL_2397469, EPI_ISL_2397473, EPI_ISL_2397481, EPI_ISL_2397483, EPI_ISL_2397488, EPI_ISL_2397492, EPI_ISL_2397497, EPI_ISL_2397502, EPI_ISL_2397505, EPI_ISL_2397510, EPI_ISL_2397514, EPI_ISL_2397520, EPI_ISL_2397523, EPI_ISL_2397524, EPI_ISL_2397526, EPI_ISL_2397529, EPI_ISL_2397532, EPI_ISL_2397537, EPI_ISL_2397541, EPI_ISL_2397548, EPI_ISL_2397550, EPI_ISL_2397638, EPI_ISL_2399476, EPI_ISL_2399478, EPI_ISL_2399480, EPI_ISL_2399484, EPI_ISL_2439320, EPI_ISL_2439365, EPI_ISL_2439384, EPI_ISL_2439409, EPI_ISL_2439415, EPI_ISL_2439425, EPI_ISL_2439428, EPI_ISL_2439430, EPI_ISL_2439431, EPI_ISL_2439440, EPI_ISL_2439442, EPI_ISL_2439443, EPI_ISL_2439444, EPI_ISL_2439451, EPI_ISL_2439452, EPI_ISL_2439454, EPI_ISL_2439462, EPI_ISL_2439464, EPI_ISL_2439465, EPI_ISL_2439466, EPI_ISL_2439467, EPI_ISL_2439468, EPI_ISL_2439469, EPI_ISL_2439471, EPI_ISL_2439474, EPI_ISL_2439475, EPI_ISL_2439481, EPI_ISL_2439486, EPI_ISL_2439488, EPI_ISL_2439490, EPI_ISL_2439495, EPI_ISL_2439497, EPI_ISL_2439501, EPI_ISL_2439502, EPI_ISL_2439503, EPI_ISL_2439514, EPI_ISL_2439516, EPI_ISL_2439521, EPI_ISL_2439522, EPI_ISL_2439523, EPI_ISL_2439524, EPI_ISL_2439525, EPI_ISL_2439528, EPI_ISL_2439529, EPI_ISL_2439531, EPI_ISL_2439534, EPI_ISL_2439539, EPI_ISL_2439541, EPI_ISL_2439542, EPI_ISL_2439544, EPI_ISL_2439545, EPI_ISL_2439549, EPI_ISL_2439551, EPI_ISL_2439552, EPI_ISL_2439556, EPI_ISL_2439559, EPI_ISL_2439560, EPI_ISL_2439564, EPI_ISL_2439700, EPI_ISL_2439712, EPI_ISL_2439719, EPI_ISL_2439732, EPI_ISL_2439743, EPI_ISL_2439744, EPI_ISL_2439746, EPI_ISL_2439755, EPI_ISL_2439757, EPI_ISL_2439762, EPI_ISL_2439770, EPI_ISL_2439773, EPI_ISL_2439777, EPI_ISL_2439778, EPI_ISL_2439787, EPI_ISL_2439801, EPI_ISL_2439813, EPI_ISL_2439814, EPI_ISL_2439818, EPI_ISL_2439827, EPI_ISL_2440687, EPI_ISL_2440777, EPI_ISL_2440781, EPI_ISL_2440794, EPI_ISL_2440815, EPI_ISL_2440820, EPI_ISL_2440862, EPI_ISL_2440876, EPI_ISL_2451278, EPI_ISL_2451279, EPI_ISL_2451282, EPI_ISL_2451292, EPI_ISL_2451294, EPI_ISL_2451360, EPI_ISL_2451386, EPI_ISL_2451423, EPI_ISL_2451428, EPI_ISL_2451429, EPI_ISL_2451434, EPI_ISL_2451436, EPI_ISL_2451439, EPI_ISL_2463023, EPI_ISL_2463037, EPI_ISL_2463049, EPI_ISL_2463055, EPI_ISL_2463097, EPI_ISL_2463102, EPI_ISL_2463105, EPI_ISL_2463125, EPI_ISL_2463126, EPI_ISL_2463135, EPI_ISL_2463147, EPI_ISL_2463149, EPI_ISL_2463152, EPI_ISL_2463154, EPI_ISL_2463157, EPI_ISL_2463160, EPI_ISL_2463161, EPI_ISL_2463168, EPI_ISL_2463170, EPI_ISL_2463171, EPI_ISL_2463172, EPI_ISL_2463174, EPI_ISL_2463175, EPI_ISL_2463176, EPI_ISL_2463180, EPI_ISL_2463181, EPI_ISL_2463184, EPI_ISL_2463186, EPI_ISL_2463187, EPI_ISL_2463188, EPI_ISL_2463190, EPI_ISL_2463192, EPI_ISL_2463197, EPI_ISL_2463199, EPI_ISL_2463200, EPI_ISL_2463201, EPI_ISL_2463205, EPI_ISL_2463209, EPI_ISL_2463211, EPI_ISL_2463213, EPI_ISL_2463214, EPI_ISL_2463215, EPI_ISL_2463220, EPI_ISL_2463221, EPI_ISL_2463225, EPI_ISL_2463226, EPI_ISL_2463227, EPI_ISL_2463229, EPI_ISL_2463230, EPI_ISL_2463232, EPI_ISL_2463235, EPI_ISL_2463237, EPI_ISL_2463239, EPI_ISL_2463241, EPI_ISL_2463243, EPI_ISL_2463244, EPI_ISL_2463246, EPI_ISL_2463248, EPI_ISL_2463251, EPI_ISL_2463253, EPI_ISL_2463254, EPI_ISL_2463256, EPI_ISL_2463258, EPI_ISL_2463262, EPI_ISL_2463265, EPI_ISL_2463266, EPI_ISL_2463268, EPI_ISL_2463269, EPI_ISL_2463270, EPI_ISL_2463281, EPI_ISL_2488824, EPI_ISL_2488833, EPI_ISL_2488835, EPI_ISL_2488845, EPI_ISL_2488846, EPI_ISL_2488847, EPI_ISL_2488849, EPI_ISL_2488853, EPI_ISL_2488854, EPI_ISL_2488855, EPI_ISL_2526803, EPI_ISL_2526859, EPI_ISL_2526931, EPI_ISL_2527016, EPI_ISL_2529091, EPI_ISL_2529127, EPI_ISL_2529155, EPI_ISL_2529159, EPI_ISL_2529183, EPI_ISL_2529295, EPI_ISL_2529299, EPI_ISL_2529312, EPI_ISL_2529314, EPI_ISL_2529331, EPI_ISL_2529333, EPI_ISL_2534529, EPI_ISL_2534554, EPI_ISL_2534564, EPI_ISL_2599782, EPI_ISL_2599858, EPI_ISL_2599886, EPI_ISL_2611825, EPI_ISL_2611835, EPI_ISL_2611836, EPI_ISL_2611839, EPI_ISL_2611853, EPI_ISL_2649976, EPI_ISL_2649978, EPI_ISL_2649985, EPI_ISL_2649990, EPI_ISL_2652368, EPI_ISL_2652376, EPI_ISL_2652377, EPI_ISL_2652382, EPI_ISL_2652401, EPI_ISL_2686378, EPI_ISL_2868689, EPI_ISL_2868691, EPI_ISL_2868695, EPI_ISL_2868702, EPI_ISL_2868709, EPI_ISL_2868716, EPI_ISL_2868717, EPI_ISL_2868722, EPI_ISL_2868730, EPI_ISL_2868731, EPI_ISL_2868734, EPI_ISL_2868737, EPI_ISL_2868740, EPI_ISL_2868742, EPI_ISL_2868743, EPI_ISL_2868748, EPI_ISL_2868752, EPI_ISL_2868756, EPI_ISL_2868761, EPI_ISL_2868763, EPI_ISL_2868767, EPI_ISL_2868769, EPI_ISL_2868772, EPI_ISL_2868773, EPI_ISL_2868775, EPI_ISL_2868776, EPI_ISL_2868778, EPI_ISL_2868780, EPI_ISL_2868782, EPI_ISL_2868787, EPI_ISL_2868789, EPI_ISL_2868795, EPI_ISL_2868797, EPI_ISL_2868802, EPI_ISL_2868804, EPI_ISL_2868806, EPI_ISL_2868810, EPI_ISL_2868812, EPI_ISL_2868814, EPI_ISL_2868823, EPI_ISL_2868824, EPI_ISL_2868826, EPI_ISL_2868830, EPI_ISL_2868832, EPI_ISL_2868834, EPI_ISL_2868836, EPI_ISL_2868839, EPI_ISL_2868841, EPI_ISL_2868842, EPI_ISL_2868846, EPI_ISL_2868858, EPI_ISL_2868863, EPI_ISL_2868870, EPI_ISL_2868871, EPI_ISL_2868872, EPI_ISL_2868885, EPI_ISL_2868898, EPI_ISL_2868903, EPI_ISL_2868908, EPI_ISL_2868924, EPI_ISL_2868948, EPI_ISL_2868951, EPI_ISL_2868952, EPI_ISL_2868953, EPI_ISL_2868960, EPI_ISL_2868965, EPI_ISL_2868970, EPI_ISL_2868972, EPI_ISL_2868976, EPI_ISL_2868983, EPI_ISL_2868985, EPI_ISL_2868988, EPI_ISL_2868994, EPI_ISL_2868999, EPI_ISL_2869004, EPI_ISL_2869005, EPI_ISL_2869011, EPI_ISL_2869012, EPI_ISL_2869013, EPI_ISL_2869017, EPI_ISL_2869025, EPI_ISL_2869033, EPI_ISL_2869039, EPI_ISL_2869042, EPI_ISL_2869044, EPI_ISL_2869048, EPI_ISL_2869058, EPI_ISL_2869061, EPI_ISL_2869070, EPI_ISL_2869074, EPI_ISL_2869081, EPI_ISL_2869085, EPI_ISL_2869086, EPI_ISL_2869091, EPI_ISL_2869093, EPI_ISL_2869094, EPI_ISL_2869095, EPI_ISL_2869100, EPI_ISL_2869103, EPI_ISL_2869105, EPI_ISL_2869106, EPI_ISL_2869110, EPI_ISL_2869111, EPI_ISL_2869114, EPI_ISL_2869136, EPI_ISL_2869139, EPI_ISL_2869142, EPI_ISL_2869145, EPI_ISL_2869154, EPI_ISL_2869163, EPI_ISL_2869164, EPI_ISL_2869177, EPI_ISL_2869178, EPI_ISL_2869182, EPI_ISL_2869183, EPI_ISL_2869192, EPI_ISL_2869194, EPI_ISL_2869195, EPI_ISL_2869207, EPI_ISL_2869210, EPI_ISL_2869212, EPI_ISL_2869221, EPI_ISL_2869231, EPI_ISL_2869243, EPI_ISL_2869245, EPI_ISL_2869250, EPI_ISL_2869258, EPI_ISL_2869265, EPI_ISL_2869276, EPI_ISL_2869280, EPI_ISL_2869284, EPI_ISL_2869289, EPI_ISL_2869313, EPI_ISL_2869322, EPI_ISL_2869331, EPI_ISL_2869337, EPI_ISL_2869357, EPI_ISL_2869358, EPI_ISL_2869368, EPI_ISL_2869369, EPI_ISL_2869371, EPI_ISL_2869374, EPI_ISL_2869383, EPI_ISL_2869386, EPI_ISL_2869392, EPI_ISL_2869397, EPI_ISL_2869398, EPI_ISL_2869405, EPI_ISL_2869411, EPI_ISL_2869417, EPI_ISL_2869419, EPI_ISL_2869424, EPI_ISL_2869425, EPI_ISL_2869428, EPI_ISL_2869437, EPI_ISL_2869438, EPI_ISL_2869449, EPI_ISL_2869451, EPI_ISL_2869454, EPI_ISL_2869465, EPI_ISL_2869476, EPI_ISL_2869487, EPI_ISL_2869495, EPI_ISL_2869496, EPI_ISL_2869502, EPI_ISL_2869503, EPI_ISL_2869509, EPI_ISL_2869519, EPI_ISL_2869528, EPI_ISL_2869529, EPI_ISL_2869536, EPI_ISL_2869540, EPI_ISL_2869569, EPI_ISL_2869573, EPI_ISL_2869578, EPI_ISL_2869585, EPI_ISL_2869587, EPI_ISL_2869589, EPI_ISL_2869594, EPI_ISL_2869600, EPI_ISL_2869602, EPI_ISL_2869608, EPI_ISL_2869638, EPI_ISL_2869646, EPI_ISL_2869652, EPI_ISL_2869654, EPI_ISL_2869655, EPI_ISL_2869659, EPI_ISL_2869660, EPI_ISL_2869661, EPI_ISL_2869662, EPI_ISL_2869665, EPI_ISL_2869670, EPI_ISL_2869675, EPI_ISL_2869676, EPI_ISL_2869679, EPI_ISL_2869680, EPI_ISL_2869681, EPI_ISL_2869687, EPI_ISL_2869688, EPI_ISL_2869696, EPI_ISL_2869700, EPI_ISL_2869705, EPI_ISL_2869707, EPI_ISL_2869712, EPI_ISL_2869714, EPI_ISL_2869716, EPI_ISL_2869719, EPI_ISL_2869725, EPI_ISL_2869727, EPI_ISL_2869729, EPI_ISL_2869732, EPI_ISL_2869733, EPI_ISL_2869739, EPI_ISL_2869748, EPI_ISL_2869760, EPI_ISL_2869763, EPI_ISL_2869773, EPI_ISL_2869789, EPI_ISL_2869792, EPI_ISL_2869795, EPI_ISL_2869810, EPI_ISL_2869817, EPI_ISL_2869833, EPI_ISL_2869839, EPI_ISL_2869842, EPI_ISL_2869854, EPI_ISL_2869857, EPI_ISL_2869863, EPI_ISL_2869866, EPI_ISL_2869881, EPI_ISL_2869892, EPI_ISL_2869898, EPI_ISL_2869905, EPI_ISL_2869916, EPI_ISL_2869958, EPI_ISL_2869967, EPI_ISL_2869979, EPI_ISL_2869982, EPI_ISL_2869987, EPI_ISL_2869993, EPI_ISL_2870016, EPI_ISL_2870027, EPI_ISL_2870044, EPI_ISL_2870052, EPI_ISL_2870070, EPI_ISL_2870072, EPI_ISL_2870086, EPI_ISL_2870089, EPI_ISL_2870092, EPI_ISL_2870098, EPI_ISL_2870106, EPI_ISL_2870120, EPI_ISL_2870127, EPI_ISL_2870138, EPI_ISL_2870164, EPI_ISL_2870173, EPI_ISL_2870176, EPI_ISL_2870199, EPI_ISL_2870205, EPI_ISL_2870208, EPI_ISL_2870217, EPI_ISL_2870240, EPI_ISL_2870246, EPI_ISL_2870254, EPI_ISL_2870263, EPI_ISL_2870266, EPI_ISL_2870269, EPI_ISL_2870274, EPI_ISL_2870277, EPI_ISL_2870286, EPI_ISL_2870295, EPI_ISL_2870307, EPI_ISL_2870711, EPI_ISL_2870856, EPI_ISL_2870945, EPI_ISL_2870969, EPI_ISL_2871039, EPI_ISL_2871148, EPI_ISL_2871169, EPI_ISL_2871231, EPI_ISL_2871243, EPI_ISL_2871254, EPI_ISL_2871296, EPI_ISL_2871329, EPI_ISL_2871433, EPI_ISL_2871448, EPI_ISL_2871521, EPI_ISL_2871535, EPI_ISL_2871661, EPI_ISL_2871684, EPI_ISL_2871693, EPI_ISL_2871710, EPI_ISL_2871890, EPI_ISL_2872001, EPI_ISL_2872115, EPI_ISL_2873888, EPI_ISL_2873901, EPI_ISL_2873903, EPI_ISL_2873911, EPI_ISL_2873987, EPI_ISL_2874006, EPI_ISL_2874034, EPI_ISL_2874069, EPI_ISL_2874076, EPI_ISL_2874096, EPI_ISL_2874112, EPI_ISL_2874117, EPI_ISL_2874147, EPI_ISL_2874154, EPI_ISL_2874162, EPI_ISL_2874163, EPI_ISL_2874165, EPI_ISL_2874219, EPI_ISL_2874221, EPI_ISL_2874238, EPI_ISL_2874258, EPI_ISL_2874273, EPI_ISL_2874281, EPI_ISL_2874304, EPI_ISL_2874313, EPI_ISL_2874317, EPI_ISL_2874327, EPI_ISL_2874329, EPI_ISL_2874349, EPI_ISL_2874350, EPI_ISL_2874373, EPI_ISL_2874377, EPI_ISL_2874384, EPI_ISL_2874395, EPI_ISL_2874401, EPI_ISL_2874404, EPI_ISL_2874408, EPI_ISL_2874411, EPI_ISL_2874418, EPI_ISL_2874437, EPI_ISL_2874445, EPI_ISL_2874450, EPI_ISL_2874455, EPI_ISL_2874463, EPI_ISL_2874466, EPI_ISL_2874469, | | | |

| | | | |
|---|---|---|---|
| EPI_ISL_2874484, EPI_ISL_2874500, EPI_ISL_2874501, EPI_ISL_2874514, EPI_ISL_2874515, EPI_ISL_2874518, EPI_ISL_2874528, EPI_ISL_2874529, EPI_ISL_2874531, EPI_ISL_2874533, EPI_ISL_2874538, EPI_ISL_2874549, EPI_ISL_2874554, EPI_ISL_2874562, EPI_ISL_2874564, EPI_ISL_2874571, EPI_ISL_2874572, EPI_ISL_2874573, EPI_ISL_2874598, EPI_ISL_2874599, EPI_ISL_2874607, EPI_ISL_2874611, EPI_ISL_2874622, EPI_ISL_2874632, EPI_ISL_2874636, EPI_ISL_2874641, EPI_ISL_2874648, EPI_ISL_2874649, EPI_ISL_2874678, EPI_ISL_2874685, EPI_ISL_2874687, EPI_ISL_2874690, EPI_ISL_2874691, EPI_ISL_2874693, EPI_ISL_2874695, EPI_ISL_2874703, EPI_ISL_2874707, EPI_ISL_2874708, EPI_ISL_2874711, EPI_ISL_2874718, EPI_ISL_2874743, EPI_ISL_2874756, EPI_ISL_2874758, EPI_ISL_2874760, EPI_ISL_2874767, EPI_ISL_2874768, EPI_ISL_2874769, EPI_ISL_2874776, EPI_ISL_2874779, EPI_ISL_2874784, EPI_ISL_2874786, EPI_ISL_2874787, EPI_ISL_2874792, EPI_ISL_2874796, EPI_ISL_2874802, EPI_ISL_2874806, EPI_ISL_2874809, EPI_ISL_2874810, EPI_ISL_2874811, EPI_ISL_2874813, EPI_ISL_2874814, EPI_ISL_2874822, EPI_ISL_2874826, EPI_ISL_2874827, EPI_ISL_2874831, EPI_ISL_2874832, EPI_ISL_2874842, EPI_ISL_2874843, EPI_ISL_2874847, EPI_ISL_2874868, EPI_ISL_2874881, EPI_ISL_2874888, EPI_ISL_2874889, EPI_ISL_2874892, EPI_ISL_2874895, EPI_ISL_2874899, EPI_ISL_2874904, EPI_ISL_2874908, EPI_ISL_2874909, EPI_ISL_2874918, EPI_ISL_2874919, EPI_ISL_2874925, EPI_ISL_2874927, EPI_ISL_2874929, EPI_ISL_2874930, EPI_ISL_2874939, EPI_ISL_2874941, EPI_ISL_2874943, EPI_ISL_2874946, EPI_ISL_2874948, EPI_ISL_2874956, EPI_ISL_2874957, EPI_ISL_2874962, EPI_ISL_2874964, EPI_ISL_2874972, EPI_ISL_2874973, EPI_ISL_2874974, EPI_ISL_2874975, EPI_ISL_2874982, EPI_ISL_2874983, EPI_ISL_2875000, EPI_ISL_2875006, EPI_ISL_2875012, EPI_ISL_2875013, EPI_ISL_2875021, EPI_ISL_2875027, EPI_ISL_2875028, EPI_ISL_2875034, EPI_ISL_2875036, EPI_ISL_2875040, EPI_ISL_2875043, EPI_ISL_2875053, EPI_ISL_2875057, EPI_ISL_2875059, EPI_ISL_2875062, EPI_ISL_2875065, EPI_ISL_2875072, EPI_ISL_2875078, EPI_ISL_2875082, EPI_ISL_2875084, EPI_ISL_2875088, EPI_ISL_2875091, EPI_ISL_2875095, EPI_ISL_2875103, EPI_ISL_2875104, EPI_ISL_2875105, EPI_ISL_2875106, EPI_ISL_2875119, EPI_ISL_2875123, EPI_ISL_2875124, EPI_ISL_2875129, EPI_ISL_2875130, EPI_ISL_2875135, EPI_ISL_2875144, EPI_ISL_2875151, EPI_ISL_2875152, EPI_ISL_2875155, EPI_ISL_2875165, EPI_ISL_2876105, EPI_ISL_2876117, EPI_ISL_2876124, EPI_ISL_2876126, EPI_ISL_2876136, EPI_ISL_2876138, EPI_ISL_2876142, EPI_ISL_2876146, EPI_ISL_2876149, EPI_ISL_2876150, EPI_ISL_2876152, EPI_ISL_2876158, EPI_ISL_2876165, EPI_ISL_2876167, EPI_ISL_2876643, EPI_ISL_2876667, EPI_ISL_2876674, EPI_ISL_2876678, EPI_ISL_2876680, EPI_ISL_2876685, EPI_ISL_2876686, EPI_ISL_2876690, EPI_ISL_2876691, EPI_ISL_2876692, EPI_ISL_2876699, EPI_ISL_2876703, EPI_ISL_2876716, EPI_ISL_2876728, EPI_ISL_2876736, EPI_ISL_2876737, EPI_ISL_2876742, EPI_ISL_2876746, EPI_ISL_2876748, EPI_ISL_2876754, EPI_ISL_2876759, EPI_ISL_2876763, EPI_ISL_2876766, EPI_ISL_2876767, EPI_ISL_2876777, EPI_ISL_2876785, EPI_ISL_2876787, EPI_ISL_2876794, EPI_ISL_2876796, EPI_ISL_2876800, EPI_ISL_2876802, EPI_ISL_2876809, EPI_ISL_2876812, EPI_ISL_2876820, EPI_ISL_2876821, EPI_ISL_2876836, EPI_ISL_2876840, EPI_ISL_2876850, EPI_ISL_2876854, EPI_ISL_2876863, EPI_ISL_2876869, EPI_ISL_2876871, EPI_ISL_2876873, EPI_ISL_2876897, EPI_ISL_2876900, EPI_ISL_2876903, EPI_ISL_2876911, EPI_ISL_2876913, EPI_ISL_2876924, EPI_ISL_2876926, EPI_ISL_2876928, EPI_ISL_2876931, EPI_ISL_2929168, EPI_ISL_2929172, EPI_ISL_2929173, EPI_ISL_2929178, EPI_ISL_2929180, EPI_ISL_2929181, EPI_ISL_2929186, EPI_ISL_2929187, EPI_ISL_2929189, EPI_ISL_2929192, EPI_ISL_2929212, EPI_ISL_2929217, EPI_ISL_2929222, EPI_ISL_2929223, EPI_ISL_2929232, EPI_ISL_2929239, EPI_ISL_2929242, EPI_ISL_2929243, EPI_ISL_2929246, EPI_ISL_2929247, EPI_ISL_2929248, EPI_ISL_2929256, EPI_ISL_2929268, EPI_ISL_2929269, EPI_ISL_2929270, EPI_ISL_2929272, EPI_ISL_2929273, EPI_ISL_2929274, EPI_ISL_2929284, EPI_ISL_2929289, EPI_ISL_2929292, EPI_ISL_2929293, EPI_ISL_2929297, EPI_ISL_2929308, EPI_ISL_2929310, EPI_ISL_2929311, EPI_ISL_2929314, EPI_ISL_2929317, EPI_ISL_2929319, EPI_ISL_2929323, EPI_ISL_2929324, EPI_ISL_2929325, EPI_ISL_2929327, EPI_ISL_2929331, EPI_ISL_2929332, EPI_ISL_2929334, EPI_ISL_2929340, EPI_ISL_2929343, EPI_ISL_2929355, EPI_ISL_2929362, EPI_ISL_2929369, EPI_ISL_2929377, EPI_ISL_2929378, EPI_ISL_2929381, EPI_ISL_2929382, EPI_ISL_2929385, EPI_ISL_2929388, EPI_ISL_2929394, EPI_ISL_2929395, EPI_ISL_2929396, EPI_ISL_2929402, EPI_ISL_2929414, EPI_ISL_2929415, EPI_ISL_2929417, EPI_ISL_2929419, EPI_ISL_2929422, EPI_ISL_2929423, EPI_ISL_2929425, EPI_ISL_2929433, EPI_ISL_2929435, EPI_ISL_2929440, EPI_ISL_2929442, EPI_ISL_2929445, EPI_ISL_2929448, EPI_ISL_2929456, EPI_ISL_2929461, EPI_ISL_2929463, EPI_ISL_2929468, EPI_ISL_2929472, EPI_ISL_2929478, EPI_ISL_2929486, EPI_ISL_2929494, EPI_ISL_2929499, EPI_ISL_2929502, EPI_ISL_2929503, EPI_ISL_2929507, EPI_ISL_2929514, EPI_ISL_2929519, EPI_ISL_2929520, EPI_ISL_2929521, EPI_ISL_2929531, EPI_ISL_2929532, EPI_ISL_2929538, EPI_ISL_2929539, EPI_ISL_2929540, EPI_ISL_2929550, EPI_ISL_2929554, EPI_ISL_2929555, EPI_ISL_2929557, EPI_ISL_2929559, EPI_ISL_2929564, EPI_ISL_2929565, EPI_ISL_2929566, EPI_ISL_2929570, EPI_ISL_2929572, EPI_ISL_2929574, EPI_ISL_2929577, EPI_ISL_2929578, EPI_ISL_2929579, EPI_ISL_2929580, EPI_ISL_2929585, EPI_ISL_2929586, EPI_ISL_2929589, EPI_ISL_2929591, EPI_ISL_2929593, EPI_ISL_2929604, EPI_ISL_2929605, EPI_ISL_2929609, EPI_ISL_2929611, EPI_ISL_2929615, EPI_ISL_2929620, EPI_ISL_2929622, EPI_ISL_2929623, EPI_ISL_2929630, EPI_ISL_2929631, EPI_ISL_2929657, EPI_ISL_2929686, EPI_ISL_2929702, EPI_ISL_2929716, EPI_ISL_2929723, EPI_ISL_2929735, EPI_ISL_2929754, EPI_ISL_2929784, EPI_ISL_2929800, EPI_ISL_2929804, EPI_ISL_2929806, EPI_ISL_2929813, EPI_ISL_2929814, EPI_ISL_2929873, EPI_ISL_2929890, EPI_ISL_2929898, EPI_ISL_2929902, EPI_ISL_2929903, EPI_ISL_2929904, EPI_ISL_2929928, EPI_ISL_2929953, EPI_ISL_2929976, EPI_ISL_2929981, EPI_ISL_2929982, EPI_ISL_2930000, EPI_ISL_2930002, EPI_ISL_2930006, EPI_ISL_2930016, EPI_ISL_2930034, EPI_ISL_2930090, EPI_ISL_2930099, EPI_ISL_2930130, EPI_ISL_2930151, EPI_ISL_2930173, EPI_ISL_2930203, EPI_ISL_2930206, EPI_ISL_2930217, EPI_ISL_2930250, EPI_ISL_2930265, EPI_ISL_2930279, EPI_ISL_2930309, EPI_ISL_2930311 | | | |
| see above | Quest Diagnostics Incorporated | Centers for Disease Control and Prevention Division of Viral Diseases, Pathogen Discovery | A. Gerasimova; A. Perez; Adrian Paskey; B. Anderson; Benjamin Rambo-Martin; Christopher Gulvick; Clinton R. Paden; Dakota Howard; Darlene Wagner; Dhwani Batra; Duncan MacCannell; F. Lacbawan; I. A. Shlyakhter; Jason Caravas; K.E. Livingston; Kara Moser; L.E. Bernstein; M. Hua; Matthew Schmerer; P. Tanpaiboon; Peter W. Cook; R. M. Kagan; R. Owen; R. V. Rolando; S. H. Rosenthal; Scott Sammons; Shatavia Morrison; Y. Liu; Yvette Unoarumhi |
| EPI_ISL_2628498 | Rady Children's Hospital - San Diego | Andersen lab at Scripps Research | Christina Clarke; David Dimmock; Denise Malicki; Kathryn Bouic; Linda Luo; SEARCH Alliance San Diego with Nanda Radamchar; Teresa Mueller |
| EPI_ISL_2934052 | Regional Medical Center | Santa Clara County Public Health Laboratory | Santa Clara County Public Health Department |
| EPI_ISL_2253203, EPI_ISL_3091564, EPI_ISL_3091570, EPI_ISL_3091571, EPI_ISL_3091577, EPI_ISL_3091588, EPI_ISL_3091589, EPI_ISL_3091597, EPI_ISL_3091598, EPI_ISL_3091603, EPI_ISL_3091604, EPI_ISL_3091610, EPI_ISL_3091611, EPI_ISL_3091615, EPI_ISL_3091620, EPI_ISL_3091640, EPI_ISL_3091646, EPI_ISL_3091651, EPI_ISL_3091653, EPI_ISL_3091655, EPI_ISL_3091665, EPI_ISL_3091668, EPI_ISL_3091669, EPI_ISL_3091673, EPI_ISL_3091695, EPI_ISL_3091698, EPI_ISL_3091706, EPI_ISL_3091710, EPI_ISL_3091711, EPI_ISL_3091713, EPI_ISL_3091717, EPI_ISL_3091720, EPI_ISL_3091724, EPI_ISL_3091732, EPI_ISL_3091749, EPI_ISL_3091762, EPI_ISL_3091763, EPI_ISL_3091777, EPI_ISL_3091781, EPI_ISL_3091783, EPI_ISL_3091797, EPI_ISL_3091798, EPI_ISL_3091802, EPI_ISL_3091806, EPI_ISL_3091813, EPI_ISL_3091819, EPI_ISL_3091823, EPI_ISL_3091830, EPI_ISL_3091837, EPI_ISL_3091852, EPI_ISL_3091854, EPI_ISL_3091867, EPI_ISL_3091872, EPI_ISL_3091875, EPI_ISL_3091878, EPI_ISL_3091879, EPI_ISL_3091887, EPI_ISL_3091891, EPI_ISL_3091903, EPI_ISL_3091908, EPI_ISL_3091913, EPI_ISL_3091917, EPI_ISL_3091931, EPI_ISL_3091935, EPI_ISL_3091979, EPI_ISL_3091980, EPI_ISL_3091981, EPI_ISL_3091982, EPI_ISL_3091996, EPI_ISL_3092008, EPI_ISL_3092012, EPI_ISL_3092013, EPI_ISL_3092016, EPI_ISL_3092017, EPI_ISL_3092020, EPI_ISL_3092027, EPI_ISL_3092039, EPI_ISL_3092045 | | | |
| see above | Respiratory Viruses Branch, Centers for Disease Control and Prevention | Respiratory Viruses Branch, Centers for Disease Control and Prevention | Anderson, B.; Batra, D.; Bernstein, L.; C.R.; Caravas, J.; Cook; Fierro, J.; Gao, H.; Gao, J.; Gerasimova, A.; Gulvick, C.; Howard, D.; Hua, M.; I.A.; Kagan; Lacbawan, F.; Li, M.; Liu, Y.; Livingston, K.; MacCannell, D.; Meng, Y.; Morrison, S.; Moser, K.; Ng, D.; Owen, R.; P.W.; Paden; Paskey, A.; Perez, A.; R.M.; R.V.; Rambo-Martin, B.; Rolando; Rosenthal; S.H.; Sammons, S.; Sapra, B.; Schmerer, M.; Shlyakhter; Tanpaiboon, P.; Tsai, B.; Unoarumhi, Y.; Wagner, D.; Xie, J. |
| EPI_ISL_2812005 | Saint Francis Medical Center | Los Angeles County PHL | P. Hemarajata et al. |
| EPI_ISL_2907565, EPI_ISL_2907566 | Saint Francis Medical Center | Los Angeles County Public Health Laboratories | P. Hemarajata et al. |
| EPI_ISL_2774497 | San Diego County Public Health Lab | San Diego County Public Health Lab | Ashleigh Murphy Schafer; Brett Austin |
| EPI_ISL_2329357, EPI_ISL_2329364, EPI_ISL_2329370, EPI_ISL_2329381, EPI_ISL_2329470, EPI_ISL_2329479, EPI_ISL_2329492, EPI_ISL_2329503, EPI_ISL_2329509, EPI_ISL_2329522, EPI_ISL_2329523, EPI_ISL_2329529, EPI_ISL_2329530, EPI_ISL_2329536, EPI_ISL_2329537, EPI_ISL_2423012, EPI_ISL_2423013, EPI_ISL_2423014, EPI_ISL_2423015, EPI_ISL_2423016, EPI_ISL_2423017, EPI_ISL_2628329, EPI_ISL_2628334, EPI_ISL_2628346, EPI_ISL_2628350, EPI_ISL_2628439, EPI_ISL_2628440, EPI_ISL_2628443, EPI_ISL_2628444, EPI_ISL_2628446, EPI_ISL_2628452, EPI_ISL_2628457, EPI_ISL_2628460, EPI_ISL_2628461, EPI_ISL_2628465, EPI_ISL_2628466, EPI_ISL_2628471, EPI_ISL_2628476, EPI_ISL_2628478, EPI_ISL_2628481, EPI_ISL_2628484, EPI_ISL_2628486, EPI_ISL_2628488, EPI_ISL_2628494, EPI_ISL_2628496, EPI_ISL_2628510, EPI_ISL_2628550, EPI_ISL_2628583, EPI_ISL_2628588, EPI_ISL_2628598, EPI_ISL_2628610, EPI_ISL_2628611, EPI_ISL_2628612, EPI_ISL_2628614, EPI_ISL_2628723, EPI_ISL_2628739, EPI_ISL_2628742, EPI_ISL_2628745, EPI_ISL_2628749, EPI_ISL_2628753, EPI_ISL_2628757, EPI_ISL_2628760, EPI_ISL_2628768, EPI_ISL_2628771, EPI_ISL_2628775, EPI_ISL_2628779, EPI_ISL_2628782, EPI_ISL_2628786, EPI_ISL_2628790, EPI_ISL_2628793, EPI_ISL_2628797, EPI_ISL_2628801, EPI_ISL_2628804, EPI_ISL_2628808, EPI_ISL_2628812, EPI_ISL_2628815, EPI_ISL_2628819, EPI_ISL_2628822, EPI_ISL_2628826, EPI_ISL_2628830, EPI_ISL_2628833, EPI_ISL_2628837, EPI_ISL_2628841, EPI_ISL_2628844, EPI_ISL_2628848, EPI_ISL_2628851, EPI_ISL_2628855, EPI_ISL_2628859, EPI_ISL_2628863, EPI_ISL_2628866, EPI_ISL_2628870, EPI_ISL_2628874, EPI_ISL_2628877, EPI_ISL_2628881, EPI_ISL_2628884, EPI_ISL_2628887, EPI_ISL_2628891, EPI_ISL_2628895, EPI_ISL_2628897, EPI_ISL_2628901, EPI_ISL_2628904, EPI_ISL_2628908, EPI_ISL_2628911, EPI_ISL_2628915, EPI_ISL_2628918, EPI_ISL_2628922, EPI_ISL_2628926, EPI_ISL_2628929, EPI_ISL_2628933, EPI_ISL_2628937, EPI_ISL_2628940, EPI_ISL_2628944, EPI_ISL_2628948, EPI_ISL_2628951, EPI_ISL_2628955, EPI_ISL_2712831 | | | |
| see above | San Diego County Public Health Laboratory | Andersen lab at Scripps Research | Brett Austin; Jovan Shephard; SEARCH Alliance San Diego with Ashleigh Murphy |
| EPI_ISL_2834946, EPI_ISL_2834947, EPI_ISL_2834948, EPI_ISL_2834949, EPI_ISL_2834950, EPI_ISL_2834951, EPI_ISL_2834952, EPI_ISL_2834953, EPI_ISL_2834954, EPI_ISL_2834955, EPI_ISL_2834956, EPI_ISL_2834957, EPI_ISL_2840592, EPI_ISL_2840593, EPI_ISL_2840594, EPI_ISL_2840606, EPI_ISL_2840607, EPI_ISL_2840608, EPI_ISL_2840638, EPI_ISL_2840852, EPI_ISL_2840861, EPI_ISL_2840862, EPI_ISL_2840863, EPI_ISL_2840864, EPI_ISL_2840867, EPI_ISL_2840895, EPI_ISL_2840896, EPI_ISL_2840897, EPI_ISL_2840898, EPI_ISL_2840899, EPI_ISL_2840900, EPI_ISL_2840901, EPI_ISL_2840902, EPI_ISL_2840903, EPI_ISL_2840917, EPI_ISL_2840918, EPI_ISL_2840920, EPI_ISL_2840921, EPI_ISL_2840922, EPI_ISL_2840926, EPI_ISL_2840927, EPI_ISL_2840928, EPI_ISL_2840930, EPI_ISL_2841488, EPI_ISL_2841489, EPI_ISL_2841490, EPI_ISL_2841534, EPI_ISL_2841536, EPI_ISL_2841537, EPI_ISL_2841538, EPI_ISL_2841540, EPI_ISL_2841603, EPI_ISL_2841604, EPI_ISL_2841613, EPI_ISL_2841614, EPI_ISL_2841615, EPI_ISL_2842692, EPI_ISL_2842693, EPI_ISL_2842694, EPI_ISL_2842695, EPI_ISL_2842696, EPI_ISL_2842697, EPI_ISL_2842698, EPI_ISL_2842699, EPI_ISL_2842700, EPI_ISL_2842701, EPI_ISL_2842702, EPI_ISL_2842703, EPI_ISL_2842704, EPI_ISL_2842705, EPI_ISL_2842706, EPI_ISL_2873837 | | | |
| see above | San Diego County Public Health Laboratory | San Diego County Public Health Laboratory | Ashleigh Murphy Schafer; Brett Austin; San Diego County Public Health Laboratory |
| EPI_ISL_2496776, EPI_ISL_2496777, EPI_ISL_2496787, EPI_ISL_2496788, EPI_ISL_2496845, EPI_ISL_2496892, EPI_ISL_2496895, EPI_ISL_2496923, EPI_ISL_2496931, EPI_ISL_2496941, EPI_ISL_2496976, EPI_ISL_2500085, EPI_ISL_2500156, EPI_ISL_2500199, EPI_ISL_2658589, EPI_ISL_2658592, EPI_ISL_2658598, EPI_ISL_2658735 | | | |
| see above | Santa Clara County Public Health Laboratory | Chan-Zuckerberg Biohub | CZB Cliahub Consortium |
| EPI_ISL_2631202, EPI_ISL_2631203, EPI_ISL_2631204, EPI_ISL_2631205, EPI_ISL_2631206, EPI_ISL_2631207, EPI_ISL_2650563, EPI_ISL_2659057, EPI_ISL_2659064 | | | |
| see above | Santa Clara County Public Health Laboratory | Santa Clara County Public Health Laboratory | Santa Clara County Public Health Department |
| EPI_ISL_2631208, EPI_ISL_2631209, EPI_ISL_2631211, EPI_ISL_2631212, EPI_ISL_2631213, EPI_ISL_2631214, EPI_ISL_2631215, EPI_ISL_2631216, EPI_ISL_2631217, EPI_ISL_2631218, EPI_ISL_2650546, EPI_ISL_2650547, EPI_ISL_2650548, EPI_ISL_2650549, EPI_ISL_2650550, EPI_ISL_2650551, EPI_ISL_2650552, EPI_ISL_2650553, EPI_ISL_2650554, EPI_ISL_2650555, EPI_ISL_2650556, EPI_ISL_2650557, EPI_ISL_2650558, EPI_ISL_2650559, EPI_ISL_2650560, EPI_ISL_2650561, EPI_ISL_2650564, EPI_ISL_2650565, EPI_ISL_2650567, EPI_ISL_2650568, EPI_ISL_2650569, EPI_ISL_2650570, EPI_ISL_2650571, EPI_ISL_2650572, EPI_ISL_2650573, EPI_ISL_2650574, EPI_ISL_2650575, EPI_ISL_2650576, EPI_ISL_2650577, EPI_ISL_2650578, EPI_ISL_2650579, EPI_ISL_2650580, EPI_ISL_2650582, EPI_ISL_2650583, EPI_ISL_2691299, EPI_ISL_2691300, EPI_ISL_2691302, EPI_ISL_2691303, EPI_ISL_2691304, EPI_ISL_2691305, EPI_ISL_2691306, EPI_ISL_2691307, EPI_ISL_2691308, EPI_ISL_2691309, EPI_ISL_2691310, EPI_ISL_2691311, EPI_ISL_2691312, EPI_ISL_2691313, EPI_ISL_2691314, EPI_ISL_2753976, EPI_ISL_2774591, EPI_ISL_2774592, EPI_ISL_2774593, EPI_ISL_2774594, EPI_ISL_2774595, EPI_ISL_2774596, EPI_ISL_2774597, EPI_ISL_2774598, EPI_ISL_2774599, EPI_ISL_2774600, EPI_ISL_2774601, EPI_ISL_2774602, EPI_ISL_2774603, EPI_ISL_2774604, EPI_ISL_2774605, EPI_ISL_2774606, EPI_ISL_2774607, EPI_ISL_2774608, EPI_ISL_2774609, EPI_ISL_2774610, EPI_ISL_2774611, EPI_ISL_2774612, EPI_ISL_2774613, EPI_ISL_2774614, EPI_ISL_2774615, EPI_ISL_2774616, EPI_ISL_2774617, EPI_ISL_2774618, EPI_ISL_2774619, EPI_ISL_2774620, EPI_ISL_2774624, EPI_ISL_2774625, EPI_ISL_2774626, EPI_ISL_2774627, EPI_ISL_2774628, EPI_ISL_2774629, EPI_ISL_2774630, EPI_ISL_2774631, EPI_ISL_2774632, EPI_ISL_2774633, EPI_ISL_2774634, EPI_ISL_2933987, EPI_ISL_2933989, EPI_ISL_2933993, EPI_ISL_2933994, EPI_ISL_2933995, EPI_ISL_2933997, EPI_ISL_2933998, EPI_ISL_2934000, EPI_ISL_2934006, EPI_ISL_2934021, EPI_ISL_2934022, EPI_ISL_2934029, EPI_ISL_2934030, EPI_ISL_2934031, EPI_ISL_2934032, EPI_ISL_2934034, EPI_ISL_2934035, EPI_ISL_2934036, EPI_ISL_2934037, EPI_ISL_2934039, EPI_ISL_2934041, EPI_ISL_2934043, EPI_ISL_2934046, EPI_ISL_2934051, EPI_ISL_2934055, EPI_ISL_2934056, EPI_ISL_2934059, EPI_ISL_2934060, EPI_ISL_2934062, EPI_ISL_2934063, EPI_ISL_2934064, EPI_ISL_2934069, EPI_ISL_2934070, EPI_ISL_2934072, EPI_ISL_2934081, EPI_ISL_2934082, EPI_ISL_2934083, EPI_ISL_2934084 | | | |
| see above | Santa Clara Valley Medical Center | Santa Clara County Public Health Laboratory | Santa Clara County Public Health Department |
| EPI_ISL_2231131, EPI_ISL_2231137, EPI_ISL_2231139, EPI_ISL_2231157, EPI_ISL_2231162, EPI_ISL_2231163, EPI_ISL_2231164, EPI_ISL_2231526, EPI_ISL_2329427, EPI_ISL_2329428, EPI_ISL_2329429, EPI_ISL_2329430, EPI_ISL_2329431, EPI_ISL_2329432, EPI_ISL_2329433, EPI_ISL_2329434, EPI_ISL_2329435, EPI_ISL_2329436, EPI_ISL_2329437, EPI_ISL_2329438, EPI_ISL_2329439, EPI_ISL_2329440, EPI_ISL_2329441, EPI_ISL_2329442, EPI_ISL_2329443, EPI_ISL_2329444, EPI_ISL_2329445, EPI_ISL_2329446, EPI_ISL_2329447, EPI_ISL_2329450, EPI_ISL_2329451, EPI_ISL_2329452, EPI_ISL_2329453, EPI_ISL_2329454, EPI_ISL_2329455, EPI_ISL_2329456, EPI_ISL_2329457, EPI_ISL_2329458, EPI_ISL_2329459, EPI_ISL_2329460, EPI_ISL_2329461, EPI_ISL_2329462, EPI_ISL_2329463, EPI_ISL_2329464, EPI_ISL_2422815, EPI_ISL_2422816, EPI_ISL_2422817, EPI_ISL_2422818, EPI_ISL_2422819, EPI_ISL_2422820, EPI_ISL_2422821, EPI_ISL_2422822, EPI_ISL_2422823, EPI_ISL_2422824, EPI_ISL_2422825, EPI_ISL_2422826, EPI_ISL_2422827, EPI_ISL_2422831, EPI_ISL_2422832, EPI_ISL_2422833, EPI_ISL_2628380, EPI_ISL_2628381, EPI_ISL_2628382, EPI_ISL_2628383, EPI_ISL_2628384, EPI_ISL_2628385, EPI_ISL_2628386, EPI_ISL_2628387, EPI_ISL_2628388, EPI_ISL_2628389, EPI_ISL_2628390, EPI_ISL_2628391, EPI_ISL_2628392, EPI_ISL_2628393, EPI_ISL_2628394, EPI_ISL_2628395, EPI_ISL_2628536, EPI_ISL_2628537, EPI_ISL_2628538, EPI_ISL_2628539, EPI_ISL_2628540, EPI_ISL_2628541, EPI_ISL_2628542, EPI_ISL_2628543, EPI_ISL_2628544, EPI_ISL_2712535, EPI_ISL_2712536, EPI_ISL_2712537, EPI_ISL_2712538, EPI_ISL_2712539, EPI_ISL_2712540, EPI_ISL_2712541, EPI_ISL_2712542, EPI_ISL_2712543, EPI_ISL_2712544, EPI_ISL_2712545, EPI_ISL_2712546, EPI_ISL_2712547, EPI_ISL_2712548, EPI_ISL_2712549, EPI_ISL_2712550, EPI_ISL_2712551, EPI_ISL_2712552, EPI_ISL_2712553, EPI_ISL_2712554, EPI_ISL_2712555, EPI_ISL_2835552, EPI_ISL_2835553, EPI_ISL_2835554, EPI_ISL_2835555, EPI_ISL_2835556, EPI_ISL_2835557, EPI_ISL_2835558, EPI_ISL_2835559, EPI_ISL_2835560, EPI_ISL_2835561, EPI_ISL_2835562, EPI_ISL_2835563, EPI_ISL_2835564, EPI_ISL_2835565, EPI_ISL_2920432, EPI_ISL_2920433, EPI_ISL_2920434, EPI_ISL_2920435, EPI_ISL_2920436, EPI_ISL_2920437, EPI_ISL_2920438, EPI_ISL_2920439, EPI_ISL_2920440, EPI_ISL_2920441, EPI_ISL_2920442, EPI_ISL_2920443 | | | |
| see above | Sharp HealthCare Laboratory | Andersen lab at Scripps Research | Art Mendoza; Cathy Woerle; Jacquelyn Berumen; Liam McGinnis; Omid Bakhtar; SEARCH Alliance San Diego with Aaron Harding |
| EPI_ISL_2457061 | Stanford Health Care | Stanford University School of Medicine, Clinical Virology Laboratory | Becky Jiang; Bernadette Troung; ChunHong Huang; Daniel Solis; James Zehnder; Malaya K. Sahoo; Mamdouh Sibai; Michelle Verghese; and Benjamin A. Pinsky |
| EPI_ISL_2391790, EPI_ISL_2391791 | Sun Clinical Laboratory | Los Angeles County PHL | P. Hemarajata et al. |
| EPI_ISL_2620779, EPI_ISL_2774572 | Torrance Memorial Medical Center | Los Angeles County PHL | P. Hemarajata et al. |
| EPI_ISL_2426985, EPI_ISL_2426986, EPI_ISL_2426987, EPI_ISL_2426988, EPI_ISL_2426989, EPI_ISL_2426990, EPI_ISL_2426991, EPI_ISL_2426992, EPI_ISL_2426993, EPI_ISL_2426994 | | | |
| see above | UC Davis Genome Center and Healthy Davis Together | UC Davis Genome Center | UC Davis |
| EPI_ISL_2391761, EPI_ISL_2391762, EPI_ISL_2391763, EPI_ISL_2391810, EPI_ISL_2391811, EPI_ISL_2391812, EPI_ISL_2392172, EPI_ISL_2392174, EPI_ISL_2392177, EPI_ISL_2392179, EPI_ISL_2392180, EPI_ISL_2392191, EPI_ISL_2392193, EPI_ISL_2392194, EPI_ISL_2399612, EPI_ISL_2399613, EPI_ISL_2399616, EPI_ISL_2399619, EPI_ISL_2399620, EPI_ISL_2404855, EPI_ISL_2404857, EPI_ISL_2404860, EPI_ISL_2404861, EPI_ISL_2404862, EPI_ISL_2404863, EPI_ISL_2404864, EPI_ISL_2404866, EPI_ISL_2404867, EPI_ISL_2405167, EPI_ISL_2405183, EPI_ISL_2424170, EPI_ISL_2424172, EPI_ISL_2424175, EPI_ISL_2424183, EPI_ISL_2424189, EPI_ISL_2424190, EPI_ISL_2424193, EPI_ISL_2424195, EPI_ISL_2424198, EPI_ISL_2424199, EPI_ISL_2424202, EPI_ISL_2424204, EPI_ISL_2424205, EPI_ISL_2424206, EPI_ISL_2424208, EPI_ISL_2424234, EPI_ISL_2617943, EPI_ISL_2617944, EPI_ISL_2617945, EPI_ISL_2617946, EPI_ISL_2617947, EPI_ISL_2617948, EPI_ISL_2617949, EPI_ISL_2617950, EPI_ISL_2617951, EPI_ISL_2617952, EPI_ISL_2617953, EPI_ISL_2617954, EPI_ISL_2617955, EPI_ISL_2617956, EPI_ISL_2617957, EPI_ISL_2617958, EPI_ISL_2620750, EPI_ISL_2772506, EPI_ISL_2772507, EPI_ISL_2772508, | | | |

| | | | |
|---|---|---|---|
| EPI_ISL_2772509, EPI_ISL_2772510, EPI_ISL_2772511, EPI_ISL_2772512, EPI_ISL_2772513, EPI_ISL_2772514, EPI_ISL_2772515, EPI_ISL_2772516, EPI_ISL_2772517, EPI_ISL_2772518, EPI_ISL_2772519, EPI_ISL_2772520, EPI_ISL_2772521, EPI_ISL_2811920, EPI_ISL_2811921 | | | |
| see above | UCLA Clinical Micro Lab | Los Angeles County PHL | P. Hemarajata et al. |
| EPI_ISL_2179217, EPI_ISL_2179218, EPI_ISL_2179270, EPI_ISL_2179271, EPI_ISL_2179272, EPI_ISL_2295456, EPI_ISL_2295457, EPI_ISL_2378755, EPI_ISL_2378756, EPI_ISL_2494632, EPI_ISL_2494633, EPI_ISL_2494634, EPI_ISL_2494635 | | | |
| see above | UCSC Genomics Institute | UCSC Genomics Institute | A. Marm Kilpatrick; Angie Hinrichs; Beth Shapiro; Bryan Thornlow; Ciara Wanket; David Haussler; Eric Beraut; Hugh Olsen; Ikenna Anigbogu; Isabel Bjork; Jakob McBroome; Jeremy Sanford; Joshua Kapp; Mark Akeson; Maximilian Haeussler; Michael Stone; Miten Jain; Molly Cassatt-Johnstone; Namrita Dhillon; Russell Corbett-Detig; Terren Chang; Yatish Turakhia |
| EPI_ISL_2426732, EPI_ISL_2450384, EPI_ISL_2715313 | US Air Force School of Aerospace Medicine | US Air Force School of Aerospace Medicine | Amanda Javorina; Anthony Fries; Carol Garrett; Clarise Starr; Elizabeth Macias; Jennifer Meyer; Sarah Purves; William Gruner |
| EPI_ISL_2391782, EPI_ISL_2391783, EPI_ISL_2391784, EPI_ISL_2391785, EPI_ISL_2391786, EPI_ISL_2391787, EPI_ISL_2405180, EPI_ISL_2405181, EPI_ISL_2620783, EPI_ISL_2811942, EPI_ISL_2811943, EPI_ISL_2811966, EPI_ISL_2812004, EPI_ISL_2894587, EPI_ISL_2894588, EPI_ISL_2894589 | | | |
| see above | USC Clinical Lab | Los Angeles County PHL | P. Hemarajata et al. |
| EPI_ISL_2907573, EPI_ISL_2907574, EPI_ISL_2907575 | USC Clinical Lab | Los Angeles County Public Health Laboratories | P. Hemarajata et al. |
| EPI_ISL_2404811, EPI_ISL_2404812, EPI_ISL_2404813, EPI_ISL_2404814, EPI_ISL_2404816, EPI_ISL_2404817, EPI_ISL_2404818, EPI_ISL_2404819, EPI_ISL_2404820, EPI_ISL_2404821, EPI_ISL_2404822, EPI_ISL_2404823, EPI_ISL_2404824, EPI_ISL_2404825, EPI_ISL_2404828, EPI_ISL_2404829, EPI_ISL_2404830, EPI_ISL_2404831, EPI_ISL_2404832, EPI_ISL_2404833, EPI_ISL_2404834, EPI_ISL_2404835, EPI_ISL_2404836, EPI_ISL_2404837, EPI_ISL_2404838, EPI_ISL_2404839 | | | |
| see above | VMC | Santa Clara County public Health Laboratory | Santa Clara County Public Health Department |

We gratefully acknowledge the following Authors from the Originating laboratories responsible for obtaining the specimens, as well as the Submitting laboratories where the genome data were generated and shared via GISAID, on which this research is based.

All Submitters of data may be contacted directly via www.gisaid.org

Authors are sorted alphabetically.

| Accession ID | Originating Laboratory | Submitting Laboratory | Authors |
|---|---|---|---|
| EPI_ISL_1354236, EPI_ISL_1433094, EPI_ISL_1433134, EPI_ISL_1433219, EPI_ISL_1433260, EPI_ISL_1438687 | ALB FILS KLINIKEN GmbH - Institut für Laboratoriumsmedizin Klinik am Eichert Göppingen | Robert Koch Institute | |
| EPI_ISL_1287284, EPI_ISL_1287286, EPI_ISL_1287288, EPI_ISL_1287289, EPI_ISL_1287290, EPI_ISL_1287291, EPI_ISL_1287293, EPI_ISL_1287294, EPI_ISL_1287295, EPI_ISL_1287296, EPI_ISL_1287298, EPI_ISL_1287299, EPI_ISL_1287300, EPI_ISL_1287301, EPI_ISL_1287302, EPI_ISL_1287303, EPI_ISL_1287304, EPI_ISL_1287307, EPI_ISL_1287309, EPI_ISL_1287310, EPI_ISL_1287311, EPI_ISL_1287312, EPI_ISL_1287313, EPI_ISL_1287315, EPI_ISL_1287317, EPI_ISL_1287318, EPI_ISL_1287319, EPI_ISL_1354475, EPI_ISL_1354476, EPI_ISL_1354477, EPI_ISL_1354479, EPI_ISL_1354480, EPI_ISL_1354481, EPI_ISL_1354482, EPI_ISL_1354483, EPI_ISL_1354485, EPI_ISL_1354488, EPI_ISL_1354490, EPI_ISL_1354491, EPI_ISL_1354499 | | | |
| see above | Alexianer DaKS GmbH (eigenes IT-Tochterunternehmen der Alexianer Krankenhäuser) | Robert Koch Institute | |
| EPI_ISL_1570333, EPI_ISL_1570339, EPI_ISL_1570355, EPI_ISL_1570360, EPI_ISL_1570368, EPI_ISL_1570377, EPI_ISL_1640830, EPI_ISL_1725276, EPI_ISL_1725279, EPI_ISL_1725280, EPI_ISL_1725281, EPI_ISL_1725282, EPI_ISL_1725284, EPI_ISL_1725286, EPI_ISL_1725287, EPI_ISL_1725288, EPI_ISL_1725291, EPI_ISL_1725292, EPI_ISL_1725294, EPI_ISL_1725296, EPI_ISL_1725298, EPI_ISL_1725300, EPI_ISL_1725304, EPI_ISL_1725306, EPI_ISL_1725307, EPI_ISL_1725315, EPI_ISL_1725316, EPI_ISL_1725318, EPI_ISL_1725323, EPI_ISL_1725324, EPI_ISL_1725326, EPI_ISL_1725327, EPI_ISL_1725329, EPI_ISL_1725331, EPI_ISL_1725332, EPI_ISL_1725335, EPI_ISL_1725336, EPI_ISL_1725341, EPI_ISL_1725342, EPI_ISL_1725343, EPI_ISL_1725344, EPI_ISL_1725346, EPI_ISL_1725347, EPI_ISL_1725349, EPI_ISL_1725350, EPI_ISL_1725352, EPI_ISL_1725355, EPI_ISL_1725356, EPI_ISL_1725357, EPI_ISL_1725358, EPI_ISL_1725361, EPI_ISL_1725362, EPI_ISL_1725363, EPI_ISL_1725364, EPI_ISL_1725365, EPI_ISL_1725366, EPI_ISL_1725368, EPI_ISL_1725369, EPI_ISL_1725372, EPI_ISL_1725374, EPI_ISL_1725377, EPI_ISL_1725378, EPI_ISL_1725379, EPI_ISL_1725380, EPI_ISL_1725382, EPI_ISL_1725383, EPI_ISL_1725384, EPI_ISL_1725386, EPI_ISL_1725387, EPI_ISL_1725389, EPI_ISL_1725390, EPI_ISL_1725391, EPI_ISL_1725393, EPI_ISL_1725394, EPI_ISL_1725395, EPI_ISL_1725398, EPI_ISL_1725399, EPI_ISL_1725402, EPI_ISL_1725405, EPI_ISL_1725408, EPI_ISL_1725410, EPI_ISL_1725411, EPI_ISL_1725412, EPI_ISL_1725414, EPI_ISL_1725415, EPI_ISL_1725416, EPI_ISL_1725421, EPI_ISL_1725423, EPI_ISL_1725424, EPI_ISL_1725426, EPI_ISL_1725427, EPI_ISL_1725430, EPI_ISL_1725432, EPI_ISL_1725434, EPI_ISL_1725437, EPI_ISL_1725439, EPI_ISL_1725440, EPI_ISL_1725441, EPI_ISL_1725442, EPI_ISL_1725443, EPI_ISL_1725444, EPI_ISL_1725447, EPI_ISL_1725448, EPI_ISL_1725449, EPI_ISL_1725451, EPI_ISL_1725452, EPI_ISL_1725455, EPI_ISL_1725458, EPI_ISL_1725462, EPI_ISL_1725463, EPI_ISL_1725464, EPI_ISL_1725469, EPI_ISL_1725470, EPI_ISL_1725471, EPI_ISL_1725472, EPI_ISL_1725474, EPI_ISL_1725475, EPI_ISL_1725477, EPI_ISL_1725478, EPI_ISL_1725479, EPI_ISL_1725482, EPI_ISL_1725483, EPI_ISL_1725484, EPI_ISL_1725487, EPI_ISL_1725488, EPI_ISL_1725489, EPI_ISL_1725490, EPI_ISL_1725491, EPI_ISL_1725494, EPI_ISL_1725496, EPI_ISL_1725497, EPI_ISL_1725499, EPI_ISL_1725500, EPI_ISL_1725501, EPI_ISL_1725503, EPI_ISL_1725504, EPI_ISL_1725506, EPI_ISL_1725507, EPI_ISL_1725508, EPI_ISL_1725509, EPI_ISL_1725511, EPI_ISL_1725512, EPI_ISL_1725513, EPI_ISL_1725514, EPI_ISL_1725515, EPI_ISL_1725518, EPI_ISL_1725519, EPI_ISL_1725520, EPI_ISL_1725522, EPI_ISL_1725523, EPI_ISL_1725525, EPI_ISL_1725526, EPI_ISL_1725529, EPI_ISL_2110936, EPI_ISL_2110961, EPI_ISL_2110966 | | | |
| see above | Bayerisches Landesamt fÃ¼r Gesundheit und Lebensmittelsicherheit (LGL) | Robert Koch Institute | |
| EPI_ISL_1349119, EPI_ISL_1357072, EPI_ISL_1357085, EPI_ISL_1357091, EPI_ISL_1357094, EPI_ISL_1357099, EPI_ISL_1357100, EPI_ISL_1357101, EPI_ISL_1357105, EPI_ISL_1357107, EPI_ISL_1357111, EPI_ISL_1357114, EPI_ISL_1357120, EPI_ISL_1357121, EPI_ISL_1357123, EPI_ISL_1357125, EPI_ISL_1357127, EPI_ISL_1357129, EPI_ISL_1357139, EPI_ISL_1357141, EPI_ISL_1357142, EPI_ISL_1357148, EPI_ISL_1357154, EPI_ISL_1357155, EPI_ISL_1357159, EPI_ISL_1357160, EPI_ISL_1357165, EPI_ISL_1357167, EPI_ISL_1357170, EPI_ISL_1357174, EPI_ISL_1357175, EPI_ISL_1357177, EPI_ISL_1357181, EPI_ISL_1357184, EPI_ISL_1357191, EPI_ISL_1357204, EPI_ISL_1357205, EPI_ISL_1357206, EPI_ISL_1357210, EPI_ISL_1357214, EPI_ISL_1357215, EPI_ISL_1357217, EPI_ISL_1357219, EPI_ISL_1357221, EPI_ISL_1357222, EPI_ISL_1357223, EPI_ISL_1357225 | | | |
| see above | Bayerisches Landesamt für Gesundheit und Lebensmittelsicherheit (LGL) | Robert Koch Institute | |
| EPI_ISL_1216467, EPI_ISL_1216479, EPI_ISL_1216527, EPI_ISL_1281996, EPI_ISL_1282001, EPI_ISL_1282005, EPI_ISL_1282007, EPI_ISL_1282018, EPI_ISL_1282019, EPI_ISL_1282021, EPI_ISL_1282023, EPI_ISL_1282036, EPI_ISL_1282037, EPI_ISL_1282044, EPI_ISL_1282045, EPI_ISL_1282046, EPI_ISL_1282050, EPI_ISL_1282051, EPI_ISL_1282060, EPI_ISL_1282062, EPI_ISL_1282071, EPI_ISL_1282072, EPI_ISL_1282077, EPI_ISL_1282081, EPI_ISL_1282091, EPI_ISL_1282092, EPI_ISL_1282098, EPI_ISL_1282101, EPI_ISL_1282103, EPI_ISL_1282110, EPI_ISL_1282113, EPI_ISL_1282114, EPI_ISL_1282115, EPI_ISL_1282118, EPI_ISL_1282123, EPI_ISL_1282125, EPI_ISL_1282133, EPI_ISL_1282134, EPI_ISL_1282137, EPI_ISL_1282162, EPI_ISL_1282167, EPI_ISL_1282170, EPI_ISL_1282180, EPI_ISL_1282182, EPI_ISL_1282191, EPI_ISL_1282193, EPI_ISL_1282200, EPI_ISL_1282201, EPI_ISL_1282203, EPI_ISL_1282220, EPI_ISL_1282221, EPI_ISL_1282222, EPI_ISL_1282223, EPI_ISL_1282225, EPI_ISL_1282226, EPI_ISL_1282232, EPI_ISL_1282237, EPI_ISL_1282239, EPI_ISL_1282240, EPI_ISL_1282247, EPI_ISL_1282249, EPI_ISL_1282250, EPI_ISL_1282251, EPI_ISL_1282254, EPI_ISL_1282259, EPI_ISL_1282260, EPI_ISL_1282266, EPI_ISL_1282267, EPI_ISL_1282268, EPI_ISL_1282274, EPI_ISL_1282278, EPI_ISL_1282292, EPI_ISL_1282295, EPI_ISL_1282309, EPI_ISL_1284554, EPI_ISL_1284555, EPI_ISL_1284556, EPI_ISL_1284559, EPI_ISL_1284560, EPI_ISL_1284565, EPI_ISL_1284566, EPI_ISL_1284572, EPI_ISL_1284573, EPI_ISL_1284585, EPI_ISL_1284589, EPI_ISL_1284592, EPI_ISL_1284593, EPI_ISL_1284598, EPI_ISL_1284601, EPI_ISL_1284602, EPI_ISL_1284607, EPI_ISL_1284609, EPI_ISL_1284610, EPI_ISL_1284611, EPI_ISL_1284612, EPI_ISL_1284629, EPI_ISL_1284636, EPI_ISL_1284638, EPI_ISL_1284647, EPI_ISL_1284651, EPI_ISL_1284652, EPI_ISL_1284657, EPI_ISL_1284658, EPI_ISL_1284662, EPI_ISL_1284667, EPI_ISL_1284668, EPI_ISL_1284671, EPI_ISL_1284679, EPI_ISL_1284684, EPI_ISL_1284689, EPI_ISL_1284692, EPI_ISL_1284693, EPI_ISL_1284695, EPI_ISL_1284702, EPI_ISL_1284710, EPI_ISL_1284713, EPI_ISL_1284714, EPI_ISL_1284717, EPI_ISL_1284722, EPI_ISL_1284723, EPI_ISL_1284725, EPI_ISL_1284726, EPI_ISL_1284728, EPI_ISL_1284729, EPI_ISL_1284730, EPI_ISL_1284734, EPI_ISL_1284738, EPI_ISL_1284743, EPI_ISL_1284750, EPI_ISL_1284754, EPI_ISL_1284755, EPI_ISL_1284757, EPI_ISL_1284758, EPI_ISL_1284759, EPI_ISL_1284761, EPI_ISL_1284765, EPI_ISL_1284767, EPI_ISL_1284770, EPI_ISL_1284772, EPI_ISL_1284773, EPI_ISL_1284774, EPI_ISL_1284775, EPI_ISL_1284776, EPI_ISL_1284778, EPI_ISL_1284780, EPI_ISL_1284781, EPI_ISL_1284783, EPI_ISL_1284784, EPI_ISL_1284785, EPI_ISL_1284787, EPI_ISL_1284792, EPI_ISL_1284793, EPI_ISL_1284796, EPI_ISL_1284802, EPI_ISL_1284803, EPI_ISL_1284805, EPI_ISL_1284810, EPI_ISL_1284817, EPI_ISL_1284818, EPI_ISL_1284820, EPI_ISL_1284825, EPI_ISL_1284826, EPI_ISL_1284829, EPI_ISL_1284830, EPI_ISL_1284834, EPI_ISL_1284838, EPI_ISL_1284839, EPI_ISL_1284841, EPI_ISL_1284845, EPI_ISL_1284846, EPI_ISL_1284854, EPI_ISL_1284856, EPI_ISL_1284857, EPI_ISL_1284861, EPI_ISL_1284863, EPI_ISL_1284864, EPI_ISL_1284870, EPI_ISL_1284878, EPI_ISL_1284879, EPI_ISL_1285446, EPI_ISL_1285485, EPI_ISL_1285534, EPI_ISL_1285556, EPI_ISL_1285565, EPI_ISL_1285590, EPI_ISL_1285612, EPI_ISL_1285623, EPI_ISL_1285632, EPI_ISL_1285674, EPI_ISL_1349728, EPI_ISL_1349730, EPI_ISL_1349732, EPI_ISL_1349737, EPI_ISL_1349739, EPI_ISL_1349740, EPI_ISL_1349743, EPI_ISL_1349744, EPI_ISL_1349745, EPI_ISL_1349746, EPI_ISL_1349747, EPI_ISL_1349748, EPI_ISL_1349749, EPI_ISL_1349751, EPI_ISL_1349753, EPI_ISL_1349755, EPI_ISL_1349756, EPI_ISL_1349757, EPI_ISL_1349758, EPI_ISL_1349760, EPI_ISL_1349763, EPI_ISL_1349767, EPI_ISL_1349768, EPI_ISL_1349769, EPI_ISL_1349770, EPI_ISL_1349771, EPI_ISL_1349775, EPI_ISL_1349778, EPI_ISL_1349780, EPI_ISL_1349781, EPI_ISL_1349782, EPI_ISL_1349784, EPI_ISL_1349785, EPI_ISL_1349788, EPI_ISL_1349789, EPI_ISL_1349790, EPI_ISL_1349796, EPI_ISL_1349799, EPI_ISL_1349801, EPI_ISL_1349802, EPI_ISL_1349804, EPI_ISL_1349805, EPI_ISL_1349806, EPI_ISL_1349807, EPI_ISL_1349808, EPI_ISL_1349810, EPI_ISL_1349811, EPI_ISL_1349813, EPI_ISL_1349814, EPI_ISL_1349815, EPI_ISL_1349816, EPI_ISL_1349817, EPI_ISL_1349818, EPI_ISL_1349823, EPI_ISL_1349825, EPI_ISL_1349826, EPI_ISL_1349827, EPI_ISL_1349828, EPI_ISL_1349829, EPI_ISL_1349833, EPI_ISL_1349837, EPI_ISL_1349838, EPI_ISL_1349842, EPI_ISL_1349843, EPI_ISL_1349845, EPI_ISL_1349849, EPI_ISL_1349852, EPI_ISL_1349853, EPI_ISL_1349854, EPI_ISL_1349855, EPI_ISL_1349856, EPI_ISL_1349857, EPI_ISL_1349860, EPI_ISL_1349861, EPI_ISL_1349865, EPI_ISL_1349866, EPI_ISL_1349867, EPI_ISL_1349869, EPI_ISL_1349872, EPI_ISL_1349873, EPI_ISL_1349876, EPI_ISL_1349878, EPI_ISL_1349880, EPI_ISL_1349882, EPI_ISL_1349884, EPI_ISL_1349885, EPI_ISL_1349886, EPI_ISL_1349889, EPI_ISL_1349891, EPI_ISL_1349892, EPI_ISL_1349893, EPI_ISL_1349897, EPI_ISL_1349902, EPI_ISL_1349904, EPI_ISL_1349905, EPI_ISL_1349907, EPI_ISL_1349908, EPI_ISL_1349911, EPI_ISL_1349913, EPI_ISL_1349915, EPI_ISL_1349916, EPI_ISL_1349918, EPI_ISL_1349920, EPI_ISL_1349921, EPI_ISL_1349922, EPI_ISL_1349923, EPI_ISL_1349924, EPI_ISL_1349925, EPI_ISL_1349927, EPI_ISL_1349929, EPI_ISL_1349930, EPI_ISL_1349932, EPI_ISL_1349934, EPI_ISL_1349935, EPI_ISL_1349936, EPI_ISL_1349939, EPI_ISL_1349940, EPI_ISL_1349941, EPI_ISL_1349942, EPI_ISL_1349944, EPI_ISL_1349945, EPI_ISL_1349946, EPI_ISL_1349947, EPI_ISL_1349948, EPI_ISL_1349949, EPI_ISL_1349952, EPI_ISL_1349954, EPI_ISL_1349955, EPI_ISL_1349956, EPI_ISL_1349958, EPI_ISL_1349959, EPI_ISL_1349970, EPI_ISL_1349972, EPI_ISL_1349979, EPI_ISL_1349980, EPI_ISL_1349982, EPI_ISL_1349984, EPI_ISL_1349985, EPI_ISL_1349989, EPI_ISL_1349990, EPI_ISL_1349993, EPI_ISL_1349995, EPI_ISL_1349997, EPI_ISL_1350000, EPI_ISL_1350002, EPI_ISL_1350003, EPI_ISL_1350006, EPI_ISL_1350007, EPI_ISL_1350008, EPI_ISL_1350009, EPI_ISL_1350010, EPI_ISL_1350014, EPI_ISL_1350015, EPI_ISL_1350018, EPI_ISL_1350021, EPI_ISL_1350023, EPI_ISL_1350025, EPI_ISL_1350027, EPI_ISL_1350028, EPI_ISL_1350029, EPI_ISL_1350036, EPI_ISL_1350038, EPI_ISL_1350039, EPI_ISL_1350041, EPI_ISL_1350042, EPI_ISL_1350043, EPI_ISL_1350045, EPI_ISL_1350047, EPI_ISL_1350048, EPI_ISL_1350049, EPI_ISL_1350050, EPI_ISL_1350051, EPI_ISL_1350053, EPI_ISL_1350054, EPI_ISL_1350056, EPI_ISL_1350058, EPI_ISL_1350059, EPI_ISL_1350060, EPI_ISL_1350063, EPI_ISL_1350064, EPI_ISL_1350065, EPI_ISL_1350066, EPI_ISL_1350068, EPI_ISL_1350069, EPI_ISL_1350070, EPI_ISL_1350071, EPI_ISL_1350073, EPI_ISL_1350076, EPI_ISL_1350078, EPI_ISL_1350080, EPI_ISL_1353642, EPI_ISL_1353643, EPI_ISL_1353645, EPI_ISL_1353649, EPI_ISL_1353651, EPI_ISL_1353657, EPI_ISL_1353662, EPI_ISL_1353663, EPI_ISL_1353664, EPI_ISL_1353665, EPI_ISL_1353677, EPI_ISL_1353688, EPI_ISL_1353690, EPI_ISL_1353693, EPI_ISL_1353707, EPI_ISL_1353711, EPI_ISL_1353721, EPI_ISL_1353722, EPI_ISL_1353727, EPI_ISL_1353733, EPI_ISL_1353734, EPI_ISL_1353737, EPI_ISL_1353743, EPI_ISL_1353746, EPI_ISL_1353747, EPI_ISL_1353751, EPI_ISL_1353755, EPI_ISL_1353757, EPI_ISL_1353773, EPI_ISL_1353775, EPI_ISL_1353789, EPI_ISL_1353794, EPI_ISL_1353811, EPI_ISL_1353812, EPI_ISL_1353813, EPI_ISL_1353816, EPI_ISL_1353818, EPI_ISL_1353824, EPI_ISL_1353825, EPI_ISL_1353831, EPI_ISL_1353835, EPI_ISL_1353847, EPI_ISL_1353848, EPI_ISL_1353849, EPI_ISL_1353858, EPI_ISL_1353864, EPI_ISL_1353866, EPI_ISL_1353872, EPI_ISL_1353874, EPI_ISL_1353878, EPI_ISL_1353886, EPI_ISL_1353887, EPI_ISL_1353892, EPI_ISL_1353895, EPI_ISL_1353897, EPI_ISL_1353898, EPI_ISL_1353901, EPI_ISL_1353902, EPI_ISL_1353903, EPI_ISL_1353921, EPI_ISL_1353935, EPI_ISL_1353942, EPI_ISL_1353948, EPI_ISL_1353950, EPI_ISL_1353951, EPI_ISL_1353953, EPI_ISL_1353956, EPI_ISL_1353967, EPI_ISL_1353971, EPI_ISL_1353974, EPI_ISL_1353978, EPI_ISL_1353987, EPI_ISL_1354015, EPI_ISL_1354017, EPI_ISL_1354026, EPI_ISL_1354061, EPI_ISL_1354128, EPI_ISL_1354157, EPI_ISL_1354161, EPI_ISL_1354187, EPI_ISL_1354228, EPI_ISL_1354230, EPI_ISL_1354250, EPI_ISL_1432945, EPI_ISL_1432991, EPI_ISL_1433002, EPI_ISL_1433048, EPI_ISL_1433062, EPI_ISL_1433071, EPI_ISL_1433088, EPI_ISL_1433090, EPI_ISL_1433106, EPI_ISL_1433119, EPI_ISL_1433133, EPI_ISL_1433145, EPI_ISL_1433149, EPI_ISL_1433161, EPI_ISL_1433173, EPI_ISL_1433181, EPI_ISL_1433218, EPI_ISL_1433249, EPI_ISL_1433250, EPI_ISL_1433267, EPI_ISL_1433273, EPI_ISL_1433274, EPI_ISL_1433282, EPI_ISL_1433300, EPI_ISL_1433360, EPI_ISL_1433372, EPI_ISL_1438977, EPI_ISL_1439179, EPI_ISL_1569268 | | | |
| see above | Bioscientia Labor Wermsdorf | Robert Koch Institute | |
| EPI_ISL_1284558, EPI_ISL_1284562, EPI_ISL_1284569, EPI_ISL_1284578, EPI_ISL_1284614, EPI_ISL_1284620, EPI_ISL_1284623, EPI_ISL_1284624, EPI_ISL_1284635, EPI_ISL_1284663, EPI_ISL_1284670, EPI_ISL_1284674, EPI_ISL_1284698, EPI_ISL_1284708, EPI_ISL_1284711, EPI_ISL_1284712, EPI_ISL_1284719, EPI_ISL_1284740, EPI_ISL_1284763, EPI_ISL_1284788, EPI_ISL_1284816, EPI_ISL_1284822, EPI_ISL_1284823, EPI_ISL_1284837, EPI_ISL_1284849, EPI_ISL_1353652, EPI_ISL_1353654, EPI_ISL_1353655, EPI_ISL_1353660, EPI_ISL_1353668, EPI_ISL_1353683, EPI_ISL_1353687, EPI_ISL_1353708, EPI_ISL_1353731, EPI_ISL_1353732, EPI_ISL_1353736, EPI_ISL_1353776, EPI_ISL_1353784, EPI_ISL_1353795, EPI_ISL_1353796, EPI_ISL_1353797, EPI_ISL_1353805, EPI_ISL_1353806, EPI_ISL_1353823, EPI_ISL_1353834, EPI_ISL_1353852, EPI_ISL_1353862, EPI_ISL_1353883, EPI_ISL_1353890, EPI_ISL_1353891, EPI_ISL_1353907, EPI_ISL_1353913, EPI_ISL_1353928, EPI_ISL_1353938, EPI_ISL_1353954, EPI_ISL_1353960, EPI_ISL_1353962, EPI_ISL_1353963, EPI_ISL_1353965, EPI_ISL_1353976, EPI_ISL_1353982 | | | |
| see above | Bioscientia MVZ Labor Karlsruhe GmbH | Robert Koch Institute | |
| EPI_ISL_1364967, EPI_ISL_1364987, EPI_ISL_1364988, EPI_ISL_1365014 | Bundeswehr Institute of Microbiology | Bundeswehr Institute of Microbiology | Alexandra Rehn; Enrico Georgi; Malena Bestehorn-Willmann; Markus Antwerpen; Mathias Walter; Mike Pillukat; Roman Wölfel; Sabine Zange |
| EPI_ISL_1364974, EPI_ISL_1364996, EPI_ISL_1365006, EPI_ISL_1365007, EPI_ISL_1365008 | Bundeswehrkrankenhaus Berlin | Bundeswehr Institute of Microbiology | Alexandra Rehn; Enrico Georgi; Malena Bestehorn-Willmann; Markus Antwerpen; Mathias Walter; Mike Pillukat; Roman Wölfel; Sabine Zange |
| EPI_ISL_1364992, EPI_ISL_1364993, EPI_ISL_1364994, EPI_ISL_1364995, EPI_ISL_1365020 | Bundeswehrkrankenhaus Hamburg | Bundeswehr Institute of Microbiology | Alexandra Rehn; Enrico Georgi; Malena Bestehorn-Willmann; Markus Antwerpen; Mathias Walter; Mike Pillukat; Roman Wölfel; Sabine Zange |
| EPI_ISL_1364979, EPI_ISL_1364989, EPI_ISL_1364990, EPI_ISL_1365005 | Bundeswehrkrankenhaus Ulm | Bundeswehr Institute of Microbiology | Alexandra Rehn; Enrico Georgi; Malena Bestehorn-Willmann; Markus Antwerpen; Mathias Walter; Mike Pillukat; Roman Wölfel; Sabine Zange |
| EPI_ISL_1364997, EPI_ISL_1364998, EPI_ISL_1364999, EPI_ISL_1365000, EPI_ISL_1365001, EPI_ISL_1365002, EPI_ISL_1365003, EPI_ISL_1365004, EPI_ISL_1365011, EPI_ISL_1365012, EPI_ISL_1365015, EPI_ISL_1365017 | | | |
| see above | Bundeswehrzentralkrankenhaus Koblenz | Bundeswehr Institute of Microbiology | Alexandra Rehn; Enrico Georgi; Malena Bestehorn-Willmann; Markus Antwerpen; Mathias Walter; Mike Pillukat; Roman Wölfel; Sabine Zange |
| EPI_ISL_1285343, EPI_ISL_1285346, EPI_ISL_1285351, EPI_ISL_1285352, EPI_ISL_1285354, EPI_ISL_1285359, EPI_ISL_1285362, EPI_ISL_1285364 | | | |
| see above | CeGaT Management | Robert Koch Institute | |
| EPI_ISL_1347451, EPI_ISL_1347452, EPI_ISL_1347453, EPI_ISL_1347454, EPI_ISL_1347455, EPI_ISL_1347456, EPI_ISL_1347457, EPI_ISL_1347458, EPI_ISL_1347459, EPI_ISL_1347460, EPI_ISL_1347461, EPI_ISL_1347462, EPI_ISL_1347464, EPI_ISL_1347465, EPI_ISL_1347466, EPI_ISL_1347467, EPI_ISL_1347468, EPI_ISL_1347469, EPI_ISL_1347470, EPI_ISL_1347711, EPI_ISL_1347713, EPI_ISL_1347714, EPI_ISL_1347716, EPI_ISL_1347717, EPI_ISL_1347720, EPI_ISL_1347721, EPI_ISL_1347722, EPI_ISL_1347723, EPI_ISL_1347724, EPI_ISL_1347725, EPI_ISL_1347726, EPI_ISL_1347727, EPI_ISL_1347729, EPI_ISL_1347731, EPI_ISL_1347732, EPI_ISL_1347734, EPI_ISL_1347755, EPI_ISL_1347756, EPI_ISL_1347757, EPI_ISL_1347758, EPI_ISL_1347759, EPI_ISL_1347760, EPI_ISL_1347761, EPI_ISL_1347762, EPI_ISL_1347763, EPI_ISL_1347764, EPI_ISL_1347765, EPI_ISL_1347766, EPI_ISL_1347767, EPI_ISL_1347768, EPI_ISL_1347769, EPI_ISL_1347770, EPI_ISL_1347771, EPI_ISL_1347772, EPI_ISL_1347773, EPI_ISL_1347774, EPI_ISL_1347775, EPI_ISL_1347776, EPI_ISL_1347790, EPI_ISL_1347791, EPI_ISL_1347792, EPI_ISL_1347793, EPI_ISL_1347794, EPI_ISL_1347795, EPI_ISL_1347796, EPI_ISL_1347797, EPI_ISL_1347798, EPI_ISL_1347799, EPI_ISL_1347800, EPI_ISL_1347801, EPI_ISL_1347802, EPI_ISL_1347803, EPI_ISL_1347804, EPI_ISL_1347805, EPI_ISL_1347806, EPI_ISL_1347807, EPI_ISL_1347808, EPI_ISL_1347809, EPI_ISL_1347810, EPI_ISL_1347811, EPI_ISL_1347812, EPI_ISL_1347813, EPI_ISL_1347814, EPI_ISL_1347815, EPI_ISL_1347816, EPI_ISL_1347817, EPI_ISL_1347818, EPI_ISL_1347819, EPI_ISL_1347820, EPI_ISL_1347951, EPI_ISL_1347952, EPI_ISL_1347953, EPI_ISL_1347954, EPI_ISL_1347955, EPI_ISL_1347956, EPI_ISL_1347957, EPI_ISL_1347958, EPI_ISL_1347960, EPI_ISL_1347961, EPI_ISL_1347962, EPI_ISL_1347964, EPI_ISL_1347965, EPI_ISL_1347966, EPI_ISL_1347967, EPI_ISL_1347993, EPI_ISL_1347994, EPI_ISL_1347995, EPI_ISL_1347996, EPI_ISL_1347997, EPI_ISL_1347998, EPI_ISL_1347999, EPI_ISL_1348000, EPI_ISL_1348002, EPI_ISL_1348003, EPI_ISL_1348004, EPI_ISL_1348005, EPI_ISL_1348006, EPI_ISL_1348007, EPI_ISL_1348008, EPI_ISL_1348009, EPI_ISL_1348010, EPI_ISL_1348011, EPI_ISL_1348012, EPI_ISL_1348013, EPI_ISL_1348014, EPI_ISL_1348015, EPI_ISL_1348016, EPI_ISL_1348017, EPI_ISL_1348018, EPI_ISL_1348019, EPI_ISL_1348020, EPI_ISL_1348021, EPI_ISL_1348022, EPI_ISL_1348023, EPI_ISL_1348024, EPI_ISL_1348025, EPI_ISL_1348026, EPI_ISL_1348027, EPI_ISL_1348028, EPI_ISL_1348029, EPI_ISL_1348030, EPI_ISL_1348031, EPI_ISL_1348032, EPI_ISL_1348033, EPI_ISL_1988683, EPI_ISL_1988687, EPI_ISL_1988688, EPI_ISL_1989635, EPI_ISL_1989636, EPI_ISL_1989637, EPI_ISL_1989638, EPI_ISL_1989639, EPI_ISL_1989640, EPI_ISL_1990068, EPI_ISL_1990069, EPI_ISL_1993606 | | | |
| see above | Center of Medical Microbiology, Virology, and Hospital Hygiene, University of Duesseldorf | Center of Medical Microbiology, Virology, and Hospital Hygiene, University of Duesseldorf | Alexander Dilthey; Andreas Walker; Daniel Strelow; Jessica Nicolai; Jörg Timm; Klaus Pfeffer; Lisanna Hülse; Malte Kohns Vasconcelos; Maximilian Damagnez; Nadine Lübke; Tobias Wienemann; Torsten Houwaart |
| EPI_ISL_1843813, EPI_ISL_1843829, EPI_ISL_1843830, EPI_ISL_1843859, EPI_ISL_1843959, EPI_ISL_1843970, EPI_ISL_2108503, EPI_ISL_2108509, EPI_ISL_2108515, EPI_ISL_2108563, EPI_ISL_2108568, EPI_ISL_2108582, EPI_ISL_2108600, EPI_ISL_2108601, EPI_ISL_2108614, EPI_ISL_2108628, EPI_ISL_2108695, EPI_ISL_2108702 | | | |
| see above | Centogene; Dr. Bauer Laboratoriums GmbH | Robert Koch Institute | |
| EPI_ISL_1280590, EPI_ISL_1280591, EPI_ISL_1280592, EPI_ISL_1280593, EPI_ISL_1280594, EPI_ISL_1280595, EPI_ISL_1280596, EPI_ISL_1280597, EPI_ISL_1280598, EPI_ISL_1280599, EPI_ISL_1280600, EPI_ISL_1349415, EPI_ISL_1349418, EPI_ISL_1349420, EPI_ISL_1349422, EPI_ISL_1349424 | | | |
| see above | Chemische und Veterinäruntersuchungsamt Münsterland-Emscher-Lippe | Robert Koch Institute | |
| EPI_ISL_1571971, EPI_ISL_1571972, EPI_ISL_1571973, EPI_ISL_1571977, EPI_ISL_1571987, EPI_ISL_1849040 | Clemenshospital - Alexianer Clemenshospital MÃ¼nster | Robert Koch Institute | |
| EPI_ISL_1563935, EPI_ISL_1563936, EPI_ISL_1563937, EPI_ISL_1563939, EPI_ISL_1563941, EPI_ISL_1563942, EPI_ISL_1563944, EPI_ISL_1563946, EPI_ISL_1563948, EPI_ISL_1563949, EPI_ISL_1563951, EPI_ISL_1563953, EPI_ISL_1563954, EPI_ISL_1563955, EPI_ISL_1563956, EPI_ISL_1563958, EPI_ISL_1563959, EPI_ISL_1642750 | | | |
| see above | Corona-Testzentrum ifp Institut fÃ¼r ProduktqualitÃ¤t GmbH | Robert Koch Institute | |
| EPI_ISL_1642766, EPI_ISL_1642772, EPI_ISL_1642775, EPI_ISL_1642783, EPI_ISL_1642785, EPI_ISL_1642794, EPI_ISL_1642798, EPI_ISL_1642799, EPI_ISL_1642802, EPI_ISL_1642808, EPI_ISL_1642815, EPI_ISL_1642821 | | | |
| see above | Department fÃ¼r Labormedizin Abteilung III Bereich Molekulare Diagnostik UniversitÃ¤tsklinikum Halle(Saale) | Robert Koch Institute | |
| EPI_ISL_1284948, EPI_ISL_1284949, EPI_ISL_1284950, EPI_ISL_1284951, EPI_ISL_1284952, EPI_ISL_1284953, EPI_ISL_1284954, EPI_ISL_1284955, EPI_ISL_1284956, EPI_ISL_1284957, EPI_ISL_1284958, EPI_ISL_1284959, EPI_ISL_1284960, EPI_ISL_1284961, EPI_ISL_1284962, EPI_ISL_1284963, EPI_ISL_1284964, EPI_ISL_1284965, EPI_ISL_1284966, EPI_ISL_1284967, EPI_ISL_1284968, EPI_ISL_1284969, EPI_ISL_1284970, EPI_ISL_1284971, EPI_ISL_1284972, EPI_ISL_1284973, EPI_ISL_1284974, EPI_ISL_1284976, EPI_ISL_1284977, EPI_ISL_1284978, EPI_ISL_1284979, EPI_ISL_1284980, EPI_ISL_1284981, EPI_ISL_1284982, EPI_ISL_1284983, EPI_ISL_1284984, EPI_ISL_1284985, EPI_ISL_1284986, EPI_ISL_1284987, EPI_ISL_1284988, EPI_ISL_1284989, EPI_ISL_1284991, EPI_ISL_1284992, EPI_ISL_1284993, EPI_ISL_1284994, EPI_ISL_1284995, EPI_ISL_1284996, EPI_ISL_1284997, EPI_ISL_1284998, EPI_ISL_1284999, EPI_ISL_1285000, EPI_ISL_1285002, EPI_ISL_1285003, EPI_ISL_1285004, EPI_ISL_1285005, EPI_ISL_1285007, EPI_ISL_1285008, EPI_ISL_1285009, EPI_ISL_1285010, EPI_ISL_1285011, EPI_ISL_1285012, EPI_ISL_1285013, EPI_ISL_1285014, EPI_ISL_1285015, EPI_ISL_1285016, EPI_ISL_1285017, EPI_ISL_1285018, EPI_ISL_1285019, EPI_ISL_1285020, EPI_ISL_1285021, EPI_ISL_1285022, EPI_ISL_1285023, EPI_ISL_1285024, EPI_ISL_1285025, EPI_ISL_1285026, EPI_ISL_1285027, EPI_ISL_1285028, EPI_ISL_1285029, EPI_ISL_1285031, EPI_ISL_1285032, EPI_ISL_1285033, EPI_ISL_1285036, EPI_ISL_1285037, EPI_ISL_1285038, EPI_ISL_1285040, EPI_ISL_1285041, EPI_ISL_1285042, EPI_ISL_1356975, EPI_ISL_1356976, EPI_ISL_1356977, EPI_ISL_1356978, EPI_ISL_1356979, EPI_ISL_1356981, EPI_ISL_1356982, EPI_ISL_1356983, EPI_ISL_1356985, EPI_ISL_1356986, EPI_ISL_1356987, EPI_ISL_1356989, EPI_ISL_1356991, EPI_ISL_1356993, EPI_ISL_1356995, EPI_ISL_1356996, EPI_ISL_1356997, EPI_ISL_1356999, EPI_ISL_1357000, EPI_ISL_1357001, EPI_ISL_1357002, EPI_ISL_1357005, EPI_ISL_1357007, EPI_ISL_1357008, EPI_ISL_1357009, EPI_ISL_1357010, EPI_ISL_1357012, EPI_ISL_1357014, EPI_ISL_1357015, EPI_ISL_1357016, EPI_ISL_1357017, EPI_ISL_1357019, EPI_ISL_1357020, EPI_ISL_1357021, EPI_ISL_1357022, EPI_ISL_1357023, EPI_ISL_1357024, EPI_ISL_1357026, EPI_ISL_1357027, EPI_ISL_1357030, EPI_ISL_1357031, EPI_ISL_1357033, EPI_ISL_1357036, EPI_ISL_1357037, EPI_ISL_1357038, EPI_ISL_1357039, EPI_ISL_1357040, EPI_ISL_1357041, EPI_ISL_1357042, EPI_ISL_1357044, EPI_ISL_1357046, EPI_ISL_1357047, EPI_ISL_1357048, EPI_ISL_1357050, EPI_ISL_1357051, EPI_ISL_1357053, EPI_ISL_1357054, EPI_ISL_1357055, EPI_ISL_1357056, EPI_ISL_1357058, EPI_ISL_1357059, EPI_ISL_1357060, EPI_ISL_1357061, EPI_ISL_1357063, EPI_ISL_1357065, EPI_ISL_1357067, EPI_ISL_1439526, EPI_ISL_1439527, EPI_ISL_1439528, EPI_ISL_1439529, EPI_ISL_1439530, EPI_ISL_1439532, EPI_ISL_1439533, EPI_ISL_1439534, EPI_ISL_1439536, EPI_ISL_1439537, EPI_ISL_1439538, EPI_ISL_1439539, EPI_ISL_1439540, EPI_ISL_1439541, EPI_ISL_1439542, EPI_ISL_1439543, EPI_ISL_1439544, EPI_ISL_1439545, EPI_ISL_1439546, EPI_ISL_1439547, EPI_ISL_1439548, EPI_ISL_1439549, EPI_ISL_1439550, EPI_ISL_1439551, EPI_ISL_1439552, EPI_ISL_1439553, EPI_ISL_1439554, EPI_ISL_1439555, EPI_ISL_1439556, EPI_ISL_1439557, EPI_ISL_1439558, EPI_ISL_1439560, EPI_ISL_1439561, EPI_ISL_1439562, EPI_ISL_1439564, EPI_ISL_1439565, EPI_ISL_1439566, EPI_ISL_1439567, EPI_ISL_1439568, EPI_ISL_1439569, EPI_ISL_1439570, EPI_ISL_1439571, EPI_ISL_1439572, EPI_ISL_1439573, EPI_ISL_1439574, | | | |

| | | | |
|---|---|---|---|
| EPI_ISL_1439576, EPI_ISL_1439577, EPI_ISL_1439579, EPI_ISL_1439581, EPI_ISL_1439582, EPI_ISL_1439583, EPI_ISL_1439585, EPI_ISL_1439586, EPI_ISL_1439587, EPI_ISL_1439591, EPI_ISL_1439592, EPI_ISL_1439594, EPI_ISL_1439595, EPI_ISL_1439596, EPI_ISL_1439598, EPI_ISL_1439599, EPI_ISL_1439600, EPI_ISL_1439601, EPI_ISL_1439602, EPI_ISL_1439603, EPI_ISL_1439604, EPI_ISL_1439605, EPI_ISL_1439606, EPI_ISL_1439607, EPI_ISL_1439608, EPI_ISL_1439609, EPI_ISL_1439610, EPI_ISL_1439611, EPI_ISL_1439612, EPI_ISL_1439613, EPI_ISL_1439615, EPI_ISL_1439616, EPI_ISL_1439617, EPI_ISL_1439618, EPI_ISL_1439619, EPI_ISL_1439620, EPI_ISL_1439621, EPI_ISL_1439622, EPI_ISL_1439623, EPI_ISL_1439624, EPI_ISL_1440972, EPI_ISL_1440997, EPI_ISL_1572525, EPI_ISL_1572527, EPI_ISL_1572536 | | | |
| see above | Diagnosticum - Labor Neukirchen | Robert Koch Institute | |
| EPI_ISL_1285797, EPI_ISL_1285798, EPI_ISL_1285801, EPI_ISL_1285802, EPI_ISL_1285803, EPI_ISL_1285804, EPI_ISL_1285805, EPI_ISL_1285808, EPI_ISL_1285809, EPI_ISL_1285810, EPI_ISL_1285811, EPI_ISL_1285814, EPI_ISL_1285815, EPI_ISL_1285818, EPI_ISL_1285820, EPI_ISL_1285821, EPI_ISL_1285822, EPI_ISL_1285823, EPI_ISL_1285824, EPI_ISL_1285825, EPI_ISL_1285826, EPI_ISL_1285828, EPI_ISL_1285829, EPI_ISL_1285831, EPI_ISL_1285832, EPI_ISL_1285834, EPI_ISL_1285836, EPI_ISL_1285837, EPI_ISL_1285838, EPI_ISL_1285839, EPI_ISL_1435285, EPI_ISL_1435286, EPI_ISL_1435287, EPI_ISL_1435288, EPI_ISL_1435289, EPI_ISL_1435290, EPI_ISL_1435291, EPI_ISL_1568104, EPI_ISL_1568114, EPI_ISL_1568124, EPI_ISL_1568132 | | | |
| see above | Dianovis GmbH Greiz | Robert Koch Institute | |
| EPI_ISL_1285044, EPI_ISL_1285046, EPI_ISL_1285048, EPI_ISL_1285053, EPI_ISL_1285061, EPI_ISL_1285062, EPI_ISL_1285063, EPI_ISL_1285065, EPI_ISL_1285071, EPI_ISL_1285076, EPI_ISL_1285083, EPI_ISL_1285086, EPI_ISL_1285089, EPI_ISL_1285092, EPI_ISL_1285095, EPI_ISL_1285100, EPI_ISL_1285108, EPI_ISL_1285109, EPI_ISL_1285113, EPI_ISL_1285122, EPI_ISL_1285123, EPI_ISL_1285131, EPI_ISL_1285133, EPI_ISL_1285138, EPI_ISL_1285142, EPI_ISL_1285150, EPI_ISL_1285155, EPI_ISL_1285157 | | | |
| see above | ELBLAB GmbH Zentrum für LaborMedizin | Robert Koch Institute | |
| EPI_ISL_1355172, EPI_ISL_1355173, EPI_ISL_1355174, EPI_ISL_1355175, EPI_ISL_1355176, EPI_ISL_1355177, EPI_ISL_1355178, EPI_ISL_1355179, EPI_ISL_1355180, EPI_ISL_1355181, EPI_ISL_1355182, EPI_ISL_1355183, EPI_ISL_1355184, EPI_ISL_1355185, EPI_ISL_1355186, EPI_ISL_1355187, EPI_ISL_1355188, EPI_ISL_1355189, EPI_ISL_1355190, EPI_ISL_1355192, EPI_ISL_1355193, EPI_ISL_1355194, EPI_ISL_1355195, EPI_ISL_1355196, EPI_ISL_1355197, EPI_ISL_1355198, EPI_ISL_1355199, EPI_ISL_1355200, EPI_ISL_1355201, EPI_ISL_1355202, EPI_ISL_1355203, EPI_ISL_1355204, EPI_ISL_1355205, EPI_ISL_1355206, EPI_ISL_1355207, EPI_ISL_1355208, EPI_ISL_1355209, EPI_ISL_1355210, EPI_ISL_1355211, EPI_ISL_1355212, EPI_ISL_1355213, EPI_ISL_1355214, EPI_ISL_1355215, EPI_ISL_1355216, EPI_ISL_1355217, EPI_ISL_1355218, EPI_ISL_1355219, EPI_ISL_1355220, EPI_ISL_1355221, EPI_ISL_1355222, EPI_ISL_1355223, EPI_ISL_1355225, EPI_ISL_1355226, EPI_ISL_1355227, EPI_ISL_1355228, EPI_ISL_1355229, EPI_ISL_1355230, EPI_ISL_1355231, EPI_ISL_1355232, EPI_ISL_1355233, EPI_ISL_1356534, EPI_ISL_1356535, EPI_ISL_1356536, EPI_ISL_1356537, EPI_ISL_1356538, EPI_ISL_1356540, EPI_ISL_1356541, EPI_ISL_1356542, EPI_ISL_1356543, EPI_ISL_1356544, EPI_ISL_1356546, EPI_ISL_1356547, EPI_ISL_1356548, EPI_ISL_1356549, EPI_ISL_1356550, EPI_ISL_1356551, EPI_ISL_1356552, EPI_ISL_1356553, EPI_ISL_1356554, EPI_ISL_1356555, EPI_ISL_1356557, EPI_ISL_1356558, EPI_ISL_1356559, EPI_ISL_1356560, EPI_ISL_1356561, EPI_ISL_1356562, EPI_ISL_1356563, EPI_ISL_1356564, EPI_ISL_1356565, EPI_ISL_1356566, EPI_ISL_1356567, EPI_ISL_1356568, EPI_ISL_1356570, EPI_ISL_1356571, EPI_ISL_1356572, EPI_ISL_1356573, EPI_ISL_1356574, EPI_ISL_1356575, EPI_ISL_1356576, EPI_ISL_1356577, EPI_ISL_1356578, EPI_ISL_1356579, EPI_ISL_1356580, EPI_ISL_1356581, EPI_ISL_1356582, EPI_ISL_1356583, EPI_ISL_1356584, EPI_ISL_1356585, EPI_ISL_1356586, EPI_ISL_1356587, EPI_ISL_1356588, EPI_ISL_1356589, EPI_ISL_1356590, EPI_ISL_1356591, EPI_ISL_1356592, EPI_ISL_1356593, EPI_ISL_1356594, EPI_ISL_1356595, EPI_ISL_1356596, EPI_ISL_1356597, EPI_ISL_1356598, EPI_ISL_1356599, EPI_ISL_1356600, EPI_ISL_1356601, EPI_ISL_1356602, EPI_ISL_1356603, EPI_ISL_1356604, EPI_ISL_1356605, EPI_ISL_1356606, EPI_ISL_1356607, EPI_ISL_1356608, EPI_ISL_1356609, EPI_ISL_1356610, EPI_ISL_1356611, EPI_ISL_1356612, EPI_ISL_1356613, EPI_ISL_1356614, EPI_ISL_1356615, EPI_ISL_1432751, EPI_ISL_1432753, EPI_ISL_1432755, EPI_ISL_1432757, EPI_ISL_1432760, EPI_ISL_1432762, EPI_ISL_1432764, EPI_ISL_1432766, EPI_ISL_1432768, EPI_ISL_1432770, EPI_ISL_1432772, EPI_ISL_1432774, EPI_ISL_1432777, EPI_ISL_1432779, EPI_ISL_1432781, EPI_ISL_1432783, EPI_ISL_1432785, EPI_ISL_1432787, EPI_ISL_1432790, EPI_ISL_1432792, EPI_ISL_1432794, EPI_ISL_1432796, EPI_ISL_1432799, EPI_ISL_1432800, EPI_ISL_1432803, EPI_ISL_1432805, EPI_ISL_1432807, EPI_ISL_1432809, EPI_ISL_1432814, EPI_ISL_1432816, EPI_ISL_1432818, EPI_ISL_1432820, EPI_ISL_1432822, EPI_ISL_1432824, EPI_ISL_1432827, EPI_ISL_1432829, EPI_ISL_1432831, EPI_ISL_1432833, EPI_ISL_1432836, EPI_ISL_1432838, EPI_ISL_1432840, EPI_ISL_1432843, EPI_ISL_1432844, EPI_ISL_1432847, EPI_ISL_1432849, EPI_ISL_1432851, EPI_ISL_1432853, EPI_ISL_1432855, EPI_ISL_1432857, EPI_ISL_1432859, EPI_ISL_1432861, EPI_ISL_1432864, EPI_ISL_1432866, EPI_ISL_1432869, EPI_ISL_1432871, EPI_ISL_1432873, EPI_ISL_1432875, EPI_ISL_1432878, EPI_ISL_1432880, EPI_ISL_1432882, EPI_ISL_1432884, EPI_ISL_1432886, EPI_ISL_1432889, EPI_ISL_1432891, EPI_ISL_1432893, EPI_ISL_1432895, EPI_ISL_1432898, EPI_ISL_1432902, EPI_ISL_1432905, EPI_ISL_1432907, EPI_ISL_1432909, EPI_ISL_1432911, EPI_ISL_1432915, EPI_ISL_1432918, EPI_ISL_1432920, EPI_ISL_1432922, EPI_ISL_1432924, EPI_ISL_1570076, EPI_ISL_1570078, EPI_ISL_1570085, EPI_ISL_1570089, EPI_ISL_1570098, EPI_ISL_1570106, EPI_ISL_1570116, EPI_ISL_1570117, EPI_ISL_1570124, EPI_ISL_1570128, EPI_ISL_1570131, EPI_ISL_1570139, EPI_ISL_1570149, EPI_ISL_1570150, EPI_ISL_1570151, EPI_ISL_1570158, EPI_ISL_1570161, EPI_ISL_1570162, EPI_ISL_1570163, EPI_ISL_1570164, EPI_ISL_1570165, EPI_ISL_1570167, EPI_ISL_1570172, EPI_ISL_1570174, EPI_ISL_1570177, EPI_ISL_1570178, EPI_ISL_1570179, EPI_ISL_1570182, EPI_ISL_1570185, EPI_ISL_1570187, EPI_ISL_1570191, EPI_ISL_1570193, EPI_ISL_1570194, EPI_ISL_1570195, EPI_ISL_1570197, EPI_ISL_1570198, EPI_ISL_1570199, EPI_ISL_1570200, EPI_ISL_1570201, EPI_ISL_1570203, EPI_ISL_1570205, EPI_ISL_1570208, EPI_ISL_1570212, EPI_ISL_1570213, EPI_ISL_1570216, EPI_ISL_1570219, EPI_ISL_1570226, EPI_ISL_1570228, EPI_ISL_1570229, EPI_ISL_1570231, EPI_ISL_1570232, EPI_ISL_1570235, EPI_ISL_1570236, EPI_ISL_1570237, EPI_ISL_1570240, EPI_ISL_1570241, EPI_ISL_1570242, EPI_ISL_1570243, EPI_ISL_1570244, EPI_ISL_1570245, EPI_ISL_1570246, EPI_ISL_1570247, EPI_ISL_1570248, EPI_ISL_1570249, EPI_ISL_1570251, EPI_ISL_1570252, EPI_ISL_1570253, EPI_ISL_1570254, EPI_ISL_1570255, EPI_ISL_1570256, EPI_ISL_1570257, EPI_ISL_1570259, EPI_ISL_1570260, EPI_ISL_1570261, EPI_ISL_1570262, EPI_ISL_1570263, EPI_ISL_1570264, EPI_ISL_1570265, EPI_ISL_1570266, EPI_ISL_1570267, EPI_ISL_1570268, EPI_ISL_1570269, EPI_ISL_1570270, EPI_ISL_1570271, EPI_ISL_1570272, EPI_ISL_1570273, EPI_ISL_1570274, EPI_ISL_1570277, EPI_ISL_1570278, EPI_ISL_1570279, EPI_ISL_1570280, EPI_ISL_1570281, EPI_ISL_1570283, EPI_ISL_1570284, EPI_ISL_1570285, EPI_ISL_1570286, EPI_ISL_1570288, EPI_ISL_1570289, EPI_ISL_1570290, EPI_ISL_1570291, EPI_ISL_1570292, EPI_ISL_1570293, EPI_ISL_1570294, EPI_ISL_1570295, EPI_ISL_1570296, EPI_ISL_1570297, EPI_ISL_1570298, EPI_ISL_1570299, EPI_ISL_1570300, EPI_ISL_1570301, EPI_ISL_1570302, EPI_ISL_1570303, EPI_ISL_1570304, EPI_ISL_1570305, EPI_ISL_1570307, EPI_ISL_1570308, EPI_ISL_1570309, EPI_ISL_1570311, EPI_ISL_1570312 | | | |
| see above | Eurofins LifeCodexx GmbH | Robert Koch Institute | |
| EPI_ISL_1563759, EPI_ISL_1563762, EPI_ISL_1563763, EPI_ISL_1563769, EPI_ISL_1563770, EPI_ISL_1563771, EPI_ISL_1563773, EPI_ISL_1563774, EPI_ISL_1563806, EPI_ISL_1563807, EPI_ISL_1563808, EPI_ISL_1563809, EPI_ISL_1563810, EPI_ISL_1563811, EPI_ISL_1563812, EPI_ISL_1563813, EPI_ISL_1563814, EPI_ISL_1563815, EPI_ISL_1563816, EPI_ISL_1563817 | | | |
| see above | Fraunhofer-Institut fÃ¼r Zelltherapie und Immunologie | Robert Koch Institute | |
| EPI_ISL_1268661, EPI_ISL_1524890, EPI_ISL_1524892, EPI_ISL_1524894, EPI_ISL_1524895 | Hannover Medical School, Institute of Virology | Hannover Medical School, Institute of Virology | Lars Steinbrück |
| EPI_ISL_1563744, EPI_ISL_1563745, EPI_ISL_1563746, EPI_ISL_1563747 | Hufeland Klinikum GmbH | Robert Koch Institute | |
| EPI_ISL_1725530, EPI_ISL_1725533, EPI_ISL_1725538, EPI_ISL_1725539, EPI_ISL_1725540, EPI_ISL_1725546, EPI_ISL_1725548, EPI_ISL_1725555, EPI_ISL_1725556, EPI_ISL_1725557, EPI_ISL_1725560, EPI_ISL_1725561, EPI_ISL_1725564, EPI_ISL_1725566, EPI_ISL_1725567, EPI_ISL_1725568, EPI_ISL_1725569, EPI_ISL_1725571, EPI_ISL_1725572, EPI_ISL_1725573, EPI_ISL_1725576, EPI_ISL_1725579, EPI_ISL_1725581, EPI_ISL_1725583, EPI_ISL_1725585, EPI_ISL_1725587, EPI_ISL_1725588, EPI_ISL_1725596 | | | |
| see above | IFLb Laboratoriumsmedizin Berlin GmbH | Robert Koch Institute | |
| EPI_ISL_1282310, EPI_ISL_1282312, EPI_ISL_1282313, EPI_ISL_1282319, EPI_ISL_1282320, EPI_ISL_1282325, EPI_ISL_1282326, EPI_ISL_1282330, EPI_ISL_1282331, EPI_ISL_1282339, EPI_ISL_1282341, EPI_ISL_1282364, EPI_ISL_1282371, EPI_ISL_1282373, EPI_ISL_1350107, EPI_ISL_1350110, EPI_ISL_1350118, EPI_ISL_1350120, EPI_ISL_1350124, EPI_ISL_1350127, EPI_ISL_1350129, EPI_ISL_1350131, EPI_ISL_1350132, EPI_ISL_1350134, EPI_ISL_1350135, EPI_ISL_1350144, EPI_ISL_1350146, EPI_ISL_1350152, EPI_ISL_1350159 | | | |
| see above | IMD - Institut für Medizinische Diagnostik Berlin-Potsdam | Robert Koch Institute | |
| EPI_ISL_1282332, EPI_ISL_1282335, EPI_ISL_1282337, EPI_ISL_1282356, EPI_ISL_1282365, EPI_ISL_1282366, EPI_ISL_1282369, EPI_ISL_1285734, EPI_ISL_1285736, EPI_ISL_1285739, EPI_ISL_1285741, EPI_ISL_1285744, EPI_ISL_1285746, EPI_ISL_1285747, EPI_ISL_1285748, EPI_ISL_1285750, EPI_ISL_1285758, EPI_ISL_1285760, EPI_ISL_1285764, EPI_ISL_1285766, EPI_ISL_1285768, EPI_ISL_1285769, EPI_ISL_1285770, EPI_ISL_1285772, EPI_ISL_1285773, EPI_ISL_1285774, EPI_ISL_1285775, EPI_ISL_1285779, EPI_ISL_1285780, EPI_ISL_1285787, EPI_ISL_1285788, EPI_ISL_1285790, EPI_ISL_1285794, EPI_ISL_1285795, EPI_ISL_1354283, EPI_ISL_1354284, EPI_ISL_1354286, EPI_ISL_1354287, EPI_ISL_1354290, EPI_ISL_1354291, EPI_ISL_1354292, EPI_ISL_1354295, EPI_ISL_1354296, EPI_ISL_1354297, EPI_ISL_1354298, EPI_ISL_1354301, EPI_ISL_1354302, EPI_ISL_1354307, EPI_ISL_1354308, EPI_ISL_1354309, EPI_ISL_1354310, EPI_ISL_1354312, EPI_ISL_1354313, EPI_ISL_1354317, EPI_ISL_1354318, EPI_ISL_1354320, EPI_ISL_1354321, EPI_ISL_1354322, EPI_ISL_1354323, EPI_ISL_1354328, EPI_ISL_1354330, EPI_ISL_1354333, EPI_ISL_1354334, EPI_ISL_1354338, EPI_ISL_1354341, EPI_ISL_1354342, EPI_ISL_1354343, EPI_ISL_1354344, EPI_ISL_1354345, EPI_ISL_1354346, EPI_ISL_1354347, EPI_ISL_1354348, EPI_ISL_1354350, EPI_ISL_1354351, EPI_ISL_1354354, EPI_ISL_1354355, EPI_ISL_1354356, EPI_ISL_1354357, EPI_ISL_1354358, EPI_ISL_1354360, EPI_ISL_1354361, EPI_ISL_1354363 | | | |
| see above | IMD - Labor Oderland | Robert Koch Institute | |
| EPI_ISL_1282317, EPI_ISL_1282379, EPI_ISL_1350117, EPI_ISL_1350119, EPI_ISL_1350137, EPI_ISL_1350138, EPI_ISL_1350156, EPI_ISL_1350157, EPI_ISL_1435854, EPI_ISL_1435876, EPI_ISL_1435891 | | | |
| see above | IMD - MVZ Labor Greifswald | Robert Koch Institute | |
| EPI_ISL_1350109, EPI_ISL_1350111, EPI_ISL_1350112, EPI_ISL_1350114, EPI_ISL_1350121, EPI_ISL_1350123, EPI_ISL_1350126, EPI_ISL_1350133, EPI_ISL_1350139, EPI_ISL_1350142, EPI_ISL_1350147, EPI_ISL_1350149, EPI_ISL_1350151, EPI_ISL_1350154, EPI_ISL_1354288, EPI_ISL_1354303, EPI_ISL_1354311, EPI_ISL_1354332 | | | |
| see above | IMD - MVZ Labor Martinsried | Robert Koch Institute | |
| EPI_ISL_1285738, EPI_ISL_1285749, EPI_ISL_1285751, EPI_ISL_1285752, EPI_ISL_1285753, EPI_ISL_1285765, EPI_ISL_1285771, EPI_ISL_1285776, EPI_ISL_1285777, EPI_ISL_1285786, EPI_ISL_1285791, EPI_ISL_1285796, EPI_ISL_1354282, EPI_ISL_1354299, EPI_ISL_1354306, EPI_ISL_1354316, EPI_ISL_1354339, EPI_ISL_1354353 | | | |
| see above | IMD - Medizinisches Labor Rostock | Robert Koch Institute | |
| EPI_ISL_1216870, EPI_ISL_1216905, EPI_ISL_1216909, EPI_ISL_1216920, EPI_ISL_1216924, EPI_ISL_1216929, EPI_ISL_1216983, EPI_ISL_1216986, EPI_ISL_1216989, EPI_ISL_1280680, EPI_ISL_1280683, EPI_ISL_1280693, EPI_ISL_1280694, EPI_ISL_1280696, EPI_ISL_1280717, EPI_ISL_1284516, EPI_ISL_1284518, EPI_ISL_1284520, EPI_ISL_1284523, EPI_ISL_1284528, EPI_ISL_1284538, EPI_ISL_1284541, EPI_ISL_1284547, EPI_ISL_1284549, EPI_ISL_1284553, EPI_ISL_1349436, EPI_ISL_1349437, EPI_ISL_1349439, EPI_ISL_1349440, EPI_ISL_1349441, EPI_ISL_1349442, EPI_ISL_1349443, EPI_ISL_1349444, EPI_ISL_1349445, EPI_ISL_1349446, EPI_ISL_1349447, EPI_ISL_1349448, EPI_ISL_1349449, EPI_ISL_1349450, EPI_ISL_1349451, EPI_ISL_1349452, EPI_ISL_1349453, EPI_ISL_1349454, EPI_ISL_1349455, EPI_ISL_1349456, EPI_ISL_1349457, EPI_ISL_1349458, EPI_ISL_1349459, EPI_ISL_1349460, EPI_ISL_1349461, EPI_ISL_1349462, EPI_ISL_1349463, EPI_ISL_1349464, EPI_ISL_1349465, EPI_ISL_1349466, EPI_ISL_1349468, EPI_ISL_1349469, EPI_ISL_1349470, EPI_ISL_1349471, EPI_ISL_1349472, EPI_ISL_1349473, EPI_ISL_1349474, EPI_ISL_1349475, EPI_ISL_1349476, EPI_ISL_1349477, EPI_ISL_1349478, EPI_ISL_1349479, EPI_ISL_1349480, EPI_ISL_1349481, EPI_ISL_1349482, EPI_ISL_1349483, EPI_ISL_1349484, EPI_ISL_1349485, EPI_ISL_1349486, EPI_ISL_1349487, EPI_ISL_1349488, EPI_ISL_1349489, EPI_ISL_1349490, EPI_ISL_1349491, EPI_ISL_1349492, EPI_ISL_1349493 | | | |
| see above | IMD Labor Frankfurt | Robert Koch Institute | |
| EPI_ISL_1568526, EPI_ISL_1639262 | Institut fÃ¼r med. Mikrobiologie u. Virologie Klinikum Oldenburg | Robert Koch Institute | |
| EPI_ISL_1217519, EPI_ISL_1217520, EPI_ISL_1284532, EPI_ISL_1284544 | Institut für Immunologie und Genetik Kaiserslautern: Medizinisches Labor Dr. med. B. Thiele | Robert Koch Institute | |
| EPI_ISL_1214986 | Institut für Klinische Chemie Transfusions- und LaboratoriumsmedizinBerufsgenossenschaftliches Universitätsklinikum Bergmannsheil | Robert Koch Institute | |
| EPI_ISL_1351183, EPI_ISL_1439748 | Institut für Medizinische Mikrobiologie - Universitätsmedizin Göttingen | Robert Koch Institute | |
| EPI_ISL_1281287 | Institut für med. Mikrobiologie u. Virologie Klinikum Oldenburg | Robert Koch Institute | |
| EPI_ISL_1279618, EPI_ISL_1279619, EPI_ISL_1279625, EPI_ISL_1279671, EPI_ISL_1279672, EPI_ISL_1279673, EPI_ISL_1279674, EPI_ISL_1279675, EPI_ISL_1279676, EPI_ISL_1279677, EPI_ISL_1279685, EPI_ISL_1279686, EPI_ISL_1279687, EPI_ISL_1279688, EPI_ISL_1279689, EPI_ISL_1279690, EPI_ISL_1279698, EPI_ISL_1279699, EPI_ISL_1279706, EPI_ISL_1279707, EPI_ISL_1279708, EPI_ISL_1279713, EPI_ISL_1279714, EPI_ISL_1279715, EPI_ISL_1279716, EPI_ISL_1279717, EPI_ISL_1279718, EPI_ISL_1279766, EPI_ISL_1279767, EPI_ISL_1279768, EPI_ISL_1279771, EPI_ISL_1279772, EPI_ISL_1279773, EPI_ISL_1279775, EPI_ISL_1279776, EPI_ISL_1279777, EPI_ISL_1279778, EPI_ISL_1279779, EPI_ISL_1279780, EPI_ISL_1279781, EPI_ISL_1279782, EPI_ISL_1279784, EPI_ISL_1279785, EPI_ISL_1279791, EPI_ISL_1279792, EPI_ISL_1279805, EPI_ISL_1279813, EPI_ISL_1279815, EPI_ISL_1279828, EPI_ISL_1279851, EPI_ISL_1279852, EPI_ISL_1279853, EPI_ISL_1279854, EPI_ISL_1279855, EPI_ISL_1279856, EPI_ISL_1279865, EPI_ISL_1279866, EPI_ISL_1279867, EPI_ISL_1279868, EPI_ISL_1279869, EPI_ISL_1279875, EPI_ISL_1279876, EPI_ISL_1279877, EPI_ISL_1279878, EPI_ISL_1279879, EPI_ISL_1361693, EPI_ISL_1361694, EPI_ISL_1361695, EPI_ISL_1361696, EPI_ISL_1361697, EPI_ISL_1361698, EPI_ISL_1361699, EPI_ISL_1361700, EPI_ISL_1361701, EPI_ISL_1361702, EPI_ISL_1361709, EPI_ISL_1361710, EPI_ISL_1361711, EPI_ISL_1361712, EPI_ISL_1361751 | | | |
| see above | Institute of Medical Microbiology and Hospital Hygiene | Institute of Medical Microbiology and Hospital Hygiene | Aljoscha Tersteegen; Prof. Dr. Achim Kaasch |
| EPI_ISL_1357648, EPI_ISL_1357658, EPI_ISL_1357659, EPI_ISL_1357660, EPI_ISL_1357661, EPI_ISL_1357662, EPI_ISL_1357663, EPI_ISL_1357664, EPI_ISL_1357665, EPI_ISL_1357667, EPI_ISL_1357668, EPI_ISL_1357669, EPI_ISL_1357670, EPI_ISL_1357671, EPI_ISL_1357672, EPI_ISL_1357677, EPI_ISL_1357678, EPI_ISL_1357679, EPI_ISL_1357680, EPI_ISL_1357681, EPI_ISL_1357682, EPI_ISL_1357689, EPI_ISL_1357690, EPI_ISL_1588600, EPI_ISL_1588601, EPI_ISL_1588602, EPI_ISL_1588603, EPI_ISL_1588604, EPI_ISL_1588605, EPI_ISL_1588606, EPI_ISL_1588607, EPI_ISL_1588608, EPI_ISL_1588609, EPI_ISL_1588610, EPI_ISL_1588611, EPI_ISL_1804997, EPI_ISL_1804998, EPI_ISL_1804999, EPI_ISL_1805002 | | | |
| see above | Institute of Virology, Medical Center, University of Freiburg, Freiburg, Germany | Institute of Virology, Clinial Virus Genomics, Medical Center, University of Freiburg, Freiburg, Germany | Hajo Grundmann; Jonas Fuchs; Lena Jaki; Lisa Kern; Marcus Panning; Sandra Reuter |
| EPI_ISL_1715869 | Johannes Wesling Klinikum Minden | Heinrich Pette Institute, Leibniz Institute for Experimental Virology | Adam Grundhoff; Alexis Robitaille; Johannes Knobloch; Martin Aepfelbacher; Nicole Fischer; Thomas Günther |
| EPI_ISL_1284924, EPI_ISL_1284926, EPI_ISL_1284927, EPI_ISL_1284928, EPI_ISL_1284929, EPI_ISL_1284930, EPI_ISL_1284931, EPI_ISL_1284932, EPI_ISL_1284933, EPI_ISL_1284934, EPI_ISL_1284935, EPI_ISL_1284936, EPI_ISL_1284937, EPI_ISL_1284938, EPI_ISL_1284939, EPI_ISL_1284941, EPI_ISL_1284943, EPI_ISL_1284945, EPI_ISL_1284946 | | | |
| see above | KRH Klinikum Region Hannover: Institut für Med. Mikrobiologie und Krankenhaushygiene | Robert Koch Institute | |
| EPI_ISL_1464269, EPI_ISL_1715872, EPI_ISL_1715873, EPI_ISL_1715874 | Kath. Marienkrankenhaus | Heinrich Pette Institute, Leibniz Institute for Experimental Virology | Adam Grundhoff; Alexis Robitaille; Johannes Knobloch; Martin Aepfelbacher; Nicole Fischer; Thomas Günther |
| EPI_ISL_1217522, EPI_ISL_1217530, EPI_ISL_1217559, EPI_ISL_1281972 | Kath. St.-Johannes-Gesellschaft Dortmund gGmbH | Robert Koch Institute | |
| EPI_ISL_1355235, EPI_ISL_1438666, EPI_ISL_1441213 | Klinikum Darmstadt GmbH | Robert Koch Institute | |
| EPI_ISL_1218189, EPI_ISL_1218190, EPI_ISL_1218191, EPI_ISL_1218193, EPI_ISL_1218195, EPI_ISL_1218197, EPI_ISL_1218198, EPI_ISL_1282754, EPI_ISL_1282755, EPI_ISL_1282756, EPI_ISL_1282757, EPI_ISL_1282758, EPI_ISL_1282759, EPI_ISL_1282760, EPI_ISL_1282761, EPI_ISL_1282762, EPI_ISL_1282763, EPI_ISL_1282764, EPI_ISL_1285166, EPI_ISL_1285167, EPI_ISL_1285168, EPI_ISL_1285169, EPI_ISL_1285170, EPI_ISL_1285171, EPI_ISL_1285172, EPI_ISL_1285173, EPI_ISL_1285174, EPI_ISL_1285175, EPI_ISL_1285176, EPI_ISL_1349227, EPI_ISL_1349228, EPI_ISL_1349229, EPI_ISL_1349230, EPI_ISL_1349232, EPI_ISL_1349234, EPI_ISL_1349235, EPI_ISL_1349236, EPI_ISL_1349239, EPI_ISL_1349240, EPI_ISL_1349241 | | | |
| see above | Klinikum Ernst von Bergmann gemeinnützige GmbH - stationärer Bereich | Robert Koch Institute | |
| EPI_ISL_1282146, EPI_ISL_1353786, EPI_ISL_1441195, EPI_ISL_1441197 | Klinikum Fulda gAG | Robert Koch Institute | |
| EPI_ISL_1569997, EPI_ISL_1569998, EPI_ISL_1569999, EPI_ISL_1570000, EPI_ISL_1570002, EPI_ISL_1570003, EPI_ISL_1570004, EPI_ISL_1570005, EPI_ISL_1570006, EPI_ISL_1570007, EPI_ISL_1570008, EPI_ISL_1570010 | | | |
| see above | Klinikum Stuttgart | Robert Koch Institute | |
| EPI_ISL_1280666, EPI_ISL_1280667, EPI_ISL_1280669, EPI_ISL_1280671, EPI_ISL_1280673, EPI_ISL_1280676, EPI_ISL_1280678, EPI_ISL_1280679, EPI_ISL_1280685, EPI_ISL_1280686, EPI_ISL_1280688, EPI_ISL_1280689, EPI_ISL_1280692, EPI_ISL_1280698, EPI_ISL_1280701, EPI_ISL_1280702, EPI_ISL_1280704, EPI_ISL_1280705, EPI_ISL_1280706, EPI_ISL_1280708, EPI_ISL_1280711, EPI_ISL_1280712, EPI_ISL_1280716, EPI_ISL_1280720, EPI_ISL_1280722, EPI_ISL_1280724, EPI_ISL_1280726, EPI_ISL_1280727, EPI_ISL_1280728, EPI_ISL_1280729, EPI_ISL_1280730, EPI_ISL_1280731, EPI_ISL_1280732, EPI_ISL_1280733, EPI_ISL_1280734 | | | |
| see above | Klinikum der Stadt Ludwigshafen - Institut für Labordiagnostik Hygiene und Transfusionsmedizin | Robert Koch Institute | |
| EPI_ISL_1217618, EPI_ISL_1217652, EPI_ISL_1217702, EPI_ISL_1285088, EPI_ISL_1285090, EPI_ISL_1285105 | LABOR CHEMNITZ – Zentrum für Diagnostik am Klinikum Chemnitz | Robert Koch Institute | |

EPI_ISL_1217374, EPI_ISL_1217392, EPI_ISL_1282321, EPI_ISL_1282340, EPI_ISL_1282347, EPI_ISL_1282351, EPI_ISL_1282368, EPI_ISL_1282380, EPI_ISL_1285737, EPI_ISL_1285740, EPI_ISL_1285743, EPI_ISL_1285755, EPI_ISL_1285756, EPI_ISL_1285759, EPI_ISL_1285761, EPI_ISL_1285762, EPI_ISL_1285767, EPI_ISL_1285781, EPI_ISL_1285783, EPI_ISL_1285785, EPI_ISL_1285793

see above — LADR MVZ Zweigpraxis Bernau — Robert Koch Institute

EPI_ISL_1281992, EPI_ISL_1281993, EPI_ISL_1281994, EPI_ISL_1281999, EPI_ISL_1282006, EPI_ISL_1282010, EPI_ISL_1282012, EPI_ISL_1282013, EPI_ISL_1282016, EPI_ISL_1282022, EPI_ISL_1282026, EPI_ISL_1282029, EPI_ISL_1282033, EPI_ISL_1282038, EPI_ISL_1282039, EPI_ISL_1282040, EPI_ISL_1282041, EPI_ISL_1282048, EPI_ISL_1282049, EPI_ISL_1282053, EPI_ISL_1282054, EPI_ISL_1282056, EPI_ISL_1282057, EPI_ISL_1282061, EPI_ISL_1282063, EPI_ISL_1282065, EPI_ISL_1282070, EPI_ISL_1282075, EPI_ISL_1282079, EPI_ISL_1282080, EPI_ISL_1282087, EPI_ISL_1282097, EPI_ISL_1282100, EPI_ISL_1282106, EPI_ISL_1282107, EPI_ISL_1282109, EPI_ISL_1282111, EPI_ISL_1282120, EPI_ISL_1282121, EPI_ISL_1282126, EPI_ISL_1282127, EPI_ISL_1282130, EPI_ISL_1282131, EPI_ISL_1282142, EPI_ISL_1282144, EPI_ISL_1282145, EPI_ISL_1282149, EPI_ISL_1282151, EPI_ISL_1282153, EPI_ISL_1282154, EPI_ISL_1282156, EPI_ISL_1282157, EPI_ISL_1282161, EPI_ISL_1282165, EPI_ISL_1282169, EPI_ISL_1282172, EPI_ISL_1282173, EPI_ISL_1282174, EPI_ISL_1282175, EPI_ISL_1282178, EPI_ISL_1282185, EPI_ISL_1282186, EPI_ISL_1282192, EPI_ISL_1282198, EPI_ISL_1282199, EPI_ISL_1282202, EPI_ISL_1282207, EPI_ISL_1282212, EPI_ISL_1282214, EPI_ISL_1282216, EPI_ISL_1282219, EPI_ISL_1282227, EPI_ISL_1282230, EPI_ISL_1282233, EPI_ISL_1282235, EPI_ISL_1282241, EPI_ISL_1282242, EPI_ISL_1282243, EPI_ISL_1282255, EPI_ISL_1282256, EPI_ISL_1282262, EPI_ISL_1282264, EPI_ISL_1282269, EPI_ISL_1282271, EPI_ISL_1282272, EPI_ISL_1282276, EPI_ISL_1282277, EPI_ISL_1282282, EPI_ISL_1282283, EPI_ISL_1282288, EPI_ISL_1282290, EPI_ISL_1282291, EPI_ISL_1282293, EPI_ISL_1282294, EPI_ISL_1282302, EPI_ISL_1282303, EPI_ISL_1282306, EPI_ISL_1282307, EPI_ISL_1285432, EPI_ISL_1285442, EPI_ISL_1285447, EPI_ISL_1285450, EPI_ISL_1285451, EPI_ISL_1285453, EPI_ISL_1285454, EPI_ISL_1285455, EPI_ISL_1285460, EPI_ISL_1285462, EPI_ISL_1285463, EPI_ISL_1285465, EPI_ISL_1285467, EPI_ISL_1285469, EPI_ISL_1285470, EPI_ISL_1285471, EPI_ISL_1285472, EPI_ISL_1285476, EPI_ISL_1285478, EPI_ISL_1285480, EPI_ISL_1285481, EPI_ISL_1285483, EPI_ISL_1285486, EPI_ISL_1285492, EPI_ISL_1285495, EPI_ISL_1285498, EPI_ISL_1285500, EPI_ISL_1285502, EPI_ISL_1285511, EPI_ISL_1285513, EPI_ISL_1285518, EPI_ISL_1285523, EPI_ISL_1285528, EPI_ISL_1285529, EPI_ISL_1285533, EPI_ISL_1285537, EPI_ISL_1285543, EPI_ISL_1285549, EPI_ISL_1285553, EPI_ISL_1285559, EPI_ISL_1285563, EPI_ISL_1285575, EPI_ISL_1285577, EPI_ISL_1285580, EPI_ISL_1285581, EPI_ISL_1285582, EPI_ISL_1285585, EPI_ISL_1285591, EPI_ISL_1285593, EPI_ISL_1285597, EPI_ISL_1285601, EPI_ISL_1285603, EPI_ISL_1285606, EPI_ISL_1285608, EPI_ISL_1285609, EPI_ISL_1285613, EPI_ISL_1285616, EPI_ISL_1285617, EPI_ISL_1285618, EPI_ISL_1285625, EPI_ISL_1285628, EPI_ISL_1285631, EPI_ISL_1285634, EPI_ISL_1285636, EPI_ISL_1285637, EPI_ISL_1285640, EPI_ISL_1285651, EPI_ISL_1285652, EPI_ISL_1285654, EPI_ISL_1285655, EPI_ISL_1285656, EPI_ISL_1285657, EPI_ISL_1285659, EPI_ISL_1285662, EPI_ISL_1285667, EPI_ISL_1285679, EPI_ISL_1285682, EPI_ISL_1285685, EPI_ISL_1285694, EPI_ISL_1285698, EPI_ISL_1285699, EPI_ISL_1285700, EPI_ISL_1285701, EPI_ISL_1285702, EPI_ISL_1285705, EPI_ISL_1285706, EPI_ISL_1285709, EPI_ISL_1285710, EPI_ISL_1285716, EPI_ISL_1349727, EPI_ISL_1349792, EPI_ISL_1349820, EPI_ISL_1349831, EPI_ISL_1349832, EPI_ISL_1349840, EPI_ISL_1349846, EPI_ISL_1349847, EPI_ISL_1349895, EPI_ISL_1349919, EPI_ISL_1349951, EPI_ISL_1349966, EPI_ISL_1349971, EPI_ISL_1349981, EPI_ISL_1349991, EPI_ISL_1349994, EPI_ISL_1350001, EPI_ISL_1350017, EPI_ISL_1350077, EPI_ISL_1353666, EPI_ISL_1353888

see above — LabKom - Labor Augsburg MVZ GmbH — Robert Koch Institute

EPI_ISL_1285435, EPI_ISL_1285487, EPI_ISL_1285550, EPI_ISL_1285554, EPI_ISL_1285598 — LabKom - Labor Hamburg-Lübeck — Robert Koch Institute

EPI_ISL_1349750, EPI_ISL_1349754, EPI_ISL_1349762, EPI_ISL_1349793, EPI_ISL_1349800, EPI_ISL_1349803, EPI_ISL_1349839, EPI_ISL_1349844, EPI_ISL_1349851, EPI_ISL_1349863, EPI_ISL_1349864, EPI_ISL_1349868, EPI_ISL_1349870, EPI_ISL_1349874, EPI_ISL_1349883, EPI_ISL_1349894, EPI_ISL_1349896, EPI_ISL_1349906, EPI_ISL_1349909, EPI_ISL_1349917, EPI_ISL_1349931, EPI_ISL_1349933, EPI_ISL_1349938, EPI_ISL_1349962, EPI_ISL_1349976, EPI_ISL_1349983, EPI_ISL_1349999, EPI_ISL_1350005, EPI_ISL_1350013, EPI_ISL_1350032, EPI_ISL_1350033, EPI_ISL_1350040, EPI_ISL_1350046, EPI_ISL_1350055, EPI_ISL_1350062, EPI_ISL_1350067

see above — LabKom - Labor Hannover MVZ GmbH — Robert Koch Institute

EPI_ISL_1282117, EPI_ISL_1349726, EPI_ISL_1349764, EPI_ISL_1349809, EPI_ISL_1349821, EPI_ISL_1349830, EPI_ISL_1349836, EPI_ISL_1349850, EPI_ISL_1349875, EPI_ISL_1349901, EPI_ISL_1349937, EPI_ISL_1349943, EPI_ISL_1349953, EPI_ISL_1349957, EPI_ISL_1349960, EPI_ISL_1349965, EPI_ISL_1349967, EPI_ISL_1349975, EPI_ISL_1349998, EPI_ISL_1350012, EPI_ISL_1350044, EPI_ISL_1350052, EPI_ISL_1350061, EPI_ISL_1350072, EPI_ISL_1350079, EPI_ISL_1353787

see above — LabKom - Labor Mainz MVZ GmbH — Robert Koch Institute

EPI_ISL_1281995, EPI_ISL_1282011, EPI_ISL_1282014, EPI_ISL_1282020, EPI_ISL_1282024, EPI_ISL_1282027, EPI_ISL_1282032, EPI_ISL_1282034, EPI_ISL_1282035, EPI_ISL_1282042, EPI_ISL_1282052, EPI_ISL_1282055, EPI_ISL_1282058, EPI_ISL_1282059, EPI_ISL_1282067, EPI_ISL_1282068, EPI_ISL_1282069, EPI_ISL_1282076, EPI_ISL_1282078, EPI_ISL_1282083, EPI_ISL_1282084, EPI_ISL_1282085, EPI_ISL_1282093, EPI_ISL_1282094, EPI_ISL_1282096, EPI_ISL_1282099, EPI_ISL_1282112, EPI_ISL_1282116, EPI_ISL_1282119, EPI_ISL_1282122, EPI_ISL_1282124, EPI_ISL_1282135, EPI_ISL_1282140, EPI_ISL_1282150, EPI_ISL_1282176, EPI_ISL_1282177, EPI_ISL_1282183, EPI_ISL_1282184, EPI_ISL_1282187, EPI_ISL_1282188, EPI_ISL_1282190, EPI_ISL_1282194, EPI_ISL_1282195, EPI_ISL_1282204, EPI_ISL_1282205, EPI_ISL_1282206, EPI_ISL_1282208, EPI_ISL_1282213, EPI_ISL_1282217, EPI_ISL_1282218, EPI_ISL_1282229, EPI_ISL_1282231, EPI_ISL_1282304, EPI_ISL_1282305, EPI_ISL_1282308, EPI_ISL_1349731, EPI_ISL_1349736, EPI_ISL_1349741, EPI_ISL_1349752, EPI_ISL_1349779, EPI_ISL_1349795, EPI_ISL_1349877, EPI_ISL_1349898, EPI_ISL_1349900, EPI_ISL_1349964, EPI_ISL_1349969, EPI_ISL_1350022, EPI_ISL_1350026

see above — LabKom - Labor an der Salzbrücke MVZ GmbH — Robert Koch Institute

EPI_ISL_1285431, EPI_ISL_1285433, EPI_ISL_1285436, EPI_ISL_1285440, EPI_ISL_1285441, EPI_ISL_1285444, EPI_ISL_1285452, EPI_ISL_1285456, EPI_ISL_1285458, EPI_ISL_1285459, EPI_ISL_1285466, EPI_ISL_1285479, EPI_ISL_1285482, EPI_ISL_1285491, EPI_ISL_1285503, EPI_ISL_1285505, EPI_ISL_1285506, EPI_ISL_1285508, EPI_ISL_1285514, EPI_ISL_1285515, EPI_ISL_1285519, EPI_ISL_1285527, EPI_ISL_1285530, EPI_ISL_1285536, EPI_ISL_1285538, EPI_ISL_1285540, EPI_ISL_1285541, EPI_ISL_1285542, EPI_ISL_1285544, EPI_ISL_1285548, EPI_ISL_1285558, EPI_ISL_1285567, EPI_ISL_1285569, EPI_ISL_1285578, EPI_ISL_1285584, EPI_ISL_1285586, EPI_ISL_1285589, EPI_ISL_1285592, EPI_ISL_1285607, EPI_ISL_1285611, EPI_ISL_1285620, EPI_ISL_1285622, EPI_ISL_1285627, EPI_ISL_1285629, EPI_ISL_1285635, EPI_ISL_1285645, EPI_ISL_1285647, EPI_ISL_1285649, EPI_ISL_1285660, EPI_ISL_1285661, EPI_ISL_1285665, EPI_ISL_1285668, EPI_ISL_1285676, EPI_ISL_1285677, EPI_ISL_1285680, EPI_ISL_1285683, EPI_ISL_1285684, EPI_ISL_1285687, EPI_ISL_1285688, EPI_ISL_1285691, EPI_ISL_1285693, EPI_ISL_1285695, EPI_ISL_1285703, EPI_ISL_1285708, EPI_ISL_1285712, EPI_ISL_1285713, EPI_ISL_1285714, EPI_ISL_1285715, EPI_ISL_1285717

see above — LabKom - MVZ Labor Bochum MLB GmbH — Robert Koch Institute

EPI_ISL_1349729, EPI_ISL_1349733, EPI_ISL_1349734, EPI_ISL_1349735, EPI_ISL_1349759, EPI_ISL_1349761, EPI_ISL_1349772, EPI_ISL_1349773, EPI_ISL_1349774, EPI_ISL_1349834, EPI_ISL_1349835, EPI_ISL_1349862, EPI_ISL_1349881, EPI_ISL_1349887, EPI_ISL_1349899, EPI_ISL_1349912, EPI_ISL_1349914, EPI_ISL_1349973, EPI_ISL_1350004, EPI_ISL_1350031, EPI_ISL_1350034, EPI_ISL_1353673, EPI_ISL_1353678, EPI_ISL_1353705, EPI_ISL_1353709, EPI_ISL_1353728, EPI_ISL_1353767, EPI_ISL_1353785, EPI_ISL_1353865, EPI_ISL_1353877, EPI_ISL_1353899, EPI_ISL_1353914, EPI_ISL_1353929, EPI_ISL_1353937, EPI_ISL_1353955, EPI_ISL_1353959, EPI_ISL_1353988, EPI_ISL_1432930, EPI_ISL_1432965, EPI_ISL_1432987, EPI_ISL_1432989, EPI_ISL_1432993, EPI_ISL_1432998, EPI_ISL_1433033, EPI_ISL_1433068, EPI_ISL_1433075, EPI_ISL_1433076, EPI_ISL_1433079, EPI_ISL_1433093, EPI_ISL_1433099, EPI_ISL_1433110, EPI_ISL_1433114, EPI_ISL_1433123, EPI_ISL_1433204, EPI_ISL_1433241, EPI_ISL_1433272, EPI_ISL_1433332

see above — Labor 28 MVZ GmbH — Robert Koch Institute

EPI_ISL_1568532, EPI_ISL_1568533, EPI_ISL_1568535, EPI_ISL_1568541, EPI_ISL_1568543, EPI_ISL_1568545, EPI_ISL_1568546, EPI_ISL_1568547, EPI_ISL_1568550, EPI_ISL_1568551, EPI_ISL_1568556, EPI_ISL_1568557, EPI_ISL_1568558, EPI_ISL_1568559, EPI_ISL_1568560, EPI_ISL_1568561, EPI_ISL_1568562, EPI_ISL_1568563, EPI_ISL_1568564, EPI_ISL_1568565, EPI_ISL_1568566, EPI_ISL_1568567, EPI_ISL_1568568, EPI_ISL_1568569, EPI_ISL_1568570, EPI_ISL_1568572, EPI_ISL_1568573, EPI_ISL_1568574, EPI_ISL_1640624, EPI_ISL_1640625, EPI_ISL_1640626, EPI_ISL_1640627, EPI_ISL_1640628, EPI_ISL_1640629, EPI_ISL_1640630, EPI_ISL_1640631, EPI_ISL_1640632, EPI_ISL_1640633, EPI_ISL_1640636, EPI_ISL_1640637, EPI_ISL_1640638, EPI_ISL_1640639, EPI_ISL_1640640, EPI_ISL_1640641, EPI_ISL_1640642, EPI_ISL_1640644, EPI_ISL_1640646, EPI_ISL_1640647, EPI_ISL_1640648, EPI_ISL_1640649, EPI_ISL_1640651, EPI_ISL_1640652, EPI_ISL_1640653, EPI_ISL_1640654, EPI_ISL_1640655, EPI_ISL_1640657, EPI_ISL_1640658, EPI_ISL_1640659, EPI_ISL_1640660, EPI_ISL_1640661, EPI_ISL_1640662, EPI_ISL_1640663, EPI_ISL_1640664, EPI_ISL_1640665, EPI_ISL_1640666, EPI_ISL_1640667, EPI_ISL_1640668, EPI_ISL_1640669, EPI_ISL_1846774, EPI_ISL_1846777, EPI_ISL_1846783, EPI_ISL_1846785, EPI_ISL_1846791, EPI_ISL_1846792, EPI_ISL_1846802, EPI_ISL_1846804

see above — Labor Becker & Kollegen (Standort MÃ¼nchen) — Robert Koch Institute

EPI_ISL_1982493, EPI_ISL_1982495, EPI_ISL_1982516, EPI_ISL_1982531, EPI_ISL_1982533, EPI_ISL_1982543, EPI_ISL_1982552, EPI_ISL_1982560, EPI_ISL_1982570, EPI_ISL_1982575, EPI_ISL_1982581, EPI_ISL_1982587, EPI_ISL_1982593, EPI_ISL_1982597, EPI_ISL_1982599, EPI_ISL_1982602, EPI_ISL_1982611, EPI_ISL_1982614, EPI_ISL_1984770, EPI_ISL_1986328, EPI_ISL_1986343, EPI_ISL_1986351, EPI_ISL_1986357, EPI_ISL_1986364, EPI_ISL_1986371, EPI_ISL_1986394, EPI_ISL_1986406, EPI_ISL_1986408, EPI_ISL_1986437, EPI_ISL_1986439, EPI_ISL_1986445, EPI_ISL_1986447, EPI_ISL_1986448, EPI_ISL_1986450, EPI_ISL_1986452, EPI_ISL_1986456, EPI_ISL_1986462, EPI_ISL_1986477, EPI_ISL_1986487, EPI_ISL_1988173, EPI_ISL_1988178, EPI_ISL_1988179, EPI_ISL_1988180, EPI_ISL_1988181, EPI_ISL_1988183, EPI_ISL_1988184, EPI_ISL_1988185, EPI_ISL_1988186, EPI_ISL_1988187, EPI_ISL_1988188, EPI_ISL_1988189, EPI_ISL_1988190, EPI_ISL_1988191, EPI_ISL_1988192, EPI_ISL_1988193, EPI_ISL_1988194, EPI_ISL_1988196, EPI_ISL_1988198, EPI_ISL_1988199, EPI_ISL_1988200, EPI_ISL_1988201, EPI_ISL_1988202, EPI_ISL_1988203, EPI_ISL_1988204, EPI_ISL_1988205, EPI_ISL_1988208, EPI_ISL_1988210, EPI_ISL_1988211, EPI_ISL_1988212, EPI_ISL_1988213, EPI_ISL_1988214, EPI_ISL_1988215, EPI_ISL_1988216, EPI_ISL_1988217, EPI_ISL_1988218, EPI_ISL_1988219, EPI_ISL_1988220, EPI_ISL_1988221, EPI_ISL_1988222, EPI_ISL_1988223, EPI_ISL_1988224, EPI_ISL_1988225, EPI_ISL_1988226, EPI_ISL_1988227, EPI_ISL_1988228, EPI_ISL_1988229, EPI_ISL_1988230, EPI_ISL_1988231, EPI_ISL_1988233, EPI_ISL_1988234, EPI_ISL_1988236, EPI_ISL_1988237, EPI_ISL_1988238, EPI_ISL_1988239, EPI_ISL_1988240, EPI_ISL_1988241, EPI_ISL_1988242, EPI_ISL_1988243, EPI_ISL_1988244, EPI_ISL_1988245, EPI_ISL_1988246, EPI_ISL_1989272, EPI_ISL_1989282, EPI_ISL_1989290, EPI_ISL_1989297, EPI_ISL_1989298, EPI_ISL_1989301, EPI_ISL_1989317, EPI_ISL_1989334, EPI_ISL_1989336

see above — Labor Berlin Charité Vivantes GmbH / Institut für Virologie — Charité Universitätsmedizin Berlin, Institut für Virologie/Labor Berlin — Barbara Mühlemann; Christian Drosten; Christine Stephan; Peter Menzel; Rolf Schwarzer; Terry Jones; Victor M Corman

EPI_ISL_1570496, EPI_ISL_1570508, EPI_ISL_1570511, EPI_ISL_1570512, EPI_ISL_1570524, EPI_ISL_1570529, EPI_ISL_1570530, EPI_ISL_1570534, EPI_ISL_1570538, EPI_ISL_1570543, EPI_ISL_1570553, EPI_ISL_1570555, EPI_ISL_1570559, EPI_ISL_1570563, EPI_ISL_1570572, EPI_ISL_1570575, EPI_ISL_1570576, EPI_ISL_1570577, EPI_ISL_1570600, EPI_ISL_1570615, EPI_ISL_1570632, EPI_ISL_1570643, EPI_ISL_1570644

see above — Labor Blackholm MVZ — Robert Koch Institute

EPI_ISL_1464417, EPI_ISL_1464418, EPI_ISL_1464447, EPI_ISL_1464461, EPI_ISL_1464462, EPI_ISL_1464463, EPI_ISL_1464464, EPI_ISL_1464465, EPI_ISL_1464466, EPI_ISL_1464467, EPI_ISL_1464469, EPI_ISL_1464470, EPI_ISL_1464471, EPI_ISL_1464472, EPI_ISL_1464473, EPI_ISL_1464474, EPI_ISL_1464475, EPI_ISL_1464476, EPI_ISL_1464477, EPI_ISL_1464478, EPI_ISL_1464479, EPI_ISL_1464480, EPI_ISL_1464481, EPI_ISL_1464482, EPI_ISL_1464483, EPI_ISL_1464484, EPI_ISL_1464485, EPI_ISL_1464486, EPI_ISL_1464487, EPI_ISL_1464488, EPI_ISL_1464489, EPI_ISL_1464490, EPI_ISL_1464491, EPI_ISL_1464492, EPI_ISL_1464493, EPI_ISL_1464494, EPI_ISL_1464495, EPI_ISL_1464496, EPI_ISL_1464497, EPI_ISL_1464498, EPI_ISL_1464499, EPI_ISL_1464500, EPI_ISL_1464501, EPI_ISL_1464502, EPI_ISL_1464503, EPI_ISL_1464504, EPI_ISL_1464505, EPI_ISL_1464506, EPI_ISL_1464507, EPI_ISL_1464508, EPI_ISL_1464509, EPI_ISL_1464510, EPI_ISL_1464511, EPI_ISL_1464512, EPI_ISL_1464513, EPI_ISL_1464514, EPI_ISL_1464515, EPI_ISL_1464516, EPI_ISL_1464517, EPI_ISL_1464518, EPI_ISL_1464519, EPI_ISL_1464520, EPI_ISL_1464521, EPI_ISL_1464522, EPI_ISL_1464523, EPI_ISL_1464524, EPI_ISL_1464525, EPI_ISL_1464526, EPI_ISL_1464527, EPI_ISL_1464528, EPI_ISL_1464529, EPI_ISL_1464530, EPI_ISL_1464531, EPI_ISL_1464532, EPI_ISL_1464533, EPI_ISL_1464534, EPI_ISL_1464535, EPI_ISL_1464536, EPI_ISL_1464537, EPI_ISL_1464538, EPI_ISL_1464539, EPI_ISL_1464540, EPI_ISL_1464541, EPI_ISL_1464542, EPI_ISL_1464543, EPI_ISL_1464544, EPI_ISL_1464545, EPI_ISL_1464546, EPI_ISL_1464547, EPI_ISL_1464548, EPI_ISL_1464549, EPI_ISL_1464550, EPI_ISL_1464551, EPI_ISL_1464552, EPI_ISL_1464553, EPI_ISL_1464554, EPI_ISL_1464555, EPI_ISL_1464556, EPI_ISL_1464557, EPI_ISL_1464558, EPI_ISL_1464559, EPI_ISL_1464560, EPI_ISL_1464561, EPI_ISL_1464562, EPI_ISL_1464563, EPI_ISL_1464564, EPI_ISL_1464565, EPI_ISL_1464566, EPI_ISL_1464567, EPI_ISL_1464568, EPI_ISL_1464569, EPI_ISL_1464570, EPI_ISL_1464571, EPI_ISL_1464572, EPI_ISL_1464573, EPI_ISL_1464574, EPI_ISL_1464575, EPI_ISL_1464576, EPI_ISL_1464577, EPI_ISL_1464578, EPI_ISL_1464579, EPI_ISL_1464580, EPI_ISL_1464581, EPI_ISL_1464582, EPI_ISL_1715875, EPI_ISL_1715876, EPI_ISL_1715877, EPI_ISL_1715878, EPI_ISL_1715879, EPI_ISL_1715880, EPI_ISL_1715881, EPI_ISL_1715882, EPI_ISL_1715883, EPI_ISL_1715884, EPI_ISL_1715885, EPI_ISL_1715886, EPI_ISL_1715887, EPI_ISL_1715888, EPI_ISL_1715889, EPI_ISL_1715890, EPI_ISL_1715891, EPI_ISL_1715892, EPI_ISL_1715893, EPI_ISL_1715894, EPI_ISL_1715895, EPI_ISL_1715896, EPI_ISL_1715897, EPI_ISL_1715898, EPI_ISL_1715899, EPI_ISL_1715900, EPI_ISL_1715901, EPI_ISL_1715902, EPI_ISL_1715903, EPI_ISL_1715904, EPI_ISL_1715905, EPI_ISL_1715906, EPI_ISL_1715907, EPI_ISL_1715908, EPI_ISL_1715909, EPI_ISL_1715910, EPI_ISL_1715911, EPI_ISL_1715912, EPI_ISL_1715913, EPI_ISL_1715914, EPI_ISL_1715915, EPI_ISL_1715916, EPI_ISL_1715917, EPI_ISL_1715918, EPI_ISL_1715919, EPI_ISL_1715920, EPI_ISL_1715921, EPI_ISL_1715922, EPI_ISL_1715923, EPI_ISL_1715924, EPI_ISL_1715925, EPI_ISL_1715926, EPI_ISL_1715927, EPI_ISL_1715928, EPI_ISL_1715929, EPI_ISL_1715930, EPI_ISL_1715931, EPI_ISL_1715932, EPI_ISL_1715933, EPI_ISL_1715934, EPI_ISL_1715935, EPI_ISL_1715936, EPI_ISL_1715937, EPI_ISL_1715938, EPI_ISL_1715939, EPI_ISL_1715940, EPI_ISL_1715941, EPI_ISL_1715942, EPI_ISL_1715943, EPI_ISL_1715944, EPI_ISL_1715945, EPI_ISL_1715946, EPI_ISL_1715947, EPI_ISL_1715948, EPI_ISL_1715949, EPI_ISL_1715950, EPI_ISL_1715951, EPI_ISL_1715952, EPI_ISL_1715953, EPI_ISL_1715954, EPI_ISL_1715955, EPI_ISL_1715956, EPI_ISL_1715957, EPI_ISL_1715958, EPI_ISL_1715959, EPI_ISL_1715960, EPI_ISL_1715961, EPI_ISL_1715962, EPI_ISL_1715963, EPI_ISL_1715964, EPI_ISL_1715965, EPI_ISL_1715966, EPI_ISL_1715967, EPI_ISL_1715968, EPI_ISL_1715969, EPI_ISL_1715970, EPI_ISL_1715971, EPI_ISL_1715972, EPI_ISL_1715973, EPI_ISL_1715975, EPI_ISL_1715976, EPI_ISL_1715977, EPI_ISL_1715978, EPI_ISL_1715979, EPI_ISL_1715980, EPI_ISL_1715981, EPI_ISL_1715982, EPI_ISL_1715983, EPI_ISL_1715984, EPI_ISL_1715986, EPI_ISL_1715987, EPI_ISL_1715988, EPI_ISL_1715989, EPI_ISL_1715990, EPI_ISL_1715991, EPI_ISL_1715992, EPI_ISL_1715993, EPI_ISL_1715994, EPI_ISL_1715995, EPI_ISL_1715996, EPI_ISL_1715997, EPI_ISL_1715998, EPI_ISL_1715999, EPI_ISL_1716000, EPI_ISL_1716001, EPI_ISL_1716002, EPI_ISL_1716003, EPI_ISL_1716004, EPI_ISL_1716005, EPI_ISL_1716006, EPI_ISL_1716007, EPI_ISL_1716008, EPI_ISL_1716009, EPI_ISL_1716010, EPI_ISL_1716011, EPI_ISL_1716012, EPI_ISL_1716013, EPI_ISL_1716014, EPI_ISL_1716015, EPI_ISL_1716016, EPI_ISL_1716017, EPI_ISL_1716018, EPI_ISL_1716019, EPI_ISL_1716020, EPI_ISL_1716021, EPI_ISL_1716022, EPI_ISL_1716023, EPI_ISL_1716024, EPI_ISL_1716025, EPI_ISL_1716026, EPI_ISL_1716027, EPI_ISL_1716028, EPI_ISL_1716029, EPI_ISL_1716030, EPI_ISL_1716031, EPI_ISL_1716032, EPI_ISL_1716033, EPI_ISL_1716034, EPI_ISL_1716035, EPI_ISL_1716036, EPI_ISL_1716037, EPI_ISL_1716038, EPI_ISL_1716039, EPI_ISL_1716040, EPI_ISL_1716041, EPI_ISL_1716042, EPI_ISL_1716043, EPI_ISL_1716044, EPI_ISL_1716045, EPI_ISL_1716046, EPI_ISL_1716047, EPI_ISL_1716048, EPI_ISL_1716049, EPI_ISL_1716050, EPI_ISL_1716051, EPI_ISL_1716052, EPI_ISL_1716053, EPI_ISL_1716054, EPI_ISL_1716055, EPI_ISL_1716215

see above — Labor Dr. Fenner und Kollegen — Heinrich Pette Institute, Leibniz Institute for Experimental Virology — Adam Grundhoff; Alexis Robitaille; Johannes Knobloch; Martin Aepfelbacher; Nicole Fischer; Thomas Günther

EPI_ISL_1217125, EPI_ISL_1217137, EPI_ISL_1217144, EPI_ISL_1217145, EPI_ISL_1217146, EPI_ISL_1217156, EPI_ISL_1217161, EPI_ISL_1217163, EPI_ISL_1217175, EPI_ISL_1217176, EPI_ISL_1286094, EPI_ISL_1286095, EPI_ISL_1286098, EPI_ISL_1286099, EPI_ISL_1286100, EPI_ISL_1286101, EPI_ISL_1286103, EPI_ISL_1286104, EPI_ISL_1286105, EPI_ISL_1286107, EPI_ISL_1286110, EPI_ISL_1286112, EPI_ISL_1286113, EPI_ISL_1286115, EPI_ISL_1286117, EPI_ISL_1286119, EPI_ISL_1286123, EPI_ISL_1286124, EPI_ISL_1286127, EPI_ISL_1286128, EPI_ISL_1286129, EPI_ISL_1286130, EPI_ISL_1286131, EPI_ISL_1286132, EPI_ISL_1286133, EPI_ISL_1354687, EPI_ISL_1354689, EPI_ISL_1354690, EPI_ISL_1354691, EPI_ISL_1354692, EPI_ISL_1354696, EPI_ISL_1354697, EPI_ISL_1354700, EPI_ISL_1354703, EPI_ISL_1354704, EPI_ISL_1354708, EPI_ISL_1354710, EPI_ISL_1354711, EPI_ISL_1354712, EPI_ISL_1354714, EPI_ISL_1354718, EPI_ISL_1354725, EPI_ISL_1354726, EPI_ISL_1354728, EPI_ISL_1354731, EPI_ISL_1354733, EPI_ISL_1354738, EPI_ISL_1354739, EPI_ISL_1354740, EPI_ISL_1354745, EPI_ISL_1354752

see above — Labor Dr. Heidrich & Kollegen MVZ GmbH Hamburg — Robert Koch Institute

EPI_ISL_1284570, EPI_ISL_1284599, EPI_ISL_1284616, EPI_ISL_1284628, EPI_ISL_1284653, EPI_ISL_1284664, EPI_ISL_1284687, EPI_ISL_1284701, EPI_ISL_1284709, EPI_ISL_1284724, EPI_ISL_1284764, EPI_ISL_1284768, EPI_ISL_1284799, EPI_ISL_1284801, EPI_ISL_1284806, EPI_ISL_1284844, EPI_ISL_1284860, EPI_ISL_1284872, EPI_ISL_1284874, EPI_ISL_1285525, EPI_ISL_1285600, EPI_ISL_1285678, EPI_ISL_1351804, EPI_ISL_1351834, EPI_ISL_1351874, EPI_ISL_1351881, EPI_ISL_1351889, EPI_ISL_1351945, EPI_ISL_1351968, EPI_ISL_1351994, EPI_ISL_1351996, EPI_ISL_1351998, EPI_ISL_1352014, EPI_ISL_1352021, EPI_ISL_1352030, EPI_ISL_1352056, EPI_ISL_1352134, EPI_ISL_1352142, EPI_ISL_1352196, EPI_ISL_1354203, EPI_ISL_1354225, EPI_ISL_1354260, EPI_ISL_1432961, EPI_ISL_1433125, EPI_ISL_1433162, EPI_ISL_1433200, EPI_ISL_1433228, EPI_ISL_1433292, EPI_ISL_1438691, EPI_ISL_1438716, EPI_ISL_1438954, EPI_ISL_1439017

see above — Labor Dr. Schumacher MVZ — Robert Koch Institute

EPI_ISL_1570403, EPI_ISL_1570405, EPI_ISL_1570409, EPI_ISL_1570416, EPI_ISL_1570420, EPI_ISL_1570422, EPI_ISL_1570424, EPI_ISL_1570435, EPI_ISL_1570441, EPI_ISL_1570442, EPI_ISL_1570443, EPI_ISL_1570444, EPI_ISL_1570445, EPI_ISL_1570448, EPI_ISL_1570456

see above — Labor Dr. Spranger — Robert Koch Institute

EPI_ISL_1215816, EPI_ISL_1215817, EPI_ISL_1215820, EPI_ISL_1215822, EPI_ISL_1215824, EPI_ISL_1215828, EPI_ISL_1215834, EPI_ISL_1215836, EPI_ISL_1215837, EPI_ISL_1215839, EPI_ISL_1215840, EPI_ISL_1215843, EPI_ISL_1215845, EPI_ISL_1215846, EPI_ISL_1215855, EPI_ISL_1215856, EPI_ISL_1215860, EPI_ISL_1215861, EPI_ISL_1215862, EPI_ISL_1215863, EPI_ISL_1215864, EPI_ISL_1215865, EPI_ISL_1215866, EPI_ISL_1215867, EPI_ISL_1215872, EPI_ISL_1215873, EPI_ISL_1215877, EPI_ISL_1215880, EPI_ISL_1215883, EPI_ISL_1215891, EPI_ISL_1215892, EPI_ISL_1215895, EPI_ISL_1215897, EPI_ISL_1215900, EPI_ISL_1215904, EPI_ISL_1215906, EPI_ISL_1215907, EPI_ISL_1215909, EPI_ISL_1215913, EPI_ISL_1215916, EPI_ISL_1215920, EPI_ISL_1215921, EPI_ISL_1215922, EPI_ISL_1215925, EPI_ISL_1215927, EPI_ISL_1215928, EPI_ISL_1215933, EPI_ISL_1215934, EPI_ISL_1215939, EPI_ISL_1215942, EPI_ISL_1215949, EPI_ISL_1215956, EPI_ISL_1215964, EPI_ISL_1215965, EPI_ISL_1215966, EPI_ISL_1215967, EPI_ISL_1215970, EPI_ISL_1215973, EPI_ISL_1215975, EPI_ISL_1215978, EPI_ISL_1215979, EPI_ISL_1215989, EPI_ISL_1215992, EPI_ISL_1215996, EPI_ISL_1215997, EPI_ISL_1215999, EPI_ISL_1216004, EPI_ISL_1216005, EPI_ISL_1216007, EPI_ISL_1216010, EPI_ISL_1216012, EPI_ISL_1216016, EPI_ISL_1216019, EPI_ISL_1216023, EPI_ISL_1216026, EPI_ISL_1216027, EPI_ISL_1216038, EPI_ISL_1216041, EPI_ISL_1282384, EPI_ISL_1282391, EPI_ISL_1282392, EPI_ISL_1282400, EPI_ISL_1282401, EPI_ISL_1282404, EPI_ISL_1282406, EPI_ISL_1282407, EPI_ISL_1282408, EPI_ISL_1282410, EPI_ISL_1282416, EPI_ISL_1282417, EPI_ISL_1282425, EPI_ISL_1282432, EPI_ISL_1282436, EPI_ISL_1282437, EPI_ISL_1282439, EPI_ISL_1282442, EPI_ISL_1282443, EPI_ISL_1282448, EPI_ISL_1282451, EPI_ISL_1282455, EPI_ISL_1282456, EPI_ISL_1282457, EPI_ISL_1282459, EPI_ISL_1282460, EPI_ISL_1282461, EPI_ISL_1282463, EPI_ISL_1282471, EPI_ISL_1282472, EPI_ISL_1282473, EPI_ISL_1282475, EPI_ISL_1282476, EPI_ISL_1282481, EPI_ISL_1282484, EPI_ISL_1282485, EPI_ISL_1282486, EPI_ISL_1282490, EPI_ISL_1282491, EPI_ISL_1282493, EPI_ISL_1282498, EPI_ISL_1282501, EPI_ISL_1282502, EPI_ISL_1282503, EPI_ISL_1282507, EPI_ISL_1282508, EPI_ISL_1282511, EPI_ISL_1282514, EPI_ISL_1282519, EPI_ISL_1282520, EPI_ISL_1282524, EPI_ISL_1282525, EPI_ISL_1282527, EPI_ISL_1282529, EPI_ISL_1282539, EPI_ISL_1282540, EPI_ISL_1282541, EPI_ISL_1282547, EPI_ISL_1282548, EPI_ISL_1282550, EPI_ISL_1282551, EPI_ISL_1282552, EPI_ISL_1282553, EPI_ISL_1282555, EPI_ISL_1282556, EPI_ISL_1282557, EPI_ISL_1282560, EPI_ISL_1282562, EPI_ISL_1282563, EPI_ISL_1282566, EPI_ISL_1282569, EPI_ISL_1282571, EPI_ISL_1282572, EPI_ISL_1282578, EPI_ISL_1282581, EPI_ISL_1282584, EPI_ISL_1282585, EPI_ISL_1282592, EPI_ISL_1282593, EPI_ISL_1282597, EPI_ISL_1282598, EPI_ISL_1282599, EPI_ISL_1282600, EPI_ISL_1282602, EPI_ISL_1282605, EPI_ISL_1282606, EPI_ISL_1282607, EPI_ISL_1282608, EPI_ISL_1282609, EPI_ISL_1282611, EPI_ISL_1282612, EPI_ISL_1282616, EPI_ISL_1282619, EPI_ISL_1282624, EPI_ISL_1282628, EPI_ISL_1282629, EPI_ISL_1282630, EPI_ISL_1282631, EPI_ISL_1282632, EPI_ISL_1282635, EPI_ISL_1282636, EPI_ISL_1282638, EPI_ISL_1282640, EPI_ISL_1282642, EPI_ISL_1282643, EPI_ISL_1282654, EPI_ISL_1282657, EPI_ISL_1282659, EPI_ISL_1282661, EPI_ISL_1282663, EPI_ISL_1282665, EPI_ISL_1282669, EPI_ISL_1282672, EPI_ISL_1282674, EPI_ISL_1282675, EPI_ISL_1282680, EPI_ISL_1282688, EPI_ISL_1282689, EPI_ISL_1282692, EPI_ISL_1282693, EPI_ISL_1282694, EPI_ISL_1282695, EPI_ISL_1282696, EPI_ISL_1282697, EPI_ISL_1282698, EPI_ISL_1282700, EPI_ISL_1282704, EPI_ISL_1282707, EPI_ISL_1282709, EPI_ISL_1282710, EPI_ISL_1282714, EPI_ISL_1282718, EPI_ISL_1282720, EPI_ISL_1282721, EPI_ISL_1282723, EPI_ISL_1282726, EPI_ISL_1282728, EPI_ISL_1282731, EPI_ISL_1282733, EPI_ISL_1282734, EPI_ISL_1282737, EPI_ISL_1282738, EPI_ISL_1282740, EPI_ISL_1282741, EPI_ISL_1282743, EPI_ISL_1282744, EPI_ISL_1282747, EPI_ISL_1282748, EPI_ISL_1282752, EPI_ISL_1286909, EPI_ISL_1286910, EPI_ISL_1286911, EPI_ISL_1286913, EPI_ISL_1286920, EPI_ISL_1286941, EPI_ISL_1286962, EPI_ISL_1286964, EPI_ISL_1286967, EPI_ISL_1286974, EPI_ISL_1286979, EPI_ISL_1286980, EPI_ISL_1286981, EPI_ISL_1286987, EPI_ISL_1287001, EPI_ISL_1287030, EPI_ISL_1287040, EPI_ISL_1287045, EPI_ISL_1287046, EPI_ISL_1287048, EPI_ISL_1287053, EPI_ISL_1287060, EPI_ISL_1287063, EPI_ISL_1287066, EPI_ISL_1287069, EPI_ISL_1287075, EPI_ISL_1287077, EPI_ISL_1287078, EPI_ISL_1287080, EPI_ISL_1287082, EPI_ISL_1287108, EPI_ISL_1287112, EPI_ISL_1287115, EPI_ISL_1287120, EPI_ISL_1287126, EPI_ISL_1287145, EPI_ISL_1287165, EPI_ISL_1287198, EPI_ISL_1287199, EPI_ISL_1287203, EPI_ISL_1287210, EPI_ISL_1287216, EPI_ISL_1287220, EPI_ISL_1287225, EPI_ISL_1287235, EPI_ISL_1287242, EPI_ISL_1287248, EPI_ISL_1287256, EPI_ISL_1287262, EPI_ISL_1287263, EPI_ISL_1354473, EPI_ISL_1433990

see above — Labor Dr. Wisplinghoff - Köln — Robert Koch Institute

EPI_ISL_1281777, EPI_ISL_1281778, EPI_ISL_1281779, EPI_ISL_1281780, EPI_ISL_1281781, EPI_ISL_1281782, EPI_ISL_1281783, EPI_ISL_1281784, EPI_ISL_1350950, EPI_ISL_1350957

see above — Labor Kneißler GmbH & Co. KG — Robert Koch Institute

EPI_ISL_1281915, EPI_ISL_1281923, EPI_ISL_1281964, EPI_ISL_1281965, EPI_ISL_1281968, EPI_ISL_1281976, EPI_ISL_1285290, EPI_ISL_1285306, EPI_ISL_1285317, EPI_ISL_1285319, EPI_ISL_1285340, EPI_ISL_1350264, EPI_ISL_1350276, EPI_ISL_1350286, EPI_ISL_1350295, EPI_ISL_1350305, EPI_ISL_1350315, EPI_ISL_1350317

see above — Labor Lübeck bzw. Laborärztliche Gemeinschaftspraxis Lübeck — Robert Koch Institute

EPI_ISL_1283131, EPI_ISL_1283133, EPI_ISL_1283134, EPI_ISL_1283135, EPI_ISL_1283136, EPI_ISL_1283138, EPI_ISL_1283142, EPI_ISL_1283149, EPI_ISL_1283150, EPI_ISL_1283154, EPI_ISL_1283166, EPI_ISL_1283175, EPI_ISL_1283177, EPI_ISL_1283182, EPI_ISL_1283189, EPI_ISL_1283193, EPI_ISL_1283201, EPI_ISL_1283203, EPI_ISL_1283206, EPI_ISL_1283209, EPI_ISL_1283211, EPI_ISL_1283214, EPI_ISL_1283227, EPI_ISL_1283242, EPI_ISL_1283705, EPI_ISL_1283733, EPI_ISL_1283739, EPI_ISL_1283758, EPI_ISL_1283770, EPI_ISL_1283787, EPI_ISL_1283805, EPI_ISL_1283825, EPI_ISL_1283836, EPI_ISL_1283852, EPI_ISL_1286136, EPI_ISL_1286137, EPI_ISL_1353070, EPI_ISL_1353072, EPI_ISL_1353076, EPI_ISL_1353078, EPI_ISL_1353085, EPI_ISL_1353086, EPI_ISL_1353087, EPI_ISL_1353091, EPI_ISL_1353092, EPI_ISL_1353093, EPI_ISL_1353101, EPI_ISL_1353113, EPI_ISL_1353127, EPI_ISL_1353130, EPI_ISL_1353132, EPI_ISL_1353134, EPI_ISL_1353137, EPI_ISL_1353139

| | | | |
|---|---|---|---|
| see above | Labor MönchengladbachMVZ Dr. Stein + Kollegen GbR | Robert Koch Institute | |
| EPI_ISL_1285347, EPI_ISL_1285350, EPI_ISL_1285353, EPI_ISL_1285357, EPI_ISL_1285360, EPI_ISL_1285365, EPI_ISL_1285369, EPI_ISL_1285370, EPI_ISL_1285375, EPI_ISL_1285376, EPI_ISL_1285377, EPI_ISL_1285379, EPI_ISL_1285380, EPI_ISL_1285381, EPI_ISL_1285382, EPI_ISL_1285384, EPI_ISL_1564010, EPI_ISL_1564014, EPI_ISL_1564015, EPI_ISL_1564016, EPI_ISL_1564017, EPI_ISL_1564021, EPI_ISL_1564022, EPI_ISL_1569716, EPI_ISL_1571935, EPI_ISL_1571939, EPI_ISL_1571943, EPI_ISL_1638356 | | | |
| see above | Labor Prof. Dr. G. Enders MVZ GbR | Robert Koch Institute | |
| EPI_ISL_1573270 | Labor ZOTZ\|KLIMAS; MVZ DÃ¼sseldorf-Centrum | Robert Koch Institute | |
| EPI_ISL_1285234, EPI_ISL_1350322, EPI_ISL_1350323, EPI_ISL_1350325, EPI_ISL_1350327, EPI_ISL_1350329, EPI_ISL_1350330, EPI_ISL_1350332, EPI_ISL_1350333, EPI_ISL_1350339, EPI_ISL_1350344, EPI_ISL_1350347, EPI_ISL_1350351, EPI_ISL_1350352, EPI_ISL_1350353, EPI_ISL_1350356, EPI_ISL_1350357, EPI_ISL_1350360, EPI_ISL_1350366, EPI_ISL_1350368, EPI_ISL_1350370, EPI_ISL_1350373, EPI_ISL_1350374, EPI_ISL_1436832, EPI_ISL_1436841, EPI_ISL_1436849, EPI_ISL_1436873, EPI_ISL_1436881, EPI_ISL_1436919, EPI_ISL_1441779 | | | |
| see above | Labor ZOTZ\|KLIMAS; MVZ Düsseldorf-Centrum | Robert Koch Institute | |
| EPI_ISL_1218301, EPI_ISL_1218307, EPI_ISL_1218314, EPI_ISL_1218332, EPI_ISL_1280789, EPI_ISL_1280807, EPI_ISL_1280830, EPI_ISL_1280867, EPI_ISL_1280871, EPI_ISL_1280879, EPI_ISL_1280897, EPI_ISL_1352071, EPI_ISL_1352078, EPI_ISL_1352129, EPI_ISL_1352131, EPI_ISL_1352215 | | | |
| see above | Labor für medizinische Mikrobiologie Bakteriologie Mykologie Virologie & Infektionsserologie | Robert Koch Institute | |
| EPI_ISL_1569712, EPI_ISL_1572158, EPI_ISL_1572174 | Laborarztpraxis Jochen HÃ¼ter | Robert Koch Institute | |
| EPI_ISL_1356620, EPI_ISL_1356635, EPI_ISL_1438676 | Laborarztpraxis Jochen Hüter | Robert Koch Institute | |
| EPI_ISL_1353354, EPI_ISL_1353363, EPI_ISL_1353387, EPI_ISL_1353397, EPI_ISL_1353406, EPI_ISL_1353409, EPI_ISL_1353410, EPI_ISL_1353413, EPI_ISL_1433736, EPI_ISL_1433737 | | | |
| see above | Labormedizin Darmstadt | Robert Koch Institute | |
| EPI_ISL_1279617, EPI_ISL_1279620, EPI_ISL_1279621, EPI_ISL_1279623, EPI_ISL_1279624, EPI_ISL_1279678, EPI_ISL_1279679, EPI_ISL_1279680, EPI_ISL_1279681, EPI_ISL_1279682, EPI_ISL_1279683, EPI_ISL_1279684, EPI_ISL_1279691, EPI_ISL_1279692, EPI_ISL_1279693, EPI_ISL_1279694, EPI_ISL_1279695, EPI_ISL_1279696, EPI_ISL_1279697, EPI_ISL_1279704, EPI_ISL_1279705, EPI_ISL_1279709, EPI_ISL_1279710, EPI_ISL_1279711, EPI_ISL_1279712, EPI_ISL_1279719, EPI_ISL_1279720, EPI_ISL_1279721, EPI_ISL_1279722, EPI_ISL_1279723, EPI_ISL_1279724, EPI_ISL_1279725, EPI_ISL_1279726, EPI_ISL_1279727, EPI_ISL_1279728, EPI_ISL_1279729, EPI_ISL_1279730, EPI_ISL_1279731, EPI_ISL_1279769, EPI_ISL_1279770, EPI_ISL_1279774, EPI_ISL_1279786, EPI_ISL_1279787, EPI_ISL_1279788, EPI_ISL_1279789, EPI_ISL_1279790, EPI_ISL_1279793, EPI_ISL_1279794, EPI_ISL_1279795, EPI_ISL_1279796, EPI_ISL_1279797, EPI_ISL_1279798, EPI_ISL_1279799, EPI_ISL_1279802, EPI_ISL_1279803, EPI_ISL_1279804, EPI_ISL_1279812, EPI_ISL_1279816, EPI_ISL_1279817, EPI_ISL_1279818, EPI_ISL_1279822, EPI_ISL_1279823, EPI_ISL_1279824, EPI_ISL_1279825, EPI_ISL_1279826, EPI_ISL_1279827, EPI_ISL_1279829, EPI_ISL_1279830, EPI_ISL_1279857, EPI_ISL_1279858, EPI_ISL_1279859, EPI_ISL_1279860, EPI_ISL_1279861, EPI_ISL_1279862, EPI_ISL_1279863, EPI_ISL_1279864, EPI_ISL_1279874, EPI_ISL_1279880, EPI_ISL_1279881, EPI_ISL_1279882, EPI_ISL_1279883, EPI_ISL_1279884, EPI_ISL_1279885, EPI_ISL_1279886, EPI_ISL_1279887, EPI_ISL_1279888, EPI_ISL_1279889, EPI_ISL_1279890, EPI_ISL_1361657, EPI_ISL_1361658, EPI_ISL_1361659, EPI_ISL_1361660, EPI_ISL_1361661, EPI_ISL_1361662, EPI_ISL_1361663, EPI_ISL_1361664, EPI_ISL_1361665, EPI_ISL_1361666, EPI_ISL_1361667, EPI_ISL_1361668, EPI_ISL_1361669, EPI_ISL_1361670, EPI_ISL_1361671, EPI_ISL_1361672, EPI_ISL_1361673, EPI_ISL_1361674, EPI_ISL_1361675, EPI_ISL_1361676, EPI_ISL_1361677, EPI_ISL_1361678, EPI_ISL_1361679, EPI_ISL_1361680, EPI_ISL_1361681, EPI_ISL_1361682, EPI_ISL_1361683, EPI_ISL_1361684, EPI_ISL_1361685, EPI_ISL_1361686, EPI_ISL_1361687, EPI_ISL_1361688, EPI_ISL_1361689, EPI_ISL_1361690, EPI_ISL_1361691, EPI_ISL_1361692, EPI_ISL_1361703, EPI_ISL_1361704, EPI_ISL_1361705, EPI_ISL_1361706, EPI_ISL_1361707, EPI_ISL_1361708, EPI_ISL_1361713, EPI_ISL_1361714, EPI_ISL_1361715, EPI_ISL_1361716, EPI_ISL_1361717, EPI_ISL_1361718, EPI_ISL_1361719, EPI_ISL_1361720, EPI_ISL_1361721, EPI_ISL_1361722, EPI_ISL_1361723, EPI_ISL_1361724, EPI_ISL_1361725, EPI_ISL_1361726, EPI_ISL_1361727, EPI_ISL_1361728, EPI_ISL_1361729, EPI_ISL_1361730, EPI_ISL_1361731, EPI_ISL_1361732, EPI_ISL_1361733, EPI_ISL_1361734, EPI_ISL_1361735, EPI_ISL_1361736, EPI_ISL_1361737, EPI_ISL_1361738, EPI_ISL_1361739, EPI_ISL_1361740, EPI_ISL_1361741, EPI_ISL_1361742, EPI_ISL_1361743, EPI_ISL_1361744, EPI_ISL_1361745, EPI_ISL_1361746, EPI_ISL_1361747, EPI_ISL_1361748, EPI_ISL_1361749, EPI_ISL_1361750 | | | |
| see above | Landesamt für Verbraucherschutz Sachsen Anhalt, Magdeburg | Institute of Medical Microbiology and Hospital Hygiene | Aljoscha Tersteegen; Prof. Dr. Achim Kaasch |
| EPI_ISL_1638644, EPI_ISL_1638645, EPI_ISL_1638649, EPI_ISL_1638650, EPI_ISL_1638651, EPI_ISL_1638652, EPI_ISL_1638655, EPI_ISL_1638658, EPI_ISL_1638659, EPI_ISL_1638661, EPI_ISL_1638662, EPI_ISL_1638663, EPI_ISL_1638664, EPI_ISL_1638665 | | | |
| see above | Landesgesundheitsamt Baden-WÃ¼rttemberg | Robert Koch Institute | |
| EPI_ISL_1350202 | Landesgesundheitsamt Baden-Württemberg | Robert Koch Institute | |
| EPI_ISL_1217620, EPI_ISL_1217623, EPI_ISL_1217630, EPI_ISL_1217641, EPI_ISL_1217643, EPI_ISL_1217648, EPI_ISL_1217658, EPI_ISL_1217659, EPI_ISL_1217661, EPI_ISL_1217668, EPI_ISL_1217669, EPI_ISL_1217672, EPI_ISL_1217675, EPI_ISL_1217679, EPI_ISL_1217692, EPI_ISL_1217694, EPI_ISL_1217695, EPI_ISL_1217703, EPI_ISL_1285060, EPI_ISL_1285069, EPI_ISL_1285079, EPI_ISL_1285080, EPI_ISL_1285093, EPI_ISL_1285094, EPI_ISL_1285096, EPI_ISL_1285110, EPI_ISL_1285115, EPI_ISL_1285117, EPI_ISL_1285120, EPI_ISL_1285127, EPI_ISL_1285128, EPI_ISL_1285129, EPI_ISL_1285130, EPI_ISL_1285134, EPI_ISL_1285135, EPI_ISL_1285143, EPI_ISL_1285146, EPI_ISL_1285147, EPI_ISL_1285149, EPI_ISL_1285152, EPI_ISL_1285154, EPI_ISL_1285156, EPI_ISL_1285158, EPI_ISL_1285163, EPI_ISL_1285164, EPI_ISL_1285165 | | | |
| see above | Landesuntersuchungsanstalt für das Gesundheits- und Veterinärwesen (LUA) Sachsen | Robert Koch Institute | |
| EPI_ISL_1372974, EPI_ISL_1531531, EPI_ISL_1531532, EPI_ISL_1711184, EPI_ISL_1711239 | Landstuhl Regional Medical Center | US Air Force School of Aerospace Medicine | Amanda Javorina; Anthony Fries; Clarise Starr; Cole Anderson; Elizabeth Macias; Fritz Castillo; Jennifer Meyer; Sarah Purves; William Buggele; William Gruner |
| EPI_ISL_1281177, EPI_ISL_1281193, EPI_ISL_1281212, EPI_ISL_1281239, EPI_ISL_1281244, EPI_ISL_1281809, EPI_ISL_1281812, EPI_ISL_1281815, EPI_ISL_1281818, EPI_ISL_1283170, EPI_ISL_1283213, EPI_ISL_1283713, EPI_ISL_1283777, EPI_ISL_1283842, EPI_ISL_1283847, EPI_ISL_1284479, EPI_ISL_1284480, EPI_ISL_1284491, EPI_ISL_1284495, EPI_ISL_1284502, EPI_ISL_1284503, EPI_ISL_1284511, EPI_ISL_1352309, EPI_ISL_1352312, EPI_ISL_1353221 | | | |
| see above | Limbach - MVZ Gemeinschaftslabor Suhl | Robert Koch Institute | |
| EPI_ISL_1281583, EPI_ISL_1281584, EPI_ISL_1281585, EPI_ISL_1281586, EPI_ISL_1281587, EPI_ISL_1281588, EPI_ISL_1281589, EPI_ISL_1281590, EPI_ISL_1281591, EPI_ISL_1281592, EPI_ISL_1281593, EPI_ISL_1281594, EPI_ISL_1281596, EPI_ISL_1281597, EPI_ISL_1281598, EPI_ISL_1281599, EPI_ISL_1281600, EPI_ISL_1281601, EPI_ISL_1281602, EPI_ISL_1281603, EPI_ISL_1281604, EPI_ISL_1281605, EPI_ISL_1281606, EPI_ISL_1281607, EPI_ISL_1281608, EPI_ISL_1281609, EPI_ISL_1281610, EPI_ISL_1281611, EPI_ISL_1281612, EPI_ISL_1281613, EPI_ISL_1281614, EPI_ISL_1281615, EPI_ISL_1281616, EPI_ISL_1281617, EPI_ISL_1281618, EPI_ISL_1281619, EPI_ISL_1281620, EPI_ISL_1281621, EPI_ISL_1281622, EPI_ISL_1281623, EPI_ISL_1281624, EPI_ISL_1281625, EPI_ISL_1281626, EPI_ISL_1281627, EPI_ISL_1281628, EPI_ISL_1281629, EPI_ISL_1281630, EPI_ISL_1281631, EPI_ISL_1281632, EPI_ISL_1281633, EPI_ISL_1281634, EPI_ISL_1281635, EPI_ISL_1281636, EPI_ISL_1281637, EPI_ISL_1281638, EPI_ISL_1281639, EPI_ISL_1281640, EPI_ISL_1281641, EPI_ISL_1281642, EPI_ISL_1281643, EPI_ISL_1281644, EPI_ISL_1281645, EPI_ISL_1281646, EPI_ISL_1281647, EPI_ISL_1281648, EPI_ISL_1281649, EPI_ISL_1281650, EPI_ISL_1281651, EPI_ISL_1281652, EPI_ISL_1281653, EPI_ISL_1281654, EPI_ISL_1281655, EPI_ISL_1281656, EPI_ISL_1281657, EPI_ISL_1281658, EPI_ISL_1281659, EPI_ISL_1281660, EPI_ISL_1281661, EPI_ISL_1281662, EPI_ISL_1281663, EPI_ISL_1281664, EPI_ISL_1281665, EPI_ISL_1281666, EPI_ISL_1281667, EPI_ISL_1281668, EPI_ISL_1281669, EPI_ISL_1281670, EPI_ISL_1281671, EPI_ISL_1281672, EPI_ISL_1281673, EPI_ISL_1281674, EPI_ISL_1281675, EPI_ISL_1281677, EPI_ISL_1281678, EPI_ISL_1281679, EPI_ISL_1281681, EPI_ISL_1281684, EPI_ISL_1281687, EPI_ISL_1281689, EPI_ISL_1281691, EPI_ISL_1281694, EPI_ISL_1281695, EPI_ISL_1281696, EPI_ISL_1281697, EPI_ISL_1281698, EPI_ISL_1281699, EPI_ISL_1281700, EPI_ISL_1281701, EPI_ISL_1281702, EPI_ISL_1281703, EPI_ISL_1281704, EPI_ISL_1281705, EPI_ISL_1281706, EPI_ISL_1281707, EPI_ISL_1281708, EPI_ISL_1281709, EPI_ISL_1281710, EPI_ISL_1281711, EPI_ISL_1281712, EPI_ISL_1281713, EPI_ISL_1281714, EPI_ISL_1281715, EPI_ISL_1281716, EPI_ISL_1281717, EPI_ISL_1281719, EPI_ISL_1281720, EPI_ISL_1281721, EPI_ISL_1281722, EPI_ISL_1281723, EPI_ISL_1281724, EPI_ISL_1281725, EPI_ISL_1281726, EPI_ISL_1281727, EPI_ISL_1281728, EPI_ISL_1281729, EPI_ISL_1281730, EPI_ISL_1281731, EPI_ISL_1281732, EPI_ISL_1281733, EPI_ISL_1281734, EPI_ISL_1281735, EPI_ISL_1281736, EPI_ISL_1281737, EPI_ISL_1281738, EPI_ISL_1281739, EPI_ISL_1281740, EPI_ISL_1281741, EPI_ISL_1281742, EPI_ISL_1281743, EPI_ISL_1281744, EPI_ISL_1281745, EPI_ISL_1281746, EPI_ISL_1281747, EPI_ISL_1281749, EPI_ISL_1281750, EPI_ISL_1281751, EPI_ISL_1281752, EPI_ISL_1281753, EPI_ISL_1281754, EPI_ISL_1281755, EPI_ISL_1281756, EPI_ISL_1281757, EPI_ISL_1281758, EPI_ISL_1281760, EPI_ISL_1281761, EPI_ISL_1281762, EPI_ISL_1281763, EPI_ISL_1281764, EPI_ISL_1281765, EPI_ISL_1281766, EPI_ISL_1281767, EPI_ISL_1281768, EPI_ISL_1281769, EPI_ISL_1281770, EPI_ISL_1281771, EPI_ISL_1281772, EPI_ISL_1281773, EPI_ISL_1281774, EPI_ISL_1281775, EPI_ISL_1281776, EPI_ISL_1350466, EPI_ISL_1350502 | | | |
| see above | Limbach - MVZ Humangenetik Ulm | Robert Koch Institute | |
| EPI_ISL_1280197, EPI_ISL_1280205, EPI_ISL_1280208, EPI_ISL_1280217, EPI_ISL_1283128, EPI_ISL_1283144, EPI_ISL_1283153, EPI_ISL_1283155, EPI_ISL_1283158, EPI_ISL_1283160, EPI_ISL_1283163, EPI_ISL_1283169, EPI_ISL_1283183, EPI_ISL_1283202, EPI_ISL_1283224, EPI_ISL_1283237, EPI_ISL_1283680, EPI_ISL_1283683, EPI_ISL_1283686, EPI_ISL_1283691, EPI_ISL_1283700, EPI_ISL_1283706, EPI_ISL_1283720, EPI_ISL_1283748, EPI_ISL_1283768, EPI_ISL_1283775, EPI_ISL_1283780, EPI_ISL_1283782, EPI_ISL_1283793, EPI_ISL_1283795, EPI_ISL_1283803, EPI_ISL_1283812, EPI_ISL_1283824, EPI_ISL_1283827, EPI_ISL_1283830, EPI_ISL_1283849, EPI_ISL_1283854, EPI_ISL_1352249, EPI_ISL_1352261, EPI_ISL_1352289, EPI_ISL_1352333, EPI_ISL_1353073, EPI_ISL_1353075, EPI_ISL_1353080, EPI_ISL_1353088, EPI_ISL_1353090, EPI_ISL_1353100, EPI_ISL_1353104, EPI_ISL_1353107, EPI_ISL_1353109, EPI_ISL_1353111, EPI_ISL_1353119, EPI_ISL_1353120, EPI_ISL_1353124, EPI_ISL_1353125, EPI_ISL_1353129, EPI_ISL_1353131, EPI_ISL_1441663, EPI_ISL_1441676 | | | |
| see above | Limbach - MVZ Labor Eveld & Kollegen Essen | Robert Koch Institute | |
| EPI_ISL_1352252, EPI_ISL_1352284, EPI_ISL_1441650 | Limbach - MVZ Labor Ludwigsburg | Robert Koch Institute | |
| EPI_ISL_1281196, EPI_ISL_1281199, EPI_ISL_1281786, EPI_ISL_1281789, EPI_ISL_1281790, EPI_ISL_1281794, EPI_ISL_1281797, EPI_ISL_1281798, EPI_ISL_1281806, EPI_ISL_1281814, EPI_ISL_1281817, EPI_ISL_1283684, EPI_ISL_1283829, EPI_ISL_1284510, EPI_ISL_1284512, EPI_ISL_1350221, EPI_ISL_1350225, EPI_ISL_1350231, EPI_ISL_1350240, EPI_ISL_1352241, EPI_ISL_1352242, EPI_ISL_1352248, EPI_ISL_1352257, EPI_ISL_1352258, EPI_ISL_1352281, EPI_ISL_1352282, EPI_ISL_1352288, EPI_ISL_1352290, EPI_ISL_1352294, EPI_ISL_1352300, EPI_ISL_1352308, EPI_ISL_1352313, EPI_ISL_1352323, EPI_ISL_1352334, EPI_ISL_1352336, EPI_ISL_1352340, EPI_ISL_1352342, EPI_ISL_1352346, EPI_ISL_1353023, EPI_ISL_1353030, EPI_ISL_1353034, EPI_ISL_1353040, EPI_ISL_1353051, EPI_ISL_1353160, EPI_ISL_1353218, EPI_ISL_1434450, EPI_ISL_1435305, EPI_ISL_1435454, EPI_ISL_1435978, EPI_ISL_1435982, EPI_ISL_1435986, EPI_ISL_1435993, EPI_ISL_1435998, EPI_ISL_1441158, EPI_ISL_1441159, EPI_ISL_1441160, EPI_ISL_1441161, EPI_ISL_1441162, EPI_ISL_1441164, EPI_ISL_1441165, EPI_ISL_1441166, EPI_ISL_1441172, EPI_ISL_1441173, EPI_ISL_1441174, EPI_ISL_1441175, EPI_ISL_1441176, EPI_ISL_1441177, EPI_ISL_1441178, EPI_ISL_1441179, EPI_ISL_1441180, EPI_ISL_1441181, EPI_ISL_1441183, EPI_ISL_1441185, EPI_ISL_1441187, EPI_ISL_1441188, EPI_ISL_1441189, EPI_ISL_1441190, EPI_ISL_1441192, EPI_ISL_1441194, EPI_ISL_1441199, EPI_ISL_1441204, EPI_ISL_1441205, EPI_ISL_1441208, EPI_ISL_1441209, EPI_ISL_1441212, EPI_ISL_1441223, EPI_ISL_1441229, EPI_ISL_1441666, EPI_ISL_1441669, EPI_ISL_1441671, EPI_ISL_1441682, EPI_ISL_1441683, EPI_ISL_1441684, EPI_ISL_1441685, EPI_ISL_1441688, EPI_ISL_1441689, EPI_ISL_1441690, EPI_ISL_1441691, EPI_ISL_1441692, EPI_ISL_1441693, EPI_ISL_1441694, EPI_ISL_1441695, EPI_ISL_1441698, EPI_ISL_1441702, EPI_ISL_1441703, EPI_ISL_1441707, EPI_ISL_1441710, EPI_ISL_1441711, EPI_ISL_1441712, EPI_ISL_1441713, EPI_ISL_1441715, EPI_ISL_1441716 | | | |
| see above | MDI Limbach Berlin GmbH | Robert Koch Institute | |
| EPI_ISL_1464263, EPI_ISL_1464264, EPI_ISL_1464265, EPI_ISL_1464266, EPI_ISL_1464267, EPI_ISL_1464268, EPI_ISL_1715870, EPI_ISL_1715871 | | | |
| see above | MEDILYS Laborgesellschaft mbH | Heinrich Pette Institute, Leibniz Institute for Experimental Virology | Adam Grundhoff; Alexis Robitaille; Johannes Knobloch; Martin Aepfelbacher; Nicole Fischer; Thomas Günther |
| EPI_ISL_1524900, EPI_ISL_1524903, EPI_ISL_1524904, EPI_ISL_1524905, EPI_ISL_1586898 | MKK Mühlenkreiskliniken, Johannes Wesling Klinikum Minden | Hannover Medical School, Institute of Virology | Lars Steinbrück |
| EPI_ISL_1214834, EPI_ISL_1214842, EPI_ISL_1214877, EPI_ISL_1214898, EPI_ISL_1214936, EPI_ISL_1214945, EPI_ISL_1214950, EPI_ISL_1214955, EPI_ISL_1214982, EPI_ISL_1217523, EPI_ISL_1217524, EPI_ISL_1217526, EPI_ISL_1217529, EPI_ISL_1217531, EPI_ISL_1217536, EPI_ISL_1217538, EPI_ISL_1217544, EPI_ISL_1217548, EPI_ISL_1217550, EPI_ISL_1217551, EPI_ISL_1217557, EPI_ISL_1217560, EPI_ISL_1281911, EPI_ISL_1281912, EPI_ISL_1281913, EPI_ISL_1281914, EPI_ISL_1281917, EPI_ISL_1281918, EPI_ISL_1281919, EPI_ISL_1281921, EPI_ISL_1281922, EPI_ISL_1281924, EPI_ISL_1281926, EPI_ISL_1281927, EPI_ISL_1281928, EPI_ISL_1281929, EPI_ISL_1281930, EPI_ISL_1281931, EPI_ISL_1281932, EPI_ISL_1281933, EPI_ISL_1281934, EPI_ISL_1281935, EPI_ISL_1281936, EPI_ISL_1281937, EPI_ISL_1281938, EPI_ISL_1281939, EPI_ISL_1281940, EPI_ISL_1281941, EPI_ISL_1281942, EPI_ISL_1281944, EPI_ISL_1281945, EPI_ISL_1281946, EPI_ISL_1281947, EPI_ISL_1281948, EPI_ISL_1281950, EPI_ISL_1281951, EPI_ISL_1281952, EPI_ISL_1281954, EPI_ISL_1281955, EPI_ISL_1281956, EPI_ISL_1281957, EPI_ISL_1281958, EPI_ISL_1281961, EPI_ISL_1281962, EPI_ISL_1281963, EPI_ISL_1281967, EPI_ISL_1281969, EPI_ISL_1281970, EPI_ISL_1281971, EPI_ISL_1281973, EPI_ISL_1281974, EPI_ISL_1281975, EPI_ISL_1281977, EPI_ISL_1281978, EPI_ISL_1281979, EPI_ISL_1281980, EPI_ISL_1281981, EPI_ISL_1281984, EPI_ISL_1281985, EPI_ISL_1281986, EPI_ISL_1281987, EPI_ISL_1281988, EPI_ISL_1281989, EPI_ISL_1281990, EPI_ISL_1281991, EPI_ISL_1285284, EPI_ISL_1285285, EPI_ISL_1285286, EPI_ISL_1285287, EPI_ISL_1285289, EPI_ISL_1285291, EPI_ISL_1285293, EPI_ISL_1285296, EPI_ISL_1285298, EPI_ISL_1285301, EPI_ISL_1285302, EPI_ISL_1285304, EPI_ISL_1285307, EPI_ISL_1285309, EPI_ISL_1285310, EPI_ISL_1285312, EPI_ISL_1285313, EPI_ISL_1285314, EPI_ISL_1285316, EPI_ISL_1285318, EPI_ISL_1285320, EPI_ISL_1285321, EPI_ISL_1285322, EPI_ISL_1285323, EPI_ISL_1285324, EPI_ISL_1285325, EPI_ISL_1285326, EPI_ISL_1285327, EPI_ISL_1285328, EPI_ISL_1285329, EPI_ISL_1285331, EPI_ISL_1285332, EPI_ISL_1285333, EPI_ISL_1285334, EPI_ISL_1285337, EPI_ISL_1285338, EPI_ISL_1285339, EPI_ISL_1285341, EPI_ISL_1285342, EPI_ISL_1350243, EPI_ISL_1350244, EPI_ISL_1350245, EPI_ISL_1350247, EPI_ISL_1350248, EPI_ISL_1350249, EPI_ISL_1350250, EPI_ISL_1350252, EPI_ISL_1350253, EPI_ISL_1350254, EPI_ISL_1350255, EPI_ISL_1350257, EPI_ISL_1350259, EPI_ISL_1350261, EPI_ISL_1350262, EPI_ISL_1350263, EPI_ISL_1350266, EPI_ISL_1350268, EPI_ISL_1350270, EPI_ISL_1350277, EPI_ISL_1350280, EPI_ISL_1350281, EPI_ISL_1350283, EPI_ISL_1350284, EPI_ISL_1350285, EPI_ISL_1350287, EPI_ISL_1350289, EPI_ISL_1350290, EPI_ISL_1350292, EPI_ISL_1350293, EPI_ISL_1350294, EPI_ISL_1350296, EPI_ISL_1350297, EPI_ISL_1350298, EPI_ISL_1350299, EPI_ISL_1350300, EPI_ISL_1350301, EPI_ISL_1350302, EPI_ISL_1350303, EPI_ISL_1350304, EPI_ISL_1350307, EPI_ISL_1350308, EPI_ISL_1350310, EPI_ISL_1350311, EPI_ISL_1350313 | | | |
| see above | MVZ Dr. Eberhard & Partner Dortmund | Robert Koch Institute | |
| EPI_ISL_1280230, EPI_ISL_1280240, EPI_ISL_1280241, EPI_ISL_1280242, EPI_ISL_1280244, EPI_ISL_1280248, EPI_ISL_1280249, EPI_ISL_1280254, EPI_ISL_1280257, EPI_ISL_1280259, EPI_ISL_1280262, EPI_ISL_1280263, EPI_ISL_1280264, EPI_ISL_1280268, EPI_ISL_1281266, EPI_ISL_1281278, EPI_ISL_1348953, EPI_ISL_1348976, EPI_ISL_1348977 | | | |
| see above | MVZ Labor Dr. Fenner und Kollegen (Standort Hamburg) | Robert Koch Institute | |
| EPI_ISL_1281788, EPI_ISL_1281792, EPI_ISL_1281796, EPI_ISL_1281799, EPI_ISL_1281805, EPI_ISL_1281810, EPI_ISL_1281819, EPI_ISL_1281820, EPI_ISL_1283122, EPI_ISL_1283123, EPI_ISL_1283125, EPI_ISL_1283126, EPI_ISL_1283129, EPI_ISL_1283141, EPI_ISL_1283146, EPI_ISL_1283147, EPI_ISL_1283151, EPI_ISL_1283156, EPI_ISL_1283162, EPI_ISL_1283165, EPI_ISL_1283168, EPI_ISL_1283172, EPI_ISL_1283174, EPI_ISL_1283185, EPI_ISL_1283199, EPI_ISL_1283212, EPI_ISL_1283217, EPI_ISL_1283218, EPI_ISL_1283219, EPI_ISL_1283221, EPI_ISL_1283223, EPI_ISL_1283225, EPI_ISL_1283226, EPI_ISL_1283228, EPI_ISL_1283229, EPI_ISL_1283230, EPI_ISL_1283232, EPI_ISL_1283234, EPI_ISL_1283235, EPI_ISL_1283239, EPI_ISL_1283240, EPI_ISL_1283244, EPI_ISL_1283675, EPI_ISL_1283678, EPI_ISL_1283681, EPI_ISL_1283688, EPI_ISL_1283693, EPI_ISL_1283694, EPI_ISL_1283696, EPI_ISL_1283698, EPI_ISL_1283703, EPI_ISL_1283708, EPI_ISL_1283711, EPI_ISL_1283715, EPI_ISL_1283716, EPI_ISL_1283718, EPI_ISL_1283721, EPI_ISL_1283723, EPI_ISL_1283725, EPI_ISL_1283726, EPI_ISL_1283728, EPI_ISL_1283729, EPI_ISL_1283731, EPI_ISL_1283734, EPI_ISL_1283736, EPI_ISL_1283738, EPI_ISL_1283741, EPI_ISL_1283744, EPI_ISL_1283746, EPI_ISL_1283751, EPI_ISL_1283753, EPI_ISL_1283755, EPI_ISL_1283757, EPI_ISL_1283760, EPI_ISL_1283761, EPI_ISL_1283763, EPI_ISL_1283766, EPI_ISL_1283769, EPI_ISL_1283774, EPI_ISL_1283779, EPI_ISL_1283785, EPI_ISL_1283789, EPI_ISL_1283790, EPI_ISL_1283797, EPI_ISL_1283800, EPI_ISL_1283802, EPI_ISL_1283809, EPI_ISL_1283810, EPI_ISL_1283814, EPI_ISL_1283815, EPI_ISL_1283817, EPI_ISL_1283818, EPI_ISL_1283820, EPI_ISL_1283832, EPI_ISL_1283834, EPI_ISL_1283837, EPI_ISL_1283839, EPI_ISL_1283850, EPI_ISL_1284470, EPI_ISL_1284471, EPI_ISL_1284474, EPI_ISL_1284476, EPI_ISL_1284477, EPI_ISL_1284478, EPI_ISL_1284484, EPI_ISL_1284485, EPI_ISL_1284486, EPI_ISL_1284489, EPI_ISL_1284490, EPI_ISL_1284492, EPI_ISL_1284493, EPI_ISL_1284494, EPI_ISL_1284497, EPI_ISL_1284498, EPI_ISL_1284499, EPI_ISL_1284501, EPI_ISL_1284504, EPI_ISL_1284505, EPI_ISL_1284507, EPI_ISL_1284508, EPI_ISL_1284509, EPI_ISL_1284513, EPI_ISL_1285718, EPI_ISL_1286135, EPI_ISL_1349513, EPI_ISL_1349515, EPI_ISL_1349516, EPI_ISL_1349517, EPI_ISL_1349518, EPI_ISL_1349519, EPI_ISL_1349520, EPI_ISL_1349521, EPI_ISL_1349522, EPI_ISL_1349523, EPI_ISL_1349524, EPI_ISL_1349526, EPI_ISL_1349527, EPI_ISL_1349529, EPI_ISL_1349530, EPI_ISL_1349532, EPI_ISL_1349533, EPI_ISL_1349534, EPI_ISL_1349535, EPI_ISL_1349536, EPI_ISL_1349537, EPI_ISL_1349538, EPI_ISL_1349540, EPI_ISL_1349541, EPI_ISL_1349542, EPI_ISL_1349543, EPI_ISL_1349545, EPI_ISL_1349547, EPI_ISL_1349548, EPI_ISL_1349549, EPI_ISL_1349551, EPI_ISL_1349552, EPI_ISL_1349553, EPI_ISL_1349554, EPI_ISL_1349555, EPI_ISL_1349556, EPI_ISL_1349558, EPI_ISL_1349559, EPI_ISL_1349560, EPI_ISL_1349562, EPI_ISL_1349563, EPI_ISL_1349564, EPI_ISL_1349565, EPI_ISL_1349566, EPI_ISL_1349567, EPI_ISL_1349568, EPI_ISL_1349572, EPI_ISL_1349573, EPI_ISL_1349574, EPI_ISL_1349575, EPI_ISL_1349577, EPI_ISL_1349579, EPI_ISL_1349586, EPI_ISL_1349589, EPI_ISL_1349593, EPI_ISL_1349599, EPI_ISL_1350227, EPI_ISL_1350232, EPI_ISL_1350235, EPI_ISL_1350238, EPI_ISL_1350422, EPI_ISL_1350423, EPI_ISL_1350424, EPI_ISL_1350426, EPI_ISL_1350427, EPI_ISL_1350428, EPI_ISL_1350429, EPI_ISL_1350430, EPI_ISL_1350431, EPI_ISL_1350432, EPI_ISL_1350433, EPI_ISL_1350435, EPI_ISL_1350436, EPI_ISL_1350437, EPI_ISL_1350438, EPI_ISL_1350439, EPI_ISL_1350440, EPI_ISL_1351356, EPI_ISL_1438594, EPI_ISL_1438641, EPI_ISL_1441648, EPI_ISL_1441652, EPI_ISL_1569726 | | | |
| see above | MVZ Labor Dr. Limbach & Kollegen GbR | Robert Koch Institute | |
| EPI_ISL_1570462, EPI_ISL_1570464, EPI_ISL_1570565, EPI_ISL_1570591, EPI_ISL_1570593, EPI_ISL_1570596, EPI_ISL_1570605, EPI_ISL_1570606, EPI_ISL_1570610, EPI_ISL_1570629, EPI_ISL_1570630, EPI_ISL_1570647, EPI_ISL_1570650, EPI_ISL_1570666, EPI_ISL_1570677 | | | |
| see above | MVZ Labor Dr. Quade & Kollegen GmbH | Robert Koch Institute | |
| EPI_ISL_1280185, EPI_ISL_1280201, EPI_ISL_1283127, EPI_ISL_1283241, EPI_ISL_1283243, EPI_ISL_1283676, EPI_ISL_1283701, EPI_ISL_1283792, EPI_ISL_1283841, EPI_ISL_1352251, EPI_ISL_1352345, EPI_ISL_1353099, EPI_ISL_1353128 | | | |
| see above | MVZ Labor Dr. Reising-Ackermann und Kollegen | Robert Koch Institute | |
| EPI_ISL_1218551, EPI_ISL_1218553, EPI_ISL_1218555, EPI_ISL_1218563, EPI_ISL_1218565, EPI_ISL_1218567, EPI_ISL_1218572, EPI_ISL_1218574, EPI_ISL_1218580, EPI_ISL_1218581, EPI_ISL_1218583, EPI_ISL_1218584, EPI_ISL_1218585, EPI_ISL_1218586, EPI_ISL_1218587, EPI_ISL_1218591, EPI_ISL_1218592, EPI_ISL_1218593, EPI_ISL_1218594, EPI_ISL_1218595, EPI_ISL_1218598, EPI_ISL_1218599, EPI_ISL_1218601, EPI_ISL_1218604, EPI_ISL_1218605, EPI_ISL_1218607, EPI_ISL_1218609, EPI_ISL_1218612, EPI_ISL_1218615, EPI_ISL_1218616, EPI_ISL_1218618, EPI_ISL_1218620, EPI_ISL_1218621, EPI_ISL_1218622, EPI_ISL_1218624, EPI_ISL_1218626, EPI_ISL_1218628, EPI_ISL_1218629, EPI_ISL_1218630, EPI_ISL_1218632, EPI_ISL_1218633, EPI_ISL_1218634, EPI_ISL_1218636, EPI_ISL_1218638, | | | |

| | | | |
|---|---|---|---|
| EPI_ISL_1218642, EPI_ISL_1218644, EPI_ISL_1281822, EPI_ISL_1281823, EPI_ISL_1281824, EPI_ISL_1281825, EPI_ISL_1281826, EPI_ISL_1281827, EPI_ISL_1281828, EPI_ISL_1281829, EPI_ISL_1281830, EPI_ISL_1281831, EPI_ISL_1281832, EPI_ISL_1281833, EPI_ISL_1281834, EPI_ISL_1281835, EPI_ISL_1281836, EPI_ISL_1281837, EPI_ISL_1281838, EPI_ISL_1281839, EPI_ISL_1281840, EPI_ISL_1281842, EPI_ISL_1281843, EPI_ISL_1281844, EPI_ISL_1281845, EPI_ISL_1281846, EPI_ISL_1281847, EPI_ISL_1281848, EPI_ISL_1281849, EPI_ISL_1281850, EPI_ISL_1281851, EPI_ISL_1281852, EPI_ISL_1281853, EPI_ISL_1281854, EPI_ISL_1281855, EPI_ISL_1281856, EPI_ISL_1281857, EPI_ISL_1281858, EPI_ISL_1281859, EPI_ISL_1281860, EPI_ISL_1281861, EPI_ISL_1281862, EPI_ISL_1281863, EPI_ISL_1281864, EPI_ISL_1281865, EPI_ISL_1281866, EPI_ISL_1281867, EPI_ISL_1281868, EPI_ISL_1281869, EPI_ISL_1281870, EPI_ISL_1281871, EPI_ISL_1281872, EPI_ISL_1281873, EPI_ISL_1281874, EPI_ISL_1281875, EPI_ISL_1281876, EPI_ISL_1281877, EPI_ISL_1281878, EPI_ISL_1281879, EPI_ISL_1281880, EPI_ISL_1281881, EPI_ISL_1281882, EPI_ISL_1281883, EPI_ISL_1281884, EPI_ISL_1281885, EPI_ISL_1281886, EPI_ISL_1281887, EPI_ISL_1281888, EPI_ISL_1281889, EPI_ISL_1281890, EPI_ISL_1281891, EPI_ISL_1281892, EPI_ISL_1281893, EPI_ISL_1281894, EPI_ISL_1281895, EPI_ISL_1281896, EPI_ISL_1281897, EPI_ISL_1281898, EPI_ISL_1281899, EPI_ISL_1281900, EPI_ISL_1281901, EPI_ISL_1281902, EPI_ISL_1281903, EPI_ISL_1281904, EPI_ISL_1281905, EPI_ISL_1281906, EPI_ISL_1281907, EPI_ISL_1281908, EPI_ISL_1281909, EPI_ISL_1281910, EPI_ISL_1284529, EPI_ISL_1285385, EPI_ISL_1285386, EPI_ISL_1285387, EPI_ISL_1285388, EPI_ISL_1285389, EPI_ISL_1285390, EPI_ISL_1285391, EPI_ISL_1285392, EPI_ISL_1285393, EPI_ISL_1285394, EPI_ISL_1285395, EPI_ISL_1285396, EPI_ISL_1285397, EPI_ISL_1285398, EPI_ISL_1285400, EPI_ISL_1285402, EPI_ISL_1285403, EPI_ISL_1285404, EPI_ISL_1285406, EPI_ISL_1285407, EPI_ISL_1285409, EPI_ISL_1285412, EPI_ISL_1285413, EPI_ISL_1285414, EPI_ISL_1285415, EPI_ISL_1285416, EPI_ISL_1285418, EPI_ISL_1285419, EPI_ISL_1285420, EPI_ISL_1285421, EPI_ISL_1285422, EPI_ISL_1285423, EPI_ISL_1285426, EPI_ISL_1285427, EPI_ISL_1285428, EPI_ISL_1349149, EPI_ISL_1349155, EPI_ISL_1349159, EPI_ISL_1349162, EPI_ISL_1349163, EPI_ISL_1349167, EPI_ISL_1349172, EPI_ISL_1349173, EPI_ISL_1349174, EPI_ISL_1349175, EPI_ISL_1349179, EPI_ISL_1349181, EPI_ISL_1349182, EPI_ISL_1349183, EPI_ISL_1349184, EPI_ISL_1349188, EPI_ISL_1349192, EPI_ISL_1349193, EPI_ISL_1349194, EPI_ISL_1349197, EPI_ISL_1349201, EPI_ISL_1349206, EPI_ISL_1349209, EPI_ISL_1349210, EPI_ISL_1349211, EPI_ISL_1349217, EPI_ISL_1349222, EPI_ISL_1349223, EPI_ISL_1349224, EPI_ISL_1351270, EPI_ISL_1351271, EPI_ISL_1351272, EPI_ISL_1351273, EPI_ISL_1351274, EPI_ISL_1351275, EPI_ISL_1351276, EPI_ISL_1351277, EPI_ISL_1351278, EPI_ISL_1351279, EPI_ISL_1351280, EPI_ISL_1351281, EPI_ISL_1351282, EPI_ISL_1351283, EPI_ISL_1351284, EPI_ISL_1351285, EPI_ISL_1351286, EPI_ISL_1351287, EPI_ISL_1351289, EPI_ISL_1351290, EPI_ISL_1351291, EPI_ISL_1351292, EPI_ISL_1351293, EPI_ISL_1351294, EPI_ISL_1351295, EPI_ISL_1351296, EPI_ISL_1351297, EPI_ISL_1351298, EPI_ISL_1351299, EPI_ISL_1351300, EPI_ISL_1351301, EPI_ISL_1351302, EPI_ISL_1351303, EPI_ISL_1351304, EPI_ISL_1351305, EPI_ISL_1351306, EPI_ISL_1351307, EPI_ISL_1351308, EPI_ISL_1351309, EPI_ISL_1351310, EPI_ISL_1351311, EPI_ISL_1351312, EPI_ISL_1351313, EPI_ISL_1351314, EPI_ISL_1351315, EPI_ISL_1351316, EPI_ISL_1351317, EPI_ISL_1351318, EPI_ISL_1351319, EPI_ISL_1351320, EPI_ISL_1351321, EPI_ISL_1351322, EPI_ISL_1351323, EPI_ISL_1351324, EPI_ISL_1351326, EPI_ISL_1351327, EPI_ISL_1351328, EPI_ISL_1351329, EPI_ISL_1351330, EPI_ISL_1351331, EPI_ISL_1351332, EPI_ISL_1351333, EPI_ISL_1351334, EPI_ISL_1351335, EPI_ISL_1351336, EPI_ISL_1351337, EPI_ISL_1351338, EPI_ISL_1351339, EPI_ISL_1351340, EPI_ISL_1351341, EPI_ISL_1351342, EPI_ISL_1351343, EPI_ISL_1351344, EPI_ISL_1351345, EPI_ISL_1351346, EPI_ISL_1351347, EPI_ISL_1351348, EPI_ISL_1351349, EPI_ISL_1351350, EPI_ISL_1351351, EPI_ISL_1351352, EPI_ISL_1351353, EPI_ISL_1351354, EPI_ISL_1354634, EPI_ISL_1354636, EPI_ISL_1354637, EPI_ISL_1354644, EPI_ISL_1354662, EPI_ISL_1354663, EPI_ISL_1354666, EPI_ISL_1354676 | | | |
| see above | MVZ Labor Krone GbR | Robert Koch Institute | |
| EPI_ISL_1281791, EPI_ISL_1284481, EPI_ISL_1284482, EPI_ISL_1284483, EPI_ISL_1353144, EPI_ISL_1353146, EPI_ISL_1353150, EPI_ISL_1353152, EPI_ISL_1353172, EPI_ISL_1353184, EPI_ISL_1353191, EPI_ISL_1353197, EPI_ISL_1353202, EPI_ISL_1353207, EPI_ISL_1353211, EPI_ISL_1353215, EPI_ISL_1434454, EPI_ISL_1434463, EPI_ISL_1434471, EPI_ISL_1434479, EPI_ISL_1434485, EPI_ISL_1435293, EPI_ISL_1435296 | | | |
| see above | MVZ Medizinisches Labor Hannover GmbH | Robert Koch Institute | |
| EPI_ISL_1280674, EPI_ISL_1353372, EPI_ISL_1353384, EPI_ISL_1353396, EPI_ISL_1433535, EPI_ISL_1433543 | MVZ für Laboratoriumsmedizin und Mikrobiologie Koblenz-Mittelrhein (Labor Koblenz) | Robert Koch Institute | |
| EPI_ISL_1285299, EPI_ISL_1285308 | MVZ für Laboratoriumsmedizin und Mikrobiologie Würzburg | Robert Koch Institute | |
| EPI_ISL_1751059, EPI_ISL_1752098, EPI_ISL_1752108, EPI_ISL_1752109, EPI_ISL_1752110, EPI_ISL_1752118, EPI_ISL_1752175, EPI_ISL_1752176, EPI_ISL_1752177, EPI_ISL_1752233, EPI_ISL_1752234, EPI_ISL_1752237, EPI_ISL_1752323, EPI_ISL_1752324, EPI_ISL_1752325, EPI_ISL_1752326, EPI_ISL_1752327, EPI_ISL_1752334, EPI_ISL_1752335, EPI_ISL_1752336, EPI_ISL_1752347, EPI_ISL_1752351, EPI_ISL_1752353, EPI_ISL_1752355, EPI_ISL_1752356, EPI_ISL_1752357, EPI_ISL_1752358, EPI_ISL_1752359, EPI_ISL_1752360, EPI_ISL_1752361, EPI_ISL_1752362, EPI_ISL_1752363, EPI_ISL_1752364, EPI_ISL_1752365, EPI_ISL_1752366, EPI_ISL_1752367, EPI_ISL_1752402, EPI_ISL_1752403, EPI_ISL_1752404, EPI_ISL_1752405, EPI_ISL_1752406, EPI_ISL_1752407, EPI_ISL_1752408, EPI_ISL_1752410, EPI_ISL_1752411, EPI_ISL_1752412, EPI_ISL_1752413, EPI_ISL_1752414, EPI_ISL_2094538, EPI_ISL_2094616, EPI_ISL_2094632, EPI_ISL_2094649, EPI_ISL_2094667, EPI_ISL_2094668, EPI_ISL_2094669, EPI_ISL_2094673, EPI_ISL_2094674, EPI_ISL_2094675, EPI_ISL_2094676, EPI_ISL_2094677, EPI_ISL_2094837, EPI_ISL_2094842, EPI_ISL_2094843, EPI_ISL_2094844, EPI_ISL_2094845, EPI_ISL_2094846, EPI_ISL_2094847, EPI_ISL_2094848, EPI_ISL_2094849, EPI_ISL_2094850, EPI_ISL_2094974, EPI_ISL_2094994, EPI_ISL_2095149, EPI_ISL_2095156, EPI_ISL_2095162, EPI_ISL_2095230, EPI_ISL_2095231, EPI_ISL_2095232, EPI_ISL_2095234, EPI_ISL_2095266, EPI_ISL_2095267, EPI_ISL_2095301 | | | |
| see above | Max von Pettenkofer Institute, Virology, National Reference Center for Retroviruses, LMU Munich | Laboratory for Functional Genome Analysis; Dept. Genomics; Gene Center of the LMU Munich | Alexander Graf; Helmut Blum; Max Muenchhoff; Oliver Keppler; Stefan Krebs |
| EPI_ISL_1217576, EPI_ISL_1217577, EPI_ISL_1217584, EPI_ISL_1217590, EPI_ISL_1217593, EPI_ISL_1217595, EPI_ISL_1217601, EPI_ISL_1284899 | | | |
| see above | Med. Labor Prof. Schenk Dr. Ansorge & Kollegen | Robert Koch Institute | |
| EPI_ISL_1347821, EPI_ISL_1347822, EPI_ISL_1347823, EPI_ISL_1347824, EPI_ISL_1347825, EPI_ISL_1347827, EPI_ISL_1347828, EPI_ISL_1347829, EPI_ISL_1347830, EPI_ISL_1347831, EPI_ISL_1347832, EPI_ISL_1347835, EPI_ISL_1347836, EPI_ISL_1347837, EPI_ISL_1347838, EPI_ISL_1347839, EPI_ISL_1347841, EPI_ISL_1347842, EPI_ISL_1347847, EPI_ISL_1347848, EPI_ISL_1347849, EPI_ISL_1347850, EPI_ISL_1347852, EPI_ISL_1347853, EPI_ISL_1347854, EPI_ISL_1347855, EPI_ISL_1347856, EPI_ISL_1347857, EPI_ISL_1347859, EPI_ISL_1347860, EPI_ISL_1347861, EPI_ISL_1347862, EPI_ISL_1347863, EPI_ISL_1347864, EPI_ISL_1347866, EPI_ISL_1347867, EPI_ISL_1347868, EPI_ISL_1347870, EPI_ISL_1347871, EPI_ISL_1347872, EPI_ISL_1347873, EPI_ISL_1347874, EPI_ISL_1347875, EPI_ISL_1347876, EPI_ISL_1347877, EPI_ISL_1347949, EPI_ISL_1347950, EPI_ISL_1347968, EPI_ISL_1347971, EPI_ISL_1347972, EPI_ISL_1347973, EPI_ISL_1347974, EPI_ISL_1347975, EPI_ISL_1347976, EPI_ISL_1347977, EPI_ISL_1347978, EPI_ISL_1347979, EPI_ISL_1347980, EPI_ISL_1347981, EPI_ISL_1347982, EPI_ISL_1347983, EPI_ISL_1347984, EPI_ISL_1347985, EPI_ISL_1347986 | | | |
| see above | Medical Laboratories Duesseldorf | Center of Medical Microbiology, Virology, and Hospital Hygiene, University of Duesseldorf | Alexander Dilthey; Andreas Walker; Angelika Helmer; Christian Lange; Daniel Strelow; Jessica Nicolai; Jörg Timm; Klaus Pfeffer; Lisanna Hülse; Malte Kohns Vasconcelos; Maximilian Damagnez; Nadine Lübke; Tobias Wienemann; Torsten Houwaart |
| EPI_ISL_1351357, EPI_ISL_1351366, EPI_ISL_1354699 | Medizinisch-Diagnostisches Labor Kempten allgäulab | Robert Koch Institute | |
| EPI_ISL_1350081, EPI_ISL_1350082, EPI_ISL_1350083, EPI_ISL_1350084, EPI_ISL_1350085, EPI_ISL_1350086, EPI_ISL_1350087, EPI_ISL_1350088, EPI_ISL_1350089, EPI_ISL_1350090, EPI_ISL_1350091, EPI_ISL_1350092, EPI_ISL_1350093, EPI_ISL_1350094, EPI_ISL_1350095, EPI_ISL_1350096, EPI_ISL_1350097, EPI_ISL_1350098, EPI_ISL_1350099, EPI_ISL_1350100, EPI_ISL_1350101, EPI_ISL_1350102, EPI_ISL_1350103, EPI_ISL_1350104, EPI_ISL_1350105, EPI_ISL_1350106 | | | |
| see above | Medizinische Laboratorien Düsseldorf | Robert Koch Institute | |
| EPI_ISL_1217616, EPI_ISL_1217621, EPI_ISL_1217622, EPI_ISL_1217624, EPI_ISL_1217625, EPI_ISL_1217626, EPI_ISL_1217628, EPI_ISL_1217631, EPI_ISL_1217632, EPI_ISL_1217633, EPI_ISL_1217635, EPI_ISL_1217636, EPI_ISL_1217638, EPI_ISL_1217640, EPI_ISL_1217644, EPI_ISL_1217646, EPI_ISL_1217649, EPI_ISL_1217650, EPI_ISL_1217655, EPI_ISL_1217657, EPI_ISL_1217660, EPI_ISL_1217662, EPI_ISL_1217665, EPI_ISL_1217671, EPI_ISL_1217676, EPI_ISL_1217677, EPI_ISL_1217678, EPI_ISL_1217685, EPI_ISL_1217687, EPI_ISL_1217688, EPI_ISL_1217696, EPI_ISL_1217699, EPI_ISL_1217700, EPI_ISL_1217701, EPI_ISL_1281800, EPI_ISL_1281802, EPI_ISL_1281803, EPI_ISL_1281804, EPI_ISL_1281808, EPI_ISL_1283139, EPI_ISL_1283148, EPI_ISL_1283173, EPI_ISL_1283176, EPI_ISL_1283179, EPI_ISL_1283180, EPI_ISL_1283188, EPI_ISL_1283190, EPI_ISL_1283192, EPI_ISL_1283195, EPI_ISL_1283197, EPI_ISL_1283204, EPI_ISL_1283205, EPI_ISL_1283207, EPI_ISL_1283208, EPI_ISL_1283210, EPI_ISL_1283215, EPI_ISL_1283231, EPI_ISL_1283233, EPI_ISL_1285043, EPI_ISL_1285047, EPI_ISL_1285049, EPI_ISL_1285052, EPI_ISL_1285054, EPI_ISL_1285055, EPI_ISL_1285058, EPI_ISL_1285059, EPI_ISL_1285066, EPI_ISL_1285067, EPI_ISL_1285068, EPI_ISL_1285070, EPI_ISL_1285073, EPI_ISL_1285077, EPI_ISL_1285081, EPI_ISL_1285084, EPI_ISL_1285085, EPI_ISL_1285087, EPI_ISL_1285091, EPI_ISL_1285099, EPI_ISL_1285102, EPI_ISL_1285111, EPI_ISL_1285114, EPI_ISL_1285119, EPI_ISL_1285140, EPI_ISL_1285141, EPI_ISL_1285145, EPI_ISL_1285148, EPI_ISL_1285153, EPI_ISL_1285159, EPI_ISL_1353166, EPI_ISL_1353173, EPI_ISL_1353186, EPI_ISL_1353213, EPI_ISL_1353220 | | | |
| see above | Medizinisches Labor Ostsachsen MVZ GbRNebenbetriebsstätte Dresden | Robert Koch Institute | |
| EPI_ISL_1361639, EPI_ISL_1361640, EPI_ISL_1361641, EPI_ISL_1361642, EPI_ISL_1361643, EPI_ISL_1361644, EPI_ISL_1361645, EPI_ISL_1361646, EPI_ISL_1361647, EPI_ISL_1361648, EPI_ISL_1361649, EPI_ISL_1361650, EPI_ISL_1361651, EPI_ISL_1361652, EPI_ISL_1361653, EPI_ISL_1361654, EPI_ISL_1361655, EPI_ISL_1361656 | | | |
| see above | Medizinisches Labor Prof. Dr. Schenk/ Dr. Ansorge & Kollegen, Magdeburg | Institute of Medical Microbiology and Hospital Hygiene | Aljoscha Tersteegen; Prof. Dr. Achim Kaasch |
| EPI_ISL_1214875, EPI_ISL_1214937, EPI_ISL_1214947, EPI_ISL_1214972, EPI_ISL_1285283, EPI_ISL_1285288, EPI_ISL_1285292, EPI_ISL_1285294, EPI_ISL_1285295, EPI_ISL_1285297, EPI_ISL_1285300, EPI_ISL_1285303, EPI_ISL_1285311, EPI_ISL_1285336 | | | |
| see above | Medizinisches Labor Wahl Lüdenscheid | Robert Koch Institute | |
| EPI_ISL_1281916 | Medizinisches Versorgungszentrum für Labormedizin und Mikrobiologie Ruhr GmbH - mvzlm RUHR GmbH | Robert Koch Institute | |
| EPI_ISL_1213870, EPI_ISL_1213918, EPI_ISL_1213923, EPI_ISL_1214077, EPI_ISL_1215651, EPI_ISL_1215653, EPI_ISL_1215658, EPI_ISL_1215660, EPI_ISL_1215663, EPI_ISL_1215666, EPI_ISL_1215675, EPI_ISL_1215677, EPI_ISL_1215684, EPI_ISL_1215687, EPI_ISL_1215689, EPI_ISL_1215690, EPI_ISL_1215691, EPI_ISL_1215695, EPI_ISL_1215697, EPI_ISL_1215701, EPI_ISL_1215705, EPI_ISL_1215706, EPI_ISL_1215712, EPI_ISL_1215713, EPI_ISL_1215723, EPI_ISL_1215734, EPI_ISL_1215735, EPI_ISL_1218217, EPI_ISL_1218226, EPI_ISL_1218249, EPI_ISL_1218253, EPI_ISL_1218260, EPI_ISL_1218263, EPI_ISL_1218266, EPI_ISL_1218271, EPI_ISL_1218275, EPI_ISL_1218295, EPI_ISL_1218302, EPI_ISL_1218306, EPI_ISL_1218313, EPI_ISL_1218317, EPI_ISL_1218327, EPI_ISL_1218331, EPI_ISL_1218336, EPI_ISL_1218342, EPI_ISL_1218346, EPI_ISL_1218354, EPI_ISL_1218359, EPI_ISL_1218366, EPI_ISL_1218374, EPI_ISL_1218388, EPI_ISL_1218393, EPI_ISL_1218397, EPI_ISL_1218403, EPI_ISL_1218408, EPI_ISL_1218411, EPI_ISL_1218417, EPI_ISL_1218420, EPI_ISL_1218422, EPI_ISL_1218430, EPI_ISL_1218434, EPI_ISL_1218436, EPI_ISL_1218451, EPI_ISL_1218455, EPI_ISL_1218460, EPI_ISL_1218466, EPI_ISL_1218471, EPI_ISL_1218473, EPI_ISL_1218475, EPI_ISL_1218491, EPI_ISL_1218499, EPI_ISL_1218506, EPI_ISL_1218520, EPI_ISL_1218521, EPI_ISL_1218522, EPI_ISL_1218531, EPI_ISL_1218533, EPI_ISL_1218535, EPI_ISL_1218546, EPI_ISL_1280799, EPI_ISL_1280826, EPI_ISL_1280839, EPI_ISL_1280844, EPI_ISL_1280849, EPI_ISL_1280857, EPI_ISL_1280861, EPI_ISL_1280863, EPI_ISL_1280872, EPI_ISL_1280878, EPI_ISL_1280885, EPI_ISL_1280889, EPI_ISL_1280894, EPI_ISL_1280901, EPI_ISL_1280903, EPI_ISL_1280905, EPI_ISL_1283913, EPI_ISL_1283918, EPI_ISL_1283951, EPI_ISL_1283966, EPI_ISL_1284081, EPI_ISL_1284153, EPI_ISL_1284159, EPI_ISL_1348034, EPI_ISL_1348035, EPI_ISL_1348037, EPI_ISL_1348040, EPI_ISL_1348046, EPI_ISL_1348065, EPI_ISL_1348069, EPI_ISL_1348075, EPI_ISL_1348077, EPI_ISL_1348078, EPI_ISL_1348080, EPI_ISL_1348083, EPI_ISL_1348088, EPI_ISL_1348090, EPI_ISL_1348093, EPI_ISL_1348100, EPI_ISL_1348102, EPI_ISL_1348111, EPI_ISL_1348122, EPI_ISL_1348126, EPI_ISL_1348133, EPI_ISL_1348143, EPI_ISL_1348183, EPI_ISL_1348190, EPI_ISL_1348309, EPI_ISL_1348312, EPI_ISL_1348318, EPI_ISL_1348325, EPI_ISL_1348342, EPI_ISL_1348346, EPI_ISL_1348347, EPI_ISL_1348348, EPI_ISL_1348356, EPI_ISL_1348362, EPI_ISL_1348371, EPI_ISL_1348374, EPI_ISL_1348375, EPI_ISL_1348379, EPI_ISL_1348386, EPI_ISL_1348397, EPI_ISL_1348412, EPI_ISL_1348415, EPI_ISL_1348416, EPI_ISL_1348418, EPI_ISL_1348420, EPI_ISL_1348422, EPI_ISL_1348430, EPI_ISL_1348434, EPI_ISL_1348437, EPI_ISL_1348446, EPI_ISL_1348463, EPI_ISL_1348465, EPI_ISL_1348469, EPI_ISL_1348473, EPI_ISL_1348474, EPI_ISL_1348476, EPI_ISL_1348477, EPI_ISL_1348478, EPI_ISL_1348480, EPI_ISL_1348481, EPI_ISL_1348485, EPI_ISL_1348487, EPI_ISL_1348488, EPI_ISL_1348489, EPI_ISL_1348490, EPI_ISL_1351859, EPI_ISL_1351892, EPI_ISL_1352063, EPI_ISL_1352101, EPI_ISL_1352183 | | | |
| see above | Medizinisches Zentrallabor Altenburg GmbH & Co.KG | Robert Koch Institute | |
| EPI_ISL_1350924, EPI_ISL_1350925, EPI_ISL_1350926, EPI_ISL_1350927, EPI_ISL_1350928, EPI_ISL_1350929, EPI_ISL_1350930, EPI_ISL_1350931, EPI_ISL_1350932, EPI_ISL_1350933, EPI_ISL_1350934, EPI_ISL_1350935, EPI_ISL_1350936, EPI_ISL_1350937, EPI_ISL_1350939, EPI_ISL_1350940, EPI_ISL_1350941, EPI_ISL_1350947 | | | |
| see above | Niedersächsisches Landesgesundheitsamt (NLGA) | Robert Koch Institute | |
| EPI_ISL_1217681 | Oberlausitz Kliniken gGmBH | Robert Koch Institute | |
| EPI_ISL_1211934, EPI_ISL_1211936, EPI_ISL_1211937, EPI_ISL_1211941, EPI_ISL_1211943, EPI_ISL_1284881, EPI_ISL_1284884, EPI_ISL_1284887 | | | |
| see above | Philipps-Universität Marburg Fachbereich Medizin Institut für Virologie | Robert Koch Institute | |
| EPI_ISL_1214089, EPI_ISL_1280896, EPI_ISL_1280916, EPI_ISL_1283969, EPI_ISL_1284050, EPI_ISL_1284115, EPI_ISL_1284228, EPI_ISL_1284238, EPI_ISL_1348453, EPI_ISL_1350669, EPI_ISL_1350705, EPI_ISL_1350708 | | | |
| see above | Praxisgemeinschaft für Laboratoriumsmedizin Labor Blumenstraße; Praxis Dr. E. Wietschel & Dr. F. Wietschel | Robert Koch Institute | |
| EPI_ISL_1214082, EPI_ISL_1214092, EPI_ISL_1280920, EPI_ISL_1280940, EPI_ISL_1284102, EPI_ISL_1348055, EPI_ISL_1348154, EPI_ISL_1348459, EPI_ISL_1348468, EPI_ISL_1350629, EPI_ISL_1350642, EPI_ISL_1350728 | | | |
| see above | Praxisgemeinschaft für Laboratoriumsmedizin Labor Blumenstraße; Praxis Dr. Sel und Dr. Späte | Robert Koch Institute | |
| EPI_ISL_1281925, EPI_ISL_1281953, EPI_ISL_1281960, EPI_ISL_1350258, EPI_ISL_1350269, EPI_ISL_1350278, EPI_ISL_1350306, EPI_ISL_1350309 | | | |
| see above | Procomcure Biotech Germany GmbH | Robert Koch Institute | |
| EPI_ISL_1285547, EPI_ISL_1354075 | Robert-Bosch-Krankenhaus | Robert Koch Institute | |
| EPI_ISL_1281221, EPI_ISL_1284473 | SLK-Kliniken Heilbronn GmbH - Institut für Laboratoriumsmedizin | Robert Koch Institute | |
| EPI_ISL_1213786, EPI_ISL_1213789, EPI_ISL_1213812, EPI_ISL_1213813, EPI_ISL_1213821, EPI_ISL_1213828, EPI_ISL_1213833, EPI_ISL_1213834, EPI_ISL_1213838, EPI_ISL_1213853, EPI_ISL_1213874, EPI_ISL_1213876, EPI_ISL_1213877, EPI_ISL_1213880, EPI_ISL_1213883, EPI_ISL_1213887, EPI_ISL_1213891, EPI_ISL_1213898, EPI_ISL_1213899, EPI_ISL_1213910, EPI_ISL_1213921, EPI_ISL_1213935, EPI_ISL_1213936, EPI_ISL_1213939, EPI_ISL_1213940, EPI_ISL_1213948, EPI_ISL_1213953, EPI_ISL_1213960, EPI_ISL_1213970, EPI_ISL_1214003, EPI_ISL_1214034, EPI_ISL_1214053, EPI_ISL_1214061, EPI_ISL_1214096, EPI_ISL_1214102, EPI_ISL_1215654, EPI_ISL_1215665, EPI_ISL_1215670, EPI_ISL_1215671, EPI_ISL_1215674, EPI_ISL_1215693, EPI_ISL_1215700, EPI_ISL_1215709, EPI_ISL_1215725, EPI_ISL_1215727, EPI_ISL_1215740, EPI_ISL_1218202, EPI_ISL_1218207, EPI_ISL_1218210, EPI_ISL_1218212, EPI_ISL_1218213, EPI_ISL_1218214, EPI_ISL_1218218, EPI_ISL_1218221, EPI_ISL_1218222, EPI_ISL_1218223, EPI_ISL_1218224, EPI_ISL_1218225, EPI_ISL_1218227, EPI_ISL_1218228, EPI_ISL_1218230, EPI_ISL_1218232, EPI_ISL_1218234, EPI_ISL_1218236, EPI_ISL_1218238, EPI_ISL_1218242, EPI_ISL_1218244, EPI_ISL_1218245, EPI_ISL_1218255, EPI_ISL_1218257, EPI_ISL_1218259, EPI_ISL_1218265, EPI_ISL_1218268, EPI_ISL_1218269, EPI_ISL_1218273, EPI_ISL_1218277, EPI_ISL_1218282, EPI_ISL_1218291, EPI_ISL_1218315, EPI_ISL_1218320, EPI_ISL_1218322, EPI_ISL_1218323, EPI_ISL_1218328, EPI_ISL_1218337, EPI_ISL_1218340, EPI_ISL_1218350, EPI_ISL_1218361, EPI_ISL_1218362, EPI_ISL_1218363, EPI_ISL_1218365, EPI_ISL_1218369, EPI_ISL_1218371, EPI_ISL_1218373, EPI_ISL_1218376, EPI_ISL_1218382, EPI_ISL_1218387, EPI_ISL_1218391, EPI_ISL_1218399, EPI_ISL_1218401, EPI_ISL_1218404, EPI_ISL_1218406, EPI_ISL_1218407, EPI_ISL_1218409, EPI_ISL_1218412, EPI_ISL_1218414, EPI_ISL_1218416, EPI_ISL_1218418, EPI_ISL_1218419, EPI_ISL_1218423, EPI_ISL_1218425, EPI_ISL_1218426, EPI_ISL_1218427, EPI_ISL_1218428, EPI_ISL_1218429, EPI_ISL_1218433, EPI_ISL_1218438, EPI_ISL_1218439, EPI_ISL_1218440, EPI_ISL_1218442, EPI_ISL_1218444, EPI_ISL_1218447, EPI_ISL_1218453, EPI_ISL_1218456, EPI_ISL_1218459, EPI_ISL_1218462, EPI_ISL_1218467, EPI_ISL_1218468, EPI_ISL_1218472, EPI_ISL_1218474, EPI_ISL_1218478, EPI_ISL_1218479, EPI_ISL_1218483, EPI_ISL_1218484, EPI_ISL_1218485, EPI_ISL_1218486, EPI_ISL_1218492, EPI_ISL_1218496, EPI_ISL_1218501, EPI_ISL_1218502, EPI_ISL_1218503, EPI_ISL_1218507, EPI_ISL_1218508, EPI_ISL_1218509, EPI_ISL_1218510, EPI_ISL_1218512, EPI_ISL_1218513, EPI_ISL_1218514, EPI_ISL_1218519, EPI_ISL_1218523, EPI_ISL_1218524, EPI_ISL_1218525, EPI_ISL_1218526, EPI_ISL_1218527, EPI_ISL_1218528, EPI_ISL_1218529, EPI_ISL_1218534, EPI_ISL_1218537, EPI_ISL_1218538, EPI_ISL_1218540, EPI_ISL_1218541, EPI_ISL_1218543, EPI_ISL_1218547, EPI_ISL_1218548, EPI_ISL_1218549, EPI_ISL_1280784, EPI_ISL_1280804, EPI_ISL_1280815, EPI_ISL_1280820, EPI_ISL_1280821, EPI_ISL_1280833, EPI_ISL_1280835, EPI_ISL_1280838, EPI_ISL_1280840, EPI_ISL_1280846, EPI_ISL_1280848, EPI_ISL_1280855, EPI_ISL_1280858, EPI_ISL_1280865, EPI_ISL_1280866, EPI_ISL_1280870, EPI_ISL_1280874, EPI_ISL_1280880, EPI_ISL_1280884, EPI_ISL_1280886, EPI_ISL_1280887, EPI_ISL_1280898, EPI_ISL_1280906, EPI_ISL_1280907, EPI_ISL_1280913, EPI_ISL_1280914, EPI_ISL_1280915, EPI_ISL_1280930, EPI_ISL_1280931, EPI_ISL_1280934, EPI_ISL_1280936, EPI_ISL_1280937, EPI_ISL_1280939, EPI_ISL_1280941, EPI_ISL_1280943, EPI_ISL_1280944, EPI_ISL_1280945, EPI_ISL_1282988, EPI_ISL_1282990, EPI_ISL_1282992, EPI_ISL_1282993, EPI_ISL_1282994, EPI_ISL_1282996, EPI_ISL_1282997, EPI_ISL_1282999, EPI_ISL_1283001, EPI_ISL_1283002, EPI_ISL_1283004, EPI_ISL_1283006, EPI_ISL_1283007, EPI_ISL_1283009, EPI_ISL_1283010, EPI_ISL_1283013, EPI_ISL_1283014, EPI_ISL_1283015, EPI_ISL_1283017, EPI_ISL_1283018, EPI_ISL_1283019, EPI_ISL_1283021, EPI_ISL_1283023, EPI_ISL_1283024, EPI_ISL_1283025, EPI_ISL_1283027, EPI_ISL_1283028, EPI_ISL_1283029, EPI_ISL_1283032, EPI_ISL_1283033, EPI_ISL_1283034, EPI_ISL_1283043, EPI_ISL_1283044, EPI_ISL_1283046, EPI_ISL_1283048, EPI_ISL_1283049, EPI_ISL_1283050, EPI_ISL_1283052, EPI_ISL_1283053, EPI_ISL_1283059, EPI_ISL_1283060, EPI_ISL_1283064, EPI_ISL_1283065, EPI_ISL_1283070, EPI_ISL_1283071, EPI_ISL_1283072, EPI_ISL_1283073, EPI_ISL_1283074, EPI_ISL_1283075, EPI_ISL_1283076, EPI_ISL_1283078, EPI_ISL_1283079, EPI_ISL_1283082, EPI_ISL_1283083, EPI_ISL_1283084, EPI_ISL_1283085, EPI_ISL_1283089, EPI_ISL_1283093, EPI_ISL_1283094, EPI_ISL_1283096, EPI_ISL_1283098, EPI_ISL_1283100, EPI_ISL_1283102, EPI_ISL_1283103, EPI_ISL_1283104, EPI_ISL_1283105, EPI_ISL_1283106, EPI_ISL_1283107, EPI_ISL_1283111, EPI_ISL_1283112, EPI_ISL_1283114, EPI_ISL_1283115, EPI_ISL_1283117, EPI_ISL_1283119, EPI_ISL_1283120, EPI_ISL_1283121, EPI_ISL_1283857, EPI_ISL_1283860, EPI_ISL_1283861, EPI_ISL_1283865, EPI_ISL_1283871, EPI_ISL_1283876, EPI_ISL_1283887, EPI_ISL_1283890, EPI_ISL_1283892, EPI_ISL_1283894, EPI_ISL_1283899, EPI_ISL_1283903, EPI_ISL_1283907, EPI_ISL_1283908, EPI_ISL_1283910, EPI_ISL_1283917, EPI_ISL_1283925, EPI_ISL_1283931, EPI_ISL_1283934, EPI_ISL_1283939, EPI_ISL_1283942, EPI_ISL_1283944, EPI_ISL_1283950, EPI_ISL_1283954, EPI_ISL_1283958, EPI_ISL_1283963, EPI_ISL_1283974, EPI_ISL_1283980, EPI_ISL_1283989, EPI_ISL_1283999, EPI_ISL_1284003, EPI_ISL_1284005, EPI_ISL_1284009, EPI_ISL_1284018, EPI_ISL_1284020, EPI_ISL_1284022, EPI_ISL_1284030, EPI_ISL_1284037, EPI_ISL_1284038, EPI_ISL_1284041, EPI_ISL_1284045, EPI_ISL_1284053, EPI_ISL_1284055, EPI_ISL_1284066, EPI_ISL_1284068, EPI_ISL_1284071, EPI_ISL_1284073, EPI_ISL_1284080, EPI_ISL_1284089, EPI_ISL_1284095, EPI_ISL_1284098, EPI_ISL_1284104, EPI_ISL_1284106, EPI_ISL_1284122, EPI_ISL_1284126, EPI_ISL_1284130, EPI_ISL_1284132, EPI_ISL_1284135, EPI_ISL_1284137, EPI_ISL_1284147, EPI_ISL_1284150, EPI_ISL_1284157, EPI_ISL_1284162, EPI_ISL_1284180, EPI_ISL_1284208, EPI_ISL_1284216, EPI_ISL_1284225, EPI_ISL_1284232, EPI_ISL_1284235, EPI_ISL_1285933, EPI_ISL_1285945, EPI_ISL_1285949, EPI_ISL_1285950, EPI_ISL_1285952, EPI_ISL_1285966, EPI_ISL_1285998, EPI_ISL_1286002, EPI_ISL_1286007, EPI_ISL_1286010, EPI_ISL_1286014, EPI_ISL_1286020, EPI_ISL_1286022, EPI_ISL_1286024, EPI_ISL_1286025, EPI_ISL_1286030, EPI_ISL_1286031, EPI_ISL_1286032, EPI_ISL_1286034, EPI_ISL_1286035, EPI_ISL_1286036, EPI_ISL_1286039, EPI_ISL_1286042, EPI_ISL_1286047, EPI_ISL_1286055, EPI_ISL_1286056, EPI_ISL_1286058, EPI_ISL_1286059, EPI_ISL_1286060, EPI_ISL_1286063, EPI_ISL_1286065, EPI_ISL_1286068, EPI_ISL_1286069, EPI_ISL_1286072, EPI_ISL_1286074, EPI_ISL_1286077, EPI_ISL_1286080, EPI_ISL_1286083, EPI_ISL_1286088, EPI_ISL_1286089, EPI_ISL_1286092, EPI_ISL_1286093, EPI_ISL_1348039, EPI_ISL_1348042, EPI_ISL_1348043, EPI_ISL_1348044, EPI_ISL_1348047, EPI_ISL_1348048, EPI_ISL_1348052, EPI_ISL_1348053, EPI_ISL_1348056, EPI_ISL_1348058, EPI_ISL_1348061, EPI_ISL_1348062, EPI_ISL_1348063, EPI_ISL_1348066, EPI_ISL_1348067, EPI_ISL_1348079, EPI_ISL_1348081, EPI_ISL_1348084, EPI_ISL_1348086, EPI_ISL_1348087, EPI_ISL_1348092, EPI_ISL_1348095, EPI_ISL_1348096, EPI_ISL_1348097, EPI_ISL_1348101, EPI_ISL_1348104, EPI_ISL_1348115, EPI_ISL_1348117, EPI_ISL_1348119, EPI_ISL_1348124, EPI_ISL_1348128, EPI_ISL_1348131, EPI_ISL_1348147, EPI_ISL_1348150, EPI_ISL_1348159, EPI_ISL_1348161, EPI_ISL_1348169, EPI_ISL_1348181, EPI_ISL_1348196, EPI_ISL_1348199, EPI_ISL_1348203, EPI_ISL_1348212, EPI_ISL_1348219, EPI_ISL_1348223, EPI_ISL_1348227, EPI_ISL_1348230, EPI_ISL_1348232, EPI_ISL_1348237, EPI_ISL_1348239, EPI_ISL_1348241, EPI_ISL_1348246, EPI_ISL_1348255, EPI_ISL_1348262, EPI_ISL_1348266, EPI_ISL_1348269, EPI_ISL_1348277, EPI_ISL_1348280, EPI_ISL_1348287, EPI_ISL_1348289, EPI_ISL_1348291, EPI_ISL_1348296, EPI_ISL_1348321, EPI_ISL_1348341, EPI_ISL_1348343, EPI_ISL_1348345, EPI_ISL_1348349, EPI_ISL_1348357, EPI_ISL_1348360, EPI_ISL_1348361, EPI_ISL_1348364, EPI_ISL_1348367, EPI_ISL_1348368, EPI_ISL_1348378, EPI_ISL_1348380, EPI_ISL_1348383, EPI_ISL_1348390, EPI_ISL_1348391, EPI_ISL_1348393, EPI_ISL_1348408, EPI_ISL_1348426, EPI_ISL_1348427, EPI_ISL_1348439, EPI_ISL_1348443, EPI_ISL_1348450, EPI_ISL_1348456, EPI_ISL_1348466, EPI_ISL_1348470, EPI_ISL_1348486, EPI_ISL_1350643, EPI_ISL_1350656, EPI_ISL_1350710, EPI_ISL_1350714, EPI_ISL_1350756, EPI_ISL_1350759, EPI_ISL_1350767 | | | |

see above SYNLAB Jena Oncoscreen Robert Koch Institute

EPI_ISL_1215652, EPI_ISL_1215656, EPI_ISL_1215685, EPI_ISL_1215710, EPI_ISL_1215714, EPI_ISL_1215717, EPI_ISL_1215722, EPI_ISL_1215724, EPI_ISL_1215726, EPI_ISL_1215729, EPI_ISL_1215730, EPI_ISL_1215732, EPI_ISL_1215733, EPI_ISL_1215739, EPI_ISL_1280788, EPI_ISL_1280793, EPI_ISL_1280794, EPI_ISL_1280796, EPI_ISL_1280801, EPI_ISL_1280813, EPI_ISL_1280817, EPI_ISL_1280822, EPI_ISL_1280834, EPI_ISL_1280854, EPI_ISL_1280881, EPI_ISL_1280890, EPI_ISL_1280893, EPI_ISL_1280895, EPI_ISL_1280926, EPI_ISL_1280929, EPI_ISL_1348041, EPI_ISL_1348372, EPI_ISL_1348410, EPI_ISL_1348413, EPI_ISL_1348417, EPI_ISL_1348441, EPI_ISL_1348451, EPI_ISL_1348457, EPI_ISL_1348458, EPI_ISL_1348460, EPI_ISL_1348461, EPI_ISL_1348483, EPI_ISL_1350651, EPI_ISL_1350665, EPI_ISL_1350698, EPI_ISL_1351918, EPI_ISL_1352016

see above SYNLAB Labor München Zentrum LMZ Robert Koch Institute

EPI_ISL_1215037, EPI_ISL_1215144, EPI_ISL_1215659, EPI_ISL_1215668, EPI_ISL_1215672, EPI_ISL_1215673, EPI_ISL_1215678, EPI_ISL_1215680, EPI_ISL_1215681, EPI_ISL_1215682, EPI_ISL_1215683, EPI_ISL_1215694, EPI_ISL_1215698, EPI_ISL_1215699, EPI_ISL_1215702, EPI_ISL_1215703, EPI_ISL_1215704, EPI_ISL_1215707, EPI_ISL_1215708, EPI_ISL_1215715, EPI_ISL_1215721, EPI_ISL_1215736, EPI_ISL_1215737, EPI_ISL_1215738, EPI_ISL_1218385, EPI_ISL_1218390, EPI_ISL_1218394, EPI_ISL_1218449, EPI_ISL_1218454, EPI_ISL_1218465, EPI_ISL_1218476, EPI_ISL_1218480, EPI_ISL_1218497, EPI_ISL_1218518, EPI_ISL_1218532, EPI_ISL_1218545, EPI_ISL_1280923, EPI_ISL_1280933, EPI_ISL_1282991, EPI_ISL_1283000, EPI_ISL_1283005, EPI_ISL_1283008, EPI_ISL_1283030, EPI_ISL_1283036, EPI_ISL_1283039, EPI_ISL_1283045, EPI_ISL_1283054, EPI_ISL_1283057, EPI_ISL_1283058, EPI_ISL_1283062, EPI_ISL_1283068, EPI_ISL_1283090, EPI_ISL_1283101, EPI_ISL_1285934, EPI_ISL_1285937, EPI_ISL_1285943, EPI_ISL_1285979, EPI_ISL_1285983, EPI_ISL_1285991, EPI_ISL_1285997, EPI_ISL_1286003, EPI_ISL_1286013, EPI_ISL_1286027, EPI_ISL_1286037, EPI_ISL_1286050, EPI_ISL_1286052, EPI_ISL_1286053, EPI_ISL_1286070, EPI_ISL_1286078, EPI_ISL_1286081, EPI_ISL_1286087, EPI_ISL_1348068, EPI_ISL_1348085, EPI_ISL_1348089, EPI_ISL_1348106, EPI_ISL_1348185, EPI_ISL_1348257, EPI_ISL_1348285, EPI_ISL_1348339, EPI_ISL_1348373, EPI_ISL_1348376, EPI_ISL_1348377, EPI_ISL_1348394, EPI_ISL_1348398, EPI_ISL_1348400, EPI_ISL_1348401, EPI_ISL_1348404, EPI_ISL_1348423, EPI_ISL_1348424, EPI_ISL_1348433, EPI_ISL_1348436, EPI_ISL_1348444, EPI_ISL_1350650, EPI_ISL_1356141

see above SYNLAB MVZ Berlin Robert Koch Institute

EPI_ISL_1351023, EPI_ISL_1351115, EPI_ISL_1354531, EPI_ISL_1354619, EPI_ISL_1432055, EPI_ISL_1432089, EPI_ISL_1432125, EPI_ISL_1432155, EPI_ISL_1432265, EPI_ISL_1432312, EPI_ISL_1432432, EPI_ISL_1432678, EPI_ISL_1432686, EPI_ISL_1432706, EPI_ISL_1432713, EPI_ISL_1432717, EPI_ISL_1437434, EPI_ISL_1437440, EPI_ISL_1437456, EPI_ISL_1437460, EPI_ISL_1437483, EPI_ISL_1437487, EPI_ISL_1437590, EPI_ISL_1568847, EPI_ISL_1720051

see above SYNLAB MVZ Bonn Robert Koch Institute

EPI_ISL_1281018, EPI_ISL_1281025, EPI_ISL_1284371, EPI_ISL_1284430 SYNLAB MVZ Dachau Robert Koch Institute

EPI_ISL_1217424, EPI_ISL_1217425, EPI_ISL_1217426, EPI_ISL_1217427, EPI_ISL_1217432, EPI_ISL_1217434, EPI_ISL_1217435, EPI_ISL_1217436, EPI_ISL_1217439, EPI_ISL_1217442, EPI_ISL_1217448, EPI_ISL_1217449, EPI_ISL_1217453, EPI_ISL_1217454, EPI_ISL_1217456, EPI_ISL_1217457, EPI_ISL_1217459, EPI_ISL_1217462, EPI_ISL_1217463, EPI_ISL_1217464, EPI_ISL_1217465, EPI_ISL_1217467, EPI_ISL_1217469, EPI_ISL_1217471, EPI_ISL_1217472, EPI_ISL_1217473, EPI_ISL_1217474, EPI_ISL_1217475, EPI_ISL_1217477, EPI_ISL_1217479, EPI_ISL_1217483, EPI_ISL_1217485, EPI_ISL_1217486, EPI_ISL_1217493, EPI_ISL_1217494, EPI_ISL_1217495, EPI_ISL_1217497, EPI_ISL_1217498, EPI_ISL_1217500, EPI_ISL_1217501, EPI_ISL_1217503, EPI_ISL_1217504, EPI_ISL_1217505, EPI_ISL_1217506, EPI_ISL_1281085, EPI_ISL_1281089, EPI_ISL_1281092, EPI_ISL_1281096, EPI_ISL_1281103, EPI_ISL_1281106, EPI_ISL_1281107, EPI_ISL_1281108, EPI_ISL_1281111, EPI_ISL_1281113, EPI_ISL_1281115, EPI_ISL_1281116, EPI_ISL_1281117, EPI_ISL_1281120, EPI_ISL_1281124, EPI_ISL_1281126, EPI_ISL_1281129, EPI_ISL_1281131, EPI_ISL_1281141, EPI_ISL_1281144, EPI_ISL_1281146, EPI_ISL_1281152, EPI_ISL_1281156, EPI_ISL_1281158, EPI_ISL_1281159, EPI_ISL_1281161, EPI_ISL_1281164, EPI_ISL_1281165, EPI_ISL_1281166, EPI_ISL_1281167, EPI_ISL_1281169, EPI_ISL_1350962, EPI_ISL_1350964, EPI_ISL_1350967, EPI_ISL_1350969, EPI_ISL_1350979, EPI_ISL_1350984, EPI_ISL_1350988, EPI_ISL_1350990, EPI_ISL_1350992, EPI_ISL_1350995, EPI_ISL_1350999, EPI_ISL_1351009, EPI_ISL_1351033, EPI_ISL_1351037, EPI_ISL_1351040, EPI_ISL_1351044, EPI_ISL_1351045, EPI_ISL_1351048, EPI_ISL_1351049, EPI_ISL_1351053, EPI_ISL_1351056, EPI_ISL_1351058, EPI_ISL_1351065, EPI_ISL_1351069, EPI_ISL_1351072, EPI_ISL_1351073, EPI_ISL_1351077, EPI_ISL_1351082, EPI_ISL_1351083, EPI_ISL_1351086, EPI_ISL_1351090, EPI_ISL_1351094, EPI_ISL_1351097, EPI_ISL_1351107, EPI_ISL_1351111, EPI_ISL_1351119, EPI_ISL_1351120, EPI_ISL_1351123, EPI_ISL_1351125, EPI_ISL_1351128, EPI_ISL_1351132, EPI_ISL_1351133, EPI_ISL_1351136, EPI_ISL_1351137, EPI_ISL_1351145, EPI_ISL_1351150, EPI_ISL_1351151, EPI_ISL_1351154, EPI_ISL_1351159, EPI_ISL_1351161, EPI_ISL_1351163, EPI_ISL_1351168, EPI_ISL_1351171, EPI_ISL_1351172, EPI_ISL_1351176, EPI_ISL_1354503, EPI_ISL_1354506, EPI_ISL_1354507, EPI_ISL_1354508, EPI_ISL_1354510, EPI_ISL_1354513, EPI_ISL_1354514, EPI_ISL_1354516, EPI_ISL_1354518, EPI_ISL_1354520, EPI_ISL_1354521, EPI_ISL_1354523, EPI_ISL_1354532, EPI_ISL_1354535, EPI_ISL_1354536, EPI_ISL_1354538, EPI_ISL_1354539, EPI_ISL_1354540, EPI_ISL_1354542, EPI_ISL_1354544, EPI_ISL_1354546, EPI_ISL_1354547, EPI_ISL_1354548, EPI_ISL_1354549, EPI_ISL_1354551, EPI_ISL_1354552, EPI_ISL_1354556, EPI_ISL_1354559, EPI_ISL_1354560, EPI_ISL_1354561, EPI_ISL_1354563, EPI_ISL_1354564, EPI_ISL_1354565, EPI_ISL_1354566, EPI_ISL_1354568, EPI_ISL_1354569, EPI_ISL_1354570, EPI_ISL_1354572, EPI_ISL_1354573, EPI_ISL_1354574, EPI_ISL_1354575, EPI_ISL_1354576, EPI_ISL_1354577, EPI_ISL_1354578, EPI_ISL_1354579, EPI_ISL_1354580, EPI_ISL_1354584, EPI_ISL_1354585, EPI_ISL_1354589, EPI_ISL_1354593, EPI_ISL_1354594, EPI_ISL_1354595, EPI_ISL_1354596, EPI_ISL_1354597, EPI_ISL_1354599, EPI_ISL_1354600, EPI_ISL_1354603, EPI_ISL_1354605, EPI_ISL_1354608, EPI_ISL_1354609, EPI_ISL_1354610, EPI_ISL_1354611, EPI_ISL_1354612, EPI_ISL_1354613, EPI_ISL_1354614, EPI_ISL_1354616, EPI_ISL_1354618, EPI_ISL_1354622, EPI_ISL_1354624, EPI_ISL_1354628, EPI_ISL_1354629, EPI_ISL_1356245, EPI_ISL_1356247, EPI_ISL_1356251, EPI_ISL_1356256, EPI_ISL_1356258, EPI_ISL_1356260, EPI_ISL_1356262, EPI_ISL_1356264, EPI_ISL_1356266, EPI_ISL_1356268, EPI_ISL_1356271, EPI_ISL_1356274, EPI_ISL_1356278, EPI_ISL_1356281, EPI_ISL_1356284, EPI_ISL_1356288, EPI_ISL_1356293, EPI_ISL_1356297, EPI_ISL_1356303, EPI_ISL_1356316, EPI_ISL_1356320, EPI_ISL_1356323, EPI_ISL_1356327, EPI_ISL_1356339, EPI_ISL_1356343, EPI_ISL_1356355, EPI_ISL_1356359, EPI_ISL_1356361, EPI_ISL_1356368, EPI_ISL_1356370, EPI_ISL_1356374, EPI_ISL_1356384, EPI_ISL_1356386, EPI_ISL_1356390, EPI_ISL_1356391, EPI_ISL_1356399, EPI_ISL_1356401, EPI_ISL_1356403, EPI_ISL_1356405, EPI_ISL_1356408, EPI_ISL_1356410, EPI_ISL_1356414, EPI_ISL_1356415, EPI_ISL_1356419, EPI_ISL_1356429, EPI_ISL_1356430, EPI_ISL_1356433, EPI_ISL_1356434, EPI_ISL_1356437, EPI_ISL_1356439, EPI_ISL_1356442, EPI_ISL_1356444, EPI_ISL_1356448, EPI_ISL_1356449, EPI_ISL_1356452, EPI_ISL_1356453, EPI_ISL_1356455, EPI_ISL_1356458, EPI_ISL_1356461, EPI_ISL_1356471, EPI_ISL_1356483, EPI_ISL_1356487, EPI_ISL_1356490, EPI_ISL_1356493, EPI_ISL_1356498, EPI_ISL_1356501, EPI_ISL_1356506, EPI_ISL_1356509, EPI_ISL_1356515, EPI_ISL_1356517, EPI_ISL_1356520, EPI_ISL_1356524, EPI_ISL_1356529, EPI_ISL_1356533, EPI_ISL_1432053, EPI_ISL_1432066, EPI_ISL_1432074, EPI_ISL_1432084, EPI_ISL_1432086, EPI_ISL_1432098, EPI_ISL_1432114, EPI_ISL_1432118, EPI_ISL_1432128, EPI_ISL_1432138, EPI_ISL_1432145, EPI_ISL_1432153, EPI_ISL_1432166, EPI_ISL_1432175, EPI_ISL_1432181, EPI_ISL_1432190, EPI_ISL_1432201, EPI_ISL_1432209, EPI_ISL_1432211, EPI_ISL_1432217, EPI_ISL_1432221, EPI_ISL_1432227, EPI_ISL_1432229, EPI_ISL_1432239, EPI_ISL_1432246, EPI_ISL_1432292, EPI_ISL_1432338, EPI_ISL_1432341, EPI_ISL_1432345, EPI_ISL_1432349, EPI_ISL_1432351, EPI_ISL_1432365, EPI_ISL_1432390, EPI_ISL_1432392, EPI_ISL_1432397, EPI_ISL_1432402, EPI_ISL_1432421, EPI_ISL_1432430, EPI_ISL_1432436, EPI_ISL_1432438, EPI_ISL_1432453, EPI_ISL_1432457, EPI_ISL_1432479, EPI_ISL_1432489, EPI_ISL_1432494, EPI_ISL_1432539, EPI_ISL_1432573, EPI_ISL_1432583, EPI_ISL_1432592, EPI_ISL_1432609, EPI_ISL_1432637, EPI_ISL_1432645, EPI_ISL_1432654, EPI_ISL_1432663, EPI_ISL_1432669, EPI_ISL_1432676, EPI_ISL_1432688, EPI_ISL_1432692, EPI_ISL_1432698, EPI_ISL_1432705, EPI_ISL_1432715, EPI_ISL_1432731, EPI_ISL_1437455, EPI_ISL_1437468, EPI_ISL_1437488, EPI_ISL_1437492, EPI_ISL_1437542, EPI_ISL_1437556, EPI_ISL_1437577, EPI_ISL_1437598, EPI_ISL_1437604, EPI_ISL_1442377, EPI_ISL_1442382, EPI_ISL_1442420, EPI_ISL_1442424, EPI_ISL_1442428, EPI_ISL_1442431, EPI_ISL_1442452, EPI_ISL_1442470, EPI_ISL_1442477, EPI_ISL_1565790, EPI_ISL_1565791, EPI_ISL_1565858, EPI_ISL_1565876, EPI_ISL_1565948, EPI_ISL_1565952, EPI_ISL_1567714, EPI_ISL_1567720, EPI_ISL_1567724, EPI_ISL_1567727, EPI_ISL_1567731, EPI_ISL_1567735, EPI_ISL_1567737, EPI_ISL_1567743, EPI_ISL_1567746, EPI_ISL_1567751, EPI_ISL_1568656, EPI_ISL_1568737, EPI_ISL_1568811, EPI_ISL_1646095, EPI_ISL_1646876, EPI_ISL_1646884, EPI_ISL_1719924, EPI_ISL_1719926, EPI_ISL_1720021, EPI_ISL_1720063, EPI_ISL_1720067, EPI_ISL_1720127, EPI_ISL_1720148

see above SYNLAB MVZ Ettlingen Robert Koch Institute

EPI_ISL_1217438, EPI_ISL_1217447, EPI_ISL_1217460, EPI_ISL_1217478, EPI_ISL_1281093, EPI_ISL_1281095, EPI_ISL_1281119, EPI_ISL_1350963, EPI_ISL_1350970, EPI_ISL_1350974, EPI_ISL_1350985, EPI_ISL_1350989, EPI_ISL_1350993, EPI_ISL_1350996, EPI_ISL_1351002, EPI_ISL_1351027, EPI_ISL_1351063, EPI_ISL_1351085, EPI_ISL_1351091, EPI_ISL_1351100, EPI_ISL_1351109, EPI_ISL_1351114, EPI_ISL_1351118, EPI_ISL_1351122, EPI_ISL_1351126, EPI_ISL_1351130, EPI_ISL_1351139, EPI_ISL_1351141, EPI_ISL_1351146, EPI_ISL_1351178, EPI_ISL_1354502, EPI_ISL_1354522, EPI_ISL_1354530, EPI_ISL_1354534, EPI_ISL_1354553, EPI_ISL_1354567, EPI_ISL_1354571, EPI_ISL_1354621, EPI_ISL_1354623, EPI_ISL_1354625, EPI_ISL_1354627, EPI_ISL_1356254, EPI_ISL_1356298, EPI_ISL_1356315, EPI_ISL_1356363, EPI_ISL_1356532, EPI_ISL_1432070, EPI_ISL_1440587, EPI_ISL_1440616, EPI_ISL_1440619, EPI_ISL_1440647, EPI_ISL_1565852, EPI_ISL_1565869, EPI_ISL_1565971, EPI_ISL_1567745, EPI_ISL_1568684, EPI_ISL_1720034, EPI_ISL_1720040

see above SYNLAB MVZ Hamburg Robert Koch Institute

EPI_ISL_1217430, EPI_ISL_1217431, EPI_ISL_1217445, EPI_ISL_1217446, EPI_ISL_1217451, EPI_ISL_1217455, EPI_ISL_1217458, EPI_ISL_1217470, EPI_ISL_1217481, EPI_ISL_1217490, EPI_ISL_1281088, EPI_ISL_1281091, EPI_ISL_1281094, EPI_ISL_1281098, EPI_ISL_1281101, EPI_ISL_1281105, EPI_ISL_1281114, EPI_ISL_1281121, EPI_ISL_1281122, EPI_ISL_1281125, EPI_ISL_1281135, EPI_ISL_1281139, EPI_ISL_1281140, EPI_ISL_1281143, EPI_ISL_1281145, EPI_ISL_1281150, EPI_ISL_1281151, EPI_ISL_1281155, EPI_ISL_1281163, EPI_ISL_1281170, EPI_ISL_1281171, EPI_ISL_1282766, EPI_ISL_1282768, EPI_ISL_1282769, EPI_ISL_1282770, EPI_ISL_1282772, EPI_ISL_1282773, EPI_ISL_1282774, EPI_ISL_1282776, EPI_ISL_1282777, EPI_ISL_1282778, EPI_ISL_1282779, EPI_ISL_1282781, EPI_ISL_1282783, EPI_ISL_1282784, EPI_ISL_1282785, EPI_ISL_1282786, EPI_ISL_1282787, EPI_ISL_1282788, EPI_ISL_1282789, EPI_ISL_1282790, EPI_ISL_1282792, EPI_ISL_1282793, EPI_ISL_1282794, EPI_ISL_1282797, EPI_ISL_1282798, EPI_ISL_1282800, EPI_ISL_1282805, EPI_ISL_1282806, EPI_ISL_1282807, EPI_ISL_1282808, EPI_ISL_1282812, EPI_ISL_1282815, EPI_ISL_1282816, EPI_ISL_1282817, EPI_ISL_1282818, EPI_ISL_1282819, EPI_ISL_1282821, EPI_ISL_1282822, EPI_ISL_1282823, EPI_ISL_1282824, EPI_ISL_1282826, EPI_ISL_1282827, EPI_ISL_1282828, EPI_ISL_1282829, EPI_ISL_1282830, EPI_ISL_1282831, EPI_ISL_1282832, EPI_ISL_1282833, EPI_ISL_1282834, EPI_ISL_1282835, EPI_ISL_1354512, EPI_ISL_1354517, EPI_ISL_1354526, EPI_ISL_1354541, EPI_ISL_1354550, EPI_ISL_1354554, EPI_ISL_1354586, EPI_ISL_1354588, EPI_ISL_1354590, EPI_ISL_1354592, EPI_ISL_1354601, EPI_ISL_1354602, EPI_ISL_1354606, EPI_ISL_1354617, EPI_ISL_1354626, EPI_ISL_1356259, EPI_ISL_1356261, EPI_ISL_1356263, EPI_ISL_1356265, EPI_ISL_1356267, EPI_ISL_1356269, EPI_ISL_1356270, EPI_ISL_1356272, EPI_ISL_1356277, EPI_ISL_1356279, EPI_ISL_1356282, EPI_ISL_1356283, EPI_ISL_1356286, EPI_ISL_1356287, EPI_ISL_1356290, EPI_ISL_1356296, EPI_ISL_1356300, EPI_ISL_1356304, EPI_ISL_1356305, EPI_ISL_1356307, EPI_ISL_1356309, EPI_ISL_1356314, EPI_ISL_1356317, EPI_ISL_1356324, EPI_ISL_1356328, EPI_ISL_1356336, EPI_ISL_1356337, EPI_ISL_1356340, EPI_ISL_1356346, EPI_ISL_1356348, EPI_ISL_1356356, EPI_ISL_1356358, EPI_ISL_1356373, EPI_ISL_1356375, EPI_ISL_1356376, EPI_ISL_1356377, EPI_ISL_1356382, EPI_ISL_1356397, EPI_ISL_1356406, EPI_ISL_1356409, EPI_ISL_1356418, EPI_ISL_1356420, EPI_ISL_1356425, EPI_ISL_1356428, EPI_ISL_1356432, EPI_ISL_1356436, EPI_ISL_1356440, EPI_ISL_1356445, EPI_ISL_1356462, EPI_ISL_1356463, EPI_ISL_1356466, EPI_ISL_1356473, EPI_ISL_1356476, EPI_ISL_1356479, EPI_ISL_1356481, EPI_ISL_1356484, EPI_ISL_1356500, EPI_ISL_1432044, EPI_ISL_1432132, EPI_ISL_1432280, EPI_ISL_1432647, EPI_ISL_1432720, EPI_ISL_1437311, EPI_ISL_1437314, EPI_ISL_1437316, EPI_ISL_1437322, EPI_ISL_1437341, EPI_ISL_1437373, EPI_ISL_1437426, EPI_ISL_1437428, EPI_ISL_1440582, EPI_ISL_1440601, EPI_ISL_1440620, EPI_ISL_1440685, EPI_ISL_1440700, EPI_ISL_1565834, EPI_ISL_1565838, EPI_ISL_1565842, EPI_ISL_1565845, EPI_ISL_1565866, EPI_ISL_1567728, EPI_ISL_1568826, EPI_ISL_1568846, EPI_ISL_1568859, EPI_ISL_1568866, EPI_ISL_1568934, EPI_ISL_1568941, EPI_ISL_1572279, EPI_ISL_1719939, EPI_ISL_1720109, EPI_ISL_1720131

see above SYNLAB MVZ Heidelberg Robert Koch Institute

EPI_ISL_1213794, EPI_ISL_1213857, EPI_ISL_1213866, EPI_ISL_1213893, EPI_ISL_1213897, EPI_ISL_1213911, EPI_ISL_1213926, EPI_ISL_1218201, EPI_ISL_1218203, EPI_ISL_1218205, EPI_ISL_1218208, EPI_ISL_1218211, EPI_ISL_1218216, EPI_ISL_1218220, EPI_ISL_1218229, EPI_ISL_1218231, EPI_ISL_1218233, EPI_ISL_1218235, EPI_ISL_1218237, EPI_ISL_1218240, EPI_ISL_1218241, EPI_ISL_1218243, EPI_ISL_1218247, EPI_ISL_1218248, EPI_ISL_1218251, EPI_ISL_1218252, EPI_ISL_1218254, EPI_ISL_1218256, EPI_ISL_1218258, EPI_ISL_1218262, EPI_ISL_1218267, EPI_ISL_1218270, EPI_ISL_1218274, EPI_ISL_1218278, EPI_ISL_1218279, EPI_ISL_1218280, EPI_ISL_1218283, EPI_ISL_1218284, EPI_ISL_1218286, EPI_ISL_1218287, EPI_ISL_1218288, EPI_ISL_1218290, EPI_ISL_1218292, EPI_ISL_1218293, EPI_ISL_1218296, EPI_ISL_1218297, EPI_ISL_1218298, EPI_ISL_1218299, EPI_ISL_1218300, EPI_ISL_1218303, EPI_ISL_1218304, EPI_ISL_1218308, EPI_ISL_1218309, EPI_ISL_1218316, EPI_ISL_1218321, EPI_ISL_1218324, EPI_ISL_1218329, EPI_ISL_1218330, EPI_ISL_1218333, EPI_ISL_1218334, EPI_ISL_1218335, EPI_ISL_1218339, EPI_ISL_1218343, EPI_ISL_1218347, EPI_ISL_1218348, EPI_ISL_1218352, EPI_ISL_1218353, EPI_ISL_1218355, EPI_ISL_1218357, EPI_ISL_1218358, EPI_ISL_1218360, EPI_ISL_1218364, EPI_ISL_1218368, EPI_ISL_1218370, EPI_ISL_1218372, EPI_ISL_1218375, EPI_ISL_1218377, EPI_ISL_1218378, EPI_ISL_1218379, EPI_ISL_1218381, EPI_ISL_1218383, EPI_ISL_1218386, EPI_ISL_1218389, EPI_ISL_1218392, EPI_ISL_1218396, EPI_ISL_1218400, EPI_ISL_1218402, EPI_ISL_1218410, EPI_ISL_1218413, EPI_ISL_1218424, EPI_ISL_1218431, EPI_ISL_1218432, EPI_ISL_1218441, EPI_ISL_1218445, EPI_ISL_1218450, EPI_ISL_1218452, EPI_ISL_1218457, EPI_ISL_1218458, EPI_ISL_1218464, EPI_ISL_1218469, EPI_ISL_1218470, EPI_ISL_1218488, EPI_ISL_1218489, EPI_ISL_1218494, EPI_ISL_1218498, EPI_ISL_1218500, EPI_ISL_1218504, EPI_ISL_1218505, EPI_ISL_1218515, EPI_ISL_1218516, EPI_ISL_1218530, EPI_ISL_1218536, EPI_ISL_1218539, EPI_ISL_1280783, EPI_ISL_1280787, EPI_ISL_1280795, EPI_ISL_1280797, EPI_ISL_1280798, EPI_ISL_1280800, EPI_ISL_1280802, EPI_ISL_1280805, EPI_ISL_1280806, EPI_ISL_1280808, EPI_ISL_1280811, EPI_ISL_1280812, EPI_ISL_1280818, EPI_ISL_1280823, EPI_ISL_1280828, EPI_ISL_1280841, EPI_ISL_1280843, EPI_ISL_1280845, EPI_ISL_1280847, EPI_ISL_1280851, EPI_ISL_1280852, EPI_ISL_1280856, EPI_ISL_1280859, EPI_ISL_1280862, EPI_ISL_1280864, EPI_ISL_1280869, EPI_ISL_1280873, EPI_ISL_1280875, EPI_ISL_1280876, EPI_ISL_1280882, EPI_ISL_1280883, EPI_ISL_1280888, EPI_ISL_1280892, EPI_ISL_1280899, EPI_ISL_1280900, EPI_ISL_1280902, EPI_ISL_1280904, EPI_ISL_1280908, EPI_ISL_1280910, EPI_ISL_1280912, EPI_ISL_1280917, EPI_ISL_1280918, EPI_ISL_1280919, EPI_ISL_1280921, EPI_ISL_1280922, EPI_ISL_1280925, EPI_ISL_1280932, EPI_ISL_1280935, EPI_ISL_1280942, EPI_ISL_1282989, EPI_ISL_1282995, EPI_ISL_1282998, EPI_ISL_1283003, EPI_ISL_1283011, EPI_ISL_1283012, EPI_ISL_1283016, EPI_ISL_1283020, EPI_ISL_1283022, EPI_ISL_1283026, EPI_ISL_1283031, EPI_ISL_1283037, EPI_ISL_1283038, EPI_ISL_1283040, EPI_ISL_1283041, EPI_ISL_1283042, EPI_ISL_1283051, EPI_ISL_1283055, EPI_ISL_1283056, EPI_ISL_1283061, EPI_ISL_1283063, EPI_ISL_1283066, EPI_ISL_1283067, EPI_ISL_1283069, EPI_ISL_1283077, EPI_ISL_1283080, EPI_ISL_1283081, EPI_ISL_1283087, EPI_ISL_1283088, EPI_ISL_1283091, EPI_ISL_1283092, EPI_ISL_1283095, EPI_ISL_1283097, EPI_ISL_1283099, EPI_ISL_1283109, EPI_ISL_1283110, EPI_ISL_1283113, EPI_ISL_1283116, EPI_ISL_1283118, EPI_ISL_1283855, EPI_ISL_1283858, EPI_ISL_1283863, EPI_ISL_1283866, EPI_ISL_1283868, EPI_ISL_1283870, EPI_ISL_1283873, EPI_ISL_1283874, EPI_ISL_1283881, EPI_ISL_1283883, EPI_ISL_1283885, EPI_ISL_1283888, EPI_ISL_1283895, EPI_ISL_1283897, EPI_ISL_1283900, EPI_ISL_1283902, EPI_ISL_1283905, EPI_ISL_1283912, EPI_ISL_1283915, EPI_ISL_1283920, EPI_ISL_1283937, EPI_ISL_1283947, EPI_ISL_1283955, EPI_ISL_1283961, EPI_ISL_1283976, EPI_ISL_1283978, EPI_ISL_1283983, EPI_ISL_1283986, EPI_ISL_1283992, EPI_ISL_1283995, EPI_ISL_1284011, EPI_ISL_1284015, EPI_ISL_1284026, EPI_ISL_1284042, EPI_ISL_1284059, EPI_ISL_1284062, EPI_ISL_1284077, EPI_ISL_1284084, EPI_ISL_1284086, EPI_ISL_1284092, EPI_ISL_1284099, EPI_ISL_1284111, EPI_ISL_1284112, EPI_ISL_1284117, EPI_ISL_1284119, EPI_ISL_1284124, EPI_ISL_1284139, EPI_ISL_1284142, EPI_ISL_1284143, EPI_ISL_1284146, EPI_ISL_1284155, EPI_ISL_1284164, EPI_ISL_1284167, EPI_ISL_1284170, EPI_ISL_1284173, EPI_ISL_1284177, EPI_ISL_1284181, EPI_ISL_1284184, EPI_ISL_1284187, EPI_ISL_1284188, EPI_ISL_1284191, EPI_ISL_1284194, EPI_ISL_1284196, EPI_ISL_1284198, EPI_ISL_1284200, EPI_ISL_1284203, EPI_ISL_1284206, EPI_ISL_1284211, EPI_ISL_1284214, EPI_ISL_1284218, EPI_ISL_1284221, EPI_ISL_1284229, EPI_ISL_1284241, EPI_ISL_1285927, EPI_ISL_1285928, EPI_ISL_1285929, EPI_ISL_1285930, EPI_ISL_1285932, EPI_ISL_1285935, EPI_ISL_1285936, EPI_ISL_1285939, EPI_ISL_1285942, EPI_ISL_1285944, EPI_ISL_1285946, EPI_ISL_1285951, EPI_ISL_1285953, EPI_ISL_1285954, EPI_ISL_1285955, EPI_ISL_1285956, EPI_ISL_1285957, EPI_ISL_1285959, EPI_ISL_1285960, EPI_ISL_1285961, EPI_ISL_1285963, EPI_ISL_1285964, EPI_ISL_1285965, EPI_ISL_1285967, EPI_ISL_1285968, EPI_ISL_1285969, EPI_ISL_1285970, EPI_ISL_1285971, EPI_ISL_1285972, EPI_ISL_1285973, EPI_ISL_1285974, EPI_ISL_1285975, EPI_ISL_1285976, EPI_ISL_1285977, EPI_ISL_1285978, EPI_ISL_1285980, EPI_ISL_1285981, EPI_ISL_1285982, EPI_ISL_1285984, EPI_ISL_1285985, EPI_ISL_1285986, EPI_ISL_1285987, EPI_ISL_1285988, EPI_ISL_1285989, EPI_ISL_1285992, EPI_ISL_1285994, EPI_ISL_1285995, EPI_ISL_1285996, EPI_ISL_1285999, EPI_ISL_1286000, EPI_ISL_1286001, EPI_ISL_1286004, EPI_ISL_1286005, EPI_ISL_1286006, EPI_ISL_1286008, EPI_ISL_1286009, EPI_ISL_1286011, EPI_ISL_1286012, EPI_ISL_1286015, EPI_ISL_1286016, EPI_ISL_1286017, EPI_ISL_1286018, EPI_ISL_1286019, EPI_ISL_1286021, EPI_ISL_1286023, EPI_ISL_1286028, EPI_ISL_1286029, EPI_ISL_1286033, EPI_ISL_1286038, EPI_ISL_1286041, EPI_ISL_1286043, EPI_ISL_1286045, EPI_ISL_1286046, EPI_ISL_1286048, EPI_ISL_1286049, EPI_ISL_1286051, EPI_ISL_1286061, EPI_ISL_1286064, EPI_ISL_1286066, EPI_ISL_1286067, EPI_ISL_1286073, EPI_ISL_1286075, EPI_ISL_1286076, EPI_ISL_1286079, EPI_ISL_1286082, EPI_ISL_1286084, EPI_ISL_1286085, EPI_ISL_1348036, EPI_ISL_1348038, EPI_ISL_1348045, EPI_ISL_1348049, EPI_ISL_1348057, EPI_ISL_1348059, EPI_ISL_1348060, EPI_ISL_1348064, EPI_ISL_1348070, EPI_ISL_1348071, EPI_ISL_1348072, EPI_ISL_1348073, EPI_ISL_1348074, EPI_ISL_1348076, EPI_ISL_1348082, EPI_ISL_1348091, EPI_ISL_1348094, EPI_ISL_1348098, EPI_ISL_1348103, EPI_ISL_1348105, EPI_ISL_1348108, EPI_ISL_1348137, EPI_ISL_1348139, EPI_ISL_1348141, EPI_ISL_1348152, EPI_ISL_1348156, EPI_ISL_1348163, EPI_ISL_1348165, EPI_ISL_1348167, EPI_ISL_1348172, EPI_ISL_1348176, EPI_ISL_1348178, EPI_ISL_1348207, EPI_ISL_1348209, EPI_ISL_1348216, EPI_ISL_1348243, EPI_ISL_1348248, EPI_ISL_1348260, EPI_ISL_1348264, EPI_ISL_1348282, EPI_ISL_1348302, EPI_ISL_1348305, EPI_ISL_1348307, EPI_ISL_1348323, EPI_ISL_1348327, EPI_ISL_1348332, EPI_ISL_1348334, EPI_ISL_1348337, EPI_ISL_1348344, EPI_ISL_1348352, EPI_ISL_1348355, EPI_ISL_1348358, EPI_ISL_1348363, EPI_ISL_1348366, EPI_ISL_1348370, EPI_ISL_1348381, EPI_ISL_1348382, EPI_ISL_1348384, EPI_ISL_1348387, EPI_ISL_1348395, EPI_ISL_1348405, EPI_ISL_1348407, EPI_ISL_1348411, EPI_ISL_1348419, EPI_ISL_1348421, EPI_ISL_1348428, EPI_ISL_1348431, EPI_ISL_1348435, EPI_ISL_1348440, EPI_ISL_1348448, EPI_ISL_1348454, EPI_ISL_1348455, EPI_ISL_1348462, EPI_ISL_1348464, EPI_ISL_1348467, EPI_ISL_1348471, EPI_ISL_1348472, EPI_ISL_1348479, EPI_ISL_1348482, EPI_ISL_1350618, EPI_ISL_1350620, EPI_ISL_1350622, EPI_ISL_1350624, EPI_ISL_1350625, EPI_ISL_1350627, EPI_ISL_1350633, EPI_ISL_1350634, EPI_ISL_1350640, EPI_ISL_1350649, EPI_ISL_1350654, EPI_ISL_1350661, EPI_ISL_1350662, EPI_ISL_1350668, EPI_ISL_1350673, EPI_ISL_1350677, EPI_ISL_1350685, EPI_ISL_1350690, EPI_ISL_1350694, EPI_ISL_1350695, EPI_ISL_1350697, EPI_ISL_1350703, EPI_ISL_1350704, EPI_ISL_1350707, EPI_ISL_1350713, EPI_ISL_1350720, EPI_ISL_1350721, EPI_ISL_1350724, EPI_ISL_1350727, EPI_ISL_1350731, EPI_ISL_1350737, EPI_ISL_1350746, EPI_ISL_1350747, EPI_ISL_1350753, EPI_ISL_1350760, EPI_ISL_1350764, EPI_ISL_1350768, EPI_ISL_1351781, EPI_ISL_1351816, EPI_ISL_1351835, EPI_ISL_1351838, EPI_ISL_1351841, EPI_ISL_1351844, EPI_ISL_1351853, EPI_ISL_1351858, EPI_ISL_1351865, EPI_ISL_1351885, EPI_ISL_1351909, EPI_ISL_1351910, EPI_ISL_1351919, EPI_ISL_1351925, EPI_ISL_1351943, EPI_ISL_1351979, EPI_ISL_1351985, EPI_ISL_1351992, EPI_ISL_1351995, EPI_ISL_1352019, EPI_ISL_1352034, EPI_ISL_1352036, EPI_ISL_1352065, EPI_ISL_1352091, EPI_ISL_1352097, EPI_ISL_1352100, EPI_ISL_1352116, EPI_ISL_1352122, EPI_ISL_1352124, EPI_ISL_1352153, EPI_ISL_1352156, EPI_ISL_1352160, EPI_ISL_1352173, EPI_ISL_1352175, EPI_ISL_1352181, EPI_ISL_1352189, EPI_ISL_1352200, EPI_ISL_1352207, EPI_ISL_1352214, EPI_ISL_1352222, EPI_ISL_1352237, EPI_ISL_1352739, EPI_ISL_1352770, EPI_ISL_1352786, EPI_ISL_1352806, EPI_ISL_1352815, EPI_ISL_1352856, EPI_ISL_1356152

see above SYNLAB MVZ Leinfelden-Echterdingen Robert Koch Institute

EPI_ISL_1213937, EPI_ISL_1217433, EPI_ISL_1217437, EPI_ISL_1217441, EPI_ISL_1217450, EPI_ISL_1217468, EPI_ISL_1217487, EPI_ISL_1217491, EPI_ISL_1217499, EPI_ISL_1281097, EPI_ISL_1281100, EPI_ISL_1281104, EPI_ISL_1281109, EPI_ISL_1281112, EPI_ISL_1281133, EPI_ISL_1281136, EPI_ISL_1282796, EPI_ISL_1282799, EPI_ISL_1282803, EPI_ISL_1282804, EPI_ISL_1282809, EPI_ISL_1282820, EPI_ISL_1350965, EPI_ISL_1350966, EPI_ISL_1350968, EPI_ISL_1350971, EPI_ISL_1350972, EPI_ISL_1350973, EPI_ISL_1350975, EPI_ISL_1350976, EPI_ISL_1350977, EPI_ISL_1350978, EPI_ISL_1350980, EPI_ISL_1350981, EPI_ISL_1350982, EPI_ISL_1350983, EPI_ISL_1350986, EPI_ISL_1350987, EPI_ISL_1350991, EPI_ISL_1350994, EPI_ISL_1350997, EPI_ISL_1350998, EPI_ISL_1351000, EPI_ISL_1351001, EPI_ISL_1351003, EPI_ISL_1351004, EPI_ISL_1351005, EPI_ISL_1351006, EPI_ISL_1351007, EPI_ISL_1351010, EPI_ISL_1351011, EPI_ISL_1351012, EPI_ISL_1351013, EPI_ISL_1351014, EPI_ISL_1351015, EPI_ISL_1351016, EPI_ISL_1351017, EPI_ISL_1351018, EPI_ISL_1351019, EPI_ISL_1351020, EPI_ISL_1351021, EPI_ISL_1351022, EPI_ISL_1351024, EPI_ISL_1351025, EPI_ISL_1351028, EPI_ISL_1351029, EPI_ISL_1351030, EPI_ISL_1351031, EPI_ISL_1351034, EPI_ISL_1351035, EPI_ISL_1351036, EPI_ISL_1351038, EPI_ISL_1351039, EPI_ISL_1351041, EPI_ISL_1351042, EPI_ISL_1351043, EPI_ISL_1351046, EPI_ISL_1351047, EPI_ISL_1351050, EPI_ISL_1351051, EPI_ISL_1351052, EPI_ISL_1351054, EPI_ISL_1351055, EPI_ISL_1351057, EPI_ISL_1351059, EPI_ISL_1351060, EPI_ISL_1351061, EPI_ISL_1351062, EPI_ISL_1351064, EPI_ISL_1351066, EPI_ISL_1351067, EPI_ISL_1351068, EPI_ISL_1351070, EPI_ISL_1351071, EPI_ISL_1351074, EPI_ISL_1351075, EPI_ISL_1351076, EPI_ISL_1351079, EPI_ISL_1351080, EPI_ISL_1351084, EPI_ISL_1351087, EPI_ISL_1351088, EPI_ISL_1351089, EPI_ISL_1351092, EPI_ISL_1351093, EPI_ISL_1351095, EPI_ISL_1351096, EPI_ISL_1351098, EPI_ISL_1351099, EPI_ISL_1351101, EPI_ISL_1351102, EPI_ISL_1351103, EPI_ISL_1351104, EPI_ISL_1351105, EPI_ISL_1351106, EPI_ISL_1351108, EPI_ISL_1351110, EPI_ISL_1351112, EPI_ISL_1351113, EPI_ISL_1351116, EPI_ISL_1351117, EPI_ISL_1351121, EPI_ISL_1351127, EPI_ISL_1351129, EPI_ISL_1351131, EPI_ISL_1351134, EPI_ISL_1351135, EPI_ISL_1351138, EPI_ISL_1351140, EPI_ISL_1351142, EPI_ISL_1351143, EPI_ISL_1351144, EPI_ISL_1351147, EPI_ISL_1351148, EPI_ISL_1351149, EPI_ISL_1351152, EPI_ISL_1351153, EPI_ISL_1351155, EPI_ISL_1351156, EPI_ISL_1351157, EPI_ISL_1351158, EPI_ISL_1351160, EPI_ISL_1351162, EPI_ISL_1351164, EPI_ISL_1351166, EPI_ISL_1351167, EPI_ISL_1351169, EPI_ISL_1351170, EPI_ISL_1351173, EPI_ISL_1351174, EPI_ISL_1351175, EPI_ISL_1351177, EPI_ISL_1351179, EPI_ISL_1351180, EPI_ISL_1351181, EPI_ISL_1351182, EPI_ISL_1354501, EPI_ISL_1354504, EPI_ISL_1354505, EPI_ISL_1354511, EPI_ISL_1354519, EPI_ISL_1354528, EPI_ISL_1354555, EPI_ISL_1354558, EPI_ISL_1354562, EPI_ISL_1354582, EPI_ISL_1354587, EPI_ISL_1354598, EPI_ISL_1354615, EPI_ISL_1354630, EPI_ISL_1354631, EPI_ISL_1356249, EPI_ISL_1356275, EPI_ISL_1356276, EPI_ISL_1356280, EPI_ISL_1356285, EPI_ISL_1356289, EPI_ISL_1356291, EPI_ISL_1356292, EPI_ISL_1356299, EPI_ISL_1356301, EPI_ISL_1356302, EPI_ISL_1356306, EPI_ISL_1356313, EPI_ISL_1356318, EPI_ISL_1356319, EPI_ISL_1356321, EPI_ISL_1356322, EPI_ISL_1356325, EPI_ISL_1356326, EPI_ISL_1356329, EPI_ISL_1356330, EPI_ISL_1356332, EPI_ISL_1356333, EPI_ISL_1356335, EPI_ISL_1356338, EPI_ISL_1356342, EPI_ISL_1356345, EPI_ISL_1356347, EPI_ISL_1356349, EPI_ISL_1356350, EPI_ISL_1356352, EPI_ISL_1356353, EPI_ISL_1356354, EPI_ISL_1356357, EPI_ISL_1356364, EPI_ISL_1356366, EPI_ISL_1356367, EPI_ISL_1356371, EPI_ISL_1356372, EPI_ISL_1356378, EPI_ISL_1356379, EPI_ISL_1356381, EPI_ISL_1356383, EPI_ISL_1356385, EPI_ISL_1356387, EPI_ISL_1356388, EPI_ISL_1356392, EPI_ISL_1356393, EPI_ISL_1356394, EPI_ISL_1356395, EPI_ISL_1356396, EPI_ISL_1356398, EPI_ISL_1356400, EPI_ISL_1356402, EPI_ISL_1356404, EPI_ISL_1356407, EPI_ISL_1356411, EPI_ISL_1356412, EPI_ISL_1356413, EPI_ISL_1356416, EPI_ISL_1356417, EPI_ISL_1356421, EPI_ISL_1356422, EPI_ISL_1356423, EPI_ISL_1356426, EPI_ISL_1356427, EPI_ISL_1356431, EPI_ISL_1356435, EPI_ISL_1356438, EPI_ISL_1356441, EPI_ISL_1356446, EPI_ISL_1356450, EPI_ISL_1356451, EPI_ISL_1356454, EPI_ISL_1356457, EPI_ISL_1356460, EPI_ISL_1356469, EPI_ISL_1356470, EPI_ISL_1356472, EPI_ISL_1356474, EPI_ISL_1356477, EPI_ISL_1356478, EPI_ISL_1356480, EPI_ISL_1356482, EPI_ISL_1356485, EPI_ISL_1356486, EPI_ISL_1356488, EPI_ISL_1356489, EPI_ISL_1356491, EPI_ISL_1356492, EPI_ISL_1356494, EPI_ISL_1356495, EPI_ISL_1356496, EPI_ISL_1356497, EPI_ISL_1356499, EPI_ISL_1356502, EPI_ISL_1356503, EPI_ISL_1356504, EPI_ISL_1356505, EPI_ISL_1356507, EPI_ISL_1356510, EPI_ISL_1356511, EPI_ISL_1356513, EPI_ISL_1356514, EPI_ISL_1356516, EPI_ISL_1356518, EPI_ISL_1356519, EPI_ISL_1356521, EPI_ISL_1356522, EPI_ISL_1356523, EPI_ISL_1356525, EPI_ISL_1356526, EPI_ISL_1356527, EPI_ISL_1356528, EPI_ISL_1356530, EPI_ISL_1356531, EPI_ISL_1432059, EPI_ISL_1432065, EPI_ISL_1432068, EPI_ISL_1432075, EPI_ISL_1432082, EPI_ISL_1432122, EPI_ISL_1432151, EPI_ISL_1432170, EPI_ISL_1432179, EPI_ISL_1432194, EPI_ISL_1432196, EPI_ISL_1432213, EPI_ISL_1432236, EPI_ISL_1432250, EPI_ISL_1432255, EPI_ISL_1432269, EPI_ISL_1432271, EPI_ISL_1432284, EPI_ISL_1432290, EPI_ISL_1432296, EPI_ISL_1432302, EPI_ISL_1432307, EPI_ISL_1432317, EPI_ISL_1432321, EPI_ISL_1432323, EPI_ISL_1432325, EPI_ISL_1432334, EPI_ISL_1432343, EPI_ISL_1432347, EPI_ISL_1432371, EPI_ISL_1432375, EPI_ISL_1432378, EPI_ISL_1432395, EPI_ISL_1432405, EPI_ISL_1432407, EPI_ISL_1432409, EPI_ISL_1432415, EPI_ISL_1432449, EPI_ISL_1432451, EPI_ISL_1432462, EPI_ISL_1432503, EPI_ISL_1432512, EPI_ISL_1432514, EPI_ISL_1432525, EPI_ISL_1432545, EPI_ISL_1432564, EPI_ISL_1432569, EPI_ISL_1432590, EPI_ISL_1432599, EPI_ISL_1432603, EPI_ISL_1432607, EPI_ISL_1432616, EPI_ISL_1432618, EPI_ISL_1432624, EPI_ISL_1432628, EPI_ISL_1432643, EPI_ISL_1432650, EPI_ISL_1432658, EPI_ISL_1432665, EPI_ISL_1432672, EPI_ISL_1432674, EPI_ISL_1432679, EPI_ISL_1432682, EPI_ISL_1432696, EPI_ISL_1432700, EPI_ISL_1432708, EPI_ISL_1432737, EPI_ISL_1437283, EPI_ISL_1437284, EPI_ISL_1437287, EPI_ISL_1437290, EPI_ISL_1437297, EPI_ISL_1437298, EPI_ISL_1437300, EPI_ISL_1437306, EPI_ISL_1437310, EPI_ISL_1437312, EPI_ISL_1437315, EPI_ISL_1437317, EPI_ISL_1437318, EPI_ISL_1437320, EPI_ISL_1437324, EPI_ISL_1437328, EPI_ISL_1437333, EPI_ISL_1437338, EPI_ISL_1437350, EPI_ISL_1437355, EPI_ISL_1437357, EPI_ISL_1437359, EPI_ISL_1437361, EPI_ISL_1437367, EPI_ISL_1437369, EPI_ISL_1437372, EPI_ISL_1437376, EPI_ISL_1437377, EPI_ISL_1437378, EPI_ISL_1437380, EPI_ISL_1437381, EPI_ISL_1437383, EPI_ISL_1437385, EPI_ISL_1437386, EPI_ISL_1437388, EPI_ISL_1437390, EPI_ISL_1437395, EPI_ISL_1437400, EPI_ISL_1437403, EPI_ISL_1437404, EPI_ISL_1437405, EPI_ISL_1437407, EPI_ISL_1437408, EPI_ISL_1437410, EPI_ISL_1437412, EPI_ISL_1437413, EPI_ISL_1437416, EPI_ISL_1437419, EPI_ISL_1437423, EPI_ISL_1437424, EPI_ISL_1437427, EPI_ISL_1437432, EPI_ISL_1437436, EPI_ISL_1437441, EPI_ISL_1437442, EPI_ISL_1437445, EPI_ISL_1437447, EPI_ISL_1437452, EPI_ISL_1437465, EPI_ISL_1437466, EPI_ISL_1437470, EPI_ISL_1437473, EPI_ISL_1437475, EPI_ISL_1437477, EPI_ISL_1437479, EPI_ISL_1437481, EPI_ISL_1437485, EPI_ISL_1437489, EPI_ISL_1437491, EPI_ISL_1437497, EPI_ISL_1437509, EPI_ISL_1437519, EPI_ISL_1437528, EPI_ISL_1437565, EPI_ISL_1437574, EPI_ISL_1437578, EPI_ISL_1437581, EPI_ISL_1437588, EPI_ISL_1437589, EPI_ISL_1437591, EPI_ISL_1437606, EPI_ISL_1440580, EPI_ISL_1440583, EPI_ISL_1440588, EPI_ISL_1440590, EPI_ISL_1440593, EPI_ISL_1440595, EPI_ISL_1440598, EPI_ISL_1440615, EPI_ISL_1440622, EPI_ISL_1440629, EPI_ISL_1440650, EPI_ISL_1440651, EPI_ISL_1440661, EPI_ISL_1440674, EPI_ISL_1440711, EPI_ISL_1440718, EPI_ISL_1440730, EPI_ISL_1441936, EPI_ISL_1441942, EPI_ISL_1441948, EPI_ISL_1441949, EPI_ISL_1441950, EPI_ISL_1441954, EPI_ISL_1441955, EPI_ISL_1441959, EPI_ISL_1441961, EPI_ISL_1441962, EPI_ISL_1441967, EPI_ISL_1441968, EPI_ISL_1441972, EPI_ISL_1441973, EPI_ISL_1441975, EPI_ISL_1441977, EPI_ISL_1441979, EPI_ISL_1441983, EPI_ISL_1441989, EPI_ISL_1441990, EPI_ISL_1442375, EPI_ISL_1442378, EPI_ISL_1442383, EPI_ISL_1442384, EPI_ISL_1442402, EPI_ISL_1442406, EPI_ISL_1442411, EPI_ISL_1442421, EPI_ISL_1442439, EPI_ISL_1442441, EPI_ISL_1442460, EPI_ISL_1442480, EPI_ISL_1565868, EPI_ISL_1565894, EPI_ISL_1565903, EPI_ISL_1568626, EPI_ISL_1568812, EPI_ISL_1568820, EPI_ISL_1568828, EPI_ISL_1568836, EPI_ISL_1568851, EPI_ISL_1568857, EPI_ISL_1568862, EPI_ISL_1568879, EPI_ISL_1568882, EPI_ISL_1568890, EPI_ISL_1568892, EPI_ISL_1568904, EPI_ISL_1568910, EPI_ISL_1568915, EPI_ISL_1568927, EPI_ISL_1568929, EPI_ISL_1568932, EPI_ISL_1568938, EPI_ISL_1568944, EPI_ISL_1645990, EPI_ISL_1646045, EPI_ISL_1646105, EPI_ISL_1646235, EPI_ISL_1646276, EPI_ISL_1646470, EPI_ISL_1646601, EPI_ISL_1646749, EPI_ISL_1646809, EPI_ISL_1646841, EPI_ISL_1646859, EPI_ISL_1646863, EPI_ISL_1719975

see above SYNLAB MVZ Leverkusen Robert Koch Institute

EPI_ISL_1565809, EPI_ISL_1567713, EPI_ISL_1567723, EPI_ISL_1567739 SYNLAB MVZ Neustadt a. d. WeinstraÃŸe Robert Koch Institute

EPI_ISL_1217428, EPI_ISL_1217488, EPI_ISL_1217489, EPI_ISL_1281154, EPI_ISL_1281168, EPI_ISL_1282765, EPI_ISL_1282775, EPI_ISL_1282782, EPI_ISL_1282791, EPI_ISL_1282810, EPI_ISL_1282814, EPI_ISL_1354524, EPI_ISL_1354525, EPI_ISL_1354529, EPI_ISL_1354533, EPI_ISL_1354537, EPI_ISL_1354545, EPI_ISL_1354583, EPI_ISL_1354620, EPI_ISL_1356246, EPI_ISL_1356248, EPI_ISL_1356250, EPI_ISL_1356252, EPI_ISL_1356253, EPI_ISL_1356255, EPI_ISL_1356257, EPI_ISL_1356334, EPI_ISL_1356344, EPI_ISL_1356351, EPI_ISL_1356389, EPI_ISL_1432656, EPI_ISL_1440129, EPI_ISL_1440215, EPI_ISL_1440596, EPI_ISL_1440628, EPI_ISL_1440649, EPI_ISL_1440675, EPI_ISL_1440713

| | | | |
|---|---|---|---|
| see above | SYNLAB MVZ Neustadt a. d. Weinstraße | Robert Koch Institute | |
| EPI_ISL_1213901, EPI_ISL_1214992, EPI_ISL_1214995, EPI_ISL_1214999, EPI_ISL_1215014, EPI_ISL_1215017, EPI_ISL_1215023, EPI_ISL_1215025, EPI_ISL_1215027, EPI_ISL_1215029, EPI_ISL_1215033, EPI_ISL_1215035, EPI_ISL_1215038, EPI_ISL_1215040, EPI_ISL_1215042, EPI_ISL_1215043, EPI_ISL_1215050, EPI_ISL_1215064, EPI_ISL_1215080, EPI_ISL_1215082, EPI_ISL_1215093, EPI_ISL_1215103, EPI_ISL_1215106, EPI_ISL_1215114, EPI_ISL_1215129, EPI_ISL_1215147, EPI_ISL_1215165, EPI_ISL_1215166, EPI_ISL_1215168, EPI_ISL_1215170, EPI_ISL_1215172, EPI_ISL_1215177, EPI_ISL_1215178, EPI_ISL_1215180, EPI_ISL_1215183, EPI_ISL_1215193, EPI_ISL_1215197, EPI_ISL_1215201, EPI_ISL_1215206, EPI_ISL_1215211, EPI_ISL_1215213, EPI_ISL_1215220, EPI_ISL_1215225, EPI_ISL_1215226, EPI_ISL_1215231, EPI_ISL_1215241, EPI_ISL_1215243, EPI_ISL_1215245, EPI_ISL_1215248, EPI_ISL_1215258, EPI_ISL_1280947, EPI_ISL_1280949, EPI_ISL_1280951, EPI_ISL_1280952, EPI_ISL_1280953, EPI_ISL_1280954, EPI_ISL_1280955, EPI_ISL_1280956, EPI_ISL_1280959, EPI_ISL_1280964, EPI_ISL_1280965, EPI_ISL_1280971, EPI_ISL_1280976, EPI_ISL_1280980, EPI_ISL_1280985, EPI_ISL_1280989, EPI_ISL_1280992, EPI_ISL_1280994, EPI_ISL_1280998, EPI_ISL_1281007, EPI_ISL_1281011, EPI_ISL_1281014, EPI_ISL_1281019, EPI_ISL_1281021, EPI_ISL_1281022, EPI_ISL_1281027, EPI_ISL_1281028, EPI_ISL_1281030, EPI_ISL_1281032, EPI_ISL_1281033, EPI_ISL_1281034, EPI_ISL_1281038, EPI_ISL_1281039, EPI_ISL_1281042, EPI_ISL_1281046, EPI_ISL_1281048, EPI_ISL_1281051, EPI_ISL_1281052, EPI_ISL_1281053, EPI_ISL_1281055, EPI_ISL_1281057, EPI_ISL_1281065, EPI_ISL_1281068, EPI_ISL_1281070, EPI_ISL_1281073, EPI_ISL_1281075, EPI_ISL_1281078, EPI_ISL_1281079, EPI_ISL_1281080, EPI_ISL_1281081, EPI_ISL_1281083, EPI_ISL_1284252, EPI_ISL_1284261, EPI_ISL_1284265, EPI_ISL_1284270, EPI_ISL_1284272, EPI_ISL_1284276, EPI_ISL_1284280, EPI_ISL_1284286, EPI_ISL_1284287, EPI_ISL_1284293, EPI_ISL_1284294, EPI_ISL_1284297, EPI_ISL_1284301, EPI_ISL_1284305, EPI_ISL_1284308, EPI_ISL_1284311, EPI_ISL_1284328, EPI_ISL_1284330, EPI_ISL_1284332, EPI_ISL_1284334, EPI_ISL_1284342, EPI_ISL_1284348, EPI_ISL_1284353, EPI_ISL_1284355, EPI_ISL_1284357, EPI_ISL_1284360, EPI_ISL_1284364, EPI_ISL_1284376, EPI_ISL_1284378, EPI_ISL_1284379, EPI_ISL_1284381, EPI_ISL_1284383, EPI_ISL_1284386, EPI_ISL_1284390, EPI_ISL_1284391, EPI_ISL_1284393, EPI_ISL_1284399, EPI_ISL_1284401, EPI_ISL_1284411, EPI_ISL_1284413, EPI_ISL_1284414, EPI_ISL_1284415, EPI_ISL_1284417, EPI_ISL_1284418, EPI_ISL_1284420, EPI_ISL_1284421, EPI_ISL_1284427, EPI_ISL_1284431, EPI_ISL_1284435, EPI_ISL_1284436, EPI_ISL_1284438, EPI_ISL_1284441, EPI_ISL_1284446, EPI_ISL_1284452, EPI_ISL_1284453, EPI_ISL_1284457, EPI_ISL_1284460, EPI_ISL_1284467, EPI_ISL_1284469, EPI_ISL_1286760, EPI_ISL_1286761, EPI_ISL_1286766, EPI_ISL_1286772, EPI_ISL_1286775, EPI_ISL_1286778, EPI_ISL_1286780, EPI_ISL_1286784, EPI_ISL_1286786, EPI_ISL_1286787, EPI_ISL_1286795, EPI_ISL_1286802, EPI_ISL_1286807, EPI_ISL_1286808, EPI_ISL_1286809, EPI_ISL_1286810, EPI_ISL_1286814, EPI_ISL_1286817, EPI_ISL_1286819, EPI_ISL_1286820, EPI_ISL_1286822, EPI_ISL_1286824, EPI_ISL_1286830, EPI_ISL_1286836, EPI_ISL_1286837, EPI_ISL_1286839, EPI_ISL_1286842, EPI_ISL_1286845, EPI_ISL_1286850, EPI_ISL_1286852, EPI_ISL_1286855, EPI_ISL_1286856, EPI_ISL_1286857, EPI_ISL_1286858, EPI_ISL_1286860, EPI_ISL_1286861, EPI_ISL_1286862, EPI_ISL_1286863, EPI_ISL_1286864, EPI_ISL_1286865, EPI_ISL_1286866, EPI_ISL_1286868, EPI_ISL_1286872, EPI_ISL_1286874, EPI_ISL_1286880, EPI_ISL_1286881, EPI_ISL_1286885, EPI_ISL_1286886, EPI_ISL_1286887, EPI_ISL_1286888, EPI_ISL_1286889, EPI_ISL_1286891, EPI_ISL_1286892, EPI_ISL_1350709, EPI_ISL_1350741, EPI_ISL_1351797, EPI_ISL_1352723, EPI_ISL_1355977, EPI_ISL_1356104 | | | |
| see above | SYNLAB MVZ Trier | Robert Koch Institute | |
| EPI_ISL_1214990, EPI_ISL_1214997, EPI_ISL_1215000, EPI_ISL_1215005, EPI_ISL_1215009, EPI_ISL_1215020, EPI_ISL_1215030, EPI_ISL_1215052, EPI_ISL_1215057, EPI_ISL_1215059, EPI_ISL_1215061, EPI_ISL_1215066, EPI_ISL_1215067, EPI_ISL_1215069, EPI_ISL_1215077, EPI_ISL_1215086, EPI_ISL_1215090, EPI_ISL_1215099, EPI_ISL_1215109, EPI_ISL_1215111, EPI_ISL_1215124, EPI_ISL_1215127, EPI_ISL_1215134, EPI_ISL_1215141, EPI_ISL_1215152, EPI_ISL_1215154, EPI_ISL_1215159, EPI_ISL_1215161, EPI_ISL_1215173, EPI_ISL_1215175, EPI_ISL_1215190, EPI_ISL_1215195, EPI_ISL_1215200, EPI_ISL_1215203, EPI_ISL_1215205, EPI_ISL_1215208, EPI_ISL_1215216, EPI_ISL_1215221, EPI_ISL_1215228, EPI_ISL_1215230, EPI_ISL_1215233, EPI_ISL_1215234, EPI_ISL_1215240, EPI_ISL_1215250, EPI_ISL_1215253, EPI_ISL_1280946, EPI_ISL_1280948, EPI_ISL_1280950, EPI_ISL_1280958, EPI_ISL_1280961, EPI_ISL_1280962, EPI_ISL_1280966, EPI_ISL_1280970, EPI_ISL_1280972, EPI_ISL_1280975, EPI_ISL_1280977, EPI_ISL_1280979, EPI_ISL_1280984, EPI_ISL_1280987, EPI_ISL_1280988, EPI_ISL_1280990, EPI_ISL_1280991, EPI_ISL_1280993, EPI_ISL_1280995, EPI_ISL_1280996, EPI_ISL_1280997, EPI_ISL_1280999, EPI_ISL_1281001, EPI_ISL_1281002, EPI_ISL_1281003, EPI_ISL_1281004, EPI_ISL_1281008, EPI_ISL_1281009, EPI_ISL_1281013, EPI_ISL_1281016, EPI_ISL_1281017, EPI_ISL_1281023, EPI_ISL_1281024, EPI_ISL_1281029, EPI_ISL_1281031, EPI_ISL_1281035, EPI_ISL_1281036, EPI_ISL_1281040, EPI_ISL_1281041, EPI_ISL_1281043, EPI_ISL_1281044, EPI_ISL_1281047, EPI_ISL_1281050, EPI_ISL_1281054, EPI_ISL_1281056, EPI_ISL_1281058, EPI_ISL_1281059, EPI_ISL_1281060, EPI_ISL_1281062, EPI_ISL_1281063, EPI_ISL_1281064, EPI_ISL_1281069, EPI_ISL_1281071, EPI_ISL_1281074, EPI_ISL_1281076, EPI_ISL_1281077, EPI_ISL_1284242, EPI_ISL_1284248, EPI_ISL_1284255, EPI_ISL_1284257, EPI_ISL_1284259, EPI_ISL_1284268, EPI_ISL_1284279, EPI_ISL_1284283, EPI_ISL_1284300, EPI_ISL_1284304, EPI_ISL_1284319, EPI_ISL_1284321, EPI_ISL_1284324, EPI_ISL_1284325, EPI_ISL_1284337, EPI_ISL_1284338, EPI_ISL_1284347, EPI_ISL_1284358, EPI_ISL_1284362, EPI_ISL_1284367, EPI_ISL_1284384, EPI_ISL_1284388, EPI_ISL_1284395, EPI_ISL_1284396, EPI_ISL_1284398, EPI_ISL_1284403, EPI_ISL_1284406, EPI_ISL_1284416, EPI_ISL_1284423, EPI_ISL_1284425, EPI_ISL_1284428, EPI_ISL_1284440, EPI_ISL_1284443, EPI_ISL_1284445, EPI_ISL_1284448, EPI_ISL_1284455, EPI_ISL_1284458, EPI_ISL_1284459, EPI_ISL_1284462, EPI_ISL_1284463, EPI_ISL_1284464, EPI_ISL_1284468, EPI_ISL_1286762, EPI_ISL_1286764, EPI_ISL_1286773, EPI_ISL_1286776, EPI_ISL_1286777, EPI_ISL_1286783, EPI_ISL_1286788, EPI_ISL_1286791, EPI_ISL_1286793, EPI_ISL_1286794, EPI_ISL_1286800, EPI_ISL_1286803, EPI_ISL_1286813, EPI_ISL_1286818, EPI_ISL_1286821, EPI_ISL_1286825, EPI_ISL_1286826, EPI_ISL_1286827, EPI_ISL_1286828, EPI_ISL_1286832, EPI_ISL_1286833, EPI_ISL_1286835, EPI_ISL_1286838, EPI_ISL_1286843, EPI_ISL_1286844, EPI_ISL_1286847, EPI_ISL_1286851, EPI_ISL_1286853, EPI_ISL_1286854, EPI_ISL_1286859, EPI_ISL_1286871, EPI_ISL_1286875, EPI_ISL_1286884, EPI_ISL_1286890 | | | |
| see above | SYNLAB MVZ Weiden | Robert Koch Institute | |
| EPI_ISL_1284630, EPI_ISL_1284669, EPI_ISL_1285438, EPI_ISL_1285449, EPI_ISL_1285493, EPI_ISL_1285521, EPI_ISL_1285551, EPI_ISL_1285564, EPI_ISL_1285610, EPI_ISL_1285633, EPI_ISL_1285642, EPI_ISL_1285648, EPI_ISL_1285653, EPI_ISL_1285671, EPI_ISL_1285681, EPI_ISL_1285689, EPI_ISL_1349859, EPI_ISL_1350024, EPI_ISL_1353648, EPI_ISL_1353702, EPI_ISL_1353703, EPI_ISL_1353719, EPI_ISL_1353730, EPI_ISL_1353735, EPI_ISL_1353749, EPI_ISL_1353760, EPI_ISL_1353766, EPI_ISL_1353778, EPI_ISL_1353779, EPI_ISL_1353782, EPI_ISL_1353807, EPI_ISL_1353819, EPI_ISL_1353829, EPI_ISL_1353833, EPI_ISL_1353856, EPI_ISL_1353859, EPI_ISL_1353868, EPI_ISL_1353870, EPI_ISL_1353873, EPI_ISL_1353939, EPI_ISL_1353980, EPI_ISL_1353990, EPI_ISL_1353999, EPI_ISL_1354000, EPI_ISL_1354069, EPI_ISL_1354079, EPI_ISL_1354091, EPI_ISL_1354095, EPI_ISL_1354131, EPI_ISL_1354153, EPI_ISL_1354160, EPI_ISL_1354205, EPI_ISL_1354209, EPI_ISL_1432941, EPI_ISL_1432972, EPI_ISL_1433070, EPI_ISL_1433104, EPI_ISL_1433109, EPI_ISL_1433126, EPI_ISL_1433169, EPI_ISL_1433187, EPI_ISL_1433188, EPI_ISL_1433201, EPI_ISL_1433222, EPI_ISL_1433225, EPI_ISL_1433237, EPI_ISL_1433243, EPI_ISL_1433246, EPI_ISL_1433264, EPI_ISL_1433269, EPI_ISL_1433270, EPI_ISL_1433290, EPI_ISL_1433296, EPI_ISL_1433313 | | | |
| see above | Sonic - Bioscientia - MVZ Labor Saar GmbH | Robert Koch Institute | |
| EPI_ISL_1282015, EPI_ISL_1282090, EPI_ISL_1282128, EPI_ISL_1282129, EPI_ISL_1282152, EPI_ISL_1282181, EPI_ISL_1282215, EPI_ISL_1282265, EPI_ISL_1282285, EPI_ISL_1282298, EPI_ISL_1282301, EPI_ISL_1354016, EPI_ISL_1354039, EPI_ISL_1354090, EPI_ISL_1354099, EPI_ISL_1354112, EPI_ISL_1354159, EPI_ISL_1354168, EPI_ISL_1354192, EPI_ISL_1354229, EPI_ISL_1354263, EPI_ISL_1436053, EPI_ISL_1436110, EPI_ISL_1436127 | | | |
| see above | Sonic - Dr. Staber & Kollegen GmbH München | Robert Koch Institute | |
| EPI_ISL_1284561, EPI_ISL_1284564, EPI_ISL_1284567, EPI_ISL_1284568, EPI_ISL_1284574, EPI_ISL_1284582, EPI_ISL_1284584, EPI_ISL_1284586, EPI_ISL_1284590, EPI_ISL_1284597, EPI_ISL_1284604, EPI_ISL_1284606, EPI_ISL_1284613, EPI_ISL_1284617, EPI_ISL_1284621, EPI_ISL_1284622, EPI_ISL_1284626, EPI_ISL_1284627, EPI_ISL_1284631, EPI_ISL_1284632, EPI_ISL_1284633, EPI_ISL_1284639, EPI_ISL_1284644, EPI_ISL_1284648, EPI_ISL_1284654, EPI_ISL_1284666, EPI_ISL_1284673, EPI_ISL_1284676, EPI_ISL_1284677, EPI_ISL_1284681, EPI_ISL_1284683, EPI_ISL_1284685, EPI_ISL_1284686, EPI_ISL_1284691, EPI_ISL_1284703, EPI_ISL_1284705, EPI_ISL_1284707, EPI_ISL_1284720, EPI_ISL_1284733, EPI_ISL_1284737, EPI_ISL_1284739, EPI_ISL_1284741, EPI_ISL_1284742, EPI_ISL_1284744, EPI_ISL_1284745, EPI_ISL_1284748, EPI_ISL_1284753, EPI_ISL_1284756, EPI_ISL_1284762, EPI_ISL_1284766, EPI_ISL_1284786, EPI_ISL_1284791, EPI_ISL_1284804, EPI_ISL_1284809, EPI_ISL_1284812, EPI_ISL_1284815, EPI_ISL_1284819, EPI_ISL_1284824, EPI_ISL_1284835, EPI_ISL_1284847, EPI_ISL_1284852, EPI_ISL_1284859, EPI_ISL_1284862, EPI_ISL_1284865, EPI_ISL_1284866, EPI_ISL_1284868, EPI_ISL_1284873, EPI_ISL_1284875, EPI_ISL_1284880, EPI_ISL_1353998, EPI_ISL_1354007, EPI_ISL_1354033, EPI_ISL_1354047, EPI_ISL_1354053, EPI_ISL_1354088, EPI_ISL_1354106, EPI_ISL_1354114, EPI_ISL_1354125, EPI_ISL_1354162, EPI_ISL_1354212, EPI_ISL_1354217, EPI_ISL_1354233, EPI_ISL_1354246, EPI_ISL_1354251, EPI_ISL_1354252, EPI_ISL_1438763, EPI_ISL_1438784, EPI_ISL_1438785, EPI_ISL_1438819, EPI_ISL_1438846, EPI_ISL_1438912, EPI_ISL_1439003 | | | |
| see above | Sonic - Labor Dr. von Froreich GmbH | Robert Koch Institute | |
| EPI_ISL_1285594, EPI_ISL_1285666 | Sonic - Labor Lademannbogen MVZ GmbH | Robert Koch Institute | |
| EPI_ISL_1281998, EPI_ISL_1282043, EPI_ISL_1282047, EPI_ISL_1282074, EPI_ISL_1282136, EPI_ISL_1282236, EPI_ISL_1282252, EPI_ISL_1353992, EPI_ISL_1354014, EPI_ISL_1354034, EPI_ISL_1354046, EPI_ISL_1354049, EPI_ISL_1354050, EPI_ISL_1354077, EPI_ISL_1354081, EPI_ISL_1354116, EPI_ISL_1354130, EPI_ISL_1354133, EPI_ISL_1354190, EPI_ISL_1354221 | | | |
| see above | Sonic - Labor Staber Bayreuth | Robert Koch Institute | |
| EPI_ISL_1282009, EPI_ISL_1282066, EPI_ISL_1282086, EPI_ISL_1282104, EPI_ISL_1282196, EPI_ISL_1282209, EPI_ISL_1282261, EPI_ISL_1282297, EPI_ISL_1353995, EPI_ISL_1354028, EPI_ISL_1354029, EPI_ISL_1354032, EPI_ISL_1354063, EPI_ISL_1354082, EPI_ISL_1354121, EPI_ISL_1354129, EPI_ISL_1354163, EPI_ISL_1354176, EPI_ISL_1354191, EPI_ISL_1354199, EPI_ISL_1354234, EPI_ISL_1354249, EPI_ISL_1354253, EPI_ISL_1354261 | | | |
| see above | Sonic - Labor Staber Dresden (Klipphausen) | Robert Koch Institute | |
| EPI_ISL_1282234, EPI_ISL_1354087, EPI_ISL_1354182 | Sonic - Labor Staber Heilbronn | Robert Koch Institute | |
| EPI_ISL_1282108, EPI_ISL_1282286, EPI_ISL_1353808, EPI_ISL_1353893, EPI_ISL_1353908, EPI_ISL_1353920, EPI_ISL_1353927 | | | |
| see above | Sonic - Labor Staber MVZ Kassel | Robert Koch Institute | |
| EPI_ISL_1282148, EPI_ISL_1282159, EPI_ISL_1282248, EPI_ISL_1282263, EPI_ISL_1282270, EPI_ISL_1354003, EPI_ISL_1354073, EPI_ISL_1354085, EPI_ISL_1354146, EPI_ISL_1354148, EPI_ISL_1354158, EPI_ISL_1354166, EPI_ISL_1354175, EPI_ISL_1354179, EPI_ISL_1354186, EPI_ISL_1354245, EPI_ISL_1436126, EPI_ISL_1436156 | | | |
| see above | Sonic - Labor Staber Nürnberg | Robert Koch Institute | |
| EPI_ISL_1284557, EPI_ISL_1284563, EPI_ISL_1284571, EPI_ISL_1284575, EPI_ISL_1284581, EPI_ISL_1284591, EPI_ISL_1284594, EPI_ISL_1284596, EPI_ISL_1284605, EPI_ISL_1284619, EPI_ISL_1284637, EPI_ISL_1284640, EPI_ISL_1284641, EPI_ISL_1284645, EPI_ISL_1284655, EPI_ISL_1284660, EPI_ISL_1284680, EPI_ISL_1284688, EPI_ISL_1284690, EPI_ISL_1284694, EPI_ISL_1284699, EPI_ISL_1284700, EPI_ISL_1284704, EPI_ISL_1284706, EPI_ISL_1284715, EPI_ISL_1284716, EPI_ISL_1284718, EPI_ISL_1284721, EPI_ISL_1284727, EPI_ISL_1284731, EPI_ISL_1284732, EPI_ISL_1284735, EPI_ISL_1284736, EPI_ISL_1284746, EPI_ISL_1284769, EPI_ISL_1284771, EPI_ISL_1284789, EPI_ISL_1284790, EPI_ISL_1284795, EPI_ISL_1284798, EPI_ISL_1284807, EPI_ISL_1284813, EPI_ISL_1284814, EPI_ISL_1284827, EPI_ISL_1284840, EPI_ISL_1284850, EPI_ISL_1284851, EPI_ISL_1284853, EPI_ISL_1284855, EPI_ISL_1284858, EPI_ISL_1284867, EPI_ISL_1284869, EPI_ISL_1284871, EPI_ISL_1284877, EPI_ISL_1285430, EPI_ISL_1285437, EPI_ISL_1285439, EPI_ISL_1285445, EPI_ISL_1285488, EPI_ISL_1285490, EPI_ISL_1285516, EPI_ISL_1285531, EPI_ISL_1285539, EPI_ISL_1285562, EPI_ISL_1285568, EPI_ISL_1285570, EPI_ISL_1285574, EPI_ISL_1285596, EPI_ISL_1285604, EPI_ISL_1285605, EPI_ISL_1285621, EPI_ISL_1285643, EPI_ISL_1285646, EPI_ISL_1285650, EPI_ISL_1285675, EPI_ISL_1285697, EPI_ISL_1285704, EPI_ISL_1285707, EPI_ISL_1353989, EPI_ISL_1353991, EPI_ISL_1353993, EPI_ISL_1353994, EPI_ISL_1354009, EPI_ISL_1354019, EPI_ISL_1354021, EPI_ISL_1354024, EPI_ISL_1354025, EPI_ISL_1354035, EPI_ISL_1354037, EPI_ISL_1354040, EPI_ISL_1354042, EPI_ISL_1354058, EPI_ISL_1354059, EPI_ISL_1354071, EPI_ISL_1354072, EPI_ISL_1354076, EPI_ISL_1354083, EPI_ISL_1354086, EPI_ISL_1354094, EPI_ISL_1354096, EPI_ISL_1354098, EPI_ISL_1354101, EPI_ISL_1354103, EPI_ISL_1354108, EPI_ISL_1354113, EPI_ISL_1354120, EPI_ISL_1354124, EPI_ISL_1354126, EPI_ISL_1354136, EPI_ISL_1354137, EPI_ISL_1354145, EPI_ISL_1354147, EPI_ISL_1354177, EPI_ISL_1354181, EPI_ISL_1354184, EPI_ISL_1354204, EPI_ISL_1354206, EPI_ISL_1354207, EPI_ISL_1354208, EPI_ISL_1354214, EPI_ISL_1354220, EPI_ISL_1354237, EPI_ISL_1354240, EPI_ISL_1354241, EPI_ISL_1354243, EPI_ISL_1354256, EPI_ISL_1354259, EPI_ISL_1438694 | | | |
| see above | Sonic - MVZ Medizinisches Labor Bremen GmbH | Robert Koch Institute | |
| EPI_ISL_1284808, EPI_ISL_1285696 | Sonic - MVZ Medizinisches Labor Nord GmbH Kiel | Robert Koch Institute | |
| EPI_ISL_1284580, EPI_ISL_1284588, EPI_ISL_1284615, EPI_ISL_1284618, EPI_ISL_1284643, EPI_ISL_1284649, EPI_ISL_1284682, EPI_ISL_1284696, EPI_ISL_1284697, EPI_ISL_1284779, EPI_ISL_1284828, EPI_ISL_1284833, EPI_ISL_1284843, EPI_ISL_1284876, EPI_ISL_1285443, EPI_ISL_1285686 | | | |
| see above | Sonic - MVZ Medizinisches Labor Oldenburg | Robert Koch Institute | |
| EPI_ISL_1285330 | St. Marien-Hospital Hamm | Robert Koch Institute | |
| EPI_ISL_1285305, EPI_ISL_1285335, EPI_ISL_1353293 | Stiftung Krankenhaus Bethanien für die Grafschaft Moers | Robert Koch Institute | |
| EPI_ISL_1217627, EPI_ISL_1217653, EPI_ISL_1217656, EPI_ISL_1217666, EPI_ISL_1217667, EPI_ISL_1217670, EPI_ISL_1217683, EPI_ISL_1217684, EPI_ISL_1217689, EPI_ISL_1217690, EPI_ISL_1217691, EPI_ISL_1217693, EPI_ISL_1217697, EPI_ISL_1217698, EPI_ISL_1285056, EPI_ISL_1285072, EPI_ISL_1285103, EPI_ISL_1285112, EPI_ISL_1285116, EPI_ISL_1285118, EPI_ISL_1285121, EPI_ISL_1285124, EPI_ISL_1285126, EPI_ISL_1285132, EPI_ISL_1285136, EPI_ISL_1285144, EPI_ISL_1285151, EPI_ISL_1285160, EPI_ISL_1285161, EPI_ISL_1285162 | | | |
| see above | Städtisches Klinikum Dresden | Robert Koch Institute | |
| EPI_ISL_1215007, EPI_ISL_1215054, EPI_ISL_1215085, EPI_ISL_1215088, EPI_ISL_1215095, EPI_ISL_1215101, EPI_ISL_1215116, EPI_ISL_1215119, EPI_ISL_1215122, EPI_ISL_1215126, EPI_ISL_1215136, EPI_ISL_1215139, EPI_ISL_1215188, EPI_ISL_1215257, EPI_ISL_1215263, EPI_ISL_1280957, EPI_ISL_1280960, EPI_ISL_1280963, EPI_ISL_1280967, EPI_ISL_1280968, EPI_ISL_1280969, EPI_ISL_1280973, EPI_ISL_1280974, EPI_ISL_1280978, EPI_ISL_1280981, EPI_ISL_1280982, EPI_ISL_1280983, EPI_ISL_1280986, EPI_ISL_1281000, EPI_ISL_1281005, EPI_ISL_1281006, EPI_ISL_1281010, EPI_ISL_1281012, EPI_ISL_1281015, EPI_ISL_1281020, EPI_ISL_1281026, EPI_ISL_1281037, EPI_ISL_1281049, EPI_ISL_1281061, EPI_ISL_1281066, EPI_ISL_1284245, EPI_ISL_1284250, EPI_ISL_1284263, EPI_ISL_1284274, EPI_ISL_1284290, EPI_ISL_1284313, EPI_ISL_1284315, EPI_ISL_1284345, EPI_ISL_1284350, EPI_ISL_1284352, EPI_ISL_1284365, EPI_ISL_1284369, EPI_ISL_1284374, EPI_ISL_1284404, EPI_ISL_1284450, EPI_ISL_1284461, EPI_ISL_1284466, EPI_ISL_1286770, EPI_ISL_1286779, EPI_ISL_1286805, EPI_ISL_1286823, EPI_ISL_1286876, EPI_ISL_1286878, EPI_ISL_1286882 | | | |
| see above | Synlab MVZ Augsburg | Robert Koch Institute | |
| EPI_ISL_1217570, EPI_ISL_1285719, EPI_ISL_1285720, EPI_ISL_1285721, EPI_ISL_1285723, EPI_ISL_1285724, EPI_ISL_1353266, EPI_ISL_1353267, EPI_ISL_1353268, EPI_ISL_1353269, EPI_ISL_1353270, EPI_ISL_1353271, EPI_ISL_1353272, EPI_ISL_1353273, EPI_ISL_1353274, EPI_ISL_1353275, EPI_ISL_1353277 | | | |
| see above | Thüringer Landesamtes für Verbraucherschutz | Robert Koch Institute | |
| EPI_ISL_1217617, EPI_ISL_1217629, EPI_ISL_1217634, EPI_ISL_1217637, EPI_ISL_1217639, EPI_ISL_1217642, EPI_ISL_1217647, EPI_ISL_1217651, EPI_ISL_1217654, EPI_ISL_1217663, EPI_ISL_1217674, EPI_ISL_1217680, EPI_ISL_1217682, EPI_ISL_1285045, EPI_ISL_1285050, EPI_ISL_1285057, EPI_ISL_1285074, EPI_ISL_1285078, EPI_ISL_1285104, EPI_ISL_1285106, EPI_ISL_1285107, EPI_ISL_1285125, EPI_ISL_1285137, EPI_ISL_1285139 | | | |
| see above | Uniklinikum Carl Gustav Carus an der TU Dresden; Institut für Virologie | Robert Koch Institute | |
| EPI_ISL_1353265, EPI_ISL_1353423, EPI_ISL_1355240, EPI_ISL_1355241, EPI_ISL_1355242, EPI_ISL_1355286, EPI_ISL_1355287, EPI_ISL_1355288, EPI_ISL_1355289, EPI_ISL_1355290, EPI_ISL_1355293, EPI_ISL_1638637, EPI_ISL_1638639 | | | |
| see above | Uniklinikum Jena | Robert Koch Institute | |
| EPI_ISL_1217510, EPI_ISL_1217512, EPI_ISL_1217513, EPI_ISL_1217515, EPI_ISL_1217516, EPI_ISL_1285728, EPI_ISL_1285729, EPI_ISL_1285730, EPI_ISL_1285731, EPI_ISL_1285732, EPI_ISL_1357266, EPI_ISL_1357271 | | | |
| see above | Uniklinikum Regensburg; Institut für Mikrobiologie | Robert Koch Institute | |
| EPI_ISL_2123895, EPI_ISL_2123898, EPI_ISL_2123899, EPI_ISL_2123906, EPI_ISL_2123954 | Universitatsklinikum Koln; Institut fur Virologie | Robert Koch Institute | |
| EPI_ISL_1464591, EPI_ISL_1464597, EPI_ISL_1464599, EPI_ISL_1716059, EPI_ISL_1716060, EPI_ISL_1716061, EPI_ISL_1716070, EPI_ISL_1716075, EPI_ISL_1716076, EPI_ISL_1716077, EPI_ISL_1716078, EPI_ISL_1716085, EPI_ISL_1716090 | | | |
| see above | University Medical Center Hamburg Eppendorf | Heinrich Pette Institute, Leibniz Institute for Experimental Virology | Adam Grundhoff; Alexis Robitaille; Johannes Knobloch; Martin Aepfelbacher; Nicole Fischer; Thomas Günther |
| EPI_ISL_1573218, EPI_ISL_1573219, EPI_ISL_1573220, EPI_ISL_1573221, EPI_ISL_1573223, EPI_ISL_1573225, EPI_ISL_1573226, EPI_ISL_1573227, EPI_ISL_1573228 | | | |
| see above | UniversitÃ¤tsmedizin Mannheim | Robert Koch Institute | |
| EPI_ISL_1284519, EPI_ISL_1284521, EPI_ISL_1284533, EPI_ISL_1284535, EPI_ISL_1284537, EPI_ISL_1353373, EPI_ISL_1353381 | | | |
| see above | Universitätsklinikum Bonn - Institut für Medizinische Mikrobiologie Immunologie und Parasitologie + Institut für Virologie | Robert Koch Institute | |
| EPI_ISL_1217795, EPI_ISL_1217797, EPI_ISL_1217798, EPI_ISL_1217802, EPI_ISL_1217804, EPI_ISL_1217806 | Universitätsklinikum Essen | Robert Koch Institute | |
| EPI_ISL_1354939, EPI_ISL_1354942, EPI_ISL_1354943, EPI_ISL_1354944, EPI_ISL_1354951, EPI_ISL_1354954, EPI_ISL_1354962, EPI_ISL_1354968, EPI_ISL_1354986, EPI_ISL_1354990, EPI_ISL_1354991, EPI_ISL_1355043, EPI_ISL_1355045, EPI_ISL_1355058 | | | |
| see above | Universitätsklinikum Heidelberg | Robert Koch Institute | |

| | | | |
|---|---|---|---|
| EPI_ISL_1364976, EPI_ISL_1364977, EPI_ISL_1364978, EPI_ISL_1365009, EPI_ISL_1365016 | Universitätsklinikum Knappschaftskrankenhaus Bochum | Bundeswehr Institute of Microbiology | Alexandra Rehn; Enrico Georgi; Malena Bestehorn-Willmann; Markus Antwerpen; Mathias Walter; Mike Pillukat; Roman Wölfel; Sabine Zange |
| EPI_ISL_1217844, EPI_ISL_1217847, EPI_ISL_1217849, EPI_ISL_1217850, EPI_ISL_1217852, EPI_ISL_1217853, EPI_ISL_1217855, EPI_ISL_1217857, EPI_ISL_1217858, EPI_ISL_1217860, EPI_ISL_1217862, EPI_ISL_1217863, EPI_ISL_1217865, EPI_ISL_1217866, EPI_ISL_1217870, EPI_ISL_1217872, EPI_ISL_1217873, EPI_ISL_1217875, EPI_ISL_1217877, EPI_ISL_1217878, EPI_ISL_1217880, EPI_ISL_1217881, EPI_ISL_1217883, EPI_ISL_1217884, EPI_ISL_1217886, EPI_ISL_1217887, EPI_ISL_1217889, EPI_ISL_1217890, EPI_ISL_1217892, EPI_ISL_1217893, EPI_ISL_1217895, EPI_ISL_1217896, EPI_ISL_1217898, EPI_ISL_1217901, EPI_ISL_1217904, EPI_ISL_1217906, EPI_ISL_1217907, EPI_ISL_1217909, EPI_ISL_1217910, EPI_ISL_1217912, EPI_ISL_1217913, EPI_ISL_1217915, EPI_ISL_1217917, EPI_ISL_1217918, EPI_ISL_1217920, EPI_ISL_1217921, EPI_ISL_1217925, EPI_ISL_1217926, EPI_ISL_1217927, EPI_ISL_1217929, EPI_ISL_1217931, EPI_ISL_1217932, EPI_ISL_1217933, EPI_ISL_1217934, EPI_ISL_1217935, EPI_ISL_1217936, EPI_ISL_1217937, EPI_ISL_1217938, EPI_ISL_1217939, EPI_ISL_1217940, EPI_ISL_1217941, EPI_ISL_1217942, EPI_ISL_1217943, EPI_ISL_1217944, EPI_ISL_1217945, EPI_ISL_1217946, EPI_ISL_1217947, EPI_ISL_1217949, EPI_ISL_1217950, EPI_ISL_1217951, EPI_ISL_1217952, EPI_ISL_1217953, EPI_ISL_1217954, EPI_ISL_1217956, EPI_ISL_1217957, EPI_ISL_1217960, EPI_ISL_1217961, EPI_ISL_1217962, EPI_ISL_1217963 | | | |
| see above | Universitätsklinikum Leipzig - Institut für Medizinische Mikrobiologie und Virologie virologisches Labor | Robert Koch Institute | |
| EPI_ISL_1353439, EPI_ISL_1353452, EPI_ISL_1353464, EPI_ISL_1353473 | Universitätsklinikum Münster Institut für Virologie | Robert Koch Institute | |
| EPI_ISL_1435459 | Universitätsklinikum Schleswig-Holstein (standortübergreifend inkl. Kiel) - Institut für Klinische Chemie und Laboratoriumsdiagnostik | Robert Koch Institute | |
| EPI_ISL_1355743, EPI_ISL_1355745, EPI_ISL_1355826, EPI_ISL_1355830, EPI_ISL_1355832, EPI_ISL_1355841, EPI_ISL_1355842, EPI_ISL_1355843, EPI_ISL_1355846, EPI_ISL_1355852, EPI_ISL_1355853, EPI_ISL_1355854, EPI_ISL_1355859, EPI_ISL_1355860, EPI_ISL_1355866, EPI_ISL_1355867, EPI_ISL_1355868, EPI_ISL_1355869, EPI_ISL_1355876, EPI_ISL_1355887, EPI_ISL_1355891, EPI_ISL_1355892, EPI_ISL_1355896 | | | |
| see above | Universitätsmedizin Mannheim | Robert Koch Institute | |
| EPI_ISL_1354265, EPI_ISL_1354270, EPI_ISL_1354273, EPI_ISL_1354276, EPI_ISL_1354280 | Virologisches Institut des Universitätsklinikums Erlangen | Robert Koch Institute | |
| EPI_ISL_1366971, EPI_ISL_1366972, EPI_ISL_1366973, EPI_ISL_1366974, EPI_ISL_1366992, EPI_ISL_1367000, EPI_ISL_1367001, EPI_ISL_1367002, EPI_ISL_1367003, EPI_ISL_1367004, EPI_ISL_1367018, EPI_ISL_1367019, EPI_ISL_1367023, EPI_ISL_1367024, EPI_ISL_1367025, EPI_ISL_1367060, EPI_ISL_1367061, EPI_ISL_1367062, EPI_ISL_1367063, EPI_ISL_1367064, EPI_ISL_1367065, EPI_ISL_1367066, EPI_ISL_1367067, EPI_ISL_1367093, EPI_ISL_1367094, EPI_ISL_1367095, EPI_ISL_1367096, EPI_ISL_1367097, EPI_ISL_1367098, EPI_ISL_1367153, EPI_ISL_1367154, EPI_ISL_1367155, EPI_ISL_1367156, EPI_ISL_1367157, EPI_ISL_1367158 | | | |
| see above | Virology, Universitätsklinikum des Saarlandes | Epigenetics, Saarland University | Jörn Walter; Kathrin Kattler; Markus Vogelgesang; Sascha Tierling; Sigrun Smola; Stefan Lohse; Thorsten Pfuhl |
| EPI_ISL_1159348, EPI_ISL_1159349, EPI_ISL_1159350, EPI_ISL_1159351, EPI_ISL_1159352, EPI_ISL_1159353, EPI_ISL_1159354, EPI_ISL_1159355, EPI_ISL_1159356, EPI_ISL_1159357, EPI_ISL_1159358, EPI_ISL_1159359, EPI_ISL_1159360, EPI_ISL_1159361, EPI_ISL_1159362, EPI_ISL_1159363, EPI_ISL_1159364, EPI_ISL_1159365, EPI_ISL_1988340, EPI_ISL_1988342, EPI_ISL_1988343, EPI_ISL_1988344, EPI_ISL_1988345, EPI_ISL_1988346, EPI_ISL_1988348, EPI_ISL_1988349, EPI_ISL_1988351, EPI_ISL_1988352, EPI_ISL_1988354, EPI_ISL_1988355, EPI_ISL_1988357, EPI_ISL_1988358, EPI_ISL_1988360, EPI_ISL_1988361, EPI_ISL_1988362, EPI_ISL_1988363, EPI_ISL_1988364, EPI_ISL_1988366, EPI_ISL_1988367, EPI_ISL_1988368, EPI_ISL_1988369, EPI_ISL_1988370, EPI_ISL_1988697, EPI_ISL_1988698, EPI_ISL_1988699, EPI_ISL_1988700, EPI_ISL_1988702, EPI_ISL_1988703, EPI_ISL_1988704, EPI_ISL_1988705, EPI_ISL_1988706, EPI_ISL_1990026, EPI_ISL_1990027, EPI_ISL_1990028, EPI_ISL_1990030, EPI_ISL_1990031, EPI_ISL_1990032, EPI_ISL_1990034, EPI_ISL_1990035 | | | |
| see above | ZOTZ KLIMAS MVZ Düsseldorf-Centrum GbR ÜBAG für Labormedizin, Genetik, Zytologie, Pathologie | Center of Medical Microbiology, Virology, and Hospital Hygiene, University of Duesseldorf | Alexander Dilthey; Andreas Walker; Daniel Strelow; Jessica Nicolai; Jörg Timm; Katrin Hoffmann; Klaus Pfeffer; Lisanna Hülse; Malte Kohns Vasconcelos; Maximilian Damagnez; Nadine Lübke; Patrick Finzer; Rainer Zotz; Tobias Wienemann; Torsten Houwaart |
| EPI_ISL_1281315, EPI_ISL_1281317, EPI_ISL_1281329, EPI_ISL_1281347, EPI_ISL_1281356, EPI_ISL_1281361, EPI_ISL_1281370, EPI_ISL_1281373, EPI_ISL_1281377, EPI_ISL_1281392, EPI_ISL_1281400, EPI_ISL_1281406, EPI_ISL_1281411, EPI_ISL_1281430, EPI_ISL_1281435, EPI_ISL_1281480, EPI_ISL_1281482, EPI_ISL_1281483, EPI_ISL_1281487, EPI_ISL_1281492, EPI_ISL_1281504, EPI_ISL_1281505, EPI_ISL_1281509, EPI_ISL_1281526, EPI_ISL_1281532, EPI_ISL_1281537, EPI_ISL_1281541, EPI_ISL_1281552, EPI_ISL_1281560, EPI_ISL_1281564, EPI_ISL_1281565, EPI_ISL_1281578, EPI_ISL_1281582, EPI_ISL_1351381, EPI_ISL_1351383, EPI_ISL_1351391, EPI_ISL_1351394, EPI_ISL_1351399, EPI_ISL_1351403, EPI_ISL_1351409, EPI_ISL_1351413, EPI_ISL_1351425, EPI_ISL_1351456, EPI_ISL_1351458, EPI_ISL_1351469, EPI_ISL_1351491, EPI_ISL_1351496, EPI_ISL_1351499, EPI_ISL_1351522, EPI_ISL_1351539, EPI_ISL_1351563, EPI_ISL_1351588, EPI_ISL_1351600, EPI_ISL_1351607, EPI_ISL_1351621, EPI_ISL_1351640, EPI_ISL_1351641, EPI_ISL_1351650, EPI_ISL_1351657, EPI_ISL_1351666, EPI_ISL_1351695, EPI_ISL_1351715, EPI_ISL_1351741, EPI_ISL_1351745, EPI_ISL_1351758, EPI_ISL_1436775 | | | |
| see above | amedes Göttingen | Robert Koch Institute | |
| EPI_ISL_1281311, EPI_ISL_1281335, EPI_ISL_1281339, EPI_ISL_1281341, EPI_ISL_1281344, EPI_ISL_1281349, EPI_ISL_1281368, EPI_ISL_1281379, EPI_ISL_1281385, EPI_ISL_1281387, EPI_ISL_1281391, EPI_ISL_1281409, EPI_ISL_1281437, EPI_ISL_1281443, EPI_ISL_1281451, EPI_ISL_1281502, EPI_ISL_1281512, EPI_ISL_1281516, EPI_ISL_1281520, EPI_ISL_1281525, EPI_ISL_1281527, EPI_ISL_1281531, EPI_ISL_1281533, EPI_ISL_1281534, EPI_ISL_1281543, EPI_ISL_1281556, EPI_ISL_1281561, EPI_ISL_1281566, EPI_ISL_1281579, EPI_ISL_1351395, EPI_ISL_1351479, EPI_ISL_1351484, EPI_ISL_1351490, EPI_ISL_1351519, EPI_ISL_1351553, EPI_ISL_1351568, EPI_ISL_1351587, EPI_ISL_1351628, EPI_ISL_1351646, EPI_ISL_1351684, EPI_ISL_1351707, EPI_ISL_1351711, EPI_ISL_1351723, EPI_ISL_1351736, EPI_ISL_1351742 | | | |
| see above | amedes MVZ DIAMEDIS Sennestadt | Robert Koch Institute | |
| EPI_ISL_1281308, EPI_ISL_1281312, EPI_ISL_1281316, EPI_ISL_1281322, EPI_ISL_1281323, EPI_ISL_1281324, EPI_ISL_1281327, EPI_ISL_1281332, EPI_ISL_1281364, EPI_ISL_1281365, EPI_ISL_1281374, EPI_ISL_1281389, EPI_ISL_1281395, EPI_ISL_1281397, EPI_ISL_1281401, EPI_ISL_1281414, EPI_ISL_1281415, EPI_ISL_1281436, EPI_ISL_1281441, EPI_ISL_1281446, EPI_ISL_1281463, EPI_ISL_1281472, EPI_ISL_1281488, EPI_ISL_1281489, EPI_ISL_1281496, EPI_ISL_1281501, EPI_ISL_1281506, EPI_ISL_1281510, EPI_ISL_1281514, EPI_ISL_1281530, EPI_ISL_1281539, EPI_ISL_1281542, EPI_ISL_1281549, EPI_ISL_1351386, EPI_ISL_1351530, EPI_ISL_1351534 | | | |
| see above | amedes MVZ Halle/Leipzig | Robert Koch Institute | |
| EPI_ISL_1212933, EPI_ISL_1212934, EPI_ISL_1212939, EPI_ISL_1212942, EPI_ISL_1212966, EPI_ISL_1212996, EPI_ISL_1213035, EPI_ISL_1213044, EPI_ISL_1213070, EPI_ISL_1213095, EPI_ISL_1213122, EPI_ISL_1213124, EPI_ISL_1281350, EPI_ISL_1281359, EPI_ISL_1281381, EPI_ISL_1281470, EPI_ISL_1281555, EPI_ISL_1281563, EPI_ISL_1351400, EPI_ISL_1351402, EPI_ISL_1351407, EPI_ISL_1351408, EPI_ISL_1351412, EPI_ISL_1351415, EPI_ISL_1351432, EPI_ISL_1351488, EPI_ISL_1351504, EPI_ISL_1351524, EPI_ISL_1351581, EPI_ISL_1351591, EPI_ISL_1351604, EPI_ISL_1351613, EPI_ISL_1351630, EPI_ISL_1351639, EPI_ISL_1351658, EPI_ISL_1351663, EPI_ISL_1351669, EPI_ISL_1351673, EPI_ISL_1351676, EPI_ISL_1351681, EPI_ISL_1351687, EPI_ISL_1351692, EPI_ISL_1351714 | | | |
| see above | amedes MVZ Hannover | Robert Koch Institute | |
| EPI_ISL_1281328, EPI_ISL_1281334, EPI_ISL_1281345, EPI_ISL_1281348, EPI_ISL_1281353, EPI_ISL_1281369, EPI_ISL_1281398, EPI_ISL_1281404, EPI_ISL_1281408, EPI_ISL_1281412, EPI_ISL_1281417, EPI_ISL_1281421, EPI_ISL_1281424, EPI_ISL_1281442, EPI_ISL_1281444, EPI_ISL_1281453, EPI_ISL_1281461, EPI_ISL_1281474, EPI_ISL_1281477, EPI_ISL_1281481, EPI_ISL_1281500, EPI_ISL_1281515, EPI_ISL_1281518, EPI_ISL_1281519, EPI_ISL_1281545, EPI_ISL_1281548, EPI_ISL_1281554, EPI_ISL_1281562, EPI_ISL_1281572, EPI_ISL_1281580 | | | |
| see above | amedes MVZ Jena | Robert Koch Institute | |
| EPI_ISL_1281493, EPI_ISL_1281550, EPI_ISL_1281575 | amedes MVZ Zentrum für Pathologie und Zytodiagnostik GmbH (Köln) | Robert Koch Institute | |
| EPI_ISL_1212929, EPI_ISL_1212937, EPI_ISL_1212945, EPI_ISL_1212991, EPI_ISL_1213000, EPI_ISL_1213027, EPI_ISL_1213086, EPI_ISL_1281330, EPI_ISL_1281331, EPI_ISL_1281354, EPI_ISL_1281393, EPI_ISL_1281396, EPI_ISL_1281402, EPI_ISL_1281429, EPI_ISL_1281452, EPI_ISL_1281455, EPI_ISL_1281456, EPI_ISL_1281458, EPI_ISL_1281497, EPI_ISL_1281546, EPI_ISL_1281574, EPI_ISL_1351388, EPI_ISL_1351393, EPI_ISL_1351423, EPI_ISL_1351455, EPI_ISL_1351464, EPI_ISL_1351472, EPI_ISL_1351501, EPI_ISL_1351533, EPI_ISL_1351544, EPI_ISL_1351556, EPI_ISL_1351562, EPI_ISL_1351570, EPI_ISL_1351573, EPI_ISL_1351586, EPI_ISL_1351610, EPI_ISL_1351627, EPI_ISL_1351708 | | | |
| see above | amedes MVZ für Laboratoriumsdiagnostik Raubling GmbH | Robert Koch Institute | |
| EPI_ISL_1281325, EPI_ISL_1281352, EPI_ISL_1281499, EPI_ISL_1351390, EPI_ISL_1351398, EPI_ISL_1436648, EPI_ISL_1436777 | | | |
| see above | amedes MVZ für Laboratoriumsmedizin Fürstenfeldbruck | Robert Koch Institute | |
| EPI_ISL_1351396, EPI_ISL_1351404, EPI_ISL_1351701, EPI_ISL_1436782 | amedes MVZ für Laboratoriumsmedizin München | Robert Koch Institute | |
| EPI_ISL_1281448, EPI_ISL_1281544 | amedes MVZ für Laboratoriumsmedizin Rhein/Ruhr | Robert Koch Institute | |
| EPI_ISL_1213042, EPI_ISL_1281310, EPI_ISL_1281336, EPI_ISL_1281342, EPI_ISL_1281357, EPI_ISL_1281362, EPI_ISL_1281367, EPI_ISL_1281439, EPI_ISL_1281524, EPI_ISL_1351387, EPI_ISL_1351572, EPI_ISL_1351634, EPI_ISL_1351727, EPI_ISL_1351747, EPI_ISL_1351750 | | | |
| see above | amedes MVZ für Labordiagnostik Rhein-Main | Robert Koch Institute | |
| EPI_ISL_1281399, EPI_ISL_1281517, EPI_ISL_1281557, EPI_ISL_1281577 | amedes Medizinisches Labor Mühlhausen | Robert Koch Institute | |
| EPI_ISL_1281319, EPI_ISL_1281321, EPI_ISL_1281343, EPI_ISL_1281428, EPI_ISL_1281449, EPI_ISL_1281486, EPI_ISL_1281529 | | | |
| see above | amedes Medlab Arnold Analytik MVZ GmbH | Robert Koch Institute | |
| EPI_ISL_1282968, EPI_ISL_1282980, EPI_ISL_1282982, EPI_ISL_1351772, EPI_ISL_1351773 | labopart - Medizinische Laboratorien Dresden | Robert Koch Institute | |
| EPI_ISL_1281309, EPI_ISL_1281313, EPI_ISL_1281333, EPI_ISL_1281346, EPI_ISL_1281351, EPI_ISL_1281355, EPI_ISL_1281358, EPI_ISL_1281360, EPI_ISL_1281363, EPI_ISL_1281366, EPI_ISL_1281371, EPI_ISL_1281380, EPI_ISL_1281383, EPI_ISL_1281386, EPI_ISL_1281390, EPI_ISL_1281394, EPI_ISL_1281403, EPI_ISL_1281407, EPI_ISL_1281413, EPI_ISL_1281418, EPI_ISL_1281425, EPI_ISL_1281431, EPI_ISL_1281432, EPI_ISL_1281433, EPI_ISL_1281447, EPI_ISL_1281459, EPI_ISL_1281465, EPI_ISL_1281466, EPI_ISL_1281468, EPI_ISL_1281473, EPI_ISL_1281476, EPI_ISL_1281478, EPI_ISL_1281485, EPI_ISL_1281490, EPI_ISL_1281495, EPI_ISL_1281508, EPI_ISL_1281511, EPI_ISL_1281513, EPI_ISL_1281522, EPI_ISL_1281528, EPI_ISL_1281535, EPI_ISL_1281536, EPI_ISL_1281540, EPI_ISL_1281551, EPI_ISL_1281558, EPI_ISL_1281559, EPI_ISL_1281567, EPI_ISL_1281570, EPI_ISL_1281581, EPI_ISL_1351375, EPI_ISL_1351377, EPI_ISL_1351379, EPI_ISL_1351382, EPI_ISL_1351385, EPI_ISL_1351411, EPI_ISL_1351414, EPI_ISL_1351466, EPI_ISL_1351637, EPI_ISL_1351693, EPI_ISL_1351712, EPI_ISL_1351738, EPI_ISL_1351748 | | | |
| see above | nordlab - Partnerschaftspraxis fu?r Laboratoriumsmedizin | Robert Koch Institute | |

We gratefully acknowledge the following Authors from the Originating laboratories responsible for obtaining the specimens, as well as the Submitting laboratories where the genome data were generated and shared via GISAID, on which this research is based.

All Submitters of data may be contacted directly via www.gisaid.org

Authors are sorted alphabetically.

| Accession ID | Originating Laboratory | Submitting Laboratory | Authors |
|---|---|---|---|
| EPI_ISL_1433035, EPI_ISL_1433037, EPI_ISL_1433248, EPI_ISL_1433276, EPI_ISL_1438712, EPI_ISL_1438726, EPI_ISL_1438731, EPI_ISL_1438778, EPI_ISL_1438797, EPI_ISL_1438798, EPI_ISL_1438799, EPI_ISL_1438815, EPI_ISL_1438843, EPI_ISL_1438865, EPI_ISL_1438880, EPI_ISL_1438929, EPI_ISL_1438958, EPI_ISL_1438974, EPI_ISL_1438983, EPI_ISL_1438985, EPI_ISL_1438995, EPI_ISL_1439007, EPI_ISL_1439018 | | | |
| see above | ALB FILS KLINIKEN GmbH - Institut für Laboratoriumsmedizin Klinik am Eichert Göppingen | Robert Koch Institute | |
| EPI_ISL_1354484, EPI_ISL_1354492, EPI_ISL_1354493, EPI_ISL_1354494, EPI_ISL_1354495, EPI_ISL_1354496, EPI_ISL_1354497, EPI_ISL_1354498, EPI_ISL_1354500 | | | |
| see above | Alexianer DaKS GmbH (eigenes IT-Tochterunternehmen der Alexianer Krankenhäuser) | Robert Koch Institute | |
| EPI_ISL_2111429 | Bayerisches Landesamt fur Gesundheit und Lebensmittelsicherheit (LGL) | Robert Koch Institute | |
| EPI_ISL_1570335, EPI_ISL_1570336, EPI_ISL_1570337, EPI_ISL_1570340, EPI_ISL_1570341, EPI_ISL_1570342, EPI_ISL_1570343, EPI_ISL_1570344, EPI_ISL_1570346, EPI_ISL_1570348, EPI_ISL_1570349, EPI_ISL_1570350, EPI_ISL_1570352, EPI_ISL_1570353, EPI_ISL_1570354, EPI_ISL_1570356, EPI_ISL_1570357, EPI_ISL_1570358, EPI_ISL_1570359, EPI_ISL_1570361, EPI_ISL_1570362, EPI_ISL_1570363, EPI_ISL_1570364, EPI_ISL_1570365, EPI_ISL_1570366, EPI_ISL_1570367, EPI_ISL_1570369, EPI_ISL_1570370, EPI_ISL_1570371, EPI_ISL_1570372, EPI_ISL_1570373, EPI_ISL_1570374, EPI_ISL_1570375, EPI_ISL_1570376, EPI_ISL_1570378, EPI_ISL_1570379, EPI_ISL_1570380, EPI_ISL_1570381, EPI_ISL_1570382, EPI_ISL_1570383, EPI_ISL_1570384, EPI_ISL_1570386, EPI_ISL_1570387, EPI_ISL_1570388, EPI_ISL_1570389, EPI_ISL_1570390, EPI_ISL_1570391, EPI_ISL_1570393, EPI_ISL_1570394, EPI_ISL_1570395, EPI_ISL_1570396, EPI_ISL_1570397, EPI_ISL_1570398, EPI_ISL_1570399, EPI_ISL_1570400, EPI_ISL_1638666, EPI_ISL_1638667, EPI_ISL_1638668, EPI_ISL_1638669, EPI_ISL_1638670, EPI_ISL_1638671, EPI_ISL_1638672, EPI_ISL_1638673, EPI_ISL_1638674, EPI_ISL_1638675, EPI_ISL_1638676, EPI_ISL_1638677, EPI_ISL_1638678, EPI_ISL_1638679, EPI_ISL_1638680, EPI_ISL_1638681, EPI_ISL_1638682, EPI_ISL_1640831, EPI_ISL_1640832, EPI_ISL_1640833, EPI_ISL_1640834, EPI_ISL_1640835, EPI_ISL_1640836, EPI_ISL_1640837, EPI_ISL_1640838, EPI_ISL_1640839, EPI_ISL_1640840, EPI_ISL_1640841, EPI_ISL_1640842, EPI_ISL_1640843, EPI_ISL_1640844, EPI_ISL_1640845, EPI_ISL_1640848, EPI_ISL_1640849, EPI_ISL_1640850, EPI_ISL_1640851, EPI_ISL_1640852, EPI_ISL_1640854, EPI_ISL_1640855, EPI_ISL_1640856, EPI_ISL_1640857, EPI_ISL_1640858, EPI_ISL_1640859, EPI_ISL_1640860, EPI_ISL_1640861, EPI_ISL_1640862, EPI_ISL_1640863, EPI_ISL_1640864, EPI_ISL_1640865, EPI_ISL_1640866, EPI_ISL_1640868, EPI_ISL_1640869, EPI_ISL_1640871, EPI_ISL_1640872, EPI_ISL_1640873, EPI_ISL_1640875, EPI_ISL_1640876, EPI_ISL_1640877, EPI_ISL_1640878, EPI_ISL_1640879, EPI_ISL_1640880, EPI_ISL_1640881, EPI_ISL_1640882, EPI_ISL_1640883, EPI_ISL_1640885, EPI_ISL_1640886, EPI_ISL_1640887, EPI_ISL_1640888, EPI_ISL_1640889, EPI_ISL_1640890, EPI_ISL_1640891, EPI_ISL_1640892, EPI_ISL_1640893, EPI_ISL_1640894, EPI_ISL_1640895, EPI_ISL_1640896, EPI_ISL_1640897, EPI_ISL_1640898, EPI_ISL_1640899, EPI_ISL_1640900, EPI_ISL_1640901, EPI_ISL_1640902, EPI_ISL_1640903, EPI_ISL_1725285, EPI_ISL_1725360, EPI_ISL_1725381, EPI_ISL_2110879, EPI_ISL_2110922 | | | |
| see above | Bayerisches Landesamt fÃ¼r Gesundheit und Lebensmittelsicherheit (LGL) | Robert Koch Institute | |
| EPI_ISL_2129723, EPI_ISL_2129725, EPI_ISL_2129727, EPI_ISL_2129728, EPI_ISL_2129730 | Bernhard-Nocht-Institut fÃ¼r Tropenmedizin | Robert Koch Institute | |
| EPI_ISL_1353644, EPI_ISL_1353650, EPI_ISL_1353659, EPI_ISL_1353661, EPI_ISL_1353676, EPI_ISL_1353680, EPI_ISL_1353689, EPI_ISL_1353692, EPI_ISL_1353694, EPI_ISL_1353697, EPI_ISL_1353700, EPI_ISL_1353706, EPI_ISL_1353715, EPI_ISL_1353724, EPI_ISL_1353725, EPI_ISL_1353741, EPI_ISL_1353744, EPI_ISL_1353753, EPI_ISL_1353756, EPI_ISL_1353764, EPI_ISL_1353768, EPI_ISL_1353774, EPI_ISL_1353783, EPI_ISL_1353792, EPI_ISL_1353799, EPI_ISL_1353804, EPI_ISL_1353814, EPI_ISL_1353817, EPI_ISL_1353821, EPI_ISL_1353840, EPI_ISL_1353841, EPI_ISL_1353842, EPI_ISL_1353845, EPI_ISL_1353846, EPI_ISL_1353854, EPI_ISL_1353855, EPI_ISL_1353867, EPI_ISL_1353871, EPI_ISL_1353879, EPI_ISL_1353881, EPI_ISL_1353900, EPI_ISL_1353904, EPI_ISL_1353910, EPI_ISL_1353912, EPI_ISL_1353915, EPI_ISL_1353916, EPI_ISL_1353917, EPI_ISL_1353922, EPI_ISL_1353926, EPI_ISL_1353934, EPI_ISL_1353944, EPI_ISL_1353945, EPI_ISL_1353958, EPI_ISL_1353966, EPI_ISL_1353969, EPI_ISL_1353970, EPI_ISL_1353981, EPI_ISL_1353984, EPI_ISL_1432939, EPI_ISL_1432943, EPI_ISL_1432952, EPI_ISL_1432954, EPI_ISL_1432958, EPI_ISL_1432963, EPI_ISL_1432970, EPI_ISL_1432978, EPI_ISL_1433000, EPI_ISL_1433004, EPI_ISL_1433015, EPI_ISL_1433017, EPI_ISL_1433019, EPI_ISL_1433023, EPI_ISL_1433050, EPI_ISL_1433052, EPI_ISL_1433059, EPI_ISL_1433061, EPI_ISL_1433063, EPI_ISL_1433064, EPI_ISL_1433065, EPI_ISL_1433066, EPI_ISL_1433069, EPI_ISL_1433073, EPI_ISL_1433074, EPI_ISL_1433080, EPI_ISL_1433086, EPI_ISL_1433097, EPI_ISL_1433101, EPI_ISL_1433115, EPI_ISL_1433116, EPI_ISL_1433128, EPI_ISL_1433137, EPI_ISL_1433138, EPI_ISL_1433139, EPI_ISL_1433150, EPI_ISL_1433151, EPI_ISL_1433152, EPI_ISL_1433155, EPI_ISL_1433156, EPI_ISL_1433157, EPI_ISL_1433160, EPI_ISL_1433163, EPI_ISL_1433172, EPI_ISL_1433177, EPI_ISL_1433178, EPI_ISL_1433180, EPI_ISL_1433182, EPI_ISL_1433183, EPI_ISL_1433185, EPI_ISL_1433186, EPI_ISL_1433194, EPI_ISL_1433196, EPI_ISL_1433198, EPI_ISL_1433199, EPI_ISL_1433205, EPI_ISL_1433210, EPI_ISL_1433211, EPI_ISL_1433212, EPI_ISL_1433217, EPI_ISL_1433221, EPI_ISL_1433224, EPI_ISL_1433232, EPI_ISL_1433234, EPI_ISL_1433236, EPI_ISL_1433240, EPI_ISL_1433244, EPI_ISL_1433245, EPI_ISL_1433247, EPI_ISL_1433253, EPI_ISL_1433255, EPI_ISL_1433256, EPI_ISL_1433262, EPI_ISL_1433271, EPI_ISL_1433278, EPI_ISL_1433280, EPI_ISL_1433283, EPI_ISL_1433305, EPI_ISL_1433308, EPI_ISL_1433309, EPI_ISL_1433311, EPI_ISL_1433321, EPI_ISL_1433323, EPI_ISL_1433325, EPI_ISL_1433327, EPI_ISL_1433329, EPI_ISL_1433334, EPI_ISL_1433340, EPI_ISL_1433362, EPI_ISL_1433374, EPI_ISL_1433376, EPI_ISL_1436034, EPI_ISL_1436305, EPI_ISL_1436307, EPI_ISL_1438686, EPI_ISL_1438693, EPI_ISL_1438696, EPI_ISL_1438697, EPI_ISL_1438698, EPI_ISL_1438699, EPI_ISL_1438704, EPI_ISL_1438705, EPI_ISL_1438710, EPI_ISL_1438711, EPI_ISL_1438717, EPI_ISL_1438721, EPI_ISL_1438723, EPI_ISL_1438724, EPI_ISL_1438729, EPI_ISL_1438730, EPI_ISL_1438732, EPI_ISL_1438739, EPI_ISL_1438740, EPI_ISL_1438741, EPI_ISL_1438743, EPI_ISL_1438744, EPI_ISL_1438746, EPI_ISL_1438747, EPI_ISL_1438749, EPI_ISL_1438750, EPI_ISL_1438751, EPI_ISL_1438753, EPI_ISL_1438754, EPI_ISL_1438755, EPI_ISL_1438756, EPI_ISL_1438757, EPI_ISL_1438758, EPI_ISL_1438764, EPI_ISL_1438765, EPI_ISL_1438767, EPI_ISL_1438768, EPI_ISL_1438770, EPI_ISL_1438773, EPI_ISL_1438774, EPI_ISL_1438775, EPI_ISL_1438776, EPI_ISL_1438777, EPI_ISL_1438781, EPI_ISL_1438782, EPI_ISL_1438788, EPI_ISL_1438790, EPI_ISL_1438792, EPI_ISL_1438793, EPI_ISL_1438794, EPI_ISL_1438796, EPI_ISL_1438800, EPI_ISL_1438801, EPI_ISL_1438802, EPI_ISL_1438803, EPI_ISL_1438804, EPI_ISL_1438807, EPI_ISL_1438810, EPI_ISL_1438816, EPI_ISL_1438818, EPI_ISL_1438820, EPI_ISL_1438822, EPI_ISL_1438827, EPI_ISL_1438828, EPI_ISL_1438830, EPI_ISL_1438832, EPI_ISL_1438837, EPI_ISL_1438839, EPI_ISL_1438840, EPI_ISL_1438844, EPI_ISL_1438848, EPI_ISL_1438850, EPI_ISL_1438853, EPI_ISL_1438857, EPI_ISL_1438858, EPI_ISL_1438860, EPI_ISL_1438864, EPI_ISL_1438866, EPI_ISL_1438868, EPI_ISL_1438873, EPI_ISL_1438875, EPI_ISL_1438876, EPI_ISL_1438877, EPI_ISL_1438881, EPI_ISL_1438883, EPI_ISL_1438885, EPI_ISL_1438886, EPI_ISL_1438888, EPI_ISL_1438890, EPI_ISL_1438891, EPI_ISL_1438895, EPI_ISL_1438898, EPI_ISL_1438899, EPI_ISL_1438902, EPI_ISL_1438906, EPI_ISL_1438911, EPI_ISL_1438913, EPI_ISL_1438915, EPI_ISL_1438916, EPI_ISL_1438921, EPI_ISL_1438922, EPI_ISL_1438923, EPI_ISL_1438930, EPI_ISL_1438933, EPI_ISL_1438936, EPI_ISL_1438942, EPI_ISL_1438944, EPI_ISL_1438945, EPI_ISL_1438948, EPI_ISL_1438949, EPI_ISL_1438950, EPI_ISL_1438951, EPI_ISL_1438952, EPI_ISL_1438953, EPI_ISL_1438956, EPI_ISL_1438957, EPI_ISL_1438960, EPI_ISL_1438962, EPI_ISL_1438963, EPI_ISL_1438964, EPI_ISL_1438965, EPI_ISL_1438966, EPI_ISL_1438967, EPI_ISL_1438968, EPI_ISL_1438970, EPI_ISL_1438975, EPI_ISL_1438978, EPI_ISL_1438979, EPI_ISL_1438981, EPI_ISL_1438982, EPI_ISL_1438987, EPI_ISL_1438988, EPI_ISL_1438990, EPI_ISL_1438993, EPI_ISL_1438994, EPI_ISL_1438996, EPI_ISL_1438999, EPI_ISL_1439000, EPI_ISL_1439006, EPI_ISL_1439013, EPI_ISL_1439015, EPI_ISL_1439023, EPI_ISL_1439026, EPI_ISL_1439035, EPI_ISL_1439036, EPI_ISL_1439037, EPI_ISL_1439039, EPI_ISL_1439047, EPI_ISL_1439057, EPI_ISL_1439058, EPI_ISL_1439067, EPI_ISL_1439076, EPI_ISL_1439081, EPI_ISL_1439082, EPI_ISL_1439086, EPI_ISL_1439089, EPI_ISL_1439090, EPI_ISL_1439094, EPI_ISL_1439105, EPI_ISL_1439111, EPI_ISL_1439115, EPI_ISL_1439117, EPI_ISL_1439123, EPI_ISL_1439132, EPI_ISL_1439139, EPI_ISL_1439140, EPI_ISL_1439143, EPI_ISL_1439150, EPI_ISL_1439151, EPI_ISL_1439153, EPI_ISL_1439156, EPI_ISL_1439166, EPI_ISL_1439167, EPI_ISL_1439168, EPI_ISL_1439169, EPI_ISL_1439181, EPI_ISL_1439182, EPI_ISL_1439185, EPI_ISL_1439187, EPI_ISL_1439188, EPI_ISL_1439190, EPI_ISL_1439191, EPI_ISL_1439196, EPI_ISL_1439198, EPI_ISL_1439205, EPI_ISL_1439206, EPI_ISL_1439214, EPI_ISL_1439215, EPI_ISL_1439223, EPI_ISL_1439227, EPI_ISL_1439231, EPI_ISL_1439242, EPI_ISL_1439251, EPI_ISL_1439255, EPI_ISL_1439257, EPI_ISL_1439258, EPI_ISL_1439259, EPI_ISL_1439261, EPI_ISL_1439262, EPI_ISL_1439263, EPI_ISL_1439274, EPI_ISL_1439276, EPI_ISL_1439280, EPI_ISL_1439295, EPI_ISL_1439309, EPI_ISL_1439311, EPI_ISL_1439319, EPI_ISL_1439325, EPI_ISL_1439326, EPI_ISL_1439330, EPI_ISL_1439336, EPI_ISL_1439337, EPI_ISL_1439338, EPI_ISL_1439350, EPI_ISL_1439358, EPI_ISL_1439359, EPI_ISL_1439360, EPI_ISL_1439364, EPI_ISL_1568946, EPI_ISL_1568947, EPI_ISL_1568948, EPI_ISL_1568951, EPI_ISL_1568954, EPI_ISL_1568955, EPI_ISL_1568956, EPI_ISL_1568957, EPI_ISL_1568959, EPI_ISL_1568960, EPI_ISL_1568966, EPI_ISL_1568967, EPI_ISL_1568968, EPI_ISL_1568970, EPI_ISL_1568971, EPI_ISL_1568973, EPI_ISL_1568977, EPI_ISL_1568979, EPI_ISL_1568982, EPI_ISL_1568985, EPI_ISL_1568987, EPI_ISL_1568989, EPI_ISL_1568990, EPI_ISL_1568993, EPI_ISL_1568994, EPI_ISL_1568996, EPI_ISL_1569000, EPI_ISL_1569002, EPI_ISL_1569004, EPI_ISL_1569006, EPI_ISL_1569009, EPI_ISL_1569010, EPI_ISL_1569016, EPI_ISL_1569019, EPI_ISL_1569020, EPI_ISL_1569022, EPI_ISL_1569026, EPI_ISL_1569028, EPI_ISL_1569031, EPI_ISL_1569039, EPI_ISL_1569040, EPI_ISL_1569042, EPI_ISL_1569044, EPI_ISL_1569048, EPI_ISL_1569051, EPI_ISL_1569053, EPI_ISL_1569055, EPI_ISL_1569056, EPI_ISL_1569057, EPI_ISL_1569060, EPI_ISL_1569061, EPI_ISL_1569063, EPI_ISL_1569064, EPI_ISL_1569065, EPI_ISL_1569067, EPI_ISL_1569068, EPI_ISL_1569069, EPI_ISL_1569070, EPI_ISL_1569071, EPI_ISL_1569072, EPI_ISL_1569073, EPI_ISL_1569074, EPI_ISL_1569075, EPI_ISL_1569077, EPI_ISL_1569078, EPI_ISL_1569079, EPI_ISL_1569084, EPI_ISL_1569085, EPI_ISL_1569086, EPI_ISL_1569087, EPI_ISL_1569088, EPI_ISL_1569090, EPI_ISL_1569092, EPI_ISL_1569093, EPI_ISL_1569094, EPI_ISL_1569095, EPI_ISL_1569096, EPI_ISL_1569097, EPI_ISL_1569106, EPI_ISL_1569107, EPI_ISL_1569116, EPI_ISL_1569119, EPI_ISL_1569120, EPI_ISL_1569121, EPI_ISL_1569122, EPI_ISL_1569123, EPI_ISL_1569124, EPI_ISL_1569127, EPI_ISL_1569129, EPI_ISL_1569130, EPI_ISL_1569132, EPI_ISL_1569134, EPI_ISL_1569135, EPI_ISL_1569136, EPI_ISL_1569138, EPI_ISL_1569140, EPI_ISL_1569141, EPI_ISL_1569143, EPI_ISL_1569144, EPI_ISL_1569146, EPI_ISL_1569147, EPI_ISL_1569149, EPI_ISL_1569150, EPI_ISL_1569151, EPI_ISL_1569152, EPI_ISL_1569154, EPI_ISL_1569157, EPI_ISL_1569158, EPI_ISL_1569160, EPI_ISL_1569161, EPI_ISL_1569162, EPI_ISL_1569164, EPI_ISL_1569165, EPI_ISL_1569168, EPI_ISL_1569169, EPI_ISL_1569171, EPI_ISL_1569174, EPI_ISL_1569175, EPI_ISL_1569180, EPI_ISL_1569181, EPI_ISL_1569182, EPI_ISL_1569184, EPI_ISL_1569188, EPI_ISL_1569192, EPI_ISL_1569195, EPI_ISL_1569196, EPI_ISL_1569197, EPI_ISL_1569198, EPI_ISL_1569199, EPI_ISL_1569200, EPI_ISL_1569201, EPI_ISL_1569202, EPI_ISL_1569203, EPI_ISL_1569206, EPI_ISL_1569207, EPI_ISL_1569210, EPI_ISL_1569211, EPI_ISL_1569213, EPI_ISL_1569217, EPI_ISL_1569218, EPI_ISL_1569221, EPI_ISL_1569224, EPI_ISL_1569230, EPI_ISL_1569231, EPI_ISL_1569232, EPI_ISL_1569233, EPI_ISL_1569234, EPI_ISL_1569236, EPI_ISL_1569240, EPI_ISL_1569243, EPI_ISL_1569246, EPI_ISL_1569247, EPI_ISL_1569249, EPI_ISL_1569250, EPI_ISL_1569251, EPI_ISL_1569252, EPI_ISL_1569259, EPI_ISL_1569260, EPI_ISL_1569269, EPI_ISL_1569272, EPI_ISL_1569273, EPI_ISL_1569274, EPI_ISL_1569286, EPI_ISL_1569318, EPI_ISL_1569323, EPI_ISL_1569325, EPI_ISL_1569344, EPI_ISL_1569453, EPI_ISL_1569618, EPI_ISL_1569647, EPI_ISL_1569687, EPI_ISL_1638721, EPI_ISL_1638754, EPI_ISL_1638773, EPI_ISL_1638778, EPI_ISL_1638783, EPI_ISL_1638784, EPI_ISL_1638801, EPI_ISL_1638802, EPI_ISL_1638804, EPI_ISL_1638810, EPI_ISL_1638834, EPI_ISL_1638850, EPI_ISL_1638857, EPI_ISL_1638893, EPI_ISL_1638907, EPI_ISL_1638910, EPI_ISL_1638914, EPI_ISL_1638921, EPI_ISL_1638946, EPI_ISL_1638949, EPI_ISL_1638964, EPI_ISL_1638968, EPI_ISL_1639000, EPI_ISL_1639067, EPI_ISL_1639074, EPI_ISL_1639140, EPI_ISL_1639151, EPI_ISL_1639163, EPI_ISL_1639169, EPI_ISL_1642511 | | | |
| see above | Bioscientia Labor Wermsdorf | Robert Koch Institute | |
| EPI_ISL_1353780, EPI_ISL_1353875, EPI_ISL_1432947, EPI_ISL_1432982, EPI_ISL_1432995, EPI_ISL_1433067, EPI_ISL_1433087, EPI_ISL_1433092, EPI_ISL_1433117, EPI_ISL_1433121, EPI_ISL_1433129, EPI_ISL_1433136, EPI_ISL_1433144, EPI_ISL_1433179, EPI_ISL_1433192, EPI_ISL_1433209, EPI_ISL_1433213, EPI_ISL_1433230, EPI_ISL_1433286, EPI_ISL_1433338, EPI_ISL_1439158, EPI_ISL_1439237, EPI_ISL_1439243, EPI_ISL_1439327, EPI_ISL_1439340, EPI_ISL_1439344, EPI_ISL_1568953, EPI_ISL_1568961, EPI_ISL_1568963, EPI_ISL_1568972, EPI_ISL_1568978, EPI_ISL_1568980, EPI_ISL_1568981, EPI_ISL_1568983, EPI_ISL_1568988, EPI_ISL_1568991, EPI_ISL_1568992, EPI_ISL_1568995, EPI_ISL_1568998, EPI_ISL_1568999, EPI_ISL_1569003, EPI_ISL_1569011, EPI_ISL_1569012, EPI_ISL_1569030, EPI_ISL_1569041, EPI_ISL_1569045, EPI_ISL_1569046, EPI_ISL_1569109, EPI_ISL_1569110, EPI_ISL_1569112, EPI_ISL_1569117, EPI_ISL_1569128, EPI_ISL_1569131, EPI_ISL_1569137, EPI_ISL_1569145, EPI_ISL_1569156, EPI_ISL_1569159, EPI_ISL_1569163, EPI_ISL_1569166, EPI_ISL_1569167, EPI_ISL_1569172, EPI_ISL_1569176, EPI_ISL_1569178, EPI_ISL_1569183, EPI_ISL_1569186, EPI_ISL_1569189, EPI_ISL_1569193, EPI_ISL_1569194, EPI_ISL_1569205, EPI_ISL_1569209, EPI_ISL_1569212, EPI_ISL_1569216, EPI_ISL_1569220, EPI_ISL_1569228, EPI_ISL_1569237, EPI_ISL_1569241, EPI_ISL_1569242, EPI_ISL_1569244, EPI_ISL_1569253, EPI_ISL_1569255, EPI_ISL_1569256, EPI_ISL_1569258, EPI_ISL_1569263, EPI_ISL_1569265, EPI_ISL_1569270 | | | |
| see above | Bioscientia MVZ Labor Karlsruhe GmbH | Robert Koch Institute | |
| EPI_ISL_1365019 | Bundeswehr Institute of Microbiology | Bundeswehr Institute of Microbiology | Alexandra Rehn; Enrico Georgi; Malena Bestehorn-Willmann; Markus Antwerpen; Mathias Walter; Mike Pillukat; Roman Wölfel; Sabine Zange |
| EPI_ISL_1833954, EPI_ISL_1833955, EPI_ISL_1833956, EPI_ISL_1833957, EPI_ISL_1833958, EPI_ISL_1833959, EPI_ISL_1833960, EPI_ISL_1833961, EPI_ISL_1833986 | | | |
| see above | Bundeswehrkrankenhaus Berlin | Bundeswehr Institute of Microbiology | Alexandra Rehn; Enrico Georgi; Malena Bestehorn-Willmann; Markus Antwerpen; Mathias Walter; Mike Pillukat; Roman Wölfel; Sabine Zange |
| EPI_ISL_1365021, EPI_ISL_1833964, EPI_ISL_1833965, EPI_ISL_1833966, EPI_ISL_1833967 | Bundeswehrkrankenhaus Hamburg | Bundeswehr Institute of Microbiology | Alexandra Rehn; Enrico Georgi; Malena Bestehorn-Willmann; Markus Antwerpen; Mathias Walter; Mike Pillukat; Roman Wölfel; Sabine Zange |
| EPI_ISL_1833949 | Bundeswehrkrankenhaus Ulm | Bundeswehr Institute of Microbiology | Alexandra Rehn; Enrico Georgi; Malena Bestehorn-Willmann; Markus Antwerpen; Mathias Walter; Mike Pillukat; Roman Wölfel; Sabine Zange |
| EPI_ISL_1365018, EPI_ISL_1833951, EPI_ISL_1833968, EPI_ISL_1833969, EPI_ISL_1833970, EPI_ISL_1833971, EPI_ISL_1833972 | | | |
| see above | Bundeswehrzentralkrankenhaus Koblenz | Bundeswehr Institute of Microbiology | Alexandra Rehn; Enrico Georgi; Malena Bestehorn-Willmann; Markus Antwerpen; Mathias Walter; Mike Pillukat; Roman Wölfel; Sabine Zange |
| EPI_ISL_1990004, EPI_ISL_1990005, EPI_ISL_1990006, EPI_ISL_1990007, EPI_ISL_1990008, EPI_ISL_1990009, EPI_ISL_1990010, EPI_ISL_1990011, EPI_ISL_1990012, EPI_ISL_1990014, EPI_ISL_1990015, EPI_ISL_1990016, EPI_ISL_1990017, EPI_ISL_1990018, EPI_ISL_1990019, EPI_ISL_1990020, EPI_ISL_1990070, EPI_ISL_1990071, EPI_ISL_1990072, EPI_ISL_1990073, EPI_ISL_1990074, EPI_ISL_1990075, EPI_ISL_1990076, EPI_ISL_1990077, EPI_ISL_1990078, EPI_ISL_1990080, EPI_ISL_1990081 | | | |
| see above | Center of Medical Microbiology, Virology, and Hospital Hygiene, University of Duesseldorf | Center of Medical Microbiology, Virology, and Hospital Hygiene, University of Duesseldorf | Alexander Dilthey; Andreas Walker; Daniel Strelow; Jessica Nicolai; Jörg Timm; Klaus Pfeffer; Lisanna Hülse; Malte Kohns Vasconcelos; Maximilian Damagnez; Nadine Lübke; Tobias Wienemann; Torsten Houwaart |
| EPI_ISL_2108542, EPI_ISL_2108546, EPI_ISL_2108562, EPI_ISL_2108564, EPI_ISL_2108575, EPI_ISL_2108579, EPI_ISL_2108591, EPI_ISL_2108612, EPI_ISL_2108649, EPI_ISL_2108661, EPI_ISL_2108671, EPI_ISL_2108679, EPI_ISL_2108692 | | | |
| see above | Centogene; Dr. Bauer Laboratoriums GmbH | Robert Koch Institute | |
| EPI_ISL_1349416, EPI_ISL_1349417, EPI_ISL_1349419, EPI_ISL_1349421, EPI_ISL_1349423, EPI_ISL_1349425, EPI_ISL_1349426, EPI_ISL_1349427, EPI_ISL_1435333, EPI_ISL_1435334, EPI_ISL_1435335, EPI_ISL_1435336, EPI_ISL_1435337, EPI_ISL_1435338, EPI_ISL_1435339, EPI_ISL_1435340 | | | |
| see above | Chemische und Veterinäruntersuchungsamt Münsterland-Emscher-Lippe | Robert Koch Institute | |
| EPI_ISL_1571964, EPI_ISL_1571965, EPI_ISL_1571966, EPI_ISL_1571968, EPI_ISL_1571969, EPI_ISL_1571970, EPI_ISL_1571974, EPI_ISL_1571975, EPI_ISL_1571976, EPI_ISL_1571978, EPI_ISL_1571979, EPI_ISL_1571980, EPI_ISL_1571981, EPI_ISL_1571982, EPI_ISL_1571983, EPI_ISL_1571984, EPI_ISL_1571985, EPI_ISL_1571986, EPI_ISL_1571989, EPI_ISL_1571991, EPI_ISL_1571992, EPI_ISL_1571993, EPI_ISL_1571995, EPI_ISL_1571996, EPI_ISL_1571997, EPI_ISL_1571998, EPI_ISL_1571999, EPI_ISL_1572000, EPI_ISL_1572001, EPI_ISL_1572003, EPI_ISL_1572004, EPI_ISL_1572005, EPI_ISL_1572006, EPI_ISL_1572007, EPI_ISL_1572008, EPI_ISL_1639347, EPI_ISL_1639348, EPI_ISL_1722971, EPI_ISL_1724293, EPI_ISL_1724294, EPI_ISL_1724299 | | | |
| see above | Clemenshospital - Alexianer Clemenshospital MÃ¼nster | Robert Koch Institute | |
| EPI_ISL_1563938, EPI_ISL_1563940, EPI_ISL_1563943, EPI_ISL_1563945, EPI_ISL_1563947, EPI_ISL_1563950, EPI_ISL_1563952, EPI_ISL_1563957, EPI_ISL_1642736, EPI_ISL_1642737, EPI_ISL_1642738, EPI_ISL_1642742, EPI_ISL_1642744, EPI_ISL_1642746, EPI_ISL_1642747, EPI_ISL_1642751, EPI_ISL_1642753, EPI_ISL_1642754, EPI_ISL_1642755, EPI_ISL_1642756, EPI_ISL_1642757, EPI_ISL_1642758, EPI_ISL_1642759, EPI_ISL_1642760, EPI_ISL_1642761 | | | |
| see above | Corona-Testzentrum ifp Institut fÃ¼r ProduktqualitÃ¤t GmbH | Robert Koch Institute | |
| EPI_ISL_1356992, EPI_ISL_1356994, EPI_ISL_1357003, EPI_ISL_1357004, EPI_ISL_1357006, EPI_ISL_1357011, EPI_ISL_1357013, EPI_ISL_1357018, EPI_ISL_1357028, EPI_ISL_1357034, EPI_ISL_1357043, EPI_ISL_1357049, EPI_ISL_1357057, EPI_ISL_1439575, EPI_ISL_1439588, EPI_ISL_1439614, EPI_ISL_1440967, EPI_ISL_1440968, EPI_ISL_1440969, EPI_ISL_1440970, EPI_ISL_1440971, EPI_ISL_1440973, EPI_ISL_1440974, EPI_ISL_1440975, EPI_ISL_1440976, EPI_ISL_1440977, EPI_ISL_1440978, EPI_ISL_1440979, EPI_ISL_1440981, EPI_ISL_1440982, EPI_ISL_1440984, EPI_ISL_1440985, EPI_ISL_1440986, EPI_ISL_1440988, EPI_ISL_1440989, EPI_ISL_1440990, EPI_ISL_1440991, EPI_ISL_1440992, EPI_ISL_1440993, EPI_ISL_1440994, EPI_ISL_1440995, EPI_ISL_1440996, EPI_ISL_1440998, EPI_ISL_1440999, EPI_ISL_1441000, EPI_ISL_1441001, EPI_ISL_1441002, EPI_ISL_1441003, EPI_ISL_1441004, EPI_ISL_1441005, EPI_ISL_1441006, EPI_ISL_1441007, EPI_ISL_1441008, EPI_ISL_1441009, EPI_ISL_1441010, EPI_ISL_1441011, EPI_ISL_1441012, EPI_ISL_1441013, EPI_ISL_1441014, EPI_ISL_1441015, EPI_ISL_1441016, EPI_ISL_1441017, EPI_ISL_1441018, EPI_ISL_1441019, EPI_ISL_1441020, EPI_ISL_1441021, EPI_ISL_1441022, EPI_ISL_1441023, EPI_ISL_1441024, EPI_ISL_1441025, EPI_ISL_1441026, EPI_ISL_1441027, EPI_ISL_1441028, EPI_ISL_1441029, EPI_ISL_1441030, EPI_ISL_1441031, EPI_ISL_1441032, EPI_ISL_1441033, EPI_ISL_1441034, EPI_ISL_1441036, EPI_ISL_1441037, EPI_ISL_1441038, EPI_ISL_1441039, EPI_ISL_1441040, EPI_ISL_1441042, EPI_ISL_1441043, EPI_ISL_1441044, EPI_ISL_1441045, EPI_ISL_1441046, EPI_ISL_1441047, EPI_ISL_1441048, EPI_ISL_1441049, EPI_ISL_1441050, EPI_ISL_1441051, EPI_ISL_1441053, EPI_ISL_1441054, EPI_ISL_1441055, EPI_ISL_1441056, EPI_ISL_1441057, EPI_ISL_1441058, EPI_ISL_1441060, EPI_ISL_1441062, EPI_ISL_1441063, EPI_ISL_1441064, EPI_ISL_1441065, EPI_ISL_1441066, EPI_ISL_1441067, EPI_ISL_1441068, EPI_ISL_1441069, EPI_ISL_1441070, EPI_ISL_1441071, EPI_ISL_1441072, EPI_ISL_1441073, EPI_ISL_1441075, EPI_ISL_1441077, EPI_ISL_1441078, EPI_ISL_1441079, EPI_ISL_1441080, EPI_ISL_1441082, EPI_ISL_1441083, EPI_ISL_1441084, EPI_ISL_1441085, EPI_ISL_1441086, EPI_ISL_1441087, EPI_ISL_1441089, EPI_ISL_1441090, EPI_ISL_1441091, EPI_ISL_1441092, EPI_ISL_1441093, EPI_ISL_1441094, EPI_ISL_1441095, EPI_ISL_1441096, EPI_ISL_1441097, EPI_ISL_1441098, EPI_ISL_1441101, EPI_ISL_1441102, EPI_ISL_1441103, EPI_ISL_1441104, EPI_ISL_1441105, EPI_ISL_1441106, EPI_ISL_1441107, EPI_ISL_1441108, EPI_ISL_1441109, EPI_ISL_1441110, EPI_ISL_1441113, EPI_ISL_1441114, EPI_ISL_1441116, EPI_ISL_1441117, EPI_ISL_1441118, EPI_ISL_1441119, EPI_ISL_1441120, EPI_ISL_1441121, EPI_ISL_1441122, EPI_ISL_1441123, EPI_ISL_1441124, EPI_ISL_1441125, EPI_ISL_1441128, EPI_ISL_1441129, EPI_ISL_1441130, EPI_ISL_1441132, EPI_ISL_1441134, EPI_ISL_1441135, EPI_ISL_1441136, EPI_ISL_1441138, EPI_ISL_1441139, EPI_ISL_1441141, EPI_ISL_1441142, EPI_ISL_1441143, EPI_ISL_1441144, EPI_ISL_1441145, EPI_ISL_1441146, EPI_ISL_1441148, EPI_ISL_1441149, EPI_ISL_1441150, EPI_ISL_1441151, EPI_ISL_1441152, EPI_ISL_1441153, EPI_ISL_1441154, EPI_ISL_1441155, EPI_ISL_1441156, EPI_ISL_1441157, EPI_ISL_1568158, EPI_ISL_1568178, EPI_ISL_1568186, EPI_ISL_1568196, EPI_ISL_1568237, EPI_ISL_1572526, EPI_ISL_1572529, EPI_ISL_1572530, EPI_ISL_1572539, EPI_ISL_1572540, EPI_ISL_1572541, EPI_ISL_1572545, EPI_ISL_1572546, EPI_ISL_1572549, EPI_ISL_1572550, EPI_ISL_1572552, EPI_ISL_1572554, EPI_ISL_1640189, EPI_ISL_1640255, EPI_ISL_1640285 | | | |
| see above | Diagnosticum - Labor Neukirchen | Robert Koch Institute | |
| EPI_ISL_1568103, EPI_ISL_1568105, EPI_ISL_1568107, EPI_ISL_1568109, EPI_ISL_1568116, EPI_ISL_1568117, EPI_ISL_1568118, EPI_ISL_1568119, EPI_ISL_1568122, EPI_ISL_1568123, EPI_ISL_1568125, EPI_ISL_1568126, EPI_ISL_1568128, EPI_ISL_1568129, EPI_ISL_1568131, EPI_ISL_1568133 | | | |
| see above | Dianovis GmbH Greiz | Robert Koch Institute | |
| EPI_ISL_1354366, EPI_ISL_1354371, EPI_ISL_1354379, EPI_ISL_1354384, EPI_ISL_1354388, EPI_ISL_1354389, EPI_ISL_1354400, EPI_ISL_1354431, EPI_ISL_1354436, EPI_ISL_1354437, EPI_ISL_1354438, EPI_ISL_1354443, EPI_ISL_1354463 | | | |

| | | | |
|---|---|---|---|
| see above | ELBLAB GmbH Zentrum für LaborMedizin | Robert Koch Institute | |
| EPI_ISL_1570011, EPI_ISL_1570012, EPI_ISL_1570013, EPI_ISL_1570014, EPI_ISL_1570015, EPI_ISL_1570016, EPI_ISL_1570017, EPI_ISL_1570018, EPI_ISL_1570019, EPI_ISL_1570020, EPI_ISL_1570021, EPI_ISL_1570022, EPI_ISL_1570024, EPI_ISL_1570025, EPI_ISL_1570026, EPI_ISL_1570027, EPI_ISL_1570028, EPI_ISL_1570029, EPI_ISL_1570030, EPI_ISL_1570031, EPI_ISL_1570032, EPI_ISL_1570034, EPI_ISL_1570035, EPI_ISL_1570036, EPI_ISL_1570037, EPI_ISL_1570038, EPI_ISL_1570039, EPI_ISL_1570040, EPI_ISL_1570041, EPI_ISL_1570042, EPI_ISL_1570043, EPI_ISL_1570044, EPI_ISL_1570045, EPI_ISL_1570046, EPI_ISL_1570047, EPI_ISL_1570048, EPI_ISL_1570049, EPI_ISL_1570050, EPI_ISL_1570051, EPI_ISL_1570052, EPI_ISL_1570053, EPI_ISL_1570054, EPI_ISL_1570055, EPI_ISL_1570056, EPI_ISL_1570057, EPI_ISL_1570058, EPI_ISL_1570059, EPI_ISL_1570060, EPI_ISL_1570061, EPI_ISL_1570062, EPI_ISL_1570063, EPI_ISL_1570064, EPI_ISL_1570065, EPI_ISL_1570066, EPI_ISL_1570067, EPI_ISL_1570068, EPI_ISL_1570070, EPI_ISL_1570071, EPI_ISL_1570072, EPI_ISL_1570073, EPI_ISL_1570074, EPI_ISL_1570075, EPI_ISL_1570077, EPI_ISL_1570079, EPI_ISL_1570080, EPI_ISL_1570081, EPI_ISL_1570082, EPI_ISL_1570083, EPI_ISL_1570084, EPI_ISL_1570086, EPI_ISL_1570087, EPI_ISL_1570088, EPI_ISL_1570090, EPI_ISL_1570091, EPI_ISL_1570092, EPI_ISL_1570093, EPI_ISL_1570094, EPI_ISL_1570095, EPI_ISL_1570096, EPI_ISL_1570097, EPI_ISL_1570099, EPI_ISL_1570100, EPI_ISL_1570102, EPI_ISL_1570103, EPI_ISL_1570104, EPI_ISL_1570105, EPI_ISL_1570107, EPI_ISL_1570108, EPI_ISL_1570109, EPI_ISL_1570110, EPI_ISL_1570111, EPI_ISL_1570112, EPI_ISL_1570113, EPI_ISL_1570114, EPI_ISL_1570115, EPI_ISL_1570118, EPI_ISL_1570119, EPI_ISL_1570120, EPI_ISL_1570121, EPI_ISL_1570122, EPI_ISL_1570123, EPI_ISL_1570125, EPI_ISL_1570126, EPI_ISL_1570127, EPI_ISL_1570129, EPI_ISL_1570130, EPI_ISL_1570132, EPI_ISL_1570133, EPI_ISL_1570134, EPI_ISL_1570135, EPI_ISL_1570136, EPI_ISL_1570137, EPI_ISL_1570138, EPI_ISL_1570141, EPI_ISL_1570142, EPI_ISL_1570143, EPI_ISL_1570144, EPI_ISL_1570145, EPI_ISL_1570146, EPI_ISL_1570147, EPI_ISL_1570148, EPI_ISL_1570152, EPI_ISL_1570153, EPI_ISL_1570154, EPI_ISL_1570155, EPI_ISL_1570156, EPI_ISL_1570157, EPI_ISL_1570159, EPI_ISL_1570160, EPI_ISL_1570166, EPI_ISL_1570168, EPI_ISL_1570169, EPI_ISL_1570170, EPI_ISL_1570173, EPI_ISL_1570176, EPI_ISL_1570180, EPI_ISL_1570181, EPI_ISL_1570183, EPI_ISL_1570184, EPI_ISL_1570186, EPI_ISL_1570188, EPI_ISL_1570189, EPI_ISL_1570190, EPI_ISL_1570192, EPI_ISL_1570196, EPI_ISL_1570204, EPI_ISL_1570206, EPI_ISL_1570207, EPI_ISL_1570209, EPI_ISL_1570210, EPI_ISL_1570211, EPI_ISL_1570214, EPI_ISL_1570215, EPI_ISL_1570217, EPI_ISL_1570218, EPI_ISL_1570220, EPI_ISL_1570221, EPI_ISL_1570222, EPI_ISL_1570223, EPI_ISL_1570224, EPI_ISL_1570225, EPI_ISL_1570227, EPI_ISL_1570230, EPI_ISL_1570233, EPI_ISL_1570234, EPI_ISL_1570238, EPI_ISL_1570239, EPI_ISL_1570250, EPI_ISL_1570275, EPI_ISL_1570276, EPI_ISL_1570282, EPI_ISL_1570287, EPI_ISL_1570306, EPI_ISL_1570310, EPI_ISL_1571206, EPI_ISL_1571207, EPI_ISL_1571209, EPI_ISL_1571210, EPI_ISL_1571211, EPI_ISL_1571212, EPI_ISL_1571213, EPI_ISL_1571214, EPI_ISL_1571215, EPI_ISL_1571216, EPI_ISL_1571217, EPI_ISL_1571218, EPI_ISL_1571219, EPI_ISL_1571220, EPI_ISL_1571221, EPI_ISL_1571222, EPI_ISL_1571223, EPI_ISL_1571224, EPI_ISL_1571225, EPI_ISL_1571226, EPI_ISL_1571228, EPI_ISL_1571229, EPI_ISL_1571230, EPI_ISL_1571231, EPI_ISL_1571233, EPI_ISL_1571234, EPI_ISL_1571235, EPI_ISL_1571236, EPI_ISL_1571240, EPI_ISL_1571242, EPI_ISL_1571245, EPI_ISL_1571247, EPI_ISL_1571250, EPI_ISL_1571252, EPI_ISL_1571254, EPI_ISL_1571257, EPI_ISL_1571260, EPI_ISL_1571262, EPI_ISL_1571263, EPI_ISL_1571264, EPI_ISL_1571265, EPI_ISL_1571266, EPI_ISL_1571267, EPI_ISL_1571268, EPI_ISL_1571269, EPI_ISL_1571270, EPI_ISL_1571271, EPI_ISL_1571272, EPI_ISL_1571273, EPI_ISL_1571274, EPI_ISL_1571275, EPI_ISL_1571276, EPI_ISL_1571277, EPI_ISL_1571279, EPI_ISL_1571280, EPI_ISL_1571281 | | | |
| see above | Eurofins LifeCodexx GmbH | Robert Koch Institute | |
| EPI_ISL_1563754, EPI_ISL_1563755, EPI_ISL_1563756, EPI_ISL_1563757, EPI_ISL_1563758, EPI_ISL_1563760, EPI_ISL_1563761, EPI_ISL_1563764, EPI_ISL_1563765, EPI_ISL_1563766, EPI_ISL_1563767, EPI_ISL_1563768, EPI_ISL_1563772, EPI_ISL_1563775, EPI_ISL_1563776, EPI_ISL_1563777, EPI_ISL_1563779, EPI_ISL_1563780, EPI_ISL_1563781, EPI_ISL_1563782, EPI_ISL_1563783, EPI_ISL_1563784, EPI_ISL_1563785, EPI_ISL_1563787, EPI_ISL_1563788, EPI_ISL_1563790, EPI_ISL_1563791, EPI_ISL_1563792, EPI_ISL_1563793, EPI_ISL_1563794, EPI_ISL_1563795, EPI_ISL_1563796, EPI_ISL_1563797, EPI_ISL_1563798, EPI_ISL_1563799, EPI_ISL_1563800, EPI_ISL_1563801, EPI_ISL_1563803, EPI_ISL_1563804, EPI_ISL_1563805 | | | |
| see above | Fraunhofer-Institut fÃ¼r Zelltherapie und Immunologie | Robert Koch Institute | |
| EPI_ISL_1524889, EPI_ISL_1524891 | Hannover Medical School, Institute of Virology | Hannover Medical School, Institute of Virology | Lars Steinbrück |
| EPI_ISL_1567776, EPI_ISL_1567833 | Helios UniversitÃ¤tsklinikum Wuppertal: UniversitÃ¤t Witten/Herdecke: Institut fÃ¼r Medizinische Labordiagnostik | Robert Koch Institute | |
| EPI_ISL_1439466 | Helios Universitätsklinikum Wuppertal: Universität Witten/Herdecke: Institut für Medizinische Labordiagnostik | Robert Koch Institute | |
| EPI_ISL_1725537, EPI_ISL_1725541, EPI_ISL_1725542, EPI_ISL_1725549, EPI_ISL_1725558, EPI_ISL_1725575 | IFLb Laboratoriumsmedizin Berlin GmbH | Robert Koch Institute | |
| EPI_ISL_1350108, EPI_ISL_1350122, EPI_ISL_1350136, EPI_ISL_1350141, EPI_ISL_1350145, EPI_ISL_1350148, EPI_ISL_1354293, EPI_ISL_1354304, EPI_ISL_1354319, EPI_ISL_1354331, EPI_ISL_1435843, EPI_ISL_1435858, EPI_ISL_1435864, EPI_ISL_1435868, EPI_ISL_1435871, EPI_ISL_1435872, EPI_ISL_1435874, EPI_ISL_1435877, EPI_ISL_1435890, EPI_ISL_1435895, EPI_ISL_1435897, EPI_ISL_1435901, EPI_ISL_1439897 | | | |
| see above | IMD - Institut für Medizinische Diagnostik Berlin-Potsdam | Robert Koch Institute | |
| EPI_ISL_1354327, EPI_ISL_1354362, EPI_ISL_1435841, EPI_ISL_1435844, EPI_ISL_1435845, EPI_ISL_1435847, EPI_ISL_1435849, EPI_ISL_1435850, EPI_ISL_1435852, EPI_ISL_1435855, EPI_ISL_1435856, EPI_ISL_1435857, EPI_ISL_1435859, EPI_ISL_1435860, EPI_ISL_1435861, EPI_ISL_1435862, EPI_ISL_1435863, EPI_ISL_1435866, EPI_ISL_1435867, EPI_ISL_1435870, EPI_ISL_1435875, EPI_ISL_1435880, EPI_ISL_1435882, EPI_ISL_1435883, EPI_ISL_1435885, EPI_ISL_1435887, EPI_ISL_1435889, EPI_ISL_1435894, EPI_ISL_1435899, EPI_ISL_1439879, EPI_ISL_1439881, EPI_ISL_1439888, EPI_ISL_1439900, EPI_ISL_1565417, EPI_ISL_1565433, EPI_ISL_1565435, EPI_ISL_1565442, EPI_ISL_1565460, EPI_ISL_1565466, EPI_ISL_1565491 | | | |
| see above | IMD - Labor Oderland | Robert Koch Institute | |
| EPI_ISL_1435873, EPI_ISL_1435893, EPI_ISL_1565413, EPI_ISL_1565415, EPI_ISL_1565416, EPI_ISL_1565420, EPI_ISL_1565439, EPI_ISL_1565445, EPI_ISL_1565452, EPI_ISL_1565457, EPI_ISL_1565458, EPI_ISL_1565465, EPI_ISL_1565468, EPI_ISL_1565473, EPI_ISL_1565487, EPI_ISL_1565489 | | | |
| see above | IMD - MVZ Labor Greifswald | Robert Koch Institute | |
| EPI_ISL_1354285, EPI_ISL_1354324, EPI_ISL_1354337, EPI_ISL_1354340, EPI_ISL_1354349, EPI_ISL_1354352, EPI_ISL_1354359, EPI_ISL_1435842, EPI_ISL_1435846, EPI_ISL_1435848, EPI_ISL_1435851, EPI_ISL_1435853, EPI_ISL_1435865, EPI_ISL_1435869, EPI_ISL_1435878, EPI_ISL_1435879, EPI_ISL_1435881, EPI_ISL_1435884, EPI_ISL_1435886, EPI_ISL_1435892, EPI_ISL_1439870, EPI_ISL_1439871, EPI_ISL_1439872, EPI_ISL_1439876, EPI_ISL_1439882, EPI_ISL_1439884, EPI_ISL_1439885, EPI_ISL_1439890, EPI_ISL_1439891, EPI_ISL_1439894, EPI_ISL_1439895, EPI_ISL_1439898, EPI_ISL_1439899, EPI_ISL_1565419 | | | |
| see above | IMD - MVZ Labor Martinsried | Robert Koch Institute | |
| EPI_ISL_1354326, EPI_ISL_1439873, EPI_ISL_1439874, EPI_ISL_1439875, EPI_ISL_1439878, EPI_ISL_1439887, EPI_ISL_1439893 | | | |
| see above | IMD - Medizinisches Labor Rostock | Robert Koch Institute | |
| EPI_ISL_1353357, EPI_ISL_1353360, EPI_ISL_1353364, EPI_ISL_1353370, EPI_ISL_1353374, EPI_ISL_1353382, EPI_ISL_1353383, EPI_ISL_1353385, EPI_ISL_1353386, EPI_ISL_1353388, EPI_ISL_1353389, EPI_ISL_1353391, EPI_ISL_1353392, EPI_ISL_1353393, EPI_ISL_1353395, EPI_ISL_1353398, EPI_ISL_1353399, EPI_ISL_1353403, EPI_ISL_1353407, EPI_ISL_1353408, EPI_ISL_1353412, EPI_ISL_1353416, EPI_ISL_1353417, EPI_ISL_1353418, EPI_ISL_1353419, EPI_ISL_1353420, EPI_ISL_1353421, EPI_ISL_1433505, EPI_ISL_1433507, EPI_ISL_1433511, EPI_ISL_1433513, EPI_ISL_1433515, EPI_ISL_1433533, EPI_ISL_1433537, EPI_ISL_1433549, EPI_ISL_1433553, EPI_ISL_1433574, EPI_ISL_1433579, EPI_ISL_1433580, EPI_ISL_1433582, EPI_ISL_1433586, EPI_ISL_1433590, EPI_ISL_1433592, EPI_ISL_1439372, EPI_ISL_1439378, EPI_ISL_1439382, EPI_ISL_1439385, EPI_ISL_1439386, EPI_ISL_1439391, EPI_ISL_1439396, EPI_ISL_1439403, EPI_ISL_1439404, EPI_ISL_1439406, EPI_ISL_1439409, EPI_ISL_1439413, EPI_ISL_1439414, EPI_ISL_1439418, EPI_ISL_1439421, EPI_ISL_1439424, EPI_ISL_1439426, EPI_ISL_1439427, EPI_ISL_1439428, EPI_ISL_1439430, EPI_ISL_1439436 | | | |
| see above | IMD Labor Frankfurt | Robert Koch Institute | |
| EPI_ISL_1563749, EPI_ISL_1563750, EPI_ISL_1563751 | Institut fÃ¼r Klinische Transfusionsmedizin und Immungenetik DRK Blutspendedienst Baden-WÃ¼rttemberg | Robert Koch Institute | |
| EPI_ISL_1571552, EPI_ISL_1571553, EPI_ISL_1571558, EPI_ISL_1571559, EPI_ISL_1571561 | Institut fÃ¼r Medizinische Laboratoriumsdiagnostik Bochum GmbH | Robert Koch Institute | |
| EPI_ISL_1565365, EPI_ISL_1565366, EPI_ISL_1565367, EPI_ISL_1565368 | Institut fÃ¼r Medizinische Mikrobiologie - UniversitÃ¤tsmedizin GÃ¶ttingen | Robert Koch Institute | |
| EPI_ISL_1568524, EPI_ISL_1568525, EPI_ISL_1568527, EPI_ISL_1568528, EPI_ISL_1568529, EPI_ISL_1639263 | Institut fÃ¼r med. Mikrobiologie u. Virologie Klinikum Oldenburg | Robert Koch Institute | |
| EPI_ISL_1353328 | Institut für Klinische Chemie Transfusions- und LaboratoriumsmedizinBerufsgenossenschaftliches Universitätsklinikum Bergmannsheil | Robert Koch Institute | |
| EPI_ISL_1361774, EPI_ISL_1361775, EPI_ISL_1361776, EPI_ISL_1361777, EPI_ISL_1361778, EPI_ISL_1361779, EPI_ISL_1361780, EPI_ISL_1361781, EPI_ISL_1361782, EPI_ISL_1361823, EPI_ISL_1361824, EPI_ISL_1361825, EPI_ISL_1361826, EPI_ISL_1361827, EPI_ISL_1361828, EPI_ISL_1361829, EPI_ISL_1361830, EPI_ISL_1361831, EPI_ISL_1361832, EPI_ISL_1361833, EPI_ISL_1361869, EPI_ISL_1361870, EPI_ISL_1361871, EPI_ISL_1361872, EPI_ISL_1361873, EPI_ISL_1361874, EPI_ISL_1361908, EPI_ISL_1670007, EPI_ISL_1670008, EPI_ISL_1670009, EPI_ISL_1670010, EPI_ISL_1670048, EPI_ISL_1670049, EPI_ISL_1670050, EPI_ISL_1670051, EPI_ISL_1670052, EPI_ISL_1670053, EPI_ISL_1670054, EPI_ISL_1670055, EPI_ISL_1670056 | | | |
| see above | Institute of Medical Microbiology and Hospital Hygiene | Institute of Medical Microbiology and Hospital Hygiene | Aljoscha Tersteegen; Prof. Dr. Achim Kaasch |
| EPI_ISL_1588594, EPI_ISL_1588595, EPI_ISL_1588596, EPI_ISL_1588597, EPI_ISL_1588598, EPI_ISL_1588599, EPI_ISL_1588612, EPI_ISL_1588613, EPI_ISL_1588614, EPI_ISL_1588615, EPI_ISL_1588618, EPI_ISL_1588622, EPI_ISL_1588623, EPI_ISL_1588624, EPI_ISL_1588625, EPI_ISL_1588626, EPI_ISL_1805000, EPI_ISL_1805001, EPI_ISL_1805003, EPI_ISL_1805009 | | | |
| see above | Institute of Virology, Medical Center, University of Freiburg, Freiburg, Germany | Institute of Virology, Clinial Virus Genomics, Medical Center, University of Freiburg, Freiburg, Germany | Hajo Grundmann; Jonas Fuchs; Lena Jaki; Lisa Kern; Marcus Panning; Sandra Reuter |
| EPI_ISL_1716056, EPI_ISL_1716057, EPI_ISL_1716058 | Kath. Marienkrankenhaus | Heinrich Pette Institute, Leibniz Institute for Experimental Virology | Adam Grundhoff; Alexis Robitaille; Johannes Knobloch; Martin Aepfelbacher; Nicole Fischer; Thomas Günther |
| EPI_ISL_1353333 | Kath. St.-Johannes-Gesellschaft Dortmund gGmbH | Robert Koch Institute | |
| EPI_ISL_1563748, EPI_ISL_1563752, EPI_ISL_1563753, EPI_ISL_1563778 | Kerckhoff-Klinik GmbH | Robert Koch Institute | |
| EPI_ISL_1441225, EPI_ISL_1569706 | Klinikum Darmstadt GmbH | Robert Koch Institute | |
| EPI_ISL_1349231, EPI_ISL_1349233, EPI_ISL_1349237, EPI_ISL_1349238, EPI_ISL_1357239, EPI_ISL_1357240, EPI_ISL_1357241, EPI_ISL_1357242, EPI_ISL_1357243, EPI_ISL_1357244, EPI_ISL_1357245, EPI_ISL_1357246, EPI_ISL_1357247, EPI_ISL_1357248, EPI_ISL_1357249, EPI_ISL_1357250, EPI_ISL_1357251, EPI_ISL_1357252, EPI_ISL_1357253, EPI_ISL_1357254, EPI_ISL_1357255, EPI_ISL_1357256, EPI_ISL_1357257, EPI_ISL_1357258, EPI_ISL_1357259, EPI_ISL_1357260, EPI_ISL_1357261, EPI_ISL_1357262, EPI_ISL_1357263, EPI_ISL_1357264, EPI_ISL_1442531, EPI_ISL_1442532, EPI_ISL_1442538, EPI_ISL_1442540 | | | |
| see above | Klinikum Ernst von Bergmann gemeinnützige GmbH - stationärer Bereich | Robert Koch Institute | |
| EPI_ISL_1435441, EPI_ISL_1438761 | Klinikum Fulda gAG | Robert Koch Institute | |
| EPI_ISL_1286893, EPI_ISL_1286895, EPI_ISL_1286897, EPI_ISL_1286900, EPI_ISL_1286902, EPI_ISL_1286904, EPI_ISL_1570001, EPI_ISL_1570703, EPI_ISL_1722847 | | | |
| see above | Klinikum Stuttgart | Robert Koch Institute | |
| EPI_ISL_1572136, EPI_ISL_1572139 | Krankenhausgesellschaft St. Vincenz mbH | Robert Koch Institute | |
| EPI_ISL_1354368, EPI_ISL_1354387, EPI_ISL_1354393, EPI_ISL_1354409, EPI_ISL_1354416, EPI_ISL_1354469 | LABOR CHEMNITZ – Zentrum für Diagnostik am Klinikum Chemnitz | Robert Koch Institute | |
| EPI_ISL_1353674, EPI_ISL_1432934, EPI_ISL_1432949, EPI_ISL_1432974, EPI_ISL_1432976, EPI_ISL_1432985, EPI_ISL_1433006, EPI_ISL_1433029, EPI_ISL_1433054, EPI_ISL_1433058, EPI_ISL_1433060, EPI_ISL_1433072, EPI_ISL_1433083, EPI_ISL_1433084, EPI_ISL_1433085, EPI_ISL_1433095, EPI_ISL_1433098, EPI_ISL_1433100, EPI_ISL_1433103, EPI_ISL_1433107, EPI_ISL_1433111, EPI_ISL_1433120, EPI_ISL_1433124, EPI_ISL_1433135, EPI_ISL_1433147, EPI_ISL_1433153, EPI_ISL_1433159, EPI_ISL_1433166, EPI_ISL_1433170, EPI_ISL_1433176, EPI_ISL_1433195, EPI_ISL_1433207, EPI_ISL_1433214, EPI_ISL_1433215, EPI_ISL_1433216, EPI_ISL_1433220, EPI_ISL_1433229, EPI_ISL_1433231, EPI_ISL_1433235, EPI_ISL_1433238, EPI_ISL_1433239, EPI_ISL_1433252, EPI_ISL_1433257, EPI_ISL_1433258, EPI_ISL_1433259, EPI_ISL_1433279, EPI_ISL_1433281, EPI_ISL_1433284, EPI_ISL_1433288, EPI_ISL_1433302, EPI_ISL_1433319, EPI_ISL_1433336, EPI_ISL_1433346, EPI_ISL_1433356, EPI_ISL_1433366, EPI_ISL_1436010, EPI_ISL_1436021, EPI_ISL_1436031, EPI_ISL_1436040, EPI_ISL_1436051, EPI_ISL_1436075, EPI_ISL_1436076, EPI_ISL_1436077, EPI_ISL_1436078, EPI_ISL_1436079, EPI_ISL_1436082, EPI_ISL_1436083, EPI_ISL_1436087, EPI_ISL_1436095, EPI_ISL_1436105, EPI_ISL_1436111, EPI_ISL_1436120, EPI_ISL_1436124, EPI_ISL_1436131, EPI_ISL_1436132, EPI_ISL_1436133, EPI_ISL_1436141, EPI_ISL_1436154, EPI_ISL_1436157, EPI_ISL_1436170, EPI_ISL_1436171, EPI_ISL_1436172, EPI_ISL_1436177, EPI_ISL_1436187, EPI_ISL_1436191, EPI_ISL_1436194, EPI_ISL_1436196, EPI_ISL_1436205, EPI_ISL_1436206, EPI_ISL_1436207, EPI_ISL_1436209, EPI_ISL_1436210, EPI_ISL_1436218, EPI_ISL_1436219, EPI_ISL_1436220, EPI_ISL_1436221, EPI_ISL_1436222, EPI_ISL_1436227, EPI_ISL_1436235, EPI_ISL_1436236, EPI_ISL_1436238, EPI_ISL_1436239, EPI_ISL_1436248, EPI_ISL_1436250, EPI_ISL_1436253, EPI_ISL_1436260, EPI_ISL_1436265, EPI_ISL_1436274, EPI_ISL_1436277, EPI_ISL_1436278, EPI_ISL_1436290, EPI_ISL_1436295, EPI_ISL_1436306, EPI_ISL_1436310, EPI_ISL_1436312, EPI_ISL_1436318, EPI_ISL_1436327, EPI_ISL_1436328, EPI_ISL_1436334, EPI_ISL_1436341, EPI_ISL_1436348, EPI_ISL_1436356, EPI_ISL_1436363, EPI_ISL_1436365, EPI_ISL_1436366, EPI_ISL_1436369, EPI_ISL_1436374, EPI_ISL_1436375, EPI_ISL_1436379, EPI_ISL_1436385, EPI_ISL_1436393, EPI_ISL_1438791, EPI_ISL_1439038, EPI_ISL_1439119, EPI_ISL_1439142, EPI_ISL_1439283, EPI_ISL_1439298 | | | |
| see above | LabKom - Labor Augsburg MVZ GmbH | Robert Koch Institute | |
| EPI_ISL_1436064, EPI_ISL_1436072, EPI_ISL_1436136, EPI_ISL_1436163, EPI_ISL_1436176, EPI_ISL_1436245, EPI_ISL_1436361 | | | |
| see above | LabKom - Labor Hamburg-Lübeck | Robert Koch Institute | |
| EPI_ISL_1436003, EPI_ISL_1436049, EPI_ISL_1436066, EPI_ISL_1436068, EPI_ISL_1436080, EPI_ISL_1436081, EPI_ISL_1436084, EPI_ISL_1436085, EPI_ISL_1436088, EPI_ISL_1436096, EPI_ISL_1436099, EPI_ISL_1436104, EPI_ISL_1436112, EPI_ISL_1436116, EPI_ISL_1436146, EPI_ISL_1436148, EPI_ISL_1436149, EPI_ISL_1436150, EPI_ISL_1436151, EPI_ISL_1436153, EPI_ISL_1436159, EPI_ISL_1436161, EPI_ISL_1436168, EPI_ISL_1436183, EPI_ISL_1436195, EPI_ISL_1436213, EPI_ISL_1436223, EPI_ISL_1436224, EPI_ISL_1436234, EPI_ISL_1436241, EPI_ISL_1436246, EPI_ISL_1436259, EPI_ISL_1436273, EPI_ISL_1436276, EPI_ISL_1436279, EPI_ISL_1436283, EPI_ISL_1436284, EPI_ISL_1436289, EPI_ISL_1436297, EPI_ISL_1436299, EPI_ISL_1436302, EPI_ISL_1436316, EPI_ISL_1436317, EPI_ISL_1436319, EPI_ISL_1436330, EPI_ISL_1436331, EPI_ISL_1436362 | | | |
| see above | LabKom - Labor Hannover MVZ GmbH | Robert Koch Institute | |
| EPI_ISL_1432980, EPI_ISL_1436262, EPI_ISL_1439032, EPI_ISL_1439034, EPI_ISL_1439040, EPI_ISL_1439055, EPI_ISL_1439062, EPI_ISL_1439063, EPI_ISL_1439064, EPI_ISL_1439071, EPI_ISL_1439083, EPI_ISL_1439100, EPI_ISL_1439113, EPI_ISL_1439121, EPI_ISL_1439137, EPI_ISL_1439138, EPI_ISL_1439157, EPI_ISL_1439171, EPI_ISL_1439180, EPI_ISL_1439189, EPI_ISL_1439199, EPI_ISL_1439200, EPI_ISL_1439211, EPI_ISL_1439232, EPI_ISL_1439235, EPI_ISL_1439238, EPI_ISL_1439244, EPI_ISL_1439253, EPI_ISL_1439256, EPI_ISL_1439264, EPI_ISL_1439268, EPI_ISL_1439273, EPI_ISL_1439287, EPI_ISL_1439293, EPI_ISL_1439296, EPI_ISL_1439308, EPI_ISL_1439323, EPI_ISL_1439345, EPI_ISL_1439365 | | | |
| see above | LabKom - Labor Mainz MVZ GmbH | Robert Koch Institute | |
| EPI_ISL_1436029, EPI_ISL_1436070, EPI_ISL_1436073, EPI_ISL_1436074, EPI_ISL_1436086, EPI_ISL_1436090, EPI_ISL_1436113, EPI_ISL_1436128, EPI_ISL_1436135, EPI_ISL_1436184, EPI_ISL_1436199, EPI_ISL_1436200, EPI_ISL_1436214, EPI_ISL_1436216, EPI_ISL_1436247, EPI_ISL_1436255, EPI_ISL_1436258, EPI_ISL_1436261, EPI_ISL_1436275, EPI_ISL_1436281, EPI_ISL_1436314, EPI_ISL_1436336, EPI_ISL_1436359, EPI_ISL_1436373, EPI_ISL_1436380, EPI_ISL_1436388, EPI_ISL_1438695, EPI_ISL_1438709, EPI_ISL_1438715, EPI_ISL_1438720, EPI_ISL_1438733, EPI_ISL_1438734, EPI_ISL_1438735, EPI_ISL_1438748, EPI_ISL_1438806, EPI_ISL_1438825, EPI_ISL_1438833, EPI_ISL_1438841, EPI_ISL_1438856, EPI_ISL_1438869, EPI_ISL_1438878, EPI_ISL_1438893, EPI_ISL_1438939, EPI_ISL_1438941, EPI_ISL_1438946, EPI_ISL_1438971, EPI_ISL_1439009, EPI_ISL_1439011, EPI_ISL_1439012, EPI_ISL_1439022, EPI_ISL_1439041, EPI_ISL_1439049, EPI_ISL_1439068, EPI_ISL_1439096, EPI_ISL_1439097, EPI_ISL_1439102, EPI_ISL_1439103, EPI_ISL_1439110, EPI_ISL_1439120, EPI_ISL_1439130, EPI_ISL_1439148, EPI_ISL_1439154, EPI_ISL_1439155, EPI_ISL_1439162, EPI_ISL_1439184, EPI_ISL_1439192, | | | |

EPI_ISL_1439204, EPI_ISL_1439207, EPI_ISL_1439210, EPI_ISL_1439221, EPI_ISL_1439226, EPI_ISL_1439269, EPI_ISL_1439275, EPI_ISL_1439284, EPI_ISL_1439301, EPI_ISL_1439304, EPI_ISL_1439305, EPI_ISL_1439313, EPI_ISL_1439316, EPI_ISL_1439317, EPI_ISL_1439333, EPI_ISL_1439341, EPI_ISL_1439346, EPI_ISL_1439348, EPI_ISL_1439352, EPI_ISL_1439362, EPI_ISL_1439363, EPI_ISL_1439366

see above | LabKom - Labor an der Salzbrücke MVZ GmbH | Robert Koch Institute

EPI_ISL_1439033, EPI_ISL_1439042, EPI_ISL_1439050, EPI_ISL_1439052, EPI_ISL_1439056, EPI_ISL_1439060, EPI_ISL_1439061, EPI_ISL_1439066, EPI_ISL_1439072, EPI_ISL_1439074, EPI_ISL_1439075, EPI_ISL_1439079, EPI_ISL_1439085, EPI_ISL_1439087, EPI_ISL_1439091, EPI_ISL_1439093, EPI_ISL_1439104, EPI_ISL_1439107, EPI_ISL_1439112, EPI_ISL_1439126, EPI_ISL_1439129, EPI_ISL_1439131, EPI_ISL_1439144, EPI_ISL_1439146, EPI_ISL_1439160, EPI_ISL_1439163, EPI_ISL_1439165, EPI_ISL_1439173, EPI_ISL_1439175, EPI_ISL_1439178, EPI_ISL_1439183, EPI_ISL_1439186, EPI_ISL_1439194, EPI_ISL_1439195, EPI_ISL_1439203, EPI_ISL_1439216, EPI_ISL_1439222, EPI_ISL_1439225, EPI_ISL_1439229, EPI_ISL_1439230, EPI_ISL_1439234, EPI_ISL_1439240, EPI_ISL_1439249, EPI_ISL_1439250, EPI_ISL_1439254, EPI_ISL_1439265, EPI_ISL_1439266, EPI_ISL_1439271, EPI_ISL_1439279, EPI_ISL_1439286, EPI_ISL_1439288, EPI_ISL_1439290, EPI_ISL_1439292, EPI_ISL_1439297, EPI_ISL_1439299, EPI_ISL_1439303, EPI_ISL_1439306, EPI_ISL_1439307, EPI_ISL_1439310, EPI_ISL_1439312, EPI_ISL_1439324, EPI_ISL_1439328, EPI_ISL_1439329, EPI_ISL_1439331, EPI_ISL_1439334, EPI_ISL_1439349, EPI_ISL_1439351, EPI_ISL_1439354, EPI_ISL_1439356

see above | LabKom - MVZ Labor Bochum MLB GmbH | Robert Koch Institute

EPI_ISL_1353669, EPI_ISL_1353698, EPI_ISL_1353740, EPI_ISL_1353761, EPI_ISL_1353772, EPI_ISL_1353790, EPI_ISL_1353798, EPI_ISL_1353827, EPI_ISL_1353828, EPI_ISL_1353931, EPI_ISL_1353949, EPI_ISL_1353964, EPI_ISL_1432926, EPI_ISL_1433013, EPI_ISL_1433021, EPI_ISL_1433105, EPI_ISL_1433113, EPI_ISL_1433122, EPI_ISL_1433130, EPI_ISL_1433154, EPI_ISL_1433158, EPI_ISL_1433167, EPI_ISL_1433175, EPI_ISL_1433191, EPI_ISL_1433193, EPI_ISL_1433208, EPI_ISL_1433233, EPI_ISL_1433266, EPI_ISL_1433268, EPI_ISL_1433285, EPI_ISL_1433344, EPI_ISL_1433368

see above | Labor 28 MVZ GmbH | Robert Koch Institute

EPI_ISL_2123981, EPI_ISL_2123982, EPI_ISL_2123986, EPI_ISL_2123988, EPI_ISL_2123989, EPI_ISL_2123991, EPI_ISL_2123994, EPI_ISL_2123995, EPI_ISL_2124000, EPI_ISL_2124001, EPI_ISL_2124005, EPI_ISL_2124006, EPI_ISL_2124008, EPI_ISL_2124009, EPI_ISL_2124010, EPI_ISL_2124011, EPI_ISL_2124015

see above | Labor Becker & Kollegen (Standort Munchen) | Robert Koch Institute

EPI_ISL_1640645, EPI_ISL_1640650, EPI_ISL_1846773, EPI_ISL_1846775, EPI_ISL_1846776, EPI_ISL_1846779, EPI_ISL_1846781, EPI_ISL_1846782, EPI_ISL_1846784, EPI_ISL_1846788, EPI_ISL_1846789, EPI_ISL_1846790, EPI_ISL_1846793, EPI_ISL_1846794, EPI_ISL_1846795, EPI_ISL_1846796, EPI_ISL_1846797, EPI_ISL_1846798, EPI_ISL_1846799, EPI_ISL_1846800, EPI_ISL_1846801, EPI_ISL_1846803, EPI_ISL_1846805, EPI_ISL_1846806, EPI_ISL_1846807, EPI_ISL_1846808, EPI_ISL_1846810, EPI_ISL_1846811, EPI_ISL_1846812, EPI_ISL_1846813, EPI_ISL_1846814

see above | Labor Becker & Kollegen (Standort MÃ¼nchen) | Robert Koch Institute

EPI_ISL_1840804, EPI_ISL_1905151, EPI_ISL_1905152, EPI_ISL_1905153, EPI_ISL_1905170, EPI_ISL_1972149, EPI_ISL_1972154, EPI_ISL_1972157, EPI_ISL_1972158, EPI_ISL_1972160, EPI_ISL_1972162, EPI_ISL_1972171, EPI_ISL_1972172, EPI_ISL_1972173, EPI_ISL_1972180, EPI_ISL_1972187, EPI_ISL_1972188, EPI_ISL_1972190, EPI_ISL_1972191, EPI_ISL_1972199, EPI_ISL_1972202, EPI_ISL_1972203, EPI_ISL_1972206, EPI_ISL_1982488, EPI_ISL_1982496, EPI_ISL_1982498, EPI_ISL_1982500, EPI_ISL_1982502, EPI_ISL_1982504, EPI_ISL_1982511, EPI_ISL_1982513, EPI_ISL_1982514, EPI_ISL_1982518, EPI_ISL_1982522, EPI_ISL_1982524, EPI_ISL_1982535, EPI_ISL_1982536, EPI_ISL_1982544, EPI_ISL_1982546, EPI_ISL_1982550, EPI_ISL_1982558, EPI_ISL_1982562, EPI_ISL_1982568, EPI_ISL_1982571, EPI_ISL_1982573, EPI_ISL_1982577, EPI_ISL_1982583, EPI_ISL_1982586, EPI_ISL_1982589, EPI_ISL_1982595, EPI_ISL_1982606, EPI_ISL_1982616, EPI_ISL_1982618, EPI_ISL_1986377, EPI_ISL_1986385, EPI_ISL_1986392, EPI_ISL_1986396, EPI_ISL_1986398, EPI_ISL_1986400, EPI_ISL_1986402, EPI_ISL_1986404, EPI_ISL_1986410, EPI_ISL_1986412, EPI_ISL_1986414, EPI_ISL_1986417, EPI_ISL_1986419, EPI_ISL_1986421, EPI_ISL_1986423, EPI_ISL_1986425, EPI_ISL_1986426, EPI_ISL_1986428, EPI_ISL_1986430, EPI_ISL_1986432, EPI_ISL_1986434, EPI_ISL_1986435, EPI_ISL_1986441, EPI_ISL_1986443, EPI_ISL_1986454, EPI_ISL_1986458, EPI_ISL_1986460, EPI_ISL_1986464, EPI_ISL_1986465, EPI_ISL_1986468, EPI_ISL_1986469, EPI_ISL_1986471, EPI_ISL_1986473, EPI_ISL_1986475, EPI_ISL_1986479, EPI_ISL_1986480, EPI_ISL_1986482, EPI_ISL_1986484, EPI_ISL_1986486, EPI_ISL_1986489, EPI_ISL_1986491

see above | Labor Berlin Charité Vivantes GmbH / Institut für Virologie | Charité Universitätsmedizin Berlin, Institut für Virologie/Labor Berlin | Barbara Mühlemann; Christian Drosten; Christine Stephan; Peter Menzel; Rolf Schwarzer; Terry Jones; Victor M Corman

EPI_ISL_1570470, EPI_ISL_1570473, EPI_ISL_1570480, EPI_ISL_1570500, EPI_ISL_1570507, EPI_ISL_1570514, EPI_ISL_1570515, EPI_ISL_1570517, EPI_ISL_1570519, EPI_ISL_1570521, EPI_ISL_1570525, EPI_ISL_1570528, EPI_ISL_1570532, EPI_ISL_1570537, EPI_ISL_1570539, EPI_ISL_1570540, EPI_ISL_1570545, EPI_ISL_1570546, EPI_ISL_1570549, EPI_ISL_1570551, EPI_ISL_1570552, EPI_ISL_1570562, EPI_ISL_1570566, EPI_ISL_1570568, EPI_ISL_1570569, EPI_ISL_1570571, EPI_ISL_1570573, EPI_ISL_1570574, EPI_ISL_1570578, EPI_ISL_1570580, EPI_ISL_1570583, EPI_ISL_1570585, EPI_ISL_1570586, EPI_ISL_1570589, EPI_ISL_1570590, EPI_ISL_1570592, EPI_ISL_1570595, EPI_ISL_1570607, EPI_ISL_1570611, EPI_ISL_1570613, EPI_ISL_1570614, EPI_ISL_1570617, EPI_ISL_1570618, EPI_ISL_1570627, EPI_ISL_1570633, EPI_ISL_1570634, EPI_ISL_1570638, EPI_ISL_1570641, EPI_ISL_1570645, EPI_ISL_1570646, EPI_ISL_1570657, EPI_ISL_1570659, EPI_ISL_1570663, EPI_ISL_1570665, EPI_ISL_1570667, EPI_ISL_1570672, EPI_ISL_1570674, EPI_ISL_1570679, EPI_ISL_1570681, EPI_ISL_1570686, EPI_ISL_1570687, EPI_ISL_1570693, EPI_ISL_1570699, EPI_ISL_1570701

see above | Labor Blackholm MVZ | Robert Koch Institute

EPI_ISL_1715802, EPI_ISL_1715803, EPI_ISL_1715804, EPI_ISL_1715974, EPI_ISL_1715985, EPI_ISL_1716092, EPI_ISL_1716093, EPI_ISL_1716094, EPI_ISL_1716095, EPI_ISL_1716096, EPI_ISL_1716097, EPI_ISL_1716098, EPI_ISL_1716099, EPI_ISL_1716101, EPI_ISL_1716102, EPI_ISL_1716103, EPI_ISL_1716104, EPI_ISL_1716105, EPI_ISL_1716106, EPI_ISL_1716107, EPI_ISL_1716109, EPI_ISL_1716110, EPI_ISL_1716111, EPI_ISL_1716112, EPI_ISL_1716113, EPI_ISL_1716114, EPI_ISL_1716115, EPI_ISL_1716116, EPI_ISL_1716117, EPI_ISL_1716118, EPI_ISL_1716119, EPI_ISL_1716120, EPI_ISL_1716121, EPI_ISL_1716122, EPI_ISL_1716123, EPI_ISL_1716124, EPI_ISL_1716125, EPI_ISL_1716126, EPI_ISL_1716127, EPI_ISL_1716128, EPI_ISL_1716129, EPI_ISL_1716130, EPI_ISL_1716131, EPI_ISL_1716132, EPI_ISL_1716133, EPI_ISL_1716134, EPI_ISL_1716135, EPI_ISL_1716136, EPI_ISL_1716137, EPI_ISL_1716138, EPI_ISL_1716139, EPI_ISL_1716140, EPI_ISL_1716141, EPI_ISL_1716142, EPI_ISL_1716143, EPI_ISL_1716144, EPI_ISL_1716145, EPI_ISL_1716146, EPI_ISL_1716147, EPI_ISL_1716148, EPI_ISL_1716149, EPI_ISL_1716150, EPI_ISL_1716151, EPI_ISL_1716152, EPI_ISL_1716153, EPI_ISL_1716154, EPI_ISL_1716155, EPI_ISL_1716156, EPI_ISL_1716157, EPI_ISL_1716158, EPI_ISL_1716159, EPI_ISL_1716160, EPI_ISL_1716161, EPI_ISL_1716162, EPI_ISL_1716163, EPI_ISL_1716164, EPI_ISL_1716165, EPI_ISL_1716166, EPI_ISL_1716167, EPI_ISL_1716168, EPI_ISL_1716169, EPI_ISL_1716170, EPI_ISL_1716171, EPI_ISL_1716172, EPI_ISL_1716173, EPI_ISL_1716174, EPI_ISL_1716175, EPI_ISL_1716176, EPI_ISL_1716177, EPI_ISL_1716178, EPI_ISL_1716179, EPI_ISL_1716180, EPI_ISL_1716181, EPI_ISL_1716182, EPI_ISL_1716183, EPI_ISL_1716184, EPI_ISL_1716185, EPI_ISL_1716186, EPI_ISL_1716187, EPI_ISL_1716188, EPI_ISL_1716189, EPI_ISL_1716190, EPI_ISL_1716191, EPI_ISL_1716192, EPI_ISL_1716193, EPI_ISL_1716194, EPI_ISL_1716195, EPI_ISL_1716196, EPI_ISL_1716197, EPI_ISL_1716198, EPI_ISL_1716199, EPI_ISL_1716200, EPI_ISL_1716201, EPI_ISL_1716202, EPI_ISL_1716203, EPI_ISL_1716204, EPI_ISL_1716205, EPI_ISL_1716206, EPI_ISL_1716207, EPI_ISL_1716208, EPI_ISL_1716209, EPI_ISL_1716210, EPI_ISL_1716211, EPI_ISL_1716212, EPI_ISL_1716213, EPI_ISL_1716214, EPI_ISL_1716216, EPI_ISL_1716217, EPI_ISL_1716218, EPI_ISL_1716219, EPI_ISL_1716220, EPI_ISL_1716221, EPI_ISL_1716222, EPI_ISL_1716223, EPI_ISL_1716224, EPI_ISL_1716225, EPI_ISL_1716226, EPI_ISL_1716227, EPI_ISL_1716228, EPI_ISL_1716229, EPI_ISL_1716230, EPI_ISL_1716231, EPI_ISL_1716232, EPI_ISL_1716233, EPI_ISL_1716234, EPI_ISL_1716235, EPI_ISL_1716236, EPI_ISL_1716237, EPI_ISL_1716238, EPI_ISL_1716239, EPI_ISL_1716240, EPI_ISL_1716241, EPI_ISL_1716242, EPI_ISL_1716243, EPI_ISL_1716244, EPI_ISL_1716245, EPI_ISL_1716246, EPI_ISL_1716247, EPI_ISL_1716248, EPI_ISL_1716249, EPI_ISL_1716250, EPI_ISL_1716251, EPI_ISL_1716252, EPI_ISL_1716253, EPI_ISL_1716254, EPI_ISL_1716255, EPI_ISL_1716256, EPI_ISL_1716257, EPI_ISL_1716258, EPI_ISL_1716259, EPI_ISL_1716260, EPI_ISL_1716261, EPI_ISL_1716262, EPI_ISL_1716263, EPI_ISL_1716264, EPI_ISL_1716387, EPI_ISL_1716389, EPI_ISL_1716390, EPI_ISL_1716391, EPI_ISL_1861707, EPI_ISL_1861709

see above | Labor Dr. Fenner und Kollegen | Heinrich Pette Institute, Leibniz Institute for Experimental Virology | Adam Grundhoff; Alexis Robitaille; Johannes Knobloch; Martin Aepfelbacher; Nicole Fischer; Thomas Günther

EPI_ISL_1439798, EPI_ISL_1439804, EPI_ISL_1439813, EPI_ISL_1439820, EPI_ISL_1439831, EPI_ISL_1439856, EPI_ISL_1439863, EPI_ISL_1442546, EPI_ISL_1442551, EPI_ISL_1442554, EPI_ISL_1442556, EPI_ISL_1442557, EPI_ISL_1442559, EPI_ISL_1442560, EPI_ISL_1442561, EPI_ISL_1442562, EPI_ISL_1442567, EPI_ISL_1442570

see above | Labor Dr. Heidrich & Kollegen MVZ GmbH Hamburg | Robert Koch Institute

EPI_ISL_1353933, EPI_ISL_1354045, EPI_ISL_1354048, EPI_ISL_1354070, EPI_ISL_1354092, EPI_ISL_1354104, EPI_ISL_1354178, EPI_ISL_1354223, EPI_ISL_1354232, EPI_ISL_1354248, EPI_ISL_1432936, EPI_ISL_1432967, EPI_ISL_1433025, EPI_ISL_1433081, EPI_ISL_1433127, EPI_ISL_1433142, EPI_ISL_1433265, EPI_ISL_1433364, EPI_ISL_1438700, EPI_ISL_1438722, EPI_ISL_1438759, EPI_ISL_1438762, EPI_ISL_1438854, EPI_ISL_1438859, EPI_ISL_1438867, EPI_ISL_1438905, EPI_ISL_1438917, EPI_ISL_1438925, EPI_ISL_1438931, EPI_ISL_1438938, EPI_ISL_1438947, EPI_ISL_1438955, EPI_ISL_1438989, EPI_ISL_1439004, EPI_ISL_1439014, EPI_ISL_1847097, EPI_ISL_1847107, EPI_ISL_1847123, EPI_ISL_1847132, EPI_ISL_1847155

see above | Labor Dr. Schumacher MVZ | Robert Koch Institute

EPI_ISL_1570417, EPI_ISL_1643834, EPI_ISL_1643835, EPI_ISL_1643836, EPI_ISL_1643838, EPI_ISL_1643839, EPI_ISL_1643840, EPI_ISL_1643846, EPI_ISL_1643850, EPI_ISL_1643851, EPI_ISL_1643852, EPI_ISL_1643853, EPI_ISL_1643854, EPI_ISL_1643855, EPI_ISL_1643856, EPI_ISL_1847416, EPI_ISL_1847417

see above | Labor Dr. Spranger | Robert Koch Institute

EPI_ISL_1286907, EPI_ISL_1286972, EPI_ISL_1287027, EPI_ISL_1287113, EPI_ISL_1287156, EPI_ISL_1287177, EPI_ISL_1287190, EPI_ISL_1349244, EPI_ISL_1349264, EPI_ISL_1349282, EPI_ISL_1349289, EPI_ISL_1349322, EPI_ISL_1349325, EPI_ISL_1349345, EPI_ISL_1349357, EPI_ISL_1349360, EPI_ISL_1433741, EPI_ISL_1433764, EPI_ISL_1433780, EPI_ISL_1433844, EPI_ISL_1433908, EPI_ISL_1433965, EPI_ISL_1433971, EPI_ISL_1434003, EPI_ISL_1434011, EPI_ISL_1434015, EPI_ISL_1434074, EPI_ISL_1434125, EPI_ISL_1434130, EPI_ISL_1434170, EPI_ISL_1434185, EPI_ISL_1434214, EPI_ISL_1434295, EPI_ISL_1434309, EPI_ISL_1434325, EPI_ISL_1434351

see above | Labor Dr. Wisplinghoff - Köln | Robert Koch Institute

EPI_ISL_1350949, EPI_ISL_1350951, EPI_ISL_1350952, EPI_ISL_1350953, EPI_ISL_1350954, EPI_ISL_1350955, EPI_ISL_1350956, EPI_ISL_1350958, EPI_ISL_1350959, EPI_ISL_1350960, EPI_ISL_1350961, EPI_ISL_1435456, EPI_ISL_1435457, EPI_ISL_1435458, EPI_ISL_1435461, EPI_ISL_1435462, EPI_ISL_1435463, EPI_ISL_1435464, EPI_ISL_1435465, EPI_ISL_1435466

see above | Labor Kneißler GmbH & Co. KG | Robert Koch Institute

EPI_ISL_1353284, EPI_ISL_1353298, EPI_ISL_1353300, EPI_ISL_1353317, EPI_ISL_1353323, EPI_ISL_1353334, EPI_ISL_1353337, EPI_ISL_1353343, EPI_ISL_1353351, EPI_ISL_1435344, EPI_ISL_1435357, EPI_ISL_1435393, EPI_ISL_1435394, EPI_ISL_1435395, EPI_ISL_1435407, EPI_ISL_1435413, EPI_ISL_1435420, EPI_ISL_1435423, EPI_ISL_1439462, EPI_ISL_1439510, EPI_ISL_1439513

see above | Labor Lübeck bzw. Laborärztliche Gemeinschaftspraxis Lübeck | Robert Koch Institute

EPI_ISL_1571201, EPI_ISL_1571955 | Labor MÃ¶nchengladbach MVZ Dr. Stein + Kollegen GbR | Robert Koch Institute

EPI_ISL_1349569, EPI_ISL_1349570, EPI_ISL_1349571, EPI_ISL_1349576, EPI_ISL_1349578, EPI_ISL_1349580, EPI_ISL_1349581, EPI_ISL_1349583, EPI_ISL_1349584, EPI_ISL_1349585, EPI_ISL_1349587, EPI_ISL_1349588, EPI_ISL_1349591, EPI_ISL_1349592, EPI_ISL_1349595, EPI_ISL_1349596, EPI_ISL_1349597, EPI_ISL_1349598, EPI_ISL_1349600, EPI_ISL_1349601, EPI_ISL_1349602, EPI_ISL_1349603, EPI_ISL_1350222, EPI_ISL_1350224, EPI_ISL_1350229, EPI_ISL_1350230, EPI_ISL_1350234, EPI_ISL_1350237, EPI_ISL_1350239, EPI_ISL_1350241, EPI_ISL_1351269, EPI_ISL_1353058, EPI_ISL_1353059, EPI_ISL_1353060, EPI_ISL_1353062, EPI_ISL_1353065, EPI_ISL_1353067, EPI_ISL_1353068, EPI_ISL_1353071, EPI_ISL_1353077, EPI_ISL_1353081, EPI_ISL_1353096, EPI_ISL_1353103, EPI_ISL_1353116, EPI_ISL_1353133, EPI_ISL_1354281, EPI_ISL_1433599, EPI_ISL_1433603, EPI_ISL_1433610, EPI_ISL_1433619, EPI_ISL_1433621, EPI_ISL_1433627, EPI_ISL_1433629, EPI_ISL_1433631, EPI_ISL_1433633, EPI_ISL_1433634, EPI_ISL_1433637, EPI_ISL_1433640, EPI_ISL_1433642, EPI_ISL_1433644, EPI_ISL_1433649, EPI_ISL_1433653, EPI_ISL_1433655, EPI_ISL_1433659, EPI_ISL_1433661, EPI_ISL_1433662, EPI_ISL_1433668, EPI_ISL_1433670, EPI_ISL_1433671, EPI_ISL_1433672, EPI_ISL_1433673, EPI_ISL_1433675, EPI_ISL_1433677, EPI_ISL_1433678, EPI_ISL_1434439, EPI_ISL_1434446, EPI_ISL_1434465, EPI_ISL_1434467, EPI_ISL_1434489, EPI_ISL_1434494, EPI_ISL_1434496, EPI_ISL_1434510, EPI_ISL_1434514, EPI_ISL_1434518

see above | Labor MönchengladbachMVZ Dr. Stein + Kollegen GbR | Robert Koch Institute

EPI_ISL_1564011, EPI_ISL_1564012, EPI_ISL_1564013, EPI_ISL_1564018, EPI_ISL_1564020, EPI_ISL_1573239, EPI_ISL_1573244, EPI_ISL_1573247, EPI_ISL_1573258, EPI_ISL_1573260, EPI_ISL_1638281

see above | Labor Prof. Dr. G. Enders MVZ GbR | Robert Koch Institute

EPI_ISL_1573269, EPI_ISL_1573275 | Labor ZOTZ|KLIMAS; MVZ DÃ¼sseldorf-Centrum | Robert Koch Institute

EPI_ISL_1436889, EPI_ISL_1436896, EPI_ISL_1436932, EPI_ISL_1436934, EPI_ISL_1436947, EPI_ISL_1441730, EPI_ISL_1441731, EPI_ISL_1441762, EPI_ISL_1441764, EPI_ISL_1441781

see above | Labor ZOTZ|KLIMAS; MVZ Düsseldorf-Centrum | Robert Koch Institute

EPI_ISL_1357399, EPI_ISL_1357421, EPI_ISL_1357430, EPI_ISL_1357438, EPI_ISL_1357455, EPI_ISL_1357503, EPI_ISL_1357523, EPI_ISL_1357546, EPI_ISL_1357572, EPI_ISL_1438400, EPI_ISL_1438491

see above | Labor für medizinische Mikrobiologie Bakteriologie Mykologie Virologie & Infektionsserologie | Robert Koch Institute

EPI_ISL_1643969, EPI_ISL_1643970, EPI_ISL_1643971, EPI_ISL_1643972, EPI_ISL_1643973, EPI_ISL_1643974, EPI_ISL_1643975, EPI_ISL_1643976, EPI_ISL_1643977, EPI_ISL_1643978, EPI_ISL_1643979, EPI_ISL_1643980, EPI_ISL_1643981, EPI_ISL_1643982, EPI_ISL_1643983, EPI_ISL_1643984, EPI_ISL_1643985, EPI_ISL_1643986, EPI_ISL_1643987, EPI_ISL_1643988, EPI_ISL_1643989, EPI_ISL_1643990, EPI_ISL_1643991, EPI_ISL_1643992, EPI_ISL_1643993, EPI_ISL_1643994, EPI_ISL_1643995

see above | Laborarztpraxis Dres. med. Walther Weindel & Kollegen | Robert Koch Institute

EPI_ISL_1563859, EPI_ISL_1563880, EPI_ISL_1569708, EPI_ISL_1569724, EPI_ISL_1572144, EPI_ISL_1572153, EPI_ISL_1572155, EPI_ISL_1572167, EPI_ISL_1572175, EPI_ISL_1572176

see above | Laborarztpraxis Jochen HÃ¼ter | Robert Koch Institute

EPI_ISL_1356618, EPI_ISL_1356619, EPI_ISL_1356622, EPI_ISL_1356623, EPI_ISL_1356624, EPI_ISL_1356626, EPI_ISL_1356627, EPI_ISL_1356629, EPI_ISL_1356630, EPI_ISL_1356631, EPI_ISL_1356632, EPI_ISL_1356633, EPI_ISL_1356634, EPI_ISL_1356636, EPI_ISL_1356637, EPI_ISL_1356638, EPI_ISL_1434452, EPI_ISL_1439523, EPI_ISL_1439525

see above | Laborarztpraxis Jochen Hüter | Robert Koch Institute

EPI_ISL_1353362, EPI_ISL_1353369, EPI_ISL_1353400, EPI_ISL_1433518, EPI_ISL_1433526, EPI_ISL_1433563, EPI_ISL_1433567, EPI_ISL_1433569, EPI_ISL_1433576, EPI_ISL_1433594, EPI_ISL_1439380, EPI_ISL_1439398, EPI_ISL_1439416, EPI_ISL_1439432, EPI_ISL_1563821, EPI_ISL_1563824, EPI_ISL_1563830, EPI_ISL_1563836, EPI_ISL_1563838, EPI_ISL_1563847, EPI_ISL_1563848, EPI_ISL_1563850

see above | Labormedizin Darmstadt | Robert Koch Institute

EPI_ISL_1361752, EPI_ISL_1361753, EPI_ISL_1361754, EPI_ISL_1361755, EPI_ISL_1361756, EPI_ISL_1361757, EPI_ISL_1361758, EPI_ISL_1361759, EPI_ISL_1361760, EPI_ISL_1361761, EPI_ISL_1361762, EPI_ISL_1361763, EPI_ISL_1361764, EPI_ISL_1361765, EPI_ISL_1361766, EPI_ISL_1361767, EPI_ISL_1361768, EPI_ISL_1361769, EPI_ISL_1361770, EPI_ISL_1361771, EPI_ISL_1361772, EPI_ISL_1361773, EPI_ISL_1361783, EPI_ISL_1361784, EPI_ISL_1361785, EPI_ISL_1361786, EPI_ISL_1361787, EPI_ISL_1361788, EPI_ISL_1361789, EPI_ISL_1361790, EPI_ISL_1361791, EPI_ISL_1361792, EPI_ISL_1361793, EPI_ISL_1361794, EPI_ISL_1361795, EPI_ISL_1361796, EPI_ISL_1361797, EPI_ISL_1361798, EPI_ISL_1361799, EPI_ISL_1361800, EPI_ISL_1361801, EPI_ISL_1361802, EPI_ISL_1361803, EPI_ISL_1361804, EPI_ISL_1361805, EPI_ISL_1361806, EPI_ISL_1361807, EPI_ISL_1361808, EPI_ISL_1361809, EPI_ISL_1361810, EPI_ISL_1361811, EPI_ISL_1361812, EPI_ISL_1361813, EPI_ISL_1361814, EPI_ISL_1361815, EPI_ISL_1361816, EPI_ISL_1361817, EPI_ISL_1361818, EPI_ISL_1361819, EPI_ISL_1361820, EPI_ISL_1361821, EPI_ISL_1361822, EPI_ISL_1361836, EPI_ISL_1361837, EPI_ISL_1361838, EPI_ISL_1361839, EPI_ISL_1361840, EPI_ISL_1361841, EPI_ISL_1361842, EPI_ISL_1361843, EPI_ISL_1361844, EPI_ISL_1361845, EPI_ISL_1361846, EPI_ISL_1361847, EPI_ISL_1361848, EPI_ISL_1361849, EPI_ISL_1361850, EPI_ISL_1361851, EPI_ISL_1361852, EPI_ISL_1361853, EPI_ISL_1361854, EPI_ISL_1361855, EPI_ISL_1361856, EPI_ISL_1361857, EPI_ISL_1361858, EPI_ISL_1361859, EPI_ISL_1361860, EPI_ISL_1361861, EPI_ISL_1361862, EPI_ISL_1361863, EPI_ISL_1361864, EPI_ISL_1361865, EPI_ISL_1361866, EPI_ISL_1361867, EPI_ISL_1361868, EPI_ISL_1361875, EPI_ISL_1361876, EPI_ISL_1361877, EPI_ISL_1361878, EPI_ISL_1361879, EPI_ISL_1361880, EPI_ISL_1361881, EPI_ISL_1361882, EPI_ISL_1361883, EPI_ISL_1361884, EPI_ISL_1361885, EPI_ISL_1361886, EPI_ISL_1361887, EPI_ISL_1361888, EPI_ISL_1361889, EPI_ISL_1361890, EPI_ISL_1361891, EPI_ISL_1361892, EPI_ISL_1361893, EPI_ISL_1361894, EPI_ISL_1361895, EPI_ISL_1361896, EPI_ISL_1361897, EPI_ISL_1361898, EPI_ISL_1361899, EPI_ISL_1361900, EPI_ISL_1361901, EPI_ISL_1361902, EPI_ISL_1361903, EPI_ISL_1361904, EPI_ISL_1361905, EPI_ISL_1361909, EPI_ISL_1361910, EPI_ISL_1361911, EPI_ISL_1361912, EPI_ISL_1361913, EPI_ISL_1361914, EPI_ISL_1361915, EPI_ISL_1361916, EPI_ISL_1361917, EPI_ISL_1361918, EPI_ISL_1361919, EPI_ISL_1361920, EPI_ISL_1361921, EPI_ISL_1361922, EPI_ISL_1361923, EPI_ISL_1361924, EPI_ISL_1361925, EPI_ISL_1361926, EPI_ISL_1361927, EPI_ISL_1361928, EPI_ISL_1361929, EPI_ISL_1361930, EPI_ISL_1361931, EPI_ISL_1361932, EPI_ISL_1361933, EPI_ISL_1670002, EPI_ISL_1670003, EPI_ISL_1670004, EPI_ISL_1670005, EPI_ISL_1670006, EPI_ISL_1670011, EPI_ISL_1670012, EPI_ISL_1670013, EPI_ISL_1670014, EPI_ISL_1670015, EPI_ISL_1670016, EPI_ISL_1670017, EPI_ISL_1670018, EPI_ISL_1670019, EPI_ISL_1670020, EPI_ISL_1670021, EPI_ISL_1670022, EPI_ISL_1670023, EPI_ISL_1670024, EPI_ISL_1670025, EPI_ISL_1670026, EPI_ISL_1670027, EPI_ISL_1670028, EPI_ISL_1670029, EPI_ISL_1670030, EPI_ISL_1670031, EPI_ISL_1670032, EPI_ISL_1670033, EPI_ISL_1670034, EPI_ISL_1670035, EPI_ISL_1670036, EPI_ISL_1670037, EPI_ISL_1670038, EPI_ISL_1670039, EPI_ISL_1670040, EPI_ISL_1670041, EPI_ISL_1670042, EPI_ISL_1670043, EPI_ISL_1670044, EPI_ISL_1670045, EPI_ISL_1670046, EPI_ISL_1670047, EPI_ISL_1670101, EPI_ISL_1670195

see above | Landesamt für Verbraucherschutz Sachsen Anhalt, Magdeburg | Institute of Medical Microbiology and Hospital Hygiene | Aljoscha Tersteegen; Prof. Dr. Achim Kaasch

EPI_ISL_2126401, EPI_ISL_2126404 | Landesgesundheitsamt Baden-Wurttemberg | Robert Koch Institute

EPI_ISL_1638646, EPI_ISL_1638647, EPI_ISL_1638648, EPI_ISL_1638653, EPI_ISL_1638654, EPI_ISL_1638660 | Landesgesundheitsamt Baden-WÃ¼rttemberg | Robert Koch Institute

EPI_ISL_1354365, EPI_ISL_1354378, EPI_ISL_1354383, EPI_ISL_1354386, EPI_ISL_1354390, EPI_ISL_1354401, EPI_ISL_1354402, EPI_ISL_1354406, EPI_ISL_1354412, EPI_ISL_1354413, EPI_ISL_1354418, EPI_ISL_1354421, EPI_ISL_1354424, EPI_ISL_1354427, EPI_ISL_1354428, EPI_ISL_1354430, EPI_ISL_1354435, EPI_ISL_1354452, EPI_ISL_1354455

see above | Landesuntersuchungsanstalt für das Gesundheits- und Veterinärwesen (LUA) Sachsen | Robert Koch Institute

EPI_ISL_1711163, EPI_ISL_1711171, EPI_ISL_1711172, EPI_ISL_1711183, EPI_ISL_1711185, EPI_ISL_1711186, EPI_ISL_1711187, EPI_ISL_1711188, EPI_ISL_1711203, EPI_ISL_1711209, EPI_ISL_1711210, EPI_ISL_1711211, EPI_ISL_1711212, EPI_ISL_1711213, EPI_ISL_1711224, EPI_ISL_1711226, EPI_ISL_1711228, EPI_ISL_1711240, EPI_ISL_1711241

| | | | |
|---|---|---|---|
| see above | Landstuhl Regional Medical Center | US Air Force School of Aerospace Medicine | Amanda Javorina; Anthony Fries; Clarise Starr; Cole Anderson; Elizabeth Macias; Fritz Castillo; Jennifer Meyer; Sarah Purves; William Buggele; William Gruner |
| EPI_ISL_1440951, EPI_ISL_1440952, EPI_ISL_1440953, EPI_ISL_1440954 | Limbach - Ganzimmun Diagnostics AG Mainz | Robert Koch Institute | |
| EPI_ISL_1352297, EPI_ISL_1353148, EPI_ISL_1353155, EPI_ISL_1353170, EPI_ISL_1353178, EPI_ISL_1353189, EPI_ISL_1353200, EPI_ISL_1435303, EPI_ISL_1435308, EPI_ISL_1435313, EPI_ISL_1435315, EPI_ISL_1435316, EPI_ISL_1435321, EPI_ISL_1435324, EPI_ISL_1435325, EPI_ISL_1435327, EPI_ISL_1435332, EPI_ISL_1441196, EPI_ISL_1441206, EPI_ISL_1441214, EPI_ISL_1441218, EPI_ISL_1441220, EPI_ISL_1567645, EPI_ISL_1567648, EPI_ISL_1567656, EPI_ISL_1571958 | | | |
| see above | Limbach - MVZ Gemeinschaftslabor Suhl | Robert Koch Institute | |
| EPI_ISL_1350441, EPI_ISL_1350442, EPI_ISL_1350443, EPI_ISL_1350444, EPI_ISL_1350445, EPI_ISL_1350446, EPI_ISL_1350447, EPI_ISL_1350448, EPI_ISL_1350449, EPI_ISL_1350450, EPI_ISL_1350451, EPI_ISL_1350452, EPI_ISL_1350453, EPI_ISL_1350454, EPI_ISL_1350455, EPI_ISL_1350456, EPI_ISL_1350457, EPI_ISL_1350458, EPI_ISL_1350459, EPI_ISL_1350460, EPI_ISL_1350461, EPI_ISL_1350462, EPI_ISL_1350463, EPI_ISL_1350464, EPI_ISL_1350465, EPI_ISL_1350467, EPI_ISL_1350468, EPI_ISL_1350469, EPI_ISL_1350470, EPI_ISL_1350471, EPI_ISL_1350472, EPI_ISL_1350473, EPI_ISL_1350474, EPI_ISL_1350475, EPI_ISL_1350476, EPI_ISL_1350477, EPI_ISL_1350478, EPI_ISL_1350479, EPI_ISL_1350480, EPI_ISL_1350481, EPI_ISL_1350482, EPI_ISL_1350484, EPI_ISL_1350485, EPI_ISL_1350486, EPI_ISL_1350487, EPI_ISL_1350488, EPI_ISL_1350489, EPI_ISL_1350490, EPI_ISL_1350491, EPI_ISL_1350492, EPI_ISL_1350493, EPI_ISL_1350494, EPI_ISL_1350495, EPI_ISL_1350496, EPI_ISL_1350497, EPI_ISL_1350498, EPI_ISL_1350499, EPI_ISL_1350500, EPI_ISL_1350501, EPI_ISL_1350503, EPI_ISL_1350504, EPI_ISL_1350505, EPI_ISL_1350506, EPI_ISL_1350507, EPI_ISL_1350508, EPI_ISL_1350509, EPI_ISL_1350510, EPI_ISL_1350511, EPI_ISL_1350512, EPI_ISL_1350513, EPI_ISL_1350514, EPI_ISL_1350515, EPI_ISL_1350516, EPI_ISL_1350517, EPI_ISL_1350518, EPI_ISL_1350519, EPI_ISL_1350520, EPI_ISL_1350521, EPI_ISL_1350522, EPI_ISL_1350523, EPI_ISL_1350524, EPI_ISL_1350525, EPI_ISL_1350526, EPI_ISL_1350527, EPI_ISL_1350528, EPI_ISL_1350529, EPI_ISL_1350530, EPI_ISL_1350532, EPI_ISL_1350533, EPI_ISL_1350534, EPI_ISL_1350535, EPI_ISL_1350536, EPI_ISL_1350537, EPI_ISL_1350538, EPI_ISL_1350539, EPI_ISL_1350540, EPI_ISL_1350541, EPI_ISL_1350542, EPI_ISL_1350543, EPI_ISL_1350544, EPI_ISL_1350545, EPI_ISL_1350546, EPI_ISL_1350547, EPI_ISL_1350548, EPI_ISL_1350549, EPI_ISL_1350550, EPI_ISL_1350551, EPI_ISL_1350552, EPI_ISL_1350553, EPI_ISL_1350554, EPI_ISL_1350555, EPI_ISL_1350556, EPI_ISL_1350557, EPI_ISL_1350558, EPI_ISL_1350559, EPI_ISL_1350560, EPI_ISL_1350561, EPI_ISL_1350562, EPI_ISL_1350563, EPI_ISL_1350564, EPI_ISL_1350565, EPI_ISL_1350566, EPI_ISL_1350567, EPI_ISL_1350568, EPI_ISL_1350569, EPI_ISL_1350570, EPI_ISL_1350571, EPI_ISL_1350572, EPI_ISL_1350573, EPI_ISL_1350574, EPI_ISL_1350575, EPI_ISL_1350576, EPI_ISL_1350577, EPI_ISL_1350578, EPI_ISL_1350579, EPI_ISL_1350581, EPI_ISL_1350582, EPI_ISL_1350583, EPI_ISL_1350584, EPI_ISL_1350585, EPI_ISL_1350586, EPI_ISL_1350587, EPI_ISL_1350588, EPI_ISL_1350589, EPI_ISL_1350590, EPI_ISL_1350591, EPI_ISL_1350592, EPI_ISL_1350593, EPI_ISL_1350594, EPI_ISL_1350595, EPI_ISL_1350596, EPI_ISL_1350597, EPI_ISL_1350598, EPI_ISL_1350599, EPI_ISL_1350600, EPI_ISL_1350601, EPI_ISL_1350602, EPI_ISL_1350603, EPI_ISL_1350604, EPI_ISL_1350605, EPI_ISL_1350606, EPI_ISL_1350607, EPI_ISL_1350608, EPI_ISL_1350609, EPI_ISL_1350610, EPI_ISL_1350611, EPI_ISL_1350612, EPI_ISL_1350613, EPI_ISL_1350614, EPI_ISL_1350615, EPI_ISL_1350616, EPI_ISL_1436395, EPI_ISL_1436399, EPI_ISL_1436401, EPI_ISL_1436403, EPI_ISL_1436405, EPI_ISL_1436408, EPI_ISL_1436409, EPI_ISL_1436410, EPI_ISL_1436412, EPI_ISL_1436413, EPI_ISL_1436414, EPI_ISL_1436415, EPI_ISL_1436416, EPI_ISL_1436417, EPI_ISL_1436420, EPI_ISL_1436423, EPI_ISL_1436424, EPI_ISL_1436427, EPI_ISL_1436428, EPI_ISL_1436431, EPI_ISL_1436433, EPI_ISL_1436435, EPI_ISL_1436436, EPI_ISL_1436437, EPI_ISL_1436440, EPI_ISL_1436443, EPI_ISL_1436444, EPI_ISL_1436447, EPI_ISL_1436448, EPI_ISL_1436451, EPI_ISL_1436455, EPI_ISL_1436456, EPI_ISL_1436457, EPI_ISL_1436461, EPI_ISL_1436463, EPI_ISL_1436464, EPI_ISL_1436465, EPI_ISL_1436468, EPI_ISL_1436471, EPI_ISL_1436473, EPI_ISL_1436475, EPI_ISL_1436476, EPI_ISL_1436478, EPI_ISL_1436480, EPI_ISL_1436481, EPI_ISL_1436482, EPI_ISL_1436483, EPI_ISL_1436484, EPI_ISL_1436485, EPI_ISL_1436487, EPI_ISL_1436488, EPI_ISL_1436489, EPI_ISL_1436490, EPI_ISL_1436492, EPI_ISL_1436493, EPI_ISL_1436494, EPI_ISL_1436496, EPI_ISL_1436499, EPI_ISL_1436501, EPI_ISL_1436503, EPI_ISL_1436504, EPI_ISL_1436507, EPI_ISL_1436508, EPI_ISL_1436510, EPI_ISL_1436512, EPI_ISL_1436514, EPI_ISL_1436515, EPI_ISL_1436516, EPI_ISL_1436517, EPI_ISL_1436520, EPI_ISL_1436521, EPI_ISL_1436522, EPI_ISL_1436524, EPI_ISL_1436526, EPI_ISL_1436527, EPI_ISL_1436530, EPI_ISL_1436532, EPI_ISL_1436534, EPI_ISL_1436536, EPI_ISL_1436538, EPI_ISL_1436539, EPI_ISL_1436543, EPI_ISL_1436544, EPI_ISL_1436545, EPI_ISL_1436547, EPI_ISL_1436551, EPI_ISL_1436552, EPI_ISL_1436554, EPI_ISL_1436555, EPI_ISL_1436556, EPI_ISL_1436558, EPI_ISL_1436559, EPI_ISL_1436560, EPI_ISL_1436562, EPI_ISL_1436563, EPI_ISL_1436566, EPI_ISL_1436569, EPI_ISL_1567239 | | | |
| see above | Limbach - MVZ Humangenetik Ulm | Robert Koch Institute | |
| EPI_ISL_1433597, EPI_ISL_1433608, EPI_ISL_1433612, EPI_ISL_1433614, EPI_ISL_1433617, EPI_ISL_1433625, EPI_ISL_1433638, EPI_ISL_1433646, EPI_ISL_1433651, EPI_ISL_1433657, EPI_ISL_1438605, EPI_ISL_1438639, EPI_ISL_1438664, EPI_ISL_1438669, EPI_ISL_1439520, EPI_ISL_1439522, EPI_ISL_1439524, EPI_ISL_1441649, EPI_ISL_1441651, EPI_ISL_1441653, EPI_ISL_1441654, EPI_ISL_1441655, EPI_ISL_1441657, EPI_ISL_1441658, EPI_ISL_1441661, EPI_ISL_1441664, EPI_ISL_1441665, EPI_ISL_1441667, EPI_ISL_1441668, EPI_ISL_1441673, EPI_ISL_1441674, EPI_ISL_1441677, EPI_ISL_1441679, EPI_ISL_1441680, EPI_ISL_1441681, EPI_ISL_1563852, EPI_ISL_1563867, EPI_ISL_1563881, EPI_ISL_1563882, EPI_ISL_1571163, EPI_ISL_1571165, EPI_ISL_1571168, EPI_ISL_1571174, EPI_ISL_1571179, EPI_ISL_1571186, EPI_ISL_1571190 | | | |
| see above | Limbach - MVZ Labor Eveld & Kollegen Essen | Robert Koch Institute | |
| EPI_ISL_1350228, EPI_ISL_1350236, EPI_ISL_1352238, EPI_ISL_1352244, EPI_ISL_1352246, EPI_ISL_1352256, EPI_ISL_1352259, EPI_ISL_1352260, EPI_ISL_1352270, EPI_ISL_1352271, EPI_ISL_1352273, EPI_ISL_1352276, EPI_ISL_1352283, EPI_ISL_1352302, EPI_ISL_1352304, EPI_ISL_1352305, EPI_ISL_1352315, EPI_ISL_1352319, EPI_ISL_1352320, EPI_ISL_1352326, EPI_ISL_1352327, EPI_ISL_1352331, EPI_ISL_1352337, EPI_ISL_1352344, EPI_ISL_1353026, EPI_ISL_1353027, EPI_ISL_1353041, EPI_ISL_1353043, EPI_ISL_1353044, EPI_ISL_1353046, EPI_ISL_1353063, EPI_ISL_1353094, EPI_ISL_1353095, EPI_ISL_1353097, EPI_ISL_1353110, EPI_ISL_1353112, EPI_ISL_1353121, EPI_ISL_1353126, EPI_ISL_1353135, EPI_ISL_1353136, EPI_ISL_1353169, EPI_ISL_1353179, EPI_ISL_1353185, EPI_ISL_1353206, EPI_ISL_1434481, EPI_ISL_1435292, EPI_ISL_1435294, EPI_ISL_1435295, EPI_ISL_1435297, EPI_ISL_1435299, EPI_ISL_1435301, EPI_ISL_1435302, EPI_ISL_1435304, EPI_ISL_1435307, EPI_ISL_1435309, EPI_ISL_1435311, EPI_ISL_1435312, EPI_ISL_1435314, EPI_ISL_1435317, EPI_ISL_1435318, EPI_ISL_1435319, EPI_ISL_1435322, EPI_ISL_1435323, EPI_ISL_1435326, EPI_ISL_1435329, EPI_ISL_1435330, EPI_ISL_1435331, EPI_ISL_1435428, EPI_ISL_1435429, EPI_ISL_1435430, EPI_ISL_1435431, EPI_ISL_1435432, EPI_ISL_1435433, EPI_ISL_1435434, EPI_ISL_1435435, EPI_ISL_1435436, EPI_ISL_1435437, EPI_ISL_1435438, EPI_ISL_1435439, EPI_ISL_1435442, EPI_ISL_1435443, EPI_ISL_1435444, EPI_ISL_1435445, EPI_ISL_1435446, EPI_ISL_1435447, EPI_ISL_1435448, EPI_ISL_1435449, EPI_ISL_1435450, EPI_ISL_1435451, EPI_ISL_1435452, EPI_ISL_1435453, EPI_ISL_1435455, EPI_ISL_1438598, EPI_ISL_1438606, EPI_ISL_1438609, EPI_ISL_1438611, EPI_ISL_1438614, EPI_ISL_1438620, EPI_ISL_1438634, EPI_ISL_1438644, EPI_ISL_1438652, EPI_ISL_1438658, EPI_ISL_1438662, EPI_ISL_1438667, EPI_ISL_1438671, EPI_ISL_1439521, EPI_ISL_1441163, EPI_ISL_1441167, EPI_ISL_1441168, EPI_ISL_1441169, EPI_ISL_1441170, EPI_ISL_1441171, EPI_ISL_1441182, EPI_ISL_1441184, EPI_ISL_1441186, EPI_ISL_1441191, EPI_ISL_1441193, EPI_ISL_1441198, EPI_ISL_1441201, EPI_ISL_1441203, EPI_ISL_1441210, EPI_ISL_1441211, EPI_ISL_1441215, EPI_ISL_1441216, EPI_ISL_1441217, EPI_ISL_1441222, EPI_ISL_1441224, EPI_ISL_1441226, EPI_ISL_1441228, EPI_ISL_1441230, EPI_ISL_1441660, EPI_ISL_1441686, EPI_ISL_1441696, EPI_ISL_1441697, EPI_ISL_1441699, EPI_ISL_1441700, EPI_ISL_1441701, EPI_ISL_1441704, EPI_ISL_1441705, EPI_ISL_1441706, EPI_ISL_1441708, EPI_ISL_1441714 | | | |
| see above | MDI Limbach Berlin GmbH | Robert Koch Institute | |
| EPI_ISL_1563857, EPI_ISL_1563883, EPI_ISL_1563885, EPI_ISL_1563888, EPI_ISL_1563890, EPI_ISL_1563892, EPI_ISL_1563894, EPI_ISL_1563899, EPI_ISL_1563903, EPI_ISL_1563906, EPI_ISL_1563908, EPI_ISL_1563909, EPI_ISL_1563912, EPI_ISL_1563914, EPI_ISL_1563918, EPI_ISL_1563920, EPI_ISL_1563921, EPI_ISL_1563923, EPI_ISL_1563926, EPI_ISL_1565390, EPI_ISL_1569714, EPI_ISL_1569719, EPI_ISL_1570707, EPI_ISL_1571931, EPI_ISL_1571934, EPI_ISL_1571940, EPI_ISL_1571942, EPI_ISL_1571950, EPI_ISL_1571952, EPI_ISL_1571954, EPI_ISL_1571957, EPI_ISL_1572142, EPI_ISL_1572146, EPI_ISL_1572148, EPI_ISL_1572151, EPI_ISL_1572157, EPI_ISL_1572165, EPI_ISL_1572173, EPI_ISL_1573255, EPI_ISL_1573257, EPI_ISL_1573261, EPI_ISL_1573262, EPI_ISL_1638321, EPI_ISL_1638362 | | | |
| see above | MDI Limbach Berlin GmbH; MVZ Labor Berlin | Robert Koch Institute | |
| EPI_ISL_1464586, EPI_ISL_1464587, EPI_ISL_1464590 | MEDILYS Laborgesellschaft mbH | Heinrich Pette Institute, Leibniz Institute for Experimental Virology | Adam Grundhoff; Alexis Robitaille; Johannes Knobloch; Martin Aepfelbacher; Nicole Fischer; Thomas Günther |
| EPI_ISL_1524896, EPI_ISL_1524897, EPI_ISL_1524898, EPI_ISL_1524899, EPI_ISL_1524901, EPI_ISL_1524902 | MKK Mühlenkreiskliniken, Johannes Wesling Klinikum Minden | Hannover Medical School, Institute of Virology | Lars Steinbrück |
| EPI_ISL_1350242, EPI_ISL_1350246, EPI_ISL_1350251, EPI_ISL_1350256, EPI_ISL_1350260, EPI_ISL_1350267, EPI_ISL_1350271, EPI_ISL_1350272, EPI_ISL_1350273, EPI_ISL_1350274, EPI_ISL_1350275, EPI_ISL_1350279, EPI_ISL_1350282, EPI_ISL_1350288, EPI_ISL_1350291, EPI_ISL_1350312, EPI_ISL_1350314, EPI_ISL_1350316, EPI_ISL_1350318, EPI_ISL_1350319, EPI_ISL_1353278, EPI_ISL_1353279, EPI_ISL_1353280, EPI_ISL_1353281, EPI_ISL_1353282, EPI_ISL_1353283, EPI_ISL_1353285, EPI_ISL_1353286, EPI_ISL_1353287, EPI_ISL_1353288, EPI_ISL_1353289, EPI_ISL_1353290, EPI_ISL_1353291, EPI_ISL_1353292, EPI_ISL_1353294, EPI_ISL_1353295, EPI_ISL_1353296, EPI_ISL_1353297, EPI_ISL_1353299, EPI_ISL_1353303, EPI_ISL_1353304, EPI_ISL_1353305, EPI_ISL_1353306, EPI_ISL_1353307, EPI_ISL_1353308, EPI_ISL_1353310, EPI_ISL_1353311, EPI_ISL_1353312, EPI_ISL_1353313, EPI_ISL_1353314, EPI_ISL_1353315, EPI_ISL_1353316, EPI_ISL_1353319, EPI_ISL_1353321, EPI_ISL_1353324, EPI_ISL_1353326, EPI_ISL_1353327, EPI_ISL_1353329, EPI_ISL_1353331, EPI_ISL_1353332, EPI_ISL_1353335, EPI_ISL_1353336, EPI_ISL_1353338, EPI_ISL_1353339, EPI_ISL_1353340, EPI_ISL_1353341, EPI_ISL_1353342, EPI_ISL_1353344, EPI_ISL_1353345, EPI_ISL_1353347, EPI_ISL_1353349, EPI_ISL_1353352, EPI_ISL_1435341, EPI_ISL_1435342, EPI_ISL_1435343, EPI_ISL_1435346, EPI_ISL_1435347, EPI_ISL_1435348, EPI_ISL_1435349, EPI_ISL_1435350, EPI_ISL_1435351, EPI_ISL_1435352, EPI_ISL_1435353, EPI_ISL_1435355, EPI_ISL_1435356, EPI_ISL_1435358, EPI_ISL_1435359, EPI_ISL_1435360, EPI_ISL_1435361, EPI_ISL_1435363, EPI_ISL_1435364, EPI_ISL_1435365, EPI_ISL_1435366, EPI_ISL_1435367, EPI_ISL_1435368, EPI_ISL_1435369, EPI_ISL_1435370, EPI_ISL_1435371, EPI_ISL_1435372, EPI_ISL_1435373, EPI_ISL_1435374, EPI_ISL_1435375, EPI_ISL_1435376, EPI_ISL_1435377, EPI_ISL_1435378, EPI_ISL_1435380, EPI_ISL_1435381, EPI_ISL_1435383, EPI_ISL_1435386, EPI_ISL_1435387, EPI_ISL_1435389, EPI_ISL_1435390, EPI_ISL_1435396, EPI_ISL_1435397, EPI_ISL_1435398, EPI_ISL_1435399, EPI_ISL_1435401, EPI_ISL_1435402, EPI_ISL_1435403, EPI_ISL_1435404, EPI_ISL_1435405, EPI_ISL_1435408, EPI_ISL_1435409, EPI_ISL_1435410, EPI_ISL_1435411, EPI_ISL_1435412, EPI_ISL_1435414, EPI_ISL_1435415, EPI_ISL_1435416, EPI_ISL_1435417, EPI_ISL_1435418, EPI_ISL_1435419, EPI_ISL_1435421, EPI_ISL_1435422, EPI_ISL_1435424, EPI_ISL_1435425, EPI_ISL_1435426, EPI_ISL_1435427 | | | |
| see above | MVZ Dr. Eberhard & Partner Dortmund | Robert Koch Institute | |
| EPI_ISL_1434506, EPI_ISL_1435320, EPI_ISL_1435328, EPI_ISL_1441202 | MVZ Labor Dortmund | Robert Koch Institute | |
| EPI_ISL_1350380, EPI_ISL_1350388, EPI_ISL_1350409, EPI_ISL_1350416, EPI_ISL_1350421, EPI_ISL_1357279, EPI_ISL_1357282, EPI_ISL_1357286, EPI_ISL_1357288, EPI_ISL_1357306, EPI_ISL_1357314, EPI_ISL_1357315, EPI_ISL_1357326 | | | |
| see above | MVZ Labor Dr. Fenner und Kollegen (Standort Hamburg) | Robert Koch Institute | |
| EPI_ISL_1353069, EPI_ISL_1353106, EPI_ISL_1353142, EPI_ISL_1353180, EPI_ISL_1433601, EPI_ISL_1433676, EPI_ISL_1438584, EPI_ISL_1438590, EPI_ISL_1438595, EPI_ISL_1438596, EPI_ISL_1438599, EPI_ISL_1438600, EPI_ISL_1438602, EPI_ISL_1438607, EPI_ISL_1438610, EPI_ISL_1438617, EPI_ISL_1438625, EPI_ISL_1438626, EPI_ISL_1438629, EPI_ISL_1438630, EPI_ISL_1438637, EPI_ISL_1438646, EPI_ISL_1438647, EPI_ISL_1438653, EPI_ISL_1438656, EPI_ISL_1438663, EPI_ISL_1438665, EPI_ISL_1438670, EPI_ISL_1438674, EPI_ISL_1438675, EPI_ISL_1438677, EPI_ISL_1438678, EPI_ISL_1438681, EPI_ISL_1438682, EPI_ISL_1438683, EPI_ISL_1438684, EPI_ISL_1438685, EPI_ISL_1441227, EPI_ISL_1638319, EPI_ISL_1638375, EPI_ISL_1638491 | | | |
| see above | MVZ Labor Dr. Limbach & Kollegen GbR | Robert Koch Institute | |
| EPI_ISL_1570463, EPI_ISL_1570467, EPI_ISL_1570495, EPI_ISL_1570497, EPI_ISL_1570499, EPI_ISL_1570502, EPI_ISL_1570503, EPI_ISL_1570504, EPI_ISL_1570523, EPI_ISL_1570531, EPI_ISL_1570533, EPI_ISL_1570535, EPI_ISL_1570541, EPI_ISL_1570542, EPI_ISL_1570544, EPI_ISL_1570547, EPI_ISL_1570548, EPI_ISL_1570556, EPI_ISL_1570557, EPI_ISL_1570581, EPI_ISL_1570587, EPI_ISL_1570598, EPI_ISL_1570604, EPI_ISL_1570612, EPI_ISL_1570619, EPI_ISL_1570620, EPI_ISL_1570621, EPI_ISL_1570622, EPI_ISL_1570623, EPI_ISL_1570635, EPI_ISL_1570636, EPI_ISL_1570637, EPI_ISL_1570639, EPI_ISL_1570642, EPI_ISL_1570652, EPI_ISL_1570655, EPI_ISL_1570662, EPI_ISL_1570664, EPI_ISL_1570668, EPI_ISL_1570669, EPI_ISL_1570670, EPI_ISL_1570671, EPI_ISL_1570673, EPI_ISL_1570676, EPI_ISL_1570683, EPI_ISL_1570685, EPI_ISL_1570688, EPI_ISL_1570689, EPI_ISL_1570696 | | | |
| see above | MVZ Labor Dr. Quade & Kollegen GmbH | Robert Koch Institute | |
| EPI_ISL_1353061, EPI_ISL_1353108, EPI_ISL_1433607, EPI_ISL_1433623, EPI_ISL_1433665, EPI_ISL_1433669, EPI_ISL_1441656, EPI_ISL_1441659, EPI_ISL_1441662, EPI_ISL_1441672, EPI_ISL_1563886 | | | |
| see above | MVZ Labor Dr. Reising-Ackermann und Kollegen | Robert Koch Institute | |
| EPI_ISL_1351325, EPI_ISL_1354632, EPI_ISL_1354633, EPI_ISL_1354635, EPI_ISL_1354638, EPI_ISL_1354639, EPI_ISL_1354640, EPI_ISL_1354641, EPI_ISL_1354642, EPI_ISL_1354643, EPI_ISL_1354645, EPI_ISL_1354646, EPI_ISL_1354647, EPI_ISL_1354648, EPI_ISL_1354649, EPI_ISL_1354650, EPI_ISL_1354651, EPI_ISL_1354652, EPI_ISL_1354653, EPI_ISL_1354654, EPI_ISL_1354655, EPI_ISL_1354656, EPI_ISL_1354657, EPI_ISL_1354658, EPI_ISL_1354659, EPI_ISL_1354660, EPI_ISL_1354661, EPI_ISL_1354664, EPI_ISL_1354665, EPI_ISL_1354667, EPI_ISL_1354668, EPI_ISL_1354669, EPI_ISL_1354670, EPI_ISL_1354671, EPI_ISL_1354672, EPI_ISL_1354673, EPI_ISL_1354674, EPI_ISL_1354675, EPI_ISL_1354677, EPI_ISL_1354678, EPI_ISL_1354679, EPI_ISL_1354680, EPI_ISL_1354681, EPI_ISL_1354682, EPI_ISL_1354683, EPI_ISL_1354684, EPI_ISL_1354685, EPI_ISL_1354686, EPI_ISL_1434520, EPI_ISL_1434521, EPI_ISL_1434524, EPI_ISL_1434526, EPI_ISL_1434528, EPI_ISL_1434530, EPI_ISL_1434532, EPI_ISL_1434534, EPI_ISL_1434536, EPI_ISL_1434538, EPI_ISL_1434540, EPI_ISL_1434542, EPI_ISL_1434544, EPI_ISL_1434546, EPI_ISL_1434548, EPI_ISL_1434550, EPI_ISL_1434552, EPI_ISL_1434554, EPI_ISL_1434556, EPI_ISL_1434559, EPI_ISL_1434561, EPI_ISL_1434563, EPI_ISL_1434567, EPI_ISL_1434568, EPI_ISL_1434570, EPI_ISL_1434572, EPI_ISL_1434574, EPI_ISL_1434576, EPI_ISL_1434578, EPI_ISL_1434581, EPI_ISL_1434582, EPI_ISL_1434584, EPI_ISL_1434586, EPI_ISL_1434588, EPI_ISL_1434590, EPI_ISL_1434592, EPI_ISL_1434594, EPI_ISL_1434596, EPI_ISL_1434598, EPI_ISL_1434600, EPI_ISL_1434601, EPI_ISL_1434603, EPI_ISL_1434605, EPI_ISL_1434607, EPI_ISL_1434609, EPI_ISL_1434611, EPI_ISL_1434613, EPI_ISL_1439438, EPI_ISL_1439625, EPI_ISL_1439629, EPI_ISL_1439631, EPI_ISL_1439633, EPI_ISL_1439635, EPI_ISL_1439637, EPI_ISL_1439641, EPI_ISL_1439642, EPI_ISL_1439646, EPI_ISL_1440873, EPI_ISL_1440874, EPI_ISL_1440875, EPI_ISL_1440876, EPI_ISL_1440877, EPI_ISL_1440878, EPI_ISL_1440881, EPI_ISL_1440882, EPI_ISL_1440883, EPI_ISL_1440884, EPI_ISL_1440886, EPI_ISL_1440887, EPI_ISL_1440888, EPI_ISL_1440889, EPI_ISL_1440890, EPI_ISL_1440891, EPI_ISL_1440892, EPI_ISL_1440893, EPI_ISL_1440894, EPI_ISL_1440895, EPI_ISL_1440896, EPI_ISL_1440897, EPI_ISL_1440898, EPI_ISL_1440899, EPI_ISL_1440901, EPI_ISL_1440902, EPI_ISL_1440903, EPI_ISL_1440905, EPI_ISL_1440906, EPI_ISL_1440907, EPI_ISL_1440908, EPI_ISL_1440909, EPI_ISL_1440911, EPI_ISL_1440912, EPI_ISL_1440913, EPI_ISL_1440914, EPI_ISL_1440915, EPI_ISL_1440916, EPI_ISL_1440917, EPI_ISL_1440918, EPI_ISL_1440919, EPI_ISL_1440920, EPI_ISL_1440921, EPI_ISL_1440922, EPI_ISL_1440923, EPI_ISL_1440924, EPI_ISL_1440925, EPI_ISL_1440926, EPI_ISL_1440927, EPI_ISL_1440928, EPI_ISL_1440929, EPI_ISL_1440930, EPI_ISL_1440931, EPI_ISL_1440932, EPI_ISL_1440933, EPI_ISL_1440934, EPI_ISL_1440936, EPI_ISL_1440937, EPI_ISL_1440938, EPI_ISL_1440939, EPI_ISL_1440940, EPI_ISL_1440941, EPI_ISL_1440942, EPI_ISL_1440943, EPI_ISL_1440944, EPI_ISL_1440945, EPI_ISL_1440946, EPI_ISL_1440947, EPI_ISL_1440948, EPI_ISL_1440949, EPI_ISL_1440950, EPI_ISL_1566009, EPI_ISL_1640675, EPI_ISL_1640711 | | | |
| see above | MVZ Labor Krone GbR | Robert Koch Institute | |
| EPI_ISL_1722068, EPI_ISL_1722069 | MVZ Labor Passau GbR | Robert Koch Institute | |
| EPI_ISL_1435298, EPI_ISL_1435306, EPI_ISL_1435310, EPI_ISL_1441219, EPI_ISL_1563889, EPI_ISL_1563893, EPI_ISL_1563896, EPI_ISL_1563897, EPI_ISL_1563898, EPI_ISL_1563904, EPI_ISL_1563905, EPI_ISL_1563916, EPI_ISL_1563925 | | | |
| see above | MVZ Medizinisches Labor Hannover GmbH | Robert Koch Institute | |
| EPI_ISL_1567562, EPI_ISL_1567574, EPI_ISL_1567655, EPI_ISL_1567658, EPI_ISL_1567660, EPI_ISL_1567661, EPI_ISL_1567663, EPI_ISL_1567684, EPI_ISL_1570824, EPI_ISL_1570826, EPI_ISL_1570830, EPI_ISL_1570835, EPI_ISL_1570838, EPI_ISL_1570885, EPI_ISL_1643770, EPI_ISL_1643776 | | | |
| see above | MVZ fÃ¼r Laboratoriumsmedizin und Mikrobiologie Koblenz-Mittelrhein (Labor Koblenz) | Robert Koch Institute | |
| EPI_ISL_1353365, EPI_ISL_1353380, EPI_ISL_1433501, EPI_ISL_1433503, EPI_ISL_1433524, EPI_ISL_1433559, EPI_ISL_1433565 | | | |
| see above | MVZ für Laboratoriumsmedizin und Mikrobiologie Koblenz-Mittelrhein (Labor Koblenz) | Robert Koch Institute | |
| EPI_ISL_1435354 | MVZ für Laboratoriumsmedizin und Mikrobiologie Würzburg | Robert Koch Institute | |
| EPI_ISL_1752352, EPI_ISL_1752354, EPI_ISL_1752368, EPI_ISL_1752369, EPI_ISL_1752370, EPI_ISL_1752371, EPI_ISL_1752372, EPI_ISL_1752373, EPI_ISL_1752374, EPI_ISL_1752375, EPI_ISL_1752376, EPI_ISL_1752393, EPI_ISL_1752394, EPI_ISL_1752395, EPI_ISL_1752396, EPI_ISL_1752397, EPI_ISL_1752409, EPI_ISL_1752416, EPI_ISL_1752417, EPI_ISL_1752418, EPI_ISL_1752419, EPI_ISL_1752421, EPI_ISL_1752423, EPI_ISL_1752425, EPI_ISL_1752426, EPI_ISL_1752427, EPI_ISL_1752428, EPI_ISL_1752429, EPI_ISL_1752430, EPI_ISL_1752431, EPI_ISL_1752432, EPI_ISL_1752433, EPI_ISL_1752434, EPI_ISL_2006675, EPI_ISL_2006676, EPI_ISL_2006677, EPI_ISL_2094526, EPI_ISL_2094583, EPI_ISL_2094623, EPI_ISL_2094650, EPI_ISL_2094678, EPI_ISL_2094679, EPI_ISL_2094680, EPI_ISL_2094693, EPI_ISL_2094694, EPI_ISL_2094695, EPI_ISL_2094696, EPI_ISL_2094769, EPI_ISL_2094774, EPI_ISL_2094785, EPI_ISL_2094786, EPI_ISL_2094787, EPI_ISL_2094788, EPI_ISL_2094789, EPI_ISL_2094790, EPI_ISL_2094791, EPI_ISL_2094793, EPI_ISL_2094800, EPI_ISL_2094801, EPI_ISL_2094802, EPI_ISL_2094803, EPI_ISL_2094804, EPI_ISL_2094805, EPI_ISL_2094806, EPI_ISL_2094851, EPI_ISL_2094852, EPI_ISL_2094853, EPI_ISL_2094854, EPI_ISL_2094855, EPI_ISL_2094856, EPI_ISL_2094889, EPI_ISL_2094891, EPI_ISL_2095002, EPI_ISL_2095003, EPI_ISL_2095004, EPI_ISL_2095005, EPI_ISL_2095006, EPI_ISL_2095007, EPI_ISL_2095008, EPI_ISL_2095009, EPI_ISL_2095010, EPI_ISL_2095011, EPI_ISL_2095013, EPI_ISL_2095014, EPI_ISL_2095022, EPI_ISL_2095023, EPI_ISL_2095024, EPI_ISL_2095025, EPI_ISL_2095026, EPI_ISL_2095070, EPI_ISL_2095072, EPI_ISL_2095075, EPI_ISL_2095083, EPI_ISL_2095090, EPI_ISL_2095100, EPI_ISL_2095111, EPI_ISL_2095112, EPI_ISL_2095113, EPI_ISL_2095114, EPI_ISL_2095122, EPI_ISL_2095134, EPI_ISL_2095140, EPI_ISL_2095143, EPI_ISL_2095154, EPI_ISL_2095167, EPI_ISL_2095184, EPI_ISL_2095188, EPI_ISL_2095189, EPI_ISL_2095235, EPI_ISL_2095261, EPI_ISL_2095268, EPI_ISL_2095285, EPI_ISL_2095287, EPI_ISL_2095288, EPI_ISL_2095292, EPI_ISL_2095302, EPI_ISL_2095304 | | | |
| see above | Max von Pettenkofer Institute, Virology, National Reference Center for Retroviruses, LMU Munich | Laboratory for Functional Genome Analysis; Dept. Genomics; Gene Center of the LMU Munich | Alexander Graf; Helmut Blum; Max Muenchhoff; Oliver Keppler; Stefan Krebs |
| EPI_ISL_1353597, EPI_ISL_1353600, EPI_ISL_1353602, EPI_ISL_1353610, EPI_ISL_1353613, EPI_ISL_1353614, EPI_ISL_1353617, EPI_ISL_1353620, EPI_ISL_1353634, EPI_ISL_1353635 | | | |
| see above | Med. Labor Prof. Schenk Dr. Ansorge & Kollegen | Robert Koch Institute | |
| EPI_ISL_1989341, EPI_ISL_1989342, EPI_ISL_1989343, EPI_ISL_1989344, EPI_ISL_1989345, EPI_ISL_1989346, EPI_ISL_1989347, EPI_ISL_1989348, EPI_ISL_1989349, EPI_ISL_1989350, EPI_ISL_1989351, EPI_ISL_1989352, EPI_ISL_1989353, EPI_ISL_1989355, EPI_ISL_1989356, EPI_ISL_1989357, EPI_ISL_1989358, EPI_ISL_1989359, EPI_ISL_1989360, EPI_ISL_1989361, EPI_ISL_1989362, EPI_ISL_1989365, EPI_ISL_1989366, EPI_ISL_1989367, EPI_ISL_1989368, EPI_ISL_1989369, EPI_ISL_1989370, EPI_ISL_1989371, EPI_ISL_1989372, EPI_ISL_1989373, EPI_ISL_1989374, EPI_ISL_1989375, EPI_ISL_1989376, EPI_ISL_1989377, EPI_ISL_1989378, EPI_ISL_1989379, EPI_ISL_1989380, EPI_ISL_1989381, EPI_ISL_1989382, EPI_ISL_1989383, EPI_ISL_1989461, EPI_ISL_1989462, EPI_ISL_1989463, EPI_ISL_1989464, EPI_ISL_1989465, EPI_ISL_1989466, EPI_ISL_1989467, EPI_ISL_1989468, EPI_ISL_1989469, EPI_ISL_1989470, EPI_ISL_1989471, EPI_ISL_1989472, EPI_ISL_1989473, EPI_ISL_1989474, EPI_ISL_1989475, EPI_ISL_1989476, EPI_ISL_1989477, EPI_ISL_1989478, EPI_ISL_1989479, EPI_ISL_1989480, EPI_ISL_1989481, EPI_ISL_1989482, EPI_ISL_1989483, EPI_ISL_1989484, EPI_ISL_1989485, EPI_ISL_1989486, EPI_ISL_1989487, EPI_ISL_1989488, EPI_ISL_1989489, EPI_ISL_1989490, EPI_ISL_1989491, EPI_ISL_1989493, EPI_ISL_1989494 | | | |
| see above | Medical Laboratories Duesseldorf | Center of Medical Microbiology, Virology, and Hospital Hygiene, University of Duesseldorf | Alexander Dilthey; Andreas Walker; Angelika Helmer; Christian Lange; Daniel Strelow; Jessica Nicolai; Jörg Timm; Klaus Pfeffer; Lisanna Hülse; Malte Kohns Vasconcelos; Maximilian Damagnez; Nadine Lübke; Tobias Wienemann; Torsten Houwaart |
| EPI_ISL_1439821, EPI_ISL_1439824, EPI_ISL_1439826, EPI_ISL_1439841, EPI_ISL_1439847, EPI_ISL_1439850, EPI_ISL_1439867 | | | |

see above | Medizinisch-Diagnostisches Labor Kempten allgäulab | Robert Koch Institute

EPI_ISL_1436580, EPI_ISL_1436581, EPI_ISL_1436582, EPI_ISL_1436583, EPI_ISL_1436584, EPI_ISL_1436585, EPI_ISL_1436586, EPI_ISL_1436587, EPI_ISL_1436588, EPI_ISL_1436589, EPI_ISL_1436590, EPI_ISL_1436591, EPI_ISL_1436592, EPI_ISL_1436593, EPI_ISL_1436594, EPI_ISL_1436595, EPI_ISL_1436596, EPI_ISL_1436597, EPI_ISL_1436598, EPI_ISL_1436599, EPI_ISL_1436600, EPI_ISL_1436601, EPI_ISL_1436602, EPI_ISL_1436603, EPI_ISL_1436604, EPI_ISL_1436605, EPI_ISL_1436606, EPI_ISL_1436607, EPI_ISL_1436608, EPI_ISL_1436609, EPI_ISL_1436610, EPI_ISL_1436611, EPI_ISL_1436612, EPI_ISL_1436613, EPI_ISL_1436614, EPI_ISL_1436615, EPI_ISL_1436616, EPI_ISL_1436617, EPI_ISL_1436618, EPI_ISL_1436619

see above | Medizinische Laboratorien Düsseldorf | Robert Koch Institute

EPI_ISL_1353143, EPI_ISL_1353145, EPI_ISL_1353149, EPI_ISL_1353151, EPI_ISL_1353153, EPI_ISL_1353154, EPI_ISL_1353156, EPI_ISL_1353157, EPI_ISL_1353158, EPI_ISL_1353159, EPI_ISL_1353162, EPI_ISL_1353163, EPI_ISL_1353165, EPI_ISL_1353168, EPI_ISL_1353171, EPI_ISL_1353175, EPI_ISL_1353176, EPI_ISL_1353183, EPI_ISL_1353187, EPI_ISL_1353188, EPI_ISL_1353192, EPI_ISL_1353193, EPI_ISL_1353194, EPI_ISL_1353195, EPI_ISL_1353198, EPI_ISL_1353201, EPI_ISL_1353203, EPI_ISL_1353204, EPI_ISL_1353205, EPI_ISL_1353208, EPI_ISL_1353209, EPI_ISL_1353210, EPI_ISL_1353216, EPI_ISL_1354364, EPI_ISL_1354367, EPI_ISL_1354369, EPI_ISL_1354370, EPI_ISL_1354372, EPI_ISL_1354373, EPI_ISL_1354375, EPI_ISL_1354376, EPI_ISL_1354377, EPI_ISL_1354380, EPI_ISL_1354381, EPI_ISL_1354382, EPI_ISL_1354385, EPI_ISL_1354396, EPI_ISL_1354398, EPI_ISL_1354405, EPI_ISL_1354407, EPI_ISL_1354414, EPI_ISL_1354415, EPI_ISL_1354417, EPI_ISL_1354422, EPI_ISL_1354423, EPI_ISL_1354426, EPI_ISL_1354433, EPI_ISL_1354434, EPI_ISL_1354439, EPI_ISL_1354447, EPI_ISL_1354448, EPI_ISL_1354449, EPI_ISL_1354451, EPI_ISL_1354456, EPI_ISL_1354459, EPI_ISL_1434437, EPI_ISL_1434441, EPI_ISL_1434456, EPI_ISL_1434458, EPI_ISL_1434461, EPI_ISL_1434473, EPI_ISL_1434475, EPI_ISL_1434477, EPI_ISL_1434483, EPI_ISL_1434487, EPI_ISL_1434492, EPI_ISL_1434498, EPI_ISL_1434500, EPI_ISL_1434502, EPI_ISL_1434504, EPI_ISL_1434508, EPI_ISL_1434512, EPI_ISL_1438586, EPI_ISL_1438587, EPI_ISL_1438588, EPI_ISL_1438589, EPI_ISL_1438592, EPI_ISL_1438593, EPI_ISL_1438597, EPI_ISL_1438601, EPI_ISL_1438603, EPI_ISL_1438608, EPI_ISL_1438615, EPI_ISL_1438619, EPI_ISL_1438621, EPI_ISL_1438622, EPI_ISL_1438623, EPI_ISL_1438627, EPI_ISL_1438628, EPI_ISL_1438632, EPI_ISL_1438633, EPI_ISL_1438636, EPI_ISL_1438638, EPI_ISL_1438640, EPI_ISL_1438643, EPI_ISL_1438645, EPI_ISL_1438648, EPI_ISL_1438651, EPI_ISL_1438654, EPI_ISL_1438657, EPI_ISL_1438659, EPI_ISL_1438661, EPI_ISL_1438668, EPI_ISL_1438679

see above | Medizinisches Labor Ostsachsen MVZ GbRNebenbetriebsstätte Dresden | Robert Koch Institute

EPI_ISL_1353302, EPI_ISL_1353309, EPI_ISL_1353320, EPI_ISL_1353322, EPI_ISL_1353325, EPI_ISL_1353346, EPI_ISL_1353348, EPI_ISL_1353350, EPI_ISL_1353353

see above | Medizinisches Labor Wahl Lüdenscheid | Robert Koch Institute

EPI_ISL_1435384 | Medizinisches Versorgungszentrum für Labormedizin und Mikrobiologie Ruhr GmbH - mvzlm RUHR GmbH | Robert Koch Institute

EPI_ISL_1357342, EPI_ISL_1357381, EPI_ISL_1357384, EPI_ISL_1357393, EPI_ISL_1357396, EPI_ISL_1357406, EPI_ISL_1357424, EPI_ISL_1357426, EPI_ISL_1357427, EPI_ISL_1357432, EPI_ISL_1357444, EPI_ISL_1357452, EPI_ISL_1357457, EPI_ISL_1357459, EPI_ISL_1357463, EPI_ISL_1357466, EPI_ISL_1357486, EPI_ISL_1357487, EPI_ISL_1357490, EPI_ISL_1357492, EPI_ISL_1357494, EPI_ISL_1357498, EPI_ISL_1357499, EPI_ISL_1357500, EPI_ISL_1357501, EPI_ISL_1357502, EPI_ISL_1357506, EPI_ISL_1357508, EPI_ISL_1357514, EPI_ISL_1357517, EPI_ISL_1357530, EPI_ISL_1357532, EPI_ISL_1357533, EPI_ISL_1357538, EPI_ISL_1357540, EPI_ISL_1357543, EPI_ISL_1357545, EPI_ISL_1357548, EPI_ISL_1357552, EPI_ISL_1357554, EPI_ISL_1357560, EPI_ISL_1357561, EPI_ISL_1357565, EPI_ISL_1357567, EPI_ISL_1357569, EPI_ISL_1357570, EPI_ISL_1357571, EPI_ISL_1357575, EPI_ISL_1357577, EPI_ISL_1357578, EPI_ISL_1357581, EPI_ISL_1357584, EPI_ISL_1357592, EPI_ISL_1357594, EPI_ISL_1434843, EPI_ISL_1434853, EPI_ISL_1434861, EPI_ISL_1435255, EPI_ISL_1435275, EPI_ISL_1435277

see above | Medizinisches Zentrallabor Altenburg GmbH & Co.KG | Robert Koch Institute

EPI_ISL_1350938, EPI_ISL_1350942, EPI_ISL_1350943, EPI_ISL_1350944, EPI_ISL_1350945, EPI_ISL_1350946, EPI_ISL_1350948, EPI_ISL_1436808, EPI_ISL_1436809, EPI_ISL_1436810, EPI_ISL_1436811, EPI_ISL_1436812, EPI_ISL_1436813, EPI_ISL_1436814, EPI_ISL_1436815, EPI_ISL_1436816, EPI_ISL_1436818, EPI_ISL_1436819, EPI_ISL_1436820, EPI_ISL_1436821, EPI_ISL_1436822, EPI_ISL_1436823, EPI_ISL_1436824, EPI_ISL_1436825, EPI_ISL_1436826, EPI_ISL_1436827, EPI_ISL_1436828

see above | Niedersächsisches Landesgesundheitsamt (NLGA) | Robert Koch Institute

EPI_ISL_1354395 | Oberlausitz Kliniken gGmBH | Robert Koch Institute

EPI_ISL_1436971, EPI_ISL_1437017, EPI_ISL_1437279 | Praxisgemeinschaft für Laboratoriumsmedizin Labor Blumenstraße; Praxis Dr. E. Wietschel & Dr. F. Wietschel | Robert Koch Institute

EPI_ISL_1350692, EPI_ISL_1352218, EPI_ISL_1352232, EPI_ISL_1352736, EPI_ISL_1352809, EPI_ISL_1352827, EPI_ISL_1352833, EPI_ISL_1436976, EPI_ISL_1436987

see above | Praxisgemeinschaft für Laboratoriumsmedizin Labor Blumenstraße; Praxis Dr. Sel und Dr. Späte | Robert Koch Institute

EPI_ISL_1350265, EPI_ISL_1435362, EPI_ISL_1435379, EPI_ISL_1435382, EPI_ISL_1435388, EPI_ISL_1435400, EPI_ISL_1439454, EPI_ISL_1439458, EPI_ISL_1439460, EPI_ISL_1439485, EPI_ISL_1439489

see above | Procomcure Biotech Germany GmbH | Robert Koch Institute

EPI_ISL_1353647, EPI_ISL_1353704, EPI_ISL_1353748, EPI_ISL_1353843, EPI_ISL_1353911, EPI_ISL_1353918, EPI_ISL_1353946, EPI_ISL_1439128, EPI_ISL_1642434, EPI_ISL_1642579, EPI_ISL_1642608, EPI_ISL_1642691, EPI_ISL_1642696

see above | Robert-Bosch-Krankenhaus | Robert Koch Institute

EPI_ISL_1286091, EPI_ISL_1348050, EPI_ISL_1348054, EPI_ISL_1348113, EPI_ISL_1348187, EPI_ISL_1348192, EPI_ISL_1348214, EPI_ISL_1348221, EPI_ISL_1348253, EPI_ISL_1348273, EPI_ISL_1348298, EPI_ISL_1348316, EPI_ISL_1348330, EPI_ISL_1348350, EPI_ISL_1348354, EPI_ISL_1348359, EPI_ISL_1348385, EPI_ISL_1348388, EPI_ISL_1348392, EPI_ISL_1348399, EPI_ISL_1348402, EPI_ISL_1348403, EPI_ISL_1348409, EPI_ISL_1348442, EPI_ISL_1348445, EPI_ISL_1350617, EPI_ISL_1350619, EPI_ISL_1350621, EPI_ISL_1350623, EPI_ISL_1350626, EPI_ISL_1350628, EPI_ISL_1350630, EPI_ISL_1350635, EPI_ISL_1350636, EPI_ISL_1350637, EPI_ISL_1350638, EPI_ISL_1350639, EPI_ISL_1350644, EPI_ISL_1350645, EPI_ISL_1350647, EPI_ISL_1350652, EPI_ISL_1350653, EPI_ISL_1350657, EPI_ISL_1350658, EPI_ISL_1350659, EPI_ISL_1350667, EPI_ISL_1350670, EPI_ISL_1350672, EPI_ISL_1350675, EPI_ISL_1350679, EPI_ISL_1350682, EPI_ISL_1350684, EPI_ISL_1350686, EPI_ISL_1350691, EPI_ISL_1350696, EPI_ISL_1350699, EPI_ISL_1350700, EPI_ISL_1350701, EPI_ISL_1350702, EPI_ISL_1350706, EPI_ISL_1350711, EPI_ISL_1350715, EPI_ISL_1350716, EPI_ISL_1350722, EPI_ISL_1350725, EPI_ISL_1350729, EPI_ISL_1350730, EPI_ISL_1350732, EPI_ISL_1350733, EPI_ISL_1350735, EPI_ISL_1350736, EPI_ISL_1350738, EPI_ISL_1350739, EPI_ISL_1350740, EPI_ISL_1350742, EPI_ISL_1350743, EPI_ISL_1350744, EPI_ISL_1350745, EPI_ISL_1350748, EPI_ISL_1350749, EPI_ISL_1350750, EPI_ISL_1350751, EPI_ISL_1350752, EPI_ISL_1350754, EPI_ISL_1350755, EPI_ISL_1350757, EPI_ISL_1350761, EPI_ISL_1350762, EPI_ISL_1350763, EPI_ISL_1350766, EPI_ISL_1351776, EPI_ISL_1351779, EPI_ISL_1351782, EPI_ISL_1351785, EPI_ISL_1351788, EPI_ISL_1351794, EPI_ISL_1351800, EPI_ISL_1351801, EPI_ISL_1351806, EPI_ISL_1351812, EPI_ISL_1351814, EPI_ISL_1351819, EPI_ISL_1351820, EPI_ISL_1351825, EPI_ISL_1351828, EPI_ISL_1351840, EPI_ISL_1351847, EPI_ISL_1351849, EPI_ISL_1351856, EPI_ISL_1351861, EPI_ISL_1351864, EPI_ISL_1351871, EPI_ISL_1351891, EPI_ISL_1351897, EPI_ISL_1351898, EPI_ISL_1351903, EPI_ISL_1351907, EPI_ISL_1351915, EPI_ISL_1351920, EPI_ISL_1351939, EPI_ISL_1351942, EPI_ISL_1351955, EPI_ISL_1351959, EPI_ISL_1351970, EPI_ISL_1351986, EPI_ISL_1351988, EPI_ISL_1352003, EPI_ISL_1352017, EPI_ISL_1352022, EPI_ISL_1352028, EPI_ISL_1352040, EPI_ISL_1352042, EPI_ISL_1352044, EPI_ISL_1352046, EPI_ISL_1352064, EPI_ISL_1352067, EPI_ISL_1352069, EPI_ISL_1352074, EPI_ISL_1352077, EPI_ISL_1352084, EPI_ISL_1352086, EPI_ISL_1352087, EPI_ISL_1352089, EPI_ISL_1352095, EPI_ISL_1352096, EPI_ISL_1352102, EPI_ISL_1352103, EPI_ISL_1352105, EPI_ISL_1352111, EPI_ISL_1352113, EPI_ISL_1352117, EPI_ISL_1352120, EPI_ISL_1352126, EPI_ISL_1352127, EPI_ISL_1352130, EPI_ISL_1352132, EPI_ISL_1352135, EPI_ISL_1352138, EPI_ISL_1352139, EPI_ISL_1352144, EPI_ISL_1352145, EPI_ISL_1352147, EPI_ISL_1352148, EPI_ISL_1352149, EPI_ISL_1352152, EPI_ISL_1352154, EPI_ISL_1352155, EPI_ISL_1352159, EPI_ISL_1352162, EPI_ISL_1352164, EPI_ISL_1352167, EPI_ISL_1352169, EPI_ISL_1352170, EPI_ISL_1352172, EPI_ISL_1352174, EPI_ISL_1352178, EPI_ISL_1352180, EPI_ISL_1352182, EPI_ISL_1352184, EPI_ISL_1352185, EPI_ISL_1352186, EPI_ISL_1352187, EPI_ISL_1352188, EPI_ISL_1352190, EPI_ISL_1352192, EPI_ISL_1352194, EPI_ISL_1352197, EPI_ISL_1352198, EPI_ISL_1352199, EPI_ISL_1352202, EPI_ISL_1352208, EPI_ISL_1352210, EPI_ISL_1352213, EPI_ISL_1352216, EPI_ISL_1352217, EPI_ISL_1352221, EPI_ISL_1352228, EPI_ISL_1352229, EPI_ISL_1352231, EPI_ISL_1352233, EPI_ISL_1352235, EPI_ISL_1352695, EPI_ISL_1352706, EPI_ISL_1352712, EPI_ISL_1352716, EPI_ISL_1352719, EPI_ISL_1352721, EPI_ISL_1352749, EPI_ISL_1352755, EPI_ISL_1352757, EPI_ISL_1352758, EPI_ISL_1352761, EPI_ISL_1352764, EPI_ISL_1352766, EPI_ISL_1352768, EPI_ISL_1352782, EPI_ISL_1352787, EPI_ISL_1352792, EPI_ISL_1352795, EPI_ISL_1352804, EPI_ISL_1352810, EPI_ISL_1352812, EPI_ISL_1352816, EPI_ISL_1352822, EPI_ISL_1352823, EPI_ISL_1352831, EPI_ISL_1352836, EPI_ISL_1352838, EPI_ISL_1352839, EPI_ISL_1352844, EPI_ISL_1352845, EPI_ISL_1352846, EPI_ISL_1352848, EPI_ISL_1352850, EPI_ISL_1352851, EPI_ISL_1352852, EPI_ISL_1352853, EPI_ISL_1352854, EPI_ISL_1352858, EPI_ISL_1352860, EPI_ISL_1352861, EPI_ISL_1352862, EPI_ISL_1355899, EPI_ISL_1355901, EPI_ISL_1355902, EPI_ISL_1355903, EPI_ISL_1355904, EPI_ISL_1355906, EPI_ISL_1355908, EPI_ISL_1355909, EPI_ISL_1355910, EPI_ISL_1355911, EPI_ISL_1355912, EPI_ISL_1355921, EPI_ISL_1355922, EPI_ISL_1355923, EPI_ISL_1355926, EPI_ISL_1355927, EPI_ISL_1355934, EPI_ISL_1355938, EPI_ISL_1355943, EPI_ISL_1355948, EPI_ISL_1355950, EPI_ISL_1355952, EPI_ISL_1355957, EPI_ISL_1355959, EPI_ISL_1355966, EPI_ISL_1355967, EPI_ISL_1355972, EPI_ISL_1355973, EPI_ISL_1355978, EPI_ISL_1355980, EPI_ISL_1355983, EPI_ISL_1355986, EPI_ISL_1355988, EPI_ISL_1355989, EPI_ISL_1355991, EPI_ISL_1355993, EPI_ISL_1355994, EPI_ISL_1355995, EPI_ISL_1356000, EPI_ISL_1356004, EPI_ISL_1356007, EPI_ISL_1356011, EPI_ISL_1356014, EPI_ISL_1356015, EPI_ISL_1356019, EPI_ISL_1356020, EPI_ISL_1356022, EPI_ISL_1356028, EPI_ISL_1356034, EPI_ISL_1356036, EPI_ISL_1356038, EPI_ISL_1356042, EPI_ISL_1356043, EPI_ISL_1356048, EPI_ISL_1356050, EPI_ISL_1356052, EPI_ISL_1356054, EPI_ISL_1356055, EPI_ISL_1356058, EPI_ISL_1356059, EPI_ISL_1356063, EPI_ISL_1356072, EPI_ISL_1356073, EPI_ISL_1356075, EPI_ISL_1356081, EPI_ISL_1356083, EPI_ISL_1356085, EPI_ISL_1356110, EPI_ISL_1356115, EPI_ISL_1356119, EPI_ISL_1356123, EPI_ISL_1356125, EPI_ISL_1356126, EPI_ISL_1356130, EPI_ISL_1356133, EPI_ISL_1356135, EPI_ISL_1356140, EPI_ISL_1356142, EPI_ISL_1356143, EPI_ISL_1356147, EPI_ISL_1356150, EPI_ISL_1356155, EPI_ISL_1356161, EPI_ISL_1356172, EPI_ISL_1356176, EPI_ISL_1356182, EPI_ISL_1356183, EPI_ISL_1356184, EPI_ISL_1356186, EPI_ISL_1356188, EPI_ISL_1356189, EPI_ISL_1356191, EPI_ISL_1356196, EPI_ISL_1356204, EPI_ISL_1356209, EPI_ISL_1356210, EPI_ISL_1356214, EPI_ISL_1356215, EPI_ISL_1356218, EPI_ISL_1356220, EPI_ISL_1356227, EPI_ISL_1356229, EPI_ISL_1356230, EPI_ISL_1356231, EPI_ISL_1356233, EPI_ISL_1356234, EPI_ISL_1357337, EPI_ISL_1357338, EPI_ISL_1357339, EPI_ISL_1357340, EPI_ISL_1357341, EPI_ISL_1357344, EPI_ISL_1357345, EPI_ISL_1357347, EPI_ISL_1357348, EPI_ISL_1357350, EPI_ISL_1357355, EPI_ISL_1357358, EPI_ISL_1357360, EPI_ISL_1357364, EPI_ISL_1357365, EPI_ISL_1357368, EPI_ISL_1357371, EPI_ISL_1357376, EPI_ISL_1357382, EPI_ISL_1357383, EPI_ISL_1357386, EPI_ISL_1357391, EPI_ISL_1357398, EPI_ISL_1357401, EPI_ISL_1357402, EPI_ISL_1357404, EPI_ISL_1357405, EPI_ISL_1357409, EPI_ISL_1357410, EPI_ISL_1357411, EPI_ISL_1357415, EPI_ISL_1357416, EPI_ISL_1357418, EPI_ISL_1357420, EPI_ISL_1357425, EPI_ISL_1357428, EPI_ISL_1357429, EPI_ISL_1357431, EPI_ISL_1357433, EPI_ISL_1357437, EPI_ISL_1357440, EPI_ISL_1357442, EPI_ISL_1357443, EPI_ISL_1357447, EPI_ISL_1357448, EPI_ISL_1357449, EPI_ISL_1357451, EPI_ISL_1357453, EPI_ISL_1357460, EPI_ISL_1357465, EPI_ISL_1357473, EPI_ISL_1357474, EPI_ISL_1357475, EPI_ISL_1357476, EPI_ISL_1357478, EPI_ISL_1357480, EPI_ISL_1357481, EPI_ISL_1357483, EPI_ISL_1357491, EPI_ISL_1357497, EPI_ISL_1357505, EPI_ISL_1357509, EPI_ISL_1357510, EPI_ISL_1357512, EPI_ISL_1357513, EPI_ISL_1357515, EPI_ISL_1357518, EPI_ISL_1357520, EPI_ISL_1357522, EPI_ISL_1357524, EPI_ISL_1357526, EPI_ISL_1357531, EPI_ISL_1357539, EPI_ISL_1357542, EPI_ISL_1357555, EPI_ISL_1357558, EPI_ISL_1357563, EPI_ISL_1357564, EPI_ISL_1357566, EPI_ISL_1357568, EPI_ISL_1357574, EPI_ISL_1357580, EPI_ISL_1357588, EPI_ISL_1357590, EPI_ISL_1357591, EPI_ISL_1434617, EPI_ISL_1434619, EPI_ISL_1434626, EPI_ISL_1434637, EPI_ISL_1434639, EPI_ISL_1434645, EPI_ISL_1434647, EPI_ISL_1434649, EPI_ISL_1434655, EPI_ISL_1434659, EPI_ISL_1434661, EPI_ISL_1434664, EPI_ISL_1434668, EPI_ISL_1434672, EPI_ISL_1434674, EPI_ISL_1434678, EPI_ISL_1434685, EPI_ISL_1434686, EPI_ISL_1434692, EPI_ISL_1434694, EPI_ISL_1434698, EPI_ISL_1434706, EPI_ISL_1434712, EPI_ISL_1434716, EPI_ISL_1434720, EPI_ISL_1434721, EPI_ISL_1434723, EPI_ISL_1434725, EPI_ISL_1434729, EPI_ISL_1434742, EPI_ISL_1434746, EPI_ISL_1434750, EPI_ISL_1434752, EPI_ISL_1434755, EPI_ISL_1434757, EPI_ISL_1434761, EPI_ISL_1434771, EPI_ISL_1434775, EPI_ISL_1434778, EPI_ISL_1434780, EPI_ISL_1434785, EPI_ISL_1434787, EPI_ISL_1434789, EPI_ISL_1434791, EPI_ISL_1434795, EPI_ISL_1434796, EPI_ISL_1434800, EPI_ISL_1434802, EPI_ISL_1434805, EPI_ISL_1434810, EPI_ISL_1434814, EPI_ISL_1434820, EPI_ISL_1434822, EPI_ISL_1434824, EPI_ISL_1434826, EPI_ISL_1434828, EPI_ISL_1434829, EPI_ISL_1434831, EPI_ISL_1434835, EPI_ISL_1434837, EPI_ISL_1434847, EPI_ISL_1434851, EPI_ISL_1434856, EPI_ISL_1434857, EPI_ISL_1434859, EPI_ISL_1434863, EPI_ISL_1434865, EPI_ISL_1434867, EPI_ISL_1434874, EPI_ISL_1434877, EPI_ISL_1434886, EPI_ISL_1434890, EPI_ISL_1434896, EPI_ISL_1434900, EPI_ISL_1434904, EPI_ISL_1434906, EPI_ISL_1434911, EPI_ISL_1434913, EPI_ISL_1434915, EPI_ISL_1434919, EPI_ISL_1434923, EPI_ISL_1434925, EPI_ISL_1434927, EPI_ISL_1434931, EPI_ISL_1434933, EPI_ISL_1434935, EPI_ISL_1434937, EPI_ISL_1434941, EPI_ISL_1434943, EPI_ISL_1434945, EPI_ISL_1434947, EPI_ISL_1434951, EPI_ISL_1434953, EPI_ISL_1434955, EPI_ISL_1434957, EPI_ISL_1434958, EPI_ISL_1434960, EPI_ISL_1434962, EPI_ISL_1434964, EPI_ISL_1434966, EPI_ISL_1434968, EPI_ISL_1434972, EPI_ISL_1434974, EPI_ISL_1434976, EPI_ISL_1434978, EPI_ISL_1434984, EPI_ISL_1434988, EPI_ISL_1434999, EPI_ISL_1435003, EPI_ISL_1435005, EPI_ISL_1435009, EPI_ISL_1435011, EPI_ISL_1435016, EPI_ISL_1435018, EPI_ISL_1435020, EPI_ISL_1435025, EPI_ISL_1435031, EPI_ISL_1435033, EPI_ISL_1435037, EPI_ISL_1435039, EPI_ISL_1435044, EPI_ISL_1435049, EPI_ISL_1435050, EPI_ISL_1435052, EPI_ISL_1435054, EPI_ISL_1435055, EPI_ISL_1435059, EPI_ISL_1435061, EPI_ISL_1435063, EPI_ISL_1435072, EPI_ISL_1435074, EPI_ISL_1435080, EPI_ISL_1435086, EPI_ISL_1435087, EPI_ISL_1435089, EPI_ISL_1435091, EPI_ISL_1435093, EPI_ISL_1435095, EPI_ISL_1435099, EPI_ISL_1435101, EPI_ISL_1435107, EPI_ISL_1435109, EPI_ISL_1435111, EPI_ISL_1435117, EPI_ISL_1435120, EPI_ISL_1435124, EPI_ISL_1435126, EPI_ISL_1435128, EPI_ISL_1435136, EPI_ISL_1435140, EPI_ISL_1435146, EPI_ISL_1435148, EPI_ISL_1435154, EPI_ISL_1435157, EPI_ISL_1435164, EPI_ISL_1435166, EPI_ISL_1435168, EPI_ISL_1435180, EPI_ISL_1435184, EPI_ISL_1435186, EPI_ISL_1435188, EPI_ISL_1435192, EPI_ISL_1435201, EPI_ISL_1435203, EPI_ISL_1435205, EPI_ISL_1435209, EPI_ISL_1435211, EPI_ISL_1435215, EPI_ISL_1435216, EPI_ISL_1435221, EPI_ISL_1435222, EPI_ISL_1435225, EPI_ISL_1435226, EPI_ISL_1435230, EPI_ISL_1435234, EPI_ISL_1435240, EPI_ISL_1435248, EPI_ISL_1435250, EPI_ISL_1435252, EPI_ISL_1435257, EPI_ISL_1435261, EPI_ISL_1435263, EPI_ISL_1435265, EPI_ISL_1435269, EPI_ISL_1435272, EPI_ISL_1435274, EPI_ISL_1435276, EPI_ISL_1435278, EPI_ISL_1435279, EPI_ISL_1435280, EPI_ISL_1435281, EPI_ISL_1435282, EPI_ISL_1435283, EPI_ISL_1435284, EPI_ISL_1436952, EPI_ISL_1436959, EPI_ISL_1436960, EPI_ISL_1436961, EPI_ISL_1436963, EPI_ISL_1436966, EPI_ISL_1436978, EPI_ISL_1436982, EPI_ISL_1436983, EPI_ISL_1436986, EPI_ISL_1437000, EPI_ISL_1437002, EPI_ISL_1437005, EPI_ISL_1437012, EPI_ISL_1437020, EPI_ISL_1437028, EPI_ISL_1437029, EPI_ISL_1437048, EPI_ISL_1437056, EPI_ISL_1437062, EPI_ISL_1437073, EPI_ISL_1437076, EPI_ISL_1437085, EPI_ISL_1437093, EPI_ISL_1437097, EPI_ISL_1437099, EPI_ISL_1437102, EPI_ISL_1437103, EPI_ISL_1437109, EPI_ISL_1437118, EPI_ISL_1437121, EPI_ISL_1437126, EPI_ISL_1437134, EPI_ISL_1437137, EPI_ISL_1437138, EPI_ISL_1437141, EPI_ISL_1437165, EPI_ISL_1437172, EPI_ISL_1437180, EPI_ISL_1437185, EPI_ISL_1437187, EPI_ISL_1437188, EPI_ISL_1437192, EPI_ISL_1437198, EPI_ISL_1437200, EPI_ISL_1437201, EPI_ISL_1437204, EPI_ISL_1437213, EPI_ISL_1437221, EPI_ISL_1437223, EPI_ISL_1437225, EPI_ISL_1437260, EPI_ISL_1437263, EPI_ISL_1437268, EPI_ISL_1438308, EPI_ISL_1438310, EPI_ISL_1438319, EPI_ISL_1438321, EPI_ISL_1438325, EPI_ISL_1438326, EPI_ISL_1438327, EPI_ISL_1438328, EPI_ISL_1438329, EPI_ISL_1438336, EPI_ISL_1438347, EPI_ISL_1438349, EPI_ISL_1438351, EPI_ISL_1438369, EPI_ISL_1438370, EPI_ISL_1438378, EPI_ISL_1438384, EPI_ISL_1438385, EPI_ISL_1438406, EPI_ISL_1438408, EPI_ISL_1438415, EPI_ISL_1438418, EPI_ISL_1438426, EPI_ISL_1438431, EPI_ISL_1438432, EPI_ISL_1438433, EPI_ISL_1438440, EPI_ISL_1438450, EPI_ISL_1438453, EPI_ISL_1438459, EPI_ISL_1438465, EPI_ISL_1438469, EPI_ISL_1438472, EPI_ISL_1438481, EPI_ISL_1438484, EPI_ISL_1438487, EPI_ISL_1438490, EPI_ISL_1438494, EPI_ISL_1438495, EPI_ISL_1438497, EPI_ISL_1438505, EPI_ISL_1438514, EPI_ISL_1438518, EPI_ISL_1438530, EPI_ISL_1438535, EPI_ISL_1438536, EPI_ISL_1438540, EPI_ISL_1438544, EPI_ISL_1438549, EPI_ISL_1438553, EPI_ISL_1438572, EPI_ISL_1438575, EPI_ISL_1438578

see above | SYNLAB Jena Oncoscreen | Robert Koch Institute

EPI_ISL_1350683, EPI_ISL_1350689, EPI_ISL_1350723, EPI_ISL_1351877, EPI_ISL_1351906, EPI_ISL_1351974, EPI_ISL_1352054, EPI_ISL_1352143

see above | SYNLAB Labor München Zentrum LMZ | Robert Koch Institute

EPI_ISL_1348432, EPI_ISL_1348438, EPI_ISL_1348449, EPI_ISL_1350632, EPI_ISL_1350641, EPI_ISL_1350664, EPI_ISL_1350676, EPI_ISL_1350681, EPI_ISL_1350687, EPI_ISL_1350688, EPI_ISL_1350712, EPI_ISL_1351798, EPI_ISL_1351808, EPI_ISL_1351817, EPI_ISL_1351832, EPI_ISL_1351850, EPI_ISL_1351867, EPI_ISL_1351868, EPI_ISL_1351879, EPI_ISL_1351882, EPI_ISL_1351894, EPI_ISL_1351900, EPI_ISL_1351904, EPI_ISL_1351913, EPI_ISL_1351926, EPI_ISL_1351989, EPI_ISL_1352104, EPI_ISL_1352109, EPI_ISL_1352121, EPI_ISL_1352128, EPI_ISL_1352140, EPI_ISL_1352150, EPI_ISL_1352161, EPI_ISL_1352163, EPI_ISL_1352176, EPI_ISL_1352193, EPI_ISL_1352204, EPI_ISL_1352205, EPI_ISL_1352212, EPI_ISL_1352220, EPI_ISL_1352223, EPI_ISL_1352227, EPI_ISL_1352776, EPI_ISL_1352779, EPI_ISL_1352781, EPI_ISL_1352789, EPI_ISL_1352794, EPI_ISL_1352805, EPI_ISL_1352814, EPI_ISL_1352818, EPI_ISL_1352824, EPI_ISL_1352829, EPI_ISL_1352832, EPI_ISL_1352841, EPI_ISL_1352857, EPI_ISL_1355954, EPI_ISL_1355996, EPI_ISL_1356017, EPI_ISL_1356067, EPI_ISL_1356168, EPI_ISL_1356187, EPI_ISL_1356211, EPI_ISL_1356225, EPI_ISL_1356226, EPI_ISL_1356236, EPI_ISL_1356237, EPI_ISL_1356239, EPI_ISL_1356241, EPI_ISL_1434615, EPI_ISL_1434657, EPI_ISL_1434666, EPI_ISL_1434683, EPI_ISL_1434688, EPI_ISL_1434714, EPI_ISL_1434731, EPI_ISL_1434737, EPI_ISL_1434744, EPI_ISL_1434765, EPI_ISL_1434818, EPI_ISL_1434888, EPI_ISL_1434892, EPI_ISL_1434898, EPI_ISL_1434970, EPI_ISL_1435023, EPI_ISL_1435070, EPI_ISL_1435076, EPI_ISL_1435084, EPI_ISL_1435115, EPI_ISL_1435130, EPI_ISL_1435134, EPI_ISL_1435160, EPI_ISL_1435172, EPI_ISL_1435178, EPI_ISL_1435190, EPI_ISL_1435197, EPI_ISL_1435213, EPI_ISL_1435228, EPI_ISL_1435242, EPI_ISL_1435254, EPI_ISL_1435259, EPI_ISL_1435267, EPI_ISL_1435271, EPI_ISL_1435273, EPI_ISL_1436974, EPI_ISL_1436979, EPI_ISL_1436992, EPI_ISL_1436996, EPI_ISL_1437038, EPI_ISL_1437039, EPI_ISL_1437110, EPI_ISL_1437119, EPI_ISL_1437135, EPI_ISL_1437147, EPI_ISL_1437151, EPI_ISL_1437156, EPI_ISL_1437171, EPI_ISL_1437179, EPI_ISL_1437184, EPI_ISL_1437207, EPI_ISL_1437216, EPI_ISL_1437227, EPI_ISL_1437228, EPI_ISL_1437231, EPI_ISL_1437232, EPI_ISL_1437238, EPI_ISL_1437240, EPI_ISL_1437247, EPI_ISL_1437249, EPI_ISL_1437252, EPI_ISL_1437255, EPI_ISL_1437259, EPI_ISL_1438364, EPI_ISL_1438524, EPI_ISL_1438539, EPI_ISL_1438558, EPI_ISL_1566905

see above | SYNLAB MVZ Berlin | Robert Koch Institute

EPI_ISL_1437496, EPI_ISL_1437517, EPI_ISL_1437563, EPI_ISL_1439972, EPI_ISL_1440218, EPI_ISL_1442368, EPI_ISL_1442374, EPI_ISL_1442376, EPI_ISL_1442381, EPI_ISL_1442449, EPI_ISL_1442456, EPI_ISL_1565829, EPI_ISL_1565918, EPI_ISL_1566133, EPI_ISL_1566164, EPI_ISL_1566167, EPI_ISL_1566195, EPI_ISL_1566213, EPI_ISL_1566217, EPI_ISL_1566242, EPI_ISL_1566321, EPI_ISL_1566325, EPI_ISL_1566362, EPI_ISL_1568687, EPI_ISL_1572246, EPI_ISL_1572415, EPI_ISL_1572493

see above | SYNLAB MVZ Bonn | Robert Koch Institute

EPI_ISL_1352954 | SYNLAB MVZ Dachau | Robert Koch Institute

EPI_ISL_1432026, EPI_ISL_1432031, EPI_ISL_1432038, EPI_ISL_1432042, EPI_ISL_1432046, EPI_ISL_1432050, EPI_ISL_1432057, EPI_ISL_1432062, EPI_ISL_1432091, EPI_ISL_1432097, EPI_ISL_1432107, EPI_ISL_1432109, EPI_ISL_1432110, EPI_ISL_1432123, EPI_ISL_1432134, EPI_ISL_1432161, EPI_ISL_1432163, EPI_ISL_1432173, EPI_ISL_1432186, EPI_ISL_1432205, EPI_ISL_1432225, EPI_ISL_1432231, EPI_ISL_1432234, EPI_ISL_1432241, EPI_ISL_1432248, EPI_ISL_1432252, EPI_ISL_1432257, EPI_ISL_1432260, EPI_ISL_1432278, EPI_ISL_1432281, EPI_ISL_1432286, EPI_ISL_1432300, EPI_ISL_1432305, EPI_ISL_1432309, EPI_ISL_1432315, EPI_ISL_1432328, EPI_ISL_1432355, EPI_ISL_1432362, EPI_ISL_1432373, EPI_ISL_1432382, EPI_ISL_1432399, EPI_ISL_1432411, EPI_ISL_1432417, EPI_ISL_1432423, EPI_ISL_1432445, EPI_ISL_1432466, EPI_ISL_1432475, EPI_ISL_1432496, EPI_ISL_1432516, EPI_ISL_1432523, EPI_ISL_1432528, EPI_ISL_1432534, EPI_ISL_1432537, EPI_ISL_1432543, EPI_ISL_1432549, EPI_ISL_1432554, EPI_ISL_1432558, EPI_ISL_1432562, EPI_ISL_1432566, EPI_ISL_1432571, EPI_ISL_1432575, EPI_ISL_1432581, EPI_ISL_1432586, EPI_ISL_1432594, EPI_ISL_1432601, EPI_ISL_1432611, EPI_ISL_1432620, EPI_ISL_1432626, EPI_ISL_1432635, EPI_ISL_1432652, EPI_ISL_1432661, EPI_ISL_1432667, EPI_ISL_1432690, EPI_ISL_1432694, EPI_ISL_1432702, EPI_ISL_1432711, EPI_ISL_1432722, EPI_ISL_1432729, EPI_ISL_1432735, EPI_ISL_1432740, EPI_ISL_1432748, EPI_ISL_1437429, EPI_ISL_1437435, EPI_ISL_1437437, EPI_ISL_1437438, EPI_ISL_1437439, EPI_ISL_1437444, EPI_ISL_1437448, EPI_ISL_1437457, EPI_ISL_1437458, EPI_ISL_1437459, EPI_ISL_1437461, EPI_ISL_1437463, EPI_ISL_1437467, EPI_ISL_1437471, EPI_ISL_1437478, EPI_ISL_1437482, EPI_ISL_1437486, EPI_ISL_1437490, EPI_ISL_1437493, EPI_ISL_1437495, EPI_ISL_1437500, EPI_ISL_1437501, EPI_ISL_1437507, EPI_ISL_1437511, EPI_ISL_1437521, EPI_ISL_1437524, EPI_ISL_1437526, EPI_ISL_1437531, EPI_ISL_1437533, EPI_ISL_1437535, EPI_ISL_1437547, EPI_ISL_1437549, EPI_ISL_1437551, EPI_ISL_1437560, EPI_ISL_1437567, EPI_ISL_1437570, EPI_ISL_1437572, EPI_ISL_1437575, EPI_ISL_1437576, EPI_ISL_1437580, EPI_ISL_1437582, EPI_ISL_1437583, EPI_ISL_1437587, EPI_ISL_1437600, EPI_ISL_1437601, EPI_ISL_1437605, EPI_ISL_1437607, EPI_ISL_1437609, EPI_ISL_1439902, EPI_ISL_1439904, EPI_ISL_1439905, EPI_ISL_1439909, EPI_ISL_1439911, EPI_ISL_1439915, EPI_ISL_1439929, EPI_ISL_1439933, EPI_ISL_1439935, EPI_ISL_1439936, EPI_ISL_1439940, EPI_ISL_1439946, EPI_ISL_1439951, EPI_ISL_1439957, EPI_ISL_1439966, EPI_ISL_1439977, EPI_ISL_1439980, EPI_ISL_1439981, EPI_ISL_1439986, EPI_ISL_1439988, EPI_ISL_1439994, EPI_ISL_1439997, EPI_ISL_1439999, EPI_ISL_1440008, EPI_ISL_1440012, EPI_ISL_1440014, EPI_ISL_1440018, EPI_ISL_1440022, EPI_ISL_1440026, EPI_ISL_1440028, EPI_ISL_1440034, EPI_ISL_1440038, EPI_ISL_1440040, EPI_ISL_1440044, EPI_ISL_1440047, EPI_ISL_1440049, EPI_ISL_1440054, EPI_ISL_1440059, EPI_ISL_1440061, EPI_ISL_1440063, EPI_ISL_1440067, EPI_ISL_1440072, EPI_ISL_1440074, EPI_ISL_1440079, EPI_ISL_1440085, EPI_ISL_1440097, EPI_ISL_1440101, EPI_ISL_1440107, EPI_ISL_1440111, EPI_ISL_1440117, EPI_ISL_1440123, EPI_ISL_1440124, EPI_ISL_1440138, EPI_ISL_1440159, EPI_ISL_1440166, EPI_ISL_1440169, EPI_ISL_1440173, EPI_ISL_1440187, EPI_ISL_1440189, EPI_ISL_1440194, EPI_ISL_1440195, EPI_ISL_1440198, EPI_ISL_1440200, EPI_ISL_1440201, EPI_ISL_1440204, EPI_ISL_1440213, EPI_ISL_1440224, EPI_ISL_1440225, EPI_ISL_1440231, EPI_ISL_1440235, EPI_ISL_1440237, EPI_ISL_1440242, EPI_ISL_1440247, EPI_ISL_1440250, EPI_ISL_1440255, EPI_ISL_1440258, EPI_ISL_1440259, EPI_ISL_1440262, EPI_ISL_1440263, EPI_ISL_1440265, EPI_ISL_1440267, EPI_ISL_1440268, EPI_ISL_1440270, EPI_ISL_1440271, EPI_ISL_1440273, EPI_ISL_1442371, EPI_ISL_1442379, EPI_ISL_1442380, EPI_ISL_1442386, EPI_ISL_1442387, EPI_ISL_1442390, EPI_ISL_1442391, EPI_ISL_1442394, EPI_ISL_1442395, EPI_ISL_1442396, EPI_ISL_1442398, EPI_ISL_1442399, EPI_ISL_1442400, EPI_ISL_1442401, EPI_ISL_1442404, EPI_ISL_1442407, EPI_ISL_1442410, EPI_ISL_1442415, EPI_ISL_1442419, EPI_ISL_1442423, EPI_ISL_1442425, EPI_ISL_1442432, EPI_ISL_1442435, EPI_ISL_1442437, EPI_ISL_1442443, EPI_ISL_1442446, EPI_ISL_1442450, EPI_ISL_1442454, EPI_ISL_1442457, EPI_ISL_1442458, EPI_ISL_1442459, EPI_ISL_1442461, EPI_ISL_1442467, EPI_ISL_1442468, EPI_ISL_1442472, EPI_ISL_1442486, EPI_ISL_1565795, EPI_ISL_1565797, EPI_ISL_1565800, EPI_ISL_1565811, EPI_ISL_1565816, EPI_ISL_1565835, EPI_ISL_1565839, EPI_ISL_1565843, EPI_ISL_1565850, EPI_ISL_1565863, EPI_ISL_1565909, EPI_ISL_1565920, EPI_ISL_1565926, EPI_ISL_1565944, EPI_ISL_1565959, EPI_ISL_1565967, EPI_ISL_1565987, EPI_ISL_1565992, EPI_ISL_1565993, EPI_ISL_1566102, EPI_ISL_1566104, EPI_ISL_1566106, EPI_ISL_1566108, EPI_ISL_1566110, EPI_ISL_1566116, EPI_ISL_1566130, EPI_ISL_1566139, EPI_ISL_1566142, EPI_ISL_1566148, EPI_ISL_1566163, EPI_ISL_1566174, EPI_ISL_1566178, EPI_ISL_1566199, EPI_ISL_1566203, EPI_ISL_1566208, EPI_ISL_1566222, EPI_ISL_1566225, EPI_ISL_1566228, EPI_ISL_1566231, EPI_ISL_1566237, EPI_ISL_1566238, EPI_ISL_1566247, EPI_ISL_1566251, EPI_ISL_1566253, EPI_ISL_1566258, EPI_ISL_1566265, EPI_ISL_1566270, EPI_ISL_1566271, EPI_ISL_1566275, EPI_ISL_1566276, EPI_ISL_1566278, EPI_ISL_1566282, EPI_ISL_1566284, EPI_ISL_1566285, EPI_ISL_1566288, EPI_ISL_1566292, EPI_ISL_1566293, EPI_ISL_1566297, EPI_ISL_1566300, EPI_ISL_1566304, EPI_ISL_1566307, EPI_ISL_1566314, EPI_ISL_1566331, EPI_ISL_1566333, EPI_ISL_1566336, EPI_ISL_1566338, EPI_ISL_1566341, EPI_ISL_1566343, EPI_ISL_1566346, EPI_ISL_1566348, EPI_ISL_1566351, EPI_ISL_1566354, EPI_ISL_1566356, EPI_ISL_1566359, EPI_ISL_1566361, EPI_ISL_1566364, EPI_ISL_1566365, EPI_ISL_1566368, EPI_ISL_1566371, EPI_ISL_1566373,

EPI_ISL_1566377, EPI_ISL_1566379, EPI_ISL_1566381, EPI_ISL_1566383, EPI_ISL_1566384, EPI_ISL_1566386, EPI_ISL_1566387, EPI_ISL_1566389, EPI_ISL_1567706, EPI_ISL_1567716, EPI_ISL_1567721, EPI_ISL_1567736, EPI_ISL_1567747, EPI_ISL_1567748, EPI_ISL_1567753, EPI_ISL_1568578, EPI_ISL_1568584, EPI_ISL_1568600, EPI_ISL_1568645, EPI_ISL_1568674, EPI_ISL_1568709, EPI_ISL_1568717,
EPI_ISL_1568730, EPI_ISL_1568731, EPI_ISL_1568733, EPI_ISL_1568747, EPI_ISL_1568784, EPI_ISL_1568794, EPI_ISL_1568804, EPI_ISL_1568813, EPI_ISL_1572234, EPI_ISL_1572236, EPI_ISL_1572238, EPI_ISL_1572239, EPI_ISL_1572240, EPI_ISL_1572241, EPI_ISL_1572242, EPI_ISL_1572247, EPI_ISL_1572248, EPI_ISL_1572252, EPI_ISL_1572255, EPI_ISL_1572257, EPI_ISL_1572259, EPI_ISL_1572261,
EPI_ISL_1572262, EPI_ISL_1572274, EPI_ISL_1572280, EPI_ISL_1572283, EPI_ISL_1572286, EPI_ISL_1572290, EPI_ISL_1572296, EPI_ISL_1572300, EPI_ISL_1572302, EPI_ISL_1572305, EPI_ISL_1572306, EPI_ISL_1572315, EPI_ISL_1572318, EPI_ISL_1572330, EPI_ISL_1572331, EPI_ISL_1572334, EPI_ISL_1572335, EPI_ISL_1572338, EPI_ISL_1572340, EPI_ISL_1572343, EPI_ISL_1572346, EPI_ISL_1572351,
EPI_ISL_1572353, EPI_ISL_1572357, EPI_ISL_1572361, EPI_ISL_1572363, EPI_ISL_1572367, EPI_ISL_1572370, EPI_ISL_1572372, EPI_ISL_1572375, EPI_ISL_1572382, EPI_ISL_1572386, EPI_ISL_1572389, EPI_ISL_1572398, EPI_ISL_1572400, EPI_ISL_1572404, EPI_ISL_1572407, EPI_ISL_1572412, EPI_ISL_1572420, EPI_ISL_1572432, EPI_ISL_1572434, EPI_ISL_1572439, EPI_ISL_1572444, EPI_ISL_1572449,
EPI_ISL_1572451, EPI_ISL_1572459, EPI_ISL_1572461, EPI_ISL_1572468, EPI_ISL_1572470, EPI_ISL_1572474, EPI_ISL_1572476, EPI_ISL_1572479, EPI_ISL_1572480, EPI_ISL_1572485, EPI_ISL_1572487, EPI_ISL_1572489, EPI_ISL_1572490, EPI_ISL_1572491, EPI_ISL_1572494, EPI_ISL_1572495, EPI_ISL_1572496, EPI_ISL_1572497, EPI_ISL_1572500, EPI_ISL_1572501, EPI_ISL_1572503, EPI_ISL_1572507,
EPI_ISL_1572511, EPI_ISL_1572513, EPI_ISL_1572515, EPI_ISL_1572517, EPI_ISL_1572519, EPI_ISL_1572521, EPI_ISL_1639616, EPI_ISL_1639686, EPI_ISL_1639865, EPI_ISL_1639871, EPI_ISL_1645960, EPI_ISL_1645971, EPI_ISL_1646017, EPI_ISL_1646082, EPI_ISL_1646890, EPI_ISL_1719923, EPI_ISL_1719953, EPI_ISL_1720000, EPI_ISL_1720059, EPI_ISL_1720062, EPI_ISL_1720106, EPI_ISL_1720129,
EPI_ISL_1849725, EPI_ISL_1849730, EPI_ISL_1849731, EPI_ISL_1849745, EPI_ISL_1849760, EPI_ISL_1849800, EPI_ISL_1849837, EPI_ISL_1849865, EPI_ISL_1849870, EPI_ISL_1849888, EPI_ISL_1849897, EPI_ISL_1849911, EPI_ISL_1849915, EPI_ISL_2108163, EPI_ISL_2119627

see above SYNLAB MVZ Ettlingen Robert Koch Institute

EPI_ISL_1356295, EPI_ISL_1356312, EPI_ISL_1356360, EPI_ISL_1356369, EPI_ISL_1356380, EPI_ISL_1356456, EPI_ISL_1356459, EPI_ISL_1356468, EPI_ISL_1432093, EPI_ISL_1432192, EPI_ISL_1432360, EPI_ISL_1432386, EPI_ISL_1432428, EPI_ISL_1432440, EPI_ISL_1432447, EPI_ISL_1432487, EPI_ISL_1432491, EPI_ISL_1432500, EPI_ISL_1432556, EPI_ISL_1432588, EPI_ISL_1432596, EPI_ISL_1432622,
EPI_ISL_1432742, EPI_ISL_1437286, EPI_ISL_1437329, EPI_ISL_1437331, EPI_ISL_1437365, EPI_ISL_1437366, EPI_ISL_1437371, EPI_ISL_1437379, EPI_ISL_1437384, EPI_ISL_1437595, EPI_ISL_1439922, EPI_ISL_1439971, EPI_ISL_1439993, EPI_ISL_1440000, EPI_ISL_1440005, EPI_ISL_1440015, EPI_ISL_1440020, EPI_ISL_1440042, EPI_ISL_1440121, EPI_ISL_1440130, EPI_ISL_1440163, EPI_ISL_1440175,
EPI_ISL_1440185, EPI_ISL_1440253, EPI_ISL_1440579, EPI_ISL_1440585, EPI_ISL_1440610, EPI_ISL_1440621, EPI_ISL_1440623, EPI_ISL_1440644, EPI_ISL_1440652, EPI_ISL_1440657, EPI_ISL_1440660, EPI_ISL_1440666, EPI_ISL_1440690, EPI_ISL_1440699, EPI_ISL_1442367, EPI_ISL_1442385, EPI_ISL_1442389, EPI_ISL_1442392, EPI_ISL_1442409, EPI_ISL_1442412, EPI_ISL_1442414, EPI_ISL_1442418,
EPI_ISL_1442463, EPI_ISL_1565792, EPI_ISL_1565794, EPI_ISL_1565806, EPI_ISL_1565808, EPI_ISL_1565818, EPI_ISL_1565824, EPI_ISL_1565832, EPI_ISL_1565836, EPI_ISL_1565856, EPI_ISL_1565877, EPI_ISL_1565887, EPI_ISL_1565901, EPI_ISL_1565914, EPI_ISL_1565931, EPI_ISL_1565958, EPI_ISL_1565974, EPI_ISL_1566117, EPI_ISL_1566138, EPI_ISL_1566158, EPI_ISL_1566170, EPI_ISL_1566182,
EPI_ISL_1566183, EPI_ISL_1566206, EPI_ISL_1566241, EPI_ISL_1566248, EPI_ISL_1566254, EPI_ISL_1566256, EPI_ISL_1566272, EPI_ISL_1566274, EPI_ISL_1566286, EPI_ISL_1566287, EPI_ISL_1566327, EPI_ISL_1566330, EPI_ISL_1567762, EPI_ISL_1568669, EPI_ISL_1568701, EPI_ISL_1568706, EPI_ISL_1568814, EPI_ISL_1568835, EPI_ISL_1568837, EPI_ISL_1572243, EPI_ISL_1572275, EPI_ISL_1572293,
EPI_ISL_1572310, EPI_ISL_1572339, EPI_ISL_1572341, EPI_ISL_1572403, EPI_ISL_1572427, EPI_ISL_1572477, EPI_ISL_1572486, EPI_ISL_1572488, EPI_ISL_1572492, EPI_ISL_1572506, EPI_ISL_1572508, EPI_ISL_1572512, EPI_ISL_1646562, EPI_ISL_1646583, EPI_ISL_1720026, EPI_ISL_1720056, EPI_ISL_1720086, EPI_ISL_1729849, EPI_ISL_1729915, EPI_ISL_1729964

see above SYNLAB MVZ Hamburg Robert Koch Institute

EPI_ISL_1432029, EPI_ISL_1432033, EPI_ISL_1432036, EPI_ISL_1432040, EPI_ISL_1432048, EPI_ISL_1432052, EPI_ISL_1432061, EPI_ISL_1432072, EPI_ISL_1432080, EPI_ISL_1432095, EPI_ISL_1432100, EPI_ISL_1432103, EPI_ISL_1432120, EPI_ISL_1432130, EPI_ISL_1432136, EPI_ISL_1432140, EPI_ISL_1432143, EPI_ISL_1432147, EPI_ISL_1432149, EPI_ISL_1432157, EPI_ISL_1432159, EPI_ISL_1432168,
EPI_ISL_1432177, EPI_ISL_1432183, EPI_ISL_1432188, EPI_ISL_1432203, EPI_ISL_1432207, EPI_ISL_1432219, EPI_ISL_1432244, EPI_ISL_1432259, EPI_ISL_1432267, EPI_ISL_1432274, EPI_ISL_1432275, EPI_ISL_1432294, EPI_ISL_1432330, EPI_ISL_1432336, EPI_ISL_1432353, EPI_ISL_1432358, EPI_ISL_1432369, EPI_ISL_1432380, EPI_ISL_1432388, EPI_ISL_1432400, EPI_ISL_1432413, EPI_ISL_1432419,
EPI_ISL_1432426, EPI_ISL_1432434, EPI_ISL_1432455, EPI_ISL_1432468, EPI_ISL_1432470, EPI_ISL_1432473, EPI_ISL_1432481, EPI_ISL_1432498, EPI_ISL_1432508, EPI_ISL_1432519, EPI_ISL_1432521, EPI_ISL_1432530, EPI_ISL_1432532, EPI_ISL_1432541, EPI_ISL_1432547, EPI_ISL_1432560, EPI_ISL_1432605, EPI_ISL_1432613, EPI_ISL_1432630, EPI_ISL_1432633, EPI_ISL_1432641, EPI_ISL_1432724,
EPI_ISL_1432733, EPI_ISL_1432746, EPI_ISL_1439913, EPI_ISL_1439918, EPI_ISL_1439928, EPI_ISL_1439931, EPI_ISL_1439962, EPI_ISL_1440037, EPI_ISL_1440140, EPI_ISL_1440146, EPI_ISL_1440589, EPI_ISL_1440608, EPI_ISL_1440609, EPI_ISL_1440618, EPI_ISL_1440624, EPI_ISL_1440633, EPI_ISL_1440639, EPI_ISL_1440640, EPI_ISL_1440643, EPI_ISL_1440646, EPI_ISL_1440656, EPI_ISL_1440681,
EPI_ISL_1440691, EPI_ISL_1440695, EPI_ISL_1440696, EPI_ISL_1440698, EPI_ISL_1440709, EPI_ISL_1440714, EPI_ISL_1440728, EPI_ISL_1565804, EPI_ISL_1565812, EPI_ISL_1565817, EPI_ISL_1565847, EPI_ISL_1565862, EPI_ISL_1566101, EPI_ISL_1566107, EPI_ISL_1566109, EPI_ISL_1566115, EPI_ISL_1566122, EPI_ISL_1566126, EPI_ISL_1566128, EPI_ISL_1566129, EPI_ISL_1566131, EPI_ISL_1566134,
EPI_ISL_1566135, EPI_ISL_1566141, EPI_ISL_1566144, EPI_ISL_1566145, EPI_ISL_1566149, EPI_ISL_1566150, EPI_ISL_1566151, EPI_ISL_1566153, EPI_ISL_1566154, EPI_ISL_1566157, EPI_ISL_1566161, EPI_ISL_1566165, EPI_ISL_1566169, EPI_ISL_1566172, EPI_ISL_1566175, EPI_ISL_1566176, EPI_ISL_1566179, EPI_ISL_1566180, EPI_ISL_1566184, EPI_ISL_1566187, EPI_ISL_1566188, EPI_ISL_1566189,
EPI_ISL_1566190, EPI_ISL_1566191, EPI_ISL_1566192, EPI_ISL_1566193, EPI_ISL_1566194, EPI_ISL_1566196, EPI_ISL_1566197, EPI_ISL_1566198, EPI_ISL_1566201, EPI_ISL_1566209, EPI_ISL_1566210, EPI_ISL_1566211, EPI_ISL_1566220, EPI_ISL_1566224, EPI_ISL_1566229, EPI_ISL_1566230, EPI_ISL_1566232, EPI_ISL_1566234, EPI_ISL_1566239, EPI_ISL_1566243, EPI_ISL_1566246, EPI_ISL_1566252,
EPI_ISL_1566257, EPI_ISL_1566260, EPI_ISL_1566264, EPI_ISL_1566267, EPI_ISL_1566269, EPI_ISL_1566273, EPI_ISL_1566291, EPI_ISL_1566295, EPI_ISL_1566298, EPI_ISL_1566303, EPI_ISL_1566309, EPI_ISL_1566311, EPI_ISL_1566313, EPI_ISL_1566316, EPI_ISL_1566319, EPI_ISL_1566322, EPI_ISL_1566324, EPI_ISL_1566329, EPI_ISL_1566334, EPI_ISL_1566335, EPI_ISL_1566339, EPI_ISL_1566340,
EPI_ISL_1566344, EPI_ISL_1566349, EPI_ISL_1566353, EPI_ISL_1566357, EPI_ISL_1566363, EPI_ISL_1566367, EPI_ISL_1566370, EPI_ISL_1566374, EPI_ISL_1566376, EPI_ISL_1566378, EPI_ISL_1567744, EPI_ISL_1567750, EPI_ISL_1567752, EPI_ISL_1567764, EPI_ISL_1568821, EPI_ISL_1568822, EPI_ISL_1568823, EPI_ISL_1568824, EPI_ISL_1568825, EPI_ISL_1568831, EPI_ISL_1568833, EPI_ISL_1568838,
EPI_ISL_1568841, EPI_ISL_1568849, EPI_ISL_1568850, EPI_ISL_1568852, EPI_ISL_1568855, EPI_ISL_1568858, EPI_ISL_1568860, EPI_ISL_1568863, EPI_ISL_1568865, EPI_ISL_1568867, EPI_ISL_1568868, EPI_ISL_1568870, EPI_ISL_1568872, EPI_ISL_1568873, EPI_ISL_1568876, EPI_ISL_1568878, EPI_ISL_1568880, EPI_ISL_1568881, EPI_ISL_1568883, EPI_ISL_1568886, EPI_ISL_1568888, EPI_ISL_1568889,
EPI_ISL_1568891, EPI_ISL_1568895, EPI_ISL_1568900, EPI_ISL_1568906, EPI_ISL_1568907, EPI_ISL_1568908, EPI_ISL_1568911, EPI_ISL_1568912, EPI_ISL_1568913, EPI_ISL_1568917, EPI_ISL_1568919, EPI_ISL_1568921, EPI_ISL_1568924, EPI_ISL_1568930, EPI_ISL_1568931, EPI_ISL_1568933, EPI_ISL_1568935, EPI_ISL_1568936, EPI_ISL_1568937, EPI_ISL_1568942, EPI_ISL_1568943, EPI_ISL_1572235,
EPI_ISL_1572237, EPI_ISL_1572244, EPI_ISL_1572249, EPI_ISL_1572250, EPI_ISL_1572253, EPI_ISL_1572258, EPI_ISL_1572260, EPI_ISL_1572263, EPI_ISL_1572264, EPI_ISL_1572266, EPI_ISL_1572268, EPI_ISL_1572269, EPI_ISL_1572270, EPI_ISL_1572272, EPI_ISL_1572273, EPI_ISL_1572277, EPI_ISL_1572278, EPI_ISL_1572282, EPI_ISL_1572285, EPI_ISL_1572287, EPI_ISL_1572289, EPI_ISL_1572292,
EPI_ISL_1572295, EPI_ISL_1572298, EPI_ISL_1572303, EPI_ISL_1572307, EPI_ISL_1572308, EPI_ISL_1572313, EPI_ISL_1572320, EPI_ISL_1572322, EPI_ISL_1572323, EPI_ISL_1572328, EPI_ISL_1572336, EPI_ISL_1572342, EPI_ISL_1572349, EPI_ISL_1572356, EPI_ISL_1572360, EPI_ISL_1572377, EPI_ISL_1572379, EPI_ISL_1572384, EPI_ISL_1572391, EPI_ISL_1572401, EPI_ISL_1572410, EPI_ISL_1572422,
EPI_ISL_1572430, EPI_ISL_1572437, EPI_ISL_1572441, EPI_ISL_1572456, EPI_ISL_1572463, EPI_ISL_1572473, EPI_ISL_1572483, EPI_ISL_1572484, EPI_ISL_1572499, EPI_ISL_1572502, EPI_ISL_1572504, EPI_ISL_1572514, EPI_ISL_1572516, EPI_ISL_1572523, EPI_ISL_1572524, EPI_ISL_1639549, EPI_ISL_1639571, EPI_ISL_1639575, EPI_ISL_1639593, EPI_ISL_1639598, EPI_ISL_1639605, EPI_ISL_1639610,
EPI_ISL_1639614, EPI_ISL_1639623, EPI_ISL_1639629, EPI_ISL_1639632, EPI_ISL_1639635, EPI_ISL_1639640, EPI_ISL_1639648, EPI_ISL_1639649, EPI_ISL_1639651, EPI_ISL_1639656, EPI_ISL_1639664, EPI_ISL_1639668, EPI_ISL_1639672, EPI_ISL_1639687, EPI_ISL_1639691, EPI_ISL_1639696, EPI_ISL_1639709, EPI_ISL_1639734, EPI_ISL_1639737, EPI_ISL_1639744, EPI_ISL_1639757, EPI_ISL_1639765,
EPI_ISL_1639772, EPI_ISL_1639776, EPI_ISL_1639778, EPI_ISL_1639789, EPI_ISL_1639794, EPI_ISL_1639799, EPI_ISL_1639808, EPI_ISL_1639814, EPI_ISL_1639818, EPI_ISL_1639831, EPI_ISL_1639833, EPI_ISL_1639891, EPI_ISL_1639947, EPI_ISL_1646504, EPI_ISL_1646513, EPI_ISL_1646522, EPI_ISL_1646530, EPI_ISL_1646552, EPI_ISL_1720091

see above SYNLAB MVZ Heidelberg Robert Koch Institute

EPI_ISL_1351778, EPI_ISL_1351784, EPI_ISL_1351786, EPI_ISL_1351789, EPI_ISL_1351791, EPI_ISL_1351792, EPI_ISL_1351795, EPI_ISL_1351803, EPI_ISL_1351807, EPI_ISL_1351813, EPI_ISL_1351822, EPI_ISL_1351823, EPI_ISL_1351826, EPI_ISL_1351843, EPI_ISL_1351846, EPI_ISL_1351855, EPI_ISL_1351862, EPI_ISL_1351870, EPI_ISL_1351887, EPI_ISL_1351888, EPI_ISL_1351912, EPI_ISL_1351916,
EPI_ISL_1351922, EPI_ISL_1351928, EPI_ISL_1351936, EPI_ISL_1351940, EPI_ISL_1351946, EPI_ISL_1351948, EPI_ISL_1351949, EPI_ISL_1351951, EPI_ISL_1351962, EPI_ISL_1351964, EPI_ISL_1351971, EPI_ISL_1351973, EPI_ISL_1351977, EPI_ISL_1351980, EPI_ISL_1351982, EPI_ISL_1351983, EPI_ISL_1351991, EPI_ISL_1351999, EPI_ISL_1352001, EPI_ISL_1352004, EPI_ISL_1352005, EPI_ISL_1352007,
EPI_ISL_1352008, EPI_ISL_1352010, EPI_ISL_1352011, EPI_ISL_1352013, EPI_ISL_1352024, EPI_ISL_1352027, EPI_ISL_1352031, EPI_ISL_1352033, EPI_ISL_1352037, EPI_ISL_1352039, EPI_ISL_1352045, EPI_ISL_1352049, EPI_ISL_1352058, EPI_ISL_1352061, EPI_ISL_1352073, EPI_ISL_1352075, EPI_ISL_1352080, EPI_ISL_1352081, EPI_ISL_1352090, EPI_ISL_1352092, EPI_ISL_1352093, EPI_ISL_1352094,
EPI_ISL_1352099, EPI_ISL_1352106, EPI_ISL_1352107, EPI_ISL_1352108, EPI_ISL_1352112, EPI_ISL_1352115, EPI_ISL_1352118, EPI_ISL_1352119, EPI_ISL_1352123, EPI_ISL_1352125, EPI_ISL_1352136, EPI_ISL_1352137, EPI_ISL_1352141, EPI_ISL_1352146, EPI_ISL_1352151, EPI_ISL_1352158, EPI_ISL_1352165, EPI_ISL_1352166, EPI_ISL_1352168, EPI_ISL_1352171, EPI_ISL_1352177, EPI_ISL_1352179,
EPI_ISL_1352191, EPI_ISL_1352195, EPI_ISL_1352201, EPI_ISL_1352211, EPI_ISL_1352224, EPI_ISL_1352225, EPI_ISL_1352226, EPI_ISL_1352230, EPI_ISL_1352234, EPI_ISL_1352236, EPI_ISL_1352694, EPI_ISL_1352696, EPI_ISL_1352697, EPI_ISL_1352698, EPI_ISL_1352699, EPI_ISL_1352700, EPI_ISL_1352702, EPI_ISL_1352703, EPI_ISL_1352704, EPI_ISL_1352705, EPI_ISL_1352708, EPI_ISL_1352709,
EPI_ISL_1352710, EPI_ISL_1352711, EPI_ISL_1352713, EPI_ISL_1352714, EPI_ISL_1352715, EPI_ISL_1352720, EPI_ISL_1352722, EPI_ISL_1352724, EPI_ISL_1352726, EPI_ISL_1352728, EPI_ISL_1352729, EPI_ISL_1352730, EPI_ISL_1352731, EPI_ISL_1352732, EPI_ISL_1352733, EPI_ISL_1352734, EPI_ISL_1352735, EPI_ISL_1352737, EPI_ISL_1352741, EPI_ISL_1352742, EPI_ISL_1352744,
EPI_ISL_1352746, EPI_ISL_1352747, EPI_ISL_1352748, EPI_ISL_1352750, EPI_ISL_1352751, EPI_ISL_1352752, EPI_ISL_1352753, EPI_ISL_1352756, EPI_ISL_1352759, EPI_ISL_1352760, EPI_ISL_1352762, EPI_ISL_1352763, EPI_ISL_1352769, EPI_ISL_1352771, EPI_ISL_1352773, EPI_ISL_1352774, EPI_ISL_1352777, EPI_ISL_1352778, EPI_ISL_1352780, EPI_ISL_1352783, EPI_ISL_1352784, EPI_ISL_1352785,
EPI_ISL_1352788, EPI_ISL_1352790, EPI_ISL_1352791, EPI_ISL_1352793, EPI_ISL_1352797, EPI_ISL_1352798, EPI_ISL_1352800, EPI_ISL_1352801, EPI_ISL_1352802, EPI_ISL_1352803, EPI_ISL_1352807, EPI_ISL_1352808, EPI_ISL_1352811, EPI_ISL_1352813, EPI_ISL_1352819, EPI_ISL_1352820, EPI_ISL_1352821, EPI_ISL_1352825, EPI_ISL_1352826, EPI_ISL_1352828, EPI_ISL_1352830, EPI_ISL_1352837,
EPI_ISL_1352840, EPI_ISL_1352842, EPI_ISL_1352847, EPI_ISL_1352849, EPI_ISL_1352859, EPI_ISL_1352863, EPI_ISL_1355898, EPI_ISL_1355900, EPI_ISL_1355914, EPI_ISL_1355915, EPI_ISL_1355916, EPI_ISL_1355917, EPI_ISL_1355918, EPI_ISL_1355919, EPI_ISL_1355920, EPI_ISL_1355924, EPI_ISL_1355925, EPI_ISL_1355928, EPI_ISL_1355929, EPI_ISL_1355930, EPI_ISL_1355931, EPI_ISL_1355932,
EPI_ISL_1355933, EPI_ISL_1355936, EPI_ISL_1355937, EPI_ISL_1355939, EPI_ISL_1355940, EPI_ISL_1355942, EPI_ISL_1355944, EPI_ISL_1355945, EPI_ISL_1355946, EPI_ISL_1355947, EPI_ISL_1355949, EPI_ISL_1355951, EPI_ISL_1355953, EPI_ISL_1355955, EPI_ISL_1355956, EPI_ISL_1355960, EPI_ISL_1355961, EPI_ISL_1355962, EPI_ISL_1355964, EPI_ISL_1355965, EPI_ISL_1355968,
EPI_ISL_1355969, EPI_ISL_1355970, EPI_ISL_1355971, EPI_ISL_1355974, EPI_ISL_1355975, EPI_ISL_1355976, EPI_ISL_1355979, EPI_ISL_1355981, EPI_ISL_1355985, EPI_ISL_1355987, EPI_ISL_1355990, EPI_ISL_1355992, EPI_ISL_1355997, EPI_ISL_1355998, EPI_ISL_1355999, EPI_ISL_1356001, EPI_ISL_1356002, EPI_ISL_1356003, EPI_ISL_1356005, EPI_ISL_1356006, EPI_ISL_1356008, EPI_ISL_1356009,
EPI_ISL_1356010, EPI_ISL_1356012, EPI_ISL_1356016, EPI_ISL_1356021, EPI_ISL_1356023, EPI_ISL_1356024, EPI_ISL_1356025, EPI_ISL_1356026, EPI_ISL_1356027, EPI_ISL_1356029, EPI_ISL_1356030, EPI_ISL_1356031, EPI_ISL_1356032, EPI_ISL_1356033, EPI_ISL_1356035, EPI_ISL_1356037, EPI_ISL_1356039, EPI_ISL_1356040, EPI_ISL_1356041, EPI_ISL_1356044, EPI_ISL_1356046, EPI_ISL_1356047,
EPI_ISL_1356049, EPI_ISL_1356051, EPI_ISL_1356057, EPI_ISL_1356060, EPI_ISL_1356061, EPI_ISL_1356064, EPI_ISL_1356065, EPI_ISL_1356068, EPI_ISL_1356070, EPI_ISL_1356074, EPI_ISL_1356076, EPI_ISL_1356077, EPI_ISL_1356079, EPI_ISL_1356080, EPI_ISL_1356084, EPI_ISL_1356086, EPI_ISL_1356087, EPI_ISL_1356088, EPI_ISL_1356090, EPI_ISL_1356091, EPI_ISL_1356095, EPI_ISL_1356096,
EPI_ISL_1356097, EPI_ISL_1356098, EPI_ISL_1356099, EPI_ISL_1356100, EPI_ISL_1356102, EPI_ISL_1356106, EPI_ISL_1356108, EPI_ISL_1356109, EPI_ISL_1356111, EPI_ISL_1356112, EPI_ISL_1356114, EPI_ISL_1356116, EPI_ISL_1356118, EPI_ISL_1356120, EPI_ISL_1356121, EPI_ISL_1356122, EPI_ISL_1356124, EPI_ISL_1356127, EPI_ISL_1356128, EPI_ISL_1356129, EPI_ISL_1356131, EPI_ISL_1356132,
EPI_ISL_1356134, EPI_ISL_1356136, EPI_ISL_1356137, EPI_ISL_1356138, EPI_ISL_1356139, EPI_ISL_1356144, EPI_ISL_1356145, EPI_ISL_1356146, EPI_ISL_1356149, EPI_ISL_1356153, EPI_ISL_1356154, EPI_ISL_1356156, EPI_ISL_1356157, EPI_ISL_1356158, EPI_ISL_1356160, EPI_ISL_1356162, EPI_ISL_1356163, EPI_ISL_1356164, EPI_ISL_1356165, EPI_ISL_1356166, EPI_ISL_1356167, EPI_ISL_1356169,
EPI_ISL_1356170, EPI_ISL_1356171, EPI_ISL_1356173, EPI_ISL_1356174, EPI_ISL_1356175, EPI_ISL_1356177, EPI_ISL_1356178, EPI_ISL_1356179, EPI_ISL_1356180, EPI_ISL_1356185, EPI_ISL_1356190, EPI_ISL_1356192, EPI_ISL_1356194, EPI_ISL_1356195, EPI_ISL_1356197, EPI_ISL_1356198, EPI_ISL_1356199, EPI_ISL_1356200, EPI_ISL_1356201, EPI_ISL_1356202, EPI_ISL_1356203, EPI_ISL_1356205,
EPI_ISL_1356206, EPI_ISL_1356207, EPI_ISL_1356208, EPI_ISL_1356212, EPI_ISL_1356213, EPI_ISL_1356216, EPI_ISL_1356217, EPI_ISL_1356221, EPI_ISL_1356222, EPI_ISL_1356223, EPI_ISL_1356224, EPI_ISL_1356228, EPI_ISL_1356232, EPI_ISL_1356235, EPI_ISL_1356238, EPI_ISL_1356240, EPI_ISL_1357346, EPI_ISL_1357349, EPI_ISL_1357352, EPI_ISL_1357353, EPI_ISL_1357357, EPI_ISL_1357359,
EPI_ISL_1357362, EPI_ISL_1357363, EPI_ISL_1357366, EPI_ISL_1357369, EPI_ISL_1357372, EPI_ISL_1357373, EPI_ISL_1357377, EPI_ISL_1357378, EPI_ISL_1357379, EPI_ISL_1357380, EPI_ISL_1357385, EPI_ISL_1357387, EPI_ISL_1357388, EPI_ISL_1357389, EPI_ISL_1357390, EPI_ISL_1357392, EPI_ISL_1357397, EPI_ISL_1357400, EPI_ISL_1357403, EPI_ISL_1357407, EPI_ISL_1357408, EPI_ISL_1357412,
EPI_ISL_1357413, EPI_ISL_1357414, EPI_ISL_1357417, EPI_ISL_1357419, EPI_ISL_1357422, EPI_ISL_1357423, EPI_ISL_1357434, EPI_ISL_1357436, EPI_ISL_1357439, EPI_ISL_1357441, EPI_ISL_1357446, EPI_ISL_1357450, EPI_ISL_1357454, EPI_ISL_1357462, EPI_ISL_1357464, EPI_ISL_1357467, EPI_ISL_1357468, EPI_ISL_1357470, EPI_ISL_1357471, EPI_ISL_1357472, EPI_ISL_1357477, EPI_ISL_1357479,
EPI_ISL_1357485, EPI_ISL_1357488, EPI_ISL_1357489, EPI_ISL_1357493, EPI_ISL_1357504, EPI_ISL_1357519, EPI_ISL_1357521, EPI_ISL_1357529, EPI_ISL_1357534, EPI_ISL_1357535, EPI_ISL_1357537, EPI_ISL_1357541, EPI_ISL_1357549, EPI_ISL_1357551, EPI_ISL_1357553, EPI_ISL_1357556, EPI_ISL_1357576, EPI_ISL_1357579, EPI_ISL_1357582, EPI_ISL_1357583, EPI_ISL_1357585,
EPI_ISL_1357586, EPI_ISL_1357587, EPI_ISL_1357589, EPI_ISL_1357593, EPI_ISL_1434620, EPI_ISL_1434622, EPI_ISL_1434624, EPI_ISL_1434631, EPI_ISL_1434633, EPI_ISL_1434635, EPI_ISL_1434651, EPI_ISL_1434653, EPI_ISL_1434663, EPI_ISL_1434676, EPI_ISL_1434681, EPI_ISL_1434696, EPI_ISL_1434700, EPI_ISL_1434702, EPI_ISL_1434704, EPI_ISL_1434708, EPI_ISL_1434710, EPI_ISL_1434718,
EPI_ISL_1434727, EPI_ISL_1434733, EPI_ISL_1434735, EPI_ISL_1434748, EPI_ISL_1434758, EPI_ISL_1434767, EPI_ISL_1434769, EPI_ISL_1434777, EPI_ISL_1434783, EPI_ISL_1434793, EPI_ISL_1434798, EPI_ISL_1434809, EPI_ISL_1434816, EPI_ISL_1434834, EPI_ISL_1434839, EPI_ISL_1434841, EPI_ISL_1434845, EPI_ISL_1434849, EPI_ISL_1434864, EPI_ISL_1434871, EPI_ISL_1434873, EPI_ISL_1434879,
EPI_ISL_1434881, EPI_ISL_1434883, EPI_ISL_1434884, EPI_ISL_1434894, EPI_ISL_1434902, EPI_ISL_1434908, EPI_ISL_1434910, EPI_ISL_1434917, EPI_ISL_1434921, EPI_ISL_1434929, EPI_ISL_1434939, EPI_ISL_1434948, EPI_ISL_1434980, EPI_ISL_1434982, EPI_ISL_1434986, EPI_ISL_1434993, EPI_ISL_1434997, EPI_ISL_1435001, EPI_ISL_1435007, EPI_ISL_1435013, EPI_ISL_1435014, EPI_ISL_1435022,
EPI_ISL_1435027, EPI_ISL_1435029, EPI_ISL_1435041, EPI_ISL_1435042, EPI_ISL_1435046, EPI_ISL_1435065, EPI_ISL_1435078, EPI_ISL_1435082, EPI_ISL_1435097, EPI_ISL_1435105, EPI_ISL_1435113, EPI_ISL_1435122, EPI_ISL_1435142, EPI_ISL_1435144, EPI_ISL_1435153, EPI_ISL_1435159, EPI_ISL_1435162, EPI_ISL_1435170, EPI_ISL_1435174, EPI_ISL_1435176, EPI_ISL_1435182, EPI_ISL_1435194,
EPI_ISL_1435196, EPI_ISL_1435207, EPI_ISL_1435232, EPI_ISL_1435236, EPI_ISL_1435244, EPI_ISL_1435246, EPI_ISL_1436949, EPI_ISL_1436950, EPI_ISL_1436951, EPI_ISL_1436953, EPI_ISL_1436954, EPI_ISL_1436955, EPI_ISL_1436956, EPI_ISL_1436957, EPI_ISL_1436958, EPI_ISL_1436962, EPI_ISL_1436964, EPI_ISL_1436965, EPI_ISL_1436967, EPI_ISL_1436968, EPI_ISL_1436969, EPI_ISL_1436970,
EPI_ISL_1436972, EPI_ISL_1436973, EPI_ISL_1436980, EPI_ISL_1436984, EPI_ISL_1436985, EPI_ISL_1436991, EPI_ISL_1436994, EPI_ISL_1436995, EPI_ISL_1436997, EPI_ISL_1436998, EPI_ISL_1436999, EPI_ISL_1437001, EPI_ISL_1437003, EPI_ISL_1437006, EPI_ISL_1437007, EPI_ISL_1437008, EPI_ISL_1437009, EPI_ISL_1437010, EPI_ISL_1437011, EPI_ISL_1437014, EPI_ISL_1437015, EPI_ISL_1437016,
EPI_ISL_1437018, EPI_ISL_1437019, EPI_ISL_1437021, EPI_ISL_1437022, EPI_ISL_1437023, EPI_ISL_1437024, EPI_ISL_1437025, EPI_ISL_1437026, EPI_ISL_1437027, EPI_ISL_1437030, EPI_ISL_1437032, EPI_ISL_1437033, EPI_ISL_1437034, EPI_ISL_1437035, EPI_ISL_1437036, EPI_ISL_1437037, EPI_ISL_1437040, EPI_ISL_1437042, EPI_ISL_1437043, EPI_ISL_1437044, EPI_ISL_1437046, EPI_ISL_1437049,
EPI_ISL_1437050, EPI_ISL_1437051, EPI_ISL_1437054, EPI_ISL_1437055, EPI_ISL_1437058, EPI_ISL_1437064, EPI_ISL_1437066, EPI_ISL_1437067, EPI_ISL_1437068, EPI_ISL_1437072, EPI_ISL_1437074, EPI_ISL_1437078, EPI_ISL_1437080, EPI_ISL_1437083, EPI_ISL_1437086, EPI_ISL_1437087, EPI_ISL_1437088, EPI_ISL_1437090, EPI_ISL_1437092, EPI_ISL_1437094, EPI_ISL_1437095, EPI_ISL_1437096,
EPI_ISL_1437098, EPI_ISL_1437100, EPI_ISL_1437101, EPI_ISL_1437104, EPI_ISL_1437105, EPI_ISL_1437107, EPI_ISL_1437108, EPI_ISL_1437111, EPI_ISL_1437112, EPI_ISL_1437113, EPI_ISL_1437114, EPI_ISL_1437115, EPI_ISL_1437117, EPI_ISL_1437120, EPI_ISL_1437122, EPI_ISL_1437123, EPI_ISL_1437127, EPI_ISL_1437128, EPI_ISL_1437129, EPI_ISL_1437130, EPI_ISL_1437131, EPI_ISL_1437132,
EPI_ISL_1437133, EPI_ISL_1437139, EPI_ISL_1437142, EPI_ISL_1437143, EPI_ISL_1437144, EPI_ISL_1437146, EPI_ISL_1437148, EPI_ISL_1437149, EPI_ISL_1437150, EPI_ISL_1437153, EPI_ISL_1437154, EPI_ISL_1437155, EPI_ISL_1437158, EPI_ISL_1437159, EPI_ISL_1437161, EPI_ISL_1437162, EPI_ISL_1437163, EPI_ISL_1437164, EPI_ISL_1437166, EPI_ISL_1437167, EPI_ISL_1437168, EPI_ISL_1437169,
EPI_ISL_1437170, EPI_ISL_1437173, EPI_ISL_1437174, EPI_ISL_1437175, EPI_ISL_1437177, EPI_ISL_1437178, EPI_ISL_1437181, EPI_ISL_1437182, EPI_ISL_1437183, EPI_ISL_1437186, EPI_ISL_1437190, EPI_ISL_1437191, EPI_ISL_1437193, EPI_ISL_1437194, EPI_ISL_1437195, EPI_ISL_1437196, EPI_ISL_1437197, EPI_ISL_1437199, EPI_ISL_1437202, EPI_ISL_1437205, EPI_ISL_1437206, EPI_ISL_1437209,
EPI_ISL_1437210, EPI_ISL_1437211, EPI_ISL_1437212, EPI_ISL_1437214, EPI_ISL_1437217, EPI_ISL_1437218, EPI_ISL_1437222, EPI_ISL_1437226, EPI_ISL_1437229, EPI_ISL_1437230, EPI_ISL_1437234, EPI_ISL_1437235, EPI_ISL_1437236, EPI_ISL_1437237, EPI_ISL_1437241, EPI_ISL_1437242, EPI_ISL_1437243, EPI_ISL_1437244, EPI_ISL_1437245, EPI_ISL_1437246, EPI_ISL_1437248, EPI_ISL_1437250,
EPI_ISL_1437251, EPI_ISL_1437253, EPI_ISL_1437254, EPI_ISL_1437256, EPI_ISL_1437258, EPI_ISL_1437261, EPI_ISL_1437262, EPI_ISL_1437264, EPI_ISL_1437265, EPI_ISL_1437266, EPI_ISL_1437267, EPI_ISL_1437269, EPI_ISL_1437270, EPI_ISL_1437271, EPI_ISL_1437272, EPI_ISL_1437273, EPI_ISL_1437274, EPI_ISL_1437275, EPI_ISL_1437277, EPI_ISL_1437278, EPI_ISL_1437280, EPI_ISL_1438296,
EPI_ISL_1438300, EPI_ISL_1438302, EPI_ISL_1438306, EPI_ISL_1438309, EPI_ISL_1438313, EPI_ISL_1438314, EPI_ISL_1438317, EPI_ISL_1438318, EPI_ISL_1438323, EPI_ISL_1438338, EPI_ISL_1438350, EPI_ISL_1438359, EPI_ISL_1438383, EPI_ISL_1438389, EPI_ISL_1438392, EPI_ISL_1438396, EPI_ISL_1438402, EPI_ISL_1438409, EPI_ISL_1438414, EPI_ISL_1438419, EPI_ISL_1438422, EPI_ISL_1438423,
EPI_ISL_1438437, EPI_ISL_1438442, EPI_ISL_1438448, EPI_ISL_1438451, EPI_ISL_1438476, EPI_ISL_1438480, EPI_ISL_1438483, EPI_ISL_1438493, EPI_ISL_1438498, EPI_ISL_1438503, EPI_ISL_1438504, EPI_ISL_1438507, EPI_ISL_1438508, EPI_ISL_1438510, EPI_ISL_1438511, EPI_ISL_1438513, EPI_ISL_1438515, EPI_ISL_1438520, EPI_ISL_1438529, EPI_ISL_1438538, EPI_ISL_1438543, EPI_ISL_1438556,
EPI_ISL_1438565, EPI_ISL_1438568, EPI_ISL_1438571, EPI_ISL_1438576

see above SYNLAB MVZ Leinfelden-Echterdingen Robert Koch Institute

EPI_ISL_1356362, EPI_ISL_1356464, EPI_ISL_1432510, EPI_ISL_1432579, EPI_ISL_1432684, EPI_ISL_1437285, EPI_ISL_1437288, EPI_ISL_1437289, EPI_ISL_1437291, EPI_ISL_1437292, EPI_ISL_1437293, EPI_ISL_1437295, EPI_ISL_1437299, EPI_ISL_1437301, EPI_ISL_1437303, EPI_ISL_1437304, EPI_ISL_1437305, EPI_ISL_1437308, EPI_ISL_1437309, EPI_ISL_1437313, EPI_ISL_1437319, EPI_ISL_1437321,
EPI_ISL_1437323, EPI_ISL_1437325, EPI_ISL_1437326, EPI_ISL_1437327, EPI_ISL_1437332, EPI_ISL_1437334, EPI_ISL_1437335, EPI_ISL_1437339, EPI_ISL_1437340, EPI_ISL_1437342, EPI_ISL_1437343, EPI_ISL_1437344, EPI_ISL_1437345, EPI_ISL_1437346, EPI_ISL_1437348, EPI_ISL_1437349, EPI_ISL_1437351, EPI_ISL_1437352, EPI_ISL_1437353, EPI_ISL_1437354, EPI_ISL_1437356, EPI_ISL_1437358,
EPI_ISL_1437360, EPI_ISL_1437363, EPI_ISL_1437364, EPI_ISL_1437370, EPI_ISL_1437374, EPI_ISL_1437382, EPI_ISL_1437389, EPI_ISL_1437391, EPI_ISL_1437392, EPI_ISL_1437393, EPI_ISL_1437394, EPI_ISL_1437396, EPI_ISL_1437399, EPI_ISL_1437401, EPI_ISL_1437402, EPI_ISL_1437411, EPI_ISL_1437414, EPI_ISL_1437415, EPI_ISL_1437418, EPI_ISL_1437420, EPI_ISL_1437421, EPI_ISL_1437422,
EPI_ISL_1437425, EPI_ISL_1437430, EPI_ISL_1437433, EPI_ISL_1437446, EPI_ISL_1437449, EPI_ISL_1437450, EPI_ISL_1437451, EPI_ISL_1437453, EPI_ISL_1437454, EPI_ISL_1437462, EPI_ISL_1437464, EPI_ISL_1437469, EPI_ISL_1437472, EPI_ISL_1437474, EPI_ISL_1437476, EPI_ISL_1437480, EPI_ISL_1437484, EPI_ISL_1437494, EPI_ISL_1437498, EPI_ISL_1437499, EPI_ISL_1437502, EPI_ISL_1437503,
EPI_ISL_1437505, EPI_ISL_1437513, EPI_ISL_1437515, EPI_ISL_1437537, EPI_ISL_1437544, EPI_ISL_1437554, EPI_ISL_1437558, EPI_ISL_1437579, EPI_ISL_1437584, EPI_ISL_1437585, EPI_ISL_1437586, EPI_ISL_1437592, EPI_ISL_1437593, EPI_ISL_1437594, EPI_ISL_1437596, EPI_ISL_1437597, EPI_ISL_1437599, EPI_ISL_1437602, EPI_ISL_1437603, EPI_ISL_1437608, EPI_ISL_1437610, EPI_ISL_1437611,
EPI_ISL_1439901, EPI_ISL_1439903, EPI_ISL_1439906, EPI_ISL_1439907, EPI_ISL_1439908, EPI_ISL_1439910, EPI_ISL_1439912, EPI_ISL_1439914, EPI_ISL_1439916, EPI_ISL_1439917, EPI_ISL_1439919, EPI_ISL_1439921, EPI_ISL_1439923, EPI_ISL_1439925, EPI_ISL_1439926, EPI_ISL_1439930, EPI_ISL_1439932, EPI_ISL_1439937, EPI_ISL_1439938, EPI_ISL_1439939, EPI_ISL_1439941, EPI_ISL_1439942,
EPI_ISL_1439943, EPI_ISL_1439945, EPI_ISL_1439947, EPI_ISL_1439949, EPI_ISL_1439950, EPI_ISL_1439952, EPI_ISL_1439953, EPI_ISL_1439954, EPI_ISL_1439955, EPI_ISL_1439956, EPI_ISL_1439958, EPI_ISL_1439959, EPI_ISL_1439960, EPI_ISL_1439963, EPI_ISL_1439964, EPI_ISL_1439965, EPI_ISL_1439967, EPI_ISL_1439968, EPI_ISL_1439970, EPI_ISL_1439973, EPI_ISL_1439974, EPI_ISL_1439976,
EPI_ISL_1439978, EPI_ISL_1439979, EPI_ISL_1439982, EPI_ISL_1439984, EPI_ISL_1439985, EPI_ISL_1439989, EPI_ISL_1439995, EPI_ISL_1439996, EPI_ISL_1439998, EPI_ISL_1440001, EPI_ISL_1440002, EPI_ISL_1440003, EPI_ISL_1440004, EPI_ISL_1440007, EPI_ISL_1440009, EPI_ISL_1440010, EPI_ISL_1440011, EPI_ISL_1440013, EPI_ISL_1440016, EPI_ISL_1440017, EPI_ISL_1440019,
EPI_ISL_1440021, EPI_ISL_1440023, EPI_ISL_1440024, EPI_ISL_1440025, EPI_ISL_1440027, EPI_ISL_1440029, EPI_ISL_1440030, EPI_ISL_1440031, EPI_ISL_1440032, EPI_ISL_1440033, EPI_ISL_1440035, EPI_ISL_1440036, EPI_ISL_1440039, EPI_ISL_1440041, EPI_ISL_1440043, EPI_ISL_1440045, EPI_ISL_1440046, EPI_ISL_1440048, EPI_ISL_1440050, EPI_ISL_1440051, EPI_ISL_1440052, EPI_ISL_1440053,
EPI_ISL_1440055, EPI_ISL_1440056, EPI_ISL_1440057, EPI_ISL_1440058, EPI_ISL_1440060, EPI_ISL_1440062, EPI_ISL_1440064, EPI_ISL_1440065, EPI_ISL_1440066, EPI_ISL_1440068, EPI_ISL_1440069, EPI_ISL_1440071, EPI_ISL_1440073, EPI_ISL_1440076, EPI_ISL_1440077, EPI_ISL_1440080, EPI_ISL_1440082, EPI_ISL_1440086, EPI_ISL_1440088, EPI_ISL_1440089, EPI_ISL_1440092, EPI_ISL_1440094,
EPI_ISL_1440095, EPI_ISL_1440098, EPI_ISL_1440100, EPI_ISL_1440103, EPI_ISL_1440104, EPI_ISL_1440105, EPI_ISL_1440108, EPI_ISL_1440110, EPI_ISL_1440113, EPI_ISL_1440114, EPI_ISL_1440120, EPI_ISL_1440126, EPI_ISL_1440127, EPI_ISL_1440132, EPI_ISL_1440133, EPI_ISL_1440135, EPI_ISL_1440137, EPI_ISL_1440141, EPI_ISL_1440143, EPI_ISL_1440144, EPI_ISL_1440147, EPI_ISL_1440149,
EPI_ISL_1440150, EPI_ISL_1440152, EPI_ISL_1440153, EPI_ISL_1440155, EPI_ISL_1440156, EPI_ISL_1440158, EPI_ISL_1440161, EPI_ISL_1440162, EPI_ISL_1440165, EPI_ISL_1440168, EPI_ISL_1440170, EPI_ISL_1440176, EPI_ISL_1440177, EPI_ISL_1440180, EPI_ISL_1440182, EPI_ISL_1440183, EPI_ISL_1440186, EPI_ISL_1440191, EPI_ISL_1440192, EPI_ISL_1440197, EPI_ISL_1440203, EPI_ISL_1440205,
EPI_ISL_1440216, EPI_ISL_1440219, EPI_ISL_1440221, EPI_ISL_1440222, EPI_ISL_1440227, EPI_ISL_1440228, EPI_ISL_1440230, EPI_ISL_1440233, EPI_ISL_1440234, EPI_ISL_1440238, EPI_ISL_1440239, EPI_ISL_1440241, EPI_ISL_1440244, EPI_ISL_1440246, EPI_ISL_1440249, EPI_ISL_1440251, EPI_ISL_1440256, EPI_ISL_1440260, EPI_ISL_1440274, EPI_ISL_1440578, EPI_ISL_1440581, EPI_ISL_1440584,
EPI_ISL_1440586, EPI_ISL_1440591, EPI_ISL_1440594, EPI_ISL_1440597, EPI_ISL_1440599, EPI_ISL_1440600, EPI_ISL_1440602, EPI_ISL_1440603, EPI_ISL_1440604, EPI_ISL_1440605, EPI_ISL_1440606, EPI_ISL_1440607, EPI_ISL_1440611, EPI_ISL_1440612, EPI_ISL_1440613, EPI_ISL_1440614, EPI_ISL_1440617, EPI_ISL_1440625, EPI_ISL_1440626, EPI_ISL_1440627, EPI_ISL_1440630, EPI_ISL_1440631,
EPI_ISL_1440632, EPI_ISL_1440634, EPI_ISL_1440635, EPI_ISL_1440636, EPI_ISL_1440637, EPI_ISL_1440638, EPI_ISL_1440641, EPI_ISL_1440642, EPI_ISL_1440645, EPI_ISL_1440648, EPI_ISL_1440653, EPI_ISL_1440654, EPI_ISL_1440658, EPI_ISL_1440659, EPI_ISL_1440662, EPI_ISL_1440663, EPI_ISL_1440664, EPI_ISL_1440665, EPI_ISL_1440667, EPI_ISL_1440668, EPI_ISL_1440670, EPI_ISL_1440671,
EPI_ISL_1440672, EPI_ISL_1440673, EPI_ISL_1440676, EPI_ISL_1440678, EPI_ISL_1440679, EPI_ISL_1440680, EPI_ISL_1440682, EPI_ISL_1440683, EPI_ISL_1440684, EPI_ISL_1440686, EPI_ISL_1440687, EPI_ISL_1440688, EPI_ISL_1440689, EPI_ISL_1440692, EPI_ISL_1440694, EPI_ISL_1440697, EPI_ISL_1440701, EPI_ISL_1440702, EPI_ISL_1440703, EPI_ISL_1440704, EPI_ISL_1440705, EPI_ISL_1440706,
EPI_ISL_1440707, EPI_ISL_1440708, EPI_ISL_1440710, EPI_ISL_1440712, EPI_ISL_1440715, EPI_ISL_1440716, EPI_ISL_1440717, EPI_ISL_1440719, EPI_ISL_1440720, EPI_ISL_1440721, EPI_ISL_1440722, EPI_ISL_1440723, EPI_ISL_1440724, EPI_ISL_1440725, EPI_ISL_1440726, EPI_ISL_1440727, EPI_ISL_1440729, EPI_ISL_1440731, EPI_ISL_1441934, EPI_ISL_1441935, EPI_ISL_1441937, EPI_ISL_1441938,
EPI_ISL_1441939, EPI_ISL_1441940, EPI_ISL_1441941, EPI_ISL_1441943, EPI_ISL_1441944, EPI_ISL_1441945, EPI_ISL_1441946, EPI_ISL_1441947, EPI_ISL_1441951, EPI_ISL_1441952, EPI_ISL_1441953, EPI_ISL_1441956, EPI_ISL_1441957, EPI_ISL_1441958, EPI_ISL_1441960, EPI_ISL_1441963, EPI_ISL_1441964, EPI_ISL_1441965, EPI_ISL_1441966, EPI_ISL_1441969, EPI_ISL_1441970, EPI_ISL_1441971,
EPI_ISL_1441974, EPI_ISL_1441976, EPI_ISL_1441978, EPI_ISL_1441980, EPI_ISL_1441981, EPI_ISL_1441982, EPI_ISL_1441984, EPI_ISL_1441985, EPI_ISL_1441986, EPI_ISL_1441987, EPI_ISL_1441988, EPI_ISL_1442369, EPI_ISL_1442370, EPI_ISL_1442372, EPI_ISL_1442373, EPI_ISL_1442388, EPI_ISL_1442393, EPI_ISL_1442397, EPI_ISL_1442403, EPI_ISL_1442405, EPI_ISL_1442413, EPI_ISL_1442416,
EPI_ISL_1442417, EPI_ISL_1442422, EPI_ISL_1442429, EPI_ISL_1442434, EPI_ISL_1442436, EPI_ISL_1442444, EPI_ISL_1442465, EPI_ISL_1442474, EPI_ISL_1442478, EPI_ISL_1442482, EPI_ISL_1442484, EPI_ISL_1565793, EPI_ISL_1565796, EPI_ISL_1565798, EPI_ISL_1565799, EPI_ISL_1565801, EPI_ISL_1565802, EPI_ISL_1565803, EPI_ISL_1565805, EPI_ISL_1565807, EPI_ISL_1565810,
EPI_ISL_1565814, EPI_ISL_1565815, EPI_ISL_1565819, EPI_ISL_1565820, EPI_ISL_1565821, EPI_ISL_1565822, EPI_ISL_1565823, EPI_ISL_1565825, EPI_ISL_1565826, EPI_ISL_1565827, EPI_ISL_1565828, EPI_ISL_1565831, EPI_ISL_1565833, EPI_ISL_1565837, EPI_ISL_1565840, EPI_ISL_1565841, EPI_ISL_1565844, EPI_ISL_1565846, EPI_ISL_1565848, EPI_ISL_1565849, EPI_ISL_1565851, EPI_ISL_1565853,
EPI_ISL_1565854, EPI_ISL_1565857, EPI_ISL_1565859, EPI_ISL_1565860, EPI_ISL_1565861, EPI_ISL_1565864, EPI_ISL_1565865, EPI_ISL_1565867, EPI_ISL_1565870, EPI_ISL_1565871, EPI_ISL_1565872, EPI_ISL_1565873, EPI_ISL_1565874, EPI_ISL_1565875, EPI_ISL_1565878, EPI_ISL_1565879, EPI_ISL_1565880, EPI_ISL_1565881, EPI_ISL_1565882, EPI_ISL_1565883, EPI_ISL_1565884, EPI_ISL_1565885,
EPI_ISL_1565886, EPI_ISL_1565888, EPI_ISL_1565889, EPI_ISL_1565890, EPI_ISL_1565891, EPI_ISL_1565892, EPI_ISL_1565893, EPI_ISL_1565895, EPI_ISL_1565897, EPI_ISL_1565898, EPI_ISL_1565899, EPI_ISL_1565900, EPI_ISL_1565902, EPI_ISL_1565904, EPI_ISL_1565905, EPI_ISL_1565906, EPI_ISL_1565907, EPI_ISL_1565908, EPI_ISL_1565910, EPI_ISL_1565912, EPI_ISL_1565913, EPI_ISL_1565915,
EPI_ISL_1565916, EPI_ISL_1565917, EPI_ISL_1565919, EPI_ISL_1565922, EPI_ISL_1565923, EPI_ISL_1565924, EPI_ISL_1565925, EPI_ISL_1565927, EPI_ISL_1565928, EPI_ISL_1565929, EPI_ISL_1565930, EPI_ISL_1565932, EPI_ISL_1565933, EPI_ISL_1565934, EPI_ISL_1565935, EPI_ISL_1565936, EPI_ISL_1565937, EPI_ISL_1565938, EPI_ISL_1565939, EPI_ISL_1565940, EPI_ISL_1565941, EPI_ISL_1565942,
EPI_ISL_1565943, EPI_ISL_1565945, EPI_ISL_1565946, EPI_ISL_1565947, EPI_ISL_1565949, EPI_ISL_1565950, EPI_ISL_1565951, EPI_ISL_1565953, EPI_ISL_1565954, EPI_ISL_1565955, EPI_ISL_1565956, EPI_ISL_1565957, EPI_ISL_1565960, EPI_ISL_1565961, EPI_ISL_1565962, EPI_ISL_1565963, EPI_ISL_1565964, EPI_ISL_1565965, EPI_ISL_1565966, EPI_ISL_1565968, EPI_ISL_1565969, EPI_ISL_1565970,
EPI_ISL_1565972, EPI_ISL_1565973, EPI_ISL_1565975, EPI_ISL_1565976, EPI_ISL_1565977, EPI_ISL_1565978, EPI_ISL_1565979, EPI_ISL_1565980, EPI_ISL_1565981, EPI_ISL_1565982, EPI_ISL_1565983, EPI_ISL_1565984, EPI_ISL_1565985, EPI_ISL_1565986, EPI_ISL_1565988, EPI_ISL_1565989, EPI_ISL_1565990, EPI_ISL_1565991, EPI_ISL_1566111, EPI_ISL_1566114, EPI_ISL_1566119, EPI_ISL_1566123,
EPI_ISL_1566127, EPI_ISL_1566137, EPI_ISL_1566140, EPI_ISL_1566143, EPI_ISL_1566146, EPI_ISL_1566147, EPI_ISL_1566152, EPI_ISL_1566155, EPI_ISL_1566156, EPI_ISL_1566159, EPI_ISL_1566160, EPI_ISL_1566162, EPI_ISL_1566168, EPI_ISL_1566171, EPI_ISL_1566173, EPI_ISL_1566177, EPI_ISL_1566181, EPI_ISL_1566185, EPI_ISL_1566186, EPI_ISL_1566200, EPI_ISL_1566202, EPI_ISL_1566205,
EPI_ISL_1566207, EPI_ISL_1566212, EPI_ISL_1566214, EPI_ISL_1566218, EPI_ISL_1566221, EPI_ISL_1566223, EPI_ISL_1566226, EPI_ISL_1566227, EPI_ISL_1566233, EPI_ISL_1566235, EPI_ISL_1566236, EPI_ISL_1566240, EPI_ISL_1566244, EPI_ISL_1566245, EPI_ISL_1566249, EPI_ISL_1566250, EPI_ISL_1566259, EPI_ISL_1566261, EPI_ISL_1566262, EPI_ISL_1566263, EPI_ISL_1566266, EPI_ISL_1566268,
EPI_ISL_1566277, EPI_ISL_1566281, EPI_ISL_1566283, EPI_ISL_1566289, EPI_ISL_1566290, EPI_ISL_1566294, EPI_ISL_1566296, EPI_ISL_1566299, EPI_ISL_1566301, EPI_ISL_1566302, EPI_ISL_1566305, EPI_ISL_1566308, EPI_ISL_1566312, EPI_ISL_1566315, EPI_ISL_1566317, EPI_ISL_1566318, EPI_ISL_1566326, EPI_ISL_1566328, EPI_ISL_1566332, EPI_ISL_1566337, EPI_ISL_1566342, EPI_ISL_1566345,
EPI_ISL_1566347, EPI_ISL_1566350, EPI_ISL_1566352, EPI_ISL_1566355, EPI_ISL_1566358, EPI_ISL_1566360, EPI_ISL_1566369, EPI_ISL_1566372, EPI_ISL_1566375, EPI_ISL_1566380, EPI_ISL_1566382, EPI_ISL_1566385, EPI_ISL_1566388, EPI_ISL_1568581, EPI_ISL_1568591, EPI_ISL_1568594, EPI_ISL_1568595, EPI_ISL_1568603, EPI_ISL_1568615, EPI_ISL_1568619, EPI_ISL_1568631, EPI_ISL_1568643,
EPI_ISL_1568644, EPI_ISL_1568657, EPI_ISL_1568660, EPI_ISL_1568662, EPI_ISL_1568671, EPI_ISL_1568673, EPI_ISL_1568675, EPI_ISL_1568695, EPI_ISL_1568697, EPI_ISL_1568703, EPI_ISL_1568714, EPI_ISL_1568719, EPI_ISL_1568723, EPI_ISL_1568725, EPI_ISL_1568735, EPI_ISL_1568738, EPI_ISL_1568745, EPI_ISL_1568752, EPI_ISL_1568788, EPI_ISL_1568818, EPI_ISL_1572233, EPI_ISL_1572245,
EPI_ISL_1572254, EPI_ISL_1572256, EPI_ISL_1572265, EPI_ISL_1572267, EPI_ISL_1572271, EPI_ISL_1572276, EPI_ISL_1572281, EPI_ISL_1572284, EPI_ISL_1572288, EPI_ISL_1572294, EPI_ISL_1572297, EPI_ISL_1572299, EPI_ISL_1572301, EPI_ISL_1572304, EPI_ISL_1572309, EPI_ISL_1572314, EPI_ISL_1572317, EPI_ISL_1572319, EPI_ISL_1572324, EPI_ISL_1572325, EPI_ISL_1572327, EPI_ISL_1572332,
EPI_ISL_1572333, EPI_ISL_1572337, EPI_ISL_1572344, EPI_ISL_1572345, EPI_ISL_1572347, EPI_ISL_1572348, EPI_ISL_1572350, EPI_ISL_1572352, EPI_ISL_1572358, EPI_ISL_1572359, EPI_ISL_1572365, EPI_ISL_1572397, EPI_ISL_1572402, EPI_ISL_1572405, EPI_ISL_1572418, EPI_ISL_1572425, EPI_ISL_1572446, EPI_ISL_1572453, EPI_ISL_1572464, EPI_ISL_1572472, EPI_ISL_1572475, EPI_ISL_1572478,
EPI_ISL_1572482, EPI_ISL_1572510, EPI_ISL_1572518, EPI_ISL_1572522, EPI_ISL_1646053, EPI_ISL_1646324, EPI_ISL_1646368, EPI_ISL_1646609, EPI_ISL_1646674, EPI_ISL_1646847, EPI_ISL_1646867, EPI_ISL_1719927, EPI_ISL_1719928, EPI_ISL_1719934, EPI_ISL_1719935, EPI_ISL_1719938, EPI_ISL_1719943, EPI_ISL_1719948, EPI_ISL_1719950, EPI_ISL_1719951, EPI_ISL_1719957, EPI_ISL_1719958,
EPI_ISL_1719960, EPI_ISL_1719966, EPI_ISL_1719969, EPI_ISL_1719974, EPI_ISL_1719988, EPI_ISL_1719990, EPI_ISL_1719997, EPI_ISL_1719999, EPI_ISL_1720002, EPI_ISL_1720003, EPI_ISL_1720004, EPI_ISL_1720005, EPI_ISL_1720006, EPI_ISL_1720010, EPI_ISL_1720013, EPI_ISL_1720015, EPI_ISL_1720018, EPI_ISL_1720020, EPI_ISL_1720022, EPI_ISL_1720023, EPI_ISL_1720024, EPI_ISL_1720028,
EPI_ISL_1720030, EPI_ISL_1720033, EPI_ISL_1720036, EPI_ISL_1720037, EPI_ISL_1720041, EPI_ISL_1720043, EPI_ISL_1720044, EPI_ISL_1720045, EPI_ISL_1720047, EPI_ISL_1720048, EPI_ISL_1720049, EPI_ISL_1720050, EPI_ISL_1720054, EPI_ISL_1720057, EPI_ISL_1720058, EPI_ISL_1720061, EPI_ISL_1720066, EPI_ISL_1720071, EPI_ISL_1720075, EPI_ISL_1720076, EPI_ISL_1720080, EPI_ISL_1720082,
EPI_ISL_1720087, EPI_ISL_1720088, EPI_ISL_1720093, EPI_ISL_1720104, EPI_ISL_1720108, EPI_ISL_1720111, EPI_ISL_1720119, EPI_ISL_1720123, EPI_ISL_1720124, EPI_ISL_1720128, EPI_ISL_1720133, EPI_ISL_1720137, EPI_ISL_1720138, EPI_ISL_1720140, EPI_ISL_1720141, EPI_ISL_1720142, EPI_ISL_1720146, EPI_ISL_1720153, EPI_ISL_1720154, EPI_ISL_1720157, EPI_ISL_1849766, EPI_ISL_1849798

see above SYNLAB MVZ Leverkusen Robert Koch Institute

EPI_ISL_1566103, EPI_ISL_1566105, EPI_ISL_1566112, EPI_ISL_1566118, EPI_ISL_1566121, EPI_ISL_1566125, EPI_ISL_1566136, EPI_ISL_1566204, EPI_ISL_1566215, EPI_ISL_1566219, EPI_ISL_1566320, EPI_ISL_1566323, EPI_ISL_1567707, EPI_ISL_1567708, EPI_ISL_1567710, EPI_ISL_1567711, EPI_ISL_1567722, EPI_ISL_1567725, EPI_ISL_1567726, EPI_ISL_1567729, EPI_ISL_1567730, EPI_ISL_1567733, EPI_ISL_1567734, EPI_ISL_1567742, EPI_ISL_1567749, EPI_ISL_1567758, EPI_ISL_1567760, EPI_ISL_1567761, EPI_ISL_1567763, EPI_ISL_1572291, EPI_ISL_1572329, EPI_ISL_1572354, EPI_ISL_1572355, EPI_ISL_1572395, EPI_ISL_1572399, EPI_ISL_1729939

see above SYNLAB MVZ Neustadt a. d. WeinstraÃŸe Robert Koch Institute

EPI_ISL_1432078, EPI_ISL_1432113, EPI_ISL_1432116, EPI_ISL_1432263, EPI_ISL_1432288, EPI_ISL_1432639, EPI_ISL_1440592, EPI_ISL_1440655, EPI_ISL_1440669, EPI_ISL_1440693

see above SYNLAB MVZ Neustadt a. d. Weinstraße Robert Koch Institute

EPI_ISL_1286767, EPI_ISL_1286768, EPI_ISL_1286769, EPI_ISL_1286771, EPI_ISL_1286774, EPI_ISL_1286781, EPI_ISL_1286782, EPI_ISL_1286785, EPI_ISL_1286789, EPI_ISL_1286792, EPI_ISL_1286797, EPI_ISL_1286801, EPI_ISL_1286811, EPI_ISL_1286812, EPI_ISL_1286816, EPI_ISL_1286829, EPI_ISL_1286834, EPI_ISL_1286840, EPI_ISL_1286846, EPI_ISL_1286849, EPI_ISL_1286869, EPI_ISL_1286870, EPI_ISL_1286879, EPI_ISL_1350646, EPI_ISL_1350655, EPI_ISL_1350660, EPI_ISL_1350693, EPI_ISL_1350765, EPI_ISL_1350769, EPI_ISL_1350770, EPI_ISL_1350772, EPI_ISL_1350773, EPI_ISL_1350774, EPI_ISL_1350775, EPI_ISL_1350776, EPI_ISL_1350777, EPI_ISL_1350778, EPI_ISL_1350779, EPI_ISL_1350780, EPI_ISL_1350781, EPI_ISL_1350782, EPI_ISL_1350783, EPI_ISL_1350784, EPI_ISL_1350786, EPI_ISL_1350787, EPI_ISL_1350788, EPI_ISL_1350789, EPI_ISL_1350790, EPI_ISL_1350791, EPI_ISL_1350792, EPI_ISL_1350794, EPI_ISL_1350796, EPI_ISL_1350797, EPI_ISL_1350798, EPI_ISL_1350801, EPI_ISL_1350802, EPI_ISL_1350803, EPI_ISL_1350804, EPI_ISL_1350807, EPI_ISL_1350808, EPI_ISL_1350809, EPI_ISL_1350811, EPI_ISL_1350812, EPI_ISL_1350814, EPI_ISL_1350816, EPI_ISL_1350819, EPI_ISL_1350821, EPI_ISL_1350823, EPI_ISL_1350824, EPI_ISL_1350827, EPI_ISL_1350828, EPI_ISL_1350829, EPI_ISL_1350830, EPI_ISL_1350831, EPI_ISL_1350833, EPI_ISL_1350834, EPI_ISL_1350835, EPI_ISL_1350837, EPI_ISL_1350838, EPI_ISL_1350840, EPI_ISL_1350843, EPI_ISL_1350844, EPI_ISL_1350846, EPI_ISL_1350847, EPI_ISL_1350848, EPI_ISL_1350849, EPI_ISL_1350850, EPI_ISL_1350851, EPI_ISL_1350852, EPI_ISL_1350853, EPI_ISL_1350854, EPI_ISL_1350855, EPI_ISL_1350856, EPI_ISL_1350857, EPI_ISL_1350858, EPI_ISL_1350859, EPI_ISL_1350860, EPI_ISL_1350861, EPI_ISL_1350862, EPI_ISL_1350864, EPI_ISL_1350868, EPI_ISL_1350869, EPI_ISL_1350872, EPI_ISL_1350873, EPI_ISL_1350874, EPI_ISL_1350875, EPI_ISL_1350876, EPI_ISL_1350877, EPI_ISL_1350879, EPI_ISL_1350880, EPI_ISL_1350886, EPI_ISL_1350887, EPI_ISL_1350889, EPI_ISL_1350890, EPI_ISL_1350891, EPI_ISL_1350892, EPI_ISL_1350893, EPI_ISL_1350894, EPI_ISL_1350896, EPI_ISL_1350898, EPI_ISL_1350899, EPI_ISL_1350900, EPI_ISL_1350901, EPI_ISL_1350903, EPI_ISL_1350904, EPI_ISL_1350905, EPI_ISL_1350907, EPI_ISL_1350908, EPI_ISL_1350909, EPI_ISL_1350910, EPI_ISL_1350911, EPI_ISL_1350912, EPI_ISL_1350916, EPI_ISL_1350917, EPI_ISL_1350918, EPI_ISL_1350919, EPI_ISL_1350920, EPI_ISL_1350921, EPI_ISL_1350922, EPI_ISL_1351965, EPI_ISL_1352738, EPI_ISL_1352745, EPI_ISL_1352865, EPI_ISL_1352867, EPI_ISL_1352869, EPI_ISL_1352877, EPI_ISL_1352879, EPI_ISL_1352880, EPI_ISL_1352884, EPI_ISL_1352885, EPI_ISL_1352889, EPI_ISL_1352890, EPI_ISL_1352891, EPI_ISL_1352892, EPI_ISL_1352894, EPI_ISL_1352895, EPI_ISL_1352896, EPI_ISL_1352897, EPI_ISL_1352898, EPI_ISL_1352899, EPI_ISL_1352900, EPI_ISL_1352901, EPI_ISL_1352902, EPI_ISL_1352903, EPI_ISL_1352904, EPI_ISL_1352905, EPI_ISL_1352907, EPI_ISL_1352908, EPI_ISL_1352909, EPI_ISL_1352911, EPI_ISL_1352912, EPI_ISL_1352913, EPI_ISL_1352915, EPI_ISL_1352916, EPI_ISL_1352917, EPI_ISL_1352921, EPI_ISL_1352922, EPI_ISL_1352923, EPI_ISL_1352925, EPI_ISL_1352926, EPI_ISL_1352928, EPI_ISL_1352929, EPI_ISL_1352930, EPI_ISL_1352931, EPI_ISL_1352933, EPI_ISL_1352934, EPI_ISL_1352935, EPI_ISL_1352936, EPI_ISL_1352938, EPI_ISL_1352939, EPI_ISL_1352940, EPI_ISL_1352941, EPI_ISL_1352942, EPI_ISL_1352945, EPI_ISL_1352946, EPI_ISL_1352947, EPI_ISL_1352948, EPI_ISL_1352950, EPI_ISL_1352951, EPI_ISL_1352953, EPI_ISL_1352956, EPI_ISL_1352957, EPI_ISL_1352958, EPI_ISL_1352959, EPI_ISL_1352960, EPI_ISL_1352961, EPI_ISL_1352963, EPI_ISL_1352964, EPI_ISL_1352967, EPI_ISL_1352969, EPI_ISL_1352975, EPI_ISL_1352978, EPI_ISL_1352979, EPI_ISL_1352980, EPI_ISL_1352981, EPI_ISL_1352983, EPI_ISL_1352984, EPI_ISL_1352985, EPI_ISL_1352986, EPI_ISL_1352987, EPI_ISL_1352989, EPI_ISL_1352990, EPI_ISL_1352991, EPI_ISL_1352998, EPI_ISL_1353003, EPI_ISL_1353004, EPI_ISL_1353008, EPI_ISL_1353009, EPI_ISL_1353012, EPI_ISL_1355982, EPI_ISL_1356053, EPI_ISL_1356078, EPI_ISL_1356107, EPI_ISL_1356193, EPI_ISL_1437614, EPI_ISL_1437615, EPI_ISL_1437616, EPI_ISL_1437617, EPI_ISL_1437619, EPI_ISL_1437622, EPI_ISL_1437623, EPI_ISL_1437624, EPI_ISL_1437625, EPI_ISL_1437626, EPI_ISL_1437627, EPI_ISL_1437628, EPI_ISL_1437629, EPI_ISL_1437630, EPI_ISL_1437631, EPI_ISL_1437632, EPI_ISL_1437633, EPI_ISL_1437635, EPI_ISL_1437636, EPI_ISL_1437637, EPI_ISL_1437638, EPI_ISL_1437639, EPI_ISL_1437640, EPI_ISL_1437641, EPI_ISL_1437643, EPI_ISL_1437644, EPI_ISL_1437646, EPI_ISL_1437648, EPI_ISL_1437651, EPI_ISL_1437652, EPI_ISL_1437654, EPI_ISL_1437655, EPI_ISL_1437656, EPI_ISL_1437657, EPI_ISL_1437658, EPI_ISL_1437659, EPI_ISL_1437660, EPI_ISL_1437661, EPI_ISL_1437662, EPI_ISL_1437665, EPI_ISL_1437666, EPI_ISL_1437669, EPI_ISL_1437670, EPI_ISL_1437671, EPI_ISL_1437672, EPI_ISL_1437673, EPI_ISL_1437674, EPI_ISL_1437675, EPI_ISL_1437676, EPI_ISL_1437677, EPI_ISL_1437678, EPI_ISL_1437679, EPI_ISL_1437680, EPI_ISL_1437682, EPI_ISL_1437683, EPI_ISL_1437684, EPI_ISL_1437685, EPI_ISL_1437687, EPI_ISL_1437689, EPI_ISL_1437691, EPI_ISL_1437693, EPI_ISL_1437695, EPI_ISL_1437697, EPI_ISL_1437700, EPI_ISL_1437702, EPI_ISL_1437705, EPI_ISL_1437708, EPI_ISL_1437710, EPI_ISL_1437712, EPI_ISL_1437714, EPI_ISL_1437716, EPI_ISL_1437721, EPI_ISL_1437724, EPI_ISL_1437726, EPI_ISL_1437731, EPI_ISL_1437732, EPI_ISL_1437733, EPI_ISL_1437735, EPI_ISL_1437738, EPI_ISL_1437739, EPI_ISL_1437740, EPI_ISL_1437741, EPI_ISL_1437743, EPI_ISL_1437744, EPI_ISL_1437745, EPI_ISL_1437747, EPI_ISL_1437752, EPI_ISL_1437753, EPI_ISL_1437755, EPI_ISL_1437756, EPI_ISL_1437758, EPI_ISL_1437759, EPI_ISL_1437760, EPI_ISL_1437761, EPI_ISL_1437763, EPI_ISL_1437766, EPI_ISL_1437768, EPI_ISL_1437769, EPI_ISL_1437770, EPI_ISL_1437771, EPI_ISL_1437773, EPI_ISL_1437775, EPI_ISL_1437778, EPI_ISL_1437779, EPI_ISL_1437782, EPI_ISL_1437783, EPI_ISL_1437784, EPI_ISL_1437785, EPI_ISL_1437787, EPI_ISL_1437789, EPI_ISL_1437790, EPI_ISL_1437791, EPI_ISL_1437792, EPI_ISL_1437793, EPI_ISL_1437794, EPI_ISL_1437795, EPI_ISL_1437796, EPI_ISL_1437797, EPI_ISL_1437798, EPI_ISL_1437799, EPI_ISL_1437800, EPI_ISL_1437801, EPI_ISL_1437802, EPI_ISL_1437803, EPI_ISL_1437804, EPI_ISL_1437805, EPI_ISL_1437806, EPI_ISL_1437807, EPI_ISL_1437809, EPI_ISL_1437811, EPI_ISL_1437812, EPI_ISL_1437813, EPI_ISL_1437817, EPI_ISL_1437818, EPI_ISL_1437820, EPI_ISL_1437821, EPI_ISL_1437822, EPI_ISL_1437823, EPI_ISL_1437824, EPI_ISL_1437825, EPI_ISL_1437826, EPI_ISL_1437827, EPI_ISL_1437828, EPI_ISL_1437829, EPI_ISL_1437830, EPI_ISL_1437831, EPI_ISL_1437832, EPI_ISL_1437833, EPI_ISL_1437834, EPI_ISL_1437835, EPI_ISL_1437836, EPI_ISL_1437838, EPI_ISL_1437839, EPI_ISL_1437840, EPI_ISL_1437841, EPI_ISL_1437842, EPI_ISL_1437844, EPI_ISL_1437849, EPI_ISL_1437850, EPI_ISL_1437851, EPI_ISL_1437852, EPI_ISL_1437853, EPI_ISL_1437854, EPI_ISL_1437855, EPI_ISL_1437856, EPI_ISL_1437857, EPI_ISL_1437858, EPI_ISL_1437860, EPI_ISL_1437862, EPI_ISL_1437864, EPI_ISL_1437865, EPI_ISL_1437866, EPI_ISL_1437867, EPI_ISL_1437868, EPI_ISL_1437870, EPI_ISL_1437871, EPI_ISL_1437872, EPI_ISL_1437873, EPI_ISL_1437874, EPI_ISL_1437875, EPI_ISL_1437876, EPI_ISL_1437877, EPI_ISL_1437878, EPI_ISL_1437879, EPI_ISL_1437880, EPI_ISL_1437883, EPI_ISL_1437884, EPI_ISL_1437885, EPI_ISL_1437886, EPI_ISL_1437887, EPI_ISL_1437888, EPI_ISL_1437889, EPI_ISL_1437891, EPI_ISL_1437894, EPI_ISL_1437895, EPI_ISL_1437896, EPI_ISL_1437897, EPI_ISL_1437898, EPI_ISL_1437899, EPI_ISL_1437900, EPI_ISL_1437902, EPI_ISL_1437903, EPI_ISL_1437906, EPI_ISL_1437907, EPI_ISL_1437908, EPI_ISL_1437910, EPI_ISL_1437912, EPI_ISL_1437914, EPI_ISL_1437916, EPI_ISL_1437917, EPI_ISL_1437918, EPI_ISL_1437919, EPI_ISL_1437920, EPI_ISL_1437921, EPI_ISL_1437922, EPI_ISL_1437924, EPI_ISL_1437926, EPI_ISL_1437928, EPI_ISL_1437929, EPI_ISL_1437930, EPI_ISL_1437931, EPI_ISL_1440299, EPI_ISL_1440302, EPI_ISL_1440315, EPI_ISL_1440321, EPI_ISL_1440328, EPI_ISL_1440331, EPI_ISL_1440333, EPI_ISL_1440334, EPI_ISL_1440343, EPI_ISL_1440344, EPI_ISL_1440348, EPI_ISL_1440362, EPI_ISL_1440365, EPI_ISL_1440372, EPI_ISL_1440374, EPI_ISL_1440378, EPI_ISL_1440383, EPI_ISL_1440385, EPI_ISL_1440390, EPI_ISL_1440395, EPI_ISL_1440398, EPI_ISL_1440419, EPI_ISL_1440430, EPI_ISL_1440445, EPI_ISL_1440447, EPI_ISL_1440455, EPI_ISL_1440456, EPI_ISL_1440461, EPI_ISL_1440482, EPI_ISL_1440484, EPI_ISL_1440497, EPI_ISL_1440501, EPI_ISL_1440502, EPI_ISL_1440508, EPI_ISL_1440512, EPI_ISL_1440517, EPI_ISL_1440522, EPI_ISL_1440539, EPI_ISL_1440549, EPI_ISL_1440564, EPI_ISL_1440575

see above SYNLAB MVZ Trier Robert Koch Institute

EPI_ISL_1286798, EPI_ISL_1286831, EPI_ISL_1286841, EPI_ISL_1286867, EPI_ISL_1286873, EPI_ISL_1350800, EPI_ISL_1350806, EPI_ISL_1350817, EPI_ISL_1350820, EPI_ISL_1350839, EPI_ISL_1350841, EPI_ISL_1350865, EPI_ISL_1350867, EPI_ISL_1350882, EPI_ISL_1350884, EPI_ISL_1350897, EPI_ISL_1350906, EPI_ISL_1350913, EPI_ISL_1350915, EPI_ISL_1352871, EPI_ISL_1352873, EPI_ISL_1352886, EPI_ISL_1352887, EPI_ISL_1352893, EPI_ISL_1352914, EPI_ISL_1352918, EPI_ISL_1352924, EPI_ISL_1352932, EPI_ISL_1352937, EPI_ISL_1352944, EPI_ISL_1352952, EPI_ISL_1352955, EPI_ISL_1352962, EPI_ISL_1352966, EPI_ISL_1352970, EPI_ISL_1352971, EPI_ISL_1352972, EPI_ISL_1352974, EPI_ISL_1352976, EPI_ISL_1352982, EPI_ISL_1352988, EPI_ISL_1352992, EPI_ISL_1352994, EPI_ISL_1353005, EPI_ISL_1353011, EPI_ISL_1353013, EPI_ISL_1437645, EPI_ISL_1437650, EPI_ISL_1437664, EPI_ISL_1437681, EPI_ISL_1437719, EPI_ISL_1437777, EPI_ISL_1437780, EPI_ISL_1437786, EPI_ISL_1437816, EPI_ISL_1437843, EPI_ISL_1437881, EPI_ISL_1437882, EPI_ISL_1437904, EPI_ISL_1440318, EPI_ISL_1440550

see above SYNLAB MVZ Weiden Robert Koch Institute

EPI_ISL_1433057, EPI_ISL_1433164, EPI_ISL_1433165, EPI_ISL_1433294, EPI_ISL_1433347, EPI_ISL_1438701, EPI_ISL_1438738, EPI_ISL_1438771, EPI_ISL_1438772, EPI_ISL_1438861, EPI_ISL_1438870, EPI_ISL_1438887, EPI_ISL_1438900, EPI_ISL_1438904, EPI_ISL_1438986, EPI_ISL_1439019, EPI_ISL_1439025, EPI_ISL_1439127, EPI_ISL_1439135, EPI_ISL_1439141, EPI_ISL_1439218, EPI_ISL_1568949, EPI_ISL_1568958, EPI_ISL_1568984, EPI_ISL_1569333, EPI_ISL_1638714, EPI_ISL_1638817, EPI_ISL_1639083, EPI_ISL_1639249, EPI_ISL_1643211, EPI_ISL_1643295, EPI_ISL_1721254, EPI_ISL_1721261, EPI_ISL_1721279, EPI_ISL_1721305

see above Sonic - Bioscientia - MVZ Labor Saar GmbH Robert Koch Institute

EPI_ISL_1354008, EPI_ISL_1436016, EPI_ISL_1436092, EPI_ISL_1436108, EPI_ISL_1436117, EPI_ISL_1436119, EPI_ISL_1436140, EPI_ISL_1436142, EPI_ISL_1436155, EPI_ISL_1436165, EPI_ISL_1436211, EPI_ISL_1436212, EPI_ISL_1436226, EPI_ISL_1436300, EPI_ISL_1436377, EPI_ISL_1439065, EPI_ISL_1439070, EPI_ISL_1439098, EPI_ISL_1439114, EPI_ISL_1439177, EPI_ISL_1439202, EPI_ISL_1439209, EPI_ISL_1439220, EPI_ISL_1439272, EPI_ISL_1439282, EPI_ISL_1439357

see above Sonic - Dr. Staber & Kollegen GmbH München Robert Koch Institute

EPI_ISL_1354001, EPI_ISL_1354006, EPI_ISL_1354010, EPI_ISL_1354011, EPI_ISL_1354013, EPI_ISL_1354020, EPI_ISL_1354043, EPI_ISL_1354044, EPI_ISL_1354051, EPI_ISL_1354066, EPI_ISL_1354068, EPI_ISL_1354074, EPI_ISL_1354084, EPI_ISL_1354105, EPI_ISL_1354110, EPI_ISL_1354111, EPI_ISL_1354115, EPI_ISL_1354119, EPI_ISL_1354123, EPI_ISL_1354132, EPI_ISL_1354138, EPI_ISL_1354140, EPI_ISL_1354142, EPI_ISL_1354155, EPI_ISL_1354169, EPI_ISL_1354171, EPI_ISL_1354172, EPI_ISL_1354180, EPI_ISL_1354194, EPI_ISL_1354200, EPI_ISL_1354201, EPI_ISL_1354202, EPI_ISL_1354222, EPI_ISL_1354244, EPI_ISL_1436019, EPI_ISL_1436023, EPI_ISL_1436027, EPI_ISL_1436042, EPI_ISL_1436044, EPI_ISL_1436055, EPI_ISL_1436059, EPI_ISL_1436065, EPI_ISL_1436069, EPI_ISL_1436091, EPI_ISL_1436115, EPI_ISL_1436118, EPI_ISL_1436125, EPI_ISL_1436138, EPI_ISL_1436152, EPI_ISL_1436166, EPI_ISL_1436185, EPI_ISL_1436188, EPI_ISL_1436197, EPI_ISL_1436202, EPI_ISL_1436204, EPI_ISL_1436217, EPI_ISL_1436229, EPI_ISL_1436240, EPI_ISL_1436251, EPI_ISL_1436252, EPI_ISL_1436266, EPI_ISL_1436267, EPI_ISL_1436291, EPI_ISL_1436308, EPI_ISL_1436309, EPI_ISL_1436321, EPI_ISL_1436325, EPI_ISL_1436335, EPI_ISL_1436339, EPI_ISL_1436346, EPI_ISL_1436350, EPI_ISL_1436352, EPI_ISL_1436364, EPI_ISL_1436368, EPI_ISL_1436372, EPI_ISL_1436376, EPI_ISL_1436378, EPI_ISL_1436381, EPI_ISL_1436383, EPI_ISL_1436387, EPI_ISL_1436392, EPI_ISL_1438689, EPI_ISL_1438690, EPI_ISL_1438707, EPI_ISL_1438708, EPI_ISL_1438725, EPI_ISL_1438728, EPI_ISL_1438766, EPI_ISL_1438779, EPI_ISL_1438789, EPI_ISL_1438795, EPI_ISL_1438805, EPI_ISL_1438823, EPI_ISL_1438826, EPI_ISL_1438831, EPI_ISL_1438834, EPI_ISL_1438851, EPI_ISL_1438862, EPI_ISL_1438872, EPI_ISL_1438884, EPI_ISL_1438901, EPI_ISL_1438907, EPI_ISL_1438908, EPI_ISL_1438926, EPI_ISL_1438928, EPI_ISL_1438934, EPI_ISL_1438935, EPI_ISL_1438959, EPI_ISL_1438976, EPI_ISL_1439001, EPI_ISL_1439002, EPI_ISL_1439005, EPI_ISL_1439008, EPI_ISL_1439010, EPI_ISL_1439024, EPI_ISL_1439030, EPI_ISL_1568964, EPI_ISL_1568969, EPI_ISL_1569013, EPI_ISL_1569017, EPI_ISL_1569035, EPI_ISL_1569037, EPI_ISL_1569047, EPI_ISL_1569049, EPI_ISL_1569054, EPI_ISL_1569058, EPI_ISL_1569076, EPI_ISL_1569080, EPI_ISL_1569082, EPI_ISL_1569089, EPI_ISL_1569104, EPI_ISL_1569105, EPI_ISL_1569113, EPI_ISL_1569115, EPI_ISL_1569118, EPI_ISL_1569125, EPI_ISL_1569142, EPI_ISL_1569185, EPI_ISL_1569187, EPI_ISL_1569215, EPI_ISL_1569229, EPI_ISL_1569238, EPI_ISL_1569245, EPI_ISL_1569261, EPI_ISL_1569262, EPI_ISL_1569266, EPI_ISL_1569271

see above Sonic - Labor Dr. von Froreich GmbH Robert Koch Institute

EPI_ISL_1568950, EPI_ISL_1569008, EPI_ISL_1569081, EPI_ISL_1569114, EPI_ISL_1569148, EPI_ISL_1569226 Sonic - Labor Lademannbogen MVZ GmbH Robert Koch Institute

EPI_ISL_1354195, EPI_ISL_1436062, EPI_ISL_1436071, EPI_ISL_1436098, EPI_ISL_1436167, EPI_ISL_1436268, EPI_ISL_1436270, EPI_ISL_1436286, EPI_ISL_1436292, EPI_ISL_1436322, EPI_ISL_1436324, EPI_ISL_1436332, EPI_ISL_1436337, EPI_ISL_1436389, EPI_ISL_1439043, EPI_ISL_1439077, EPI_ISL_1439172, EPI_ISL_1439201, EPI_ISL_1439224, EPI_ISL_1439246, EPI_ISL_1439252, EPI_ISL_1439302, EPI_ISL_1439321, EPI_ISL_1439361

see above Sonic - Labor Staber Bayreuth Robert Koch Institute

EPI_ISL_1436012, EPI_ISL_1436025, EPI_ISL_1436093, EPI_ISL_1436121, EPI_ISL_1436137, EPI_ISL_1436143, EPI_ISL_1436147, EPI_ISL_1436160, EPI_ISL_1436164, EPI_ISL_1436179, EPI_ISL_1436242, EPI_ISL_1436243, EPI_ISL_1436254, EPI_ISL_1436263, EPI_ISL_1436329, EPI_ISL_1436384, EPI_ISL_1436391, EPI_ISL_1439048, EPI_ISL_1439078, EPI_ISL_1439101, EPI_ISL_1439124, EPI_ISL_1439136, EPI_ISL_1439147, EPI_ISL_1439213, EPI_ISL_1439241, EPI_ISL_1439291, EPI_ISL_1439347

see above Sonic - Labor Staber Dresden (Klipphausen) Robert Koch Institute

EPI_ISL_1436114, EPI_ISL_1436182, EPI_ISL_1436189, EPI_ISL_1436208, EPI_ISL_1436271, EPI_ISL_1439125, EPI_ISL_1439193, EPI_ISL_1439320

see above Sonic - Labor Staber Heilbronn Robert Koch Institute

EPI_ISL_1353781, EPI_ISL_1353861, EPI_ISL_1353882, EPI_ISL_1433197, EPI_ISL_1433277, EPI_ISL_1438909, EPI_ISL_1438969

see above Sonic - Labor Staber MVZ Kassel Robert Koch Institute

EPI_ISL_1436007, EPI_ISL_1436060, EPI_ISL_1436061, EPI_ISL_1436175, EPI_ISL_1436178, EPI_ISL_1436186, EPI_ISL_1436228, EPI_ISL_1436280, EPI_ISL_1439046, EPI_ISL_1439059, EPI_ISL_1439080, EPI_ISL_1439095, EPI_ISL_1439228, EPI_ISL_1439353

see above Sonic - Labor Staber Nürnberg Robert Koch Institute

EPI_ISL_1353653, EPI_ISL_1353656, EPI_ISL_1353658, EPI_ISL_1353671, EPI_ISL_1353672, EPI_ISL_1353675, EPI_ISL_1353679, EPI_ISL_1353681, EPI_ISL_1353682, EPI_ISL_1353686, EPI_ISL_1353696, EPI_ISL_1353699, EPI_ISL_1353701, EPI_ISL_1353710, EPI_ISL_1353712, EPI_ISL_1353714, EPI_ISL_1353716, EPI_ISL_1353717, EPI_ISL_1353720, EPI_ISL_1353723, EPI_ISL_1353726, EPI_ISL_1353729, EPI_ISL_1353738, EPI_ISL_1353742, EPI_ISL_1353752, EPI_ISL_1353754, EPI_ISL_1353762, EPI_ISL_1353770, EPI_ISL_1353771, EPI_ISL_1353800, EPI_ISL_1353801, EPI_ISL_1353802, EPI_ISL_1353815, EPI_ISL_1353820, EPI_ISL_1353822, EPI_ISL_1353826, EPI_ISL_1353830, EPI_ISL_1353832, EPI_ISL_1353836, EPI_ISL_1353837, EPI_ISL_1353844, EPI_ISL_1353853, EPI_ISL_1353857, EPI_ISL_1353860, EPI_ISL_1353876, EPI_ISL_1353880, EPI_ISL_1353884, EPI_ISL_1353885, EPI_ISL_1353896, EPI_ISL_1353923, EPI_ISL_1353924, EPI_ISL_1353930, EPI_ISL_1353932, EPI_ISL_1353943, EPI_ISL_1353947, EPI_ISL_1353957, EPI_ISL_1353961, EPI_ISL_1353977, EPI_ISL_1353979, EPI_ISL_1353986, EPI_ISL_1353997, EPI_ISL_1354002, EPI_ISL_1354005, EPI_ISL_1354018, EPI_ISL_1354022, EPI_ISL_1354030, EPI_ISL_1354036, EPI_ISL_1354038, EPI_ISL_1354041, EPI_ISL_1354052, EPI_ISL_1354054, EPI_ISL_1354055, EPI_ISL_1354057, EPI_ISL_1354060, EPI_ISL_1354062, EPI_ISL_1354064, EPI_ISL_1354065, EPI_ISL_1354067, EPI_ISL_1354078, EPI_ISL_1354080, EPI_ISL_1354089, EPI_ISL_1354093, EPI_ISL_1354100, EPI_ISL_1354109, EPI_ISL_1354118, EPI_ISL_1354122, EPI_ISL_1354135, EPI_ISL_1354141, EPI_ISL_1354143, EPI_ISL_1354150, EPI_ISL_1354151, EPI_ISL_1354152, EPI_ISL_1354154, EPI_ISL_1354165, EPI_ISL_1354170, EPI_ISL_1354173, EPI_ISL_1354174, EPI_ISL_1354183, EPI_ISL_1354185, EPI_ISL_1354188, EPI_ISL_1354198, EPI_ISL_1354210, EPI_ISL_1354213, EPI_ISL_1354218, EPI_ISL_1354219, EPI_ISL_1354224, EPI_ISL_1354231, EPI_ISL_1354239, EPI_ISL_1354242, EPI_ISL_1354247, EPI_ISL_1354254, EPI_ISL_1354255, EPI_ISL_1354257, EPI_ISL_1354258, EPI_ISL_1436014, EPI_ISL_1436036, EPI_ISL_1436057, EPI_ISL_1436089, EPI_ISL_1436097, EPI_ISL_1436100, EPI_ISL_1436103, EPI_ISL_1436107, EPI_ISL_1436109, EPI_ISL_1436129, EPI_ISL_1436130, EPI_ISL_1436174, EPI_ISL_1436180, EPI_ISL_1436181, EPI_ISL_1436193, EPI_ISL_1436203, EPI_ISL_1436232, EPI_ISL_1436249, EPI_ISL_1436257, EPI_ISL_1436269, EPI_ISL_1436272, EPI_ISL_1436296, EPI_ISL_1436298, EPI_ISL_1436301, EPI_ISL_1436303, EPI_ISL_1436304, EPI_ISL_1436315, EPI_ISL_1436344, EPI_ISL_1436370, EPI_ISL_1436371, EPI_ISL_1436382, EPI_ISL_1436394, EPI_ISL_1438688, EPI_ISL_1438706, EPI_ISL_1438714, EPI_ISL_1438719, EPI_ISL_1438727, EPI_ISL_1438736, EPI_ISL_1438737, EPI_ISL_1438745, EPI_ISL_1438760, EPI_ISL_1438783, EPI_ISL_1438787, EPI_ISL_1438808, EPI_ISL_1438809, EPI_ISL_1438812, EPI_ISL_1438813, EPI_ISL_1438814, EPI_ISL_1438817, EPI_ISL_1438821, EPI_ISL_1438829, EPI_ISL_1438835, EPI_ISL_1438836, EPI_ISL_1438838, EPI_ISL_1438842, EPI_ISL_1438847, EPI_ISL_1438855, EPI_ISL_1438871, EPI_ISL_1438874, EPI_ISL_1438879, EPI_ISL_1438889, EPI_ISL_1438894, EPI_ISL_1438897, EPI_ISL_1438910, EPI_ISL_1438914, EPI_ISL_1438918, EPI_ISL_1438919, EPI_ISL_1438924, EPI_ISL_1438972, EPI_ISL_1438973, EPI_ISL_1438991, EPI_ISL_1438997, EPI_ISL_1438998, EPI_ISL_1439016, EPI_ISL_1439020, EPI_ISL_1439021, EPI_ISL_1439028, EPI_ISL_1569582

see above Sonic - MVZ Medizinisches Labor Bremen GmbH Robert Koch Institute

EPI_ISL_1353996, EPI_ISL_1354144, EPI_ISL_1354156, EPI_ISL_1354211, EPI_ISL_1354264, EPI_ISL_1436230, EPI_ISL_1438692, EPI_ISL_1438713, EPI_ISL_1439027, EPI_ISL_1439029, EPI_ISL_1568962, EPI_ISL_1569025, EPI_ISL_1569036, EPI_ISL_1569264

see above Sonic - MVZ Medizinisches Labor Oldenburg Robert Koch Institute

EPI_ISL_1353301, EPI_ISL_1435385, EPI_ISL_1435391, EPI_ISL_1435392, EPI_ISL_1435406 Stiftung Krankenhaus Bethanien für die Grafschaft Moers Robert Koch Institute

EPI_ISL_1354391, EPI_ISL_1354403, EPI_ISL_1354410, EPI_ISL_1354432, EPI_ISL_1354442, EPI_ISL_1354450, EPI_ISL_1354454, EPI_ISL_1354458, EPI_ISL_1354462, EPI_ISL_1354465

see above Städtisches Klinikum Dresden Robert Koch Institute

EPI_ISL_1286796, EPI_ISL_1286799, EPI_ISL_1286815, EPI_ISL_1350785, EPI_ISL_1350793, EPI_ISL_1350795, EPI_ISL_1350799, EPI_ISL_1350805, EPI_ISL_1350818, EPI_ISL_1350822, EPI_ISL_1350826, EPI_ISL_1350832, EPI_ISL_1350842, EPI_ISL_1350870, EPI_ISL_1350878, EPI_ISL_1350883, EPI_ISL_1350895, EPI_ISL_1350914, EPI_ISL_1352866, EPI_ISL_1352868, EPI_ISL_1352870, EPI_ISL_1352872, EPI_ISL_1352874, EPI_ISL_1352876, EPI_ISL_1352878, EPI_ISL_1352882, EPI_ISL_1352888, EPI_ISL_1352910, EPI_ISL_1352927, EPI_ISL_1352943, EPI_ISL_1352949, EPI_ISL_1352968, EPI_ISL_1352973, EPI_ISL_1352977, EPI_ISL_1352993, EPI_ISL_1352995, EPI_ISL_1352999, EPI_ISL_1353006, EPI_ISL_1353007, EPI_ISL_1437612, EPI_ISL_1437621, EPI_ISL_1437653, EPI_ISL_1437667, EPI_ISL_1437668, EPI_ISL_1437699, EPI_ISL_1437715, EPI_ISL_1437728, EPI_ISL_1437742, EPI_ISL_1437746, EPI_ISL_1437748, EPI_ISL_1437750, EPI_ISL_1437757, EPI_ISL_1437762, EPI_ISL_1437764, EPI_ISL_1437767, EPI_ISL_1437772, EPI_ISL_1437776, EPI_ISL_1437810, EPI_ISL_1437815, EPI_ISL_1437819, EPI_ISL_1437845, EPI_ISL_1437859, EPI_ISL_1437861, EPI_ISL_1437863, EPI_ISL_1437892, EPI_ISL_1437901, EPI_ISL_1437905, EPI_ISL_1437913, EPI_ISL_1437915, EPI_ISL_1437927, EPI_ISL_1440338, EPI_ISL_1440346, EPI_ISL_1440352, EPI_ISL_1440355, EPI_ISL_1440359, EPI_ISL_1440394, EPI_ISL_1440460, EPI_ISL_1440462, EPI_ISL_1440492, EPI_ISL_1440524, EPI_ISL_1440531

see above Synlab MVZ Augsburg Robert Koch Institute

EPI_ISL_1565994, EPI_ISL_1565995, EPI_ISL_1565996, EPI_ISL_1565997, EPI_ISL_1572839 ThÃ¼ringer Landesamtes fÃ¼r Verbraucherschutz Robert Koch Institute

EPI_ISL_1354392, EPI_ISL_1354394, EPI_ISL_1354399, EPI_ISL_1354404, EPI_ISL_1354408, EPI_ISL_1354411, EPI_ISL_1354419, EPI_ISL_1354425, EPI_ISL_1354441, EPI_ISL_1354444, EPI_ISL_1354445, EPI_ISL_1354453, EPI_ISL_1354460, EPI_ISL_1354461, EPI_ISL_1354464

see above Uniklinikum Carl Gustav Carus an der TU Dresden; Institut für Virologie Robert Koch Institute

EPI_ISL_1355236, EPI_ISL_1355237, EPI_ISL_1355238, EPI_ISL_1355239, EPI_ISL_1565998, EPI_ISL_1565999, EPI_ISL_1566001, EPI_ISL_1566003, EPI_ISL_1566004, EPI_ISL_1566005, EPI_ISL_1566006, EPI_ISL_1638628, EPI_ISL_1638632

see above Uniklinikum Jena Robert Koch Institute

EPI_ISL_1569756, EPI_ISL_1569758 Uniklinikum Regensburg; Institut fÃ¼r klinische Mikrobiologie und Hygiene Robert Koch Institute

EPI_ISL_1357265, EPI_ISL_1357267, EPI_ISL_1357268, EPI_ISL_1357269, EPI_ISL_1357270, EPI_ISL_1357272, EPI_ISL_1357273, EPI_ISL_1357274, EPI_ISL_1357276

| | | | |
|---|---|---|---|
| see above | Uniklinikum Regensburg; Institut für Mikrobiologie | Robert Koch Institute | |
| EPI_ISL_2123863, EPI_ISL_2123872, EPI_ISL_2123920, EPI_ISL_2123938, EPI_ISL_2123944, EPI_ISL_2123945 | Universitatsklinikum Koln; Institut fur Virologie | Robert Koch Institute | |
| EPI_ISL_1464588, EPI_ISL_1464589, EPI_ISL_1464592, EPI_ISL_1464593, EPI_ISL_1464594, EPI_ISL_1464595, EPI_ISL_1464596, EPI_ISL_1464598, EPI_ISL_1464600, EPI_ISL_1716062, EPI_ISL_1716063, EPI_ISL_1716064, EPI_ISL_1716065, EPI_ISL_1716066, EPI_ISL_1716067, EPI_ISL_1716068, EPI_ISL_1716069, EPI_ISL_1716079, EPI_ISL_1716080, EPI_ISL_1716081, EPI_ISL_1716083, EPI_ISL_1716084, EPI_ISL_1716086, EPI_ISL_1716087, EPI_ISL_1716088, EPI_ISL_1716089, EPI_ISL_1716091, EPI_ISL_1716270 | | | |
| see above | University Medical Center Hamburg Eppendorf | Heinrich Pette Institute, Leibniz Institute for Experimental Virology | Adam Grundhoff; Alexis Robitaille; Johannes Knobloch; Martin Aepfelbacher; Nicole Fischer; Thomas Günther |
| EPI_ISL_1572026, EPI_ISL_1572047, EPI_ISL_1572051, EPI_ISL_1572053, EPI_ISL_1572057, EPI_ISL_1572063, EPI_ISL_1572093, EPI_ISL_1572101 | | | |
| see above | UniversitÃ¤tsklinikum Bonn - Institut fÃ¼r Virologie | Robert Koch Institute | |
| EPI_ISL_1569091, EPI_ISL_1569153, EPI_ISL_1569235, EPI_ISL_1569254 | UniversitÃ¤tsklinikum Frankfurt - Institut fÃ¼r Medizinische Virologie | Robert Koch Institute | |
| EPI_ISL_1570313, EPI_ISL_1570314, EPI_ISL_1570315, EPI_ISL_1570316, EPI_ISL_1570317, EPI_ISL_1570318, EPI_ISL_1570319, EPI_ISL_1570320, EPI_ISL_1570321, EPI_ISL_1570322, EPI_ISL_1570326, EPI_ISL_1570327 | | | |
| see above | UniversitÃ¤tsklinikum Heidelberg | Robert Koch Institute | |
| EPI_ISL_1726859, EPI_ISL_1726864, EPI_ISL_1851137, EPI_ISL_1851139 | UniversitÃ¤tsklinikum Schleswig-Holstein (standortÃ¼bergreifend inkl. Kiel) - Institut fÃ¼r Klinische Chemie und Laboratoriumsdiagnostik | Robert Koch Institute | |
| EPI_ISL_1573222, EPI_ISL_1573224, EPI_ISL_1573229, EPI_ISL_1573230, EPI_ISL_1573231, EPI_ISL_1573232, EPI_ISL_1640150, EPI_ISL_1640151, EPI_ISL_1640152, EPI_ISL_1640153, EPI_ISL_1640154, EPI_ISL_1640155, EPI_ISL_1640156 | | | |
| see above | UniversitÃ¤tsmedizin Mannheim | Robert Koch Institute | |
| EPI_ISL_1353358, EPI_ISL_1353361, EPI_ISL_1353368, EPI_ISL_1353371, EPI_ISL_1353377, EPI_ISL_1353401, EPI_ISL_1353404, EPI_ISL_1439369, EPI_ISL_1439371, EPI_ISL_1439377, EPI_ISL_1439381, EPI_ISL_1439383, EPI_ISL_1439384, EPI_ISL_1439400 | | | |
| see above | Universitätsklinikum Bonn - Institut für Medizinische Mikrobiologie Immunologie und Parasitologie + Institut für Virologie | Robert Koch Institute | |
| EPI_ISL_1355006, EPI_ISL_1355024, EPI_ISL_1355085, EPI_ISL_1355093, EPI_ISL_1355095, EPI_ISL_1355097, EPI_ISL_1355123, EPI_ISL_1355134, EPI_ISL_1355135, EPI_ISL_1355141, EPI_ISL_1355142, EPI_ISL_1355146, EPI_ISL_1355149, EPI_ISL_1355158, EPI_ISL_1355165 | | | |
| see above | Universitätsklinikum Heidelberg | Robert Koch Institute | |
| EPI_ISL_1833953 | Universitätsklinikum Knappschaftskrankenhaus Bochum | Bundeswehr Institute of Microbiology | Alexandra Rehn; Enrico Georgi; Malena Bestehorn-Willmann; Markus Antwerpen; Mathias Walter; Mike Pillukat; Roman Wölfel; Sabine Zange |
| EPI_ISL_1349647, EPI_ISL_1349648, EPI_ISL_1349649, EPI_ISL_1349650, EPI_ISL_1349651, EPI_ISL_1349652, EPI_ISL_1349653, EPI_ISL_1349654, EPI_ISL_1349655, EPI_ISL_1349656, EPI_ISL_1349657, EPI_ISL_1349659, EPI_ISL_1349660, EPI_ISL_1349661, EPI_ISL_1349662, EPI_ISL_1349663, EPI_ISL_1349664, EPI_ISL_1349665, EPI_ISL_1349666, EPI_ISL_1349667, EPI_ISL_1349668, EPI_ISL_1349669, EPI_ISL_1349670, EPI_ISL_1349671, EPI_ISL_1349672, EPI_ISL_1349673, EPI_ISL_1349674, EPI_ISL_1349675, EPI_ISL_1349677, EPI_ISL_1349678, EPI_ISL_1349679, EPI_ISL_1349680, EPI_ISL_1349681, EPI_ISL_1349682, EPI_ISL_1349683, EPI_ISL_1349684, EPI_ISL_1349685, EPI_ISL_1349686, EPI_ISL_1349687, EPI_ISL_1349688, EPI_ISL_1349689, EPI_ISL_1349690, EPI_ISL_1349691, EPI_ISL_1349692, EPI_ISL_1349693, EPI_ISL_1349694, EPI_ISL_1349695, EPI_ISL_1349696, EPI_ISL_1349697, EPI_ISL_1349698, EPI_ISL_1349700, EPI_ISL_1349701, EPI_ISL_1349702, EPI_ISL_1349703, EPI_ISL_1349704, EPI_ISL_1349705, EPI_ISL_1349706, EPI_ISL_1349707, EPI_ISL_1349708, EPI_ISL_1349709, EPI_ISL_1349710, EPI_ISL_1349712, EPI_ISL_1349713, EPI_ISL_1349714, EPI_ISL_1349715, EPI_ISL_1349716, EPI_ISL_1349717, EPI_ISL_1349719, EPI_ISL_1349720, EPI_ISL_1349721, EPI_ISL_1349722, EPI_ISL_1349723, EPI_ISL_1349724, EPI_ISL_1349725 | | | |
| see above | Universitätsklinikum Leipzig - Institut für Medizinische Mikrobiologie und Virologie virologisches Labor | Robert Koch Institute | |
| EPI_ISL_1353426, EPI_ISL_1353432, EPI_ISL_1353435, EPI_ISL_1353436, EPI_ISL_1353450, EPI_ISL_1353455, EPI_ISL_1353456, EPI_ISL_1353462, EPI_ISL_1353465, EPI_ISL_1353467, EPI_ISL_1353469, EPI_ISL_1353470, EPI_ISL_1353472, EPI_ISL_1435905, EPI_ISL_1435907, EPI_ISL_1435910, EPI_ISL_1435912, EPI_ISL_1435914, EPI_ISL_1435916, EPI_ISL_1435918, EPI_ISL_1435921, EPI_ISL_1435923, EPI_ISL_1435925, EPI_ISL_1435927, EPI_ISL_1435929, EPI_ISL_1435932, EPI_ISL_1435934, EPI_ISL_1435936, EPI_ISL_1435938, EPI_ISL_1435940, EPI_ISL_1435945, EPI_ISL_1435947, EPI_ISL_1435949, EPI_ISL_1435951, EPI_ISL_1435953, EPI_ISL_1435955, EPI_ISL_1435957, EPI_ISL_1435959 | | | |
| see above | Universitätsklinikum Münster Institut für Virologie | Robert Koch Institute | |
| EPI_ISL_1439401, EPI_ISL_1439405 | Universitätsklinikum des Saarlandes - Institut für Virologie | Robert Koch Institute | |
| EPI_ISL_1563962, EPI_ISL_1563965, EPI_ISL_1563967 | Virologisches Institut des UniversitÃ¤tsklinikums Erlangen | Robert Koch Institute | |
| EPI_ISL_1354267 | Virologisches Institut des Universitätsklinikums Erlangen | Robert Koch Institute | |
| EPI_ISL_1366975, EPI_ISL_1366993, EPI_ISL_1367005, EPI_ISL_1367006, EPI_ISL_1367007, EPI_ISL_1367008, EPI_ISL_1367009, EPI_ISL_1367010, EPI_ISL_1367011, EPI_ISL_1367017, EPI_ISL_1367020, EPI_ISL_1367021, EPI_ISL_1367022, EPI_ISL_1367026, EPI_ISL_1367027, EPI_ISL_1367028, EPI_ISL_1367029, EPI_ISL_1367030, EPI_ISL_1367031, EPI_ISL_1367032, EPI_ISL_1367033, EPI_ISL_1367034, EPI_ISL_1367035, EPI_ISL_1367036, EPI_ISL_1367037, EPI_ISL_1367038, EPI_ISL_1367039, EPI_ISL_1367040, EPI_ISL_1367041, EPI_ISL_1367042, EPI_ISL_1367043, EPI_ISL_1367099, EPI_ISL_1367100, EPI_ISL_1367101, EPI_ISL_1367102, EPI_ISL_1367103, EPI_ISL_1367104, EPI_ISL_1367105, EPI_ISL_1367159, EPI_ISL_1367160, EPI_ISL_1367161 | | | |
| see above | Virology, Universitätsklinikum des Saarlandes | Epigenetics, Saarland University | Jörn Walter; Kathrin Kattler; Markus Vogelgesang; Sascha Tierling; Sigrun Smola; Stefan Lohse; Thorsten Pfuhl |
| EPI_ISL_1435963, EPI_ISL_1435965, EPI_ISL_1435967, EPI_ISL_1435969, EPI_ISL_1435974, EPI_ISL_1435976 | alphaomega Labor GbR (Medizinische KooperationsgemeinschaftDres. Ackermann/Tennert/Zaiß) | Robert Koch Institute | |
| EPI_ISL_1564621, EPI_ISL_1564641 | amedes GÃ¶ttingen | Robert Koch Institute | |
| EPI_ISL_1351427, EPI_ISL_1351438, EPI_ISL_1351508, EPI_ISL_1351680, EPI_ISL_1436640, EPI_ISL_1436694, EPI_ISL_1436703, EPI_ISL_1436704, EPI_ISL_1436708, EPI_ISL_1436711, EPI_ISL_1436718, EPI_ISL_1436726, EPI_ISL_1436729, EPI_ISL_1436731, EPI_ISL_1436736, EPI_ISL_1436738, EPI_ISL_1436740, EPI_ISL_1436747, EPI_ISL_1436750, EPI_ISL_1436756, EPI_ISL_1436760, EPI_ISL_1436763, EPI_ISL_1436773 | | | |
| see above | amedes Göttingen | Robert Koch Institute | |
| EPI_ISL_1351450, EPI_ISL_1351475, EPI_ISL_1351476, EPI_ISL_1351507, EPI_ISL_1351582, EPI_ISL_1351594, EPI_ISL_1351622, EPI_ISL_1351624, EPI_ISL_1351625, EPI_ISL_1351636, EPI_ISL_1351644, EPI_ISL_1351652, EPI_ISL_1351667, EPI_ISL_1351720, EPI_ISL_1436696, EPI_ISL_1436700, EPI_ISL_1436710, EPI_ISL_1436716, EPI_ISL_1436719, EPI_ISL_1436724, EPI_ISL_1436735, EPI_ISL_1436741, EPI_ISL_1436743, EPI_ISL_1436748, EPI_ISL_1436753, EPI_ISL_1436758, EPI_ISL_1436765, EPI_ISL_1436770, EPI_ISL_1436774, EPI_ISL_1436786, EPI_ISL_1436796, EPI_ISL_1436803, EPI_ISL_1436805, EPI_ISL_1564487 | | | |
| see above | amedes MVZ DIAMEDIS Sennestadt | Robert Koch Institute | |
| EPI_ISL_1351389, EPI_ISL_1351406, EPI_ISL_1351410, EPI_ISL_1351417, EPI_ISL_1351436, EPI_ISL_1351463, EPI_ISL_1351470, EPI_ISL_1351473, EPI_ISL_1351495, EPI_ISL_1351512, EPI_ISL_1351521, EPI_ISL_1351528, EPI_ISL_1351541, EPI_ISL_1351557, EPI_ISL_1351565, EPI_ISL_1351575, EPI_ISL_1351576, EPI_ISL_1351578, EPI_ISL_1351599, EPI_ISL_1351618, EPI_ISL_1351633, EPI_ISL_1351649, EPI_ISL_1351660, EPI_ISL_1351696, EPI_ISL_1351733, EPI_ISL_1436620, EPI_ISL_1436625, EPI_ISL_1436686, EPI_ISL_1436687, EPI_ISL_1436688, EPI_ISL_1436693, EPI_ISL_1436705, EPI_ISL_1436706, EPI_ISL_1436709, EPI_ISL_1436713, EPI_ISL_1436734, EPI_ISL_1436739, EPI_ISL_1436742, EPI_ISL_1436745, EPI_ISL_1436800 | | | |
| see above | amedes MVZ Halle/Leipzig | Robert Koch Institute | |
| EPI_ISL_1351444, EPI_ISL_1351548, EPI_ISL_1351551, EPI_ISL_1351584, EPI_ISL_1351643, EPI_ISL_1436621, EPI_ISL_1436623, EPI_ISL_1436627, EPI_ISL_1436632, EPI_ISL_1436644, EPI_ISL_1436650, EPI_ISL_1436657, EPI_ISL_1436672, EPI_ISL_1436679, EPI_ISL_1436681, EPI_ISL_1436689, EPI_ISL_1436692, EPI_ISL_1436697, EPI_ISL_1436717, EPI_ISL_1436723, EPI_ISL_1436757, EPI_ISL_1436768, EPI_ISL_1436779, EPI_ISL_1436784 | | | |
| see above | amedes MVZ Hannover | Robert Koch Institute | |
| EPI_ISL_1436630, EPI_ISL_1436634, EPI_ISL_1436685, EPI_ISL_1436690, EPI_ISL_1436707, EPI_ISL_1436715, EPI_ISL_1436730, EPI_ISL_1436746, EPI_ISL_1436755, EPI_ISL_1436762, EPI_ISL_1436771, EPI_ISL_1436780, EPI_ISL_1436783, EPI_ISL_1436787, EPI_ISL_1436802 | | | |
| see above | amedes MVZ Jena | Robert Koch Institute | |
| EPI_ISL_1351467, EPI_ISL_1436654, EPI_ISL_1436669, EPI_ISL_1436776 | amedes MVZ Zentrum für Pathologie und Zytodiagnostik GmbH (Köln) | Robert Koch Institute | |
| EPI_ISL_1564349, EPI_ISL_1565361 | amedes MVZ fÃ¼r Laboratoriumsmedizin FÃ¼rstenfeldbruck | Robert Koch Institute | |
| EPI_ISL_1564725 | amedes MVZ fÃ¼r Laboratoriumsmedizin Hamburg | Robert Koch Institute | |
| EPI_ISL_1351440, EPI_ISL_1351452, EPI_ISL_1351461, EPI_ISL_1351478, EPI_ISL_1351481, EPI_ISL_1351482, EPI_ISL_1351487, EPI_ISL_1351505, EPI_ISL_1351510, EPI_ISL_1351537, EPI_ISL_1351569, EPI_ISL_1351597, EPI_ISL_1351664, EPI_ISL_1351724, EPI_ISL_1351735, EPI_ISL_1351744, EPI_ISL_1436638, EPI_ISL_1436642, EPI_ISL_1436646, EPI_ISL_1436652, EPI_ISL_1436655, EPI_ISL_1436665, EPI_ISL_1436722, EPI_ISL_1436728, EPI_ISL_1436737, EPI_ISL_1436744, EPI_ISL_1436749, EPI_ISL_1436752, EPI_ISL_1436781, EPI_ISL_1436785, EPI_ISL_1436789, EPI_ISL_1436792, EPI_ISL_1436794, EPI_ISL_1436795, EPI_ISL_1436799 | | | |
| see above | amedes MVZ für Laboratoriumsdiagnostik Raubling GmbH | Robert Koch Institute | |
| EPI_ISL_1436674, EPI_ISL_1436684, EPI_ISL_1436691, EPI_ISL_1436797, EPI_ISL_1436798, EPI_ISL_1436806 | amedes MVZ für Laboratoriumsmedizin Fürstenfeldbruck | Robert Koch Institute | |
| EPI_ISL_1351515, EPI_ISL_1351518, EPI_ISL_1436732, EPI_ISL_1436759, EPI_ISL_1436778, EPI_ISL_1436791 | amedes MVZ für Laboratoriumsmedizin Rhein/Ruhr | Robert Koch Institute | |
| EPI_ISL_1351446, EPI_ISL_1351449, EPI_ISL_1351536, EPI_ISL_1351542, EPI_ISL_1351647, EPI_ISL_1351674, EPI_ISL_1351704, EPI_ISL_1436678, EPI_ISL_1436682 | | | |
| see above | amedes MVZ für Labordiagnostik Rhein-Main | Robert Koch Institute | |
| EPI_ISL_1436712 | amedes Medizinisches Labor Mühlhausen | Robert Koch Institute | |
| EPI_ISL_1351392, EPI_ISL_1351421, EPI_ISL_1351442, EPI_ISL_1351459 | amedes Medlab Arnold Analytik MVZ GmbH | Robert Koch Institute | |
| EPI_ISL_1351768, EPI_ISL_1439675 | labopart - Medizinische Laboratorien Dresden | Robert Koch Institute | |
| EPI_ISL_1351373, EPI_ISL_1351374, EPI_ISL_1351378, EPI_ISL_1351397, EPI_ISL_1351401, EPI_ISL_1351405, EPI_ISL_1351431, EPI_ISL_1351434, EPI_ISL_1351453, EPI_ISL_1351485, EPI_ISL_1351497, EPI_ISL_1351502, EPI_ISL_1351513, EPI_ISL_1351516, EPI_ISL_1351527, EPI_ISL_1351531, EPI_ISL_1351545, EPI_ISL_1351559, EPI_ISL_1351560, EPI_ISL_1351566, EPI_ISL_1351579, EPI_ISL_1351593, EPI_ISL_1351596, EPI_ISL_1351602, EPI_ISL_1351603, EPI_ISL_1351609, EPI_ISL_1351612, EPI_ISL_1351615, EPI_ISL_1351616, EPI_ISL_1351619, EPI_ISL_1351631, EPI_ISL_1351653, EPI_ISL_1351661, EPI_ISL_1351671, EPI_ISL_1351677, EPI_ISL_1351678, EPI_ISL_1351683, EPI_ISL_1351686, EPI_ISL_1351690, EPI_ISL_1351699, EPI_ISL_1351702, EPI_ISL_1351705, EPI_ISL_1351710, EPI_ISL_1351717, EPI_ISL_1351718, EPI_ISL_1351721, EPI_ISL_1351722, EPI_ISL_1351726, EPI_ISL_1351729, EPI_ISL_1351730, EPI_ISL_1351732, EPI_ISL_1351740, EPI_ISL_1351751, EPI_ISL_1351753, EPI_ISL_1351755, EPI_ISL_1351756, EPI_ISL_1436695, EPI_ISL_1436761, EPI_ISL_1436764, EPI_ISL_1436804 | | | |
| see above | nordlab - Partnerschaftspraxis fu?r Laboratoriumsmedizin | Robert Koch Institute | |

We gratefully acknowledge the following Authors from the Originating laboratories responsible for obtaining the specimens, as well as the Submitting laboratories where the genome data were generated and shared via GISAID, on which this research is based.

All Submitters of data may be contacted directly via www.gisaid.org

Authors are sorted alphabetically.

| Accession ID | Originating Laboratory | Submitting Laboratory | Authors |
|---|---|---|---|
| EPI_ISL_2110901, EPI_ISL_2110902, EPI_ISL_2111000, EPI_ISL_2111388 | Bayerisches Landesamt fur Gesundheit und Lebensmittelsicherheit (LGL) | Robert Koch Institute | |
| EPI_ISL_1570347, EPI_ISL_1640846, EPI_ISL_1640847, EPI_ISL_1640853, EPI_ISL_1640867, EPI_ISL_1640870, EPI_ISL_1640874, EPI_ISL_1640884, EPI_ISL_1725275, EPI_ISL_1725289, EPI_ISL_1725293, EPI_ISL_1725295, EPI_ISL_1725301, EPI_ISL_1725302, EPI_ISL_1725303, EPI_ISL_1725311, EPI_ISL_1725313, EPI_ISL_1725325, EPI_ISL_1725328, EPI_ISL_1725330, EPI_ISL_1725334, EPI_ISL_1725345, EPI_ISL_1725348, EPI_ISL_1725354, EPI_ISL_1725373, EPI_ISL_1725375, EPI_ISL_1725385, EPI_ISL_1725388, EPI_ISL_1725392, EPI_ISL_1725396, EPI_ISL_1725397, EPI_ISL_1725400, EPI_ISL_1725404, EPI_ISL_1725407, EPI_ISL_1725409, EPI_ISL_1725418, EPI_ISL_1725419, EPI_ISL_1725425, EPI_ISL_1725429, EPI_ISL_1725433, EPI_ISL_1725450, EPI_ISL_1725453, EPI_ISL_1725454, EPI_ISL_1725460, EPI_ISL_1725461, EPI_ISL_1725465, EPI_ISL_1725466, EPI_ISL_1725467, EPI_ISL_1725468, EPI_ISL_1725473, EPI_ISL_1725481, EPI_ISL_1725485, EPI_ISL_1725493, EPI_ISL_1725510, EPI_ISL_1725516, EPI_ISL_1725521, EPI_ISL_1725524, EPI_ISL_1725528, EPI_ISL_2110864, EPI_ISL_2110865, EPI_ISL_2110868, EPI_ISL_2110869, EPI_ISL_2110870, EPI_ISL_2110882, EPI_ISL_2110883, EPI_ISL_2110887, EPI_ISL_2110889, EPI_ISL_2110893, EPI_ISL_2110895, EPI_ISL_2110898, EPI_ISL_2110899, EPI_ISL_2110904, EPI_ISL_2110912, EPI_ISL_2110919, EPI_ISL_2110920, EPI_ISL_2110923, EPI_ISL_2110924, EPI_ISL_2110925, EPI_ISL_2110930, EPI_ISL_2110931, EPI_ISL_2110934, EPI_ISL_2110939, EPI_ISL_2110943, EPI_ISL_2110952, EPI_ISL_2110954, EPI_ISL_2110959, EPI_ISL_2110963, EPI_ISL_2110965, EPI_ISL_2110968, EPI_ISL_2110970, EPI_ISL_2110971, EPI_ISL_2110979, EPI_ISL_2110980, EPI_ISL_2110983, EPI_ISL_2110991, EPI_ISL_2110992, EPI_ISL_2110995, EPI_ISL_2110996, EPI_ISL_2110998 | | | |
| see above | Bayerisches Landesamt fÃ¼r Gesundheit und Lebensmittelsicherheit (LGL) | Robert Koch Institute | |
| EPI_ISL_1568952, EPI_ISL_1569018, EPI_ISL_1569108, EPI_ISL_1569320, EPI_ISL_1569379, EPI_ISL_1569445, EPI_ISL_1569469, EPI_ISL_1569521, EPI_ISL_1569573, EPI_ISL_1569616, EPI_ISL_1638683, EPI_ISL_1638685, EPI_ISL_1638689, EPI_ISL_1638691, EPI_ISL_1638695, EPI_ISL_1638696, EPI_ISL_1638698, EPI_ISL_1638702, EPI_ISL_1638704, EPI_ISL_1638705, EPI_ISL_1638706, EPI_ISL_1638707, EPI_ISL_1638708, EPI_ISL_1638709, EPI_ISL_1638710, EPI_ISL_1638712, EPI_ISL_1638716, EPI_ISL_1638717, EPI_ISL_1638718, EPI_ISL_1638720, EPI_ISL_1638724, EPI_ISL_1638725, EPI_ISL_1638727, EPI_ISL_1638728, EPI_ISL_1638729, EPI_ISL_1638731, EPI_ISL_1638732, EPI_ISL_1638734, EPI_ISL_1638735, EPI_ISL_1638738, EPI_ISL_1638739, EPI_ISL_1638740, EPI_ISL_1638741, EPI_ISL_1638744, EPI_ISL_1638745, EPI_ISL_1638747, EPI_ISL_1638751, EPI_ISL_1638752, EPI_ISL_1638753, EPI_ISL_1638756, EPI_ISL_1638758, EPI_ISL_1638759, EPI_ISL_1638761, EPI_ISL_1638762, EPI_ISL_1638766, EPI_ISL_1638767, EPI_ISL_1638769, EPI_ISL_1638770, EPI_ISL_1638771, EPI_ISL_1638772, EPI_ISL_1638774, EPI_ISL_1638775, EPI_ISL_1638779, EPI_ISL_1638780, EPI_ISL_1638782, EPI_ISL_1638785, EPI_ISL_1638786, EPI_ISL_1638788, EPI_ISL_1638795, EPI_ISL_1638798, EPI_ISL_1638799, EPI_ISL_1638800, EPI_ISL_1638805, EPI_ISL_1638811, EPI_ISL_1638812, EPI_ISL_1638813, EPI_ISL_1638814, EPI_ISL_1638818, EPI_ISL_1638819, EPI_ISL_1638820, EPI_ISL_1638821, EPI_ISL_1638822, EPI_ISL_1638823, EPI_ISL_1638826, EPI_ISL_1638827, EPI_ISL_1638828, EPI_ISL_1638832, EPI_ISL_1638833, EPI_ISL_1638835, EPI_ISL_1638836, EPI_ISL_1638837, EPI_ISL_1638841, EPI_ISL_1638842, EPI_ISL_1638843, EPI_ISL_1638844, EPI_ISL_1638846, EPI_ISL_1638847, EPI_ISL_1638848, EPI_ISL_1638849, EPI_ISL_1638851, EPI_ISL_1638852, EPI_ISL_1638853, EPI_ISL_1638854, EPI_ISL_1638855, EPI_ISL_1638856, EPI_ISL_1638858, EPI_ISL_1638860, EPI_ISL_1638862, EPI_ISL_1638864, EPI_ISL_1638865, EPI_ISL_1638868, EPI_ISL_1638869, EPI_ISL_1638870, EPI_ISL_1638873, EPI_ISL_1638874, EPI_ISL_1638875, EPI_ISL_1638876, EPI_ISL_1638877, EPI_ISL_1638878, EPI_ISL_1638880, EPI_ISL_1638882, EPI_ISL_1638883, EPI_ISL_1638884, EPI_ISL_1638885, EPI_ISL_1638886, EPI_ISL_1638889, EPI_ISL_1638892, EPI_ISL_1638894, EPI_ISL_1638899, EPI_ISL_1638900, EPI_ISL_1638901, EPI_ISL_1638903, EPI_ISL_1638904, EPI_ISL_1638906, EPI_ISL_1638909, EPI_ISL_1638911, EPI_ISL_1638912, EPI_ISL_1638913, EPI_ISL_1638915, EPI_ISL_1638916, EPI_ISL_1638917, EPI_ISL_1638919, EPI_ISL_1638925, EPI_ISL_1638928, EPI_ISL_1638929, EPI_ISL_1638930, EPI_ISL_1638931, EPI_ISL_1638932, EPI_ISL_1638933, EPI_ISL_1638935, EPI_ISL_1638936, EPI_ISL_1638938, EPI_ISL_1638939, EPI_ISL_1638940, EPI_ISL_1638942, EPI_ISL_1638943, EPI_ISL_1638944, EPI_ISL_1638947, EPI_ISL_1638948, EPI_ISL_1638950, EPI_ISL_1638951, EPI_ISL_1638953, EPI_ISL_1638954, EPI_ISL_1638958, EPI_ISL_1638959, EPI_ISL_1638961, EPI_ISL_1638965, EPI_ISL_1638966, EPI_ISL_1638967, EPI_ISL_1638972, EPI_ISL_1638975, EPI_ISL_1638976, EPI_ISL_1638977, EPI_ISL_1638981, EPI_ISL_1638982, EPI_ISL_1638983, EPI_ISL_1638984, EPI_ISL_1638985, EPI_ISL_1638986, EPI_ISL_1638987, EPI_ISL_1638997, EPI_ISL_1639003, EPI_ISL_1639004, EPI_ISL_1639005, EPI_ISL_1639016, EPI_ISL_1639023, EPI_ISL_1639028, EPI_ISL_1639029, EPI_ISL_1639033, EPI_ISL_1639034, EPI_ISL_1639038, EPI_ISL_1639044, EPI_ISL_1639048, EPI_ISL_1639050, EPI_ISL_1639051, EPI_ISL_1639052, EPI_ISL_1639056, EPI_ISL_1639058, EPI_ISL_1639060, EPI_ISL_1639062, EPI_ISL_1639063, EPI_ISL_1639065, EPI_ISL_1639066, EPI_ISL_1639072, EPI_ISL_1639073, EPI_ISL_1639075, EPI_ISL_1639078, EPI_ISL_1639079, EPI_ISL_1639080, EPI_ISL_1639086, EPI_ISL_1639088, EPI_ISL_1639091, EPI_ISL_1639092, EPI_ISL_1639095, EPI_ISL_1639102, EPI_ISL_1639103, EPI_ISL_1639104, EPI_ISL_1639105, EPI_ISL_1639106, EPI_ISL_1639108, EPI_ISL_1639109, EPI_ISL_1639110, EPI_ISL_1639111, EPI_ISL_1639113, EPI_ISL_1639114, EPI_ISL_1639118, EPI_ISL_1639119, EPI_ISL_1639124, EPI_ISL_1639125, EPI_ISL_1639128, EPI_ISL_1639129, EPI_ISL_1639130, EPI_ISL_1639133, EPI_ISL_1639135, EPI_ISL_1639139, EPI_ISL_1639141, EPI_ISL_1639142, EPI_ISL_1639144, EPI_ISL_1639147, EPI_ISL_1639152, EPI_ISL_1639155, EPI_ISL_1639158, EPI_ISL_1639160, EPI_ISL_1639161, EPI_ISL_1639167, EPI_ISL_1639168, EPI_ISL_1639170, EPI_ISL_1639173, EPI_ISL_1639174, EPI_ISL_1639175, EPI_ISL_1639177, EPI_ISL_1639179, EPI_ISL_1639180, EPI_ISL_1639181, EPI_ISL_1639184, EPI_ISL_1639191, EPI_ISL_1639192, EPI_ISL_1639197, EPI_ISL_1639198, EPI_ISL_1639199, EPI_ISL_1639200, EPI_ISL_1639203, EPI_ISL_1639207, EPI_ISL_1639209, EPI_ISL_1639212, EPI_ISL_1639213, EPI_ISL_1639218, EPI_ISL_1639221, EPI_ISL_1639222, EPI_ISL_1639223, EPI_ISL_1639225, EPI_ISL_1639226, EPI_ISL_1639228, EPI_ISL_1639229, EPI_ISL_1639230, EPI_ISL_1639233, EPI_ISL_1639237, EPI_ISL_1639239, EPI_ISL_1639241, EPI_ISL_1639242, EPI_ISL_1639243, EPI_ISL_1639244, EPI_ISL_1639245, EPI_ISL_1639246, EPI_ISL_1639247, EPI_ISL_1642421, EPI_ISL_1642445, EPI_ISL_1642453, EPI_ISL_1642470, EPI_ISL_1642477, EPI_ISL_1642484, EPI_ISL_1642486, EPI_ISL_1642494, EPI_ISL_1642514, EPI_ISL_1642517, EPI_ISL_1642520, EPI_ISL_1642539, EPI_ISL_1642547, EPI_ISL_1642576, EPI_ISL_1642581, EPI_ISL_1642610, EPI_ISL_1642620, EPI_ISL_1642626, EPI_ISL_1642628, EPI_ISL_1642636, EPI_ISL_1642639, EPI_ISL_1642642, EPI_ISL_1642654, EPI_ISL_1643118, EPI_ISL_1643125, EPI_ISL_1643131, EPI_ISL_1643142, EPI_ISL_1643149, EPI_ISL_1643152, EPI_ISL_1643162, EPI_ISL_1643164, EPI_ISL_1643177, EPI_ISL_1643193, EPI_ISL_1643210, EPI_ISL_1643218, EPI_ISL_1643221, EPI_ISL_1643228, EPI_ISL_1643238, EPI_ISL_1643259, EPI_ISL_1643263, EPI_ISL_1643274, EPI_ISL_1643281, EPI_ISL_1643285, EPI_ISL_1643288, EPI_ISL_1643294, EPI_ISL_1643300, EPI_ISL_1643314, EPI_ISL_1643329, EPI_ISL_1643356, EPI_ISL_1643362, EPI_ISL_1643364, EPI_ISL_1643387, EPI_ISL_1721123, EPI_ISL_1721202, EPI_ISL_1721275, EPI_ISL_1721384, EPI_ISL_1721385, EPI_ISL_1721419, EPI_ISL_1721454, EPI_ISL_1721483, EPI_ISL_1721516, EPI_ISL_1721525, EPI_ISL_1721632, EPI_ISL_1721655, EPI_ISL_1848447 | | | |
| see above | Bioscientia Labor Wermsdorf | Robert Koch Institute | |
| EPI_ISL_1638713, EPI_ISL_1638737, EPI_ISL_1638763, EPI_ISL_1638787, EPI_ISL_1638809, EPI_ISL_1638969, EPI_ISL_1638979, EPI_ISL_1638989, EPI_ISL_1638994, EPI_ISL_1638998, EPI_ISL_1639001, EPI_ISL_1639007, EPI_ISL_1639018, EPI_ISL_1639035, EPI_ISL_1639043, EPI_ISL_1639046, EPI_ISL_1639053, EPI_ISL_1639055, EPI_ISL_1639069, EPI_ISL_1639071, EPI_ISL_1639076, EPI_ISL_1639087, EPI_ISL_1639090, EPI_ISL_1639097, EPI_ISL_1639100, EPI_ISL_1639131, EPI_ISL_1639132, EPI_ISL_1639136, EPI_ISL_1639149, EPI_ISL_1639162, EPI_ISL_1639164, EPI_ISL_1639187, EPI_ISL_1639208, EPI_ISL_1639231, EPI_ISL_1639232, EPI_ISL_1639234, EPI_ISL_1642425, EPI_ISL_1642446, EPI_ISL_1642459, EPI_ISL_1642472, EPI_ISL_1642475, EPI_ISL_1642507, EPI_ISL_1642515, EPI_ISL_1642532, EPI_ISL_1642588, EPI_ISL_1642647, EPI_ISL_1643146, EPI_ISL_1643165, EPI_ISL_1643173, EPI_ISL_1643197, EPI_ISL_1643207, EPI_ISL_1643241, EPI_ISL_1643242, EPI_ISL_1643249, EPI_ISL_1643292, EPI_ISL_1643320, EPI_ISL_1643357, EPI_ISL_1643386, EPI_ISL_1721230, EPI_ISL_1721257, EPI_ISL_1721380 | | | |
| see above | Bioscientia MVZ Labor Karlsruhe GmbH | Robert Koch Institute | |
| EPI_ISL_1833950, EPI_ISL_1833974 | Bundeswehr Institute of Microbiology | Bundeswehr Institute of Microbiology | Alexandra Rehn; Enrico Georgi; Malena Bestehorn-Willmann; Markus Antwerpen; Mathias Walter; Mike Pillukat; Roman Wölfel; Sabine Zange |
| EPI_ISL_1833983, EPI_ISL_1833984, EPI_ISL_1833985, EPI_ISL_1833987, EPI_ISL_1833988, EPI_ISL_1833989, EPI_ISL_1833990, EPI_ISL_1834011, EPI_ISL_1834017 | | | |
| see above | Bundeswehrkrankenhaus Berlin | Bundeswehr Institute of Microbiology | Alexandra Rehn; Enrico Georgi; Malena Bestehorn-Willmann; Markus Antwerpen; Mathias Walter; Mike Pillukat; Roman Wölfel; Sabine Zange |
| EPI_ISL_1833962, EPI_ISL_1833963, EPI_ISL_1833991, EPI_ISL_1833992, EPI_ISL_1833993 | Bundeswehrkrankenhaus Hamburg | Bundeswehr Institute of Microbiology | Alexandra Rehn; Enrico Georgi; Malena Bestehorn-Willmann; Markus Antwerpen; Mathias Walter; Mike Pillukat; Roman Wölfel; Sabine Zange |
| EPI_ISL_1833973, EPI_ISL_1833975, EPI_ISL_1833978, EPI_ISL_1833980 | Bundeswehrzentralkrankenhaus Koblenz | Bundeswehr Institute of Microbiology | Alexandra Rehn; Enrico Georgi; Malena Bestehorn-Willmann; Markus Antwerpen; Mathias Walter; Mike Pillukat; Roman Wölfel; Sabine Zange |
| EPI_ISL_1990021, EPI_ISL_1990079, EPI_ISL_1990082, EPI_ISL_1990084, EPI_ISL_1990085, EPI_ISL_1990086, EPI_ISL_1990087, EPI_ISL_1991611, EPI_ISL_1991613, EPI_ISL_1993607, EPI_ISL_1993608 | | | |
| see above | Center of Medical Microbiology, Virology, and Hospital Hygiene, University of Duesseldorf | Center of Medical Microbiology, Virology, and Hospital Hygiene, University of Duesseldorf | Alexander Dilthey; Andreas Walker; Daniel Strelow; Jessica Nicolai; Jörg Timm; Klaus Pfeffer; Lisanna Hülse; Malte Kohns Vasconcelos; Maximilian Damagnez; Nadine Lübke; Tobias Wienemann; Torsten Houwaart |
| EPI_ISL_2108484, EPI_ISL_2108486, EPI_ISL_2108488, EPI_ISL_2108489, EPI_ISL_2108499, EPI_ISL_2108500, EPI_ISL_2108511, EPI_ISL_2108514, EPI_ISL_2108524, EPI_ISL_2108525, EPI_ISL_2108526, EPI_ISL_2108527, EPI_ISL_2108528, EPI_ISL_2108530, EPI_ISL_2108535, EPI_ISL_2108537, EPI_ISL_2108538, EPI_ISL_2108550, EPI_ISL_2108553, EPI_ISL_2108555, EPI_ISL_2108557, EPI_ISL_2108571, EPI_ISL_2108573, EPI_ISL_2108577, EPI_ISL_2108584, EPI_ISL_2108587, EPI_ISL_2108606, EPI_ISL_2108608, EPI_ISL_2108615, EPI_ISL_2108620, EPI_ISL_2108621, EPI_ISL_2108631, EPI_ISL_2108633, EPI_ISL_2108634, EPI_ISL_2108636, EPI_ISL_2108639, EPI_ISL_2108645, EPI_ISL_2108646, EPI_ISL_2108653, EPI_ISL_2108664, EPI_ISL_2108674, EPI_ISL_2108675, EPI_ISL_2108676, EPI_ISL_2108678, EPI_ISL_2108680, EPI_ISL_2108682, EPI_ISL_2108683, EPI_ISL_2108686, EPI_ISL_2108687, EPI_ISL_2108688, EPI_ISL_2108703, EPI_ISL_2108704 | | | |
| see above | Centogene; Dr. Bauer Laboratoriums GmbH | Robert Koch Institute | |
| EPI_ISL_1569800, EPI_ISL_1569801, EPI_ISL_1569802, EPI_ISL_1569803, EPI_ISL_1569804, EPI_ISL_1569805, EPI_ISL_1569806, EPI_ISL_1569807, EPI_ISL_1569808 | | | |
| see above | Chemische und VeterinÃ¤runtersuchungsamt MÃ¼nsterland-Emscher-Lippe (CVUA-MEL) | Robert Koch Institute | |
| EPI_ISL_1571498, EPI_ISL_1571499, EPI_ISL_1571500, EPI_ISL_1571501, EPI_ISL_1571502, EPI_ISL_1571503, EPI_ISL_1571504, EPI_ISL_1571505, EPI_ISL_1571506, EPI_ISL_1571507, EPI_ISL_1571508, EPI_ISL_1571509, EPI_ISL_1571510, EPI_ISL_1571511, EPI_ISL_1571512, EPI_ISL_1571513, EPI_ISL_1571514, EPI_ISL_1571516, EPI_ISL_1571517, EPI_ISL_1571518, EPI_ISL_1571519, EPI_ISL_1571521, EPI_ISL_1571522, EPI_ISL_1571523, EPI_ISL_1571524, EPI_ISL_1571525, EPI_ISL_1571526, EPI_ISL_1571533, EPI_ISL_1571534, EPI_ISL_1571535, EPI_ISL_1571536, EPI_ISL_1571537, EPI_ISL_1571538, EPI_ISL_1571967, EPI_ISL_1571988, EPI_ISL_1571994, EPI_ISL_1639335, EPI_ISL_1639340, EPI_ISL_1639349, EPI_ISL_1639355, EPI_ISL_1639358, EPI_ISL_1639364, EPI_ISL_1639366, EPI_ISL_1639367, EPI_ISL_1639369, EPI_ISL_1639372, EPI_ISL_1639373, EPI_ISL_1639374, EPI_ISL_1639378, EPI_ISL_1639379, EPI_ISL_1639380, EPI_ISL_1639383, EPI_ISL_1639387, EPI_ISL_1639390, EPI_ISL_1639392, EPI_ISL_1639393, EPI_ISL_1639396, EPI_ISL_1639399, EPI_ISL_1639400, EPI_ISL_1639401, EPI_ISL_1639405, EPI_ISL_1639408, EPI_ISL_1639411, EPI_ISL_1639412, EPI_ISL_1639413, EPI_ISL_1639415, EPI_ISL_1639416, EPI_ISL_1639417, EPI_ISL_1639418, EPI_ISL_1639423, EPI_ISL_1639425, EPI_ISL_1639426, EPI_ISL_1639428, EPI_ISL_1639429, EPI_ISL_1639430, EPI_ISL_1639435, EPI_ISL_1639436, EPI_ISL_1722937, EPI_ISL_1722938, EPI_ISL_1722939, EPI_ISL_1722966, EPI_ISL_1722967, EPI_ISL_1722968, EPI_ISL_1722969, EPI_ISL_1722970, EPI_ISL_1722972, EPI_ISL_1844407 | | | |
| see above | Clemenshospital - Alexianer Clemenshospital MÃ¼nster | Robert Koch Institute | |
| EPI_ISL_1642739, EPI_ISL_1642740, EPI_ISL_1642741, EPI_ISL_1642743, EPI_ISL_1642745, EPI_ISL_1642748, EPI_ISL_1642749, EPI_ISL_1642752, EPI_ISL_1642762, EPI_ISL_1642763, EPI_ISL_1843180, EPI_ISL_1843185, EPI_ISL_1843187, EPI_ISL_1843189, EPI_ISL_1843191, EPI_ISL_1843192, EPI_ISL_1843193, EPI_ISL_1843195, EPI_ISL_1843196 | | | |
| see above | Corona-Testzentrum ifp Institut fÃ¼r ProduktqualitÃ¤t GmbH | Robert Koch Institute | |
| EPI_ISL_1435840, EPI_ISL_1568154, EPI_ISL_1568155, EPI_ISL_1568156, EPI_ISL_1568157, EPI_ISL_1568159, EPI_ISL_1568160, EPI_ISL_1568161, EPI_ISL_1568162, EPI_ISL_1568163, EPI_ISL_1568164, EPI_ISL_1568165, EPI_ISL_1568166, EPI_ISL_1568167, EPI_ISL_1568168, EPI_ISL_1568169, EPI_ISL_1568170, EPI_ISL_1568171, EPI_ISL_1568172, EPI_ISL_1568173, EPI_ISL_1568174, EPI_ISL_1568175, EPI_ISL_1568176, EPI_ISL_1568177, EPI_ISL_1568179, EPI_ISL_1568180, EPI_ISL_1568181, EPI_ISL_1568182, EPI_ISL_1568183, EPI_ISL_1568184, EPI_ISL_1568185, EPI_ISL_1568187, EPI_ISL_1568188, EPI_ISL_1568189, EPI_ISL_1568190, EPI_ISL_1568191, EPI_ISL_1568192, EPI_ISL_1568193, EPI_ISL_1568194, EPI_ISL_1568195, EPI_ISL_1568197, EPI_ISL_1568198, EPI_ISL_1568199, EPI_ISL_1568200, EPI_ISL_1568201, EPI_ISL_1568202, EPI_ISL_1568203, EPI_ISL_1568204, EPI_ISL_1568205, EPI_ISL_1568206, EPI_ISL_1568207, EPI_ISL_1568208, EPI_ISL_1568209, EPI_ISL_1568210, EPI_ISL_1568211, EPI_ISL_1568212, EPI_ISL_1568213, EPI_ISL_1568214, EPI_ISL_1568215, EPI_ISL_1568216, EPI_ISL_1568217, EPI_ISL_1568218, EPI_ISL_1568219, EPI_ISL_1568220, EPI_ISL_1568221, EPI_ISL_1568222, EPI_ISL_1568223, EPI_ISL_1568224, EPI_ISL_1568225, EPI_ISL_1568226, EPI_ISL_1568227, EPI_ISL_1568228, EPI_ISL_1568229, EPI_ISL_1568230, EPI_ISL_1568231, EPI_ISL_1568232, EPI_ISL_1568233, EPI_ISL_1568234, EPI_ISL_1568235, EPI_ISL_1568236, EPI_ISL_1568238, EPI_ISL_1568239, EPI_ISL_1568240, EPI_ISL_1568241, EPI_ISL_1572531, EPI_ISL_1572533, EPI_ISL_1572534, EPI_ISL_1572535, EPI_ISL_1572537, EPI_ISL_1572538, EPI_ISL_1572542, EPI_ISL_1572547, EPI_ISL_1572548, EPI_ISL_1572551, EPI_ISL_1572553, EPI_ISL_1640171, EPI_ISL_1640174, EPI_ISL_1640176, EPI_ISL_1640182, EPI_ISL_1640185, EPI_ISL_1640190, EPI_ISL_1640193, EPI_ISL_1640200, EPI_ISL_1640206, EPI_ISL_1640241, EPI_ISL_1640249, EPI_ISL_1640265, EPI_ISL_1640268, EPI_ISL_1640272, EPI_ISL_1640273, EPI_ISL_1640274, EPI_ISL_1640277, EPI_ISL_1640286, EPI_ISL_1640288, EPI_ISL_1640293, EPI_ISL_1640302, EPI_ISL_1640306, EPI_ISL_1640317, EPI_ISL_2116223, EPI_ISL_2116236 | | | |
| see above | Diagnosticum - Labor Neukirchen | Robert Koch Institute | |
| EPI_ISL_1568102, EPI_ISL_1568106, EPI_ISL_1568108, EPI_ISL_1568110, EPI_ISL_1568111, EPI_ISL_1568112, EPI_ISL_1568113, EPI_ISL_1568115, EPI_ISL_1568120, EPI_ISL_1568121, EPI_ISL_1568127, EPI_ISL_1568130, EPI_ISL_1644921, EPI_ISL_1644923, EPI_ISL_1644924, EPI_ISL_1644926, EPI_ISL_1644928, EPI_ISL_1644930 | | | |
| see above | Dianovis GmbH Greiz | Robert Koch Institute | |
| EPI_ISL_1569893, EPI_ISL_1569897, EPI_ISL_1569918, EPI_ISL_1569921, EPI_ISL_1569924, EPI_ISL_1569929, EPI_ISL_1569940, EPI_ISL_1569943, EPI_ISL_1569971, EPI_ISL_1569982, EPI_ISL_1569983 | | | |
| see above | ELBLAB GmbH Zentrum fÃ¼r LaborMedizin | Robert Koch Institute | |
| EPI_ISL_1571337, EPI_ISL_1571338, EPI_ISL_1571339, EPI_ISL_1571340, EPI_ISL_1571341, EPI_ISL_1571342, EPI_ISL_1571343, EPI_ISL_1571344, EPI_ISL_1571345, EPI_ISL_1571346, EPI_ISL_1571347, EPI_ISL_1571348, EPI_ISL_1571349, EPI_ISL_1571350, EPI_ISL_1571351, EPI_ISL_1571352, EPI_ISL_1571353, EPI_ISL_1571354, EPI_ISL_1571355, EPI_ISL_1571356, EPI_ISL_1571357, EPI_ISL_1571358, EPI_ISL_1571359, EPI_ISL_1571360, EPI_ISL_1571361, EPI_ISL_1571362, EPI_ISL_1571363, EPI_ISL_1571364, EPI_ISL_1571365, EPI_ISL_1571366, EPI_ISL_1571367, EPI_ISL_1571368, EPI_ISL_1571369, EPI_ISL_1571370, EPI_ISL_1571371, EPI_ISL_1571372, EPI_ISL_1571373, EPI_ISL_1571374, EPI_ISL_1571375, EPI_ISL_1571376, EPI_ISL_1571377, EPI_ISL_1571378, EPI_ISL_1571379, EPI_ISL_1571380, EPI_ISL_1571381, EPI_ISL_1571382, EPI_ISL_1571383, EPI_ISL_1571384, EPI_ISL_1571385, EPI_ISL_1571386, EPI_ISL_1571387, EPI_ISL_1571388, EPI_ISL_1571389, EPI_ISL_1571390, EPI_ISL_1571391, EPI_ISL_1571392, EPI_ISL_1571393, EPI_ISL_1571394, EPI_ISL_1571395, EPI_ISL_1571396, EPI_ISL_1571397, EPI_ISL_1571398, EPI_ISL_1571399, EPI_ISL_1571400, EPI_ISL_1571401, EPI_ISL_1571402, EPI_ISL_1571403, EPI_ISL_1571404, EPI_ISL_1571405, EPI_ISL_1571406, EPI_ISL_1571407, EPI_ISL_1571408, EPI_ISL_1571409, EPI_ISL_1571410, EPI_ISL_1571411, EPI_ISL_1571412, EPI_ISL_1571413, EPI_ISL_1571414, EPI_ISL_1571415, EPI_ISL_1571416, EPI_ISL_1571417, EPI_ISL_1571418, EPI_ISL_1571419, EPI_ISL_1571420, EPI_ISL_1571421, EPI_ISL_1571422, EPI_ISL_1571423, EPI_ISL_1571424, EPI_ISL_1571425, EPI_ISL_1571426, EPI_ISL_1571427, EPI_ISL_1571428, EPI_ISL_1571429, EPI_ISL_1571430, EPI_ISL_1571431, EPI_ISL_1571432, EPI_ISL_1571433, EPI_ISL_1571434, EPI_ISL_1571435, EPI_ISL_1571436, EPI_ISL_1571438, EPI_ISL_1571439, EPI_ISL_1571440, EPI_ISL_1571441, EPI_ISL_1571442, EPI_ISL_1571443, EPI_ISL_1571444, EPI_ISL_1571445, EPI_ISL_1571446, EPI_ISL_1571447, EPI_ISL_1571448, EPI_ISL_1571449, EPI_ISL_1571450, EPI_ISL_1571451, EPI_ISL_1571452, EPI_ISL_1571453, EPI_ISL_1571454, EPI_ISL_1571455, EPI_ISL_1571456, EPI_ISL_1571457, EPI_ISL_1571458, EPI_ISL_1571459, EPI_ISL_1571460, EPI_ISL_1571461, EPI_ISL_1571462, EPI_ISL_1571463, EPI_ISL_1571464, EPI_ISL_1571465 | | | |
| see above | Eurofins LifeCodexx GmbH | Robert Koch Institute | |
| EPI_ISL_1563786, EPI_ISL_1563789, EPI_ISL_1563802, EPI_ISL_1570850, EPI_ISL_1570851, EPI_ISL_1570852, EPI_ISL_1570853, EPI_ISL_1570854, EPI_ISL_1570855, EPI_ISL_1570856, EPI_ISL_1570857, EPI_ISL_1570858, EPI_ISL_1570859, EPI_ISL_1570860, EPI_ISL_1570861, EPI_ISL_1570862, EPI_ISL_1570863, EPI_ISL_1570864, EPI_ISL_1570866, EPI_ISL_1570867, EPI_ISL_1570868, EPI_ISL_1570869, EPI_ISL_1570870, EPI_ISL_1570871, EPI_ISL_1570872, EPI_ISL_1570873, EPI_ISL_1570874, EPI_ISL_1570875, EPI_ISL_1570876, EPI_ISL_1570877, EPI_ISL_1570878, EPI_ISL_1570879, EPI_ISL_1570880, EPI_ISL_1570881, EPI_ISL_1570882, EPI_ISL_1570883, EPI_ISL_1642302 | | | |
| see above | Fraunhofer-Institut fÃ¼r Zelltherapie und Immunologie | Robert Koch Institute | |
| EPI_ISL_1524906 | Hannover Medical School, Institute of Virology | Hannover Medical School, Institute of Virology | Lars Steinbrück |
| EPI_ISL_1571635, EPI_ISL_1571704, EPI_ISL_1571800, EPI_ISL_1571860 | Helios Klinikum Niederberg | Robert Koch Institute | |
| EPI_ISL_1565412, EPI_ISL_1565418, EPI_ISL_1565421, EPI_ISL_1565429, EPI_ISL_1565437, EPI_ISL_1565438, EPI_ISL_1565456, EPI_ISL_1565463, EPI_ISL_1565469, EPI_ISL_1565472, EPI_ISL_1565481, EPI_ISL_1565482, EPI_ISL_1565490, EPI_ISL_1565495, EPI_ISL_1565497, EPI_ISL_1639288, EPI_ISL_1639289, EPI_ISL_1639291, EPI_ISL_1639298, EPI_ISL_1639299, EPI_ISL_1639300, EPI_ISL_1639302, EPI_ISL_1639303, EPI_ISL_1639305, EPI_ISL_1639306, EPI_ISL_1639317, EPI_ISL_1639320, EPI_ISL_1639321, EPI_ISL_1639323, EPI_ISL_1639324 | | | |
| see above | IMD - Institut fÃ¼r Medizinische Diagnostik Berlin-Potsdam | Robert Koch Institute | |
| EPI_ISL_1565414, EPI_ISL_1565423, EPI_ISL_1565426, EPI_ISL_1565427, EPI_ISL_1565432, EPI_ISL_1565434, EPI_ISL_1565440, EPI_ISL_1565441, EPI_ISL_1565444, EPI_ISL_1565447, EPI_ISL_1565449, EPI_ISL_1565450, EPI_ISL_1565451, EPI_ISL_1565454, EPI_ISL_1565461, EPI_ISL_1565478, EPI_ISL_1565484, EPI_ISL_1565485, EPI_ISL_1565486, EPI_ISL_1565488, EPI_ISL_1565493, EPI_ISL_1565501, EPI_ISL_1639277, EPI_ISL_1639296, EPI_ISL_1639307, EPI_ISL_1639311, EPI_ISL_1639312, EPI_ISL_1639314, EPI_ISL_1639315, EPI_ISL_1639322, EPI_ISL_1643685, EPI_ISL_1643686, EPI_ISL_1643689, EPI_ISL_1643759 | | | |
| see above | IMD - Labor Oderland | Robert Koch Institute | |
| EPI_ISL_1565443, EPI_ISL_1565459, EPI_ISL_1565479 | IMD - MVZ Labor Greifswald | Robert Koch Institute | |

| | | | |
|---|---|---|---|
| EPI_ISL_1439883, EPI_ISL_1439889, EPI_ISL_1439892, EPI_ISL_1565422, EPI_ISL_1565424, EPI_ISL_1565425, EPI_ISL_1565428, EPI_ISL_1565430, EPI_ISL_1565431, EPI_ISL_1565436, EPI_ISL_1565446, EPI_ISL_1565448, EPI_ISL_1565453, EPI_ISL_1565455, EPI_ISL_1565462, EPI_ISL_1565464, EPI_ISL_1565467, EPI_ISL_1565470, EPI_ISL_1565471, EPI_ISL_1565474, EPI_ISL_1565475, EPI_ISL_1565476, EPI_ISL_1565477, EPI_ISL_1565480, EPI_ISL_1565483, EPI_ISL_1565492, EPI_ISL_1565494, EPI_ISL_1565496, EPI_ISL_1565498, EPI_ISL_1565499, EPI_ISL_1565500, EPI_ISL_1639275, EPI_ISL_1639278, EPI_ISL_1639279, EPI_ISL_1639284, EPI_ISL_1639286, EPI_ISL_1639292, EPI_ISL_1639294, EPI_ISL_1639295, EPI_ISL_1639301, EPI_ISL_1639308, EPI_ISL_1639310, EPI_ISL_1639316, EPI_ISL_1639318, EPI_ISL_1639440, EPI_ISL_1639443, EPI_ISL_1639448, EPI_ISL_1639453, EPI_ISL_1639456, EPI_ISL_1639470, EPI_ISL_1639495 | | | |
| see above | IMD - MVZ Labor Martinsried | Robert Koch Institute | |
| EPI_ISL_1354336, EPI_ISL_1639280, EPI_ISL_1639281, EPI_ISL_1639285, EPI_ISL_1639290, EPI_ISL_1639293, EPI_ISL_1639297, EPI_ISL_1639309, EPI_ISL_1639313, EPI_ISL_1639325 | | | |
| see above | IMD - Medizinisches Labor Rostock | Robert Koch Institute | |
| EPI_ISL_1439368, EPI_ISL_1439370, EPI_ISL_1439374, EPI_ISL_1439376, EPI_ISL_1439379, EPI_ISL_1439393, EPI_ISL_1439402, EPI_ISL_1439407, EPI_ISL_1439410, EPI_ISL_1439412, EPI_ISL_1439415, EPI_ISL_1439417, EPI_ISL_1439419, EPI_ISL_1439420, EPI_ISL_1439422, EPI_ISL_1439429, EPI_ISL_1439431, EPI_ISL_1439434, EPI_ISL_1563822, EPI_ISL_1563823, EPI_ISL_1563826, EPI_ISL_1563827, EPI_ISL_1563828, EPI_ISL_1563829, EPI_ISL_1563831, EPI_ISL_1563832, EPI_ISL_1563833, EPI_ISL_1563834, EPI_ISL_1563837, EPI_ISL_1563839, EPI_ISL_1563840, EPI_ISL_1563841, EPI_ISL_1563843, EPI_ISL_1563844, EPI_ISL_1563845, EPI_ISL_1563846, EPI_ISL_1563849, EPI_ISL_1572009, EPI_ISL_1572012, EPI_ISL_1572014, EPI_ISL_1572017, EPI_ISL_1572018, EPI_ISL_1572020, EPI_ISL_1572022, EPI_ISL_1572023, EPI_ISL_1572025, EPI_ISL_1572029, EPI_ISL_1572031, EPI_ISL_1572032, EPI_ISL_1572033, EPI_ISL_1572038, EPI_ISL_1572044, EPI_ISL_1572048, EPI_ISL_1572052, EPI_ISL_1572054, EPI_ISL_1572056, EPI_ISL_1572061, EPI_ISL_1572064, EPI_ISL_1572065, EPI_ISL_1572071, EPI_ISL_1572075, EPI_ISL_1572076, EPI_ISL_1572078, EPI_ISL_1572079, EPI_ISL_1572080, EPI_ISL_1572081, EPI_ISL_1572083, EPI_ISL_1572084, EPI_ISL_1572085, EPI_ISL_1572086, EPI_ISL_1572087, EPI_ISL_1572088, EPI_ISL_1572090, EPI_ISL_1572091, EPI_ISL_1572094, EPI_ISL_1572095, EPI_ISL_1572097, EPI_ISL_1572099, EPI_ISL_1572100, EPI_ISL_1572102, EPI_ISL_1572104, EPI_ISL_1572106 | | | |
| see above | IMD Labor Frankfurt | Robert Koch Institute | |
| EPI_ISL_1563835, EPI_ISL_1563842 | Institut fÃ¼r Immunologie und Genetik Kaiserslautern: Medizinisches Labor Dr. med. B. Thiele | Robert Koch Institute | |
| EPI_ISL_1569789, EPI_ISL_1569790, EPI_ISL_1569791, EPI_ISL_1569792, EPI_ISL_1569793, EPI_ISL_1569794, EPI_ISL_1569795 | | | |
| see above | Institut fÃ¼r Klinische Transfusionsmedizin und Immungenetik DRK Blutspendedienst Baden-WÃ¼rttemberg | Robert Koch Institute | |
| EPI_ISL_1571549, EPI_ISL_1571550, EPI_ISL_1571551, EPI_ISL_1571554, EPI_ISL_1571557, EPI_ISL_1571560, EPI_ISL_1724238 | | | |
| see above | Institut fÃ¼r Medizinische Laboratoriumsdiagnostik Bochum GmbH | Robert Koch Institute | |
| EPI_ISL_1565369 | Institut fÃ¼r Medizinische Mikrobiologie - UniversitÃ¤tsmedizin GÃ¶ttingen | Robert Koch Institute | |
| EPI_ISL_1639260, EPI_ISL_1639261 | Institut fÃ¼r med. Mikrobiologie u. Virologie Klinikum Oldenburg | Robert Koch Institute | |
| EPI_ISL_1670093, EPI_ISL_1670094, EPI_ISL_1670095, EPI_ISL_1670096, EPI_ISL_1670097, EPI_ISL_1670098, EPI_ISL_1670099, EPI_ISL_1670100, EPI_ISL_1670144, EPI_ISL_1670145, EPI_ISL_1670146, EPI_ISL_1670147, EPI_ISL_1670148, EPI_ISL_1670179, EPI_ISL_1670180, EPI_ISL_1670181, EPI_ISL_1670182, EPI_ISL_1670183, EPI_ISL_1670184, EPI_ISL_1670185, EPI_ISL_1670186, EPI_ISL_1670187, EPI_ISL_1670188, EPI_ISL_1670189, EPI_ISL_1670190, EPI_ISL_1670191, EPI_ISL_1670192, EPI_ISL_1670193, EPI_ISL_1670194, EPI_ISL_1670276, EPI_ISL_1670277 | | | |
| see above | Institute of Medical Microbiology and Hospital Hygiene | Institute of Medical Microbiology and Hospital Hygiene | Aljoscha Tersteegen; Prof. Dr. Achim Kaasch |
| EPI_ISL_1648924, EPI_ISL_1648925, EPI_ISL_1648926, EPI_ISL_1648927, EPI_ISL_1648928, EPI_ISL_1648929, EPI_ISL_1648930, EPI_ISL_1648931, EPI_ISL_1653691, EPI_ISL_1653692 | | | |
| see above | Institute of Tropical Medicine | Institute of Tropical Medicine | Prof. Dr. Thirumalaisamy P. Velavan and Prof. Dr. Peter Kremsner |
| EPI_ISL_1588619, EPI_ISL_1588620, EPI_ISL_1588627, EPI_ISL_1588628, EPI_ISL_1588629, EPI_ISL_1588630, EPI_ISL_1588631, EPI_ISL_1588632, EPI_ISL_1588633, EPI_ISL_1588634, EPI_ISL_1805004, EPI_ISL_1805005, EPI_ISL_1805006, EPI_ISL_1805008, EPI_ISL_1805011, EPI_ISL_1805012, EPI_ISL_1805018, EPI_ISL_1805019, EPI_ISL_1805020, EPI_ISL_1805021, EPI_ISL_1805022, EPI_ISL_1805023, EPI_ISL_1805024, EPI_ISL_1805026, EPI_ISL_1805027, EPI_ISL_1805028, EPI_ISL_1805029, EPI_ISL_1805030, EPI_ISL_1805031, EPI_ISL_1805032, EPI_ISL_1805033, EPI_ISL_1805034, EPI_ISL_1805035 | | | |
| see above | Institute of Virology, Medical Center, University of Freiburg, Freiburg, Germany | Institute of Virology, Clinial Virus Genomics, Medical Center, University of Freiburg, Freiburg, Germany | Hajo Grundmann; Jonas Fuchs; Lena Jaki; Lisa Kern; Marcus Panning; Sandra Reuter |
| EPI_ISL_1640904, EPI_ISL_1640907, EPI_ISL_1640908, EPI_ISL_1640910, EPI_ISL_1640911, EPI_ISL_1640912, EPI_ISL_1640915, EPI_ISL_1640918, EPI_ISL_1640920, EPI_ISL_1640921, EPI_ISL_1640924, EPI_ISL_1640926, EPI_ISL_1640927, EPI_ISL_1640928, EPI_ISL_1640931 | | | |
| see above | KRH Klinikum Region Hannover: Institut fÃ¼r Med. Mikrobiologie und Krankenhaushygiene | Robert Koch Institute | |
| EPI_ISL_1437281, EPI_ISL_1437282 | KRH Klinikum Region Hannover: Institut für Med. Mikrobiologie und Krankenhaushygiene | Robert Koch Institute | |
| EPI_ISL_1715844, EPI_ISL_1715847, EPI_ISL_1716268, EPI_ISL_1716269 | Kath. Marienkrankenhaus | Heinrich Pette Institute, Leibniz Institute for Experimental Virology | Adam Grundhoff; Alexis Robitaille; Johannes Knobloch; Martin Aepfelbacher; Nicole Fischer; Thomas Günther |
| EPI_ISL_1439487 | Kath. St.-Johannes-Gesellschaft Dortmund gGmbH | Robert Koch Institute | |
| EPI_ISL_1563907, EPI_ISL_1567615, EPI_ISL_1573242 | Kerckhoff-Klinik GmbH | Robert Koch Institute | |
| EPI_ISL_1438660, EPI_ISL_1570705 | Klinikum Darmstadt GmbH | Robert Koch Institute | |
| EPI_ISL_1570751, EPI_ISL_1570753, EPI_ISL_1570755, EPI_ISL_1570757, EPI_ISL_1570762, EPI_ISL_1570765, EPI_ISL_1570766, EPI_ISL_1570767, EPI_ISL_1570771, EPI_ISL_1570773, EPI_ISL_1570776, EPI_ISL_1570777, EPI_ISL_1570780, EPI_ISL_1570781, EPI_ISL_1570788, EPI_ISL_1570792, EPI_ISL_1570795, EPI_ISL_1570798, EPI_ISL_1570806, EPI_ISL_1570809, EPI_ISL_1570811, EPI_ISL_1570814, EPI_ISL_1573063, EPI_ISL_1722266, EPI_ISL_1722275 | | | |
| see above | Klinikum Ernst von Bergmann gemeinnÃ¼tzige GmbH - stationÃ¤rer Bereich | Robert Koch Institute | |
| EPI_ISL_1442533, EPI_ISL_1442534, EPI_ISL_1442535, EPI_ISL_1442537, EPI_ISL_1442539, EPI_ISL_1442541, EPI_ISL_1442542 | | | |
| see above | Klinikum Ernst von Bergmann gemeinnützige GmbH - stationärer Bereich | Robert Koch Institute | |
| EPI_ISL_1569295, EPI_ISL_1569458 | Klinikum Fulda gAG | Robert Koch Institute | |
| EPI_ISL_1722843, EPI_ISL_1722844, EPI_ISL_1722845, EPI_ISL_1722846 | Klinikum Stuttgart | Robert Koch Institute | |
| EPI_ISL_1572035, EPI_ISL_1572037, EPI_ISL_1572042, EPI_ISL_1572058, EPI_ISL_1572072 | Klinikum der Stadt Ludwigshafen - Institut fÃ¼r Labordiagnostik Hygiene und Transfusionsmedizin | Robert Koch Institute | |
| EPI_ISL_1572137, EPI_ISL_1572138 | Krankenhausgesellschaft St. Vincenz mbH | Robert Koch Institute | |
| EPI_ISL_1569951, EPI_ISL_1569964, EPI_ISL_1569967, EPI_ISL_1569991 | LABOR CHEMNITZ â€" Zentrum fÃ¼r Diagnostik am Klinikum Chemnitz | Robert Koch Institute | |
| EPI_ISL_1568965, EPI_ISL_1569033, EPI_ISL_1569100, EPI_ISL_1569103, EPI_ISL_1569173, EPI_ISL_1569208, EPI_ISL_1569223, EPI_ISL_1569248, EPI_ISL_1569257, EPI_ISL_1569267, EPI_ISL_1569276, EPI_ISL_1569278, EPI_ISL_1569279, EPI_ISL_1569285, EPI_ISL_1569289, EPI_ISL_1569290, EPI_ISL_1569297, EPI_ISL_1569302, EPI_ISL_1569307, EPI_ISL_1569313, EPI_ISL_1569316, EPI_ISL_1569317, EPI_ISL_1569322, EPI_ISL_1569324, EPI_ISL_1569327, EPI_ISL_1569331, EPI_ISL_1569336, EPI_ISL_1569339, EPI_ISL_1569342, EPI_ISL_1569345, EPI_ISL_1569347, EPI_ISL_1569348, EPI_ISL_1569351, EPI_ISL_1569356, EPI_ISL_1569359, EPI_ISL_1569363, EPI_ISL_1569367, EPI_ISL_1569368, EPI_ISL_1569370, EPI_ISL_1569377, EPI_ISL_1569378, EPI_ISL_1569389, EPI_ISL_1569390, EPI_ISL_1569391, EPI_ISL_1569392, EPI_ISL_1569396, EPI_ISL_1569401, EPI_ISL_1569403, EPI_ISL_1569410, EPI_ISL_1569417, EPI_ISL_1569419, EPI_ISL_1569423, EPI_ISL_1569424, EPI_ISL_1569428, EPI_ISL_1569429, EPI_ISL_1569430, EPI_ISL_1569436, EPI_ISL_1569441, EPI_ISL_1569450, EPI_ISL_1569452, EPI_ISL_1569457, EPI_ISL_1569464, EPI_ISL_1569465, EPI_ISL_1569474, EPI_ISL_1569481, EPI_ISL_1569490, EPI_ISL_1569503, EPI_ISL_1569505, EPI_ISL_1569510, EPI_ISL_1569524, EPI_ISL_1569527, EPI_ISL_1569537, EPI_ISL_1569555, EPI_ISL_1569561, EPI_ISL_1569570, EPI_ISL_1569594, EPI_ISL_1569607, EPI_ISL_1569627, EPI_ISL_1569690, EPI_ISL_1569698, EPI_ISL_1569703, EPI_ISL_1638748, EPI_ISL_1639205 | | | |
| see above | LabKom - Labor Augsburg MVZ GmbH | Robert Koch Institute | |
| EPI_ISL_1638872, EPI_ISL_1638898, EPI_ISL_1638924 | LabKom - Labor Hannover MVZ GmbH | Robert Koch Institute | |
| EPI_ISL_1638684, EPI_ISL_1638686, EPI_ISL_1638693, EPI_ISL_1638694, EPI_ISL_1638701, EPI_ISL_1638703, EPI_ISL_1638723, EPI_ISL_1638726, EPI_ISL_1638742, EPI_ISL_1638755, EPI_ISL_1638789, EPI_ISL_1638790, EPI_ISL_1638793, EPI_ISL_1638824, EPI_ISL_1638830, EPI_ISL_1638840, EPI_ISL_1638859, EPI_ISL_1638871, EPI_ISL_1638879, EPI_ISL_1638887, EPI_ISL_1638891, EPI_ISL_1638905, EPI_ISL_1638908, EPI_ISL_1638920, EPI_ISL_1638923, EPI_ISL_1638927, EPI_ISL_1638934, EPI_ISL_1638937, EPI_ISL_1642431, EPI_ISL_1642438, EPI_ISL_1642452, EPI_ISL_1642463, EPI_ISL_1642489, EPI_ISL_1642577, EPI_ISL_1642606, EPI_ISL_1642619, EPI_ISL_1642632, EPI_ISL_1642646, EPI_ISL_1642677, EPI_ISL_1643196, EPI_ISL_1643202, EPI_ISL_1643252, EPI_ISL_1643277, EPI_ISL_1643291, EPI_ISL_1643318, EPI_ISL_1643321, EPI_ISL_1643371, EPI_ISL_1643380, EPI_ISL_1643388, EPI_ISL_1643389 | | | |
| see above | Labor 28 MVZ GmbH | Robert Koch Institute | |
| EPI_ISL_2123979, EPI_ISL_2123985, EPI_ISL_2123996, EPI_ISL_2123998, EPI_ISL_2123999, EPI_ISL_2124002, EPI_ISL_2124004, EPI_ISL_2124007, EPI_ISL_2124012, EPI_ISL_2124014 | | | |
| see above | Labor Becker & Kollegen (Standort Munchen) | Robert Koch Institute | |
| EPI_ISL_1640635, EPI_ISL_1846778, EPI_ISL_1846780, EPI_ISL_1846787 | Labor Becker & Kollegen (Standort MÃ¼nchen) | Robert Koch Institute | |
| EPI_ISL_1905063, EPI_ISL_1905064, EPI_ISL_1905071, EPI_ISL_1905075, EPI_ISL_1905080, EPI_ISL_1905085, EPI_ISL_1905089, EPI_ISL_1905107, EPI_ISL_1905115, EPI_ISL_1905134, EPI_ISL_1905138, EPI_ISL_1905155, EPI_ISL_1905159, EPI_ISL_1905161, EPI_ISL_1905162, EPI_ISL_1905163, EPI_ISL_1905164, EPI_ISL_1905167, EPI_ISL_1905168, EPI_ISL_1972148, EPI_ISL_1972150, EPI_ISL_1972151, EPI_ISL_1972153, EPI_ISL_1972155, EPI_ISL_1972156, EPI_ISL_1972159, EPI_ISL_1972161, EPI_ISL_1972163, EPI_ISL_1972165, EPI_ISL_1972166, EPI_ISL_1972167, EPI_ISL_1972168, EPI_ISL_1972174, EPI_ISL_1972175, EPI_ISL_1972176, EPI_ISL_1972177, EPI_ISL_1972178, EPI_ISL_1972179, EPI_ISL_1972181, EPI_ISL_1972182, EPI_ISL_1972183, EPI_ISL_1972184, EPI_ISL_1972185, EPI_ISL_1972186, EPI_ISL_1972189, EPI_ISL_1972192, EPI_ISL_1972193, EPI_ISL_1972194, EPI_ISL_1972195, EPI_ISL_1972196, EPI_ISL_1972197, EPI_ISL_1972198, EPI_ISL_1972201, EPI_ISL_1972204, EPI_ISL_1972205, EPI_ISL_1972207, EPI_ISL_1972208, EPI_ISL_1972209, EPI_ISL_1972210, EPI_ISL_1972211, EPI_ISL_1982486, EPI_ISL_1982491, EPI_ISL_1982506, EPI_ISL_1982508, EPI_ISL_1982520, EPI_ISL_1982526, EPI_ISL_1982528, EPI_ISL_1982539, EPI_ISL_1982541, EPI_ISL_1982548, EPI_ISL_1982555, EPI_ISL_1982564, EPI_ISL_1982566, EPI_ISL_1982579, EPI_ISL_1982591, EPI_ISL_1982601, EPI_ISL_1982604, EPI_ISL_1982612, EPI_ISL_1984772 | | | |
| see above | Labor Berlin Charité Vivantes GmbH / Institut für Virologie | Charité Universitätsmedizin Berlin, Institut für Virologie/Labor Berlin | Barbara Mühlemann; Christian Drosten; Christine Stephan; Peter Menzel; Rolf Schwarzer; Terry Jones; Victor M Corman |
| EPI_ISL_1570475, EPI_ISL_1570484, EPI_ISL_1570487, EPI_ISL_1570498, EPI_ISL_1570501, EPI_ISL_1570506, EPI_ISL_1570509, EPI_ISL_1570510, EPI_ISL_1570513, EPI_ISL_1570520, EPI_ISL_1570522, EPI_ISL_1570526, EPI_ISL_1570550, EPI_ISL_1570554, EPI_ISL_1570558, EPI_ISL_1570560, EPI_ISL_1570561, EPI_ISL_1570564, EPI_ISL_1570570, EPI_ISL_1570579, EPI_ISL_1570582, EPI_ISL_1570584, EPI_ISL_1570594, EPI_ISL_1570597, EPI_ISL_1570603, EPI_ISL_1570609, EPI_ISL_1570616, EPI_ISL_1570624, EPI_ISL_1570626, EPI_ISL_1570628, EPI_ISL_1570631, EPI_ISL_1570649, EPI_ISL_1570651, EPI_ISL_1570653, EPI_ISL_1570656, EPI_ISL_1570660, EPI_ISL_1570661, EPI_ISL_1570675, EPI_ISL_1570680, EPI_ISL_1570682, EPI_ISL_1570690, EPI_ISL_1570691, EPI_ISL_1570694, EPI_ISL_1570697, EPI_ISL_1570698 | | | |
| see above | Labor Blackholm MVZ | Robert Koch Institute | |
| EPI_ISL_1715769, EPI_ISL_1715770, EPI_ISL_1715771, EPI_ISL_1715772, EPI_ISL_1715773, EPI_ISL_1715774, EPI_ISL_1715775, EPI_ISL_1715776, EPI_ISL_1715777, EPI_ISL_1715778, EPI_ISL_1715779, EPI_ISL_1715780, EPI_ISL_1715781, EPI_ISL_1715782, EPI_ISL_1715783, EPI_ISL_1715784, EPI_ISL_1715785, EPI_ISL_1715786, EPI_ISL_1715787, EPI_ISL_1715788, EPI_ISL_1715789, EPI_ISL_1715790, EPI_ISL_1715791, EPI_ISL_1715792, EPI_ISL_1715793, EPI_ISL_1715794, EPI_ISL_1715795, EPI_ISL_1715796, EPI_ISL_1715797, EPI_ISL_1715798, EPI_ISL_1715799, EPI_ISL_1715800, EPI_ISL_1715801, EPI_ISL_1715805, EPI_ISL_1715806, EPI_ISL_1715807, EPI_ISL_1715808, EPI_ISL_1715809, EPI_ISL_1715810, EPI_ISL_1715811, EPI_ISL_1715812, EPI_ISL_1715813, EPI_ISL_1715814, EPI_ISL_1715815, EPI_ISL_1715816, EPI_ISL_1715817, EPI_ISL_1715818, EPI_ISL_1715819, EPI_ISL_1715820, EPI_ISL_1715821, EPI_ISL_1715822, EPI_ISL_1715823, EPI_ISL_1715824, EPI_ISL_1715825, EPI_ISL_1715826, EPI_ISL_1715827, EPI_ISL_1716100, EPI_ISL_1716280, EPI_ISL_1716281, EPI_ISL_1716282, EPI_ISL_1716283, EPI_ISL_1716284, EPI_ISL_1716285, EPI_ISL_1716286, EPI_ISL_1716287, EPI_ISL_1716288, EPI_ISL_1716289, EPI_ISL_1716290, EPI_ISL_1716291, EPI_ISL_1716292, EPI_ISL_1716293, EPI_ISL_1716294, EPI_ISL_1716295, EPI_ISL_1716296, EPI_ISL_1716297, EPI_ISL_1716298, EPI_ISL_1716299, EPI_ISL_1716300, EPI_ISL_1716301, EPI_ISL_1716302, EPI_ISL_1716303, EPI_ISL_1716304, EPI_ISL_1716305, EPI_ISL_1716306, EPI_ISL_1716307, EPI_ISL_1716308, EPI_ISL_1716309, EPI_ISL_1716310, EPI_ISL_1716311, EPI_ISL_1716312, EPI_ISL_1716313, EPI_ISL_1716314, EPI_ISL_1716315, EPI_ISL_1716316, EPI_ISL_1716317, EPI_ISL_1716318, EPI_ISL_1716319, EPI_ISL_1716320, EPI_ISL_1716321, EPI_ISL_1716322, EPI_ISL_1716323, EPI_ISL_1716324, EPI_ISL_1716325, EPI_ISL_1716326, EPI_ISL_1716327, EPI_ISL_1716328, EPI_ISL_1716329, EPI_ISL_1716330, EPI_ISL_1716331, EPI_ISL_1716332, EPI_ISL_1716333, EPI_ISL_1716334, EPI_ISL_1716335, EPI_ISL_1716336, EPI_ISL_1716337, EPI_ISL_1716338, EPI_ISL_1716339, EPI_ISL_1716340, EPI_ISL_1716341, EPI_ISL_1716342, EPI_ISL_1716343, EPI_ISL_1716344, EPI_ISL_1716345, EPI_ISL_1716346, EPI_ISL_1716347, EPI_ISL_1716348, EPI_ISL_1716349, EPI_ISL_1716350, EPI_ISL_1716351, EPI_ISL_1716352, EPI_ISL_1716353, EPI_ISL_1716354, EPI_ISL_1716355, EPI_ISL_1716356, EPI_ISL_1716357, EPI_ISL_1716358, EPI_ISL_1716359, EPI_ISL_1716360, EPI_ISL_1716361, EPI_ISL_1716362, EPI_ISL_1716363, EPI_ISL_1716364, EPI_ISL_1716365, EPI_ISL_1716366, EPI_ISL_1716367, EPI_ISL_1716368, EPI_ISL_1716369, EPI_ISL_1716370, EPI_ISL_1716371, EPI_ISL_1716372, EPI_ISL_1716373, EPI_ISL_1716374, EPI_ISL_1716375, EPI_ISL_1716376, EPI_ISL_1716377, EPI_ISL_1716378, EPI_ISL_1716379, EPI_ISL_1716380, EPI_ISL_1716381, EPI_ISL_1716382, EPI_ISL_1716383, EPI_ISL_1716384, EPI_ISL_1716385, EPI_ISL_1716386, EPI_ISL_1716388, EPI_ISL_1716392, EPI_ISL_1716393, EPI_ISL_1716394, EPI_ISL_1716395, EPI_ISL_1716396, EPI_ISL_1861722, EPI_ISL_1861724, EPI_ISL_1861726, EPI_ISL_1861727, EPI_ISL_1861729, EPI_ISL_1861731, EPI_ISL_1861744, EPI_ISL_1861746, EPI_ISL_1861748, EPI_ISL_1861750, EPI_ISL_1861752, EPI_ISL_1861754, EPI_ISL_1861759, EPI_ISL_1861765, EPI_ISL_1861767, EPI_ISL_1861768, EPI_ISL_1861770, EPI_ISL_1861772, EPI_ISL_1861774, EPI_ISL_1861776, EPI_ISL_1861786, EPI_ISL_1861788, EPI_ISL_1861790, EPI_ISL_1861792, EPI_ISL_1861793, EPI_ISL_1861795, EPI_ISL_1861806, EPI_ISL_1861808, EPI_ISL_1861810, EPI_ISL_1861812, EPI_ISL_1861813, EPI_ISL_1861814, EPI_ISL_1861818, EPI_ISL_1861822, EPI_ISL_1861824, EPI_ISL_1861826, EPI_ISL_1861828, EPI_ISL_1861830, EPI_ISL_1861832, EPI_ISL_1861833, EPI_ISL_1861835, EPI_ISL_1861845, EPI_ISL_1861847, EPI_ISL_1861848, EPI_ISL_1861850, EPI_ISL_1861852, EPI_ISL_1861854, EPI_ISL_1861856, EPI_ISL_1861865, EPI_ISL_1861867, EPI_ISL_1861869, EPI_ISL_1861871, EPI_ISL_1861873, EPI_ISL_1861875, EPI_ISL_1861877, EPI_ISL_1861878, EPI_ISL_1861880, EPI_ISL_1861891, EPI_ISL_1861897, EPI_ISL_1861901, EPI_ISL_1861922 | | | |
| see above | Labor Dr. Fenner und Kollegen | Heinrich Pette Institute, Leibniz Institute for Experimental Virology | Adam Grundhoff; Alexis Robitaille; Johannes Knobloch; Martin Aepfelbacher; Nicole Fischer; Thomas Günther |
| EPI_ISL_1439805, EPI_ISL_1439832, EPI_ISL_1439845, EPI_ISL_1439857, EPI_ISL_1439865, EPI_ISL_1442548, EPI_ISL_1442553, EPI_ISL_1442555, EPI_ISL_1442566, EPI_ISL_1442568, EPI_ISL_1442569, EPI_ISL_1442571, EPI_ISL_1566728, EPI_ISL_1566729, EPI_ISL_1566730, EPI_ISL_1566731, EPI_ISL_1566732, EPI_ISL_1566733, EPI_ISL_1566734, EPI_ISL_1566735, EPI_ISL_1566736, EPI_ISL_1566737, EPI_ISL_1566738, EPI_ISL_1566739, EPI_ISL_1566740, EPI_ISL_1566741, EPI_ISL_1566742, EPI_ISL_1566743, EPI_ISL_1566744, EPI_ISL_1566745, EPI_ISL_1566746, EPI_ISL_1566747, EPI_ISL_1566752, EPI_ISL_1566753, EPI_ISL_1566755, EPI_ISL_1566756, EPI_ISL_1566757, EPI_ISL_1566759, EPI_ISL_1566760, EPI_ISL_1566761, EPI_ISL_1566762, EPI_ISL_1566764, EPI_ISL_1566765, EPI_ISL_1566767, EPI_ISL_1566768, EPI_ISL_1566771, EPI_ISL_1566772, EPI_ISL_1566773, EPI_ISL_1566774, EPI_ISL_1566775, EPI_ISL_1566776, EPI_ISL_1566778, EPI_ISL_1566780, EPI_ISL_1566782, EPI_ISL_1566783, EPI_ISL_1566784 | | | |
| see above | Labor Dr. Heidrich & Kollegen MVZ GmbH Hamburg | Robert Koch Institute | |

| | | | |
|---|---|---|---|
| EPI_ISL_1569298, EPI_ISL_1569334, EPI_ISL_1569376, EPI_ISL_1569386, EPI_ISL_1569388, EPI_ISL_1569393, EPI_ISL_1569394, EPI_ISL_1569397, EPI_ISL_1569399, EPI_ISL_1569411, EPI_ISL_1569461, EPI_ISL_1569480, EPI_ISL_1569487, EPI_ISL_1569497, EPI_ISL_1569501, EPI_ISL_1569605, EPI_ISL_1569674, EPI_ISL_1721127, EPI_ISL_1721158, EPI_ISL_1721160, EPI_ISL_1721189, EPI_ISL_1721299, EPI_ISL_1842838, EPI_ISL_1842856, EPI_ISL_1842857, EPI_ISL_1842905, EPI_ISL_1843628, EPI_ISL_1847099, EPI_ISL_1847102, EPI_ISL_1847114, EPI_ISL_1847117, EPI_ISL_1847135, EPI_ISL_1847154, EPI_ISL_1847165 | | | |
| see above | Labor Dr. Schumacher MVZ | Robert Koch Institute | |
| EPI_ISL_1643832, EPI_ISL_1643833, EPI_ISL_1643837, EPI_ISL_1643841, EPI_ISL_1643842, EPI_ISL_1643843, EPI_ISL_1643848, EPI_ISL_1643849, EPI_ISL_1643857 | | | |
| see above | Labor Dr. Spranger | Robert Koch Institute | |
| EPI_ISL_1563927, EPI_ISL_1563928, EPI_ISL_1563929, EPI_ISL_1563930, EPI_ISL_1563931, EPI_ISL_1571544 | Labor Dr. Wisplinghoff - KÃ¶ln | Robert Koch Institute | |
| EPI_ISL_1433950 | Labor Dr. Wisplinghoff - Köln | Robert Koch Institute | |
| EPI_ISL_1569796, EPI_ISL_1569797, EPI_ISL_1569798, EPI_ISL_1569799 | Labor KneiÃŸler GmbH & Co. KG | Robert Koch Institute | |
| EPI_ISL_1567781, EPI_ISL_1567817, EPI_ISL_1567830 | Labor LÃ¼beck bzw. LaborÃ¤rztliche Gemeinschaftspraxis LÃ¼beck | Robert Koch Institute | |
| EPI_ISL_1567565, EPI_ISL_1567570, EPI_ISL_1567573, EPI_ISL_1567575, EPI_ISL_1567576, EPI_ISL_1567579, EPI_ISL_1567582, EPI_ISL_1567584, EPI_ISL_1567586, EPI_ISL_1567596, EPI_ISL_1567598, EPI_ISL_1567601, EPI_ISL_1567603, EPI_ISL_1567605, EPI_ISL_1567609, EPI_ISL_1567612, EPI_ISL_1567613, EPI_ISL_1567617, EPI_ISL_1567620, EPI_ISL_1567623, EPI_ISL_1567624, EPI_ISL_1567625, EPI_ISL_1567628, EPI_ISL_1567632, EPI_ISL_1567640, EPI_ISL_1567659, EPI_ISL_1567664, EPI_ISL_1567666, EPI_ISL_1567671, EPI_ISL_1567680, EPI_ISL_1567686, EPI_ISL_1567690, EPI_ISL_1570746, EPI_ISL_1570816, EPI_ISL_1571164, EPI_ISL_1571180, EPI_ISL_1571185, EPI_ISL_1571191, EPI_ISL_1571196, EPI_ISL_1571199, EPI_ISL_1571204, EPI_ISL_1571941 | | | |
| see above | Labor MÃ¶nchengladbach MVZ Dr. Stein + Kollegen GbR | Robert Koch Institute | |
| EPI_ISL_1570839, EPI_ISL_1570842, EPI_ISL_1570846, EPI_ISL_1573245, EPI_ISL_1573264 | Labor Prof. Dr. G. Enders MVZ GbR | Robert Koch Institute | |
| EPI_ISL_1573266, EPI_ISL_1573267, EPI_ISL_1573268, EPI_ISL_1573271, EPI_ISL_1573272, EPI_ISL_1573273, EPI_ISL_1573274, EPI_ISL_1573276, EPI_ISL_1573277, EPI_ISL_1573278, EPI_ISL_1573279, EPI_ISL_1573280, EPI_ISL_1573281, EPI_ISL_1573282, EPI_ISL_1573283, EPI_ISL_1573284, EPI_ISL_1573285, EPI_ISL_1573286, EPI_ISL_1573287, EPI_ISL_1573288, EPI_ISL_1573290 | | | |
| see above | Labor ZOTZ\|KLIMAS; MVZ DÃ¼sseldorf-Centrum | Robert Koch Institute | |
| EPI_ISL_1567288, EPI_ISL_1567291, EPI_ISL_1567293, EPI_ISL_1567298, EPI_ISL_1567307, EPI_ISL_1567330, EPI_ISL_1567342, EPI_ISL_1567350, EPI_ISL_1567442, EPI_ISL_1567483, EPI_ISL_1567530, EPI_ISL_1567544 | | | |
| see above | Labor fÃ¼r medizinische Mikrobiologie Bakteriologie Mykologie Virologie & Infektionsserologie - Labor MÃ¶lbis | Robert Koch Institute | |
| EPI_ISL_1438427, EPI_ISL_1438509 | Labor für medizinische Mikrobiologie Bakteriologie Mykologie Virologie & Infektionsserologie | Robert Koch Institute | |
| EPI_ISL_1643996, EPI_ISL_1643997, EPI_ISL_1643998, EPI_ISL_1643999, EPI_ISL_1644000, EPI_ISL_1644001, EPI_ISL_1644002, EPI_ISL_1644003 | | | |
| see above | Laborarztpraxis Dres. med. Walther Weindel & Kollegen | Robert Koch Institute | |
| EPI_ISL_1570844 | Laborarztpraxis Jochen HÃ¼ter | Robert Koch Institute | |
| EPI_ISL_1438616, EPI_ISL_1438631 | Laborarztpraxis Jochen Hüter | Robert Koch Institute | |
| EPI_ISL_1563820, EPI_ISL_1563825, EPI_ISL_1572013, EPI_ISL_1572015, EPI_ISL_1572016, EPI_ISL_1572027, EPI_ISL_1572034, EPI_ISL_1572039, EPI_ISL_1572040, EPI_ISL_1572055, EPI_ISL_1572059, EPI_ISL_1572077, EPI_ISL_1572082, EPI_ISL_1572092, EPI_ISL_1572105, EPI_ISL_1638572, EPI_ISL_1638577, EPI_ISL_1638584, EPI_ISL_1638589, EPI_ISL_1638614, EPI_ISL_1642318, EPI_ISL_1642332, EPI_ISL_1642338, EPI_ISL_1642342, EPI_ISL_1642349 | | | |
| see above | Labormedizin Darmstadt | Robert Koch Institute | |
| EPI_ISL_1361934, EPI_ISL_1670057, EPI_ISL_1670058, EPI_ISL_1670059, EPI_ISL_1670060, EPI_ISL_1670061, EPI_ISL_1670062, EPI_ISL_1670063, EPI_ISL_1670064, EPI_ISL_1670065, EPI_ISL_1670066, EPI_ISL_1670067, EPI_ISL_1670068, EPI_ISL_1670069, EPI_ISL_1670070, EPI_ISL_1670071, EPI_ISL_1670072, EPI_ISL_1670073, EPI_ISL_1670074, EPI_ISL_1670075, EPI_ISL_1670076, EPI_ISL_1670077, EPI_ISL_1670078, EPI_ISL_1670079, EPI_ISL_1670080, EPI_ISL_1670081, EPI_ISL_1670082, EPI_ISL_1670083, EPI_ISL_1670084, EPI_ISL_1670085, EPI_ISL_1670086, EPI_ISL_1670087, EPI_ISL_1670088, EPI_ISL_1670089, EPI_ISL_1670090, EPI_ISL_1670091, EPI_ISL_1670092, EPI_ISL_1670102, EPI_ISL_1670103, EPI_ISL_1670104, EPI_ISL_1670105, EPI_ISL_1670106, EPI_ISL_1670107, EPI_ISL_1670108, EPI_ISL_1670109, EPI_ISL_1670110, EPI_ISL_1670111, EPI_ISL_1670112, EPI_ISL_1670113, EPI_ISL_1670114, EPI_ISL_1670115, EPI_ISL_1670116, EPI_ISL_1670117, EPI_ISL_1670118, EPI_ISL_1670119, EPI_ISL_1670120, EPI_ISL_1670121, EPI_ISL_1670122, EPI_ISL_1670123, EPI_ISL_1670124, EPI_ISL_1670125, EPI_ISL_1670126, EPI_ISL_1670127, EPI_ISL_1670128, EPI_ISL_1670129, EPI_ISL_1670130, EPI_ISL_1670131, EPI_ISL_1670132, EPI_ISL_1670133, EPI_ISL_1670134, EPI_ISL_1670135, EPI_ISL_1670136, EPI_ISL_1670137, EPI_ISL_1670138, EPI_ISL_1670139, EPI_ISL_1670140, EPI_ISL_1670141, EPI_ISL_1670142, EPI_ISL_1670143, EPI_ISL_1670149, EPI_ISL_1670150, EPI_ISL_1670151, EPI_ISL_1670152, EPI_ISL_1670153, EPI_ISL_1670154, EPI_ISL_1670155, EPI_ISL_1670156, EPI_ISL_1670157, EPI_ISL_1670158, EPI_ISL_1670159, EPI_ISL_1670160, EPI_ISL_1670161, EPI_ISL_1670162, EPI_ISL_1670163, EPI_ISL_1670164, EPI_ISL_1670165, EPI_ISL_1670166, EPI_ISL_1670167, EPI_ISL_1670168, EPI_ISL_1670169, EPI_ISL_1670170, EPI_ISL_1670171, EPI_ISL_1670172, EPI_ISL_1670173, EPI_ISL_1670174, EPI_ISL_1670175, EPI_ISL_1670176, EPI_ISL_1670177, EPI_ISL_1670178, EPI_ISL_1670196, EPI_ISL_1670197, EPI_ISL_1670198, EPI_ISL_1670199, EPI_ISL_1670200, EPI_ISL_1670201, EPI_ISL_1670202, EPI_ISL_1670203, EPI_ISL_1670204, EPI_ISL_1670205, EPI_ISL_1670206, EPI_ISL_1670207, EPI_ISL_1670208, EPI_ISL_1670209, EPI_ISL_1670210, EPI_ISL_1670211, EPI_ISL_1670212, EPI_ISL_1670213, EPI_ISL_1670214, EPI_ISL_1670215, EPI_ISL_1670216, EPI_ISL_1670217, EPI_ISL_1670218, EPI_ISL_1670219, EPI_ISL_1670220, EPI_ISL_1670221, EPI_ISL_1670222, EPI_ISL_1670223, EPI_ISL_1670224, EPI_ISL_1670225, EPI_ISL_1670226, EPI_ISL_1670227, EPI_ISL_1670228, EPI_ISL_1670229, EPI_ISL_1670230, EPI_ISL_1670231, EPI_ISL_1670232, EPI_ISL_1670233, EPI_ISL_1670234, EPI_ISL_1670235, EPI_ISL_1670236, EPI_ISL_1670237, EPI_ISL_1670238, EPI_ISL_1670239, EPI_ISL_1670240, EPI_ISL_1670241, EPI_ISL_1670242, EPI_ISL_1670243, EPI_ISL_1670244, EPI_ISL_1670245, EPI_ISL_1670246, EPI_ISL_1670247, EPI_ISL_1670248, EPI_ISL_1670249, EPI_ISL_1670250, EPI_ISL_1670251, EPI_ISL_1670252, EPI_ISL_1670253, EPI_ISL_1670254, EPI_ISL_1670255, EPI_ISL_1670256, EPI_ISL_1670257, EPI_ISL_1670258, EPI_ISL_1670259, EPI_ISL_1670260, EPI_ISL_1670261, EPI_ISL_1670262, EPI_ISL_1670263, EPI_ISL_1670264, EPI_ISL_1670265, EPI_ISL_1670266, EPI_ISL_1670267, EPI_ISL_1670268, EPI_ISL_1670269, EPI_ISL_1670270, EPI_ISL_1670427, EPI_ISL_1670428, EPI_ISL_1670524, EPI_ISL_1940857, EPI_ISL_1941171 | | | |
| see above | Landesamt für Verbraucherschutz Sachsen Anhalt, Magdeburg | Institute of Medical Microbiology and Hospital Hygiene | Aljoscha Tersteegen; Prof. Dr. Achim Kaasch |
| EPI_ISL_1441242, EPI_ISL_1441244, EPI_ISL_1441245, EPI_ISL_1441250, EPI_ISL_1441251, EPI_ISL_1441255, EPI_ISL_1441257, EPI_ISL_1441258, EPI_ISL_1441260, EPI_ISL_1441261, EPI_ISL_1441263, EPI_ISL_1441265, EPI_ISL_1441268, EPI_ISL_1441270, EPI_ISL_1441271, EPI_ISL_1441272, EPI_ISL_1441273, EPI_ISL_1441274, EPI_ISL_1441275, EPI_ISL_1441278, EPI_ISL_1441281, EPI_ISL_1441284, EPI_ISL_1441287, EPI_ISL_1441292, EPI_ISL_1441294, EPI_ISL_1441295, EPI_ISL_1441298, EPI_ISL_1441300, EPI_ISL_1441306, EPI_ISL_1441308 | | | |
| see above | Landesuntersuchungsanstalt für das Gesundheits- und Veterinärwesen (LUA) Sachsen | Robert Koch Institute | |
| EPI_ISL_1711173, EPI_ISL_1711189, EPI_ISL_1711190, EPI_ISL_1711191, EPI_ISL_1711192, EPI_ISL_1711204, EPI_ISL_1711220, EPI_ISL_1711222 | | | |
| see above | Landstuhl Regional Medical Center | US Air Force School of Aerospace Medicine | Amanda Javorina; Anthony Fries; Clarise Starr; Cole Anderson; Elizabeth Macias; Fritz Castillo; Jennifer Meyer; Sarah Purves; William Buggele; William Gruner |
| EPI_ISL_1440955, EPI_ISL_1440956, EPI_ISL_1440957, EPI_ISL_1440958, EPI_ISL_1440959, EPI_ISL_1440961, EPI_ISL_1440962, EPI_ISL_1440963, EPI_ISL_1440964, EPI_ISL_1440965, EPI_ISL_1440966, EPI_ISL_1572177, EPI_ISL_1572178, EPI_ISL_1572179, EPI_ISL_1572181, EPI_ISL_1572183 | | | |
| see above | Limbach - Ganzimmun Diagnostics AG Mainz | Robert Koch Institute | |
| EPI_ISL_1438635, EPI_ISL_1438672, EPI_ISL_1563895, EPI_ISL_1563900, EPI_ISL_1563913, EPI_ISL_1565370, EPI_ISL_1565384, EPI_ISL_1565395, EPI_ISL_1567569, EPI_ISL_1567572, EPI_ISL_1567595, EPI_ISL_1567604, EPI_ISL_1567618, EPI_ISL_1567619, EPI_ISL_1567637, EPI_ISL_1567646, EPI_ISL_1567669, EPI_ISL_1567695, EPI_ISL_1572150 | | | |
| see above | Limbach - MVZ Gemeinschaftslabor Suhl | Robert Koch Institute | |
| EPI_ISL_1436396, EPI_ISL_1436397, EPI_ISL_1436398, EPI_ISL_1436400, EPI_ISL_1436402, EPI_ISL_1436404, EPI_ISL_1436406, EPI_ISL_1436407, EPI_ISL_1436411, EPI_ISL_1436418, EPI_ISL_1436419, EPI_ISL_1436421, EPI_ISL_1436422, EPI_ISL_1436425, EPI_ISL_1436426, EPI_ISL_1436429, EPI_ISL_1436430, EPI_ISL_1436432, EPI_ISL_1436438, EPI_ISL_1436439, EPI_ISL_1436441, EPI_ISL_1436442, EPI_ISL_1436445, EPI_ISL_1436446, EPI_ISL_1436449, EPI_ISL_1436450, EPI_ISL_1436452, EPI_ISL_1436453, EPI_ISL_1436454, EPI_ISL_1436458, EPI_ISL_1436459, EPI_ISL_1436460, EPI_ISL_1436462, EPI_ISL_1436466, EPI_ISL_1436467, EPI_ISL_1436469, EPI_ISL_1436470, EPI_ISL_1436474, EPI_ISL_1436477, EPI_ISL_1436479, EPI_ISL_1436486, EPI_ISL_1436491, EPI_ISL_1436497, EPI_ISL_1436498, EPI_ISL_1436500, EPI_ISL_1436505, EPI_ISL_1436506, EPI_ISL_1436509, EPI_ISL_1436511, EPI_ISL_1436513, EPI_ISL_1436518, EPI_ISL_1436519, EPI_ISL_1436523, EPI_ISL_1436525, EPI_ISL_1436528, EPI_ISL_1436531, EPI_ISL_1436533, EPI_ISL_1436535, EPI_ISL_1436537, EPI_ISL_1436540, EPI_ISL_1436541, EPI_ISL_1436542, EPI_ISL_1436546, EPI_ISL_1436548, EPI_ISL_1436549, EPI_ISL_1436550, EPI_ISL_1436553, EPI_ISL_1436557, EPI_ISL_1436561, EPI_ISL_1436564, EPI_ISL_1436565, EPI_ISL_1436567, EPI_ISL_1436568, EPI_ISL_1436570, EPI_ISL_1436571, EPI_ISL_1436572, EPI_ISL_1436573, EPI_ISL_1436574, EPI_ISL_1436575, EPI_ISL_1436576, EPI_ISL_1436577, EPI_ISL_1436578, EPI_ISL_1436579, EPI_ISL_1567118, EPI_ISL_1567120, EPI_ISL_1567124, EPI_ISL_1567125, EPI_ISL_1567126, EPI_ISL_1567127, EPI_ISL_1567129, EPI_ISL_1567130, EPI_ISL_1567131, EPI_ISL_1567134, EPI_ISL_1567137, EPI_ISL_1567141, EPI_ISL_1567142, EPI_ISL_1567146, EPI_ISL_1567149, EPI_ISL_1567152, EPI_ISL_1567158, EPI_ISL_1567161, EPI_ISL_1567162, EPI_ISL_1567163, EPI_ISL_1567166, EPI_ISL_1567167, EPI_ISL_1567169, EPI_ISL_1567172, EPI_ISL_1567173, EPI_ISL_1567175, EPI_ISL_1567176, EPI_ISL_1567179, EPI_ISL_1567181, EPI_ISL_1567186, EPI_ISL_1567188, EPI_ISL_1567192, EPI_ISL_1567193, EPI_ISL_1567194, EPI_ISL_1567195, EPI_ISL_1567197, EPI_ISL_1567199, EPI_ISL_1567204, EPI_ISL_1567208, EPI_ISL_1567212, EPI_ISL_1567216, EPI_ISL_1567218, EPI_ISL_1567220, EPI_ISL_1567221, EPI_ISL_1567226, EPI_ISL_1567228, EPI_ISL_1567229, EPI_ISL_1567230, EPI_ISL_1567232, EPI_ISL_1567233, EPI_ISL_1567234, EPI_ISL_1567235, EPI_ISL_1567237, EPI_ISL_1567238, EPI_ISL_1567240, EPI_ISL_1567241, EPI_ISL_1567242, EPI_ISL_1567244, EPI_ISL_1567245, EPI_ISL_1567247, EPI_ISL_1567248, EPI_ISL_1567252, EPI_ISL_1567253, EPI_ISL_1567256, EPI_ISL_1567258, EPI_ISL_1567259, EPI_ISL_1567260, EPI_ISL_1567261, EPI_ISL_1567262, EPI_ISL_1567265, EPI_ISL_1567267, EPI_ISL_1567268, EPI_ISL_1567269, EPI_ISL_1567270, EPI_ISL_1567271, EPI_ISL_1567273, EPI_ISL_1567274, EPI_ISL_1567275, EPI_ISL_1567277, EPI_ISL_1567279, EPI_ISL_1567280, EPI_ISL_1567281, EPI_ISL_1567282, EPI_ISL_1567283, EPI_ISL_1567284, EPI_ISL_1567285 | | | |
| see above | Limbach - MVZ Humangenetik Ulm | Robert Koch Institute | |
| EPI_ISL_1438624, EPI_ISL_1563858, EPI_ISL_1563861, EPI_ISL_1563862, EPI_ISL_1563864, EPI_ISL_1563865, EPI_ISL_1563873, EPI_ISL_1563874, EPI_ISL_1563875, EPI_ISL_1563884, EPI_ISL_1567553, EPI_ISL_1567556, EPI_ISL_1567558, EPI_ISL_1567560, EPI_ISL_1567564, EPI_ISL_1567587, EPI_ISL_1567607, EPI_ISL_1567614, EPI_ISL_1567616, EPI_ISL_1567627, EPI_ISL_1570708, EPI_ISL_1570761, EPI_ISL_1570778, EPI_ISL_1643397 | | | |
| see above | Limbach - MVZ Labor Eveld & Kollegen Essen | Robert Koch Institute | |
| EPI_ISL_1563851, EPI_ISL_1563863, EPI_ISL_1563866, EPI_ISL_1563868, EPI_ISL_1563887, EPI_ISL_1563891, EPI_ISL_1563911, EPI_ISL_1563915, EPI_ISL_1563919, EPI_ISL_1565372, EPI_ISL_1565374, EPI_ISL_1565375, EPI_ISL_1565378, EPI_ISL_1565380, EPI_ISL_1565381, EPI_ISL_1565383, EPI_ISL_1565386, EPI_ISL_1565391, EPI_ISL_1565392, EPI_ISL_1565394, EPI_ISL_1565396, EPI_ISL_1565398, EPI_ISL_1565402, EPI_ISL_1565403, EPI_ISL_1565405, EPI_ISL_1565407, EPI_ISL_1565409, EPI_ISL_1567551, EPI_ISL_1567552, EPI_ISL_1567554, EPI_ISL_1567557, EPI_ISL_1567559, EPI_ISL_1567561, EPI_ISL_1567563, EPI_ISL_1567568, EPI_ISL_1567577, EPI_ISL_1567578, EPI_ISL_1567591, EPI_ISL_1567592, EPI_ISL_1567597, EPI_ISL_1567599, EPI_ISL_1567600, EPI_ISL_1567608, EPI_ISL_1567611, EPI_ISL_1567626, EPI_ISL_1567630, EPI_ISL_1567634, EPI_ISL_1567636, EPI_ISL_1567638, EPI_ISL_1567641, EPI_ISL_1567642, EPI_ISL_1567643, EPI_ISL_1567644, EPI_ISL_1567647, EPI_ISL_1567649, EPI_ISL_1567651, EPI_ISL_1567652, EPI_ISL_1567654, EPI_ISL_1567657, EPI_ISL_1567662, EPI_ISL_1567677, EPI_ISL_1567678, EPI_ISL_1567682, EPI_ISL_1567683, EPI_ISL_1567689, EPI_ISL_1567691, EPI_ISL_1567693, EPI_ISL_1567694, EPI_ISL_1567696, EPI_ISL_1567697, EPI_ISL_1567698, EPI_ISL_1567699, EPI_ISL_1567700, EPI_ISL_1567702, EPI_ISL_1567703, EPI_ISL_1567705, EPI_ISL_1569996, EPI_ISL_1570704, EPI_ISL_1570706, EPI_ISL_1570819, EPI_ISL_1570825, EPI_ISL_1570829, EPI_ISL_1570840, EPI_ISL_1570845, EPI_ISL_1570848, EPI_ISL_1570887, EPI_ISL_1570890, EPI_ISL_1570932, EPI_ISL_1570944, EPI_ISL_1570947, EPI_ISL_1571121, EPI_ISL_1571126, EPI_ISL_1571128, EPI_ISL_1571135, EPI_ISL_1571137, EPI_ISL_1571139, EPI_ISL_1571143, EPI_ISL_1571148, EPI_ISL_1571149, EPI_ISL_1571154, EPI_ISL_1573240, EPI_ISL_1573241, EPI_ISL_1573246, EPI_ISL_1573248, EPI_ISL_1573249, EPI_ISL_1573250, EPI_ISL_1573251, EPI_ISL_1573252, EPI_ISL_1573256, EPI_ISL_1573263, EPI_ISL_1573265, EPI_ISL_1638280, EPI_ISL_1638289, EPI_ISL_1638296, EPI_ISL_1638335, EPI_ISL_1638358, EPI_ISL_1638397, EPI_ISL_1638406, EPI_ISL_1638409, EPI_ISL_1638411, EPI_ISL_1638415, EPI_ISL_1638434, EPI_ISL_1638440, EPI_ISL_1638441, EPI_ISL_1638466, EPI_ISL_1638468, EPI_ISL_1638473, EPI_ISL_1638478, EPI_ISL_1638494 | | | |
| see above | MDI Limbach Berlin GmbH; MVZ Labor Berlin | Robert Koch Institute | |
| EPI_ISL_1716273, EPI_ISL_1716274, EPI_ISL_1716275, EPI_ISL_1716276 | MEDILYS Laborgesellschaft mbH | Heinrich Pette Institute, Leibniz Institute for Experimental Virology | Adam Grundhoff; Alexis Robitaille; Johannes Knobloch; Martin Aepfelbacher; Nicole Fischer; Thomas Günther |
| EPI_ISL_1439442, EPI_ISL_1439443, EPI_ISL_1439447, EPI_ISL_1439450, EPI_ISL_1439456, EPI_ISL_1439469, EPI_ISL_1439474, EPI_ISL_1439478, EPI_ISL_1439480, EPI_ISL_1439481, EPI_ISL_1439488, EPI_ISL_1439491, EPI_ISL_1439492, EPI_ISL_1439493, EPI_ISL_1439494, EPI_ISL_1439495, EPI_ISL_1439497, EPI_ISL_1439498, EPI_ISL_1439499, EPI_ISL_1439500, EPI_ISL_1439502, EPI_ISL_1439503, EPI_ISL_1439505, EPI_ISL_1439506, EPI_ISL_1439507, EPI_ISL_1439508, EPI_ISL_1439509, EPI_ISL_1439512, EPI_ISL_1439514, EPI_ISL_1567766, EPI_ISL_1567773, EPI_ISL_1567777, EPI_ISL_1567782, EPI_ISL_1567784, EPI_ISL_1567787, EPI_ISL_1567788, EPI_ISL_1567790, EPI_ISL_1567792, EPI_ISL_1567793, EPI_ISL_1567796, EPI_ISL_1567804, EPI_ISL_1567808, EPI_ISL_1567810, EPI_ISL_1567811, EPI_ISL_1567815, EPI_ISL_1567816, EPI_ISL_1567819, EPI_ISL_1567828, EPI_ISL_1572108, EPI_ISL_1638506, EPI_ISL_1638523 | | | |
| see above | MVZ Dr. Eberhard & Partner Dortmund | Robert Koch Institute | |
| EPI_ISL_1565373, EPI_ISL_1567571 | MVZ Labor Dortmund | Robert Koch Institute | |
| EPI_ISL_1442509, EPI_ISL_1727069, EPI_ISL_1727070, EPI_ISL_1727071, EPI_ISL_1727073, EPI_ISL_1727074, EPI_ISL_1727076, EPI_ISL_1727079, EPI_ISL_1727080, EPI_ISL_1727083, EPI_ISL_1727084, EPI_ISL_1727085, EPI_ISL_1727086, EPI_ISL_1727088, EPI_ISL_1727089, EPI_ISL_1727091, EPI_ISL_1727092 | | | |
| see above | MVZ Labor Dr. Fenner und Kollegen (Standort Hamburg) | Robert Koch Institute | |
| EPI_ISL_1563854, EPI_ISL_1563876, EPI_ISL_1563877, EPI_ISL_1563879, EPI_ISL_1567631, EPI_ISL_1569995, EPI_ISL_1570709, EPI_ISL_1570722, EPI_ISL_1570726, EPI_ISL_1570727, EPI_ISL_1570730, EPI_ISL_1570731, EPI_ISL_1570735, EPI_ISL_1570738, EPI_ISL_1570748, EPI_ISL_1570750, EPI_ISL_1570752, EPI_ISL_1570754, EPI_ISL_1570758, EPI_ISL_1570759, EPI_ISL_1570808, EPI_ISL_1570812, EPI_ISL_1570831, EPI_ISL_1570893, EPI_ISL_1570895, EPI_ISL_1570905, EPI_ISL_1570918, EPI_ISL_1570934, EPI_ISL_1570939, EPI_ISL_1570948, EPI_ISL_1570949, EPI_ISL_1570950, EPI_ISL_1570951, EPI_ISL_1570953, EPI_ISL_1570955, EPI_ISL_1570957, EPI_ISL_1570958, EPI_ISL_1638460, EPI_ISL_1643410 | | | |
| see above | MVZ Labor Dr. Limbach & Kollegen GbR | Robert Koch Institute | |
| EPI_ISL_1570491, EPI_ISL_1570527, EPI_ISL_1570536, EPI_ISL_1570599, EPI_ISL_1570602, EPI_ISL_1570625, EPI_ISL_1570640, EPI_ISL_1570678, EPI_ISL_1570692 | | | |
| see above | MVZ Labor Dr. Quade & Kollegen GmbH | Robert Koch Institute | |
| EPI_ISL_1563853, EPI_ISL_1563855, EPI_ISL_1563856, EPI_ISL_1563869, EPI_ISL_1567567, EPI_ISL_1567594, EPI_ISL_1567650, EPI_ISL_1567673, EPI_ISL_1567674, EPI_ISL_1570774, EPI_ISL_1570787, EPI_ISL_1570813 | | | |
| see above | MVZ Labor Dr. Reising-Ackermann und Kollegen | Robert Koch Institute | |
| EPI_ISL_1439626, EPI_ISL_1439627, EPI_ISL_1439628, EPI_ISL_1439630, EPI_ISL_1439632, EPI_ISL_1439634, EPI_ISL_1439636, EPI_ISL_1439638, EPI_ISL_1439639, EPI_ISL_1439640, EPI_ISL_1439643, EPI_ISL_1439644, EPI_ISL_1439645, EPI_ISL_1439648, EPI_ISL_1439649, EPI_ISL_1439650, EPI_ISL_1439651, EPI_ISL_1439652, EPI_ISL_1439653, EPI_ISL_1439654, EPI_ISL_1439655, EPI_ISL_1439656, EPI_ISL_1439657, EPI_ISL_1439658, EPI_ISL_1439659, EPI_ISL_1439660, EPI_ISL_1439661, EPI_ISL_1439662, EPI_ISL_1439663, EPI_ISL_1439664, EPI_ISL_1439665, EPI_ISL_1439666, EPI_ISL_1439667, EPI_ISL_1439668, EPI_ISL_1439669, EPI_ISL_1439670, EPI_ISL_1440880, EPI_ISL_1440904, EPI_ISL_1442896, EPI_ISL_1442897, EPI_ISL_1442898, EPI_ISL_1442899, EPI_ISL_1442900, EPI_ISL_1442901, EPI_ISL_1442902, EPI_ISL_1442903, EPI_ISL_1442904, EPI_ISL_1442905, EPI_ISL_1442906, EPI_ISL_1442907, EPI_ISL_1442908, EPI_ISL_1442909, EPI_ISL_1442910, EPI_ISL_1442911, EPI_ISL_1442912, EPI_ISL_1442914, EPI_ISL_1442915, EPI_ISL_1442916, EPI_ISL_1442917, EPI_ISL_1442918, EPI_ISL_1442919, EPI_ISL_1442920, EPI_ISL_1442921, EPI_ISL_1442922, EPI_ISL_1442923, EPI_ISL_1442924, EPI_ISL_1442925, EPI_ISL_1442927, EPI_ISL_1442928, EPI_ISL_1442929, EPI_ISL_1442930, EPI_ISL_1442931, EPI_ISL_1442932, EPI_ISL_1442933, EPI_ISL_1442934, EPI_ISL_1442935, EPI_ISL_1442936, EPI_ISL_1442937, EPI_ISL_1442938, EPI_ISL_1442939, EPI_ISL_1442940, EPI_ISL_1442941, EPI_ISL_1563978, EPI_ISL_1563980, EPI_ISL_1563981, EPI_ISL_1563983, EPI_ISL_1563995, EPI_ISL_1563996, EPI_ISL_1564009, EPI_ISL_1566007, EPI_ISL_1566010, EPI_ISL_1566012, EPI_ISL_1566013, EPI_ISL_1566014, EPI_ISL_1566015, EPI_ISL_1566016, EPI_ISL_1566019, EPI_ISL_1566020, EPI_ISL_1566021, EPI_ISL_1566023, EPI_ISL_1566024, EPI_ISL_1566026, EPI_ISL_1566030, EPI_ISL_1566031, EPI_ISL_1566034, EPI_ISL_1566035, EPI_ISL_1566036, EPI_ISL_1566037, EPI_ISL_1566038, EPI_ISL_1566041, | | | |

| | | | |
|---|---|---|---|
| EPI_ISL_1566042, EPI_ISL_1566045, EPI_ISL_1566046, EPI_ISL_1566047, EPI_ISL_1566049, EPI_ISL_1566050, EPI_ISL_1566051, EPI_ISL_1566052, EPI_ISL_1566054, EPI_ISL_1566055, EPI_ISL_1566057, EPI_ISL_1566058, EPI_ISL_1566061, EPI_ISL_1566062, EPI_ISL_1566064, EPI_ISL_1566065, EPI_ISL_1566066, EPI_ISL_1566067, EPI_ISL_1566068, EPI_ISL_1566069, EPI_ISL_1566070, EPI_ISL_1566071, EPI_ISL_1566072, EPI_ISL_1566073, EPI_ISL_1566076, EPI_ISL_1566077, EPI_ISL_1566078, EPI_ISL_1566079, EPI_ISL_1566080, EPI_ISL_1566083, EPI_ISL_1566084, EPI_ISL_1566086, EPI_ISL_1566087, EPI_ISL_1566088, EPI_ISL_1566089, EPI_ISL_1566090, EPI_ISL_1566091, EPI_ISL_1566095, EPI_ISL_1566096, EPI_ISL_1566097, EPI_ISL_1566098, EPI_ISL_1566099, EPI_ISL_1566100, EPI_ISL_1569810, EPI_ISL_1569811, EPI_ISL_1569813, EPI_ISL_1569814, EPI_ISL_1569818, EPI_ISL_1569819, EPI_ISL_1569821, EPI_ISL_1569822, EPI_ISL_1569824, EPI_ISL_1569825, EPI_ISL_1569827, EPI_ISL_1569829, EPI_ISL_1569830, EPI_ISL_1569831, EPI_ISL_1569833, EPI_ISL_1569834, EPI_ISL_1569835, EPI_ISL_1569837, EPI_ISL_1569838, EPI_ISL_1569839, EPI_ISL_1569840, EPI_ISL_1569842, EPI_ISL_1569846, EPI_ISL_1569848, EPI_ISL_1569849, EPI_ISL_1569851, EPI_ISL_1569854, EPI_ISL_1569858, EPI_ISL_1569859, EPI_ISL_1569861, EPI_ISL_1569862, EPI_ISL_1569863, EPI_ISL_1569869, EPI_ISL_1569872, EPI_ISL_1569874, EPI_ISL_1569875, EPI_ISL_1569876, EPI_ISL_1569879, EPI_ISL_1569882, EPI_ISL_1640708, EPI_ISL_1640712, EPI_ISL_1640714, EPI_ISL_1640720, EPI_ISL_1640721, EPI_ISL_1640727, EPI_ISL_1640740 | | | |
| see above | MVZ Labor Krone GbR | Robert Koch Institute | |
| EPI_ISL_1720906, EPI_ISL_1722066, EPI_ISL_1722067, EPI_ISL_1846335 | MVZ Labor Passau GbR | Robert Koch Institute | |
| EPI_ISL_1572145, EPI_ISL_1572152, EPI_ISL_1572156, EPI_ISL_1572159, EPI_ISL_1572160, EPI_ISL_1572161, EPI_ISL_1572162, EPI_ISL_1572163, EPI_ISL_1572168, EPI_ISL_1572170, EPI_ISL_1572171, EPI_ISL_1572172, EPI_ISL_1643772 | | | |
| see above | MVZ Medizinisches Labor Hannover GmbH | Robert Koch Institute | |
| EPI_ISL_1567629, EPI_ISL_1567670, EPI_ISL_1567672, EPI_ISL_1567676, EPI_ISL_1570832, EPI_ISL_1570884, EPI_ISL_1572010, EPI_ISL_1638581, EPI_ISL_1638587, EPI_ISL_1638588, EPI_ISL_1638606 | | | |
| see above | MVZ fÃ¼r Laboratoriumsmedizin und Mikrobiologie Koblenz-Mittelrhein (Labor Koblenz) | Robert Koch Institute | |
| EPI_ISL_1439452 | MVZ für Laboratoriumsmedizin und Mikrobiologie Würzburg | Robert Koch Institute | |
| EPI_ISL_1752377, EPI_ISL_1752378, EPI_ISL_1752379, EPI_ISL_1752380, EPI_ISL_1752381, EPI_ISL_1752382, EPI_ISL_1752383, EPI_ISL_1752384, EPI_ISL_1752385, EPI_ISL_1752420, EPI_ISL_1752422, EPI_ISL_1752424, EPI_ISL_1752435, EPI_ISL_1752436, EPI_ISL_1752437, EPI_ISL_1752438, EPI_ISL_1752439, EPI_ISL_1752440, EPI_ISL_1752441, EPI_ISL_1752442, EPI_ISL_1752443, EPI_ISL_1752444, EPI_ISL_1752445, EPI_ISL_1752446, EPI_ISL_1752447, EPI_ISL_1752448, EPI_ISL_1752449, EPI_ISL_1752450, EPI_ISL_1752451, EPI_ISL_1752452, EPI_ISL_1752453, EPI_ISL_1752454, EPI_ISL_1752455, EPI_ISL_1752459, EPI_ISL_2094544, EPI_ISL_2094550, EPI_ISL_2094590, EPI_ISL_2094625, EPI_ISL_2094629, EPI_ISL_2094640, EPI_ISL_2094646, EPI_ISL_2094654, EPI_ISL_2094681, EPI_ISL_2094682, EPI_ISL_2094683, EPI_ISL_2094684, EPI_ISL_2094741, EPI_ISL_2094742, EPI_ISL_2094743, EPI_ISL_2094744, EPI_ISL_2094745, EPI_ISL_2094746, EPI_ISL_2094766, EPI_ISL_2094767, EPI_ISL_2094768, EPI_ISL_2094770, EPI_ISL_2094771, EPI_ISL_2094828, EPI_ISL_2094857, EPI_ISL_2094858, EPI_ISL_2094859, EPI_ISL_2094860, EPI_ISL_2094861, EPI_ISL_2094862, EPI_ISL_2094863, EPI_ISL_2094864, EPI_ISL_2094932, EPI_ISL_2094933, EPI_ISL_2094934, EPI_ISL_2094938, EPI_ISL_2094954, EPI_ISL_2094956, EPI_ISL_2094962, EPI_ISL_2094965, EPI_ISL_2094966, EPI_ISL_2094967, EPI_ISL_2094977, EPI_ISL_2094978, EPI_ISL_2094987, EPI_ISL_2094988, EPI_ISL_2094989, EPI_ISL_2095012, EPI_ISL_2095066, EPI_ISL_2095094, EPI_ISL_2095104, EPI_ISL_2095109, EPI_ISL_2095115, EPI_ISL_2095116, EPI_ISL_2095133, EPI_ISL_2095139, EPI_ISL_2095146, EPI_ISL_2095163, EPI_ISL_2095166, EPI_ISL_2095171, EPI_ISL_2095185, EPI_ISL_2095233, EPI_ISL_2095236, EPI_ISL_2095258, EPI_ISL_2095286, EPI_ISL_2095293, EPI_ISL_2095294, EPI_ISL_2095295 | | | |
| see above | Max von Pettenkofer Institute, Virology, National Reference Center for Retroviruses, LMU Munich | Laboratory for Functional Genome Analysis; Dept. Genomics; Gene Center of the LMU Munich | Alexander Graf; Helmut Blum; Max Muenchhoff; Oliver Keppler; Stefan Krebs |
| EPI_ISL_1439758, EPI_ISL_1439763, EPI_ISL_1439769 | Med. Labor Prof. Schenk Dr. Ansorge & Kollegen | Robert Koch Institute | |
| EPI_ISL_1989501, EPI_ISL_1989502, EPI_ISL_1989503, EPI_ISL_1989548, EPI_ISL_1989550, EPI_ISL_1989552, EPI_ISL_1989553, EPI_ISL_1989555, EPI_ISL_1989557, EPI_ISL_1989558, EPI_ISL_1989560, EPI_ISL_1989562, EPI_ISL_1989564, EPI_ISL_1989565, EPI_ISL_1989566, EPI_ISL_1989567, EPI_ISL_1989568, EPI_ISL_1989569, EPI_ISL_1989570, EPI_ISL_1989571, EPI_ISL_1989572, EPI_ISL_1989573, EPI_ISL_1989574, EPI_ISL_1989575, EPI_ISL_1989576, EPI_ISL_1989577, EPI_ISL_1989578, EPI_ISL_1989579, EPI_ISL_1989580, EPI_ISL_1989581, EPI_ISL_1989582, EPI_ISL_1989583, EPI_ISL_1989584, EPI_ISL_1989585, EPI_ISL_1989586, EPI_ISL_1989587, EPI_ISL_1989588, EPI_ISL_1989589, EPI_ISL_1989590, EPI_ISL_1989591, EPI_ISL_1989593, EPI_ISL_1989594, EPI_ISL_1989596, EPI_ISL_1989598, EPI_ISL_1989599, EPI_ISL_1989600, EPI_ISL_1989602, EPI_ISL_1989603, EPI_ISL_1989604, EPI_ISL_1989605, EPI_ISL_1989606, EPI_ISL_1989607, EPI_ISL_1989608, EPI_ISL_1989609, EPI_ISL_1989610, EPI_ISL_1989611, EPI_ISL_1989612, EPI_ISL_1989613, EPI_ISL_1989614, EPI_ISL_1989615, EPI_ISL_1989616, EPI_ISL_1989619, EPI_ISL_1989620, EPI_ISL_1989621, EPI_ISL_1989622, EPI_ISL_1989623, EPI_ISL_1989624, EPI_ISL_1990039, EPI_ISL_1990040, EPI_ISL_1990044, EPI_ISL_1990045, EPI_ISL_1990046, EPI_ISL_1990053, EPI_ISL_1990054, EPI_ISL_1990055, EPI_ISL_1990056, EPI_ISL_1990057, EPI_ISL_1990059, EPI_ISL_1990060, EPI_ISL_1990061, EPI_ISL_1990062, EPI_ISL_1990063, EPI_ISL_1990064, EPI_ISL_1990065, EPI_ISL_1990066, EPI_ISL_1990067, EPI_ISL_1990108, EPI_ISL_1990112, EPI_ISL_1990114, EPI_ISL_1990115, EPI_ISL_1990117, EPI_ISL_1990118, EPI_ISL_1990119, EPI_ISL_1990120, EPI_ISL_1990122, EPI_ISL_1990123, EPI_ISL_1990125, EPI_ISL_1990128, EPI_ISL_1990129, EPI_ISL_1990130, EPI_ISL_1990131, EPI_ISL_1990132, EPI_ISL_1990133, EPI_ISL_1990134, EPI_ISL_1990138, EPI_ISL_1990651, EPI_ISL_1990653, EPI_ISL_1990655, EPI_ISL_1990658, EPI_ISL_1990659, EPI_ISL_1990662, EPI_ISL_1990663 | | | |
| see above | Medical Laboratories Duesseldorf | Center of Medical Microbiology, Virology, and Hospital Hygiene, University of Duesseldorf | Alexander Dilthey; Andreas Walker; Angelika Helmer; Christian Lange; Daniel Strelow; Jessica Nicolai; Jörg Timm; Klaus Pfeffer; Lisanna Hülse; Malte Kohns Vasconcelos; Maximilian Damagnez; Nadine Lübke; Tobias Wienemann; Torsten Houwaart |
| EPI_ISL_1439797, EPI_ISL_1439799, EPI_ISL_1439800, EPI_ISL_1439803, EPI_ISL_1439809, EPI_ISL_1439810, EPI_ISL_1439819, EPI_ISL_1439822, EPI_ISL_1439825, EPI_ISL_1439827, EPI_ISL_1439828, EPI_ISL_1439830, EPI_ISL_1439834, EPI_ISL_1439835, EPI_ISL_1439836, EPI_ISL_1439839, EPI_ISL_1439858, EPI_ISL_1439861, EPI_ISL_1439869, EPI_ISL_1442543 | | | |
| see above | Medizinisch-Diagnostisches Labor Kempten allgäulab | Robert Koch Institute | |
| EPI_ISL_1566697, EPI_ISL_1566698, EPI_ISL_1566699, EPI_ISL_1566701, EPI_ISL_1566702, EPI_ISL_1566703, EPI_ISL_1566704, EPI_ISL_1566705, EPI_ISL_1566706, EPI_ISL_1566707, EPI_ISL_1566708, EPI_ISL_1566709, EPI_ISL_1566710, EPI_ISL_1566711, EPI_ISL_1566713, EPI_ISL_1566714, EPI_ISL_1566715, EPI_ISL_1566716, EPI_ISL_1566717, EPI_ISL_1566718, EPI_ISL_1566719, EPI_ISL_1566720, EPI_ISL_1566721, EPI_ISL_1566722, EPI_ISL_1566723, EPI_ISL_1566724, EPI_ISL_1566725, EPI_ISL_1566726, EPI_ISL_1566727 | | | |
| see above | Medizinische Laboratorien DÃ¼sseldorf | Robert Koch Institute | |
| EPI_ISL_1563902, EPI_ISL_1563910, EPI_ISL_1565371, EPI_ISL_1565376, EPI_ISL_1565377, EPI_ISL_1565379, EPI_ISL_1565382, EPI_ISL_1565385, EPI_ISL_1565388, EPI_ISL_1565389, EPI_ISL_1565393, EPI_ISL_1565397, EPI_ISL_1565399, EPI_ISL_1565400, EPI_ISL_1565401, EPI_ISL_1565404, EPI_ISL_1565406, EPI_ISL_1565408, EPI_ISL_1565411, EPI_ISL_1567580, EPI_ISL_1567581, EPI_ISL_1567583, EPI_ISL_1567590, EPI_ISL_1567621, EPI_ISL_1567633, EPI_ISL_1567635, EPI_ISL_1567665, EPI_ISL_1567667, EPI_ISL_1567668, EPI_ISL_1567679, EPI_ISL_1567701, EPI_ISL_1569923, EPI_ISL_1569972, EPI_ISL_1569988 | | | |
| see above | Medizinisches Labor Ostsachsen MVZ GbR NebenbetriebsstÃ¤tte Dresden | Robert Koch Institute | |
| EPI_ISL_1441234, EPI_ISL_1441236, EPI_ISL_1441238, EPI_ISL_1441246, EPI_ISL_1441249, EPI_ISL_1441252, EPI_ISL_1441253, EPI_ISL_1441256, EPI_ISL_1441259, EPI_ISL_1441279, EPI_ISL_1441282, EPI_ISL_1441285, EPI_ISL_1441286, EPI_ISL_1441288, EPI_ISL_1441289, EPI_ISL_1441296, EPI_ISL_1441301, EPI_ISL_1441303, EPI_ISL_1441309 | | | |
| see above | Medizinisches Labor Ostsachsen MVZ GbRNebenbetriebsstätte Dresden | Robert Koch Institute | |
| EPI_ISL_1439445, EPI_ISL_1439464, EPI_ISL_1439468, EPI_ISL_1439471, EPI_ISL_1439475, EPI_ISL_1439483, EPI_ISL_1439490, EPI_ISL_1439496, EPI_ISL_1439501, EPI_ISL_1439504, EPI_ISL_1439511, EPI_ISL_1439515, EPI_ISL_1439516, EPI_ISL_1439517, EPI_ISL_1439518, EPI_ISL_1439519 | | | |
| see above | Medizinisches Labor Wahl Lüdenscheid | Robert Koch Institute | |
| EPI_ISL_1439440 | Medizinisches Versorgungszentrum für Labormedizin und Mikrobiologie Ruhr GmbH - mvzlm RUHR GmbH | Robert Koch Institute | |
| EPI_ISL_1438298, EPI_ISL_1438299, EPI_ISL_1438316, EPI_ISL_1438344, EPI_ISL_1438348, EPI_ISL_1438352, EPI_ISL_1438371, EPI_ISL_1438401, EPI_ISL_1438502, EPI_ISL_1438537, EPI_ISL_1441316, EPI_ISL_1441332, EPI_ISL_1441349, EPI_ISL_1441357, EPI_ISL_1441364, EPI_ISL_1441365, EPI_ISL_1441380, EPI_ISL_1441384, EPI_ISL_1441387, EPI_ISL_1441389, EPI_ISL_1441397, EPI_ISL_1441399, EPI_ISL_1441401, EPI_ISL_1441414, EPI_ISL_1441417, EPI_ISL_1441422, EPI_ISL_1441432, EPI_ISL_1441488, EPI_ISL_1441513, EPI_ISL_1441517, EPI_ISL_1441519, EPI_ISL_1441525, EPI_ISL_1441530, EPI_ISL_1441544, EPI_ISL_1441555, EPI_ISL_1441570, EPI_ISL_1441581, EPI_ISL_1441585, EPI_ISL_1441608, EPI_ISL_1441645, EPI_ISL_1441646, EPI_ISL_1441647, EPI_ISL_1565515, EPI_ISL_1565525, EPI_ISL_1565527, EPI_ISL_1565530, EPI_ISL_1565534, EPI_ISL_1565562, EPI_ISL_1565578, EPI_ISL_1565582, EPI_ISL_1565589, EPI_ISL_1565596, EPI_ISL_1565599, EPI_ISL_1565614, EPI_ISL_1565618, EPI_ISL_1565634, EPI_ISL_1565640, EPI_ISL_1565667, EPI_ISL_1565681, EPI_ISL_1565685, EPI_ISL_1565696, EPI_ISL_1565728, EPI_ISL_1565737, EPI_ISL_1565742, EPI_ISL_1565748, EPI_ISL_1565751, EPI_ISL_1565757, EPI_ISL_1565772, EPI_ISL_1565774, EPI_ISL_1565776, EPI_ISL_1570963, EPI_ISL_1570969, EPI_ISL_1570973, EPI_ISL_1570974, EPI_ISL_1570977, EPI_ISL_1570985, EPI_ISL_1570996, EPI_ISL_1571005, EPI_ISL_1571013, EPI_ISL_1571032, EPI_ISL_1571034, EPI_ISL_1571037, EPI_ISL_1571038, EPI_ISL_1571046, EPI_ISL_1571074, EPI_ISL_1571075, EPI_ISL_1571078, EPI_ISL_1571080, EPI_ISL_1571089, EPI_ISL_1571105, EPI_ISL_1571108, EPI_ISL_1571109, EPI_ISL_1571115, EPI_ISL_1571638, EPI_ISL_1571671, EPI_ISL_1571680, EPI_ISL_1571738, EPI_ISL_1571743, EPI_ISL_1571770, EPI_ISL_1571837 | | | |
| see above | Medizinisches Zentrallabor Altenburg GmbH & Co.KG | Robert Koch Institute | |
| EPI_ISL_1441264, EPI_ISL_1441266, EPI_ISL_1441277 | Oberlausitz Kliniken gGmBH | Robert Koch Institute | |
| EPI_ISL_1563932, EPI_ISL_1563933 | Philipps-UniversitÃ¤t Marburg Fachbereich Medizin Institut fÃ¼r Virologie | Robert Koch Institute | |
| EPI_ISL_1566987, EPI_ISL_1567004, EPI_ISL_1567013, EPI_ISL_1567018 | Praxisgemeinschaft fÃ¼r Laboratoriumsmedizin Labor BlumenstraÃŸe; Praxis Dr. Sel und Dr. SpÃ¤te | Robert Koch Institute | |
| EPI_ISL_1436993, EPI_ISL_1437089, EPI_ISL_1438311, EPI_ISL_1441366, EPI_ISL_1442846 | Praxisgemeinschaft für Laboratoriumsmedizin Labor Blumenstraße; Praxis Dr. E. Wietschel & Dr. F. Wietschel | Robert Koch Institute | |
| EPI_ISL_1436989, EPI_ISL_1438479, EPI_ISL_1438546, EPI_ISL_1441486, EPI_ISL_1441631, EPI_ISL_1441636, EPI_ISL_1442831, EPI_ISL_1442833 | | | |
| see above | Praxisgemeinschaft für Laboratoriumsmedizin Labor Blumenstraße; Praxis Dr. Sel und Dr. Späte | Robert Koch Institute | |
| EPI_ISL_1642429, EPI_ISL_1642537, EPI_ISL_1642561, EPI_ISL_1642641, EPI_ISL_1642674, EPI_ISL_1642680 | Robert-Bosch-Krankenhaus | Robert Koch Institute | |
| EPI_ISL_1438583, EPI_ISL_1438642, EPI_ISL_1438655 | SLK-Kliniken Heilbronn GmbH - Institut für Laboratoriumsmedizin | Robert Koch Institute | |
| EPI_ISL_1437004, EPI_ISL_1437140, EPI_ISL_1437160, EPI_ISL_1437215, EPI_ISL_1438315, EPI_ISL_1438320, EPI_ISL_1438322, EPI_ISL_1438333, EPI_ISL_1438334, EPI_ISL_1438337, EPI_ISL_1438339, EPI_ISL_1438340, EPI_ISL_1438346, EPI_ISL_1438358, EPI_ISL_1438362, EPI_ISL_1438375, EPI_ISL_1438379, EPI_ISL_1438380, EPI_ISL_1438391, EPI_ISL_1438399, EPI_ISL_1438407, EPI_ISL_1438438, EPI_ISL_1438446, EPI_ISL_1438475, EPI_ISL_1438477, EPI_ISL_1438500, EPI_ISL_1438519, EPI_ISL_1438557, EPI_ISL_1438561, EPI_ISL_1438566, EPI_ISL_1441310, EPI_ISL_1441313, EPI_ISL_1441318, EPI_ISL_1441321, EPI_ISL_1441323, EPI_ISL_1441325, EPI_ISL_1441326, EPI_ISL_1441328, EPI_ISL_1441330, EPI_ISL_1441334, EPI_ISL_1441336, EPI_ISL_1441337, EPI_ISL_1441338, EPI_ISL_1441339, EPI_ISL_1441343, EPI_ISL_1441348, EPI_ISL_1441353, EPI_ISL_1441356, EPI_ISL_1441359, EPI_ISL_1441362, EPI_ISL_1441374, EPI_ISL_1441376, EPI_ISL_1441379, EPI_ISL_1441383, EPI_ISL_1441391, EPI_ISL_1441409, EPI_ISL_1441411, EPI_ISL_1441416, EPI_ISL_1441418, EPI_ISL_1441427, EPI_ISL_1441428, EPI_ISL_1441430, EPI_ISL_1441434, EPI_ISL_1441437, EPI_ISL_1441440, EPI_ISL_1441455, EPI_ISL_1441459, EPI_ISL_1441472, EPI_ISL_1441473, EPI_ISL_1441496, EPI_ISL_1441500, EPI_ISL_1441502, EPI_ISL_1441510, EPI_ISL_1441521, EPI_ISL_1441524, EPI_ISL_1441526, EPI_ISL_1441528, EPI_ISL_1441531, EPI_ISL_1441535, EPI_ISL_1441537, EPI_ISL_1441542, EPI_ISL_1441551, EPI_ISL_1441553, EPI_ISL_1441559, EPI_ISL_1441561, EPI_ISL_1441565, EPI_ISL_1441567, EPI_ISL_1441568, EPI_ISL_1441577, EPI_ISL_1441582, EPI_ISL_1441586, EPI_ISL_1441589, EPI_ISL_1441591, EPI_ISL_1441592, EPI_ISL_1441604, EPI_ISL_1441610, EPI_ISL_1441615, EPI_ISL_1441622, EPI_ISL_1441624, EPI_ISL_1441626, EPI_ISL_1441635, EPI_ISL_1441638, EPI_ISL_1441641, EPI_ISL_1441642, EPI_ISL_1441643, EPI_ISL_1441644, EPI_ISL_1442766, EPI_ISL_1442768, EPI_ISL_1442769, EPI_ISL_1442771, EPI_ISL_1442773, EPI_ISL_1442776, EPI_ISL_1442785, EPI_ISL_1442786, EPI_ISL_1442791, EPI_ISL_1442797, EPI_ISL_1442798, EPI_ISL_1442801, EPI_ISL_1442805, EPI_ISL_1442812, EPI_ISL_1442815, EPI_ISL_1442818, EPI_ISL_1442819, EPI_ISL_1442826, EPI_ISL_1442827, EPI_ISL_1442828, EPI_ISL_1442834, EPI_ISL_1442836, EPI_ISL_1442839, EPI_ISL_1442840, EPI_ISL_1442842, EPI_ISL_1442845, EPI_ISL_1442849, EPI_ISL_1442854, EPI_ISL_1442855, EPI_ISL_1442864, EPI_ISL_1442865, EPI_ISL_1442866, EPI_ISL_1442871, EPI_ISL_1442873, EPI_ISL_1442874, EPI_ISL_1442876, EPI_ISL_1442877, EPI_ISL_1442879, EPI_ISL_1442884, EPI_ISL_1442885, EPI_ISL_1442887, EPI_ISL_1565502, EPI_ISL_1565504, EPI_ISL_1565505, EPI_ISL_1565506, EPI_ISL_1565507, EPI_ISL_1565512, EPI_ISL_1565513, EPI_ISL_1565514, EPI_ISL_1565517, EPI_ISL_1565518, EPI_ISL_1565523, EPI_ISL_1565529, EPI_ISL_1565533, EPI_ISL_1565536, EPI_ISL_1565538, EPI_ISL_1565541, EPI_ISL_1565543, EPI_ISL_1565547, EPI_ISL_1565548, EPI_ISL_1565550, EPI_ISL_1565551, EPI_ISL_1565554, EPI_ISL_1565558, EPI_ISL_1565563, EPI_ISL_1565565, EPI_ISL_1565566, EPI_ISL_1565569, EPI_ISL_1565571, EPI_ISL_1565574, EPI_ISL_1565575, EPI_ISL_1565580, EPI_ISL_1565585, EPI_ISL_1565587, EPI_ISL_1565588, EPI_ISL_1565591, EPI_ISL_1565592, EPI_ISL_1565600, EPI_ISL_1565602, EPI_ISL_1565603, EPI_ISL_1565606, EPI_ISL_1565608, EPI_ISL_1565609, EPI_ISL_1565610, EPI_ISL_1565612, EPI_ISL_1565613, EPI_ISL_1565616, EPI_ISL_1565617, EPI_ISL_1565619, EPI_ISL_1565620, EPI_ISL_1565622, EPI_ISL_1565623, EPI_ISL_1565624, EPI_ISL_1565625, EPI_ISL_1565627, EPI_ISL_1565629, EPI_ISL_1565631, EPI_ISL_1565632, EPI_ISL_1565635, EPI_ISL_1565636, EPI_ISL_1565637, EPI_ISL_1565638, EPI_ISL_1565641, EPI_ISL_1565642, EPI_ISL_1565645, EPI_ISL_1565646, EPI_ISL_1565648, EPI_ISL_1565649, EPI_ISL_1565650, EPI_ISL_1565652, EPI_ISL_1565653, EPI_ISL_1565654, EPI_ISL_1565656, EPI_ISL_1565657, EPI_ISL_1565658, EPI_ISL_1565661, EPI_ISL_1565664, EPI_ISL_1565666, EPI_ISL_1565668, EPI_ISL_1565671, EPI_ISL_1565675, EPI_ISL_1565677, EPI_ISL_1565678, EPI_ISL_1565680, EPI_ISL_1565688, EPI_ISL_1565690, EPI_ISL_1565692, EPI_ISL_1565693, EPI_ISL_1565694, EPI_ISL_1565695, EPI_ISL_1565699, EPI_ISL_1565702, EPI_ISL_1565709, EPI_ISL_1565714, EPI_ISL_1565715, EPI_ISL_1565717, EPI_ISL_1565719, EPI_ISL_1565724, EPI_ISL_1565726, EPI_ISL_1565727, EPI_ISL_1565729, EPI_ISL_1565731, EPI_ISL_1565732, EPI_ISL_1565734, EPI_ISL_1565735, EPI_ISL_1565736, EPI_ISL_1565739, EPI_ISL_1565741, EPI_ISL_1565744, EPI_ISL_1565745, EPI_ISL_1565746, EPI_ISL_1565747, EPI_ISL_1565750, EPI_ISL_1565752, EPI_ISL_1565756, EPI_ISL_1565758, EPI_ISL_1565759, EPI_ISL_1565760, EPI_ISL_1565763, EPI_ISL_1565766, EPI_ISL_1565767, EPI_ISL_1565768, EPI_ISL_1565771, EPI_ISL_1565777, EPI_ISL_1565778, EPI_ISL_1565779, EPI_ISL_1566785, EPI_ISL_1566789, EPI_ISL_1566792, EPI_ISL_1566793, EPI_ISL_1566798, EPI_ISL_1566799, EPI_ISL_1566801, EPI_ISL_1566803, EPI_ISL_1566808, EPI_ISL_1566811, EPI_ISL_1566814, EPI_ISL_1566815, EPI_ISL_1566823, EPI_ISL_1566825, EPI_ISL_1566829, EPI_ISL_1566831, EPI_ISL_1566835, EPI_ISL_1566836, EPI_ISL_1566837, EPI_ISL_1566841, EPI_ISL_1566843, EPI_ISL_1566845, EPI_ISL_1566854, EPI_ISL_1566856, EPI_ISL_1566859, EPI_ISL_1566860, EPI_ISL_1566861, EPI_ISL_1566867, EPI_ISL_1566870, EPI_ISL_1566873, EPI_ISL_1566877, EPI_ISL_1566878, EPI_ISL_1566881, EPI_ISL_1566885, EPI_ISL_1566889, EPI_ISL_1566891, EPI_ISL_1566892, EPI_ISL_1566893, EPI_ISL_1566894, EPI_ISL_1566898, EPI_ISL_1566903, EPI_ISL_1566904, EPI_ISL_1566906, EPI_ISL_1566907, EPI_ISL_1566908, EPI_ISL_1566909, EPI_ISL_1566913, EPI_ISL_1566914, EPI_ISL_1566916, EPI_ISL_1566917, EPI_ISL_1566918, EPI_ISL_1566919, EPI_ISL_1566920, EPI_ISL_1566921, EPI_ISL_1566922, EPI_ISL_1566923, EPI_ISL_1566924, EPI_ISL_1566925, EPI_ISL_1566926, EPI_ISL_1566928, EPI_ISL_1566930, EPI_ISL_1566932, EPI_ISL_1566934, EPI_ISL_1566936, EPI_ISL_1566938, EPI_ISL_1566940, EPI_ISL_1566941, EPI_ISL_1566944, EPI_ISL_1566948, EPI_ISL_1566949, EPI_ISL_1566950, EPI_ISL_1566953, EPI_ISL_1566955, EPI_ISL_1566956, EPI_ISL_1566959, EPI_ISL_1566960, EPI_ISL_1566963, EPI_ISL_1566965, EPI_ISL_1566967, EPI_ISL_1566968, EPI_ISL_1566971, EPI_ISL_1566972, EPI_ISL_1566973, EPI_ISL_1566976, EPI_ISL_1566978, EPI_ISL_1566980, EPI_ISL_1566982, EPI_ISL_1566984, EPI_ISL_1566986, EPI_ISL_1566988, EPI_ISL_1566989, EPI_ISL_1566991, EPI_ISL_1566992, EPI_ISL_1566993, EPI_ISL_1566995, EPI_ISL_1566998, EPI_ISL_1567000, EPI_ISL_1567001, EPI_ISL_1567002, EPI_ISL_1567003, EPI_ISL_1567006, EPI_ISL_1567007, EPI_ISL_1567009, EPI_ISL_1567012, EPI_ISL_1567017, EPI_ISL_1567021, EPI_ISL_1567022, EPI_ISL_1567023, EPI_ISL_1567025, EPI_ISL_1567031, EPI_ISL_1567033, EPI_ISL_1567034, EPI_ISL_1567038, EPI_ISL_1567040, EPI_ISL_1567042, EPI_ISL_1567043, EPI_ISL_1567044, EPI_ISL_1567045, EPI_ISL_1567048, EPI_ISL_1567051, EPI_ISL_1567053, EPI_ISL_1567056, EPI_ISL_1567057, EPI_ISL_1567059, EPI_ISL_1567060, EPI_ISL_1567061, EPI_ISL_1567064, EPI_ISL_1567065, EPI_ISL_1567068, EPI_ISL_1567072, EPI_ISL_1567074, EPI_ISL_1567080, EPI_ISL_1567082, EPI_ISL_1567083, EPI_ISL_1567287, EPI_ISL_1567289, EPI_ISL_1567292, EPI_ISL_1567295, EPI_ISL_1567300, EPI_ISL_1567302, EPI_ISL_1567304, EPI_ISL_1567305, EPI_ISL_1567306, EPI_ISL_1567308, EPI_ISL_1567310, EPI_ISL_1567311, EPI_ISL_1567312, EPI_ISL_1567314, EPI_ISL_1567316, EPI_ISL_1567318, EPI_ISL_1567321, EPI_ISL_1567332, EPI_ISL_1567345, EPI_ISL_1567353, EPI_ISL_1567357, EPI_ISL_1567361, EPI_ISL_1567364, EPI_ISL_1567365, EPI_ISL_1567371, EPI_ISL_1567374, EPI_ISL_1567375, EPI_ISL_1567385, EPI_ISL_1567386, EPI_ISL_1567392, EPI_ISL_1567396, EPI_ISL_1567398, EPI_ISL_1567399, EPI_ISL_1567400, EPI_ISL_1567401, EPI_ISL_1567402, EPI_ISL_1567405, EPI_ISL_1567407, EPI_ISL_1567408, EPI_ISL_1567410, EPI_ISL_1567411, EPI_ISL_1567412, EPI_ISL_1567414, EPI_ISL_1567418, EPI_ISL_1567419, EPI_ISL_1567422, EPI_ISL_1567424, EPI_ISL_1567425, EPI_ISL_1567426, EPI_ISL_1567429, EPI_ISL_1567431, EPI_ISL_1567433, EPI_ISL_1567434, EPI_ISL_1567436, EPI_ISL_1567437, EPI_ISL_1567438, EPI_ISL_1567441, EPI_ISL_1567443, EPI_ISL_1567444, EPI_ISL_1567448, EPI_ISL_1567449, EPI_ISL_1567451, EPI_ISL_1567456, EPI_ISL_1567458, EPI_ISL_1567459, EPI_ISL_1567462, EPI_ISL_1567463, EPI_ISL_1567466, EPI_ISL_1567467, EPI_ISL_1567468, EPI_ISL_1567470, EPI_ISL_1567471, EPI_ISL_1567472, EPI_ISL_1567475, EPI_ISL_1567476, EPI_ISL_1567477, EPI_ISL_1567480, EPI_ISL_1567486, EPI_ISL_1567487, EPI_ISL_1567489, EPI_ISL_1567494, EPI_ISL_1567500, EPI_ISL_1567502, EPI_ISL_1567504, EPI_ISL_1567505, EPI_ISL_1567506, EPI_ISL_1567507, EPI_ISL_1567511, EPI_ISL_1567516, EPI_ISL_1567519, EPI_ISL_1567520, EPI_ISL_1567523, EPI_ISL_1567535, EPI_ISL_1567537, EPI_ISL_1567542, EPI_ISL_1567543, EPI_ISL_1567545, EPI_ISL_1567838, EPI_ISL_1567844, EPI_ISL_1567845, EPI_ISL_1567846, EPI_ISL_1567849, EPI_ISL_1567852, EPI_ISL_1567854, EPI_ISL_1567858, EPI_ISL_1567861, EPI_ISL_1567863, EPI_ISL_1567866, EPI_ISL_1567870, EPI_ISL_1567874, EPI_ISL_1567875, EPI_ISL_1567879, EPI_ISL_1567880, EPI_ISL_1567882, EPI_ISL_1567886, EPI_ISL_1567889, EPI_ISL_1567892, EPI_ISL_1567893, EPI_ISL_1567894, EPI_ISL_1567895, EPI_ISL_1567896, EPI_ISL_1567900, EPI_ISL_1567901, EPI_ISL_1567902, EPI_ISL_1567904, EPI_ISL_1567905, EPI_ISL_1567908, EPI_ISL_1567909, EPI_ISL_1567912, EPI_ISL_1567913, EPI_ISL_1567916, EPI_ISL_1567917, EPI_ISL_1567922, EPI_ISL_1567923, EPI_ISL_1567924, EPI_ISL_1567925, EPI_ISL_1567926, EPI_ISL_1567930, EPI_ISL_1567934, EPI_ISL_1567936, EPI_ISL_1567937, EPI_ISL_1567938, EPI_ISL_1567940, EPI_ISL_1567944, EPI_ISL_1567946, EPI_ISL_1567956, EPI_ISL_1567957, EPI_ISL_1567959, EPI_ISL_1567961, EPI_ISL_1567967, EPI_ISL_1567971, EPI_ISL_1567973, EPI_ISL_1567977, EPI_ISL_1567981, EPI_ISL_1567986, EPI_ISL_1567989, EPI_ISL_1567990, EPI_ISL_1567993, EPI_ISL_1567994, EPI_ISL_1567995, EPI_ISL_1567997, EPI_ISL_1567999, EPI_ISL_1568001, EPI_ISL_1568002, EPI_ISL_1568008, EPI_ISL_1568014, EPI_ISL_1568017, EPI_ISL_1568018, EPI_ISL_1568019, EPI_ISL_1568020, EPI_ISL_1568022, EPI_ISL_1568030, EPI_ISL_1568035, EPI_ISL_1568037, EPI_ISL_1568038, EPI_ISL_1568039, EPI_ISL_1568040, EPI_ISL_1568042, EPI_ISL_1568043, EPI_ISL_1568045, EPI_ISL_1568048, EPI_ISL_1568049, EPI_ISL_1568052, EPI_ISL_1568055, EPI_ISL_1568057, EPI_ISL_1568062, EPI_ISL_1568063, EPI_ISL_1568065, EPI_ISL_1568066, EPI_ISL_1568071, EPI_ISL_1568072, EPI_ISL_1568076, EPI_ISL_1568077, EPI_ISL_1568078, EPI_ISL_1568080, EPI_ISL_1568081, EPI_ISL_1568082, EPI_ISL_1568084, EPI_ISL_1568085, EPI_ISL_1568087, EPI_ISL_1568088, EPI_ISL_1568089, EPI_ISL_1568090, EPI_ISL_1568092, EPI_ISL_1568093, EPI_ISL_1568095, EPI_ISL_1568096, EPI_ISL_1568098, EPI_ISL_1568099, EPI_ISL_1568101, EPI_ISL_1570968, EPI_ISL_1570970, EPI_ISL_1570972, EPI_ISL_1570975, EPI_ISL_1570984, EPI_ISL_1570990, EPI_ISL_1570991, EPI_ISL_1570993, EPI_ISL_1570994, EPI_ISL_1570997, EPI_ISL_1570998, EPI_ISL_1571001, EPI_ISL_1571012, EPI_ISL_1571018, EPI_ISL_1571020, EPI_ISL_1571021, EPI_ISL_1571023, EPI_ISL_1571027, EPI_ISL_1571039, EPI_ISL_1571042, EPI_ISL_1571048, EPI_ISL_1571054, EPI_ISL_1571058, EPI_ISL_1571065, EPI_ISL_1571070, EPI_ISL_1571077, EPI_ISL_1571079, EPI_ISL_1571082, EPI_ISL_1571088, EPI_ISL_1571093, EPI_ISL_1571110, EPI_ISL_1571610, EPI_ISL_1571643, EPI_ISL_1571648, EPI_ISL_1571654, EPI_ISL_1571664, EPI_ISL_1571665, EPI_ISL_1571724, EPI_ISL_1571725, EPI_ISL_1571726, EPI_ISL_1571733, EPI_ISL_1571737, EPI_ISL_1571741, EPI_ISL_1571750, EPI_ISL_1571763, EPI_ISL_1571772, EPI_ISL_1571802, EPI_ISL_1571840, EPI_ISL_1571841, EPI_ISL_1571850, EPI_ISL_1571853, EPI_ISL_1571863, EPI_ISL_1571876, EPI_ISL_1571878, EPI_ISL_1571889, EPI_ISL_1571905, EPI_ISL_1571924 | | | |
| see above | SYNLAB Jena Oncoscreen | Robert Koch Institute | |

EPI_ISL_1357395, EPI_ISL_1357435, EPI_ISL_1357511, EPI_ISL_1357516, EPI_ISL_1357527, EPI_ISL_1357536, EPI_ISL_1357562, EPI_ISL_1438381, EPI_ISL_1438499, EPI_ISL_1438574

see above SYNLAB Labor München Zentrum LMZ Robert Koch Institute

EPI_ISL_1437031, EPI_ISL_1437145, EPI_ISL_1437176, EPI_ISL_1437219, EPI_ISL_1441312, EPI_ISL_1441320, EPI_ISL_1441324, EPI_ISL_1441331, EPI_ISL_1441350, EPI_ISL_1441360, EPI_ISL_1441370, EPI_ISL_1441375, EPI_ISL_1441382, EPI_ISL_1441386, EPI_ISL_1441388, EPI_ISL_1441394, EPI_ISL_1441396, EPI_ISL_1441400, EPI_ISL_1441405, EPI_ISL_1441410, EPI_ISL_1441413, EPI_ISL_1441415, EPI_ISL_1441420, EPI_ISL_1441425, EPI_ISL_1441439, EPI_ISL_1441465, EPI_ISL_1441467, EPI_ISL_1441476, EPI_ISL_1441506, EPI_ISL_1441508, EPI_ISL_1441515, EPI_ISL_1441529, EPI_ISL_1441534, EPI_ISL_1441540, EPI_ISL_1441546, EPI_ISL_1441557, EPI_ISL_1441563, EPI_ISL_1441571, EPI_ISL_1441572, EPI_ISL_1441574, EPI_ISL_1441576, EPI_ISL_1441579, EPI_ISL_1441587, EPI_ISL_1441599, EPI_ISL_1441606, EPI_ISL_1441612, EPI_ISL_1441623, EPI_ISL_1441630, EPI_ISL_1441632, EPI_ISL_1441634, EPI_ISL_1565511, EPI_ISL_1565516, EPI_ISL_1565520, EPI_ISL_1565537, EPI_ISL_1565597, EPI_ISL_1565626, EPI_ISL_1565639, EPI_ISL_1565643, EPI_ISL_1565644, EPI_ISL_1565647, EPI_ISL_1565655, EPI_ISL_1565672, EPI_ISL_1565683, EPI_ISL_1565684, EPI_ISL_1565689, EPI_ISL_1565697, EPI_ISL_1565701, EPI_ISL_1565704, EPI_ISL_1565706, EPI_ISL_1565707, EPI_ISL_1565710, EPI_ISL_1565720, EPI_ISL_1565723, EPI_ISL_1565730, EPI_ISL_1565761, EPI_ISL_1565764, EPI_ISL_1565769, EPI_ISL_1566818, EPI_ISL_1566871, EPI_ISL_1566985, EPI_ISL_1567019, EPI_ISL_1567077, EPI_ISL_1567317, EPI_ISL_1567320, EPI_ISL_1567323, EPI_ISL_1567324, EPI_ISL_1567325, EPI_ISL_1567329, EPI_ISL_1567336, EPI_ISL_1567338, EPI_ISL_1567341, EPI_ISL_1567346, EPI_ISL_1567370, EPI_ISL_1567378, EPI_ISL_1567382, EPI_ISL_1567384, EPI_ISL_1567387, EPI_ISL_1567389, EPI_ISL_1567394, EPI_ISL_1567415, EPI_ISL_1567416, EPI_ISL_1567421, EPI_ISL_1567428, EPI_ISL_1567432, EPI_ISL_1567439, EPI_ISL_1567445, EPI_ISL_1567450, EPI_ISL_1567452, EPI_ISL_1567454, EPI_ISL_1567457, EPI_ISL_1567478, EPI_ISL_1567484, EPI_ISL_1567488, EPI_ISL_1567497, EPI_ISL_1567503, EPI_ISL_1567508, EPI_ISL_1567509, EPI_ISL_1567515, EPI_ISL_1567526, EPI_ISL_1567531, EPI_ISL_1567546, EPI_ISL_1567548, EPI_ISL_1567847, EPI_ISL_1567857, EPI_ISL_1567906, EPI_ISL_1567921, EPI_ISL_1567929, EPI_ISL_1567939, EPI_ISL_1567949, EPI_ISL_1567992, EPI_ISL_1568007, EPI_ISL_1568034, EPI_ISL_1568046, EPI_ISL_1568061, EPI_ISL_1568075, EPI_ISL_1568079, EPI_ISL_1568083, EPI_ISL_1568091, EPI_ISL_1570965, EPI_ISL_1570980, EPI_ISL_1570982, EPI_ISL_1570987, EPI_ISL_1571002, EPI_ISL_1571007, EPI_ISL_1571011, EPI_ISL_1571019, EPI_ISL_1571024, EPI_ISL_1571053, EPI_ISL_1571055, EPI_ISL_1571057, EPI_ISL_1571073, EPI_ISL_1571085, EPI_ISL_1571095, EPI_ISL_1571104, EPI_ISL_1571111, EPI_ISL_1571112, EPI_ISL_1571114, EPI_ISL_1571117, EPI_ISL_1571614, EPI_ISL_1571620, EPI_ISL_1571625, EPI_ISL_1571645, EPI_ISL_1571656, EPI_ISL_1571662, EPI_ISL_1571669, EPI_ISL_1571674, EPI_ISL_1571692, EPI_ISL_1571697, EPI_ISL_1571701, EPI_ISL_1571746, EPI_ISL_1571755, EPI_ISL_1571854, EPI_ISL_1571875, EPI_ISL_1571904, EPI_ISL_1571921, EPI_ISL_1573537, EPI_ISL_1573546, EPI_ISL_1573632, EPI_ISL_1573653, EPI_ISL_1573656, EPI_ISL_1573662, EPI_ISL_1573665, EPI_ISL_1573836

see above SYNLAB MVZ Berlin Robert Koch Institute

EPI_ISL_1567712, EPI_ISL_1567719, EPI_ISL_1567732, EPI_ISL_1567740, EPI_ISL_1567755, EPI_ISL_1567759, EPI_ISL_1568679, EPI_ISL_1568845, EPI_ISL_1639542, EPI_ISL_1639553, EPI_ISL_1639570, EPI_ISL_1639591, EPI_ISL_1639608, EPI_ISL_1639806, EPI_ISL_1639840, EPI_ISL_1639844, EPI_ISL_1639967, EPI_ISL_1640010, EPI_ISL_1641909, EPI_ISL_1641944, EPI_ISL_1641949, EPI_ISL_1641966, EPI_ISL_1642016, EPI_ISL_1642083, EPI_ISL_1642122, EPI_ISL_1642207, EPI_ISL_1646074, EPI_ISL_1646099, EPI_ISL_1646341, EPI_ISL_1646597, EPI_ISL_1646724, EPI_ISL_1646733, EPI_ISL_1646794, EPI_ISL_1722923

see above SYNLAB MVZ Bonn Robert Koch Institute

EPI_ISL_1567756, EPI_ISL_1568576, EPI_ISL_1568580, EPI_ISL_1568582, EPI_ISL_1568585, EPI_ISL_1568586, EPI_ISL_1568588, EPI_ISL_1568589, EPI_ISL_1568593, EPI_ISL_1568597, EPI_ISL_1568602, EPI_ISL_1568606, EPI_ISL_1568607, EPI_ISL_1568610, EPI_ISL_1568613, EPI_ISL_1568614, EPI_ISL_1568618, EPI_ISL_1568622, EPI_ISL_1568623, EPI_ISL_1568625, EPI_ISL_1568627, EPI_ISL_1568629, EPI_ISL_1568634, EPI_ISL_1568637, EPI_ISL_1568641, EPI_ISL_1568647, EPI_ISL_1568648, EPI_ISL_1568650, EPI_ISL_1568663, EPI_ISL_1568664, EPI_ISL_1568667, EPI_ISL_1568668, EPI_ISL_1568670, EPI_ISL_1568672, EPI_ISL_1568689, EPI_ISL_1568692, EPI_ISL_1568694, EPI_ISL_1568696, EPI_ISL_1568704, EPI_ISL_1568708, EPI_ISL_1568711, EPI_ISL_1568712, EPI_ISL_1568715, EPI_ISL_1568716, EPI_ISL_1568718, EPI_ISL_1568721, EPI_ISL_1568724, EPI_ISL_1568727, EPI_ISL_1568736, EPI_ISL_1568739, EPI_ISL_1568741, EPI_ISL_1568742, EPI_ISL_1568744, EPI_ISL_1568746, EPI_ISL_1568750, EPI_ISL_1568751, EPI_ISL_1568754, EPI_ISL_1568755, EPI_ISL_1568756, EPI_ISL_1568758, EPI_ISL_1568759, EPI_ISL_1568760, EPI_ISL_1568761, EPI_ISL_1568763, EPI_ISL_1568765, EPI_ISL_1568766, EPI_ISL_1568768, EPI_ISL_1568770, EPI_ISL_1568771, EPI_ISL_1568775, EPI_ISL_1568776, EPI_ISL_1568777, EPI_ISL_1568779, EPI_ISL_1568780, EPI_ISL_1568781, EPI_ISL_1568782, EPI_ISL_1568785, EPI_ISL_1568786, EPI_ISL_1568787, EPI_ISL_1568789, EPI_ISL_1568790, EPI_ISL_1568791, EPI_ISL_1568792, EPI_ISL_1568793, EPI_ISL_1568795, EPI_ISL_1568796, EPI_ISL_1568797, EPI_ISL_1568798, EPI_ISL_1568799, EPI_ISL_1568800, EPI_ISL_1568801, EPI_ISL_1568803, EPI_ISL_1568805, EPI_ISL_1568806, EPI_ISL_1568807, EPI_ISL_1568808, EPI_ISL_1568809, EPI_ISL_1568810, EPI_ISL_1568816, EPI_ISL_1568817, EPI_ISL_1572311, EPI_ISL_1572321, EPI_ISL_1639541, EPI_ISL_1639554, EPI_ISL_1639577, EPI_ISL_1639580, EPI_ISL_1639596, EPI_ISL_1639601, EPI_ISL_1639609, EPI_ISL_1639618, EPI_ISL_1639622, EPI_ISL_1639654, EPI_ISL_1639657, EPI_ISL_1639670, EPI_ISL_1639673, EPI_ISL_1639678, EPI_ISL_1639705, EPI_ISL_1639706, EPI_ISL_1639711, EPI_ISL_1639724, EPI_ISL_1639736, EPI_ISL_1639739, EPI_ISL_1639742, EPI_ISL_1639753, EPI_ISL_1639754, EPI_ISL_1639758, EPI_ISL_1639762, EPI_ISL_1639784, EPI_ISL_1639816, EPI_ISL_1639827, EPI_ISL_1639829, EPI_ISL_1639835, EPI_ISL_1639851, EPI_ISL_1639854, EPI_ISL_1639885, EPI_ISL_1639890, EPI_ISL_1639893, EPI_ISL_1639898, EPI_ISL_1639902, EPI_ISL_1639903, EPI_ISL_1639913, EPI_ISL_1639918, EPI_ISL_1639921, EPI_ISL_1639927, EPI_ISL_1639929, EPI_ISL_1639930, EPI_ISL_1639931, EPI_ISL_1639933, EPI_ISL_1639943, EPI_ISL_1639949, EPI_ISL_1639950, EPI_ISL_1639952, EPI_ISL_1639954, EPI_ISL_1639956, EPI_ISL_1639958, EPI_ISL_1639962, EPI_ISL_1639963, EPI_ISL_1639966, EPI_ISL_1639970, EPI_ISL_1639971, EPI_ISL_1639973, EPI_ISL_1639975, EPI_ISL_1639977, EPI_ISL_1639978, EPI_ISL_1639981, EPI_ISL_1639986, EPI_ISL_1639987, EPI_ISL_1639991, EPI_ISL_1639993, EPI_ISL_1639996, EPI_ISL_1639997, EPI_ISL_1640000, EPI_ISL_1640001, EPI_ISL_1640004, EPI_ISL_1640007, EPI_ISL_1640011, EPI_ISL_1640014, EPI_ISL_1640015, EPI_ISL_1640018, EPI_ISL_1640021, EPI_ISL_1640023, EPI_ISL_1640027, EPI_ISL_1640030, EPI_ISL_1640036, EPI_ISL_1640040, EPI_ISL_1640041, EPI_ISL_1640049, EPI_ISL_1640056, EPI_ISL_1640060, EPI_ISL_1640062, EPI_ISL_1641880, EPI_ISL_1641890, EPI_ISL_1641895, EPI_ISL_1641917, EPI_ISL_1641932, EPI_ISL_1641971, EPI_ISL_1641972, EPI_ISL_1641977, EPI_ISL_1641985, EPI_ISL_1642002, EPI_ISL_1642030, EPI_ISL_1642075, EPI_ISL_1642077, EPI_ISL_1642131, EPI_ISL_1642135, EPI_ISL_1642140, EPI_ISL_1642163, EPI_ISL_1642164, EPI_ISL_1642170, EPI_ISL_1642175, EPI_ISL_1642180, EPI_ISL_1642191, EPI_ISL_1642199, EPI_ISL_1642210, EPI_ISL_1642219, EPI_ISL_1642223, EPI_ISL_1642227, EPI_ISL_1642229, EPI_ISL_1642240, EPI_ISL_1642247, EPI_ISL_1642252, EPI_ISL_1642266, EPI_ISL_1642268, EPI_ISL_1642270, EPI_ISL_1645965, EPI_ISL_1645975, EPI_ISL_1645999, EPI_ISL_1646007, EPI_ISL_1646021, EPI_ISL_1646030, EPI_ISL_1646051, EPI_ISL_1646055, EPI_ISL_1646057, EPI_ISL_1646063, EPI_ISL_1646084, EPI_ISL_1646088, EPI_ISL_1646092, EPI_ISL_1646096, EPI_ISL_1646101, EPI_ISL_1646103, EPI_ISL_1646107, EPI_ISL_1646113, EPI_ISL_1646115, EPI_ISL_1646117, EPI_ISL_1646128, EPI_ISL_1646130, EPI_ISL_1646132, EPI_ISL_1646136, EPI_ISL_1646138, EPI_ISL_1646165, EPI_ISL_1646169, EPI_ISL_1646171, EPI_ISL_1646177, EPI_ISL_1646181, EPI_ISL_1646185, EPI_ISL_1646190, EPI_ISL_1646194, EPI_ISL_1646196, EPI_ISL_1646200, EPI_ISL_1646209, EPI_ISL_1646211, EPI_ISL_1646217, EPI_ISL_1646219, EPI_ISL_1646225, EPI_ISL_1646229, EPI_ISL_1646247, EPI_ISL_1646262, EPI_ISL_1646270, EPI_ISL_1646286, EPI_ISL_1646292, EPI_ISL_1646301, EPI_ISL_1646307, EPI_ISL_1646328, EPI_ISL_1646333, EPI_ISL_1646352, EPI_ISL_1646384, EPI_ISL_1646386, EPI_ISL_1646397, EPI_ISL_1646410, EPI_ISL_1646413, EPI_ISL_1646416, EPI_ISL_1646426, EPI_ISL_1646433, EPI_ISL_1646451, EPI_ISL_1646463, EPI_ISL_1646476, EPI_ISL_1646478, EPI_ISL_1646482, EPI_ISL_1646484, EPI_ISL_1646493, EPI_ISL_1646501, EPI_ISL_1646542, EPI_ISL_1646591, EPI_ISL_1646616, EPI_ISL_1646620, EPI_ISL_1646639, EPI_ISL_1646670, EPI_ISL_1646678, EPI_ISL_1646680, EPI_ISL_1646696, EPI_ISL_1646710, EPI_ISL_1646712, EPI_ISL_1646714, EPI_ISL_1646720, EPI_ISL_1646729, EPI_ISL_1646739, EPI_ISL_1646743, EPI_ISL_1646745, EPI_ISL_1646751, EPI_ISL_1646770, EPI_ISL_1646778, EPI_ISL_1646792, EPI_ISL_1646801, EPI_ISL_1646807, EPI_ISL_1646818, EPI_ISL_1646869, EPI_ISL_1719918, EPI_ISL_1719933, EPI_ISL_1719936, EPI_ISL_1719962, EPI_ISL_1719985, EPI_ISL_1719989, EPI_ISL_1719995, EPI_ISL_1720077, EPI_ISL_1720103, EPI_ISL_1720105, EPI_ISL_1720135, EPI_ISL_1720151, EPI_ISL_1720159, EPI_ISL_1720160, EPI_ISL_1720165, EPI_ISL_1722860, EPI_ISL_1722861, EPI_ISL_1722862, EPI_ISL_1722864, EPI_ISL_1722865, EPI_ISL_1722867, EPI_ISL_1722870, EPI_ISL_1722871, EPI_ISL_1722874, EPI_ISL_1722875, EPI_ISL_1722876, EPI_ISL_1722880, EPI_ISL_1722888, EPI_ISL_1722892, EPI_ISL_1722894, EPI_ISL_1722895, EPI_ISL_1722901, EPI_ISL_1722902, EPI_ISL_1722903, EPI_ISL_1722905, EPI_ISL_1722906, EPI_ISL_1722908, EPI_ISL_1722909, EPI_ISL_1722911, EPI_ISL_1722914, EPI_ISL_1722915, EPI_ISL_1722916, EPI_ISL_1722917, EPI_ISL_1722919, EPI_ISL_1722921, EPI_ISL_1722922, EPI_ISL_1729721, EPI_ISL_1729732, EPI_ISL_1729758, EPI_ISL_1729802, EPI_ISL_1729810, EPI_ISL_1729889, EPI_ISL_1729895, EPI_ISL_1729914, EPI_ISL_1729918, EPI_ISL_2108009, EPI_ISL_2108016, EPI_ISL_2108077, EPI_ISL_2108079, EPI_ISL_2108197, EPI_ISL_2108216, EPI_ISL_2119035, EPI_ISL_2119616

see above SYNLAB MVZ Ettlingen Robert Koch Institute

EPI_ISL_1565813, EPI_ISL_1565830, EPI_ISL_1565896, EPI_ISL_1565911, EPI_ISL_1567715, EPI_ISL_1568601, EPI_ISL_1568611, EPI_ISL_1568620, EPI_ISL_1568621, EPI_ISL_1568624, EPI_ISL_1568628, EPI_ISL_1568632, EPI_ISL_1568635, EPI_ISL_1568636, EPI_ISL_1568639, EPI_ISL_1568642, EPI_ISL_1568649, EPI_ISL_1568654, EPI_ISL_1568691, EPI_ISL_1568702, EPI_ISL_1568705, EPI_ISL_1568710, EPI_ISL_1568722, EPI_ISL_1568740, EPI_ISL_1568743, EPI_ISL_1568783, EPI_ISL_1568925, EPI_ISL_1568928, EPI_ISL_1572312, EPI_ISL_1572481, EPI_ISL_1572498, EPI_ISL_1639520, EPI_ISL_1639540, EPI_ISL_1639677, EPI_ISL_1639695, EPI_ISL_1639702, EPI_ISL_1639712, EPI_ISL_1639718, EPI_ISL_1639732, EPI_ISL_1639788, EPI_ISL_1639800, EPI_ISL_1639832, EPI_ISL_1639875, EPI_ISL_1639923, EPI_ISL_1639946, EPI_ISL_1640033, EPI_ISL_1640035, EPI_ISL_1640038, EPI_ISL_1640045, EPI_ISL_1641915, EPI_ISL_1641947, EPI_ISL_1641999, EPI_ISL_1642004, EPI_ISL_1642182, EPI_ISL_1642213, EPI_ISL_1642244, EPI_ISL_1645969, EPI_ISL_1645994, EPI_ISL_1646070, EPI_ISL_1646120, EPI_ISL_1646150, EPI_ISL_1646163, EPI_ISL_1646180, EPI_ISL_1646188, EPI_ISL_1646213, EPI_ISL_1646223, EPI_ISL_1646252, EPI_ISL_1646254, EPI_ISL_1646274, EPI_ISL_1646284, EPI_ISL_1646335, EPI_ISL_1646395, EPI_ISL_1646440, EPI_ISL_1646453, EPI_ISL_1646459, EPI_ISL_1646508, EPI_ISL_1646575, EPI_ISL_1646635, EPI_ISL_1646659, EPI_ISL_1646661, EPI_ISL_1646668, EPI_ISL_1646676, EPI_ISL_1646706, EPI_ISL_1646726, EPI_ISL_1646754, EPI_ISL_1646788, EPI_ISL_1646851, EPI_ISL_1646874, EPI_ISL_1646880, EPI_ISL_1719922, EPI_ISL_1720009, EPI_ISL_1720121, EPI_ISL_1722863, EPI_ISL_1722887, EPI_ISL_1722898, EPI_ISL_1722900, EPI_ISL_1722904, EPI_ISL_1728764, EPI_ISL_1729722, EPI_ISL_1729776, EPI_ISL_1729852, EPI_ISL_1729884, EPI_ISL_1729924, EPI_ISL_1729947

see above SYNLAB MVZ Hamburg Robert Koch Institute

EPI_ISL_1567741, EPI_ISL_1568575, EPI_ISL_1568577, EPI_ISL_1568592, EPI_ISL_1568598, EPI_ISL_1568599, EPI_ISL_1568604, EPI_ISL_1568605, EPI_ISL_1568608, EPI_ISL_1568612, EPI_ISL_1568676, EPI_ISL_1568678, EPI_ISL_1568690, EPI_ISL_1568707, EPI_ISL_1568726, EPI_ISL_1568728, EPI_ISL_1568748, EPI_ISL_1568753, EPI_ISL_1568767, EPI_ISL_1568772, EPI_ISL_1568778, EPI_ISL_1568843, EPI_ISL_1568896, EPI_ISL_1568916, EPI_ISL_1639521, EPI_ISL_1639528, EPI_ISL_1639532, EPI_ISL_1639536, EPI_ISL_1639543, EPI_ISL_1639560, EPI_ISL_1639561, EPI_ISL_1639563, EPI_ISL_1639564, EPI_ISL_1639565, EPI_ISL_1639566, EPI_ISL_1639572, EPI_ISL_1639573, EPI_ISL_1639581, EPI_ISL_1639582, EPI_ISL_1639583, EPI_ISL_1639588, EPI_ISL_1639589, EPI_ISL_1639597, EPI_ISL_1639602, EPI_ISL_1639603, EPI_ISL_1639607, EPI_ISL_1639611, EPI_ISL_1639617, EPI_ISL_1639619, EPI_ISL_1639620, EPI_ISL_1639621, EPI_ISL_1639625, EPI_ISL_1639626, EPI_ISL_1639627, EPI_ISL_1639639, EPI_ISL_1639643, EPI_ISL_1639644, EPI_ISL_1639647, EPI_ISL_1639652, EPI_ISL_1639653, EPI_ISL_1639655, EPI_ISL_1639659, EPI_ISL_1639660, EPI_ISL_1639663, EPI_ISL_1639666, EPI_ISL_1639671, EPI_ISL_1639674, EPI_ISL_1639675, EPI_ISL_1639676, EPI_ISL_1639683, EPI_ISL_1639684, EPI_ISL_1639685, EPI_ISL_1639688, EPI_ISL_1639692, EPI_ISL_1639693, EPI_ISL_1639697, EPI_ISL_1639699, EPI_ISL_1639701, EPI_ISL_1639703, EPI_ISL_1639704, EPI_ISL_1639708, EPI_ISL_1639710, EPI_ISL_1639713, EPI_ISL_1639714, EPI_ISL_1639715, EPI_ISL_1639716, EPI_ISL_1639719, EPI_ISL_1639720, EPI_ISL_1639721, EPI_ISL_1639722, EPI_ISL_1639723, EPI_ISL_1639726, EPI_ISL_1639728, EPI_ISL_1639730, EPI_ISL_1639735, EPI_ISL_1639738, EPI_ISL_1639740, EPI_ISL_1639741, EPI_ISL_1639743, EPI_ISL_1639745, EPI_ISL_1639746, EPI_ISL_1639748, EPI_ISL_1639749, EPI_ISL_1639750, EPI_ISL_1639751, EPI_ISL_1639752, EPI_ISL_1639760, EPI_ISL_1639761, EPI_ISL_1639764, EPI_ISL_1639770, EPI_ISL_1639771, EPI_ISL_1639774, EPI_ISL_1639777, EPI_ISL_1639779, EPI_ISL_1639781, EPI_ISL_1639782, EPI_ISL_1639785, EPI_ISL_1639786, EPI_ISL_1639792, EPI_ISL_1639793, EPI_ISL_1639803, EPI_ISL_1639804, EPI_ISL_1639805, EPI_ISL_1639807, EPI_ISL_1639809, EPI_ISL_1639817, EPI_ISL_1639820, EPI_ISL_1639822, EPI_ISL_1639824, EPI_ISL_1639826, EPI_ISL_1639828, EPI_ISL_1639830, EPI_ISL_1639836, EPI_ISL_1639837, EPI_ISL_1639839, EPI_ISL_1639842, EPI_ISL_1639843, EPI_ISL_1639846, EPI_ISL_1639847, EPI_ISL_1639848, EPI_ISL_1639850, EPI_ISL_1639853, EPI_ISL_1639855, EPI_ISL_1639856, EPI_ISL_1639859, EPI_ISL_1639861, EPI_ISL_1639862, EPI_ISL_1639864, EPI_ISL_1639868, EPI_ISL_1639869, EPI_ISL_1639870, EPI_ISL_1639872, EPI_ISL_1639877, EPI_ISL_1639879, EPI_ISL_1639880, EPI_ISL_1639882, EPI_ISL_1639883, EPI_ISL_1639884, EPI_ISL_1639886, EPI_ISL_1639889, EPI_ISL_1639896, EPI_ISL_1639900, EPI_ISL_1639901, EPI_ISL_1639904, EPI_ISL_1639905, EPI_ISL_1639906, EPI_ISL_1639907, EPI_ISL_1639908, EPI_ISL_1639909, EPI_ISL_1639910, EPI_ISL_1639915, EPI_ISL_1639917, EPI_ISL_1639919, EPI_ISL_1639920, EPI_ISL_1639922, EPI_ISL_1639928, EPI_ISL_1639932, EPI_ISL_1639934, EPI_ISL_1639935, EPI_ISL_1639937, EPI_ISL_1639938, EPI_ISL_1639940, EPI_ISL_1639942, EPI_ISL_1639945, EPI_ISL_1639948, EPI_ISL_1639960, EPI_ISL_1639964, EPI_ISL_1639968, EPI_ISL_1639972, EPI_ISL_1639976, EPI_ISL_1639980, EPI_ISL_1639982, EPI_ISL_1639985, EPI_ISL_1639988, EPI_ISL_1639989, EPI_ISL_1639990, EPI_ISL_1639992, EPI_ISL_1639994, EPI_ISL_1639995, EPI_ISL_1639998, EPI_ISL_1639999, EPI_ISL_1640002, EPI_ISL_1640005, EPI_ISL_1640009, EPI_ISL_1640012, EPI_ISL_1640042, EPI_ISL_1640044, EPI_ISL_1640046, EPI_ISL_1640048, EPI_ISL_1640055, EPI_ISL_1640061, EPI_ISL_1640063, EPI_ISL_1640064, EPI_ISL_1640065, EPI_ISL_1640067, EPI_ISL_1640068, EPI_ISL_1640069, EPI_ISL_1641857, EPI_ISL_1641868, EPI_ISL_1641870, EPI_ISL_1641873, EPI_ISL_1641879, EPI_ISL_1641882, EPI_ISL_1641885, EPI_ISL_1641887, EPI_ISL_1641892, EPI_ISL_1641916, EPI_ISL_1641926, EPI_ISL_1641928, EPI_ISL_1641930, EPI_ISL_1641933, EPI_ISL_1641936, EPI_ISL_1641941, EPI_ISL_1641954, EPI_ISL_1641955, EPI_ISL_1641960, EPI_ISL_1641967, EPI_ISL_1641973, EPI_ISL_1641989, EPI_ISL_1641990, EPI_ISL_1641994, EPI_ISL_1641997, EPI_ISL_1642000, EPI_ISL_1642005, EPI_ISL_1642008, EPI_ISL_1642021, EPI_ISL_1642023, EPI_ISL_1642025, EPI_ISL_1642028, EPI_ISL_1642031, EPI_ISL_1642033, EPI_ISL_1642035, EPI_ISL_1642038, EPI_ISL_1642040, EPI_ISL_1642042, EPI_ISL_1642046, EPI_ISL_1642050, EPI_ISL_1642052, EPI_ISL_1642054, EPI_ISL_1642059, EPI_ISL_1642064, EPI_ISL_1642069, EPI_ISL_1642072, EPI_ISL_1642079, EPI_ISL_1642081, EPI_ISL_1642086, EPI_ISL_1642089, EPI_ISL_1642093, EPI_ISL_1642097, EPI_ISL_1642101, EPI_ISL_1642102, EPI_ISL_1642103, EPI_ISL_1642110, EPI_ISL_1642111, EPI_ISL_1642112, EPI_ISL_1642114, EPI_ISL_1642123, EPI_ISL_1642124, EPI_ISL_1642143, EPI_ISL_1642144, EPI_ISL_1642150, EPI_ISL_1642154, EPI_ISL_1642155, EPI_ISL_1642158, EPI_ISL_1642160, EPI_ISL_1642162, EPI_ISL_1642167, EPI_ISL_1642168, EPI_ISL_1642176, EPI_ISL_1642177, EPI_ISL_1642190, EPI_ISL_1642197, EPI_ISL_1642204, EPI_ISL_1642208, EPI_ISL_1642214, EPI_ISL_1642215, EPI_ISL_1642216, EPI_ISL_1642221, EPI_ISL_1642226, EPI_ISL_1642231, EPI_ISL_1642232, EPI_ISL_1642236, EPI_ISL_1642242, EPI_ISL_1642246, EPI_ISL_1642255, EPI_ISL_1642272, EPI_ISL_1642274, EPI_ISL_1646320, EPI_ISL_1646382, EPI_ISL_1646422, EPI_ISL_1646491, EPI_ISL_1646506, EPI_ISL_1646515, EPI_ISL_1646526, EPI_ISL_1646533, EPI_ISL_1646554, EPI_ISL_1646835, EPI_ISL_1646849, EPI_ISL_1646872, EPI_ISL_1646878, EPI_ISL_1646882, EPI_ISL_1719977, EPI_ISL_1719993, EPI_ISL_1719998, EPI_ISL_1722881, EPI_ISL_1729714, EPI_ISL_1729779, EPI_ISL_1729783, EPI_ISL_1729890, EPI_ISL_1729921, EPI_ISL_1729923, EPI_ISL_1729934, EPI_ISL_2119070, EPI_ISL_2119156, EPI_ISL_2119198, EPI_ISL_2119257, EPI_ISL_2119374, EPI_ISL_2119407, EPI_ISL_2119716, EPI_ISL_2119744

see above SYNLAB MVZ Heidelberg Robert Koch Institute

EPI_ISL_1438297, EPI_ISL_1438301, EPI_ISL_1438303, EPI_ISL_1438304, EPI_ISL_1438305, EPI_ISL_1438307, EPI_ISL_1438312, EPI_ISL_1438324, EPI_ISL_1438331, EPI_ISL_1438332, EPI_ISL_1438341, EPI_ISL_1438342, EPI_ISL_1438345, EPI_ISL_1438353, EPI_ISL_1438355, EPI_ISL_1438356, EPI_ISL_1438357, EPI_ISL_1438360, EPI_ISL_1438363, EPI_ISL_1438366, EPI_ISL_1438367, EPI_ISL_1438368, EPI_ISL_1438372, EPI_ISL_1438373, EPI_ISL_1438374, EPI_ISL_1438376, EPI_ISL_1438377, EPI_ISL_1438382, EPI_ISL_1438386, EPI_ISL_1438387, EPI_ISL_1438388, EPI_ISL_1438390, EPI_ISL_1438394, EPI_ISL_1438395, EPI_ISL_1438397, EPI_ISL_1438398, EPI_ISL_1438403, EPI_ISL_1438404, EPI_ISL_1438410, EPI_ISL_1438411, EPI_ISL_1438412, EPI_ISL_1438413, EPI_ISL_1438420, EPI_ISL_1438421, EPI_ISL_1438424, EPI_ISL_1438425, EPI_ISL_1438428, EPI_ISL_1438429, EPI_ISL_1438430, EPI_ISL_1438435, EPI_ISL_1438436, EPI_ISL_1438439, EPI_ISL_1438444, EPI_ISL_1438447, EPI_ISL_1438452, EPI_ISL_1438455, EPI_ISL_1438457, EPI_ISL_1438461, EPI_ISL_1438463, EPI_ISL_1438467, EPI_ISL_1438468, EPI_ISL_1438470, EPI_ISL_1438471, EPI_ISL_1438473, EPI_ISL_1438478, EPI_ISL_1438482, EPI_ISL_1438485, EPI_ISL_1438488, EPI_ISL_1438489, EPI_ISL_1438492, EPI_ISL_1438496, EPI_ISL_1438501, EPI_ISL_1438506, EPI_ISL_1438512, EPI_ISL_1438517, EPI_ISL_1438521, EPI_ISL_1438522, EPI_ISL_1438523, EPI_ISL_1438525, EPI_ISL_1438526, EPI_ISL_1438527, EPI_ISL_1438528, EPI_ISL_1438531, EPI_ISL_1438532, EPI_ISL_1438534, EPI_ISL_1438541, EPI_ISL_1438545, EPI_ISL_1438547, EPI_ISL_1438548, EPI_ISL_1438550, EPI_ISL_1438552, EPI_ISL_1438554, EPI_ISL_1438555, EPI_ISL_1438560, EPI_ISL_1438562, EPI_ISL_1438563, EPI_ISL_1438570, EPI_ISL_1438577, EPI_ISL_1438579, EPI_ISL_1438580, EPI_ISL_1438581, EPI_ISL_1441311, EPI_ISL_1441315, EPI_ISL_1441317, EPI_ISL_1441319, EPI_ISL_1441329, EPI_ISL_1441333, EPI_ISL_1441335, EPI_ISL_1441342, EPI_ISL_1441344, EPI_ISL_1441345, EPI_ISL_1441346, EPI_ISL_1441347, EPI_ISL_1441351, EPI_ISL_1441352, EPI_ISL_1441354, EPI_ISL_1441361, EPI_ISL_1441363, EPI_ISL_1441367, EPI_ISL_1441369, EPI_ISL_1441371, EPI_ISL_1441372, EPI_ISL_1441373, EPI_ISL_1441377, EPI_ISL_1441381, EPI_ISL_1441385, EPI_ISL_1441390, EPI_ISL_1441392, EPI_ISL_1441393, EPI_ISL_1441395, EPI_ISL_1441398, EPI_ISL_1441404, EPI_ISL_1441406, EPI_ISL_1441407, EPI_ISL_1441408, EPI_ISL_1441412, EPI_ISL_1441443, EPI_ISL_1441444, EPI_ISL_1441446, EPI_ISL_1441448, EPI_ISL_1441451, EPI_ISL_1441453, EPI_ISL_1441457, EPI_ISL_1441461, EPI_ISL_1441463, EPI_ISL_1441469, EPI_ISL_1441478, EPI_ISL_1441479, EPI_ISL_1441481, EPI_ISL_1441483, EPI_ISL_1441494, EPI_ISL_1441498, EPI_ISL_1441505, EPI_ISL_1441507, EPI_ISL_1441516, EPI_ISL_1441520, EPI_ISL_1441532, EPI_ISL_1441536, EPI_ISL_1441548, EPI_ISL_1441550, EPI_ISL_1441575, EPI_ISL_1441584, EPI_ISL_1441593, EPI_ISL_1441597, EPI_ISL_1441601, EPI_ISL_1441602, EPI_ISL_1441605, EPI_ISL_1441607, EPI_ISL_1441611, EPI_ISL_1441613, EPI_ISL_1441614, EPI_ISL_1441617, EPI_ISL_1441618, EPI_ISL_1441619, EPI_ISL_1441620, EPI_ISL_1441621, EPI_ISL_1441625, EPI_ISL_1441627, EPI_ISL_1441628, EPI_ISL_1441629, EPI_ISL_1441633, EPI_ISL_1441637, EPI_ISL_1441639, EPI_ISL_1442751, EPI_ISL_1442752, EPI_ISL_1442753, EPI_ISL_1442754, EPI_ISL_1442755, EPI_ISL_1442756, EPI_ISL_1442757, EPI_ISL_1442758, EPI_ISL_1442759, EPI_ISL_1442760, EPI_ISL_1442761, EPI_ISL_1442762, EPI_ISL_1442763, EPI_ISL_1442764, EPI_ISL_1442765, EPI_ISL_1442767, EPI_ISL_1442770, EPI_ISL_1442772, EPI_ISL_1442775, EPI_ISL_1442778, EPI_ISL_1442779, EPI_ISL_1442780, EPI_ISL_1442781, EPI_ISL_1442782, EPI_ISL_1442783, EPI_ISL_1442784, EPI_ISL_1442787, EPI_ISL_1442788, EPI_ISL_1442789, EPI_ISL_1442790, EPI_ISL_1442792, EPI_ISL_1442793, EPI_ISL_1442794, EPI_ISL_1442795, EPI_ISL_1442796, EPI_ISL_1442799, EPI_ISL_1442800, EPI_ISL_1442802, EPI_ISL_1442803, EPI_ISL_1442804, EPI_ISL_1442806, EPI_ISL_1442807, EPI_ISL_1442809, EPI_ISL_1442810, EPI_ISL_1442811, EPI_ISL_1442813, EPI_ISL_1442816, EPI_ISL_1442820, EPI_ISL_1442821, EPI_ISL_1442822, EPI_ISL_1442823, EPI_ISL_1442824, EPI_ISL_1442825, EPI_ISL_1442829, EPI_ISL_1442830, EPI_ISL_1442832, EPI_ISL_1442835, EPI_ISL_1442837, EPI_ISL_1442838, EPI_ISL_1442841, EPI_ISL_1442843, EPI_ISL_1442844, EPI_ISL_1442847, EPI_ISL_1442850, EPI_ISL_1442851, EPI_ISL_1442852, EPI_ISL_1442853, EPI_ISL_1442856, EPI_ISL_1442857, EPI_ISL_1442858, EPI_ISL_1442859, EPI_ISL_1442861, EPI_ISL_1442862, EPI_ISL_1442863, EPI_ISL_1442867, EPI_ISL_1442869, EPI_ISL_1442870, EPI_ISL_1442872, EPI_ISL_1442875, EPI_ISL_1442880, EPI_ISL_1442882, EPI_ISL_1442883, EPI_ISL_1442886, EPI_ISL_1442888, EPI_ISL_1442889, EPI_ISL_1442890, EPI_ISL_1442891, EPI_ISL_1442892, EPI_ISL_1442893, EPI_ISL_1442894, EPI_ISL_1442895, EPI_ISL_1565508, EPI_ISL_1565509, EPI_ISL_1565510, EPI_ISL_1565519, EPI_ISL_1565521, EPI_ISL_1565522, EPI_ISL_1565524, EPI_ISL_1565526, EPI_ISL_1565528, EPI_ISL_1565531, EPI_ISL_1565532, EPI_ISL_1565535, EPI_ISL_1565539, EPI_ISL_1565540, EPI_ISL_1565542, EPI_ISL_1565544, EPI_ISL_1565545, EPI_ISL_1565546, EPI_ISL_1565549, EPI_ISL_1565552, EPI_ISL_1565553, EPI_ISL_1565555, EPI_ISL_1565556, EPI_ISL_1565557, EPI_ISL_1565559, EPI_ISL_1565560, EPI_ISL_1565561, EPI_ISL_1565564, EPI_ISL_1565567, EPI_ISL_1565568, EPI_ISL_1565570, EPI_ISL_1565572, EPI_ISL_1565573, EPI_ISL_1565576, EPI_ISL_1565577, EPI_ISL_1565579, EPI_ISL_1565581, EPI_ISL_1565583, EPI_ISL_1565584, EPI_ISL_1565586, EPI_ISL_1565590, EPI_ISL_1565594, EPI_ISL_1565598, EPI_ISL_1565601, EPI_ISL_1565604, EPI_ISL_1565605, EPI_ISL_1565607, EPI_ISL_1565615, EPI_ISL_1565621, EPI_ISL_1565628, EPI_ISL_1565630, EPI_ISL_1565633, EPI_ISL_1565651, EPI_ISL_1565659, EPI_ISL_1565660, EPI_ISL_1565662, EPI_ISL_1565663, EPI_ISL_1565665, EPI_ISL_1565669, EPI_ISL_1565670, EPI_ISL_1565673, EPI_ISL_1565674, EPI_ISL_1565676, EPI_ISL_1565679, EPI_ISL_1565682, EPI_ISL_1565686, EPI_ISL_1565687, EPI_ISL_1565691, EPI_ISL_1565698, EPI_ISL_1565700, EPI_ISL_1565703, EPI_ISL_1565705, EPI_ISL_1565708, EPI_ISL_1565711, EPI_ISL_1565712, EPI_ISL_1565713, EPI_ISL_1565716, EPI_ISL_1565718, EPI_ISL_1565721, EPI_ISL_1565722, EPI_ISL_1565725, EPI_ISL_1565738, EPI_ISL_1565740, EPI_ISL_1565743, EPI_ISL_1565749, EPI_ISL_1565753, EPI_ISL_1565754, EPI_ISL_1565755, EPI_ISL_1565765, EPI_ISL_1565773, EPI_ISL_1565775, EPI_ISL_1566786, EPI_ISL_1566787, EPI_ISL_1566788, EPI_ISL_1566790, EPI_ISL_1566791, EPI_ISL_1566794, EPI_ISL_1566795, EPI_ISL_1566796, EPI_ISL_1566797, EPI_ISL_1566800, EPI_ISL_1566802, EPI_ISL_1566804, EPI_ISL_1566805, EPI_ISL_1566806, EPI_ISL_1566807, EPI_ISL_1566809, EPI_ISL_1566810, EPI_ISL_1566812, EPI_ISL_1566813, EPI_ISL_1566816, EPI_ISL_1566817, EPI_ISL_1566819, EPI_ISL_1566820, EPI_ISL_1566821, EPI_ISL_1566827, EPI_ISL_1566828, EPI_ISL_1566830, EPI_ISL_1566833, EPI_ISL_1566834, EPI_ISL_1566839, EPI_ISL_1566840, EPI_ISL_1566842, EPI_ISL_1566844, EPI_ISL_1566846, EPI_ISL_1566848, EPI_ISL_1566849, EPI_ISL_1566850, EPI_ISL_1566851, EPI_ISL_1566852, EPI_ISL_1566853, EPI_ISL_1566855, EPI_ISL_1566857, EPI_ISL_1566858, EPI_ISL_1566862, EPI_ISL_1566863, EPI_ISL_1566864, EPI_ISL_1566866, EPI_ISL_1566868, EPI_ISL_1566869, EPI_ISL_1566872, EPI_ISL_1566874, EPI_ISL_1566876, EPI_ISL_1566880, EPI_ISL_1566882, EPI_ISL_1566884, EPI_ISL_1566886, EPI_ISL_1566887, EPI_ISL_1566888, EPI_ISL_1566890, EPI_ISL_1566895, EPI_ISL_1566897, EPI_ISL_1566899, EPI_ISL_1566900, EPI_ISL_1566901, EPI_ISL_1566902, EPI_ISL_1566910, EPI_ISL_1566911, EPI_ISL_1566912, EPI_ISL_1566915, EPI_ISL_1566927, EPI_ISL_1566929, EPI_ISL_1566931, EPI_ISL_1566933, EPI_ISL_1566935, EPI_ISL_1566937, EPI_ISL_1566939, EPI_ISL_1566942, EPI_ISL_1566943, EPI_ISL_1566945, EPI_ISL_1566946, EPI_ISL_1566947, EPI_ISL_1566951, EPI_ISL_1566952, EPI_ISL_1566957, EPI_ISL_1566958, EPI_ISL_1566961, EPI_ISL_1566962, EPI_ISL_1566964, EPI_ISL_1566966, EPI_ISL_1566969, EPI_ISL_1566970, EPI_ISL_1566974, EPI_ISL_1566975, EPI_ISL_1566977, EPI_ISL_1566979, EPI_ISL_1566981, EPI_ISL_1566990, EPI_ISL_1566996, EPI_ISL_1566997, EPI_ISL_1567005, EPI_ISL_1567008, EPI_ISL_1567011, EPI_ISL_1567014, EPI_ISL_1567015, EPI_ISL_1567020, EPI_ISL_1567024, EPI_ISL_1567026, EPI_ISL_1567028, EPI_ISL_1567029, EPI_ISL_1567030, EPI_ISL_1567032, EPI_ISL_1567035, EPI_ISL_1567036, EPI_ISL_1567037, EPI_ISL_1567039, EPI_ISL_1567041, EPI_ISL_1567046, EPI_ISL_1567047, EPI_ISL_1567049, EPI_ISL_1567050, EPI_ISL_1567052, EPI_ISL_1567054, EPI_ISL_1567055, EPI_ISL_1567058, EPI_ISL_1567062, EPI_ISL_1567063, EPI_ISL_1567066, EPI_ISL_1567067, EPI_ISL_1567070, EPI_ISL_1567071, EPI_ISL_1567073, EPI_ISL_1567075, EPI_ISL_1567076, EPI_ISL_1567078, EPI_ISL_1567079, EPI_ISL_1567081, EPI_ISL_1567085, EPI_ISL_1567086, EPI_ISL_1567087, EPI_ISL_1567088, EPI_ISL_1567089, EPI_ISL_1567090, EPI_ISL_1567091, EPI_ISL_1567290, EPI_ISL_1567294, EPI_ISL_1567296, EPI_ISL_1567297, EPI_ISL_1567299, EPI_ISL_1567301, EPI_ISL_1567303, EPI_ISL_1567309, EPI_ISL_1567313, EPI_ISL_1567315, EPI_ISL_1567319, EPI_ISL_1567322, EPI_ISL_1567328, EPI_ISL_1567331, EPI_ISL_1567334, EPI_ISL_1567335, EPI_ISL_1567337, EPI_ISL_1567339, EPI_ISL_1567340, EPI_ISL_1567343, EPI_ISL_1567344, EPI_ISL_1567348, EPI_ISL_1567349, EPI_ISL_1567351, EPI_ISL_1567352, EPI_ISL_1567354, EPI_ISL_1567355, EPI_ISL_1567356, EPI_ISL_1567358, EPI_ISL_1567359, EPI_ISL_1567360, EPI_ISL_1567362, EPI_ISL_1567363, EPI_ISL_1567366, EPI_ISL_1567367, EPI_ISL_1567368, EPI_ISL_1567369, EPI_ISL_1567372, EPI_ISL_1567373, EPI_ISL_1567376, EPI_ISL_1567379, EPI_ISL_1567380, EPI_ISL_1567383, EPI_ISL_1567388, EPI_ISL_1567390, EPI_ISL_1567391, EPI_ISL_1567393, EPI_ISL_1567397, EPI_ISL_1567404, EPI_ISL_1567406, EPI_ISL_1567409, EPI_ISL_1567413, EPI_ISL_1567417, EPI_ISL_1567420, EPI_ISL_1567423, EPI_ISL_1567427, EPI_ISL_1567430, EPI_ISL_1567435, EPI_ISL_1567440, EPI_ISL_1567446, EPI_ISL_1567447, EPI_ISL_1567453, EPI_ISL_1567455, EPI_ISL_1567460, EPI_ISL_1567464, EPI_ISL_1567469, EPI_ISL_1567473, EPI_ISL_1567474, EPI_ISL_1567479, EPI_ISL_1567481, EPI_ISL_1567482, EPI_ISL_1567485, EPI_ISL_1567490, EPI_ISL_1567491, EPI_ISL_1567492, EPI_ISL_1567493, EPI_ISL_1567495, EPI_ISL_1567496, EPI_ISL_1567498, EPI_ISL_1567499, EPI_ISL_1567501, EPI_ISL_1567510, EPI_ISL_1567512, EPI_ISL_1567513, EPI_ISL_1567514, EPI_ISL_1567517, EPI_ISL_1567518, EPI_ISL_1567521, EPI_ISL_1567522, EPI_ISL_1567524, EPI_ISL_1567525, EPI_ISL_1567527, EPI_ISL_1567528, EPI_ISL_1567529, EPI_ISL_1567532, EPI_ISL_1567533, EPI_ISL_1567536, EPI_ISL_1567538, EPI_ISL_1567539, EPI_ISL_1567540, EPI_ISL_1567541, EPI_ISL_1567549, EPI_ISL_1567550, EPI_ISL_1567834, EPI_ISL_1567835, EPI_ISL_1567836, EPI_ISL_1567837, EPI_ISL_1567839, EPI_ISL_1567840, EPI_ISL_1567841, EPI_ISL_1567842, EPI_ISL_1567843, EPI_ISL_1567848, EPI_ISL_1567850, EPI_ISL_1567851, EPI_ISL_1567853, EPI_ISL_1567855, EPI_ISL_1567856, EPI_ISL_1567859, EPI_ISL_1567860, EPI_ISL_1567864, EPI_ISL_1567867, EPI_ISL_1567868, EPI_ISL_1567869, EPI_ISL_1567871, EPI_ISL_1567872, EPI_ISL_1567878, EPI_ISL_1567881, EPI_ISL_1567883, EPI_ISL_1567884, EPI_ISL_1567885, EPI_ISL_1567888, EPI_ISL_1567890, EPI_ISL_1567891, EPI_ISL_1567897, EPI_ISL_1567898, EPI_ISL_1567899, EPI_ISL_1567903, EPI_ISL_1567907, EPI_ISL_1567910, EPI_ISL_1567911, EPI_ISL_1567914, EPI_ISL_1567915, EPI_ISL_1567918, EPI_ISL_1567919, EPI_ISL_1567920, EPI_ISL_1567927, EPI_ISL_1567928, EPI_ISL_1567931, EPI_ISL_1567932, EPI_ISL_1567933, EPI_ISL_1567935, EPI_ISL_1567941, EPI_ISL_1567942, EPI_ISL_1567943, EPI_ISL_1567945, EPI_ISL_1567947, EPI_ISL_1567948, EPI_ISL_1567950, EPI_ISL_1567951, EPI_ISL_1567952, EPI_ISL_1567953, EPI_ISL_1567954, EPI_ISL_1567955, EPI_ISL_1567958, EPI_ISL_1567962, EPI_ISL_1567963, EPI_ISL_1567964, EPI_ISL_1567965, EPI_ISL_1567966, EPI_ISL_1567968, EPI_ISL_1567969, EPI_ISL_1567970, EPI_ISL_1567972, EPI_ISL_1567974, EPI_ISL_1567975, EPI_ISL_1567976, EPI_ISL_1567978, EPI_ISL_1567979, EPI_ISL_1567980, EPI_ISL_1567982, EPI_ISL_1567983, EPI_ISL_1567984, EPI_ISL_1567985, EPI_ISL_1567987, EPI_ISL_1567988, EPI_ISL_1567991, EPI_ISL_1567996, EPI_ISL_1567998, EPI_ISL_1568000, EPI_ISL_1568003, EPI_ISL_1568004, EPI_ISL_1568005, EPI_ISL_1568006, EPI_ISL_1568009, EPI_ISL_1568010, EPI_ISL_1568011, EPI_ISL_1568012, EPI_ISL_1568013, EPI_ISL_1568015, EPI_ISL_1568016, EPI_ISL_1568021, EPI_ISL_1568023, EPI_ISL_1568024, EPI_ISL_1568025, EPI_ISL_1568026, EPI_ISL_1568028, EPI_ISL_1568029, EPI_ISL_1568031, EPI_ISL_1568032, EPI_ISL_1568033, EPI_ISL_1568035, EPI_ISL_1568041, EPI_ISL_1568044, EPI_ISL_1568047, EPI_ISL_1568051, EPI_ISL_1568053, EPI_ISL_1568054, EPI_ISL_1568056, EPI_ISL_1568058, EPI_ISL_1568059, EPI_ISL_1568060, EPI_ISL_1568064, EPI_ISL_1568067, EPI_ISL_1568068, EPI_ISL_1568069, EPI_ISL_1568070, EPI_ISL_1568073, EPI_ISL_1568074, EPI_ISL_1568086, EPI_ISL_1568094, EPI_ISL_1568097, EPI_ISL_1568100, EPI_ISL_1570962, EPI_ISL_1570964, EPI_ISL_1570966, EPI_ISL_1570979, EPI_ISL_1570983, EPI_ISL_1570988, EPI_ISL_1570992, EPI_ISL_1570999, EPI_ISL_1571009, EPI_ISL_1571010, EPI_ISL_1571014, EPI_ISL_1571015, EPI_ISL_1571017, EPI_ISL_1571028, EPI_ISL_1571029, EPI_ISL_1571031, EPI_ISL_1571036, EPI_ISL_1571040, EPI_ISL_1571041, EPI_ISL_1571045, EPI_ISL_1571050, EPI_ISL_1571051, EPI_ISL_1571056, EPI_ISL_1571059, EPI_ISL_1571061, EPI_ISL_1571064, EPI_ISL_1571069, EPI_ISL_1571091, EPI_ISL_1571106, EPI_ISL_1571107, EPI_ISL_1571113, EPI_ISL_1571116

see above SYNLAB MVZ Leinfelden-Echterdingen Robert Koch Institute

EPI_ISL_1566310, EPI_ISL_1566366, EPI_ISL_1567709, EPI_ISL_1567717, EPI_ISL_1567738, EPI_ISL_1567754, EPI_ISL_1567757, EPI_ISL_1568579, EPI_ISL_1568583, EPI_ISL_1568587, EPI_ISL_1568590, EPI_ISL_1568596, EPI_ISL_1568609, EPI_ISL_1568616, EPI_ISL_1568617, EPI_ISL_1568630, EPI_ISL_1568633, EPI_ISL_1568638, EPI_ISL_1568640, EPI_ISL_1568646, EPI_ISL_1568651, EPI_ISL_1568652, EPI_ISL_1568653, EPI_ISL_1568655, EPI_ISL_1568658, EPI_ISL_1568659, EPI_ISL_1568661, EPI_ISL_1568665, EPI_ISL_1568666, EPI_ISL_1568677, EPI_ISL_1568680, EPI_ISL_1568681, EPI_ISL_1568682, EPI_ISL_1568686, EPI_ISL_1568693, EPI_ISL_1568698, EPI_ISL_1568699, EPI_ISL_1568700, EPI_ISL_1568713, EPI_ISL_1568720, EPI_ISL_1568729, EPI_ISL_1568732, EPI_ISL_1568734, EPI_ISL_1568749, EPI_ISL_1568764, EPI_ISL_1568769, EPI_ISL_1568773, EPI_ISL_1568774, EPI_ISL_1568802, EPI_ISL_1568815, EPI_ISL_1568819, EPI_ISL_1568827, EPI_ISL_1568829, EPI_ISL_1568830, EPI_ISL_1568832, EPI_ISL_1568834, EPI_ISL_1568839, EPI_ISL_1568840, EPI_ISL_1568842, EPI_ISL_1568844, EPI_ISL_1568848, EPI_ISL_1568853, EPI_ISL_1568854, EPI_ISL_1568856, EPI_ISL_1568861, EPI_ISL_1568868, EPI_ISL_1568869, EPI_ISL_1568871, EPI_ISL_1568874, EPI_ISL_1568875, EPI_ISL_1568877, EPI_ISL_1568884, EPI_ISL_1568885, EPI_ISL_1568887, EPI_ISL_1568893, EPI_ISL_1568894, EPI_ISL_1568897, EPI_ISL_1568898, EPI_ISL_1568899, EPI_ISL_1568901, EPI_ISL_1568902, EPI_ISL_1568903, EPI_ISL_1568905, EPI_ISL_1568909, EPI_ISL_1568914, EPI_ISL_1568918, EPI_ISL_1568920, EPI_ISL_1568922, EPI_ISL_1568923, EPI_ISL_1568926, EPI_ISL_1568940, EPI_ISL_1568945, EPI_ISL_1572251, EPI_ISL_1572520, EPI_ISL_1639519, EPI_ISL_1639522, EPI_ISL_1639523, EPI_ISL_1639524, EPI_ISL_1639526, EPI_ISL_1639527, EPI_ISL_1639529, EPI_ISL_1639530, EPI_ISL_1639534, EPI_ISL_1639535, EPI_ISL_1639537, EPI_ISL_1639539, EPI_ISL_1639545, EPI_ISL_1639546, EPI_ISL_1639548, EPI_ISL_1639551, EPI_ISL_1639556, EPI_ISL_1639557, EPI_ISL_1639559, EPI_ISL_1639562, EPI_ISL_1639568, EPI_ISL_1639569, EPI_ISL_1639574, EPI_ISL_1639576, EPI_ISL_1639578, EPI_ISL_1639579, EPI_ISL_1639587, EPI_ISL_1639590, EPI_ISL_1639595, EPI_ISL_1639600, EPI_ISL_1639604, EPI_ISL_1639612, EPI_ISL_1639615, EPI_ISL_1639624, EPI_ISL_1639630, EPI_ISL_1639631, EPI_ISL_1639633, EPI_ISL_1639634, EPI_ISL_1639637, EPI_ISL_1639641, EPI_ISL_1639645, EPI_ISL_1639650, EPI_ISL_1639658, EPI_ISL_1639661, EPI_ISL_1639665, EPI_ISL_1639667, EPI_ISL_1639669, EPI_ISL_1639679, EPI_ISL_1639681, EPI_ISL_1639694, EPI_ISL_1639698, EPI_ISL_1639700, EPI_ISL_1639707, EPI_ISL_1639729, EPI_ISL_1639731, EPI_ISL_1639733, EPI_ISL_1639747, EPI_ISL_1639755, EPI_ISL_1639756, EPI_ISL_1639759, EPI_ISL_1639763, EPI_ISL_1639766, EPI_ISL_1639767, EPI_ISL_1639768, EPI_ISL_1639769, EPI_ISL_1639773, EPI_ISL_1639775, EPI_ISL_1639780, EPI_ISL_1639787, EPI_ISL_1639791, EPI_ISL_1639795, EPI_ISL_1639796, EPI_ISL_1639797, EPI_ISL_1639798, EPI_ISL_1639801, EPI_ISL_1639802, EPI_ISL_1639810, EPI_ISL_1639811, EPI_ISL_1639812, EPI_ISL_1639813, EPI_ISL_1639819, EPI_ISL_1639821, EPI_ISL_1639823, EPI_ISL_1639825, EPI_ISL_1639834, EPI_ISL_1639838, EPI_ISL_1639841, EPI_ISL_1639845, EPI_ISL_1639849, EPI_ISL_1639852, EPI_ISL_1639857, EPI_ISL_1639858, EPI_ISL_1639860, EPI_ISL_1639863, EPI_ISL_1639866, EPI_ISL_1639867, EPI_ISL_1639873, EPI_ISL_1639874, EPI_ISL_1639876, EPI_ISL_1639878, EPI_ISL_1639881, EPI_ISL_1639887, EPI_ISL_1639892, EPI_ISL_1639895, EPI_ISL_1639899, EPI_ISL_1639911, EPI_ISL_1639912, EPI_ISL_1639914, EPI_ISL_1639916, EPI_ISL_1639925, EPI_ISL_1639926, EPI_ISL_1639936, EPI_ISL_1639939, EPI_ISL_1639941, EPI_ISL_1639944, EPI_ISL_1639951, EPI_ISL_1639953, EPI_ISL_1639957, EPI_ISL_1639959, EPI_ISL_1639961, EPI_ISL_1639965, EPI_ISL_1639969, EPI_ISL_1639974, EPI_ISL_1639979, EPI_ISL_1639983,

| | | |
|---|---|---|
| EPI_ISL_1639984, EPI_ISL_1640003, EPI_ISL_1640006, EPI_ISL_1640008, EPI_ISL_1640013, EPI_ISL_1640016, EPI_ISL_1640017, EPI_ISL_1640019, EPI_ISL_1640020, EPI_ISL_1640022, EPI_ISL_1640024, EPI_ISL_1640026, EPI_ISL_1640028, EPI_ISL_1640031, EPI_ISL_1640032, EPI_ISL_1640034, EPI_ISL_1640037, EPI_ISL_1640039, EPI_ISL_1640047, EPI_ISL_1640050, EPI_ISL_1640052, EPI_ISL_1640054, EPI_ISL_1640057, EPI_ISL_1640058, EPI_ISL_1640066, EPI_ISL_1641856, EPI_ISL_1641858, EPI_ISL_1641859, EPI_ISL_1641860, EPI_ISL_1641862, EPI_ISL_1641864, EPI_ISL_1641865, EPI_ISL_1641866, EPI_ISL_1641867, EPI_ISL_1641871, EPI_ISL_1641874, EPI_ISL_1641877, EPI_ISL_1641881, EPI_ISL_1641886, EPI_ISL_1641891, EPI_ISL_1641894, EPI_ISL_1641898, EPI_ISL_1641901, EPI_ISL_1641904, EPI_ISL_1641906, EPI_ISL_1641908, EPI_ISL_1641910, EPI_ISL_1641911, EPI_ISL_1641912, EPI_ISL_1641913, EPI_ISL_1641918, EPI_ISL_1641919, EPI_ISL_1641920, EPI_ISL_1641921, EPI_ISL_1641923, EPI_ISL_1641924, EPI_ISL_1641927, EPI_ISL_1641929, EPI_ISL_1641931, EPI_ISL_1641934, EPI_ISL_1641935, EPI_ISL_1641937, EPI_ISL_1641938, EPI_ISL_1641940, EPI_ISL_1641942, EPI_ISL_1641945, EPI_ISL_1641946, EPI_ISL_1641950, EPI_ISL_1641951, EPI_ISL_1641953, EPI_ISL_1641956, EPI_ISL_1641957, EPI_ISL_1641959, EPI_ISL_1641963, EPI_ISL_1641964, EPI_ISL_1641969, EPI_ISL_1641970, EPI_ISL_1641974, EPI_ISL_1641975, EPI_ISL_1641978, EPI_ISL_1641979, EPI_ISL_1641982, EPI_ISL_1641984, EPI_ISL_1641986, EPI_ISL_1641988, EPI_ISL_1641991, EPI_ISL_1641993, EPI_ISL_1641995, EPI_ISL_1641996, EPI_ISL_1641998, EPI_ISL_1642001, EPI_ISL_1642006, EPI_ISL_1642007, EPI_ISL_1642009, EPI_ISL_1642011, EPI_ISL_1642012, EPI_ISL_1642014, EPI_ISL_1642017, EPI_ISL_1642019, EPI_ISL_1642020, EPI_ISL_1642022, EPI_ISL_1642024, EPI_ISL_1642026, EPI_ISL_1642027, EPI_ISL_1642029, EPI_ISL_1642032, EPI_ISL_1642034, EPI_ISL_1642036, EPI_ISL_1642037, EPI_ISL_1642039, EPI_ISL_1642041, EPI_ISL_1642043, EPI_ISL_1642044, EPI_ISL_1642047, EPI_ISL_1642048, EPI_ISL_1642051, EPI_ISL_1642053, EPI_ISL_1642055, EPI_ISL_1642056, EPI_ISL_1642058, EPI_ISL_1642061, EPI_ISL_1642063, EPI_ISL_1642065, EPI_ISL_1642066, EPI_ISL_1642067, EPI_ISL_1642068, EPI_ISL_1642070, EPI_ISL_1642071, EPI_ISL_1642073, EPI_ISL_1642074, EPI_ISL_1642078, EPI_ISL_1642080, EPI_ISL_1642082, EPI_ISL_1642084, EPI_ISL_1642085, EPI_ISL_1642087, EPI_ISL_1642088, EPI_ISL_1642091, EPI_ISL_1642092, EPI_ISL_1642094, EPI_ISL_1642095, EPI_ISL_1642096, EPI_ISL_1642098, EPI_ISL_1642099, EPI_ISL_1642100, EPI_ISL_1642104, EPI_ISL_1642105, EPI_ISL_1642107, EPI_ISL_1642109, EPI_ISL_1642113, EPI_ISL_1642115, EPI_ISL_1642117, EPI_ISL_1642118, EPI_ISL_1642119, EPI_ISL_1642120, EPI_ISL_1642127, EPI_ISL_1642128, EPI_ISL_1642132, EPI_ISL_1642133, EPI_ISL_1642136, EPI_ISL_1642137, EPI_ISL_1642138, EPI_ISL_1642139, EPI_ISL_1642146, EPI_ISL_1642147, EPI_ISL_1642148, EPI_ISL_1642149, EPI_ISL_1642166, EPI_ISL_1642169, EPI_ISL_1642172, EPI_ISL_1642174, EPI_ISL_1642178, EPI_ISL_1642181, EPI_ISL_1642183, EPI_ISL_1642185, EPI_ISL_1642186, EPI_ISL_1642188, EPI_ISL_1642192, EPI_ISL_1642194, EPI_ISL_1642196, EPI_ISL_1642198, EPI_ISL_1642200, EPI_ISL_1642202, EPI_ISL_1642203, EPI_ISL_1642205, EPI_ISL_1642206, EPI_ISL_1642212, EPI_ISL_1642217, EPI_ISL_1642218, EPI_ISL_1642220, EPI_ISL_1642224, EPI_ISL_1642228, EPI_ISL_1642235, EPI_ISL_1642239, EPI_ISL_1642251, EPI_ISL_1642254, EPI_ISL_1642257, EPI_ISL_1642260, EPI_ISL_1642261, EPI_ISL_1642262, EPI_ISL_1642263, EPI_ISL_1642273, EPI_ISL_1642275, EPI_ISL_1642276, EPI_ISL_1645935, EPI_ISL_1645944, EPI_ISL_1645948, EPI_ISL_1645950, EPI_ISL_1645952, EPI_ISL_1645956, EPI_ISL_1645963, EPI_ISL_1645977, EPI_ISL_1645981, EPI_ISL_1645986, EPI_ISL_1645988, EPI_ISL_1645995, EPI_ISL_1645998, EPI_ISL_1646000, EPI_ISL_1646003, EPI_ISL_1646005, EPI_ISL_1646011, EPI_ISL_1646013, EPI_ISL_1646019, EPI_ISL_1646025, EPI_ISL_1646027, EPI_ISL_1646031, EPI_ISL_1646034, EPI_ISL_1646039, EPI_ISL_1646047, EPI_ISL_1646066, EPI_ISL_1646068, EPI_ISL_1646072, EPI_ISL_1646076, EPI_ISL_1646077, EPI_ISL_1646080, EPI_ISL_1646086, EPI_ISL_1646093, EPI_ISL_1646111, EPI_ISL_1646124, EPI_ISL_1646134, EPI_ISL_1646142, EPI_ISL_1646155, EPI_ISL_1646156, EPI_ISL_1646161, EPI_ISL_1646167, EPI_ISL_1646173, EPI_ISL_1646175, EPI_ISL_1646183, EPI_ISL_1646202, EPI_ISL_1646205, EPI_ISL_1646206, EPI_ISL_1646215, EPI_ISL_1646221, EPI_ISL_1646227, EPI_ISL_1646233, EPI_ISL_1646239, EPI_ISL_1646241, EPI_ISL_1646245, EPI_ISL_1646255, EPI_ISL_1646260, EPI_ISL_1646268, EPI_ISL_1646272, EPI_ISL_1646278, EPI_ISL_1646288, EPI_ISL_1646290, EPI_ISL_1646295, EPI_ISL_1646298, EPI_ISL_1646305, EPI_ISL_1646309, EPI_ISL_1646316, EPI_ISL_1646322, EPI_ISL_1646330, EPI_ISL_1646337, EPI_ISL_1646339, EPI_ISL_1646344, EPI_ISL_1646346, EPI_ISL_1646350, EPI_ISL_1646359, EPI_ISL_1646361, EPI_ISL_1646364, EPI_ISL_1646372, EPI_ISL_1646374, EPI_ISL_1646376, EPI_ISL_1646389, EPI_ISL_1646402, EPI_ISL_1646403, EPI_ISL_1646405, EPI_ISL_1646424, EPI_ISL_1646428, EPI_ISL_1646444, EPI_ISL_1646457, EPI_ISL_1646465, EPI_ISL_1646467, EPI_ISL_1646487, EPI_ISL_1646495, EPI_ISL_1646510, EPI_ISL_1646519, EPI_ISL_1646524, EPI_ISL_1646528, EPI_ISL_1646535, EPI_ISL_1646538, EPI_ISL_1646544, EPI_ISL_1646546, EPI_ISL_1646548, EPI_ISL_1646556, EPI_ISL_1646558, EPI_ISL_1646569, EPI_ISL_1646571, EPI_ISL_1646579, EPI_ISL_1646581, EPI_ISL_1646587, EPI_ISL_1646590, EPI_ISL_1646599, EPI_ISL_1646603, EPI_ISL_1646607, EPI_ISL_1646612, EPI_ISL_1646614, EPI_ISL_1646618, EPI_ISL_1646624, EPI_ISL_1646626, EPI_ISL_1646628, EPI_ISL_1646630, EPI_ISL_1646632, EPI_ISL_1646641, EPI_ISL_1646643, EPI_ISL_1646645, EPI_ISL_1646647, EPI_ISL_1646649, EPI_ISL_1646653, EPI_ISL_1646655, EPI_ISL_1646657, EPI_ISL_1646664, EPI_ISL_1646666, EPI_ISL_1646685, EPI_ISL_1646687, EPI_ISL_1646689, EPI_ISL_1646691, EPI_ISL_1646699, EPI_ISL_1646704, EPI_ISL_1646708, EPI_ISL_1646716, EPI_ISL_1646718, EPI_ISL_1646737, EPI_ISL_1646756, EPI_ISL_1646758, EPI_ISL_1646763, EPI_ISL_1646765, EPI_ISL_1646767, EPI_ISL_1646776, EPI_ISL_1646782, EPI_ISL_1646784, EPI_ISL_1646786, EPI_ISL_1646796, EPI_ISL_1646803, EPI_ISL_1646813, EPI_ISL_1646815, EPI_ISL_1646820, EPI_ISL_1646822, EPI_ISL_1646824, EPI_ISL_1646829, EPI_ISL_1646833, EPI_ISL_1646837, EPI_ISL_1646843, EPI_ISL_1646845, EPI_ISL_1646853, EPI_ISL_1646855, EPI_ISL_1646857, EPI_ISL_1646861, EPI_ISL_1646865, EPI_ISL_1646886, EPI_ISL_1719919, EPI_ISL_1719920, EPI_ISL_1719929, EPI_ISL_1719942, EPI_ISL_1719947, EPI_ISL_1719952, EPI_ISL_1719963, EPI_ISL_1719971, EPI_ISL_1719973, EPI_ISL_1719979, EPI_ISL_1719981, EPI_ISL_1719982, EPI_ISL_1719986, EPI_ISL_1719992, EPI_ISL_1719994, EPI_ISL_1720007, EPI_ISL_1720011, EPI_ISL_1720019, EPI_ISL_1720025, EPI_ISL_1720027, EPI_ISL_1720029, EPI_ISL_1720032, EPI_ISL_1720035, EPI_ISL_1720038, EPI_ISL_1720039, EPI_ISL_1720042, EPI_ISL_1720046, EPI_ISL_1720060, EPI_ISL_1720064, EPI_ISL_1720065, EPI_ISL_1720069, EPI_ISL_1720074, EPI_ISL_1720078, EPI_ISL_1720081, EPI_ISL_1720083, EPI_ISL_1720085, EPI_ISL_1720089, EPI_ISL_1720090, EPI_ISL_1720095, EPI_ISL_1720096, EPI_ISL_1720101, EPI_ISL_1720107, EPI_ISL_1720115, EPI_ISL_1720116, EPI_ISL_1720118, EPI_ISL_1720122, EPI_ISL_1720125, EPI_ISL_1720126, EPI_ISL_1720134, EPI_ISL_1720147, EPI_ISL_1720149, EPI_ISL_1720158, EPI_ISL_1720166, EPI_ISL_1720167, EPI_ISL_1720168, EPI_ISL_1720169, EPI_ISL_1720171, EPI_ISL_1722866, EPI_ISL_1722868, EPI_ISL_1722869, EPI_ISL_1722872, EPI_ISL_1722873, EPI_ISL_1722877, EPI_ISL_1722878, EPI_ISL_1722879, EPI_ISL_1722882, EPI_ISL_1722883, EPI_ISL_1722884, EPI_ISL_1722886, EPI_ISL_1722889, EPI_ISL_1722890, EPI_ISL_1722891, EPI_ISL_1722893, EPI_ISL_1722896, EPI_ISL_1722897, EPI_ISL_1722899, EPI_ISL_1722907, EPI_ISL_1722910, EPI_ISL_1722912, EPI_ISL_1722913, EPI_ISL_1722918, EPI_ISL_1722920, EPI_ISL_1728662, EPI_ISL_1728667, EPI_ISL_1729752, EPI_ISL_1729756, EPI_ISL_1729807, EPI_ISL_1729819, EPI_ISL_1729858, EPI_ISL_1729902, EPI_ISL_1729909, EPI_ISL_1729928, EPI_ISL_2108013, EPI_ISL_2108130, EPI_ISL_2108137, EPI_ISL_2108154, EPI_ISL_2108158, EPI_ISL_2108182, EPI_ISL_2108206, EPI_ISL_2108222, EPI_ISL_2119310, EPI_ISL_2119866, EPI_ISL_2119885 | | |
| see above | SYNLAB MVZ Leverkusen | Robert Koch Institute |
| EPI_ISL_1567718, EPI_ISL_1568683, EPI_ISL_1639531, EPI_ISL_1639533, EPI_ISL_1639538, EPI_ISL_1639544, EPI_ISL_1639547, EPI_ISL_1639550, EPI_ISL_1639552, EPI_ISL_1639555, EPI_ISL_1639558, EPI_ISL_1639567, EPI_ISL_1639584, EPI_ISL_1639586, EPI_ISL_1639594, EPI_ISL_1639599, EPI_ISL_1639606, EPI_ISL_1639613, EPI_ISL_1639628, EPI_ISL_1639636, EPI_ISL_1639638, EPI_ISL_1639642, EPI_ISL_1639646, EPI_ISL_1639662, EPI_ISL_1639682, EPI_ISL_1639689, EPI_ISL_1639690, EPI_ISL_1639717, EPI_ISL_1639725, EPI_ISL_1639727, EPI_ISL_1639783, EPI_ISL_1639790, EPI_ISL_1639815, EPI_ISL_1639888, EPI_ISL_1639897, EPI_ISL_1639924, EPI_ISL_1639955, EPI_ISL_1640025, EPI_ISL_1640029, EPI_ISL_1640043, EPI_ISL_1640051, EPI_ISL_1640059, EPI_ISL_1641869, EPI_ISL_1641872, EPI_ISL_1641875, EPI_ISL_1641878, EPI_ISL_1641883, EPI_ISL_1641884, EPI_ISL_1641889, EPI_ISL_1641896, EPI_ISL_1641899, EPI_ISL_1641902, EPI_ISL_1641905, EPI_ISL_1641907, EPI_ISL_1641914, EPI_ISL_1641965, EPI_ISL_1641968, EPI_ISL_1641976, EPI_ISL_1641980, EPI_ISL_1641987, EPI_ISL_1642062, EPI_ISL_1642121, EPI_ISL_1642126, EPI_ISL_1642129, EPI_ISL_1642142, EPI_ISL_1642152, EPI_ISL_1642157, EPI_ISL_1642159, EPI_ISL_1642161, EPI_ISL_1642171, EPI_ISL_1642187, EPI_ISL_1642195, EPI_ISL_1642211, EPI_ISL_1642225, EPI_ISL_1642230, EPI_ISL_1642233, EPI_ISL_1642245, EPI_ISL_1642249, EPI_ISL_1642259, EPI_ISL_1646231, EPI_ISL_1646393, EPI_ISL_1646474, EPI_ISL_1646499 | | |
| see above | SYNLAB MVZ Neustadt a. d. WeinstraÃŸe | Robert Koch Institute |
| EPI_ISL_2128039 | SYNLAB MVZ Neustadt a. d. Weinstraße | Robert Koch Institute |
| EPI_ISL_1440280, EPI_ISL_1440288, EPI_ISL_1440293, EPI_ISL_1440300, EPI_ISL_1440306, EPI_ISL_1440311, EPI_ISL_1440329, EPI_ISL_1440330, EPI_ISL_1440342, EPI_ISL_1440347, EPI_ISL_1440353, EPI_ISL_1440368, EPI_ISL_1440380, EPI_ISL_1440381, EPI_ISL_1440387, EPI_ISL_1440388, EPI_ISL_1440391, EPI_ISL_1440397, EPI_ISL_1440405, EPI_ISL_1440407, EPI_ISL_1440408, EPI_ISL_1440410, EPI_ISL_1440415, EPI_ISL_1440420, EPI_ISL_1440431, EPI_ISL_1440433, EPI_ISL_1440435, EPI_ISL_1440448, EPI_ISL_1440450, EPI_ISL_1440453, EPI_ISL_1440466, EPI_ISL_1440473, EPI_ISL_1440476, EPI_ISL_1440491, EPI_ISL_1440495, EPI_ISL_1440499, EPI_ISL_1440503, EPI_ISL_1440504, EPI_ISL_1440505, EPI_ISL_1440506, EPI_ISL_1440509, EPI_ISL_1440511, EPI_ISL_1440513, EPI_ISL_1440514, EPI_ISL_1440519, EPI_ISL_1440526, EPI_ISL_1440528, EPI_ISL_1440541, EPI_ISL_1440543, EPI_ISL_1440547, EPI_ISL_1440552, EPI_ISL_1440553, EPI_ISL_1440555, EPI_ISL_1440565, EPI_ISL_1442586, EPI_ISL_1442587, EPI_ISL_1442589, EPI_ISL_1442590, EPI_ISL_1442591, EPI_ISL_1442592, EPI_ISL_1442594, EPI_ISL_1442595, EPI_ISL_1442596, EPI_ISL_1442597, EPI_ISL_1442599, EPI_ISL_1442601, EPI_ISL_1442602, EPI_ISL_1442603, EPI_ISL_1442605, EPI_ISL_1442607, EPI_ISL_1442610, EPI_ISL_1442615, EPI_ISL_1442618, EPI_ISL_1442619, EPI_ISL_1442620, EPI_ISL_1442621, EPI_ISL_1442623, EPI_ISL_1442624, EPI_ISL_1442625, EPI_ISL_1442627, EPI_ISL_1442629, EPI_ISL_1442630, EPI_ISL_1442632, EPI_ISL_1442635, EPI_ISL_1442637, EPI_ISL_1442638, EPI_ISL_1442640, EPI_ISL_1442642, EPI_ISL_1442643, EPI_ISL_1442645, EPI_ISL_1442647, EPI_ISL_1442648, EPI_ISL_1442649, EPI_ISL_1442651, EPI_ISL_1442652, EPI_ISL_1442653, EPI_ISL_1442655, EPI_ISL_1442657, EPI_ISL_1442659, EPI_ISL_1442665, EPI_ISL_1442668, EPI_ISL_1442669, EPI_ISL_1442670, EPI_ISL_1442672, EPI_ISL_1442673, EPI_ISL_1442674, EPI_ISL_1442675, EPI_ISL_1442677, EPI_ISL_1442679, EPI_ISL_1442681, EPI_ISL_1442683, EPI_ISL_1442684, EPI_ISL_1442685, EPI_ISL_1442686, EPI_ISL_1442687, EPI_ISL_1442688, EPI_ISL_1442689, EPI_ISL_1442690, EPI_ISL_1442691, EPI_ISL_1442692, EPI_ISL_1442693, EPI_ISL_1442694, EPI_ISL_1442695, EPI_ISL_1442696, EPI_ISL_1442698, EPI_ISL_1442699, EPI_ISL_1442700, EPI_ISL_1442701, EPI_ISL_1442702, EPI_ISL_1442703, EPI_ISL_1442704, EPI_ISL_1442705, EPI_ISL_1442706, EPI_ISL_1442708, EPI_ISL_1442710, EPI_ISL_1442713, EPI_ISL_1442716, EPI_ISL_1442717, EPI_ISL_1442718, EPI_ISL_1442719, EPI_ISL_1442720, EPI_ISL_1442721, EPI_ISL_1442722, EPI_ISL_1442723, EPI_ISL_1442724, EPI_ISL_1442726, EPI_ISL_1442729, EPI_ISL_1442730, EPI_ISL_1442733, EPI_ISL_1442734, EPI_ISL_1442735, EPI_ISL_1442736, EPI_ISL_1442737, EPI_ISL_1442738, EPI_ISL_1442739, EPI_ISL_1442740, EPI_ISL_1442741, EPI_ISL_1442742, EPI_ISL_1442743, EPI_ISL_1442745, EPI_ISL_1442746, EPI_ISL_1442747, EPI_ISL_1442748, EPI_ISL_1442749, EPI_ISL_1442750, EPI_ISL_1566390, EPI_ISL_1566391, EPI_ISL_1566392, EPI_ISL_1566393, EPI_ISL_1566394, EPI_ISL_1566395, EPI_ISL_1566396, EPI_ISL_1566397, EPI_ISL_1566398, EPI_ISL_1566400, EPI_ISL_1566401, EPI_ISL_1566402, EPI_ISL_1566403, EPI_ISL_1566404, EPI_ISL_1566405, EPI_ISL_1566406, EPI_ISL_1566407, EPI_ISL_1566409, EPI_ISL_1566410, EPI_ISL_1566411, EPI_ISL_1566412, EPI_ISL_1566413, EPI_ISL_1566414, EPI_ISL_1566415, EPI_ISL_1566418, EPI_ISL_1566419, EPI_ISL_1566420, EPI_ISL_1566421, EPI_ISL_1566422, EPI_ISL_1566423, EPI_ISL_1566424, EPI_ISL_1566427, EPI_ISL_1566428, EPI_ISL_1566430, EPI_ISL_1566431, EPI_ISL_1566432, EPI_ISL_1566433, EPI_ISL_1566434, EPI_ISL_1566435, EPI_ISL_1566436, EPI_ISL_1566438, EPI_ISL_1566440, EPI_ISL_1566441, EPI_ISL_1566442, EPI_ISL_1566443, EPI_ISL_1566444, EPI_ISL_1566445, EPI_ISL_1566446, EPI_ISL_1566447, EPI_ISL_1566448, EPI_ISL_1566451, EPI_ISL_1566452, EPI_ISL_1566453, EPI_ISL_1566454, EPI_ISL_1566455, EPI_ISL_1566456, EPI_ISL_1566457, EPI_ISL_1566458, EPI_ISL_1566459, EPI_ISL_1566461, EPI_ISL_1566463, EPI_ISL_1566464, EPI_ISL_1566466, EPI_ISL_1566468, EPI_ISL_1566471, EPI_ISL_1566472, EPI_ISL_1566474, EPI_ISL_1566475, EPI_ISL_1566477, EPI_ISL_1566478, EPI_ISL_1566479, EPI_ISL_1566480, EPI_ISL_1566481, EPI_ISL_1566482, EPI_ISL_1566484, EPI_ISL_1566485, EPI_ISL_1566489, EPI_ISL_1566490, EPI_ISL_1566491, EPI_ISL_1566492, EPI_ISL_1566494, EPI_ISL_1566495, EPI_ISL_1566496, EPI_ISL_1566499, EPI_ISL_1566500, EPI_ISL_1566501, EPI_ISL_1566502, EPI_ISL_1566503, EPI_ISL_1566504, EPI_ISL_1566505, EPI_ISL_1566506, EPI_ISL_1566507, EPI_ISL_1566509, EPI_ISL_1566510, EPI_ISL_1566511, EPI_ISL_1566512, EPI_ISL_1566513, EPI_ISL_1566515, EPI_ISL_1566516, EPI_ISL_1566517, EPI_ISL_1566518, EPI_ISL_1566519, EPI_ISL_1566520, EPI_ISL_1566521, EPI_ISL_1566522, EPI_ISL_1566523, EPI_ISL_1566524, EPI_ISL_1566527, EPI_ISL_1566531, EPI_ISL_1566532, EPI_ISL_1566533, EPI_ISL_1566534, EPI_ISL_1566535, EPI_ISL_1566536, EPI_ISL_1566537, EPI_ISL_1566538, EPI_ISL_1566540, EPI_ISL_1566542, EPI_ISL_1566543, EPI_ISL_1566544, EPI_ISL_1566545, EPI_ISL_1566546, EPI_ISL_1566547, EPI_ISL_1566548, EPI_ISL_1566549, EPI_ISL_1566551, EPI_ISL_1566552, EPI_ISL_1566553, EPI_ISL_1566554, EPI_ISL_1566555, EPI_ISL_1566557, EPI_ISL_1566558, EPI_ISL_1566559, EPI_ISL_1566560, EPI_ISL_1566561, EPI_ISL_1566563, EPI_ISL_1566564, EPI_ISL_1566567, EPI_ISL_1566568, EPI_ISL_1566569, EPI_ISL_1566571, EPI_ISL_1566572, EPI_ISL_1566573, EPI_ISL_1566574, EPI_ISL_1566575, EPI_ISL_1566576, EPI_ISL_1566579, EPI_ISL_1566580, EPI_ISL_1566581, EPI_ISL_1566582, EPI_ISL_1566583, EPI_ISL_1566584, EPI_ISL_1566585, EPI_ISL_1566586, EPI_ISL_1566589, EPI_ISL_1566590, EPI_ISL_1566591, EPI_ISL_1566592, EPI_ISL_1566593, EPI_ISL_1566596, EPI_ISL_1566597, EPI_ISL_1566599, EPI_ISL_1566600, EPI_ISL_1566601, EPI_ISL_1566602, EPI_ISL_1566605, EPI_ISL_1566606, EPI_ISL_1566607, EPI_ISL_1566610, EPI_ISL_1566612, EPI_ISL_1566613, EPI_ISL_1566614, EPI_ISL_1566615, EPI_ISL_1566616, EPI_ISL_1566618, EPI_ISL_1566619, EPI_ISL_1566620, EPI_ISL_1566622, EPI_ISL_1566624, EPI_ISL_1566626, EPI_ISL_1566627, EPI_ISL_1566628, EPI_ISL_1566629, EPI_ISL_1566630, EPI_ISL_1566632, EPI_ISL_1566633, EPI_ISL_1566634, EPI_ISL_1566635, EPI_ISL_1566637, EPI_ISL_1566638, EPI_ISL_1566641, EPI_ISL_1566643, EPI_ISL_1566644, EPI_ISL_1566645, EPI_ISL_1566646, EPI_ISL_1566648, EPI_ISL_1566649, EPI_ISL_1566650, EPI_ISL_1566652, EPI_ISL_1566653, EPI_ISL_1566655, EPI_ISL_1566656, EPI_ISL_1566657, EPI_ISL_1566660, EPI_ISL_1566661, EPI_ISL_1566662, EPI_ISL_1566663, EPI_ISL_1566664, EPI_ISL_1566665, EPI_ISL_1566666, EPI_ISL_1566667, EPI_ISL_1566668, EPI_ISL_1566669, EPI_ISL_1566670, EPI_ISL_1566672, EPI_ISL_1566673, EPI_ISL_1566674, EPI_ISL_1566675, EPI_ISL_1566676, EPI_ISL_1566678, EPI_ISL_1566679, EPI_ISL_1566680, EPI_ISL_1566681, EPI_ISL_1566682, EPI_ISL_1566683, EPI_ISL_1566684, EPI_ISL_1566685, EPI_ISL_1566686, EPI_ISL_1566687, EPI_ISL_1566688, EPI_ISL_1566689, EPI_ISL_1566691, EPI_ISL_1566692, EPI_ISL_1566693, EPI_ISL_1566695, EPI_ISL_1568348 | | |
| see above | SYNLAB MVZ Trier | Robert Koch Institute |
| EPI_ISL_1440277, EPI_ISL_1440285, EPI_ISL_1440286, EPI_ISL_1440291, EPI_ISL_1440317, EPI_ISL_1440325, EPI_ISL_1440326, EPI_ISL_1440340, EPI_ISL_1440341, EPI_ISL_1440360, EPI_ISL_1440363, EPI_ISL_1440375, EPI_ISL_1440376, EPI_ISL_1440377, EPI_ISL_1440392, EPI_ISL_1440417, EPI_ISL_1440423, EPI_ISL_1440424, EPI_ISL_1440427, EPI_ISL_1440428, EPI_ISL_1440434, EPI_ISL_1440436, EPI_ISL_1440437, EPI_ISL_1440441, EPI_ISL_1440449, EPI_ISL_1440457, EPI_ISL_1440468, EPI_ISL_1440469, EPI_ISL_1440472, EPI_ISL_1440481, EPI_ISL_1440483, EPI_ISL_1440489, EPI_ISL_1440493, EPI_ISL_1440556, EPI_ISL_1440570, EPI_ISL_1440576, EPI_ISL_1442604, EPI_ISL_1442609, EPI_ISL_1442613, EPI_ISL_1442614, EPI_ISL_1442633, EPI_ISL_1442663, EPI_ISL_1442711, EPI_ISL_1442732, EPI_ISL_1442744, EPI_ISL_1566399, EPI_ISL_1566416, EPI_ISL_1566429, EPI_ISL_1566437, EPI_ISL_1566449, EPI_ISL_1566450, EPI_ISL_1566462, EPI_ISL_1566465, EPI_ISL_1566498, EPI_ISL_1566514, EPI_ISL_1566530, EPI_ISL_1566541, EPI_ISL_1566550, EPI_ISL_1566556, EPI_ISL_1566562, EPI_ISL_1566565, EPI_ISL_1566570, EPI_ISL_1566578, EPI_ISL_1566587, EPI_ISL_1566594, EPI_ISL_1566609, EPI_ISL_1566611, EPI_ISL_1566625, EPI_ISL_1566631, EPI_ISL_1566639, EPI_ISL_1566640, EPI_ISL_1566647, EPI_ISL_1566654, EPI_ISL_1566671, EPI_ISL_1566694, EPI_ISL_1566696, EPI_ISL_1568259, EPI_ISL_1568264, EPI_ISL_1568271, EPI_ISL_1568276, EPI_ISL_1568286, EPI_ISL_1568292, EPI_ISL_1568295, EPI_ISL_1568299, EPI_ISL_1568302, EPI_ISL_1568306, EPI_ISL_1568314, EPI_ISL_1568318, EPI_ISL_1568321, EPI_ISL_1568327, EPI_ISL_1568338, EPI_ISL_1568342, EPI_ISL_1568345, EPI_ISL_1568350, EPI_ISL_1568351, EPI_ISL_1568357, EPI_ISL_1568361, EPI_ISL_1568363, EPI_ISL_1568372, EPI_ISL_1568379, EPI_ISL_1568385, EPI_ISL_1568389, EPI_ISL_1568390, EPI_ISL_1568399, EPI_ISL_1568413, EPI_ISL_1568414, EPI_ISL_1568415, EPI_ISL_1568419, EPI_ISL_1568425, EPI_ISL_1568439, EPI_ISL_1568449, EPI_ISL_1568453, EPI_ISL_1568456, EPI_ISL_1568462, EPI_ISL_1568466, EPI_ISL_1568503, EPI_ISL_1568507, EPI_ISL_1568511 | | |
| see above | SYNLAB MVZ Weiden | Robert Koch Institute |
| EPI_ISL_1569299, EPI_ISL_1638749, EPI_ISL_1638777, EPI_ISL_1638803, EPI_ISL_1638807, EPI_ISL_1638867, EPI_ISL_1638897, EPI_ISL_1638902, EPI_ISL_1638945, EPI_ISL_1639099, EPI_ISL_1639101, EPI_ISL_1639127, EPI_ISL_1639219, EPI_ISL_1639248, EPI_ISL_1642424, EPI_ISL_1642504, EPI_ISL_1642549, EPI_ISL_1642554, EPI_ISL_1642566, EPI_ISL_1642575, EPI_ISL_1642583, EPI_ISL_1642671, EPI_ISL_1643240, EPI_ISL_1643334, EPI_ISL_1643366, EPI_ISL_1721129, EPI_ISL_1721218, EPI_ISL_1721248, EPI_ISL_1721295, EPI_ISL_1721587, EPI_ISL_1847286 | | |
| see above | Sonic - Bioscientia - MVZ Labor Saar GmbH | Robert Koch Institute |
| EPI_ISL_1638687, EPI_ISL_1638690, EPI_ISL_1638692, EPI_ISL_1638719, EPI_ISL_1638730, EPI_ISL_1638733, EPI_ISL_1638746, EPI_ISL_1638768, EPI_ISL_1638794, EPI_ISL_1638831, EPI_ISL_1638866, EPI_ISL_1638895, EPI_ISL_1638952, EPI_ISL_1638962 | | |
| see above | Sonic - Dr. Staber & Kollegen GmbH MÃ¼nchen | Robert Koch Institute |
| EPI_ISL_1439233, EPI_ISL_1439342 | Sonic - Dr. Staber & Kollegen GmbH München | Robert Koch Institute |
| EPI_ISL_1569275, EPI_ISL_1569291, EPI_ISL_1569294, EPI_ISL_1569303, EPI_ISL_1569308, EPI_ISL_1569309, EPI_ISL_1569310, EPI_ISL_1569328, EPI_ISL_1569330, EPI_ISL_1569335, EPI_ISL_1569337, EPI_ISL_1569346, EPI_ISL_1569350, EPI_ISL_1569354, EPI_ISL_1569365, EPI_ISL_1569369, EPI_ISL_1569372, EPI_ISL_1569380, EPI_ISL_1569383, EPI_ISL_1569406, EPI_ISL_1569407, EPI_ISL_1569408, EPI_ISL_1569414, EPI_ISL_1569422, EPI_ISL_1569434, EPI_ISL_1569437, EPI_ISL_1569440, EPI_ISL_1569449, EPI_ISL_1569451, EPI_ISL_1569454, EPI_ISL_1569455, EPI_ISL_1569468, EPI_ISL_1569471, EPI_ISL_1569472, EPI_ISL_1569473, EPI_ISL_1569475, EPI_ISL_1569479, EPI_ISL_1569482, EPI_ISL_1569493, EPI_ISL_1569506, EPI_ISL_1569509, EPI_ISL_1569512, EPI_ISL_1569513, EPI_ISL_1569518, EPI_ISL_1569535, EPI_ISL_1569542, EPI_ISL_1569545, EPI_ISL_1569551, EPI_ISL_1569553, EPI_ISL_1569563, EPI_ISL_1569567, EPI_ISL_1569577, EPI_ISL_1569579, EPI_ISL_1569609, EPI_ISL_1569630, EPI_ISL_1569658, EPI_ISL_1569668, EPI_ISL_1569670, EPI_ISL_1569692, EPI_ISL_1569694, EPI_ISL_1569700, EPI_ISL_1638970, EPI_ISL_1639012, EPI_ISL_1639057, EPI_ISL_1639120, EPI_ISL_1639143, EPI_ISL_1639153, EPI_ISL_1639194, EPI_ISL_1639215, EPI_ISL_1639217, EPI_ISL_1639235 | | |
| see above | Sonic - Labor Dr. von Froreich GmbH | Robert Koch Institute |
| EPI_ISL_1439197, EPI_ISL_1439285, EPI_ISL_1638700, EPI_ISL_1638743, EPI_ISL_1638750, EPI_ISL_1638757, EPI_ISL_1638791, EPI_ISL_1638796, EPI_ISL_1638839, EPI_ISL_1638845, EPI_ISL_1638861, EPI_ISL_1638863, EPI_ISL_1638881, EPI_ISL_1638918, EPI_ISL_1638922, EPI_ISL_1638960, EPI_ISL_1642483, EPI_ISL_1642529 | | |
| see above | Sonic - Labor Staber Bayreuth | Robert Koch Institute |
| EPI_ISL_1439245, EPI_ISL_1638688, EPI_ISL_1638715, EPI_ISL_1638760, EPI_ISL_1638764, EPI_ISL_1638765, EPI_ISL_1638792, EPI_ISL_1638797, EPI_ISL_1638808, EPI_ISL_1638825, EPI_ISL_1638888, EPI_ISL_1638890, EPI_ISL_1638955, EPI_ISL_1638956, EPI_ISL_1638963, EPI_ISL_1642528 | | |
| see above | Sonic - Labor Staber Dresden (Klipphausen) | Robert Koch Institute |
| EPI_ISL_1638699, EPI_ISL_1638711, EPI_ISL_1638722, EPI_ISL_1638736, EPI_ISL_1638815, EPI_ISL_1638816, EPI_ISL_1638838, EPI_ISL_1638896, EPI_ISL_1638926 | | |
| see above | Sonic - Labor Staber Heilbronn | Robert Koch Institute |
| EPI_ISL_1569304, EPI_ISL_1569427, EPI_ISL_1569500, EPI_ISL_1638776, EPI_ISL_1643187 | Sonic - Labor Staber MVZ Kassel | Robert Koch Institute |
| EPI_ISL_1638781, EPI_ISL_1638806, EPI_ISL_1638941, EPI_ISL_1642428, EPI_ISL_1721197, EPI_ISL_1721378 | Sonic - Labor Staber NÃ¼rnberg | Robert Koch Institute |
| EPI_ISL_1439281 | Sonic - Labor Staber Nürnberg | Robert Koch Institute |
| EPI_ISL_1568974, EPI_ISL_1568975, EPI_ISL_1568976, EPI_ISL_1568997, EPI_ISL_1569001, EPI_ISL_1569005, EPI_ISL_1569007, EPI_ISL_1569014, EPI_ISL_1569015, EPI_ISL_1569023, EPI_ISL_1569024, EPI_ISL_1569034, EPI_ISL_1569038, EPI_ISL_1569050, EPI_ISL_1569052, EPI_ISL_1569059, EPI_ISL_1569062, EPI_ISL_1569066, EPI_ISL_1569083, EPI_ISL_1569098, EPI_ISL_1569102, EPI_ISL_1569139, EPI_ISL_1569179, EPI_ISL_1569191, EPI_ISL_1569214, EPI_ISL_1569219, EPI_ISL_1569222, EPI_ISL_1569225, EPI_ISL_1569227, EPI_ISL_1569281, EPI_ISL_1569282, EPI_ISL_1569283, EPI_ISL_1569284, EPI_ISL_1569287, EPI_ISL_1569288, EPI_ISL_1569292, EPI_ISL_1569293, EPI_ISL_1569301, EPI_ISL_1569305, EPI_ISL_1569306, EPI_ISL_1569311, EPI_ISL_1569312, EPI_ISL_1569314, EPI_ISL_1569319, EPI_ISL_1569321, EPI_ISL_1569326, EPI_ISL_1569329, EPI_ISL_1569338, EPI_ISL_1569340, EPI_ISL_1569341, EPI_ISL_1569343, EPI_ISL_1569349, EPI_ISL_1569352, EPI_ISL_1569355, EPI_ISL_1569357, EPI_ISL_1569358, EPI_ISL_1569360, EPI_ISL_1569361, EPI_ISL_1569362, EPI_ISL_1569364, EPI_ISL_1569366, EPI_ISL_1569373, EPI_ISL_1569374, EPI_ISL_1569381, EPI_ISL_1569382, EPI_ISL_1569384, EPI_ISL_1569385, EPI_ISL_1569387, EPI_ISL_1569395, EPI_ISL_1569398, EPI_ISL_1569400, EPI_ISL_1569404, EPI_ISL_1569412, EPI_ISL_1569413, EPI_ISL_1569415, EPI_ISL_1569416, EPI_ISL_1569418, EPI_ISL_1569420, EPI_ISL_1569421, EPI_ISL_1569425, EPI_ISL_1569426, EPI_ISL_1569431, EPI_ISL_1569432, EPI_ISL_1569433, EPI_ISL_1569435, EPI_ISL_1569438, EPI_ISL_1569439, EPI_ISL_1569443, EPI_ISL_1569444, EPI_ISL_1569446, EPI_ISL_1569447, EPI_ISL_1569448, EPI_ISL_1569460, EPI_ISL_1569462, EPI_ISL_1569463, EPI_ISL_1569466, EPI_ISL_1569467, EPI_ISL_1569476, EPI_ISL_1569477, EPI_ISL_1569478, EPI_ISL_1569483, EPI_ISL_1569484, EPI_ISL_1569485, EPI_ISL_1569486, EPI_ISL_1569488, EPI_ISL_1569489, EPI_ISL_1569492, EPI_ISL_1569495, EPI_ISL_1569496, EPI_ISL_1569502, EPI_ISL_1569508, EPI_ISL_1569511, EPI_ISL_1569515, EPI_ISL_1569531, EPI_ISL_1569540, EPI_ISL_1569547, EPI_ISL_1569565, EPI_ISL_1569585, EPI_ISL_1569596, EPI_ISL_1569599, EPI_ISL_1569612, EPI_ISL_1569621, EPI_ISL_1569624, EPI_ISL_1569632, EPI_ISL_1569636, EPI_ISL_1569639, EPI_ISL_1569642, EPI_ISL_1569651, EPI_ISL_1569653, EPI_ISL_1569655, EPI_ISL_1569662, EPI_ISL_1569665, EPI_ISL_1569672, EPI_ISL_1569679, EPI_ISL_1569682, EPI_ISL_1569684, EPI_ISL_1569696, EPI_ISL_1638990, EPI_ISL_1638999, EPI_ISL_1639006, EPI_ISL_1639011, EPI_ISL_1639014, EPI_ISL_1639024, EPI_ISL_1639025, EPI_ISL_1639026, EPI_ISL_1639027, EPI_ISL_1639030, EPI_ISL_1639036, EPI_ISL_1639037, EPI_ISL_1639041, EPI_ISL_1639082, EPI_ISL_1639107, EPI_ISL_1639145, EPI_ISL_1639148, EPI_ISL_1639154, EPI_ISL_1639166, EPI_ISL_1639171, EPI_ISL_1639182, EPI_ISL_1639183, EPI_ISL_1639190, EPI_ISL_1639201, EPI_ISL_1639216, EPI_ISL_1639224, EPI_ISL_1639236, EPI_ISL_1639240 | | |
| see above | Sonic - MVZ Medizinisches Labor Bremen GmbH | Robert Koch Institute |
| EPI_ISL_1569375, EPI_ISL_1569456, EPI_ISL_1569504, EPI_ISL_1642578, EPI_ISL_1642609, EPI_ISL_1642625 | Sonic - MVZ Medizinisches Labor Oldenburg | Robert Koch Institute |
| EPI_ISL_1441247, EPI_ISL_1441254, EPI_ISL_1441290, EPI_ISL_1441305 | Städtisches Klinikum Dresden | Robert Koch Institute |
| EPI_ISL_1440305, EPI_ISL_1440312, EPI_ISL_1440345, EPI_ISL_1440350, EPI_ISL_1440416, EPI_ISL_1440421, EPI_ISL_1440429, EPI_ISL_1440446, EPI_ISL_1440452, EPI_ISL_1440475, EPI_ISL_1440518, EPI_ISL_1440527, EPI_ISL_1440551, EPI_ISL_1440568, EPI_ISL_1442588, EPI_ISL_1442598, EPI_ISL_1442608, EPI_ISL_1442616, EPI_ISL_1442697, EPI_ISL_1442707, EPI_ISL_1442709, EPI_ISL_1442714, EPI_ISL_1442715, EPI_ISL_1442728, EPI_ISL_1566408, EPI_ISL_1566417, EPI_ISL_1566425, EPI_ISL_1566426, EPI_ISL_1566460, EPI_ISL_1566467, EPI_ISL_1566469, EPI_ISL_1566470, EPI_ISL_1566473, EPI_ISL_1566476, EPI_ISL_1566483, EPI_ISL_1566486, EPI_ISL_1566487, EPI_ISL_1566488, EPI_ISL_1566493, EPI_ISL_1566497, EPI_ISL_1566508, EPI_ISL_1566525, EPI_ISL_1566529, EPI_ISL_1566566, EPI_ISL_1566577, EPI_ISL_1566588, EPI_ISL_1566595, EPI_ISL_1566598, EPI_ISL_1566604, EPI_ISL_1566608, EPI_ISL_1566617, EPI_ISL_1566621, EPI_ISL_1566623, EPI_ISL_1566636, EPI_ISL_1566642, EPI_ISL_1566658, EPI_ISL_1566677, EPI_ISL_1568282, EPI_ISL_1568341, EPI_ISL_1568344, EPI_ISL_1568347, EPI_ISL_1568360, EPI_ISL_1568375, EPI_ISL_1568398, EPI_ISL_1568438, EPI_ISL_1568445, EPI_ISL_1568480, EPI_ISL_1568494, EPI_ISL_1568500 | | |
| see above | Synlab MVZ Augsburg | Robert Koch Institute |
| EPI_ISL_1572836, EPI_ISL_1572837, EPI_ISL_1572838, EPI_ISL_1722838 | ThÃ¼ringer Landesamtes fÃ¼r Verbraucherschutz | Robert Koch Institute |

| | | | |
|---|---|---|---|
| EPI_ISL_1441231, EPI_ISL_1441233, EPI_ISL_1441235, EPI_ISL_1441237, EPI_ISL_1441239, EPI_ISL_1441241, EPI_ISL_1441248, EPI_ISL_1441299 | | | |
| see above | Uniklinikum Carl Gustav Carus an der TU Dresden; Institut für Virologie | Robert Koch Institute | |
| EPI_ISL_1566000, EPI_ISL_1639254, EPI_ISL_1639255, EPI_ISL_1639256, EPI_ISL_1639257, EPI_ISL_1639258 | Uniklinikum Jena | Robert Koch Institute | |
| EPI_ISL_1569729, EPI_ISL_1569731, EPI_ISL_1569733, EPI_ISL_1569735, EPI_ISL_1569738, EPI_ISL_1569743, EPI_ISL_1569746, EPI_ISL_1569749, EPI_ISL_1569751, EPI_ISL_1569753, EPI_ISL_1638271, EPI_ISL_1722063, EPI_ISL_1722064, EPI_ISL_1722065 | | | |
| see above | Uniklinikum Regensburg; Institut fÃ¼r klinische Mikrobiologie und Hygiene | Robert Koch Institute | |
| EPI_ISL_1716071, EPI_ISL_1716072, EPI_ISL_1716073, EPI_ISL_1716074, EPI_ISL_1716265, EPI_ISL_1716266, EPI_ISL_1716271, EPI_ISL_1716272 | | | |
| see above | University Medical Center Hamburg Eppendorf | Heinrich Pette Institute, Leibniz Institute for Experimental Virology | Adam Grundhoff; Alexis Robitaille; Johannes Knobloch; Martin Aepfelbacher; Nicole Fischer; Thomas Günther |
| EPI_ISL_1572045, EPI_ISL_1572050, EPI_ISL_1572074, EPI_ISL_1572089, EPI_ISL_1638578, EPI_ISL_1638585, EPI_ISL_1638592, EPI_ISL_1638599, EPI_ISL_1638602, EPI_ISL_1638605, EPI_ISL_1638608, EPI_ISL_1638613, EPI_ISL_1638619, EPI_ISL_1638622, EPI_ISL_1638624 | | | |
| see above | UniversitÃ¤tsklinikum Bonn - Institut fÃ¼r Virologie | Robert Koch Institute | |
| EPI_ISL_1569280, EPI_ISL_1569296, EPI_ISL_1569300, EPI_ISL_1569371, EPI_ISL_1569442, EPI_ISL_1569494, EPI_ISL_1569602 | | | |
| see above | UniversitÃ¤tsklinikum Frankfurt - Institut fÃ¼r Medizinische Virologie | Robert Koch Institute | |
| EPI_ISL_1563934 | UniversitÃ¤tsklinikum GieÃŸen - Institut fÃ¼r Medizinische Virologie | Robert Koch Institute | |
| EPI_ISL_1570323, EPI_ISL_1570324, EPI_ISL_1570325, EPI_ISL_1570328, EPI_ISL_1570329, EPI_ISL_1570330, EPI_ISL_1640141, EPI_ISL_1640142, EPI_ISL_1640143, EPI_ISL_1640144, EPI_ISL_1640145, EPI_ISL_1640146, EPI_ISL_1640147, EPI_ISL_1640148, EPI_ISL_1640149 | | | |
| see above | UniversitÃ¤tsklinikum Heidelberg | Robert Koch Institute | |
| EPI_ISL_1567092, EPI_ISL_1567093, EPI_ISL_1567094, EPI_ISL_1567095, EPI_ISL_1567096, EPI_ISL_1567097, EPI_ISL_1567098, EPI_ISL_1567099, EPI_ISL_1567100, EPI_ISL_1567102, EPI_ISL_1567103, EPI_ISL_1567104, EPI_ISL_1567105, EPI_ISL_1567106, EPI_ISL_1567107, EPI_ISL_1567108, EPI_ISL_1567109, EPI_ISL_1567111, EPI_ISL_1567112, EPI_ISL_1567113, EPI_ISL_1567114, EPI_ISL_1567115, EPI_ISL_1567116 | | | |
| see above | UniversitÃ¤tsklinikum MÃ¼nster Institut fÃ¼r Virologie | Robert Koch Institute | |
| EPI_ISL_1572021, EPI_ISL_1572028, EPI_ISL_1572043, EPI_ISL_1572049, EPI_ISL_1572070, EPI_ISL_1572096, EPI_ISL_1572098 | | | |
| see above | UniversitÃ¤tsklinikum des Saarlandes - Institut fÃ¼r Virologie | Robert Koch Institute | |
| EPI_ISL_1640157, EPI_ISL_1640158, EPI_ISL_1640159, EPI_ISL_1640160, EPI_ISL_1640161, EPI_ISL_1725177, EPI_ISL_1725178, EPI_ISL_1725179, EPI_ISL_1725180, EPI_ISL_1725181 | | | |
| see above | UniversitÃ¤tsmedizin Mannheim | Robert Koch Institute | |
| EPI_ISL_1833952, EPI_ISL_1833994 | Universitätsklinikum Knappschaftskrankenhaus Bochum | Bundeswehr Institute of Microbiology | Alexandra Rehn; Enrico Georgi; Malena Bestehorn-Willmann; Markus Antwerpen; Mathias Walter; Mike Pillukat; Roman Wölfel; Sabine Zange |
| EPI_ISL_1355243, EPI_ISL_1355244, EPI_ISL_1355245, EPI_ISL_1355247, EPI_ISL_1355248, EPI_ISL_1355250, EPI_ISL_1355251, EPI_ISL_1355252, EPI_ISL_1355253, EPI_ISL_1355254, EPI_ISL_1355255, EPI_ISL_1355256, EPI_ISL_1355257, EPI_ISL_1355258, EPI_ISL_1355260, EPI_ISL_1355261, EPI_ISL_1355262, EPI_ISL_1355263, EPI_ISL_1355264, EPI_ISL_1355265, EPI_ISL_1355266, EPI_ISL_1355267, EPI_ISL_1355268, EPI_ISL_1355269, EPI_ISL_1355270, EPI_ISL_1355273, EPI_ISL_1355274, EPI_ISL_1355275, EPI_ISL_1355276, EPI_ISL_1355277, EPI_ISL_1355278, EPI_ISL_1355279, EPI_ISL_1355280, EPI_ISL_1355281, EPI_ISL_1355282, EPI_ISL_1355283, EPI_ISL_1355284, EPI_ISL_1356616, EPI_ISL_1356617, EPI_ISL_1434366, EPI_ISL_1434367, EPI_ISL_1434368, EPI_ISL_1434369, EPI_ISL_1434371, EPI_ISL_1434372, EPI_ISL_1434373, EPI_ISL_1434374, EPI_ISL_1434375, EPI_ISL_1434376, EPI_ISL_1434377, EPI_ISL_1434378, EPI_ISL_1434379, EPI_ISL_1434380, EPI_ISL_1434381, EPI_ISL_1434382, EPI_ISL_1434383, EPI_ISL_1434384, EPI_ISL_1434385, EPI_ISL_1434386, EPI_ISL_1434387, EPI_ISL_1434388, EPI_ISL_1434389, EPI_ISL_1434390, EPI_ISL_1434391, EPI_ISL_1434392, EPI_ISL_1434393, EPI_ISL_1434394, EPI_ISL_1434395, EPI_ISL_1434396, EPI_ISL_1434397, EPI_ISL_1434398, EPI_ISL_1434399, EPI_ISL_1434400, EPI_ISL_1434402, EPI_ISL_1434403, EPI_ISL_1434404, EPI_ISL_1434406, EPI_ISL_1434408, EPI_ISL_1434409, EPI_ISL_1434410, EPI_ISL_1434411, EPI_ISL_1434412, EPI_ISL_1434413, EPI_ISL_1434414, EPI_ISL_1434415, EPI_ISL_1434416, EPI_ISL_1434417, EPI_ISL_1434419, EPI_ISL_1434423, EPI_ISL_1434429, EPI_ISL_1434431, EPI_ISL_1434433, EPI_ISL_1434435 | | | |
| see above | Universitätsklinikum Leipzig - Institut für Medizinische Mikrobiologie und Virologie virologisches Labor | Robert Koch Institute | |
| EPI_ISL_1435943, EPI_ISL_1435961 | Universitätsklinikum Münster Institut für Virologie | Robert Koch Institute | |
| EPI_ISL_1439387, EPI_ISL_1439388, EPI_ISL_1439390, EPI_ISL_1439395, EPI_ISL_1439397, EPI_ISL_1439399, EPI_ISL_1439408, EPI_ISL_1439423, EPI_ISL_1439425 | | | |
| see above | Universitätsklinikum des Saarlandes - Institut für Virologie | Robert Koch Institute | |
| EPI_ISL_1563960, EPI_ISL_1563961, EPI_ISL_1563964, EPI_ISL_1563966, EPI_ISL_1563968, EPI_ISL_1563969, EPI_ISL_1563970, EPI_ISL_1563971, EPI_ISL_1563972 | | | |
| see above | Virologisches Institut des UniversitÃ¤tsklinikums Erlangen | Robert Koch Institute | |
| EPI_ISL_1394506, EPI_ISL_1394507, EPI_ISL_1394508, EPI_ISL_1394509, EPI_ISL_1394510, EPI_ISL_1394511, EPI_ISL_1394512, EPI_ISL_1394513, EPI_ISL_1394514, EPI_ISL_1394515, EPI_ISL_1394516, EPI_ISL_1394517, EPI_ISL_1394518, EPI_ISL_1394519, EPI_ISL_1394520, EPI_ISL_1394521, EPI_ISL_1394522, EPI_ISL_1394523, EPI_ISL_1394524, EPI_ISL_1394525, EPI_ISL_1394526, EPI_ISL_1394527, EPI_ISL_1394528, EPI_ISL_1394529, EPI_ISL_1394530, EPI_ISL_1394531, EPI_ISL_1394532, EPI_ISL_1394533, EPI_ISL_1394534, EPI_ISL_1394535, EPI_ISL_1394536, EPI_ISL_1394537, EPI_ISL_1394538, EPI_ISL_1394539, EPI_ISL_1577823, EPI_ISL_1577826, EPI_ISL_1577833, EPI_ISL_1577834, EPI_ISL_1577835, EPI_ISL_1577841, EPI_ISL_1577842, EPI_ISL_1577845, EPI_ISL_1577846, EPI_ISL_1577847, EPI_ISL_1577848, EPI_ISL_1577853, EPI_ISL_1577864, EPI_ISL_1577865, EPI_ISL_1577884, EPI_ISL_1577885, EPI_ISL_1577890, EPI_ISL_1577893, EPI_ISL_1577894, EPI_ISL_1577895, EPI_ISL_1577896, EPI_ISL_1577897, EPI_ISL_1577898, EPI_ISL_1577899 | | | |
| see above | Virology, Universitätsklinikum des Saarlandes | Epigenetics, Saarland University | Jörn Walter; Kathrin Kattler; Markus Vogelgesang; Sascha Tierling; Sigrun Smola; Stefan Lohse; Thorsten Pfuhl |
| EPI_ISL_1990088, EPI_ISL_1990090, EPI_ISL_1990091, EPI_ISL_1990092, EPI_ISL_1990094, EPI_ISL_1990095, EPI_ISL_1990096, EPI_ISL_1990097, EPI_ISL_1990098, EPI_ISL_1990099, EPI_ISL_1990100, EPI_ISL_1990101, EPI_ISL_1990102, EPI_ISL_1990103, EPI_ISL_1990104, EPI_ISL_1990105, EPI_ISL_1990106, EPI_ISL_1990107, EPI_ISL_1991618, EPI_ISL_1991621, EPI_ISL_1991634, EPI_ISL_1991637, EPI_ISL_1991638, EPI_ISL_1991640, EPI_ISL_1991641, EPI_ISL_1991643, EPI_ISL_1991644 | | | |
| see above | ZOTZ KLIMAS MVZ Düsseldorf-Centrum GbR ÜBAG für Labormedizin, Genetik, Zytologie, Pathologie | Center of Medical Microbiology, Virology, and Hospital Hygiene, University of Duesseldorf | Alexander Dilthey; Andreas Walker; Daniel Strelow; Jessica Nicolai; Jörg Timm; Katrin Hoffmann; Klaus Pfeffer; Lisanna Hülse; Malte Kohns Vasconcelos; Maximilian Damagnez; Nadine Lübke; Patrick Finzer; Rainer Zotz; Tobias Wienemann; Torsten Houwaart |
| EPI_ISL_1569781 | alphaomega Labor GbR (Medizinische Kooperationsgemeinschaft Dres. Ackermann/Tennert/ZaiÃŸ) | Robert Koch Institute | |
| EPI_ISL_1442577, EPI_ISL_1442579, EPI_ISL_1442580, EPI_ISL_1442581, EPI_ISL_1442582, EPI_ISL_1442584, EPI_ISL_1442585 | | | |
| see above | alphaomega Labor GbR (Medizinische KooperationsgemeinschaftDres. Ackermann/Tennert/Zaiß) | Robert Koch Institute | |
| EPI_ISL_1564030, EPI_ISL_1564127, EPI_ISL_1564197, EPI_ISL_1564211, EPI_ISL_1564280, EPI_ISL_1564407, EPI_ISL_1564436, EPI_ISL_1564441, EPI_ISL_1564465, EPI_ISL_1564516, EPI_ISL_1564610, EPI_ISL_1564739, EPI_ISL_1564774, EPI_ISL_1564874, EPI_ISL_1565005, EPI_ISL_1565154, EPI_ISL_1565214, EPI_ISL_1565225, EPI_ISL_1565307, EPI_ISL_1565334, EPI_ISL_1565347, EPI_ISL_1565351, EPI_ISL_1565356, EPI_ISL_1565358, EPI_ISL_1572848, EPI_ISL_1572857, EPI_ISL_1572904, EPI_ISL_1572941, EPI_ISL_1573007, EPI_ISL_1573014, EPI_ISL_1573018, EPI_ISL_1573024, EPI_ISL_1573034, EPI_ISL_1573035 | | | |
| see above | amedes GÃ¶ttingen | Robert Koch Institute | |
| EPI_ISL_1436751, EPI_ISL_1436772 | amedes Göttingen | Robert Koch Institute | |
| EPI_ISL_1564068, EPI_ISL_1564136, EPI_ISL_1564166, EPI_ISL_1564176, EPI_ISL_1564182, EPI_ISL_1564430, EPI_ISL_1564498, EPI_ISL_1564532, EPI_ISL_1564543, EPI_ISL_1564595, EPI_ISL_1564617, EPI_ISL_1564667, EPI_ISL_1564790, EPI_ISL_1564854, EPI_ISL_1564921, EPI_ISL_1565211, EPI_ISL_1565346, EPI_ISL_1565362 | | | |
| see above | amedes MVZ DIAMEDIS Sennestadt | Robert Koch Institute | |
| EPI_ISL_1436636, EPI_ISL_1436659, EPI_ISL_1436663, EPI_ISL_1436677, EPI_ISL_1436698, EPI_ISL_1436702, EPI_ISL_1436714, EPI_ISL_1436721, EPI_ISL_1436725, EPI_ISL_1436727, EPI_ISL_1436754, EPI_ISL_1436788, EPI_ISL_1436793, EPI_ISL_1564080, EPI_ISL_1564119, EPI_ISL_1564144, EPI_ISL_1564295, EPI_ISL_1564315, EPI_ISL_1564378, EPI_ISL_1564470, EPI_ISL_1564537, EPI_ISL_1564600, EPI_ISL_1564606, EPI_ISL_1564616, EPI_ISL_1564618, EPI_ISL_1564825, EPI_ISL_1564859, EPI_ISL_1564867, EPI_ISL_1564897, EPI_ISL_1564913, EPI_ISL_1565014, EPI_ISL_1565045, EPI_ISL_1565051, EPI_ISL_1565136, EPI_ISL_1565194, EPI_ISL_1565202, EPI_ISL_1565205, EPI_ISL_1565208, EPI_ISL_1565212, EPI_ISL_1565268, EPI_ISL_1565292, EPI_ISL_1565348, EPI_ISL_1565349, EPI_ISL_1565352, EPI_ISL_1565354 | | | |
| see above | amedes MVZ Halle/Leipzig | Robert Koch Institute | |
| EPI_ISL_1564024, EPI_ISL_1564323, EPI_ISL_1564460, EPI_ISL_1564658, EPI_ISL_1564746, EPI_ISL_1564754, EPI_ISL_1564837, EPI_ISL_1564928, EPI_ISL_1564953, EPI_ISL_1564965, EPI_ISL_1564977, EPI_ISL_1564985, EPI_ISL_1565029, EPI_ISL_1565187, EPI_ISL_1565210, EPI_ISL_1565237, EPI_ISL_1565357 | | | |
| see above | amedes MVZ Hannover | Robert Koch Institute | |
| EPI_ISL_1564064, EPI_ISL_1564253, EPI_ISL_1564373, EPI_ISL_1564397, EPI_ISL_1564479, EPI_ISL_1564510, EPI_ISL_1564611, EPI_ISL_1564624, EPI_ISL_1564649, EPI_ISL_1564699, EPI_ISL_1564731, EPI_ISL_1564781, EPI_ISL_1564809, EPI_ISL_1565038, EPI_ISL_1565072, EPI_ISL_1565213, EPI_ISL_1565218, EPI_ISL_1565230, EPI_ISL_1565259, EPI_ISL_1565280, EPI_ISL_1565303, EPI_ISL_1565325, EPI_ISL_1565353, EPI_ISL_1565363 | | | |
| see above | amedes MVZ Jena | Robert Koch Institute | |
| EPI_ISL_1564247, EPI_ISL_1564550, EPI_ISL_1564881, EPI_ISL_1564946, EPI_ISL_1564970 | amedes MVZ Zentrum fÃ¼r Pathologie und Zytodiagnostik GmbH (KÃ¶ln) | Robert Koch Institute | |
| EPI_ISL_1564095, EPI_ISL_1564219, EPI_ISL_1564265, EPI_ISL_1564277, EPI_ISL_1564289, EPI_ISL_1564301, EPI_ISL_1564423, EPI_ISL_1564475, EPI_ISL_1564503, EPI_ISL_1564614, EPI_ISL_1564619, EPI_ISL_1564626, EPI_ISL_1564674, EPI_ISL_1564693, EPI_ISL_1564721, EPI_ISL_1565085, EPI_ISL_1565203, EPI_ISL_1565242, EPI_ISL_1565253 | | | |
| see above | amedes MVZ fÃ¼r Laboratoriumsdiagnostik Raubling GmbH | Robert Koch Institute | |
| EPI_ISL_1564035, EPI_ISL_1564084, EPI_ISL_1564231, EPI_ISL_1564590, EPI_ISL_1565364 | amedes MVZ fÃ¼r Laboratoriumsmedizin FÃ¼rstenfeldbruck | Robert Koch Institute | |
| EPI_ISL_1564025, EPI_ISL_1564076, EPI_ISL_1564089, EPI_ISL_1564113, EPI_ISL_1564152, EPI_ISL_1564239, EPI_ISL_1564259, EPI_ISL_1564341, EPI_ISL_1564362, EPI_ISL_1564449, EPI_ISL_1564561, EPI_ISL_1564582, EPI_ISL_1564622, EPI_ISL_1564625, EPI_ISL_1564707, EPI_ISL_1564817, EPI_ISL_1564907, EPI_ISL_1565106, EPI_ISL_1565112, EPI_ISL_1565142, EPI_ISL_1565248, EPI_ISL_1565287, EPI_ISL_1565359, EPI_ISL_1565360 | | | |
| see above | amedes MVZ fÃ¼r Laboratoriumsmedizin Hamburg | Robert Koch Institute | |
| EPI_ISL_1564051, EPI_ISL_1564389 | amedes MVZ fÃ¼r Laboratoriumsmedizin MÃ¼nchen | Robert Koch Institute | |
| EPI_ISL_1564107, EPI_ISL_1564902 | amedes MVZ fÃ¼r Laboratoriumsmedizin Rhein/Ruhr | Robert Koch Institute | |
| EPI_ISL_1564849, EPI_ISL_1572846 | amedes MVZ fÃ¼r Labordiagnostik Rhein-Main | Robert Koch Institute | |
| EPI_ISL_1436661, EPI_ISL_1436667, EPI_ISL_1436676, EPI_ISL_1436680, EPI_ISL_1436683, EPI_ISL_1436769, EPI_ISL_1436790, EPI_ISL_1436801, EPI_ISL_1436807 | | | |
| see above | amedes MVZ für Labordiagnostik Rhein-Main | Robert Koch Institute | |
| EPI_ISL_1436701, EPI_ISL_1436733, EPI_ISL_1436767 | amedes Medlab Arnold Analytik MVZ GmbH | Robert Koch Institute | |
| EPI_ISL_1439683 | labopart - Medizinische Laboratorien Dresden | Robert Koch Institute | |
| EPI_ISL_1564057, EPI_ISL_1564101, EPI_ISL_1564171, EPI_ISL_1564204, EPI_ISL_1564454, EPI_ISL_1564493, EPI_ISL_1564557, EPI_ISL_1564567, EPI_ISL_1564575, EPI_ISL_1564612, EPI_ISL_1564615, EPI_ISL_1564620, EPI_ISL_1564623, EPI_ISL_1564633, EPI_ISL_1564683, EPI_ISL_1564759, EPI_ISL_1564842, EPI_ISL_1564887, EPI_ISL_1564932, EPI_ISL_1564938, EPI_ISL_1565092, EPI_ISL_1565121, EPI_ISL_1565172, EPI_ISL_1565178, EPI_ISL_1565204, EPI_ISL_1565206, EPI_ISL_1565209, EPI_ISL_1565313, EPI_ISL_1565343, EPI_ISL_1565345, EPI_ISL_1565355 | | | |
| see above | nordlab - Partnerschaftspraxis fuÌˆr Laboratoriumsmedizin | Robert Koch Institute | |

We gratefully acknowledge the following Authors from the Originating laboratories responsible for obtaining the specimens, as well as the Submitting laboratories where the genome data were generated and shared via GISAID, on which this research is based.

All Submitters of data may be contacted directly via www.gisaid.org

Authors are sorted alphabetically.

| Accession ID | Originating Laboratory | Submitting Laboratory | Authors |
|---|---|---|---|
| EPI_ISL_2115592 | ALB FILS KLINIKEN GmbH - Institut fur Laboratoriumsmedizin Klinik am Eichert Gappingen | Robert Koch Institute | |
| EPI_ISL_1721135, EPI_ISL_1721381, EPI_ISL_1842827, EPI_ISL_1842829, EPI_ISL_2115660, EPI_ISL_2115748 | ALB FILS KLINIKEN GmbH - Institut fÃ¼r Laboratoriumsmedizin Klinik am Eichert GÃ¶ppingen | Robert Koch Institute | |
| EPI_ISL_2111001, EPI_ISL_2111002, EPI_ISL_2111280, EPI_ISL_2111284, EPI_ISL_2111289, EPI_ISL_2111293, EPI_ISL_2111294, EPI_ISL_2111296, EPI_ISL_2111298, EPI_ISL_2111303, EPI_ISL_2111304, EPI_ISL_2111305, EPI_ISL_2111310, EPI_ISL_2111311, EPI_ISL_2111314, EPI_ISL_2111317, EPI_ISL_2111318, EPI_ISL_2111319, EPI_ISL_2111321, EPI_ISL_2111323, EPI_ISL_2111326, EPI_ISL_2111328, EPI_ISL_2111330, EPI_ISL_2111336, EPI_ISL_2111337, EPI_ISL_2111339, EPI_ISL_2111342, EPI_ISL_2111345, EPI_ISL_2111347, EPI_ISL_2111348, EPI_ISL_2111349, EPI_ISL_2111350, EPI_ISL_2111351, EPI_ISL_2111353, EPI_ISL_2111356, EPI_ISL_2111358, EPI_ISL_2111360, EPI_ISL_2111363, EPI_ISL_2111367, EPI_ISL_2111369, EPI_ISL_2111371, EPI_ISL_2111377, EPI_ISL_2111379, EPI_ISL_2111380, EPI_ISL_2111387, EPI_ISL_2111392, EPI_ISL_2111395, EPI_ISL_2111397, EPI_ISL_2111398, EPI_ISL_2111401, EPI_ISL_2111402, EPI_ISL_2111403, EPI_ISL_2111404, EPI_ISL_2111408, EPI_ISL_2111409, EPI_ISL_2111411, EPI_ISL_2111413, EPI_ISL_2111414, EPI_ISL_2111416, EPI_ISL_2111422, EPI_ISL_2111423, EPI_ISL_2111424, EPI_ISL_2111426, EPI_ISL_2111427, EPI_ISL_2111428, EPI_ISL_2111431, EPI_ISL_2111432 | | | |
| see above | Bayerisches Landesamt fur Gesundheit und Lebensmittelsicherheit (LGL) | Robert Koch Institute | |
| EPI_ISL_1725277, EPI_ISL_1725278, EPI_ISL_1725283, EPI_ISL_1725290, EPI_ISL_1725297, EPI_ISL_1725305, EPI_ISL_1725310, EPI_ISL_1725312, EPI_ISL_1725314, EPI_ISL_1725317, EPI_ISL_1725319, EPI_ISL_1725321, EPI_ISL_1725333, EPI_ISL_1725337, EPI_ISL_1725351, EPI_ISL_1725353, EPI_ISL_1725359, EPI_ISL_1725367, EPI_ISL_1725370, EPI_ISL_1725376, EPI_ISL_1725403, EPI_ISL_1725413, EPI_ISL_1725417, EPI_ISL_1725420, EPI_ISL_1725422, EPI_ISL_1725431, EPI_ISL_1725435, EPI_ISL_1725436, EPI_ISL_1725438, EPI_ISL_1725445, EPI_ISL_1725457, EPI_ISL_1725459, EPI_ISL_1725476, EPI_ISL_1725480, EPI_ISL_1725486, EPI_ISL_1725492, EPI_ISL_1725495, EPI_ISL_1725502, EPI_ISL_1725505, EPI_ISL_1725527, EPI_ISL_2110863, EPI_ISL_2110866, EPI_ISL_2110867, EPI_ISL_2110871, EPI_ISL_2110872, EPI_ISL_2110873, EPI_ISL_2110874, EPI_ISL_2110875, EPI_ISL_2110876, EPI_ISL_2110877, EPI_ISL_2110878, EPI_ISL_2110880, EPI_ISL_2110884, EPI_ISL_2110885, EPI_ISL_2110886, EPI_ISL_2110888, EPI_ISL_2110890, EPI_ISL_2110891, EPI_ISL_2110894, EPI_ISL_2110896, EPI_ISL_2110897, EPI_ISL_2110903, EPI_ISL_2110907, EPI_ISL_2110908, EPI_ISL_2110909, EPI_ISL_2110911, EPI_ISL_2110913, EPI_ISL_2110914, EPI_ISL_2110915, EPI_ISL_2110916, EPI_ISL_2110917, EPI_ISL_2110918, EPI_ISL_2110921, EPI_ISL_2110926, EPI_ISL_2110927, EPI_ISL_2110932, EPI_ISL_2110935, EPI_ISL_2110938, EPI_ISL_2110940, EPI_ISL_2110941, EPI_ISL_2110942, EPI_ISL_2110944, EPI_ISL_2110945, EPI_ISL_2110946, EPI_ISL_2110947, EPI_ISL_2110948, EPI_ISL_2110949, EPI_ISL_2110950, EPI_ISL_2110951, EPI_ISL_2110953, EPI_ISL_2110957, EPI_ISL_2110958, EPI_ISL_2110960, EPI_ISL_2110962, EPI_ISL_2110964, EPI_ISL_2110967, EPI_ISL_2110969, EPI_ISL_2110972, EPI_ISL_2110973, EPI_ISL_2110974, EPI_ISL_2110975, EPI_ISL_2110976, EPI_ISL_2110977, EPI_ISL_2110981, EPI_ISL_2110982, EPI_ISL_2110985, EPI_ISL_2110986, EPI_ISL_2110987, EPI_ISL_2110988, EPI_ISL_2110989, EPI_ISL_2110990, EPI_ISL_2110993, EPI_ISL_2110994, EPI_ISL_2110997, EPI_ISL_2110999 | | | |
| see above | Bayerisches Landesamt fÃ¼r Gesundheit und Lebensmittelsicherheit (LGL) | Robert Koch Institute | |
| EPI_ISL_1642449, EPI_ISL_1642464, EPI_ISL_1642469, EPI_ISL_1642481, EPI_ISL_1642485, EPI_ISL_1642493, EPI_ISL_1642497, EPI_ISL_1642509, EPI_ISL_1642518, EPI_ISL_1642563, EPI_ISL_1642634, EPI_ISL_1642637, EPI_ISL_1642662, EPI_ISL_1642672, EPI_ISL_1642685, EPI_ISL_1642692, EPI_ISL_1643119, EPI_ISL_1643180, EPI_ISL_1643188, EPI_ISL_1643232, EPI_ISL_1643239, EPI_ISL_1643333, EPI_ISL_1643335, EPI_ISL_1643354, EPI_ISL_1643360, EPI_ISL_1643378, EPI_ISL_1643390, EPI_ISL_1721108, EPI_ISL_1721110, EPI_ISL_1721113, EPI_ISL_1721115, EPI_ISL_1721117, EPI_ISL_1721126, EPI_ISL_1721138, EPI_ISL_1721139, EPI_ISL_1721142, EPI_ISL_1721149, EPI_ISL_1721151, EPI_ISL_1721162, EPI_ISL_1721168, EPI_ISL_1721170, EPI_ISL_1721172, EPI_ISL_1721175, EPI_ISL_1721178, EPI_ISL_1721186, EPI_ISL_1721194, EPI_ISL_1721199, EPI_ISL_1721200, EPI_ISL_1721201, EPI_ISL_1721206, EPI_ISL_1721209, EPI_ISL_1721215, EPI_ISL_1721221, EPI_ISL_1721222, EPI_ISL_1721231, EPI_ISL_1721235, EPI_ISL_1721239, EPI_ISL_1721263, EPI_ISL_1721274, EPI_ISL_1721287, EPI_ISL_1721293, EPI_ISL_1721308, EPI_ISL_1721312, EPI_ISL_1721318, EPI_ISL_1721322, EPI_ISL_1721324, EPI_ISL_1721331, EPI_ISL_1721336, EPI_ISL_1721344, EPI_ISL_1721348, EPI_ISL_1721350, EPI_ISL_1721351, EPI_ISL_1721355, EPI_ISL_1721359, EPI_ISL_1721360, EPI_ISL_1721373, EPI_ISL_1721376, EPI_ISL_1721383, EPI_ISL_1721388, EPI_ISL_1721389, EPI_ISL_1721391, EPI_ISL_1721392, EPI_ISL_1721394, EPI_ISL_1721395, EPI_ISL_1721396, EPI_ISL_1721397, EPI_ISL_1721398, EPI_ISL_1721402, EPI_ISL_1721403, EPI_ISL_1721404, EPI_ISL_1721406, EPI_ISL_1721411, EPI_ISL_1721416, EPI_ISL_1721417, EPI_ISL_1721420, EPI_ISL_1721423, EPI_ISL_1721424, EPI_ISL_1721426, EPI_ISL_1721428, EPI_ISL_1721433, EPI_ISL_1721434, EPI_ISL_1721437, EPI_ISL_1721438, EPI_ISL_1721439, EPI_ISL_1721444, EPI_ISL_1721445, EPI_ISL_1721447, EPI_ISL_1721448, EPI_ISL_1721450, EPI_ISL_1721455, EPI_ISL_1721457, EPI_ISL_1721458, EPI_ISL_1721459, EPI_ISL_1721460, EPI_ISL_1721461, EPI_ISL_1721462, EPI_ISL_1721463, EPI_ISL_1721468, EPI_ISL_1721469, EPI_ISL_1721470, EPI_ISL_1721472, EPI_ISL_1721474, EPI_ISL_1721476, EPI_ISL_1721477, EPI_ISL_1721480, EPI_ISL_1721482, EPI_ISL_1721485, EPI_ISL_1721486, EPI_ISL_1721487, EPI_ISL_1721488, EPI_ISL_1721489, EPI_ISL_1721490, EPI_ISL_1721491, EPI_ISL_1721494, EPI_ISL_1721495, EPI_ISL_1721497, EPI_ISL_1721498, EPI_ISL_1721501, EPI_ISL_1721503, EPI_ISL_1721504, EPI_ISL_1721505, EPI_ISL_1721506, EPI_ISL_1721507, EPI_ISL_1721512, EPI_ISL_1721515, EPI_ISL_1721519, EPI_ISL_1721520, EPI_ISL_1721521, EPI_ISL_1721526, EPI_ISL_1721527, EPI_ISL_1721528, EPI_ISL_1721531, EPI_ISL_1721533, EPI_ISL_1721535, EPI_ISL_1721538, EPI_ISL_1721541, EPI_ISL_1721544, EPI_ISL_1721545, EPI_ISL_1721547, EPI_ISL_1721548, EPI_ISL_1721549, EPI_ISL_1721550, EPI_ISL_1721551, EPI_ISL_1721552, EPI_ISL_1721553, EPI_ISL_1721557, EPI_ISL_1721558, EPI_ISL_1721560, EPI_ISL_1721562, EPI_ISL_1721563, EPI_ISL_1721564, EPI_ISL_1721565, EPI_ISL_1721566, EPI_ISL_1721567, EPI_ISL_1721568, EPI_ISL_1721569, EPI_ISL_1721570, EPI_ISL_1721571, EPI_ISL_1721575, EPI_ISL_1721579, EPI_ISL_1721581, EPI_ISL_1721583, EPI_ISL_1721584, EPI_ISL_1721585, EPI_ISL_1721586, EPI_ISL_1721588, EPI_ISL_1721590, EPI_ISL_1721592, EPI_ISL_1721597, EPI_ISL_1721598, EPI_ISL_1721599, EPI_ISL_1721600, EPI_ISL_1721601, EPI_ISL_1721602, EPI_ISL_1721604, EPI_ISL_1721607, EPI_ISL_1721611, EPI_ISL_1721612, EPI_ISL_1721613, EPI_ISL_1721615, EPI_ISL_1721619, EPI_ISL_1721621, EPI_ISL_1721623, EPI_ISL_1721624, EPI_ISL_1721625, EPI_ISL_1721626, EPI_ISL_1721627, EPI_ISL_1721628, EPI_ISL_1721629, EPI_ISL_1721635, EPI_ISL_1721637, EPI_ISL_1721638, EPI_ISL_1721641, EPI_ISL_1721642, EPI_ISL_1721643, EPI_ISL_1721644, EPI_ISL_1721645, EPI_ISL_1721647, EPI_ISL_1721648, EPI_ISL_1721649, EPI_ISL_1721650, EPI_ISL_1721656, EPI_ISL_1721657, EPI_ISL_1721658, EPI_ISL_1721659, EPI_ISL_1721660, EPI_ISL_1721662, EPI_ISL_1721667, EPI_ISL_1721668, EPI_ISL_1721669, EPI_ISL_1721670, EPI_ISL_1721671, EPI_ISL_1721672, EPI_ISL_1721673, EPI_ISL_1721675, EPI_ISL_1721678, EPI_ISL_1721679, EPI_ISL_1721681, EPI_ISL_1721682, EPI_ISL_1721683, EPI_ISL_1721684, EPI_ISL_1721685, EPI_ISL_1721686, EPI_ISL_1721688, EPI_ISL_1721690, EPI_ISL_1721699, EPI_ISL_1721706, EPI_ISL_1721707, EPI_ISL_1721710, EPI_ISL_1721714, EPI_ISL_1721721, EPI_ISL_1721727, EPI_ISL_1721733, EPI_ISL_1721734, EPI_ISL_1721735, EPI_ISL_1721738, EPI_ISL_1721747, EPI_ISL_1721748, EPI_ISL_1721750, EPI_ISL_1721751, EPI_ISL_1721756, EPI_ISL_1721757, EPI_ISL_1721762, EPI_ISL_1721764, EPI_ISL_1721774, EPI_ISL_1721775, EPI_ISL_1721787, EPI_ISL_1721790, EPI_ISL_1721792, EPI_ISL_1721795, EPI_ISL_1721796, EPI_ISL_1721799, EPI_ISL_1721806, EPI_ISL_1721810, EPI_ISL_1721811, EPI_ISL_1721817, EPI_ISL_1721820, EPI_ISL_1721821, EPI_ISL_1721823, EPI_ISL_1721824, EPI_ISL_1721832, EPI_ISL_1721833, EPI_ISL_1721835, EPI_ISL_1721836, EPI_ISL_1721838, EPI_ISL_1721847, EPI_ISL_1721849, EPI_ISL_1721853, EPI_ISL_1721857, EPI_ISL_1721862, EPI_ISL_1721868, EPI_ISL_1721870, EPI_ISL_1721871, EPI_ISL_1721874, EPI_ISL_1721876, EPI_ISL_1721886, EPI_ISL_1721894, EPI_ISL_1721895, EPI_ISL_1721896, EPI_ISL_1721899, EPI_ISL_1721901, EPI_ISL_1721921, EPI_ISL_1721943, EPI_ISL_1721945, EPI_ISL_1721947, EPI_ISL_1721951, EPI_ISL_1721956, EPI_ISL_1721969, EPI_ISL_1721977, EPI_ISL_1721978, EPI_ISL_1721979, EPI_ISL_1842999, EPI_ISL_1843018, EPI_ISL_1843144, EPI_ISL_1843682, EPI_ISL_1844046, EPI_ISL_1844089, EPI_ISL_1844090, EPI_ISL_1844116, EPI_ISL_1844133, EPI_ISL_1844139, EPI_ISL_1844151, EPI_ISL_1844161, EPI_ISL_1844163, EPI_ISL_1844166, EPI_ISL_1844191, EPI_ISL_1844196, EPI_ISL_1844206, EPI_ISL_1844227, EPI_ISL_1844265, EPI_ISL_1847129, EPI_ISL_1847131, EPI_ISL_1847276, EPI_ISL_1847300 | | | |
| see above | Bioscientia Labor Wermsdorf | Robert Koch Institute | |
| EPI_ISL_1642498, EPI_ISL_1643330, EPI_ISL_1721103, EPI_ISL_1721106, EPI_ISL_1721114, EPI_ISL_1721133, EPI_ISL_1721140, EPI_ISL_1721141, EPI_ISL_1721145, EPI_ISL_1721146, EPI_ISL_1721153, EPI_ISL_1721156, EPI_ISL_1721169, EPI_ISL_1721173, EPI_ISL_1721174, EPI_ISL_1721177, EPI_ISL_1721185, EPI_ISL_1721190, EPI_ISL_1721211, EPI_ISL_1721213, EPI_ISL_1721223, EPI_ISL_1721224, EPI_ISL_1721226, EPI_ISL_1721229, EPI_ISL_1721244, EPI_ISL_1721258, EPI_ISL_1721264, EPI_ISL_1721265, EPI_ISL_1721266, EPI_ISL_1721281, EPI_ISL_1721288, EPI_ISL_1721289, EPI_ISL_1721294, EPI_ISL_1721296, EPI_ISL_1721303, EPI_ISL_1721309, EPI_ISL_1721310, EPI_ISL_1721317, EPI_ISL_1721319, EPI_ISL_1721320, EPI_ISL_1721321, EPI_ISL_1721323, EPI_ISL_1721353, EPI_ISL_1721358, EPI_ISL_1721361, EPI_ISL_1721362, EPI_ISL_1721365, EPI_ISL_1721370, EPI_ISL_1721676, EPI_ISL_1721746, EPI_ISL_1721749, EPI_ISL_1721785, EPI_ISL_1721800, EPI_ISL_1721812, EPI_ISL_1721905, EPI_ISL_1721961, EPI_ISL_1721973, EPI_ISL_1842977, EPI_ISL_1842982, EPI_ISL_1843010, EPI_ISL_1843034, EPI_ISL_1843041, EPI_ISL_1843057, EPI_ISL_1843107, EPI_ISL_1843114, EPI_ISL_1843127, EPI_ISL_1843160, EPI_ISL_1847309 | | | |
| see above | Bioscientia MVZ Labor Karlsruhe GmbH | Robert Koch Institute | |
| EPI_ISL_1834008, EPI_ISL_1834009, EPI_ISL_1834010, EPI_ISL_1834012, EPI_ISL_1834013, EPI_ISL_1834014, EPI_ISL_1834015, EPI_ISL_1834016, EPI_ISL_1834018, EPI_ISL_1834019, EPI_ISL_1834020, EPI_ISL_1834021, EPI_ISL_1834022, EPI_ISL_1834023, EPI_ISL_1834024, EPI_ISL_1834025, EPI_ISL_1834026, EPI_ISL_1834027, EPI_ISL_1834028, EPI_ISL_1834030, EPI_ISL_1834031, EPI_ISL_1834043 | | | |
| see above | Bundeswehrkrankenhaus Berlin | Bundeswehr Institute of Microbiology | Alexandra Rehn; Enrico Georgi; Malena Bestehorn-Willmann; Markus Antwerpen; Mathias Walter; Mike Pillukat; Roman Wölfel; Sabine Zange |
| EPI_ISL_1834006 | Bundeswehrkrankenhaus Hamburg | Bundeswehr Institute of Microbiology | Alexandra Rehn; Enrico Georgi; Malena Bestehorn-Willmann; Markus Antwerpen; Mathias Walter; Mike Pillukat; Roman Wölfel; Sabine Zange |
| EPI_ISL_1833981, EPI_ISL_1833982, EPI_ISL_1834032 | Bundeswehrkrankenhaus Ulm | Bundeswehr Institute of Microbiology | Alexandra Rehn; Enrico Georgi; Malena Bestehorn-Willmann; Markus Antwerpen; Mathias Walter; Mike Pillukat; Roman Wölfel; Sabine Zange |
| EPI_ISL_1834033 | Bundeswehrkrankenhaus Westerstede | Bundeswehr Institute of Microbiology | Alexandra Rehn; Enrico Georgi; Malena Bestehorn-Willmann; Markus Antwerpen; Mathias Walter; Mike Pillukat; Roman Wölfel; Sabine Zange |
| EPI_ISL_1833976, EPI_ISL_1833977, EPI_ISL_1833979, EPI_ISL_1833998, EPI_ISL_1834000, EPI_ISL_1834001, EPI_ISL_1834002, EPI_ISL_1834005 | | | |
| see above | Bundeswehrzentralkrankenhaus Koblenz | Bundeswehr Institute of Microbiology | Alexandra Rehn; Enrico Georgi; Malena Bestehorn-Willmann; Markus Antwerpen; Mathias Walter; Mike Pillukat; Roman Wölfel; Sabine Zange |
| EPI_ISL_1721981, EPI_ISL_1843730, EPI_ISL_1843743, EPI_ISL_1843753, EPI_ISL_1843810, EPI_ISL_1843827, EPI_ISL_1843831, EPI_ISL_1843842, EPI_ISL_1843860, EPI_ISL_1843890, EPI_ISL_1843917, EPI_ISL_1843920, EPI_ISL_1843930, EPI_ISL_1843932, EPI_ISL_1843936, EPI_ISL_1843939, EPI_ISL_1843944, EPI_ISL_1843962, EPI_ISL_1843964, EPI_ISL_1843965, EPI_ISL_1843997, EPI_ISL_1844009, EPI_ISL_1844018 | | | |
| see above | Centogene; Dr. Bauer Laboratoriums GmbH | Robert Koch Institute | |
| EPI_ISL_1639326, EPI_ISL_1639327, EPI_ISL_1639328, EPI_ISL_1639329, EPI_ISL_1639330, EPI_ISL_1639331 | Chemische und VeterinÃ¤runtersuchungsamt MÃ¼nsterland-Emscher-Lippe (CVUA-MEL) | Robert Koch Institute | |
| EPI_ISL_1571515, EPI_ISL_1571520, EPI_ISL_1571527, EPI_ISL_1571529, EPI_ISL_1571530, EPI_ISL_1571531, EPI_ISL_1571532, EPI_ISL_1639333, EPI_ISL_1639334, EPI_ISL_1639336, EPI_ISL_1639337, EPI_ISL_1639338, EPI_ISL_1639339, EPI_ISL_1639341, EPI_ISL_1639342, EPI_ISL_1639343, EPI_ISL_1639344, EPI_ISL_1639345, EPI_ISL_1639346, EPI_ISL_1639350, EPI_ISL_1639352, EPI_ISL_1639353, EPI_ISL_1639354, EPI_ISL_1639356, EPI_ISL_1639357, EPI_ISL_1639360, EPI_ISL_1639361, EPI_ISL_1639362, EPI_ISL_1639363, EPI_ISL_1639365, EPI_ISL_1639368, EPI_ISL_1639371, EPI_ISL_1639375, EPI_ISL_1639376, EPI_ISL_1639377, EPI_ISL_1639381, EPI_ISL_1639382, EPI_ISL_1639384, EPI_ISL_1639385, EPI_ISL_1639386, EPI_ISL_1639388, EPI_ISL_1639389, EPI_ISL_1639391, EPI_ISL_1639394, EPI_ISL_1639395, EPI_ISL_1639397, EPI_ISL_1639398, EPI_ISL_1639402, EPI_ISL_1639404, EPI_ISL_1639406, EPI_ISL_1639407, EPI_ISL_1639409, EPI_ISL_1639410, EPI_ISL_1639414, EPI_ISL_1639419, EPI_ISL_1639420, EPI_ISL_1639421, EPI_ISL_1639422, EPI_ISL_1639427, EPI_ISL_1639431, EPI_ISL_1639432, EPI_ISL_1639433, EPI_ISL_1639434, EPI_ISL_1639437, EPI_ISL_1722941, EPI_ISL_1722942, EPI_ISL_1722943, EPI_ISL_1722944, EPI_ISL_1722945, EPI_ISL_1722946, EPI_ISL_1722950, EPI_ISL_1722951, EPI_ISL_1722952 | | | |
| see above | Clemenshospital - Alexianer Clemenshospital MÃ¼nster | Robert Koch Institute | |
| EPI_ISL_1843175, EPI_ISL_1843178, EPI_ISL_1843179, EPI_ISL_1843181, EPI_ISL_1843183, EPI_ISL_1843186, EPI_ISL_1843188, EPI_ISL_1843190, EPI_ISL_1843194, EPI_ISL_1843198 | | | |
| see above | Corona-Testzentrum ifp Institut fÃ¼r ProduktqualitÃ¤t GmbH | Robert Koch Institute | |
| EPI_ISL_1640162, EPI_ISL_1640163, EPI_ISL_1640164, EPI_ISL_1640165, EPI_ISL_1640166, EPI_ISL_1640167, EPI_ISL_1640168, EPI_ISL_1640169, EPI_ISL_1640170, EPI_ISL_1640172, EPI_ISL_1640173, EPI_ISL_1640175, EPI_ISL_1640177, EPI_ISL_1640178, EPI_ISL_1640179, EPI_ISL_1640180, EPI_ISL_1640181, EPI_ISL_1640183, EPI_ISL_1640184, EPI_ISL_1640186, EPI_ISL_1640187, EPI_ISL_1640188, EPI_ISL_1640191, EPI_ISL_1640192, EPI_ISL_1640194, EPI_ISL_1640195, EPI_ISL_1640196, EPI_ISL_1640197, EPI_ISL_1640198, EPI_ISL_1640199, EPI_ISL_1640201, EPI_ISL_1640202, EPI_ISL_1640203, EPI_ISL_1640204, EPI_ISL_1640205, EPI_ISL_1640207, EPI_ISL_1640208, EPI_ISL_1640209, EPI_ISL_1640210, EPI_ISL_1640211, EPI_ISL_1640212, EPI_ISL_1640213, EPI_ISL_1640214, EPI_ISL_1640215, EPI_ISL_1640216, EPI_ISL_1640217, EPI_ISL_1640218, EPI_ISL_1640219, EPI_ISL_1640220, EPI_ISL_1640221, EPI_ISL_1640222, EPI_ISL_1640223, EPI_ISL_1640224, EPI_ISL_1640225, EPI_ISL_1640226, EPI_ISL_1640227, EPI_ISL_1640228, EPI_ISL_1640229, EPI_ISL_1640230, EPI_ISL_1640231, EPI_ISL_1640232, EPI_ISL_1640233, EPI_ISL_1640234, EPI_ISL_1640235, EPI_ISL_1640236, EPI_ISL_1640237, EPI_ISL_1640238, EPI_ISL_1640239, EPI_ISL_1640240, EPI_ISL_1640242, EPI_ISL_1640243, EPI_ISL_1640244, EPI_ISL_1640245, EPI_ISL_1640246, EPI_ISL_1640247, EPI_ISL_1640248, EPI_ISL_1640250, EPI_ISL_1640251, EPI_ISL_1640252, EPI_ISL_1640253, EPI_ISL_1640254, EPI_ISL_1640256, EPI_ISL_1640257, EPI_ISL_1640258, EPI_ISL_1640259, EPI_ISL_1640260, EPI_ISL_1640261, EPI_ISL_1640262, EPI_ISL_1640263, EPI_ISL_1640264, EPI_ISL_1640266, EPI_ISL_1640267, EPI_ISL_1640269, EPI_ISL_1640270, EPI_ISL_1640271, EPI_ISL_1640275, EPI_ISL_1640276, EPI_ISL_1640278, EPI_ISL_1640279, EPI_ISL_1640280, EPI_ISL_1640281, EPI_ISL_1640282, EPI_ISL_1640283, EPI_ISL_1640284, EPI_ISL_1640287, EPI_ISL_1640289, EPI_ISL_1640290, EPI_ISL_1640291, EPI_ISL_1640292, EPI_ISL_1640294, EPI_ISL_1640295, EPI_ISL_1640296, EPI_ISL_1640297, EPI_ISL_1640298, EPI_ISL_1640299, EPI_ISL_1640300, EPI_ISL_1640301, EPI_ISL_1640303, EPI_ISL_1640304, EPI_ISL_1640305, EPI_ISL_1640307, EPI_ISL_1640308, EPI_ISL_1640309, EPI_ISL_1640310, EPI_ISL_1640311, EPI_ISL_1640312, EPI_ISL_1640313, EPI_ISL_1640314, EPI_ISL_1640315, EPI_ISL_1640316, EPI_ISL_1640318, EPI_ISL_1640319, EPI_ISL_1722279, EPI_ISL_1722281, EPI_ISL_1722283, EPI_ISL_1722285, EPI_ISL_1722286, EPI_ISL_1722291, EPI_ISL_1722296, EPI_ISL_1722307, EPI_ISL_1722308, EPI_ISL_1722314, EPI_ISL_1722316, EPI_ISL_1722320, EPI_ISL_1722323, EPI_ISL_1722328, EPI_ISL_1722337, EPI_ISL_1722342, EPI_ISL_1722343, EPI_ISL_1722348, EPI_ISL_1722349, EPI_ISL_1722356, EPI_ISL_1725241, EPI_ISL_1725246, EPI_ISL_1725247 | | | |
| see above | Diagnosticum - Labor Neukirchen | Robert Koch Institute | |
| EPI_ISL_1644915, EPI_ISL_1644916, EPI_ISL_1644917, EPI_ISL_1644918, EPI_ISL_1644919, EPI_ISL_1644920, EPI_ISL_1644922, EPI_ISL_1644925, EPI_ISL_1644931 | | | |
| see above | Dianovis GmbH Greiz | Robert Koch Institute | |
| EPI_ISL_1569900, EPI_ISL_1569914, EPI_ISL_1569917, EPI_ISL_1569919, EPI_ISL_1569925, EPI_ISL_1569936, EPI_ISL_1569937, EPI_ISL_1569941, EPI_ISL_1569959, EPI_ISL_1569963, EPI_ISL_1569968, EPI_ISL_1569974, EPI_ISL_1569990 | | | |
| see above | ELBLAB GmbH Zentrum fÃ¼r LaborMedizin | Robert Koch Institute | |
| EPI_ISL_1571282, EPI_ISL_1571283, EPI_ISL_1571284, EPI_ISL_1571285, EPI_ISL_1571286, EPI_ISL_1571287, EPI_ISL_1571288, EPI_ISL_1571289, EPI_ISL_1571290, EPI_ISL_1571291, EPI_ISL_1571292, EPI_ISL_1571293, EPI_ISL_1571294, EPI_ISL_1571295, EPI_ISL_1571296, EPI_ISL_1571297, EPI_ISL_1571298, EPI_ISL_1571299, EPI_ISL_1571300, EPI_ISL_1571301, EPI_ISL_1571302, EPI_ISL_1571303, EPI_ISL_1571304, EPI_ISL_1571305, EPI_ISL_1571306, EPI_ISL_1571307, EPI_ISL_1571308, EPI_ISL_1571309, EPI_ISL_1571310, EPI_ISL_1571311, EPI_ISL_1571312, EPI_ISL_1571313, EPI_ISL_1571314, EPI_ISL_1571315, EPI_ISL_1571316, EPI_ISL_1571317, EPI_ISL_1571318, EPI_ISL_1571319, EPI_ISL_1571320, EPI_ISL_1571321, EPI_ISL_1571322, EPI_ISL_1571323, EPI_ISL_1571324, EPI_ISL_1571325, EPI_ISL_1571326, EPI_ISL_1571327, EPI_ISL_1571328, EPI_ISL_1571329, EPI_ISL_1571330, EPI_ISL_1571331, EPI_ISL_1571332, EPI_ISL_1571333, EPI_ISL_1571334, EPI_ISL_1571335, EPI_ISL_1571336, EPI_ISL_1643884, EPI_ISL_1643885, EPI_ISL_1643886, EPI_ISL_1643887, EPI_ISL_1643888, EPI_ISL_1643889, EPI_ISL_1643890, EPI_ISL_1643891, EPI_ISL_1643892, EPI_ISL_1643893, EPI_ISL_1643894, EPI_ISL_1643895, EPI_ISL_1643896, EPI_ISL_1643897, EPI_ISL_1643898, EPI_ISL_1643899, EPI_ISL_1643900, EPI_ISL_1643901, EPI_ISL_1643902, EPI_ISL_1643903, EPI_ISL_1643904, EPI_ISL_1643905, EPI_ISL_1643906, EPI_ISL_1643907, EPI_ISL_1643908, EPI_ISL_1643909, EPI_ISL_1643910, EPI_ISL_1643911, EPI_ISL_1643912, EPI_ISL_1643913, EPI_ISL_1643914, EPI_ISL_1643915, EPI_ISL_1643916, EPI_ISL_1643917, EPI_ISL_1643918, EPI_ISL_1643919, EPI_ISL_1643920, EPI_ISL_1643921, EPI_ISL_1643922, EPI_ISL_1643923, EPI_ISL_1643924, EPI_ISL_1643925, EPI_ISL_1643926, EPI_ISL_1643927, EPI_ISL_1643928, EPI_ISL_1643929, EPI_ISL_1643930, EPI_ISL_1643931, EPI_ISL_1643932, EPI_ISL_1643933, EPI_ISL_1643934, EPI_ISL_1643935, EPI_ISL_1643936, EPI_ISL_1643937, EPI_ISL_1643938, EPI_ISL_1643939, EPI_ISL_1643940, EPI_ISL_1643941, EPI_ISL_1643942, EPI_ISL_1643943, EPI_ISL_1643944, EPI_ISL_1643945, EPI_ISL_1643946, EPI_ISL_1643947, EPI_ISL_1643948, EPI_ISL_1643949, EPI_ISL_1643950, EPI_ISL_1643951, EPI_ISL_1643952, EPI_ISL_1643953, EPI_ISL_1643954, EPI_ISL_1643955, EPI_ISL_1643956, EPI_ISL_1643957, EPI_ISL_1643958, EPI_ISL_1643959, EPI_ISL_1643960, EPI_ISL_1643961, EPI_ISL_1643962, EPI_ISL_1643963, EPI_ISL_1643964, EPI_ISL_1643965, EPI_ISL_1722673, EPI_ISL_1722674, EPI_ISL_1722675, EPI_ISL_1722676, EPI_ISL_1722677, EPI_ISL_1722678, EPI_ISL_1722679, EPI_ISL_1722680, EPI_ISL_1722682, EPI_ISL_1722683, EPI_ISL_1722684, EPI_ISL_1722685, EPI_ISL_1722686, EPI_ISL_1722687, EPI_ISL_1722688, EPI_ISL_1722689, EPI_ISL_1722690, EPI_ISL_1722691, EPI_ISL_1722692, EPI_ISL_1722693, EPI_ISL_1722694, EPI_ISL_1722695, EPI_ISL_1722696, EPI_ISL_1722697, EPI_ISL_1722698, EPI_ISL_1722699, EPI_ISL_1722700, EPI_ISL_1722701, EPI_ISL_1722702, EPI_ISL_1722703, EPI_ISL_1722704, EPI_ISL_1722705, EPI_ISL_1722706, EPI_ISL_1722707, EPI_ISL_1722708, EPI_ISL_1722709, EPI_ISL_1722710, EPI_ISL_1722711, EPI_ISL_1722712, EPI_ISL_1722713, EPI_ISL_1722714, EPI_ISL_1722715, EPI_ISL_1722716, EPI_ISL_1722717, EPI_ISL_1722718, EPI_ISL_1722719, EPI_ISL_1722720, EPI_ISL_1722721, EPI_ISL_1722722, EPI_ISL_1722723, EPI_ISL_1722724, EPI_ISL_1722725, EPI_ISL_1722726, EPI_ISL_1722727, EPI_ISL_1722728, EPI_ISL_1722729, EPI_ISL_1722730, EPI_ISL_1722731, EPI_ISL_1722732, EPI_ISL_1722733, EPI_ISL_1722734, EPI_ISL_1722735, EPI_ISL_1722736, EPI_ISL_1722737, EPI_ISL_1722738, EPI_ISL_1722739, EPI_ISL_1722740, EPI_ISL_1722741, EPI_ISL_1722742, EPI_ISL_1722743, EPI_ISL_1722744, EPI_ISL_1722745, EPI_ISL_1722746, EPI_ISL_1722747, EPI_ISL_1722748, EPI_ISL_1722749, EPI_ISL_1722750, EPI_ISL_1722751, EPI_ISL_1722752, EPI_ISL_1722753, EPI_ISL_1722754, EPI_ISL_1722755, EPI_ISL_1722756, EPI_ISL_1722757, EPI_ISL_1722758, EPI_ISL_1722759, EPI_ISL_1722760, EPI_ISL_1722761, EPI_ISL_1722762, EPI_ISL_1722763, EPI_ISL_1722764, EPI_ISL_1722765, EPI_ISL_1722766, EPI_ISL_1722767, EPI_ISL_1722768, EPI_ISL_1722769, EPI_ISL_1722770, EPI_ISL_1722771, EPI_ISL_1722772, EPI_ISL_1722773, EPI_ISL_1722774, EPI_ISL_1722775, EPI_ISL_1722776, EPI_ISL_1722777, EPI_ISL_1722778, EPI_ISL_1722779, EPI_ISL_1722780, EPI_ISL_1722781, EPI_ISL_1722782, EPI_ISL_1722783, EPI_ISL_1722784, EPI_ISL_1722785, EPI_ISL_1722786, EPI_ISL_1722787, EPI_ISL_1722788, EPI_ISL_1722789, EPI_ISL_1722790, EPI_ISL_1722791, EPI_ISL_1722792, EPI_ISL_1722793, EPI_ISL_1722794, EPI_ISL_1722795, EPI_ISL_1722796, EPI_ISL_1722797, EPI_ISL_1722798, EPI_ISL_1722799, EPI_ISL_1722800, EPI_ISL_1722801, EPI_ISL_1722802, EPI_ISL_1722803, EPI_ISL_1722804, EPI_ISL_1722805, EPI_ISL_1722806, EPI_ISL_1722807, EPI_ISL_1722808, EPI_ISL_1722809, EPI_ISL_1722810, EPI_ISL_1722811, EPI_ISL_1722812, EPI_ISL_1722813, EPI_ISL_1722814, EPI_ISL_1722815, EPI_ISL_1722816, EPI_ISL_1722817, EPI_ISL_1722818, EPI_ISL_1722819, EPI_ISL_1722820, EPI_ISL_1722821, EPI_ISL_1722822, EPI_ISL_1722823, EPI_ISL_1722824, EPI_ISL_1722825, EPI_ISL_1722826, EPI_ISL_1722827, EPI_ISL_1722828, EPI_ISL_1722829, EPI_ISL_1845802, EPI_ISL_1845803, EPI_ISL_1845804, EPI_ISL_1845805, EPI_ISL_1845806, EPI_ISL_1845807, EPI_ISL_1845808, EPI_ISL_1845809, EPI_ISL_1845810, EPI_ISL_1845811, EPI_ISL_1845812, EPI_ISL_1845813, EPI_ISL_1845814, EPI_ISL_1845815, EPI_ISL_1845816, EPI_ISL_1845817, EPI_ISL_1845818, EPI_ISL_1845819, EPI_ISL_1845820, EPI_ISL_1845821, EPI_ISL_1845822, EPI_ISL_1845823, EPI_ISL_1845824, EPI_ISL_1845825, EPI_ISL_1845826, EPI_ISL_1845827, EPI_ISL_1845828, EPI_ISL_1845829, EPI_ISL_1845830, EPI_ISL_1845831, EPI_ISL_1845832, EPI_ISL_1845833, EPI_ISL_1845834, EPI_ISL_1845835, EPI_ISL_1845836, EPI_ISL_1845837, EPI_ISL_1845838, EPI_ISL_1845839, EPI_ISL_1845840, EPI_ISL_1845841, EPI_ISL_1845842, EPI_ISL_1845843, EPI_ISL_1845844, EPI_ISL_1845845, EPI_ISL_1845846, EPI_ISL_1845847, EPI_ISL_1845848, EPI_ISL_1845849, EPI_ISL_1845850, EPI_ISL_1845851, EPI_ISL_1845852, EPI_ISL_1845853, EPI_ISL_1845854, EPI_ISL_1845855, EPI_ISL_1845856, EPI_ISL_1845857, EPI_ISL_1845858, EPI_ISL_1845859, EPI_ISL_1845860, | | | |

| | | | |
|---|---|---|---|
| EPI_ISL_1845861, EPI_ISL_1845862, EPI_ISL_1845863, EPI_ISL_1845864, EPI_ISL_1845865, EPI_ISL_1845866, EPI_ISL_1845867, EPI_ISL_1845868, EPI_ISL_1845869, EPI_ISL_1845870, EPI_ISL_1845871, EPI_ISL_1845872, EPI_ISL_1845873, EPI_ISL_1845874, EPI_ISL_1845875, EPI_ISL_1845876, EPI_ISL_1845877, EPI_ISL_1845878, EPI_ISL_1845879, EPI_ISL_1845880, EPI_ISL_1845881, EPI_ISL_1845882, EPI_ISL_1845883, EPI_ISL_1845884, EPI_ISL_1845885, EPI_ISL_1845886, EPI_ISL_1845887, EPI_ISL_1845888, EPI_ISL_1845889, EPI_ISL_1845890, EPI_ISL_1845891, EPI_ISL_1845892, EPI_ISL_1845893, EPI_ISL_1845894, EPI_ISL_1845895, EPI_ISL_1845896, EPI_ISL_1845897, EPI_ISL_1845898, EPI_ISL_1845899, EPI_ISL_1845900, EPI_ISL_1845901, EPI_ISL_1845902, EPI_ISL_1845903, EPI_ISL_1845904, EPI_ISL_1845905, EPI_ISL_1845906, EPI_ISL_1845907, EPI_ISL_1845908, EPI_ISL_1845909, EPI_ISL_1845910, EPI_ISL_1845911, EPI_ISL_1845912, EPI_ISL_1845913, EPI_ISL_1845914, EPI_ISL_1845915, EPI_ISL_1845916, EPI_ISL_1845917, EPI_ISL_1845918, EPI_ISL_1845919, EPI_ISL_1845920, EPI_ISL_1845921, EPI_ISL_1845922, EPI_ISL_1845923, EPI_ISL_1845924, EPI_ISL_1845925, EPI_ISL_1845926, EPI_ISL_1845927, EPI_ISL_1845928, EPI_ISL_1845929, EPI_ISL_1845930, EPI_ISL_1845931, EPI_ISL_1845932, EPI_ISL_1845933, EPI_ISL_1845934, EPI_ISL_1845935, EPI_ISL_1845936, EPI_ISL_1845937, EPI_ISL_1845938, EPI_ISL_1845939, EPI_ISL_1845940, EPI_ISL_1845941, EPI_ISL_1845942, EPI_ISL_1845943, EPI_ISL_1845944, EPI_ISL_1845945, EPI_ISL_1845946, EPI_ISL_1845947, EPI_ISL_1845948, EPI_ISL_1845949, EPI_ISL_1845950, EPI_ISL_1845951, EPI_ISL_1845952, EPI_ISL_1845953, EPI_ISL_1845954, EPI_ISL_1845955, EPI_ISL_1845956, EPI_ISL_1845957, EPI_ISL_1845958, EPI_ISL_1845959, EPI_ISL_1845960, EPI_ISL_1845961, EPI_ISL_1845962, EPI_ISL_1845963, EPI_ISL_1845964, EPI_ISL_1845965, EPI_ISL_1845966, EPI_ISL_1845967, EPI_ISL_1845968, EPI_ISL_1845969, EPI_ISL_1845970, EPI_ISL_1845971, EPI_ISL_1845972, EPI_ISL_1845973, EPI_ISL_1845974, EPI_ISL_1845975, EPI_ISL_1845976, EPI_ISL_1845977, EPI_ISL_1845978, EPI_ISL_1845979, EPI_ISL_1845980, EPI_ISL_1845981, EPI_ISL_1845982, EPI_ISL_1845983, EPI_ISL_1845984, EPI_ISL_1845985, EPI_ISL_1845986, EPI_ISL_1845987, EPI_ISL_1845988, EPI_ISL_1845989, EPI_ISL_1845990, EPI_ISL_1845991, EPI_ISL_1845992, EPI_ISL_1845993, EPI_ISL_1845994, EPI_ISL_1845995, EPI_ISL_1845996, EPI_ISL_1845997, EPI_ISL_1845998, EPI_ISL_1845999, EPI_ISL_1846000, EPI_ISL_1846001, EPI_ISL_1846002, EPI_ISL_1846003, EPI_ISL_1846004, EPI_ISL_1846005, EPI_ISL_1846006, EPI_ISL_1846007, EPI_ISL_1846008, EPI_ISL_1846009, EPI_ISL_1846010, EPI_ISL_1846011, EPI_ISL_1846012, EPI_ISL_1846013, EPI_ISL_1846014, EPI_ISL_1846015, EPI_ISL_1846016, EPI_ISL_1846017, EPI_ISL_1846018, EPI_ISL_1846019, EPI_ISL_1846020, EPI_ISL_1846021, EPI_ISL_1846022, EPI_ISL_1846023, EPI_ISL_1846024, EPI_ISL_1846025, EPI_ISL_1846026, EPI_ISL_1846027, EPI_ISL_1846028, EPI_ISL_1846029, EPI_ISL_1846030, EPI_ISL_1846031, EPI_ISL_1846032, EPI_ISL_1846033, EPI_ISL_1846034, EPI_ISL_1846035, EPI_ISL_1846036, EPI_ISL_1846037, EPI_ISL_1846038, EPI_ISL_1846039, EPI_ISL_1846040, EPI_ISL_1846041, EPI_ISL_1846042, EPI_ISL_1846043, EPI_ISL_1846044, EPI_ISL_1846045, EPI_ISL_1846046, EPI_ISL_1846047, EPI_ISL_1846048, EPI_ISL_1846049, EPI_ISL_1846050, EPI_ISL_1846051, EPI_ISL_1846052, EPI_ISL_1846053, EPI_ISL_1846054, EPI_ISL_1846055, EPI_ISL_1846056, EPI_ISL_1846057, EPI_ISL_1846058, EPI_ISL_1846059, EPI_ISL_1846060, EPI_ISL_1846061, EPI_ISL_1846062, EPI_ISL_1846063, EPI_ISL_1846064, EPI_ISL_1846065, EPI_ISL_1846066, EPI_ISL_1846067, EPI_ISL_1846068, EPI_ISL_1846069, EPI_ISL_1846070, EPI_ISL_1846071, EPI_ISL_1846072, EPI_ISL_1846073, EPI_ISL_1846074, EPI_ISL_1846075, EPI_ISL_1846076, EPI_ISL_1846077, EPI_ISL_1846078, EPI_ISL_1846079, EPI_ISL_1846080, EPI_ISL_1846081, EPI_ISL_1846082, EPI_ISL_1846083, EPI_ISL_1846084, EPI_ISL_1846085, EPI_ISL_1846086, EPI_ISL_1846087, EPI_ISL_1846088, EPI_ISL_1846089, EPI_ISL_1846090, EPI_ISL_1846091, EPI_ISL_1846092, EPI_ISL_1846093, EPI_ISL_1846094, EPI_ISL_1846095, EPI_ISL_1846096, EPI_ISL_1846097, EPI_ISL_1846098, EPI_ISL_1846099, EPI_ISL_1846100, EPI_ISL_1846101, EPI_ISL_1846102, EPI_ISL_1846103, EPI_ISL_1846104, EPI_ISL_1846105, EPI_ISL_1846106, EPI_ISL_1846107, EPI_ISL_1846108, EPI_ISL_1846109, EPI_ISL_1846110, EPI_ISL_1846111, EPI_ISL_1846112, EPI_ISL_1846113, EPI_ISL_1846114, EPI_ISL_1846115, EPI_ISL_1846116, EPI_ISL_1846117, EPI_ISL_1846118, EPI_ISL_1846119, EPI_ISL_1846120, EPI_ISL_1846121, EPI_ISL_1846122, EPI_ISL_1846123, EPI_ISL_1846124, EPI_ISL_1846125, EPI_ISL_1846126, EPI_ISL_1846127, EPI_ISL_1846128, EPI_ISL_1846129, EPI_ISL_1846130, EPI_ISL_1846131, EPI_ISL_1846132, EPI_ISL_1846133, EPI_ISL_1846134, EPI_ISL_1846135, EPI_ISL_1846136, EPI_ISL_1846137, EPI_ISL_1846138, EPI_ISL_1846139, EPI_ISL_1846140, EPI_ISL_1846141, EPI_ISL_1846142, EPI_ISL_1846143, EPI_ISL_1846144, EPI_ISL_1846145, EPI_ISL_1846146, EPI_ISL_1846147, EPI_ISL_1846148, EPI_ISL_1846149, EPI_ISL_1846150, EPI_ISL_1846151, EPI_ISL_1846152, EPI_ISL_1846153, EPI_ISL_1846154, EPI_ISL_1846155, EPI_ISL_1846156, EPI_ISL_1846157, EPI_ISL_1846158, EPI_ISL_1846159, EPI_ISL_1846160, EPI_ISL_1846161, EPI_ISL_1846162, EPI_ISL_1846163, EPI_ISL_1846164, EPI_ISL_1846165, EPI_ISL_1846166, EPI_ISL_1846167, EPI_ISL_1846168, EPI_ISL_1846169, EPI_ISL_1846170, EPI_ISL_1846171, EPI_ISL_1846172, EPI_ISL_1846173, EPI_ISL_1846174, EPI_ISL_1846175, EPI_ISL_1846176, EPI_ISL_1846177, EPI_ISL_1846178, EPI_ISL_1846179, EPI_ISL_1846180, EPI_ISL_1846181, EPI_ISL_1846182, EPI_ISL_1846183, EPI_ISL_1846184, EPI_ISL_1846185, EPI_ISL_1846186, EPI_ISL_1846187, EPI_ISL_1846188, EPI_ISL_1846189, EPI_ISL_1846190, EPI_ISL_1846191, EPI_ISL_1846192, EPI_ISL_1846193, EPI_ISL_1846194, EPI_ISL_1846195, EPI_ISL_1846196, EPI_ISL_1846197, EPI_ISL_1846198, EPI_ISL_1846199, EPI_ISL_1846200, EPI_ISL_1846201, EPI_ISL_1846202, EPI_ISL_1846203, EPI_ISL_1846204, EPI_ISL_1846205, EPI_ISL_1846206, EPI_ISL_1846207, EPI_ISL_1846208, EPI_ISL_1846209, EPI_ISL_1846210, EPI_ISL_1846211, EPI_ISL_1846212, EPI_ISL_1846213 | | | |
| see above | Eurofins LifeCodexx GmbH | Robert Koch Institute | |
| EPI_ISL_1642277, EPI_ISL_1642278, EPI_ISL_1642279, EPI_ISL_1642280, EPI_ISL_1642281, EPI_ISL_1642282, EPI_ISL_1642283, EPI_ISL_1642284, EPI_ISL_1642285, EPI_ISL_1642286, EPI_ISL_1642287, EPI_ISL_1642288, EPI_ISL_1642289, EPI_ISL_1642290, EPI_ISL_1642291, EPI_ISL_1642292, EPI_ISL_1642293, EPI_ISL_1642294, EPI_ISL_1642295, EPI_ISL_1642296, EPI_ISL_1642297, EPI_ISL_1642298, EPI_ISL_1642299, EPI_ISL_1642300, EPI_ISL_1642301, EPI_ISL_1642303, EPI_ISL_1642304, EPI_ISL_1642305, EPI_ISL_1642306, EPI_ISL_1642307, EPI_ISL_1642308, EPI_ISL_1642309 | | | |
| see above | Fraunhofer-Institut fÃ¼r Zelltherapie und Immunologie | Robert Koch Institute | |
| EPI_ISL_1720952, EPI_ISL_1720957, EPI_ISL_1721005 | Helios UniversitÃ¤tsklinikum Wuppertal: UniversitÃ¤t Witten/Herdecke: Institut fÃ¼r Medizinische Labordiagnostik | Robert Koch Institute | |
| EPI_ISL_1725565 | IFLb Laboratoriumsmedizin Berlin GmbH | Robert Koch Institute | |
| EPI_ISL_1760614 | ILTM | ILTM | Martin Farr |
| EPI_ISL_1639447, EPI_ISL_1639454, EPI_ISL_1639455, EPI_ISL_1639458, EPI_ISL_1639459, EPI_ISL_1639466, EPI_ISL_1639474, EPI_ISL_1639475, EPI_ISL_1639477, EPI_ISL_1639483, EPI_ISL_1639487, EPI_ISL_1639493, EPI_ISL_1639499, EPI_ISL_1639505, EPI_ISL_1639508, EPI_ISL_1639511, EPI_ISL_1639515, EPI_ISL_1722182 | | | |
| see above | IMD - Institut fÃ¼r Medizinische Diagnostik Berlin-Potsdam | Robert Koch Institute | |
| EPI_ISL_1643683, EPI_ISL_1643688, EPI_ISL_1643690, EPI_ISL_1643695, EPI_ISL_1643698, EPI_ISL_1643699, EPI_ISL_1643701, EPI_ISL_1643702, EPI_ISL_1643703, EPI_ISL_1643704, EPI_ISL_1643706, EPI_ISL_1643709, EPI_ISL_1643712, EPI_ISL_1643713, EPI_ISL_1643715, EPI_ISL_1643716, EPI_ISL_1643718, EPI_ISL_1643720, EPI_ISL_1643723, EPI_ISL_1643727, EPI_ISL_1643728, EPI_ISL_1643731, EPI_ISL_1643732, EPI_ISL_1643733, EPI_ISL_1643734, EPI_ISL_1643738, EPI_ISL_1643739, EPI_ISL_1643740, EPI_ISL_1643741, EPI_ISL_1643742, EPI_ISL_1643743, EPI_ISL_1643744, EPI_ISL_1643746, EPI_ISL_1643747, EPI_ISL_1643751, EPI_ISL_1643752, EPI_ISL_1643754, EPI_ISL_1643755, EPI_ISL_1643756, EPI_ISL_1722181, EPI_ISL_1722190, EPI_ISL_1722236 | | | |
| see above | IMD - Labor Oderland | Robert Koch Institute | |
| EPI_ISL_1639472, EPI_ISL_1639476, EPI_ISL_1639479, EPI_ISL_1639481, EPI_ISL_1639491, EPI_ISL_1639514, EPI_ISL_1643711, EPI_ISL_1643730, EPI_ISL_1643748, EPI_ISL_1643758 | | | |
| see above | IMD - MVZ Labor Greifswald | Robert Koch Institute | |
| EPI_ISL_1639276, EPI_ISL_1639282, EPI_ISL_1639283, EPI_ISL_1639287, EPI_ISL_1639304, EPI_ISL_1639319, EPI_ISL_1639438, EPI_ISL_1639441, EPI_ISL_1639442, EPI_ISL_1639444, EPI_ISL_1639445, EPI_ISL_1639446, EPI_ISL_1639449, EPI_ISL_1639450, EPI_ISL_1639451, EPI_ISL_1639452, EPI_ISL_1639457, EPI_ISL_1639460, EPI_ISL_1639461, EPI_ISL_1639462, EPI_ISL_1639463, EPI_ISL_1639464, EPI_ISL_1639465, EPI_ISL_1639467, EPI_ISL_1639468, EPI_ISL_1639469, EPI_ISL_1639471, EPI_ISL_1639473, EPI_ISL_1639478, EPI_ISL_1639482, EPI_ISL_1639484, EPI_ISL_1639486, EPI_ISL_1639488, EPI_ISL_1639489, EPI_ISL_1639490, EPI_ISL_1639492, EPI_ISL_1639496, EPI_ISL_1639497, EPI_ISL_1639498, EPI_ISL_1639501, EPI_ISL_1639502, EPI_ISL_1639503, EPI_ISL_1639507, EPI_ISL_1639509, EPI_ISL_1639510, EPI_ISL_1639512, EPI_ISL_1639513, EPI_ISL_1639516, EPI_ISL_1639517, EPI_ISL_1639518, EPI_ISL_1643682, EPI_ISL_1643697, EPI_ISL_1643717, EPI_ISL_1643726 | | | |
| see above | IMD - MVZ Labor Martinsried | Robert Koch Institute | |
| EPI_ISL_1639439, EPI_ISL_1639480, EPI_ISL_1639485, EPI_ISL_1639494, EPI_ISL_1639500, EPI_ISL_1639504, EPI_ISL_1639506 | | | |
| see above | IMD - Medizinisches Labor Rostock | Robert Koch Institute | |
| EPI_ISL_1572019, EPI_ISL_1572024, EPI_ISL_1572030, EPI_ISL_1572036, EPI_ISL_1572041, EPI_ISL_1572060, EPI_ISL_1572062, EPI_ISL_1572066, EPI_ISL_1572068, EPI_ISL_1572069, EPI_ISL_1572103, EPI_ISL_1638533, EPI_ISL_1638534, EPI_ISL_1638535, EPI_ISL_1638536, EPI_ISL_1638537, EPI_ISL_1638538, EPI_ISL_1638539, EPI_ISL_1638540, EPI_ISL_1638541, EPI_ISL_1638542, EPI_ISL_1638543, EPI_ISL_1638544, EPI_ISL_1638545, EPI_ISL_1638546, EPI_ISL_1638548, EPI_ISL_1638549, EPI_ISL_1638550, EPI_ISL_1638551, EPI_ISL_1638552, EPI_ISL_1638553, EPI_ISL_1638554, EPI_ISL_1638555, EPI_ISL_1638556, EPI_ISL_1638557, EPI_ISL_1638558, EPI_ISL_1638559, EPI_ISL_1638560, EPI_ISL_1638561, EPI_ISL_1638562, EPI_ISL_1638563, EPI_ISL_1638564, EPI_ISL_1638565, EPI_ISL_1638566, EPI_ISL_1638567, EPI_ISL_1638568, EPI_ISL_1638569, EPI_ISL_1638570, EPI_ISL_1638571, EPI_ISL_1638575, EPI_ISL_1638576, EPI_ISL_1638579, EPI_ISL_1638582, EPI_ISL_1638586, EPI_ISL_1638591, EPI_ISL_1638593, EPI_ISL_1638594, EPI_ISL_1638600, EPI_ISL_1638601, EPI_ISL_1638603, EPI_ISL_1638609, EPI_ISL_1638610, EPI_ISL_1638611, EPI_ISL_1638612, EPI_ISL_1638615, EPI_ISL_1638616, EPI_ISL_1638617, EPI_ISL_1638618, EPI_ISL_1638620, EPI_ISL_1638621, EPI_ISL_1642316, EPI_ISL_1642321, EPI_ISL_1642326, EPI_ISL_1642335 | | | |
| see above | IMD Labor Frankfurt | Robert Koch Institute | |
| EPI_ISL_2108477 | Institut fur Medizinische Mikrobiologie - Universitatsmedizin Gottingen | Robert Koch Institute | |
| EPI_ISL_1642733, EPI_ISL_1642734, EPI_ISL_1642735 | Institut fÃ¼r Klinische Transfusionsmedizin und Immungenetik DRK Blutspendedienst Baden-WÃ¼rttemberg | Robert Koch Institute | |
| EPI_ISL_1724239, EPI_ISL_1724240, EPI_ISL_1724241, EPI_ISL_1724242, EPI_ISL_1724243 | Institut fÃ¼r Medizinische Laboratoriumsdiagnostik Bochum GmbH | Robert Koch Institute | |
| EPI_ISL_1643882 | Institut fÃ¼r Medizinische Mikrobiologie - UniversitÃ¤tsmedizin GÃ¶ttingen | Robert Koch Institute | |
| EPI_ISL_1724442, EPI_ISL_1724443, EPI_ISL_1724444, EPI_ISL_1724445, EPI_ISL_1724446, EPI_ISL_1724447, EPI_ISL_1846341 | | | |
| see above | Institut fÃ¼r med. Mikrobiologie u. Virologie Klinikum Oldenburg | Robert Koch Institute | |
| EPI_ISL_1670272, EPI_ISL_1670273, EPI_ISL_1670274, EPI_ISL_1670275, EPI_ISL_1670327, EPI_ISL_1670328, EPI_ISL_1670329, EPI_ISL_1670330, EPI_ISL_1670331, EPI_ISL_1670332, EPI_ISL_1670333, EPI_ISL_1670334, EPI_ISL_1670335, EPI_ISL_1670336, EPI_ISL_1670337, EPI_ISL_1670376, EPI_ISL_1670377, EPI_ISL_1670378 | | | |
| see above | Institute of Medical Microbiology and Hospital Hygiene | Institute of Medical Microbiology and Hospital Hygiene | Aljoscha Tersteegen; Prof. Dr. Achim Kaasch |
| EPI_ISL_1634481, EPI_ISL_1648932, EPI_ISL_1648933, EPI_ISL_1648936, EPI_ISL_1648937, EPI_ISL_1653693 | Institute of Tropical Medicine | Institute of Tropical Medicine | Prof. Dr. Thirumalaisamy P. Velavan and Prof. Dr. Peter Kremsner |
| EPI_ISL_1804995, EPI_ISL_1804996, EPI_ISL_1805007, EPI_ISL_1805010, EPI_ISL_1805013, EPI_ISL_1805014, EPI_ISL_1805015, EPI_ISL_1805016, EPI_ISL_1805017, EPI_ISL_1805051, EPI_ISL_1805052, EPI_ISL_1805053, EPI_ISL_1805054 | | | |
| see above | Institute of Virology, Medical Center, University of Freiburg, Freiburg, Germany | Institute of Virology, Clinial Virus Genomics, Medical Center, University of Freiburg, Freiburg, Germany | Hajo Grundmann; Jonas Fuchs; Lena Jaki; Lisa Kern; Marcus Panning; Sandra Reuter |
| EPI_ISL_1640905, EPI_ISL_1640906, EPI_ISL_1640909, EPI_ISL_1640913, EPI_ISL_1640914, EPI_ISL_1640916, EPI_ISL_1640917, EPI_ISL_1640919, EPI_ISL_1640922, EPI_ISL_1640923, EPI_ISL_1640925, EPI_ISL_1640929, EPI_ISL_1640930, EPI_ISL_1722359, EPI_ISL_1722361, EPI_ISL_1722365, EPI_ISL_1722368 | | | |
| see above | KRH Klinikum Region Hannover: Institut fÃ¼r Med. Mikrobiologie und Krankenhaushygiene | Robert Koch Institute | |
| EPI_ISL_1715843, EPI_ISL_1715845, EPI_ISL_1715846, EPI_ISL_1715850, EPI_ISL_1715852, EPI_ISL_1715853 | Kath. Marienkrankenhaus | Heinrich Pette Institute, Leibniz Institute for Experimental Virology | Adam Grundhoff; Alexis Robitaille; Johannes Knobloch; Martin Aepfelbacher; Nicole Fischer; Thomas Günther |
| EPI_ISL_1567681, EPI_ISL_1570833, EPI_ISL_1570847, EPI_ISL_1571141, EPI_ISL_1571144, EPI_ISL_1572166, EPI_ISL_1638342, EPI_ISL_1638352 | | | |
| see above | Kerckhoff-Klinik GmbH | Robert Koch Institute | |
| EPI_ISL_1638463 | Klinikum Darmstadt GmbH | Robert Koch Institute | |
| EPI_ISL_1570747, EPI_ISL_1570760, EPI_ISL_1570764, EPI_ISL_1570769, EPI_ISL_1570772, EPI_ISL_1570783, EPI_ISL_1570785, EPI_ISL_1570791, EPI_ISL_1570799, EPI_ISL_1570801, EPI_ISL_1570803, EPI_ISL_1573057, EPI_ISL_1573058, EPI_ISL_1573059, EPI_ISL_1573060, EPI_ISL_1573061, EPI_ISL_1573062, EPI_ISL_1573064, EPI_ISL_1573065, EPI_ISL_1573066, EPI_ISL_1573067, EPI_ISL_1573068, EPI_ISL_1573069, EPI_ISL_1573070, EPI_ISL_1573071, EPI_ISL_1573072, EPI_ISL_1573073, EPI_ISL_1573074, EPI_ISL_1573075, EPI_ISL_1573076 | | | |
| see above | Klinikum Ernst von Bergmann gemeinnÃ¼tzige GmbH - stationÃ¤rer Bereich | Robert Koch Institute | |
| EPI_ISL_1571145, EPI_ISL_1639126, EPI_ISL_1642580, EPI_ISL_1642615 | Klinikum Fulda gAG | Robert Koch Institute | |
| EPI_ISL_1722842 | Klinikum Stuttgart | Robert Koch Institute | |
| EPI_ISL_1569976, EPI_ISL_1569979, EPI_ISL_1569984 | LABOR CHEMNITZ â€“ Zentrum fÃ¼r Diagnostik am Klinikum Chemnitz | Robert Koch Institute | |
| EPI_ISL_1642441, EPI_ISL_1642457, EPI_ISL_1642508, EPI_ISL_1642531, EPI_ISL_1642536, EPI_ISL_1642584, EPI_ISL_1642597, EPI_ISL_1642611, EPI_ISL_1643120, EPI_ISL_1643122, EPI_ISL_1643123, EPI_ISL_1643124, EPI_ISL_1643126, EPI_ISL_1643127, EPI_ISL_1643128, EPI_ISL_1643129, EPI_ISL_1643132, EPI_ISL_1643134, EPI_ISL_1643135, EPI_ISL_1643136, EPI_ISL_1643137, EPI_ISL_1643138, EPI_ISL_1643139, EPI_ISL_1643140, EPI_ISL_1643141, EPI_ISL_1643143, EPI_ISL_1643145, EPI_ISL_1643147, EPI_ISL_1643148, EPI_ISL_1643150, EPI_ISL_1643151, EPI_ISL_1643154, EPI_ISL_1643156, EPI_ISL_1643157, EPI_ISL_1643158, EPI_ISL_1643160, EPI_ISL_1643163, EPI_ISL_1643166, EPI_ISL_1643167, EPI_ISL_1643169, EPI_ISL_1643170, EPI_ISL_1643171, EPI_ISL_1643172, EPI_ISL_1643175, EPI_ISL_1643176, EPI_ISL_1643178, EPI_ISL_1643181, EPI_ISL_1643182, EPI_ISL_1643183, EPI_ISL_1643185, EPI_ISL_1643190, EPI_ISL_1643192, EPI_ISL_1643194, EPI_ISL_1643195, EPI_ISL_1643198, EPI_ISL_1643201, EPI_ISL_1643203, EPI_ISL_1643206, EPI_ISL_1643208, EPI_ISL_1643209, EPI_ISL_1643213, EPI_ISL_1643214, EPI_ISL_1643217, EPI_ISL_1643219, EPI_ISL_1643224, EPI_ISL_1643225, EPI_ISL_1643227, EPI_ISL_1643230, EPI_ISL_1643231, EPI_ISL_1643233, EPI_ISL_1643234, EPI_ISL_1643235, EPI_ISL_1643236, EPI_ISL_1643243, EPI_ISL_1643244, EPI_ISL_1643245, EPI_ISL_1643246, EPI_ISL_1643248, EPI_ISL_1643250, EPI_ISL_1643251, EPI_ISL_1643256, EPI_ISL_1643258, EPI_ISL_1643261, EPI_ISL_1643262, EPI_ISL_1643264, EPI_ISL_1643265, EPI_ISL_1643266, EPI_ISL_1643268, EPI_ISL_1643269, EPI_ISL_1643270, EPI_ISL_1643271, EPI_ISL_1643272, EPI_ISL_1643273, EPI_ISL_1643278, EPI_ISL_1643279, EPI_ISL_1643282, EPI_ISL_1643283, EPI_ISL_1643286, EPI_ISL_1643289, EPI_ISL_1643290, EPI_ISL_1643296, EPI_ISL_1643297, EPI_ISL_1643299, EPI_ISL_1643301, EPI_ISL_1643302, EPI_ISL_1643303, EPI_ISL_1643304, EPI_ISL_1643305, EPI_ISL_1643306, EPI_ISL_1643307, EPI_ISL_1643308, EPI_ISL_1643309, EPI_ISL_1643310, EPI_ISL_1643316, EPI_ISL_1643319, EPI_ISL_1643323, EPI_ISL_1643324, EPI_ISL_1643325, EPI_ISL_1643326, EPI_ISL_1643328, EPI_ISL_1643338, EPI_ISL_1643339, EPI_ISL_1643340, EPI_ISL_1643341, EPI_ISL_1643342, EPI_ISL_1643343, EPI_ISL_1643345, EPI_ISL_1643348, EPI_ISL_1643349, EPI_ISL_1643350, EPI_ISL_1643351, EPI_ISL_1643352, EPI_ISL_1643355, EPI_ISL_1643358, EPI_ISL_1643359, EPI_ISL_1643365, EPI_ISL_1643368, EPI_ISL_1643369, EPI_ISL_1643370, EPI_ISL_1643372, EPI_ISL_1643373, EPI_ISL_1643374, EPI_ISL_1643381, EPI_ISL_1643382, EPI_ISL_1643383, EPI_ISL_1643384, EPI_ISL_1643391, EPI_ISL_1643392 | | | |
| see above | LabKom - Labor Augsburg MVZ GmbH | Robert Koch Institute | |
| EPI_ISL_1642422, EPI_ISL_1642430, EPI_ISL_1642553, EPI_ISL_1721453, EPI_ISL_1721652 | LabKom - Labor Hamburg-LÃ¼beck | Robert Koch Institute | |
| EPI_ISL_1642427, EPI_ISL_1642433, EPI_ISL_1642440, EPI_ISL_1642444, EPI_ISL_1642450, EPI_ISL_1642465, EPI_ISL_1642466, EPI_ISL_1642473, EPI_ISL_1642480, EPI_ISL_1642491, EPI_ISL_1642492, EPI_ISL_1642501, EPI_ISL_1642510, EPI_ISL_1642513, EPI_ISL_1642521, EPI_ISL_1642542, EPI_ISL_1642551, EPI_ISL_1642560, EPI_ISL_1642568, EPI_ISL_1642601, EPI_ISL_1642605, EPI_ISL_1642612, EPI_ISL_1642621, EPI_ISL_1642635, EPI_ISL_1642645, EPI_ISL_1642651, EPI_ISL_1642652, EPI_ISL_1642653, EPI_ISL_1642656, EPI_ISL_1642658, EPI_ISL_1642661, EPI_ISL_1642663, EPI_ISL_1642679, EPI_ISL_1642681, EPI_ISL_1642689 | | | |
| see above | LabKom - Labor Hannover MVZ GmbH | Robert Koch Institute | |
| EPI_ISL_1642420, EPI_ISL_1642423, EPI_ISL_1642426, EPI_ISL_1642435, EPI_ISL_1642437, EPI_ISL_1642442, EPI_ISL_1642451, EPI_ISL_1642456, EPI_ISL_1642458, EPI_ISL_1642461, EPI_ISL_1642468, EPI_ISL_1642479, EPI_ISL_1642482, EPI_ISL_1642487, EPI_ISL_1642495, EPI_ISL_1642502, EPI_ISL_1642503, EPI_ISL_1642512, EPI_ISL_1642522, EPI_ISL_1642526, EPI_ISL_1642533, EPI_ISL_1642535, EPI_ISL_1642546, EPI_ISL_1642548, EPI_ISL_1642557, EPI_ISL_1642559, EPI_ISL_1642565, EPI_ISL_1642567, EPI_ISL_1642569, EPI_ISL_1642585, EPI_ISL_1642586, EPI_ISL_1642589, EPI_ISL_1642592, EPI_ISL_1642598, EPI_ISL_1642602, EPI_ISL_1642616, EPI_ISL_1642623, EPI_ISL_1642629, EPI_ISL_1642630, EPI_ISL_1642631, EPI_ISL_1642633, EPI_ISL_1642648, EPI_ISL_1642649, EPI_ISL_1642655, EPI_ISL_1642660, EPI_ISL_1642673, EPI_ISL_1642684, EPI_ISL_1642686, EPI_ISL_1721393, EPI_ISL_1721408, EPI_ISL_1721409, EPI_ISL_1721412, EPI_ISL_1721418, EPI_ISL_1721431, EPI_ISL_1721442, EPI_ISL_1721465, EPI_ISL_1721478, EPI_ISL_1721479, EPI_ISL_1721534, EPI_ISL_1721537, EPI_ISL_1721546, EPI_ISL_1721595, EPI_ISL_1721603, EPI_ISL_1721609, EPI_ISL_1721633, EPI_ISL_1721640, EPI_ISL_1721661, EPI_ISL_1721664 | | | |

| | | | |
|---|---|---|---|
| see above | LabKom - Labor an der SalzbrÃ¼cke MVZ GmbH | Robert Koch Institute | |
| EPI_ISL_1721104, EPI_ISL_1721107, EPI_ISL_1721116, EPI_ISL_1721122, EPI_ISL_1721130, EPI_ISL_1721131, EPI_ISL_1721132, EPI_ISL_1721134, EPI_ISL_1721137, EPI_ISL_1721148, EPI_ISL_1721155, EPI_ISL_1721157, EPI_ISL_1721159, EPI_ISL_1721161, EPI_ISL_1721166, EPI_ISL_1721171, EPI_ISL_1721176, EPI_ISL_1721179, EPI_ISL_1721182, EPI_ISL_1721184, EPI_ISL_1721203, EPI_ISL_1721212, EPI_ISL_1721216, EPI_ISL_1721217, EPI_ISL_1721227, EPI_ISL_1721237, EPI_ISL_1721238, EPI_ISL_1721245, EPI_ISL_1721247, EPI_ISL_1721249, EPI_ISL_1721253, EPI_ISL_1721255, EPI_ISL_1721262, EPI_ISL_1721267, EPI_ISL_1721269, EPI_ISL_1721271, EPI_ISL_1721280, EPI_ISL_1721282, EPI_ISL_1721285, EPI_ISL_1721291, EPI_ISL_1721298, EPI_ISL_1721304, EPI_ISL_1721311, EPI_ISL_1721314, EPI_ISL_1721325, EPI_ISL_1721328, EPI_ISL_1721330, EPI_ISL_1721333, EPI_ISL_1721334, EPI_ISL_1721335, EPI_ISL_1721338, EPI_ISL_1721340, EPI_ISL_1721341, EPI_ISL_1721342, EPI_ISL_1721347, EPI_ISL_1721349, EPI_ISL_1721352, EPI_ISL_1721363, EPI_ISL_1721367, EPI_ISL_1721382 | | | |
| see above | LabKom - MVZ Labor Bochum MLB GmbH | Robert Koch Institute | |
| EPI_ISL_1642462, EPI_ISL_1642474, EPI_ISL_1642530, EPI_ISL_1642556, EPI_ISL_1642582, EPI_ISL_1642599, EPI_ISL_1642622, EPI_ISL_1643226, EPI_ISL_1721889, EPI_ISL_1843012, EPI_ISL_1843017, EPI_ISL_1843089, EPI_ISL_1843116, EPI_ISL_1843120, EPI_ISL_1843134, EPI_ISL_1847100, EPI_ISL_1847109, EPI_ISL_1847136, EPI_ISL_1847150 | | | |
| see above | Labor 28 MVZ GmbH | Robert Koch Institute | |
| EPI_ISL_2123980, EPI_ISL_2123983, EPI_ISL_2123984, EPI_ISL_2123987, EPI_ISL_2123990, EPI_ISL_2123992, EPI_ISL_2123993, EPI_ISL_2123997, EPI_ISL_2124003, EPI_ISL_2124013 | | | |
| see above | Labor Becker & Kollegen (Standort Munchen) | Robert Koch Institute | |
| EPI_ISL_1846786, EPI_ISL_1846809 | Labor Becker & Kollegen (Standort MÃ¼nchen) | Robert Koch Institute | |
| EPI_ISL_1905067, EPI_ISL_1905077, EPI_ISL_1905093, EPI_ISL_1905097, EPI_ISL_1905101, EPI_ISL_1905103, EPI_ISL_1905104, EPI_ISL_1905108, EPI_ISL_1905109, EPI_ISL_1905132, EPI_ISL_1905133, EPI_ISL_1905135, EPI_ISL_1905137, EPI_ISL_1905139, EPI_ISL_1905140, EPI_ISL_1905141, EPI_ISL_1905142, EPI_ISL_1905143, EPI_ISL_1905144, EPI_ISL_1905146, EPI_ISL_1905147, EPI_ISL_1905148, EPI_ISL_1905149, EPI_ISL_1905150, EPI_ISL_1905156, EPI_ISL_1905157, EPI_ISL_1905158, EPI_ISL_1905160, EPI_ISL_1905165, EPI_ISL_1905166, EPI_ISL_1905169, EPI_ISL_1905171, EPI_ISL_1905172, EPI_ISL_1905174, EPI_ISL_1905175, EPI_ISL_1905176, EPI_ISL_1905177, EPI_ISL_1905178, EPI_ISL_1905181, EPI_ISL_1905182, EPI_ISL_1972152, EPI_ISL_1972164, EPI_ISL_1972169, EPI_ISL_1972170, EPI_ISL_1972200 | | | |
| see above | Labor Berlin Charité Vivantes GmbH / Institut für Virologie | Charité Universitätsmedizin Berlin, Institut für Virologie/Labor Berlin | Barbara Mühlemann; Christian Drosten; Christine Stephan; Peter Menzel; Rolf Schwarzer; Terry Jones; Victor M Corman |
| EPI_ISL_1861710, EPI_ISL_1861712, EPI_ISL_1861714, EPI_ISL_1861716, EPI_ISL_1861718, EPI_ISL_1861720, EPI_ISL_1861733, EPI_ISL_1861735, EPI_ISL_1861737, EPI_ISL_1861739, EPI_ISL_1861741, EPI_ISL_1861743, EPI_ISL_1861755, EPI_ISL_1861757, EPI_ISL_1861761, EPI_ISL_1861763, EPI_ISL_1861778, EPI_ISL_1861780, EPI_ISL_1861782, EPI_ISL_1861784, EPI_ISL_1861797, EPI_ISL_1861799, EPI_ISL_1861801, EPI_ISL_1861803, EPI_ISL_1861805, EPI_ISL_1861815, EPI_ISL_1861816, EPI_ISL_1861820, EPI_ISL_1861837, EPI_ISL_1861839, EPI_ISL_1861841, EPI_ISL_1861843, EPI_ISL_1861857, EPI_ISL_1861859, EPI_ISL_1861861, EPI_ISL_1861863, EPI_ISL_1861882, EPI_ISL_1861884, EPI_ISL_1861886, EPI_ISL_1861888, EPI_ISL_1861889, EPI_ISL_1861893, EPI_ISL_1861895, EPI_ISL_1861899, EPI_ISL_1861903, EPI_ISL_1861905, EPI_ISL_1861906, EPI_ISL_1861908, EPI_ISL_1861910, EPI_ISL_1861912, EPI_ISL_1861914, EPI_ISL_1861916, EPI_ISL_1861918, EPI_ISL_1861920, EPI_ISL_1861924, EPI_ISL_1861926, EPI_ISL_1861928, EPI_ISL_1861930, EPI_ISL_1861931, EPI_ISL_1861933, EPI_ISL_1861935, EPI_ISL_1861937, EPI_ISL_1861939, EPI_ISL_1861941, EPI_ISL_1861943, EPI_ISL_1861944, EPI_ISL_1861946, EPI_ISL_1861948, EPI_ISL_1861950, EPI_ISL_1861952, EPI_ISL_1861954, EPI_ISL_1861956, EPI_ISL_1861958, EPI_ISL_1861960, EPI_ISL_1861962, EPI_ISL_1861964, EPI_ISL_1861966, EPI_ISL_1861968, EPI_ISL_1861971, EPI_ISL_1861973, EPI_ISL_1861975, EPI_ISL_1861977, EPI_ISL_1861979, EPI_ISL_1861981, EPI_ISL_1861984, EPI_ISL_1861986, EPI_ISL_1861988, EPI_ISL_1861990, EPI_ISL_1861992, EPI_ISL_1861994, EPI_ISL_1861996, EPI_ISL_1861998, EPI_ISL_1862001, EPI_ISL_1862003, EPI_ISL_1862005, EPI_ISL_1862007, EPI_ISL_1862009, EPI_ISL_1862011, EPI_ISL_1862013, EPI_ISL_1862015, EPI_ISL_1862017, EPI_ISL_1862019, EPI_ISL_1862021, EPI_ISL_1862023, EPI_ISL_1862025, EPI_ISL_1862028, EPI_ISL_1862030, EPI_ISL_1862032, EPI_ISL_1862034, EPI_ISL_1862037, EPI_ISL_1862039, EPI_ISL_1862041, EPI_ISL_1862044, EPI_ISL_1862046, EPI_ISL_1862048, EPI_ISL_1862050, EPI_ISL_1862063, EPI_ISL_1862064, EPI_ISL_1862078, EPI_ISL_1862080, EPI_ISL_1862082, EPI_ISL_1862086, EPI_ISL_1862096, EPI_ISL_1862098, EPI_ISL_1862103, EPI_ISL_1862107, EPI_ISL_1862119, EPI_ISL_1862123, EPI_ISL_1862125, EPI_ISL_1862131, EPI_ISL_1862144, EPI_ISL_1862165, EPI_ISL_1862169, EPI_ISL_1862171, EPI_ISL_1862173, EPI_ISL_1862175, EPI_ISL_1862183, EPI_ISL_1862185, EPI_ISL_1862189, EPI_ISL_1862210, EPI_ISL_1862214, EPI_ISL_1862216, EPI_ISL_1862218, EPI_ISL_1862222, EPI_ISL_1862224, EPI_ISL_1862226, EPI_ISL_1862228, EPI_ISL_1862231, EPI_ISL_1862233, EPI_ISL_1862235, EPI_ISL_1862237, EPI_ISL_1862239, EPI_ISL_1862249, EPI_ISL_1862254, EPI_ISL_1862256, EPI_ISL_1862264, EPI_ISL_1862267, EPI_ISL_1862269, EPI_ISL_1862271, EPI_ISL_1862273, EPI_ISL_1862283, EPI_ISL_1862287, EPI_ISL_1862289, EPI_ISL_1862291, EPI_ISL_1862293, EPI_ISL_1862295, EPI_ISL_1862297, EPI_ISL_1862299, EPI_ISL_1862307, EPI_ISL_1862309, EPI_ISL_1862311, EPI_ISL_1862313, EPI_ISL_1862316, EPI_ISL_1862318, EPI_ISL_1862320, EPI_ISL_1862322, EPI_ISL_1862330, EPI_ISL_1862332, EPI_ISL_1862333, EPI_ISL_1862336, EPI_ISL_1862338, EPI_ISL_1862345, EPI_ISL_1862347, EPI_ISL_1862352, EPI_ISL_1862354, EPI_ISL_1862356, EPI_ISL_1862368, EPI_ISL_1862371, EPI_ISL_1862373, EPI_ISL_1862375, EPI_ISL_1862378, EPI_ISL_1862380 | | | |
| see above | Labor Dr. Fenner und Kollegen | Heinrich Pette Institute, Leibniz Institute for Experimental Virology | Adam Grundhoff; Alexis Robitaille; Johannes Knobloch; Martin Aepfelbacher; Nicole Fischer; Thomas Günther |
| EPI_ISL_1566769, EPI_ISL_1640070, EPI_ISL_1640071, EPI_ISL_1640072, EPI_ISL_1640073, EPI_ISL_1640074, EPI_ISL_1640075, EPI_ISL_1640076, EPI_ISL_1640077, EPI_ISL_1640078, EPI_ISL_1640080, EPI_ISL_1640081, EPI_ISL_1640082, EPI_ISL_1640084, EPI_ISL_1640085, EPI_ISL_1640086, EPI_ISL_1640087, EPI_ISL_1640088, EPI_ISL_1640089, EPI_ISL_1640090, EPI_ISL_1640092, EPI_ISL_1640093, EPI_ISL_1640094, EPI_ISL_1640095, EPI_ISL_1640096, EPI_ISL_1640097, EPI_ISL_1640098, EPI_ISL_1640099, EPI_ISL_1640100, EPI_ISL_1640101, EPI_ISL_1640102, EPI_ISL_1640103, EPI_ISL_1640104, EPI_ISL_1640105, EPI_ISL_1640106, EPI_ISL_1640107, EPI_ISL_1640108, EPI_ISL_1640109, EPI_ISL_1640111, EPI_ISL_1640112, EPI_ISL_1640113, EPI_ISL_1640116, EPI_ISL_1640117, EPI_ISL_1640118, EPI_ISL_1640119, EPI_ISL_1640120, EPI_ISL_1723037 | | | |
| see above | Labor Dr. Heidrich & Kollegen MVZ GmbH Hamburg | Robert Koch Institute | |
| EPI_ISL_1643844, EPI_ISL_1643845, EPI_ISL_1643847, EPI_ISL_1847405, EPI_ISL_1847409, EPI_ISL_1847411, EPI_ISL_1847412, EPI_ISL_1847413, EPI_ISL_1847414 | | | |
| see above | Labor Dr. Spranger | Robert Koch Institute | |
| EPI_ISL_1571539, EPI_ISL_1571540, EPI_ISL_1571541, EPI_ISL_1571542, EPI_ISL_1571543, EPI_ISL_1571545, EPI_ISL_1571546, EPI_ISL_1571547, EPI_ISL_1571548 | | | |
| see above | Labor Dr. Wisplinghoff - KÃ¶ln | Robert Koch Institute | |
| EPI_ISL_1639264, EPI_ISL_1639265, EPI_ISL_1639266, EPI_ISL_1639267, EPI_ISL_1639268, EPI_ISL_1639269, EPI_ISL_1639270, EPI_ISL_1639271, EPI_ISL_1639272, EPI_ISL_1639273, EPI_ISL_1639274, EPI_ISL_1643966, EPI_ISL_1643967 | | | |
| see above | Labor KneiÃŸler GmbH & Co. KG | Robert Koch Institute | |
| EPI_ISL_1643416, EPI_ISL_1643423, EPI_ISL_1643424, EPI_ISL_1643435, EPI_ISL_1643437, EPI_ISL_1643439, EPI_ISL_1643450, EPI_ISL_1643451, EPI_ISL_1643459, EPI_ISL_1643468, EPI_ISL_1643474, EPI_ISL_1643492 | | | |
| see above | Labor LÃ¼beck bzw. LaborÃ¤rztliche Gemeinschaftspraxis LÃ¼beck | Robert Koch Institute | |
| EPI_ISL_1571172, EPI_ISL_1571178, EPI_ISL_1571181, EPI_ISL_1571182, EPI_ISL_1571189, EPI_ISL_1571195, EPI_ISL_1571197, EPI_ISL_1571202, EPI_ISL_1571930, EPI_ISL_1571932, EPI_ISL_1571933, EPI_ISL_1571936, EPI_ISL_1571937, EPI_ISL_1571938, EPI_ISL_1571944, EPI_ISL_1571945, EPI_ISL_1571947, EPI_ISL_1571948, EPI_ISL_1571951, EPI_ISL_1571953, EPI_ISL_1571956, EPI_ISL_1571959, EPI_ISL_1571960, EPI_ISL_1571961, EPI_ISL_1571962, EPI_ISL_1571963, EPI_ISL_1638298, EPI_ISL_1643765, EPI_ISL_1643767, EPI_ISL_1643769, EPI_ISL_1643773, EPI_ISL_1643784, EPI_ISL_1643801, EPI_ISL_1643812 | | | |
| see above | Labor MÃ¶nchengladbach MVZ Dr. Stein + Kollegen GbR | Robert Koch Institute | |
| EPI_ISL_1638309, EPI_ISL_1638318, EPI_ISL_1638390, EPI_ISL_1638418, EPI_ISL_1643785, EPI_ISL_1643786 | Labor Prof. Dr. G. Enders MVZ GbR | Robert Koch Institute | |
| EPI_ISL_1644844, EPI_ISL_1644845, EPI_ISL_1644846, EPI_ISL_1644847, EPI_ISL_1644848, EPI_ISL_1644849, EPI_ISL_1644850, EPI_ISL_1644851, EPI_ISL_1644852, EPI_ISL_1644853, EPI_ISL_1644854, EPI_ISL_1644855, EPI_ISL_1725072, EPI_ISL_1725105, EPI_ISL_1725109, EPI_ISL_1725112, EPI_ISL_1725113, EPI_ISL_1725117, EPI_ISL_1725144, EPI_ISL_1725146, EPI_ISL_1725154, EPI_ISL_1851148, EPI_ISL_1851174, EPI_ISL_1851192, EPI_ISL_1851196, EPI_ISL_1851200, EPI_ISL_1851206, EPI_ISL_1851224, EPI_ISL_1851227, EPI_ISL_1851235, EPI_ISL_1851252, EPI_ISL_1851259, EPI_ISL_1851263, EPI_ISL_1851271, EPI_ISL_1851274, EPI_ISL_1851288, EPI_ISL_1851290, EPI_ISL_1851310, EPI_ISL_1851326, EPI_ISL_1851331, EPI_ISL_1851337, EPI_ISL_1851366, EPI_ISL_2129293 | | | |
| see above | Labor ZOTZ\|KLIMAS; MVZ DÃ¼sseldorf-Centrum | Robert Koch Institute | |
| EPI_ISL_1570967, EPI_ISL_1570981, EPI_ISL_1570989, EPI_ISL_1571022, EPI_ISL_1571062, EPI_ISL_1571063, EPI_ISL_1571067, EPI_ISL_1571071, EPI_ISL_1571083, EPI_ISL_1571092, EPI_ISL_1571094, EPI_ISL_1571100, EPI_ISL_1573614, EPI_ISL_1573647, EPI_ISL_1573764, EPI_ISL_1573845, EPI_ISL_1574006, EPI_ISL_1574039, EPI_ISL_1574755, EPI_ISL_1574790, EPI_ISL_1574921, EPI_ISL_1574936, EPI_ISL_1574939 | | | |
| see above | Labor fÃ¼r medizinische Mikrobiologie Bakteriologie Mykologie Virologie & Infektionsserologie - Labor MÃ¶lbis | Robert Koch Institute | |
| EPI_ISL_1570823, EPI_ISL_1570849, EPI_ISL_1570886, EPI_ISL_1570892, EPI_ISL_1570894, EPI_ISL_1570896, EPI_ISL_1570901, EPI_ISL_1570922 | | | |
| see above | Laborarztpraxis Jochen HÃ¼ter | Robert Koch Institute | |
| EPI_ISL_1642310, EPI_ISL_1642317, EPI_ISL_1642319, EPI_ISL_1642320, EPI_ISL_1642324, EPI_ISL_1642336, EPI_ISL_1642344, EPI_ISL_1642345, EPI_ISL_1642346, EPI_ISL_1642347, EPI_ISL_1642348, EPI_ISL_1721064 | | | |
| see above | Labormedizin Darmstadt | Robert Koch Institute | |
| EPI_ISL_1670271, EPI_ISL_1670278, EPI_ISL_1670279, EPI_ISL_1670280, EPI_ISL_1670281, EPI_ISL_1670282, EPI_ISL_1670283, EPI_ISL_1670284, EPI_ISL_1670285, EPI_ISL_1670286, EPI_ISL_1670287, EPI_ISL_1670288, EPI_ISL_1670289, EPI_ISL_1670290, EPI_ISL_1670291, EPI_ISL_1670292, EPI_ISL_1670295, EPI_ISL_1670296, EPI_ISL_1670297, EPI_ISL_1670298, EPI_ISL_1670299, EPI_ISL_1670300, EPI_ISL_1670301, EPI_ISL_1670302, EPI_ISL_1670303, EPI_ISL_1670304, EPI_ISL_1670305, EPI_ISL_1670306, EPI_ISL_1670307, EPI_ISL_1670308, EPI_ISL_1670309, EPI_ISL_1670310, EPI_ISL_1670311, EPI_ISL_1670312, EPI_ISL_1670313, EPI_ISL_1670314, EPI_ISL_1670315, EPI_ISL_1670316, EPI_ISL_1670317, EPI_ISL_1670318, EPI_ISL_1670319, EPI_ISL_1670320, EPI_ISL_1670321, EPI_ISL_1670322, EPI_ISL_1670323, EPI_ISL_1670324, EPI_ISL_1670325, EPI_ISL_1670326, EPI_ISL_1670338, EPI_ISL_1670339, EPI_ISL_1670340, EPI_ISL_1670341, EPI_ISL_1670342, EPI_ISL_1670343, EPI_ISL_1670344, EPI_ISL_1670345, EPI_ISL_1670346, EPI_ISL_1670347, EPI_ISL_1670348, EPI_ISL_1670349, EPI_ISL_1670350, EPI_ISL_1670351, EPI_ISL_1670352, EPI_ISL_1670353, EPI_ISL_1670354, EPI_ISL_1670355, EPI_ISL_1670356, EPI_ISL_1670357, EPI_ISL_1670358, EPI_ISL_1670359, EPI_ISL_1670360, EPI_ISL_1670361, EPI_ISL_1670362, EPI_ISL_1670363, EPI_ISL_1670364, EPI_ISL_1670365, EPI_ISL_1670366, EPI_ISL_1670367, EPI_ISL_1670368, EPI_ISL_1670369, EPI_ISL_1670370, EPI_ISL_1670371, EPI_ISL_1670372, EPI_ISL_1670373, EPI_ISL_1670374, EPI_ISL_1670375, EPI_ISL_1670379, EPI_ISL_1670380, EPI_ISL_1670381, EPI_ISL_1670382, EPI_ISL_1670383, EPI_ISL_1670384, EPI_ISL_1670385, EPI_ISL_1670386, EPI_ISL_1670387, EPI_ISL_1670388, EPI_ISL_1670389, EPI_ISL_1670390, EPI_ISL_1670391, EPI_ISL_1670392, EPI_ISL_1670393, EPI_ISL_1670394, EPI_ISL_1670395, EPI_ISL_1670396, EPI_ISL_1670397, EPI_ISL_1670398, EPI_ISL_1670399, EPI_ISL_1670400, EPI_ISL_1670401, EPI_ISL_1670402, EPI_ISL_1670403, EPI_ISL_1670404, EPI_ISL_1670405, EPI_ISL_1670406, EPI_ISL_1670407, EPI_ISL_1670408, EPI_ISL_1670409, EPI_ISL_1670410, EPI_ISL_1670411, EPI_ISL_1670412, EPI_ISL_1670413, EPI_ISL_1670414, EPI_ISL_1670415, EPI_ISL_1670416, EPI_ISL_1670417, EPI_ISL_1940849, EPI_ISL_1940850, EPI_ISL_1940851, EPI_ISL_1940852, EPI_ISL_1940853, EPI_ISL_1940854, EPI_ISL_1940855, EPI_ISL_1940856, EPI_ISL_1941170 | | | |
| see above | Landesamt für Verbraucherschutz Sachsen Anhalt, Magdeburg | Institute of Medical Microbiology and Hospital Hygiene | Aljoscha Tersteegen; Prof. Dr. Achim Kaasch |
| EPI_ISL_2110766, EPI_ISL_2110771, EPI_ISL_2110772, EPI_ISL_2110775, EPI_ISL_2110778, EPI_ISL_2110779, EPI_ISL_2110780, EPI_ISL_2110783 | | | |
| see above | Landesgesundheitsamt Baden-Wurttemberg | Robert Koch Institute | |
| EPI_ISL_1846318, EPI_ISL_1846319, EPI_ISL_1846320, EPI_ISL_1846321, EPI_ISL_1846322, EPI_ISL_1846326, EPI_ISL_1846328, EPI_ISL_1846329, EPI_ISL_1846330, EPI_ISL_1846331, EPI_ISL_1846356 | | | |
| see above | Landesgesundheitsamt Baden-WÃ¼rttemberg | Robert Koch Institute | |
| EPI_ISL_1569885, EPI_ISL_1569887, EPI_ISL_1569890, EPI_ISL_1569898, EPI_ISL_1569901, EPI_ISL_1569904, EPI_ISL_1569907, EPI_ISL_1569909, EPI_ISL_1569913, EPI_ISL_1569926, EPI_ISL_1569931, EPI_ISL_1569932, EPI_ISL_1569933, EPI_ISL_1569934, EPI_ISL_1569938, EPI_ISL_1569944, EPI_ISL_1569947, EPI_ISL_1569949, EPI_ISL_1569952, EPI_ISL_1569953, EPI_ISL_1569954, EPI_ISL_1569960, EPI_ISL_1569965, EPI_ISL_1569980, EPI_ISL_1569981, EPI_ISL_1569994 | | | |
| see above | Landesuntersuchungsanstalt fÃ¼r das Gesundheits- und VeterinÃ¤rwesen (LUA) Sachsen | Robert Koch Institute | |
| EPI_ISL_1572180, EPI_ISL_1572182, EPI_ISL_1572184, EPI_ISL_1572185, EPI_ISL_1572186 | Limbach - Ganzimmun Diagnostics AG Mainz | Robert Koch Institute | |
| EPI_ISL_1570817, EPI_ISL_1570828, EPI_ISL_1570836, EPI_ISL_1570936, EPI_ISL_1570938, EPI_ISL_1571136, EPI_ISL_1571158, EPI_ISL_1571159, EPI_ISL_1572140, EPI_ISL_1572141, EPI_ISL_1572143, EPI_ISL_1572147, EPI_ISL_1572149, EPI_ISL_1572169 | | | |
| see above | Limbach - MVZ Gemeinschaftslabor Suhl | Robert Koch Institute | |
| EPI_ISL_1567117, EPI_ISL_1567119, EPI_ISL_1567121, EPI_ISL_1567122, EPI_ISL_1567123, EPI_ISL_1567128, EPI_ISL_1567132, EPI_ISL_1567133, EPI_ISL_1567135, EPI_ISL_1567136, EPI_ISL_1567138, EPI_ISL_1567139, EPI_ISL_1567140, EPI_ISL_1567143, EPI_ISL_1567144, EPI_ISL_1567145, EPI_ISL_1567147, EPI_ISL_1567148, EPI_ISL_1567150, EPI_ISL_1567151, EPI_ISL_1567153, EPI_ISL_1567154, EPI_ISL_1567155, EPI_ISL_1567156, EPI_ISL_1567157, EPI_ISL_1567159, EPI_ISL_1567160, EPI_ISL_1567165, EPI_ISL_1567168, EPI_ISL_1567171, EPI_ISL_1567174, EPI_ISL_1567177, EPI_ISL_1567178, EPI_ISL_1567180, EPI_ISL_1567182, EPI_ISL_1567183, EPI_ISL_1567184, EPI_ISL_1567185, EPI_ISL_1567187, EPI_ISL_1567189, EPI_ISL_1567190, EPI_ISL_1567191, EPI_ISL_1567196, EPI_ISL_1567200, EPI_ISL_1567201, EPI_ISL_1567202, EPI_ISL_1567203, EPI_ISL_1567205, EPI_ISL_1567206, EPI_ISL_1567207, EPI_ISL_1567209, EPI_ISL_1567210, EPI_ISL_1567211, EPI_ISL_1567213, EPI_ISL_1567215, EPI_ISL_1567217, EPI_ISL_1567219, EPI_ISL_1567222, EPI_ISL_1567223, EPI_ISL_1567224, EPI_ISL_1567225, EPI_ISL_1567227, EPI_ISL_1567231, EPI_ISL_1567243, EPI_ISL_1567246, EPI_ISL_1567249, EPI_ISL_1567250, EPI_ISL_1567254, EPI_ISL_1567255, EPI_ISL_1567257, EPI_ISL_1567263, EPI_ISL_1567264, EPI_ISL_1567266, EPI_ISL_1567272, EPI_ISL_1567276, EPI_ISL_1567278, EPI_ISL_1573298, EPI_ISL_1573301, EPI_ISL_1573302, EPI_ISL_1573304, EPI_ISL_1573306, EPI_ISL_1573308, EPI_ISL_1573311, EPI_ISL_1573312, EPI_ISL_1573314, EPI_ISL_1573318, EPI_ISL_1573319, EPI_ISL_1573322, EPI_ISL_1573323, EPI_ISL_1573324, EPI_ISL_1573325, EPI_ISL_1573327, EPI_ISL_1573328, EPI_ISL_1573330, EPI_ISL_1573332, EPI_ISL_1573335, EPI_ISL_1573336, EPI_ISL_1573339, EPI_ISL_1573340, EPI_ISL_1573343, EPI_ISL_1573344, EPI_ISL_1573345, EPI_ISL_1573348, EPI_ISL_1573350, EPI_ISL_1573353, EPI_ISL_1573354, EPI_ISL_1573356, EPI_ISL_1573357, EPI_ISL_1573359, EPI_ISL_1573361, EPI_ISL_1573363, EPI_ISL_1573366, EPI_ISL_1573368, EPI_ISL_1573369, EPI_ISL_1573370, EPI_ISL_1573373, EPI_ISL_1573375, EPI_ISL_1573378, EPI_ISL_1573379, EPI_ISL_1573386, EPI_ISL_1573394, EPI_ISL_1573398, EPI_ISL_1573400, EPI_ISL_1573401, EPI_ISL_1573403, EPI_ISL_1573405, EPI_ISL_1573408, EPI_ISL_1573409, EPI_ISL_1573411, EPI_ISL_1573412, EPI_ISL_1573414, EPI_ISL_1573417, EPI_ISL_1573419, EPI_ISL_1573420, EPI_ISL_1573422, EPI_ISL_1573423, EPI_ISL_1573425, EPI_ISL_1573427, EPI_ISL_1573429, EPI_ISL_1573436, EPI_ISL_1573438, EPI_ISL_1573439, EPI_ISL_1573442, EPI_ISL_1573443, EPI_ISL_1573445, EPI_ISL_1573448, EPI_ISL_1573450, EPI_ISL_1573453, EPI_ISL_1573455, EPI_ISL_1573458, EPI_ISL_1573459, EPI_ISL_1573460, EPI_ISL_1573461, EPI_ISL_1573462, EPI_ISL_1573463, EPI_ISL_1573464, EPI_ISL_1573465 | | | |
| see above | Limbach - MVZ Humangenetik Ulm | Robert Koch Institute | |
| EPI_ISL_1567566, EPI_ISL_1570790, EPI_ISL_1570800, EPI_ISL_1570810, EPI_ISL_1570815, EPI_ISL_1571160, EPI_ISL_1571161, EPI_ISL_1571169, EPI_ISL_1571170, EPI_ISL_1571171, EPI_ISL_1571173, EPI_ISL_1571175, EPI_ISL_1571176, EPI_ISL_1571184, EPI_ISL_1571187, EPI_ISL_1571188, EPI_ISL_1571192, EPI_ISL_1571193, EPI_ISL_1571200, EPI_ISL_1571203, EPI_ISL_1638291, EPI_ISL_1638293, EPI_ISL_1638300, EPI_ISL_1638331, EPI_ISL_1638355 | | | |
| see above | Limbach - MVZ Labor Eveld & Kollegen Essen | Robert Koch Institute | |
| EPI_ISL_1571132, EPI_ISL_1571133, EPI_ISL_1571142, EPI_ISL_1571150, EPI_ISL_1571156, EPI_ISL_1571157 | Limbach - MVZ Labor Ludwigsburg | Robert Koch Institute | |
| EPI_ISL_1570960 | Limbach - MVZ Labor Main-Taunus GbR | Robert Koch Institute | |

| | | | |
|---|---|---|---|
| EPI_ISL_1567555, EPI_ISL_1567606, EPI_ISL_1567653, EPI_ISL_1567675, EPI_ISL_1567685, EPI_ISL_1567688, EPI_ISL_1567704, EPI_ISL_1570837, EPI_ISL_1571120, EPI_ISL_1571122, EPI_ISL_1571123, EPI_ISL_1571124, EPI_ISL_1571125, EPI_ISL_1571127, EPI_ISL_1571129, EPI_ISL_1571131, EPI_ISL_1571134, EPI_ISL_1571138, EPI_ISL_1571140, EPI_ISL_1571146, EPI_ISL_1571147, EPI_ISL_1571151, EPI_ISL_1571152, EPI_ISL_1571153, EPI_ISL_1571155, EPI_ISL_1638285, EPI_ISL_1638287, EPI_ISL_1638290, EPI_ISL_1638301, EPI_ISL_1638304, EPI_ISL_1638306, EPI_ISL_1638308, EPI_ISL_1638312, EPI_ISL_1638315, EPI_ISL_1638320, EPI_ISL_1638325, EPI_ISL_1638326, EPI_ISL_1638327, EPI_ISL_1638328, EPI_ISL_1638329, EPI_ISL_1638346, EPI_ISL_1638347, EPI_ISL_1638349, EPI_ISL_1638353, EPI_ISL_1638361, EPI_ISL_1638364, EPI_ISL_1638366, EPI_ISL_1638371, EPI_ISL_1638376, EPI_ISL_1638378, EPI_ISL_1638381, EPI_ISL_1638382, EPI_ISL_1638383, EPI_ISL_1638387, EPI_ISL_1638398, EPI_ISL_1638403, EPI_ISL_1638408, EPI_ISL_1638416, EPI_ISL_1638419, EPI_ISL_1638421, EPI_ISL_1638424, EPI_ISL_1638425, EPI_ISL_1638427, EPI_ISL_1638430, EPI_ISL_1638431, EPI_ISL_1638436, EPI_ISL_1638447, EPI_ISL_1638449, EPI_ISL_1638453, EPI_ISL_1638459, EPI_ISL_1638461, EPI_ISL_1638464, EPI_ISL_1638474, EPI_ISL_1638475, EPI_ISL_1638476, EPI_ISL_1639253, EPI_ISL_1643863, EPI_ISL_1643876, EPI_ISL_1643877, EPI_ISL_1643878, EPI_ISL_1720817, EPI_ISL_1720818, EPI_ISL_1720819, EPI_ISL_1720821, EPI_ISL_1720829, EPI_ISL_1720836, EPI_ISL_1720839, EPI_ISL_1720841, EPI_ISL_1720842, EPI_ISL_1720843, EPI_ISL_1720845, EPI_ISL_1725624, EPI_ISL_1846226 | | | |
| see above | MDI Limbach Berlin GmbH; MVZ Labor Berlin | Robert Koch Institute | |
| EPI_ISL_1715842, EPI_ISL_1716267, EPI_ISL_1716277, EPI_ISL_1716278, EPI_ISL_1716279 | MEDILYS Laborgesellschaft mbH | Heinrich Pette Institute, Leibniz Institute for Experimental Virology | Adam Grundhoff; Alexis Robitaille; Johannes Knobloch; Martin Aepfelbacher; Nicole Fischer; Thomas Günther |
| EPI_ISL_1567767, EPI_ISL_1567768, EPI_ISL_1567769, EPI_ISL_1567770, EPI_ISL_1567771, EPI_ISL_1567774, EPI_ISL_1567778, EPI_ISL_1567780, EPI_ISL_1567783, EPI_ISL_1567786, EPI_ISL_1567789, EPI_ISL_1567798, EPI_ISL_1567800, EPI_ISL_1567801, EPI_ISL_1567802, EPI_ISL_1567803, EPI_ISL_1567805, EPI_ISL_1567809, EPI_ISL_1567814, EPI_ISL_1567820, EPI_ISL_1567822, EPI_ISL_1567824, EPI_ISL_1567825, EPI_ISL_1567826, EPI_ISL_1567827, EPI_ISL_1567831, EPI_ISL_1567832, EPI_ISL_1572107, EPI_ISL_1572109, EPI_ISL_1572111, EPI_ISL_1572112, EPI_ISL_1572113, EPI_ISL_1572114, EPI_ISL_1572115, EPI_ISL_1572116, EPI_ISL_1572117, EPI_ISL_1572118, EPI_ISL_1572119, EPI_ISL_1572120, EPI_ISL_1572121, EPI_ISL_1572122, EPI_ISL_1572123, EPI_ISL_1572124, EPI_ISL_1572125, EPI_ISL_1572126, EPI_ISL_1572127, EPI_ISL_1572128, EPI_ISL_1572129, EPI_ISL_1572130, EPI_ISL_1572131, EPI_ISL_1572132, EPI_ISL_1572133, EPI_ISL_1572134, EPI_ISL_1572135, EPI_ISL_1638497, EPI_ISL_1638511, EPI_ISL_1638513, EPI_ISL_1638515, EPI_ISL_1638517, EPI_ISL_1638519, EPI_ISL_1638522, EPI_ISL_1638524, EPI_ISL_1638526, EPI_ISL_1638531, EPI_ISL_1643417, EPI_ISL_1643422 | | | |
| see above | MVZ Dr. Eberhard & Partner Dortmund | Robert Koch Institute | |
| EPI_ISL_1571119, EPI_ISL_1572164, EPI_ISL_1638367 | MVZ Labor Dortmund | Robert Koch Institute | |
| EPI_ISL_1571118 | MVZ Labor Dr. Fenner und Kollegen (Standort Hamburg) | Robert Koch Institute | |
| EPI_ISL_1570710, EPI_ISL_1570711, EPI_ISL_1570712, EPI_ISL_1570713, EPI_ISL_1570714, EPI_ISL_1570715, EPI_ISL_1570716, EPI_ISL_1570718, EPI_ISL_1570719, EPI_ISL_1570720, EPI_ISL_1570721, EPI_ISL_1570723, EPI_ISL_1570724, EPI_ISL_1570725, EPI_ISL_1570729, EPI_ISL_1570732, EPI_ISL_1570733, EPI_ISL_1570734, EPI_ISL_1570736, EPI_ISL_1570737, EPI_ISL_1570739, EPI_ISL_1570740, EPI_ISL_1570741, EPI_ISL_1570742, EPI_ISL_1570743, EPI_ISL_1570745, EPI_ISL_1570756, EPI_ISL_1570763, EPI_ISL_1570768, EPI_ISL_1570770, EPI_ISL_1570779, EPI_ISL_1570782, EPI_ISL_1570784, EPI_ISL_1570789, EPI_ISL_1570793, EPI_ISL_1570794, EPI_ISL_1570796, EPI_ISL_1570802, EPI_ISL_1570805, EPI_ISL_1570807, EPI_ISL_1570818, EPI_ISL_1570820, EPI_ISL_1570821, EPI_ISL_1570834, EPI_ISL_1570914, EPI_ISL_1570925, EPI_ISL_1570928, EPI_ISL_1570937, EPI_ISL_1570941, EPI_ISL_1570942, EPI_ISL_1570943, EPI_ISL_1570945, EPI_ISL_1570946, EPI_ISL_1570952, EPI_ISL_1570954, EPI_ISL_1570956, EPI_ISL_1570959, EPI_ISL_1570961, EPI_ISL_1572187, EPI_ISL_1572188, EPI_ISL_1572189, EPI_ISL_1572190, EPI_ISL_1572191, EPI_ISL_1572192, EPI_ISL_1572193, EPI_ISL_1572194, EPI_ISL_1572195, EPI_ISL_1572196, EPI_ISL_1572197, EPI_ISL_1572198, EPI_ISL_1572199, EPI_ISL_1572200, EPI_ISL_1572201, EPI_ISL_1572202, EPI_ISL_1572203, EPI_ISL_1572206, EPI_ISL_1572207, EPI_ISL_1572208, EPI_ISL_1572209, EPI_ISL_1572210, EPI_ISL_1572211, EPI_ISL_1572212, EPI_ISL_1572213, EPI_ISL_1572214, EPI_ISL_1572216, EPI_ISL_1572217, EPI_ISL_1572218, EPI_ISL_1572219, EPI_ISL_1572220, EPI_ISL_1572222, EPI_ISL_1572223, EPI_ISL_1572224, EPI_ISL_1572226, EPI_ISL_1572227, EPI_ISL_1572228, EPI_ISL_1572230, EPI_ISL_1572231, EPI_ISL_1573182, EPI_ISL_1573183, EPI_ISL_1573184, EPI_ISL_1573185, EPI_ISL_1573186, EPI_ISL_1573187, EPI_ISL_1573189, EPI_ISL_1573190, EPI_ISL_1573191, EPI_ISL_1573192, EPI_ISL_1573193, EPI_ISL_1573195, EPI_ISL_1573196, EPI_ISL_1573197, EPI_ISL_1573198, EPI_ISL_1573200, EPI_ISL_1573201, EPI_ISL_1573202, EPI_ISL_1573204, EPI_ISL_1573205, EPI_ISL_1573206, EPI_ISL_1573207, EPI_ISL_1573208, EPI_ISL_1573209, EPI_ISL_1573210, EPI_ISL_1573211, EPI_ISL_1573212, EPI_ISL_1573213, EPI_ISL_1573214, EPI_ISL_1573216, EPI_ISL_1573217, EPI_ISL_1573236, EPI_ISL_1573237, EPI_ISL_1573238, EPI_ISL_1638279, EPI_ISL_1638283, EPI_ISL_1638295, EPI_ISL_1638299, EPI_ISL_1638305, EPI_ISL_1638310, EPI_ISL_1638314, EPI_ISL_1638324, EPI_ISL_1638345, EPI_ISL_1638348, EPI_ISL_1638350, EPI_ISL_1638351, EPI_ISL_1638354, EPI_ISL_1638359, EPI_ISL_1638363, EPI_ISL_1638365, EPI_ISL_1638368, EPI_ISL_1638377, EPI_ISL_1638380, EPI_ISL_1638391, EPI_ISL_1638395, EPI_ISL_1638405, EPI_ISL_1638420, EPI_ISL_1638426, EPI_ISL_1638433, EPI_ISL_1638439, EPI_ISL_1638442, EPI_ISL_1638444, EPI_ISL_1638462, EPI_ISL_1638469, EPI_ISL_1638472, EPI_ISL_1638477, EPI_ISL_1638483, EPI_ISL_1638487, EPI_ISL_1638488, EPI_ISL_1638489, EPI_ISL_1641175, EPI_ISL_1642724, EPI_ISL_1643403, EPI_ISL_1643865, EPI_ISL_1722672 | | | |
| see above | MVZ Labor Dr. Limbach & Kollegen GbR | Robert Koch Institute | |
| EPI_ISL_1571162, EPI_ISL_1571166, EPI_ISL_1571167, EPI_ISL_1571177, EPI_ISL_1571194, EPI_ISL_1571198, EPI_ISL_1571205 | | | |
| see above | MVZ Labor Dr. Reising-Ackermann und Kollegen | Robert Koch Institute | |
| EPI_ISL_1563973, EPI_ISL_1563974, EPI_ISL_1563975, EPI_ISL_1563976, EPI_ISL_1563977, EPI_ISL_1563979, EPI_ISL_1563982, EPI_ISL_1563984, EPI_ISL_1563985, EPI_ISL_1563986, EPI_ISL_1563987, EPI_ISL_1563989, EPI_ISL_1563990, EPI_ISL_1563991, EPI_ISL_1563992, EPI_ISL_1563993, EPI_ISL_1563994, EPI_ISL_1563997, EPI_ISL_1563998, EPI_ISL_1563999, EPI_ISL_1564000, EPI_ISL_1564002, EPI_ISL_1564003, EPI_ISL_1564004, EPI_ISL_1564005, EPI_ISL_1564006, EPI_ISL_1564007, EPI_ISL_1564008, EPI_ISL_1640670, EPI_ISL_1640671, EPI_ISL_1640672, EPI_ISL_1640673, EPI_ISL_1640674, EPI_ISL_1640676, EPI_ISL_1640677, EPI_ISL_1640678, EPI_ISL_1640679, EPI_ISL_1640680, EPI_ISL_1640681, EPI_ISL_1640682, EPI_ISL_1640683, EPI_ISL_1640684, EPI_ISL_1640685, EPI_ISL_1640686, EPI_ISL_1640687, EPI_ISL_1640688, EPI_ISL_1640689, EPI_ISL_1640690, EPI_ISL_1640692, EPI_ISL_1640693, EPI_ISL_1640694, EPI_ISL_1640695, EPI_ISL_1640696, EPI_ISL_1640697, EPI_ISL_1640698, EPI_ISL_1640699, EPI_ISL_1640700, EPI_ISL_1640701, EPI_ISL_1640702, EPI_ISL_1640703, EPI_ISL_1640704, EPI_ISL_1640705, EPI_ISL_1640706, EPI_ISL_1640707, EPI_ISL_1640709, EPI_ISL_1640710, EPI_ISL_1640713, EPI_ISL_1640716, EPI_ISL_1640717, EPI_ISL_1640718, EPI_ISL_1640719, EPI_ISL_1640722, EPI_ISL_1640723, EPI_ISL_1640724, EPI_ISL_1640726, EPI_ISL_1640731, EPI_ISL_1640732, EPI_ISL_1640733, EPI_ISL_1640734, EPI_ISL_1640735, EPI_ISL_1640737, EPI_ISL_1640738, EPI_ISL_1640739 | | | |
| see above | MVZ Labor Krone GbR | Robert Koch Institute | |
| EPI_ISL_1720869, EPI_ISL_1720887, EPI_ISL_1720901, EPI_ISL_1720904, EPI_ISL_1720905, EPI_ISL_1720908, EPI_ISL_1720909, EPI_ISL_1720910, EPI_ISL_1720911, EPI_ISL_1720912, EPI_ISL_1720913, EPI_ISL_1720914, EPI_ISL_1720915, EPI_ISL_1720916, EPI_ISL_1720918, EPI_ISL_1720919, EPI_ISL_1720924, EPI_ISL_1720926, EPI_ISL_1720928, EPI_ISL_1720929, EPI_ISL_1720934, EPI_ISL_1720935, EPI_ISL_1720937, EPI_ISL_1720938, EPI_ISL_1843340, EPI_ISL_1846338 | | | |
| see above | MVZ Labor Passau GbR | Robert Koch Institute | |
| EPI_ISL_1572154, EPI_ISL_1725649 | MVZ Medizinisches Labor Hannover GmbH | Robert Koch Institute | |
| EPI_ISL_1642343, EPI_ISL_1724307, EPI_ISL_1724422, EPI_ISL_1724436 | MVZ fÃ¼r Laboratoriumsmedizin und Mikrobiologie Koblenz-Mittelrhein (Labor Koblenz) | Robert Koch Institute | |
| EPI_ISL_1752386, EPI_ISL_1752387, EPI_ISL_1752388, EPI_ISL_1752389, EPI_ISL_1752456, EPI_ISL_1752457, EPI_ISL_1752460, EPI_ISL_1752461, EPI_ISL_1752462, EPI_ISL_1752463, EPI_ISL_1752464, EPI_ISL_1752465, EPI_ISL_1752466, EPI_ISL_1752467, EPI_ISL_2006681, EPI_ISL_2094515, EPI_ISL_2094523, EPI_ISL_2094536, EPI_ISL_2094552, EPI_ISL_2094564, EPI_ISL_2094568, EPI_ISL_2094576, EPI_ISL_2094588, EPI_ISL_2094602, EPI_ISL_2094612, EPI_ISL_2094630, EPI_ISL_2094631, EPI_ISL_2094638, EPI_ISL_2094685, EPI_ISL_2094686, EPI_ISL_2094687, EPI_ISL_2094727, EPI_ISL_2094728, EPI_ISL_2094729, EPI_ISL_2094730, EPI_ISL_2094731, EPI_ISL_2094737, EPI_ISL_2094747, EPI_ISL_2094748, EPI_ISL_2094749, EPI_ISL_2094750, EPI_ISL_2094751, EPI_ISL_2094752, EPI_ISL_2094753, EPI_ISL_2094763, EPI_ISL_2094772, EPI_ISL_2094776, EPI_ISL_2094809, EPI_ISL_2094817, EPI_ISL_2094821, EPI_ISL_2094825, EPI_ISL_2094865, EPI_ISL_2094866, EPI_ISL_2094867, EPI_ISL_2094868, EPI_ISL_2094869, EPI_ISL_2094870, EPI_ISL_2094871, EPI_ISL_2094872, EPI_ISL_2094873, EPI_ISL_2094874, EPI_ISL_2094935, EPI_ISL_2094936, EPI_ISL_2094937, EPI_ISL_2094939, EPI_ISL_2094940, EPI_ISL_2094941, EPI_ISL_2094968, EPI_ISL_2094969, EPI_ISL_2094979, EPI_ISL_2094982, EPI_ISL_2094983, EPI_ISL_2094985, EPI_ISL_2094986, EPI_ISL_2094991, EPI_ISL_2094992, EPI_ISL_2095030, EPI_ISL_2095107, EPI_ISL_2095164, EPI_ISL_2095180, EPI_ISL_2095187, EPI_ISL_2095271 | | | |
| see above | Max von Pettenkofer Institute, Virology, National Reference Center for Retroviruses, LMU Munich | Laboratory for Functional Genome Analysis; Dept. Genomics; Gene Center of the LMU Munich | Alexander Graf; Helmut Blum; Max Muenchhoff; Oliver Keppler; Stefan Krebs |
| EPI_ISL_1990665, EPI_ISL_1990666, EPI_ISL_1990667, EPI_ISL_1990669, EPI_ISL_1990670, EPI_ISL_1990672, EPI_ISL_1990673, EPI_ISL_1990674, EPI_ISL_1990677, EPI_ISL_1990679, EPI_ISL_1990680, EPI_ISL_1990681, EPI_ISL_1990683, EPI_ISL_1990684, EPI_ISL_1990685, EPI_ISL_1990688, EPI_ISL_1990690, EPI_ISL_1990691, EPI_ISL_1990692, EPI_ISL_1990694, EPI_ISL_1990695, EPI_ISL_1990699, EPI_ISL_1990702, EPI_ISL_1990703, EPI_ISL_1990705, EPI_ISL_1990706, EPI_ISL_1990708, EPI_ISL_1990709, EPI_ISL_1991597, EPI_ISL_1991599, EPI_ISL_1991600, EPI_ISL_1991602 | | | |
| see above | Medical Laboratories Duesseldorf | Center of Medical Microbiology, Virology, and Hospital Hygiene, University of Duesseldorf | Alexander Dilthey; Andreas Walker; Angelika Helmer; Christian Lange; Daniel Strelow; Jessica Nicolai; Jörg Timm; Klaus Pfeffer; Lisanna Hülse; Malte Kohns Vasconcelos; Maximilian Damagnez; Nadine Lübke; Tobias Wienemann; Torsten Houwaart |
| EPI_ISL_1566749, EPI_ISL_1566750, EPI_ISL_1566751, EPI_ISL_1566763, EPI_ISL_1566770, EPI_ISL_1566777, EPI_ISL_1566779, EPI_ISL_1566781 | | | |
| see above | Medizinisch-Diagnostisches Labor Kempten allgÃ¤ulab | Robert Koch Institute | |
| EPI_ISL_1573132, EPI_ISL_1573133, EPI_ISL_1573134, EPI_ISL_1573135, EPI_ISL_1573136, EPI_ISL_1573137, EPI_ISL_1573139, EPI_ISL_1573140, EPI_ISL_1573141, EPI_ISL_1573142, EPI_ISL_1573143, EPI_ISL_1573144, EPI_ISL_1573145, EPI_ISL_1573146, EPI_ISL_1573147, EPI_ISL_1573148, EPI_ISL_1573149, EPI_ISL_1573150, EPI_ISL_1573151, EPI_ISL_1573152, EPI_ISL_1573153, EPI_ISL_1573154, EPI_ISL_1573155, EPI_ISL_1573156, EPI_ISL_1573157, EPI_ISL_1573158, EPI_ISL_1573159, EPI_ISL_1573160, EPI_ISL_1573161, EPI_ISL_1573162, EPI_ISL_1573163, EPI_ISL_1573164, EPI_ISL_1573165, EPI_ISL_1573167, EPI_ISL_1573168, EPI_ISL_1573169, EPI_ISL_1573171, EPI_ISL_1573172, EPI_ISL_1573173, EPI_ISL_1573174, EPI_ISL_1573175, EPI_ISL_1573176, EPI_ISL_1573177, EPI_ISL_1573178, EPI_ISL_1573179, EPI_ISL_1573180, EPI_ISL_1573181 | | | |
| see above | Medizinische Laboratorien DÃ¼sseldorf | Robert Koch Institute | |
| EPI_ISL_1569886, EPI_ISL_1569889, EPI_ISL_1569892, EPI_ISL_1569896, EPI_ISL_1569903, EPI_ISL_1569908, EPI_ISL_1569920, EPI_ISL_1569922, EPI_ISL_1569935, EPI_ISL_1569942, EPI_ISL_1569950, EPI_ISL_1569956, EPI_ISL_1569958, EPI_ISL_1569961, EPI_ISL_1569970, EPI_ISL_1569978, EPI_ISL_1569987, EPI_ISL_1569989, EPI_ISL_1569992, EPI_ISL_1569993, EPI_ISL_1638278, EPI_ISL_1638284, EPI_ISL_1638286, EPI_ISL_1638307, EPI_ISL_1638311, EPI_ISL_1638316, EPI_ISL_1638330, EPI_ISL_1638332, EPI_ISL_1638336, EPI_ISL_1638337, EPI_ISL_1638339, EPI_ISL_1638341, EPI_ISL_1638344, EPI_ISL_1638372, EPI_ISL_1638373, EPI_ISL_1638385, EPI_ISL_1638389, EPI_ISL_1638392, EPI_ISL_1638396, EPI_ISL_1638400, EPI_ISL_1638407, EPI_ISL_1638413, EPI_ISL_1638417, EPI_ISL_1638422, EPI_ISL_1638428, EPI_ISL_1638429, EPI_ISL_1638437, EPI_ISL_1638445, EPI_ISL_1638446, EPI_ISL_1638448, EPI_ISL_1638458, EPI_ISL_1638465, EPI_ISL_1638467, EPI_ISL_1638470, EPI_ISL_1638479, EPI_ISL_1638480, EPI_ISL_1638481, EPI_ISL_1638486, EPI_ISL_1638490, EPI_ISL_1638493, EPI_ISL_1642383, EPI_ISL_1642699, EPI_ISL_1642700, EPI_ISL_1642706, EPI_ISL_1642710, EPI_ISL_1642728, EPI_ISL_1643768 | | | |
| see above | Medizinisches Labor Ostsachsen MVZ GbR NebenbetriebsstÃ¤tte Dresden | Robert Koch Institute | |
| EPI_ISL_1567765, EPI_ISL_1567772, EPI_ISL_1567775, EPI_ISL_1567779, EPI_ISL_1567785, EPI_ISL_1567791, EPI_ISL_1567794, EPI_ISL_1567795, EPI_ISL_1567797, EPI_ISL_1567799, EPI_ISL_1567807, EPI_ISL_1567818, EPI_ISL_1567821, EPI_ISL_1567823 | | | |
| see above | Medizinisches Labor Wahl LÃ¼denscheid | Robert Koch Institute | |
| EPI_ISL_1643456 | Medizinisches Versorgungszentrum fÃ¼r Labormedizin und Mikrobiologie Ruhr GmbH - mvzlm RUHR GmbH | Robert Koch Institute | |
| EPI_ISL_1570976, EPI_ISL_1571003, EPI_ISL_1571008, EPI_ISL_1571016, EPI_ISL_1571044, EPI_ISL_1571076, EPI_ISL_1571081, EPI_ISL_1571102, EPI_ISL_1571605, EPI_ISL_1571612, EPI_ISL_1571621, EPI_ISL_1571630, EPI_ISL_1571658, EPI_ISL_1571661, EPI_ISL_1571667, EPI_ISL_1571679, EPI_ISL_1571695, EPI_ISL_1571705, EPI_ISL_1571713, EPI_ISL_1571714, EPI_ISL_1571719, EPI_ISL_1571730, EPI_ISL_1571731, EPI_ISL_1571740, EPI_ISL_1571745, EPI_ISL_1571748, EPI_ISL_1571777, EPI_ISL_1571779, EPI_ISL_1571791, EPI_ISL_1571793, EPI_ISL_1571809, EPI_ISL_1571816, EPI_ISL_1571821, EPI_ISL_1571826, EPI_ISL_1571844, EPI_ISL_1571851, EPI_ISL_1571859, EPI_ISL_1571864, EPI_ISL_1571877, EPI_ISL_1571879, EPI_ISL_1571883, EPI_ISL_1571896, EPI_ISL_1571903, EPI_ISL_1571911, EPI_ISL_1571916, EPI_ISL_1573544, EPI_ISL_1573560, EPI_ISL_1573633, EPI_ISL_1573643, EPI_ISL_1573658, EPI_ISL_1573667, EPI_ISL_1573672, EPI_ISL_1573676, EPI_ISL_1573690, EPI_ISL_1573694, EPI_ISL_1573695, EPI_ISL_1573696, EPI_ISL_1573699, EPI_ISL_1573703, EPI_ISL_1573707, EPI_ISL_1573712, EPI_ISL_1573719, EPI_ISL_1573723, EPI_ISL_1573725, EPI_ISL_1573744, EPI_ISL_1573750, EPI_ISL_1573754, EPI_ISL_1573757, EPI_ISL_1573760, EPI_ISL_1573773, EPI_ISL_1573775, EPI_ISL_1573777, EPI_ISL_1573780, EPI_ISL_1573789, EPI_ISL_1573791, EPI_ISL_1573819, EPI_ISL_1573834, EPI_ISL_1573838, EPI_ISL_1573854, EPI_ISL_1573977, EPI_ISL_1573979, EPI_ISL_1573980, EPI_ISL_1573982, EPI_ISL_1573999, EPI_ISL_1574002, EPI_ISL_1574007, EPI_ISL_1574014, EPI_ISL_1574020, EPI_ISL_1574024, EPI_ISL_1574032, EPI_ISL_1574033, EPI_ISL_1574053, EPI_ISL_1574055, EPI_ISL_1574057, EPI_ISL_1645250, EPI_ISL_1645270, EPI_ISL_1645277, EPI_ISL_1645278, EPI_ISL_1645288, EPI_ISL_1645289, EPI_ISL_1645325, EPI_ISL_1645331, EPI_ISL_1645342, EPI_ISL_1645348, EPI_ISL_1645353, EPI_ISL_1645355, EPI_ISL_1645361, EPI_ISL_1645471, EPI_ISL_1645492 | | | |
| see above | Medizinisches Zentrallabor Altenburg GmbH & Co.KG | Robert Koch Institute | |
| EPI_ISL_1569916, EPI_ISL_1569969, EPI_ISL_1569977 | Oberlausitz Kliniken gGmbH: Standort Bautzen | Robert Koch Institute | |
| EPI_ISL_1566983, EPI_ISL_1567327, EPI_ISL_1567347, EPI_ISL_1567377, EPI_ISL_1567381, EPI_ISL_1574121, EPI_ISL_1574272, EPI_ISL_1574275, EPI_ISL_1574333 | | | |
| see above | Praxisgemeinschaft fÃ¼r Laboratoriumsmedizin Labor BlumenstraÃŸe; Praxis Dr. E. Wietschel & Dr. F. Wietschel | Robert Koch Institute | |
| EPI_ISL_1566832, EPI_ISL_1571004, EPI_ISL_1571006, EPI_ISL_1571049, EPI_ISL_1574103, EPI_ISL_1574172, EPI_ISL_1574258, EPI_ISL_1574267, EPI_ISL_1574299, EPI_ISL_1574342, EPI_ISL_1641037, EPI_ISL_1641064, EPI_ISL_1641073, EPI_ISL_1641075, EPI_ISL_1641137, EPI_ISL_1641143 | | | |
| see above | Praxisgemeinschaft fÃ¼r Laboratoriumsmedizin Labor BlumenstraÃŸe; Praxis Dr. Sel und Dr. SpÃ¤te | Robert Koch Institute | |
| EPI_ISL_1720953, EPI_ISL_1720990, EPI_ISL_1721010 | Procomcure Biotech Germany GmbH | Robert Koch Institute | |
| EPI_ISL_1642538, EPI_ISL_1642603, EPI_ISL_1642664, EPI_ISL_1721377, EPI_ISL_1721399, EPI_ISL_2108718, EPI_ISL_2108739, EPI_ISL_2108751, EPI_ISL_2108816, EPI_ISL_2108832, EPI_ISL_2108942 | | | |
| see above | Robert-Bosch-Krankenhaus | Robert Koch Institute | |
| EPI_ISL_1567593, EPI_ISL_1567692 | SLK-Kliniken Heilbronn GmbH - Institut fÃ¼r Laboratoriumsmedizin | Robert Koch Institute | |
| EPI_ISL_1570971, EPI_ISL_1570978, EPI_ISL_1570986, EPI_ISL_1571000, EPI_ISL_1571025, EPI_ISL_1571026, EPI_ISL_1571030, EPI_ISL_1571035, EPI_ISL_1571043, EPI_ISL_1571047, EPI_ISL_1571052, EPI_ISL_1571066, EPI_ISL_1571068, EPI_ISL_1571072, EPI_ISL_1571084, EPI_ISL_1571086, EPI_ISL_1571087, EPI_ISL_1571090, EPI_ISL_1571097, EPI_ISL_1571098, EPI_ISL_1571099, EPI_ISL_1571101, EPI_ISL_1571599, EPI_ISL_1571608, EPI_ISL_1571615, EPI_ISL_1571619, EPI_ISL_1571622, EPI_ISL_1571626, EPI_ISL_1571629, EPI_ISL_1571633, EPI_ISL_1571640, EPI_ISL_1571649, EPI_ISL_1571657, EPI_ISL_1571668, EPI_ISL_1571672, EPI_ISL_1571682, EPI_ISL_1571686, EPI_ISL_1571690, EPI_ISL_1571694, EPI_ISL_1571698, EPI_ISL_1571699, EPI_ISL_1571702, EPI_ISL_1571707, EPI_ISL_1571710, EPI_ISL_1571712, EPI_ISL_1571715, EPI_ISL_1571718, EPI_ISL_1571721, EPI_ISL_1571734, EPI_ISL_1571742, EPI_ISL_1571749, EPI_ISL_1571761, EPI_ISL_1571766, EPI_ISL_1571786, EPI_ISL_1571796, EPI_ISL_1571804, EPI_ISL_1571811, EPI_ISL_1571824, EPI_ISL_1571828, EPI_ISL_1571833, EPI_ISL_1571835, EPI_ISL_1571836, EPI_ISL_1571843, EPI_ISL_1571845, EPI_ISL_1571846, EPI_ISL_1571847, EPI_ISL_1571852, EPI_ISL_1571855, EPI_ISL_1571857, EPI_ISL_1571862, EPI_ISL_1571865, EPI_ISL_1571868, EPI_ISL_1571870, EPI_ISL_1571873, EPI_ISL_1571874, EPI_ISL_1571882, EPI_ISL_1571884, EPI_ISL_1571885, EPI_ISL_1571886, EPI_ISL_1571887, EPI_ISL_1571888, EPI_ISL_1571890, EPI_ISL_1571892, EPI_ISL_1571895, EPI_ISL_1571897, EPI_ISL_1571898, EPI_ISL_1571899, EPI_ISL_1571900, EPI_ISL_1571907, EPI_ISL_1571909, EPI_ISL_1571910, EPI_ISL_1571913, EPI_ISL_1571914, EPI_ISL_1571923, EPI_ISL_1571926, EPI_ISL_1571928, EPI_ISL_1572557, EPI_ISL_1572558, EPI_ISL_1572560, EPI_ISL_1572561, EPI_ISL_1572565, EPI_ISL_1572571, EPI_ISL_1572574, EPI_ISL_1572576, EPI_ISL_1572582, EPI_ISL_1572584, EPI_ISL_1572585, EPI_ISL_1572586, EPI_ISL_1572587, EPI_ISL_1572589, EPI_ISL_1572590, EPI_ISL_1572592, EPI_ISL_1572597, EPI_ISL_1572599, EPI_ISL_1572600, EPI_ISL_1572603, EPI_ISL_1572604, EPI_ISL_1572606, EPI_ISL_1572610, EPI_ISL_1572611, EPI_ISL_1572612, EPI_ISL_1572614, EPI_ISL_1572616, EPI_ISL_1572618, EPI_ISL_1572620, EPI_ISL_1572621, EPI_ISL_1572623, EPI_ISL_1572625, EPI_ISL_1572626, EPI_ISL_1572628, EPI_ISL_1572631, EPI_ISL_1572632, EPI_ISL_1572633, EPI_ISL_1572636, EPI_ISL_1572637, EPI_ISL_1572638, EPI_ISL_1572639, EPI_ISL_1572640, EPI_ISL_1572642, EPI_ISL_1572644, EPI_ISL_1572645, EPI_ISL_1572647, EPI_ISL_1572649, EPI_ISL_1572650, EPI_ISL_1572652, EPI_ISL_1572653, EPI_ISL_1572654, EPI_ISL_1572655, EPI_ISL_1572657, EPI_ISL_1572658, EPI_ISL_1572660, EPI_ISL_1572661, EPI_ISL_1572662, EPI_ISL_1572663, EPI_ISL_1572665, EPI_ISL_1572667, EPI_ISL_1572669, EPI_ISL_1572670, EPI_ISL_1572671, EPI_ISL_1572672, EPI_ISL_1572673, EPI_ISL_1572674, EPI_ISL_1572676, EPI_ISL_1572679, EPI_ISL_1572680, EPI_ISL_1572682, EPI_ISL_1572683, EPI_ISL_1572685, EPI_ISL_1572687, EPI_ISL_1572688, EPI_ISL_1572689, EPI_ISL_1572691, EPI_ISL_1572692, EPI_ISL_1572694, EPI_ISL_1572695, EPI_ISL_1572697, EPI_ISL_1572698, EPI_ISL_1572700, EPI_ISL_1572701, EPI_ISL_1572702, EPI_ISL_1572703, EPI_ISL_1572704, EPI_ISL_1572706, EPI_ISL_1572707, EPI_ISL_1572708, EPI_ISL_1572711, EPI_ISL_1572712, EPI_ISL_1572713, EPI_ISL_1572714, EPI_ISL_1572716, EPI_ISL_1572717, EPI_ISL_1572719, EPI_ISL_1572720, EPI_ISL_1572722, EPI_ISL_1572723, EPI_ISL_1572724, EPI_ISL_1572725, EPI_ISL_1572727, EPI_ISL_1572728, EPI_ISL_1572729, EPI_ISL_1572730, EPI_ISL_1572731, EPI_ISL_1572732, EPI_ISL_1572733, EPI_ISL_1572735, EPI_ISL_1572736, EPI_ISL_1572737, EPI_ISL_1572738, EPI_ISL_1572739, EPI_ISL_1572740, EPI_ISL_1572742, EPI_ISL_1572743, EPI_ISL_1572746, EPI_ISL_1572747, EPI_ISL_1572749, EPI_ISL_1572750, EPI_ISL_1572751, EPI_ISL_1572752, EPI_ISL_1572753, EPI_ISL_1572754, EPI_ISL_1572755, EPI_ISL_1572756, EPI_ISL_1572758, EPI_ISL_1572759, EPI_ISL_1572760, EPI_ISL_1572762, EPI_ISL_1572763, EPI_ISL_1572764, EPI_ISL_1572765, EPI_ISL_1572768, EPI_ISL_1572769, EPI_ISL_1572770, EPI_ISL_1572771, EPI_ISL_1572772, EPI_ISL_1572774, EPI_ISL_1572775, EPI_ISL_1572776, EPI_ISL_1572777, EPI_ISL_1572779, EPI_ISL_1572780, EPI_ISL_1572781, EPI_ISL_1572783, EPI_ISL_1572786, EPI_ISL_1572787, EPI_ISL_1572788, EPI_ISL_1572789, EPI_ISL_1572790, EPI_ISL_1572792, EPI_ISL_1572793, EPI_ISL_1572794, EPI_ISL_1572796, EPI_ISL_1572798, EPI_ISL_1572800, EPI_ISL_1572801, EPI_ISL_1572804, EPI_ISL_1572807, EPI_ISL_1572808, EPI_ISL_1572809, EPI_ISL_1572811, EPI_ISL_1572812, EPI_ISL_1572813, EPI_ISL_1572817, EPI_ISL_1572819, EPI_ISL_1572820, EPI_ISL_1572821, EPI_ISL_1572823, EPI_ISL_1572825, EPI_ISL_1572826, EPI_ISL_1572827, EPI_ISL_1572828, EPI_ISL_1572829, EPI_ISL_1572830, EPI_ISL_1572831, EPI_ISL_1572833, EPI_ISL_1572835, EPI_ISL_1573466, EPI_ISL_1573468, EPI_ISL_1573470, EPI_ISL_1573476, EPI_ISL_1573483, EPI_ISL_1573491, EPI_ISL_1573494, EPI_ISL_1573498, EPI_ISL_1573503, EPI_ISL_1573505, EPI_ISL_1573506, EPI_ISL_1573509, EPI_ISL_1573512, EPI_ISL_1573515, EPI_ISL_1573516, EPI_ISL_1573518, EPI_ISL_1573519, EPI_ISL_1573521, EPI_ISL_1573523, EPI_ISL_1573526, EPI_ISL_1573527, EPI_ISL_1573530, EPI_ISL_1573532, EPI_ISL_1573534, EPI_ISL_1573535, EPI_ISL_1573540, EPI_ISL_1573542, EPI_ISL_1573553, EPI_ISL_1573555, EPI_ISL_1573558, | | | |

EPI_ISL_1573561, EPI_ISL_1573562, EPI_ISL_1573565, EPI_ISL_1573566, EPI_ISL_1573571, EPI_ISL_1573576, EPI_ISL_1573581, EPI_ISL_1573586, EPI_ISL_1573590, EPI_ISL_1573591, EPI_ISL_1573594, EPI_ISL_1573597, EPI_ISL_1573598, EPI_ISL_1573599, EPI_ISL_1573600, EPI_ISL_1573602, EPI_ISL_1573604, EPI_ISL_1573606, EPI_ISL_1573607, EPI_ISL_1573609, EPI_ISL_1573613, EPI_ISL_1573618, EPI_ISL_1573619, EPI_ISL_1573622, EPI_ISL_1573623, EPI_ISL_1573626, EPI_ISL_1573628, EPI_ISL_1573629, EPI_ISL_1573639, EPI_ISL_1573645, EPI_ISL_1573646, EPI_ISL_1573650, EPI_ISL_1573654, EPI_ISL_1573660, EPI_ISL_1573669, EPI_ISL_1573670, EPI_ISL_1573671, EPI_ISL_1573673, EPI_ISL_1573674, EPI_ISL_1573678, EPI_ISL_1573682, EPI_ISL_1573684, EPI_ISL_1573686, EPI_ISL_1573692, EPI_ISL_1573697, EPI_ISL_1573698, EPI_ISL_1573709, EPI_ISL_1573715, EPI_ISL_1573724, EPI_ISL_1573727, EPI_ISL_1573731, EPI_ISL_1573733, EPI_ISL_1573736, EPI_ISL_1573740, EPI_ISL_1573745, EPI_ISL_1573751, EPI_ISL_1573756, EPI_ISL_1573761, EPI_ISL_1573762, EPI_ISL_1573768, EPI_ISL_1573781, EPI_ISL_1573784, EPI_ISL_1573788, EPI_ISL_1573792, EPI_ISL_1573794, EPI_ISL_1573796, EPI_ISL_1573798, EPI_ISL_1573805, EPI_ISL_1573808, EPI_ISL_1573809, EPI_ISL_1573810, EPI_ISL_1573812, EPI_ISL_1573813, EPI_ISL_1573814, EPI_ISL_1573822, EPI_ISL_1573823, EPI_ISL_1573824, EPI_ISL_1573825, EPI_ISL_1573826, EPI_ISL_1573829, EPI_ISL_1573830, EPI_ISL_1573831, EPI_ISL_1573833, EPI_ISL_1573842, EPI_ISL_1573846, EPI_ISL_1573850, EPI_ISL_1573853, EPI_ISL_1573855, EPI_ISL_1573858, EPI_ISL_1573860, EPI_ISL_1573867, EPI_ISL_1573870, EPI_ISL_1573874, EPI_ISL_1573876, EPI_ISL_1573880, EPI_ISL_1573882, EPI_ISL_1573883, EPI_ISL_1573884, EPI_ISL_1573886, EPI_ISL_1573888, EPI_ISL_1573890, EPI_ISL_1573891, EPI_ISL_1573893, EPI_ISL_1573900, EPI_ISL_1573905, EPI_ISL_1573913, EPI_ISL_1573914, EPI_ISL_1573916, EPI_ISL_1573917, EPI_ISL_1573919, EPI_ISL_1573922, EPI_ISL_1573923, EPI_ISL_1573924, EPI_ISL_1573925, EPI_ISL_1573926, EPI_ISL_1573928, EPI_ISL_1573929, EPI_ISL_1573931, EPI_ISL_1573932, EPI_ISL_1573934, EPI_ISL_1573935, EPI_ISL_1573937, EPI_ISL_1573939, EPI_ISL_1573941, EPI_ISL_1573942, EPI_ISL_1573945, EPI_ISL_1573947, EPI_ISL_1573949, EPI_ISL_1573950, EPI_ISL_1573953, EPI_ISL_1573954, EPI_ISL_1573956, EPI_ISL_1573959, EPI_ISL_1573962, EPI_ISL_1573963, EPI_ISL_1573964, EPI_ISL_1573965, EPI_ISL_1573974, EPI_ISL_1573978, EPI_ISL_1573985, EPI_ISL_1573986, EPI_ISL_1573991, EPI_ISL_1573993, EPI_ISL_1573995, EPI_ISL_1574000, EPI_ISL_1574003, EPI_ISL_1574009, EPI_ISL_1574012, EPI_ISL_1574017, EPI_ISL_1574022, EPI_ISL_1574027, EPI_ISL_1574037, EPI_ISL_1574041, EPI_ISL_1574042, EPI_ISL_1574045, EPI_ISL_1574047, EPI_ISL_1574049, EPI_ISL_1574051, EPI_ISL_1574061, EPI_ISL_1574062, EPI_ISL_1574064, EPI_ISL_1574065, EPI_ISL_1574066, EPI_ISL_1574068, EPI_ISL_1574070, EPI_ISL_1574074, EPI_ISL_1574076, EPI_ISL_1574077, EPI_ISL_1574078, EPI_ISL_1574080, EPI_ISL_1574082, EPI_ISL_1574084, EPI_ISL_1574088, EPI_ISL_1574092, EPI_ISL_1574093, EPI_ISL_1574094, EPI_ISL_1574095, EPI_ISL_1574106, EPI_ISL_1574111, EPI_ISL_1574113, EPI_ISL_1574115, EPI_ISL_1574119, EPI_ISL_1574120, EPI_ISL_1574123, EPI_ISL_1574124, EPI_ISL_1574125, EPI_ISL_1574132, EPI_ISL_1574133, EPI_ISL_1574135, EPI_ISL_1574137, EPI_ISL_1574138, EPI_ISL_1574140, EPI_ISL_1574141, EPI_ISL_1574144, EPI_ISL_1574145, EPI_ISL_1574149, EPI_ISL_1574154, EPI_ISL_1574159, EPI_ISL_1574162, EPI_ISL_1574163, EPI_ISL_1574168, EPI_ISL_1574171, EPI_ISL_1574177, EPI_ISL_1574179, EPI_ISL_1574182, EPI_ISL_1574184, EPI_ISL_1574185, EPI_ISL_1574186, EPI_ISL_1574187, EPI_ISL_1574194, EPI_ISL_1574195, EPI_ISL_1574200, EPI_ISL_1574203, EPI_ISL_1574204, EPI_ISL_1574205, EPI_ISL_1574207, EPI_ISL_1574209, EPI_ISL_1574210, EPI_ISL_1574213, EPI_ISL_1574214, EPI_ISL_1574217, EPI_ISL_1574220, EPI_ISL_1574221, EPI_ISL_1574223, EPI_ISL_1574229, EPI_ISL_1574233, EPI_ISL_1574239, EPI_ISL_1574243, EPI_ISL_1574248, EPI_ISL_1574249, EPI_ISL_1574250, EPI_ISL_1574253, EPI_ISL_1574254, EPI_ISL_1574255, EPI_ISL_1574257, EPI_ISL_1574266, EPI_ISL_1574271, EPI_ISL_1574276, EPI_ISL_1574288, EPI_ISL_1574291, EPI_ISL_1574298, EPI_ISL_1574302, EPI_ISL_1574311, EPI_ISL_1574320, EPI_ISL_1574321, EPI_ISL_1574323, EPI_ISL_1574336, EPI_ISL_1574339, EPI_ISL_1574343, EPI_ISL_1574345, EPI_ISL_1574346, EPI_ISL_1574357, EPI_ISL_1574358, EPI_ISL_1574690, EPI_ISL_1574691, EPI_ISL_1574692, EPI_ISL_1574695, EPI_ISL_1574699, EPI_ISL_1574700, EPI_ISL_1574703, EPI_ISL_1574704, EPI_ISL_1574712, EPI_ISL_1574719, EPI_ISL_1574720, EPI_ISL_1574731, EPI_ISL_1574733, EPI_ISL_1574738, EPI_ISL_1574741, EPI_ISL_1574743, EPI_ISL_1574746, EPI_ISL_1574753, EPI_ISL_1574754, EPI_ISL_1574758, EPI_ISL_1574765, EPI_ISL_1574768, EPI_ISL_1574771, EPI_ISL_1574772, EPI_ISL_1574776, EPI_ISL_1574780, EPI_ISL_1574782, EPI_ISL_1574783, EPI_ISL_1574785, EPI_ISL_1574789, EPI_ISL_1574793, EPI_ISL_1574796, EPI_ISL_1574797, EPI_ISL_1574803, EPI_ISL_1574805, EPI_ISL_1574807, EPI_ISL_1574809, EPI_ISL_1574814, EPI_ISL_1574816, EPI_ISL_1574817, EPI_ISL_1574822, EPI_ISL_1574825, EPI_ISL_1574833, EPI_ISL_1574837, EPI_ISL_1574838, EPI_ISL_1574841, EPI_ISL_1574842, EPI_ISL_1574844, EPI_ISL_1574846, EPI_ISL_1574848, EPI_ISL_1574850, EPI_ISL_1574852, EPI_ISL_1574853, EPI_ISL_1574854, EPI_ISL_1574856, EPI_ISL_1574857, EPI_ISL_1574859, EPI_ISL_1574869, EPI_ISL_1574870, EPI_ISL_1574872, EPI_ISL_1574876, EPI_ISL_1574878, EPI_ISL_1574882, EPI_ISL_1574884, EPI_ISL_1574888, EPI_ISL_1574889, EPI_ISL_1574891, EPI_ISL_1574894, EPI_ISL_1574895, EPI_ISL_1574896, EPI_ISL_1574897, EPI_ISL_1574900, EPI_ISL_1574903, EPI_ISL_1574905, EPI_ISL_1574913, EPI_ISL_1574915, EPI_ISL_1574920, EPI_ISL_1574924, EPI_ISL_1574925, EPI_ISL_1574930, EPI_ISL_1574933, EPI_ISL_1574938, EPI_ISL_1574950, EPI_ISL_1574955, EPI_ISL_1574958, EPI_ISL_1574960, EPI_ISL_1574964, EPI_ISL_1640339, EPI_ISL_1640341, EPI_ISL_1640342, EPI_ISL_1640347, EPI_ISL_1640363, EPI_ISL_1640367, EPI_ISL_1640369, EPI_ISL_1640371, EPI_ISL_1640372, EPI_ISL_1640377, EPI_ISL_1640378, EPI_ISL_1640381, EPI_ISL_1640384, EPI_ISL_1640394, EPI_ISL_1640396, EPI_ISL_1640400, EPI_ISL_1640401, EPI_ISL_1640406, EPI_ISL_1640410, EPI_ISL_1640412, EPI_ISL_1640414, EPI_ISL_1640415, EPI_ISL_1640417, EPI_ISL_1640418, EPI_ISL_1640420, EPI_ISL_1640424, EPI_ISL_1640428, EPI_ISL_1640432, EPI_ISL_1640434, EPI_ISL_1640435, EPI_ISL_1640442, EPI_ISL_1640448, EPI_ISL_1640451, EPI_ISL_1640455, EPI_ISL_1640458, EPI_ISL_1640460, EPI_ISL_1640466, EPI_ISL_1640467, EPI_ISL_1640469, EPI_ISL_1640473, EPI_ISL_1640475, EPI_ISL_1640479, EPI_ISL_1640482, EPI_ISL_1640485, EPI_ISL_1640486, EPI_ISL_1640487, EPI_ISL_1640492, EPI_ISL_1640493, EPI_ISL_1640507, EPI_ISL_1640515, EPI_ISL_1640519, EPI_ISL_1640523, EPI_ISL_1640533, EPI_ISL_1640539, EPI_ISL_1640550, EPI_ISL_1640576, EPI_ISL_1640587, EPI_ISL_1640593, EPI_ISL_1640610, EPI_ISL_1640940, EPI_ISL_1640946, EPI_ISL_1640959, EPI_ISL_1640964, EPI_ISL_1640966, EPI_ISL_1640967, EPI_ISL_1640971, EPI_ISL_1640972, EPI_ISL_1640981, EPI_ISL_1640982, EPI_ISL_1640986, EPI_ISL_1640993, EPI_ISL_1640995, EPI_ISL_1641012, EPI_ISL_1641021, EPI_ISL_1641022, EPI_ISL_1641024, EPI_ISL_1641026, EPI_ISL_1641028, EPI_ISL_1641035, EPI_ISL_1641038, EPI_ISL_1641041, EPI_ISL_1641042, EPI_ISL_1641045, EPI_ISL_1641048, EPI_ISL_1641049, EPI_ISL_1641054, EPI_ISL_1641055, EPI_ISL_1641057, EPI_ISL_1641058, EPI_ISL_1641067, EPI_ISL_1641077, EPI_ISL_1641080, EPI_ISL_1641082, EPI_ISL_1641086, EPI_ISL_1641092, EPI_ISL_1641112, EPI_ISL_1641114, EPI_ISL_1641118, EPI_ISL_1641121, EPI_ISL_1641126, EPI_ISL_1641130, EPI_ISL_1641135, EPI_ISL_1641156, EPI_ISL_1644055, EPI_ISL_1644058, EPI_ISL_1644066, EPI_ISL_1644092, EPI_ISL_1644094, EPI_ISL_1644096, EPI_ISL_1644103, EPI_ISL_1644118, EPI_ISL_1644126, EPI_ISL_1644130, EPI_ISL_1644228, EPI_ISL_1644282, EPI_ISL_1644379, EPI_ISL_1644464, EPI_ISL_1645319, EPI_ISL_1645354, EPI_ISL_1645460, EPI_ISL_1645463, EPI_ISL_1645489, EPI_ISL_1645494, EPI_ISL_1645495, EPI_ISL_1645498, EPI_ISL_1645527

see above | SYNLAB Jena Oncoscreen | Robert Koch Institute

EPI_ISL_1571611, EPI_ISL_1571617, EPI_ISL_1571623, EPI_ISL_1571632, EPI_ISL_1571637, EPI_ISL_1571653, EPI_ISL_1571678, EPI_ISL_1571684, EPI_ISL_1571685, EPI_ISL_1571687, EPI_ISL_1571709, EPI_ISL_1571717, EPI_ISL_1571722, EPI_ISL_1573592, EPI_ISL_1574831, EPI_ISL_1574883

see above | SYNLAB Labor MÃ¼nchen Zentrum LMZ | Robert Koch Institute

EPI_ISL_1572562, EPI_ISL_1572567, EPI_ISL_1572569, EPI_ISL_1572577, EPI_ISL_1572578, EPI_ISL_1572580, EPI_ISL_1572591, EPI_ISL_1572595, EPI_ISL_1572598, EPI_ISL_1572601, EPI_ISL_1572605, EPI_ISL_1572608, EPI_ISL_1572609, EPI_ISL_1572613, EPI_ISL_1572617, EPI_ISL_1572619, EPI_ISL_1572622, EPI_ISL_1572624, EPI_ISL_1572627, EPI_ISL_1572641, EPI_ISL_1572646, EPI_ISL_1572648, EPI_ISL_1572651, EPI_ISL_1572656, EPI_ISL_1572659, EPI_ISL_1572666, EPI_ISL_1572677, EPI_ISL_1572681, EPI_ISL_1572690, EPI_ISL_1572693, EPI_ISL_1572699, EPI_ISL_1572705, EPI_ISL_1572767, EPI_ISL_1572779, EPI_ISL_1572782, EPI_ISL_1572797, EPI_ISL_1572803, EPI_ISL_1572822, EPI_ISL_1572824, EPI_ISL_1572832, EPI_ISL_1573477, EPI_ISL_1573480, EPI_ISL_1573485, EPI_ISL_1573490, EPI_ISL_1573495, EPI_ISL_1573508, EPI_ISL_1573513, EPI_ISL_1573541, EPI_ISL_1573545, EPI_ISL_1573549, EPI_ISL_1573605, EPI_ISL_1573610, EPI_ISL_1573637, EPI_ISL_1573668, EPI_ISL_1573683, EPI_ISL_1573811, EPI_ISL_1573840, EPI_ISL_1573887, EPI_ISL_1573895, EPI_ISL_1573903, EPI_ISL_1573909, EPI_ISL_1573912, EPI_ISL_1573915, EPI_ISL_1573918, EPI_ISL_1573920, EPI_ISL_1573927, EPI_ISL_1573930, EPI_ISL_1573936, EPI_ISL_1573990, EPI_ISL_1574011, EPI_ISL_1574083, EPI_ISL_1574105, EPI_ISL_1574112, EPI_ISL_1574146, EPI_ISL_1574173, EPI_ISL_1574180, EPI_ISL_1574192, EPI_ISL_1574198, EPI_ISL_1574206, EPI_ISL_1574212, EPI_ISL_1574226, EPI_ISL_1574232, EPI_ISL_1574247, EPI_ISL_1574265, EPI_ISL_1574286, EPI_ISL_1574306, EPI_ISL_1574316, EPI_ISL_1574334, EPI_ISL_1574716, EPI_ISL_1574718, EPI_ISL_1574722, EPI_ISL_1574724, EPI_ISL_1574728, EPI_ISL_1574732, EPI_ISL_1574734, EPI_ISL_1574735, EPI_ISL_1574737, EPI_ISL_1574747, EPI_ISL_1574748, EPI_ISL_1574752, EPI_ISL_1574756, EPI_ISL_1574760, EPI_ISL_1574764, EPI_ISL_1574767, EPI_ISL_1574778, EPI_ISL_1574792, EPI_ISL_1574801, EPI_ISL_1574818, EPI_ISL_1574836, EPI_ISL_1574843, EPI_ISL_1574864, EPI_ISL_1574877, EPI_ISL_1574890, EPI_ISL_1574893, EPI_ISL_1574899, EPI_ISL_1574906, EPI_ISL_1574907, EPI_ISL_1574918, EPI_ISL_1574931, EPI_ISL_1574935, EPI_ISL_1574937, EPI_ISL_1574956, EPI_ISL_1574959, EPI_ISL_1640329, EPI_ISL_1640338, EPI_ISL_1640348, EPI_ISL_1640359, EPI_ISL_1640457, EPI_ISL_1640470, EPI_ISL_1640494, EPI_ISL_1640513, EPI_ISL_1640517, EPI_ISL_1640536, EPI_ISL_1640541, EPI_ISL_1640559, EPI_ISL_1640570, EPI_ISL_1640574, EPI_ISL_1640583, EPI_ISL_1640585, EPI_ISL_1640941, EPI_ISL_1640960, EPI_ISL_1640978, EPI_ISL_1641034, EPI_ISL_1641089, EPI_ISL_1641095, EPI_ISL_1641099, EPI_ISL_1641103, EPI_ISL_1641117, EPI_ISL_1641120, EPI_ISL_1641127, EPI_ISL_1641129, EPI_ISL_1641133, EPI_ISL_1641144, EPI_ISL_1641146, EPI_ISL_1641149, EPI_ISL_1642975, EPI_ISL_1645237, EPI_ISL_1645307, EPI_ISL_1645334, EPI_ISL_1645338, EPI_ISL_1645349, EPI_ISL_1645365, EPI_ISL_1645384, EPI_ISL_1645455

see above | SYNLAB MVZ Berlin | Robert Koch Institute

EPI_ISL_1728734, EPI_ISL_1728739, EPI_ISL_1728771, EPI_ISL_1728804, EPI_ISL_1728811, EPI_ISL_1728857, EPI_ISL_1729719, EPI_ISL_1729727, EPI_ISL_1729764, EPI_ISL_1729769, EPI_ISL_1729820, EPI_ISL_1729863, EPI_ISL_1729866, EPI_ISL_1729876, EPI_ISL_1729877, EPI_ISL_1849515, EPI_ISL_1849599, EPI_ISL_2108140, EPI_ISL_2128733

see above | SYNLAB MVZ Bonn | Robert Koch Institute

EPI_ISL_1574460, EPI_ISL_1574650 | SYNLAB MVZ Dachau | Robert Koch Institute

EPI_ISL_1726538, EPI_ISL_1726569, EPI_ISL_1726622, EPI_ISL_1726629, EPI_ISL_1726630, EPI_ISL_1726633, EPI_ISL_1726649, EPI_ISL_1726664, EPI_ISL_1726689, EPI_ISL_1726710, EPI_ISL_1726735, EPI_ISL_1726746, EPI_ISL_1726764, EPI_ISL_1728731, EPI_ISL_1728733, EPI_ISL_1728735, EPI_ISL_1728736, EPI_ISL_1728740, EPI_ISL_1728743, EPI_ISL_1728746, EPI_ISL_1728749, EPI_ISL_1728750, EPI_ISL_1728751, EPI_ISL_1728752, EPI_ISL_1728754, EPI_ISL_1728755, EPI_ISL_1728756, EPI_ISL_1728757, EPI_ISL_1728758, EPI_ISL_1728760, EPI_ISL_1728762, EPI_ISL_1728765, EPI_ISL_1728767, EPI_ISL_1728768, EPI_ISL_1728770, EPI_ISL_1728773, EPI_ISL_1728774, EPI_ISL_1728779, EPI_ISL_1728780, EPI_ISL_1728783, EPI_ISL_1728785, EPI_ISL_1728786, EPI_ISL_1728787, EPI_ISL_1728788, EPI_ISL_1728789, EPI_ISL_1728790, EPI_ISL_1728794, EPI_ISL_1728795, EPI_ISL_1728796, EPI_ISL_1728797, EPI_ISL_1728798, EPI_ISL_1728799, EPI_ISL_1728800, EPI_ISL_1728803, EPI_ISL_1728805, EPI_ISL_1728807, EPI_ISL_1728809, EPI_ISL_1728810, EPI_ISL_1728812, EPI_ISL_1728813, EPI_ISL_1728814, EPI_ISL_1728816, EPI_ISL_1728817, EPI_ISL_1728819, EPI_ISL_1728820, EPI_ISL_1728822, EPI_ISL_1728823, EPI_ISL_1728824, EPI_ISL_1728826, EPI_ISL_1728827, EPI_ISL_1728828, EPI_ISL_1728829, EPI_ISL_1728830, EPI_ISL_1728831, EPI_ISL_1728833, EPI_ISL_1728834, EPI_ISL_1728835, EPI_ISL_1728836, EPI_ISL_1728838, EPI_ISL_1728839, EPI_ISL_1728841, EPI_ISL_1728842, EPI_ISL_1728844, EPI_ISL_1728845, EPI_ISL_1728846, EPI_ISL_1728847, EPI_ISL_1728848, EPI_ISL_1728849, EPI_ISL_1728850, EPI_ISL_1728851, EPI_ISL_1728852, EPI_ISL_1728853, EPI_ISL_1728854, EPI_ISL_1728855, EPI_ISL_1728856, EPI_ISL_1728858, EPI_ISL_1728859, EPI_ISL_1728861, EPI_ISL_1728862, EPI_ISL_1728863, EPI_ISL_1729707, EPI_ISL_1729710, EPI_ISL_1729713, EPI_ISL_1729715, EPI_ISL_1729718, EPI_ISL_1729723, EPI_ISL_1729728, EPI_ISL_1729729, EPI_ISL_1729734, EPI_ISL_1729735, EPI_ISL_1729738, EPI_ISL_1729739, EPI_ISL_1729741, EPI_ISL_1729742, EPI_ISL_1729743, EPI_ISL_1729749, EPI_ISL_1729750, EPI_ISL_1729754, EPI_ISL_1729761, EPI_ISL_1729762, EPI_ISL_1729763, EPI_ISL_1729767, EPI_ISL_1729768, EPI_ISL_1729770, EPI_ISL_1729771, EPI_ISL_1729774, EPI_ISL_1729775, EPI_ISL_1729782, EPI_ISL_1729784, EPI_ISL_1729786, EPI_ISL_1729789, EPI_ISL_1729790, EPI_ISL_1729793, EPI_ISL_1729801, EPI_ISL_1729803, EPI_ISL_1729804, EPI_ISL_1729805, EPI_ISL_1729812, EPI_ISL_1729816, EPI_ISL_1729817, EPI_ISL_1729824, EPI_ISL_1729827, EPI_ISL_1729828, EPI_ISL_1729830, EPI_ISL_1729834, EPI_ISL_1729836, EPI_ISL_1729838, EPI_ISL_1729839, EPI_ISL_1729843, EPI_ISL_1729848, EPI_ISL_1729850, EPI_ISL_1729855, EPI_ISL_1729860, EPI_ISL_1729862, EPI_ISL_1729865, EPI_ISL_1729867, EPI_ISL_1729870, EPI_ISL_1729871, EPI_ISL_1729873, EPI_ISL_1729875, EPI_ISL_1729881, EPI_ISL_1729882, EPI_ISL_1729886, EPI_ISL_1729887, EPI_ISL_1729892, EPI_ISL_1729896, EPI_ISL_1729898, EPI_ISL_1729899, EPI_ISL_1729910, EPI_ISL_1729916, EPI_ISL_1729917, EPI_ISL_1729919, EPI_ISL_1729937, EPI_ISL_1729940, EPI_ISL_1729942, EPI_ISL_1729944, EPI_ISL_1729945, EPI_ISL_1729946, EPI_ISL_1729948, EPI_ISL_1729951, EPI_ISL_1729954, EPI_ISL_1729956, EPI_ISL_1729957, EPI_ISL_1729958, EPI_ISL_1729960, EPI_ISL_1729963, EPI_ISL_1849517, EPI_ISL_1849524, EPI_ISL_1849531, EPI_ISL_1849534, EPI_ISL_1849535, EPI_ISL_1849537, EPI_ISL_1849551, EPI_ISL_1849567, EPI_ISL_1849568, EPI_ISL_1849572, EPI_ISL_1849574, EPI_ISL_1849576, EPI_ISL_1849578, EPI_ISL_1849584, EPI_ISL_1849585, EPI_ISL_1849593, EPI_ISL_1849608, EPI_ISL_1849616, EPI_ISL_1849633, EPI_ISL_1849644, EPI_ISL_1849646, EPI_ISL_1849667, EPI_ISL_1849670, EPI_ISL_1849681, EPI_ISL_1849682, EPI_ISL_1849684, EPI_ISL_1849687, EPI_ISL_1849691, EPI_ISL_2108012, EPI_ISL_2108032, EPI_ISL_2108034, EPI_ISL_2108045, EPI_ISL_2108100, EPI_ISL_2108115, EPI_ISL_2108124, EPI_ISL_2108171, EPI_ISL_2108209, EPI_ISL_2108229, EPI_ISL_2108230, EPI_ISL_2108240, EPI_ISL_2108256, EPI_ISL_2108261, EPI_ISL_2114517, EPI_ISL_2114521, EPI_ISL_2114522, EPI_ISL_2114524, EPI_ISL_2114525, EPI_ISL_2114529, EPI_ISL_2114534, EPI_ISL_2114538, EPI_ISL_2114549, EPI_ISL_2114551, EPI_ISL_2114554, EPI_ISL_2114559, EPI_ISL_2114576, EPI_ISL_2114578, EPI_ISL_2114579, EPI_ISL_2114584, EPI_ISL_2114596, EPI_ISL_2114602, EPI_ISL_2114605, EPI_ISL_2114619, EPI_ISL_2114621, EPI_ISL_2114626, EPI_ISL_2114632, EPI_ISL_2114637, EPI_ISL_2114643, EPI_ISL_2114649, EPI_ISL_2114660, EPI_ISL_2114663, EPI_ISL_2114664, EPI_ISL_2114674, EPI_ISL_2114675, EPI_ISL_2114677, EPI_ISL_2114686, EPI_ISL_2114688, EPI_ISL_2114702, EPI_ISL_2114714, EPI_ISL_2114715, EPI_ISL_2114722, EPI_ISL_2114725, EPI_ISL_2114728, EPI_ISL_2114732, EPI_ISL_2114745, EPI_ISL_2114756, EPI_ISL_2114766, EPI_ISL_2114767, EPI_ISL_2114779, EPI_ISL_2114783, EPI_ISL_2114784, EPI_ISL_2114797, EPI_ISL_2114799, EPI_ISL_2114828, EPI_ISL_2114829, EPI_ISL_2114834, EPI_ISL_2114835, EPI_ISL_2114836, EPI_ISL_2114837, EPI_ISL_2114841, EPI_ISL_2114848, EPI_ISL_2114857, EPI_ISL_2114858, EPI_ISL_2114863, EPI_ISL_2114873, EPI_ISL_2114877, EPI_ISL_2114879, EPI_ISL_2114884, EPI_ISL_2114899, EPI_ISL_2114900, EPI_ISL_2114901, EPI_ISL_2114914, EPI_ISL_2114916, EPI_ISL_2114920, EPI_ISL_2114921, EPI_ISL_2114924, EPI_ISL_2114940, EPI_ISL_2114941, EPI_ISL_2114942, EPI_ISL_2114943, EPI_ISL_2114947, EPI_ISL_2114948, EPI_ISL_2114950, EPI_ISL_2114956, EPI_ISL_2114957, EPI_ISL_2114980, EPI_ISL_2115004, EPI_ISL_2115013, EPI_ISL_2115030, EPI_ISL_2115031, EPI_ISL_2115034, EPI_ISL_2115036, EPI_ISL_2115049, EPI_ISL_2115061, EPI_ISL_2115071, EPI_ISL_2115074, EPI_ISL_2115089, EPI_ISL_2115092, EPI_ISL_2115098, EPI_ISL_2115101, EPI_ISL_2115104, EPI_ISL_2115107, EPI_ISL_2115113, EPI_ISL_2115122, EPI_ISL_2119010, EPI_ISL_2119173, EPI_ISL_2119176, EPI_ISL_2119180, EPI_ISL_2119451, EPI_ISL_2119937, EPI_ISL_2128677, EPI_ISL_2128696, EPI_ISL_2128717, EPI_ISL_2128754, EPI_ISL_2128858, EPI_ISL_2128914, EPI_ISL_2128946, EPI_ISL_2128948

see above | SYNLAB MVZ Ettlingen | Robert Koch Institute

EPI_ISL_1645973, EPI_ISL_1645984, EPI_ISL_1646015, EPI_ISL_1646024, EPI_ISL_1646059, EPI_ISL_1646075, EPI_ISL_1646573, EPI_ISL_1719921, EPI_ISL_1719931, EPI_ISL_1719932, EPI_ISL_1719937, EPI_ISL_1719941, EPI_ISL_1719944, EPI_ISL_1719945, EPI_ISL_1719946, EPI_ISL_1719949, EPI_ISL_1719955, EPI_ISL_1719961, EPI_ISL_1719965, EPI_ISL_1719970, EPI_ISL_1720008, EPI_ISL_1720017, EPI_ISL_1720052, EPI_ISL_1720099, EPI_ISL_1720161, EPI_ISL_1720164, EPI_ISL_1726761, EPI_ISL_1728618, EPI_ISL_1728687, EPI_ISL_1728696, EPI_ISL_1728711, EPI_ISL_1728713, EPI_ISL_1728782, EPI_ISL_1728784, EPI_ISL_1728832, EPI_ISL_1729780, EPI_ISL_1729826, EPI_ISL_1729853, EPI_ISL_1729859, EPI_ISL_1729869, EPI_ISL_1729885, EPI_ISL_1729888, EPI_ISL_1729893, EPI_ISL_1729959, EPI_ISL_1849523, EPI_ISL_1849623, EPI_ISL_1849631, EPI_ISL_1849668, EPI_ISL_1849674, EPI_ISL_2108075, EPI_ISL_2108098, EPI_ISL_2108134, EPI_ISL_2108141, EPI_ISL_2108155, EPI_ISL_2108189, EPI_ISL_2108262, EPI_ISL_2114515, EPI_ISL_2114533, EPI_ISL_2114555, EPI_ISL_2114562, EPI_ISL_2114592, EPI_ISL_2114593, EPI_ISL_2114638, EPI_ISL_2114648, EPI_ISL_2114658, EPI_ISL_2114694, EPI_ISL_2114733, EPI_ISL_2114777, EPI_ISL_2114781, EPI_ISL_2114815, EPI_ISL_2114832, EPI_ISL_2114882, EPI_ISL_2114933, EPI_ISL_2114955, EPI_ISL_2114999, EPI_ISL_2115001, EPI_ISL_2115021, EPI_ISL_2115052, EPI_ISL_2115066, EPI_ISL_2115111, EPI_ISL_2119608, EPI_ISL_2119653, EPI_ISL_2119887, EPI_ISL_2128694, EPI_ISL_2128703, EPI_ISL_2128704, EPI_ISL_2128705, EPI_ISL_2128749, EPI_ISL_2128751, EPI_ISL_2128787

see above | SYNLAB MVZ Hamburg | Robert Koch Institute

EPI_ISL_1641861, EPI_ISL_1641863, EPI_ISL_1641876, EPI_ISL_1641888, EPI_ISL_1641893, EPI_ISL_1641897, EPI_ISL_1641900, EPI_ISL_1641903, EPI_ISL_1641922, EPI_ISL_1641925, EPI_ISL_1641939, EPI_ISL_1641943, EPI_ISL_1641948, EPI_ISL_1641952, EPI_ISL_1641958, EPI_ISL_1641962, EPI_ISL_1641981, EPI_ISL_1641983, EPI_ISL_1641992, EPI_ISL_1642003, EPI_ISL_1642010, EPI_ISL_1642013, EPI_ISL_1642015, EPI_ISL_1642018, EPI_ISL_1642045, EPI_ISL_1642049, EPI_ISL_1642057, EPI_ISL_1642060, EPI_ISL_1642076, EPI_ISL_1642090, EPI_ISL_1642106, EPI_ISL_1642108, EPI_ISL_1642116, EPI_ISL_1642125, EPI_ISL_1642130, EPI_ISL_1642134, EPI_ISL_1642141, EPI_ISL_1642145, EPI_ISL_1642151, EPI_ISL_1642153, EPI_ISL_1642156, EPI_ISL_1642165, EPI_ISL_1642173, EPI_ISL_1642179, EPI_ISL_1642184, EPI_ISL_1642189, EPI_ISL_1642193, EPI_ISL_1642201, EPI_ISL_1642222, EPI_ISL_1642234, EPI_ISL_1642237, EPI_ISL_1642238, EPI_ISL_1642241, EPI_ISL_1642243, EPI_ISL_1642248, EPI_ISL_1642250, EPI_ISL_1642253, EPI_ISL_1642256, EPI_ISL_1642258, EPI_ISL_1642264, EPI_ISL_1642265, EPI_ISL_1642267, EPI_ISL_1642269, EPI_ISL_1642271, EPI_ISL_1645937, EPI_ISL_1645939, EPI_ISL_1645941, EPI_ISL_1645946, EPI_ISL_1645954, EPI_ISL_1645958, EPI_ISL_1645967, EPI_ISL_1645979, EPI_ISL_1645992, EPI_ISL_1645997, EPI_ISL_1646001, EPI_ISL_1646009, EPI_ISL_1646036, EPI_ISL_1646040, EPI_ISL_1646043, EPI_ISL_1646049, EPI_ISL_1646061, EPI_ISL_1646078, EPI_ISL_1646091, EPI_ISL_1646094, EPI_ISL_1646109, EPI_ISL_1646122, EPI_ISL_1646126, EPI_ISL_1646140, EPI_ISL_1646144, EPI_ISL_1646146, EPI_ISL_1646148, EPI_ISL_1646152, EPI_ISL_1646159, EPI_ISL_1646192, EPI_ISL_1646198, EPI_ISL_1646237, EPI_ISL_1646243, EPI_ISL_1646249, EPI_ISL_1646257, EPI_ISL_1646266, EPI_ISL_1646280, EPI_ISL_1646282, EPI_ISL_1646303, EPI_ISL_1646312, EPI_ISL_1646314, EPI_ISL_1646318, EPI_ISL_1646326, EPI_ISL_1646348, EPI_ISL_1646355, EPI_ISL_1646366, EPI_ISL_1646370, EPI_ISL_1646379, EPI_ISL_1646391, EPI_ISL_1646399, EPI_ISL_1646407, EPI_ISL_1646412, EPI_ISL_1646418, EPI_ISL_1646420, EPI_ISL_1646435, EPI_ISL_1646437, EPI_ISL_1646441, EPI_ISL_1646446, EPI_ISL_1646455, EPI_ISL_1646461, EPI_ISL_1646472, EPI_ISL_1646480, EPI_ISL_1646489, EPI_ISL_1646497, EPI_ISL_1646540, EPI_ISL_1646550, EPI_ISL_1646560, EPI_ISL_1646567, EPI_ISL_1646577, EPI_ISL_1646585, EPI_ISL_1646593, EPI_ISL_1646595, EPI_ISL_1646605, EPI_ISL_1646622, EPI_ISL_1646637, EPI_ISL_1646651, EPI_ISL_1646672, EPI_ISL_1646683, EPI_ISL_1646694, EPI_ISL_1646698, EPI_ISL_1646702, EPI_ISL_1646722, EPI_ISL_1646730, EPI_ISL_1646735, EPI_ISL_1646741, EPI_ISL_1646747, EPI_ISL_1646772, EPI_ISL_1646779, EPI_ISL_1646790, EPI_ISL_1646799, EPI_ISL_1646805, EPI_ISL_1646811, EPI_ISL_1646826, EPI_ISL_1646831, EPI_ISL_1646839, EPI_ISL_1726473, EPI_ISL_1726481, EPI_ISL_1726487, EPI_ISL_1726491, EPI_ISL_1726494, EPI_ISL_1726501, EPI_ISL_1726504, EPI_ISL_1726509, EPI_ISL_1726511, EPI_ISL_1726513, EPI_ISL_1726515, EPI_ISL_1726520, EPI_ISL_1726522, EPI_ISL_1726524, EPI_ISL_1726527, EPI_ISL_1726532, EPI_ISL_1726547, EPI_ISL_1726551, EPI_ISL_1726553, EPI_ISL_1726554, EPI_ISL_1726556, EPI_ISL_1726557, EPI_ISL_1726561, EPI_ISL_1726563, EPI_ISL_1726564, EPI_ISL_1726566, EPI_ISL_1726572, EPI_ISL_1726575, EPI_ISL_1726580, EPI_ISL_1726583, EPI_ISL_1726584, EPI_ISL_1726585, EPI_ISL_1726588, EPI_ISL_1726592, EPI_ISL_1726598, EPI_ISL_1726599, EPI_ISL_1726601, EPI_ISL_1726603, EPI_ISL_1726609, EPI_ISL_1726631, EPI_ISL_1726634, EPI_ISL_1726635, EPI_ISL_1726653, EPI_ISL_1726658, EPI_ISL_1726666, EPI_ISL_1726671, EPI_ISL_1726678, EPI_ISL_1726691, EPI_ISL_1726694, EPI_ISL_1726708, EPI_ISL_1726709, EPI_ISL_1726713, EPI_ISL_1726715, EPI_ISL_1726717, EPI_ISL_1728600, EPI_ISL_1849499, EPI_ISL_1849501, EPI_ISL_1849503, EPI_ISL_1849507, EPI_ISL_1849508, EPI_ISL_1849509, EPI_ISL_1849510, EPI_ISL_1849512, EPI_ISL_1849514, EPI_ISL_1849518, EPI_ISL_1849519, EPI_ISL_1849520, EPI_ISL_1849525, EPI_ISL_1849527, EPI_ISL_1849528, EPI_ISL_1849529, EPI_ISL_1849533, EPI_ISL_1849538, EPI_ISL_1849539, EPI_ISL_1849540, EPI_ISL_1849542, EPI_ISL_1849543, EPI_ISL_1849544, EPI_ISL_1849548, EPI_ISL_1849549, EPI_ISL_1849556, EPI_ISL_1849558, EPI_ISL_1849561, EPI_ISL_1849565, EPI_ISL_1849579, EPI_ISL_1849580, EPI_ISL_1849581, EPI_ISL_1849583, EPI_ISL_1849587, EPI_ISL_1849588, EPI_ISL_1849598, EPI_ISL_1849601, EPI_ISL_1849602, EPI_ISL_1849603, EPI_ISL_1849604, EPI_ISL_1849607, EPI_ISL_1849609, EPI_ISL_1849611, EPI_ISL_1849612, EPI_ISL_1849617, EPI_ISL_1849618, EPI_ISL_1849621, EPI_ISL_1849622, EPI_ISL_1849626, EPI_ISL_1849627, EPI_ISL_1849628, EPI_ISL_1849632, EPI_ISL_1849636, EPI_ISL_1849637, EPI_ISL_1849638, EPI_ISL_1849641, EPI_ISL_1849648, EPI_ISL_1849650, EPI_ISL_1849651, EPI_ISL_1849655, EPI_ISL_1849663, EPI_ISL_1849664, EPI_ISL_1849665, EPI_ISL_1849677, EPI_ISL_1849679, EPI_ISL_1849686, EPI_ISL_1849688, EPI_ISL_1849696, EPI_ISL_1849697, EPI_ISL_1849890, EPI_ISL_2128676, EPI_ISL_2128690, EPI_ISL_2128931, EPI_ISL_2128947

see above | SYNLAB MVZ Heidelberg | Robert Koch Institute

EPI_ISL_1571596, EPI_ISL_1571597, EPI_ISL_1571598, EPI_ISL_1571600, EPI_ISL_1571601, EPI_ISL_1571602, EPI_ISL_1571603, EPI_ISL_1571604, EPI_ISL_1571606, EPI_ISL_1571607, EPI_ISL_1571609, EPI_ISL_1571613, EPI_ISL_1571616, EPI_ISL_1571618, EPI_ISL_1571624, EPI_ISL_1571631, EPI_ISL_1571634, EPI_ISL_1571636, EPI_ISL_1571641, EPI_ISL_1571642, EPI_ISL_1571644, EPI_ISL_1571646, EPI_ISL_1571650, EPI_ISL_1571652, EPI_ISL_1571655, EPI_ISL_1571659, EPI_ISL_1571663, EPI_ISL_1571666, EPI_ISL_1571670, EPI_ISL_1571673, EPI_ISL_1571675, EPI_ISL_1571676, EPI_ISL_1571677, EPI_ISL_1571681, EPI_ISL_1571683, EPI_ISL_1571688, EPI_ISL_1571689, EPI_ISL_1571693, EPI_ISL_1571696, EPI_ISL_1571700, EPI_ISL_1571703, EPI_ISL_1571706, EPI_ISL_1571708, EPI_ISL_1571711, EPI_ISL_1571716, EPI_ISL_1571720, EPI_ISL_1571727, EPI_ISL_1571728, EPI_ISL_1571729, EPI_ISL_1571732, EPI_ISL_1571735, EPI_ISL_1571736, EPI_ISL_1571739, EPI_ISL_1571744, EPI_ISL_1571747, EPI_ISL_1571751, EPI_ISL_1571752, EPI_ISL_1571753, EPI_ISL_1571754, EPI_ISL_1571756, EPI_ISL_1571757, EPI_ISL_1571758, EPI_ISL_1571759, EPI_ISL_1571760, EPI_ISL_1571762, EPI_ISL_1571764, EPI_ISL_1571765, EPI_ISL_1571767, EPI_ISL_1571774, EPI_ISL_1571789, EPI_ISL_1571798, EPI_ISL_1571814, EPI_ISL_1571818, EPI_ISL_1571831, EPI_ISL_1571832, EPI_ISL_1571834, EPI_ISL_1571838, EPI_ISL_1571839, EPI_ISL_1571848, EPI_ISL_1571849, EPI_ISL_1571856, EPI_ISL_1571858, EPI_ISL_1571866, EPI_ISL_1571867, EPI_ISL_1571869, EPI_ISL_1571871, EPI_ISL_1571872, EPI_ISL_1571880, EPI_ISL_1571881, EPI_ISL_1571891, EPI_ISL_1571893, EPI_ISL_1571894, EPI_ISL_1571902, EPI_ISL_1571906, EPI_ISL_1571912, EPI_ISL_1571915, EPI_ISL_1571917, EPI_ISL_1571918, EPI_ISL_1571919, EPI_ISL_1571920, EPI_ISL_1571922, EPI_ISL_1571923, EPI_ISL_1571927, EPI_ISL_1572555, EPI_ISL_1572556, EPI_ISL_1572559, EPI_ISL_1572563, EPI_ISL_1572564, EPI_ISL_1572566, EPI_ISL_1572568, EPI_ISL_1572570, EPI_ISL_1572572, EPI_ISL_1572573, EPI_ISL_1572575, EPI_ISL_1572579, EPI_ISL_1572583, EPI_ISL_1572593, EPI_ISL_1572594, EPI_ISL_1572596, EPI_ISL_1572602, EPI_ISL_1572607, EPI_ISL_1572615, EPI_ISL_1572629, EPI_ISL_1572630, EPI_ISL_1572634, EPI_ISL_1572635, EPI_ISL_1572643, EPI_ISL_1572664, EPI_ISL_1572668, EPI_ISL_1572675, EPI_ISL_1572678, EPI_ISL_1572684, EPI_ISL_1572686, EPI_ISL_1572696, EPI_ISL_1572710, EPI_ISL_1572715, EPI_ISL_1572718, EPI_ISL_1572721, EPI_ISL_1572726, EPI_ISL_1572734, EPI_ISL_1572741, EPI_ISL_1572744, EPI_ISL_1572748, EPI_ISL_1572757, EPI_ISL_1572761, EPI_ISL_1572766, EPI_ISL_1572773, EPI_ISL_1572784, EPI_ISL_1572785, EPI_ISL_1572791, EPI_ISL_1572795, EPI_ISL_1572799, EPI_ISL_1572802, EPI_ISL_1572805, EPI_ISL_1572806, EPI_ISL_1572810, EPI_ISL_1572814, EPI_ISL_1572815, EPI_ISL_1572816, EPI_ISL_1572818, EPI_ISL_1572834, EPI_ISL_1573467, EPI_ISL_1573469, EPI_ISL_1573472, EPI_ISL_1573473, EPI_ISL_1573474, EPI_ISL_1573475, EPI_ISL_1573478, EPI_ISL_1573479, EPI_ISL_1573481, EPI_ISL_1573482, EPI_ISL_1573484, EPI_ISL_1573486, EPI_ISL_1573487, EPI_ISL_1573488, EPI_ISL_1573489, EPI_ISL_1573492, EPI_ISL_1573493, EPI_ISL_1573496, EPI_ISL_1573497, EPI_ISL_1573499, EPI_ISL_1573500, EPI_ISL_1573501, EPI_ISL_1573502, EPI_ISL_1573504, EPI_ISL_1573507, EPI_ISL_1573510, EPI_ISL_1573511, EPI_ISL_1573514, EPI_ISL_1573517, EPI_ISL_1573520, EPI_ISL_1573522, EPI_ISL_1573524, EPI_ISL_1573525, EPI_ISL_1573528, EPI_ISL_1573529, EPI_ISL_1573531, EPI_ISL_1573533, EPI_ISL_1573536, EPI_ISL_1573538, EPI_ISL_1573539, EPI_ISL_1573543, EPI_ISL_1573547, EPI_ISL_1573548, EPI_ISL_1573550, EPI_ISL_1573551, EPI_ISL_1573552, EPI_ISL_1573554, EPI_ISL_1573556, EPI_ISL_1573557, EPI_ISL_1573559, EPI_ISL_1573563, EPI_ISL_1573564, EPI_ISL_1573567, EPI_ISL_1573569, EPI_ISL_1573570, EPI_ISL_1573573, EPI_ISL_1573574, EPI_ISL_1573575, EPI_ISL_1573577, EPI_ISL_1573578, EPI_ISL_1573579, EPI_ISL_1573580, EPI_ISL_1573582, EPI_ISL_1573583, EPI_ISL_1573584, EPI_ISL_1573585, EPI_ISL_1573587, EPI_ISL_1573588, EPI_ISL_1573589, EPI_ISL_1573593, EPI_ISL_1573595, EPI_ISL_1573596, EPI_ISL_1573601, EPI_ISL_1573603, EPI_ISL_1573608, EPI_ISL_1573611, EPI_ISL_1573612, EPI_ISL_1573615, EPI_ISL_1573616, EPI_ISL_1573617, EPI_ISL_1573620, EPI_ISL_1573621, EPI_ISL_1573624, EPI_ISL_1573625, EPI_ISL_1573627, EPI_ISL_1573630, EPI_ISL_1573631, EPI_ISL_1573634, EPI_ISL_1573635, EPI_ISL_1573636, EPI_ISL_1573638, EPI_ISL_1573640, EPI_ISL_1573641, EPI_ISL_1573642, EPI_ISL_1573644, EPI_ISL_1573648, EPI_ISL_1573649, EPI_ISL_1573651, EPI_ISL_1573655, EPI_ISL_1573657, EPI_ISL_1573659, EPI_ISL_1573661, EPI_ISL_1573663, EPI_ISL_1573664, EPI_ISL_1573666, EPI_ISL_1573677, EPI_ISL_1573679, EPI_ISL_1573680, EPI_ISL_1573685, EPI_ISL_1573687, EPI_ISL_1573688, EPI_ISL_1573689, EPI_ISL_1573691, EPI_ISL_1573693, EPI_ISL_1573700, EPI_ISL_1573701, EPI_ISL_1573702, EPI_ISL_1573704, EPI_ISL_1573705, EPI_ISL_1573706, EPI_ISL_1573708, EPI_ISL_1573710, EPI_ISL_1573711, EPI_ISL_1573713, EPI_ISL_1573714, EPI_ISL_1573716, EPI_ISL_1573717, EPI_ISL_1573718, EPI_ISL_1573720, EPI_ISL_1573721, EPI_ISL_1573722, EPI_ISL_1573726, EPI_ISL_1573729, EPI_ISL_1573730, EPI_ISL_1573732, EPI_ISL_1573734, EPI_ISL_1573735, EPI_ISL_1573737, EPI_ISL_1573738, EPI_ISL_1573739, EPI_ISL_1573741, EPI_ISL_1573742, EPI_ISL_1573743, EPI_ISL_1573746, EPI_ISL_1573747, EPI_ISL_1573748, EPI_ISL_1573749, EPI_ISL_1573752, EPI_ISL_1573753, EPI_ISL_1573755, EPI_ISL_1573758, EPI_ISL_1573759, EPI_ISL_1573763, EPI_ISL_1573765, EPI_ISL_1573766, EPI_ISL_1573767, EPI_ISL_1573769, EPI_ISL_1573771, EPI_ISL_1573772, EPI_ISL_1573774, EPI_ISL_1573776, EPI_ISL_1573778, EPI_ISL_1573779, EPI_ISL_1573782, EPI_ISL_1573783, EPI_ISL_1573786, EPI_ISL_1573787, EPI_ISL_1573790, EPI_ISL_1573793, EPI_ISL_1573795, EPI_ISL_1573797, EPI_ISL_1573799, EPI_ISL_1573800, EPI_ISL_1573801, EPI_ISL_1573802, EPI_ISL_1573803, EPI_ISL_1573804, EPI_ISL_1573806, EPI_ISL_1573807, EPI_ISL_1573815, EPI_ISL_1573816, EPI_ISL_1573817, EPI_ISL_1573818, EPI_ISL_1573820, EPI_ISL_1573821, EPI_ISL_1573827, EPI_ISL_1573828, EPI_ISL_1573832, EPI_ISL_1573835, EPI_ISL_1573837, EPI_ISL_1573839, EPI_ISL_1573841, EPI_ISL_1573843, EPI_ISL_1573844, EPI_ISL_1573847, EPI_ISL_1573848, EPI_ISL_1573849, EPI_ISL_1573851, EPI_ISL_1573852, EPI_ISL_1573856, EPI_ISL_1573857, EPI_ISL_1573859, EPI_ISL_1573861, EPI_ISL_1573862, EPI_ISL_1573863, EPI_ISL_1573864, EPI_ISL_1573865, EPI_ISL_1573866, EPI_ISL_1573868, EPI_ISL_1573869, EPI_ISL_1573871, EPI_ISL_1573872, EPI_ISL_1573873, EPI_ISL_1573875, EPI_ISL_1573877, EPI_ISL_1573878, EPI_ISL_1573879, EPI_ISL_1573881, EPI_ISL_1573885, EPI_ISL_1573889, EPI_ISL_1573898, EPI_ISL_1573907, EPI_ISL_1573911, EPI_ISL_1573921, EPI_ISL_1573933, EPI_ISL_1573938, EPI_ISL_1573940, EPI_ISL_1573943, EPI_ISL_1573944, EPI_ISL_1573946, EPI_ISL_1573948, EPI_ISL_1573951, EPI_ISL_1573952, EPI_ISL_1573957, EPI_ISL_1573958, EPI_ISL_1573960, EPI_ISL_1573961, EPI_ISL_1573966, EPI_ISL_1573967, EPI_ISL_1573968, EPI_ISL_1573969, EPI_ISL_1573970, EPI_ISL_1573971, EPI_ISL_1573972, EPI_ISL_1573973, EPI_ISL_1573975, EPI_ISL_1573976, EPI_ISL_1573981, EPI_ISL_1573983, EPI_ISL_1573984, EPI_ISL_1573987, EPI_ISL_1573988, EPI_ISL_1573989, EPI_ISL_1573992, EPI_ISL_1573994, EPI_ISL_1573996, EPI_ISL_1573997, EPI_ISL_1573998, EPI_ISL_1574001, EPI_ISL_1574004, EPI_ISL_1574005, EPI_ISL_1574008, EPI_ISL_1574010, EPI_ISL_1574013, EPI_ISL_1574015, EPI_ISL_1574016, EPI_ISL_1574018, EPI_ISL_1574019, EPI_ISL_1574021, EPI_ISL_1574023, EPI_ISL_1574025, EPI_ISL_1574026, EPI_ISL_1574028, EPI_ISL_1574029, EPI_ISL_1574030, EPI_ISL_1574031, EPI_ISL_1574034, EPI_ISL_1574035, EPI_ISL_1574036, EPI_ISL_1574038, EPI_ISL_1574040, EPI_ISL_1574043, EPI_ISL_1574046, EPI_ISL_1574048, EPI_ISL_1574050, EPI_ISL_1574052, EPI_ISL_1574054, EPI_ISL_1574056, EPI_ISL_1574058, EPI_ISL_1574059, EPI_ISL_1574060, EPI_ISL_1574063, EPI_ISL_1574067, EPI_ISL_1574069, EPI_ISL_1574071, EPI_ISL_1574072, EPI_ISL_1574073, EPI_ISL_1574075, EPI_ISL_1574079, EPI_ISL_1574081, EPI_ISL_1574085, EPI_ISL_1574086, EPI_ISL_1574087, EPI_ISL_1574089, EPI_ISL_1574090, EPI_ISL_1574091, EPI_ISL_1574096, EPI_ISL_1574097, EPI_ISL_1574098, EPI_ISL_1574099, EPI_ISL_1574100, EPI_ISL_1574101, EPI_ISL_1574102, EPI_ISL_1574104, EPI_ISL_1574107, EPI_ISL_1574108, EPI_ISL_1574109, EPI_ISL_1574110, EPI_ISL_1574114, EPI_ISL_1574116, EPI_ISL_1574117, EPI_ISL_1574118, EPI_ISL_1574122, EPI_ISL_1574126, EPI_ISL_1574127, EPI_ISL_1574128, EPI_ISL_1574129, EPI_ISL_1574130, EPI_ISL_1574131, EPI_ISL_1574134, EPI_ISL_1574136, EPI_ISL_1574139, EPI_ISL_1574142, EPI_ISL_1574143, EPI_ISL_1574147, EPI_ISL_1574148, EPI_ISL_1574150, EPI_ISL_1574151, EPI_ISL_1574152, EPI_ISL_1574153, EPI_ISL_1574155, EPI_ISL_1574156, EPI_ISL_1574157, EPI_ISL_1574158, EPI_ISL_1574160, EPI_ISL_1574161, EPI_ISL_1574164, EPI_ISL_1574165, EPI_ISL_1574166, EPI_ISL_1574167, EPI_ISL_1574169, EPI_ISL_1574170, EPI_ISL_1574174, EPI_ISL_1574175, EPI_ISL_1574176, EPI_ISL_1574178, EPI_ISL_1574181, EPI_ISL_1574183, EPI_ISL_1574188, EPI_ISL_1574189, EPI_ISL_1574190, EPI_ISL_1574191, EPI_ISL_1574193, EPI_ISL_1574196, EPI_ISL_1574197, EPI_ISL_1574199, EPI_ISL_1574201, EPI_ISL_1574202, EPI_ISL_1574208, EPI_ISL_1574211, EPI_ISL_1574215, EPI_ISL_1574216, EPI_ISL_1574219, EPI_ISL_1574222, EPI_ISL_1574224, EPI_ISL_1574225, EPI_ISL_1574227, EPI_ISL_1574228, EPI_ISL_1574230, EPI_ISL_1574231, EPI_ISL_1574234, EPI_ISL_1574235, EPI_ISL_1574236, EPI_ISL_1574237, EPI_ISL_1574238, EPI_ISL_1574240, EPI_ISL_1574241, EPI_ISL_1574242, EPI_ISL_1574245, EPI_ISL_1574246, EPI_ISL_1574251, EPI_ISL_1574252, EPI_ISL_1574256, EPI_ISL_1574259, EPI_ISL_1574260, EPI_ISL_1574261, EPI_ISL_1574262, EPI_ISL_1574263, EPI_ISL_1574264, EPI_ISL_1574268, EPI_ISL_1574269, EPI_ISL_1574270, EPI_ISL_1574273, EPI_ISL_1574274, EPI_ISL_1574277, EPI_ISL_1574278, EPI_ISL_1574279, EPI_ISL_1574280, EPI_ISL_1574281, EPI_ISL_1574282, EPI_ISL_1574283, EPI_ISL_1574284, EPI_ISL_1574285, EPI_ISL_1574287, EPI_ISL_1574289, EPI_ISL_1574290, EPI_ISL_1574292, EPI_ISL_1574293, EPI_ISL_1574294, EPI_ISL_1574295, EPI_ISL_1574296, EPI_ISL_1574297, EPI_ISL_1574300, EPI_ISL_1574301, EPI_ISL_1574303, EPI_ISL_1574304, EPI_ISL_1574305, EPI_ISL_1574307, EPI_ISL_1574308, EPI_ISL_1574309, EPI_ISL_1574310, EPI_ISL_1574312, EPI_ISL_1574313, EPI_ISL_1574314, EPI_ISL_1574315, EPI_ISL_1574317, EPI_ISL_1574318, EPI_ISL_1574319, EPI_ISL_1574322, EPI_ISL_1574324, EPI_ISL_1574325, EPI_ISL_1574326, EPI_ISL_1574327, EPI_ISL_1574329, EPI_ISL_1574330, EPI_ISL_1574331, EPI_ISL_1574332, EPI_ISL_1574335, EPI_ISL_1574337, EPI_ISL_1574340, EPI_ISL_1574341, EPI_ISL_1574344, EPI_ISL_1574347, EPI_ISL_1574348, EPI_ISL_1574349, EPI_ISL_1574350, EPI_ISL_1574351, EPI_ISL_1574352, EPI_ISL_1574353, EPI_ISL_1574354, EPI_ISL_1574355, EPI_ISL_1574356, EPI_ISL_1574359, EPI_ISL_1574360, EPI_ISL_1574361, EPI_ISL_1574362, EPI_ISL_1574687, EPI_ISL_1574688, EPI_ISL_1574689, EPI_ISL_1574693, EPI_ISL_1574694, EPI_ISL_1574696, EPI_ISL_1574697,

EPI_ISL_1574698, EPI_ISL_1574701, EPI_ISL_1574702, EPI_ISL_1574705, EPI_ISL_1574706, EPI_ISL_1574707, EPI_ISL_1574708, EPI_ISL_1574709, EPI_ISL_1574710, EPI_ISL_1574711, EPI_ISL_1574713, EPI_ISL_1574714, EPI_ISL_1574715, EPI_ISL_1574717, EPI_ISL_1574721, EPI_ISL_1574723, EPI_ISL_1574725, EPI_ISL_1574726, EPI_ISL_1574727, EPI_ISL_1574729, EPI_ISL_1574730, EPI_ISL_1574736, EPI_ISL_1574740, EPI_ISL_1574742, EPI_ISL_1574744, EPI_ISL_1574745, EPI_ISL_1574749, EPI_ISL_1574750, EPI_ISL_1574751, EPI_ISL_1574757, EPI_ISL_1574759, EPI_ISL_1574761, EPI_ISL_1574762, EPI_ISL_1574763, EPI_ISL_1574766, EPI_ISL_1574769, EPI_ISL_1574770, EPI_ISL_1574773, EPI_ISL_1574774, EPI_ISL_1574775, EPI_ISL_1574777, EPI_ISL_1574779, EPI_ISL_1574781, EPI_ISL_1574784, EPI_ISL_1574786, EPI_ISL_1574787, EPI_ISL_1574788, EPI_ISL_1574791, EPI_ISL_1574794, EPI_ISL_1574795, EPI_ISL_1574798, EPI_ISL_1574799, EPI_ISL_1574800, EPI_ISL_1574802, EPI_ISL_1574804, EPI_ISL_1574806, EPI_ISL_1574808, EPI_ISL_1574810, EPI_ISL_1574811, EPI_ISL_1574812, EPI_ISL_1574813, EPI_ISL_1574815, EPI_ISL_1574819, EPI_ISL_1574820, EPI_ISL_1574821, EPI_ISL_1574823, EPI_ISL_1574824, EPI_ISL_1574826, EPI_ISL_1574827, EPI_ISL_1574828, EPI_ISL_1574829, EPI_ISL_1574830, EPI_ISL_1574832, EPI_ISL_1574834, EPI_ISL_1574835, EPI_ISL_1574839, EPI_ISL_1574840, EPI_ISL_1574845, EPI_ISL_1574847, EPI_ISL_1574849, EPI_ISL_1574851, EPI_ISL_1574855, EPI_ISL_1574858, EPI_ISL_1574860, EPI_ISL_1574861, EPI_ISL_1574862, EPI_ISL_1574863, EPI_ISL_1574865, EPI_ISL_1574866, EPI_ISL_1574867, EPI_ISL_1574868, EPI_ISL_1574871, EPI_ISL_1574873, EPI_ISL_1574874, EPI_ISL_1574875, EPI_ISL_1574879, EPI_ISL_1574880, EPI_ISL_1574881, EPI_ISL_1574885, EPI_ISL_1574886, EPI_ISL_1574887, EPI_ISL_1574892, EPI_ISL_1574898, EPI_ISL_1574901, EPI_ISL_1574902, EPI_ISL_1574904, EPI_ISL_1574908, EPI_ISL_1574909, EPI_ISL_1574911, EPI_ISL_1574912, EPI_ISL_1574914, EPI_ISL_1574916, EPI_ISL_1574917, EPI_ISL_1574919, EPI_ISL_1574922, EPI_ISL_1574923, EPI_ISL_1574926, EPI_ISL_1574927, EPI_ISL_1574928, EPI_ISL_1574929, EPI_ISL_1574932, EPI_ISL_1574934, EPI_ISL_1574940, EPI_ISL_1574941, EPI_ISL_1574942, EPI_ISL_1574943, EPI_ISL_1574944, EPI_ISL_1574945, EPI_ISL_1574946, EPI_ISL_1574947, EPI_ISL_1574948, EPI_ISL_1574951, EPI_ISL_1574952, EPI_ISL_1574953, EPI_ISL_1574954, EPI_ISL_1574961, EPI_ISL_1574962, EPI_ISL_1574963, EPI_ISL_1574965, EPI_ISL_1640321, EPI_ISL_1640322, EPI_ISL_1640323, EPI_ISL_1640324, EPI_ISL_1640325, EPI_ISL_1640326, EPI_ISL_1640327, EPI_ISL_1640328, EPI_ISL_1640330, EPI_ISL_1640331, EPI_ISL_1640332, EPI_ISL_1640333, EPI_ISL_1640334, EPI_ISL_1640335, EPI_ISL_1640336, EPI_ISL_1640337, EPI_ISL_1640340, EPI_ISL_1640343, EPI_ISL_1640344, EPI_ISL_1640345, EPI_ISL_1640346, EPI_ISL_1640349, EPI_ISL_1640350, EPI_ISL_1640351, EPI_ISL_1640352, EPI_ISL_1640353, EPI_ISL_1640354, EPI_ISL_1640355, EPI_ISL_1640356, EPI_ISL_1640357, EPI_ISL_1640358, EPI_ISL_1640360, EPI_ISL_1640361, EPI_ISL_1640362, EPI_ISL_1640364, EPI_ISL_1640365, EPI_ISL_1640366, EPI_ISL_1640368, EPI_ISL_1640370, EPI_ISL_1640373, EPI_ISL_1640374, EPI_ISL_1640375, EPI_ISL_1640376, EPI_ISL_1640379, EPI_ISL_1640380, EPI_ISL_1640382, EPI_ISL_1640383, EPI_ISL_1640385, EPI_ISL_1640386, EPI_ISL_1640387, EPI_ISL_1640388, EPI_ISL_1640389, EPI_ISL_1640390, EPI_ISL_1640391, EPI_ISL_1640392, EPI_ISL_1640393, EPI_ISL_1640395, EPI_ISL_1640397, EPI_ISL_1640398, EPI_ISL_1640399, EPI_ISL_1640402, EPI_ISL_1640403, EPI_ISL_1640404, EPI_ISL_1640405, EPI_ISL_1640407, EPI_ISL_1640408, EPI_ISL_1640409, EPI_ISL_1640411, EPI_ISL_1640413, EPI_ISL_1640416, EPI_ISL_1640419, EPI_ISL_1640421, EPI_ISL_1640422, EPI_ISL_1640423, EPI_ISL_1640425, EPI_ISL_1640426, EPI_ISL_1640427, EPI_ISL_1640429, EPI_ISL_1640430, EPI_ISL_1640431, EPI_ISL_1640433, EPI_ISL_1640436, EPI_ISL_1640437, EPI_ISL_1640438, EPI_ISL_1640439, EPI_ISL_1640440, EPI_ISL_1640441, EPI_ISL_1640443, EPI_ISL_1640444, EPI_ISL_1640445, EPI_ISL_1640446, EPI_ISL_1640447, EPI_ISL_1640449, EPI_ISL_1640450, EPI_ISL_1640452, EPI_ISL_1640453, EPI_ISL_1640454, EPI_ISL_1640456, EPI_ISL_1640459, EPI_ISL_1640461, EPI_ISL_1640462, EPI_ISL_1640463, EPI_ISL_1640464, EPI_ISL_1640465, EPI_ISL_1640468, EPI_ISL_1640471, EPI_ISL_1640472, EPI_ISL_1640474, EPI_ISL_1640476, EPI_ISL_1640477, EPI_ISL_1640478, EPI_ISL_1640480, EPI_ISL_1640481, EPI_ISL_1640483, EPI_ISL_1640484, EPI_ISL_1640488, EPI_ISL_1640489, EPI_ISL_1640490, EPI_ISL_1640491, EPI_ISL_1640495, EPI_ISL_1640496, EPI_ISL_1640497, EPI_ISL_1640498, EPI_ISL_1640499, EPI_ISL_1640500, EPI_ISL_1640501, EPI_ISL_1640502, EPI_ISL_1640503, EPI_ISL_1640504, EPI_ISL_1640505, EPI_ISL_1640506, EPI_ISL_1640508, EPI_ISL_1640509, EPI_ISL_1640510, EPI_ISL_1640511, EPI_ISL_1640512, EPI_ISL_1640514, EPI_ISL_1640516, EPI_ISL_1640518, EPI_ISL_1640520, EPI_ISL_1640521, EPI_ISL_1640522, EPI_ISL_1640524, EPI_ISL_1640525, EPI_ISL_1640526, EPI_ISL_1640527, EPI_ISL_1640528, EPI_ISL_1640529, EPI_ISL_1640530, EPI_ISL_1640531, EPI_ISL_1640532, EPI_ISL_1640534, EPI_ISL_1640535, EPI_ISL_1640537, EPI_ISL_1640538, EPI_ISL_1640540, EPI_ISL_1640542, EPI_ISL_1640543, EPI_ISL_1640544, EPI_ISL_1640545, EPI_ISL_1640546, EPI_ISL_1640547, EPI_ISL_1640548, EPI_ISL_1640549, EPI_ISL_1640551, EPI_ISL_1640552, EPI_ISL_1640553, EPI_ISL_1640554, EPI_ISL_1640555, EPI_ISL_1640556, EPI_ISL_1640557, EPI_ISL_1640558, EPI_ISL_1640560, EPI_ISL_1640561, EPI_ISL_1640562, EPI_ISL_1640563, EPI_ISL_1640564, EPI_ISL_1640565, EPI_ISL_1640566, EPI_ISL_1640567, EPI_ISL_1640568, EPI_ISL_1640569, EPI_ISL_1640571, EPI_ISL_1640572, EPI_ISL_1640573, EPI_ISL_1640575, EPI_ISL_1640577, EPI_ISL_1640578, EPI_ISL_1640579, EPI_ISL_1640580, EPI_ISL_1640581, EPI_ISL_1640582, EPI_ISL_1640584, EPI_ISL_1640586, EPI_ISL_1640588, EPI_ISL_1640589, EPI_ISL_1640590, EPI_ISL_1640591, EPI_ISL_1640592, EPI_ISL_1640594, EPI_ISL_1640595, EPI_ISL_1640596, EPI_ISL_1640597, EPI_ISL_1640598, EPI_ISL_1640599, EPI_ISL_1640600, EPI_ISL_1640601, EPI_ISL_1640602, EPI_ISL_1640603, EPI_ISL_1640604, EPI_ISL_1640605, EPI_ISL_1640606, EPI_ISL_1640607, EPI_ISL_1640608, EPI_ISL_1640609, EPI_ISL_1640611, EPI_ISL_1640612, EPI_ISL_1640613, EPI_ISL_1640614, EPI_ISL_1640615, EPI_ISL_1640616, EPI_ISL_1640617, EPI_ISL_1640618, EPI_ISL_1640619, EPI_ISL_1640620, EPI_ISL_1640621, EPI_ISL_1640622, EPI_ISL_1640623, EPI_ISL_1640932, EPI_ISL_1640933, EPI_ISL_1640935, EPI_ISL_1640938, EPI_ISL_1640939, EPI_ISL_1640942, EPI_ISL_1640943, EPI_ISL_1640944, EPI_ISL_1640945, EPI_ISL_1640947, EPI_ISL_1640948, EPI_ISL_1640950, EPI_ISL_1640951, EPI_ISL_1640952, EPI_ISL_1640953, EPI_ISL_1640954, EPI_ISL_1640956, EPI_ISL_1640957, EPI_ISL_1640958, EPI_ISL_1640961, EPI_ISL_1640962, EPI_ISL_1640963, EPI_ISL_1640965, EPI_ISL_1640968, EPI_ISL_1640969, EPI_ISL_1640970, EPI_ISL_1640973, EPI_ISL_1640974, EPI_ISL_1640975, EPI_ISL_1640979, EPI_ISL_1640980, EPI_ISL_1640983, EPI_ISL_1640984, EPI_ISL_1640985, EPI_ISL_1640987, EPI_ISL_1640988, EPI_ISL_1640989, EPI_ISL_1640990, EPI_ISL_1640991, EPI_ISL_1640994, EPI_ISL_1640996, EPI_ISL_1640998, EPI_ISL_1640999, EPI_ISL_1641000, EPI_ISL_1641001, EPI_ISL_1641002, EPI_ISL_1641003, EPI_ISL_1641004, EPI_ISL_1641005, EPI_ISL_1641007, EPI_ISL_1641008, EPI_ISL_1641009, EPI_ISL_1641010, EPI_ISL_1641011, EPI_ISL_1641013, EPI_ISL_1641014, EPI_ISL_1641015, EPI_ISL_1641016, EPI_ISL_1641017, EPI_ISL_1641018, EPI_ISL_1641019, EPI_ISL_1641020, EPI_ISL_1641023, EPI_ISL_1641025, EPI_ISL_1641027, EPI_ISL_1641029, EPI_ISL_1641030, EPI_ISL_1641031, EPI_ISL_1641033, EPI_ISL_1641036, EPI_ISL_1641040, EPI_ISL_1641043, EPI_ISL_1641044, EPI_ISL_1641046, EPI_ISL_1641047, EPI_ISL_1641051, EPI_ISL_1641052, EPI_ISL_1641053, EPI_ISL_1641056, EPI_ISL_1641059, EPI_ISL_1641061, EPI_ISL_1641062, EPI_ISL_1641063, EPI_ISL_1641065, EPI_ISL_1641069, EPI_ISL_1641070, EPI_ISL_1641072, EPI_ISL_1641074, EPI_ISL_1641076, EPI_ISL_1641078, EPI_ISL_1641079, EPI_ISL_1641081, EPI_ISL_1641083, EPI_ISL_1641084, EPI_ISL_1641085, EPI_ISL_1641087, EPI_ISL_1641088, EPI_ISL_1641091, EPI_ISL_1641093, EPI_ISL_1641094, EPI_ISL_1641096, EPI_ISL_1641097, EPI_ISL_1641098, EPI_ISL_1641100, EPI_ISL_1641101, EPI_ISL_1641102, EPI_ISL_1641104, EPI_ISL_1641105, EPI_ISL_1641106, EPI_ISL_1641107, EPI_ISL_1641108, EPI_ISL_1641110, EPI_ISL_1641111, EPI_ISL_1641113, EPI_ISL_1641116, EPI_ISL_1641119, EPI_ISL_1641122, EPI_ISL_1641124, EPI_ISL_1641128, EPI_ISL_1641131, EPI_ISL_1641132, EPI_ISL_1641134, EPI_ISL_1641136, EPI_ISL_1641138, EPI_ISL_1641139, EPI_ISL_1641140, EPI_ISL_1641141, EPI_ISL_1641142, EPI_ISL_1641145, EPI_ISL_1641147, EPI_ISL_1641148, EPI_ISL_1641150, EPI_ISL_1641151, EPI_ISL_1641152, EPI_ISL_1641153, EPI_ISL_1641154, EPI_ISL_1641155, EPI_ISL_1641158, EPI_ISL_1645247, EPI_ISL_1645271, EPI_ISL_1645298, EPI_ISL_1645345, EPI_ISL_1645358, EPI_ISL_1645385, EPI_ISL_1645453, EPI_ISL_1645474, EPI_ISL_1645479, EPI_ISL_1645484, EPI_ISL_1645487, EPI_ISL_1645497, EPI_ISL_1645499, EPI_ISL_1645500

see above | SYNLAB MVZ Leinfelden-Echterdingen | Robert Koch Institute

EPI_ISL_1639525, EPI_ISL_1646264, EPI_ISL_1646356, EPI_ISL_1646430, EPI_ISL_1646517, EPI_ISL_1646774, EPI_ISL_1646888, EPI_ISL_1719925, EPI_ISL_1719930, EPI_ISL_1719940, EPI_ISL_1719954, EPI_ISL_1719956, EPI_ISL_1719959, EPI_ISL_1719964, EPI_ISL_1719967, EPI_ISL_1719968, EPI_ISL_1719972, EPI_ISL_1719976, EPI_ISL_1719978, EPI_ISL_1719980, EPI_ISL_1719983, EPI_ISL_1719984, EPI_ISL_1719987, EPI_ISL_1719991, EPI_ISL_1719996, EPI_ISL_1720001, EPI_ISL_1720012, EPI_ISL_1720014, EPI_ISL_1720016, EPI_ISL_1720031, EPI_ISL_1720053, EPI_ISL_1720055, EPI_ISL_1720068, EPI_ISL_1720070, EPI_ISL_1720072, EPI_ISL_1720073, EPI_ISL_1720079, EPI_ISL_1720084, EPI_ISL_1720092, EPI_ISL_1720094, EPI_ISL_1720097, EPI_ISL_1720098, EPI_ISL_1720100, EPI_ISL_1720102, EPI_ISL_1720110, EPI_ISL_1720112, EPI_ISL_1720113, EPI_ISL_1720114, EPI_ISL_1720117, EPI_ISL_1720120, EPI_ISL_1720130, EPI_ISL_1720132, EPI_ISL_1720139, EPI_ISL_1720143, EPI_ISL_1720144, EPI_ISL_1720145, EPI_ISL_1720150, EPI_ISL_1720152, EPI_ISL_1720155, EPI_ISL_1720156, EPI_ISL_1720162, EPI_ISL_1720163, EPI_ISL_1720170, EPI_ISL_1726468, EPI_ISL_1726470, EPI_ISL_1726471, EPI_ISL_1726472, EPI_ISL_1726474, EPI_ISL_1726475, EPI_ISL_1726476, EPI_ISL_1726478, EPI_ISL_1726479, EPI_ISL_1726480, EPI_ISL_1726482, EPI_ISL_1726485, EPI_ISL_1726486, EPI_ISL_1726489, EPI_ISL_1726490, EPI_ISL_1726493, EPI_ISL_1726495, EPI_ISL_1726496, EPI_ISL_1726497, EPI_ISL_1726499, EPI_ISL_1726500, EPI_ISL_1726502, EPI_ISL_1726503, EPI_ISL_1726505, EPI_ISL_1726506, EPI_ISL_1726507, EPI_ISL_1726508, EPI_ISL_1726510, EPI_ISL_1726512, EPI_ISL_1726514, EPI_ISL_1726517, EPI_ISL_1726518, EPI_ISL_1726519, EPI_ISL_1726521, EPI_ISL_1726523, EPI_ISL_1726525, EPI_ISL_1726526, EPI_ISL_1726528, EPI_ISL_1726530, EPI_ISL_1726531, EPI_ISL_1726533, EPI_ISL_1726535, EPI_ISL_1726537, EPI_ISL_1726540, EPI_ISL_1726541, EPI_ISL_1726543, EPI_ISL_1726545, EPI_ISL_1726546, EPI_ISL_1726548, EPI_ISL_1726549, EPI_ISL_1726558, EPI_ISL_1726559, EPI_ISL_1726562, EPI_ISL_1726565, EPI_ISL_1726567, EPI_ISL_1726568, EPI_ISL_1726571, EPI_ISL_1726573, EPI_ISL_1726574, EPI_ISL_1726576, EPI_ISL_1726577, EPI_ISL_1726578, EPI_ISL_1726579, EPI_ISL_1726582, EPI_ISL_1726589, EPI_ISL_1726591, EPI_ISL_1726594, EPI_ISL_1726595, EPI_ISL_1726597, EPI_ISL_1726600, EPI_ISL_1726605, EPI_ISL_1726607, EPI_ISL_1726608, EPI_ISL_1726610, EPI_ISL_1726612, EPI_ISL_1726613, EPI_ISL_1726614, EPI_ISL_1726616, EPI_ISL_1726617, EPI_ISL_1726618, EPI_ISL_1726619, EPI_ISL_1726620, EPI_ISL_1726621, EPI_ISL_1726624, EPI_ISL_1726625, EPI_ISL_1726626, EPI_ISL_1726627, EPI_ISL_1726632, EPI_ISL_1726637, EPI_ISL_1726638, EPI_ISL_1726640, EPI_ISL_1726641, EPI_ISL_1726643, EPI_ISL_1726644, EPI_ISL_1726645, EPI_ISL_1726647, EPI_ISL_1726650, EPI_ISL_1726657, EPI_ISL_1726659, EPI_ISL_1726662, EPI_ISL_1726665, EPI_ISL_1726667, EPI_ISL_1726669, EPI_ISL_1726673, EPI_ISL_1726675, EPI_ISL_1726676, EPI_ISL_1726677, EPI_ISL_1726680, EPI_ISL_1726684, EPI_ISL_1726685, EPI_ISL_1726688, EPI_ISL_1726692, EPI_ISL_1726693, EPI_ISL_1726696, EPI_ISL_1726697, EPI_ISL_1726699, EPI_ISL_1726701, EPI_ISL_1726702, EPI_ISL_1726704, EPI_ISL_1726705, EPI_ISL_1726707, EPI_ISL_1726712, EPI_ISL_1726716, EPI_ISL_1726719, EPI_ISL_1726720, EPI_ISL_1726725, EPI_ISL_1726731, EPI_ISL_1726733, EPI_ISL_1726738, EPI_ISL_1726739, EPI_ISL_1726741, EPI_ISL_1726743, EPI_ISL_1726744, EPI_ISL_1726747, EPI_ISL_1726749, EPI_ISL_1726750, EPI_ISL_1726753, EPI_ISL_1726756, EPI_ISL_1726757, EPI_ISL_1726758, EPI_ISL_1726759, EPI_ISL_1726760, EPI_ISL_1726762, EPI_ISL_1726763, EPI_ISL_1726765, EPI_ISL_1726767, EPI_ISL_1726768, EPI_ISL_1726770, EPI_ISL_1726771, EPI_ISL_1726772, EPI_ISL_1726773, EPI_ISL_1726774, EPI_ISL_1726775, EPI_ISL_1726776, EPI_ISL_1726777, EPI_ISL_1728591, EPI_ISL_1728592, EPI_ISL_1728593, EPI_ISL_1728594, EPI_ISL_1728595, EPI_ISL_1728596, EPI_ISL_1728597, EPI_ISL_1728598, EPI_ISL_1728599, EPI_ISL_1728601, EPI_ISL_1728602, EPI_ISL_1728603, EPI_ISL_1728604, EPI_ISL_1728605, EPI_ISL_1728606, EPI_ISL_1728607, EPI_ISL_1728608, EPI_ISL_1728609, EPI_ISL_1728610, EPI_ISL_1728612, EPI_ISL_1728613, EPI_ISL_1728614, EPI_ISL_1728615, EPI_ISL_1728616, EPI_ISL_1728617, EPI_ISL_1728619, EPI_ISL_1728620, EPI_ISL_1728621, EPI_ISL_1728622, EPI_ISL_1728623, EPI_ISL_1728624, EPI_ISL_1728625, EPI_ISL_1728626, EPI_ISL_1728627, EPI_ISL_1728628, EPI_ISL_1728629, EPI_ISL_1728630, EPI_ISL_1728631, EPI_ISL_1728632, EPI_ISL_1728633, EPI_ISL_1728634, EPI_ISL_1728635, EPI_ISL_1728636, EPI_ISL_1728637, EPI_ISL_1728638, EPI_ISL_1728639, EPI_ISL_1728640, EPI_ISL_1728641, EPI_ISL_1728642, EPI_ISL_1728643, EPI_ISL_1728645, EPI_ISL_1728646, EPI_ISL_1728647, EPI_ISL_1728648, EPI_ISL_1728649, EPI_ISL_1728650, EPI_ISL_1728651, EPI_ISL_1728652, EPI_ISL_1728653, EPI_ISL_1728654, EPI_ISL_1728655, EPI_ISL_1728656, EPI_ISL_1728657, EPI_ISL_1728658, EPI_ISL_1728659, EPI_ISL_1728660, EPI_ISL_1728661, EPI_ISL_1728663, EPI_ISL_1728664, EPI_ISL_1728665, EPI_ISL_1728666, EPI_ISL_1728668, EPI_ISL_1728669, EPI_ISL_1728670, EPI_ISL_1728671, EPI_ISL_1728672, EPI_ISL_1728673, EPI_ISL_1728674, EPI_ISL_1728675, EPI_ISL_1728676, EPI_ISL_1728677, EPI_ISL_1728678, EPI_ISL_1728679, EPI_ISL_1728680, EPI_ISL_1728681, EPI_ISL_1728682, EPI_ISL_1728683, EPI_ISL_1728684, EPI_ISL_1728685, EPI_ISL_1728686, EPI_ISL_1728688, EPI_ISL_1728689, EPI_ISL_1728690, EPI_ISL_1728691, EPI_ISL_1728692, EPI_ISL_1728693, EPI_ISL_1728694, EPI_ISL_1728695, EPI_ISL_1728697, EPI_ISL_1728698, EPI_ISL_1728699, EPI_ISL_1728700, EPI_ISL_1728701, EPI_ISL_1728702, EPI_ISL_1728703, EPI_ISL_1728704, EPI_ISL_1728705, EPI_ISL_1728706, EPI_ISL_1728707, EPI_ISL_1728708, EPI_ISL_1728709, EPI_ISL_1728710, EPI_ISL_1728712, EPI_ISL_1728714, EPI_ISL_1728715, EPI_ISL_1728716, EPI_ISL_1728717, EPI_ISL_1728718, EPI_ISL_1728719, EPI_ISL_1728720, EPI_ISL_1728721, EPI_ISL_1728722, EPI_ISL_1728723, EPI_ISL_1728724, EPI_ISL_1728725, EPI_ISL_1728726, EPI_ISL_1728727, EPI_ISL_1728728, EPI_ISL_1728729, EPI_ISL_1728730, EPI_ISL_1728732, EPI_ISL_1728737, EPI_ISL_1728738, EPI_ISL_1728741, EPI_ISL_1728742, EPI_ISL_1728744, EPI_ISL_1728745, EPI_ISL_1728747, EPI_ISL_1728748, EPI_ISL_1728753, EPI_ISL_1728759, EPI_ISL_1728761, EPI_ISL_1728763, EPI_ISL_1728766, EPI_ISL_1728769, EPI_ISL_1728772, EPI_ISL_1728775, EPI_ISL_1728776, EPI_ISL_1728777, EPI_ISL_1728778, EPI_ISL_1728781, EPI_ISL_1728791, EPI_ISL_1728792, EPI_ISL_1728793, EPI_ISL_1728801, EPI_ISL_1728802, EPI_ISL_1728806, EPI_ISL_1728808, EPI_ISL_1728815, EPI_ISL_1728818, EPI_ISL_1728821, EPI_ISL_1728825, EPI_ISL_1728837, EPI_ISL_1728840, EPI_ISL_1728843, EPI_ISL_1728860, EPI_ISL_1728864, EPI_ISL_1729708, EPI_ISL_1729709, EPI_ISL_1729711, EPI_ISL_1729712, EPI_ISL_1729716, EPI_ISL_1729717, EPI_ISL_1729720, EPI_ISL_1729724, EPI_ISL_1729725, EPI_ISL_1729726, EPI_ISL_1729730, EPI_ISL_1729731, EPI_ISL_1729733, EPI_ISL_1729736, EPI_ISL_1729737, EPI_ISL_1729740, EPI_ISL_1729744, EPI_ISL_1729745, EPI_ISL_1729746, EPI_ISL_1729747, EPI_ISL_1729748, EPI_ISL_1729751, EPI_ISL_1729753, EPI_ISL_1729755, EPI_ISL_1729757, EPI_ISL_1729759, EPI_ISL_1729760, EPI_ISL_1729765, EPI_ISL_1729766, EPI_ISL_1729772, EPI_ISL_1729773, EPI_ISL_1729777, EPI_ISL_1729778, EPI_ISL_1729781, EPI_ISL_1729785, EPI_ISL_1729787, EPI_ISL_1729788, EPI_ISL_1729791, EPI_ISL_1729792, EPI_ISL_1729794, EPI_ISL_1729795, EPI_ISL_1729796, EPI_ISL_1729797, EPI_ISL_1729798, EPI_ISL_1729799, EPI_ISL_1729800, EPI_ISL_1729806, EPI_ISL_1729808, EPI_ISL_1729809, EPI_ISL_1729811, EPI_ISL_1729813, EPI_ISL_1729814, EPI_ISL_1729815, EPI_ISL_1729818, EPI_ISL_1729821, EPI_ISL_1729822, EPI_ISL_1729823, EPI_ISL_1729825, EPI_ISL_1729829, EPI_ISL_1729831, EPI_ISL_1729832, EPI_ISL_1729833, EPI_ISL_1729835, EPI_ISL_1729837, EPI_ISL_1729840, EPI_ISL_1729841, EPI_ISL_1729842, EPI_ISL_1729844, EPI_ISL_1729845, EPI_ISL_1729846, EPI_ISL_1729847, EPI_ISL_1729854, EPI_ISL_1729856, EPI_ISL_1729857, EPI_ISL_1729861, EPI_ISL_1729864, EPI_ISL_1729868, EPI_ISL_1729874, EPI_ISL_1729878, EPI_ISL_1729879, EPI_ISL_1729880, EPI_ISL_1729883, EPI_ISL_1729891, EPI_ISL_1729894, EPI_ISL_1729897, EPI_ISL_1729900, EPI_ISL_1729901, EPI_ISL_1729903, EPI_ISL_1729904, EPI_ISL_1729905, EPI_ISL_1729906, EPI_ISL_1729907, EPI_ISL_1729908, EPI_ISL_1729911, EPI_ISL_1729912, EPI_ISL_1729913, EPI_ISL_1729920, EPI_ISL_1729922, EPI_ISL_1729925, EPI_ISL_1729926, EPI_ISL_1729927, EPI_ISL_1729929, EPI_ISL_1729930, EPI_ISL_1729931, EPI_ISL_1729932, EPI_ISL_1729933, EPI_ISL_1729935, EPI_ISL_1729936, EPI_ISL_1729938, EPI_ISL_1729941, EPI_ISL_1729943, EPI_ISL_1729949, EPI_ISL_1729950, EPI_ISL_1729952, EPI_ISL_1729953, EPI_ISL_1729955, EPI_ISL_1729961, EPI_ISL_1729962, EPI_ISL_1729965, EPI_ISL_1729966, EPI_ISL_1849513, EPI_ISL_1849545, EPI_ISL_1849547, EPI_ISL_1849550, EPI_ISL_1849560, EPI_ISL_1849562, EPI_ISL_1849563, EPI_ISL_1849566, EPI_ISL_1849570, EPI_ISL_1849589, EPI_ISL_1849592, EPI_ISL_1849596, EPI_ISL_1849597, EPI_ISL_1849606, EPI_ISL_1849613, EPI_ISL_1849614, EPI_ISL_1849624, EPI_ISL_1849629, EPI_ISL_1849634, EPI_ISL_1849639, EPI_ISL_1849642, EPI_ISL_1849643, EPI_ISL_1849647, EPI_ISL_1849652, EPI_ISL_1849658, EPI_ISL_1849660, EPI_ISL_1849662, EPI_ISL_1849669, EPI_ISL_1849672, EPI_ISL_1849673, EPI_ISL_1849675, EPI_ISL_1849678, EPI_ISL_1849683, EPI_ISL_1849689, EPI_ISL_1849692, EPI_ISL_1849693, EPI_ISL_2108004, EPI_ISL_2108014, EPI_ISL_2108015, EPI_ISL_2108019, EPI_ISL_2108020, EPI_ISL_2108021, EPI_ISL_2108023, EPI_ISL_2108029, EPI_ISL_2108030, EPI_ISL_2108031, EPI_ISL_2108037, EPI_ISL_2108039, EPI_ISL_2108047, EPI_ISL_2108048, EPI_ISL_2108049, EPI_ISL_2108051, EPI_ISL_2108057, EPI_ISL_2108060, EPI_ISL_2108062, EPI_ISL_2108063, EPI_ISL_2108064, EPI_ISL_2108066, EPI_ISL_2108072, EPI_ISL_2108076, EPI_ISL_2108080, EPI_ISL_2108091, EPI_ISL_2108096, EPI_ISL_2108099, EPI_ISL_2108106, EPI_ISL_2108111, EPI_ISL_2108112, EPI_ISL_2108113, EPI_ISL_2108119, EPI_ISL_2108122, EPI_ISL_2108125, EPI_ISL_2108128, EPI_ISL_2108133, EPI_ISL_2108135, EPI_ISL_2108139, EPI_ISL_2108149, EPI_ISL_2108152, EPI_ISL_2108156, EPI_ISL_2108160, EPI_ISL_2108161, EPI_ISL_2108162, EPI_ISL_2108166, EPI_ISL_2108179, EPI_ISL_2108193, EPI_ISL_2108194, EPI_ISL_2108205, EPI_ISL_2108208, EPI_ISL_2108210, EPI_ISL_2108212, EPI_ISL_2108224, EPI_ISL_2108225, EPI_ISL_2108231, EPI_ISL_2108232, EPI_ISL_2108236, EPI_ISL_2108237, EPI_ISL_2108238, EPI_ISL_2108241, EPI_ISL_2108246, EPI_ISL_2108247, EPI_ISL_2108248, EPI_ISL_2108250, EPI_ISL_2108251, EPI_ISL_2108254, EPI_ISL_2114516, EPI_ISL_2114520, EPI_ISL_2114527, EPI_ISL_2114530, EPI_ISL_2114545, EPI_ISL_2114547, EPI_ISL_2114553, EPI_ISL_2114564, EPI_ISL_2114567, EPI_ISL_2114575, EPI_ISL_2114577, EPI_ISL_2114586, EPI_ISL_2114595, EPI_ISL_2114606, EPI_ISL_2114608, EPI_ISL_2114611, EPI_ISL_2114615, EPI_ISL_2114616, EPI_ISL_2114628, EPI_ISL_2114641, EPI_ISL_2114644, EPI_ISL_2114652, EPI_ISL_2114665, EPI_ISL_2114666, EPI_ISL_2114668, EPI_ISL_2114676, EPI_ISL_2114679, EPI_ISL_2114697, EPI_ISL_2114699, EPI_ISL_2114703, EPI_ISL_2114704, EPI_ISL_2114711, EPI_ISL_2114712, EPI_ISL_2114721, EPI_ISL_2114737, EPI_ISL_2114738, EPI_ISL_2114757, EPI_ISL_2114760, EPI_ISL_2114769, EPI_ISL_2114786, EPI_ISL_2114794, EPI_ISL_2114812, EPI_ISL_2114851, EPI_ISL_2114856, EPI_ISL_2114867, EPI_ISL_2114871, EPI_ISL_2114878, EPI_ISL_2114886, EPI_ISL_2114893, EPI_ISL_2114904, EPI_ISL_2114917, EPI_ISL_2114926, EPI_ISL_2114929, EPI_ISL_2114931, EPI_ISL_2114951, EPI_ISL_2114953, EPI_ISL_2114970, EPI_ISL_2114982, EPI_ISL_2114989, EPI_ISL_2114990, EPI_ISL_2114991, EPI_ISL_2114993, EPI_ISL_2114995, EPI_ISL_2114998, EPI_ISL_2115006, EPI_ISL_2115008, EPI_ISL_2115012, EPI_ISL_2115019, EPI_ISL_2115026, EPI_ISL_2115029, EPI_ISL_2115032, EPI_ISL_2115033, EPI_ISL_2115040, EPI_ISL_2115055, EPI_ISL_2115081, EPI_ISL_2115082, EPI_ISL_2115094, EPI_ISL_2115102, EPI_ISL_2115108, EPI_ISL_2115115, EPI_ISL_2115117, EPI_ISL_2115123, EPI_ISL_2115124, EPI_ISL_2118857, EPI_ISL_2118904, EPI_ISL_2118916, EPI_ISL_2118981, EPI_ISL_2118990, EPI_ISL_2118999, EPI_ISL_2119026, EPI_ISL_2119060, EPI_ISL_2119067, EPI_ISL_2119069, EPI_ISL_2119093, EPI_ISL_2119117, EPI_ISL_2119120, EPI_ISL_2119167, EPI_ISL_2119184, EPI_ISL_2119187, EPI_ISL_2119245, EPI_ISL_2119248, EPI_ISL_2119249, EPI_ISL_2119264, EPI_ISL_2119282, EPI_ISL_2119286, EPI_ISL_2119290, EPI_ISL_2119295, EPI_ISL_2119313, EPI_ISL_2119321, EPI_ISL_2119323, EPI_ISL_2119329, EPI_ISL_2119337, EPI_ISL_2119347, EPI_ISL_2119355, EPI_ISL_2119366, EPI_ISL_2119380, EPI_ISL_2119391, EPI_ISL_2119394, EPI_ISL_2119399, EPI_ISL_2119481, EPI_ISL_2119486, EPI_ISL_2119521, EPI_ISL_2119524, EPI_ISL_2119581, EPI_ISL_2119614, EPI_ISL_2119645, EPI_ISL_2119665, EPI_ISL_2119714, EPI_ISL_2119722, EPI_ISL_2119758, EPI_ISL_2119783, EPI_ISL_2119788, EPI_ISL_2119837, EPI_ISL_2119842, EPI_ISL_2119878, EPI_ISL_2119879, EPI_ISL_2119881, EPI_ISL_2119892, EPI_ISL_2119893, EPI_ISL_2119894, EPI_ISL_2119897, EPI_ISL_2119953, EPI_ISL_2119989, EPI_ISL_2119998, EPI_ISL_2120001, EPI_ISL_2120023, EPI_ISL_2120029, EPI_ISL_2120041, EPI_ISL_2128638, EPI_ISL_2128644, EPI_ISL_2128663, EPI_ISL_2128666, EPI_ISL_2128675, EPI_ISL_2128679, EPI_ISL_2128684, EPI_ISL_2128706, EPI_ISL_2128718, EPI_ISL_2128746, EPI_ISL_2128752, EPI_ISL_2128770, EPI_ISL_2128776, EPI_ISL_2128865, EPI_ISL_2128939, EPI_ISL_2128940, EPI_ISL_2128950

see above | SYNLAB MVZ Leverkusen | Robert Koch Institute

EPI_ISL_1642209, EPI_ISL_1646761, EPI_ISL_1726469, EPI_ISL_1726484, EPI_ISL_1726498, EPI_ISL_1726542, EPI_ISL_1726648, EPI_ISL_1726663, EPI_ISL_1726722, EPI_ISL_1849505, EPI_ISL_1849522, EPI_ISL_1849532, EPI_ISL_1849554, EPI_ISL_1849590, EPI_ISL_1849594, EPI_ISL_1849619, EPI_ISL_1849653, EPI_ISL_1849695

see above | SYNLAB MVZ Neustadt a. d. WeinstraÃŸe | Robert Koch Institute

EPI_ISL_1568242, EPI_ISL_1568243, EPI_ISL_1568244, EPI_ISL_1568246, EPI_ISL_1568247, EPI_ISL_1568250, EPI_ISL_1568251, EPI_ISL_1568252, EPI_ISL_1568254, EPI_ISL_1568255, EPI_ISL_1568256, EPI_ISL_1568258, EPI_ISL_1568260, EPI_ISL_1568261, EPI_ISL_1568262, EPI_ISL_1568263, EPI_ISL_1568265, EPI_ISL_1568267, EPI_ISL_1568268, EPI_ISL_1568269, EPI_ISL_1568273, EPI_ISL_1568275, EPI_ISL_1568277, EPI_ISL_1568278, EPI_ISL_1568279, EPI_ISL_1568280, EPI_ISL_1568281, EPI_ISL_1568283, EPI_ISL_1568284, EPI_ISL_1568285, EPI_ISL_1568288, EPI_ISL_1568290, EPI_ISL_1568291, EPI_ISL_1568293, EPI_ISL_1568294, EPI_ISL_1568296, EPI_ISL_1568297, EPI_ISL_1568300, EPI_ISL_1568303, EPI_ISL_1568305, EPI_ISL_1568310, EPI_ISL_1568311, EPI_ISL_1568312, EPI_ISL_1568315, EPI_ISL_1568316, EPI_ISL_1568317, EPI_ISL_1568319, EPI_ISL_1568320, EPI_ISL_1568323, EPI_ISL_1568326, EPI_ISL_1568329, EPI_ISL_1568331, EPI_ISL_1568332, EPI_ISL_1568333, EPI_ISL_1568334, EPI_ISL_1568335, EPI_ISL_1568336, EPI_ISL_1568337, EPI_ISL_1568339, EPI_ISL_1568340, EPI_ISL_1568349, EPI_ISL_1568352, EPI_ISL_1568354, EPI_ISL_1568355, EPI_ISL_1568356, EPI_ISL_1568358, EPI_ISL_1568359, EPI_ISL_1568364, EPI_ISL_1568365, EPI_ISL_1568366, EPI_ISL_1568368, EPI_ISL_1568370, EPI_ISL_1568371, EPI_ISL_1568373, EPI_ISL_1568376, EPI_ISL_1568377, EPI_ISL_1568378, EPI_ISL_1568380, EPI_ISL_1568381, EPI_ISL_1568382, EPI_ISL_1568383, EPI_ISL_1568384, EPI_ISL_1568386, EPI_ISL_1568387, EPI_ISL_1568388, EPI_ISL_1568391, EPI_ISL_1568393, EPI_ISL_1568394, EPI_ISL_1568395, EPI_ISL_1568396, EPI_ISL_1568397, EPI_ISL_1568400, EPI_ISL_1568402, EPI_ISL_1568403, EPI_ISL_1568405, EPI_ISL_1568406, EPI_ISL_1568407, EPI_ISL_1568408, EPI_ISL_1568409, EPI_ISL_1568410, EPI_ISL_1568411, EPI_ISL_1568412, EPI_ISL_1568416, EPI_ISL_1568417, EPI_ISL_1568418, EPI_ISL_1568421, EPI_ISL_1568422, EPI_ISL_1568423, EPI_ISL_1568426, EPI_ISL_1568427, EPI_ISL_1568429, EPI_ISL_1568431, EPI_ISL_1568433, EPI_ISL_1568435, EPI_ISL_1568436, EPI_ISL_1568437, EPI_ISL_1568440, EPI_ISL_1568441, EPI_ISL_1568443, EPI_ISL_1568444, EPI_ISL_1568446, EPI_ISL_1568447, EPI_ISL_1568448, EPI_ISL_1568450, EPI_ISL_1568451, EPI_ISL_1568452, EPI_ISL_1568454, EPI_ISL_1568455, EPI_ISL_1568457, EPI_ISL_1568458, EPI_ISL_1568459, EPI_ISL_1568460, EPI_ISL_1568461, EPI_ISL_1568463, EPI_ISL_1568464, EPI_ISL_1568465, EPI_ISL_1568467, EPI_ISL_1568468, EPI_ISL_1568469, EPI_ISL_1568470, EPI_ISL_1568473, EPI_ISL_1568474, EPI_ISL_1568475, EPI_ISL_1568476, EPI_ISL_1568477, EPI_ISL_1568478, EPI_ISL_1568479, EPI_ISL_1568481, EPI_ISL_1568482, EPI_ISL_1568484, EPI_ISL_1568485, EPI_ISL_1568486, EPI_ISL_1568487, EPI_ISL_1568490, EPI_ISL_1568492, EPI_ISL_1568493, EPI_ISL_1568497, EPI_ISL_1568498, EPI_ISL_1568499, EPI_ISL_1568501, EPI_ISL_1568504, EPI_ISL_1568505, EPI_ISL_1568506, EPI_ISL_1568508, EPI_ISL_1568510, EPI_ISL_1568512, EPI_ISL_1568513, EPI_ISL_1568514, EPI_ISL_1568515, EPI_ISL_1568516, EPI_ISL_1568517, EPI_ISL_1568518, EPI_ISL_1568521, EPI_ISL_1568522, EPI_ISL_1568523, EPI_ISL_1573077, EPI_ISL_1573079, EPI_ISL_1573080, EPI_ISL_1573081, EPI_ISL_1573082, EPI_ISL_1573083, EPI_ISL_1573084, EPI_ISL_1573085, EPI_ISL_1573087, EPI_ISL_1573088, EPI_ISL_1573089, EPI_ISL_1573091, EPI_ISL_1573092, EPI_ISL_1573093, EPI_ISL_1573094, EPI_ISL_1573095, EPI_ISL_1573098, EPI_ISL_1573099, EPI_ISL_1573103, EPI_ISL_1573104, EPI_ISL_1573105, EPI_ISL_1573108, EPI_ISL_1573109, EPI_ISL_1573111, EPI_ISL_1573113, EPI_ISL_1573114, EPI_ISL_1573115, EPI_ISL_1573116, EPI_ISL_1573117, EPI_ISL_1573122, EPI_ISL_1573124, EPI_ISL_1573126, EPI_ISL_1573128, EPI_ISL_1573129, EPI_ISL_1573130, EPI_ISL_1573131, EPI_ISL_1574363, EPI_ISL_1574365, EPI_ISL_1574367, EPI_ISL_1574369, EPI_ISL_1574370, EPI_ISL_1574372, EPI_ISL_1574374, EPI_ISL_1574375, EPI_ISL_1574378, EPI_ISL_1574380, EPI_ISL_1574381, EPI_ISL_1574383, EPI_ISL_1574385, EPI_ISL_1574386, EPI_ISL_1574387, EPI_ISL_1574388, EPI_ISL_1574389, EPI_ISL_1574390, EPI_ISL_1574391, EPI_ISL_1574392, EPI_ISL_1574393, EPI_ISL_1574394, EPI_ISL_1574395, EPI_ISL_1574396, EPI_ISL_1574397, EPI_ISL_1574398, EPI_ISL_1574399, EPI_ISL_1574400, EPI_ISL_1574401, EPI_ISL_1574402, EPI_ISL_1574403, EPI_ISL_1574404, EPI_ISL_1574405, EPI_ISL_1574406, EPI_ISL_1574408, EPI_ISL_1574409, EPI_ISL_1574411, EPI_ISL_1574412, EPI_ISL_1574414, EPI_ISL_1574416, EPI_ISL_1574417, EPI_ISL_1574418, EPI_ISL_1574419, EPI_ISL_1574420, EPI_ISL_1574422, EPI_ISL_1574423, EPI_ISL_1574424, EPI_ISL_1574425, EPI_ISL_1574427, EPI_ISL_1574428, EPI_ISL_1574429, EPI_ISL_1574430, EPI_ISL_1574431, EPI_ISL_1574432, EPI_ISL_1574433, EPI_ISL_1574435, EPI_ISL_1574436, EPI_ISL_1574437, EPI_ISL_1574439, EPI_ISL_1574442, EPI_ISL_1574443, EPI_ISL_1574444, EPI_ISL_1574445, EPI_ISL_1574448, EPI_ISL_1574449, EPI_ISL_1574450, EPI_ISL_1574451, EPI_ISL_1574452, EPI_ISL_1574456, EPI_ISL_1574457, EPI_ISL_1574461, EPI_ISL_1574462, EPI_ISL_1574463, EPI_ISL_1574466, EPI_ISL_1574467, EPI_ISL_1574468, EPI_ISL_1574470, EPI_ISL_1574471, EPI_ISL_1574472, EPI_ISL_1574473, EPI_ISL_1574476, EPI_ISL_1574478, EPI_ISL_1574480, EPI_ISL_1574481, EPI_ISL_1574484, EPI_ISL_1574486, EPI_ISL_1574487, EPI_ISL_1574488, EPI_ISL_1574489, EPI_ISL_1574490, EPI_ISL_1574492, EPI_ISL_1574493, EPI_ISL_1574494, EPI_ISL_1574495, EPI_ISL_1574496, EPI_ISL_1574497, EPI_ISL_1574498, EPI_ISL_1574500, EPI_ISL_1574503, EPI_ISL_1574504, EPI_ISL_1574505, EPI_ISL_1574507, EPI_ISL_1574508, EPI_ISL_1574509, EPI_ISL_1574510, EPI_ISL_1574511, EPI_ISL_1574512, EPI_ISL_1574513, EPI_ISL_1574514, EPI_ISL_1574515, EPI_ISL_1574516, EPI_ISL_1574517, EPI_ISL_1574518, EPI_ISL_1574519, EPI_ISL_1574520, EPI_ISL_1574521, EPI_ISL_1574522, EPI_ISL_1574523, EPI_ISL_1574524, EPI_ISL_1574525, EPI_ISL_1574527, EPI_ISL_1574528, EPI_ISL_1574529, EPI_ISL_1574530, EPI_ISL_1574531, EPI_ISL_1574532, EPI_ISL_1574533, EPI_ISL_1574534, EPI_ISL_1574535, EPI_ISL_1574536, EPI_ISL_1574539, EPI_ISL_1574540, EPI_ISL_1574541, EPI_ISL_1574543, EPI_ISL_1574544, EPI_ISL_1574545, EPI_ISL_1574546, EPI_ISL_1574547, EPI_ISL_1574548, EPI_ISL_1574549, EPI_ISL_1574552, EPI_ISL_1574554, EPI_ISL_1574557, EPI_ISL_1574558, EPI_ISL_1574559, EPI_ISL_1574560, EPI_ISL_1574561, EPI_ISL_1574563, EPI_ISL_1574564, EPI_ISL_1574565, EPI_ISL_1574566, EPI_ISL_1574567, EPI_ISL_1574568, EPI_ISL_1574569, EPI_ISL_1574570, EPI_ISL_1574571, EPI_ISL_1574573, EPI_ISL_1574574, EPI_ISL_1574576, EPI_ISL_1574577, EPI_ISL_1574578, EPI_ISL_1574579, EPI_ISL_1574580, EPI_ISL_1574581, EPI_ISL_1574583, EPI_ISL_1574584, EPI_ISL_1574585, EPI_ISL_1574587, EPI_ISL_1574588, EPI_ISL_1574590, EPI_ISL_1574591, EPI_ISL_1574593, EPI_ISL_1574595, EPI_ISL_1574596, EPI_ISL_1574598, EPI_ISL_1574599, EPI_ISL_1574600, EPI_ISL_1574601, EPI_ISL_1574603, EPI_ISL_1574604, EPI_ISL_1574606, EPI_ISL_1574607, EPI_ISL_1574608, EPI_ISL_1574609, EPI_ISL_1574611, EPI_ISL_1574612, EPI_ISL_1574613, EPI_ISL_1574614, EPI_ISL_1574615, EPI_ISL_1574618, EPI_ISL_1574619, EPI_ISL_1574623, EPI_ISL_1574625, EPI_ISL_1574626, EPI_ISL_1574627, EPI_ISL_1574628, EPI_ISL_1574629, EPI_ISL_1574630, EPI_ISL_1574632, EPI_ISL_1574634, EPI_ISL_1574636, EPI_ISL_1574637, EPI_ISL_1574639, EPI_ISL_1574640, EPI_ISL_1574642, EPI_ISL_1574643, EPI_ISL_1574644, EPI_ISL_1574645, EPI_ISL_1574646, EPI_ISL_1574648, EPI_ISL_1574649, EPI_ISL_1574651, EPI_ISL_1574653, EPI_ISL_1574654, EPI_ISL_1574655, EPI_ISL_1574656, EPI_ISL_1574657, EPI_ISL_1574658, EPI_ISL_1574659, EPI_ISL_1574661, EPI_ISL_1574662, EPI_ISL_1574663, EPI_ISL_1574665, EPI_ISL_1574666, EPI_ISL_1574667, EPI_ISL_1574668, EPI_ISL_1574670, EPI_ISL_1574671, EPI_ISL_1574672, EPI_ISL_1574673, EPI_ISL_1574674, EPI_ISL_1574675, EPI_ISL_1574676, EPI_ISL_1574677, EPI_ISL_1574678, EPI_ISL_1574679, EPI_ISL_1574681, EPI_ISL_1574683, EPI_ISL_1574684, EPI_ISL_1574685, EPI_ISL_1574686, EPI_ISL_1641272, EPI_ISL_1641277, EPI_ISL_1641281, EPI_ISL_1641293, EPI_ISL_1641308, EPI_ISL_1641310, EPI_ISL_1641312, EPI_ISL_1641315, EPI_ISL_1641330, EPI_ISL_1641354, EPI_ISL_1641360, EPI_ISL_1641361, EPI_ISL_1641365, EPI_ISL_1641367, EPI_ISL_1641370, EPI_ISL_1641373, EPI_ISL_1641375, EPI_ISL_1641379, EPI_ISL_1641380, EPI_ISL_1641384, EPI_ISL_1641395, EPI_ISL_1641396, EPI_ISL_1641397, EPI_ISL_1641398, EPI_ISL_1641399, EPI_ISL_1641400, EPI_ISL_1641401, EPI_ISL_1641402, EPI_ISL_1641409, EPI_ISL_1641412, EPI_ISL_1641416, EPI_ISL_1641418, EPI_ISL_1641421, EPI_ISL_1641424, EPI_ISL_1641426, EPI_ISL_1641439, EPI_ISL_1641440, EPI_ISL_1641445, EPI_ISL_1641450, EPI_ISL_1641453, EPI_ISL_1641464, EPI_ISL_1641467, EPI_ISL_1641470, EPI_ISL_1641475, EPI_ISL_1641487, EPI_ISL_1641511, EPI_ISL_1641512, EPI_ISL_1641530, EPI_ISL_1641536, EPI_ISL_1641541

see above | SYNLAB MVZ Trier | Robert Koch Institute

EPI_ISL_1568245, EPI_ISL_1568274, EPI_ISL_1568304, EPI_ISL_1568308, EPI_ISL_1568322, EPI_ISL_1568325, EPI_ISL_1568330, EPI_ISL_1568369, EPI_ISL_1568374, EPI_ISL_1568404, EPI_ISL_1568483, EPI_ISL_1568489, EPI_ISL_1568496, EPI_ISL_1568519, EPI_ISL_1573086, EPI_ISL_1573100, EPI_ISL_1573107, EPI_ISL_1574373, EPI_ISL_1574377, EPI_ISL_1574410, EPI_ISL_1574434, EPI_ISL_1574446, EPI_ISL_1574458, EPI_ISL_1574464, EPI_ISL_1574469, EPI_ISL_1574474, EPI_ISL_1574477, EPI_ISL_1574483, EPI_ISL_1574502, EPI_ISL_1574526, EPI_ISL_1574538, EPI_ISL_1574551, EPI_ISL_1574556, EPI_ISL_1574572, EPI_ISL_1574575, EPI_ISL_1574624, EPI_ISL_1574635, EPI_ISL_1574664, EPI_ISL_1574669

see above | SYNLAB MVZ Weiden | Robert Koch Institute

EPI_ISL_1721214, EPI_ISL_1721233, EPI_ISL_1721241, EPI_ISL_1721301, EPI_ISL_1721357, EPI_ISL_1721475, EPI_ISL_1721496, EPI_ISL_1721511, EPI_ISL_1721556, EPI_ISL_1721559, EPI_ISL_1721574, EPI_ISL_1721576, EPI_ISL_1721605, EPI_ISL_1721610, EPI_ISL_1721616, EPI_ISL_1721663, EPI_ISL_1843035, EPI_ISL_1843446, EPI_ISL_1847281

see above | Sonic - Bioscientia - MVZ Labor Saar GmbH | Robert Koch Institute

EPI_ISL_1642523, EPI_ISL_1642524, EPI_ISL_1642562, EPI_ISL_1642590, EPI_ISL_1642591, EPI_ISL_1642650, EPI_ISL_1642675, EPI_ISL_1642678, EPI_ISL_1642687, EPI_ISL_1642688, EPI_ISL_1721136, EPI_ISL_1721243, EPI_ISL_1721302, EPI_ISL_1842793

see above | Sonic - Dr. Staber & Kollegen GmbH MÃ¼nchen | Robert Koch Institute

EPI_ISL_1721109, EPI_ISL_1721111, EPI_ISL_1721112, EPI_ISL_1721119, EPI_ISL_1721120, EPI_ISL_1721121, EPI_ISL_1721128, EPI_ISL_1721143, EPI_ISL_1721144, EPI_ISL_1721152, EPI_ISL_1721163, EPI_ISL_1721164, EPI_ISL_1721183, EPI_ISL_1721188, EPI_ISL_1721192, EPI_ISL_1721195, EPI_ISL_1721196, EPI_ISL_1721204, EPI_ISL_1721210, EPI_ISL_1721220, EPI_ISL_1721225, EPI_ISL_1721228, EPI_ISL_1721240, EPI_ISL_1721242, EPI_ISL_1721246, EPI_ISL_1721252, EPI_ISL_1721256, EPI_ISL_1721259, EPI_ISL_1721268, EPI_ISL_1721270, EPI_ISL_1721272, EPI_ISL_1721273, EPI_ISL_1721277, EPI_ISL_1721297, EPI_ISL_1721306, EPI_ISL_1721307, EPI_ISL_1721316, EPI_ISL_1721329, EPI_ISL_1721332, EPI_ISL_1721339, EPI_ISL_1721343, EPI_ISL_1721366, EPI_ISL_1721368, EPI_ISL_1721371, EPI_ISL_1721372, EPI_ISL_1721379, EPI_ISL_1721386, EPI_ISL_1721425, EPI_ISL_1721429, EPI_ISL_1721436, EPI_ISL_1721473, EPI_ISL_1721572, EPI_ISL_1721620, EPI_ISL_1721622, EPI_ISL_1721630, EPI_ISL_1721631, EPI_ISL_1721677, EPI_ISL_1843473

see above | Sonic - Labor Dr. von Froreich GmbH | Robert Koch Institute

EPI_ISL_1642436, EPI_ISL_1642455, EPI_ISL_1642476, EPI_ISL_1642499, EPI_ISL_1642550, EPI_ISL_1642558, EPI_ISL_1642573, EPI_ISL_1642587, EPI_ISL_1642594, EPI_ISL_1642617, EPI_ISL_1642643, EPI_ISL_1721181, EPI_ISL_1721205, EPI_ISL_1721250, EPI_ISL_1721260, EPI_ISL_1842821, EPI_ISL_1842833

see above | Sonic - Labor Staber Bayreuth | Robert Koch Institute

EPI_ISL_1642419, EPI_ISL_1642439, EPI_ISL_1642471, EPI_ISL_1642505, EPI_ISL_1642541, EPI_ISL_1642543, EPI_ISL_1642572, EPI_ISL_1642600, EPI_ISL_1642627, EPI_ISL_1642676, EPI_ISL_1721193, EPI_ISL_1721276, EPI_ISL_1721356

| | | | |
|---|---|---|---|
| see above | Sonic - Labor Staber Dresden (Klipphausen) | Robert Koch Institute | |
| EPI_ISL_1642448, EPI_ISL_1642496, EPI_ISL_1642527, EPI_ISL_1642540, EPI_ISL_1721147, EPI_ISL_1721369 | Sonic - Labor Staber Heilbronn | Robert Koch Institute | |
| EPI_ISL_1847111 | Sonic - Labor Staber MVZ Kassel | Robert Koch Institute | |
| EPI_ISL_1642432, EPI_ISL_1642443, EPI_ISL_1642488, EPI_ISL_1642564, EPI_ISL_1642596, EPI_ISL_1642683 | Sonic - Labor Staber NÃ¼rnberg | Robert Koch Institute | |
| EPI_ISL_1638971, EPI_ISL_1638973, EPI_ISL_1638974, EPI_ISL_1638978, EPI_ISL_1638980, EPI_ISL_1638988, EPI_ISL_1638991, EPI_ISL_1638992, EPI_ISL_1638993, EPI_ISL_1638995, EPI_ISL_1638996, EPI_ISL_1639002, EPI_ISL_1639008, EPI_ISL_1639009, EPI_ISL_1639010, EPI_ISL_1639013, EPI_ISL_1639015, EPI_ISL_1639017, EPI_ISL_1639019, EPI_ISL_1639020, EPI_ISL_1639021, EPI_ISL_1639022, EPI_ISL_1639031, EPI_ISL_1639032, EPI_ISL_1639039, EPI_ISL_1639040, EPI_ISL_1639042, EPI_ISL_1639045, EPI_ISL_1639047, EPI_ISL_1639049, EPI_ISL_1639054, EPI_ISL_1639059, EPI_ISL_1639061, EPI_ISL_1639064, EPI_ISL_1639068, EPI_ISL_1639077, EPI_ISL_1639081, EPI_ISL_1639085, EPI_ISL_1639089, EPI_ISL_1639094, EPI_ISL_1639096, EPI_ISL_1639098, EPI_ISL_1639112, EPI_ISL_1639115, EPI_ISL_1639116, EPI_ISL_1639117, EPI_ISL_1639121, EPI_ISL_1639122, EPI_ISL_1639123, EPI_ISL_1639134, EPI_ISL_1639137, EPI_ISL_1639138, EPI_ISL_1639146, EPI_ISL_1639150, EPI_ISL_1639156, EPI_ISL_1639157, EPI_ISL_1639159, EPI_ISL_1639165, EPI_ISL_1639172, EPI_ISL_1639176, EPI_ISL_1639178, EPI_ISL_1639185, EPI_ISL_1639186, EPI_ISL_1639188, EPI_ISL_1639189, EPI_ISL_1639193, EPI_ISL_1639195, EPI_ISL_1639196, EPI_ISL_1639202, EPI_ISL_1639204, EPI_ISL_1639206, EPI_ISL_1639210, EPI_ISL_1639211, EPI_ISL_1639214, EPI_ISL_1639220, EPI_ISL_1639227, EPI_ISL_1639238, EPI_ISL_1639250, EPI_ISL_1642447, EPI_ISL_1642467, EPI_ISL_1642478, EPI_ISL_1642490, EPI_ISL_1642500, EPI_ISL_1642506, EPI_ISL_1642516, EPI_ISL_1642519, EPI_ISL_1642525, EPI_ISL_1642534, EPI_ISL_1642544, EPI_ISL_1642552, EPI_ISL_1642555, EPI_ISL_1642570, EPI_ISL_1642593, EPI_ISL_1642595, EPI_ISL_1642604, EPI_ISL_1642607, EPI_ISL_1642613, EPI_ISL_1642618, EPI_ISL_1642624, EPI_ISL_1642640, EPI_ISL_1642644, EPI_ISL_1642657, EPI_ISL_1642659, EPI_ISL_1642666, EPI_ISL_1642667, EPI_ISL_1642668, EPI_ISL_1642669, EPI_ISL_1642670, EPI_ISL_1642693, EPI_ISL_1642694, EPI_ISL_1642695, EPI_ISL_1643121, EPI_ISL_1643130, EPI_ISL_1643133, EPI_ISL_1643144, EPI_ISL_1643153, EPI_ISL_1643155, EPI_ISL_1643159, EPI_ISL_1643161, EPI_ISL_1643168, EPI_ISL_1643174, EPI_ISL_1643179, EPI_ISL_1643184, EPI_ISL_1643186, EPI_ISL_1643191, EPI_ISL_1643199, EPI_ISL_1643200, EPI_ISL_1643204, EPI_ISL_1643205, EPI_ISL_1643212, EPI_ISL_1643215, EPI_ISL_1643216, EPI_ISL_1643220, EPI_ISL_1643222, EPI_ISL_1643223, EPI_ISL_1643229, EPI_ISL_1643237, EPI_ISL_1643247, EPI_ISL_1643253, EPI_ISL_1643254, EPI_ISL_1643255, EPI_ISL_1643257, EPI_ISL_1643260, EPI_ISL_1643267, EPI_ISL_1643275, EPI_ISL_1643284, EPI_ISL_1643287, EPI_ISL_1643293, EPI_ISL_1643298, EPI_ISL_1643311, EPI_ISL_1643312, EPI_ISL_1643313, EPI_ISL_1643317, EPI_ISL_1643322, EPI_ISL_1643327, EPI_ISL_1643331, EPI_ISL_1643332, EPI_ISL_1643336, EPI_ISL_1643337, EPI_ISL_1643344, EPI_ISL_1643346, EPI_ISL_1643347, EPI_ISL_1643361, EPI_ISL_1643363, EPI_ISL_1643367, EPI_ISL_1643375, EPI_ISL_1643376, EPI_ISL_1643377, EPI_ISL_1643379, EPI_ISL_1643393, EPI_ISL_1721105, EPI_ISL_1721118, EPI_ISL_1721124, EPI_ISL_1721125, EPI_ISL_1721150, EPI_ISL_1721154, EPI_ISL_1721165, EPI_ISL_1721167, EPI_ISL_1721180, EPI_ISL_1721187, EPI_ISL_1721191, EPI_ISL_1721198, EPI_ISL_1721208, EPI_ISL_1721219, EPI_ISL_1721232, EPI_ISL_1721234, EPI_ISL_1721236, EPI_ISL_1721251, EPI_ISL_1721278, EPI_ISL_1721283, EPI_ISL_1721284, EPI_ISL_1721286, EPI_ISL_1721290, EPI_ISL_1721292, EPI_ISL_1721300, EPI_ISL_1721313, EPI_ISL_1721315, EPI_ISL_1721326, EPI_ISL_1721327, EPI_ISL_1721337, EPI_ISL_1721345, EPI_ISL_1721354, EPI_ISL_1721364, EPI_ISL_1721374, EPI_ISL_1721375, EPI_ISL_1721387 | | | |
| see above | Sonic - MVZ Medizinisches Labor Bremen GmbH | Robert Koch Institute | |
| EPI_ISL_1642454, EPI_ISL_1642571, EPI_ISL_1642614, EPI_ISL_1642638, EPI_ISL_1642690, EPI_ISL_1721207, EPI_ISL_1721608 | | | |
| see above | Sonic - MVZ Medizinisches Labor Oldenburg | Robert Koch Institute | |
| EPI_ISL_1567829, EPI_ISL_1638512 | St. Marien-Hospital Hamm | Robert Koch Institute | |
| EPI_ISL_1567813, EPI_ISL_1572110, EPI_ISL_1643482, EPI_ISL_1643483 | Stiftung Krankenhaus Bethanien fÃ¼r die Grafschaft Moers | Robert Koch Institute | |
| EPI_ISL_1569891, EPI_ISL_1569906, EPI_ISL_1569911, EPI_ISL_1569912, EPI_ISL_1569927, EPI_ISL_1569945, EPI_ISL_1569946, EPI_ISL_1569966, EPI_ISL_1569973, EPI_ISL_1569975, EPI_ISL_1569986 | | | |
| see above | StÃ¤dtisches Klinikum Dresden Institut fÃ¼r Klinische Chemie und Laboratoriumsmedizin | Robert Koch Institute | |
| EPI_ISL_1568272, EPI_ISL_1568287, EPI_ISL_1568305, EPI_ISL_1568328, EPI_ISL_1568343, EPI_ISL_1568346, EPI_ISL_1568362, EPI_ISL_1568424, EPI_ISL_1568428, EPI_ISL_1568442, EPI_ISL_1568491, EPI_ISL_1573120, EPI_ISL_1573123, EPI_ISL_1574364, EPI_ISL_1574371, EPI_ISL_1574379, EPI_ISL_1574382, EPI_ISL_1574384, EPI_ISL_1574407, EPI_ISL_1574413, EPI_ISL_1574415, EPI_ISL_1574421, EPI_ISL_1574426, EPI_ISL_1574441, EPI_ISL_1574447, EPI_ISL_1574459, EPI_ISL_1574465, EPI_ISL_1574475, EPI_ISL_1574482, EPI_ISL_1574485, EPI_ISL_1574499, EPI_ISL_1574501, EPI_ISL_1574537, EPI_ISL_1574542, EPI_ISL_1574550, EPI_ISL_1574553, EPI_ISL_1574555, EPI_ISL_1574562, EPI_ISL_1574582, EPI_ISL_1574586, EPI_ISL_1574589, EPI_ISL_1574594, EPI_ISL_1574597, EPI_ISL_1574602, EPI_ISL_1574610, EPI_ISL_1574616, EPI_ISL_1574617, EPI_ISL_1574620, EPI_ISL_1574621, EPI_ISL_1574622, EPI_ISL_1574631, EPI_ISL_1574633, EPI_ISL_1574638, EPI_ISL_1574641, EPI_ISL_1574652, EPI_ISL_1574660, EPI_ISL_1574680, EPI_ISL_1574682, EPI_ISL_1641393 | | | |
| see above | Synlab MVZ Augsburg | Robert Koch Institute | |
| EPI_ISL_1722831, EPI_ISL_1722832, EPI_ISL_1722833, EPI_ISL_1722834, EPI_ISL_1722836 | ThÃ¼ringer Landesamtes fÃ¼r Verbraucherschutz | Robert Koch Institute | |
| EPI_ISL_1569884, EPI_ISL_1569888, EPI_ISL_1569895, EPI_ISL_1569899, EPI_ISL_1569902, EPI_ISL_1569905, EPI_ISL_1569910, EPI_ISL_1569915, EPI_ISL_1569930, EPI_ISL_1569939, EPI_ISL_1569948, EPI_ISL_1569955, EPI_ISL_1569957, EPI_ISL_1569962, EPI_ISL_1569985 | | | |
| see above | Uniklinikum Carl Gustav Carus an der TU Dresden; Institut fÃ¼r Virologie | Robert Koch Institute | |
| EPI_ISL_1722852, EPI_ISL_1722858, EPI_ISL_1722859 | Uniklinikum Jena | Robert Koch Institute | |
| EPI_ISL_1638269, EPI_ISL_1638270, EPI_ISL_1638272, EPI_ISL_1638273, EPI_ISL_1638274, EPI_ISL_1638275, EPI_ISL_1638276, EPI_ISL_1638277 | | | |
| see above | Uniklinikum Regensburg; Institut fÃ¼r klinische Mikrobiologie und Hygiene | Robert Koch Institute | |
| EPI_ISL_1715828, EPI_ISL_1715829, EPI_ISL_1715858, EPI_ISL_1862392 | University Medical Center Hamburg Eppendorf | Heinrich Pette Institute, Leibniz Institute for Experimental Virology | Adam Grundhoff; Alexis Robitaille; Johannes Knobloch; Martin Aepfelbacher; Nicole Fischer; Thomas Günther |
| EPI_ISL_1638573, EPI_ISL_1638580, EPI_ISL_1638583, EPI_ISL_1638590, EPI_ISL_1638595, EPI_ISL_1638597, EPI_ISL_1638625, EPI_ISL_1638626, EPI_ISL_1721033, EPI_ISL_1721043, EPI_ISL_1721050, EPI_ISL_1721054, EPI_ISL_1721067, EPI_ISL_1721071, EPI_ISL_1721085, EPI_ISL_1721087, EPI_ISL_1721088, EPI_ISL_1721091, EPI_ISL_1721096 | | | |
| see above | UniversitÃ¤tsklinikum Bonn - Institut fÃ¼r Virologie | Robert Koch Institute | |
| EPI_ISL_1722848, EPI_ISL_1722849, EPI_ISL_1722850, EPI_ISL_1722851 | UniversitÃ¤tsklinikum Heidelberg | Robert Koch Institute | |
| EPI_ISL_1567101, EPI_ISL_1571562, EPI_ISL_1571563, EPI_ISL_1571564, EPI_ISL_1571565, EPI_ISL_1571566, EPI_ISL_1571567, EPI_ISL_1571568, EPI_ISL_1571569, EPI_ISL_1571570, EPI_ISL_1571571, EPI_ISL_1571572, EPI_ISL_1571573, EPI_ISL_1571574, EPI_ISL_1571575, EPI_ISL_1571576, EPI_ISL_1571577, EPI_ISL_1571578, EPI_ISL_1571579, EPI_ISL_1571580, EPI_ISL_1571581, EPI_ISL_1571582, EPI_ISL_1571583, EPI_ISL_1571584, EPI_ISL_1571585, EPI_ISL_1571586, EPI_ISL_1571587, EPI_ISL_1571588, EPI_ISL_1571589, EPI_ISL_1571590, EPI_ISL_1571591, EPI_ISL_1571592, EPI_ISL_1571593, EPI_ISL_1571594, EPI_ISL_1571595, EPI_ISL_1643829, EPI_ISL_1643831 | | | |
| see above | UniversitÃ¤tsklinikum MÃ¼nster Institut fÃ¼r Virologie | Robert Koch Institute | |
| EPI_ISL_1638574, EPI_ISL_1638596, EPI_ISL_1638598, EPI_ISL_1638604, EPI_ISL_1638607, EPI_ISL_1638623 | UniversitÃ¤tsklinikum des Saarlandes - Institut fÃ¼r Virologie | Robert Koch Institute | |
| EPI_ISL_1725182, EPI_ISL_1725183, EPI_ISL_1725184 | UniversitÃ¤tsmedizin Mannheim | Robert Koch Institute | |
| EPI_ISL_1833995, EPI_ISL_1833996, EPI_ISL_1834055 | Universitätsklinikum Knappschaftskrankenhaus Bochum | Bundeswehr Institute of Microbiology | Alexandra Rehn; Enrico Georgi; Malena Bestehorn-Willmann; Markus Antwerpen; Mathias Walter; Mike Pillukat; Roman Wölfel; Sabine Zange |
| EPI_ISL_1439708, EPI_ISL_1439709, EPI_ISL_1439710, EPI_ISL_1439711, EPI_ISL_1439712, EPI_ISL_1439713, EPI_ISL_1439714, EPI_ISL_1439715, EPI_ISL_1439716, EPI_ISL_1439717, EPI_ISL_1439718, EPI_ISL_1439719, EPI_ISL_1439720, EPI_ISL_1439721, EPI_ISL_1439722, EPI_ISL_1439723, EPI_ISL_1439724, EPI_ISL_1439725, EPI_ISL_1439727, EPI_ISL_1439728, EPI_ISL_1439729, EPI_ISL_1439730, EPI_ISL_1439731, EPI_ISL_1439735, EPI_ISL_1439736, EPI_ISL_1439738, EPI_ISL_1439739, EPI_ISL_1439740, EPI_ISL_1439742, EPI_ISL_1439743, EPI_ISL_1439745, EPI_ISL_1439746, EPI_ISL_1440732, EPI_ISL_1440733, EPI_ISL_1440734, EPI_ISL_1440735, EPI_ISL_1440736, EPI_ISL_1440737, EPI_ISL_1440738, EPI_ISL_1440739, EPI_ISL_1440741, EPI_ISL_1440742, EPI_ISL_1440743, EPI_ISL_1440744, EPI_ISL_1440745, EPI_ISL_1440746, EPI_ISL_1440747, EPI_ISL_1440748, EPI_ISL_1440749, EPI_ISL_1440750, EPI_ISL_1440751, EPI_ISL_1440752, EPI_ISL_1440753, EPI_ISL_1440754, EPI_ISL_1440755, EPI_ISL_1440756, EPI_ISL_1440758, EPI_ISL_1440759, EPI_ISL_1440760, EPI_ISL_1440761, EPI_ISL_1440762, EPI_ISL_1440763, EPI_ISL_1440764, EPI_ISL_1440765, EPI_ISL_1440766, EPI_ISL_1440767, EPI_ISL_1440768, EPI_ISL_1440769, EPI_ISL_1440770, EPI_ISL_1440772, EPI_ISL_1440773, EPI_ISL_1440775, EPI_ISL_1440776, EPI_ISL_1440777, EPI_ISL_1440779, EPI_ISL_1440780, EPI_ISL_1440782, EPI_ISL_1440783, EPI_ISL_1440784, EPI_ISL_1440785, EPI_ISL_1440786, EPI_ISL_1440787, EPI_ISL_1440788, EPI_ISL_1440789, EPI_ISL_1440790, EPI_ISL_1440792, EPI_ISL_1440793, EPI_ISL_1440794, EPI_ISL_1440795, EPI_ISL_1440796, EPI_ISL_1440797, EPI_ISL_1440798, EPI_ISL_1440799, EPI_ISL_1440800, EPI_ISL_1440801, EPI_ISL_1440802, EPI_ISL_1440803, EPI_ISL_1440804, EPI_ISL_1440805, EPI_ISL_1440807, EPI_ISL_1440808, EPI_ISL_1440809, EPI_ISL_1440811, EPI_ISL_1440812, EPI_ISL_1440813, EPI_ISL_1440814, EPI_ISL_1440815, EPI_ISL_1440816, EPI_ISL_1440817, EPI_ISL_1440818, EPI_ISL_1440819, EPI_ISL_1440820, EPI_ISL_1440821, EPI_ISL_1440822, EPI_ISL_1440823, EPI_ISL_1440824, EPI_ISL_1440825, EPI_ISL_1440826, EPI_ISL_1440827, EPI_ISL_1440828, EPI_ISL_1440829, EPI_ISL_1440830, EPI_ISL_1440831, EPI_ISL_1440832, EPI_ISL_1440833, EPI_ISL_1440834, EPI_ISL_1440835, EPI_ISL_1440836, EPI_ISL_1440837, EPI_ISL_1440838, EPI_ISL_1440839, EPI_ISL_1440840, EPI_ISL_1440841, EPI_ISL_1440842, EPI_ISL_1440843, EPI_ISL_1440844, EPI_ISL_1440845, EPI_ISL_1440847, EPI_ISL_1440848, EPI_ISL_1440849, EPI_ISL_1440850, EPI_ISL_1440851, EPI_ISL_1440852, EPI_ISL_1440854, EPI_ISL_1440856, EPI_ISL_1440857, EPI_ISL_1440858, EPI_ISL_1440859, EPI_ISL_1440860, EPI_ISL_1440861, EPI_ISL_1440862, EPI_ISL_1440863, EPI_ISL_1440864, EPI_ISL_1440865, EPI_ISL_1440866, EPI_ISL_1440867, EPI_ISL_1440868, EPI_ISL_1440869, EPI_ISL_1440870, EPI_ISL_1440871 | | | |
| see above | Universitätsklinikum Leipzig - Institut für Medizinische Mikrobiologie und Virologie virologisches Labor | Robert Koch Institute | |
| EPI_ISL_2115504, EPI_ISL_2115507, EPI_ISL_2115510 | Virologisches Institut des Universitatsklinikums Erlangen | Robert Koch Institute | |
| EPI_ISL_1786465, EPI_ISL_1786466, EPI_ISL_1786467 | Virology, Universitätsklinikum des Saarlandes | Epigenetics, Saarland University | Jörn Walter; Kathrin Kattler; Sascha Tierling; Sigrun Smola; Stefan Lohse; Thorsten Pfuhl |
| EPI_ISL_1991617, EPI_ISL_1991620, EPI_ISL_1991623, EPI_ISL_1991624, EPI_ISL_1991626, EPI_ISL_1991627, EPI_ISL_1991628 | | | |
| see above | ZOTZ KLIMAS MVZ Düsseldorf-Centrum GbR ÜBAG für Labormedizin, Genetik, Zytologie, Pathologie | Center of Medical Microbiology, Virology, and Hospital Hygiene, University of Duesseldorf | Alexander Dilthey; Andreas Walker; Daniel Strelow; Jessica Nicolai; Jörg Timm; Katrin Hoffmann; Klaus Pfeffer; Lisanna Hülse; Malte Kohns Vasconcelos; Maximilian Damagnez; Nadine Lübke; Patrick Finzer; Rainer Zotz; Tobias Wienemann; Torsten Houwaart |
| EPI_ISL_1569761, EPI_ISL_1569764, EPI_ISL_1569768, EPI_ISL_1569770, EPI_ISL_1569772, EPI_ISL_1569779, EPI_ISL_1569786 | | | |
| see above | alphaomega Labor GbR (Medizinische Kooperationsgemeinschaft Dres. Ackermann/Tennert/ZaiÃŸ) | Robert Koch Institute | |
| EPI_ISL_1572864, EPI_ISL_1572866, EPI_ISL_1572869, EPI_ISL_1572873, EPI_ISL_1572879, EPI_ISL_1572883, EPI_ISL_1572884, EPI_ISL_1572890, EPI_ISL_1572893, EPI_ISL_1572895, EPI_ISL_1572908, EPI_ISL_1572911, EPI_ISL_1572936, EPI_ISL_1572953, EPI_ISL_1572967, EPI_ISL_1572976, EPI_ISL_1572990, EPI_ISL_1573040, EPI_ISL_1573042, EPI_ISL_1573051, EPI_ISL_1644987, EPI_ISL_1644994, EPI_ISL_1645005, EPI_ISL_1645010, EPI_ISL_1645014, EPI_ISL_1645021, EPI_ISL_1645024, EPI_ISL_1645028, EPI_ISL_1645030, EPI_ISL_1645037, EPI_ISL_1645086, EPI_ISL_1645134, EPI_ISL_1645135, EPI_ISL_1645176, EPI_ISL_1645178, EPI_ISL_1645191, EPI_ISL_1645204, EPI_ISL_1645227 | | | |
| see above | amedes GÃ¶ttingen | Robert Koch Institute | |
| EPI_ISL_1572851, EPI_ISL_1572855, EPI_ISL_1572856, EPI_ISL_1572865, EPI_ISL_1572870, EPI_ISL_1572899, EPI_ISL_1572901, EPI_ISL_1572902, EPI_ISL_1572938, EPI_ISL_1572942, EPI_ISL_1572952, EPI_ISL_1572956, EPI_ISL_1572960, EPI_ISL_1572962, EPI_ISL_1572965, EPI_ISL_1572968, EPI_ISL_1572970, EPI_ISL_1572979, EPI_ISL_1572980, EPI_ISL_1572989, EPI_ISL_1572992, EPI_ISL_1572994, EPI_ISL_1572997, EPI_ISL_1572998, EPI_ISL_1573008, EPI_ISL_1573013, EPI_ISL_1573015, EPI_ISL_1573020, EPI_ISL_1573036, EPI_ISL_1573043 | | | |
| see above | amedes MVZ DIAMEDIS Sennestadt | Robert Koch Institute | |
| EPI_ISL_1572840, EPI_ISL_1572842, EPI_ISL_1572844, EPI_ISL_1572849, EPI_ISL_1572852, EPI_ISL_1572876, EPI_ISL_1572882, EPI_ISL_1572887, EPI_ISL_1572889, EPI_ISL_1572894, EPI_ISL_1572900, EPI_ISL_1572916, EPI_ISL_1572920, EPI_ISL_1572926, EPI_ISL_1572934, EPI_ISL_1572937, EPI_ISL_1572940, EPI_ISL_1572944, EPI_ISL_1572945, EPI_ISL_1572948, EPI_ISL_1572964, EPI_ISL_1572971, EPI_ISL_1572973, EPI_ISL_1572974, EPI_ISL_1572988, EPI_ISL_1572996, EPI_ISL_1573005, EPI_ISL_1573006, EPI_ISL_1573027, EPI_ISL_1573032, EPI_ISL_1573039, EPI_ISL_1573049 | | | |
| see above | amedes MVZ Halle/Leipzig | Robert Koch Institute | |
| EPI_ISL_1572858, EPI_ISL_1572886, EPI_ISL_1572896, EPI_ISL_1572930, EPI_ISL_1572951, EPI_ISL_1572955, EPI_ISL_1572958, EPI_ISL_1572975, EPI_ISL_1572977, EPI_ISL_1572981, EPI_ISL_1572986, EPI_ISL_1573019, EPI_ISL_1573022, EPI_ISL_1573025, EPI_ISL_1573033, EPI_ISL_1573047, EPI_ISL_1573053, EPI_ISL_1573055, EPI_ISL_1850481 | | | |
| see above | amedes MVZ Hannover | Robert Koch Institute | |
| EPI_ISL_1572841, EPI_ISL_1572845, EPI_ISL_1572847, EPI_ISL_1572850, EPI_ISL_1572861, EPI_ISL_1572888, EPI_ISL_1572914, EPI_ISL_1572924, EPI_ISL_1572939, EPI_ISL_1572961, EPI_ISL_1572963, EPI_ISL_1572978, EPI_ISL_1572983, EPI_ISL_1572999, EPI_ISL_1573000, EPI_ISL_1573004, EPI_ISL_1573011, EPI_ISL_1573016, EPI_ISL_1573017, EPI_ISL_1573021, EPI_ISL_1573030, EPI_ISL_1573031, EPI_ISL_1573038, EPI_ISL_1573044 | | | |
| see above | amedes MVZ Jena | Robert Koch Institute | |
| EPI_ISL_1572943 | amedes MVZ Zentrum fÃ¼r Pathologie und Zytodiagnostik GmbH (KÃ¶ln) | Robert Koch Institute | |
| EPI_ISL_1572854, EPI_ISL_1572863, EPI_ISL_1572909, EPI_ISL_1572913, EPI_ISL_1572921, EPI_ISL_1572928, EPI_ISL_1572991, EPI_ISL_1573028, EPI_ISL_1573029, EPI_ISL_1573056 | | | |
| see above | amedes MVZ fÃ¼r Laboratoriumsdiagnostik Raubling GmbH | Robert Koch Institute | |
| EPI_ISL_1572891, EPI_ISL_1572935, EPI_ISL_1572949, EPI_ISL_1572959, EPI_ISL_1573010, EPI_ISL_1644984, EPI_ISL_1645034, EPI_ISL_1645088, EPI_ISL_1645101, EPI_ISL_1645133, EPI_ISL_1645160, EPI_ISL_1645174 | | | |
| see above | amedes MVZ fÃ¼r Laboratoriumsmedizin FÃ¼rstenfeldbruck | Robert Koch Institute | |
| EPI_ISL_1572966 | amedes MVZ fÃ¼r Laboratoriumsmedizin MÃ¼nchen | Robert Koch Institute | |
| EPI_ISL_1572867, EPI_ISL_1572946 | amedes MVZ fÃ¼r Laboratoriumsmedizin Rhein/Ruhr | Robert Koch Institute | |
| EPI_ISL_1572871, EPI_ISL_1572918, EPI_ISL_1572925, EPI_ISL_1572933, EPI_ISL_1573002, EPI_ISL_1573009, EPI_ISL_1573041 | | | |

| | | |
|---|---|---|
| see above | amedes MVZ fÃ¼r Labordiagnostik Rhein-Main | Robert Koch Institute |
| EPI_ISL_1572853, EPI_ISL_1572907, EPI_ISL_1572917 | amedes Medlab Arnold Analytik MVZ GmbH | Robert Koch Institute |
| EPI_ISL_1638641, EPI_ISL_1638642 | labopart - Medizinische Laboratorien Dresden | Robert Koch Institute |
| EPI_ISL_1564043, EPI_ISL_1564072, EPI_ISL_1564272, EPI_ISL_1564332, EPI_ISL_1564355, EPI_ISL_1564522, EPI_ISL_1564613, EPI_ISL_1564715, EPI_ISL_1564801, EPI_ISL_1564832, EPI_ISL_1564959, EPI_ISL_1565100, EPI_ISL_1565215, EPI_ISL_1565273, EPI_ISL_1565297, EPI_ISL_1565319, EPI_ISL_1565341, EPI_ISL_1572843, EPI_ISL_1572859, EPI_ISL_1572860, EPI_ISL_1572862, EPI_ISL_1572868, EPI_ISL_1572872, EPI_ISL_1572874, EPI_ISL_1572875, EPI_ISL_1572877, EPI_ISL_1572878, EPI_ISL_1572880, EPI_ISL_1572881, EPI_ISL_1572885, EPI_ISL_1572892, EPI_ISL_1572897, EPI_ISL_1572898, EPI_ISL_1572903, EPI_ISL_1572905, EPI_ISL_1572915, EPI_ISL_1572919, EPI_ISL_1572922, EPI_ISL_1572923, EPI_ISL_1572929, EPI_ISL_1572947, EPI_ISL_1572954, EPI_ISL_1572969, EPI_ISL_1572972, EPI_ISL_1572984, EPI_ISL_1572985, EPI_ISL_1572987, EPI_ISL_1572993, EPI_ISL_1572995, EPI_ISL_1573003, EPI_ISL_1573012, EPI_ISL_1573023, EPI_ISL_1573037, EPI_ISL_1573045, EPI_ISL_1573046, EPI_ISL_1573048, EPI_ISL_1573052, EPI_ISL_1573054 | | |
| see above | nordlab - Partnerschaftspraxis fuÌˆr Laboratoriumsmedizin | Robert Koch Institute |

We gratefully acknowledge the following Authors from the Originating laboratories responsible for obtaining the specimens, as well as the Submitting laboratories where the genome data were generated and shared via GISAID, on which this research is based.

All Submitters of data may be contacted directly via www.gisaid.org

Authors are sorted alphabetically.

| Accession ID | Originating Laboratory | Submitting Laboratory | Authors |
|---|---|---|---|
| EPI_ISL_2115594 | ALB FILS KLINIKEN GmbH - Institut fur Laboratoriumsmedizin Klinik am Eichert Gappingen | Robert Koch Institute | |
| EPI_ISL_2115712, EPI_ISL_2115754 | ALB FILS KLINIKEN GmbH - Institut fÃ¼r Laboratoriumsmedizin Klinik am Eichert GÃ¶ppingen | Robert Koch Institute | |
| EPI_ISL_2110900, EPI_ISL_2111003, EPI_ISL_2111281, EPI_ISL_2111282, EPI_ISL_2111285, EPI_ISL_2111286, EPI_ISL_2111288, EPI_ISL_2111291, EPI_ISL_2111295, EPI_ISL_2111297, EPI_ISL_2111299, EPI_ISL_2111300, EPI_ISL_2111306, EPI_ISL_2111309, EPI_ISL_2111312, EPI_ISL_2111313, EPI_ISL_2111315, EPI_ISL_2111316, EPI_ISL_2111324, EPI_ISL_2111327, EPI_ISL_2111329, EPI_ISL_2111331, EPI_ISL_2111332, EPI_ISL_2111334, EPI_ISL_2111338, EPI_ISL_2111341, EPI_ISL_2111346, EPI_ISL_2111352, EPI_ISL_2111354, EPI_ISL_2111355, EPI_ISL_2111362, EPI_ISL_2111364, EPI_ISL_2111366, EPI_ISL_2111368, EPI_ISL_2111370, EPI_ISL_2111372, EPI_ISL_2111373, EPI_ISL_2111378, EPI_ISL_2111381, EPI_ISL_2111391, EPI_ISL_2111393, EPI_ISL_2111396, EPI_ISL_2111399, EPI_ISL_2111405, EPI_ISL_2111407, EPI_ISL_2111412, EPI_ISL_2111415, EPI_ISL_2111417, EPI_ISL_2111418, EPI_ISL_2111419, EPI_ISL_2111420, EPI_ISL_2111425, EPI_ISL_2111433 | | | |
| see above | Bayerisches Landesamt fur Gesundheit und Lebensmittelsicherheit (LGL) | Robert Koch Institute | |
| EPI_ISL_2110881, EPI_ISL_2110892, EPI_ISL_2110905, EPI_ISL_2110906, EPI_ISL_2110910, EPI_ISL_2110928, EPI_ISL_2110929, EPI_ISL_2110933, EPI_ISL_2110937, EPI_ISL_2110955, EPI_ISL_2110956, EPI_ISL_2110978, EPI_ISL_2110984 | | | |
| see above | Bayerisches Landesamt fÃ¼r Gesundheit und Lebensmittelsicherheit (LGL) | Robert Koch Institute | |
| EPI_ISL_1721725, EPI_ISL_1721850, EPI_ISL_1721931, EPI_ISL_1842966, EPI_ISL_1842968, EPI_ISL_1842969, EPI_ISL_1842970, EPI_ISL_1842973, EPI_ISL_1842975, EPI_ISL_1842976, EPI_ISL_1842981, EPI_ISL_1842983, EPI_ISL_1842984, EPI_ISL_1842986, EPI_ISL_1842987, EPI_ISL_1842988, EPI_ISL_1842989, EPI_ISL_1842990, EPI_ISL_1842991, EPI_ISL_1842993, EPI_ISL_1842994, EPI_ISL_1842995, EPI_ISL_1842996, EPI_ISL_1842997, EPI_ISL_1842998, EPI_ISL_1843000, EPI_ISL_1843001, EPI_ISL_1843005, EPI_ISL_1843007, EPI_ISL_1843009, EPI_ISL_1843011, EPI_ISL_1843013, EPI_ISL_1843014, EPI_ISL_1843016, EPI_ISL_1843019, EPI_ISL_1843021, EPI_ISL_1843022, EPI_ISL_1843025, EPI_ISL_1843026, EPI_ISL_1843028, EPI_ISL_1843030, EPI_ISL_1843031, EPI_ISL_1843033, EPI_ISL_1843039, EPI_ISL_1843040, EPI_ISL_1843043, EPI_ISL_1843044, EPI_ISL_1843045, EPI_ISL_1843046, EPI_ISL_1843048, EPI_ISL_1843051, EPI_ISL_1843052, EPI_ISL_1843053, EPI_ISL_1843054, EPI_ISL_1843055, EPI_ISL_1843056, EPI_ISL_1843060, EPI_ISL_1843061, EPI_ISL_1843062, EPI_ISL_1843063, EPI_ISL_1843067, EPI_ISL_1843068, EPI_ISL_1843069, EPI_ISL_1843070, EPI_ISL_1843078, EPI_ISL_1843079, EPI_ISL_1843081, EPI_ISL_1843085, EPI_ISL_1843087, EPI_ISL_1843092, EPI_ISL_1843094, EPI_ISL_1843095, EPI_ISL_1843096, EPI_ISL_1843097, EPI_ISL_1843100, EPI_ISL_1843101, EPI_ISL_1843106, EPI_ISL_1843110, EPI_ISL_1843111, EPI_ISL_1843113, EPI_ISL_1843115, EPI_ISL_1843118, EPI_ISL_1843123, EPI_ISL_1843125, EPI_ISL_1843128, EPI_ISL_1843129, EPI_ISL_1843132, EPI_ISL_1843133, EPI_ISL_1843135, EPI_ISL_1843138, EPI_ISL_1843139, EPI_ISL_1843140, EPI_ISL_1843142, EPI_ISL_1843145, EPI_ISL_1843146, EPI_ISL_1843148, EPI_ISL_1843150, EPI_ISL_1843151, EPI_ISL_1843157, EPI_ISL_1843158, EPI_ISL_1843163, EPI_ISL_1843165, EPI_ISL_1843166, EPI_ISL_1843168, EPI_ISL_1843172, EPI_ISL_1843173, EPI_ISL_1843174, EPI_ISL_1843445, EPI_ISL_1843449, EPI_ISL_1843452, EPI_ISL_1843457, EPI_ISL_1843474, EPI_ISL_1843489, EPI_ISL_1843491, EPI_ISL_1843492, EPI_ISL_1843494, EPI_ISL_1843495, EPI_ISL_1843499, EPI_ISL_1843502, EPI_ISL_1843508, EPI_ISL_1843510, EPI_ISL_1843511, EPI_ISL_1843513, EPI_ISL_1843520, EPI_ISL_1843527, EPI_ISL_1843528, EPI_ISL_1843530, EPI_ISL_1843534, EPI_ISL_1843535, EPI_ISL_1843538, EPI_ISL_1843539, EPI_ISL_1843542, EPI_ISL_1843546, EPI_ISL_1843547, EPI_ISL_1843551, EPI_ISL_1843553, EPI_ISL_1843561, EPI_ISL_1843562, EPI_ISL_1843563, EPI_ISL_1843566, EPI_ISL_1843570, EPI_ISL_1843571, EPI_ISL_1843572, EPI_ISL_1843573, EPI_ISL_1843577, EPI_ISL_1843578, EPI_ISL_1843582, EPI_ISL_1843583, EPI_ISL_1843584, EPI_ISL_1843586, EPI_ISL_1843588, EPI_ISL_1843590, EPI_ISL_1843592, EPI_ISL_1843593, EPI_ISL_1843595, EPI_ISL_1843597, EPI_ISL_1843598, EPI_ISL_1843599, EPI_ISL_1843600, EPI_ISL_1843601, EPI_ISL_1843602, EPI_ISL_1843605, EPI_ISL_1843606, EPI_ISL_1843608, EPI_ISL_1843613, EPI_ISL_1843614, EPI_ISL_1843619, EPI_ISL_1843620, EPI_ISL_1843623, EPI_ISL_1843625, EPI_ISL_1843626, EPI_ISL_1843632, EPI_ISL_1843636, EPI_ISL_1843638, EPI_ISL_1843642, EPI_ISL_1843644, EPI_ISL_1843645, EPI_ISL_1843650, EPI_ISL_1843655, EPI_ISL_1843659, EPI_ISL_1843661, EPI_ISL_1843663, EPI_ISL_1843665, EPI_ISL_1843667, EPI_ISL_1843669, EPI_ISL_1843670, EPI_ISL_1843675, EPI_ISL_1843683, EPI_ISL_1843685, EPI_ISL_1843688, EPI_ISL_1843689, EPI_ISL_1843690, EPI_ISL_1843693, EPI_ISL_1843694, EPI_ISL_1843695, EPI_ISL_1843698, EPI_ISL_1843701, EPI_ISL_1843703, EPI_ISL_1843704, EPI_ISL_1843706, EPI_ISL_1843707, EPI_ISL_1843708, EPI_ISL_1843709, EPI_ISL_1843710, EPI_ISL_1843712, EPI_ISL_1843714, EPI_ISL_1843717, EPI_ISL_1843719, EPI_ISL_1844047, EPI_ISL_1844061, EPI_ISL_1844083, EPI_ISL_1844087, EPI_ISL_1844102, EPI_ISL_1844104, EPI_ISL_1844108, EPI_ISL_1844112, EPI_ISL_1844113, EPI_ISL_1844119, EPI_ISL_1844121, EPI_ISL_1844122, EPI_ISL_1844127, EPI_ISL_1844129, EPI_ISL_1844140, EPI_ISL_1844142, EPI_ISL_1844143, EPI_ISL_1844154, EPI_ISL_1844170, EPI_ISL_1844171, EPI_ISL_1844173, EPI_ISL_1844176, EPI_ISL_1844183, EPI_ISL_1844189, EPI_ISL_1844197, EPI_ISL_1844198, EPI_ISL_1844210, EPI_ISL_1844211, EPI_ISL_1844218, EPI_ISL_1844225, EPI_ISL_1844229, EPI_ISL_1844237, EPI_ISL_1844238, EPI_ISL_1844248, EPI_ISL_1844250, EPI_ISL_1844251, EPI_ISL_1844255, EPI_ISL_1844260, EPI_ISL_1844267, EPI_ISL_1846239, EPI_ISL_1846241, EPI_ISL_1846242, EPI_ISL_1846243, EPI_ISL_1846245, EPI_ISL_1846248, EPI_ISL_1846249, EPI_ISL_1846250, EPI_ISL_1846252, EPI_ISL_1846253, EPI_ISL_1846254, EPI_ISL_1846255, EPI_ISL_1846257, EPI_ISL_1846259, EPI_ISL_1846260, EPI_ISL_1846261, EPI_ISL_1846262, EPI_ISL_1846265, EPI_ISL_1846268, EPI_ISL_1846269, EPI_ISL_1846270, EPI_ISL_1846272, EPI_ISL_1846273, EPI_ISL_1846277, EPI_ISL_1846278, EPI_ISL_1846279, EPI_ISL_1846282, EPI_ISL_1846285, EPI_ISL_1846286, EPI_ISL_1846287, EPI_ISL_1846289, EPI_ISL_1846290, EPI_ISL_1846291, EPI_ISL_1846292, EPI_ISL_1846294, EPI_ISL_1846295, EPI_ISL_1846302, EPI_ISL_1846303, EPI_ISL_1846306, EPI_ISL_1846308, EPI_ISL_1846309, EPI_ISL_1846310, EPI_ISL_1846311, EPI_ISL_1846312, EPI_ISL_1846313, EPI_ISL_1846314, EPI_ISL_1846315, EPI_ISL_1847121, EPI_ISL_1847122, EPI_ISL_1847137, EPI_ISL_1847138, EPI_ISL_1847147, EPI_ISL_1847235, EPI_ISL_1847236, EPI_ISL_1847238, EPI_ISL_1847239, EPI_ISL_1847242, EPI_ISL_1847243, EPI_ISL_1847244, EPI_ISL_1847245, EPI_ISL_1847246, EPI_ISL_1847247, EPI_ISL_1847249, EPI_ISL_1847250, EPI_ISL_1847251, EPI_ISL_1847252, EPI_ISL_1847253, EPI_ISL_1847254, EPI_ISL_1847255, EPI_ISL_1847256, EPI_ISL_1847257, EPI_ISL_1847258, EPI_ISL_1847259, EPI_ISL_1847260, EPI_ISL_1847261, EPI_ISL_1847263, EPI_ISL_1847264, EPI_ISL_1847265, EPI_ISL_1847266, EPI_ISL_1847267, EPI_ISL_1847268, EPI_ISL_1847269, EPI_ISL_1847270, EPI_ISL_1847272, EPI_ISL_1847273, EPI_ISL_1847274, EPI_ISL_1847275, EPI_ISL_1847278, EPI_ISL_1847280, EPI_ISL_1847282, EPI_ISL_1847283, EPI_ISL_1847284, EPI_ISL_1847285, EPI_ISL_1847287, EPI_ISL_1847288, EPI_ISL_1847289, EPI_ISL_1847291, EPI_ISL_1847292, EPI_ISL_1847293, EPI_ISL_1847295, EPI_ISL_1847296, EPI_ISL_1847297, EPI_ISL_1847298, EPI_ISL_1847299, EPI_ISL_1847301, EPI_ISL_1847304, EPI_ISL_1847305, EPI_ISL_1847306, EPI_ISL_1847307, EPI_ISL_1847308, EPI_ISL_1847311, EPI_ISL_1847485, EPI_ISL_1847514, EPI_ISL_1848406, EPI_ISL_1848409, EPI_ISL_1848411, EPI_ISL_1848413, EPI_ISL_1848416, EPI_ISL_1848425, EPI_ISL_1848430, EPI_ISL_1848450, EPI_ISL_1848452, EPI_ISL_1848456, EPI_ISL_1848462, EPI_ISL_1848466, EPI_ISL_1848471, EPI_ISL_1848474, EPI_ISL_1848482, EPI_ISL_1848490, EPI_ISL_1848497, EPI_ISL_1848503, EPI_ISL_1848506, EPI_ISL_1848512, EPI_ISL_1848515, EPI_ISL_1848519, EPI_ISL_1848521, EPI_ISL_1848522, EPI_ISL_1848527, EPI_ISL_1848528, EPI_ISL_1848530, EPI_ISL_1848538, EPI_ISL_1848546, EPI_ISL_1848549, EPI_ISL_1848554, EPI_ISL_1848556, EPI_ISL_1848574, EPI_ISL_1848575, EPI_ISL_1848577, EPI_ISL_1848580, EPI_ISL_1848583, EPI_ISL_1848584, EPI_ISL_1848586, EPI_ISL_1848592, EPI_ISL_1848593, EPI_ISL_1848595, EPI_ISL_1848600, EPI_ISL_1848611, EPI_ISL_1848614, EPI_ISL_1848620, EPI_ISL_1848626, EPI_ISL_1848636, EPI_ISL_1848640, EPI_ISL_1848648, EPI_ISL_1848653, EPI_ISL_1848655, EPI_ISL_1848657, EPI_ISL_1848665, EPI_ISL_1848677, EPI_ISL_1848681, EPI_ISL_1848683, EPI_ISL_1848699, EPI_ISL_1848702, EPI_ISL_1848705, EPI_ISL_1848706, EPI_ISL_1848708, EPI_ISL_1848709, EPI_ISL_1848711, EPI_ISL_1848712, EPI_ISL_1848717, EPI_ISL_1848721, EPI_ISL_1848723, EPI_ISL_1848725, EPI_ISL_1848731, EPI_ISL_1848740, EPI_ISL_2108733, EPI_ISL_2108736, EPI_ISL_2108744, EPI_ISL_2108752, EPI_ISL_2108762, EPI_ISL_2108764, EPI_ISL_2108768, EPI_ISL_2108788, EPI_ISL_2108794, EPI_ISL_2108812, EPI_ISL_2108834, EPI_ISL_2108841, EPI_ISL_2108846, EPI_ISL_2108850, EPI_ISL_2108861, EPI_ISL_2108863, EPI_ISL_2108879, EPI_ISL_2108882, EPI_ISL_2108895, EPI_ISL_2108901, EPI_ISL_2108908, EPI_ISL_2108913, EPI_ISL_2108920, EPI_ISL_2108930, EPI_ISL_2108944, EPI_ISL_2109045, EPI_ISL_2109055, EPI_ISL_2109078, EPI_ISL_2109133, EPI_ISL_2109143, EPI_ISL_2109186, EPI_ISL_2109193, EPI_ISL_2109196, EPI_ISL_2109211, EPI_ISL_2109283, EPI_ISL_2109288, EPI_ISL_2109342, EPI_ISL_2109759, EPI_ISL_2109790, EPI_ISL_2115799 | | | |
| see above | Bioscientia Labor Wermsdorf | Robert Koch Institute | |
| EPI_ISL_1842980, EPI_ISL_1843002, EPI_ISL_1843006, EPI_ISL_1843015, EPI_ISL_1843032, EPI_ISL_1843036, EPI_ISL_1843038, EPI_ISL_1843049, EPI_ISL_1843050, EPI_ISL_1843071, EPI_ISL_1843075, EPI_ISL_1843076, EPI_ISL_1843084, EPI_ISL_1843086, EPI_ISL_1843090, EPI_ISL_1843093, EPI_ISL_1843098, EPI_ISL_1843103, EPI_ISL_1843112, EPI_ISL_1843117, EPI_ISL_1843121, EPI_ISL_1843124, EPI_ISL_1843136, EPI_ISL_1843143, EPI_ISL_1843152, EPI_ISL_1843162, EPI_ISL_1843469, EPI_ISL_1843487, EPI_ISL_1843536, EPI_ISL_1843558, EPI_ISL_1843567, EPI_ISL_1843635, EPI_ISL_1843637, EPI_ISL_1843640, EPI_ISL_1843672, EPI_ISL_1843691, EPI_ISL_1843716, EPI_ISL_1843729, EPI_ISL_1847237, EPI_ISL_1847240, EPI_ISL_1847241, EPI_ISL_1847248, EPI_ISL_1847262, EPI_ISL_1847271, EPI_ISL_1847277, EPI_ISL_1847279, EPI_ISL_1847290, EPI_ISL_1847294, EPI_ISL_1847302, EPI_ISL_1847303, EPI_ISL_1847310, EPI_ISL_1848428, EPI_ISL_1848444, EPI_ISL_1848448, EPI_ISL_1848457, EPI_ISL_1848523, EPI_ISL_1848567, EPI_ISL_1848570, EPI_ISL_1848587, EPI_ISL_1848608, EPI_ISL_1848619, EPI_ISL_1848638, EPI_ISL_1848671, EPI_ISL_1848673, EPI_ISL_1848675, EPI_ISL_1848685, EPI_ISL_1848730 | | | |
| see above | Bioscientia MVZ Labor Karlsruhe GmbH | Robert Koch Institute | |
| EPI_ISL_1834007 | Bundeswehr Institute of Microbiology | Bundeswehr Institute of Microbiology | Alexandra Rehn; Enrico Georgi; Malena Bestehorn-Willmann; Markus Antwerpen; Mathias Walter; Mike Pillukat; Roman Wölfel; Sabine Zange |
| EPI_ISL_1834029, EPI_ISL_1834037, EPI_ISL_1834038, EPI_ISL_1834040, EPI_ISL_1834041, EPI_ISL_1834042, EPI_ISL_1834062, EPI_ISL_1834066 | | | |
| see above | Bundeswehrkrankenhaus Berlin | Bundeswehr Institute of Microbiology | Alexandra Rehn; Enrico Georgi; Malena Bestehorn-Willmann; Markus Antwerpen; Mathias Walter; Mike Pillukat; Roman Wölfel; Sabine Zange |
| EPI_ISL_1834047 | Bundeswehrkrankenhaus Hamburg | Bundeswehr Institute of Microbiology | Alexandra Rehn; Enrico Georgi; Malena Bestehorn-Willmann; Markus Antwerpen; Mathias Walter; Mike Pillukat; Roman Wölfel; Sabine Zange |
| EPI_ISL_1833999, EPI_ISL_1834035, EPI_ISL_1834036 | Bundeswehrkrankenhaus Ulm | Bundeswehr Institute of Microbiology | Alexandra Rehn; Enrico Georgi; Malena Bestehorn-Willmann; Markus Antwerpen; Mathias Walter; Mike Pillukat; Roman Wölfel; Sabine Zange |
| EPI_ISL_1833997, EPI_ISL_1834003, EPI_ISL_1834004, EPI_ISL_1834034, EPI_ISL_1834044, EPI_ISL_1834045, EPI_ISL_1834046 | | | |
| see above | Bundeswehrzentralkrankenhaus Koblenz | Bundeswehr Institute of Microbiology | Alexandra Rehn; Enrico Georgi; Malena Bestehorn-Willmann; Markus Antwerpen; Mathias Walter; Mike Pillukat; Roman Wölfel; Sabine Zange |
| EPI_ISL_1843732, EPI_ISL_1843737, EPI_ISL_1843741, EPI_ISL_1843748, EPI_ISL_1843757, EPI_ISL_1843760, EPI_ISL_1843764, EPI_ISL_1843770, EPI_ISL_1843780, EPI_ISL_1843781, EPI_ISL_1843785, EPI_ISL_1843790, EPI_ISL_1843793, EPI_ISL_1843794, EPI_ISL_1843796, EPI_ISL_1843797, EPI_ISL_1843799, EPI_ISL_1843801, EPI_ISL_1843802, EPI_ISL_1843803, EPI_ISL_1843805, EPI_ISL_1843806, EPI_ISL_1843812, EPI_ISL_1843819, EPI_ISL_1843820, EPI_ISL_1843822, EPI_ISL_1843824, EPI_ISL_1843825, EPI_ISL_1843826, EPI_ISL_1843828, EPI_ISL_1843832, EPI_ISL_1843833, EPI_ISL_1843836, EPI_ISL_1843839, EPI_ISL_1843841, EPI_ISL_1843843, EPI_ISL_1843844, EPI_ISL_1843845, EPI_ISL_1843848, EPI_ISL_1843850, EPI_ISL_1843852, EPI_ISL_1843853, EPI_ISL_1843854, EPI_ISL_1843855, EPI_ISL_1843857, EPI_ISL_1843858, EPI_ISL_1843861, EPI_ISL_1843863, EPI_ISL_1843864, EPI_ISL_1843865, EPI_ISL_1843866, EPI_ISL_1843867, EPI_ISL_1843869, EPI_ISL_1843872, EPI_ISL_1843873, EPI_ISL_1843874, EPI_ISL_1843875, EPI_ISL_1843876, EPI_ISL_1843880, EPI_ISL_1843884, EPI_ISL_1843886, EPI_ISL_1843887, EPI_ISL_1843888, EPI_ISL_1843889, EPI_ISL_1843892, EPI_ISL_1843893, EPI_ISL_1843897, EPI_ISL_1843898, EPI_ISL_1843900, EPI_ISL_1843901, EPI_ISL_1843902, EPI_ISL_1843903, EPI_ISL_1843904, EPI_ISL_1843906, EPI_ISL_1843907, EPI_ISL_1843908, EPI_ISL_1843909, EPI_ISL_1843910, EPI_ISL_1843913, EPI_ISL_1843915, EPI_ISL_1843916, EPI_ISL_1843919, EPI_ISL_1843921, EPI_ISL_1843922, EPI_ISL_1843923, EPI_ISL_1843924, EPI_ISL_1843925, EPI_ISL_1843926, EPI_ISL_1843927, EPI_ISL_1843929, EPI_ISL_1843931, EPI_ISL_1843933, EPI_ISL_1843934, EPI_ISL_1843935, EPI_ISL_1843937, EPI_ISL_1843940, EPI_ISL_1843941, EPI_ISL_1843942, EPI_ISL_1843945, EPI_ISL_1843947, EPI_ISL_1843949, EPI_ISL_1843950, EPI_ISL_1843952, EPI_ISL_1843953, EPI_ISL_1843955, EPI_ISL_1843957, EPI_ISL_1843958, EPI_ISL_1843960, EPI_ISL_1843961, EPI_ISL_1843963, EPI_ISL_1843966, EPI_ISL_1843967, EPI_ISL_1843968, EPI_ISL_1843974, EPI_ISL_1843975, EPI_ISL_1843976, EPI_ISL_1843977, EPI_ISL_1843978, EPI_ISL_1843981, EPI_ISL_1843983, EPI_ISL_1843985, EPI_ISL_1843987, EPI_ISL_1843993, EPI_ISL_1843995, EPI_ISL_1843998, EPI_ISL_1843999, EPI_ISL_1844002, EPI_ISL_1844003, EPI_ISL_1844004, EPI_ISL_1844006, EPI_ISL_1844007, EPI_ISL_1844008, EPI_ISL_1844010, EPI_ISL_1844013, EPI_ISL_1844015, EPI_ISL_1844017, EPI_ISL_1844021, EPI_ISL_1844022, EPI_ISL_1844025, EPI_ISL_1844026, EPI_ISL_2123141 | | | |
| see above | Centogene; Dr. Bauer Laboratoriums GmbH | Robert Koch Institute | |
| EPI_ISL_1643111, EPI_ISL_1643112, EPI_ISL_1643113, EPI_ISL_1643114, EPI_ISL_1643115, EPI_ISL_1643116, EPI_ISL_1643117, EPI_ISL_1725601 | | | |
| see above | Chemische und VeterinÃ¤runtersuchungsamt MÃ¼nsterland-Emscher-Lippe (CVUA-MEL) | Robert Koch Institute | |
| EPI_ISL_1639332, EPI_ISL_1639351, EPI_ISL_1639359, EPI_ISL_1639370, EPI_ISL_1639403, EPI_ISL_1639424, EPI_ISL_1722940, EPI_ISL_1722947, EPI_ISL_1722948, EPI_ISL_1722949, EPI_ISL_1722953, EPI_ISL_1722954, EPI_ISL_1722955, EPI_ISL_1722956, EPI_ISL_1722957, EPI_ISL_1722958, EPI_ISL_1722959, EPI_ISL_1722960, EPI_ISL_1722961, EPI_ISL_1722962, EPI_ISL_1722963, EPI_ISL_1722964, EPI_ISL_1722965, EPI_ISL_1722973, EPI_ISL_1722974, EPI_ISL_1722975, EPI_ISL_1722976, EPI_ISL_1722977, EPI_ISL_1722978, EPI_ISL_1722979, EPI_ISL_1722980, EPI_ISL_1722981, EPI_ISL_1722982, EPI_ISL_1722983, EPI_ISL_1722984, EPI_ISL_1722985, EPI_ISL_1722986, EPI_ISL_1722987, EPI_ISL_1722988, EPI_ISL_1722989, EPI_ISL_1722990, EPI_ISL_1722991, EPI_ISL_1722992, EPI_ISL_1722993, EPI_ISL_1722994, EPI_ISL_1722995, EPI_ISL_1722996, EPI_ISL_1722997, EPI_ISL_1722998, EPI_ISL_1722999, EPI_ISL_1723000, EPI_ISL_1723001, EPI_ISL_1723002, EPI_ISL_1723003, EPI_ISL_1723004, EPI_ISL_1723005, EPI_ISL_1723006, EPI_ISL_1723007, EPI_ISL_1723008, EPI_ISL_1723009, EPI_ISL_1723010, EPI_ISL_1723011, EPI_ISL_1723012, EPI_ISL_1723013, EPI_ISL_1723014, EPI_ISL_1723015, EPI_ISL_1724278, EPI_ISL_1724279, EPI_ISL_1724280, EPI_ISL_1724281, EPI_ISL_1724282, EPI_ISL_1724283, EPI_ISL_1724284, EPI_ISL_1724285, EPI_ISL_1724286, EPI_ISL_1724287, EPI_ISL_1724288, EPI_ISL_1724289, EPI_ISL_1724290, EPI_ISL_1724291, EPI_ISL_1724292, EPI_ISL_1724295, EPI_ISL_1724296, EPI_ISL_1724297, EPI_ISL_1724298, EPI_ISL_1724300, EPI_ISL_1724301, EPI_ISL_1724302, EPI_ISL_1724303, EPI_ISL_1724304, EPI_ISL_1724305, EPI_ISL_1724306, EPI_ISL_1844415, EPI_ISL_1844434, EPI_ISL_1844435 | | | |
| see above | Clemenshospital - Alexianer Clemenshospital MÃ¼nster | Robert Koch Institute | |
| EPI_ISL_2112746, EPI_ISL_2112748, EPI_ISL_2112759, EPI_ISL_2112761 | Corona-Testzentrum ifp Institut fur Produktqualitat GmbH | Robert Koch Institute | |
| EPI_ISL_1843176, EPI_ISL_1843177, EPI_ISL_1843182, EPI_ISL_1843184, EPI_ISL_1843197 | Corona-Testzentrum ifp Institut fÃ¼r ProduktqualitÃ¤t GmbH | Robert Koch Institute | |
| EPI_ISL_2108983, EPI_ISL_2108990, EPI_ISL_2108993, EPI_ISL_2108994, EPI_ISL_2108999, EPI_ISL_2109003, EPI_ISL_2109004, EPI_ISL_2109010, EPI_ISL_2109016, EPI_ISL_2109017 | | | |
| see above | Department fur Labormedizin Abteilung III Bereich Molekulare Diagnostik Universitatsklinikum Halle(Saale) | Robert Koch Institute | |
| EPI_ISL_1722278, EPI_ISL_1722280, EPI_ISL_1722282, EPI_ISL_1722284, EPI_ISL_1722287, EPI_ISL_1722288, EPI_ISL_1722289, EPI_ISL_1722290, EPI_ISL_1722292, EPI_ISL_1722293, EPI_ISL_1722294, EPI_ISL_1722295, EPI_ISL_1722297, EPI_ISL_1722298, EPI_ISL_1722299, EPI_ISL_1722300, EPI_ISL_1722301, EPI_ISL_1722302, EPI_ISL_1722303, EPI_ISL_1722304, EPI_ISL_1722305, EPI_ISL_1722309, EPI_ISL_1722310, EPI_ISL_1722311, EPI_ISL_1722312, EPI_ISL_1722313, EPI_ISL_1722315, EPI_ISL_1722317, EPI_ISL_1722318, EPI_ISL_1722319, EPI_ISL_1722321, EPI_ISL_1722322, EPI_ISL_1722324, EPI_ISL_1722325, EPI_ISL_1722326, EPI_ISL_1722327, EPI_ISL_1722329, EPI_ISL_1722330, EPI_ISL_1722331, EPI_ISL_1722332, EPI_ISL_1722333, EPI_ISL_1722334, EPI_ISL_1722335, EPI_ISL_1722336, EPI_ISL_1722338, EPI_ISL_1722339, EPI_ISL_1722340, EPI_ISL_1722341, EPI_ISL_1722344, EPI_ISL_1722345, EPI_ISL_1722346, EPI_ISL_1722347, EPI_ISL_1722350, EPI_ISL_1722351, EPI_ISL_1722352, EPI_ISL_1722353, EPI_ISL_1722354, EPI_ISL_1722355, EPI_ISL_1722357, EPI_ISL_1722358, EPI_ISL_1725239, EPI_ISL_1725240, EPI_ISL_1725242, EPI_ISL_1725243, EPI_ISL_1725244, EPI_ISL_1725245, EPI_ISL_1848843 | | | |
| see above | Diagnosticum - Labor Neukirchen | Robert Koch Institute | |
| EPI_ISL_1644927, EPI_ISL_1644929, EPI_ISL_1644932, EPI_ISL_1644933, EPI_ISL_1724448, EPI_ISL_1724449, EPI_ISL_1724450, EPI_ISL_1724451, EPI_ISL_1724452, EPI_ISL_1724453, EPI_ISL_1724454, EPI_ISL_1724455, EPI_ISL_1724456, EPI_ISL_1724457, EPI_ISL_1724458, EPI_ISL_1724459, EPI_ISL_1727065 | | | |
| see above | Dianovis GmbH Greiz | Robert Koch Institute | |
| EPI_ISL_1724338, EPI_ISL_1724349, EPI_ISL_1724353, EPI_ISL_1724357, EPI_ISL_1724359, EPI_ISL_1724363, EPI_ISL_1724364, EPI_ISL_1724366, EPI_ISL_1724369, EPI_ISL_1724372, EPI_ISL_1724375, EPI_ISL_1724377, EPI_ISL_1724380, EPI_ISL_1724384, EPI_ISL_1724385, EPI_ISL_1724390 | | | |
| see above | ELBLAB GmbH Zentrum fÃ¼r LaborMedizin | Robert Koch Institute | |
| EPI_ISL_2110417, EPI_ISL_2110420, EPI_ISL_2110423, EPI_ISL_2110426, EPI_ISL_2110427, EPI_ISL_2110428, EPI_ISL_2110429, EPI_ISL_2110430, EPI_ISL_2110431, EPI_ISL_2110432, EPI_ISL_2110433, EPI_ISL_2110434, EPI_ISL_2110435, EPI_ISL_2110436, EPI_ISL_2110437, EPI_ISL_2110438, EPI_ISL_2110439, EPI_ISL_2110440, EPI_ISL_2110441, EPI_ISL_2110442, EPI_ISL_2110443, EPI_ISL_2110444, EPI_ISL_2110445, EPI_ISL_2110446, EPI_ISL_2110447, EPI_ISL_2110448, EPI_ISL_2110449, EPI_ISL_2110450, EPI_ISL_2110451, EPI_ISL_2110452, EPI_ISL_2110453, EPI_ISL_2110454, EPI_ISL_2110455, EPI_ISL_2110456, EPI_ISL_2110457, EPI_ISL_2110458, EPI_ISL_2110459, EPI_ISL_2110460, EPI_ISL_2110461, EPI_ISL_2110462, EPI_ISL_2110463, EPI_ISL_2110464, EPI_ISL_2110465, EPI_ISL_2110466, EPI_ISL_2110467, EPI_ISL_2110468, EPI_ISL_2110469, EPI_ISL_2110470, EPI_ISL_2110471, EPI_ISL_2110472, EPI_ISL_2110473, EPI_ISL_2110474, EPI_ISL_2110475, EPI_ISL_2110476, EPI_ISL_2110477, EPI_ISL_2110478, EPI_ISL_2110479, EPI_ISL_2110480, EPI_ISL_2110481, EPI_ISL_2110482, EPI_ISL_2110483, EPI_ISL_2110484, EPI_ISL_2110485, EPI_ISL_2110486, EPI_ISL_2110487, EPI_ISL_2110488, EPI_ISL_2110489, EPI_ISL_2110490, EPI_ISL_2110491, EPI_ISL_2110492, EPI_ISL_2110493, EPI_ISL_2110494, EPI_ISL_2110495, EPI_ISL_2110496, EPI_ISL_2110497, EPI_ISL_2110498, EPI_ISL_2110499, EPI_ISL_2110500, EPI_ISL_2110501, EPI_ISL_2110502, EPI_ISL_2110503, EPI_ISL_2110504, EPI_ISL_2110505, EPI_ISL_2110506, EPI_ISL_2110507, EPI_ISL_2110508, EPI_ISL_2110509, EPI_ISL_2110510, EPI_ISL_2110511, EPI_ISL_2110512, EPI_ISL_2110513, EPI_ISL_2110514, EPI_ISL_2110515, EPI_ISL_2110516, EPI_ISL_2110517, EPI_ISL_2110518, EPI_ISL_2110519, EPI_ISL_2110520, EPI_ISL_2110521, EPI_ISL_2110522, EPI_ISL_2110523, EPI_ISL_2110524, EPI_ISL_2110525, EPI_ISL_2110526, EPI_ISL_2110527, EPI_ISL_2110528, EPI_ISL_2110529, EPI_ISL_2110530, EPI_ISL_2110531, EPI_ISL_2110532, EPI_ISL_2110533, EPI_ISL_2110534, EPI_ISL_2110535, EPI_ISL_2110536, EPI_ISL_2110537, EPI_ISL_2110538, EPI_ISL_2110539, EPI_ISL_2110540, EPI_ISL_2110541, EPI_ISL_2110542, EPI_ISL_2110543, EPI_ISL_2110544, EPI_ISL_2110545, EPI_ISL_2110546, EPI_ISL_2110547, EPI_ISL_2110548, EPI_ISL_2110549, EPI_ISL_2110550, EPI_ISL_2110551, EPI_ISL_2110552, EPI_ISL_2110553, EPI_ISL_2110554, EPI_ISL_2110555, EPI_ISL_2110556, EPI_ISL_2110557, EPI_ISL_2110558, EPI_ISL_2110559, EPI_ISL_2110560, EPI_ISL_2110561, EPI_ISL_2110562, EPI_ISL_2110563, EPI_ISL_2110564, EPI_ISL_2110565, EPI_ISL_2110566, EPI_ISL_2110567, EPI_ISL_2110568, EPI_ISL_2110569, EPI_ISL_2110570, EPI_ISL_2110571, EPI_ISL_2110572, EPI_ISL_2110573, EPI_ISL_2110574, EPI_ISL_2110575, EPI_ISL_2110576, EPI_ISL_2110577, EPI_ISL_2110578, EPI_ISL_2110579, EPI_ISL_2110580, EPI_ISL_2110581, EPI_ISL_2110582, EPI_ISL_2110583, EPI_ISL_2110584, EPI_ISL_2110585, EPI_ISL_2110586, EPI_ISL_2110587, EPI_ISL_2110588, EPI_ISL_2110589, EPI_ISL_2110590, EPI_ISL_2110591, EPI_ISL_2110592, EPI_ISL_2110593, EPI_ISL_2110594, EPI_ISL_2110595, EPI_ISL_2110596, EPI_ISL_2110597, EPI_ISL_2110598, EPI_ISL_2110599, EPI_ISL_2110600, EPI_ISL_2110601, EPI_ISL_2110602, EPI_ISL_2110603, EPI_ISL_2110604, EPI_ISL_2110605, EPI_ISL_2110606, EPI_ISL_2110607, EPI_ISL_2110608, EPI_ISL_2110609, EPI_ISL_2110610, EPI_ISL_2110611, EPI_ISL_2110612, EPI_ISL_2110613, EPI_ISL_2110614, EPI_ISL_2110615, EPI_ISL_2110616, EPI_ISL_2110617, EPI_ISL_2110618, EPI_ISL_2110619, EPI_ISL_2110620, EPI_ISL_2110621, EPI_ISL_2110622, EPI_ISL_2110623, EPI_ISL_2110624, EPI_ISL_2110625, EPI_ISL_2110626, EPI_ISL_2110627, EPI_ISL_2110628, EPI_ISL_2110629, EPI_ISL_2110630, EPI_ISL_2110631, EPI_ISL_2110632, EPI_ISL_2110633, EPI_ISL_2110634, EPI_ISL_2110635, EPI_ISL_2110636, EPI_ISL_2110637, EPI_ISL_2110638, EPI_ISL_2110639, EPI_ISL_2110640, EPI_ISL_2110641, EPI_ISL_2110642, | | | |

| | | | |
|---|---|---|---|
| EPI_ISL_2110644, EPI_ISL_2110645, EPI_ISL_2110646, EPI_ISL_2110647, EPI_ISL_2110648, EPI_ISL_2110649, EPI_ISL_2110650, EPI_ISL_2110651, EPI_ISL_2110652, EPI_ISL_2110653, EPI_ISL_2110654, EPI_ISL_2110655, EPI_ISL_2110656, EPI_ISL_2110657, EPI_ISL_2110658, EPI_ISL_2110659, EPI_ISL_2110660, EPI_ISL_2110661, EPI_ISL_2110662, EPI_ISL_2110663, EPI_ISL_2110664, EPI_ISL_2110665, EPI_ISL_2110666, EPI_ISL_2110667, EPI_ISL_2110668, EPI_ISL_2110669, EPI_ISL_2110670, EPI_ISL_2110671, EPI_ISL_2110672, EPI_ISL_2110673, EPI_ISL_2110674, EPI_ISL_2110675, EPI_ISL_2110676, EPI_ISL_2110677, EPI_ISL_2110678, EPI_ISL_2110679, EPI_ISL_2110680, EPI_ISL_2110681, EPI_ISL_2110682, EPI_ISL_2110683, EPI_ISL_2110684, EPI_ISL_2110685, EPI_ISL_2110686, EPI_ISL_2110687, EPI_ISL_2110688, EPI_ISL_2110689, EPI_ISL_2110690, EPI_ISL_2110691, EPI_ISL_2110692, EPI_ISL_2110693, EPI_ISL_2110694, EPI_ISL_2110695, EPI_ISL_2110696, EPI_ISL_2110697, EPI_ISL_2110698, EPI_ISL_2110699, EPI_ISL_2110700, EPI_ISL_2110701, EPI_ISL_2110702, EPI_ISL_2110703, EPI_ISL_2110704, EPI_ISL_2110705, EPI_ISL_2110706, EPI_ISL_2110707, EPI_ISL_2110708, EPI_ISL_2110709, EPI_ISL_2110710, EPI_ISL_2110711, EPI_ISL_2110712, EPI_ISL_2110713, EPI_ISL_2110714, EPI_ISL_2110715, EPI_ISL_2110716, EPI_ISL_2110717, EPI_ISL_2110718, EPI_ISL_2110719, EPI_ISL_2110720, EPI_ISL_2110721, EPI_ISL_2110722, EPI_ISL_2110723, EPI_ISL_2110724, EPI_ISL_2110725, EPI_ISL_2110726, EPI_ISL_2110727, EPI_ISL_2110728, EPI_ISL_2110729, EPI_ISL_2110730, EPI_ISL_2110731, EPI_ISL_2110732, EPI_ISL_2110733, EPI_ISL_2110734, EPI_ISL_2110735, EPI_ISL_2110736, EPI_ISL_2110737, EPI_ISL_2110738, EPI_ISL_2110739, EPI_ISL_2110740, EPI_ISL_2110741, EPI_ISL_2110742, EPI_ISL_2110743, EPI_ISL_2110744, EPI_ISL_2110745, EPI_ISL_2110746, EPI_ISL_2110747, EPI_ISL_2110748, EPI_ISL_2110749, EPI_ISL_2110750, EPI_ISL_2110751, EPI_ISL_2110752, EPI_ISL_2110753, EPI_ISL_2110754, EPI_ISL_2110755, EPI_ISL_2110756, EPI_ISL_2110757, EPI_ISL_2110758, EPI_ISL_2110759, EPI_ISL_2110760 | | | |
| see above | Eurofins LifeCodexx GmbH | Robert Koch Institute | |
| EPI_ISL_1571466, EPI_ISL_1571467, EPI_ISL_1571468, EPI_ISL_1571469, EPI_ISL_1571470, EPI_ISL_1571471, EPI_ISL_1571472, EPI_ISL_1571473, EPI_ISL_1571474, EPI_ISL_1571475, EPI_ISL_1571476, EPI_ISL_1571477, EPI_ISL_1571478, EPI_ISL_1571479, EPI_ISL_1571480, EPI_ISL_1571481, EPI_ISL_1571482, EPI_ISL_1571483, EPI_ISL_1571484, EPI_ISL_1571485, EPI_ISL_1571486, EPI_ISL_1571487, EPI_ISL_1571488, EPI_ISL_1571489, EPI_ISL_1571490, EPI_ISL_1571491, EPI_ISL_1571492, EPI_ISL_1571493, EPI_ISL_1571494, EPI_ISL_1571495, EPI_ISL_1571497 | | | |
| see above | Eurofins MVZ Labor Gelsenkirchen | Robert Koch Institute | |
| EPI_ISL_1847342, EPI_ISL_1847346, EPI_ISL_1847358, EPI_ISL_1847361, EPI_ISL_1847363, EPI_ISL_1847367, EPI_ISL_1847372, EPI_ISL_1847376 | | | |
| see above | Fraunhofer-Institut fur Zelltherapie und Immunologie IZI AG Next-Generation Diagnostics | Robert Koch Institute | |
| EPI_ISL_1725156, EPI_ISL_1725157, EPI_ISL_1725158, EPI_ISL_1725159, EPI_ISL_1725160, EPI_ISL_1725161, EPI_ISL_1725162, EPI_ISL_1725163, EPI_ISL_1725164, EPI_ISL_1725165, EPI_ISL_1725166, EPI_ISL_1725167, EPI_ISL_1725168, EPI_ISL_1725169, EPI_ISL_1725170, EPI_ISL_1725171, EPI_ISL_1725172, EPI_ISL_1725173, EPI_ISL_1725174, EPI_ISL_1725175, EPI_ISL_1725176 | | | |
| see above | Fraunhofer-Institut fÃ¼r Zelltherapie und Immunologie | Robert Koch Institute | |
| EPI_ISL_1720967, EPI_ISL_1721001, EPI_ISL_1721002, EPI_ISL_1721003 | Helios UniversitÃ¤tsklinikum Wuppertal: UniversitÃ¤t Witten/Herdecke: Institut fÃ¼r Medizinische Labordiagnostik | Robert Koch Institute | |
| EPI_ISL_1643687, EPI_ISL_1643691, EPI_ISL_1643692, EPI_ISL_1643693, EPI_ISL_1643694, EPI_ISL_1643700, EPI_ISL_1643707, EPI_ISL_1643708, EPI_ISL_1643710, EPI_ISL_1643719, EPI_ISL_1643721, EPI_ISL_1643735, EPI_ISL_1643736, EPI_ISL_1643749, EPI_ISL_1722186, EPI_ISL_1722229, EPI_ISL_1722230, EPI_ISL_1722259, EPI_ISL_1844461, EPI_ISL_1844468, EPI_ISL_1844477, EPI_ISL_1844485, EPI_ISL_1844492, EPI_ISL_1844515, EPI_ISL_1844517, EPI_ISL_1844523 | | | |
| see above | IMD - Institut fÃ¼r Medizinische Diagnostik Berlin-Potsdam | Robert Koch Institute | |
| EPI_ISL_1722174, EPI_ISL_1722175, EPI_ISL_1722177, EPI_ISL_1722178, EPI_ISL_1722180, EPI_ISL_1722183, EPI_ISL_1722184, EPI_ISL_1722187, EPI_ISL_1722188, EPI_ISL_1722192, EPI_ISL_1722193, EPI_ISL_1722194, EPI_ISL_1722195, EPI_ISL_1722197, EPI_ISL_1722198, EPI_ISL_1722199, EPI_ISL_1722203, EPI_ISL_1722205, EPI_ISL_1722206, EPI_ISL_1722207, EPI_ISL_1722212, EPI_ISL_1722213, EPI_ISL_1722216, EPI_ISL_1722218, EPI_ISL_1722219, EPI_ISL_1722220, EPI_ISL_1722224, EPI_ISL_1722225, EPI_ISL_1722226, EPI_ISL_1722232, EPI_ISL_1722235, EPI_ISL_1722240, EPI_ISL_1722246, EPI_ISL_1722247, EPI_ISL_1722251, EPI_ISL_1722252, EPI_ISL_1722253, EPI_ISL_1722254, EPI_ISL_1722255, EPI_ISL_1722256, EPI_ISL_1722258, EPI_ISL_1844525, EPI_ISL_1844527, EPI_ISL_1844537, EPI_ISL_1844543, EPI_ISL_1844548, EPI_ISL_1844551, EPI_ISL_1844555, EPI_ISL_1844556, EPI_ISL_1844558, EPI_ISL_1844563, EPI_ISL_1844566, EPI_ISL_1844570, EPI_ISL_1844571, EPI_ISL_1844573, EPI_ISL_1844574, EPI_ISL_1844576, EPI_ISL_1844578, EPI_ISL_1844586, EPI_ISL_1844587, EPI_ISL_1844589, EPI_ISL_1844590, EPI_ISL_1844596, EPI_ISL_1844598, EPI_ISL_1844601, EPI_ISL_1844603, EPI_ISL_1844604, EPI_ISL_1844608 | | | |
| see above | IMD - Labor Oderland | Robert Koch Institute | |
| EPI_ISL_1643684, EPI_ISL_1643696, EPI_ISL_1643705, EPI_ISL_1643714, EPI_ISL_1643724, EPI_ISL_1643725, EPI_ISL_1643745, EPI_ISL_1643750, EPI_ISL_1844471, EPI_ISL_1844479, EPI_ISL_1844486, EPI_ISL_1844489, EPI_ISL_1844490, EPI_ISL_1844493, EPI_ISL_1844494, EPI_ISL_1844495, EPI_ISL_1844496, EPI_ISL_1844501, EPI_ISL_1844502, EPI_ISL_1844512, EPI_ISL_1844514, EPI_ISL_1844516, EPI_ISL_1844520 | | | |
| see above | IMD - MVZ Labor Greifswald | Robert Koch Institute | |
| EPI_ISL_1643729, EPI_ISL_1643737, EPI_ISL_1643753, EPI_ISL_1643757, EPI_ISL_1722176, EPI_ISL_1722179, EPI_ISL_1722185, EPI_ISL_1722191, EPI_ISL_1722200, EPI_ISL_1722201, EPI_ISL_1722202, EPI_ISL_1722208, EPI_ISL_1722211, EPI_ISL_1722217, EPI_ISL_1722233, EPI_ISL_1722241, EPI_ISL_1722242, EPI_ISL_1722243, EPI_ISL_1722248, EPI_ISL_1722249, EPI_ISL_1722250, EPI_ISL_1722257, EPI_ISL_1844462, EPI_ISL_1844464, EPI_ISL_1844466, EPI_ISL_1844467, EPI_ISL_1844473, EPI_ISL_1844478, EPI_ISL_1844484, EPI_ISL_1844508, EPI_ISL_1844510, EPI_ISL_1844513, EPI_ISL_1844521, EPI_ISL_1844522 | | | |
| see above | IMD - MVZ Labor Martinsried | Robert Koch Institute | |
| EPI_ISL_1722189, EPI_ISL_1722204, EPI_ISL_1722210, EPI_ISL_1722214, EPI_ISL_1722221, EPI_ISL_1722223, EPI_ISL_1722227, EPI_ISL_1722239, EPI_ISL_1722245, EPI_ISL_1844591 | | | |
| see above | IMD - Medizinisches Labor Rostock | Robert Koch Institute | |
| EPI_ISL_1642311, EPI_ISL_1642312, EPI_ISL_1642315, EPI_ISL_1642322, EPI_ISL_1642323, EPI_ISL_1642325, EPI_ISL_1642327, EPI_ISL_1642331, EPI_ISL_1642339, EPI_ISL_1642340, EPI_ISL_1642341, EPI_ISL_1721014, EPI_ISL_1721016, EPI_ISL_1721017, EPI_ISL_1721018, EPI_ISL_1721019, EPI_ISL_1721020, EPI_ISL_1721021, EPI_ISL_1721022, EPI_ISL_1721023, EPI_ISL_1721024, EPI_ISL_1721025, EPI_ISL_1721027, EPI_ISL_1721028, EPI_ISL_1721029, EPI_ISL_1721030, EPI_ISL_1721032, EPI_ISL_1721034, EPI_ISL_1721036, EPI_ISL_1721038, EPI_ISL_1721041, EPI_ISL_1721042, EPI_ISL_1721046, EPI_ISL_1721051, EPI_ISL_1721056, EPI_ISL_1721057, EPI_ISL_1721058, EPI_ISL_1721059, EPI_ISL_1721060, EPI_ISL_1721061, EPI_ISL_1721063, EPI_ISL_1721066, EPI_ISL_1721068, EPI_ISL_1721069, EPI_ISL_1721073, EPI_ISL_1721074, EPI_ISL_1721077, EPI_ISL_1721081, EPI_ISL_1721082, EPI_ISL_1721083, EPI_ISL_1721084, EPI_ISL_1721090, EPI_ISL_1721093, EPI_ISL_1721095, EPI_ISL_1721097, EPI_ISL_1721098, EPI_ISL_1721099, EPI_ISL_1725219, EPI_ISL_1725223, EPI_ISL_1725226, EPI_ISL_1725227, EPI_ISL_1725230, EPI_ISL_1725233, EPI_ISL_1725235, EPI_ISL_1843321 | | | |
| see above | IMD Labor Frankfurt | Robert Koch Institute | |
| EPI_ISL_2122762, EPI_ISL_2122770 | Institut fur Medizinische Mikrobiologie - Universitatsmedizin Gottingen | Robert Koch Institute | |
| EPI_ISL_1638547 | Institut fÃ¼r Immunologie und Genetik Kaiserslautern: Medizinisches Labor Dr. med. B. Thiele | Robert Koch Institute | |
| EPI_ISL_1643425 | Institut fÃ¼r Klinische Chemie Transfusions- und Laboratoriumsmedizin Berufsgenossenschaftliches UniversitÃ¤tsklinikum Bergmannsheil | Robert Koch Institute | |
| EPI_ISL_1643818 | Institut fÃ¼r Klinische Transfusionsmedizin und Immungenetik DRK Blutspendedienst Baden-WÃ¼rttemberg | Robert Koch Institute | |
| EPI_ISL_1724237, EPI_ISL_1843433, EPI_ISL_1843434, EPI_ISL_1843435, EPI_ISL_1843436, EPI_ISL_1843438, EPI_ISL_1843439, EPI_ISL_1843440, EPI_ISL_1843441, EPI_ISL_1843442 | | | |
| see above | Institut fÃ¼r Medizinische Laboratoriumsdiagnostik Bochum GmbH | Robert Koch Institute | |
| EPI_ISL_1643883 | Institut fÃ¼r Medizinische Mikrobiologie - UniversitÃ¤tsmedizin GÃ¶ttingen | Robert Koch Institute | |
| EPI_ISL_1724441, EPI_ISL_1846339, EPI_ISL_1846340, EPI_ISL_1846342 | Institut fÃ¼r med. Mikrobiologie u. Virologie Klinikum Oldenburg | Robert Koch Institute | |
| EPI_ISL_1670418, EPI_ISL_1670419, EPI_ISL_1670420, EPI_ISL_1670421, EPI_ISL_1670422, EPI_ISL_1670423, EPI_ISL_1670424, EPI_ISL_1670425, EPI_ISL_1670426, EPI_ISL_1670465, EPI_ISL_1670466, EPI_ISL_1670467, EPI_ISL_1670468, EPI_ISL_1670469, EPI_ISL_1670470, EPI_ISL_1670471, EPI_ISL_1670472, EPI_ISL_1670473, EPI_ISL_1670474, EPI_ISL_1670475, EPI_ISL_1670476, EPI_ISL_1670477, EPI_ISL_1670478, EPI_ISL_1670479, EPI_ISL_1670480, EPI_ISL_1670481, EPI_ISL_1670482, EPI_ISL_1670483, EPI_ISL_1670484, EPI_ISL_1670485, EPI_ISL_1670486, EPI_ISL_1670487, EPI_ISL_1670490, EPI_ISL_1670491, EPI_ISL_1670499, EPI_ISL_1670537, EPI_ISL_1670538, EPI_ISL_1670539, EPI_ISL_1670540, EPI_ISL_1670541, EPI_ISL_1670542, EPI_ISL_1670543, EPI_ISL_1670544, EPI_ISL_1670545, EPI_ISL_1670546, EPI_ISL_1670547, EPI_ISL_1670548, EPI_ISL_1940757, EPI_ISL_1940758, EPI_ISL_1940759, EPI_ISL_1940760, EPI_ISL_1940761, EPI_ISL_1940762 | | | |
| see above | Institute of Medical Microbiology and Hospital Hygiene | Institute of Medical Microbiology and Hospital Hygiene | Aljoscha Tersteegen; Prof. Dr. Achim Kaasch |
| EPI_ISL_1648934, EPI_ISL_1648935, EPI_ISL_1648938, EPI_ISL_1648939, EPI_ISL_1653694 | Institute of Tropical Medicine | Institute of Tropical Medicine | Prof. Dr. Thirumalaisamy P. Velavan and Prof. Dr. Peter Kremsner |
| EPI_ISL_1805025, EPI_ISL_1805036, EPI_ISL_1805038, EPI_ISL_1805039, EPI_ISL_1805040, EPI_ISL_1805041, EPI_ISL_1805055, EPI_ISL_1805056 | | | |
| see above | Institute of Virology, Medical Center, University of Freiburg, Freiburg, Germany | Institute of Virology, Clinial Virus Genomics, Medical Center, University of Freiburg, Freiburg, Germany | Hajo Grundmann; Jonas Fuchs; Lena Jaki; Lisa Kern; Marcus Panning; Sandra Reuter |
| EPI_ISL_1722360, EPI_ISL_1722363, EPI_ISL_1722364, EPI_ISL_1722367 | KRH Klinikum Region Hannover: Institut fÃ¼r Med. Mikrobiologie und Krankenhaushygiene | Robert Koch Institute | |
| EPI_ISL_1715849, EPI_ISL_1715851, EPI_ISL_1715854, EPI_ISL_1715855, EPI_ISL_1715856, EPI_ISL_1715857, EPI_ISL_1862397, EPI_ISL_1862399, EPI_ISL_1862401, EPI_ISL_1862406, EPI_ISL_1862410 | | | |
| see above | Kath. Marienkrankenhaus | Heinrich Pette Institute, Leibniz Institute for Experimental Virology | Adam Grundhoff; Alexis Robitaille; Johannes Knobloch; Martin Aepfelbacher; Nicole Fischer; Thomas Günther |
| EPI_ISL_1643484 | Kath. St.-Johannes-Gesellschaft Dortmund gGmbH | Robert Koch Institute | |
| EPI_ISL_1638393, EPI_ISL_1643763, EPI_ISL_1643859, EPI_ISL_1643866, EPI_ISL_1724423, EPI_ISL_1724428 | Kerckhoff-Klinik GmbH | Robert Koch Institute | |
| EPI_ISL_1724316 | Klinikum Darmstadt GmbH | Robert Koch Institute | |
| EPI_ISL_1722261, EPI_ISL_1722262, EPI_ISL_1722263, EPI_ISL_1722265, EPI_ISL_1722267, EPI_ISL_1722268, EPI_ISL_1722269, EPI_ISL_1722271, EPI_ISL_1722273, EPI_ISL_1722274, EPI_ISL_1722276, EPI_ISL_1722277, EPI_ISL_1722931, EPI_ISL_1722932, EPI_ISL_1722933, EPI_ISL_1722934, EPI_ISL_1722935, EPI_ISL_1722936, EPI_ISL_1724460, EPI_ISL_1724461, EPI_ISL_1724462, EPI_ISL_1724464, EPI_ISL_1724465, EPI_ISL_1724466, EPI_ISL_1724468, EPI_ISL_1724469, EPI_ISL_1724470, EPI_ISL_1724472, EPI_ISL_1724477 | | | |
| see above | Klinikum Ernst von Bergmann gemeinnÃ¼tzige GmbH - stationÃ¤rer Bereich | Robert Koch Institute | |
| EPI_ISL_1847158 | Klinikum Fulda gAG | Robert Koch Institute | |
| EPI_ISL_1643880, EPI_ISL_1643881 | Krankenhausgesellschaft St. Vincenz mbH | Robert Koch Institute | |
| EPI_ISL_1722108, EPI_ISL_1722109, EPI_ISL_1722111, EPI_ISL_1722117, EPI_ISL_1722119, EPI_ISL_1724332, EPI_ISL_1724334, EPI_ISL_1724335, EPI_ISL_1724348, EPI_ISL_1724354, EPI_ISL_1724362, EPI_ISL_1724370, EPI_ISL_1724388, EPI_ISL_1846364, EPI_ISL_1846367, EPI_ISL_1846373, EPI_ISL_1846377 | | | |
| see above | LABOR CHEMNITZ â€“ Zentrum fÃ¼r Diagnostik am Klinikum Chemnitz | Robert Koch Institute | |
| EPI_ISL_1721390, EPI_ISL_1721400, EPI_ISL_1721405, EPI_ISL_1721407, EPI_ISL_1721410, EPI_ISL_1721413, EPI_ISL_1721414, EPI_ISL_1721415, EPI_ISL_1721421, EPI_ISL_1721422, EPI_ISL_1721427, EPI_ISL_1721430, EPI_ISL_1721432, EPI_ISL_1721435, EPI_ISL_1721440, EPI_ISL_1721441, EPI_ISL_1721443, EPI_ISL_1721446, EPI_ISL_1721449, EPI_ISL_1721451, EPI_ISL_1721452, EPI_ISL_1721456, EPI_ISL_1721464, EPI_ISL_1721466, EPI_ISL_1721467, EPI_ISL_1721471, EPI_ISL_1721481, EPI_ISL_1721484, EPI_ISL_1721492, EPI_ISL_1721493, EPI_ISL_1721499, EPI_ISL_1721500, EPI_ISL_1721502, EPI_ISL_1721508, EPI_ISL_1721509, EPI_ISL_1721510, EPI_ISL_1721513, EPI_ISL_1721514, EPI_ISL_1721517, EPI_ISL_1721518, EPI_ISL_1721522, EPI_ISL_1721523, EPI_ISL_1721529, EPI_ISL_1721530, EPI_ISL_1721532, EPI_ISL_1721536, EPI_ISL_1721539, EPI_ISL_1721540, EPI_ISL_1721542, EPI_ISL_1721543, EPI_ISL_1721554, EPI_ISL_1721555, EPI_ISL_1721561, EPI_ISL_1721573, EPI_ISL_1721577, EPI_ISL_1721578, EPI_ISL_1721580, EPI_ISL_1721582, EPI_ISL_1721593, EPI_ISL_1721594, EPI_ISL_1721596, EPI_ISL_1721606, EPI_ISL_1721617, EPI_ISL_1721618, EPI_ISL_1721634, EPI_ISL_1721636, EPI_ISL_1721639, EPI_ISL_1721646, EPI_ISL_1721651, EPI_ISL_1721653, EPI_ISL_1721654, EPI_ISL_1721665, EPI_ISL_1721666, EPI_ISL_1721674, EPI_ISL_1721680, EPI_ISL_1721687, EPI_ISL_1721689, EPI_ISL_1721691, EPI_ISL_1721692, EPI_ISL_1721694, EPI_ISL_1721696, EPI_ISL_1721701, EPI_ISL_1721703, EPI_ISL_1721704, EPI_ISL_1721715, EPI_ISL_1721719, EPI_ISL_1721723, EPI_ISL_1721726, EPI_ISL_1721728, EPI_ISL_1721736, EPI_ISL_1721737, EPI_ISL_1721739, EPI_ISL_1721743, EPI_ISL_1721744, EPI_ISL_1721745, EPI_ISL_1721752, EPI_ISL_1721754, EPI_ISL_1721759, EPI_ISL_1721760, EPI_ISL_1721763, EPI_ISL_1721768, EPI_ISL_1721769, EPI_ISL_1721780, EPI_ISL_1721786, EPI_ISL_1721788, EPI_ISL_1721798, EPI_ISL_1721801, EPI_ISL_1721803, EPI_ISL_1721808, EPI_ISL_1721816, EPI_ISL_1721822, EPI_ISL_1721827, EPI_ISL_1721834, EPI_ISL_1721837, EPI_ISL_1721841, EPI_ISL_1721842, EPI_ISL_1721844, EPI_ISL_1721852, EPI_ISL_1721856, EPI_ISL_1721861, EPI_ISL_1721863, EPI_ISL_1721864, EPI_ISL_1721866, EPI_ISL_1721873, EPI_ISL_1721881, EPI_ISL_1721883, EPI_ISL_1721887, EPI_ISL_1721888, EPI_ISL_1721892, EPI_ISL_1721897, EPI_ISL_1721903, EPI_ISL_1721904, EPI_ISL_1721906, EPI_ISL_1721908, EPI_ISL_1721915, EPI_ISL_1721924, EPI_ISL_1721925, EPI_ISL_1721929, EPI_ISL_1721932, EPI_ISL_1721938, EPI_ISL_1721940, EPI_ISL_1721942, EPI_ISL_1721948, EPI_ISL_1721955, EPI_ISL_1721957, EPI_ISL_1721958, EPI_ISL_1721959, EPI_ISL_1721964, EPI_ISL_1721968, EPI_ISL_1721972, EPI_ISL_1721974, EPI_ISL_1847157, EPI_ISL_1847162 | | | |
| see above | LabKom - Labor Augsburg MVZ GmbH | Robert Koch Institute | |
| EPI_ISL_1721815, EPI_ISL_1721859, EPI_ISL_1721934, EPI_ISL_1721946 | LabKom - Labor Hamburg-LÃ¼beck | Robert Koch Institute | |
| EPI_ISL_1721401, EPI_ISL_1721524, EPI_ISL_1721591, EPI_ISL_1721614, EPI_ISL_1721698, EPI_ISL_1721705, EPI_ISL_1721741, EPI_ISL_1721742, EPI_ISL_1721771, EPI_ISL_1721777, EPI_ISL_1721783, EPI_ISL_1721797, EPI_ISL_1721813, EPI_ISL_1721830, EPI_ISL_1721848, EPI_ISL_1721865, EPI_ISL_1721879, EPI_ISL_1721880, EPI_ISL_1721882, EPI_ISL_1721911, EPI_ISL_1721912, EPI_ISL_1721916, EPI_ISL_1721919, EPI_ISL_1721922, EPI_ISL_1721933, EPI_ISL_1721936, EPI_ISL_1721937, EPI_ISL_1721971, EPI_ISL_1721976, EPI_ISL_2108763, EPI_ISL_2108854, EPI_ISL_2108923 | | | |
| see above | LabKom - Labor Mainz MVZ GmbH | Robert Koch Institute | |
| EPI_ISL_1721695, EPI_ISL_1721697, EPI_ISL_1721708, EPI_ISL_1721711, EPI_ISL_1721712, EPI_ISL_1721713, EPI_ISL_1721720, EPI_ISL_1721722, EPI_ISL_1721724, EPI_ISL_1721730, EPI_ISL_1721731, EPI_ISL_1721758, EPI_ISL_1721779, EPI_ISL_1721781, EPI_ISL_1721805, EPI_ISL_1721831, EPI_ISL_1721846, EPI_ISL_1721851, EPI_ISL_1721855, EPI_ISL_1721872, EPI_ISL_1721878, EPI_ISL_1721884, EPI_ISL_1721885, EPI_ISL_1721909, EPI_ISL_1721910, EPI_ISL_1721913, EPI_ISL_1721914, EPI_ISL_1721917, EPI_ISL_1721920, EPI_ISL_1721928, EPI_ISL_1721949, EPI_ISL_1721962, EPI_ISL_1721965, EPI_ISL_1721966, EPI_ISL_1721967, EPI_ISL_1721970, EPI_ISL_1721975, EPI_ISL_1842967, EPI_ISL_1843105, EPI_ISL_1843154, EPI_ISL_1843164, EPI_ISL_1844049, EPI_ISL_1844069, EPI_ISL_1844096, EPI_ISL_1844101, EPI_ISL_1844134, EPI_ISL_1844141, EPI_ISL_1844167, EPI_ISL_1844172 | | | |
| see above | LabKom - Labor an der SalzbrÃ¼cke MVZ GmbH | Robert Koch Institute | |
| EPI_ISL_1842781, EPI_ISL_1842782, EPI_ISL_1842784, EPI_ISL_1842786, EPI_ISL_1842800, EPI_ISL_1842803, EPI_ISL_1842804, EPI_ISL_1842805, EPI_ISL_1842810, EPI_ISL_1842811, EPI_ISL_1842826, EPI_ISL_1842830, EPI_ISL_1842831, EPI_ISL_1842834, EPI_ISL_1842839, EPI_ISL_1842843, EPI_ISL_1842844, EPI_ISL_1842848, EPI_ISL_1842851, EPI_ISL_1842852, EPI_ISL_1842853, EPI_ISL_1842854, EPI_ISL_1842858, EPI_ISL_1842859, EPI_ISL_1842864, EPI_ISL_1842880, EPI_ISL_1842887, EPI_ISL_1842894, EPI_ISL_1842896, EPI_ISL_1842897, EPI_ISL_1842898, EPI_ISL_1842901, EPI_ISL_1842912, EPI_ISL_1842914, EPI_ISL_1842915, EPI_ISL_1842917, EPI_ISL_1842922, EPI_ISL_1842923, EPI_ISL_1842924, EPI_ISL_1842932, EPI_ISL_1842936, EPI_ISL_1842942, EPI_ISL_1842954, EPI_ISL_1842955, | | | |

| | | | |
|---|---|---|---|
| EPI_ISL_1842959, EPI_ISL_1842964, EPI_ISL_1844256 | | | |
| see above | LabKom - MVZ Labor Bochum MLB GmbH | Robert Koch Institute | |
| EPI_ISL_1843042, EPI_ISL_1843088, EPI_ISL_1843099, EPI_ISL_1843137, EPI_ISL_1843171, EPI_ISL_1843464, EPI_ISL_1843484, EPI_ISL_1843496, EPI_ISL_1843509, EPI_ISL_1843522, EPI_ISL_1843531, EPI_ISL_1843543, EPI_ISL_1843607, EPI_ISL_1843674, EPI_ISL_1843677, EPI_ISL_1843713, EPI_ISL_1843724, EPI_ISL_1847105, EPI_ISL_1847116, EPI_ISL_1847127, EPI_ISL_2109052, EPI_ISL_2109118, EPI_ISL_2109251, EPI_ISL_2109291, EPI_ISL_2109303, EPI_ISL_2109316, EPI_ISL_2109338, EPI_ISL_2109352 | | | |
| see above | Labor 28 MVZ GmbH | Robert Koch Institute | |
| EPI_ISL_2123978 | Labor Becker & Kollegen (Standort Munchen) | Robert Koch Institute | |
| EPI_ISL_1885867, EPI_ISL_1885870, EPI_ISL_1885876, EPI_ISL_1885882, EPI_ISL_1885885, EPI_ISL_1885887, EPI_ISL_1885902, EPI_ISL_1885927, EPI_ISL_1885930, EPI_ISL_1885956, EPI_ISL_1885961, EPI_ISL_1885964, EPI_ISL_1886001, EPI_ISL_1886101, EPI_ISL_1886114, EPI_ISL_1886130, EPI_ISL_1886140, EPI_ISL_1886163, EPI_ISL_1886193, EPI_ISL_1886196, EPI_ISL_1886198, EPI_ISL_1905061, EPI_ISL_1905062, EPI_ISL_1905065, EPI_ISL_1905066, EPI_ISL_1905068, EPI_ISL_1905069, EPI_ISL_1905070, EPI_ISL_1905072, EPI_ISL_1905073, EPI_ISL_1905074, EPI_ISL_1905076, EPI_ISL_1905078, EPI_ISL_1905081, EPI_ISL_1905082, EPI_ISL_1905083, EPI_ISL_1905084, EPI_ISL_1905086, EPI_ISL_1905087, EPI_ISL_1905088, EPI_ISL_1905090, EPI_ISL_1905091, EPI_ISL_1905092, EPI_ISL_1905094, EPI_ISL_1905095, EPI_ISL_1905096, EPI_ISL_1905098, EPI_ISL_1905099, EPI_ISL_1905100, EPI_ISL_1905102, EPI_ISL_1905105, EPI_ISL_1905106, EPI_ISL_1905110, EPI_ISL_1905111, EPI_ISL_1905112, EPI_ISL_1905113, EPI_ISL_1905114, EPI_ISL_1905116, EPI_ISL_1905117, EPI_ISL_1905118, EPI_ISL_1905119, EPI_ISL_1905120, EPI_ISL_1905136, EPI_ISL_1905145, EPI_ISL_1905154, EPI_ISL_1905173, EPI_ISL_1905179, EPI_ISL_1905180 | | | |
| see above | Labor Berlin Charité Vivantes GmbH / Institut für Virologie | Charité Universitätsmedizin Berlin, Institut für Virologie/Labor Berlin | Barbara Mühlemann; Christian Drosten; Christine Stephan; Peter Menzel; Rolf Schwarzer; Terry Jones; Victor M Corman |
| EPI_ISL_1862052, EPI_ISL_1862054, EPI_ISL_1862056, EPI_ISL_1862058, EPI_ISL_1862060, EPI_ISL_1862067, EPI_ISL_1862069, EPI_ISL_1862071, EPI_ISL_1862073, EPI_ISL_1862075, EPI_ISL_1862077, EPI_ISL_1862084, EPI_ISL_1862088, EPI_ISL_1862090, EPI_ISL_1862092, EPI_ISL_1862094, EPI_ISL_1862101, EPI_ISL_1862105, EPI_ISL_1862109, EPI_ISL_1862111, EPI_ISL_1862113, EPI_ISL_1862115, EPI_ISL_1862117, EPI_ISL_1862121, EPI_ISL_1862127, EPI_ISL_1862129, EPI_ISL_1862133, EPI_ISL_1862136, EPI_ISL_1862137, EPI_ISL_1862140, EPI_ISL_1862141, EPI_ISL_1862146, EPI_ISL_1862148, EPI_ISL_1862150, EPI_ISL_1862152, EPI_ISL_1862154, EPI_ISL_1862156, EPI_ISL_1862158, EPI_ISL_1862161, EPI_ISL_1862162, EPI_ISL_1862167, EPI_ISL_1862177, EPI_ISL_1862179, EPI_ISL_1862181, EPI_ISL_1862187, EPI_ISL_1862192, EPI_ISL_1862193, EPI_ISL_1862196, EPI_ISL_1862197, EPI_ISL_1862199, EPI_ISL_1862201, EPI_ISL_1862203, EPI_ISL_1862206, EPI_ISL_1862208, EPI_ISL_1862212, EPI_ISL_1862220, EPI_ISL_1862241, EPI_ISL_1862244, EPI_ISL_1862245, EPI_ISL_1862248, EPI_ISL_1862252, EPI_ISL_1862258, EPI_ISL_1862260, EPI_ISL_1862262, EPI_ISL_1862275, EPI_ISL_1862278, EPI_ISL_1862280, EPI_ISL_1862285, EPI_ISL_1862301, EPI_ISL_1862303, EPI_ISL_1862305, EPI_ISL_1862323, EPI_ISL_1862325, EPI_ISL_1862327, EPI_ISL_1862340, EPI_ISL_1862342, EPI_ISL_1862349, EPI_ISL_1862358, EPI_ISL_1862360, EPI_ISL_1862362, EPI_ISL_1862364, EPI_ISL_1862366 | | | |
| see above | Labor Dr. Fenner und Kollegen | Heinrich Pette Institute, Leibniz Institute for Experimental Virology | Adam Grundhoff; Alexis Robitaille; Johannes Knobloch; Martin Aepfelbacher; Nicole Fischer; Thomas Günther |
| EPI_ISL_1723016, EPI_ISL_1723017, EPI_ISL_1723018, EPI_ISL_1723019, EPI_ISL_1723020, EPI_ISL_1723021, EPI_ISL_1723022, EPI_ISL_1723023, EPI_ISL_1723024, EPI_ISL_1723026, EPI_ISL_1723027, EPI_ISL_1723028, EPI_ISL_1723029, EPI_ISL_1723030, EPI_ISL_1723031, EPI_ISL_1723032, EPI_ISL_1723033, EPI_ISL_1723034, EPI_ISL_1723036, EPI_ISL_1723038, EPI_ISL_1723039, EPI_ISL_1723040, EPI_ISL_1723041, EPI_ISL_1723042, EPI_ISL_1723043, EPI_ISL_1723044, EPI_ISL_1723045, EPI_ISL_1723046, EPI_ISL_1723047, EPI_ISL_1723048, EPI_ISL_1723049, EPI_ISL_1723050, EPI_ISL_1723051, EPI_ISL_1723052, EPI_ISL_1723054, EPI_ISL_1723055, EPI_ISL_1723056, EPI_ISL_1723057, EPI_ISL_1723058, EPI_ISL_1723059, EPI_ISL_1723060, EPI_ISL_1723062, EPI_ISL_1727103, EPI_ISL_1727107 | | | |
| see above | Labor Dr. Heidrich & Kollegen MVZ GmbH Hamburg | Robert Koch Institute | |
| EPI_ISL_1842795, EPI_ISL_1842798, EPI_ISL_1842799, EPI_ISL_1842801, EPI_ISL_1842808, EPI_ISL_1842809, EPI_ISL_1842823, EPI_ISL_1842840, EPI_ISL_1842882, EPI_ISL_1842889, EPI_ISL_1842895, EPI_ISL_1842911, EPI_ISL_1842913, EPI_ISL_1842925, EPI_ISL_1842927, EPI_ISL_1842937, EPI_ISL_1842947, EPI_ISL_1842952, EPI_ISL_1842962, EPI_ISL_1847095, EPI_ISL_1847106, EPI_ISL_1847113, EPI_ISL_1847115, EPI_ISL_1847140, EPI_ISL_1847145, EPI_ISL_1847148, EPI_ISL_1847149, EPI_ISL_1847151, EPI_ISL_1847152, EPI_ISL_1847159, EPI_ISL_1847160, EPI_ISL_1847161, EPI_ISL_2115683, EPI_ISL_2115779, EPI_ISL_2120678, EPI_ISL_2121339 | | | |
| see above | Labor Dr. Schumacher MVZ | Robert Koch Institute | |
| EPI_ISL_1847403, EPI_ISL_1847404, EPI_ISL_1847406, EPI_ISL_1847415, EPI_ISL_1847418, EPI_ISL_1847419, EPI_ISL_1847420, EPI_ISL_1847421, EPI_ISL_1847423, EPI_ISL_1847424 | | | |
| see above | Labor Dr. Spranger | Robert Koch Institute | |
| EPI_ISL_1643494, EPI_ISL_1643495, EPI_ISL_1643496, EPI_ISL_1643497, EPI_ISL_1643498, EPI_ISL_1643499, EPI_ISL_1643500, EPI_ISL_1643501, EPI_ISL_1643502, EPI_ISL_1643503, EPI_ISL_1643504, EPI_ISL_1643505, EPI_ISL_1643506, EPI_ISL_1643507, EPI_ISL_1643508, EPI_ISL_1643509, EPI_ISL_1643510, EPI_ISL_1643511, EPI_ISL_1725186, EPI_ISL_1725189 | | | |
| see above | Labor Dr. Wisplinghoff - KÃ¶ln | Robert Koch Institute | |
| EPI_ISL_1643968, EPI_ISL_1726865, EPI_ISL_1851136, EPI_ISL_1851138 | Labor KneiÃŸler GmbH & Co. KG | Robert Koch Institute | |
| EPI_ISL_1643418, EPI_ISL_1643420, EPI_ISL_1643421, EPI_ISL_1643429, EPI_ISL_1643473, EPI_ISL_1643477, EPI_ISL_1720946, EPI_ISL_1720948, EPI_ISL_1720979, EPI_ISL_1720981, EPI_ISL_1720992, EPI_ISL_1726788, EPI_ISL_1726792, EPI_ISL_1726807, EPI_ISL_1726828, EPI_ISL_1726834, EPI_ISL_1726836 | | | |
| see above | Labor LÃ¼beck bzw. LaborÃ¤rztliche Gemeinschaftspraxis LÃ¼beck | Robert Koch Institute | |
| EPI_ISL_1643775, EPI_ISL_1643788, EPI_ISL_1643789, EPI_ISL_1643790, EPI_ISL_1643791, EPI_ISL_1643792, EPI_ISL_1643793, EPI_ISL_1643794, EPI_ISL_1643795, EPI_ISL_1643796, EPI_ISL_1643797, EPI_ISL_1643798, EPI_ISL_1643799, EPI_ISL_1643800, EPI_ISL_1643802, EPI_ISL_1643803, EPI_ISL_1643804, EPI_ISL_1643805, EPI_ISL_1643806, EPI_ISL_1643807, EPI_ISL_1643808, EPI_ISL_1643809, EPI_ISL_1643810, EPI_ISL_1643811, EPI_ISL_1643813, EPI_ISL_1643814, EPI_ISL_1643815, EPI_ISL_1643816, EPI_ISL_1643817, EPI_ISL_1724262, EPI_ISL_1725190, EPI_ISL_1725191, EPI_ISL_1725199, EPI_ISL_1725208, EPI_ISL_1725209, EPI_ISL_1725211, EPI_ISL_1725213, EPI_ISL_1725216 | | | |
| see above | Labor MÃ¶nchengladbach MVZ Dr. Stein + Kollegen GbR | Robert Koch Institute | |
| EPI_ISL_1724308, EPI_ISL_1724310, EPI_ISL_1724313, EPI_ISL_1724314, EPI_ISL_1724325, EPI_ISL_1843336, EPI_ISL_1843342 | | | |
| see above | Labor Prof. Dr. G. Enders MVZ GbR | Robert Koch Institute | |
| EPI_ISL_2111579 | Labor ZOTZ\|KLIMAS; MVZ Dusseldorf-Centrum | Robert Koch Institute | |
| EPI_ISL_1725070, EPI_ISL_1725071, EPI_ISL_1725073, EPI_ISL_1725074, EPI_ISL_1725075, EPI_ISL_1725076, EPI_ISL_1725077, EPI_ISL_1725078, EPI_ISL_1725079, EPI_ISL_1725080, EPI_ISL_1725081, EPI_ISL_1725082, EPI_ISL_1725083, EPI_ISL_1725084, EPI_ISL_1725085, EPI_ISL_1725086, EPI_ISL_1725087, EPI_ISL_1725088, EPI_ISL_1725089, EPI_ISL_1725090, EPI_ISL_1725091, EPI_ISL_1725092, EPI_ISL_1725093, EPI_ISL_1725094, EPI_ISL_1725095, EPI_ISL_1725096, EPI_ISL_1725097, EPI_ISL_1725098, EPI_ISL_1725099, EPI_ISL_1725100, EPI_ISL_1725101, EPI_ISL_1725102, EPI_ISL_1725103, EPI_ISL_1725104, EPI_ISL_1725106, EPI_ISL_1725107, EPI_ISL_1725108, EPI_ISL_1725110, EPI_ISL_1725111, EPI_ISL_1725114, EPI_ISL_1725115, EPI_ISL_1725116, EPI_ISL_1725118, EPI_ISL_1725119, EPI_ISL_1725120, EPI_ISL_1725121, EPI_ISL_1725122, EPI_ISL_1725123, EPI_ISL_1725124, EPI_ISL_1725125, EPI_ISL_1725126, EPI_ISL_1725127, EPI_ISL_1725128, EPI_ISL_1725129, EPI_ISL_1725130, EPI_ISL_1725131, EPI_ISL_1725132, EPI_ISL_1725133, EPI_ISL_1725134, EPI_ISL_1725135, EPI_ISL_1725136, EPI_ISL_1725137, EPI_ISL_1725138, EPI_ISL_1725139, EPI_ISL_1725140, EPI_ISL_1725141, EPI_ISL_1725142, EPI_ISL_1725143, EPI_ISL_1725145, EPI_ISL_1725147, EPI_ISL_1725148, EPI_ISL_1725149, EPI_ISL_1725150, EPI_ISL_1725151, EPI_ISL_1725152, EPI_ISL_1725153, EPI_ISL_1725155, EPI_ISL_1851152, EPI_ISL_1851155, EPI_ISL_1851156, EPI_ISL_1851158, EPI_ISL_1851160, EPI_ISL_1851161, EPI_ISL_1851162, EPI_ISL_1851163, EPI_ISL_1851166, EPI_ISL_1851168, EPI_ISL_1851170, EPI_ISL_1851173, EPI_ISL_1851179, EPI_ISL_1851180, EPI_ISL_1851181, EPI_ISL_1851184, EPI_ISL_1851187, EPI_ISL_1851191, EPI_ISL_1851193, EPI_ISL_1851197, EPI_ISL_1851202, EPI_ISL_1851203, EPI_ISL_1851204, EPI_ISL_1851205, EPI_ISL_1851211, EPI_ISL_1851212, EPI_ISL_1851214, EPI_ISL_1851216, EPI_ISL_1851218, EPI_ISL_1851219, EPI_ISL_1851221, EPI_ISL_1851228, EPI_ISL_1851231, EPI_ISL_1851232, EPI_ISL_1851236, EPI_ISL_1851237, EPI_ISL_1851240, EPI_ISL_1851242, EPI_ISL_1851243, EPI_ISL_1851244, EPI_ISL_1851245, EPI_ISL_1851246, EPI_ISL_1851247, EPI_ISL_1851249, EPI_ISL_1851250, EPI_ISL_1851255, EPI_ISL_1851258, EPI_ISL_1851260, EPI_ISL_1851264, EPI_ISL_1851265, EPI_ISL_1851266, EPI_ISL_1851267, EPI_ISL_1851270, EPI_ISL_1851272, EPI_ISL_1851275, EPI_ISL_1851278, EPI_ISL_1851279, EPI_ISL_1851283, EPI_ISL_1851287, EPI_ISL_1851289, EPI_ISL_1851292, EPI_ISL_1851299, EPI_ISL_1851302, EPI_ISL_1851303, EPI_ISL_1851304, EPI_ISL_1851307, EPI_ISL_1851311, EPI_ISL_1851316, EPI_ISL_1851318, EPI_ISL_1851321, EPI_ISL_1851323, EPI_ISL_1851325, EPI_ISL_1851329, EPI_ISL_1851330, EPI_ISL_1851332, EPI_ISL_1851334, EPI_ISL_1851335, EPI_ISL_1851339, EPI_ISL_1851340, EPI_ISL_1851341, EPI_ISL_1851346, EPI_ISL_1851347, EPI_ISL_1851350, EPI_ISL_1851355, EPI_ISL_1851358, EPI_ISL_1851359, EPI_ISL_1851360, EPI_ISL_1851362, EPI_ISL_1851363, EPI_ISL_1851365, EPI_ISL_1851368, EPI_ISL_1851369, EPI_ISL_1851372, EPI_ISL_1851374, EPI_ISL_1851375, EPI_ISL_1851377, EPI_ISL_1851379, EPI_ISL_1851380, EPI_ISL_1851383, EPI_ISL_1851386 | | | |
| see above | Labor ZOTZ\|KLIMAS; MVZ DÃ¼sseldorf-Centrum | Robert Koch Institute | |
| EPI_ISL_1644500, EPI_ISL_1644505, EPI_ISL_1722515, EPI_ISL_1722588, EPI_ISL_1722598, EPI_ISL_1722664 | Labor fÃ¼r medizinische Mikrobiologie Bakteriologie Mykologie Virologie & Infektionsserologie - Labor MÃ¶lbis | Robert Koch Institute | |
| EPI_ISL_1644004, EPI_ISL_1644005, EPI_ISL_1644006, EPI_ISL_1644007, EPI_ISL_1644008, EPI_ISL_1644009, EPI_ISL_1644010, EPI_ISL_1644011, EPI_ISL_1644012, EPI_ISL_1644021, EPI_ISL_1644022, EPI_ISL_1644024, EPI_ISL_1644025, EPI_ISL_1644026, EPI_ISL_1644027, EPI_ISL_1644028, EPI_ISL_1644029, EPI_ISL_1644030, EPI_ISL_1644031, EPI_ISL_1644032, EPI_ISL_1644033, EPI_ISL_1644034, EPI_ISL_1644035, EPI_ISL_1644036, EPI_ISL_1644037, EPI_ISL_1644038, EPI_ISL_1644039, EPI_ISL_1644040, EPI_ISL_1644041, EPI_ISL_1644043, EPI_ISL_1644044 | | | |
| see above | Laborarztpraxis Dres. med. Walther Weindel & Kollegen | Robert Koch Institute | |
| EPI_ISL_1638282, EPI_ISL_1638334, EPI_ISL_1638338, EPI_ISL_1638340, EPI_ISL_1638357, EPI_ISL_1638360, EPI_ISL_1638414, EPI_ISL_1638485 | | | |
| see above | Laborarztpraxis Jochen HÃ¼ter | Robert Koch Institute | |
| EPI_ISL_1642313, EPI_ISL_1642314, EPI_ISL_1642328, EPI_ISL_1642329, EPI_ISL_1642330, EPI_ISL_1642333, EPI_ISL_1642334, EPI_ISL_1642337, EPI_ISL_1642350, EPI_ISL_1721013, EPI_ISL_1721026, EPI_ISL_1721039, EPI_ISL_1721048, EPI_ISL_1721055, EPI_ISL_1721062, EPI_ISL_1721072, EPI_ISL_1721080, EPI_ISL_1721086, EPI_ISL_1721101, EPI_ISL_1721102, EPI_ISL_1725222, EPI_ISL_1725232 | | | |
| see above | Labormedizin Darmstadt | Robert Koch Institute | |
| EPI_ISL_1670429, EPI_ISL_1670430, EPI_ISL_1670431, EPI_ISL_1670432, EPI_ISL_1670433, EPI_ISL_1670434, EPI_ISL_1670435, EPI_ISL_1670436, EPI_ISL_1670437, EPI_ISL_1670438, EPI_ISL_1670439, EPI_ISL_1670440, EPI_ISL_1670441, EPI_ISL_1670442, EPI_ISL_1670443, EPI_ISL_1670444, EPI_ISL_1670445, EPI_ISL_1670446, EPI_ISL_1670447, EPI_ISL_1670448, EPI_ISL_1670449, EPI_ISL_1670450, EPI_ISL_1670451, EPI_ISL_1670452, EPI_ISL_1670453, EPI_ISL_1670454, EPI_ISL_1670455, EPI_ISL_1670456, EPI_ISL_1670457, EPI_ISL_1670458, EPI_ISL_1670459, EPI_ISL_1670460, EPI_ISL_1670461, EPI_ISL_1670462, EPI_ISL_1670463, EPI_ISL_1670464, EPI_ISL_1670488, EPI_ISL_1670489, EPI_ISL_1670492, EPI_ISL_1670493, EPI_ISL_1670494, EPI_ISL_1670495, EPI_ISL_1670496, EPI_ISL_1670497, EPI_ISL_1670498, EPI_ISL_1670500, EPI_ISL_1670501, EPI_ISL_1670502, EPI_ISL_1670503, EPI_ISL_1670504, EPI_ISL_1670505, EPI_ISL_1670506, EPI_ISL_1670507, EPI_ISL_1670508, EPI_ISL_1670509, EPI_ISL_1670510, EPI_ISL_1670511, EPI_ISL_1670512, EPI_ISL_1670513, EPI_ISL_1670514, EPI_ISL_1670515, EPI_ISL_1670516, EPI_ISL_1670517, EPI_ISL_1670518, EPI_ISL_1670519, EPI_ISL_1670520, EPI_ISL_1670521, EPI_ISL_1670522, EPI_ISL_1670523, EPI_ISL_1670525, EPI_ISL_1670526, EPI_ISL_1670527, EPI_ISL_1670528, EPI_ISL_1670529, EPI_ISL_1670530, EPI_ISL_1670531, EPI_ISL_1670532, EPI_ISL_1670533, EPI_ISL_1670534, EPI_ISL_1670535, EPI_ISL_1670536, EPI_ISL_1940738, EPI_ISL_1940739, EPI_ISL_1940740, EPI_ISL_1940741, EPI_ISL_1940742, EPI_ISL_1940743, EPI_ISL_1940744, EPI_ISL_1940745, EPI_ISL_1940746, EPI_ISL_1940747, EPI_ISL_1940748, EPI_ISL_1940749, EPI_ISL_1940750, EPI_ISL_1940751, EPI_ISL_1940752, EPI_ISL_1940753, EPI_ISL_1940754, EPI_ISL_1940755, EPI_ISL_1940756, EPI_ISL_1940763, EPI_ISL_1940764, EPI_ISL_1940765, EPI_ISL_1940766, EPI_ISL_1940767, EPI_ISL_1940768, EPI_ISL_1940769, EPI_ISL_1940770, EPI_ISL_1940771, EPI_ISL_1940772, EPI_ISL_1940773, EPI_ISL_1940774, EPI_ISL_1940775, EPI_ISL_1940776, EPI_ISL_1940777, EPI_ISL_1940778, EPI_ISL_1940779, EPI_ISL_1940780, EPI_ISL_1940781, EPI_ISL_1940782, EPI_ISL_1940783, EPI_ISL_1940784, EPI_ISL_1940785, EPI_ISL_1940786, EPI_ISL_1940787, EPI_ISL_1940788, EPI_ISL_1940789, EPI_ISL_1940790, EPI_ISL_1940791, EPI_ISL_1940792, EPI_ISL_1940793, EPI_ISL_1940794, EPI_ISL_1940795, EPI_ISL_1940796, EPI_ISL_1940797, EPI_ISL_1940798, EPI_ISL_1940799, EPI_ISL_1940800, EPI_ISL_1940801 | | | |
| see above | Landesamt für Verbraucherschutz Sachsen Anhalt, Magdeburg | Institute of Medical Microbiology and Hospital Hygiene | Aljoscha Tersteegen; Prof. Dr. Achim Kaasch |
| EPI_ISL_2110770 | Landesgesundheitsamt Baden-Wurttemberg | Robert Koch Institute | |
| EPI_ISL_1846323, EPI_ISL_1846324, EPI_ISL_1846325, EPI_ISL_1846327, EPI_ISL_1846332, EPI_ISL_1846357 | Landesgesundheitsamt Baden-WÃ¼rttemberg | Robert Koch Institute | |
| EPI_ISL_1640123, EPI_ISL_1640125, EPI_ISL_1640129, EPI_ISL_1640133, EPI_ISL_1640137, EPI_ISL_1640139, EPI_ISL_1640140, EPI_ISL_1722087, EPI_ISL_1722088, EPI_ISL_1722089, EPI_ISL_1722100, EPI_ISL_1722113, EPI_ISL_1724356, EPI_ISL_1724376, EPI_ISL_1724387 | | | |
| see above | Landesuntersuchungsanstalt fÃ¼r das Gesundheits- und VeterinÃ¤rwesen (LUA) Sachsen | Robert Koch Institute | |
| EPI_ISL_1722924, EPI_ISL_1722925, EPI_ISL_1722926, EPI_ISL_1722927, EPI_ISL_1722928, EPI_ISL_1722929, EPI_ISL_1722930 | | | |
| see above | Limbach - Ganzimmun Diagnostics AG Mainz | Robert Koch Institute | |
| EPI_ISL_1638369, EPI_ISL_1638374, EPI_ISL_1638456, EPI_ISL_1638482, EPI_ISL_1638492, EPI_ISL_1643761, EPI_ISL_1643764, EPI_ISL_1643777, EPI_ISL_1643787, EPI_ISL_1643860, EPI_ISL_1643874 | | | |
| see above | Limbach - MVZ Gemeinschaftslabor Suhl | Robert Koch Institute | |
| EPI_ISL_1573299, EPI_ISL_1573300, EPI_ISL_1573303, EPI_ISL_1573305, EPI_ISL_1573307, EPI_ISL_1573309, EPI_ISL_1573310, EPI_ISL_1573315, EPI_ISL_1573316, EPI_ISL_1573317, EPI_ISL_1573320, EPI_ISL_1573326, EPI_ISL_1573329, EPI_ISL_1573331, EPI_ISL_1573333, EPI_ISL_1573334, EPI_ISL_1573337, EPI_ISL_1573338, EPI_ISL_1573341, EPI_ISL_1573342, EPI_ISL_1573346, EPI_ISL_1573347, EPI_ISL_1573351, EPI_ISL_1573352, EPI_ISL_1573355, EPI_ISL_1573358, EPI_ISL_1573360, EPI_ISL_1573362, EPI_ISL_1573364, EPI_ISL_1573365, EPI_ISL_1573367, EPI_ISL_1573371, EPI_ISL_1573372, EPI_ISL_1573374, EPI_ISL_1573376, EPI_ISL_1573377, EPI_ISL_1573380, EPI_ISL_1573381, EPI_ISL_1573383, EPI_ISL_1573387, EPI_ISL_1573388, EPI_ISL_1573389, EPI_ISL_1573390, EPI_ISL_1573391, EPI_ISL_1573392, EPI_ISL_1573393, EPI_ISL_1573395, EPI_ISL_1573396, EPI_ISL_1573397, EPI_ISL_1573399, EPI_ISL_1573402, EPI_ISL_1573404, EPI_ISL_1573406, EPI_ISL_1573407, EPI_ISL_1573410, EPI_ISL_1573415, EPI_ISL_1573416, EPI_ISL_1573418, EPI_ISL_1573421, EPI_ISL_1573424, EPI_ISL_1573426, EPI_ISL_1573428, EPI_ISL_1573430, EPI_ISL_1573431, EPI_ISL_1573432, EPI_ISL_1573433, EPI_ISL_1573434, EPI_ISL_1573437, EPI_ISL_1573440, EPI_ISL_1573441, EPI_ISL_1573444, EPI_ISL_1573446, EPI_ISL_1573447, EPI_ISL_1573449, EPI_ISL_1573451, EPI_ISL_1573452, EPI_ISL_1573454, EPI_ISL_1573456, EPI_ISL_1573457, EPI_ISL_1643512, EPI_ISL_1643515, EPI_ISL_1643517, EPI_ISL_1643518, EPI_ISL_1643520, EPI_ISL_1643521, EPI_ISL_1643522, EPI_ISL_1643524, EPI_ISL_1643525, EPI_ISL_1643527, EPI_ISL_1643528, EPI_ISL_1643531, EPI_ISL_1643532, EPI_ISL_1643534, EPI_ISL_1643535, EPI_ISL_1643536, EPI_ISL_1643539, EPI_ISL_1643542, EPI_ISL_1643544, EPI_ISL_1643545, EPI_ISL_1643546, EPI_ISL_1643547, EPI_ISL_1643548, EPI_ISL_1643551, EPI_ISL_1643552, EPI_ISL_1643555, EPI_ISL_1643556, EPI_ISL_1643558, EPI_ISL_1643559, EPI_ISL_1643560, EPI_ISL_1643561, EPI_ISL_1643563, EPI_ISL_1643568, EPI_ISL_1643572, EPI_ISL_1643574, EPI_ISL_1643576, EPI_ISL_1643578, EPI_ISL_1643579, EPI_ISL_1643584, EPI_ISL_1643587, EPI_ISL_1643590, EPI_ISL_1643592, EPI_ISL_1643594, EPI_ISL_1643596, EPI_ISL_1643597, EPI_ISL_1643601, EPI_ISL_1643603, EPI_ISL_1643606, EPI_ISL_1643607, EPI_ISL_1643608, EPI_ISL_1643609, EPI_ISL_1643611, EPI_ISL_1643612, EPI_ISL_1643614, EPI_ISL_1643615, EPI_ISL_1643616, EPI_ISL_1643617, EPI_ISL_1643622, EPI_ISL_1643623, EPI_ISL_1643626, EPI_ISL_1643627, EPI_ISL_1643628, EPI_ISL_1643630, EPI_ISL_1643631, EPI_ISL_1643632, EPI_ISL_1643633, EPI_ISL_1643635, EPI_ISL_1643636, EPI_ISL_1643637, EPI_ISL_1643642, EPI_ISL_1643643, EPI_ISL_1643648, EPI_ISL_1643650, EPI_ISL_1643651, EPI_ISL_1643654, EPI_ISL_1643655, EPI_ISL_1643657, EPI_ISL_1643658, EPI_ISL_1643664, EPI_ISL_1643666, EPI_ISL_1643667, EPI_ISL_1643668, EPI_ISL_1643670, EPI_ISL_1643671, EPI_ISL_1643674, EPI_ISL_1643675, EPI_ISL_1643676, EPI_ISL_1643677, EPI_ISL_1643679, EPI_ISL_1643680, EPI_ISL_1643681 | | | |
| see above | Limbach - MVZ Humangenetik Ulm | Robert Koch Institute | |
| EPI_ISL_1638288, EPI_ISL_1643396, EPI_ISL_1643398, EPI_ISL_1643399, EPI_ISL_1643400, EPI_ISL_1643401, EPI_ISL_1643402, EPI_ISL_1643404, EPI_ISL_1643405, EPI_ISL_1643407, EPI_ISL_1643408, EPI_ISL_1643409, EPI_ISL_1724249, EPI_ISL_1724250, EPI_ISL_1724251, EPI_ISL_1724253, EPI_ISL_1724254, EPI_ISL_1724255, EPI_ISL_1724257, EPI_ISL_1724259, EPI_ISL_1724260, EPI_ISL_1724261, EPI_ISL_1724265, EPI_ISL_1724269, EPI_ISL_1724273, EPI_ISL_1724274, EPI_ISL_1724276, EPI_ISL_1725194, EPI_ISL_1725207, EPI_ISL_1846216, EPI_ISL_1846219, EPI_ISL_1848363 | | | |
| see above | Limbach - MVZ Labor Eveld & Kollegen Essen | Robert Koch Institute | |
| EPI_ISL_1643864, EPI_ISL_1643868 | Limbach - MVZ Labor Ludwigsburg | Robert Koch Institute | |
| EPI_ISL_1638294, EPI_ISL_1638297, EPI_ISL_1638302, EPI_ISL_1638343, EPI_ISL_1638379, EPI_ISL_1638388, EPI_ISL_1638394, EPI_ISL_1638399, EPI_ISL_1638402, EPI_ISL_1638423, EPI_ISL_1638432, EPI_ISL_1638450, EPI_ISL_1638454, EPI_ISL_1638471, EPI_ISL_1638484, EPI_ISL_1639251, EPI_ISL_1643766, EPI_ISL_1643771, EPI_ISL_1643774, EPI_ISL_1643779, EPI_ISL_1643780, EPI_ISL_1643782, EPI_ISL_1643858, EPI_ISL_1643861, EPI_ISL_1643867, EPI_ISL_1643869, EPI_ISL_1643870, EPI_ISL_1643871, EPI_ISL_1643872, EPI_ISL_1643873, EPI_ISL_1643875, EPI_ISL_1720816, EPI_ISL_1720830, EPI_ISL_1720838, EPI_ISL_1720847, EPI_ISL_1720849, EPI_ISL_1720850, EPI_ISL_1720851, EPI_ISL_1720921, EPI_ISL_1724309, EPI_ISL_1724312, EPI_ISL_1724318, EPI_ISL_1724319, EPI_ISL_1724322, EPI_ISL_1725248, EPI_ISL_1725265, EPI_ISL_1725603, EPI_ISL_1725604, EPI_ISL_1725606, EPI_ISL_1725607, EPI_ISL_1725608, EPI_ISL_1725609, EPI_ISL_1725610, EPI_ISL_1725611, EPI_ISL_1725613, EPI_ISL_1725614, EPI_ISL_1725615, EPI_ISL_1725617, EPI_ISL_1725620, EPI_ISL_1725622, EPI_ISL_1725628, EPI_ISL_1725629, EPI_ISL_1725630, EPI_ISL_1725631, EPI_ISL_1725632, EPI_ISL_1846223, EPI_ISL_1847204, EPI_ISL_1847324, EPI_ISL_1847330, EPI_ISL_1847331 | | | |

| | | | |
|---|---|---|---|
| see above | MDI Limbach Berlin GmbH; MVZ Labor Berlin | Robert Koch Institute | |
| EPI_ISL_1715848 | MEDILYS Laborgesellschaft mbH | Heinrich Pette Institute, Leibniz Institute for Experimental Virology | Adam Grundhoff; Alexis Robitaille; Johannes Knobloch; Martin Aepfelbacher; Nicole Fischer; Thomas Günther |
| EPI_ISL_1638496, EPI_ISL_1638510, EPI_ISL_1638516, EPI_ISL_1638529, EPI_ISL_1643419, EPI_ISL_1643427, EPI_ISL_1643428, EPI_ISL_1643431, EPI_ISL_1643433, EPI_ISL_1643434, EPI_ISL_1643436, EPI_ISL_1643438, EPI_ISL_1643440, EPI_ISL_1643441, EPI_ISL_1643443, EPI_ISL_1643445, EPI_ISL_1643446, EPI_ISL_1643448, EPI_ISL_1643449, EPI_ISL_1643452, EPI_ISL_1643453, EPI_ISL_1643457, EPI_ISL_1643458, EPI_ISL_1643460, EPI_ISL_1643461, EPI_ISL_1643462, EPI_ISL_1643463, EPI_ISL_1643464, EPI_ISL_1643465, EPI_ISL_1643466, EPI_ISL_1643467, EPI_ISL_1643469, EPI_ISL_1643471, EPI_ISL_1643472, EPI_ISL_1643475, EPI_ISL_1643476, EPI_ISL_1643478, EPI_ISL_1643480, EPI_ISL_1643481, EPI_ISL_1643485, EPI_ISL_1643486, EPI_ISL_1643487, EPI_ISL_1643488, EPI_ISL_1643489, EPI_ISL_1643490, EPI_ISL_1643491, EPI_ISL_1643493, EPI_ISL_1720939, EPI_ISL_1720942, EPI_ISL_1720943, EPI_ISL_1720947, EPI_ISL_1720949, EPI_ISL_1720954, EPI_ISL_1720960, EPI_ISL_1720961, EPI_ISL_1720962, EPI_ISL_1720964, EPI_ISL_1720965, EPI_ISL_1720966, EPI_ISL_1720968, EPI_ISL_1720970, EPI_ISL_1720971, EPI_ISL_1720972, EPI_ISL_1720976, EPI_ISL_1720977, EPI_ISL_1720978, EPI_ISL_1720980, EPI_ISL_1720983, EPI_ISL_1720984, EPI_ISL_1720985, EPI_ISL_1720987, EPI_ISL_1720989, EPI_ISL_1720991, EPI_ISL_1720995, EPI_ISL_1720998, EPI_ISL_1720999, EPI_ISL_1721000, EPI_ISL_1721004, EPI_ISL_1726781, EPI_ISL_1726783, EPI_ISL_1726799, EPI_ISL_1726801, EPI_ISL_1726810, EPI_ISL_1726816, EPI_ISL_1726826, EPI_ISL_1726829, EPI_ISL_1726849, EPI_ISL_1726852 | | | |
| see above | MVZ Dr. Eberhard & Partner Dortmund | Robert Koch Institute | |
| EPI_ISL_1725644 | MVZ Labor Dortmund | Robert Koch Institute | |
| EPI_ISL_1722070, EPI_ISL_1722071, EPI_ISL_1722072, EPI_ISL_1722073, EPI_ISL_1722074, EPI_ISL_1722075, EPI_ISL_1727072, EPI_ISL_1727075, EPI_ISL_1727077, EPI_ISL_1727078, EPI_ISL_1727081, EPI_ISL_1727082, EPI_ISL_1727087, EPI_ISL_1727090, EPI_ISL_1727093, EPI_ISL_1849357, EPI_ISL_1849371 | | | |
| see above | MVZ Labor Dr. Fenner und Kollegen (Standort Hamburg) | Robert Koch Institute | |
| EPI_ISL_1638401, EPI_ISL_1641159, EPI_ISL_1641160, EPI_ISL_1641161, EPI_ISL_1641162, EPI_ISL_1641163, EPI_ISL_1641164, EPI_ISL_1641165, EPI_ISL_1641166, EPI_ISL_1641167, EPI_ISL_1641168, EPI_ISL_1641169, EPI_ISL_1641170, EPI_ISL_1641171, EPI_ISL_1641172, EPI_ISL_1641173, EPI_ISL_1641174, EPI_ISL_1641177, EPI_ISL_1641178, EPI_ISL_1641179, EPI_ISL_1641180, EPI_ISL_1641181, EPI_ISL_1641182, EPI_ISL_1641183, EPI_ISL_1641184, EPI_ISL_1641185, EPI_ISL_1641186, EPI_ISL_1641187, EPI_ISL_1641188, EPI_ISL_1641189, EPI_ISL_1641190, EPI_ISL_1641191, EPI_ISL_1641192, EPI_ISL_1641193, EPI_ISL_1641194, EPI_ISL_1641195, EPI_ISL_1641196, EPI_ISL_1642351, EPI_ISL_1642352, EPI_ISL_1642353, EPI_ISL_1642354, EPI_ISL_1642355, EPI_ISL_1642357, EPI_ISL_1642358, EPI_ISL_1642359, EPI_ISL_1642360, EPI_ISL_1642361, EPI_ISL_1642362, EPI_ISL_1642363, EPI_ISL_1642364, EPI_ISL_1642365, EPI_ISL_1642366, EPI_ISL_1642367, EPI_ISL_1642368, EPI_ISL_1642369, EPI_ISL_1642371, EPI_ISL_1642372, EPI_ISL_1642373, EPI_ISL_1642375, EPI_ISL_1642376, EPI_ISL_1642377, EPI_ISL_1642380, EPI_ISL_1642381, EPI_ISL_1642382, EPI_ISL_1642384, EPI_ISL_1642385, EPI_ISL_1642386, EPI_ISL_1642387, EPI_ISL_1642388, EPI_ISL_1642389, EPI_ISL_1642390, EPI_ISL_1642391, EPI_ISL_1642392, EPI_ISL_1642393, EPI_ISL_1642394, EPI_ISL_1642395, EPI_ISL_1642396, EPI_ISL_1642397, EPI_ISL_1642398, EPI_ISL_1642399, EPI_ISL_1642400, EPI_ISL_1642401, EPI_ISL_1642402, EPI_ISL_1642403, EPI_ISL_1642404, EPI_ISL_1642405, EPI_ISL_1642406, EPI_ISL_1642407, EPI_ISL_1642408, EPI_ISL_1642409, EPI_ISL_1642410, EPI_ISL_1642412, EPI_ISL_1642413, EPI_ISL_1642414, EPI_ISL_1642415, EPI_ISL_1642416, EPI_ISL_1642417, EPI_ISL_1642418, EPI_ISL_1642697, EPI_ISL_1642698, EPI_ISL_1642701, EPI_ISL_1642702, EPI_ISL_1642703, EPI_ISL_1642704, EPI_ISL_1642705, EPI_ISL_1642707, EPI_ISL_1642708, EPI_ISL_1642709, EPI_ISL_1642712, EPI_ISL_1642713, EPI_ISL_1642715, EPI_ISL_1642716, EPI_ISL_1642717, EPI_ISL_1642718, EPI_ISL_1642719, EPI_ISL_1642720, EPI_ISL_1642721, EPI_ISL_1642722, EPI_ISL_1642723, EPI_ISL_1642726, EPI_ISL_1642727, EPI_ISL_1642729, EPI_ISL_1642730, EPI_ISL_1642731, EPI_ISL_1642732, EPI_ISL_1643862, EPI_ISL_1720775, EPI_ISL_1720776, EPI_ISL_1720777, EPI_ISL_1720778, EPI_ISL_1720779, EPI_ISL_1720780, EPI_ISL_1720781, EPI_ISL_1720782, EPI_ISL_1720783, EPI_ISL_1720784, EPI_ISL_1720785, EPI_ISL_1720786, EPI_ISL_1720787, EPI_ISL_1720788, EPI_ISL_1720789, EPI_ISL_1720790, EPI_ISL_1720791, EPI_ISL_1720792, EPI_ISL_1720793, EPI_ISL_1720794, EPI_ISL_1720795, EPI_ISL_1720796, EPI_ISL_1720797, EPI_ISL_1720798, EPI_ISL_1720799, EPI_ISL_1720800, EPI_ISL_1720801, EPI_ISL_1720802, EPI_ISL_1720803, EPI_ISL_1720804, EPI_ISL_1720805, EPI_ISL_1720806, EPI_ISL_1720807, EPI_ISL_1720808, EPI_ISL_1720809, EPI_ISL_1720810, EPI_ISL_1720811, EPI_ISL_1720812, EPI_ISL_1720814, EPI_ISL_1720820, EPI_ISL_1720822, EPI_ISL_1720823, EPI_ISL_1720831, EPI_ISL_1720833, EPI_ISL_1720840, EPI_ISL_1720846, EPI_ISL_1720848, EPI_ISL_1720852, EPI_ISL_1720853, EPI_ISL_1720854, EPI_ISL_1720855, EPI_ISL_1720856, EPI_ISL_1720857, EPI_ISL_1720858, EPI_ISL_1720859, EPI_ISL_1720860, EPI_ISL_1720861, EPI_ISL_1720862, EPI_ISL_1720863, EPI_ISL_1720864, EPI_ISL_1720865, EPI_ISL_1720866, EPI_ISL_1720867, EPI_ISL_1720868, EPI_ISL_1720870, EPI_ISL_1720871, EPI_ISL_1720872, EPI_ISL_1720873, EPI_ISL_1720874, EPI_ISL_1720877, EPI_ISL_1720879, EPI_ISL_1720880, EPI_ISL_1720881, EPI_ISL_1720882, EPI_ISL_1720883, EPI_ISL_1720884, EPI_ISL_1720885, EPI_ISL_1720886, EPI_ISL_1720888, EPI_ISL_1720889, EPI_ISL_1720890, EPI_ISL_1720891, EPI_ISL_1720892, EPI_ISL_1720893, EPI_ISL_1720894, EPI_ISL_1720895, EPI_ISL_1720896, EPI_ISL_1720897, EPI_ISL_1720898, EPI_ISL_1720899, EPI_ISL_1720900, EPI_ISL_1723063, EPI_ISL_1725205, EPI_ISL_1727384, EPI_ISL_1727385, EPI_ISL_1727386, EPI_ISL_1727387, EPI_ISL_1727389, EPI_ISL_1727390, EPI_ISL_1727391, EPI_ISL_1727392, EPI_ISL_1727393, EPI_ISL_1727394, EPI_ISL_1727395, EPI_ISL_1727396, EPI_ISL_1727397, EPI_ISL_1727398, EPI_ISL_1727399, EPI_ISL_1727400, EPI_ISL_1727401, EPI_ISL_1727402, EPI_ISL_1727403, EPI_ISL_1727404, EPI_ISL_1727405, EPI_ISL_1727406, EPI_ISL_1727407, EPI_ISL_1727408, EPI_ISL_1727409, EPI_ISL_1727410, EPI_ISL_1727411, EPI_ISL_1727412, EPI_ISL_1727414, EPI_ISL_1727415, EPI_ISL_1727416, EPI_ISL_1727417, EPI_ISL_1727418, EPI_ISL_1727419, EPI_ISL_1727420, EPI_ISL_1727554, EPI_ISL_1727555, EPI_ISL_1727558, EPI_ISL_1727561, EPI_ISL_1727562, EPI_ISL_1727567, EPI_ISL_1727571, EPI_ISL_1727573, EPI_ISL_1727576, EPI_ISL_1727577, EPI_ISL_1727579, EPI_ISL_1843383, EPI_ISL_1843384, EPI_ISL_1843385, EPI_ISL_1843387, EPI_ISL_1843389, EPI_ISL_1843393, EPI_ISL_1843395, EPI_ISL_1843397, EPI_ISL_1843398, EPI_ISL_1843399, EPI_ISL_1843401, EPI_ISL_1843403, EPI_ISL_1843404, EPI_ISL_1843405, EPI_ISL_1843409, EPI_ISL_1843411, EPI_ISL_1843414, EPI_ISL_1843419, EPI_ISL_1843420, EPI_ISL_1843423, EPI_ISL_1843424, EPI_ISL_1843426, EPI_ISL_1843427, EPI_ISL_1843429, EPI_ISL_1843430, EPI_ISL_1844269, EPI_ISL_1846224, EPI_ISL_1846233, EPI_ISL_1846234, EPI_ISL_1848764, EPI_ISL_1848780, EPI_ISL_1852046, EPI_ISL_1852107 | | | |
| see above | MVZ Labor Dr. Limbach & Kollegen GbR | Robert Koch Institute | |
| EPI_ISL_1638323, EPI_ISL_1638404, EPI_ISL_1638435, EPI_ISL_1643395, EPI_ISL_1643411, EPI_ISL_1643412, EPI_ISL_1643413, EPI_ISL_1643414, EPI_ISL_1643415, EPI_ISL_1724248, EPI_ISL_1724267 | | | |
| see above | MVZ Labor Dr. Reising-Ackermann und Kollegen | Robert Koch Institute | |
| EPI_ISL_1640691, EPI_ISL_1640715, EPI_ISL_1640725, EPI_ISL_1640728, EPI_ISL_1640729, EPI_ISL_1640736, EPI_ISL_1641197, EPI_ISL_1641198, EPI_ISL_1641199, EPI_ISL_1641200, EPI_ISL_1641201, EPI_ISL_1641202, EPI_ISL_1641203, EPI_ISL_1641204, EPI_ISL_1641205, EPI_ISL_1641206, EPI_ISL_1641207, EPI_ISL_1641208, EPI_ISL_1641209, EPI_ISL_1641210, EPI_ISL_1641211, EPI_ISL_1641212, EPI_ISL_1641213, EPI_ISL_1641214, EPI_ISL_1641215, EPI_ISL_1641216, EPI_ISL_1641217, EPI_ISL_1641218, EPI_ISL_1641219, EPI_ISL_1641220, EPI_ISL_1641221, EPI_ISL_1641222, EPI_ISL_1641223, EPI_ISL_1641224, EPI_ISL_1641225, EPI_ISL_1641226, EPI_ISL_1641227, EPI_ISL_1641228, EPI_ISL_1641229, EPI_ISL_1641230, EPI_ISL_1641231, EPI_ISL_1641232, EPI_ISL_1641233, EPI_ISL_1641234, EPI_ISL_1641235, EPI_ISL_1641236, EPI_ISL_1641237, EPI_ISL_1642828, EPI_ISL_1642829, EPI_ISL_1642830, EPI_ISL_1642831, EPI_ISL_1642832, EPI_ISL_1642833, EPI_ISL_1642834, EPI_ISL_1642835, EPI_ISL_1642836, EPI_ISL_1642837, EPI_ISL_1642838, EPI_ISL_1642839, EPI_ISL_1642840, EPI_ISL_1642841, EPI_ISL_1642842, EPI_ISL_1642843, EPI_ISL_1642844, EPI_ISL_1642845, EPI_ISL_1642846, EPI_ISL_1642847, EPI_ISL_1642848, EPI_ISL_1642849, EPI_ISL_1642850, EPI_ISL_1642851, EPI_ISL_1642852, EPI_ISL_1642853, EPI_ISL_1642854, EPI_ISL_1642855, EPI_ISL_1642856, EPI_ISL_1642857, EPI_ISL_1642858, EPI_ISL_1642859, EPI_ISL_1642860, EPI_ISL_1642861, EPI_ISL_1642862, EPI_ISL_1642863, EPI_ISL_1642864, EPI_ISL_1642865, EPI_ISL_1642866, EPI_ISL_1642867, EPI_ISL_1642868, EPI_ISL_1642869, EPI_ISL_1642870, EPI_ISL_1644934, EPI_ISL_1644935, EPI_ISL_1644936, EPI_ISL_1644937, EPI_ISL_1644938, EPI_ISL_1644939, EPI_ISL_1644940, EPI_ISL_1644941, EPI_ISL_1644943, EPI_ISL_1644944, EPI_ISL_1644945, EPI_ISL_1644946, EPI_ISL_1644947, EPI_ISL_1644948, EPI_ISL_1644949, EPI_ISL_1644950, EPI_ISL_1644951, EPI_ISL_1644952, EPI_ISL_1644953, EPI_ISL_1644954, EPI_ISL_1644955, EPI_ISL_1644956, EPI_ISL_1644957, EPI_ISL_1644959, EPI_ISL_1644960, EPI_ISL_1644961, EPI_ISL_1644962, EPI_ISL_1644963, EPI_ISL_1644964, EPI_ISL_1644965, EPI_ISL_1644966, EPI_ISL_1644967, EPI_ISL_1644968, EPI_ISL_1644969, EPI_ISL_1644970, EPI_ISL_1644971, EPI_ISL_1644972, EPI_ISL_1644973, EPI_ISL_1644974, EPI_ISL_1644975, EPI_ISL_1644976, EPI_ISL_1644977, EPI_ISL_1644978, EPI_ISL_1721982, EPI_ISL_1721983, EPI_ISL_1721984, EPI_ISL_1721985, EPI_ISL_1721986, EPI_ISL_1721987, EPI_ISL_1721988, EPI_ISL_1721989, EPI_ISL_1721990, EPI_ISL_1721991, EPI_ISL_1721992, EPI_ISL_1721993, EPI_ISL_1721994, EPI_ISL_1721995, EPI_ISL_1721996, EPI_ISL_1721997, EPI_ISL_1721998, EPI_ISL_1721999, EPI_ISL_1722000, EPI_ISL_1722001, EPI_ISL_1722002, EPI_ISL_1722003, EPI_ISL_1722004, EPI_ISL_1722005, EPI_ISL_1722006, EPI_ISL_1722007, EPI_ISL_1722008, EPI_ISL_1722009, EPI_ISL_1722010, EPI_ISL_1722011, EPI_ISL_1722012, EPI_ISL_1722013, EPI_ISL_1722014, EPI_ISL_1722015, EPI_ISL_1722016, EPI_ISL_1722017, EPI_ISL_1722018, EPI_ISL_1722019, EPI_ISL_1722020, EPI_ISL_1722021, EPI_ISL_1722022, EPI_ISL_1722023, EPI_ISL_1722024, EPI_ISL_1722025, EPI_ISL_1722026, EPI_ISL_1722027, EPI_ISL_1722028, EPI_ISL_1722029, EPI_ISL_1722030, EPI_ISL_1722031, EPI_ISL_1722032, EPI_ISL_1722033, EPI_ISL_1722034, EPI_ISL_1722035, EPI_ISL_1722036, EPI_ISL_1722037, EPI_ISL_1722038, EPI_ISL_1722039, EPI_ISL_1722040, EPI_ISL_1722041, EPI_ISL_1722042, EPI_ISL_1722043, EPI_ISL_1722044, EPI_ISL_1722045, EPI_ISL_1722046, EPI_ISL_1722047, EPI_ISL_1722048, EPI_ISL_1722049, EPI_ISL_1722050, EPI_ISL_1722051, EPI_ISL_1722052, EPI_ISL_1722053, EPI_ISL_1722054, EPI_ISL_1722055, EPI_ISL_1722056, EPI_ISL_1722057, EPI_ISL_1722058, EPI_ISL_1722059, EPI_ISL_1722060, EPI_ISL_1722061, EPI_ISL_1722062, EPI_ISL_1726937, EPI_ISL_1726943, EPI_ISL_1726951, EPI_ISL_1726972 | | | |
| see above | MVZ Labor Krone GbR | Robert Koch Institute | |
| EPI_ISL_1720875, EPI_ISL_1720878, EPI_ISL_1720903, EPI_ISL_1720907, EPI_ISL_1720917, EPI_ISL_1720920, EPI_ISL_1720922, EPI_ISL_1720923, EPI_ISL_1720925, EPI_ISL_1720927, EPI_ISL_1720930, EPI_ISL_1720932, EPI_ISL_1720933 | | | |
| see above | MVZ Labor Passau GbR | Robert Koch Institute | |
| EPI_ISL_1725634, EPI_ISL_1725643, EPI_ISL_1725646, EPI_ISL_1725656, EPI_ISL_1725658, EPI_ISL_1843332, EPI_ISL_1843343, EPI_ISL_1843349, EPI_ISL_1848798, EPI_ISL_1848799, EPI_ISL_1852460 | | | |
| see above | MVZ Medizinisches Labor Hannover GmbH | Robert Koch Institute | |
| EPI_ISL_1724424, EPI_ISL_1725214, EPI_ISL_1725249, EPI_ISL_1725250, EPI_ISL_1725255, EPI_ISL_1725261, EPI_ISL_1725263, EPI_ISL_1725266, EPI_ISL_1725273, EPI_ISL_1725612, EPI_ISL_1725618, EPI_ISL_1725635, EPI_ISL_1725639, EPI_ISL_1725641, EPI_ISL_1725645, EPI_ISL_1843345, EPI_ISL_1846333, EPI_ISL_1847211, EPI_ISL_1847212, EPI_ISL_1847319 | | | |
| see above | MVZ fÃ¼r Laboratoriumsmedizin und Mikrobiologie Koblenz-Mittelrhein (Labor Koblenz) | Robert Koch Institute | |
| EPI_ISL_1850384 | MVZ fÃ¼r Laboratoriumsmedizin und Mikrobiologie WÃ¼rzburg | Robert Koch Institute | |
| EPI_ISL_1752390, EPI_ISL_1752391, EPI_ISL_1752398, EPI_ISL_1752399, EPI_ISL_1752400, EPI_ISL_1752458, EPI_ISL_1752468, EPI_ISL_1752469, EPI_ISL_1752470, EPI_ISL_2006679, EPI_ISL_2006680, EPI_ISL_2094513, EPI_ISL_2094520, EPI_ISL_2094522, EPI_ISL_2094530, EPI_ISL_2094539, EPI_ISL_2094540, EPI_ISL_2094541, EPI_ISL_2094542, EPI_ISL_2094560, EPI_ISL_2094563, EPI_ISL_2094566, EPI_ISL_2094569, EPI_ISL_2094570, EPI_ISL_2094573, EPI_ISL_2094577, EPI_ISL_2094587, EPI_ISL_2094592, EPI_ISL_2094593, EPI_ISL_2094597, EPI_ISL_2094601, EPI_ISL_2094608, EPI_ISL_2094614, EPI_ISL_2094621, EPI_ISL_2094622, EPI_ISL_2094626, EPI_ISL_2094627, EPI_ISL_2094639, EPI_ISL_2094644, EPI_ISL_2094648, EPI_ISL_2094651, EPI_ISL_2094656, EPI_ISL_2094688, EPI_ISL_2094689, EPI_ISL_2094690, EPI_ISL_2094691, EPI_ISL_2094692, EPI_ISL_2094732, EPI_ISL_2094733, EPI_ISL_2094734, EPI_ISL_2094735, EPI_ISL_2094754, EPI_ISL_2094807, EPI_ISL_2094808, EPI_ISL_2094810, EPI_ISL_2094811, EPI_ISL_2094812, EPI_ISL_2094813, EPI_ISL_2094814, EPI_ISL_2094815, EPI_ISL_2094816, EPI_ISL_2094818, EPI_ISL_2094819, EPI_ISL_2094820, EPI_ISL_2094823, EPI_ISL_2094824, EPI_ISL_2094875, EPI_ISL_2094876, EPI_ISL_2094877, EPI_ISL_2094878, EPI_ISL_2094879, EPI_ISL_2094880, EPI_ISL_2094881, EPI_ISL_2094882, EPI_ISL_2094883, EPI_ISL_2094884, EPI_ISL_2094885, EPI_ISL_2094886, EPI_ISL_2094887, EPI_ISL_2094888, EPI_ISL_2094890, EPI_ISL_2094942, EPI_ISL_2094943, EPI_ISL_2094944, EPI_ISL_2094945, EPI_ISL_2094957, EPI_ISL_2094970, EPI_ISL_2094993, EPI_ISL_2095027, EPI_ISL_2095028, EPI_ISL_2095029, EPI_ISL_2095031, EPI_ISL_2095032, EPI_ISL_2095033, EPI_ISL_2095034, EPI_ISL_2095048, EPI_ISL_2095051, EPI_ISL_2095052, EPI_ISL_2095056, EPI_ISL_2095057, EPI_ISL_2095060, EPI_ISL_2095085, EPI_ISL_2095086, EPI_ISL_2095087, EPI_ISL_2095095, EPI_ISL_2095102, EPI_ISL_2095103, EPI_ISL_2095118, EPI_ISL_2095121, EPI_ISL_2095153, EPI_ISL_2095155, EPI_ISL_2095160, EPI_ISL_2095179, EPI_ISL_2095186, EPI_ISL_2095190, EPI_ISL_2095289, EPI_ISL_2095290, EPI_ISL_2095299, EPI_ISL_2095303 | | | |
| see above | Max von Pettenkofer Institute, Virology, National Reference Center for Retroviruses, LMU Munich | Laboratory for Functional Genome Analysis; Dept. Genomics; Gene Center of the LMU Munich | Alexander Graf; Helmut Blum; Max Muenchhoff; Oliver Keppler; Stefan Krebs |
| EPI_ISL_1722076, EPI_ISL_1722077, EPI_ISL_1722078, EPI_ISL_1722079, EPI_ISL_1722080, EPI_ISL_1722081, EPI_ISL_1722082, EPI_ISL_1722083, EPI_ISL_1722084 | | | |
| see above | Med. Labor Prof. Schenk Dr. Ansorge & Kollegen | Robert Koch Institute | |
| EPI_ISL_1640079, EPI_ISL_1640091, EPI_ISL_1640114, EPI_ISL_1640115 | Medizinisch-Diagnostisches Labor Kempten allgÃ¤ulab | Robert Koch Institute | |
| EPI_ISL_1644856, EPI_ISL_1644857, EPI_ISL_1644858, EPI_ISL_1644859, EPI_ISL_1644860, EPI_ISL_1644861, EPI_ISL_1644862, EPI_ISL_1644863, EPI_ISL_1644864, EPI_ISL_1644865, EPI_ISL_1644866, EPI_ISL_1644867, EPI_ISL_1644868, EPI_ISL_1644869, EPI_ISL_1644870, EPI_ISL_1644871, EPI_ISL_1644872, EPI_ISL_1644873, EPI_ISL_1644874, EPI_ISL_1644875, EPI_ISL_1644876, EPI_ISL_1644877, EPI_ISL_1644878, EPI_ISL_1644879, EPI_ISL_1644880, EPI_ISL_1644881, EPI_ISL_1644882, EPI_ISL_1644883, EPI_ISL_1644884, EPI_ISL_1644885, EPI_ISL_1644886, EPI_ISL_1644887, EPI_ISL_1644888, EPI_ISL_1644889, EPI_ISL_1644890, EPI_ISL_1644891, EPI_ISL_1644892, EPI_ISL_1644893, EPI_ISL_1644894, EPI_ISL_1644895, EPI_ISL_1644896, EPI_ISL_1644897, EPI_ISL_1644898, EPI_ISL_1644899, EPI_ISL_1644900, EPI_ISL_1644901, EPI_ISL_1644902, EPI_ISL_1644903, EPI_ISL_1644904, EPI_ISL_1644905, EPI_ISL_1644906, EPI_ISL_1644907, EPI_ISL_1644908, EPI_ISL_1644909, EPI_ISL_1644910, EPI_ISL_1644911, EPI_ISL_1644912, EPI_ISL_1644913, EPI_ISL_1644914, EPI_ISL_1727028, EPI_ISL_1727037, EPI_ISL_1727049 | | | |
| see above | Medizinische Laboratorien DÃ¼sseldorf | Robert Koch Institute | |
| EPI_ISL_1638384, EPI_ISL_1638386, EPI_ISL_1638410, EPI_ISL_1638457, EPI_ISL_1642370, EPI_ISL_1642374, EPI_ISL_1642378, EPI_ISL_1642379, EPI_ISL_1643760, EPI_ISL_1643762, EPI_ISL_1643778, EPI_ISL_1643781, EPI_ISL_1643783, EPI_ISL_1722091, EPI_ISL_1722094, EPI_ISL_1722095, EPI_ISL_1722096, EPI_ISL_1722101, EPI_ISL_1722102, EPI_ISL_1722104, EPI_ISL_1722105, EPI_ISL_1722110, EPI_ISL_1722114, EPI_ISL_1722115, EPI_ISL_1722116, EPI_ISL_1722120, EPI_ISL_1722121, EPI_ISL_1724328, EPI_ISL_1724330, EPI_ISL_1724337, EPI_ISL_1724341, EPI_ISL_1724350, EPI_ISL_1724361, EPI_ISL_1724365, EPI_ISL_1724368, EPI_ISL_1724374, EPI_ISL_1724378, EPI_ISL_1724379, EPI_ISL_1724389, EPI_ISL_1725269 | | | |
| see above | Medizinisches Labor Ostsachsen MVZ GbR NebenbetriebsstÃ¤tte Dresden | Robert Koch Institute | |
| EPI_ISL_1638495, EPI_ISL_1638498, EPI_ISL_1638499, EPI_ISL_1638500, EPI_ISL_1638501, EPI_ISL_1638502, EPI_ISL_1638503, EPI_ISL_1638504, EPI_ISL_1638505, EPI_ISL_1638507, EPI_ISL_1638514, EPI_ISL_1638518, EPI_ISL_1638520, EPI_ISL_1638521, EPI_ISL_1638525, EPI_ISL_1638527, EPI_ISL_1638528, EPI_ISL_1638530, EPI_ISL_1638532, EPI_ISL_1643426, EPI_ISL_1643432, EPI_ISL_1643442, EPI_ISL_1643444, EPI_ISL_1643447, EPI_ISL_1643454, EPI_ISL_1643455, EPI_ISL_1643470, EPI_ISL_1643479, EPI_ISL_1720950, EPI_ISL_1720951, EPI_ISL_1720993, EPI_ISL_1720996, EPI_ISL_1721011, EPI_ISL_1850411 | | | |
| see above | Medizinisches Labor Wahl LÃ¼denscheid | Robert Koch Institute | |
| EPI_ISL_1638508, EPI_ISL_1638509 | Medizinisches Versorgungszentrum fÃ¼r Labormedizin und Mikrobiologie Ruhr GmbH - mvzlm RUHR GmbH | Robert Koch Institute | |
| EPI_ISL_1645240, EPI_ISL_1645242, EPI_ISL_1645245, EPI_ISL_1645249, EPI_ISL_1645252, EPI_ISL_1645328, EPI_ISL_1645362, EPI_ISL_1645369, EPI_ISL_1645373, EPI_ISL_1645378, EPI_ISL_1645380, EPI_ISL_1645398, EPI_ISL_1645439, EPI_ISL_1645440, EPI_ISL_1645443, EPI_ISL_1645444, EPI_ISL_1645445, EPI_ISL_1645450, EPI_ISL_1645472, EPI_ISL_1645496, EPI_ISL_1645536, EPI_ISL_1645549, EPI_ISL_1645571, EPI_ISL_1645589, EPI_ISL_1645591, EPI_ISL_1645594, EPI_ISL_1645599, EPI_ISL_1645604, EPI_ISL_1645611, EPI_ISL_1645614, EPI_ISL_1645615, EPI_ISL_1645618, EPI_ISL_1645629, EPI_ISL_1645634, EPI_ISL_1645636, EPI_ISL_1645645, EPI_ISL_1645647, EPI_ISL_1645660, EPI_ISL_1645686, EPI_ISL_1645819, EPI_ISL_1645836, EPI_ISL_1645848, EPI_ISL_1645850, EPI_ISL_1645863, EPI_ISL_1722630, EPI_ISL_1722650 | | | |
| see above | Medizinisches Zentrallabor Altenburg GmbH & Co.KG | Robert Koch Institute | |
| EPI_ISL_1721037, EPI_ISL_1721052 | NA | Robert Koch Institute | |
| EPI_ISL_1722123, EPI_ISL_1722124, EPI_ISL_1722125, EPI_ISL_1722126, EPI_ISL_1722127, EPI_ISL_1722128 | Philipps-UniversitÃ¤t Marburg Fachbereich Medizin Institut fÃ¼r Virologie | Robert Koch Institute | |
| EPI_ISL_1640934, EPI_ISL_1640976, EPI_ISL_1641050, EPI_ISL_1641060, EPI_ISL_1641068, EPI_ISL_1641071, EPI_ISL_1641109, EPI_ISL_1643043, EPI_ISL_1645452, EPI_ISL_1645464 | | | |
| see above | Praxisgemeinschaft fÃ¼r Laboratoriumsmedizin Labor BlumenstraÃŸe; Praxis Dr. E. Wietschel & Dr. F. Wietschel | Robert Koch Institute | |
| EPI_ISL_1641066, EPI_ISL_1641115, EPI_ISL_1644083, EPI_ISL_1645282, EPI_ISL_1645388, EPI_ISL_1722414, EPI_ISL_1722551, EPI_ISL_1722579, EPI_ISL_1722590, EPI_ISL_1722609 | | | |
| see above | Praxisgemeinschaft fÃ¼r Laboratoriumsmedizin Labor BlumenstraÃŸe; Praxis Dr. Sel und Dr. SpÃ¤te | Robert Koch Institute | |
| EPI_ISL_1720955, EPI_ISL_1720956, EPI_ISL_1720969, EPI_ISL_1720974 | Procomcure Biotech Germany GmbH | Robert Koch Institute | |
| EPI_ISL_1842855, EPI_ISL_2108811 | Robert-Bosch-Krankenhaus | Robert Koch Institute | |
| EPI_ISL_1638317, EPI_ISL_1638322, EPI_ISL_1638333, EPI_ISL_1638370, EPI_ISL_1638438 | SLK-Kliniken Heilbronn GmbH - Institut fÃ¼r Laboratoriumsmedizin | Robert Koch Institute | |
| EPI_ISL_1640936, EPI_ISL_1640937, EPI_ISL_1640949, EPI_ISL_1640955, EPI_ISL_1640977, EPI_ISL_1640992, EPI_ISL_1640997, EPI_ISL_1641006, EPI_ISL_1641039, EPI_ISL_1641090, EPI_ISL_1641123, EPI_ISL_1641125, EPI_ISL_1641157, EPI_ISL_1642877, EPI_ISL_1642879, EPI_ISL_1642880, EPI_ISL_1642895, EPI_ISL_1642899, EPI_ISL_1642905, EPI_ISL_1642907, EPI_ISL_1642911, EPI_ISL_1642915, EPI_ISL_1642916, EPI_ISL_1642918, EPI_ISL_1642919, EPI_ISL_1642920, EPI_ISL_1642921, EPI_ISL_1642922, EPI_ISL_1642924, EPI_ISL_1642927, EPI_ISL_1642929, EPI_ISL_1642932, EPI_ISL_1642934, EPI_ISL_1642937, EPI_ISL_1642941, EPI_ISL_1642942, EPI_ISL_1642943, EPI_ISL_1642945, EPI_ISL_1642946, EPI_ISL_1642947, EPI_ISL_1642952, EPI_ISL_1642956, EPI_ISL_1642962, EPI_ISL_1642966, EPI_ISL_1642967, EPI_ISL_1642970, EPI_ISL_1642971, EPI_ISL_1642972, EPI_ISL_1642976, EPI_ISL_1642977, EPI_ISL_1642978, EPI_ISL_1642979, EPI_ISL_1642981, EPI_ISL_1642982, EPI_ISL_1642990, EPI_ISL_1642994, EPI_ISL_1642999, EPI_ISL_1643002, EPI_ISL_1643007, EPI_ISL_1643011, EPI_ISL_1643020, EPI_ISL_1643023, EPI_ISL_1643024, EPI_ISL_1643036, EPI_ISL_1643039, EPI_ISL_1643049, EPI_ISL_1643061, EPI_ISL_1643068, EPI_ISL_1643080, EPI_ISL_1643084, EPI_ISL_1643088, EPI_ISL_1643089, EPI_ISL_1643090, EPI_ISL_1643092, EPI_ISL_1643093, EPI_ISL_1643101, EPI_ISL_1643105, EPI_ISL_1643106, EPI_ISL_1643109, EPI_ISL_1644045, EPI_ISL_1644046, EPI_ISL_1644047, EPI_ISL_1644048, EPI_ISL_1644049, EPI_ISL_1644050, EPI_ISL_1644051, EPI_ISL_1644052, EPI_ISL_1644054, EPI_ISL_1644060, EPI_ISL_1644061, EPI_ISL_1644065, EPI_ISL_1644068, EPI_ISL_1644069, EPI_ISL_1644072, EPI_ISL_1644076, EPI_ISL_1644079, EPI_ISL_1644081, EPI_ISL_1644086, EPI_ISL_1644088, EPI_ISL_1644093, EPI_ISL_1644095, EPI_ISL_1644098, EPI_ISL_1644101, EPI_ISL_1644104, EPI_ISL_1644105, EPI_ISL_1644107, EPI_ISL_1644109, EPI_ISL_1644111, EPI_ISL_1644113, EPI_ISL_1644115, EPI_ISL_1644116, EPI_ISL_1644117, EPI_ISL_1644119, EPI_ISL_1644122, EPI_ISL_1644123, EPI_ISL_1644124, EPI_ISL_1644132, EPI_ISL_1644133, EPI_ISL_1644137, EPI_ISL_1644138, EPI_ISL_1644139, EPI_ISL_1644142, EPI_ISL_1644145, EPI_ISL_1644146, EPI_ISL_1644149, EPI_ISL_1644154, EPI_ISL_1644158, EPI_ISL_1644159, EPI_ISL_1644162, EPI_ISL_1644164, EPI_ISL_1644167, EPI_ISL_1644171, | | | |

EPI_ISL_1644173, EPI_ISL_1644181, EPI_ISL_1644183, EPI_ISL_1644184, EPI_ISL_1644186, EPI_ISL_1644192, EPI_ISL_1644194, EPI_ISL_1644198, EPI_ISL_1644202, EPI_ISL_1644203, EPI_ISL_1644206, EPI_ISL_1644208, EPI_ISL_1644209, EPI_ISL_1644211, EPI_ISL_1644212, EPI_ISL_1644213, EPI_ISL_1644214, EPI_ISL_1644217, EPI_ISL_1644218, EPI_ISL_1644219, EPI_ISL_1644220, EPI_ISL_1644222, EPI_ISL_1644223, EPI_ISL_1644225, EPI_ISL_1644227, EPI_ISL_1644229, EPI_ISL_1644233, EPI_ISL_1644237, EPI_ISL_1644239, EPI_ISL_1644242, EPI_ISL_1644243, EPI_ISL_1644247, EPI_ISL_1644250, EPI_ISL_1644251, EPI_ISL_1644253, EPI_ISL_1644258, EPI_ISL_1644259, EPI_ISL_1644262, EPI_ISL_1644263, EPI_ISL_1644265, EPI_ISL_1644268, EPI_ISL_1644269, EPI_ISL_1644270, EPI_ISL_1644273, EPI_ISL_1644277, EPI_ISL_1644284, EPI_ISL_1644286, EPI_ISL_1644287, EPI_ISL_1644290, EPI_ISL_1644291, EPI_ISL_1644297, EPI_ISL_1644298, EPI_ISL_1644300, EPI_ISL_1644301, EPI_ISL_1644302, EPI_ISL_1644304, EPI_ISL_1644305, EPI_ISL_1644307, EPI_ISL_1644313, EPI_ISL_1644316, EPI_ISL_1644318, EPI_ISL_1644319, EPI_ISL_1644320, EPI_ISL_1644322, EPI_ISL_1644323, EPI_ISL_1644324, EPI_ISL_1644327, EPI_ISL_1644328, EPI_ISL_1644330, EPI_ISL_1644331, EPI_ISL_1644332, EPI_ISL_1644335, EPI_ISL_1644343, EPI_ISL_1644344, EPI_ISL_1644345, EPI_ISL_1644352, EPI_ISL_1644353, EPI_ISL_1644355, EPI_ISL_1644357, EPI_ISL_1644371, EPI_ISL_1644374, EPI_ISL_1644384, EPI_ISL_1644388, EPI_ISL_1644399, EPI_ISL_1644412, EPI_ISL_1644417, EPI_ISL_1644420, EPI_ISL_1644422, EPI_ISL_1644428, EPI_ISL_1644429, EPI_ISL_1644432, EPI_ISL_1644439, EPI_ISL_1644442, EPI_ISL_1644443, EPI_ISL_1644444, EPI_ISL_1644445, EPI_ISL_1644446, EPI_ISL_1644447, EPI_ISL_1644449, EPI_ISL_1644450, EPI_ISL_1644454, EPI_ISL_1644456, EPI_ISL_1644458, EPI_ISL_1644459, EPI_ISL_1644462, EPI_ISL_1644463, EPI_ISL_1644466, EPI_ISL_1644469, EPI_ISL_1644470, EPI_ISL_1644473, EPI_ISL_1644474, EPI_ISL_1644475, EPI_ISL_1644476, EPI_ISL_1644477, EPI_ISL_1644478, EPI_ISL_1644480, EPI_ISL_1644481, EPI_ISL_1644482, EPI_ISL_1644484, EPI_ISL_1644485, EPI_ISL_1644489, EPI_ISL_1644493, EPI_ISL_1644495, EPI_ISL_1644496, EPI_ISL_1644497, EPI_ISL_1644498, EPI_ISL_1644499, EPI_ISL_1644501, EPI_ISL_1644502, EPI_ISL_1644503, EPI_ISL_1644510, EPI_ISL_1644530, EPI_ISL_1644532, EPI_ISL_1644534, EPI_ISL_1644536, EPI_ISL_1644537, EPI_ISL_1644539, EPI_ISL_1645232, EPI_ISL_1645233, EPI_ISL_1645239, EPI_ISL_1645244, EPI_ISL_1645261, EPI_ISL_1645274, EPI_ISL_1645279, EPI_ISL_1645281, EPI_ISL_1645300, EPI_ISL_1645302, EPI_ISL_1645311, EPI_ISL_1645314, EPI_ISL_1645315, EPI_ISL_1645320, EPI_ISL_1645322, EPI_ISL_1645327, EPI_ISL_1645329, EPI_ISL_1645330, EPI_ISL_1645332, EPI_ISL_1645333, EPI_ISL_1645339, EPI_ISL_1645340, EPI_ISL_1645343, EPI_ISL_1645344, EPI_ISL_1645346, EPI_ISL_1645350, EPI_ISL_1645357, EPI_ISL_1645359, EPI_ISL_1645364, EPI_ISL_1645383, EPI_ISL_1645387, EPI_ISL_1645389, EPI_ISL_1645390, EPI_ISL_1645407, EPI_ISL_1645416, EPI_ISL_1645432, EPI_ISL_1645436, EPI_ISL_1645446, EPI_ISL_1645447, EPI_ISL_1645449, EPI_ISL_1645451, EPI_ISL_1645457, EPI_ISL_1645469, EPI_ISL_1645473, EPI_ISL_1645475, EPI_ISL_1645476, EPI_ISL_1645478, EPI_ISL_1645481, EPI_ISL_1645483, EPI_ISL_1645485, EPI_ISL_1645501, EPI_ISL_1645503, EPI_ISL_1645506, EPI_ISL_1645507, EPI_ISL_1645510, EPI_ISL_1645512, EPI_ISL_1645515, EPI_ISL_1645516, EPI_ISL_1645518, EPI_ISL_1645519, EPI_ISL_1645521, EPI_ISL_1645522, EPI_ISL_1645526, EPI_ISL_1645531, EPI_ISL_1645533, EPI_ISL_1645541, EPI_ISL_1645543, EPI_ISL_1645546, EPI_ISL_1645547, EPI_ISL_1645551, EPI_ISL_1645555, EPI_ISL_1645557, EPI_ISL_1645560, EPI_ISL_1645563, EPI_ISL_1645565, EPI_ISL_1645566, EPI_ISL_1645567, EPI_ISL_1645569, EPI_ISL_1645570, EPI_ISL_1645572, EPI_ISL_1645573, EPI_ISL_1645574, EPI_ISL_1645577, EPI_ISL_1645578, EPI_ISL_1645579, EPI_ISL_1645581, EPI_ISL_1645583, EPI_ISL_1645584, EPI_ISL_1645587, EPI_ISL_1645592, EPI_ISL_1645598, EPI_ISL_1645602, EPI_ISL_1645608, EPI_ISL_1645609, EPI_ISL_1645610, EPI_ISL_1645619, EPI_ISL_1645623, EPI_ISL_1645626, EPI_ISL_1645627, EPI_ISL_1645631, EPI_ISL_1645632, EPI_ISL_1645633, EPI_ISL_1645638, EPI_ISL_1645642, EPI_ISL_1645646, EPI_ISL_1645651, EPI_ISL_1645654, EPI_ISL_1645655, EPI_ISL_1645658, EPI_ISL_1645659, EPI_ISL_1645662, EPI_ISL_1645664, EPI_ISL_1645668, EPI_ISL_1645669, EPI_ISL_1645673, EPI_ISL_1645675, EPI_ISL_1645677, EPI_ISL_1645678, EPI_ISL_1645679, EPI_ISL_1645681, EPI_ISL_1645694, EPI_ISL_1645696, EPI_ISL_1645703, EPI_ISL_1645707, EPI_ISL_1645711, EPI_ISL_1645713, EPI_ISL_1645717, EPI_ISL_1645721, EPI_ISL_1645731, EPI_ISL_1645740, EPI_ISL_1645742, EPI_ISL_1645744, EPI_ISL_1645749, EPI_ISL_1645750, EPI_ISL_1645752, EPI_ISL_1645753, EPI_ISL_1645757, EPI_ISL_1645759, EPI_ISL_1645769, EPI_ISL_1645772, EPI_ISL_1645774, EPI_ISL_1645776, EPI_ISL_1645778, EPI_ISL_1645784, EPI_ISL_1645786, EPI_ISL_1645788, EPI_ISL_1645790, EPI_ISL_1645791, EPI_ISL_1645793, EPI_ISL_1645796, EPI_ISL_1645801, EPI_ISL_1645809, EPI_ISL_1645813, EPI_ISL_1645817, EPI_ISL_1645821, EPI_ISL_1645823, EPI_ISL_1645827, EPI_ISL_1645829, EPI_ISL_1645831, EPI_ISL_1645834, EPI_ISL_1645838, EPI_ISL_1645840, EPI_ISL_1645842, EPI_ISL_1645846, EPI_ISL_1645853, EPI_ISL_1645856, EPI_ISL_1645859, EPI_ISL_1645860, EPI_ISL_1645862, EPI_ISL_1645873, EPI_ISL_1645878, EPI_ISL_1645880, EPI_ISL_1645882, EPI_ISL_1645890, EPI_ISL_1645894, EPI_ISL_1645896, EPI_ISL_1645901, EPI_ISL_1645911, EPI_ISL_1645915, EPI_ISL_1645922, EPI_ISL_1645924, EPI_ISL_1645926, EPI_ISL_1645929, EPI_ISL_1645933, EPI_ISL_1722387, EPI_ISL_1722389, EPI_ISL_1722398, EPI_ISL_1722400, EPI_ISL_1722402, EPI_ISL_1722405, EPI_ISL_1722406, EPI_ISL_1722407, EPI_ISL_1722419, EPI_ISL_1722429, EPI_ISL_1722432, EPI_ISL_1722433, EPI_ISL_1722452, EPI_ISL_1722465, EPI_ISL_1722466, EPI_ISL_1722470, EPI_ISL_1722477, EPI_ISL_1722480, EPI_ISL_1722486, EPI_ISL_1722496, EPI_ISL_1722499, EPI_ISL_1722502, EPI_ISL_1722505, EPI_ISL_1722509, EPI_ISL_1722510, EPI_ISL_1722518, EPI_ISL_1722522, EPI_ISL_1722526, EPI_ISL_1722531, EPI_ISL_1722535, EPI_ISL_1722537, EPI_ISL_1722539, EPI_ISL_1722541, EPI_ISL_1722542, EPI_ISL_1722544, EPI_ISL_1722549, EPI_ISL_1722554, EPI_ISL_1722560, EPI_ISL_1722563, EPI_ISL_1722567, EPI_ISL_1722568, EPI_ISL_1722572, EPI_ISL_1722577, EPI_ISL_1722578, EPI_ISL_1722580, EPI_ISL_1722581, EPI_ISL_1722582, EPI_ISL_1722587, EPI_ISL_1722592, EPI_ISL_1722616, EPI_ISL_1722617, EPI_ISL_1722618, EPI_ISL_1722619, EPI_ISL_1722622, EPI_ISL_1722623, EPI_ISL_1722627, EPI_ISL_1722632, EPI_ISL_1722633, EPI_ISL_1722637, EPI_ISL_1722640, EPI_ISL_1722644, EPI_ISL_1722645, EPI_ISL_1722649, EPI_ISL_1722656, EPI_ISL_1722659, EPI_ISL_1722660, EPI_ISL_1722665, EPI_ISL_1723065, EPI_ISL_1723067, EPI_ISL_1723068, EPI_ISL_1723072, EPI_ISL_1723077, EPI_ISL_1723080, EPI_ISL_1723085, EPI_ISL_1723093, EPI_ISL_1723100, EPI_ISL_1723105, EPI_ISL_1723109, EPI_ISL_1723113, EPI_ISL_1723120, EPI_ISL_1723124, EPI_ISL_1723135, EPI_ISL_1723143, EPI_ISL_1723144, EPI_ISL_1723157, EPI_ISL_1723160, EPI_ISL_1723163, EPI_ISL_1723166, EPI_ISL_1723168, EPI_ISL_1723178, EPI_ISL_1723181, EPI_ISL_1723184, EPI_ISL_1723185, EPI_ISL_1723190, EPI_ISL_1723193, EPI_ISL_1723195, EPI_ISL_1723198, EPI_ISL_1723200, EPI_ISL_1723203, EPI_ISL_1723205, EPI_ISL_1723207, EPI_ISL_1723209, EPI_ISL_1723210, EPI_ISL_1723213, EPI_ISL_1723214, EPI_ISL_1723218, EPI_ISL_1723224, EPI_ISL_1723225, EPI_ISL_1723227, EPI_ISL_1723230, EPI_ISL_1723232, EPI_ISL_1723233, EPI_ISL_1723236, EPI_ISL_1723237, EPI_ISL_1723240, EPI_ISL_1723241, EPI_ISL_1723243, EPI_ISL_1723244, EPI_ISL_1723247, EPI_ISL_1723249, EPI_ISL_1723253, EPI_ISL_1723254, EPI_ISL_1723257, EPI_ISL_1723259, EPI_ISL_1723262, EPI_ISL_1723263, EPI_ISL_1723267, EPI_ISL_1723271, EPI_ISL_1723272, EPI_ISL_1723275, EPI_ISL_1723280, EPI_ISL_1723283, EPI_ISL_1723287, EPI_ISL_1723296, EPI_ISL_1723301, EPI_ISL_1723310, EPI_ISL_1723312, EPI_ISL_1723316, EPI_ISL_1723319, EPI_ISL_1723322, EPI_ISL_1723325, EPI_ISL_1723328, EPI_ISL_1723329, EPI_ISL_1723335, EPI_ISL_1723337, EPI_ISL_1723344, EPI_ISL_1723346, EPI_ISL_1724492, EPI_ISL_1724501, EPI_ISL_1724507, EPI_ISL_1724514, EPI_ISL_1724515, EPI_ISL_1724516, EPI_ISL_1724522, EPI_ISL_1724548, EPI_ISL_1724551, EPI_ISL_1724554, EPI_ISL_1724557, EPI_ISL_1724559, EPI_ISL_1724568, EPI_ISL_1724570, EPI_ISL_1724571, EPI_ISL_1724576, EPI_ISL_1724579, EPI_ISL_1724582, EPI_ISL_1724585, EPI_ISL_1724617, EPI_ISL_1724622, EPI_ISL_1724627, EPI_ISL_1724631, EPI_ISL_1724643, EPI_ISL_1724649, EPI_ISL_1724651, EPI_ISL_1724652, EPI_ISL_1724654, EPI_ISL_1724659, EPI_ISL_1724662, EPI_ISL_1724672, EPI_ISL_1724681, EPI_ISL_1724684, EPI_ISL_1724691, EPI_ISL_1724692, EPI_ISL_1724700, EPI_ISL_1724718, EPI_ISL_1724737, EPI_ISL_1724740, EPI_ISL_1724755, EPI_ISL_1725970, EPI_ISL_1726006, EPI_ISL_1726082, EPI_ISL_1727600, EPI_ISL_1727609

see above SYNLAB Jena Oncoscreen Robert Koch Institute

EPI_ISL_1645517, EPI_ISL_1645558, EPI_ISL_1645576, EPI_ISL_1645607, EPI_ISL_1645621, EPI_ISL_1645656, EPI_ISL_1645698, EPI_ISL_1645709, EPI_ISL_1722485, EPI_ISL_1722545, EPI_ISL_1722624, EPI_ISL_1722658, EPI_ISL_1722661

see above SYNLAB Labor MÃ¼nchen Zentrum LMZ Robert Koch Institute

EPI_ISL_1642873, EPI_ISL_1642875, EPI_ISL_1642876, EPI_ISL_1642878, EPI_ISL_1642881, EPI_ISL_1642884, EPI_ISL_1643012, EPI_ISL_1643033, EPI_ISL_1643074, EPI_ISL_1643097, EPI_ISL_1644056, EPI_ISL_1644063, EPI_ISL_1644071, EPI_ISL_1644074, EPI_ISL_1644078, EPI_ISL_1644080, EPI_ISL_1644082, EPI_ISL_1644085, EPI_ISL_1644087, EPI_ISL_1644089, EPI_ISL_1644091, EPI_ISL_1644108, EPI_ISL_1644112, EPI_ISL_1644114, EPI_ISL_1644166, EPI_ISL_1644169, EPI_ISL_1644170, EPI_ISL_1644188, EPI_ISL_1644189, EPI_ISL_1644201, EPI_ISL_1644224, EPI_ISL_1644241, EPI_ISL_1644254, EPI_ISL_1644256, EPI_ISL_1644267, EPI_ISL_1644271, EPI_ISL_1644295, EPI_ISL_1644348, EPI_ISL_1644368, EPI_ISL_1644416, EPI_ISL_1644421, EPI_ISL_1644427, EPI_ISL_1644518, EPI_ISL_1644521, EPI_ISL_1644524, EPI_ISL_1644526, EPI_ISL_1644528, EPI_ISL_1645234, EPI_ISL_1645238, EPI_ISL_1645260, EPI_ISL_1645267, EPI_ISL_1645269, EPI_ISL_1645335, EPI_ISL_1645371, EPI_ISL_1645421, EPI_ISL_1645424, EPI_ISL_1645441, EPI_ISL_1645461, EPI_ISL_1645488, EPI_ISL_1645493, EPI_ISL_1645509, EPI_ISL_1645520, EPI_ISL_1645525, EPI_ISL_1645538, EPI_ISL_1645544, EPI_ISL_1645553, EPI_ISL_1645559, EPI_ISL_1645585, EPI_ISL_1645586, EPI_ISL_1645596, EPI_ISL_1645613, EPI_ISL_1645637, EPI_ISL_1645640, EPI_ISL_1645641, EPI_ISL_1645653, EPI_ISL_1645663, EPI_ISL_1645671, EPI_ISL_1645692, EPI_ISL_1645854, EPI_ISL_1645879, EPI_ISL_1645907, EPI_ISL_1722417, EPI_ISL_1722434, EPI_ISL_1722439, EPI_ISL_1722448, EPI_ISL_1722454, EPI_ISL_1722457, EPI_ISL_1722460, EPI_ISL_1722463, EPI_ISL_1722468, EPI_ISL_1722474, EPI_ISL_1722478, EPI_ISL_1722481, EPI_ISL_1722487, EPI_ISL_1722493, EPI_ISL_1722495, EPI_ISL_1722504, EPI_ISL_1722517, EPI_ISL_1722521, EPI_ISL_1722524, EPI_ISL_1722552, EPI_ISL_1722557, EPI_ISL_1722574, EPI_ISL_1722595, EPI_ISL_1722599, EPI_ISL_1722605, EPI_ISL_1722610, EPI_ISL_1722611, EPI_ISL_1722613, EPI_ISL_1722615, EPI_ISL_1722621, EPI_ISL_1722625, EPI_ISL_1722628, EPI_ISL_1722639, EPI_ISL_1722641, EPI_ISL_1722643, EPI_ISL_1722647, EPI_ISL_1722663, EPI_ISL_1723116, EPI_ISL_1723127, EPI_ISL_1723148, EPI_ISL_1723189, EPI_ISL_1723202, EPI_ISL_1723221, EPI_ISL_1723252, EPI_ISL_1723256, EPI_ISL_1723260, EPI_ISL_1723266, EPI_ISL_1723285, EPI_ISL_1723290, EPI_ISL_1723294, EPI_ISL_1723327, EPI_ISL_1723330, EPI_ISL_1723333, EPI_ISL_1724625, EPI_ISL_1724686, EPI_ISL_1725984, EPI_ISL_1725988, EPI_ISL_1725992, EPI_ISL_1725993, EPI_ISL_1726004, EPI_ISL_1726011, EPI_ISL_1726018, EPI_ISL_1726022, EPI_ISL_1726037, EPI_ISL_1726048, EPI_ISL_1726053, EPI_ISL_1726084, EPI_ISL_1726086, EPI_ISL_1726089, EPI_ISL_1726097, EPI_ISL_1726107, EPI_ISL_1726115, EPI_ISL_1726118, EPI_ISL_1726120, EPI_ISL_1726128, EPI_ISL_1726137, EPI_ISL_1726138, EPI_ISL_1726146, EPI_ISL_1726151, EPI_ISL_1726152, EPI_ISL_1727584, EPI_ISL_1727586, EPI_ISL_1727594, EPI_ISL_1727598, EPI_ISL_1727602, EPI_ISL_1727624, EPI_ISL_1727628, EPI_ISL_1727635, EPI_ISL_1728880, EPI_ISL_1728882, EPI_ISL_1728883, EPI_ISL_1728911, EPI_ISL_1728970, EPI_ISL_1728979, EPI_ISL_1728985

see above SYNLAB MVZ Berlin Robert Koch Institute

EPI_ISL_1645158, EPI_ISL_1645171, EPI_ISL_1645184, EPI_ISL_1645188, EPI_ISL_1645192, EPI_ISL_1645202, EPI_ISL_1645207, EPI_ISL_1645210

see above SYNLAB MVZ Gera Robert Koch Institute

EPI_ISL_1726477, EPI_ISL_1726483, EPI_ISL_1726488, EPI_ISL_1726492, EPI_ISL_1726516, EPI_ISL_1726529, EPI_ISL_1726534, EPI_ISL_1726536, EPI_ISL_1726539, EPI_ISL_1726544, EPI_ISL_1726550, EPI_ISL_1726552, EPI_ISL_1726555, EPI_ISL_1726560, EPI_ISL_1726570, EPI_ISL_1726581, EPI_ISL_1726586, EPI_ISL_1726587, EPI_ISL_1726590, EPI_ISL_1726593, EPI_ISL_1726596, EPI_ISL_1726602, EPI_ISL_1726604, EPI_ISL_1726606, EPI_ISL_1726611, EPI_ISL_1726615, EPI_ISL_1726623, EPI_ISL_1726636, EPI_ISL_1726639, EPI_ISL_1726646, EPI_ISL_1726651, EPI_ISL_1726652, EPI_ISL_1726654, EPI_ISL_1726655, EPI_ISL_1726656, EPI_ISL_1726660, EPI_ISL_1726661, EPI_ISL_1726670, EPI_ISL_1726672, EPI_ISL_1726679, EPI_ISL_1726681, EPI_ISL_1726683, EPI_ISL_1726686, EPI_ISL_1726687, EPI_ISL_1726690, EPI_ISL_1726695, EPI_ISL_1726698, EPI_ISL_1726700, EPI_ISL_1726703, EPI_ISL_1726706, EPI_ISL_1726711, EPI_ISL_1726714, EPI_ISL_1726718, EPI_ISL_1726721, EPI_ISL_1726724, EPI_ISL_1726726, EPI_ISL_1726727, EPI_ISL_1726728, EPI_ISL_1726729, EPI_ISL_1726730, EPI_ISL_1726732, EPI_ISL_1726734, EPI_ISL_1726737, EPI_ISL_1726740, EPI_ISL_1726742, EPI_ISL_1726745, EPI_ISL_1726748, EPI_ISL_1726751, EPI_ISL_1726752, EPI_ISL_1726754, EPI_ISL_1726755, EPI_ISL_1726766, EPI_ISL_1726769, EPI_ISL_1849698, EPI_ISL_1849700, EPI_ISL_1849701, EPI_ISL_1849703, EPI_ISL_1849705, EPI_ISL_1849706, EPI_ISL_1849707, EPI_ISL_1849709, EPI_ISL_1849710, EPI_ISL_1849711, EPI_ISL_1849714, EPI_ISL_1849715, EPI_ISL_1849716, EPI_ISL_1849721, EPI_ISL_1849722, EPI_ISL_1849724, EPI_ISL_1849728, EPI_ISL_1849729, EPI_ISL_1849732, EPI_ISL_1849734, EPI_ISL_1849735, EPI_ISL_1849738, EPI_ISL_1849740, EPI_ISL_1849742, EPI_ISL_1849747, EPI_ISL_1849749, EPI_ISL_1849750, EPI_ISL_1849752, EPI_ISL_1849753, EPI_ISL_1849754, EPI_ISL_1849755, EPI_ISL_1849758, EPI_ISL_1849762, EPI_ISL_1849764, EPI_ISL_1849765, EPI_ISL_1849770, EPI_ISL_1849771, EPI_ISL_1849772, EPI_ISL_1849774, EPI_ISL_1849776, EPI_ISL_1849778, EPI_ISL_1849780, EPI_ISL_1849783, EPI_ISL_1849785, EPI_ISL_1849787, EPI_ISL_1849788, EPI_ISL_1849789, EPI_ISL_1849790, EPI_ISL_1849791, EPI_ISL_1849793, EPI_ISL_1849794, EPI_ISL_1849795, EPI_ISL_1849797, EPI_ISL_1849799, EPI_ISL_1849803, EPI_ISL_1849804, EPI_ISL_1849806, EPI_ISL_1849807, EPI_ISL_1849809, EPI_ISL_1849811, EPI_ISL_1849812, EPI_ISL_1849813, EPI_ISL_1849814, EPI_ISL_1849816, EPI_ISL_1849818, EPI_ISL_1849819, EPI_ISL_1849821, EPI_ISL_1849823, EPI_ISL_1849824, EPI_ISL_1849826, EPI_ISL_1849828, EPI_ISL_1849830, EPI_ISL_1849832, EPI_ISL_1849834, EPI_ISL_1849836, EPI_ISL_1849839, EPI_ISL_1849840, EPI_ISL_1849842, EPI_ISL_1849845, EPI_ISL_1849846, EPI_ISL_1849848, EPI_ISL_1849849, EPI_ISL_1849850, EPI_ISL_1849852, EPI_ISL_1849853, EPI_ISL_1849855, EPI_ISL_1849856, EPI_ISL_1849858, EPI_ISL_1849859, EPI_ISL_1849860, EPI_ISL_1849862, EPI_ISL_1849863, EPI_ISL_1849864, EPI_ISL_1849867, EPI_ISL_1849868, EPI_ISL_1849869, EPI_ISL_1849872, EPI_ISL_1849873, EPI_ISL_1849874, EPI_ISL_1849876, EPI_ISL_1849877, EPI_ISL_1849878, EPI_ISL_1849879, EPI_ISL_1849882, EPI_ISL_1849883, EPI_ISL_1849884, EPI_ISL_1849886, EPI_ISL_1849887, EPI_ISL_1849891, EPI_ISL_1849892, EPI_ISL_1849893, EPI_ISL_1849895, EPI_ISL_1849896, EPI_ISL_1849899, EPI_ISL_1849900, EPI_ISL_1849903, EPI_ISL_1849905, EPI_ISL_1849907, EPI_ISL_1849909, EPI_ISL_1849910, EPI_ISL_1849914, EPI_ISL_1849916, EPI_ISL_1849917, EPI_ISL_2108010, EPI_ISL_2108018, EPI_ISL_2108026, EPI_ISL_2108028, EPI_ISL_2108040, EPI_ISL_2108043, EPI_ISL_2108046, EPI_ISL_2108052, EPI_ISL_2108053, EPI_ISL_2108056, EPI_ISL_2108061, EPI_ISL_2108071, EPI_ISL_2108073, EPI_ISL_2108074, EPI_ISL_2108078, EPI_ISL_2108081, EPI_ISL_2108083, EPI_ISL_2108085, EPI_ISL_2108087, EPI_ISL_2108088, EPI_ISL_2108092, EPI_ISL_2108093, EPI_ISL_2108094, EPI_ISL_2108095, EPI_ISL_2108102, EPI_ISL_2108103, EPI_ISL_2108105, EPI_ISL_2108109, EPI_ISL_2108118, EPI_ISL_2108120, EPI_ISL_2108123, EPI_ISL_2108131, EPI_ISL_2108136, EPI_ISL_2108138, EPI_ISL_2108144, EPI_ISL_2108145, EPI_ISL_2108147, EPI_ISL_2108150, EPI_ISL_2108153, EPI_ISL_2108164, EPI_ISL_2108167, EPI_ISL_2108169, EPI_ISL_2108170, EPI_ISL_2108175, EPI_ISL_2108176, EPI_ISL_2108177, EPI_ISL_2108180, EPI_ISL_2108181, EPI_ISL_2108183, EPI_ISL_2108184, EPI_ISL_2108186, EPI_ISL_2108190, EPI_ISL_2108191, EPI_ISL_2108192, EPI_ISL_2108195, EPI_ISL_2108198, EPI_ISL_2108200, EPI_ISL_2108203, EPI_ISL_2108213, EPI_ISL_2108218, EPI_ISL_2108219, EPI_ISL_2108221, EPI_ISL_2108227, EPI_ISL_2108234, EPI_ISL_2108239, EPI_ISL_2108242, EPI_ISL_2108253, EPI_ISL_2108259, EPI_ISL_2108260, EPI_ISL_2114514, EPI_ISL_2114528, EPI_ISL_2114537, EPI_ISL_2114566, EPI_ISL_2114614, EPI_ISL_2114624, EPI_ISL_2114642, EPI_ISL_2114647, EPI_ISL_2114655, EPI_ISL_2114661, EPI_ISL_2114680, EPI_ISL_2114689, EPI_ISL_2114696, EPI_ISL_2114708, EPI_ISL_2114726, EPI_ISL_2114740, EPI_ISL_2114741, EPI_ISL_2114764, EPI_ISL_2114768, EPI_ISL_2114789, EPI_ISL_2114806, EPI_ISL_2114842, EPI_ISL_2114861, EPI_ISL_2114866, EPI_ISL_2114887, EPI_ISL_2114907, EPI_ISL_2114928, EPI_ISL_2114938, EPI_ISL_2114985, EPI_ISL_2114992, EPI_ISL_2115014, EPI_ISL_2115022, EPI_ISL_2115027, EPI_ISL_2115051, EPI_ISL_2115078, EPI_ISL_2115086, EPI_ISL_2118842, EPI_ISL_2118867, EPI_ISL_2118884, EPI_ISL_2118889, EPI_ISL_2118901, EPI_ISL_2118907, EPI_ISL_2118937, EPI_ISL_2118971, EPI_ISL_2118993, EPI_ISL_2118997, EPI_ISL_2119023, EPI_ISL_2119032, EPI_ISL_2119044, EPI_ISL_2119049, EPI_ISL_2119052, EPI_ISL_2119055, EPI_ISL_2119063, EPI_ISL_2119064, EPI_ISL_2119065, EPI_ISL_2119068, EPI_ISL_2119073, EPI_ISL_2119074, EPI_ISL_2119079, EPI_ISL_2119129, EPI_ISL_2119132, EPI_ISL_2119135, EPI_ISL_2119140, EPI_ISL_2119153, EPI_ISL_2119162, EPI_ISL_2119165, EPI_ISL_2119190, EPI_ISL_2119201, EPI_ISL_2119215, EPI_ISL_2119224, EPI_ISL_2119229, EPI_ISL_2119267, EPI_ISL_2119269, EPI_ISL_2119288, EPI_ISL_2119340, EPI_ISL_2119360, EPI_ISL_2119415, EPI_ISL_2119423, EPI_ISL_2119436, EPI_ISL_2119446, EPI_ISL_2119455, EPI_ISL_2119464, EPI_ISL_2119475, EPI_ISL_2119505, EPI_ISL_2119511, EPI_ISL_2119516, EPI_ISL_2119533, EPI_ISL_2119536, EPI_ISL_2119563, EPI_ISL_2119575, EPI_ISL_2119594, EPI_ISL_2119603, EPI_ISL_2119634, EPI_ISL_2119663, EPI_ISL_2119670, EPI_ISL_2119719, EPI_ISL_2119760, EPI_ISL_2119763, EPI_ISL_2119791, EPI_ISL_2119802, EPI_ISL_2119810, EPI_ISL_2119840, EPI_ISL_2119871, EPI_ISL_2119880, EPI_ISL_2119888, EPI_ISL_2119895, EPI_ISL_2119898, EPI_ISL_2119899, EPI_ISL_2119903, EPI_ISL_2119905, EPI_ISL_2119913, EPI_ISL_2119929, EPI_ISL_2119932, EPI_ISL_2119940, EPI_ISL_2119956, EPI_ISL_2119959, EPI_ISL_2119962, EPI_ISL_2119971, EPI_ISL_2120034, EPI_ISL_2128637, EPI_ISL_2128660, EPI_ISL_2128662, EPI_ISL_2128685, EPI_ISL_2128691, EPI_ISL_2128698, EPI_ISL_2128700, EPI_ISL_2128708, EPI_ISL_2128713, EPI_ISL_2128725, EPI_ISL_2128731, EPI_ISL_2128734, EPI_ISL_2128737, EPI_ISL_2128757, EPI_ISL_2128778, EPI_ISL_2128782, EPI_ISL_2128789, EPI_ISL_2128790, EPI_ISL_2128794, EPI_ISL_2128814, EPI_ISL_2128817, EPI_ISL_2128831, EPI_ISL_2128832, EPI_ISL_2128834, EPI_ISL_2128836, EPI_ISL_2128837, EPI_ISL_2128848, EPI_ISL_2128855, EPI_ISL_2128860, EPI_ISL_2128870, EPI_ISL_2128877, EPI_ISL_2128886, EPI_ISL_2128889, EPI_ISL_2128899, EPI_ISL_2128907, EPI_ISL_2128915, EPI_ISL_2128916, EPI_ISL_2128920, EPI_ISL_2128929

see above SYNLAB MVZ Heidelberg Robert Koch Institute

EPI_ISL_1642871, EPI_ISL_1642872, EPI_ISL_1642874, EPI_ISL_1642882, EPI_ISL_1642883, EPI_ISL_1642885, EPI_ISL_1642886, EPI_ISL_1642887, EPI_ISL_1642888, EPI_ISL_1642889, EPI_ISL_1642890, EPI_ISL_1642891, EPI_ISL_1642892, EPI_ISL_1642893, EPI_ISL_1642894, EPI_ISL_1642896, EPI_ISL_1642897, EPI_ISL_1642898, EPI_ISL_1642900, EPI_ISL_1642901, EPI_ISL_1642902, EPI_ISL_1642903, EPI_ISL_1642904, EPI_ISL_1642906, EPI_ISL_1642908, EPI_ISL_1642909, EPI_ISL_1642910, EPI_ISL_1642912, EPI_ISL_1642913, EPI_ISL_1642914, EPI_ISL_1642917, EPI_ISL_1642923, EPI_ISL_1642925, EPI_ISL_1642926, EPI_ISL_1642928, EPI_ISL_1642930, EPI_ISL_1642931, EPI_ISL_1642933, EPI_ISL_1642935, EPI_ISL_1642936, EPI_ISL_1642938, EPI_ISL_1642939, EPI_ISL_1642940, EPI_ISL_1642944, EPI_ISL_1642948, EPI_ISL_1642949, EPI_ISL_1642950, EPI_ISL_1642951, EPI_ISL_1642953, EPI_ISL_1642954, EPI_ISL_1642955, EPI_ISL_1642957, EPI_ISL_1642958, EPI_ISL_1642959, EPI_ISL_1642960, EPI_ISL_1642961, EPI_ISL_1642963, EPI_ISL_1642964, EPI_ISL_1642965, EPI_ISL_1642968, EPI_ISL_1642969, EPI_ISL_1642973, EPI_ISL_1642974, EPI_ISL_1642980, EPI_ISL_1642983, EPI_ISL_1642984, EPI_ISL_1642985, EPI_ISL_1642986, EPI_ISL_1642987, EPI_ISL_1642988, EPI_ISL_1642989, EPI_ISL_1642991, EPI_ISL_1642992, EPI_ISL_1642993, EPI_ISL_1642995, EPI_ISL_1642996, EPI_ISL_1642997, EPI_ISL_1642998, EPI_ISL_1643000, EPI_ISL_1643001, EPI_ISL_1643003, EPI_ISL_1643004, EPI_ISL_1643005, EPI_ISL_1643006, EPI_ISL_1643008, EPI_ISL_1643009, EPI_ISL_1643010, EPI_ISL_1643013, EPI_ISL_1643014, EPI_ISL_1643015, EPI_ISL_1643016, EPI_ISL_1643017, EPI_ISL_1643018, EPI_ISL_1643019, EPI_ISL_1643021, EPI_ISL_1643022, EPI_ISL_1643025, EPI_ISL_1643026, EPI_ISL_1643027, EPI_ISL_1643028, EPI_ISL_1643029, EPI_ISL_1643030, EPI_ISL_1643031, EPI_ISL_1643032, EPI_ISL_1643034, EPI_ISL_1643035, EPI_ISL_1643037, EPI_ISL_1643038, EPI_ISL_1643040, EPI_ISL_1643041, EPI_ISL_1643042, EPI_ISL_1643044, EPI_ISL_1643045, EPI_ISL_1643046, EPI_ISL_1643047, EPI_ISL_1643048, EPI_ISL_1643050, EPI_ISL_1643051, EPI_ISL_1643052, EPI_ISL_1643053, EPI_ISL_1643054, EPI_ISL_1643055, EPI_ISL_1643056, EPI_ISL_1643057, EPI_ISL_1643058, EPI_ISL_1643059, EPI_ISL_1643060, EPI_ISL_1643062, EPI_ISL_1643063, EPI_ISL_1643064, EPI_ISL_1643065, EPI_ISL_1643066, EPI_ISL_1643067, EPI_ISL_1643069, EPI_ISL_1643070, EPI_ISL_1643071, EPI_ISL_1643073, EPI_ISL_1643075, EPI_ISL_1643076, EPI_ISL_1643077, EPI_ISL_1643078, EPI_ISL_1643079, EPI_ISL_1643082, EPI_ISL_1643083, EPI_ISL_1643085, EPI_ISL_1643086, EPI_ISL_1643087, EPI_ISL_1643091, EPI_ISL_1643094, EPI_ISL_1643095, EPI_ISL_1643096, EPI_ISL_1643098, EPI_ISL_1643099, EPI_ISL_1643100, EPI_ISL_1643102, EPI_ISL_1643103, EPI_ISL_1643104, EPI_ISL_1643107, EPI_ISL_1643108, EPI_ISL_1643110, EPI_ISL_1644053, EPI_ISL_1644057, EPI_ISL_1644059, EPI_ISL_1644062, EPI_ISL_1644064, EPI_ISL_1644067, EPI_ISL_1644070, EPI_ISL_1644073, EPI_ISL_1644075, EPI_ISL_1644077, EPI_ISL_1644084, EPI_ISL_1644090, EPI_ISL_1644097, EPI_ISL_1644099, EPI_ISL_1644100, EPI_ISL_1644102, EPI_ISL_1644106, EPI_ISL_1644110, EPI_ISL_1644120, EPI_ISL_1644121, EPI_ISL_1644125, EPI_ISL_1644127, EPI_ISL_1644128, EPI_ISL_1644129, EPI_ISL_1644131, EPI_ISL_1644134, EPI_ISL_1644135, EPI_ISL_1644136, EPI_ISL_1644140, EPI_ISL_1644141, EPI_ISL_1644143, EPI_ISL_1644144, EPI_ISL_1644147, EPI_ISL_1644148, EPI_ISL_1644150, EPI_ISL_1644151, EPI_ISL_1644152, EPI_ISL_1644153, EPI_ISL_1644155, EPI_ISL_1644156, EPI_ISL_1644157, EPI_ISL_1644160, EPI_ISL_1644161, EPI_ISL_1644163, EPI_ISL_1644165, EPI_ISL_1644168, EPI_ISL_1644172, EPI_ISL_1644174, EPI_ISL_1644175, EPI_ISL_1644176, EPI_ISL_1644177, EPI_ISL_1644178, EPI_ISL_1644179, EPI_ISL_1644180, EPI_ISL_1644182, EPI_ISL_1644185, EPI_ISL_1644187, EPI_ISL_1644190, EPI_ISL_1644191, EPI_ISL_1644193, EPI_ISL_1644195, EPI_ISL_1644196, EPI_ISL_1644197, EPI_ISL_1644199, EPI_ISL_1644200, EPI_ISL_1644204, EPI_ISL_1644205, EPI_ISL_1644207, EPI_ISL_1644210, EPI_ISL_1644215, EPI_ISL_1644216, EPI_ISL_1644221, EPI_ISL_1644226, EPI_ISL_1644230, EPI_ISL_1644231, EPI_ISL_1644232, EPI_ISL_1644234, EPI_ISL_1644235, EPI_ISL_1644236, EPI_ISL_1644238, EPI_ISL_1644240, EPI_ISL_1644244, EPI_ISL_1644245, EPI_ISL_1644246, EPI_ISL_1644248, EPI_ISL_1644249, EPI_ISL_1644252, EPI_ISL_1644255, EPI_ISL_1644257, EPI_ISL_1644260, EPI_ISL_1644261, EPI_ISL_1644264, EPI_ISL_1644266, EPI_ISL_1644272, EPI_ISL_1644274, EPI_ISL_1644275, EPI_ISL_1644276, EPI_ISL_1644278, EPI_ISL_1644279, EPI_ISL_1644280, EPI_ISL_1644281, EPI_ISL_1644283, EPI_ISL_1644285, EPI_ISL_1644288, EPI_ISL_1644289, EPI_ISL_1644292, EPI_ISL_1644293, EPI_ISL_1644294, EPI_ISL_1644296, EPI_ISL_1644299, EPI_ISL_1644303, EPI_ISL_1644306, EPI_ISL_1644308, EPI_ISL_1644309, EPI_ISL_1644310, EPI_ISL_1644311, EPI_ISL_1644312, EPI_ISL_1644314, EPI_ISL_1644315, EPI_ISL_1644317, EPI_ISL_1644325, EPI_ISL_1644326, EPI_ISL_1644333, EPI_ISL_1644334, EPI_ISL_1644336, EPI_ISL_1644337, EPI_ISL_1644338, EPI_ISL_1644339, EPI_ISL_1644340, EPI_ISL_1644341, EPI_ISL_1644342, EPI_ISL_1644346, EPI_ISL_1644349, EPI_ISL_1644350, EPI_ISL_1644351, EPI_ISL_1644354, EPI_ISL_1644356, EPI_ISL_1644358, EPI_ISL_1644359, EPI_ISL_1644360, EPI_ISL_1644361, EPI_ISL_1644362, EPI_ISL_1644363, EPI_ISL_1644364, EPI_ISL_1644365, EPI_ISL_1644366, EPI_ISL_1644367, EPI_ISL_1644369, EPI_ISL_1644370, EPI_ISL_1644372, EPI_ISL_1644375, EPI_ISL_1644376, EPI_ISL_1644377, EPI_ISL_1644378, EPI_ISL_1644380, EPI_ISL_1644381, EPI_ISL_1644382, EPI_ISL_1644383, EPI_ISL_1644385, EPI_ISL_1644386, EPI_ISL_1644387, EPI_ISL_1644389, EPI_ISL_1644390, EPI_ISL_1644391, EPI_ISL_1644392, EPI_ISL_1644393, EPI_ISL_1644394, EPI_ISL_1644395, EPI_ISL_1644396, EPI_ISL_1644397, EPI_ISL_1644398, EPI_ISL_1644400, EPI_ISL_1644401, EPI_ISL_1644402, EPI_ISL_1644403, EPI_ISL_1644404, EPI_ISL_1644405, EPI_ISL_1644406, EPI_ISL_1644407, EPI_ISL_1644408, EPI_ISL_1644409, EPI_ISL_1644410, EPI_ISL_1644411, EPI_ISL_1644413, EPI_ISL_1644414, EPI_ISL_1644415, EPI_ISL_1644418, EPI_ISL_1644419, EPI_ISL_1644423, EPI_ISL_1644424, EPI_ISL_1644425, EPI_ISL_1644426, EPI_ISL_1644430, EPI_ISL_1644431, EPI_ISL_1644434, EPI_ISL_1644435, EPI_ISL_1644436, EPI_ISL_1644437, EPI_ISL_1644438, EPI_ISL_1644440, EPI_ISL_1644441, EPI_ISL_1644448, EPI_ISL_1644451, EPI_ISL_1644452, EPI_ISL_1644453, EPI_ISL_1644455, EPI_ISL_1644457, EPI_ISL_1644460, EPI_ISL_1644461, EPI_ISL_1644465, EPI_ISL_1644467, EPI_ISL_1644468, EPI_ISL_1644471, EPI_ISL_1644472, EPI_ISL_1644479, EPI_ISL_1644483, EPI_ISL_1644486, EPI_ISL_1644487, EPI_ISL_1644488, EPI_ISL_1644490, EPI_ISL_1644491, EPI_ISL_1644492, EPI_ISL_1644494, EPI_ISL_1644506, EPI_ISL_1644507, EPI_ISL_1644508, EPI_ISL_1644509, EPI_ISL_1644511, EPI_ISL_1644512, EPI_ISL_1644513, EPI_ISL_1644514, EPI_ISL_1644515, EPI_ISL_1644516, EPI_ISL_1644517, EPI_ISL_1644519, EPI_ISL_1644520, EPI_ISL_1644522, EPI_ISL_1644523, EPI_ISL_1644525, EPI_ISL_1644527, EPI_ISL_1644529, EPI_ISL_1644531, EPI_ISL_1644533, EPI_ISL_1644535, EPI_ISL_1644538, EPI_ISL_1644540, EPI_ISL_1645235, EPI_ISL_1645236, EPI_ISL_1645241, EPI_ISL_1645243, EPI_ISL_1645246, EPI_ISL_1645248, EPI_ISL_1645251, EPI_ISL_1645253, EPI_ISL_1645254, EPI_ISL_1645255, EPI_ISL_1645256, EPI_ISL_1645257, EPI_ISL_1645258, EPI_ISL_1645259, EPI_ISL_1645262, EPI_ISL_1645263, EPI_ISL_1645264, EPI_ISL_1645265, EPI_ISL_1645266, EPI_ISL_1645268, EPI_ISL_1645272, EPI_ISL_1645273, EPI_ISL_1645275, EPI_ISL_1645276, EPI_ISL_1645280, EPI_ISL_1645283, EPI_ISL_1645284, EPI_ISL_1645285, EPI_ISL_1645286, EPI_ISL_1645287, EPI_ISL_1645290, EPI_ISL_1645291, EPI_ISL_1645292, EPI_ISL_1645293, EPI_ISL_1645294, EPI_ISL_1645295, EPI_ISL_1645296, EPI_ISL_1645297, EPI_ISL_1645299, EPI_ISL_1645301, EPI_ISL_1645303, EPI_ISL_1645304, EPI_ISL_1645305, EPI_ISL_1645306, EPI_ISL_1645308, EPI_ISL_1645309, EPI_ISL_1645310, EPI_ISL_1645312, EPI_ISL_1645313, EPI_ISL_1645316, EPI_ISL_1645317, EPI_ISL_1645318, EPI_ISL_1645321, EPI_ISL_1645323, EPI_ISL_1645324, EPI_ISL_1645326, EPI_ISL_1645336, EPI_ISL_1645337, EPI_ISL_1645341, EPI_ISL_1645347, EPI_ISL_1645351, EPI_ISL_1645352, EPI_ISL_1645356, EPI_ISL_1645363, EPI_ISL_1645366, EPI_ISL_1645367, EPI_ISL_1645368, EPI_ISL_1645370, EPI_ISL_1645372, EPI_ISL_1645374, EPI_ISL_1645375, EPI_ISL_1645376, EPI_ISL_1645377, EPI_ISL_1645379, EPI_ISL_1645381, EPI_ISL_1645382, EPI_ISL_1645386, EPI_ISL_1645391, EPI_ISL_1645392, EPI_ISL_1645393, EPI_ISL_1645394, EPI_ISL_1645395, EPI_ISL_1645396, EPI_ISL_1645397, EPI_ISL_1645399, EPI_ISL_1645400, EPI_ISL_1645401, EPI_ISL_1645402, EPI_ISL_1645403, EPI_ISL_1645404, EPI_ISL_1645405, EPI_ISL_1645406, EPI_ISL_1645408, EPI_ISL_1645409, EPI_ISL_1645410, EPI_ISL_1645411, EPI_ISL_1645412, EPI_ISL_1645414, EPI_ISL_1645415, EPI_ISL_1645417, EPI_ISL_1645418, EPI_ISL_1645419, EPI_ISL_1645420, EPI_ISL_1645422, EPI_ISL_1645423, EPI_ISL_1645425, EPI_ISL_1645426, EPI_ISL_1645427, EPI_ISL_1645428, EPI_ISL_1645429, EPI_ISL_1645430, EPI_ISL_1645431, EPI_ISL_1645433, EPI_ISL_1645434, EPI_ISL_1645435, EPI_ISL_1645437, EPI_ISL_1645438, EPI_ISL_1645442, EPI_ISL_1645448, EPI_ISL_1645454, EPI_ISL_1645456, EPI_ISL_1645458, EPI_ISL_1645459, EPI_ISL_1645462, EPI_ISL_1645465, EPI_ISL_1645466, EPI_ISL_1645467, EPI_ISL_1645468, EPI_ISL_1645470, EPI_ISL_1645477, EPI_ISL_1645480, EPI_ISL_1645482, EPI_ISL_1645486, EPI_ISL_1645490, EPI_ISL_1645491, EPI_ISL_1645502, EPI_ISL_1645504, EPI_ISL_1645505, EPI_ISL_1645508, EPI_ISL_1645511, EPI_ISL_1645513, EPI_ISL_1645514, EPI_ISL_1645523, EPI_ISL_1645524, EPI_ISL_1645528, EPI_ISL_1645529, EPI_ISL_1645530, EPI_ISL_1645532, EPI_ISL_1645534, EPI_ISL_1645535, EPI_ISL_1645537, EPI_ISL_1645539, EPI_ISL_1645540, EPI_ISL_1645542, EPI_ISL_1645545, EPI_ISL_1645548, EPI_ISL_1645550, EPI_ISL_1645552, EPI_ISL_1645554, EPI_ISL_1645556, EPI_ISL_1645561, EPI_ISL_1645562, EPI_ISL_1645564, EPI_ISL_1645568, EPI_ISL_1645575, EPI_ISL_1645580, EPI_ISL_1645582, EPI_ISL_1645590, EPI_ISL_1645593, EPI_ISL_1645595, EPI_ISL_1645597, EPI_ISL_1645600, EPI_ISL_1645601, EPI_ISL_1645603, EPI_ISL_1645605, EPI_ISL_1645606, EPI_ISL_1645612, EPI_ISL_1645616, EPI_ISL_1645617, EPI_ISL_1645620, EPI_ISL_1645622, EPI_ISL_1645624, EPI_ISL_1645625, EPI_ISL_1645628, EPI_ISL_1645630, EPI_ISL_1645635, EPI_ISL_1645639, EPI_ISL_1645643, EPI_ISL_1645644, EPI_ISL_1645648, EPI_ISL_1645649, EPI_ISL_1645650, EPI_ISL_1645652, EPI_ISL_1645657, EPI_ISL_1645661, EPI_ISL_1645665, EPI_ISL_1645666, EPI_ISL_1645667, EPI_ISL_1645670, EPI_ISL_1645672, EPI_ISL_1645674, EPI_ISL_1645680, EPI_ISL_1645682, EPI_ISL_1645684, EPI_ISL_1645688, EPI_ISL_1645690, EPI_ISL_1645700, EPI_ISL_1645705, EPI_ISL_1645708, EPI_ISL_1645710, EPI_ISL_1645715, EPI_ISL_1645719, EPI_ISL_1645723, EPI_ISL_1645725, EPI_ISL_1645727, EPI_ISL_1645729, EPI_ISL_1645733, EPI_ISL_1645736, EPI_ISL_1645738, EPI_ISL_1645746, EPI_ISL_1645748, EPI_ISL_1645751, EPI_ISL_1645755, EPI_ISL_1645762, EPI_ISL_1645764, EPI_ISL_1645767, EPI_ISL_1645780, EPI_ISL_1645782, EPI_ISL_1645789, EPI_ISL_1645792, EPI_ISL_1645794, EPI_ISL_1645799, EPI_ISL_1645803, EPI_ISL_1645805, EPI_ISL_1645807, EPI_ISL_1645811, EPI_ISL_1645815, EPI_ISL_1645844, EPI_ISL_1645858, EPI_ISL_1645861, EPI_ISL_1645865, EPI_ISL_1645871, EPI_ISL_1645875, EPI_ISL_1645876, EPI_ISL_1645877, EPI_ISL_1645884, EPI_ISL_1645886, EPI_ISL_1645888, EPI_ISL_1645892, EPI_ISL_1645899, EPI_ISL_1645902, EPI_ISL_1645904, EPI_ISL_1645909, EPI_ISL_1645913, EPI_ISL_1645917, EPI_ISL_1645919, EPI_ISL_1645931, EPI_ISL_1722388, EPI_ISL_1722390, EPI_ISL_1722391, EPI_ISL_1722392, EPI_ISL_1722393, EPI_ISL_1722394, EPI_ISL_1722395, EPI_ISL_1722396, EPI_ISL_1722397, EPI_ISL_1722401, EPI_ISL_1722403, EPI_ISL_1722404, EPI_ISL_1722408, EPI_ISL_1722409, EPI_ISL_1722410, EPI_ISL_1722411, EPI_ISL_1722412, EPI_ISL_1722413, EPI_ISL_1722415, EPI_ISL_1722416, EPI_ISL_1722418, EPI_ISL_1722420, EPI_ISL_1722421, EPI_ISL_1722422, EPI_ISL_1722423, EPI_ISL_1722424, EPI_ISL_1722425, EPI_ISL_1722426, EPI_ISL_1722427, EPI_ISL_1722428, EPI_ISL_1722430, EPI_ISL_1722431, EPI_ISL_1722435, EPI_ISL_1722436, EPI_ISL_1722437, EPI_ISL_1722438, EPI_ISL_1722440, EPI_ISL_1722441, EPI_ISL_1722442, EPI_ISL_1722443, EPI_ISL_1722444, EPI_ISL_1722445, EPI_ISL_1722446, EPI_ISL_1722447, EPI_ISL_1722449, EPI_ISL_1722450, EPI_ISL_1722451, EPI_ISL_1722453, EPI_ISL_1722455, EPI_ISL_1722456, EPI_ISL_1722458, EPI_ISL_1722459, EPI_ISL_1722461, EPI_ISL_1722462, EPI_ISL_1722464, EPI_ISL_1722467, EPI_ISL_1722469, EPI_ISL_1722471, EPI_ISL_1722472, EPI_ISL_1722473, EPI_ISL_1722475, EPI_ISL_1722476, EPI_ISL_1722479, EPI_ISL_1722482, EPI_ISL_1722483, EPI_ISL_1722484, EPI_ISL_1722488, EPI_ISL_1722489, EPI_ISL_1722490, EPI_ISL_1722491, EPI_ISL_1722492, EPI_ISL_1722494, EPI_ISL_1722497, EPI_ISL_1722498, EPI_ISL_1722500, EPI_ISL_1722501, EPI_ISL_1722503, EPI_ISL_1722506, EPI_ISL_1722507, EPI_ISL_1722508, EPI_ISL_1722511, EPI_ISL_1722512, EPI_ISL_1722513, EPI_ISL_1722514, EPI_ISL_1722516, EPI_ISL_1722519, EPI_ISL_1722520, EPI_ISL_1722523, EPI_ISL_1722525, EPI_ISL_1722527, EPI_ISL_1722528, EPI_ISL_1722529, EPI_ISL_1722530, EPI_ISL_1722532, EPI_ISL_1722533, EPI_ISL_1722534, EPI_ISL_1722536, EPI_ISL_1722538, EPI_ISL_1722540, EPI_ISL_1722543, EPI_ISL_1722546, EPI_ISL_1722547, EPI_ISL_1722548, EPI_ISL_1722550, EPI_ISL_1722553, EPI_ISL_1722555, EPI_ISL_1722556, EPI_ISL_1722558, EPI_ISL_1722559, EPI_ISL_1722561, EPI_ISL_1722562, EPI_ISL_1722564, EPI_ISL_1722565, EPI_ISL_1722566, EPI_ISL_1722569, EPI_ISL_1722570, EPI_ISL_1722571, EPI_ISL_1722573, EPI_ISL_1722575, EPI_ISL_1722576, EPI_ISL_1722583, EPI_ISL_1722584, EPI_ISL_1722585, EPI_ISL_1722586, EPI_ISL_1722589, EPI_ISL_1722591, EPI_ISL_1722593, EPI_ISL_1722594, EPI_ISL_1722596, EPI_ISL_1722597, EPI_ISL_1722600, EPI_ISL_1722601, EPI_ISL_1722602, EPI_ISL_1722603, EPI_ISL_1722604, EPI_ISL_1722606, EPI_ISL_1722607, EPI_ISL_1722608, EPI_ISL_1722612, EPI_ISL_1722614, EPI_ISL_1722620, EPI_ISL_1722626, EPI_ISL_1722629, EPI_ISL_1722631, EPI_ISL_1722634, EPI_ISL_1722635, EPI_ISL_1722636, EPI_ISL_1722638, EPI_ISL_1722642, EPI_ISL_1722646, EPI_ISL_1722648, EPI_ISL_1722651, EPI_ISL_1722652, EPI_ISL_1722653, EPI_ISL_1722654, EPI_ISL_1722655, EPI_ISL_1722657, EPI_ISL_1722662, EPI_ISL_1722666, EPI_ISL_1722667, EPI_ISL_1722668, EPI_ISL_1722669, EPI_ISL_1722670, EPI_ISL_1722671, EPI_ISL_1723064, EPI_ISL_1723066, EPI_ISL_1723069, EPI_ISL_1723070, EPI_ISL_1723071, EPI_ISL_1723073, EPI_ISL_1723074, EPI_ISL_1723075, EPI_ISL_1723076, EPI_ISL_1723078, EPI_ISL_1723079, EPI_ISL_1723081, EPI_ISL_1723082, EPI_ISL_1723083, EPI_ISL_1723084, EPI_ISL_1723086, EPI_ISL_1723087, EPI_ISL_1723088, EPI_ISL_1723089, EPI_ISL_1723090, EPI_ISL_1723091, EPI_ISL_1723092, EPI_ISL_1723094, EPI_ISL_1723095, EPI_ISL_1723096, EPI_ISL_1723097, EPI_ISL_1723099, EPI_ISL_1723101, EPI_ISL_1723102, EPI_ISL_1723103, EPI_ISL_1723104, EPI_ISL_1723106, EPI_ISL_1723107, EPI_ISL_1723108, EPI_ISL_1723110, EPI_ISL_1723111, EPI_ISL_1723112, EPI_ISL_1723114, EPI_ISL_1723115, EPI_ISL_1723117, EPI_ISL_1723118, EPI_ISL_1723119, EPI_ISL_1723121, EPI_ISL_1723122, EPI_ISL_1723123, EPI_ISL_1723125, EPI_ISL_1723126, EPI_ISL_1723128, EPI_ISL_1723129, EPI_ISL_1723130, EPI_ISL_1723131, EPI_ISL_1723132, EPI_ISL_1723133, EPI_ISL_1723134, EPI_ISL_1723136, EPI_ISL_1723137, EPI_ISL_1723138, EPI_ISL_1723139, EPI_ISL_1723140, EPI_ISL_1723141, EPI_ISL_1723142, EPI_ISL_1723145, EPI_ISL_1723146, EPI_ISL_1723147, EPI_ISL_1723149, EPI_ISL_1723150, EPI_ISL_1723151, EPI_ISL_1723152, EPI_ISL_1723153, EPI_ISL_1723155, EPI_ISL_1723156, EPI_ISL_1723158, EPI_ISL_1723159, EPI_ISL_1723161, EPI_ISL_1723162, EPI_ISL_1723164, EPI_ISL_1723165, EPI_ISL_1723167, EPI_ISL_1723169, EPI_ISL_1723170, EPI_ISL_1723171, EPI_ISL_1723173, EPI_ISL_1723174, EPI_ISL_1723175, EPI_ISL_1723176, EPI_ISL_1723179, EPI_ISL_1723180, EPI_ISL_1723182, EPI_ISL_1723183, EPI_ISL_1723186, EPI_ISL_1723187, EPI_ISL_1723188, EPI_ISL_1723191, EPI_ISL_1723192, EPI_ISL_1723194, EPI_ISL_1723196, EPI_ISL_1723197, EPI_ISL_1723199, EPI_ISL_1723201, EPI_ISL_1723204, EPI_ISL_1723206, EPI_ISL_1723208, EPI_ISL_1723211, EPI_ISL_1723212, EPI_ISL_1723215, EPI_ISL_1723216, EPI_ISL_1723217, EPI_ISL_1723219, EPI_ISL_1723220, EPI_ISL_1723222, EPI_ISL_1723223, EPI_ISL_1723226, EPI_ISL_1723228, EPI_ISL_1723229, EPI_ISL_1723231, EPI_ISL_1723234, EPI_ISL_1723235, EPI_ISL_1723238, EPI_ISL_1723239, EPI_ISL_1723242, EPI_ISL_1723245, EPI_ISL_1723246, EPI_ISL_1723248, EPI_ISL_1723250, EPI_ISL_1723251, EPI_ISL_1723255, EPI_ISL_1723258, EPI_ISL_1723261, EPI_ISL_1723264, EPI_ISL_1723265, EPI_ISL_1723268, EPI_ISL_1723269, EPI_ISL_1723270, EPI_ISL_1723273, EPI_ISL_1723274, EPI_ISL_1723276, EPI_ISL_1723277, EPI_ISL_1723278, EPI_ISL_1723279, EPI_ISL_1723281, EPI_ISL_1723282, EPI_ISL_1723284, EPI_ISL_1723286, EPI_ISL_1723288, EPI_ISL_1723289, EPI_ISL_1723291, EPI_ISL_1723292, EPI_ISL_1723293, EPI_ISL_1723295, EPI_ISL_1723297, EPI_ISL_1723298, EPI_ISL_1723299, EPI_ISL_1723300, EPI_ISL_1723302, EPI_ISL_1723303, EPI_ISL_1723304, EPI_ISL_1723305, EPI_ISL_1723306, EPI_ISL_1723307, EPI_ISL_1723308, EPI_ISL_1723309, EPI_ISL_1723311, EPI_ISL_1723313, EPI_ISL_1723314, EPI_ISL_1723315, EPI_ISL_1723317, EPI_ISL_1723318, EPI_ISL_1723320, EPI_ISL_1723321, EPI_ISL_1723323, EPI_ISL_1723324, EPI_ISL_1723326, EPI_ISL_1723331, EPI_ISL_1723332, EPI_ISL_1723334, EPI_ISL_1723336, EPI_ISL_1723338, EPI_ISL_1723339, EPI_ISL_1723340, EPI_ISL_1723341, EPI_ISL_1723342, EPI_ISL_1723343, EPI_ISL_1723345, EPI_ISL_1723347, EPI_ISL_1723348, EPI_ISL_1723349, EPI_ISL_1723350, EPI_ISL_1724485, EPI_ISL_1724487, EPI_ISL_1724489, EPI_ISL_1724491, EPI_ISL_1724493, EPI_ISL_1724494, EPI_ISL_1724496, EPI_ISL_1724497, EPI_ISL_1724498, EPI_ISL_1724500, EPI_ISL_1724503, EPI_ISL_1724504, EPI_ISL_1724505, EPI_ISL_1724508, EPI_ISL_1724509, EPI_ISL_1724510, EPI_ISL_1724511,

EPI_ISL_1724512, EPI_ISL_1724513, EPI_ISL_1724517, EPI_ISL_1724518, EPI_ISL_1724519, EPI_ISL_1724520, EPI_ISL_1724521, EPI_ISL_1724523, EPI_ISL_1724524, EPI_ISL_1724525, EPI_ISL_1724526, EPI_ISL_1724527, EPI_ISL_1724528, EPI_ISL_1724529, EPI_ISL_1724530, EPI_ISL_1724531, EPI_ISL_1724533, EPI_ISL_1724534, EPI_ISL_1724535, EPI_ISL_1724537, EPI_ISL_1724538, EPI_ISL_1724539, EPI_ISL_1724540, EPI_ISL_1724541, EPI_ISL_1724542, EPI_ISL_1724543, EPI_ISL_1724545, EPI_ISL_1724546, EPI_ISL_1724549, EPI_ISL_1724550, EPI_ISL_1724553, EPI_ISL_1724555, EPI_ISL_1724556, EPI_ISL_1724558, EPI_ISL_1724560, EPI_ISL_1724561, EPI_ISL_1724563, EPI_ISL_1724564, EPI_ISL_1724565, EPI_ISL_1724566, EPI_ISL_1724567, EPI_ISL_1724569, EPI_ISL_1724572, EPI_ISL_1724573, EPI_ISL_1724574, EPI_ISL_1724577, EPI_ISL_1724578, EPI_ISL_1724580, EPI_ISL_1724581, EPI_ISL_1724583, EPI_ISL_1724586, EPI_ISL_1724587, EPI_ISL_1724588, EPI_ISL_1724589, EPI_ISL_1724590, EPI_ISL_1724591, EPI_ISL_1724592, EPI_ISL_1724594, EPI_ISL_1724595, EPI_ISL_1724596, EPI_ISL_1724597, EPI_ISL_1724598, EPI_ISL_1724600, EPI_ISL_1724601, EPI_ISL_1724602, EPI_ISL_1724603, EPI_ISL_1724605, EPI_ISL_1724606, EPI_ISL_1724607, EPI_ISL_1724608, EPI_ISL_1724609, EPI_ISL_1724610, EPI_ISL_1724611, EPI_ISL_1724612, EPI_ISL_1724613, EPI_ISL_1724614, EPI_ISL_1724615, EPI_ISL_1724616, EPI_ISL_1724618, EPI_ISL_1724619, EPI_ISL_1724620, EPI_ISL_1724621, EPI_ISL_1724623, EPI_ISL_1724624, EPI_ISL_1724626, EPI_ISL_1724628, EPI_ISL_1724629, EPI_ISL_1724630, EPI_ISL_1724632, EPI_ISL_1724633, EPI_ISL_1724634, EPI_ISL_1724635, EPI_ISL_1724636, EPI_ISL_1724637, EPI_ISL_1724638, EPI_ISL_1724640, EPI_ISL_1724641, EPI_ISL_1724642, EPI_ISL_1724644, EPI_ISL_1724645, EPI_ISL_1724646, EPI_ISL_1724647, EPI_ISL_1724648, EPI_ISL_1724650, EPI_ISL_1724653, EPI_ISL_1724655, EPI_ISL_1724656, EPI_ISL_1724657, EPI_ISL_1724658, EPI_ISL_1724660, EPI_ISL_1724665, EPI_ISL_1724666, EPI_ISL_1724667, EPI_ISL_1724669, EPI_ISL_1724670, EPI_ISL_1724671, EPI_ISL_1724673, EPI_ISL_1724674, EPI_ISL_1724675, EPI_ISL_1724676, EPI_ISL_1724678, EPI_ISL_1724679, EPI_ISL_1724680, EPI_ISL_1724682, EPI_ISL_1724687, EPI_ISL_1724688, EPI_ISL_1724689, EPI_ISL_1724693, EPI_ISL_1724694, EPI_ISL_1724695, EPI_ISL_1724696, EPI_ISL_1724698, EPI_ISL_1724699, EPI_ISL_1724701, EPI_ISL_1724703, EPI_ISL_1724704, EPI_ISL_1724705, EPI_ISL_1724706, EPI_ISL_1724707, EPI_ISL_1724708, EPI_ISL_1724709, EPI_ISL_1724710, EPI_ISL_1724712, EPI_ISL_1724713, EPI_ISL_1724714, EPI_ISL_1724715, EPI_ISL_1724716, EPI_ISL_1724719, EPI_ISL_1724720, EPI_ISL_1724722, EPI_ISL_1724723, EPI_ISL_1724724, EPI_ISL_1724727, EPI_ISL_1724728, EPI_ISL_1724729, EPI_ISL_1724731, EPI_ISL_1724733, EPI_ISL_1724734, EPI_ISL_1724735, EPI_ISL_1724736, EPI_ISL_1724738, EPI_ISL_1724739, EPI_ISL_1724741, EPI_ISL_1724742, EPI_ISL_1724745, EPI_ISL_1724746, EPI_ISL_1724749, EPI_ISL_1724750, EPI_ISL_1724753, EPI_ISL_1724754, EPI_ISL_1724756, EPI_ISL_1724757, EPI_ISL_1724759, EPI_ISL_1724760, EPI_ISL_1725966, EPI_ISL_1725968, EPI_ISL_1725976, EPI_ISL_1725979, EPI_ISL_1725987, EPI_ISL_1725996, EPI_ISL_1725999, EPI_ISL_1726000, EPI_ISL_1726007, EPI_ISL_1726008, EPI_ISL_1726013, EPI_ISL_1726014, EPI_ISL_1726015, EPI_ISL_1726023, EPI_ISL_1726026, EPI_ISL_1726027, EPI_ISL_1726031, EPI_ISL_1726036, EPI_ISL_1726043, EPI_ISL_1726045, EPI_ISL_1726052, EPI_ISL_1726058, EPI_ISL_1726060, EPI_ISL_1726062, EPI_ISL_1726076, EPI_ISL_1726078, EPI_ISL_1726079, EPI_ISL_1726081, EPI_ISL_1726087, EPI_ISL_1726091, EPI_ISL_1726093, EPI_ISL_1726101, EPI_ISL_1726106, EPI_ISL_1726111, EPI_ISL_1726114, EPI_ISL_1726119, EPI_ISL_1726124, EPI_ISL_1726126, EPI_ISL_1726127, EPI_ISL_1726129, EPI_ISL_1726140, EPI_ISL_1726142, EPI_ISL_1726144, EPI_ISL_1726145, EPI_ISL_1726149, EPI_ISL_1726150, EPI_ISL_1726153, EPI_ISL_1727588, EPI_ISL_1727592, EPI_ISL_1727596, EPI_ISL_1727597, EPI_ISL_1727599, EPI_ISL_1727601, EPI_ISL_1727603, EPI_ISL_1727608, EPI_ISL_1727610, EPI_ISL_1727612, EPI_ISL_1727613, EPI_ISL_1727614, EPI_ISL_1727620, EPI_ISL_1727622, EPI_ISL_1727623, EPI_ISL_1727629, EPI_ISL_1727632, EPI_ISL_1727633, EPI_ISL_1727634, EPI_ISL_1727642, EPI_ISL_1727643, EPI_ISL_1727644, EPI_ISL_1727647, EPI_ISL_1727651, EPI_ISL_1727653, EPI_ISL_1846822, EPI_ISL_1846837, EPI_ISL_1846899, EPI_ISL_1847012

see above | SYNLAB MVZ Leinfelden-Echterdingen | Robert Koch Institute

EPI_ISL_1641260, EPI_ISL_1641262, EPI_ISL_1641264, EPI_ISL_1641266, EPI_ISL_1641268, EPI_ISL_1641269, EPI_ISL_1641270, EPI_ISL_1641273, EPI_ISL_1641274, EPI_ISL_1641275, EPI_ISL_1641276, EPI_ISL_1641278, EPI_ISL_1641279, EPI_ISL_1641280, EPI_ISL_1641282, EPI_ISL_1641283, EPI_ISL_1641284, EPI_ISL_1641285, EPI_ISL_1641286, EPI_ISL_1641287, EPI_ISL_1641288, EPI_ISL_1641289, EPI_ISL_1641290, EPI_ISL_1641291, EPI_ISL_1641292, EPI_ISL_1641294, EPI_ISL_1641295, EPI_ISL_1641296, EPI_ISL_1641297, EPI_ISL_1641298, EPI_ISL_1641299, EPI_ISL_1641300, EPI_ISL_1641301, EPI_ISL_1641303, EPI_ISL_1641304, EPI_ISL_1641305, EPI_ISL_1641306, EPI_ISL_1641307, EPI_ISL_1641309, EPI_ISL_1641311, EPI_ISL_1641313, EPI_ISL_1641314, EPI_ISL_1641316, EPI_ISL_1641317, EPI_ISL_1641318, EPI_ISL_1641319, EPI_ISL_1641320, EPI_ISL_1641321, EPI_ISL_1641322, EPI_ISL_1641323, EPI_ISL_1641325, EPI_ISL_1641326, EPI_ISL_1641327, EPI_ISL_1641328, EPI_ISL_1641329, EPI_ISL_1641331, EPI_ISL_1641332, EPI_ISL_1641333, EPI_ISL_1641334, EPI_ISL_1641335, EPI_ISL_1641337, EPI_ISL_1641338, EPI_ISL_1641339, EPI_ISL_1641340, EPI_ISL_1641341, EPI_ISL_1641342, EPI_ISL_1641343, EPI_ISL_1641344, EPI_ISL_1641345, EPI_ISL_1641346, EPI_ISL_1641347, EPI_ISL_1641348, EPI_ISL_1641349, EPI_ISL_1641350, EPI_ISL_1641351, EPI_ISL_1641352, EPI_ISL_1641355, EPI_ISL_1641357, EPI_ISL_1641358, EPI_ISL_1641359, EPI_ISL_1641362, EPI_ISL_1641363, EPI_ISL_1641364, EPI_ISL_1641366, EPI_ISL_1641368, EPI_ISL_1641369, EPI_ISL_1641371, EPI_ISL_1641372, EPI_ISL_1641374, EPI_ISL_1641376, EPI_ISL_1641377, EPI_ISL_1641378, EPI_ISL_1641381, EPI_ISL_1641382, EPI_ISL_1641383, EPI_ISL_1641385, EPI_ISL_1641387, EPI_ISL_1641388, EPI_ISL_1641389, EPI_ISL_1641391, EPI_ISL_1641392, EPI_ISL_1641403, EPI_ISL_1641404, EPI_ISL_1641405, EPI_ISL_1641406, EPI_ISL_1641407, EPI_ISL_1641408, EPI_ISL_1641410, EPI_ISL_1641411, EPI_ISL_1641414, EPI_ISL_1641415, EPI_ISL_1641417, EPI_ISL_1641419, EPI_ISL_1641422, EPI_ISL_1641423, EPI_ISL_1641425, EPI_ISL_1641427, EPI_ISL_1641428, EPI_ISL_1641429, EPI_ISL_1641431, EPI_ISL_1641432, EPI_ISL_1641433, EPI_ISL_1641434, EPI_ISL_1641435, EPI_ISL_1641436, EPI_ISL_1641437, EPI_ISL_1641438, EPI_ISL_1641441, EPI_ISL_1641442, EPI_ISL_1641443, EPI_ISL_1641444, EPI_ISL_1641446, EPI_ISL_1641447, EPI_ISL_1641448, EPI_ISL_1641449, EPI_ISL_1641451, EPI_ISL_1641452, EPI_ISL_1641454, EPI_ISL_1641455, EPI_ISL_1641456, EPI_ISL_1641457, EPI_ISL_1641458, EPI_ISL_1641459, EPI_ISL_1641460, EPI_ISL_1641461, EPI_ISL_1641462, EPI_ISL_1641463, EPI_ISL_1641465, EPI_ISL_1641466, EPI_ISL_1641468, EPI_ISL_1641469, EPI_ISL_1641471, EPI_ISL_1641472, EPI_ISL_1641473, EPI_ISL_1641474, EPI_ISL_1641476, EPI_ISL_1641477, EPI_ISL_1641478, EPI_ISL_1641479, EPI_ISL_1641480, EPI_ISL_1641481, EPI_ISL_1641482, EPI_ISL_1641483, EPI_ISL_1641484, EPI_ISL_1641485, EPI_ISL_1641486, EPI_ISL_1641488, EPI_ISL_1641489, EPI_ISL_1641491, EPI_ISL_1641492, EPI_ISL_1641493, EPI_ISL_1641495, EPI_ISL_1641497, EPI_ISL_1641498, EPI_ISL_1641499, EPI_ISL_1641500, EPI_ISL_1641501, EPI_ISL_1641503, EPI_ISL_1641504, EPI_ISL_1641505, EPI_ISL_1641507, EPI_ISL_1641508, EPI_ISL_1641509, EPI_ISL_1641510, EPI_ISL_1641513, EPI_ISL_1641515, EPI_ISL_1641517, EPI_ISL_1641518, EPI_ISL_1641519, EPI_ISL_1641520, EPI_ISL_1641521, EPI_ISL_1641522, EPI_ISL_1641523, EPI_ISL_1641524, EPI_ISL_1641525, EPI_ISL_1641526, EPI_ISL_1641527, EPI_ISL_1641528, EPI_ISL_1641529, EPI_ISL_1641531, EPI_ISL_1641532, EPI_ISL_1641533, EPI_ISL_1641534, EPI_ISL_1641535, EPI_ISL_1641537, EPI_ISL_1641538, EPI_ISL_1641539, EPI_ISL_1641540, EPI_ISL_1641542, EPI_ISL_1641543, EPI_ISL_1641544, EPI_ISL_1641545, EPI_ISL_1641546, EPI_ISL_1641547, EPI_ISL_1641548, EPI_ISL_1641549, EPI_ISL_1641550, EPI_ISL_1641551, EPI_ISL_1641552, EPI_ISL_1641553, EPI_ISL_1641554, EPI_ISL_1641555, EPI_ISL_1641556, EPI_ISL_1641557, EPI_ISL_1641558, EPI_ISL_1641560, EPI_ISL_1641561, EPI_ISL_1641562, EPI_ISL_1641564, EPI_ISL_1641565, EPI_ISL_1641566, EPI_ISL_1641567, EPI_ISL_1641568, EPI_ISL_1641569, EPI_ISL_1641570, EPI_ISL_1641572, EPI_ISL_1641573, EPI_ISL_1641576, EPI_ISL_1641578, EPI_ISL_1641582, EPI_ISL_1641583, EPI_ISL_1641584, EPI_ISL_1641585, EPI_ISL_1641586, EPI_ISL_1641587, EPI_ISL_1641588, EPI_ISL_1641590, EPI_ISL_1641591, EPI_ISL_1641593, EPI_ISL_1641595, EPI_ISL_1641597, EPI_ISL_1641599, EPI_ISL_1641600, EPI_ISL_1641601, EPI_ISL_1641602, EPI_ISL_1641603, EPI_ISL_1641605, EPI_ISL_1641608, EPI_ISL_1641610, EPI_ISL_1641611, EPI_ISL_1641613, EPI_ISL_1641614, EPI_ISL_1641615, EPI_ISL_1641616, EPI_ISL_1641617, EPI_ISL_1641618, EPI_ISL_1641620, EPI_ISL_1641621, EPI_ISL_1641623, EPI_ISL_1641624, EPI_ISL_1641629, EPI_ISL_1641631, EPI_ISL_1641633, EPI_ISL_1641634, EPI_ISL_1641635, EPI_ISL_1641636, EPI_ISL_1641637, EPI_ISL_1641638, EPI_ISL_1641640, EPI_ISL_1641643, EPI_ISL_1641645, EPI_ISL_1641646, EPI_ISL_1641648, EPI_ISL_1641649, EPI_ISL_1641650, EPI_ISL_1641651, EPI_ISL_1641652, EPI_ISL_1641653, EPI_ISL_1641654, EPI_ISL_1641656, EPI_ISL_1641657, EPI_ISL_1641659, EPI_ISL_1641660, EPI_ISL_1641662, EPI_ISL_1641663, EPI_ISL_1641664, EPI_ISL_1641665, EPI_ISL_1641666, EPI_ISL_1641667, EPI_ISL_1641669, EPI_ISL_1641670, EPI_ISL_1641673, EPI_ISL_1641675, EPI_ISL_1641676, EPI_ISL_1641678, EPI_ISL_1641682, EPI_ISL_1641683, EPI_ISL_1641685, EPI_ISL_1641690, EPI_ISL_1641691, EPI_ISL_1641694, EPI_ISL_1641695, EPI_ISL_1641697, EPI_ISL_1641698, EPI_ISL_1641700, EPI_ISL_1641702, EPI_ISL_1641704, EPI_ISL_1641705, EPI_ISL_1641707, EPI_ISL_1641708, EPI_ISL_1641709, EPI_ISL_1641711, EPI_ISL_1641712, EPI_ISL_1641713, EPI_ISL_1641715, EPI_ISL_1641716, EPI_ISL_1641717, EPI_ISL_1641718, EPI_ISL_1641719, EPI_ISL_1641721, EPI_ISL_1641722, EPI_ISL_1641723, EPI_ISL_1641725, EPI_ISL_1641726, EPI_ISL_1641727, EPI_ISL_1641729, EPI_ISL_1641730, EPI_ISL_1641731, EPI_ISL_1641733, EPI_ISL_1641734, EPI_ISL_1641735, EPI_ISL_1641737, EPI_ISL_1641738, EPI_ISL_1641739, EPI_ISL_1641742, EPI_ISL_1641744, EPI_ISL_1641745, EPI_ISL_1641746, EPI_ISL_1641748, EPI_ISL_1641749, EPI_ISL_1641750, EPI_ISL_1641753, EPI_ISL_1641754, EPI_ISL_1641757, EPI_ISL_1641758, EPI_ISL_1641759, EPI_ISL_1641760, EPI_ISL_1641762, EPI_ISL_1641763, EPI_ISL_1641764, EPI_ISL_1641765, EPI_ISL_1641767, EPI_ISL_1641768, EPI_ISL_1641773, EPI_ISL_1641774, EPI_ISL_1641776, EPI_ISL_1641777, EPI_ISL_1641778, EPI_ISL_1641779, EPI_ISL_1641789, EPI_ISL_1641790, EPI_ISL_1641791, EPI_ISL_1641792, EPI_ISL_1641795, EPI_ISL_1641796, EPI_ISL_1641799, EPI_ISL_1641800, EPI_ISL_1641802, EPI_ISL_1641805, EPI_ISL_1641806, EPI_ISL_1641807, EPI_ISL_1641808, EPI_ISL_1641810, EPI_ISL_1641811, EPI_ISL_1641814, EPI_ISL_1641815, EPI_ISL_1641816, EPI_ISL_1641817, EPI_ISL_1641818, EPI_ISL_1641820, EPI_ISL_1641822, EPI_ISL_1641825, EPI_ISL_1641826, EPI_ISL_1641828, EPI_ISL_1641829, EPI_ISL_1641832, EPI_ISL_1641835, EPI_ISL_1641836, EPI_ISL_1641839, EPI_ISL_1641840, EPI_ISL_1641841, EPI_ISL_1641842, EPI_ISL_1641843, EPI_ISL_1641844, EPI_ISL_1641845, EPI_ISL_1641846, EPI_ISL_1641848, EPI_ISL_1641850, EPI_ISL_1641851, EPI_ISL_1641852, EPI_ISL_1641854, EPI_ISL_1641855, EPI_ISL_1644541, EPI_ISL_1644542, EPI_ISL_1644543, EPI_ISL_1644544, EPI_ISL_1644546, EPI_ISL_1644547, EPI_ISL_1644548, EPI_ISL_1644549, EPI_ISL_1644550, EPI_ISL_1644551, EPI_ISL_1644552, EPI_ISL_1644554, EPI_ISL_1644555, EPI_ISL_1644556, EPI_ISL_1644557, EPI_ISL_1644560, EPI_ISL_1644561, EPI_ISL_1644563, EPI_ISL_1644564, EPI_ISL_1644566, EPI_ISL_1644567, EPI_ISL_1644568, EPI_ISL_1644570, EPI_ISL_1644572, EPI_ISL_1644573, EPI_ISL_1644575, EPI_ISL_1644576, EPI_ISL_1644577, EPI_ISL_1644578, EPI_ISL_1644579, EPI_ISL_1644580, EPI_ISL_1644581, EPI_ISL_1644584, EPI_ISL_1644585, EPI_ISL_1644587, EPI_ISL_1644589, EPI_ISL_1644590, EPI_ISL_1644593, EPI_ISL_1644594, EPI_ISL_1644595, EPI_ISL_1644598, EPI_ISL_1644599, EPI_ISL_1644601, EPI_ISL_1644602, EPI_ISL_1644605, EPI_ISL_1644609, EPI_ISL_1644610, EPI_ISL_1644613, EPI_ISL_1644614, EPI_ISL_1644615, EPI_ISL_1644618, EPI_ISL_1644620, EPI_ISL_1644623, EPI_ISL_1644625, EPI_ISL_1644626, EPI_ISL_1644627, EPI_ISL_1644628, EPI_ISL_1644630, EPI_ISL_1644631, EPI_ISL_1644632, EPI_ISL_1644633, EPI_ISL_1644635, EPI_ISL_1644638, EPI_ISL_1644639, EPI_ISL_1644641, EPI_ISL_1644642, EPI_ISL_1644643, EPI_ISL_1644644, EPI_ISL_1644645, EPI_ISL_1644646, EPI_ISL_1644647, EPI_ISL_1644648, EPI_ISL_1644649, EPI_ISL_1644650, EPI_ISL_1644652, EPI_ISL_1644657, EPI_ISL_1644659, EPI_ISL_1644660, EPI_ISL_1644662, EPI_ISL_1644663, EPI_ISL_1644664, EPI_ISL_1644666, EPI_ISL_1644667, EPI_ISL_1644668, EPI_ISL_1644671, EPI_ISL_1644672, EPI_ISL_1644674, EPI_ISL_1644675, EPI_ISL_1644677, EPI_ISL_1644679, EPI_ISL_1644680, EPI_ISL_1644681, EPI_ISL_1644683, EPI_ISL_1644684, EPI_ISL_1644685, EPI_ISL_1644686, EPI_ISL_1644688, EPI_ISL_1644689, EPI_ISL_1644690, EPI_ISL_1644692, EPI_ISL_1644693, EPI_ISL_1644695, EPI_ISL_1644696, EPI_ISL_1644697, EPI_ISL_1644698, EPI_ISL_1644699, EPI_ISL_1644700, EPI_ISL_1644704, EPI_ISL_1644705, EPI_ISL_1644707, EPI_ISL_1644708, EPI_ISL_1644709, EPI_ISL_1644712, EPI_ISL_1644713, EPI_ISL_1644715, EPI_ISL_1644716, EPI_ISL_1644717, EPI_ISL_1644719, EPI_ISL_1644720, EPI_ISL_1644721, EPI_ISL_1644722, EPI_ISL_1644723, EPI_ISL_1644724, EPI_ISL_1644725, EPI_ISL_1644726, EPI_ISL_1644727, EPI_ISL_1644728, EPI_ISL_1644729, EPI_ISL_1644730, EPI_ISL_1644731, EPI_ISL_1644732, EPI_ISL_1644733, EPI_ISL_1644734, EPI_ISL_1644735, EPI_ISL_1644736, EPI_ISL_1644737, EPI_ISL_1644738, EPI_ISL_1644739, EPI_ISL_1644741, EPI_ISL_1644742, EPI_ISL_1644743, EPI_ISL_1644744, EPI_ISL_1644745, EPI_ISL_1644746, EPI_ISL_1644748, EPI_ISL_1644749, EPI_ISL_1644751, EPI_ISL_1644752, EPI_ISL_1644753, EPI_ISL_1644754, EPI_ISL_1644755, EPI_ISL_1644756, EPI_ISL_1644758, EPI_ISL_1644759, EPI_ISL_1644760, EPI_ISL_1644763, EPI_ISL_1644764, EPI_ISL_1644766, EPI_ISL_1644767, EPI_ISL_1644768, EPI_ISL_1644770, EPI_ISL_1644771, EPI_ISL_1644772, EPI_ISL_1644773, EPI_ISL_1644774, EPI_ISL_1644775, EPI_ISL_1644776, EPI_ISL_1644777, EPI_ISL_1644779, EPI_ISL_1644780, EPI_ISL_1644781, EPI_ISL_1644782, EPI_ISL_1644783, EPI_ISL_1644784, EPI_ISL_1644785, EPI_ISL_1644786, EPI_ISL_1644788, EPI_ISL_1644790, EPI_ISL_1644791, EPI_ISL_1644792, EPI_ISL_1644793, EPI_ISL_1644794, EPI_ISL_1644795, EPI_ISL_1644796, EPI_ISL_1644797, EPI_ISL_1644798, EPI_ISL_1644800, EPI_ISL_1644801, EPI_ISL_1644802, EPI_ISL_1644803, EPI_ISL_1644804, EPI_ISL_1644805, EPI_ISL_1644806, EPI_ISL_1644807, EPI_ISL_1644808, EPI_ISL_1644809, EPI_ISL_1644810, EPI_ISL_1644811, EPI_ISL_1644812, EPI_ISL_1644813, EPI_ISL_1644814, EPI_ISL_1644815, EPI_ISL_1644816, EPI_ISL_1644817, EPI_ISL_1644819, EPI_ISL_1644820, EPI_ISL_1644821, EPI_ISL_1644822, EPI_ISL_1644823, EPI_ISL_1644824, EPI_ISL_1644825, EPI_ISL_1644827, EPI_ISL_1644828, EPI_ISL_1644829, EPI_ISL_1644830, EPI_ISL_1644831, EPI_ISL_1644832, EPI_ISL_1644833, EPI_ISL_1644834, EPI_ISL_1644835, EPI_ISL_1644836, EPI_ISL_1644837, EPI_ISL_1644838, EPI_ISL_1644839, EPI_ISL_1644840, EPI_ISL_1644841, EPI_ISL_1644842, EPI_ISL_1644843, EPI_ISL_1720172, EPI_ISL_1720173, EPI_ISL_1720174, EPI_ISL_1720175, EPI_ISL_1720176, EPI_ISL_1720177, EPI_ISL_1720178, EPI_ISL_1720179, EPI_ISL_1720180, EPI_ISL_1720181, EPI_ISL_1720182, EPI_ISL_1720183, EPI_ISL_1720186, EPI_ISL_1720189, EPI_ISL_1720193, EPI_ISL_1720194, EPI_ISL_1720195, EPI_ISL_1720196, EPI_ISL_1720197, EPI_ISL_1720198, EPI_ISL_1720200, EPI_ISL_1720201, EPI_ISL_1720202, EPI_ISL_1720203, EPI_ISL_1720204, EPI_ISL_1720207, EPI_ISL_1720210, EPI_ISL_1720212, EPI_ISL_1720213, EPI_ISL_1720215, EPI_ISL_1720216, EPI_ISL_1720217, EPI_ISL_1720220, EPI_ISL_1720221, EPI_ISL_1720223, EPI_ISL_1720226, EPI_ISL_1720228, EPI_ISL_1720229, EPI_ISL_1720231, EPI_ISL_1720233, EPI_ISL_1720236, EPI_ISL_1720238, EPI_ISL_1720239, EPI_ISL_1720242, EPI_ISL_1720243, EPI_ISL_1720246, EPI_ISL_1720247, EPI_ISL_1720248, EPI_ISL_1720249, EPI_ISL_1720250, EPI_ISL_1720252, EPI_ISL_1720253, EPI_ISL_1720255, EPI_ISL_1720256, EPI_ISL_1720258, EPI_ISL_1720260, EPI_ISL_1720261, EPI_ISL_1720262, EPI_ISL_1720263, EPI_ISL_1720265, EPI_ISL_1720266, EPI_ISL_1720267, EPI_ISL_1720269, EPI_ISL_1720270, EPI_ISL_1720271, EPI_ISL_1720273, EPI_ISL_1720274, EPI_ISL_1720275, EPI_ISL_1720276, EPI_ISL_1720277, EPI_ISL_1720279, EPI_ISL_1720280, EPI_ISL_1720281, EPI_ISL_1720283, EPI_ISL_1720284, EPI_ISL_1720286, EPI_ISL_1720287, EPI_ISL_1720289, EPI_ISL_1720290, EPI_ISL_1720292, EPI_ISL_1720293, EPI_ISL_1720294, EPI_ISL_1720296, EPI_ISL_1720297, EPI_ISL_1720298, EPI_ISL_1720300, EPI_ISL_1720301, EPI_ISL_1720302, EPI_ISL_1720305, EPI_ISL_1720307, EPI_ISL_1720308, EPI_ISL_1720309, EPI_ISL_1720311, EPI_ISL_1720312, EPI_ISL_1720314, EPI_ISL_1720315, EPI_ISL_1720316, EPI_ISL_1720319, EPI_ISL_1720320, EPI_ISL_1720322, EPI_ISL_1720323, EPI_ISL_1720324, EPI_ISL_1720326, EPI_ISL_1720327, EPI_ISL_1720328, EPI_ISL_1720329, EPI_ISL_1720330, EPI_ISL_1720331, EPI_ISL_1720332, EPI_ISL_1720333, EPI_ISL_1720334, EPI_ISL_1720335, EPI_ISL_1720336, EPI_ISL_1720337, EPI_ISL_1720338, EPI_ISL_1720339, EPI_ISL_1720341, EPI_ISL_1720344, EPI_ISL_1720346, EPI_ISL_1720348, EPI_ISL_1720351, EPI_ISL_1720352, EPI_ISL_1720353, EPI_ISL_1720354, EPI_ISL_1720355, EPI_ISL_1720356, EPI_ISL_1720357, EPI_ISL_1720359, EPI_ISL_1720360, EPI_ISL_1720362, EPI_ISL_1720363, EPI_ISL_1720364, EPI_ISL_1720367, EPI_ISL_1720369, EPI_ISL_1720370, EPI_ISL_1720371, EPI_ISL_1720373, EPI_ISL_1720374, EPI_ISL_1720375, EPI_ISL_1720376, EPI_ISL_1720377, EPI_ISL_1720378, EPI_ISL_1720379, EPI_ISL_1720381, EPI_ISL_1720382, EPI_ISL_1720383, EPI_ISL_1720384, EPI_ISL_1720385, EPI_ISL_1720386, EPI_ISL_1720388, EPI_ISL_1720389, EPI_ISL_1720391, EPI_ISL_1720392, EPI_ISL_1720393, EPI_ISL_1720394, EPI_ISL_1720395, EPI_ISL_1720396, EPI_ISL_1720397, EPI_ISL_1720398, EPI_ISL_1720399, EPI_ISL_1720400, EPI_ISL_1720401, EPI_ISL_1720403, EPI_ISL_1720404, EPI_ISL_1720405, EPI_ISL_1720406, EPI_ISL_1720407, EPI_ISL_1720409, EPI_ISL_1720410, EPI_ISL_1720411, EPI_ISL_1720412, EPI_ISL_1720413, EPI_ISL_1720414, EPI_ISL_1720416, EPI_ISL_1720417, EPI_ISL_1720418, EPI_ISL_1720419, EPI_ISL_1720420, EPI_ISL_1720421, EPI_ISL_1720424, EPI_ISL_1720425, EPI_ISL_1720426, EPI_ISL_1720427, EPI_ISL_1720428, EPI_ISL_1720430, EPI_ISL_1720431, EPI_ISL_1720432, EPI_ISL_1720433, EPI_ISL_1720434, EPI_ISL_1720435, EPI_ISL_1720436, EPI_ISL_1720437, EPI_ISL_1720438, EPI_ISL_1720439, EPI_ISL_1720440, EPI_ISL_1720441, EPI_ISL_1720442, EPI_ISL_1720443, EPI_ISL_1720444, EPI_ISL_1720445, EPI_ISL_1720446, EPI_ISL_1720447, EPI_ISL_1720448, EPI_ISL_1720450, EPI_ISL_1720452, EPI_ISL_1720456, EPI_ISL_1720458, EPI_ISL_1720475, EPI_ISL_1720508, EPI_ISL_1720518, EPI_ISL_1720539, EPI_ISL_1720550, EPI_ISL_1720557, EPI_ISL_1720561, EPI_ISL_1720563, EPI_ISL_1720598, EPI_ISL_1720608, EPI_ISL_1720612, EPI_ISL_1720616, EPI_ISL_1720621, EPI_ISL_1720637, EPI_ISL_1720640, EPI_ISL_1720652, EPI_ISL_1720657, EPI_ISL_1720662, EPI_ISL_1720663, EPI_ISL_1720674, EPI_ISL_1720680, EPI_ISL_1720687, EPI_ISL_1720689, EPI_ISL_1720691, EPI_ISL_1720694, EPI_ISL_1720703, EPI_ISL_1720713, EPI_ISL_1720719, EPI_ISL_1720752, EPI_ISL_1720753, EPI_ISL_1720754, EPI_ISL_1720755, EPI_ISL_1720756, EPI_ISL_1720757, EPI_ISL_1720758, EPI_ISL_1720759, EPI_ISL_1720760, EPI_ISL_1720761, EPI_ISL_1720762, EPI_ISL_1720763, EPI_ISL_1720764, EPI_ISL_1720765, EPI_ISL_1720766, EPI_ISL_1720767, EPI_ISL_1720768, EPI_ISL_1720769, EPI_ISL_1720770, EPI_ISL_1720771, EPI_ISL_1720772, EPI_ISL_1720773, EPI_ISL_1720774, EPI_ISL_2115110, EPI_ISL_2119544

see above | SYNLAB MVZ Leverkusen | Robert Koch Institute

EPI_ISL_2108011, EPI_ISL_2108035, EPI_ISL_2108038, EPI_ISL_2108068, EPI_ISL_2108082, EPI_ISL_2108127, EPI_ISL_2108146, EPI_ISL_2108148, EPI_ISL_2108173, EPI_ISL_2108220, EPI_ISL_2108228, EPI_ISL_2114542, EPI_ISL_2114684, EPI_ISL_2114724, EPI_ISL_2114934

see above | SYNLAB MVZ Neustadt a. d. Weinstrasse | Robert Koch Institute

EPI_ISL_1726628, EPI_ISL_1726642, EPI_ISL_1726668, EPI_ISL_1726674, EPI_ISL_1726682, EPI_ISL_1726723, EPI_ISL_1849702, EPI_ISL_1849712, EPI_ISL_1849718, EPI_ISL_1849719, EPI_ISL_1849720, EPI_ISL_1849726, EPI_ISL_1849736, EPI_ISL_1849748, EPI_ISL_1849767, EPI_ISL_1849769, EPI_ISL_1849782, EPI_ISL_1849784, EPI_ISL_1849802, EPI_ISL_1849808, EPI_ISL_1849817, EPI_ISL_1849822, EPI_ISL_1849835, EPI_ISL_1849844, EPI_ISL_1849854, EPI_ISL_1849881, EPI_ISL_1849901, EPI_ISL_1849919

see above | SYNLAB MVZ Neustadt a. d. WeinstraÃŸe | Robert Koch Institute

EPI_ISL_2118833, EPI_ISL_2118850, EPI_ISL_2118887, EPI_ISL_2119029, EPI_ISL_2119047, EPI_ISL_2119057, EPI_ISL_2119151, EPI_ISL_2119195, EPI_ISL_2119205, EPI_ISL_2119232, EPI_ISL_2119279, EPI_ISL_2119284, EPI_ISL_2119297, EPI_ISL_2119305, EPI_ISL_2119345, EPI_ISL_2119350, EPI_ISL_2119404, EPI_ISL_2119409, EPI_ISL_2119469, EPI_ISL_2119488, EPI_ISL_2119490, EPI_ISL_2119493, EPI_ISL_2119540, EPI_ISL_2119566, EPI_ISL_2119598, EPI_ISL_2119632, EPI_ISL_2119636, EPI_ISL_2119648, EPI_ISL_2119650, EPI_ISL_2119656, EPI_ISL_2119698, EPI_ISL_2119708, EPI_ISL_2119742, EPI_ISL_2119755, EPI_ISL_2119796, EPI_ISL_2119829, EPI_ISL_2119831, EPI_ISL_2119834, EPI_ISL_2119877, EPI_ISL_2119889, EPI_ISL_2119926, EPI_ISL_2119992, EPI_ISL_2119995, EPI_ISL_2120004, EPI_ISL_2128771, EPI_ISL_2128828, EPI_ISL_2128883

see above | SYNLAB MVZ Neustadt a. d. Weinstraße | Robert Koch Institute

EPI_ISL_1573090, EPI_ISL_1573096, EPI_ISL_1573101, EPI_ISL_1573102, EPI_ISL_1573106, EPI_ISL_1573110, EPI_ISL_1573112, EPI_ISL_1573121, EPI_ISL_1573127, EPI_ISL_1641261, EPI_ISL_1641263, EPI_ISL_1641265, EPI_ISL_1641267, EPI_ISL_1641271, EPI_ISL_1641336, EPI_ISL_1641353, EPI_ISL_1641356, EPI_ISL_1641386, EPI_ISL_1641394, EPI_ISL_1641413, EPI_ISL_1641420, EPI_ISL_1641430, EPI_ISL_1641490, EPI_ISL_1641494, EPI_ISL_1641496, EPI_ISL_1641502, EPI_ISL_1641514, EPI_ISL_1641516, EPI_ISL_1641559, EPI_ISL_1641563, EPI_ISL_1641571, EPI_ISL_1641575, EPI_ISL_1641577, EPI_ISL_1641579, EPI_ISL_1641580, EPI_ISL_1641581, EPI_ISL_1641592, EPI_ISL_1641594, EPI_ISL_1641596, EPI_ISL_1641598, EPI_ISL_1641604, EPI_ISL_1641606, EPI_ISL_1641609, EPI_ISL_1641612, EPI_ISL_1641619, EPI_ISL_1641622, EPI_ISL_1641626, EPI_ISL_1641627, EPI_ISL_1641628, EPI_ISL_1641630, EPI_ISL_1641639, EPI_ISL_1641641, EPI_ISL_1641642, EPI_ISL_1641644, EPI_ISL_1641647, EPI_ISL_1641655, EPI_ISL_1641658, EPI_ISL_1641661, EPI_ISL_1641668, EPI_ISL_1641671, EPI_ISL_1641672, EPI_ISL_1641674, EPI_ISL_1641677, EPI_ISL_1641679, EPI_ISL_1641680, EPI_ISL_1641681, EPI_ISL_1641686, EPI_ISL_1641687, EPI_ISL_1641688, EPI_ISL_1641689, EPI_ISL_1641692, EPI_ISL_1641693, EPI_ISL_1641696, EPI_ISL_1641699, EPI_ISL_1641701, EPI_ISL_1641703, EPI_ISL_1641706, EPI_ISL_1641710, EPI_ISL_1641714, EPI_ISL_1641720, EPI_ISL_1641724, EPI_ISL_1641728, EPI_ISL_1641732, EPI_ISL_1641736, EPI_ISL_1641740, EPI_ISL_1641741, EPI_ISL_1641743, EPI_ISL_1641747, EPI_ISL_1641751, EPI_ISL_1641752, EPI_ISL_1641755, EPI_ISL_1641756, EPI_ISL_1641761, EPI_ISL_1641766, EPI_ISL_1641769, EPI_ISL_1641770, EPI_ISL_1641771, EPI_ISL_1641772, EPI_ISL_1641775, EPI_ISL_1641780, EPI_ISL_1641781, EPI_ISL_1641783, EPI_ISL_1641784, EPI_ISL_1641785, EPI_ISL_1641786, EPI_ISL_1641787, EPI_ISL_1641788, EPI_ISL_1641793, EPI_ISL_1641794, EPI_ISL_1641797, EPI_ISL_1641798, EPI_ISL_1641801, EPI_ISL_1641803, EPI_ISL_1641804, EPI_ISL_1641809, EPI_ISL_1641812, EPI_ISL_1641813, EPI_ISL_1641819, EPI_ISL_1641821, EPI_ISL_1641823, EPI_ISL_1641824, EPI_ISL_1641827, EPI_ISL_1641830, EPI_ISL_1641833, EPI_ISL_1641834, EPI_ISL_1641837, EPI_ISL_1641838, EPI_ISL_1641847, EPI_ISL_1641849, EPI_ISL_1641853, EPI_ISL_1644545, EPI_ISL_1644559, EPI_ISL_1644565, EPI_ISL_1644569, EPI_ISL_1644571, EPI_ISL_1644574, EPI_ISL_1644582, EPI_ISL_1644586, EPI_ISL_1644591, EPI_ISL_1644592, EPI_ISL_1644596, EPI_ISL_1644597, EPI_ISL_1644600, EPI_ISL_1644604, EPI_ISL_1644606, EPI_ISL_1644607, EPI_ISL_1644608, EPI_ISL_1644611, EPI_ISL_1644616, EPI_ISL_1644617, EPI_ISL_1644619, EPI_ISL_1644621, EPI_ISL_1644622, EPI_ISL_1644624, EPI_ISL_1644629, EPI_ISL_1644634, EPI_ISL_1644636, EPI_ISL_1644637, EPI_ISL_1644640, EPI_ISL_1644651, EPI_ISL_1644653, EPI_ISL_1644654, EPI_ISL_1644655, EPI_ISL_1644656, EPI_ISL_1644658, EPI_ISL_1644661, EPI_ISL_1644665, EPI_ISL_1644669, EPI_ISL_1644670, EPI_ISL_1644673, EPI_ISL_1644676, EPI_ISL_1644678, EPI_ISL_1644682, EPI_ISL_1644687, EPI_ISL_1644691, EPI_ISL_1644694, EPI_ISL_1644701, EPI_ISL_1644702, EPI_ISL_1644703, EPI_ISL_1644706, EPI_ISL_1644710, EPI_ISL_1644711, EPI_ISL_1644714, EPI_ISL_1644718, EPI_ISL_1644740, EPI_ISL_1644747, EPI_ISL_1644750, EPI_ISL_1644761, EPI_ISL_1644762, EPI_ISL_1644765, EPI_ISL_1644769, EPI_ISL_1644778, EPI_ISL_1644787, EPI_ISL_1644789, EPI_ISL_1644799, EPI_ISL_1644818, EPI_ISL_1644826, EPI_ISL_1720184, EPI_ISL_1720187, EPI_ISL_1720188, EPI_ISL_1720191, EPI_ISL_1720192, EPI_ISL_1720199, EPI_ISL_1720205, EPI_ISL_1720206, EPI_ISL_1720208, EPI_ISL_1720209, EPI_ISL_1720214, EPI_ISL_1720218, EPI_ISL_1720219, EPI_ISL_1720222, EPI_ISL_1720224, EPI_ISL_1720225, EPI_ISL_1720227, EPI_ISL_1720230, EPI_ISL_1720232, EPI_ISL_1720234, EPI_ISL_1720235, EPI_ISL_1720237, EPI_ISL_1720240, EPI_ISL_1720241, EPI_ISL_1720244, EPI_ISL_1720245, EPI_ISL_1720251, EPI_ISL_1720254, EPI_ISL_1720257, EPI_ISL_1720264, EPI_ISL_1720268, EPI_ISL_1720272, EPI_ISL_1720278, EPI_ISL_1720282, EPI_ISL_1720285, EPI_ISL_1720288, EPI_ISL_1720303, EPI_ISL_1720304, EPI_ISL_1720306, EPI_ISL_1720310, EPI_ISL_1720313, EPI_ISL_1720321, EPI_ISL_1720340, EPI_ISL_1720342, EPI_ISL_1720343, EPI_ISL_1720345, EPI_ISL_1720347, EPI_ISL_1720349, EPI_ISL_1720350, EPI_ISL_1720361, EPI_ISL_1720365, EPI_ISL_1720366, EPI_ISL_1720368, EPI_ISL_1720372, EPI_ISL_1720380, EPI_ISL_1720387, EPI_ISL_1720390, EPI_ISL_1720402, EPI_ISL_1720408, EPI_ISL_1720415, EPI_ISL_1720422, EPI_ISL_1720423, EPI_ISL_1720429, EPI_ISL_1720451, EPI_ISL_1720593, EPI_ISL_1720702

see above | SYNLAB MVZ Trier | Robert Koch Institute

EPI_ISL_1573097, EPI_ISL_1641632, EPI_ISL_1641684, EPI_ISL_1641782, EPI_ISL_1641831, EPI_ISL_1720453, EPI_ISL_1720597, EPI_ISL_1724115, EPI_ISL_1724151, EPI_ISL_1724161, EPI_ISL_1724222

see above | SYNLAB MVZ Weiden | Robert Koch Institute

EPI_ISL_1843517, EPI_ISL_1847595, EPI_ISL_1847631, EPI_ISL_1848433, EPI_ISL_1848459, EPI_ISL_1848479, EPI_ISL_1848643, EPI_ISL_1848696, EPI_ISL_1848704, EPI_ISL_2108740, EPI_ISL_2108798, EPI_ISL_2108857, EPI_ISL_2108886, EPI_ISL_2108899, EPI_ISL_2108912, EPI_ISL_2109085, EPI_ISL_2109137, EPI_ISL_2109141, EPI_ISL_2109153, EPI_ISL_2109176, EPI_ISL_2109182, EPI_ISL_2109226, EPI_ISL_2109238, EPI_ISL_2109266, EPI_ISL_2109279, EPI_ISL_2109525, EPI_ISL_2109543, EPI_ISL_2109568, EPI_ISL_2109579, EPI_ISL_2109587, EPI_ISL_2109617, EPI_ISL_2109670, EPI_ISL_2109708, EPI_ISL_2109715, EPI_ISL_2109751, EPI_ISL_2109784, EPI_ISL_2109827, EPI_ISL_2109838

see above | Sonic - Bioscientia - MVZ Labor Saar GmbH | Robert Koch Institute

EPI_ISL_1842789, EPI_ISL_1842819, EPI_ISL_1842832, EPI_ISL_1842862, EPI_ISL_1842875, EPI_ISL_1842877, EPI_ISL_1842900, EPI_ISL_1842921, EPI_ISL_1842945, EPI_ISL_1843059, EPI_ISL_1843073, EPI_ISL_1843109

see above | Sonic - Dr. Staber & Kollegen GmbH MÃ¼nchen | Robert Koch Institute

EPI_ISL_1721700, EPI_ISL_1721702, EPI_ISL_1721709, EPI_ISL_1721717, EPI_ISL_1721718, EPI_ISL_1721729, EPI_ISL_1721753, EPI_ISL_1721755, EPI_ISL_1721765, EPI_ISL_1721770, EPI_ISL_1721772, EPI_ISL_1721776, EPI_ISL_1721784, EPI_ISL_1721789, EPI_ISL_1721793, EPI_ISL_1721802, EPI_ISL_1721814, EPI_ISL_1721818, EPI_ISL_1721845, EPI_ISL_1721854, EPI_ISL_1721867, EPI_ISL_1721875, EPI_ISL_1721877, EPI_ISL_1721890, EPI_ISL_1721900, EPI_ISL_1721902, EPI_ISL_1721918, EPI_ISL_1721923, EPI_ISL_1721926, EPI_ISL_1721927, EPI_ISL_1721930, EPI_ISL_1721939, EPI_ISL_1721941, EPI_ISL_1721944, EPI_ISL_1721950, EPI_ISL_1721953, EPI_ISL_1721954, EPI_ISL_1721963, EPI_ISL_1721980, EPI_ISL_1842796, EPI_ISL_1842797, EPI_ISL_1842802, EPI_ISL_1842813, EPI_ISL_1842815, EPI_ISL_1842817, EPI_ISL_1842820, EPI_ISL_1842824, EPI_ISL_1842836, EPI_ISL_1842837, EPI_ISL_1842841, EPI_ISL_1842842, EPI_ISL_1842847, EPI_ISL_1842868, EPI_ISL_1842876, EPI_ISL_1842899, EPI_ISL_1842902, EPI_ISL_1842908, EPI_ISL_1842919, EPI_ISL_1842920, EPI_ISL_1842926, EPI_ISL_1842928, EPI_ISL_1842930, EPI_ISL_1842938, EPI_ISL_1842943, EPI_ISL_1842946, EPI_ISL_1842948, EPI_ISL_1842956

see above | Sonic - Labor Dr. von Froreich GmbH | Robert Koch Institute

EPI_ISL_1842785, EPI_ISL_1842806, EPI_ISL_1842861, EPI_ISL_1842869, EPI_ISL_1842873, EPI_ISL_1842886, EPI_ISL_1842965, EPI_ISL_1843108, EPI_ISL_1843523

see above | Sonic - Labor Staber Bayreuth | Robert Koch Institute

EPI_ISL_1842807, EPI_ISL_1842814, EPI_ISL_1842850, EPI_ISL_1842878, EPI_ISL_1842941, EPI_ISL_1842944, EPI_ISL_1842949

see above | Sonic - Labor Staber Dresden (Klipphausen) | Robert Koch Institute

EPI_ISL_1842780, EPI_ISL_1842879, EPI_ISL_1842881, EPI_ISL_1842885, EPI_ISL_1842891, EPI_ISL_1842953, EPI_ISL_1843091

see above | Sonic - Labor Staber Heilbronn | Robert Koch Institute

EPI_ISL_1842825, EPI_ISL_1843451, EPI_ISL_1847139, EPI_ISL_1847146, EPI_ISL_1847612, EPI_ISL_1847620 | Sonic - Labor Staber MVZ Kassel | Robert Koch Institute

| | | | |
|---|---|---|---|
| EPI_ISL_1842783, EPI_ISL_1842791, EPI_ISL_1842849, EPI_ISL_1842910, EPI_ISL_1842933, EPI_ISL_1842960, EPI_ISL_1842978, EPI_ISL_1842992, EPI_ISL_1843119 | | | |
| see above | Sonic - Labor Staber NÃ¼rnberg | Robert Koch Institute | |
| EPI_ISL_1721693, EPI_ISL_1721716, EPI_ISL_1721732, EPI_ISL_1721740, EPI_ISL_1721761, EPI_ISL_1721766, EPI_ISL_1721767, EPI_ISL_1721773, EPI_ISL_1721778, EPI_ISL_1721782, EPI_ISL_1721791, EPI_ISL_1721794, EPI_ISL_1721804, EPI_ISL_1721807, EPI_ISL_1721809, EPI_ISL_1721819, EPI_ISL_1721826, EPI_ISL_1721828, EPI_ISL_1721829, EPI_ISL_1721839, EPI_ISL_1721840, EPI_ISL_1721843, EPI_ISL_1721858, EPI_ISL_1721860, EPI_ISL_1721869, EPI_ISL_1721891, EPI_ISL_1721893, EPI_ISL_1721898, EPI_ISL_1721907, EPI_ISL_1721935, EPI_ISL_1721952, EPI_ISL_1721960, EPI_ISL_1842778, EPI_ISL_1842779, EPI_ISL_1842787, EPI_ISL_1842788, EPI_ISL_1842790, EPI_ISL_1842792, EPI_ISL_1842794, EPI_ISL_1842812, EPI_ISL_1842816, EPI_ISL_1842818, EPI_ISL_1842822, EPI_ISL_1842828, EPI_ISL_1842835, EPI_ISL_1842845, EPI_ISL_1842846, EPI_ISL_1842860, EPI_ISL_1842863, EPI_ISL_1842865, EPI_ISL_1842866, EPI_ISL_1842867, EPI_ISL_1842870, EPI_ISL_1842871, EPI_ISL_1842872, EPI_ISL_1842874, EPI_ISL_1842883, EPI_ISL_1842884, EPI_ISL_1842888, EPI_ISL_1842890, EPI_ISL_1842892, EPI_ISL_1842893, EPI_ISL_1842903, EPI_ISL_1842904, EPI_ISL_1842906, EPI_ISL_1842907, EPI_ISL_1842909, EPI_ISL_1842916, EPI_ISL_1842918, EPI_ISL_1842929, EPI_ISL_1842931, EPI_ISL_1842934, EPI_ISL_1842935, EPI_ISL_1842939, EPI_ISL_1842940, EPI_ISL_1842950, EPI_ISL_1842951, EPI_ISL_1842957, EPI_ISL_1842958, EPI_ISL_1842961, EPI_ISL_1842963, EPI_ISL_1842971, EPI_ISL_1842972, EPI_ISL_1842974, EPI_ISL_1842979, EPI_ISL_1843003, EPI_ISL_1843004, EPI_ISL_1843008, EPI_ISL_1843020, EPI_ISL_1843023, EPI_ISL_1843024, EPI_ISL_1843027, EPI_ISL_1843029, EPI_ISL_1843037, EPI_ISL_1843047, EPI_ISL_1843058, EPI_ISL_1843064, EPI_ISL_1843065, EPI_ISL_1843066, EPI_ISL_1843072, EPI_ISL_1843074, EPI_ISL_1843077, EPI_ISL_1843080, EPI_ISL_1843082, EPI_ISL_1843083, EPI_ISL_1843102, EPI_ISL_1843104, EPI_ISL_1843122, EPI_ISL_1843126, EPI_ISL_1843130, EPI_ISL_1843141, EPI_ISL_1843147, EPI_ISL_1843149, EPI_ISL_1843153, EPI_ISL_1843155, EPI_ISL_1843156, EPI_ISL_1843159, EPI_ISL_1843161, EPI_ISL_1843167, EPI_ISL_1843169, EPI_ISL_1843170, EPI_ISL_1847093, EPI_ISL_1847094, EPI_ISL_1847096, EPI_ISL_1847098, EPI_ISL_1847101, EPI_ISL_1847103, EPI_ISL_1847104, EPI_ISL_1847108, EPI_ISL_1847110, EPI_ISL_1847112, EPI_ISL_1847118, EPI_ISL_1847119, EPI_ISL_1847120, EPI_ISL_1847124, EPI_ISL_1847125, EPI_ISL_1847126, EPI_ISL_1847128, EPI_ISL_1847130, EPI_ISL_1847133, EPI_ISL_1847134, EPI_ISL_1847141, EPI_ISL_1847142, EPI_ISL_1847143, EPI_ISL_1847144, EPI_ISL_1847153, EPI_ISL_1847156, EPI_ISL_1847163, EPI_ISL_1847164, EPI_ISL_1847454, EPI_ISL_1847487, EPI_ISL_1847489, EPI_ISL_1847497, EPI_ISL_1847541, EPI_ISL_1847608, EPI_ISL_1847651, EPI_ISL_1847689 | | | |
| see above | Sonic - MVZ Medizinisches Labor Bremen GmbH | Robert Koch Institute | |
| EPI_ISL_1842985, EPI_ISL_1843131 | Sonic - MVZ Medizinisches Labor Oldenburg | Robert Koch Institute | |
| EPI_ISL_2109418 | Stadtisches Klinikum Dresden Institut fur Klinische Chemie und Laboratoriumsmedizin | Robert Koch Institute | |
| EPI_ISL_1720963 | Stiftung Krankenhaus Bethanien fÃ¼r die Grafschaft Moers | Robert Koch Institute | |
| EPI_ISL_1722085, EPI_ISL_1722090, EPI_ISL_1722092, EPI_ISL_1722093, EPI_ISL_1722097, EPI_ISL_1722098, EPI_ISL_1722099, EPI_ISL_1722103, EPI_ISL_1722106, EPI_ISL_1722107, EPI_ISL_1722118, EPI_ISL_1722122, EPI_ISL_1724340, EPI_ISL_1724342, EPI_ISL_1724343, EPI_ISL_1724344, EPI_ISL_1724345, EPI_ISL_1724346, EPI_ISL_1724347, EPI_ISL_1724351, EPI_ISL_1724352, EPI_ISL_1724355, EPI_ISL_1724360, EPI_ISL_1724371, EPI_ISL_1724373, EPI_ISL_1724381, EPI_ISL_1724382 | | | |
| see above | StÃ¤dtisches Klinikum Dresden Institut fÃ¼r Klinische Chemie und Laboratoriumsmedizin | Robert Koch Institute | |
| EPI_ISL_1573078, EPI_ISL_1641390, EPI_ISL_1641574, EPI_ISL_1641589, EPI_ISL_1641607, EPI_ISL_1644553, EPI_ISL_1644558, EPI_ISL_1644562, EPI_ISL_1644583, EPI_ISL_1644588, EPI_ISL_1644603, EPI_ISL_1720185, EPI_ISL_1720190, EPI_ISL_1720211, EPI_ISL_1720259, EPI_ISL_1720291, EPI_ISL_1720295, EPI_ISL_1720299, EPI_ISL_1720318, EPI_ISL_1720358, EPI_ISL_1720449, EPI_ISL_1720469, EPI_ISL_1720478, EPI_ISL_1720481, EPI_ISL_1720485, EPI_ISL_1720487, EPI_ISL_1720501, EPI_ISL_1720513, EPI_ISL_1720551, EPI_ISL_1720587, EPI_ISL_1720592, EPI_ISL_1720664, EPI_ISL_1720731, EPI_ISL_1720736, EPI_ISL_1720748, EPI_ISL_1723887, EPI_ISL_1723951, EPI_ISL_1723972, EPI_ISL_1724072, EPI_ISL_1724075 | | | |
| see above | Synlab MVZ Augsburg | Robert Koch Institute | |
| EPI_ISL_1722830, EPI_ISL_1722835, EPI_ISL_1722837, EPI_ISL_1722839, EPI_ISL_1722840, EPI_ISL_1844034, EPI_ISL_1844039, EPI_ISL_1844040, EPI_ISL_1844041, EPI_ISL_1844043 | | | |
| see above | ThÃ¼ringer Landesamtes fÃ¼r Verbraucherschutz | Robert Koch Institute | |
| EPI_ISL_2109374 | Uniklinikum Carl Gustav Carus an der TU Dresden; Institut fur Virologie | Robert Koch Institute | |
| EPI_ISL_1640121, EPI_ISL_1640122, EPI_ISL_1640124, EPI_ISL_1640126, EPI_ISL_1640127, EPI_ISL_1640128, EPI_ISL_1640130, EPI_ISL_1640131, EPI_ISL_1640132, EPI_ISL_1640134, EPI_ISL_1640136, EPI_ISL_1640138, EPI_ISL_1722086, EPI_ISL_1722112, EPI_ISL_1724329, EPI_ISL_1724331, EPI_ISL_1724333, EPI_ISL_1724336, EPI_ISL_1724339, EPI_ISL_1724358, EPI_ISL_1724367, EPI_ISL_1724383, EPI_ISL_1724386, EPI_ISL_1724391, EPI_ISL_1724392 | | | |
| see above | Uniklinikum Carl Gustav Carus an der TU Dresden; Institut fÃ¼r Virologie | Robert Koch Institute | |
| EPI_ISL_1722853, EPI_ISL_1722854, EPI_ISL_1722855, EPI_ISL_1722856, EPI_ISL_1722857, EPI_ISL_1844029, EPI_ISL_1844032 | | | |
| see above | Uniklinikum Jena | Robert Koch Institute | |
| EPI_ISL_1843374, EPI_ISL_1843375, EPI_ISL_1843378, EPI_ISL_1843379, EPI_ISL_1843380, EPI_ISL_1843381 | Uniklinikum Regensburg; Institut fÃ¼r klinische Mikrobiologie und Hygiene | Robert Koch Institute | |
| EPI_ISL_2109852, EPI_ISL_2109853, EPI_ISL_2109854, EPI_ISL_2109855, EPI_ISL_2109857, EPI_ISL_2109860, EPI_ISL_2109861, EPI_ISL_2109862, EPI_ISL_2109863, EPI_ISL_2109864 | | | |
| see above | Universitatsklinikum Bonn - Institut fur Virologie | Robert Koch Institute | |
| EPI_ISL_1715830, EPI_ISL_1715831, EPI_ISL_1715832, EPI_ISL_1715833, EPI_ISL_1715834, EPI_ISL_1715835, EPI_ISL_1715836, EPI_ISL_1715837, EPI_ISL_1715838, EPI_ISL_1715840, EPI_ISL_1715841, EPI_ISL_1862390, EPI_ISL_1862395 | | | |
| see above | University Medical Center Hamburg Eppendorf | Heinrich Pette Institute, Leibniz Institute for Experimental Virology | Adam Grundhoff; Alexis Robitaille; Johannes Knobloch; Martin Aepfelbacher; Nicole Fischer; Thomas Günther |
| EPI_ISL_1721070 | UniversitÃ¤tsklinikum Bonn - Institut fÃ¼r Virologie | Robert Koch Institute | |
| EPI_ISL_1843498, EPI_ISL_1843501, EPI_ISL_1843519, EPI_ISL_1843541, EPI_ISL_1843545, EPI_ISL_1843565, EPI_ISL_1843591, EPI_ISL_1843603, EPI_ISL_1843612, EPI_ISL_1843658, EPI_ISL_1843662, EPI_ISL_1843705 | | | |
| see above | UniversitÃ¤tsklinikum Frankfurt - Institut fÃ¼r Medizinische Virologie | Robert Koch Institute | |
| EPI_ISL_1722129, EPI_ISL_1722130 | UniversitÃ¤tsklinikum GieÃŸen - Institut fÃ¼r Medizinische Virologie | Robert Koch Institute | |
| EPI_ISL_1847166, EPI_ISL_1848365, EPI_ISL_1848366, EPI_ISL_1848367 | UniversitÃ¤tsklinikum Heidelberg | Robert Koch Institute | |
| EPI_ISL_1643819, EPI_ISL_1643820, EPI_ISL_1643821, EPI_ISL_1643822, EPI_ISL_1643823, EPI_ISL_1643824, EPI_ISL_1643825, EPI_ISL_1643826, EPI_ISL_1643827, EPI_ISL_1643828, EPI_ISL_1643830, EPI_ISL_1726856 | | | |
| see above | UniversitÃ¤tsklinikum MÃ¼nster Institut fÃ¼r Virologie | Robert Koch Institute | |
| EPI_ISL_1721015, EPI_ISL_1721035, EPI_ISL_1721040, EPI_ISL_1721044, EPI_ISL_1721045, EPI_ISL_1721047, EPI_ISL_1721049, EPI_ISL_1721053, EPI_ISL_1721075, EPI_ISL_1721076, EPI_ISL_1721078, EPI_ISL_1721079, EPI_ISL_1721089, EPI_ISL_1721092, EPI_ISL_1721094, EPI_ISL_1721100 | | | |
| see above | UniversitÃ¤tsklinikum des Saarlandes - Institut fÃ¼r Virologie | Robert Koch Institute | |
| EPI_ISL_1725185, EPI_ISL_1848347 | UniversitÃ¤tsmedizin Mannheim | Robert Koch Institute | |
| EPI_ISL_1834054 | Universitätsklinikum Knappschaftskrankenhaus Bochum | Bundeswehr Institute of Microbiology | Alexandra Rehn; Enrico Georgi; Malena Bestehorn-Willmann; Markus Antwerpen; Mathias Walter; Mike Pillukat; Roman Wölfel; Sabine Zange |
| EPI_ISL_2115500, EPI_ISL_2115505 | Virologisches Institut des Universitatsklinikums Erlangen | Robert Koch Institute | |
| EPI_ISL_1647971, EPI_ISL_1647976, EPI_ISL_1647978, EPI_ISL_1647979, EPI_ISL_1647980, EPI_ISL_1647981, EPI_ISL_1647982, EPI_ISL_1647983, EPI_ISL_1647985, EPI_ISL_1647986, EPI_ISL_1647987, EPI_ISL_1647988, EPI_ISL_1647990, EPI_ISL_1647991, EPI_ISL_1647992, EPI_ISL_1647993, EPI_ISL_1647994, EPI_ISL_1647999, EPI_ISL_1648000, EPI_ISL_1648002, EPI_ISL_1648004, EPI_ISL_1648007, EPI_ISL_1648017, EPI_ISL_1648018, EPI_ISL_1648019, EPI_ISL_1648020, EPI_ISL_1648031, EPI_ISL_1648032, EPI_ISL_1648037, EPI_ISL_1648038, EPI_ISL_1648039, EPI_ISL_1648042 | | | |
| see above | Virology, Universitätsklinikum des Saarlandes | Epigenetics, Saarland University | Jörn Walter; Kathrin Kattler; Sascha Tierling; Sigrun Smola; Stefan Lohse; Thorsten Pfuhl |
| EPI_ISL_1573291, EPI_ISL_1573292, EPI_ISL_1573293, EPI_ISL_1573294, EPI_ISL_1573295, EPI_ISL_1573296, EPI_ISL_1573297, EPI_ISL_1644013, EPI_ISL_1644014, EPI_ISL_1644015, EPI_ISL_1644016, EPI_ISL_1644017, EPI_ISL_1644018, EPI_ISL_1644019, EPI_ISL_1644020 | | | |
| see above | alphaomega Labor GbR (Medizinische Kooperationsgemeinschaft Dres. Ackermann/Tennert/ZaiÃŸ) | Robert Koch Institute | |
| EPI_ISL_1645008, EPI_ISL_1645018, EPI_ISL_1645047, EPI_ISL_1645053, EPI_ISL_1645108, EPI_ISL_1645167, EPI_ISL_1645193, EPI_ISL_1645213, EPI_ISL_1645230, EPI_ISL_1727153, EPI_ISL_1727177, EPI_ISL_1727197, EPI_ISL_1727217, EPI_ISL_1727238, EPI_ISL_1727243, EPI_ISL_1727248, EPI_ISL_1727258, EPI_ISL_1727262, EPI_ISL_1727275, EPI_ISL_1727280, EPI_ISL_1727285, EPI_ISL_1727306, EPI_ISL_1727311, EPI_ISL_1727319, EPI_ISL_1727321, EPI_ISL_1727332, EPI_ISL_1727333, EPI_ISL_1727352, EPI_ISL_1727354, EPI_ISL_1727357, EPI_ISL_1727359, EPI_ISL_1727361 | | | |
| see above | amedes GÃ¶ttingen | Robert Koch Institute | |
| EPI_ISL_1644998, EPI_ISL_1645002, EPI_ISL_1645012, EPI_ISL_1645013, EPI_ISL_1645019, EPI_ISL_1645023, EPI_ISL_1645025, EPI_ISL_1645029, EPI_ISL_1645032, EPI_ISL_1645038, EPI_ISL_1645043, EPI_ISL_1645049, EPI_ISL_1645071, EPI_ISL_1645075, EPI_ISL_1645082, EPI_ISL_1645090, EPI_ISL_1645102, EPI_ISL_1645104, EPI_ISL_1645106, EPI_ISL_1645107, EPI_ISL_1645112, EPI_ISL_1645118, EPI_ISL_1645122, EPI_ISL_1645128, EPI_ISL_1645141, EPI_ISL_1645145, EPI_ISL_1645148, EPI_ISL_1645150, EPI_ISL_1645156, EPI_ISL_1645164, EPI_ISL_1645165, EPI_ISL_1645195, EPI_ISL_1727192, EPI_ISL_1727212, EPI_ISL_1727221, EPI_ISL_1727225, EPI_ISL_1727235, EPI_ISL_1727269, EPI_ISL_1727281, EPI_ISL_1727310, EPI_ISL_1727312, EPI_ISL_1727325, EPI_ISL_1727330, EPI_ISL_1727376 | | | |
| see above | amedes MVZ DIAMEDIS Sennestadt | Robert Koch Institute | |
| EPI_ISL_1644997, EPI_ISL_1645004, EPI_ISL_1645009, EPI_ISL_1645016, EPI_ISL_1645020, EPI_ISL_1645026, EPI_ISL_1645031, EPI_ISL_1645036, EPI_ISL_1645041, EPI_ISL_1645046, EPI_ISL_1645050, EPI_ISL_1645057, EPI_ISL_1645063, EPI_ISL_1645066, EPI_ISL_1645089, EPI_ISL_1645099, EPI_ISL_1645105, EPI_ISL_1645110, EPI_ISL_1645115, EPI_ISL_1645120, EPI_ISL_1645121, EPI_ISL_1645126, EPI_ISL_1645138, EPI_ISL_1645142, EPI_ISL_1645144, EPI_ISL_1645152, EPI_ISL_1645155, EPI_ISL_1645157, EPI_ISL_1645162, EPI_ISL_1645169, EPI_ISL_1645173, EPI_ISL_1645180, EPI_ISL_1645186, EPI_ISL_1645194, EPI_ISL_1645198, EPI_ISL_1645209, EPI_ISL_1645226, EPI_ISL_1727135 | | | |
| see above | amedes MVZ Halle/Leipzig | Robert Koch Institute | |
| EPI_ISL_1644979, EPI_ISL_1644980, EPI_ISL_1644985, EPI_ISL_1644986, EPI_ISL_1644990, EPI_ISL_1644991, EPI_ISL_1644992, EPI_ISL_1644993, EPI_ISL_1644996, EPI_ISL_1645001, EPI_ISL_1645027, EPI_ISL_1645035, EPI_ISL_1645040, EPI_ISL_1645055, EPI_ISL_1645056, EPI_ISL_1645062, EPI_ISL_1645068, EPI_ISL_1645073, EPI_ISL_1645077, EPI_ISL_1645078, EPI_ISL_1645083, EPI_ISL_1645119, EPI_ISL_1645137, EPI_ISL_1645140, EPI_ISL_1645151, EPI_ISL_1645197, EPI_ISL_1645203, EPI_ISL_1645216, EPI_ISL_1645222, EPI_ISL_1645223 | | | |
| see above | amedes MVZ Hannover | Robert Koch Institute | |
| EPI_ISL_1645048, EPI_ISL_1645051, EPI_ISL_1645058, EPI_ISL_1645067, EPI_ISL_1645069, EPI_ISL_1645072, EPI_ISL_1645076, EPI_ISL_1645079, EPI_ISL_1645084, EPI_ISL_1645087, EPI_ISL_1645096, EPI_ISL_1645103, EPI_ISL_1645109, EPI_ISL_1645114, EPI_ISL_1645117, EPI_ISL_1645124, EPI_ISL_1645129, EPI_ISL_1645154, EPI_ISL_1645163, EPI_ISL_1645166, EPI_ISL_1645172, EPI_ISL_1645175, EPI_ISL_1645199, EPI_ISL_1645211, EPI_ISL_1645217 | | | |
| see above | amedes MVZ Jena | Robert Koch Institute | |
| EPI_ISL_1645044, EPI_ISL_1645181 | amedes MVZ Zentrum fÃ¼r Pathologie und Zytodiagnostik GmbH (KÃ¶ln) | Robert Koch Institute | |
| EPI_ISL_1644988, EPI_ISL_1645007, EPI_ISL_1645022, EPI_ISL_1645052, EPI_ISL_1645065, EPI_ISL_1645080, EPI_ISL_1645085, EPI_ISL_1645091, EPI_ISL_1645123, EPI_ISL_1645130, EPI_ISL_1645136, EPI_ISL_1645153, EPI_ISL_1645168, EPI_ISL_1645177, EPI_ISL_1645189, EPI_ISL_1645200, EPI_ISL_1727126 | | | |
| see above | amedes MVZ fÃ¼r Laboratoriumsdiagnostik Raubling GmbH | Robert Koch Institute | |
| EPI_ISL_1644989, EPI_ISL_1644999, EPI_ISL_1645059, EPI_ISL_1645070, EPI_ISL_1645081, EPI_ISL_1645092, EPI_ISL_1645094, EPI_ISL_1645132, EPI_ISL_1645143, EPI_ISL_1645149, EPI_ISL_1645170, EPI_ISL_1645187, EPI_ISL_1645205, EPI_ISL_1645215 | | | |
| see above | amedes MVZ fÃ¼r Laboratoriumsmedizin FÃ¼rstenfeldbruck | Robert Koch Institute | |
| EPI_ISL_1645161, EPI_ISL_1727139 | amedes MVZ fÃ¼r Laboratoriumsmedizin MÃ¼nchen | Robert Koch Institute | |
| EPI_ISL_1644982, EPI_ISL_1645093, EPI_ISL_1645098 | amedes MVZ fÃ¼r Laboratoriumsmedizin Rhein/Ruhr | Robert Koch Institute | |
| EPI_ISL_1572910, EPI_ISL_1572927, EPI_ISL_1572931, EPI_ISL_1572932, EPI_ISL_1572950, EPI_ISL_1572957, EPI_ISL_1573026, EPI_ISL_1573050, EPI_ISL_1645006, EPI_ISL_1645017 | | | |
| see above | amedes MVZ fÃ¼r Labordiagnostik Rhein-Main | Robert Koch Institute | |
| EPI_ISL_1727203, EPI_ISL_1727210, EPI_ISL_1727255 | amedes Medlab Arnold Analytik MVZ GmbH | Robert Koch Institute | |
| EPI_ISL_1638627, EPI_ISL_1638643 | labopart - Medizinische Laboratorien Dresden | Robert Koch Institute | |
| EPI_ISL_1644981, EPI_ISL_1644983, EPI_ISL_1644995, EPI_ISL_1645000, EPI_ISL_1645003, EPI_ISL_1645011, EPI_ISL_1645015, EPI_ISL_1645033, EPI_ISL_1645039, EPI_ISL_1645042, EPI_ISL_1645045, EPI_ISL_1645054, EPI_ISL_1645060, EPI_ISL_1645061, EPI_ISL_1645064, EPI_ISL_1645074, EPI_ISL_1645095, EPI_ISL_1645097, EPI_ISL_1645100, EPI_ISL_1645111, EPI_ISL_1645113, EPI_ISL_1645116, EPI_ISL_1645125, EPI_ISL_1645127, EPI_ISL_1645131, EPI_ISL_1645139, EPI_ISL_1645146, EPI_ISL_1645147, EPI_ISL_1645159, EPI_ISL_1645179, EPI_ISL_1645182, EPI_ISL_1645185, EPI_ISL_1645190, EPI_ISL_1645196, EPI_ISL_1645201, EPI_ISL_1645206, EPI_ISL_1645208, EPI_ISL_1645212, EPI_ISL_1645214, EPI_ISL_1645218, EPI_ISL_1645219, EPI_ISL_1645220, EPI_ISL_1645221, EPI_ISL_1645224, EPI_ISL_1645225, EPI_ISL_1645228, EPI_ISL_1645229, EPI_ISL_1645231 | | | |
| see above | nordlab - Partnerschaftspraxis fuÌˆr Laboratoriumsmedizin | Robert Koch Institute | |

We gratefully acknowledge the following Authors from the Originating laboratories responsible for obtaining the specimens, as well as the Submitting laboratories where the genome data were generated and shared via GISAID, on which this research is based.

All Submitters of data may be contacted directly via www.gisaid.org

Authors are sorted alphabetically.

| Accession ID | Originating Laboratory | Submitting Laboratory | Authors |
|---|---|---|---|
| EPI_ISL_2115522, EPI_ISL_2115538 | ALB FILS KLINIKEN GmbH - Institut fur Laboratoriumsmedizin Klinik am Eichert Gappingen | Robert Koch Institute | |
| EPI_ISL_2115691 | ALB FILS KLINIKEN GmbH - Institut fÃ¼r Laboratoriumsmedizin Klinik am Eichert GÃ¶ppingen | Robert Koch Institute | |
| EPI_ISL_2111283, EPI_ISL_2111287, EPI_ISL_2111292, EPI_ISL_2111301, EPI_ISL_2111302, EPI_ISL_2111307, EPI_ISL_2111308, EPI_ISL_2111320, EPI_ISL_2111322, EPI_ISL_2111325, EPI_ISL_2111333, EPI_ISL_2111335, EPI_ISL_2111343, EPI_ISL_2111344, EPI_ISL_2111357, EPI_ISL_2111359, EPI_ISL_2111361, EPI_ISL_2111365, EPI_ISL_2111374, EPI_ISL_2111375, EPI_ISL_2111376, EPI_ISL_2111382, EPI_ISL_2111383, EPI_ISL_2111384, EPI_ISL_2111385, EPI_ISL_2111386, EPI_ISL_2111389, EPI_ISL_2111390, EPI_ISL_2111394, EPI_ISL_2111400, EPI_ISL_2111406, EPI_ISL_2111410, EPI_ISL_2111421 | | | |
| see above | Bayerisches Landesamt fur Gesundheit und Lebensmittelsicherheit (LGL) | Robert Koch Institute | |
| EPI_ISL_2129726, EPI_ISL_2129729 | Bernhard-Nocht-Institut fÃ¼r Tropenmedizin | Robert Koch Institute | |
| EPI_ISL_1843459, EPI_ISL_1843472, EPI_ISL_1843497, EPI_ISL_1843507, EPI_ISL_1843516, EPI_ISL_1843533, EPI_ISL_1843548, EPI_ISL_1843560, EPI_ISL_1843574, EPI_ISL_1843576, EPI_ISL_1843585, EPI_ISL_1843615, EPI_ISL_1843648, EPI_ISL_1843649, EPI_ISL_1843656, EPI_ISL_1843666, EPI_ISL_1844062, EPI_ISL_1844135, EPI_ISL_1844145, EPI_ISL_1844157, EPI_ISL_1844190, EPI_ISL_1844193, EPI_ISL_1844201, EPI_ISL_1844207, EPI_ISL_1844226, EPI_ISL_1844232, EPI_ISL_1844240, EPI_ISL_1844262, EPI_ISL_1844263, EPI_ISL_1846274, EPI_ISL_1846288, EPI_ISL_1846298, EPI_ISL_1846299, EPI_ISL_1846307, EPI_ISL_1848404, EPI_ISL_1848412, EPI_ISL_1848414, EPI_ISL_1848418, EPI_ISL_1848419, EPI_ISL_1848421, EPI_ISL_1848423, EPI_ISL_1848424, EPI_ISL_1848427, EPI_ISL_1848431, EPI_ISL_1848441, EPI_ISL_1848443, EPI_ISL_1848445, EPI_ISL_1848449, EPI_ISL_1848453, EPI_ISL_1848454, EPI_ISL_1848461, EPI_ISL_1848463, EPI_ISL_1848465, EPI_ISL_1848468, EPI_ISL_1848470, EPI_ISL_1848472, EPI_ISL_1848473, EPI_ISL_1848475, EPI_ISL_1848477, EPI_ISL_1848481, EPI_ISL_1848487, EPI_ISL_1848492, EPI_ISL_1848493, EPI_ISL_1848500, EPI_ISL_1848502, EPI_ISL_1848504, EPI_ISL_1848505, EPI_ISL_1848507, EPI_ISL_1848511, EPI_ISL_1848518, EPI_ISL_1848525, EPI_ISL_1848533, EPI_ISL_1848534, EPI_ISL_1848537, EPI_ISL_1848540, EPI_ISL_1848541, EPI_ISL_1848542, EPI_ISL_1848545, EPI_ISL_1848548, EPI_ISL_1848550, EPI_ISL_1848553, EPI_ISL_1848555, EPI_ISL_1848558, EPI_ISL_1848559, EPI_ISL_1848560, EPI_ISL_1848562, EPI_ISL_1848563, EPI_ISL_1848564, EPI_ISL_1848566, EPI_ISL_1848572, EPI_ISL_1848579, EPI_ISL_1848581, EPI_ISL_1848585, EPI_ISL_1848588, EPI_ISL_1848589, EPI_ISL_1848590, EPI_ISL_1848594, EPI_ISL_1848597, EPI_ISL_1848598, EPI_ISL_1848599, EPI_ISL_1848601, EPI_ISL_1848602, EPI_ISL_1848603, EPI_ISL_1848605, EPI_ISL_1848607, EPI_ISL_1848612, EPI_ISL_1848613, EPI_ISL_1848615, EPI_ISL_1848616, EPI_ISL_1848617, EPI_ISL_1848621, EPI_ISL_1848624, EPI_ISL_1848627, EPI_ISL_1848628, EPI_ISL_1848632, EPI_ISL_1848633, EPI_ISL_1848634, EPI_ISL_1848635, EPI_ISL_1848641, EPI_ISL_1848645, EPI_ISL_1848646, EPI_ISL_1848647, EPI_ISL_1848649, EPI_ISL_1848650, EPI_ISL_1848651, EPI_ISL_1848652, EPI_ISL_1848654, EPI_ISL_1848656, EPI_ISL_1848659, EPI_ISL_1848660, EPI_ISL_1848661, EPI_ISL_1848662, EPI_ISL_1848663, EPI_ISL_1848667, EPI_ISL_1848669, EPI_ISL_1848670, EPI_ISL_1848676, EPI_ISL_1848678, EPI_ISL_1848679, EPI_ISL_1848680, EPI_ISL_1848684, EPI_ISL_1848686, EPI_ISL_1848687, EPI_ISL_1848688, EPI_ISL_1848689, EPI_ISL_1848691, EPI_ISL_1848692, EPI_ISL_1848693, EPI_ISL_1848694, EPI_ISL_1848695, EPI_ISL_1848698, EPI_ISL_1848700, EPI_ISL_1848701, EPI_ISL_1848703, EPI_ISL_1848707, EPI_ISL_1848710, EPI_ISL_1848715, EPI_ISL_1848718, EPI_ISL_1848719, EPI_ISL_1848720, EPI_ISL_1848722, EPI_ISL_1848724, EPI_ISL_1848727, EPI_ISL_1848728, EPI_ISL_1848733, EPI_ISL_1848734, EPI_ISL_1848735, EPI_ISL_1848736, EPI_ISL_1848738, EPI_ISL_1848739, EPI_ISL_1848741, EPI_ISL_1848742, EPI_ISL_1848743, EPI_ISL_2108717, EPI_ISL_2108720, EPI_ISL_2108723, EPI_ISL_2108726, EPI_ISL_2108727, EPI_ISL_2108734, EPI_ISL_2108737, EPI_ISL_2108745, EPI_ISL_2108746, EPI_ISL_2108749, EPI_ISL_2108757, EPI_ISL_2108770, EPI_ISL_2108777, EPI_ISL_2108779, EPI_ISL_2108780, EPI_ISL_2108781, EPI_ISL_2108786, EPI_ISL_2108790, EPI_ISL_2108792, EPI_ISL_2108797, EPI_ISL_2108799, EPI_ISL_2108801, EPI_ISL_2108810, EPI_ISL_2108817, EPI_ISL_2108818, EPI_ISL_2108824, EPI_ISL_2108829, EPI_ISL_2108840, EPI_ISL_2108844, EPI_ISL_2108848, EPI_ISL_2108853, EPI_ISL_2108855, EPI_ISL_2108864, EPI_ISL_2108869, EPI_ISL_2108871, EPI_ISL_2108875, EPI_ISL_2108881, EPI_ISL_2108888, EPI_ISL_2108894, EPI_ISL_2108898, EPI_ISL_2108902, EPI_ISL_2108903, EPI_ISL_2108909, EPI_ISL_2108925, EPI_ISL_2108927, EPI_ISL_2108936, EPI_ISL_2108949, EPI_ISL_2108955, EPI_ISL_2108957, EPI_ISL_2108963, EPI_ISL_2108969, EPI_ISL_2108970, EPI_ISL_2108975, EPI_ISL_2108977, EPI_ISL_2108978, EPI_ISL_2109054, EPI_ISL_2109067, EPI_ISL_2109094, EPI_ISL_2109157, EPI_ISL_2109215, EPI_ISL_2109234, EPI_ISL_2109324, EPI_ISL_2109340, EPI_ISL_2109351, EPI_ISL_2109540, EPI_ISL_2109554, EPI_ISL_2109696, EPI_ISL_2109728, EPI_ISL_2109783, EPI_ISL_2115639 | | | |
| see above | Bioscientia Labor Wermsdorf | Robert Koch Institute | |
| EPI_ISL_1843575, EPI_ISL_1848437, EPI_ISL_1848438, EPI_ISL_1848460, EPI_ISL_1848486, EPI_ISL_1848495, EPI_ISL_1848516, EPI_ISL_1848543, EPI_ISL_1848544, EPI_ISL_1848571, EPI_ISL_1848622, EPI_ISL_1848668, EPI_ISL_1848690, EPI_ISL_1848713, EPI_ISL_1848726, EPI_ISL_2108783, EPI_ISL_2108821, EPI_ISL_2108890, EPI_ISL_2109061, EPI_ISL_2109069, EPI_ISL_2109071, EPI_ISL_2109080, EPI_ISL_2109093, EPI_ISL_2109114, EPI_ISL_2109146, EPI_ISL_2109149, EPI_ISL_2109166, EPI_ISL_2109218, EPI_ISL_2109228, EPI_ISL_2109240, EPI_ISL_2109306, EPI_ISL_2109322, EPI_ISL_2109326, EPI_ISL_2109344 | | | |
| see above | Bioscientia MVZ Labor Karlsruhe GmbH | Robert Koch Institute | |
| EPI_ISL_1834048, EPI_ISL_1834049 | Bundeswehr Institute of Microbiology | Bundeswehr Institute of Microbiology | Alexandra Rehn; Enrico Georgi; Malena Bestehorn-Willmann; Markus Antwerpen; Mathias Walter; Mike Pillukat; Roman Wölfel; Sabine Zange |
| EPI_ISL_1834039, EPI_ISL_1834057, EPI_ISL_1834058, EPI_ISL_1834059, EPI_ISL_1834060, EPI_ISL_1834061, EPI_ISL_1834064, EPI_ISL_1834065, EPI_ISL_1834067, EPI_ISL_1834068, EPI_ISL_1834069 | | | |
| see above | Bundeswehrkrankenhaus Berlin | Bundeswehr Institute of Microbiology | Alexandra Rehn; Enrico Georgi; Malena Bestehorn-Willmann; Markus Antwerpen; Mathias Walter; Mike Pillukat; Roman Wölfel; Sabine Zange |
| EPI_ISL_1834071, EPI_ISL_1834073, EPI_ISL_1834074, EPI_ISL_1834075 | Bundeswehrzentralkrankenhaus Koblenz | Bundeswehr Institute of Microbiology | Alexandra Rehn; Enrico Georgi; Malena Bestehorn-Willmann; Markus Antwerpen; Mathias Walter; Mike Pillukat; Roman Wölfel; Sabine Zange |
| EPI_ISL_1991859, EPI_ISL_1991860 | Center of Medical Microbiology, Virology, and Hospital Hygiene, University of Duesseldorf | Center of Medical Microbiology, Virology, and Hospital Hygiene, University of Duesseldorf | Alexander Dilthey; Andreas Walker; Daniel Strelow; Jessica Nicolai; Jörg Timm; Klaus Pfeffer; Lisanna Hülse; Malte Kohns Vasconcelos; Maximilian Damagnez; Nadine Lübke; Tobias Wienemann; Torsten Houwaart |
| EPI_ISL_1843735, EPI_ISL_1843739, EPI_ISL_1843746, EPI_ISL_1843750, EPI_ISL_1843755, EPI_ISL_1843762, EPI_ISL_1843766, EPI_ISL_1843768, EPI_ISL_1843773, EPI_ISL_1843775, EPI_ISL_1843777, EPI_ISL_1843782, EPI_ISL_1843783, EPI_ISL_1843784, EPI_ISL_1843786, EPI_ISL_1843787, EPI_ISL_1843788, EPI_ISL_1843789, EPI_ISL_1843791, EPI_ISL_1843792, EPI_ISL_1843795, EPI_ISL_1843798, EPI_ISL_1843800, EPI_ISL_1843804, EPI_ISL_1843807, EPI_ISL_1843808, EPI_ISL_1843809, EPI_ISL_1843811, EPI_ISL_1843814, EPI_ISL_1843815, EPI_ISL_1843816, EPI_ISL_1843817, EPI_ISL_1843818, EPI_ISL_1843821, EPI_ISL_1843823, EPI_ISL_1843834, EPI_ISL_1843835, EPI_ISL_1843837, EPI_ISL_1843838, EPI_ISL_1843840, EPI_ISL_1843846, EPI_ISL_1843847, EPI_ISL_1843849, EPI_ISL_1843851, EPI_ISL_1843856, EPI_ISL_1843862, EPI_ISL_1843868, EPI_ISL_1843870, EPI_ISL_1843871, EPI_ISL_1843877, EPI_ISL_1843878, EPI_ISL_1843879, EPI_ISL_1843881, EPI_ISL_1843882, EPI_ISL_1843883, EPI_ISL_1843885, EPI_ISL_1843891, EPI_ISL_1843894, EPI_ISL_1843895, EPI_ISL_1843896, EPI_ISL_1843899, EPI_ISL_1843905, EPI_ISL_1843911, EPI_ISL_1843912, EPI_ISL_1843914, EPI_ISL_1843918, EPI_ISL_1843928, EPI_ISL_1843938, EPI_ISL_1843943, EPI_ISL_1843946, EPI_ISL_1843948, EPI_ISL_1843951, EPI_ISL_1843954, EPI_ISL_1843956, EPI_ISL_1843969, EPI_ISL_1843971, EPI_ISL_1843972, EPI_ISL_1843973, EPI_ISL_1843979, EPI_ISL_1843980, EPI_ISL_1843982, EPI_ISL_1843984, EPI_ISL_1843986, EPI_ISL_1843988, EPI_ISL_1843989, EPI_ISL_1843990, EPI_ISL_1843991, EPI_ISL_1843992, EPI_ISL_1843994, EPI_ISL_1843996, EPI_ISL_1844000, EPI_ISL_1844001, EPI_ISL_1844005, EPI_ISL_1844011, EPI_ISL_1844012, EPI_ISL_1844014, EPI_ISL_1844016, EPI_ISL_1844019, EPI_ISL_1844020, EPI_ISL_1844023, EPI_ISL_1844024, EPI_ISL_2108480, EPI_ISL_2108481, EPI_ISL_2108482, EPI_ISL_2108483, EPI_ISL_2108485, EPI_ISL_2108487, EPI_ISL_2108490, EPI_ISL_2108491, EPI_ISL_2108492, EPI_ISL_2108493, EPI_ISL_2108494, EPI_ISL_2108495, EPI_ISL_2108496, EPI_ISL_2108497, EPI_ISL_2108498, EPI_ISL_2108501, EPI_ISL_2108502, EPI_ISL_2108504, EPI_ISL_2108505, EPI_ISL_2108506, EPI_ISL_2108507, EPI_ISL_2108508, EPI_ISL_2108510, EPI_ISL_2108512, EPI_ISL_2108516, EPI_ISL_2108517, EPI_ISL_2108518, EPI_ISL_2108519, EPI_ISL_2108520, EPI_ISL_2108521, EPI_ISL_2108523, EPI_ISL_2108529, EPI_ISL_2108531, EPI_ISL_2108532, EPI_ISL_2108533, EPI_ISL_2108534, EPI_ISL_2108536, EPI_ISL_2108539, EPI_ISL_2108541, EPI_ISL_2108543, EPI_ISL_2108544, EPI_ISL_2108545, EPI_ISL_2108547, EPI_ISL_2108548, EPI_ISL_2108549, EPI_ISL_2108551, EPI_ISL_2108552, EPI_ISL_2108554, EPI_ISL_2108556, EPI_ISL_2108558, EPI_ISL_2108559, EPI_ISL_2108560, EPI_ISL_2108561, EPI_ISL_2108565, EPI_ISL_2108567, EPI_ISL_2108569, EPI_ISL_2108570, EPI_ISL_2108572, EPI_ISL_2108574, EPI_ISL_2108576, EPI_ISL_2108578, EPI_ISL_2108580, EPI_ISL_2108581, EPI_ISL_2108583, EPI_ISL_2108585, EPI_ISL_2108586, EPI_ISL_2108588, EPI_ISL_2108589, EPI_ISL_2108590, EPI_ISL_2108592, EPI_ISL_2108593, EPI_ISL_2108594, EPI_ISL_2108595, EPI_ISL_2108596, EPI_ISL_2108597, EPI_ISL_2108598, EPI_ISL_2108599, EPI_ISL_2108602, EPI_ISL_2108603, EPI_ISL_2108604, EPI_ISL_2108605, EPI_ISL_2108607, EPI_ISL_2108609, EPI_ISL_2108610, EPI_ISL_2108611, EPI_ISL_2108613, EPI_ISL_2108616, EPI_ISL_2108617, EPI_ISL_2108618, EPI_ISL_2108619, EPI_ISL_2108622, EPI_ISL_2108623, EPI_ISL_2108624, EPI_ISL_2108625, EPI_ISL_2108626, EPI_ISL_2108627, EPI_ISL_2108629, EPI_ISL_2108630, EPI_ISL_2108632, EPI_ISL_2108635, EPI_ISL_2108637, EPI_ISL_2108638, EPI_ISL_2108640, EPI_ISL_2108641, EPI_ISL_2108642, EPI_ISL_2108643, EPI_ISL_2108644, EPI_ISL_2108647, EPI_ISL_2108648, EPI_ISL_2108650, EPI_ISL_2108651, EPI_ISL_2108652, EPI_ISL_2108654, EPI_ISL_2108655, EPI_ISL_2108656, EPI_ISL_2108657, EPI_ISL_2108658, EPI_ISL_2108659, EPI_ISL_2108660, EPI_ISL_2108662, EPI_ISL_2108663, EPI_ISL_2108665, EPI_ISL_2108666, EPI_ISL_2108667, EPI_ISL_2108668, EPI_ISL_2108669, EPI_ISL_2108670, EPI_ISL_2108672, EPI_ISL_2108673, EPI_ISL_2108677, EPI_ISL_2108681, EPI_ISL_2108684, EPI_ISL_2108685, EPI_ISL_2108689, EPI_ISL_2108690, EPI_ISL_2108691, EPI_ISL_2108693, EPI_ISL_2108694, EPI_ISL_2108696, EPI_ISL_2108697, EPI_ISL_2108698, EPI_ISL_2108699, EPI_ISL_2108700, EPI_ISL_2108701, EPI_ISL_2108705, EPI_ISL_2108706, EPI_ISL_2108707, EPI_ISL_2108708, EPI_ISL_2108709, EPI_ISL_2108710, EPI_ISL_2108711, EPI_ISL_2123081, EPI_ISL_2123083, EPI_ISL_2123084, EPI_ISL_2123087, EPI_ISL_2123089, EPI_ISL_2123090, EPI_ISL_2123091, EPI_ISL_2123092, EPI_ISL_2123093, EPI_ISL_2123094, EPI_ISL_2123095, EPI_ISL_2123096, EPI_ISL_2123097, EPI_ISL_2123098, EPI_ISL_2123099, EPI_ISL_2123100, EPI_ISL_2123101, EPI_ISL_2123105, EPI_ISL_2123106, EPI_ISL_2123110, EPI_ISL_2123113, EPI_ISL_2123115, EPI_ISL_2123116, EPI_ISL_2123117, EPI_ISL_2123118, EPI_ISL_2123120, EPI_ISL_2123121, EPI_ISL_2123123, EPI_ISL_2123124, EPI_ISL_2123126, EPI_ISL_2123127, EPI_ISL_2123129, EPI_ISL_2123131, EPI_ISL_2123132, EPI_ISL_2123134, EPI_ISL_2123135, EPI_ISL_2123136, EPI_ISL_2123139, EPI_ISL_2123140, EPI_ISL_2123145, EPI_ISL_2123146, EPI_ISL_2123152, EPI_ISL_2123154, EPI_ISL_2123155, EPI_ISL_2123156, EPI_ISL_2123161, EPI_ISL_2123162, EPI_ISL_2123163, EPI_ISL_2123165, EPI_ISL_2123166, EPI_ISL_2123167, EPI_ISL_2123168, EPI_ISL_2123169, EPI_ISL_2123170, EPI_ISL_2123172, EPI_ISL_2123173, EPI_ISL_2123174, EPI_ISL_2123175, EPI_ISL_2123179, EPI_ISL_2123180, EPI_ISL_2123182, EPI_ISL_2123183, EPI_ISL_2123184, EPI_ISL_2123185, EPI_ISL_2123186, EPI_ISL_2123194, EPI_ISL_2123196, EPI_ISL_2123197, EPI_ISL_2123198, EPI_ISL_2123199, EPI_ISL_2123200, EPI_ISL_2123201, EPI_ISL_2123207, EPI_ISL_2123209, EPI_ISL_2123210, EPI_ISL_2123212, EPI_ISL_2123214, EPI_ISL_2123216, EPI_ISL_2123217, EPI_ISL_2123221, EPI_ISL_2123222, EPI_ISL_2123225, EPI_ISL_2123228, EPI_ISL_2123229, EPI_ISL_2123231, EPI_ISL_2123232, EPI_ISL_2123233, EPI_ISL_2123234, EPI_ISL_2123235, EPI_ISL_2123238, EPI_ISL_2123240, EPI_ISL_2123241, EPI_ISL_2123242, EPI_ISL_2123243, EPI_ISL_2123244, EPI_ISL_2123246, EPI_ISL_2123248, EPI_ISL_2123250, EPI_ISL_2123251, EPI_ISL_2123252, EPI_ISL_2123253, EPI_ISL_2123254, EPI_ISL_2123257, EPI_ISL_2123258, EPI_ISL_2123259, EPI_ISL_2123261, EPI_ISL_2123262, EPI_ISL_2123263, EPI_ISL_2123264, EPI_ISL_2123268, EPI_ISL_2123269, EPI_ISL_2123271, EPI_ISL_2123273, EPI_ISL_2123274, EPI_ISL_2123275, EPI_ISL_2123276, EPI_ISL_2123280, EPI_ISL_2123283, EPI_ISL_2123285, EPI_ISL_2123287, EPI_ISL_2123288, EPI_ISL_2123289, EPI_ISL_2123290, EPI_ISL_2123291, EPI_ISL_2123295, EPI_ISL_2123296, EPI_ISL_2123297, EPI_ISL_2123298, EPI_ISL_2123299, EPI_ISL_2123302, EPI_ISL_2123303, EPI_ISL_2123305, EPI_ISL_2123306, EPI_ISL_2123307, EPI_ISL_2123308, EPI_ISL_2123310, EPI_ISL_2123311, EPI_ISL_2123312, EPI_ISL_2123314, EPI_ISL_2123315, EPI_ISL_2123317, EPI_ISL_2123318, EPI_ISL_2123319, EPI_ISL_2123321, EPI_ISL_2123325, EPI_ISL_2123328, EPI_ISL_2123330, EPI_ISL_2123331, EPI_ISL_2123332, EPI_ISL_2123333, EPI_ISL_2123334, EPI_ISL_2123336, EPI_ISL_2123337, EPI_ISL_2123339, EPI_ISL_2123340, EPI_ISL_2123343, EPI_ISL_2123344, EPI_ISL_2123345, EPI_ISL_2123347, EPI_ISL_2123348, EPI_ISL_2123349, EPI_ISL_2123351, EPI_ISL_2123352, EPI_ISL_2123354, EPI_ISL_2123355, EPI_ISL_2123356, EPI_ISL_2123358, EPI_ISL_2123359, EPI_ISL_2123361, EPI_ISL_2123362, EPI_ISL_2123364, EPI_ISL_2123365, EPI_ISL_2123366 | | | |
| see above | Centogene; Dr. Bauer Laboratoriums GmbH | Robert Koch Institute | |
| EPI_ISL_1725597, EPI_ISL_1725598, EPI_ISL_1725599, EPI_ISL_1725600, EPI_ISL_1725602 | Chemische und VeterinÃ¤runtersuchungsamt MÃ¼nsterland-Emscher-Lippe (CVUA-MEL) | Robert Koch Institute | |
| EPI_ISL_1844374, EPI_ISL_1844377, EPI_ISL_1844378, EPI_ISL_1844379, EPI_ISL_1844380, EPI_ISL_1844382, EPI_ISL_1844384, EPI_ISL_1844385, EPI_ISL_1844388, EPI_ISL_1844390, EPI_ISL_1844391, EPI_ISL_1844393, EPI_ISL_1844394, EPI_ISL_1844395, EPI_ISL_1844397, EPI_ISL_1844398, EPI_ISL_1844399, EPI_ISL_1844401, EPI_ISL_1844403, EPI_ISL_1844404, EPI_ISL_1844405, EPI_ISL_1844408, EPI_ISL_1844409, EPI_ISL_1844410, EPI_ISL_1844411, EPI_ISL_1844413, EPI_ISL_1844414, EPI_ISL_1844417, EPI_ISL_1844419, EPI_ISL_1844420, EPI_ISL_1844421, EPI_ISL_1844422, EPI_ISL_1844423, EPI_ISL_1844425, EPI_ISL_1844427, EPI_ISL_1844428, EPI_ISL_1844429, EPI_ISL_1844430, EPI_ISL_1844431, EPI_ISL_1844432, EPI_ISL_1844436, EPI_ISL_1844438, EPI_ISL_1844439, EPI_ISL_1844440, EPI_ISL_1844442, EPI_ISL_1844443, EPI_ISL_1844444, EPI_ISL_1844445, EPI_ISL_1844447, EPI_ISL_1844448, EPI_ISL_1844449, EPI_ISL_1844450, EPI_ISL_1844451, EPI_ISL_1844453, EPI_ISL_1844454, EPI_ISL_1844455, EPI_ISL_1844456, EPI_ISL_1844457, EPI_ISL_1844458, EPI_ISL_1844460 | | | |
| see above | Clemenshospital - Alexianer Clemenshospital MÃ¼nster | Robert Koch Institute | |
| EPI_ISL_2112751, EPI_ISL_2112754, EPI_ISL_2112756, EPI_ISL_2112758, EPI_ISL_2112760 | Corona-Testzentrum ifp Institut fur Produktqualitat GmbH | Robert Koch Institute | |
| EPI_ISL_2108986, EPI_ISL_2108987, EPI_ISL_2108997, EPI_ISL_2109014 | Department fur Labormedizin Abteilung III Bereich Molekulare Diagnostik Universitatsklinikum Halle(Saale) | Robert Koch Institute | |
| EPI_ISL_1848817, EPI_ISL_1848818, EPI_ISL_1848819, EPI_ISL_1848820, EPI_ISL_1848821, EPI_ISL_1848822, EPI_ISL_1848823, EPI_ISL_1848824, EPI_ISL_1848825, EPI_ISL_1848826, EPI_ISL_1848827, EPI_ISL_1848828, EPI_ISL_1848829, EPI_ISL_1848830, EPI_ISL_1848831, EPI_ISL_1848832, EPI_ISL_1848833, EPI_ISL_1848834, EPI_ISL_1848835, EPI_ISL_1848836, EPI_ISL_1848837, EPI_ISL_1848838, EPI_ISL_1848839, EPI_ISL_1848840, EPI_ISL_1848841, EPI_ISL_1848842, EPI_ISL_1848844, EPI_ISL_1848845, EPI_ISL_1848846, EPI_ISL_1848847, EPI_ISL_1848848, EPI_ISL_1848849, EPI_ISL_1848850, EPI_ISL_1848851, EPI_ISL_1848852, EPI_ISL_1848853, EPI_ISL_1848854, EPI_ISL_1848855, EPI_ISL_1848856, EPI_ISL_1848857, EPI_ISL_1848858, EPI_ISL_1848859, EPI_ISL_1848860, EPI_ISL_1848861, EPI_ISL_1848862, EPI_ISL_1848863, EPI_ISL_1848864, EPI_ISL_1848865, EPI_ISL_1848866, EPI_ISL_1848867, EPI_ISL_1848868, EPI_ISL_1848869, EPI_ISL_1848870, EPI_ISL_1848871, EPI_ISL_1848872, EPI_ISL_1848873, EPI_ISL_1848874, EPI_ISL_1848875, EPI_ISL_1848876, EPI_ISL_1848877, EPI_ISL_1848878, EPI_ISL_1848879, EPI_ISL_1848880, EPI_ISL_1848881, EPI_ISL_1848882, EPI_ISL_1848883, EPI_ISL_1848884, EPI_ISL_1848885, EPI_ISL_1848886, EPI_ISL_1848887, EPI_ISL_1848888, EPI_ISL_1848889, EPI_ISL_1848890, EPI_ISL_1848891, EPI_ISL_1848892, EPI_ISL_1848893, EPI_ISL_1848894, EPI_ISL_1848895, EPI_ISL_1848896, EPI_ISL_1848897, EPI_ISL_1848898, EPI_ISL_1848899, EPI_ISL_1848900, EPI_ISL_1848901, EPI_ISL_1848902, EPI_ISL_1848903, EPI_ISL_1848904, EPI_ISL_1848905, EPI_ISL_1848906, EPI_ISL_1848907, EPI_ISL_1848908, EPI_ISL_1848909, EPI_ISL_1848910, EPI_ISL_1848911, EPI_ISL_1848912, EPI_ISL_1848913, EPI_ISL_1848914, EPI_ISL_1848915, EPI_ISL_1848916, EPI_ISL_1848917, EPI_ISL_1848918, EPI_ISL_1848919, EPI_ISL_1848920, EPI_ISL_1848921, EPI_ISL_1848922, EPI_ISL_1848923, EPI_ISL_1848924, EPI_ISL_1848925, EPI_ISL_1848926, EPI_ISL_1848927, EPI_ISL_1848928, EPI_ISL_1848929, EPI_ISL_1848930, EPI_ISL_1848931, EPI_ISL_1848932, EPI_ISL_1848933, EPI_ISL_1848934, EPI_ISL_1848935, EPI_ISL_1848936, EPI_ISL_1848937, EPI_ISL_1848938, EPI_ISL_1848939, EPI_ISL_1848940, EPI_ISL_1848941, EPI_ISL_1848942, EPI_ISL_1848943, EPI_ISL_1848944, EPI_ISL_1848945, EPI_ISL_1848946, EPI_ISL_1848947, EPI_ISL_1848948, EPI_ISL_1848949, EPI_ISL_1848950, EPI_ISL_1848951, EPI_ISL_1848952, EPI_ISL_1848953, EPI_ISL_1848954, EPI_ISL_1848955, EPI_ISL_1848956, EPI_ISL_1848957, EPI_ISL_1848958, EPI_ISL_1848959, EPI_ISL_1848960, EPI_ISL_1848961, EPI_ISL_1848962, EPI_ISL_1848963, EPI_ISL_1848964, EPI_ISL_1848965, EPI_ISL_1848966, EPI_ISL_2116152, EPI_ISL_2116153, EPI_ISL_2116156, EPI_ISL_2116157, EPI_ISL_2116161, EPI_ISL_2116164, EPI_ISL_2116165, EPI_ISL_2116168, EPI_ISL_2116175, EPI_ISL_2116177, EPI_ISL_2116184, EPI_ISL_2116186, EPI_ISL_2116187, EPI_ISL_2116189, EPI_ISL_2116190, EPI_ISL_2116193, EPI_ISL_2116194, EPI_ISL_2116198, EPI_ISL_2116205, EPI_ISL_2116207, EPI_ISL_2116213, EPI_ISL_2116214, EPI_ISL_2116216, EPI_ISL_2116219, EPI_ISL_2116225, EPI_ISL_2116227, EPI_ISL_2116229, EPI_ISL_2116230, EPI_ISL_2116237, EPI_ISL_2116241, EPI_ISL_2116245, EPI_ISL_2116246, EPI_ISL_2116249, EPI_ISL_2116250, EPI_ISL_2116251, EPI_ISL_2116255, EPI_ISL_2116259, EPI_ISL_2116268, EPI_ISL_2116270, EPI_ISL_2116273, EPI_ISL_2116275, EPI_ISL_2116295, EPI_ISL_2116300, EPI_ISL_2116301, EPI_ISL_2116312 | | | |
| see above | Diagnosticum - Labor Neukirchen | Robert Koch Institute | |
| EPI_ISL_1727052, EPI_ISL_1727053, EPI_ISL_1727054, EPI_ISL_1727055, EPI_ISL_1727056, EPI_ISL_1727057, EPI_ISL_1727058, EPI_ISL_1727059, EPI_ISL_1727060, EPI_ISL_1727061, EPI_ISL_1727062, EPI_ISL_1727063, EPI_ISL_1727064, EPI_ISL_1727066, EPI_ISL_1727067, EPI_ISL_1727068 | | | |
| see above | Dianovis GmbH Greiz | Robert Koch Institute | |
| EPI_ISL_2111435, EPI_ISL_2111436, EPI_ISL_2111437, EPI_ISL_2111438 | Eurofins LifeCodexx GmbH | Robert Koch Institute | |
| EPI_ISL_1640741, EPI_ISL_1640742, EPI_ISL_1640743, EPI_ISL_1640744, EPI_ISL_1640745, EPI_ISL_1640746, EPI_ISL_1640747, EPI_ISL_1640748, EPI_ISL_1640749, EPI_ISL_1640750, EPI_ISL_1640751, EPI_ISL_1640752, EPI_ISL_1640753, EPI_ISL_1640754, EPI_ISL_1640755, EPI_ISL_1640756, EPI_ISL_1640757, EPI_ISL_1640758, EPI_ISL_1640759, EPI_ISL_1640760, EPI_ISL_1640761, EPI_ISL_1640762, EPI_ISL_1640763, EPI_ISL_1640764, EPI_ISL_1640765, EPI_ISL_1640766, EPI_ISL_1640767, EPI_ISL_1640768, EPI_ISL_1640769, EPI_ISL_1640770, EPI_ISL_1640771, EPI_ISL_1640772, EPI_ISL_1640773, EPI_ISL_1640774, EPI_ISL_1640775, EPI_ISL_1640776, EPI_ISL_1640777, EPI_ISL_1640778, EPI_ISL_1640779, EPI_ISL_1640780, EPI_ISL_1640781, EPI_ISL_1640782, EPI_ISL_1640783, EPI_ISL_1640784, EPI_ISL_1640785, EPI_ISL_1640786, EPI_ISL_1640787, EPI_ISL_1640788, EPI_ISL_1640789, EPI_ISL_1640790, EPI_ISL_1640791, EPI_ISL_1640792, EPI_ISL_1640793, EPI_ISL_1640794, EPI_ISL_1640795, EPI_ISL_1640796, EPI_ISL_1640797, EPI_ISL_1640798, EPI_ISL_1640799, EPI_ISL_1640800, EPI_ISL_1640801, EPI_ISL_1640802, EPI_ISL_1640803, EPI_ISL_1640804, EPI_ISL_1640805, EPI_ISL_1640806, EPI_ISL_1640807, EPI_ISL_1640808, EPI_ISL_1640809, EPI_ISL_1640810, EPI_ISL_1640811, EPI_ISL_1640812, EPI_ISL_1640813, EPI_ISL_1640814, EPI_ISL_1640815, EPI_ISL_1640816, EPI_ISL_1640817, EPI_ISL_1640818, EPI_ISL_1640819, EPI_ISL_1640820, EPI_ISL_1640821, EPI_ISL_1640822, EPI_ISL_1640823, EPI_ISL_1640824, EPI_ISL_1640825, EPI_ISL_1640826, EPI_ISL_1640827, EPI_ISL_1640828, EPI_ISL_1640829 | | | |
| see above | Eurofins MVZ Labor Gelsenkirchen | Robert Koch Institute | |
| EPI_ISL_1847337, EPI_ISL_1847338, EPI_ISL_1847339, EPI_ISL_1847340, EPI_ISL_1847341, EPI_ISL_1847343, EPI_ISL_1847344, EPI_ISL_1847345, EPI_ISL_1847347, EPI_ISL_1847348, EPI_ISL_1847349, EPI_ISL_1847350, EPI_ISL_1847351, EPI_ISL_1847352, EPI_ISL_1847353, EPI_ISL_1847354, EPI_ISL_1847355, EPI_ISL_1847356, EPI_ISL_1847357, EPI_ISL_1847359, EPI_ISL_1847360, EPI_ISL_1847362, EPI_ISL_1847364, EPI_ISL_1847365, EPI_ISL_1847366, EPI_ISL_1847368, EPI_ISL_1847369, EPI_ISL_1847370, EPI_ISL_1847371, EPI_ISL_1847373, EPI_ISL_1847374, EPI_ISL_1847375, EPI_ISL_1847377, EPI_ISL_1847378, EPI_ISL_1847379, EPI_ISL_1847380, EPI_ISL_1847381, EPI_ISL_1847382 | | | |
| see above | Fraunhofer-Institut fur Zelltherapie und Immunologie IZI AG Next-Generation Diagnostics | Robert Koch Institute | |
| EPI_ISL_1720973, EPI_ISL_1721006, EPI_ISL_1843219, EPI_ISL_1843231, EPI_ISL_1843234, EPI_ISL_1843239, EPI_ISL_1843246, EPI_ISL_1843258, EPI_ISL_1843277, EPI_ISL_1843287 | | | |
| see above | Helios UniversitÃ¤tsklinikum Wuppertal: UniversitÃ¤t Witten/Herdecke: Institut fÃ¼r | Robert Koch Institute | |

| | | | |
|---|---|---|---|
| | Medizinische Labordiagnostik | | |
| EPI_ISL_1844469, EPI_ISL_1844472, EPI_ISL_1844474, EPI_ISL_1844475, EPI_ISL_1844476, EPI_ISL_1844487, EPI_ISL_1844488, EPI_ISL_1844491, EPI_ISL_1844497, EPI_ISL_1844499, EPI_ISL_1844505, EPI_ISL_1844506, EPI_ISL_1844511, EPI_ISL_1844519, EPI_ISL_1844524, EPI_ISL_1844544, EPI_ISL_1844552, EPI_ISL_1844561, EPI_ISL_1844585 | | | |
| see above | IMD - Institut fÃ¼r Medizinische Diagnostik Berlin-Potsdam | Robert Koch Institute | |
| EPI_ISL_1844528, EPI_ISL_1844531, EPI_ISL_1844534, EPI_ISL_1844541, EPI_ISL_1844545, EPI_ISL_1844546, EPI_ISL_1844549, EPI_ISL_1844554, EPI_ISL_1844565, EPI_ISL_1844567, EPI_ISL_1844572, EPI_ISL_1844575, EPI_ISL_1844579, EPI_ISL_1844583, EPI_ISL_1844584, EPI_ISL_1844588, EPI_ISL_1844599, EPI_ISL_1844600, EPI_ISL_1844605, EPI_ISL_1844606, EPI_ISL_1844607, EPI_ISL_2109427, EPI_ISL_2109437, EPI_ISL_2109447, EPI_ISL_2109469, EPI_ISL_2109471, EPI_ISL_2109482, EPI_ISL_2109494 | | | |
| see above | IMD - Labor Oderland | Robert Koch Institute | |
| EPI_ISL_1844498, EPI_ISL_2108436, EPI_ISL_2108443, EPI_ISL_2108444, EPI_ISL_2108455, EPI_ISL_2108468 | IMD - MVZ Labor Greifswald | Robert Koch Institute | |
| EPI_ISL_1722196, EPI_ISL_1722209, EPI_ISL_1722215, EPI_ISL_1722222, EPI_ISL_1722228, EPI_ISL_1722231, EPI_ISL_1722234, EPI_ISL_1722237, EPI_ISL_1722238, EPI_ISL_1722244, EPI_ISL_1844463, EPI_ISL_1844465, EPI_ISL_1844470, EPI_ISL_1844480, EPI_ISL_1844482, EPI_ISL_1844483, EPI_ISL_1844500, EPI_ISL_1844504, EPI_ISL_1844507, EPI_ISL_1844509, EPI_ISL_1844526, EPI_ISL_1844530, EPI_ISL_1844532, EPI_ISL_1844533, EPI_ISL_1844538, EPI_ISL_1844550, EPI_ISL_1844557, EPI_ISL_1844560, EPI_ISL_1844562, EPI_ISL_1844564, EPI_ISL_1844568, EPI_ISL_1844569, EPI_ISL_1844581, EPI_ISL_1844582, EPI_ISL_1844592, EPI_ISL_1844593, EPI_ISL_1844597 | | | |
| see above | IMD - MVZ Labor Martinsried | Robert Koch Institute | |
| EPI_ISL_1844529, EPI_ISL_1844535, EPI_ISL_1844536, EPI_ISL_1844539, EPI_ISL_1844540, EPI_ISL_1844542, EPI_ISL_1844547, EPI_ISL_1844553, EPI_ISL_1844559, EPI_ISL_1844577, EPI_ISL_1844580, EPI_ISL_1844594, EPI_ISL_1844595, EPI_ISL_1844602 | | | |
| see above | IMD - Medizinisches Labor Rostock | Robert Koch Institute | |
| EPI_ISL_1725221, EPI_ISL_1725224, EPI_ISL_1725225, EPI_ISL_1725228, EPI_ISL_1725229, EPI_ISL_1725231, EPI_ISL_1725234, EPI_ISL_1725236, EPI_ISL_1725237, EPI_ISL_1843322, EPI_ISL_1848373, EPI_ISL_2108263 | | | |
| see above | IMD Labor Frankfurt | Robert Koch Institute | |
| EPI_ISL_2110862 | Institut fur Klinische Transfusionsmedizin und Immungenetik DRK Blutspendedienst Baden-Wurttemberg | Robert Koch Institute | |
| EPI_ISL_2122768 | Institut fur Medizinische Mikrobiologie - Universitatsmedizin Gottingen | Robert Koch Institute | |
| EPI_ISL_1721031, EPI_ISL_1843328 | Institut fÃ¼r Immunologie und Genetik Kaiserslautern: Medizinisches Labor Dr. med. B. Thiele | Robert Koch Institute | |
| EPI_ISL_1726800 | Institut fÃ¼r Klinische Chemie Transfusions- und Laboratoriumsmedizin Berufsgenossenschaftliches UniversitÃ¤tsklinikum Bergmannsheil | Robert Koch Institute | |
| EPI_ISL_1843371, EPI_ISL_1843372, EPI_ISL_1843373 | Institut fÃ¼r Klinische Transfusionsmedizin und Immungenetik DRK Blutspendedienst Baden-WÃ¼rttemberg | Robert Koch Institute | |
| EPI_ISL_1843432, EPI_ISL_1843437 | Institut fÃ¼r Medizinische Laboratoriumsdiagnostik Bochum GmbH | Robert Koch Institute | |
| EPI_ISL_2112719 | Institut fÃ¼r med. Mikrobiologie u. Virologie Klinikum Oldenburg | Robert Koch Institute | |
| EPI_ISL_1940802, EPI_ISL_1940803, EPI_ISL_1940804, EPI_ISL_1940805, EPI_ISL_1940806, EPI_ISL_1940807, EPI_ISL_1940808, EPI_ISL_1940809, EPI_ISL_1940858, EPI_ISL_1940859, EPI_ISL_1940899, EPI_ISL_1940900, EPI_ISL_1940901, EPI_ISL_1940902, EPI_ISL_1940903, EPI_ISL_1940904, EPI_ISL_1940948, EPI_ISL_1940949, EPI_ISL_1940950, EPI_ISL_1940951, EPI_ISL_1940952, EPI_ISL_1940953 | | | |
| see above | Institute of Medical Microbiology and Hospital Hygiene | Institute of Medical Microbiology and Hospital Hygiene | Aljoscha Tersteegen; Prof. Dr. Achim Kaasch |
| EPI_ISL_1648940, EPI_ISL_1648941, EPI_ISL_1648942, EPI_ISL_1648943, EPI_ISL_1653695, EPI_ISL_1653696, EPI_ISL_1805414, EPI_ISL_1805415, EPI_ISL_1805416, EPI_ISL_1805417 | | | |
| see above | Institute of Tropical Medicine | Institute of Tropical Medicine | Prof. Dr. Thirumalaisamy P. Velavan and Prof. Dr. Peter Kremsner |
| EPI_ISL_1805037, EPI_ISL_1805042, EPI_ISL_1805043, EPI_ISL_1805044, EPI_ISL_1805045, EPI_ISL_1805046, EPI_ISL_1805047, EPI_ISL_1805050 | | | |
| see above | Institute of Virology, Medical Center, University of Freiburg, Freiburg, Germany | Institute of Virology, Clinial Virus Genomics, Medical Center, University of Freiburg, Freiburg, Germany | Hajo Grundmann; Jonas Fuchs; Lena Jaki; Lisa Kern; Marcus Panning; Sandra Reuter |
| EPI_ISL_1722362, EPI_ISL_1722366 | KRH Klinikum Region Hannover: Institut fÃ¼r Med. Mikrobiologie und Krankenhaushygiene | Robert Koch Institute | |
| EPI_ISL_1862403, EPI_ISL_1862408, EPI_ISL_1862412 | Kath. Marienkrankenhaus | Heinrich Pette Institute, Leibniz Institute for Experimental Virology | Adam Grundhoff; Alexis Robitaille; Johannes Knobloch; Martin Aepfelbacher; Nicole Fischer; Thomas Günther |
| EPI_ISL_1725251, EPI_ISL_1725267, EPI_ISL_1725274, EPI_ISL_1843331, EPI_ISL_1852092 | Kerckhoff-Klinik GmbH | Robert Koch Institute | |
| EPI_ISL_1843368 | Klinikum Darmstadt GmbH | Robert Koch Institute | |
| EPI_ISL_1722260, EPI_ISL_1722264, EPI_ISL_1722270, EPI_ISL_1722272, EPI_ISL_1724463, EPI_ISL_1724467, EPI_ISL_1724471, EPI_ISL_1724473, EPI_ISL_1724474, EPI_ISL_1724475, EPI_ISL_1724476, EPI_ISL_1724478, EPI_ISL_1724479, EPI_ISL_1724480, EPI_ISL_1724481, EPI_ISL_1724482, EPI_ISL_1724483, EPI_ISL_1726925, EPI_ISL_1726926, EPI_ISL_1726927, EPI_ISL_1726934, EPI_ISL_1844277 | | | |
| see above | Klinikum Ernst von Bergmann gemeinnÃ¼tzige GmbH - stationÃ¤rer Bereich | Robert Koch Institute | |
| EPI_ISL_1847531, EPI_ISL_1847572 | Klinikum Fulda gAG | Robert Koch Institute | |
| EPI_ISL_1725238 | Krankenhausgesellschaft St. Vincenz mbH | Robert Koch Institute | |
| EPI_ISL_1843537, EPI_ISL_1843720, EPI_ISL_1846246, EPI_ISL_1846256, EPI_ISL_1847447, EPI_ISL_1847460, EPI_ISL_1847463, EPI_ISL_1847466, EPI_ISL_1847467, EPI_ISL_1847472, EPI_ISL_1847475, EPI_ISL_1847481, EPI_ISL_1847484, EPI_ISL_1847488, EPI_ISL_1847490, EPI_ISL_1847496, EPI_ISL_1847500, EPI_ISL_1847503, EPI_ISL_1847506, EPI_ISL_1847513, EPI_ISL_1847526, EPI_ISL_1847528, EPI_ISL_1847529, EPI_ISL_1847530, EPI_ISL_1847533, EPI_ISL_1847537, EPI_ISL_1847543, EPI_ISL_1847544, EPI_ISL_1847546, EPI_ISL_1847552, EPI_ISL_1847556, EPI_ISL_1847557, EPI_ISL_1847559, EPI_ISL_1847560, EPI_ISL_1847561, EPI_ISL_1847563, EPI_ISL_1847568, EPI_ISL_1847570, EPI_ISL_1847578, EPI_ISL_1847579, EPI_ISL_1847581, EPI_ISL_1847585, EPI_ISL_1847586, EPI_ISL_1847589, EPI_ISL_1847594, EPI_ISL_1847598, EPI_ISL_1847599, EPI_ISL_1847603, EPI_ISL_1847604, EPI_ISL_1847606, EPI_ISL_1847611, EPI_ISL_1847613, EPI_ISL_1847614, EPI_ISL_1847621, EPI_ISL_1847623, EPI_ISL_1847627, EPI_ISL_1847628, EPI_ISL_1847630, EPI_ISL_1847634, EPI_ISL_1847635, EPI_ISL_1847636, EPI_ISL_1847641, EPI_ISL_1847646, EPI_ISL_1847647, EPI_ISL_1847648, EPI_ISL_1847654, EPI_ISL_1847655, EPI_ISL_1847658, EPI_ISL_1847659, EPI_ISL_1847660, EPI_ISL_1847665, EPI_ISL_1847666, EPI_ISL_1847675, EPI_ISL_1847677, EPI_ISL_1847678, EPI_ISL_1847679, EPI_ISL_1847681, EPI_ISL_1847682, EPI_ISL_1847683, EPI_ISL_1847690, EPI_ISL_1847691, EPI_ISL_1847695, EPI_ISL_1847697, EPI_ISL_1848417, EPI_ISL_1848420, EPI_ISL_1848432, EPI_ISL_1848434, EPI_ISL_1848435, EPI_ISL_1848440, EPI_ISL_1848467, EPI_ISL_1848478, EPI_ISL_1848483, EPI_ISL_1848485, EPI_ISL_1848489, EPI_ISL_1848499, EPI_ISL_1848509, EPI_ISL_1848510, EPI_ISL_1848514, EPI_ISL_1848532, EPI_ISL_1848536, EPI_ISL_1848551, EPI_ISL_1848569, EPI_ISL_1848573, EPI_ISL_1848576, EPI_ISL_1848578, EPI_ISL_1848582, EPI_ISL_1848591, EPI_ISL_1848596, EPI_ISL_1848604, EPI_ISL_1848609, EPI_ISL_1848610, EPI_ISL_1848618, EPI_ISL_1848623, EPI_ISL_1848625, EPI_ISL_1848629, EPI_ISL_1848631, EPI_ISL_1848639, EPI_ISL_1848642, EPI_ISL_1848644, EPI_ISL_1848658, EPI_ISL_1848664, EPI_ISL_1848666, EPI_ISL_1848672, EPI_ISL_1848674, EPI_ISL_1848682, EPI_ISL_1848697, EPI_ISL_1848714, EPI_ISL_1848716, EPI_ISL_1848729, EPI_ISL_1848732, EPI_ISL_1848737 | | | |
| see above | LabKom - Labor Augsburg MVZ GmbH | Robert Koch Institute | |
| EPI_ISL_1844052, EPI_ISL_1844064, EPI_ISL_1844084, EPI_ISL_1844100, EPI_ISL_1844125, EPI_ISL_1844138, EPI_ISL_1844149, EPI_ISL_1844186, EPI_ISL_1844208, EPI_ISL_1844249, EPI_ISL_1846240, EPI_ISL_1846244, EPI_ISL_1846251, EPI_ISL_1846258, EPI_ISL_1846263, EPI_ISL_1846264, EPI_ISL_1846266, EPI_ISL_1846271, EPI_ISL_1846275, EPI_ISL_1846276, EPI_ISL_1846281, EPI_ISL_1846283, EPI_ISL_1846293, EPI_ISL_1846297, EPI_ISL_1846300, EPI_ISL_1846301, EPI_ISL_1846304, EPI_ISL_1846305, EPI_ISL_1846317, EPI_ISL_2115521 | | | |
| see above | LabKom - Labor Hannover MVZ GmbH | Robert Koch Institute | |
| EPI_ISL_1844080, EPI_ISL_1844110, EPI_ISL_1844159, EPI_ISL_1844257, EPI_ISL_1846247, EPI_ISL_1846267, EPI_ISL_1846280, EPI_ISL_1846284, EPI_ISL_1846296, EPI_ISL_1846316, EPI_ISL_2108778, EPI_ISL_2108831, EPI_ISL_2108860, EPI_ISL_2108943 | | | |
| see above | LabKom - Labor Mainz MVZ GmbH | Robert Koch Institute | |
| EPI_ISL_1843455, EPI_ISL_1843458, EPI_ISL_1843460, EPI_ISL_1843477, EPI_ISL_1843481, EPI_ISL_1843504, EPI_ISL_1843506, EPI_ISL_1843526, EPI_ISL_1843532, EPI_ISL_1843557, EPI_ISL_1843587, EPI_ISL_1843611, EPI_ISL_1843627, EPI_ISL_1843631, EPI_ISL_1843668, EPI_ISL_1843681, EPI_ISL_1843684, EPI_ISL_1843700, EPI_ISL_2109092, EPI_ISL_2109122, EPI_ISL_2109136, EPI_ISL_2109139, EPI_ISL_2109169, EPI_ISL_2109177, EPI_ISL_2109225, EPI_ISL_2109235, EPI_ISL_2109276, EPI_ISL_2109278, EPI_ISL_2109311, EPI_ISL_2109317, EPI_ISL_2109337 | | | |
| see above | Labor 28 MVZ GmbH | Robert Koch Institute | |
| EPI_ISL_1885872, EPI_ISL_1885879, EPI_ISL_1885892, EPI_ISL_1885895, EPI_ISL_1885897, EPI_ISL_1885899, EPI_ISL_1885904, EPI_ISL_1885917, EPI_ISL_1885920, EPI_ISL_1885922, EPI_ISL_1885932, EPI_ISL_1885934, EPI_ISL_1885937, EPI_ISL_1885939, EPI_ISL_1885949, EPI_ISL_1885951, EPI_ISL_1885953, EPI_ISL_1885958, EPI_ISL_1885967, EPI_ISL_1885976, EPI_ISL_1885978, EPI_ISL_1885981, EPI_ISL_1885983, EPI_ISL_1885986, EPI_ISL_1885989, EPI_ISL_1886104, EPI_ISL_1886116, EPI_ISL_1886119, EPI_ISL_1886122, EPI_ISL_1886125, EPI_ISL_1886132, EPI_ISL_1886137, EPI_ISL_1886143, EPI_ISL_1886146, EPI_ISL_1886148, EPI_ISL_1886151, EPI_ISL_1886153, EPI_ISL_1886155, EPI_ISL_1886158, EPI_ISL_1886161, EPI_ISL_1886168, EPI_ISL_1886173, EPI_ISL_1886179, EPI_ISL_1886182, EPI_ISL_1886188, EPI_ISL_1886191, EPI_ISL_1886203 | | | |
| see above | Labor Berlin Charité Vivantes GmbH / Institut für Virologie | Charité Universitätsmedizin Berlin, Institut für Virologie/Labor Berlin | Barbara Mühlemann; Christian Drosten; Christine Stephan; Peter Menzel; Rolf Schwarzer; Terry Jones; Victor M Corman |
| EPI_ISL_1727094, EPI_ISL_1727095, EPI_ISL_1727096, EPI_ISL_1727097, EPI_ISL_1727098, EPI_ISL_1727099, EPI_ISL_1727100, EPI_ISL_1727101, EPI_ISL_1727102, EPI_ISL_1727104, EPI_ISL_1727105, EPI_ISL_1727106, EPI_ISL_1727108, EPI_ISL_1727109, EPI_ISL_1727110, EPI_ISL_1727111, EPI_ISL_1727112, EPI_ISL_1727113, EPI_ISL_1727114, EPI_ISL_1727115, EPI_ISL_1727116, EPI_ISL_1727117, EPI_ISL_1727118, EPI_ISL_1727119, EPI_ISL_1727120, EPI_ISL_1727121, EPI_ISL_1727122, EPI_ISL_1844609, EPI_ISL_1844611, EPI_ISL_1844615, EPI_ISL_1844621, EPI_ISL_1844625, EPI_ISL_1844627, EPI_ISL_1844628, EPI_ISL_1844632, EPI_ISL_1844633, EPI_ISL_1844635 | | | |
| see above | Labor Dr. Heidrich & Kollegen MVZ GmbH Hamburg | Robert Koch Institute | |
| EPI_ISL_1843448, EPI_ISL_1843454, EPI_ISL_1843456, EPI_ISL_1843470, EPI_ISL_1843478, EPI_ISL_1843485, EPI_ISL_1843514, EPI_ISL_1843515, EPI_ISL_1843549, EPI_ISL_1843552, EPI_ISL_1843564, EPI_ISL_1843581, EPI_ISL_1843604, EPI_ISL_1843616, EPI_ISL_1843618, EPI_ISL_1843646, EPI_ISL_1843660, EPI_ISL_1843664, EPI_ISL_1843673, EPI_ISL_1843679, EPI_ISL_1843687, EPI_ISL_1843702, EPI_ISL_1843728, EPI_ISL_1844056, EPI_ISL_1844057, EPI_ISL_1844067, EPI_ISL_1844074, EPI_ISL_1844077, EPI_ISL_1844082, EPI_ISL_1844118, EPI_ISL_1844120, EPI_ISL_1844124, EPI_ISL_1844126, EPI_ISL_1844146, EPI_ISL_1844153, EPI_ISL_1844165, EPI_ISL_1844168, EPI_ISL_1844175, EPI_ISL_1844177, EPI_ISL_1844178, EPI_ISL_1844185, EPI_ISL_1844192, EPI_ISL_1844204, EPI_ISL_1844220, EPI_ISL_1844222, EPI_ISL_1844228, EPI_ISL_1844243, EPI_ISL_1844268, EPI_ISL_2120621, EPI_ISL_2120629, EPI_ISL_2120639, EPI_ISL_2120663, EPI_ISL_2120682, EPI_ISL_2120686, EPI_ISL_2120725, EPI_ISL_2120734, EPI_ISL_2120826, EPI_ISL_2120914, EPI_ISL_2120988, EPI_ISL_2121071, EPI_ISL_2121124, EPI_ISL_2121212, EPI_ISL_2121260, EPI_ISL_2121438 | | | |
| see above | Labor Dr. Schumacher MVZ | Robert Koch Institute | |
| EPI_ISL_1847408, EPI_ISL_1847410, EPI_ISL_1847422, EPI_ISL_2125116, EPI_ISL_2125117, EPI_ISL_2125120, EPI_ISL_2125123, EPI_ISL_2125125, EPI_ISL_2125130, EPI_ISL_2125131, EPI_ISL_2125133 | | | |
| see above | Labor Dr. Spranger | Robert Koch Institute | |
| EPI_ISL_1724327, EPI_ISL_1725187, EPI_ISL_1725188 | Labor Dr. Wisplinghoff - KÃ¶ln | Robert Koch Institute | |
| EPI_ISL_1726860, EPI_ISL_1726861, EPI_ISL_1726862, EPI_ISL_1726863, EPI_ISL_1726866, EPI_ISL_1726867, EPI_ISL_1851131, EPI_ISL_1851132, EPI_ISL_1851133, EPI_ISL_1851134, EPI_ISL_1851135 | | | |
| see above | Labor KneiÃŸler GmbH & Co. KG | Robert Koch Institute | |
| EPI_ISL_1726785, EPI_ISL_1726814, EPI_ISL_1850381, EPI_ISL_1850405, EPI_ISL_1850416, EPI_ISL_1850425 | Labor LÃ¼beck bzw. LaborÃ¤rztliche Gemeinschaftspraxis LÃ¼beck | Robert Koch Institute | |
| EPI_ISL_1724252, EPI_ISL_1724256, EPI_ISL_1724258, EPI_ISL_1724263, EPI_ISL_1724264, EPI_ISL_1724266, EPI_ISL_1724268, EPI_ISL_1724270, EPI_ISL_1724275, EPI_ISL_1724277, EPI_ISL_1724326, EPI_ISL_1724393, EPI_ISL_1724394, EPI_ISL_1724395, EPI_ISL_1724396, EPI_ISL_1724397, EPI_ISL_1724398, EPI_ISL_1724399, EPI_ISL_1724400, EPI_ISL_1724401, EPI_ISL_1724402, EPI_ISL_1724403, EPI_ISL_1724404, EPI_ISL_1724405, EPI_ISL_1724406, EPI_ISL_1724407, EPI_ISL_1724408, EPI_ISL_1724409, EPI_ISL_1724410, EPI_ISL_1724411, EPI_ISL_1724412, EPI_ISL_1724413, EPI_ISL_1724414, EPI_ISL_1724415, EPI_ISL_1724416, EPI_ISL_1724417, EPI_ISL_1724419, EPI_ISL_1725193, EPI_ISL_1725195, EPI_ISL_1725217, EPI_ISL_1846231, EPI_ISL_1846232, EPI_ISL_1846235, EPI_ISL_1846237, EPI_ISL_1846334, EPI_ISL_1846336, EPI_ISL_1847188, EPI_ISL_1847190, EPI_ISL_1847191, EPI_ISL_1847193, EPI_ISL_1847314, EPI_ISL_1847321, EPI_ISL_1847323, EPI_ISL_1847325, EPI_ISL_1847327 | | | |
| see above | Labor MÃ¶nchengladbach MVZ Dr. Stein + Kollegen GbR | Robert Koch Institute | |
| EPI_ISL_1843356, EPI_ISL_1843359, EPI_ISL_1843367, EPI_ISL_1846772 | Labor Prof. Dr. G. Enders MVZ GbR | Robert Koch Institute | |
| EPI_ISL_2111442, EPI_ISL_2111448, EPI_ISL_2111454, EPI_ISL_2111477, EPI_ISL_2111498, EPI_ISL_2111561, EPI_ISL_2111593, EPI_ISL_2111603 | | | |
| see above | Labor ZOTZ\|KLIMAS; MVZ Dusseldorf-Centrum | Robert Koch Institute | |
| EPI_ISL_1851149, EPI_ISL_1851150, EPI_ISL_1851151, EPI_ISL_1851153, EPI_ISL_1851154, EPI_ISL_1851157, EPI_ISL_1851159, EPI_ISL_1851164, EPI_ISL_1851165, EPI_ISL_1851167, EPI_ISL_1851169, EPI_ISL_1851171, EPI_ISL_1851172, EPI_ISL_1851175, EPI_ISL_1851176, EPI_ISL_1851177, EPI_ISL_1851178, EPI_ISL_1851182, EPI_ISL_1851183, EPI_ISL_1851185, EPI_ISL_1851186, EPI_ISL_1851188, EPI_ISL_1851189, EPI_ISL_1851190, EPI_ISL_1851194, EPI_ISL_1851195, EPI_ISL_1851198, EPI_ISL_1851199, EPI_ISL_1851201, EPI_ISL_1851207, EPI_ISL_1851208, EPI_ISL_1851209, EPI_ISL_1851210, EPI_ISL_1851213, EPI_ISL_1851215, EPI_ISL_1851217, EPI_ISL_1851220, EPI_ISL_1851222, EPI_ISL_1851223, EPI_ISL_1851225, EPI_ISL_1851226, EPI_ISL_1851229, EPI_ISL_1851230, EPI_ISL_1851233, | | | |

| | | | |
|---|---|---|---|
| EPI_ISL_1851234, EPI_ISL_1851238, EPI_ISL_1851239, EPI_ISL_1851241, EPI_ISL_1851248, EPI_ISL_1851251, EPI_ISL_1851253, EPI_ISL_1851254, EPI_ISL_1851256, EPI_ISL_1851257, EPI_ISL_1851261, EPI_ISL_1851262, EPI_ISL_1851268, EPI_ISL_1851269, EPI_ISL_1851273, EPI_ISL_1851276, EPI_ISL_1851277, EPI_ISL_1851280, EPI_ISL_1851281, EPI_ISL_1851282, EPI_ISL_1851284, EPI_ISL_1851285, EPI_ISL_1851286, EPI_ISL_1851291, EPI_ISL_1851293, EPI_ISL_1851294, EPI_ISL_1851295, EPI_ISL_1851296, EPI_ISL_1851297, EPI_ISL_1851298, EPI_ISL_1851300, EPI_ISL_1851301, EPI_ISL_1851305, EPI_ISL_1851306, EPI_ISL_1851308, EPI_ISL_1851309, EPI_ISL_1851312, EPI_ISL_1851313, EPI_ISL_1851314, EPI_ISL_1851315, EPI_ISL_1851317, EPI_ISL_1851319, EPI_ISL_1851320, EPI_ISL_1851322, EPI_ISL_1851324, EPI_ISL_1851327, EPI_ISL_1851328, EPI_ISL_1851333, EPI_ISL_1851336, EPI_ISL_1851338, EPI_ISL_1851342, EPI_ISL_1851343, EPI_ISL_1851344, EPI_ISL_1851345, EPI_ISL_1851348, EPI_ISL_1851349, EPI_ISL_1851351, EPI_ISL_1851352, EPI_ISL_1851353, EPI_ISL_1851354, EPI_ISL_1851356, EPI_ISL_1851357, EPI_ISL_1851361, EPI_ISL_1851364, EPI_ISL_1851367, EPI_ISL_1851370, EPI_ISL_1851371, EPI_ISL_1851373, EPI_ISL_1851376, EPI_ISL_1851378, EPI_ISL_1851381, EPI_ISL_1851382, EPI_ISL_1851384, EPI_ISL_1851385 | | | |
| see above | Labor ZOTZ\|KLIMAS; MVZ DÃ¼sseldorf-Centrum | Robert Koch Institute | |
| EPI_ISL_1729041, EPI_ISL_1729052, EPI_ISL_1729061 | Labor fÃ¼r medizinische Mikrobiologie Bakteriologie Mykologie Virologie & Infektionsserologie - Labor MÃ¶lbis | Robert Koch Institute | |
| EPI_ISL_1843295, EPI_ISL_1843296, EPI_ISL_1843298, EPI_ISL_1843302, EPI_ISL_1843307, EPI_ISL_1843309, EPI_ISL_1843311, EPI_ISL_1843317, EPI_ISL_1843319, EPI_ISL_1843326, EPI_ISL_1843327, EPI_ISL_1843330, EPI_ISL_1848370, EPI_ISL_1848385, EPI_ISL_1848389, EPI_ISL_1848396, EPI_ISL_1848397, EPI_ISL_1848399 | | | |
| see above | Labormedizin Darmstadt | Robert Koch Institute | |
| EPI_ISL_1940810, EPI_ISL_1940811, EPI_ISL_1940812, EPI_ISL_1940813, EPI_ISL_1940814, EPI_ISL_1940815, EPI_ISL_1940816, EPI_ISL_1940817, EPI_ISL_1940818, EPI_ISL_1940819, EPI_ISL_1940820, EPI_ISL_1940821, EPI_ISL_1940822, EPI_ISL_1940823, EPI_ISL_1940824, EPI_ISL_1940825, EPI_ISL_1940826, EPI_ISL_1940827, EPI_ISL_1940828, EPI_ISL_1940829, EPI_ISL_1940830, EPI_ISL_1940831, EPI_ISL_1940832, EPI_ISL_1940833, EPI_ISL_1940834, EPI_ISL_1940835, EPI_ISL_1940836, EPI_ISL_1940837, EPI_ISL_1940838, EPI_ISL_1940839, EPI_ISL_1940840, EPI_ISL_1940841, EPI_ISL_1940842, EPI_ISL_1940843, EPI_ISL_1940844, EPI_ISL_1940845, EPI_ISL_1940846, EPI_ISL_1940847, EPI_ISL_1940848, EPI_ISL_1940860, EPI_ISL_1940861, EPI_ISL_1940862, EPI_ISL_1940863, EPI_ISL_1940864, EPI_ISL_1940865, EPI_ISL_1940866, EPI_ISL_1940867, EPI_ISL_1940868, EPI_ISL_1940869, EPI_ISL_1940870, EPI_ISL_1940871, EPI_ISL_1940872, EPI_ISL_1940873, EPI_ISL_1940874, EPI_ISL_1940875, EPI_ISL_1940876, EPI_ISL_1940877, EPI_ISL_1940878, EPI_ISL_1940879, EPI_ISL_1940880, EPI_ISL_1940881, EPI_ISL_1940882, EPI_ISL_1940883, EPI_ISL_1940884, EPI_ISL_1940885, EPI_ISL_1940886, EPI_ISL_1940887, EPI_ISL_1940888, EPI_ISL_1940889, EPI_ISL_1940890, EPI_ISL_1940891, EPI_ISL_1940892, EPI_ISL_1940893, EPI_ISL_1940894, EPI_ISL_1940895, EPI_ISL_1940896, EPI_ISL_1940897, EPI_ISL_1940898, EPI_ISL_1940905, EPI_ISL_1940906, EPI_ISL_1940907, EPI_ISL_1940908, EPI_ISL_1940909, EPI_ISL_1940910, EPI_ISL_1940911, EPI_ISL_1940912, EPI_ISL_1940913, EPI_ISL_1940914, EPI_ISL_1940915, EPI_ISL_1940916, EPI_ISL_1940917, EPI_ISL_1940918, EPI_ISL_1940919, EPI_ISL_1940920, EPI_ISL_1940921, EPI_ISL_1940922, EPI_ISL_1940923, EPI_ISL_1940924, EPI_ISL_1940925, EPI_ISL_1940926, EPI_ISL_1940927, EPI_ISL_1940928, EPI_ISL_1940929, EPI_ISL_1940930, EPI_ISL_1940931, EPI_ISL_1940932, EPI_ISL_1940933, EPI_ISL_1940934, EPI_ISL_1940935, EPI_ISL_1940936, EPI_ISL_1940937, EPI_ISL_1940938, EPI_ISL_1940939, EPI_ISL_1940940, EPI_ISL_1940941, EPI_ISL_1940942, EPI_ISL_1940943, EPI_ISL_1940944, EPI_ISL_1940945, EPI_ISL_1940946, EPI_ISL_1940947, EPI_ISL_1940954, EPI_ISL_1940955, EPI_ISL_1940956, EPI_ISL_1940957, EPI_ISL_1940958, EPI_ISL_1940959, EPI_ISL_1940960, EPI_ISL_1940961, EPI_ISL_1940962, EPI_ISL_1940963, EPI_ISL_1940964, EPI_ISL_1940965, EPI_ISL_1940966, EPI_ISL_1940967, EPI_ISL_1940968, EPI_ISL_1940969, EPI_ISL_1940970, EPI_ISL_1940971, EPI_ISL_1940972, EPI_ISL_1940973, EPI_ISL_1940974, EPI_ISL_1940975, EPI_ISL_1940976, EPI_ISL_1940977, EPI_ISL_1940978, EPI_ISL_1940979, EPI_ISL_1940980, EPI_ISL_1940981, EPI_ISL_1940982, EPI_ISL_1940983, EPI_ISL_1940984, EPI_ISL_1940985, EPI_ISL_1940986, EPI_ISL_1940987, EPI_ISL_1940988, EPI_ISL_1940989, EPI_ISL_1940990, EPI_ISL_1940991, EPI_ISL_1940992, EPI_ISL_1940993, EPI_ISL_1940994, EPI_ISL_1940995, EPI_ISL_1940996, EPI_ISL_1940997, EPI_ISL_1940998, EPI_ISL_1940999, EPI_ISL_1941000, EPI_ISL_1941001 | | | |
| see above | Landesamt für Verbraucherschutz Sachsen Anhalt, Magdeburg | Institute of Medical Microbiology and Hospital Hygiene | Aljoscha Tersteegen; Prof. Dr. Achim Kaasch |
| EPI_ISL_2110767, EPI_ISL_2110768, EPI_ISL_2110774 | Landesgesundheitsamt Baden-Wurttemberg | Robert Koch Institute | |
| EPI_ISL_1846346, EPI_ISL_1846347, EPI_ISL_1846348, EPI_ISL_1846349, EPI_ISL_1846350, EPI_ISL_1846351, EPI_ISL_1846352, EPI_ISL_1846353, EPI_ISL_1846354, EPI_ISL_1846355, EPI_ISL_1846358, EPI_ISL_1846359, EPI_ISL_1846360, EPI_ISL_1846361, EPI_ISL_1846362 | | | |
| see above | Landesgesundheitsamt Baden-WÃ¼rttemberg | Robert Koch Institute | |
| EPI_ISL_1846756 | Limbach - Ganzimmun Diagnostics AG Mainz | Robert Koch Institute | |
| EPI_ISL_1720876, EPI_ISL_1725258, EPI_ISL_1725260, EPI_ISL_1725262, EPI_ISL_1725650, EPI_ISL_1725654, EPI_ISL_1852117 | | | |
| see above | Limbach - MVZ Gemeinschaftslabor Suhl | Robert Koch Institute | |
| EPI_ISL_1643513, EPI_ISL_1643514, EPI_ISL_1643516, EPI_ISL_1643519, EPI_ISL_1643523, EPI_ISL_1643526, EPI_ISL_1643529, EPI_ISL_1643530, EPI_ISL_1643533, EPI_ISL_1643537, EPI_ISL_1643538, EPI_ISL_1643540, EPI_ISL_1643541, EPI_ISL_1643543, EPI_ISL_1643549, EPI_ISL_1643550, EPI_ISL_1643553, EPI_ISL_1643554, EPI_ISL_1643557, EPI_ISL_1643562, EPI_ISL_1643564, EPI_ISL_1643565, EPI_ISL_1643566, EPI_ISL_1643567, EPI_ISL_1643569, EPI_ISL_1643570, EPI_ISL_1643571, EPI_ISL_1643573, EPI_ISL_1643575, EPI_ISL_1643577, EPI_ISL_1643580, EPI_ISL_1643581, EPI_ISL_1643582, EPI_ISL_1643583, EPI_ISL_1643585, EPI_ISL_1643586, EPI_ISL_1643588, EPI_ISL_1643589, EPI_ISL_1643591, EPI_ISL_1643593, EPI_ISL_1643595, EPI_ISL_1643598, EPI_ISL_1643599, EPI_ISL_1643600, EPI_ISL_1643602, EPI_ISL_1643604, EPI_ISL_1643605, EPI_ISL_1643610, EPI_ISL_1643613, EPI_ISL_1643618, EPI_ISL_1643619, EPI_ISL_1643620, EPI_ISL_1643621, EPI_ISL_1643624, EPI_ISL_1643625, EPI_ISL_1643629, EPI_ISL_1643634, EPI_ISL_1643638, EPI_ISL_1643639, EPI_ISL_1643640, EPI_ISL_1643641, EPI_ISL_1643644, EPI_ISL_1643645, EPI_ISL_1643646, EPI_ISL_1643647, EPI_ISL_1643649, EPI_ISL_1643652, EPI_ISL_1643653, EPI_ISL_1643656, EPI_ISL_1643659, EPI_ISL_1643660, EPI_ISL_1643661, EPI_ISL_1643662, EPI_ISL_1643663, EPI_ISL_1643665, EPI_ISL_1643669, EPI_ISL_1643672, EPI_ISL_1643673, EPI_ISL_1643678, EPI_ISL_1727422, EPI_ISL_1727424, EPI_ISL_1727425, EPI_ISL_1727426, EPI_ISL_1727427, EPI_ISL_1727428, EPI_ISL_1727431, EPI_ISL_1727433, EPI_ISL_1727434, EPI_ISL_1727435, EPI_ISL_1727436, EPI_ISL_1727437, EPI_ISL_1727438, EPI_ISL_1727439, EPI_ISL_1727440, EPI_ISL_1727442, EPI_ISL_1727443, EPI_ISL_1727444, EPI_ISL_1727445, EPI_ISL_1727446, EPI_ISL_1727447, EPI_ISL_1727448, EPI_ISL_1727449, EPI_ISL_1727450, EPI_ISL_1727451, EPI_ISL_1727452, EPI_ISL_1727453, EPI_ISL_1727455, EPI_ISL_1727458, EPI_ISL_1727459, EPI_ISL_1727460, EPI_ISL_1727461, EPI_ISL_1727462, EPI_ISL_1727464, EPI_ISL_1727465, EPI_ISL_1727466, EPI_ISL_1727468, EPI_ISL_1727469, EPI_ISL_1727470, EPI_ISL_1727472, EPI_ISL_1727474, EPI_ISL_1727475, EPI_ISL_1727477, EPI_ISL_1727479, EPI_ISL_1727480, EPI_ISL_1727481, EPI_ISL_1727483, EPI_ISL_1727485, EPI_ISL_1727486, EPI_ISL_1727487, EPI_ISL_1727488, EPI_ISL_1727489, EPI_ISL_1727490, EPI_ISL_1727491, EPI_ISL_1727492, EPI_ISL_1727493, EPI_ISL_1727494, EPI_ISL_1727495, EPI_ISL_1727496, EPI_ISL_1727498, EPI_ISL_1727499, EPI_ISL_1727502, EPI_ISL_1727504, EPI_ISL_1727505, EPI_ISL_1727506, EPI_ISL_1727508, EPI_ISL_1727509, EPI_ISL_1727512, EPI_ISL_1727514, EPI_ISL_1727516, EPI_ISL_1727517, EPI_ISL_1727518, EPI_ISL_1727519, EPI_ISL_1727520, EPI_ISL_1727523, EPI_ISL_1727524, EPI_ISL_1727527, EPI_ISL_1727528, EPI_ISL_1727530, EPI_ISL_1727532, EPI_ISL_1727533, EPI_ISL_1727534, EPI_ISL_1727537, EPI_ISL_1727538, EPI_ISL_1727542, EPI_ISL_1727545, EPI_ISL_1727550, EPI_ISL_1727551 | | | |
| see above | Limbach - MVZ Humangenetik Ulm | Robert Koch Institute | |
| EPI_ISL_1725192, EPI_ISL_1725196, EPI_ISL_1725198, EPI_ISL_1725200, EPI_ISL_1725201, EPI_ISL_1725202, EPI_ISL_1725203, EPI_ISL_1725204, EPI_ISL_1725210, EPI_ISL_1725212, EPI_ISL_1725215, EPI_ISL_1725218, EPI_ISL_1846217, EPI_ISL_1846218, EPI_ISL_1846220, EPI_ISL_1846221, EPI_ISL_1846222, EPI_ISL_1846225, EPI_ISL_1846227, EPI_ISL_1846228, EPI_ISL_1846229, EPI_ISL_1852078 | | | |
| see above | Limbach - MVZ Labor Eveld & Kollegen Essen | Robert Koch Institute | |
| EPI_ISL_1725638, EPI_ISL_1725653 | Limbach - MVZ Labor Ludwigsburg | Robert Koch Institute | |
| EPI_ISL_1725633, EPI_ISL_1725648, EPI_ISL_1725651, EPI_ISL_1725652, EPI_ISL_1725657 | Limbach - MVZ Labor MÃ¼nster (Cullen) | Robert Koch Institute | |
| EPI_ISL_1720813, EPI_ISL_1720815, EPI_ISL_1720824, EPI_ISL_1720825, EPI_ISL_1720826, EPI_ISL_1720827, EPI_ISL_1720828, EPI_ISL_1720832, EPI_ISL_1720834, EPI_ISL_1720835, EPI_ISL_1720837, EPI_ISL_1720844, EPI_ISL_1720902, EPI_ISL_1720936, EPI_ISL_1724323, EPI_ISL_1724418, EPI_ISL_1724420, EPI_ISL_1724421, EPI_ISL_1724425, EPI_ISL_1724426, EPI_ISL_1724427, EPI_ISL_1724429, EPI_ISL_1724430, EPI_ISL_1724431, EPI_ISL_1724432, EPI_ISL_1724433, EPI_ISL_1724434, EPI_ISL_1724435, EPI_ISL_1724437, EPI_ISL_1724438, EPI_ISL_1724439, EPI_ISL_1724440, EPI_ISL_1725605, EPI_ISL_1725616, EPI_ISL_1725619, EPI_ISL_1725621, EPI_ISL_1725623, EPI_ISL_1725625, EPI_ISL_1725626, EPI_ISL_1725627, EPI_ISL_1725637, EPI_ISL_1725642, EPI_ISL_1725647, EPI_ISL_1725655, EPI_ISL_1843339, EPI_ISL_1843348, EPI_ISL_1843355, EPI_ISL_1843358, EPI_ISL_1843362, EPI_ISL_1843363, EPI_ISL_1843364, EPI_ISL_1843369, EPI_ISL_1846337, EPI_ISL_1847198, EPI_ISL_1847199, EPI_ISL_1847202, EPI_ISL_1847203, EPI_ISL_1847206, EPI_ISL_1847210, EPI_ISL_1847213, EPI_ISL_1847215, EPI_ISL_1847221, EPI_ISL_1847233, EPI_ISL_1847234, EPI_ISL_1847425, EPI_ISL_1847428, EPI_ISL_1847429, EPI_ISL_1847441, EPI_ISL_1847728, EPI_ISL_1847729, EPI_ISL_1847734, EPI_ISL_1852124 | | | |
| see above | MDI Limbach Berlin GmbH; MVZ Labor Berlin | Robert Koch Institute | |
| EPI_ISL_1715859 | MEDILYS Laborgesellschaft mbH | Heinrich Pette Institute, Leibniz Institute for Experimental Virology | Adam Grundhoff; Alexis Robitaille; Johannes Knobloch; Martin Aepfelbacher; Nicole Fischer; Thomas Günther |
| EPI_ISL_1726778, EPI_ISL_1726780, EPI_ISL_1726784, EPI_ISL_1726786, EPI_ISL_1726787, EPI_ISL_1726789, EPI_ISL_1726790, EPI_ISL_1726793, EPI_ISL_1726794, EPI_ISL_1726795, EPI_ISL_1726797, EPI_ISL_1726798, EPI_ISL_1726802, EPI_ISL_1726804, EPI_ISL_1726805, EPI_ISL_1726806, EPI_ISL_1726808, EPI_ISL_1726809, EPI_ISL_1726811, EPI_ISL_1726812, EPI_ISL_1726813, EPI_ISL_1726815, EPI_ISL_1726818, EPI_ISL_1726820, EPI_ISL_1726821, EPI_ISL_1726822, EPI_ISL_1726823, EPI_ISL_1726824, EPI_ISL_1726825, EPI_ISL_1726827, EPI_ISL_1726830, EPI_ISL_1726831, EPI_ISL_1726832, EPI_ISL_1726833, EPI_ISL_1726835, EPI_ISL_1726837, EPI_ISL_1726838, EPI_ISL_1726839, EPI_ISL_1726840, EPI_ISL_1726841, EPI_ISL_1726843, EPI_ISL_1726844, EPI_ISL_1726845, EPI_ISL_1726846, EPI_ISL_1726847, EPI_ISL_1726848, EPI_ISL_1726853, EPI_ISL_1843216, EPI_ISL_1843217, EPI_ISL_1843220, EPI_ISL_1843221, EPI_ISL_1843222, EPI_ISL_1843225, EPI_ISL_1843228, EPI_ISL_1843232, EPI_ISL_1843240, EPI_ISL_1843241, EPI_ISL_1843242, EPI_ISL_1843243, EPI_ISL_1843244, EPI_ISL_1843247, EPI_ISL_1843248, EPI_ISL_1843250, EPI_ISL_1843251, EPI_ISL_1843253, EPI_ISL_1843254, EPI_ISL_1843256, EPI_ISL_1843257, EPI_ISL_1843259, EPI_ISL_1843261, EPI_ISL_1843263, EPI_ISL_1843265, EPI_ISL_1843267, EPI_ISL_1843269, EPI_ISL_1843270, EPI_ISL_1843271, EPI_ISL_1843272, EPI_ISL_1843276, EPI_ISL_1843280, EPI_ISL_1843281, EPI_ISL_1843282, EPI_ISL_1843283, EPI_ISL_1843285, EPI_ISL_1843286, EPI_ISL_1843288, EPI_ISL_1843292 | | | |
| see above | MVZ Dr. Eberhard & Partner Dortmund | Robert Koch Institute | |
| EPI_ISL_1722131, EPI_ISL_1722132, EPI_ISL_1722133, EPI_ISL_1722134, EPI_ISL_1722135, EPI_ISL_1722136, EPI_ISL_1722137, EPI_ISL_1722138, EPI_ISL_1722139, EPI_ISL_1722140, EPI_ISL_1722141, EPI_ISL_1722142, EPI_ISL_1722143, EPI_ISL_1722144, EPI_ISL_1722145, EPI_ISL_1722146, EPI_ISL_1722147, EPI_ISL_1722148, EPI_ISL_1722149, EPI_ISL_1722150, EPI_ISL_1722151, EPI_ISL_1722152, EPI_ISL_1722153, EPI_ISL_1722154, EPI_ISL_1722155, EPI_ISL_1722156, EPI_ISL_1722157, EPI_ISL_1722158, EPI_ISL_1722159, EPI_ISL_1722160, EPI_ISL_1722161, EPI_ISL_1722162, EPI_ISL_1722163, EPI_ISL_1722164, EPI_ISL_1722165, EPI_ISL_1722166, EPI_ISL_1722167, EPI_ISL_1722168, EPI_ISL_1722169, EPI_ISL_1722170, EPI_ISL_1722171, EPI_ISL_1722172, EPI_ISL_1722173, EPI_ISL_1849350, EPI_ISL_1849356, EPI_ISL_1849360, EPI_ISL_1849364, EPI_ISL_1849372, EPI_ISL_1849375, EPI_ISL_1849387, EPI_ISL_1849394, EPI_ISL_1849396, EPI_ISL_1849397, EPI_ISL_1849401, EPI_ISL_1849411 | | | |
| see above | MVZ Labor Dr. Fenner und Kollegen (Standort Hamburg) | Robert Koch Institute | |
| EPI_ISL_1722841, EPI_ISL_1727383, EPI_ISL_1727388, EPI_ISL_1727413, EPI_ISL_1727565, EPI_ISL_1843335, EPI_ISL_1843346, EPI_ISL_1843360, EPI_ISL_1843382, EPI_ISL_1843386, EPI_ISL_1843388, EPI_ISL_1843390, EPI_ISL_1843391, EPI_ISL_1843392, EPI_ISL_1843394, EPI_ISL_1843396, EPI_ISL_1843400, EPI_ISL_1843402, EPI_ISL_1843406, EPI_ISL_1843407, EPI_ISL_1843408, EPI_ISL_1843410, EPI_ISL_1843415, EPI_ISL_1843417, EPI_ISL_1843421, EPI_ISL_1843422, EPI_ISL_1843428, EPI_ISL_1843431, EPI_ISL_1846363 | | | |
| see above | MVZ Labor Dr. Limbach & Kollegen GbR | Robert Koch Institute | |
| EPI_ISL_1643406, EPI_ISL_1724271, EPI_ISL_1724272, EPI_ISL_1725197, EPI_ISL_1725206, EPI_ISL_1846236, EPI_ISL_1846238, EPI_ISL_1847192 | | | |
| see above | MVZ Labor Dr. Reising-Ackermann und Kollegen | Robert Koch Institute | |
| EPI_ISL_1644942, EPI_ISL_1644958, EPI_ISL_1726868, EPI_ISL_1726869, EPI_ISL_1726870, EPI_ISL_1726871, EPI_ISL_1726872, EPI_ISL_1726873, EPI_ISL_1726874, EPI_ISL_1726875, EPI_ISL_1726876, EPI_ISL_1726877, EPI_ISL_1726878, EPI_ISL_1726879, EPI_ISL_1726880, EPI_ISL_1726881, EPI_ISL_1726882, EPI_ISL_1726883, EPI_ISL_1726884, EPI_ISL_1726885, EPI_ISL_1726886, EPI_ISL_1726887, EPI_ISL_1726888, EPI_ISL_1726889, EPI_ISL_1726890, EPI_ISL_1726891, EPI_ISL_1726892, EPI_ISL_1726893, EPI_ISL_1726894, EPI_ISL_1726895, EPI_ISL_1726896, EPI_ISL_1726897, EPI_ISL_1726898, EPI_ISL_1726899, EPI_ISL_1726900, EPI_ISL_1726901, EPI_ISL_1726902, EPI_ISL_1726903, EPI_ISL_1726904, EPI_ISL_1726905, EPI_ISL_1726906, EPI_ISL_1726907, EPI_ISL_1726908, EPI_ISL_1726909, EPI_ISL_1726910, EPI_ISL_1726911, EPI_ISL_1726912, EPI_ISL_1726913, EPI_ISL_1726914, EPI_ISL_1726915, EPI_ISL_1726916, EPI_ISL_1726917, EPI_ISL_1726918, EPI_ISL_1726919, EPI_ISL_1726920, EPI_ISL_1726921, EPI_ISL_1726938, EPI_ISL_1726940, EPI_ISL_1726942, EPI_ISL_1726944, EPI_ISL_1726945, EPI_ISL_1726946, EPI_ISL_1726947, EPI_ISL_1726948, EPI_ISL_1726949, EPI_ISL_1726950, EPI_ISL_1726952, EPI_ISL_1726954, EPI_ISL_1726955, EPI_ISL_1726956, EPI_ISL_1726959, EPI_ISL_1726960, EPI_ISL_1726963, EPI_ISL_1726966, EPI_ISL_1726967, EPI_ISL_1726968, EPI_ISL_1726969, EPI_ISL_1726970, EPI_ISL_1726973, EPI_ISL_1726974, EPI_ISL_1844283, EPI_ISL_1844284, EPI_ISL_1844285, EPI_ISL_1844286, EPI_ISL_1844287, EPI_ISL_1844291, EPI_ISL_1844293, EPI_ISL_1844294, EPI_ISL_1844295, EPI_ISL_1844297, EPI_ISL_1844298, EPI_ISL_1844300, EPI_ISL_1844304, EPI_ISL_1844307, EPI_ISL_1844308, EPI_ISL_1844309, EPI_ISL_1844310, EPI_ISL_1844311, EPI_ISL_1844313, EPI_ISL_1844317, EPI_ISL_1844318, EPI_ISL_1844319, EPI_ISL_1844320, EPI_ISL_1844321, EPI_ISL_1844323, EPI_ISL_1844325, EPI_ISL_1844328, EPI_ISL_1844329, EPI_ISL_1844332, EPI_ISL_1844333, EPI_ISL_1844334, EPI_ISL_1844336, EPI_ISL_1844338, EPI_ISL_1844339, EPI_ISL_1844340, EPI_ISL_1844341, EPI_ISL_1844345, EPI_ISL_1844346, EPI_ISL_1844350, EPI_ISL_1844352, EPI_ISL_1844355, EPI_ISL_1844356, EPI_ISL_1844357, EPI_ISL_1844358, EPI_ISL_1844359, EPI_ISL_1844360, EPI_ISL_1844361, EPI_ISL_1844363, EPI_ISL_1844364, EPI_ISL_1844365, EPI_ISL_1844366, EPI_ISL_1844367, EPI_ISL_1844368, EPI_ISL_1844369, EPI_ISL_1844370, EPI_ISL_1844371, EPI_ISL_1844372, EPI_ISL_1849428 | | | |
| see above | MVZ Labor Krone GbR | Robert Koch Institute | |
| EPI_ISL_1725640, EPI_ISL_1847430, EPI_ISL_1847442, EPI_ISL_1848797, EPI_ISL_1852461 | MVZ Medizinisches Labor Hannover GmbH | Robert Koch Institute | |
| EPI_ISL_1725272, EPI_ISL_1725636, EPI_ISL_1847182, EPI_ISL_1847183, EPI_ISL_1847196, EPI_ISL_1847197, EPI_ISL_1847200, EPI_ISL_1847207, EPI_ISL_1847214, EPI_ISL_1847216, EPI_ISL_1847217, EPI_ISL_1847218, EPI_ISL_1847226, EPI_ISL_1847227, EPI_ISL_1847228, EPI_ISL_1847231, EPI_ISL_1847232, EPI_ISL_1847720, EPI_ISL_1847721, EPI_ISL_1847722, EPI_ISL_1847724, EPI_ISL_1847732 | | | |
| see above | MVZ fÃ¼r Laboratoriumsmedizin und Mikrobiologie Koblenz-Mittelrhein (Labor Koblenz) | Robert Koch Institute | |
| EPI_ISL_1850404 | MVZ fÃ¼r Laboratoriumsmedizin und Mikrobiologie WÃ¼rzburg | Robert Koch Institute | |
| EPI_ISL_1752415, EPI_ISL_2006682, EPI_ISL_2094527, EPI_ISL_2094529, EPI_ISL_2094532, EPI_ISL_2094535, EPI_ISL_2094548, EPI_ISL_2094556, EPI_ISL_2094559, EPI_ISL_2094567, EPI_ISL_2094574, EPI_ISL_2094578, EPI_ISL_2094581, EPI_ISL_2094584, EPI_ISL_2094605, EPI_ISL_2094607, EPI_ISL_2094609, EPI_ISL_2094610, EPI_ISL_2094615, EPI_ISL_2094617, EPI_ISL_2094624, EPI_ISL_2094641, EPI_ISL_2094657, EPI_ISL_2094697, EPI_ISL_2094892, EPI_ISL_2094893, EPI_ISL_2094894, EPI_ISL_2094895, EPI_ISL_2094896, EPI_ISL_2094897, EPI_ISL_2094946, EPI_ISL_2094947, EPI_ISL_2094948, EPI_ISL_2094949, EPI_ISL_2094958, EPI_ISL_2094981, EPI_ISL_2095044, EPI_ISL_2095046, EPI_ISL_2095058, EPI_ISL_2095062, EPI_ISL_2095068, EPI_ISL_2095098, EPI_ISL_2095144, EPI_ISL_2095151, EPI_ISL_2095277 | | | |
| see above | Max von Pettenkofer Institute, Virology, National Reference Center for Retroviruses, LMU Munich | Laboratory for Functional Genome Analysis; Dept. Genomics; Gene Center of the LMU Munich | Alexander Graf; Helmut Blum; Max Muenchhoff; Oliver Keppler; Stefan Krebs |
| EPI_ISL_1843199, EPI_ISL_1843201, EPI_ISL_1843203, EPI_ISL_1843211, EPI_ISL_1843212, EPI_ISL_1843215 | Med. Labor Prof. Schenk Dr. Ansorge & Kollegen | Robert Koch Institute | |
| EPI_ISL_1991648, EPI_ISL_1991650, EPI_ISL_1991651, EPI_ISL_1991652, EPI_ISL_1991655, EPI_ISL_1991656, EPI_ISL_1991845, EPI_ISL_1991846, EPI_ISL_1991847, EPI_ISL_1991848, EPI_ISL_1991849, EPI_ISL_1991852, EPI_ISL_1991853, EPI_ISL_1991854, EPI_ISL_1991855, EPI_ISL_1991857, EPI_ISL_1991858 | | | |
| see above | Medical Laboratories Duesseldorf | Center of Medical Microbiology, Virology, and Hospital Hygiene, University of Duesseldorf | Alexander Dilthey; Andreas Walker; Angelika Helmer; Christian Lange; Daniel Strelow; Jessica Nicolai; Jörg Timm; Klaus Pfeffer; Lisanna Hülse; Malte Kohns Vasconcelos; Maximilian Damagnez; Nadine Lübke; Tobias Wienemann; Torsten Houwaart |
| EPI_ISL_1723025, EPI_ISL_1723035, EPI_ISL_1723053, EPI_ISL_1723061 | Medizinisch-Diagnostisches Labor Kempten allgÃ¤ulab | Robert Koch Institute | |
| EPI_ISL_1726986, EPI_ISL_1726987, EPI_ISL_1726988, EPI_ISL_1726989, EPI_ISL_1726990, EPI_ISL_1726991, EPI_ISL_1726992, EPI_ISL_1726993, EPI_ISL_1726994, EPI_ISL_1726995, EPI_ISL_1726996, EPI_ISL_1726997, EPI_ISL_1726998, EPI_ISL_1726999, EPI_ISL_1727000, EPI_ISL_1727001, EPI_ISL_1727002, EPI_ISL_1727003, EPI_ISL_1727004, EPI_ISL_1727005, EPI_ISL_1727006, EPI_ISL_1727007, EPI_ISL_1727008, EPI_ISL_1727009, EPI_ISL_1727010, EPI_ISL_1727011, EPI_ISL_1727012, EPI_ISL_1727013, EPI_ISL_1727014, EPI_ISL_1727015, EPI_ISL_1727016, EPI_ISL_1727017, EPI_ISL_1727018, EPI_ISL_1727019, EPI_ISL_1727020, EPI_ISL_1727021, EPI_ISL_1727022, EPI_ISL_1727023, EPI_ISL_1727024, EPI_ISL_1727025, EPI_ISL_1727026, EPI_ISL_1727027, EPI_ISL_1727029, EPI_ISL_1727030, EPI_ISL_1727031, EPI_ISL_1727032, EPI_ISL_1727033, EPI_ISL_1727034, EPI_ISL_1727035, EPI_ISL_1727036, EPI_ISL_1727038, EPI_ISL_1727039, EPI_ISL_1727040, EPI_ISL_1727041, EPI_ISL_1727042, EPI_ISL_1727043, EPI_ISL_1727044, EPI_ISL_1727045, EPI_ISL_1727046, EPI_ISL_1727047, EPI_ISL_1727048, EPI_ISL_1727050, EPI_ISL_1727051 | | | |
| see above | Medizinische Laboratorien DÃ¼sseldorf | Robert Koch Institute | |
| EPI_ISL_1725252, EPI_ISL_1725253, EPI_ISL_1725254, EPI_ISL_1725256, EPI_ISL_1725257, EPI_ISL_1725259, EPI_ISL_1725264, EPI_ISL_1725268, EPI_ISL_1725270, EPI_ISL_1725271, EPI_ISL_1727552, EPI_ISL_1727553, EPI_ISL_1727556, EPI_ISL_1727557, EPI_ISL_1727559, EPI_ISL_1727560, EPI_ISL_1727563, EPI_ISL_1727564, EPI_ISL_1727566, EPI_ISL_1727568, EPI_ISL_1727569, EPI_ISL_1727570, EPI_ISL_1727572, EPI_ISL_1727574, EPI_ISL_1727575, EPI_ISL_1727578, EPI_ISL_1727580, EPI_ISL_1727581, EPI_ISL_1843412, EPI_ISL_1843413, EPI_ISL_1843416, EPI_ISL_1843418, EPI_ISL_1843425, EPI_ISL_1847317 | | | |
| see above | Medizinisches Labor Ostsachsen MVZ GbR NebenbetriebsstÃ¤tte Dresden | Robert Koch Institute | |

EPI_ISL_1720941, EPI_ISL_1720988, EPI_ISL_1721009, EPI_ISL_1726779, EPI_ISL_1726782, EPI_ISL_1726791, EPI_ISL_1726796, EPI_ISL_1726803, EPI_ISL_1726817, EPI_ISL_1726819, EPI_ISL_1726842, EPI_ISL_1726850, EPI_ISL_1726851

| | | |
|---|---|---|
| see above | Medizinisches Labor Wahl LÃ¼denscheid | Robert Koch Institute |
| EPI_ISL_1725997, EPI_ISL_1726056, EPI_ISL_1726074, EPI_ISL_1726104 | Praxisgemeinschaft fÃ¼r Laboratoriumsmedizin Labor BlumenstraÃŸe; Praxis Dr. E. Wietschel & Dr. F. Wietschel | Robert Koch Institute |

EPI_ISL_1724547, EPI_ISL_1724599, EPI_ISL_1724683, EPI_ISL_1724732, EPI_ISL_1724747, EPI_ISL_1725990, EPI_ISL_1725994, EPI_ISL_1726001, EPI_ISL_1726034, EPI_ISL_1726039

| | | |
|---|---|---|
| see above | Praxisgemeinschaft fÃ¼r Laboratoriumsmedizin Labor BlumenstraÃŸe; Praxis Dr. Sel und Dr. SpÃ¤te | Robert Koch Institute |

EPI_ISL_1720940, EPI_ISL_1720944, EPI_ISL_1720945, EPI_ISL_1720958, EPI_ISL_1720959, EPI_ISL_1720975, EPI_ISL_1720982, EPI_ISL_1720986, EPI_ISL_1720994, EPI_ISL_1720997, EPI_ISL_1721007, EPI_ISL_1721008, EPI_ISL_1721012

| | | |
|---|---|---|
| see above | Procomcure Biotech Germany GmbH | Robert Koch Institute |

EPI_ISL_1843480, EPI_ISL_1843486, EPI_ISL_1843521, EPI_ISL_1844214, EPI_ISL_2109522, EPI_ISL_2109622, EPI_ISL_2109625, EPI_ISL_2115626

| | | |
|---|---|---|
| see above | Robert-Bosch-Krankenhaus | Robert Koch Institute |
| EPI_ISL_1724311, EPI_ISL_1724315, EPI_ISL_1724317, EPI_ISL_1724320, EPI_ISL_1724321, EPI_ISL_1724324 | SLK-Kliniken Heilbronn GmbH - Institut fÃ¼r Laboratoriumsmedizin | Robert Koch Institute |

EPI_ISL_1722399, EPI_ISL_1723172, EPI_ISL_1724484, EPI_ISL_1724486, EPI_ISL_1724488, EPI_ISL_1724490, EPI_ISL_1724495, EPI_ISL_1724499, EPI_ISL_1724502, EPI_ISL_1724506, EPI_ISL_1724532, EPI_ISL_1724536, EPI_ISL_1724544, EPI_ISL_1724552, EPI_ISL_1724562, EPI_ISL_1724575, EPI_ISL_1724584, EPI_ISL_1724593, EPI_ISL_1724604, EPI_ISL_1724639, EPI_ISL_1724661, EPI_ISL_1724663, EPI_ISL_1724664, EPI_ISL_1724668, EPI_ISL_1724677, EPI_ISL_1724685, EPI_ISL_1724690, EPI_ISL_1724697, EPI_ISL_1724702, EPI_ISL_1724711, EPI_ISL_1724717, EPI_ISL_1724721, EPI_ISL_1724725, EPI_ISL_1724726, EPI_ISL_1724730, EPI_ISL_1724743, EPI_ISL_1724744, EPI_ISL_1724748, EPI_ISL_1724751, EPI_ISL_1724752, EPI_ISL_1724758, EPI_ISL_1724761, EPI_ISL_1724762, EPI_ISL_1724763, EPI_ISL_1724764, EPI_ISL_1725967, EPI_ISL_1725985, EPI_ISL_1725998, EPI_ISL_1726002, EPI_ISL_1726016, EPI_ISL_1726019, EPI_ISL_1726021, EPI_ISL_1726025, EPI_ISL_1726033, EPI_ISL_1726040, EPI_ISL_1726044, EPI_ISL_1726046, EPI_ISL_1726051, EPI_ISL_1726063, EPI_ISL_1726064, EPI_ISL_1726065, EPI_ISL_1726066, EPI_ISL_1726067, EPI_ISL_1726073, EPI_ISL_1726075, EPI_ISL_1726077, EPI_ISL_1726092, EPI_ISL_1726094, EPI_ISL_1726103, EPI_ISL_1726117, EPI_ISL_1726130, EPI_ISL_1726135, EPI_ISL_1727582, EPI_ISL_1727583, EPI_ISL_1727585, EPI_ISL_1727587, EPI_ISL_1727589, EPI_ISL_1727590, EPI_ISL_1727591, EPI_ISL_1727593, EPI_ISL_1727595, EPI_ISL_1727604, EPI_ISL_1727605, EPI_ISL_1727606, EPI_ISL_1727607, EPI_ISL_1727611, EPI_ISL_1727615, EPI_ISL_1727616, EPI_ISL_1727617, EPI_ISL_1727618, EPI_ISL_1727619, EPI_ISL_1727621, EPI_ISL_1727625, EPI_ISL_1727627, EPI_ISL_1727630, EPI_ISL_1727631, EPI_ISL_1727636, EPI_ISL_1727637, EPI_ISL_1727638, EPI_ISL_1727639, EPI_ISL_1727640, EPI_ISL_1727645, EPI_ISL_1727648, EPI_ISL_1727649, EPI_ISL_1727650, EPI_ISL_1727652, EPI_ISL_1727654, EPI_ISL_1729018, EPI_ISL_1729019, EPI_ISL_1729021, EPI_ISL_1729022, EPI_ISL_1729023, EPI_ISL_1729029, EPI_ISL_1729033, EPI_ISL_1729034, EPI_ISL_1729035, EPI_ISL_1729051, EPI_ISL_1729092, EPI_ISL_1729095, EPI_ISL_1729130, EPI_ISL_1729134, EPI_ISL_1729141, EPI_ISL_1729150, EPI_ISL_1729151, EPI_ISL_1729156, EPI_ISL_1729157, EPI_ISL_1729171, EPI_ISL_1729175, EPI_ISL_1729183, EPI_ISL_1729193, EPI_ISL_1729196, EPI_ISL_1729197, EPI_ISL_1729199, EPI_ISL_1729201, EPI_ISL_1729203, EPI_ISL_1729204, EPI_ISL_1729206, EPI_ISL_1729208, EPI_ISL_1729209, EPI_ISL_1729211, EPI_ISL_1729214, EPI_ISL_1729225, EPI_ISL_1729228, EPI_ISL_1729229, EPI_ISL_1729232, EPI_ISL_1729233, EPI_ISL_1729235, EPI_ISL_1729246, EPI_ISL_1729249, EPI_ISL_1729250, EPI_ISL_1729251, EPI_ISL_1729253, EPI_ISL_1729254, EPI_ISL_1729255, EPI_ISL_1729256, EPI_ISL_1729257, EPI_ISL_1729258, EPI_ISL_1729259, EPI_ISL_1729261, EPI_ISL_1729262, EPI_ISL_1729264, EPI_ISL_1729265, EPI_ISL_1729266, EPI_ISL_1729267, EPI_ISL_1729269, EPI_ISL_1729270, EPI_ISL_1729271, EPI_ISL_1729272, EPI_ISL_1729273, EPI_ISL_1729274, EPI_ISL_1729275, EPI_ISL_1729276, EPI_ISL_1729277, EPI_ISL_1729279, EPI_ISL_1729280, EPI_ISL_1729281, EPI_ISL_1729282, EPI_ISL_1729283, EPI_ISL_1729284, EPI_ISL_1729285, EPI_ISL_1729286, EPI_ISL_1729287, EPI_ISL_1729288, EPI_ISL_1729289, EPI_ISL_1729290, EPI_ISL_1729291, EPI_ISL_1729292, EPI_ISL_1729293, EPI_ISL_1729294, EPI_ISL_1729295, EPI_ISL_1729296, EPI_ISL_1729298, EPI_ISL_1729299, EPI_ISL_1729300, EPI_ISL_1729301, EPI_ISL_1729302, EPI_ISL_1729303, EPI_ISL_1729304, EPI_ISL_1729305, EPI_ISL_1729306, EPI_ISL_1729307, EPI_ISL_1729308, EPI_ISL_1729309, EPI_ISL_1729310, EPI_ISL_1729311, EPI_ISL_1729313, EPI_ISL_1729314, EPI_ISL_1729317, EPI_ISL_1729318, EPI_ISL_1729319, EPI_ISL_1729320, EPI_ISL_1729321, EPI_ISL_1729322, EPI_ISL_1729323, EPI_ISL_1729324, EPI_ISL_1729325, EPI_ISL_1729326, EPI_ISL_1729327, EPI_ISL_1729328, EPI_ISL_1729329, EPI_ISL_1729330, EPI_ISL_1729331, EPI_ISL_1729332, EPI_ISL_1729333, EPI_ISL_1729334, EPI_ISL_1729335, EPI_ISL_1729336, EPI_ISL_1729339, EPI_ISL_1729340, EPI_ISL_1729341, EPI_ISL_1729342, EPI_ISL_1729343, EPI_ISL_1729344, EPI_ISL_1729345, EPI_ISL_1729346, EPI_ISL_1729347, EPI_ISL_1729348, EPI_ISL_1729349, EPI_ISL_1729350, EPI_ISL_1729351, EPI_ISL_1729352, EPI_ISL_1729354, EPI_ISL_1729356, EPI_ISL_1729357, EPI_ISL_1729358, EPI_ISL_1729359, EPI_ISL_1729362, EPI_ISL_1729363, EPI_ISL_1729364, EPI_ISL_1729365, EPI_ISL_1729367, EPI_ISL_1729371, EPI_ISL_1729372, EPI_ISL_1729376, EPI_ISL_1729377, EPI_ISL_1729378, EPI_ISL_1729379, EPI_ISL_1729380, EPI_ISL_1729382, EPI_ISL_1729385, EPI_ISL_1729389, EPI_ISL_1729390, EPI_ISL_1729391, EPI_ISL_1846816, EPI_ISL_1846817, EPI_ISL_1846838, EPI_ISL_1846841, EPI_ISL_1846842, EPI_ISL_1846843, EPI_ISL_1846844, EPI_ISL_1846846, EPI_ISL_1846851, EPI_ISL_1846852, EPI_ISL_1846859, EPI_ISL_1846861, EPI_ISL_1846864, EPI_ISL_1846870, EPI_ISL_1846873, EPI_ISL_1846875, EPI_ISL_1846878, EPI_ISL_1846886, EPI_ISL_1846888, EPI_ISL_1846894, EPI_ISL_1846896, EPI_ISL_1846901, EPI_ISL_1846902, EPI_ISL_1846904, EPI_ISL_1846912, EPI_ISL_1846914, EPI_ISL_1846916, EPI_ISL_1846919, EPI_ISL_1846924, EPI_ISL_1846926, EPI_ISL_1846927, EPI_ISL_1846939, EPI_ISL_1846946, EPI_ISL_1846948, EPI_ISL_1846949, EPI_ISL_1846952, EPI_ISL_1846959, EPI_ISL_1846960, EPI_ISL_1846962, EPI_ISL_1846965, EPI_ISL_1846966, EPI_ISL_1846988, EPI_ISL_1846998, EPI_ISL_1847000, EPI_ISL_1847001, EPI_ISL_1847006, EPI_ISL_1847008, EPI_ISL_1847017, EPI_ISL_1847020, EPI_ISL_1847021, EPI_ISL_1847025, EPI_ISL_1847028, EPI_ISL_1847037, EPI_ISL_1847038, EPI_ISL_1847041, EPI_ISL_1847046, EPI_ISL_1847047, EPI_ISL_1847052, EPI_ISL_1847053, EPI_ISL_1847064, EPI_ISL_1847067, EPI_ISL_1847070, EPI_ISL_1847072, EPI_ISL_1847073, EPI_ISL_1847081, EPI_ISL_1847083, EPI_ISL_1847089, EPI_ISL_1847747, EPI_ISL_1847749, EPI_ISL_1847755, EPI_ISL_1847759, EPI_ISL_1847760, EPI_ISL_1847764, EPI_ISL_1847766, EPI_ISL_1847771, EPI_ISL_1847782, EPI_ISL_1847785, EPI_ISL_1847786, EPI_ISL_1847795, EPI_ISL_1847798, EPI_ISL_1847799, EPI_ISL_1847803, EPI_ISL_1847806, EPI_ISL_1847808, EPI_ISL_1847811, EPI_ISL_1847819, EPI_ISL_1847820, EPI_ISL_1847823, EPI_ISL_1847825, EPI_ISL_1847827, EPI_ISL_1847829, EPI_ISL_1847837, EPI_ISL_1847838, EPI_ISL_1847843, EPI_ISL_1847851, EPI_ISL_1847853, EPI_ISL_1847854, EPI_ISL_1847859, EPI_ISL_1847860, EPI_ISL_1847862, EPI_ISL_1847865, EPI_ISL_1847869, EPI_ISL_1847870, EPI_ISL_1847872, EPI_ISL_1847873, EPI_ISL_1847874, EPI_ISL_1847877, EPI_ISL_1847888, EPI_ISL_1847889, EPI_ISL_1847892, EPI_ISL_1847896, EPI_ISL_1847901, EPI_ISL_1847913, EPI_ISL_1847915, EPI_ISL_1847917, EPI_ISL_1847923, EPI_ISL_1847925, EPI_ISL_1847931, EPI_ISL_1847933, EPI_ISL_1847936, EPI_ISL_1847940, EPI_ISL_1847941, EPI_ISL_1847942, EPI_ISL_1847944, EPI_ISL_1847945, EPI_ISL_1847946, EPI_ISL_1847949, EPI_ISL_1847951, EPI_ISL_1847953, EPI_ISL_1847956, EPI_ISL_1847958, EPI_ISL_1847983, EPI_ISL_1847987, EPI_ISL_1847988, EPI_ISL_1847994, EPI_ISL_1847995, EPI_ISL_1847998, EPI_ISL_1847999, EPI_ISL_1848014, EPI_ISL_1848016, EPI_ISL_1848017, EPI_ISL_1848019, EPI_ISL_1848030, EPI_ISL_1848033, EPI_ISL_1848040, EPI_ISL_1848042, EPI_ISL_1848043, EPI_ISL_1848049, EPI_ISL_1848052, EPI_ISL_1848062, EPI_ISL_1848076, EPI_ISL_1848079, EPI_ISL_1848080, EPI_ISL_1848083, EPI_ISL_1848084, EPI_ISL_1848085, EPI_ISL_1848090, EPI_ISL_1848098, EPI_ISL_1848102, EPI_ISL_1848103, EPI_ISL_1848110, EPI_ISL_1848115, EPI_ISL_1848119, EPI_ISL_1848120, EPI_ISL_1848124, EPI_ISL_1848130, EPI_ISL_1848133, EPI_ISL_1848135, EPI_ISL_1848136, EPI_ISL_1848140, EPI_ISL_1848150, EPI_ISL_1848162, EPI_ISL_1848165, EPI_ISL_1848170, EPI_ISL_1848174, EPI_ISL_1848181, EPI_ISL_1848188, EPI_ISL_1848190, EPI_ISL_1848197, EPI_ISL_1848198, EPI_ISL_1848214, EPI_ISL_1848218, EPI_ISL_1848220, EPI_ISL_1848225, EPI_ISL_1848227, EPI_ISL_1848233, EPI_ISL_1848239, EPI_ISL_1848241, EPI_ISL_1848252, EPI_ISL_1848255, EPI_ISL_1848258, EPI_ISL_1848259, EPI_ISL_1848264, EPI_ISL_1848269, EPI_ISL_1848275, EPI_ISL_1848281, EPI_ISL_1848282, EPI_ISL_1848284, EPI_ISL_1848285, EPI_ISL_1848286, EPI_ISL_1848290, EPI_ISL_1848291, EPI_ISL_1848293, EPI_ISL_1848296, EPI_ISL_1848297, EPI_ISL_1848299, EPI_ISL_1848301, EPI_ISL_1848304, EPI_ISL_1848305, EPI_ISL_1848309, EPI_ISL_1848311, EPI_ISL_1848312, EPI_ISL_1848318, EPI_ISL_1848320, EPI_ISL_1848325, EPI_ISL_1848328, EPI_ISL_1848332, EPI_ISL_1848340, EPI_ISL_1848341, EPI_ISL_1848346, EPI_ISL_1850808, EPI_ISL_1850831, EPI_ISL_1850836, EPI_ISL_1850845, EPI_ISL_1850850, EPI_ISL_1850852, EPI_ISL_1850863, EPI_ISL_1850876, EPI_ISL_1850894, EPI_ISL_1850905, EPI_ISL_1850929, EPI_ISL_1850935, EPI_ISL_1850960, EPI_ISL_1850979, EPI_ISL_1850990, EPI_ISL_1851080

| | | |
|---|---|---|
| see above | SYNLAB Jena Oncoscreen | Robert Koch Institute |
| EPI_ISL_1725983, EPI_ISL_1726003, EPI_ISL_1726041, EPI_ISL_1726070, EPI_ISL_1726105, EPI_ISL_1726110 | SYNLAB Labor MÃ¼nchen Zentrum LMZ | Robert Koch Institute |

EPI_ISL_1729000, EPI_ISL_1729012, EPI_ISL_1729036, EPI_ISL_1729045, EPI_ISL_1729048, EPI_ISL_1729057, EPI_ISL_1729062, EPI_ISL_1729066, EPI_ISL_1729069, EPI_ISL_1729084, EPI_ISL_1729087, EPI_ISL_1729088, EPI_ISL_1729096, EPI_ISL_1729102, EPI_ISL_1729112, EPI_ISL_1729118, EPI_ISL_1729218, EPI_ISL_1729227, EPI_ISL_1729237, EPI_ISL_1729248, EPI_ISL_1729252, EPI_ISL_1729260, EPI_ISL_1729312, EPI_ISL_1729353, EPI_ISL_1729361, EPI_ISL_1729366, EPI_ISL_1729368, EPI_ISL_1729373, EPI_ISL_1729374, EPI_ISL_1729381, EPI_ISL_1729383, EPI_ISL_1729384, EPI_ISL_1847050, EPI_ISL_1847741, EPI_ISL_1847753, EPI_ISL_1847756, EPI_ISL_1847770, EPI_ISL_1847810, EPI_ISL_1847815, EPI_ISL_1847842, EPI_ISL_1847903, EPI_ISL_1847920, EPI_ISL_1847928, EPI_ISL_1847968, EPI_ISL_1847991, EPI_ISL_1848001, EPI_ISL_1848002, EPI_ISL_1848004, EPI_ISL_1848025, EPI_ISL_1848026, EPI_ISL_1848045, EPI_ISL_1848046, EPI_ISL_1848071, EPI_ISL_1848077, EPI_ISL_1848081, EPI_ISL_1848108, EPI_ISL_1848154, EPI_ISL_1848166, EPI_ISL_1848173, EPI_ISL_1848196, EPI_ISL_1848221, EPI_ISL_1848323, EPI_ISL_1850722, EPI_ISL_1850751, EPI_ISL_1850783, EPI_ISL_1850793, EPI_ISL_1850798, EPI_ISL_1850803, EPI_ISL_1850820, EPI_ISL_1850827, EPI_ISL_1850833, EPI_ISL_1850835, EPI_ISL_1850840, EPI_ISL_1850841, EPI_ISL_1850843, EPI_ISL_1850853, EPI_ISL_1850855, EPI_ISL_1850861, EPI_ISL_1850862, EPI_ISL_1850874, EPI_ISL_1850878, EPI_ISL_1850886, EPI_ISL_1850887, EPI_ISL_1850904, EPI_ISL_1850913, EPI_ISL_1850916, EPI_ISL_1850918, EPI_ISL_1850920, EPI_ISL_1850930, EPI_ISL_1850943, EPI_ISL_1850947, EPI_ISL_1850948, EPI_ISL_1850949, EPI_ISL_1850969, EPI_ISL_1850973, EPI_ISL_1850977, EPI_ISL_1850996, EPI_ISL_1851001, EPI_ISL_1851002, EPI_ISL_1851009, EPI_ISL_1851018, EPI_ISL_1851033, EPI_ISL_1851035, EPI_ISL_1851037, EPI_ISL_1851044, EPI_ISL_1851045, EPI_ISL_1851057, EPI_ISL_1851074, EPI_ISL_1851075, EPI_ISL_1851076, EPI_ISL_1851085

| | | |
|---|---|---|
| see above | SYNLAB MVZ Berlin | Robert Koch Institute |

EPI_ISL_2108005, EPI_ISL_2108006, EPI_ISL_2108007, EPI_ISL_2108008, EPI_ISL_2108017, EPI_ISL_2108022, EPI_ISL_2108024, EPI_ISL_2108025, EPI_ISL_2108027, EPI_ISL_2108033, EPI_ISL_2108036, EPI_ISL_2108041, EPI_ISL_2108042, EPI_ISL_2108044, EPI_ISL_2108050, EPI_ISL_2108054, EPI_ISL_2108055, EPI_ISL_2108058, EPI_ISL_2108059, EPI_ISL_2108065, EPI_ISL_2108067, EPI_ISL_2108069, EPI_ISL_2108070, EPI_ISL_2108084, EPI_ISL_2108086, EPI_ISL_2108090, EPI_ISL_2108097, EPI_ISL_2108101, EPI_ISL_2108104, EPI_ISL_2108107, EPI_ISL_2108108, EPI_ISL_2108114, EPI_ISL_2108116, EPI_ISL_2108121, EPI_ISL_2108126, EPI_ISL_2108129, EPI_ISL_2108132, EPI_ISL_2108141, EPI_ISL_2108151, EPI_ISL_2108157, EPI_ISL_2108159, EPI_ISL_2108165, EPI_ISL_2108174, EPI_ISL_2108178, EPI_ISL_2108185, EPI_ISL_2108187, EPI_ISL_2108188, EPI_ISL_2108196, EPI_ISL_2108199, EPI_ISL_2108201, EPI_ISL_2108202, EPI_ISL_2108204, EPI_ISL_2108207, EPI_ISL_2108211, EPI_ISL_2108214, EPI_ISL_2108215, EPI_ISL_2108217, EPI_ISL_2108223, EPI_ISL_2108226, EPI_ISL_2108235, EPI_ISL_2108243, EPI_ISL_2108244, EPI_ISL_2108245, EPI_ISL_2108249, EPI_ISL_2108252, EPI_ISL_2108255, EPI_ISL_2108257, EPI_ISL_2108258, EPI_ISL_2114526, EPI_ISL_2114535, EPI_ISL_2114536, EPI_ISL_2114539, EPI_ISL_2114540, EPI_ISL_2114543, EPI_ISL_2114548, EPI_ISL_2114550, EPI_ISL_2114560, EPI_ISL_2114568, EPI_ISL_2114569, EPI_ISL_2114571, EPI_ISL_2114573, EPI_ISL_2114581, EPI_ISL_2114582, EPI_ISL_2114585, EPI_ISL_2114588, EPI_ISL_2114590, EPI_ISL_2114599, EPI_ISL_2114607, EPI_ISL_2114610, EPI_ISL_2114612, EPI_ISL_2114613, EPI_ISL_2114617, EPI_ISL_2114620, EPI_ISL_2114622, EPI_ISL_2114627, EPI_ISL_2114630, EPI_ISL_2114634, EPI_ISL_2114635, EPI_ISL_2114636, EPI_ISL_2114646, EPI_ISL_2114650, EPI_ISL_2114653, EPI_ISL_2114654, EPI_ISL_2114656, EPI_ISL_2114662, EPI_ISL_2114669, EPI_ISL_2114678, EPI_ISL_2114691, EPI_ISL_2114698, EPI_ISL_2114709, EPI_ISL_2114718, EPI_ISL_2114720, EPI_ISL_2114723, EPI_ISL_2114731, EPI_ISL_2114746, EPI_ISL_2114750, EPI_ISL_2114752, EPI_ISL_2114759, EPI_ISL_2114762, EPI_ISL_2114765, EPI_ISL_2114771, EPI_ISL_2114776, EPI_ISL_2114785, EPI_ISL_2114787, EPI_ISL_2114791, EPI_ISL_2114796, EPI_ISL_2114801, EPI_ISL_2114807, EPI_ISL_2114809, EPI_ISL_2114814, EPI_ISL_2114817, EPI_ISL_2114821, EPI_ISL_2114822, EPI_ISL_2114823, EPI_ISL_2114824, EPI_ISL_2114825, EPI_ISL_2114826, EPI_ISL_2114830, EPI_ISL_2114838, EPI_ISL_2114843, EPI_ISL_2114845, EPI_ISL_2114850, EPI_ISL_2114855, EPI_ISL_2114860, EPI_ISL_2114870, EPI_ISL_2114880, EPI_ISL_2114883, EPI_ISL_2114895, EPI_ISL_2114903, EPI_ISL_2114910, EPI_ISL_2114915, EPI_ISL_2114919, EPI_ISL_2114922, EPI_ISL_2114925, EPI_ISL_2114930, EPI_ISL_2114932, EPI_ISL_2114936, EPI_ISL_2114944, EPI_ISL_2114952, EPI_ISL_2114954, EPI_ISL_2114958, EPI_ISL_2114961, EPI_ISL_2114963, EPI_ISL_2114966, EPI_ISL_2114968, EPI_ISL_2114979, EPI_ISL_2114986, EPI_ISL_2114988, EPI_ISL_2114994, EPI_ISL_2114996, EPI_ISL_2115000, EPI_ISL_2115003, EPI_ISL_2115005, EPI_ISL_2115007, EPI_ISL_2115010, EPI_ISL_2115016, EPI_ISL_2115020, EPI_ISL_2115025, EPI_ISL_2115037, EPI_ISL_2115039, EPI_ISL_2115042, EPI_ISL_2115047, EPI_ISL_2115048, EPI_ISL_2115053, EPI_ISL_2115057, EPI_ISL_2115063, EPI_ISL_2115068, EPI_ISL_2115070, EPI_ISL_2115077, EPI_ISL_2115079, EPI_ISL_2115095, EPI_ISL_2115096, EPI_ISL_2115105, EPI_ISL_2115109, EPI_ISL_2115118, EPI_ISL_2115121, EPI_ISL_2118848, EPI_ISL_2118860, EPI_ISL_2118872, EPI_ISL_2118873, EPI_ISL_2118875, EPI_ISL_2118881, EPI_ISL_2118895, EPI_ISL_2118943, EPI_ISL_2118946, EPI_ISL_2118948, EPI_ISL_2118961, EPI_ISL_2118963, EPI_ISL_2119002, EPI_ISL_2119016, EPI_ISL_2119038, EPI_ISL_2119042, EPI_ISL_2119061, EPI_ISL_2119066, EPI_ISL_2119072, EPI_ISL_2119076, EPI_ISL_2119077, EPI_ISL_2119087, EPI_ISL_2119096, EPI_ISL_2119101, EPI_ISL_2119104, EPI_ISL_2119108, EPI_ISL_2119126, EPI_ISL_2119159, EPI_ISL_2119212, EPI_ISL_2119236, EPI_ISL_2119241, EPI_ISL_2119253, EPI_ISL_2119287, EPI_ISL_2119299, EPI_ISL_2119300, EPI_ISL_2119308, EPI_ISL_2119318, EPI_ISL_2119357, EPI_ISL_2119365, EPI_ISL_2119369, EPI_ISL_2119382, EPI_ISL_2119384, EPI_ISL_2119401, EPI_ISL_2119418, EPI_ISL_2119426, EPI_ISL_2119461, EPI_ISL_2119483, EPI_ISL_2119494, EPI_ISL_2119496, EPI_ISL_2119530, EPI_ISL_2119553, EPI_ISL_2119558, EPI_ISL_2119568, EPI_ISL_2119570, EPI_ISL_2119588, EPI_ISL_2119619, EPI_ISL_2119624, EPI_ISL_2119629, EPI_ISL_2119637, EPI_ISL_2119668, EPI_ISL_2119733, EPI_ISL_2119737, EPI_ISL_2119747, EPI_ISL_2119750, EPI_ISL_2119752, EPI_ISL_2119766, EPI_ISL_2119804, EPI_ISL_2119821, EPI_ISL_2119824, EPI_ISL_2119847, EPI_ISL_2119856, EPI_ISL_2119874, EPI_ISL_2119876, EPI_ISL_2119883, EPI_ISL_2119886, EPI_ISL_2119900, EPI_ISL_2119901, EPI_ISL_2119949, EPI_ISL_2119978, EPI_ISL_2120020, EPI_ISL_2120026, EPI_ISL_2120032, EPI_ISL_2128074

| | | |
|---|---|---|
| see above | SYNLAB MVZ Heidelberg | Robert Koch Institute |

EPI_ISL_1725965, EPI_ISL_1725969, EPI_ISL_1725971, EPI_ISL_1725972, EPI_ISL_1725973, EPI_ISL_1725974, EPI_ISL_1725975, EPI_ISL_1725977, EPI_ISL_1725978, EPI_ISL_1725980, EPI_ISL_1725981, EPI_ISL_1725982, EPI_ISL_1725986, EPI_ISL_1725989, EPI_ISL_1725991, EPI_ISL_1725995, EPI_ISL_1726005, EPI_ISL_1726009, EPI_ISL_1726010, EPI_ISL_1726012, EPI_ISL_1726017, EPI_ISL_1726020, EPI_ISL_1726024, EPI_ISL_1726028, EPI_ISL_1726029, EPI_ISL_1726030, EPI_ISL_1726032, EPI_ISL_1726035, EPI_ISL_1726038, EPI_ISL_1726042, EPI_ISL_1726047, EPI_ISL_1726049, EPI_ISL_1726050, EPI_ISL_1726054, EPI_ISL_1726055, EPI_ISL_1726057, EPI_ISL_1726059, EPI_ISL_1726061, EPI_ISL_1726068, EPI_ISL_1726069, EPI_ISL_1726071, EPI_ISL_1726072, EPI_ISL_1726080, EPI_ISL_1726083, EPI_ISL_1726085, EPI_ISL_1726088, EPI_ISL_1726090, EPI_ISL_1726095, EPI_ISL_1726096, EPI_ISL_1726098, EPI_ISL_1726099, EPI_ISL_1726100, EPI_ISL_1726102, EPI_ISL_1726108, EPI_ISL_1726109, EPI_ISL_1726112, EPI_ISL_1726113, EPI_ISL_1726116, EPI_ISL_1726121, EPI_ISL_1726122, EPI_ISL_1726123, EPI_ISL_1726125, EPI_ISL_1726131, EPI_ISL_1726132, EPI_ISL_1726133, EPI_ISL_1726134, EPI_ISL_1726136, EPI_ISL_1726139, EPI_ISL_1726141, EPI_ISL_1726143, EPI_ISL_1726147, EPI_ISL_1726148, EPI_ISL_1728865, EPI_ISL_1728866, EPI_ISL_1728867, EPI_ISL_1728868, EPI_ISL_1728869, EPI_ISL_1728870, EPI_ISL_1728871, EPI_ISL_1728872, EPI_ISL_1728873, EPI_ISL_1728874, EPI_ISL_1728875, EPI_ISL_1728876, EPI_ISL_1728877, EPI_ISL_1728878, EPI_ISL_1728879, EPI_ISL_1728881, EPI_ISL_1728884, EPI_ISL_1728885, EPI_ISL_1728886, EPI_ISL_1728887, EPI_ISL_1728888, EPI_ISL_1728889, EPI_ISL_1728890, EPI_ISL_1728891, EPI_ISL_1728892, EPI_ISL_1728893, EPI_ISL_1728894, EPI_ISL_1728895, EPI_ISL_1728896, EPI_ISL_1728897, EPI_ISL_1728898, EPI_ISL_1728899, EPI_ISL_1728900, EPI_ISL_1728901, EPI_ISL_1728902, EPI_ISL_1728903, EPI_ISL_1728904, EPI_ISL_1728905, EPI_ISL_1728906, EPI_ISL_1728907, EPI_ISL_1728908, EPI_ISL_1728909, EPI_ISL_1728910, EPI_ISL_1728912, EPI_ISL_1728913, EPI_ISL_1728914, EPI_ISL_1728915, EPI_ISL_1728916, EPI_ISL_1728917, EPI_ISL_1728918, EPI_ISL_1728919, EPI_ISL_1728920, EPI_ISL_1728921, EPI_ISL_1728922, EPI_ISL_1728923, EPI_ISL_1728924, EPI_ISL_1728925, EPI_ISL_1728926, EPI_ISL_1728927, EPI_ISL_1728928, EPI_ISL_1728929, EPI_ISL_1728930, EPI_ISL_1728931, EPI_ISL_1728932, EPI_ISL_1728933, EPI_ISL_1728934, EPI_ISL_1728935, EPI_ISL_1728936, EPI_ISL_1728937, EPI_ISL_1728938, EPI_ISL_1728939, EPI_ISL_1728940, EPI_ISL_1728941, EPI_ISL_1728942, EPI_ISL_1728943, EPI_ISL_1728944, EPI_ISL_1728945, EPI_ISL_1728946, EPI_ISL_1728947, EPI_ISL_1728948, EPI_ISL_1728949, EPI_ISL_1728950, EPI_ISL_1728951, EPI_ISL_1728952, EPI_ISL_1728953, EPI_ISL_1728954, EPI_ISL_1728955, EPI_ISL_1728956, EPI_ISL_1728957, EPI_ISL_1728958, EPI_ISL_1728959, EPI_ISL_1728960, EPI_ISL_1728961, EPI_ISL_1728962, EPI_ISL_1728963, EPI_ISL_1728964, EPI_ISL_1728965, EPI_ISL_1728966, EPI_ISL_1728967, EPI_ISL_1728968, EPI_ISL_1728969, EPI_ISL_1728971, EPI_ISL_1728972, EPI_ISL_1728973, EPI_ISL_1728974, EPI_ISL_1728975, EPI_ISL_1728976, EPI_ISL_1728977, EPI_ISL_1728978, EPI_ISL_1728980, EPI_ISL_1728981, EPI_ISL_1728982, EPI_ISL_1728983, EPI_ISL_1728984, EPI_ISL_1728986, EPI_ISL_1728987, EPI_ISL_1728988, EPI_ISL_1728989, EPI_ISL_1728990, EPI_ISL_1728991, EPI_ISL_1728992, EPI_ISL_1728993, EPI_ISL_1728994, EPI_ISL_1728995, EPI_ISL_1728996, EPI_ISL_1728997, EPI_ISL_1728998, EPI_ISL_1728999, EPI_ISL_1729001, EPI_ISL_1729002, EPI_ISL_1729003, EPI_ISL_1729004, EPI_ISL_1729005, EPI_ISL_1729006, EPI_ISL_1729007, EPI_ISL_1729008, EPI_ISL_1729009, EPI_ISL_1729010, EPI_ISL_1729011, EPI_ISL_1729013, EPI_ISL_1729014, EPI_ISL_1729015, EPI_ISL_1729016, EPI_ISL_1729017, EPI_ISL_1729020, EPI_ISL_1729024, EPI_ISL_1729025, EPI_ISL_1729026, EPI_ISL_1729027, EPI_ISL_1729028, EPI_ISL_1729030, EPI_ISL_1729031, EPI_ISL_1729032, EPI_ISL_1729037, EPI_ISL_1729038, EPI_ISL_1729039, EPI_ISL_1729040, EPI_ISL_1729042, EPI_ISL_1729043, EPI_ISL_1729044, EPI_ISL_1729046, EPI_ISL_1729047, EPI_ISL_1729049, EPI_ISL_1729050, EPI_ISL_1729053, EPI_ISL_1729054, EPI_ISL_1729055, EPI_ISL_1729056, EPI_ISL_1729058, EPI_ISL_1729059, EPI_ISL_1729060, EPI_ISL_1729063, EPI_ISL_1729064, EPI_ISL_1729065, EPI_ISL_1729067, EPI_ISL_1729068, EPI_ISL_1729070, EPI_ISL_1729071, EPI_ISL_1729072, EPI_ISL_1729073, EPI_ISL_1729074, EPI_ISL_1729075, EPI_ISL_1729076, EPI_ISL_1729077, EPI_ISL_1729078, EPI_ISL_1729079, EPI_ISL_1729080, EPI_ISL_1729081, EPI_ISL_1729082, EPI_ISL_1729083, EPI_ISL_1729085, EPI_ISL_1729086, EPI_ISL_1729089, EPI_ISL_1729090, EPI_ISL_1729091, EPI_ISL_1729093, EPI_ISL_1729094, EPI_ISL_1729097, EPI_ISL_1729098, EPI_ISL_1729099, EPI_ISL_1729100, EPI_ISL_1729101, EPI_ISL_1729103, EPI_ISL_1729104, EPI_ISL_1729105, EPI_ISL_1729107, EPI_ISL_1729108, EPI_ISL_1729109, EPI_ISL_1729110, EPI_ISL_1729111, EPI_ISL_1729113, EPI_ISL_1729114, EPI_ISL_1729115, EPI_ISL_1729116, EPI_ISL_1729117, EPI_ISL_1729119, EPI_ISL_1729120, EPI_ISL_1729121, EPI_ISL_1729122, EPI_ISL_1729123, EPI_ISL_1729124, EPI_ISL_1729125, EPI_ISL_1729126, EPI_ISL_1729127, EPI_ISL_1729128, EPI_ISL_1729129, EPI_ISL_1729131, EPI_ISL_1729132, EPI_ISL_1729133, EPI_ISL_1729135, EPI_ISL_1729136, EPI_ISL_1729137, EPI_ISL_1729138, EPI_ISL_1729139, EPI_ISL_1729140, EPI_ISL_1729142, EPI_ISL_1729143, EPI_ISL_1729144, EPI_ISL_1729145, EPI_ISL_1729146, EPI_ISL_1729147, EPI_ISL_1729148, EPI_ISL_1729149, EPI_ISL_1729152, EPI_ISL_1729153, EPI_ISL_1729154, EPI_ISL_1729155, EPI_ISL_1729158, EPI_ISL_1729159, EPI_ISL_1729160, EPI_ISL_1729161, EPI_ISL_1729162, EPI_ISL_1729163, EPI_ISL_1729164, EPI_ISL_1729165, EPI_ISL_1729166, EPI_ISL_1729167, EPI_ISL_1729168, EPI_ISL_1729169, EPI_ISL_1729170, EPI_ISL_1729172, EPI_ISL_1729173, EPI_ISL_1729174, EPI_ISL_1729176, EPI_ISL_1729177, EPI_ISL_1729178, EPI_ISL_1729179, EPI_ISL_1729180, EPI_ISL_1729181, EPI_ISL_1729182, EPI_ISL_1729184, EPI_ISL_1729185, EPI_ISL_1729186, EPI_ISL_1729187, EPI_ISL_1729188, EPI_ISL_1729189, EPI_ISL_1729190, EPI_ISL_1729191, EPI_ISL_1729192, EPI_ISL_1729194, EPI_ISL_1729195, EPI_ISL_1729198, EPI_ISL_1729200, EPI_ISL_1729202, EPI_ISL_1729205, EPI_ISL_1729207, EPI_ISL_1729210, EPI_ISL_1729212, EPI_ISL_1729213, EPI_ISL_1729215, EPI_ISL_1729216, EPI_ISL_1729217, EPI_ISL_1729219, EPI_ISL_1729220, EPI_ISL_1729221, EPI_ISL_1729222, EPI_ISL_1729223, EPI_ISL_1729224, EPI_ISL_1729226, EPI_ISL_1729230, EPI_ISL_1729231, EPI_ISL_1729234, EPI_ISL_1729236, EPI_ISL_1729238, EPI_ISL_1729239, EPI_ISL_1729240, EPI_ISL_1729241, EPI_ISL_1729242, EPI_ISL_1729243, EPI_ISL_1729244, EPI_ISL_1729245, EPI_ISL_1729247, EPI_ISL_1729263, EPI_ISL_1729268, EPI_ISL_1729278, EPI_ISL_1729315, EPI_ISL_1729316, EPI_ISL_1729337, EPI_ISL_1729338, EPI_ISL_1729355, EPI_ISL_1729360, EPI_ISL_1729369, EPI_ISL_1729370, EPI_ISL_1729375, EPI_ISL_1729386, EPI_ISL_1729387, EPI_ISL_1729388, EPI_ISL_1846815, EPI_ISL_1846818, EPI_ISL_1846819, EPI_ISL_1846820, EPI_ISL_1846821, EPI_ISL_1846823, EPI_ISL_1846824, EPI_ISL_1846825, EPI_ISL_1846826, EPI_ISL_1846827, EPI_ISL_1846828, EPI_ISL_1846829, EPI_ISL_1846830, EPI_ISL_1846831, EPI_ISL_1846832, EPI_ISL_1846833, EPI_ISL_1846834, EPI_ISL_1846835, EPI_ISL_1846836, EPI_ISL_1846839, EPI_ISL_1846840, EPI_ISL_1846845, EPI_ISL_1846847, EPI_ISL_1846848, EPI_ISL_1846849, EPI_ISL_1846850, EPI_ISL_1846853, EPI_ISL_1846854, EPI_ISL_1846855, EPI_ISL_1846856, EPI_ISL_1846857, EPI_ISL_1846858, EPI_ISL_1846860, EPI_ISL_1846862, EPI_ISL_1846863, EPI_ISL_1846865, EPI_ISL_1846866, EPI_ISL_1846867, EPI_ISL_1846868, EPI_ISL_1846869, EPI_ISL_1846871, EPI_ISL_1846872, EPI_ISL_1846874, EPI_ISL_1846876, EPI_ISL_1846877, EPI_ISL_1846879, EPI_ISL_1846880, EPI_ISL_1846881, EPI_ISL_1846882, EPI_ISL_1846883, EPI_ISL_1846884, EPI_ISL_1846885, EPI_ISL_1846887, EPI_ISL_1846889, EPI_ISL_1846890, EPI_ISL_1846891, EPI_ISL_1846892, EPI_ISL_1846893, EPI_ISL_1846895, EPI_ISL_1846897, EPI_ISL_1846898, EPI_ISL_1846900, EPI_ISL_1846903, EPI_ISL_1846905, EPI_ISL_1846906, EPI_ISL_1846907, EPI_ISL_1846908, EPI_ISL_1846909, EPI_ISL_1846910, EPI_ISL_1846911, EPI_ISL_1846913, EPI_ISL_1846915, EPI_ISL_1846917, EPI_ISL_1846918, EPI_ISL_1846920, EPI_ISL_1846921, EPI_ISL_1846922, EPI_ISL_1846923, EPI_ISL_1846925, EPI_ISL_1846928, EPI_ISL_1846929, EPI_ISL_1846930, EPI_ISL_1846931, EPI_ISL_1846932, EPI_ISL_1846933, EPI_ISL_1846934, EPI_ISL_1846935, EPI_ISL_1846936, EPI_ISL_1846937, EPI_ISL_1846938, EPI_ISL_1846940, EPI_ISL_1846941, EPI_ISL_1846942, EPI_ISL_1846943, EPI_ISL_1846944, EPI_ISL_1846945, EPI_ISL_1846947, EPI_ISL_1846950, EPI_ISL_1846951, EPI_ISL_1846953, EPI_ISL_1846954, EPI_ISL_1846955, EPI_ISL_1846956, EPI_ISL_1846957, EPI_ISL_1846958, EPI_ISL_1846961, EPI_ISL_1846963, EPI_ISL_1846964, EPI_ISL_1846967, EPI_ISL_1846968, EPI_ISL_1846969, EPI_ISL_1846970, EPI_ISL_1846971, EPI_ISL_1846972, EPI_ISL_1846973, EPI_ISL_1846974, EPI_ISL_1846975, EPI_ISL_1846976, EPI_ISL_1846977, EPI_ISL_1846978, EPI_ISL_1846979, EPI_ISL_1846980, EPI_ISL_1846981, EPI_ISL_1846982, EPI_ISL_1846983, EPI_ISL_1846984, EPI_ISL_1846985, EPI_ISL_1846986, EPI_ISL_1846987, EPI_ISL_1846989, EPI_ISL_1846990, EPI_ISL_1846991, EPI_ISL_1846992, EPI_ISL_1846993, EPI_ISL_1846994, EPI_ISL_1846995, EPI_ISL_1846996, EPI_ISL_1846997, EPI_ISL_1846999, EPI_ISL_1847002, EPI_ISL_1847003, EPI_ISL_1847004, EPI_ISL_1847005, EPI_ISL_1847007, EPI_ISL_1847009, EPI_ISL_1847010, EPI_ISL_1847011, EPI_ISL_1847013, EPI_ISL_1847014, EPI_ISL_1847015, EPI_ISL_1847016, EPI_ISL_1847018, EPI_ISL_1847019, EPI_ISL_1847022, EPI_ISL_1847023, EPI_ISL_1847024, EPI_ISL_1847026, EPI_ISL_1847027, EPI_ISL_1847029, EPI_ISL_1847030, EPI_ISL_1847031, EPI_ISL_1847032, EPI_ISL_1847033, EPI_ISL_1847034, EPI_ISL_1847035, EPI_ISL_1847036, EPI_ISL_1847039, EPI_ISL_1847040, EPI_ISL_1847042, EPI_ISL_1847043, EPI_ISL_1847044, EPI_ISL_1847045, EPI_ISL_1847048, EPI_ISL_1847049, EPI_ISL_1847051, EPI_ISL_1847054, EPI_ISL_1847055, EPI_ISL_1847056, EPI_ISL_1847057, EPI_ISL_1847058, EPI_ISL_1847059, EPI_ISL_1847060, EPI_ISL_1847061, EPI_ISL_1847062, EPI_ISL_1847063, EPI_ISL_1847065, EPI_ISL_1847066, EPI_ISL_1847068, EPI_ISL_1847069, EPI_ISL_1847071, EPI_ISL_1847074, EPI_ISL_1847075, EPI_ISL_1847076, EPI_ISL_1847077, EPI_ISL_1847078, EPI_ISL_1847079, EPI_ISL_1847080, EPI_ISL_1847082, EPI_ISL_1847084, EPI_ISL_1847085, EPI_ISL_1847086, EPI_ISL_1847087, EPI_ISL_1847088, EPI_ISL_1847090, EPI_ISL_1847091, EPI_ISL_1847092, EPI_ISL_1847736, EPI_ISL_1847738, EPI_ISL_1847739, EPI_ISL_1847740, EPI_ISL_1847742, EPI_ISL_1847743, EPI_ISL_1847744, EPI_ISL_1847745, EPI_ISL_1847748, EPI_ISL_1847750, EPI_ISL_1847751, EPI_ISL_1847752, EPI_ISL_1847754, EPI_ISL_1847758, EPI_ISL_1847761, EPI_ISL_1847762, EPI_ISL_1847765, EPI_ISL_1847767, EPI_ISL_1847768, EPI_ISL_1847769, EPI_ISL_1847773, EPI_ISL_1847774, EPI_ISL_1847775, EPI_ISL_1847776, EPI_ISL_1847777, EPI_ISL_1847778, EPI_ISL_1847779, EPI_ISL_1847781, EPI_ISL_1847783, EPI_ISL_1847784, EPI_ISL_1847787, EPI_ISL_1847788, EPI_ISL_1847789, EPI_ISL_1847790, EPI_ISL_1847791, EPI_ISL_1847792, EPI_ISL_1847793, EPI_ISL_1847794, EPI_ISL_1847796, EPI_ISL_1847797, EPI_ISL_1847800, EPI_ISL_1847801, EPI_ISL_1847802, EPI_ISL_1847804, EPI_ISL_1847805, EPI_ISL_1847807, EPI_ISL_1847809, EPI_ISL_1847812, EPI_ISL_1847813, EPI_ISL_1847814, EPI_ISL_1847816, EPI_ISL_1847817, EPI_ISL_1847818, EPI_ISL_1847821, EPI_ISL_1847822, EPI_ISL_1847824, EPI_ISL_1847826, EPI_ISL_1847828, EPI_ISL_1847830, EPI_ISL_1847831, EPI_ISL_1847832, EPI_ISL_1847833, EPI_ISL_1847834, EPI_ISL_1847835, EPI_ISL_1847839, EPI_ISL_1847840, EPI_ISL_1847844, EPI_ISL_1847845, EPI_ISL_1847846, EPI_ISL_1847848, EPI_ISL_1847850, EPI_ISL_1847852, EPI_ISL_1847855, EPI_ISL_1847856, EPI_ISL_1847857, EPI_ISL_1847858, EPI_ISL_1847861, EPI_ISL_1847863, EPI_ISL_1847864, EPI_ISL_1847866, EPI_ISL_1847867, EPI_ISL_1847868, EPI_ISL_1847871, EPI_ISL_1847875, EPI_ISL_1847876, EPI_ISL_1847878, EPI_ISL_1847879, EPI_ISL_1847880, EPI_ISL_1847881, EPI_ISL_1847882, EPI_ISL_1847883, EPI_ISL_1847885, EPI_ISL_1847886, EPI_ISL_1847887, EPI_ISL_1847890, EPI_ISL_1847891, EPI_ISL_1847893, EPI_ISL_1847894, EPI_ISL_1847895, EPI_ISL_1847898, EPI_ISL_1847900, EPI_ISL_1847902, EPI_ISL_1847905, EPI_ISL_1847906, EPI_ISL_1847908, EPI_ISL_1847909, EPI_ISL_1847911, EPI_ISL_1847912, EPI_ISL_1847914, EPI_ISL_1847916, EPI_ISL_1847918, EPI_ISL_1847919, EPI_ISL_1847921, EPI_ISL_1847922, EPI_ISL_1847926, EPI_ISL_1847927, EPI_ISL_1847930, EPI_ISL_1847932, EPI_ISL_1847934, EPI_ISL_1847935, EPI_ISL_1847937, EPI_ISL_1847938, EPI_ISL_1847939, EPI_ISL_1847943, EPI_ISL_1847947, EPI_ISL_1847948, EPI_ISL_1847950, EPI_ISL_1847952, EPI_ISL_1847954, EPI_ISL_1847955, EPI_ISL_1847957, EPI_ISL_1847959, EPI_ISL_1847960, EPI_ISL_1847961, EPI_ISL_1847963, EPI_ISL_1847964, EPI_ISL_1847965, EPI_ISL_1847966, EPI_ISL_1847967, EPI_ISL_1847969, EPI_ISL_1847970, EPI_ISL_1847971, EPI_ISL_1847972, EPI_ISL_1847973, EPI_ISL_1847974, EPI_ISL_1847975, EPI_ISL_1847977, EPI_ISL_1847979, EPI_ISL_1847980, EPI_ISL_1847981, EPI_ISL_1847982, EPI_ISL_1847984, EPI_ISL_1847985, EPI_ISL_1847986, EPI_ISL_1847989, EPI_ISL_1847990, EPI_ISL_1847992, EPI_ISL_1847996, EPI_ISL_1848000, EPI_ISL_1848003, EPI_ISL_1848005, EPI_ISL_1848006, EPI_ISL_1848008, EPI_ISL_1848009, EPI_ISL_1848010, EPI_ISL_1848011, EPI_ISL_1848012, EPI_ISL_1848013, EPI_ISL_1848015, EPI_ISL_1848018, EPI_ISL_1848020, EPI_ISL_1848021, EPI_ISL_1848022, EPI_ISL_1848023, EPI_ISL_1848024, EPI_ISL_1848028, EPI_ISL_1848029, EPI_ISL_1848031, EPI_ISL_1848032, EPI_ISL_1848034, EPI_ISL_1848035, EPI_ISL_1848036, EPI_ISL_1848038, EPI_ISL_1848039, EPI_ISL_1848041, EPI_ISL_1848044, EPI_ISL_1848047, EPI_ISL_1848048, EPI_ISL_1848050, EPI_ISL_1848051, EPI_ISL_1848053, EPI_ISL_1848054, EPI_ISL_1848055, EPI_ISL_1848056, EPI_ISL_1848057, EPI_ISL_1848058, EPI_ISL_1848059, EPI_ISL_1848060, EPI_ISL_1848061, EPI_ISL_1848063, EPI_ISL_1848064, EPI_ISL_1848065, EPI_ISL_1848066, EPI_ISL_1848067, EPI_ISL_1848068, EPI_ISL_1848069, EPI_ISL_1848070, EPI_ISL_1848072, EPI_ISL_1848075, EPI_ISL_1848078, EPI_ISL_1848082, EPI_ISL_1848086, EPI_ISL_1848087, EPI_ISL_1848088, EPI_ISL_1848089, EPI_ISL_1848091, EPI_ISL_1848092, EPI_ISL_1848093, EPI_ISL_1848094, EPI_ISL_1848096, EPI_ISL_1848097, EPI_ISL_1848100, EPI_ISL_1848101, EPI_ISL_1848104, EPI_ISL_1848105, EPI_ISL_1848106, EPI_ISL_1848107, EPI_ISL_1848109, EPI_ISL_1848112, EPI_ISL_1848113, EPI_ISL_1848114, EPI_ISL_1848116, EPI_ISL_1848117, EPI_ISL_1848118, EPI_ISL_1848121, EPI_ISL_1848122, EPI_ISL_1848123, EPI_ISL_1848126, EPI_ISL_1848128, EPI_ISL_1848129, EPI_ISL_1848131, EPI_ISL_1848132, EPI_ISL_1848137, EPI_ISL_1848138, EPI_ISL_1848139, EPI_ISL_1848141, EPI_ISL_1848142, EPI_ISL_1848143, EPI_ISL_1848145, EPI_ISL_1848146, EPI_ISL_1848147, EPI_ISL_1848148, EPI_ISL_1848149, EPI_ISL_1848151, EPI_ISL_1848152, EPI_ISL_1848153, EPI_ISL_1848155, EPI_ISL_1848156, EPI_ISL_1848157, EPI_ISL_1848158, EPI_ISL_1848159, EPI_ISL_1848160, EPI_ISL_1848161, EPI_ISL_1848163, EPI_ISL_1848167, EPI_ISL_1848168, EPI_ISL_1848171, EPI_ISL_1848172, EPI_ISL_1848175, EPI_ISL_1848176, EPI_ISL_1848177, EPI_ISL_1848178, EPI_ISL_1848180, EPI_ISL_1848182, EPI_ISL_1848183, EPI_ISL_1848184, EPI_ISL_1848185, EPI_ISL_1848187, EPI_ISL_1848189, EPI_ISL_1848191, EPI_ISL_1848192, EPI_ISL_1848193, EPI_ISL_1848194, EPI_ISL_1848195, EPI_ISL_1848199, EPI_ISL_1848200, EPI_ISL_1848201, EPI_ISL_1848202, EPI_ISL_1848203, EPI_ISL_1848204, EPI_ISL_1848205, EPI_ISL_1848206, EPI_ISL_1848207, EPI_ISL_1848208, EPI_ISL_1848210, EPI_ISL_1848212, EPI_ISL_1848213, EPI_ISL_1848215, EPI_ISL_1848216, EPI_ISL_1848217, EPI_ISL_1848219, EPI_ISL_1848222, EPI_ISL_1848223, EPI_ISL_1848226, EPI_ISL_1848229, EPI_ISL_1848230, EPI_ISL_1848231, EPI_ISL_1848232, EPI_ISL_1848234, EPI_ISL_1848235, EPI_ISL_1848236, EPI_ISL_1848237, EPI_ISL_1848238, EPI_ISL_1848240, EPI_ISL_1848242, EPI_ISL_1848244, EPI_ISL_1848245, EPI_ISL_1848246, EPI_ISL_1848247, EPI_ISL_1848248, EPI_ISL_1848249, EPI_ISL_1848253, EPI_ISL_1848254, EPI_ISL_1848257, EPI_ISL_1848260, EPI_ISL_1848262, EPI_ISL_1848263, EPI_ISL_1848265, EPI_ISL_1848266, EPI_ISL_1848267, EPI_ISL_1848268, EPI_ISL_1848271, EPI_ISL_1848272, EPI_ISL_1848273, EPI_ISL_1848276, EPI_ISL_1848277, EPI_ISL_1848280, EPI_ISL_1848283, EPI_ISL_1848288, EPI_ISL_1848289,

EPI_ISL_1848292, EPI_ISL_1848294, EPI_ISL_1848295, EPI_ISL_1848298, EPI_ISL_1848300, EPI_ISL_1848302, EPI_ISL_1848303, EPI_ISL_1848306, EPI_ISL_1848307, EPI_ISL_1848308, EPI_ISL_1848310, EPI_ISL_1848313, EPI_ISL_1848314, EPI_ISL_1848315, EPI_ISL_1848316, EPI_ISL_1848317, EPI_ISL_1848319, EPI_ISL_1848321, EPI_ISL_1848322, EPI_ISL_1848324, EPI_ISL_1848326, EPI_ISL_1848329, EPI_ISL_1848331, EPI_ISL_1848334, EPI_ISL_1848335, EPI_ISL_1848336, EPI_ISL_1848337, EPI_ISL_1848338, EPI_ISL_1848339, EPI_ISL_1848342, EPI_ISL_1848345, EPI_ISL_1850724, EPI_ISL_1850802, EPI_ISL_1850804, EPI_ISL_1850807, EPI_ISL_1850810, EPI_ISL_1850811, EPI_ISL_1850816, EPI_ISL_1850819, EPI_ISL_1850821, EPI_ISL_1850823, EPI_ISL_1850837, EPI_ISL_1850866, EPI_ISL_1850872, EPI_ISL_1850883, EPI_ISL_1850896, EPI_ISL_1850897, EPI_ISL_1850903, EPI_ISL_1850909, EPI_ISL_1850915, EPI_ISL_1850919, EPI_ISL_1850940, EPI_ISL_1850954, EPI_ISL_1850955, EPI_ISL_1850957, EPI_ISL_1850962, EPI_ISL_1850963, EPI_ISL_1850972, EPI_ISL_1850976, EPI_ISL_1850981, EPI_ISL_1850983, EPI_ISL_1850988, EPI_ISL_1851000, EPI_ISL_1851008, EPI_ISL_1851010, EPI_ISL_1851013, EPI_ISL_1851017, EPI_ISL_1851022, EPI_ISL_1851025, EPI_ISL_1851027, EPI_ISL_1851039, EPI_ISL_1851040, EPI_ISL_1851063, EPI_ISL_1851066, EPI_ISL_1851078

see above | SYNLAB MVZ Leinfelden-Echterdingen | Robert Koch Institute

EPI_ISL_1720454, EPI_ISL_1720455, EPI_ISL_1720457, EPI_ISL_1720459, EPI_ISL_1720460, EPI_ISL_1720461, EPI_ISL_1720462, EPI_ISL_1720463, EPI_ISL_1720464, EPI_ISL_1720465, EPI_ISL_1720466, EPI_ISL_1720467, EPI_ISL_1720468, EPI_ISL_1720470, EPI_ISL_1720471, EPI_ISL_1720472, EPI_ISL_1720473, EPI_ISL_1720474, EPI_ISL_1720476, EPI_ISL_1720477, EPI_ISL_1720479, EPI_ISL_1720480, EPI_ISL_1720482, EPI_ISL_1720483, EPI_ISL_1720484, EPI_ISL_1720486, EPI_ISL_1720488, EPI_ISL_1720489, EPI_ISL_1720490, EPI_ISL_1720491, EPI_ISL_1720492, EPI_ISL_1720493, EPI_ISL_1720494, EPI_ISL_1720495, EPI_ISL_1720496, EPI_ISL_1720497, EPI_ISL_1720498, EPI_ISL_1720499, EPI_ISL_1720500, EPI_ISL_1720502, EPI_ISL_1720503, EPI_ISL_1720504, EPI_ISL_1720505, EPI_ISL_1720506, EPI_ISL_1720507, EPI_ISL_1720509, EPI_ISL_1720510, EPI_ISL_1720511, EPI_ISL_1720512, EPI_ISL_1720514, EPI_ISL_1720515, EPI_ISL_1720516, EPI_ISL_1720517, EPI_ISL_1720519, EPI_ISL_1720520, EPI_ISL_1720521, EPI_ISL_1720522, EPI_ISL_1720523, EPI_ISL_1720524, EPI_ISL_1720525, EPI_ISL_1720526, EPI_ISL_1720527, EPI_ISL_1720528, EPI_ISL_1720529, EPI_ISL_1720530, EPI_ISL_1720531, EPI_ISL_1720532, EPI_ISL_1720533, EPI_ISL_1720534, EPI_ISL_1720535, EPI_ISL_1720536, EPI_ISL_1720537, EPI_ISL_1720538, EPI_ISL_1720540, EPI_ISL_1720541, EPI_ISL_1720542, EPI_ISL_1720543, EPI_ISL_1720544, EPI_ISL_1720545, EPI_ISL_1720546, EPI_ISL_1720547, EPI_ISL_1720548, EPI_ISL_1720549, EPI_ISL_1720552, EPI_ISL_1720553, EPI_ISL_1720554, EPI_ISL_1720555, EPI_ISL_1720556, EPI_ISL_1720558, EPI_ISL_1720559, EPI_ISL_1720560, EPI_ISL_1720562, EPI_ISL_1720564, EPI_ISL_1720565, EPI_ISL_1720566, EPI_ISL_1720567, EPI_ISL_1720568, EPI_ISL_1720569, EPI_ISL_1720570, EPI_ISL_1720571, EPI_ISL_1720572, EPI_ISL_1720573, EPI_ISL_1720574, EPI_ISL_1720575, EPI_ISL_1720576, EPI_ISL_1720577, EPI_ISL_1720578, EPI_ISL_1720579, EPI_ISL_1720580, EPI_ISL_1720581, EPI_ISL_1720582, EPI_ISL_1720583, EPI_ISL_1720584, EPI_ISL_1720585, EPI_ISL_1720586, EPI_ISL_1720588, EPI_ISL_1720589, EPI_ISL_1720590, EPI_ISL_1720591, EPI_ISL_1720594, EPI_ISL_1720595, EPI_ISL_1720596, EPI_ISL_1720599, EPI_ISL_1720600, EPI_ISL_1720601, EPI_ISL_1720602, EPI_ISL_1720603, EPI_ISL_1720604, EPI_ISL_1720605, EPI_ISL_1720606, EPI_ISL_1720607, EPI_ISL_1720609, EPI_ISL_1720610, EPI_ISL_1720611, EPI_ISL_1720613, EPI_ISL_1720615, EPI_ISL_1720617, EPI_ISL_1720618, EPI_ISL_1720619, EPI_ISL_1720620, EPI_ISL_1720622, EPI_ISL_1720623, EPI_ISL_1720624, EPI_ISL_1720625, EPI_ISL_1720626, EPI_ISL_1720627, EPI_ISL_1720628, EPI_ISL_1720629, EPI_ISL_1720630, EPI_ISL_1720631, EPI_ISL_1720632, EPI_ISL_1720633, EPI_ISL_1720634, EPI_ISL_1720635, EPI_ISL_1720636, EPI_ISL_1720638, EPI_ISL_1720639, EPI_ISL_1720641, EPI_ISL_1720642, EPI_ISL_1720643, EPI_ISL_1720644, EPI_ISL_1720645, EPI_ISL_1720646, EPI_ISL_1720647, EPI_ISL_1720648, EPI_ISL_1720649, EPI_ISL_1720650, EPI_ISL_1720651, EPI_ISL_1720653, EPI_ISL_1720654, EPI_ISL_1720655, EPI_ISL_1720656, EPI_ISL_1720658, EPI_ISL_1720659, EPI_ISL_1720660, EPI_ISL_1720661, EPI_ISL_1720665, EPI_ISL_1720666, EPI_ISL_1720667, EPI_ISL_1720668, EPI_ISL_1720669, EPI_ISL_1720670, EPI_ISL_1720671, EPI_ISL_1720672, EPI_ISL_1720673, EPI_ISL_1720675, EPI_ISL_1720676, EPI_ISL_1720677, EPI_ISL_1720678, EPI_ISL_1720679, EPI_ISL_1720681, EPI_ISL_1720682, EPI_ISL_1720683, EPI_ISL_1720684, EPI_ISL_1720685, EPI_ISL_1720686, EPI_ISL_1720688, EPI_ISL_1720690, EPI_ISL_1720692, EPI_ISL_1720693, EPI_ISL_1720695, EPI_ISL_1720696, EPI_ISL_1720697, EPI_ISL_1720698, EPI_ISL_1720699, EPI_ISL_1720700, EPI_ISL_1720701, EPI_ISL_1720704, EPI_ISL_1720705, EPI_ISL_1720706, EPI_ISL_1720707, EPI_ISL_1720708, EPI_ISL_1720709, EPI_ISL_1720710, EPI_ISL_1720711, EPI_ISL_1720712, EPI_ISL_1720714, EPI_ISL_1720715, EPI_ISL_1720716, EPI_ISL_1720717, EPI_ISL_1720718, EPI_ISL_1720720, EPI_ISL_1720721, EPI_ISL_1720722, EPI_ISL_1720723, EPI_ISL_1720724, EPI_ISL_1720725, EPI_ISL_1720726, EPI_ISL_1720727, EPI_ISL_1720728, EPI_ISL_1720729, EPI_ISL_1720730, EPI_ISL_1720732, EPI_ISL_1720733, EPI_ISL_1720734, EPI_ISL_1720735, EPI_ISL_1720737, EPI_ISL_1720738, EPI_ISL_1720739, EPI_ISL_1720740, EPI_ISL_1720741, EPI_ISL_1720742, EPI_ISL_1720743, EPI_ISL_1720744, EPI_ISL_1720745, EPI_ISL_1720746, EPI_ISL_1720747, EPI_ISL_1720749, EPI_ISL_1720750, EPI_ISL_1720751, EPI_ISL_1723351, EPI_ISL_1723352, EPI_ISL_1723353, EPI_ISL_1723354, EPI_ISL_1723355, EPI_ISL_1723356, EPI_ISL_1723357, EPI_ISL_1723358, EPI_ISL_1723359, EPI_ISL_1723360, EPI_ISL_1723361, EPI_ISL_1723362, EPI_ISL_1723363, EPI_ISL_1723364, EPI_ISL_1723365, EPI_ISL_1723366, EPI_ISL_1723367, EPI_ISL_1723368, EPI_ISL_1723369, EPI_ISL_1723370, EPI_ISL_1723371, EPI_ISL_1723372, EPI_ISL_1723373, EPI_ISL_1723374, EPI_ISL_1723375, EPI_ISL_1723376, EPI_ISL_1723377, EPI_ISL_1723378, EPI_ISL_1723379, EPI_ISL_1723380, EPI_ISL_1723381, EPI_ISL_1723382, EPI_ISL_1723383, EPI_ISL_1723384, EPI_ISL_1723385, EPI_ISL_1723386, EPI_ISL_1723387, EPI_ISL_1723388, EPI_ISL_1723389, EPI_ISL_1723390, EPI_ISL_1723391, EPI_ISL_1723392, EPI_ISL_1723393, EPI_ISL_1723394, EPI_ISL_1723395, EPI_ISL_1723396, EPI_ISL_1723397, EPI_ISL_1723398, EPI_ISL_1723399, EPI_ISL_1723400, EPI_ISL_1723401, EPI_ISL_1723402, EPI_ISL_1723403, EPI_ISL_1723404, EPI_ISL_1723405, EPI_ISL_1723406, EPI_ISL_1723407, EPI_ISL_1723408, EPI_ISL_1723409, EPI_ISL_1723410, EPI_ISL_1723411, EPI_ISL_1723412, EPI_ISL_1723413, EPI_ISL_1723414, EPI_ISL_1723415, EPI_ISL_1723416, EPI_ISL_1723417, EPI_ISL_1723418, EPI_ISL_1723419, EPI_ISL_1723420, EPI_ISL_1723421, EPI_ISL_1723422, EPI_ISL_1723423, EPI_ISL_1723424, EPI_ISL_1723425, EPI_ISL_1723426, EPI_ISL_1723427, EPI_ISL_1723428, EPI_ISL_1723429, EPI_ISL_1723430, EPI_ISL_1723431, EPI_ISL_1723432, EPI_ISL_1723433, EPI_ISL_1723434, EPI_ISL_1723435, EPI_ISL_1723436, EPI_ISL_1723437, EPI_ISL_1723438, EPI_ISL_1723439, EPI_ISL_1723440, EPI_ISL_1723441, EPI_ISL_1723442, EPI_ISL_1723443, EPI_ISL_1723444, EPI_ISL_1723445, EPI_ISL_1723446, EPI_ISL_1723447, EPI_ISL_1723448, EPI_ISL_1723449, EPI_ISL_1723450, EPI_ISL_1723451, EPI_ISL_1723452, EPI_ISL_1723453, EPI_ISL_1723454, EPI_ISL_1723455, EPI_ISL_1723456, EPI_ISL_1723457, EPI_ISL_1723458, EPI_ISL_1723459, EPI_ISL_1723460, EPI_ISL_1723461, EPI_ISL_1723462, EPI_ISL_1723463, EPI_ISL_1723464, EPI_ISL_1723465, EPI_ISL_1723466, EPI_ISL_1723467, EPI_ISL_1723468, EPI_ISL_1723469, EPI_ISL_1723470, EPI_ISL_1723471, EPI_ISL_1723472, EPI_ISL_1723473, EPI_ISL_1723474, EPI_ISL_1723475, EPI_ISL_1723476, EPI_ISL_1723477, EPI_ISL_1723478, EPI_ISL_1723479, EPI_ISL_1723480, EPI_ISL_1723481, EPI_ISL_1723482, EPI_ISL_1723483, EPI_ISL_1723484, EPI_ISL_1723485, EPI_ISL_1723486, EPI_ISL_1723487, EPI_ISL_1723488, EPI_ISL_1723489, EPI_ISL_1723490, EPI_ISL_1723491, EPI_ISL_1723492, EPI_ISL_1723493, EPI_ISL_1723494, EPI_ISL_1723495, EPI_ISL_1723496, EPI_ISL_1723497, EPI_ISL_1723498, EPI_ISL_1723499, EPI_ISL_1723500, EPI_ISL_1723501, EPI_ISL_1723502, EPI_ISL_1723503, EPI_ISL_1723504, EPI_ISL_1723505, EPI_ISL_1723506, EPI_ISL_1723507, EPI_ISL_1723508, EPI_ISL_1723509, EPI_ISL_1723510, EPI_ISL_1723511, EPI_ISL_1723512, EPI_ISL_1723513, EPI_ISL_1723514, EPI_ISL_1723515, EPI_ISL_1723516, EPI_ISL_1723517, EPI_ISL_1723518, EPI_ISL_1723519, EPI_ISL_1723520, EPI_ISL_1723521, EPI_ISL_1723522, EPI_ISL_1723523, EPI_ISL_1723524, EPI_ISL_1723525, EPI_ISL_1723526, EPI_ISL_1723527, EPI_ISL_1723528, EPI_ISL_1723529, EPI_ISL_1723530, EPI_ISL_1723531, EPI_ISL_1723532, EPI_ISL_1723533, EPI_ISL_1723534, EPI_ISL_1723535, EPI_ISL_1723536, EPI_ISL_1723537, EPI_ISL_1723538, EPI_ISL_1723539, EPI_ISL_1723540, EPI_ISL_1723541, EPI_ISL_1723542, EPI_ISL_1723543, EPI_ISL_1723544, EPI_ISL_1723545, EPI_ISL_1723546, EPI_ISL_1723547, EPI_ISL_1723548, EPI_ISL_1723549, EPI_ISL_1723550, EPI_ISL_1723551, EPI_ISL_1723552, EPI_ISL_1723553, EPI_ISL_1723554, EPI_ISL_1723555, EPI_ISL_1723556, EPI_ISL_1723557, EPI_ISL_1723558, EPI_ISL_1723559, EPI_ISL_1723560, EPI_ISL_1723561, EPI_ISL_1723562, EPI_ISL_1723563, EPI_ISL_1723564, EPI_ISL_1723565, EPI_ISL_1723566, EPI_ISL_1723567, EPI_ISL_1723568, EPI_ISL_1723569, EPI_ISL_1723570, EPI_ISL_1723571, EPI_ISL_1723572, EPI_ISL_1723573, EPI_ISL_1723574, EPI_ISL_1723575, EPI_ISL_1723576, EPI_ISL_1723577, EPI_ISL_1723578, EPI_ISL_1723579, EPI_ISL_1723580, EPI_ISL_1723581, EPI_ISL_1723582, EPI_ISL_1723583, EPI_ISL_1723584, EPI_ISL_1723585, EPI_ISL_1723586, EPI_ISL_1723587, EPI_ISL_1723588, EPI_ISL_1723589, EPI_ISL_1723590, EPI_ISL_1723591, EPI_ISL_1723592, EPI_ISL_1723593, EPI_ISL_1723594, EPI_ISL_1723595, EPI_ISL_1723596, EPI_ISL_1723597, EPI_ISL_1723598, EPI_ISL_1723599, EPI_ISL_1723600, EPI_ISL_1723601, EPI_ISL_1723602, EPI_ISL_1723603, EPI_ISL_1723604, EPI_ISL_1723605, EPI_ISL_1723606, EPI_ISL_1723607, EPI_ISL_1723608, EPI_ISL_1723609, EPI_ISL_1723610, EPI_ISL_1723611, EPI_ISL_1723612, EPI_ISL_1723613, EPI_ISL_1723614, EPI_ISL_1723615, EPI_ISL_1723616, EPI_ISL_1723617, EPI_ISL_1723618, EPI_ISL_1723619, EPI_ISL_1723620, EPI_ISL_1723621, EPI_ISL_1723622, EPI_ISL_1723623, EPI_ISL_1723624, EPI_ISL_1723625, EPI_ISL_1723626, EPI_ISL_1723627, EPI_ISL_1723628, EPI_ISL_1723629, EPI_ISL_1723630, EPI_ISL_1723631, EPI_ISL_1723632, EPI_ISL_1723633, EPI_ISL_1723634, EPI_ISL_1723635, EPI_ISL_1723636, EPI_ISL_1723637, EPI_ISL_1723638, EPI_ISL_1723639, EPI_ISL_1723640, EPI_ISL_1723641, EPI_ISL_1723642, EPI_ISL_1723643, EPI_ISL_1723644, EPI_ISL_1723645, EPI_ISL_1723646, EPI_ISL_1723647, EPI_ISL_1723648, EPI_ISL_1723649, EPI_ISL_1723650, EPI_ISL_1723651, EPI_ISL_1723652, EPI_ISL_1723653, EPI_ISL_1723654, EPI_ISL_1723655, EPI_ISL_1723656, EPI_ISL_1723657, EPI_ISL_1723659, EPI_ISL_1723660, EPI_ISL_1723661, EPI_ISL_1723662, EPI_ISL_1723664, EPI_ISL_1723665, EPI_ISL_1723666, EPI_ISL_1723667, EPI_ISL_1723668, EPI_ISL_1723669, EPI_ISL_1723670, EPI_ISL_1723671, EPI_ISL_1723672, EPI_ISL_1723673, EPI_ISL_1723675, EPI_ISL_1723676, EPI_ISL_1723678, EPI_ISL_1723680, EPI_ISL_1723681, EPI_ISL_1723682, EPI_ISL_1723683, EPI_ISL_1723684, EPI_ISL_1723686, EPI_ISL_1723687, EPI_ISL_1723690, EPI_ISL_1723691, EPI_ISL_1723692, EPI_ISL_1723693, EPI_ISL_1723694, EPI_ISL_1723698, EPI_ISL_1723701, EPI_ISL_1723703, EPI_ISL_1723704, EPI_ISL_1723705, EPI_ISL_1723706, EPI_ISL_1723707, EPI_ISL_1723708, EPI_ISL_1723709, EPI_ISL_1723710, EPI_ISL_1723711, EPI_ISL_1723712, EPI_ISL_1723714, EPI_ISL_1723717, EPI_ISL_1723718, EPI_ISL_1723719, EPI_ISL_1723720, EPI_ISL_1723721, EPI_ISL_1723723, EPI_ISL_1723724, EPI_ISL_1723725, EPI_ISL_1723726, EPI_ISL_1723727, EPI_ISL_1723728, EPI_ISL_1723729, EPI_ISL_1723730, EPI_ISL_1723731, EPI_ISL_1723732, EPI_ISL_1723734, EPI_ISL_1723735, EPI_ISL_1723736, EPI_ISL_1723737, EPI_ISL_1723738, EPI_ISL_1723740, EPI_ISL_1723741, EPI_ISL_1723742, EPI_ISL_1723744, EPI_ISL_1723745, EPI_ISL_1723746, EPI_ISL_1723747, EPI_ISL_1723748, EPI_ISL_1723749, EPI_ISL_1723750, EPI_ISL_1723751, EPI_ISL_1723752, EPI_ISL_1723754, EPI_ISL_1723755, EPI_ISL_1723756, EPI_ISL_1723758, EPI_ISL_1723759, EPI_ISL_1723761, EPI_ISL_1723763, EPI_ISL_1723765, EPI_ISL_1723767, EPI_ISL_1723768, EPI_ISL_1723770, EPI_ISL_1723771, EPI_ISL_1723772, EPI_ISL_1723773, EPI_ISL_1723774, EPI_ISL_1723775, EPI_ISL_1723776, EPI_ISL_1723777, EPI_ISL_1723779, EPI_ISL_1723780, EPI_ISL_1723781, EPI_ISL_1723782, EPI_ISL_1723783, EPI_ISL_1723784, EPI_ISL_1723785, EPI_ISL_1723786, EPI_ISL_1723787, EPI_ISL_1723788, EPI_ISL_1723789, EPI_ISL_1723790, EPI_ISL_1723791, EPI_ISL_1723795, EPI_ISL_1723796, EPI_ISL_1723797, EPI_ISL_1723799, EPI_ISL_1723800, EPI_ISL_1723802, EPI_ISL_1723804, EPI_ISL_1723805, EPI_ISL_1723806, EPI_ISL_1723808, EPI_ISL_1723809, EPI_ISL_1723810, EPI_ISL_1723811, EPI_ISL_1723812, EPI_ISL_1723813, EPI_ISL_1723814, EPI_ISL_1723815, EPI_ISL_1723816, EPI_ISL_1723817, EPI_ISL_1723818, EPI_ISL_1723820, EPI_ISL_1723821, EPI_ISL_1723822, EPI_ISL_1723823, EPI_ISL_1723824, EPI_ISL_1723825, EPI_ISL_1723826, EPI_ISL_1723828, EPI_ISL_1723829, EPI_ISL_1723831, EPI_ISL_1723832, EPI_ISL_1723833, EPI_ISL_1723834, EPI_ISL_1723835, EPI_ISL_1723836, EPI_ISL_1723837, EPI_ISL_1723838, EPI_ISL_1723839, EPI_ISL_1723840, EPI_ISL_1723841, EPI_ISL_1723842, EPI_ISL_1723843, EPI_ISL_1723844, EPI_ISL_1723845, EPI_ISL_1723846, EPI_ISL_1723847, EPI_ISL_1723848, EPI_ISL_1723849, EPI_ISL_1723850, EPI_ISL_1723851, EPI_ISL_1723852, EPI_ISL_1723853, EPI_ISL_1723854, EPI_ISL_1723855, EPI_ISL_1723857, EPI_ISL_1723858, EPI_ISL_1723859, EPI_ISL_1723860, EPI_ISL_1723861, EPI_ISL_1723862, EPI_ISL_1723864, EPI_ISL_1723865, EPI_ISL_1723866, EPI_ISL_1723868, EPI_ISL_1723869, EPI_ISL_1723870, EPI_ISL_1723871, EPI_ISL_1723872, EPI_ISL_1723873, EPI_ISL_1723874, EPI_ISL_1723875, EPI_ISL_1723876, EPI_ISL_1723877, EPI_ISL_1723878, EPI_ISL_1723879, EPI_ISL_1723881, EPI_ISL_1723882, EPI_ISL_1723883, EPI_ISL_1723884, EPI_ISL_1723885, EPI_ISL_1723886, EPI_ISL_1723889, EPI_ISL_1723891, EPI_ISL_1723892, EPI_ISL_1723894, EPI_ISL_1723895, EPI_ISL_1723897, EPI_ISL_1723898, EPI_ISL_1723899, EPI_ISL_1723900, EPI_ISL_1723902, EPI_ISL_1723903, EPI_ISL_1723904, EPI_ISL_1723905, EPI_ISL_1723906, EPI_ISL_1723907, EPI_ISL_1723908, EPI_ISL_1723909, EPI_ISL_1723910, EPI_ISL_1723911, EPI_ISL_1723912, EPI_ISL_1723913, EPI_ISL_1723914, EPI_ISL_1723915, EPI_ISL_1723916, EPI_ISL_1723917, EPI_ISL_1723918, EPI_ISL_1723920, EPI_ISL_1723921, EPI_ISL_1723922, EPI_ISL_1723923, EPI_ISL_1723924, EPI_ISL_1723925, EPI_ISL_1723928, EPI_ISL_1723930, EPI_ISL_1723931, EPI_ISL_1723932, EPI_ISL_1723934, EPI_ISL_1723936, EPI_ISL_1723937, EPI_ISL_1723938, EPI_ISL_1723940, EPI_ISL_1723941, EPI_ISL_1723942, EPI_ISL_1723945, EPI_ISL_1723946, EPI_ISL_1723947, EPI_ISL_1723948, EPI_ISL_1723949, EPI_ISL_1723950, EPI_ISL_1723952, EPI_ISL_1723953, EPI_ISL_1723954, EPI_ISL_1723955, EPI_ISL_1723957, EPI_ISL_1723959, EPI_ISL_1723960, EPI_ISL_1723961, EPI_ISL_1723962, EPI_ISL_1723964, EPI_ISL_1723965, EPI_ISL_1723966, EPI_ISL_1723967, EPI_ISL_1723968, EPI_ISL_1723969, EPI_ISL_1723970, EPI_ISL_1723971, EPI_ISL_1723973, EPI_ISL_1723975, EPI_ISL_1723976, EPI_ISL_1723978, EPI_ISL_1723979, EPI_ISL_1723980, EPI_ISL_1723981, EPI_ISL_1723983, EPI_ISL_1723985, EPI_ISL_1723986, EPI_ISL_1723987, EPI_ISL_1723988, EPI_ISL_1723989, EPI_ISL_1723991, EPI_ISL_1723992, EPI_ISL_1723993, EPI_ISL_1723994, EPI_ISL_1723995, EPI_ISL_1723996, EPI_ISL_1723997, EPI_ISL_1723999, EPI_ISL_1724000, EPI_ISL_1724001, EPI_ISL_1724002, EPI_ISL_1724003, EPI_ISL_1724004, EPI_ISL_1724005, EPI_ISL_1724006, EPI_ISL_1724007, EPI_ISL_1724008, EPI_ISL_1724009, EPI_ISL_1724010, EPI_ISL_1724011, EPI_ISL_1724012, EPI_ISL_1724013, EPI_ISL_1724014, EPI_ISL_1724015, EPI_ISL_1724016, EPI_ISL_1724017, EPI_ISL_1724018, EPI_ISL_1724019, EPI_ISL_1724020, EPI_ISL_1724021, EPI_ISL_1724022, EPI_ISL_1724023, EPI_ISL_1724024, EPI_ISL_1724026, EPI_ISL_1724027, EPI_ISL_1724028, EPI_ISL_1724030, EPI_ISL_1724031, EPI_ISL_1724032, EPI_ISL_1724033, EPI_ISL_1724034, EPI_ISL_1724035, EPI_ISL_1724037, EPI_ISL_1724038, EPI_ISL_1724039, EPI_ISL_1724040, EPI_ISL_1724041, EPI_ISL_1724042, EPI_ISL_1724044, EPI_ISL_1724045, EPI_ISL_1724046, EPI_ISL_1724048, EPI_ISL_1724050, EPI_ISL_1724052, EPI_ISL_1724054, EPI_ISL_1724055, EPI_ISL_1724056, EPI_ISL_1724057, EPI_ISL_1724058, EPI_ISL_1724059, EPI_ISL_1724060, EPI_ISL_1724061, EPI_ISL_1724063, EPI_ISL_1724064, EPI_ISL_1724065, EPI_ISL_1724066, EPI_ISL_1724067, EPI_ISL_1724068, EPI_ISL_1724070, EPI_ISL_1724073, EPI_ISL_1724074, EPI_ISL_1724076, EPI_ISL_1724077, EPI_ISL_1724078, EPI_ISL_1724079, EPI_ISL_1724080, EPI_ISL_1724081, EPI_ISL_1724082, EPI_ISL_1724083, EPI_ISL_1724084, EPI_ISL_1724087, EPI_ISL_1724089, EPI_ISL_1724090, EPI_ISL_1724091, EPI_ISL_1724095, EPI_ISL_1724097, EPI_ISL_1724099, EPI_ISL_1724102, EPI_ISL_1724103, EPI_ISL_1724104, EPI_ISL_1724105, EPI_ISL_1724106, EPI_ISL_1724107, EPI_ISL_1724114, EPI_ISL_1724118, EPI_ISL_1724119, EPI_ISL_1724121, EPI_ISL_1724122, EPI_ISL_1724123, EPI_ISL_1724124, EPI_ISL_1724126, EPI_ISL_1724127, EPI_ISL_1724128, EPI_ISL_1724130, EPI_ISL_1724131, EPI_ISL_1724132, EPI_ISL_1724133, EPI_ISL_1724134, EPI_ISL_1724135, EPI_ISL_1724136, EPI_ISL_1724137, EPI_ISL_1724138, EPI_ISL_1724140, EPI_ISL_1724141, EPI_ISL_1724142, EPI_ISL_1724144, EPI_ISL_1724145, EPI_ISL_1724146, EPI_ISL_1724148, EPI_ISL_1724149, EPI_ISL_1724150, EPI_ISL_1724152, EPI_ISL_1724153, EPI_ISL_1724154, EPI_ISL_1724155, EPI_ISL_1724156, EPI_ISL_1724157, EPI_ISL_1724158, EPI_ISL_1724159, EPI_ISL_1724160, EPI_ISL_1724162, EPI_ISL_1724163, EPI_ISL_1724164, EPI_ISL_1724165, EPI_ISL_1724166, EPI_ISL_1724167, EPI_ISL_1724169, EPI_ISL_1724170, EPI_ISL_1724171, EPI_ISL_1724172, EPI_ISL_1724174, EPI_ISL_1724175, EPI_ISL_1724177, EPI_ISL_1724178, EPI_ISL_1724179, EPI_ISL_1724181, EPI_ISL_1724182, EPI_ISL_1724183, EPI_ISL_1724184, EPI_ISL_1724186, EPI_ISL_1724187, EPI_ISL_1724188, EPI_ISL_1724190, EPI_ISL_1724192, EPI_ISL_1724193, EPI_ISL_1724196, EPI_ISL_1724198, EPI_ISL_1724199, EPI_ISL_1724200, EPI_ISL_1724201, EPI_ISL_1724203, EPI_ISL_1724204, EPI_ISL_1724205, EPI_ISL_1724206, EPI_ISL_1724207, EPI_ISL_1724208, EPI_ISL_1724210, EPI_ISL_1724211, EPI_ISL_1724212, EPI_ISL_1724214, EPI_ISL_1724216, EPI_ISL_1724217, EPI_ISL_1724218, EPI_ISL_1724219, EPI_ISL_1724220, EPI_ISL_1724221, EPI_ISL_1724223, EPI_ISL_1724224, EPI_ISL_1724227, EPI_ISL_1724228, EPI_ISL_1724229, EPI_ISL_1724230, EPI_ISL_1724233, EPI_ISL_1724234, EPI_ISL_1724236, EPI_ISL_1724765, EPI_ISL_1724768, EPI_ISL_1724771, EPI_ISL_1724772, EPI_ISL_1724777, EPI_ISL_1724779, EPI_ISL_1724780, EPI_ISL_1724783, EPI_ISL_1724786, EPI_ISL_1724791, EPI_ISL_1724793, EPI_ISL_1724797, EPI_ISL_1724798, EPI_ISL_1724800, EPI_ISL_1724804, EPI_ISL_1724808, EPI_ISL_1724809, EPI_ISL_1724811, EPI_ISL_1724815, EPI_ISL_1724816, EPI_ISL_1724818, EPI_ISL_1724819, EPI_ISL_1724821, EPI_ISL_1724823, EPI_ISL_1724825, EPI_ISL_1724827, EPI_ISL_1724828, EPI_ISL_1724829, EPI_ISL_1724831, EPI_ISL_1724836, EPI_ISL_1724837, EPI_ISL_1724838, EPI_ISL_1724841, EPI_ISL_1724845, EPI_ISL_1724846, EPI_ISL_1724847, EPI_ISL_1724848, EPI_ISL_1724849, EPI_ISL_1724850, EPI_ISL_1724855, EPI_ISL_1724859, EPI_ISL_1724861, EPI_ISL_1724862, EPI_ISL_1724864, EPI_ISL_1724865, EPI_ISL_1724867, EPI_ISL_1724868, EPI_ISL_1724870, EPI_ISL_1724872, EPI_ISL_1724873, EPI_ISL_1724874, EPI_ISL_1724878, EPI_ISL_1724880, EPI_ISL_1724881, EPI_ISL_1724886, EPI_ISL_1724887, EPI_ISL_1724890, EPI_ISL_1724891, EPI_ISL_1724894, EPI_ISL_1724896, EPI_ISL_1724899, EPI_ISL_1724901, EPI_ISL_1724903, EPI_ISL_1724906, EPI_ISL_1724907, EPI_ISL_1724911, EPI_ISL_1724915, EPI_ISL_1724918, EPI_ISL_1724924, EPI_ISL_1724929, EPI_ISL_1724934, EPI_ISL_1724936, EPI_ISL_1724939, EPI_ISL_1724942, EPI_ISL_1724945, EPI_ISL_1724946, EPI_ISL_1724947, EPI_ISL_1724949, EPI_ISL_1724950, EPI_ISL_1724952, EPI_ISL_1724955, EPI_ISL_1724956, EPI_ISL_1724959, EPI_ISL_1724962, EPI_ISL_1724963, EPI_ISL_1724964, EPI_ISL_1724969, EPI_ISL_1724971, EPI_ISL_1724974, EPI_ISL_1724975, EPI_ISL_1724981, EPI_ISL_1724984, EPI_ISL_1724985, EPI_ISL_1724986, EPI_ISL_1724990, EPI_ISL_1724992, EPI_ISL_1724995, EPI_ISL_1725000, EPI_ISL_1725004, EPI_ISL_1725008, EPI_ISL_1725011, EPI_ISL_1725015, EPI_ISL_1725018, EPI_ISL_1725024, EPI_ISL_1725025, EPI_ISL_1725026, EPI_ISL_1725029, EPI_ISL_1725030, EPI_ISL_1725033, EPI_ISL_1725042, EPI_ISL_1725047, EPI_ISL_1725050, EPI_ISL_1725053, EPI_ISL_1725054, EPI_ISL_1725055, EPI_ISL_1725056, EPI_ISL_1725060, EPI_ISL_1725061, EPI_ISL_1725065, EPI_ISL_1725068, EPI_ISL_1725069, EPI_ISL_1725659, EPI_ISL_1725660, EPI_ISL_1725661, EPI_ISL_1725663, EPI_ISL_1725665, EPI_ISL_1725668, EPI_ISL_1725674, EPI_ISL_1725675, EPI_ISL_1725677, EPI_ISL_1725679, EPI_ISL_1725680, EPI_ISL_1725682, EPI_ISL_1725684, EPI_ISL_1725685, EPI_ISL_1725686, EPI_ISL_1725689, EPI_ISL_1725690, EPI_ISL_1725692, EPI_ISL_1725693, EPI_ISL_1725696, EPI_ISL_1725697, EPI_ISL_1725701, EPI_ISL_1725703, EPI_ISL_1725705, EPI_ISL_1725707, EPI_ISL_1725709, EPI_ISL_1725713, EPI_ISL_1725714, EPI_ISL_1725718, EPI_ISL_1725722, EPI_ISL_1725724, EPI_ISL_1725725, EPI_ISL_1725729, EPI_ISL_1725731, EPI_ISL_1725732, EPI_ISL_1725735, EPI_ISL_1725736, EPI_ISL_1725738, EPI_ISL_1725739, EPI_ISL_1725740, EPI_ISL_1725742, EPI_ISL_1725745, EPI_ISL_1725746, EPI_ISL_1725749, EPI_ISL_1725750, EPI_ISL_1725751, EPI_ISL_1725753, EPI_ISL_1725755, EPI_ISL_1725756, EPI_ISL_1725757, EPI_ISL_1725759, EPI_ISL_1725760, EPI_ISL_1725762, EPI_ISL_1725764, EPI_ISL_1725765, EPI_ISL_1725767, EPI_ISL_1725768, EPI_ISL_1725770, EPI_ISL_1725772, EPI_ISL_1725773, EPI_ISL_1725774, EPI_ISL_1725776, EPI_ISL_1725777, EPI_ISL_1725780, EPI_ISL_1725783, EPI_ISL_1725787, EPI_ISL_1725790, EPI_ISL_1725794, EPI_ISL_1725797, EPI_ISL_1725801, EPI_ISL_1725804, EPI_ISL_1725806, EPI_ISL_1725808, EPI_ISL_1725809, EPI_ISL_1725812, EPI_ISL_1725813, EPI_ISL_1725817, EPI_ISL_1725820, EPI_ISL_1725822, EPI_ISL_1725824, EPI_ISL_1725827, EPI_ISL_1725828, EPI_ISL_1725829, EPI_ISL_1725830, EPI_ISL_1725831, EPI_ISL_1725832, EPI_ISL_1725833, EPI_ISL_1725834, EPI_ISL_1725836, EPI_ISL_1725837, EPI_ISL_1725840, EPI_ISL_1725841, EPI_ISL_1725843, EPI_ISL_1725846, EPI_ISL_1725848, EPI_ISL_1725849, EPI_ISL_1725850, EPI_ISL_1725852, EPI_ISL_1725853, EPI_ISL_1725855, EPI_ISL_1725857, EPI_ISL_1725858, EPI_ISL_1725859, EPI_ISL_1725863, EPI_ISL_1725865, EPI_ISL_1725866, EPI_ISL_1725867, EPI_ISL_1725871, EPI_ISL_1725874, EPI_ISL_1725876, EPI_ISL_1725877, EPI_ISL_1725878, EPI_ISL_1725882, EPI_ISL_1725884, EPI_ISL_1725885, EPI_ISL_1725886, EPI_ISL_1725887, EPI_ISL_1725888, EPI_ISL_1725891, EPI_ISL_1725892, EPI_ISL_1725893, EPI_ISL_1725895, EPI_ISL_1725896, EPI_ISL_1725898, EPI_ISL_1725899, EPI_ISL_1725901, EPI_ISL_1725903, EPI_ISL_1725907, EPI_ISL_1725909, EPI_ISL_1725911, EPI_ISL_1725912, EPI_ISL_1725913, EPI_ISL_1725914, EPI_ISL_1725916, EPI_ISL_1725918, EPI_ISL_1725919, EPI_ISL_1725921, EPI_ISL_1725922, EPI_ISL_1725923, EPI_ISL_1725925, EPI_ISL_1725926, EPI_ISL_1725927, EPI_ISL_1725928, EPI_ISL_1725929, EPI_ISL_1725931, EPI_ISL_1725932, EPI_ISL_1725935, EPI_ISL_1725936, EPI_ISL_1725937, EPI_ISL_1725939, EPI_ISL_1725940, EPI_ISL_1725941, EPI_ISL_1725942, EPI_ISL_1725943, EPI_ISL_1725944, EPI_ISL_1725945, EPI_ISL_1725946, EPI_ISL_1725948, EPI_ISL_1725949, EPI_ISL_1725950, EPI_ISL_1725951, EPI_ISL_1725952, EPI_ISL_1725953, EPI_ISL_1725954, EPI_ISL_1725955, EPI_ISL_1725956, EPI_ISL_1725957, EPI_ISL_1725959, EPI_ISL_1725960, EPI_ISL_1725962, EPI_ISL_1725963, EPI_ISL_1725964, EPI_ISL_1726154, EPI_ISL_1726155, EPI_ISL_1726156, EPI_ISL_1726157, EPI_ISL_1726158, EPI_ISL_1726159, EPI_ISL_1726160, EPI_ISL_1726161, EPI_ISL_1726162, EPI_ISL_1726163, EPI_ISL_1726164, EPI_ISL_1726165, EPI_ISL_1726166, EPI_ISL_1726167, EPI_ISL_1726168, EPI_ISL_1726169, EPI_ISL_1726170, EPI_ISL_1726171, EPI_ISL_1726173, EPI_ISL_1726174, EPI_ISL_1726175, EPI_ISL_1726176, EPI_ISL_1726177, EPI_ISL_1726178, EPI_ISL_1726179, EPI_ISL_1726180, EPI_ISL_1726181, EPI_ISL_1726182, EPI_ISL_1726183, EPI_ISL_1726184, EPI_ISL_1726185, EPI_ISL_1726186, EPI_ISL_1726187, EPI_ISL_1726188, EPI_ISL_1726189, EPI_ISL_1726190, EPI_ISL_1726191, EPI_ISL_1726192, EPI_ISL_1726193, EPI_ISL_1726194, EPI_ISL_1726195, EPI_ISL_1726196, EPI_ISL_1726197, EPI_ISL_1726198, EPI_ISL_1726199, EPI_ISL_1726200, EPI_ISL_1726201, EPI_ISL_1726202, EPI_ISL_1726203, EPI_ISL_1726204, EPI_ISL_1726205, EPI_ISL_1726206, EPI_ISL_1726207, EPI_ISL_1726208, EPI_ISL_1726209, EPI_ISL_1726210, EPI_ISL_1726211, EPI_ISL_1726212, EPI_ISL_1726213, EPI_ISL_1726214, EPI_ISL_1726215, EPI_ISL_1726216, EPI_ISL_1726217, EPI_ISL_1726218, EPI_ISL_1726219, EPI_ISL_1726220, EPI_ISL_1726221, EPI_ISL_1726222, EPI_ISL_1726223, EPI_ISL_1726224, EPI_ISL_1726225, EPI_ISL_1726226, EPI_ISL_1726227, EPI_ISL_1726228, EPI_ISL_1726229, EPI_ISL_1726230, EPI_ISL_1726231, EPI_ISL_1726232, EPI_ISL_1726233, EPI_ISL_1726234, EPI_ISL_1726235, EPI_ISL_1726236, EPI_ISL_1726237, EPI_ISL_1726238, EPI_ISL_1726239, EPI_ISL_1726240, EPI_ISL_1726241, EPI_ISL_1726242, EPI_ISL_1726243, EPI_ISL_1726244, EPI_ISL_1726245, EPI_ISL_1726246, EPI_ISL_1726247, EPI_ISL_1726248, EPI_ISL_1726249, EPI_ISL_1726250, EPI_ISL_1726251, EPI_ISL_1726252, EPI_ISL_1726253, EPI_ISL_1726254, EPI_ISL_1726255, EPI_ISL_1726256, EPI_ISL_1726257, EPI_ISL_1726258, EPI_ISL_1726259, EPI_ISL_1726260, EPI_ISL_1726261, EPI_ISL_1726262, EPI_ISL_1726263, EPI_ISL_1726264, EPI_ISL_1726265, EPI_ISL_1726266, EPI_ISL_1726267, EPI_ISL_1726268, EPI_ISL_1726269, EPI_ISL_1726270, EPI_ISL_1726271, EPI_ISL_1726272, EPI_ISL_1726273, EPI_ISL_1726274, EPI_ISL_1726275, EPI_ISL_1726276, EPI_ISL_1726277, EPI_ISL_1726278, EPI_ISL_1726279, EPI_ISL_1726280, EPI_ISL_1726281, EPI_ISL_1726282, EPI_ISL_1726283, EPI_ISL_1726284, EPI_ISL_1726285, EPI_ISL_1726286, EPI_ISL_1726287, EPI_ISL_1726288, EPI_ISL_1726289, EPI_ISL_1726290, EPI_ISL_1726291, EPI_ISL_1726292, EPI_ISL_1726293, EPI_ISL_1726294, EPI_ISL_1726295, EPI_ISL_1726296, EPI_ISL_1726297, EPI_ISL_1726298, EPI_ISL_1726299, EPI_ISL_1726300, EPI_ISL_1726301, EPI_ISL_1726302, EPI_ISL_1726303, EPI_ISL_1726304, EPI_ISL_1726305, EPI_ISL_1726306, EPI_ISL_1726307, EPI_ISL_1726308, EPI_ISL_1726309, EPI_ISL_1726310, EPI_ISL_1726311, EPI_ISL_1726312, EPI_ISL_1726313, EPI_ISL_1726314, EPI_ISL_1726315, EPI_ISL_1726316, EPI_ISL_1726317, EPI_ISL_1726318, EPI_ISL_1726319, EPI_ISL_1726320, EPI_ISL_1726321, EPI_ISL_1726322, EPI_ISL_1726323, EPI_ISL_1726324, EPI_ISL_1726325, EPI_ISL_1726326, EPI_ISL_1726327, EPI_ISL_1726328, EPI_ISL_1726329, EPI_ISL_1726330, EPI_ISL_1726331, EPI_ISL_1726332, EPI_ISL_1726333, EPI_ISL_1726334, EPI_ISL_1726335, EPI_ISL_1726336, EPI_ISL_1726337, EPI_ISL_1726338, EPI_ISL_1726339, EPI_ISL_1726340, EPI_ISL_1726341, EPI_ISL_1726342, EPI_ISL_1726343, EPI_ISL_1726344, EPI_ISL_1726345, EPI_ISL_1726346, EPI_ISL_1726347, EPI_ISL_1726348, EPI_ISL_1726349, EPI_ISL_1726350, EPI_ISL_1726351, EPI_ISL_1726352, EPI_ISL_1726353, EPI_ISL_1726354, EPI_ISL_1726355, EPI_ISL_1726356, EPI_ISL_1726357, EPI_ISL_1726358, EPI_ISL_1726359, EPI_ISL_1726360, EPI_ISL_1726361, EPI_ISL_1726362, EPI_ISL_1726363, EPI_ISL_1726364, EPI_ISL_1726365, EPI_ISL_1726366, EPI_ISL_1726367, EPI_ISL_1726368, EPI_ISL_1726369, EPI_ISL_1726370, EPI_ISL_1726371, EPI_ISL_1726372, EPI_ISL_1726373, EPI_ISL_1726374, EPI_ISL_1726375, EPI_ISL_1726376, EPI_ISL_1726377, EPI_ISL_1726378, EPI_ISL_1726379, EPI_ISL_1726381, EPI_ISL_1726382, EPI_ISL_1726383, EPI_ISL_1726384, EPI_ISL_1726385, EPI_ISL_1726386, EPI_ISL_1726387, EPI_ISL_1726388, EPI_ISL_1726389, EPI_ISL_1726390, EPI_ISL_1726391, EPI_ISL_1726392, EPI_ISL_1726393, EPI_ISL_1726394, EPI_ISL_1726395, EPI_ISL_1726396, EPI_ISL_1726397, EPI_ISL_1726398, EPI_ISL_1726399, EPI_ISL_1726400, EPI_ISL_1726401, EPI_ISL_1726402, EPI_ISL_1726403, EPI_ISL_1726404, EPI_ISL_1726405, EPI_ISL_1726406, EPI_ISL_1726407, EPI_ISL_1726408, EPI_ISL_1726409, EPI_ISL_1726410, EPI_ISL_1726411, EPI_ISL_1726412, EPI_ISL_1726413, EPI_ISL_1726414, EPI_ISL_1726415, EPI_ISL_1726416, EPI_ISL_1726417, EPI_ISL_1726418, EPI_ISL_1726419, EPI_ISL_1726420, EPI_ISL_1726421, EPI_ISL_1726422, EPI_ISL_1726423, EPI_ISL_1726424, EPI_ISL_1726425, EPI_ISL_1726426, EPI_ISL_1726427, EPI_ISL_1726428, EPI_ISL_1726429, EPI_ISL_1726430, EPI_ISL_1726431, EPI_ISL_1726432, EPI_ISL_1726433, EPI_ISL_1726434, EPI_ISL_1726435, EPI_ISL_1726436, EPI_ISL_1726437, EPI_ISL_1726438, EPI_ISL_1726439, EPI_ISL_1726440, EPI_ISL_1726441, EPI_ISL_1726442, EPI_ISL_1726443, EPI_ISL_1726444, EPI_ISL_1726445, EPI_ISL_1726446, EPI_ISL_1726447, EPI_ISL_1726448, EPI_ISL_1726449, EPI_ISL_1726450, EPI_ISL_1726451, EPI_ISL_1726452, EPI_ISL_1726453, EPI_ISL_1726454, EPI_ISL_1726455, EPI_ISL_1726456, EPI_ISL_1726457, EPI_ISL_1726458, EPI_ISL_1726459, EPI_ISL_1726460, EPI_ISL_1726461, EPI_ISL_1726462, EPI_ISL_1726463, EPI_ISL_1726464, EPI_ISL_1726465, EPI_ISL_1726466, EPI_ISL_1726467, EPI_ISL_1727656, EPI_ISL_1727658, EPI_ISL_1727660, EPI_ISL_1727661, EPI_ISL_1727662, EPI_ISL_1727664, EPI_ISL_1727666, EPI_ISL_1727669, EPI_ISL_1727670, EPI_ISL_1727671, EPI_ISL_1727672, EPI_ISL_1727674, EPI_ISL_1727675, EPI_ISL_1727677, EPI_ISL_1727678, EPI_ISL_1727680, EPI_ISL_1727681, EPI_ISL_1727682, EPI_ISL_1727683, EPI_ISL_1727684, EPI_ISL_1727685, EPI_ISL_1727686, EPI_ISL_1727687, EPI_ISL_1727688, EPI_ISL_1727689, EPI_ISL_1727691, EPI_ISL_1727692, EPI_ISL_1727693, EPI_ISL_1727694, EPI_ISL_1727695, EPI_ISL_1727696, EPI_ISL_1727697, EPI_ISL_1727698, EPI_ISL_1727699, EPI_ISL_1727700, EPI_ISL_1727702, EPI_ISL_1727703, EPI_ISL_1727704, EPI_ISL_1727705, EPI_ISL_1727707, EPI_ISL_1727708, EPI_ISL_1727709, EPI_ISL_1727710, EPI_ISL_1727712, EPI_ISL_1727713, EPI_ISL_1727714, EPI_ISL_1727715, EPI_ISL_1727716, EPI_ISL_1727717, EPI_ISL_1727719, EPI_ISL_1727720, EPI_ISL_1727721, EPI_ISL_1727722, EPI_ISL_1727723, EPI_ISL_1727724, EPI_ISL_1727725, EPI_ISL_1727726, EPI_ISL_1727727, EPI_ISL_1727728, EPI_ISL_1727729, EPI_ISL_1727730, EPI_ISL_1727731, EPI_ISL_1727732, EPI_ISL_1727733, EPI_ISL_1727735, EPI_ISL_1727736, EPI_ISL_1727737, EPI_ISL_1727738, EPI_ISL_1727739, EPI_ISL_1727740, EPI_ISL_1727741, EPI_ISL_1727742, EPI_ISL_1727743, EPI_ISL_1727744, EPI_ISL_1727745, EPI_ISL_1727746, EPI_ISL_1727747, EPI_ISL_1727748, EPI_ISL_1727749, EPI_ISL_1727750, EPI_ISL_1727751, EPI_ISL_1727753, EPI_ISL_1727754, EPI_ISL_1727755, EPI_ISL_1727756, EPI_ISL_1727757, EPI_ISL_1727758, EPI_ISL_1727759, EPI_ISL_1727760, EPI_ISL_1727762, EPI_ISL_1727764, EPI_ISL_1727765, EPI_ISL_1727766, EPI_ISL_1727767, EPI_ISL_1727769, EPI_ISL_1727770, EPI_ISL_1727771, EPI_ISL_1727772, EPI_ISL_1727774, EPI_ISL_1727775, EPI_ISL_1727776, EPI_ISL_1727777, EPI_ISL_1727778, EPI_ISL_1727779, EPI_ISL_1727780, EPI_ISL_1727781, EPI_ISL_1727782, EPI_ISL_1727783, EPI_ISL_1727784, EPI_ISL_1727785, EPI_ISL_1727786, EPI_ISL_1727787, EPI_ISL_1727788, EPI_ISL_1727790, EPI_ISL_1727791, EPI_ISL_1727792, EPI_ISL_1727793, EPI_ISL_1727795, EPI_ISL_1727796, EPI_ISL_1727797, EPI_ISL_1727798, EPI_ISL_1727799, EPI_ISL_1727800, EPI_ISL_1727801, EPI_ISL_1727802, EPI_ISL_1727803, EPI_ISL_1727804, EPI_ISL_1727805, EPI_ISL_1727807, EPI_ISL_1727808, EPI_ISL_1727809, EPI_ISL_1727810, EPI_ISL_1727812, EPI_ISL_1727813, EPI_ISL_1727814, EPI_ISL_1727815, EPI_ISL_1727816, EPI_ISL_1727817, EPI_ISL_1727818, EPI_ISL_1727819, EPI_ISL_1727820, EPI_ISL_1727821, EPI_ISL_1727822, EPI_ISL_1727823, EPI_ISL_1727824, EPI_ISL_1727825, EPI_ISL_1727826, EPI_ISL_1727827, EPI_ISL_1727828, EPI_ISL_1727829, EPI_ISL_1727830, EPI_ISL_1727831, EPI_ISL_1727832, EPI_ISL_1727833, EPI_ISL_1727834, EPI_ISL_1727835, EPI_ISL_1727837, EPI_ISL_1727838, EPI_ISL_1727839, EPI_ISL_1727841, EPI_ISL_1727842, EPI_ISL_1727843, EPI_ISL_1727844, EPI_ISL_1727845, EPI_ISL_1727846, EPI_ISL_1727847, EPI_ISL_1727848, EPI_ISL_1727849, EPI_ISL_1727852, EPI_ISL_1727853, EPI_ISL_1727857, EPI_ISL_1727858, EPI_ISL_1727859, EPI_ISL_1727860, EPI_ISL_1727862, EPI_ISL_1727863, EPI_ISL_1727864, EPI_ISL_1727865, EPI_ISL_1727866, EPI_ISL_1727868, EPI_ISL_1727870, EPI_ISL_1727871, EPI_ISL_1727872, EPI_ISL_1727873, EPI_ISL_1727874, EPI_ISL_1727875, EPI_ISL_1727876, EPI_ISL_1727878, EPI_ISL_1727879, EPI_ISL_1727882, EPI_ISL_1727883, EPI_ISL_1727884, EPI_ISL_1727885, EPI_ISL_1727886, EPI_ISL_1727887, EPI_ISL_1727888, EPI_ISL_1727889, EPI_ISL_1727891, EPI_ISL_1727892, EPI_ISL_1727893, EPI_ISL_1727895, EPI_ISL_1727896, EPI_ISL_1727897, EPI_ISL_1727898, EPI_ISL_1727899, EPI_ISL_1727900, EPI_ISL_1727901, EPI_ISL_1727902, EPI_ISL_1727903, EPI_ISL_1727904, EPI_ISL_1727905, EPI_ISL_1727906, EPI_ISL_1727907, EPI_ISL_1727910, EPI_ISL_1727911, EPI_ISL_1727913, EPI_ISL_1727914, EPI_ISL_1727915, EPI_ISL_1727916, EPI_ISL_1727917, EPI_ISL_1727918, EPI_ISL_1727919, EPI_ISL_1727920, EPI_ISL_1727921, EPI_ISL_1727922, EPI_ISL_1727923, EPI_ISL_1727925, EPI_ISL_1727926, EPI_ISL_1727928, EPI_ISL_1727930, EPI_ISL_1727932, EPI_ISL_1727933, EPI_ISL_1727935, EPI_ISL_1727936, EPI_ISL_1727937, EPI_ISL_1727939, EPI_ISL_1727940, EPI_ISL_1727942, EPI_ISL_1727944, EPI_ISL_1727945, EPI_ISL_1727946, EPI_ISL_1727947, EPI_ISL_1727948, EPI_ISL_1727949, EPI_ISL_1727950, EPI_ISL_1727952, EPI_ISL_1727953, EPI_ISL_1727954, EPI_ISL_1727955, EPI_ISL_1727956, EPI_ISL_1727958, EPI_ISL_1727960, EPI_ISL_1727962, EPI_ISL_1727964, EPI_ISL_1727966, EPI_ISL_1727968, EPI_ISL_1727970, EPI_ISL_1727974, EPI_ISL_1727976, EPI_ISL_1727977, EPI_ISL_1727978, EPI_ISL_1727979, EPI_ISL_1727980, EPI_ISL_1727981, EPI_ISL_1727983, EPI_ISL_1727985, EPI_ISL_1727986, EPI_ISL_1727987, EPI_ISL_1727988, EPI_ISL_1727991, EPI_ISL_1727992, EPI_ISL_1727993, EPI_ISL_1727994, EPI_ISL_1727995, EPI_ISL_1727996, EPI_ISL_1727998, EPI_ISL_1727999, EPI_ISL_1728000, EPI_ISL_1728002, EPI_ISL_1728003, EPI_ISL_1728004, EPI_ISL_1728005, EPI_ISL_1728006, EPI_ISL_1728007, EPI_ISL_1728008, EPI_ISL_1728010, EPI_ISL_1728011, EPI_ISL_1728012, EPI_ISL_1728013, EPI_ISL_1728016, EPI_ISL_1728017, EPI_ISL_1728018, EPI_ISL_1728019, EPI_ISL_1728020, EPI_ISL_1728021, EPI_ISL_1728023, EPI_ISL_1728024, EPI_ISL_1728025, EPI_ISL_1728026, EPI_ISL_1728027, EPI_ISL_1728028, EPI_ISL_1728029, EPI_ISL_1728030, EPI_ISL_1728031, EPI_ISL_1728034, EPI_ISL_1728035, EPI_ISL_1728036, EPI_ISL_1728037, EPI_ISL_1728039, EPI_ISL_1728041, EPI_ISL_1728043, EPI_ISL_1728044, EPI_ISL_1728045, EPI_ISL_1728048, EPI_ISL_1728049, EPI_ISL_1728050, EPI_ISL_1728051, EPI_ISL_1728053, EPI_ISL_1728054, EPI_ISL_1728055, EPI_ISL_1728056, EPI_ISL_1728058, EPI_ISL_1728059, EPI_ISL_1728060, EPI_ISL_1728061, EPI_ISL_1728062, EPI_ISL_1728063, EPI_ISL_1728064, EPI_ISL_1728065, EPI_ISL_1728066, EPI_ISL_1728067, EPI_ISL_1728068, EPI_ISL_1728069, EPI_ISL_1728070, EPI_ISL_1728071, EPI_ISL_1728073, EPI_ISL_1728074, EPI_ISL_1728076, EPI_ISL_1728077, EPI_ISL_1728078, EPI_ISL_1728079, EPI_ISL_1728081, EPI_ISL_1728082, EPI_ISL_1728083, EPI_ISL_1728084, EPI_ISL_1728086, EPI_ISL_1728087, EPI_ISL_1728088, EPI_ISL_1728089, EPI_ISL_1728090, EPI_ISL_1728092, EPI_ISL_1728093, EPI_ISL_1728094, EPI_ISL_1728095, EPI_ISL_1728097, EPI_ISL_1728099, EPI_ISL_1728101, EPI_ISL_1728102, EPI_ISL_1728103, EPI_ISL_1728104, EPI_ISL_1728105, EPI_ISL_1728106, EPI_ISL_1728107, EPI_ISL_1728108, EPI_ISL_1728109, EPI_ISL_1728111, EPI_ISL_1728112, EPI_ISL_1728113, EPI_ISL_1728114, EPI_ISL_1728115, EPI_ISL_1728116, EPI_ISL_1728117, EPI_ISL_1728118, EPI_ISL_1728120, EPI_ISL_1728121, EPI_ISL_1728122, EPI_ISL_1728123, EPI_ISL_1728124, EPI_ISL_1728125, EPI_ISL_1728127, EPI_ISL_1728128, EPI_ISL_1728129, EPI_ISL_1728131, EPI_ISL_1728132, EPI_ISL_1728134, EPI_ISL_1728135, EPI_ISL_1728136, EPI_ISL_1728137, EPI_ISL_1728138, EPI_ISL_1728140, EPI_ISL_1728142, EPI_ISL_1728143, EPI_ISL_1728144, EPI_ISL_1728145, EPI_ISL_1728146, EPI_ISL_1728147, EPI_ISL_1728149, EPI_ISL_1728150, EPI_ISL_1728151, EPI_ISL_1728152, EPI_ISL_1728153, EPI_ISL_1728154, EPI_ISL_1728155, EPI_ISL_1728156, EPI_ISL_1728157, EPI_ISL_1728158, EPI_ISL_1728159, EPI_ISL_1728160, EPI_ISL_1728161, EPI_ISL_1728162, EPI_ISL_1728163, EPI_ISL_1728164, EPI_ISL_1728165, EPI_ISL_1728166, EPI_ISL_1728167, EPI_ISL_1728168, EPI_ISL_1728169, EPI_ISL_1728170, EPI_ISL_1728171, EPI_ISL_1728172, EPI_ISL_1728173, EPI_ISL_1728174, EPI_ISL_1728176, EPI_ISL_1728177, EPI_ISL_1728178, EPI_ISL_1728179, EPI_ISL_1728180, EPI_ISL_1728181, EPI_ISL_1728184, EPI_ISL_1728185, EPI_ISL_1728186, EPI_ISL_1728187, EPI_ISL_1728189, EPI_ISL_1728190, EPI_ISL_1728191, EPI_ISL_1728192, EPI_ISL_1728193, EPI_ISL_1728194, EPI_ISL_1728195, EPI_ISL_1728196, EPI_ISL_1728197, EPI_ISL_1728198, EPI_ISL_1728199, EPI_ISL_1728200, EPI_ISL_1728201, EPI_ISL_1728202, EPI_ISL_1728203, EPI_ISL_1728204, EPI_ISL_1728205, EPI_ISL_1728206, EPI_ISL_1728207, EPI_ISL_1728208, EPI_ISL_1728210, EPI_ISL_1728211, EPI_ISL_1728212, EPI_ISL_1728213, EPI_ISL_1728215, EPI_ISL_1728216, EPI_ISL_1728217, EPI_ISL_1728219, EPI_ISL_1728220, EPI_ISL_1728221, EPI_ISL_1728223, EPI_ISL_1728224, EPI_ISL_1728225, EPI_ISL_1728226, EPI_ISL_1728227, EPI_ISL_1728228, EPI_ISL_1728229, EPI_ISL_1728230, EPI_ISL_1728231, EPI_ISL_1728232, EPI_ISL_1728233, EPI_ISL_1728235, EPI_ISL_1728236, EPI_ISL_1728238, EPI_ISL_1728239, EPI_ISL_1728240, EPI_ISL_1728241, EPI_ISL_1728242, EPI_ISL_1728243, EPI_ISL_1728244, EPI_ISL_1728245, EPI_ISL_1728246, EPI_ISL_1728248, EPI_ISL_1728249, EPI_ISL_1728250, EPI_ISL_1728251, EPI_ISL_1728253, EPI_ISL_1728254, EPI_ISL_1728255, EPI_ISL_1728256, EPI_ISL_1728257, EPI_ISL_1728258, EPI_ISL_1728259, EPI_ISL_1728260, EPI_ISL_1728261, EPI_ISL_1728262, EPI_ISL_1728263, EPI_ISL_1728264, EPI_ISL_1728265, EPI_ISL_1728266, EPI_ISL_1728267, EPI_ISL_1728268, EPI_ISL_1728269, EPI_ISL_1728270, EPI_ISL_1728271, EPI_ISL_1728272, EPI_ISL_1728273, EPI_ISL_1728274, EPI_ISL_1728275, EPI_ISL_1728276, EPI_ISL_1728277, EPI_ISL_1728278, EPI_ISL_1728279, EPI_ISL_1728280, EPI_ISL_1728281, EPI_ISL_1728282, EPI_ISL_1728283, EPI_ISL_1728284, EPI_ISL_1728285, EPI_ISL_1729460, EPI_ISL_1729461, EPI_ISL_1729531, EPI_ISL_1729535, EPI_ISL_1729538, EPI_ISL_1729541, EPI_ISL_1729544, EPI_ISL_1729554, EPI_ISL_1729578, EPI_ISL_1729582, EPI_ISL_1729592, EPI_ISL_1729613, EPI_ISL_1729614, EPI_ISL_1729618, EPI_ISL_1729676

see above | SYNLAB MVZ Leverkusen | Robert Koch Institute

EPI_ISL_2108089, EPI_ISL_2108110, EPI_ISL_2108143, EPI_ISL_2108168, EPI_ISL_2108172, EPI_ISL_2108233, EPI_ISL_2114597, EPI_ISL_2114604, EPI_ISL_2114625, EPI_ISL_2114633, EPI_ISL_2114657, EPI_ISL_2114659, EPI_ISL_2114670, EPI_ISL_2114695, EPI_ISL_2114700, EPI_ISL_2114742, EPI_ISL_2114805, EPI_ISL_2114811, EPI_ISL_2114833, EPI_ISL_2114839, EPI_ISL_2114844, EPI_ISL_2114869, EPI_ISL_2114874, EPI_ISL_2114923, EPI_ISL_2114945, EPI_ISL_2114964

see above | SYNLAB MVZ Neustadt a. d. Weinstrasse | Robert Koch Institute

EPI_ISL_2115103, EPI_ISL_2115127
SYNLAB MVZ Neustadt a. d. WeinstraÃŸe
Robert Koch Institute

EPI_ISL_2114997, EPI_ISL_2115011, EPI_ISL_2115024, EPI_ISL_2115060, EPI_ISL_2115062, EPI_ISL_2115069, EPI_ISL_2118836, EPI_ISL_2118878, EPI_ISL_2118951, EPI_ISL_2119004, EPI_ISL_2119050, EPI_ISL_2119098, EPI_ISL_2119145, EPI_ISL_2119207, EPI_ISL_2119372, EPI_ISL_2119377, EPI_ISL_2119421, EPI_ISL_2119501, EPI_ISL_2119550, EPI_ISL_2119600, EPI_ISL_2119891, EPI_ISL_2119969, EPI_ISL_2120018

see above
SYNLAB MVZ Neustadt a. d. Weinstraße
Robert Koch Institute

EPI_ISL_1723658, EPI_ISL_1723663, EPI_ISL_1723674, EPI_ISL_1723677, EPI_ISL_1723679, EPI_ISL_1723685, EPI_ISL_1723688, EPI_ISL_1723689, EPI_ISL_1723695, EPI_ISL_1723696, EPI_ISL_1723697, EPI_ISL_1723699, EPI_ISL_1723700, EPI_ISL_1723702, EPI_ISL_1723713, EPI_ISL_1723715, EPI_ISL_1723716, EPI_ISL_1723722, EPI_ISL_1723733, EPI_ISL_1723739, EPI_ISL_1723743, EPI_ISL_1723753, EPI_ISL_1723757, EPI_ISL_1723760, EPI_ISL_1723762, EPI_ISL_1723764, EPI_ISL_1723766, EPI_ISL_1723769, EPI_ISL_1723778, EPI_ISL_1723792, EPI_ISL_1723793, EPI_ISL_1723798, EPI_ISL_1723801, EPI_ISL_1723803, EPI_ISL_1723807, EPI_ISL_1723819, EPI_ISL_1723827, EPI_ISL_1723830, EPI_ISL_1723856, EPI_ISL_1723863, EPI_ISL_1723867, EPI_ISL_1723888, EPI_ISL_1723890, EPI_ISL_1723893, EPI_ISL_1723896, EPI_ISL_1723901, EPI_ISL_1723919, EPI_ISL_1723926, EPI_ISL_1723927, EPI_ISL_1723929, EPI_ISL_1723933, EPI_ISL_1723935, EPI_ISL_1723939, EPI_ISL_1723943, EPI_ISL_1723944, EPI_ISL_1723956, EPI_ISL_1723958, EPI_ISL_1723963, EPI_ISL_1723974, EPI_ISL_1723977, EPI_ISL_1723982, EPI_ISL_1723984, EPI_ISL_1723990, EPI_ISL_1723998, EPI_ISL_1724029, EPI_ISL_1724036, EPI_ISL_1724043, EPI_ISL_1724047, EPI_ISL_1724049, EPI_ISL_1724051, EPI_ISL_1724053, EPI_ISL_1724062, EPI_ISL_1724069, EPI_ISL_1724071, EPI_ISL_1724085, EPI_ISL_1724086, EPI_ISL_1724088, EPI_ISL_1724092, EPI_ISL_1724093, EPI_ISL_1724094, EPI_ISL_1724096, EPI_ISL_1724098, EPI_ISL_1724100, EPI_ISL_1724101, EPI_ISL_1724109, EPI_ISL_1724110, EPI_ISL_1724111, EPI_ISL_1724112, EPI_ISL_1724113, EPI_ISL_1724117, EPI_ISL_1724120, EPI_ISL_1724125, EPI_ISL_1724129, EPI_ISL_1724139, EPI_ISL_1724143, EPI_ISL_1724147, EPI_ISL_1724173, EPI_ISL_1724176, EPI_ISL_1724180, EPI_ISL_1724185, EPI_ISL_1724189, EPI_ISL_1724191, EPI_ISL_1724194, EPI_ISL_1724195, EPI_ISL_1724197, EPI_ISL_1724202, EPI_ISL_1724209, EPI_ISL_1724213, EPI_ISL_1724215, EPI_ISL_1724225, EPI_ISL_1724226, EPI_ISL_1724231, EPI_ISL_1724232, EPI_ISL_1724235, EPI_ISL_1724766, EPI_ISL_1724767, EPI_ISL_1724769, EPI_ISL_1724770, EPI_ISL_1724773, EPI_ISL_1724774, EPI_ISL_1724775, EPI_ISL_1724776, EPI_ISL_1724778, EPI_ISL_1724781, EPI_ISL_1724782, EPI_ISL_1724784, EPI_ISL_1724785, EPI_ISL_1724787, EPI_ISL_1724788, EPI_ISL_1724789, EPI_ISL_1724790, EPI_ISL_1724792, EPI_ISL_1724794, EPI_ISL_1724795, EPI_ISL_1724796, EPI_ISL_1724799, EPI_ISL_1724801, EPI_ISL_1724802, EPI_ISL_1724803, EPI_ISL_1724805, EPI_ISL_1724806, EPI_ISL_1724810, EPI_ISL_1724812, EPI_ISL_1724813, EPI_ISL_1724814, EPI_ISL_1724822, EPI_ISL_1724824, EPI_ISL_1724826, EPI_ISL_1724830, EPI_ISL_1724832, EPI_ISL_1724834, EPI_ISL_1724835, EPI_ISL_1724839, EPI_ISL_1724840, EPI_ISL_1724842, EPI_ISL_1724843, EPI_ISL_1724844, EPI_ISL_1724851, EPI_ISL_1724852, EPI_ISL_1724853, EPI_ISL_1724854, EPI_ISL_1724856, EPI_ISL_1724857, EPI_ISL_1724858, EPI_ISL_1724860, EPI_ISL_1724863, EPI_ISL_1724866, EPI_ISL_1724869, EPI_ISL_1724871, EPI_ISL_1724875, EPI_ISL_1724876, EPI_ISL_1724877, EPI_ISL_1724879, EPI_ISL_1724882, EPI_ISL_1724883, EPI_ISL_1724884, EPI_ISL_1724885, EPI_ISL_1724888, EPI_ISL_1724889, EPI_ISL_1724892, EPI_ISL_1724893, EPI_ISL_1724895, EPI_ISL_1724897, EPI_ISL_1724898, EPI_ISL_1724900, EPI_ISL_1724902, EPI_ISL_1724904, EPI_ISL_1724905, EPI_ISL_1724908, EPI_ISL_1724909, EPI_ISL_1724910, EPI_ISL_1724912, EPI_ISL_1724913, EPI_ISL_1724914, EPI_ISL_1724916, EPI_ISL_1724917, EPI_ISL_1724919, EPI_ISL_1724920, EPI_ISL_1724921, EPI_ISL_1724922, EPI_ISL_1724923, EPI_ISL_1724925, EPI_ISL_1724926, EPI_ISL_1724927, EPI_ISL_1724928, EPI_ISL_1724930, EPI_ISL_1724931, EPI_ISL_1724932, EPI_ISL_1724933, EPI_ISL_1724935, EPI_ISL_1724938, EPI_ISL_1724940, EPI_ISL_1724941, EPI_ISL_1724943, EPI_ISL_1724944, EPI_ISL_1724948, EPI_ISL_1724951, EPI_ISL_1724954, EPI_ISL_1724957, EPI_ISL_1724958, EPI_ISL_1724960, EPI_ISL_1724961, EPI_ISL_1724965, EPI_ISL_1724966, EPI_ISL_1724967, EPI_ISL_1724968, EPI_ISL_1724970, EPI_ISL_1724972, EPI_ISL_1724973, EPI_ISL_1724976, EPI_ISL_1724977, EPI_ISL_1724978, EPI_ISL_1724979, EPI_ISL_1724980, EPI_ISL_1724982, EPI_ISL_1724983, EPI_ISL_1724987, EPI_ISL_1724988, EPI_ISL_1724989, EPI_ISL_1724991, EPI_ISL_1724993, EPI_ISL_1724994, EPI_ISL_1724996, EPI_ISL_1724997, EPI_ISL_1724998, EPI_ISL_1724999, EPI_ISL_1725002, EPI_ISL_1725003, EPI_ISL_1725005, EPI_ISL_1725007, EPI_ISL_1725009, EPI_ISL_1725010, EPI_ISL_1725013, EPI_ISL_1725014, EPI_ISL_1725016, EPI_ISL_1725017, EPI_ISL_1725019, EPI_ISL_1725020, EPI_ISL_1725021, EPI_ISL_1725023, EPI_ISL_1725027, EPI_ISL_1725028, EPI_ISL_1725031, EPI_ISL_1725032, EPI_ISL_1725035, EPI_ISL_1725036, EPI_ISL_1725038, EPI_ISL_1725039, EPI_ISL_1725040, EPI_ISL_1725041, EPI_ISL_1725043, EPI_ISL_1725044, EPI_ISL_1725045, EPI_ISL_1725046, EPI_ISL_1725048, EPI_ISL_1725049, EPI_ISL_1725051, EPI_ISL_1725052, EPI_ISL_1725057, EPI_ISL_1725059, EPI_ISL_1725062, EPI_ISL_1725063, EPI_ISL_1725064, EPI_ISL_1725066, EPI_ISL_1725067, EPI_ISL_1725664, EPI_ISL_1725666, EPI_ISL_1725667, EPI_ISL_1725669, EPI_ISL_1725670, EPI_ISL_1725671, EPI_ISL_1725672, EPI_ISL_1725673, EPI_ISL_1725676, EPI_ISL_1725678, EPI_ISL_1725681, EPI_ISL_1725683, EPI_ISL_1725687, EPI_ISL_1725688, EPI_ISL_1725691, EPI_ISL_1725694, EPI_ISL_1725699, EPI_ISL_1725700, EPI_ISL_1725704, EPI_ISL_1725706, EPI_ISL_1725708, EPI_ISL_1725710, EPI_ISL_1725711, EPI_ISL_1725716, EPI_ISL_1725717, EPI_ISL_1725719, EPI_ISL_1725720, EPI_ISL_1725721, EPI_ISL_1725723, EPI_ISL_1725726, EPI_ISL_1725727, EPI_ISL_1725728, EPI_ISL_1725730, EPI_ISL_1725733, EPI_ISL_1725734, EPI_ISL_1725737, EPI_ISL_1725743, EPI_ISL_1725744, EPI_ISL_1725747, EPI_ISL_1725748, EPI_ISL_1725752, EPI_ISL_1725754, EPI_ISL_1725761, EPI_ISL_1725763, EPI_ISL_1725766, EPI_ISL_1725769, EPI_ISL_1725771, EPI_ISL_1725775, EPI_ISL_1725778, EPI_ISL_1725779, EPI_ISL_1725781, EPI_ISL_1725782, EPI_ISL_1725784, EPI_ISL_1725785, EPI_ISL_1725786, EPI_ISL_1725788, EPI_ISL_1725789, EPI_ISL_1725791, EPI_ISL_1725792, EPI_ISL_1725793, EPI_ISL_1725795, EPI_ISL_1725796, EPI_ISL_1725798, EPI_ISL_1725799, EPI_ISL_1725800, EPI_ISL_1725802, EPI_ISL_1725803, EPI_ISL_1725805, EPI_ISL_1725807, EPI_ISL_1725810, EPI_ISL_1725811, EPI_ISL_1725814, EPI_ISL_1725816, EPI_ISL_1725818, EPI_ISL_1725819, EPI_ISL_1725821, EPI_ISL_1725826, EPI_ISL_1725835, EPI_ISL_1725838, EPI_ISL_1725842, EPI_ISL_1725844, EPI_ISL_1725847, EPI_ISL_1725851, EPI_ISL_1725854, EPI_ISL_1725860, EPI_ISL_1725861, EPI_ISL_1725864, EPI_ISL_1725868, EPI_ISL_1725870, EPI_ISL_1725872, EPI_ISL_1725873, EPI_ISL_1725875, EPI_ISL_1725880, EPI_ISL_1725881, EPI_ISL_1725883, EPI_ISL_1725890, EPI_ISL_1725897, EPI_ISL_1725902, EPI_ISL_1725904, EPI_ISL_1725905, EPI_ISL_1725906, EPI_ISL_1725908, EPI_ISL_1725915, EPI_ISL_1725920, EPI_ISL_1725924, EPI_ISL_1725933, EPI_ISL_1725938, EPI_ISL_1725947, EPI_ISL_1726172, EPI_ISL_1727655, EPI_ISL_1727659, EPI_ISL_1727663, EPI_ISL_1727665, EPI_ISL_1727667, EPI_ISL_1727673, EPI_ISL_1727676, EPI_ISL_1727679, EPI_ISL_1727690, EPI_ISL_1727701, EPI_ISL_1727706, EPI_ISL_1727711, EPI_ISL_1727718, EPI_ISL_1727752, EPI_ISL_1727763, EPI_ISL_1727768, EPI_ISL_1727773, EPI_ISL_1727789, EPI_ISL_1727794, EPI_ISL_1727806, EPI_ISL_1727811, EPI_ISL_1727840, EPI_ISL_1727851, EPI_ISL_1727854, EPI_ISL_1727856, EPI_ISL_1727861, EPI_ISL_1727867, EPI_ISL_1727869, EPI_ISL_1727877, EPI_ISL_1727880, EPI_ISL_1727881, EPI_ISL_1727894, EPI_ISL_1727908, EPI_ISL_1727909, EPI_ISL_1727912, EPI_ISL_1727924, EPI_ISL_1727927, EPI_ISL_1727929, EPI_ISL_1727931, EPI_ISL_1727934, EPI_ISL_1727938, EPI_ISL_1727943, EPI_ISL_1727951, EPI_ISL_1727957, EPI_ISL_1727959, EPI_ISL_1727965, EPI_ISL_1727967, EPI_ISL_1727969, EPI_ISL_1727971, EPI_ISL_1727972, EPI_ISL_1727975, EPI_ISL_1727982, EPI_ISL_1727984, EPI_ISL_1727989, EPI_ISL_1727990, EPI_ISL_1727997, EPI_ISL_1728001, EPI_ISL_1728009, EPI_ISL_1728014, EPI_ISL_1728015, EPI_ISL_1728022, EPI_ISL_1728032, EPI_ISL_1728033, EPI_ISL_1728038, EPI_ISL_1728040, EPI_ISL_1728047, EPI_ISL_1728052, EPI_ISL_1728057, EPI_ISL_1728072, EPI_ISL_1728075, EPI_ISL_1728080, EPI_ISL_1728085, EPI_ISL_1728091, EPI_ISL_1728096, EPI_ISL_1728098, EPI_ISL_1728100, EPI_ISL_1728110, EPI_ISL_1728119, EPI_ISL_1728126, EPI_ISL_1728130, EPI_ISL_1728133, EPI_ISL_1728139, EPI_ISL_1728141, EPI_ISL_1728148, EPI_ISL_1728175, EPI_ISL_1728182, EPI_ISL_1728183, EPI_ISL_1728188, EPI_ISL_1728209, EPI_ISL_1728214, EPI_ISL_1728218, EPI_ISL_1728222, EPI_ISL_1728234, EPI_ISL_1728237, EPI_ISL_1728247, EPI_ISL_1728252, EPI_ISL_1729397, EPI_ISL_1729400, EPI_ISL_1729405, EPI_ISL_1729408, EPI_ISL_1729413, EPI_ISL_1729414, EPI_ISL_1729416, EPI_ISL_1729418, EPI_ISL_1729420, EPI_ISL_1729421, EPI_ISL_1729424, EPI_ISL_1729425, EPI_ISL_1729427, EPI_ISL_1729429, EPI_ISL_1729430, EPI_ISL_1729432, EPI_ISL_1729433, EPI_ISL_1729434, EPI_ISL_1729435, EPI_ISL_1729436, EPI_ISL_1729437, EPI_ISL_1729438, EPI_ISL_1729439, EPI_ISL_1729441, EPI_ISL_1729443, EPI_ISL_1729445, EPI_ISL_1729446, EPI_ISL_1729447, EPI_ISL_1729450, EPI_ISL_1729451, EPI_ISL_1729452, EPI_ISL_1729455, EPI_ISL_1729457, EPI_ISL_1729458, EPI_ISL_1729459, EPI_ISL_1729463, EPI_ISL_1729466, EPI_ISL_1729467, EPI_ISL_1729468, EPI_ISL_1729470, EPI_ISL_1729472, EPI_ISL_1729473, EPI_ISL_1729476, EPI_ISL_1729478, EPI_ISL_1729479, EPI_ISL_1729480, EPI_ISL_1729481, EPI_ISL_1729486, EPI_ISL_1729490, EPI_ISL_1729492, EPI_ISL_1729493, EPI_ISL_1729499, EPI_ISL_1729500, EPI_ISL_1729501, EPI_ISL_1729503, EPI_ISL_1729512, EPI_ISL_1729514, EPI_ISL_1729516, EPI_ISL_1729519, EPI_ISL_1729521, EPI_ISL_1729524, EPI_ISL_1729525, EPI_ISL_1729526, EPI_ISL_1729527, EPI_ISL_1729537, EPI_ISL_1729540, EPI_ISL_1729542, EPI_ISL_1729549, EPI_ISL_1729551, EPI_ISL_1729558, EPI_ISL_1729562, EPI_ISL_1729567, EPI_ISL_1729568, EPI_ISL_1729570, EPI_ISL_1729573, EPI_ISL_1729581, EPI_ISL_1729586, EPI_ISL_1729591, EPI_ISL_1729596, EPI_ISL_1729597, EPI_ISL_1729600, EPI_ISL_1729619, EPI_ISL_1729623, EPI_ISL_1729624, EPI_ISL_1729628, EPI_ISL_1729633, EPI_ISL_1729634, EPI_ISL_1729636, EPI_ISL_1729639, EPI_ISL_1729642, EPI_ISL_1729644, EPI_ISL_1729646, EPI_ISL_1729647, EPI_ISL_1729651, EPI_ISL_1729652, EPI_ISL_1729653, EPI_ISL_1729654, EPI_ISL_1729656, EPI_ISL_1729657, EPI_ISL_1729659, EPI_ISL_1729660, EPI_ISL_1729661, EPI_ISL_1729662, EPI_ISL_1729664, EPI_ISL_1729665, EPI_ISL_1729667, EPI_ISL_1729668, EPI_ISL_1729670, EPI_ISL_1729672, EPI_ISL_1729674, EPI_ISL_1729679, EPI_ISL_1729680, EPI_ISL_1729681, EPI_ISL_1729684, EPI_ISL_1729687, EPI_ISL_1729691, EPI_ISL_1729693, EPI_ISL_1729696, EPI_ISL_1729697, EPI_ISL_1729698, EPI_ISL_1729699, EPI_ISL_1729700, EPI_ISL_1729701, EPI_ISL_1729706

see above
SYNLAB MVZ Trier
Robert Koch Institute

EPI_ISL_1724953, EPI_ISL_1725058, EPI_ISL_1725698, EPI_ISL_1725758, EPI_ISL_1725815, EPI_ISL_1725845, EPI_ISL_1725900, EPI_ISL_1725930, EPI_ISL_1725958, EPI_ISL_1727836, EPI_ISL_1727941, EPI_ISL_1727961, EPI_ISL_1727963, EPI_ISL_1728533, EPI_ISL_1729454, EPI_ISL_1729565, EPI_ISL_1729575, EPI_ISL_1729603, EPI_ISL_1729625, EPI_ISL_1729631

see above
SYNLAB MVZ Weiden
Robert Koch Institute

EPI_ISL_1848439, EPI_ISL_1848606, EPI_ISL_1848630, EPI_ISL_1848637, EPI_ISL_2108774, EPI_ISL_2108796, EPI_ISL_2108806, EPI_ISL_2108883, EPI_ISL_2108956, EPI_ISL_2108968, EPI_ISL_2109039, EPI_ISL_2109047, EPI_ISL_2109073, EPI_ISL_2109098, EPI_ISL_2109113, EPI_ISL_2109132, EPI_ISL_2109156, EPI_ISL_2109162, EPI_ISL_2109232, EPI_ISL_2109275, EPI_ISL_2109301, EPI_ISL_2109305, EPI_ISL_2109309, EPI_ISL_2109313, EPI_ISL_2109349, EPI_ISL_2109508, EPI_ISL_2109521, EPI_ISL_2109533, EPI_ISL_2109559, EPI_ISL_2109572, EPI_ISL_2109591, EPI_ISL_2109619, EPI_ISL_2109639, EPI_ISL_2109656, EPI_ISL_2109661, EPI_ISL_2109684, EPI_ISL_2109688, EPI_ISL_2109775, EPI_ISL_2109843, EPI_ISL_2115513, EPI_ISL_2115518, EPI_ISL_2115619, EPI_ISL_2115646, EPI_ISL_2115742, EPI_ISL_2115775, EPI_ISL_2115777

see above
Sonic - Bioscientia - MVZ Labor Saar GmbH
Robert Koch Institute

EPI_ISL_2108808, EPI_ISL_2108828, EPI_ISL_2108905, EPI_ISL_2108921, EPI_ISL_2108972
Sonic - Dr. Staber & Kollegen GmbH Munchen
Robert Koch Institute

EPI_ISL_1843476, EPI_ISL_1843488, EPI_ISL_1843500, EPI_ISL_1843524, EPI_ISL_1843617, EPI_ISL_1843697, EPI_ISL_2115573, EPI_ISL_2115656, EPI_ISL_2115766

see above
Sonic - Dr. Staber & Kollegen GmbH MÃ¼nchen
Robert Koch Institute

EPI_ISL_1844079, EPI_ISL_1844088, EPI_ISL_1844117, EPI_ISL_1844158, EPI_ISL_1844194, EPI_ISL_1844224, EPI_ISL_1844230, EPI_ISL_1844231, EPI_ISL_1844233, EPI_ISL_1844242, EPI_ISL_1844244, EPI_ISL_1844258, EPI_ISL_1847451, EPI_ISL_1847457, EPI_ISL_1847458, EPI_ISL_1847464, EPI_ISL_1847470, EPI_ISL_1847473, EPI_ISL_1847477, EPI_ISL_1847479, EPI_ISL_1847482, EPI_ISL_1847486, EPI_ISL_1847492, EPI_ISL_1847494, EPI_ISL_1847498, EPI_ISL_1847499, EPI_ISL_1847504, EPI_ISL_1847517, EPI_ISL_1847522, EPI_ISL_1847523, EPI_ISL_1847539, EPI_ISL_1847545, EPI_ISL_1847547, EPI_ISL_1847549, EPI_ISL_1847551, EPI_ISL_1847554, EPI_ISL_1847564, EPI_ISL_1847565, EPI_ISL_1847566, EPI_ISL_1847567, EPI_ISL_1847569, EPI_ISL_1847573, EPI_ISL_1847574, EPI_ISL_1847576, EPI_ISL_1847577, EPI_ISL_1847582, EPI_ISL_1847583, EPI_ISL_1847587, EPI_ISL_1847588, EPI_ISL_1847596, EPI_ISL_1847607, EPI_ISL_1847609, EPI_ISL_1847616, EPI_ISL_1847638, EPI_ISL_1847643, EPI_ISL_1847656, EPI_ISL_1847662, EPI_ISL_1847664, EPI_ISL_1847673, EPI_ISL_1847674, EPI_ISL_1847676, EPI_ISL_1847686, EPI_ISL_1847694

see above
Sonic - Labor Dr. von Froreich GmbH
Robert Koch Institute

EPI_ISL_1843450, EPI_ISL_1843453, EPI_ISL_1843462, EPI_ISL_1843475, EPI_ISL_1843490, EPI_ISL_1843512, EPI_ISL_1843544, EPI_ISL_1843568, EPI_ISL_2108773, EPI_ISL_2108856, EPI_ISL_2108870, EPI_ISL_2108900, EPI_ISL_2108906, EPI_ISL_2108914, EPI_ISL_2108928, EPI_ISL_2108935

see above
Sonic - Labor Staber Bayreuth
Robert Koch Institute

EPI_ISL_1843493, EPI_ISL_1843569, EPI_ISL_1843579, EPI_ISL_1843580, EPI_ISL_1843657, EPI_ISL_1843718, EPI_ISL_1843722, EPI_ISL_2108712, EPI_ISL_2108729, EPI_ISL_2108756, EPI_ISL_2108776, EPI_ISL_2108795, EPI_ISL_2108825, EPI_ISL_2108867, EPI_ISL_2108917, EPI_ISL_2108945, EPI_ISL_2108954

see above
Sonic - Labor Staber Dresden (Klipphausen)
Robert Koch Institute

EPI_ISL_1843447, EPI_ISL_1843699, EPI_ISL_2108830, EPI_ISL_2108924
Sonic - Labor Staber Heilbronn
Robert Koch Institute

EPI_ISL_1847507, EPI_ISL_1847534
Sonic - Labor Staber MVZ Kassel
Robert Koch Institute

EPI_ISL_2108804, EPI_ISL_2108843, EPI_ISL_2108862, EPI_ISL_2108877, EPI_ISL_2108937, EPI_ISL_2108950, EPI_ISL_2108953

see above
Sonic - Labor Staber Nurnberg
Robert Koch Institute

EPI_ISL_1843443, EPI_ISL_1843466, EPI_ISL_1843596, EPI_ISL_1843651
Sonic - Labor Staber NÃ¼rnberg
Robert Koch Institute

EPI_ISL_1843461, EPI_ISL_1843463, EPI_ISL_1843465, EPI_ISL_1843467, EPI_ISL_1843468, EPI_ISL_1843471, EPI_ISL_1843479, EPI_ISL_1843482, EPI_ISL_1843483, EPI_ISL_1843503, EPI_ISL_1843505, EPI_ISL_1843518, EPI_ISL_1843525, EPI_ISL_1843529, EPI_ISL_1843540, EPI_ISL_1843550, EPI_ISL_1843554, EPI_ISL_1843555, EPI_ISL_1843556, EPI_ISL_1843559, EPI_ISL_1843589, EPI_ISL_1843609, EPI_ISL_1843621, EPI_ISL_1843622, EPI_ISL_1843624, EPI_ISL_1843629, EPI_ISL_1843633, EPI_ISL_1843634, EPI_ISL_1843641, EPI_ISL_1843643, EPI_ISL_1843647, EPI_ISL_1843652, EPI_ISL_1843653, EPI_ISL_1843654, EPI_ISL_1843671, EPI_ISL_1843676, EPI_ISL_1843678, EPI_ISL_1843680, EPI_ISL_1843686, EPI_ISL_1843692, EPI_ISL_1843696, EPI_ISL_1843711, EPI_ISL_1843715, EPI_ISL_1843727, EPI_ISL_1844048, EPI_ISL_1844050, EPI_ISL_1844051, EPI_ISL_1844053, EPI_ISL_1844054, EPI_ISL_1844055, EPI_ISL_1844058, EPI_ISL_1844059, EPI_ISL_1844060, EPI_ISL_1844063, EPI_ISL_1844065, EPI_ISL_1844066, EPI_ISL_1844068, EPI_ISL_1844070, EPI_ISL_1844071, EPI_ISL_1844072, EPI_ISL_1844073, EPI_ISL_1844075, EPI_ISL_1844076, EPI_ISL_1844078, EPI_ISL_1844081, EPI_ISL_1844085, EPI_ISL_1844086, EPI_ISL_1844091, EPI_ISL_1844092, EPI_ISL_1844093, EPI_ISL_1844094, EPI_ISL_1844095, EPI_ISL_1844097, EPI_ISL_1844098, EPI_ISL_1844099, EPI_ISL_1844103, EPI_ISL_1844105, EPI_ISL_1844106, EPI_ISL_1844107, EPI_ISL_1844109, EPI_ISL_1844111, EPI_ISL_1844114, EPI_ISL_1844115, EPI_ISL_1844123, EPI_ISL_1844128, EPI_ISL_1844130, EPI_ISL_1844131, EPI_ISL_1844132, EPI_ISL_1844136, EPI_ISL_1844137, EPI_ISL_1844144, EPI_ISL_1844147, EPI_ISL_1844148, EPI_ISL_1844150, EPI_ISL_1844152, EPI_ISL_1844155, EPI_ISL_1844156, EPI_ISL_1844160, EPI_ISL_1844162, EPI_ISL_1844164, EPI_ISL_1844169, EPI_ISL_1844174, EPI_ISL_1844179, EPI_ISL_1844180, EPI_ISL_1844181, EPI_ISL_1844182, EPI_ISL_1844184, EPI_ISL_1844187, EPI_ISL_1844188, EPI_ISL_1844195, EPI_ISL_1844199, EPI_ISL_1844200, EPI_ISL_1844202, EPI_ISL_1844203, EPI_ISL_1844205, EPI_ISL_1844209, EPI_ISL_1844212, EPI_ISL_1844213, EPI_ISL_1844215, EPI_ISL_1844216, EPI_ISL_1844217, EPI_ISL_1844219, EPI_ISL_1844221, EPI_ISL_1844223, EPI_ISL_1844234, EPI_ISL_1844235, EPI_ISL_1844236, EPI_ISL_1844239, EPI_ISL_1844241, EPI_ISL_1844245, EPI_ISL_1844246, EPI_ISL_1844247, EPI_ISL_1844252, EPI_ISL_1844253, EPI_ISL_1844254, EPI_ISL_1844259, EPI_ISL_1844261, EPI_ISL_1844264, EPI_ISL_1844266, EPI_ISL_1847448, EPI_ISL_1847449, EPI_ISL_1847450, EPI_ISL_1847452, EPI_ISL_1847453, EPI_ISL_1847455, EPI_ISL_1847456, EPI_ISL_1847459, EPI_ISL_1847461, EPI_ISL_1847462, EPI_ISL_1847465, EPI_ISL_1847468, EPI_ISL_1847469, EPI_ISL_1847471, EPI_ISL_1847474, EPI_ISL_1847476, EPI_ISL_1847478, EPI_ISL_1847480, EPI_ISL_1847483, EPI_ISL_1847491, EPI_ISL_1847493, EPI_ISL_1847495, EPI_ISL_1847501, EPI_ISL_1847502, EPI_ISL_1847505, EPI_ISL_1847508, EPI_ISL_1847509, EPI_ISL_1847510, EPI_ISL_1847511, EPI_ISL_1847512, EPI_ISL_1847515, EPI_ISL_1847516, EPI_ISL_1847518, EPI_ISL_1847519, EPI_ISL_1847520, EPI_ISL_1847521, EPI_ISL_1847524, EPI_ISL_1847525, EPI_ISL_1847527, EPI_ISL_1847532, EPI_ISL_1847535, EPI_ISL_1847536, EPI_ISL_1847538, EPI_ISL_1847540, EPI_ISL_1847542, EPI_ISL_1847548, EPI_ISL_1847550, EPI_ISL_1847553, EPI_ISL_1847555, EPI_ISL_1847558, EPI_ISL_1847562, EPI_ISL_1847571, EPI_ISL_1847575, EPI_ISL_1847580, EPI_ISL_1847584, EPI_ISL_1847590, EPI_ISL_1847591, EPI_ISL_1847592, EPI_ISL_1847593, EPI_ISL_1847597, EPI_ISL_1847600, EPI_ISL_1847601, EPI_ISL_1847602, EPI_ISL_1847605, EPI_ISL_1847610, EPI_ISL_1847615, EPI_ISL_1847617, EPI_ISL_1847619, EPI_ISL_1847622, EPI_ISL_1847625, EPI_ISL_1847626, EPI_ISL_1847629, EPI_ISL_1847632, EPI_ISL_1847633, EPI_ISL_1847637, EPI_ISL_1847640, EPI_ISL_1847642, EPI_ISL_1847644, EPI_ISL_1847645, EPI_ISL_1847649, EPI_ISL_1847650, EPI_ISL_1847652, EPI_ISL_1847653, EPI_ISL_1847657, EPI_ISL_1847661, EPI_ISL_1847663, EPI_ISL_1847667, EPI_ISL_1847668, EPI_ISL_1847669, EPI_ISL_1847671, EPI_ISL_1847672, EPI_ISL_1847684, EPI_ISL_1847685, EPI_ISL_1847687, EPI_ISL_1847688, EPI_ISL_1847692, EPI_ISL_1847693, EPI_ISL_1847696, EPI_ISL_2120950

see above
Sonic - MVZ Medizinisches Labor Bremen GmbH
Robert Koch Institute

EPI_ISL_1843444, EPI_ISL_1843594, EPI_ISL_1843610, EPI_ISL_1843630, EPI_ISL_1843639, EPI_ISL_1847618, EPI_ISL_1847624, EPI_ISL_1847639, EPI_ISL_1847670, EPI_ISL_1847680, EPI_ISL_2109174, EPI_ISL_2109286

see above
Sonic - MVZ Medizinisches Labor Oldenburg
Robert Koch Institute

EPI_ISL_1843289
Stiftung Krankenhaus Bethanien fÃ¼r die Grafschaft Moers
Robert Koch Institute

EPI_ISL_1723794, EPI_ISL_1723880, EPI_ISL_1724025, EPI_ISL_1724108, EPI_ISL_1724116, EPI_ISL_1724168, EPI_ISL_1724807, EPI_ISL_1724817, EPI_ISL_1724820, EPI_ISL_1724833, EPI_ISL_1724937, EPI_ISL_1725001, EPI_ISL_1725006, EPI_ISL_1725012, EPI_ISL_1725022, EPI_ISL_1725034, EPI_ISL_1725037, EPI_ISL_1725662, EPI_ISL_1725695, EPI_ISL_1725702, EPI_ISL_1725712, EPI_ISL_1725715, EPI_ISL_1725741, EPI_ISL_1725823, EPI_ISL_1725825, EPI_ISL_1725856, EPI_ISL_1725869, EPI_ISL_1725879, EPI_ISL_1725889, EPI_ISL_1725894, EPI_ISL_1725910, EPI_ISL_1725917, EPI_ISL_1725934, EPI_ISL_1725961, EPI_ISL_1727657, EPI_ISL_1727734, EPI_ISL_1727761, EPI_ISL_1727850, EPI_ISL_1727890, EPI_ISL_1728042, EPI_ISL_1728520, EPI_ISL_1728545, EPI_ISL_1729410, EPI_ISL_1729483, EPI_ISL_1729487, EPI_ISL_1729506, EPI_ISL_1729509, EPI_ISL_1729548, EPI_ISL_1729560, EPI_ISL_1729604, EPI_ISL_1729617, EPI_ISL_1729620, EPI_ISL_1729627, EPI_ISL_1729640

see above
Synlab MVZ Augsburg
Robert Koch Institute

EPI_ISL_2109022, EPI_ISL_2109023, EPI_ISL_2109024, EPI_ISL_2109026, EPI_ISL_2109027, EPI_ISL_2109028, EPI_ISL_2109030, EPI_ISL_2109032, EPI_ISL_2109034, EPI_ISL_2109037, EPI_ISL_2123857

see above
Thuringer Landesamtes fur Verbraucherschutz
Robert Koch Institute

EPI_ISL_1844035, EPI_ISL_1844036, EPI_ISL_1844037, EPI_ISL_1844038, EPI_ISL_1844042, EPI_ISL_1844044, EPI_ISL_1844045

see above
ThÃ¼ringer Landesamtes fÃ¼r Verbraucherschutz
Robert Koch Institute

EPI_ISL_1844027, EPI_ISL_1844028, EPI_ISL_1844030, EPI_ISL_1844033
Uniklinikum Jena
Robert Koch Institute

EPI_ISL_1843376, EPI_ISL_1843377, EPI_ISL_1851145
Uniklinikum Regensburg; Institut fÃ¼r klinische Mikrobiologie und Hygiene
Robert Koch Institute

EPI_ISL_2109856, EPI_ISL_2109858, EPI_ISL_2109859, EPI_ISL_2127218, EPI_ISL_2127219, EPI_ISL_2127221, EPI_ISL_2127225, EPI_ISL_2127227, EPI_ISL_2127229, EPI_ISL_2127231

see above
Universitatsklinikum Bonn - Institut fur Virologie

EPI_ISL_2108714, EPI_ISL_2108719, EPI_ISL_2108754, EPI_ISL_2108852, EPI_ISL_2108897, EPI_ISL_2108922, EPI_ISL_2108940, EPI_ISL_2108976

see above
Universitatsklinikum Frankfurt - Institut fur Medizinische Virologie
Robert Koch Institute

EPI_ISL_2112479
Universitatsklinikum Heidelberg
Robert Koch Institute

EPI_ISL_1862382, EPI_ISL_1862384, EPI_ISL_1862386, EPI_ISL_1862388, EPI_ISL_1862427, EPI_ISL_1862429
University Medical Center Hamburg Eppendorf
Heinrich Pette Institute, Leibniz Institute for Experimental Virology
Adam Grundhoff; Alexis Robitaille; Johannes Knobloch; Martin Aepfelbacher; Nicole Fischer; Thomas Günther

EPI_ISL_1726854, EPI_ISL_1726857, EPI_ISL_1726858
UniversitÃ¤tsklinikum MÃ¼nster Institut fÃ¼r Virologie
Robert Koch Institute

EPI_ISL_1725220, EPI_ISL_1843299, EPI_ISL_1843304, EPI_ISL_1843310, EPI_ISL_1843313, EPI_ISL_1843315, EPI_ISL_1843320, EPI_ISL_1843324, EPI_ISL_1843325

see above
UniversitÃ¤tsklinikum des Saarlandes - Institut fÃ¼r Virologie
Robert Koch Institute

| | | | |
|---|---|---|---|
| EPI_ISL_1786468, EPI_ISL_1786469, EPI_ISL_1786470, EPI_ISL_1786471, EPI_ISL_1786472, EPI_ISL_1786473, EPI_ISL_1786474, EPI_ISL_1786475, EPI_ISL_1786476, EPI_ISL_1786477, EPI_ISL_1786478, EPI_ISL_1786479, EPI_ISL_1786480, EPI_ISL_1786481, EPI_ISL_1786482, EPI_ISL_1786483, EPI_ISL_1786484, EPI_ISL_1786485, EPI_ISL_1786486, EPI_ISL_1786487, EPI_ISL_1786488, EPI_ISL_2003750, EPI_ISL_2003751 | | | |
| see above | Virology, Universitätsklinikum des Saarlandes | Epigenetics, Saarland University | Jörn Walter; Kathrin Kattler; Sascha Tierling; Sigrun Smola; Stefan Lohse; Thorsten Pfuhl |
| EPI_ISL_1991861, EPI_ISL_1991862, EPI_ISL_1991863, EPI_ISL_1991864, EPI_ISL_1991865, EPI_ISL_1991866, EPI_ISL_1991870, EPI_ISL_1991871, EPI_ISL_1991872, EPI_ISL_1991873, EPI_ISL_1991874, EPI_ISL_1991875, EPI_ISL_1991876, EPI_ISL_1991877, EPI_ISL_1991880, EPI_ISL_1991881, EPI_ISL_1991884, EPI_ISL_1991885, EPI_ISL_1991886, EPI_ISL_1991887, EPI_ISL_1991888, EPI_ISL_1991890, EPI_ISL_1991892, EPI_ISL_1991893, EPI_ISL_1991894, EPI_ISL_1991895, EPI_ISL_1991896, EPI_ISL_1991934, EPI_ISL_1991935, EPI_ISL_1991936, EPI_ISL_1991937, EPI_ISL_1991938, EPI_ISL_1991939, EPI_ISL_1991940, EPI_ISL_1991941, EPI_ISL_1991942, EPI_ISL_1991943, EPI_ISL_1991944, EPI_ISL_1991947, EPI_ISL_1991948, EPI_ISL_1991949, EPI_ISL_1991950, EPI_ISL_1991951, EPI_ISL_1991952, EPI_ISL_1991954, EPI_ISL_1991955, EPI_ISL_1991956, EPI_ISL_1991957, EPI_ISL_1991958, EPI_ISL_1991959, EPI_ISL_1991962, EPI_ISL_1991963, EPI_ISL_1991964, EPI_ISL_1991965 | | | |
| see above | ZOTZ KLIMAS MVZ Düsseldorf-Centrum GbR ÜBAG für Labormedizin, Genetik, Zytologie, Pathologie | Center of Medical Microbiology, Virology, and Hospital Hygiene, University of Duesseldorf | Alexander Dilthey; Andreas Walker; Daniel Strelow; Jessica Nicolai; Jörg Timm; Katrin Hoffmann; Klaus Pfeffer; Lisanna Hülse; Malte Kohns Vasconcelos; Maximilian Damagnez; Nadine Lübke; Patrick Finzer; Rainer Zotz; Tobias Wienemann; Torsten Houwaart |
| EPI_ISL_1726982 | alphaomega Labor GbR (Medizinische Kooperationsgemeinschaft Dres. Ackermann/Tennert/ZaiÃŸ) | Robert Koch Institute | |
| EPI_ISL_1727309, EPI_ISL_1727313, EPI_ISL_1850690 | amedes GÃ¶ttingen | Robert Koch Institute | |
| EPI_ISL_1727146, EPI_ISL_1727151, EPI_ISL_1727156, EPI_ISL_1727170, EPI_ISL_1727179, EPI_ISL_1727185, EPI_ISL_1727186, EPI_ISL_1727201, EPI_ISL_1727209, EPI_ISL_1727213, EPI_ISL_1727214, EPI_ISL_1727218, EPI_ISL_1727219, EPI_ISL_1727222, EPI_ISL_1727242, EPI_ISL_1727247, EPI_ISL_1727279, EPI_ISL_1727283, EPI_ISL_1727288, EPI_ISL_1727297, EPI_ISL_1727301, EPI_ISL_1727302, EPI_ISL_1727305, EPI_ISL_1727307, EPI_ISL_1727315, EPI_ISL_1727320, EPI_ISL_1727322, EPI_ISL_1727334, EPI_ISL_1727335, EPI_ISL_1727337, EPI_ISL_1727340, EPI_ISL_1727343, EPI_ISL_1727349, EPI_ISL_1727356, EPI_ISL_1727360, EPI_ISL_1727370, EPI_ISL_1727373, EPI_ISL_1727380, EPI_ISL_1850559 | | | |
| see above | amedes MVZ DIAMEDIS Sennestadt | Robert Koch Institute | |
| EPI_ISL_1727138, EPI_ISL_1727141, EPI_ISL_1727164, EPI_ISL_1727167, EPI_ISL_1727174, EPI_ISL_1727188, EPI_ISL_1727189, EPI_ISL_1727191, EPI_ISL_1727196, EPI_ISL_1727204, EPI_ISL_1727206, EPI_ISL_1727233, EPI_ISL_1727236, EPI_ISL_1727237, EPI_ISL_1727244, EPI_ISL_1727249, EPI_ISL_1727254, EPI_ISL_1727256, EPI_ISL_1727257, EPI_ISL_1727266, EPI_ISL_1727271, EPI_ISL_1727286, EPI_ISL_1727304, EPI_ISL_1727314, EPI_ISL_1727323, EPI_ISL_1727326, EPI_ISL_1727331, EPI_ISL_1727339, EPI_ISL_1727346, EPI_ISL_1727350, EPI_ISL_1727366, EPI_ISL_1727372 | | | |
| see above | amedes MVZ Halle/Leipzig | Robert Koch Institute | |
| EPI_ISL_1727133, EPI_ISL_1727148, EPI_ISL_1727150, EPI_ISL_1727154, EPI_ISL_1727159, EPI_ISL_1727160, EPI_ISL_1727175, EPI_ISL_1727205, EPI_ISL_1727229, EPI_ISL_1727234, EPI_ISL_1727240, EPI_ISL_1727261, EPI_ISL_1727274, EPI_ISL_1727276, EPI_ISL_1727282, EPI_ISL_1727290, EPI_ISL_1727293, EPI_ISL_1727295, EPI_ISL_1727296, EPI_ISL_1727308, EPI_ISL_1727324, EPI_ISL_1727358, EPI_ISL_1727364, EPI_ISL_1727369 | | | |
| see above | amedes MVZ Hannover | Robert Koch Institute | |
| EPI_ISL_1727125, EPI_ISL_1727129, EPI_ISL_1727134, EPI_ISL_1727147, EPI_ISL_1727158, EPI_ISL_1727178, EPI_ISL_1727183, EPI_ISL_1727187, EPI_ISL_1727193, EPI_ISL_1727207, EPI_ISL_1727220, EPI_ISL_1727223, EPI_ISL_1727226, EPI_ISL_1727241, EPI_ISL_1727316, EPI_ISL_1727338, EPI_ISL_1727351, EPI_ISL_1727363, EPI_ISL_1727368, EPI_ISL_1727377, EPI_ISL_1727378, EPI_ISL_1727379 | | | |
| see above | amedes MVZ Jena | Robert Koch Institute | |
| EPI_ISL_1727136, EPI_ISL_1727264 | amedes MVZ Zentrum fÃ¼r Pathologie und Zytodiagnostik GmbH (KÃ¶ln) | Robert Koch Institute | |
| EPI_ISL_1727128, EPI_ISL_1727132, EPI_ISL_1727166, EPI_ISL_1727194, EPI_ISL_1727211, EPI_ISL_1727245, EPI_ISL_1727253, EPI_ISL_1727260, EPI_ISL_1727270, EPI_ISL_1727284, EPI_ISL_1727298, EPI_ISL_1727318, EPI_ISL_1727329, EPI_ISL_1727345 | | | |
| see above | amedes MVZ fÃ¼r Laboratoriumsdiagnostik Raubling GmbH | Robert Koch Institute | |
| EPI_ISL_1727327 | amedes MVZ fÃ¼r Laboratoriumsmedizin FÃ¼rstenfeldbruck | Robert Koch Institute | |
| EPI_ISL_1850489 | amedes MVZ fÃ¼r Laboratoriumsmedizin Hamburg | Robert Koch Institute | |
| EPI_ISL_1727130, EPI_ISL_1727157, EPI_ISL_1727169, EPI_ISL_1727181, EPI_ISL_1727199, EPI_ISL_1727200, EPI_ISL_1727227, EPI_ISL_1727232, EPI_ISL_1727250, EPI_ISL_1727362 | | | |
| see above | amedes MVZ fÃ¼r Laboratoriumsmedizin MÃ¼nchen | Robert Koch Institute | |
| EPI_ISL_1727127, EPI_ISL_1727162, EPI_ISL_1727208, EPI_ISL_1727278 | amedes MVZ fÃ¼r Laboratoriumsmedizin Rhein/Ruhr | Robert Koch Institute | |
| EPI_ISL_1727176, EPI_ISL_1727273, EPI_ISL_1727277, EPI_ISL_1727289, EPI_ISL_1727348 | amedes MVZ fÃ¼r Labordiagnostik Rhein-Main | Robert Koch Institute | |
| EPI_ISL_1727252, EPI_ISL_1727267, EPI_ISL_1727341 | amedes Medlab Arnold Analytik MVZ GmbH | Robert Koch Institute | |
| EPI_ISL_1727123, EPI_ISL_1727124 | labopart - Medizinische Laboratorien Dresden | Robert Koch Institute | |
| EPI_ISL_1727144, EPI_ISL_1727145, EPI_ISL_1727155, EPI_ISL_1727172, EPI_ISL_1727184, EPI_ISL_1727202, EPI_ISL_1727231, EPI_ISL_1727287, EPI_ISL_1727317, EPI_ISL_1727342, EPI_ISL_1727371, EPI_ISL_1727374 | | | |
| see above | nordlab - Partnerschaftspraxis fuÌˆr Laboratoriumsmedizin | Robert Koch Institute | |

We gratefully acknowledge the following Authors from the Originating laboratories responsible for obtaining the specimens, as well as the Submitting laboratories where the genome data were generated and shared via GISAID, on which this research is based.

All Submitters of data may be contacted directly via www.gisaid.org

Authors are sorted alphabetically.

| Accession ID | Originating Laboratory | Submitting Laboratory | Authors |
|---|---|---|---|
| EPI_ISL_2111290, EPI_ISL_2111340, EPI_ISL_2111430 | Bayerisches Landesamt fur Gesundheit und Lebensmittelsicherheit (LGL) | Robert Koch Institute | |
| EPI_ISL_2129724 | Bernhard-Nocht-Institut fÃ¼r Tropenmedizin | Robert Koch Institute | |
| EPI_ISL_2108724, EPI_ISL_2108725, EPI_ISL_2108735, EPI_ISL_2108743, EPI_ISL_2108771, EPI_ISL_2108772, EPI_ISL_2108775, EPI_ISL_2108787, EPI_ISL_2108791, EPI_ISL_2108807, EPI_ISL_2108827, EPI_ISL_2108836, EPI_ISL_2108839, EPI_ISL_2108845, EPI_ISL_2108896, EPI_ISL_2108919, EPI_ISL_2108966, EPI_ISL_2109042, EPI_ISL_2109049, EPI_ISL_2109050, EPI_ISL_2109051, EPI_ISL_2109053, EPI_ISL_2109062, EPI_ISL_2109064, EPI_ISL_2109065, EPI_ISL_2109072, EPI_ISL_2109082, EPI_ISL_2109086, EPI_ISL_2109087, EPI_ISL_2109088, EPI_ISL_2109097, EPI_ISL_2109106, EPI_ISL_2109109, EPI_ISL_2109115, EPI_ISL_2109116, EPI_ISL_2109117, EPI_ISL_2109124, EPI_ISL_2109138, EPI_ISL_2109142, EPI_ISL_2109144, EPI_ISL_2109145, EPI_ISL_2109154, EPI_ISL_2109159, EPI_ISL_2109160, EPI_ISL_2109163, EPI_ISL_2109164, EPI_ISL_2109167, EPI_ISL_2109172, EPI_ISL_2109179, EPI_ISL_2109183, EPI_ISL_2109194, EPI_ISL_2109197, EPI_ISL_2109198, EPI_ISL_2109204, EPI_ISL_2109206, EPI_ISL_2109210, EPI_ISL_2109212, EPI_ISL_2109227, EPI_ISL_2109237, EPI_ISL_2109239, EPI_ISL_2109247, EPI_ISL_2109253, EPI_ISL_2109256, EPI_ISL_2109259, EPI_ISL_2109268, EPI_ISL_2109272, EPI_ISL_2109280, EPI_ISL_2109290, EPI_ISL_2109296, EPI_ISL_2109297, EPI_ISL_2109298, EPI_ISL_2109323, EPI_ISL_2109329, EPI_ISL_2109330, EPI_ISL_2109336, EPI_ISL_2109343, EPI_ISL_2109345, EPI_ISL_2109346, EPI_ISL_2109347, EPI_ISL_2109350, EPI_ISL_2109353, EPI_ISL_2109356, EPI_ISL_2109503, EPI_ISL_2109504, EPI_ISL_2109507, EPI_ISL_2109509, EPI_ISL_2109510, EPI_ISL_2109511, EPI_ISL_2109513, EPI_ISL_2109515, EPI_ISL_2109518, EPI_ISL_2109520, EPI_ISL_2109530, EPI_ISL_2109535, EPI_ISL_2109536, EPI_ISL_2109538, EPI_ISL_2109541, EPI_ISL_2109548, EPI_ISL_2109549, EPI_ISL_2109553, EPI_ISL_2109557, EPI_ISL_2109563, EPI_ISL_2109567, EPI_ISL_2109569, EPI_ISL_2109576, EPI_ISL_2109580, EPI_ISL_2109583, EPI_ISL_2109584, EPI_ISL_2109588, EPI_ISL_2109590, EPI_ISL_2109595, EPI_ISL_2109598, EPI_ISL_2109600, EPI_ISL_2109601, EPI_ISL_2109603, EPI_ISL_2109604, EPI_ISL_2109605, EPI_ISL_2109614, EPI_ISL_2109621, EPI_ISL_2109624, EPI_ISL_2109626, EPI_ISL_2109627, EPI_ISL_2109628, EPI_ISL_2109630, EPI_ISL_2109634, EPI_ISL_2109636, EPI_ISL_2109637, EPI_ISL_2109638, EPI_ISL_2109640, EPI_ISL_2109641, EPI_ISL_2109644, EPI_ISL_2109649, EPI_ISL_2109650, EPI_ISL_2109652, EPI_ISL_2109653, EPI_ISL_2109659, EPI_ISL_2109662, EPI_ISL_2109664, EPI_ISL_2109665, EPI_ISL_2109668, EPI_ISL_2109669, EPI_ISL_2109672, EPI_ISL_2109678, EPI_ISL_2109698, EPI_ISL_2109700, EPI_ISL_2109710, EPI_ISL_2109714, EPI_ISL_2109716, EPI_ISL_2109719, EPI_ISL_2109721, EPI_ISL_2109725, EPI_ISL_2109727, EPI_ISL_2109729, EPI_ISL_2109738, EPI_ISL_2109740, EPI_ISL_2109741, EPI_ISL_2109745, EPI_ISL_2109754, EPI_ISL_2109762, EPI_ISL_2109766, EPI_ISL_2109767, EPI_ISL_2109770, EPI_ISL_2109773, EPI_ISL_2109782, EPI_ISL_2109786, EPI_ISL_2109788, EPI_ISL_2109794, EPI_ISL_2109795, EPI_ISL_2109796, EPI_ISL_2109799, EPI_ISL_2109800, EPI_ISL_2109802, EPI_ISL_2109803, EPI_ISL_2109805, EPI_ISL_2109807, EPI_ISL_2109811, EPI_ISL_2109818, EPI_ISL_2109820, EPI_ISL_2109822, EPI_ISL_2109828, EPI_ISL_2109834, EPI_ISL_2109835, EPI_ISL_2109836, EPI_ISL_2109839, EPI_ISL_2109840, EPI_ISL_2109842, EPI_ISL_2109845, EPI_ISL_2109846, EPI_ISL_2109847, EPI_ISL_2109848, EPI_ISL_2109849, EPI_ISL_2109850, EPI_ISL_2109851, EPI_ISL_2115516, EPI_ISL_2115520, EPI_ISL_2115527, EPI_ISL_2115535, EPI_ISL_2115544, EPI_ISL_2115546, EPI_ISL_2115577, EPI_ISL_2115597, EPI_ISL_2115627, EPI_ISL_2115628, EPI_ISL_2115636, EPI_ISL_2115717, EPI_ISL_2115729, EPI_ISL_2115738, EPI_ISL_2115767, EPI_ISL_2115776, EPI_ISL_2115786, EPI_ISL_2120644, EPI_ISL_2120658, EPI_ISL_2120669, EPI_ISL_2120696, EPI_ISL_2120701, EPI_ISL_2120741, EPI_ISL_2120747, EPI_ISL_2120765, EPI_ISL_2120779, EPI_ISL_2120823, EPI_ISL_2120829, EPI_ISL_2120844, EPI_ISL_2120864, EPI_ISL_2120896, EPI_ISL_2120927, EPI_ISL_2120967, EPI_ISL_2121043, EPI_ISL_2121090, EPI_ISL_2121130, EPI_ISL_2121155, EPI_ISL_2121173, EPI_ISL_2121357, EPI_ISL_2121363, EPI_ISL_2121375, EPI_ISL_2121433, EPI_ISL_2121453, EPI_ISL_2121455, EPI_ISL_2121456, EPI_ISL_2121460, EPI_ISL_2121463, EPI_ISL_2121464, EPI_ISL_2121467, EPI_ISL_2121468, EPI_ISL_2121471, EPI_ISL_2121477, EPI_ISL_2121486, EPI_ISL_2121488, EPI_ISL_2121491, EPI_ISL_2121499, EPI_ISL_2121502, EPI_ISL_2121505, EPI_ISL_2121510, EPI_ISL_2121515, EPI_ISL_2121518, EPI_ISL_2121520, EPI_ISL_2121523, EPI_ISL_2121526, EPI_ISL_2121529, EPI_ISL_2121544, EPI_ISL_2121550, EPI_ISL_2121553, EPI_ISL_2121558, EPI_ISL_2121565, EPI_ISL_2121573, EPI_ISL_2121578, EPI_ISL_2121583, EPI_ISL_2121589, EPI_ISL_2121595, EPI_ISL_2121597, EPI_ISL_2121599, EPI_ISL_2121602, EPI_ISL_2121610, EPI_ISL_2121612, EPI_ISL_2121616, EPI_ISL_2121621, EPI_ISL_2121627, EPI_ISL_2121630, EPI_ISL_2121633, EPI_ISL_2121635, EPI_ISL_2121638, EPI_ISL_2121641, EPI_ISL_2121644, EPI_ISL_2121649, EPI_ISL_2121652, EPI_ISL_2121664, EPI_ISL_2121669, EPI_ISL_2121672, EPI_ISL_2121675, EPI_ISL_2121689, EPI_ISL_2121698, EPI_ISL_2121703, EPI_ISL_2121706, EPI_ISL_2121709, EPI_ISL_2121712, EPI_ISL_2121717, EPI_ISL_2121720, EPI_ISL_2121726, EPI_ISL_2121728, EPI_ISL_2121730, EPI_ISL_2121734, EPI_ISL_2121736, EPI_ISL_2121742, EPI_ISL_2121744, EPI_ISL_2121746, EPI_ISL_2121749, EPI_ISL_2121752, EPI_ISL_2121757, EPI_ISL_2121760, EPI_ISL_2121768, EPI_ISL_2121789, EPI_ISL_2121792, EPI_ISL_2121794, EPI_ISL_2121803, EPI_ISL_2121818, EPI_ISL_2121821, EPI_ISL_2121827, EPI_ISL_2121832, EPI_ISL_2121836, EPI_ISL_2121839, EPI_ISL_2121848, EPI_ISL_2121860, EPI_ISL_2121862, EPI_ISL_2121868, EPI_ISL_2121871, EPI_ISL_2121879, EPI_ISL_2121882, EPI_ISL_2121884, EPI_ISL_2121892, EPI_ISL_2121900, EPI_ISL_2121908, EPI_ISL_2121910, EPI_ISL_2121921, EPI_ISL_2121924, EPI_ISL_2121926, EPI_ISL_2121929, EPI_ISL_2121935, EPI_ISL_2121937, EPI_ISL_2121940, EPI_ISL_2121945, EPI_ISL_2121953, EPI_ISL_2121961, EPI_ISL_2121967, EPI_ISL_2121981, EPI_ISL_2121987, EPI_ISL_2121990, EPI_ISL_2121993, EPI_ISL_2121996, EPI_ISL_2122001, EPI_ISL_2122006, EPI_ISL_2122008, EPI_ISL_2122020, EPI_ISL_2122025, EPI_ISL_2122033, EPI_ISL_2122041, EPI_ISL_2122047, EPI_ISL_2122054, EPI_ISL_2122057, EPI_ISL_2122064, EPI_ISL_2122067, EPI_ISL_2122072, EPI_ISL_2122083, EPI_ISL_2122090, EPI_ISL_2122099, EPI_ISL_2122101, EPI_ISL_2122112, EPI_ISL_2122132, EPI_ISL_2122141, EPI_ISL_2122154, EPI_ISL_2122168, EPI_ISL_2122172, EPI_ISL_2122173, EPI_ISL_2122175, EPI_ISL_2122177, EPI_ISL_2122179, EPI_ISL_2122188, EPI_ISL_2122201, EPI_ISL_2122207, EPI_ISL_2122209, EPI_ISL_2122214, EPI_ISL_2122216, EPI_ISL_2122225, EPI_ISL_2122228, EPI_ISL_2122230, EPI_ISL_2122239, EPI_ISL_2122247, EPI_ISL_2122252, EPI_ISL_2122255, EPI_ISL_2122258, EPI_ISL_2122261, EPI_ISL_2122262, EPI_ISL_2122263, EPI_ISL_2122264, EPI_ISL_2122266, EPI_ISL_2122268, EPI_ISL_2122271, EPI_ISL_2122277, EPI_ISL_2122283, EPI_ISL_2122299, EPI_ISL_2122302, EPI_ISL_2122307, EPI_ISL_2122311, EPI_ISL_2125220, EPI_ISL_2125225, EPI_ISL_2125304, EPI_ISL_2125413 | | | |
| see above | Bioscientia Labor Wermsdorf | Robert Koch Institute | |
| EPI_ISL_2109084, EPI_ISL_2109110, EPI_ISL_2109175, EPI_ISL_2109262, EPI_ISL_2109294, EPI_ISL_2109502, EPI_ISL_2109512, EPI_ISL_2109524, EPI_ISL_2109526, EPI_ISL_2109539, EPI_ISL_2109542, EPI_ISL_2109547, EPI_ISL_2109561, EPI_ISL_2109566, EPI_ISL_2109571, EPI_ISL_2109574, EPI_ISL_2109585, EPI_ISL_2109589, EPI_ISL_2109594, EPI_ISL_2109597, EPI_ISL_2109599, EPI_ISL_2109608, EPI_ISL_2109616, EPI_ISL_2109620, EPI_ISL_2109645, EPI_ISL_2109647, EPI_ISL_2109655, EPI_ISL_2109658, EPI_ISL_2109673, EPI_ISL_2109680, EPI_ISL_2109731, EPI_ISL_2109733, EPI_ISL_2109734, EPI_ISL_2109742, EPI_ISL_2109764, EPI_ISL_2109765, EPI_ISL_2109771, EPI_ISL_2109778, EPI_ISL_2109779, EPI_ISL_2109785, EPI_ISL_2109792, EPI_ISL_2109809, EPI_ISL_2109819, EPI_ISL_2109841, EPI_ISL_2121458, EPI_ISL_2121461, EPI_ISL_2121466, EPI_ISL_2121534, EPI_ISL_2121537, EPI_ISL_2121570, EPI_ISL_2121581, EPI_ISL_2121678, EPI_ISL_2121683, EPI_ISL_2121754, EPI_ISL_2121776, EPI_ISL_2121797, EPI_ISL_2121829, EPI_ISL_2121854, EPI_ISL_2121932, EPI_ISL_2121978, EPI_ISL_2122081, EPI_ISL_2122093, EPI_ISL_2122115, EPI_ISL_2122138, EPI_ISL_2122166, EPI_ISL_2122193, EPI_ISL_2122227, EPI_ISL_2122233, EPI_ISL_2122260, EPI_ISL_2126135 | | | |
| see above | Bioscientia MVZ Labor Karlsruhe GmbH | Robert Koch Institute | |
| EPI_ISL_1834050, EPI_ISL_1834051, EPI_ISL_1834052, EPI_ISL_1834053, EPI_ISL_1834056, EPI_ISL_1834072, EPI_ISL_1911987 | | | |
| see above | Bundeswehr Institute of Microbiology | Bundeswehr Institute of Microbiology | Alexandra Rehn; Enrico Georgi; Malena Bestehorn-Willmann; Markus Antwerpen; Mathias Walter; Mike Pillukat; Roman Wölfel; Sabine Zange |
| EPI_ISL_1834063, EPI_ISL_1911995, EPI_ISL_1911996, EPI_ISL_1911997, EPI_ISL_1911998, EPI_ISL_1911999, EPI_ISL_1912001, EPI_ISL_2131162 | | | |
| see above | Bundeswehrkrankenhaus Berlin | Bundeswehr Institute of Microbiology | Alexandra Rehn; Enrico Georgi; Malena Bestehorn-Willmann; Markus Antwerpen; Mathias Walter; Mike Pillukat; Roman Wölfel; Sabine Zange |
| EPI_ISL_1834076, EPI_ISL_1911990, EPI_ISL_1911991 | Bundeswehrkrankenhaus Ulm | Bundeswehr Institute of Microbiology | Alexandra Rehn; Enrico Georgi; Malena Bestehorn-Willmann; Markus Antwerpen; Mathias Walter; Mike Pillukat; Roman Wölfel; Sabine Zange |
| EPI_ISL_1834077 | Bundeswehrkrankenhaus Westerstede | Bundeswehr Institute of Microbiology | Alexandra Rehn; Enrico Georgi; Malena Bestehorn-Willmann; Markus Antwerpen; Mathias Walter; Mike Pillukat; Roman Wölfel; Sabine Zange |
| EPI_ISL_1834070, EPI_ISL_1911993 | Bundeswehrzentralkrankenhaus Koblenz | Bundeswehr Institute of Microbiology | Alexandra Rehn; Enrico Georgi; Malena Bestehorn-Willmann; Markus Antwerpen; Mathias Walter; Mike Pillukat; Roman Wölfel; Sabine Zange |
| EPI_ISL_1991932, EPI_ISL_1991933, EPI_ISL_1993609, EPI_ISL_1993610, EPI_ISL_1993611, EPI_ISL_1993612, EPI_ISL_1993613 | | | |
| see above | Center of Medical Microbiology, Virology, and Hospital Hygiene, University of Duesseldorf | Center of Medical Microbiology, Virology, and Hospital Hygiene, University of Duesseldorf | Alexander Dilthey; Andreas Walker; Daniel Strelow; Jessica Nicolai; Jörg Timm; Klaus Pfeffer; Lisanna Hülse; Malte Kohns Vasconcelos; Maximilian Damagnez; Nadine Lübke; Tobias Wienemann; Torsten Houwaart |
| EPI_ISL_2108513, EPI_ISL_2108522, EPI_ISL_2108540, EPI_ISL_2108566, EPI_ISL_2123082, EPI_ISL_2123085, EPI_ISL_2123086, EPI_ISL_2123088, EPI_ISL_2123102, EPI_ISL_2123103, EPI_ISL_2123104, EPI_ISL_2123107, EPI_ISL_2123109, EPI_ISL_2123111, EPI_ISL_2123112, EPI_ISL_2123114, EPI_ISL_2123122, EPI_ISL_2123125, EPI_ISL_2123128, EPI_ISL_2123130, EPI_ISL_2123133, EPI_ISL_2123137, EPI_ISL_2123143, EPI_ISL_2123144, EPI_ISL_2123147, EPI_ISL_2123148, EPI_ISL_2123149, EPI_ISL_2123150, EPI_ISL_2123151, EPI_ISL_2123153, EPI_ISL_2123157, EPI_ISL_2123158, EPI_ISL_2123160, EPI_ISL_2123164, EPI_ISL_2123171, EPI_ISL_2123176, EPI_ISL_2123178, EPI_ISL_2123181, EPI_ISL_2123187, EPI_ISL_2123188, EPI_ISL_2123189, EPI_ISL_2123190, EPI_ISL_2123192, EPI_ISL_2123193, EPI_ISL_2123195, EPI_ISL_2123202, EPI_ISL_2123203, EPI_ISL_2123204, EPI_ISL_2123205, EPI_ISL_2123206, EPI_ISL_2123208, EPI_ISL_2123211, EPI_ISL_2123213, EPI_ISL_2123215, EPI_ISL_2123218, EPI_ISL_2123219, EPI_ISL_2123220, EPI_ISL_2123223, EPI_ISL_2123227, EPI_ISL_2123230, EPI_ISL_2123236, EPI_ISL_2123245, EPI_ISL_2123247, EPI_ISL_2123249, EPI_ISL_2123255, EPI_ISL_2123256, EPI_ISL_2123260, EPI_ISL_2123265, EPI_ISL_2123266, EPI_ISL_2123267, EPI_ISL_2123270, EPI_ISL_2123272, EPI_ISL_2123277, EPI_ISL_2123278, EPI_ISL_2123279, EPI_ISL_2123281, EPI_ISL_2123282, EPI_ISL_2123284, EPI_ISL_2123286, EPI_ISL_2123292, EPI_ISL_2123294, EPI_ISL_2123300, EPI_ISL_2123301, EPI_ISL_2123304, EPI_ISL_2123309, EPI_ISL_2123313, EPI_ISL_2123316, EPI_ISL_2123320, EPI_ISL_2123322, EPI_ISL_2123323, EPI_ISL_2123324, EPI_ISL_2123326, EPI_ISL_2123327, EPI_ISL_2123329, EPI_ISL_2123335, EPI_ISL_2123338, EPI_ISL_2123341, EPI_ISL_2123342, EPI_ISL_2123346, EPI_ISL_2123350, EPI_ISL_2123353, EPI_ISL_2123357, EPI_ISL_2123360, EPI_ISL_2123363, EPI_ISL_2123367, EPI_ISL_2123368 | | | |
| see above | Centogene; Dr. Bauer Laboratoriums GmbH | Robert Koch Institute | |
| EPI_ISL_2112945, EPI_ISL_2112948, EPI_ISL_2112949, EPI_ISL_2112951, EPI_ISL_2112953 | Chemische und Veterinaruntersuchungsamt Monsterland-Emscher-Lippe (CVUA-MEL) | Robert Koch Institute | |
| EPI_ISL_1849497 | Chemische und VeterinÃ¤runtersuchungsamt MÃ¼nsterland-Emscher-Lippe (CVUA-MEL) | Robert Koch Institute | |
| EPI_ISL_2111014, EPI_ISL_2111015, EPI_ISL_2111016, EPI_ISL_2111017, EPI_ISL_2111018, EPI_ISL_2111019, EPI_ISL_2111020, EPI_ISL_2111021, EPI_ISL_2111022, EPI_ISL_2111027, EPI_ISL_2111028, EPI_ISL_2111029, EPI_ISL_2111032, EPI_ISL_2111033, EPI_ISL_2111034, EPI_ISL_2111035, EPI_ISL_2111036, EPI_ISL_2111038, EPI_ISL_2111039, EPI_ISL_2111040, EPI_ISL_2111041, EPI_ISL_2111042, EPI_ISL_2111043, EPI_ISL_2111044, EPI_ISL_2111045, EPI_ISL_2111047, EPI_ISL_2111049, EPI_ISL_2111050, EPI_ISL_2111051, EPI_ISL_2111052, EPI_ISL_2111053, EPI_ISL_2111054, EPI_ISL_2111055, EPI_ISL_2111056, EPI_ISL_2111057, EPI_ISL_2111058, EPI_ISL_2111060, EPI_ISL_2111062, EPI_ISL_2111063, EPI_ISL_2111064, EPI_ISL_2111065, EPI_ISL_2111066, EPI_ISL_2111067, EPI_ISL_2111068, EPI_ISL_2111071, EPI_ISL_2111072, EPI_ISL_2111073, EPI_ISL_2111075, EPI_ISL_2111076, EPI_ISL_2111078, EPI_ISL_2111079, EPI_ISL_2111082, EPI_ISL_2111083, EPI_ISL_2111084, EPI_ISL_2111085, EPI_ISL_2111086, EPI_ISL_2111087, EPI_ISL_2111088, EPI_ISL_2111092, EPI_ISL_2111093, EPI_ISL_2111094, EPI_ISL_2111095, EPI_ISL_2111097, EPI_ISL_2111098, EPI_ISL_2111099, EPI_ISL_2111103, EPI_ISL_2111105, EPI_ISL_2111106, EPI_ISL_2111107, EPI_ISL_2124320 | | | |
| see above | Clemenshospital - Alexianer Clemenshospital Munster | Robert Koch Institute | |
| EPI_ISL_1844373, EPI_ISL_1844375, EPI_ISL_1844376, EPI_ISL_1844381, EPI_ISL_1844383, EPI_ISL_1844386, EPI_ISL_1844387, EPI_ISL_1844389, EPI_ISL_1844392, EPI_ISL_1844396, EPI_ISL_1844400, EPI_ISL_1844402, EPI_ISL_1844406, EPI_ISL_1844412, EPI_ISL_1844416, EPI_ISL_1844418, EPI_ISL_1844424, EPI_ISL_1844426, EPI_ISL_1844433, EPI_ISL_1844437, EPI_ISL_1844441, EPI_ISL_1844446, EPI_ISL_1844452, EPI_ISL_1844459 | | | |
| see above | Clemenshospital - Alexianer Clemenshospital MÃ¼nster | Robert Koch Institute | |
| EPI_ISL_2112744, EPI_ISL_2112745, EPI_ISL_2112747, EPI_ISL_2112749, EPI_ISL_2112750, EPI_ISL_2112752, EPI_ISL_2112753, EPI_ISL_2112755, EPI_ISL_2112757 | | | |
| see above | Corona-Testzentrum ifp Institut fur Produktqualitat GmbH | Robert Koch Institute | |
| EPI_ISL_2108981, EPI_ISL_2108985, EPI_ISL_2109001, EPI_ISL_2109002, EPI_ISL_2109008, EPI_ISL_2109013 | Department fur Labormedizin Abteilung III Bereich Molekulare Diagnostik Universitatsklinikum Halle(Saale) | Robert Koch Institute | |
| EPI_ISL_2116150, EPI_ISL_2116151, EPI_ISL_2116154, EPI_ISL_2116155, EPI_ISL_2116158, EPI_ISL_2116159, EPI_ISL_2116160, EPI_ISL_2116162, EPI_ISL_2116163, EPI_ISL_2116166, EPI_ISL_2116167, EPI_ISL_2116169, EPI_ISL_2116170, EPI_ISL_2116171, EPI_ISL_2116172, EPI_ISL_2116173, EPI_ISL_2116174, EPI_ISL_2116176, EPI_ISL_2116178, EPI_ISL_2116179, EPI_ISL_2116180, EPI_ISL_2116181, EPI_ISL_2116182, EPI_ISL_2116183, EPI_ISL_2116185, EPI_ISL_2116188, EPI_ISL_2116191, EPI_ISL_2116192, EPI_ISL_2116196, EPI_ISL_2116197, EPI_ISL_2116199, EPI_ISL_2116200, EPI_ISL_2116201, EPI_ISL_2116202, EPI_ISL_2116203, EPI_ISL_2116204, EPI_ISL_2116206, EPI_ISL_2116208, EPI_ISL_2116209, EPI_ISL_2116210, EPI_ISL_2116211, EPI_ISL_2116212, EPI_ISL_2116215, EPI_ISL_2116217, EPI_ISL_2116218, EPI_ISL_2116220, EPI_ISL_2116221, EPI_ISL_2116222, EPI_ISL_2116224, EPI_ISL_2116226, EPI_ISL_2116228, EPI_ISL_2116231, EPI_ISL_2116232, EPI_ISL_2116233, EPI_ISL_2116234, EPI_ISL_2116235, EPI_ISL_2116238, EPI_ISL_2116239, EPI_ISL_2116240, EPI_ISL_2116242, EPI_ISL_2116243, EPI_ISL_2116244, EPI_ISL_2116247, EPI_ISL_2116248, EPI_ISL_2116252, EPI_ISL_2116253, EPI_ISL_2116254, EPI_ISL_2116256, EPI_ISL_2116257, EPI_ISL_2116258, EPI_ISL_2116260, EPI_ISL_2116261, EPI_ISL_2116262, EPI_ISL_2116263, EPI_ISL_2116264, EPI_ISL_2116265, EPI_ISL_2116266, EPI_ISL_2116267, EPI_ISL_2116269, EPI_ISL_2116271, EPI_ISL_2116272, EPI_ISL_2116274, EPI_ISL_2116276, EPI_ISL_2116277, EPI_ISL_2116278, EPI_ISL_2116279, EPI_ISL_2116280, EPI_ISL_2116281, EPI_ISL_2116282, EPI_ISL_2116283, EPI_ISL_2116284, EPI_ISL_2116285, EPI_ISL_2116286, EPI_ISL_2116287, EPI_ISL_2116288, EPI_ISL_2116289, EPI_ISL_2116290, EPI_ISL_2116291, EPI_ISL_2116292, EPI_ISL_2116293, EPI_ISL_2116294, EPI_ISL_2116296, EPI_ISL_2116297, EPI_ISL_2116298, EPI_ISL_2116299, EPI_ISL_2116302, EPI_ISL_2116303, EPI_ISL_2116304, EPI_ISL_2116305, EPI_ISL_2116306, EPI_ISL_2116307, EPI_ISL_2116308, EPI_ISL_2116309, EPI_ISL_2116310, EPI_ISL_2116311, EPI_ISL_2128412, EPI_ISL_2128414, EPI_ISL_2128415, EPI_ISL_2128416, EPI_ISL_2128418, EPI_ISL_2128419, EPI_ISL_2128420, EPI_ISL_2128421, EPI_ISL_2128422, EPI_ISL_2128424, EPI_ISL_2128425, EPI_ISL_2128426, EPI_ISL_2128428, EPI_ISL_2128429, EPI_ISL_2128430, EPI_ISL_2128431, EPI_ISL_2128432, EPI_ISL_2128433, EPI_ISL_2128434, EPI_ISL_2128435, EPI_ISL_2128437, EPI_ISL_2128438, EPI_ISL_2128439, EPI_ISL_2128440, EPI_ISL_2128441, EPI_ISL_2128442, EPI_ISL_2128444, EPI_ISL_2128445, EPI_ISL_2128446, EPI_ISL_2128447, EPI_ISL_2128448, EPI_ISL_2128449, EPI_ISL_2128450, EPI_ISL_2128451, EPI_ISL_2128452, EPI_ISL_2128453, EPI_ISL_2128454, EPI_ISL_2128455, EPI_ISL_2128456, EPI_ISL_2128457, EPI_ISL_2128459, EPI_ISL_2128464, EPI_ISL_2128468, EPI_ISL_2128472, EPI_ISL_2128477, EPI_ISL_2128480, EPI_ISL_2128482, EPI_ISL_2128486, EPI_ISL_2128488, EPI_ISL_2128490, EPI_ISL_2128496, EPI_ISL_2128501, EPI_ISL_2128504, EPI_ISL_2128505, EPI_ISL_2128506, EPI_ISL_2128507, EPI_ISL_2128510, EPI_ISL_2128513, EPI_ISL_2128514, EPI_ISL_2128520, EPI_ISL_2128525, EPI_ISL_2128527, EPI_ISL_2128531, EPI_ISL_2128533, EPI_ISL_2128536, EPI_ISL_2128540, EPI_ISL_2128543, EPI_ISL_2128547, EPI_ISL_2128549, EPI_ISL_2128551, EPI_ISL_2128553, EPI_ISL_2128557, EPI_ISL_2128563, EPI_ISL_2128564, EPI_ISL_2128567, EPI_ISL_2128569, EPI_ISL_2128571, EPI_ISL_2128573, EPI_ISL_2128575, EPI_ISL_2128577, EPI_ISL_2128581, EPI_ISL_2128583, EPI_ISL_2128591, EPI_ISL_2128593, EPI_ISL_2128600, EPI_ISL_2128601, EPI_ISL_2128602, EPI_ISL_2128603, EPI_ISL_2128605, EPI_ISL_2128606, EPI_ISL_2128607, EPI_ISL_2128608, EPI_ISL_2128609, EPI_ISL_2128611, EPI_ISL_2128612, EPI_ISL_2128613, EPI_ISL_2128615, EPI_ISL_2128616, EPI_ISL_2128617, EPI_ISL_2128618, EPI_ISL_2128619, EPI_ISL_2128620, EPI_ISL_2128621, EPI_ISL_2128623, EPI_ISL_2128625, EPI_ISL_2128628, EPI_ISL_2128629, EPI_ISL_2128630, EPI_ISL_2128632, EPI_ISL_2128634, EPI_ISL_2128635, EPI_ISL_2128636 | | | |
| see above | Diagnosticum - Labor Neukirchen | Robert Koch Institute | |
| EPI_ISL_1849284, EPI_ISL_1849285, EPI_ISL_1849286, EPI_ISL_1849287, EPI_ISL_1849289, EPI_ISL_1849290, EPI_ISL_1849291, EPI_ISL_1849292, EPI_ISL_1849294, EPI_ISL_1849295, EPI_ISL_1849296, EPI_ISL_1849297, EPI_ISL_1849299, EPI_ISL_1849300, EPI_ISL_1849301, EPI_ISL_1849302, EPI_ISL_1849303, EPI_ISL_1849305, EPI_ISL_1849306, EPI_ISL_1849307, EPI_ISL_1849308, EPI_ISL_1849310, EPI_ISL_1849311, EPI_ISL_1849312, EPI_ISL_1849313, EPI_ISL_1849314, EPI_ISL_1849316, EPI_ISL_1849317, EPI_ISL_1849318, EPI_ISL_1849320, EPI_ISL_1849321, EPI_ISL_1849322, EPI_ISL_1849323, EPI_ISL_1849325 | | | |
| see above | Dianovis GmbH Greiz | Robert Koch Institute | |
| EPI_ISL_1846371, EPI_ISL_1846374, EPI_ISL_1846385, EPI_ISL_1846395, EPI_ISL_1846398, EPI_ISL_1846403, EPI_ISL_1846420, EPI_ISL_1846424 | | | |
| see above | ELBLAB GmbH Zentrum fÃ¼r LaborMedizin | Robert Koch Institute | |
| EPI_ISL_2123723, EPI_ISL_2123724, EPI_ISL_2123725, EPI_ISL_2123726, EPI_ISL_2123727, EPI_ISL_2123728, EPI_ISL_2123729, EPI_ISL_2123730, EPI_ISL_2123731, EPI_ISL_2123732, EPI_ISL_2123733, EPI_ISL_2123734, EPI_ISL_2123735, EPI_ISL_2123736, EPI_ISL_2123737, EPI_ISL_2123738, EPI_ISL_2123739, EPI_ISL_2123740, EPI_ISL_2123741, EPI_ISL_2123742, EPI_ISL_2123743, EPI_ISL_2123744, EPI_ISL_2123745, EPI_ISL_2123746, EPI_ISL_2123747, EPI_ISL_2123748, EPI_ISL_2123749, EPI_ISL_2123750, EPI_ISL_2123751, EPI_ISL_2123752, EPI_ISL_2123753, EPI_ISL_2123754, EPI_ISL_2123755, EPI_ISL_2123756, EPI_ISL_2123757, EPI_ISL_2123758, EPI_ISL_2123759, EPI_ISL_2123760, EPI_ISL_2123761, EPI_ISL_2123762, EPI_ISL_2123763, EPI_ISL_2123764, EPI_ISL_2123765, EPI_ISL_2123766, EPI_ISL_2123767, EPI_ISL_2123768, EPI_ISL_2123769, EPI_ISL_2123770, EPI_ISL_2123771, EPI_ISL_2123772, EPI_ISL_2123773, EPI_ISL_2123774, EPI_ISL_2123775, EPI_ISL_2123776, EPI_ISL_2123777, EPI_ISL_2123778, EPI_ISL_2123779, EPI_ISL_2123780, EPI_ISL_2123781, EPI_ISL_2123782, EPI_ISL_2123783, EPI_ISL_2123784, EPI_ISL_2123785, EPI_ISL_2123786, EPI_ISL_2123787, EPI_ISL_2123788, EPI_ISL_2123789, EPI_ISL_2123790, EPI_ISL_2123791, EPI_ISL_2123792, EPI_ISL_2123793, EPI_ISL_2123794, EPI_ISL_2123795, EPI_ISL_2123796, EPI_ISL_2123797, EPI_ISL_2123798, EPI_ISL_2123799, EPI_ISL_2123800, EPI_ISL_2123801, EPI_ISL_2123802, EPI_ISL_2123803, EPI_ISL_2123804, EPI_ISL_2123805, EPI_ISL_2123806, EPI_ISL_2123807 | | | |
| see above | Eurofins LifeCodexx GmbH | Robert Koch Institute | |
| EPI_ISL_1722369, EPI_ISL_1722370, EPI_ISL_1722371, EPI_ISL_1722372, EPI_ISL_1722373, EPI_ISL_1722374, EPI_ISL_1722375, EPI_ISL_1722376, EPI_ISL_1722377, EPI_ISL_1722378, EPI_ISL_1722379, EPI_ISL_1722380, EPI_ISL_1722381, EPI_ISL_1722382, EPI_ISL_1722383, EPI_ISL_1722384, EPI_ISL_1722385, EPI_ISL_1722386, EPI_ISL_1724244, EPI_ISL_1724245, EPI_ISL_1724246, EPI_ISL_1724247 | | | |
| see above | Eurofins MVZ Labor Gelsenkirchen | Robert Koch Institute | |

| | | | |
|---|---|---|---|
| EPI_ISL_2112482, EPI_ISL_2112483, EPI_ISL_2112484, EPI_ISL_2112485, EPI_ISL_2112486, EPI_ISL_2112487, EPI_ISL_2112488, EPI_ISL_2112489, EPI_ISL_2125020, EPI_ISL_2125025 | | | |
| see above | Fraunhofer-Institut fur Zelltherapie und Immunologie IZI AG Next-Generation Diagnostics | Robert Koch Institute | |
| EPI_ISL_1848037, EPI_ISL_1848095, EPI_ISL_1848144, EPI_ISL_1848164, EPI_ISL_1848261, EPI_ISL_1848333 | Hufeland Klinikum GmbH | Robert Koch Institute | |
| EPI_ISL_2108418, EPI_ISL_2108421, EPI_ISL_2108422, EPI_ISL_2108423, EPI_ISL_2108427, EPI_ISL_2108428, EPI_ISL_2108437, EPI_ISL_2108442, EPI_ISL_2108445, EPI_ISL_2108446, EPI_ISL_2108449, EPI_ISL_2108451, EPI_ISL_2108452, EPI_ISL_2108453, EPI_ISL_2108456, EPI_ISL_2108461, EPI_ISL_2108462, EPI_ISL_2108463, EPI_ISL_2108465, EPI_ISL_2108466, EPI_ISL_2108469, EPI_ISL_2108475, EPI_ISL_2109450, EPI_ISL_2109472, EPI_ISL_2109475, EPI_ISL_2109488 | | | |
| see above | IMD - Institut fur Medizinische Diagnostik Berlin-Potsdam | Robert Koch Institute | |
| EPI_ISL_2109428, EPI_ISL_2109429, EPI_ISL_2109430, EPI_ISL_2109433, EPI_ISL_2109434, EPI_ISL_2109435, EPI_ISL_2109438, EPI_ISL_2109439, EPI_ISL_2109443, EPI_ISL_2109445, EPI_ISL_2109446, EPI_ISL_2109449, EPI_ISL_2109454, EPI_ISL_2109458, EPI_ISL_2109460, EPI_ISL_2109461, EPI_ISL_2109462, EPI_ISL_2109474, EPI_ISL_2109478, EPI_ISL_2109479, EPI_ISL_2109481, EPI_ISL_2109483, EPI_ISL_2109484, EPI_ISL_2109486, EPI_ISL_2109487, EPI_ISL_2109490, EPI_ISL_2109491, EPI_ISL_2109495, EPI_ISL_2109496, EPI_ISL_2109497, EPI_ISL_2109498, EPI_ISL_2109500, EPI_ISL_2109501, EPI_ISL_2123402, EPI_ISL_2123404, EPI_ISL_2123406, EPI_ISL_2123415 | | | |
| see above | IMD - Labor Oderland | Robert Koch Institute | |
| EPI_ISL_2108424, EPI_ISL_2108429, EPI_ISL_2108432, EPI_ISL_2108439, EPI_ISL_2108440, EPI_ISL_2108441, EPI_ISL_2108454, EPI_ISL_2108467, EPI_ISL_2108474, EPI_ISL_2108476, EPI_ISL_2115371 | | | |
| see above | IMD - MVZ Labor Greifswald | Robert Koch Institute | |
| EPI_ISL_1844481, EPI_ISL_1844503, EPI_ISL_1844518, EPI_ISL_2108426, EPI_ISL_2108430, EPI_ISL_2108431, EPI_ISL_2108433, EPI_ISL_2108434, EPI_ISL_2108435, EPI_ISL_2108438, EPI_ISL_2108448, EPI_ISL_2108450, EPI_ISL_2108457, EPI_ISL_2108459, EPI_ISL_2108464, EPI_ISL_2108470, EPI_ISL_2108471, EPI_ISL_2108472, EPI_ISL_2108473, EPI_ISL_2109441, EPI_ISL_2109451, EPI_ISL_2109453, EPI_ISL_2109459, EPI_ISL_2109465, EPI_ISL_2109466, EPI_ISL_2109468, EPI_ISL_2109470, EPI_ISL_2109476, EPI_ISL_2109480, EPI_ISL_2109489, EPI_ISL_2109499, EPI_ISL_2123383, EPI_ISL_2123390 | | | |
| see above | IMD - MVZ Labor Martinsried | Robert Koch Institute | |
| EPI_ISL_2109431, EPI_ISL_2109432, EPI_ISL_2109436, EPI_ISL_2109440, EPI_ISL_2109448, EPI_ISL_2109452, EPI_ISL_2109455, EPI_ISL_2109456, EPI_ISL_2109457, EPI_ISL_2109463, EPI_ISL_2109467, EPI_ISL_2109477, EPI_ISL_2109485, EPI_ISL_2109492, EPI_ISL_2109493 | | | |
| see above | IMD - Medizinisches Labor Rostock | Robert Koch Institute | |
| EPI_ISL_1843293, EPI_ISL_1843294, EPI_ISL_1843297, EPI_ISL_1843300, EPI_ISL_1843301, EPI_ISL_1843303, EPI_ISL_1843305, EPI_ISL_1843306, EPI_ISL_1843308, EPI_ISL_1843312, EPI_ISL_1843314, EPI_ISL_1843316, EPI_ISL_1843329, EPI_ISL_1848368, EPI_ISL_1848369, EPI_ISL_1848372, EPI_ISL_1848374, EPI_ISL_1848375, EPI_ISL_1848377, EPI_ISL_1848378, EPI_ISL_1848379, EPI_ISL_1848380, EPI_ISL_1848381, EPI_ISL_1848382, EPI_ISL_1848383, EPI_ISL_1848384, EPI_ISL_1848386, EPI_ISL_1848388, EPI_ISL_1848390, EPI_ISL_1848391, EPI_ISL_1848392, EPI_ISL_1848393, EPI_ISL_1848394, EPI_ISL_1848395, EPI_ISL_1848398, EPI_ISL_1848400, EPI_ISL_1848401, EPI_ISL_1848402, EPI_ISL_2108270, EPI_ISL_2108271, EPI_ISL_2108272, EPI_ISL_2108273, EPI_ISL_2108276, EPI_ISL_2108280, EPI_ISL_2108283, EPI_ISL_2108284, EPI_ISL_2108285, EPI_ISL_2108286, EPI_ISL_2108287, EPI_ISL_2108288, EPI_ISL_2108290, EPI_ISL_2108292, EPI_ISL_2108297, EPI_ISL_2108298, EPI_ISL_2108299, EPI_ISL_2108300, EPI_ISL_2108301, EPI_ISL_2108303, EPI_ISL_2108304, EPI_ISL_2108306, EPI_ISL_2108308, EPI_ISL_2108310, EPI_ISL_2108311, EPI_ISL_2108314, EPI_ISL_2108315, EPI_ISL_2108317, EPI_ISL_2108318, EPI_ISL_2108319 | | | |
| see above | IMD Labor Frankfurt | Robert Koch Institute | |
| EPI_ISL_2123421, EPI_ISL_2123422, EPI_ISL_2123423 | Institut fur Klinische Transfusionsmedizin und Immungenetik DRK Blutspendedienst Baden-Wurttemberg | Robert Koch Institute | |
| EPI_ISL_2122765, EPI_ISL_2122777, EPI_ISL_2122778, EPI_ISL_2122783 | Institut fur Medizinische Mikrobiologie - Universitatsmedizin Gottingen | Robert Koch Institute | |
| EPI_ISL_1843318, EPI_ISL_1843323, EPI_ISL_1848371, EPI_ISL_1848376 | Institut fÃ¼r Immunologie und Genetik Kaiserslautern: Medizinisches Labor Dr. med. B. Thiele | Robert Koch Institute | |
| EPI_ISL_2112718, EPI_ISL_2112720 | Institut fÃ¼r med. Mikrobiologie u. Virologie Klinikum Oldenburg | Robert Koch Institute | |
| EPI_ISL_1941002, EPI_ISL_1941003, EPI_ISL_1941004, EPI_ISL_1941005, EPI_ISL_1941006, EPI_ISL_1941007, EPI_ISL_1941008, EPI_ISL_1941009, EPI_ISL_1941010, EPI_ISL_1941011, EPI_ISL_1941012, EPI_ISL_1941013, EPI_ISL_1941014, EPI_ISL_1941015, EPI_ISL_1941016, EPI_ISL_1941063, EPI_ISL_1941064, EPI_ISL_1941065, EPI_ISL_1941112, EPI_ISL_1941113, EPI_ISL_1941160, EPI_ISL_1941161, EPI_ISL_1941162, EPI_ISL_1941163, EPI_ISL_1941164, EPI_ISL_1941165, EPI_ISL_1941166, EPI_ISL_1941167, EPI_ISL_1941168, EPI_ISL_1941169, EPI_ISL_1941218, EPI_ISL_1941219, EPI_ISL_1941220, EPI_ISL_1941221, EPI_ISL_1941222, EPI_ISL_1941223, EPI_ISL_1941224, EPI_ISL_1941225, EPI_ISL_1941226, EPI_ISL_1941227, EPI_ISL_1941228, EPI_ISL_1941229, EPI_ISL_1941230, EPI_ISL_1941275, EPI_ISL_1941276 | | | |
| see above | Institute of Medical Microbiology and Hospital Hygiene | Institute of Medical Microbiology and Hospital Hygiene | Aljoscha Tersteegen; Prof. Dr. Achim Kaasch |
| EPI_ISL_1805413, EPI_ISL_1805418, EPI_ISL_1805419, EPI_ISL_1805420, EPI_ISL_1805421, EPI_ISL_1805422, EPI_ISL_1805423, EPI_ISL_1805424, EPI_ISL_1805425, EPI_ISL_1805426, EPI_ISL_1805427, EPI_ISL_1805428, EPI_ISL_1805429, EPI_ISL_1828786, EPI_ISL_1960944, EPI_ISL_1960947 | | | |
| see above | Institute of Tropical Medicine | Institute of Tropical Medicine | Prof. Dr. Thirumalaisamy P. Velavan and Prof. Dr. Peter Kremsner |
| EPI_ISL_1805048, EPI_ISL_1805049 | Institute of Virology, Medical Center, University of Freiburg, Freiburg, Germany | Institute of Virology, Clinial Virus Genomics, Medical Center, University of Freiburg, Freiburg, Germany | Hajo Grundmann; Jonas Fuchs; Lena Jaki; Lisa Kern; Marcus Panning; Sandra Reuter |
| EPI_ISL_1862446, EPI_ISL_1862448, EPI_ISL_1862450, EPI_ISL_1862452, EPI_ISL_1862454 | Kath. Marienkrankenhaus | Heinrich Pette Institute, Leibniz Institute for Experimental Virology | Adam Grundhoff; Alexis Robitaille; Johannes Knobloch; Martin Aepfelbacher; Nicole Fischer; Thomas Günther |
| EPI_ISL_1843264 | Kath. St.-Johannes-Gesellschaft Dortmund gGmbH | Robert Koch Institute | |
| EPI_ISL_1846770, EPI_ISL_1846771, EPI_ISL_1847208, EPI_ISL_1847437 | Kerckhoff-Klinik GmbH | Robert Koch Institute | |
| EPI_ISL_1726922, EPI_ISL_1726923, EPI_ISL_1726924, EPI_ISL_1726928, EPI_ISL_1726929, EPI_ISL_1726930, EPI_ISL_1726931, EPI_ISL_1726932, EPI_ISL_1726933, EPI_ISL_1726935, EPI_ISL_1726936, EPI_ISL_1844270, EPI_ISL_1844271, EPI_ISL_1844272, EPI_ISL_1844273, EPI_ISL_1844274, EPI_ISL_1844275, EPI_ISL_1844276, EPI_ISL_1844278, EPI_ISL_1844279, EPI_ISL_1844280, EPI_ISL_1844281, EPI_ISL_1844282, EPI_ISL_1849326, EPI_ISL_1849327, EPI_ISL_1849328, EPI_ISL_1849330, EPI_ISL_1849331, EPI_ISL_1849332, EPI_ISL_1849334, EPI_ISL_1849335, EPI_ISL_1849336, EPI_ISL_1849337, EPI_ISL_1849338, EPI_ISL_1849340, EPI_ISL_1849341, EPI_ISL_1849344, EPI_ISL_1849345, EPI_ISL_1849346, EPI_ISL_1849347 | | | |
| see above | Klinikum Ernst von Bergmann gemeinnÃ¼tzige GmbH - stationÃ¤rer Bereich | Robert Koch Institute | |
| EPI_ISL_2115539, EPI_ISL_2115555, EPI_ISL_2115736, EPI_ISL_2115789 | Klinikum Fulda gAG | Robert Koch Institute | |
| EPI_ISL_2111434 | Klinikum Stuttgart | Robert Koch Institute | |
| EPI_ISL_1847181, EPI_ISL_1847431 | Krankenhausgesellschaft St. Vincenz mbH | Robert Koch Institute | |
| EPI_ISL_1846392, EPI_ISL_1846396, EPI_ISL_1846411, EPI_ISL_1846413, EPI_ISL_1846429, EPI_ISL_1846437, EPI_ISL_1846438, EPI_ISL_1846444 | | | |
| see above | LABOR CHEMNITZ â€“ Zentrum fÃ¼r Diagnostik am Klinikum Chemnitz | Robert Koch Institute | |
| EPI_ISL_2108713, EPI_ISL_2108715, EPI_ISL_2108716, EPI_ISL_2108721, EPI_ISL_2108722, EPI_ISL_2108728, EPI_ISL_2108730, EPI_ISL_2108731, EPI_ISL_2108732, EPI_ISL_2108738, EPI_ISL_2108741, EPI_ISL_2108742, EPI_ISL_2108747, EPI_ISL_2108748, EPI_ISL_2108750, EPI_ISL_2108753, EPI_ISL_2108755, EPI_ISL_2108758, EPI_ISL_2108759, EPI_ISL_2108760, EPI_ISL_2108761, EPI_ISL_2108765, EPI_ISL_2108766, EPI_ISL_2108767, EPI_ISL_2108769, EPI_ISL_2108782, EPI_ISL_2108784, EPI_ISL_2108785, EPI_ISL_2108789, EPI_ISL_2108793, EPI_ISL_2108800, EPI_ISL_2108802, EPI_ISL_2108803, EPI_ISL_2108805, EPI_ISL_2108809, EPI_ISL_2108813, EPI_ISL_2108814, EPI_ISL_2108815, EPI_ISL_2108819, EPI_ISL_2108820, EPI_ISL_2108822, EPI_ISL_2108823, EPI_ISL_2108826, EPI_ISL_2108833, EPI_ISL_2108835, EPI_ISL_2108837, EPI_ISL_2108838, EPI_ISL_2108842, EPI_ISL_2108847, EPI_ISL_2108849, EPI_ISL_2108851, EPI_ISL_2108858, EPI_ISL_2108859, EPI_ISL_2108865, EPI_ISL_2108866, EPI_ISL_2108868, EPI_ISL_2108872, EPI_ISL_2108873, EPI_ISL_2108874, EPI_ISL_2108876, EPI_ISL_2108878, EPI_ISL_2108880, EPI_ISL_2108884, EPI_ISL_2108885, EPI_ISL_2108887, EPI_ISL_2108889, EPI_ISL_2108891, EPI_ISL_2108892, EPI_ISL_2108893, EPI_ISL_2108904, EPI_ISL_2108907, EPI_ISL_2108910, EPI_ISL_2108911, EPI_ISL_2108915, EPI_ISL_2108916, EPI_ISL_2108918, EPI_ISL_2108926, EPI_ISL_2108929, EPI_ISL_2108931, EPI_ISL_2108932, EPI_ISL_2108933, EPI_ISL_2108934, EPI_ISL_2108938, EPI_ISL_2108939, EPI_ISL_2108941, EPI_ISL_2108946, EPI_ISL_2108947, EPI_ISL_2108948, EPI_ISL_2108951, EPI_ISL_2108952, EPI_ISL_2108958, EPI_ISL_2108959, EPI_ISL_2108960, EPI_ISL_2108961, EPI_ISL_2108962, EPI_ISL_2108964, EPI_ISL_2108965, EPI_ISL_2108967, EPI_ISL_2108971, EPI_ISL_2108973, EPI_ISL_2108974, EPI_ISL_2109076, EPI_ISL_2109077, EPI_ISL_2109090, EPI_ISL_2109101, EPI_ISL_2109102, EPI_ISL_2109108, EPI_ISL_2109135, EPI_ISL_2109171, EPI_ISL_2109180, EPI_ISL_2109195, EPI_ISL_2109219, EPI_ISL_2109254, EPI_ISL_2109261, EPI_ISL_2109264, EPI_ISL_2109267, EPI_ISL_2109282, EPI_ISL_2109341 | | | |
| see above | LabKom - Labor Augsburg MVZ GmbH | Robert Koch Institute | |
| EPI_ISL_2109081, EPI_ISL_2109100, EPI_ISL_2109148, EPI_ISL_2109224, EPI_ISL_2109292, EPI_ISL_2109295, EPI_ISL_2109327 | | | |
| see above | LabKom - Labor Hamburg-Lubeck | Robert Koch Institute | |
| EPI_ISL_2115524, EPI_ISL_2115525, EPI_ISL_2115532, EPI_ISL_2115543, EPI_ISL_2115547, EPI_ISL_2115548, EPI_ISL_2115570, EPI_ISL_2115583, EPI_ISL_2115596, EPI_ISL_2115599, EPI_ISL_2115610, EPI_ISL_2115615, EPI_ISL_2115631, EPI_ISL_2115637, EPI_ISL_2115638, EPI_ISL_2115640, EPI_ISL_2115642, EPI_ISL_2115643, EPI_ISL_2115658, EPI_ISL_2115665, EPI_ISL_2115688, EPI_ISL_2115692, EPI_ISL_2115698, EPI_ISL_2115702, EPI_ISL_2115707, EPI_ISL_2115709, EPI_ISL_2115715, EPI_ISL_2115719, EPI_ISL_2115739, EPI_ISL_2115743, EPI_ISL_2115759, EPI_ISL_2115764, EPI_ISL_2115768, EPI_ISL_2115771, EPI_ISL_2115774, EPI_ISL_2115784, EPI_ISL_2115800 | | | |
| see above | LabKom - Labor Hannover MVZ GmbH | Robert Koch Institute | |
| EPI_ISL_2109260 | LabKom - Labor Mainz MVZ GmbH | Robert Koch Institute | |
| EPI_ISL_2109040, EPI_ISL_2109046, EPI_ISL_2109057, EPI_ISL_2109074, EPI_ISL_2109095, EPI_ISL_2109103, EPI_ISL_2109104, EPI_ISL_2109107, EPI_ISL_2109126, EPI_ISL_2109127, EPI_ISL_2109140, EPI_ISL_2109152, EPI_ISL_2109155, EPI_ISL_2109165, EPI_ISL_2109168, EPI_ISL_2109178, EPI_ISL_2109181, EPI_ISL_2109188, EPI_ISL_2109189, EPI_ISL_2109192, EPI_ISL_2109205, EPI_ISL_2109214, EPI_ISL_2109216, EPI_ISL_2109221, EPI_ISL_2109230, EPI_ISL_2109231, EPI_ISL_2109242, EPI_ISL_2109243, EPI_ISL_2109244, EPI_ISL_2109245, EPI_ISL_2109255, EPI_ISL_2109257, EPI_ISL_2109258, EPI_ISL_2109270, EPI_ISL_2109308, EPI_ISL_2109312, EPI_ISL_2109314, EPI_ISL_2109319, EPI_ISL_2109332, EPI_ISL_2109357, EPI_ISL_2115530, EPI_ISL_2115541, EPI_ISL_2115568, EPI_ISL_2115575 | | | |
| see above | LabKom - Labor an der Salzbrucke MVZ GmbH | Robert Koch Institute | |
| EPI_ISL_2115612, EPI_ISL_2115616, EPI_ISL_2115620, EPI_ISL_2115622, EPI_ISL_2115645, EPI_ISL_2115647, EPI_ISL_2115652, EPI_ISL_2115657, EPI_ISL_2115662, EPI_ISL_2115670, EPI_ISL_2115671, EPI_ISL_2115672, EPI_ISL_2115684, EPI_ISL_2115690, EPI_ISL_2115727, EPI_ISL_2115730, EPI_ISL_2115756, EPI_ISL_2115761, EPI_ISL_2115769, EPI_ISL_2115797, EPI_ISL_2115798 | | | |
| see above | LabKom - Labor an der SalzbrÃ¼cke MVZ GmbH | Robert Koch Institute | |
| EPI_ISL_2115512, EPI_ISL_2115517, EPI_ISL_2115519, EPI_ISL_2115523, EPI_ISL_2115534, EPI_ISL_2115537, EPI_ISL_2115542, EPI_ISL_2115553, EPI_ISL_2115556, EPI_ISL_2115558, EPI_ISL_2115561, EPI_ISL_2115563, EPI_ISL_2115567, EPI_ISL_2115569, EPI_ISL_2115571, EPI_ISL_2115574, EPI_ISL_2115579, EPI_ISL_2115580, EPI_ISL_2115582, EPI_ISL_2115584, EPI_ISL_2115586, EPI_ISL_2115587, EPI_ISL_2115591, EPI_ISL_2115593, EPI_ISL_2115600, EPI_ISL_2115601, EPI_ISL_2115603, EPI_ISL_2115604, EPI_ISL_2115605, EPI_ISL_2115606, EPI_ISL_2115608, EPI_ISL_2115617, EPI_ISL_2115618, EPI_ISL_2115623, EPI_ISL_2115625, EPI_ISL_2115635, EPI_ISL_2115644, EPI_ISL_2115659, EPI_ISL_2115667, EPI_ISL_2115673, EPI_ISL_2115674, EPI_ISL_2115676, EPI_ISL_2115677, EPI_ISL_2115678, EPI_ISL_2115679, EPI_ISL_2115680, EPI_ISL_2115687, EPI_ISL_2115689, EPI_ISL_2115693, EPI_ISL_2115699, EPI_ISL_2115701, EPI_ISL_2115706, EPI_ISL_2115710, EPI_ISL_2115713, EPI_ISL_2115714, EPI_ISL_2115716, EPI_ISL_2115718, EPI_ISL_2115720, EPI_ISL_2115725, EPI_ISL_2115732, EPI_ISL_2115733, EPI_ISL_2115735, EPI_ISL_2115741, EPI_ISL_2115747, EPI_ISL_2115749, EPI_ISL_2115750, EPI_ISL_2115753, EPI_ISL_2115755, EPI_ISL_2115757, EPI_ISL_2115773, EPI_ISL_2115778, EPI_ISL_2115780, EPI_ISL_2115783, EPI_ISL_2115785, EPI_ISL_2115788, EPI_ISL_2115791, EPI_ISL_2115792, EPI_ISL_2115793, EPI_ISL_2115795, EPI_ISL_2115804, EPI_ISL_2115805, EPI_ISL_2115806, EPI_ISL_2115807 | | | |
| see above | LabKom - MVZ Labor Bochum MLB GmbH | Robert Koch Institute | |
| EPI_ISL_2109041, EPI_ISL_2109044, EPI_ISL_2109091, EPI_ISL_2109105, EPI_ISL_2109111, EPI_ISL_2109121, EPI_ISL_2109147, EPI_ISL_2109158, EPI_ISL_2109161, EPI_ISL_2109170, EPI_ISL_2109184, EPI_ISL_2109208, EPI_ISL_2109222, EPI_ISL_2109223, EPI_ISL_2109271, EPI_ISL_2109274, EPI_ISL_2109285, EPI_ISL_2109287, EPI_ISL_2109304, EPI_ISL_2109320, EPI_ISL_2109334, EPI_ISL_2109339, EPI_ISL_2121531, EPI_ISL_2121560, EPI_ISL_2121605, EPI_ISL_2121619, EPI_ISL_2121661, EPI_ISL_2121686, EPI_ISL_2121800, EPI_ISL_2121845, EPI_ISL_2121956, EPI_ISL_2121984, EPI_ISL_2122012, EPI_ISL_2122017, EPI_ISL_2122028, EPI_ISL_2122070, EPI_ISL_2122075, EPI_ISL_2122110, EPI_ISL_2122212, EPI_ISL_2122242, EPI_ISL_2122257, EPI_ISL_2122259, EPI_ISL_2122305, EPI_ISL_2125605 | | | |
| see above | Labor 28 MVZ GmbH | Robert Koch Institute | |
| EPI_ISL_1840789, EPI_ISL_1840790, EPI_ISL_1840791, EPI_ISL_1840793, EPI_ISL_1840794, EPI_ISL_1840795, EPI_ISL_1840796, EPI_ISL_1840799, EPI_ISL_1840800, EPI_ISL_1840802, EPI_ISL_1840803, EPI_ISL_1840805, EPI_ISL_1840806, EPI_ISL_1840807, EPI_ISL_1840808, EPI_ISL_1840809, EPI_ISL_1840810, EPI_ISL_1840813, EPI_ISL_1840814, EPI_ISL_1840815, EPI_ISL_1840816, EPI_ISL_1840817, EPI_ISL_1840818, EPI_ISL_1840819, EPI_ISL_1840820, EPI_ISL_1840821, EPI_ISL_1840823, EPI_ISL_1840824, EPI_ISL_1840825, EPI_ISL_1840826, EPI_ISL_1840827, EPI_ISL_1840829, EPI_ISL_1840830, EPI_ISL_1840831, EPI_ISL_1840832, EPI_ISL_1840833, EPI_ISL_1840834, EPI_ISL_1840835, EPI_ISL_1840838, EPI_ISL_1840839, EPI_ISL_1840840, EPI_ISL_1840841, EPI_ISL_1840842, EPI_ISL_1840843, EPI_ISL_1840844, EPI_ISL_1840845, EPI_ISL_1840846, EPI_ISL_1840848, EPI_ISL_1840849, EPI_ISL_1840850, EPI_ISL_1840851, EPI_ISL_1840852, EPI_ISL_1840853, EPI_ISL_1840854, EPI_ISL_1840857, EPI_ISL_1840858, EPI_ISL_1840859, EPI_ISL_1886170, EPI_ISL_2107835, EPI_ISL_2107836, EPI_ISL_2107846, EPI_ISL_2107853, EPI_ISL_2107861, EPI_ISL_2107882, EPI_ISL_2107885 | | | |
| see above | Labor Berlin Charité Vivantes GmbH / Institut für Virologie | Charité Universitätsmedizin Berlin, Institut für Virologie/Labor Berlin | Barbara Mühlemann; Christian Drosten; Christine Stephan; Peter Menzel; Rolf Schwarzer; Terry Jones; Victor M Corman |
| EPI_ISL_1862440, EPI_ISL_1862442, EPI_ISL_1862444 | Labor Dr. Fenner und Kollegen | Heinrich Pette Institute, Leibniz Institute for Experimental Virology | Adam Grundhoff; Alexis Robitaille; Johannes Knobloch; Martin Aepfelbacher; Nicole Fischer; Thomas Günther |
| EPI_ISL_1844613, EPI_ISL_1844616, EPI_ISL_1844618, EPI_ISL_1844620, EPI_ISL_1844622, EPI_ISL_1844626, EPI_ISL_1844637, EPI_ISL_1851387, EPI_ISL_1851388, EPI_ISL_1851389, EPI_ISL_1851390, EPI_ISL_1851391, EPI_ISL_1851392, EPI_ISL_1851393, EPI_ISL_1851395, EPI_ISL_1851396, EPI_ISL_1851397, EPI_ISL_1851398, EPI_ISL_1851400, EPI_ISL_1851401, EPI_ISL_1851402, EPI_ISL_1851403, EPI_ISL_1851404, EPI_ISL_1851406, EPI_ISL_1851407, EPI_ISL_1851408, EPI_ISL_1851409, EPI_ISL_1851411, EPI_ISL_1851412, EPI_ISL_1851413, EPI_ISL_1851414, EPI_ISL_1851415 | | | |
| see above | Labor Dr. Heidrich & Kollegen MVZ GmbH Hamburg | Robert Koch Institute | |
| EPI_ISL_2115557, EPI_ISL_2115578, EPI_ISL_2115629, EPI_ISL_2120672, EPI_ISL_2120676, EPI_ISL_2120680, EPI_ISL_2120687, EPI_ISL_2121006, EPI_ISL_2121018, EPI_ISL_2121136, EPI_ISL_2121189, EPI_ISL_2121214, EPI_ISL_2121226, EPI_ISL_2121272, EPI_ISL_2121298, EPI_ISL_2121304, EPI_ISL_2121328, EPI_ISL_2121366, EPI_ISL_2121389, EPI_ISL_2121435, EPI_ISL_2121442, EPI_ISL_2125464, EPI_ISL_2125626, EPI_ISL_2125660, EPI_ISL_2126009, EPI_ISL_2126143, EPI_ISL_2126182 | | | |

| | | | |
|---|---|---|---|
| see above | Labor Dr. Schumacher MVZ | Robert Koch Institute | |
| EPI_ISL_2125118, EPI_ISL_2125124, EPI_ISL_2125128, EPI_ISL_2125129, EPI_ISL_2125132, EPI_ISL_2125134, EPI_ISL_2125137 | | | |
| see above | Labor Dr. Spranger | Robert Koch Institute | |
| EPI_ISL_2111026 | Labor Dr. Wisplinghoff - Kaln | Robert Koch Institute | |
| EPI_ISL_1850332, EPI_ISL_1850334, EPI_ISL_1850336 | Labor Dr. Wisplinghoff - KÃ¶ln | Robert Koch Institute | |
| EPI_ISL_2108352, EPI_ISL_2108391, EPI_ISL_2108406, EPI_ISL_2108410 | Labor Lubeck bzw. Laborarztliche Gemeinschaftspraxis Lubeck | Robert Koch Institute | |
| EPI_ISL_1850357, EPI_ISL_1850364, EPI_ISL_1850382, EPI_ISL_1850385, EPI_ISL_1850388, EPI_ISL_1850430 | Labor LÃ¼beck bzw. LaborÃ¤rztliche Gemeinschaftspraxis LÃ¼beck | Robert Koch Institute | |
| EPI_ISL_1847312, EPI_ISL_1847315, EPI_ISL_1847316, EPI_ISL_1847320, EPI_ISL_1847328, EPI_ISL_1848744, EPI_ISL_1848745, EPI_ISL_1848746, EPI_ISL_1848748, EPI_ISL_1848750, EPI_ISL_1848751, EPI_ISL_1848753, EPI_ISL_1848754, EPI_ISL_1848755, EPI_ISL_1848756, EPI_ISL_1848757, EPI_ISL_1848759, EPI_ISL_1848760, EPI_ISL_1848761, EPI_ISL_1848763, EPI_ISL_1848767, EPI_ISL_1848768, EPI_ISL_1848769, EPI_ISL_1848772, EPI_ISL_1848774, EPI_ISL_1848776, EPI_ISL_1848777, EPI_ISL_1848778, EPI_ISL_1848782, EPI_ISL_1849241, EPI_ISL_1849243, EPI_ISL_1849244, EPI_ISL_1849245, EPI_ISL_1849246, EPI_ISL_1849248, EPI_ISL_1849249, EPI_ISL_1849250, EPI_ISL_1849251, EPI_ISL_1849253, EPI_ISL_1849254, EPI_ISL_1849255, EPI_ISL_1849256, EPI_ISL_1849258, EPI_ISL_1849259, EPI_ISL_1849260, EPI_ISL_1849261, EPI_ISL_1849263, EPI_ISL_1849264, EPI_ISL_1849265, EPI_ISL_1849266, EPI_ISL_1849268, EPI_ISL_1849269, EPI_ISL_1849270, EPI_ISL_1849271, EPI_ISL_1849273, EPI_ISL_1849274, EPI_ISL_1849275, EPI_ISL_1849276, EPI_ISL_1849278, EPI_ISL_1849279, EPI_ISL_1849280, EPI_ISL_1849281, EPI_ISL_1849283, EPI_ISL_1852087, EPI_ISL_1852129, EPI_ISL_1852486 | | | |
| see above | Labor MÃ¶nchengladbach MVZ Dr. Stein + Kollegen GbR | Robert Koch Institute | |
| EPI_ISL_2111439, EPI_ISL_2111440, EPI_ISL_2111441, EPI_ISL_2111443, EPI_ISL_2111444, EPI_ISL_2111445, EPI_ISL_2111446, EPI_ISL_2111447, EPI_ISL_2111449, EPI_ISL_2111450, EPI_ISL_2111451, EPI_ISL_2111452, EPI_ISL_2111453, EPI_ISL_2111455, EPI_ISL_2111456, EPI_ISL_2111457, EPI_ISL_2111458, EPI_ISL_2111459, EPI_ISL_2111460, EPI_ISL_2111461, EPI_ISL_2111462, EPI_ISL_2111463, EPI_ISL_2111465, EPI_ISL_2111466, EPI_ISL_2111467, EPI_ISL_2111468, EPI_ISL_2111469, EPI_ISL_2111470, EPI_ISL_2111471, EPI_ISL_2111472, EPI_ISL_2111473, EPI_ISL_2111474, EPI_ISL_2111475, EPI_ISL_2111476, EPI_ISL_2111478, EPI_ISL_2111480, EPI_ISL_2111481, EPI_ISL_2111482, EPI_ISL_2111483, EPI_ISL_2111484, EPI_ISL_2111485, EPI_ISL_2111486, EPI_ISL_2111487, EPI_ISL_2111488, EPI_ISL_2111489, EPI_ISL_2111490, EPI_ISL_2111491, EPI_ISL_2111492, EPI_ISL_2111494, EPI_ISL_2111495, EPI_ISL_2111496, EPI_ISL_2111497, EPI_ISL_2111499, EPI_ISL_2111500, EPI_ISL_2111501, EPI_ISL_2111502, EPI_ISL_2111503, EPI_ISL_2111504, EPI_ISL_2111505, EPI_ISL_2111506, EPI_ISL_2111507, EPI_ISL_2111508, EPI_ISL_2111509, EPI_ISL_2111510, EPI_ISL_2111511, EPI_ISL_2111512, EPI_ISL_2111513, EPI_ISL_2111514, EPI_ISL_2111515, EPI_ISL_2111516, EPI_ISL_2111517, EPI_ISL_2111518, EPI_ISL_2111519, EPI_ISL_2111521, EPI_ISL_2111522, EPI_ISL_2111523, EPI_ISL_2111524, EPI_ISL_2111525, EPI_ISL_2111526, EPI_ISL_2111527, EPI_ISL_2111528, EPI_ISL_2111529, EPI_ISL_2111530, EPI_ISL_2111531, EPI_ISL_2111532, EPI_ISL_2111533, EPI_ISL_2111534, EPI_ISL_2111535, EPI_ISL_2111536, EPI_ISL_2111537, EPI_ISL_2111538, EPI_ISL_2111539, EPI_ISL_2111540, EPI_ISL_2111541, EPI_ISL_2111542, EPI_ISL_2111543, EPI_ISL_2111544, EPI_ISL_2111545, EPI_ISL_2111546, EPI_ISL_2111547, EPI_ISL_2111548, EPI_ISL_2111550, EPI_ISL_2111551, EPI_ISL_2111552, EPI_ISL_2111553, EPI_ISL_2111554, EPI_ISL_2111555, EPI_ISL_2111556, EPI_ISL_2111557, EPI_ISL_2111558, EPI_ISL_2111559, EPI_ISL_2111560, EPI_ISL_2111562, EPI_ISL_2111563, EPI_ISL_2111564, EPI_ISL_2111565, EPI_ISL_2111566, EPI_ISL_2111567, EPI_ISL_2111568, EPI_ISL_2111569, EPI_ISL_2111570, EPI_ISL_2111571, EPI_ISL_2111572, EPI_ISL_2111573, EPI_ISL_2111574, EPI_ISL_2111575, EPI_ISL_2111576, EPI_ISL_2111577, EPI_ISL_2111578, EPI_ISL_2111580, EPI_ISL_2111581, EPI_ISL_2111582, EPI_ISL_2111584, EPI_ISL_2111585, EPI_ISL_2111586, EPI_ISL_2111587, EPI_ISL_2111588, EPI_ISL_2111589, EPI_ISL_2111590, EPI_ISL_2111591, EPI_ISL_2111592, EPI_ISL_2111594, EPI_ISL_2111595, EPI_ISL_2111596, EPI_ISL_2111597, EPI_ISL_2111598, EPI_ISL_2111599, EPI_ISL_2111600, EPI_ISL_2111601, EPI_ISL_2111602, EPI_ISL_2111604, EPI_ISL_2129250 | | | |
| see above | Labor ZOTZ\|KLIMAS; MVZ Dusseldorf-Centrum | Robert Koch Institute | |
| EPI_ISL_2129290 | Labor ZOTZ\|KLIMAS; MVZ DÃ¼sseldorf-Centrum | Robert Koch Institute | |
| EPI_ISL_1847746 | Labor fur medizinische Mikrobiologie Bakteriologie Mykologie Virologie & Infektionsserologie - Labor Molbis | Robert Koch Institute | |
| EPI_ISL_1847884, EPI_ISL_1847904, EPI_ISL_1848111, EPI_ISL_1848125, EPI_ISL_1848330, EPI_ISL_1852141, EPI_ISL_1852157, EPI_ISL_1852243, EPI_ISL_1852341, EPI_ISL_1852374 | | | |
| see above | Labor fÃ¼r medizinische Mikrobiologie Bakteriologie Mykologie Virologie & Infektionsserologie - Labor MÃ¶lbis | Robert Koch Institute | |
| EPI_ISL_2111110, EPI_ISL_2111111, EPI_ISL_2111112, EPI_ISL_2111113, EPI_ISL_2111114, EPI_ISL_2111115, EPI_ISL_2111116, EPI_ISL_2111117, EPI_ISL_2111118, EPI_ISL_2111119, EPI_ISL_2111120, EPI_ISL_2111121, EPI_ISL_2111122, EPI_ISL_2111123, EPI_ISL_2111124, EPI_ISL_2111125, EPI_ISL_2111126, EPI_ISL_2111127, EPI_ISL_2111128, EPI_ISL_2111129, EPI_ISL_2111130, EPI_ISL_2111131, EPI_ISL_2111132, EPI_ISL_2111133, EPI_ISL_2111134, EPI_ISL_2111135, EPI_ISL_2111136, EPI_ISL_2111137, EPI_ISL_2111138, EPI_ISL_2111139, EPI_ISL_2111140, EPI_ISL_2111141, EPI_ISL_2111142, EPI_ISL_2111143, EPI_ISL_2111144, EPI_ISL_2111145, EPI_ISL_2111146, EPI_ISL_2111147, EPI_ISL_2111148, EPI_ISL_2111149, EPI_ISL_2111150, EPI_ISL_2111151, EPI_ISL_2111152, EPI_ISL_2111153 | | | |
| see above | Laborarztpraxis Dres. med. Walther Weindel & Kollegen | Robert Koch Institute | |
| EPI_ISL_2108322, EPI_ISL_2108324, EPI_ISL_2108327, EPI_ISL_2108329, EPI_ISL_2108331, EPI_ISL_2108333, EPI_ISL_2108338, EPI_ISL_2108339, EPI_ISL_2108342, EPI_ISL_2108343, EPI_ISL_2108344, EPI_ISL_2108345, EPI_ISL_2108346, EPI_ISL_2108348, EPI_ISL_2108351, EPI_ISL_2108354, EPI_ISL_2108356, EPI_ISL_2108362, EPI_ISL_2108367, EPI_ISL_2108369, EPI_ISL_2108372, EPI_ISL_2108380, EPI_ISL_2108381, EPI_ISL_2108382, EPI_ISL_2108388, EPI_ISL_2108394, EPI_ISL_2108397, EPI_ISL_2108400, EPI_ISL_2108414 | | | |
| see above | Laborarztpraxis Osnabruck | Robert Koch Institute | |
| EPI_ISL_1848387, EPI_ISL_2108265, EPI_ISL_2108267, EPI_ISL_2108296, EPI_ISL_2108321, EPI_ISL_2112830, EPI_ISL_2112832, EPI_ISL_2112837, EPI_ISL_2112839 | | | |
| see above | Labormedizin Darmstadt | Robert Koch Institute | |
| EPI_ISL_1941017, EPI_ISL_1941018, EPI_ISL_1941019, EPI_ISL_1941020, EPI_ISL_1941021, EPI_ISL_1941022, EPI_ISL_1941023, EPI_ISL_1941024, EPI_ISL_1941025, EPI_ISL_1941026, EPI_ISL_1941027, EPI_ISL_1941028, EPI_ISL_1941029, EPI_ISL_1941030, EPI_ISL_1941031, EPI_ISL_1941032, EPI_ISL_1941033, EPI_ISL_1941034, EPI_ISL_1941035, EPI_ISL_1941036, EPI_ISL_1941037, EPI_ISL_1941038, EPI_ISL_1941039, EPI_ISL_1941040, EPI_ISL_1941041, EPI_ISL_1941042, EPI_ISL_1941043, EPI_ISL_1941044, EPI_ISL_1941045, EPI_ISL_1941046, EPI_ISL_1941047, EPI_ISL_1941048, EPI_ISL_1941049, EPI_ISL_1941050, EPI_ISL_1941051, EPI_ISL_1941052, EPI_ISL_1941053, EPI_ISL_1941054, EPI_ISL_1941055, EPI_ISL_1941056, EPI_ISL_1941057, EPI_ISL_1941058, EPI_ISL_1941059, EPI_ISL_1941060, EPI_ISL_1941061, EPI_ISL_1941062, EPI_ISL_1941066, EPI_ISL_1941067, EPI_ISL_1941068, EPI_ISL_1941069, EPI_ISL_1941070, EPI_ISL_1941071, EPI_ISL_1941072, EPI_ISL_1941073, EPI_ISL_1941074, EPI_ISL_1941075, EPI_ISL_1941076, EPI_ISL_1941077, EPI_ISL_1941078, EPI_ISL_1941079, EPI_ISL_1941080, EPI_ISL_1941081, EPI_ISL_1941082, EPI_ISL_1941083, EPI_ISL_1941084, EPI_ISL_1941085, EPI_ISL_1941086, EPI_ISL_1941087, EPI_ISL_1941088, EPI_ISL_1941089, EPI_ISL_1941090, EPI_ISL_1941091, EPI_ISL_1941092, EPI_ISL_1941093, EPI_ISL_1941094, EPI_ISL_1941095, EPI_ISL_1941096, EPI_ISL_1941097, EPI_ISL_1941098, EPI_ISL_1941099, EPI_ISL_1941100, EPI_ISL_1941101, EPI_ISL_1941102, EPI_ISL_1941103, EPI_ISL_1941104, EPI_ISL_1941105, EPI_ISL_1941106, EPI_ISL_1941107, EPI_ISL_1941108, EPI_ISL_1941109, EPI_ISL_1941110, EPI_ISL_1941111, EPI_ISL_1941114, EPI_ISL_1941115, EPI_ISL_1941116, EPI_ISL_1941117, EPI_ISL_1941118, EPI_ISL_1941119, EPI_ISL_1941120, EPI_ISL_1941121, EPI_ISL_1941122, EPI_ISL_1941123, EPI_ISL_1941124, EPI_ISL_1941125, EPI_ISL_1941126, EPI_ISL_1941127, EPI_ISL_1941128, EPI_ISL_1941129, EPI_ISL_1941130, EPI_ISL_1941131, EPI_ISL_1941132, EPI_ISL_1941133, EPI_ISL_1941134, EPI_ISL_1941135, EPI_ISL_1941136, EPI_ISL_1941137, EPI_ISL_1941138, EPI_ISL_1941139, EPI_ISL_1941140, EPI_ISL_1941141, EPI_ISL_1941142, EPI_ISL_1941143, EPI_ISL_1941144, EPI_ISL_1941145, EPI_ISL_1941146, EPI_ISL_1941147, EPI_ISL_1941148, EPI_ISL_1941149, EPI_ISL_1941150, EPI_ISL_1941151, EPI_ISL_1941152, EPI_ISL_1941153, EPI_ISL_1941154, EPI_ISL_1941155, EPI_ISL_1941156, EPI_ISL_1941157, EPI_ISL_1941158, EPI_ISL_1941159, EPI_ISL_1941172, EPI_ISL_1941173, EPI_ISL_1941174, EPI_ISL_1941175, EPI_ISL_1941176, EPI_ISL_1941177, EPI_ISL_1941178, EPI_ISL_1941179, EPI_ISL_1941180, EPI_ISL_1941181, EPI_ISL_1941182, EPI_ISL_1941183, EPI_ISL_1941184, EPI_ISL_1941185, EPI_ISL_1941186, EPI_ISL_1941187, EPI_ISL_1941188, EPI_ISL_1941189, EPI_ISL_1941190, EPI_ISL_1941191, EPI_ISL_1941192, EPI_ISL_1941193, EPI_ISL_1941194, EPI_ISL_1941195, EPI_ISL_1941196, EPI_ISL_1941197, EPI_ISL_1941198, EPI_ISL_1941199, EPI_ISL_1941200, EPI_ISL_1941201, EPI_ISL_1941202, EPI_ISL_1941203, EPI_ISL_1941204, EPI_ISL_1941205, EPI_ISL_1941206, EPI_ISL_1941207, EPI_ISL_1941208, EPI_ISL_1941209, EPI_ISL_1941210, EPI_ISL_1941211, EPI_ISL_1941212, EPI_ISL_1941213, EPI_ISL_1941214, EPI_ISL_1941215, EPI_ISL_1941216, EPI_ISL_1941217, EPI_ISL_1941231, EPI_ISL_1941232, EPI_ISL_1941233, EPI_ISL_1941234, EPI_ISL_1941235, EPI_ISL_1941236, EPI_ISL_1941237, EPI_ISL_1941238, EPI_ISL_1941239, EPI_ISL_1941240, EPI_ISL_1941241, EPI_ISL_1941242, EPI_ISL_1941243, EPI_ISL_1941244, EPI_ISL_1941245, EPI_ISL_1941246, EPI_ISL_1941247, EPI_ISL_1941248, EPI_ISL_1941249, EPI_ISL_1941250, EPI_ISL_1941251, EPI_ISL_1941252, EPI_ISL_1941253, EPI_ISL_1941254, EPI_ISL_1941255, EPI_ISL_1941256, EPI_ISL_1941257, EPI_ISL_1941258, EPI_ISL_1941259, EPI_ISL_1941260, EPI_ISL_1941261, EPI_ISL_1941263, EPI_ISL_1941264, EPI_ISL_1941265, EPI_ISL_1941266, EPI_ISL_1941267, EPI_ISL_1941268, EPI_ISL_1941269, EPI_ISL_1941270, EPI_ISL_1941271, EPI_ISL_1941272, EPI_ISL_1941273, EPI_ISL_1941274 | | | |
| see above | Landesamt für Verbraucherschutz Sachsen Anhalt, Magdeburg | Institute of Medical Microbiology and Hospital Hygiene | Aljoscha Tersteegen; Prof. Dr. Achim Kaasch |
| EPI_ISL_2110763, EPI_ISL_2110764, EPI_ISL_2110765, EPI_ISL_2110769, EPI_ISL_2110773, EPI_ISL_2110776, EPI_ISL_2110777, EPI_ISL_2110781, EPI_ISL_2110782, EPI_ISL_2126385, EPI_ISL_2126387, EPI_ISL_2126388, EPI_ISL_2126392, EPI_ISL_2126394, EPI_ISL_2126406, EPI_ISL_2126410 | | | |
| see above | Landesgesundheitsamt Baden-Wurttemberg | Robert Koch Institute | |
| EPI_ISL_1846369, EPI_ISL_1846375, EPI_ISL_1846379, EPI_ISL_1846382, EPI_ISL_1846383, EPI_ISL_1846386, EPI_ISL_1846391, EPI_ISL_1846400, EPI_ISL_1846402, EPI_ISL_1846405, EPI_ISL_1846406, EPI_ISL_1846416, EPI_ISL_1846417, EPI_ISL_1846422, EPI_ISL_1846430, EPI_ISL_1846432, EPI_ISL_1846434, EPI_ISL_1846442, EPI_ISL_1846445, EPI_ISL_1846446, EPI_ISL_1846451 | | | |
| see above | Landesuntersuchungsanstalt fÃ¼r das Gesundheits- und VeterinÃ¤rwesen (LUA) Sachsen | Robert Koch Institute | |
| EPI_ISL_1846755, EPI_ISL_1846757, EPI_ISL_1846758, EPI_ISL_1846759, EPI_ISL_1846760, EPI_ISL_1846761, EPI_ISL_1846762, EPI_ISL_1846763, EPI_ISL_1846764, EPI_ISL_1846765, EPI_ISL_1846766, EPI_ISL_1846767, EPI_ISL_1846768, EPI_ISL_1846769, EPI_ISL_2111004, EPI_ISL_2111005, EPI_ISL_2111006, EPI_ISL_2111007, EPI_ISL_2111008, EPI_ISL_2111009, EPI_ISL_2111010, EPI_ISL_2111011, EPI_ISL_2111012, EPI_ISL_2111013 | | | |
| see above | Limbach - Ganzimmun Diagnostics AG Mainz | Robert Koch Institute | |
| EPI_ISL_1843334, EPI_ISL_1843347, EPI_ISL_1843350, EPI_ISL_1847195, EPI_ISL_1847209, EPI_ISL_1847322, EPI_ISL_1847329, EPI_ISL_1847435, EPI_ISL_1848810, EPI_ISL_1850307, EPI_ISL_1850310, EPI_ISL_1850322, EPI_ISL_1850329, EPI_ISL_1852451, EPI_ISL_1852470, EPI_ISL_1852474, EPI_ISL_1852485 | | | |
| see above | Limbach - MVZ Gemeinschaftslabor Suhl | Robert Koch Institute | |
| EPI_ISL_1727421, EPI_ISL_1727423, EPI_ISL_1727429, EPI_ISL_1727430, EPI_ISL_1727432, EPI_ISL_1727441, EPI_ISL_1727454, EPI_ISL_1727456, EPI_ISL_1727457, EPI_ISL_1727463, EPI_ISL_1727467, EPI_ISL_1727471, EPI_ISL_1727473, EPI_ISL_1727476, EPI_ISL_1727478, EPI_ISL_1727484, EPI_ISL_1727497, EPI_ISL_1727500, EPI_ISL_1727501, EPI_ISL_1727503, EPI_ISL_1727507, EPI_ISL_1727510, EPI_ISL_1727511, EPI_ISL_1727513, EPI_ISL_1727515, EPI_ISL_1727521, EPI_ISL_1727522, EPI_ISL_1727526, EPI_ISL_1727529, EPI_ISL_1727531, EPI_ISL_1727535, EPI_ISL_1727536, EPI_ISL_1727539, EPI_ISL_1727540, EPI_ISL_1727541, EPI_ISL_1727543, EPI_ISL_1727544, EPI_ISL_1727546, EPI_ISL_1727547, EPI_ISL_1727548, EPI_ISL_1727549, EPI_ISL_1853111, EPI_ISL_1853121, EPI_ISL_1853127, EPI_ISL_1853132, EPI_ISL_1853138, EPI_ISL_1853139, EPI_ISL_1853147, EPI_ISL_1853194, EPI_ISL_1853230, EPI_ISL_1853241 | | | |
| see above | Limbach - MVZ Humangenetik Ulm | Robert Koch Institute | |
| EPI_ISL_1846214, EPI_ISL_1846215, EPI_ISL_1846230, EPI_ISL_1847167, EPI_ISL_1847170, EPI_ISL_1847171, EPI_ISL_1847172, EPI_ISL_1847174, EPI_ISL_1847175, EPI_ISL_1848348, EPI_ISL_1848349, EPI_ISL_1848350, EPI_ISL_1848352, EPI_ISL_1848353, EPI_ISL_1848354, EPI_ISL_1848355, EPI_ISL_1848357, EPI_ISL_1848358, EPI_ISL_1848359, EPI_ISL_1848360, EPI_ISL_1848362, EPI_ISL_1848364, EPI_ISL_1848749, EPI_ISL_1848752, EPI_ISL_1848762, EPI_ISL_1848765, EPI_ISL_1848766, EPI_ISL_1848771, EPI_ISL_1848775, EPI_ISL_1848779, EPI_ISL_1848781, EPI_ISL_1852051, EPI_ISL_1852079, EPI_ISL_1852091, EPI_ISL_1852093, EPI_ISL_1852095, EPI_ISL_1852098, EPI_ISL_1852110, EPI_ISL_1852119 | | | |
| see above | Limbach - MVZ Labor Eveld & Kollegen Essen | Robert Koch Institute | |
| EPI_ISL_1850293, EPI_ISL_1850309, EPI_ISL_1850319, EPI_ISL_1850321 | Limbach - MVZ Labor Ludwigsburg | Robert Koch Institute | |
| EPI_ISL_1850283, EPI_ISL_1850284, EPI_ISL_1850294, EPI_ISL_1850297, EPI_ISL_1850305 | Limbach - MVZ Labor MÃ¼nster (Cullen) | Robert Koch Institute | |
| EPI_ISL_1843333, EPI_ISL_1843337, EPI_ISL_1843338, EPI_ISL_1843341, EPI_ISL_1843344, EPI_ISL_1843351, EPI_ISL_1843352, EPI_ISL_1843353, EPI_ISL_1843354, EPI_ISL_1843361, EPI_ISL_1847178, EPI_ISL_1847179, EPI_ISL_1847180, EPI_ISL_1847184, EPI_ISL_1847194, EPI_ISL_1847201, EPI_ISL_1847205, EPI_ISL_1847219, EPI_ISL_1847220, EPI_ISL_1847222, EPI_ISL_1847223, EPI_ISL_1847224, EPI_ISL_1847225, EPI_ISL_1847229, EPI_ISL_1847230, EPI_ISL_1847313, EPI_ISL_1847318, EPI_ISL_1847326, EPI_ISL_1847426, EPI_ISL_1847427, EPI_ISL_1847433, EPI_ISL_1847440, EPI_ISL_1847443, EPI_ISL_1847723, EPI_ISL_1847725, EPI_ISL_1847726, EPI_ISL_1847727, EPI_ISL_1847730, EPI_ISL_1847731, EPI_ISL_1847733, EPI_ISL_1848783, EPI_ISL_1848784, EPI_ISL_1848785, EPI_ISL_1848786, EPI_ISL_1848787, EPI_ISL_1848788, EPI_ISL_1848790, EPI_ISL_1848791, EPI_ISL_1848795, EPI_ISL_1848800, EPI_ISL_1848801, EPI_ISL_1848802, EPI_ISL_1848803, EPI_ISL_1848804, EPI_ISL_1848806, EPI_ISL_1848807, EPI_ISL_1848808, EPI_ISL_1848809, EPI_ISL_1848811, EPI_ISL_1848812, EPI_ISL_1850282, EPI_ISL_1850285, EPI_ISL_1850286, EPI_ISL_1850288, EPI_ISL_1850289, EPI_ISL_1850290, EPI_ISL_1850291, EPI_ISL_1850295, EPI_ISL_1850299, EPI_ISL_1850300, EPI_ISL_1850301, EPI_ISL_1850302, EPI_ISL_1850304, EPI_ISL_1850306, EPI_ISL_1850311, EPI_ISL_1850312, EPI_ISL_1850314, EPI_ISL_1850315, EPI_ISL_1850316, EPI_ISL_1850320, EPI_ISL_1850324, EPI_ISL_1850325, EPI_ISL_1850327, EPI_ISL_1850330, EPI_ISL_1852047, EPI_ISL_1852471 | | | |
| see above | MDI Limbach Berlin GmbH; MVZ Labor Berlin | Robert Koch Institute | |
| EPI_ISL_1862456 | MEDILYS Laborgesellschaft mbH | Heinrich Pette Institute, Leibniz Institute for Experimental Virology | Adam Grundhoff; Alexis Robitaille; Johannes Knobloch; Martin Aepfelbacher; Nicole Fischer; Thomas Günther |
| EPI_ISL_1843218, EPI_ISL_1843223, EPI_ISL_1843224, EPI_ISL_1843226, EPI_ISL_1843227, EPI_ISL_1843229, EPI_ISL_1843233, EPI_ISL_1843236, EPI_ISL_1843237, EPI_ISL_1843238, EPI_ISL_1843245, EPI_ISL_1843249, EPI_ISL_1843260, EPI_ISL_1843262, EPI_ISL_1843266, EPI_ISL_1843268, EPI_ISL_1843274, EPI_ISL_1843275, EPI_ISL_1843278, EPI_ISL_1843279, EPI_ISL_1843291, EPI_ISL_1850338, EPI_ISL_1850340, EPI_ISL_1850342, EPI_ISL_1850343, EPI_ISL_1850345, EPI_ISL_1850346, EPI_ISL_1850347, EPI_ISL_1850350, EPI_ISL_1850351, EPI_ISL_1850352, EPI_ISL_1850355, EPI_ISL_1850356, EPI_ISL_1850359, EPI_ISL_1850360, EPI_ISL_1850361, EPI_ISL_1850365, EPI_ISL_1850370, EPI_ISL_1850373, EPI_ISL_1850374, EPI_ISL_1850375, EPI_ISL_1850376, EPI_ISL_1850377, EPI_ISL_1850379, EPI_ISL_1850389, EPI_ISL_1850390, EPI_ISL_1850391, EPI_ISL_1850394, EPI_ISL_1850395, EPI_ISL_1850396, EPI_ISL_1850399, EPI_ISL_1850401, EPI_ISL_1850402, EPI_ISL_1850407, EPI_ISL_1850409, EPI_ISL_1850412, EPI_ISL_1850415, EPI_ISL_1850417, EPI_ISL_1850419, EPI_ISL_1850420, EPI_ISL_1850421, EPI_ISL_1850423, EPI_ISL_1850426, EPI_ISL_1850428, EPI_ISL_1850431, EPI_ISL_1850432, EPI_ISL_1850433, EPI_ISL_1850435, EPI_ISL_1850436, EPI_ISL_2108323, EPI_ISL_2108326, EPI_ISL_2108330, EPI_ISL_2108332, EPI_ISL_2108334, EPI_ISL_2108335, EPI_ISL_2108336, EPI_ISL_2108337, EPI_ISL_2108340, EPI_ISL_2108347, EPI_ISL_2108350, EPI_ISL_2108358, EPI_ISL_2108359, EPI_ISL_2108361, EPI_ISL_2108363, EPI_ISL_2108365, EPI_ISL_2108366, EPI_ISL_2108368, EPI_ISL_2108371, EPI_ISL_2108373, EPI_ISL_2108374, EPI_ISL_2108377, EPI_ISL_2108378, EPI_ISL_2108383, EPI_ISL_2108385, EPI_ISL_2108387, EPI_ISL_2108395, EPI_ISL_2108399, EPI_ISL_2108401, EPI_ISL_2108402, EPI_ISL_2108403, EPI_ISL_2108407, EPI_ISL_2108411, EPI_ISL_2108412, EPI_ISL_2108413, EPI_ISL_2115448 | | | |
| see above | MVZ Dr. Eberhard & Partner Dortmund | Robert Koch Institute | |
| EPI_ISL_1849349, EPI_ISL_1849351, EPI_ISL_1849352, EPI_ISL_1849354, EPI_ISL_1849355, EPI_ISL_1849359, EPI_ISL_1849361, EPI_ISL_1849362, EPI_ISL_1849365, EPI_ISL_1849366, EPI_ISL_1849367, EPI_ISL_1849368, EPI_ISL_1849370, EPI_ISL_1849373, EPI_ISL_1849376, EPI_ISL_1849377, EPI_ISL_1849378, EPI_ISL_1849379, EPI_ISL_1849380, EPI_ISL_1849381, EPI_ISL_1849382, EPI_ISL_1849383, EPI_ISL_1849385, EPI_ISL_1849386, EPI_ISL_1849388, EPI_ISL_1849390, EPI_ISL_1849391, EPI_ISL_1849392, EPI_ISL_1849395, EPI_ISL_1849399, EPI_ISL_1849400, EPI_ISL_1849402, EPI_ISL_1849404, EPI_ISL_1849405, EPI_ISL_1849406, EPI_ISL_1849407, EPI_ISL_1849409, EPI_ISL_1849410, EPI_ISL_1849412, EPI_ISL_2111238, EPI_ISL_2111253, EPI_ISL_2111274 | | | |
| see above | MVZ Labor Dr. Fenner und Kollegen (Standort Hamburg) | Robert Koch Institute | |
| EPI_ISL_1847168, EPI_ISL_1847176, EPI_ISL_1848747, EPI_ISL_1850331, EPI_ISL_1852045, EPI_ISL_1852048, EPI_ISL_1852049, EPI_ISL_1852050, EPI_ISL_1852052, EPI_ISL_1852053, EPI_ISL_1852054, EPI_ISL_1852055, EPI_ISL_1852056, EPI_ISL_1852059, EPI_ISL_1852060, EPI_ISL_1852061, EPI_ISL_1852062, EPI_ISL_1852063, EPI_ISL_1852065, EPI_ISL_1852066, EPI_ISL_1852067, EPI_ISL_1852068, EPI_ISL_1852075, EPI_ISL_1852077, EPI_ISL_1852080, EPI_ISL_1852081, EPI_ISL_1852082, EPI_ISL_1852083, EPI_ISL_1852084, EPI_ISL_1852088, EPI_ISL_1852090, EPI_ISL_1852097, EPI_ISL_1852099, EPI_ISL_1852100, EPI_ISL_1852102, EPI_ISL_1852104, EPI_ISL_1852105, EPI_ISL_1852106, EPI_ISL_1852108, EPI_ISL_1852109, EPI_ISL_1852111, EPI_ISL_1852113, EPI_ISL_1852116, EPI_ISL_1852118, EPI_ISL_1852120, EPI_ISL_1852121, EPI_ISL_1852122, EPI_ISL_1852123, EPI_ISL_1852125, EPI_ISL_1852126, EPI_ISL_1852127, EPI_ISL_1852128, EPI_ISL_1852131, EPI_ISL_1852134, EPI_ISL_1852453, EPI_ISL_1852455, EPI_ISL_1852457, EPI_ISL_1852467, EPI_ISL_1852468, EPI_ISL_1852476, EPI_ISL_1852478, EPI_ISL_1852480 | | | |
| see above | MVZ Labor Dr. Limbach & Kollegen GbR | Robert Koch Institute | |
| EPI_ISL_1847169, EPI_ISL_1847173, EPI_ISL_1847177, EPI_ISL_1847186, EPI_ISL_1847187, EPI_ISL_1847189, EPI_ISL_1848356, EPI_ISL_1848361, EPI_ISL_1848758, EPI_ISL_1848770, EPI_ISL_1848773 | | | |
| see above | MVZ Labor Dr. Reising-Ackermann und Kollegen | Robert Koch Institute | |
| EPI_ISL_1726939, EPI_ISL_1726941, EPI_ISL_1726953, EPI_ISL_1726957, EPI_ISL_1726958, EPI_ISL_1726961, EPI_ISL_1726962, EPI_ISL_1726964, EPI_ISL_1726965, EPI_ISL_1726971, EPI_ISL_1844288, EPI_ISL_1844289, EPI_ISL_1844290, EPI_ISL_1844292, EPI_ISL_1844296, EPI_ISL_1844299, EPI_ISL_1844301, EPI_ISL_1844302, EPI_ISL_1844303, EPI_ISL_1844305, EPI_ISL_1844306, EPI_ISL_1844312, | | | |

| | | | |
|---|---|---|---|
| EPI_ISL_1844314, EPI_ISL_1844315, EPI_ISL_1844316, EPI_ISL_1844322, EPI_ISL_1844324, EPI_ISL_1844326, EPI_ISL_1844327, EPI_ISL_1844330, EPI_ISL_1844331, EPI_ISL_1844335, EPI_ISL_1844337, EPI_ISL_1844342, EPI_ISL_1844343, EPI_ISL_1844344, EPI_ISL_1844347, EPI_ISL_1844348, EPI_ISL_1844349, EPI_ISL_1844351, EPI_ISL_1844353, EPI_ISL_1844354, EPI_ISL_1844362, EPI_ISL_1848403, EPI_ISL_1848405, EPI_ISL_1848407, EPI_ISL_1848408, EPI_ISL_1848410, EPI_ISL_1848415, EPI_ISL_1848422, EPI_ISL_1848426, EPI_ISL_1848429, EPI_ISL_1848436, EPI_ISL_1848442, EPI_ISL_1848446, EPI_ISL_1848451, EPI_ISL_1848455, EPI_ISL_1848458, EPI_ISL_1848464, EPI_ISL_1848469, EPI_ISL_1848476, EPI_ISL_1848480, EPI_ISL_1848484, EPI_ISL_1848488, EPI_ISL_1848491, EPI_ISL_1848494, EPI_ISL_1848496, EPI_ISL_1848498, EPI_ISL_1848501, EPI_ISL_1848508, EPI_ISL_1848513, EPI_ISL_1848517, EPI_ISL_1848520, EPI_ISL_1848524, EPI_ISL_1848526, EPI_ISL_1848529, EPI_ISL_1848531, EPI_ISL_1848535, EPI_ISL_1848539, EPI_ISL_1848547, EPI_ISL_1848552, EPI_ISL_1848557, EPI_ISL_1848561, EPI_ISL_1848565, EPI_ISL_1848568, EPI_ISL_1849414, EPI_ISL_1849415, EPI_ISL_1849417, EPI_ISL_1849418, EPI_ISL_1849419, EPI_ISL_1849421, EPI_ISL_1849422, EPI_ISL_1849423, EPI_ISL_1849425, EPI_ISL_1849426, EPI_ISL_1849427, EPI_ISL_1849430, EPI_ISL_1849431, EPI_ISL_1849432, EPI_ISL_1849433, EPI_ISL_1849435, EPI_ISL_1849436, EPI_ISL_1849437, EPI_ISL_1849440, EPI_ISL_1849441, EPI_ISL_1849442, EPI_ISL_1849444, EPI_ISL_1849445, EPI_ISL_1849446, EPI_ISL_1849447, EPI_ISL_1849448, EPI_ISL_1849450, EPI_ISL_1849451, EPI_ISL_1849452, EPI_ISL_1849454, EPI_ISL_1849455, EPI_ISL_1849456, EPI_ISL_1849457, EPI_ISL_1849459, EPI_ISL_1849460, EPI_ISL_1849461, EPI_ISL_1849462, EPI_ISL_1849464, EPI_ISL_1849465, EPI_ISL_1849466, EPI_ISL_1849467, EPI_ISL_1849469, EPI_ISL_1849470, EPI_ISL_1849471, EPI_ISL_1849472, EPI_ISL_1849474, EPI_ISL_1849475, EPI_ISL_1849476, EPI_ISL_1849477, EPI_ISL_1849479, EPI_ISL_1849480, EPI_ISL_1849481, EPI_ISL_1849482, EPI_ISL_1849483, EPI_ISL_1849485, EPI_ISL_1849486, EPI_ISL_1849487, EPI_ISL_1849488, EPI_ISL_1849490, EPI_ISL_1849491, EPI_ISL_1849492, EPI_ISL_1849493, EPI_ISL_1849495, EPI_ISL_1849496, EPI_ISL_1851087, EPI_ISL_1851088, EPI_ISL_1851089, EPI_ISL_1851090, EPI_ISL_1851091, EPI_ISL_1851092, EPI_ISL_1851093, EPI_ISL_1851094, EPI_ISL_1851095, EPI_ISL_1851096, EPI_ISL_1851097, EPI_ISL_1851098, EPI_ISL_1851099, EPI_ISL_1851100, EPI_ISL_1851101, EPI_ISL_1851103, EPI_ISL_1851104, EPI_ISL_1851105, EPI_ISL_1851106, EPI_ISL_1851107, EPI_ISL_1851108, EPI_ISL_1851109, EPI_ISL_1851110, EPI_ISL_1851111, EPI_ISL_1851112, EPI_ISL_1851113, EPI_ISL_1851114, EPI_ISL_1851115, EPI_ISL_1851116, EPI_ISL_1851117, EPI_ISL_1851118, EPI_ISL_1851119, EPI_ISL_1851120, EPI_ISL_1851121, EPI_ISL_1851122, EPI_ISL_1851123, EPI_ISL_1851124, EPI_ISL_1851125, EPI_ISL_1851126, EPI_ISL_1851127, EPI_ISL_1851128, EPI_ISL_1851129, EPI_ISL_1851130, EPI_ISL_1851429, EPI_ISL_1851430, EPI_ISL_1851431, EPI_ISL_1851432, EPI_ISL_1851433, EPI_ISL_1851434, EPI_ISL_1851435, EPI_ISL_1851436, EPI_ISL_1851437, EPI_ISL_1851438, EPI_ISL_1851439, EPI_ISL_1851440, EPI_ISL_1851441, EPI_ISL_2110787, EPI_ISL_2110804, EPI_ISL_2110819, EPI_ISL_2110826, EPI_ISL_2115306, EPI_ISL_2115322 | | | |
| see above | MVZ Labor Krone GbR | Robert Koch Institute | |
| EPI_ISL_1850317, EPI_ISL_1850326 | MVZ Medizinisches Labor Hannover GmbH | Robert Koch Institute | |
| EPI_ISL_1850354, EPI_ISL_1850368, EPI_ISL_1850369, EPI_ISL_1850397, EPI_ISL_1850410 | MVZ fÃ¼r Laboratoriumsmedizin und Mikrobiologie WÃ¼rzburg | Robert Koch Institute | |
| EPI_ISL_2006683, EPI_ISL_2006684, EPI_ISL_2094518, EPI_ISL_2094549, EPI_ISL_2094572, EPI_ISL_2094595, EPI_ISL_2094628, EPI_ISL_2094634, EPI_ISL_2094645, EPI_ISL_2094653, EPI_ISL_2094698, EPI_ISL_2094699, EPI_ISL_2094700, EPI_ISL_2094701, EPI_ISL_2094702, EPI_ISL_2094703, EPI_ISL_2094704, EPI_ISL_2094705, EPI_ISL_2094706, EPI_ISL_2094707, EPI_ISL_2094708, EPI_ISL_2094709, EPI_ISL_2094710, EPI_ISL_2094711, EPI_ISL_2094712, EPI_ISL_2094713, EPI_ISL_2094714, EPI_ISL_2094755, EPI_ISL_2094756, EPI_ISL_2094757, EPI_ISL_2094758, EPI_ISL_2094759, EPI_ISL_2094760, EPI_ISL_2094761, EPI_ISL_2094762, EPI_ISL_2094773, EPI_ISL_2094775, EPI_ISL_2094792, EPI_ISL_2094826, EPI_ISL_2094829, EPI_ISL_2094898, EPI_ISL_2094899, EPI_ISL_2094900, EPI_ISL_2094901, EPI_ISL_2094902, EPI_ISL_2094903, EPI_ISL_2094904, EPI_ISL_2094905, EPI_ISL_2094906, EPI_ISL_2094907, EPI_ISL_2094908, EPI_ISL_2094909, EPI_ISL_2094910, EPI_ISL_2094911, EPI_ISL_2094912, EPI_ISL_2094913, EPI_ISL_2094914, EPI_ISL_2094915, EPI_ISL_2094916, EPI_ISL_2094917, EPI_ISL_2094950, EPI_ISL_2094951, EPI_ISL_2094971, EPI_ISL_2094972, EPI_ISL_2094973, EPI_ISL_2095041, EPI_ISL_2095053, EPI_ISL_2095065, EPI_ISL_2095071, EPI_ISL_2095076, EPI_ISL_2095079, EPI_ISL_2095080, EPI_ISL_2095093, EPI_ISL_2095110, EPI_ISL_2095127, EPI_ISL_2095136, EPI_ISL_2095141, EPI_ISL_2095158, EPI_ISL_2095175, EPI_ISL_2095176, EPI_ISL_2095181, EPI_ISL_2095257, EPI_ISL_2095259, EPI_ISL_2095269, EPI_ISL_2095274, EPI_ISL_2095296, EPI_ISL_2095297, EPI_ISL_2095300 | | | |
| see above | Max von Pettenkofer Institute, Virology, National Reference Center for Retroviruses, LMU Munich | Laboratory for Functional Genome Analysis; Dept. Genomics; Gene Center of the LMU Munich | Alexander Graf; Helmut Blum; Max Muenchhoff; Oliver Keppler; Stefan Krebs |
| EPI_ISL_1843200, EPI_ISL_1843202, EPI_ISL_1843204, EPI_ISL_1843205, EPI_ISL_1843206, EPI_ISL_1843207, EPI_ISL_1843208, EPI_ISL_1843209, EPI_ISL_1843210, EPI_ISL_1843213, EPI_ISL_1843214, EPI_ISL_2110381, EPI_ISL_2110388, EPI_ISL_2110389, EPI_ISL_2110402 | | | |
| see above | Med. Labor Prof. Schenk Dr. Ansorge & Kollegen | Robert Koch Institute | |
| EPI_ISL_1991897, EPI_ISL_1991898, EPI_ISL_1991899, EPI_ISL_1991900, EPI_ISL_1991901, EPI_ISL_1991903, EPI_ISL_1991904, EPI_ISL_1991905, EPI_ISL_1991906, EPI_ISL_1991907, EPI_ISL_1991908, EPI_ISL_1991909, EPI_ISL_1991910, EPI_ISL_1991911, EPI_ISL_1991912, EPI_ISL_1991913, EPI_ISL_1991914, EPI_ISL_1991915, EPI_ISL_1991917, EPI_ISL_1991919, EPI_ISL_1991920, EPI_ISL_1991921, EPI_ISL_1991922, EPI_ISL_1991923, EPI_ISL_1991924, EPI_ISL_1991926, EPI_ISL_1991927, EPI_ISL_1991928, EPI_ISL_1991929, EPI_ISL_1991931, EPI_ISL_1991966, EPI_ISL_1991967, EPI_ISL_1991969, EPI_ISL_1991970, EPI_ISL_1991971, EPI_ISL_1991972, EPI_ISL_1991973, EPI_ISL_1991975, EPI_ISL_1991976, EPI_ISL_1991977, EPI_ISL_1991978, EPI_ISL_1991979, EPI_ISL_1991981, EPI_ISL_1991982, EPI_ISL_1991983, EPI_ISL_1991984, EPI_ISL_1991985, EPI_ISL_1991986, EPI_ISL_1991987, EPI_ISL_1991988, EPI_ISL_1991989, EPI_ISL_1991990, EPI_ISL_1991991, EPI_ISL_1991992, EPI_ISL_1991993, EPI_ISL_1991994, EPI_ISL_1991995, EPI_ISL_1991996, EPI_ISL_1991997, EPI_ISL_1991998, EPI_ISL_1991999, EPI_ISL_1992000, EPI_ISL_1992003, EPI_ISL_1992005, EPI_ISL_1992006, EPI_ISL_1992007, EPI_ISL_1992008, EPI_ISL_1992009, EPI_ISL_1992010, EPI_ISL_1992012, EPI_ISL_1992013, EPI_ISL_1992014, EPI_ISL_1992015, EPI_ISL_1992016, EPI_ISL_1992017, EPI_ISL_1992852, EPI_ISL_1992853, EPI_ISL_1992854, EPI_ISL_1992856, EPI_ISL_1992857, EPI_ISL_1992859, EPI_ISL_1992860, EPI_ISL_1992861, EPI_ISL_1992863, EPI_ISL_1992864, EPI_ISL_1992866, EPI_ISL_1992867, EPI_ISL_1992868, EPI_ISL_1992870, EPI_ISL_1992871, EPI_ISL_1992872, EPI_ISL_1992874, EPI_ISL_1992877, EPI_ISL_1992878, EPI_ISL_1992879, EPI_ISL_1992880, EPI_ISL_1992882, EPI_ISL_1992883, EPI_ISL_1992885, EPI_ISL_1992887, EPI_ISL_1992890, EPI_ISL_1992891, EPI_ISL_1992893, EPI_ISL_1992894, EPI_ISL_1992895 | | | |
| see above | Medical Laboratories Duesseldorf | Center of Medical Microbiology, Virology, and Hospital Hygiene, University of Duesseldorf | Alexander Dilthey; Andreas Walker; Angelika Helmer; Christian Lange; Daniel Strelow; Jessica Nicolai; Jörg Timm; Klaus Pfeffer; Lisanna Hülse; Malte Kohns Vasconcelos; Maximilian Damagnez; Nadine Lübke; Tobias Wienemann; Torsten Houwaart |
| EPI_ISL_1844610, EPI_ISL_1844612, EPI_ISL_1844614, EPI_ISL_1844617, EPI_ISL_1844619, EPI_ISL_1844623, EPI_ISL_1844624, EPI_ISL_1844629, EPI_ISL_1844630, EPI_ISL_1844631, EPI_ISL_1844634, EPI_ISL_1844636, EPI_ISL_1844638 | | | |
| see above | Medizinisch-Diagnostisches Labor Kempten allgÃ¤ulab | Robert Koch Institute | |
| EPI_ISL_2116103, EPI_ISL_2116104, EPI_ISL_2116105, EPI_ISL_2116106, EPI_ISL_2116107, EPI_ISL_2116108, EPI_ISL_2116109, EPI_ISL_2116110, EPI_ISL_2116111, EPI_ISL_2116112, EPI_ISL_2116113, EPI_ISL_2116114, EPI_ISL_2116115, EPI_ISL_2116116, EPI_ISL_2116117, EPI_ISL_2116118, EPI_ISL_2116119, EPI_ISL_2116120, EPI_ISL_2116121, EPI_ISL_2116122, EPI_ISL_2116123, EPI_ISL_2116124, EPI_ISL_2116125, EPI_ISL_2116126, EPI_ISL_2116127, EPI_ISL_2116128, EPI_ISL_2116129, EPI_ISL_2116130, EPI_ISL_2116131, EPI_ISL_2116132, EPI_ISL_2116133, EPI_ISL_2116134 | | | |
| see above | Medizinische Laboratorien Dusseldorf | Robert Koch Institute | |
| EPI_ISL_1850723, EPI_ISL_1850725, EPI_ISL_1850726, EPI_ISL_1850727, EPI_ISL_1850728, EPI_ISL_1850729, EPI_ISL_1850730, EPI_ISL_1850731, EPI_ISL_1850732, EPI_ISL_1850733, EPI_ISL_1850734, EPI_ISL_1850735, EPI_ISL_1850736, EPI_ISL_1850737, EPI_ISL_1850738, EPI_ISL_1850739, EPI_ISL_1850740, EPI_ISL_1850741, EPI_ISL_1850742, EPI_ISL_1850743, EPI_ISL_1850744, EPI_ISL_1850745, EPI_ISL_1850746, EPI_ISL_1850747, EPI_ISL_1850748, EPI_ISL_1850750, EPI_ISL_1850752, EPI_ISL_1850754, EPI_ISL_1850755, EPI_ISL_1850756, EPI_ISL_1850758, EPI_ISL_1850759, EPI_ISL_1850760, EPI_ISL_1850762, EPI_ISL_1850763, EPI_ISL_1850765, EPI_ISL_1850766, EPI_ISL_1850767, EPI_ISL_1850769, EPI_ISL_1850770, EPI_ISL_1850772, EPI_ISL_1850773, EPI_ISL_1850774, EPI_ISL_1850775, EPI_ISL_1850777, EPI_ISL_1850778, EPI_ISL_1850779, EPI_ISL_1850780, EPI_ISL_1850781, EPI_ISL_1850782, EPI_ISL_1850783, EPI_ISL_1850785, EPI_ISL_1850787, EPI_ISL_1850788, EPI_ISL_1850790, EPI_ISL_1850791, EPI_ISL_1850792, EPI_ISL_1850794, EPI_ISL_1850795, EPI_ISL_1850796, EPI_ISL_1850797, EPI_ISL_1850799, EPI_ISL_2116093, EPI_ISL_2116094, EPI_ISL_2116095, EPI_ISL_2116096, EPI_ISL_2116097, EPI_ISL_2116098, EPI_ISL_2116099, EPI_ISL_2116100, EPI_ISL_2116101, EPI_ISL_2116102 | | | |
| see above | Medizinische Laboratorien DÃ¼sseldorf | Robert Koch Institute | |
| EPI_ISL_1846365, EPI_ISL_1846366, EPI_ISL_1846370, EPI_ISL_1846372, EPI_ISL_1846376, EPI_ISL_1846378, EPI_ISL_1846381, EPI_ISL_1846390, EPI_ISL_1846397, EPI_ISL_1846407, EPI_ISL_1846408, EPI_ISL_1846409, EPI_ISL_1846410, EPI_ISL_1846414, EPI_ISL_1846425, EPI_ISL_1846427, EPI_ISL_1846431, EPI_ISL_1846449, EPI_ISL_1847432, EPI_ISL_1847434, EPI_ISL_1847436, EPI_ISL_1847438, EPI_ISL_1847439, EPI_ISL_1848789, EPI_ISL_1848792, EPI_ISL_1848793, EPI_ISL_1848794, EPI_ISL_1848796, EPI_ISL_1848805, EPI_ISL_1848813, EPI_ISL_1852057, EPI_ISL_1852073, EPI_ISL_1852086, EPI_ISL_1852101, EPI_ISL_1852130, EPI_ISL_1852132, EPI_ISL_1852133, EPI_ISL_1852135, EPI_ISL_1852136, EPI_ISL_1852448, EPI_ISL_1852450, EPI_ISL_1852452, EPI_ISL_1852454, EPI_ISL_1852456, EPI_ISL_1852458, EPI_ISL_1852459, EPI_ISL_1852465, EPI_ISL_1852469, EPI_ISL_1852472, EPI_ISL_1852475, EPI_ISL_1852484 | | | |
| see above | Medizinisches Labor Ostsachsen MVZ GbR NebenbetriebsstÃ¤tte Dresden | Robert Koch Institute | |
| EPI_ISL_1843230, EPI_ISL_1843252, EPI_ISL_1843255, EPI_ISL_1843273, EPI_ISL_1843284, EPI_ISL_1843290, EPI_ISL_1850341, EPI_ISL_1850348, EPI_ISL_1850366, EPI_ISL_1850371, EPI_ISL_1850380, EPI_ISL_1850386, EPI_ISL_1850400, EPI_ISL_1850406, EPI_ISL_1850414, EPI_ISL_1850422 | | | |
| see above | Medizinisches Labor Wahl LÃ¼denscheid | Robert Koch Institute | |
| EPI_ISL_1847737, EPI_ISL_1847772, EPI_ISL_1848127, EPI_ISL_1850749, EPI_ISL_1850768, EPI_ISL_1850801, EPI_ISL_1850806, EPI_ISL_1850908, EPI_ISL_1850991, EPI_ISL_1851012, EPI_ISL_1851058 | | | |
| see above | Praxisgemeinschaft fÃ¼r Laboratoriumsmedizin Labor BlumenstraÃŸe; Praxis Dr. E. Wietschel & Dr. F. Wietschel | Robert Koch Institute | |
| EPI_ISL_1848134, EPI_ISL_1848211, EPI_ISL_1848224, EPI_ISL_1850815, EPI_ISL_1850864, EPI_ISL_1850933, EPI_ISL_1850939, EPI_ISL_1851041, EPI_ISL_1851053, EPI_ISL_1851059 | | | |
| see above | Praxisgemeinschaft fÃ¼r Laboratoriumsmedizin Labor BlumenstraÃŸe; Praxis Dr. Sel und Dr. SpÃ¤te | Robert Koch Institute | |
| EPI_ISL_2109128, EPI_ISL_2109201, EPI_ISL_2109248, EPI_ISL_2109573, EPI_ISL_2109611, EPI_ISL_2109706, EPI_ISL_2115661, EPI_ISL_2115711, EPI_ISL_2115765, EPI_ISL_2115796, EPI_ISL_2125388 | | | |
| see above | Robert-Bosch-Krankenhaus | Robert Koch Institute | |
| EPI_ISL_1843357, EPI_ISL_1843365, EPI_ISL_1843366, EPI_ISL_1843370 | SLK-Kliniken Heilbronn GmbH - Institut fÃ¼r Laboratoriumsmedizin | Robert Koch Institute | |
| EPI_ISL_1847735, EPI_ISL_1847757, EPI_ISL_1847763, EPI_ISL_1847780, EPI_ISL_1847836, EPI_ISL_1847841, EPI_ISL_1847849, EPI_ISL_1847899, EPI_ISL_1847910, EPI_ISL_1847929, EPI_ISL_1847962, EPI_ISL_1847976, EPI_ISL_1847978, EPI_ISL_1847993, EPI_ISL_1848007, EPI_ISL_1848027, EPI_ISL_1848073, EPI_ISL_1848074, EPI_ISL_1848099, EPI_ISL_1848169, EPI_ISL_1848179, EPI_ISL_1848186, EPI_ISL_1848209, EPI_ISL_1848250, EPI_ISL_1848251, EPI_ISL_1848256, EPI_ISL_1848270, EPI_ISL_1848274, EPI_ISL_1848278, EPI_ISL_1848279, EPI_ISL_1848287, EPI_ISL_1848343, EPI_ISL_1848344, EPI_ISL_1850753, EPI_ISL_1850757, EPI_ISL_1850761, EPI_ISL_1850764, EPI_ISL_1850776, EPI_ISL_1850786, EPI_ISL_1850789, EPI_ISL_1850809, EPI_ISL_1850812, EPI_ISL_1850814, EPI_ISL_1850817, EPI_ISL_1850822, EPI_ISL_1850824, EPI_ISL_1850828, EPI_ISL_1850829, EPI_ISL_1850830, EPI_ISL_1850838, EPI_ISL_1850839, EPI_ISL_1850842, EPI_ISL_1850846, EPI_ISL_1850847, EPI_ISL_1850849, EPI_ISL_1850851, EPI_ISL_1850856, EPI_ISL_1850858, EPI_ISL_1850870, EPI_ISL_1850871, EPI_ISL_1850877, EPI_ISL_1850879, EPI_ISL_1850880, EPI_ISL_1850884, EPI_ISL_1850885, EPI_ISL_1850890, EPI_ISL_1850891, EPI_ISL_1850893, EPI_ISL_1850898, EPI_ISL_1850899, EPI_ISL_1850901, EPI_ISL_1850906, EPI_ISL_1850907, EPI_ISL_1850910, EPI_ISL_1850914, EPI_ISL_1850917, EPI_ISL_1850921, EPI_ISL_1850922, EPI_ISL_1850923, EPI_ISL_1850924, EPI_ISL_1850926, EPI_ISL_1850934, EPI_ISL_1850936, EPI_ISL_1850937, EPI_ISL_1850938, EPI_ISL_1850941, EPI_ISL_1850944, EPI_ISL_1850945, EPI_ISL_1850946, EPI_ISL_1850950, EPI_ISL_1850951, EPI_ISL_1850953, EPI_ISL_1850956, EPI_ISL_1850959, EPI_ISL_1850961, EPI_ISL_1850965, EPI_ISL_1850966, EPI_ISL_1850968, EPI_ISL_1850970, EPI_ISL_1850974, EPI_ISL_1850975, EPI_ISL_1850978, EPI_ISL_1850982, EPI_ISL_1850984, EPI_ISL_1850985, EPI_ISL_1850986, EPI_ISL_1850989, EPI_ISL_1850992, EPI_ISL_1850993, EPI_ISL_1850997, EPI_ISL_1850998, EPI_ISL_1850999, EPI_ISL_1851003, EPI_ISL_1851014, EPI_ISL_1851015, EPI_ISL_1851016, EPI_ISL_1851021, EPI_ISL_1851023, EPI_ISL_1851024, EPI_ISL_1851028, EPI_ISL_1851031, EPI_ISL_1851036, EPI_ISL_1851038, EPI_ISL_1851042, EPI_ISL_1851046, EPI_ISL_1851047, EPI_ISL_1851048, EPI_ISL_1851049, EPI_ISL_1851050, EPI_ISL_1851051, EPI_ISL_1851052, EPI_ISL_1851054, EPI_ISL_1851055, EPI_ISL_1851056, EPI_ISL_1851060, EPI_ISL_1851062, EPI_ISL_1851067, EPI_ISL_1851068, EPI_ISL_1851070, EPI_ISL_1851071, EPI_ISL_1851072, EPI_ISL_1851077, EPI_ISL_1851082, EPI_ISL_1851083, EPI_ISL_1851084, EPI_ISL_1851750, EPI_ISL_1851754, EPI_ISL_1851758, EPI_ISL_1851763, EPI_ISL_1851764, EPI_ISL_1851767, EPI_ISL_1851768, EPI_ISL_1851769, EPI_ISL_1851773, EPI_ISL_1851775, EPI_ISL_1851776, EPI_ISL_1851779, EPI_ISL_1851781, EPI_ISL_1851783, EPI_ISL_1851784, EPI_ISL_1851785, EPI_ISL_1851786, EPI_ISL_1851790, EPI_ISL_1851793, EPI_ISL_1851795, EPI_ISL_1851796, EPI_ISL_1851797, EPI_ISL_1851801, EPI_ISL_1851802, EPI_ISL_1851803, EPI_ISL_1851812, EPI_ISL_1851814, EPI_ISL_1851816, EPI_ISL_1851824, EPI_ISL_1851825, EPI_ISL_1851828, EPI_ISL_1851829, EPI_ISL_1851833, EPI_ISL_1851841, EPI_ISL_1851843, EPI_ISL_1851844, EPI_ISL_1851845, EPI_ISL_1851848, EPI_ISL_1851849, EPI_ISL_1851850, EPI_ISL_1851851, EPI_ISL_1851853, EPI_ISL_1851858, EPI_ISL_1851861, EPI_ISL_1851862, EPI_ISL_1851867, EPI_ISL_1851868, EPI_ISL_1851870, EPI_ISL_1851873, EPI_ISL_1851874, EPI_ISL_1851877, EPI_ISL_1851879, EPI_ISL_1851880, EPI_ISL_1851881, EPI_ISL_1851882, EPI_ISL_1851883, EPI_ISL_1851885, EPI_ISL_1851887, EPI_ISL_1851890, EPI_ISL_1851891, EPI_ISL_1851894, EPI_ISL_1851896, EPI_ISL_1851899, EPI_ISL_1851901, EPI_ISL_1851905, EPI_ISL_1851908, EPI_ISL_1851909, EPI_ISL_1851910, EPI_ISL_1851912, EPI_ISL_1851917, EPI_ISL_1851918, EPI_ISL_1851919, EPI_ISL_1851921, EPI_ISL_1851922, EPI_ISL_1851923, EPI_ISL_1851924, EPI_ISL_1851925, EPI_ISL_1851926, EPI_ISL_1851929, EPI_ISL_1851930, EPI_ISL_1851937, EPI_ISL_1851939, EPI_ISL_1851940, EPI_ISL_1851942, EPI_ISL_1851943, EPI_ISL_1851947, EPI_ISL_1851948, EPI_ISL_1851949, EPI_ISL_1851952, EPI_ISL_1851954, EPI_ISL_1851956, EPI_ISL_1851957, EPI_ISL_1851958, EPI_ISL_1851959, EPI_ISL_1851965, EPI_ISL_1851966, EPI_ISL_1851967, EPI_ISL_1851968, EPI_ISL_1851969, EPI_ISL_1851971, EPI_ISL_1851973, EPI_ISL_1851975, EPI_ISL_1851976, EPI_ISL_1851978, EPI_ISL_1851982, EPI_ISL_1851984, EPI_ISL_1851985, EPI_ISL_1851986, EPI_ISL_1851987, EPI_ISL_1851988, EPI_ISL_1851989, EPI_ISL_1851991, EPI_ISL_1851992, EPI_ISL_1851994, EPI_ISL_1851996, EPI_ISL_1851998, EPI_ISL_1851999, EPI_ISL_1852000, EPI_ISL_1852001, EPI_ISL_1852002, EPI_ISL_1852005, EPI_ISL_1852009, EPI_ISL_1852012, EPI_ISL_1852013, EPI_ISL_1852014, EPI_ISL_1852016, EPI_ISL_1852017, EPI_ISL_1852018, EPI_ISL_1852019, EPI_ISL_1852021, EPI_ISL_1852027, EPI_ISL_1852029, EPI_ISL_1852030, EPI_ISL_1852032, EPI_ISL_1852033, EPI_ISL_1852036, EPI_ISL_1852037, EPI_ISL_1852041, EPI_ISL_1852043, EPI_ISL_1852137, EPI_ISL_1852138, EPI_ISL_1852139, EPI_ISL_1852145, EPI_ISL_1852146, EPI_ISL_1852147, EPI_ISL_1852149, EPI_ISL_1852151, EPI_ISL_1852152, EPI_ISL_1852153, EPI_ISL_1852154, EPI_ISL_1852156, EPI_ISL_1852160, EPI_ISL_1852161, EPI_ISL_1852162, EPI_ISL_1852163, EPI_ISL_1852164, EPI_ISL_1852165, EPI_ISL_1852166, EPI_ISL_1852167, EPI_ISL_1852168, EPI_ISL_1852171, EPI_ISL_1852175, EPI_ISL_1852176, EPI_ISL_1852177, EPI_ISL_1852178, EPI_ISL_1852179, EPI_ISL_1852181, EPI_ISL_1852183, EPI_ISL_1852184, EPI_ISL_1852185, EPI_ISL_1852187, EPI_ISL_1852188, EPI_ISL_1852189, EPI_ISL_1852190, EPI_ISL_1852191, EPI_ISL_1852193, EPI_ISL_1852194, EPI_ISL_1852195, EPI_ISL_1852198, EPI_ISL_1852201, EPI_ISL_1852202, EPI_ISL_1852203, EPI_ISL_1852204, EPI_ISL_1852205, EPI_ISL_1852207, EPI_ISL_1852212, EPI_ISL_1852213, EPI_ISL_1852215, EPI_ISL_1852216, EPI_ISL_1852218, EPI_ISL_1852219, EPI_ISL_1852220, EPI_ISL_1852225, EPI_ISL_1852226, EPI_ISL_1852227, EPI_ISL_1852229, EPI_ISL_1852239, EPI_ISL_1852240, EPI_ISL_1852242, EPI_ISL_1852245, EPI_ISL_1852246, EPI_ISL_1852247, EPI_ISL_1852255, EPI_ISL_1852256, EPI_ISL_1852258, EPI_ISL_1852259, EPI_ISL_1852260, EPI_ISL_1852261, EPI_ISL_1852263, EPI_ISL_1852264, EPI_ISL_1852265, EPI_ISL_1852266, EPI_ISL_1852267, EPI_ISL_1852269, EPI_ISL_1852270, EPI_ISL_1852271, EPI_ISL_1852272, EPI_ISL_1852273, EPI_ISL_1852274, EPI_ISL_1852275, EPI_ISL_1852282, EPI_ISL_1852283, EPI_ISL_1852284, EPI_ISL_1852286, EPI_ISL_1852287, EPI_ISL_1852295, EPI_ISL_1852296, EPI_ISL_1852301, EPI_ISL_1852302, EPI_ISL_1852303, EPI_ISL_1852304, EPI_ISL_1852305, EPI_ISL_1852306, EPI_ISL_1852307, EPI_ISL_1852308, EPI_ISL_1852310, EPI_ISL_1852311, EPI_ISL_1852312, EPI_ISL_1852314, EPI_ISL_1852317, EPI_ISL_1852318, EPI_ISL_1852319, EPI_ISL_1852323, EPI_ISL_1852324, EPI_ISL_1852330, EPI_ISL_1852331, EPI_ISL_1852332, EPI_ISL_1852334, EPI_ISL_1852336, EPI_ISL_1852337, EPI_ISL_1852340, EPI_ISL_1852343, EPI_ISL_1852344, EPI_ISL_1852345, EPI_ISL_1852346, EPI_ISL_1852347, EPI_ISL_1852348, EPI_ISL_1852349, EPI_ISL_1852350, EPI_ISL_1852351, EPI_ISL_1852353, EPI_ISL_1852354, EPI_ISL_1852356, EPI_ISL_1852357, EPI_ISL_1852359, EPI_ISL_1852360, EPI_ISL_1852361, EPI_ISL_1852362, EPI_ISL_1852365, EPI_ISL_1852367, EPI_ISL_1852368, EPI_ISL_1852377, EPI_ISL_1852378, EPI_ISL_1852379, EPI_ISL_1852380, EPI_ISL_1852382, EPI_ISL_1852385, EPI_ISL_1852387, EPI_ISL_1852388, EPI_ISL_1852389, EPI_ISL_1852390, EPI_ISL_1852394, EPI_ISL_1852396, EPI_ISL_1852399, EPI_ISL_1852401, EPI_ISL_1852402, EPI_ISL_1852405, EPI_ISL_1852406, EPI_ISL_1852410, EPI_ISL_1852411, EPI_ISL_1852412, EPI_ISL_1852413, EPI_ISL_1852415, EPI_ISL_1852418, EPI_ISL_1852421, EPI_ISL_1852423, EPI_ISL_1852424, EPI_ISL_1852425, EPI_ISL_1852426, EPI_ISL_1852428, EPI_ISL_1852430, EPI_ISL_1852431, EPI_ISL_1852432, EPI_ISL_1852436, EPI_ISL_1852438, EPI_ISL_1852440, EPI_ISL_1852442, EPI_ISL_1852445, EPI_ISL_2109865, EPI_ISL_2109866, EPI_ISL_2109867, EPI_ISL_2109870, EPI_ISL_2109873, EPI_ISL_2109881, EPI_ISL_2109885, EPI_ISL_2109888, EPI_ISL_2109889, EPI_ISL_2109893, EPI_ISL_2109895, EPI_ISL_2109896, EPI_ISL_2109899, EPI_ISL_2109900, EPI_ISL_2109902, EPI_ISL_2109903, EPI_ISL_2109904, EPI_ISL_2109905, EPI_ISL_2109906, EPI_ISL_2109907, EPI_ISL_2109909, EPI_ISL_2109917, EPI_ISL_2109918, EPI_ISL_2109922, EPI_ISL_2109923, EPI_ISL_2109925, EPI_ISL_2109931, EPI_ISL_2109932, EPI_ISL_2109933, EPI_ISL_2109934, EPI_ISL_2109937, EPI_ISL_2109939, EPI_ISL_2109946, EPI_ISL_2109949, EPI_ISL_2109951, EPI_ISL_2109952, EPI_ISL_2109953, EPI_ISL_2109958, EPI_ISL_2109964, EPI_ISL_2109972, EPI_ISL_2109974, EPI_ISL_2109975, EPI_ISL_2109981, EPI_ISL_2109982, EPI_ISL_2109983, EPI_ISL_2109985, EPI_ISL_2109988, EPI_ISL_2109993, EPI_ISL_2109997, EPI_ISL_2110001, EPI_ISL_2110008, EPI_ISL_2110010, EPI_ISL_2110013, EPI_ISL_2110020, EPI_ISL_2110022, EPI_ISL_2110026, EPI_ISL_2110028, EPI_ISL_2110036, EPI_ISL_2110038, EPI_ISL_2110040, EPI_ISL_2110042, EPI_ISL_2110043, EPI_ISL_2110045, EPI_ISL_2110046, EPI_ISL_2110049, EPI_ISL_2110053, EPI_ISL_2110055, EPI_ISL_2110057, EPI_ISL_2110063, EPI_ISL_2110066, EPI_ISL_2110067, EPI_ISL_2110071, EPI_ISL_2110072, EPI_ISL_2110076, EPI_ISL_2110079, EPI_ISL_2110081, EPI_ISL_2110085, EPI_ISL_2110088, EPI_ISL_2110092, EPI_ISL_2110094, EPI_ISL_2110096, EPI_ISL_2110097, EPI_ISL_2110103, EPI_ISL_2110104, EPI_ISL_2110107, EPI_ISL_2110109, EPI_ISL_2110111, EPI_ISL_2110113, EPI_ISL_2110118, EPI_ISL_2110120, EPI_ISL_2110123, EPI_ISL_2110126, EPI_ISL_2110128, EPI_ISL_2110130, EPI_ISL_2110132, EPI_ISL_2110137, EPI_ISL_2110138, EPI_ISL_2110146, EPI_ISL_2110155, EPI_ISL_2110158, EPI_ISL_2110159, EPI_ISL_2110160, EPI_ISL_2110161, EPI_ISL_2110166, EPI_ISL_2110167, EPI_ISL_2110174, EPI_ISL_2110175, EPI_ISL_2110176, EPI_ISL_2110177, EPI_ISL_2110178, EPI_ISL_2110179, EPI_ISL_2110182, EPI_ISL_2110183, EPI_ISL_2110187, EPI_ISL_2110193, EPI_ISL_2110196, EPI_ISL_2110197, EPI_ISL_2110201, EPI_ISL_2110202, EPI_ISL_2110208, EPI_ISL_2110210, EPI_ISL_2110214, EPI_ISL_2110215, EPI_ISL_2110217, EPI_ISL_2110221, EPI_ISL_2110223, EPI_ISL_2110224, EPI_ISL_2110225, EPI_ISL_2110228, EPI_ISL_2110231, EPI_ISL_2110236, EPI_ISL_2110240, EPI_ISL_2110243, EPI_ISL_2110245, EPI_ISL_2110246, EPI_ISL_2110247, EPI_ISL_2110254, EPI_ISL_2110256, EPI_ISL_2110257, EPI_ISL_2110265, EPI_ISL_2110266, EPI_ISL_2110267, EPI_ISL_2110269, EPI_ISL_2110272, EPI_ISL_2110275, EPI_ISL_2110276, EPI_ISL_2110278, EPI_ISL_2110279, EPI_ISL_2110280, EPI_ISL_2110282, EPI_ISL_2110284, EPI_ISL_2110285, EPI_ISL_2110287, EPI_ISL_2110288, EPI_ISL_2110290, EPI_ISL_2110295, EPI_ISL_2110299, EPI_ISL_2110300, EPI_ISL_2110302, EPI_ISL_2110305, EPI_ISL_2110313, EPI_ISL_2110316, EPI_ISL_2110317, EPI_ISL_2110319, EPI_ISL_2110323, EPI_ISL_2110328, EPI_ISL_2110332, EPI_ISL_2110335, EPI_ISL_2110336, EPI_ISL_2110338, EPI_ISL_2110342, EPI_ISL_2110343, EPI_ISL_2110350, EPI_ISL_2110354, EPI_ISL_2110355, EPI_ISL_2110364, EPI_ISL_2110368, EPI_ISL_2110369, EPI_ISL_2110370, EPI_ISL_2110375, EPI_ISL_2110377, EPI_ISL_2112965, EPI_ISL_2112970, EPI_ISL_2112999, EPI_ISL_2113025, EPI_ISL_2113037, EPI_ISL_2113049, EPI_ISL_2113056, EPI_ISL_2113068, EPI_ISL_2113069, EPI_ISL_2113076, EPI_ISL_2113080, EPI_ISL_2113088, EPI_ISL_2113104, EPI_ISL_2113114, EPI_ISL_2113128, EPI_ISL_2113129, EPI_ISL_2113131, EPI_ISL_2113133, EPI_ISL_2113158, EPI_ISL_2113174, EPI_ISL_2113175, EPI_ISL_2113186, EPI_ISL_2113199, EPI_ISL_2113207, EPI_ISL_2113213, EPI_ISL_2113218, EPI_ISL_2113226, EPI_ISL_2113227 | | | |
| see above | SYNLAB Jena Oncoscreen | Robert Koch Institute | |
| EPI_ISL_1847897, EPI_ISL_1847907, EPI_ISL_1847924, EPI_ISL_1847997, EPI_ISL_1848228, EPI_ISL_1848327, EPI_ISL_1852209, EPI_ISL_1852292, EPI_ISL_1852433 | | | |
| see above | SYNLAB Labor MÃ¼nchen Zentrum LMZ | Robert Koch Institute | |
| EPI_ISL_1851756, EPI_ISL_1851757, EPI_ISL_1851760, EPI_ISL_1851762, EPI_ISL_1851782, EPI_ISL_1851794, EPI_ISL_1851798, EPI_ISL_1851799, EPI_ISL_1851800, EPI_ISL_1851805, EPI_ISL_1851807, EPI_ISL_1851809, EPI_ISL_1851810, EPI_ISL_1851811, EPI_ISL_1851821, EPI_ISL_1851822, EPI_ISL_1851823, EPI_ISL_1851835, EPI_ISL_1851837, EPI_ISL_1851838, EPI_ISL_1851840, EPI_ISL_1851869, EPI_ISL_1851876, EPI_ISL_1851886, EPI_ISL_1851892, EPI_ISL_1851902, EPI_ISL_1851903, EPI_ISL_1851932, EPI_ISL_1851933, EPI_ISL_1851935, EPI_ISL_1851953, EPI_ISL_1851955, EPI_ISL_1851964, EPI_ISL_1851970, EPI_ISL_1851983, EPI_ISL_1851990, EPI_ISL_1851993, EPI_ISL_1852003, EPI_ISL_1852011, EPI_ISL_1852015, EPI_ISL_1852020, EPI_ISL_1852024, EPI_ISL_1852031, EPI_ISL_1852144, EPI_ISL_1852206, EPI_ISL_1852268, EPI_ISL_1852386, EPI_ISL_2109869, EPI_ISL_2109878, EPI_ISL_2109879, EPI_ISL_2109884, EPI_ISL_2109887, EPI_ISL_2109908, EPI_ISL_2109926, EPI_ISL_2109943, EPI_ISL_2109956, EPI_ISL_2109957, EPI_ISL_2109959, EPI_ISL_2109965, EPI_ISL_2109970, EPI_ISL_2109971, EPI_ISL_2109979, EPI_ISL_2109986, EPI_ISL_2109995, EPI_ISL_2109999, EPI_ISL_2110002, EPI_ISL_2110005, EPI_ISL_2110015, EPI_ISL_2110017, EPI_ISL_2110023, EPI_ISL_2110031, EPI_ISL_2110032, EPI_ISL_2110039, EPI_ISL_2110059, EPI_ISL_2110062, EPI_ISL_2110068, EPI_ISL_2110083, EPI_ISL_2110084, EPI_ISL_2110086, EPI_ISL_2110090, EPI_ISL_2110091, EPI_ISL_2110106, EPI_ISL_2110110, EPI_ISL_2110114, EPI_ISL_2110125, EPI_ISL_2110131, EPI_ISL_2110136, EPI_ISL_2110140, EPI_ISL_2110141, EPI_ISL_2110149, EPI_ISL_2110153, EPI_ISL_2110157, EPI_ISL_2110169, EPI_ISL_2110170, EPI_ISL_2110185, EPI_ISL_2110186, EPI_ISL_2110203, EPI_ISL_2110222, EPI_ISL_2110227, EPI_ISL_2110230, EPI_ISL_2110235, EPI_ISL_2110238, EPI_ISL_2110242, EPI_ISL_2110252, EPI_ISL_2110253, EPI_ISL_2110281, EPI_ISL_2110293, EPI_ISL_2110301, EPI_ISL_2110304, EPI_ISL_2110308, EPI_ISL_2110331, EPI_ISL_2110334, EPI_ISL_2110339, EPI_ISL_2110349, EPI_ISL_2110356, EPI_ISL_2110365, EPI_ISL_2110373, EPI_ISL_2110374, EPI_ISL_2113008, EPI_ISL_2113092, EPI_ISL_2113147, EPI_ISL_2113188, EPI_ISL_2113224 | | | |
| see above | SYNLAB MVZ Berlin | Robert Koch Institute | |
| EPI_ISL_2114518, EPI_ISL_2114519, EPI_ISL_2114523, EPI_ISL_2114531, EPI_ISL_2114532, EPI_ISL_2114546, EPI_ISL_2114552, EPI_ISL_2114556, EPI_ISL_2114557, EPI_ISL_2114561, EPI_ISL_2114563, EPI_ISL_2114565, EPI_ISL_2114570, EPI_ISL_2114572, EPI_ISL_2114574, EPI_ISL_2114583, EPI_ISL_2114589, EPI_ISL_2114591, EPI_ISL_2114594, EPI_ISL_2114601, EPI_ISL_2114603, EPI_ISL_2114609, | | | |

EPI_ISL_2114618, EPI_ISL_2114623, EPI_ISL_2114629, EPI_ISL_2114639, EPI_ISL_2114640, EPI_ISL_2114645, EPI_ISL_2114651, EPI_ISL_2114667, EPI_ISL_2114671, EPI_ISL_2114673, EPI_ISL_2114682, EPI_ISL_2114685, EPI_ISL_2114687, EPI_ISL_2114690, EPI_ISL_2114692, EPI_ISL_2114693, EPI_ISL_2114705, EPI_ISL_2114706, EPI_ISL_2114707, EPI_ISL_2114710, EPI_ISL_2114717, EPI_ISL_2114719,
EPI_ISL_2114729, EPI_ISL_2114730, EPI_ISL_2114734, EPI_ISL_2114735, EPI_ISL_2114736, EPI_ISL_2114739, EPI_ISL_2114743, EPI_ISL_2114744, EPI_ISL_2114748, EPI_ISL_2114749, EPI_ISL_2114754, EPI_ISL_2114761, EPI_ISL_2114770, EPI_ISL_2114772, EPI_ISL_2114773, EPI_ISL_2114774, EPI_ISL_2114775, EPI_ISL_2114782, EPI_ISL_2114788, EPI_ISL_2114792, EPI_ISL_2114793, EPI_ISL_2114795,
EPI_ISL_2114798, EPI_ISL_2114800, EPI_ISL_2114802, EPI_ISL_2114803, EPI_ISL_2114804, EPI_ISL_2114808, EPI_ISL_2114810, EPI_ISL_2114813, EPI_ISL_2114818, EPI_ISL_2114819, EPI_ISL_2114827, EPI_ISL_2114831, EPI_ISL_2114840, EPI_ISL_2114846, EPI_ISL_2114847, EPI_ISL_2114849, EPI_ISL_2114852, EPI_ISL_2114853, EPI_ISL_2114862, EPI_ISL_2114864, EPI_ISL_2114865, EPI_ISL_2114868,
EPI_ISL_2114872, EPI_ISL_2114875, EPI_ISL_2114876, EPI_ISL_2114881, EPI_ISL_2114885, EPI_ISL_2114888, EPI_ISL_2114889, EPI_ISL_2114890, EPI_ISL_2114892, EPI_ISL_2114896, EPI_ISL_2114902, EPI_ISL_2114906, EPI_ISL_2114908, EPI_ISL_2114909, EPI_ISL_2114911, EPI_ISL_2114912, EPI_ISL_2114935, EPI_ISL_2114937, EPI_ISL_2114939, EPI_ISL_2114946, EPI_ISL_2114959, EPI_ISL_2114960,
EPI_ISL_2114962, EPI_ISL_2114965, EPI_ISL_2114967, EPI_ISL_2114969, EPI_ISL_2114972, EPI_ISL_2114973, EPI_ISL_2114974, EPI_ISL_2114975, EPI_ISL_2114976, EPI_ISL_2114977, EPI_ISL_2114978, EPI_ISL_2114981, EPI_ISL_2114983, EPI_ISL_2114987, EPI_ISL_2115002, EPI_ISL_2115015, EPI_ISL_2115017, EPI_ISL_2115018, EPI_ISL_2115028, EPI_ISL_2115038, EPI_ISL_2115041,
EPI_ISL_2115044, EPI_ISL_2115045, EPI_ISL_2115046, EPI_ISL_2115050, EPI_ISL_2115054, EPI_ISL_2115056, EPI_ISL_2115058, EPI_ISL_2115059, EPI_ISL_2115064, EPI_ISL_2115067, EPI_ISL_2115072, EPI_ISL_2115073, EPI_ISL_2115075, EPI_ISL_2115076, EPI_ISL_2115080, EPI_ISL_2115083, EPI_ISL_2115085, EPI_ISL_2115088, EPI_ISL_2115090, EPI_ISL_2115091, EPI_ISL_2115093, EPI_ISL_2115097,
EPI_ISL_2115099, EPI_ISL_2115100, EPI_ISL_2115106, EPI_ISL_2115112, EPI_ISL_2115114, EPI_ISL_2115119, EPI_ISL_2115120, EPI_ISL_2115125, EPI_ISL_2115126, EPI_ISL_2115128, EPI_ISL_2118853, EPI_ISL_2118863, EPI_ISL_2118869, EPI_ISL_2118893, EPI_ISL_2118898, EPI_ISL_2118910, EPI_ISL_2118913, EPI_ISL_2118940, EPI_ISL_2118954, EPI_ISL_2118957, EPI_ISL_2118968, EPI_ISL_2118978,
EPI_ISL_2118984, EPI_ISL_2118987, EPI_ISL_2119005, EPI_ISL_2119008, EPI_ISL_2119014, EPI_ISL_2119019, EPI_ISL_2119051, EPI_ISL_2119054, EPI_ISL_2119056, EPI_ISL_2119058, EPI_ISL_2119059, EPI_ISL_2119062, EPI_ISL_2119071, EPI_ISL_2119078, EPI_ISL_2119081, EPI_ISL_2119111, EPI_ISL_2119114, EPI_ISL_2119123, EPI_ISL_2119137, EPI_ISL_2119143, EPI_ISL_2119147, EPI_ISL_2119170,
EPI_ISL_2119182, EPI_ISL_2119209, EPI_ISL_2119221, EPI_ISL_2119227, EPI_ISL_2119239, EPI_ISL_2119251, EPI_ISL_2119259, EPI_ISL_2119262, EPI_ISL_2119276, EPI_ISL_2119291, EPI_ISL_2119302, EPI_ISL_2119316, EPI_ISL_2119331, EPI_ISL_2119334, EPI_ISL_2119342, EPI_ISL_2119353, EPI_ISL_2119362, EPI_ISL_2119388, EPI_ISL_2119429, EPI_ISL_2119433, EPI_ISL_2119466, EPI_ISL_2119472,
EPI_ISL_2119478, EPI_ISL_2119499, EPI_ISL_2119508, EPI_ISL_2119514, EPI_ISL_2119519, EPI_ISL_2119527, EPI_ISL_2119538, EPI_ISL_2119542, EPI_ISL_2119547, EPI_ISL_2119555, EPI_ISL_2119560, EPI_ISL_2119578, EPI_ISL_2119585, EPI_ISL_2119586, EPI_ISL_2119592, EPI_ISL_2119605, EPI_ISL_2119621, EPI_ISL_2119640, EPI_ISL_2119643, EPI_ISL_2119659, EPI_ISL_2119673, EPI_ISL_2119675,
EPI_ISL_2119678, EPI_ISL_2119681, EPI_ISL_2119684, EPI_ISL_2119687, EPI_ISL_2119690, EPI_ISL_2119693, EPI_ISL_2119700, EPI_ISL_2119703, EPI_ISL_2119705, EPI_ISL_2119725, EPI_ISL_2119728, EPI_ISL_2119730, EPI_ISL_2119739, EPI_ISL_2119773, EPI_ISL_2119776, EPI_ISL_2119779, EPI_ISL_2119785, EPI_ISL_2119793, EPI_ISL_2119799, EPI_ISL_2119807, EPI_ISL_2119815, EPI_ISL_2119818,
EPI_ISL_2119827, EPI_ISL_2119862, EPI_ISL_2119882, EPI_ISL_2119884, EPI_ISL_2119890, EPI_ISL_2119896, EPI_ISL_2119909, EPI_ISL_2119916, EPI_ISL_2119920, EPI_ISL_2119923, EPI_ISL_2119947, EPI_ISL_2119966, EPI_ISL_2119983, EPI_ISL_2120007, EPI_ISL_2120014, EPI_ISL_2120038, EPI_ISL_2127704, EPI_ISL_2127705, EPI_ISL_2127707, EPI_ISL_2127712, EPI_ISL_2127713, EPI_ISL_2127715,
EPI_ISL_2127716, EPI_ISL_2127720, EPI_ISL_2127721, EPI_ISL_2127737, EPI_ISL_2127747, EPI_ISL_2127754, EPI_ISL_2127783, EPI_ISL_2127786, EPI_ISL_2127788, EPI_ISL_2127790, EPI_ISL_2127796, EPI_ISL_2127819, EPI_ISL_2127824, EPI_ISL_2127856, EPI_ISL_2127857, EPI_ISL_2127862, EPI_ISL_2127868, EPI_ISL_2127872, EPI_ISL_2127891, EPI_ISL_2127898, EPI_ISL_2127900,
EPI_ISL_2127907, EPI_ISL_2127913, EPI_ISL_2127947, EPI_ISL_2127957, EPI_ISL_2127965, EPI_ISL_2127970, EPI_ISL_2127980, EPI_ISL_2127982, EPI_ISL_2127996, EPI_ISL_2128006, EPI_ISL_2128008, EPI_ISL_2128012, EPI_ISL_2128019, EPI_ISL_2128022, EPI_ISL_2128024, EPI_ISL_2128030, EPI_ISL_2128033, EPI_ISL_2128041, EPI_ISL_2128072, EPI_ISL_2128078, EPI_ISL_2128083, EPI_ISL_2128098,
EPI_ISL_2128100, EPI_ISL_2128102, EPI_ISL_2128114, EPI_ISL_2128133, EPI_ISL_2128154, EPI_ISL_2128166, EPI_ISL_2128170, EPI_ISL_2128179, EPI_ISL_2128181, EPI_ISL_2128187, EPI_ISL_2128197, EPI_ISL_2128206, EPI_ISL_2128209, EPI_ISL_2128211, EPI_ISL_2128217, EPI_ISL_2128228, EPI_ISL_2128232, EPI_ISL_2128243, EPI_ISL_2128252, EPI_ISL_2128258, EPI_ISL_2128262, EPI_ISL_2128271,
EPI_ISL_2128276, EPI_ISL_2128283, EPI_ISL_2128296, EPI_ISL_2128303, EPI_ISL_2128309, EPI_ISL_2128313, EPI_ISL_2128318, EPI_ISL_2128322, EPI_ISL_2128326, EPI_ISL_2128336, EPI_ISL_2128340, EPI_ISL_2128344, EPI_ISL_2128355, EPI_ISL_2128367, EPI_ISL_2128640, EPI_ISL_2128642, EPI_ISL_2128645, EPI_ISL_2128649, EPI_ISL_2128654, EPI_ISL_2128668, EPI_ISL_2128672, EPI_ISL_2128674,
EPI_ISL_2128681, EPI_ISL_2128682, EPI_ISL_2128689, EPI_ISL_2128692, EPI_ISL_2128697, EPI_ISL_2128701, EPI_ISL_2128709, EPI_ISL_2128715, EPI_ISL_2128721, EPI_ISL_2128726, EPI_ISL_2128728, EPI_ISL_2128732, EPI_ISL_2128736, EPI_ISL_2128738, EPI_ISL_2128739, EPI_ISL_2128740, EPI_ISL_2128741, EPI_ISL_2128756, EPI_ISL_2128758, EPI_ISL_2128774, EPI_ISL_2128775, EPI_ISL_2128780,
EPI_ISL_2128786, EPI_ISL_2128795, EPI_ISL_2128803, EPI_ISL_2128811, EPI_ISL_2128826, EPI_ISL_2128838, EPI_ISL_2128845, EPI_ISL_2128846, EPI_ISL_2128863, EPI_ISL_2128864, EPI_ISL_2128868, EPI_ISL_2128872, EPI_ISL_2128879, EPI_ISL_2128888, EPI_ISL_2128894, EPI_ISL_2128900, EPI_ISL_2128904, EPI_ISL_2128909, EPI_ISL_2128910, EPI_ISL_2128911, EPI_ISL_2128921, EPI_ISL_2128923,
EPI_ISL_2128924, EPI_ISL_2128927, EPI_ISL_2128933, EPI_ISL_2128942, EPI_ISL_2128943, EPI_ISL_2128949

see above SYNLAB MVZ Heidelberg Robert Koch Institute

EPI_ISL_1850771, EPI_ISL_1850800, EPI_ISL_1850805, EPI_ISL_1850813, EPI_ISL_1850818, EPI_ISL_1850825, EPI_ISL_1850826, EPI_ISL_1850832, EPI_ISL_1850834, EPI_ISL_1850844, EPI_ISL_1850848, EPI_ISL_1850854, EPI_ISL_1850857, EPI_ISL_1850859, EPI_ISL_1850860, EPI_ISL_1850865, EPI_ISL_1850867, EPI_ISL_1850868, EPI_ISL_1850869, EPI_ISL_1850873, EPI_ISL_1850875, EPI_ISL_1850881,
EPI_ISL_1850882, EPI_ISL_1850888, EPI_ISL_1850889, EPI_ISL_1850892, EPI_ISL_1850895, EPI_ISL_1850900, EPI_ISL_1850902, EPI_ISL_1850911, EPI_ISL_1850912, EPI_ISL_1850925, EPI_ISL_1850927, EPI_ISL_1850928, EPI_ISL_1850931, EPI_ISL_1850932, EPI_ISL_1850942, EPI_ISL_1850952, EPI_ISL_1850958, EPI_ISL_1850964, EPI_ISL_1850967, EPI_ISL_1850971, EPI_ISL_1850980, EPI_ISL_1850987,
EPI_ISL_1850994, EPI_ISL_1850995, EPI_ISL_1851004, EPI_ISL_1851005, EPI_ISL_1851006, EPI_ISL_1851007, EPI_ISL_1851011, EPI_ISL_1851019, EPI_ISL_1851020, EPI_ISL_1851026, EPI_ISL_1851029, EPI_ISL_1851030, EPI_ISL_1851032, EPI_ISL_1851034, EPI_ISL_1851043, EPI_ISL_1851061, EPI_ISL_1851064, EPI_ISL_1851065, EPI_ISL_1851069, EPI_ISL_1851073, EPI_ISL_1851079, EPI_ISL_1851081,
EPI_ISL_1851086, EPI_ISL_1851751, EPI_ISL_1851752, EPI_ISL_1851753, EPI_ISL_1851755, EPI_ISL_1851759, EPI_ISL_1851761, EPI_ISL_1851765, EPI_ISL_1851766, EPI_ISL_1851770, EPI_ISL_1851771, EPI_ISL_1851772, EPI_ISL_1851774, EPI_ISL_1851777, EPI_ISL_1851778, EPI_ISL_1851780, EPI_ISL_1851787, EPI_ISL_1851788, EPI_ISL_1851789, EPI_ISL_1851791, EPI_ISL_1851792, EPI_ISL_1851804,
EPI_ISL_1851806, EPI_ISL_1851808, EPI_ISL_1851813, EPI_ISL_1851817, EPI_ISL_1851818, EPI_ISL_1851819, EPI_ISL_1851827, EPI_ISL_1851830, EPI_ISL_1851832, EPI_ISL_1851834, EPI_ISL_1851839, EPI_ISL_1851846, EPI_ISL_1851854, EPI_ISL_1851855, EPI_ISL_1851856, EPI_ISL_1851859, EPI_ISL_1851860, EPI_ISL_1851863, EPI_ISL_1851864, EPI_ISL_1851865, EPI_ISL_1851866, EPI_ISL_1851871,
EPI_ISL_1851872, EPI_ISL_1851875, EPI_ISL_1851878, EPI_ISL_1851884, EPI_ISL_1851888, EPI_ISL_1851889, EPI_ISL_1851893, EPI_ISL_1851895, EPI_ISL_1851897, EPI_ISL_1851898, EPI_ISL_1851900, EPI_ISL_1851904, EPI_ISL_1851906, EPI_ISL_1851907, EPI_ISL_1851911, EPI_ISL_1851913, EPI_ISL_1851914, EPI_ISL_1851915, EPI_ISL_1851916, EPI_ISL_1851920, EPI_ISL_1851927, EPI_ISL_1851928,
EPI_ISL_1851934, EPI_ISL_1851936, EPI_ISL_1851938, EPI_ISL_1851941, EPI_ISL_1851944, EPI_ISL_1851945, EPI_ISL_1851946, EPI_ISL_1851950, EPI_ISL_1851951, EPI_ISL_1851960, EPI_ISL_1851961, EPI_ISL_1851962, EPI_ISL_1851963, EPI_ISL_1851972, EPI_ISL_1851974, EPI_ISL_1851977, EPI_ISL_1851979, EPI_ISL_1851980, EPI_ISL_1851981, EPI_ISL_1851995, EPI_ISL_1851997, EPI_ISL_1852004,
EPI_ISL_1852006, EPI_ISL_1852007, EPI_ISL_1852008, EPI_ISL_1852010, EPI_ISL_1852022, EPI_ISL_1852023, EPI_ISL_1852025, EPI_ISL_1852026, EPI_ISL_1852028, EPI_ISL_1852034, EPI_ISL_1852035, EPI_ISL_1852038, EPI_ISL_1852039, EPI_ISL_1852040, EPI_ISL_1852042, EPI_ISL_1852140, EPI_ISL_1852142, EPI_ISL_1852143, EPI_ISL_1852148, EPI_ISL_1852150, EPI_ISL_1852155, EPI_ISL_1852158,
EPI_ISL_1852159, EPI_ISL_1852169, EPI_ISL_1852170, EPI_ISL_1852172, EPI_ISL_1852173, EPI_ISL_1852174, EPI_ISL_1852180, EPI_ISL_1852182, EPI_ISL_1852186, EPI_ISL_1852196, EPI_ISL_1852199, EPI_ISL_1852200, EPI_ISL_1852208, EPI_ISL_1852210, EPI_ISL_1852211, EPI_ISL_1852214, EPI_ISL_1852217, EPI_ISL_1852221, EPI_ISL_1852222, EPI_ISL_1852223, EPI_ISL_1852224, EPI_ISL_1852228,
EPI_ISL_1852230, EPI_ISL_1852231, EPI_ISL_1852232, EPI_ISL_1852233, EPI_ISL_1852234, EPI_ISL_1852235, EPI_ISL_1852236, EPI_ISL_1852237, EPI_ISL_1852238, EPI_ISL_1852241, EPI_ISL_1852244, EPI_ISL_1852248, EPI_ISL_1852249, EPI_ISL_1852250, EPI_ISL_1852251, EPI_ISL_1852252, EPI_ISL_1852253, EPI_ISL_1852254, EPI_ISL_1852257, EPI_ISL_1852262, EPI_ISL_1852276, EPI_ISL_1852277,
EPI_ISL_1852278, EPI_ISL_1852279, EPI_ISL_1852280, EPI_ISL_1852281, EPI_ISL_1852285, EPI_ISL_1852288, EPI_ISL_1852289, EPI_ISL_1852290, EPI_ISL_1852291, EPI_ISL_1852293, EPI_ISL_1852294, EPI_ISL_1852297, EPI_ISL_1852298, EPI_ISL_1852299, EPI_ISL_1852300, EPI_ISL_1852309, EPI_ISL_1852313, EPI_ISL_1852315, EPI_ISL_1852316, EPI_ISL_1852320, EPI_ISL_1852321, EPI_ISL_1852322,
EPI_ISL_1852325, EPI_ISL_1852327, EPI_ISL_1852328, EPI_ISL_1852329, EPI_ISL_1852333, EPI_ISL_1852335, EPI_ISL_1852338, EPI_ISL_1852339, EPI_ISL_1852342, EPI_ISL_1852352, EPI_ISL_1852355, EPI_ISL_1852358, EPI_ISL_1852363, EPI_ISL_1852364, EPI_ISL_1852366, EPI_ISL_1852369, EPI_ISL_1852370, EPI_ISL_1852371, EPI_ISL_1852372, EPI_ISL_1852373, EPI_ISL_1852375, EPI_ISL_1852376,
EPI_ISL_1852381, EPI_ISL_1852383, EPI_ISL_1852384, EPI_ISL_1852391, EPI_ISL_1852392, EPI_ISL_1852393, EPI_ISL_1852395, EPI_ISL_1852397, EPI_ISL_1852398, EPI_ISL_1852400, EPI_ISL_1852403, EPI_ISL_1852404, EPI_ISL_1852407, EPI_ISL_1852408, EPI_ISL_1852409, EPI_ISL_1852414, EPI_ISL_1852416, EPI_ISL_1852417, EPI_ISL_1852419, EPI_ISL_1852420, EPI_ISL_1852422, EPI_ISL_1852427,
EPI_ISL_1852429, EPI_ISL_1852434, EPI_ISL_1852435, EPI_ISL_1852437, EPI_ISL_1852439, EPI_ISL_1852441, EPI_ISL_1852443, EPI_ISL_1852444, EPI_ISL_1852446, EPI_ISL_1852447, EPI_ISL_2109920, EPI_ISL_2109928, EPI_ISL_2109929, EPI_ISL_2109936, EPI_ISL_2109940, EPI_ISL_2109954, EPI_ISL_2109961, EPI_ISL_2109962, EPI_ISL_2109966, EPI_ISL_2109976, EPI_ISL_2109990, EPI_ISL_2109992,
EPI_ISL_2109994, EPI_ISL_2109996, EPI_ISL_2110009, EPI_ISL_2110011, EPI_ISL_2110016, EPI_ISL_2110018, EPI_ISL_2110021, EPI_ISL_2110033, EPI_ISL_2110041, EPI_ISL_2110044, EPI_ISL_2110048, EPI_ISL_2110052, EPI_ISL_2110054, EPI_ISL_2110060, EPI_ISL_2110061, EPI_ISL_2110064, EPI_ISL_2110087, EPI_ISL_2110093, EPI_ISL_2110116, EPI_ISL_2110117, EPI_ISL_2110121, EPI_ISL_2110122,
EPI_ISL_2110142, EPI_ISL_2110145, EPI_ISL_2110150, EPI_ISL_2110152, EPI_ISL_2110168, EPI_ISL_2110171, EPI_ISL_2110173, EPI_ISL_2110189, EPI_ISL_2110190, EPI_ISL_2110199, EPI_ISL_2110220, EPI_ISL_2110229, EPI_ISL_2110232, EPI_ISL_2110233, EPI_ISL_2110248, EPI_ISL_2110251, EPI_ISL_2110259, EPI_ISL_2110260, EPI_ISL_2110264, EPI_ISL_2110270, EPI_ISL_2110271, EPI_ISL_2110291,
EPI_ISL_2110294, EPI_ISL_2110306, EPI_ISL_2110322, EPI_ISL_2110327, EPI_ISL_2110329, EPI_ISL_2110344, EPI_ISL_2110346, EPI_ISL_2110348, EPI_ISL_2110351, EPI_ISL_2110352, EPI_ISL_2110353, EPI_ISL_2110357, EPI_ISL_2110359, EPI_ISL_2110366, EPI_ISL_2110372

see above SYNLAB MVZ Leinfelden-Echterdingen Robert Koch Institute

EPI_ISL_1728286, EPI_ISL_1728287, EPI_ISL_1728288, EPI_ISL_1728289, EPI_ISL_1728290, EPI_ISL_1728291, EPI_ISL_1728292, EPI_ISL_1728293, EPI_ISL_1728294, EPI_ISL_1728295, EPI_ISL_1728296, EPI_ISL_1728297, EPI_ISL_1728298, EPI_ISL_1728299, EPI_ISL_1728300, EPI_ISL_1728301, EPI_ISL_1728302, EPI_ISL_1728303, EPI_ISL_1728304, EPI_ISL_1728305, EPI_ISL_1728306, EPI_ISL_1728307,
EPI_ISL_1728308, EPI_ISL_1728309, EPI_ISL_1728310, EPI_ISL_1728311, EPI_ISL_1728312, EPI_ISL_1728313, EPI_ISL_1728314, EPI_ISL_1728315, EPI_ISL_1728316, EPI_ISL_1728317, EPI_ISL_1728318, EPI_ISL_1728319, EPI_ISL_1728320, EPI_ISL_1728321, EPI_ISL_1728322, EPI_ISL_1728323, EPI_ISL_1728324, EPI_ISL_1728325, EPI_ISL_1728326, EPI_ISL_1728327, EPI_ISL_1728328, EPI_ISL_1728329,
EPI_ISL_1728330, EPI_ISL_1728331, EPI_ISL_1728332, EPI_ISL_1728333, EPI_ISL_1728334, EPI_ISL_1728335, EPI_ISL_1728336, EPI_ISL_1728337, EPI_ISL_1728338, EPI_ISL_1728339, EPI_ISL_1728340, EPI_ISL_1728341, EPI_ISL_1728342, EPI_ISL_1728343, EPI_ISL_1728344, EPI_ISL_1728345, EPI_ISL_1728346, EPI_ISL_1728347, EPI_ISL_1728348, EPI_ISL_1728349, EPI_ISL_1728350, EPI_ISL_1728351,
EPI_ISL_1728352, EPI_ISL_1728353, EPI_ISL_1728354, EPI_ISL_1728355, EPI_ISL_1728356, EPI_ISL_1728357, EPI_ISL_1728358, EPI_ISL_1728359, EPI_ISL_1728360, EPI_ISL_1728361, EPI_ISL_1728362, EPI_ISL_1728363, EPI_ISL_1728364, EPI_ISL_1728365, EPI_ISL_1728366, EPI_ISL_1728367, EPI_ISL_1728368, EPI_ISL_1728369, EPI_ISL_1728370, EPI_ISL_1728371, EPI_ISL_1728372, EPI_ISL_1728373,
EPI_ISL_1728374, EPI_ISL_1728375, EPI_ISL_1728376, EPI_ISL_1728377, EPI_ISL_1728378, EPI_ISL_1728379, EPI_ISL_1728380, EPI_ISL_1728381, EPI_ISL_1728382, EPI_ISL_1728383, EPI_ISL_1728384, EPI_ISL_1728385, EPI_ISL_1728386, EPI_ISL_1728387, EPI_ISL_1728388, EPI_ISL_1728389, EPI_ISL_1728390, EPI_ISL_1728391, EPI_ISL_1728392, EPI_ISL_1728393, EPI_ISL_1728394, EPI_ISL_1728395,
EPI_ISL_1728396, EPI_ISL_1728397, EPI_ISL_1728398, EPI_ISL_1728399, EPI_ISL_1728400, EPI_ISL_1728401, EPI_ISL_1728402, EPI_ISL_1728403, EPI_ISL_1728404, EPI_ISL_1728405, EPI_ISL_1728406, EPI_ISL_1728407, EPI_ISL_1728408, EPI_ISL_1728409, EPI_ISL_1728410, EPI_ISL_1728411, EPI_ISL_1728412, EPI_ISL_1728413, EPI_ISL_1728414, EPI_ISL_1728415, EPI_ISL_1728416, EPI_ISL_1728417,
EPI_ISL_1728418, EPI_ISL_1728419, EPI_ISL_1728420, EPI_ISL_1728421, EPI_ISL_1728422, EPI_ISL_1728423, EPI_ISL_1728424, EPI_ISL_1728425, EPI_ISL_1728426, EPI_ISL_1728427, EPI_ISL_1728428, EPI_ISL_1728429, EPI_ISL_1728430, EPI_ISL_1728431, EPI_ISL_1728432, EPI_ISL_1728433, EPI_ISL_1728434, EPI_ISL_1728435, EPI_ISL_1728436, EPI_ISL_1728437, EPI_ISL_1728438, EPI_ISL_1728439,
EPI_ISL_1728440, EPI_ISL_1728441, EPI_ISL_1728442, EPI_ISL_1728443, EPI_ISL_1728444, EPI_ISL_1728445, EPI_ISL_1728446, EPI_ISL_1728447, EPI_ISL_1728448, EPI_ISL_1728449, EPI_ISL_1728450, EPI_ISL_1728451, EPI_ISL_1728452, EPI_ISL_1728453, EPI_ISL_1728454, EPI_ISL_1728455, EPI_ISL_1728456, EPI_ISL_1728457, EPI_ISL_1728458, EPI_ISL_1728459, EPI_ISL_1728460, EPI_ISL_1728461,
EPI_ISL_1728462, EPI_ISL_1728463, EPI_ISL_1728464, EPI_ISL_1728465, EPI_ISL_1728466, EPI_ISL_1728467, EPI_ISL_1728468, EPI_ISL_1728469, EPI_ISL_1728470, EPI_ISL_1728471, EPI_ISL_1728472, EPI_ISL_1728473, EPI_ISL_1728474, EPI_ISL_1728475, EPI_ISL_1728476, EPI_ISL_1728477, EPI_ISL_1728478, EPI_ISL_1728479, EPI_ISL_1728480, EPI_ISL_1728481, EPI_ISL_1728482, EPI_ISL_1728483,
EPI_ISL_1728484, EPI_ISL_1728485, EPI_ISL_1728486, EPI_ISL_1728487, EPI_ISL_1728488, EPI_ISL_1728489, EPI_ISL_1728490, EPI_ISL_1728491, EPI_ISL_1728492, EPI_ISL_1728493, EPI_ISL_1728494, EPI_ISL_1728495, EPI_ISL_1728496, EPI_ISL_1728497, EPI_ISL_1728498, EPI_ISL_1728499, EPI_ISL_1728500, EPI_ISL_1728501, EPI_ISL_1728502, EPI_ISL_1728503, EPI_ISL_1728504, EPI_ISL_1728505,
EPI_ISL_1728506, EPI_ISL_1728507, EPI_ISL_1728508, EPI_ISL_1728509, EPI_ISL_1728510, EPI_ISL_1728511, EPI_ISL_1728512, EPI_ISL_1728513, EPI_ISL_1728514, EPI_ISL_1728515, EPI_ISL_1728516, EPI_ISL_1728517, EPI_ISL_1728518, EPI_ISL_1728519, EPI_ISL_1728521, EPI_ISL_1728522, EPI_ISL_1728523, EPI_ISL_1728524, EPI_ISL_1728525, EPI_ISL_1728526, EPI_ISL_1728527, EPI_ISL_1728528,
EPI_ISL_1728529, EPI_ISL_1728530, EPI_ISL_1728531, EPI_ISL_1728532, EPI_ISL_1728534, EPI_ISL_1728535, EPI_ISL_1728536, EPI_ISL_1728537, EPI_ISL_1728538, EPI_ISL_1728539, EPI_ISL_1728540, EPI_ISL_1728541, EPI_ISL_1728542, EPI_ISL_1728543, EPI_ISL_1728544, EPI_ISL_1728546, EPI_ISL_1728547, EPI_ISL_1728548, EPI_ISL_1728549, EPI_ISL_1728550, EPI_ISL_1728551, EPI_ISL_1728552,
EPI_ISL_1728553, EPI_ISL_1728554, EPI_ISL_1728555, EPI_ISL_1728556, EPI_ISL_1728557, EPI_ISL_1728558, EPI_ISL_1728559, EPI_ISL_1728560, EPI_ISL_1728561, EPI_ISL_1728562, EPI_ISL_1728563, EPI_ISL_1728564, EPI_ISL_1728565, EPI_ISL_1728566, EPI_ISL_1728567, EPI_ISL_1728568, EPI_ISL_1728569, EPI_ISL_1728570, EPI_ISL_1728571, EPI_ISL_1728572, EPI_ISL_1728573, EPI_ISL_1728574,
EPI_ISL_1728575, EPI_ISL_1728576, EPI_ISL_1728577, EPI_ISL_1728578, EPI_ISL_1728579, EPI_ISL_1728580, EPI_ISL_1728581, EPI_ISL_1728582, EPI_ISL_1728583, EPI_ISL_1728584, EPI_ISL_1728585, EPI_ISL_1728586, EPI_ISL_1728587, EPI_ISL_1728588, EPI_ISL_1728589, EPI_ISL_1728590, EPI_ISL_1729392, EPI_ISL_1729393, EPI_ISL_1729394, EPI_ISL_1729395, EPI_ISL_1729396, EPI_ISL_1729398,
EPI_ISL_1729399, EPI_ISL_1729401, EPI_ISL_1729402, EPI_ISL_1729403, EPI_ISL_1729404, EPI_ISL_1729406, EPI_ISL_1729407, EPI_ISL_1729409, EPI_ISL_1729411, EPI_ISL_1729412, EPI_ISL_1729415, EPI_ISL_1729417, EPI_ISL_1729419, EPI_ISL_1729422, EPI_ISL_1729423, EPI_ISL_1729426, EPI_ISL_1729428, EPI_ISL_1729431, EPI_ISL_1729440, EPI_ISL_1729442, EPI_ISL_1729444, EPI_ISL_1729448,
EPI_ISL_1729449, EPI_ISL_1729453, EPI_ISL_1729456, EPI_ISL_1729462, EPI_ISL_1729464, EPI_ISL_1729465, EPI_ISL_1729469, EPI_ISL_1729471, EPI_ISL_1729474, EPI_ISL_1729475, EPI_ISL_1729477, EPI_ISL_1729482, EPI_ISL_1729484, EPI_ISL_1729485, EPI_ISL_1729488, EPI_ISL_1729489, EPI_ISL_1729491, EPI_ISL_1729494, EPI_ISL_1729495, EPI_ISL_1729496, EPI_ISL_1729497, EPI_ISL_1729498,
EPI_ISL_1729502, EPI_ISL_1729504, EPI_ISL_1729505, EPI_ISL_1729507, EPI_ISL_1729508, EPI_ISL_1729511, EPI_ISL_1729513, EPI_ISL_1729515, EPI_ISL_1729517, EPI_ISL_1729518, EPI_ISL_1729520, EPI_ISL_1729522, EPI_ISL_1729523, EPI_ISL_1729528, EPI_ISL_1729529, EPI_ISL_1729530, EPI_ISL_1729532, EPI_ISL_1729533, EPI_ISL_1729534, EPI_ISL_1729536, EPI_ISL_1729539,
EPI_ISL_1729543, EPI_ISL_1729545, EPI_ISL_1729546, EPI_ISL_1729547, EPI_ISL_1729550, EPI_ISL_1729552, EPI_ISL_1729553, EPI_ISL_1729555, EPI_ISL_1729556, EPI_ISL_1729557, EPI_ISL_1729559, EPI_ISL_1729561, EPI_ISL_1729563, EPI_ISL_1729564, EPI_ISL_1729566, EPI_ISL_1729569, EPI_ISL_1729571, EPI_ISL_1729572, EPI_ISL_1729574, EPI_ISL_1729576, EPI_ISL_1729577, EPI_ISL_1729579,
EPI_ISL_1729580, EPI_ISL_1729583, EPI_ISL_1729584, EPI_ISL_1729585, EPI_ISL_1729588, EPI_ISL_1729589, EPI_ISL_1729590, EPI_ISL_1729593, EPI_ISL_1729594, EPI_ISL_1729595, EPI_ISL_1729599, EPI_ISL_1729601, EPI_ISL_1729602, EPI_ISL_1729605, EPI_ISL_1729606, EPI_ISL_1729607, EPI_ISL_1729608, EPI_ISL_1729609, EPI_ISL_1729610, EPI_ISL_1729611, EPI_ISL_1729612,
EPI_ISL_1729615, EPI_ISL_1729616, EPI_ISL_1729621, EPI_ISL_1729622, EPI_ISL_1729626, EPI_ISL_1729629, EPI_ISL_1729630, EPI_ISL_1729632, EPI_ISL_1729635, EPI_ISL_1729637, EPI_ISL_1729638, EPI_ISL_1729641, EPI_ISL_1729643, EPI_ISL_1729645, EPI_ISL_1729648, EPI_ISL_1729649, EPI_ISL_1729650, EPI_ISL_1729655, EPI_ISL_1729658, EPI_ISL_1729663, EPI_ISL_1729666, EPI_ISL_1729669,
EPI_ISL_1729671, EPI_ISL_1729675, EPI_ISL_1729677, EPI_ISL_1729678, EPI_ISL_1729682, EPI_ISL_1729683, EPI_ISL_1729686, EPI_ISL_1729689, EPI_ISL_1729690, EPI_ISL_1729692, EPI_ISL_1729694, EPI_ISL_1729695, EPI_ISL_1729702, EPI_ISL_1729703, EPI_ISL_1729704, EPI_ISL_1729705, EPI_ISL_1844644, EPI_ISL_1844645, EPI_ISL_1844646,
EPI_ISL_1844647, EPI_ISL_1844649, EPI_ISL_1844650, EPI_ISL_1844651, EPI_ISL_1844654, EPI_ISL_1844656, EPI_ISL_1844657, EPI_ISL_1844658, EPI_ISL_1844659, EPI_ISL_1844662, EPI_ISL_1844664, EPI_ISL_1844666, EPI_ISL_1844668, EPI_ISL_1844669, EPI_ISL_1844670, EPI_ISL_1844674, EPI_ISL_1844676, EPI_ISL_1844677, EPI_ISL_1844679, EPI_ISL_1844680, EPI_ISL_1844681, EPI_ISL_1844695,
EPI_ISL_1844696, EPI_ISL_1844697, EPI_ISL_1844699, EPI_ISL_1844700, EPI_ISL_1844701, EPI_ISL_1844705, EPI_ISL_1844708, EPI_ISL_1844712, EPI_ISL_1844713, EPI_ISL_1844717, EPI_ISL_1844718, EPI_ISL_1844719, EPI_ISL_1844722, EPI_ISL_1844723, EPI_ISL_1844726, EPI_ISL_1844730, EPI_ISL_1844732, EPI_ISL_1844735, EPI_ISL_1844737, EPI_ISL_1844739, EPI_ISL_1844740,
EPI_ISL_1844742, EPI_ISL_1844745, EPI_ISL_1844746, EPI_ISL_1844751, EPI_ISL_1844754, EPI_ISL_1844757, EPI_ISL_1844759, EPI_ISL_1844760, EPI_ISL_1844764, EPI_ISL_1844766, EPI_ISL_1844770, EPI_ISL_1844775, EPI_ISL_1844783, EPI_ISL_1844785, EPI_ISL_1844786, EPI_ISL_1844789, EPI_ISL_1844790, EPI_ISL_1844794, EPI_ISL_1844795, EPI_ISL_1844797, EPI_ISL_1844799,
EPI_ISL_1844800, EPI_ISL_1844801, EPI_ISL_1844805, EPI_ISL_1844806, EPI_ISL_1844807, EPI_ISL_1844810, EPI_ISL_1844811, EPI_ISL_1844813, EPI_ISL_1844815, EPI_ISL_1844819, EPI_ISL_1844821, EPI_ISL_1844822, EPI_ISL_1844823, EPI_ISL_1844824, EPI_ISL_1844825, EPI_ISL_1844826, EPI_ISL_1844827, EPI_ISL_1844829, EPI_ISL_1844830, EPI_ISL_1844831, EPI_ISL_1844833, EPI_ISL_1844834,
EPI_ISL_1844835, EPI_ISL_1844839, EPI_ISL_1844841, EPI_ISL_1844842, EPI_ISL_1844843, EPI_ISL_1844844, EPI_ISL_1844845, EPI_ISL_1844846, EPI_ISL_1844847, EPI_ISL_1844850, EPI_ISL_1844852, EPI_ISL_1844854, EPI_ISL_1844855, EPI_ISL_1844859, EPI_ISL_1844860, EPI_ISL_1844861, EPI_ISL_1844862, EPI_ISL_1844863, EPI_ISL_1844864, EPI_ISL_1844866, EPI_ISL_1844867, EPI_ISL_1844868,
EPI_ISL_1844872, EPI_ISL_1844874, EPI_ISL_1844875, EPI_ISL_1844877, EPI_ISL_1844878, EPI_ISL_1844879, EPI_ISL_1844881, EPI_ISL_1844882, EPI_ISL_1844883, EPI_ISL_1844884, EPI_ISL_1844889, EPI_ISL_1844891, EPI_ISL_1844892, EPI_ISL_1844893, EPI_ISL_1844895, EPI_ISL_1844897, EPI_ISL_1844899, EPI_ISL_1844900, EPI_ISL_1844901, EPI_ISL_1844902, EPI_ISL_1844903, EPI_ISL_1844904,
EPI_ISL_1844906, EPI_ISL_1844908, EPI_ISL_1844910, EPI_ISL_1844911, EPI_ISL_1844912, EPI_ISL_1844919, EPI_ISL_1844920, EPI_ISL_1844921, EPI_ISL_1844922, EPI_ISL_1844923, EPI_ISL_1844927, EPI_ISL_1844928, EPI_ISL_1844933, EPI_ISL_1844937, EPI_ISL_1844938, EPI_ISL_1844939, EPI_ISL_1844940, EPI_ISL_1844941, EPI_ISL_1844942, EPI_ISL_1844946, EPI_ISL_1844948, EPI_ISL_1844951,
EPI_ISL_1844952, EPI_ISL_1844954, EPI_ISL_1844955, EPI_ISL_1844957, EPI_ISL_1844959, EPI_ISL_1844961, EPI_ISL_1844966, EPI_ISL_1844967, EPI_ISL_1844969, EPI_ISL_1844970, EPI_ISL_1844972, EPI_ISL_1844973, EPI_ISL_1844974, EPI_ISL_1844975, EPI_ISL_1844978, EPI_ISL_1844979, EPI_ISL_1844982, EPI_ISL_1844984, EPI_ISL_1844989, EPI_ISL_1844990, EPI_ISL_1844991, EPI_ISL_1844992,
EPI_ISL_1844993, EPI_ISL_1844994, EPI_ISL_1844995, EPI_ISL_1844996, EPI_ISL_1844998, EPI_ISL_1845001, EPI_ISL_1845002, EPI_ISL_1845008, EPI_ISL_1845010, EPI_ISL_1845011, EPI_ISL_1845012, EPI_ISL_1845015, EPI_ISL_1845018, EPI_ISL_1845019, EPI_ISL_1845023, EPI_ISL_1845024, EPI_ISL_1845025, EPI_ISL_1845026, EPI_ISL_1845028, EPI_ISL_1845030, EPI_ISL_1845033, EPI_ISL_1845034,
EPI_ISL_1845036, EPI_ISL_1845038, EPI_ISL_1845041, EPI_ISL_1845044, EPI_ISL_1845045, EPI_ISL_1845046, EPI_ISL_1845047, EPI_ISL_1845049, EPI_ISL_1845051, EPI_ISL_1845052, EPI_ISL_1845054, EPI_ISL_1845055, EPI_ISL_1845056, EPI_ISL_1845057, EPI_ISL_1845059, EPI_ISL_1845060, EPI_ISL_1845061, EPI_ISL_1845062, EPI_ISL_1845064, EPI_ISL_1845065, EPI_ISL_1845067, EPI_ISL_1845068,
EPI_ISL_1845070, EPI_ISL_1845075, EPI_ISL_1845078, EPI_ISL_1845081, EPI_ISL_1845083, EPI_ISL_1845085, EPI_ISL_1845086, EPI_ISL_1845090, EPI_ISL_1845096, EPI_ISL_1845098, EPI_ISL_1845099, EPI_ISL_1845100, EPI_ISL_1845101, EPI_ISL_1845102, EPI_ISL_1845104, EPI_ISL_1845106, EPI_ISL_1845107, EPI_ISL_1845109, EPI_ISL_1845113, EPI_ISL_1845118, EPI_ISL_1845119, EPI_ISL_1845120,
EPI_ISL_1845122, EPI_ISL_1845123, EPI_ISL_1845124, EPI_ISL_1845125, EPI_ISL_1845127, EPI_ISL_1845128, EPI_ISL_1845130, EPI_ISL_1845132, EPI_ISL_1845134, EPI_ISL_1845135, EPI_ISL_1845138, EPI_ISL_1845139, EPI_ISL_1845140, EPI_ISL_1845141, EPI_ISL_1845142, EPI_ISL_1845146, EPI_ISL_1845147, EPI_ISL_1845148, EPI_ISL_1845150, EPI_ISL_1845153, EPI_ISL_1845155, EPI_ISL_1845156,
EPI_ISL_1845158, EPI_ISL_1845159, EPI_ISL_1845162, EPI_ISL_1845163, EPI_ISL_1845164, EPI_ISL_1845165, EPI_ISL_1845167, EPI_ISL_1845170, EPI_ISL_1845171, EPI_ISL_1845172, EPI_ISL_1845174, EPI_ISL_1845176, EPI_ISL_1845177, EPI_ISL_1845178, EPI_ISL_1845179, EPI_ISL_1845180, EPI_ISL_1845182, EPI_ISL_1845183, EPI_ISL_1845184, EPI_ISL_1845185, EPI_ISL_1845186, EPI_ISL_1845188,
EPI_ISL_1845190, EPI_ISL_1845191, EPI_ISL_1845192, EPI_ISL_1845193, EPI_ISL_1845194, EPI_ISL_1845195, EPI_ISL_1845196, EPI_ISL_1845199, EPI_ISL_1845201, EPI_ISL_1845202, EPI_ISL_1845203, EPI_ISL_1845205, EPI_ISL_1845208, EPI_ISL_1845210, EPI_ISL_1845211, EPI_ISL_1845212, EPI_ISL_1845213, EPI_ISL_1845214, EPI_ISL_1845217, EPI_ISL_1845220, EPI_ISL_1845222, EPI_ISL_1845223,
EPI_ISL_1845224, EPI_ISL_1845225, EPI_ISL_1845226, EPI_ISL_1845227, EPI_ISL_1845228, EPI_ISL_1845230, EPI_ISL_1845231, EPI_ISL_1845232, EPI_ISL_1845233, EPI_ISL_1845234, EPI_ISL_1845235, EPI_ISL_1845238, EPI_ISL_1845239, EPI_ISL_1845241, EPI_ISL_1845242, EPI_ISL_1845243, EPI_ISL_1845244, EPI_ISL_1845245, EPI_ISL_1845247, EPI_ISL_1845248, EPI_ISL_1845249, EPI_ISL_1845250,
EPI_ISL_1845251, EPI_ISL_1845252, EPI_ISL_1845253, EPI_ISL_1845254, EPI_ISL_1845256, EPI_ISL_1845259, EPI_ISL_1845261, EPI_ISL_1845262, EPI_ISL_1845263, EPI_ISL_1845264, EPI_ISL_1845265, EPI_ISL_1845267, EPI_ISL_1845268, EPI_ISL_1845270, EPI_ISL_1845272, EPI_ISL_1845273, EPI_ISL_1845275, EPI_ISL_1845276, EPI_ISL_1845277, EPI_ISL_1845279, EPI_ISL_1845280, EPI_ISL_1845281,
EPI_ISL_1845282, EPI_ISL_1845283, EPI_ISL_1845284, EPI_ISL_1845285, EPI_ISL_1845287, EPI_ISL_1845290, EPI_ISL_1845291, EPI_ISL_1845293, EPI_ISL_1845294, EPI_ISL_1845295, EPI_ISL_1845296, EPI_ISL_1845297, EPI_ISL_1845298, EPI_ISL_1845301, EPI_ISL_1845302, EPI_ISL_1845305, EPI_ISL_1845306, EPI_ISL_1845307, EPI_ISL_1845308, EPI_ISL_1845309, EPI_ISL_1845311, EPI_ISL_1845313,
EPI_ISL_1845314, EPI_ISL_1845315, EPI_ISL_1845317, EPI_ISL_1845318, EPI_ISL_1845319, EPI_ISL_1845322, EPI_ISL_1845323, EPI_ISL_1845324, EPI_ISL_1845325, EPI_ISL_1845326, EPI_ISL_1845327, EPI_ISL_1845328, EPI_ISL_1845329, EPI_ISL_1845331, EPI_ISL_1845332, EPI_ISL_1845333, EPI_ISL_1845334, EPI_ISL_1845335, EPI_ISL_1845336, EPI_ISL_1845337, EPI_ISL_1845339, EPI_ISL_1845340,
EPI_ISL_1845341, EPI_ISL_1845342, EPI_ISL_1845343, EPI_ISL_1845346, EPI_ISL_1845348, EPI_ISL_1845349, EPI_ISL_1845350, EPI_ISL_1845351, EPI_ISL_1845352, EPI_ISL_1845353, EPI_ISL_1845355, EPI_ISL_1845356, EPI_ISL_1845358, EPI_ISL_1845360, EPI_ISL_1845362, EPI_ISL_1845363, EPI_ISL_1845364, EPI_ISL_1845365, EPI_ISL_1845366, EPI_ISL_1845367, EPI_ISL_1845368, EPI_ISL_1845369,
EPI_ISL_1845371, EPI_ISL_1845372, EPI_ISL_1845373, EPI_ISL_1845374, EPI_ISL_1845376, EPI_ISL_1845378, EPI_ISL_1845379, EPI_ISL_1845380, EPI_ISL_1845382, EPI_ISL_1845383, EPI_ISL_1845385, EPI_ISL_1845386, EPI_ISL_1845387, EPI_ISL_1845388, EPI_ISL_1845391, EPI_ISL_1845392, EPI_ISL_1845393, EPI_ISL_1845394, EPI_ISL_1845395, EPI_ISL_1845396, EPI_ISL_1845397, EPI_ISL_1845398,
EPI_ISL_1845399, EPI_ISL_1845400, EPI_ISL_1845401, EPI_ISL_1845403, EPI_ISL_1845404, EPI_ISL_1845405, EPI_ISL_1845406, EPI_ISL_1845408, EPI_ISL_1845409, EPI_ISL_1845410, EPI_ISL_1845411, EPI_ISL_1845412, EPI_ISL_1845414, EPI_ISL_1845415, EPI_ISL_1845416, EPI_ISL_1845417, EPI_ISL_1845418, EPI_ISL_1845419, EPI_ISL_1845421, EPI_ISL_1845422, EPI_ISL_1845425,
EPI_ISL_1845426, EPI_ISL_1845427, EPI_ISL_1845428, EPI_ISL_1845429, EPI_ISL_1845430, EPI_ISL_1845431, EPI_ISL_1845432, EPI_ISL_1845434, EPI_ISL_1845435, EPI_ISL_1845436, EPI_ISL_1845437, EPI_ISL_1845438, EPI_ISL_1845439, EPI_ISL_1845440, EPI_ISL_1845441, EPI_ISL_1845442, EPI_ISL_1845443, EPI_ISL_1845444, EPI_ISL_1845446, EPI_ISL_1845447, EPI_ISL_1845448, EPI_ISL_1845450,
EPI_ISL_1845451, EPI_ISL_1845452, EPI_ISL_1845453, EPI_ISL_1845454, EPI_ISL_1845456, EPI_ISL_1845457, EPI_ISL_1845459, EPI_ISL_1845460, EPI_ISL_1845461, EPI_ISL_1845462, EPI_ISL_1845463, EPI_ISL_1845464, EPI_ISL_1845465, EPI_ISL_1845466, EPI_ISL_1845468, EPI_ISL_1845469, EPI_ISL_1845470, EPI_ISL_1845472, EPI_ISL_1845473, EPI_ISL_1845474, EPI_ISL_1845476,
EPI_ISL_1845477, EPI_ISL_1845478, EPI_ISL_1845480, EPI_ISL_1845481, EPI_ISL_1845482, EPI_ISL_1845484, EPI_ISL_1845485, EPI_ISL_1845486, EPI_ISL_1845487, EPI_ISL_1845488, EPI_ISL_1845489, EPI_ISL_1845491, EPI_ISL_1845494, EPI_ISL_1845495, EPI_ISL_1845496, EPI_ISL_1845497, EPI_ISL_1845499, EPI_ISL_1845500, EPI_ISL_1845501, EPI_ISL_1845502, EPI_ISL_1845505, EPI_ISL_1845506,
EPI_ISL_1845507, EPI_ISL_1845508, EPI_ISL_1845509, EPI_ISL_1845512, EPI_ISL_1845513, EPI_ISL_1845514, EPI_ISL_1845515, EPI_ISL_1845516, EPI_ISL_1845517, EPI_ISL_1845518, EPI_ISL_1845519, EPI_ISL_1845520, EPI_ISL_1845521, EPI_ISL_1845522, EPI_ISL_1845523, EPI_ISL_1845525, EPI_ISL_1845526, EPI_ISL_1845531, EPI_ISL_1845532, EPI_ISL_1845534, EPI_ISL_1845535, EPI_ISL_1845536,
EPI_ISL_1845537, EPI_ISL_1845540, EPI_ISL_1845541, EPI_ISL_1845542, EPI_ISL_1845543, EPI_ISL_1845545, EPI_ISL_1845546, EPI_ISL_1845547, EPI_ISL_1845548, EPI_ISL_1845550, EPI_ISL_1845552, EPI_ISL_1845553, EPI_ISL_1845554, EPI_ISL_1845555, EPI_ISL_1845556, EPI_ISL_1845557, EPI_ISL_1845558, EPI_ISL_1845559, EPI_ISL_1845561, EPI_ISL_1845562, EPI_ISL_1845564, EPI_ISL_1845565,
EPI_ISL_1845567, EPI_ISL_1845568, EPI_ISL_1845569, EPI_ISL_1845570, EPI_ISL_1845572, EPI_ISL_1845573, EPI_ISL_1845574, EPI_ISL_1845575, EPI_ISL_1845576, EPI_ISL_1845577, EPI_ISL_1845578, EPI_ISL_1845579, EPI_ISL_1845580, EPI_ISL_1845581, EPI_ISL_1845582, EPI_ISL_1845583, EPI_ISL_1845584, EPI_ISL_1845585, EPI_ISL_1845586, EPI_ISL_1845587, EPI_ISL_1845589, EPI_ISL_1845590,
EPI_ISL_1845591, EPI_ISL_1845593, EPI_ISL_1845594, EPI_ISL_1845595, EPI_ISL_1845596, EPI_ISL_1845597, EPI_ISL_1845599, EPI_ISL_1845600, EPI_ISL_1845601, EPI_ISL_1845602, EPI_ISL_1845603, EPI_ISL_1845604, EPI_ISL_1845605, EPI_ISL_1845606, EPI_ISL_1845607, EPI_ISL_1845608, EPI_ISL_1845609, EPI_ISL_1845610, EPI_ISL_1845611, EPI_ISL_1845612, EPI_ISL_1845616, EPI_ISL_1845617,
EPI_ISL_1845618, EPI_ISL_1845619, EPI_ISL_1845620, EPI_ISL_1845621, EPI_ISL_1845622, EPI_ISL_1845624, EPI_ISL_1845625, EPI_ISL_1845626, EPI_ISL_1845627, EPI_ISL_1845628, EPI_ISL_1845630, EPI_ISL_1845632, EPI_ISL_1845634, EPI_ISL_1845635, EPI_ISL_1845636, EPI_ISL_1845637, EPI_ISL_1845639, EPI_ISL_1845640, EPI_ISL_1845641, EPI_ISL_1845642, EPI_ISL_1845643, EPI_ISL_1845646,
EPI_ISL_1845647, EPI_ISL_1845648, EPI_ISL_1845649, EPI_ISL_1845651, EPI_ISL_1845652, EPI_ISL_1845653, EPI_ISL_1845654, EPI_ISL_1845655, EPI_ISL_1845656, EPI_ISL_1845657, EPI_ISL_1845658, EPI_ISL_1845659, EPI_ISL_1845660, EPI_ISL_1845661, EPI_ISL_1845662, EPI_ISL_1845663, EPI_ISL_1845664, EPI_ISL_1845666, EPI_ISL_1845667, EPI_ISL_1845668, EPI_ISL_1845669, EPI_ISL_1845670,
EPI_ISL_1845671, EPI_ISL_1845672, EPI_ISL_1845673, EPI_ISL_1845674, EPI_ISL_1845675, EPI_ISL_1845676, EPI_ISL_1845677, EPI_ISL_1845678, EPI_ISL_1845679, EPI_ISL_1845682, EPI_ISL_1845683, EPI_ISL_1845684, EPI_ISL_1845685, EPI_ISL_1845686, EPI_ISL_1845687, EPI_ISL_1845688, EPI_ISL_1845689, EPI_ISL_1845690, EPI_ISL_1845691, EPI_ISL_1845692, EPI_ISL_1845693, EPI_ISL_1845695,
EPI_ISL_1845696, EPI_ISL_1845697, EPI_ISL_1845698, EPI_ISL_1845699, EPI_ISL_1845700, EPI_ISL_1845702, EPI_ISL_1845703, EPI_ISL_1845705, EPI_ISL_1845706, EPI_ISL_1845708, EPI_ISL_1845709, EPI_ISL_1845710, EPI_ISL_1845712, EPI_ISL_1845713, EPI_ISL_1845714, EPI_ISL_1845715, EPI_ISL_1845716, EPI_ISL_1845717, EPI_ISL_1845718, EPI_ISL_1845719, EPI_ISL_1845720, EPI_ISL_1845721,
EPI_ISL_1845723, EPI_ISL_1845724, EPI_ISL_1845725, EPI_ISL_1845726, EPI_ISL_1845728, EPI_ISL_1845729, EPI_ISL_1845730, EPI_ISL_1845731, EPI_ISL_1845733, EPI_ISL_1845734, EPI_ISL_1845735, EPI_ISL_1845736, EPI_ISL_1845737, EPI_ISL_1845738, EPI_ISL_1845739, EPI_ISL_1845742, EPI_ISL_1845743, EPI_ISL_1845744, EPI_ISL_1845745, EPI_ISL_1845746, EPI_ISL_1845747, EPI_ISL_1845748,
EPI_ISL_1845749, EPI_ISL_1845750, EPI_ISL_1845751, EPI_ISL_1845752, EPI_ISL_1845754, EPI_ISL_1845755, EPI_ISL_1845756, EPI_ISL_1845758, EPI_ISL_1845761, EPI_ISL_1845762, EPI_ISL_1845763, EPI_ISL_1845764, EPI_ISL_1845765, EPI_ISL_1845766, EPI_ISL_1845767, EPI_ISL_1845768, EPI_ISL_1845769, EPI_ISL_1845770, EPI_ISL_1845772, EPI_ISL_1845773, EPI_ISL_1845774, EPI_ISL_1845775,
EPI_ISL_1845776, EPI_ISL_1845777, EPI_ISL_1845779, EPI_ISL_1845780, EPI_ISL_1845785, EPI_ISL_1845786, EPI_ISL_1845788, EPI_ISL_1845790, EPI_ISL_1845791, EPI_ISL_1845792, EPI_ISL_1845793, EPI_ISL_1845794, EPI_ISL_1845795, EPI_ISL_1845796, EPI_ISL_1845797, EPI_ISL_1845800, EPI_ISL_1845801, EPI_ISL_1846452, EPI_ISL_1846454, EPI_ISL_1846455, EPI_ISL_1846456,
EPI_ISL_1846457, EPI_ISL_1846458, EPI_ISL_1846459, EPI_ISL_1846460, EPI_ISL_1846461, EPI_ISL_1846462, EPI_ISL_1846463, EPI_ISL_1846464, EPI_ISL_1846465, EPI_ISL_1846466, EPI_ISL_1846467, EPI_ISL_1846468, EPI_ISL_1846469, EPI_ISL_1846470, EPI_ISL_1846471, EPI_ISL_1846472, EPI_ISL_1846473, EPI_ISL_1846474, EPI_ISL_1846475, EPI_ISL_1846476, EPI_ISL_1846477, EPI_ISL_1846478,
EPI_ISL_1846479, EPI_ISL_1846480, EPI_ISL_1846481, EPI_ISL_1846482, EPI_ISL_1846483, EPI_ISL_1846484, EPI_ISL_1846485, EPI_ISL_1846486, EPI_ISL_1846487, EPI_ISL_1846488, EPI_ISL_1846489, EPI_ISL_1846490, EPI_ISL_1846491, EPI_ISL_1846492, EPI_ISL_1846493, EPI_ISL_1846494, EPI_ISL_1846495, EPI_ISL_1846496, EPI_ISL_1846497, EPI_ISL_1846498, EPI_ISL_1846499, EPI_ISL_1846500,
EPI_ISL_1846501, EPI_ISL_1846502, EPI_ISL_1846503, EPI_ISL_1846504, EPI_ISL_1846505, EPI_ISL_1846506, EPI_ISL_1846507, EPI_ISL_1846508, EPI_ISL_1846509, EPI_ISL_1846510, EPI_ISL_1846511, EPI_ISL_1846512, EPI_ISL_1846513, EPI_ISL_1846514, EPI_ISL_1846515, EPI_ISL_1846516, EPI_ISL_1846517, EPI_ISL_1846518, EPI_ISL_1846519, EPI_ISL_1846520, EPI_ISL_1846521, EPI_ISL_1846522,
EPI_ISL_1846523, EPI_ISL_1846524, EPI_ISL_1846525, EPI_ISL_1846526, EPI_ISL_1846527, EPI_ISL_1846528, EPI_ISL_1846529, EPI_ISL_1846530, EPI_ISL_1846531, EPI_ISL_1846532, EPI_ISL_1846533, EPI_ISL_1846534, EPI_ISL_1846535, EPI_ISL_1846536, EPI_ISL_1846537, EPI_ISL_1846538, EPI_ISL_1846539, EPI_ISL_1846540, EPI_ISL_1846541, EPI_ISL_1846542, EPI_ISL_1846543, EPI_ISL_1846544,
EPI_ISL_1846545, EPI_ISL_1846546, EPI_ISL_1846547, EPI_ISL_1846548, EPI_ISL_1846549, EPI_ISL_1846550, EPI_ISL_1846551, EPI_ISL_1846552, EPI_ISL_1846553, EPI_ISL_1846554, EPI_ISL_1846555, EPI_ISL_1846556, EPI_ISL_1846557, EPI_ISL_1846558, EPI_ISL_1846559, EPI_ISL_1846560, EPI_ISL_1846561, EPI_ISL_1846562, EPI_ISL_1846563, EPI_ISL_1846564, EPI_ISL_1846565, EPI_ISL_1846566,
EPI_ISL_1846567, EPI_ISL_1846568, EPI_ISL_1846569, EPI_ISL_1846570, EPI_ISL_1846571, EPI_ISL_1846572, EPI_ISL_1846573, EPI_ISL_1846574, EPI_ISL_1846575, EPI_ISL_1846576, EPI_ISL_1846577, EPI_ISL_1846578, EPI_ISL_1846580, EPI_ISL_1846581, EPI_ISL_1846582, EPI_ISL_1846583, EPI_ISL_1846584, EPI_ISL_1846585, EPI_ISL_1846586, EPI_ISL_1846587, EPI_ISL_1846588,
EPI_ISL_1846589, EPI_ISL_1846590, EPI_ISL_1846591, EPI_ISL_1846592, EPI_ISL_1846593, EPI_ISL_1846594, EPI_ISL_1846595, EPI_ISL_1846596, EPI_ISL_1846597, EPI_ISL_1846598, EPI_ISL_1846599, EPI_ISL_1846600, EPI_ISL_1846601, EPI_ISL_1846602, EPI_ISL_1846603, EPI_ISL_1846604, EPI_ISL_1846605, EPI_ISL_1846606, EPI_ISL_1846607, EPI_ISL_1846608, EPI_ISL_1846609, EPI_ISL_1846610,
EPI_ISL_1846611, EPI_ISL_1846612, EPI_ISL_1846613, EPI_ISL_1846614, EPI_ISL_1846615, EPI_ISL_1846616, EPI_ISL_1846617, EPI_ISL_1846618, EPI_ISL_1846619, EPI_ISL_1846620, EPI_ISL_1846621, EPI_ISL_1846622, EPI_ISL_1846623, EPI_ISL_1846624, EPI_ISL_1846625, EPI_ISL_1846626, EPI_ISL_1846627, EPI_ISL_1846628, EPI_ISL_1846629, EPI_ISL_1846630, EPI_ISL_1846631, EPI_ISL_1846632,
EPI_ISL_1846633, EPI_ISL_1846634, EPI_ISL_1846635, EPI_ISL_1846636, EPI_ISL_1846637, EPI_ISL_1846638, EPI_ISL_1846639, EPI_ISL_1846640, EPI_ISL_1846641, EPI_ISL_1846642, EPI_ISL_1846643, EPI_ISL_1846644, EPI_ISL_1846645, EPI_ISL_1846646, EPI_ISL_1846647, EPI_ISL_1846648, EPI_ISL_1846649, EPI_ISL_1846650, EPI_ISL_1846651, EPI_ISL_1846652, EPI_ISL_1846653, EPI_ISL_1846654,
EPI_ISL_1846655, EPI_ISL_1846656, EPI_ISL_1846657, EPI_ISL_1846658, EPI_ISL_1846659, EPI_ISL_1846660, EPI_ISL_1846661, EPI_ISL_1846662, EPI_ISL_1846663, EPI_ISL_1846664, EPI_ISL_1846665, EPI_ISL_1846666, EPI_ISL_1846667, EPI_ISL_1846668, EPI_ISL_1846669, EPI_ISL_1846670, EPI_ISL_1846671, EPI_ISL_1846672, EPI_ISL_1846673, EPI_ISL_1846674, EPI_ISL_1846675, EPI_ISL_1846676,
EPI_ISL_1846677, EPI_ISL_1846678, EPI_ISL_1846679, EPI_ISL_1846680, EPI_ISL_1846681, EPI_ISL_1846682, EPI_ISL_1846683, EPI_ISL_1846684, EPI_ISL_1846685, EPI_ISL_1846686, EPI_ISL_1846687, EPI_ISL_1846688, EPI_ISL_1846689, EPI_ISL_1846690, EPI_ISL_1846691, EPI_ISL_1846692, EPI_ISL_1846693, EPI_ISL_1846694, EPI_ISL_1846695, EPI_ISL_1846696, EPI_ISL_1846697, EPI_ISL_1846698,
EPI_ISL_1846699, EPI_ISL_1846700, EPI_ISL_1846701, EPI_ISL_1846702, EPI_ISL_1846703, EPI_ISL_1846704, EPI_ISL_1846705, EPI_ISL_1846706, EPI_ISL_1846707, EPI_ISL_1846708, EPI_ISL_1846709, EPI_ISL_1846710, EPI_ISL_1846711, EPI_ISL_1846712, EPI_ISL_1846713, EPI_ISL_1846714, EPI_ISL_1846715, EPI_ISL_1846716, EPI_ISL_1846717, EPI_ISL_1846718, EPI_ISL_1846719, EPI_ISL_1846720,
EPI_ISL_1846721, EPI_ISL_1846722, EPI_ISL_1846723, EPI_ISL_1846724, EPI_ISL_1846725, EPI_ISL_1846726, EPI_ISL_1846727, EPI_ISL_1846728, EPI_ISL_1846729, EPI_ISL_1846730, EPI_ISL_1846731, EPI_ISL_1846732, EPI_ISL_1846733, EPI_ISL_1846734, EPI_ISL_1846735, EPI_ISL_1846736, EPI_ISL_1846737, EPI_ISL_1846738, EPI_ISL_1846739, EPI_ISL_1846740, EPI_ISL_1846741, EPI_ISL_1846742,
EPI_ISL_1846743, EPI_ISL_1846744, EPI_ISL_1846745, EPI_ISL_1846746, EPI_ISL_1846747, EPI_ISL_1846748, EPI_ISL_1846749, EPI_ISL_1846750, EPI_ISL_1846751, EPI_ISL_1846752, EPI_ISL_1846753, EPI_ISL_1846754, EPI_ISL_1849920, EPI_ISL_1849921, EPI_ISL_1849922, EPI_ISL_1849924, EPI_ISL_1849925, EPI_ISL_1849926, EPI_ISL_1849928, EPI_ISL_1849929, EPI_ISL_1849930, EPI_ISL_1849931,
EPI_ISL_1849933, EPI_ISL_1849934, EPI_ISL_1849935, EPI_ISL_1849936, EPI_ISL_1849938, EPI_ISL_1849939, EPI_ISL_1849940, EPI_ISL_1849941, EPI_ISL_1849942, EPI_ISL_1849944, EPI_ISL_1849945, EPI_ISL_1849946, EPI_ISL_1849947, EPI_ISL_1849948, EPI_ISL_1849949, EPI_ISL_1849950, EPI_ISL_1849952, EPI_ISL_1849953, EPI_ISL_1849954, EPI_ISL_1849955, EPI_ISL_1849958, EPI_ISL_1849959,

EPI_ISL_1849960, EPI_ISL_1849962, EPI_ISL_1849963, EPI_ISL_1849964, EPI_ISL_1849966, EPI_ISL_1849968, EPI_ISL_1849969, EPI_ISL_1849970, EPI_ISL_1849971, EPI_ISL_1849973, EPI_ISL_1849974, EPI_ISL_1849975, EPI_ISL_1849976, EPI_ISL_1849978, EPI_ISL_1849979, EPI_ISL_1849980, EPI_ISL_1849981, EPI_ISL_1849982, EPI_ISL_1849984, EPI_ISL_1849985, EPI_ISL_1849986, EPI_ISL_1849987, EPI_ISL_1849989, EPI_ISL_1849990, EPI_ISL_1849991, EPI_ISL_1849992, EPI_ISL_1849994, EPI_ISL_1849995, EPI_ISL_1849996, EPI_ISL_1849997, EPI_ISL_1849998, EPI_ISL_1850000, EPI_ISL_1850001, EPI_ISL_1850002, EPI_ISL_1850004, EPI_ISL_1850005, EPI_ISL_1850006, EPI_ISL_1850009, EPI_ISL_1850010, EPI_ISL_1850011, EPI_ISL_1850012, EPI_ISL_1850014, EPI_ISL_1850015, EPI_ISL_1850016, EPI_ISL_1850017, EPI_ISL_1850019, EPI_ISL_1850020, EPI_ISL_1850021, EPI_ISL_1850022, EPI_ISL_1850023, EPI_ISL_1850025, EPI_ISL_1850027, EPI_ISL_1850028, EPI_ISL_1850030, EPI_ISL_1850032, EPI_ISL_1850033, EPI_ISL_1850036, EPI_ISL_1850038, EPI_ISL_1850039, EPI_ISL_1850041, EPI_ISL_1850042, EPI_ISL_1850045, EPI_ISL_1850047, EPI_ISL_1850048, EPI_ISL_1850050, EPI_ISL_1850051, EPI_ISL_1850052, EPI_ISL_1850053, EPI_ISL_1850056, EPI_ISL_1850057, EPI_ISL_1850058, EPI_ISL_1850060, EPI_ISL_1850061, EPI_ISL_1850062, EPI_ISL_1850063, EPI_ISL_1850065, EPI_ISL_1850067, EPI_ISL_1850068, EPI_ISL_1850069, EPI_ISL_1850071, EPI_ISL_1850073, EPI_ISL_1850074, EPI_ISL_1850075, EPI_ISL_1850077, EPI_ISL_1850079, EPI_ISL_1850080, EPI_ISL_1850082, EPI_ISL_1850083, EPI_ISL_1850084, EPI_ISL_1850085, EPI_ISL_1850087, EPI_ISL_1850089, EPI_ISL_1850091, EPI_ISL_1850094, EPI_ISL_1850096, EPI_ISL_1850097, EPI_ISL_1850098, EPI_ISL_1850101, EPI_ISL_1850102, EPI_ISL_1850103, EPI_ISL_1850104, EPI_ISL_1850106, EPI_ISL_1850107, EPI_ISL_1850108, EPI_ISL_1850109, EPI_ISL_1850111, EPI_ISL_1850112, EPI_ISL_1850113, EPI_ISL_1850114, EPI_ISL_1850116, EPI_ISL_1850118, EPI_ISL_1850119, EPI_ISL_1850120, EPI_ISL_1850122, EPI_ISL_1850124, EPI_ISL_1850125, EPI_ISL_1850127, EPI_ISL_1850128, EPI_ISL_1850129, EPI_ISL_1850131, EPI_ISL_1850132, EPI_ISL_1850134, EPI_ISL_1850136, EPI_ISL_1850138, EPI_ISL_1850139, EPI_ISL_1850141, EPI_ISL_1850142, EPI_ISL_1850143, EPI_ISL_1850144, EPI_ISL_1850146, EPI_ISL_1850147, EPI_ISL_1850148, EPI_ISL_1850150, EPI_ISL_1850151, EPI_ISL_1850153, EPI_ISL_1850154, EPI_ISL_1850156, EPI_ISL_1850157, EPI_ISL_1850158, EPI_ISL_1850160, EPI_ISL_1850162, EPI_ISL_1850163, EPI_ISL_1850164, EPI_ISL_1850165, EPI_ISL_1850167, EPI_ISL_1850168, EPI_ISL_1850169, EPI_ISL_1850170, EPI_ISL_1850172, EPI_ISL_1850174, EPI_ISL_1850175, EPI_ISL_1850178, EPI_ISL_1850179, EPI_ISL_1850182, EPI_ISL_1850183, EPI_ISL_1850184, EPI_ISL_1850186, EPI_ISL_1850187, EPI_ISL_1850188, EPI_ISL_1850189, EPI_ISL_1850190, EPI_ISL_1850192, EPI_ISL_1850193, EPI_ISL_1850194, EPI_ISL_1850195, EPI_ISL_1850197, EPI_ISL_1850198, EPI_ISL_1850199, EPI_ISL_1850200, EPI_ISL_1850202, EPI_ISL_1850203, EPI_ISL_1850204, EPI_ISL_1850205, EPI_ISL_1850207, EPI_ISL_1850208, EPI_ISL_1850209, EPI_ISL_1850210, EPI_ISL_1850212, EPI_ISL_1850213, EPI_ISL_1850214, EPI_ISL_1850215, EPI_ISL_1850217, EPI_ISL_1850218, EPI_ISL_1850219, EPI_ISL_1850220, EPI_ISL_1850222, EPI_ISL_1850223, EPI_ISL_1850224, EPI_ISL_1850225, EPI_ISL_1850227, EPI_ISL_1850228, EPI_ISL_1850229, EPI_ISL_1850230, EPI_ISL_1850232, EPI_ISL_1850233, EPI_ISL_1850234, EPI_ISL_1850235, EPI_ISL_1850237, EPI_ISL_1850238, EPI_ISL_1850239, EPI_ISL_1850240, EPI_ISL_1850242, EPI_ISL_1850243, EPI_ISL_1850244, EPI_ISL_1850245, EPI_ISL_1850247, EPI_ISL_1850248, EPI_ISL_1850249, EPI_ISL_1850252, EPI_ISL_1850253, EPI_ISL_1850254, EPI_ISL_1850255, EPI_ISL_1850257, EPI_ISL_1850258, EPI_ISL_1850259, EPI_ISL_1850261, EPI_ISL_1850262, EPI_ISL_1850263, EPI_ISL_1850264, EPI_ISL_1850266, EPI_ISL_1850267, EPI_ISL_1850268, EPI_ISL_1850269, EPI_ISL_1850270, EPI_ISL_1850272, EPI_ISL_1850273, EPI_ISL_1850274, EPI_ISL_1850275, EPI_ISL_1850277, EPI_ISL_1850278, EPI_ISL_1850279, EPI_ISL_1850280, EPI_ISL_1851445, EPI_ISL_1851446, EPI_ISL_1851448, EPI_ISL_1851449, EPI_ISL_1851450, EPI_ISL_1851452, EPI_ISL_1851453, EPI_ISL_1851454, EPI_ISL_1851455, EPI_ISL_1851456, EPI_ISL_1851457, EPI_ISL_1851458, EPI_ISL_1851459, EPI_ISL_1851462, EPI_ISL_1851463, EPI_ISL_1851465, EPI_ISL_1851468, EPI_ISL_1851469, EPI_ISL_1851471, EPI_ISL_1851475, EPI_ISL_1851476, EPI_ISL_1851477, EPI_ISL_1851478, EPI_ISL_1851479, EPI_ISL_1851480, EPI_ISL_1851481, EPI_ISL_1851482, EPI_ISL_1851483, EPI_ISL_1851484, EPI_ISL_1851485, EPI_ISL_1851486, EPI_ISL_1851489, EPI_ISL_1851491, EPI_ISL_1851493, EPI_ISL_1851494, EPI_ISL_1851499, EPI_ISL_1851501, EPI_ISL_1851504, EPI_ISL_1851507, EPI_ISL_1851508, EPI_ISL_1851509, EPI_ISL_1851510, EPI_ISL_1851511, EPI_ISL_1851512, EPI_ISL_1851513, EPI_ISL_1851515, EPI_ISL_1851516, EPI_ISL_1851517, EPI_ISL_1851519, EPI_ISL_1851520, EPI_ISL_1851521, EPI_ISL_1851523, EPI_ISL_1851524, EPI_ISL_1851525, EPI_ISL_1851526, EPI_ISL_1851527, EPI_ISL_1851529, EPI_ISL_1851530, EPI_ISL_1851533, EPI_ISL_1851534, EPI_ISL_1851536, EPI_ISL_1851537, EPI_ISL_1851538, EPI_ISL_1851542, EPI_ISL_1851544, EPI_ISL_1851545, EPI_ISL_1851546, EPI_ISL_1851547, EPI_ISL_1851548, EPI_ISL_1851550, EPI_ISL_1851552, EPI_ISL_1851553, EPI_ISL_1851557, EPI_ISL_1851558, EPI_ISL_1851561, EPI_ISL_1851562, EPI_ISL_1851563, EPI_ISL_1851564, EPI_ISL_1851566, EPI_ISL_1851567, EPI_ISL_1851572, EPI_ISL_1851573, EPI_ISL_1851574, EPI_ISL_1851575, EPI_ISL_1851579, EPI_ISL_1851581, EPI_ISL_1851583, EPI_ISL_1851584, EPI_ISL_1851589, EPI_ISL_1851591, EPI_ISL_1851593, EPI_ISL_1851594, EPI_ISL_1851595, EPI_ISL_1851600, EPI_ISL_1851602, EPI_ISL_1851605, EPI_ISL_1851606, EPI_ISL_1851607, EPI_ISL_1851608, EPI_ISL_1851612, EPI_ISL_1851613, EPI_ISL_1851615, EPI_ISL_1851616, EPI_ISL_1851617, EPI_ISL_1851619, EPI_ISL_1851620, EPI_ISL_1851622, EPI_ISL_1851623, EPI_ISL_1851625, EPI_ISL_1851627, EPI_ISL_1851631, EPI_ISL_1851633, EPI_ISL_1851634, EPI_ISL_1851635, EPI_ISL_1851638, EPI_ISL_1851640, EPI_ISL_1851641, EPI_ISL_1851646, EPI_ISL_1851647, EPI_ISL_1851651, EPI_ISL_1851653, EPI_ISL_1851656, EPI_ISL_1851657, EPI_ISL_1851658, EPI_ISL_1851660, EPI_ISL_1851661, EPI_ISL_1851663, EPI_ISL_1851665, EPI_ISL_1851666, EPI_ISL_1851667, EPI_ISL_1851668, EPI_ISL_1851669, EPI_ISL_1851670, EPI_ISL_1851673, EPI_ISL_1851675, EPI_ISL_1851677, EPI_ISL_1851678, EPI_ISL_1851681, EPI_ISL_1851683, EPI_ISL_1851685, EPI_ISL_1851686, EPI_ISL_1851687, EPI_ISL_1851690, EPI_ISL_1851691, EPI_ISL_1851697, EPI_ISL_1851698, EPI_ISL_1851699, EPI_ISL_1851700, EPI_ISL_1851701, EPI_ISL_1851702, EPI_ISL_1851703, EPI_ISL_1851704, EPI_ISL_1851708, EPI_ISL_1851710, EPI_ISL_1851712, EPI_ISL_1851713, EPI_ISL_1851714, EPI_ISL_1851715, EPI_ISL_1851717, EPI_ISL_1851718, EPI_ISL_1851720, EPI_ISL_1851721, EPI_ISL_1851722, EPI_ISL_1851723, EPI_ISL_1851724, EPI_ISL_1851726, EPI_ISL_1851727, EPI_ISL_1851730, EPI_ISL_1851731, EPI_ISL_1851733, EPI_ISL_1851735, EPI_ISL_1851737, EPI_ISL_1851738, EPI_ISL_1851741, EPI_ISL_1851742, EPI_ISL_1851743, EPI_ISL_1851745, EPI_ISL_1851746, EPI_ISL_1851749, EPI_ISL_1852488, EPI_ISL_1852489, EPI_ISL_1852493, EPI_ISL_1852494, EPI_ISL_1852496, EPI_ISL_1852499, EPI_ISL_1852501, EPI_ISL_1852502, EPI_ISL_1852508, EPI_ISL_1852510, EPI_ISL_1852517, EPI_ISL_1852518, EPI_ISL_1852520, EPI_ISL_1852521, EPI_ISL_1852523, EPI_ISL_1852533, EPI_ISL_1852534, EPI_ISL_1852548, EPI_ISL_1852553, EPI_ISL_1852556, EPI_ISL_1852566, EPI_ISL_1852569, EPI_ISL_1852575, EPI_ISL_1852576, EPI_ISL_1852577, EPI_ISL_1852581, EPI_ISL_1852584, EPI_ISL_1852585, EPI_ISL_1852587, EPI_ISL_1852589, EPI_ISL_1852590, EPI_ISL_1852593, EPI_ISL_1852598, EPI_ISL_1852600, EPI_ISL_1852607, EPI_ISL_1852609, EPI_ISL_1852616, EPI_ISL_1852619, EPI_ISL_1852621, EPI_ISL_1852631, EPI_ISL_1852632, EPI_ISL_1852634, EPI_ISL_1852635, EPI_ISL_1852637, EPI_ISL_1852638, EPI_ISL_1852640, EPI_ISL_1852641, EPI_ISL_1852643, EPI_ISL_1852647, EPI_ISL_1852648, EPI_ISL_1852650, EPI_ISL_1852651, EPI_ISL_1852658, EPI_ISL_1852661, EPI_ISL_1852662, EPI_ISL_1852663, EPI_ISL_1852667, EPI_ISL_1852668, EPI_ISL_1852669, EPI_ISL_1852670, EPI_ISL_1852673, EPI_ISL_1852676, EPI_ISL_1852677, EPI_ISL_1852681, EPI_ISL_1852683, EPI_ISL_1852688, EPI_ISL_1852689, EPI_ISL_1852694, EPI_ISL_1852703, EPI_ISL_1852704, EPI_ISL_1852705, EPI_ISL_1852709, EPI_ISL_1852711, EPI_ISL_1852712, EPI_ISL_1852715, EPI_ISL_1852717, EPI_ISL_1852719, EPI_ISL_1852720, EPI_ISL_1852721, EPI_ISL_1852723, EPI_ISL_1852724, EPI_ISL_1852726, EPI_ISL_1852728, EPI_ISL_1852730, EPI_ISL_1852731, EPI_ISL_1852738, EPI_ISL_1852740, EPI_ISL_1852741, EPI_ISL_1852743, EPI_ISL_1852745, EPI_ISL_1852751, EPI_ISL_1852752, EPI_ISL_1852755, EPI_ISL_1852759, EPI_ISL_1852763, EPI_ISL_1852766, EPI_ISL_1852772, EPI_ISL_1852777, EPI_ISL_1852780, EPI_ISL_1852785, EPI_ISL_1852792, EPI_ISL_1852794, EPI_ISL_1852798, EPI_ISL_1852801, EPI_ISL_1852802, EPI_ISL_1852806, EPI_ISL_1852807, EPI_ISL_1852808, EPI_ISL_1852809, EPI_ISL_1852816, EPI_ISL_1852819, EPI_ISL_1852820, EPI_ISL_1852821, EPI_ISL_1852823, EPI_ISL_1852826, EPI_ISL_1852828, EPI_ISL_1852829, EPI_ISL_1852832, EPI_ISL_1852834, EPI_ISL_1852836, EPI_ISL_1852837, EPI_ISL_1852840, EPI_ISL_1852842, EPI_ISL_1852843, EPI_ISL_1852844, EPI_ISL_1852849, EPI_ISL_1852851, EPI_ISL_1852853, EPI_ISL_1852854, EPI_ISL_1852856, EPI_ISL_1852859, EPI_ISL_1852860, EPI_ISL_1852861, EPI_ISL_1852863, EPI_ISL_1852865, EPI_ISL_1852866, EPI_ISL_1852869, EPI_ISL_1852872, EPI_ISL_1852873, EPI_ISL_1852878, EPI_ISL_1852881, EPI_ISL_1852882, EPI_ISL_1852883, EPI_ISL_1852886, EPI_ISL_1852889, EPI_ISL_1852892, EPI_ISL_1852895, EPI_ISL_1852898, EPI_ISL_1852900, EPI_ISL_1852903, EPI_ISL_1852908, EPI_ISL_1852910, EPI_ISL_1852912, EPI_ISL_1852918, EPI_ISL_1852919, EPI_ISL_1852922, EPI_ISL_1852926, EPI_ISL_1852929, EPI_ISL_1852932, EPI_ISL_1852935, EPI_ISL_1852936, EPI_ISL_1852939, EPI_ISL_1852940, EPI_ISL_1852941, EPI_ISL_1852943, EPI_ISL_1852945, EPI_ISL_1852946, EPI_ISL_1852947, EPI_ISL_1852951, EPI_ISL_1852952, EPI_ISL_1852959, EPI_ISL_1852961, EPI_ISL_1852963, EPI_ISL_1852966, EPI_ISL_1852967, EPI_ISL_1852969, EPI_ISL_1852975, EPI_ISL_1852982, EPI_ISL_1852987, EPI_ISL_1852989, EPI_ISL_1852995, EPI_ISL_1852998, EPI_ISL_1853007, EPI_ISL_1853014, EPI_ISL_1853021, EPI_ISL_1853022, EPI_ISL_1853027, EPI_ISL_1853030, EPI_ISL_1853031, EPI_ISL_1853033, EPI_ISL_1853038, EPI_ISL_1853041, EPI_ISL_1853042, EPI_ISL_1853047, EPI_ISL_1853063, EPI_ISL_1853065, EPI_ISL_1853066, EPI_ISL_1853068, EPI_ISL_1853072, EPI_ISL_1853073, EPI_ISL_1853074, EPI_ISL_1853081, EPI_ISL_1853087, EPI_ISL_1853093, EPI_ISL_1853095, EPI_ISL_1853097, EPI_ISL_1853099, EPI_ISL_2111611, EPI_ISL_2111672, EPI_ISL_2111679, EPI_ISL_2111680, EPI_ISL_2111771, EPI_ISL_2111775, EPI_ISL_2111781, EPI_ISL_2111789, EPI_ISL_2111800, EPI_ISL_2111902, EPI_ISL_2111921, EPI_ISL_2111965, EPI_ISL_2111971, EPI_ISL_2111983, EPI_ISL_2112043, EPI_ISL_2112060, EPI_ISL_2112115, EPI_ISL_2112145, EPI_ISL_2113379

see above SYNLAB MVZ Leverkusen Robert Koch Institute

EPI_ISL_2114541, EPI_ISL_2114544, EPI_ISL_2114558, EPI_ISL_2114580, EPI_ISL_2114587, EPI_ISL_2114598, EPI_ISL_2114600, EPI_ISL_2114631, EPI_ISL_2114681, EPI_ISL_2114701, EPI_ISL_2114716, EPI_ISL_2114727, EPI_ISL_2114747, EPI_ISL_2114753, EPI_ISL_2114755, EPI_ISL_2114758, EPI_ISL_2114763, EPI_ISL_2114778, EPI_ISL_2114780, EPI_ISL_2114790, EPI_ISL_2114816, EPI_ISL_2114820, EPI_ISL_2114854, EPI_ISL_2114859, EPI_ISL_2114891, EPI_ISL_2114894, EPI_ISL_2114897, EPI_ISL_2114913, EPI_ISL_2114918, EPI_ISL_2114927, EPI_ISL_2114949, EPI_ISL_2114971

see above SYNLAB MVZ Neustadt a. d. Weinstrasse Robert Koch Institute

EPI_ISL_2115116 SYNLAB MVZ Neustadt a. d. WeinstraÃŸe Robert Koch Institute

EPI_ISL_2114984, EPI_ISL_2115009, EPI_ISL_2115065, EPI_ISL_2115084, EPI_ISL_2115087, EPI_ISL_2118839, EPI_ISL_2118845, EPI_ISL_2119053, EPI_ISL_2119075, EPI_ISL_2119090, EPI_ISL_2119164, EPI_ISL_2119217, EPI_ISL_2119272, EPI_ISL_2119315, EPI_ISL_2119326, EPI_ISL_2119396, EPI_ISL_2119443, EPI_ISL_2119459, EPI_ISL_2119573, EPI_ISL_2119611, EPI_ISL_2119695, EPI_ISL_2119711, EPI_ISL_2119721, EPI_ISL_2119768, EPI_ISL_2119771, EPI_ISL_2119813, EPI_ISL_2119845, EPI_ISL_2119850, EPI_ISL_2119853, EPI_ISL_2119868, EPI_ISL_2119935, EPI_ISL_2119944, EPI_ISL_2120011, EPI_ISL_2127708, EPI_ISL_2127758, EPI_ISL_2127794, EPI_ISL_2127848, EPI_ISL_2127853, EPI_ISL_2127909, EPI_ISL_2127915, EPI_ISL_2127927, EPI_ISL_2127929, EPI_ISL_2127998, EPI_ISL_2128016, EPI_ISL_2128070, EPI_ISL_2128090, EPI_ISL_2128189, EPI_ISL_2128200, EPI_ISL_2128221, EPI_ISL_2128286, EPI_ISL_2128324, EPI_ISL_2128331, EPI_ISL_2128791, EPI_ISL_2128825, EPI_ISL_2128912

see above SYNLAB MVZ Neustadt a. d. Weinstraße Robert Koch Institute

EPI_ISL_1844639, EPI_ISL_1844640, EPI_ISL_1844641, EPI_ISL_1844642, EPI_ISL_1844643, EPI_ISL_1844652, EPI_ISL_1844653, EPI_ISL_1844655, EPI_ISL_1844660, EPI_ISL_1844661, EPI_ISL_1844663, EPI_ISL_1844665, EPI_ISL_1844671, EPI_ISL_1844672, EPI_ISL_1844673, EPI_ISL_1844678, EPI_ISL_1844682, EPI_ISL_1844683, EPI_ISL_1844684, EPI_ISL_1844685, EPI_ISL_1844686, EPI_ISL_1844687, EPI_ISL_1844688, EPI_ISL_1844691, EPI_ISL_1844692, EPI_ISL_1844694, EPI_ISL_1844702, EPI_ISL_1844704, EPI_ISL_1844706, EPI_ISL_1844709, EPI_ISL_1844710, EPI_ISL_1844714, EPI_ISL_1844716, EPI_ISL_1844725, EPI_ISL_1844727, EPI_ISL_1844728, EPI_ISL_1844731, EPI_ISL_1844734, EPI_ISL_1844736, EPI_ISL_1844741, EPI_ISL_1844744, EPI_ISL_1844748, EPI_ISL_1844753, EPI_ISL_1844755, EPI_ISL_1844758, EPI_ISL_1844762, EPI_ISL_1844763, EPI_ISL_1844768, EPI_ISL_1844772, EPI_ISL_1844774, EPI_ISL_1844777, EPI_ISL_1844779, EPI_ISL_1844780, EPI_ISL_1844784, EPI_ISL_1844787, EPI_ISL_1844791, EPI_ISL_1844792, EPI_ISL_1844793, EPI_ISL_1844796, EPI_ISL_1844798, EPI_ISL_1844802, EPI_ISL_1844803, EPI_ISL_1844808, EPI_ISL_1844809, EPI_ISL_1844812, EPI_ISL_1844814, EPI_ISL_1844816, EPI_ISL_1844817, EPI_ISL_1844818, EPI_ISL_1844820, EPI_ISL_1844828, EPI_ISL_1844832, EPI_ISL_1844836, EPI_ISL_1844840, EPI_ISL_1844848, EPI_ISL_1844849, EPI_ISL_1844851, EPI_ISL_1844853, EPI_ISL_1844856, EPI_ISL_1844857, EPI_ISL_1844858, EPI_ISL_1844865, EPI_ISL_1844869, EPI_ISL_1844871, EPI_ISL_1844873, EPI_ISL_1844880, EPI_ISL_1844885, EPI_ISL_1844886, EPI_ISL_1844887, EPI_ISL_1844888, EPI_ISL_1844890, EPI_ISL_1844894, EPI_ISL_1844896, EPI_ISL_1844898, EPI_ISL_1844905, EPI_ISL_1844907, EPI_ISL_1844909, EPI_ISL_1844913, EPI_ISL_1844914, EPI_ISL_1844915, EPI_ISL_1844916, EPI_ISL_1844917, EPI_ISL_1844918, EPI_ISL_1844924, EPI_ISL_1844925, EPI_ISL_1844926, EPI_ISL_1844929, EPI_ISL_1844930, EPI_ISL_1844931, EPI_ISL_1844932, EPI_ISL_1844934, EPI_ISL_1844935, EPI_ISL_1844936, EPI_ISL_1844943, EPI_ISL_1844944, EPI_ISL_1844945, EPI_ISL_1844947, EPI_ISL_1844949, EPI_ISL_1844950, EPI_ISL_1844953, EPI_ISL_1844956, EPI_ISL_1844958, EPI_ISL_1844960, EPI_ISL_1844962, EPI_ISL_1844963, EPI_ISL_1844964, EPI_ISL_1844965, EPI_ISL_1844968, EPI_ISL_1844971, EPI_ISL_1844976, EPI_ISL_1844977, EPI_ISL_1844980, EPI_ISL_1844981, EPI_ISL_1844983, EPI_ISL_1844985, EPI_ISL_1844986, EPI_ISL_1844987, EPI_ISL_1844988, EPI_ISL_1844997, EPI_ISL_1844999, EPI_ISL_1845000, EPI_ISL_1845003, EPI_ISL_1845004, EPI_ISL_1845005, EPI_ISL_1845006, EPI_ISL_1845007, EPI_ISL_1845009, EPI_ISL_1845013, EPI_ISL_1845014, EPI_ISL_1845016, EPI_ISL_1845017, EPI_ISL_1845020, EPI_ISL_1845021, EPI_ISL_1845022, EPI_ISL_1845027, EPI_ISL_1845029, EPI_ISL_1845031, EPI_ISL_1845032, EPI_ISL_1845035, EPI_ISL_1845037, EPI_ISL_1845039, EPI_ISL_1845040, EPI_ISL_1845042, EPI_ISL_1845043, EPI_ISL_1845048, EPI_ISL_1845050, EPI_ISL_1845053, EPI_ISL_1845058, EPI_ISL_1845063, EPI_ISL_1845066, EPI_ISL_1845069, EPI_ISL_1845071, EPI_ISL_1845072, EPI_ISL_1845073, EPI_ISL_1845074, EPI_ISL_1845076, EPI_ISL_1845077, EPI_ISL_1845079, EPI_ISL_1845080, EPI_ISL_1845082, EPI_ISL_1845084, EPI_ISL_1845087, EPI_ISL_1845088, EPI_ISL_1845089, EPI_ISL_1845091, EPI_ISL_1845092, EPI_ISL_1845093, EPI_ISL_1845094, EPI_ISL_1845095, EPI_ISL_1845097, EPI_ISL_1845103, EPI_ISL_1845105, EPI_ISL_1845108, EPI_ISL_1845110, EPI_ISL_1845111, EPI_ISL_1845112, EPI_ISL_1845114, EPI_ISL_1845115, EPI_ISL_1845116, EPI_ISL_1845117, EPI_ISL_1845121, EPI_ISL_1845126, EPI_ISL_1845129, EPI_ISL_1845131, EPI_ISL_1845136, EPI_ISL_1845137, EPI_ISL_1845143, EPI_ISL_1845145, EPI_ISL_1845149, EPI_ISL_1845151, EPI_ISL_1845152, EPI_ISL_1845154, EPI_ISL_1845157, EPI_ISL_1845160, EPI_ISL_1845161, EPI_ISL_1845166, EPI_ISL_1845168, EPI_ISL_1845169, EPI_ISL_1845173, EPI_ISL_1845181, EPI_ISL_1845187, EPI_ISL_1845197, EPI_ISL_1845198, EPI_ISL_1845200, EPI_ISL_1845204, EPI_ISL_1845206, EPI_ISL_1845207, EPI_ISL_1845209, EPI_ISL_1845215, EPI_ISL_1845216, EPI_ISL_1845218, EPI_ISL_1845219, EPI_ISL_1845229, EPI_ISL_1845236, EPI_ISL_1845237, EPI_ISL_1845240, EPI_ISL_1845246, EPI_ISL_1845255, EPI_ISL_1845257, EPI_ISL_1845258, EPI_ISL_1845260, EPI_ISL_1845266, EPI_ISL_1845269, EPI_ISL_1845271, EPI_ISL_1845274, EPI_ISL_1845278, EPI_ISL_1845286, EPI_ISL_1845288, EPI_ISL_1845289, EPI_ISL_1845299, EPI_ISL_1845300, EPI_ISL_1845303, EPI_ISL_1845304, EPI_ISL_1845310, EPI_ISL_1845312, EPI_ISL_1845316, EPI_ISL_1845320, EPI_ISL_1845321, EPI_ISL_1845330, EPI_ISL_1845338, EPI_ISL_1845344, EPI_ISL_1845345, EPI_ISL_1845347, EPI_ISL_1845354, EPI_ISL_1845359, EPI_ISL_1845361, EPI_ISL_1845375, EPI_ISL_1845377, EPI_ISL_1845381, EPI_ISL_1845384, EPI_ISL_1845389, EPI_ISL_1845390, EPI_ISL_1845423, EPI_ISL_1845433, EPI_ISL_1845445, EPI_ISL_1845449, EPI_ISL_1845458, EPI_ISL_1845471, EPI_ISL_1845483, EPI_ISL_1845490, EPI_ISL_1845492, EPI_ISL_1845493, EPI_ISL_1845503, EPI_ISL_1845504, EPI_ISL_1845510, EPI_ISL_1845511, EPI_ISL_1845527, EPI_ISL_1845528, EPI_ISL_1845529, EPI_ISL_1845530, EPI_ISL_1845533, EPI_ISL_1845538, EPI_ISL_1845539, EPI_ISL_1845544, EPI_ISL_1845549, EPI_ISL_1845560, EPI_ISL_1845566, EPI_ISL_1845571, EPI_ISL_1845588, EPI_ISL_1845592, EPI_ISL_1845598, EPI_ISL_1845613, EPI_ISL_1845614, EPI_ISL_1845615, EPI_ISL_1845623, EPI_ISL_1845631, EPI_ISL_1845638, EPI_ISL_1845644, EPI_ISL_1845645, EPI_ISL_1845650, EPI_ISL_1845681, EPI_ISL_1845694, EPI_ISL_1845701, EPI_ISL_1845704, EPI_ISL_1845711, EPI_ISL_1845722, EPI_ISL_1845727, EPI_ISL_1845740, EPI_ISL_1845741, EPI_ISL_1845753, EPI_ISL_1845759, EPI_ISL_1845760, EPI_ISL_1845771, EPI_ISL_1845781, EPI_ISL_1845782, EPI_ISL_1845784, EPI_ISL_1845787, EPI_ISL_1845789, EPI_ISL_1845798, EPI_ISL_1845799, EPI_ISL_1851442, EPI_ISL_1851443, EPI_ISL_1851444, EPI_ISL_1851447, EPI_ISL_1851451, EPI_ISL_1851460, EPI_ISL_1851461, EPI_ISL_1851464, EPI_ISL_1851466, EPI_ISL_1851467, EPI_ISL_1851470, EPI_ISL_1851473, EPI_ISL_1851474, EPI_ISL_1851487, EPI_ISL_1851492, EPI_ISL_1851495, EPI_ISL_1851496, EPI_ISL_1851497, EPI_ISL_1851498, EPI_ISL_1851500, EPI_ISL_1851502, EPI_ISL_1851503, EPI_ISL_1851505, EPI_ISL_1851506, EPI_ISL_1851514, EPI_ISL_1851518, EPI_ISL_1851522, EPI_ISL_1851528, EPI_ISL_1851532, EPI_ISL_1851535, EPI_ISL_1851539, EPI_ISL_1851540, EPI_ISL_1851541, EPI_ISL_1851549, EPI_ISL_1851551, EPI_ISL_1851556, EPI_ISL_1851559, EPI_ISL_1851568, EPI_ISL_1851569, EPI_ISL_1851571, EPI_ISL_1851576, EPI_ISL_1851577, EPI_ISL_1851578, EPI_ISL_1851580, EPI_ISL_1851582, EPI_ISL_1851585, EPI_ISL_1851587, EPI_ISL_1851588, EPI_ISL_1851592, EPI_ISL_1851597, EPI_ISL_1851598, EPI_ISL_1851599, EPI_ISL_1851601, EPI_ISL_1851603, EPI_ISL_1851604, EPI_ISL_1851610, EPI_ISL_1851611, EPI_ISL_1851614, EPI_ISL_1851618, EPI_ISL_1851626, EPI_ISL_1851628, EPI_ISL_1851629, EPI_ISL_1851630, EPI_ISL_1851632, EPI_ISL_1851636, EPI_ISL_1851637, EPI_ISL_1851639, EPI_ISL_1851642, EPI_ISL_1851643, EPI_ISL_1851645, EPI_ISL_1851648, EPI_ISL_1851650, EPI_ISL_1851652, EPI_ISL_1851654, EPI_ISL_1851655, EPI_ISL_1851659, EPI_ISL_1851662, EPI_ISL_1851664, EPI_ISL_1851671, EPI_ISL_1851672, EPI_ISL_1851674, EPI_ISL_1851676, EPI_ISL_1851679, EPI_ISL_1851680, EPI_ISL_1851682, EPI_ISL_1851684, EPI_ISL_1851688, EPI_ISL_1851693, EPI_ISL_1851694, EPI_ISL_1851695, EPI_ISL_1851696, EPI_ISL_1851705, EPI_ISL_1851707, EPI_ISL_1851711, EPI_ISL_1851716, EPI_ISL_1851719, EPI_ISL_1851725, EPI_ISL_1851728, EPI_ISL_1851729, EPI_ISL_1851732, EPI_ISL_1851734, EPI_ISL_1851736, EPI_ISL_1851739, EPI_ISL_1851740, EPI_ISL_1851744, EPI_ISL_1851747, EPI_ISL_1851748, EPI_ISL_1852511, EPI_ISL_1852513, EPI_ISL_1852514, EPI_ISL_1852515, EPI_ISL_1852519, EPI_ISL_1852526, EPI_ISL_1852527, EPI_ISL_1852528, EPI_ISL_1852535, EPI_ISL_1852538, EPI_ISL_1852540, EPI_ISL_1852550, EPI_ISL_1852562, EPI_ISL_1852564, EPI_ISL_1852574, EPI_ISL_1852578, EPI_ISL_1852582, EPI_ISL_1852583, EPI_ISL_1852597, EPI_ISL_1852622, EPI_ISL_1852624, EPI_ISL_1852639, EPI_ISL_1852646, EPI_ISL_1852655, EPI_ISL_1852657, EPI_ISL_1852672, EPI_ISL_1852680, EPI_ISL_1852695, EPI_ISL_1852707, EPI_ISL_1852714, EPI_ISL_1852732, EPI_ISL_1852733, EPI_ISL_1852735, EPI_ISL_1852736, EPI_ISL_1852737, EPI_ISL_1852739, EPI_ISL_1852742, EPI_ISL_1852747, EPI_ISL_1852748, EPI_ISL_1852754, EPI_ISL_1852756, EPI_ISL_1852757, EPI_ISL_1852758, EPI_ISL_1852760, EPI_ISL_1852764, EPI_ISL_1852765, EPI_ISL_1852769, EPI_ISL_1852776, EPI_ISL_1852778, EPI_ISL_1852783, EPI_ISL_1852789, EPI_ISL_1852815, EPI_ISL_1852822, EPI_ISL_1852827, EPI_ISL_1852830, EPI_ISL_1852838, EPI_ISL_1852855, EPI_ISL_1852867, EPI_ISL_1852868, EPI_ISL_1852875, EPI_ISL_1852885, EPI_ISL_1852905, EPI_ISL_1852915, EPI_ISL_1852924, EPI_ISL_1852956, EPI_ISL_1852960, EPI_ISL_1852979, EPI_ISL_1852992, EPI_ISL_1853001, EPI_ISL_1853009, EPI_ISL_1853018, EPI_ISL_1853023, EPI_ISL_1853025, EPI_ISL_1853039, EPI_ISL_1853040, EPI_ISL_1853057, EPI_ISL_1853077, EPI_ISL_1853088, EPI_ISL_1853100, EPI_ISL_2111705, EPI_ISL_2111744, EPI_ISL_2111757, EPI_ISL_2111758, EPI_ISL_2111774, EPI_ISL_2111798, EPI_ISL_2111925, EPI_ISL_2111927, EPI_ISL_2111937, EPI_ISL_2111958, EPI_ISL_2111985, EPI_ISL_2112001, EPI_ISL_2112002, EPI_ISL_2112009, EPI_ISL_2112036, EPI_ISL_2112087, EPI_ISL_2112098, EPI_ISL_2112170, EPI_ISL_2112179

see above SYNLAB MVZ Trier Robert Koch Institute

EPI_ISL_1844749, EPI_ISL_1844776, EPI_ISL_1844804, EPI_ISL_1844870, EPI_ISL_1844876, EPI_ISL_1845221, EPI_ISL_1845402, EPI_ISL_1845407, EPI_ISL_1845413, EPI_ISL_1845524, EPI_ISL_1845629, EPI_ISL_1845633, EPI_ISL_1845665, EPI_ISL_1845707, EPI_ISL_1845732, EPI_ISL_1845757, EPI_ISL_1845778, EPI_ISL_1845783, EPI_ISL_1849965, EPI_ISL_1850007, EPI_ISL_1850026, EPI_ISL_1850031, EPI_ISL_1850035, EPI_ISL_1850037, EPI_ISL_1850043, EPI_ISL_1850046, EPI_ISL_1850055, EPI_ISL_1850066, EPI_ISL_1850088, EPI_ISL_1850093, EPI_ISL_1850099, EPI_ISL_1850117, EPI_ISL_1850133, EPI_ISL_1850137, EPI_ISL_1850173, EPI_ISL_1850177, EPI_ISL_1850180, EPI_ISL_1851490, EPI_ISL_1851531, EPI_ISL_1851543, EPI_ISL_1851586, EPI_ISL_1851590, EPI_ISL_1851609, EPI_ISL_1851621, EPI_ISL_1851644, EPI_ISL_1851692, EPI_ISL_1852799, EPI_ISL_1852817, EPI_ISL_1852847, EPI_ISL_1852907, EPI_ISL_1852964, EPI_ISL_1852980, EPI_ISL_1853070, EPI_ISL_1853101, EPI_ISL_2111641, EPI_ISL_2111654, EPI_ISL_2111736, EPI_ISL_2111978, EPI_ISL_2113481

see above SYNLAB MVZ Weiden Robert Koch Institute

EPI_ISL_2109059, EPI_ISL_2109079, EPI_ISL_2109096, EPI_ISL_2109099, EPI_ISL_2109120, EPI_ISL_2109131, EPI_ISL_2109150, EPI_ISL_2109190, EPI_ISL_2109199, EPI_ISL_2109209, EPI_ISL_2109236, EPI_ISL_2109241, EPI_ISL_2109265, EPI_ISL_2109277, EPI_ISL_2109552, EPI_ISL_2109560, EPI_ISL_2109562, EPI_ISL_2109575, EPI_ISL_2109593, EPI_ISL_2109607, EPI_ISL_2109618, EPI_ISL_2109633, EPI_ISL_2109635, EPI_ISL_2109646, EPI_ISL_2109667, EPI_ISL_2109676, EPI_ISL_2109682, EPI_ISL_2109686, EPI_ISL_2109717, EPI_ISL_2109732, EPI_ISL_2109739, EPI_ISL_2109755, EPI_ISL_2109760, EPI_ISL_2109772, EPI_ISL_2109776, EPI_ISL_2109781, EPI_ISL_2109793, EPI_ISL_2109798, EPI_ISL_2109808, EPI_ISL_2109813, EPI_ISL_2109816, EPI_ISL_2109821, EPI_ISL_2109823, EPI_ISL_2109826, EPI_ISL_2109832, EPI_ISL_2115540, EPI_ISL_2115559, EPI_ISL_2115562, EPI_ISL_2115588, EPI_ISL_2115675, EPI_ISL_2115705, EPI_ISL_2115722, EPI_ISL_2115760, EPI_ISL_2120675, EPI_ISL_2120707, EPI_ISL_2120867, EPI_ISL_2120878, EPI_ISL_2120903, EPI_ISL_2121009, EPI_ISL_2121048, EPI_ISL_2121181, EPI_ISL_2121421, EPI_ISL_2122272, EPI_ISL_2125163, EPI_ISL_2125186, EPI_ISL_2125389, EPI_ISL_2125505, EPI_ISL_2125549, EPI_ISL_2125679, EPI_ISL_2126118, EPI_ISL_2126169

see above Sonic - Bioscientia - MVZ Labor Saar GmbH Robert Koch Institute

EPI_ISL_2125197, EPI_ISL_2125253 Sonic - Dr. Staber & Kollegen GmbH Munchen Robert Koch Institute

EPI_ISL_2115514, EPI_ISL_2115526, EPI_ISL_2115566, EPI_ISL_2115572, EPI_ISL_2115581, EPI_ISL_2115607, EPI_ISL_2115641, EPI_ISL_2115668, EPI_ISL_2115669, EPI_ISL_2115694, EPI_ISL_2115724, EPI_ISL_2115731, EPI_ISL_2115737, EPI_ISL_2115751, EPI_ISL_2115758, EPI_ISL_2115790

see above Sonic - Dr. Staber & Kollegen GmbH MÃ¼nchen Robert Koch Institute

EPI_ISL_2109505, EPI_ISL_2109516, EPI_ISL_2109517, EPI_ISL_2109519, EPI_ISL_2109523, EPI_ISL_2109528, EPI_ISL_2109529, EPI_ISL_2109531, EPI_ISL_2109532, EPI_ISL_2109534, EPI_ISL_2109537, EPI_ISL_2109544, EPI_ISL_2109545, EPI_ISL_2109546, EPI_ISL_2109550, EPI_ISL_2109551, EPI_ISL_2109556, EPI_ISL_2109581, EPI_ISL_2109586, EPI_ISL_2109596, EPI_ISL_2109606, EPI_ISL_2109609, EPI_ISL_2109610, EPI_ISL_2109612, EPI_ISL_2109615, EPI_ISL_2109623, EPI_ISL_2109629, EPI_ISL_2109631, EPI_ISL_2109642, EPI_ISL_2109643, EPI_ISL_2109648, EPI_ISL_2109651, EPI_ISL_2109657, EPI_ISL_2109660, EPI_ISL_2109666, EPI_ISL_2109690, EPI_ISL_2109693, EPI_ISL_2109718, EPI_ISL_2109720, EPI_ISL_2109722, EPI_ISL_2109723, EPI_ISL_2109724, EPI_ISL_2109726, EPI_ISL_2109730, EPI_ISL_2109736, EPI_ISL_2109737, EPI_ISL_2109744, EPI_ISL_2109749, EPI_ISL_2109750, EPI_ISL_2109752, EPI_ISL_2109753, EPI_ISL_2109756, EPI_ISL_2109757, EPI_ISL_2109758, EPI_ISL_2109763, EPI_ISL_2109768, EPI_ISL_2109774, EPI_ISL_2109777, EPI_ISL_2109780, EPI_ISL_2109787, EPI_ISL_2109789, EPI_ISL_2109791, EPI_ISL_2109797, EPI_ISL_2109801, EPI_ISL_2109806, EPI_ISL_2109810, EPI_ISL_2109812, EPI_ISL_2109814, EPI_ISL_2109815, EPI_ISL_2109817, EPI_ISL_2109824, EPI_ISL_2109825, EPI_ISL_2109829, EPI_ISL_2109830, EPI_ISL_2109833, EPI_ISL_2125221, EPI_ISL_2125268

see above Sonic - Labor Dr. von Froreich GmbH Robert Koch Institute

EPI_ISL_2120719, EPI_ISL_2121100, EPI_ISL_2121289 Sonic - Labor Lademannbogen MVZ GmbH Robert Koch Institute

EPI_ISL_2115550, EPI_ISL_2115585, EPI_ISL_2115589, EPI_ISL_2115590, EPI_ISL_2115595, EPI_ISL_2115624, EPI_ISL_2115650, EPI_ISL_2115663, EPI_ISL_2115682, EPI_ISL_2115685, EPI_ISL_2115704, EPI_ISL_2115740, EPI_ISL_2115745, EPI_ISL_2115770, EPI_ISL_2115781, EPI_ISL_2115801, EPI_ISL_2125162, EPI_ISL_2125165, EPI_ISL_2125281, EPI_ISL_2125360

see above Sonic - Labor Staber Bayreuth Robert Koch Institute

EPI_ISL_2115529, EPI_ISL_2115549, EPI_ISL_2115551, EPI_ISL_2115554, EPI_ISL_2115602, EPI_ISL_2115614, EPI_ISL_2115649, EPI_ISL_2115653, EPI_ISL_2115681, EPI_ISL_2115697, EPI_ISL_2115728, EPI_ISL_2115746, EPI_ISL_2115782, EPI_ISL_2115794, EPI_ISL_2125157, EPI_ISL_2125303

see above Sonic - Labor Staber Dresden (Klipphausen) Robert Koch Institute

EPI_ISL_2115565, EPI_ISL_2115633, EPI_ISL_2115648, EPI_ISL_2115708, EPI_ISL_2115763 Sonic - Labor Staber MVZ Kassel Robert Koch Institute

EPI_ISL_2125194, EPI_ISL_2125245, EPI_ISL_2125329 Sonic - Labor Staber Nurnberg Robert Koch Institute

EPI_ISL_2115531, EPI_ISL_2115613, EPI_ISL_2115721, EPI_ISL_2115744, EPI_ISL_2115762, EPI_ISL_2115772, EPI_ISL_2115787

see above Sonic - Labor Staber NÃ¼rnberg Robert Koch Institute

EPI_ISL_2109038, EPI_ISL_2109043, EPI_ISL_2109048, EPI_ISL_2109056, EPI_ISL_2109058, EPI_ISL_2109060, EPI_ISL_2109063, EPI_ISL_2109066, EPI_ISL_2109068, EPI_ISL_2109070, EPI_ISL_2109075, EPI_ISL_2109083, EPI_ISL_2109089, EPI_ISL_2109112, EPI_ISL_2109119, EPI_ISL_2109123, EPI_ISL_2109125, EPI_ISL_2109129, EPI_ISL_2109130, EPI_ISL_2109134, EPI_ISL_2109151, EPI_ISL_2109173, EPI_ISL_2109185, EPI_ISL_2109187, EPI_ISL_2109191, EPI_ISL_2109200, EPI_ISL_2109202, EPI_ISL_2109203, EPI_ISL_2109207, EPI_ISL_2109213, EPI_ISL_2109217, EPI_ISL_2109220, EPI_ISL_2109229, EPI_ISL_2109233, EPI_ISL_2109246, EPI_ISL_2109249, EPI_ISL_2109250, EPI_ISL_2109252, EPI_ISL_2109263, EPI_ISL_2109269, EPI_ISL_2109273, EPI_ISL_2109281, EPI_ISL_2109284, EPI_ISL_2109289, EPI_ISL_2109293, EPI_ISL_2109299, EPI_ISL_2109300, EPI_ISL_2109302, EPI_ISL_2109307, EPI_ISL_2109310, EPI_ISL_2109315, EPI_ISL_2109318, EPI_ISL_2109321, EPI_ISL_2109325, EPI_ISL_2109328, EPI_ISL_2109331, EPI_ISL_2109333, EPI_ISL_2109335, EPI_ISL_2109348, EPI_ISL_2109354, EPI_ISL_2109355, EPI_ISL_2109506, EPI_ISL_2109514, EPI_ISL_2109527, EPI_ISL_2109555, EPI_ISL_2109558, EPI_ISL_2109564, EPI_ISL_2109565, EPI_ISL_2109577, EPI_ISL_2109578, EPI_ISL_2109582, EPI_ISL_2109592, EPI_ISL_2109602, EPI_ISL_2109613, EPI_ISL_2109632, EPI_ISL_2109654, EPI_ISL_2109663, EPI_ISL_2109694, EPI_ISL_2109699, EPI_ISL_2109703, EPI_ISL_2109712, EPI_ISL_2109735, EPI_ISL_2109743, EPI_ISL_2109746, EPI_ISL_2109747, EPI_ISL_2109748, EPI_ISL_2109761, EPI_ISL_2109769, EPI_ISL_2109804, EPI_ISL_2109831, EPI_ISL_2109837, EPI_ISL_2109844, EPI_ISL_2115515, EPI_ISL_2115560, EPI_ISL_2115576, EPI_ISL_2115598, EPI_ISL_2115634, EPI_ISL_2115651, EPI_ISL_2115654, EPI_ISL_2115666, EPI_ISL_2115686, EPI_ISL_2115700, EPI_ISL_2115703, EPI_ISL_2115726, EPI_ISL_2115752

see above Sonic - MVZ Medizinisches Labor Bremen GmbH Robert Koch Institute

| | | | |
|---|---|---|---|
| EPI_ISL_2115528, EPI_ISL_2115536, EPI_ISL_2115545, EPI_ISL_2115632, EPI_ISL_2115655, EPI_ISL_2115696, EPI_ISL_2115723, EPI_ISL_2115802 | | | |
| see above | Sonic - MVZ Medizinisches Labor Oldenburg | Robert Koch Institute | |
| EPI_ISL_1843235 | Stiftung Krankenhaus Bethanien fÃ¼r die Grafschaft Moers | Robert Koch Institute | |
| EPI_ISL_1846388, EPI_ISL_1846399, EPI_ISL_1846401, EPI_ISL_1846418, EPI_ISL_1846419, EPI_ISL_1846423, EPI_ISL_1846426, EPI_ISL_1846433, EPI_ISL_1846435, EPI_ISL_1846436, EPI_ISL_1846439, EPI_ISL_1846440, EPI_ISL_1846443, EPI_ISL_1846447, EPI_ISL_1846448 | | | |
| see above | StÃ¤dtisches Klinikum Dresden Institut fÃ¼r Klinische Chemie und Laboratoriumsmedizin | Robert Koch Institute | |
| EPI_ISL_1729587, EPI_ISL_1844648, EPI_ISL_1844667, EPI_ISL_1844675, EPI_ISL_1844690, EPI_ISL_1844750, EPI_ISL_1844781, EPI_ISL_1844837, EPI_ISL_1844838, EPI_ISL_1845133, EPI_ISL_1845144, EPI_ISL_1845175, EPI_ISL_1845189, EPI_ISL_1845292, EPI_ISL_1845357, EPI_ISL_1845370, EPI_ISL_1845424, EPI_ISL_1845467, EPI_ISL_1845475, EPI_ISL_1845479, EPI_ISL_1845498, EPI_ISL_1845551, EPI_ISL_1845563, EPI_ISL_1845680, EPI_ISL_1849957, EPI_ISL_1850078, EPI_ISL_1850092, EPI_ISL_1850123, EPI_ISL_1850152, EPI_ISL_1850159, EPI_ISL_1850250, EPI_ISL_1851472, EPI_ISL_1851488, EPI_ISL_1851555, EPI_ISL_1851570, EPI_ISL_1851596, EPI_ISL_1851624, EPI_ISL_1851649, EPI_ISL_1851689, EPI_ISL_1851706, EPI_ISL_1851709, EPI_ISL_1852507, EPI_ISL_1852551, EPI_ISL_1852633, EPI_ISL_1852796, EPI_ISL_1852803, EPI_ISL_1852874, EPI_ISL_1852996, EPI_ISL_1853055, EPI_ISL_1853071, EPI_ISL_1853092, EPI_ISL_2111633, EPI_ISL_2111861, EPI_ISL_2111905, EPI_ISL_2111943, EPI_ISL_2111961, EPI_ISL_2113787 | | | |
| see above | Synlab MVZ Augsburg | Robert Koch Institute | |
| EPI_ISL_2109018, EPI_ISL_2109019, EPI_ISL_2109020, EPI_ISL_2109021, EPI_ISL_2109025, EPI_ISL_2109029, EPI_ISL_2109031, EPI_ISL_2109033, EPI_ISL_2109035, EPI_ISL_2109036, EPI_ISL_2123852, EPI_ISL_2123853, EPI_ISL_2123854, EPI_ISL_2123855, EPI_ISL_2123858, EPI_ISL_2123859, EPI_ISL_2123860, EPI_ISL_2123861 | | | |
| see above | Thuringer Landesamtes fur Verbraucherschutz | Robert Koch Institute | |
| EPI_ISL_1846368, EPI_ISL_1846380, EPI_ISL_1846384, EPI_ISL_1846387, EPI_ISL_1846389, EPI_ISL_1846393, EPI_ISL_1846394, EPI_ISL_1846404, EPI_ISL_1846412, EPI_ISL_1846415, EPI_ISL_1846421, EPI_ISL_1846428, EPI_ISL_1846441, EPI_ISL_1846450 | | | |
| see above | Uniklinikum Carl Gustav Carus an der TU Dresden; Institut fÃ¼r Virologie | Robert Koch Institute | |
| EPI_ISL_1844031, EPI_ISL_2110412, EPI_ISL_2110414 | Uniklinikum Jena | Robert Koch Institute | |
| EPI_ISL_1851140, EPI_ISL_1851141, EPI_ISL_1851142, EPI_ISL_1851143, EPI_ISL_1851144, EPI_ISL_1851146, EPI_ISL_1851147 | | | |
| see above | Uniklinikum Regensburg; Institut fÃ¼r klinische Mikrobiologie und Hygiene | Robert Koch Institute | |
| EPI_ISL_2127223 | Universitatsklinikum Bonn - Institut fur Virologie | Robert Koch Institute | |
| EPI_ISL_2112480, EPI_ISL_2123976, EPI_ISL_2123977 | Universitatsklinikum Heidelberg | Robert Koch Institute | |
| EPI_ISL_2108266, EPI_ISL_2108277, EPI_ISL_2108279, EPI_ISL_2108282, EPI_ISL_2108289, EPI_ISL_2108293, EPI_ISL_2108295, EPI_ISL_2108305, EPI_ISL_2108307, EPI_ISL_2108309, EPI_ISL_2108320 | | | |
| see above | Universitatsklinikum des Saarlandes - Institut fur Virologie | Robert Koch Institute | |
| EPI_ISL_1862413, EPI_ISL_1862416, EPI_ISL_1862417, EPI_ISL_1862420, EPI_ISL_1862421, EPI_ISL_1862423, EPI_ISL_1862425, EPI_ISL_1862432, EPI_ISL_1862434, EPI_ISL_1862435, EPI_ISL_1862438 | | | |
| see above | University Medical Center Hamburg Eppendorf | Heinrich Pette Institute, Leibniz Institute for Experimental Virology | Adam Grundhoff; Alexis Robitaille; Johannes Knobloch; Martin Aepfelbacher; Nicole Fischer; Thomas Günther |
| EPI_ISL_2115533, EPI_ISL_2115552, EPI_ISL_2115564 | UniversitÃ¤tsklinikum Frankfurt - Institut fur Medizinische Virologie | Robert Koch Institute | |
| EPI_ISL_2115609, EPI_ISL_2115611, EPI_ISL_2115621, EPI_ISL_2115630, EPI_ISL_2115664, EPI_ISL_2115695, EPI_ISL_2115803 | | | |
| see above | UniversitÃ¤tsklinikum Frankfurt - Institut fÃ¼r Medizinische Virologie | Robert Koch Institute | |
| EPI_ISL_1850437, EPI_ISL_1850438, EPI_ISL_1850440, EPI_ISL_1850441, EPI_ISL_1850442, EPI_ISL_1850443, EPI_ISL_1850445 | | | |
| see above | UniversitÃ¤tsklinikum MÃ¼nster Institut fÃ¼r Virologie | Robert Koch Institute | |
| EPI_ISL_2115501, EPI_ISL_2115502, EPI_ISL_2115503, EPI_ISL_2115506, EPI_ISL_2115508, EPI_ISL_2115509 | Virologisches Institut des Universitatsklinikums Erlangen | Robert Koch Institute | |
| EPI_ISL_1864490, EPI_ISL_1864491, EPI_ISL_1864494, EPI_ISL_1864496, EPI_ISL_1864498, EPI_ISL_1864500, EPI_ISL_1864502, EPI_ISL_1864504, EPI_ISL_1864509, EPI_ISL_1864514, EPI_ISL_1864519, EPI_ISL_1864521, EPI_ISL_1864523, EPI_ISL_1864525, EPI_ISL_1864527, EPI_ISL_1864529, EPI_ISL_1864532, EPI_ISL_1864534, EPI_ISL_1864536, EPI_ISL_1864538, EPI_ISL_1864540, EPI_ISL_1864542, EPI_ISL_1864544, EPI_ISL_1864546, EPI_ISL_1864552, EPI_ISL_1864557, EPI_ISL_1864559 | | | |
| see above | Virology, Universitätsklinikum des Saarlandes | Epigenetics, Saarland University | Jörn Walter; Kathrin Kattler; Sascha Tierling; Sigrun Smola; Stefan Lohse; Thorsten Pfuhl |
| EPI_ISL_1992897, EPI_ISL_1992898, EPI_ISL_1992899, EPI_ISL_1993384, EPI_ISL_1993386, EPI_ISL_1993388, EPI_ISL_1993389, EPI_ISL_1993390, EPI_ISL_1993391, EPI_ISL_1993392, EPI_ISL_1993393, EPI_ISL_1993394, EPI_ISL_1993395, EPI_ISL_1993396, EPI_ISL_1993397, EPI_ISL_1993398, EPI_ISL_1993400, EPI_ISL_1993411, EPI_ISL_1993412, EPI_ISL_1993413, EPI_ISL_1993414, EPI_ISL_1993415, EPI_ISL_1993416, EPI_ISL_1993417, EPI_ISL_1993418, EPI_ISL_1993419, EPI_ISL_1993420, EPI_ISL_1993421, EPI_ISL_1993430, EPI_ISL_1993431, EPI_ISL_1993433, EPI_ISL_1993439, EPI_ISL_1993441, EPI_ISL_1993635, EPI_ISL_1993636, EPI_ISL_1993637, EPI_ISL_1993638, EPI_ISL_1993639, EPI_ISL_1993640, EPI_ISL_1993641, EPI_ISL_1993642, EPI_ISL_1993643, EPI_ISL_1993644, EPI_ISL_1993645, EPI_ISL_1993646, EPI_ISL_1993647, EPI_ISL_1993648, EPI_ISL_1993649, EPI_ISL_1993687, EPI_ISL_1993688, EPI_ISL_1993689, EPI_ISL_1993690, EPI_ISL_1993692, EPI_ISL_1993693, EPI_ISL_1993697, EPI_ISL_1993698, EPI_ISL_1993699, EPI_ISL_1993701, EPI_ISL_1993702, EPI_ISL_1993703, EPI_ISL_1993704, EPI_ISL_1993706, EPI_ISL_1993708, EPI_ISL_1993709, EPI_ISL_1993710, EPI_ISL_1993711, EPI_ISL_1993712, EPI_ISL_1993714, EPI_ISL_1993715, EPI_ISL_1993717, EPI_ISL_1993718, EPI_ISL_1993721, EPI_ISL_1993722, EPI_ISL_1993723, EPI_ISL_1993725, EPI_ISL_1993726, EPI_ISL_1993727, EPI_ISL_1993728, EPI_ISL_1993729, EPI_ISL_1993730 | | | |
| see above | ZOTZ KLIMAS MVZ Düsseldorf-Centrum GbR ÜBAG für Labormedizin, Genetik, Zytologie, Pathologie | Center of Medical Microbiology, Virology, and Hospital Hygiene, University of Duesseldorf | Alexander Dilthey; Andreas Walker; Daniel Strelow; Jessica Nicolai; Jörg Timm; Katrin Hoffmann; Klaus Pfeffer; Lisanna Hülse; Malte Kohns Vasconcelos; Maximilian Damagnez; Nadine Lübke; Patrick Finzer; Rainer Zotz; Tobias Wienemann; Torsten Houwaart |
| EPI_ISL_1726975, EPI_ISL_1726976, EPI_ISL_1726977, EPI_ISL_1726978, EPI_ISL_1726979, EPI_ISL_1726980, EPI_ISL_1726983, EPI_ISL_1726984, EPI_ISL_1726985, EPI_ISL_1851417, EPI_ISL_1851418, EPI_ISL_1851420, EPI_ISL_1851422, EPI_ISL_1851423, EPI_ISL_1851424, EPI_ISL_1851425, EPI_ISL_1851426, EPI_ISL_1851427 | | | |
| see above | alphaomega Labor GbR (Medizinische Kooperationsgemeinschaft Dres. Ackermann/Tennert/ZaiÃŸ) | Robert Koch Institute | |
| EPI_ISL_1727259, EPI_ISL_1727268, EPI_ISL_1727336, EPI_ISL_1727344, EPI_ISL_1727347, EPI_ISL_1850462, EPI_ISL_1850472, EPI_ISL_1850473, EPI_ISL_1850493, EPI_ISL_1850510, EPI_ISL_1850536, EPI_ISL_1850554, EPI_ISL_1850565, EPI_ISL_1850572, EPI_ISL_1850575, EPI_ISL_1850580, EPI_ISL_1850585, EPI_ISL_1850596, EPI_ISL_1850598, EPI_ISL_1850624, EPI_ISL_1850629, EPI_ISL_1850635, EPI_ISL_1850646, EPI_ISL_1850648, EPI_ISL_1850658, EPI_ISL_1850662, EPI_ISL_1850673, EPI_ISL_1850676, EPI_ISL_1850679, EPI_ISL_1850684, EPI_ISL_1850693, EPI_ISL_1850708 | | | |
| see above | amedes GÃ¶ttingen | Robert Koch Institute | |
| EPI_ISL_1850463, EPI_ISL_1850474, EPI_ISL_1850486, EPI_ISL_1850506, EPI_ISL_1850509, EPI_ISL_1850512, EPI_ISL_1850514, EPI_ISL_1850516, EPI_ISL_1850520, EPI_ISL_1850521, EPI_ISL_1850524, EPI_ISL_1850531, EPI_ISL_1850553, EPI_ISL_1850558, EPI_ISL_1850570, EPI_ISL_1850577, EPI_ISL_1850587, EPI_ISL_1850591, EPI_ISL_1850593, EPI_ISL_1850603, EPI_ISL_1850605, EPI_ISL_1850606, EPI_ISL_1850608, EPI_ISL_1850609, EPI_ISL_1850614, EPI_ISL_1850615, EPI_ISL_1850618, EPI_ISL_1850619, EPI_ISL_1850622, EPI_ISL_1850623, EPI_ISL_1850628, EPI_ISL_1850632, EPI_ISL_1850633, EPI_ISL_1850634, EPI_ISL_1850637, EPI_ISL_1850638, EPI_ISL_1850641, EPI_ISL_1850643, EPI_ISL_1850651, EPI_ISL_1850652, EPI_ISL_1850653, EPI_ISL_1850654, EPI_ISL_1850661, EPI_ISL_1850668, EPI_ISL_1850674, EPI_ISL_1850675, EPI_ISL_1850677, EPI_ISL_1850682, EPI_ISL_1850685, EPI_ISL_1850688, EPI_ISL_1850692, EPI_ISL_1850696, EPI_ISL_1850702, EPI_ISL_2115996 | | | |
| see above | amedes MVZ DIAMEDIS Sennestadt | Robert Koch Institute | |
| EPI_ISL_1850447, EPI_ISL_1850451, EPI_ISL_1850454, EPI_ISL_1850467, EPI_ISL_1850488, EPI_ISL_1850494, EPI_ISL_1850526, EPI_ISL_1850529, EPI_ISL_1850537, EPI_ISL_1850542, EPI_ISL_1850546, EPI_ISL_1850551, EPI_ISL_1850557, EPI_ISL_1850566, EPI_ISL_1850579, EPI_ISL_1850586, EPI_ISL_1850602, EPI_ISL_1850611, EPI_ISL_1850627, EPI_ISL_1850642, EPI_ISL_1850655, EPI_ISL_1850657, EPI_ISL_1850663, EPI_ISL_1850669, EPI_ISL_1850680, EPI_ISL_1850698, EPI_ISL_1850707 | | | |
| see above | amedes MVZ Halle/Leipzig | Robert Koch Institute | |
| EPI_ISL_1850448, EPI_ISL_1850452, EPI_ISL_1850456, EPI_ISL_1850457, EPI_ISL_1850461, EPI_ISL_1850465, EPI_ISL_1850483, EPI_ISL_1850490, EPI_ISL_1850495, EPI_ISL_1850498, EPI_ISL_1850501, EPI_ISL_1850519, EPI_ISL_1850525, EPI_ISL_1850528, EPI_ISL_1850534, EPI_ISL_1850539, EPI_ISL_1850571, EPI_ISL_1850582, EPI_ISL_1850584, EPI_ISL_1850590, EPI_ISL_1850616, EPI_ISL_1850621, EPI_ISL_1850665, EPI_ISL_1850670, EPI_ISL_1850714 | | | |
| see above | amedes MVZ Hannover | Robert Koch Institute | |
| EPI_ISL_1850460, EPI_ISL_1850482, EPI_ISL_1850497, EPI_ISL_1850503, EPI_ISL_1850532, EPI_ISL_1850533, EPI_ISL_1850541, EPI_ISL_1850544, EPI_ISL_1850548, EPI_ISL_1850550, EPI_ISL_1850561, EPI_ISL_1850564, EPI_ISL_1850574, EPI_ISL_1850589, EPI_ISL_1850592, EPI_ISL_1850594, EPI_ISL_1850604, EPI_ISL_1850617, EPI_ISL_1850636, EPI_ISL_1850650, EPI_ISL_1850664, EPI_ISL_1850666, EPI_ISL_1850671, EPI_ISL_1850687, EPI_ISL_1850709, EPI_ISL_1850715, EPI_ISL_1850720 | | | |
| see above | amedes MVZ Jena | Robert Koch Institute | |
| EPI_ISL_1850656, EPI_ISL_1850703 | amedes MVZ Zentrum fÃ¼r Pathologie und Zytodiagnostik GmbH (KÃ¶ln) | Robert Koch Institute | |
| EPI_ISL_1850446, EPI_ISL_1850450, EPI_ISL_1850453, EPI_ISL_1850468, EPI_ISL_1850478, EPI_ISL_1850555, EPI_ISL_1850556, EPI_ISL_1850573, EPI_ISL_1850581, EPI_ISL_1850583, EPI_ISL_1850601, EPI_ISL_1850613, EPI_ISL_1850699, EPI_ISL_1850711 | | | |
| see above | amedes MVZ fÃ¼r Laboratoriumsdiagnostik Raubling GmbH | Robert Koch Institute | |
| EPI_ISL_1850504, EPI_ISL_1850508, EPI_ISL_1850563, EPI_ISL_1850626, EPI_ISL_1850681, EPI_ISL_1850691, EPI_ISL_1850717 | | | |
| see above | amedes MVZ fÃ¼r Laboratoriumsmedizin FÃ¼rstenfeldbruck | Robert Koch Institute | |
| EPI_ISL_1850484 | amedes MVZ fÃ¼r Laboratoriumsmedizin Hamburg | Robert Koch Institute | |
| EPI_ISL_1850620 | amedes MVZ fÃ¼r Laboratoriumsmedizin MÃ¼nchen | Robert Koch Institute | |
| EPI_ISL_1850543, EPI_ISL_1850547, EPI_ISL_1850568, EPI_ISL_1850607 | amedes MVZ fÃ¼r Laboratoriumsmedizin Rhein/Ruhr | Robert Koch Institute | |
| EPI_ISL_2115810, EPI_ISL_2115888, EPI_ISL_2115913, EPI_ISL_2115956, EPI_ISL_2115986, EPI_ISL_2116006 | amedes MVZ fÃ¼r Labordiagnostik Rhein-Main | Robert Koch Institute | |
| EPI_ISL_1727131, EPI_ISL_1727137, EPI_ISL_1727140, EPI_ISL_1727142, EPI_ISL_1727143, EPI_ISL_1727149, EPI_ISL_1727152, EPI_ISL_1727161, EPI_ISL_1727163, EPI_ISL_1727165, EPI_ISL_1727168, EPI_ISL_1727171, EPI_ISL_1727173, EPI_ISL_1727180, EPI_ISL_1727182, EPI_ISL_1727190, EPI_ISL_1727195, EPI_ISL_1727198, EPI_ISL_1727215, EPI_ISL_1727216, EPI_ISL_1727224, EPI_ISL_1727228, EPI_ISL_1727230, EPI_ISL_1727246, EPI_ISL_1727251, EPI_ISL_1727263, EPI_ISL_1727265, EPI_ISL_1727272, EPI_ISL_1727291, EPI_ISL_1727292, EPI_ISL_1727294, EPI_ISL_1727299, EPI_ISL_1727300, EPI_ISL_1727303, EPI_ISL_1727328, EPI_ISL_1727353, EPI_ISL_1727355, EPI_ISL_1727365, EPI_ISL_1727367, EPI_ISL_1727375, EPI_ISL_1727381, EPI_ISL_1727382, EPI_ISL_1850466, EPI_ISL_1850471, EPI_ISL_1850476, EPI_ISL_1850487, EPI_ISL_1850492, EPI_ISL_1850499, EPI_ISL_1850505, EPI_ISL_1850515, EPI_ISL_1850517, EPI_ISL_1850523, EPI_ISL_1850538, EPI_ISL_1850578, EPI_ISL_1850588, EPI_ISL_1850599, EPI_ISL_1850625, EPI_ISL_1850630, EPI_ISL_1850645, EPI_ISL_1850647, EPI_ISL_1850667, EPI_ISL_1850686, EPI_ISL_1850694, EPI_ISL_1850697, EPI_ISL_1850700, EPI_ISL_1850704, EPI_ISL_1850716, EPI_ISL_1850718, EPI_ISL_1850719, EPI_ISL_1850721 | | | |
| see above | nordlab - Partnerschaftspraxis fuÌˆr Laboratoriumsmedizin | Robert Koch Institute | |

We gratefully acknowledge the following Authors from the Originating laboratories responsible for obtaining the specimens, as well as the Submitting laboratories where the genome data were generated and shared via GISAID, on which this research is based.

All Submitters of data may be contacted directly via www.gisaid.org

Authors are sorted alphabetically.

| Accession ID | Originating Laboratory | Submitting Laboratory | Authors |
|---|---|---|---|
| EPI_ISL_2120599, EPI_ISL_2120632, EPI_ISL_2120660, EPI_ISL_2120661, EPI_ISL_2120666, EPI_ISL_2120670, EPI_ISL_2120674, EPI_ISL_2120681, EPI_ISL_2120683, EPI_ISL_2120685, EPI_ISL_2120693, EPI_ISL_2120711, EPI_ISL_2120716, EPI_ISL_2120729, EPI_ISL_2120807, EPI_ISL_2120813, EPI_ISL_2120832, EPI_ISL_2120850, EPI_ISL_2120859, EPI_ISL_2120862, EPI_ISL_2120870, EPI_ISL_2120917, EPI_ISL_2120952, EPI_ISL_2120961, EPI_ISL_2120982, EPI_ISL_2120997, EPI_ISL_2121012, EPI_ISL_2121024, EPI_ISL_2121045, EPI_ISL_2121051, EPI_ISL_2121065, EPI_ISL_2121074, EPI_ISL_2121077, EPI_ISL_2121087, EPI_ISL_2121093, EPI_ISL_2121106, EPI_ISL_2121142, EPI_ISL_2121158, EPI_ISL_2121170, EPI_ISL_2121184, EPI_ISL_2121200, EPI_ISL_2121220, EPI_ISL_2121257, EPI_ISL_2121276, EPI_ISL_2121278, EPI_ISL_2121348, EPI_ISL_2121398, EPI_ISL_2121403, EPI_ISL_2121406, EPI_ISL_2121415, EPI_ISL_2121424, EPI_ISL_2121439, EPI_ISL_2121443, EPI_ISL_2121445, EPI_ISL_2121450, EPI_ISL_2121451, EPI_ISL_2121452, EPI_ISL_2121454, EPI_ISL_2121457, EPI_ISL_2121539, EPI_ISL_2121655, EPI_ISL_2121667, EPI_ISL_2121692, EPI_ISL_2121779, EPI_ISL_2121786, EPI_ISL_2121815, EPI_ISL_2121834, EPI_ISL_2121842, EPI_ISL_2121873, EPI_ISL_2121898, EPI_ISL_2121905, EPI_ISL_2121913, EPI_ISL_2121943, EPI_ISL_2121948, EPI_ISL_2121951, EPI_ISL_2121972, EPI_ISL_2122003, EPI_ISL_2122015, EPI_ISL_2122038, EPI_ISL_2122124, EPI_ISL_2122134, EPI_ISL_2122157, EPI_ISL_2122163, EPI_ISL_2122196, EPI_ISL_2122265, EPI_ISL_2122269, EPI_ISL_2122270, EPI_ISL_2125144, EPI_ISL_2125167, EPI_ISL_2125250, EPI_ISL_2125275, EPI_ISL_2125324, EPI_ISL_2125468, EPI_ISL_2125475, EPI_ISL_2125478, EPI_ISL_2125483, EPI_ISL_2125487, EPI_ISL_2125489, EPI_ISL_2125491, EPI_ISL_2125494, EPI_ISL_2125513, EPI_ISL_2125524, EPI_ISL_2125526, EPI_ISL_2125530, EPI_ISL_2125532, EPI_ISL_2125539, EPI_ISL_2125542, EPI_ISL_2125546, EPI_ISL_2125558, EPI_ISL_2125563, EPI_ISL_2125571, EPI_ISL_2125574, EPI_ISL_2125576, EPI_ISL_2125583, EPI_ISL_2125589, EPI_ISL_2125598, EPI_ISL_2125601, EPI_ISL_2125603, EPI_ISL_2125615, EPI_ISL_2125619, EPI_ISL_2125621, EPI_ISL_2125628, EPI_ISL_2125630, EPI_ISL_2125636, EPI_ISL_2125639, EPI_ISL_2125645, EPI_ISL_2125663, EPI_ISL_2125672, EPI_ISL_2125674, EPI_ISL_2125676, EPI_ISL_2125681, EPI_ISL_2125683, EPI_ISL_2125688, EPI_ISL_2125705, EPI_ISL_2125712, EPI_ISL_2125714, EPI_ISL_2125721, EPI_ISL_2125723, EPI_ISL_2125730, EPI_ISL_2125735, EPI_ISL_2125754, EPI_ISL_2125759, EPI_ISL_2125762, EPI_ISL_2125775, EPI_ISL_2125779, EPI_ISL_2125781, EPI_ISL_2125783, EPI_ISL_2125788, EPI_ISL_2125790, EPI_ISL_2125792, EPI_ISL_2125794, EPI_ISL_2125801, EPI_ISL_2125812, EPI_ISL_2125823, EPI_ISL_2125825, EPI_ISL_2125827, EPI_ISL_2125834, EPI_ISL_2125838, EPI_ISL_2125852, EPI_ISL_2125856, EPI_ISL_2125860, EPI_ISL_2125862, EPI_ISL_2125871, EPI_ISL_2125880, EPI_ISL_2125882, EPI_ISL_2125884, EPI_ISL_2125895, EPI_ISL_2125908, EPI_ISL_2125915, EPI_ISL_2125920, EPI_ISL_2125922, EPI_ISL_2125930, EPI_ISL_2125935, EPI_ISL_2125936, EPI_ISL_2125937, EPI_ISL_2125938, EPI_ISL_2125940, EPI_ISL_2125942, EPI_ISL_2125974, EPI_ISL_2125979, EPI_ISL_2125987, EPI_ISL_2125991, EPI_ISL_2125994, EPI_ISL_2125996, EPI_ISL_2125998, EPI_ISL_2126003, EPI_ISL_2126005, EPI_ISL_2126022, EPI_ISL_2126032, EPI_ISL_2126034, EPI_ISL_2126041, EPI_ISL_2126043, EPI_ISL_2126048, EPI_ISL_2126052, EPI_ISL_2126063, EPI_ISL_2126065, EPI_ISL_2126069, EPI_ISL_2126073, EPI_ISL_2126075, EPI_ISL_2126077, EPI_ISL_2126083, EPI_ISL_2126089, EPI_ISL_2126091, EPI_ISL_2126095, EPI_ISL_2126098, EPI_ISL_2126108, EPI_ISL_2126116, EPI_ISL_2126120, EPI_ISL_2126129, EPI_ISL_2126131, EPI_ISL_2126139, EPI_ISL_2126141, EPI_ISL_2126146, EPI_ISL_2126158, EPI_ISL_2126160, EPI_ISL_2126162, EPI_ISL_2126174, EPI_ISL_2126179, EPI_ISL_2126185 | | | |
| see above | Bioscientia Labor Wermsdorf | Robert Koch Institute | |
| EPI_ISL_2121459, EPI_ISL_2122022, EPI_ISL_2125473, EPI_ISL_2125493, EPI_ISL_2125521, EPI_ISL_2125560, EPI_ISL_2125592, EPI_ISL_2125641, EPI_ISL_2125685, EPI_ISL_2125690, EPI_ISL_2125697, EPI_ISL_2125702, EPI_ISL_2125707, EPI_ISL_2125716, EPI_ISL_2125725, EPI_ISL_2125757, EPI_ISL_2125771, EPI_ISL_2125773, EPI_ISL_2125777, EPI_ISL_2125809, EPI_ISL_2125854, EPI_ISL_2125900, EPI_ISL_2125904, EPI_ISL_2125910, EPI_ISL_2125912, EPI_ISL_2125928, EPI_ISL_2125941, EPI_ISL_2125972, EPI_ISL_2125999, EPI_ISL_2126000, EPI_ISL_2126008, EPI_ISL_2126016, EPI_ISL_2126025, EPI_ISL_2126030, EPI_ISL_2126059, EPI_ISL_2126104, EPI_ISL_2126106, EPI_ISL_2126122, EPI_ISL_2126137, EPI_ISL_2126150, EPI_ISL_2126167, EPI_ISL_2126176, EPI_ISL_2126189 | | | |
| see above | Bioscientia MVZ Labor Karlsruhe GmbH | Robert Koch Institute | |
| EPI_ISL_1834078, EPI_ISL_1834079, EPI_ISL_1834080, EPI_ISL_1834081, EPI_ISL_1911988, EPI_ISL_2131153 | Bundeswehr Institute of Microbiology | Bundeswehr Institute of Microbiology | Alexandra Rehn; Enrico Georgi; Malena Bestehorn-Willmann; Markus Antwerpen; Mathias Walter; Mike Pillukat; Roman Wölfel; Sabine Zange |
| EPI_ISL_1912000, EPI_ISL_2131156, EPI_ISL_2131157, EPI_ISL_2131158, EPI_ISL_2131159, EPI_ISL_2131160, EPI_ISL_2131161, EPI_ISL_2131164, EPI_ISL_2131165, EPI_ISL_2131166, EPI_ISL_2131167, EPI_ISL_2131168, EPI_ISL_2131170, EPI_ISL_2131171 | | | |
| see above | Bundeswehrkrankenhaus Berlin | Bundeswehr Institute of Microbiology | Alexandra Rehn; Enrico Georgi; Malena Bestehorn-Willmann; Markus Antwerpen; Mathias Walter; Mike Pillukat; Roman Wölfel; Sabine Zange |
| EPI_ISL_1911989, EPI_ISL_1911992, EPI_ISL_2131154, EPI_ISL_2131155 | Bundeswehrkrankenhaus Ulm | Bundeswehr Institute of Microbiology | Alexandra Rehn; Enrico Georgi; Malena Bestehorn-Willmann; Markus Antwerpen; Mathias Walter; Mike Pillukat; Roman Wölfel; Sabine Zange |
| EPI_ISL_1911994 | Bundeswehrzentralkrankenhaus Koblenz | Bundeswehr Institute of Microbiology | Alexandra Rehn; Enrico Georgi; Malena Bestehorn-Willmann; Markus Antwerpen; Mathias Walter; Mike Pillukat; Roman Wölfel; Sabine Zange |
| EPI_ISL_1993429, EPI_ISL_1993614, EPI_ISL_1993615, EPI_ISL_1993616, EPI_ISL_1993617, EPI_ISL_1993618, EPI_ISL_1993619, EPI_ISL_1993620, EPI_ISL_1993621, EPI_ISL_1993622, EPI_ISL_1993623, EPI_ISL_1993624, EPI_ISL_1993625, EPI_ISL_1993626, EPI_ISL_1993627, EPI_ISL_1993628, EPI_ISL_1993629, EPI_ISL_1993630, EPI_ISL_1993631 | | | |
| see above | Center of Medical Microbiology, Virology, and Hospital Hygiene, University of Duesseldorf | Center of Medical Microbiology, Virology, and Hospital Hygiene, University of Duesseldorf | Alexander Dilthey; Andreas Walker; Daniel Strelow; Jessica Nicolai; Jörg Timm; Klaus Pfeffer; Lisanna Hülse; Malte Kohns Vasconcelos; Maximilian Damagnez; Nadine Lübke; Tobias Wienemann; Torsten Houwaart |
| EPI_ISL_2123108, EPI_ISL_2123119, EPI_ISL_2123138, EPI_ISL_2123142, EPI_ISL_2123159, EPI_ISL_2123177, EPI_ISL_2123191, EPI_ISL_2123224, EPI_ISL_2123226, EPI_ISL_2123237, EPI_ISL_2123239, EPI_ISL_2123293 | | | |
| see above | Centogene; Dr. Bauer Laboratoriums GmbH | Robert Koch Institute | |
| EPI_ISL_2112942, EPI_ISL_2112943, EPI_ISL_2112946, EPI_ISL_2112947, EPI_ISL_2112950, EPI_ISL_2112952 | Chemische und Veterinaruntersuchungsamt Monsterland-Emscher-Lippe (CVUA-MEL) | Robert Koch Institute | |
| EPI_ISL_2129678 | Chemische und VeterinÃ¤runtersuchungsamt MÃ¼nsterland-Emscher-Lippe (CVUA-MEL) | Robert Koch Institute | |
| EPI_ISL_2111030, EPI_ISL_2111031, EPI_ISL_2111037, EPI_ISL_2111046, EPI_ISL_2111048, EPI_ISL_2111059, EPI_ISL_2111061, EPI_ISL_2111069, EPI_ISL_2111070, EPI_ISL_2111074, EPI_ISL_2111077, EPI_ISL_2111080, EPI_ISL_2111081, EPI_ISL_2111089, EPI_ISL_2111090, EPI_ISL_2111091, EPI_ISL_2111096, EPI_ISL_2111100, EPI_ISL_2111101, EPI_ISL_2111102, EPI_ISL_2111104, EPI_ISL_2124271, EPI_ISL_2124272, EPI_ISL_2124275, EPI_ISL_2124278, EPI_ISL_2124280, EPI_ISL_2124281, EPI_ISL_2124283, EPI_ISL_2124285, EPI_ISL_2124286, EPI_ISL_2124288, EPI_ISL_2124289, EPI_ISL_2124290, EPI_ISL_2124292, EPI_ISL_2124294, EPI_ISL_2124295, EPI_ISL_2124296, EPI_ISL_2124297, EPI_ISL_2124299, EPI_ISL_2124300, EPI_ISL_2124301, EPI_ISL_2124302, EPI_ISL_2124304, EPI_ISL_2124305, EPI_ISL_2124306, EPI_ISL_2124308, EPI_ISL_2124310, EPI_ISL_2124311, EPI_ISL_2124313, EPI_ISL_2124314, EPI_ISL_2124316, EPI_ISL_2124317, EPI_ISL_2124318, EPI_ISL_2124319, EPI_ISL_2124321, EPI_ISL_2124324, EPI_ISL_2124329, EPI_ISL_2124332, EPI_ISL_2124334, EPI_ISL_2124335, EPI_ISL_2124336, EPI_ISL_2124338 | | | |
| see above | Clemenshospital - Alexianer Clemenshospital Munster | Robert Koch Institute | |
| EPI_ISL_2108979, EPI_ISL_2108980, EPI_ISL_2108982, EPI_ISL_2108984, EPI_ISL_2108988, EPI_ISL_2108989, EPI_ISL_2108991, EPI_ISL_2108992, EPI_ISL_2108995, EPI_ISL_2108996, EPI_ISL_2108998, EPI_ISL_2109000, EPI_ISL_2109005, EPI_ISL_2109006, EPI_ISL_2109007, EPI_ISL_2109009, EPI_ISL_2109011, EPI_ISL_2109012, EPI_ISL_2109015 | | | |
| see above | Department fur Labormedizin Abteilung III Bereich Molekulare Diagnostik Universitatsklinikum Halle(Saale) | Robert Koch Institute | |
| EPI_ISL_2128413, EPI_ISL_2128417, EPI_ISL_2128423, EPI_ISL_2128427, EPI_ISL_2128436, EPI_ISL_2128443, EPI_ISL_2128458, EPI_ISL_2128462, EPI_ISL_2128466, EPI_ISL_2128470, EPI_ISL_2128475, EPI_ISL_2128484, EPI_ISL_2128489, EPI_ISL_2128493, EPI_ISL_2128498, EPI_ISL_2128500, EPI_ISL_2128508, EPI_ISL_2128509, EPI_ISL_2128511, EPI_ISL_2128512, EPI_ISL_2128515, EPI_ISL_2128516, EPI_ISL_2128522, EPI_ISL_2128529, EPI_ISL_2128537, EPI_ISL_2128545, EPI_ISL_2128555, EPI_ISL_2128559, EPI_ISL_2128561, EPI_ISL_2128580, EPI_ISL_2128585, EPI_ISL_2128589, EPI_ISL_2128596, EPI_ISL_2128604, EPI_ISL_2128610, EPI_ISL_2128614, EPI_ISL_2128622, EPI_ISL_2128624, EPI_ISL_2128626, EPI_ISL_2128627, EPI_ISL_2128631, EPI_ISL_2128633 | | | |
| see above | Diagnosticum - Labor Neukirchen | Robert Koch Institute | |
| EPI_ISL_2115129, EPI_ISL_2115130, EPI_ISL_2115131, EPI_ISL_2115132, EPI_ISL_2115133, EPI_ISL_2115134, EPI_ISL_2115135, EPI_ISL_2115136, EPI_ISL_2115137, EPI_ISL_2115138, EPI_ISL_2115139, EPI_ISL_2115140, EPI_ISL_2115141, EPI_ISL_2115142, EPI_ISL_2115143, EPI_ISL_2115144, EPI_ISL_2115145, EPI_ISL_2115146 | | | |
| see above | Dianovis GmbH Greiz | Robert Koch Institute | |
| EPI_ISL_2109362, EPI_ISL_2109370, EPI_ISL_2109377, EPI_ISL_2109401, EPI_ISL_2109413, EPI_ISL_2109414, EPI_ISL_2109426 | | | |
| see above | ELBLAB GmbH Zentrum fur LaborMedizin | Robert Koch Institute | |
| EPI_ISL_1846343, EPI_ISL_1846344, EPI_ISL_1846345 | Eurofins MVZ Labor Gelsenkirchen | Robert Koch Institute | |
| EPI_ISL_2125018, EPI_ISL_2125019, EPI_ISL_2125021, EPI_ISL_2125022, EPI_ISL_2125023, EPI_ISL_2125024, EPI_ISL_2125026, EPI_ISL_2125027 | | | |
| see above | Fraunhofer-Institut fur Zelltherapie und Immunologie IZI AG Next-Generation Diagnostics | Robert Koch Institute | |
| EPI_ISL_2108355, EPI_ISL_2108360, EPI_ISL_2108393, EPI_ISL_2108408, EPI_ISL_2108416 | Helios Universitatsklinikum Wuppertal: Universitat Witten/Herdecke: Institut fur Medizinische Labordiagnostik | Robert Koch Institute | |
| EPI_ISL_2115369, EPI_ISL_2115383 | IMD - Institut fur Medizinische Diagnostik Berlin-Potsdam | Robert Koch Institute | |
| EPI_ISL_2109444, EPI_ISL_2123384, EPI_ISL_2123386, EPI_ISL_2123387, EPI_ISL_2123388, EPI_ISL_2123393, EPI_ISL_2123394, EPI_ISL_2123395, EPI_ISL_2123396, EPI_ISL_2123397, EPI_ISL_2123400, EPI_ISL_2123409, EPI_ISL_2123411, EPI_ISL_2123419 | | | |
| see above | IMD - Labor Oderland | Robert Koch Institute | |
| EPI_ISL_2115372, EPI_ISL_2115373, EPI_ISL_2115374, EPI_ISL_2115376, EPI_ISL_2115380, EPI_ISL_2115381, EPI_ISL_2115382, EPI_ISL_2115384, EPI_ISL_2115386, EPI_ISL_2115387, EPI_ISL_2115388 | | | |
| see above | IMD - MVZ Labor Greifswald | Robert Koch Institute | |
| EPI_ISL_2108417, EPI_ISL_2108419, EPI_ISL_2108420, EPI_ISL_2108425, EPI_ISL_2108447, EPI_ISL_2108458, EPI_ISL_2108460, EPI_ISL_2109442, EPI_ISL_2109464, EPI_ISL_2109473, EPI_ISL_2115370, EPI_ISL_2115375, EPI_ISL_2115377, EPI_ISL_2115378, EPI_ISL_2115379, EPI_ISL_2115385, EPI_ISL_2123385, EPI_ISL_2123392, EPI_ISL_2123405, EPI_ISL_2123407, EPI_ISL_2123408, EPI_ISL_2123416, EPI_ISL_2123420 | | | |
| see above | IMD - MVZ Labor Martinsried | Robert Koch Institute | |
| EPI_ISL_2123389, EPI_ISL_2123391, EPI_ISL_2123398, EPI_ISL_2123401, EPI_ISL_2123403, EPI_ISL_2123410, EPI_ISL_2123412, EPI_ISL_2123413, EPI_ISL_2123414, EPI_ISL_2123417, EPI_ISL_2123418 | | | |
| see above | IMD - Medizinisches Labor Rostock | Robert Koch Institute | |
| EPI_ISL_2108264, EPI_ISL_2108268, EPI_ISL_2108269, EPI_ISL_2108274, EPI_ISL_2108281, EPI_ISL_2108291, EPI_ISL_2108294, EPI_ISL_2108312, EPI_ISL_2108313, EPI_ISL_2108316, EPI_ISL_2112810, EPI_ISL_2112811, EPI_ISL_2112812, EPI_ISL_2112813, EPI_ISL_2112814, EPI_ISL_2112815, EPI_ISL_2112816, EPI_ISL_2112817, EPI_ISL_2112818, EPI_ISL_2112819, EPI_ISL_2112820, EPI_ISL_2112821, EPI_ISL_2112822, EPI_ISL_2112823, EPI_ISL_2112824, EPI_ISL_2112825, EPI_ISL_2112826, EPI_ISL_2112827, EPI_ISL_2112828, EPI_ISL_2112829, EPI_ISL_2112833, EPI_ISL_2112834, EPI_ISL_2112835, EPI_ISL_2112836, EPI_ISL_2112838, EPI_ISL_2112840, EPI_ISL_2120131, EPI_ISL_2120151, EPI_ISL_2120153, EPI_ISL_2120159, EPI_ISL_2120174, EPI_ISL_2120177, EPI_ISL_2120187, EPI_ISL_2120194, EPI_ISL_2120200, EPI_ISL_2120212, EPI_ISL_2120219, EPI_ISL_2120223, EPI_ISL_2120225, EPI_ISL_2120241, EPI_ISL_2120246, EPI_ISL_2120249, EPI_ISL_2120251, EPI_ISL_2120260, EPI_ISL_2120263, EPI_ISL_2120266, EPI_ISL_2120269, EPI_ISL_2120272, EPI_ISL_2120281, EPI_ISL_2120284 | | | |
| see above | IMD Labor Frankfurt | Robert Koch Institute | |
| EPI_ISL_2120147 | Institut fur Immunologie und Genetik Kaiserslautern: Medizinisches Labor Dr. med. B. Thiele | Robert Koch Institute | |
| EPI_ISL_2123424 | Institut fur Klinische Transfusionsmedizin und Immungenetik DRK Blutspendedienst Baden-Wurttemberg | Robert Koch Institute | |
| EPI_ISL_2122780 | Institut fur Medizinische Mikrobiologie - Universitatsmedizin Gottingen | Robert Koch Institute | |
| EPI_ISL_1850427 | Institut fÃ¼r Klinische Chemie Transfusions- und Laboratoriumsmedizin Berufsgenossenschaftliches UniversitÃ¤tsklinikum Bergmannsheil | Robert Koch Institute | |
| EPI_ISL_1941277, EPI_ISL_1941278, EPI_ISL_1941279, EPI_ISL_1941280, EPI_ISL_1941281, EPI_ISL_1941335, EPI_ISL_1941336, EPI_ISL_1941337, EPI_ISL_1941338, EPI_ISL_1941339, EPI_ISL_1941340, EPI_ISL_1941341, EPI_ISL_1941342, EPI_ISL_1941464, EPI_ISL_1941465, EPI_ISL_1941466, EPI_ISL_1941467, EPI_ISL_1941468, EPI_ISL_1941469, EPI_ISL_1941470, EPI_ISL_1941471, EPI_ISL_1941472, EPI_ISL_1941473, EPI_ISL_1941474, EPI_ISL_1941475, EPI_ISL_1941476, EPI_ISL_1941477 | | | |
| see above | Institute of Medical Microbiology and Hospital Hygiene | Institute of Medical Microbiology and Hospital Hygiene | Aljoscha Tersteegen; Prof. Dr. Achim Kaasch |
| EPI_ISL_1805430, EPI_ISL_1960945, EPI_ISL_1960946, EPI_ISL_1969667, EPI_ISL_1969668 | Institute of Tropical Medicine | Institute of Tropical Medicine | Prof. Dr. Thirumalaisamy P. Velavan and Prof. Dr. Peter Kremsner |
| EPI_ISL_2108478, EPI_ISL_2108479 | Kath. Klinikum Bochum gGmbH | Robert Koch Institute | |
| EPI_ISL_1850363, EPI_ISL_1850393, EPI_ISL_2115393 | Kath. St.-Johannes-Gesellschaft Dortmund gGmbH | Robert Koch Institute | |
| EPI_ISL_2110827, EPI_ISL_2110828, EPI_ISL_2110829, EPI_ISL_2110830, EPI_ISL_2110831, EPI_ISL_2110832, EPI_ISL_2110833, EPI_ISL_2110834, EPI_ISL_2110835, EPI_ISL_2110837, EPI_ISL_2110839, EPI_ISL_2110842, EPI_ISL_2110844, EPI_ISL_2110849, EPI_ISL_2110852, EPI_ISL_2110855, EPI_ISL_2110857, EPI_ISL_2110860, EPI_ISL_2110861 | | | |
| see above | Klinikum Ernst von Bergmann gemeinnutzige GmbH - stationorer Bereich | Robert Koch Institute | |

| | | | |
|---|---|---|---|
| EPI_ISL_1849343, EPI_ISL_2116055, EPI_ISL_2116056, EPI_ISL_2116057, EPI_ISL_2116058, EPI_ISL_2116059, EPI_ISL_2116060, EPI_ISL_2116062, EPI_ISL_2116063, EPI_ISL_2116064, EPI_ISL_2116065, EPI_ISL_2116066, EPI_ISL_2116067, EPI_ISL_2116068, EPI_ISL_2116069, EPI_ISL_2116070, EPI_ISL_2116071, EPI_ISL_2116072, EPI_ISL_2116073, EPI_ISL_2116076, EPI_ISL_2116088 | | | |
| see above | Klinikum Ernst von Bergmann gemeinnÃ¼tzige GmbH - stationÃ¤rer Bereich | Robert Koch Institute | |
| EPI_ISL_2125166, EPI_ISL_2125168, EPI_ISL_2125205 | Klinikum Fulda gAG | Robert Koch Institute | |
| EPI_ISL_2109358, EPI_ISL_2109367, EPI_ISL_2109379, EPI_ISL_2109386, EPI_ISL_2109388, EPI_ISL_2109393, EPI_ISL_2109397, EPI_ISL_2109402, EPI_ISL_2109419, EPI_ISL_2109421 | | | |
| see above | LABOR CHEMNITZ Zentrum fur Diagnostik am Klinikum Chemnitz | Robert Koch Institute | |
| EPI_ISL_2120593, EPI_ISL_2120596, EPI_ISL_2120603, EPI_ISL_2120606, EPI_ISL_2120612, EPI_ISL_2120615, EPI_ISL_2120618, EPI_ISL_2120626, EPI_ISL_2120635, EPI_ISL_2120641, EPI_ISL_2120650, EPI_ISL_2120653, EPI_ISL_2120659, EPI_ISL_2120662, EPI_ISL_2120664, EPI_ISL_2120668, EPI_ISL_2120671, EPI_ISL_2120677, EPI_ISL_2120679, EPI_ISL_2120688, EPI_ISL_2120689, EPI_ISL_2120698, EPI_ISL_2120699, EPI_ISL_2120704, EPI_ISL_2120714, EPI_ISL_2120731, EPI_ISL_2120744, EPI_ISL_2120752, EPI_ISL_2120755, EPI_ISL_2120762, EPI_ISL_2120777, EPI_ISL_2120783, EPI_ISL_2120788, EPI_ISL_2120792, EPI_ISL_2120801, EPI_ISL_2120810, EPI_ISL_2120817, EPI_ISL_2120820, EPI_ISL_2120835, EPI_ISL_2120838, EPI_ISL_2120841, EPI_ISL_2120855, EPI_ISL_2120863, EPI_ISL_2120875, EPI_ISL_2120881, EPI_ISL_2120884, EPI_ISL_2120887, EPI_ISL_2120890, EPI_ISL_2120893, EPI_ISL_2120906, EPI_ISL_2120920, EPI_ISL_2120937, EPI_ISL_2120946, EPI_ISL_2120970, EPI_ISL_2120975, EPI_ISL_2120978, EPI_ISL_2120985, EPI_ISL_2120994, EPI_ISL_2121002, EPI_ISL_2121003, EPI_ISL_2121015, EPI_ISL_2121027, EPI_ISL_2121030, EPI_ISL_2121054, EPI_ISL_2121059, EPI_ISL_2121062, EPI_ISL_2121080, EPI_ISL_2121096, EPI_ISL_2121109, EPI_ISL_2121112, EPI_ISL_2121121, EPI_ISL_2121133, EPI_ISL_2121145, EPI_ISL_2121152, EPI_ISL_2121161, EPI_ISL_2121164, EPI_ISL_2121167, EPI_ISL_2121187, EPI_ISL_2121202, EPI_ISL_2121205, EPI_ISL_2121208, EPI_ISL_2121211, EPI_ISL_2121217, EPI_ISL_2121223, EPI_ISL_2121238, EPI_ISL_2121241, EPI_ISL_2121245, EPI_ISL_2121248, EPI_ISL_2121267, EPI_ISL_2121284, EPI_ISL_2121292, EPI_ISL_2121295, EPI_ISL_2121301, EPI_ISL_2121314, EPI_ISL_2121317, EPI_ISL_2121320, EPI_ISL_2121323, EPI_ISL_2121326, EPI_ISL_2121329, EPI_ISL_2121332, EPI_ISL_2121336, EPI_ISL_2121369, EPI_ISL_2121372, EPI_ISL_2121377, EPI_ISL_2121379, EPI_ISL_2121384, EPI_ISL_2121387, EPI_ISL_2121395, EPI_ISL_2121400, EPI_ISL_2121409, EPI_ISL_2121411, EPI_ISL_2121418, EPI_ISL_2121427, EPI_ISL_2121430, EPI_ISL_2121444, EPI_ISL_2121446, EPI_ISL_2121447, EPI_ISL_2121448, EPI_ISL_2121449, EPI_ISL_2121723, EPI_ISL_2121876, EPI_ISL_2121969, EPI_ISL_2121992, EPI_ISL_2122077, EPI_ISL_2122121, EPI_ISL_2122127, EPI_ISL_2122129, EPI_ISL_2122236, EPI_ISL_2122280, EPI_ISL_2125173, EPI_ISL_2125196, EPI_ISL_2125236, EPI_ISL_2125239, EPI_ISL_2125334, EPI_ISL_2125380 | | | |
| see above | LabKom - Labor Augsburg MVZ GmbH | Robert Koch Institute | |
| EPI_ISL_2120673, EPI_ISL_2121148, EPI_ISL_2121191, EPI_ISL_2121441, EPI_ISL_2125326 | LabKom - Labor Hamburg-Lubeck | Robert Koch Institute | |
| EPI_ISL_2125150, EPI_ISL_2125158, EPI_ISL_2125179, EPI_ISL_2125184, EPI_ISL_2125192, EPI_ISL_2125200, EPI_ISL_2125203, EPI_ISL_2125206, EPI_ISL_2125213, EPI_ISL_2125226, EPI_ISL_2125238, EPI_ISL_2125252, EPI_ISL_2125258, EPI_ISL_2125262, EPI_ISL_2125267, EPI_ISL_2125269, EPI_ISL_2125271, EPI_ISL_2125283, EPI_ISL_2125290, EPI_ISL_2125292, EPI_ISL_2125298, EPI_ISL_2125299, EPI_ISL_2125301, EPI_ISL_2125308, EPI_ISL_2125312, EPI_ISL_2125313, EPI_ISL_2125318, EPI_ISL_2125340, EPI_ISL_2125356, EPI_ISL_2125357, EPI_ISL_2125358, EPI_ISL_2125362, EPI_ISL_2125363, EPI_ISL_2125372, EPI_ISL_2125390, EPI_ISL_2125404, EPI_ISL_2125405, EPI_ISL_2125419, EPI_ISL_2125462 | | | |
| see above | LabKom - Labor Hannover MVZ GmbH | Robert Koch Institute | |
| EPI_ISL_2121462, EPI_ISL_2121480, EPI_ISL_2121494, EPI_ISL_2121608, EPI_ISL_2121624, EPI_ISL_2121646, EPI_ISL_2121695, EPI_ISL_2121808, EPI_ISL_2121814, EPI_ISL_2121857, EPI_ISL_2121865, EPI_ISL_2121890, EPI_ISL_2121959, EPI_ISL_2121975, EPI_ISL_2121998, EPI_ISL_2122030, EPI_ISL_2122052, EPI_ISL_2122061, EPI_ISL_2122087, EPI_ISL_2122107, EPI_ISL_2122118, EPI_ISL_2122137, EPI_ISL_2122145, EPI_ISL_2122182, EPI_ISL_2122185, EPI_ISL_2122219, EPI_ISL_2122244, EPI_ISL_2122249, EPI_ISL_2122273, EPI_ISL_2122285, EPI_ISL_2122288, EPI_ISL_2122291, EPI_ISL_2122294, EPI_ISL_2122296 | | | |
| see above | LabKom - Labor Mainz MVZ GmbH | Robert Koch Institute | |
| EPI_ISL_2120623, EPI_ISL_2120647, EPI_ISL_2120656, EPI_ISL_2120667, EPI_ISL_2120684, EPI_ISL_2120758, EPI_ISL_2120794, EPI_ISL_2120847, EPI_ISL_2120900, EPI_ISL_2120924, EPI_ISL_2120931, EPI_ISL_2120955, EPI_ISL_2120958, EPI_ISL_2120964, EPI_ISL_2120973, EPI_ISL_2121036, EPI_ISL_2121040, EPI_ISL_2121103, EPI_ISL_2121127, EPI_ISL_2121140, EPI_ISL_2121178, EPI_ISL_2121197, EPI_ISL_2121233, EPI_ISL_2121254, EPI_ISL_2121270, EPI_ISL_2121282, EPI_ISL_2121307, EPI_ISL_2121311, EPI_ISL_2121342, EPI_ISL_2121345, EPI_ISL_2121351, EPI_ISL_2121354, EPI_ISL_2121360, EPI_ISL_2121412, EPI_ISL_2125141, EPI_ISL_2125147, EPI_ISL_2125152, EPI_ISL_2125177, EPI_ISL_2125180, EPI_ISL_2125188, EPI_ISL_2125198, EPI_ISL_2125209, EPI_ISL_2125210, EPI_ISL_2125259, EPI_ISL_2125266, EPI_ISL_2125273, EPI_ISL_2125284, EPI_ISL_2125286, EPI_ISL_2125294, EPI_ISL_2125296, EPI_ISL_2125297, EPI_ISL_2125316, EPI_ISL_2125330, EPI_ISL_2125341, EPI_ISL_2125343, EPI_ISL_2125351, EPI_ISL_2125367, EPI_ISL_2125376, EPI_ISL_2125378, EPI_ISL_2125381, EPI_ISL_2125382, EPI_ISL_2125385, EPI_ISL_2125387, EPI_ISL_2125394, EPI_ISL_2125399, EPI_ISL_2125402, EPI_ISL_2125408, EPI_ISL_2125422, EPI_ISL_2125431, EPI_ISL_2125433, EPI_ISL_2125452, EPI_ISL_2125455 | | | |
| see above | LabKom - Labor an der Salzbrucke MVZ GmbH | Robert Koch Institute | |
| EPI_ISL_2121465, EPI_ISL_2121474, EPI_ISL_2121482, EPI_ISL_2121497, EPI_ISL_2121512, EPI_ISL_2121541, EPI_ISL_2121556, EPI_ISL_2121592, EPI_ISL_2121659, EPI_ISL_2121681, EPI_ISL_2121715, EPI_ISL_2121771, EPI_ISL_2121774, EPI_ISL_2121805, EPI_ISL_2121851, EPI_ISL_2121887, EPI_ISL_2121895, EPI_ISL_2121903, EPI_ISL_2121916, EPI_ISL_2121919, EPI_ISL_2121964, EPI_ISL_2122095, EPI_ISL_2122104, EPI_ISL_2122160, EPI_ISL_2122222, EPI_ISL_2122267, EPI_ISL_2122274, EPI_ISL_2125917 | | | |
| see above | Labor 28 MVZ GmbH | Robert Koch Institute | |
| EPI_ISL_1921855 | Labor Berlin Charite Vivantes GmbH / Institut für Virologie | Charité Universitätsmedizin Berlin, Institut für Virologie/Labor Berlin | Barbara Mühlemann; Christian Drosten; Christine Stephan; Peter Menzel; Rolf Schwarzer; Terry Jones; Victor M Corman |
| EPI_ISL_1840792, EPI_ISL_1840797, EPI_ISL_1840798, EPI_ISL_1840801, EPI_ISL_1840811, EPI_ISL_1840812, EPI_ISL_1840822, EPI_ISL_1840828, EPI_ISL_1840836, EPI_ISL_1840837, EPI_ISL_1840847, EPI_ISL_1840855, EPI_ISL_1840856, EPI_ISL_2107829, EPI_ISL_2107830, EPI_ISL_2107831, EPI_ISL_2107832, EPI_ISL_2107833, EPI_ISL_2107834, EPI_ISL_2107839, EPI_ISL_2107840, EPI_ISL_2107841, EPI_ISL_2107843, EPI_ISL_2107844, EPI_ISL_2107845, EPI_ISL_2107847, EPI_ISL_2107848, EPI_ISL_2107849, EPI_ISL_2107851, EPI_ISL_2107854, EPI_ISL_2107855, EPI_ISL_2107856, EPI_ISL_2107858, EPI_ISL_2107859, EPI_ISL_2107860, EPI_ISL_2107862, EPI_ISL_2107863, EPI_ISL_2107865, EPI_ISL_2107866, EPI_ISL_2107867, EPI_ISL_2107868, EPI_ISL_2107869, EPI_ISL_2107870, EPI_ISL_2107871, EPI_ISL_2107872, EPI_ISL_2107873, EPI_ISL_2107874, EPI_ISL_2107875, EPI_ISL_2107877, EPI_ISL_2107880, EPI_ISL_2107883, EPI_ISL_2107884, EPI_ISL_2107886 | | | |
| see above | Labor Berlin Charité Vivantes GmbH / Institut für Virologie | Charité Universitätsmedizin Berlin, Institut für Virologie/Labor Berlin | Barbara Mühlemann; Christian Drosten; Christine Stephan; Peter Menzel; Rolf Schwarzer; Terry Jones; Victor M Corman |
| EPI_ISL_2122961, EPI_ISL_2122962, EPI_ISL_2122964, EPI_ISL_2122965, EPI_ISL_2122966, EPI_ISL_2122968, EPI_ISL_2122969, EPI_ISL_2122974, EPI_ISL_2122975, EPI_ISL_2122976, EPI_ISL_2122977, EPI_ISL_2122978, EPI_ISL_2122980, EPI_ISL_2122981, EPI_ISL_2122982, EPI_ISL_2122983, EPI_ISL_2122985, EPI_ISL_2122986, EPI_ISL_2122989, EPI_ISL_2122991, EPI_ISL_2122995 | | | |
| see above | Labor Dr. Heidrich & Kollegen MVZ GmbH Hamburg | Robert Koch Institute | |
| EPI_ISL_2120722, EPI_ISL_2120749, EPI_ISL_2120786, EPI_ISL_2120934, EPI_ISL_2120940, EPI_ISL_2120974, EPI_ISL_2121000, EPI_ISL_2121115, EPI_ISL_2121118, EPI_ISL_2125466, EPI_ISL_2125496, EPI_ISL_2125503, EPI_ISL_2125507, EPI_ISL_2125509, EPI_ISL_2125519, EPI_ISL_2125565, EPI_ISL_2125624, EPI_ISL_2125643, EPI_ISL_2125658, EPI_ISL_2125667, EPI_ISL_2125695, EPI_ISL_2125699, EPI_ISL_2125747, EPI_ISL_2125797, EPI_ISL_2125836, EPI_ISL_2125845, EPI_ISL_2125967, EPI_ISL_2126001, EPI_ISL_2126037, EPI_ISL_2126079, EPI_ISL_2126081, EPI_ISL_2126085, EPI_ISL_2126126, EPI_ISL_2126187 | | | |
| see above | Labor Dr. Schumacher MVZ | Robert Koch Institute | |
| EPI_ISL_2125119, EPI_ISL_2125121, EPI_ISL_2125122, EPI_ISL_2125127, EPI_ISL_2125135, EPI_ISL_2125136 | Labor Dr. Spranger | Robert Koch Institute | |
| EPI_ISL_2111023, EPI_ISL_2111024, EPI_ISL_2111025, EPI_ISL_2111108, EPI_ISL_2111109 | Labor Dr. Wisplinghoff - Kaln | Robert Koch Institute | |
| EPI_ISL_2112903, EPI_ISL_2112904, EPI_ISL_2112905, EPI_ISL_2112906, EPI_ISL_2112907, EPI_ISL_2112908, EPI_ISL_2112909, EPI_ISL_2112910, EPI_ISL_2112911, EPI_ISL_2112912 | | | |
| see above | Labor KneiÃŸler GmbH & Co. KG | Robert Koch Institute | |
| EPI_ISL_2122353, EPI_ISL_2122364, EPI_ISL_2122391, EPI_ISL_2122580 | Labor Lobeck bzw. Laborarztliche Gemeinschaftspraxis Lobeck | Robert Koch Institute | |
| EPI_ISL_2115395, EPI_ISL_2115440, EPI_ISL_2115463, EPI_ISL_2115487, EPI_ISL_2115494 | Labor Lubeck bzw. Laborarztliche Gemeinschaftspraxis Lubeck | Robert Koch Institute | |
| EPI_ISL_2111479, EPI_ISL_2111493, EPI_ISL_2111520, EPI_ISL_2111549, EPI_ISL_2111583, EPI_ISL_2129183, EPI_ISL_2129185, EPI_ISL_2129186, EPI_ISL_2129187, EPI_ISL_2129188, EPI_ISL_2129190, EPI_ISL_2129191, EPI_ISL_2129192, EPI_ISL_2129193, EPI_ISL_2129195, EPI_ISL_2129196, EPI_ISL_2129197, EPI_ISL_2129198, EPI_ISL_2129199, EPI_ISL_2129200, EPI_ISL_2129201, EPI_ISL_2129202, EPI_ISL_2129203, EPI_ISL_2129204, EPI_ISL_2129206, EPI_ISL_2129207, EPI_ISL_2129208, EPI_ISL_2129209, EPI_ISL_2129210, EPI_ISL_2129211, EPI_ISL_2129213, EPI_ISL_2129214, EPI_ISL_2129216, EPI_ISL_2129217, EPI_ISL_2129218, EPI_ISL_2129219, EPI_ISL_2129221, EPI_ISL_2129222, EPI_ISL_2129223, EPI_ISL_2129225, EPI_ISL_2129226, EPI_ISL_2129227, EPI_ISL_2129228, EPI_ISL_2129229, EPI_ISL_2129231, EPI_ISL_2129232, EPI_ISL_2129233, EPI_ISL_2129234, EPI_ISL_2129235, EPI_ISL_2129236, EPI_ISL_2129238, EPI_ISL_2129240, EPI_ISL_2129241, EPI_ISL_2129243, EPI_ISL_2129244, EPI_ISL_2129245, EPI_ISL_2129246, EPI_ISL_2129247, EPI_ISL_2129251, EPI_ISL_2129252, EPI_ISL_2129253, EPI_ISL_2129254, EPI_ISL_2129255, EPI_ISL_2129256, EPI_ISL_2129257, EPI_ISL_2129258, EPI_ISL_2129259, EPI_ISL_2129260, EPI_ISL_2129261, EPI_ISL_2129262, EPI_ISL_2129263, EPI_ISL_2129264, EPI_ISL_2129265, EPI_ISL_2129266, EPI_ISL_2129267, EPI_ISL_2129268 | | | |
| see above | Labor ZOTZ\|KLIMAS; MVZ Dusseldorf-Centrum | Robert Koch Institute | |
| EPI_ISL_2129269, EPI_ISL_2129270, EPI_ISL_2129271, EPI_ISL_2129272, EPI_ISL_2129273, EPI_ISL_2129274, EPI_ISL_2129276, EPI_ISL_2129278, EPI_ISL_2129279, EPI_ISL_2129280, EPI_ISL_2129281, EPI_ISL_2129282, EPI_ISL_2129283, EPI_ISL_2129284, EPI_ISL_2129285, EPI_ISL_2129286, EPI_ISL_2129287, EPI_ISL_2129288, EPI_ISL_2129289, EPI_ISL_2129292, EPI_ISL_2129295, EPI_ISL_2129296, EPI_ISL_2129297, EPI_ISL_2129298, EPI_ISL_2129299, EPI_ISL_2129300, EPI_ISL_2129301, EPI_ISL_2129302, EPI_ISL_2129303, EPI_ISL_2129305, EPI_ISL_2129306, EPI_ISL_2129307, EPI_ISL_2129308, EPI_ISL_2129309, EPI_ISL_2129310, EPI_ISL_2129311, EPI_ISL_2129312, EPI_ISL_2129313, EPI_ISL_2129314, EPI_ISL_2129315, EPI_ISL_2129316, EPI_ISL_2129317, EPI_ISL_2129318 | | | |
| see above | Labor ZOTZ\|KLIMAS; MVZ DÃ¼sseldorf-Centrum | Robert Koch Institute | |
| EPI_ISL_2113171, EPI_ISL_2113184 | Labor fur medizinische Mikrobiologie Bakteriologie Mykologie Virologie & Infektionsserologie - Labor Malbis | Robert Koch Institute | |
| EPI_ISL_2112986, EPI_ISL_2113041, EPI_ISL_2113059 | Labor fÃ¼r medizinische Mikrobiologie Bakteriologie Mykologie Virologie & Infektionsserologie - Labor MÃ¶lbis | Robert Koch Institute | |
| EPI_ISL_2111154, EPI_ISL_2111155, EPI_ISL_2111156, EPI_ISL_2111157, EPI_ISL_2111159, EPI_ISL_2111160, EPI_ISL_2111161, EPI_ISL_2111162, EPI_ISL_2111163, EPI_ISL_2111164, EPI_ISL_2111165, EPI_ISL_2111166, EPI_ISL_2111167, EPI_ISL_2111168, EPI_ISL_2111169, EPI_ISL_2111170, EPI_ISL_2111171, EPI_ISL_2111172, EPI_ISL_2111173, EPI_ISL_2111174, EPI_ISL_2111175, EPI_ISL_2111176 | | | |
| see above | Laborarztpraxis Dres. med. Walther Weindel & Kollegen | Robert Koch Institute | |
| EPI_ISL_2108328, EPI_ISL_2108353, EPI_ISL_2108375, EPI_ISL_2108389, EPI_ISL_2108396, EPI_ISL_2108409, EPI_ISL_2108415, EPI_ISL_2122313, EPI_ISL_2122315, EPI_ISL_2122318, EPI_ISL_2122324, EPI_ISL_2122330, EPI_ISL_2122336, EPI_ISL_2122347, EPI_ISL_2122350, EPI_ISL_2122367, EPI_ISL_2122368, EPI_ISL_2122376, EPI_ISL_2122382, EPI_ISL_2122385, EPI_ISL_2122387, EPI_ISL_2122393, EPI_ISL_2122396, EPI_ISL_2122404, EPI_ISL_2122407, EPI_ISL_2122417, EPI_ISL_2122419, EPI_ISL_2122427, EPI_ISL_2122449, EPI_ISL_2122454, EPI_ISL_2122457, EPI_ISL_2122460, EPI_ISL_2122466, EPI_ISL_2122483, EPI_ISL_2122489, EPI_ISL_2122495, EPI_ISL_2122498, EPI_ISL_2122509, EPI_ISL_2122511, EPI_ISL_2122555, EPI_ISL_2122577, EPI_ISL_2122582, EPI_ISL_2122585, EPI_ISL_2122589, EPI_ISL_2122592 | | | |
| see above | Laborarztpraxis Osnabruck | Robert Koch Institute | |
| EPI_ISL_2108275, EPI_ISL_2108302, EPI_ISL_2120228, EPI_ISL_2120234, EPI_ISL_2120235, EPI_ISL_2120243, EPI_ISL_2120258, EPI_ISL_2120278 | | | |
| see above | Labormedizin Darmstadt | Robert Koch Institute | |
| EPI_ISL_1941282, EPI_ISL_1941283, EPI_ISL_1941284, EPI_ISL_1941285, EPI_ISL_1941286, EPI_ISL_1941287, EPI_ISL_1941288, EPI_ISL_1941289, EPI_ISL_1941290, EPI_ISL_1941291, EPI_ISL_1941292, EPI_ISL_1941293, EPI_ISL_1941294, EPI_ISL_1941295, EPI_ISL_1941296, EPI_ISL_1941297, EPI_ISL_1941298, EPI_ISL_1941299, EPI_ISL_1941300, EPI_ISL_1941301, EPI_ISL_1941302, EPI_ISL_1941303, EPI_ISL_1941304, EPI_ISL_1941305, EPI_ISL_1941306, EPI_ISL_1941307, EPI_ISL_1941308, EPI_ISL_1941309, EPI_ISL_1941310, EPI_ISL_1941311, EPI_ISL_1941312, EPI_ISL_1941313, EPI_ISL_1941314, EPI_ISL_1941315, EPI_ISL_1941316, EPI_ISL_1941317, EPI_ISL_1941318, EPI_ISL_1941319, EPI_ISL_1941320, EPI_ISL_1941321, EPI_ISL_1941322, EPI_ISL_1941323, EPI_ISL_1941324, EPI_ISL_1941325, EPI_ISL_1941326, EPI_ISL_1941327, EPI_ISL_1941328, EPI_ISL_1941329, EPI_ISL_1941330, EPI_ISL_1941331, EPI_ISL_1941332, EPI_ISL_1941333, EPI_ISL_1941334, EPI_ISL_1941343, EPI_ISL_1941344, EPI_ISL_1941345, EPI_ISL_1941346, EPI_ISL_1941347, EPI_ISL_1941348, EPI_ISL_1941349, EPI_ISL_1941350, EPI_ISL_1941352, EPI_ISL_1941353, EPI_ISL_1941354, EPI_ISL_1941355, EPI_ISL_1941356, EPI_ISL_1941357, EPI_ISL_1941358, EPI_ISL_1941359, EPI_ISL_1941360, EPI_ISL_1941361, EPI_ISL_1941362, EPI_ISL_1941363, EPI_ISL_1941364, EPI_ISL_1941366, EPI_ISL_1941367, EPI_ISL_1941368, EPI_ISL_1941369, EPI_ISL_1941370, EPI_ISL_1941371, EPI_ISL_1941372, EPI_ISL_1941373, EPI_ISL_1941374, EPI_ISL_1941376, EPI_ISL_1941377, EPI_ISL_1941378, EPI_ISL_1941379, EPI_ISL_1941380, EPI_ISL_1941381, EPI_ISL_1941382, EPI_ISL_1941383, EPI_ISL_1941384, EPI_ISL_1941385, EPI_ISL_1941386, EPI_ISL_1941387, EPI_ISL_1941388, EPI_ISL_1941389, EPI_ISL_1941390, EPI_ISL_1941391, EPI_ISL_1941392, EPI_ISL_1941393, EPI_ISL_1941394, EPI_ISL_1941395, EPI_ISL_1941396, EPI_ISL_1941397, EPI_ISL_1941398, EPI_ISL_1941399, EPI_ISL_1941400, EPI_ISL_1941401, EPI_ISL_1941402, EPI_ISL_1941403, EPI_ISL_1941404, EPI_ISL_1941405, EPI_ISL_1941406, EPI_ISL_1941407, EPI_ISL_1941408, EPI_ISL_1941409, EPI_ISL_1941410, EPI_ISL_1941411, EPI_ISL_1941412, EPI_ISL_1941413, EPI_ISL_1941414, EPI_ISL_1941415, EPI_ISL_1941416, EPI_ISL_1941418, EPI_ISL_1941419, EPI_ISL_1941420, EPI_ISL_1941421, EPI_ISL_1941422, EPI_ISL_1941423, EPI_ISL_1941424, EPI_ISL_1941425, EPI_ISL_1941426, EPI_ISL_1941427, EPI_ISL_1941428, EPI_ISL_1941429, EPI_ISL_1941430, EPI_ISL_1941431, EPI_ISL_1941432, EPI_ISL_1941433, EPI_ISL_1941434, EPI_ISL_1941435, EPI_ISL_1941436, EPI_ISL_1941437, EPI_ISL_1941438, EPI_ISL_1941439, EPI_ISL_1941440, EPI_ISL_1941441, EPI_ISL_1941442, EPI_ISL_1941443, EPI_ISL_1941445, EPI_ISL_1941446, EPI_ISL_1941447, EPI_ISL_1941448, EPI_ISL_1941449, EPI_ISL_1941450, EPI_ISL_1941451, EPI_ISL_1941452, EPI_ISL_1941453, EPI_ISL_1941454, EPI_ISL_1941455, EPI_ISL_1941456, EPI_ISL_1941457, EPI_ISL_1941458, EPI_ISL_1941459, EPI_ISL_1941460, EPI_ISL_1941461, EPI_ISL_1941462, EPI_ISL_1941463, EPI_ISL_1941478, EPI_ISL_1941479, EPI_ISL_1941481, EPI_ISL_1941482, EPI_ISL_1941483, EPI_ISL_1941484 | | | |
| see above | Landesamt für Verbraucherschutz Sachsen Anhalt, Magdeburg | Institute of Medical Microbiology and Hospital Hygiene | Aljoscha Tersteegen; Prof. Dr. Achim Kaasch |
| EPI_ISL_2126390, EPI_ISL_2126396, EPI_ISL_2126399, EPI_ISL_2126408 | Landesgesundheitsamt Baden-Wurttemberg | Robert Koch Institute | |
| EPI_ISL_2109360, EPI_ISL_2109368, EPI_ISL_2109378, EPI_ISL_2109383, EPI_ISL_2109392, EPI_ISL_2109398, EPI_ISL_2109403, EPI_ISL_2109404, EPI_ISL_2109411, EPI_ISL_2109420, EPI_ISL_2109424 | | | |
| see above | Landesuntersuchungsanstalt fur das Gesundheits- und Veterinarwesen (LUA) Sachsen | Robert Koch Institute | |
| EPI_ISL_1852473, EPI_ISL_1852477 | Limbach - MVZ Gemeinschaftslabor Suhl | Robert Koch Institute | |
| EPI_ISL_1853102, EPI_ISL_1853103, EPI_ISL_1853104, EPI_ISL_1853105, EPI_ISL_1853106, EPI_ISL_1853107, EPI_ISL_1853108, EPI_ISL_1853109, EPI_ISL_1853110, EPI_ISL_1853112, EPI_ISL_1853113, EPI_ISL_1853114, EPI_ISL_1853115, EPI_ISL_1853116, EPI_ISL_1853117, EPI_ISL_1853118, EPI_ISL_1853119, EPI_ISL_1853120, EPI_ISL_1853122, EPI_ISL_1853123, EPI_ISL_1853124, EPI_ISL_1853125, EPI_ISL_1853126, EPI_ISL_1853128, EPI_ISL_1853129, EPI_ISL_1853130, EPI_ISL_1853131, EPI_ISL_1853133, EPI_ISL_1853134, EPI_ISL_1853135, EPI_ISL_1853136, EPI_ISL_1853137, EPI_ISL_1853140, EPI_ISL_1853141, EPI_ISL_1853142, EPI_ISL_1853143, EPI_ISL_1853144, EPI_ISL_1853145, EPI_ISL_1853146, EPI_ISL_1853148, EPI_ISL_1853149, EPI_ISL_1853150, EPI_ISL_1853151, EPI_ISL_1853152, EPI_ISL_1853153, EPI_ISL_1853154, EPI_ISL_1853155, EPI_ISL_1853156, EPI_ISL_1853157, EPI_ISL_1853158, EPI_ISL_1853159, EPI_ISL_1853160, EPI_ISL_1853161, EPI_ISL_1853162, EPI_ISL_1853163, EPI_ISL_1853164, EPI_ISL_1853165, EPI_ISL_1853166, EPI_ISL_1853167, EPI_ISL_1853168, EPI_ISL_1853169, EPI_ISL_1853170, EPI_ISL_1853171, EPI_ISL_1853172, EPI_ISL_1853173, EPI_ISL_1853174, EPI_ISL_1853175, EPI_ISL_1853176, EPI_ISL_1853177, EPI_ISL_1853178, EPI_ISL_1853179, EPI_ISL_1853180, EPI_ISL_1853181, EPI_ISL_1853182, EPI_ISL_1853183, EPI_ISL_1853184, EPI_ISL_1853185, EPI_ISL_1853186, EPI_ISL_1853187, EPI_ISL_1853188, EPI_ISL_1853189, EPI_ISL_1853190, EPI_ISL_1853191, EPI_ISL_1853192, EPI_ISL_1853193, EPI_ISL_1853195, EPI_ISL_1853196, EPI_ISL_1853197, EPI_ISL_1853198, EPI_ISL_1853199, EPI_ISL_1853200, EPI_ISL_1853201, EPI_ISL_1853202, EPI_ISL_1853203, EPI_ISL_1853204, EPI_ISL_1853205, EPI_ISL_1853206, EPI_ISL_1853207, EPI_ISL_1853208, EPI_ISL_1853209, EPI_ISL_1853210, EPI_ISL_1853211, EPI_ISL_1853212, EPI_ISL_1853213, EPI_ISL_1853214, EPI_ISL_1853215, EPI_ISL_1853216, EPI_ISL_1853217, EPI_ISL_1853218, EPI_ISL_1853220, EPI_ISL_1853221, EPI_ISL_1853222, EPI_ISL_1853223, EPI_ISL_1853224, EPI_ISL_1853225, EPI_ISL_1853226, EPI_ISL_1853227, EPI_ISL_1853228, EPI_ISL_1853229, EPI_ISL_1853231, EPI_ISL_1853232, EPI_ISL_1853233, EPI_ISL_1853234, EPI_ISL_1853235, EPI_ISL_1853236, EPI_ISL_1853237, EPI_ISL_1853238, EPI_ISL_1853239, EPI_ISL_1853240, EPI_ISL_1853242, EPI_ISL_1853243, EPI_ISL_1853244, | | | |

| | | | |
|---|---|---|---|
| EPI_ISL_1853245, EPI_ISL_1853246, EPI_ISL_1853247, EPI_ISL_1853248, EPI_ISL_1853249, EPI_ISL_1853250, EPI_ISL_1853251, EPI_ISL_1853252, EPI_ISL_1853253, EPI_ISL_1853254, EPI_ISL_1853255, EPI_ISL_1853256, EPI_ISL_1853257, EPI_ISL_1853258, EPI_ISL_1853259, EPI_ISL_1853260, EPI_ISL_1853261, EPI_ISL_1853262, EPI_ISL_1853263, EPI_ISL_1853264, EPI_ISL_1853265, EPI_ISL_1853266, EPI_ISL_1853267, EPI_ISL_1853268, EPI_ISL_1853269, EPI_ISL_1853270, EPI_ISL_1853271, EPI_ISL_1853272, EPI_ISL_1853273, EPI_ISL_1853274, EPI_ISL_1853275, EPI_ISL_1853276, EPI_ISL_1853277, EPI_ISL_1853278, EPI_ISL_2117438, EPI_ISL_2117441, EPI_ISL_2117445, EPI_ISL_2117450, EPI_ISL_2117452, EPI_ISL_2117455, EPI_ISL_2117458, EPI_ISL_2117461, EPI_ISL_2117469, EPI_ISL_2117471, EPI_ISL_2117473, EPI_ISL_2117479, EPI_ISL_2117481, EPI_ISL_2117484, EPI_ISL_2117493, EPI_ISL_2117495, EPI_ISL_2117502, EPI_ISL_2117504, EPI_ISL_2117506, EPI_ISL_2117508, EPI_ISL_2117514, EPI_ISL_2117518, EPI_ISL_2117522, EPI_ISL_2117527, EPI_ISL_2117541, EPI_ISL_2117545, EPI_ISL_2117548, EPI_ISL_2117555, EPI_ISL_2117558, EPI_ISL_2117567, EPI_ISL_2117570, EPI_ISL_2117574, EPI_ISL_2117575, EPI_ISL_2117599, EPI_ISL_2117605, EPI_ISL_2117616, EPI_ISL_2117628, EPI_ISL_2117634, EPI_ISL_2117646, EPI_ISL_2117653, EPI_ISL_2117655, EPI_ISL_2117659, EPI_ISL_2117669, EPI_ISL_2117671, EPI_ISL_2117675, EPI_ISL_2117680, EPI_ISL_2117704, EPI_ISL_2117724, EPI_ISL_2117735, EPI_ISL_2117746, EPI_ISL_2117750, EPI_ISL_2117759, EPI_ISL_2117768, EPI_ISL_2117775, EPI_ISL_2117776, EPI_ISL_2117778, EPI_ISL_2117780, EPI_ISL_2117789, EPI_ISL_2117802, EPI_ISL_2117815, EPI_ISL_2117827, EPI_ISL_2117836, EPI_ISL_2117839, EPI_ISL_2117847, EPI_ISL_2117853, EPI_ISL_2117856, EPI_ISL_2117858, EPI_ISL_2117866, EPI_ISL_2117875, EPI_ISL_2117881, EPI_ISL_2117887, EPI_ISL_2117889, EPI_ISL_2117891, EPI_ISL_2117896, EPI_ISL_2117899, EPI_ISL_2117902, EPI_ISL_2117911, EPI_ISL_2117914, EPI_ISL_2117917, EPI_ISL_2117920, EPI_ISL_2117923 | | | |
| see above | Limbach - MVZ Humangenetik Ulm | Robert Koch Institute | |
| EPI_ISL_1852103 | Limbach - MVZ Labor Eveld & Kollegen Essen | Robert Koch Institute | |
| EPI_ISL_1852483 | Limbach - MVZ Labor MÃ¼nster (Cullen) | Robert Koch Institute | |
| EPI_ISL_2108325, EPI_ISL_2108357, EPI_ISL_2108364, EPI_ISL_2108370, EPI_ISL_2108376, EPI_ISL_2108379, EPI_ISL_2108390, EPI_ISL_2108398, EPI_ISL_2108405, EPI_ISL_2115389, EPI_ISL_2115391, EPI_ISL_2115392, EPI_ISL_2115394, EPI_ISL_2115396, EPI_ISL_2115397, EPI_ISL_2115398, EPI_ISL_2115399, EPI_ISL_2115400, EPI_ISL_2115401, EPI_ISL_2115402, EPI_ISL_2115403, EPI_ISL_2115404, EPI_ISL_2115405, EPI_ISL_2115406, EPI_ISL_2115407, EPI_ISL_2115409, EPI_ISL_2115410, EPI_ISL_2115411, EPI_ISL_2115412, EPI_ISL_2115414, EPI_ISL_2115415, EPI_ISL_2115416, EPI_ISL_2115417, EPI_ISL_2115418, EPI_ISL_2115419, EPI_ISL_2115420, EPI_ISL_2115421, EPI_ISL_2115422, EPI_ISL_2115423, EPI_ISL_2115425, EPI_ISL_2115427, EPI_ISL_2115428, EPI_ISL_2115430, EPI_ISL_2115431, EPI_ISL_2115433, EPI_ISL_2115434, EPI_ISL_2115436, EPI_ISL_2115437, EPI_ISL_2115438, EPI_ISL_2115439, EPI_ISL_2115441, EPI_ISL_2115442, EPI_ISL_2115443, EPI_ISL_2115445, EPI_ISL_2115447, EPI_ISL_2115449, EPI_ISL_2115450, EPI_ISL_2115451, EPI_ISL_2115453, EPI_ISL_2115454, EPI_ISL_2115455, EPI_ISL_2115456, EPI_ISL_2115457, EPI_ISL_2115458, EPI_ISL_2115459, EPI_ISL_2115460, EPI_ISL_2115461, EPI_ISL_2115462, EPI_ISL_2115464, EPI_ISL_2115465, EPI_ISL_2115466, EPI_ISL_2115467, EPI_ISL_2115468, EPI_ISL_2115470, EPI_ISL_2115471, EPI_ISL_2115472, EPI_ISL_2115473, EPI_ISL_2115474, EPI_ISL_2115475, EPI_ISL_2115477, EPI_ISL_2115478, EPI_ISL_2115479, EPI_ISL_2115480, EPI_ISL_2115481, EPI_ISL_2115482, EPI_ISL_2115483, EPI_ISL_2115484, EPI_ISL_2115485, EPI_ISL_2115486, EPI_ISL_2115488, EPI_ISL_2115489, EPI_ISL_2115490, EPI_ISL_2115491, EPI_ISL_2115492, EPI_ISL_2115493, EPI_ISL_2115495, EPI_ISL_2115496, EPI_ISL_2115497, EPI_ISL_2115498, EPI_ISL_2115499, EPI_ISL_2122327, EPI_ISL_2122333, EPI_ISL_2122361, EPI_ISL_2122370, EPI_ISL_2122401, EPI_ISL_2122413, EPI_ISL_2122422, EPI_ISL_2122425, EPI_ISL_2122440, EPI_ISL_2122464, EPI_ISL_2122481, EPI_ISL_2122542, EPI_ISL_2122567, EPI_ISL_2122570, EPI_ISL_2122595 | | | |
| see above | MVZ Dr. Eberhard & Partner Dortmund | Robert Koch Institute | |
| EPI_ISL_2111220, EPI_ISL_2111221, EPI_ISL_2111222, EPI_ISL_2111223, EPI_ISL_2111224, EPI_ISL_2111225, EPI_ISL_2111226, EPI_ISL_2111227, EPI_ISL_2111228, EPI_ISL_2111229, EPI_ISL_2111230, EPI_ISL_2111231, EPI_ISL_2111232, EPI_ISL_2111233, EPI_ISL_2111234, EPI_ISL_2111235, EPI_ISL_2111236, EPI_ISL_2111237, EPI_ISL_2111239, EPI_ISL_2111240, EPI_ISL_2111241, EPI_ISL_2111242, EPI_ISL_2111243, EPI_ISL_2111244, EPI_ISL_2111245, EPI_ISL_2111246, EPI_ISL_2111247, EPI_ISL_2111248, EPI_ISL_2111249, EPI_ISL_2111250, EPI_ISL_2111251, EPI_ISL_2111252, EPI_ISL_2111254, EPI_ISL_2111255, EPI_ISL_2111256, EPI_ISL_2111257, EPI_ISL_2111258, EPI_ISL_2111259, EPI_ISL_2111260, EPI_ISL_2111261, EPI_ISL_2111262, EPI_ISL_2111263, EPI_ISL_2111264, EPI_ISL_2111265, EPI_ISL_2111266, EPI_ISL_2111267, EPI_ISL_2111268, EPI_ISL_2111269, EPI_ISL_2111270, EPI_ISL_2111271, EPI_ISL_2111272, EPI_ISL_2111273, EPI_ISL_2111275, EPI_ISL_2111276, EPI_ISL_2111277, EPI_ISL_2111278, EPI_ISL_2111279 | | | |
| see above | MVZ Labor Dr. Fenner und Kollegen (Standort Hamburg) | Robert Koch Institute | |
| EPI_ISL_1847332, EPI_ISL_1847333, EPI_ISL_1847334, EPI_ISL_1847335, EPI_ISL_1847336, EPI_ISL_1847383, EPI_ISL_1847384, EPI_ISL_1847385, EPI_ISL_1847386, EPI_ISL_1847387, EPI_ISL_1847388, EPI_ISL_1847389, EPI_ISL_1847390, EPI_ISL_1847391, EPI_ISL_1847392, EPI_ISL_1847393, EPI_ISL_1847394, EPI_ISL_1847395, EPI_ISL_1847396, EPI_ISL_1847397, EPI_ISL_1847398, EPI_ISL_1847399, EPI_ISL_1847400, EPI_ISL_1847401, EPI_ISL_1847402, EPI_ISL_1847444, EPI_ISL_1847445, EPI_ISL_1847446, EPI_ISL_1847698, EPI_ISL_1847699, EPI_ISL_1847700, EPI_ISL_1847701, EPI_ISL_1847702, EPI_ISL_1847703, EPI_ISL_1847704, EPI_ISL_1847705, EPI_ISL_1847706, EPI_ISL_1847707, EPI_ISL_1847708, EPI_ISL_1847709, EPI_ISL_1847710, EPI_ISL_1847711, EPI_ISL_1847712, EPI_ISL_1847713, EPI_ISL_1847714, EPI_ISL_1847715, EPI_ISL_1847716, EPI_ISL_1847717, EPI_ISL_1847718, EPI_ISL_1847719, EPI_ISL_1848814, EPI_ISL_1848815, EPI_ISL_1848816, EPI_ISL_1852044, EPI_ISL_1852058, EPI_ISL_1852064, EPI_ISL_1852069, EPI_ISL_1852070, EPI_ISL_1852071, EPI_ISL_1852072, EPI_ISL_1852074, EPI_ISL_1852076, EPI_ISL_1852085, EPI_ISL_1852089, EPI_ISL_1852094, EPI_ISL_1852096, EPI_ISL_1852112, EPI_ISL_1852114, EPI_ISL_1852449, EPI_ISL_1852462, EPI_ISL_1852463, EPI_ISL_1852464, EPI_ISL_1852466, EPI_ISL_1852479, EPI_ISL_1852481, EPI_ISL_1852482, EPI_ISL_2123003, EPI_ISL_2123008, EPI_ISL_2123021, EPI_ISL_2124016, EPI_ISL_2124021 | | | |
| see above | MVZ Labor Dr. Limbach & Kollegen GbR | Robert Koch Institute | |
| EPI_ISL_1849439, EPI_ISL_1851102, EPI_ISL_2110784, EPI_ISL_2110785, EPI_ISL_2110786, EPI_ISL_2110788, EPI_ISL_2110789, EPI_ISL_2110790, EPI_ISL_2110791, EPI_ISL_2110792, EPI_ISL_2110793, EPI_ISL_2110794, EPI_ISL_2110795, EPI_ISL_2110796, EPI_ISL_2110797, EPI_ISL_2110798, EPI_ISL_2110799, EPI_ISL_2110800, EPI_ISL_2110801, EPI_ISL_2110802, EPI_ISL_2110803, EPI_ISL_2110805, EPI_ISL_2110806, EPI_ISL_2110807, EPI_ISL_2110808, EPI_ISL_2110809, EPI_ISL_2110810, EPI_ISL_2110811, EPI_ISL_2110812, EPI_ISL_2110813, EPI_ISL_2110814, EPI_ISL_2110815, EPI_ISL_2110816, EPI_ISL_2110817, EPI_ISL_2110818, EPI_ISL_2110820, EPI_ISL_2110821, EPI_ISL_2110822, EPI_ISL_2110823, EPI_ISL_2110824, EPI_ISL_2110825, EPI_ISL_2111177, EPI_ISL_2111178, EPI_ISL_2111179, EPI_ISL_2111180, EPI_ISL_2111181, EPI_ISL_2111182, EPI_ISL_2111183, EPI_ISL_2111184, EPI_ISL_2111185, EPI_ISL_2111186, EPI_ISL_2111187, EPI_ISL_2111188, EPI_ISL_2111189, EPI_ISL_2111190, EPI_ISL_2111191, EPI_ISL_2111192, EPI_ISL_2111193, EPI_ISL_2111194, EPI_ISL_2111195, EPI_ISL_2111196, EPI_ISL_2111197, EPI_ISL_2111198, EPI_ISL_2111199, EPI_ISL_2111200, EPI_ISL_2111201, EPI_ISL_2111202, EPI_ISL_2111203, EPI_ISL_2111204, EPI_ISL_2111205, EPI_ISL_2111206, EPI_ISL_2111207, EPI_ISL_2111208, EPI_ISL_2111209, EPI_ISL_2111210, EPI_ISL_2111211, EPI_ISL_2111212, EPI_ISL_2111213, EPI_ISL_2111214, EPI_ISL_2111215, EPI_ISL_2111216, EPI_ISL_2111217, EPI_ISL_2111218, EPI_ISL_2111219, EPI_ISL_2112841, EPI_ISL_2112842, EPI_ISL_2112843, EPI_ISL_2112844, EPI_ISL_2112845, EPI_ISL_2112846, EPI_ISL_2112847, EPI_ISL_2112848, EPI_ISL_2112849, EPI_ISL_2112851, EPI_ISL_2112853, EPI_ISL_2112854, EPI_ISL_2112855, EPI_ISL_2112857, EPI_ISL_2112858, EPI_ISL_2112860, EPI_ISL_2112861, EPI_ISL_2112863, EPI_ISL_2115255, EPI_ISL_2115256, EPI_ISL_2115257, EPI_ISL_2115258, EPI_ISL_2115259, EPI_ISL_2115260, EPI_ISL_2115261, EPI_ISL_2115262, EPI_ISL_2115263, EPI_ISL_2115264, EPI_ISL_2115265, EPI_ISL_2115266, EPI_ISL_2115267, EPI_ISL_2115268, EPI_ISL_2115269, EPI_ISL_2115270, EPI_ISL_2115271, EPI_ISL_2115272, EPI_ISL_2115273, EPI_ISL_2115274, EPI_ISL_2115275, EPI_ISL_2115276, EPI_ISL_2115277, EPI_ISL_2115278, EPI_ISL_2115279, EPI_ISL_2115281, EPI_ISL_2115282, EPI_ISL_2115283, EPI_ISL_2115284, EPI_ISL_2115285, EPI_ISL_2115286, EPI_ISL_2115287, EPI_ISL_2115288, EPI_ISL_2115289, EPI_ISL_2115290, EPI_ISL_2115291, EPI_ISL_2115292, EPI_ISL_2115293, EPI_ISL_2115294, EPI_ISL_2115295, EPI_ISL_2115296, EPI_ISL_2115297, EPI_ISL_2115298, EPI_ISL_2115299, EPI_ISL_2115300, EPI_ISL_2115301, EPI_ISL_2115302, EPI_ISL_2115303, EPI_ISL_2115304, EPI_ISL_2115305, EPI_ISL_2115307, EPI_ISL_2115308, EPI_ISL_2115309, EPI_ISL_2115310, EPI_ISL_2115311, EPI_ISL_2115312, EPI_ISL_2115313, EPI_ISL_2115314, EPI_ISL_2115315, EPI_ISL_2115316, EPI_ISL_2115317, EPI_ISL_2115318, EPI_ISL_2115319, EPI_ISL_2115320, EPI_ISL_2115321, EPI_ISL_2115323, EPI_ISL_2115324, EPI_ISL_2115325, EPI_ISL_2115326, EPI_ISL_2115327, EPI_ISL_2115328, EPI_ISL_2115329, EPI_ISL_2115330, EPI_ISL_2115331, EPI_ISL_2115332, EPI_ISL_2115333, EPI_ISL_2115334, EPI_ISL_2115335, EPI_ISL_2115336, EPI_ISL_2115337, EPI_ISL_2116136, EPI_ISL_2116138, EPI_ISL_2116139, EPI_ISL_2116140, EPI_ISL_2116141, EPI_ISL_2116142, EPI_ISL_2116143, EPI_ISL_2116144, EPI_ISL_2116145, EPI_ISL_2116146, EPI_ISL_2116147, EPI_ISL_2116148, EPI_ISL_2116149 | | | |
| see above | MVZ Labor Krone GbR | Robert Koch Institute | |
| EPI_ISL_2122431, EPI_ISL_2122477, EPI_ISL_2122486, EPI_ISL_2122519, EPI_ISL_2122545 | MVZ fur Laboratoriumsmedizin und Mikrobiologie Wurzburg | Robert Koch Institute | |
| EPI_ISL_2094516, EPI_ISL_2094528, EPI_ISL_2094643, EPI_ISL_2094652, EPI_ISL_2094715, EPI_ISL_2094716, EPI_ISL_2094717, EPI_ISL_2094718, EPI_ISL_2094719, EPI_ISL_2094720, EPI_ISL_2094736, EPI_ISL_2094918, EPI_ISL_2094919, EPI_ISL_2094920, EPI_ISL_2094921, EPI_ISL_2094922, EPI_ISL_2094923, EPI_ISL_2094924, EPI_ISL_2094925, EPI_ISL_2094926, EPI_ISL_2094927, EPI_ISL_2094928, EPI_ISL_2094929, EPI_ISL_2094952, EPI_ISL_2094953, EPI_ISL_2094959, EPI_ISL_2094960, EPI_ISL_2094961, EPI_ISL_2094984, EPI_ISL_2095040, EPI_ISL_2095050, EPI_ISL_2095054, EPI_ISL_2095059, EPI_ISL_2095074, EPI_ISL_2095077, EPI_ISL_2095084, EPI_ISL_2095091, EPI_ISL_2095092, EPI_ISL_2095117, EPI_ISL_2095131, EPI_ISL_2095137, EPI_ISL_2095150, EPI_ISL_2095168, EPI_ISL_2095170, EPI_ISL_2095177, EPI_ISL_2095178, EPI_ISL_2095260, EPI_ISL_2095265, EPI_ISL_2095272, EPI_ISL_2095273, EPI_ISL_2095291, EPI_ISL_2095305 | | | |
| see above | Max von Pettenkofer Institute, Virology, National Reference Center for Retroviruses, LMU Munich | Laboratory for Functional Genome Analysis; Dept. Genomics; Gene Center of the LMU Munich | Alexander Graf; Helmut Blum; Max Muenchhoff; Oliver Keppler; Stefan Krebs |
| EPI_ISL_2110378, EPI_ISL_2110379, EPI_ISL_2110380, EPI_ISL_2110382, EPI_ISL_2110383, EPI_ISL_2110384, EPI_ISL_2110385, EPI_ISL_2110386, EPI_ISL_2110387, EPI_ISL_2110391, EPI_ISL_2110393, EPI_ISL_2110396, EPI_ISL_2110397, EPI_ISL_2110399, EPI_ISL_2110406 | | | |
| see above | Med. Labor Prof. Schenk Dr. Ansorge & Kollegen | Robert Koch Institute | |
| EPI_ISL_1993422, EPI_ISL_1993423, EPI_ISL_1993426, EPI_ISL_1993481, EPI_ISL_1993482, EPI_ISL_1993485, EPI_ISL_1993487, EPI_ISL_1993488, EPI_ISL_1993489, EPI_ISL_1993490, EPI_ISL_1993491, EPI_ISL_1993492, EPI_ISL_1993493, EPI_ISL_1993494, EPI_ISL_1993496, EPI_ISL_1993497, EPI_ISL_1993498, EPI_ISL_1993499, EPI_ISL_1993500, EPI_ISL_1993501, EPI_ISL_1993502, EPI_ISL_1993505, EPI_ISL_1993506, EPI_ISL_1993507, EPI_ISL_1993508, EPI_ISL_1993509, EPI_ISL_1993510, EPI_ISL_1993511, EPI_ISL_1993512, EPI_ISL_1993513, EPI_ISL_1993514, EPI_ISL_1993515, EPI_ISL_1993516, EPI_ISL_1993517, EPI_ISL_1993518, EPI_ISL_1993519, EPI_ISL_1993520, EPI_ISL_1993521, EPI_ISL_1993522, EPI_ISL_1993523, EPI_ISL_1993524, EPI_ISL_1993525, EPI_ISL_1993577, EPI_ISL_1993579, EPI_ISL_1993580, EPI_ISL_1993581, EPI_ISL_1993582, EPI_ISL_1993583, EPI_ISL_1993584, EPI_ISL_1993585, EPI_ISL_1993586, EPI_ISL_1993587, EPI_ISL_1993589, EPI_ISL_1993590, EPI_ISL_1993591, EPI_ISL_1993592, EPI_ISL_1993593, EPI_ISL_1993594, EPI_ISL_1993595, EPI_ISL_1993596, EPI_ISL_1993597, EPI_ISL_1993598, EPI_ISL_1993599, EPI_ISL_1993600, EPI_ISL_1993601, EPI_ISL_1993602, EPI_ISL_1993603, EPI_ISL_1993604 | | | |
| see above | Medical Laboratories Duesseldorf | Center of Medical Microbiology, Virology, and Hospital Hygiene, University of Duesseldorf | Alexander Dilthey; Andreas Walker; Angelika Helmer; Christian Lange; Daniel Strelow; Jessica Nicolai; Jörg Timm; Klaus Pfeffer; Lisanna Hülse; Malte Kohns Vasconcelos; Maximilian Damagnez; Nadine Lübke; Tobias Wienemann; Torsten Houwaart |
| EPI_ISL_2109359, EPI_ISL_2109365, EPI_ISL_2109375, EPI_ISL_2109376, EPI_ISL_2109380, EPI_ISL_2109382, EPI_ISL_2109387, EPI_ISL_2109391, EPI_ISL_2109395, EPI_ISL_2109396, EPI_ISL_2109405, EPI_ISL_2109406, EPI_ISL_2109408, EPI_ISL_2109412, EPI_ISL_2109416, EPI_ISL_2109423 | | | |
| see above | Medizinisches Labor Ostsachsen MVZ GbR Nebenbetriebsstotte Dresden | Robert Koch Institute | |
| EPI_ISL_2108341, EPI_ISL_2108349, EPI_ISL_2108384, EPI_ISL_2108386, EPI_ISL_2108392, EPI_ISL_2108404, EPI_ISL_2115390, EPI_ISL_2115408, EPI_ISL_2115413, EPI_ISL_2115426, EPI_ISL_2115429, EPI_ISL_2115432, EPI_ISL_2115435, EPI_ISL_2115444, EPI_ISL_2115446, EPI_ISL_2115452, EPI_ISL_2115469, EPI_ISL_2115476 | | | |
| see above | Medizinisches Labor Wahl Ludenscheid | Robert Koch Institute | |
| EPI_ISL_2113161 | Medizinisches Zentrallabor Altenburg GmbH & Co.KG | Robert Koch Institute | |
| EPI_ISL_2110025, EPI_ISL_2110080, EPI_ISL_2112186, EPI_ISL_2112208, EPI_ISL_2112242 | Praxisgemeinschaft fur Laboratoriumsmedizin Labor Blumenstrasse; Praxis Dr. E. Wietschel & Dr. F. Wietschel | Robert Koch Institute | |
| EPI_ISL_2109886, EPI_ISL_2109912, EPI_ISL_2109967, EPI_ISL_2109969, EPI_ISL_2109978, EPI_ISL_2110003, EPI_ISL_2110006, EPI_ISL_2110030, EPI_ISL_2110089, EPI_ISL_2110098, EPI_ISL_2110144, EPI_ISL_2110194, EPI_ISL_2110206, EPI_ISL_2110262, EPI_ISL_2110312, EPI_ISL_2110347 | | | |
| see above | Praxisgemeinschaft fur Laboratoriumsmedizin Labor Blumenstrasse; Praxis Dr. Sel und Dr. Spote | Robert Koch Institute | |
| EPI_ISL_2125243, EPI_ISL_2125278, EPI_ISL_2125280, EPI_ISL_2125359, EPI_ISL_2125366 | Robert-Bosch-Krankenhaus | Robert Koch Institute | |
| EPI_ISL_2109875, EPI_ISL_2109876, EPI_ISL_2109880, EPI_ISL_2109882, EPI_ISL_2109883, EPI_ISL_2109891, EPI_ISL_2109892, EPI_ISL_2109897, EPI_ISL_2109901, EPI_ISL_2109911, EPI_ISL_2109913, EPI_ISL_2109914, EPI_ISL_2109916, EPI_ISL_2109919, EPI_ISL_2109921, EPI_ISL_2109924, EPI_ISL_2109927, EPI_ISL_2109930, EPI_ISL_2109935, EPI_ISL_2109941, EPI_ISL_2109942, EPI_ISL_2109944, EPI_ISL_2109947, EPI_ISL_2109950, EPI_ISL_2109960, EPI_ISL_2109973, EPI_ISL_2109980, EPI_ISL_2109984, EPI_ISL_2109989, EPI_ISL_2109998, EPI_ISL_2110000, EPI_ISL_2110004, EPI_ISL_2110007, EPI_ISL_2110012, EPI_ISL_2110014, EPI_ISL_2110019, EPI_ISL_2110024, EPI_ISL_2110027, EPI_ISL_2110029, EPI_ISL_2110034, EPI_ISL_2110037, EPI_ISL_2110050, EPI_ISL_2110051, EPI_ISL_2110065, EPI_ISL_2110069, EPI_ISL_2110070, EPI_ISL_2110073, EPI_ISL_2110074, EPI_ISL_2110075, EPI_ISL_2110077, EPI_ISL_2110078, EPI_ISL_2110082, EPI_ISL_2110095, EPI_ISL_2110101, EPI_ISL_2110102, EPI_ISL_2110105, EPI_ISL_2110108, EPI_ISL_2110112, EPI_ISL_2110115, EPI_ISL_2110124, EPI_ISL_2110127, EPI_ISL_2110133, EPI_ISL_2110134, EPI_ISL_2110139, EPI_ISL_2110143, EPI_ISL_2110147, EPI_ISL_2110148, EPI_ISL_2110154, EPI_ISL_2110156, EPI_ISL_2110162, EPI_ISL_2110163, EPI_ISL_2110164, EPI_ISL_2110165, EPI_ISL_2110172, EPI_ISL_2110181, EPI_ISL_2110191, EPI_ISL_2110192, EPI_ISL_2110195, EPI_ISL_2110198, EPI_ISL_2110200, EPI_ISL_2110204, EPI_ISL_2110207, EPI_ISL_2110209, EPI_ISL_2110211, EPI_ISL_2110212, EPI_ISL_2110213, EPI_ISL_2110218, EPI_ISL_2110219, EPI_ISL_2110234, EPI_ISL_2110237, EPI_ISL_2110239, EPI_ISL_2110241, EPI_ISL_2110244, EPI_ISL_2110250, EPI_ISL_2110255, EPI_ISL_2110261, EPI_ISL_2110263, EPI_ISL_2110268, EPI_ISL_2110273, EPI_ISL_2110277, EPI_ISL_2110283, EPI_ISL_2110286, EPI_ISL_2110289, EPI_ISL_2110297, EPI_ISL_2110298, EPI_ISL_2110303, EPI_ISL_2110309, EPI_ISL_2110310, EPI_ISL_2110311, EPI_ISL_2110314, EPI_ISL_2110318, EPI_ISL_2110320, EPI_ISL_2110324, EPI_ISL_2110325, EPI_ISL_2110326, EPI_ISL_2110330, EPI_ISL_2110337, EPI_ISL_2110340, EPI_ISL_2110341, EPI_ISL_2110345, EPI_ISL_2110358, EPI_ISL_2110360, EPI_ISL_2110362, EPI_ISL_2110363, EPI_ISL_2110367, EPI_ISL_2110371, EPI_ISL_2110376, EPI_ISL_2112185, EPI_ISL_2112188, EPI_ISL_2112189, EPI_ISL_2112190, EPI_ISL_2112191, EPI_ISL_2112192, EPI_ISL_2112193, EPI_ISL_2112194, EPI_ISL_2112195, EPI_ISL_2112196, EPI_ISL_2112197, EPI_ISL_2112198, EPI_ISL_2112199, EPI_ISL_2112201, EPI_ISL_2112202, EPI_ISL_2112203, EPI_ISL_2112206, EPI_ISL_2112207, EPI_ISL_2112209, EPI_ISL_2112211, EPI_ISL_2112215, EPI_ISL_2112216, EPI_ISL_2112217, EPI_ISL_2112219, EPI_ISL_2112221, EPI_ISL_2112222, EPI_ISL_2112224, EPI_ISL_2112225, EPI_ISL_2112226, EPI_ISL_2112230, EPI_ISL_2112231, EPI_ISL_2112232, EPI_ISL_2112233, EPI_ISL_2112234, EPI_ISL_2112236, EPI_ISL_2112237, EPI_ISL_2112238, EPI_ISL_2112239, EPI_ISL_2112240, EPI_ISL_2112241, EPI_ISL_2112245, EPI_ISL_2112248, EPI_ISL_2112249, EPI_ISL_2112250, EPI_ISL_2112251, EPI_ISL_2112252, EPI_ISL_2112253, EPI_ISL_2112256, EPI_ISL_2112258, EPI_ISL_2112259, EPI_ISL_2112262, EPI_ISL_2112263, EPI_ISL_2112264, EPI_ISL_2112267, EPI_ISL_2112268, EPI_ISL_2112269, EPI_ISL_2112270, EPI_ISL_2112272, EPI_ISL_2112275, EPI_ISL_2112276, EPI_ISL_2112277, EPI_ISL_2112280, EPI_ISL_2112281, EPI_ISL_2112283, EPI_ISL_2112284, EPI_ISL_2112285, EPI_ISL_2112286, EPI_ISL_2112288, EPI_ISL_2112289, EPI_ISL_2112291, EPI_ISL_2112292, EPI_ISL_2112293, EPI_ISL_2112294, EPI_ISL_2112295, EPI_ISL_2112297, EPI_ISL_2112300, EPI_ISL_2112301, EPI_ISL_2112304, EPI_ISL_2112305, EPI_ISL_2112307, EPI_ISL_2112309, EPI_ISL_2112311, EPI_ISL_2112312, EPI_ISL_2112313, EPI_ISL_2112314, EPI_ISL_2112316, EPI_ISL_2112322, EPI_ISL_2112323, EPI_ISL_2112324, EPI_ISL_2112325, EPI_ISL_2112326, EPI_ISL_2112327, EPI_ISL_2112329, EPI_ISL_2112330, EPI_ISL_2112333, EPI_ISL_2112337, EPI_ISL_2112338, EPI_ISL_2112339, EPI_ISL_2112340, EPI_ISL_2112341, EPI_ISL_2112342, EPI_ISL_2112344, EPI_ISL_2112345, EPI_ISL_2112347, EPI_ISL_2112348, EPI_ISL_2112349, EPI_ISL_2112351, EPI_ISL_2112353, EPI_ISL_2112354, EPI_ISL_2112355, EPI_ISL_2112356, EPI_ISL_2112357, EPI_ISL_2112358, EPI_ISL_2112359, EPI_ISL_2112361, EPI_ISL_2112362, EPI_ISL_2112363, EPI_ISL_2112368, EPI_ISL_2112369, EPI_ISL_2112370, EPI_ISL_2112372, EPI_ISL_2112373, EPI_ISL_2112374, EPI_ISL_2112376, EPI_ISL_2112377, EPI_ISL_2112379, EPI_ISL_2112381, EPI_ISL_2112382, EPI_ISL_2112383, EPI_ISL_2112385, EPI_ISL_2112386, EPI_ISL_2112387, EPI_ISL_2112388, EPI_ISL_2112389, EPI_ISL_2112390, EPI_ISL_2112391, EPI_ISL_2112393, EPI_ISL_2112395, EPI_ISL_2112396, EPI_ISL_2112397, EPI_ISL_2112398, EPI_ISL_2112399, EPI_ISL_2112401, EPI_ISL_2112402, EPI_ISL_2112403, EPI_ISL_2112404, EPI_ISL_2112405, EPI_ISL_2112406, EPI_ISL_2112407, EPI_ISL_2112410, EPI_ISL_2112411, EPI_ISL_2112412, EPI_ISL_2112414, EPI_ISL_2112415, EPI_ISL_2112416, EPI_ISL_2112419, EPI_ISL_2112420, EPI_ISL_2112421, EPI_ISL_2112422, EPI_ISL_2112423, EPI_ISL_2112424, EPI_ISL_2112425, EPI_ISL_2112427, EPI_ISL_2112428, EPI_ISL_2112431, EPI_ISL_2112432, EPI_ISL_2112433, EPI_ISL_2112434, EPI_ISL_2112435, EPI_ISL_2112436, EPI_ISL_2112437, EPI_ISL_2112442, EPI_ISL_2112445, EPI_ISL_2112448, EPI_ISL_2112449, EPI_ISL_2112451, EPI_ISL_2112452, EPI_ISL_2112453, EPI_ISL_2112454, EPI_ISL_2112455, EPI_ISL_2112456, EPI_ISL_2112458, EPI_ISL_2112460, EPI_ISL_2112462, EPI_ISL_2112464, EPI_ISL_2112469, EPI_ISL_2112470, EPI_ISL_2112471, EPI_ISL_2112473, EPI_ISL_2112475, EPI_ISL_2112478, EPI_ISL_2112492, EPI_ISL_2112494, EPI_ISL_2112495, EPI_ISL_2112496, EPI_ISL_2112501, EPI_ISL_2112502, EPI_ISL_2112505, EPI_ISL_2112506, EPI_ISL_2112508, EPI_ISL_2112511, EPI_ISL_2112512, EPI_ISL_2112513, EPI_ISL_2112514, EPI_ISL_2112515, EPI_ISL_2112517, EPI_ISL_2112518, EPI_ISL_2112520, EPI_ISL_2112521, EPI_ISL_2112522, EPI_ISL_2112523, EPI_ISL_2112526, EPI_ISL_2112530, EPI_ISL_2112531, EPI_ISL_2112532, EPI_ISL_2112533, EPI_ISL_2112534, EPI_ISL_2112535, EPI_ISL_2112537, EPI_ISL_2112539, EPI_ISL_2112543, EPI_ISL_2112545, EPI_ISL_2112546, EPI_ISL_2112547, EPI_ISL_2112549, EPI_ISL_2112552, EPI_ISL_2112559, EPI_ISL_2112561, EPI_ISL_2112562, EPI_ISL_2112563, EPI_ISL_2112566, EPI_ISL_2112567, EPI_ISL_2112568, EPI_ISL_2112569, EPI_ISL_2112570, EPI_ISL_2112571, EPI_ISL_2112574, EPI_ISL_2112576, EPI_ISL_2112579, EPI_ISL_2112581, EPI_ISL_2112582, EPI_ISL_2112585, EPI_ISL_2112590, EPI_ISL_2112594, EPI_ISL_2112596, EPI_ISL_2112598, EPI_ISL_2112599, EPI_ISL_2112600, EPI_ISL_2112602, EPI_ISL_2112603, EPI_ISL_2112605, EPI_ISL_2112606, EPI_ISL_2112607, EPI_ISL_2112608, EPI_ISL_2112611, EPI_ISL_2112615, EPI_ISL_2112617, EPI_ISL_2112619, EPI_ISL_2112620, EPI_ISL_2112621, EPI_ISL_2112622, EPI_ISL_2112623, EPI_ISL_2112624, EPI_ISL_2112626, EPI_ISL_2112627, EPI_ISL_2112629, EPI_ISL_2112631, EPI_ISL_2112632, EPI_ISL_2112633, EPI_ISL_2112634, EPI_ISL_2112637, EPI_ISL_2112638, EPI_ISL_2112642, EPI_ISL_2112643, EPI_ISL_2112644, EPI_ISL_2112645, EPI_ISL_2112650, EPI_ISL_2112653, EPI_ISL_2112655, EPI_ISL_2112656, EPI_ISL_2112657, EPI_ISL_2112659, EPI_ISL_2112660, EPI_ISL_2112661, EPI_ISL_2112662, EPI_ISL_2112666, EPI_ISL_2112667, EPI_ISL_2112668, EPI_ISL_2112669, EPI_ISL_2112670, EPI_ISL_2112672, EPI_ISL_2112674, EPI_ISL_2112675, EPI_ISL_2112676, EPI_ISL_2112677, EPI_ISL_2112678, EPI_ISL_2112680, EPI_ISL_2112681, EPI_ISL_2112682, EPI_ISL_2112683, EPI_ISL_2112685, EPI_ISL_2112686, EPI_ISL_2112693, EPI_ISL_2112694, EPI_ISL_2112695, EPI_ISL_2112696, EPI_ISL_2112701, EPI_ISL_2112704, EPI_ISL_2112708, EPI_ISL_2112709, EPI_ISL_2112710, EPI_ISL_2112711, EPI_ISL_2112713, EPI_ISL_2112714, EPI_ISL_2112715, EPI_ISL_2112716, EPI_ISL_2112717, EPI_ISL_2112954, EPI_ISL_2112955, EPI_ISL_2112958, EPI_ISL_2112962, EPI_ISL_2112963, EPI_ISL_2112966, EPI_ISL_2112967, EPI_ISL_2112968, EPI_ISL_2112969, EPI_ISL_2112974, EPI_ISL_2112975, EPI_ISL_2112976, EPI_ISL_2112980, EPI_ISL_2112981, EPI_ISL_2112983, EPI_ISL_2112984, EPI_ISL_2112985, EPI_ISL_2112987, EPI_ISL_2112988, EPI_ISL_2112990, EPI_ISL_2112992, EPI_ISL_2112993, EPI_ISL_2112994, EPI_ISL_2112995, EPI_ISL_2112996, EPI_ISL_2112997, EPI_ISL_2112998, EPI_ISL_2113000, EPI_ISL_2113001, EPI_ISL_2113002, EPI_ISL_2113004, EPI_ISL_2113007, EPI_ISL_2113010, EPI_ISL_2113012, EPI_ISL_2113013, EPI_ISL_2113014, EPI_ISL_2113016, EPI_ISL_2113017, EPI_ISL_2113018, EPI_ISL_2113020, EPI_ISL_2113022, EPI_ISL_2113023, EPI_ISL_2113026, EPI_ISL_2113029, EPI_ISL_2113033, EPI_ISL_2113034, EPI_ISL_2113035, EPI_ISL_2113036, EPI_ISL_2113042, EPI_ISL_2113045, EPI_ISL_2113050, EPI_ISL_2113051, EPI_ISL_2113052, EPI_ISL_2113053, EPI_ISL_2113057, EPI_ISL_2113058, EPI_ISL_2113060, EPI_ISL_2113062, EPI_ISL_2113063, EPI_ISL_2113065, EPI_ISL_2113066, EPI_ISL_2113067, EPI_ISL_2113070, EPI_ISL_2113071, EPI_ISL_2113072, EPI_ISL_2113073, EPI_ISL_2113075, EPI_ISL_2113077, EPI_ISL_2113078, EPI_ISL_2113082, EPI_ISL_2113083, EPI_ISL_2113084, EPI_ISL_2113085, EPI_ISL_2113086, EPI_ISL_2113087, EPI_ISL_2113089, EPI_ISL_2113094, EPI_ISL_2113095, EPI_ISL_2113096, EPI_ISL_2113098, EPI_ISL_2113101, EPI_ISL_2113102, EPI_ISL_2113103, EPI_ISL_2113105, EPI_ISL_2113107, EPI_ISL_2113109, EPI_ISL_2113110, EPI_ISL_2113111, EPI_ISL_2113115, EPI_ISL_2113116, EPI_ISL_2113118, EPI_ISL_2113119, EPI_ISL_2113121, EPI_ISL_2113122, EPI_ISL_2113124, EPI_ISL_2113125, EPI_ISL_2113127, EPI_ISL_2113132, EPI_ISL_2113134, EPI_ISL_2113136, EPI_ISL_2113137, EPI_ISL_2113138, EPI_ISL_2113139, EPI_ISL_2113140, EPI_ISL_2113144, EPI_ISL_2113149, EPI_ISL_2113153, EPI_ISL_2113154, EPI_ISL_2113160, EPI_ISL_2113164, EPI_ISL_2113168, EPI_ISL_2113172, EPI_ISL_2113173, EPI_ISL_2113176, EPI_ISL_2113177, EPI_ISL_2113178, EPI_ISL_2113181, EPI_ISL_2113182, EPI_ISL_2113187, EPI_ISL_2113191, EPI_ISL_2113192, EPI_ISL_2113196, EPI_ISL_2113197, EPI_ISL_2113198, EPI_ISL_2113201, EPI_ISL_2113202, EPI_ISL_2113204, EPI_ISL_2113205, EPI_ISL_2113208, EPI_ISL_2113210, EPI_ISL_2113215, EPI_ISL_2113216, EPI_ISL_2113219, EPI_ISL_2113223, EPI_ISL_2113228, EPI_ISL_2113229, EPI_ISL_2113230, EPI_ISL_2113231, EPI_ISL_2113233, EPI_ISL_2113234, EPI_ISL_2117298, EPI_ISL_2117300, EPI_ISL_2117309, EPI_ISL_2117315, EPI_ISL_2117319, EPI_ISL_2117328, EPI_ISL_2117332, EPI_ISL_2117339, EPI_ISL_2117340, EPI_ISL_2117345, EPI_ISL_2117361, EPI_ISL_2117376, EPI_ISL_2117398, EPI_ISL_2117422 | | | |
| see above | SYNLAB Jena Oncoscreen | Robert Koch Institute | |
| EPI_ISL_2109898, EPI_ISL_2109915, EPI_ISL_2110035, EPI_ISL_2110274 | SYNLAB Labor MÃ¼nchen Zentrum LMZ | Robert Koch Institute | |
| EPI_ISL_2109871, EPI_ISL_2109874, EPI_ISL_2109890, EPI_ISL_2109910, EPI_ISL_2109945, EPI_ISL_2109963, EPI_ISL_2109987, EPI_ISL_2110056, EPI_ISL_2110100, EPI_ISL_2110119, EPI_ISL_2110180, EPI_ISL_2110188, EPI_ISL_2110216, EPI_ISL_2110292, EPI_ISL_2112205, EPI_ISL_2112210, EPI_ISL_2112213, EPI_ISL_2112218, EPI_ISL_2112220, EPI_ISL_2112229, EPI_ISL_2112244, EPI_ISL_2112246, EPI_ISL_2112247, EPI_ISL_2112254, EPI_ISL_2112255, EPI_ISL_2112257, EPI_ISL_2112261, EPI_ISL_2112266, EPI_ISL_2112273, EPI_ISL_2112279, EPI_ISL_2112287, EPI_ISL_2112290, EPI_ISL_2112296, EPI_ISL_2112298, EPI_ISL_2112299, EPI_ISL_2112303, EPI_ISL_2112306, EPI_ISL_2112308, EPI_ISL_2112310, EPI_ISL_2112315, EPI_ISL_2112318, EPI_ISL_2112319, EPI_ISL_2112320, EPI_ISL_2112321, EPI_ISL_2112328, EPI_ISL_2112331, EPI_ISL_2112334, EPI_ISL_2112335, EPI_ISL_2112336, EPI_ISL_2112343, EPI_ISL_2112350, EPI_ISL_2112352, EPI_ISL_2112360, EPI_ISL_2112364, EPI_ISL_2112365, EPI_ISL_2112371, EPI_ISL_2112375, EPI_ISL_2112378, EPI_ISL_2112408, EPI_ISL_2112413, EPI_ISL_2112417, EPI_ISL_2112418, EPI_ISL_2112426, EPI_ISL_2112429, EPI_ISL_2112438, EPI_ISL_2112439, EPI_ISL_2112440, EPI_ISL_2112441, EPI_ISL_2112444, EPI_ISL_2112446, EPI_ISL_2112447, EPI_ISL_2112450, EPI_ISL_2112459, EPI_ISL_2112463, EPI_ISL_2112465, EPI_ISL_2112466, EPI_ISL_2112468, EPI_ISL_2112472, EPI_ISL_2112474, EPI_ISL_2112476, EPI_ISL_2112477, EPI_ISL_2112504, EPI_ISL_2112528, EPI_ISL_2117368, EPI_ISL_2117400 | | | |

see above SYNLAB MVZ Berlin Robert Koch Institute

EPI_ISL_2127699, EPI_ISL_2127702, EPI_ISL_2127703, EPI_ISL_2127706, EPI_ISL_2127709, EPI_ISL_2127711, EPI_ISL_2127714, EPI_ISL_2127717, EPI_ISL_2127718, EPI_ISL_2127719, EPI_ISL_2127722, EPI_ISL_2127723, EPI_ISL_2127725, EPI_ISL_2127740, EPI_ISL_2127742, EPI_ISL_2127749, EPI_ISL_2127756, EPI_ISL_2127760, EPI_ISL_2127762, EPI_ISL_2127766, EPI_ISL_2127769, EPI_ISL_2127771,
EPI_ISL_2127781, EPI_ISL_2127793, EPI_ISL_2127799, EPI_ISL_2127801, EPI_ISL_2127803, EPI_ISL_2127806, EPI_ISL_2127810, EPI_ISL_2127813, EPI_ISL_2127817, EPI_ISL_2127821, EPI_ISL_2127826, EPI_ISL_2127828, EPI_ISL_2127830, EPI_ISL_2127832, EPI_ISL_2127834, EPI_ISL_2127836, EPI_ISL_2127838, EPI_ISL_2127840, EPI_ISL_2127843, EPI_ISL_2127844, EPI_ISL_2127846, EPI_ISL_2127850,
EPI_ISL_2127855, EPI_ISL_2127859, EPI_ISL_2127864, EPI_ISL_2127866, EPI_ISL_2127870, EPI_ISL_2127877, EPI_ISL_2127879, EPI_ISL_2127881, EPI_ISL_2127884, EPI_ISL_2127886, EPI_ISL_2127887, EPI_ISL_2127896, EPI_ISL_2127901, EPI_ISL_2127902, EPI_ISL_2127905, EPI_ISL_2127917, EPI_ISL_2127919, EPI_ISL_2127921, EPI_ISL_2127925, EPI_ISL_2127937, EPI_ISL_2127941, EPI_ISL_2127943,
EPI_ISL_2127946, EPI_ISL_2127953, EPI_ISL_2127955, EPI_ISL_2127961, EPI_ISL_2127968, EPI_ISL_2127973, EPI_ISL_2127976, EPI_ISL_2127978, EPI_ISL_2127984, EPI_ISL_2127992, EPI_ISL_2127993, EPI_ISL_2128002, EPI_ISL_2128004, EPI_ISL_2128010, EPI_ISL_2128014, EPI_ISL_2128021, EPI_ISL_2128026, EPI_ISL_2128034, EPI_ISL_2128036, EPI_ISL_2128043, EPI_ISL_2128045, EPI_ISL_2128048,
EPI_ISL_2128050, EPI_ISL_2128052, EPI_ISL_2128054, EPI_ISL_2128057, EPI_ISL_2128059, EPI_ISL_2128061, EPI_ISL_2128063, EPI_ISL_2128066, EPI_ISL_2128068, EPI_ISL_2128076, EPI_ISL_2128081, EPI_ISL_2128085, EPI_ISL_2128088, EPI_ISL_2128092, EPI_ISL_2128096, EPI_ISL_2128107, EPI_ISL_2128109, EPI_ISL_2128111, EPI_ISL_2128116, EPI_ISL_2128118, EPI_ISL_2128120, EPI_ISL_2128125,
EPI_ISL_2128129, EPI_ISL_2128135, EPI_ISL_2128138, EPI_ISL_2128142, EPI_ISL_2128144, EPI_ISL_2128146, EPI_ISL_2128148, EPI_ISL_2128156, EPI_ISL_2128159, EPI_ISL_2128163, EPI_ISL_2128168, EPI_ISL_2128172, EPI_ISL_2128175, EPI_ISL_2128177, EPI_ISL_2128183, EPI_ISL_2128191, EPI_ISL_2128193, EPI_ISL_2128195, EPI_ISL_2128202, EPI_ISL_2128204, EPI_ISL_2128213, EPI_ISL_2128215,
EPI_ISL_2128220, EPI_ISL_2128223, EPI_ISL_2128230, EPI_ISL_2128239, EPI_ISL_2128241, EPI_ISL_2128247, EPI_ISL_2128249, EPI_ISL_2128254, EPI_ISL_2128256, EPI_ISL_2128260, EPI_ISL_2128267, EPI_ISL_2128269, EPI_ISL_2128273, EPI_ISL_2128278, EPI_ISL_2128287, EPI_ISL_2128290, EPI_ISL_2128292, EPI_ISL_2128293, EPI_ISL_2128299, EPI_ISL_2128301, EPI_ISL_2128311, EPI_ISL_2128315,
EPI_ISL_2128329, EPI_ISL_2128333, EPI_ISL_2128338, EPI_ISL_2128348, EPI_ISL_2128353, EPI_ISL_2128357, EPI_ISL_2128359, EPI_ISL_2128361, EPI_ISL_2128363, EPI_ISL_2128365, EPI_ISL_2128369, EPI_ISL_2128371, EPI_ISL_2128373, EPI_ISL_2128374, EPI_ISL_2128375, EPI_ISL_2128643, EPI_ISL_2128646, EPI_ISL_2128648, EPI_ISL_2128651, EPI_ISL_2128653, EPI_ISL_2128655, EPI_ISL_2128664,
EPI_ISL_2128667, EPI_ISL_2128669, EPI_ISL_2128670, EPI_ISL_2128671, EPI_ISL_2128680, EPI_ISL_2128687, EPI_ISL_2128712, EPI_ISL_2128714, EPI_ISL_2128722, EPI_ISL_2128724, EPI_ISL_2128730, EPI_ISL_2128743, EPI_ISL_2128745, EPI_ISL_2128747, EPI_ISL_2128750, EPI_ISL_2128753, EPI_ISL_2128755, EPI_ISL_2128760, EPI_ISL_2128761, EPI_ISL_2128764, EPI_ISL_2128765, EPI_ISL_2128766,
EPI_ISL_2128767, EPI_ISL_2128768, EPI_ISL_2128769, EPI_ISL_2128772, EPI_ISL_2128777, EPI_ISL_2128779, EPI_ISL_2128781, EPI_ISL_2128784, EPI_ISL_2128788, EPI_ISL_2128793, EPI_ISL_2128800, EPI_ISL_2128802, EPI_ISL_2128805, EPI_ISL_2128806, EPI_ISL_2128813, EPI_ISL_2128818, EPI_ISL_2128821, EPI_ISL_2128827, EPI_ISL_2128829, EPI_ISL_2128830, EPI_ISL_2128833, EPI_ISL_2128835,
EPI_ISL_2128839, EPI_ISL_2128840, EPI_ISL_2128843, EPI_ISL_2128844, EPI_ISL_2128850, EPI_ISL_2128854, EPI_ISL_2128859, EPI_ISL_2128861, EPI_ISL_2128862, EPI_ISL_2128866, EPI_ISL_2128867, EPI_ISL_2128874, EPI_ISL_2128875, EPI_ISL_2128878, EPI_ISL_2128880, EPI_ISL_2128885, EPI_ISL_2128887, EPI_ISL_2128891, EPI_ISL_2128892, EPI_ISL_2128901, EPI_ISL_2128903, EPI_ISL_2128928,
EPI_ISL_2128932, EPI_ISL_2128935, EPI_ISL_2128937, EPI_ISL_2128945, EPI_ISL_2128951, EPI_ISL_2128953, EPI_ISL_2130837, EPI_ISL_2130842, EPI_ISL_2130845, EPI_ISL_2130861, EPI_ISL_2130866, EPI_ISL_2130877, EPI_ISL_2130879, EPI_ISL_2130884, EPI_ISL_2130889, EPI_ISL_2130891, EPI_ISL_2130893, EPI_ISL_2130895, EPI_ISL_2130898, EPI_ISL_2130900, EPI_ISL_2130903, EPI_ISL_2130906,
EPI_ISL_2130907, EPI_ISL_2130908, EPI_ISL_2130915, EPI_ISL_2130923, EPI_ISL_2130925, EPI_ISL_2130931, EPI_ISL_2130933, EPI_ISL_2130935, EPI_ISL_2130951, EPI_ISL_2130955, EPI_ISL_2130959, EPI_ISL_2130967, EPI_ISL_2130972, EPI_ISL_2130978, EPI_ISL_2130982, EPI_ISL_2131002, EPI_ISL_2131004, EPI_ISL_2131007, EPI_ISL_2131010, EPI_ISL_2131016, EPI_ISL_2131024, EPI_ISL_2131025,
EPI_ISL_2131026, EPI_ISL_2131039, EPI_ISL_2131042, EPI_ISL_2131043, EPI_ISL_2131052, EPI_ISL_2131055, EPI_ISL_2131056, EPI_ISL_2131057, EPI_ISL_2131064, EPI_ISL_2131068, EPI_ISL_2131070, EPI_ISL_2131072, EPI_ISL_2131087, EPI_ISL_2131093, EPI_ISL_2131096, EPI_ISL_2131099, EPI_ISL_2131108, EPI_ISL_2131113, EPI_ISL_2131118, EPI_ISL_2131119, EPI_ISL_2131120, EPI_ISL_2131121,
EPI_ISL_2131123, EPI_ISL_2131124, EPI_ISL_2131128, EPI_ISL_2131137, EPI_ISL_2131138, EPI_ISL_2131144

see above SYNLAB MVZ Heidelberg Robert Koch Institute

EPI_ISL_2109868, EPI_ISL_2109877, EPI_ISL_2109894, EPI_ISL_2109938, EPI_ISL_2109977, EPI_ISL_2110047, EPI_ISL_2110058, EPI_ISL_2110099, EPI_ISL_2110129, EPI_ISL_2110135, EPI_ISL_2110151, EPI_ISL_2110184, EPI_ISL_2110205, EPI_ISL_2110249, EPI_ISL_2110258, EPI_ISL_2110296, EPI_ISL_2110321, EPI_ISL_2110361, EPI_ISL_2112187, EPI_ISL_2112200, EPI_ISL_2112214, EPI_ISL_2112223,
EPI_ISL_2112228, EPI_ISL_2112260, EPI_ISL_2112265, EPI_ISL_2112271, EPI_ISL_2112274, EPI_ISL_2112302, EPI_ISL_2112317, EPI_ISL_2112332, EPI_ISL_2112346, EPI_ISL_2112366, EPI_ISL_2112380, EPI_ISL_2112384, EPI_ISL_2112392, EPI_ISL_2112394, EPI_ISL_2112409, EPI_ISL_2112430, EPI_ISL_2112443, EPI_ISL_2112457, EPI_ISL_2112461, EPI_ISL_2112490, EPI_ISL_2112491, EPI_ISL_2112497,
EPI_ISL_2112498, EPI_ISL_2112500, EPI_ISL_2112503, EPI_ISL_2112507, EPI_ISL_2112509, EPI_ISL_2112510, EPI_ISL_2112516, EPI_ISL_2112519, EPI_ISL_2112536, EPI_ISL_2112538, EPI_ISL_2112540, EPI_ISL_2112541, EPI_ISL_2112542, EPI_ISL_2112544, EPI_ISL_2112548, EPI_ISL_2112550, EPI_ISL_2112551, EPI_ISL_2112553, EPI_ISL_2112554, EPI_ISL_2112556, EPI_ISL_2112557, EPI_ISL_2112558,
EPI_ISL_2112560, EPI_ISL_2112564, EPI_ISL_2112565, EPI_ISL_2112572, EPI_ISL_2112573, EPI_ISL_2112575, EPI_ISL_2112577, EPI_ISL_2112578, EPI_ISL_2112580, EPI_ISL_2112583, EPI_ISL_2112584, EPI_ISL_2112586, EPI_ISL_2112587, EPI_ISL_2112588, EPI_ISL_2112591, EPI_ISL_2112592, EPI_ISL_2112593, EPI_ISL_2112595, EPI_ISL_2112597, EPI_ISL_2112601, EPI_ISL_2112609, EPI_ISL_2112610,
EPI_ISL_2112612, EPI_ISL_2112613, EPI_ISL_2112614, EPI_ISL_2112616, EPI_ISL_2112618, EPI_ISL_2112628, EPI_ISL_2112630, EPI_ISL_2112635, EPI_ISL_2112636, EPI_ISL_2112639, EPI_ISL_2112640, EPI_ISL_2112641, EPI_ISL_2112646, EPI_ISL_2112647, EPI_ISL_2112649, EPI_ISL_2112651, EPI_ISL_2112654, EPI_ISL_2112658, EPI_ISL_2112663, EPI_ISL_2112664, EPI_ISL_2112671,
EPI_ISL_2112673, EPI_ISL_2112679, EPI_ISL_2112684, EPI_ISL_2112687, EPI_ISL_2112688, EPI_ISL_2112689, EPI_ISL_2112690, EPI_ISL_2112691, EPI_ISL_2112692, EPI_ISL_2112697, EPI_ISL_2112698, EPI_ISL_2112699, EPI_ISL_2112700, EPI_ISL_2112702, EPI_ISL_2112703, EPI_ISL_2112705, EPI_ISL_2112706, EPI_ISL_2112707, EPI_ISL_2112712, EPI_ISL_2112956, EPI_ISL_2112959, EPI_ISL_2112960,
EPI_ISL_2112961, EPI_ISL_2112971, EPI_ISL_2112973, EPI_ISL_2112977, EPI_ISL_2112979, EPI_ISL_2112982, EPI_ISL_2112989, EPI_ISL_2112991, EPI_ISL_2113005, EPI_ISL_2113009, EPI_ISL_2113011, EPI_ISL_2113019, EPI_ISL_2113021, EPI_ISL_2113024, EPI_ISL_2113027, EPI_ISL_2113028, EPI_ISL_2113031, EPI_ISL_2113038, EPI_ISL_2113039, EPI_ISL_2113040, EPI_ISL_2113044, EPI_ISL_2113047,
EPI_ISL_2113064, EPI_ISL_2113091, EPI_ISL_2113106, EPI_ISL_2113108, EPI_ISL_2113113, EPI_ISL_2113126, EPI_ISL_2113135, EPI_ISL_2113146, EPI_ISL_2113148, EPI_ISL_2113150, EPI_ISL_2113155, EPI_ISL_2113157, EPI_ISL_2113159, EPI_ISL_2113162, EPI_ISL_2113163, EPI_ISL_2113165, EPI_ISL_2113166, EPI_ISL_2113179, EPI_ISL_2113189, EPI_ISL_2113193, EPI_ISL_2113195,
EPI_ISL_2113200, EPI_ISL_2113203, EPI_ISL_2113206, EPI_ISL_2113212, EPI_ISL_2113214, EPI_ISL_2113221, EPI_ISL_2113222, EPI_ISL_2113232, EPI_ISL_2113235, EPI_ISL_2113305, EPI_ISL_2117307, EPI_ISL_2117324, EPI_ISL_2117344, EPI_ISL_2117347, EPI_ISL_2117362, EPI_ISL_2117370, EPI_ISL_2117416, EPI_ISL_2123491, EPI_ISL_2123497, EPI_ISL_2123524, EPI_ISL_2123574, EPI_ISL_2123601,
EPI_ISL_2123629, EPI_ISL_2123651, EPI_ISL_2123717, EPI_ISL_2124124, EPI_ISL_2127238, EPI_ISL_2127240, EPI_ISL_2127245, EPI_ISL_2127246, EPI_ISL_2127248, EPI_ISL_2127251, EPI_ISL_2127253, EPI_ISL_2127256, EPI_ISL_2127258, EPI_ISL_2127262, EPI_ISL_2127267, EPI_ISL_2127273, EPI_ISL_2127275, EPI_ISL_2127277, EPI_ISL_2127282, EPI_ISL_2127283, EPI_ISL_2127286, EPI_ISL_2127290,
EPI_ISL_2127293, EPI_ISL_2127295, EPI_ISL_2127300, EPI_ISL_2127302, EPI_ISL_2127304, EPI_ISL_2127306, EPI_ISL_2127313, EPI_ISL_2127315, EPI_ISL_2127317, EPI_ISL_2127327, EPI_ISL_2127329, EPI_ISL_2127331, EPI_ISL_2127339, EPI_ISL_2127341, EPI_ISL_2127343, EPI_ISL_2127345, EPI_ISL_2127347, EPI_ISL_2127349, EPI_ISL_2127354, EPI_ISL_2127358, EPI_ISL_2127361, EPI_ISL_2127363,
EPI_ISL_2127364, EPI_ISL_2127370, EPI_ISL_2127372, EPI_ISL_2127374, EPI_ISL_2127376, EPI_ISL_2127379, EPI_ISL_2127381, EPI_ISL_2127383, EPI_ISL_2127385, EPI_ISL_2127387, EPI_ISL_2127392, EPI_ISL_2127395, EPI_ISL_2127397, EPI_ISL_2127405, EPI_ISL_2127408, EPI_ISL_2127412, EPI_ISL_2127414, EPI_ISL_2127421, EPI_ISL_2127425, EPI_ISL_2127427, EPI_ISL_2127429, EPI_ISL_2127431,
EPI_ISL_2127433, EPI_ISL_2127436, EPI_ISL_2127441, EPI_ISL_2127445, EPI_ISL_2127447, EPI_ISL_2127450, EPI_ISL_2127457, EPI_ISL_2127461, EPI_ISL_2127466, EPI_ISL_2127472, EPI_ISL_2127474, EPI_ISL_2127475, EPI_ISL_2127477, EPI_ISL_2127479, EPI_ISL_2127481, EPI_ISL_2127487, EPI_ISL_2127489, EPI_ISL_2127492, EPI_ISL_2127494, EPI_ISL_2127496, EPI_ISL_2127500, EPI_ISL_2127502,
EPI_ISL_2127506, EPI_ISL_2127510, EPI_ISL_2127515, EPI_ISL_2127517, EPI_ISL_2127519, EPI_ISL_2127521, EPI_ISL_2127523, EPI_ISL_2127525, EPI_ISL_2127527, EPI_ISL_2127531, EPI_ISL_2127533, EPI_ISL_2127535, EPI_ISL_2127537, EPI_ISL_2127539, EPI_ISL_2127541, EPI_ISL_2127542, EPI_ISL_2127544, EPI_ISL_2127547, EPI_ISL_2127551, EPI_ISL_2127554, EPI_ISL_2127558, EPI_ISL_2127560,
EPI_ISL_2127569, EPI_ISL_2127571, EPI_ISL_2127573, EPI_ISL_2127575, EPI_ISL_2127577, EPI_ISL_2127582, EPI_ISL_2127587, EPI_ISL_2127589, EPI_ISL_2127591, EPI_ISL_2127593, EPI_ISL_2127598, EPI_ISL_2127600, EPI_ISL_2127603, EPI_ISL_2127605, EPI_ISL_2127609, EPI_ISL_2127614, EPI_ISL_2127620, EPI_ISL_2127622, EPI_ISL_2127629, EPI_ISL_2127631, EPI_ISL_2127633, EPI_ISL_2127636,
EPI_ISL_2127638, EPI_ISL_2127639, EPI_ISL_2127641, EPI_ISL_2127642, EPI_ISL_2127644, EPI_ISL_2127645, EPI_ISL_2127646, EPI_ISL_2127647, EPI_ISL_2127649, EPI_ISL_2127650, EPI_ISL_2127652, EPI_ISL_2127653, EPI_ISL_2127654, EPI_ISL_2127655, EPI_ISL_2127657, EPI_ISL_2127659, EPI_ISL_2127661, EPI_ISL_2127663, EPI_ISL_2127664, EPI_ISL_2127665, EPI_ISL_2127666, EPI_ISL_2127667,
EPI_ISL_2127669, EPI_ISL_2127670, EPI_ISL_2127671, EPI_ISL_2127672, EPI_ISL_2127673, EPI_ISL_2127674, EPI_ISL_2127675, EPI_ISL_2127677, EPI_ISL_2127680, EPI_ISL_2127683, EPI_ISL_2127686, EPI_ISL_2127690, EPI_ISL_2127691, EPI_ISL_2127692, EPI_ISL_2127693, EPI_ISL_2127694, EPI_ISL_2127695, EPI_ISL_2127696, EPI_ISL_2129731, EPI_ISL_2129732, EPI_ISL_2129733, EPI_ISL_2129740,
EPI_ISL_2129741, EPI_ISL_2129743, EPI_ISL_2129744, EPI_ISL_2129745, EPI_ISL_2129747, EPI_ISL_2129752, EPI_ISL_2129754, EPI_ISL_2129755, EPI_ISL_2129756, EPI_ISL_2129757, EPI_ISL_2129758, EPI_ISL_2129759, EPI_ISL_2129760, EPI_ISL_2129761, EPI_ISL_2129763, EPI_ISL_2129765, EPI_ISL_2129769, EPI_ISL_2129770, EPI_ISL_2129771, EPI_ISL_2129773, EPI_ISL_2129776, EPI_ISL_2129777,
EPI_ISL_2129779, EPI_ISL_2129780, EPI_ISL_2129781, EPI_ISL_2129782, EPI_ISL_2129783, EPI_ISL_2129784, EPI_ISL_2129787, EPI_ISL_2129790, EPI_ISL_2129791, EPI_ISL_2129792, EPI_ISL_2129795, EPI_ISL_2129796, EPI_ISL_2129797, EPI_ISL_2129798, EPI_ISL_2129800, EPI_ISL_2129802, EPI_ISL_2129803, EPI_ISL_2129804, EPI_ISL_2129805, EPI_ISL_2129806, EPI_ISL_2129807, EPI_ISL_2129812,
EPI_ISL_2129813, EPI_ISL_2129814, EPI_ISL_2129815, EPI_ISL_2129817, EPI_ISL_2129821, EPI_ISL_2129822, EPI_ISL_2129823, EPI_ISL_2129825, EPI_ISL_2129827, EPI_ISL_2129829, EPI_ISL_2129830, EPI_ISL_2129831, EPI_ISL_2129832, EPI_ISL_2129833, EPI_ISL_2129835, EPI_ISL_2129836, EPI_ISL_2129837, EPI_ISL_2129838, EPI_ISL_2129839, EPI_ISL_2129840, EPI_ISL_2129841, EPI_ISL_2129845,
EPI_ISL_2129847, EPI_ISL_2129849, EPI_ISL_2129850, EPI_ISL_2129852, EPI_ISL_2129853, EPI_ISL_2129854, EPI_ISL_2129858, EPI_ISL_2129859, EPI_ISL_2129861, EPI_ISL_2129862, EPI_ISL_2129863, EPI_ISL_2129864, EPI_ISL_2129867, EPI_ISL_2129868, EPI_ISL_2129871, EPI_ISL_2129872, EPI_ISL_2129876, EPI_ISL_2129877, EPI_ISL_2129878, EPI_ISL_2129879, EPI_ISL_2129880, EPI_ISL_2129881,
EPI_ISL_2129883, EPI_ISL_2129885, EPI_ISL_2129886, EPI_ISL_2129887, EPI_ISL_2129888, EPI_ISL_2129889, EPI_ISL_2129890, EPI_ISL_2129892, EPI_ISL_2129897, EPI_ISL_2129899, EPI_ISL_2129901, EPI_ISL_2129903, EPI_ISL_2129904, EPI_ISL_2129905, EPI_ISL_2129908, EPI_ISL_2129909, EPI_ISL_2129911, EPI_ISL_2129913, EPI_ISL_2129915, EPI_ISL_2129917, EPI_ISL_2129918, EPI_ISL_2129920,
EPI_ISL_2129921, EPI_ISL_2129922, EPI_ISL_2129925, EPI_ISL_2129926, EPI_ISL_2129927, EPI_ISL_2129929, EPI_ISL_2129930, EPI_ISL_2129931, EPI_ISL_2129933, EPI_ISL_2129934, EPI_ISL_2129936, EPI_ISL_2129938, EPI_ISL_2129939, EPI_ISL_2129945, EPI_ISL_2129950, EPI_ISL_2129952, EPI_ISL_2129953, EPI_ISL_2129954, EPI_ISL_2129956, EPI_ISL_2129957, EPI_ISL_2129958, EPI_ISL_2129959,
EPI_ISL_2129960, EPI_ISL_2129961, EPI_ISL_2129962, EPI_ISL_2129963, EPI_ISL_2129966, EPI_ISL_2129971, EPI_ISL_2129972, EPI_ISL_2129973, EPI_ISL_2129974, EPI_ISL_2129975, EPI_ISL_2129976, EPI_ISL_2129978

see above SYNLAB MVZ Leinfelden-Echterdingen Robert Koch Institute

EPI_ISL_1852487, EPI_ISL_1852490, EPI_ISL_1852491, EPI_ISL_1852492, EPI_ISL_1852495, EPI_ISL_1852497, EPI_ISL_1852498, EPI_ISL_1852500, EPI_ISL_1852503, EPI_ISL_1852504, EPI_ISL_1852505, EPI_ISL_1852506, EPI_ISL_1852509, EPI_ISL_1852512, EPI_ISL_1852516, EPI_ISL_1852522, EPI_ISL_1852524, EPI_ISL_1852525, EPI_ISL_1852529, EPI_ISL_1852530, EPI_ISL_1852531, EPI_ISL_1852532,
EPI_ISL_1852536, EPI_ISL_1852537, EPI_ISL_1852539, EPI_ISL_1852541, EPI_ISL_1852542, EPI_ISL_1852543, EPI_ISL_1852544, EPI_ISL_1852545, EPI_ISL_1852546, EPI_ISL_1852547, EPI_ISL_1852549, EPI_ISL_1852552, EPI_ISL_1852554, EPI_ISL_1852555, EPI_ISL_1852557, EPI_ISL_1852558, EPI_ISL_1852559, EPI_ISL_1852560, EPI_ISL_1852561, EPI_ISL_1852565, EPI_ISL_1852567, EPI_ISL_1852570,
EPI_ISL_1852571, EPI_ISL_1852572, EPI_ISL_1852573, EPI_ISL_1852579, EPI_ISL_1852580, EPI_ISL_1852586, EPI_ISL_1852588, EPI_ISL_1852591, EPI_ISL_1852592, EPI_ISL_1852594, EPI_ISL_1852595, EPI_ISL_1852596, EPI_ISL_1852599, EPI_ISL_1852601, EPI_ISL_1852602, EPI_ISL_1852603, EPI_ISL_1852604, EPI_ISL_1852605, EPI_ISL_1852606, EPI_ISL_1852608, EPI_ISL_1852610, EPI_ISL_1852611,
EPI_ISL_1852612, EPI_ISL_1852613, EPI_ISL_1852614, EPI_ISL_1852615, EPI_ISL_1852617, EPI_ISL_1852618, EPI_ISL_1852620, EPI_ISL_1852623, EPI_ISL_1852625, EPI_ISL_1852626, EPI_ISL_1852627, EPI_ISL_1852628, EPI_ISL_1852629, EPI_ISL_1852630, EPI_ISL_1852636, EPI_ISL_1852642, EPI_ISL_1852644, EPI_ISL_1852645, EPI_ISL_1852649, EPI_ISL_1852652, EPI_ISL_1852653, EPI_ISL_1852654,
EPI_ISL_1852656, EPI_ISL_1852659, EPI_ISL_1852660, EPI_ISL_1852664, EPI_ISL_1852665, EPI_ISL_1852666, EPI_ISL_1852671, EPI_ISL_1852674, EPI_ISL_1852675, EPI_ISL_1852678, EPI_ISL_1852679, EPI_ISL_1852682, EPI_ISL_1852684, EPI_ISL_1852685, EPI_ISL_1852686, EPI_ISL_1852687, EPI_ISL_1852690, EPI_ISL_1852691, EPI_ISL_1852692, EPI_ISL_1852693, EPI_ISL_1852696, EPI_ISL_1852697,
EPI_ISL_1852698, EPI_ISL_1852699, EPI_ISL_1852700, EPI_ISL_1852701, EPI_ISL_1852702, EPI_ISL_1852706, EPI_ISL_1852708, EPI_ISL_1852710, EPI_ISL_1852713, EPI_ISL_1852716, EPI_ISL_1852718, EPI_ISL_1852722, EPI_ISL_1852725, EPI_ISL_1852727, EPI_ISL_1852729, EPI_ISL_1852734, EPI_ISL_1852744, EPI_ISL_1852746, EPI_ISL_1852749, EPI_ISL_1852750, EPI_ISL_1852753, EPI_ISL_1852761,
EPI_ISL_1852762, EPI_ISL_1852767, EPI_ISL_1852768, EPI_ISL_1852770, EPI_ISL_1852771, EPI_ISL_1852773, EPI_ISL_1852774, EPI_ISL_1852775, EPI_ISL_1852779, EPI_ISL_1852781, EPI_ISL_1852782, EPI_ISL_1852784, EPI_ISL_1852786, EPI_ISL_1852787, EPI_ISL_1852788, EPI_ISL_1852790, EPI_ISL_1852791, EPI_ISL_1852793, EPI_ISL_1852795, EPI_ISL_1852797, EPI_ISL_1852800, EPI_ISL_1852804,
EPI_ISL_1852805, EPI_ISL_1852810, EPI_ISL_1852811, EPI_ISL_1852812, EPI_ISL_1852813, EPI_ISL_1852814, EPI_ISL_1852818, EPI_ISL_1852824, EPI_ISL_1852825, EPI_ISL_1852831, EPI_ISL_1852833, EPI_ISL_1852835, EPI_ISL_1852839, EPI_ISL_1852841, EPI_ISL_1852845, EPI_ISL_1852846, EPI_ISL_1852848, EPI_ISL_1852850, EPI_ISL_1852852, EPI_ISL_1852857, EPI_ISL_1852858, EPI_ISL_1852862,
EPI_ISL_1852864, EPI_ISL_1852870, EPI_ISL_1852871, EPI_ISL_1852876, EPI_ISL_1852877, EPI_ISL_1852879, EPI_ISL_1852880, EPI_ISL_1852884, EPI_ISL_1852887, EPI_ISL_1852888, EPI_ISL_1852890, EPI_ISL_1852891, EPI_ISL_1852893, EPI_ISL_1852894, EPI_ISL_1852896, EPI_ISL_1852897, EPI_ISL_1852899, EPI_ISL_1852901, EPI_ISL_1852902, EPI_ISL_1852904, EPI_ISL_1852906, EPI_ISL_1852909,
EPI_ISL_1852911, EPI_ISL_1852913, EPI_ISL_1852914, EPI_ISL_1852916, EPI_ISL_1852917, EPI_ISL_1852920, EPI_ISL_1852921, EPI_ISL_1852923, EPI_ISL_1852925, EPI_ISL_1852927, EPI_ISL_1852928, EPI_ISL_1852930, EPI_ISL_1852931, EPI_ISL_1852933, EPI_ISL_1852934, EPI_ISL_1852937, EPI_ISL_1852938, EPI_ISL_1852942, EPI_ISL_1852944, EPI_ISL_1852948, EPI_ISL_1852949, EPI_ISL_1852953,
EPI_ISL_1852954, EPI_ISL_1852955, EPI_ISL_1852957, EPI_ISL_1852958, EPI_ISL_1852962, EPI_ISL_1852965, EPI_ISL_1852968, EPI_ISL_1852970, EPI_ISL_1852971, EPI_ISL_1852972, EPI_ISL_1852973, EPI_ISL_1852974, EPI_ISL_1852976, EPI_ISL_1852977, EPI_ISL_1852978, EPI_ISL_1852981, EPI_ISL_1852983, EPI_ISL_1852984, EPI_ISL_1852986, EPI_ISL_1852988, EPI_ISL_1852990, EPI_ISL_1852993,
EPI_ISL_1852994, EPI_ISL_1852999, EPI_ISL_1853000, EPI_ISL_1853002, EPI_ISL_1853004, EPI_ISL_1853005, EPI_ISL_1853006, EPI_ISL_1853010, EPI_ISL_1853011, EPI_ISL_1853012, EPI_ISL_1853015, EPI_ISL_1853016, EPI_ISL_1853017, EPI_ISL_1853020, EPI_ISL_1853026, EPI_ISL_1853028, EPI_ISL_1853029, EPI_ISL_1853032, EPI_ISL_1853034, EPI_ISL_1853035, EPI_ISL_1853036, EPI_ISL_1853037,
EPI_ISL_1853043, EPI_ISL_1853044, EPI_ISL_1853045, EPI_ISL_1853046, EPI_ISL_1853048, EPI_ISL_1853049, EPI_ISL_1853050, EPI_ISL_1853051, EPI_ISL_1853052, EPI_ISL_1853053, EPI_ISL_1853054, EPI_ISL_1853056, EPI_ISL_1853058, EPI_ISL_1853059, EPI_ISL_1853060, EPI_ISL_1853061, EPI_ISL_1853062, EPI_ISL_1853064, EPI_ISL_1853067, EPI_ISL_1853069, EPI_ISL_1853075, EPI_ISL_1853076,
EPI_ISL_1853078, EPI_ISL_1853079, EPI_ISL_1853080, EPI_ISL_1853082, EPI_ISL_1853083, EPI_ISL_1853084, EPI_ISL_1853085, EPI_ISL_1853086, EPI_ISL_1853089, EPI_ISL_1853090, EPI_ISL_1853091, EPI_ISL_1853094, EPI_ISL_1853096, EPI_ISL_1853098, EPI_ISL_2111606, EPI_ISL_2111607, EPI_ISL_2111608, EPI_ISL_2111609, EPI_ISL_2111610, EPI_ISL_2111613, EPI_ISL_2111616, EPI_ISL_2111617,
EPI_ISL_2111618, EPI_ISL_2111619, EPI_ISL_2111620, EPI_ISL_2111621, EPI_ISL_2111622, EPI_ISL_2111623, EPI_ISL_2111624, EPI_ISL_2111627, EPI_ISL_2111629, EPI_ISL_2111630, EPI_ISL_2111632, EPI_ISL_2111634, EPI_ISL_2111635, EPI_ISL_2111636, EPI_ISL_2111638, EPI_ISL_2111639, EPI_ISL_2111640, EPI_ISL_2111642, EPI_ISL_2111643, EPI_ISL_2111646, EPI_ISL_2111648, EPI_ISL_2111649,
EPI_ISL_2111650, EPI_ISL_2111651, EPI_ISL_2111652, EPI_ISL_2111655, EPI_ISL_2111656, EPI_ISL_2111657, EPI_ISL_2111659, EPI_ISL_2111661, EPI_ISL_2111662, EPI_ISL_2111665, EPI_ISL_2111667, EPI_ISL_2111668, EPI_ISL_2111670, EPI_ISL_2111671, EPI_ISL_2111673, EPI_ISL_2111674, EPI_ISL_2111675, EPI_ISL_2111676, EPI_ISL_2111677, EPI_ISL_2111678, EPI_ISL_2111681, EPI_ISL_2111682,
EPI_ISL_2111683, EPI_ISL_2111685, EPI_ISL_2111686, EPI_ISL_2111688, EPI_ISL_2111689, EPI_ISL_2111690, EPI_ISL_2111691, EPI_ISL_2111692, EPI_ISL_2111693, EPI_ISL_2111694, EPI_ISL_2111696, EPI_ISL_2111697, EPI_ISL_2111698, EPI_ISL_2111699, EPI_ISL_2111700, EPI_ISL_2111703, EPI_ISL_2111704, EPI_ISL_2111706, EPI_ISL_2111707, EPI_ISL_2111708, EPI_ISL_2111709, EPI_ISL_2111710,
EPI_ISL_2111712, EPI_ISL_2111715, EPI_ISL_2111716, EPI_ISL_2111718, EPI_ISL_2111719, EPI_ISL_2111720, EPI_ISL_2111721, EPI_ISL_2111722, EPI_ISL_2111723, EPI_ISL_2111726, EPI_ISL_2111727, EPI_ISL_2111730, EPI_ISL_2111731, EPI_ISL_2111735, EPI_ISL_2111737, EPI_ISL_2111738, EPI_ISL_2111739, EPI_ISL_2111740, EPI_ISL_2111741, EPI_ISL_2111742, EPI_ISL_2111743, EPI_ISL_2111746,
EPI_ISL_2111747, EPI_ISL_2111748, EPI_ISL_2111749, EPI_ISL_2111751, EPI_ISL_2111753, EPI_ISL_2111755, EPI_ISL_2111759, EPI_ISL_2111760, EPI_ISL_2111761, EPI_ISL_2111762, EPI_ISL_2111763, EPI_ISL_2111764, EPI_ISL_2111765, EPI_ISL_2111768, EPI_ISL_2111769, EPI_ISL_2111770, EPI_ISL_2111773, EPI_ISL_2111776, EPI_ISL_2111777, EPI_ISL_2111779, EPI_ISL_2111780, EPI_ISL_2111782,
EPI_ISL_2111784, EPI_ISL_2111785, EPI_ISL_2111788, EPI_ISL_2111791, EPI_ISL_2111794, EPI_ISL_2111797, EPI_ISL_2111801, EPI_ISL_2111802, EPI_ISL_2111803, EPI_ISL_2111805, EPI_ISL_2111806, EPI_ISL_2111807, EPI_ISL_2111808, EPI_ISL_2111809, EPI_ISL_2111811, EPI_ISL_2111812, EPI_ISL_2111813, EPI_ISL_2111816, EPI_ISL_2111818, EPI_ISL_2111819, EPI_ISL_2111820, EPI_ISL_2111821,
EPI_ISL_2111822, EPI_ISL_2111823, EPI_ISL_2111826, EPI_ISL_2111827, EPI_ISL_2111828, EPI_ISL_2111829, EPI_ISL_2111830, EPI_ISL_2111831, EPI_ISL_2111832, EPI_ISL_2111833, EPI_ISL_2111834, EPI_ISL_2111836, EPI_ISL_2111838, EPI_ISL_2111839, EPI_ISL_2111840, EPI_ISL_2111842, EPI_ISL_2111843, EPI_ISL_2111845, EPI_ISL_2111847, EPI_ISL_2111848, EPI_ISL_2111851, EPI_ISL_2111852,
EPI_ISL_2111857, EPI_ISL_2111859, EPI_ISL_2111860, EPI_ISL_2111862, EPI_ISL_2111863, EPI_ISL_2111864, EPI_ISL_2111866, EPI_ISL_2111867, EPI_ISL_2111868, EPI_ISL_2111869, EPI_ISL_2111870, EPI_ISL_2111873, EPI_ISL_2111875, EPI_ISL_2111876, EPI_ISL_2111878, EPI_ISL_2111880, EPI_ISL_2111881, EPI_ISL_2111882, EPI_ISL_2111883, EPI_ISL_2111885, EPI_ISL_2111887, EPI_ISL_2111888,
EPI_ISL_2111892, EPI_ISL_2111893, EPI_ISL_2111894, EPI_ISL_2111895, EPI_ISL_2111896, EPI_ISL_2111897, EPI_ISL_2111898, EPI_ISL_2111901, EPI_ISL_2111904, EPI_ISL_2111907, EPI_ISL_2111908, EPI_ISL_2111909, EPI_ISL_2111910, EPI_ISL_2111911, EPI_ISL_2111912, EPI_ISL_2111914, EPI_ISL_2111916, EPI_ISL_2111917, EPI_ISL_2111919, EPI_ISL_2111922, EPI_ISL_2111923, EPI_ISL_2111926,
EPI_ISL_2111929, EPI_ISL_2111930, EPI_ISL_2111933, EPI_ISL_2111939, EPI_ISL_2111942, EPI_ISL_2111944, EPI_ISL_2111945, EPI_ISL_2111946, EPI_ISL_2111950, EPI_ISL_2111952, EPI_ISL_2111953, EPI_ISL_2111955, EPI_ISL_2111956, EPI_ISL_2111959, EPI_ISL_2111960, EPI_ISL_2111962, EPI_ISL_2111963, EPI_ISL_2111964, EPI_ISL_2111966, EPI_ISL_2111968, EPI_ISL_2111969, EPI_ISL_2111972,
EPI_ISL_2111973, EPI_ISL_2111975, EPI_ISL_2111976, EPI_ISL_2111977, EPI_ISL_2111979, EPI_ISL_2111980, EPI_ISL_2111981, EPI_ISL_2111982, EPI_ISL_2111984, EPI_ISL_2111986, EPI_ISL_2111988, EPI_ISL_2111989, EPI_ISL_2111990, EPI_ISL_2111991, EPI_ISL_2111992, EPI_ISL_2111993, EPI_ISL_2111994, EPI_ISL_2111995, EPI_ISL_2111997, EPI_ISL_2111998, EPI_ISL_2111999, EPI_ISL_2112000,
EPI_ISL_2112003, EPI_ISL_2112004, EPI_ISL_2112005, EPI_ISL_2112006, EPI_ISL_2112007, EPI_ISL_2112008, EPI_ISL_2112010, EPI_ISL_2112011, EPI_ISL_2112012, EPI_ISL_2112013, EPI_ISL_2112015, EPI_ISL_2112016, EPI_ISL_2112017, EPI_ISL_2112018, EPI_ISL_2112020, EPI_ISL_2112021, EPI_ISL_2112023, EPI_ISL_2112024, EPI_ISL_2112025, EPI_ISL_2112026, EPI_ISL_2112029, EPI_ISL_2112031,
EPI_ISL_2112034, EPI_ISL_2112037, EPI_ISL_2112039, EPI_ISL_2112041, EPI_ISL_2112042, EPI_ISL_2112047, EPI_ISL_2112048, EPI_ISL_2112049, EPI_ISL_2112050, EPI_ISL_2112051, EPI_ISL_2112052, EPI_ISL_2112054, EPI_ISL_2112057, EPI_ISL_2112058, EPI_ISL_2112059, EPI_ISL_2112062, EPI_ISL_2112063, EPI_ISL_2112065, EPI_ISL_2112067, EPI_ISL_2112069, EPI_ISL_2112071, EPI_ISL_2112072,
EPI_ISL_2112073, EPI_ISL_2112074, EPI_ISL_2112075, EPI_ISL_2112076, EPI_ISL_2112078, EPI_ISL_2112079, EPI_ISL_2112083, EPI_ISL_2112085, EPI_ISL_2112086, EPI_ISL_2112089, EPI_ISL_2112091, EPI_ISL_2112093, EPI_ISL_2112096, EPI_ISL_2112097, EPI_ISL_2112100, EPI_ISL_2112102, EPI_ISL_2112103, EPI_ISL_2112104, EPI_ISL_2112107, EPI_ISL_2112108, EPI_ISL_2112110,
EPI_ISL_2112111, EPI_ISL_2112112, EPI_ISL_2112114, EPI_ISL_2112116, EPI_ISL_2112117, EPI_ISL_2112119, EPI_ISL_2112121, EPI_ISL_2112122, EPI_ISL_2112125, EPI_ISL_2112126, EPI_ISL_2112127, EPI_ISL_2112128, EPI_ISL_2112129, EPI_ISL_2112130, EPI_ISL_2112131, EPI_ISL_2112132, EPI_ISL_2112133, EPI_ISL_2112134, EPI_ISL_2112135, EPI_ISL_2112137, EPI_ISL_2112140, EPI_ISL_2112142,
EPI_ISL_2112143, EPI_ISL_2112144, EPI_ISL_2112146, EPI_ISL_2112147, EPI_ISL_2112149, EPI_ISL_2112151, EPI_ISL_2112153, EPI_ISL_2112155, EPI_ISL_2112157, EPI_ISL_2112159, EPI_ISL_2112160, EPI_ISL_2112161, EPI_ISL_2112165, EPI_ISL_2112166, EPI_ISL_2112167, EPI_ISL_2112169, EPI_ISL_2112171, EPI_ISL_2112173, EPI_ISL_2112174, EPI_ISL_2112175, EPI_ISL_2112176, EPI_ISL_2112181,
EPI_ISL_2112182, EPI_ISL_2112183, EPI_ISL_2112184, EPI_ISL_2113236, EPI_ISL_2113237, EPI_ISL_2113238, EPI_ISL_2113239, EPI_ISL_2113240, EPI_ISL_2113242, EPI_ISL_2113244, EPI_ISL_2113245, EPI_ISL_2113247, EPI_ISL_2113248, EPI_ISL_2113249, EPI_ISL_2113250, EPI_ISL_2113251, EPI_ISL_2113253, EPI_ISL_2113254, EPI_ISL_2113255, EPI_ISL_2113256, EPI_ISL_2113257, EPI_ISL_2113258,
EPI_ISL_2113260, EPI_ISL_2113261, EPI_ISL_2113263, EPI_ISL_2113264, EPI_ISL_2113265, EPI_ISL_2113266, EPI_ISL_2113267, EPI_ISL_2113268, EPI_ISL_2113269, EPI_ISL_2113273, EPI_ISL_2113274, EPI_ISL_2113275, EPI_ISL_2113276, EPI_ISL_2113278, EPI_ISL_2113279, EPI_ISL_2113280, EPI_ISL_2113281, EPI_ISL_2113283, EPI_ISL_2113285, EPI_ISL_2113286, EPI_ISL_2113287, EPI_ISL_2113289,
EPI_ISL_2113290, EPI_ISL_2113292, EPI_ISL_2113295, EPI_ISL_2113296, EPI_ISL_2113297, EPI_ISL_2113298, EPI_ISL_2113299, EPI_ISL_2113300, EPI_ISL_2113301, EPI_ISL_2113302, EPI_ISL_2113303, EPI_ISL_2113304, EPI_ISL_2113305, EPI_ISL_2113306, EPI_ISL_2113307, EPI_ISL_2113308, EPI_ISL_2113309, EPI_ISL_2113310, EPI_ISL_2113311, EPI_ISL_2113312, EPI_ISL_2113313, EPI_ISL_2113314,
EPI_ISL_2113316, EPI_ISL_2113317, EPI_ISL_2113318, EPI_ISL_2113319, EPI_ISL_2113320, EPI_ISL_2113321, EPI_ISL_2113322, EPI_ISL_2113323, EPI_ISL_2113325, EPI_ISL_2113326, EPI_ISL_2113327, EPI_ISL_2113329, EPI_ISL_2113330, EPI_ISL_2113331, EPI_ISL_2113333, EPI_ISL_2113335, EPI_ISL_2113336, EPI_ISL_2113337, EPI_ISL_2113338, EPI_ISL_2113339, EPI_ISL_2113340, EPI_ISL_2113342,
EPI_ISL_2113343, EPI_ISL_2113344, EPI_ISL_2113345, EPI_ISL_2113346, EPI_ISL_2113347, EPI_ISL_2113348, EPI_ISL_2113349, EPI_ISL_2113350, EPI_ISL_2113353, EPI_ISL_2113355, EPI_ISL_2113357, EPI_ISL_2113358, EPI_ISL_2113359, EPI_ISL_2113360, EPI_ISL_2113361, EPI_ISL_2113362, EPI_ISL_2113363, EPI_ISL_2113364, EPI_ISL_2113365, EPI_ISL_2113367, EPI_ISL_2113368, EPI_ISL_2113369,
EPI_ISL_2113370, EPI_ISL_2113371, EPI_ISL_2113372, EPI_ISL_2113374, EPI_ISL_2113375, EPI_ISL_2113377, EPI_ISL_2113378, EPI_ISL_2113380, EPI_ISL_2113381, EPI_ISL_2113383, EPI_ISL_2113384, EPI_ISL_2113386, EPI_ISL_2113387, EPI_ISL_2113388, EPI_ISL_2113390, EPI_ISL_2113391, EPI_ISL_2113392, EPI_ISL_2113394, EPI_ISL_2113396, EPI_ISL_2113397, EPI_ISL_2113399, EPI_ISL_2113400,
EPI_ISL_2113401, EPI_ISL_2113402, EPI_ISL_2113403, EPI_ISL_2113405, EPI_ISL_2113406, EPI_ISL_2113408, EPI_ISL_2113411, EPI_ISL_2113412, EPI_ISL_2113416, EPI_ISL_2113417, EPI_ISL_2113418, EPI_ISL_2113420, EPI_ISL_2113422, EPI_ISL_2113423, EPI_ISL_2113424, EPI_ISL_2113425, EPI_ISL_2113426, EPI_ISL_2113427, EPI_ISL_2113428, EPI_ISL_2113431, EPI_ISL_2113432, EPI_ISL_2113433,
EPI_ISL_2113434, EPI_ISL_2113435, EPI_ISL_2113436, EPI_ISL_2113437, EPI_ISL_2113440, EPI_ISL_2113441, EPI_ISL_2113442, EPI_ISL_2113443, EPI_ISL_2113444, EPI_ISL_2113445, EPI_ISL_2113446, EPI_ISL_2113447, EPI_ISL_2113448, EPI_ISL_2113449, EPI_ISL_2113450, EPI_ISL_2113451, EPI_ISL_2113452, EPI_ISL_2113453, EPI_ISL_2113455, EPI_ISL_2113456, EPI_ISL_2113457, EPI_ISL_2113458,
EPI_ISL_2113459, EPI_ISL_2113461, EPI_ISL_2113462, EPI_ISL_2113463, EPI_ISL_2113465, EPI_ISL_2113466, EPI_ISL_2113469, EPI_ISL_2113470, EPI_ISL_2113471, EPI_ISL_2113472, EPI_ISL_2113474, EPI_ISL_2113475, EPI_ISL_2113476, EPI_ISL_2113477, EPI_ISL_2113478, EPI_ISL_2113480, EPI_ISL_2113482, EPI_ISL_2113483, EPI_ISL_2113484, EPI_ISL_2113486, EPI_ISL_2113487, EPI_ISL_2113488,
EPI_ISL_2113489, EPI_ISL_2113490, EPI_ISL_2113491, EPI_ISL_2113494, EPI_ISL_2113496, EPI_ISL_2113497, EPI_ISL_2113498, EPI_ISL_2113499, EPI_ISL_2113500, EPI_ISL_2113501, EPI_ISL_2113502, EPI_ISL_2113504, EPI_ISL_2113505, EPI_ISL_2113506, EPI_ISL_2113508, EPI_ISL_2113509, EPI_ISL_2113510, EPI_ISL_2113511, EPI_ISL_2113514, EPI_ISL_2113515, EPI_ISL_2113516,
EPI_ISL_2113518, EPI_ISL_2113520, EPI_ISL_2113522, EPI_ISL_2113523, EPI_ISL_2113524, EPI_ISL_2113526, EPI_ISL_2113527, EPI_ISL_2113528, EPI_ISL_2113529, EPI_ISL_2113530, EPI_ISL_2113532, EPI_ISL_2113533, EPI_ISL_2113534, EPI_ISL_2113535, EPI_ISL_2113537, EPI_ISL_2113538, EPI_ISL_2113540, EPI_ISL_2113541, EPI_ISL_2113542, EPI_ISL_2113544, EPI_ISL_2113545, EPI_ISL_2113546,
EPI_ISL_2113547, EPI_ISL_2113548, EPI_ISL_2113549, EPI_ISL_2113550, EPI_ISL_2113551, EPI_ISL_2113552, EPI_ISL_2113553, EPI_ISL_2113555, EPI_ISL_2113556, EPI_ISL_2113557, EPI_ISL_2113558, EPI_ISL_2113559, EPI_ISL_2113561, EPI_ISL_2113562, EPI_ISL_2113563, EPI_ISL_2113564, EPI_ISL_2113566, EPI_ISL_2113567, EPI_ISL_2113568, EPI_ISL_2113569, EPI_ISL_2113570,
EPI_ISL_2113571, EPI_ISL_2113572, EPI_ISL_2113573, EPI_ISL_2113574, EPI_ISL_2113575, EPI_ISL_2113576, EPI_ISL_2113577, EPI_ISL_2113580, EPI_ISL_2113581, EPI_ISL_2113583, EPI_ISL_2113584, EPI_ISL_2113586, EPI_ISL_2113587, EPI_ISL_2113589, EPI_ISL_2113591, EPI_ISL_2113592, EPI_ISL_2113593, EPI_ISL_2113594, EPI_ISL_2113595, EPI_ISL_2113597, EPI_ISL_2113598, EPI_ISL_2113599,
EPI_ISL_2113600, EPI_ISL_2113603, EPI_ISL_2113605, EPI_ISL_2113606, EPI_ISL_2113607, EPI_ISL_2113609, EPI_ISL_2113610, EPI_ISL_2113612, EPI_ISL_2113614, EPI_ISL_2113615, EPI_ISL_2113616, EPI_ISL_2113617, EPI_ISL_2113621, EPI_ISL_2113622, EPI_ISL_2113624, EPI_ISL_2113625, EPI_ISL_2113626, EPI_ISL_2113627, EPI_ISL_2113628, EPI_ISL_2113629, EPI_ISL_2113630, EPI_ISL_2113631,
EPI_ISL_2113634, EPI_ISL_2113635, EPI_ISL_2113637, EPI_ISL_2113640, EPI_ISL_2113643, EPI_ISL_2113645, EPI_ISL_2113647, EPI_ISL_2113648, EPI_ISL_2113649, EPI_ISL_2113651, EPI_ISL_2113652, EPI_ISL_2113653, EPI_ISL_2113654, EPI_ISL_2113655, EPI_ISL_2113656, EPI_ISL_2113657, EPI_ISL_2113658, EPI_ISL_2113662, EPI_ISL_2113663, EPI_ISL_2113664, EPI_ISL_2113667, EPI_ISL_2113671,
EPI_ISL_2113674, EPI_ISL_2113675, EPI_ISL_2113677, EPI_ISL_2113680, EPI_ISL_2113681, EPI_ISL_2113682, EPI_ISL_2113683, EPI_ISL_2113686, EPI_ISL_2113687, EPI_ISL_2113688, EPI_ISL_2113690, EPI_ISL_2113692, EPI_ISL_2113695, EPI_ISL_2113696, EPI_ISL_2113697, EPI_ISL_2113698, EPI_ISL_2113699, EPI_ISL_2113700, EPI_ISL_2113701, EPI_ISL_2113702, EPI_ISL_2113703, EPI_ISL_2113704,
EPI_ISL_2113707, EPI_ISL_2113708, EPI_ISL_2113710, EPI_ISL_2113711, EPI_ISL_2113712, EPI_ISL_2113713, EPI_ISL_2113714, EPI_ISL_2113715, EPI_ISL_2113716, EPI_ISL_2113717, EPI_ISL_2113718, EPI_ISL_2113719, EPI_ISL_2113721, EPI_ISL_2113723, EPI_ISL_2113724, EPI_ISL_2113725, EPI_ISL_2113726, EPI_ISL_2113727, EPI_ISL_2113728, EPI_ISL_2113729, EPI_ISL_2113730, EPI_ISL_2113732,
EPI_ISL_2113733, EPI_ISL_2113735, EPI_ISL_2113736, EPI_ISL_2113737, EPI_ISL_2113739, EPI_ISL_2113740, EPI_ISL_2113742, EPI_ISL_2113744, EPI_ISL_2113745, EPI_ISL_2113747, EPI_ISL_2113748, EPI_ISL_2113749, EPI_ISL_2113751, EPI_ISL_2113752, EPI_ISL_2113753, EPI_ISL_2113754, EPI_ISL_2113755, EPI_ISL_2113756, EPI_ISL_2113758, EPI_ISL_2113759, EPI_ISL_2113761, EPI_ISL_2113763,
EPI_ISL_2113764, EPI_ISL_2113766, EPI_ISL_2113767, EPI_ISL_2113768, EPI_ISL_2113769, EPI_ISL_2113771, EPI_ISL_2113772, EPI_ISL_2113773, EPI_ISL_2113775, EPI_ISL_2113778, EPI_ISL_2113779, EPI_ISL_2113780, EPI_ISL_2113783, EPI_ISL_2113784, EPI_ISL_2113786, EPI_ISL_2113788, EPI_ISL_2113789, EPI_ISL_2113790, EPI_ISL_2113791, EPI_ISL_2113792, EPI_ISL_2113793, EPI_ISL_2113794,
EPI_ISL_2113795, EPI_ISL_2113796, EPI_ISL_2113797, EPI_ISL_2113799, EPI_ISL_2113800, EPI_ISL_2113801, EPI_ISL_2113803, EPI_ISL_2113804, EPI_ISL_2113805, EPI_ISL_2113806, EPI_ISL_2113808, EPI_ISL_2113809, EPI_ISL_2113810, EPI_ISL_2113811, EPI_ISL_2113812, EPI_ISL_2113813, EPI_ISL_2113816, EPI_ISL_2113818, EPI_ISL_2113819, EPI_ISL_2113820, EPI_ISL_2113821, EPI_ISL_2113822,
EPI_ISL_2113824, EPI_ISL_2113825, EPI_ISL_2113827, EPI_ISL_2113828, EPI_ISL_2113831, EPI_ISL_2113832, EPI_ISL_2113833, EPI_ISL_2113834, EPI_ISL_2113835, EPI_ISL_2113838, EPI_ISL_2113840, EPI_ISL_2113841, EPI_ISL_2113844, EPI_ISL_2113845, EPI_ISL_2113846, EPI_ISL_2113847, EPI_ISL_2113850, EPI_ISL_2113854, EPI_ISL_2113855, EPI_ISL_2113856, EPI_ISL_2113860, EPI_ISL_2113863,
EPI_ISL_2113864, EPI_ISL_2113868, EPI_ISL_2113871, EPI_ISL_2113874, EPI_ISL_2113875, EPI_ISL_2113876, EPI_ISL_2113877, EPI_ISL_2113880, EPI_ISL_2113882, EPI_ISL_2113884, EPI_ISL_2113886, EPI_ISL_2113888, EPI_ISL_2113890, EPI_ISL_2113891, EPI_ISL_2113895, EPI_ISL_2113896, EPI_ISL_2113897, EPI_ISL_2113898, EPI_ISL_2113900, EPI_ISL_2113901,
EPI_ISL_2113902, EPI_ISL_2113903, EPI_ISL_2113904, EPI_ISL_2113906, EPI_ISL_2113907, EPI_ISL_2113908, EPI_ISL_2113909, EPI_ISL_2113911, EPI_ISL_2113912, EPI_ISL_2113913, EPI_ISL_2113914, EPI_ISL_2113915, EPI_ISL_2113916, EPI_ISL_2113917, EPI_ISL_2113919, EPI_ISL_2113920, EPI_ISL_2113921, EPI_ISL_2113922, EPI_ISL_2113923, EPI_ISL_2113924, EPI_ISL_2113925, EPI_ISL_2113926,
EPI_ISL_2113927, EPI_ISL_2113929, EPI_ISL_2113930, EPI_ISL_2113932, EPI_ISL_2113933, EPI_ISL_2113934, EPI_ISL_2113935, EPI_ISL_2113936, EPI_ISL_2113937, EPI_ISL_2113938, EPI_ISL_2113939, EPI_ISL_2113940, EPI_ISL_2113941, EPI_ISL_2113942, EPI_ISL_2113943, EPI_ISL_2113944, EPI_ISL_2113945, EPI_ISL_2113947, EPI_ISL_2113948, EPI_ISL_2113949, EPI_ISL_2113950, EPI_ISL_2113951,
EPI_ISL_2113952, EPI_ISL_2113953, EPI_ISL_2113954, EPI_ISL_2113955, EPI_ISL_2113956, EPI_ISL_2113957, EPI_ISL_2113958, EPI_ISL_2113959, EPI_ISL_2113960, EPI_ISL_2113961, EPI_ISL_2113962, EPI_ISL_2113963, EPI_ISL_2113964, EPI_ISL_2113965, EPI_ISL_2113966, EPI_ISL_2113967, EPI_ISL_2113968, EPI_ISL_2113969, EPI_ISL_2113970, EPI_ISL_2113971, EPI_ISL_2113972, EPI_ISL_2113973,
EPI_ISL_2113974, EPI_ISL_2113975, EPI_ISL_2113976, EPI_ISL_2113977, EPI_ISL_2113978, EPI_ISL_2113979, EPI_ISL_2113980, EPI_ISL_2113981, EPI_ISL_2113982, EPI_ISL_2113983, EPI_ISL_2113985, EPI_ISL_2113986, EPI_ISL_2113987, EPI_ISL_2113988, EPI_ISL_2113990, EPI_ISL_2113991, EPI_ISL_2113992, EPI_ISL_2113993, EPI_ISL_2113995, EPI_ISL_2113997,
EPI_ISL_2113998, EPI_ISL_2114000, EPI_ISL_2114001, EPI_ISL_2114002, EPI_ISL_2114004, EPI_ISL_2114006, EPI_ISL_2114007, EPI_ISL_2114009, EPI_ISL_2114010, EPI_ISL_2114011, EPI_ISL_2114014, EPI_ISL_2114015, EPI_ISL_2114016, EPI_ISL_2114017, EPI_ISL_2114018, EPI_ISL_2114019, EPI_ISL_2114020, EPI_ISL_2114021, EPI_ISL_2114022, EPI_ISL_2114023, EPI_ISL_2114024, EPI_ISL_2114025,
EPI_ISL_2114026, EPI_ISL_2114027, EPI_ISL_2114029, EPI_ISL_2114030, EPI_ISL_2114031, EPI_ISL_2114032, EPI_ISL_2114033, EPI_ISL_2114034, EPI_ISL_2114035, EPI_ISL_2114036, EPI_ISL_2114037, EPI_ISL_2114038, EPI_ISL_2114040, EPI_ISL_2114041, EPI_ISL_2114042, EPI_ISL_2114043, EPI_ISL_2114044, EPI_ISL_2114045, EPI_ISL_2114046, EPI_ISL_2114047, EPI_ISL_2114048, EPI_ISL_2114049,
EPI_ISL_2114050, EPI_ISL_2114052, EPI_ISL_2114053, EPI_ISL_2114054, EPI_ISL_2114055, EPI_ISL_2114056, EPI_ISL_2114057, EPI_ISL_2114058, EPI_ISL_2114059, EPI_ISL_2114060, EPI_ISL_2114061, EPI_ISL_2114062, EPI_ISL_2114063, EPI_ISL_2114064, EPI_ISL_2114065, EPI_ISL_2114066, EPI_ISL_2114067, EPI_ISL_2114068, EPI_ISL_2114069, EPI_ISL_2114070, EPI_ISL_2114071, EPI_ISL_2114072,
EPI_ISL_2114074, EPI_ISL_2114075, EPI_ISL_2114076, EPI_ISL_2114077, EPI_ISL_2114078, EPI_ISL_2114079, EPI_ISL_2114080, EPI_ISL_2114081, EPI_ISL_2114082, EPI_ISL_2114083, EPI_ISL_2114084, EPI_ISL_2114085, EPI_ISL_2114086, EPI_ISL_2114087, EPI_ISL_2114088, EPI_ISL_2114089, EPI_ISL_2114090, EPI_ISL_2114091, EPI_ISL_2114092, EPI_ISL_2114093, EPI_ISL_2114094, EPI_ISL_2114095, EPI_ISL_2114096,
EPI_ISL_2114097, EPI_ISL_2114098, EPI_ISL_2114099, EPI_ISL_2114100, EPI_ISL_2114101, EPI_ISL_2114103, EPI_ISL_2114104, EPI_ISL_2114106, EPI_ISL_2114107, EPI_ISL_2114108, EPI_ISL_2114109, EPI_ISL_2114110, EPI_ISL_2114111, EPI_ISL_2114112, EPI_ISL_2114114, EPI_ISL_2114116, EPI_ISL_2114117, EPI_ISL_2114118, EPI_ISL_2114119, EPI_ISL_2114120, EPI_ISL_2114122, EPI_ISL_2114124,
EPI_ISL_2114125, EPI_ISL_2114126, EPI_ISL_2114127, EPI_ISL_2114128, EPI_ISL_2114129, EPI_ISL_2114130, EPI_ISL_2114131, EPI_ISL_2114133, EPI_ISL_2114134, EPI_ISL_2114135, EPI_ISL_2114136, EPI_ISL_2114137, EPI_ISL_2114138, EPI_ISL_2114139, EPI_ISL_2114140, EPI_ISL_2114141, EPI_ISL_2114142, EPI_ISL_2114143, EPI_ISL_2114144, EPI_ISL_2114145, EPI_ISL_2114146,
EPI_ISL_2114147, EPI_ISL_2114148, EPI_ISL_2114149, EPI_ISL_2114150, EPI_ISL_2114151, EPI_ISL_2114152, EPI_ISL_2114153, EPI_ISL_2114154, EPI_ISL_2114155, EPI_ISL_2114156, EPI_ISL_2114157, EPI_ISL_2114158, EPI_ISL_2114159, EPI_ISL_2114160, EPI_ISL_2114161, EPI_ISL_2114162, EPI_ISL_2114163, EPI_ISL_2114164, EPI_ISL_2114165, EPI_ISL_2114166, EPI_ISL_2114167, EPI_ISL_2114168,
EPI_ISL_2114169, EPI_ISL_2114170, EPI_ISL_2114171, EPI_ISL_2114172, EPI_ISL_2114174, EPI_ISL_2114175, EPI_ISL_2114176, EPI_ISL_2114177, EPI_ISL_2114178, EPI_ISL_2114179, EPI_ISL_2114180, EPI_ISL_2114181, EPI_ISL_2114182, EPI_ISL_2114183, EPI_ISL_2114184, EPI_ISL_2114185, EPI_ISL_2114186, EPI_ISL_2114188, EPI_ISL_2114189, EPI_ISL_2114190, EPI_ISL_2114191, EPI_ISL_2114192,
EPI_ISL_2114193, EPI_ISL_2114194, EPI_ISL_2114195, EPI_ISL_2114196, EPI_ISL_2114197, EPI_ISL_2114198, EPI_ISL_2114199, EPI_ISL_2114200, EPI_ISL_2114201, EPI_ISL_2114202, EPI_ISL_2114203, EPI_ISL_2114204, EPI_ISL_2114205, EPI_ISL_2114206, EPI_ISL_2114207, EPI_ISL_2114208, EPI_ISL_2114209, EPI_ISL_2114210, EPI_ISL_2114211, EPI_ISL_2114212, EPI_ISL_2114213, EPI_ISL_2114214,
EPI_ISL_2114215, EPI_ISL_2114216, EPI_ISL_2114217, EPI_ISL_2114219, EPI_ISL_2114220, EPI_ISL_2114221, EPI_ISL_2114222, EPI_ISL_2114223, EPI_ISL_2114224, EPI_ISL_2114225, EPI_ISL_2114226, EPI_ISL_2114227, EPI_ISL_2114228, EPI_ISL_2114229, EPI_ISL_2114230, EPI_ISL_2114231, EPI_ISL_2114232, EPI_ISL_2114233, EPI_ISL_2114235, EPI_ISL_2114236, EPI_ISL_2114237, EPI_ISL_2114238,

| | | | |
|---|---|---|---|
| EPI_ISL_2114239, EPI_ISL_2114240, EPI_ISL_2114241, EPI_ISL_2114242, EPI_ISL_2114243, EPI_ISL_2114244, EPI_ISL_2114245, EPI_ISL_2114246, EPI_ISL_2114247, EPI_ISL_2114248, EPI_ISL_2114249, EPI_ISL_2114250, EPI_ISL_2114252, EPI_ISL_2114254, EPI_ISL_2114256, EPI_ISL_2114257, EPI_ISL_2114258, EPI_ISL_2114260, EPI_ISL_2114261, EPI_ISL_2114262, EPI_ISL_2114263, EPI_ISL_2114264, EPI_ISL_2114265, EPI_ISL_2114266, EPI_ISL_2114268, EPI_ISL_2114269, EPI_ISL_2114270, EPI_ISL_2114271, EPI_ISL_2114272, EPI_ISL_2114273, EPI_ISL_2114274, EPI_ISL_2114276, EPI_ISL_2114277, EPI_ISL_2114278, EPI_ISL_2114279, EPI_ISL_2114280, EPI_ISL_2114281, EPI_ISL_2114282, EPI_ISL_2114283, EPI_ISL_2114284, EPI_ISL_2114285, EPI_ISL_2114286, EPI_ISL_2114287, EPI_ISL_2114288, EPI_ISL_2114289, EPI_ISL_2114290, EPI_ISL_2114291, EPI_ISL_2114292, EPI_ISL_2114293, EPI_ISL_2114294, EPI_ISL_2114295, EPI_ISL_2114296, EPI_ISL_2114297, EPI_ISL_2114298, EPI_ISL_2114299, EPI_ISL_2114300, EPI_ISL_2114301, EPI_ISL_2114302, EPI_ISL_2114303, EPI_ISL_2114304, EPI_ISL_2114305, EPI_ISL_2114306, EPI_ISL_2114307, EPI_ISL_2114308, EPI_ISL_2114309, EPI_ISL_2114310, EPI_ISL_2114311, EPI_ISL_2114312, EPI_ISL_2114313, EPI_ISL_2114315, EPI_ISL_2114316, EPI_ISL_2114317, EPI_ISL_2114318, EPI_ISL_2114319, EPI_ISL_2114320, EPI_ISL_2114321, EPI_ISL_2114322, EPI_ISL_2114323, EPI_ISL_2114324, EPI_ISL_2114325, EPI_ISL_2114326, EPI_ISL_2114327, EPI_ISL_2114328, EPI_ISL_2114329, EPI_ISL_2114330, EPI_ISL_2114331, EPI_ISL_2114332, EPI_ISL_2114333, EPI_ISL_2114334, EPI_ISL_2114335, EPI_ISL_2114336, EPI_ISL_2114337, EPI_ISL_2114338, EPI_ISL_2114339, EPI_ISL_2114340, EPI_ISL_2114341, EPI_ISL_2114342, EPI_ISL_2114343, EPI_ISL_2114344, EPI_ISL_2114345, EPI_ISL_2114346, EPI_ISL_2114347, EPI_ISL_2114348, EPI_ISL_2114349, EPI_ISL_2114350, EPI_ISL_2114351, EPI_ISL_2114352, EPI_ISL_2114353, EPI_ISL_2114354, EPI_ISL_2114355, EPI_ISL_2114356, EPI_ISL_2114357, EPI_ISL_2114358, EPI_ISL_2114359, EPI_ISL_2114360, EPI_ISL_2114361, EPI_ISL_2114362, EPI_ISL_2114363, EPI_ISL_2114364, EPI_ISL_2114365, EPI_ISL_2114366, EPI_ISL_2114367, EPI_ISL_2114368, EPI_ISL_2114369, EPI_ISL_2114370, EPI_ISL_2114371, EPI_ISL_2114372, EPI_ISL_2114373, EPI_ISL_2114376, EPI_ISL_2114377, EPI_ISL_2114378, EPI_ISL_2114379, EPI_ISL_2114380, EPI_ISL_2114381, EPI_ISL_2114382, EPI_ISL_2114383, EPI_ISL_2114384, EPI_ISL_2114385, EPI_ISL_2114386, EPI_ISL_2114387, EPI_ISL_2114388, EPI_ISL_2114389, EPI_ISL_2114390, EPI_ISL_2114391, EPI_ISL_2114392, EPI_ISL_2114393, EPI_ISL_2114394, EPI_ISL_2114395, EPI_ISL_2114396, EPI_ISL_2114397, EPI_ISL_2114398, EPI_ISL_2114399, EPI_ISL_2114400, EPI_ISL_2114401, EPI_ISL_2114403, EPI_ISL_2114404, EPI_ISL_2114405, EPI_ISL_2114406, EPI_ISL_2114407, EPI_ISL_2114408, EPI_ISL_2114409, EPI_ISL_2114410, EPI_ISL_2114411, EPI_ISL_2114413, EPI_ISL_2114414, EPI_ISL_2114415, EPI_ISL_2114416, EPI_ISL_2114417, EPI_ISL_2114418, EPI_ISL_2114419, EPI_ISL_2114420, EPI_ISL_2114421, EPI_ISL_2114422, EPI_ISL_2114424, EPI_ISL_2114425, EPI_ISL_2114426, EPI_ISL_2114427, EPI_ISL_2114428, EPI_ISL_2114429, EPI_ISL_2114430, EPI_ISL_2114431, EPI_ISL_2114432, EPI_ISL_2114433, EPI_ISL_2114434, EPI_ISL_2114435, EPI_ISL_2114436, EPI_ISL_2114437, EPI_ISL_2114438, EPI_ISL_2114439, EPI_ISL_2114440, EPI_ISL_2114441, EPI_ISL_2114442, EPI_ISL_2114443, EPI_ISL_2114444, EPI_ISL_2114445, EPI_ISL_2114446, EPI_ISL_2114447, EPI_ISL_2114448, EPI_ISL_2114449, EPI_ISL_2114450, EPI_ISL_2114451, EPI_ISL_2114452, EPI_ISL_2114455, EPI_ISL_2114456, EPI_ISL_2114457, EPI_ISL_2114458, EPI_ISL_2114459, EPI_ISL_2114460, EPI_ISL_2114461, EPI_ISL_2114462, EPI_ISL_2114463, EPI_ISL_2114464, EPI_ISL_2114465, EPI_ISL_2114466, EPI_ISL_2114467, EPI_ISL_2114468, EPI_ISL_2114469, EPI_ISL_2114470, EPI_ISL_2114471, EPI_ISL_2114472, EPI_ISL_2114473, EPI_ISL_2114474, EPI_ISL_2114475, EPI_ISL_2114476, EPI_ISL_2114477, EPI_ISL_2114478, EPI_ISL_2114479, EPI_ISL_2114480, EPI_ISL_2114481, EPI_ISL_2114482, EPI_ISL_2114483, EPI_ISL_2114484, EPI_ISL_2114485, EPI_ISL_2114486, EPI_ISL_2114487, EPI_ISL_2114488, EPI_ISL_2114489, EPI_ISL_2114490, EPI_ISL_2114491, EPI_ISL_2114492, EPI_ISL_2114493, EPI_ISL_2114494, EPI_ISL_2114495, EPI_ISL_2114496, EPI_ISL_2114497, EPI_ISL_2114498, EPI_ISL_2114499, EPI_ISL_2114500, EPI_ISL_2114501, EPI_ISL_2114502, EPI_ISL_2114504, EPI_ISL_2114505, EPI_ISL_2114506, EPI_ISL_2114507, EPI_ISL_2114508, EPI_ISL_2114509, EPI_ISL_2114510, EPI_ISL_2114511, EPI_ISL_2114512, EPI_ISL_2114513, EPI_ISL_2116657, EPI_ISL_2116661, EPI_ISL_2116668, EPI_ISL_2116669, EPI_ISL_2116676, EPI_ISL_2116689, EPI_ISL_2116706, EPI_ISL_2116709, EPI_ISL_2116721, EPI_ISL_2116723, EPI_ISL_2116747, EPI_ISL_2116751, EPI_ISL_2116777, EPI_ISL_2116809, EPI_ISL_2116820, EPI_ISL_2116824, EPI_ISL_2116834, EPI_ISL_2116835, EPI_ISL_2116838, EPI_ISL_2116846, EPI_ISL_2116852, EPI_ISL_2116864, EPI_ISL_2116868, EPI_ISL_2116873, EPI_ISL_2116905, EPI_ISL_2116917, EPI_ISL_2116931, EPI_ISL_2116951, EPI_ISL_2116952, EPI_ISL_2116955, EPI_ISL_2116968, EPI_ISL_2116979, EPI_ISL_2116980, EPI_ISL_2116982, EPI_ISL_2117003, EPI_ISL_2117011, EPI_ISL_2117015, EPI_ISL_2117017, EPI_ISL_2117027, EPI_ISL_2117029, EPI_ISL_2117035, EPI_ISL_2117047, EPI_ISL_2117055, EPI_ISL_2117056, EPI_ISL_2117062, EPI_ISL_2117069, EPI_ISL_2117070, EPI_ISL_2117073, EPI_ISL_2117088, EPI_ISL_2117091, EPI_ISL_2117105, EPI_ISL_2117106, EPI_ISL_2117109, EPI_ISL_2117120, EPI_ISL_2117134, EPI_ISL_2117136, EPI_ISL_2117142, EPI_ISL_2117150, EPI_ISL_2117152, EPI_ISL_2117157, EPI_ISL_2117182, EPI_ISL_2117183, EPI_ISL_2117184, EPI_ISL_2117185, EPI_ISL_2117186, EPI_ISL_2117200, EPI_ISL_2117201, EPI_ISL_2117206, EPI_ISL_2117214, EPI_ISL_2117215, EPI_ISL_2117224, EPI_ISL_2117227, EPI_ISL_2117231, EPI_ISL_2117237, EPI_ISL_2117249, EPI_ISL_2117257, EPI_ISL_2117261, EPI_ISL_2117269, EPI_ISL_2117278, EPI_ISL_2117283, EPI_ISL_2117286, EPI_ISL_2117289, EPI_ISL_2117290, EPI_ISL_2117294 | | | |
| see above | SYNLAB MVZ Leverkusen | Robert Koch Institute | |
| EPI_ISL_2130835, EPI_ISL_2130838, EPI_ISL_2130844, EPI_ISL_2130856, EPI_ISL_2130940, EPI_ISL_2130971, EPI_ISL_2130981, EPI_ISL_2131001, EPI_ISL_2131011, EPI_ISL_2131020, EPI_ISL_2131067, EPI_ISL_2131136 | | | |
| see above | SYNLAB MVZ Neustadt a. d. WeinstraÃŸe | Robert Koch Institute | |
| EPI_ISL_2127697, EPI_ISL_2127698, EPI_ISL_2127700, EPI_ISL_2127701, EPI_ISL_2127710, EPI_ISL_2127724, EPI_ISL_2127727, EPI_ISL_2127744, EPI_ISL_2127751, EPI_ISL_2127775, EPI_ISL_2127779, EPI_ISL_2127808, EPI_ISL_2127890, EPI_ISL_2127894, EPI_ISL_2127911, EPI_ISL_2127923, EPI_ISL_2127933, EPI_ISL_2127935, EPI_ISL_2127939, EPI_ISL_2127944, EPI_ISL_2127949, EPI_ISL_2127951, EPI_ISL_2127959, EPI_ISL_2127962, EPI_ISL_2128000, EPI_ISL_2128029, EPI_ISL_2128105, EPI_ISL_2128122, EPI_ISL_2128140, EPI_ISL_2128152, EPI_ISL_2128161, EPI_ISL_2128185, EPI_ISL_2128212, EPI_ISL_2128226, EPI_ISL_2128235, EPI_ISL_2128237, EPI_ISL_2128245, EPI_ISL_2128264, EPI_ISL_2128280, EPI_ISL_2128285, EPI_ISL_2128294, EPI_ISL_2128305, EPI_ISL_2128320, EPI_ISL_2128335, EPI_ISL_2128342, EPI_ISL_2128350, EPI_ISL_2128641, EPI_ISL_2128647, EPI_ISL_2128688, EPI_ISL_2128699, EPI_ISL_2128720, EPI_ISL_2128723, EPI_ISL_2128742, EPI_ISL_2128773, EPI_ISL_2128799, EPI_ISL_2128808, EPI_ISL_2128816, EPI_ISL_2128819, EPI_ISL_2128822, EPI_ISL_2128841, EPI_ISL_2128842, EPI_ISL_2128851, EPI_ISL_2128853, EPI_ISL_2128857, EPI_ISL_2128873, EPI_ISL_2128876, EPI_ISL_2128905, EPI_ISL_2128906, EPI_ISL_2128919, EPI_ISL_2128938, EPI_ISL_2128952 | | | |
| see above | SYNLAB MVZ Neustadt a. d. Weinstraße | Robert Koch Institute | |
| EPI_ISL_2111605, EPI_ISL_2111614, EPI_ISL_2111615, EPI_ISL_2111626, EPI_ISL_2111628, EPI_ISL_2111631, EPI_ISL_2111637, EPI_ISL_2111644, EPI_ISL_2111645, EPI_ISL_2111647, EPI_ISL_2111653, EPI_ISL_2111658, EPI_ISL_2111660, EPI_ISL_2111663, EPI_ISL_2111664, EPI_ISL_2111666, EPI_ISL_2111669, EPI_ISL_2111684, EPI_ISL_2111687, EPI_ISL_2111695, EPI_ISL_2111701, EPI_ISL_2111702, EPI_ISL_2111713, EPI_ISL_2111714, EPI_ISL_2111717, EPI_ISL_2111724, EPI_ISL_2111729, EPI_ISL_2111732, EPI_ISL_2111733, EPI_ISL_2111734, EPI_ISL_2111745, EPI_ISL_2111752, EPI_ISL_2111754, EPI_ISL_2111756, EPI_ISL_2111766, EPI_ISL_2111767, EPI_ISL_2111772, EPI_ISL_2111783, EPI_ISL_2111786, EPI_ISL_2111787, EPI_ISL_2111790, EPI_ISL_2111792, EPI_ISL_2111793, EPI_ISL_2111796, EPI_ISL_2111799, EPI_ISL_2111804, EPI_ISL_2111810, EPI_ISL_2111814, EPI_ISL_2111815, EPI_ISL_2111824, EPI_ISL_2111825, EPI_ISL_2111835, EPI_ISL_2111837, EPI_ISL_2111841, EPI_ISL_2111844, EPI_ISL_2111846, EPI_ISL_2111850, EPI_ISL_2111854, EPI_ISL_2111855, EPI_ISL_2111856, EPI_ISL_2111858, EPI_ISL_2111865, EPI_ISL_2111871, EPI_ISL_2111872, EPI_ISL_2111874, EPI_ISL_2111877, EPI_ISL_2111879, EPI_ISL_2111884, EPI_ISL_2111886, EPI_ISL_2111889, EPI_ISL_2111890, EPI_ISL_2111891, EPI_ISL_2111899, EPI_ISL_2111900, EPI_ISL_2111906, EPI_ISL_2111913, EPI_ISL_2111915, EPI_ISL_2111918, EPI_ISL_2111920, EPI_ISL_2111924, EPI_ISL_2111928, EPI_ISL_2111931, EPI_ISL_2111934, EPI_ISL_2111935, EPI_ISL_2111938, EPI_ISL_2111940, EPI_ISL_2111941, EPI_ISL_2111947, EPI_ISL_2111948, EPI_ISL_2111949, EPI_ISL_2111951, EPI_ISL_2111954, EPI_ISL_2111957, EPI_ISL_2111967, EPI_ISL_2111970, EPI_ISL_2111974, EPI_ISL_2111987, EPI_ISL_2111996, EPI_ISL_2112014, EPI_ISL_2112019, EPI_ISL_2112022, EPI_ISL_2112027, EPI_ISL_2112028, EPI_ISL_2112030, EPI_ISL_2112032, EPI_ISL_2112033, EPI_ISL_2112035, EPI_ISL_2112038, EPI_ISL_2112040, EPI_ISL_2112044, EPI_ISL_2112045, EPI_ISL_2112046, EPI_ISL_2112053, EPI_ISL_2112055, EPI_ISL_2112056, EPI_ISL_2112061, EPI_ISL_2112064, EPI_ISL_2112066, EPI_ISL_2112068, EPI_ISL_2112070, EPI_ISL_2112077, EPI_ISL_2112080, EPI_ISL_2112081, EPI_ISL_2112084, EPI_ISL_2112088, EPI_ISL_2112090, EPI_ISL_2112092, EPI_ISL_2112094, EPI_ISL_2112095, EPI_ISL_2112105, EPI_ISL_2112109, EPI_ISL_2112113, EPI_ISL_2112123, EPI_ISL_2112124, EPI_ISL_2112136, EPI_ISL_2112139, EPI_ISL_2112141, EPI_ISL_2112148, EPI_ISL_2112150, EPI_ISL_2112152, EPI_ISL_2112154, EPI_ISL_2112156, EPI_ISL_2112158, EPI_ISL_2112162, EPI_ISL_2112163, EPI_ISL_2112164, EPI_ISL_2112168, EPI_ISL_2112172, EPI_ISL_2112177, EPI_ISL_2112178, EPI_ISL_2112180, EPI_ISL_2113241, EPI_ISL_2113243, EPI_ISL_2113246, EPI_ISL_2113252, EPI_ISL_2113259, EPI_ISL_2113262, EPI_ISL_2113270, EPI_ISL_2113271, EPI_ISL_2113272, EPI_ISL_2113277, EPI_ISL_2113282, EPI_ISL_2113288, EPI_ISL_2113291, EPI_ISL_2113293, EPI_ISL_2113294, EPI_ISL_2113315, EPI_ISL_2113324, EPI_ISL_2113328, EPI_ISL_2113332, EPI_ISL_2113334, EPI_ISL_2113341, EPI_ISL_2113351, EPI_ISL_2113352, EPI_ISL_2113354, EPI_ISL_2113373, EPI_ISL_2113382, EPI_ISL_2113385, EPI_ISL_2113393, EPI_ISL_2113395, EPI_ISL_2113397, EPI_ISL_2113404, EPI_ISL_2113407, EPI_ISL_2113409, EPI_ISL_2113410, EPI_ISL_2113413, EPI_ISL_2113414, EPI_ISL_2113415, EPI_ISL_2113419, EPI_ISL_2113421, EPI_ISL_2113429, EPI_ISL_2113430, EPI_ISL_2113438, EPI_ISL_2113439, EPI_ISL_2113454, EPI_ISL_2113460, EPI_ISL_2113464, EPI_ISL_2113467, EPI_ISL_2113473, EPI_ISL_2113479, EPI_ISL_2113485, EPI_ISL_2113492, EPI_ISL_2113503, EPI_ISL_2113507, EPI_ISL_2113510, EPI_ISL_2113513, EPI_ISL_2113517, EPI_ISL_2113521, EPI_ISL_2113525, EPI_ISL_2113531, EPI_ISL_2113536, EPI_ISL_2113539, EPI_ISL_2113560, EPI_ISL_2113565, EPI_ISL_2113578, EPI_ISL_2113579, EPI_ISL_2113582, EPI_ISL_2113585, EPI_ISL_2113588, EPI_ISL_2113590, EPI_ISL_2113596, EPI_ISL_2113601, EPI_ISL_2113602, EPI_ISL_2113604, EPI_ISL_2113608, EPI_ISL_2113611, EPI_ISL_2113613, EPI_ISL_2113618, EPI_ISL_2113619, EPI_ISL_2113620, EPI_ISL_2113623, EPI_ISL_2113632, EPI_ISL_2113633, EPI_ISL_2113636, EPI_ISL_2113638, EPI_ISL_2113639, EPI_ISL_2113641, EPI_ISL_2113642, EPI_ISL_2113646, EPI_ISL_2113650, EPI_ISL_2113659, EPI_ISL_2113660, EPI_ISL_2113661, EPI_ISL_2113665, EPI_ISL_2113666, EPI_ISL_2113668, EPI_ISL_2113669, EPI_ISL_2113670, EPI_ISL_2113672, EPI_ISL_2113673, EPI_ISL_2113676, EPI_ISL_2113678, EPI_ISL_2113679, EPI_ISL_2113684, EPI_ISL_2113685, EPI_ISL_2113689, EPI_ISL_2113691, EPI_ISL_2113693, EPI_ISL_2113694, EPI_ISL_2113705, EPI_ISL_2113706, EPI_ISL_2113720, EPI_ISL_2113722, EPI_ISL_2113731, EPI_ISL_2113738, EPI_ISL_2113743, EPI_ISL_2113746, EPI_ISL_2113750, EPI_ISL_2113757, EPI_ISL_2113760, EPI_ISL_2113762, EPI_ISL_2113770, EPI_ISL_2113774, EPI_ISL_2113777, EPI_ISL_2113781, EPI_ISL_2113782, EPI_ISL_2113785, EPI_ISL_2113798, EPI_ISL_2113802, EPI_ISL_2113807, EPI_ISL_2113814, EPI_ISL_2113815, EPI_ISL_2113817, EPI_ISL_2113823, EPI_ISL_2113826, EPI_ISL_2113829, EPI_ISL_2113830, EPI_ISL_2113836, EPI_ISL_2113837, EPI_ISL_2113839, EPI_ISL_2113842, EPI_ISL_2113848, EPI_ISL_2113849, EPI_ISL_2113851, EPI_ISL_2113852, EPI_ISL_2113853, EPI_ISL_2113857, EPI_ISL_2113859, EPI_ISL_2113861, EPI_ISL_2113862, EPI_ISL_2113865, EPI_ISL_2113867, EPI_ISL_2113869, EPI_ISL_2113870, EPI_ISL_2113873, EPI_ISL_2113878, EPI_ISL_2113879, EPI_ISL_2113883, EPI_ISL_2113885, EPI_ISL_2113887, EPI_ISL_2113889, EPI_ISL_2113892, EPI_ISL_2113894, EPI_ISL_2113905, EPI_ISL_2113910, EPI_ISL_2113918, EPI_ISL_2113928, EPI_ISL_2113931, EPI_ISL_2113946, EPI_ISL_2113996, EPI_ISL_2113999, EPI_ISL_2114003, EPI_ISL_2114005, EPI_ISL_2114008, EPI_ISL_2114012, EPI_ISL_2114028, EPI_ISL_2114039, EPI_ISL_2114051, EPI_ISL_2114073, EPI_ISL_2114105, EPI_ISL_2114113, EPI_ISL_2114121, EPI_ISL_2114123, EPI_ISL_2114173, EPI_ISL_2114187, EPI_ISL_2114218, EPI_ISL_2114234, EPI_ISL_2114251, EPI_ISL_2114253, EPI_ISL_2114255, EPI_ISL_2114259, EPI_ISL_2114314, EPI_ISL_2114374, EPI_ISL_2114375, EPI_ISL_2114402, EPI_ISL_2114412, EPI_ISL_2114453, EPI_ISL_2114454, EPI_ISL_2116642, EPI_ISL_2116646, EPI_ISL_2116652, EPI_ISL_2116673, EPI_ISL_2116675, EPI_ISL_2116686, EPI_ISL_2116710, EPI_ISL_2116722, EPI_ISL_2116727, EPI_ISL_2116737, EPI_ISL_2116738, EPI_ISL_2116740, EPI_ISL_2116742, EPI_ISL_2116749, EPI_ISL_2116753, EPI_ISL_2116754, EPI_ISL_2116759, EPI_ISL_2116762, EPI_ISL_2116766, EPI_ISL_2116786, EPI_ISL_2116788, EPI_ISL_2116791, EPI_ISL_2116799, EPI_ISL_2116819, EPI_ISL_2116823, EPI_ISL_2116825, EPI_ISL_2116830, EPI_ISL_2116832, EPI_ISL_2116836, EPI_ISL_2116842, EPI_ISL_2116845, EPI_ISL_2116848, EPI_ISL_2116855, EPI_ISL_2116860, EPI_ISL_2116865, EPI_ISL_2116872, EPI_ISL_2116893, EPI_ISL_2116897, EPI_ISL_2116902, EPI_ISL_2116903, EPI_ISL_2116906, EPI_ISL_2116909, EPI_ISL_2116913, EPI_ISL_2116914, EPI_ISL_2116932, EPI_ISL_2116938, EPI_ISL_2116978, EPI_ISL_2116990, EPI_ISL_2116995, EPI_ISL_2117000, EPI_ISL_2117005, EPI_ISL_2117034, EPI_ISL_2117036, EPI_ISL_2117037, EPI_ISL_2117040, EPI_ISL_2117048, EPI_ISL_2117049, EPI_ISL_2117066, EPI_ISL_2117090, EPI_ISL_2117093, EPI_ISL_2117097, EPI_ISL_2117101, EPI_ISL_2117114, EPI_ISL_2117119, EPI_ISL_2117123, EPI_ISL_2117126, EPI_ISL_2117131, EPI_ISL_2117149, EPI_ISL_2117153, EPI_ISL_2117156, EPI_ISL_2117166, EPI_ISL_2117172, EPI_ISL_2117173, EPI_ISL_2117174, EPI_ISL_2117178, EPI_ISL_2117181, EPI_ISL_2117189, EPI_ISL_2117192, EPI_ISL_2117193, EPI_ISL_2117194, EPI_ISL_2117198, EPI_ISL_2117205, EPI_ISL_2117207, EPI_ISL_2117210, EPI_ISL_2117211, EPI_ISL_2117218, EPI_ISL_2117219, EPI_ISL_2117228, EPI_ISL_2117229, EPI_ISL_2117232, EPI_ISL_2117265, EPI_ISL_2117280, EPI_ISL_2117281, EPI_ISL_2117282, EPI_ISL_2117284, EPI_ISL_2117291, EPI_ISL_2118731 | | | |
| see above | SYNLAB MVZ Trier | Robert Koch Institute | |
| EPI_ISL_2111711, EPI_ISL_2111725, EPI_ISL_2111750, EPI_ISL_2111817, EPI_ISL_2111903, EPI_ISL_2111932, EPI_ISL_2112118, EPI_ISL_2112120, EPI_ISL_2113376, EPI_ISL_2113398, EPI_ISL_2113776, EPI_ISL_2113858, EPI_ISL_2116743, EPI_ISL_2116933 | | | |
| see above | SYNLAB MVZ Weiden | Robert Koch Institute | |
| EPI_ISL_2121507, EPI_ISL_2121562, EPI_ISL_2121567, EPI_ISL_2121575, EPI_ISL_2121586, EPI_ISL_2121701, EPI_ISL_2121739, EPI_ISL_2121763, EPI_ISL_2121766, EPI_ISL_2121783, EPI_ISL_2121811, EPI_ISL_2121824, EPI_ISL_2122035, EPI_ISL_2122049, EPI_ISL_2122190, EPI_ISL_2122199, EPI_ISL_2122204, EPI_ISL_2125182, EPI_ISL_2125272, EPI_ISL_2125437, EPI_ISL_2125470, EPI_ISL_2125477, EPI_ISL_2125534, EPI_ISL_2125536, EPI_ISL_2125562, EPI_ISL_2125567, EPI_ISL_2125585, EPI_ISL_2125665, EPI_ISL_2125752, EPI_ISL_2125766, EPI_ISL_2125802, EPI_ISL_2125807, EPI_ISL_2125821, EPI_ISL_2125847, EPI_ISL_2125892, EPI_ISL_2125902, EPI_ISL_2125943, EPI_ISL_2125949, EPI_ISL_2125951, EPI_ISL_2125955, EPI_ISL_2125959, EPI_ISL_2125981, EPI_ISL_2126002, EPI_ISL_2126087, EPI_ISL_2126124, EPI_ISL_2126165 | | | |
| see above | Sonic - Bioscientia - MVZ Labor Saar GmbH | Robert Koch Institute | |
| EPI_ISL_2125145, EPI_ISL_2125164, EPI_ISL_2125172, EPI_ISL_2125181, EPI_ISL_2125185, EPI_ISL_2125195, EPI_ISL_2125214, EPI_ISL_2125241, EPI_ISL_2125321, EPI_ISL_2125352, EPI_ISL_2125361, EPI_ISL_2125386, EPI_ISL_2125393, EPI_ISL_2125396, EPI_ISL_2125415 | | | |
| see above | Sonic - Dr. Staber & Kollegen GmbH Munchen | Robert Koch Institute | |
| EPI_ISL_2125139, EPI_ISL_2125142, EPI_ISL_2125143, EPI_ISL_2125148, EPI_ISL_2125151, EPI_ISL_2125154, EPI_ISL_2125159, EPI_ISL_2125161, EPI_ISL_2125171, EPI_ISL_2125174, EPI_ISL_2125178, EPI_ISL_2125187, EPI_ISL_2125189, EPI_ISL_2125193, EPI_ISL_2125199, EPI_ISL_2125202, EPI_ISL_2125208, EPI_ISL_2125211, EPI_ISL_2125212, EPI_ISL_2125215, EPI_ISL_2125219, EPI_ISL_2125223, EPI_ISL_2125224, EPI_ISL_2125228, EPI_ISL_2125229, EPI_ISL_2125230, EPI_ISL_2125233, EPI_ISL_2125235, EPI_ISL_2125242, EPI_ISL_2125244, EPI_ISL_2125251, EPI_ISL_2125254, EPI_ISL_2125260, EPI_ISL_2125270, EPI_ISL_2125277, EPI_ISL_2125279, EPI_ISL_2125282, EPI_ISL_2125285, EPI_ISL_2125287, EPI_ISL_2125289, EPI_ISL_2125291, EPI_ISL_2125306, EPI_ISL_2125307, EPI_ISL_2125309, EPI_ISL_2125314, EPI_ISL_2125320, EPI_ISL_2125322, EPI_ISL_2125323, EPI_ISL_2125327, EPI_ISL_2125335, EPI_ISL_2125336, EPI_ISL_2125339, EPI_ISL_2125342, EPI_ISL_2125344, EPI_ISL_2125348, EPI_ISL_2125350, EPI_ISL_2125353, EPI_ISL_2125354, EPI_ISL_2125371, EPI_ISL_2125374, EPI_ISL_2125377, EPI_ISL_2125379, EPI_ISL_2125383, EPI_ISL_2125392, EPI_ISL_2125395, EPI_ISL_2125398, EPI_ISL_2125400, EPI_ISL_2125403, EPI_ISL_2125406, EPI_ISL_2125411, EPI_ISL_2125427, EPI_ISL_2125429, EPI_ISL_2125435, EPI_ISL_2125448, EPI_ISL_2125453, EPI_ISL_2125454, EPI_ISL_2125457, EPI_ISL_2125460 | | | |
| see above | Sonic - Labor Dr. von Froreich GmbH | Robert Koch Institute | |
| EPI_ISL_2121083, EPI_ISL_2121194, EPI_ISL_2121236, EPI_ISL_2125170, EPI_ISL_2125183, EPI_ISL_2125249, EPI_ISL_2125255, EPI_ISL_2125261, EPI_ISL_2125265, EPI_ISL_2125276, EPI_ISL_2125311, EPI_ISL_2125319, EPI_ISL_2125409 | | | |
| see above | Sonic - Labor Lademannbogen MVZ GmbH | Robert Koch Institute | |
| EPI_ISL_2125155, EPI_ISL_2125191, EPI_ISL_2125204, EPI_ISL_2125231, EPI_ISL_2125310, EPI_ISL_2125347, EPI_ISL_2125364, EPI_ISL_2125445 | | | |
| see above | Sonic - Labor Staber Bayreuth | Robert Koch Institute | |
| EPI_ISL_2125153, EPI_ISL_2125169, EPI_ISL_2125175, EPI_ISL_2125176, EPI_ISL_2125222, EPI_ISL_2125246, EPI_ISL_2125315, EPI_ISL_2125355, EPI_ISL_2125369, EPI_ISL_2125373, EPI_ISL_2125375, EPI_ISL_2125401, EPI_ISL_2125440 | | | |
| see above | Sonic - Labor Staber Dresden (Klipphausen) | Robert Koch Institute | |
| EPI_ISL_2125138, EPI_ISL_2125256 | Sonic - Labor Staber MVZ Kassel | Robert Koch Institute | |
| EPI_ISL_2125237, EPI_ISL_2125274, EPI_ISL_2125328, EPI_ISL_2125332, EPI_ISL_2125337, EPI_ISL_2125368 | Sonic - Labor Staber Nurnberg | Robert Koch Institute | |
| EPI_ISL_2120609, EPI_ISL_2120665, EPI_ISL_2120797, EPI_ISL_2120804, EPI_ISL_2120853, EPI_ISL_2120991, EPI_ISL_2121022, EPI_ISL_2121033, EPI_ISL_2121068, EPI_ISL_2121176, EPI_ISL_2121229, EPI_ISL_2121251, EPI_ISL_2121264, EPI_ISL_2121287, EPI_ISL_2121392, EPI_ISL_2121423, EPI_ISL_2121440, EPI_ISL_2125146, EPI_ISL_2125149, EPI_ISL_2125218, EPI_ISL_2125248, EPI_ISL_2125288, EPI_ISL_2125293, EPI_ISL_2125325, EPI_ISL_2125331, EPI_ISL_2125338, EPI_ISL_2125365, EPI_ISL_2125397, EPI_ISL_2125418, EPI_ISL_2125424 | | | |
| see above | Sonic - MVZ Medizinisches Labor Bremen GmbH | Robert Koch Institute | |
| EPI_ISL_2109366, EPI_ISL_2109371, EPI_ISL_2109372, EPI_ISL_2109373, EPI_ISL_2109381, EPI_ISL_2109384, EPI_ISL_2109390, EPI_ISL_2109400, EPI_ISL_2109409, EPI_ISL_2109410, EPI_ISL_2109417, EPI_ISL_2109422 | | | |
| see above | Stadtisches Klinikum Dresden Institut fur Klinische Chemie und Laboratoriumsmedizin | Robert Koch Institute | |
| EPI_ISL_1852950, EPI_ISL_2111612, EPI_ISL_2111728, EPI_ISL_2111849, EPI_ISL_2111853, EPI_ISL_2111936, EPI_ISL_2112099, EPI_ISL_2112101, EPI_ISL_2112138, EPI_ISL_2113356, EPI_ISL_2113366, EPI_ISL_2113468, EPI_ISL_2113543, EPI_ISL_2113709, EPI_ISL_2113734, EPI_ISL_2113741, EPI_ISL_2113765, EPI_ISL_2113872, EPI_ISL_2113881, EPI_ISL_2116687, EPI_ISL_2116701, EPI_ISL_2116745, EPI_ISL_2116950, EPI_ISL_2117057, EPI_ISL_2117107, EPI_ISL_2117143, EPI_ISL_2117171, EPI_ISL_2117195, EPI_ISL_2117255 | | | |
| see above | Synlab MVZ Augsburg | Robert Koch Institute | |
| EPI_ISL_2109361, EPI_ISL_2109363, EPI_ISL_2109364, EPI_ISL_2109369, EPI_ISL_2109385, EPI_ISL_2109389, EPI_ISL_2109394, EPI_ISL_2109399, EPI_ISL_2109407, EPI_ISL_2109415, EPI_ISL_2109425 | | | |
| see above | Uniklinikum Carl Gustav Carus an der TU Dresden; Institut fur Virologie | Robert Koch Institute | |
| EPI_ISL_2110409, EPI_ISL_2110415, EPI_ISL_2123856 | Uniklinikum Jena | Robert Koch Institute | |
| EPI_ISL_2116089, EPI_ISL_2116090, EPI_ISL_2116092 | Uniklinikum Regensburg; Institut fÃ¼r klinische Mikrobiologie und Hygiene | Robert Koch Institute | |
| EPI_ISL_2125140, EPI_ISL_2125190, EPI_ISL_2125216, EPI_ISL_2125240, EPI_ISL_2125257, EPI_ISL_2125263, EPI_ISL_2125295, EPI_ISL_2125300, EPI_ISL_2125302, EPI_ISL_2125317, EPI_ISL_2125333, EPI_ISL_2125345, EPI_ISL_2125370 | | | |
| see above | Universitatsklinikum Bonn - Institut fur Virologie | Robert Koch Institute | |
| EPI_ISL_2120144, EPI_ISL_2120145, EPI_ISL_2120164, EPI_ISL_2120170 | Universitatsklinikum des Saarlandes - Institut fur Virologie | Robert Koch Institute | |
| EPI_ISL_2116049 | UniversitÃ¤tsklinikum GieÃŸen - Institut fÃ¼r Medizinische Virologie | Robert Koch Institute | |
| EPI_ISL_2126192, EPI_ISL_2126379, EPI_ISL_2126382 | UniversitÃ¤tsklinikum Heidelberg | Robert Koch Institute | |
| EPI_ISL_2115338, EPI_ISL_2115339, EPI_ISL_2115340, EPI_ISL_2115341, EPI_ISL_2115342, EPI_ISL_2115343, EPI_ISL_2115344, EPI_ISL_2115345, EPI_ISL_2115346, EPI_ISL_2115347, EPI_ISL_2115348, EPI_ISL_2115349 | | | |
| see above | UniversitÃ¤tsklinikum Munster Institut fur Virologie | Robert Koch Institute | |
| EPI_ISL_2115511 | Virologisches Institut des Universitatsklinikums Erlangen | Robert Koch Institute | |
| EPI_ISL_2003752, EPI_ISL_2003753, EPI_ISL_2003754, EPI_ISL_2003755, EPI_ISL_2003756, EPI_ISL_2003757, EPI_ISL_2003758, EPI_ISL_2003759, EPI_ISL_2003760, EPI_ISL_2003761, EPI_ISL_2003762, EPI_ISL_2003763, EPI_ISL_2003764, EPI_ISL_2003765, EPI_ISL_2003766, EPI_ISL_2003767, EPI_ISL_2003768, EPI_ISL_2003769, EPI_ISL_2003770, EPI_ISL_2003771, EPI_ISL_2003772, EPI_ISL_2003773, EPI_ISL_2003774, EPI_ISL_2003775, EPI_ISL_2003776, EPI_ISL_2003777, EPI_ISL_2135730, EPI_ISL_2135731, EPI_ISL_2135732, EPI_ISL_2135733, EPI_ISL_2135734, EPI_ISL_2135738, EPI_ISL_2135751 | | | |
| see above | Virology, Universitätsklinikum des Saarlandes | Epigenetics, Saarland University | Jörn Walter; Kathrin Kattler; Nastasja Seiwert; Sascha Tierling; Sigrun Smola; Stefan Lohse; Thorsten Pfuhl |

| | | | |
|---|---|---|---|
| EPI_ISL_1993434, EPI_ISL_1993435, EPI_ISL_1993436, EPI_ISL_1993437, EPI_ISL_1993438, EPI_ISL_1993444, EPI_ISL_1993445, EPI_ISL_1993446, EPI_ISL_1993448, EPI_ISL_1993449, EPI_ISL_1993451, EPI_ISL_1993453, EPI_ISL_1993454, EPI_ISL_1993456, EPI_ISL_1993457, EPI_ISL_1993458, EPI_ISL_1993459, EPI_ISL_1993461, EPI_ISL_1993462, EPI_ISL_1993463, EPI_ISL_1993464, EPI_ISL_1993466, EPI_ISL_1993467, EPI_ISL_1993468, EPI_ISL_1993469, EPI_ISL_1993471, EPI_ISL_1993472, EPI_ISL_1993473, EPI_ISL_1993474, EPI_ISL_1993476, EPI_ISL_1993477, EPI_ISL_1993478, EPI_ISL_1993479, EPI_ISL_1993480, EPI_ISL_1993527, EPI_ISL_1993528, EPI_ISL_1993532, EPI_ISL_1993533, EPI_ISL_1993534, EPI_ISL_1993535, EPI_ISL_1993536, EPI_ISL_1993537, EPI_ISL_1993538, EPI_ISL_1993539, EPI_ISL_1993540, EPI_ISL_1993541, EPI_ISL_1993542, EPI_ISL_1993543, EPI_ISL_1993544, EPI_ISL_1993545, EPI_ISL_1993546, EPI_ISL_1993564, EPI_ISL_1993565, EPI_ISL_1993568, EPI_ISL_1993569, EPI_ISL_1993570, EPI_ISL_1993572, EPI_ISL_1993573, EPI_ISL_1993574, EPI_ISL_1993575, EPI_ISL_1993576, EPI_ISL_1993632, EPI_ISL_1993633, EPI_ISL_1993634 | | | |
| see above | ZOTZ KLIMAS MVZ Düsseldorf-Centrum GbR ÜBAG für Labormedizin, Genetik, Zytologie, Pathologie | Center of Medical Microbiology, Virology, and Hospital Hygiene, University of Duesseldorf | Alexander Dilthey; Andreas Walker; Daniel Strelow; Jessica Nicolai; Jörg Timm; Katrin Hoffmann; Klaus Pfeffer; Lisanna Hülse; Malte Kohns Vasconcelos; Maximilian Damagnez; Nadine Lübke; Patrick Finzer; Rainer Zotz; Tobias Wienemann; Torsten Houwaart |
| EPI_ISL_1851419, EPI_ISL_1851428, EPI_ISL_2116044, EPI_ISL_2116045, EPI_ISL_2116047 | alphaomega Labor GbR (Medizinische Kooperationsgemeinschaft Dres. Ackermann/Tennert/ZaiÃŸ) | Robert Koch Institute | |
| EPI_ISL_2115811, EPI_ISL_2115813, EPI_ISL_2115815, EPI_ISL_2115817, EPI_ISL_2115818, EPI_ISL_2115839, EPI_ISL_2115855, EPI_ISL_2115859, EPI_ISL_2115878, EPI_ISL_2115886, EPI_ISL_2115902, EPI_ISL_2115930, EPI_ISL_2115947, EPI_ISL_2115950, EPI_ISL_2115954, EPI_ISL_2115960, EPI_ISL_2115964, EPI_ISL_2115977, EPI_ISL_2115980, EPI_ISL_2115984, EPI_ISL_2116001, EPI_ISL_2116009, EPI_ISL_2116026 | | | |
| see above | amedes GÃ¶ttingen | Robert Koch Institute | |
| EPI_ISL_2115809, EPI_ISL_2115816, EPI_ISL_2115822, EPI_ISL_2115827, EPI_ISL_2115831, EPI_ISL_2115835, EPI_ISL_2115840, EPI_ISL_2115851, EPI_ISL_2115856, EPI_ISL_2115862, EPI_ISL_2115865, EPI_ISL_2115866, EPI_ISL_2115870, EPI_ISL_2115871, EPI_ISL_2115879, EPI_ISL_2115892, EPI_ISL_2115895, EPI_ISL_2115898, EPI_ISL_2115901, EPI_ISL_2115908, EPI_ISL_2115912, EPI_ISL_2115923, EPI_ISL_2115926, EPI_ISL_2115931, EPI_ISL_2115934, EPI_ISL_2115935, EPI_ISL_2115937, EPI_ISL_2115939, EPI_ISL_2115943, EPI_ISL_2115945, EPI_ISL_2115953, EPI_ISL_2115958, EPI_ISL_2115978, EPI_ISL_2115982, EPI_ISL_2115985, EPI_ISL_2116013, EPI_ISL_2116015, EPI_ISL_2116030, EPI_ISL_2116032, EPI_ISL_2116034, EPI_ISL_2116036, EPI_ISL_2116038 | | | |
| see above | amedes MVZ DIAMEDIS Sennestadt | Robert Koch Institute | |
| EPI_ISL_1850470, EPI_ISL_1850500, EPI_ISL_1850540, EPI_ISL_1850545, EPI_ISL_1850549, EPI_ISL_1850595, EPI_ISL_1850597, EPI_ISL_1850640, EPI_ISL_1850644, EPI_ISL_1850649, EPI_ISL_1850683, EPI_ISL_1850695, EPI_ISL_1850705, EPI_ISL_2115823, EPI_ISL_2115842, EPI_ISL_2115863, EPI_ISL_2115867, EPI_ISL_2115891, EPI_ISL_2115894, EPI_ISL_2115905, EPI_ISL_2115911, EPI_ISL_2115933, EPI_ISL_2115940, EPI_ISL_2115962, EPI_ISL_2115973, EPI_ISL_2115992, EPI_ISL_2116005, EPI_ISL_2116011, EPI_ISL_2116014, EPI_ISL_2116016, EPI_ISL_2116033, EPI_ISL_2116035, EPI_ISL_2116037 | | | |
| see above | amedes MVZ Halle/Leipzig | Robert Koch Institute | |
| EPI_ISL_1850689, EPI_ISL_2115812, EPI_ISL_2115819, EPI_ISL_2115836, EPI_ISL_2115844, EPI_ISL_2115857, EPI_ISL_2115858, EPI_ISL_2115874, EPI_ISL_2115882, EPI_ISL_2115883, EPI_ISL_2115884, EPI_ISL_2115890, EPI_ISL_2115906, EPI_ISL_2115914, EPI_ISL_2115918, EPI_ISL_2115919, EPI_ISL_2115925, EPI_ISL_2115975, EPI_ISL_2115987, EPI_ISL_2116000, EPI_ISL_2116008, EPI_ISL_2116024, EPI_ISL_2116027, EPI_ISL_2116028, EPI_ISL_2116039 | | | |
| see above | amedes MVZ Hannover | Robert Koch Institute | |
| EPI_ISL_2115821, EPI_ISL_2115846, EPI_ISL_2115864, EPI_ISL_2115903, EPI_ISL_2115915, EPI_ISL_2115921, EPI_ISL_2115932, EPI_ISL_2115942, EPI_ISL_2115952, EPI_ISL_2115957, EPI_ISL_2115961, EPI_ISL_2115968, EPI_ISL_2115971, EPI_ISL_2115974, EPI_ISL_2115983, EPI_ISL_2115989, EPI_ISL_2116002, EPI_ISL_2116007, EPI_ISL_2116010, EPI_ISL_2116017, EPI_ISL_2116018, EPI_ISL_2116023, EPI_ISL_2116031 | | | |
| see above | amedes MVZ Jena | Robert Koch Institute | |
| EPI_ISL_2115852, EPI_ISL_2115877 | amedes MVZ Zentrum fÃ¼r Pathologie und Zytodiagnostik GmbH (KÃ¶ln) | Robert Koch Institute | |
| EPI_ISL_2115814, EPI_ISL_2115837, EPI_ISL_2115849, EPI_ISL_2115853, EPI_ISL_2115861, EPI_ISL_2115880, EPI_ISL_2115917, EPI_ISL_2115920, EPI_ISL_2115949, EPI_ISL_2115959, EPI_ISL_2115972, EPI_ISL_2115991, EPI_ISL_2115999, EPI_ISL_2116004, EPI_ISL_2116019 | | | |
| see above | amedes MVZ fÃ¼r Laboratoriumsdiagnostik Raubling GmbH | Robert Koch Institute | |
| EPI_ISL_2115828, EPI_ISL_2115829, EPI_ISL_2115838, EPI_ISL_2115904, EPI_ISL_2115928, EPI_ISL_2116021 | amedes MVZ fÃ¼r Laboratoriumsmedizin FÃ¼rstenfeldbruck | Robert Koch Institute | |
| EPI_ISL_2115825, EPI_ISL_2115832 | amedes MVZ fÃ¼r Laboratoriumsmedizin MÃ¼nchen | Robert Koch Institute | |
| EPI_ISL_2115850, EPI_ISL_2115875, EPI_ISL_2115979 | amedes MVZ fÃ¼r Laboratoriumsmedizin Rhein/Ruhr | Robert Koch Institute | |
| EPI_ISL_2115897, EPI_ISL_2115899, EPI_ISL_2115922, EPI_ISL_2115929, EPI_ISL_2115966, EPI_ISL_2116025, EPI_ISL_2116029 | | | |
| see above | amedes MVZ fÃ¼r Labordiagnostik Rhein-Main | Robert Koch Institute | |
| EPI_ISL_1850458, EPI_ISL_1850610, EPI_ISL_1850660, EPI_ISL_1850710, EPI_ISL_1850712, EPI_ISL_2115907, EPI_ISL_2116020, EPI_ISL_2116022 | | | |
| see above | amedes Medlab Arnold Analytik MVZ GmbH | Robert Koch Institute | |
| EPI_ISL_1850477, EPI_ISL_1850479, EPI_ISL_1850511, EPI_ISL_1850552, EPI_ISL_1850560, EPI_ISL_1850562, EPI_ISL_1850567, EPI_ISL_1850569, EPI_ISL_1850576, EPI_ISL_1850600, EPI_ISL_1850612, EPI_ISL_1850631, EPI_ISL_1850639, EPI_ISL_1850659, EPI_ISL_1850672, EPI_ISL_1850678, EPI_ISL_1850701, EPI_ISL_1850706, EPI_ISL_1850713 | | | |
| see above | nordlab - Partnerschaftspraxis fuÌˆr Laboratoriumsmedizin | Robert Koch Institute | |

We gratefully acknowledge the following Authors from the Originating laboratories responsible for obtaining the specimens, as well as the Submitting laboratories where the genome data were generated and shared via GISAID, on which this research is based.

All Submitters of data may be contacted directly via www.gisaid.org

Authors are sorted alphabetically.

| Accession ID | Originating Laboratory | Submitting Laboratory | Authors |
|---|---|---|---|
| EPI_ISL_2260602, EPI_ISL_2260622, EPI_ISL_2260714, EPI_ISL_2260748, EPI_ISL_2260816, EPI_ISL_2260861, EPI_ISL_2260896, EPI_ISL_2388384, EPI_ISL_2388654, EPI_ISL_2388703 | | | |
| see above | ALB FILS KLINIKEN GmbH - Institut für Laboratoriumsmedizin Klinik am Eichert Göppingen | Robert Koch Institute | |
| EPI_ISL_2262662, EPI_ISL_2262692, EPI_ISL_2262696, EPI_ISL_2262697, EPI_ISL_2262700, EPI_ISL_2262712, EPI_ISL_2262717, EPI_ISL_2262718, EPI_ISL_2262732, EPI_ISL_2262739, EPI_ISL_2262741, EPI_ISL_2262788, EPI_ISL_2262794, EPI_ISL_2262811, EPI_ISL_2315669, EPI_ISL_2315670, EPI_ISL_2315673, EPI_ISL_2315684, EPI_ISL_2315688, EPI_ISL_2315690, EPI_ISL_2315691, EPI_ISL_2315692, EPI_ISL_2315693, EPI_ISL_2315694, EPI_ISL_2315701, EPI_ISL_2315716, EPI_ISL_2315723, EPI_ISL_2315731, EPI_ISL_2315733, EPI_ISL_2315744, EPI_ISL_2315748, EPI_ISL_2315753, EPI_ISL_2315754, EPI_ISL_2315755, EPI_ISL_2315757, EPI_ISL_2315759, EPI_ISL_2315761, EPI_ISL_2315764, EPI_ISL_2315766, EPI_ISL_2315770, EPI_ISL_2315775, EPI_ISL_2315776, EPI_ISL_2315784, EPI_ISL_2315787, EPI_ISL_2315788, EPI_ISL_2315794, EPI_ISL_2315795, EPI_ISL_2315796, EPI_ISL_2315797, EPI_ISL_2315801, EPI_ISL_2315802, EPI_ISL_2315807, EPI_ISL_2315808, EPI_ISL_2315809, EPI_ISL_2315811, EPI_ISL_2315815, EPI_ISL_2315816, EPI_ISL_2315818, EPI_ISL_2390475, EPI_ISL_2390478, EPI_ISL_2390480, EPI_ISL_2390481, EPI_ISL_2390482, EPI_ISL_2390485, EPI_ISL_2390486, EPI_ISL_2390487, EPI_ISL_2390489, EPI_ISL_2390490, EPI_ISL_2390495, EPI_ISL_2390496, EPI_ISL_2390497, EPI_ISL_2390498, EPI_ISL_2390499, EPI_ISL_2390502, EPI_ISL_2390503, EPI_ISL_2390504, EPI_ISL_2390508, EPI_ISL_2390511, EPI_ISL_2390512, EPI_ISL_2390514, EPI_ISL_2390517, EPI_ISL_2390521, EPI_ISL_2390522, EPI_ISL_2390523, EPI_ISL_2390525, EPI_ISL_2390526, EPI_ISL_2390527, EPI_ISL_2390528, EPI_ISL_2390530, EPI_ISL_2390531, EPI_ISL_2390533, EPI_ISL_2390534, EPI_ISL_2390535, EPI_ISL_2390536, EPI_ISL_2390537, EPI_ISL_2390538, EPI_ISL_2390539, EPI_ISL_2390540, EPI_ISL_2390542, EPI_ISL_2390543, EPI_ISL_2390544, EPI_ISL_2390545, EPI_ISL_2390547, EPI_ISL_2390549, EPI_ISL_2390552, EPI_ISL_2390553, EPI_ISL_2390554, EPI_ISL_2390560, EPI_ISL_2390561, EPI_ISL_2390562, EPI_ISL_2390564, EPI_ISL_2390566, EPI_ISL_2390570, EPI_ISL_2390572, EPI_ISL_2390573, EPI_ISL_2390576, EPI_ISL_2390577, EPI_ISL_2390579, EPI_ISL_2472551, EPI_ISL_2472553, EPI_ISL_2472554, EPI_ISL_2472563, EPI_ISL_2472564, EPI_ISL_2472566, EPI_ISL_2472569, EPI_ISL_2472573, EPI_ISL_2472574, EPI_ISL_2472575, EPI_ISL_2472578, EPI_ISL_2472579, EPI_ISL_2472585, EPI_ISL_2472588, EPI_ISL_2472591, EPI_ISL_2472592, EPI_ISL_2472596, EPI_ISL_2472602, EPI_ISL_2472604, EPI_ISL_2472606, EPI_ISL_2472610, EPI_ISL_2472617, EPI_ISL_2472621, EPI_ISL_2472622, EPI_ISL_2472626, EPI_ISL_2472628, EPI_ISL_2472629, EPI_ISL_2472632, EPI_ISL_2472634, EPI_ISL_2472635, EPI_ISL_2472638, EPI_ISL_2634978, EPI_ISL_2635003 | | | |
| see above | Bayerisches Landesamt für Gesundheit und Lebensmittelsicherheit (LGL) | Robert Koch Institute | |
| EPI_ISL_2125485, EPI_ISL_2125492, EPI_ISL_2125499, EPI_ISL_2125544, EPI_ISL_2125551, EPI_ISL_2125596, EPI_ISL_2125613, EPI_ISL_2125616, EPI_ISL_2125656, EPI_ISL_2125800, EPI_ISL_2125842, EPI_ISL_2125873, EPI_ISL_2125929, EPI_ISL_2126128, EPI_ISL_2126180, EPI_ISL_2260090, EPI_ISL_2260099, EPI_ISL_2260117, EPI_ISL_2260118, EPI_ISL_2260119, EPI_ISL_2260132, EPI_ISL_2260135, EPI_ISL_2260147, EPI_ISL_2260156, EPI_ISL_2260159, EPI_ISL_2260167, EPI_ISL_2260180, EPI_ISL_2260193, EPI_ISL_2260213, EPI_ISL_2260220, EPI_ISL_2260226, EPI_ISL_2260241, EPI_ISL_2260256, EPI_ISL_2260258, EPI_ISL_2260264, EPI_ISL_2260275, EPI_ISL_2260323, EPI_ISL_2260337, EPI_ISL_2260360, EPI_ISL_2260363, EPI_ISL_2260369, EPI_ISL_2260380, EPI_ISL_2260400, EPI_ISL_2260412, EPI_ISL_2260418, EPI_ISL_2260601, EPI_ISL_2260606, EPI_ISL_2260609, EPI_ISL_2260611, EPI_ISL_2260614, EPI_ISL_2260620, EPI_ISL_2260623, EPI_ISL_2260636, EPI_ISL_2260641, EPI_ISL_2260643, EPI_ISL_2260658, EPI_ISL_2260661, EPI_ISL_2260669, EPI_ISL_2260678, EPI_ISL_2260680, EPI_ISL_2260682, EPI_ISL_2260684, EPI_ISL_2260687, EPI_ISL_2260690, EPI_ISL_2260691, EPI_ISL_2260693, EPI_ISL_2260695, EPI_ISL_2260699, EPI_ISL_2260704, EPI_ISL_2260705, EPI_ISL_2260710, EPI_ISL_2260720, EPI_ISL_2260728, EPI_ISL_2260731, EPI_ISL_2260739, EPI_ISL_2260754, EPI_ISL_2260755, EPI_ISL_2260758, EPI_ISL_2260761, EPI_ISL_2260762, EPI_ISL_2260767, EPI_ISL_2260768, EPI_ISL_2260769, EPI_ISL_2260771, EPI_ISL_2260778, EPI_ISL_2260782, EPI_ISL_2260785, EPI_ISL_2260792, EPI_ISL_2260805, EPI_ISL_2260812, EPI_ISL_2260813, EPI_ISL_2260819, EPI_ISL_2260826, EPI_ISL_2260830, EPI_ISL_2260832, EPI_ISL_2260840, EPI_ISL_2260842, EPI_ISL_2260848, EPI_ISL_2260853, EPI_ISL_2260856, EPI_ISL_2260860, EPI_ISL_2260868, EPI_ISL_2260872, EPI_ISL_2260877, EPI_ISL_2260880, EPI_ISL_2260884, EPI_ISL_2260886, EPI_ISL_2260889, EPI_ISL_2260895, EPI_ISL_2260897, EPI_ISL_2260903, EPI_ISL_2260906, EPI_ISL_2260909, EPI_ISL_2265535, EPI_ISL_2265536, EPI_ISL_2265538, EPI_ISL_2265539, EPI_ISL_2265540, EPI_ISL_2265544, EPI_ISL_2265546, EPI_ISL_2265547, EPI_ISL_2265548, EPI_ISL_2265549, EPI_ISL_2265551, EPI_ISL_2265557, EPI_ISL_2265558, EPI_ISL_2265559, EPI_ISL_2265560, EPI_ISL_2265563, EPI_ISL_2265564, EPI_ISL_2265565, EPI_ISL_2265570, EPI_ISL_2265571, EPI_ISL_2265572, EPI_ISL_2265574, EPI_ISL_2265579, EPI_ISL_2265581, EPI_ISL_2265582, EPI_ISL_2265583, EPI_ISL_2265584, EPI_ISL_2265586, EPI_ISL_2265587, EPI_ISL_2265589, EPI_ISL_2265590, EPI_ISL_2265598, EPI_ISL_2265599, EPI_ISL_2265601, EPI_ISL_2265602, EPI_ISL_2265604, EPI_ISL_2265605, EPI_ISL_2265607, EPI_ISL_2265610, EPI_ISL_2265611, EPI_ISL_2265614, EPI_ISL_2265615, EPI_ISL_2265618, EPI_ISL_2265621, EPI_ISL_2265623, EPI_ISL_2265625, EPI_ISL_2265629, EPI_ISL_2265632, EPI_ISL_2265637, EPI_ISL_2265643, EPI_ISL_2265644, EPI_ISL_2265646, EPI_ISL_2265649, EPI_ISL_2265650, EPI_ISL_2265652, EPI_ISL_2265656, EPI_ISL_2265659, EPI_ISL_2265660, EPI_ISL_2265661, EPI_ISL_2265662, EPI_ISL_2265665, EPI_ISL_2265667, EPI_ISL_2265668, EPI_ISL_2265674, EPI_ISL_2265675, EPI_ISL_2265676, EPI_ISL_2265677, EPI_ISL_2265678, EPI_ISL_2265684, EPI_ISL_2265687, EPI_ISL_2265688, EPI_ISL_2265689, EPI_ISL_2265690, EPI_ISL_2265692, EPI_ISL_2265693, EPI_ISL_2265696, EPI_ISL_2265698, EPI_ISL_2265699, EPI_ISL_2265700, EPI_ISL_2265704, EPI_ISL_2265706, EPI_ISL_2265707, EPI_ISL_2265708, EPI_ISL_2265709, EPI_ISL_2265712, EPI_ISL_2265714, EPI_ISL_2265716, EPI_ISL_2265717, EPI_ISL_2265718, EPI_ISL_2265722, EPI_ISL_2265724, EPI_ISL_2265725, EPI_ISL_2265727, EPI_ISL_2265728, EPI_ISL_2265730, EPI_ISL_2265733, EPI_ISL_2265734, EPI_ISL_2265735, EPI_ISL_2265739, EPI_ISL_2265742, EPI_ISL_2265743, EPI_ISL_2265744, EPI_ISL_2265745, EPI_ISL_2265746, EPI_ISL_2265748, EPI_ISL_2265750, EPI_ISL_2265751, EPI_ISL_2265752, EPI_ISL_2265757, EPI_ISL_2265760, EPI_ISL_2265761, EPI_ISL_2265764, EPI_ISL_2265765, EPI_ISL_2265766, EPI_ISL_2265769, EPI_ISL_2265770, EPI_ISL_2265773, EPI_ISL_2265774, EPI_ISL_2265775, EPI_ISL_2265776, EPI_ISL_2265777, EPI_ISL_2265778, EPI_ISL_2265779, EPI_ISL_2265780, EPI_ISL_2265781, EPI_ISL_2265782, EPI_ISL_2265783, EPI_ISL_2265785, EPI_ISL_2265787, EPI_ISL_2265788, EPI_ISL_2265791, EPI_ISL_2265794, EPI_ISL_2265797, EPI_ISL_2265800, EPI_ISL_2265801, EPI_ISL_2265802, EPI_ISL_2265803, EPI_ISL_2265806, EPI_ISL_2265808, EPI_ISL_2265809, EPI_ISL_2265811, EPI_ISL_2265814, EPI_ISL_2265815, EPI_ISL_2265816, EPI_ISL_2265817, EPI_ISL_2265818, EPI_ISL_2265819, EPI_ISL_2265821, EPI_ISL_2265823, EPI_ISL_2265824, EPI_ISL_2265825, EPI_ISL_2265826, EPI_ISL_2265829, EPI_ISL_2265831, EPI_ISL_2265834, EPI_ISL_2265835, EPI_ISL_2265837, EPI_ISL_2265839, EPI_ISL_2265840, EPI_ISL_2265841, EPI_ISL_2265842, EPI_ISL_2265843, EPI_ISL_2265850, EPI_ISL_2265851, EPI_ISL_2265852, EPI_ISL_2265854, EPI_ISL_2265855, EPI_ISL_2265856, EPI_ISL_2265859, EPI_ISL_2265861, EPI_ISL_2265864, EPI_ISL_2265865, EPI_ISL_2265867, EPI_ISL_2265868, EPI_ISL_2313457, EPI_ISL_2313505, EPI_ISL_2313811, EPI_ISL_2314111, EPI_ISL_2314195, EPI_ISL_2314222, EPI_ISL_2314297 | | | |
| see above | Bioscientia Labor Wermsdorf | Robert Koch Institute | |
| EPI_ISL_2260109, EPI_ISL_2260141, EPI_ISL_2260262, EPI_ISL_2260272, EPI_ISL_2265542, EPI_ISL_2265562, EPI_ISL_2265567, EPI_ISL_2265568, EPI_ISL_2265578, EPI_ISL_2265588, EPI_ISL_2265594, EPI_ISL_2265603, EPI_ISL_2265606, EPI_ISL_2265608, EPI_ISL_2265612, EPI_ISL_2265613, EPI_ISL_2265626, EPI_ISL_2265630, EPI_ISL_2265636, EPI_ISL_2265642, EPI_ISL_2265654, EPI_ISL_2265671, EPI_ISL_2265681, EPI_ISL_2265691, EPI_ISL_2265701, EPI_ISL_2265721, EPI_ISL_2265723, EPI_ISL_2265758, EPI_ISL_2265768, EPI_ISL_2265790, EPI_ISL_2265805, EPI_ISL_2265807, EPI_ISL_2265828, EPI_ISL_2265836, EPI_ISL_2265849, EPI_ISL_2265853, EPI_ISL_2265862 | | | |
| see above | Bioscientia MVZ Labor Karlsruhe GmbH | Robert Koch Institute | |
| EPI_ISL_2131152, EPI_ISL_2131163 | Bundeswehr Institute of Microbiology | Bundeswehr Institute of Microbiology | Alexandra Rehn; Enrico Georgi; Malena Bestehorn-Willmann; Markus Antwerpen; Mathias Walter; Mike Pillukat; Roman Wölfel; Sabine Zange |
| EPI_ISL_2131169, EPI_ISL_2536134, EPI_ISL_2536135, EPI_ISL_2536136, EPI_ISL_2536137, EPI_ISL_2536138 | Bundeswehrkrankenhaus Berlin | Bundeswehr Institute of Microbiology | Alexandra Rehn; Enrico Georgi; Malena Bestehorn-Willmann; Markus Antwerpen; Mathias Walter; Mike Pillukat; Roman Wölfel; Sabine Zange |
| EPI_ISL_2536131 | Bundeswehrkrankenhaus Ulm | Bundeswehr Institute of Microbiology | Alexandra Rehn; Enrico Georgi; Malena Bestehorn-Willmann; Markus Antwerpen; Mathias Walter; Mike Pillukat; Roman Wölfel; Sabine Zange |
| EPI_ISL_2131172, EPI_ISL_2131173, EPI_ISL_2131174 | Bundeswehrzentralkrankenhaus Koblenz | Bundeswehr Institute of Microbiology | Alexandra Rehn; Enrico Georgi; Malena Bestehorn-Willmann; Markus Antwerpen; Mathias Walter; Mike Pillukat; Roman Wölfel; Sabine Zange |
| EPI_ISL_2259810, EPI_ISL_2259811, EPI_ISL_2259812, EPI_ISL_2259813, EPI_ISL_2259814, EPI_ISL_2259816, EPI_ISL_2259817, EPI_ISL_2259818, EPI_ISL_2259819, EPI_ISL_2259820, EPI_ISL_2259821, EPI_ISL_2259822, EPI_ISL_2259823, EPI_ISL_2259824, EPI_ISL_2259825, EPI_ISL_2259826, EPI_ISL_2259827, EPI_ISL_2259828, EPI_ISL_2259829, EPI_ISL_2259830, EPI_ISL_2259831, EPI_ISL_2259832, EPI_ISL_2259833, EPI_ISL_2259835, EPI_ISL_2259836, EPI_ISL_2259837, EPI_ISL_2259838, EPI_ISL_2259839, EPI_ISL_2259840, EPI_ISL_2259841, EPI_ISL_2259842, EPI_ISL_2259843, EPI_ISL_2259844, EPI_ISL_2259846, EPI_ISL_2259847, EPI_ISL_2259848, EPI_ISL_2259849, EPI_ISL_2259850, EPI_ISL_2259851, EPI_ISL_2259852, EPI_ISL_2259853, EPI_ISL_2259854, EPI_ISL_2259855, EPI_ISL_2259856, EPI_ISL_2259857, EPI_ISL_2259858, EPI_ISL_2259860, EPI_ISL_2259861, EPI_ISL_2259862, EPI_ISL_2259863, EPI_ISL_2259864, EPI_ISL_2259865, EPI_ISL_2259866, EPI_ISL_2259867, EPI_ISL_2259868, EPI_ISL_2259869, EPI_ISL_2259871, EPI_ISL_2259872, EPI_ISL_2259874, EPI_ISL_2259875, EPI_ISL_2259876, EPI_ISL_2259877, EPI_ISL_2259878, EPI_ISL_2259879, EPI_ISL_2259880, EPI_ISL_2259881, EPI_ISL_2259882, EPI_ISL_2259883, EPI_ISL_2259884, EPI_ISL_2259885, EPI_ISL_2259886, EPI_ISL_2259888, EPI_ISL_2259889, EPI_ISL_2259890, EPI_ISL_2259891, EPI_ISL_2259892, EPI_ISL_2259893, EPI_ISL_2259894, EPI_ISL_2259895, EPI_ISL_2259896, EPI_ISL_2259897, EPI_ISL_2259898, EPI_ISL_2259900, EPI_ISL_2259901, EPI_ISL_2259902, EPI_ISL_2259903, EPI_ISL_2259904, EPI_ISL_2259905, EPI_ISL_2259906, EPI_ISL_2259907, EPI_ISL_2259908, EPI_ISL_2259909, EPI_ISL_2259910, EPI_ISL_2259911, EPI_ISL_2259913, EPI_ISL_2259914, EPI_ISL_2259915, EPI_ISL_2259916, EPI_ISL_2259917, EPI_ISL_2259918, EPI_ISL_2259919, EPI_ISL_2259920, EPI_ISL_2259921, EPI_ISL_2259922, EPI_ISL_2259923, EPI_ISL_2259924, EPI_ISL_2259926, EPI_ISL_2259927, EPI_ISL_2259928, EPI_ISL_2259929, EPI_ISL_2259930, EPI_ISL_2259931, EPI_ISL_2259932, EPI_ISL_2259934, EPI_ISL_2259935, EPI_ISL_2259936, EPI_ISL_2259937, EPI_ISL_2259938, EPI_ISL_2259940, EPI_ISL_2259941, EPI_ISL_2259942, EPI_ISL_2259943, EPI_ISL_2259944, EPI_ISL_2259945, EPI_ISL_2259946, EPI_ISL_2259947, EPI_ISL_2259948, EPI_ISL_2259949, EPI_ISL_2259951, EPI_ISL_2259952, EPI_ISL_2259953, EPI_ISL_2259954, EPI_ISL_2259955, EPI_ISL_2259956, EPI_ISL_2259957, EPI_ISL_2259958, EPI_ISL_2259959, EPI_ISL_2259960, EPI_ISL_2259961, EPI_ISL_2259962, EPI_ISL_2259965, EPI_ISL_2259966, EPI_ISL_2259967, EPI_ISL_2259968, EPI_ISL_2259969, EPI_ISL_2259970, EPI_ISL_2259971, EPI_ISL_2259973, EPI_ISL_2259974, EPI_ISL_2259975, EPI_ISL_2259976, EPI_ISL_2259977, EPI_ISL_2259978, EPI_ISL_2259979, EPI_ISL_2259980, EPI_ISL_2259981, EPI_ISL_2259982, EPI_ISL_2259983, EPI_ISL_2259985, EPI_ISL_2259986, EPI_ISL_2259987, EPI_ISL_2259988, EPI_ISL_2259989, EPI_ISL_2259990, EPI_ISL_2259991, EPI_ISL_2259992, EPI_ISL_2259993, EPI_ISL_2259994, EPI_ISL_2259996, EPI_ISL_2259997, EPI_ISL_2259998, EPI_ISL_2259999, EPI_ISL_2260000, EPI_ISL_2260001, EPI_ISL_2260002, EPI_ISL_2260003, EPI_ISL_2260004, EPI_ISL_2260005, EPI_ISL_2260006, EPI_ISL_2260007, EPI_ISL_2260009, EPI_ISL_2260010, EPI_ISL_2260011, EPI_ISL_2260012, EPI_ISL_2260013, EPI_ISL_2260014, EPI_ISL_2260015, EPI_ISL_2260017, EPI_ISL_2260018, EPI_ISL_2260019, EPI_ISL_2260020, EPI_ISL_2260021, EPI_ISL_2260022, EPI_ISL_2260024, EPI_ISL_2260025, EPI_ISL_2260026, EPI_ISL_2260028, EPI_ISL_2260029, EPI_ISL_2260030, EPI_ISL_2260031, EPI_ISL_2260032, EPI_ISL_2260033, EPI_ISL_2260034, EPI_ISL_2260036, EPI_ISL_2260038, EPI_ISL_2260039, EPI_ISL_2260040, EPI_ISL_2267954, EPI_ISL_2268200, EPI_ISL_2386887, EPI_ISL_2386916, EPI_ISL_2386936, EPI_ISL_2386980, EPI_ISL_2386985, EPI_ISL_2386993, EPI_ISL_2386995, EPI_ISL_2387009, EPI_ISL_2387010, EPI_ISL_2387019, EPI_ISL_2387031, EPI_ISL_2387037, EPI_ISL_2387041, EPI_ISL_2387055, EPI_ISL_2387073, EPI_ISL_2387084, EPI_ISL_2387104, EPI_ISL_2387114, EPI_ISL_2387116, EPI_ISL_2387117, EPI_ISL_2387136, EPI_ISL_2387141, EPI_ISL_2387165, EPI_ISL_2387178, EPI_ISL_2387181, EPI_ISL_2471185, EPI_ISL_2471186, EPI_ISL_2471191, EPI_ISL_2471199, EPI_ISL_2471206, EPI_ISL_2471217, EPI_ISL_2471220, EPI_ISL_2471224, EPI_ISL_2471235, EPI_ISL_2471238, EPI_ISL_2471243, EPI_ISL_2471245, EPI_ISL_2471259, EPI_ISL_2471274, EPI_ISL_2471297, EPI_ISL_2471311, EPI_ISL_2471318, EPI_ISL_2471337, EPI_ISL_2471350, EPI_ISL_2471352, EPI_ISL_2471359, EPI_ISL_2471360, EPI_ISL_2471361, EPI_ISL_2471368, EPI_ISL_2471369, EPI_ISL_2471376, EPI_ISL_2471380, EPI_ISL_2471383, EPI_ISL_2471384, EPI_ISL_2471387, EPI_ISL_2471398, EPI_ISL_2471411, EPI_ISL_2471413, EPI_ISL_2471415, EPI_ISL_2471423, EPI_ISL_2471451, EPI_ISL_2471455 | | | |
| see above | CENTOGENE Frankfurt Laboratory: Niederlassung Industriepark Höchst | Robert Koch Institute | |
| EPI_ISL_2678682, EPI_ISL_2678685 | Carl-Thiem-Klinikum Cottbus - Zentrum für Laboratoriumsmedizin Mikrobiologie und Krankenhaushygiene | Robert Koch Institute | |
| EPI_ISL_2380589, EPI_ISL_2380590, EPI_ISL_2380591, EPI_ISL_2380592, EPI_ISL_2380593, EPI_ISL_2380735, EPI_ISL_2380736, EPI_ISL_2380868, EPI_ISL_2380869, EPI_ISL_2380941, EPI_ISL_2380942, EPI_ISL_2380944, EPI_ISL_2381463, EPI_ISL_2381464, EPI_ISL_2381465 | | | |
| see above | Center of Medical Microbiology, Virology, and Hospital Hygiene, University of Duesseldorf | Center of Medical Microbiology, Virology, and Hospital Hygiene, University of Duesseldorf | Alexander Dilthey; Andreas Walker; Daniel Strelow; Jessica Nicolai; Jörg Timm; Klaus Pfeffer; Lisanna Hülse; Malte Kohns Vasconcelos; Maximilian Damagnez; Nadine Lübke; Tobias Wienemann; Torsten Houwaart |
| EPI_ISL_2112944 | Chemische und Veterinaruntersuchungsamt Monsterland-Emscher-Lippe (CVUA-MEL) | Robert Koch Institute | |
| EPI_ISL_2129673, EPI_ISL_2129674, EPI_ISL_2129675, EPI_ISL_2129676, EPI_ISL_2129677, EPI_ISL_2129679 | Chemische und VeterinÃ¤runtersuchungsamt MÃ¼nsterland-Emscher-Lippe (CVUA-MEL) | Robert Koch Institute | |
| EPI_ISL_2264645 | Chemische und Veterinäruntersuchungsamt Münsterland-Emscher-Lippe (CVUA-MEL) | Robert Koch Institute | |
| EPI_ISL_2124269, EPI_ISL_2124270, EPI_ISL_2124273, EPI_ISL_2124274, EPI_ISL_2124276, EPI_ISL_2124277, EPI_ISL_2124279, EPI_ISL_2124282, EPI_ISL_2124284, EPI_ISL_2124287, EPI_ISL_2124291, EPI_ISL_2124293, EPI_ISL_2124298, EPI_ISL_2124303, EPI_ISL_2124307, EPI_ISL_2124309, EPI_ISL_2124312, EPI_ISL_2124315, EPI_ISL_2124322, EPI_ISL_2124323, EPI_ISL_2124325, EPI_ISL_2124326, EPI_ISL_2124327, EPI_ISL_2124328, EPI_ISL_2124330, EPI_ISL_2124331, EPI_ISL_2124333, EPI_ISL_2124337 | | | |
| see above | Clemenshospital - Alexianer Clemenshospital Munster | Robert Koch Institute | |
| EPI_ISL_2263201, EPI_ISL_2263202, EPI_ISL_2263203, EPI_ISL_2263204, EPI_ISL_2263205, EPI_ISL_2263206, EPI_ISL_2263207, EPI_ISL_2263208, EPI_ISL_2264589, EPI_ISL_2264590, EPI_ISL_2264591, EPI_ISL_2264593, EPI_ISL_2264595, EPI_ISL_2264596, EPI_ISL_2264598, EPI_ISL_2264599, EPI_ISL_2264601, EPI_ISL_2264602, EPI_ISL_2264605, EPI_ISL_2264607, EPI_ISL_2264608, EPI_ISL_2264609, EPI_ISL_2264610, EPI_ISL_2264612, EPI_ISL_2264613, EPI_ISL_2264614, EPI_ISL_2264615, EPI_ISL_2264616, EPI_ISL_2264635, EPI_ISL_2264636, EPI_ISL_2264637, EPI_ISL_2264638, EPI_ISL_2264639, EPI_ISL_2264640, EPI_ISL_2264641, EPI_ISL_2264642, EPI_ISL_2315842, EPI_ISL_2315851, EPI_ISL_2315863, EPI_ISL_2315864 | | | |
| see above | Clemenshospital - Alexianer Clemenshospital Münster | Robert Koch Institute | |
| EPI_ISL_2469762, EPI_ISL_2469766, EPI_ISL_2469770, EPI_ISL_2469771, EPI_ISL_2469774, EPI_ISL_2469776, EPI_ISL_2469777, EPI_ISL_2469780, EPI_ISL_2469781, EPI_ISL_2469782, EPI_ISL_2632124, EPI_ISL_2632128, EPI_ISL_2632133, EPI_ISL_2632135 | | | |
| see above | Corona-Testzentrum ifp Institut für Produktqualität GmbH | Robert Koch Institute | |
| EPI_ISL_2261492, EPI_ISL_2261493, EPI_ISL_2261494, EPI_ISL_2261495, EPI_ISL_2261496, EPI_ISL_2261497, EPI_ISL_2261498, EPI_ISL_2261499, EPI_ISL_2261501, EPI_ISL_2261502, EPI_ISL_2261503, EPI_ISL_2261504, EPI_ISL_2261505, EPI_ISL_2261506, EPI_ISL_2261507, EPI_ISL_2261509, EPI_ISL_2261511, EPI_ISL_2261512, EPI_ISL_2261513, EPI_ISL_2261514, EPI_ISL_2261515, EPI_ISL_2261516, EPI_ISL_2261517, EPI_ISL_2261518, EPI_ISL_2261519, EPI_ISL_2261521, EPI_ISL_2261522, EPI_ISL_2261523, EPI_ISL_2261524, EPI_ISL_2261525, EPI_ISL_2261526, EPI_ISL_2261527, EPI_ISL_2261528, EPI_ISL_2261531, EPI_ISL_2261532, EPI_ISL_2261533, EPI_ISL_2261534, EPI_ISL_2261536, EPI_ISL_2261537, EPI_ISL_2261538, EPI_ISL_2261539, EPI_ISL_2261541, EPI_ISL_2261542, EPI_ISL_2261544, EPI_ISL_2261545, EPI_ISL_2261549, EPI_ISL_2266073, EPI_ISL_2266074, EPI_ISL_2266075, EPI_ISL_2266076, EPI_ISL_2266077, EPI_ISL_2266078, EPI_ISL_2266079, EPI_ISL_2266081, EPI_ISL_2266082, EPI_ISL_2266083, EPI_ISL_2266084, EPI_ISL_2266085, EPI_ISL_2266086, EPI_ISL_2266087, EPI_ISL_2266088, EPI_ISL_2266089, EPI_ISL_2266090, EPI_ISL_2266091, EPI_ISL_2266092, EPI_ISL_2266093, EPI_ISL_2266095, EPI_ISL_2266097, EPI_ISL_2313894, EPI_ISL_2313895, EPI_ISL_2313896, EPI_ISL_2313897, EPI_ISL_2313898, EPI_ISL_2313899 | | | |
| see above | Department für Labormedizin Abteilung III Bereich Molekulare Diagnostik Universitätsklinikum Halle(Saale) | Robert Koch Institute | |
| EPI_ISL_2314886, EPI_ISL_2314887, EPI_ISL_2314888, EPI_ISL_2314889, EPI_ISL_2314890, EPI_ISL_2314891, EPI_ISL_2314892, EPI_ISL_2314893, EPI_ISL_2314894, EPI_ISL_2314895, EPI_ISL_2314896, EPI_ISL_2314897, EPI_ISL_2314898, EPI_ISL_2314899, EPI_ISL_2314900, EPI_ISL_2314901, EPI_ISL_2314902, EPI_ISL_2314903, EPI_ISL_2314904, EPI_ISL_2314905, EPI_ISL_2314906, EPI_ISL_2314907, EPI_ISL_2314908, EPI_ISL_2314909, EPI_ISL_2314910, EPI_ISL_2314911, EPI_ISL_2314912, EPI_ISL_2314913, EPI_ISL_2314914, EPI_ISL_2314915, EPI_ISL_2314916, EPI_ISL_2314917, EPI_ISL_2314918, EPI_ISL_2314919, EPI_ISL_2314920, EPI_ISL_2314921, EPI_ISL_2314922, EPI_ISL_2314923, EPI_ISL_2314924, EPI_ISL_2314925, EPI_ISL_2314926, EPI_ISL_2314927, EPI_ISL_2314928, EPI_ISL_2314929, EPI_ISL_2314930, EPI_ISL_2314931, EPI_ISL_2314932, EPI_ISL_2314933, EPI_ISL_2314934, EPI_ISL_2314935, EPI_ISL_2314936, EPI_ISL_2314937, EPI_ISL_2314938, EPI_ISL_2314939, EPI_ISL_2314940, EPI_ISL_2314941, EPI_ISL_2314942, EPI_ISL_2314943, EPI_ISL_2314944, EPI_ISL_2314945, EPI_ISL_2314946, EPI_ISL_2314947, EPI_ISL_2314948, EPI_ISL_2314950, EPI_ISL_2314951, EPI_ISL_2314952, EPI_ISL_2314953, EPI_ISL_2314954, EPI_ISL_2314955, EPI_ISL_2314956, EPI_ISL_2314957, EPI_ISL_2314958, EPI_ISL_2314959, EPI_ISL_2314960, EPI_ISL_2314961, EPI_ISL_2314962, EPI_ISL_2314963, EPI_ISL_2314964, EPI_ISL_2314966, EPI_ISL_2314967, EPI_ISL_2314968, EPI_ISL_2314969, EPI_ISL_2314970, EPI_ISL_2314971, EPI_ISL_2314972, EPI_ISL_2314973, EPI_ISL_2314974, EPI_ISL_2314975, EPI_ISL_2314976, EPI_ISL_2314977, EPI_ISL_2314978, EPI_ISL_2314979, EPI_ISL_2314980, EPI_ISL_2314981, EPI_ISL_2314982, EPI_ISL_2314983, EPI_ISL_2314984, EPI_ISL_2314985, EPI_ISL_2314986, EPI_ISL_2314987, EPI_ISL_2314988, EPI_ISL_2314989, EPI_ISL_2314990, EPI_ISL_2314991, EPI_ISL_2314992, EPI_ISL_2314993, EPI_ISL_2314994, EPI_ISL_2314995, EPI_ISL_2314996, EPI_ISL_2314997, EPI_ISL_2314998, EPI_ISL_2314999, EPI_ISL_2315000, EPI_ISL_2315001, EPI_ISL_2315002, EPI_ISL_2315003, EPI_ISL_2315004, EPI_ISL_2315006, EPI_ISL_2315007, EPI_ISL_2315008, EPI_ISL_2315010, EPI_ISL_2315011, EPI_ISL_2315012, EPI_ISL_2315013, EPI_ISL_2315014, EPI_ISL_2315015, EPI_ISL_2315016, EPI_ISL_2315017, EPI_ISL_2315018, EPI_ISL_2315019, EPI_ISL_2315021, EPI_ISL_2315022, EPI_ISL_2315023, EPI_ISL_2315024, EPI_ISL_2315025, EPI_ISL_2315026, EPI_ISL_2315027, EPI_ISL_2315028, EPI_ISL_2315030, EPI_ISL_2315032, EPI_ISL_2315033, EPI_ISL_2315034, EPI_ISL_2315035, EPI_ISL_2315036, EPI_ISL_2315037, EPI_ISL_2315038, EPI_ISL_2315039, EPI_ISL_2315040, EPI_ISL_2315041, EPI_ISL_2315042, EPI_ISL_2315583, EPI_ISL_2315584, EPI_ISL_2315585, EPI_ISL_2315586, EPI_ISL_2315587, EPI_ISL_2315588, EPI_ISL_2315589, EPI_ISL_2315590, EPI_ISL_2315591, EPI_ISL_2315592, EPI_ISL_2315593, EPI_ISL_2315594, EPI_ISL_2470540 | | | |
| see above | Diagnosticum - Labor Neukirchen | Robert Koch Institute | |
| EPI_ISL_2129319, EPI_ISL_2129320, EPI_ISL_2129321, EPI_ISL_2129322, EPI_ISL_2129323, EPI_ISL_2129324, EPI_ISL_2129325, EPI_ISL_2129326, EPI_ISL_2129327, EPI_ISL_2129328, EPI_ISL_2129329, EPI_ISL_2129330, EPI_ISL_2129331, EPI_ISL_2129332, EPI_ISL_2129333, EPI_ISL_2129334, EPI_ISL_2129335, EPI_ISL_2129336, EPI_ISL_2129337, EPI_ISL_2129338, EPI_ISL_2129339, EPI_ISL_2129340, EPI_ISL_2129341, EPI_ISL_2129342, EPI_ISL_2129343, EPI_ISL_2129344, EPI_ISL_2129345, EPI_ISL_2129346, EPI_ISL_2129347, EPI_ISL_2129348, EPI_ISL_2129349, EPI_ISL_2129350, EPI_ISL_2129351, EPI_ISL_2129352, EPI_ISL_2129353, EPI_ISL_2129354, EPI_ISL_2129355, EPI_ISL_2129356, EPI_ISL_2129357, EPI_ISL_2129358, EPI_ISL_2129359, EPI_ISL_2129360 | | | |
| see above | Dianovis GmbH Greiz | Robert Koch Institute | |
| EPI_ISL_2122786, EPI_ISL_2122791, EPI_ISL_2122795, EPI_ISL_2122808, EPI_ISL_2122845, EPI_ISL_2122880, EPI_ISL_2122892, EPI_ISL_2122955 | | | |

| | | | |
|---|---|---|---|
| see above | ELBLAB GmbH Zentrum fur LaborMedizin | Robert Koch Institute | |
| EPI_ISL_2314349, EPI_ISL_2314350, EPI_ISL_2314351, EPI_ISL_2314352, EPI_ISL_2314353, EPI_ISL_2314354, EPI_ISL_2314355, EPI_ISL_2314356, EPI_ISL_2314357, EPI_ISL_2314358, EPI_ISL_2314359, EPI_ISL_2314360, EPI_ISL_2314361, EPI_ISL_2314362, EPI_ISL_2314363, EPI_ISL_2314364, EPI_ISL_2314365, EPI_ISL_2314366, EPI_ISL_2314367, EPI_ISL_2314368, EPI_ISL_2314369, EPI_ISL_2314370, EPI_ISL_2314371, EPI_ISL_2314372, EPI_ISL_2314373, EPI_ISL_2314374, EPI_ISL_2314375, EPI_ISL_2314376, EPI_ISL_2314377, EPI_ISL_2314378, EPI_ISL_2314379, EPI_ISL_2314380, EPI_ISL_2314381, EPI_ISL_2314382, EPI_ISL_2314383, EPI_ISL_2314384, EPI_ISL_2314385, EPI_ISL_2314386, EPI_ISL_2314387, EPI_ISL_2314388, EPI_ISL_2314389, EPI_ISL_2314390, EPI_ISL_2314391, EPI_ISL_2314392, EPI_ISL_2314393, EPI_ISL_2314394, EPI_ISL_2314395, EPI_ISL_2314396, EPI_ISL_2314397, EPI_ISL_2314398, EPI_ISL_2314399, EPI_ISL_2314400, EPI_ISL_2314401, EPI_ISL_2314402, EPI_ISL_2314403, EPI_ISL_2314404, EPI_ISL_2314405, EPI_ISL_2314406, EPI_ISL_2314407, EPI_ISL_2314408, EPI_ISL_2314409, EPI_ISL_2314410, EPI_ISL_2315595, EPI_ISL_2315596, EPI_ISL_2315597, EPI_ISL_2315598, EPI_ISL_2315599, EPI_ISL_2315600, EPI_ISL_2315601, EPI_ISL_2315602, EPI_ISL_2315603, EPI_ISL_2315604, EPI_ISL_2315605, EPI_ISL_2315606, EPI_ISL_2315607, EPI_ISL_2315608, EPI_ISL_2315609, EPI_ISL_2315610, EPI_ISL_2315611, EPI_ISL_2315612, EPI_ISL_2315613, EPI_ISL_2315614, EPI_ISL_2315615, EPI_ISL_2315616, EPI_ISL_2315617, EPI_ISL_2315618, EPI_ISL_2315619, EPI_ISL_2315620, EPI_ISL_2315621, EPI_ISL_2315622, EPI_ISL_2315623, EPI_ISL_2315624, EPI_ISL_2315625, EPI_ISL_2315626, EPI_ISL_2315627, EPI_ISL_2315628, EPI_ISL_2315629, EPI_ISL_2315630, EPI_ISL_2315631, EPI_ISL_2315632, EPI_ISL_2315633, EPI_ISL_2315634, EPI_ISL_2315635, EPI_ISL_2315636, EPI_ISL_2315637, EPI_ISL_2315638, EPI_ISL_2315639, EPI_ISL_2315640, EPI_ISL_2315641, EPI_ISL_2315642, EPI_ISL_2315643, EPI_ISL_2315644, EPI_ISL_2315645, EPI_ISL_2315646, EPI_ISL_2315647, EPI_ISL_2315648, EPI_ISL_2315649, EPI_ISL_2315650, EPI_ISL_2315651, EPI_ISL_2315652, EPI_ISL_2315653, EPI_ISL_2315654, EPI_ISL_2315655, EPI_ISL_2315656, EPI_ISL_2315657, EPI_ISL_2315658, EPI_ISL_2315659, EPI_ISL_2315660, EPI_ISL_2315661, EPI_ISL_2315662, EPI_ISL_2315663, EPI_ISL_2315664, EPI_ISL_2315665, EPI_ISL_2315666, EPI_ISL_2316829, EPI_ISL_2316833, EPI_ISL_2316836, EPI_ISL_2316840, EPI_ISL_2316843, EPI_ISL_2316847, EPI_ISL_2316849, EPI_ISL_2316853, EPI_ISL_2316856, EPI_ISL_2316858, EPI_ISL_2316861, EPI_ISL_2316863, EPI_ISL_2316871, EPI_ISL_2316874, EPI_ISL_2316877, EPI_ISL_2316880, EPI_ISL_2316884, EPI_ISL_2316888, EPI_ISL_2316891, EPI_ISL_2316894, EPI_ISL_2316898, EPI_ISL_2316901, EPI_ISL_2316905, EPI_ISL_2316909, EPI_ISL_2316911, EPI_ISL_2316915, EPI_ISL_2316918, EPI_ISL_2316921, EPI_ISL_2316925, EPI_ISL_2316928, EPI_ISL_2316931, EPI_ISL_2316934, EPI_ISL_2316937, EPI_ISL_2316938, EPI_ISL_2316939, EPI_ISL_2316942, EPI_ISL_2316945, EPI_ISL_2316950, EPI_ISL_2316952, EPI_ISL_2316955, EPI_ISL_2316959, EPI_ISL_2316962, EPI_ISL_2316966, EPI_ISL_2316969, EPI_ISL_2316974, EPI_ISL_2316977, EPI_ISL_2316981, EPI_ISL_2316984, EPI_ISL_2316988, EPI_ISL_2316990, EPI_ISL_2316994, EPI_ISL_2316997, EPI_ISL_2317001, EPI_ISL_2317003, EPI_ISL_2317004, EPI_ISL_2317005, EPI_ISL_2317006, EPI_ISL_2317007, EPI_ISL_2317008, EPI_ISL_2317009, EPI_ISL_2317010, EPI_ISL_2317011, EPI_ISL_2317012, EPI_ISL_2317013, EPI_ISL_2317014, EPI_ISL_2317015, EPI_ISL_2317016, EPI_ISL_2317017, EPI_ISL_2317018, EPI_ISL_2317019, EPI_ISL_2317020, EPI_ISL_2317021, EPI_ISL_2317022, EPI_ISL_2317023, EPI_ISL_2317024, EPI_ISL_2317025, EPI_ISL_2317026, EPI_ISL_2317027, EPI_ISL_2317028, EPI_ISL_2317029, EPI_ISL_2317030, EPI_ISL_2317031, EPI_ISL_2317032, EPI_ISL_2317033, EPI_ISL_2317034, EPI_ISL_2317035, EPI_ISL_2317036 | | | |
| see above | Eurofins LifeCodexx GmbH | Robert Koch Institute | |
| EPI_ISL_2110761, EPI_ISL_2110762 | Eurofins MVZ Labor Gelsenkirchen | Robert Koch Institute | |
| EPI_ISL_2262306, EPI_ISL_2262307, EPI_ISL_2262308, EPI_ISL_2262309, EPI_ISL_2262310, EPI_ISL_2262311, EPI_ISL_2262314, EPI_ISL_2262315, EPI_ISL_2262316, EPI_ISL_2262318, EPI_ISL_2262319, EPI_ISL_2262321, EPI_ISL_2262322, EPI_ISL_2262323 | | | |
| see above | Fraunhofer-Institut für Zelltherapie und Immunologie IZI AG Next-Generation Diagnostics | Robert Koch Institute | |
| EPI_ISL_2260964, EPI_ISL_2260967, EPI_ISL_2260974, EPI_ISL_2261003, EPI_ISL_2261015, EPI_ISL_2261017, EPI_ISL_2261038, EPI_ISL_2261042 | | | |
| see above | Helios Universitätsklinikum Wuppertal: Universität Witten/Herdecke: Institut für Medizinische Labordiagnostik | Robert Koch Institute | |
| EPI_ISL_2112589, EPI_ISL_2112652, EPI_ISL_2129964 | Hufeland Klinikum GmbH | Robert Koch Institute | |
| EPI_ISL_2262405, EPI_ISL_2262410, EPI_ISL_2262429, EPI_ISL_2314414, EPI_ISL_2314421, EPI_ISL_2314433 | IMD - Institut für Medizinische Diagnostik Berlin-Potsdam | Robert Koch Institute | |
| EPI_ISL_2262403, EPI_ISL_2262404, EPI_ISL_2262408, EPI_ISL_2262409, EPI_ISL_2262412, EPI_ISL_2262413, EPI_ISL_2262415, EPI_ISL_2262416, EPI_ISL_2262419, EPI_ISL_2262422, EPI_ISL_2262424, EPI_ISL_2262426, EPI_ISL_2262430, EPI_ISL_2262431, EPI_ISL_2262432, EPI_ISL_2262434, EPI_ISL_2262435, EPI_ISL_2314070 | | | |
| see above | IMD - Labor Oderland | Robert Koch Institute | |
| EPI_ISL_2266151, EPI_ISL_2266165, EPI_ISL_2314413, EPI_ISL_2314415, EPI_ISL_2314418, EPI_ISL_2314422, EPI_ISL_2314423, EPI_ISL_2314427, EPI_ISL_2314430, EPI_ISL_2314431, EPI_ISL_2314436, EPI_ISL_2314437, EPI_ISL_2314438 | | | |
| see above | IMD - MVZ Labor Greifswald | Robert Koch Institute | |
| EPI_ISL_2123399, EPI_ISL_2262407, EPI_ISL_2262411, EPI_ISL_2262417, EPI_ISL_2262433, EPI_ISL_2262436, EPI_ISL_2262437, EPI_ISL_2262439, EPI_ISL_2266137, EPI_ISL_2266144, EPI_ISL_2314416, EPI_ISL_2314417, EPI_ISL_2314419, EPI_ISL_2314420, EPI_ISL_2314424, EPI_ISL_2314426, EPI_ISL_2314428, EPI_ISL_2314429, EPI_ISL_2314432, EPI_ISL_2314434, EPI_ISL_2314435, EPI_ISL_2314439 | | | |
| see above | IMD - MVZ Labor Martinsried | Robert Koch Institute | |
| EPI_ISL_2262406, EPI_ISL_2262414, EPI_ISL_2262418, EPI_ISL_2262421, EPI_ISL_2262423, EPI_ISL_2262425, EPI_ISL_2262427, EPI_ISL_2262438, EPI_ISL_2262440 | | | |
| see above | IMD - Medizinisches Labor Rostock | Robert Koch Institute | |
| EPI_ISL_2120139, EPI_ISL_2120156, EPI_ISL_2120162, EPI_ISL_2120167, EPI_ISL_2120203, EPI_ISL_2120231, EPI_ISL_2120237, EPI_ISL_2120255, EPI_ISL_2120275, EPI_ISL_2260062, EPI_ISL_2260063, EPI_ISL_2260065, EPI_ISL_2260066, EPI_ISL_2260067, EPI_ISL_2260068, EPI_ISL_2260069, EPI_ISL_2260071, EPI_ISL_2260072, EPI_ISL_2260073, EPI_ISL_2260075, EPI_ISL_2260076, EPI_ISL_2260078, EPI_ISL_2260079, EPI_ISL_2260080, EPI_ISL_2260081, EPI_ISL_2260082, EPI_ISL_2260083, EPI_ISL_2260084, EPI_ISL_2260085, EPI_ISL_2260086, EPI_ISL_2260087, EPI_ISL_2260088, EPI_ISL_2260089, EPI_ISL_2264486, EPI_ISL_2264487, EPI_ISL_2264503, EPI_ISL_2264510, EPI_ISL_2264512, EPI_ISL_2264517, EPI_ISL_2264520, EPI_ISL_2264522, EPI_ISL_2264528, EPI_ISL_2264529, EPI_ISL_2264532, EPI_ISL_2264536, EPI_ISL_2264537, EPI_ISL_2264538, EPI_ISL_2264540 | | | |
| see above | IMD Labor Frankfurt | Robert Koch Institute | |
| EPI_ISL_2112831 | Institut fÃ¼r Immunologie und Genetik Kaiserslautern: Medizinisches Labor Dr. med. B. Thiele | Robert Koch Institute | |
| EPI_ISL_2260997, EPI_ISL_2261000 | Institut für Klinische Chemie Transfusions- und Laboratoriumsmedizin Berufsgenossenschaftliches Universitätsklinikum Bergmannsheil | Robert Koch Institute | |
| EPI_ISL_2262442, EPI_ISL_2262443, EPI_ISL_2262446, EPI_ISL_2262448 | Institut für Klinische Transfusionsmedizin und Immungenetik DRK Blutspendedienst Baden-Württemberg | Robert Koch Institute | |
| EPI_ISL_2262632, EPI_ISL_2262639, EPI_ISL_2262640, EPI_ISL_2265461 | Institut für Medizinische Laboratoriumsdiagnostik Bochum GmbH | Robert Koch Institute | |
| EPI_ISL_2261487, EPI_ISL_2266415, EPI_ISL_2266416, EPI_ISL_2266417, EPI_ISL_2266421, EPI_ISL_2631862, EPI_ISL_2631867 | | | |
| see above | Institut für Medizinische Mikrobiologie - Universitätsmedizin Göttingen | Robert Koch Institute | |
| EPI_ISL_2262216, EPI_ISL_2262217, EPI_ISL_2262219, EPI_ISL_2262220, EPI_ISL_2262221 | Institut für med. Mikrobiologie u. Virologie Klinikum Oldenburg | Robert Koch Institute | |
| EPI_ISL_1941537, EPI_ISL_2285386, EPI_ISL_2285387, EPI_ISL_2285431, EPI_ISL_2285432, EPI_ISL_2285476, EPI_ISL_2285477, EPI_ISL_2285513, EPI_ISL_2285514, EPI_ISL_2285515, EPI_ISL_2285516 | | | |
| see above | Institute of Medical Microbiology and Hospital Hygiene | Institute of Medical Microbiology and Hospital Hygiene | Aljoscha Tersteegen; Prof. Dr. Achim Kaasch |
| EPI_ISL_2535303, EPI_ISL_2535304, EPI_ISL_2535305, EPI_ISL_2535327, EPI_ISL_2535337, EPI_ISL_2535338, EPI_ISL_2535340, EPI_ISL_2535341, EPI_ISL_2535345, EPI_ISL_2535346, EPI_ISL_2535351, EPI_ISL_2535352, EPI_ISL_2535354, EPI_ISL_2535355, EPI_ISL_2535374, EPI_ISL_2535375, EPI_ISL_2535376, EPI_ISL_2535377, EPI_ISL_2535399, EPI_ISL_2535401, EPI_ISL_2535406 | | | |
| see above | Institute of Virology, Medical Center, University of Freiburg, Freiburg, Germany | Institute of Virology, Clinial Virus Genomics, Medical Center, University of Freiburg, Freiburg, Germany | Hajo Grundmann; Jonas Fuchs; Lena Jaki; Lisa Kern; Marcus Panning; Sandra Reuter |
| EPI_ISL_2385065, EPI_ISL_2385066, EPI_ISL_2385067, EPI_ISL_2385068, EPI_ISL_2385069 | Kath. Marienkrankenhaus | Heinrich Pette Institute, Leibniz Institute for Experimental Virology | Adam Grundhoff; Alexis Robitaille; Johannes Knobloch; Martin Aepfelbacher; Nicole Fischer; Thomas Günther |
| EPI_ISL_2120589, EPI_ISL_2122605, EPI_ISL_2129112, EPI_ISL_2259751, EPI_ISL_2260935 | Kerckhoff-Klinik GmbH | Robert Koch Institute | |
| EPI_ISL_2120441, EPI_ISL_2120550, EPI_ISL_2120577 | Klinikum Darmstadt GmbH | Robert Koch Institute | |
| EPI_ISL_2116074, EPI_ISL_2116075, EPI_ISL_2116077, EPI_ISL_2116078, EPI_ISL_2116079, EPI_ISL_2116080, EPI_ISL_2116081, EPI_ISL_2116082, EPI_ISL_2116083, EPI_ISL_2116084, EPI_ISL_2116085, EPI_ISL_2116086, EPI_ISL_2116087 | | | |
| see above | Klinikum Ernst von Bergmann gemeinnÃ¼tzige GmbH - stationÃ¤rer Bereich | Robert Koch Institute | |
| EPI_ISL_2264741, EPI_ISL_2264742, EPI_ISL_2264744, EPI_ISL_2264745, EPI_ISL_2264747, EPI_ISL_2264748, EPI_ISL_2264749, EPI_ISL_2264754, EPI_ISL_2264755, EPI_ISL_2264759, EPI_ISL_2264761, EPI_ISL_2264798 | | | |
| see above | Klinikum Ernst von Bergmann gemeinnützige GmbH - stationärer Bereich | Robert Koch Institute | |
| EPI_ISL_2260610, EPI_ISL_2260721, EPI_ISL_2260725 | Klinikum Fulda gAG | Robert Koch Institute | |
| EPI_ISL_2386751, EPI_ISL_2386752 | Klinikum Osnabrück Medizinisches Labor | Robert Koch Institute | |
| EPI_ISL_2389457, EPI_ISL_2389458 | Klinikum Stuttgart | Robert Koch Institute | |
| EPI_ISL_2123808, EPI_ISL_2124349, EPI_ISL_2262827, EPI_ISL_2264543, EPI_ISL_2264553, EPI_ISL_2264738 | Krankenhausgesellschaft St. Vincenz mbH | Robert Koch Institute | |
| EPI_ISL_2122793, EPI_ISL_2122799, EPI_ISL_2122821, EPI_ISL_2122838, EPI_ISL_2122877, EPI_ISL_2122890, EPI_ISL_2122898, EPI_ISL_2122924, EPI_ISL_2122936, EPI_ISL_2122944, EPI_ISL_2122953 | | | |
| see above | LABOR CHEMNITZ Zentrum fur Diagnostik am Klinikum Chemnitz | Robert Koch Institute | |
| EPI_ISL_2260612, EPI_ISL_2260655, EPI_ISL_2260734, EPI_ISL_2260834 | LabKom - Labor Hamburg-Lübeck | Robert Koch Institute | |
| EPI_ISL_2260616, EPI_ISL_2260625, EPI_ISL_2260631, EPI_ISL_2260640, EPI_ISL_2260647, EPI_ISL_2260657, EPI_ISL_2260672, EPI_ISL_2260713, EPI_ISL_2260722, EPI_ISL_2260723, EPI_ISL_2260746, EPI_ISL_2260759, EPI_ISL_2260773, EPI_ISL_2260780, EPI_ISL_2260787, EPI_ISL_2260793, EPI_ISL_2260797, EPI_ISL_2260800, EPI_ISL_2260801, EPI_ISL_2260836, EPI_ISL_2260841, EPI_ISL_2260852, EPI_ISL_2260858, EPI_ISL_2260859, EPI_ISL_2260862, EPI_ISL_2260869, EPI_ISL_2260890, EPI_ISL_2260902, EPI_ISL_2260907, EPI_ISL_2260910, EPI_ISL_2260915 | | | |
| see above | LabKom - Labor Hannover MVZ GmbH | Robert Koch Institute | |
| EPI_ISL_2260098, EPI_ISL_2260110, EPI_ISL_2260120, EPI_ISL_2260121, EPI_ISL_2260145, EPI_ISL_2260164, EPI_ISL_2260181, EPI_ISL_2260190, EPI_ISL_2260191, EPI_ISL_2260194, EPI_ISL_2260217, EPI_ISL_2260236, EPI_ISL_2260237, EPI_ISL_2260239, EPI_ISL_2260252, EPI_ISL_2260273, EPI_ISL_2260276, EPI_ISL_2260283, EPI_ISL_2260288, EPI_ISL_2260291, EPI_ISL_2260292, EPI_ISL_2260293, EPI_ISL_2260296, EPI_ISL_2260297, EPI_ISL_2260304, EPI_ISL_2260311, EPI_ISL_2260326, EPI_ISL_2260335, EPI_ISL_2260352, EPI_ISL_2260358, EPI_ISL_2260374, EPI_ISL_2260389, EPI_ISL_2260410, EPI_ISL_2260415, EPI_ISL_2260688, EPI_ISL_2260703 | | | |
| see above | LabKom - Labor Mainz MVZ GmbH | Robert Koch Institute | |
| EPI_ISL_2260608, EPI_ISL_2260615, EPI_ISL_2260617, EPI_ISL_2260624, EPI_ISL_2260626, EPI_ISL_2260633, EPI_ISL_2260639, EPI_ISL_2260644, EPI_ISL_2260645, EPI_ISL_2260648, EPI_ISL_2260652, EPI_ISL_2260671, EPI_ISL_2260679, EPI_ISL_2260685, EPI_ISL_2260686, EPI_ISL_2260696, EPI_ISL_2260697, EPI_ISL_2260708, EPI_ISL_2260715, EPI_ISL_2260719, EPI_ISL_2260727, EPI_ISL_2260729, EPI_ISL_2260732, EPI_ISL_2260741, EPI_ISL_2260747, EPI_ISL_2260749, EPI_ISL_2260751, EPI_ISL_2260753, EPI_ISL_2260757, EPI_ISL_2260760, EPI_ISL_2260763, EPI_ISL_2260772, EPI_ISL_2260786, EPI_ISL_2260788, EPI_ISL_2260804, EPI_ISL_2260821, EPI_ISL_2260824, EPI_ISL_2260825, EPI_ISL_2260828, EPI_ISL_2260837, EPI_ISL_2260838, EPI_ISL_2260843, EPI_ISL_2260849, EPI_ISL_2260850, EPI_ISL_2260851, EPI_ISL_2260854, EPI_ISL_2260857, EPI_ISL_2260864, EPI_ISL_2260875, EPI_ISL_2260892, EPI_ISL_2260894, EPI_ISL_2260898, EPI_ISL_2260899, EPI_ISL_2260900, EPI_ISL_2260905, EPI_ISL_2260912, EPI_ISL_2265591, EPI_ISL_2265609, EPI_ISL_2265622, EPI_ISL_2265658 | | | |
| see above | LabKom - Labor an der Salzbrücke MVZ GmbH | Robert Koch Institute | |
| EPI_ISL_2125488, EPI_ISL_2125490, EPI_ISL_2125508, EPI_ISL_2125515, EPI_ISL_2125517, EPI_ISL_2125553, EPI_ISL_2125555, EPI_ISL_2125569, EPI_ISL_2125594, EPI_ISL_2125608, EPI_ISL_2125611, EPI_ISL_2125650, EPI_ISL_2125653, EPI_ISL_2125670, EPI_ISL_2125692, EPI_ISL_2125718, EPI_ISL_2125741, EPI_ISL_2125764, EPI_ISL_2125769, EPI_ISL_2125785, EPI_ISL_2125805, EPI_ISL_2125811, EPI_ISL_2125832, EPI_ISL_2125840, EPI_ISL_2125849, EPI_ISL_2125858, EPI_ISL_2125864, EPI_ISL_2125877, EPI_ISL_2125889, EPI_ISL_2125897, EPI_ISL_2125906, EPI_ISL_2125924, EPI_ISL_2125939, EPI_ISL_2125945, EPI_ISL_2125947, EPI_ISL_2125963, EPI_ISL_2125970, EPI_ISL_2125976, EPI_ISL_2125985, EPI_ISL_2126004, EPI_ISL_2126012, EPI_ISL_2126014, EPI_ISL_2126039, EPI_ISL_2126045, EPI_ISL_2126050, EPI_ISL_2126056, EPI_ISL_2126067, EPI_ISL_2126071, EPI_ISL_2126100, EPI_ISL_2126112, EPI_ISL_2126172, EPI_ISL_2126178, EPI_ISL_2260107, EPI_ISL_2260124, EPI_ISL_2260136, EPI_ISL_2260140, EPI_ISL_2260155, EPI_ISL_2260160, EPI_ISL_2260165, EPI_ISL_2260174, EPI_ISL_2260178, EPI_ISL_2260179, EPI_ISL_2260187, EPI_ISL_2260198, EPI_ISL_2260204, EPI_ISL_2260214, EPI_ISL_2260229, EPI_ISL_2260242, EPI_ISL_2260246, EPI_ISL_2260247, EPI_ISL_2260269, EPI_ISL_2260274, EPI_ISL_2260282, EPI_ISL_2260328, EPI_ISL_2260354, EPI_ISL_2260402, EPI_ISL_2260403, EPI_ISL_2260409, EPI_ISL_2260413, EPI_ISL_2260419, EPI_ISL_2260421, EPI_ISL_2265543, EPI_ISL_2265545, EPI_ISL_2265550, EPI_ISL_2265553, EPI_ISL_2265555, EPI_ISL_2265556, EPI_ISL_2265566, EPI_ISL_2265569, EPI_ISL_2265576, EPI_ISL_2265577, EPI_ISL_2265580, EPI_ISL_2265592, EPI_ISL_2265593, EPI_ISL_2265596, EPI_ISL_2265597, EPI_ISL_2265600, EPI_ISL_2265620, EPI_ISL_2265624, EPI_ISL_2265628, EPI_ISL_2265633, EPI_ISL_2265635, EPI_ISL_2265638, EPI_ISL_2265639, EPI_ISL_2265640, EPI_ISL_2265641, EPI_ISL_2265647, EPI_ISL_2265651, EPI_ISL_2265666, EPI_ISL_2265672, EPI_ISL_2265673, EPI_ISL_2265682, EPI_ISL_2265683, EPI_ISL_2265686, EPI_ISL_2265695, EPI_ISL_2265697, EPI_ISL_2265702, EPI_ISL_2265703, EPI_ISL_2265705, EPI_ISL_2265711, EPI_ISL_2265713, EPI_ISL_2265715, EPI_ISL_2265719, EPI_ISL_2265731, EPI_ISL_2265737, EPI_ISL_2265741, EPI_ISL_2265747, EPI_ISL_2265749, EPI_ISL_2265755, EPI_ISL_2265756, EPI_ISL_2265763, EPI_ISL_2265767, EPI_ISL_2265771, EPI_ISL_2265784, EPI_ISL_2265789, EPI_ISL_2265792, EPI_ISL_2265795, EPI_ISL_2265798, EPI_ISL_2265822, EPI_ISL_2265827, EPI_ISL_2265832, EPI_ISL_2265833, EPI_ISL_2265838, EPI_ISL_2265846, EPI_ISL_2265848, EPI_ISL_2265857, EPI_ISL_2265866, EPI_ISL_2265869 | | | |
| see above | LabKom - MVZ Labor Bochum MLB GmbH | Robert Koch Institute | |
| EPI_ISL_2125829, EPI_ISL_2260093, EPI_ISL_2260101, EPI_ISL_2260153, EPI_ISL_2260161, EPI_ISL_2260200, EPI_ISL_2260248, EPI_ISL_2260306, EPI_ISL_2260361, EPI_ISL_2260362, EPI_ISL_2260367, EPI_ISL_2260417 | | | |

| | | | |
|---|---|---|---|
| see above | Labor 28 MVZ GmbH | Robert Koch Institute | |
| EPI_ISL_2386818, EPI_ISL_2678386, EPI_ISL_2678387, EPI_ISL_2678388, EPI_ISL_2678389, EPI_ISL_2678390, EPI_ISL_2678391, EPI_ISL_2678392, EPI_ISL_2678393, EPI_ISL_2678394, EPI_ISL_2678395, EPI_ISL_2678396, EPI_ISL_2678397, EPI_ISL_2678398, EPI_ISL_2678399, EPI_ISL_2678400, EPI_ISL_2678401, EPI_ISL_2678402, EPI_ISL_2678403, EPI_ISL_2678404, EPI_ISL_2678405, EPI_ISL_2678406, EPI_ISL_2678407, EPI_ISL_2678408, EPI_ISL_2678409, EPI_ISL_2678410, EPI_ISL_2678411, EPI_ISL_2678412, EPI_ISL_2678413, EPI_ISL_2678414, EPI_ISL_2678415, EPI_ISL_2678416, EPI_ISL_2678417, EPI_ISL_2678418, EPI_ISL_2678420, EPI_ISL_2678421, EPI_ISL_2678422, EPI_ISL_2678423, EPI_ISL_2678424, EPI_ISL_2678425, EPI_ISL_2678426, EPI_ISL_2678427, EPI_ISL_2678428, EPI_ISL_2678429, EPI_ISL_2678430, EPI_ISL_2678431, EPI_ISL_2678432, EPI_ISL_2678433, EPI_ISL_2678434, EPI_ISL_2678435, EPI_ISL_2678436, EPI_ISL_2678437, EPI_ISL_2678438 | | | |
| see above | Labor Becker & Kollegen (Standort München) | Robert Koch Institute | |
| EPI_ISL_2107828, EPI_ISL_2107837, EPI_ISL_2107838, EPI_ISL_2107842, EPI_ISL_2107850, EPI_ISL_2107852, EPI_ISL_2107857, EPI_ISL_2107864, EPI_ISL_2107876, EPI_ISL_2107878, EPI_ISL_2107879, EPI_ISL_2107881, EPI_ISL_2227353, EPI_ISL_2227354, EPI_ISL_2227355, EPI_ISL_2227356, EPI_ISL_2227357, EPI_ISL_2227358, EPI_ISL_2227359, EPI_ISL_2227360, EPI_ISL_2227361, EPI_ISL_2227362, EPI_ISL_2227363, EPI_ISL_2227364, EPI_ISL_2227366, EPI_ISL_2227367, EPI_ISL_2227368, EPI_ISL_2227369, EPI_ISL_2227370, EPI_ISL_2227371, EPI_ISL_2227372, EPI_ISL_2227373, EPI_ISL_2339436, EPI_ISL_2339437, EPI_ISL_2339438, EPI_ISL_2339440, EPI_ISL_2339441, EPI_ISL_2339442, EPI_ISL_2339443, EPI_ISL_2339444, EPI_ISL_2339445, EPI_ISL_2339446, EPI_ISL_2339449, EPI_ISL_2339450, EPI_ISL_2339465, EPI_ISL_2339476 | | | |
| see above | Labor Berlin Charité Vivantes GmbH / Institut für Virologie | Charité Universitätsmedizin Berlin, Institut für Virologie/Labor Berlin | Barbara Mühlemann; Christian Drosten; Christine Stephan; Peter Menzel; Rolf Schwarzer; Terry Jones; Victor M Corman |
| EPI_ISL_2261058, EPI_ISL_2261061, EPI_ISL_2261065, EPI_ISL_2261070, EPI_ISL_2261091, EPI_ISL_2261099, EPI_ISL_2261103, EPI_ISL_2261107, EPI_ISL_2261126, EPI_ISL_2261129, EPI_ISL_2261158, EPI_ISL_2261167, EPI_ISL_2261171, EPI_ISL_2261176, EPI_ISL_2261183, EPI_ISL_2261194, EPI_ISL_2261200, EPI_ISL_2261203, EPI_ISL_2261204, EPI_ISL_2261209, EPI_ISL_2261212, EPI_ISL_2261217, EPI_ISL_2261221, EPI_ISL_2261226, EPI_ISL_2261231, EPI_ISL_2261239, EPI_ISL_2261252, EPI_ISL_2261254, EPI_ISL_2261255, EPI_ISL_2261263, EPI_ISL_2261268, EPI_ISL_2261273, EPI_ISL_2261284, EPI_ISL_2261288, EPI_ISL_2261294, EPI_ISL_2261299, EPI_ISL_2261308, EPI_ISL_2261318, EPI_ISL_2261326, EPI_ISL_2261334, EPI_ISL_2261347, EPI_ISL_2261356, EPI_ISL_2261373, EPI_ISL_2261389, EPI_ISL_2261396, EPI_ISL_2261401, EPI_ISL_2261402, EPI_ISL_2261405, EPI_ISL_2261407, EPI_ISL_2261409, EPI_ISL_2261410, EPI_ISL_2261418, EPI_ISL_2261423, EPI_ISL_2261431, EPI_ISL_2261436, EPI_ISL_2261440, EPI_ISL_2261445, EPI_ISL_2261448, EPI_ISL_2261451, EPI_ISL_2261453, EPI_ISL_2261456, EPI_ISL_2261457, EPI_ISL_2261459, EPI_ISL_2261462, EPI_ISL_2261463, EPI_ISL_2261465, EPI_ISL_2261467, EPI_ISL_2261468, EPI_ISL_2261470, EPI_ISL_2261472, EPI_ISL_2261473, EPI_ISL_2261478, EPI_ISL_2261479, EPI_ISL_2261480, EPI_ISL_2261482, EPI_ISL_2261484, EPI_ISL_2262485, EPI_ISL_2262487, EPI_ISL_2262520, EPI_ISL_2262560, EPI_ISL_2262565 | | | |
| see above | Labor Blackholm MVZ | Robert Koch Institute | |
| EPI_ISL_2384917, EPI_ISL_2384918, EPI_ISL_2384919, EPI_ISL_2384920, EPI_ISL_2384921, EPI_ISL_2384922, EPI_ISL_2384923, EPI_ISL_2384924, EPI_ISL_2384925, EPI_ISL_2384926, EPI_ISL_2384927, EPI_ISL_2384928, EPI_ISL_2384929, EPI_ISL_2384930, EPI_ISL_2384931, EPI_ISL_2384932, EPI_ISL_2384933, EPI_ISL_2384934, EPI_ISL_2384935, EPI_ISL_2384936, EPI_ISL_2384937, EPI_ISL_2384938, EPI_ISL_2384939, EPI_ISL_2384940, EPI_ISL_2384941, EPI_ISL_2384943, EPI_ISL_2384944, EPI_ISL_2384945, EPI_ISL_2384946, EPI_ISL_2384947, EPI_ISL_2384948, EPI_ISL_2384949, EPI_ISL_2384950, EPI_ISL_2384951, EPI_ISL_2384952, EPI_ISL_2384953, EPI_ISL_2384954, EPI_ISL_2384955, EPI_ISL_2384956, EPI_ISL_2384957, EPI_ISL_2384958, EPI_ISL_2384959, EPI_ISL_2384960, EPI_ISL_2384961, EPI_ISL_2384962, EPI_ISL_2384963, EPI_ISL_2384964, EPI_ISL_2384965, EPI_ISL_2384966, EPI_ISL_2384967, EPI_ISL_2384968, EPI_ISL_2384969, EPI_ISL_2384970, EPI_ISL_2384971, EPI_ISL_2384972, EPI_ISL_2384973, EPI_ISL_2384974, EPI_ISL_2384975, EPI_ISL_2384976, EPI_ISL_2384977, EPI_ISL_2384978, EPI_ISL_2384979, EPI_ISL_2384980, EPI_ISL_2384981, EPI_ISL_2384982, EPI_ISL_2384983, EPI_ISL_2384984, EPI_ISL_2384985, EPI_ISL_2384986, EPI_ISL_2384987, EPI_ISL_2384988, EPI_ISL_2384989, EPI_ISL_2384990, EPI_ISL_2384991, EPI_ISL_2384992, EPI_ISL_2384993, EPI_ISL_2384994, EPI_ISL_2384995, EPI_ISL_2384996, EPI_ISL_2384997, EPI_ISL_2384998, EPI_ISL_2384999, EPI_ISL_2385000, EPI_ISL_2385001, EPI_ISL_2385002, EPI_ISL_2385003, EPI_ISL_2385004, EPI_ISL_2385005, EPI_ISL_2385006, EPI_ISL_2385007, EPI_ISL_2385008, EPI_ISL_2385009, EPI_ISL_2385010, EPI_ISL_2385011, EPI_ISL_2385012, EPI_ISL_2385013, EPI_ISL_2385014, EPI_ISL_2385015, EPI_ISL_2385016, EPI_ISL_2385017, EPI_ISL_2385018, EPI_ISL_2385019, EPI_ISL_2385020, EPI_ISL_2385021, EPI_ISL_2385022, EPI_ISL_2385023, EPI_ISL_2385024, EPI_ISL_2385025, EPI_ISL_2385026, EPI_ISL_2385027, EPI_ISL_2385028, EPI_ISL_2385029, EPI_ISL_2385030, EPI_ISL_2385031, EPI_ISL_2385032, EPI_ISL_2385033, EPI_ISL_2385034, EPI_ISL_2385035, EPI_ISL_2385036, EPI_ISL_2385037, EPI_ISL_2385038, EPI_ISL_2385039, EPI_ISL_2385040, EPI_ISL_2385041, EPI_ISL_2385042, EPI_ISL_2385043, EPI_ISL_2385044, EPI_ISL_2385045, EPI_ISL_2385046, EPI_ISL_2385047, EPI_ISL_2385048, EPI_ISL_2385049, EPI_ISL_2385050, EPI_ISL_2385051, EPI_ISL_2385052, EPI_ISL_2385053, EPI_ISL_2385054, EPI_ISL_2385055, EPI_ISL_2385056, EPI_ISL_2385057, EPI_ISL_2385058, EPI_ISL_2385059, EPI_ISL_2385060, EPI_ISL_2385061, EPI_ISL_2385062, EPI_ISL_2385063, EPI_ISL_2385064 | | | |
| see above | Labor Dr. Fenner und Kollegen | Heinrich Pette Institute, Leibniz Institute for Experimental Virology | Adam Grundhoff; Alexis Robitaille; Johannes Knobloch; Martin Aepfelbacher; Nicole Fischer; Thomas Günther |
| EPI_ISL_2122958, EPI_ISL_2122959, EPI_ISL_2122960, EPI_ISL_2122970, EPI_ISL_2122984, EPI_ISL_2122992, EPI_ISL_2122994, EPI_ISL_2122996, EPI_ISL_2129681, EPI_ISL_2129683, EPI_ISL_2129684, EPI_ISL_2129685, EPI_ISL_2129687, EPI_ISL_2129694, EPI_ISL_2129696, EPI_ISL_2129698, EPI_ISL_2129699, EPI_ISL_2129700, EPI_ISL_2129701, EPI_ISL_2129702, EPI_ISL_2129703, EPI_ISL_2129704, EPI_ISL_2129705, EPI_ISL_2129706, EPI_ISL_2129707, EPI_ISL_2129710, EPI_ISL_2129711, EPI_ISL_2129712, EPI_ISL_2129714, EPI_ISL_2129715, EPI_ISL_2129716, EPI_ISL_2129717, EPI_ISL_2129720, EPI_ISL_2129721, EPI_ISL_2129722 | | | |
| see above | Labor Dr. Heidrich & Kollegen MVZ GmbH Hamburg | Robert Koch Institute | |
| EPI_ISL_2125481, EPI_ISL_2125501, EPI_ISL_2125528, EPI_ISL_2125578, EPI_ISL_2125580, EPI_ISL_2125633, EPI_ISL_2125648, EPI_ISL_2125709, EPI_ISL_2125727, EPI_ISL_2125732, EPI_ISL_2125743, EPI_ISL_2125746, EPI_ISL_2125750, EPI_ISL_2125814, EPI_ISL_2125816, EPI_ISL_2125819, EPI_ISL_2125869, EPI_ISL_2125886, EPI_ISL_2125953, EPI_ISL_2125956, EPI_ISL_2125961, EPI_ISL_2125965, EPI_ISL_2125983, EPI_ISL_2126020, EPI_ISL_2126027, EPI_ISL_2126054, EPI_ISL_2126060, EPI_ISL_2126093, EPI_ISL_2126102, EPI_ISL_2126114, EPI_ISL_2126133, EPI_ISL_2126148, EPI_ISL_2126153, EPI_ISL_2126156, EPI_ISL_2126183, EPI_ISL_2316030, EPI_ISL_2316156, EPI_ISL_2316191, EPI_ISL_2316200 | | | |
| see above | Labor Dr. Schumacher MVZ | Robert Koch Institute | |
| EPI_ISL_2265963, EPI_ISL_2265964, EPI_ISL_2265966, EPI_ISL_2265968, EPI_ISL_2265969 | Labor Dr. Spranger | Robert Koch Institute | |
| EPI_ISL_2125015, EPI_ISL_2125016, EPI_ISL_2125017 | Labor Dr. Wisplinghoff - Koln | Robert Koch Institute | |
| EPI_ISL_2263199, EPI_ISL_2263200, EPI_ISL_2266929 | Labor Dr. Wisplinghoff - Köln | Robert Koch Institute | |
| EPI_ISL_2260581, EPI_ISL_2260582, EPI_ISL_2260583, EPI_ISL_2260584, EPI_ISL_2260585, EPI_ISL_2260586, EPI_ISL_2260587 | | | |
| see above | Labor Kneißler GmbH & Co. KG | Robert Koch Institute | |
| EPI_ISL_2122434 | Labor Lobeck bzw. Laborarztliche Gemeinschaftspraxis Lobeck | Robert Koch Institute | |
| EPI_ISL_2129547, EPI_ISL_2129550, EPI_ISL_2129557 | Labor LÃ¼beck bzw. LaborÃ¤rztliche Gemeinschaftspraxis LÃ¼beck | Robert Koch Institute | |
| EPI_ISL_2260979, EPI_ISL_2261014, EPI_ISL_2261025, EPI_ISL_2261049, EPI_ISL_2261053 | Labor Lübeck bzw. Laborärztliche Gemeinschaftspraxis Lübeck | Robert Koch Institute | |
| EPI_ISL_2112767, EPI_ISL_2112807, EPI_ISL_2112864, EPI_ISL_2112865, EPI_ISL_2112866, EPI_ISL_2112867, EPI_ISL_2112868, EPI_ISL_2112869, EPI_ISL_2112870, EPI_ISL_2112871, EPI_ISL_2112872, EPI_ISL_2112873, EPI_ISL_2112874, EPI_ISL_2112875, EPI_ISL_2112876, EPI_ISL_2112877, EPI_ISL_2112878, EPI_ISL_2112879, EPI_ISL_2112880, EPI_ISL_2112881, EPI_ISL_2112882, EPI_ISL_2112883, EPI_ISL_2112884, EPI_ISL_2112885, EPI_ISL_2112886, EPI_ISL_2112887, EPI_ISL_2112888, EPI_ISL_2112889, EPI_ISL_2112890, EPI_ISL_2112891, EPI_ISL_2112892, EPI_ISL_2112893, EPI_ISL_2112894, EPI_ISL_2112895, EPI_ISL_2112896, EPI_ISL_2112897, EPI_ISL_2112898, EPI_ISL_2112899, EPI_ISL_2112900, EPI_ISL_2112901, EPI_ISL_2112902, EPI_ISL_2112913, EPI_ISL_2112914, EPI_ISL_2112915, EPI_ISL_2112916, EPI_ISL_2112917, EPI_ISL_2112918, EPI_ISL_2112919, EPI_ISL_2112920, EPI_ISL_2112921, EPI_ISL_2112922, EPI_ISL_2112923, EPI_ISL_2112924, EPI_ISL_2112925, EPI_ISL_2112926, EPI_ISL_2112927, EPI_ISL_2112928, EPI_ISL_2112929, EPI_ISL_2112930, EPI_ISL_2112931, EPI_ISL_2112932, EPI_ISL_2112933, EPI_ISL_2112934, EPI_ISL_2112935, EPI_ISL_2112936, EPI_ISL_2112937, EPI_ISL_2112938, EPI_ISL_2112939, EPI_ISL_2112940, EPI_ISL_2112941, EPI_ISL_2115353, EPI_ISL_2115354, EPI_ISL_2115357, EPI_ISL_2115364, EPI_ISL_2123065, EPI_ISL_2124343, EPI_ISL_2124347, EPI_ISL_2125071, EPI_ISL_2125074, EPI_ISL_2125077, EPI_ISL_2128993, EPI_ISL_2128994, EPI_ISL_2128997, EPI_ISL_2128999, EPI_ISL_2129000, EPI_ISL_2129001, EPI_ISL_2129003, EPI_ISL_2129008, EPI_ISL_2129011, EPI_ISL_2129012, EPI_ISL_2129074, EPI_ISL_2129075, EPI_ISL_2129076, EPI_ISL_2129077, EPI_ISL_2129078, EPI_ISL_2129079, EPI_ISL_2129080, EPI_ISL_2129081, EPI_ISL_2129082, EPI_ISL_2129083, EPI_ISL_2129084, EPI_ISL_2129085, EPI_ISL_2129086, EPI_ISL_2129087, EPI_ISL_2129088, EPI_ISL_2129089, EPI_ISL_2129090, EPI_ISL_2129091, EPI_ISL_2129092, EPI_ISL_2129093, EPI_ISL_2129094, EPI_ISL_2129095, EPI_ISL_2129096, EPI_ISL_2129097, EPI_ISL_2129103, EPI_ISL_2129104, EPI_ISL_2129105, EPI_ISL_2129106, EPI_ISL_2129117, EPI_ISL_2129118, EPI_ISL_2129122, EPI_ISL_2129123, EPI_ISL_2129124, EPI_ISL_2129127, EPI_ISL_2129129, EPI_ISL_2129134, EPI_ISL_2129135, EPI_ISL_2129137, EPI_ISL_2129138, EPI_ISL_2129139, EPI_ISL_2129140, EPI_ISL_2129141, EPI_ISL_2129142, EPI_ISL_2129143, EPI_ISL_2129144, EPI_ISL_2129145, EPI_ISL_2129146, EPI_ISL_2129147, EPI_ISL_2129148, EPI_ISL_2129149, EPI_ISL_2129150, EPI_ISL_2129151, EPI_ISL_2129152, EPI_ISL_2129153, EPI_ISL_2129154, EPI_ISL_2129155, EPI_ISL_2129156, EPI_ISL_2129157, EPI_ISL_2129158, EPI_ISL_2129159, EPI_ISL_2129160, EPI_ISL_2129161, EPI_ISL_2129162, EPI_ISL_2129163, EPI_ISL_2129164, EPI_ISL_2129165, EPI_ISL_2129166, EPI_ISL_2129167, EPI_ISL_2129168, EPI_ISL_2129169, EPI_ISL_2129170, EPI_ISL_2129171, EPI_ISL_2129172, EPI_ISL_2129173, EPI_ISL_2129174, EPI_ISL_2129175, EPI_ISL_2129176, EPI_ISL_2129177, EPI_ISL_2129178, EPI_ISL_2129179, EPI_ISL_2129180 | | | |
| see above | Labor Monchengladbach MVZ Dr. Stein + Kollegen GbR | Robert Koch Institute | |
| EPI_ISL_2129535, EPI_ISL_2129536, EPI_ISL_2129537, EPI_ISL_2129538, EPI_ISL_2129539, EPI_ISL_2129540, EPI_ISL_2129541, EPI_ISL_2129542, EPI_ISL_2129543, EPI_ISL_2129544, EPI_ISL_2129545, EPI_ISL_2129546, EPI_ISL_2129638, EPI_ISL_2129639, EPI_ISL_2129642, EPI_ISL_2129646, EPI_ISL_2129647, EPI_ISL_2129648, EPI_ISL_2129649, EPI_ISL_2129650, EPI_ISL_2129651, EPI_ISL_2129652, EPI_ISL_2129653, EPI_ISL_2129654, EPI_ISL_2129655, EPI_ISL_2129656, EPI_ISL_2129657, EPI_ISL_2129658, EPI_ISL_2129659, EPI_ISL_2129660, EPI_ISL_2129661, EPI_ISL_2129662 | | | |
| see above | Labor MÃ¶nchengladbach MVZ Dr. Stein + Kollegen GbR | Robert Koch Institute | |
| EPI_ISL_2259744, EPI_ISL_2259745, EPI_ISL_2259747, EPI_ISL_2259753, EPI_ISL_2259763, EPI_ISL_2259775, EPI_ISL_2259782, EPI_ISL_2264547, EPI_ISL_2264564, EPI_ISL_2264577, EPI_ISL_2264580, EPI_ISL_2264581, EPI_ISL_2264582, EPI_ISL_2264587 | | | |
| see above | Labor Mönchengladbach MVZ Dr. Stein + Kollegen GbR | Robert Koch Institute | |
| EPI_ISL_2123371, EPI_ISL_2123372, EPI_ISL_2123373, EPI_ISL_2123374, EPI_ISL_2123380, EPI_ISL_2125029, EPI_ISL_2125103, EPI_ISL_2125105, EPI_ISL_2125109, EPI_ISL_2125112, EPI_ISL_2125114, EPI_ISL_2125115 | | | |
| see above | Labor Prof. Dr. G. Enders MVZ GbR | Robert Koch Institute | |
| EPI_ISL_2129184, EPI_ISL_2129189, EPI_ISL_2129194, EPI_ISL_2129205, EPI_ISL_2129212, EPI_ISL_2129215, EPI_ISL_2129220, EPI_ISL_2129224, EPI_ISL_2129230, EPI_ISL_2129237, EPI_ISL_2129239, EPI_ISL_2129242, EPI_ISL_2129248, EPI_ISL_2129249 | | | |
| see above | Labor ZOTZ\|KLIMAS; MVZ Dusseldorf-Centrum | Robert Koch Institute | |
| EPI_ISL_2129275, EPI_ISL_2129277, EPI_ISL_2129291, EPI_ISL_2129294, EPI_ISL_2129304 | Labor ZOTZ\|KLIMAS; MVZ DÃ¼sseldorf-Centrum | Robert Koch Institute | |
| EPI_ISL_2264813, EPI_ISL_2264818, EPI_ISL_2264820, EPI_ISL_2264822, EPI_ISL_2264827, EPI_ISL_2264829, EPI_ISL_2264830, EPI_ISL_2264832, EPI_ISL_2264837, EPI_ISL_2264839, EPI_ISL_2264841, EPI_ISL_2264843, EPI_ISL_2264845, EPI_ISL_2264847, EPI_ISL_2264849, EPI_ISL_2264851, EPI_ISL_2264853, EPI_ISL_2264855, EPI_ISL_2264857, EPI_ISL_2264860, EPI_ISL_2264862, EPI_ISL_2264864, EPI_ISL_2264866, EPI_ISL_2264868, EPI_ISL_2264870, EPI_ISL_2264872, EPI_ISL_2264874, EPI_ISL_2264876, EPI_ISL_2264878, EPI_ISL_2264881, EPI_ISL_2264883, EPI_ISL_2264885, EPI_ISL_2264887, EPI_ISL_2264889, EPI_ISL_2264891, EPI_ISL_2264892, EPI_ISL_2264894, EPI_ISL_2264895, EPI_ISL_2264896, EPI_ISL_2264897, EPI_ISL_2264898, EPI_ISL_2264899, EPI_ISL_2264900, EPI_ISL_2264901, EPI_ISL_2264902, EPI_ISL_2264903, EPI_ISL_2264904, EPI_ISL_2264905, EPI_ISL_2264906, EPI_ISL_2264907, EPI_ISL_2264908, EPI_ISL_2264909, EPI_ISL_2264910, EPI_ISL_2264911, EPI_ISL_2264912, EPI_ISL_2264913, EPI_ISL_2264914, EPI_ISL_2264915, EPI_ISL_2264916, EPI_ISL_2264917, EPI_ISL_2264918, EPI_ISL_2264919, EPI_ISL_2264920, EPI_ISL_2264921, EPI_ISL_2264922, EPI_ISL_2264924, EPI_ISL_2264925, EPI_ISL_2264926, EPI_ISL_2264927, EPI_ISL_2264928, EPI_ISL_2264929, EPI_ISL_2264930, EPI_ISL_2264931, EPI_ISL_2264933, EPI_ISL_2264934, EPI_ISL_2264935, EPI_ISL_2264936, EPI_ISL_2264937, EPI_ISL_2264938, EPI_ISL_2264939, EPI_ISL_2264940, EPI_ISL_2264941, EPI_ISL_2264942, EPI_ISL_2264943, EPI_ISL_2264944, EPI_ISL_2264945, EPI_ISL_2264946, EPI_ISL_2264947, EPI_ISL_2264948, EPI_ISL_2264949, EPI_ISL_2264950, EPI_ISL_2264951, EPI_ISL_2264952, EPI_ISL_2264953, EPI_ISL_2264954, EPI_ISL_2264955, EPI_ISL_2264956, EPI_ISL_2264957, EPI_ISL_2264958, EPI_ISL_2264959, EPI_ISL_2264960, EPI_ISL_2264961, EPI_ISL_2264962, EPI_ISL_2264963, EPI_ISL_2264964, EPI_ISL_2264965, EPI_ISL_2264966, EPI_ISL_2264967, EPI_ISL_2264968, EPI_ISL_2264969, EPI_ISL_2264970, EPI_ISL_2264971, EPI_ISL_2264972, EPI_ISL_2264973, EPI_ISL_2264975, EPI_ISL_2264976, EPI_ISL_2264977, EPI_ISL_2264978, EPI_ISL_2264979, EPI_ISL_2264980, EPI_ISL_2264981, EPI_ISL_2264982, EPI_ISL_2264983, EPI_ISL_2264984 | | | |
| see above | Labor ZOTZ\|KLIMAS; MVZ Düsseldorf-Centrum | Robert Koch Institute | |
| EPI_ISL_2112204, EPI_ISL_2112282, EPI_ISL_2112467, EPI_ISL_2117337, EPI_ISL_2117342, EPI_ISL_2117393, EPI_ISL_2117427, EPI_ISL_2123810, EPI_ISL_2123811, EPI_ISL_2124024, EPI_ISL_2124194, EPI_ISL_2124233 | | | |
| see above | Labor fur medizinische Mikrobiologie Bakteriologie Mykologie Virologie & Infektionsserologie - Labor Malbis | Robert Koch Institute | |
| EPI_ISL_2263188, EPI_ISL_2263189, EPI_ISL_2263190, EPI_ISL_2263191, EPI_ISL_2263192 | Laborarztpraxis Dres. med. Walther Weindel & Kollegen | Robert Koch Institute | |
| EPI_ISL_2260919, EPI_ISL_2260920, EPI_ISL_2260924, EPI_ISL_2260927, EPI_ISL_2260931, EPI_ISL_2260933, EPI_ISL_2260939, EPI_ISL_2260944, EPI_ISL_2260947, EPI_ISL_2260949 | | | |
| see above | Laborarztpraxis Jochen Hüter | Robert Koch Institute | |
| EPI_ISL_2122446, EPI_ISL_2122458, EPI_ISL_2122501, EPI_ISL_2122549 | Laborarztpraxis Osnabruck | Robert Koch Institute | |
| EPI_ISL_2260965, EPI_ISL_2260966, EPI_ISL_2260972, EPI_ISL_2260977, EPI_ISL_2260981, EPI_ISL_2260982, EPI_ISL_2260983, EPI_ISL_2260985, EPI_ISL_2260986, EPI_ISL_2260989, EPI_ISL_2260992, EPI_ISL_2260995, EPI_ISL_2260996, EPI_ISL_2260998, EPI_ISL_2261002, EPI_ISL_2261004, EPI_ISL_2261005, EPI_ISL_2261007, EPI_ISL_2261008, EPI_ISL_2261009, EPI_ISL_2261016, EPI_ISL_2261018, EPI_ISL_2261020, EPI_ISL_2261021, EPI_ISL_2261024, EPI_ISL_2261026, EPI_ISL_2261029, EPI_ISL_2261033, EPI_ISL_2261035, EPI_ISL_2261036, EPI_ISL_2261037, EPI_ISL_2261040, EPI_ISL_2261043, EPI_ISL_2261050, EPI_ISL_2261051 | | | |
| see above | Laborarztpraxis Osnabrück | Robert Koch Institute | |
| EPI_ISL_2120180, EPI_ISL_2120190, EPI_ISL_2260074, EPI_ISL_2264518, EPI_ISL_2264526, EPI_ISL_2313965, EPI_ISL_2314023, EPI_ISL_2314031 | | | |
| see above | Labormedizin Darmstadt | Robert Koch Institute | |
| EPI_ISL_1941485, EPI_ISL_1941486, EPI_ISL_1941487, EPI_ISL_1941488, EPI_ISL_1941489, EPI_ISL_1941490, EPI_ISL_1941491, EPI_ISL_1941492, EPI_ISL_1941493, EPI_ISL_1941494, EPI_ISL_1941495, EPI_ISL_1941496, EPI_ISL_1941497, EPI_ISL_1941498, EPI_ISL_1941499, EPI_ISL_1941500, EPI_ISL_1941501, EPI_ISL_1941502, EPI_ISL_1941503, EPI_ISL_1941504, EPI_ISL_1941505, EPI_ISL_1941506, EPI_ISL_1941507, EPI_ISL_1941508, EPI_ISL_1941509, EPI_ISL_1941511, EPI_ISL_1941512, EPI_ISL_1941513, EPI_ISL_1941514, EPI_ISL_1941518, EPI_ISL_1941520, EPI_ISL_1941522, EPI_ISL_1941524, EPI_ISL_1941526, EPI_ISL_1941528, EPI_ISL_1941530, EPI_ISL_1941535, EPI_ISL_2285376, EPI_ISL_2285377, EPI_ISL_2285378, EPI_ISL_2285379, EPI_ISL_2285380, EPI_ISL_2285381, EPI_ISL_2285382, EPI_ISL_2285383, EPI_ISL_2285384, EPI_ISL_2285385, EPI_ISL_2285388, EPI_ISL_2285389, EPI_ISL_2285390, EPI_ISL_2285391, EPI_ISL_2285392, EPI_ISL_2285393, EPI_ISL_2285394, EPI_ISL_2285395, EPI_ISL_2285396, EPI_ISL_2285397, EPI_ISL_2285398, EPI_ISL_2285399, EPI_ISL_2285400, EPI_ISL_2285401, EPI_ISL_2285402, EPI_ISL_2285403, EPI_ISL_2285404, EPI_ISL_2285405, EPI_ISL_2285406, EPI_ISL_2285407, EPI_ISL_2285408, EPI_ISL_2285409, EPI_ISL_2285410, EPI_ISL_2285411, EPI_ISL_2285412, EPI_ISL_2285413, EPI_ISL_2285414, EPI_ISL_2285415, EPI_ISL_2285416, EPI_ISL_2285417, EPI_ISL_2285418, EPI_ISL_2285419, EPI_ISL_2285420, EPI_ISL_2285421, EPI_ISL_2285422, EPI_ISL_2285423, EPI_ISL_2285424, EPI_ISL_2285425, EPI_ISL_2285426, EPI_ISL_2285427, EPI_ISL_2285428, EPI_ISL_2285429, EPI_ISL_2285430, EPI_ISL_2285433, EPI_ISL_2285434, EPI_ISL_2285435, EPI_ISL_2285436, EPI_ISL_2285437, EPI_ISL_2285438, EPI_ISL_2285439, EPI_ISL_2285440, EPI_ISL_2285441, EPI_ISL_2285442, EPI_ISL_2285443, EPI_ISL_2285444, EPI_ISL_2285445, EPI_ISL_2285446, EPI_ISL_2285447, EPI_ISL_2285448, EPI_ISL_2285449, EPI_ISL_2285450, EPI_ISL_2285451, EPI_ISL_2285452, EPI_ISL_2285453, EPI_ISL_2285454, EPI_ISL_2285455, EPI_ISL_2285456, EPI_ISL_2285457, EPI_ISL_2285458, EPI_ISL_2285459, EPI_ISL_2285460, EPI_ISL_2285461, EPI_ISL_2285462, EPI_ISL_2285463, EPI_ISL_2285464, EPI_ISL_2285465, EPI_ISL_2285466, EPI_ISL_2285467, EPI_ISL_2285468, EPI_ISL_2285469, EPI_ISL_2285470, EPI_ISL_2285471, EPI_ISL_2285472, EPI_ISL_2285473, EPI_ISL_2285474, EPI_ISL_2285475, EPI_ISL_2285478, EPI_ISL_2285479, EPI_ISL_2285480, EPI_ISL_2285481, EPI_ISL_2285482, EPI_ISL_2285483, EPI_ISL_2285484, EPI_ISL_2285485, EPI_ISL_2285486, EPI_ISL_2285487, EPI_ISL_2285488, EPI_ISL_2285489, EPI_ISL_2285490, EPI_ISL_2285491, EPI_ISL_2285492, EPI_ISL_2285493, EPI_ISL_2285494, EPI_ISL_2285495, EPI_ISL_2285496, EPI_ISL_2285497, EPI_ISL_2285498, EPI_ISL_2285499, EPI_ISL_2285500, EPI_ISL_2285501, EPI_ISL_2285502, EPI_ISL_2285503, EPI_ISL_2285504, EPI_ISL_2285505, EPI_ISL_2285506, EPI_ISL_2285507, EPI_ISL_2285508, EPI_ISL_2285509, EPI_ISL_2285510, EPI_ISL_2285511, EPI_ISL_2285517, EPI_ISL_2285518, EPI_ISL_2285519, EPI_ISL_2285520, EPI_ISL_2285521, EPI_ISL_2285522, EPI_ISL_2285523, EPI_ISL_2285524, EPI_ISL_2285525, EPI_ISL_2285526, EPI_ISL_2285527, EPI_ISL_2285528, EPI_ISL_2285529, EPI_ISL_2285530, EPI_ISL_2285531, EPI_ISL_2285532, EPI_ISL_2285533, EPI_ISL_2285534, EPI_ISL_2285535, EPI_ISL_2285536, EPI_ISL_2285537, EPI_ISL_2285538, EPI_ISL_2285539, EPI_ISL_2285540, EPI_ISL_2285541, EPI_ISL_2285542, EPI_ISL_2285543, EPI_ISL_2285544, EPI_ISL_2285545, EPI_ISL_2285546, EPI_ISL_2285547, EPI_ISL_2285548, EPI_ISL_2285549, EPI_ISL_2285550 | | | |
| see above | Landesamt für Verbraucherschutz Sachsen Anhalt, Magdeburg | Institute of Medical Microbiology and Hospital Hygiene | Aljoscha Tersteegen; Prof. Dr. Achim Kaasch |
| EPI_ISL_2259442, EPI_ISL_2259443, EPI_ISL_2259444, EPI_ISL_2259445, EPI_ISL_2259446, EPI_ISL_2259447, EPI_ISL_2259448, EPI_ISL_2259449, EPI_ISL_2259450, EPI_ISL_2259451, EPI_ISL_2259452, EPI_ISL_2259453, EPI_ISL_2259454, EPI_ISL_2259455, EPI_ISL_2259457, EPI_ISL_2259458, EPI_ISL_2259459, EPI_ISL_2259460, EPI_ISL_2259461, EPI_ISL_2259463, EPI_ISL_2259464, EPI_ISL_2259465, | | | |

| | | | |
|---|---|---|---|
| EPI_ISL_2259466, EPI_ISL_2259467, EPI_ISL_2259468, EPI_ISL_2259469, EPI_ISL_2259471, EPI_ISL_2259472, EPI_ISL_2259473, EPI_ISL_2259474, EPI_ISL_2259475, EPI_ISL_2259476, EPI_ISL_2259477, EPI_ISL_2259478, EPI_ISL_2259479, EPI_ISL_2259482, EPI_ISL_2259485, EPI_ISL_2259486, EPI_ISL_2259487, EPI_ISL_2259488, EPI_ISL_2259489, EPI_ISL_2259490, EPI_ISL_2259491, EPI_ISL_2259492, EPI_ISL_2259493, EPI_ISL_2314834, EPI_ISL_2314836, EPI_ISL_2314840, EPI_ISL_2314841, EPI_ISL_2314842, EPI_ISL_2314844, EPI_ISL_2314845, EPI_ISL_2314846, EPI_ISL_2314851, EPI_ISL_2314852, EPI_ISL_2314855, EPI_ISL_2314859, EPI_ISL_2314860, EPI_ISL_2314861, EPI_ISL_2314862, EPI_ISL_2314863, EPI_ISL_2314864, EPI_ISL_2314865, EPI_ISL_2389875, EPI_ISL_2389881, EPI_ISL_2389896 | | | |
| see above | Landesgesundheitsamt Baden-Württemberg | Robert Koch Institute | |
| EPI_ISL_2122785, EPI_ISL_2122787, EPI_ISL_2122790, EPI_ISL_2122812, EPI_ISL_2122816, EPI_ISL_2122824, EPI_ISL_2122828, EPI_ISL_2122848, EPI_ISL_2122852, EPI_ISL_2122871, EPI_ISL_2122895, EPI_ISL_2122911, EPI_ISL_2122914, EPI_ISL_2122918, EPI_ISL_2122937, EPI_ISL_2122942, EPI_ISL_2122945, EPI_ISL_2122948, EPI_ISL_2122952, EPI_ISL_2122957 | | | |
| see above | Landesuntersuchungsanstalt fur das Gesundheits- und VeterinÃ¤rwesen (LUA) Sachsen | Robert Koch Institute | |
| EPI_ISL_2263144, EPI_ISL_2263145, EPI_ISL_2263146, EPI_ISL_2263147, EPI_ISL_2263148, EPI_ISL_2263149, EPI_ISL_2263150, EPI_ISL_2263151 | | | |
| see above | Limbach - Ganzimmun Diagnostics AG Mainz | Robert Koch Institute | |
| EPI_ISL_2115222, EPI_ISL_2115249, EPI_ISL_2115252, EPI_ISL_2115355, EPI_ISL_2115368, EPI_ISL_2120325, EPI_ISL_2120344, EPI_ISL_2120354, EPI_ISL_2120404, EPI_ISL_2120416, EPI_ISL_2120462, EPI_ISL_2120553, EPI_ISL_2120571, EPI_ISL_2120576, EPI_ISL_2120583, EPI_ISL_2124350, EPI_ISL_2125031, EPI_ISL_2125037, EPI_ISL_2125089, EPI_ISL_2125093, EPI_ISL_2128957, EPI_ISL_2128977, EPI_ISL_2128985, EPI_ISL_2259746, EPI_ISL_2259760, EPI_ISL_2259773 | | | |
| see above | Limbach - MVZ Gemeinschaftslabor Suhl | Robert Koch Institute | |
| EPI_ISL_2117436, EPI_ISL_2117443, EPI_ISL_2117447, EPI_ISL_2117453, EPI_ISL_2117463, EPI_ISL_2117467, EPI_ISL_2117475, EPI_ISL_2117486, EPI_ISL_2117488, EPI_ISL_2117491, EPI_ISL_2117497, EPI_ISL_2117499, EPI_ISL_2117510, EPI_ISL_2117531, EPI_ISL_2117535, EPI_ISL_2117551, EPI_ISL_2117561, EPI_ISL_2117564, EPI_ISL_2117576, EPI_ISL_2117583, EPI_ISL_2117587, EPI_ISL_2117589, EPI_ISL_2117593, EPI_ISL_2117596, EPI_ISL_2117602, EPI_ISL_2117609, EPI_ISL_2117612, EPI_ISL_2117618, EPI_ISL_2117625, EPI_ISL_2117631, EPI_ISL_2117640, EPI_ISL_2117642, EPI_ISL_2117662, EPI_ISL_2117666, EPI_ISL_2117677, EPI_ISL_2117683, EPI_ISL_2117686, EPI_ISL_2117692, EPI_ISL_2117695, EPI_ISL_2117698, EPI_ISL_2117701, EPI_ISL_2117707, EPI_ISL_2117710, EPI_ISL_2117712, EPI_ISL_2117715, EPI_ISL_2117718, EPI_ISL_2117721, EPI_ISL_2117732, EPI_ISL_2117738, EPI_ISL_2117741, EPI_ISL_2117744, EPI_ISL_2117753, EPI_ISL_2117757, EPI_ISL_2117763, EPI_ISL_2117766, EPI_ISL_2117771, EPI_ISL_2117774, EPI_ISL_2117777, EPI_ISL_2117782, EPI_ISL_2117786, EPI_ISL_2117792, EPI_ISL_2117795, EPI_ISL_2117798, EPI_ISL_2117805, EPI_ISL_2117807, EPI_ISL_2117810, EPI_ISL_2117813, EPI_ISL_2117818, EPI_ISL_2117821, EPI_ISL_2117824, EPI_ISL_2117826, EPI_ISL_2117830, EPI_ISL_2117833, EPI_ISL_2117841, EPI_ISL_2117844, EPI_ISL_2117849, EPI_ISL_2117852, EPI_ISL_2117861, EPI_ISL_2117863, EPI_ISL_2117869, EPI_ISL_2117872, EPI_ISL_2117878, EPI_ISL_2117884, EPI_ISL_2117890, EPI_ISL_2117892, EPI_ISL_2129361, EPI_ISL_2129364, EPI_ISL_2129366, EPI_ISL_2129368, EPI_ISL_2129371, EPI_ISL_2129373, EPI_ISL_2129374, EPI_ISL_2129378, EPI_ISL_2129380, EPI_ISL_2129381, EPI_ISL_2129383, EPI_ISL_2129385, EPI_ISL_2129386, EPI_ISL_2129387, EPI_ISL_2129388, EPI_ISL_2129389, EPI_ISL_2129391, EPI_ISL_2129392, EPI_ISL_2129393, EPI_ISL_2129394, EPI_ISL_2129395, EPI_ISL_2129397, EPI_ISL_2129398, EPI_ISL_2129399, EPI_ISL_2129401, EPI_ISL_2129403, EPI_ISL_2129405, EPI_ISL_2129406, EPI_ISL_2129407, EPI_ISL_2129409, EPI_ISL_2129411, EPI_ISL_2129412, EPI_ISL_2129413, EPI_ISL_2129416, EPI_ISL_2129417, EPI_ISL_2129419, EPI_ISL_2129421, EPI_ISL_2129422, EPI_ISL_2129423, EPI_ISL_2129424, EPI_ISL_2129425, EPI_ISL_2129428, EPI_ISL_2129429, EPI_ISL_2129433, EPI_ISL_2129434, EPI_ISL_2129436, EPI_ISL_2129437, EPI_ISL_2129439, EPI_ISL_2129440, EPI_ISL_2129441, EPI_ISL_2129443, EPI_ISL_2129444, EPI_ISL_2129448, EPI_ISL_2129449, EPI_ISL_2129450, EPI_ISL_2129453, EPI_ISL_2129456, EPI_ISL_2129459, EPI_ISL_2129464, EPI_ISL_2129466, EPI_ISL_2129467, EPI_ISL_2129470, EPI_ISL_2129473, EPI_ISL_2129475, EPI_ISL_2129477, EPI_ISL_2129479, EPI_ISL_2129480, EPI_ISL_2129484, EPI_ISL_2129487, EPI_ISL_2129490, EPI_ISL_2129491, EPI_ISL_2129492, EPI_ISL_2129495, EPI_ISL_2129496, EPI_ISL_2129497, EPI_ISL_2129499, EPI_ISL_2129502, EPI_ISL_2129503, EPI_ISL_2129504, EPI_ISL_2129506, EPI_ISL_2129508, EPI_ISL_2129509, EPI_ISL_2129510, EPI_ISL_2129512, EPI_ISL_2129513, EPI_ISL_2129516, EPI_ISL_2129525, EPI_ISL_2129529, EPI_ISL_2129530 | | | |
| see above | Limbach - MVZ Humangenetik Ulm | Robert Koch Institute | |
| EPI_ISL_2112725, EPI_ISL_2112732, EPI_ISL_2112733, EPI_ISL_2112738, EPI_ISL_2112800, EPI_ISL_2112809, EPI_ISL_2115147, EPI_ISL_2115148, EPI_ISL_2115151, EPI_ISL_2115154, EPI_ISL_2115156, EPI_ISL_2115157, EPI_ISL_2115158, EPI_ISL_2115160, EPI_ISL_2115165, EPI_ISL_2115166, EPI_ISL_2115167, EPI_ISL_2115168, EPI_ISL_2115169, EPI_ISL_2115170, EPI_ISL_2115171, EPI_ISL_2115172, EPI_ISL_2115173, EPI_ISL_2115174, EPI_ISL_2115175, EPI_ISL_2115182, EPI_ISL_2115183, EPI_ISL_2115184, EPI_ISL_2115185, EPI_ISL_2120090, EPI_ISL_2120093, EPI_ISL_2120097, EPI_ISL_2120099, EPI_ISL_2120102, EPI_ISL_2120105, EPI_ISL_2120108, EPI_ISL_2120111, EPI_ISL_2120118, EPI_ISL_2120122, EPI_ISL_2120125, EPI_ISL_2120128, EPI_ISL_2120288, EPI_ISL_2120300, EPI_ISL_2122670, EPI_ISL_2122673, EPI_ISL_2122675, EPI_ISL_2122678, EPI_ISL_2122686, EPI_ISL_2122688, EPI_ISL_2122694, EPI_ISL_2122700, EPI_ISL_2122709, EPI_ISL_2122720, EPI_ISL_2124358, EPI_ISL_2124361, EPI_ISL_2124364, EPI_ISL_2124365, EPI_ISL_2124369, EPI_ISL_2124370, EPI_ISL_2124371, EPI_ISL_2124373, EPI_ISL_2124375, EPI_ISL_2124377, EPI_ISL_2128990, EPI_ISL_2128991, EPI_ISL_2128995, EPI_ISL_2128996, EPI_ISL_2128998, EPI_ISL_2129002, EPI_ISL_2129004, EPI_ISL_2129005, EPI_ISL_2129006, EPI_ISL_2129007, EPI_ISL_2129009, EPI_ISL_2129010, EPI_ISL_2129637, EPI_ISL_2129640, EPI_ISL_2129641, EPI_ISL_2129643, EPI_ISL_2129644, EPI_ISL_2129645, EPI_ISL_2264546, EPI_ISL_2264556, EPI_ISL_2264559, EPI_ISL_2264563, EPI_ISL_2264566, EPI_ISL_2264568, EPI_ISL_2264570, EPI_ISL_2264574, EPI_ISL_2264585 | | | |
| see above | Limbach - MVZ Labor Eveld & Kollegen Essen | Robert Koch Institute | |
| EPI_ISL_2120565, EPI_ISL_2125057, EPI_ISL_2129125, EPI_ISL_2262237 | Limbach - MVZ Labor Ludwigsburg | Robert Koch Institute | |
| EPI_ISL_2112722, EPI_ISL_2112723, EPI_ISL_2112727, EPI_ISL_2112728, EPI_ISL_2112729, EPI_ISL_2112735, EPI_ISL_2112736, EPI_ISL_2112742, EPI_ISL_2112763, EPI_ISL_2112766, EPI_ISL_2112771, EPI_ISL_2112772, EPI_ISL_2112774, EPI_ISL_2112775, EPI_ISL_2112776, EPI_ISL_2112778, EPI_ISL_2112780, EPI_ISL_2112781, EPI_ISL_2112782, EPI_ISL_2112783, EPI_ISL_2112788, EPI_ISL_2112789, EPI_ISL_2112793, EPI_ISL_2112794, EPI_ISL_2112795, EPI_ISL_2112796, EPI_ISL_2112797, EPI_ISL_2112798, EPI_ISL_2112804, EPI_ISL_2112806, EPI_ISL_2115187, EPI_ISL_2115188, EPI_ISL_2115189, EPI_ISL_2115192, EPI_ISL_2115194, EPI_ISL_2115195, EPI_ISL_2115196, EPI_ISL_2115198, EPI_ISL_2115199, EPI_ISL_2115201, EPI_ISL_2115202, EPI_ISL_2115203, EPI_ISL_2115204, EPI_ISL_2115205, EPI_ISL_2115206, EPI_ISL_2115208, EPI_ISL_2115210, EPI_ISL_2115212, EPI_ISL_2115213, EPI_ISL_2115214, EPI_ISL_2115215, EPI_ISL_2115216, EPI_ISL_2115217, EPI_ISL_2115218, EPI_ISL_2115219, EPI_ISL_2115220, EPI_ISL_2115221, EPI_ISL_2115224, EPI_ISL_2115225, EPI_ISL_2115226, EPI_ISL_2115227, EPI_ISL_2115228, EPI_ISL_2115229, EPI_ISL_2115230, EPI_ISL_2115231, EPI_ISL_2115232, EPI_ISL_2115233, EPI_ISL_2115234, EPI_ISL_2115235, EPI_ISL_2115236, EPI_ISL_2115237, EPI_ISL_2115238, EPI_ISL_2115239, EPI_ISL_2115240, EPI_ISL_2115241, EPI_ISL_2115242, EPI_ISL_2115243, EPI_ISL_2115244, EPI_ISL_2115245, EPI_ISL_2115246, EPI_ISL_2115248, EPI_ISL_2115250, EPI_ISL_2115251, EPI_ISL_2115253, EPI_ISL_2115254, EPI_ISL_2115351, EPI_ISL_2115356, EPI_ISL_2115358, EPI_ISL_2115359, EPI_ISL_2115360, EPI_ISL_2115361, EPI_ISL_2115363, EPI_ISL_2115365, EPI_ISL_2115367, EPI_ISL_2120302, EPI_ISL_2120303, EPI_ISL_2120306, EPI_ISL_2120309, EPI_ISL_2120315, EPI_ISL_2120338, EPI_ISL_2120348, EPI_ISL_2120398, EPI_ISL_2120399, EPI_ISL_2120411, EPI_ISL_2120435, EPI_ISL_2120438, EPI_ISL_2120454, EPI_ISL_2120456, EPI_ISL_2122682, EPI_ISL_2122684, EPI_ISL_2122690, EPI_ISL_2122703, EPI_ISL_2122706, EPI_ISL_2122711, EPI_ISL_2122715, EPI_ISL_2122717, EPI_ISL_2122723, EPI_ISL_2122743, EPI_ISL_2122759, EPI_ISL_2123370, EPI_ISL_2123377, EPI_ISL_2123809, EPI_ISL_2123815, EPI_ISL_2123816, EPI_ISL_2123817, EPI_ISL_2123818, EPI_ISL_2123819, EPI_ISL_2123820, EPI_ISL_2123821, EPI_ISL_2123822, EPI_ISL_2123823, EPI_ISL_2123824, EPI_ISL_2123825, EPI_ISL_2123826, EPI_ISL_2123827, EPI_ISL_2123828, EPI_ISL_2123829, EPI_ISL_2123830, EPI_ISL_2123831, EPI_ISL_2123833, EPI_ISL_2123834, EPI_ISL_2123835, EPI_ISL_2123836, EPI_ISL_2123837, EPI_ISL_2123838, EPI_ISL_2123839, EPI_ISL_2123840, EPI_ISL_2123841, EPI_ISL_2123842, EPI_ISL_2123843, EPI_ISL_2124339, EPI_ISL_2124341, EPI_ISL_2124342, EPI_ISL_2124345, EPI_ISL_2124346, EPI_ISL_2124348, EPI_ISL_2124351, EPI_ISL_2124352, EPI_ISL_2124353, EPI_ISL_2124357, EPI_ISL_2124360, EPI_ISL_2124367, EPI_ISL_2124374, EPI_ISL_2125028, EPI_ISL_2125030, EPI_ISL_2125033, EPI_ISL_2125034, EPI_ISL_2125035, EPI_ISL_2125036, EPI_ISL_2125039, EPI_ISL_2125040, EPI_ISL_2125041, EPI_ISL_2125042, EPI_ISL_2125044, EPI_ISL_2125046, EPI_ISL_2125047, EPI_ISL_2125048, EPI_ISL_2125049, EPI_ISL_2125051, EPI_ISL_2125052, EPI_ISL_2125054, EPI_ISL_2125067, EPI_ISL_2125072, EPI_ISL_2125073, EPI_ISL_2125075, EPI_ISL_2125082, EPI_ISL_2125083, EPI_ISL_2125086, EPI_ISL_2125088, EPI_ISL_2125090, EPI_ISL_2125091, EPI_ISL_2125092, EPI_ISL_2125095, EPI_ISL_2125097, EPI_ISL_2125099, EPI_ISL_2128979, EPI_ISL_2128984, EPI_ISL_2129098, EPI_ISL_2129099, EPI_ISL_2129100, EPI_ISL_2129101, EPI_ISL_2129102, EPI_ISL_2129107, EPI_ISL_2129108, EPI_ISL_2129109, EPI_ISL_2129110, EPI_ISL_2129111, EPI_ISL_2129113, EPI_ISL_2129114, EPI_ISL_2129115, EPI_ISL_2129116, EPI_ISL_2129119, EPI_ISL_2129120, EPI_ISL_2129121, EPI_ISL_2129126, EPI_ISL_2129128, EPI_ISL_2129130, EPI_ISL_2129131, EPI_ISL_2129132, EPI_ISL_2129133, EPI_ISL_2129136 | | | |
| see above | Limbach - MVZ Labor Ravensburg Labor Dr. Gartner | Robert Koch Institute | |
| EPI_ISL_2260042, EPI_ISL_2260048, EPI_ISL_2260052, EPI_ISL_2260056, EPI_ISL_2260922, EPI_ISL_2260923, EPI_ISL_2260925, EPI_ISL_2260926, EPI_ISL_2260928, EPI_ISL_2260929, EPI_ISL_2260934, EPI_ISL_2260937, EPI_ISL_2260940, EPI_ISL_2260941, EPI_ISL_2260942, EPI_ISL_2260943, EPI_ISL_2260945, EPI_ISL_2260948, EPI_ISL_2262441, EPI_ISL_2263056, EPI_ISL_2263185, EPI_ISL_2264576, EPI_ISL_2264579, EPI_ISL_2264583, EPI_ISL_2264584, EPI_ISL_2264586, EPI_ISL_2264715 | | | |
| see above | Limbach - MVZ Labor Ravensburg Labor Dr. Gärtner | Robert Koch Institute | |
| EPI_ISL_2112731, EPI_ISL_2112734, EPI_ISL_2112739, EPI_ISL_2112740, EPI_ISL_2112741, EPI_ISL_2112743, EPI_ISL_2112764, EPI_ISL_2112768, EPI_ISL_2112769, EPI_ISL_2112770, EPI_ISL_2112773, EPI_ISL_2112777, EPI_ISL_2112779, EPI_ISL_2112784, EPI_ISL_2112785, EPI_ISL_2112786, EPI_ISL_2112787, EPI_ISL_2112790, EPI_ISL_2112792, EPI_ISL_2112799, EPI_ISL_2112803, EPI_ISL_2115193, EPI_ISL_2115200, EPI_ISL_2115223, EPI_ISL_2115362, EPI_ISL_2120294, EPI_ISL_2120297, EPI_ISL_2120308, EPI_ISL_2120312, EPI_ISL_2120318, EPI_ISL_2120321, EPI_ISL_2120327, EPI_ISL_2120330, EPI_ISL_2120333, EPI_ISL_2120335, EPI_ISL_2120341, EPI_ISL_2120351, EPI_ISL_2120414, EPI_ISL_2120421, EPI_ISL_2120427, EPI_ISL_2120444, EPI_ISL_2120451, EPI_ISL_2120459, EPI_ISL_2120465, EPI_ISL_2120475, EPI_ISL_2120478, EPI_ISL_2120481, EPI_ISL_2120488, EPI_ISL_2120490, EPI_ISL_2120493, EPI_ISL_2120496, EPI_ISL_2120499, EPI_ISL_2120501, EPI_ISL_2120502, EPI_ISL_2120504, EPI_ISL_2120507, EPI_ISL_2120510, EPI_ISL_2120513, EPI_ISL_2120516, EPI_ISL_2120519, EPI_ISL_2120521, EPI_ISL_2120524, EPI_ISL_2120527, EPI_ISL_2120531, EPI_ISL_2120537, EPI_ISL_2120540, EPI_ISL_2120543, EPI_ISL_2120546, EPI_ISL_2120568, EPI_ISL_2120580, EPI_ISL_2122601, EPI_ISL_2122697, EPI_ISL_2122729, EPI_ISL_2122732, EPI_ISL_2122734, EPI_ISL_2122737, EPI_ISL_2122740, EPI_ISL_2122746, EPI_ISL_2122749, EPI_ISL_2122753, EPI_ISL_2122755, EPI_ISL_2122757, EPI_ISL_2123064, EPI_ISL_2123376, EPI_ISL_2123426, EPI_ISL_2124366, EPI_ISL_2125043, EPI_ISL_2125050, EPI_ISL_2125068, EPI_ISL_2125069, EPI_ISL_2125070, EPI_ISL_2125076, EPI_ISL_2125078, EPI_ISL_2125079, EPI_ISL_2125080, EPI_ISL_2125081, EPI_ISL_2125084, EPI_ISL_2125085, EPI_ISL_2125087, EPI_ISL_2125096, EPI_ISL_2125098, EPI_ISL_2125100, EPI_ISL_2125101, EPI_ISL_2128955, EPI_ISL_2128960, EPI_ISL_2128961, EPI_ISL_2128962, EPI_ISL_2128963, EPI_ISL_2128964, EPI_ISL_2128965, EPI_ISL_2128968, EPI_ISL_2128969, EPI_ISL_2128971, EPI_ISL_2128975, EPI_ISL_2128976, EPI_ISL_2128978, EPI_ISL_2128988, EPI_ISL_2260041, EPI_ISL_2260043, EPI_ISL_2260044, EPI_ISL_2260046, EPI_ISL_2260049, EPI_ISL_2260051, EPI_ISL_2260053, EPI_ISL_2260054, EPI_ISL_2260055, EPI_ISL_2260058, EPI_ISL_2260059, EPI_ISL_2260061, EPI_ISL_2260921, EPI_ISL_2260930, EPI_ISL_2260932, EPI_ISL_2260936, EPI_ISL_2260938, EPI_ISL_2260946, EPI_ISL_2262824, EPI_ISL_2263184, EPI_ISL_2264569, EPI_ISL_2264706, EPI_ISL_2264713, EPI_ISL_2264714, EPI_ISL_2264717, EPI_ISL_2264718, EPI_ISL_2264723, EPI_ISL_2314664, EPI_ISL_2314672 | | | |
| see above | MDI Limbach Berlin GmbH; MVZ Labor Berlin | Robert Koch Institute | |
| EPI_ISL_2772714, EPI_ISL_2772717 | MKK Mühlenkreiskliniken, Johannes Wesling Klinikum Minden | Hannover Medical School, Institute of Virology | Lars Steinbrück |
| EPI_ISL_2122321, EPI_ISL_2122329, EPI_ISL_2122339, EPI_ISL_2122342, EPI_ISL_2122344, EPI_ISL_2122373, EPI_ISL_2122379, EPI_ISL_2122411, EPI_ISL_2122436, EPI_ISL_2122469, EPI_ISL_2122472, EPI_ISL_2122492, EPI_ISL_2122493, EPI_ISL_2122506, EPI_ISL_2122514, EPI_ISL_2122517, EPI_ISL_2122522, EPI_ISL_2122526, EPI_ISL_2122527, EPI_ISL_2122528, EPI_ISL_2122531, EPI_ISL_2122534, EPI_ISL_2122536, EPI_ISL_2122539, EPI_ISL_2122544, EPI_ISL_2122552, EPI_ISL_2122558, EPI_ISL_2122561, EPI_ISL_2122564, EPI_ISL_2122574, EPI_ISL_2129548, EPI_ISL_2129549, EPI_ISL_2129551, EPI_ISL_2129552, EPI_ISL_2129553, EPI_ISL_2129555, EPI_ISL_2129556, EPI_ISL_2129559, EPI_ISL_2129560, EPI_ISL_2129561, EPI_ISL_2129562, EPI_ISL_2129563, EPI_ISL_2129564, EPI_ISL_2129566, EPI_ISL_2129567, EPI_ISL_2129568, EPI_ISL_2129569, EPI_ISL_2129570, EPI_ISL_2129571, EPI_ISL_2129572, EPI_ISL_2129573, EPI_ISL_2129575, EPI_ISL_2129576, EPI_ISL_2129577, EPI_ISL_2129578, EPI_ISL_2129579, EPI_ISL_2129580, EPI_ISL_2129581, EPI_ISL_2129582, EPI_ISL_2129583, EPI_ISL_2129584, EPI_ISL_2129585, EPI_ISL_2129587, EPI_ISL_2129588, EPI_ISL_2129590, EPI_ISL_2129591, EPI_ISL_2129592, EPI_ISL_2129593, EPI_ISL_2129594, EPI_ISL_2129595, EPI_ISL_2129596, EPI_ISL_2129597, EPI_ISL_2129598, EPI_ISL_2129599, EPI_ISL_2129600, EPI_ISL_2129601, EPI_ISL_2129602, EPI_ISL_2129603, EPI_ISL_2129604, EPI_ISL_2129606, EPI_ISL_2129607, EPI_ISL_2129608, EPI_ISL_2129609, EPI_ISL_2129610, EPI_ISL_2129611, EPI_ISL_2129612, EPI_ISL_2129613, EPI_ISL_2129614, EPI_ISL_2129615, EPI_ISL_2129616, EPI_ISL_2129617, EPI_ISL_2129618, EPI_ISL_2129619, EPI_ISL_2129620, EPI_ISL_2129621, EPI_ISL_2129623, EPI_ISL_2129625, EPI_ISL_2129626, EPI_ISL_2129627, EPI_ISL_2129628, EPI_ISL_2129629, EPI_ISL_2129630, EPI_ISL_2129631, EPI_ISL_2129632, EPI_ISL_2129633, EPI_ISL_2129634, EPI_ISL_2129635, EPI_ISL_2129636, EPI_ISL_2260980, EPI_ISL_2260999, EPI_ISL_2261010, EPI_ISL_2261011, EPI_ISL_2261019, EPI_ISL_2261023, EPI_ISL_2261034, EPI_ISL_2261045, EPI_ISL_2313256, EPI_ISL_2387404, EPI_ISL_2388183, EPI_ISL_2388304, EPI_ISL_2469897 | | | |
| see above | MVZ Dr. Eberhard & Partner Dortmund | Robert Koch Institute | |
| EPI_ISL_2127101, EPI_ISL_2127103, EPI_ISL_2127106, EPI_ISL_2127107, EPI_ISL_2127110, EPI_ISL_2127112, EPI_ISL_2127114, EPI_ISL_2127119, EPI_ISL_2127121, EPI_ISL_2127123, EPI_ISL_2127125, EPI_ISL_2127127, EPI_ISL_2127129, EPI_ISL_2127132, EPI_ISL_2127134, EPI_ISL_2127136, EPI_ISL_2127138, EPI_ISL_2127140, EPI_ISL_2127143, EPI_ISL_2127145, EPI_ISL_2127148, EPI_ISL_2127150, EPI_ISL_2127152, EPI_ISL_2127155, EPI_ISL_2127157, EPI_ISL_2127159, EPI_ISL_2127161, EPI_ISL_2127163, EPI_ISL_2127166, EPI_ISL_2127168, EPI_ISL_2127170, EPI_ISL_2127172, EPI_ISL_2127174, EPI_ISL_2127176, EPI_ISL_2127178, EPI_ISL_2127180, EPI_ISL_2127182, EPI_ISL_2127185, EPI_ISL_2127186, EPI_ISL_2127188, EPI_ISL_2127190, EPI_ISL_2127192, EPI_ISL_2127194, EPI_ISL_2127196, EPI_ISL_2127198, EPI_ISL_2127200, EPI_ISL_2127203, EPI_ISL_2127204, EPI_ISL_2127207, EPI_ISL_2127209, EPI_ISL_2127211, EPI_ISL_2127213, EPI_ISL_2127215, EPI_ISL_2127216, EPI_ISL_2264097, EPI_ISL_2264121, EPI_ISL_2264138 | | | |
| see above | MVZ Labor Dr. Fenner und Kollegen (Standort Hamburg) | Robert Koch Institute | |
| EPI_ISL_2120044, EPI_ISL_2120047, EPI_ISL_2120050, EPI_ISL_2120055, EPI_ISL_2120056, EPI_ISL_2120058, EPI_ISL_2120063, EPI_ISL_2120065, EPI_ISL_2120070, EPI_ISL_2120073, EPI_ISL_2120076, EPI_ISL_2120079, EPI_ISL_2120081, EPI_ISL_2120083, EPI_ISL_2120086, EPI_ISL_2120586, EPI_ISL_2122599, EPI_ISL_2122608, EPI_ISL_2122610, EPI_ISL_2122614, EPI_ISL_2122615, EPI_ISL_2122617, EPI_ISL_2122620, EPI_ISL_2122622, EPI_ISL_2122624, EPI_ISL_2122628, EPI_ISL_2122631, EPI_ISL_2122634, EPI_ISL_2122637, EPI_ISL_2122640, EPI_ISL_2122643, EPI_ISL_2122645, EPI_ISL_2122648, EPI_ISL_2122650, EPI_ISL_2122651, EPI_ISL_2122653, EPI_ISL_2122657, EPI_ISL_2122660, EPI_ISL_2122663, EPI_ISL_2122666, EPI_ISL_2122726, EPI_ISL_2122997, EPI_ISL_2122998, EPI_ISL_2122999, EPI_ISL_2123000, EPI_ISL_2123001, EPI_ISL_2123002, EPI_ISL_2123004, EPI_ISL_2123005, EPI_ISL_2123006, EPI_ISL_2123007, EPI_ISL_2123009, EPI_ISL_2123010, EPI_ISL_2123011, EPI_ISL_2123012, EPI_ISL_2123013, EPI_ISL_2123014, EPI_ISL_2123015, EPI_ISL_2123016, EPI_ISL_2123017, EPI_ISL_2123018, EPI_ISL_2123019, EPI_ISL_2123020, EPI_ISL_2123067, EPI_ISL_2123068, EPI_ISL_2123069, EPI_ISL_2123070, EPI_ISL_2123071, EPI_ISL_2123072, EPI_ISL_2123073, EPI_ISL_2123074, EPI_ISL_2123075, EPI_ISL_2123076, EPI_ISL_2123077, EPI_ISL_2123078, EPI_ISL_2123079, EPI_ISL_2123080, EPI_ISL_2123812, EPI_ISL_2123813, EPI_ISL_2123814, EPI_ISL_2123844, EPI_ISL_2123845, EPI_ISL_2123846, EPI_ISL_2123847, EPI_ISL_2123848, EPI_ISL_2123849, EPI_ISL_2123850, EPI_ISL_2123851, EPI_ISL_2124017, EPI_ISL_2124018, EPI_ISL_2124019, EPI_ISL_2124020, EPI_ISL_2124022, EPI_ISL_2259639, EPI_ISL_2259641, EPI_ISL_2259642, EPI_ISL_2259643, EPI_ISL_2259644, EPI_ISL_2259655, EPI_ISL_2259657, EPI_ISL_2259660, EPI_ISL_2259663, EPI_ISL_2259665, EPI_ISL_2259666, EPI_ISL_2259668, EPI_ISL_2259669, EPI_ISL_2259672, EPI_ISL_2259674, EPI_ISL_2259676, EPI_ISL_2259678, EPI_ISL_2259680, EPI_ISL_2259681, EPI_ISL_2259682, EPI_ISL_2259683, EPI_ISL_2259684, EPI_ISL_2259685, EPI_ISL_2259688, EPI_ISL_2259689, EPI_ISL_2259690, EPI_ISL_2259691, EPI_ISL_2259692, EPI_ISL_2259693, EPI_ISL_2259695, EPI_ISL_2259696, EPI_ISL_2259754, EPI_ISL_2259770, EPI_ISL_2259779, EPI_ISL_2259780, EPI_ISL_2259788, EPI_ISL_2259789, EPI_ISL_2259796, EPI_ISL_2259797, EPI_ISL_2259798, EPI_ISL_2259799, EPI_ISL_2259800, EPI_ISL_2259802, EPI_ISL_2259805, EPI_ISL_2259806, EPI_ISL_2259808, EPI_ISL_2259809, EPI_ISL_2260917, EPI_ISL_2262223, EPI_ISL_2262226, EPI_ISL_2262227, EPI_ISL_2262233, EPI_ISL_2262240, EPI_ISL_2262247, EPI_ISL_2262249, EPI_ISL_2262250, EPI_ISL_2262251, EPI_ISL_2262253, EPI_ISL_2262258, EPI_ISL_2262275, EPI_ISL_2264558, EPI_ISL_2264562, EPI_ISL_2264578, EPI_ISL_2264588, EPI_ISL_2265887, EPI_ISL_2313170, EPI_ISL_2313188, EPI_ISL_2314440, EPI_ISL_2314680 | | | |
| see above | MVZ Labor Dr. Limbach & Kollegen GbR | Robert Koch Institute | |
| EPI_ISL_2261056, EPI_ISL_2261063, EPI_ISL_2261084, EPI_ISL_2261086, EPI_ISL_2261116, EPI_ISL_2261120, EPI_ISL_2261133, EPI_ISL_2261151, EPI_ISL_2261155, EPI_ISL_2261236, EPI_ISL_2261293, EPI_ISL_2261298, EPI_ISL_2261303, EPI_ISL_2261309, EPI_ISL_2261312, EPI_ISL_2261345, EPI_ISL_2261357, EPI_ISL_2261368, EPI_ISL_2261393, EPI_ISL_2261397, EPI_ISL_2261403, EPI_ISL_2261406, EPI_ISL_2261415, EPI_ISL_2261419, EPI_ISL_2261426, EPI_ISL_2261446, EPI_ISL_2261450, EPI_ISL_2261454, EPI_ISL_2261475, EPI_ISL_2261476, EPI_ISL_2262455, EPI_ISL_2262460, EPI_ISL_2262465, EPI_ISL_2262470, EPI_ISL_2262472, EPI_ISL_2262480, EPI_ISL_2262483, EPI_ISL_2262496, EPI_ISL_2262501, EPI_ISL_2262503, EPI_ISL_2262523, EPI_ISL_2262532, EPI_ISL_2262534, EPI_ISL_2262547, EPI_ISL_2262549, EPI_ISL_2262551, EPI_ISL_2262558, EPI_ISL_2262569, EPI_ISL_2262574, EPI_ISL_2262587, EPI_ISL_2262603, EPI_ISL_2262608, EPI_ISL_2262627 | | | |
| see above | MVZ Labor Dr. Quade & Kollegen GmbH | Robert Koch Institute | |
| EPI_ISL_2115149, EPI_ISL_2115150, EPI_ISL_2115152, EPI_ISL_2115153, EPI_ISL_2115155, EPI_ISL_2115159, EPI_ISL_2115164, EPI_ISL_2115176, EPI_ISL_2115178, EPI_ISL_2115180, EPI_ISL_2115181, EPI_ISL_2124363, EPI_ISL_2124368, EPI_ISL_2124372, EPI_ISL_2128989, EPI_ISL_2128992, EPI_ISL_2259756, EPI_ISL_2259771, EPI_ISL_2264565 | | | |
| see above | MVZ Labor Dr. Reising-Ackermann und Kollegen | Robert Koch Institute | |
| EPI_ISL_2112850, EPI_ISL_2112852, EPI_ISL_2112856, EPI_ISL_2112859, EPI_ISL_2112862, EPI_ISL_2115280, EPI_ISL_2116135, EPI_ISL_2116137, EPI_ISL_2123022, EPI_ISL_2123023, EPI_ISL_2123024, EPI_ISL_2123025, EPI_ISL_2123026, EPI_ISL_2123027, EPI_ISL_2123028, EPI_ISL_2123029, EPI_ISL_2123030, EPI_ISL_2123031, EPI_ISL_2123032, EPI_ISL_2123033, EPI_ISL_2123034, EPI_ISL_2123035, EPI_ISL_2123036, EPI_ISL_2123037, EPI_ISL_2123038, EPI_ISL_2123039, EPI_ISL_2123040, EPI_ISL_2123041, EPI_ISL_2123042, EPI_ISL_2123043, EPI_ISL_2123044, EPI_ISL_2123045, EPI_ISL_2123046, EPI_ISL_2123047, EPI_ISL_2123048, EPI_ISL_2123049, EPI_ISL_2123050, EPI_ISL_2123051, EPI_ISL_2123052, EPI_ISL_2123053, EPI_ISL_2123054, EPI_ISL_2123055, EPI_ISL_2123056, EPI_ISL_2123057, EPI_ISL_2123058, EPI_ISL_2123059, EPI_ISL_2123060, EPI_ISL_2123061, EPI_ISL_2123062, EPI_ISL_2123063, EPI_ISL_2126194, EPI_ISL_2126196, EPI_ISL_2126198, EPI_ISL_2126200, EPI_ISL_2126202, EPI_ISL_2126204, EPI_ISL_2126206, EPI_ISL_2126208, EPI_ISL_2126210, EPI_ISL_2126213, EPI_ISL_2126215, EPI_ISL_2126217, EPI_ISL_2126219, EPI_ISL_2126221, EPI_ISL_2126223, EPI_ISL_2126225, EPI_ISL_2126227, EPI_ISL_2126229, EPI_ISL_2126231, EPI_ISL_2126234, EPI_ISL_2126236, EPI_ISL_2126238, EPI_ISL_2126239, EPI_ISL_2126241, EPI_ISL_2126243, EPI_ISL_2126245, EPI_ISL_2126247, EPI_ISL_2126249, EPI_ISL_2126252, EPI_ISL_2126254, EPI_ISL_2126257, EPI_ISL_2126259, EPI_ISL_2126261, EPI_ISL_2126264, EPI_ISL_2126266, EPI_ISL_2126268, EPI_ISL_2126271, EPI_ISL_2126273, EPI_ISL_2126275, EPI_ISL_2126277, EPI_ISL_2126279, EPI_ISL_2126281, EPI_ISL_2126284, EPI_ISL_2126286, EPI_ISL_2126288, EPI_ISL_2126290, EPI_ISL_2126293, EPI_ISL_2126295, EPI_ISL_2126297, EPI_ISL_2126299, EPI_ISL_2126301, EPI_ISL_2126304, EPI_ISL_2126308, EPI_ISL_2126310, EPI_ISL_2126312, EPI_ISL_2126313, EPI_ISL_2126315, EPI_ISL_2126318, EPI_ISL_2126320, EPI_ISL_2126322, EPI_ISL_2126324, EPI_ISL_2126328, EPI_ISL_2126330, EPI_ISL_2126332, EPI_ISL_2126335, EPI_ISL_2126337, EPI_ISL_2126339, EPI_ISL_2126341, EPI_ISL_2126343, EPI_ISL_2126346, EPI_ISL_2126352, EPI_ISL_2126354, EPI_ISL_2126357, EPI_ISL_2126359, EPI_ISL_2126362, EPI_ISL_2126364, EPI_ISL_2126366, EPI_ISL_2126368, EPI_ISL_2126371, EPI_ISL_2126373, EPI_ISL_2126375, EPI_ISL_2126377, EPI_ISL_2126378, EPI_ISL_2128390, EPI_ISL_2128391, EPI_ISL_2128392, EPI_ISL_2128393, EPI_ISL_2128394, EPI_ISL_2128395, EPI_ISL_2128396, EPI_ISL_2128397, EPI_ISL_2128398, EPI_ISL_2128399, EPI_ISL_2128400, EPI_ISL_2128401, EPI_ISL_2128402, EPI_ISL_2128403, EPI_ISL_2128404, EPI_ISL_2128405, EPI_ISL_2128406, EPI_ISL_2128407, EPI_ISL_2128408, EPI_ISL_2128409, EPI_ISL_2128410, EPI_ISL_2128411, EPI_ISL_2260950, EPI_ISL_2260951, EPI_ISL_2260952, EPI_ISL_2260953, EPI_ISL_2260954, EPI_ISL_2260955, EPI_ISL_2260956, EPI_ISL_2260957, EPI_ISL_2260958, EPI_ISL_2260959, EPI_ISL_2260960, EPI_ISL_2260961, EPI_ISL_2260962, EPI_ISL_2260963, EPI_ISL_2264665, EPI_ISL_2264674, EPI_ISL_2264697 | | | |
| see above | MVZ Labor Krone GbR | Robert Koch Institute | |
| EPI_ISL_2112762, EPI_ISL_2115161, EPI_ISL_2115162, EPI_ISL_2115163, EPI_ISL_2115177, EPI_ISL_2115179, EPI_ISL_2115186, EPI_ISL_2115247, EPI_ISL_2124354, EPI_ISL_2124359, EPI_ISL_2124362, EPI_ISL_2124376, EPI_ISL_2125102, EPI_ISL_2125104, EPI_ISL_2125106, EPI_ISL_2125107, EPI_ISL_2125108, EPI_ISL_2125111, EPI_ISL_2125113 | | | |
| see above | MVZ Medizinisches Labor Hannover GmbH | Robert Koch Institute | |
| EPI_ISL_2120548, EPI_ISL_2120555, EPI_ISL_2120561, EPI_ISL_2120573, EPI_ISL_2120575, EPI_ISL_2123425, EPI_ISL_2124355, EPI_ISL_2124356, EPI_ISL_2125094, EPI_ISL_2125110, EPI_ISL_2128956, EPI_ISL_2128958, EPI_ISL_2128959, EPI_ISL_2128966, EPI_ISL_2128967, EPI_ISL_2128970, EPI_ISL_2128972, EPI_ISL_2128973, EPI_ISL_2128974, EPI_ISL_2128980, EPI_ISL_2128981, EPI_ISL_2128982, EPI_ISL_2128983 | | | |
| see above | MVZ fur Laboratoriumsmedizin und Mikrobiologie Koblenz-Mittelrhein (Labor Koblenz) | Robert Koch Institute | |
| EPI_ISL_2260045, EPI_ISL_2260057, EPI_ISL_2260060, EPI_ISL_2263187, EPI_ISL_2264712, EPI_ISL_2264722 | MVZ für Laboratoriumsmedizin und Mikrobiologie Koblenz-Mittelrhein (Labor Koblenz) | Robert Koch Institute | |
| EPI_ISL_2261039 | MVZ für Laboratoriumsmedizin und Mikrobiologie Würzburg | Robert Koch Institute | |

| | | | |
|---|---|---|---|
| EPI_ISL_2094517, EPI_ISL_2094647, EPI_ISL_2094721, EPI_ISL_2094722, EPI_ISL_2094723, EPI_ISL_2094724, EPI_ISL_2094725, EPI_ISL_2094726, EPI_ISL_2094827, EPI_ISL_2094930, EPI_ISL_2094931, EPI_ISL_2094955, EPI_ISL_2095055, EPI_ISL_2095106, EPI_ISL_2095120, EPI_ISL_2095135, EPI_ISL_2095275, EPI_ISL_2095276, EPI_ISL_2095278, EPI_ISL_2095279, EPI_ISL_2095280, EPI_ISL_2095281, EPI_ISL_2095282, EPI_ISL_2450223, EPI_ISL_2450224, EPI_ISL_2450225, EPI_ISL_2450226, EPI_ISL_2450227, EPI_ISL_2450228, EPI_ISL_2450229, EPI_ISL_2450230, EPI_ISL_2450285, EPI_ISL_2450286, EPI_ISL_2450288, EPI_ISL_2450289, EPI_ISL_2450290, EPI_ISL_2450291, EPI_ISL_2450292, EPI_ISL_2450293, EPI_ISL_2450294, EPI_ISL_2450295, EPI_ISL_2450296, EPI_ISL_2450297, EPI_ISL_2450298, EPI_ISL_2450299, EPI_ISL_2450300, EPI_ISL_2450301, EPI_ISL_2450302, EPI_ISL_2450303, EPI_ISL_2450304, EPI_ISL_2450309, EPI_ISL_2450313, EPI_ISL_2450314, EPI_ISL_2450315, EPI_ISL_2450316, EPI_ISL_2772674, EPI_ISL_2772675, EPI_ISL_2772690, EPI_ISL_2772691 | | | |
| see above | Max von Pettenkofer Institute, Virology, National Reference Center for Retroviruses, LMU Munich | Laboratory for Functional Genome Analysis; Dept. Genomics; Gene Center of the LMU Munich | Alexander Graf; Helmut Blum; Max Muenchhoff; Oliver Keppler; Stefan Krebs |
| EPI_ISL_2120356, EPI_ISL_2120359, EPI_ISL_2120363, EPI_ISL_2120366, EPI_ISL_2120368, EPI_ISL_2120372, EPI_ISL_2120375, EPI_ISL_2120377, EPI_ISL_2120380, EPI_ISL_2120383, EPI_ISL_2120386, EPI_ISL_2120389, EPI_ISL_2120392, EPI_ISL_2120396, EPI_ISL_2260591, EPI_ISL_2260595 | | | |
| see above | Med. Labor Prof. Schenk Dr. Ansorge & Kollegen | Robert Koch Institute | |
| EPI_ISL_2381582 | Medical Laboratories Duesseldorf | Center of Medical Microbiology, Virology, and Hospital Hygiene, University of Duesseldorf | Alexander Dilthey; Andreas Walker; Angelika Helmer; Christian Lange; Daniel Strelow; Jessica Nicolai; Jörg Timm; Klaus Pfeffer; Lisanna Hülse; Malte Kohns Vasconcelos; Maximilian Damagnez; Nadine Lübke; Tobias Wienemann; Torsten Houwaart |
| EPI_ISL_2122963, EPI_ISL_2122967, EPI_ISL_2122971, EPI_ISL_2122972, EPI_ISL_2122973, EPI_ISL_2122979, EPI_ISL_2122987, EPI_ISL_2122988, EPI_ISL_2122990, EPI_ISL_2122993 | | | |
| see above | Medizinisch-Diagnostisches Labor Kempten allgaulab | Robert Koch Institute | |
| EPI_ISL_2315831, EPI_ISL_2315832, EPI_ISL_2471751, EPI_ISL_2471752, EPI_ISL_2471761, EPI_ISL_2471763, EPI_ISL_2471765, EPI_ISL_2471771, EPI_ISL_2471772, EPI_ISL_2471773, EPI_ISL_2471774, EPI_ISL_2471775, EPI_ISL_2471776, EPI_ISL_2471777, EPI_ISL_2471784, EPI_ISL_2472779, EPI_ISL_2472783, EPI_ISL_2635981, EPI_ISL_2635985, EPI_ISL_2635986, EPI_ISL_2635992, EPI_ISL_2635997, EPI_ISL_2636078, EPI_ISL_2636097, EPI_ISL_2636870, EPI_ISL_2636871 | | | |
| see above | Medizinisch-Diagnostisches Labor Kempten allgäulab | Robert Koch Institute | |
| EPI_ISL_2129013, EPI_ISL_2129014, EPI_ISL_2129015, EPI_ISL_2129016, EPI_ISL_2129017, EPI_ISL_2129018, EPI_ISL_2129019, EPI_ISL_2129021, EPI_ISL_2129022, EPI_ISL_2129023, EPI_ISL_2129025, EPI_ISL_2129026, EPI_ISL_2129027, EPI_ISL_2129028, EPI_ISL_2129029, EPI_ISL_2129030, EPI_ISL_2129031, EPI_ISL_2129032, EPI_ISL_2129033, EPI_ISL_2129034, EPI_ISL_2129035, EPI_ISL_2129036, EPI_ISL_2129037, EPI_ISL_2129039, EPI_ISL_2129040, EPI_ISL_2129041, EPI_ISL_2129042, EPI_ISL_2129043, EPI_ISL_2129044, EPI_ISL_2129045, EPI_ISL_2129046, EPI_ISL_2129047, EPI_ISL_2129048, EPI_ISL_2129049, EPI_ISL_2129051, EPI_ISL_2129052, EPI_ISL_2129053, EPI_ISL_2129054, EPI_ISL_2129056, EPI_ISL_2129057, EPI_ISL_2129058, EPI_ISL_2129059, EPI_ISL_2129060, EPI_ISL_2129061, EPI_ISL_2129062, EPI_ISL_2129063, EPI_ISL_2129064, EPI_ISL_2129065, EPI_ISL_2129067, EPI_ISL_2129068, EPI_ISL_2129069, EPI_ISL_2129070, EPI_ISL_2129071, EPI_ISL_2129072, EPI_ISL_2129073 | | | |
| see above | Medizinische Laboratorien Dusseldorf | Robert Koch Institute | |
| EPI_ISL_2112721, EPI_ISL_2112724, EPI_ISL_2112726, EPI_ISL_2112730, EPI_ISL_2112737, EPI_ISL_2112765, EPI_ISL_2112791, EPI_ISL_2112801, EPI_ISL_2112802, EPI_ISL_2112805, EPI_ISL_2112808, EPI_ISL_2120401, EPI_ISL_2120408, EPI_ISL_2120424, EPI_ISL_2120432, EPI_ISL_2120447, EPI_ISL_2120467, EPI_ISL_2120470, EPI_ISL_2120473, EPI_ISL_2123369, EPI_ISL_2123375, EPI_ISL_2123378, EPI_ISL_2123379, EPI_ISL_2123381, EPI_ISL_2123382, EPI_ISL_2124340, EPI_ISL_2124344 | | | |
| see above | Medizinisches Labor Ostsachsen MVZ GbR Nebenbetriebsstatte Dresden | Robert Koch Institute | |
| EPI_ISL_2115190, EPI_ISL_2115191, EPI_ISL_2115197, EPI_ISL_2115207, EPI_ISL_2115209, EPI_ISL_2115211, EPI_ISL_2122788, EPI_ISL_2122815, EPI_ISL_2122832, EPI_ISL_2122834, EPI_ISL_2122855, EPI_ISL_2122861, EPI_ISL_2122863, EPI_ISL_2122867, EPI_ISL_2122904, EPI_ISL_2122927, EPI_ISL_2122935, EPI_ISL_2122940, EPI_ISL_2122941, EPI_ISL_2122943, EPI_ISL_2122946, EPI_ISL_2122949, EPI_ISL_2122950, EPI_ISL_2123832 | | | |
| see above | Medizinisches Labor Ostsachsen MVZ GbR NebenbetriebsstÃ¤tte Dresden | Robert Koch Institute | |
| EPI_ISL_2263044, EPI_ISL_2263058, EPI_ISL_2263186 | Medizinisches Labor Ostsachsen MVZ GbR Nebenbetriebsstätte Dresden | Robert Koch Institute | |
| EPI_ISL_2125032, EPI_ISL_2125038, EPI_ISL_2125045, EPI_ISL_2125053, EPI_ISL_2125055, EPI_ISL_2125056, EPI_ISL_2125058, EPI_ISL_2125059, EPI_ISL_2125060, EPI_ISL_2125061, EPI_ISL_2125062, EPI_ISL_2125063, EPI_ISL_2125064, EPI_ISL_2125065, EPI_ISL_2125066 | | | |
| see above | Medizinisches Labor Ostsachsen MVZ GbR Nebenbetriebsstatte Dresden | Robert Koch Institute | |
| EPI_ISL_2122356, EPI_ISL_2122389, EPI_ISL_2122399, EPI_ISL_2122443, EPI_ISL_2122451 | Medizinisches Labor Wahl Ludenscheid | Robert Koch Institute | |
| EPI_ISL_2129554, EPI_ISL_2129558, EPI_ISL_2129565, EPI_ISL_2129574, EPI_ISL_2129586, EPI_ISL_2129589, EPI_ISL_2129605, EPI_ISL_2129622, EPI_ISL_2129624 | | | |
| see above | Medizinisches Labor Wahl LÃ¼denscheid | Robert Koch Institute | |
| EPI_ISL_2122475 | Medizinisches Versorgungszentrum fur Labormedizin und Mikrobiologie Ruhr GmbH - mvzlm RUHR GmbH | Robert Koch Institute | |
| EPI_ISL_2112278, EPI_ISL_2112367, EPI_ISL_2112400, EPI_ISL_2123589, EPI_ISL_2263015 | Medizinisches Zentrallabor Altenburg GmbH & Co.KG | Robert Koch Institute | |
| EPI_ISL_2259725, EPI_ISL_2259726, EPI_ISL_2259727, EPI_ISL_2259728, EPI_ISL_2259729, EPI_ISL_2259731, EPI_ISL_2259732, EPI_ISL_2259733, EPI_ISL_2259734, EPI_ISL_2259735, EPI_ISL_2259736, EPI_ISL_2259738, EPI_ISL_2259739, EPI_ISL_2259741, EPI_ISL_2266012, EPI_ISL_2390750, EPI_ISL_2390751, EPI_ISL_2632240 | | | |
| see above | Niedersächsisches Landesgesundheitsamt (NLGA) | Robert Koch Institute | |
| EPI_ISL_2116048, EPI_ISL_2116050, EPI_ISL_2116052, EPI_ISL_2116054 | Philipps-UniversitÃ¤t Marburg Fachbereich Medizin Institut fÃ¼r Virologie | Robert Koch Institute | |
| EPI_ISL_2112212, EPI_ISL_2112235, EPI_ISL_2123441, EPI_ISL_2123501, EPI_ISL_2123538, EPI_ISL_2123610 | Praxisgemeinschaft fur Laboratoriumsmedizin Labor Blumenstrasse; Praxis Dr. E. Wietschel & Dr. F. Wietschel | Robert Koch Institute | |
| EPI_ISL_2123448, EPI_ISL_2123492, EPI_ISL_2123533, EPI_ISL_2123595, EPI_ISL_2123621 | Praxisgemeinschaft fur Laboratoriumsmedizin Labor Blumenstrasse; Praxis Dr. Sel und Dr. SpÃ¤te | Robert Koch Institute | |
| EPI_ISL_2123646, EPI_ISL_2123663, EPI_ISL_2123671, EPI_ISL_2123718, EPI_ISL_2123720 | Praxisgemeinschaft fur Laboratoriumsmedizin Labor BlumenstraÃŸe; Praxis Dr. E. Wietschel & Dr. F. Wietschel | Robert Koch Institute | |
| EPI_ISL_2117317, EPI_ISL_2117325, EPI_ISL_2117358 | Praxisgemeinschaft fÃ¼r Laboratoriumsmedizin Labor BlumenstraÃŸe; Praxis Dr. Sel und Dr. SpÃ¤te | Robert Koch Institute | |
| EPI_ISL_2263436, EPI_ISL_2263588, EPI_ISL_2264160 | Praxisgemeinschaft für Laboratoriumsmedizin Labor Blumenstraße; Praxis Dr. E. Wietschel & Dr. F. Wietschel | Robert Koch Institute | |
| EPI_ISL_2263325, EPI_ISL_2263328, EPI_ISL_2264315, EPI_ISL_2264323 | Praxisgemeinschaft für Laboratoriumsmedizin Labor Blumenstraße; Praxis Dr. Sel und Dr. Späte | Robert Koch Institute | |
| EPI_ISL_2260794, EPI_ISL_2260817, EPI_ISL_2260914 | Robert-Bosch-Krankenhaus | Robert Koch Institute | |
| EPI_ISL_2120291, EPI_ISL_2120429, EPI_ISL_2120485, EPI_ISL_2120534, EPI_ISL_2120558, EPI_ISL_2120574 | SLK-Kliniken Heilbronn GmbH - Institut fur Laboratoriumsmedizin | Robert Koch Institute | |
| EPI_ISL_2115350, EPI_ISL_2115366 | SLK-Kliniken Heilbronn GmbH - Institut fÃ¼r Laboratoriumsmedizin | Robert Koch Institute | |
| EPI_ISL_2112499, EPI_ISL_2112555, EPI_ISL_2112604, EPI_ISL_2112625, EPI_ISL_2112665, EPI_ISL_2112957, EPI_ISL_2112964, EPI_ISL_2112972, EPI_ISL_2113003, EPI_ISL_2113006, EPI_ISL_2113015, EPI_ISL_2113030, EPI_ISL_2113032, EPI_ISL_2113046, EPI_ISL_2113054, EPI_ISL_2113055, EPI_ISL_2113061, EPI_ISL_2113074, EPI_ISL_2113079, EPI_ISL_2113081, EPI_ISL_2113090, EPI_ISL_2113093, EPI_ISL_2113097, EPI_ISL_2113100, EPI_ISL_2113112, EPI_ISL_2113117, EPI_ISL_2113120, EPI_ISL_2113130, EPI_ISL_2113141, EPI_ISL_2113142, EPI_ISL_2113143, EPI_ISL_2113145, EPI_ISL_2113169, EPI_ISL_2113170, EPI_ISL_2113180, EPI_ISL_2113183, EPI_ISL_2113185, EPI_ISL_2113190, EPI_ISL_2113194, EPI_ISL_2113209, EPI_ISL_2113211, EPI_ISL_2113217, EPI_ISL_2113220, EPI_ISL_2113225, EPI_ISL_2117301, EPI_ISL_2117303, EPI_ISL_2117304, EPI_ISL_2117306, EPI_ISL_2117310, EPI_ISL_2117320, EPI_ISL_2117321, EPI_ISL_2117323, EPI_ISL_2117327, EPI_ISL_2117329, EPI_ISL_2117330, EPI_ISL_2117331, EPI_ISL_2117334, EPI_ISL_2117335, EPI_ISL_2117338, EPI_ISL_2117343, EPI_ISL_2117348, EPI_ISL_2117355, EPI_ISL_2117357, EPI_ISL_2117360, EPI_ISL_2117363, EPI_ISL_2117364, EPI_ISL_2117366, EPI_ISL_2117374, EPI_ISL_2117379, EPI_ISL_2117381, EPI_ISL_2117389, EPI_ISL_2117391, EPI_ISL_2117395, EPI_ISL_2117402, EPI_ISL_2117407, EPI_ISL_2117414, EPI_ISL_2117420, EPI_ISL_2117429, EPI_ISL_2117431, EPI_ISL_2123427, EPI_ISL_2123428, EPI_ISL_2123429, EPI_ISL_2123431, EPI_ISL_2123432, EPI_ISL_2123434, EPI_ISL_2123436, EPI_ISL_2123438, EPI_ISL_2123439, EPI_ISL_2123442, EPI_ISL_2123444, EPI_ISL_2123452, EPI_ISL_2123454, EPI_ISL_2123455, EPI_ISL_2123457, EPI_ISL_2123461, EPI_ISL_2123462, EPI_ISL_2123464, EPI_ISL_2123466, EPI_ISL_2123467, EPI_ISL_2123468, EPI_ISL_2123469, EPI_ISL_2123471, EPI_ISL_2123472, EPI_ISL_2123473, EPI_ISL_2123474, EPI_ISL_2123476, EPI_ISL_2123478, EPI_ISL_2123479, EPI_ISL_2123480, EPI_ISL_2123481, EPI_ISL_2123482, EPI_ISL_2123484, EPI_ISL_2123485, EPI_ISL_2123486, EPI_ISL_2123487, EPI_ISL_2123488, EPI_ISL_2123493, EPI_ISL_2123495, EPI_ISL_2123496, EPI_ISL_2123498, EPI_ISL_2123499, EPI_ISL_2123500, EPI_ISL_2123502, EPI_ISL_2123504, EPI_ISL_2123508, EPI_ISL_2123509, EPI_ISL_2123510, EPI_ISL_2123511, EPI_ISL_2123512, EPI_ISL_2123513, EPI_ISL_2123514, EPI_ISL_2123515, EPI_ISL_2123518, EPI_ISL_2123520, EPI_ISL_2123522, EPI_ISL_2123525, EPI_ISL_2123527, EPI_ISL_2123528, EPI_ISL_2123529, EPI_ISL_2123531, EPI_ISL_2123536, EPI_ISL_2123537, EPI_ISL_2123539, EPI_ISL_2123541, EPI_ISL_2123543, EPI_ISL_2123544, EPI_ISL_2123546, EPI_ISL_2123548, EPI_ISL_2123550, EPI_ISL_2123551, EPI_ISL_2123552, EPI_ISL_2123553, EPI_ISL_2123554, EPI_ISL_2123555, EPI_ISL_2123556, EPI_ISL_2123557, EPI_ISL_2123559, EPI_ISL_2123561, EPI_ISL_2123562, EPI_ISL_2123565, EPI_ISL_2123569, EPI_ISL_2123570, EPI_ISL_2123571, EPI_ISL_2123572, EPI_ISL_2123575, EPI_ISL_2123576, EPI_ISL_2123580, EPI_ISL_2123581, EPI_ISL_2123584, EPI_ISL_2123585, EPI_ISL_2123588, EPI_ISL_2123590, EPI_ISL_2123593, EPI_ISL_2123596, EPI_ISL_2123597, EPI_ISL_2123603, EPI_ISL_2123605, EPI_ISL_2123606, EPI_ISL_2123607, EPI_ISL_2123608, EPI_ISL_2123609, EPI_ISL_2123611, EPI_ISL_2123612, EPI_ISL_2123613, EPI_ISL_2123615, EPI_ISL_2123617, EPI_ISL_2123618, EPI_ISL_2123619, EPI_ISL_2123620, EPI_ISL_2123622, EPI_ISL_2123623, EPI_ISL_2123624, EPI_ISL_2123625, EPI_ISL_2123626, EPI_ISL_2123627, EPI_ISL_2123628, EPI_ISL_2123630, EPI_ISL_2123632, EPI_ISL_2123633, EPI_ISL_2123635, EPI_ISL_2123636, EPI_ISL_2123637, EPI_ISL_2123639, EPI_ISL_2123641, EPI_ISL_2123643, EPI_ISL_2123644, EPI_ISL_2123645, EPI_ISL_2123647, EPI_ISL_2123652, EPI_ISL_2123655, EPI_ISL_2123656, EPI_ISL_2123657, EPI_ISL_2123658, EPI_ISL_2123659, EPI_ISL_2123662, EPI_ISL_2123666, EPI_ISL_2123668, EPI_ISL_2123672, EPI_ISL_2123674, EPI_ISL_2123676, EPI_ISL_2123677, EPI_ISL_2123678, EPI_ISL_2123679, EPI_ISL_2123680, EPI_ISL_2123681, EPI_ISL_2123684, EPI_ISL_2123685, EPI_ISL_2123686, EPI_ISL_2123687, EPI_ISL_2123688, EPI_ISL_2123689, EPI_ISL_2123690, EPI_ISL_2123691, EPI_ISL_2123693, EPI_ISL_2123697, EPI_ISL_2123703, EPI_ISL_2123704, EPI_ISL_2123706, EPI_ISL_2123707, EPI_ISL_2123708, EPI_ISL_2123709, EPI_ISL_2123710, EPI_ISL_2123711, EPI_ISL_2123715, EPI_ISL_2123716, EPI_ISL_2123721, EPI_ISL_2124023, EPI_ISL_2124026, EPI_ISL_2124027, EPI_ISL_2124029, EPI_ISL_2124030, EPI_ISL_2124031, EPI_ISL_2124033, EPI_ISL_2124035, EPI_ISL_2124036, EPI_ISL_2124038, EPI_ISL_2124039, EPI_ISL_2124040, EPI_ISL_2124046, EPI_ISL_2124052, EPI_ISL_2124053, EPI_ISL_2124055, EPI_ISL_2124056, EPI_ISL_2124057, EPI_ISL_2124058, EPI_ISL_2124059, EPI_ISL_2124072, EPI_ISL_2124074, EPI_ISL_2124075, EPI_ISL_2124077, EPI_ISL_2124078, EPI_ISL_2124082, EPI_ISL_2124083, EPI_ISL_2124087, EPI_ISL_2124089, EPI_ISL_2124090, EPI_ISL_2124092, EPI_ISL_2124093, EPI_ISL_2124098, EPI_ISL_2124101, EPI_ISL_2124104, EPI_ISL_2124107, EPI_ISL_2124108, EPI_ISL_2124110, EPI_ISL_2124114, EPI_ISL_2124115, EPI_ISL_2124116, EPI_ISL_2124117, EPI_ISL_2124118, EPI_ISL_2124119, EPI_ISL_2124120, EPI_ISL_2124122, EPI_ISL_2124126, EPI_ISL_2124136, EPI_ISL_2124139, EPI_ISL_2124140, EPI_ISL_2124146, EPI_ISL_2124147, EPI_ISL_2124153, EPI_ISL_2124155, EPI_ISL_2124156, EPI_ISL_2124157, EPI_ISL_2124158, EPI_ISL_2124159, EPI_ISL_2124160, EPI_ISL_2124167, EPI_ISL_2124171, EPI_ISL_2124172, EPI_ISL_2124173, EPI_ISL_2124175, EPI_ISL_2124178, EPI_ISL_2124183, EPI_ISL_2124185, EPI_ISL_2124186, EPI_ISL_2124189, EPI_ISL_2124191, EPI_ISL_2124198, EPI_ISL_2124203, EPI_ISL_2124205, EPI_ISL_2124206, EPI_ISL_2124212, EPI_ISL_2124213, EPI_ISL_2124222, EPI_ISL_2124224, EPI_ISL_2124227, EPI_ISL_2124228, EPI_ISL_2124241, EPI_ISL_2124247, EPI_ISL_2124249, EPI_ISL_2124251, EPI_ISL_2124258, EPI_ISL_2124261, EPI_ISL_2124264, EPI_ISL_2128376, EPI_ISL_2128378, EPI_ISL_2128381, EPI_ISL_2128382, EPI_ISL_2128383, EPI_ISL_2128384, EPI_ISL_2128386, EPI_ISL_2128387, EPI_ISL_2129734, EPI_ISL_2129742, EPI_ISL_2129764, EPI_ISL_2129775, EPI_ISL_2129809, EPI_ISL_2129816, EPI_ISL_2129819, EPI_ISL_2129828, EPI_ISL_2129842, EPI_ISL_2129844, EPI_ISL_2129848, EPI_ISL_2129857, EPI_ISL_2129865, EPI_ISL_2129869, EPI_ISL_2129874, EPI_ISL_2129875, EPI_ISL_2129882, EPI_ISL_2129891, EPI_ISL_2129894, EPI_ISL_2129895, EPI_ISL_2129902, EPI_ISL_2129906, EPI_ISL_2129942, EPI_ISL_2129949, EPI_ISL_2129955, EPI_ISL_2129970, EPI_ISL_2129979, EPI_ISL_2261630, EPI_ISL_2261632, EPI_ISL_2261634, EPI_ISL_2261643, EPI_ISL_2261651, EPI_ISL_2261654, EPI_ISL_2261663, EPI_ISL_2261675, EPI_ISL_2261676, EPI_ISL_2261677, EPI_ISL_2261679, EPI_ISL_2261687, EPI_ISL_2261690, EPI_ISL_2261698, EPI_ISL_2261700, EPI_ISL_2261707, EPI_ISL_2261708, EPI_ISL_2261713, EPI_ISL_2261714, EPI_ISL_2261716, EPI_ISL_2261737, EPI_ISL_2261741, EPI_ISL_2261747, EPI_ISL_2261755, EPI_ISL_2261766, EPI_ISL_2261775, EPI_ISL_2261777, EPI_ISL_2261781, EPI_ISL_2261800, EPI_ISL_2261817, EPI_ISL_2261820, EPI_ISL_2261824, EPI_ISL_2261841, EPI_ISL_2261848, EPI_ISL_2261853, EPI_ISL_2261860, EPI_ISL_2261866, EPI_ISL_2261867, EPI_ISL_2261882, EPI_ISL_2261888, EPI_ISL_2261904, EPI_ISL_2261905, EPI_ISL_2261906, EPI_ISL_2261914, EPI_ISL_2261915, EPI_ISL_2261916, EPI_ISL_2261918, EPI_ISL_2261927, EPI_ISL_2261934, EPI_ISL_2261935, EPI_ISL_2261938, EPI_ISL_2261939, EPI_ISL_2261941, EPI_ISL_2261949, EPI_ISL_2261955, EPI_ISL_2261957, EPI_ISL_2261958, EPI_ISL_2261959, EPI_ISL_2261963, EPI_ISL_2261970, EPI_ISL_2261971, EPI_ISL_2261977, EPI_ISL_2261983, EPI_ISL_2261987, EPI_ISL_2261994, EPI_ISL_2261998, EPI_ISL_2262002, EPI_ISL_2262005, EPI_ISL_2262031, EPI_ISL_2262032, EPI_ISL_2262037, EPI_ISL_2262044, EPI_ISL_2262048, EPI_ISL_2262052, EPI_ISL_2262058, EPI_ISL_2262061, EPI_ISL_2262064, EPI_ISL_2262067, EPI_ISL_2262068, EPI_ISL_2262069, EPI_ISL_2262072, EPI_ISL_2262080, EPI_ISL_2262096, EPI_ISL_2262897, EPI_ISL_2263215, EPI_ISL_2263220, EPI_ISL_2263223, EPI_ISL_2263237, EPI_ISL_2263239, EPI_ISL_2263243, EPI_ISL_2263246, EPI_ISL_2263253, EPI_ISL_2263259, EPI_ISL_2263260, EPI_ISL_2263269, EPI_ISL_2263271, EPI_ISL_2263278, EPI_ISL_2263280, EPI_ISL_2263282, EPI_ISL_2263289, EPI_ISL_2263290, EPI_ISL_2263292, EPI_ISL_2263293, EPI_ISL_2263296, EPI_ISL_2263297, EPI_ISL_2263302, EPI_ISL_2263305, EPI_ISL_2263313, EPI_ISL_2263316, EPI_ISL_2263317, EPI_ISL_2263321, EPI_ISL_2263324, EPI_ISL_2263331, EPI_ISL_2263332, EPI_ISL_2263337, EPI_ISL_2263338, EPI_ISL_2263354, EPI_ISL_2263357, EPI_ISL_2263359, EPI_ISL_2263365, EPI_ISL_2263368, EPI_ISL_2263380, EPI_ISL_2263381, EPI_ISL_2263383, EPI_ISL_2263387, EPI_ISL_2263389, EPI_ISL_2263390, EPI_ISL_2263395, EPI_ISL_2263396, EPI_ISL_2263414, EPI_ISL_2263426, EPI_ISL_2263432, EPI_ISL_2263437, EPI_ISL_2263438, EPI_ISL_2263440, EPI_ISL_2263445, EPI_ISL_2263446, EPI_ISL_2263451, EPI_ISL_2263456, EPI_ISL_2263475, EPI_ISL_2263478, EPI_ISL_2263481, EPI_ISL_2263486, EPI_ISL_2263493, EPI_ISL_2263495, EPI_ISL_2263498, EPI_ISL_2263503, EPI_ISL_2263507, EPI_ISL_2263511, EPI_ISL_2263513, EPI_ISL_2263516, EPI_ISL_2263521, EPI_ISL_2263522, EPI_ISL_2263523, EPI_ISL_2263526, EPI_ISL_2263527, EPI_ISL_2263530, EPI_ISL_2263531, EPI_ISL_2263533, EPI_ISL_2263535, EPI_ISL_2263540, EPI_ISL_2263542, EPI_ISL_2263543, EPI_ISL_2263554, EPI_ISL_2263556, EPI_ISL_2263567, EPI_ISL_2263568, EPI_ISL_2263569, EPI_ISL_2263571, EPI_ISL_2263577, EPI_ISL_2263579, EPI_ISL_2263583, EPI_ISL_2263585, EPI_ISL_2263586, EPI_ISL_2263587, EPI_ISL_2263591, EPI_ISL_2263598, EPI_ISL_2263600, EPI_ISL_2263602, EPI_ISL_2263608, EPI_ISL_2263611, EPI_ISL_2263620, EPI_ISL_2263626, EPI_ISL_2263629, EPI_ISL_2263639, EPI_ISL_2263652, EPI_ISL_2263661, EPI_ISL_2263667, EPI_ISL_2263669, EPI_ISL_2263671, EPI_ISL_2263677, EPI_ISL_2263689, EPI_ISL_2263693, EPI_ISL_2263697, EPI_ISL_2263698, EPI_ISL_2263699, EPI_ISL_2263702, EPI_ISL_2263704, EPI_ISL_2263705, EPI_ISL_2263708, EPI_ISL_2263712, EPI_ISL_2263713, EPI_ISL_2263726, EPI_ISL_2263732, EPI_ISL_2263734, EPI_ISL_2263737, EPI_ISL_2263749, EPI_ISL_2263756, EPI_ISL_2263757, EPI_ISL_2263759, EPI_ISL_2263763, EPI_ISL_2263775, EPI_ISL_2263777, EPI_ISL_2263779, EPI_ISL_2263780 | | | |
| see above | SYNLAB Jena Oncoscreen | Robert Koch Institute | |
| EPI_ISL_2124152, EPI_ISL_2124170, EPI_ISL_2124195, EPI_ISL_2124202, EPI_ISL_2124226, EPI_ISL_2124236 | SYNLAB Labor Munchen Zentrum LMZ | Robert Koch Institute | |
| EPI_ISL_2112227, EPI_ISL_2123430, EPI_ISL_2123440, EPI_ISL_2123475, EPI_ISL_2123483, EPI_ISL_2123489, EPI_ISL_2123503, EPI_ISL_2123526, EPI_ISL_2123534, EPI_ISL_2123560, EPI_ISL_2123564, EPI_ISL_2123573, EPI_ISL_2123579, EPI_ISL_2123587, EPI_ISL_2123592, EPI_ISL_2123616, EPI_ISL_2123713, EPI_ISL_2123722, EPI_ISL_2124051, EPI_ISL_2124105 | | | |
| see above | SYNLAB Labor MÃ¼nchen Zentrum LMZ | Robert Koch Institute | |
| EPI_ISL_2262828, EPI_ISL_2262950 | SYNLAB Labor München Zentrum LMZ | Robert Koch Institute | |
| EPI_ISL_2112978, EPI_ISL_2113043, EPI_ISL_2113048, EPI_ISL_2113099, EPI_ISL_2113152, EPI_ISL_2117297, EPI_ISL_2117299, EPI_ISL_2117302, EPI_ISL_2117308, EPI_ISL_2117312, EPI_ISL_2117313, EPI_ISL_2117314, EPI_ISL_2117318, EPI_ISL_2117322, EPI_ISL_2117333, EPI_ISL_2117336, EPI_ISL_2117350, EPI_ISL_2117351, EPI_ISL_2117354, EPI_ISL_2117356, EPI_ISL_2117385, EPI_ISL_2117387, EPI_ISL_2117411, EPI_ISL_2117418, EPI_ISL_2123456, EPI_ISL_2123530, EPI_ISL_2123535, EPI_ISL_2123549, EPI_ISL_2123566, EPI_ISL_2123567, EPI_ISL_2123591, EPI_ISL_2123598, EPI_ISL_2123599, EPI_ISL_2123650, EPI_ISL_2123665, EPI_ISL_2123694, EPI_ISL_2123698, EPI_ISL_2123699, EPI_ISL_2123700, EPI_ISL_2124037, EPI_ISL_2124041, EPI_ISL_2124054, EPI_ISL_2124060, EPI_ISL_2124062, EPI_ISL_2124067, EPI_ISL_2124068, EPI_ISL_2124088, EPI_ISL_2124099, EPI_ISL_2124103, EPI_ISL_2124121, EPI_ISL_2124162, EPI_ISL_2124176, EPI_ISL_2124180, EPI_ISL_2124182, EPI_ISL_2124187, EPI_ISL_2124192, EPI_ISL_2124193, EPI_ISL_2124196, EPI_ISL_2124197, EPI_ISL_2124209, EPI_ISL_2124215, EPI_ISL_2124216, EPI_ISL_2124230, EPI_ISL_2124235, EPI_ISL_2124238, EPI_ISL_2124266, EPI_ISL_2124268, EPI_ISL_2129789, EPI_ISL_2129826, EPI_ISL_2129846, EPI_ISL_2262042 | | | |

| | | |
|---|---|---|
| see above | SYNLAB MVZ Berlin | Robert Koch Institute |
| EPI_ISL_2124700, EPI_ISL_2126904 | SYNLAB MVZ Dachau | Robert Koch Institute |

EPI_ISL_2128639, EPI_ISL_2128650, EPI_ISL_2128656, EPI_ISL_2128657, EPI_ISL_2128658, EPI_ISL_2128659, EPI_ISL_2128661, EPI_ISL_2128665, EPI_ISL_2128673, EPI_ISL_2128678, EPI_ISL_2128683, EPI_ISL_2128693, EPI_ISL_2128695, EPI_ISL_2128702, EPI_ISL_2128707, EPI_ISL_2128710, EPI_ISL_2128711, EPI_ISL_2128716, EPI_ISL_2128719, EPI_ISL_2128727, EPI_ISL_2128735, EPI_ISL_2128744,
EPI_ISL_2128748, EPI_ISL_2128762, EPI_ISL_2128763, EPI_ISL_2128783, EPI_ISL_2128796, EPI_ISL_2128797, EPI_ISL_2128798, EPI_ISL_2128801, EPI_ISL_2128804, EPI_ISL_2128807, EPI_ISL_2128809, EPI_ISL_2128810, EPI_ISL_2128812, EPI_ISL_2128815, EPI_ISL_2128820, EPI_ISL_2128823, EPI_ISL_2128824, EPI_ISL_2128847, EPI_ISL_2128849, EPI_ISL_2128852, EPI_ISL_2128856,
EPI_ISL_2128869, EPI_ISL_2128871, EPI_ISL_2128881, EPI_ISL_2128884, EPI_ISL_2128890, EPI_ISL_2128893, EPI_ISL_2128895, EPI_ISL_2128896, EPI_ISL_2128897, EPI_ISL_2128902, EPI_ISL_2128908, EPI_ISL_2128913, EPI_ISL_2128917, EPI_ISL_2128918, EPI_ISL_2128922, EPI_ISL_2128925, EPI_ISL_2128926, EPI_ISL_2128934, EPI_ISL_2128936, EPI_ISL_2128941, EPI_ISL_2128944, EPI_ISL_2128954,
EPI_ISL_2130826, EPI_ISL_2130827, EPI_ISL_2130828, EPI_ISL_2130829, EPI_ISL_2130830, EPI_ISL_2130831, EPI_ISL_2130833, EPI_ISL_2130834, EPI_ISL_2130836, EPI_ISL_2130840, EPI_ISL_2130841, EPI_ISL_2130843, EPI_ISL_2130846, EPI_ISL_2130847, EPI_ISL_2130848, EPI_ISL_2130850, EPI_ISL_2130851, EPI_ISL_2130853, EPI_ISL_2130854, EPI_ISL_2130859, EPI_ISL_2130860, EPI_ISL_2130864,
EPI_ISL_2130865, EPI_ISL_2130867, EPI_ISL_2130868, EPI_ISL_2130869, EPI_ISL_2130870, EPI_ISL_2130871, EPI_ISL_2130872, EPI_ISL_2130873, EPI_ISL_2130874, EPI_ISL_2130875, EPI_ISL_2130876, EPI_ISL_2130878, EPI_ISL_2130880, EPI_ISL_2130881, EPI_ISL_2130882, EPI_ISL_2130883, EPI_ISL_2130885, EPI_ISL_2130886, EPI_ISL_2130888, EPI_ISL_2130890, EPI_ISL_2130892, EPI_ISL_2130894,
EPI_ISL_2130896, EPI_ISL_2130897, EPI_ISL_2130899, EPI_ISL_2130901, EPI_ISL_2130902, EPI_ISL_2130904, EPI_ISL_2130905, EPI_ISL_2130909, EPI_ISL_2130910, EPI_ISL_2130911, EPI_ISL_2130912, EPI_ISL_2130913, EPI_ISL_2130914, EPI_ISL_2130916, EPI_ISL_2130917, EPI_ISL_2130918, EPI_ISL_2130919, EPI_ISL_2130920, EPI_ISL_2130921, EPI_ISL_2130922, EPI_ISL_2130924, EPI_ISL_2130926,
EPI_ISL_2130928, EPI_ISL_2130929, EPI_ISL_2130930, EPI_ISL_2130932, EPI_ISL_2130936, EPI_ISL_2130937, EPI_ISL_2130939, EPI_ISL_2130941, EPI_ISL_2130942, EPI_ISL_2130944, EPI_ISL_2130945, EPI_ISL_2130946, EPI_ISL_2130947, EPI_ISL_2130950, EPI_ISL_2130952, EPI_ISL_2130954, EPI_ISL_2130956, EPI_ISL_2130957, EPI_ISL_2130960, EPI_ISL_2130962, EPI_ISL_2130963, EPI_ISL_2130965,
EPI_ISL_2130966, EPI_ISL_2130968, EPI_ISL_2130969, EPI_ISL_2130970, EPI_ISL_2130973, EPI_ISL_2130974, EPI_ISL_2130976, EPI_ISL_2130977, EPI_ISL_2130979, EPI_ISL_2130980, EPI_ISL_2130983, EPI_ISL_2130984, EPI_ISL_2130985, EPI_ISL_2130986, EPI_ISL_2130987, EPI_ISL_2130988, EPI_ISL_2130989, EPI_ISL_2130990, EPI_ISL_2130991, EPI_ISL_2130992, EPI_ISL_2130993, EPI_ISL_2130994,
EPI_ISL_2130995, EPI_ISL_2130996, EPI_ISL_2130997, EPI_ISL_2130999, EPI_ISL_2131003, EPI_ISL_2131006, EPI_ISL_2131008, EPI_ISL_2131009, EPI_ISL_2131012, EPI_ISL_2131013, EPI_ISL_2131014, EPI_ISL_2131015, EPI_ISL_2131018, EPI_ISL_2131019, EPI_ISL_2131021, EPI_ISL_2131023, EPI_ISL_2131027, EPI_ISL_2131028, EPI_ISL_2131029, EPI_ISL_2131030, EPI_ISL_2131031, EPI_ISL_2131035,
EPI_ISL_2131038, EPI_ISL_2131040, EPI_ISL_2131041, EPI_ISL_2131044, EPI_ISL_2131045, EPI_ISL_2131046, EPI_ISL_2131047, EPI_ISL_2131049, EPI_ISL_2131050, EPI_ISL_2131053, EPI_ISL_2131054, EPI_ISL_2131058, EPI_ISL_2131059, EPI_ISL_2131060, EPI_ISL_2131061, EPI_ISL_2131062, EPI_ISL_2131063, EPI_ISL_2131065, EPI_ISL_2131066, EPI_ISL_2131069, EPI_ISL_2131071, EPI_ISL_2131073,
EPI_ISL_2131074, EPI_ISL_2131075, EPI_ISL_2131076, EPI_ISL_2131077, EPI_ISL_2131078, EPI_ISL_2131079, EPI_ISL_2131080, EPI_ISL_2131081, EPI_ISL_2131082, EPI_ISL_2131083, EPI_ISL_2131084, EPI_ISL_2131085, EPI_ISL_2131086, EPI_ISL_2131088, EPI_ISL_2131089, EPI_ISL_2131091, EPI_ISL_2131092, EPI_ISL_2131094, EPI_ISL_2131097, EPI_ISL_2131098, EPI_ISL_2131100, EPI_ISL_2131101,
EPI_ISL_2131102, EPI_ISL_2131103, EPI_ISL_2131104, EPI_ISL_2131105, EPI_ISL_2131106, EPI_ISL_2131107, EPI_ISL_2131109, EPI_ISL_2131110, EPI_ISL_2131114, EPI_ISL_2131116, EPI_ISL_2131122, EPI_ISL_2131125, EPI_ISL_2131126, EPI_ISL_2131127, EPI_ISL_2131130, EPI_ISL_2131131, EPI_ISL_2131132, EPI_ISL_2131135, EPI_ISL_2131139, EPI_ISL_2131141, EPI_ISL_2131142, EPI_ISL_2131145,
EPI_ISL_2131146, EPI_ISL_2131147, EPI_ISL_2131148, EPI_ISL_2131149, EPI_ISL_2131150, EPI_ISL_2131151, EPI_ISL_2267296, EPI_ISL_2267302, EPI_ISL_2267328, EPI_ISL_2267334, EPI_ISL_2267362, EPI_ISL_2267363, EPI_ISL_2267366, EPI_ISL_2267368, EPI_ISL_2267379, EPI_ISL_2267383, EPI_ISL_2267388, EPI_ISL_2267389, EPI_ISL_2267406, EPI_ISL_2267429, EPI_ISL_2267439, EPI_ISL_2267450,
EPI_ISL_2267455, EPI_ISL_2267462, EPI_ISL_2267487, EPI_ISL_2267508, EPI_ISL_2267513, EPI_ISL_2267521, EPI_ISL_2267540, EPI_ISL_2267568, EPI_ISL_2267573, EPI_ISL_2267578, EPI_ISL_2267590, EPI_ISL_2267594

| | | |
|---|---|---|
| see above | SYNLAB MVZ Heidelberg | Robert Koch Institute |

EPI_ISL_2117311, EPI_ISL_2117316, EPI_ISL_2117326, EPI_ISL_2117341, EPI_ISL_2117346, EPI_ISL_2117349, EPI_ISL_2117352, EPI_ISL_2117353, EPI_ISL_2117359, EPI_ISL_2117372, EPI_ISL_2117383, EPI_ISL_2117403, EPI_ISL_2117405, EPI_ISL_2117409, EPI_ISL_2117425, EPI_ISL_2117433, EPI_ISL_2123433, EPI_ISL_2123435, EPI_ISL_2123437, EPI_ISL_2123443, EPI_ISL_2123445, EPI_ISL_2123446,
EPI_ISL_2123447, EPI_ISL_2123449, EPI_ISL_2123450, EPI_ISL_2123451, EPI_ISL_2123453, EPI_ISL_2123458, EPI_ISL_2123459, EPI_ISL_2123460, EPI_ISL_2123463, EPI_ISL_2123465, EPI_ISL_2123470, EPI_ISL_2123477, EPI_ISL_2123490, EPI_ISL_2123494, EPI_ISL_2123505, EPI_ISL_2123506, EPI_ISL_2123507, EPI_ISL_2123516, EPI_ISL_2123521, EPI_ISL_2123523, EPI_ISL_2123532,
EPI_ISL_2123540, EPI_ISL_2123542, EPI_ISL_2123545, EPI_ISL_2123547, EPI_ISL_2123558, EPI_ISL_2123563, EPI_ISL_2123568, EPI_ISL_2123577, EPI_ISL_2123578, EPI_ISL_2123582, EPI_ISL_2123583, EPI_ISL_2123586, EPI_ISL_2123594, EPI_ISL_2123600, EPI_ISL_2123602, EPI_ISL_2123604, EPI_ISL_2123614, EPI_ISL_2123631, EPI_ISL_2123634, EPI_ISL_2123638, EPI_ISL_2123640, EPI_ISL_2123642,
EPI_ISL_2123648, EPI_ISL_2123649, EPI_ISL_2123653, EPI_ISL_2123654, EPI_ISL_2123660, EPI_ISL_2123661, EPI_ISL_2123664, EPI_ISL_2123669, EPI_ISL_2123670, EPI_ISL_2123673, EPI_ISL_2123675, EPI_ISL_2123682, EPI_ISL_2123683, EPI_ISL_2123692, EPI_ISL_2123695, EPI_ISL_2123696, EPI_ISL_2123701, EPI_ISL_2123702, EPI_ISL_2123705, EPI_ISL_2123712, EPI_ISL_2123714, EPI_ISL_2123719,
EPI_ISL_2124025, EPI_ISL_2124028, EPI_ISL_2124032, EPI_ISL_2124038, EPI_ISL_2124042, EPI_ISL_2124043, EPI_ISL_2124044, EPI_ISL_2124045, EPI_ISL_2124047, EPI_ISL_2124048, EPI_ISL_2124049, EPI_ISL_2124050, EPI_ISL_2124061, EPI_ISL_2124063, EPI_ISL_2124064, EPI_ISL_2124065, EPI_ISL_2124066, EPI_ISL_2124069, EPI_ISL_2124070, EPI_ISL_2124071, EPI_ISL_2124073, EPI_ISL_2124076,
EPI_ISL_2124079, EPI_ISL_2124080, EPI_ISL_2124081, EPI_ISL_2124084, EPI_ISL_2124085, EPI_ISL_2124086, EPI_ISL_2124091, EPI_ISL_2124094, EPI_ISL_2124095, EPI_ISL_2124096, EPI_ISL_2124097, EPI_ISL_2124100, EPI_ISL_2124102, EPI_ISL_2124106, EPI_ISL_2124109, EPI_ISL_2124111, EPI_ISL_2124112, EPI_ISL_2124113, EPI_ISL_2124123, EPI_ISL_2124125, EPI_ISL_2124127, EPI_ISL_2124128,
EPI_ISL_2124129, EPI_ISL_2124130, EPI_ISL_2124131, EPI_ISL_2124132, EPI_ISL_2124134, EPI_ISL_2124135, EPI_ISL_2124137, EPI_ISL_2124138, EPI_ISL_2124141, EPI_ISL_2124142, EPI_ISL_2124143, EPI_ISL_2124144, EPI_ISL_2124145, EPI_ISL_2124148, EPI_ISL_2124149, EPI_ISL_2124150, EPI_ISL_2124151, EPI_ISL_2124154, EPI_ISL_2124161, EPI_ISL_2124163, EPI_ISL_2124164, EPI_ISL_2124165,
EPI_ISL_2124166, EPI_ISL_2124168, EPI_ISL_2124169, EPI_ISL_2124174, EPI_ISL_2124177, EPI_ISL_2124179, EPI_ISL_2124181, EPI_ISL_2124184, EPI_ISL_2124188, EPI_ISL_2124190, EPI_ISL_2124199, EPI_ISL_2124200, EPI_ISL_2124201, EPI_ISL_2124204, EPI_ISL_2124207, EPI_ISL_2124208, EPI_ISL_2124211, EPI_ISL_2124214, EPI_ISL_2124217, EPI_ISL_2124218, EPI_ISL_2124219, EPI_ISL_2124220,
EPI_ISL_2124221, EPI_ISL_2124223, EPI_ISL_2124225, EPI_ISL_2124229, EPI_ISL_2124231, EPI_ISL_2124232, EPI_ISL_2124234, EPI_ISL_2124237, EPI_ISL_2124239, EPI_ISL_2124240, EPI_ISL_2124242, EPI_ISL_2124243, EPI_ISL_2124244, EPI_ISL_2124245, EPI_ISL_2124246, EPI_ISL_2124248, EPI_ISL_2124250, EPI_ISL_2124252, EPI_ISL_2124253, EPI_ISL_2124254, EPI_ISL_2124255, EPI_ISL_2124256,
EPI_ISL_2124257, EPI_ISL_2124259, EPI_ISL_2124260, EPI_ISL_2124262, EPI_ISL_2124263, EPI_ISL_2124265, EPI_ISL_2124267, EPI_ISL_2127234, EPI_ISL_2127236, EPI_ISL_2127242, EPI_ISL_2127255, EPI_ISL_2127260, EPI_ISL_2127264, EPI_ISL_2127269, EPI_ISL_2127271, EPI_ISL_2127279, EPI_ISL_2127288, EPI_ISL_2127297, EPI_ISL_2127309, EPI_ISL_2127311, EPI_ISL_2127319, EPI_ISL_2127321,
EPI_ISL_2127323, EPI_ISL_2127325, EPI_ISL_2127333, EPI_ISL_2127335, EPI_ISL_2127337, EPI_ISL_2127352, EPI_ISL_2127356, EPI_ISL_2127389, EPI_ISL_2127399, EPI_ISL_2127401, EPI_ISL_2127403, EPI_ISL_2127411, EPI_ISL_2127416, EPI_ISL_2127419, EPI_ISL_2127423, EPI_ISL_2127434, EPI_ISL_2127439, EPI_ISL_2127443, EPI_ISL_2127452, EPI_ISL_2127454, EPI_ISL_2127459, EPI_ISL_2127463,
EPI_ISL_2127468, EPI_ISL_2127470, EPI_ISL_2127484, EPI_ISL_2127486, EPI_ISL_2127498, EPI_ISL_2127504, EPI_ISL_2127508, EPI_ISL_2127512, EPI_ISL_2127549, EPI_ISL_2127555, EPI_ISL_2127562, EPI_ISL_2127564, EPI_ISL_2127566, EPI_ISL_2127579, EPI_ISL_2127584, EPI_ISL_2127596, EPI_ISL_2127607, EPI_ISL_2127612, EPI_ISL_2127616, EPI_ISL_2127618, EPI_ISL_2127625, EPI_ISL_2127627,
EPI_ISL_2127637, EPI_ISL_2127640, EPI_ISL_2127643, EPI_ISL_2127648, EPI_ISL_2127651, EPI_ISL_2127656, EPI_ISL_2127658, EPI_ISL_2127660, EPI_ISL_2127662, EPI_ISL_2127668, EPI_ISL_2127676, EPI_ISL_2127678, EPI_ISL_2127682, EPI_ISL_2127684, EPI_ISL_2127689, EPI_ISL_2128377, EPI_ISL_2128379, EPI_ISL_2128380, EPI_ISL_2128385, EPI_ISL_2128388, EPI_ISL_2128389, EPI_ISL_2129735,
EPI_ISL_2129736, EPI_ISL_2129737, EPI_ISL_2129738, EPI_ISL_2129739, EPI_ISL_2129746, EPI_ISL_2129748, EPI_ISL_2129749, EPI_ISL_2129750, EPI_ISL_2129751, EPI_ISL_2129753, EPI_ISL_2129762, EPI_ISL_2129766, EPI_ISL_2129767, EPI_ISL_2129768, EPI_ISL_2129772, EPI_ISL_2129774, EPI_ISL_2129778, EPI_ISL_2129785, EPI_ISL_2129788, EPI_ISL_2129793, EPI_ISL_2129794,
EPI_ISL_2129799, EPI_ISL_2129801, EPI_ISL_2129808, EPI_ISL_2129810, EPI_ISL_2129811, EPI_ISL_2129818, EPI_ISL_2129820, EPI_ISL_2129824, EPI_ISL_2129834, EPI_ISL_2129843, EPI_ISL_2129851, EPI_ISL_2129855, EPI_ISL_2129856, EPI_ISL_2129860, EPI_ISL_2129866, EPI_ISL_2129870, EPI_ISL_2129873, EPI_ISL_2129884, EPI_ISL_2129893, EPI_ISL_2129896, EPI_ISL_2129898, EPI_ISL_2129900,
EPI_ISL_2129907, EPI_ISL_2129910, EPI_ISL_2129912, EPI_ISL_2129914, EPI_ISL_2129916, EPI_ISL_2129919, EPI_ISL_2129923, EPI_ISL_2129924, EPI_ISL_2129928, EPI_ISL_2129932, EPI_ISL_2129935, EPI_ISL_2129937, EPI_ISL_2129940, EPI_ISL_2129941, EPI_ISL_2129943, EPI_ISL_2129944, EPI_ISL_2129946, EPI_ISL_2129947, EPI_ISL_2129948, EPI_ISL_2129951, EPI_ISL_2129965, EPI_ISL_2129967,
EPI_ISL_2129968, EPI_ISL_2129969, EPI_ISL_2129977, EPI_ISL_2261631, EPI_ISL_2261668, EPI_ISL_2261692, EPI_ISL_2261697, EPI_ISL_2261701, EPI_ISL_2261710, EPI_ISL_2261731, EPI_ISL_2261739, EPI_ISL_2261745, EPI_ISL_2261762, EPI_ISL_2261783, EPI_ISL_2261804, EPI_ISL_2261827, EPI_ISL_2261830, EPI_ISL_2261840, EPI_ISL_2261878, EPI_ISL_2261911, EPI_ISL_2261919, EPI_ISL_2261920,
EPI_ISL_2261923, EPI_ISL_2261929, EPI_ISL_2261930, EPI_ISL_2261936, EPI_ISL_2261967, EPI_ISL_2261982, EPI_ISL_2261989, EPI_ISL_2261992, EPI_ISL_2262012, EPI_ISL_2262019, EPI_ISL_2262024, EPI_ISL_2262028, EPI_ISL_2262038, EPI_ISL_2262074, EPI_ISL_2262095, EPI_ISL_2262829, EPI_ISL_2262830, EPI_ISL_2262834, EPI_ISL_2262835, EPI_ISL_2262837, EPI_ISL_2262838, EPI_ISL_2262839,
EPI_ISL_2262840, EPI_ISL_2262841, EPI_ISL_2262843, EPI_ISL_2262846, EPI_ISL_2262849, EPI_ISL_2262850, EPI_ISL_2262852, EPI_ISL_2262855, EPI_ISL_2262856, EPI_ISL_2262857, EPI_ISL_2262859, EPI_ISL_2262860, EPI_ISL_2262861, EPI_ISL_2262862, EPI_ISL_2262863, EPI_ISL_2262864, EPI_ISL_2262865, EPI_ISL_2262867, EPI_ISL_2262868, EPI_ISL_2262872, EPI_ISL_2262873, EPI_ISL_2262874,
EPI_ISL_2262875, EPI_ISL_2262876, EPI_ISL_2262878, EPI_ISL_2262879, EPI_ISL_2262881, EPI_ISL_2262882, EPI_ISL_2262885, EPI_ISL_2262886, EPI_ISL_2262887, EPI_ISL_2262888, EPI_ISL_2262889, EPI_ISL_2262890, EPI_ISL_2262892, EPI_ISL_2262893, EPI_ISL_2262895, EPI_ISL_2262898, EPI_ISL_2262899, EPI_ISL_2262900, EPI_ISL_2262902, EPI_ISL_2262903, EPI_ISL_2262905, EPI_ISL_2262906,
EPI_ISL_2262907, EPI_ISL_2262908, EPI_ISL_2262911, EPI_ISL_2262917, EPI_ISL_2262918, EPI_ISL_2262919, EPI_ISL_2262920, EPI_ISL_2262921, EPI_ISL_2262922, EPI_ISL_2262924, EPI_ISL_2262925, EPI_ISL_2262926, EPI_ISL_2262927, EPI_ISL_2262928, EPI_ISL_2262929, EPI_ISL_2262933, EPI_ISL_2262934, EPI_ISL_2262935, EPI_ISL_2262936, EPI_ISL_2262937, EPI_ISL_2262938, EPI_ISL_2262940,
EPI_ISL_2262941, EPI_ISL_2262942, EPI_ISL_2262943, EPI_ISL_2262944, EPI_ISL_2262945, EPI_ISL_2262946, EPI_ISL_2262951, EPI_ISL_2262952, EPI_ISL_2262953, EPI_ISL_2262955, EPI_ISL_2262956, EPI_ISL_2262957, EPI_ISL_2262959, EPI_ISL_2262960, EPI_ISL_2262963, EPI_ISL_2262964, EPI_ISL_2262965, EPI_ISL_2262966, EPI_ISL_2262967, EPI_ISL_2262968, EPI_ISL_2262969, EPI_ISL_2262970,
EPI_ISL_2262972, EPI_ISL_2262973, EPI_ISL_2262975, EPI_ISL_2262976, EPI_ISL_2262980, EPI_ISL_2262981, EPI_ISL_2262983, EPI_ISL_2262984, EPI_ISL_2262985, EPI_ISL_2262987, EPI_ISL_2262988, EPI_ISL_2262989, EPI_ISL_2262990, EPI_ISL_2262991, EPI_ISL_2262992, EPI_ISL_2262993, EPI_ISL_2262994, EPI_ISL_2262996, EPI_ISL_2262997, EPI_ISL_2263000, EPI_ISL_2263001, EPI_ISL_2263002,
EPI_ISL_2263004, EPI_ISL_2263006, EPI_ISL_2263007, EPI_ISL_2263008, EPI_ISL_2263009, EPI_ISL_2263011, EPI_ISL_2263012, EPI_ISL_2263014, EPI_ISL_2263016, EPI_ISL_2263017, EPI_ISL_2263018, EPI_ISL_2263019, EPI_ISL_2263020, EPI_ISL_2263021, EPI_ISL_2263022, EPI_ISL_2263024, EPI_ISL_2263025, EPI_ISL_2263026, EPI_ISL_2263027, EPI_ISL_2263029, EPI_ISL_2263030, EPI_ISL_2263031,
EPI_ISL_2263033, EPI_ISL_2263035, EPI_ISL_2263036, EPI_ISL_2263214, EPI_ISL_2263216, EPI_ISL_2263217, EPI_ISL_2263218, EPI_ISL_2263219, EPI_ISL_2263222, EPI_ISL_2263224, EPI_ISL_2263226, EPI_ISL_2263229, EPI_ISL_2263230, EPI_ISL_2263232, EPI_ISL_2263235, EPI_ISL_2263236, EPI_ISL_2263238, EPI_ISL_2263241, EPI_ISL_2263244, EPI_ISL_2263245, EPI_ISL_2263249, EPI_ISL_2263255,
EPI_ISL_2263257, EPI_ISL_2263258, EPI_ISL_2263262, EPI_ISL_2263265, EPI_ISL_2263267, EPI_ISL_2263270, EPI_ISL_2263273, EPI_ISL_2263274, EPI_ISL_2263275, EPI_ISL_2263279, EPI_ISL_2263283, EPI_ISL_2263291, EPI_ISL_2263294, EPI_ISL_2263295, EPI_ISL_2263298, EPI_ISL_2263301, EPI_ISL_2263304, EPI_ISL_2263306, EPI_ISL_2263309, EPI_ISL_2263312, EPI_ISL_2263315, EPI_ISL_2263319,
EPI_ISL_2263320, EPI_ISL_2263330, EPI_ISL_2263335, EPI_ISL_2263339, EPI_ISL_2263341, EPI_ISL_2263342, EPI_ISL_2263343, EPI_ISL_2263344, EPI_ISL_2263345, EPI_ISL_2263348, EPI_ISL_2263349, EPI_ISL_2263350, EPI_ISL_2263351, EPI_ISL_2263353, EPI_ISL_2263355, EPI_ISL_2263360, EPI_ISL_2263362, EPI_ISL_2263363, EPI_ISL_2263367, EPI_ISL_2263369, EPI_ISL_2263371, EPI_ISL_2263372,
EPI_ISL_2263373, EPI_ISL_2263378, EPI_ISL_2263382, EPI_ISL_2263385, EPI_ISL_2263386, EPI_ISL_2263391, EPI_ISL_2263392, EPI_ISL_2263398, EPI_ISL_2263399, EPI_ISL_2263401, EPI_ISL_2263403, EPI_ISL_2263405, EPI_ISL_2263408, EPI_ISL_2263409, EPI_ISL_2263416, EPI_ISL_2263417, EPI_ISL_2263418, EPI_ISL_2263422, EPI_ISL_2263423, EPI_ISL_2263430, EPI_ISL_2263431,
EPI_ISL_2263434, EPI_ISL_2263435, EPI_ISL_2263439, EPI_ISL_2263441, EPI_ISL_2263442, EPI_ISL_2263443, EPI_ISL_2263444, EPI_ISL_2263447, EPI_ISL_2263449, EPI_ISL_2263455, EPI_ISL_2263460, EPI_ISL_2263461, EPI_ISL_2263463, EPI_ISL_2263466, EPI_ISL_2263470, EPI_ISL_2263471, EPI_ISL_2263483, EPI_ISL_2263489, EPI_ISL_2263491, EPI_ISL_2263492, EPI_ISL_2263494, EPI_ISL_2263496,
EPI_ISL_2263499, EPI_ISL_2263500, EPI_ISL_2263508, EPI_ISL_2263512, EPI_ISL_2263514, EPI_ISL_2263515, EPI_ISL_2263517, EPI_ISL_2263519, EPI_ISL_2263524, EPI_ISL_2263534, EPI_ISL_2263537, EPI_ISL_2263539, EPI_ISL_2263544, EPI_ISL_2263545, EPI_ISL_2263550, EPI_ISL_2263552, EPI_ISL_2263555, EPI_ISL_2263558, EPI_ISL_2263559, EPI_ISL_2263560, EPI_ISL_2263561, EPI_ISL_2263563,
EPI_ISL_2263565, EPI_ISL_2263566, EPI_ISL_2263570, EPI_ISL_2263572, EPI_ISL_2263573, EPI_ISL_2263574, EPI_ISL_2263575, EPI_ISL_2263578, EPI_ISL_2263580, EPI_ISL_2263581, EPI_ISL_2263584, EPI_ISL_2263590, EPI_ISL_2263593, EPI_ISL_2263594, EPI_ISL_2263596, EPI_ISL_2263599, EPI_ISL_2263601, EPI_ISL_2263604, EPI_ISL_2263605, EPI_ISL_2263606, EPI_ISL_2263607, EPI_ISL_2263612,
EPI_ISL_2263613, EPI_ISL_2263614, EPI_ISL_2263617, EPI_ISL_2263618, EPI_ISL_2263624, EPI_ISL_2263625, EPI_ISL_2263628, EPI_ISL_2263630, EPI_ISL_2263631, EPI_ISL_2263632, EPI_ISL_2263634, EPI_ISL_2263638, EPI_ISL_2263640, EPI_ISL_2263641, EPI_ISL_2263642, EPI_ISL_2263643, EPI_ISL_2263644, EPI_ISL_2263646, EPI_ISL_2263649, EPI_ISL_2263655, EPI_ISL_2263656,
EPI_ISL_2263659, EPI_ISL_2263660, EPI_ISL_2263663, EPI_ISL_2263665, EPI_ISL_2263666, EPI_ISL_2263672, EPI_ISL_2263674, EPI_ISL_2263678, EPI_ISL_2263679, EPI_ISL_2263680, EPI_ISL_2263681, EPI_ISL_2263682, EPI_ISL_2263683, EPI_ISL_2263686, EPI_ISL_2263687, EPI_ISL_2263690, EPI_ISL_2263691, EPI_ISL_2263692, EPI_ISL_2263694, EPI_ISL_2263695, EPI_ISL_2263700, EPI_ISL_2263703,
EPI_ISL_2263706, EPI_ISL_2263715, EPI_ISL_2263716, EPI_ISL_2263717, EPI_ISL_2263719, EPI_ISL_2263720, EPI_ISL_2263721, EPI_ISL_2263723, EPI_ISL_2263725, EPI_ISL_2263729, EPI_ISL_2263730, EPI_ISL_2263731, EPI_ISL_2263733, EPI_ISL_2263735, EPI_ISL_2263736, EPI_ISL_2263742, EPI_ISL_2263743, EPI_ISL_2263744, EPI_ISL_2263745, EPI_ISL_2263746, EPI_ISL_2263747, EPI_ISL_2263748,
EPI_ISL_2263752, EPI_ISL_2263758, EPI_ISL_2263760, EPI_ISL_2263761, EPI_ISL_2263766, EPI_ISL_2263768, EPI_ISL_2263771, EPI_ISL_2263772, EPI_ISL_2263774, EPI_ISL_2263778, EPI_ISL_2263784, EPI_ISL_2264144, EPI_ISL_2264146, EPI_ISL_2264148, EPI_ISL_2264149, EPI_ISL_2264150, EPI_ISL_2264152, EPI_ISL_2264153, EPI_ISL_2264154, EPI_ISL_2264157, EPI_ISL_2264158, EPI_ISL_2264159,
EPI_ISL_2264161, EPI_ISL_2264163, EPI_ISL_2264164, EPI_ISL_2264165, EPI_ISL_2264166, EPI_ISL_2264168, EPI_ISL_2264170, EPI_ISL_2264171, EPI_ISL_2264172, EPI_ISL_2264173, EPI_ISL_2264174, EPI_ISL_2264175, EPI_ISL_2264177, EPI_ISL_2264178, EPI_ISL_2264179, EPI_ISL_2264181, EPI_ISL_2264183, EPI_ISL_2264184, EPI_ISL_2264187, EPI_ISL_2264190, EPI_ISL_2264192, EPI_ISL_2264193,
EPI_ISL_2264195, EPI_ISL_2264197, EPI_ISL_2264199, EPI_ISL_2264200, EPI_ISL_2264201, EPI_ISL_2264202, EPI_ISL_2264204, EPI_ISL_2264205, EPI_ISL_2264206, EPI_ISL_2264207, EPI_ISL_2264208, EPI_ISL_2264210, EPI_ISL_2264212, EPI_ISL_2264213, EPI_ISL_2264214, EPI_ISL_2264217, EPI_ISL_2264218, EPI_ISL_2264219, EPI_ISL_2264221, EPI_ISL_2264222, EPI_ISL_2264223, EPI_ISL_2264224,
EPI_ISL_2264225, EPI_ISL_2264226, EPI_ISL_2264227, EPI_ISL_2264228, EPI_ISL_2264229, EPI_ISL_2264230, EPI_ISL_2264231, EPI_ISL_2264232, EPI_ISL_2264234, EPI_ISL_2264236, EPI_ISL_2264237, EPI_ISL_2264241, EPI_ISL_2264243, EPI_ISL_2264244, EPI_ISL_2264245, EPI_ISL_2264246, EPI_ISL_2264248, EPI_ISL_2264253, EPI_ISL_2264255, EPI_ISL_2264259, EPI_ISL_2264261, EPI_ISL_2264263,
EPI_ISL_2264265, EPI_ISL_2264267, EPI_ISL_2264272, EPI_ISL_2264273, EPI_ISL_2264274, EPI_ISL_2264275, EPI_ISL_2264277, EPI_ISL_2264282, EPI_ISL_2264284, EPI_ISL_2264286, EPI_ISL_2264288, EPI_ISL_2264289, EPI_ISL_2264290, EPI_ISL_2264291, EPI_ISL_2264293, EPI_ISL_2264295, EPI_ISL_2264296, EPI_ISL_2264300, EPI_ISL_2264301, EPI_ISL_2264302, EPI_ISL_2264304, EPI_ISL_2264306,
EPI_ISL_2264309, EPI_ISL_2264312, EPI_ISL_2264313, EPI_ISL_2264317, EPI_ISL_2264318, EPI_ISL_2264319, EPI_ISL_2264320, EPI_ISL_2264321, EPI_ISL_2264325, EPI_ISL_2264328, EPI_ISL_2264330, EPI_ISL_2264331, EPI_ISL_2264332, EPI_ISL_2264333, EPI_ISL_2264335, EPI_ISL_2264337, EPI_ISL_2264339, EPI_ISL_2264340, EPI_ISL_2264343, EPI_ISL_2264344, EPI_ISL_2264345, EPI_ISL_2264346,
EPI_ISL_2264349, EPI_ISL_2264350, EPI_ISL_2264361, EPI_ISL_2264367, EPI_ISL_2264370, EPI_ISL_2264375, EPI_ISL_2264380, EPI_ISL_2264382, EPI_ISL_2264387, EPI_ISL_2264442, EPI_ISL_2314459, EPI_ISL_2314506, EPI_ISL_2314539, EPI_ISL_2314553, EPI_ISL_2314614, EPI_ISL_2314618

| | | |
|---|---|---|
| see above | SYNLAB MVZ Leinfelden-Echterdingen | Robert Koch Institute |

EPI_ISL_2116313, EPI_ISL_2116314, EPI_ISL_2116315, EPI_ISL_2116316, EPI_ISL_2116317, EPI_ISL_2116318, EPI_ISL_2116319, EPI_ISL_2116320, EPI_ISL_2116321, EPI_ISL_2116322, EPI_ISL_2116323, EPI_ISL_2116324, EPI_ISL_2116325, EPI_ISL_2116326, EPI_ISL_2116327, EPI_ISL_2116328, EPI_ISL_2116329, EPI_ISL_2116330, EPI_ISL_2116331, EPI_ISL_2116332, EPI_ISL_2116333, EPI_ISL_2116334,
EPI_ISL_2116335, EPI_ISL_2116336, EPI_ISL_2116337, EPI_ISL_2116338, EPI_ISL_2116340, EPI_ISL_2116341, EPI_ISL_2116342, EPI_ISL_2116343, EPI_ISL_2116344, EPI_ISL_2116345, EPI_ISL_2116346, EPI_ISL_2116347, EPI_ISL_2116348, EPI_ISL_2116349, EPI_ISL_2116350, EPI_ISL_2116351, EPI_ISL_2116352, EPI_ISL_2116353, EPI_ISL_2116354, EPI_ISL_2116355, EPI_ISL_2116356, EPI_ISL_2116357,
EPI_ISL_2116358, EPI_ISL_2116359, EPI_ISL_2116360, EPI_ISL_2116361, EPI_ISL_2116362, EPI_ISL_2116363, EPI_ISL_2116364, EPI_ISL_2116365, EPI_ISL_2116367, EPI_ISL_2116368, EPI_ISL_2116369, EPI_ISL_2116370, EPI_ISL_2116372, EPI_ISL_2116373, EPI_ISL_2116374, EPI_ISL_2116375, EPI_ISL_2116376, EPI_ISL_2116377, EPI_ISL_2116378, EPI_ISL_2116379, EPI_ISL_2116380, EPI_ISL_2116381,
EPI_ISL_2116382, EPI_ISL_2116383, EPI_ISL_2116384, EPI_ISL_2116385, EPI_ISL_2116386, EPI_ISL_2116387, EPI_ISL_2116388, EPI_ISL_2116390, EPI_ISL_2116391, EPI_ISL_2116393, EPI_ISL_2116394, EPI_ISL_2116395, EPI_ISL_2116396, EPI_ISL_2116397, EPI_ISL_2116398, EPI_ISL_2116400, EPI_ISL_2116401, EPI_ISL_2116402, EPI_ISL_2116403, EPI_ISL_2116404, EPI_ISL_2116405, EPI_ISL_2116406,
EPI_ISL_2116407, EPI_ISL_2116408, EPI_ISL_2116409, EPI_ISL_2116410, EPI_ISL_2116411, EPI_ISL_2116412, EPI_ISL_2116413, EPI_ISL_2116416, EPI_ISL_2116417, EPI_ISL_2116419, EPI_ISL_2116420, EPI_ISL_2116421, EPI_ISL_2116422, EPI_ISL_2116423, EPI_ISL_2116424, EPI_ISL_2116425, EPI_ISL_2116426, EPI_ISL_2116427, EPI_ISL_2116428, EPI_ISL_2116430, EPI_ISL_2116431,
EPI_ISL_2116433, EPI_ISL_2116434, EPI_ISL_2116435, EPI_ISL_2116436, EPI_ISL_2116437, EPI_ISL_2116438, EPI_ISL_2116439, EPI_ISL_2116440, EPI_ISL_2116441, EPI_ISL_2116442, EPI_ISL_2116443, EPI_ISL_2116444, EPI_ISL_2116445, EPI_ISL_2116446, EPI_ISL_2116447, EPI_ISL_2116448, EPI_ISL_2116449, EPI_ISL_2116450, EPI_ISL_2116451, EPI_ISL_2116452, EPI_ISL_2116453, EPI_ISL_2116454,
EPI_ISL_2116455, EPI_ISL_2116456, EPI_ISL_2116458, EPI_ISL_2116459, EPI_ISL_2116460, EPI_ISL_2116461, EPI_ISL_2116462, EPI_ISL_2116463, EPI_ISL_2116464, EPI_ISL_2116465, EPI_ISL_2116466, EPI_ISL_2116467, EPI_ISL_2116468, EPI_ISL_2116469, EPI_ISL_2116470, EPI_ISL_2116471, EPI_ISL_2116472, EPI_ISL_2116473, EPI_ISL_2116474, EPI_ISL_2116475, EPI_ISL_2116476, EPI_ISL_2116477,
EPI_ISL_2116478, EPI_ISL_2116479, EPI_ISL_2116480, EPI_ISL_2116481, EPI_ISL_2116482, EPI_ISL_2116483, EPI_ISL_2116484, EPI_ISL_2116485, EPI_ISL_2116486, EPI_ISL_2116487, EPI_ISL_2116488, EPI_ISL_2116489, EPI_ISL_2116490, EPI_ISL_2116491, EPI_ISL_2116492, EPI_ISL_2116493, EPI_ISL_2116494, EPI_ISL_2116495, EPI_ISL_2116496, EPI_ISL_2116497, EPI_ISL_2116498, EPI_ISL_2116500,
EPI_ISL_2116501, EPI_ISL_2116502, EPI_ISL_2116503, EPI_ISL_2116504, EPI_ISL_2116505, EPI_ISL_2116506, EPI_ISL_2116507, EPI_ISL_2116508, EPI_ISL_2116509, EPI_ISL_2116510, EPI_ISL_2116511, EPI_ISL_2116512, EPI_ISL_2116513, EPI_ISL_2116514, EPI_ISL_2116515, EPI_ISL_2116516, EPI_ISL_2116517, EPI_ISL_2116518, EPI_ISL_2116519, EPI_ISL_2116520, EPI_ISL_2116521, EPI_ISL_2116522,
EPI_ISL_2116523, EPI_ISL_2116524, EPI_ISL_2116525, EPI_ISL_2116526, EPI_ISL_2116527, EPI_ISL_2116528, EPI_ISL_2116529, EPI_ISL_2116530, EPI_ISL_2116531, EPI_ISL_2116532, EPI_ISL_2116534, EPI_ISL_2116535, EPI_ISL_2116536, EPI_ISL_2116537, EPI_ISL_2116538, EPI_ISL_2116540, EPI_ISL_2116541, EPI_ISL_2116542, EPI_ISL_2116543, EPI_ISL_2116545, EPI_ISL_2116546, EPI_ISL_2116547,
EPI_ISL_2116548, EPI_ISL_2116549, EPI_ISL_2116550, EPI_ISL_2116551, EPI_ISL_2116552, EPI_ISL_2116553, EPI_ISL_2116554, EPI_ISL_2116555, EPI_ISL_2116556, EPI_ISL_2116557, EPI_ISL_2116558, EPI_ISL_2116560, EPI_ISL_2116561, EPI_ISL_2116562, EPI_ISL_2116563, EPI_ISL_2116564, EPI_ISL_2116565, EPI_ISL_2116566, EPI_ISL_2116567, EPI_ISL_2116568, EPI_ISL_2116569, EPI_ISL_2116570,
EPI_ISL_2116571, EPI_ISL_2116572, EPI_ISL_2116573, EPI_ISL_2116574, EPI_ISL_2116575, EPI_ISL_2116576, EPI_ISL_2116577, EPI_ISL_2116578, EPI_ISL_2116579, EPI_ISL_2116580, EPI_ISL_2116581, EPI_ISL_2116582, EPI_ISL_2116583, EPI_ISL_2116584, EPI_ISL_2116585, EPI_ISL_2116586, EPI_ISL_2116587, EPI_ISL_2116588, EPI_ISL_2116589, EPI_ISL_2116590, EPI_ISL_2116591, EPI_ISL_2116592,
EPI_ISL_2116593, EPI_ISL_2116594, EPI_ISL_2116596, EPI_ISL_2116597, EPI_ISL_2116598, EPI_ISL_2116599, EPI_ISL_2116600, EPI_ISL_2116601, EPI_ISL_2116602, EPI_ISL_2116603, EPI_ISL_2116604, EPI_ISL_2116605, EPI_ISL_2116606, EPI_ISL_2116607, EPI_ISL_2116608, EPI_ISL_2116609, EPI_ISL_2116610, EPI_ISL_2116612, EPI_ISL_2116614, EPI_ISL_2116615, EPI_ISL_2116616, EPI_ISL_2116617,
EPI_ISL_2116618, EPI_ISL_2116619, EPI_ISL_2116620, EPI_ISL_2116621, EPI_ISL_2116622, EPI_ISL_2116623, EPI_ISL_2116624, EPI_ISL_2116625, EPI_ISL_2116626, EPI_ISL_2116627, EPI_ISL_2116628, EPI_ISL_2116629, EPI_ISL_2116630, EPI_ISL_2116631, EPI_ISL_2116632, EPI_ISL_2116633, EPI_ISL_2116634, EPI_ISL_2116635, EPI_ISL_2116636, EPI_ISL_2116638, EPI_ISL_2116639, EPI_ISL_2116641,
EPI_ISL_2116644, EPI_ISL_2116645, EPI_ISL_2116648, EPI_ISL_2116649, EPI_ISL_2116650, EPI_ISL_2116651, EPI_ISL_2116653, EPI_ISL_2116654, EPI_ISL_2116658, EPI_ISL_2116659, EPI_ISL_2116663, EPI_ISL_2116664, EPI_ISL_2116665, EPI_ISL_2116666, EPI_ISL_2116667, EPI_ISL_2116670, EPI_ISL_2116671, EPI_ISL_2116672, EPI_ISL_2116674, EPI_ISL_2116677, EPI_ISL_2116679, EPI_ISL_2116680,
EPI_ISL_2116681, EPI_ISL_2116683, EPI_ISL_2116684, EPI_ISL_2116685, EPI_ISL_2116688, EPI_ISL_2116691, EPI_ISL_2116692, EPI_ISL_2116693, EPI_ISL_2116694, EPI_ISL_2116695, EPI_ISL_2116696, EPI_ISL_2116697, EPI_ISL_2116698, EPI_ISL_2116699, EPI_ISL_2116700, EPI_ISL_2116703, EPI_ISL_2116704, EPI_ISL_2116705, EPI_ISL_2116707, EPI_ISL_2116708, EPI_ISL_2116712, EPI_ISL_2116713,
EPI_ISL_2116714, EPI_ISL_2116716, EPI_ISL_2116717, EPI_ISL_2116718, EPI_ISL_2116719, EPI_ISL_2116720, EPI_ISL_2116724, EPI_ISL_2116725, EPI_ISL_2116726, EPI_ISL_2116728, EPI_ISL_2116730, EPI_ISL_2116731, EPI_ISL_2116732, EPI_ISL_2116733, EPI_ISL_2116734, EPI_ISL_2116735, EPI_ISL_2116739, EPI_ISL_2116741, EPI_ISL_2116748, EPI_ISL_2116750, EPI_ISL_2116752, EPI_ISL_2116756,
EPI_ISL_2116757, EPI_ISL_2116760, EPI_ISL_2116761, EPI_ISL_2116763, EPI_ISL_2116764, EPI_ISL_2116765, EPI_ISL_2116767, EPI_ISL_2116770, EPI_ISL_2116771, EPI_ISL_2116773, EPI_ISL_2116775, EPI_ISL_2116779, EPI_ISL_2116780, EPI_ISL_2116781, EPI_ISL_2116782, EPI_ISL_2116785, EPI_ISL_2116787, EPI_ISL_2116789, EPI_ISL_2116790, EPI_ISL_2116792, EPI_ISL_2116795, EPI_ISL_2116796,
EPI_ISL_2116797, EPI_ISL_2116801, EPI_ISL_2116806, EPI_ISL_2116808, EPI_ISL_2116812, EPI_ISL_2116816, EPI_ISL_2116817, EPI_ISL_2116821, EPI_ISL_2116826, EPI_ISL_2116827, EPI_ISL_2116828, EPI_ISL_2116829, EPI_ISL_2116831, EPI_ISL_2116833, EPI_ISL_2116837, EPI_ISL_2116839, EPI_ISL_2116840, EPI_ISL_2116841, EPI_ISL_2116847, EPI_ISL_2116849,
EPI_ISL_2116850, EPI_ISL_2116854, EPI_ISL_2116858, EPI_ISL_2116859, EPI_ISL_2116861, EPI_ISL_2116863, EPI_ISL_2116866, EPI_ISL_2116867, EPI_ISL_2116869, EPI_ISL_2116870, EPI_ISL_2116871, EPI_ISL_2116875, EPI_ISL_2116878, EPI_ISL_2116879, EPI_ISL_2116880, EPI_ISL_2116881, EPI_ISL_2116884, EPI_ISL_2116885, EPI_ISL_2116886, EPI_ISL_2116887, EPI_ISL_2116888, EPI_ISL_2116889,
EPI_ISL_2116894, EPI_ISL_2116895, EPI_ISL_2116896, EPI_ISL_2116900, EPI_ISL_2116901, EPI_ISL_2116904, EPI_ISL_2116907, EPI_ISL_2116908, EPI_ISL_2116910, EPI_ISL_2116915, EPI_ISL_2116920, EPI_ISL_2116921, EPI_ISL_2116922, EPI_ISL_2116924, EPI_ISL_2116926, EPI_ISL_2116927, EPI_ISL_2116929, EPI_ISL_2116930, EPI_ISL_2116934, EPI_ISL_2116936, EPI_ISL_2116937, EPI_ISL_2116941,
EPI_ISL_2116944, EPI_ISL_2116946, EPI_ISL_2116947, EPI_ISL_2116948, EPI_ISL_2116949, EPI_ISL_2116953, EPI_ISL_2116954, EPI_ISL_2116956, EPI_ISL_2116957, EPI_ISL_2116958, EPI_ISL_2116959, EPI_ISL_2116960, EPI_ISL_2116961, EPI_ISL_2116964, EPI_ISL_2116965, EPI_ISL_2116966, EPI_ISL_2116967, EPI_ISL_2116970, EPI_ISL_2116971, EPI_ISL_2116973, EPI_ISL_2116974, EPI_ISL_2116975,
EPI_ISL_2116976, EPI_ISL_2116977, EPI_ISL_2116981, EPI_ISL_2116984, EPI_ISL_2116985, EPI_ISL_2116987, EPI_ISL_2116989, EPI_ISL_2116991, EPI_ISL_2116992, EPI_ISL_2116993, EPI_ISL_2116994, EPI_ISL_2116996, EPI_ISL_2116998, EPI_ISL_2117002, EPI_ISL_2117004, EPI_ISL_2117009, EPI_ISL_2117010, EPI_ISL_2117012, EPI_ISL_2117014, EPI_ISL_2117016, EPI_ISL_2117018, EPI_ISL_2117019,
EPI_ISL_2117021, EPI_ISL_2117022, EPI_ISL_2117023, EPI_ISL_2117024, EPI_ISL_2117025, EPI_ISL_2117026, EPI_ISL_2117028, EPI_ISL_2117030, EPI_ISL_2117031, EPI_ISL_2117033, EPI_ISL_2117039, EPI_ISL_2117041, EPI_ISL_2117043, EPI_ISL_2117044, EPI_ISL_2117045, EPI_ISL_2117050, EPI_ISL_2117051, EPI_ISL_2117052, EPI_ISL_2117053, EPI_ISL_2117054, EPI_ISL_2117058, EPI_ISL_2117059,
EPI_ISL_2117061, EPI_ISL_2117063, EPI_ISL_2117067, EPI_ISL_2117072, EPI_ISL_2117079, EPI_ISL_2117080, EPI_ISL_2117082, EPI_ISL_2117087, EPI_ISL_2117094, EPI_ISL_2117103, EPI_ISL_2117104, EPI_ISL_2117108, EPI_ISL_2117116, EPI_ISL_2117117, EPI_ISL_2117124, EPI_ISL_2117125, EPI_ISL_2117127, EPI_ISL_2117128, EPI_ISL_2117129, EPI_ISL_2117135, EPI_ISL_2117138,
EPI_ISL_2117139, EPI_ISL_2117145, EPI_ISL_2117146, EPI_ISL_2117147, EPI_ISL_2117151, EPI_ISL_2117154, EPI_ISL_2117155, EPI_ISL_2117158, EPI_ISL_2117159, EPI_ISL_2117160, EPI_ISL_2117161, EPI_ISL_2117164, EPI_ISL_2117169, EPI_ISL_2117170, EPI_ISL_2117175, EPI_ISL_2117176, EPI_ISL_2117177, EPI_ISL_2117180, EPI_ISL_2117190, EPI_ISL_2117196, EPI_ISL_2117199,
EPI_ISL_2117208, EPI_ISL_2117212, EPI_ISL_2117213, EPI_ISL_2117217, EPI_ISL_2117222, EPI_ISL_2117223, EPI_ISL_2117225, EPI_ISL_2117226, EPI_ISL_2117230, EPI_ISL_2117235, EPI_ISL_2117236, EPI_ISL_2117238, EPI_ISL_2117239, EPI_ISL_2117242, EPI_ISL_2117244, EPI_ISL_2117245, EPI_ISL_2117246, EPI_ISL_2117247, EPI_ISL_2117248, EPI_ISL_2117250, EPI_ISL_2117251, EPI_ISL_2117252,
EPI_ISL_2117253, EPI_ISL_2117256, EPI_ISL_2117259, EPI_ISL_2117260, EPI_ISL_2117262, EPI_ISL_2117266, EPI_ISL_2117268, EPI_ISL_2117271, EPI_ISL_2117274, EPI_ISL_2117275, EPI_ISL_2117276, EPI_ISL_2117279, EPI_ISL_2117285, EPI_ISL_2117287, EPI_ISL_2117288, EPI_ISL_2117292, EPI_ISL_2117295, EPI_ISL_2117296, EPI_ISL_2117926, EPI_ISL_2117937, EPI_ISL_2117940,
EPI_ISL_2117944, EPI_ISL_2117950, EPI_ISL_2117953, EPI_ISL_2117956, EPI_ISL_2117959, EPI_ISL_2117965, EPI_ISL_2117968, EPI_ISL_2117973, EPI_ISL_2117982, EPI_ISL_2117986, EPI_ISL_2117989, EPI_ISL_2117992, EPI_ISL_2117999, EPI_ISL_2118003, EPI_ISL_2118006, EPI_ISL_2118009, EPI_ISL_2118012, EPI_ISL_2118015, EPI_ISL_2118018, EPI_ISL_2118024, EPI_ISL_2118028, EPI_ISL_2118031,
EPI_ISL_2118039, EPI_ISL_2118043, EPI_ISL_2118047, EPI_ISL_2118049, EPI_ISL_2118051, EPI_ISL_2118057, EPI_ISL_2118063, EPI_ISL_2118069, EPI_ISL_2118071, EPI_ISL_2118075, EPI_ISL_2118079, EPI_ISL_2118082, EPI_ISL_2118085, EPI_ISL_2118092, EPI_ISL_2118098, EPI_ISL_2118101, EPI_ISL_2118107, EPI_ISL_2118110, EPI_ISL_2118112, EPI_ISL_2118115, EPI_ISL_2118121,
EPI_ISL_2118128, EPI_ISL_2118131, EPI_ISL_2118136, EPI_ISL_2118138, EPI_ISL_2118141, EPI_ISL_2118144, EPI_ISL_2118147, EPI_ISL_2118160, EPI_ISL_2118163, EPI_ISL_2118166, EPI_ISL_2118172, EPI_ISL_2118175, EPI_ISL_2118178, EPI_ISL_2118187, EPI_ISL_2118190, EPI_ISL_2118198, EPI_ISL_2118202, EPI_ISL_2118203, EPI_ISL_2118213, EPI_ISL_2118218, EPI_ISL_2118221, EPI_ISL_2118224,
EPI_ISL_2118227, EPI_ISL_2118239, EPI_ISL_2118242, EPI_ISL_2118245, EPI_ISL_2118248, EPI_ISL_2118253, EPI_ISL_2118257, EPI_ISL_2118260, EPI_ISL_2118266, EPI_ISL_2118273, EPI_ISL_2118275, EPI_ISL_2118276, EPI_ISL_2118277, EPI_ISL_2118280, EPI_ISL_2118281, EPI_ISL_2118282, EPI_ISL_2118284, EPI_ISL_2118285, EPI_ISL_2118287, EPI_ISL_2118289, EPI_ISL_2118290, EPI_ISL_2118292,
EPI_ISL_2118293, EPI_ISL_2118295, EPI_ISL_2118296, EPI_ISL_2118297, EPI_ISL_2118298, EPI_ISL_2118299, EPI_ISL_2118301, EPI_ISL_2118302, EPI_ISL_2118303, EPI_ISL_2118304, EPI_ISL_2118305, EPI_ISL_2118307, EPI_ISL_2118308, EPI_ISL_2118309, EPI_ISL_2118313, EPI_ISL_2118315, EPI_ISL_2118323, EPI_ISL_2118328, EPI_ISL_2118332, EPI_ISL_2118335, EPI_ISL_2118340, EPI_ISL_2118344,
EPI_ISL_2118347, EPI_ISL_2118349, EPI_ISL_2118352, EPI_ISL_2118355, EPI_ISL_2118358, EPI_ISL_2118361, EPI_ISL_2118364, EPI_ISL_2118371, EPI_ISL_2118373, EPI_ISL_2118376, EPI_ISL_2118383, EPI_ISL_2118389, EPI_ISL_2118392, EPI_ISL_2118395, EPI_ISL_2118400, EPI_ISL_2118405, EPI_ISL_2118409, EPI_ISL_2118417, EPI_ISL_2118423, EPI_ISL_2118442, EPI_ISL_2118449,
EPI_ISL_2118454, EPI_ISL_2118458, EPI_ISL_2118461, EPI_ISL_2118464, EPI_ISL_2118477, EPI_ISL_2118480, EPI_ISL_2118493, EPI_ISL_2118495, EPI_ISL_2118498, EPI_ISL_2118524, EPI_ISL_2118533, EPI_ISL_2118536, EPI_ISL_2118539, EPI_ISL_2118542, EPI_ISL_2118545, EPI_ISL_2118547, EPI_ISL_2118550, EPI_ISL_2118559, EPI_ISL_2118563, EPI_ISL_2118565, EPI_ISL_2118568, EPI_ISL_2118571,
EPI_ISL_2118580, EPI_ISL_2118584, EPI_ISL_2118594, EPI_ISL_2118597, EPI_ISL_2118600, EPI_ISL_2118608, EPI_ISL_2118611, EPI_ISL_2118614, EPI_ISL_2118629, EPI_ISL_2118637, EPI_ISL_2118641, EPI_ISL_2118646, EPI_ISL_2118653, EPI_ISL_2118656, EPI_ISL_2118663, EPI_ISL_2118666, EPI_ISL_2118677, EPI_ISL_2118683, EPI_ISL_2118688, EPI_ISL_2118691, EPI_ISL_2118696,
EPI_ISL_2118697, EPI_ISL_2118699, EPI_ISL_2118710, EPI_ISL_2118714, EPI_ISL_2118717, EPI_ISL_2118734, EPI_ISL_2118737, EPI_ISL_2118743, EPI_ISL_2118746, EPI_ISL_2118752, EPI_ISL_2118755, EPI_ISL_2118757, EPI_ISL_2118761, EPI_ISL_2118764, EPI_ISL_2118768, EPI_ISL_2118771, EPI_ISL_2118774, EPI_ISL_2118784, EPI_ISL_2118792, EPI_ISL_2118795, EPI_ISL_2118799, EPI_ISL_2118802,
EPI_ISL_2118815, EPI_ISL_2118818, EPI_ISL_2118821, EPI_ISL_2118824, EPI_ISL_2118827, EPI_ISL_2124378, EPI_ISL_2124379, EPI_ISL_2124380, EPI_ISL_2124382, EPI_ISL_2124384, EPI_ISL_2124385, EPI_ISL_2124389, EPI_ISL_2124391, EPI_ISL_2124392, EPI_ISL_2124397, EPI_ISL_2124401, EPI_ISL_2124402, EPI_ISL_2124404, EPI_ISL_2124407, EPI_ISL_2124408, EPI_ISL_2124409, EPI_ISL_2124410,
EPI_ISL_2124411, EPI_ISL_2124413, EPI_ISL_2124414, EPI_ISL_2124415, EPI_ISL_2124416, EPI_ISL_2124417, EPI_ISL_2124419, EPI_ISL_2124420, EPI_ISL_2124423, EPI_ISL_2124424, EPI_ISL_2124425, EPI_ISL_2124426, EPI_ISL_2124427, EPI_ISL_2124428, EPI_ISL_2124429, EPI_ISL_2124431, EPI_ISL_2124432, EPI_ISL_2124433, EPI_ISL_2124436, EPI_ISL_2124437, EPI_ISL_2124439, EPI_ISL_2124440,
EPI_ISL_2124441, EPI_ISL_2124442, EPI_ISL_2124443, EPI_ISL_2124447, EPI_ISL_2124449, EPI_ISL_2124450, EPI_ISL_2124451, EPI_ISL_2124452, EPI_ISL_2124453, EPI_ISL_2124454, EPI_ISL_2124455, EPI_ISL_2124456, EPI_ISL_2124458, EPI_ISL_2124459, EPI_ISL_2124460, EPI_ISL_2124461, EPI_ISL_2124463, EPI_ISL_2124466, EPI_ISL_2124468, EPI_ISL_2124469, EPI_ISL_2124471, EPI_ISL_2124472,
EPI_ISL_2124474, EPI_ISL_2124475, EPI_ISL_2124477, EPI_ISL_2124478, EPI_ISL_2124479, EPI_ISL_2124480, EPI_ISL_2124481, EPI_ISL_2124482, EPI_ISL_2124485, EPI_ISL_2124486, EPI_ISL_2124487, EPI_ISL_2124488, EPI_ISL_2124490, EPI_ISL_2124491, EPI_ISL_2124492, EPI_ISL_2124493, EPI_ISL_2124494, EPI_ISL_2124495, EPI_ISL_2124496, EPI_ISL_2124497, EPI_ISL_2124498, EPI_ISL_2124499,
EPI_ISL_2124500, EPI_ISL_2124501, EPI_ISL_2124502, EPI_ISL_2124503, EPI_ISL_2124504, EPI_ISL_2124505, EPI_ISL_2124507, EPI_ISL_2124508, EPI_ISL_2124510, EPI_ISL_2124511, EPI_ISL_2124512, EPI_ISL_2124513, EPI_ISL_2124514, EPI_ISL_2124515, EPI_ISL_2124518, EPI_ISL_2124520, EPI_ISL_2124521, EPI_ISL_2124522, EPI_ISL_2124523, EPI_ISL_2124525, EPI_ISL_2124527, EPI_ISL_2124528,
EPI_ISL_2124529, EPI_ISL_2124530, EPI_ISL_2124531, EPI_ISL_2124536, EPI_ISL_2124537, EPI_ISL_2124539, EPI_ISL_2124541, EPI_ISL_2124542, EPI_ISL_2124543, EPI_ISL_2124544, EPI_ISL_2124545, EPI_ISL_2124546, EPI_ISL_2124548, EPI_ISL_2124549, EPI_ISL_2124550, EPI_ISL_2124551, EPI_ISL_2124552, EPI_ISL_2124554, EPI_ISL_2124556, EPI_ISL_2124557, EPI_ISL_2124558, EPI_ISL_2124559,
EPI_ISL_2124560, EPI_ISL_2124561, EPI_ISL_2124562, EPI_ISL_2124563, EPI_ISL_2124565, EPI_ISL_2124566, EPI_ISL_2124567, EPI_ISL_2124568, EPI_ISL_2124569, EPI_ISL_2124570, EPI_ISL_2124571, EPI_ISL_2124572, EPI_ISL_2124574, EPI_ISL_2124575, EPI_ISL_2124576, EPI_ISL_2124577, EPI_ISL_2124578, EPI_ISL_2124579, EPI_ISL_2124580, EPI_ISL_2124582, EPI_ISL_2124584, EPI_ISL_2124585,
EPI_ISL_2124586, EPI_ISL_2124587, EPI_ISL_2124588, EPI_ISL_2124590, EPI_ISL_2124591, EPI_ISL_2124592, EPI_ISL_2124593, EPI_ISL_2124594, EPI_ISL_2124595, EPI_ISL_2124597, EPI_ISL_2124598, EPI_ISL_2124599, EPI_ISL_2124600, EPI_ISL_2124602, EPI_ISL_2124603, EPI_ISL_2124605, EPI_ISL_2124606, EPI_ISL_2124607, EPI_ISL_2124608, EPI_ISL_2124609, EPI_ISL_2124611, EPI_ISL_2124612,
EPI_ISL_2124613, EPI_ISL_2124616, EPI_ISL_2124617, EPI_ISL_2124619, EPI_ISL_2124622, EPI_ISL_2124624, EPI_ISL_2124625, EPI_ISL_2124626, EPI_ISL_2124627, EPI_ISL_2124629, EPI_ISL_2124631, EPI_ISL_2124636, EPI_ISL_2124637, EPI_ISL_2124638, EPI_ISL_2124639, EPI_ISL_2124640, EPI_ISL_2124641, EPI_ISL_2124642, EPI_ISL_2124644, EPI_ISL_2124645, EPI_ISL_2124646, EPI_ISL_2124647,
EPI_ISL_2124649, EPI_ISL_2124652, EPI_ISL_2124653, EPI_ISL_2124654, EPI_ISL_2124655, EPI_ISL_2124656, EPI_ISL_2124658, EPI_ISL_2124659, EPI_ISL_2124660, EPI_ISL_2124662, EPI_ISL_2124664, EPI_ISL_2124667, EPI_ISL_2124668, EPI_ISL_2124669, EPI_ISL_2124670, EPI_ISL_2124671, EPI_ISL_2124672, EPI_ISL_2124673, EPI_ISL_2124674, EPI_ISL_2124677, EPI_ISL_2124679, EPI_ISL_2124680,
EPI_ISL_2124681, EPI_ISL_2124683, EPI_ISL_2124686, EPI_ISL_2124687, EPI_ISL_2124688, EPI_ISL_2124689, EPI_ISL_2124691, EPI_ISL_2124692, EPI_ISL_2124694, EPI_ISL_2124696, EPI_ISL_2124697, EPI_ISL_2124698, EPI_ISL_2124701, EPI_ISL_2124702, EPI_ISL_2124703, EPI_ISL_2124704, EPI_ISL_2124705, EPI_ISL_2124707, EPI_ISL_2124708,
EPI_ISL_2124709, EPI_ISL_2124710, EPI_ISL_2124711, EPI_ISL_2124712, EPI_ISL_2124713, EPI_ISL_2124714, EPI_ISL_2124715, EPI_ISL_2124716, EPI_ISL_2124717, EPI_ISL_2124718, EPI_ISL_2124719, EPI_ISL_2124720, EPI_ISL_2124721, EPI_ISL_2124722, EPI_ISL_2124723, EPI_ISL_2124725, EPI_ISL_2124726, EPI_ISL_2124727, EPI_ISL_2124728, EPI_ISL_2124729, EPI_ISL_2124730, EPI_ISL_2124733,

EPI_ISL_2124734, EPI_ISL_2124735, EPI_ISL_2124736, EPI_ISL_2124737, EPI_ISL_2124738, EPI_ISL_2124739, EPI_ISL_2124741, EPI_ISL_2124742, EPI_ISL_2124743, EPI_ISL_2124744, EPI_ISL_2124745, EPI_ISL_2124746, EPI_ISL_2124748, EPI_ISL_2124749, EPI_ISL_2124750, EPI_ISL_2124751, EPI_ISL_2124753, EPI_ISL_2124754, EPI_ISL_2124755, EPI_ISL_2124756, EPI_ISL_2124757, EPI_ISL_2124758, EPI_ISL_2124759, EPI_ISL_2124760, EPI_ISL_2124761, EPI_ISL_2124762, EPI_ISL_2124763, EPI_ISL_2124764, EPI_ISL_2124765, EPI_ISL_2124766, EPI_ISL_2124768, EPI_ISL_2124770, EPI_ISL_2124771, EPI_ISL_2124772, EPI_ISL_2124773, EPI_ISL_2124774, EPI_ISL_2124775, EPI_ISL_2124776, EPI_ISL_2124777, EPI_ISL_2124778, EPI_ISL_2124780, EPI_ISL_2124781, EPI_ISL_2124782, EPI_ISL_2124783, EPI_ISL_2124784, EPI_ISL_2124785, EPI_ISL_2124786, EPI_ISL_2124787, EPI_ISL_2124788, EPI_ISL_2124789, EPI_ISL_2124790, EPI_ISL_2124791, EPI_ISL_2124792, EPI_ISL_2124793, EPI_ISL_2124794, EPI_ISL_2124797, EPI_ISL_2124798, EPI_ISL_2124799, EPI_ISL_2124801, EPI_ISL_2124802, EPI_ISL_2124803, EPI_ISL_2124804, EPI_ISL_2124805, EPI_ISL_2124806, EPI_ISL_2124807, EPI_ISL_2124809, EPI_ISL_2124810, EPI_ISL_2124811, EPI_ISL_2124812, EPI_ISL_2124813, EPI_ISL_2124814, EPI_ISL_2124815, EPI_ISL_2124817, EPI_ISL_2124818, EPI_ISL_2124819, EPI_ISL_2124820, EPI_ISL_2124821, EPI_ISL_2124822, EPI_ISL_2124823, EPI_ISL_2124824, EPI_ISL_2124826, EPI_ISL_2124827, EPI_ISL_2124828, EPI_ISL_2124829, EPI_ISL_2124830, EPI_ISL_2124831, EPI_ISL_2124833, EPI_ISL_2124834, EPI_ISL_2124837, EPI_ISL_2124838, EPI_ISL_2124839, EPI_ISL_2124841, EPI_ISL_2124842, EPI_ISL_2124843, EPI_ISL_2124844, EPI_ISL_2124845, EPI_ISL_2124846, EPI_ISL_2124848, EPI_ISL_2124849, EPI_ISL_2124850, EPI_ISL_2124851, EPI_ISL_2124852, EPI_ISL_2124854, EPI_ISL_2124855, EPI_ISL_2124856, EPI_ISL_2124857, EPI_ISL_2124858, EPI_ISL_2124859, EPI_ISL_2124860, EPI_ISL_2124862, EPI_ISL_2124863, EPI_ISL_2124864, EPI_ISL_2124865, EPI_ISL_2124867, EPI_ISL_2124869, EPI_ISL_2124870, EPI_ISL_2124871, EPI_ISL_2124872, EPI_ISL_2124873, EPI_ISL_2124876, EPI_ISL_2124877, EPI_ISL_2124880, EPI_ISL_2124881, EPI_ISL_2124882, EPI_ISL_2124883, EPI_ISL_2124884, EPI_ISL_2124886, EPI_ISL_2124888, EPI_ISL_2124889, EPI_ISL_2124890, EPI_ISL_2124892, EPI_ISL_2124894, EPI_ISL_2124895, EPI_ISL_2124896, EPI_ISL_2124899, EPI_ISL_2124901, EPI_ISL_2124902, EPI_ISL_2124905, EPI_ISL_2124906, EPI_ISL_2124907, EPI_ISL_2124908, EPI_ISL_2124909, EPI_ISL_2124911, EPI_ISL_2124913, EPI_ISL_2124915, EPI_ISL_2124916, EPI_ISL_2124918, EPI_ISL_2124919, EPI_ISL_2124920, EPI_ISL_2124921, EPI_ISL_2124922, EPI_ISL_2124923, EPI_ISL_2124925, EPI_ISL_2124926, EPI_ISL_2124928, EPI_ISL_2124929, EPI_ISL_2124930, EPI_ISL_2124932, EPI_ISL_2124934, EPI_ISL_2124935, EPI_ISL_2124936, EPI_ISL_2124937, EPI_ISL_2124939, EPI_ISL_2124940, EPI_ISL_2124942, EPI_ISL_2124943, EPI_ISL_2124945, EPI_ISL_2124946, EPI_ISL_2124947, EPI_ISL_2124948, EPI_ISL_2124949, EPI_ISL_2124951, EPI_ISL_2124952, EPI_ISL_2124953, EPI_ISL_2124954, EPI_ISL_2124955, EPI_ISL_2124956, EPI_ISL_2124958, EPI_ISL_2124959, EPI_ISL_2124961, EPI_ISL_2124962, EPI_ISL_2124963, EPI_ISL_2124964, EPI_ISL_2124965, EPI_ISL_2124966, EPI_ISL_2124968, EPI_ISL_2124969, EPI_ISL_2124970, EPI_ISL_2124971, EPI_ISL_2124974, EPI_ISL_2124975, EPI_ISL_2124976, EPI_ISL_2124978, EPI_ISL_2124980, EPI_ISL_2124981, EPI_ISL_2124982, EPI_ISL_2124983, EPI_ISL_2124984, EPI_ISL_2124986, EPI_ISL_2124987, EPI_ISL_2124988, EPI_ISL_2124989, EPI_ISL_2124990, EPI_ISL_2124991, EPI_ISL_2124993, EPI_ISL_2124994, EPI_ISL_2124995, EPI_ISL_2124996, EPI_ISL_2124997, EPI_ISL_2124998, EPI_ISL_2124999, EPI_ISL_2125000, EPI_ISL_2125001, EPI_ISL_2125002, EPI_ISL_2125003, EPI_ISL_2125004, EPI_ISL_2125005, EPI_ISL_2125006, EPI_ISL_2125007, EPI_ISL_2125008, EPI_ISL_2125009, EPI_ISL_2125011, EPI_ISL_2125013, EPI_ISL_2126413, EPI_ISL_2126415, EPI_ISL_2126417, EPI_ISL_2126420, EPI_ISL_2126422, EPI_ISL_2126424, EPI_ISL_2126426, EPI_ISL_2126428, EPI_ISL_2126430, EPI_ISL_2126432, EPI_ISL_2126438, EPI_ISL_2126443, EPI_ISL_2126445, EPI_ISL_2126447, EPI_ISL_2126449, EPI_ISL_2126455, EPI_ISL_2126457, EPI_ISL_2126461, EPI_ISL_2126463, EPI_ISL_2126468, EPI_ISL_2126470, EPI_ISL_2126476, EPI_ISL_2126479, EPI_ISL_2126485, EPI_ISL_2126487, EPI_ISL_2126489, EPI_ISL_2126494, EPI_ISL_2126496, EPI_ISL_2126498, EPI_ISL_2126500, EPI_ISL_2126503, EPI_ISL_2126505, EPI_ISL_2126507, EPI_ISL_2126509, EPI_ISL_2126512, EPI_ISL_2126514, EPI_ISL_2126521, EPI_ISL_2126523, EPI_ISL_2126526, EPI_ISL_2126528, EPI_ISL_2126529, EPI_ISL_2126531, EPI_ISL_2126536, EPI_ISL_2126540, EPI_ISL_2126542, EPI_ISL_2126543, EPI_ISL_2126545, EPI_ISL_2126547, EPI_ISL_2126549, EPI_ISL_2126552, EPI_ISL_2126554, EPI_ISL_2126556, EPI_ISL_2126558, EPI_ISL_2126561, EPI_ISL_2126563, EPI_ISL_2126565, EPI_ISL_2126566, EPI_ISL_2126568, EPI_ISL_2126571, EPI_ISL_2126573, EPI_ISL_2126575, EPI_ISL_2126577, EPI_ISL_2126579, EPI_ISL_2126583, EPI_ISL_2126585, EPI_ISL_2126587, EPI_ISL_2126589, EPI_ISL_2126593, EPI_ISL_2126595, EPI_ISL_2126597, EPI_ISL_2126599, EPI_ISL_2126601, EPI_ISL_2126604, EPI_ISL_2126606, EPI_ISL_2126608, EPI_ISL_2126612, EPI_ISL_2126614, EPI_ISL_2126616, EPI_ISL_2126618, EPI_ISL_2126619, EPI_ISL_2126621, EPI_ISL_2126623, EPI_ISL_2126625, EPI_ISL_2126627, EPI_ISL_2126629, EPI_ISL_2126632, EPI_ISL_2126633, EPI_ISL_2126636, EPI_ISL_2126638, EPI_ISL_2126640, EPI_ISL_2126642, EPI_ISL_2126644, EPI_ISL_2126647, EPI_ISL_2126649, EPI_ISL_2126651, EPI_ISL_2126653, EPI_ISL_2126655, EPI_ISL_2126658, EPI_ISL_2126661, EPI_ISL_2126663, EPI_ISL_2126665, EPI_ISL_2126667, EPI_ISL_2126669, EPI_ISL_2126675, EPI_ISL_2126676, EPI_ISL_2126678, EPI_ISL_2126680, EPI_ISL_2126682, EPI_ISL_2126684, EPI_ISL_2126687, EPI_ISL_2126688, EPI_ISL_2126690, EPI_ISL_2126693, EPI_ISL_2126694, EPI_ISL_2126697, EPI_ISL_2126700, EPI_ISL_2126701, EPI_ISL_2126703, EPI_ISL_2126705, EPI_ISL_2126708, EPI_ISL_2126710, EPI_ISL_2126711, EPI_ISL_2126713, EPI_ISL_2126716, EPI_ISL_2126717, EPI_ISL_2126719, EPI_ISL_2126721, EPI_ISL_2126723, EPI_ISL_2126725, EPI_ISL_2126727, EPI_ISL_2126729, EPI_ISL_2126731, EPI_ISL_2126734, EPI_ISL_2126736, EPI_ISL_2126738, EPI_ISL_2126740, EPI_ISL_2126741, EPI_ISL_2126743, EPI_ISL_2126746, EPI_ISL_2126748, EPI_ISL_2126750, EPI_ISL_2126752, EPI_ISL_2126754, EPI_ISL_2126755, EPI_ISL_2126756, EPI_ISL_2126758, EPI_ISL_2126761, EPI_ISL_2126763, EPI_ISL_2126765, EPI_ISL_2126770, EPI_ISL_2126772, EPI_ISL_2126774, EPI_ISL_2126776, EPI_ISL_2126778, EPI_ISL_2126780, EPI_ISL_2126782, EPI_ISL_2126785, EPI_ISL_2126787, EPI_ISL_2126789, EPI_ISL_2126791, EPI_ISL_2126793, EPI_ISL_2126795, EPI_ISL_2126797, EPI_ISL_2126799, EPI_ISL_2126801, EPI_ISL_2126804, EPI_ISL_2126808, EPI_ISL_2126814, EPI_ISL_2126816, EPI_ISL_2126818, EPI_ISL_2126821, EPI_ISL_2126823, EPI_ISL_2126826, EPI_ISL_2126828, EPI_ISL_2126830, EPI_ISL_2126832, EPI_ISL_2126834, EPI_ISL_2126836, EPI_ISL_2126839, EPI_ISL_2126841, EPI_ISL_2126845, EPI_ISL_2126847, EPI_ISL_2126848, EPI_ISL_2126851, EPI_ISL_2126852, EPI_ISL_2126855, EPI_ISL_2126857, EPI_ISL_2126858, EPI_ISL_2126862, EPI_ISL_2126864, EPI_ISL_2126866, EPI_ISL_2126868, EPI_ISL_2126870, EPI_ISL_2126872, EPI_ISL_2126874, EPI_ISL_2126876, EPI_ISL_2126878, EPI_ISL_2126880, EPI_ISL_2126883, EPI_ISL_2126885, EPI_ISL_2126887, EPI_ISL_2126891, EPI_ISL_2126895, EPI_ISL_2126897, EPI_ISL_2126900, EPI_ISL_2126902, EPI_ISL_2126906, EPI_ISL_2126908, EPI_ISL_2126910, EPI_ISL_2126914, EPI_ISL_2126916, EPI_ISL_2126918, EPI_ISL_2126919, EPI_ISL_2126920, EPI_ISL_2126921, EPI_ISL_2126923, EPI_ISL_2126925, EPI_ISL_2126929, EPI_ISL_2126935, EPI_ISL_2126937, EPI_ISL_2126939, EPI_ISL_2126941, EPI_ISL_2126943, EPI_ISL_2126945, EPI_ISL_2126947, EPI_ISL_2126949, EPI_ISL_2126951, EPI_ISL_2126953, EPI_ISL_2126955, EPI_ISL_2126957, EPI_ISL_2126961, EPI_ISL_2126963, EPI_ISL_2126965, EPI_ISL_2126967, EPI_ISL_2126969, EPI_ISL_2126971, EPI_ISL_2126974, EPI_ISL_2126987, EPI_ISL_2126989, EPI_ISL_2126991, EPI_ISL_2126993, EPI_ISL_2126997, EPI_ISL_2126999, EPI_ISL_2127002, EPI_ISL_2127004, EPI_ISL_2127008, EPI_ISL_2127010, EPI_ISL_2127014, EPI_ISL_2127017, EPI_ISL_2127020, EPI_ISL_2127022, EPI_ISL_2127025, EPI_ISL_2127027, EPI_ISL_2127028, EPI_ISL_2127030, EPI_ISL_2127033, EPI_ISL_2127035, EPI_ISL_2127037, EPI_ISL_2127040, EPI_ISL_2127043, EPI_ISL_2127045, EPI_ISL_2127046, EPI_ISL_2127051, EPI_ISL_2127053, EPI_ISL_2127056, EPI_ISL_2127058, EPI_ISL_2127060, EPI_ISL_2127061, EPI_ISL_2127062, EPI_ISL_2127064, EPI_ISL_2127067, EPI_ISL_2127069, EPI_ISL_2127075, EPI_ISL_2127076, EPI_ISL_2127080, EPI_ISL_2127083, EPI_ISL_2127085, EPI_ISL_2127088, EPI_ISL_2127089, EPI_ISL_2127091, EPI_ISL_2127094, EPI_ISL_2127096, EPI_ISL_2129980, EPI_ISL_2129982, EPI_ISL_2129984, EPI_ISL_2129987, EPI_ISL_2129991, EPI_ISL_2129992, EPI_ISL_2129999, EPI_ISL_2130005, EPI_ISL_2130008, EPI_ISL_2130009, EPI_ISL_2130010, EPI_ISL_2130012, EPI_ISL_2130014, EPI_ISL_2130016, EPI_ISL_2130018, EPI_ISL_2130019, EPI_ISL_2130020, EPI_ISL_2130025, EPI_ISL_2130027, EPI_ISL_2130033, EPI_ISL_2130034, EPI_ISL_2130036, EPI_ISL_2130039, EPI_ISL_2130040, EPI_ISL_2130041, EPI_ISL_2130048, EPI_ISL_2130050, EPI_ISL_2130051, EPI_ISL_2130053, EPI_ISL_2130055, EPI_ISL_2130066, EPI_ISL_2130067, EPI_ISL_2130074, EPI_ISL_2130076, EPI_ISL_2130081, EPI_ISL_2130087, EPI_ISL_2130092, EPI_ISL_2130096, EPI_ISL_2130097, EPI_ISL_2130098, EPI_ISL_2130101, EPI_ISL_2130102, EPI_ISL_2130103, EPI_ISL_2130104, EPI_ISL_2130111, EPI_ISL_2130116, EPI_ISL_2130125, EPI_ISL_2130129, EPI_ISL_2130132, EPI_ISL_2130141, EPI_ISL_2130145, EPI_ISL_2130149, EPI_ISL_2130150, EPI_ISL_2130151, EPI_ISL_2130154, EPI_ISL_2130158, EPI_ISL_2130159, EPI_ISL_2130160, EPI_ISL_2130162, EPI_ISL_2130163, EPI_ISL_2130166, EPI_ISL_2130172, EPI_ISL_2130176, EPI_ISL_2130181, EPI_ISL_2130185, EPI_ISL_2130186, EPI_ISL_2130190, EPI_ISL_2130193, EPI_ISL_2130195, EPI_ISL_2130196, EPI_ISL_2130198, EPI_ISL_2130199, EPI_ISL_2130201, EPI_ISL_2130202, EPI_ISL_2130203, EPI_ISL_2130204, EPI_ISL_2130205, EPI_ISL_2130206, EPI_ISL_2130208, EPI_ISL_2130210, EPI_ISL_2130212, EPI_ISL_2130214, EPI_ISL_2130215, EPI_ISL_2130216, EPI_ISL_2130219, EPI_ISL_2130220, EPI_ISL_2130223, EPI_ISL_2130224, EPI_ISL_2130225, EPI_ISL_2130229, EPI_ISL_2130231, EPI_ISL_2130232, EPI_ISL_2130233, EPI_ISL_2130235, EPI_ISL_2130237, EPI_ISL_2130241, EPI_ISL_2130244, EPI_ISL_2130245, EPI_ISL_2130247, EPI_ISL_2130248, EPI_ISL_2130252, EPI_ISL_2130256, EPI_ISL_2130259, EPI_ISL_2130261, EPI_ISL_2130262, EPI_ISL_2130264, EPI_ISL_2130265, EPI_ISL_2130270, EPI_ISL_2130272, EPI_ISL_2130277, EPI_ISL_2130278, EPI_ISL_2130281, EPI_ISL_2130282, EPI_ISL_2130283, EPI_ISL_2130286, EPI_ISL_2130289, EPI_ISL_2130292, EPI_ISL_2130293, EPI_ISL_2130296, EPI_ISL_2130298, EPI_ISL_2130301, EPI_ISL_2130303, EPI_ISL_2130307, EPI_ISL_2130309, EPI_ISL_2130310, EPI_ISL_2130313, EPI_ISL_2130317, EPI_ISL_2130319, EPI_ISL_2130321, EPI_ISL_2130322, EPI_ISL_2130325, EPI_ISL_2130333, EPI_ISL_2130340, EPI_ISL_2130343, EPI_ISL_2130352, EPI_ISL_2130359, EPI_ISL_2130361, EPI_ISL_2130365, EPI_ISL_2130367, EPI_ISL_2130368, EPI_ISL_2130369, EPI_ISL_2130372, EPI_ISL_2130373, EPI_ISL_2130374, EPI_ISL_2130377, EPI_ISL_2130381, EPI_ISL_2130385, EPI_ISL_2130388, EPI_ISL_2130389, EPI_ISL_2130390, EPI_ISL_2130399, EPI_ISL_2130401, EPI_ISL_2130403, EPI_ISL_2130410, EPI_ISL_2130414, EPI_ISL_2130419, EPI_ISL_2130420, EPI_ISL_2130421, EPI_ISL_2130422, EPI_ISL_2130427, EPI_ISL_2130430, EPI_ISL_2130433, EPI_ISL_2130434, EPI_ISL_2130438, EPI_ISL_2130445, EPI_ISL_2130446, EPI_ISL_2130447, EPI_ISL_2130449, EPI_ISL_2130450, EPI_ISL_2130454, EPI_ISL_2130458, EPI_ISL_2130460, EPI_ISL_2130464, EPI_ISL_2130465, EPI_ISL_2130466, EPI_ISL_2130476, EPI_ISL_2130477, EPI_ISL_2130478, EPI_ISL_2130482, EPI_ISL_2130486, EPI_ISL_2130488, EPI_ISL_2130493, EPI_ISL_2130498, EPI_ISL_2130499, EPI_ISL_2130502, EPI_ISL_2130505, EPI_ISL_2130507, EPI_ISL_2130510, EPI_ISL_2130513, EPI_ISL_2130515, EPI_ISL_2130521, EPI_ISL_2130524, EPI_ISL_2130526, EPI_ISL_2130528, EPI_ISL_2130531, EPI_ISL_2130535, EPI_ISL_2130540, EPI_ISL_2130541, EPI_ISL_2130546, EPI_ISL_2130547, EPI_ISL_2130549, EPI_ISL_2130550, EPI_ISL_2130551, EPI_ISL_2130552, EPI_ISL_2130556, EPI_ISL_2130557, EPI_ISL_2130561, EPI_ISL_2130573, EPI_ISL_2130575, EPI_ISL_2130576, EPI_ISL_2130577, EPI_ISL_2130579, EPI_ISL_2130581, EPI_ISL_2130583, EPI_ISL_2130584, EPI_ISL_2130588, EPI_ISL_2130589

see above SYNLAB MVZ Leverkusen Robert Koch Institute

EPI_ISL_2130825, EPI_ISL_2130832, EPI_ISL_2130839, EPI_ISL_2130849, EPI_ISL_2130852, EPI_ISL_2130855, EPI_ISL_2130857, EPI_ISL_2130858, EPI_ISL_2130862, EPI_ISL_2130863, EPI_ISL_2130887, EPI_ISL_2130927, EPI_ISL_2130934, EPI_ISL_2130938, EPI_ISL_2130943, EPI_ISL_2130948, EPI_ISL_2130949, EPI_ISL_2130953, EPI_ISL_2130958, EPI_ISL_2130961, EPI_ISL_2130964, EPI_ISL_2130975, EPI_ISL_2130998, EPI_ISL_2131000, EPI_ISL_2131005, EPI_ISL_2131017, EPI_ISL_2131022, EPI_ISL_2131032, EPI_ISL_2131033, EPI_ISL_2131036, EPI_ISL_2131037, EPI_ISL_2131048, EPI_ISL_2131051, EPI_ISL_2131090, EPI_ISL_2131095, EPI_ISL_2131111, EPI_ISL_2131112, EPI_ISL_2131115, EPI_ISL_2131117, EPI_ISL_2131129, EPI_ISL_2131133, EPI_ISL_2131134, EPI_ISL_2131140, EPI_ISL_2131143

see above SYNLAB MVZ Neustadt a. d. WeinstraÃŸe Robert Koch Institute

EPI_ISL_2128652, EPI_ISL_2128686, EPI_ISL_2128729, EPI_ISL_2128759, EPI_ISL_2128792, EPI_ISL_2128882, EPI_ISL_2128898, EPI_ISL_2128930, EPI_ISL_2267404, EPI_ISL_2267475, EPI_ISL_2267511, EPI_ISL_2267525

see above SYNLAB MVZ Neustadt a. d. Weinstraße Robert Koch Institute

EPI_ISL_2116366, EPI_ISL_2116389, EPI_ISL_2116392, EPI_ISL_2116399, EPI_ISL_2116415, EPI_ISL_2116418, EPI_ISL_2116429, EPI_ISL_2116432, EPI_ISL_2116457, EPI_ISL_2116544, EPI_ISL_2116559, EPI_ISL_2116595, EPI_ISL_2116611, EPI_ISL_2116613, EPI_ISL_2116643, EPI_ISL_2116647, EPI_ISL_2116655, EPI_ISL_2116656, EPI_ISL_2116660, EPI_ISL_2116662, EPI_ISL_2116678, EPI_ISL_2116682, EPI_ISL_2116690, EPI_ISL_2116702, EPI_ISL_2116711, EPI_ISL_2116715, EPI_ISL_2116729, EPI_ISL_2116736, EPI_ISL_2116744, EPI_ISL_2116746, EPI_ISL_2116755, EPI_ISL_2116768, EPI_ISL_2116769, EPI_ISL_2116774, EPI_ISL_2116776, EPI_ISL_2116778, EPI_ISL_2116783, EPI_ISL_2116784, EPI_ISL_2116793, EPI_ISL_2116794, EPI_ISL_2116798, EPI_ISL_2116800, EPI_ISL_2116803, EPI_ISL_2116804, EPI_ISL_2116805, EPI_ISL_2116807, EPI_ISL_2116810, EPI_ISL_2116811, EPI_ISL_2116815, EPI_ISL_2116818, EPI_ISL_2116822, EPI_ISL_2116844, EPI_ISL_2116851, EPI_ISL_2116853, EPI_ISL_2116856, EPI_ISL_2116857, EPI_ISL_2116862, EPI_ISL_2116874, EPI_ISL_2116882, EPI_ISL_2116883, EPI_ISL_2116890, EPI_ISL_2116891, EPI_ISL_2116892, EPI_ISL_2116898, EPI_ISL_2116899, EPI_ISL_2116911, EPI_ISL_2116912, EPI_ISL_2116916, EPI_ISL_2116918, EPI_ISL_2116919, EPI_ISL_2116923, EPI_ISL_2116925, EPI_ISL_2116928, EPI_ISL_2116935, EPI_ISL_2116939, EPI_ISL_2116940, EPI_ISL_2116942, EPI_ISL_2116943, EPI_ISL_2116945, EPI_ISL_2116962, EPI_ISL_2116963, EPI_ISL_2116969, EPI_ISL_2116972, EPI_ISL_2116983, EPI_ISL_2116986, EPI_ISL_2116988, EPI_ISL_2116997, EPI_ISL_2116999, EPI_ISL_2117001, EPI_ISL_2117006, EPI_ISL_2117007, EPI_ISL_2117008, EPI_ISL_2117020, EPI_ISL_2117032, EPI_ISL_2117042, EPI_ISL_2117046, EPI_ISL_2117064, EPI_ISL_2117068, EPI_ISL_2117074, EPI_ISL_2117075, EPI_ISL_2117077, EPI_ISL_2117085, EPI_ISL_2117096, EPI_ISL_2117099, EPI_ISL_2117110, EPI_ISL_2117111, EPI_ISL_2117112, EPI_ISL_2117113, EPI_ISL_2117115, EPI_ISL_2117118, EPI_ISL_2117121, EPI_ISL_2117122, EPI_ISL_2117130, EPI_ISL_2117132, EPI_ISL_2117133, EPI_ISL_2117137, EPI_ISL_2117140, EPI_ISL_2117141, EPI_ISL_2117144, EPI_ISL_2117148, EPI_ISL_2117162, EPI_ISL_2117163, EPI_ISL_2117165, EPI_ISL_2117167, EPI_ISL_2117179, EPI_ISL_2117187, EPI_ISL_2117197, EPI_ISL_2117202, EPI_ISL_2117204, EPI_ISL_2117209, EPI_ISL_2117216, EPI_ISL_2117220, EPI_ISL_2117221, EPI_ISL_2117233, EPI_ISL_2117234, EPI_ISL_2117240, EPI_ISL_2117241, EPI_ISL_2117243, EPI_ISL_2117254, EPI_ISL_2117258, EPI_ISL_2117263, EPI_ISL_2117267, EPI_ISL_2117270, EPI_ISL_2117272, EPI_ISL_2117273, EPI_ISL_2117277, EPI_ISL_2117293, EPI_ISL_2117929, EPI_ISL_2117932, EPI_ISL_2117947, EPI_ISL_2117962, EPI_ISL_2117971, EPI_ISL_2117976, EPI_ISL_2117979, EPI_ISL_2117995, EPI_ISL_2118034, EPI_ISL_2118037, EPI_ISL_2118055, EPI_ISL_2118066, EPI_ISL_2118077, EPI_ISL_2118095, EPI_ISL_2118104, EPI_ISL_2118117, EPI_ISL_2118124, EPI_ISL_2118151, EPI_ISL_2118154, EPI_ISL_2118157, EPI_ISL_2118165, EPI_ISL_2118169, EPI_ISL_2118181, EPI_ISL_2118194, EPI_ISL_2118196, EPI_ISL_2118204, EPI_ISL_2118207, EPI_ISL_2118209, EPI_ISL_2118216, EPI_ISL_2118230, EPI_ISL_2118236, EPI_ISL_2118251, EPI_ISL_2118263, EPI_ISL_2118269, EPI_ISL_2118278, EPI_ISL_2118279, EPI_ISL_2118286, EPI_ISL_2118288, EPI_ISL_2118291, EPI_ISL_2118294, EPI_ISL_2118300, EPI_ISL_2118306, EPI_ISL_2118319, EPI_ISL_2118326, EPI_ISL_2118337, EPI_ISL_2118367, EPI_ISL_2118379, EPI_ISL_2118386, EPI_ISL_2118399, EPI_ISL_2118403, EPI_ISL_2118412, EPI_ISL_2118415, EPI_ISL_2118418, EPI_ISL_2118420, EPI_ISL_2118426, EPI_ISL_2118429, EPI_ISL_2118432, EPI_ISL_2118439, EPI_ISL_2118445, EPI_ISL_2118451, EPI_ISL_2118455, EPI_ISL_2118467, EPI_ISL_2118470, EPI_ISL_2118474, EPI_ISL_2118483, EPI_ISL_2118486, EPI_ISL_2118490, EPI_ISL_2118505, EPI_ISL_2118508, EPI_ISL_2118511, EPI_ISL_2118514, EPI_ISL_2118517, EPI_ISL_2118520, EPI_ISL_2118527, EPI_ISL_2118530, EPI_ISL_2118548, EPI_ISL_2118553, EPI_ISL_2118555, EPI_ISL_2118574, EPI_ISL_2118591, EPI_ISL_2118602, EPI_ISL_2118606, EPI_ISL_2118617, EPI_ISL_2118620, EPI_ISL_2118623, EPI_ISL_2118626, EPI_ISL_2118632, EPI_ISL_2118635, EPI_ISL_2118644, EPI_ISL_2118650, EPI_ISL_2118660, EPI_ISL_2118669, EPI_ISL_2118672, EPI_ISL_2118674, EPI_ISL_2118680, EPI_ISL_2118686, EPI_ISL_2118720, EPI_ISL_2118723, EPI_ISL_2118726, EPI_ISL_2118729, EPI_ISL_2118741, EPI_ISL_2118749, EPI_ISL_2118777, EPI_ISL_2118790, EPI_ISL_2118805, EPI_ISL_2118808, EPI_ISL_2118812, EPI_ISL_2124381, EPI_ISL_2124383, EPI_ISL_2124386, EPI_ISL_2124387, EPI_ISL_2124393, EPI_ISL_2124394, EPI_ISL_2124395, EPI_ISL_2124398, EPI_ISL_2124399, EPI_ISL_2124400, EPI_ISL_2124403, EPI_ISL_2124405, EPI_ISL_2124406, EPI_ISL_2124412, EPI_ISL_2124421, EPI_ISL_2124422, EPI_ISL_2124430, EPI_ISL_2124434, EPI_ISL_2124438, EPI_ISL_2124444, EPI_ISL_2124445, EPI_ISL_2124446, EPI_ISL_2124448, EPI_ISL_2124457, EPI_ISL_2124462, EPI_ISL_2124464, EPI_ISL_2124465, EPI_ISL_2124467, EPI_ISL_2124470, EPI_ISL_2124473, EPI_ISL_2124476, EPI_ISL_2124483, EPI_ISL_2124484, EPI_ISL_2124489, EPI_ISL_2124509, EPI_ISL_2124516, EPI_ISL_2124517, EPI_ISL_2124519, EPI_ISL_2124524, EPI_ISL_2124526, EPI_ISL_2124532, EPI_ISL_2124533, EPI_ISL_2124534, EPI_ISL_2124535, EPI_ISL_2124538, EPI_ISL_2124540, EPI_ISL_2124547, EPI_ISL_2124553, EPI_ISL_2124555, EPI_ISL_2124564, EPI_ISL_2124581, EPI_ISL_2124583, EPI_ISL_2124589, EPI_ISL_2124596, EPI_ISL_2124601, EPI_ISL_2124604, EPI_ISL_2124610, EPI_ISL_2124614, EPI_ISL_2124615, EPI_ISL_2124618, EPI_ISL_2124620, EPI_ISL_2124621, EPI_ISL_2124623, EPI_ISL_2124628, EPI_ISL_2124630, EPI_ISL_2124632, EPI_ISL_2124633, EPI_ISL_2124635, EPI_ISL_2124643, EPI_ISL_2124648, EPI_ISL_2124650, EPI_ISL_2124651, EPI_ISL_2124657, EPI_ISL_2124661, EPI_ISL_2124663, EPI_ISL_2124665, EPI_ISL_2124675, EPI_ISL_2124676, EPI_ISL_2124678, EPI_ISL_2124685, EPI_ISL_2124690, EPI_ISL_2124695, EPI_ISL_2124699, EPI_ISL_2124724, EPI_ISL_2124731, EPI_ISL_2124732, EPI_ISL_2124740, EPI_ISL_2124747, EPI_ISL_2124752, EPI_ISL_2124769, EPI_ISL_2124779, EPI_ISL_2124796, EPI_ISL_2124800, EPI_ISL_2124816, EPI_ISL_2124825, EPI_ISL_2124835, EPI_ISL_2124836, EPI_ISL_2124840, EPI_ISL_2124847, EPI_ISL_2124853, EPI_ISL_2124866, EPI_ISL_2124868, EPI_ISL_2124874, EPI_ISL_2124875, EPI_ISL_2124878, EPI_ISL_2124879, EPI_ISL_2124885, EPI_ISL_2124887, EPI_ISL_2124891, EPI_ISL_2124893, EPI_ISL_2124897, EPI_ISL_2124898, EPI_ISL_2124900, EPI_ISL_2124903, EPI_ISL_2124904, EPI_ISL_2124910, EPI_ISL_2124912, EPI_ISL_2124914, EPI_ISL_2124917, EPI_ISL_2124924, EPI_ISL_2124927, EPI_ISL_2124931, EPI_ISL_2124933, EPI_ISL_2124938, EPI_ISL_2124941, EPI_ISL_2124944, EPI_ISL_2124950, EPI_ISL_2124957, EPI_ISL_2124960, EPI_ISL_2124967, EPI_ISL_2124972, EPI_ISL_2124973, EPI_ISL_2124977, EPI_ISL_2124979, EPI_ISL_2124985, EPI_ISL_2124992, EPI_ISL_2125010, EPI_ISL_2125012, EPI_ISL_2125014, EPI_ISL_2126434, EPI_ISL_2126441, EPI_ISL_2126451, EPI_ISL_2126459, EPI_ISL_2126475, EPI_ISL_2126481, EPI_ISL_2126516, EPI_ISL_2126518, EPI_ISL_2126533, EPI_ISL_2126538, EPI_ISL_2126671, EPI_ISL_2126674, EPI_ISL_2126695, EPI_ISL_2126810, EPI_ISL_2126812, EPI_ISL_2126843, EPI_ISL_2126860, EPI_ISL_2126889, EPI_ISL_2126894, EPI_ISL_2126912, EPI_ISL_2126932, EPI_ISL_2126959, EPI_ISL_2126978, EPI_ISL_2126980, EPI_ISL_2126985, EPI_ISL_2126995, EPI_ISL_2127071, EPI_ISL_2127078, EPI_ISL_2129985, EPI_ISL_2130023, EPI_ISL_2130054, EPI_ISL_2130064, EPI_ISL_2130147, EPI_ISL_2130179, EPI_ISL_2130222, EPI_ISL_2130246, EPI_ISL_2130250, EPI_ISL_2130295, EPI_ISL_2130380, EPI_ISL_2130471, EPI_ISL_2130473, EPI_ISL_2130489, EPI_ISL_2130543, EPI_ISL_2263906, EPI_ISL_2263922, EPI_ISL_2263967, EPI_ISL_2264027

see above SYNLAB MVZ Trier Robert Koch Institute

EPI_ISL_2116371, EPI_ISL_2116772, EPI_ISL_2116876, EPI_ISL_2117084, EPI_ISL_2124390, EPI_ISL_2124506, EPI_ISL_2124634, EPI_ISL_2124666, EPI_ISL_2124682, EPI_ISL_2124767, EPI_ISL_2126453, EPI_ISL_2126465, EPI_ISL_2126472, EPI_ISL_2126506, EPI_ISL_2126767, EPI_ISL_2127006, EPI_ISL_2127039, EPI_ISL_2130280, EPI_ISL_2130437, EPI_ISL_2130440, EPI_ISL_2130451, EPI_ISL_2130455, EPI_ISL_2130463, EPI_ISL_2130472, EPI_ISL_2130485, EPI_ISL_2130519, EPI_ISL_2130542

see above SYNLAB MVZ Weiden Robert Koch Institute

EPI_ISL_2260096, EPI_ISL_2260203, EPI_ISL_2260234, EPI_ISL_2265537, EPI_ISL_2265541, EPI_ISL_2265561, EPI_ISL_2265585, EPI_ISL_2265595, EPI_ISL_2265616, EPI_ISL_2265627, EPI_ISL_2265634, EPI_ISL_2265653, EPI_ISL_2265655, EPI_ISL_2265663, EPI_ISL_2265670, EPI_ISL_2265680, EPI_ISL_2265685, EPI_ISL_2265694, EPI_ISL_2265726, EPI_ISL_2265759, EPI_ISL_2265772, EPI_ISL_2265793, EPI_ISL_2265810, EPI_ISL_2265820, EPI_ISL_2265830, EPI_ISL_2265844, EPI_ISL_2265847, EPI_ISL_2265860, EPI_ISL_2313489, EPI_ISL_2313627, EPI_ISL_2313659, EPI_ISL_2313829, EPI_ISL_2314085, EPI_ISL_2314093, EPI_ISL_2314311, EPI_ISL_2388347, EPI_ISL_2388557, EPI_ISL_2388743

see above Sonic - Bioscientia - MVZ Labor Saar GmbH Robert Koch Institute

EPI_ISL_2260184, EPI_ISL_2260221, EPI_ISL_2260245, EPI_ISL_2260260, EPI_ISL_2260267, EPI_ISL_2313377, EPI_ISL_2313417, EPI_ISL_2313445, EPI_ISL_2313540, EPI_ISL_2313782

see above Sonic - Dr. Staber & Kollegen GmbH München Robert Koch Institute

EPI_ISL_2260604, EPI_ISL_2260605, EPI_ISL_2260607, EPI_ISL_2260618, EPI_ISL_2260619, EPI_ISL_2260621, EPI_ISL_2260628, EPI_ISL_2260635, EPI_ISL_2260637, EPI_ISL_2260649, EPI_ISL_2260650, EPI_ISL_2260653, EPI_ISL_2260664, EPI_ISL_2260665, EPI_ISL_2260667, EPI_ISL_2260673, EPI_ISL_2260674, EPI_ISL_2260681, EPI_ISL_2260683, EPI_ISL_2260692, EPI_ISL_2260694, EPI_ISL_2260700, EPI_ISL_2260707, EPI_ISL_2260717, EPI_ISL_2260718, EPI_ISL_2260730, EPI_ISL_2260733, EPI_ISL_2260740, EPI_ISL_2260745, EPI_ISL_2260750, EPI_ISL_2260752, EPI_ISL_2260756, EPI_ISL_2260764, EPI_ISL_2260765, EPI_ISL_2260781, EPI_ISL_2260783, EPI_ISL_2260790, EPI_ISL_2260795, EPI_ISL_2260796, EPI_ISL_2260799, EPI_ISL_2260802, EPI_ISL_2260803, EPI_ISL_2260806, EPI_ISL_2260810, EPI_ISL_2260814, EPI_ISL_2260815, EPI_ISL_2260822, EPI_ISL_2260823, EPI_ISL_2260831, EPI_ISL_2260833, EPI_ISL_2260839, EPI_ISL_2260847, EPI_ISL_2260863, EPI_ISL_2260871, EPI_ISL_2260876, EPI_ISL_2260878, EPI_ISL_2260882, EPI_ISL_2260887, EPI_ISL_2260893, EPI_ISL_2260911, EPI_ISL_2260913, EPI_ISL_2260916

see above Sonic - Labor Dr. von Froreich GmbH Robert Koch Institute

EPI_ISL_2260638, EPI_ISL_2260656, EPI_ISL_2260663, EPI_ISL_2260666, EPI_ISL_2260709, EPI_ISL_2260716, EPI_ISL_2260724, EPI_ISL_2260726, EPI_ISL_2260738, EPI_ISL_2260743, EPI_ISL_2260770, EPI_ISL_2260784, EPI_ISL_2260835, EPI_ISL_2260865

see above Sonic - Labor Lademannbogen MVZ GmbH Robert Koch Institute

EPI_ISL_2260102, EPI_ISL_2260108, EPI_ISL_2260122, EPI_ISL_2260137, EPI_ISL_2260154, EPI_ISL_2260173, EPI_ISL_2260177, EPI_ISL_2260183, EPI_ISL_2260268, EPI_ISL_2260285, EPI_ISL_2260321, EPI_ISL_2260347, EPI_ISL_2260372, EPI_ISL_2260382, EPI_ISL_2260387, EPI_ISL_2313405, EPI_ISL_2313413, EPI_ISL_2313441, EPI_ISL_2313450, EPI_ISL_2313454, EPI_ISL_2313622, EPI_ISL_2313676, EPI_ISL_2313735, EPI_ISL_2313763, EPI_ISL_2313768

see above Sonic - Labor Staber Bayreuth Robert Koch Institute

EPI_ISL_2260094, EPI_ISL_2260126, EPI_ISL_2260128, EPI_ISL_2260175, EPI_ISL_2260176, EPI_ISL_2260265, EPI_ISL_2260287, EPI_ISL_2260303, EPI_ISL_2260333, EPI_ISL_2260355, EPI_ISL_2260406, EPI_ISL_2260420, EPI_ISL_2313406, EPI_ISL_2313472, EPI_ISL_2313536, EPI_ISL_2313583, EPI_ISL_2313625, EPI_ISL_2313770

see above Sonic - Labor Staber Dresden (Klipphausen) Robert Koch Institute

EPI_ISL_2260232, EPI_ISL_2260395, EPI_ISL_2260407, EPI_ISL_2260411, EPI_ISL_2313381, EPI_ISL_2313392, EPI_ISL_2313394, EPI_ISL_2313533, EPI_ISL_2313666

see above Sonic - Labor Staber Heilbronn Robert Koch Institute

EPI_ISL_2260811, EPI_ISL_2260908 Sonic - Labor Staber MVZ Kassel Robert Koch Institute

EPI_ISL_2260095, EPI_ISL_2260169, EPI_ISL_2260233, EPI_ISL_2260309, EPI_ISL_2260312, EPI_ISL_2260324, EPI_ISL_2260327, EPI_ISL_2260339, EPI_ISL_2260348, EPI_ISL_2260416, EPI_ISL_2313379, EPI_ISL_2313449, EPI_ISL_2313740, EPI_ISL_2313759, EPI_ISL_2313786

see above Sonic - Labor Staber Nürnberg Robert Koch Institute

EPI_ISL_2125201, EPI_ISL_2125217, EPI_ISL_2125234, EPI_ISL_2125264, EPI_ISL_2125305, EPI_ISL_2125384, EPI_ISL_2125407, EPI_ISL_2260600, EPI_ISL_2260613, EPI_ISL_2260627, EPI_ISL_2260629, EPI_ISL_2260659, EPI_ISL_2260662, EPI_ISL_2260668, EPI_ISL_2260675, EPI_ISL_2260676, EPI_ISL_2260689, EPI_ISL_2260702, EPI_ISL_2260706, EPI_ISL_2260712, EPI_ISL_2260735, EPI_ISL_2260742, EPI_ISL_2260744, EPI_ISL_2260774, EPI_ISL_2260776, EPI_ISL_2260779, EPI_ISL_2260807, EPI_ISL_2260809, EPI_ISL_2260818, EPI_ISL_2260827, EPI_ISL_2260829, EPI_ISL_2260845, EPI_ISL_2260855, EPI_ISL_2260870, EPI_ISL_2260874, EPI_ISL_2260879, EPI_ISL_2260881, EPI_ISL_2260883, EPI_ISL_2260885, EPI_ISL_2260888, EPI_ISL_2260901, EPI_ISL_2260904

see above Sonic - MVZ Medizinisches Labor Bremen GmbH Robert Koch Institute

EPI_ISL_2260660 Sonic - MVZ Medizinisches Labor Nord GmbH Kiel Robert Koch Institute

EPI_ISL_2125156, EPI_ISL_2125160, EPI_ISL_2125227, EPI_ISL_2125232, EPI_ISL_2125247, EPI_ISL_2125346, EPI_ISL_2125349, EPI_ISL_2125391, EPI_ISL_2125410, EPI_ISL_2125442, EPI_ISL_2260603, EPI_ISL_2260798, EPI_ISL_2314285

see above Sonic - MVZ Medizinisches Labor Oldenburg Robert Koch Institute

EPI_ISL_2123066 St. Elisabeth-Gruppe GmbH - Katholische Kliniken Rhein-Ruhr Robert Koch Institute

EPI_ISL_2122802, EPI_ISL_2122818, EPI_ISL_2122831, EPI_ISL_2122870, EPI_ISL_2122889, EPI_ISL_2122901, EPI_ISL_2122907, EPI_ISL_2122929, EPI_ISL_2122939, EPI_ISL_2122947, EPI_ISL_2122951, EPI_ISL_2122954, EPI_ISL_2122956

see above Stadtisches Klinikum Dresden Institut fur Klinische Chemie und Laboratoriumsmedizin Robert Koch Institute

EPI_ISL_2116533, EPI_ISL_2116637, EPI_ISL_2116640, EPI_ISL_2116758, EPI_ISL_2116802, EPI_ISL_2116814, EPI_ISL_2116877, EPI_ISL_2117013, EPI_ISL_2117038, EPI_ISL_2117060, EPI_ISL_2117071, EPI_ISL_2117191, EPI_ISL_2117203, EPI_ISL_2118021, EPI_ISL_2118044, EPI_ISL_2118184, EPI_ISL_2118283, EPI_ISL_2118501, EPI_ISL_2118577, EPI_ISL_2118703, EPI_ISL_2124388, EPI_ISL_2124435,

| | | | |
|---|---|---|---|
| EPI_ISL_2124795, EPI_ISL_2124808, EPI_ISL_2124832, EPI_ISL_2126436, EPI_ISL_2126483, EPI_ISL_2126492, EPI_ISL_2126806, EPI_ISL_2126927, EPI_ISL_2126934, EPI_ISL_2127098, EPI_ISL_2129989, EPI_ISL_2130003, EPI_ISL_2130171, EPI_ISL_2130364, EPI_ISL_2130511 | | | |
| see above | Synlab MVZ Augsburg | Robert Koch Institute | |
| EPI_ISL_2262286, EPI_ISL_2262287, EPI_ISL_2262290, EPI_ISL_2262293, EPI_ISL_2262296, EPI_ISL_2262297 | Thüringer Landesamtes für Verbraucherschutz | Robert Koch Institute | |
| EPI_ISL_2450419, EPI_ISL_2450420, EPI_ISL_2450421, EPI_ISL_2450422, EPI_ISL_2450425, EPI_ISL_2450426, EPI_ISL_2450427, EPI_ISL_2450428, EPI_ISL_2450429, EPI_ISL_2450431, EPI_ISL_2450432, EPI_ISL_2450433, EPI_ISL_2450434, EPI_ISL_2450435, EPI_ISL_2450436, EPI_ISL_2450437, EPI_ISL_2450438, EPI_ISL_2450439, EPI_ISL_2450442, EPI_ISL_2450443, EPI_ISL_2450444, EPI_ISL_2450445, EPI_ISL_2450446 | | | |
| see above | US Air Force School of Aerospace Medicine | US Air Force School of Aerospace Medicine | Amanda Javorina; Anthony Fries; Carol Garrett; Clarise Starr; Cole Anderson; Elizabeth Macias; Fritz Castillo; Jennifer Meyer; Sarah Purves; William Gruner |
| EPI_ISL_2122805, EPI_ISL_2122843, EPI_ISL_2122858, EPI_ISL_2122873, EPI_ISL_2122883, EPI_ISL_2122886, EPI_ISL_2122921, EPI_ISL_2122932, EPI_ISL_2122938 | | | |
| see above | Uniklinikum Carl Gustav Carus an der TU Dresden; Institut fÃ¼r Virologie | Robert Koch Institute | |
| EPI_ISL_2123862, EPI_ISL_2262299, EPI_ISL_2262300, EPI_ISL_2262301, EPI_ISL_2262302, EPI_ISL_2262303, EPI_ISL_2262304, EPI_ISL_2262305, EPI_ISL_2314338, EPI_ISL_2314348 | | | |
| see above | Uniklinikum Jena | Robert Koch Institute | |
| EPI_ISL_2116091 | Uniklinikum Regensburg; Institut fÃ¼r klinische Mikrobiologie und Hygiene | Robert Koch Institute | |
| EPI_ISL_2259697, EPI_ISL_2262097, EPI_ISL_2262098, EPI_ISL_2262099, EPI_ISL_2262100 | Uniklinikum Regensburg; Institut für klinische Mikrobiologie und Hygiene | Robert Koch Institute | |
| EPI_ISL_2129182 | Universitatsklinikum Munster Institut fur Virologie | Robert Koch Institute | |
| EPI_ISL_2120134, EPI_ISL_2120137, EPI_ISL_2120142, EPI_ISL_2120183, EPI_ISL_2120197, EPI_ISL_2120205, EPI_ISL_2120215 | | | |
| see above | Universitatsklinikum des Saarlandes - Institut fur Virologie | Robert Koch Institute | |
| EPI_ISL_2384725, EPI_ISL_2384729, EPI_ISL_2384730, EPI_ISL_2384731, EPI_ISL_2384740, EPI_ISL_2689151, EPI_ISL_2689158, EPI_ISL_2689160, EPI_ISL_2689161, EPI_ISL_2689162 | | | |
| see above | University Medical Center Hamburg Eppendorf | Heinrich Pette Institute, Leibniz Institute for Experimental Virology | Adam Grundhoff; Alexis Robitaille; Johannes Knobloch; Martin Aepfelbacher; Nicole Fischer; Thomas Günther |
| EPI_ISL_2116053 | UniversitÃ¤tsklinikum GieÃŸen - Institut fÃ¼r Medizinische Virologie | Robert Koch Institute | |
| EPI_ISL_2264489, EPI_ISL_2264507, EPI_ISL_2264511, EPI_ISL_2264534, EPI_ISL_2264539, EPI_ISL_2390182, EPI_ISL_2390183, EPI_ISL_2390184, EPI_ISL_2390193, EPI_ISL_2390194 | | | |
| see above | Universitätsklinikum Bonn - Institut für Virologie | Robert Koch Institute | |
| EPI_ISL_2262325, EPI_ISL_2262327, EPI_ISL_2262328, EPI_ISL_2262329, EPI_ISL_2262330, EPI_ISL_2262331, EPI_ISL_2262332 | | | |
| see above | Universitätsklinikum Essen | Robert Koch Institute | |
| EPI_ISL_2469924, EPI_ISL_2469926 | Universitätsklinikum Heidelberg | Robert Koch Institute | |
| EPI_ISL_2845585, EPI_ISL_2845590, EPI_ISL_2845593, EPI_ISL_2845594, EPI_ISL_2845595, EPI_ISL_2845598, EPI_ISL_2845599, EPI_ISL_2845600, EPI_ISL_2845601, EPI_ISL_2845603, EPI_ISL_2845606, EPI_ISL_2845609, EPI_ISL_2845626, EPI_ISL_2845628, EPI_ISL_2845635, EPI_ISL_2845637, EPI_ISL_2845647, EPI_ISL_2845658, EPI_ISL_2845659, EPI_ISL_2845661, EPI_ISL_2845662, EPI_ISL_2845664, EPI_ISL_2845666, EPI_ISL_2845667, EPI_ISL_2845670, EPI_ISL_2845672, EPI_ISL_2845673, EPI_ISL_2845674, EPI_ISL_2845678 | | | |
| see above | Universitätsklinikum Köln; Institut für Virologie | Robert Koch Institute | |
| EPI_ISL_2264484, EPI_ISL_2264498, EPI_ISL_2264501, EPI_ISL_2264515, EPI_ISL_2264523, EPI_ISL_2264524, EPI_ISL_2264531, EPI_ISL_2264533 | | | |
| see above | Universitätsklinikum des Saarlandes - Institut für Virologie | Robert Koch Institute | |
| EPI_ISL_2388092, EPI_ISL_2633735 | Universitätsmedizin Mannheim | Robert Koch Institute | |
| EPI_ISL_2135735, EPI_ISL_2135736, EPI_ISL_2135737, EPI_ISL_2135739, EPI_ISL_2135740, EPI_ISL_2135741, EPI_ISL_2135742, EPI_ISL_2135743, EPI_ISL_2135744, EPI_ISL_2135745, EPI_ISL_2135746, EPI_ISL_2135747, EPI_ISL_2135748, EPI_ISL_2135749, EPI_ISL_2135750, EPI_ISL_2135752, EPI_ISL_2135753, EPI_ISL_2135754, EPI_ISL_2135755, EPI_ISL_2135756, EPI_ISL_2135757, EPI_ISL_2135758, EPI_ISL_2135759, EPI_ISL_2135760, EPI_ISL_2447909, EPI_ISL_2447917 | | | |
| see above | Virology, Universitätsklinikum des Saarlandes | Epigenetics, Saarland University | Jörn Walter; Kathrin Kattler; Nastasja Seiwert; Sascha Tierling; Sigrun Smola; Stefan Lohse; Thorsten Pfuhl |
| EPI_ISL_2380594, EPI_ISL_2380595, EPI_ISL_2380596, EPI_ISL_2380597, EPI_ISL_2380599, EPI_ISL_2380600, EPI_ISL_2380601, EPI_ISL_2380602, EPI_ISL_2380603, EPI_ISL_2380605, EPI_ISL_2380606, EPI_ISL_2380607, EPI_ISL_2380608, EPI_ISL_2380610, EPI_ISL_2380611, EPI_ISL_2380612, EPI_ISL_2380613, EPI_ISL_2380614, EPI_ISL_2380615, EPI_ISL_2380616, EPI_ISL_2380617, EPI_ISL_2380618, EPI_ISL_2380619, EPI_ISL_2380620, EPI_ISL_2380621, EPI_ISL_2380622, EPI_ISL_2380623, EPI_ISL_2380624, EPI_ISL_2380637, EPI_ISL_2380638, EPI_ISL_2380639, EPI_ISL_2380640, EPI_ISL_2380641, EPI_ISL_2380642, EPI_ISL_2380643, EPI_ISL_2380644, EPI_ISL_2380645, EPI_ISL_2380646, EPI_ISL_2380647, EPI_ISL_2380648, EPI_ISL_2380653, EPI_ISL_2380654, EPI_ISL_2380655, EPI_ISL_2380656, EPI_ISL_2380657, EPI_ISL_2380658, EPI_ISL_2380660, EPI_ISL_2380661, EPI_ISL_2380662, EPI_ISL_2380663, EPI_ISL_2380664, EPI_ISL_2380665, EPI_ISL_2380667, EPI_ISL_2380668, EPI_ISL_2380669, EPI_ISL_2380670, EPI_ISL_2380671, EPI_ISL_2380672, EPI_ISL_2380674, EPI_ISL_2380675, EPI_ISL_2380676, EPI_ISL_2380677, EPI_ISL_2380678, EPI_ISL_2380679, EPI_ISL_2380681, EPI_ISL_2380682, EPI_ISL_2380683, EPI_ISL_2380684, EPI_ISL_2380685, EPI_ISL_2380686, EPI_ISL_2380687, EPI_ISL_2380688, EPI_ISL_2380689, EPI_ISL_2380690, EPI_ISL_2380691, EPI_ISL_2380692, EPI_ISL_2380693, EPI_ISL_2380694, EPI_ISL_2380695, EPI_ISL_2380696, EPI_ISL_2380697, EPI_ISL_2380698, EPI_ISL_2380699, EPI_ISL_2380700, EPI_ISL_2380701, EPI_ISL_2380702, EPI_ISL_2380703, EPI_ISL_2380704, EPI_ISL_2380705, EPI_ISL_2380706, EPI_ISL_2380707, EPI_ISL_2380708, EPI_ISL_2380709, EPI_ISL_2380710, EPI_ISL_2380711, EPI_ISL_2380712, EPI_ISL_2380713, EPI_ISL_2380714, EPI_ISL_2380715, EPI_ISL_2380716, EPI_ISL_2380717, EPI_ISL_2380718, EPI_ISL_2380719, EPI_ISL_2380720, EPI_ISL_2380721, EPI_ISL_2380722, EPI_ISL_2380723, EPI_ISL_2380726, EPI_ISL_2380727, EPI_ISL_2380728, EPI_ISL_2380729, EPI_ISL_2380730, EPI_ISL_2380731, EPI_ISL_2380746, EPI_ISL_2380747, EPI_ISL_2380748, EPI_ISL_2380749, EPI_ISL_2380750, EPI_ISL_2380751, EPI_ISL_2380752, EPI_ISL_2380753, EPI_ISL_2380754, EPI_ISL_2380755, EPI_ISL_2380756, EPI_ISL_2380757, EPI_ISL_2380758, EPI_ISL_2380759, EPI_ISL_2380760, EPI_ISL_2380761, EPI_ISL_2380762, EPI_ISL_2380764, EPI_ISL_2380765, EPI_ISL_2380766, EPI_ISL_2380767, EPI_ISL_2380768, EPI_ISL_2380859, EPI_ISL_2380861, EPI_ISL_2380862, EPI_ISL_2380863, EPI_ISL_2380864, EPI_ISL_2380865, EPI_ISL_2380866, EPI_ISL_2380867, EPI_ISL_2380927, EPI_ISL_2380928, EPI_ISL_2380930, EPI_ISL_2381468, EPI_ISL_2381469, EPI_ISL_2381470, EPI_ISL_2381471, EPI_ISL_2381472, EPI_ISL_2381473, EPI_ISL_2381474, EPI_ISL_2381475, EPI_ISL_2381476, EPI_ISL_2381477, EPI_ISL_2381478, EPI_ISL_2381479, EPI_ISL_2381480, EPI_ISL_2381481, EPI_ISL_2381482, EPI_ISL_2381483, EPI_ISL_2381484, EPI_ISL_2381485, EPI_ISL_2381486, EPI_ISL_2381487, EPI_ISL_2381488, EPI_ISL_2381489, EPI_ISL_2381490, EPI_ISL_2381491, EPI_ISL_2381492, EPI_ISL_2381494, EPI_ISL_2381495, EPI_ISL_2381496, EPI_ISL_2381497, EPI_ISL_2381499, EPI_ISL_2381500, EPI_ISL_2381501, EPI_ISL_2381502, EPI_ISL_2381505, EPI_ISL_2381506, EPI_ISL_2381507, EPI_ISL_2381508, EPI_ISL_2381509, EPI_ISL_2381510, EPI_ISL_2381511, EPI_ISL_2381512, EPI_ISL_2381514, EPI_ISL_2381515, EPI_ISL_2381516, EPI_ISL_2381517, EPI_ISL_2381519, EPI_ISL_2381520, EPI_ISL_2381521, EPI_ISL_2381583, EPI_ISL_2381584, EPI_ISL_2381585, EPI_ISL_2381586, EPI_ISL_2381607, EPI_ISL_2381608 | | | |
| see above | ZOTZ KLIMAS MVZ Düsseldorf-Centrum GbR ÜBAG für Labormedizin, Genetik, Zytologie, Pathologie | Center of Medical Microbiology, Virology, and Hospital Hygiene, University of Duesseldorf | Alexander Dilthey; Andreas Walker; Daniel Strelow; Jessica Nicolai; Jörg Timm; Katrin Hoffmann; Klaus Pfeffer; Lisanna Hülse; Malte Kohns Vasconcelos; Maximilian Damagnez; Nadine Lübke; Patrick Finzer; Rainer Zotz; Tobias Wienemann; Torsten Houwaart |
| EPI_ISL_2116040, EPI_ISL_2116041, EPI_ISL_2116042, EPI_ISL_2116043, EPI_ISL_2116046 | alphaomega Labor GbR (Medizinische Kooperationsgemeinschaft Dres. Ackermann/Tennert/ZaiÃŸ) | Robert Koch Institute | |
| EPI_ISL_2262278, EPI_ISL_2262282 | alphaomega Labor GbR (Medizinische Kooperationsgemeinschaft Dres. Ackermann/Tennert/Zaiß) | Robert Koch Institute | |
| EPI_ISL_2130592, EPI_ISL_2130595, EPI_ISL_2130599, EPI_ISL_2130604, EPI_ISL_2130621, EPI_ISL_2130639, EPI_ISL_2130642, EPI_ISL_2130650, EPI_ISL_2130704, EPI_ISL_2130716, EPI_ISL_2130736, EPI_ISL_2130740, EPI_ISL_2130749, EPI_ISL_2130756, EPI_ISL_2130759, EPI_ISL_2130786, EPI_ISL_2130790, EPI_ISL_2130796, EPI_ISL_2130797, EPI_ISL_2130800, EPI_ISL_2130802, EPI_ISL_2130813, EPI_ISL_2130815, EPI_ISL_2130818, EPI_ISL_2130820, EPI_ISL_2130821, EPI_ISL_2130822, EPI_ISL_2130824 | | | |
| see above | amedes GÃ¶ttingen | Robert Koch Institute | |
| EPI_ISL_2130593, EPI_ISL_2130594, EPI_ISL_2130600, EPI_ISL_2130606, EPI_ISL_2130611, EPI_ISL_2130635, EPI_ISL_2130644, EPI_ISL_2130648, EPI_ISL_2130651, EPI_ISL_2130655, EPI_ISL_2130657, EPI_ISL_2130661, EPI_ISL_2130664, EPI_ISL_2130666, EPI_ISL_2130671, EPI_ISL_2130679, EPI_ISL_2130687, EPI_ISL_2130691, EPI_ISL_2130693, EPI_ISL_2130694, EPI_ISL_2130699, EPI_ISL_2130707, EPI_ISL_2130710, EPI_ISL_2130714, EPI_ISL_2130718, EPI_ISL_2130727, EPI_ISL_2130731, EPI_ISL_2130733, EPI_ISL_2130742, EPI_ISL_2130753, EPI_ISL_2130763, EPI_ISL_2130767, EPI_ISL_2130768, EPI_ISL_2130782, EPI_ISL_2130789, EPI_ISL_2130810 | | | |
| see above | amedes MVZ DIAMEDIS Sennestadt | Robert Koch Institute | |
| EPI_ISL_2115808, EPI_ISL_2115833, EPI_ISL_2115847, EPI_ISL_2115885, EPI_ISL_2115887, EPI_ISL_2115941, EPI_ISL_2115944, EPI_ISL_2115981, EPI_ISL_2116012, EPI_ISL_2130596, EPI_ISL_2130597, EPI_ISL_2130620, EPI_ISL_2130631, EPI_ISL_2130647, EPI_ISL_2130668, EPI_ISL_2130672, EPI_ISL_2130681, EPI_ISL_2130682, EPI_ISL_2130708, EPI_ISL_2130734, EPI_ISL_2130737, EPI_ISL_2130745, EPI_ISL_2130750, EPI_ISL_2130760, EPI_ISL_2130774, EPI_ISL_2130798, EPI_ISL_2130808, EPI_ISL_2130823 | | | |
| see above | amedes MVZ Halle/Leipzig | Robert Koch Institute | |
| EPI_ISL_2130598, EPI_ISL_2130608, EPI_ISL_2130616, EPI_ISL_2130619, EPI_ISL_2130623, EPI_ISL_2130624, EPI_ISL_2130625, EPI_ISL_2130637, EPI_ISL_2130641, EPI_ISL_2130643, EPI_ISL_2130660, EPI_ISL_2130688, EPI_ISL_2130690, EPI_ISL_2130701, EPI_ISL_2130703, EPI_ISL_2130721, EPI_ISL_2130757, EPI_ISL_2130788, EPI_ISL_2130793 | | | |
| see above | amedes MVZ Hannover | Robert Koch Institute | |
| EPI_ISL_2130615, EPI_ISL_2130617, EPI_ISL_2130622, EPI_ISL_2130628, EPI_ISL_2130629, EPI_ISL_2130646, EPI_ISL_2130649, EPI_ISL_2130653, EPI_ISL_2130663, EPI_ISL_2130669, EPI_ISL_2130673, EPI_ISL_2130675, EPI_ISL_2130677, EPI_ISL_2130684, EPI_ISL_2130702, EPI_ISL_2130705, EPI_ISL_2130709, EPI_ISL_2130711, EPI_ISL_2130713, EPI_ISL_2130719, EPI_ISL_2130722, EPI_ISL_2130726, EPI_ISL_2130729, EPI_ISL_2130732, EPI_ISL_2130739, EPI_ISL_2130746, EPI_ISL_2130747, EPI_ISL_2130755, EPI_ISL_2130758, EPI_ISL_2130764, EPI_ISL_2130766, EPI_ISL_2130772, EPI_ISL_2130776, EPI_ISL_2130779, EPI_ISL_2130781, EPI_ISL_2130783, EPI_ISL_2130803, EPI_ISL_2130804, EPI_ISL_2130806, EPI_ISL_2130807 | | | |
| see above | amedes MVZ Jena | Robert Koch Institute | |
| EPI_ISL_2130610, EPI_ISL_2130613, EPI_ISL_2130630, EPI_ISL_2130632, EPI_ISL_2130633, EPI_ISL_2130645, EPI_ISL_2130652, EPI_ISL_2130667, EPI_ISL_2130683, EPI_ISL_2130715, EPI_ISL_2130720, EPI_ISL_2130819 | | | |
| see above | amedes MVZ fÃ¼r Laboratoriumsdiagnostik Raubling GmbH | Robert Koch Institute | |
| EPI_ISL_2130601, EPI_ISL_2130612, EPI_ISL_2130706, EPI_ISL_2130730, EPI_ISL_2130748, EPI_ISL_2130780, EPI_ISL_2130801 | | | |
| see above | amedes MVZ fÃ¼r Laboratoriumsmedizin FÃ¼rstenfeldbruck | Robert Koch Institute | |
| EPI_ISL_2130761 | amedes MVZ fÃ¼r Laboratoriumsmedizin Hamburg | Robert Koch Institute | |
| EPI_ISL_2130607 | amedes MVZ fÃ¼r Laboratoriumsmedizin MÃ¼nchen | Robert Koch Institute | |
| EPI_ISL_2130603, EPI_ISL_2130656, EPI_ISL_2130724 | amedes MVZ fÃ¼r Laboratoriumsmedizin Rhein/Ruhr | Robert Koch Institute | |
| EPI_ISL_2130723 | amedes MVZ fÃ¼r Labordiagnostik Rhein-Main | Robert Koch Institute | |
| EPI_ISL_2263152, EPI_ISL_2263153, EPI_ISL_2263155, EPI_ISL_2263156, EPI_ISL_2263157, EPI_ISL_2263158, EPI_ISL_2263160, EPI_ISL_2263166, EPI_ISL_2263168, EPI_ISL_2263172, EPI_ISL_2263173, EPI_ISL_2263174, EPI_ISL_2263175, EPI_ISL_2263177, EPI_ISL_2263181 | | | |
| see above | labopart - Medizinische Laboratorien Dresden | Robert Koch Institute | |
| EPI_ISL_2115820, EPI_ISL_2115824, EPI_ISL_2115826, EPI_ISL_2115830, EPI_ISL_2115834, EPI_ISL_2115841, EPI_ISL_2115843, EPI_ISL_2115845, EPI_ISL_2115848, EPI_ISL_2115854, EPI_ISL_2115860, EPI_ISL_2115868, EPI_ISL_2115869, EPI_ISL_2115872, EPI_ISL_2115873, EPI_ISL_2115876, EPI_ISL_2115881, EPI_ISL_2115889, EPI_ISL_2115893, EPI_ISL_2115896, EPI_ISL_2115900, EPI_ISL_2115910, EPI_ISL_2115916, EPI_ISL_2115924, EPI_ISL_2115927, EPI_ISL_2115936, EPI_ISL_2115938, EPI_ISL_2115946, EPI_ISL_2115948, EPI_ISL_2115951, EPI_ISL_2115955, EPI_ISL_2115963, EPI_ISL_2115965, EPI_ISL_2115967, EPI_ISL_2115969, EPI_ISL_2115970, EPI_ISL_2115976, EPI_ISL_2115988, EPI_ISL_2115990, EPI_ISL_2115993, EPI_ISL_2115994, EPI_ISL_2115995, EPI_ISL_2115997, EPI_ISL_2115998, EPI_ISL_2116003 | | | |
| see above | nordlab - Partnerschaftspraxis fuÌˆr Laboratoriumsmedizin | Robert Koch Institute | |

We gratefully acknowledge the following Authors from the Originating laboratories responsible for obtaining the specimens, as well as the Submitting laboratories where the genome data were generated and shared via GISAID, on which this research is based.

All Submitters of data may be contacted directly via www.gisaid.org

Authors are sorted alphabetically.

| Accession ID | Originating Laboratory | Submitting Laboratory | Authors |
|---|---|---|---|
| EPI_ISL_2634051, EPI_ISL_2634354 | ALB FILS KLINIKEN GmbH - Institut für Laboratoriumsmedizin Klinik am Eichert Göppingen | Robert Koch Institute | |
| EPI_ISL_2390477, EPI_ISL_2390483, EPI_ISL_2390484, EPI_ISL_2390491, EPI_ISL_2390492, EPI_ISL_2390493, EPI_ISL_2390494, EPI_ISL_2390500, EPI_ISL_2390501, EPI_ISL_2390505, EPI_ISL_2390506, EPI_ISL_2390507, EPI_ISL_2390509, EPI_ISL_2390510, EPI_ISL_2390513, EPI_ISL_2390515, EPI_ISL_2390516, EPI_ISL_2390518, EPI_ISL_2390520, EPI_ISL_2390524, EPI_ISL_2390529, EPI_ISL_2390532, EPI_ISL_2390541, EPI_ISL_2390546, EPI_ISL_2390548, EPI_ISL_2390550, EPI_ISL_2390555, EPI_ISL_2390556, EPI_ISL_2390557, EPI_ISL_2390558, EPI_ISL_2390559, EPI_ISL_2390565, EPI_ISL_2390567, EPI_ISL_2390568, EPI_ISL_2390569, EPI_ISL_2390571, EPI_ISL_2390575, EPI_ISL_2472550, EPI_ISL_2472552, EPI_ISL_2472555, EPI_ISL_2472556, EPI_ISL_2472557, EPI_ISL_2472559, EPI_ISL_2472560, EPI_ISL_2472561, EPI_ISL_2472562, EPI_ISL_2472565, EPI_ISL_2472567, EPI_ISL_2472568, EPI_ISL_2472570, EPI_ISL_2472571, EPI_ISL_2472572, EPI_ISL_2472576, EPI_ISL_2472577, EPI_ISL_2472580, EPI_ISL_2472581, EPI_ISL_2472582, EPI_ISL_2472583, EPI_ISL_2472586, EPI_ISL_2472587, EPI_ISL_2472589, EPI_ISL_2472590, EPI_ISL_2472593, EPI_ISL_2472595, EPI_ISL_2472597, EPI_ISL_2472598, EPI_ISL_2472600, EPI_ISL_2472603, EPI_ISL_2472605, EPI_ISL_2472607, EPI_ISL_2472608, EPI_ISL_2472609, EPI_ISL_2472611, EPI_ISL_2472612, EPI_ISL_2472613, EPI_ISL_2472616, EPI_ISL_2472618, EPI_ISL_2472619, EPI_ISL_2472620, EPI_ISL_2472623, EPI_ISL_2472624, EPI_ISL_2472625, EPI_ISL_2472627, EPI_ISL_2472630, EPI_ISL_2472631, EPI_ISL_2472633, EPI_ISL_2472636, EPI_ISL_2472637, EPI_ISL_2472639, EPI_ISL_2634946, EPI_ISL_2634948, EPI_ISL_2634950, EPI_ISL_2634962, EPI_ISL_2634968, EPI_ISL_2634972, EPI_ISL_2634980, EPI_ISL_2634982, EPI_ISL_2634985, EPI_ISL_2634994, EPI_ISL_2634997, EPI_ISL_2634998, EPI_ISL_2635001, EPI_ISL_2635006, EPI_ISL_2635011, EPI_ISL_2635012, EPI_ISL_2635020, EPI_ISL_2635022, EPI_ISL_2635026, EPI_ISL_2635028, EPI_ISL_2635030, EPI_ISL_2635038, EPI_ISL_2635040, EPI_ISL_2635053, EPI_ISL_2635060, EPI_ISL_2635071, EPI_ISL_2635089, EPI_ISL_2635094, EPI_ISL_2635096, EPI_ISL_2635101, EPI_ISL_2635112, EPI_ISL_2635121 | | | |
| see above | Bayerisches Landesamt für Gesundheit und Lebensmittelsicherheit (LGL) | Robert Koch Institute | |
| EPI_ISL_2313371, EPI_ISL_2313380, EPI_ISL_2313384, EPI_ISL_2313385, EPI_ISL_2313386, EPI_ISL_2313387, EPI_ISL_2313388, EPI_ISL_2313389, EPI_ISL_2313397, EPI_ISL_2313403, EPI_ISL_2313411, EPI_ISL_2313415, EPI_ISL_2313416, EPI_ISL_2313418, EPI_ISL_2313422, EPI_ISL_2313423, EPI_ISL_2313426, EPI_ISL_2313427, EPI_ISL_2313430, EPI_ISL_2313431, EPI_ISL_2313432, EPI_ISL_2313435, EPI_ISL_2313443, EPI_ISL_2313444, EPI_ISL_2313447, EPI_ISL_2313460, EPI_ISL_2313461, EPI_ISL_2313463, EPI_ISL_2313465, EPI_ISL_2313468, EPI_ISL_2313469, EPI_ISL_2313470, EPI_ISL_2313473, EPI_ISL_2313475, EPI_ISL_2313478, EPI_ISL_2313487, EPI_ISL_2313496, EPI_ISL_2313500, EPI_ISL_2313503, EPI_ISL_2313509, EPI_ISL_2313529, EPI_ISL_2313531, EPI_ISL_2313538, EPI_ISL_2313543, EPI_ISL_2313545, EPI_ISL_2313547, EPI_ISL_2313549, EPI_ISL_2313556, EPI_ISL_2313559, EPI_ISL_2313569, EPI_ISL_2313581, EPI_ISL_2313585, EPI_ISL_2313588, EPI_ISL_2313590, EPI_ISL_2313595, EPI_ISL_2313598, EPI_ISL_2313603, EPI_ISL_2313605, EPI_ISL_2313615, EPI_ISL_2313616, EPI_ISL_2313620, EPI_ISL_2313629, EPI_ISL_2313641, EPI_ISL_2313645, EPI_ISL_2313648, EPI_ISL_2313661, EPI_ISL_2313669, EPI_ISL_2313673, EPI_ISL_2313678, EPI_ISL_2313685, EPI_ISL_2313688, EPI_ISL_2313692, EPI_ISL_2313693, EPI_ISL_2313694, EPI_ISL_2313697, EPI_ISL_2313704, EPI_ISL_2313706, EPI_ISL_2313708, EPI_ISL_2313731, EPI_ISL_2313733, EPI_ISL_2313745, EPI_ISL_2313750, EPI_ISL_2313752, EPI_ISL_2313754, EPI_ISL_2313757, EPI_ISL_2313769, EPI_ISL_2313772, EPI_ISL_2313774, EPI_ISL_2313776, EPI_ISL_2313779, EPI_ISL_2313781, EPI_ISL_2313807, EPI_ISL_2313831, EPI_ISL_2313833, EPI_ISL_2313839, EPI_ISL_2313842, EPI_ISL_2313845, EPI_ISL_2313853, EPI_ISL_2313860, EPI_ISL_2313863, EPI_ISL_2313872, EPI_ISL_2313875, EPI_ISL_2313880, EPI_ISL_2313884, EPI_ISL_2313885, EPI_ISL_2313886, EPI_ISL_2314136, EPI_ISL_2314160, EPI_ISL_2314161, EPI_ISL_2314162, EPI_ISL_2314210, EPI_ISL_2314216, EPI_ISL_2314218, EPI_ISL_2314227, EPI_ISL_2314238, EPI_ISL_2314240, EPI_ISL_2314247, EPI_ISL_2314248, EPI_ISL_2314265, EPI_ISL_2314268, EPI_ISL_2314282, EPI_ISL_2387197, EPI_ISL_2387199, EPI_ISL_2387204, EPI_ISL_2387206, EPI_ISL_2387207, EPI_ISL_2387209, EPI_ISL_2387213, EPI_ISL_2387231, EPI_ISL_2387233, EPI_ISL_2387235, EPI_ISL_2387238, EPI_ISL_2387241, EPI_ISL_2387242, EPI_ISL_2387245, EPI_ISL_2387249, EPI_ISL_2387250, EPI_ISL_2387258, EPI_ISL_2387265, EPI_ISL_2387270, EPI_ISL_2387273, EPI_ISL_2387275, EPI_ISL_2387289, EPI_ISL_2387295, EPI_ISL_2387301, EPI_ISL_2387340, EPI_ISL_2388335, EPI_ISL_2388368, EPI_ISL_2388372, EPI_ISL_2388380, EPI_ISL_2388422, EPI_ISL_2388432, EPI_ISL_2388457, EPI_ISL_2388459, EPI_ISL_2388466, EPI_ISL_2388477, EPI_ISL_2388498, EPI_ISL_2388502, EPI_ISL_2388526, EPI_ISL_2388541, EPI_ISL_2388553, EPI_ISL_2388559, EPI_ISL_2388599, EPI_ISL_2388613, EPI_ISL_2388617, EPI_ISL_2388626, EPI_ISL_2388628, EPI_ISL_2388638, EPI_ISL_2388658, EPI_ISL_2388660, EPI_ISL_2388670, EPI_ISL_2388677, EPI_ISL_2388683, EPI_ISL_2388684, EPI_ISL_2388696, EPI_ISL_2388700, EPI_ISL_2388701, EPI_ISL_2388720, EPI_ISL_2388723, EPI_ISL_2388726, EPI_ISL_2388729, EPI_ISL_2388732, EPI_ISL_2388733, EPI_ISL_2388741, EPI_ISL_2388768, EPI_ISL_2388776, EPI_ISL_2388784, EPI_ISL_2388786, EPI_ISL_2388788, EPI_ISL_2388801, EPI_ISL_2388804, EPI_ISL_2388805, EPI_ISL_2388807, EPI_ISL_2388812, EPI_ISL_2388813, EPI_ISL_2388814, EPI_ISL_2388820, EPI_ISL_2388822, EPI_ISL_2388823, EPI_ISL_2388824, EPI_ISL_2388825, EPI_ISL_2388826, EPI_ISL_2388827, EPI_ISL_2388829, EPI_ISL_2388831, EPI_ISL_2388834, EPI_ISL_2388835, EPI_ISL_2388836, EPI_ISL_2388838, EPI_ISL_2388840, EPI_ISL_2388844, EPI_ISL_2388849, EPI_ISL_2388853, EPI_ISL_2388856, EPI_ISL_2388861, EPI_ISL_2388869, EPI_ISL_2388870, EPI_ISL_2388871, EPI_ISL_2388872, EPI_ISL_2388873, EPI_ISL_2388874, EPI_ISL_2388876, EPI_ISL_2388877, EPI_ISL_2388878, EPI_ISL_2388881, EPI_ISL_2388882, EPI_ISL_2388883, EPI_ISL_2388884, EPI_ISL_2388885, EPI_ISL_2388888, EPI_ISL_2388889, EPI_ISL_2388890, EPI_ISL_2388892, EPI_ISL_2388894, EPI_ISL_2388895, EPI_ISL_2388896, EPI_ISL_2388897, EPI_ISL_2388899, EPI_ISL_2388901, EPI_ISL_2388902, EPI_ISL_2388903, EPI_ISL_2388904, EPI_ISL_2388908, EPI_ISL_2388910, EPI_ISL_2388911, EPI_ISL_2388912, EPI_ISL_2388913, EPI_ISL_2388915, EPI_ISL_2388917, EPI_ISL_2388918, EPI_ISL_2388921, EPI_ISL_2388922, EPI_ISL_2388924, EPI_ISL_2388925, EPI_ISL_2388926, EPI_ISL_2388927, EPI_ISL_2388928, EPI_ISL_2388929, EPI_ISL_2388930, EPI_ISL_2388931, EPI_ISL_2388932, EPI_ISL_2388933, EPI_ISL_2388934, EPI_ISL_2388938, EPI_ISL_2388939, EPI_ISL_2388941, EPI_ISL_2388942, EPI_ISL_2388943, EPI_ISL_2388944, EPI_ISL_2388946, EPI_ISL_2388947, EPI_ISL_2388951, EPI_ISL_2388952, EPI_ISL_2388953, EPI_ISL_2388954, EPI_ISL_2388955, EPI_ISL_2388961, EPI_ISL_2388962, EPI_ISL_2388964, EPI_ISL_2388965, EPI_ISL_2388966, EPI_ISL_2388967, EPI_ISL_2388968, EPI_ISL_2388969, EPI_ISL_2388972, EPI_ISL_2388973, EPI_ISL_2388974, EPI_ISL_2388976, EPI_ISL_2388979, EPI_ISL_2388980, EPI_ISL_2388983, EPI_ISL_2388984, EPI_ISL_2388985, EPI_ISL_2388986, EPI_ISL_2388988, EPI_ISL_2388991, EPI_ISL_2388992, EPI_ISL_2388993, EPI_ISL_2388997, EPI_ISL_2388999, EPI_ISL_2389000, EPI_ISL_2389001, EPI_ISL_2389004, EPI_ISL_2389006, EPI_ISL_2389007, EPI_ISL_2389010, EPI_ISL_2389011, EPI_ISL_2389014, EPI_ISL_2389015, EPI_ISL_2389017, EPI_ISL_2389020, EPI_ISL_2389021, EPI_ISL_2389023, EPI_ISL_2389024, EPI_ISL_2389025, EPI_ISL_2389026, EPI_ISL_2389028, EPI_ISL_2389029, EPI_ISL_2389031, EPI_ISL_2389032, EPI_ISL_2389042, EPI_ISL_2389043, EPI_ISL_2389044, EPI_ISL_2389045, EPI_ISL_2389047, EPI_ISL_2389048, EPI_ISL_2389049, EPI_ISL_2389050, EPI_ISL_2389051, EPI_ISL_2389052, EPI_ISL_2389054, EPI_ISL_2389057, EPI_ISL_2389058, EPI_ISL_2389059, EPI_ISL_2389060, EPI_ISL_2389061, EPI_ISL_2389063, EPI_ISL_2389064, EPI_ISL_2389068, EPI_ISL_2389071, EPI_ISL_2389072, EPI_ISL_2389073, EPI_ISL_2389074, EPI_ISL_2389075, EPI_ISL_2389078, EPI_ISL_2389079, EPI_ISL_2389370, EPI_ISL_2469963, EPI_ISL_2470007, EPI_ISL_2470032, EPI_ISL_2470132, EPI_ISL_2470181, EPI_ISL_2470207, EPI_ISL_2470209, EPI_ISL_2470210, EPI_ISL_2470215, EPI_ISL_2470225, EPI_ISL_2470235, EPI_ISL_2470236, EPI_ISL_2470237, EPI_ISL_2470241, EPI_ISL_2470245, EPI_ISL_2470255, EPI_ISL_2470256, EPI_ISL_2470260, EPI_ISL_2470263, EPI_ISL_2470269, EPI_ISL_2470272, EPI_ISL_2470276, EPI_ISL_2470281, EPI_ISL_2470283, EPI_ISL_2470300, EPI_ISL_2470302, EPI_ISL_2470316, EPI_ISL_2470319, EPI_ISL_2470325, EPI_ISL_2470339, EPI_ISL_2470341, EPI_ISL_2470348, EPI_ISL_2470357, EPI_ISL_2470360, EPI_ISL_2470362, EPI_ISL_2470366, EPI_ISL_2470368, EPI_ISL_2470370, EPI_ISL_2470375, EPI_ISL_2470381, EPI_ISL_2470385, EPI_ISL_2470389, EPI_ISL_2470400, EPI_ISL_2470403, EPI_ISL_2470407, EPI_ISL_2470413, EPI_ISL_2470419, EPI_ISL_2470428, EPI_ISL_2470429, EPI_ISL_2470431, EPI_ISL_2470434, EPI_ISL_2470439, EPI_ISL_2470443, EPI_ISL_2470446, EPI_ISL_2470449, EPI_ISL_2470454, EPI_ISL_2470456, EPI_ISL_2470457, EPI_ISL_2470462, EPI_ISL_2470464, EPI_ISL_2470474, EPI_ISL_2470477, EPI_ISL_2470478, EPI_ISL_2470480, EPI_ISL_2632469 | | | |
| see above | Bioscientia Labor Wermsdorf | Robert Koch Institute | |
| EPI_ISL_2313374, EPI_ISL_2313390, EPI_ISL_2313398, EPI_ISL_2313399, EPI_ISL_2313412, EPI_ISL_2313414, EPI_ISL_2313429, EPI_ISL_2313476, EPI_ISL_2313566, EPI_ISL_2313579, EPI_ISL_2313593, EPI_ISL_2313607, EPI_ISL_2313636, EPI_ISL_2313638, EPI_ISL_2313683, EPI_ISL_2313690, EPI_ISL_2313729, EPI_ISL_2313747, EPI_ISL_2313817, EPI_ISL_2313866, EPI_ISL_2313868, EPI_ISL_2313877, EPI_ISL_2313879, EPI_ISL_2313883, EPI_ISL_2314084, EPI_ISL_2314089, EPI_ISL_2314094, EPI_ISL_2314095, EPI_ISL_2314096, EPI_ISL_2314097, EPI_ISL_2314100, EPI_ISL_2314108, EPI_ISL_2314123, EPI_ISL_2314131, EPI_ISL_2314141, EPI_ISL_2314142, EPI_ISL_2314146, EPI_ISL_2314154, EPI_ISL_2314166, EPI_ISL_2314167, EPI_ISL_2314168, EPI_ISL_2314190, EPI_ISL_2314204, EPI_ISL_2314219, EPI_ISL_2314220, EPI_ISL_2314237, EPI_ISL_2314245, EPI_ISL_2314250, EPI_ISL_2314252, EPI_ISL_2314262, EPI_ISL_2314263, EPI_ISL_2314267, EPI_ISL_2314275, EPI_ISL_2314283, EPI_ISL_2314284, EPI_ISL_2314288, EPI_ISL_2388364, EPI_ISL_2388439, EPI_ISL_2388482, EPI_ISL_2388620, EPI_ISL_2388711, EPI_ISL_2388735 | | | |
| see above | Bioscientia MVZ Labor Karlsruhe GmbH | Robert Koch Institute | |
| EPI_ISL_2536133, EPI_ISL_2536139, EPI_ISL_2536140, EPI_ISL_2536141, EPI_ISL_2536143 | Bundeswehr Institute of Microbiology | Bundeswehr Institute of Microbiology | Alexandra Rehn; Enrico Georgi; Malena Bestehorn-Willmann; Markus Antwerpen; Mathias Walter; Mike Pillukat; Roman Wölfel; Sabine Zange |
| EPI_ISL_2536145, EPI_ISL_2536146, EPI_ISL_2536147, EPI_ISL_2536148, EPI_ISL_2536149, EPI_ISL_2536150 | Bundeswehrkrankenhaus Berlin | Bundeswehr Institute of Microbiology | Alexandra Rehn; Enrico Georgi; Malena Bestehorn-Willmann; Markus Antwerpen; Mathias Walter; Mike Pillukat; Roman Wölfel; Sabine Zange |
| EPI_ISL_2536142, EPI_ISL_2536144 | Bundeswehrkrankenhaus Ulm | Bundeswehr Institute of Microbiology | Alexandra Rehn; Enrico Georgi; Malena Bestehorn-Willmann; Markus Antwerpen; Mathias Walter; Mike Pillukat; Roman Wölfel; Sabine Zange |
| EPI_ISL_2259815, EPI_ISL_2260027, EPI_ISL_2267951, EPI_ISL_2267952, EPI_ISL_2267955, EPI_ISL_2267956, EPI_ISL_2267957, EPI_ISL_2267960, EPI_ISL_2267961, EPI_ISL_2267962, EPI_ISL_2267964, EPI_ISL_2267965, EPI_ISL_2267966, EPI_ISL_2267968, EPI_ISL_2267969, EPI_ISL_2267971, EPI_ISL_2267972, EPI_ISL_2267973, EPI_ISL_2267975, EPI_ISL_2267977, EPI_ISL_2267978, EPI_ISL_2267979, EPI_ISL_2267981, EPI_ISL_2267984, EPI_ISL_2267985, EPI_ISL_2267986, EPI_ISL_2267987, EPI_ISL_2267992, EPI_ISL_2267994, EPI_ISL_2267995, EPI_ISL_2267999, EPI_ISL_2268000, EPI_ISL_2268002, EPI_ISL_2268004, EPI_ISL_2268006, EPI_ISL_2268008, EPI_ISL_2268010, EPI_ISL_2268015, EPI_ISL_2268018, EPI_ISL_2268020, EPI_ISL_2268022, EPI_ISL_2268025, EPI_ISL_2268027, EPI_ISL_2268028, EPI_ISL_2268029, EPI_ISL_2268036, EPI_ISL_2268039, EPI_ISL_2268041, EPI_ISL_2268043, EPI_ISL_2268044, EPI_ISL_2268046, EPI_ISL_2268051, EPI_ISL_2268052, EPI_ISL_2268054, EPI_ISL_2268056, EPI_ISL_2268059, EPI_ISL_2268061, EPI_ISL_2268062, EPI_ISL_2268066, EPI_ISL_2268067, EPI_ISL_2268069, EPI_ISL_2268071, EPI_ISL_2268072, EPI_ISL_2268075, EPI_ISL_2268077, EPI_ISL_2268081, EPI_ISL_2268083, EPI_ISL_2268086, EPI_ISL_2268088, EPI_ISL_2268090, EPI_ISL_2268091, EPI_ISL_2268093, EPI_ISL_2268095, EPI_ISL_2268096, EPI_ISL_2268098, EPI_ISL_2268100, EPI_ISL_2268102, EPI_ISL_2268103, EPI_ISL_2268105, EPI_ISL_2268108, EPI_ISL_2268110, EPI_ISL_2268112, EPI_ISL_2268114, EPI_ISL_2268116, EPI_ISL_2268121, EPI_ISL_2268122, EPI_ISL_2268126, EPI_ISL_2268129, EPI_ISL_2268131, EPI_ISL_2268132, EPI_ISL_2268134, EPI_ISL_2268135, EPI_ISL_2268136, EPI_ISL_2268138, EPI_ISL_2268139, EPI_ISL_2268140, EPI_ISL_2268141, EPI_ISL_2268143, EPI_ISL_2268147, EPI_ISL_2268150, EPI_ISL_2268153, EPI_ISL_2268155, EPI_ISL_2268157, EPI_ISL_2268158, EPI_ISL_2268160, EPI_ISL_2268162, EPI_ISL_2268163, EPI_ISL_2268165, EPI_ISL_2268167, EPI_ISL_2268168, EPI_ISL_2268170, EPI_ISL_2268172, EPI_ISL_2268175, EPI_ISL_2268177, EPI_ISL_2268179, EPI_ISL_2268180, EPI_ISL_2268183, EPI_ISL_2268185, EPI_ISL_2268192, EPI_ISL_2268193, EPI_ISL_2268195, EPI_ISL_2268196, EPI_ISL_2268197, EPI_ISL_2268199, EPI_ISL_2268202, EPI_ISL_2268204, EPI_ISL_2268205, EPI_ISL_2268207, EPI_ISL_2268209, EPI_ISL_2268210, EPI_ISL_2268212, EPI_ISL_2268214, EPI_ISL_2268215, EPI_ISL_2268217, EPI_ISL_2268222, EPI_ISL_2268224, EPI_ISL_2268226, EPI_ISL_2268227, EPI_ISL_2268229, EPI_ISL_2268231, EPI_ISL_2268232, EPI_ISL_2268234, EPI_ISL_2268237, EPI_ISL_2268239, EPI_ISL_2268244, EPI_ISL_2268246, EPI_ISL_2268248, EPI_ISL_2268250, EPI_ISL_2268251, EPI_ISL_2268253, EPI_ISL_2268258, EPI_ISL_2268260, EPI_ISL_2268261, EPI_ISL_2268262, EPI_ISL_2268263, EPI_ISL_2268265, EPI_ISL_2268266, EPI_ISL_2268268, EPI_ISL_2268269, EPI_ISL_2268270, EPI_ISL_2268272, EPI_ISL_2268274, EPI_ISL_2268277, EPI_ISL_2268279, EPI_ISL_2268283, EPI_ISL_2268284, EPI_ISL_2268285, EPI_ISL_2268286, EPI_ISL_2268288, EPI_ISL_2268292, EPI_ISL_2268294, EPI_ISL_2268296, EPI_ISL_2268301, EPI_ISL_2268303, EPI_ISL_2268305, EPI_ISL_2268306, EPI_ISL_2268308, EPI_ISL_2268310, EPI_ISL_2268312, EPI_ISL_2268313, EPI_ISL_2268315, EPI_ISL_2268318, EPI_ISL_2268320, EPI_ISL_2268322, EPI_ISL_2268324, EPI_ISL_2268330, EPI_ISL_2268332, EPI_ISL_2268334, EPI_ISL_2386889, EPI_ISL_2386891, EPI_ISL_2386898, EPI_ISL_2386903, EPI_ISL_2386904, EPI_ISL_2386906, EPI_ISL_2386909, EPI_ISL_2386912, EPI_ISL_2386913, EPI_ISL_2386917, EPI_ISL_2386919, EPI_ISL_2386928, EPI_ISL_2386929, EPI_ISL_2386930, EPI_ISL_2386932, EPI_ISL_2386938, EPI_ISL_2386941, EPI_ISL_2386942, EPI_ISL_2386944, EPI_ISL_2386948, EPI_ISL_2386951, EPI_ISL_2386952, EPI_ISL_2386954, EPI_ISL_2386957, EPI_ISL_2386958, EPI_ISL_2386970, EPI_ISL_2386971, EPI_ISL_2386972, EPI_ISL_2386975, EPI_ISL_2386976, EPI_ISL_2386981, EPI_ISL_2386982, EPI_ISL_2386983, EPI_ISL_2386987, EPI_ISL_2386990, EPI_ISL_2387000, EPI_ISL_2387004, EPI_ISL_2387005, EPI_ISL_2387006, EPI_ISL_2387013, EPI_ISL_2387015, EPI_ISL_2387018, EPI_ISL_2387020, EPI_ISL_2387023, EPI_ISL_2387028, EPI_ISL_2387032, EPI_ISL_2387038, EPI_ISL_2387039, EPI_ISL_2387049, EPI_ISL_2387064, EPI_ISL_2387066, EPI_ISL_2387077, EPI_ISL_2387081, EPI_ISL_2387086, EPI_ISL_2387090, EPI_ISL_2387093, EPI_ISL_2387101, EPI_ISL_2387103, EPI_ISL_2387105, EPI_ISL_2387106, EPI_ISL_2387108, EPI_ISL_2387109, EPI_ISL_2387110, EPI_ISL_2387112, EPI_ISL_2387115, EPI_ISL_2387120, EPI_ISL_2387121, EPI_ISL_2387122, EPI_ISL_2387131, EPI_ISL_2387146, EPI_ISL_2387149, EPI_ISL_2387154, EPI_ISL_2387167, EPI_ISL_2387169, EPI_ISL_2387172, EPI_ISL_2387173, EPI_ISL_2387177, EPI_ISL_2387180, EPI_ISL_2387185, EPI_ISL_2387186, EPI_ISL_2387188, EPI_ISL_2387189, EPI_ISL_2471192, EPI_ISL_2471234, EPI_ISL_2471261, EPI_ISL_2471267, EPI_ISL_2471283, EPI_ISL_2471291, EPI_ISL_2471298, EPI_ISL_2471338, EPI_ISL_2471340, EPI_ISL_2471358, EPI_ISL_2471426, EPI_ISL_2471430, EPI_ISL_2471445, EPI_ISL_2471450 | | | |
| see above | CENTOGENE Frankfurt Laboratory: Niederlassung Industriepark Höchst | Robert Koch Institute | |
| EPI_ISL_2471822, EPI_ISL_2471826, EPI_ISL_2471829, EPI_ISL_2471831, EPI_ISL_2471832, EPI_ISL_2471833, EPI_ISL_2471838, EPI_ISL_2635968, EPI_ISL_2635969, EPI_ISL_2635970, EPI_ISL_2635972, EPI_ISL_2678683, EPI_ISL_2678684, EPI_ISL_2678710 | | | |
| see above | Carl-Thiem-Klinikum Cottbus - Zentrum für Laboratoriumsmedizin Mikrobiologie und Krankenhaushygiene | Robert Koch Institute | |
| EPI_ISL_2380737, EPI_ISL_2380738, EPI_ISL_2380739, EPI_ISL_2380741, EPI_ISL_2380742, EPI_ISL_2380743, EPI_ISL_2380744, EPI_ISL_2380745, EPI_ISL_2380870, EPI_ISL_2380871, EPI_ISL_2381456, EPI_ISL_2381457, EPI_ISL_2381460, EPI_ISL_2381462, EPI_ISL_2381467, EPI_ISL_2381650, EPI_ISL_2381652, EPI_ISL_2381653, EPI_ISL_2381777, EPI_ISL_2381812, EPI_ISL_2381813 | | | |
| see above | Center of Medical Microbiology, Virology, and Hospital Hygiene, University of Duesseldorf | Center of Medical Microbiology, Virology, and Hospital Hygiene, University of Duesseldorf | Alexander Dilthey; Andreas Walker; Daniel Strelow; Jessica Nicolai; Jörg Timm; Klaus Pfeffer; Lisanna Hülse; Malte Kohns Vasconcelos; Maximilian Damagnez; Nadine Lübke; Tobias Wienemann; Torsten Houwaart |
| EPI_ISL_2447991 | Charité Universitätsmedizin Berlin, Institut für Virologie/Labor Berlin | Charité Universitätsmedizin Berlin, Institut für Virologie | Barbara Mühlemann; Christian Drosten; Julia Schneider; Jörn Beheim-Schwarzbach; Talitha Veith; Terry Jones; Victor M Corman |
| EPI_ISL_2264643, EPI_ISL_2264644, EPI_ISL_2264646, EPI_ISL_2264647, EPI_ISL_2264648, EPI_ISL_2760267 | Chemische und Veterinäruntersuchungsamt Münsterland-Emscher-Lippe (CVUA-MEL) | Robert Koch Institute | |
| EPI_ISL_2263209, EPI_ISL_2263210, EPI_ISL_2263211, EPI_ISL_2264592, EPI_ISL_2264594, EPI_ISL_2264597, EPI_ISL_2264600, EPI_ISL_2264603, EPI_ISL_2264604, EPI_ISL_2264606, EPI_ISL_2264611, EPI_ISL_2315835, EPI_ISL_2315836, EPI_ISL_2315837, EPI_ISL_2315838, EPI_ISL_2315839, EPI_ISL_2315840, EPI_ISL_2315841, EPI_ISL_2315843, EPI_ISL_2315844, EPI_ISL_2315845, EPI_ISL_2315846, EPI_ISL_2315847, EPI_ISL_2315848, EPI_ISL_2315849, EPI_ISL_2315850, EPI_ISL_2315852, EPI_ISL_2315853, EPI_ISL_2315854, EPI_ISL_2315855, EPI_ISL_2315856, EPI_ISL_2315857, EPI_ISL_2315858, EPI_ISL_2315859, EPI_ISL_2315860, EPI_ISL_2315861, EPI_ISL_2315862, EPI_ISL_2315865, EPI_ISL_2315866, EPI_ISL_2315867, EPI_ISL_2315868, EPI_ISL_2315869, EPI_ISL_2315870, EPI_ISL_2315871, EPI_ISL_2390114, EPI_ISL_2390115, EPI_ISL_2390375, EPI_ISL_2390376, EPI_ISL_2390377, EPI_ISL_2390378, EPI_ISL_2390383, EPI_ISL_2390384, EPI_ISL_2390386, EPI_ISL_2390387, EPI_ISL_2390388, EPI_ISL_2390391, EPI_ISL_2390393, EPI_ISL_2390402, EPI_ISL_2390413 | | | |
| see above | Clemenshospital - Alexianer Clemenshospital Münster | Robert Koch Institute | |
| EPI_ISL_2632122, EPI_ISL_2632126, EPI_ISL_2632129, EPI_ISL_2632130, EPI_ISL_2632131, EPI_ISL_2632132, EPI_ISL_2632136, EPI_ISL_2632138, EPI_ISL_2632139, EPI_ISL_2632140 | | | |
| see above | Corona-Testzentrum ifp Institut für Produktqualität GmbH | Robert Koch Institute | |
| EPI_ISL_2266080, EPI_ISL_2266094, EPI_ISL_2266096, EPI_ISL_2266098, EPI_ISL_2471132, EPI_ISL_2471133, EPI_ISL_2471134, EPI_ISL_2471135, EPI_ISL_2471136, EPI_ISL_2471137, EPI_ISL_2471138, EPI_ISL_2471139, EPI_ISL_2471140, EPI_ISL_2471141, EPI_ISL_2471142, EPI_ISL_2471143, EPI_ISL_2471144, EPI_ISL_2471145, EPI_ISL_2471146, EPI_ISL_2471147, EPI_ISL_2471148, EPI_ISL_2471149, EPI_ISL_2471150, EPI_ISL_2471151, EPI_ISL_2471152, EPI_ISL_2471153, EPI_ISL_2471154, EPI_ISL_2471155, EPI_ISL_2471156, EPI_ISL_2471157, EPI_ISL_2471158, EPI_ISL_2471159, EPI_ISL_2471160, EPI_ISL_2471161, EPI_ISL_2471162, EPI_ISL_2471163, EPI_ISL_2471164, EPI_ISL_2471165, EPI_ISL_2471166, EPI_ISL_2471167, EPI_ISL_2471168, EPI_ISL_2471169, EPI_ISL_2471170, EPI_ISL_2471171, EPI_ISL_2471172, EPI_ISL_2471173, EPI_ISL_2471174, EPI_ISL_2471175, EPI_ISL_2471176, EPI_ISL_2471177, EPI_ISL_2471178, EPI_ISL_2471179, EPI_ISL_2471180, EPI_ISL_2471181, EPI_ISL_2471182, EPI_ISL_2759786 | | | |
| see above | Department für Labormedizin Abteilung III Bereich Molekulare Diagnostik Universitätsklinikum Halle(Saale) | Robert Koch Institute | |
| EPI_ISL_2314885, EPI_ISL_2314949, EPI_ISL_2314965, EPI_ISL_2315005, EPI_ISL_2315009, EPI_ISL_2315020, EPI_ISL_2315029, EPI_ISL_2315031, EPI_ISL_2470497, EPI_ISL_2470498, EPI_ISL_2470499, EPI_ISL_2470500, EPI_ISL_2470501, EPI_ISL_2470502, EPI_ISL_2470503, EPI_ISL_2470504, EPI_ISL_2470505, EPI_ISL_2470506, EPI_ISL_2470507, EPI_ISL_2470508, EPI_ISL_2470509, EPI_ISL_2470510, EPI_ISL_2470511, EPI_ISL_2470512, EPI_ISL_2470513, EPI_ISL_2470514, EPI_ISL_2470515, EPI_ISL_2470516, EPI_ISL_2470517, EPI_ISL_2470518, EPI_ISL_2470519, EPI_ISL_2470520, EPI_ISL_2470521, EPI_ISL_2470522, EPI_ISL_2470523, EPI_ISL_2470524, EPI_ISL_2470525, EPI_ISL_2470526, EPI_ISL_2470527, EPI_ISL_2470528, EPI_ISL_2470529, EPI_ISL_2470530, EPI_ISL_2470531, EPI_ISL_2470532, EPI_ISL_2470533, EPI_ISL_2470534, EPI_ISL_2470535, EPI_ISL_2470536, EPI_ISL_2470537, EPI_ISL_2470538, EPI_ISL_2470539, EPI_ISL_2470541, EPI_ISL_2470542, EPI_ISL_2470543, EPI_ISL_2470544, EPI_ISL_2470545, EPI_ISL_2470546, EPI_ISL_2470547, EPI_ISL_2470548, EPI_ISL_2470549, EPI_ISL_2470550, EPI_ISL_2470551, EPI_ISL_2470552, EPI_ISL_2470553, EPI_ISL_2470554, EPI_ISL_2470555, EPI_ISL_2470556, EPI_ISL_2470557, EPI_ISL_2470558, EPI_ISL_2470559, EPI_ISL_2470560, EPI_ISL_2470561, EPI_ISL_2470562, EPI_ISL_2470563, EPI_ISL_2470564, EPI_ISL_2470565, EPI_ISL_2470566, EPI_ISL_2470567, EPI_ISL_2470568, EPI_ISL_2470569, EPI_ISL_2470570, EPI_ISL_2470571, EPI_ISL_2470572, EPI_ISL_2470573, EPI_ISL_2470574, EPI_ISL_2470575, EPI_ISL_2470576, EPI_ISL_2470577, EPI_ISL_2470578, EPI_ISL_2470579, EPI_ISL_2470580, EPI_ISL_2470581, EPI_ISL_2470582, EPI_ISL_2470583, EPI_ISL_2470584, EPI_ISL_2470585, EPI_ISL_2470586, EPI_ISL_2470587, EPI_ISL_2470588, EPI_ISL_2470589, EPI_ISL_2470590, EPI_ISL_2470591, EPI_ISL_2470592, EPI_ISL_2470593, EPI_ISL_2470594, EPI_ISL_2470595, EPI_ISL_2470596, EPI_ISL_2470597, EPI_ISL_2470598, EPI_ISL_2470599, EPI_ISL_2470600, EPI_ISL_2470601, EPI_ISL_2470602, EPI_ISL_2470603, EPI_ISL_2470604, EPI_ISL_2470605, EPI_ISL_2470606, EPI_ISL_2470607, EPI_ISL_2470608, EPI_ISL_2470609, EPI_ISL_2470610, EPI_ISL_2470611, EPI_ISL_2470612, EPI_ISL_2470613, EPI_ISL_2470614, EPI_ISL_2470615, EPI_ISL_2470616, EPI_ISL_2470617, EPI_ISL_2470618, EPI_ISL_2470619, EPI_ISL_2470620, EPI_ISL_2470621, EPI_ISL_2470622, EPI_ISL_2470623, EPI_ISL_2470624, EPI_ISL_2470625, EPI_ISL_2470626, EPI_ISL_2470627, EPI_ISL_2470628, EPI_ISL_2470629, EPI_ISL_2470630, EPI_ISL_2470631, EPI_ISL_2470632, EPI_ISL_2470633, EPI_ISL_2470634, EPI_ISL_2470635, EPI_ISL_2470636, EPI_ISL_2470637, EPI_ISL_2470638, EPI_ISL_2470639, EPI_ISL_2470640, EPI_ISL_2470641, EPI_ISL_2470642, EPI_ISL_2470643, EPI_ISL_2470644, EPI_ISL_2470645, EPI_ISL_2470646, EPI_ISL_2470647, EPI_ISL_2470648, EPI_ISL_2470649, EPI_ISL_2470650, EPI_ISL_2636232, EPI_ISL_2636234, EPI_ISL_2636236, EPI_ISL_2636238, EPI_ISL_2636240, EPI_ISL_2636243, EPI_ISL_2636245, EPI_ISL_2636247, EPI_ISL_2636250, EPI_ISL_2636252, EPI_ISL_2636254, EPI_ISL_2636257, EPI_ISL_2636259, EPI_ISL_2636261, EPI_ISL_2636264, EPI_ISL_2636266, EPI_ISL_2636268, EPI_ISL_2636270, EPI_ISL_2636273, EPI_ISL_2636275, EPI_ISL_2636277, EPI_ISL_2636280, EPI_ISL_2636282, EPI_ISL_2636284, EPI_ISL_2636286, EPI_ISL_2636288, EPI_ISL_2636291, EPI_ISL_2636294, EPI_ISL_2636296, EPI_ISL_2636299, EPI_ISL_2636301, EPI_ISL_2636303, EPI_ISL_2636306, EPI_ISL_2636308, EPI_ISL_2636311, EPI_ISL_2636313, EPI_ISL_2636315, EPI_ISL_2636318, EPI_ISL_2636320, EPI_ISL_2636322, EPI_ISL_2636324, EPI_ISL_2636327, EPI_ISL_2636330, EPI_ISL_2636332, EPI_ISL_2636335, EPI_ISL_2636337, EPI_ISL_2636339, EPI_ISL_2636342, EPI_ISL_2636344, EPI_ISL_2636348, EPI_ISL_2636350, EPI_ISL_2636352, EPI_ISL_2636355, EPI_ISL_2636357, EPI_ISL_2636359, EPI_ISL_2636362, EPI_ISL_2636364, EPI_ISL_2636366, EPI_ISL_2636369, EPI_ISL_2636372, EPI_ISL_2636374, EPI_ISL_2636376, EPI_ISL_2636378, EPI_ISL_2636381, EPI_ISL_2636384, EPI_ISL_2636388, EPI_ISL_2636390, EPI_ISL_2636393, EPI_ISL_2636395, EPI_ISL_2636397, EPI_ISL_2636400, EPI_ISL_2636402, EPI_ISL_2636405, EPI_ISL_2636407, EPI_ISL_2636409, EPI_ISL_2636412, EPI_ISL_2636413, EPI_ISL_2636414, EPI_ISL_2636415, EPI_ISL_2636417, EPI_ISL_2636418, EPI_ISL_2636422, EPI_ISL_2636427, EPI_ISL_2636430, EPI_ISL_2636433, EPI_ISL_2636434, EPI_ISL_2636438, EPI_ISL_2636439, EPI_ISL_2636440, EPI_ISL_2636445, EPI_ISL_2636446, EPI_ISL_2636449, EPI_ISL_2636450, EPI_ISL_2636452, EPI_ISL_2636455, EPI_ISL_2636463, EPI_ISL_2636465, EPI_ISL_2636466, EPI_ISL_2636467, EPI_ISL_2636468, EPI_ISL_2636469, EPI_ISL_2636471, EPI_ISL_2636472 | | | |
| see above | Diagnosticum - Labor Neukirchen | Robert Koch Institute | |
| EPI_ISL_2264649, EPI_ISL_2264650, EPI_ISL_2264651, EPI_ISL_2264652, EPI_ISL_2264653, EPI_ISL_2264654, EPI_ISL_2264655, EPI_ISL_2264656, EPI_ISL_2264657, EPI_ISL_2264658, EPI_ISL_2264659, EPI_ISL_2264660, EPI_ISL_2264661, EPI_ISL_2264662, EPI_ISL_2264663, EPI_ISL_2264664, EPI_ISL_2386859, EPI_ISL_2386862, EPI_ISL_2386864, EPI_ISL_2386869, EPI_ISL_2386872, EPI_ISL_2386873, EPI_ISL_2386874 | | | |

| | | | |
|---|---|---|---|
| see above | Dianovis GmbH Greiz | Robert Koch Institute | |
| EPI_ISL_2262357, EPI_ISL_2262375, EPI_ISL_2262381, EPI_ISL_2262387, EPI_ISL_2313906, EPI_ISL_2313922, EPI_ISL_2313939, EPI_ISL_2313945, EPI_ISL_2313960, EPI_ISL_2313961, EPI_ISL_2313962 | | | |
| see above | ELBLAB GmbH Zentrum für LaborMedizin | Robert Koch Institute | |
| EPI_ISL_2471609, EPI_ISL_2471610, EPI_ISL_2471611, EPI_ISL_2471612, EPI_ISL_2471613, EPI_ISL_2471614, EPI_ISL_2471615, EPI_ISL_2471616, EPI_ISL_2471617, EPI_ISL_2471618, EPI_ISL_2471619, EPI_ISL_2471620, EPI_ISL_2471621, EPI_ISL_2471622, EPI_ISL_2471623, EPI_ISL_2471624, EPI_ISL_2471625, EPI_ISL_2471626, EPI_ISL_2471627, EPI_ISL_2471628, EPI_ISL_2471629, EPI_ISL_2471630, EPI_ISL_2471631, EPI_ISL_2471632, EPI_ISL_2471633, EPI_ISL_2471634, EPI_ISL_2471635, EPI_ISL_2471636, EPI_ISL_2471637, EPI_ISL_2471638, EPI_ISL_2471639, EPI_ISL_2471640, EPI_ISL_2471641, EPI_ISL_2471642, EPI_ISL_2471643, EPI_ISL_2471644, EPI_ISL_2471645, EPI_ISL_2471646, EPI_ISL_2471647, EPI_ISL_2471648, EPI_ISL_2471649, EPI_ISL_2471650, EPI_ISL_2471651, EPI_ISL_2471652, EPI_ISL_2471653, EPI_ISL_2471654, EPI_ISL_2471655, EPI_ISL_2471656, EPI_ISL_2471657, EPI_ISL_2471658, EPI_ISL_2471659, EPI_ISL_2471660, EPI_ISL_2471661, EPI_ISL_2471662, EPI_ISL_2471663, EPI_ISL_2471664, EPI_ISL_2471665, EPI_ISL_2471666, EPI_ISL_2471667, EPI_ISL_2471668, EPI_ISL_2471669, EPI_ISL_2471670, EPI_ISL_2471671, EPI_ISL_2471672, EPI_ISL_2471673, EPI_ISL_2471674, EPI_ISL_2471675, EPI_ISL_2471676, EPI_ISL_2471677, EPI_ISL_2471678, EPI_ISL_2471679, EPI_ISL_2471680, EPI_ISL_2471681, EPI_ISL_2471682, EPI_ISL_2471683, EPI_ISL_2471684, EPI_ISL_2471685, EPI_ISL_2471686, EPI_ISL_2471687, EPI_ISL_2471688, EPI_ISL_2471689, EPI_ISL_2471690, EPI_ISL_2472040, EPI_ISL_2472043, EPI_ISL_2472050, EPI_ISL_2472053, EPI_ISL_2472057, EPI_ISL_2472060, EPI_ISL_2472066, EPI_ISL_2472070, EPI_ISL_2472072, EPI_ISL_2472077, EPI_ISL_2472081, EPI_ISL_2472087, EPI_ISL_2472090, EPI_ISL_2472095, EPI_ISL_2472112, EPI_ISL_2472118, EPI_ISL_2472120, EPI_ISL_2472122, EPI_ISL_2472125, EPI_ISL_2472128, EPI_ISL_2472132, EPI_ISL_2472134, EPI_ISL_2472136, EPI_ISL_2472140, EPI_ISL_2472648, EPI_ISL_2472649, EPI_ISL_2472650, EPI_ISL_2472651, EPI_ISL_2472652, EPI_ISL_2472653, EPI_ISL_2472654, EPI_ISL_2472655, EPI_ISL_2472656, EPI_ISL_2472657, EPI_ISL_2472658, EPI_ISL_2472659, EPI_ISL_2472660, EPI_ISL_2472661, EPI_ISL_2472662, EPI_ISL_2472663, EPI_ISL_2472664, EPI_ISL_2472665, EPI_ISL_2472666, EPI_ISL_2472667, EPI_ISL_2472668, EPI_ISL_2472669, EPI_ISL_2472670, EPI_ISL_2472671, EPI_ISL_2472672, EPI_ISL_2472673, EPI_ISL_2472674, EPI_ISL_2472675, EPI_ISL_2472676, EPI_ISL_2472677, EPI_ISL_2472678, EPI_ISL_2472679, EPI_ISL_2472680, EPI_ISL_2472681, EPI_ISL_2472682, EPI_ISL_2472683, EPI_ISL_2472684, EPI_ISL_2472685, EPI_ISL_2472686, EPI_ISL_2472687, EPI_ISL_2472688, EPI_ISL_2472689, EPI_ISL_2472690, EPI_ISL_2472691, EPI_ISL_2472692, EPI_ISL_2472693, EPI_ISL_2472694, EPI_ISL_2472695, EPI_ISL_2631894, EPI_ISL_2631895, EPI_ISL_2631896, EPI_ISL_2631897, EPI_ISL_2631898, EPI_ISL_2631899, EPI_ISL_2631900, EPI_ISL_2631901, EPI_ISL_2631902, EPI_ISL_2631903, EPI_ISL_2631904, EPI_ISL_2631905, EPI_ISL_2631906, EPI_ISL_2631907, EPI_ISL_2631908, EPI_ISL_2631909, EPI_ISL_2631910, EPI_ISL_2631911, EPI_ISL_2631912, EPI_ISL_2631913, EPI_ISL_2631914, EPI_ISL_2631915, EPI_ISL_2631916, EPI_ISL_2631917, EPI_ISL_2631918, EPI_ISL_2631919, EPI_ISL_2631920, EPI_ISL_2631921, EPI_ISL_2631922, EPI_ISL_2631923, EPI_ISL_2631924, EPI_ISL_2631925, EPI_ISL_2631926, EPI_ISL_2631927, EPI_ISL_2631928, EPI_ISL_2631929, EPI_ISL_2631930, EPI_ISL_2631931 | | | |
| see above | Eurofins LifeCodexx GmbH | Robert Koch Institute | |
| EPI_ISL_2259784, EPI_ISL_2259785, EPI_ISL_2259786 | Eurofins MVZ Labor Gelsenkirchen | Robert Koch Institute | |
| EPI_ISL_2386289, EPI_ISL_2386290, EPI_ISL_2386291, EPI_ISL_2386292, EPI_ISL_2386293, EPI_ISL_2386294, EPI_ISL_2386295, EPI_ISL_2386296, EPI_ISL_2386297, EPI_ISL_2386298, EPI_ISL_2386299, EPI_ISL_2390444, EPI_ISL_2390445, EPI_ISL_2390446, EPI_ISL_2390448, EPI_ISL_2390451, EPI_ISL_2390454, EPI_ISL_2390455, EPI_ISL_2390458, EPI_ISL_2390460 | | | |
| see above | Fraunhofer-Institut für Zelltherapie und Immunologie IZI AG Next-Generation Diagnostics | Robert Koch Institute | |
| EPI_ISL_2471817, EPI_ISL_2471818, EPI_ISL_2471823, EPI_ISL_2471827, EPI_ISL_2471830 | Gesundheitszentrum Brandenburg an der Havel GmbH bzw. Institut für Laboratoriumsmedizin Brandenburg an der Havel | Robert Koch Institute | |
| EPI_ISL_2635926, EPI_ISL_2635927, EPI_ISL_2635928, EPI_ISL_2635929, EPI_ISL_2635930, EPI_ISL_2635931, EPI_ISL_2635932, EPI_ISL_2635933, EPI_ISL_2635934, EPI_ISL_2635935, EPI_ISL_2635936 | | | |
| see above | Helios Universitätsklinikum Wuppertal: Universität Witten/Herdecke: Institut für Medizinische Labordiagnostik | Robert Koch Institute | |
| EPI_ISL_2265053, EPI_ISL_2265109 | Hufeland Klinikum GmbH | Robert Koch Institute | |
| EPI_ISL_2266136, EPI_ISL_2266139, EPI_ISL_2266145, EPI_ISL_2266148, EPI_ISL_2266154, EPI_ISL_2266157, EPI_ISL_2266158, EPI_ISL_2266161, EPI_ISL_2266163, EPI_ISL_2389995 | | | |
| see above | IMD - Institut für Medizinische Diagnostik Berlin-Potsdam | Robert Koch Institute | |
| EPI_ISL_2314040, EPI_ISL_2314046, EPI_ISL_2314049, EPI_ISL_2314050, EPI_ISL_2314051, EPI_ISL_2314054, EPI_ISL_2314055, EPI_ISL_2314056, EPI_ISL_2314057, EPI_ISL_2314058, EPI_ISL_2314059, EPI_ISL_2314061, EPI_ISL_2314062, EPI_ISL_2314063, EPI_ISL_2314064, EPI_ISL_2314065, EPI_ISL_2314066, EPI_ISL_2314067, EPI_ISL_2314068, EPI_ISL_2314069, EPI_ISL_2314072, EPI_ISL_2314073, EPI_ISL_2314074, EPI_ISL_2314076, EPI_ISL_2389969, EPI_ISL_2389970, EPI_ISL_2389971, EPI_ISL_2389973, EPI_ISL_2389977, EPI_ISL_2389979, EPI_ISL_2389981, EPI_ISL_2389985, EPI_ISL_2389986, EPI_ISL_2389987, EPI_ISL_2389988, EPI_ISL_2471992, EPI_ISL_2472004, EPI_ISL_2472007 | | | |
| see above | IMD - Labor Oderland | Robert Koch Institute | |
| EPI_ISL_2266138, EPI_ISL_2266140, EPI_ISL_2266143, EPI_ISL_2266146, EPI_ISL_2266147, EPI_ISL_2266149, EPI_ISL_2266150, EPI_ISL_2266155, EPI_ISL_2266156, EPI_ISL_2266159, EPI_ISL_2266160, EPI_ISL_2266162, EPI_ISL_2266164, EPI_ISL_2266166, EPI_ISL_2389991, EPI_ISL_2389993, EPI_ISL_2389996, EPI_ISL_2389998, EPI_ISL_2390001, EPI_ISL_2390002, EPI_ISL_2390004, EPI_ISL_2390008, EPI_ISL_2390010 | | | |
| see above | IMD - MVZ Labor Greifswald | Robert Koch Institute | |
| EPI_ISL_2262420, EPI_ISL_2262428, EPI_ISL_2266141, EPI_ISL_2266142, EPI_ISL_2266152, EPI_ISL_2266153, EPI_ISL_2314041, EPI_ISL_2314052, EPI_ISL_2314075, EPI_ISL_2314077, EPI_ISL_2314425, EPI_ISL_2389992, EPI_ISL_2389994, EPI_ISL_2389997, EPI_ISL_2390000, EPI_ISL_2390003, EPI_ISL_2390005, EPI_ISL_2390006, EPI_ISL_2390007, EPI_ISL_2390009 | | | |
| see above | IMD - MVZ Labor Martinsried | Robert Koch Institute | |
| EPI_ISL_2314039, EPI_ISL_2314042, EPI_ISL_2314043, EPI_ISL_2314044, EPI_ISL_2314045, EPI_ISL_2314047, EPI_ISL_2314048, EPI_ISL_2314053, EPI_ISL_2314060, EPI_ISL_2314071, EPI_ISL_2389975, EPI_ISL_2389976, EPI_ISL_2389982, EPI_ISL_2389984, EPI_ISL_2389990 | | | |
| see above | IMD - Medizinisches Labor Rostock | Robert Koch Institute | |
| EPI_ISL_2264490, EPI_ISL_2264495, EPI_ISL_2264497, EPI_ISL_2264500, EPI_ISL_2264504, EPI_ISL_2264506, EPI_ISL_2264508, EPI_ISL_2264509, EPI_ISL_2264513, EPI_ISL_2264514, EPI_ISL_2264516, EPI_ISL_2264521, EPI_ISL_2264527, EPI_ISL_2264530, EPI_ISL_2264535, EPI_ISL_2313966, EPI_ISL_2313968, EPI_ISL_2313969, EPI_ISL_2313970, EPI_ISL_2313971, EPI_ISL_2313972, EPI_ISL_2313973, EPI_ISL_2313975, EPI_ISL_2313976, EPI_ISL_2313977, EPI_ISL_2313979, EPI_ISL_2313980, EPI_ISL_2313981, EPI_ISL_2313983, EPI_ISL_2313985, EPI_ISL_2313986, EPI_ISL_2313987, EPI_ISL_2313988, EPI_ISL_2313989, EPI_ISL_2313990, EPI_ISL_2313992, EPI_ISL_2313993, EPI_ISL_2313994, EPI_ISL_2313995, EPI_ISL_2313996, EPI_ISL_2313997, EPI_ISL_2313998, EPI_ISL_2314000, EPI_ISL_2314001, EPI_ISL_2314002, EPI_ISL_2314003, EPI_ISL_2314004, EPI_ISL_2314006, EPI_ISL_2314008, EPI_ISL_2314010, EPI_ISL_2314015, EPI_ISL_2314016, EPI_ISL_2314018, EPI_ISL_2314019, EPI_ISL_2314021, EPI_ISL_2314024, EPI_ISL_2314025, EPI_ISL_2314027, EPI_ISL_2314029, EPI_ISL_2314030, EPI_ISL_2314032, EPI_ISL_2314033, EPI_ISL_2314034, EPI_ISL_2314037, EPI_ISL_2389384, EPI_ISL_2389385, EPI_ISL_2389388, EPI_ISL_2389389, EPI_ISL_2389391, EPI_ISL_2389402, EPI_ISL_2389408, EPI_ISL_2389409, EPI_ISL_2389412, EPI_ISL_2389413, EPI_ISL_2389414, EPI_ISL_2389415, EPI_ISL_2389423, EPI_ISL_2389424, EPI_ISL_2389432, EPI_ISL_2389433, EPI_ISL_2389435 | | | |
| see above | IMD Labor Frankfurt | Robert Koch Institute | |
| EPI_ISL_2267932, EPI_ISL_2267949 | Institut für Klinische Chemie Transfusions- und Laboratoriumsmedizin Berufsgenossenschaftliches Universitätsklinikum Bergmannsheil | Robert Koch Institute | |
| EPI_ISL_2314317, EPI_ISL_2386875, EPI_ISL_2471750 | Institut für Klinische Transfusionsmedizin und Immungenetik DRK Blutspendedienst Baden-Württemberg | Robert Koch Institute | |
| EPI_ISL_2266414, EPI_ISL_2631860, EPI_ISL_2631861, EPI_ISL_2631866, EPI_ISL_2631868, EPI_ISL_2631869, EPI_ISL_2635797, EPI_ISL_2844881 | | | |
| see above | Institut für Medizinische Mikrobiologie - Universitätsmedizin Göttingen | Robert Koch Institute | |
| EPI_ISL_2389378, EPI_ISL_2389379, EPI_ISL_2389380, EPI_ISL_2389381, EPI_ISL_2389382, EPI_ISL_2472540 | Institut für med. Mikrobiologie u. Virologie Klinikum Oldenburg | Robert Koch Institute | |
| EPI_ISL_2285551, EPI_ISL_2285552, EPI_ISL_2285553, EPI_ISL_2285596, EPI_ISL_2285604, EPI_ISL_2285605, EPI_ISL_2285606, EPI_ISL_2285624, EPI_ISL_2285625, EPI_ISL_2285626, EPI_ISL_2285627, EPI_ISL_2285628, EPI_ISL_2285629, EPI_ISL_2285630, EPI_ISL_2285631, EPI_ISL_2285632, EPI_ISL_2285648, EPI_ISL_2285649, EPI_ISL_2285650, EPI_ISL_2285651, EPI_ISL_2285689, EPI_ISL_2285690, EPI_ISL_2285691, EPI_ISL_2285692, EPI_ISL_2285693, EPI_ISL_2285694, EPI_ISL_2285695, EPI_ISL_2285696 | | | |
| see above | Institute of Medical Microbiology and Hospital Hygiene | Institute of Medical Microbiology and Hospital Hygiene | Aljoscha Tersteegen; Prof. Dr. Achim Kaasch |
| EPI_ISL_2535306, EPI_ISL_2535307, EPI_ISL_2535308, EPI_ISL_2535339, EPI_ISL_2535342, EPI_ISL_2535343, EPI_ISL_2535358, EPI_ISL_2535359, EPI_ISL_2535360, EPI_ISL_2535361, EPI_ISL_2535362, EPI_ISL_2535363, EPI_ISL_2535364, EPI_ISL_2535365, EPI_ISL_2535366, EPI_ISL_2535367, EPI_ISL_2535368, EPI_ISL_2535369, EPI_ISL_2535378, EPI_ISL_2535379, EPI_ISL_2535380, EPI_ISL_2535381, EPI_ISL_2535382, EPI_ISL_2535383, EPI_ISL_2535384, EPI_ISL_2535385, EPI_ISL_2535388, EPI_ISL_2535389, EPI_ISL_2535390, EPI_ISL_2535391, EPI_ISL_2535392, EPI_ISL_2535393, EPI_ISL_2535394, EPI_ISL_2535395, EPI_ISL_2535396, EPI_ISL_2535407, EPI_ISL_2535408, EPI_ISL_2535409, EPI_ISL_2535410, EPI_ISL_2535411, EPI_ISL_2535412, EPI_ISL_2535434, EPI_ISL_2535435 | | | |
| see above | Institute of Virology, Medical Center, University of Freiburg, Freiburg, Germany | Institute of Virology, Clinial Virus Genomics, Medical Center, University of Freiburg, Freiburg, Germany | Hajo Grundmann; Jonas Fuchs; Lena Jaki; Lisa Kern; Marcus Panning; Sandra Reuter |
| EPI_ISL_2678804 | KRH Klinikum Region Hannover: Institut für Med. Mikrobiologie und Krankenhaushygiene | Robert Koch Institute | |
| EPI_ISL_2385072 | Kath. Marienkrankenhaus | Heinrich Pette Institute, Leibniz Institute for Experimental Virology | Adam Grundhoff; Alexis Robitaille; Johannes Knobloch; Martin Aepfelbacher; Nicole Fischer; Thomas Günther |
| EPI_ISL_2388253 | Kath. St.-Johannes-Gesellschaft Dortmund gGmbH | Robert Koch Institute | |
| EPI_ISL_2313359 | Kerckhoff-Klinik GmbH | Robert Koch Institute | |
| EPI_ISL_2263115, EPI_ISL_2313219, EPI_ISL_2313220 | Klinikum Darmstadt GmbH | Robert Koch Institute | |
| EPI_ISL_2264743, EPI_ISL_2264746, EPI_ISL_2264750, EPI_ISL_2264751, EPI_ISL_2264752, EPI_ISL_2264753, EPI_ISL_2264756, EPI_ISL_2264757, EPI_ISL_2264758, EPI_ISL_2264760, EPI_ISL_2264762, EPI_ISL_2264763, EPI_ISL_2264764, EPI_ISL_2264765, EPI_ISL_2264768, EPI_ISL_2264770, EPI_ISL_2264772, EPI_ISL_2264774, EPI_ISL_2264776, EPI_ISL_2264778, EPI_ISL_2264780, EPI_ISL_2264783, EPI_ISL_2264788, EPI_ISL_2264790, EPI_ISL_2264792, EPI_ISL_2264794, EPI_ISL_2264796, EPI_ISL_2264800, EPI_ISL_2264803, EPI_ISL_2264805, EPI_ISL_2264807, EPI_ISL_2264809, EPI_ISL_2316615, EPI_ISL_2316617, EPI_ISL_2316620, EPI_ISL_2316622, EPI_ISL_2316625, EPI_ISL_2316628, EPI_ISL_2316631, EPI_ISL_2316635, EPI_ISL_2316638, EPI_ISL_2316642, EPI_ISL_2316644 | | | |
| see above | Klinikum Ernst von Bergmann gemeinnützige GmbH - stationärer Bereich | Robert Koch Institute | |
| EPI_ISL_2314087, EPI_ISL_2388319, EPI_ISL_2388488 | Klinikum Fulda gAG | Robert Koch Institute | |
| EPI_ISL_2386719, EPI_ISL_2386758, EPI_ISL_2386763, EPI_ISL_2386782, EPI_ISL_2390735, EPI_ISL_2390745, EPI_ISL_2390747 | | | |
| see above | Klinikum Osnabrück Medizinisches Labor | Robert Koch Institute | |
| EPI_ISL_2264730 | Krankenhausgesellschaft St. Vincenz mbH | Robert Koch Institute | |
| EPI_ISL_2262340, EPI_ISL_2262342, EPI_ISL_2262351, EPI_ISL_2262352, EPI_ISL_2262368, EPI_ISL_2262380, EPI_ISL_2262389, EPI_ISL_2313931, EPI_ISL_2313935, EPI_ISL_2313936, EPI_ISL_2313942, EPI_ISL_2313944, EPI_ISL_2313948, EPI_ISL_2313953 | | | |
| see above | LABOR CHEMNITZ – Zentrum für Diagnostik am Klinikum Chemnitz | Robert Koch Institute | |
| EPI_ISL_2388321, EPI_ISL_2388323, EPI_ISL_2388328, EPI_ISL_2388333, EPI_ISL_2388337, EPI_ISL_2388352, EPI_ISL_2388353, EPI_ISL_2388360, EPI_ISL_2388362, EPI_ISL_2388376, EPI_ISL_2388378, EPI_ISL_2388388, EPI_ISL_2388392, EPI_ISL_2388396, EPI_ISL_2388398, EPI_ISL_2388402, EPI_ISL_2388404, EPI_ISL_2388408, EPI_ISL_2388413, EPI_ISL_2388420, EPI_ISL_2388426, EPI_ISL_2388443, EPI_ISL_2388445, EPI_ISL_2388447, EPI_ISL_2388449, EPI_ISL_2388453, EPI_ISL_2388464, EPI_ISL_2388470, EPI_ISL_2388473, EPI_ISL_2388475, EPI_ISL_2388490, EPI_ISL_2388496, EPI_ISL_2388500, EPI_ISL_2388506, EPI_ISL_2388513, EPI_ISL_2388515, EPI_ISL_2388523, EPI_ISL_2388530, EPI_ISL_2388532, EPI_ISL_2388534, EPI_ISL_2388538, EPI_ISL_2388543, EPI_ISL_2388545, EPI_ISL_2388547, EPI_ISL_2388549, EPI_ISL_2388555, EPI_ISL_2388563, EPI_ISL_2388565, EPI_ISL_2388567, EPI_ISL_2388571, EPI_ISL_2388575, EPI_ISL_2388580, EPI_ISL_2388588, EPI_ISL_2388594, EPI_ISL_2388597, EPI_ISL_2388601, EPI_ISL_2388603, EPI_ISL_2388605, EPI_ISL_2388607, EPI_ISL_2388609, EPI_ISL_2388615, EPI_ISL_2388640, EPI_ISL_2388649, EPI_ISL_2388653, EPI_ISL_2388657, EPI_ISL_2388663, EPI_ISL_2388664, EPI_ISL_2388668, EPI_ISL_2388669, EPI_ISL_2388673, EPI_ISL_2388675, EPI_ISL_2388678, EPI_ISL_2388679, EPI_ISL_2388680, EPI_ISL_2388693, EPI_ISL_2388694, EPI_ISL_2388702, EPI_ISL_2388704, EPI_ISL_2388705, EPI_ISL_2388707, EPI_ISL_2388712, EPI_ISL_2388714, EPI_ISL_2388715, EPI_ISL_2388716, EPI_ISL_2388717, EPI_ISL_2388719, EPI_ISL_2388721, EPI_ISL_2388722, EPI_ISL_2388724, EPI_ISL_2388730, EPI_ISL_2388736, EPI_ISL_2388739, EPI_ISL_2388747, EPI_ISL_2388749, EPI_ISL_2388751, EPI_ISL_2388754, EPI_ISL_2388755, EPI_ISL_2388756, EPI_ISL_2388760, EPI_ISL_2388762, EPI_ISL_2388763, EPI_ISL_2388766, EPI_ISL_2388767, EPI_ISL_2388775, EPI_ISL_2388778, EPI_ISL_2388779, EPI_ISL_2388780, EPI_ISL_2388781, EPI_ISL_2388787, EPI_ISL_2388789, EPI_ISL_2388790, EPI_ISL_2388792, EPI_ISL_2388794, EPI_ISL_2388795, EPI_ISL_2388797, EPI_ISL_2388799, EPI_ISL_2388802 | | | |
| see above | LabKom - Labor Augsburg MVZ GmbH | Robert Koch Institute | |
| EPI_ISL_2388434, EPI_ISL_2388480, EPI_ISL_2388528, EPI_ISL_2388655, EPI_ISL_2388759 | LabKom - Labor Hamburg-Lübeck | Robert Koch Institute | |
| EPI_ISL_2314086, EPI_ISL_2314105, EPI_ISL_2314107, EPI_ISL_2314114, EPI_ISL_2314115, EPI_ISL_2314126, EPI_ISL_2314139, EPI_ISL_2314147, EPI_ISL_2314149, EPI_ISL_2314200, EPI_ISL_2314208, EPI_ISL_2314217, EPI_ISL_2314225, EPI_ISL_2314229, EPI_ISL_2314243, EPI_ISL_2314259, EPI_ISL_2314269, EPI_ISL_2314277, EPI_ISL_2314290, EPI_ISL_2314294, EPI_ISL_2314296, EPI_ISL_2314302 | | | |
| see above | LabKom - Labor Hannover MVZ GmbH | Robert Koch Institute | |
| EPI_ISL_2314082, EPI_ISL_2314098, EPI_ISL_2314099, EPI_ISL_2314112, EPI_ISL_2314120, EPI_ISL_2314129, EPI_ISL_2314137, EPI_ISL_2314152, EPI_ISL_2314155, EPI_ISL_2314164, EPI_ISL_2314179, EPI_ISL_2314189, EPI_ISL_2314191, EPI_ISL_2314192, EPI_ISL_2314203, EPI_ISL_2314232, EPI_ISL_2314242, EPI_ISL_2314254, EPI_ISL_2314255, EPI_ISL_2314264, EPI_ISL_2314295, EPI_ISL_2314299, EPI_ISL_2388394, EPI_ISL_2388468, EPI_ISL_2388484, EPI_ISL_2388573, EPI_ISL_2388710, EPI_ISL_2388734, EPI_ISL_2388796, EPI_ISL_2388828, EPI_ISL_2388832, EPI_ISL_2388845, EPI_ISL_2388864, EPI_ISL_2388971, EPI_ISL_2388987, EPI_ISL_2388995, EPI_ISL_2389027, EPI_ISL_2389041, EPI_ISL_2389067 | | | |
| see above | LabKom - Labor Mainz MVZ GmbH | Robert Koch Institute | |
| EPI_ISL_2313373, EPI_ISL_2313382, EPI_ISL_2313383, EPI_ISL_2313391, EPI_ISL_2313393, EPI_ISL_2313400, EPI_ISL_2313402, EPI_ISL_2313404, EPI_ISL_2313407, EPI_ISL_2313409, EPI_ISL_2313420, EPI_ISL_2313425, EPI_ISL_2313428, EPI_ISL_2313433, EPI_ISL_2313436, EPI_ISL_2313440, EPI_ISL_2313442, EPI_ISL_2313455, EPI_ISL_2313456, EPI_ISL_2313459, EPI_ISL_2313464, EPI_ISL_2313471, EPI_ISL_2313480, EPI_ISL_2313482, EPI_ISL_2313484, EPI_ISL_2313493, EPI_ISL_2313498, EPI_ISL_2313512, EPI_ISL_2313522, EPI_ISL_2313524, EPI_ISL_2313526, EPI_ISL_2313552, EPI_ISL_2313554, EPI_ISL_2313561, EPI_ISL_2313571, EPI_ISL_2313574, EPI_ISL_2313577, EPI_ISL_2313600, EPI_ISL_2313609, EPI_ISL_2313614, EPI_ISL_2313618, EPI_ISL_2313631, EPI_ISL_2313643, EPI_ISL_2313652, EPI_ISL_2313656, EPI_ISL_2313700, EPI_ISL_2313712, EPI_ISL_2313714, EPI_ISL_2313761, EPI_ISL_2313766, EPI_ISL_2313784, EPI_ISL_2313785, EPI_ISL_2313790, EPI_ISL_2313804, EPI_ISL_2313809, EPI_ISL_2313815, EPI_ISL_2313837, EPI_ISL_2313851, EPI_ISL_2313882 | | | |
| see above | LabKom - Labor an der Salzbrücke MVZ GmbH | Robert Koch Institute | |

| | | | |
|---|---|---|---|
| EPI_ISL_2265552, EPI_ISL_2314083, EPI_ISL_2314088, EPI_ISL_2314092, EPI_ISL_2314101, EPI_ISL_2314106, EPI_ISL_2314110, EPI_ISL_2314113, EPI_ISL_2314118, EPI_ISL_2314119, EPI_ISL_2314121, EPI_ISL_2314132, EPI_ISL_2314134, EPI_ISL_2314140, EPI_ISL_2314143, EPI_ISL_2314157, EPI_ISL_2314158, EPI_ISL_2314159, EPI_ISL_2314165, EPI_ISL_2314171, EPI_ISL_2314173, EPI_ISL_2314174, EPI_ISL_2314178, EPI_ISL_2314180, EPI_ISL_2314184, EPI_ISL_2314188, EPI_ISL_2314193, EPI_ISL_2314201, EPI_ISL_2314205, EPI_ISL_2314207, EPI_ISL_2314209, EPI_ISL_2314212, EPI_ISL_2314213, EPI_ISL_2314214, EPI_ISL_2314215, EPI_ISL_2314221, EPI_ISL_2314228, EPI_ISL_2314234, EPI_ISL_2314236, EPI_ISL_2314241, EPI_ISL_2314244, EPI_ISL_2314246, EPI_ISL_2314249, EPI_ISL_2314253, EPI_ISL_2314257, EPI_ISL_2314260, EPI_ISL_2314261, EPI_ISL_2314270, EPI_ISL_2314276, EPI_ISL_2314293, EPI_ISL_2314298, EPI_ISL_2314300, EPI_ISL_2314301, EPI_ISL_2314306 | | | |
| see above | LabKom - MVZ Labor Bochum MLB GmbH | Robert Koch Institute | |
| EPI_ISL_2313375, EPI_ISL_2313376, EPI_ISL_2313378, EPI_ISL_2313421, EPI_ISL_2313424, EPI_ISL_2313446, EPI_ISL_2313453, EPI_ISL_2313458, EPI_ISL_2313466, EPI_ISL_2313474, EPI_ISL_2313507, EPI_ISL_2313516, EPI_ISL_2313519, EPI_ISL_2313563, EPI_ISL_2313611, EPI_ISL_2313634, EPI_ISL_2313650, EPI_ISL_2313702, EPI_ISL_2313710, EPI_ISL_2313717, EPI_ISL_2313721, EPI_ISL_2313788, EPI_ISL_2313798, EPI_ISL_2313822, EPI_ISL_2313849, EPI_ISL_2313881, EPI_ISL_2388374, EPI_ISL_2388417, EPI_ISL_2388451, EPI_ISL_2388462, EPI_ISL_2388611, EPI_ISL_2388671, EPI_ISL_2388672, EPI_ISL_2388690, EPI_ISL_2388697, EPI_ISL_2388698, EPI_ISL_2388742, EPI_ISL_2388744, EPI_ISL_2388748, EPI_ISL_2469951, EPI_ISL_2469965, EPI_ISL_2469971, EPI_ISL_2469990, EPI_ISL_2470000, EPI_ISL_2470001, EPI_ISL_2470023, EPI_ISL_2470045, EPI_ISL_2470136, EPI_ISL_2470137, EPI_ISL_2470151 | | | |
| see above | Labor 28 MVZ GmbH | Robert Koch Institute | |
| EPI_ISL_2635527, EPI_ISL_2635528, EPI_ISL_2635529, EPI_ISL_2635530, EPI_ISL_2635531, EPI_ISL_2635532, EPI_ISL_2635533, EPI_ISL_2635534, EPI_ISL_2635535, EPI_ISL_2635536, EPI_ISL_2635537, EPI_ISL_2635538, EPI_ISL_2635539, EPI_ISL_2635540, EPI_ISL_2635541, EPI_ISL_2635542, EPI_ISL_2635543, EPI_ISL_2635544, EPI_ISL_2635545, EPI_ISL_2635546, EPI_ISL_2635547, EPI_ISL_2635548, EPI_ISL_2635549, EPI_ISL_2635550, EPI_ISL_2635551, EPI_ISL_2635552, EPI_ISL_2635553, EPI_ISL_2635554, EPI_ISL_2635555, EPI_ISL_2635556, EPI_ISL_2635557, EPI_ISL_2635558, EPI_ISL_2635559, EPI_ISL_2635560, EPI_ISL_2635561, EPI_ISL_2635562, EPI_ISL_2635563, EPI_ISL_2635908, EPI_ISL_2635911, EPI_ISL_2635912, EPI_ISL_2635913, EPI_ISL_2635914, EPI_ISL_2635916, EPI_ISL_2635917, EPI_ISL_2635919, EPI_ISL_2635920, EPI_ISL_2635922, EPI_ISL_2635923, EPI_ISL_2635924, EPI_ISL_2678419 | | | |
| see above | Labor Becker & Kollegen (Standort München) | Robert Koch Institute | |
| EPI_ISL_2227365, EPI_ISL_2339452, EPI_ISL_2339453, EPI_ISL_2339454, EPI_ISL_2339455, EPI_ISL_2339456, EPI_ISL_2339457, EPI_ISL_2339458, EPI_ISL_2339459, EPI_ISL_2339460, EPI_ISL_2339461, EPI_ISL_2339462, EPI_ISL_2339463, EPI_ISL_2339464, EPI_ISL_2339466, EPI_ISL_2339467, EPI_ISL_2339468, EPI_ISL_2339469, EPI_ISL_2339470, EPI_ISL_2339471, EPI_ISL_2339472, EPI_ISL_2339473, EPI_ISL_2339474, EPI_ISL_2339475, EPI_ISL_2339477, EPI_ISL_2339478, EPI_ISL_2339479, EPI_ISL_2339480, EPI_ISL_2339481, EPI_ISL_2339482, EPI_ISL_2339483, EPI_ISL_2339484, EPI_ISL_2339485, EPI_ISL_2339486, EPI_ISL_2339487, EPI_ISL_2339488, EPI_ISL_2339490, EPI_ISL_2339491, EPI_ISL_2339492, EPI_ISL_2339493, EPI_ISL_2339494, EPI_ISL_2339495, EPI_ISL_2339496, EPI_ISL_2339497, EPI_ISL_2339498, EPI_ISL_2339500, EPI_ISL_2339501, EPI_ISL_2382570, EPI_ISL_2382571, EPI_ISL_2382572, EPI_ISL_2382575, EPI_ISL_2382580, EPI_ISL_2382581, EPI_ISL_2382584, EPI_ISL_2382587, EPI_ISL_2382588, EPI_ISL_2382589, EPI_ISL_2382590, EPI_ISL_2382591, EPI_ISL_2382595, EPI_ISL_2382597, EPI_ISL_2382598, EPI_ISL_2382603, EPI_ISL_2382605, EPI_ISL_2382606, EPI_ISL_2382607, EPI_ISL_2382609, EPI_ISL_2382610, EPI_ISL_2382613, EPI_ISL_2382614, EPI_ISL_2382617, EPI_ISL_2382618, EPI_ISL_2382620, EPI_ISL_2382623, EPI_ISL_2382624, EPI_ISL_2382625, EPI_ISL_2382629, EPI_ISL_2382631, EPI_ISL_2382633, EPI_ISL_2382636, EPI_ISL_2382637, EPI_ISL_2382640, EPI_ISL_2382644, EPI_ISL_2382645, EPI_ISL_2382648, EPI_ISL_2466772 | | | |
| see above | Labor Berlin Charité Vivantes GmbH / Institut für Virologie | Charité Universitätsmedizin Berlin, Institut für Virologie/Labor Berlin | Barbara Mühlemann; Christian Drosten; Christine Stephan; Peter Menzel; Rolf Schwarzer; Terry Jones; Victor M Corman |
| EPI_ISL_2262450, EPI_ISL_2262451, EPI_ISL_2262452, EPI_ISL_2262453, EPI_ISL_2262454, EPI_ISL_2262457, EPI_ISL_2262458, EPI_ISL_2262459, EPI_ISL_2262461, EPI_ISL_2262462, EPI_ISL_2262463, EPI_ISL_2262464, EPI_ISL_2262466, EPI_ISL_2262467, EPI_ISL_2262468, EPI_ISL_2262469, EPI_ISL_2262471, EPI_ISL_2262475, EPI_ISL_2262477, EPI_ISL_2262478, EPI_ISL_2262479, EPI_ISL_2262481, EPI_ISL_2262482, EPI_ISL_2262484, EPI_ISL_2262488, EPI_ISL_2262489, EPI_ISL_2262490, EPI_ISL_2262491, EPI_ISL_2262494, EPI_ISL_2262498, EPI_ISL_2262505, EPI_ISL_2262512, EPI_ISL_2262514, EPI_ISL_2262516, EPI_ISL_2262525, EPI_ISL_2262527, EPI_ISL_2262530, EPI_ISL_2262536, EPI_ISL_2262538, EPI_ISL_2262541, EPI_ISL_2262543, EPI_ISL_2262545, EPI_ISL_2262553, EPI_ISL_2262556, EPI_ISL_2262562, EPI_ISL_2262567, EPI_ISL_2262571, EPI_ISL_2262576, EPI_ISL_2262578, EPI_ISL_2262580, EPI_ISL_2262583, EPI_ISL_2262585, EPI_ISL_2262589, EPI_ISL_2262593, EPI_ISL_2262596, EPI_ISL_2262598, EPI_ISL_2262600, EPI_ISL_2262605, EPI_ISL_2262610, EPI_ISL_2262611, EPI_ISL_2262616, EPI_ISL_2262619, EPI_ISL_2262621, EPI_ISL_2262623, EPI_ISL_2262625, EPI_ISL_2262630 | | | |
| see above | Labor Blackholm MVZ | Robert Koch Institute | |
| EPI_ISL_2689167, EPI_ISL_2689168, EPI_ISL_2689169, EPI_ISL_2689170, EPI_ISL_2689171, EPI_ISL_2689172, EPI_ISL_2689173, EPI_ISL_2689174, EPI_ISL_2689175, EPI_ISL_2689176, EPI_ISL_2689177, EPI_ISL_2689178, EPI_ISL_2689179, EPI_ISL_2689180, EPI_ISL_2689181, EPI_ISL_2689182, EPI_ISL_2689183, EPI_ISL_2689184, EPI_ISL_2689185, EPI_ISL_2689186, EPI_ISL_2689187, EPI_ISL_2689188, EPI_ISL_2689189, EPI_ISL_2689190, EPI_ISL_2689191, EPI_ISL_2689192, EPI_ISL_2689193, EPI_ISL_2689194, EPI_ISL_2689195, EPI_ISL_2689196, EPI_ISL_2689197, EPI_ISL_2689198, EPI_ISL_2689199, EPI_ISL_2689200, EPI_ISL_2689201, EPI_ISL_2689202, EPI_ISL_2689203, EPI_ISL_2689204, EPI_ISL_2689205, EPI_ISL_2689206, EPI_ISL_2689207, EPI_ISL_2689208, EPI_ISL_2689209, EPI_ISL_2689210, EPI_ISL_2689211, EPI_ISL_2689212, EPI_ISL_2689213, EPI_ISL_2689214, EPI_ISL_2689215, EPI_ISL_2689216, EPI_ISL_2689217, EPI_ISL_2689218, EPI_ISL_2689219, EPI_ISL_2689220, EPI_ISL_2689221, EPI_ISL_2689222, EPI_ISL_2689223, EPI_ISL_2689224, EPI_ISL_2689225, EPI_ISL_2689226, EPI_ISL_2689227, EPI_ISL_2689228, EPI_ISL_2689229, EPI_ISL_2689230, EPI_ISL_2689231, EPI_ISL_2689232, EPI_ISL_2689233, EPI_ISL_2689234, EPI_ISL_2689235, EPI_ISL_2689236, EPI_ISL_2689237, EPI_ISL_2689238, EPI_ISL_2689239, EPI_ISL_2689240, EPI_ISL_2689241, EPI_ISL_2689242, EPI_ISL_2689243, EPI_ISL_2689244, EPI_ISL_2689245, EPI_ISL_2689246, EPI_ISL_2689247, EPI_ISL_2689248, EPI_ISL_2689249, EPI_ISL_2689250, EPI_ISL_2689251, EPI_ISL_2689252, EPI_ISL_2689253, EPI_ISL_2689254, EPI_ISL_2689255, EPI_ISL_2689256, EPI_ISL_2689257, EPI_ISL_2689258, EPI_ISL_2689259, EPI_ISL_2689260, EPI_ISL_2689261, EPI_ISL_2689262, EPI_ISL_2689263, EPI_ISL_2689264, EPI_ISL_2689265, EPI_ISL_2689266, EPI_ISL_2689267, EPI_ISL_2689268, EPI_ISL_2689269, EPI_ISL_2689270, EPI_ISL_2689271, EPI_ISL_2689272, EPI_ISL_2689273, EPI_ISL_2689274, EPI_ISL_2689275, EPI_ISL_2689276, EPI_ISL_2689277, EPI_ISL_2689278, EPI_ISL_2689279, EPI_ISL_2689280, EPI_ISL_2689281, EPI_ISL_2689282, EPI_ISL_2689283, EPI_ISL_2689284, EPI_ISL_2689285, EPI_ISL_2689286, EPI_ISL_2689287, EPI_ISL_2689288, EPI_ISL_2689289, EPI_ISL_2689290, EPI_ISL_2689291, EPI_ISL_2689292, EPI_ISL_2689293, EPI_ISL_2689294, EPI_ISL_2689295, EPI_ISL_2689296, EPI_ISL_2689297, EPI_ISL_2689298, EPI_ISL_2689299, EPI_ISL_2689300, EPI_ISL_2689301, EPI_ISL_2689302, EPI_ISL_2689303, EPI_ISL_2689304, EPI_ISL_2689305, EPI_ISL_2689306, EPI_ISL_2689307, EPI_ISL_2689308, EPI_ISL_2689309, EPI_ISL_2689310, EPI_ISL_2689311, EPI_ISL_2689312, EPI_ISL_2689313, EPI_ISL_2689314, EPI_ISL_2689315, EPI_ISL_2689316, EPI_ISL_2689317, EPI_ISL_2689320, EPI_ISL_2689321, EPI_ISL_2689326, EPI_ISL_2689327, EPI_ISL_2689330, EPI_ISL_2689331, EPI_ISL_2689335, EPI_ISL_2689336, EPI_ISL_2689338, EPI_ISL_2689339, EPI_ISL_2689340, EPI_ISL_2689346, EPI_ISL_2689347, EPI_ISL_2689350, EPI_ISL_2689351, EPI_ISL_2689356, EPI_ISL_2689357, EPI_ISL_2689360, EPI_ISL_2689361, EPI_ISL_2689362, EPI_ISL_2689373, EPI_ISL_2689376, EPI_ISL_2689377, EPI_ISL_2689378, EPI_ISL_2689383, EPI_ISL_2689386, EPI_ISL_2689387, EPI_ISL_2689392, EPI_ISL_2689393, EPI_ISL_2689394, EPI_ISL_2689395, EPI_ISL_2689396, EPI_ISL_2689397, EPI_ISL_2689398, EPI_ISL_2689399, EPI_ISL_2689400, EPI_ISL_2689401, EPI_ISL_2689402, EPI_ISL_2689403, EPI_ISL_2689404, EPI_ISL_2689405, EPI_ISL_2689406, EPI_ISL_2689407, EPI_ISL_2689409, EPI_ISL_2689410, EPI_ISL_2689411, EPI_ISL_2689412, EPI_ISL_2689413, EPI_ISL_2689414, EPI_ISL_2689415, EPI_ISL_2689416, EPI_ISL_2689417, EPI_ISL_2689418, EPI_ISL_2689419, EPI_ISL_2689420, EPI_ISL_2689421, EPI_ISL_2689422, EPI_ISL_2689423, EPI_ISL_2689424, EPI_ISL_2689425, EPI_ISL_2689428, EPI_ISL_2689429, EPI_ISL_2689430, EPI_ISL_2689431, EPI_ISL_2689432, EPI_ISL_2689433, EPI_ISL_2689434, EPI_ISL_2689435, EPI_ISL_2689437, EPI_ISL_2689438, EPI_ISL_2689439, EPI_ISL_2689440, EPI_ISL_2689441, EPI_ISL_2689442, EPI_ISL_2689443, EPI_ISL_2689444, EPI_ISL_2689445, EPI_ISL_2689446, EPI_ISL_2689447, EPI_ISL_2689448, EPI_ISL_2689449, EPI_ISL_2689450, EPI_ISL_2689451, EPI_ISL_2689452, EPI_ISL_2689453, EPI_ISL_2689454, EPI_ISL_2689455, EPI_ISL_2689456, EPI_ISL_2689457, EPI_ISL_2689459 | | | |
| see above | Labor Dr. Fenner und Kollegen | Heinrich Pette Institute, Leibniz Institute for Experimental Virology | Adam Grundhoff; Alexis Robitaille; Johannes Knobloch; Martin Aepfelbacher; Nicole Fischer; Thomas Günther |
| EPI_ISL_2129682, EPI_ISL_2129688, EPI_ISL_2129689, EPI_ISL_2129695, EPI_ISL_2129708, EPI_ISL_2129713, EPI_ISL_2129718, EPI_ISL_2266099, EPI_ISL_2266100, EPI_ISL_2266101, EPI_ISL_2266103, EPI_ISL_2266106, EPI_ISL_2266108, EPI_ISL_2266109, EPI_ISL_2266110, EPI_ISL_2266111, EPI_ISL_2266112, EPI_ISL_2266113, EPI_ISL_2266114, EPI_ISL_2266117, EPI_ISL_2266118, EPI_ISL_2266120, EPI_ISL_2266121, EPI_ISL_2266122, EPI_ISL_2266123, EPI_ISL_2266124, EPI_ISL_2266126, EPI_ISL_2266128, EPI_ISL_2266130, EPI_ISL_2266131, EPI_ISL_2266132, EPI_ISL_2266133, EPI_ISL_2266134, EPI_ISL_2266135, EPI_ISL_2389326, EPI_ISL_2389327, EPI_ISL_2389329, EPI_ISL_2389330, EPI_ISL_2389332, EPI_ISL_2389334, EPI_ISL_2389335, EPI_ISL_2389336, EPI_ISL_2389337, EPI_ISL_2389340, EPI_ISL_2389341, EPI_ISL_2389343, EPI_ISL_2389344, EPI_ISL_2389347, EPI_ISL_2389350, EPI_ISL_2389352, EPI_ISL_2389353, EPI_ISL_2389355, EPI_ISL_2389356, EPI_ISL_2389359 | | | |
| see above | Labor Dr. Heidrich & Kollegen MVZ GmbH Hamburg | Robert Koch Institute | |
| EPI_ISL_2265554, EPI_ISL_2265573, EPI_ISL_2265575, EPI_ISL_2265617, EPI_ISL_2265619, EPI_ISL_2265631, EPI_ISL_2265645, EPI_ISL_2265657, EPI_ISL_2265664, EPI_ISL_2265679, EPI_ISL_2265710, EPI_ISL_2265720, EPI_ISL_2265729, EPI_ISL_2265732, EPI_ISL_2265736, EPI_ISL_2265740, EPI_ISL_2265753, EPI_ISL_2265762, EPI_ISL_2265786, EPI_ISL_2265796, EPI_ISL_2265799, EPI_ISL_2265804, EPI_ISL_2265812, EPI_ISL_2265813, EPI_ISL_2265845, EPI_ISL_2265858, EPI_ISL_2265863, EPI_ISL_2316055, EPI_ISL_2316058, EPI_ISL_2316078, EPI_ISL_2316099, EPI_ISL_2316100, EPI_ISL_2316123, EPI_ISL_2316135, EPI_ISL_2316152, EPI_ISL_2316160, EPI_ISL_2316179, EPI_ISL_2316188, EPI_ISL_2316228, EPI_ISL_2316238, EPI_ISL_2316261, EPI_ISL_2316282, EPI_ISL_2316321, EPI_ISL_2316388, EPI_ISL_2634108 | | | |
| see above | Labor Dr. Schumacher MVZ | Robert Koch Institute | |
| EPI_ISL_2265965, EPI_ISL_2265967, EPI_ISL_2472544, EPI_ISL_2472547, EPI_ISL_2472548 | Labor Dr. Spranger | Robert Koch Institute | |
| EPI_ISL_2128986, EPI_ISL_2128987 | Labor Dr. Wisplinghoff - Koln | Robert Koch Institute | |
| EPI_ISL_2263037, EPI_ISL_2386546, EPI_ISL_2386548, EPI_ISL_2386549 | Labor Dr. Wisplinghoff - Köln | Robert Koch Institute | |
| EPI_ISL_2314312, EPI_ISL_2314313, EPI_ISL_2314314, EPI_ISL_2314315, EPI_ISL_2314316, EPI_ISL_2386593, EPI_ISL_2386594, EPI_ISL_2386595, EPI_ISL_2386596, EPI_ISL_2386600, EPI_ISL_2386603, EPI_ISL_2386604 | | | |
| see above | Labor Kneißler GmbH & Co. KG | Robert Koch Institute | |
| EPI_ISL_2265000, EPI_ISL_2265005, EPI_ISL_2265007, EPI_ISL_2265008, EPI_ISL_2265048, EPI_ISL_2267929, EPI_ISL_2267943, EPI_ISL_2267948, EPI_ISL_2387349, EPI_ISL_2387385, EPI_ISL_2387413, EPI_ISL_2387425, EPI_ISL_2387439, EPI_ISL_2387446 | | | |
| see above | Labor Lübeck bzw. Laborärztliche Gemeinschaftspraxis Lübeck | Robert Koch Institute | |
| EPI_ISL_2265875, EPI_ISL_2265880, EPI_ISL_2265884, EPI_ISL_2265924, EPI_ISL_2265925, EPI_ISL_2265926, EPI_ISL_2265927, EPI_ISL_2265928, EPI_ISL_2265929, EPI_ISL_2265930, EPI_ISL_2265931, EPI_ISL_2265932, EPI_ISL_2265933, EPI_ISL_2265934, EPI_ISL_2265935, EPI_ISL_2265936, EPI_ISL_2265937, EPI_ISL_2265938, EPI_ISL_2265939, EPI_ISL_2265940, EPI_ISL_2265941, EPI_ISL_2265942, EPI_ISL_2265943, EPI_ISL_2265944, EPI_ISL_2265945, EPI_ISL_2265946, EPI_ISL_2265947, EPI_ISL_2265948, EPI_ISL_2265949, EPI_ISL_2265950, EPI_ISL_2265951, EPI_ISL_2265952, EPI_ISL_2265953, EPI_ISL_2265954, EPI_ISL_2265955, EPI_ISL_2265956, EPI_ISL_2265957, EPI_ISL_2265958, EPI_ISL_2265959, EPI_ISL_2265960, EPI_ISL_2265961, EPI_ISL_2265962, EPI_ISL_2265970, EPI_ISL_2265972, EPI_ISL_2265973, EPI_ISL_2265974, EPI_ISL_2265975, EPI_ISL_2265976, EPI_ISL_2265977, EPI_ISL_2265978, EPI_ISL_2265979, EPI_ISL_2265980, EPI_ISL_2265981, EPI_ISL_2265982, EPI_ISL_2265983, EPI_ISL_2265984, EPI_ISL_2265985, EPI_ISL_2265986, EPI_ISL_2265987, EPI_ISL_2265988, EPI_ISL_2265989, EPI_ISL_2265990, EPI_ISL_2265991, EPI_ISL_2265992, EPI_ISL_2265994, EPI_ISL_2265995, EPI_ISL_2265996, EPI_ISL_2265997, EPI_ISL_2265998, EPI_ISL_2265999, EPI_ISL_2266000, EPI_ISL_2266015, EPI_ISL_2266016, EPI_ISL_2266017, EPI_ISL_2266018, EPI_ISL_2266019, EPI_ISL_2266020, EPI_ISL_2266021, EPI_ISL_2266022, EPI_ISL_2266023, EPI_ISL_2266024, EPI_ISL_2266025, EPI_ISL_2266026, EPI_ISL_2266027, EPI_ISL_2266028, EPI_ISL_2266029, EPI_ISL_2266030, EPI_ISL_2266031, EPI_ISL_2313191, EPI_ISL_2313192, EPI_ISL_2313193, EPI_ISL_2313194, EPI_ISL_2313195, EPI_ISL_2313196, EPI_ISL_2313197, EPI_ISL_2313198, EPI_ISL_2313200, EPI_ISL_2313201, EPI_ISL_2313203, EPI_ISL_2313206, EPI_ISL_2313208, EPI_ISL_2313210, EPI_ISL_2313212, EPI_ISL_2313213, EPI_ISL_2314647, EPI_ISL_2314648, EPI_ISL_2314650, EPI_ISL_2314652, EPI_ISL_2314653, EPI_ISL_2314655, EPI_ISL_2314656, EPI_ISL_2314660, EPI_ISL_2314662, EPI_ISL_2314665, EPI_ISL_2314666, EPI_ISL_2314667, EPI_ISL_2314668, EPI_ISL_2314669, EPI_ISL_2314670, EPI_ISL_2314673, EPI_ISL_2314675, EPI_ISL_2314677, EPI_ISL_2314678, EPI_ISL_2314679, EPI_ISL_2314682, EPI_ISL_2314683, EPI_ISL_2386216, EPI_ISL_2386222 | | | |
| see above | Labor Mönchengladbach MVZ Dr. Stein + Kollegen GbR | Robert Koch Institute | |
| EPI_ISL_2264550, EPI_ISL_2264552, EPI_ISL_2265519, EPI_ISL_2265521, EPI_ISL_2265523, EPI_ISL_2265526, EPI_ISL_2265532, EPI_ISL_2265533, EPI_ISL_2314742, EPI_ISL_2314753 | | | |
| see above | Labor Prof. Dr. G. Enders MVZ GbR | Robert Koch Institute | |
| EPI_ISL_2264811, EPI_ISL_2264816, EPI_ISL_2264835, EPI_ISL_2264974, EPI_ISL_2386605, EPI_ISL_2386606, EPI_ISL_2386607, EPI_ISL_2386608, EPI_ISL_2386609, EPI_ISL_2386610, EPI_ISL_2386611, EPI_ISL_2386612, EPI_ISL_2386613, EPI_ISL_2386614, EPI_ISL_2386615, EPI_ISL_2386616, EPI_ISL_2386617, EPI_ISL_2386618, EPI_ISL_2386619, EPI_ISL_2386620, EPI_ISL_2386621, EPI_ISL_2386622, EPI_ISL_2386623, EPI_ISL_2386624, EPI_ISL_2386625, EPI_ISL_2386626, EPI_ISL_2386627, EPI_ISL_2386628, EPI_ISL_2386629, EPI_ISL_2386630, EPI_ISL_2386631, EPI_ISL_2386632, EPI_ISL_2386633, EPI_ISL_2386634, EPI_ISL_2386635, EPI_ISL_2386636, EPI_ISL_2386637, EPI_ISL_2386638, EPI_ISL_2386639, EPI_ISL_2386640, EPI_ISL_2386641, EPI_ISL_2386642, EPI_ISL_2386643, EPI_ISL_2386644, EPI_ISL_2386645, EPI_ISL_2386646, EPI_ISL_2386647, EPI_ISL_2386648, EPI_ISL_2386649, EPI_ISL_2386650, EPI_ISL_2386651, EPI_ISL_2386652, EPI_ISL_2386653, EPI_ISL_2386654, EPI_ISL_2386655, EPI_ISL_2386656, EPI_ISL_2386657, EPI_ISL_2386658, EPI_ISL_2386659, EPI_ISL_2386660, EPI_ISL_2386661, EPI_ISL_2386662, EPI_ISL_2386663, EPI_ISL_2386664, EPI_ISL_2386665, EPI_ISL_2386666, EPI_ISL_2386667, EPI_ISL_2386668, EPI_ISL_2386669, EPI_ISL_2386670, EPI_ISL_2386671, EPI_ISL_2386672, EPI_ISL_2386673, EPI_ISL_2386674, EPI_ISL_2386675, EPI_ISL_2386676, EPI_ISL_2386677, EPI_ISL_2386678, EPI_ISL_2386679, EPI_ISL_2386680, EPI_ISL_2386681, EPI_ISL_2386682, EPI_ISL_2386683, EPI_ISL_2386684, EPI_ISL_2386685, EPI_ISL_2386686, EPI_ISL_2386687, EPI_ISL_2386688, EPI_ISL_2386689, EPI_ISL_2386690, EPI_ISL_2386691, EPI_ISL_2386692, EPI_ISL_2386693, EPI_ISL_2386694, EPI_ISL_2386695, EPI_ISL_2386696, EPI_ISL_2386697, EPI_ISL_2386698, EPI_ISL_2386699, EPI_ISL_2386700, EPI_ISL_2386701, EPI_ISL_2386702, EPI_ISL_2386703, EPI_ISL_2386704, EPI_ISL_2386705, EPI_ISL_2386706, EPI_ISL_2386707, EPI_ISL_2386708, EPI_ISL_2386709, EPI_ISL_2386710, EPI_ISL_2386711, EPI_ISL_2386712, EPI_ISL_2386713, EPI_ISL_2386714, EPI_ISL_2386715, EPI_ISL_2386716, EPI_ISL_2386717, EPI_ISL_2386718, EPI_ISL_2386720, EPI_ISL_2386721, EPI_ISL_2386722, EPI_ISL_2386723, EPI_ISL_2386724, EPI_ISL_2386725, EPI_ISL_2386726, EPI_ISL_2386727, EPI_ISL_2386728, EPI_ISL_2386729, EPI_ISL_2386730, EPI_ISL_2386731, EPI_ISL_2386732, EPI_ISL_2386733, EPI_ISL_2386734, EPI_ISL_2386735, EPI_ISL_2386736, EPI_ISL_2386737, EPI_ISL_2386738, EPI_ISL_2386739, EPI_ISL_2386740, EPI_ISL_2386741, EPI_ISL_2386742, EPI_ISL_2386743, EPI_ISL_2386744, EPI_ISL_2386745, EPI_ISL_2386746, EPI_ISL_2386747, EPI_ISL_2386748, EPI_ISL_2386749, EPI_ISL_2386750, EPI_ISL_2386753, EPI_ISL_2386754, EPI_ISL_2386755, EPI_ISL_2386756, EPI_ISL_2386757, EPI_ISL_2386759, EPI_ISL_2386760, EPI_ISL_2386761, EPI_ISL_2386762, EPI_ISL_2386764, EPI_ISL_2386765, EPI_ISL_2386766, EPI_ISL_2386767, EPI_ISL_2386768, EPI_ISL_2386769, EPI_ISL_2386770, EPI_ISL_2386771, EPI_ISL_2386772, EPI_ISL_2386773, EPI_ISL_2386774, EPI_ISL_2386775, EPI_ISL_2386776, EPI_ISL_2386777, EPI_ISL_2386778, EPI_ISL_2386779, EPI_ISL_2386780, EPI_ISL_2386781, EPI_ISL_2386783, EPI_ISL_2386784, EPI_ISL_2386785, EPI_ISL_2386786, EPI_ISL_2386787, EPI_ISL_2386788, EPI_ISL_2386789, EPI_ISL_2386790, EPI_ISL_2386791, EPI_ISL_2386792, EPI_ISL_2386793, EPI_ISL_2386794, EPI_ISL_2386795, EPI_ISL_2386796, EPI_ISL_2386797, EPI_ISL_2386798, EPI_ISL_2390580, EPI_ISL_2390582, EPI_ISL_2390583, EPI_ISL_2390584, EPI_ISL_2390585, EPI_ISL_2390586, EPI_ISL_2390590, EPI_ISL_2390591, EPI_ISL_2390593, EPI_ISL_2390594, EPI_ISL_2390595, EPI_ISL_2390598, EPI_ISL_2390600, EPI_ISL_2390601, EPI_ISL_2390605, EPI_ISL_2390606, EPI_ISL_2390607, EPI_ISL_2390610, EPI_ISL_2390612, EPI_ISL_2390613, EPI_ISL_2390614, EPI_ISL_2390615, EPI_ISL_2390617, EPI_ISL_2390618, EPI_ISL_2390619, EPI_ISL_2390620, EPI_ISL_2390622, EPI_ISL_2390623, EPI_ISL_2390624, EPI_ISL_2390627, EPI_ISL_2390628, EPI_ISL_2390629, EPI_ISL_2390630, EPI_ISL_2390631, EPI_ISL_2390632, EPI_ISL_2390633, EPI_ISL_2390634, EPI_ISL_2390636, EPI_ISL_2390637, EPI_ISL_2390638, EPI_ISL_2390639, EPI_ISL_2390640, EPI_ISL_2390641, EPI_ISL_2390642, EPI_ISL_2390643, EPI_ISL_2390644, EPI_ISL_2390645, EPI_ISL_2390646, EPI_ISL_2390648, EPI_ISL_2390651, EPI_ISL_2390652, EPI_ISL_2390653, EPI_ISL_2390654, EPI_ISL_2390656, EPI_ISL_2390657, EPI_ISL_2390658, EPI_ISL_2390659, EPI_ISL_2390660, EPI_ISL_2390661, EPI_ISL_2390662, EPI_ISL_2390663, EPI_ISL_2390665, EPI_ISL_2390667, EPI_ISL_2390670, EPI_ISL_2390673, EPI_ISL_2390675, EPI_ISL_2390676, EPI_ISL_2390678, EPI_ISL_2390679, EPI_ISL_2390681, EPI_ISL_2390682, EPI_ISL_2390685, EPI_ISL_2390686, EPI_ISL_2390687, EPI_ISL_2390689, EPI_ISL_2390690, EPI_ISL_2390693, EPI_ISL_2390694, EPI_ISL_2390695, EPI_ISL_2390698, EPI_ISL_2390701, EPI_ISL_2390702, EPI_ISL_2390704, EPI_ISL_2390706, EPI_ISL_2390707, EPI_ISL_2390709, EPI_ISL_2390710, EPI_ISL_2390714, EPI_ISL_2390716, EPI_ISL_2390718, EPI_ISL_2390719, EPI_ISL_2390720, EPI_ISL_2390721, EPI_ISL_2390723, EPI_ISL_2390724, EPI_ISL_2390725, EPI_ISL_2390726, EPI_ISL_2390729, EPI_ISL_2390731, EPI_ISL_2390732, EPI_ISL_2390733, EPI_ISL_2390734, EPI_ISL_2390738, EPI_ISL_2390739, EPI_ISL_2390741, EPI_ISL_2390743, EPI_ISL_2390744, EPI_ISL_2390746, EPI_ISL_2390748 | | | |
| see above | Labor ZOTZ\|KLIMAS; MVZ Düsseldorf-Centrum | Robert Koch Institute | |
| EPI_ISL_2262279, EPI_ISL_2262284, EPI_ISL_2314470, EPI_ISL_2314499, EPI_ISL_2314608, EPI_ISL_2314609, EPI_ISL_2316147, EPI_ISL_2316203, EPI_ISL_2316348, EPI_ISL_2389325 | | | |
| see above | Labor für medizinische Mikrobiologie Bakteriologie Mykologie Virologie & Infektionsserologie - Labor Mölbis | Robert Koch Institute | |
| EPI_ISL_2263193, EPI_ISL_2263194, EPI_ISL_2263195, EPI_ISL_2263196, EPI_ISL_2263197, EPI_ISL_2263198, EPI_ISL_2314318, EPI_ISL_2314319, EPI_ISL_2314320, EPI_ISL_2314321, EPI_ISL_2314322, EPI_ISL_2314323, EPI_ISL_2314324, EPI_ISL_2314325, EPI_ISL_2314326, EPI_ISL_2314327, EPI_ISL_2314328, EPI_ISL_2314329, EPI_ISL_2314330, EPI_ISL_2314331, EPI_ISL_2314332, EPI_ISL_2314333, EPI_ISL_2314334, EPI_ISL_2314335, EPI_ISL_2314336, EPI_ISL_2314337 | | | |
| see above | Laborarztpraxis Dres. med. Walther Weindel & Kollegen | Robert Koch Institute | |
| EPI_ISL_2267931, EPI_ISL_2267940, EPI_ISL_2313249, EPI_ISL_2313250, EPI_ISL_2313252, EPI_ISL_2313254, EPI_ISL_2313255, EPI_ISL_2313257, EPI_ISL_2313258, EPI_ISL_2313259, EPI_ISL_2313260, EPI_ISL_2313261, EPI_ISL_2313263, EPI_ISL_2313264, EPI_ISL_2313269, EPI_ISL_2313272, EPI_ISL_2313273, EPI_ISL_2313274, EPI_ISL_2313275, EPI_ISL_2313277, EPI_ISL_2313279, EPI_ISL_2313280, EPI_ISL_2313282, EPI_ISL_2313283, EPI_ISL_2313284, EPI_ISL_2313291, EPI_ISL_2313295, EPI_ISL_2313302, EPI_ISL_2313307, EPI_ISL_2313308, EPI_ISL_2313312, EPI_ISL_2313314, EPI_ISL_2313317, EPI_ISL_2313318, EPI_ISL_2313319, EPI_ISL_2313323, EPI_ISL_2313326, EPI_ISL_2313336, EPI_ISL_2313337, EPI_ISL_2313893, EPI_ISL_2388191, EPI_ISL_2388199, EPI_ISL_2388202, EPI_ISL_2388207, EPI_ISL_2388219, EPI_ISL_2388228, EPI_ISL_2388242, EPI_ISL_2388246, EPI_ISL_2388248, EPI_ISL_2388250, EPI_ISL_2388261, EPI_ISL_2388263, EPI_ISL_2388267, EPI_ISL_2388277, EPI_ISL_2388289, EPI_ISL_2388295 | | | |
| see above | Laborarztpraxis Osnabrück | Robert Koch Institute | |
| EPI_ISL_2313888, EPI_ISL_2313963, EPI_ISL_2313978, EPI_ISL_2313991, EPI_ISL_2314011, EPI_ISL_2314012, EPI_ISL_2314013, EPI_ISL_2314020, EPI_ISL_2389386, EPI_ISL_2389396, EPI_ISL_2389403, EPI_ISL_2389407, EPI_ISL_2389417, EPI_ISL_2389421, EPI_ISL_2389436 | | | |
| see above | Labormedizin Darmstadt | Robert Koch Institute | |
| EPI_ISL_2285554, EPI_ISL_2285555, EPI_ISL_2285556, EPI_ISL_2285557, EPI_ISL_2285558, EPI_ISL_2285559, EPI_ISL_2285560, EPI_ISL_2285561, EPI_ISL_2285562, EPI_ISL_2285563, EPI_ISL_2285564, EPI_ISL_2285565, EPI_ISL_2285566, EPI_ISL_2285567, EPI_ISL_2285568, EPI_ISL_2285569, EPI_ISL_2285570, EPI_ISL_2285571, EPI_ISL_2285572, EPI_ISL_2285573, EPI_ISL_2285574, EPI_ISL_2285575, EPI_ISL_2285576, EPI_ISL_2285577, EPI_ISL_2285578, EPI_ISL_2285579, EPI_ISL_2285580, EPI_ISL_2285581, EPI_ISL_2285582, EPI_ISL_2285583, EPI_ISL_2285584, EPI_ISL_2285585, EPI_ISL_2285586, EPI_ISL_2285587, EPI_ISL_2285588, EPI_ISL_2285589, EPI_ISL_2285590, EPI_ISL_2285591, EPI_ISL_2285592, EPI_ISL_2285593, EPI_ISL_2285594, EPI_ISL_2285595, EPI_ISL_2285597, EPI_ISL_2285598, EPI_ISL_2285599, EPI_ISL_2285600, EPI_ISL_2285601, EPI_ISL_2285602, EPI_ISL_2285603, EPI_ISL_2285607, EPI_ISL_2285608, EPI_ISL_2285609, EPI_ISL_2285610, EPI_ISL_2285611, EPI_ISL_2285612, EPI_ISL_2285613, EPI_ISL_2285614, EPI_ISL_2285615, EPI_ISL_2285616, EPI_ISL_2285617, EPI_ISL_2285618, EPI_ISL_2285619, EPI_ISL_2285620, EPI_ISL_2285621, EPI_ISL_2285622, EPI_ISL_2285623, EPI_ISL_2285633, EPI_ISL_2285634, EPI_ISL_2285635, EPI_ISL_2285636, EPI_ISL_2285637, EPI_ISL_2285638, EPI_ISL_2285639, EPI_ISL_2285640, EPI_ISL_2285641, EPI_ISL_2285642, EPI_ISL_2285643, EPI_ISL_2285644, EPI_ISL_2285645, EPI_ISL_2285646, EPI_ISL_2285647, EPI_ISL_2285652, EPI_ISL_2285653, EPI_ISL_2285654, EPI_ISL_2285655, EPI_ISL_2285656, EPI_ISL_2285657, EPI_ISL_2285658, EPI_ISL_2285659, EPI_ISL_2285660, EPI_ISL_2285661, EPI_ISL_2285662, EPI_ISL_2285663, EPI_ISL_2285664, EPI_ISL_2285665, EPI_ISL_2285666, EPI_ISL_2285667, EPI_ISL_2285668, EPI_ISL_2285669, EPI_ISL_2285670, EPI_ISL_2285671, EPI_ISL_2285672, EPI_ISL_2285673, EPI_ISL_2285674, EPI_ISL_2285675, EPI_ISL_2285676, EPI_ISL_2285677, EPI_ISL_2285678, EPI_ISL_2285679, EPI_ISL_2285680, EPI_ISL_2285681, EPI_ISL_2285682, EPI_ISL_2285683, EPI_ISL_2285684, EPI_ISL_2285685, EPI_ISL_2285686, EPI_ISL_2285687, EPI_ISL_2285688, EPI_ISL_2285697, EPI_ISL_2285698, EPI_ISL_2285699, EPI_ISL_2285700, EPI_ISL_2285701, EPI_ISL_2285702, EPI_ISL_2285703, EPI_ISL_2285704, EPI_ISL_2285705, EPI_ISL_2285706, EPI_ISL_2285707, EPI_ISL_2285708, EPI_ISL_2285709, EPI_ISL_2285710, EPI_ISL_2285711, EPI_ISL_2285712, EPI_ISL_2285713, EPI_ISL_2285714, EPI_ISL_2285715, EPI_ISL_2285716, EPI_ISL_2285717, EPI_ISL_2285718, EPI_ISL_2285719, EPI_ISL_2285720, EPI_ISL_2285721, EPI_ISL_2285722 | | | |
| see above | Landesamt für Verbraucherschutz Sachsen Anhalt, Magdeburg | Institute of Medical Microbiology and Hospital Hygiene | Aljoscha Tersteegen; Prof. Dr. Achim Kaasch |

| | | | |
|---|---|---|---|
| EPI_ISL_2314833, EPI_ISL_2314835, EPI_ISL_2314837, EPI_ISL_2314838, EPI_ISL_2314839, EPI_ISL_2314843, EPI_ISL_2314848, EPI_ISL_2314849, EPI_ISL_2314850, EPI_ISL_2314853, EPI_ISL_2314856, EPI_ISL_2314857, EPI_ISL_2314858, EPI_ISL_2314866, EPI_ISL_2314867, EPI_ISL_2314868, EPI_ISL_2314870, EPI_ISL_2389865, EPI_ISL_2389866, EPI_ISL_2389867, EPI_ISL_2389868, EPI_ISL_2389869, EPI_ISL_2389870, EPI_ISL_2389871, EPI_ISL_2389872, EPI_ISL_2389873, EPI_ISL_2389874, EPI_ISL_2389876, EPI_ISL_2389877, EPI_ISL_2389878, EPI_ISL_2389879, EPI_ISL_2389880, EPI_ISL_2389882, EPI_ISL_2389883, EPI_ISL_2389884, EPI_ISL_2389885, EPI_ISL_2389886, EPI_ISL_2389887, EPI_ISL_2389888, EPI_ISL_2389889, EPI_ISL_2389890, EPI_ISL_2389891, EPI_ISL_2389892, EPI_ISL_2389893, EPI_ISL_2389894, EPI_ISL_2389895, EPI_ISL_2389897, EPI_ISL_2389898 | | | |
| see above | Landesgesundheitsamt Baden-Württemberg | Robert Koch Institute | |
| EPI_ISL_2262335, EPI_ISL_2262344, EPI_ISL_2262345, EPI_ISL_2262350, EPI_ISL_2262360, EPI_ISL_2262361, EPI_ISL_2262369, EPI_ISL_2262370, EPI_ISL_2262373, EPI_ISL_2262374, EPI_ISL_2262378, EPI_ISL_2262390, EPI_ISL_2313900, EPI_ISL_2313902, EPI_ISL_2313904, EPI_ISL_2313905, EPI_ISL_2313909, EPI_ISL_2313926, EPI_ISL_2313929, EPI_ISL_2313933, EPI_ISL_2313949, EPI_ISL_2313956, EPI_ISL_2313957 | | | |
| see above | Landesuntersuchungsanstalt für das Gesundheits- und Veterinärwesen (LUA) Sachsen | Robert Koch Institute | |
| EPI_ISL_2386242, EPI_ISL_2386243, EPI_ISL_2386244, EPI_ISL_2386245, EPI_ISL_2386246, EPI_ISL_2386247, EPI_ISL_2386248, EPI_ISL_2386249, EPI_ISL_2386877, EPI_ISL_2386879 | | | |
| see above | Limbach - Ganzimmun Diagnostics AG Mainz | Robert Koch Institute | |
| EPI_ISL_2259765, EPI_ISL_2264621, EPI_ISL_2264629, EPI_ISL_2265971, EPI_ISL_2265993, EPI_ISL_2266594, EPI_ISL_2266595, EPI_ISL_2266597, EPI_ISL_2266602, EPI_ISL_2266610, EPI_ISL_2266614, EPI_ISL_2314686, EPI_ISL_2314697, EPI_ISL_2314729, EPI_ISL_2314757, EPI_ISL_2314767, EPI_ISL_2386288 | | | |
| see above | Limbach - MVZ Gemeinschaftslabor Suhl | Robert Koch Institute | |
| EPI_ISL_2129362, EPI_ISL_2129363, EPI_ISL_2129365, EPI_ISL_2129367, EPI_ISL_2129369, EPI_ISL_2129370, EPI_ISL_2129372, EPI_ISL_2129375, EPI_ISL_2129376, EPI_ISL_2129377, EPI_ISL_2129379, EPI_ISL_2129382, EPI_ISL_2129384, EPI_ISL_2129390, EPI_ISL_2129396, EPI_ISL_2129400, EPI_ISL_2129402, EPI_ISL_2129404, EPI_ISL_2129408, EPI_ISL_2129410, EPI_ISL_2129414, EPI_ISL_2129415, EPI_ISL_2129418, EPI_ISL_2129420, EPI_ISL_2129426, EPI_ISL_2129427, EPI_ISL_2129430, EPI_ISL_2129431, EPI_ISL_2129432, EPI_ISL_2129435, EPI_ISL_2129438, EPI_ISL_2129442, EPI_ISL_2129445, EPI_ISL_2129446, EPI_ISL_2129447, EPI_ISL_2129451, EPI_ISL_2129452, EPI_ISL_2129454, EPI_ISL_2129455, EPI_ISL_2129457, EPI_ISL_2129458, EPI_ISL_2129460, EPI_ISL_2129461, EPI_ISL_2129462, EPI_ISL_2129463, EPI_ISL_2129465, EPI_ISL_2129468, EPI_ISL_2129469, EPI_ISL_2129471, EPI_ISL_2129472, EPI_ISL_2129474, EPI_ISL_2129476, EPI_ISL_2129478, EPI_ISL_2129481, EPI_ISL_2129482, EPI_ISL_2129483, EPI_ISL_2129485, EPI_ISL_2129486, EPI_ISL_2129488, EPI_ISL_2129489, EPI_ISL_2129493, EPI_ISL_2129494, EPI_ISL_2129498, EPI_ISL_2129500, EPI_ISL_2129501, EPI_ISL_2129505, EPI_ISL_2129507, EPI_ISL_2129511, EPI_ISL_2129514, EPI_ISL_2129515, EPI_ISL_2129517, EPI_ISL_2129518, EPI_ISL_2129519, EPI_ISL_2129520, EPI_ISL_2129521, EPI_ISL_2129522, EPI_ISL_2129523, EPI_ISL_2129524, EPI_ISL_2129526, EPI_ISL_2129527, EPI_ISL_2129528, EPI_ISL_2129531, EPI_ISL_2129532, EPI_ISL_2129533, EPI_ISL_2129534, EPI_ISL_2266422, EPI_ISL_2266423, EPI_ISL_2266424, EPI_ISL_2266425, EPI_ISL_2266426, EPI_ISL_2266427, EPI_ISL_2266428, EPI_ISL_2266429, EPI_ISL_2266430, EPI_ISL_2266431, EPI_ISL_2266432, EPI_ISL_2266433, EPI_ISL_2266434, EPI_ISL_2266435, EPI_ISL_2266436, EPI_ISL_2266437, EPI_ISL_2266438, EPI_ISL_2266439, EPI_ISL_2266440, EPI_ISL_2266441, EPI_ISL_2266442, EPI_ISL_2266443, EPI_ISL_2266444, EPI_ISL_2266445, EPI_ISL_2266446, EPI_ISL_2266447, EPI_ISL_2266448, EPI_ISL_2266449, EPI_ISL_2266450, EPI_ISL_2266451, EPI_ISL_2266452, EPI_ISL_2266453, EPI_ISL_2266454, EPI_ISL_2266455, EPI_ISL_2266456, EPI_ISL_2266457, EPI_ISL_2266458, EPI_ISL_2266459, EPI_ISL_2266460, EPI_ISL_2266461, EPI_ISL_2266462, EPI_ISL_2266463, EPI_ISL_2266464, EPI_ISL_2266465, EPI_ISL_2266466, EPI_ISL_2266467, EPI_ISL_2266468, EPI_ISL_2266469, EPI_ISL_2266470, EPI_ISL_2266471, EPI_ISL_2266472, EPI_ISL_2266473, EPI_ISL_2266474, EPI_ISL_2266475, EPI_ISL_2266476, EPI_ISL_2266477, EPI_ISL_2266478, EPI_ISL_2266479, EPI_ISL_2266480, EPI_ISL_2266481, EPI_ISL_2266482, EPI_ISL_2266483, EPI_ISL_2266484, EPI_ISL_2266485, EPI_ISL_2266486, EPI_ISL_2266487, EPI_ISL_2266488, EPI_ISL_2266489, EPI_ISL_2266490, EPI_ISL_2266491, EPI_ISL_2266492, EPI_ISL_2266493, EPI_ISL_2266494, EPI_ISL_2266495, EPI_ISL_2266496, EPI_ISL_2266497, EPI_ISL_2266498, EPI_ISL_2266499, EPI_ISL_2266500, EPI_ISL_2266501, EPI_ISL_2266502, EPI_ISL_2266503, EPI_ISL_2266504, EPI_ISL_2266505, EPI_ISL_2266506, EPI_ISL_2266507, EPI_ISL_2266508, EPI_ISL_2266509, EPI_ISL_2266510, EPI_ISL_2266511, EPI_ISL_2266512, EPI_ISL_2266513, EPI_ISL_2266514, EPI_ISL_2266515, EPI_ISL_2266516, EPI_ISL_2266517, EPI_ISL_2266518, EPI_ISL_2266519, EPI_ISL_2266520, EPI_ISL_2266521, EPI_ISL_2266522, EPI_ISL_2266523, EPI_ISL_2266524, EPI_ISL_2266525, EPI_ISL_2266526, EPI_ISL_2266527, EPI_ISL_2266528, EPI_ISL_2266529, EPI_ISL_2266530, EPI_ISL_2266531, EPI_ISL_2266532, EPI_ISL_2266533, EPI_ISL_2266534, EPI_ISL_2266535, EPI_ISL_2266536, EPI_ISL_2266537, EPI_ISL_2266538, EPI_ISL_2266539, EPI_ISL_2266540, EPI_ISL_2266541, EPI_ISL_2266542, EPI_ISL_2266543, EPI_ISL_2266544, EPI_ISL_2266545, EPI_ISL_2266546, EPI_ISL_2266547, EPI_ISL_2266548, EPI_ISL_2266549, EPI_ISL_2266550, EPI_ISL_2266551, EPI_ISL_2266552, EPI_ISL_2266553, EPI_ISL_2266554, EPI_ISL_2266555, EPI_ISL_2266556, EPI_ISL_2266557, EPI_ISL_2266558, EPI_ISL_2266559, EPI_ISL_2266560, EPI_ISL_2266561, EPI_ISL_2266562, EPI_ISL_2266563, EPI_ISL_2266564, EPI_ISL_2266565, EPI_ISL_2266566, EPI_ISL_2266567, EPI_ISL_2266568, EPI_ISL_2266569, EPI_ISL_2266570, EPI_ISL_2266571, EPI_ISL_2266572, EPI_ISL_2266573, EPI_ISL_2266574, EPI_ISL_2266575, EPI_ISL_2266576, EPI_ISL_2266577, EPI_ISL_2266578, EPI_ISL_2266579, EPI_ISL_2266580, EPI_ISL_2266581, EPI_ISL_2266582, EPI_ISL_2266583, EPI_ISL_2266584, EPI_ISL_2266585, EPI_ISL_2266586, EPI_ISL_2266587, EPI_ISL_2266588, EPI_ISL_2266589, EPI_ISL_2266590, EPI_ISL_2266591, EPI_ISL_2266592 | | | |
| see above | Limbach - MVZ Humangenetik Ulm | Robert Koch Institute | |
| EPI_ISL_2263039, EPI_ISL_2263040, EPI_ISL_2263041, EPI_ISL_2263042, EPI_ISL_2263043, EPI_ISL_2263045, EPI_ISL_2263046, EPI_ISL_2263047, EPI_ISL_2263048, EPI_ISL_2263050, EPI_ISL_2263051, EPI_ISL_2263052, EPI_ISL_2263054, EPI_ISL_2263055, EPI_ISL_2263057, EPI_ISL_2264481, EPI_ISL_2264554, EPI_ISL_2264555, EPI_ISL_2264567, EPI_ISL_2264571, EPI_ISL_2264572, EPI_ISL_2264575, EPI_ISL_2265870, EPI_ISL_2265872, EPI_ISL_2265873, EPI_ISL_2265874, EPI_ISL_2265877, EPI_ISL_2265878, EPI_ISL_2265879, EPI_ISL_2265881, EPI_ISL_2265883, EPI_ISL_2265885, EPI_ISL_2265888, EPI_ISL_2313118, EPI_ISL_2313142, EPI_ISL_2313149, EPI_ISL_2313174, EPI_ISL_2313181, EPI_ISL_2313184, EPI_ISL_2313187, EPI_ISL_2314644, EPI_ISL_2314645, EPI_ISL_2314646, EPI_ISL_2314651, EPI_ISL_2314658, EPI_ISL_2314661, EPI_ISL_2314663 | | | |
| see above | Limbach - MVZ Labor Eveld & Kollegen Essen | Robert Koch Institute | |
| EPI_ISL_2262225, EPI_ISL_2262238, EPI_ISL_2262242, EPI_ISL_2262274, EPI_ISL_2264620, EPI_ISL_2264627, EPI_ISL_2264628, EPI_ISL_2265534, EPI_ISL_2314773, EPI_ISL_2314802, EPI_ISL_2314805, EPI_ISL_2386277 | | | |
| see above | Limbach - MVZ Labor Ludwigsburg | Robert Koch Institute | |
| EPI_ISL_2259750, EPI_ISL_2259755, EPI_ISL_2259758, EPI_ISL_2259761, EPI_ISL_2259764, EPI_ISL_2259772, EPI_ISL_2259776, EPI_ISL_2259778, EPI_ISL_2259783, EPI_ISL_2262224, EPI_ISL_2262230, EPI_ISL_2262234, EPI_ISL_2262236, EPI_ISL_2262266, EPI_ISL_2262823, EPI_ISL_2262826, EPI_ISL_2263049, EPI_ISL_2263060, EPI_ISL_2263063, EPI_ISL_2263064, EPI_ISL_2263066, EPI_ISL_2263068, EPI_ISL_2263073, EPI_ISL_2263077, EPI_ISL_2263078, EPI_ISL_2263080, EPI_ISL_2263081, EPI_ISL_2263082, EPI_ISL_2263083, EPI_ISL_2263084, EPI_ISL_2263085, EPI_ISL_2263086, EPI_ISL_2263087, EPI_ISL_2263088, EPI_ISL_2263090, EPI_ISL_2263091, EPI_ISL_2263092, EPI_ISL_2263093, EPI_ISL_2263094, EPI_ISL_2263095, EPI_ISL_2263096, EPI_ISL_2263097, EPI_ISL_2263098, EPI_ISL_2263099, EPI_ISL_2263100, EPI_ISL_2263104, EPI_ISL_2263105, EPI_ISL_2263106, EPI_ISL_2263108, EPI_ISL_2263113, EPI_ISL_2264548, EPI_ISL_2264551, EPI_ISL_2264707, EPI_ISL_2264708, EPI_ISL_2264709, EPI_ISL_2264710, EPI_ISL_2264711, EPI_ISL_2264716, EPI_ISL_2264720, EPI_ISL_2265514, EPI_ISL_2265515, EPI_ISL_2265516, EPI_ISL_2265517, EPI_ISL_2265518, EPI_ISL_2265520, EPI_ISL_2265522, EPI_ISL_2265524, EPI_ISL_2265525, EPI_ISL_2265527, EPI_ISL_2265528, EPI_ISL_2265529, EPI_ISL_2265530, EPI_ISL_2265531, EPI_ISL_2266593, EPI_ISL_2266596, EPI_ISL_2266598, EPI_ISL_2266599, EPI_ISL_2266601, EPI_ISL_2266603, EPI_ISL_2266604, EPI_ISL_2266606, EPI_ISL_2266609, EPI_ISL_2266611, EPI_ISL_2266612, EPI_ISL_2266613, EPI_ISL_2266615, EPI_ISL_2266616, EPI_ISL_2266618, EPI_ISL_2266619, EPI_ISL_2266620, EPI_ISL_2266621, EPI_ISL_2266622, EPI_ISL_2266623, EPI_ISL_2266624, EPI_ISL_2266625, EPI_ISL_2266626, EPI_ISL_2266628, EPI_ISL_2313349, EPI_ISL_2313350, EPI_ISL_2313351, EPI_ISL_2313352, EPI_ISL_2313353, EPI_ISL_2313354, EPI_ISL_2313355, EPI_ISL_2313356, EPI_ISL_2313357, EPI_ISL_2313358, EPI_ISL_2313360, EPI_ISL_2313361, EPI_ISL_2313362, EPI_ISL_2313363, EPI_ISL_2313365, EPI_ISL_2313366, EPI_ISL_2313367, EPI_ISL_2313368, EPI_ISL_2313370, EPI_ISL_2314412, EPI_ISL_2314676, EPI_ISL_2314681, EPI_ISL_2314684, EPI_ISL_2314693, EPI_ISL_2314694, EPI_ISL_2314695, EPI_ISL_2314696, EPI_ISL_2314700, EPI_ISL_2314702, EPI_ISL_2314703, EPI_ISL_2314704, EPI_ISL_2314706, EPI_ISL_2314707, EPI_ISL_2314708, EPI_ISL_2314709, EPI_ISL_2314710, EPI_ISL_2314711, EPI_ISL_2314712, EPI_ISL_2314713, EPI_ISL_2314714, EPI_ISL_2314715, EPI_ISL_2314716, EPI_ISL_2314717, EPI_ISL_2314718, EPI_ISL_2314719, EPI_ISL_2314720, EPI_ISL_2314721, EPI_ISL_2314722, EPI_ISL_2314724, EPI_ISL_2314725, EPI_ISL_2314726, EPI_ISL_2314727, EPI_ISL_2314728, EPI_ISL_2314730, EPI_ISL_2314731, EPI_ISL_2314732, EPI_ISL_2314733, EPI_ISL_2314734, EPI_ISL_2314736, EPI_ISL_2314737, EPI_ISL_2314739, EPI_ISL_2314740, EPI_ISL_2314741, EPI_ISL_2314743, EPI_ISL_2314744, EPI_ISL_2314747, EPI_ISL_2314748, EPI_ISL_2314749, EPI_ISL_2314750, EPI_ISL_2314751, EPI_ISL_2314752, EPI_ISL_2314754, EPI_ISL_2314755, EPI_ISL_2314756, EPI_ISL_2314758, EPI_ISL_2314760, EPI_ISL_2314762, EPI_ISL_2314764, EPI_ISL_2314765, EPI_ISL_2314766, EPI_ISL_2314770, EPI_ISL_2314771, EPI_ISL_2386259 | | | |
| see above | Limbach - MVZ Labor Ravensburg Labor Dr. Gärtner | Robert Koch Institute | |
| EPI_ISL_2262241, EPI_ISL_2262264, EPI_ISL_2262268, EPI_ISL_2262825, EPI_ISL_2263061, EPI_ISL_2263062, EPI_ISL_2263065, EPI_ISL_2263067, EPI_ISL_2263069, EPI_ISL_2263070, EPI_ISL_2263071, EPI_ISL_2263072, EPI_ISL_2263074, EPI_ISL_2263075, EPI_ISL_2263076, EPI_ISL_2263101, EPI_ISL_2263102, EPI_ISL_2263103, EPI_ISL_2263107, EPI_ISL_2263109, EPI_ISL_2263110, EPI_ISL_2263111, EPI_ISL_2263112, EPI_ISL_2263114, EPI_ISL_2263116, EPI_ISL_2263117, EPI_ISL_2263118, EPI_ISL_2263119, EPI_ISL_2263120, EPI_ISL_2263121, EPI_ISL_2263122, EPI_ISL_2263123, EPI_ISL_2263124, EPI_ISL_2263125, EPI_ISL_2263126, EPI_ISL_2263127, EPI_ISL_2263128, EPI_ISL_2263129, EPI_ISL_2263130, EPI_ISL_2263131, EPI_ISL_2263132, EPI_ISL_2263133, EPI_ISL_2263134, EPI_ISL_2263135, EPI_ISL_2263136, EPI_ISL_2263137, EPI_ISL_2263138, EPI_ISL_2263139, EPI_ISL_2263140, EPI_ISL_2263141, EPI_ISL_2263142, EPI_ISL_2263143, EPI_ISL_2264480, EPI_ISL_2264541, EPI_ISL_2264542, EPI_ISL_2264544, EPI_ISL_2264549, EPI_ISL_2264719, EPI_ISL_2264721, EPI_ISL_2264724, EPI_ISL_2264725, EPI_ISL_2264728, EPI_ISL_2264729, EPI_ISL_2264731, EPI_ISL_2264733, EPI_ISL_2264734, EPI_ISL_2264737, EPI_ISL_2264740, EPI_ISL_2266600, EPI_ISL_2266605, EPI_ISL_2266608, EPI_ISL_2266617, EPI_ISL_2266627, EPI_ISL_2314411, EPI_ISL_2314674, EPI_ISL_2314687, EPI_ISL_2314689, EPI_ISL_2314691, EPI_ISL_2314692, EPI_ISL_2314698, EPI_ISL_2314761, EPI_ISL_2386250, EPI_ISL_2386256, EPI_ISL_2386263, EPI_ISL_2386270, EPI_ISL_2386284 | | | |
| see above | MDI Limbach Berlin GmbH; MVZ Labor Berlin | Robert Koch Institute | |
| EPI_ISL_2772710 | MKK Mühlenkreiskliniken, Johannes Wesling Klinikum Minden | Hannover Medical School, Institute of Virology | Lars Steinbrück |
| EPI_ISL_2260968, EPI_ISL_2260970, EPI_ISL_2260971, EPI_ISL_2260975, EPI_ISL_2260976, EPI_ISL_2260978, EPI_ISL_2260984, EPI_ISL_2260988, EPI_ISL_2260990, EPI_ISL_2260993, EPI_ISL_2260994, EPI_ISL_2261001, EPI_ISL_2261006, EPI_ISL_2261013, EPI_ISL_2261022, EPI_ISL_2261027, EPI_ISL_2261028, EPI_ISL_2261030, EPI_ISL_2261031, EPI_ISL_2261032, EPI_ISL_2261041, EPI_ISL_2261044, EPI_ISL_2261046, EPI_ISL_2261047, EPI_ISL_2261048, EPI_ISL_2261052, EPI_ISL_2261055, EPI_ISL_2264985, EPI_ISL_2264986, EPI_ISL_2264987, EPI_ISL_2264988, EPI_ISL_2264989, EPI_ISL_2264990, EPI_ISL_2264991, EPI_ISL_2264992, EPI_ISL_2264993, EPI_ISL_2264994, EPI_ISL_2264995, EPI_ISL_2264996, EPI_ISL_2264998, EPI_ISL_2264999, EPI_ISL_2265001, EPI_ISL_2265002, EPI_ISL_2265004, EPI_ISL_2265006, EPI_ISL_2265009, EPI_ISL_2265011, EPI_ISL_2265012, EPI_ISL_2265013, EPI_ISL_2265016, EPI_ISL_2265017, EPI_ISL_2265018, EPI_ISL_2265020, EPI_ISL_2265021, EPI_ISL_2265022, EPI_ISL_2265023, EPI_ISL_2265024, EPI_ISL_2265025, EPI_ISL_2265026, EPI_ISL_2265027, EPI_ISL_2265028, EPI_ISL_2265029, EPI_ISL_2265030, EPI_ISL_2265031, EPI_ISL_2265032, EPI_ISL_2265033, EPI_ISL_2265034, EPI_ISL_2265035, EPI_ISL_2265036, EPI_ISL_2265037, EPI_ISL_2265038, EPI_ISL_2265039, EPI_ISL_2265040, EPI_ISL_2265041, EPI_ISL_2265042, EPI_ISL_2265043, EPI_ISL_2265044, EPI_ISL_2265046, EPI_ISL_2265047, EPI_ISL_2267934, EPI_ISL_2267935, EPI_ISL_2267939, EPI_ISL_2267941, EPI_ISL_2267942, EPI_ISL_2267946, EPI_ISL_2267947, EPI_ISL_2267950, EPI_ISL_2313251, EPI_ISL_2313253, EPI_ISL_2313262, EPI_ISL_2313265, EPI_ISL_2313266, EPI_ISL_2313267, EPI_ISL_2313268, EPI_ISL_2313270, EPI_ISL_2313271, EPI_ISL_2313276, EPI_ISL_2313278, EPI_ISL_2313281, EPI_ISL_2313285, EPI_ISL_2313286, EPI_ISL_2313287, EPI_ISL_2313288, EPI_ISL_2313289, EPI_ISL_2313290, EPI_ISL_2313292, EPI_ISL_2313293, EPI_ISL_2313294, EPI_ISL_2313296, EPI_ISL_2313297, EPI_ISL_2313298, EPI_ISL_2313299, EPI_ISL_2313300, EPI_ISL_2313301, EPI_ISL_2313303, EPI_ISL_2313304, EPI_ISL_2313305, EPI_ISL_2313306, EPI_ISL_2313309, EPI_ISL_2313310, EPI_ISL_2313311, EPI_ISL_2313313, EPI_ISL_2313315, EPI_ISL_2313316, EPI_ISL_2313320, EPI_ISL_2313321, EPI_ISL_2313322, EPI_ISL_2313324, EPI_ISL_2313325, EPI_ISL_2313327, EPI_ISL_2313328, EPI_ISL_2313329, EPI_ISL_2313330, EPI_ISL_2313331, EPI_ISL_2313332, EPI_ISL_2313333, EPI_ISL_2313334, EPI_ISL_2313335, EPI_ISL_2313338, EPI_ISL_2387346, EPI_ISL_2387350, EPI_ISL_2387352, EPI_ISL_2387354, EPI_ISL_2387360, EPI_ISL_2387362, EPI_ISL_2387364, EPI_ISL_2387365, EPI_ISL_2387366, EPI_ISL_2387367, EPI_ISL_2387368, EPI_ISL_2387369, EPI_ISL_2387372, EPI_ISL_2387373, EPI_ISL_2387375, EPI_ISL_2387376, EPI_ISL_2387377, EPI_ISL_2387378, EPI_ISL_2387379, EPI_ISL_2387380, EPI_ISL_2387383, EPI_ISL_2387384, EPI_ISL_2387387, EPI_ISL_2387390, EPI_ISL_2387391, EPI_ISL_2387393, EPI_ISL_2387394, EPI_ISL_2387395, EPI_ISL_2387397, EPI_ISL_2387401, EPI_ISL_2387402, EPI_ISL_2387403, EPI_ISL_2387405, EPI_ISL_2387406, EPI_ISL_2387407, EPI_ISL_2387408, EPI_ISL_2387410, EPI_ISL_2387412, EPI_ISL_2387415, EPI_ISL_2387416, EPI_ISL_2387422, EPI_ISL_2387424, EPI_ISL_2387428, EPI_ISL_2387430, EPI_ISL_2387435, EPI_ISL_2387438, EPI_ISL_2387440, EPI_ISL_2387442, EPI_ISL_2387443, EPI_ISL_2387449, EPI_ISL_2387450, EPI_ISL_2387451, EPI_ISL_2387453 | | | |
| see above | MVZ Dr. Eberhard & Partner Dortmund | Robert Koch Institute | |
| EPI_ISL_2264094, EPI_ISL_2264095, EPI_ISL_2264096, EPI_ISL_2264098, EPI_ISL_2264099, EPI_ISL_2264100, EPI_ISL_2264101, EPI_ISL_2264102, EPI_ISL_2264103, EPI_ISL_2264104, EPI_ISL_2264105, EPI_ISL_2264106, EPI_ISL_2264107, EPI_ISL_2264108, EPI_ISL_2264109, EPI_ISL_2264110, EPI_ISL_2264111, EPI_ISL_2264112, EPI_ISL_2264113, EPI_ISL_2264114, EPI_ISL_2264115, EPI_ISL_2264116, EPI_ISL_2264117, EPI_ISL_2264118, EPI_ISL_2264119, EPI_ISL_2264120, EPI_ISL_2264122, EPI_ISL_2264123, EPI_ISL_2264124, EPI_ISL_2264125, EPI_ISL_2264126, EPI_ISL_2264127, EPI_ISL_2264128, EPI_ISL_2264129, EPI_ISL_2264130, EPI_ISL_2264131, EPI_ISL_2264132, EPI_ISL_2264133, EPI_ISL_2264134, EPI_ISL_2264135, EPI_ISL_2264136, EPI_ISL_2264137, EPI_ISL_2264139, EPI_ISL_2264140, EPI_ISL_2264141, EPI_ISL_2314806, EPI_ISL_2314807, EPI_ISL_2314808, EPI_ISL_2314810, EPI_ISL_2314811, EPI_ISL_2314812, EPI_ISL_2314813, EPI_ISL_2314814, EPI_ISL_2314815, EPI_ISL_2314816, EPI_ISL_2314817, EPI_ISL_2314818, EPI_ISL_2314819, EPI_ISL_2314820, EPI_ISL_2314821, EPI_ISL_2314822, EPI_ISL_2314823, EPI_ISL_2314824, EPI_ISL_2314825, EPI_ISL_2314826, EPI_ISL_2314827, EPI_ISL_2314828, EPI_ISL_2314829, EPI_ISL_2314830, EPI_ISL_2314831, EPI_ISL_2314832, EPI_ISL_2390156, EPI_ISL_2390158, EPI_ISL_2390167, EPI_ISL_2390175 | | | |
| see above | MVZ Labor Dr. Fenner und Kollegen (Standort Hamburg) | Robert Koch Institute | |
| EPI_ISL_2259638, EPI_ISL_2259645, EPI_ISL_2259646, EPI_ISL_2259647, EPI_ISL_2259648, EPI_ISL_2259649, EPI_ISL_2259651, EPI_ISL_2259652, EPI_ISL_2259653, EPI_ISL_2259654, EPI_ISL_2259656, EPI_ISL_2259659, EPI_ISL_2259661, EPI_ISL_2259662, EPI_ISL_2259664, EPI_ISL_2259670, EPI_ISL_2259671, EPI_ISL_2259673, EPI_ISL_2259675, EPI_ISL_2259679, EPI_ISL_2259687, EPI_ISL_2259742, EPI_ISL_2259743, EPI_ISL_2259748, EPI_ISL_2259752, EPI_ISL_2259762, EPI_ISL_2259766, EPI_ISL_2259767, EPI_ISL_2259769, EPI_ISL_2259774, EPI_ISL_2259777, EPI_ISL_2259790, EPI_ISL_2259791, EPI_ISL_2259792, EPI_ISL_2259793, EPI_ISL_2259794, EPI_ISL_2259795, EPI_ISL_2259803, EPI_ISL_2259804, EPI_ISL_2259807, EPI_ISL_2260918, EPI_ISL_2262222, EPI_ISL_2262229, EPI_ISL_2262231, EPI_ISL_2262232, EPI_ISL_2262239, EPI_ISL_2262243, EPI_ISL_2262244, EPI_ISL_2262245, EPI_ISL_2262246, EPI_ISL_2262256, EPI_ISL_2262259, EPI_ISL_2262272, EPI_ISL_2262449, EPI_ISL_2265871, EPI_ISL_2266607, EPI_ISL_2313117, EPI_ISL_2313119, EPI_ISL_2313120, EPI_ISL_2313121, EPI_ISL_2313122, EPI_ISL_2313123, EPI_ISL_2313124, EPI_ISL_2313125, EPI_ISL_2313126, EPI_ISL_2313127, EPI_ISL_2313128, EPI_ISL_2313129, EPI_ISL_2313130, EPI_ISL_2313131, EPI_ISL_2313132, EPI_ISL_2313133, EPI_ISL_2313134, EPI_ISL_2313135, EPI_ISL_2313136, EPI_ISL_2313137, EPI_ISL_2313138, EPI_ISL_2313139, EPI_ISL_2313140, EPI_ISL_2313141, EPI_ISL_2313143, EPI_ISL_2313144, EPI_ISL_2313145, EPI_ISL_2313146, EPI_ISL_2313147, EPI_ISL_2313148, EPI_ISL_2313150, EPI_ISL_2313151, EPI_ISL_2313152, EPI_ISL_2313153, EPI_ISL_2313154, EPI_ISL_2313155, EPI_ISL_2313156, EPI_ISL_2313157, EPI_ISL_2313158, EPI_ISL_2313159, EPI_ISL_2313160, EPI_ISL_2313161, EPI_ISL_2313162, EPI_ISL_2313163, EPI_ISL_2313164, EPI_ISL_2313165, EPI_ISL_2313166, EPI_ISL_2313167, EPI_ISL_2313168, EPI_ISL_2313169, EPI_ISL_2313171, EPI_ISL_2313172, EPI_ISL_2313173, EPI_ISL_2313175, EPI_ISL_2313177, EPI_ISL_2313178, EPI_ISL_2313179, EPI_ISL_2313180, EPI_ISL_2313182, EPI_ISL_2313183, EPI_ISL_2313185, EPI_ISL_2313186, EPI_ISL_2313189, EPI_ISL_2313190, EPI_ISL_2313199, EPI_ISL_2313202, EPI_ISL_2313204, EPI_ISL_2313205, EPI_ISL_2313207, EPI_ISL_2313209, EPI_ISL_2313211, EPI_ISL_2313214, EPI_ISL_2313215, EPI_ISL_2313216, EPI_ISL_2313217, EPI_ISL_2313218, EPI_ISL_2313221, EPI_ISL_2313223, EPI_ISL_2313224, EPI_ISL_2313225, EPI_ISL_2313227, EPI_ISL_2313228, EPI_ISL_2313229, EPI_ISL_2313230, EPI_ISL_2313232, EPI_ISL_2313233, EPI_ISL_2313234, EPI_ISL_2313235, EPI_ISL_2313236, EPI_ISL_2313237, EPI_ISL_2313239, EPI_ISL_2313364, EPI_ISL_2313369, EPI_ISL_2314038, EPI_ISL_2386165, EPI_ISL_2386171, EPI_ISL_2386177, EPI_ISL_2386237, EPI_ISL_2388011, EPI_ISL_2388014, EPI_ISL_2388016, EPI_ISL_2388017, EPI_ISL_2388020, EPI_ISL_2388021, EPI_ISL_2388022, EPI_ISL_2388024, EPI_ISL_2388025, EPI_ISL_2388026, EPI_ISL_2388027, EPI_ISL_2388030, EPI_ISL_2388032, EPI_ISL_2388068, EPI_ISL_2388072, EPI_ISL_2388080, EPI_ISL_2388082, EPI_ISL_2388084, EPI_ISL_2388086, EPI_ISL_2388089, EPI_ISL_2388097, EPI_ISL_2388102, EPI_ISL_2388108, EPI_ISL_2388114, EPI_ISL_2388115, EPI_ISL_2388118, EPI_ISL_2388119, EPI_ISL_2388121, EPI_ISL_2388122, EPI_ISL_2388125, EPI_ISL_2388127, EPI_ISL_2388130, EPI_ISL_2388135, EPI_ISL_2388136, EPI_ISL_2388137, EPI_ISL_2388139, EPI_ISL_2388142, EPI_ISL_2388147, EPI_ISL_2388157, EPI_ISL_2388162, EPI_ISL_2388163, EPI_ISL_2388166, EPI_ISL_2388176, EPI_ISL_2388181, EPI_ISL_2390092, EPI_ISL_2390095, EPI_ISL_2390102, EPI_ISL_2390302, EPI_ISL_2390307, EPI_ISL_2390312, EPI_ISL_2390314, EPI_ISL_2390317, EPI_ISL_2471466, EPI_ISL_2471471 | | | |
| see above | MVZ Labor Dr. Limbach & Kollegen GbR | Robert Koch Institute | |
| EPI_ISL_2262456, EPI_ISL_2262473, EPI_ISL_2262474, EPI_ISL_2262476, EPI_ISL_2262486, EPI_ISL_2262492, EPI_ISL_2262507, EPI_ISL_2262509, EPI_ISL_2262518, EPI_ISL_2262591, EPI_ISL_2262614, EPI_ISL_2315456, EPI_ISL_2315457, EPI_ISL_2315458, EPI_ISL_2315460, EPI_ISL_2315461, EPI_ISL_2315463, EPI_ISL_2315466, EPI_ISL_2315467, EPI_ISL_2315468, EPI_ISL_2315469, EPI_ISL_2315470, EPI_ISL_2315471, EPI_ISL_2315472, EPI_ISL_2315473, EPI_ISL_2315474, EPI_ISL_2315475, EPI_ISL_2315476, EPI_ISL_2315477, EPI_ISL_2315479, EPI_ISL_2315480, EPI_ISL_2315482, EPI_ISL_2315483, EPI_ISL_2315484, EPI_ISL_2315486, EPI_ISL_2315487, EPI_ISL_2315488, EPI_ISL_2315489, EPI_ISL_2315490, EPI_ISL_2315491, EPI_ISL_2315492, EPI_ISL_2315493, EPI_ISL_2315494, EPI_ISL_2315495, EPI_ISL_2315496, EPI_ISL_2315499, EPI_ISL_2315500, EPI_ISL_2315502, EPI_ISL_2315504, EPI_ISL_2315505, EPI_ISL_2315507, EPI_ISL_2315508, EPI_ISL_2315510, EPI_ISL_2315511, EPI_ISL_2315512, EPI_ISL_2315513, EPI_ISL_2315514, EPI_ISL_2315516, EPI_ISL_2315517, EPI_ISL_2315518, EPI_ISL_2315520, EPI_ISL_2315521, EPI_ISL_2315523, EPI_ISL_2315524, EPI_ISL_2315525, EPI_ISL_2315526, EPI_ISL_2315527, EPI_ISL_2315528, EPI_ISL_2315529, EPI_ISL_2315532, EPI_ISL_2315536, EPI_ISL_2315538, EPI_ISL_2315540, EPI_ISL_2315541, EPI_ISL_2315542, EPI_ISL_2315543, EPI_ISL_2315547, EPI_ISL_2315548, EPI_ISL_2315551, EPI_ISL_2315553, EPI_ISL_2315555, EPI_ISL_2315558, EPI_ISL_2315561, EPI_ISL_2315562, EPI_ISL_2315563, EPI_ISL_2315564, EPI_ISL_2315566, EPI_ISL_2315567, EPI_ISL_2315568, EPI_ISL_2315569, EPI_ISL_2315570, EPI_ISL_2315571, EPI_ISL_2315572, EPI_ISL_2315574, EPI_ISL_2315576, EPI_ISL_2315577, EPI_ISL_2315578, EPI_ISL_2315580, EPI_ISL_2390023, EPI_ISL_2390038, EPI_ISL_2390040, EPI_ISL_2390048, EPI_ISL_2390063, EPI_ISL_2390068, EPI_ISL_2390077, EPI_ISL_2390089, EPI_ISL_2678546 | | | |
| see above | MVZ Labor Dr. Quade & Kollegen GmbH | Robert Koch Institute | |
| EPI_ISL_2263038, EPI_ISL_2263053, EPI_ISL_2263059, EPI_ISL_2264557, EPI_ISL_2264560, EPI_ISL_2264561, EPI_ISL_2264573, EPI_ISL_2265876, EPI_ISL_2265882, EPI_ISL_2265886, EPI_ISL_2314643, EPI_ISL_2314649, EPI_ISL_2314654, EPI_ISL_2314657 | | | |
| see above | MVZ Labor Dr. Reising-Ackermann und Kollegen | Robert Koch Institute | |
| EPI_ISL_2126306, EPI_ISL_2264666, EPI_ISL_2264667, EPI_ISL_2264668, EPI_ISL_2264669, EPI_ISL_2264670, EPI_ISL_2264671, EPI_ISL_2264672, EPI_ISL_2264673, EPI_ISL_2264675, EPI_ISL_2264676, EPI_ISL_2264677, EPI_ISL_2264678, EPI_ISL_2264679, EPI_ISL_2264680, EPI_ISL_2264681, EPI_ISL_2264682, EPI_ISL_2264683, EPI_ISL_2264684, EPI_ISL_2264685, EPI_ISL_2264686, EPI_ISL_2264687, EPI_ISL_2264688, EPI_ISL_2264689, EPI_ISL_2264690, EPI_ISL_2264691, EPI_ISL_2264692, EPI_ISL_2264693, EPI_ISL_2264694, EPI_ISL_2264695, EPI_ISL_2264696, EPI_ISL_2264698, EPI_ISL_2264699, EPI_ISL_2264700, EPI_ISL_2264701, EPI_ISL_2264702, EPI_ISL_2264703, EPI_ISL_2264704, EPI_ISL_2264705, EPI_ISL_2265889, EPI_ISL_2265890, EPI_ISL_2265891, EPI_ISL_2265892, EPI_ISL_2265893, EPI_ISL_2265894, EPI_ISL_2265895, EPI_ISL_2265896, EPI_ISL_2265897, EPI_ISL_2265898, EPI_ISL_2265899, EPI_ISL_2265900, EPI_ISL_2265901, EPI_ISL_2265902, EPI_ISL_2265903, EPI_ISL_2265904, EPI_ISL_2265905, EPI_ISL_2265906, EPI_ISL_2265907, EPI_ISL_2265908, EPI_ISL_2265909, EPI_ISL_2265910, EPI_ISL_2265911, EPI_ISL_2265912, EPI_ISL_2265913, EPI_ISL_2265914, EPI_ISL_2265915, EPI_ISL_2265916, EPI_ISL_2265917, EPI_ISL_2265918, EPI_ISL_2265919, EPI_ISL_2265920, EPI_ISL_2265921, EPI_ISL_2265922, EPI_ISL_2265923, EPI_ISL_2266032, EPI_ISL_2266033, EPI_ISL_2266034, EPI_ISL_2266035, EPI_ISL_2266036, EPI_ISL_2266037, EPI_ISL_2266038, EPI_ISL_2266039, EPI_ISL_2266040, EPI_ISL_2266041, EPI_ISL_2266042, EPI_ISL_2266043, EPI_ISL_2266044, EPI_ISL_2266045, EPI_ISL_2266046, EPI_ISL_2266047, EPI_ISL_2266048, EPI_ISL_2266049, EPI_ISL_2266050, EPI_ISL_2266051, EPI_ISL_2266052, EPI_ISL_2266053, EPI_ISL_2266054, EPI_ISL_2266055, EPI_ISL_2266056, EPI_ISL_2266057, EPI_ISL_2266058, EPI_ISL_2266059, EPI_ISL_2266060, EPI_ISL_2266061, EPI_ISL_2266062, EPI_ISL_2266063, EPI_ISL_2266064, EPI_ISL_2266065, EPI_ISL_2266066, EPI_ISL_2266067, EPI_ISL_2266068, EPI_ISL_2266069, EPI_ISL_2266070, EPI_ISL_2266071, EPI_ISL_2266072, EPI_ISL_2266352, EPI_ISL_2266353, EPI_ISL_2266354, EPI_ISL_2266355, EPI_ISL_2266356, EPI_ISL_2266357, EPI_ISL_2266358, EPI_ISL_2266359, EPI_ISL_2266360, EPI_ISL_2266361, EPI_ISL_2266362, EPI_ISL_2266363, EPI_ISL_2266364, EPI_ISL_2266365, EPI_ISL_2266366, EPI_ISL_2266367, EPI_ISL_2266368, EPI_ISL_2266369, EPI_ISL_2266370, EPI_ISL_2266371, EPI_ISL_2266372, EPI_ISL_2266373, EPI_ISL_2266374, EPI_ISL_2266375, EPI_ISL_2266376, EPI_ISL_2266377, EPI_ISL_2266378, EPI_ISL_2266379, EPI_ISL_2266380, EPI_ISL_2266381, EPI_ISL_2266382, EPI_ISL_2266383, EPI_ISL_2266384, EPI_ISL_2266385, EPI_ISL_2266386, EPI_ISL_2266387, EPI_ISL_2266388, EPI_ISL_2266389, EPI_ISL_2266390, EPI_ISL_2266391, EPI_ISL_2266392, EPI_ISL_2266393, EPI_ISL_2266394, EPI_ISL_2266395, EPI_ISL_2266396, EPI_ISL_2266397, EPI_ISL_2266398, EPI_ISL_2266399, EPI_ISL_2266400, EPI_ISL_2266401, EPI_ISL_2266402, EPI_ISL_2266403, EPI_ISL_2266404, EPI_ISL_2266405, EPI_ISL_2266406, EPI_ISL_2266407, EPI_ISL_2266408, EPI_ISL_2266409, EPI_ISL_2266410, EPI_ISL_2266411, EPI_ISL_2266412, EPI_ISL_2266413, EPI_ISL_2266629, EPI_ISL_2266630, EPI_ISL_2266631, EPI_ISL_2266632, EPI_ISL_2266633, EPI_ISL_2266634, EPI_ISL_2266635, EPI_ISL_2266636, EPI_ISL_2266637, EPI_ISL_2266638, EPI_ISL_2266639, EPI_ISL_2266640, EPI_ISL_2266641, EPI_ISL_2266642, EPI_ISL_2266643, EPI_ISL_2266644, EPI_ISL_2266645, EPI_ISL_2266646, EPI_ISL_2315876, EPI_ISL_2315877, EPI_ISL_2315878, EPI_ISL_2315879, EPI_ISL_2315880, EPI_ISL_2315881, EPI_ISL_2315882, EPI_ISL_2315883, EPI_ISL_2315884, EPI_ISL_2315885, EPI_ISL_2315886, EPI_ISL_2315887, EPI_ISL_2315888, EPI_ISL_2315889, EPI_ISL_2315890, EPI_ISL_2315891, EPI_ISL_2315892, EPI_ISL_2315893, EPI_ISL_2315894, EPI_ISL_2315895, EPI_ISL_2315896, EPI_ISL_2315897, EPI_ISL_2315898, EPI_ISL_2315899, EPI_ISL_2315900, EPI_ISL_2315901, EPI_ISL_2315902, EPI_ISL_2315903, EPI_ISL_2315904, EPI_ISL_2315905, EPI_ISL_2315906, EPI_ISL_2315907, EPI_ISL_2315908, EPI_ISL_2315909, EPI_ISL_2315910, EPI_ISL_2315911, EPI_ISL_2315912, EPI_ISL_2315913, EPI_ISL_2315914, EPI_ISL_2315915, EPI_ISL_2315916, EPI_ISL_2315917, EPI_ISL_2316648, EPI_ISL_2316650, EPI_ISL_2316654, EPI_ISL_2316657, EPI_ISL_2316661, EPI_ISL_2316664, EPI_ISL_2316667, EPI_ISL_2316670, EPI_ISL_2316672, EPI_ISL_2316675, EPI_ISL_2316678, EPI_ISL_2316681, EPI_ISL_2316685, EPI_ISL_2316688, EPI_ISL_2316691, EPI_ISL_2316694, EPI_ISL_2316698, EPI_ISL_2316702, EPI_ISL_2316705, EPI_ISL_2316708, EPI_ISL_2316711, EPI_ISL_2316714, EPI_ISL_2316718, EPI_ISL_2316722, EPI_ISL_2316728, EPI_ISL_2316731, EPI_ISL_2316733, EPI_ISL_2316735, EPI_ISL_2316738, EPI_ISL_2316741, EPI_ISL_2316744, EPI_ISL_2316748, EPI_ISL_2316751, EPI_ISL_2316754, EPI_ISL_2316757, EPI_ISL_2316760, EPI_ISL_2316768, EPI_ISL_2316772, EPI_ISL_2316775, EPI_ISL_2316779, EPI_ISL_2316782, EPI_ISL_2316785, EPI_ISL_2316788, EPI_ISL_2316792, EPI_ISL_2316795, EPI_ISL_2316798, EPI_ISL_2316799, EPI_ISL_2316804, EPI_ISL_2316807, EPI_ISL_2316811, EPI_ISL_2316814, EPI_ISL_2316817, EPI_ISL_2316823, EPI_ISL_2316826, EPI_ISL_2389459, EPI_ISL_2389460, EPI_ISL_2389462, EPI_ISL_2389463, EPI_ISL_2389465, EPI_ISL_2389466, EPI_ISL_2389467, EPI_ISL_2389468, EPI_ISL_2389469, EPI_ISL_2389471, EPI_ISL_2389472, EPI_ISL_2389474, EPI_ISL_2389475, EPI_ISL_2389476, EPI_ISL_2389477, EPI_ISL_2389478, EPI_ISL_2389480, EPI_ISL_2389481, EPI_ISL_2389483, EPI_ISL_2389484, EPI_ISL_2389485, EPI_ISL_2389486, EPI_ISL_2389487, EPI_ISL_2389488, EPI_ISL_2389489, EPI_ISL_2389490, EPI_ISL_2389491, EPI_ISL_2389492, EPI_ISL_2389493, EPI_ISL_2389494, EPI_ISL_2389495, EPI_ISL_2389496, EPI_ISL_2389497, EPI_ISL_2389498, EPI_ISL_2389499, EPI_ISL_2389501, EPI_ISL_2389502, EPI_ISL_2389503, EPI_ISL_2389504, EPI_ISL_2389505, EPI_ISL_2389506, EPI_ISL_2389508, EPI_ISL_2389509, EPI_ISL_2389510 | | | |

| | | | |
|---|---|---|---|
| see above | MVZ Labor Krone GbR | Robert Koch Institute | |
| EPI_ISL_2263079, EPI_ISL_2263089, EPI_ISL_2314688 | MVZ Labor Limbach Erfurt GmbH | Robert Koch Institute | |
| EPI_ISL_2264545, EPI_ISL_2264618, EPI_ISL_2264726, EPI_ISL_2264727, EPI_ISL_2264735, EPI_ISL_2264736, EPI_ISL_2314685, EPI_ISL_2314690, EPI_ISL_2314699, EPI_ISL_2314701, EPI_ISL_2314705, EPI_ISL_2314723, EPI_ISL_2314763, EPI_ISL_2314768 | | | |
| see above | MVZ Medizinisches Labor Hannover GmbH | Robert Koch Institute | |
| EPI_ISL_2313964, EPI_ISL_2313967, EPI_ISL_2313974, EPI_ISL_2313982, EPI_ISL_2313984, EPI_ISL_2313999, EPI_ISL_2314005, EPI_ISL_2314007, EPI_ISL_2314009, EPI_ISL_2314014, EPI_ISL_2314017, EPI_ISL_2314022, EPI_ISL_2314026, EPI_ISL_2314028, EPI_ISL_2314035, EPI_ISL_2314036, EPI_ISL_2314735, EPI_ISL_2314738, EPI_ISL_2314745, EPI_ISL_2314769, EPI_ISL_2386188, EPI_ISL_2386275, EPI_ISL_2386276, EPI_ISL_2390109 | | | |
| see above | MVZ für Laboratoriumsmedizin und Mikrobiologie Koblenz-Mittelrhein (Labor Koblenz) | Robert Koch Institute | |
| EPI_ISL_2450231, EPI_ISL_2450232, EPI_ISL_2450233, EPI_ISL_2450234, EPI_ISL_2450235, EPI_ISL_2450236, EPI_ISL_2450237, EPI_ISL_2450238, EPI_ISL_2450239, EPI_ISL_2450240, EPI_ISL_2450241, EPI_ISL_2450242, EPI_ISL_2450243, EPI_ISL_2450244, EPI_ISL_2450245, EPI_ISL_2450246, EPI_ISL_2450247, EPI_ISL_2450248, EPI_ISL_2450249, EPI_ISL_2450250, EPI_ISL_2450251, EPI_ISL_2450252, EPI_ISL_2450253, EPI_ISL_2450254, EPI_ISL_2450255, EPI_ISL_2450256, EPI_ISL_2450257, EPI_ISL_2450258, EPI_ISL_2450259, EPI_ISL_2450260, EPI_ISL_2450261, EPI_ISL_2450262, EPI_ISL_2450263, EPI_ISL_2450264, EPI_ISL_2450265, EPI_ISL_2450266, EPI_ISL_2450287, EPI_ISL_2450305, EPI_ISL_2450306, EPI_ISL_2450307, EPI_ISL_2450308, EPI_ISL_2450310, EPI_ISL_2450311, EPI_ISL_2450312, EPI_ISL_2450317, EPI_ISL_2450318, EPI_ISL_2450319, EPI_ISL_2450320, EPI_ISL_2450326 | | | |
| see above | Max von Pettenkofer Institute, Virology, National Reference Center for Retroviruses, LMU Munich | Laboratory for Functional Genome Analysis; Dept. Genomics; Gene Center of the LMU Munich | Alexander Graf; Helmut Blum; Max Muenchhoff; Oliver Keppler; Stefan Krebs |
| EPI_ISL_2260588, EPI_ISL_2260589, EPI_ISL_2260590, EPI_ISL_2260592, EPI_ISL_2260593, EPI_ISL_2260594, EPI_ISL_2260596, EPI_ISL_2260597, EPI_ISL_2260598, EPI_ISL_2260599, EPI_ISL_2389360, EPI_ISL_2389368 | | | |
| see above | Med. Labor Prof. Schenk Dr. Ansorge & Kollegen | Robert Koch Institute | |
| EPI_ISL_2380534, EPI_ISL_2380535, EPI_ISL_2380536, EPI_ISL_2380537, EPI_ISL_2380538, EPI_ISL_2380539, EPI_ISL_2380540, EPI_ISL_2380542, EPI_ISL_2380584, EPI_ISL_2380585, EPI_ISL_2380586, EPI_ISL_2380587, EPI_ISL_2380588, EPI_ISL_2638049, EPI_ISL_2638055, EPI_ISL_2638056, EPI_ISL_2638060, EPI_ISL_2638061, EPI_ISL_2638062, EPI_ISL_2638064, EPI_ISL_2638065, EPI_ISL_2638067, EPI_ISL_2638068, EPI_ISL_2638069, EPI_ISL_2638070, EPI_ISL_2638071, EPI_ISL_2638072, EPI_ISL_2638073, EPI_ISL_2638074, EPI_ISL_2638075, EPI_ISL_2638076, EPI_ISL_2638078, EPI_ISL_2638079, EPI_ISL_2638080, EPI_ISL_2638081, EPI_ISL_2638083, EPI_ISL_2638084, EPI_ISL_2638086, EPI_ISL_2638087, EPI_ISL_2638088, EPI_ISL_2638089, EPI_ISL_2638091 | | | |
| see above | Medical Laboratories Duesseldorf | Center of Medical Microbiology, Virology, and Hospital Hygiene, University of Duesseldorf | Alexander Dilthey; Andreas Walker; Angelika Helmer; Christian Lange; Daniel Strelow; Jessica Nicolai; Jörg Timm; Klaus Pfeffer; Lisanna Hülse; Malte Kohns Vasconcelos; Maximilian Damagnez; Nadine Lübke; Tobias Wienemann; Torsten Houwaart |
| EPI_ISL_2129680, EPI_ISL_2129686, EPI_ISL_2129690, EPI_ISL_2129691, EPI_ISL_2129692, EPI_ISL_2129693, EPI_ISL_2129697, EPI_ISL_2129709, EPI_ISL_2129719 | | | |
| see above | Medizinisch-Diagnostisches Labor Kempten allgÃ¤ulab | Robert Koch Institute | |
| EPI_ISL_2266102, EPI_ISL_2266104, EPI_ISL_2266105, EPI_ISL_2266115, EPI_ISL_2266116, EPI_ISL_2266119, EPI_ISL_2266125, EPI_ISL_2266127, EPI_ISL_2266129, EPI_ISL_2471768, EPI_ISL_2471782, EPI_ISL_2471783, EPI_ISL_2472771, EPI_ISL_2472773, EPI_ISL_2472774, EPI_ISL_2472775, EPI_ISL_2472778, EPI_ISL_2472780, EPI_ISL_2472781, EPI_ISL_2472782, EPI_ISL_2635976, EPI_ISL_2635978, EPI_ISL_2635979, EPI_ISL_2635982, EPI_ISL_2635984, EPI_ISL_2635988, EPI_ISL_2635989, EPI_ISL_2635990, EPI_ISL_2635991, EPI_ISL_2635995, EPI_ISL_2636077, EPI_ISL_2636082, EPI_ISL_2636085, EPI_ISL_2636087, EPI_ISL_2636089, EPI_ISL_2636092, EPI_ISL_2636096, EPI_ISL_2636100, EPI_ISL_2636102, EPI_ISL_2636103, EPI_ISL_2636112, EPI_ISL_2636113, EPI_ISL_2636116, EPI_ISL_2636121, EPI_ISL_2636788, EPI_ISL_2636789, EPI_ISL_2636790, EPI_ISL_2636797, EPI_ISL_2636802, EPI_ISL_2636803, EPI_ISL_2636804, EPI_ISL_2636805, EPI_ISL_2636806, EPI_ISL_2636807, EPI_ISL_2636810, EPI_ISL_2636812, EPI_ISL_2636819, EPI_ISL_2636823, EPI_ISL_2636826, EPI_ISL_2636829, EPI_ISL_2636831, EPI_ISL_2636836, EPI_ISL_2636838, EPI_ISL_2636840, EPI_ISL_2636844, EPI_ISL_2636845, EPI_ISL_2636846, EPI_ISL_2636851, EPI_ISL_2636852, EPI_ISL_2636854, EPI_ISL_2636856, EPI_ISL_2636860, EPI_ISL_2636862, EPI_ISL_2636868, EPI_ISL_2636869, EPI_ISL_2636874, EPI_ISL_2636875 | | | |
| see above | Medizinisch-Diagnostisches Labor Kempten allgäulab | Robert Koch Institute | |
| EPI_ISL_2129020, EPI_ISL_2129024, EPI_ISL_2129038, EPI_ISL_2129050, EPI_ISL_2129055, EPI_ISL_2129066 | Medizinische Laboratorien Dusseldorf | Robert Koch Institute | |
| EPI_ISL_2265477, EPI_ISL_2265478, EPI_ISL_2265479, EPI_ISL_2265480, EPI_ISL_2265481, EPI_ISL_2265482, EPI_ISL_2265483, EPI_ISL_2265484, EPI_ISL_2265485, EPI_ISL_2265486, EPI_ISL_2265487, EPI_ISL_2265488, EPI_ISL_2265489, EPI_ISL_2265490, EPI_ISL_2265491, EPI_ISL_2265492, EPI_ISL_2265493, EPI_ISL_2265494, EPI_ISL_2265495, EPI_ISL_2265496, EPI_ISL_2265497, EPI_ISL_2265498, EPI_ISL_2265499, EPI_ISL_2265500, EPI_ISL_2265501, EPI_ISL_2265502, EPI_ISL_2265503, EPI_ISL_2265504, EPI_ISL_2265505, EPI_ISL_2265506, EPI_ISL_2265507, EPI_ISL_2265508, EPI_ISL_2265509, EPI_ISL_2265510, EPI_ISL_2265511, EPI_ISL_2265512, EPI_ISL_2265513, EPI_ISL_2266647, EPI_ISL_2266648, EPI_ISL_2266649, EPI_ISL_2266650, EPI_ISL_2266651, EPI_ISL_2266652, EPI_ISL_2266653, EPI_ISL_2266654, EPI_ISL_2266655, EPI_ISL_2266656, EPI_ISL_2266657, EPI_ISL_2266658, EPI_ISL_2266659, EPI_ISL_2266660, EPI_ISL_2266661, EPI_ISL_2266662, EPI_ISL_2266663, EPI_ISL_2389080, EPI_ISL_2389082, EPI_ISL_2389084, EPI_ISL_2389085, EPI_ISL_2389087, EPI_ISL_2389088, EPI_ISL_2389089, EPI_ISL_2389092, EPI_ISL_2389094, EPI_ISL_2389095, EPI_ISL_2389096, EPI_ISL_2389097, EPI_ISL_2389098, EPI_ISL_2389099, EPI_ISL_2389102, EPI_ISL_2389104, EPI_ISL_2389105, EPI_ISL_2389106, EPI_ISL_2389107, EPI_ISL_2389108, EPI_ISL_2389109, EPI_ISL_2389111, EPI_ISL_2389112, EPI_ISL_2389115, EPI_ISL_2389116, EPI_ISL_2389117, EPI_ISL_2389118, EPI_ISL_2389119, EPI_ISL_2389120, EPI_ISL_2389126 | | | |
| see above | Medizinische Laboratorien Düsseldorf | Robert Koch Institute | |
| EPI_ISL_2262336, EPI_ISL_2262339, EPI_ISL_2262346, EPI_ISL_2262348, EPI_ISL_2262349, EPI_ISL_2262355, EPI_ISL_2262356, EPI_ISL_2262358, EPI_ISL_2262362, EPI_ISL_2262366, EPI_ISL_2262372, EPI_ISL_2262376, EPI_ISL_2262382, EPI_ISL_2262384, EPI_ISL_2264617, EPI_ISL_2264619, EPI_ISL_2264622, EPI_ISL_2264623, EPI_ISL_2264624, EPI_ISL_2264625, EPI_ISL_2264626, EPI_ISL_2264630, EPI_ISL_2264631, EPI_ISL_2264632, EPI_ISL_2264633, EPI_ISL_2264634, EPI_ISL_2264732, EPI_ISL_2264739, EPI_ISL_2313907, EPI_ISL_2313913, EPI_ISL_2313915, EPI_ISL_2313916, EPI_ISL_2313917, EPI_ISL_2313918, EPI_ISL_2313920, EPI_ISL_2313921, EPI_ISL_2313924, EPI_ISL_2313938, EPI_ISL_2313941, EPI_ISL_2313947, EPI_ISL_2313952, EPI_ISL_2313954, EPI_ISL_2313959 | | | |
| see above | Medizinisches Labor Ostsachsen MVZ GbR Nebenbetriebsstätte Dresden | Robert Koch Institute | |
| EPI_ISL_2260969, EPI_ISL_2260973, EPI_ISL_2260991, EPI_ISL_2261012, EPI_ISL_2261054, EPI_ISL_2264997, EPI_ISL_2265003, EPI_ISL_2265010, EPI_ISL_2265014, EPI_ISL_2265015, EPI_ISL_2265019, EPI_ISL_2267930, EPI_ISL_2267933, EPI_ISL_2267936, EPI_ISL_2267937, EPI_ISL_2267938, EPI_ISL_2267944, EPI_ISL_2387355, EPI_ISL_2387419, EPI_ISL_2387431 | | | |
| see above | Medizinisches Labor Wahl Lüdenscheid | Robert Koch Institute | |
| EPI_ISL_2267945 | Medizinisches Versorgungszentrum für Labormedizin und Mikrobiologie Ruhr GmbH - mvzlm RUHR GmbH | Robert Koch Institute | |
| EPI_ISL_2266001, EPI_ISL_2266002, EPI_ISL_2266003, EPI_ISL_2266004, EPI_ISL_2266005, EPI_ISL_2266006, EPI_ISL_2266007, EPI_ISL_2266008, EPI_ISL_2266009, EPI_ISL_2266010, EPI_ISL_2266011, EPI_ISL_2266013, EPI_ISL_2266014, EPI_ISL_2390795, EPI_ISL_2390796, EPI_ISL_2390797, EPI_ISL_2390798, EPI_ISL_2390799, EPI_ISL_2632323, EPI_ISL_2632324, EPI_ISL_2632325, EPI_ISL_2632326, EPI_ISL_2632327, EPI_ISL_2632328, EPI_ISL_2632329, EPI_ISL_2632821, EPI_ISL_2635803 | | | |
| see above | Niedersächsisches Landesgesundheitsamt (NLGA) | Robert Koch Institute | |
| EPI_ISL_2129663, EPI_ISL_2129664, EPI_ISL_2129668, EPI_ISL_2129672 | Philipps-UniversitÃ¤t Marburg Fachbereich Medizin Institut fÃ¼r Virologie | Robert Koch Institute | |
| EPI_ISL_2265465, EPI_ISL_2265467, EPI_ISL_2265468 | Philipps-Universität Marburg Fachbereich Medizin Institut für Virologie | Robert Koch Institute | |
| EPI_ISL_2263645, EPI_ISL_2263755, EPI_ISL_2264384, EPI_ISL_2265206, EPI_ISL_2314636 | Praxisgemeinschaft für Laboratoriumsmedizin Labor Blumenstraße; Praxis Dr. E. Wietschel & Dr. F. Wietschel | Robert Koch Institute | |
| EPI_ISL_2263287, EPI_ISL_2263352, EPI_ISL_2263429, EPI_ISL_2263722, EPI_ISL_2264409, EPI_ISL_2265065, EPI_ISL_2265171, EPI_ISL_2265280, EPI_ISL_2316114, EPI_ISL_2316246 | | | |
| see above | Praxisgemeinschaft für Laboratoriumsmedizin Labor Blumenstraße; Praxis Dr. Sel und Dr. Späte | Robert Koch Institute | |
| EPI_ISL_2260987, EPI_ISL_2387370 | Procomcure Biotech Germany GmbH | Robert Koch Institute | |
| EPI_ISL_2314117, EPI_ISL_2314196, EPI_ISL_2314272, EPI_ISL_2314286, EPI_ISL_2388472, EPI_ISL_2388830, EPI_ISL_2388848, EPI_ISL_2388857, EPI_ISL_2389012 | | | |
| see above | Robert-Bosch-Krankenhaus | Robert Koch Institute | |
| EPI_ISL_2262228, EPI_ISL_2262235, EPI_ISL_2262248, EPI_ISL_2262270, EPI_ISL_2313222, EPI_ISL_2313226, EPI_ISL_2313231, EPI_ISL_2313238 | | | |
| see above | SLK-Kliniken Heilbronn GmbH - Institut für Laboratoriumsmedizin | Robert Koch Institute | |
| EPI_ISL_2261894, EPI_ISL_2262832, EPI_ISL_2262844, EPI_ISL_2262845, EPI_ISL_2262912, EPI_ISL_2262948, EPI_ISL_2262979, EPI_ISL_2263003, EPI_ISL_2263013, EPI_ISL_2263034, EPI_ISL_2263228, EPI_ISL_2263326, EPI_ISL_2263340, EPI_ISL_2263424, EPI_ISL_2263541, EPI_ISL_2263714, EPI_ISL_2263765, EPI_ISL_2264357, EPI_ISL_2264400, EPI_ISL_2264405, EPI_ISL_2264451, EPI_ISL_2264453, EPI_ISL_2264465, EPI_ISL_2265133, EPI_ISL_2265300, EPI_ISL_2265315, EPI_ISL_2314468, EPI_ISL_2314488, EPI_ISL_2314626, EPI_ISL_2315053, EPI_ISL_2315058, EPI_ISL_2315073, EPI_ISL_2315085, EPI_ISL_2315094 | | | |
| see above | SYNLAB Jena Oncoscreen | Robert Koch Institute | |
| EPI_ISL_2265125, EPI_ISL_2265153, EPI_ISL_2315050, EPI_ISL_2315071, EPI_ISL_2315110 | SYNLAB Labor München Zentrum LMZ | Robert Koch Institute | |
| EPI_ISL_2263816, EPI_ISL_2264079, EPI_ISL_2264084, EPI_ISL_2266766, EPI_ISL_2266818, EPI_ISL_2267012, EPI_ISL_2267114, EPI_ISL_2267145, EPI_ISL_2267199, EPI_ISL_2267695, EPI_ISL_2267823, EPI_ISL_2267854, EPI_ISL_2315330 | | | |
| see above | SYNLAB MVZ Berlin | Robert Koch Institute | |
| EPI_ISL_2267615, EPI_ISL_2267893 | SYNLAB MVZ Dachau | Robert Koch Institute | |
| EPI_ISL_2315217 | SYNLAB MVZ Hamburg | Robert Koch Institute | |
| EPI_ISL_2131034, EPI_ISL_2267292, EPI_ISL_2267293, EPI_ISL_2267294, EPI_ISL_2267295, EPI_ISL_2267297, EPI_ISL_2267298, EPI_ISL_2267299, EPI_ISL_2267300, EPI_ISL_2267301, EPI_ISL_2267303, EPI_ISL_2267304, EPI_ISL_2267305, EPI_ISL_2267306, EPI_ISL_2267307, EPI_ISL_2267308, EPI_ISL_2267309, EPI_ISL_2267310, EPI_ISL_2267311, EPI_ISL_2267312, EPI_ISL_2267313, EPI_ISL_2267314, EPI_ISL_2267315, EPI_ISL_2267316, EPI_ISL_2267317, EPI_ISL_2267318, EPI_ISL_2267319, EPI_ISL_2267320, EPI_ISL_2267321, EPI_ISL_2267322, EPI_ISL_2267323, EPI_ISL_2267324, EPI_ISL_2267325, EPI_ISL_2267326, EPI_ISL_2267329, EPI_ISL_2267330, EPI_ISL_2267331, EPI_ISL_2267332, EPI_ISL_2267333, EPI_ISL_2267335, EPI_ISL_2267338, EPI_ISL_2267339, EPI_ISL_2267342, EPI_ISL_2267343, EPI_ISL_2267345, EPI_ISL_2267348, EPI_ISL_2267350, EPI_ISL_2267351, EPI_ISL_2267352, EPI_ISL_2267353, EPI_ISL_2267354, EPI_ISL_2267355, EPI_ISL_2267356, EPI_ISL_2267357, EPI_ISL_2267358, EPI_ISL_2267359, EPI_ISL_2267361, EPI_ISL_2267364, EPI_ISL_2267365, EPI_ISL_2267367, EPI_ISL_2267369, EPI_ISL_2267370, EPI_ISL_2267371, EPI_ISL_2267372, EPI_ISL_2267373, EPI_ISL_2267374, EPI_ISL_2267375, EPI_ISL_2267376, EPI_ISL_2267377, EPI_ISL_2267378, EPI_ISL_2267380, EPI_ISL_2267382, EPI_ISL_2267384, EPI_ISL_2267385, EPI_ISL_2267386, EPI_ISL_2267387, EPI_ISL_2267390, EPI_ISL_2267391, EPI_ISL_2267392, EPI_ISL_2267394, EPI_ISL_2267395, EPI_ISL_2267396, EPI_ISL_2267397, EPI_ISL_2267398, EPI_ISL_2267399, EPI_ISL_2267400, EPI_ISL_2267401, EPI_ISL_2267402, EPI_ISL_2267403, EPI_ISL_2267405, EPI_ISL_2267407, EPI_ISL_2267408, EPI_ISL_2267409, EPI_ISL_2267410, EPI_ISL_2267411, EPI_ISL_2267412, EPI_ISL_2267415, EPI_ISL_2267416, EPI_ISL_2267417, EPI_ISL_2267418, EPI_ISL_2267419, EPI_ISL_2267420, EPI_ISL_2267421, EPI_ISL_2267423, EPI_ISL_2267424, EPI_ISL_2267425, EPI_ISL_2267426, EPI_ISL_2267427, EPI_ISL_2267428, EPI_ISL_2267430, EPI_ISL_2267431, EPI_ISL_2267432, EPI_ISL_2267433, EPI_ISL_2267434, EPI_ISL_2267435, EPI_ISL_2267436, EPI_ISL_2267437, EPI_ISL_2267438, EPI_ISL_2267441, EPI_ISL_2267442, EPI_ISL_2267443, EPI_ISL_2267444, EPI_ISL_2267446, EPI_ISL_2267447, EPI_ISL_2267448, EPI_ISL_2267449, EPI_ISL_2267451, EPI_ISL_2267452, EPI_ISL_2267453, EPI_ISL_2267456, EPI_ISL_2267457, EPI_ISL_2267458, EPI_ISL_2267459, EPI_ISL_2267460, EPI_ISL_2267461, EPI_ISL_2267463, EPI_ISL_2267464, EPI_ISL_2267465, EPI_ISL_2267466, EPI_ISL_2267468, EPI_ISL_2267469, EPI_ISL_2267470, EPI_ISL_2267471, EPI_ISL_2267472, EPI_ISL_2267473, EPI_ISL_2267474, EPI_ISL_2267476, EPI_ISL_2267478, EPI_ISL_2267479, EPI_ISL_2267480, EPI_ISL_2267481, EPI_ISL_2267482, EPI_ISL_2267483, EPI_ISL_2267484, EPI_ISL_2267485, EPI_ISL_2267486, EPI_ISL_2267488, EPI_ISL_2267489, EPI_ISL_2267490, EPI_ISL_2267491, EPI_ISL_2267492, EPI_ISL_2267493, EPI_ISL_2267494, EPI_ISL_2267495, EPI_ISL_2267496, EPI_ISL_2267497, EPI_ISL_2267498, EPI_ISL_2267499, EPI_ISL_2267500, EPI_ISL_2267503, EPI_ISL_2267505, EPI_ISL_2267506, EPI_ISL_2267507, EPI_ISL_2267509, EPI_ISL_2267510, EPI_ISL_2267512, EPI_ISL_2267514, EPI_ISL_2267515, EPI_ISL_2267516, EPI_ISL_2267517, EPI_ISL_2267518, EPI_ISL_2267519, EPI_ISL_2267520, EPI_ISL_2267522, EPI_ISL_2267523, EPI_ISL_2267524, EPI_ISL_2267526, EPI_ISL_2267527, EPI_ISL_2267528, EPI_ISL_2267529, EPI_ISL_2267530, EPI_ISL_2267531, EPI_ISL_2267533, EPI_ISL_2267534, EPI_ISL_2267535, EPI_ISL_2267536, EPI_ISL_2267537, EPI_ISL_2267538, EPI_ISL_2267539, EPI_ISL_2267542, EPI_ISL_2267543, EPI_ISL_2267544, EPI_ISL_2267545, EPI_ISL_2267546, EPI_ISL_2267547, EPI_ISL_2267548, EPI_ISL_2267549, EPI_ISL_2267550, EPI_ISL_2267551, EPI_ISL_2267552, EPI_ISL_2267553, EPI_ISL_2267554, EPI_ISL_2267555, EPI_ISL_2267556, EPI_ISL_2267557, EPI_ISL_2267559, EPI_ISL_2267560, EPI_ISL_2267561, EPI_ISL_2267562, EPI_ISL_2267563, EPI_ISL_2267564, EPI_ISL_2267565, EPI_ISL_2267566, EPI_ISL_2267567, EPI_ISL_2267570, EPI_ISL_2267572, EPI_ISL_2267574, EPI_ISL_2267575, EPI_ISL_2267576, EPI_ISL_2267577, EPI_ISL_2267579, EPI_ISL_2267580, EPI_ISL_2267581, EPI_ISL_2267582, EPI_ISL_2267583, EPI_ISL_2267584, EPI_ISL_2267585, EPI_ISL_2267586, EPI_ISL_2267587, EPI_ISL_2267588, EPI_ISL_2267589, EPI_ISL_2267591, EPI_ISL_2267592, EPI_ISL_2267593, EPI_ISL_2267595, EPI_ISL_2267596, EPI_ISL_2267597, EPI_ISL_2267598 | | | |
| see above | SYNLAB MVZ Heidelberg | Robert Koch Institute | |
| EPI_ISL_2262831, EPI_ISL_2262833, EPI_ISL_2262836, EPI_ISL_2262842, EPI_ISL_2262847, EPI_ISL_2262848, EPI_ISL_2262851, EPI_ISL_2262853, EPI_ISL_2262854, EPI_ISL_2262858, EPI_ISL_2262866, EPI_ISL_2262869, EPI_ISL_2262870, EPI_ISL_2262871, EPI_ISL_2262877, EPI_ISL_2262880, EPI_ISL_2262883, EPI_ISL_2262884, EPI_ISL_2262891, EPI_ISL_2262894, EPI_ISL_2262896, EPI_ISL_2262901, EPI_ISL_2262904, EPI_ISL_2262909, EPI_ISL_2262910, EPI_ISL_2262913, EPI_ISL_2262914, EPI_ISL_2262915, EPI_ISL_2262916, EPI_ISL_2262923, EPI_ISL_2262930, EPI_ISL_2262931, EPI_ISL_2262932, EPI_ISL_2262939, EPI_ISL_2262947, EPI_ISL_2262949, EPI_ISL_2262954, EPI_ISL_2262958, EPI_ISL_2262961, EPI_ISL_2262962, EPI_ISL_2262971, EPI_ISL_2262974, EPI_ISL_2262977, EPI_ISL_2262978, EPI_ISL_2262982, EPI_ISL_2262986, EPI_ISL_2262995, EPI_ISL_2262998, EPI_ISL_2262999, EPI_ISL_2263005, EPI_ISL_2263010, EPI_ISL_2263023, EPI_ISL_2263028, EPI_ISL_2263032, EPI_ISL_2263227, EPI_ISL_2263240, EPI_ISL_2263247, EPI_ISL_2263251, EPI_ISL_2263284, EPI_ISL_2263286, EPI_ISL_2263299, EPI_ISL_2263311, EPI_ISL_2263314, EPI_ISL_2263323, EPI_ISL_2263334, EPI_ISL_2263346, EPI_ISL_2263356, EPI_ISL_2263384, EPI_ISL_2263394, EPI_ISL_2263400, EPI_ISL_2263404, EPI_ISL_2263406, EPI_ISL_2263407, EPI_ISL_2263411, EPI_ISL_2263412, EPI_ISL_2263420, EPI_ISL_2263448, EPI_ISL_2263458, EPI_ISL_2263465, EPI_ISL_2263473, EPI_ISL_2263488, EPI_ISL_2263501, EPI_ISL_2263505, EPI_ISL_2263525, EPI_ISL_2263528, EPI_ISL_2263546, EPI_ISL_2263551, EPI_ISL_2263562, EPI_ISL_2263592, EPI_ISL_2263595, EPI_ISL_2263603, EPI_ISL_2263615, EPI_ISL_2263621, EPI_ISL_2263622, EPI_ISL_2263633, EPI_ISL_2263635, EPI_ISL_2263636, EPI_ISL_2263647, EPI_ISL_2263650, EPI_ISL_2263651, EPI_ISL_2263654, EPI_ISL_2263668, EPI_ISL_2263673, EPI_ISL_2263684, EPI_ISL_2263685, EPI_ISL_2263688, EPI_ISL_2263709, EPI_ISL_2263711, EPI_ISL_2263724, EPI_ISL_2263727, EPI_ISL_2263738, EPI_ISL_2263750, EPI_ISL_2263751, EPI_ISL_2263753, EPI_ISL_2263773, EPI_ISL_2263781, EPI_ISL_2264351, EPI_ISL_2264352, EPI_ISL_2264353, EPI_ISL_2264354, EPI_ISL_2264355, EPI_ISL_2264356, EPI_ISL_2264358, EPI_ISL_2264359, EPI_ISL_2264360, EPI_ISL_2264362, EPI_ISL_2264363, EPI_ISL_2264364, EPI_ISL_2264365, EPI_ISL_2264366, EPI_ISL_2264368, EPI_ISL_2264369, EPI_ISL_2264371, EPI_ISL_2264372, EPI_ISL_2264373, EPI_ISL_2264374, EPI_ISL_2264376, EPI_ISL_2264377, EPI_ISL_2264378, EPI_ISL_2264379, EPI_ISL_2264381, EPI_ISL_2264383, EPI_ISL_2264385, EPI_ISL_2264386, EPI_ISL_2264388, EPI_ISL_2264389, EPI_ISL_2264390, EPI_ISL_2264391, EPI_ISL_2264392, EPI_ISL_2264393, EPI_ISL_2264394, EPI_ISL_2264395, EPI_ISL_2264396, EPI_ISL_2264397, EPI_ISL_2264398, EPI_ISL_2264399, EPI_ISL_2264401, EPI_ISL_2264402, EPI_ISL_2264403, EPI_ISL_2264404, EPI_ISL_2264406, EPI_ISL_2264407, EPI_ISL_2264411, EPI_ISL_2264413, EPI_ISL_2264414, EPI_ISL_2264415, EPI_ISL_2264416, EPI_ISL_2264418, EPI_ISL_2264420, EPI_ISL_2264422, EPI_ISL_2264425, EPI_ISL_2264426, EPI_ISL_2264427, EPI_ISL_2264429, EPI_ISL_2264430, EPI_ISL_2264436, EPI_ISL_2264438, EPI_ISL_2264440, EPI_ISL_2264441, EPI_ISL_2264444, EPI_ISL_2264445, EPI_ISL_2264447, EPI_ISL_2264448, EPI_ISL_2264450, EPI_ISL_2264455, EPI_ISL_2264457, EPI_ISL_2264459, EPI_ISL_2264460, EPI_ISL_2264462, EPI_ISL_2264463, EPI_ISL_2264466, EPI_ISL_2264468, EPI_ISL_2264469, EPI_ISL_2264471, EPI_ISL_2264472, EPI_ISL_2264473, EPI_ISL_2264475, EPI_ISL_2264477, EPI_ISL_2264478, EPI_ISL_2265049, EPI_ISL_2265050, EPI_ISL_2265051, EPI_ISL_2265052, EPI_ISL_2265054, EPI_ISL_2265055, EPI_ISL_2265056, EPI_ISL_2265057, EPI_ISL_2265058, EPI_ISL_2265059, EPI_ISL_2265060, EPI_ISL_2265061, EPI_ISL_2265062, EPI_ISL_2265063, EPI_ISL_2265064, EPI_ISL_2265066, EPI_ISL_2265067, EPI_ISL_2265068, EPI_ISL_2265069, EPI_ISL_2265070, EPI_ISL_2265071, EPI_ISL_2265072, EPI_ISL_2265073, EPI_ISL_2265074, EPI_ISL_2265075, EPI_ISL_2265076, EPI_ISL_2265077, EPI_ISL_2265078, EPI_ISL_2265079, EPI_ISL_2265080, EPI_ISL_2265081, EPI_ISL_2265082, EPI_ISL_2265083, EPI_ISL_2265084, EPI_ISL_2265085, EPI_ISL_2265086, EPI_ISL_2265087, EPI_ISL_2265088, EPI_ISL_2265089, EPI_ISL_2265090, EPI_ISL_2265091, EPI_ISL_2265092, EPI_ISL_2265093, EPI_ISL_2265094, EPI_ISL_2265095, EPI_ISL_2265096, EPI_ISL_2265097, EPI_ISL_2265098, EPI_ISL_2265099, EPI_ISL_2265100, EPI_ISL_2265101, EPI_ISL_2265102, EPI_ISL_2265103, EPI_ISL_2265104, EPI_ISL_2265105, EPI_ISL_2265106, EPI_ISL_2265107, EPI_ISL_2265108, EPI_ISL_2265110, EPI_ISL_2265111, EPI_ISL_2265112, EPI_ISL_2265113, EPI_ISL_2265114, EPI_ISL_2265115, EPI_ISL_2265116, EPI_ISL_2265117, EPI_ISL_2265118, EPI_ISL_2265119, EPI_ISL_2265120, EPI_ISL_2265121, EPI_ISL_2265122, EPI_ISL_2265123, EPI_ISL_2265124, EPI_ISL_2265126, EPI_ISL_2265127, EPI_ISL_2265128, EPI_ISL_2265129, EPI_ISL_2265130, EPI_ISL_2265131, EPI_ISL_2265132, EPI_ISL_2265134, EPI_ISL_2265135, EPI_ISL_2265136, EPI_ISL_2265137, EPI_ISL_2265138, EPI_ISL_2265139, EPI_ISL_2265140, EPI_ISL_2265141, EPI_ISL_2265142, EPI_ISL_2265143, EPI_ISL_2265144, EPI_ISL_2265145, EPI_ISL_2265146, EPI_ISL_2265147, EPI_ISL_2265148, EPI_ISL_2265149, EPI_ISL_2265150, EPI_ISL_2265151, EPI_ISL_2265152, EPI_ISL_2265154, EPI_ISL_2265155, EPI_ISL_2265156, EPI_ISL_2265157, EPI_ISL_2265158, EPI_ISL_2265159, EPI_ISL_2265160, EPI_ISL_2265161, EPI_ISL_2265162, EPI_ISL_2265163, | | | |

EPI_ISL_2265164, EPI_ISL_2265165, EPI_ISL_2265166, EPI_ISL_2265167, EPI_ISL_2265168, EPI_ISL_2265169, EPI_ISL_2265170, EPI_ISL_2265172, EPI_ISL_2265173, EPI_ISL_2265174, EPI_ISL_2265175, EPI_ISL_2265176, EPI_ISL_2265177, EPI_ISL_2265178, EPI_ISL_2265179, EPI_ISL_2265180, EPI_ISL_2265181, EPI_ISL_2265182, EPI_ISL_2265183, EPI_ISL_2265184, EPI_ISL_2265185, EPI_ISL_2265186,
EPI_ISL_2265187, EPI_ISL_2265188, EPI_ISL_2265189, EPI_ISL_2265190, EPI_ISL_2265191, EPI_ISL_2265192, EPI_ISL_2265193, EPI_ISL_2265194, EPI_ISL_2265195, EPI_ISL_2265196, EPI_ISL_2265197, EPI_ISL_2265198, EPI_ISL_2265199, EPI_ISL_2265200, EPI_ISL_2265202, EPI_ISL_2265203, EPI_ISL_2265204, EPI_ISL_2265205, EPI_ISL_2265207, EPI_ISL_2265208, EPI_ISL_2265209, EPI_ISL_2265210,
EPI_ISL_2265211, EPI_ISL_2265212, EPI_ISL_2265213, EPI_ISL_2265214, EPI_ISL_2265215, EPI_ISL_2265216, EPI_ISL_2265217, EPI_ISL_2265218, EPI_ISL_2265219, EPI_ISL_2265220, EPI_ISL_2265221, EPI_ISL_2265222, EPI_ISL_2265223, EPI_ISL_2265224, EPI_ISL_2265225, EPI_ISL_2265226, EPI_ISL_2265227, EPI_ISL_2265228, EPI_ISL_2265229, EPI_ISL_2265230, EPI_ISL_2265231, EPI_ISL_2265232,
EPI_ISL_2265233, EPI_ISL_2265234, EPI_ISL_2265235, EPI_ISL_2265236, EPI_ISL_2265237, EPI_ISL_2265238, EPI_ISL_2265239, EPI_ISL_2265240, EPI_ISL_2265241, EPI_ISL_2265242, EPI_ISL_2265243, EPI_ISL_2265244, EPI_ISL_2265245, EPI_ISL_2265246, EPI_ISL_2265247, EPI_ISL_2265248, EPI_ISL_2265249, EPI_ISL_2265250, EPI_ISL_2265251, EPI_ISL_2265252, EPI_ISL_2265253, EPI_ISL_2265254,
EPI_ISL_2265255, EPI_ISL_2265256, EPI_ISL_2265257, EPI_ISL_2265258, EPI_ISL_2265259, EPI_ISL_2265260, EPI_ISL_2265261, EPI_ISL_2265262, EPI_ISL_2265263, EPI_ISL_2265264, EPI_ISL_2265265, EPI_ISL_2265266, EPI_ISL_2265267, EPI_ISL_2265268, EPI_ISL_2265269, EPI_ISL_2265270, EPI_ISL_2265271, EPI_ISL_2265272, EPI_ISL_2265273, EPI_ISL_2265274, EPI_ISL_2265275, EPI_ISL_2265276,
EPI_ISL_2265277, EPI_ISL_2265278, EPI_ISL_2265279, EPI_ISL_2265281, EPI_ISL_2265282, EPI_ISL_2265283, EPI_ISL_2265284, EPI_ISL_2265285, EPI_ISL_2265286, EPI_ISL_2265287, EPI_ISL_2265288, EPI_ISL_2265289, EPI_ISL_2265290, EPI_ISL_2265291, EPI_ISL_2265292, EPI_ISL_2265293, EPI_ISL_2265294, EPI_ISL_2265295, EPI_ISL_2265296, EPI_ISL_2265297, EPI_ISL_2265298, EPI_ISL_2265299,
EPI_ISL_2265301, EPI_ISL_2265302, EPI_ISL_2265303, EPI_ISL_2265304, EPI_ISL_2265305, EPI_ISL_2265306, EPI_ISL_2265307, EPI_ISL_2265308, EPI_ISL_2265309, EPI_ISL_2265310, EPI_ISL_2265311, EPI_ISL_2265313, EPI_ISL_2265314, EPI_ISL_2265316, EPI_ISL_2265317, EPI_ISL_2265318, EPI_ISL_2265319, EPI_ISL_2265320, EPI_ISL_2265321, EPI_ISL_2265322, EPI_ISL_2265323, EPI_ISL_2265324,
EPI_ISL_2265325, EPI_ISL_2265326, EPI_ISL_2265327, EPI_ISL_2265328, EPI_ISL_2265329, EPI_ISL_2265330, EPI_ISL_2265331, EPI_ISL_2265332, EPI_ISL_2265333, EPI_ISL_2265334, EPI_ISL_2265335, EPI_ISL_2265336, EPI_ISL_2265337, EPI_ISL_2265338, EPI_ISL_2265339, EPI_ISL_2265340, EPI_ISL_2265341, EPI_ISL_2265342, EPI_ISL_2265343, EPI_ISL_2265344, EPI_ISL_2265345, EPI_ISL_2265346,
EPI_ISL_2265347, EPI_ISL_2265348, EPI_ISL_2265349, EPI_ISL_2265350, EPI_ISL_2265351, EPI_ISL_2265352, EPI_ISL_2265353, EPI_ISL_2265354, EPI_ISL_2265355, EPI_ISL_2265356, EPI_ISL_2265357, EPI_ISL_2265358, EPI_ISL_2265359, EPI_ISL_2265360, EPI_ISL_2265361, EPI_ISL_2265362, EPI_ISL_2265363, EPI_ISL_2265364, EPI_ISL_2265365, EPI_ISL_2265366, EPI_ISL_2265367, EPI_ISL_2265368,
EPI_ISL_2265369, EPI_ISL_2265370, EPI_ISL_2265371, EPI_ISL_2265372, EPI_ISL_2265373, EPI_ISL_2265374, EPI_ISL_2265375, EPI_ISL_2265376, EPI_ISL_2265377, EPI_ISL_2265378, EPI_ISL_2265379, EPI_ISL_2265380, EPI_ISL_2265381, EPI_ISL_2265382, EPI_ISL_2265383, EPI_ISL_2265384, EPI_ISL_2265385, EPI_ISL_2265386, EPI_ISL_2265387, EPI_ISL_2265388, EPI_ISL_2265389, EPI_ISL_2265390,
EPI_ISL_2265391, EPI_ISL_2265392, EPI_ISL_2265393, EPI_ISL_2265394, EPI_ISL_2265395, EPI_ISL_2265396, EPI_ISL_2265397, EPI_ISL_2265398, EPI_ISL_2265399, EPI_ISL_2265400, EPI_ISL_2265401, EPI_ISL_2265402, EPI_ISL_2265403, EPI_ISL_2265404, EPI_ISL_2265405, EPI_ISL_2265406, EPI_ISL_2265407, EPI_ISL_2265408, EPI_ISL_2265409, EPI_ISL_2265410, EPI_ISL_2265411, EPI_ISL_2265412,
EPI_ISL_2265413, EPI_ISL_2265414, EPI_ISL_2265415, EPI_ISL_2265416, EPI_ISL_2265417, EPI_ISL_2265418, EPI_ISL_2265419, EPI_ISL_2265420, EPI_ISL_2265421, EPI_ISL_2265422, EPI_ISL_2265423, EPI_ISL_2265424, EPI_ISL_2265425, EPI_ISL_2265426, EPI_ISL_2265427, EPI_ISL_2265428, EPI_ISL_2265429, EPI_ISL_2265430, EPI_ISL_2265431, EPI_ISL_2265432, EPI_ISL_2265433, EPI_ISL_2265434,
EPI_ISL_2265435, EPI_ISL_2265436, EPI_ISL_2265437, EPI_ISL_2265438, EPI_ISL_2265439, EPI_ISL_2265440, EPI_ISL_2265441, EPI_ISL_2265442, EPI_ISL_2265443, EPI_ISL_2265444, EPI_ISL_2265445, EPI_ISL_2265446, EPI_ISL_2265447, EPI_ISL_2265448, EPI_ISL_2265449, EPI_ISL_2265450, EPI_ISL_2265451, EPI_ISL_2265452, EPI_ISL_2265453, EPI_ISL_2265454, EPI_ISL_2265455, EPI_ISL_2265456,
EPI_ISL_2265457, EPI_ISL_2265458, EPI_ISL_2265459, EPI_ISL_2265460, EPI_ISL_2314441, EPI_ISL_2314442, EPI_ISL_2314443, EPI_ISL_2314444, EPI_ISL_2314445, EPI_ISL_2314446, EPI_ISL_2314447, EPI_ISL_2314448, EPI_ISL_2314449, EPI_ISL_2314450, EPI_ISL_2314451, EPI_ISL_2314452, EPI_ISL_2314453, EPI_ISL_2314454, EPI_ISL_2314455, EPI_ISL_2314456, EPI_ISL_2314457, EPI_ISL_2314458,
EPI_ISL_2314460, EPI_ISL_2314461, EPI_ISL_2314462, EPI_ISL_2314463, EPI_ISL_2314464, EPI_ISL_2314465, EPI_ISL_2314466, EPI_ISL_2314467, EPI_ISL_2314469, EPI_ISL_2314471, EPI_ISL_2314472, EPI_ISL_2314473, EPI_ISL_2314474, EPI_ISL_2314475, EPI_ISL_2314476, EPI_ISL_2314477, EPI_ISL_2314478, EPI_ISL_2314479, EPI_ISL_2314480, EPI_ISL_2314481, EPI_ISL_2314482, EPI_ISL_2314483,
EPI_ISL_2314484, EPI_ISL_2314485, EPI_ISL_2314486, EPI_ISL_2314487, EPI_ISL_2314489, EPI_ISL_2314490, EPI_ISL_2314491, EPI_ISL_2314492, EPI_ISL_2314493, EPI_ISL_2314494, EPI_ISL_2314495, EPI_ISL_2314496, EPI_ISL_2314497, EPI_ISL_2314498, EPI_ISL_2314500, EPI_ISL_2314501, EPI_ISL_2314502, EPI_ISL_2314503, EPI_ISL_2314504, EPI_ISL_2314505, EPI_ISL_2314507, EPI_ISL_2314508,
EPI_ISL_2314509, EPI_ISL_2314510, EPI_ISL_2314511, EPI_ISL_2314512, EPI_ISL_2314513, EPI_ISL_2314514, EPI_ISL_2314515, EPI_ISL_2314516, EPI_ISL_2314517, EPI_ISL_2314518, EPI_ISL_2314519, EPI_ISL_2314520, EPI_ISL_2314521, EPI_ISL_2314522, EPI_ISL_2314523, EPI_ISL_2314524, EPI_ISL_2314525, EPI_ISL_2314526, EPI_ISL_2314527, EPI_ISL_2314528, EPI_ISL_2314529, EPI_ISL_2314530,
EPI_ISL_2314531, EPI_ISL_2314532, EPI_ISL_2314533, EPI_ISL_2314534, EPI_ISL_2314535, EPI_ISL_2314536, EPI_ISL_2314537, EPI_ISL_2314538, EPI_ISL_2314540, EPI_ISL_2314541, EPI_ISL_2314542, EPI_ISL_2314543, EPI_ISL_2314544, EPI_ISL_2314545, EPI_ISL_2314546, EPI_ISL_2314547, EPI_ISL_2314548, EPI_ISL_2314549, EPI_ISL_2314550, EPI_ISL_2314551, EPI_ISL_2314552, EPI_ISL_2314554,
EPI_ISL_2314555, EPI_ISL_2314556, EPI_ISL_2314557, EPI_ISL_2314558, EPI_ISL_2314559, EPI_ISL_2314560, EPI_ISL_2314561, EPI_ISL_2314562, EPI_ISL_2314563, EPI_ISL_2314564, EPI_ISL_2314565, EPI_ISL_2314566, EPI_ISL_2314567, EPI_ISL_2314568, EPI_ISL_2314569, EPI_ISL_2314570, EPI_ISL_2314571, EPI_ISL_2314572, EPI_ISL_2314573, EPI_ISL_2314574, EPI_ISL_2314575, EPI_ISL_2314576,
EPI_ISL_2314577, EPI_ISL_2314578, EPI_ISL_2314579, EPI_ISL_2314580, EPI_ISL_2314581, EPI_ISL_2314582, EPI_ISL_2314583, EPI_ISL_2314584, EPI_ISL_2314585, EPI_ISL_2314586, EPI_ISL_2314587, EPI_ISL_2314588, EPI_ISL_2314589, EPI_ISL_2314590, EPI_ISL_2314591, EPI_ISL_2314592, EPI_ISL_2314593, EPI_ISL_2314594, EPI_ISL_2314595, EPI_ISL_2314596, EPI_ISL_2314597, EPI_ISL_2314598,
EPI_ISL_2314599, EPI_ISL_2314600, EPI_ISL_2314601, EPI_ISL_2314602, EPI_ISL_2314603, EPI_ISL_2314604, EPI_ISL_2314605, EPI_ISL_2314606, EPI_ISL_2314607, EPI_ISL_2314610, EPI_ISL_2314611, EPI_ISL_2314612, EPI_ISL_2314613, EPI_ISL_2314615, EPI_ISL_2314616, EPI_ISL_2314617, EPI_ISL_2314619, EPI_ISL_2314620, EPI_ISL_2314621, EPI_ISL_2314622, EPI_ISL_2314623, EPI_ISL_2314624,
EPI_ISL_2314625, EPI_ISL_2314628, EPI_ISL_2314629, EPI_ISL_2314630, EPI_ISL_2314631, EPI_ISL_2314632, EPI_ISL_2314633, EPI_ISL_2314634, EPI_ISL_2314635, EPI_ISL_2314637, EPI_ISL_2314638, EPI_ISL_2314639, EPI_ISL_2314640, EPI_ISL_2314641, EPI_ISL_2314642, EPI_ISL_2315043, EPI_ISL_2315044, EPI_ISL_2315045, EPI_ISL_2315046, EPI_ISL_2315047, EPI_ISL_2315048, EPI_ISL_2315049,
EPI_ISL_2315051, EPI_ISL_2315052, EPI_ISL_2315054, EPI_ISL_2315055, EPI_ISL_2315056, EPI_ISL_2315057, EPI_ISL_2315059, EPI_ISL_2315060, EPI_ISL_2315061, EPI_ISL_2315062, EPI_ISL_2315063, EPI_ISL_2315064, EPI_ISL_2315065, EPI_ISL_2315066, EPI_ISL_2315067, EPI_ISL_2315068, EPI_ISL_2315069, EPI_ISL_2315070, EPI_ISL_2315072, EPI_ISL_2315074, EPI_ISL_2315075, EPI_ISL_2315076,
EPI_ISL_2315077, EPI_ISL_2315078, EPI_ISL_2315079, EPI_ISL_2315080, EPI_ISL_2315081, EPI_ISL_2315082, EPI_ISL_2315083, EPI_ISL_2315084, EPI_ISL_2315086, EPI_ISL_2315087, EPI_ISL_2315088, EPI_ISL_2315089, EPI_ISL_2315090, EPI_ISL_2315091, EPI_ISL_2315092, EPI_ISL_2315093, EPI_ISL_2315095, EPI_ISL_2315096, EPI_ISL_2315097, EPI_ISL_2315098, EPI_ISL_2315099, EPI_ISL_2315100,
EPI_ISL_2315101, EPI_ISL_2315102, EPI_ISL_2315103, EPI_ISL_2315104, EPI_ISL_2315105, EPI_ISL_2315106, EPI_ISL_2315107, EPI_ISL_2315108, EPI_ISL_2315109, EPI_ISL_2315111, EPI_ISL_2315112, EPI_ISL_2315113, EPI_ISL_2315114, EPI_ISL_2315115, EPI_ISL_2315116, EPI_ISL_2316027, EPI_ISL_2316028, EPI_ISL_2316029, EPI_ISL_2316031, EPI_ISL_2316032, EPI_ISL_2316033, EPI_ISL_2316034,
EPI_ISL_2316035, EPI_ISL_2316036, EPI_ISL_2316037, EPI_ISL_2316038, EPI_ISL_2316040, EPI_ISL_2316041, EPI_ISL_2316043, EPI_ISL_2316044, EPI_ISL_2316045, EPI_ISL_2316046, EPI_ISL_2316047, EPI_ISL_2316048, EPI_ISL_2316049, EPI_ISL_2316050, EPI_ISL_2316052, EPI_ISL_2316053, EPI_ISL_2316054, EPI_ISL_2316056, EPI_ISL_2316057, EPI_ISL_2316059, EPI_ISL_2316060, EPI_ISL_2316061,
EPI_ISL_2316062, EPI_ISL_2316063, EPI_ISL_2316064, EPI_ISL_2316065, EPI_ISL_2316066, EPI_ISL_2316067, EPI_ISL_2316068, EPI_ISL_2316069, EPI_ISL_2316070, EPI_ISL_2316072, EPI_ISL_2316073, EPI_ISL_2316074, EPI_ISL_2316075, EPI_ISL_2316076, EPI_ISL_2316077, EPI_ISL_2316079, EPI_ISL_2316080, EPI_ISL_2316081, EPI_ISL_2316082, EPI_ISL_2316083, EPI_ISL_2316085, EPI_ISL_2316086,
EPI_ISL_2316087, EPI_ISL_2316088, EPI_ISL_2316089, EPI_ISL_2316090, EPI_ISL_2316092, EPI_ISL_2316093, EPI_ISL_2316094, EPI_ISL_2316095, EPI_ISL_2316096, EPI_ISL_2316097, EPI_ISL_2316098, EPI_ISL_2316101, EPI_ISL_2316102, EPI_ISL_2316103, EPI_ISL_2316104, EPI_ISL_2316105, EPI_ISL_2316106, EPI_ISL_2316107, EPI_ISL_2316108, EPI_ISL_2316109, EPI_ISL_2316111, EPI_ISL_2316112,
EPI_ISL_2316113, EPI_ISL_2316115, EPI_ISL_2316116, EPI_ISL_2316117, EPI_ISL_2316118, EPI_ISL_2316119, EPI_ISL_2316120, EPI_ISL_2316121, EPI_ISL_2316122, EPI_ISL_2316124, EPI_ISL_2316125, EPI_ISL_2316126, EPI_ISL_2316127, EPI_ISL_2316128, EPI_ISL_2316129, EPI_ISL_2316130, EPI_ISL_2316131, EPI_ISL_2316132, EPI_ISL_2316133, EPI_ISL_2316134, EPI_ISL_2316136, EPI_ISL_2316137,
EPI_ISL_2316138, EPI_ISL_2316139, EPI_ISL_2316140, EPI_ISL_2316141, EPI_ISL_2316142, EPI_ISL_2316143, EPI_ISL_2316144, EPI_ISL_2316145, EPI_ISL_2316146, EPI_ISL_2316148, EPI_ISL_2316149, EPI_ISL_2316150, EPI_ISL_2316151, EPI_ISL_2316153, EPI_ISL_2316154, EPI_ISL_2316155, EPI_ISL_2316157, EPI_ISL_2316158, EPI_ISL_2316159, EPI_ISL_2316161, EPI_ISL_2316162, EPI_ISL_2316163,
EPI_ISL_2316164, EPI_ISL_2316165, EPI_ISL_2316166, EPI_ISL_2316167, EPI_ISL_2316168, EPI_ISL_2316169, EPI_ISL_2316170, EPI_ISL_2316171, EPI_ISL_2316172, EPI_ISL_2316173, EPI_ISL_2316174, EPI_ISL_2316175, EPI_ISL_2316176, EPI_ISL_2316177, EPI_ISL_2316178, EPI_ISL_2316180, EPI_ISL_2316181, EPI_ISL_2316182, EPI_ISL_2316183, EPI_ISL_2316184, EPI_ISL_2316185, EPI_ISL_2316186,
EPI_ISL_2316189, EPI_ISL_2316190, EPI_ISL_2316192, EPI_ISL_2316193, EPI_ISL_2316195, EPI_ISL_2316196, EPI_ISL_2316197, EPI_ISL_2316198, EPI_ISL_2316199, EPI_ISL_2316201, EPI_ISL_2316202, EPI_ISL_2316205, EPI_ISL_2316206, EPI_ISL_2316207, EPI_ISL_2316208, EPI_ISL_2316209, EPI_ISL_2316210, EPI_ISL_2316211, EPI_ISL_2316212, EPI_ISL_2316213, EPI_ISL_2316214, EPI_ISL_2316215,
EPI_ISL_2316216, EPI_ISL_2316217, EPI_ISL_2316219, EPI_ISL_2316220, EPI_ISL_2316222, EPI_ISL_2316223, EPI_ISL_2316224, EPI_ISL_2316225, EPI_ISL_2316226, EPI_ISL_2316227, EPI_ISL_2316229, EPI_ISL_2316231, EPI_ISL_2316232, EPI_ISL_2316233, EPI_ISL_2316234, EPI_ISL_2316235, EPI_ISL_2316236, EPI_ISL_2316237, EPI_ISL_2316239, EPI_ISL_2316240, EPI_ISL_2316241, EPI_ISL_2316243,
EPI_ISL_2316244, EPI_ISL_2316245, EPI_ISL_2316247, EPI_ISL_2316248, EPI_ISL_2316249, EPI_ISL_2316250, EPI_ISL_2316251, EPI_ISL_2316252, EPI_ISL_2316253, EPI_ISL_2316255, EPI_ISL_2316256, EPI_ISL_2316257, EPI_ISL_2316258, EPI_ISL_2316259, EPI_ISL_2316260, EPI_ISL_2316262, EPI_ISL_2316263, EPI_ISL_2316264, EPI_ISL_2316266, EPI_ISL_2316267, EPI_ISL_2316268, EPI_ISL_2316269,
EPI_ISL_2316270, EPI_ISL_2316271, EPI_ISL_2316272, EPI_ISL_2316273, EPI_ISL_2316275, EPI_ISL_2316276, EPI_ISL_2316277, EPI_ISL_2316278, EPI_ISL_2316280, EPI_ISL_2316286, EPI_ISL_2316289, EPI_ISL_2316292, EPI_ISL_2316296, EPI_ISL_2316301, EPI_ISL_2316305, EPI_ISL_2316308, EPI_ISL_2316315, EPI_ISL_2316319, EPI_ISL_2316324, EPI_ISL_2316328, EPI_ISL_2316334, EPI_ISL_2316338,
EPI_ISL_2316341, EPI_ISL_2316344, EPI_ISL_2316352, EPI_ISL_2316355, EPI_ISL_2316357, EPI_ISL_2316360, EPI_ISL_2316363, EPI_ISL_2316367, EPI_ISL_2316370, EPI_ISL_2316374, EPI_ISL_2316377, EPI_ISL_2316380, EPI_ISL_2316382, EPI_ISL_2316383, EPI_ISL_2316385, EPI_ISL_2316391, EPI_ISL_2316394, EPI_ISL_2316396

see above SYNLAB MVZ Leinfelden-Echterdingen Robert Koch Institute

EPI_ISL_2129981, EPI_ISL_2129983, EPI_ISL_2129986, EPI_ISL_2129988, EPI_ISL_2129990, EPI_ISL_2129993, EPI_ISL_2129994, EPI_ISL_2129995, EPI_ISL_2129996, EPI_ISL_2129997, EPI_ISL_2129998, EPI_ISL_2130000, EPI_ISL_2130001, EPI_ISL_2130002, EPI_ISL_2130004, EPI_ISL_2130006, EPI_ISL_2130007, EPI_ISL_2130011, EPI_ISL_2130013, EPI_ISL_2130015, EPI_ISL_2130017, EPI_ISL_2130021,
EPI_ISL_2130022, EPI_ISL_2130024, EPI_ISL_2130026, EPI_ISL_2130028, EPI_ISL_2130029, EPI_ISL_2130030, EPI_ISL_2130031, EPI_ISL_2130032, EPI_ISL_2130035, EPI_ISL_2130037, EPI_ISL_2130038, EPI_ISL_2130042, EPI_ISL_2130043, EPI_ISL_2130044, EPI_ISL_2130045, EPI_ISL_2130046, EPI_ISL_2130047, EPI_ISL_2130049, EPI_ISL_2130052, EPI_ISL_2130056, EPI_ISL_2130057, EPI_ISL_2130058,
EPI_ISL_2130059, EPI_ISL_2130060, EPI_ISL_2130061, EPI_ISL_2130062, EPI_ISL_2130063, EPI_ISL_2130065, EPI_ISL_2130068, EPI_ISL_2130069, EPI_ISL_2130070, EPI_ISL_2130071, EPI_ISL_2130072, EPI_ISL_2130073, EPI_ISL_2130075, EPI_ISL_2130077, EPI_ISL_2130078, EPI_ISL_2130079, EPI_ISL_2130080, EPI_ISL_2130082, EPI_ISL_2130083, EPI_ISL_2130084, EPI_ISL_2130085, EPI_ISL_2130086,
EPI_ISL_2130088, EPI_ISL_2130089, EPI_ISL_2130090, EPI_ISL_2130091, EPI_ISL_2130093, EPI_ISL_2130094, EPI_ISL_2130095, EPI_ISL_2130099, EPI_ISL_2130100, EPI_ISL_2130105, EPI_ISL_2130106, EPI_ISL_2130107, EPI_ISL_2130108, EPI_ISL_2130109, EPI_ISL_2130110, EPI_ISL_2130112, EPI_ISL_2130113, EPI_ISL_2130114, EPI_ISL_2130115, EPI_ISL_2130117, EPI_ISL_2130118, EPI_ISL_2130119,
EPI_ISL_2130120, EPI_ISL_2130121, EPI_ISL_2130122, EPI_ISL_2130123, EPI_ISL_2130124, EPI_ISL_2130126, EPI_ISL_2130127, EPI_ISL_2130128, EPI_ISL_2130130, EPI_ISL_2130131, EPI_ISL_2130133, EPI_ISL_2130134, EPI_ISL_2130136, EPI_ISL_2130137, EPI_ISL_2130138, EPI_ISL_2130139, EPI_ISL_2130140, EPI_ISL_2130142, EPI_ISL_2130143, EPI_ISL_2130144, EPI_ISL_2130146, EPI_ISL_2130148,
EPI_ISL_2130152, EPI_ISL_2130153, EPI_ISL_2130155, EPI_ISL_2130156, EPI_ISL_2130157, EPI_ISL_2130161, EPI_ISL_2130164, EPI_ISL_2130165, EPI_ISL_2130167, EPI_ISL_2130168, EPI_ISL_2130169, EPI_ISL_2130170, EPI_ISL_2130173, EPI_ISL_2130174, EPI_ISL_2130175, EPI_ISL_2130177, EPI_ISL_2130178, EPI_ISL_2130180, EPI_ISL_2130182, EPI_ISL_2130183, EPI_ISL_2130184, EPI_ISL_2130187,
EPI_ISL_2130188, EPI_ISL_2130189, EPI_ISL_2130191, EPI_ISL_2130192, EPI_ISL_2130194, EPI_ISL_2130197, EPI_ISL_2130200, EPI_ISL_2130207, EPI_ISL_2130209, EPI_ISL_2130211, EPI_ISL_2130213, EPI_ISL_2130217, EPI_ISL_2130218, EPI_ISL_2130221, EPI_ISL_2130226, EPI_ISL_2130227, EPI_ISL_2130228, EPI_ISL_2130230, EPI_ISL_2130234, EPI_ISL_2130236, EPI_ISL_2130238, EPI_ISL_2130239,
EPI_ISL_2130240, EPI_ISL_2130242, EPI_ISL_2130243, EPI_ISL_2130249, EPI_ISL_2130251, EPI_ISL_2130253, EPI_ISL_2130254, EPI_ISL_2130255, EPI_ISL_2130257, EPI_ISL_2130258, EPI_ISL_2130260, EPI_ISL_2130263, EPI_ISL_2130266, EPI_ISL_2130267, EPI_ISL_2130268, EPI_ISL_2130269, EPI_ISL_2130271, EPI_ISL_2130273, EPI_ISL_2130274, EPI_ISL_2130275, EPI_ISL_2130276, EPI_ISL_2130279,
EPI_ISL_2130284, EPI_ISL_2130285, EPI_ISL_2130287, EPI_ISL_2130288, EPI_ISL_2130290, EPI_ISL_2130291, EPI_ISL_2130294, EPI_ISL_2130297, EPI_ISL_2130299, EPI_ISL_2130300, EPI_ISL_2130302, EPI_ISL_2130304, EPI_ISL_2130305, EPI_ISL_2130306, EPI_ISL_2130308, EPI_ISL_2130311, EPI_ISL_2130312, EPI_ISL_2130314, EPI_ISL_2130315, EPI_ISL_2130316, EPI_ISL_2130318, EPI_ISL_2130320,
EPI_ISL_2130323, EPI_ISL_2130324, EPI_ISL_2130326, EPI_ISL_2130327, EPI_ISL_2130329, EPI_ISL_2130330, EPI_ISL_2130331, EPI_ISL_2130332, EPI_ISL_2130334, EPI_ISL_2130335, EPI_ISL_2130336, EPI_ISL_2130337, EPI_ISL_2130338, EPI_ISL_2130339, EPI_ISL_2130341, EPI_ISL_2130342, EPI_ISL_2130344, EPI_ISL_2130345, EPI_ISL_2130346, EPI_ISL_2130347, EPI_ISL_2130348, EPI_ISL_2130349,
EPI_ISL_2130350, EPI_ISL_2130351, EPI_ISL_2130353, EPI_ISL_2130354, EPI_ISL_2130355, EPI_ISL_2130356, EPI_ISL_2130357, EPI_ISL_2130358, EPI_ISL_2130360, EPI_ISL_2130362, EPI_ISL_2130363, EPI_ISL_2130366, EPI_ISL_2130370, EPI_ISL_2130371, EPI_ISL_2130375, EPI_ISL_2130376, EPI_ISL_2130378, EPI_ISL_2130379, EPI_ISL_2130382, EPI_ISL_2130383, EPI_ISL_2130384, EPI_ISL_2130386,
EPI_ISL_2130387, EPI_ISL_2130391, EPI_ISL_2130392, EPI_ISL_2130393, EPI_ISL_2130394, EPI_ISL_2130395, EPI_ISL_2130396, EPI_ISL_2130397, EPI_ISL_2130398, EPI_ISL_2130400, EPI_ISL_2130402, EPI_ISL_2130404, EPI_ISL_2130405, EPI_ISL_2130406, EPI_ISL_2130407, EPI_ISL_2130408, EPI_ISL_2130409, EPI_ISL_2130411, EPI_ISL_2130412, EPI_ISL_2130413, EPI_ISL_2130415, EPI_ISL_2130416,
EPI_ISL_2130417, EPI_ISL_2130418, EPI_ISL_2130423, EPI_ISL_2130424, EPI_ISL_2130425, EPI_ISL_2130426, EPI_ISL_2130428, EPI_ISL_2130429, EPI_ISL_2130431, EPI_ISL_2130432, EPI_ISL_2130435, EPI_ISL_2130436, EPI_ISL_2130439, EPI_ISL_2130441, EPI_ISL_2130442, EPI_ISL_2130443, EPI_ISL_2130444, EPI_ISL_2130448, EPI_ISL_2130452, EPI_ISL_2130453, EPI_ISL_2130456, EPI_ISL_2130457,
EPI_ISL_2130459, EPI_ISL_2130461, EPI_ISL_2130462, EPI_ISL_2130467, EPI_ISL_2130468, EPI_ISL_2130469, EPI_ISL_2130470, EPI_ISL_2130474, EPI_ISL_2130475, EPI_ISL_2130479, EPI_ISL_2130480, EPI_ISL_2130481, EPI_ISL_2130483, EPI_ISL_2130484, EPI_ISL_2130487, EPI_ISL_2130490, EPI_ISL_2130491, EPI_ISL_2130492, EPI_ISL_2130494, EPI_ISL_2130495, EPI_ISL_2130496, EPI_ISL_2130497,
EPI_ISL_2130500, EPI_ISL_2130501, EPI_ISL_2130503, EPI_ISL_2130504, EPI_ISL_2130506, EPI_ISL_2130508, EPI_ISL_2130509, EPI_ISL_2130512, EPI_ISL_2130514, EPI_ISL_2130516, EPI_ISL_2130517, EPI_ISL_2130518, EPI_ISL_2130520, EPI_ISL_2130522, EPI_ISL_2130523, EPI_ISL_2130525, EPI_ISL_2130527, EPI_ISL_2130529, EPI_ISL_2130530, EPI_ISL_2130532, EPI_ISL_2130533, EPI_ISL_2130534,
EPI_ISL_2130536, EPI_ISL_2130537, EPI_ISL_2130538, EPI_ISL_2130539, EPI_ISL_2130544, EPI_ISL_2130545, EPI_ISL_2130548, EPI_ISL_2130553, EPI_ISL_2130554, EPI_ISL_2130555, EPI_ISL_2130558, EPI_ISL_2130559, EPI_ISL_2130560, EPI_ISL_2130562, EPI_ISL_2130563, EPI_ISL_2130564, EPI_ISL_2130565, EPI_ISL_2130566, EPI_ISL_2130567, EPI_ISL_2130568, EPI_ISL_2130569, EPI_ISL_2130570,
EPI_ISL_2130571, EPI_ISL_2130572, EPI_ISL_2130574, EPI_ISL_2130578, EPI_ISL_2130580, EPI_ISL_2130582, EPI_ISL_2130585, EPI_ISL_2130586, EPI_ISL_2130587, EPI_ISL_2130590, EPI_ISL_2130591, EPI_ISL_2263785, EPI_ISL_2263786, EPI_ISL_2263787, EPI_ISL_2263788, EPI_ISL_2263789, EPI_ISL_2263791, EPI_ISL_2263792, EPI_ISL_2263793, EPI_ISL_2263794, EPI_ISL_2263795, EPI_ISL_2263796,
EPI_ISL_2263797, EPI_ISL_2263798, EPI_ISL_2263799, EPI_ISL_2263800, EPI_ISL_2263801, EPI_ISL_2263802, EPI_ISL_2263803, EPI_ISL_2263804, EPI_ISL_2263805, EPI_ISL_2263806, EPI_ISL_2263807, EPI_ISL_2263808, EPI_ISL_2263809, EPI_ISL_2263810, EPI_ISL_2263811, EPI_ISL_2263812, EPI_ISL_2263813, EPI_ISL_2263814, EPI_ISL_2263815, EPI_ISL_2263817, EPI_ISL_2263818, EPI_ISL_2263819,
EPI_ISL_2263820, EPI_ISL_2263821, EPI_ISL_2263822, EPI_ISL_2263823, EPI_ISL_2263824, EPI_ISL_2263825, EPI_ISL_2263826, EPI_ISL_2263827, EPI_ISL_2263828, EPI_ISL_2263829, EPI_ISL_2263831, EPI_ISL_2263833, EPI_ISL_2263834, EPI_ISL_2263835, EPI_ISL_2263836, EPI_ISL_2263837, EPI_ISL_2263838, EPI_ISL_2263839, EPI_ISL_2263840, EPI_ISL_2263841, EPI_ISL_2263842, EPI_ISL_2263843,
EPI_ISL_2263845, EPI_ISL_2263846, EPI_ISL_2263847, EPI_ISL_2263848, EPI_ISL_2263849, EPI_ISL_2263850, EPI_ISL_2263851, EPI_ISL_2263852, EPI_ISL_2263853, EPI_ISL_2263854, EPI_ISL_2263855, EPI_ISL_2263856, EPI_ISL_2263857, EPI_ISL_2263858, EPI_ISL_2263859, EPI_ISL_2263860, EPI_ISL_2263861, EPI_ISL_2263862, EPI_ISL_2263863, EPI_ISL_2263864, EPI_ISL_2263865, EPI_ISL_2263866,
EPI_ISL_2263867, EPI_ISL_2263868, EPI_ISL_2263869, EPI_ISL_2263870, EPI_ISL_2263871, EPI_ISL_2263872, EPI_ISL_2263873, EPI_ISL_2263874, EPI_ISL_2263875, EPI_ISL_2263876, EPI_ISL_2263877, EPI_ISL_2263879, EPI_ISL_2263880, EPI_ISL_2263881, EPI_ISL_2263882, EPI_ISL_2263883, EPI_ISL_2263884, EPI_ISL_2263885, EPI_ISL_2263887, EPI_ISL_2263888, EPI_ISL_2263890, EPI_ISL_2263892,
EPI_ISL_2263893, EPI_ISL_2263895, EPI_ISL_2263896, EPI_ISL_2263897, EPI_ISL_2263900, EPI_ISL_2263901, EPI_ISL_2263903, EPI_ISL_2263904, EPI_ISL_2263907, EPI_ISL_2263908, EPI_ISL_2263909, EPI_ISL_2263910, EPI_ISL_2263911, EPI_ISL_2263912, EPI_ISL_2263913, EPI_ISL_2263914, EPI_ISL_2263915, EPI_ISL_2263916, EPI_ISL_2263917, EPI_ISL_2263919, EPI_ISL_2263920, EPI_ISL_2263921,
EPI_ISL_2263923, EPI_ISL_2263924, EPI_ISL_2263925, EPI_ISL_2263926, EPI_ISL_2263927, EPI_ISL_2263928, EPI_ISL_2263929, EPI_ISL_2263930, EPI_ISL_2263932, EPI_ISL_2263933, EPI_ISL_2263934, EPI_ISL_2263935, EPI_ISL_2263936, EPI_ISL_2263937, EPI_ISL_2263938, EPI_ISL_2263939, EPI_ISL_2263942, EPI_ISL_2263943, EPI_ISL_2263945, EPI_ISL_2263946, EPI_ISL_2263947, EPI_ISL_2263948,
EPI_ISL_2263949, EPI_ISL_2263950, EPI_ISL_2263951, EPI_ISL_2263952, EPI_ISL_2263953, EPI_ISL_2263954, EPI_ISL_2263955, EPI_ISL_2263956, EPI_ISL_2263957, EPI_ISL_2263958, EPI_ISL_2263959, EPI_ISL_2263960, EPI_ISL_2263961, EPI_ISL_2263962, EPI_ISL_2263963, EPI_ISL_2263964, EPI_ISL_2263965, EPI_ISL_2263966, EPI_ISL_2263968, EPI_ISL_2263969, EPI_ISL_2263970, EPI_ISL_2263971,
EPI_ISL_2263972, EPI_ISL_2263973, EPI_ISL_2263974, EPI_ISL_2263975, EPI_ISL_2263976, EPI_ISL_2263977, EPI_ISL_2263978, EPI_ISL_2263979, EPI_ISL_2263980, EPI_ISL_2263981, EPI_ISL_2263982, EPI_ISL_2263983, EPI_ISL_2263984, EPI_ISL_2263985, EPI_ISL_2263986, EPI_ISL_2263987, EPI_ISL_2263988, EPI_ISL_2263989, EPI_ISL_2263990, EPI_ISL_2263991, EPI_ISL_2263992, EPI_ISL_2263993,
EPI_ISL_2263994, EPI_ISL_2263995, EPI_ISL_2263996, EPI_ISL_2263997, EPI_ISL_2263998, EPI_ISL_2263999, EPI_ISL_2264000, EPI_ISL_2264001, EPI_ISL_2264002, EPI_ISL_2264003, EPI_ISL_2264004, EPI_ISL_2264005, EPI_ISL_2264006, EPI_ISL_2264007, EPI_ISL_2264008, EPI_ISL_2264010, EPI_ISL_2264011, EPI_ISL_2264012, EPI_ISL_2264013, EPI_ISL_2264015, EPI_ISL_2264016, EPI_ISL_2264018,
EPI_ISL_2264019, EPI_ISL_2264020, EPI_ISL_2264021, EPI_ISL_2264022, EPI_ISL_2264023, EPI_ISL_2264024, EPI_ISL_2264025, EPI_ISL_2264026, EPI_ISL_2264028, EPI_ISL_2264029, EPI_ISL_2264030, EPI_ISL_2264031, EPI_ISL_2264032, EPI_ISL_2264033, EPI_ISL_2264034, EPI_ISL_2264035, EPI_ISL_2264036, EPI_ISL_2264037, EPI_ISL_2264038, EPI_ISL_2264039, EPI_ISL_2264041, EPI_ISL_2264042,
EPI_ISL_2264043, EPI_ISL_2264044, EPI_ISL_2264045, EPI_ISL_2264046, EPI_ISL_2264047, EPI_ISL_2264048, EPI_ISL_2264049, EPI_ISL_2264050, EPI_ISL_2264051, EPI_ISL_2264054, EPI_ISL_2264055, EPI_ISL_2264056, EPI_ISL_2264057, EPI_ISL_2264058, EPI_ISL_2264059, EPI_ISL_2264061, EPI_ISL_2264062, EPI_ISL_2264063, EPI_ISL_2264065, EPI_ISL_2264066, EPI_ISL_2264067, EPI_ISL_2264068,
EPI_ISL_2264069, EPI_ISL_2264070, EPI_ISL_2264071, EPI_ISL_2264072, EPI_ISL_2264073, EPI_ISL_2264074, EPI_ISL_2264075, EPI_ISL_2264076, EPI_ISL_2264080, EPI_ISL_2264081, EPI_ISL_2264082, EPI_ISL_2264083, EPI_ISL_2264085, EPI_ISL_2264086, EPI_ISL_2264087, EPI_ISL_2264088, EPI_ISL_2264089, EPI_ISL_2264090, EPI_ISL_2264091, EPI_ISL_2264092, EPI_ISL_2264093, EPI_ISL_2266664,
EPI_ISL_2266665, EPI_ISL_2266666, EPI_ISL_2266667, EPI_ISL_2266669, EPI_ISL_2266670, EPI_ISL_2266671, EPI_ISL_2266672, EPI_ISL_2266673, EPI_ISL_2266674, EPI_ISL_2266675, EPI_ISL_2266676, EPI_ISL_2266677, EPI_ISL_2266678, EPI_ISL_2266679, EPI_ISL_2266680, EPI_ISL_2266681, EPI_ISL_2266682, EPI_ISL_2266683, EPI_ISL_2266684, EPI_ISL_2266685, EPI_ISL_2266686, EPI_ISL_2266687,
EPI_ISL_2266688, EPI_ISL_2266689, EPI_ISL_2266690, EPI_ISL_2266691, EPI_ISL_2266692, EPI_ISL_2266693, EPI_ISL_2266694, EPI_ISL_2266695, EPI_ISL_2266696, EPI_ISL_2266697, EPI_ISL_2266698, EPI_ISL_2266699, EPI_ISL_2266700, EPI_ISL_2266701, EPI_ISL_2266702, EPI_ISL_2266703, EPI_ISL_2266704, EPI_ISL_2266705, EPI_ISL_2266706, EPI_ISL_2266707, EPI_ISL_2266708, EPI_ISL_2266709,
EPI_ISL_2266710, EPI_ISL_2266711, EPI_ISL_2266712, EPI_ISL_2266713, EPI_ISL_2266714, EPI_ISL_2266715, EPI_ISL_2266716, EPI_ISL_2266717, EPI_ISL_2266718, EPI_ISL_2266719, EPI_ISL_2266720, EPI_ISL_2266721, EPI_ISL_2266722, EPI_ISL_2266723, EPI_ISL_2266724, EPI_ISL_2266725, EPI_ISL_2266726, EPI_ISL_2266727, EPI_ISL_2266728, EPI_ISL_2266729, EPI_ISL_2266730, EPI_ISL_2266731,
EPI_ISL_2266732, EPI_ISL_2266733, EPI_ISL_2266734, EPI_ISL_2266735, EPI_ISL_2266736, EPI_ISL_2266737, EPI_ISL_2266738, EPI_ISL_2266739, EPI_ISL_2266740, EPI_ISL_2266741, EPI_ISL_2266742, EPI_ISL_2266743, EPI_ISL_2266744, EPI_ISL_2266745, EPI_ISL_2266746, EPI_ISL_2266747, EPI_ISL_2266748, EPI_ISL_2266749, EPI_ISL_2266750, EPI_ISL_2266751, EPI_ISL_2266752, EPI_ISL_2266753,
EPI_ISL_2266755, EPI_ISL_2266756, EPI_ISL_2266757, EPI_ISL_2266758, EPI_ISL_2266759, EPI_ISL_2266760, EPI_ISL_2266762, EPI_ISL_2266763, EPI_ISL_2266765, EPI_ISL_2266767, EPI_ISL_2266768, EPI_ISL_2266769, EPI_ISL_2266770, EPI_ISL_2266771, EPI_ISL_2266772, EPI_ISL_2266773, EPI_ISL_2266774, EPI_ISL_2266775, EPI_ISL_2266776, EPI_ISL_2266777, EPI_ISL_2266778, EPI_ISL_2266779,
EPI_ISL_2266780, EPI_ISL_2266781, EPI_ISL_2266782, EPI_ISL_2266783, EPI_ISL_2266784, EPI_ISL_2266785, EPI_ISL_2266786, EPI_ISL_2266787, EPI_ISL_2266788, EPI_ISL_2266789, EPI_ISL_2266790, EPI_ISL_2266791, EPI_ISL_2266792, EPI_ISL_2266793, EPI_ISL_2266794, EPI_ISL_2266795, EPI_ISL_2266796, EPI_ISL_2266797, EPI_ISL_2266798, EPI_ISL_2266799, EPI_ISL_2266800, EPI_ISL_2266801,
EPI_ISL_2266802, EPI_ISL_2266803, EPI_ISL_2266804, EPI_ISL_2266805, EPI_ISL_2266806, EPI_ISL_2266807, EPI_ISL_2266808, EPI_ISL_2266809, EPI_ISL_2266810, EPI_ISL_2266811, EPI_ISL_2266812, EPI_ISL_2266813, EPI_ISL_2266814, EPI_ISL_2266815, EPI_ISL_2266816, EPI_ISL_2266817, EPI_ISL_2266819, EPI_ISL_2266820, EPI_ISL_2266821, EPI_ISL_2266822, EPI_ISL_2266823, EPI_ISL_2266824,
EPI_ISL_2266825, EPI_ISL_2266826, EPI_ISL_2266827, EPI_ISL_2266828, EPI_ISL_2266829, EPI_ISL_2266830, EPI_ISL_2266831, EPI_ISL_2266832, EPI_ISL_2266833, EPI_ISL_2266834, EPI_ISL_2266835, EPI_ISL_2266836, EPI_ISL_2266837, EPI_ISL_2266838, EPI_ISL_2266839, EPI_ISL_2266840, EPI_ISL_2266841, EPI_ISL_2266842, EPI_ISL_2266844, EPI_ISL_2266845, EPI_ISL_2266846, EPI_ISL_2266847,
EPI_ISL_2266848, EPI_ISL_2266849, EPI_ISL_2266850, EPI_ISL_2266852, EPI_ISL_2266853, EPI_ISL_2266854, EPI_ISL_2266855, EPI_ISL_2266856, EPI_ISL_2266857, EPI_ISL_2266858, EPI_ISL_2266859, EPI_ISL_2266861, EPI_ISL_2266862, EPI_ISL_2266863, EPI_ISL_2266864, EPI_ISL_2266865, EPI_ISL_2266866, EPI_ISL_2266867, EPI_ISL_2266868, EPI_ISL_2266869, EPI_ISL_2266870, EPI_ISL_2266871,
EPI_ISL_2266872, EPI_ISL_2266873, EPI_ISL_2266874, EPI_ISL_2266875, EPI_ISL_2266876, EPI_ISL_2266878, EPI_ISL_2266879, EPI_ISL_2266880, EPI_ISL_2266881, EPI_ISL_2266882, EPI_ISL_2266883, EPI_ISL_2266884, EPI_ISL_2266885, EPI_ISL_2266886, EPI_ISL_2266887, EPI_ISL_2266888, EPI_ISL_2266889, EPI_ISL_2266890, EPI_ISL_2266891, EPI_ISL_2266892, EPI_ISL_2266894, EPI_ISL_2266895,
EPI_ISL_2266896, EPI_ISL_2266897, EPI_ISL_2266898, EPI_ISL_2266899, EPI_ISL_2266900, EPI_ISL_2266901, EPI_ISL_2266902, EPI_ISL_2266903, EPI_ISL_2266904, EPI_ISL_2266905, EPI_ISL_2266906, EPI_ISL_2266907, EPI_ISL_2266908, EPI_ISL_2266909, EPI_ISL_2266910, EPI_ISL_2266911, EPI_ISL_2266912, EPI_ISL_2266913, EPI_ISL_2266914, EPI_ISL_2266915, EPI_ISL_2266916, EPI_ISL_2266917,
EPI_ISL_2266919, EPI_ISL_2266920, EPI_ISL_2266921, EPI_ISL_2266922, EPI_ISL_2266923, EPI_ISL_2266924, EPI_ISL_2266926, EPI_ISL_2266927, EPI_ISL_2266928, EPI_ISL_2266930, EPI_ISL_2266932, EPI_ISL_2266933, EPI_ISL_2266934, EPI_ISL_2266935, EPI_ISL_2266936, EPI_ISL_2266937, EPI_ISL_2266938, EPI_ISL_2266939, EPI_ISL_2266940, EPI_ISL_2266941, EPI_ISL_2266942, EPI_ISL_2266943,
EPI_ISL_2266944, EPI_ISL_2266945, EPI_ISL_2266946, EPI_ISL_2266947, EPI_ISL_2266948, EPI_ISL_2266949, EPI_ISL_2266950, EPI_ISL_2266951, EPI_ISL_2266952, EPI_ISL_2266953, EPI_ISL_2266954, EPI_ISL_2266955, EPI_ISL_2266956, EPI_ISL_2266957, EPI_ISL_2266958, EPI_ISL_2266959, EPI_ISL_2266960, EPI_ISL_2266961, EPI_ISL_2266962, EPI_ISL_2266963, EPI_ISL_2266964, EPI_ISL_2266965,
EPI_ISL_2266966, EPI_ISL_2266967, EPI_ISL_2266968, EPI_ISL_2266969, EPI_ISL_2266970, EPI_ISL_2266971, EPI_ISL_2266972, EPI_ISL_2266973, EPI_ISL_2266974, EPI_ISL_2266975, EPI_ISL_2266976, EPI_ISL_2266977, EPI_ISL_2266978, EPI_ISL_2266980, EPI_ISL_2266981, EPI_ISL_2266982, EPI_ISL_2266983, EPI_ISL_2266984, EPI_ISL_2266985, EPI_ISL_2266986, EPI_ISL_2266987, EPI_ISL_2266988,
EPI_ISL_2266990, EPI_ISL_2266991, EPI_ISL_2266992, EPI_ISL_2266994, EPI_ISL_2266995, EPI_ISL_2266996, EPI_ISL_2266997, EPI_ISL_2266998, EPI_ISL_2266999, EPI_ISL_2267000, EPI_ISL_2267001, EPI_ISL_2267002, EPI_ISL_2267003, EPI_ISL_2267004, EPI_ISL_2267005, EPI_ISL_2267006, EPI_ISL_2267007, EPI_ISL_2267008, EPI_ISL_2267009, EPI_ISL_2267010, EPI_ISL_2267011, EPI_ISL_2267013,
EPI_ISL_2267014, EPI_ISL_2267015, EPI_ISL_2267016, EPI_ISL_2267017, EPI_ISL_2267018, EPI_ISL_2267019, EPI_ISL_2267020, EPI_ISL_2267021, EPI_ISL_2267022, EPI_ISL_2267024, EPI_ISL_2267025, EPI_ISL_2267026, EPI_ISL_2267027, EPI_ISL_2267028, EPI_ISL_2267029, EPI_ISL_2267031, EPI_ISL_2267032, EPI_ISL_2267033, EPI_ISL_2267034, EPI_ISL_2267035, EPI_ISL_2267036, EPI_ISL_2267037,
EPI_ISL_2267038, EPI_ISL_2267039, EPI_ISL_2267040, EPI_ISL_2267041, EPI_ISL_2267042, EPI_ISL_2267043, EPI_ISL_2267044, EPI_ISL_2267045, EPI_ISL_2267046, EPI_ISL_2267047, EPI_ISL_2267048, EPI_ISL_2267049, EPI_ISL_2267050, EPI_ISL_2267051, EPI_ISL_2267052, EPI_ISL_2267053, EPI_ISL_2267054, EPI_ISL_2267055, EPI_ISL_2267056, EPI_ISL_2267057, EPI_ISL_2267058, EPI_ISL_2267059,
EPI_ISL_2267060, EPI_ISL_2267061, EPI_ISL_2267062, EPI_ISL_2267063, EPI_ISL_2267064, EPI_ISL_2267065, EPI_ISL_2267066, EPI_ISL_2267067, EPI_ISL_2267068, EPI_ISL_2267069, EPI_ISL_2267070, EPI_ISL_2267071, EPI_ISL_2267072, EPI_ISL_2267073, EPI_ISL_2267074, EPI_ISL_2267075, EPI_ISL_2267076, EPI_ISL_2267077, EPI_ISL_2267078, EPI_ISL_2267079, EPI_ISL_2267080, EPI_ISL_2267081,
EPI_ISL_2267082, EPI_ISL_2267083, EPI_ISL_2267084, EPI_ISL_2267085, EPI_ISL_2267086, EPI_ISL_2267087, EPI_ISL_2267088, EPI_ISL_2267089, EPI_ISL_2267090, EPI_ISL_2267091, EPI_ISL_2267092, EPI_ISL_2267093, EPI_ISL_2267094, EPI_ISL_2267095, EPI_ISL_2267096, EPI_ISL_2267097, EPI_ISL_2267098, EPI_ISL_2267099, EPI_ISL_2267100, EPI_ISL_2267101, EPI_ISL_2267102, EPI_ISL_2267103,
EPI_ISL_2267104, EPI_ISL_2267105, EPI_ISL_2267106, EPI_ISL_2267107, EPI_ISL_2267108, EPI_ISL_2267109, EPI_ISL_2267110, EPI_ISL_2267111, EPI_ISL_2267112, EPI_ISL_2267113, EPI_ISL_2267115, EPI_ISL_2267116, EPI_ISL_2267117, EPI_ISL_2267118, EPI_ISL_2267119, EPI_ISL_2267120, EPI_ISL_2267121, EPI_ISL_2267122, EPI_ISL_2267123, EPI_ISL_2267124, EPI_ISL_2267125, EPI_ISL_2267126,
EPI_ISL_2267127, EPI_ISL_2267129, EPI_ISL_2267130, EPI_ISL_2267131, EPI_ISL_2267132, EPI_ISL_2267133, EPI_ISL_2267134, EPI_ISL_2267135, EPI_ISL_2267137, EPI_ISL_2267138, EPI_ISL_2267139, EPI_ISL_2267140, EPI_ISL_2267141, EPI_ISL_2267143, EPI_ISL_2267144, EPI_ISL_2267146, EPI_ISL_2267147, EPI_ISL_2267148, EPI_ISL_2267149, EPI_ISL_2267150, EPI_ISL_2267151, EPI_ISL_2267152,
EPI_ISL_2267153, EPI_ISL_2267154, EPI_ISL_2267155, EPI_ISL_2267156, EPI_ISL_2267157, EPI_ISL_2267158, EPI_ISL_2267159, EPI_ISL_2267160, EPI_ISL_2267161, EPI_ISL_2267162, EPI_ISL_2267163, EPI_ISL_2267164, EPI_ISL_2267165, EPI_ISL_2267166, EPI_ISL_2267167, EPI_ISL_2267168, EPI_ISL_2267169, EPI_ISL_2267170, EPI_ISL_2267171, EPI_ISL_2267172, EPI_ISL_2267173, EPI_ISL_2267174,
EPI_ISL_2267175, EPI_ISL_2267176, EPI_ISL_2267177, EPI_ISL_2267178, EPI_ISL_2267179, EPI_ISL_2267180, EPI_ISL_2267181, EPI_ISL_2267182, EPI_ISL_2267183, EPI_ISL_2267184, EPI_ISL_2267185, EPI_ISL_2267186, EPI_ISL_2267187, EPI_ISL_2267188, EPI_ISL_2267189, EPI_ISL_2267190, EPI_ISL_2267191, EPI_ISL_2267192, EPI_ISL_2267193, EPI_ISL_2267194, EPI_ISL_2267195, EPI_ISL_2267196,
EPI_ISL_2267197, EPI_ISL_2267198, EPI_ISL_2267201, EPI_ISL_2267202, EPI_ISL_2267204, EPI_ISL_2267205, EPI_ISL_2267206, EPI_ISL_2267207, EPI_ISL_2267208, EPI_ISL_2267209, EPI_ISL_2267210, EPI_ISL_2267211, EPI_ISL_2267212, EPI_ISL_2267213, EPI_ISL_2267214, EPI_ISL_2267215, EPI_ISL_2267216, EPI_ISL_2267217, EPI_ISL_2267218, EPI_ISL_2267219, EPI_ISL_2267220, EPI_ISL_2267221,
EPI_ISL_2267222, EPI_ISL_2267223, EPI_ISL_2267224, EPI_ISL_2267225, EPI_ISL_2267226, EPI_ISL_2267227, EPI_ISL_2267228, EPI_ISL_2267231, EPI_ISL_2267233, EPI_ISL_2267234, EPI_ISL_2267235, EPI_ISL_2267236, EPI_ISL_2267237, EPI_ISL_2267238, EPI_ISL_2267239, EPI_ISL_2267240, EPI_ISL_2267241, EPI_ISL_2267242, EPI_ISL_2267244, EPI_ISL_2267245, EPI_ISL_2267246, EPI_ISL_2267247,
EPI_ISL_2267248, EPI_ISL_2267249, EPI_ISL_2267250, EPI_ISL_2267251, EPI_ISL_2267252, EPI_ISL_2267253, EPI_ISL_2267254, EPI_ISL_2267255, EPI_ISL_2267256, EPI_ISL_2267257, EPI_ISL_2267258, EPI_ISL_2267259, EPI_ISL_2267260, EPI_ISL_2267261, EPI_ISL_2267262, EPI_ISL_2267263, EPI_ISL_2267264, EPI_ISL_2267265, EPI_ISL_2267266, EPI_ISL_2267267, EPI_ISL_2267268, EPI_ISL_2267269,
EPI_ISL_2267270, EPI_ISL_2267271, EPI_ISL_2267272, EPI_ISL_2267273, EPI_ISL_2267274, EPI_ISL_2267275, EPI_ISL_2267276, EPI_ISL_2267277, EPI_ISL_2267278, EPI_ISL_2267279, EPI_ISL_2267280, EPI_ISL_2267281, EPI_ISL_2267282, EPI_ISL_2267283, EPI_ISL_2267284, EPI_ISL_2267285, EPI_ISL_2267286, EPI_ISL_2267287, EPI_ISL_2267289, EPI_ISL_2267290, EPI_ISL_2267291, EPI_ISL_2267600,
EPI_ISL_2267601, EPI_ISL_2267602, EPI_ISL_2267603, EPI_ISL_2267604, EPI_ISL_2267605, EPI_ISL_2267606, EPI_ISL_2267607, EPI_ISL_2267608, EPI_ISL_2267609, EPI_ISL_2267611, EPI_ISL_2267612, EPI_ISL_2267613, EPI_ISL_2267614, EPI_ISL_2267616, EPI_ISL_2267617, EPI_ISL_2267619, EPI_ISL_2267620, EPI_ISL_2267621, EPI_ISL_2267622, EPI_ISL_2267624, EPI_ISL_2267625, EPI_ISL_2267626,
EPI_ISL_2267627, EPI_ISL_2267628, EPI_ISL_2267629, EPI_ISL_2267630, EPI_ISL_2267632, EPI_ISL_2267633, EPI_ISL_2267635, EPI_ISL_2267636, EPI_ISL_2267637, EPI_ISL_2267638, EPI_ISL_2267639, EPI_ISL_2267640, EPI_ISL_2267641, EPI_ISL_2267642, EPI_ISL_2267643, EPI_ISL_2267644, EPI_ISL_2267645, EPI_ISL_2267646, EPI_ISL_2267649, EPI_ISL_2267650, EPI_ISL_2267651, EPI_ISL_2267652,
EPI_ISL_2267653, EPI_ISL_2267654, EPI_ISL_2267655, EPI_ISL_2267656, EPI_ISL_2267657, EPI_ISL_2267658, EPI_ISL_2267660, EPI_ISL_2267661, EPI_ISL_2267663, EPI_ISL_2267664, EPI_ISL_2267665, EPI_ISL_2267666, EPI_ISL_2267667, EPI_ISL_2267668, EPI_ISL_2267669, EPI_ISL_2267670, EPI_ISL_2267671, EPI_ISL_2267672, EPI_ISL_2267673, EPI_ISL_2267674, EPI_ISL_2267678, EPI_ISL_2267679,
EPI_ISL_2267680, EPI_ISL_2267682, EPI_ISL_2267683, EPI_ISL_2267685, EPI_ISL_2267686, EPI_ISL_2267687, EPI_ISL_2267688, EPI_ISL_2267691, EPI_ISL_2267694, EPI_ISL_2267699, EPI_ISL_2267700, EPI_ISL_2267701, EPI_ISL_2267702, EPI_ISL_2267703, EPI_ISL_2267704, EPI_ISL_2267705, EPI_ISL_2267706, EPI_ISL_2267707, EPI_ISL_2267708, EPI_ISL_2267709, EPI_ISL_2267710, EPI_ISL_2267712,
EPI_ISL_2267713, EPI_ISL_2267714, EPI_ISL_2267715, EPI_ISL_2267717, EPI_ISL_2267718, EPI_ISL_2267720, EPI_ISL_2267721, EPI_ISL_2267722, EPI_ISL_2267724, EPI_ISL_2267727, EPI_ISL_2267728, EPI_ISL_2267729, EPI_ISL_2267730, EPI_ISL_2267731, EPI_ISL_2267732, EPI_ISL_2267734, EPI_ISL_2267736, EPI_ISL_2267737, EPI_ISL_2267738, EPI_ISL_2267739, EPI_ISL_2267740, EPI_ISL_2267741,
EPI_ISL_2267742, EPI_ISL_2267744, EPI_ISL_2267745, EPI_ISL_2267746, EPI_ISL_2267747, EPI_ISL_2267748, EPI_ISL_2267749, EPI_ISL_2267750, EPI_ISL_2267751, EPI_ISL_2267752, EPI_ISL_2267753, EPI_ISL_2267754, EPI_ISL_2267755, EPI_ISL_2267757, EPI_ISL_2267758, EPI_ISL_2267759, EPI_ISL_2267760, EPI_ISL_2267761, EPI_ISL_2267762, EPI_ISL_2267763, EPI_ISL_2267765, EPI_ISL_2267766,
EPI_ISL_2267767, EPI_ISL_2267768, EPI_ISL_2267769, EPI_ISL_2267770, EPI_ISL_2267773, EPI_ISL_2267774, EPI_ISL_2267775, EPI_ISL_2267777, EPI_ISL_2267778, EPI_ISL_2267779, EPI_ISL_2267780, EPI_ISL_2267781, EPI_ISL_2267783, EPI_ISL_2267784, EPI_ISL_2267785, EPI_ISL_2267786, EPI_ISL_2267793, EPI_ISL_2267794, EPI_ISL_2267795, EPI_ISL_2267796, EPI_ISL_2267797, EPI_ISL_2267798,
EPI_ISL_2267799, EPI_ISL_2267801, EPI_ISL_2267802, EPI_ISL_2267803, EPI_ISL_2267804, EPI_ISL_2267805, EPI_ISL_2267806, EPI_ISL_2267807, EPI_ISL_2267809, EPI_ISL_2267810, EPI_ISL_2267811, EPI_ISL_2267812, EPI_ISL_2267813, EPI_ISL_2267815, EPI_ISL_2267817, EPI_ISL_2267819, EPI_ISL_2267820, EPI_ISL_2267821, EPI_ISL_2267822, EPI_ISL_2267824, EPI_ISL_2267827, EPI_ISL_2267828,
EPI_ISL_2267829, EPI_ISL_2267832, EPI_ISL_2267833, EPI_ISL_2267835, EPI_ISL_2267836, EPI_ISL_2267837, EPI_ISL_2267840, EPI_ISL_2267841, EPI_ISL_2267843, EPI_ISL_2267844, EPI_ISL_2267845, EPI_ISL_2267846, EPI_ISL_2267847, EPI_ISL_2267848, EPI_ISL_2267851, EPI_ISL_2267852, EPI_ISL_2267853, EPI_ISL_2267855, EPI_ISL_2267856, EPI_ISL_2267857, EPI_ISL_2267858, EPI_ISL_2267859,
EPI_ISL_2267860, EPI_ISL_2267861, EPI_ISL_2267863, EPI_ISL_2267865, EPI_ISL_2267867, EPI_ISL_2267868, EPI_ISL_2267869, EPI_ISL_2267870, EPI_ISL_2267871, EPI_ISL_2267873, EPI_ISL_2267874, EPI_ISL_2267875, EPI_ISL_2267876, EPI_ISL_2267877, EPI_ISL_2267878, EPI_ISL_2267879, EPI_ISL_2267881, EPI_ISL_2267882, EPI_ISL_2267883, EPI_ISL_2267884, EPI_ISL_2267885, EPI_ISL_2267886,
EPI_ISL_2267888, EPI_ISL_2267890, EPI_ISL_2267892, EPI_ISL_2267895, EPI_ISL_2267897, EPI_ISL_2267898, EPI_ISL_2267900, EPI_ISL_2267901, EPI_ISL_2267903, EPI_ISL_2315285, EPI_ISL_2386460

see above SYNLAB MVZ Leverkusen Robert Koch Institute

EPI_ISL_2267327, EPI_ISL_2267336, EPI_ISL_2267337, EPI_ISL_2267340, EPI_ISL_2267341, EPI_ISL_2267344, EPI_ISL_2267346, EPI_ISL_2267349, EPI_ISL_2267360, EPI_ISL_2267381, EPI_ISL_2267393, EPI_ISL_2267413, EPI_ISL_2267414, EPI_ISL_2267440, EPI_ISL_2267445, EPI_ISL_2267477, EPI_ISL_2267501, EPI_ISL_2267502, EPI_ISL_2267504, EPI_ISL_2267532, EPI_ISL_2267558, EPI_ISL_2267569,
EPI_ISL_2267571

| | | | |
|---|---|---|---|
| see above | SYNLAB MVZ Neustadt a. d. Weinstraße | Robert Koch Institute | |
| EPI_ISL_2263790, EPI_ISL_2263889, EPI_ISL_2263894, EPI_ISL_2263899, EPI_ISL_2263902, EPI_ISL_2263905, EPI_ISL_2263918, EPI_ISL_2263931, EPI_ISL_2263941, EPI_ISL_2264009, EPI_ISL_2264014, EPI_ISL_2264017, EPI_ISL_2264040, EPI_ISL_2264052, EPI_ISL_2264053, EPI_ISL_2264060, EPI_ISL_2266668, EPI_ISL_2266754, EPI_ISL_2266761, EPI_ISL_2266764, EPI_ISL_2266843, EPI_ISL_2266851, EPI_ISL_2266893, EPI_ISL_2266925, EPI_ISL_2266931, EPI_ISL_2266979, EPI_ISL_2266989, EPI_ISL_2266993, EPI_ISL_2267023, EPI_ISL_2267136, EPI_ISL_2267142, EPI_ISL_2267200, EPI_ISL_2267232, EPI_ISL_2267243, EPI_ISL_2267610, EPI_ISL_2267659, EPI_ISL_2267662, EPI_ISL_2267675, EPI_ISL_2267690, EPI_ISL_2267692, EPI_ISL_2267693, EPI_ISL_2267711, EPI_ISL_2267716, EPI_ISL_2267719, EPI_ISL_2267723, EPI_ISL_2267756, EPI_ISL_2267771, EPI_ISL_2267782, EPI_ISL_2267800, EPI_ISL_2267814, EPI_ISL_2267816, EPI_ISL_2267825, EPI_ISL_2267830, EPI_ISL_2267838, EPI_ISL_2267849, EPI_ISL_2267880, EPI_ISL_2267894, EPI_ISL_2267896, EPI_ISL_2267902 | | | |
| see above | SYNLAB MVZ Trier | Robert Koch Institute | |
| EPI_ISL_2130328, EPI_ISL_2263844, EPI_ISL_2263878, EPI_ISL_2263886, EPI_ISL_2263891, EPI_ISL_2263940, EPI_ISL_2263944, EPI_ISL_2264064, EPI_ISL_2264077, EPI_ISL_2264078, EPI_ISL_2267229, EPI_ISL_2267618, EPI_ISL_2267623, EPI_ISL_2267676, EPI_ISL_2267677, EPI_ISL_2267689, EPI_ISL_2267696, EPI_ISL_2267697, EPI_ISL_2267834, EPI_ISL_2267842, EPI_ISL_2267850, EPI_ISL_2386346, EPI_ISL_2386403 | | | |
| see above | SYNLAB MVZ Weiden | Robert Koch Institute | |
| EPI_ISL_2313372, EPI_ISL_2313410, EPI_ISL_2313419, EPI_ISL_2313434, EPI_ISL_2313437, EPI_ISL_2313438, EPI_ISL_2313451, EPI_ISL_2313452, EPI_ISL_2313671, EPI_ISL_2313726, EPI_ISL_2313738, EPI_ISL_2313742, EPI_ISL_2313813, EPI_ISL_2313820, EPI_ISL_2313824, EPI_ISL_2313826, EPI_ISL_2313847, EPI_ISL_2313856, EPI_ISL_2313858, EPI_ISL_2314079, EPI_ISL_2314130, EPI_ISL_2314135, EPI_ISL_2314144, EPI_ISL_2314206, EPI_ISL_2314226, EPI_ISL_2314230, EPI_ISL_2314258, EPI_ISL_2314266, EPI_ISL_2314289, EPI_ISL_2314291, EPI_ISL_2388339, EPI_ISL_2388341, EPI_ISL_2388350, EPI_ISL_2388382, EPI_ISL_2388400, EPI_ISL_2388406, EPI_ISL_2388415, EPI_ISL_2388424, EPI_ISL_2388437, EPI_ISL_2388486, EPI_ISL_2388494, EPI_ISL_2388551, EPI_ISL_2388622, EPI_ISL_2388624, EPI_ISL_2388632, EPI_ISL_2388643, EPI_ISL_2388647, EPI_ISL_2388651, EPI_ISL_2388652, EPI_ISL_2388661, EPI_ISL_2388662, EPI_ISL_2388666, EPI_ISL_2388667, EPI_ISL_2388676, EPI_ISL_2388681, EPI_ISL_2388685, EPI_ISL_2388686, EPI_ISL_2388689, EPI_ISL_2388691, EPI_ISL_2388692, EPI_ISL_2388695, EPI_ISL_2388699, EPI_ISL_2388708, EPI_ISL_2388728, EPI_ISL_2388750, EPI_ISL_2388752, EPI_ISL_2388757, EPI_ISL_2388761, EPI_ISL_2388764, EPI_ISL_2388771, EPI_ISL_2388772, EPI_ISL_2388782, EPI_ISL_2388783, EPI_ISL_2388785, EPI_ISL_2388798, EPI_ISL_2388808, EPI_ISL_2388816, EPI_ISL_2388818, EPI_ISL_2388841, EPI_ISL_2388843, EPI_ISL_2388846, EPI_ISL_2388847, EPI_ISL_2388858, EPI_ISL_2388862, EPI_ISL_2388868, EPI_ISL_2388898, EPI_ISL_2388900, EPI_ISL_2388905, EPI_ISL_2388907, EPI_ISL_2388923, EPI_ISL_2388957, EPI_ISL_2388958, EPI_ISL_2388975, EPI_ISL_2388977, EPI_ISL_2388978, EPI_ISL_2388981, EPI_ISL_2388982, EPI_ISL_2388996, EPI_ISL_2389005, EPI_ISL_2389009, EPI_ISL_2389030, EPI_ISL_2389035, EPI_ISL_2389036, EPI_ISL_2389039, EPI_ISL_2389040, EPI_ISL_2389062, EPI_ISL_2470205, EPI_ISL_2470208, EPI_ISL_2470228, EPI_ISL_2470249, EPI_ISL_2470274, EPI_ISL_2470311, EPI_ISL_2470359, EPI_ISL_2470376, EPI_ISL_2470465, EPI_ISL_2470469, EPI_ISL_2470484 | | | |
| see above | Sonic - Bioscientia - MVZ Labor Saar GmbH | Robert Koch Institute | |
| EPI_ISL_2313462, EPI_ISL_2313491, EPI_ISL_2313654, EPI_ISL_2313719, EPI_ISL_2313801, EPI_ISL_2313835, EPI_ISL_2388865, EPI_ISL_2388879, EPI_ISL_2388914, EPI_ISL_2388919, EPI_ISL_2388960, EPI_ISL_2388989, EPI_ISL_2389022, EPI_ISL_2389033, EPI_ISL_2389065 | | | |
| see above | Sonic - Dr. Staber & Kollegen GmbH München | Robert Koch Institute | |
| EPI_ISL_2314078, EPI_ISL_2314103, EPI_ISL_2314104, EPI_ISL_2314109, EPI_ISL_2314116, EPI_ISL_2314124, EPI_ISL_2314125, EPI_ISL_2314127, EPI_ISL_2314128, EPI_ISL_2314133, EPI_ISL_2314145, EPI_ISL_2314148, EPI_ISL_2314151, EPI_ISL_2314153, EPI_ISL_2314156, EPI_ISL_2314175, EPI_ISL_2314177, EPI_ISL_2314187, EPI_ISL_2314194, EPI_ISL_2314211, EPI_ISL_2314239, EPI_ISL_2314251, EPI_ISL_2314271, EPI_ISL_2314278, EPI_ISL_2314280, EPI_ISL_2314308, EPI_ISL_2314309, EPI_ISL_2388358, EPI_ISL_2388370, EPI_ISL_2388411, EPI_ISL_2388430, EPI_ISL_2388441, EPI_ISL_2388508, EPI_ISL_2388510, EPI_ISL_2388519, EPI_ISL_2388521, EPI_ISL_2388536, EPI_ISL_2388561, EPI_ISL_2388578, EPI_ISL_2388586, EPI_ISL_2388590, EPI_ISL_2388630, EPI_ISL_2388634, EPI_ISL_2388636, EPI_ISL_2388656, EPI_ISL_2388665, EPI_ISL_2388674, EPI_ISL_2388687, EPI_ISL_2388688, EPI_ISL_2388706, EPI_ISL_2388713, EPI_ISL_2388725, EPI_ISL_2388727, EPI_ISL_2388731, EPI_ISL_2388745, EPI_ISL_2388746, EPI_ISL_2388753, EPI_ISL_2388758, EPI_ISL_2388765, EPI_ISL_2388770, EPI_ISL_2388773, EPI_ISL_2388774, EPI_ISL_2388791, EPI_ISL_2388800, EPI_ISL_2469960, EPI_ISL_2470219, EPI_ISL_2470271, EPI_ISL_2470314, EPI_ISL_2470317, EPI_ISL_2470322, EPI_ISL_2470411, EPI_ISL_2470481 | | | |
| see above | Sonic - Labor Dr. von Froreich GmbH | Robert Koch Institute | |
| EPI_ISL_2314091, EPI_ISL_2314172, EPI_ISL_2314176, EPI_ISL_2314181, EPI_ISL_2314182, EPI_ISL_2314231, EPI_ISL_2314274, EPI_ISL_2314307, EPI_ISL_2314310, EPI_ISL_2634414 | | | |
| see above | Sonic - Labor Lademannbogen MVZ GmbH | Robert Koch Institute | |
| EPI_ISL_2313408, EPI_ISL_2313448, EPI_ISL_2313467, EPI_ISL_2388817, EPI_ISL_2388837, EPI_ISL_2388852, EPI_ISL_2388854, EPI_ISL_2388859, EPI_ISL_2388886, EPI_ISL_2388916, EPI_ISL_2388948, EPI_ISL_2388956, EPI_ISL_2388970, EPI_ISL_2389018, EPI_ISL_2389034, EPI_ISL_2389046, EPI_ISL_2389056, EPI_ISL_2389070 | | | |
| see above | Sonic - Labor Staber Dresden (Klipphausen) | Robert Koch Institute | |
| EPI_ISL_2313395, EPI_ISL_2313401, EPI_ISL_2313439, EPI_ISL_2313514, EPI_ISL_2313724, EPI_ISL_2388863, EPI_ISL_2388875, EPI_ISL_2388994, EPI_ISL_2388998, EPI_ISL_2389066 | | | |
| see above | Sonic - Labor Staber Heilbronn | Robert Koch Institute | |
| EPI_ISL_2313664, EPI_ISL_2388345, EPI_ISL_2470248, EPI_ISL_2470404 | Sonic - Labor Staber MVZ Kassel | Robert Koch Institute | |
| EPI_ISL_2313396, EPI_ISL_2388891, EPI_ISL_2388909 | Sonic - Labor Staber Nürnberg | Robert Koch Institute | |
| EPI_ISL_2314080, EPI_ISL_2314102, EPI_ISL_2314138, EPI_ISL_2314197, EPI_ISL_2314199, EPI_ISL_2314224, EPI_ISL_2314292, EPI_ISL_2314304, EPI_ISL_2388326, EPI_ISL_2388343, EPI_ISL_2388366, EPI_ISL_2388386, EPI_ISL_2388428, EPI_ISL_2388455, EPI_ISL_2388492, EPI_ISL_2388504, EPI_ISL_2388569, EPI_ISL_2388581, EPI_ISL_2388583, EPI_ISL_2388592, EPI_ISL_2388659, EPI_ISL_2388709, EPI_ISL_2388718, EPI_ISL_2388737, EPI_ISL_2388738, EPI_ISL_2388740, EPI_ISL_2388769, EPI_ISL_2388803 | | | |
| see above | Sonic - MVZ Medizinisches Labor Bremen GmbH | Robert Koch Institute | |
| EPI_ISL_2389008 | Sonic - MVZ Medizinisches Labor Nord GmbH Kiel | Robert Koch Institute | |
| EPI_ISL_2314122, EPI_ISL_2314163, EPI_ISL_2314170, EPI_ISL_2314198, EPI_ISL_2314223, EPI_ISL_2314235, EPI_ISL_2388355, EPI_ISL_2388517, EPI_ISL_2388793 | | | |
| see above | Sonic - MVZ Medizinisches Labor Oldenburg | Robert Koch Institute | |
| EPI_ISL_2265045 | St. Marien-Hospital Hamm | Robert Koch Institute | |
| EPI_ISL_2262337, EPI_ISL_2262338, EPI_ISL_2262341, EPI_ISL_2262343, EPI_ISL_2262359, EPI_ISL_2262363, EPI_ISL_2262371, EPI_ISL_2262383, EPI_ISL_2262386, EPI_ISL_2262391, EPI_ISL_2313908, EPI_ISL_2313910, EPI_ISL_2313912, EPI_ISL_2313925, EPI_ISL_2313927, EPI_ISL_2313932, EPI_ISL_2313937, EPI_ISL_2313940, EPI_ISL_2313946, EPI_ISL_2313950, EPI_ISL_2313951, EPI_ISL_2313955, EPI_ISL_2313958 | | | |
| see above | Städtisches Klinikum Dresden Institut für Klinische Chemie und Laboratoriumsmedizin | Robert Koch Institute | |
| EPI_ISL_2263830, EPI_ISL_2263832, EPI_ISL_2266860, EPI_ISL_2266877, EPI_ISL_2266918, EPI_ISL_2267030, EPI_ISL_2267128, EPI_ISL_2267203, EPI_ISL_2267230, EPI_ISL_2267288, EPI_ISL_2267634, EPI_ISL_2267648, EPI_ISL_2267698, EPI_ISL_2267764, EPI_ISL_2267818 | | | |
| see above | Synlab MVZ Augsburg | Robert Koch Institute | |
| EPI_ISL_2314339, EPI_ISL_2314340, EPI_ISL_2314341, EPI_ISL_2314342, EPI_ISL_2314343, EPI_ISL_2314344, EPI_ISL_2314345, EPI_ISL_2314346, EPI_ISL_2389854, EPI_ISL_2389855, EPI_ISL_2389856, EPI_ISL_2389857, EPI_ISL_2389859, EPI_ISL_2389861, EPI_ISL_2389862, EPI_ISL_2389863, EPI_ISL_2389864 | | | |
| see above | Thüringer Landesamtes für Verbraucherschutz | Robert Koch Institute | |
| EPI_ISL_2450449, EPI_ISL_2450450, EPI_ISL_2450451, EPI_ISL_2450452, EPI_ISL_2450453, EPI_ISL_2661498, EPI_ISL_2661500, EPI_ISL_2661503, EPI_ISL_2661504, EPI_ISL_2661505, EPI_ISL_2661506, EPI_ISL_2661509, EPI_ISL_2661510, EPI_ISL_2661511, EPI_ISL_2661512, EPI_ISL_2661513 | | | |
| see above | US Air Force School of Aerospace Medicine | US Air Force School of Aerospace Medicine | Amanda Javorina; Anthony Fries; Carol Garrett; Clarise Starr; Cole Anderson; Elizabeth Macias; Fritz Castillo; Jennifer Meyer; Sarah Purves; William Gruner |
| EPI_ISL_2262347, EPI_ISL_2262353, EPI_ISL_2262354, EPI_ISL_2262364, EPI_ISL_2262365, EPI_ISL_2262367, EPI_ISL_2262377, EPI_ISL_2262379, EPI_ISL_2262385, EPI_ISL_2262388, EPI_ISL_2313901, EPI_ISL_2313903, EPI_ISL_2313911, EPI_ISL_2313914, EPI_ISL_2313919, EPI_ISL_2313923, EPI_ISL_2313928, EPI_ISL_2313930, EPI_ISL_2313934, EPI_ISL_2313943 | | | |
| see above | Uniklinikum Carl Gustav Carus an der TU Dresden; Institut für Virologie | Robert Koch Institute | |
| EPI_ISL_2262298, EPI_ISL_2314879, EPI_ISL_2314880, EPI_ISL_2314884, EPI_ISL_2389900, EPI_ISL_2389901, EPI_ISL_2389902, EPI_ISL_2389905, EPI_ISL_2389911 | | | |
| see above | Uniklinikum Jena | Robert Koch Institute | |
| EPI_ISL_2472020, EPI_ISL_2472022, EPI_ISL_2472024, EPI_ISL_2472026, EPI_ISL_2472027 | Uniklinikum Regensburg; Institut für klinische Mikrobiologie und Hygiene | Robert Koch Institute | |
| EPI_ISL_2129181 | Universitatsklinikum Munster Institut fur Virologie | Robert Koch Institute | |
| EPI_ISL_2384911, EPI_ISL_2384912, EPI_ISL_2384913, EPI_ISL_2384914, EPI_ISL_2384915, EPI_ISL_2384916, EPI_ISL_2689150, EPI_ISL_2689152, EPI_ISL_2689153, EPI_ISL_2689154, EPI_ISL_2689155, EPI_ISL_2689156, EPI_ISL_2689157, EPI_ISL_2689159, EPI_ISL_2689163, EPI_ISL_2689164, EPI_ISL_2689165, EPI_ISL_2689166 | | | |
| see above | University Medical Center Hamburg Eppendorf | Heinrich Pette Institute, Leibniz Institute for Experimental Virology | Adam Grundhoff; Alexis Robitaille; Johannes Knobloch; Martin Aepfelbacher; Nicole Fischer; Thomas Günther |
| EPI_ISL_2129665, EPI_ISL_2129666, EPI_ISL_2129667, EPI_ISL_2129669, EPI_ISL_2129670, EPI_ISL_2129671 | UniversitÃ¤tsklinikum GieÃŸen - Institut fÃ¼r Medizinische Virologie | Robert Koch Institute | |
| EPI_ISL_2390181, EPI_ISL_2390185, EPI_ISL_2390186, EPI_ISL_2390187, EPI_ISL_2390188, EPI_ISL_2390189, EPI_ISL_2390190, EPI_ISL_2390191, EPI_ISL_2390192, EPI_ISL_2390195, EPI_ISL_2390196, EPI_ISL_2390197 | | | |
| see above | Universitätsklinikum Bonn - Institut für Virologie | Robert Koch Institute | |
| EPI_ISL_2262326, EPI_ISL_2262333 | Universitätsklinikum Essen | Robert Koch Institute | |
| EPI_ISL_2314081, EPI_ISL_2314090, EPI_ISL_2314150, EPI_ISL_2314169, EPI_ISL_2314183, EPI_ISL_2314186, EPI_ISL_2314202, EPI_ISL_2314233, EPI_ISL_2314256, EPI_ISL_2314273, EPI_ISL_2314279, EPI_ISL_2314281, EPI_ISL_2314287, EPI_ISL_2314303, EPI_ISL_2314305, EPI_ISL_2388682 | | | |
| see above | Universitätsklinikum Frankfurt - Institut für Medizinische Virologie | Robert Koch Institute | |
| EPI_ISL_2265462, EPI_ISL_2265463, EPI_ISL_2265464, EPI_ISL_2265466, EPI_ISL_2678373 | Universitätsklinikum Gießen - Institut für Medizinische Virologie | Robert Koch Institute | |
| EPI_ISL_2469925, EPI_ISL_2469927, EPI_ISL_2470203, EPI_ISL_2470204 | Universitätsklinikum Heidelberg | Robert Koch Institute | |
| EPI_ISL_2635959, EPI_ISL_2635960 | Universitätsklinikum Schleswig-Holstein (standortübergreifend inkl. Kiel) - Institut für Klinische Chemie und Laboratoriumsdiagnostik | Robert Koch Institute | |
| EPI_ISL_2264483, EPI_ISL_2264492, EPI_ISL_2313887, EPI_ISL_2313889, EPI_ISL_2313890, EPI_ISL_2313891, EPI_ISL_2313892, EPI_ISL_2389398, EPI_ISL_2389410, EPI_ISL_2389411, EPI_ISL_2389429, EPI_ISL_2389430 | | | |
| see above | Universitätsklinikum des Saarlandes - Institut für Virologie | Robert Koch Institute | |
| EPI_ISL_2633737, EPI_ISL_2633738, EPI_ISL_2633739 | Universitätsmedizin Mannheim | Robert Koch Institute | |
| EPI_ISL_2678378, EPI_ISL_2678382, EPI_ISL_2678385 | Virologisches Institut des Universitätsklinikums Erlangen | Robert Koch Institute | |
| EPI_ISL_2447903, EPI_ISL_2447904, EPI_ISL_2447905, EPI_ISL_2447907, EPI_ISL_2447908, EPI_ISL_2447913, EPI_ISL_2447916, EPI_ISL_2447918, EPI_ISL_2447919, EPI_ISL_2447921, EPI_ISL_2447924, EPI_ISL_2447925, EPI_ISL_2447930, EPI_ISL_2447931, EPI_ISL_2447932, EPI_ISL_2447934, EPI_ISL_2447935, EPI_ISL_2447936, EPI_ISL_2447937, EPI_ISL_2447938, EPI_ISL_2447939, EPI_ISL_2447940, EPI_ISL_2447941, EPI_ISL_2447942, EPI_ISL_2447944, EPI_ISL_2447946, EPI_ISL_2447949, EPI_ISL_2447950, EPI_ISL_2447952, EPI_ISL_2447960, EPI_ISL_2447965, EPI_ISL_2447968, EPI_ISL_2447969, EPI_ISL_2447970, EPI_ISL_2447984 | | | |
| see above | Virology, Universitätsklinikum des Saarlandes | Epigenetics, Saarland University | Jörn Walter; Kathrin Kattler; Nastasja Seiwert; Sascha Tierling; Sigrun Smola; Stefan Lohse; Thorsten Pfuhl |
| EPI_ISL_2448369, EPI_ISL_2448370, EPI_ISL_2448371, EPI_ISL_2448372, EPI_ISL_2448373, EPI_ISL_2448374, EPI_ISL_2448375 | | | |
| see above | Virology, Universitätsklinikum des Saarlandes | Epigenetics, Saarland University | Jörn Walter; Kathrin Kattler; Sascha Tierling; Sigrun Smola; Stefan Lohse; Thorsten Pfuhl |
| EPI_ISL_2380872, EPI_ISL_2380873, EPI_ISL_2380874, EPI_ISL_2380875, EPI_ISL_2380876, EPI_ISL_2380877, EPI_ISL_2380878, EPI_ISL_2380879, EPI_ISL_2380880, EPI_ISL_2380881, EPI_ISL_2380882, EPI_ISL_2380883, EPI_ISL_2380884, EPI_ISL_2380885, EPI_ISL_2380886, EPI_ISL_2380887, EPI_ISL_2380888, EPI_ISL_2380889, EPI_ISL_2380890, EPI_ISL_2380891, EPI_ISL_2380892, EPI_ISL_2380893, EPI_ISL_2380894, EPI_ISL_2380895, EPI_ISL_2380897, EPI_ISL_2380898, EPI_ISL_2380899, EPI_ISL_2380900, EPI_ISL_2380901, EPI_ISL_2380904, EPI_ISL_2380906, EPI_ISL_2380907, EPI_ISL_2380908, EPI_ISL_2380909, EPI_ISL_2380910, EPI_ISL_2380911, EPI_ISL_2380912, EPI_ISL_2380913, EPI_ISL_2380917, EPI_ISL_2380920, EPI_ISL_2380922, EPI_ISL_2380925, EPI_ISL_2380932, EPI_ISL_2380934, EPI_ISL_2380946, EPI_ISL_2380949, EPI_ISL_2380950, EPI_ISL_2380952, EPI_ISL_2380954, EPI_ISL_2380956, EPI_ISL_2380958, EPI_ISL_2380961, EPI_ISL_2380963, EPI_ISL_2380967, EPI_ISL_2380969, EPI_ISL_2380971, EPI_ISL_2380973, EPI_ISL_2380976, EPI_ISL_2380979, EPI_ISL_2380981, EPI_ISL_2381416, EPI_ISL_2381417, EPI_ISL_2381418, EPI_ISL_2381419, EPI_ISL_2381420, EPI_ISL_2381421, EPI_ISL_2381422, EPI_ISL_2381423, EPI_ISL_2381424, EPI_ISL_2381425, EPI_ISL_2381426, EPI_ISL_2381430, EPI_ISL_2381431, EPI_ISL_2381432, EPI_ISL_2381437, EPI_ISL_2381438, EPI_ISL_2381439, EPI_ISL_2381440, EPI_ISL_2381441, EPI_ISL_2381442, EPI_ISL_2381443, EPI_ISL_2381444, EPI_ISL_2381445, EPI_ISL_2381446, EPI_ISL_2381447, EPI_ISL_2381448, EPI_ISL_2381449, EPI_ISL_2381450, EPI_ISL_2381451, EPI_ISL_2381452, EPI_ISL_2381453, EPI_ISL_2381522, EPI_ISL_2381523, EPI_ISL_2381525, EPI_ISL_2381526, EPI_ISL_2381529, EPI_ISL_2381530, EPI_ISL_2381531, EPI_ISL_2381532, EPI_ISL_2381533, EPI_ISL_2381534, EPI_ISL_2381535, EPI_ISL_2381536, EPI_ISL_2381537, EPI_ISL_2381538, EPI_ISL_2381539, EPI_ISL_2381540, EPI_ISL_2381541, EPI_ISL_2381542, EPI_ISL_2381543, EPI_ISL_2381544, EPI_ISL_2381545, EPI_ISL_2381546, EPI_ISL_2381547, EPI_ISL_2381550, EPI_ISL_2381551, EPI_ISL_2381552, EPI_ISL_2381554, EPI_ISL_2381555, EPI_ISL_2381556, EPI_ISL_2381557, EPI_ISL_2381558, EPI_ISL_2381559, EPI_ISL_2381560, EPI_ISL_2381561, EPI_ISL_2381562, EPI_ISL_2381563, EPI_ISL_2381564, EPI_ISL_2381566, EPI_ISL_2381567, EPI_ISL_2381568, EPI_ISL_2381569, EPI_ISL_2381570, EPI_ISL_2381571, EPI_ISL_2381572, EPI_ISL_2381573, EPI_ISL_2381574, EPI_ISL_2381575, EPI_ISL_2381576, EPI_ISL_2381577, EPI_ISL_2381578, EPI_ISL_2381579, EPI_ISL_2381580, EPI_ISL_2381587, EPI_ISL_2381588, EPI_ISL_2381589, EPI_ISL_2381590, EPI_ISL_2381592, EPI_ISL_2381593, EPI_ISL_2381594, EPI_ISL_2381595, EPI_ISL_2381596, EPI_ISL_2381597, EPI_ISL_2381598, EPI_ISL_2381599, EPI_ISL_2381600, EPI_ISL_2381601, EPI_ISL_2381602, EPI_ISL_2381603, EPI_ISL_2381604, EPI_ISL_2381605, EPI_ISL_2381609, EPI_ISL_2381610, EPI_ISL_2381611, EPI_ISL_2381612, EPI_ISL_2381613, EPI_ISL_2381614, EPI_ISL_2381615, EPI_ISL_2381616, EPI_ISL_2381617, EPI_ISL_2381618, EPI_ISL_2381619, EPI_ISL_2381620, EPI_ISL_2381621, EPI_ISL_2381622, EPI_ISL_2381623, | | | |

| | | | |
|---|---|---|---|
| EPI_ISL_2381626, EPI_ISL_2381627, EPI_ISL_2381628, EPI_ISL_2381629, EPI_ISL_2381630, EPI_ISL_2381631, EPI_ISL_2381632, EPI_ISL_2381633, EPI_ISL_2381634, EPI_ISL_2381635, EPI_ISL_2381636, EPI_ISL_2381637, EPI_ISL_2381638, EPI_ISL_2381639, EPI_ISL_2381640, EPI_ISL_2381641, EPI_ISL_2381642, EPI_ISL_2381643, EPI_ISL_2381644, EPI_ISL_2381646, EPI_ISL_2381647, EPI_ISL_2381649, EPI_ISL_2381654, EPI_ISL_2381655, EPI_ISL_2381656, EPI_ISL_2381658, EPI_ISL_2381659, EPI_ISL_2381660, EPI_ISL_2381661, EPI_ISL_2381662, EPI_ISL_2381663, EPI_ISL_2381664, EPI_ISL_2381665, EPI_ISL_2381666, EPI_ISL_2381667, EPI_ISL_2381669, EPI_ISL_2381670, EPI_ISL_2381671, EPI_ISL_2381672, EPI_ISL_2381673, EPI_ISL_2381674, EPI_ISL_2381675, EPI_ISL_2381676, EPI_ISL_2381677 | | | |
| see above | ZOTZ KLIMAS MVZ Düsseldorf-Centrum GbR ÜBAG für Labormedizin, Genetik, Zytologie, Pathologie | Center of Medical Microbiology, Virology, and Hospital Hygiene, University of Duesseldorf | Alexander Dilthey; Andreas Walker; Daniel Strelow; Jessica Nicolai; Jörg Timm; Katrin Hoffmann; Klaus Pfeffer; Lisanna Hülse; Malte Kohns Vasconcelos; Maximilian Damagnez; Nadine Lübke; Patrick Finzer; Rainer Zotz; Tobias Wienemann; Torsten Houwaart |
| EPI_ISL_2313240, EPI_ISL_2313241, EPI_ISL_2313242, EPI_ISL_2313243, EPI_ISL_2313244, EPI_ISL_2313245, EPI_ISL_2313246, EPI_ISL_2313247, EPI_ISL_2313248 | | | |
| see above | alphaomega Labor GbR (Medizinische Kooperationsgemeinschaft Dres. Ackermann/Tennert/Zaiß) | Robert Koch Institute | |
| EPI_ISL_2266170, EPI_ISL_2266180, EPI_ISL_2266186, EPI_ISL_2266188, EPI_ISL_2266191, EPI_ISL_2266205, EPI_ISL_2266220, EPI_ISL_2266221, EPI_ISL_2266231, EPI_ISL_2266239, EPI_ISL_2266240, EPI_ISL_2266244, EPI_ISL_2266248, EPI_ISL_2266249, EPI_ISL_2266251, EPI_ISL_2266255, EPI_ISL_2266261, EPI_ISL_2266262, EPI_ISL_2266270, EPI_ISL_2266272, EPI_ISL_2266278, EPI_ISL_2266300, EPI_ISL_2266302, EPI_ISL_2266308, EPI_ISL_2266311, EPI_ISL_2266314, EPI_ISL_2266320, EPI_ISL_2266327, EPI_ISL_2266329, EPI_ISL_2266330, EPI_ISL_2266333, EPI_ISL_2266335, EPI_ISL_2266338, EPI_ISL_2266340, EPI_ISL_2266342 | | | |
| see above | amedes Göttingen | Robert Koch Institute | |
| EPI_ISL_2266168, EPI_ISL_2266177, EPI_ISL_2266185, EPI_ISL_2266189, EPI_ISL_2266214, EPI_ISL_2266227, EPI_ISL_2266230, EPI_ISL_2266232, EPI_ISL_2266252, EPI_ISL_2266260, EPI_ISL_2266274, EPI_ISL_2266280, EPI_ISL_2266297, EPI_ISL_2266309, EPI_ISL_2266313, EPI_ISL_2266316, EPI_ISL_2266326, EPI_ISL_2387196, EPI_ISL_2387198, EPI_ISL_2387200, EPI_ISL_2387205, EPI_ISL_2387212, EPI_ISL_2387216, EPI_ISL_2387219, EPI_ISL_2387224, EPI_ISL_2387228, EPI_ISL_2387229, EPI_ISL_2387237, EPI_ISL_2387239, EPI_ISL_2387252, EPI_ISL_2387257, EPI_ISL_2387262, EPI_ISL_2387264, EPI_ISL_2387269, EPI_ISL_2387276, EPI_ISL_2387280, EPI_ISL_2387282, EPI_ISL_2387285, EPI_ISL_2387287, EPI_ISL_2387290, EPI_ISL_2387294, EPI_ISL_2387297, EPI_ISL_2387300, EPI_ISL_2387304, EPI_ISL_2387306, EPI_ISL_2387317, EPI_ISL_2387318, EPI_ISL_2387319, EPI_ISL_2387320, EPI_ISL_2387323, EPI_ISL_2387324, EPI_ISL_2387327, EPI_ISL_2387333, EPI_ISL_2387345, EPI_ISL_2632261, EPI_ISL_2632299 | | | |
| see above | amedes MVZ DIAMEDIS Sennestadt | Robert Koch Institute | |
| EPI_ISL_2130634, EPI_ISL_2130665, EPI_ISL_2130754, EPI_ISL_2130778, EPI_ISL_2130785, EPI_ISL_2130799, EPI_ISL_2130814, EPI_ISL_2130817, EPI_ISL_2266174, EPI_ISL_2266178, EPI_ISL_2266194, EPI_ISL_2266215, EPI_ISL_2266218, EPI_ISL_2266219, EPI_ISL_2266223, EPI_ISL_2266224, EPI_ISL_2266229, EPI_ISL_2266234, EPI_ISL_2266235, EPI_ISL_2266236, EPI_ISL_2266238, EPI_ISL_2266242, EPI_ISL_2266247, EPI_ISL_2266256, EPI_ISL_2266257, EPI_ISL_2266259, EPI_ISL_2266267, EPI_ISL_2266269, EPI_ISL_2266271, EPI_ISL_2266277, EPI_ISL_2266281, EPI_ISL_2266284, EPI_ISL_2266286, EPI_ISL_2266303, EPI_ISL_2266304, EPI_ISL_2266306, EPI_ISL_2266328, EPI_ISL_2266343, EPI_ISL_2470865, EPI_ISL_2470872, EPI_ISL_2470875, EPI_ISL_2470898, EPI_ISL_2470933, EPI_ISL_2470947, EPI_ISL_2470963, EPI_ISL_2470998, EPI_ISL_2471008 | | | |
| see above | amedes MVZ Halle/Leipzig | Robert Koch Institute | |
| EPI_ISL_2266181, EPI_ISL_2266192, EPI_ISL_2266195, EPI_ISL_2266198, EPI_ISL_2266199, EPI_ISL_2266201, EPI_ISL_2266204, EPI_ISL_2266207, EPI_ISL_2266211, EPI_ISL_2266212, EPI_ISL_2266226, EPI_ISL_2266241, EPI_ISL_2266246, EPI_ISL_2266275, EPI_ISL_2266290, EPI_ISL_2266325, EPI_ISL_2266331, EPI_ISL_2266348, EPI_ISL_2387217, EPI_ISL_2387247, EPI_ISL_2387274, EPI_ISL_2387283, EPI_ISL_2387293, EPI_ISL_2387305, EPI_ISL_2387344 | | | |
| see above | amedes MVZ Hannover | Robert Koch Institute | |
| EPI_ISL_2130602, EPI_ISL_2130626, EPI_ISL_2130640, EPI_ISL_2130771, EPI_ISL_2130816 | amedes MVZ fÃ¼r Labordiagnostik Rhein-Main | Robert Koch Institute | |
| EPI_ISL_2266171, EPI_ISL_2266175, EPI_ISL_2266190, EPI_ISL_2266203, EPI_ISL_2266209, EPI_ISL_2266216, EPI_ISL_2266217, EPI_ISL_2266228, EPI_ISL_2266233, EPI_ISL_2266295, EPI_ISL_2266336, EPI_ISL_2266337, EPI_ISL_2387202, EPI_ISL_2387222, EPI_ISL_2387255, EPI_ISL_2387286, EPI_ISL_2387302, EPI_ISL_2387321, EPI_ISL_2387334, EPI_ISL_2387339 | | | |
| see above | amedes MVZ für Laboratoriumsdiagnostik Raubling GmbH | Robert Koch Institute | |
| EPI_ISL_2266197, EPI_ISL_2266254, EPI_ISL_2266349 | amedes MVZ für Laboratoriumsmedizin Fürstenfeldbruck | Robert Koch Institute | |
| EPI_ISL_2387277 | amedes MVZ für Laboratoriumsmedizin München | Robert Koch Institute | |
| EPI_ISL_2266202, EPI_ISL_2266291, EPI_ISL_2266322 | amedes MVZ für Laboratoriumsmedizin Rhein/Ruhr | Robert Koch Institute | |
| EPI_ISL_2266169, EPI_ISL_2266176, EPI_ISL_2266210, EPI_ISL_2266213, EPI_ISL_2266266, EPI_ISL_2266285, EPI_ISL_2266289, EPI_ISL_2266298, EPI_ISL_2266312, EPI_ISL_2266339, EPI_ISL_2266344, EPI_ISL_2470934, EPI_ISL_2470943, EPI_ISL_2470968, EPI_ISL_2470983 | | | |
| see above | amedes MVZ für Labordiagnostik Rhein-Main | Robert Koch Institute | |
| EPI_ISL_2130680, EPI_ISL_2130697, EPI_ISL_2130743, EPI_ISL_2130809, EPI_ISL_2266279, EPI_ISL_2266283 | amedes Medlab Arnold Analytik MVZ GmbH | Robert Koch Institute | |
| EPI_ISL_2263154, EPI_ISL_2263159, EPI_ISL_2263161, EPI_ISL_2263162, EPI_ISL_2263163, EPI_ISL_2263164, EPI_ISL_2263165, EPI_ISL_2263167, EPI_ISL_2263169, EPI_ISL_2263170, EPI_ISL_2263171, EPI_ISL_2263176, EPI_ISL_2263178, EPI_ISL_2263179, EPI_ISL_2263180, EPI_ISL_2263182, EPI_ISL_2263183, EPI_ISL_2267905, EPI_ISL_2267906, EPI_ISL_2267907, EPI_ISL_2267908, EPI_ISL_2267909, EPI_ISL_2267910, EPI_ISL_2267911, EPI_ISL_2267912, EPI_ISL_2267913, EPI_ISL_2267914, EPI_ISL_2267915, EPI_ISL_2267916, EPI_ISL_2267917, EPI_ISL_2267918, EPI_ISL_2267919, EPI_ISL_2267920, EPI_ISL_2267921, EPI_ISL_2267922, EPI_ISL_2267923, EPI_ISL_2267924, EPI_ISL_2267925, EPI_ISL_2267926, EPI_ISL_2267927, EPI_ISL_2267928 | | | |
| see above | labopart - Medizinische Laboratorien Dresden | Robert Koch Institute | |
| EPI_ISL_2130605, EPI_ISL_2130609, EPI_ISL_2130614, EPI_ISL_2130618, EPI_ISL_2130627, EPI_ISL_2130636, EPI_ISL_2130638, EPI_ISL_2130654, EPI_ISL_2130658, EPI_ISL_2130659, EPI_ISL_2130662, EPI_ISL_2130670, EPI_ISL_2130674, EPI_ISL_2130676, EPI_ISL_2130678, EPI_ISL_2130685, EPI_ISL_2130686, EPI_ISL_2130689, EPI_ISL_2130692, EPI_ISL_2130695, EPI_ISL_2130696, EPI_ISL_2130698, EPI_ISL_2130700, EPI_ISL_2130712, EPI_ISL_2130717, EPI_ISL_2130725, EPI_ISL_2130728, EPI_ISL_2130735, EPI_ISL_2130738, EPI_ISL_2130741, EPI_ISL_2130744, EPI_ISL_2130751, EPI_ISL_2130752, EPI_ISL_2130762, EPI_ISL_2130765, EPI_ISL_2130769, EPI_ISL_2130770, EPI_ISL_2130773, EPI_ISL_2130775, EPI_ISL_2130777, EPI_ISL_2130784, EPI_ISL_2130787, EPI_ISL_2130791, EPI_ISL_2130792, EPI_ISL_2130794, EPI_ISL_2130795, EPI_ISL_2130805, EPI_ISL_2130811, EPI_ISL_2130812 | | | |
| see above | nordlab - Partnerschaftspraxis fuÌˆr Laboratoriumsmedizin | Robert Koch Institute | |
| EPI_ISL_2266172, EPI_ISL_2266173, EPI_ISL_2266179, EPI_ISL_2266182, EPI_ISL_2266183, EPI_ISL_2266184, EPI_ISL_2266187, EPI_ISL_2266193, EPI_ISL_2266196, EPI_ISL_2266200, EPI_ISL_2266206, EPI_ISL_2266208, EPI_ISL_2266222, EPI_ISL_2266225, EPI_ISL_2266237, EPI_ISL_2266243, EPI_ISL_2266245, EPI_ISL_2266250, EPI_ISL_2266253, EPI_ISL_2266258, EPI_ISL_2266263, EPI_ISL_2266264, EPI_ISL_2266265, EPI_ISL_2266268, EPI_ISL_2266273, EPI_ISL_2266276, EPI_ISL_2266282, EPI_ISL_2266287, EPI_ISL_2266288, EPI_ISL_2266292, EPI_ISL_2266293, EPI_ISL_2266294, EPI_ISL_2266296, EPI_ISL_2266299, EPI_ISL_2266301, EPI_ISL_2266305, EPI_ISL_2266307, EPI_ISL_2266310, EPI_ISL_2266318, EPI_ISL_2266324, EPI_ISL_2266332, EPI_ISL_2266334, EPI_ISL_2266341, EPI_ISL_2266345, EPI_ISL_2266346, EPI_ISL_2266347 | | | |
| see above | nordlab - Partnerschaftspraxis für Laboratoriumsmedizin | Robert Koch Institute | |

We gratefully acknowledge the following Authors from the Originating laboratories responsible for obtaining the specimens, as well as the Submitting laboratories where the genome data were generated and shared via GISAID, on which this research is based.

All Submitters of data may be contacted directly via www.gisaid.org

Authors are sorted alphabetically.

| Accession ID | Originating Laboratory | Submitting Laboratory | Authors |
|---|---|---|---|
| EPI_ISL_2447992, EPI_ISL_2447993, EPI_ISL_2447994, EPI_ISL_2447995, EPI_ISL_2447996, EPI_ISL_2447997, EPI_ISL_2447998, EPI_ISL_2447999, EPI_ISL_2448000, EPI_ISL_2448001, EPI_ISL_2448002, EPI_ISL_2448003, EPI_ISL_2448004, EPI_ISL_2448005, EPI_ISL_2448006, EPI_ISL_2448007, EPI_ISL_2448008, EPI_ISL_2448009, EPI_ISL_2500363, EPI_ISL_2500364, EPI_ISL_2500365, EPI_ISL_2500366, EPI_ISL_2500367, EPI_ISL_2500368, EPI_ISL_2500369, EPI_ISL_2500370 | | | |
| see above | A. Krumbholz, Labor Dr. Krause und Kollegen MVZ GmbH, Kiel | Charité Universitätsmedizin Berlin, Institut für Virologie | Barbara Mühlemann; Christian Drosten; Julia Schneider; Julia Tesch; Jörn Beheim-Schwarzbach; Talitha Veith; Terry Jones; Tobias Bleicker; Victor M Corman |
| EPI_ISL_2504051, EPI_ISL_2504052, EPI_ISL_2504053, EPI_ISL_2504054, EPI_ISL_2504055, EPI_ISL_2504056, EPI_ISL_2504057, EPI_ISL_2504062, EPI_ISL_2504064, EPI_ISL_2504065 | | | |
| see above | A. Krumbholz, Labor Dr. Krause und Kollegen MVZ GmbH, Kiel | Charité Universitätsmedizin Berlin, Institut für Virologie/Labor Berlin | Andi Krumbholz; Barbara Muehlemann; Christian Drosten; Christine Stephan; Joern Beheim-Schwarzbach; Julia Schneider; Julia Tesch; Peter Menzel; Rolf Schwarzer; Talitha Veith; Terry Jones; Tobias Bleicker; Victor M Corman |
| EPI_ISL_2633918, EPI_ISL_2633955, EPI_ISL_2634053, EPI_ISL_2634101, EPI_ISL_2634113, EPI_ISL_2634179, EPI_ISL_2634244, EPI_ISL_2634328, EPI_ISL_2634348, EPI_ISL_2636966 | | | |
| see above | ALB FILS KLINIKEN GmbH - Institut für Laboratoriumsmedizin Klinik am Eichert Göppingen | Robert Koch Institute | |
| EPI_ISL_2472558, EPI_ISL_2472584, EPI_ISL_2472594, EPI_ISL_2472599, EPI_ISL_2472614, EPI_ISL_2472615, EPI_ISL_2634941, EPI_ISL_2634944, EPI_ISL_2634953, EPI_ISL_2634955, EPI_ISL_2634957, EPI_ISL_2634960, EPI_ISL_2634964, EPI_ISL_2634966, EPI_ISL_2634970, EPI_ISL_2634975, EPI_ISL_2634976, EPI_ISL_2634977, EPI_ISL_2634979, EPI_ISL_2634981, EPI_ISL_2634983, EPI_ISL_2634984, EPI_ISL_2634986, EPI_ISL_2634987, EPI_ISL_2634988, EPI_ISL_2634989, EPI_ISL_2634990, EPI_ISL_2634991, EPI_ISL_2634992, EPI_ISL_2634993, EPI_ISL_2634995, EPI_ISL_2634996, EPI_ISL_2634999, EPI_ISL_2635000, EPI_ISL_2635002, EPI_ISL_2635004, EPI_ISL_2635005, EPI_ISL_2635007, EPI_ISL_2635008, EPI_ISL_2635009, EPI_ISL_2635010, EPI_ISL_2635013, EPI_ISL_2635014, EPI_ISL_2635015, EPI_ISL_2635016, EPI_ISL_2635017, EPI_ISL_2635018, EPI_ISL_2635019, EPI_ISL_2635021, EPI_ISL_2635023, EPI_ISL_2635024, EPI_ISL_2635025, EPI_ISL_2635027, EPI_ISL_2635029, EPI_ISL_2635031, EPI_ISL_2635032, EPI_ISL_2635033, EPI_ISL_2635034, EPI_ISL_2635035, EPI_ISL_2635036, EPI_ISL_2635042, EPI_ISL_2635044, EPI_ISL_2635046, EPI_ISL_2635049, EPI_ISL_2635051, EPI_ISL_2635056, EPI_ISL_2635058, EPI_ISL_2635062, EPI_ISL_2635064, EPI_ISL_2635067, EPI_ISL_2635069, EPI_ISL_2635074, EPI_ISL_2635076, EPI_ISL_2635078, EPI_ISL_2635081, EPI_ISL_2635084, EPI_ISL_2635087, EPI_ISL_2635092, EPI_ISL_2635099, EPI_ISL_2635103, EPI_ISL_2635106, EPI_ISL_2635108, EPI_ISL_2635110, EPI_ISL_2635114, EPI_ISL_2635117, EPI_ISL_2635119, EPI_ISL_2635123, EPI_ISL_2760020, EPI_ISL_2760021, EPI_ISL_2760024, EPI_ISL_2760026, EPI_ISL_2760027, EPI_ISL_2760028, EPI_ISL_2760029, EPI_ISL_2760030, EPI_ISL_2760031, EPI_ISL_2760032, EPI_ISL_2760033, EPI_ISL_2760034, EPI_ISL_2760035, EPI_ISL_2760036, EPI_ISL_2760037, EPI_ISL_2760040, EPI_ISL_2760041, EPI_ISL_2760044, EPI_ISL_2760045, EPI_ISL_2760046, EPI_ISL_2760047, EPI_ISL_2760048, EPI_ISL_2760049, EPI_ISL_2760050, EPI_ISL_2760051, EPI_ISL_2760052, EPI_ISL_2760053, EPI_ISL_2760057, EPI_ISL_2760059, EPI_ISL_2760060, EPI_ISL_2760061, EPI_ISL_2760062, EPI_ISL_2760063, EPI_ISL_2760064, EPI_ISL_2760065, EPI_ISL_2760066, EPI_ISL_2760069, EPI_ISL_2760072, EPI_ISL_2760073, EPI_ISL_2760076, EPI_ISL_2760081, EPI_ISL_2760086, EPI_ISL_2760089, EPI_ISL_2760091, EPI_ISL_2760093, EPI_ISL_2760096, EPI_ISL_2760103, EPI_ISL_2760106, EPI_ISL_2760108, EPI_ISL_2760111, EPI_ISL_2760114, EPI_ISL_2760117, EPI_ISL_2760122, EPI_ISL_2760124, EPI_ISL_2760126, EPI_ISL_2760129, EPI_ISL_2760133, EPI_ISL_2760136, EPI_ISL_2760138, EPI_ISL_2760143, EPI_ISL_2760146, EPI_ISL_2760149, EPI_ISL_2762211, EPI_ISL_2762213, EPI_ISL_2762216, EPI_ISL_2762227, EPI_ISL_2762238, EPI_ISL_2762241, EPI_ISL_2762243, EPI_ISL_2762270 | | | |
| see above | Bayerisches Landesamt für Gesundheit und Lebensmittelsicherheit (LGL) | Robert Koch Institute | |
| EPI_ISL_2387214, EPI_ISL_2387236, EPI_ISL_2469930, EPI_ISL_2469933, EPI_ISL_2469934, EPI_ISL_2469935, EPI_ISL_2469937, EPI_ISL_2469939, EPI_ISL_2469945, EPI_ISL_2469947, EPI_ISL_2469959, EPI_ISL_2469962, EPI_ISL_2469966, EPI_ISL_2469970, EPI_ISL_2469973, EPI_ISL_2469974, EPI_ISL_2469978, EPI_ISL_2469987, EPI_ISL_2469999, EPI_ISL_2470009, EPI_ISL_2470012, EPI_ISL_2470018, EPI_ISL_2470028, EPI_ISL_2470034, EPI_ISL_2470050, EPI_ISL_2470051, EPI_ISL_2470056, EPI_ISL_2470058, EPI_ISL_2470061, EPI_ISL_2470062, EPI_ISL_2470065, EPI_ISL_2470066, EPI_ISL_2470069, EPI_ISL_2470076, EPI_ISL_2470078, EPI_ISL_2470085, EPI_ISL_2470087, EPI_ISL_2470089, EPI_ISL_2470090, EPI_ISL_2470091, EPI_ISL_2470095, EPI_ISL_2470110, EPI_ISL_2470115, EPI_ISL_2470119, EPI_ISL_2470124, EPI_ISL_2470126, EPI_ISL_2470127, EPI_ISL_2470131, EPI_ISL_2470134, EPI_ISL_2470138, EPI_ISL_2470144, EPI_ISL_2470148, EPI_ISL_2470149, EPI_ISL_2470153, EPI_ISL_2470154, EPI_ISL_2470157, EPI_ISL_2470158, EPI_ISL_2470161, EPI_ISL_2470162, EPI_ISL_2470163, EPI_ISL_2470164, EPI_ISL_2470165, EPI_ISL_2470167, EPI_ISL_2470174, EPI_ISL_2470175, EPI_ISL_2470185, EPI_ISL_2470187, EPI_ISL_2470188, EPI_ISL_2470190, EPI_ISL_2470193, EPI_ISL_2470197, EPI_ISL_2470199, EPI_ISL_2470206, EPI_ISL_2470218, EPI_ISL_2470221, EPI_ISL_2470222, EPI_ISL_2470224, EPI_ISL_2470242, EPI_ISL_2470243, EPI_ISL_2470259, EPI_ISL_2470264, EPI_ISL_2470267, EPI_ISL_2470284, EPI_ISL_2470285, EPI_ISL_2470289, EPI_ISL_2470292, EPI_ISL_2470310, EPI_ISL_2470331, EPI_ISL_2470334, EPI_ISL_2470337, EPI_ISL_2470340, EPI_ISL_2470345, EPI_ISL_2470350, EPI_ISL_2470369, EPI_ISL_2470377, EPI_ISL_2470394, EPI_ISL_2470424, EPI_ISL_2470430, EPI_ISL_2470437, EPI_ISL_2470448, EPI_ISL_2470450, EPI_ISL_2470452, EPI_ISL_2470459, EPI_ISL_2470461, EPI_ISL_2470472, EPI_ISL_2470473, EPI_ISL_2470475, EPI_ISL_2470476, EPI_ISL_2470479, EPI_ISL_2470485, EPI_ISL_2470486, EPI_ISL_2632338, EPI_ISL_2632339, EPI_ISL_2632344, EPI_ISL_2632347, EPI_ISL_2632348, EPI_ISL_2632349, EPI_ISL_2632350, EPI_ISL_2632351, EPI_ISL_2632352, EPI_ISL_2632353, EPI_ISL_2632354, EPI_ISL_2632355, EPI_ISL_2632356, EPI_ISL_2632359, EPI_ISL_2632360, EPI_ISL_2632361, EPI_ISL_2632367, EPI_ISL_2632368, EPI_ISL_2632369, EPI_ISL_2632370, EPI_ISL_2632371, EPI_ISL_2632372, EPI_ISL_2632375, EPI_ISL_2632376, EPI_ISL_2632378, EPI_ISL_2632385, EPI_ISL_2632386, EPI_ISL_2632387, EPI_ISL_2632388, EPI_ISL_2632390, EPI_ISL_2632392, EPI_ISL_2632393, EPI_ISL_2632394, EPI_ISL_2632397, EPI_ISL_2632398, EPI_ISL_2632401, EPI_ISL_2632402, EPI_ISL_2632403, EPI_ISL_2632405, EPI_ISL_2632408, EPI_ISL_2632409, EPI_ISL_2632411, EPI_ISL_2632413, EPI_ISL_2632415, EPI_ISL_2632419, EPI_ISL_2632421, EPI_ISL_2632422, EPI_ISL_2632423, EPI_ISL_2632424, EPI_ISL_2632426, EPI_ISL_2632427, EPI_ISL_2632429, EPI_ISL_2632430, EPI_ISL_2632433, EPI_ISL_2632436, EPI_ISL_2632438, EPI_ISL_2632445, EPI_ISL_2632446, EPI_ISL_2632449, EPI_ISL_2632451, EPI_ISL_2632453, EPI_ISL_2632455, EPI_ISL_2632456, EPI_ISL_2632462, EPI_ISL_2632465, EPI_ISL_2632467, EPI_ISL_2632472, EPI_ISL_2632473, EPI_ISL_2632474, EPI_ISL_2632478, EPI_ISL_2632482, EPI_ISL_2632483, EPI_ISL_2632484, EPI_ISL_2632485, EPI_ISL_2632486, EPI_ISL_2632487, EPI_ISL_2632488, EPI_ISL_2632492, EPI_ISL_2632494, EPI_ISL_2632495, EPI_ISL_2632496, EPI_ISL_2632497, EPI_ISL_2632499, EPI_ISL_2632500, EPI_ISL_2632502, EPI_ISL_2632504, EPI_ISL_2632505, EPI_ISL_2632506, EPI_ISL_2632510, EPI_ISL_2632514, EPI_ISL_2632516, EPI_ISL_2632517, EPI_ISL_2632521, EPI_ISL_2632525, EPI_ISL_2632528, EPI_ISL_2632529, EPI_ISL_2632530, EPI_ISL_2632532, EPI_ISL_2632535, EPI_ISL_2632536, EPI_ISL_2632540, EPI_ISL_2632542, EPI_ISL_2632543, EPI_ISL_2632545, EPI_ISL_2632548, EPI_ISL_2632550, EPI_ISL_2632552, EPI_ISL_2632558, EPI_ISL_2632560, EPI_ISL_2632562, EPI_ISL_2632567, EPI_ISL_2632568, EPI_ISL_2632571, EPI_ISL_2632575, EPI_ISL_2632577, EPI_ISL_2632578, EPI_ISL_2632586, EPI_ISL_2632589, EPI_ISL_2632593, EPI_ISL_2632594, EPI_ISL_2632596, EPI_ISL_2632600, EPI_ISL_2632603, EPI_ISL_2632607, EPI_ISL_2632608, EPI_ISL_2632611, EPI_ISL_2632614, EPI_ISL_2632615, EPI_ISL_2632616, EPI_ISL_2632619, EPI_ISL_2632620, EPI_ISL_2632621, EPI_ISL_2632622, EPI_ISL_2632623, EPI_ISL_2632624, EPI_ISL_2632626, EPI_ISL_2632628, EPI_ISL_2632629, EPI_ISL_2632630, EPI_ISL_2632633, EPI_ISL_2632636, EPI_ISL_2632637, EPI_ISL_2632638, EPI_ISL_2632639, EPI_ISL_2632640, EPI_ISL_2632641, EPI_ISL_2632644, EPI_ISL_2633901, EPI_ISL_2633904, EPI_ISL_2633920, EPI_ISL_2633925, EPI_ISL_2633927, EPI_ISL_2633934, EPI_ISL_2633936, EPI_ISL_2633951, EPI_ISL_2633964, EPI_ISL_2633966, EPI_ISL_2633969, EPI_ISL_2633972, EPI_ISL_2633973, EPI_ISL_2633986, EPI_ISL_2634012, EPI_ISL_2634018, EPI_ISL_2634019, EPI_ISL_2634020, EPI_ISL_2634029, EPI_ISL_2634030, EPI_ISL_2634031, EPI_ISL_2634033, EPI_ISL_2634039, EPI_ISL_2634041, EPI_ISL_2634045, EPI_ISL_2634052, EPI_ISL_2634058, EPI_ISL_2634073, EPI_ISL_2634077, EPI_ISL_2634081, EPI_ISL_2634082, EPI_ISL_2634087, EPI_ISL_2634097, EPI_ISL_2634102, EPI_ISL_2634107, EPI_ISL_2634115, EPI_ISL_2634117, EPI_ISL_2634119, EPI_ISL_2634126, EPI_ISL_2634131, EPI_ISL_2634142, EPI_ISL_2634146, EPI_ISL_2634151, EPI_ISL_2634161, EPI_ISL_2634162, EPI_ISL_2634169, EPI_ISL_2634174, EPI_ISL_2634190, EPI_ISL_2634192, EPI_ISL_2634199, EPI_ISL_2634200, EPI_ISL_2634202, EPI_ISL_2634207, EPI_ISL_2634215, EPI_ISL_2634237, EPI_ISL_2634257, EPI_ISL_2634266, EPI_ISL_2634281, EPI_ISL_2634283, EPI_ISL_2634294, EPI_ISL_2634299, EPI_ISL_2634312, EPI_ISL_2634315, EPI_ISL_2634321, EPI_ISL_2634335, EPI_ISL_2634339, EPI_ISL_2634359, EPI_ISL_2634376, EPI_ISL_2634411, EPI_ISL_2634421, EPI_ISL_2634443, EPI_ISL_2634448, EPI_ISL_2634452, EPI_ISL_2636910, EPI_ISL_2636917, EPI_ISL_2636932, EPI_ISL_2636940, EPI_ISL_2636943, EPI_ISL_2636944, EPI_ISL_2636946, EPI_ISL_2636951, EPI_ISL_2636953, EPI_ISL_2636956, EPI_ISL_2636961, EPI_ISL_2636969, EPI_ISL_2636970, EPI_ISL_2636971, EPI_ISL_2636974, EPI_ISL_2636975, EPI_ISL_2636989, EPI_ISL_2636992, EPI_ISL_2636999, EPI_ISL_2637005, EPI_ISL_2637007, EPI_ISL_2637008, EPI_ISL_2637011, EPI_ISL_2637016, EPI_ISL_2637018, EPI_ISL_2637022, EPI_ISL_2637039, EPI_ISL_2637045, EPI_ISL_2637048, EPI_ISL_2637049, EPI_ISL_2637055, EPI_ISL_2637061, EPI_ISL_2637064, EPI_ISL_2637068, EPI_ISL_2637082, EPI_ISL_2637085, EPI_ISL_2637094, EPI_ISL_2637095, EPI_ISL_2637096, EPI_ISL_2637097, EPI_ISL_2637102, EPI_ISL_2637106, EPI_ISL_2637114, EPI_ISL_2637127, EPI_ISL_2637142, EPI_ISL_2637147, EPI_ISL_2637149, EPI_ISL_2637166, EPI_ISL_2637168, EPI_ISL_2637171, EPI_ISL_2637176, EPI_ISL_2762089 | | | |
| see above | Bioscientia Labor Wermsdorf | Robert Koch Institute | |
| EPI_ISL_2469967, EPI_ISL_2469980, EPI_ISL_2470021, EPI_ISL_2470030, EPI_ISL_2470105, EPI_ISL_2470139, EPI_ISL_2470159, EPI_ISL_2470183, EPI_ISL_2470200, EPI_ISL_2470202, EPI_ISL_2470211, EPI_ISL_2470223, EPI_ISL_2470230, EPI_ISL_2470275, EPI_ISL_2470295, EPI_ISL_2470299, EPI_ISL_2470306, EPI_ISL_2470312, EPI_ISL_2470320, EPI_ISL_2470321, EPI_ISL_2470323, EPI_ISL_2470327, EPI_ISL_2470328, EPI_ISL_2470329, EPI_ISL_2470354, EPI_ISL_2470373, EPI_ISL_2470396, EPI_ISL_2470406, EPI_ISL_2470436, EPI_ISL_2632395, EPI_ISL_2632447, EPI_ISL_2632460, EPI_ISL_2632554, EPI_ISL_2633914, EPI_ISL_2633929, EPI_ISL_2633958, EPI_ISL_2634023, EPI_ISL_2634062, EPI_ISL_2634069, EPI_ISL_2634089, EPI_ISL_2634096, EPI_ISL_2634098, EPI_ISL_2634172, EPI_ISL_2634188, EPI_ISL_2634225, EPI_ISL_2634249, EPI_ISL_2634253, EPI_ISL_2634310, EPI_ISL_2634352, EPI_ISL_2634395, EPI_ISL_2634424, EPI_ISL_2634428, EPI_ISL_2637148 | | | |
| see above | Bioscientia MVZ Labor Karlsruhe GmbH | Robert Koch Institute | |
| EPI_ISL_2536151 | Bundeswehr Institute of Microbiology | Bundeswehr Institute of Microbiology | Alexandra Rehn; Enrico Georgi; Malena Bestehorn-Willmann; Markus Antwerpen; Mathias Walter; Mike Pillukat; Roman Wölfel; Sabine Zange |
| EPI_ISL_2536152, EPI_ISL_2727658, EPI_ISL_2727659, EPI_ISL_2727660, EPI_ISL_2727661, EPI_ISL_2727666 | Bundeswehrkrankenhaus Berlin | Bundeswehr Institute of Microbiology | Alexandra Rehn; Enrico Georgi; Malena Bestehorn-Willmann; Markus Antwerpen; Mathias Walter; Mike Pillukat; Roman Wölfel; Sabine Zange |
| EPI_ISL_2727663 | Bundeswehrkrankenhaus Hamburg | Bundeswehr Institute of Microbiology | Alexandra Rehn; Enrico Georgi; Malena Bestehorn-Willmann; Markus Antwerpen; Mathias Walter; Mike Pillukat; Roman Wölfel; Sabine Zange |
| EPI_ISL_2536154, EPI_ISL_2727662 | Bundeswehrkrankenhaus Ulm | Bundeswehr Institute of Microbiology | Alexandra Rehn; Enrico Georgi; Malena Bestehorn-Willmann; Markus Antwerpen; Mathias Walter; Mike Pillukat; Roman Wölfel; Sabine Zange |
| EPI_ISL_2536153 | Bundeswehrzentralkrankenhaus Koblenz | Bundeswehr Institute of Microbiology | Alexandra Rehn; Enrico Georgi; Malena Bestehorn-Willmann; Markus Antwerpen; Mathias Walter; Mike Pillukat; Roman Wölfel; Sabine Zange |
| EPI_ISL_2386884, EPI_ISL_2386886, EPI_ISL_2386888, EPI_ISL_2386890, EPI_ISL_2386894, EPI_ISL_2386895, EPI_ISL_2386896, EPI_ISL_2386900, EPI_ISL_2386902, EPI_ISL_2386905, EPI_ISL_2386907, EPI_ISL_2386915, EPI_ISL_2386918, EPI_ISL_2386920, EPI_ISL_2386925, EPI_ISL_2386933, EPI_ISL_2386937, EPI_ISL_2386939, EPI_ISL_2386940, EPI_ISL_2386943, EPI_ISL_2386950, EPI_ISL_2386956, EPI_ISL_2386959, EPI_ISL_2386962, EPI_ISL_2386963, EPI_ISL_2386968, EPI_ISL_2386969, EPI_ISL_2386973, EPI_ISL_2386974, EPI_ISL_2386977, EPI_ISL_2386978, EPI_ISL_2386979, EPI_ISL_2386984, EPI_ISL_2386988, EPI_ISL_2386991, EPI_ISL_2386992, EPI_ISL_2386994, EPI_ISL_2386998, EPI_ISL_2386999, EPI_ISL_2387002, EPI_ISL_2387003, EPI_ISL_2387007, EPI_ISL_2387011, EPI_ISL_2387012, EPI_ISL_2387016, EPI_ISL_2387022, EPI_ISL_2387025, EPI_ISL_2387026, EPI_ISL_2387029, EPI_ISL_2387030, EPI_ISL_2387033, EPI_ISL_2387034, EPI_ISL_2387035, EPI_ISL_2387036, EPI_ISL_2387042, EPI_ISL_2387043, EPI_ISL_2387045, EPI_ISL_2387046, EPI_ISL_2387048, EPI_ISL_2387052, EPI_ISL_2387056, EPI_ISL_2387057, EPI_ISL_2387059, EPI_ISL_2387062, EPI_ISL_2387065, EPI_ISL_2387067, EPI_ISL_2387068, EPI_ISL_2387070, EPI_ISL_2387071, EPI_ISL_2387074, EPI_ISL_2387075, EPI_ISL_2387076, EPI_ISL_2387078, EPI_ISL_2387082, EPI_ISL_2387083, EPI_ISL_2387085, EPI_ISL_2387087, EPI_ISL_2387088, EPI_ISL_2387091, EPI_ISL_2387092, EPI_ISL_2387094, EPI_ISL_2387095, EPI_ISL_2387096, EPI_ISL_2387097, EPI_ISL_2387098, EPI_ISL_2387099, EPI_ISL_2387107, EPI_ISL_2387113, EPI_ISL_2387124, EPI_ISL_2387125, EPI_ISL_2387126, EPI_ISL_2387128, EPI_ISL_2387129, EPI_ISL_2387132, EPI_ISL_2387133, EPI_ISL_2387134, EPI_ISL_2387135, EPI_ISL_2387137, EPI_ISL_2387138, EPI_ISL_2387140, EPI_ISL_2387142, EPI_ISL_2387148, EPI_ISL_2387150, EPI_ISL_2387152, EPI_ISL_2387159, EPI_ISL_2387161, EPI_ISL_2387163, EPI_ISL_2387166, EPI_ISL_2387170, EPI_ISL_2387174, EPI_ISL_2387179, EPI_ISL_2387187, EPI_ISL_2471183, EPI_ISL_2471184, EPI_ISL_2471187, EPI_ISL_2471188, EPI_ISL_2471189, EPI_ISL_2471190, EPI_ISL_2471194, EPI_ISL_2471195, EPI_ISL_2471196, EPI_ISL_2471197, EPI_ISL_2471198, EPI_ISL_2471200, EPI_ISL_2471201, EPI_ISL_2471202, EPI_ISL_2471203, EPI_ISL_2471204, EPI_ISL_2471205, EPI_ISL_2471207, EPI_ISL_2471208, EPI_ISL_2471209, EPI_ISL_2471210, EPI_ISL_2471211, EPI_ISL_2471212, EPI_ISL_2471213, EPI_ISL_2471214, EPI_ISL_2471215, EPI_ISL_2471216, EPI_ISL_2471218, EPI_ISL_2471219, EPI_ISL_2471221, EPI_ISL_2471222, EPI_ISL_2471223, EPI_ISL_2471225, EPI_ISL_2471226, EPI_ISL_2471227, EPI_ISL_2471228, EPI_ISL_2471229, EPI_ISL_2471230, EPI_ISL_2471231, EPI_ISL_2471232, EPI_ISL_2471233, EPI_ISL_2471236, EPI_ISL_2471237, EPI_ISL_2471239, EPI_ISL_2471240, EPI_ISL_2471241, EPI_ISL_2471242, EPI_ISL_2471244, EPI_ISL_2471246, EPI_ISL_2471247, EPI_ISL_2471248, EPI_ISL_2471249, EPI_ISL_2471250, EPI_ISL_2471251, EPI_ISL_2471252, EPI_ISL_2471253, EPI_ISL_2471254, EPI_ISL_2471255, EPI_ISL_2471256, EPI_ISL_2471257, EPI_ISL_2471258, EPI_ISL_2471260, EPI_ISL_2471262, EPI_ISL_2471263, EPI_ISL_2471264, EPI_ISL_2471265, EPI_ISL_2471266, EPI_ISL_2471268, EPI_ISL_2471269, EPI_ISL_2471270, EPI_ISL_2471271, EPI_ISL_2471272, EPI_ISL_2471273, EPI_ISL_2471275, EPI_ISL_2471276, EPI_ISL_2471277, EPI_ISL_2471278, EPI_ISL_2471279, EPI_ISL_2471280, EPI_ISL_2471281, EPI_ISL_2471282, EPI_ISL_2471284, EPI_ISL_2471285, EPI_ISL_2471286, EPI_ISL_2471287, EPI_ISL_2471288, EPI_ISL_2471289, EPI_ISL_2471290, EPI_ISL_2471292, EPI_ISL_2471293, EPI_ISL_2471294, EPI_ISL_2471295, EPI_ISL_2471296, EPI_ISL_2471299, EPI_ISL_2471300, EPI_ISL_2471301, EPI_ISL_2471302, EPI_ISL_2471303, EPI_ISL_2471304, EPI_ISL_2471305, EPI_ISL_2471306, EPI_ISL_2471307, EPI_ISL_2471308, EPI_ISL_2471309, EPI_ISL_2471310, EPI_ISL_2471312, EPI_ISL_2471313, EPI_ISL_2471314, EPI_ISL_2471315, EPI_ISL_2471316, EPI_ISL_2471317, EPI_ISL_2471319, EPI_ISL_2471320, EPI_ISL_2471321, EPI_ISL_2471322, EPI_ISL_2471323, EPI_ISL_2471324, EPI_ISL_2471325, EPI_ISL_2471326, EPI_ISL_2471327, EPI_ISL_2471328, EPI_ISL_2471329, EPI_ISL_2471330, EPI_ISL_2471331, EPI_ISL_2471332, EPI_ISL_2471333, EPI_ISL_2471334, EPI_ISL_2471335, EPI_ISL_2471336, EPI_ISL_2471339, EPI_ISL_2471341, EPI_ISL_2471342, EPI_ISL_2471343, EPI_ISL_2471344, EPI_ISL_2471345, EPI_ISL_2471346, EPI_ISL_2471347, EPI_ISL_2471348, EPI_ISL_2471349, EPI_ISL_2471351, EPI_ISL_2471353, EPI_ISL_2471354, EPI_ISL_2471355, EPI_ISL_2471356, EPI_ISL_2471357, EPI_ISL_2471362, EPI_ISL_2471363, EPI_ISL_2471364, EPI_ISL_2471365, EPI_ISL_2471366, EPI_ISL_2471367, EPI_ISL_2471370, EPI_ISL_2471371, EPI_ISL_2471372, EPI_ISL_2471373, EPI_ISL_2471374, EPI_ISL_2471375, EPI_ISL_2471377, EPI_ISL_2471378, EPI_ISL_2471379, EPI_ISL_2471381, EPI_ISL_2471382, EPI_ISL_2471385, EPI_ISL_2471386, EPI_ISL_2471388, EPI_ISL_2471389, EPI_ISL_2471390, EPI_ISL_2471391, EPI_ISL_2471392, EPI_ISL_2471393, EPI_ISL_2471394, EPI_ISL_2471395, EPI_ISL_2471396, EPI_ISL_2471397, EPI_ISL_2471399, EPI_ISL_2471400, EPI_ISL_2471401, EPI_ISL_2471402, EPI_ISL_2471403, EPI_ISL_2471404, EPI_ISL_2471405, EPI_ISL_2471406, EPI_ISL_2471407, EPI_ISL_2471408, EPI_ISL_2471409, EPI_ISL_2471410, EPI_ISL_2471412, EPI_ISL_2471414, EPI_ISL_2471416, EPI_ISL_2471417, EPI_ISL_2471418, EPI_ISL_2471419, EPI_ISL_2471420, EPI_ISL_2471421, EPI_ISL_2471422, EPI_ISL_2471424, EPI_ISL_2471425, EPI_ISL_2471427, EPI_ISL_2471428, EPI_ISL_2471429, EPI_ISL_2471431, EPI_ISL_2471432, EPI_ISL_2471433, EPI_ISL_2471434, EPI_ISL_2471435, EPI_ISL_2471436, EPI_ISL_2471437, EPI_ISL_2471438, EPI_ISL_2471439, EPI_ISL_2471440, EPI_ISL_2471441, EPI_ISL_2471442, EPI_ISL_2471443, EPI_ISL_2471444, EPI_ISL_2471446, EPI_ISL_2471447, EPI_ISL_2471448, EPI_ISL_2471449, EPI_ISL_2471452, EPI_ISL_2471453, EPI_ISL_2471454, EPI_ISL_2471456, EPI_ISL_2471457, EPI_ISL_2471458, EPI_ISL_2471459, EPI_ISL_2471461, EPI_ISL_2471462, EPI_ISL_2633465, EPI_ISL_2633470, EPI_ISL_2633477, EPI_ISL_2633478, EPI_ISL_2633482, EPI_ISL_2633483, EPI_ISL_2633485, EPI_ISL_2633493, EPI_ISL_2633495, EPI_ISL_2633496, EPI_ISL_2633499, EPI_ISL_2633500, EPI_ISL_2633504, EPI_ISL_2633507, EPI_ISL_2633510, EPI_ISL_2633512, EPI_ISL_2633514, EPI_ISL_2633516, EPI_ISL_2633518, EPI_ISL_2633521, EPI_ISL_2633523, EPI_ISL_2633524, EPI_ISL_2633526, EPI_ISL_2633529, EPI_ISL_2633531, EPI_ISL_2633532, EPI_ISL_2633537, EPI_ISL_2633539, EPI_ISL_2633540, EPI_ISL_2633542, EPI_ISL_2633545, EPI_ISL_2633547, EPI_ISL_2633549, EPI_ISL_2633559, EPI_ISL_2633561, EPI_ISL_2633563, EPI_ISL_2633566, EPI_ISL_2633568, EPI_ISL_2633569, EPI_ISL_2633570, EPI_ISL_2633576, EPI_ISL_2633578, EPI_ISL_2633585, EPI_ISL_2633591, EPI_ISL_2633592, EPI_ISL_2633594, EPI_ISL_2633598, EPI_ISL_2633599, EPI_ISL_2633601, EPI_ISL_2633602, EPI_ISL_2633603, EPI_ISL_2633605, EPI_ISL_2633607, EPI_ISL_2633616, EPI_ISL_2633620, EPI_ISL_2633621, EPI_ISL_2633623, EPI_ISL_2633630, EPI_ISL_2633639, EPI_ISL_2633641, EPI_ISL_2633643, EPI_ISL_2633648, EPI_ISL_2633665, EPI_ISL_2633667, EPI_ISL_2633668, EPI_ISL_2633676, EPI_ISL_2633683, EPI_ISL_2633686, EPI_ISL_2633692, EPI_ISL_2633693, EPI_ISL_2633699, EPI_ISL_2633701, EPI_ISL_2633702, EPI_ISL_2633703, EPI_ISL_2633705, EPI_ISL_2633708, EPI_ISL_2633709, EPI_ISL_2633711, EPI_ISL_2633712, EPI_ISL_2633713, EPI_ISL_2633715, EPI_ISL_2633718, EPI_ISL_2633726, EPI_ISL_2633733, EPI_ISL_2633734 | | | |
| see above | CENTOGENE Frankfurt Laboratory: Niederlassung Industriepark Höchst | Robert Koch Institute | |
| EPI_ISL_2471820, EPI_ISL_2471824, EPI_ISL_2471834, EPI_ISL_2471835, EPI_ISL_2471840, EPI_ISL_2635973, EPI_ISL_2678681, EPI_ISL_2760193, EPI_ISL_2760196, EPI_ISL_2760198, EPI_ISL_2760201, EPI_ISL_2760203, EPI_ISL_2760206, EPI_ISL_2760209, EPI_ISL_2760211, EPI_ISL_2760217, EPI_ISL_2760219, EPI_ISL_2760221, EPI_ISL_2760223, EPI_ISL_2760226 | | | |
| see above | Carl-Thiem-Klinikum Cottbus - Zentrum für Laboratoriumsmedizin Mikrobiologie und Krankenhaushygiene | Robert Koch Institute | |
| EPI_ISL_2381770, EPI_ISL_2381771, EPI_ISL_2381772, EPI_ISL_2381773, EPI_ISL_2381775, EPI_ISL_2381776, EPI_ISL_2381814, EPI_ISL_2381815 | | | |
| see above | Center of Medical Microbiology, Virology, and Hospital Hygiene, University of Duesseldorf | Center of Medical Microbiology, Virology, and Hospital Hygiene, University of Duesseldorf | Alexander Dilthey; Andreas Walker; Daniel Strelow; Jessica Nicolai; Jörg Timm; Klaus Pfeffer; Lisanna Hülse; Malte Kohns Vasconcelos; Maximilian Damagnez; Nadine Lübke; Tobias Wienemann; Torsten Houwaart |
| EPI_ISL_2389383, EPI_ISL_2632154, EPI_ISL_2632155, EPI_ISL_2632156, EPI_ISL_2760270 | Chemische und Veterinäruntersuchungsamt Münsterland-Emscher-Lippe (CVUA-MEL) | Robert Koch Institute | |
| EPI_ISL_2390110, EPI_ISL_2390111, EPI_ISL_2390112, EPI_ISL_2390113, EPI_ISL_2390116, EPI_ISL_2390379, EPI_ISL_2390380, EPI_ISL_2390381, EPI_ISL_2390382, EPI_ISL_2390385, EPI_ISL_2390389, EPI_ISL_2390390, EPI_ISL_2390392, EPI_ISL_2390394, EPI_ISL_2390395, EPI_ISL_2390396, EPI_ISL_2390397, EPI_ISL_2390398, EPI_ISL_2390399, EPI_ISL_2390400, EPI_ISL_2390401, EPI_ISL_2390403, EPI_ISL_2390404, EPI_ISL_2390405, EPI_ISL_2390406, EPI_ISL_2390407, EPI_ISL_2390408, EPI_ISL_2390409, EPI_ISL_2390410, EPI_ISL_2390411, EPI_ISL_2390412, EPI_ISL_2390414, EPI_ISL_2390415, EPI_ISL_2390416, EPI_ISL_2631870, EPI_ISL_2631871, EPI_ISL_2631872, EPI_ISL_2631873, EPI_ISL_2631874, EPI_ISL_2631875, EPI_ISL_2631876, EPI_ISL_2631877, EPI_ISL_2631878, EPI_ISL_2631880, EPI_ISL_2631881, EPI_ISL_2631882, EPI_ISL_2631883, EPI_ISL_2631884, EPI_ISL_2631885, EPI_ISL_2631886, EPI_ISL_2631887, EPI_ISL_2631888, EPI_ISL_2631889, EPI_ISL_2631890, EPI_ISL_2631891, EPI_ISL_2631892, EPI_ISL_2631893, EPI_ISL_2634689, EPI_ISL_2634690, EPI_ISL_2634693, EPI_ISL_2634695, EPI_ISL_2634697, EPI_ISL_2634700, EPI_ISL_2634702, EPI_ISL_2634704, EPI_ISL_2634707, EPI_ISL_2634709, EPI_ISL_2634711, EPI_ISL_2634717, EPI_ISL_2634719, EPI_ISL_2634722, EPI_ISL_2634723 | | | |
| see above | Clemenshospital - Alexianer Clemenshospital Münster | Robert Koch Institute | |
| EPI_ISL_2632119, EPI_ISL_2632120, EPI_ISL_2632121, EPI_ISL_2632123, EPI_ISL_2632125, EPI_ISL_2632127, EPI_ISL_2632134, EPI_ISL_2632137, EPI_ISL_2636125, EPI_ISL_2636127, EPI_ISL_2636128, EPI_ISL_2636129, EPI_ISL_2636130, EPI_ISL_2636131, EPI_ISL_2636132, EPI_ISL_2636134, EPI_ISL_2636137, EPI_ISL_2636141, EPI_ISL_2636144, EPI_ISL_2636146, EPI_ISL_2636149, EPI_ISL_2636151, EPI_ISL_2636153, EPI_ISL_2636156, EPI_ISL_2636158, EPI_ISL_2636160, EPI_ISL_2636162 | | | |
| see above | Corona-Testzentrum ifp Institut für Produktqualität GmbH | Robert Koch Institute | |
| EPI_ISL_2759779, EPI_ISL_2759780, EPI_ISL_2759782, EPI_ISL_2759783, EPI_ISL_2759784, EPI_ISL_2759785, EPI_ISL_2759787, EPI_ISL_2759788, EPI_ISL_2759790, EPI_ISL_2759791, EPI_ISL_2759792, EPI_ISL_2759794, EPI_ISL_2759795, EPI_ISL_2759796, EPI_ISL_2759797, EPI_ISL_2759799, EPI_ISL_2759800, EPI_ISL_2759801, EPI_ISL_2759803, EPI_ISL_2759804, EPI_ISL_2759805, EPI_ISL_2759806, EPI_ISL_2759807, EPI_ISL_2759808, EPI_ISL_2759809, EPI_ISL_2759810, EPI_ISL_2759811, EPI_ISL_2759813 | | | |
| see above | Department für Labormedizin Abteilung III Bereich Molekulare Diagnostik | Robert Koch Institute | |

| | | | |
|---|---|---|---|
| | Universitätsklinikum Halle(Saale) | | |
| EPI_ISL_2636416, EPI_ISL_2636419, EPI_ISL_2636420, EPI_ISL_2636421, EPI_ISL_2636423, EPI_ISL_2636424, EPI_ISL_2636425, EPI_ISL_2636426, EPI_ISL_2636428, EPI_ISL_2636429, EPI_ISL_2636431, EPI_ISL_2636432, EPI_ISL_2636435, EPI_ISL_2636436, EPI_ISL_2636437, EPI_ISL_2636441, EPI_ISL_2636442, EPI_ISL_2636443, EPI_ISL_2636444, EPI_ISL_2636447, EPI_ISL_2636448, EPI_ISL_2636451, EPI_ISL_2636453, EPI_ISL_2636454, EPI_ISL_2636456, EPI_ISL_2636457, EPI_ISL_2636458, EPI_ISL_2636459, EPI_ISL_2636460, EPI_ISL_2636461, EPI_ISL_2636462, EPI_ISL_2636464, EPI_ISL_2636470, EPI_ISL_2636480, EPI_ISL_2636531, EPI_ISL_2636532, EPI_ISL_2636533, EPI_ISL_2636534, EPI_ISL_2636535, EPI_ISL_2636536, EPI_ISL_2636537, EPI_ISL_2636538, EPI_ISL_2636539, EPI_ISL_2636540, EPI_ISL_2636541, EPI_ISL_2636542, EPI_ISL_2636543, EPI_ISL_2636544, EPI_ISL_2636545, EPI_ISL_2636546, EPI_ISL_2636547, EPI_ISL_2636548, EPI_ISL_2636549, EPI_ISL_2636550, EPI_ISL_2636551, EPI_ISL_2636552, EPI_ISL_2636553, EPI_ISL_2636554, EPI_ISL_2636555, EPI_ISL_2636556, EPI_ISL_2636557, EPI_ISL_2636558, EPI_ISL_2636559, EPI_ISL_2636560, EPI_ISL_2636561, EPI_ISL_2636562, EPI_ISL_2636563, EPI_ISL_2636564, EPI_ISL_2636565, EPI_ISL_2636566, EPI_ISL_2636567, EPI_ISL_2636568, EPI_ISL_2636569, EPI_ISL_2636570, EPI_ISL_2636571, EPI_ISL_2636572, EPI_ISL_2636573, EPI_ISL_2636574, EPI_ISL_2636575, EPI_ISL_2636576, EPI_ISL_2636577, EPI_ISL_2636578, EPI_ISL_2636579, EPI_ISL_2636580, EPI_ISL_2636581, EPI_ISL_2636582, EPI_ISL_2636583, EPI_ISL_2636584, EPI_ISL_2636585, EPI_ISL_2636586, EPI_ISL_2636587, EPI_ISL_2636588, EPI_ISL_2636589, EPI_ISL_2636590, EPI_ISL_2636591, EPI_ISL_2636592, EPI_ISL_2636593, EPI_ISL_2636594, EPI_ISL_2636595, EPI_ISL_2636596, EPI_ISL_2636597, EPI_ISL_2636598, EPI_ISL_2636599, EPI_ISL_2636600, EPI_ISL_2636601, EPI_ISL_2636602, EPI_ISL_2636603, EPI_ISL_2636604, EPI_ISL_2636605, EPI_ISL_2636606, EPI_ISL_2636607, EPI_ISL_2636608, EPI_ISL_2636609, EPI_ISL_2636610, EPI_ISL_2636611, EPI_ISL_2636612, EPI_ISL_2636613, EPI_ISL_2636614, EPI_ISL_2636615, EPI_ISL_2636616, EPI_ISL_2636617, EPI_ISL_2636618, EPI_ISL_2636619, EPI_ISL_2636620, EPI_ISL_2636621, EPI_ISL_2636622, EPI_ISL_2636623, EPI_ISL_2636624, EPI_ISL_2636625, EPI_ISL_2636626, EPI_ISL_2636627, EPI_ISL_2636628, EPI_ISL_2636629, EPI_ISL_2636630, EPI_ISL_2636631, EPI_ISL_2636632, EPI_ISL_2636633, EPI_ISL_2636634, EPI_ISL_2636635, EPI_ISL_2636636, EPI_ISL_2636637, EPI_ISL_2636638, EPI_ISL_2636639, EPI_ISL_2636640, EPI_ISL_2636641, EPI_ISL_2636642, EPI_ISL_2636643, EPI_ISL_2636644, EPI_ISL_2636645, EPI_ISL_2636646, EPI_ISL_2636647, EPI_ISL_2636648, EPI_ISL_2636649, EPI_ISL_2636650, EPI_ISL_2636651, EPI_ISL_2636652, EPI_ISL_2636653, EPI_ISL_2636654, EPI_ISL_2636655, EPI_ISL_2636656, EPI_ISL_2636657, EPI_ISL_2636658, EPI_ISL_2636659, EPI_ISL_2636660, EPI_ISL_2636661, EPI_ISL_2636662, EPI_ISL_2636663, EPI_ISL_2636664, EPI_ISL_2636665, EPI_ISL_2636666, EPI_ISL_2636667, EPI_ISL_2636668, EPI_ISL_2636669, EPI_ISL_2636670, EPI_ISL_2636671, EPI_ISL_2636672, EPI_ISL_2636673, EPI_ISL_2636674, EPI_ISL_2636675, EPI_ISL_2636676, EPI_ISL_2636677, EPI_ISL_2636678, EPI_ISL_2636679, EPI_ISL_2636680, EPI_ISL_2636681, EPI_ISL_2636682, EPI_ISL_2636683, EPI_ISL_2636684, EPI_ISL_2636685, EPI_ISL_2636686, EPI_ISL_2636687, EPI_ISL_2636688, EPI_ISL_2636689, EPI_ISL_2636690, EPI_ISL_2636691, EPI_ISL_2636692, EPI_ISL_2636693, EPI_ISL_2636694, EPI_ISL_2636695, EPI_ISL_2636696, EPI_ISL_2636697, EPI_ISL_2636698, EPI_ISL_2636699, EPI_ISL_2636700, EPI_ISL_2636701, EPI_ISL_2636702, EPI_ISL_2636703, EPI_ISL_2636704, EPI_ISL_2636705, EPI_ISL_2636706, EPI_ISL_2636707, EPI_ISL_2636708, EPI_ISL_2636709, EPI_ISL_2636710, EPI_ISL_2636711, EPI_ISL_2636712, EPI_ISL_2636713, EPI_ISL_2636714, EPI_ISL_2636715, EPI_ISL_2636716, EPI_ISL_2636717, EPI_ISL_2636718, EPI_ISL_2636719, EPI_ISL_2636720, EPI_ISL_2636721, EPI_ISL_2636722, EPI_ISL_2636723, EPI_ISL_2636724, EPI_ISL_2636725, EPI_ISL_2636726, EPI_ISL_2636727, EPI_ISL_2636728, EPI_ISL_2636729, EPI_ISL_2636730, EPI_ISL_2636731, EPI_ISL_2636760, EPI_ISL_2636761, EPI_ISL_2636762, EPI_ISL_2636763, EPI_ISL_2636764, EPI_ISL_2636765, EPI_ISL_2636766, EPI_ISL_2636767, EPI_ISL_2636768, EPI_ISL_2636769, EPI_ISL_2636770, EPI_ISL_2636771, EPI_ISL_2762313, EPI_ISL_2762315, EPI_ISL_2762347, EPI_ISL_2762368, EPI_ISL_2762388, EPI_ISL_2762426 | | | |
| see above | Diagnosticum - Labor Neukirchen | Robert Koch Institute | |
| EPI_ISL_2386856, EPI_ISL_2386857, EPI_ISL_2386858, EPI_ISL_2386860, EPI_ISL_2386861, EPI_ISL_2386863, EPI_ISL_2386865, EPI_ISL_2386866, EPI_ISL_2386867, EPI_ISL_2386868, EPI_ISL_2386870, EPI_ISL_2386871, EPI_ISL_2390461, EPI_ISL_2390462, EPI_ISL_2469786, EPI_ISL_2469787, EPI_ISL_2469788, EPI_ISL_2469789, EPI_ISL_2469790, EPI_ISL_2469791, EPI_ISL_2469792, EPI_ISL_2469793, EPI_ISL_2469794, EPI_ISL_2469795, EPI_ISL_2469796, EPI_ISL_2469797, EPI_ISL_2469798, EPI_ISL_2469799, EPI_ISL_2469800, EPI_ISL_2469801, EPI_ISL_2469802, EPI_ISL_2469803, EPI_ISL_2469804, EPI_ISL_2469805, EPI_ISL_2469806, EPI_ISL_2472510, EPI_ISL_2472511, EPI_ISL_2472512, EPI_ISL_2472513, EPI_ISL_2472514, EPI_ISL_2472515, EPI_ISL_2472516, EPI_ISL_2472517, EPI_ISL_2632824, EPI_ISL_2632825, EPI_ISL_2632826, EPI_ISL_2632827, EPI_ISL_2632828, EPI_ISL_2632829, EPI_ISL_2635963, EPI_ISL_2636014 | | | |
| see above | Dianovis GmbH Greiz | Robert Koch Institute | |
| EPI_ISL_2390322, EPI_ISL_2390348, EPI_ISL_2390351, EPI_ISL_2390361, EPI_ISL_2390366, EPI_ISL_2471850, EPI_ISL_2471858, EPI_ISL_2471866, EPI_ISL_2471868, EPI_ISL_2471878 | | | |
| see above | ELBLAB GmbH Zentrum für LaborMedizin | Robert Koch Institute | |
| EPI_ISL_2634506, EPI_ISL_2634508, EPI_ISL_2634510, EPI_ISL_2634513, EPI_ISL_2634515, EPI_ISL_2634518, EPI_ISL_2634520, EPI_ISL_2634522, EPI_ISL_2634524, EPI_ISL_2634526, EPI_ISL_2634529, EPI_ISL_2634531, EPI_ISL_2634534, EPI_ISL_2634536, EPI_ISL_2634538, EPI_ISL_2634540, EPI_ISL_2634543, EPI_ISL_2634545, EPI_ISL_2634551, EPI_ISL_2634553, EPI_ISL_2634556, EPI_ISL_2634558, EPI_ISL_2634561, EPI_ISL_2634563, EPI_ISL_2634565, EPI_ISL_2634567, EPI_ISL_2634569, EPI_ISL_2634572, EPI_ISL_2634574, EPI_ISL_2634577, EPI_ISL_2634579, EPI_ISL_2634581, EPI_ISL_2634583, EPI_ISL_2634585, EPI_ISL_2634588, EPI_ISL_2634590, EPI_ISL_2634592, EPI_ISL_2634594, EPI_ISL_2634596, EPI_ISL_2634599, EPI_ISL_2634601, EPI_ISL_2634603, EPI_ISL_2634605, EPI_ISL_2634608, EPI_ISL_2634610, EPI_ISL_2634612, EPI_ISL_2634614, EPI_ISL_2634617, EPI_ISL_2634619, EPI_ISL_2634621, EPI_ISL_2634623, EPI_ISL_2634625, EPI_ISL_2634628, EPI_ISL_2634630, EPI_ISL_2634632, EPI_ISL_2634634, EPI_ISL_2634637, EPI_ISL_2634639, EPI_ISL_2634641, EPI_ISL_2634643, EPI_ISL_2634645, EPI_ISL_2634648, EPI_ISL_2634650, EPI_ISL_2634652, EPI_ISL_2634654, EPI_ISL_2634656, EPI_ISL_2634658, EPI_ISL_2634661, EPI_ISL_2634664, EPI_ISL_2634667, EPI_ISL_2634669, EPI_ISL_2634671, EPI_ISL_2634674, EPI_ISL_2634676, EPI_ISL_2634678, EPI_ISL_2634680, EPI_ISL_2634683, EPI_ISL_2634685, EPI_ISL_2634687, EPI_ISL_2678472, EPI_ISL_2678473, EPI_ISL_2678474, EPI_ISL_2678475, EPI_ISL_2678476, EPI_ISL_2678477, EPI_ISL_2678478, EPI_ISL_2678479, EPI_ISL_2678480, EPI_ISL_2678481, EPI_ISL_2678482, EPI_ISL_2678483, EPI_ISL_2678484, EPI_ISL_2678485, EPI_ISL_2678486, EPI_ISL_2678487, EPI_ISL_2678488, EPI_ISL_2678489, EPI_ISL_2678490, EPI_ISL_2678491, EPI_ISL_2678492, EPI_ISL_2678493, EPI_ISL_2678494, EPI_ISL_2678495, EPI_ISL_2678496, EPI_ISL_2678497, EPI_ISL_2678498 | | | |
| see above | Eurofins LifeCodexx GmbH | Robert Koch Institute | |
| EPI_ISL_2316397, EPI_ISL_2316399, EPI_ISL_2316402, EPI_ISL_2316406, EPI_ISL_2316409, EPI_ISL_2316412, EPI_ISL_2316415, EPI_ISL_2389852, EPI_ISL_2389853 | | | |
| see above | Eurofins MVZ Labor Gelsenkirchen | Robert Koch Institute | |
| EPI_ISL_2390447, EPI_ISL_2390449, EPI_ISL_2390450, EPI_ISL_2390452, EPI_ISL_2390453, EPI_ISL_2390456, EPI_ISL_2390457, EPI_ISL_2390459, EPI_ISL_2472640, EPI_ISL_2472641, EPI_ISL_2472642, EPI_ISL_2472643, EPI_ISL_2472644, EPI_ISL_2472645, EPI_ISL_2472646, EPI_ISL_2472647, EPI_ISL_2634937, EPI_ISL_2634939 | | | |
| see above | Fraunhofer-Institut für Zelltherapie und Immunologie IZI AG Next-Generation Diagnostics | Robert Koch Institute | |
| EPI_ISL_2635971 | Gesundheitszentrum Brandenburg an der Havel GmbH bzw. Institut für Laboratoriumsmedizin Brandenburg an der Havel | Robert Koch Institute | |
| EPI_ISL_2635937, EPI_ISL_2635938, EPI_ISL_2635939, EPI_ISL_2635940, EPI_ISL_2635941, EPI_ISL_2635942, EPI_ISL_2635943, EPI_ISL_2635944, EPI_ISL_2635945, EPI_ISL_2635946, EPI_ISL_2635947, EPI_ISL_2635948 | | | |
| see above | Helios Universitätsklinikum Wuppertal: Universität Witten/Herdecke: Institut für Medizinische Labordiagnostik | Robert Koch Institute | |
| EPI_ISL_2635503 | IMD - Institut für Medizinische Diagnostik Berlin-Potsdam | Robert Koch Institute | |
| EPI_ISL_2389978, EPI_ISL_2471985, EPI_ISL_2471987, EPI_ISL_2471989, EPI_ISL_2471996, EPI_ISL_2471999, EPI_ISL_2472000, EPI_ISL_2472001, EPI_ISL_2472003, EPI_ISL_2472005, EPI_ISL_2472008, EPI_ISL_2472012, EPI_ISL_2635491, EPI_ISL_2635493, EPI_ISL_2635494, EPI_ISL_2635495, EPI_ISL_2635496, EPI_ISL_2635497, EPI_ISL_2635498, EPI_ISL_2635499, EPI_ISL_2635500, EPI_ISL_2635501, EPI_ISL_2635502, EPI_ISL_2635504, EPI_ISL_2635506, EPI_ISL_2635509 | | | |
| see above | IMD - Labor Oderland | Robert Koch Institute | |
| EPI_ISL_2471986, EPI_ISL_2471988, EPI_ISL_2471991, EPI_ISL_2471993, EPI_ISL_2471994, EPI_ISL_2471997, EPI_ISL_2471998, EPI_ISL_2472002, EPI_ISL_2472009, EPI_ISL_2472010, EPI_ISL_2472013, EPI_ISL_2472014, EPI_ISL_2472016, EPI_ISL_2472017, EPI_ISL_2472018, EPI_ISL_2472019, EPI_ISL_2632143, EPI_ISL_2632144, EPI_ISL_2632145, EPI_ISL_2632146, EPI_ISL_2632148, EPI_ISL_2632149, EPI_ISL_2632150, EPI_ISL_2632152, EPI_ISL_2632153, EPI_ISL_2637202, EPI_ISL_2637204 | | | |
| see above | IMD - MVZ Labor Greifswald | Robert Koch Institute | |
| EPI_ISL_2389999, EPI_ISL_2471990, EPI_ISL_2471995, EPI_ISL_2472006, EPI_ISL_2472011, EPI_ISL_2632141, EPI_ISL_2632142, EPI_ISL_2632147, EPI_ISL_2632151, EPI_ISL_2635492, EPI_ISL_2637196, EPI_ISL_2637197, EPI_ISL_2637203 | | | |
| see above | IMD - MVZ Labor Martinsried | Robert Koch Institute | |
| EPI_ISL_2389968, EPI_ISL_2389972, EPI_ISL_2389974, EPI_ISL_2389980, EPI_ISL_2389983, EPI_ISL_2389989, EPI_ISL_2472015, EPI_ISL_2635505, EPI_ISL_2635507, EPI_ISL_2635508, EPI_ISL_2635510 | | | |
| see above | IMD - Medizinisches Labor Rostock | Robert Koch Institute | |
| EPI_ISL_2389390, EPI_ISL_2389393, EPI_ISL_2389397, EPI_ISL_2389405, EPI_ISL_2389406, EPI_ISL_2389418, EPI_ISL_2389419, EPI_ISL_2389425, EPI_ISL_2389428, EPI_ISL_2389438, EPI_ISL_2471020, EPI_ISL_2471021, EPI_ISL_2471022, EPI_ISL_2471023, EPI_ISL_2471025, EPI_ISL_2471026, EPI_ISL_2471028, EPI_ISL_2471029, EPI_ISL_2471031, EPI_ISL_2471032, EPI_ISL_2471033, EPI_ISL_2471035, EPI_ISL_2633758, EPI_ISL_2633771, EPI_ISL_2633779, EPI_ISL_2633790, EPI_ISL_2633791, EPI_ISL_2633793, EPI_ISL_2633798, EPI_ISL_2633819, EPI_ISL_2633821, EPI_ISL_2633829, EPI_ISL_2633835, EPI_ISL_2633847, EPI_ISL_2633850, EPI_ISL_2633867, EPI_ISL_2633868, EPI_ISL_2633870, EPI_ISL_2633871, EPI_ISL_2633877, EPI_ISL_2633882, EPI_ISL_2633885, EPI_ISL_2633886, EPI_ISL_2633888, EPI_ISL_2633892 | | | |
| see above | IMD Labor Frankfurt | Robert Koch Institute | |
| EPI_ISL_2388182, EPI_ISL_2388196, EPI_ISL_2471699, EPI_ISL_2633444 | Institut für Klinische Chemie Transfusions- und Laboratoriumsmedizin Berufsgenossenschaftliches Universitätsklinikum Bergmannsheil | Robert Koch Institute | |
| EPI_ISL_2386540, EPI_ISL_2386542, EPI_ISL_2386544, EPI_ISL_2386545 | Institut für Labormedizin Mikrobiologie und Hygiene | Robert Koch Institute | |
| EPI_ISL_2631864, EPI_ISL_2631865, EPI_ISL_2635795, EPI_ISL_2635796, EPI_ISL_2635799, EPI_ISL_2635800 | Institut für Medizinische Mikrobiologie - Universitätsmedizin Göttingen | Robert Koch Institute | |
| EPI_ISL_2472538, EPI_ISL_2472539, EPI_ISL_2635125, EPI_ISL_2635128, EPI_ISL_2635129 | Institut für med. Mikrobiologie u. Virologie Klinikum Oldenburg | Robert Koch Institute | |
| EPI_ISL_2535371, EPI_ISL_2535372, EPI_ISL_2535373, EPI_ISL_2535386, EPI_ISL_2535413, EPI_ISL_2535414, EPI_ISL_2535415, EPI_ISL_2535416, EPI_ISL_2535417, EPI_ISL_2535418, EPI_ISL_2535419, EPI_ISL_2535420, EPI_ISL_2535421, EPI_ISL_2535422, EPI_ISL_2535423, EPI_ISL_2535424, EPI_ISL_2535425, EPI_ISL_2535426, EPI_ISL_2535427, EPI_ISL_2535428, EPI_ISL_2535429, EPI_ISL_2535430, EPI_ISL_2535431, EPI_ISL_2535432, EPI_ISL_2535433 | | | |
| see above | Institute of Virology, Medical Center, University of Freiburg, Freiburg, Germany | Institute of Virology, Clinial Virus Genomics, Medical Center, University of Freiburg, Freiburg, Germany | Hajo Grundmann; Jonas Fuchs; Lena Jaki; Lisa Kern; Marcus Panning; Sandra Reuter |
| EPI_ISL_2678805 | KRH Klinikum Region Hannover: Institut für Med. Mikrobiologie und Krankenhaushygiene | Robert Koch Institute | |
| EPI_ISL_2385070, EPI_ISL_2385071, EPI_ISL_2689462, EPI_ISL_2689463, EPI_ISL_2689466, EPI_ISL_2689475, EPI_ISL_2689476 | | | |
| see above | Kath. Marienkrankenhaus | Heinrich Pette Institute, Leibniz Institute for Experimental Virology | Adam Grundhoff; Alexis Robitaille; Johannes Knobloch; Martin Aepfelbacher; Nicole Fischer; Thomas Günther |
| EPI_ISL_2388198, EPI_ISL_2388301 | Kath. St.-Johannes-Gesellschaft Dortmund gGmbH | Robert Koch Institute | |
| EPI_ISL_2472463, EPI_ISL_2472481, EPI_ISL_2472504 | Kerckhoff-Klinik GmbH | Robert Koch Institute | |
| EPI_ISL_2390306, EPI_ISL_2471899 | Klinikum Darmstadt GmbH | Robert Koch Institute | |
| EPI_ISL_2386576, EPI_ISL_2386577, EPI_ISL_2386578, EPI_ISL_2386579, EPI_ISL_2386580, EPI_ISL_2386581, EPI_ISL_2386582, EPI_ISL_2386583, EPI_ISL_2386584, EPI_ISL_2386585, EPI_ISL_2386586, EPI_ISL_2386587, EPI_ISL_2386588, EPI_ISL_2386589, EPI_ISL_2386590, EPI_ISL_2386591, EPI_ISL_2386592, EPI_ISL_2470487, EPI_ISL_2470488, EPI_ISL_2470489, EPI_ISL_2470490, EPI_ISL_2470491, EPI_ISL_2470492, EPI_ISL_2470493, EPI_ISL_2470494, EPI_ISL_2470495, EPI_ISL_2470496, EPI_ISL_2471972, EPI_ISL_2471973, EPI_ISL_2471974, EPI_ISL_2471975, EPI_ISL_2471976, EPI_ISL_2471977, EPI_ISL_2471978, EPI_ISL_2471979, EPI_ISL_2471980, EPI_ISL_2471981, EPI_ISL_2471982, EPI_ISL_2471983, EPI_ISL_2678731, EPI_ISL_2678732, EPI_ISL_2678733, EPI_ISL_2678734, EPI_ISL_2678735, EPI_ISL_2678736, EPI_ISL_2678737, EPI_ISL_2678738, EPI_ISL_2678740 | | | |
| see above | Klinikum Ernst von Bergmann gemeinnützige GmbH - stationärer Bereich | Robert Koch Institute | |
| EPI_ISL_2470191, EPI_ISL_2634011 | Klinikum Fulda gAG | Robert Koch Institute | |
| EPI_ISL_2390680, EPI_ISL_2390742, EPI_ISL_2678761, EPI_ISL_2678764 | Klinikum Osnabrück Medizinisches Labor | Robert Koch Institute | |
| EPI_ISL_2761963, EPI_ISL_2761966 | Klinikum Stuttgart | Robert Koch Institute | |
| EPI_ISL_2631840 | Krankenhausgesellschaft St. Vincenz mbH | Robert Koch Institute | |
| EPI_ISL_2390320, EPI_ISL_2390323, EPI_ISL_2390334, EPI_ISL_2390365, EPI_ISL_2390367, EPI_ISL_2390372, EPI_ISL_2471869, EPI_ISL_2471874, EPI_ISL_2471883 | | | |
| see above | LABOR CHEMNITZ – Zentrum für Diagnostik am Klinikum Chemnitz | Robert Koch Institute | |
| EPI_ISL_2469928, EPI_ISL_2469931, EPI_ISL_2469932, EPI_ISL_2469940, EPI_ISL_2469944, EPI_ISL_2469946, EPI_ISL_2469948, EPI_ISL_2469949, EPI_ISL_2469952, EPI_ISL_2469953, EPI_ISL_2469955, EPI_ISL_2469957, EPI_ISL_2469958, EPI_ISL_2469976, EPI_ISL_2469981, EPI_ISL_2469982, EPI_ISL_2469983, EPI_ISL_2469984, EPI_ISL_2469985, EPI_ISL_2469988, EPI_ISL_2469989, EPI_ISL_2469992, EPI_ISL_2469994, EPI_ISL_2469995, EPI_ISL_2469996, EPI_ISL_2470002, EPI_ISL_2470003, EPI_ISL_2470008, EPI_ISL_2470014, EPI_ISL_2470016, EPI_ISL_2470022, EPI_ISL_2470027, EPI_ISL_2470031, EPI_ISL_2470033, EPI_ISL_2470036, EPI_ISL_2470038, EPI_ISL_2470040, EPI_ISL_2470041, EPI_ISL_2470042, EPI_ISL_2470047, EPI_ISL_2470048, EPI_ISL_2470049, EPI_ISL_2470052, EPI_ISL_2470053, EPI_ISL_2470054, EPI_ISL_2470055, EPI_ISL_2470059, EPI_ISL_2470060, EPI_ISL_2470063, EPI_ISL_2470064, EPI_ISL_2470070, EPI_ISL_2470073, EPI_ISL_2470074, EPI_ISL_2470075, EPI_ISL_2470081, EPI_ISL_2470083, EPI_ISL_2470088, EPI_ISL_2470092, EPI_ISL_2470094, EPI_ISL_2470097, EPI_ISL_2470099, EPI_ISL_2470101, EPI_ISL_2470102, EPI_ISL_2470103, EPI_ISL_2470106, EPI_ISL_2470108, EPI_ISL_2470111, EPI_ISL_2470114, EPI_ISL_2470117, EPI_ISL_2470123, EPI_ISL_2470125, EPI_ISL_2470129, EPI_ISL_2470130, EPI_ISL_2470133, EPI_ISL_2470142, EPI_ISL_2470143, EPI_ISL_2470145, EPI_ISL_2470150, EPI_ISL_2470152, EPI_ISL_2470155, EPI_ISL_2470160, EPI_ISL_2470168, EPI_ISL_2470169, EPI_ISL_2470172, EPI_ISL_2470176, EPI_ISL_2470177, EPI_ISL_2470189, EPI_ISL_2470192, EPI_ISL_2470195, EPI_ISL_2470198, EPI_ISL_2470201, EPI_ISL_2470342, EPI_ISL_2470379, EPI_ISL_2632346, EPI_ISL_2632364, EPI_ISL_2632380, EPI_ISL_2632406, EPI_ISL_2632410, EPI_ISL_2632417, EPI_ISL_2632428, EPI_ISL_2632435, EPI_ISL_2632443, EPI_ISL_2632444, EPI_ISL_2632452, EPI_ISL_2632461, EPI_ISL_2632463, EPI_ISL_2632464, EPI_ISL_2632475, EPI_ISL_2632479, EPI_ISL_2632513, EPI_ISL_2632522, EPI_ISL_2632533, EPI_ISL_2632537, EPI_ISL_2632551, EPI_ISL_2632556, EPI_ISL_2632585, EPI_ISL_2632588, EPI_ISL_2632631, EPI_ISL_2633910, EPI_ISL_2633915, EPI_ISL_2633923, EPI_ISL_2633926, EPI_ISL_2633930, EPI_ISL_2633933, EPI_ISL_2633943, EPI_ISL_2633968, EPI_ISL_2633977, EPI_ISL_2633980, EPI_ISL_2633982, EPI_ISL_2633984, EPI_ISL_2634000, EPI_ISL_2634025, EPI_ISL_2634056, EPI_ISL_2634104, EPI_ISL_2634110, EPI_ISL_2634138, EPI_ISL_2634144, EPI_ISL_2634153, EPI_ISL_2634167, EPI_ISL_2634186, EPI_ISL_2634222, EPI_ISL_2634297, EPI_ISL_2634323, EPI_ISL_2634345, EPI_ISL_2634361, EPI_ISL_2634390, EPI_ISL_2634410, EPI_ISL_2634415, EPI_ISL_2634417, EPI_ISL_2634432, EPI_ISL_2634446, EPI_ISL_2634450 | | | |
| see above | LabKom - Labor Augsburg MVZ GmbH | Robert Koch Institute | |
| EPI_ISL_2470072, EPI_ISL_2470438, EPI_ISL_2470460, EPI_ISL_2632481, EPI_ISL_2632583, EPI_ISL_2632599 | LabKom - Labor Hamburg-Lübeck | Robert Koch Institute | |
| EPI_ISL_2388317, EPI_ISL_2470226, EPI_ISL_2470233, EPI_ISL_2470239, EPI_ISL_2470246, EPI_ISL_2470247, EPI_ISL_2470258, EPI_ISL_2470270, EPI_ISL_2470280, EPI_ISL_2470290, EPI_ISL_2470291, EPI_ISL_2470294, EPI_ISL_2470372, EPI_ISL_2470380, EPI_ISL_2470388, EPI_ISL_2470415, EPI_ISL_2470420, EPI_ISL_2470421, EPI_ISL_2470433, EPI_ISL_2470445, EPI_ISL_2470468, EPI_ISL_2632335, | | | |

EPI_ISL_2632341, EPI_ISL_2632358, EPI_ISL_2632362, EPI_ISL_2632379, EPI_ISL_2632425, EPI_ISL_2632448, EPI_ISL_2632468, EPI_ISL_2632470, EPI_ISL_2632549, EPI_ISL_2632563, EPI_ISL_2632564, EPI_ISL_2632581, EPI_ISL_2632587, EPI_ISL_2632592, EPI_ISL_2632595, EPI_ISL_2632606, EPI_ISL_2632612, EPI_ISL_2634164, EPI_ISL_2636918, EPI_ISL_2636998, EPI_ISL_2637029, EPI_ISL_2637033, EPI_ISL_2637051, EPI_ISL_2637076, EPI_ISL_2637128, EPI_ISL_2637187

see above — LabKom - Labor Hannover MVZ GmbH — Robert Koch Institute

EPI_ISL_2470231, EPI_ISL_2470238, EPI_ISL_2470262, EPI_ISL_2470268, EPI_ISL_2470282, EPI_ISL_2470286, EPI_ISL_2470288, EPI_ISL_2470296, EPI_ISL_2470297, EPI_ISL_2470301, EPI_ISL_2470308, EPI_ISL_2470324, EPI_ISL_2470326, EPI_ISL_2470344, EPI_ISL_2470347, EPI_ISL_2470351, EPI_ISL_2470361, EPI_ISL_2470374, EPI_ISL_2470382, EPI_ISL_2470383, EPI_ISL_2470390, EPI_ISL_2470409, EPI_ISL_2470423, EPI_ISL_2470427, EPI_ISL_2470432, EPI_ISL_2470441, EPI_ISL_2470444, EPI_ISL_2470455, EPI_ISL_2470458, EPI_ISL_2470483, EPI_ISL_2632365, EPI_ISL_2632454, EPI_ISL_2632459, EPI_ISL_2632493, EPI_ISL_2632520, EPI_ISL_2632527, EPI_ISL_2632547, EPI_ISL_2632561, EPI_ISL_2632604, EPI_ISL_2632618, EPI_ISL_2632625

see above — LabKom - Labor Mainz MVZ GmbH — Robert Koch Institute

EPI_ISL_2388806, EPI_ISL_2388809, EPI_ISL_2388810, EPI_ISL_2388811, EPI_ISL_2388815, EPI_ISL_2388819, EPI_ISL_2388821, EPI_ISL_2388833, EPI_ISL_2388839, EPI_ISL_2388842, EPI_ISL_2388850, EPI_ISL_2388851, EPI_ISL_2388860, EPI_ISL_2388866, EPI_ISL_2388893, EPI_ISL_2388920, EPI_ISL_2388936, EPI_ISL_2388940, EPI_ISL_2388945, EPI_ISL_2388950, EPI_ISL_2388959, EPI_ISL_2388963, EPI_ISL_2389002, EPI_ISL_2389003, EPI_ISL_2389013, EPI_ISL_2389019, EPI_ISL_2389037, EPI_ISL_2389038, EPI_ISL_2389069, EPI_ISL_2389077, EPI_ISL_2469936, EPI_ISL_2469942, EPI_ISL_2469943, EPI_ISL_2469950, EPI_ISL_2469956, EPI_ISL_2469972, EPI_ISL_2469991, EPI_ISL_2470006, EPI_ISL_2470013, EPI_ISL_2470019, EPI_ISL_2470020, EPI_ISL_2470024, EPI_ISL_2470026, EPI_ISL_2470080, EPI_ISL_2470084, EPI_ISL_2470107, EPI_ISL_2470118, EPI_ISL_2470166, EPI_ISL_2470173, EPI_ISL_2470178, EPI_ISL_2470182, EPI_ISL_2470186, EPI_ISL_2470217, EPI_ISL_2470261, EPI_ISL_2470278, EPI_ISL_2470352, EPI_ISL_2470364, EPI_ISL_2470410, EPI_ISL_2470414, EPI_ISL_2470453, EPI_ISL_2632373, EPI_ISL_2632434, EPI_ISL_2632490, EPI_ISL_2632498, EPI_ISL_2632501, EPI_ISL_2632508, EPI_ISL_2632523, EPI_ISL_2632531, EPI_ISL_2632538, EPI_ISL_2632557, EPI_ISL_2632570, EPI_ISL_2632576, EPI_ISL_2632584, EPI_ISL_2632591, EPI_ISL_2632627, EPI_ISL_2632632, EPI_ISL_2633961, EPI_ISL_2634070, EPI_ISL_2634078, EPI_ISL_2634133, EPI_ISL_2634157, EPI_ISL_2634184, EPI_ISL_2634285, EPI_ISL_2634343, EPI_ISL_2634430, EPI_ISL_2637077, EPI_ISL_2637158

see above — LabKom - Labor an der Salzbrücke MVZ GmbH — Robert Koch Institute

EPI_ISL_2470213, EPI_ISL_2470216, EPI_ISL_2470220, EPI_ISL_2470250, EPI_ISL_2470252, EPI_ISL_2470254, EPI_ISL_2470266, EPI_ISL_2470273, EPI_ISL_2470277, EPI_ISL_2470287, EPI_ISL_2470293, EPI_ISL_2470298, EPI_ISL_2470303, EPI_ISL_2470346, EPI_ISL_2470356, EPI_ISL_2470358, EPI_ISL_2470367, EPI_ISL_2470378, EPI_ISL_2470384, EPI_ISL_2470386, EPI_ISL_2470393, EPI_ISL_2470397, EPI_ISL_2470398, EPI_ISL_2470399, EPI_ISL_2470401, EPI_ISL_2470426, EPI_ISL_2470440, EPI_ISL_2470466, EPI_ISL_2470470, EPI_ISL_2632340, EPI_ISL_2632343, EPI_ISL_2632363, EPI_ISL_2632377, EPI_ISL_2632389, EPI_ISL_2632391, EPI_ISL_2632418, EPI_ISL_2632431, EPI_ISL_2632437, EPI_ISL_2632476, EPI_ISL_2632507, EPI_ISL_2632511, EPI_ISL_2632512, EPI_ISL_2632524, EPI_ISL_2632541, EPI_ISL_2632544, EPI_ISL_2632553, EPI_ISL_2632582, EPI_ISL_2632609

see above — LabKom - MVZ Labor Bochum MLB GmbH — Robert Koch Institute

EPI_ISL_2469941, EPI_ISL_2470004, EPI_ISL_2470011, EPI_ISL_2470015, EPI_ISL_2470035, EPI_ISL_2470068, EPI_ISL_2470071, EPI_ISL_2470077, EPI_ISL_2470112, EPI_ISL_2470121, EPI_ISL_2470194, EPI_ISL_2632357, EPI_ISL_2632366, EPI_ISL_2632383, EPI_ISL_2632399, EPI_ISL_2632440, EPI_ISL_2632450, EPI_ISL_2632480, EPI_ISL_2632489, EPI_ISL_2632491, EPI_ISL_2632526, EPI_ISL_2632559, EPI_ISL_2632579, EPI_ISL_2632597, EPI_ISL_2632635, EPI_ISL_2632643, EPI_ISL_2637026, EPI_ISL_2637184, EPI_ISL_2637190

see above — Labor 28 MVZ GmbH — Robert Koch Institute

EPI_ISL_2635907, EPI_ISL_2635909, EPI_ISL_2635910, EPI_ISL_2635915, EPI_ISL_2635918, EPI_ISL_2635921, EPI_ISL_2635925, EPI_ISL_2845694, EPI_ISL_2845695, EPI_ISL_2845696, EPI_ISL_2845697, EPI_ISL_2845698, EPI_ISL_2845699, EPI_ISL_2845700, EPI_ISL_2845701, EPI_ISL_2845702, EPI_ISL_2845703, EPI_ISL_2845704, EPI_ISL_2845705, EPI_ISL_2845706, EPI_ISL_2845707, EPI_ISL_2845708, EPI_ISL_2845709, EPI_ISL_2845710, EPI_ISL_2845711, EPI_ISL_2845712, EPI_ISL_2845713, EPI_ISL_2845714, EPI_ISL_2845715, EPI_ISL_2845716, EPI_ISL_2845717, EPI_ISL_2845718, EPI_ISL_2845719, EPI_ISL_2845720, EPI_ISL_2845790, EPI_ISL_2845792, EPI_ISL_2845793, EPI_ISL_2845794, EPI_ISL_2845795, EPI_ISL_2845796, EPI_ISL_2845800, EPI_ISL_2845802, EPI_ISL_2845803, EPI_ISL_2845805, EPI_ISL_2845806, EPI_ISL_2845808, EPI_ISL_2845809, EPI_ISL_2845811, EPI_ISL_2845812

see above — Labor Becker & Kollegen (Standort München) — Robert Koch Institute

EPI_ISL_2382573, EPI_ISL_2382574, EPI_ISL_2382576, EPI_ISL_2382577, EPI_ISL_2382578, EPI_ISL_2382579, EPI_ISL_2382582, EPI_ISL_2382583, EPI_ISL_2382585, EPI_ISL_2382586, EPI_ISL_2382592, EPI_ISL_2382593, EPI_ISL_2382594, EPI_ISL_2382596, EPI_ISL_2382599, EPI_ISL_2382600, EPI_ISL_2382601, EPI_ISL_2382602, EPI_ISL_2382604, EPI_ISL_2382608, EPI_ISL_2382611, EPI_ISL_2382612, EPI_ISL_2382615, EPI_ISL_2382616, EPI_ISL_2382619, EPI_ISL_2382621, EPI_ISL_2382622, EPI_ISL_2382626, EPI_ISL_2382627, EPI_ISL_2382628, EPI_ISL_2382630, EPI_ISL_2382632, EPI_ISL_2382634, EPI_ISL_2382635, EPI_ISL_2382638, EPI_ISL_2382639, EPI_ISL_2382641, EPI_ISL_2382642, EPI_ISL_2382643, EPI_ISL_2382646, EPI_ISL_2382647, EPI_ISL_2466773, EPI_ISL_2466774, EPI_ISL_2466775, EPI_ISL_2466776, EPI_ISL_2466777, EPI_ISL_2466778, EPI_ISL_2466779, EPI_ISL_2466780, EPI_ISL_2466781, EPI_ISL_2466782, EPI_ISL_2466783, EPI_ISL_2466784, EPI_ISL_2466785, EPI_ISL_2466786, EPI_ISL_2466787, EPI_ISL_2466788, EPI_ISL_2466789, EPI_ISL_2466790, EPI_ISL_2466791, EPI_ISL_2466792, EPI_ISL_2466793, EPI_ISL_2466794, EPI_ISL_2466795, EPI_ISL_2466796, EPI_ISL_2466797, EPI_ISL_2466798, EPI_ISL_2466799, EPI_ISL_2466800, EPI_ISL_2466801, EPI_ISL_2466802, EPI_ISL_2466803, EPI_ISL_2466804, EPI_ISL_2466805, EPI_ISL_2466806, EPI_ISL_2466807, EPI_ISL_2466808, EPI_ISL_2466809, EPI_ISL_2466810, EPI_ISL_2466811, EPI_ISL_2466812, EPI_ISL_2466813, EPI_ISL_2466814, EPI_ISL_2466815, EPI_ISL_2466816, EPI_ISL_2466817, EPI_ISL_2466818, EPI_ISL_2466819, EPI_ISL_2466820, EPI_ISL_2466821, EPI_ISL_2466823, EPI_ISL_2466824, EPI_ISL_2466825, EPI_ISL_2466826, EPI_ISL_2466827, EPI_ISL_2466828, EPI_ISL_2466829, EPI_ISL_2504014, EPI_ISL_2504015, EPI_ISL_2504016, EPI_ISL_2504017, EPI_ISL_2504018, EPI_ISL_2504019, EPI_ISL_2504022, EPI_ISL_2504023, EPI_ISL_2504024, EPI_ISL_2504026, EPI_ISL_2504027, EPI_ISL_2504028, EPI_ISL_2504030, EPI_ISL_2504031, EPI_ISL_2504032, EPI_ISL_2504033, EPI_ISL_2504034, EPI_ISL_2504035, EPI_ISL_2504036, EPI_ISL_2504037, EPI_ISL_2504038, EPI_ISL_2504039, EPI_ISL_2504040, EPI_ISL_2504041, EPI_ISL_2504042, EPI_ISL_2504043, EPI_ISL_2504044, EPI_ISL_2504045, EPI_ISL_2504046, EPI_ISL_2504047, EPI_ISL_2504048, EPI_ISL_2504049, EPI_ISL_2504050, EPI_ISL_2694799, EPI_ISL_2694800, EPI_ISL_2833532

see above — Labor Berlin Charité Vivantes GmbH / Institut für Virologie — Charité Universitätsmedizin Berlin, Institut für Virologie/Labor Berlin — Barbara Mühlemann; Christian Drosten; Christine Stephan; Peter Menzel; Rolf Schwarzer; Terry Jones; Victor M Corman

EPI_ISL_2315459, EPI_ISL_2315462, EPI_ISL_2315464, EPI_ISL_2315465, EPI_ISL_2315478, EPI_ISL_2315481, EPI_ISL_2315485, EPI_ISL_2315497, EPI_ISL_2315498, EPI_ISL_2315501, EPI_ISL_2315503, EPI_ISL_2315506, EPI_ISL_2315509, EPI_ISL_2315515, EPI_ISL_2315519, EPI_ISL_2315522, EPI_ISL_2315530, EPI_ISL_2315531, EPI_ISL_2315533, EPI_ISL_2315534, EPI_ISL_2315535, EPI_ISL_2315537, EPI_ISL_2315539, EPI_ISL_2315544, EPI_ISL_2315545, EPI_ISL_2315546, EPI_ISL_2315549, EPI_ISL_2315550, EPI_ISL_2315552, EPI_ISL_2315554, EPI_ISL_2315556, EPI_ISL_2315557, EPI_ISL_2315559, EPI_ISL_2315560, EPI_ISL_2315565, EPI_ISL_2315573, EPI_ISL_2315579, EPI_ISL_2315581, EPI_ISL_2315582, EPI_ISL_2390012, EPI_ISL_2390014, EPI_ISL_2390015, EPI_ISL_2390016, EPI_ISL_2390017, EPI_ISL_2390018, EPI_ISL_2390019, EPI_ISL_2390020, EPI_ISL_2390021, EPI_ISL_2390022, EPI_ISL_2390026, EPI_ISL_2390027, EPI_ISL_2390028, EPI_ISL_2390029, EPI_ISL_2390031, EPI_ISL_2390032, EPI_ISL_2390033, EPI_ISL_2390034, EPI_ISL_2390035, EPI_ISL_2390039, EPI_ISL_2390041, EPI_ISL_2390043, EPI_ISL_2390046, EPI_ISL_2390047, EPI_ISL_2390049, EPI_ISL_2390051, EPI_ISL_2390052, EPI_ISL_2390053, EPI_ISL_2390054, EPI_ISL_2390055, EPI_ISL_2390056, EPI_ISL_2390057, EPI_ISL_2390058, EPI_ISL_2390059, EPI_ISL_2390060, EPI_ISL_2390061, EPI_ISL_2390062, EPI_ISL_2390064, EPI_ISL_2390066, EPI_ISL_2390069, EPI_ISL_2390070, EPI_ISL_2390071, EPI_ISL_2390072, EPI_ISL_2390074, EPI_ISL_2390075, EPI_ISL_2390076, EPI_ISL_2390078, EPI_ISL_2390079, EPI_ISL_2390080, EPI_ISL_2390081, EPI_ISL_2390082, EPI_ISL_2390083, EPI_ISL_2390084, EPI_ISL_2390085, EPI_ISL_2390086, EPI_ISL_2390087, EPI_ISL_2390088, EPI_ISL_2390090, EPI_ISL_2678504, EPI_ISL_2678508, EPI_ISL_2678513, EPI_ISL_2678515, EPI_ISL_2678517, EPI_ISL_2678520, EPI_ISL_2678521, EPI_ISL_2678522, EPI_ISL_2678523, EPI_ISL_2678525, EPI_ISL_2678526, EPI_ISL_2678527, EPI_ISL_2678530, EPI_ISL_2678531, EPI_ISL_2678532, EPI_ISL_2678535, EPI_ISL_2678536, EPI_ISL_2678537, EPI_ISL_2678539, EPI_ISL_2678540, EPI_ISL_2678544, EPI_ISL_2678547, EPI_ISL_2678549, EPI_ISL_2678552, EPI_ISL_2678553, EPI_ISL_2678554, EPI_ISL_2678558, EPI_ISL_2678561, EPI_ISL_2678576, EPI_ISL_2678577, EPI_ISL_2678586, EPI_ISL_2678596, EPI_ISL_2678597, EPI_ISL_2678612, EPI_ISL_2678615, EPI_ISL_2678616, EPI_ISL_2678621, EPI_ISL_2678622, EPI_ISL_2678623, EPI_ISL_2678625, EPI_ISL_2678631, EPI_ISL_2678641

see above — Labor Blackholm MVZ — Robert Koch Institute

EPI_ISL_2689318, EPI_ISL_2689319, EPI_ISL_2689322, EPI_ISL_2689323, EPI_ISL_2689324, EPI_ISL_2689325, EPI_ISL_2689328, EPI_ISL_2689329, EPI_ISL_2689332, EPI_ISL_2689333, EPI_ISL_2689334, EPI_ISL_2689337, EPI_ISL_2689341, EPI_ISL_2689342, EPI_ISL_2689343, EPI_ISL_2689344, EPI_ISL_2689345, EPI_ISL_2689348, EPI_ISL_2689349, EPI_ISL_2689352, EPI_ISL_2689353, EPI_ISL_2689354, EPI_ISL_2689355, EPI_ISL_2689358, EPI_ISL_2689359, EPI_ISL_2689363, EPI_ISL_2689364, EPI_ISL_2689365, EPI_ISL_2689366, EPI_ISL_2689367, EPI_ISL_2689368, EPI_ISL_2689369, EPI_ISL_2689370, EPI_ISL_2689371, EPI_ISL_2689372, EPI_ISL_2689374, EPI_ISL_2689375, EPI_ISL_2689379, EPI_ISL_2689380, EPI_ISL_2689381, EPI_ISL_2689382, EPI_ISL_2689384, EPI_ISL_2689385, EPI_ISL_2689388, EPI_ISL_2689389, EPI_ISL_2689390, EPI_ISL_2689391, EPI_ISL_2689408, EPI_ISL_2689426, EPI_ISL_2689427, EPI_ISL_2689436, EPI_ISL_2689458, EPI_ISL_2689480, EPI_ISL_2689481, EPI_ISL_2689482, EPI_ISL_2689483, EPI_ISL_2689484, EPI_ISL_2689485, EPI_ISL_2689486, EPI_ISL_2689487, EPI_ISL_2689488, EPI_ISL_2689489, EPI_ISL_2689490, EPI_ISL_2689491, EPI_ISL_2689492, EPI_ISL_2689493, EPI_ISL_2689494, EPI_ISL_2689495, EPI_ISL_2689496, EPI_ISL_2689497, EPI_ISL_2689498, EPI_ISL_2689499, EPI_ISL_2689500, EPI_ISL_2689501, EPI_ISL_2689502, EPI_ISL_2689503, EPI_ISL_2689504, EPI_ISL_2689505, EPI_ISL_2689506, EPI_ISL_2689507, EPI_ISL_2689508, EPI_ISL_2689509, EPI_ISL_2689510, EPI_ISL_2689511, EPI_ISL_2689512, EPI_ISL_2689513, EPI_ISL_2689514, EPI_ISL_2689515, EPI_ISL_2689516, EPI_ISL_2689517, EPI_ISL_2689518, EPI_ISL_2689519, EPI_ISL_2689520, EPI_ISL_2689521, EPI_ISL_2689522, EPI_ISL_2689523, EPI_ISL_2689524, EPI_ISL_2689525, EPI_ISL_2689526, EPI_ISL_2689527, EPI_ISL_2689528, EPI_ISL_2689529, EPI_ISL_2689530, EPI_ISL_2689531, EPI_ISL_2689532, EPI_ISL_2689533, EPI_ISL_2689534, EPI_ISL_2689535, EPI_ISL_2689536, EPI_ISL_2689537, EPI_ISL_2689538, EPI_ISL_2689539, EPI_ISL_2689540, EPI_ISL_2689541, EPI_ISL_2689542, EPI_ISL_2689543, EPI_ISL_2689544, EPI_ISL_2689545, EPI_ISL_2689546, EPI_ISL_2689547, EPI_ISL_2689548, EPI_ISL_2689549, EPI_ISL_2689550, EPI_ISL_2689551, EPI_ISL_2689552, EPI_ISL_2689553, EPI_ISL_2689554, EPI_ISL_2689555, EPI_ISL_2689556, EPI_ISL_2689557, EPI_ISL_2689558, EPI_ISL_2689559, EPI_ISL_2689560, EPI_ISL_2689561, EPI_ISL_2689562, EPI_ISL_2689563, EPI_ISL_2689564, EPI_ISL_2689565, EPI_ISL_2689566, EPI_ISL_2689567, EPI_ISL_2689568, EPI_ISL_2689569, EPI_ISL_2689570, EPI_ISL_2689571, EPI_ISL_2689572, EPI_ISL_2689573, EPI_ISL_2689574, EPI_ISL_2689575, EPI_ISL_2689576, EPI_ISL_2689577, EPI_ISL_2689578, EPI_ISL_2689579, EPI_ISL_2689580, EPI_ISL_2689581, EPI_ISL_2689582, EPI_ISL_2689583, EPI_ISL_2689584, EPI_ISL_2689585, EPI_ISL_2689586, EPI_ISL_2689587, EPI_ISL_2689588, EPI_ISL_2689589, EPI_ISL_2689590, EPI_ISL_2689591, EPI_ISL_2689592

see above — Labor Dr. Fenner und Kollegen — Heinrich Pette Institute, Leibniz Institute for Experimental Virology — Adam Grundhoff; Alexis Robitaille; Johannes Knobloch; Martin Aepfelbacher; Nicole Fischer; Thomas Günther

EPI_ISL_2389328, EPI_ISL_2389331, EPI_ISL_2389333, EPI_ISL_2389338, EPI_ISL_2389339, EPI_ISL_2389342, EPI_ISL_2389345, EPI_ISL_2389346, EPI_ISL_2389348, EPI_ISL_2389349, EPI_ISL_2389351, EPI_ISL_2389354, EPI_ISL_2389357, EPI_ISL_2389358, EPI_ISL_2471041, EPI_ISL_2471042, EPI_ISL_2471043, EPI_ISL_2471044, EPI_ISL_2471045, EPI_ISL_2471046, EPI_ISL_2471047, EPI_ISL_2471048, EPI_ISL_2471049, EPI_ISL_2471050, EPI_ISL_2471051, EPI_ISL_2471052, EPI_ISL_2471053, EPI_ISL_2471054, EPI_ISL_2471055, EPI_ISL_2471056, EPI_ISL_2471057, EPI_ISL_2471058, EPI_ISL_2471059, EPI_ISL_2471060, EPI_ISL_2632157, EPI_ISL_2632158, EPI_ISL_2632159, EPI_ISL_2632160, EPI_ISL_2632161, EPI_ISL_2632162, EPI_ISL_2632163, EPI_ISL_2632164, EPI_ISL_2632165, EPI_ISL_2632166, EPI_ISL_2632167, EPI_ISL_2632168, EPI_ISL_2632169, EPI_ISL_2632170, EPI_ISL_2632171, EPI_ISL_2632172, EPI_ISL_2632173, EPI_ISL_2632174, EPI_ISL_2632175, EPI_ISL_2632176, EPI_ISL_2635846, EPI_ISL_2635848, EPI_ISL_2635855

see above — Labor Dr. Heidrich & Kollegen MVZ GmbH Hamburg — Robert Koch Institute

EPI_ISL_2632336, EPI_ISL_2632342, EPI_ISL_2632374, EPI_ISL_2632396, EPI_ISL_2632404, EPI_ISL_2632407, EPI_ISL_2632420, EPI_ISL_2632432, EPI_ISL_2632439, EPI_ISL_2632503, EPI_ISL_2632509, EPI_ISL_2632518, EPI_ISL_2632519, EPI_ISL_2632534, EPI_ISL_2632539, EPI_ISL_2632546, EPI_ISL_2632555, EPI_ISL_2632572, EPI_ISL_2632573, EPI_ISL_2632574, EPI_ISL_2632580, EPI_ISL_2632601, EPI_ISL_2632605, EPI_ISL_2632610, EPI_ISL_2633916, EPI_ISL_2633991, EPI_ISL_2634002, EPI_ISL_2634026, EPI_ISL_2634042, EPI_ISL_2634083, EPI_ISL_2634085, EPI_ISL_2634086, EPI_ISL_2634100, EPI_ISL_2634105, EPI_ISL_2634121, EPI_ISL_2634128, EPI_ISL_2634149, EPI_ISL_2634204, EPI_ISL_2634227, EPI_ISL_2634234, EPI_ISL_2634241, EPI_ISL_2634319, EPI_ISL_2634332, EPI_ISL_2634337, EPI_ISL_2634426, EPI_ISL_2636911, EPI_ISL_2636923, EPI_ISL_2636958, EPI_ISL_2637069, EPI_ISL_2637105, EPI_ISL_2637119, EPI_ISL_2637123, EPI_ISL_2637137, EPI_ISL_2637180, EPI_ISL_2637181

see above — Labor Dr. Schumacher MVZ — Robert Koch Institute

EPI_ISL_2472545, EPI_ISL_2472546, EPI_ISL_2472549 — Labor Dr. Spranger — Robert Koch Institute

EPI_ISL_2386547, EPI_ISL_2469752, EPI_ISL_2472543, EPI_ISL_2631828, EPI_ISL_2631829, EPI_ISL_2631831, EPI_ISL_2631832, EPI_ISL_2631833, EPI_ISL_2631834, EPI_ISL_2631835

see above — Labor Dr. Wisplinghoff - Köln — Robert Koch Institute

EPI_ISL_2389395, EPI_ISL_2389401, EPI_ISL_2389431 — Labor Dr. med. Knechten — Robert Koch Institute

EPI_ISL_2386597, EPI_ISL_2386598, EPI_ISL_2386599, EPI_ISL_2386601, EPI_ISL_2386602, EPI_ISL_2471916, EPI_ISL_2471917, EPI_ISL_2471918, EPI_ISL_2471919, EPI_ISL_2471920, EPI_ISL_2471921, EPI_ISL_2631968, EPI_ISL_2631969, EPI_ISL_2631971, EPI_ISL_2631972

see above — Labor Kneißler GmbH & Co. KG — Robert Koch Institute

EPI_ISL_2387434, EPI_ISL_2469858, EPI_ISL_2469859, EPI_ISL_2469861, EPI_ISL_2469863, EPI_ISL_2469867, EPI_ISL_2469868, EPI_ISL_2469872, EPI_ISL_2469885, EPI_ISL_2469892, EPI_ISL_2469906, EPI_ISL_2632039, EPI_ISL_2632058, EPI_ISL_2632064, EPI_ISL_2632071, EPI_ISL_2632072, EPI_ISL_2632075, EPI_ISL_2632082, EPI_ISL_2632093, EPI_ISL_2632097, EPI_ISL_2632101, EPI_ISL_2632104, EPI_ISL_2632111, EPI_ISL_2632113, EPI_ISL_2635475, EPI_ISL_2635487

see above — Labor Lübeck bzw. Laborärztliche Gemeinschaftspraxis Lübeck — Robert Koch Institute

EPI_ISL_2314671, EPI_ISL_2386213, EPI_ISL_2386215, EPI_ISL_2386217, EPI_ISL_2386218, EPI_ISL_2386220, EPI_ISL_2386223, EPI_ISL_2386225, EPI_ISL_2386228, EPI_ISL_2386229, EPI_ISL_2386231, EPI_ISL_2386233, EPI_ISL_2386234, EPI_ISL_2386235, EPI_ISL_2386236, EPI_ISL_2386238, EPI_ISL_2386239, EPI_ISL_2386241, EPI_ISL_2386251, EPI_ISL_2386252, EPI_ISL_2386253, EPI_ISL_2386254, EPI_ISL_2386255, EPI_ISL_2386257, EPI_ISL_2386258, EPI_ISL_2386260, EPI_ISL_2386261, EPI_ISL_2386262, EPI_ISL_2386264, EPI_ISL_2386265, EPI_ISL_2386266, EPI_ISL_2386267, EPI_ISL_2386268, EPI_ISL_2386269, EPI_ISL_2386271, EPI_ISL_2386272, EPI_ISL_2386273, EPI_ISL_2386274, EPI_ISL_2386278, EPI_ISL_2386279, EPI_ISL_2386280, EPI_ISL_2386281, EPI_ISL_2386282, EPI_ISL_2386283, EPI_ISL_2386285, EPI_ISL_2386286, EPI_ISL_2386287, EPI_ISL_2471476, EPI_ISL_2471787, EPI_ISL_2471788, EPI_ISL_2471789, EPI_ISL_2471790, EPI_ISL_2471791, EPI_ISL_2471792, EPI_ISL_2471793, EPI_ISL_2471794, EPI_ISL_2471795, EPI_ISL_2471796, EPI_ISL_2471797, EPI_ISL_2471798, EPI_ISL_2471799, EPI_ISL_2471800, EPI_ISL_2471801, EPI_ISL_2471802, EPI_ISL_2471803, EPI_ISL_2471804, EPI_ISL_2471805, EPI_ISL_2471806, EPI_ISL_2471807, EPI_ISL_2471808, EPI_ISL_2471809, EPI_ISL_2471812, EPI_ISL_2471814, EPI_ISL_2471815, EPI_ISL_2472485, EPI_ISL_2472486, EPI_ISL_2472487, EPI_ISL_2472489, EPI_ISL_2472490, EPI_ISL_2472492, EPI_ISL_2472493, EPI_ISL_2472495, EPI_ISL_2472496, EPI_ISL_2472497, EPI_ISL_2472498, EPI_ISL_2472500, EPI_ISL_2472501, EPI_ISL_2472502, EPI_ISL_2472503, EPI_ISL_2472505, EPI_ISL_2472506, EPI_ISL_2472507, EPI_ISL_2472508, EPI_ISL_2472705, EPI_ISL_2472723, EPI_ISL_2472725

see above — Labor Mönchengladbach MVZ Dr. Stein + Kollegen GbR — Robert Koch Institute

EPI_ISL_2472731, EPI_ISL_2635835 — Labor Prof. Dr. G. Enders MVZ GbR — Robert Koch Institute

EPI_ISL_2390581, EPI_ISL_2390587, EPI_ISL_2390588, EPI_ISL_2390589, EPI_ISL_2390592, EPI_ISL_2390596, EPI_ISL_2390597, EPI_ISL_2390599, EPI_ISL_2390602, EPI_ISL_2390603, EPI_ISL_2390604, EPI_ISL_2390608, EPI_ISL_2390609, EPI_ISL_2390611, EPI_ISL_2390616, EPI_ISL_2390621, EPI_ISL_2390625, EPI_ISL_2390626, EPI_ISL_2390635, EPI_ISL_2390647, EPI_ISL_2390649, EPI_ISL_2390650, EPI_ISL_2390655, EPI_ISL_2390664, EPI_ISL_2390666, EPI_ISL_2390668, EPI_ISL_2390669, EPI_ISL_2390671, EPI_ISL_2390672, EPI_ISL_2390676, EPI_ISL_2390677, EPI_ISL_2390683, EPI_ISL_2390684, EPI_ISL_2390688, EPI_ISL_2390691, EPI_ISL_2390692, EPI_ISL_2390696, EPI_ISL_2390697, EPI_ISL_2390699, EPI_ISL_2390700, EPI_ISL_2390703, EPI_ISL_2390705, EPI_ISL_2390708, EPI_ISL_2390711, EPI_ISL_2390712, EPI_ISL_2390713, EPI_ISL_2390715, EPI_ISL_2390717, EPI_ISL_2390722, EPI_ISL_2390727, EPI_ISL_2390728, EPI_ISL_2390730, EPI_ISL_2390736, EPI_ISL_2390737, EPI_ISL_2390740, EPI_ISL_2472784, EPI_ISL_2472785, EPI_ISL_2472786, EPI_ISL_2472787, EPI_ISL_2472788, EPI_ISL_2472789, EPI_ISL_2472790, EPI_ISL_2472791, EPI_ISL_2472792, EPI_ISL_2472793, EPI_ISL_2472794, EPI_ISL_2472795, EPI_ISL_2472796, EPI_ISL_2472797, EPI_ISL_2472798, EPI_ISL_2472799, EPI_ISL_2472800, EPI_ISL_2472801, EPI_ISL_2472802, EPI_ISL_2472803, EPI_ISL_2472804, EPI_ISL_2472805, EPI_ISL_2472806, EPI_ISL_2472807, EPI_ISL_2472808, EPI_ISL_2472809, EPI_ISL_2472810, EPI_ISL_2472811, EPI_ISL_2472812, EPI_ISL_2472813, EPI_ISL_2472814, EPI_ISL_2472815, EPI_ISL_2472816, EPI_ISL_2472817, EPI_ISL_2472818, EPI_ISL_2472819, EPI_ISL_2472820, EPI_ISL_2472821, EPI_ISL_2472822, EPI_ISL_2472823, EPI_ISL_2472824, EPI_ISL_2472825, EPI_ISL_2472826, EPI_ISL_2472827, EPI_ISL_2472828, EPI_ISL_2472829, EPI_ISL_2472830, EPI_ISL_2472831, EPI_ISL_2472832, EPI_ISL_2472833, EPI_ISL_2472834, EPI_ISL_2472835, EPI_ISL_2472836, EPI_ISL_2472837, EPI_ISL_2472838, EPI_ISL_2472839, EPI_ISL_2472840, EPI_ISL_2472841, EPI_ISL_2472842, EPI_ISL_2472843, EPI_ISL_2472844, EPI_ISL_2472845, EPI_ISL_2472846, EPI_ISL_2472847, EPI_ISL_2472848, EPI_ISL_2472849, EPI_ISL_2472850, EPI_ISL_2472851, EPI_ISL_2472852, EPI_ISL_2472853, EPI_ISL_2472854, EPI_ISL_2472855, EPI_ISL_2472856, EPI_ISL_2472857, EPI_ISL_2472858, EPI_ISL_2472859, EPI_ISL_2472860, EPI_ISL_2472861, EPI_ISL_2472862, EPI_ISL_2472863, EPI_ISL_2472864, EPI_ISL_2472865, EPI_ISL_2472866, EPI_ISL_2472867, EPI_ISL_2472868, EPI_ISL_2472869, EPI_ISL_2472870, EPI_ISL_2472871, EPI_ISL_2472872, EPI_ISL_2472873, EPI_ISL_2472874, EPI_ISL_2472875, EPI_ISL_2472876, EPI_ISL_2472877, EPI_ISL_2472878, EPI_ISL_2472879, EPI_ISL_2472880, EPI_ISL_2472881, EPI_ISL_2472882, EPI_ISL_2472883, EPI_ISL_2472884, EPI_ISL_2472885, EPI_ISL_2472886, EPI_ISL_2636017, EPI_ISL_2636018, EPI_ISL_2636019, EPI_ISL_2636020, EPI_ISL_2636021, EPI_ISL_2636022, EPI_ISL_2636023, EPI_ISL_2636024, EPI_ISL_2636025, EPI_ISL_2636026, EPI_ISL_2636027, EPI_ISL_2636028, EPI_ISL_2636029, EPI_ISL_2636030, EPI_ISL_2636031, EPI_ISL_2636032, EPI_ISL_2636033, EPI_ISL_2636034, EPI_ISL_2636035, EPI_ISL_2636036, EPI_ISL_2636037, EPI_ISL_2636038, EPI_ISL_2636039, EPI_ISL_2636040, EPI_ISL_2636041, EPI_ISL_2636042, EPI_ISL_2636043, EPI_ISL_2636044, EPI_ISL_2636045, EPI_ISL_2636046, EPI_ISL_2636047, EPI_ISL_2636048, EPI_ISL_2636049, EPI_ISL_2636050, EPI_ISL_2636051, EPI_ISL_2636052, EPI_ISL_2636053, EPI_ISL_2636054, EPI_ISL_2636055, EPI_ISL_2636056, EPI_ISL_2636057, EPI_ISL_2636058, EPI_ISL_2636059, EPI_ISL_2636060, EPI_ISL_2636061, EPI_ISL_2636062, EPI_ISL_2636063, EPI_ISL_2636064, EPI_ISL_2636065, EPI_ISL_2636066, EPI_ISL_2636067, EPI_ISL_2636068, EPI_ISL_2636069, EPI_ISL_2636070, EPI_ISL_2636071, EPI_ISL_2636072, EPI_ISL_2636073, EPI_ISL_2636074, EPI_ISL_2636075, EPI_ISL_2636076, EPI_ISL_2678751, EPI_ISL_2678792, EPI_ISL_2678796

see above — Labor ZOTZ|KLIMAS; MVZ Düsseldorf-Centrum — Robert Koch Institute

EPI_ISL_2389322, EPI_ISL_2389324, EPI_ISL_2470807, EPI_ISL_2470812, EPI_ISL_2632901 — Labor für medizinische Mikrobiologie Bakteriologie Mykologie Virologie & Infektionsserologie - Labor Mölbis — Robert Koch Institute

EPI_ISL_2389442, EPI_ISL_2389443, EPI_ISL_2389444, EPI_ISL_2389445, EPI_ISL_2389446, EPI_ISL_2389447, EPI_ISL_2389448, EPI_ISL_2389449, EPI_ISL_2389451, EPI_ISL_2389452, EPI_ISL_2389453, EPI_ISL_2389455, EPI_ISL_2389456, EPI_ISL_2635999, EPI_ISL_2636000, EPI_ISL_2636001, EPI_ISL_2636007

see above — Laborarztpraxis Dres. med. Walther Weindel & Kollegen — Robert Koch Institute

EPI_ISL_2471505, EPI_ISL_2471508, EPI_ISL_2471510, EPI_ISL_2471512, EPI_ISL_2471515, EPI_ISL_2471522, EPI_ISL_2472519, EPI_ISL_2472521, EPI_ISL_2472526, EPI_ISL_2472530, EPI_ISL_2472531, EPI_ISL_2472533

see above — Laborarztpraxis Jochen Hüter — Robert Koch Institute

EPI_ISL_2388197, EPI_ISL_2388200, EPI_ISL_2388206, EPI_ISL_2388210, EPI_ISL_2388211, EPI_ISL_2388217, EPI_ISL_2388218, EPI_ISL_2388220, EPI_ISL_2388221, EPI_ISL_2388222, EPI_ISL_2388227, EPI_ISL_2388231, EPI_ISL_2388255, EPI_ISL_2388257, EPI_ISL_2388259, EPI_ISL_2388271, EPI_ISL_2388282, EPI_ISL_2388291, EPI_ISL_2388293, EPI_ISL_2388297, EPI_ISL_2388310, EPI_ISL_2388315, EPI_ISL_2469904, EPI_ISL_2632029, EPI_ISL_2632030, EPI_ISL_2632037, EPI_ISL_2632038, EPI_ISL_2632041, EPI_ISL_2632043, EPI_ISL_2632044, EPI_ISL_2632049, EPI_ISL_2632051, EPI_ISL_2632052, EPI_ISL_2632054, EPI_ISL_2632056, EPI_ISL_2632059, EPI_ISL_2632060, EPI_ISL_2632063, EPI_ISL_2632065, EPI_ISL_2632067, EPI_ISL_2632074, EPI_ISL_2632076, EPI_ISL_2632077, EPI_ISL_2632078, EPI_ISL_2632079, EPI_ISL_2632080, EPI_ISL_2632081, EPI_ISL_2632083, EPI_ISL_2632084, EPI_ISL_2632085, EPI_ISL_2632086, EPI_ISL_2632090, EPI_ISL_2632091, EPI_ISL_2632092, EPI_ISL_2632094, EPI_ISL_2632095, EPI_ISL_2632096, EPI_ISL_2632099, EPI_ISL_2632100, EPI_ISL_2632108, EPI_ISL_2632110, EPI_ISL_2633397, EPI_ISL_2633398, EPI_ISL_2633399, EPI_ISL_2633401,

EPI_ISL_2633406, EPI_ISL_2633408, EPI_ISL_2633413, EPI_ISL_2633420, EPI_ISL_2633421, EPI_ISL_2633423, EPI_ISL_2633428, EPI_ISL_2633429, EPI_ISL_2633437, EPI_ISL_2633440, EPI_ISL_2633441, EPI_ISL_2633443, EPI_ISL_2633452, EPI_ISL_2633455, EPI_ISL_2633456, EPI_ISL_2633457, EPI_ISL_2633460, EPI_ISL_2633462, EPI_ISL_2636886, EPI_ISL_2636891

see above    Laborarztpraxis Osnabrück    Robert Koch Institute

EPI_ISL_2389394, EPI_ISL_2389434, EPI_ISL_2471017, EPI_ISL_2471018, EPI_ISL_2471019, EPI_ISL_2471024, EPI_ISL_2471027, EPI_ISL_2471030, EPI_ISL_2471034, EPI_ISL_2633742, EPI_ISL_2633744, EPI_ISL_2633747, EPI_ISL_2633754, EPI_ISL_2633756, EPI_ISL_2633760, EPI_ISL_2633762, EPI_ISL_2633765, EPI_ISL_2633767, EPI_ISL_2633775, EPI_ISL_2633777, EPI_ISL_2633782, EPI_ISL_2633784, EPI_ISL_2633787, EPI_ISL_2633789, EPI_ISL_2633794, EPI_ISL_2633795, EPI_ISL_2633797, EPI_ISL_2633800, EPI_ISL_2633801, EPI_ISL_2633802, EPI_ISL_2633804, EPI_ISL_2633805, EPI_ISL_2633806, EPI_ISL_2633808, EPI_ISL_2633811, EPI_ISL_2633812, EPI_ISL_2633813, EPI_ISL_2633816, EPI_ISL_2633817, EPI_ISL_2633818, EPI_ISL_2633824, EPI_ISL_2633827, EPI_ISL_2633828, EPI_ISL_2633831, EPI_ISL_2633832, EPI_ISL_2633834, EPI_ISL_2633836, EPI_ISL_2633838, EPI_ISL_2633841, EPI_ISL_2633843, EPI_ISL_2633846, EPI_ISL_2633852, EPI_ISL_2633853, EPI_ISL_2633857, EPI_ISL_2633860, EPI_ISL_2633862, EPI_ISL_2633863, EPI_ISL_2633866, EPI_ISL_2633873, EPI_ISL_2633876, EPI_ISL_2633880, EPI_ISL_2633881, EPI_ISL_2633891, EPI_ISL_2633893, EPI_ISL_2633895, EPI_ISL_2633898, EPI_ISL_2633899

see above    Labormedizin Darmstadt    Robert Koch Institute

EPI_ISL_2634858, EPI_ISL_2634860, EPI_ISL_2634862, EPI_ISL_2634864, EPI_ISL_2634866, EPI_ISL_2634869, EPI_ISL_2634876, EPI_ISL_2634878, EPI_ISL_2634880, EPI_ISL_2634882, EPI_ISL_2634884, EPI_ISL_2634886, EPI_ISL_2634888, EPI_ISL_2634891, EPI_ISL_2634893, EPI_ISL_2634895, EPI_ISL_2634898, EPI_ISL_2634905, EPI_ISL_2634908, EPI_ISL_2634910, EPI_ISL_2634912, EPI_ISL_2634914, EPI_ISL_2634919, EPI_ISL_2634921, EPI_ISL_2634926, EPI_ISL_2634928, EPI_ISL_2634930, EPI_ISL_2634932, EPI_ISL_2634935, EPI_ISL_2760155, EPI_ISL_2760164, EPI_ISL_2760168, EPI_ISL_2760171, EPI_ISL_2760174, EPI_ISL_2760176, EPI_ISL_2760178, EPI_ISL_2760189, EPI_ISL_2761646, EPI_ISL_2761647, EPI_ISL_2761651, EPI_ISL_2761656, EPI_ISL_2761660

see above    Landesgesundheitsamt Baden-Württemberg    Robert Koch Institute

EPI_ISL_2390355, EPI_ISL_2390359, EPI_ISL_2390364, EPI_ISL_2390368, EPI_ISL_2390369, EPI_ISL_2471842, EPI_ISL_2471844, EPI_ISL_2471847, EPI_ISL_2471848, EPI_ISL_2471851, EPI_ISL_2471852, EPI_ISL_2471861, EPI_ISL_2471877, EPI_ISL_2471887

see above    Landesuntersuchungsanstalt für das Gesundheits- und Veterinärwesen (LUA) Sachsen    Robert Koch Institute

EPI_ISL_2386876, EPI_ISL_2386878, EPI_ISL_2386880, EPI_ISL_2386881, EPI_ISL_2386882, EPI_ISL_2636518, EPI_ISL_2636519, EPI_ISL_2636520, EPI_ISL_2636521, EPI_ISL_2636522, EPI_ISL_2636523, EPI_ISL_2636524, EPI_ISL_2636525, EPI_ISL_2636526, EPI_ISL_2636527, EPI_ISL_2636528, EPI_ISL_2636529, EPI_ISL_2636530, EPI_ISL_2762169

see above    Limbach - Ganzimmun Diagnostics AG Mainz    Robert Koch Institute

EPI_ISL_2314746, EPI_ISL_2314759, EPI_ISL_2316446, EPI_ISL_2316466, EPI_ISL_2316470, EPI_ISL_2316478, EPI_ISL_2386172, EPI_ISL_2386173, EPI_ISL_2386180, EPI_ISL_2386189, EPI_ISL_2390222, EPI_ISL_2390233, EPI_ISL_2390234, EPI_ISL_2390261, EPI_ISL_2390282, EPI_ISL_2390286, EPI_ISL_2471493, EPI_ISL_2471495, EPI_ISL_2471499, EPI_ISL_2471557, EPI_ISL_2471559, EPI_ISL_2471561, EPI_ISL_2472744, EPI_ISL_2472746, EPI_ISL_2472750, EPI_ISL_2472759, EPI_ISL_2472761, EPI_ISL_2472764, EPI_ISL_2632941, EPI_ISL_2632942, EPI_ISL_2632945, EPI_ISL_2632955, EPI_ISL_2632959

see above    Limbach - MVZ Gemeinschaftslabor Suhl    Robert Koch Institute

EPI_ISL_2389128, EPI_ISL_2389129, EPI_ISL_2389130, EPI_ISL_2389131, EPI_ISL_2389132, EPI_ISL_2389133, EPI_ISL_2389134, EPI_ISL_2389135, EPI_ISL_2389136, EPI_ISL_2389137, EPI_ISL_2389138, EPI_ISL_2389139, EPI_ISL_2389140, EPI_ISL_2389141, EPI_ISL_2389142, EPI_ISL_2389143, EPI_ISL_2389144, EPI_ISL_2389145, EPI_ISL_2389146, EPI_ISL_2389147, EPI_ISL_2389148, EPI_ISL_2389149, EPI_ISL_2389150, EPI_ISL_2389151, EPI_ISL_2389152, EPI_ISL_2389153, EPI_ISL_2389154, EPI_ISL_2389155, EPI_ISL_2389156, EPI_ISL_2389157, EPI_ISL_2389158, EPI_ISL_2389159, EPI_ISL_2389160, EPI_ISL_2389161, EPI_ISL_2389162, EPI_ISL_2389163, EPI_ISL_2389164, EPI_ISL_2389165, EPI_ISL_2389166, EPI_ISL_2389167, EPI_ISL_2389168, EPI_ISL_2389169, EPI_ISL_2389170, EPI_ISL_2389171, EPI_ISL_2389172, EPI_ISL_2389173, EPI_ISL_2389174, EPI_ISL_2389175, EPI_ISL_2389176, EPI_ISL_2389177, EPI_ISL_2389178, EPI_ISL_2389179, EPI_ISL_2389180, EPI_ISL_2389181, EPI_ISL_2389183, EPI_ISL_2389185, EPI_ISL_2389187, EPI_ISL_2389189, EPI_ISL_2389191, EPI_ISL_2389193, EPI_ISL_2389197, EPI_ISL_2389199, EPI_ISL_2389201, EPI_ISL_2389204, EPI_ISL_2389206, EPI_ISL_2389208, EPI_ISL_2389210, EPI_ISL_2389212, EPI_ISL_2389214, EPI_ISL_2389216, EPI_ISL_2389217, EPI_ISL_2389218, EPI_ISL_2389219, EPI_ISL_2389220, EPI_ISL_2389221, EPI_ISL_2389222, EPI_ISL_2389223, EPI_ISL_2389224, EPI_ISL_2389225, EPI_ISL_2389226, EPI_ISL_2389227, EPI_ISL_2389228, EPI_ISL_2389229, EPI_ISL_2389230, EPI_ISL_2389231, EPI_ISL_2389232, EPI_ISL_2389233, EPI_ISL_2389234, EPI_ISL_2389235, EPI_ISL_2389236, EPI_ISL_2389237, EPI_ISL_2389238, EPI_ISL_2389239, EPI_ISL_2389240, EPI_ISL_2389241, EPI_ISL_2389242, EPI_ISL_2389243, EPI_ISL_2389244, EPI_ISL_2389245, EPI_ISL_2389246, EPI_ISL_2389247, EPI_ISL_2389248, EPI_ISL_2389249, EPI_ISL_2389250, EPI_ISL_2389251, EPI_ISL_2389252, EPI_ISL_2389253, EPI_ISL_2389254, EPI_ISL_2389255, EPI_ISL_2389256, EPI_ISL_2389257, EPI_ISL_2389258, EPI_ISL_2389259, EPI_ISL_2389260, EPI_ISL_2389261, EPI_ISL_2389262, EPI_ISL_2389263, EPI_ISL_2389264, EPI_ISL_2389265, EPI_ISL_2389266, EPI_ISL_2389267, EPI_ISL_2389268, EPI_ISL_2389269, EPI_ISL_2389270, EPI_ISL_2389271, EPI_ISL_2389272, EPI_ISL_2389273, EPI_ISL_2389274, EPI_ISL_2389275, EPI_ISL_2389276, EPI_ISL_2389277, EPI_ISL_2389278, EPI_ISL_2389279, EPI_ISL_2389280, EPI_ISL_2389281, EPI_ISL_2389282, EPI_ISL_2389283, EPI_ISL_2389284, EPI_ISL_2389285, EPI_ISL_2389286, EPI_ISL_2389287, EPI_ISL_2389288, EPI_ISL_2389289, EPI_ISL_2389290, EPI_ISL_2389291, EPI_ISL_2389292, EPI_ISL_2389293, EPI_ISL_2389294, EPI_ISL_2389295, EPI_ISL_2389296, EPI_ISL_2389297, EPI_ISL_2389298, EPI_ISL_2389299, EPI_ISL_2389300, EPI_ISL_2389301, EPI_ISL_2389302, EPI_ISL_2389303, EPI_ISL_2389304, EPI_ISL_2389305, EPI_ISL_2389306, EPI_ISL_2389307, EPI_ISL_2389308, EPI_ISL_2389309, EPI_ISL_2389310, EPI_ISL_2389311, EPI_ISL_2389312, EPI_ISL_2389313, EPI_ISL_2389314, EPI_ISL_2389315, EPI_ISL_2470651, EPI_ISL_2470652, EPI_ISL_2470653, EPI_ISL_2470654, EPI_ISL_2470655, EPI_ISL_2470656, EPI_ISL_2470657, EPI_ISL_2470658, EPI_ISL_2470659, EPI_ISL_2470660, EPI_ISL_2470661, EPI_ISL_2470662, EPI_ISL_2470663, EPI_ISL_2470664, EPI_ISL_2470665, EPI_ISL_2470666, EPI_ISL_2470667, EPI_ISL_2470668, EPI_ISL_2470669, EPI_ISL_2470670, EPI_ISL_2470671, EPI_ISL_2470672, EPI_ISL_2470673, EPI_ISL_2470674, EPI_ISL_2470675, EPI_ISL_2470676, EPI_ISL_2470677, EPI_ISL_2470678, EPI_ISL_2470679, EPI_ISL_2470680, EPI_ISL_2470681, EPI_ISL_2470682, EPI_ISL_2470683, EPI_ISL_2470684, EPI_ISL_2470685, EPI_ISL_2470686, EPI_ISL_2470687, EPI_ISL_2470688, EPI_ISL_2470689, EPI_ISL_2470690, EPI_ISL_2470691, EPI_ISL_2470692, EPI_ISL_2470693, EPI_ISL_2470694, EPI_ISL_2470695, EPI_ISL_2470696, EPI_ISL_2470697, EPI_ISL_2470698, EPI_ISL_2470699, EPI_ISL_2470700, EPI_ISL_2470701, EPI_ISL_2470702, EPI_ISL_2470703, EPI_ISL_2470704, EPI_ISL_2470705, EPI_ISL_2470706, EPI_ISL_2470707, EPI_ISL_2470708, EPI_ISL_2470709, EPI_ISL_2470710, EPI_ISL_2470711, EPI_ISL_2470712, EPI_ISL_2470713, EPI_ISL_2470714, EPI_ISL_2470715, EPI_ISL_2470716, EPI_ISL_2470717, EPI_ISL_2470718, EPI_ISL_2470719, EPI_ISL_2470720, EPI_ISL_2470721, EPI_ISL_2470722, EPI_ISL_2470723, EPI_ISL_2470724, EPI_ISL_2470725, EPI_ISL_2470726, EPI_ISL_2470728, EPI_ISL_2470729, EPI_ISL_2470730, EPI_ISL_2470731, EPI_ISL_2470732, EPI_ISL_2470733, EPI_ISL_2470734, EPI_ISL_2470735, EPI_ISL_2470736, EPI_ISL_2470737, EPI_ISL_2470738, EPI_ISL_2470739, EPI_ISL_2470740, EPI_ISL_2470741, EPI_ISL_2470742, EPI_ISL_2470743, EPI_ISL_2470744, EPI_ISL_2470745, EPI_ISL_2470746, EPI_ISL_2470747, EPI_ISL_2470748, EPI_ISL_2470749, EPI_ISL_2470750, EPI_ISL_2470751, EPI_ISL_2470752, EPI_ISL_2470753, EPI_ISL_2470754, EPI_ISL_2470755, EPI_ISL_2470756, EPI_ISL_2470757, EPI_ISL_2470758, EPI_ISL_2470759, EPI_ISL_2470760, EPI_ISL_2470761, EPI_ISL_2470762, EPI_ISL_2470763, EPI_ISL_2470764, EPI_ISL_2470765, EPI_ISL_2470766, EPI_ISL_2470767, EPI_ISL_2470768, EPI_ISL_2470769, EPI_ISL_2470770, EPI_ISL_2470771, EPI_ISL_2470772, EPI_ISL_2470773, EPI_ISL_2470774, EPI_ISL_2470775, EPI_ISL_2470776, EPI_ISL_2470777, EPI_ISL_2470778, EPI_ISL_2470779, EPI_ISL_2470780, EPI_ISL_2470781, EPI_ISL_2470782, EPI_ISL_2470783, EPI_ISL_2470784, EPI_ISL_2470785, EPI_ISL_2470786, EPI_ISL_2470787, EPI_ISL_2470788, EPI_ISL_2470789, EPI_ISL_2470790, EPI_ISL_2470791, EPI_ISL_2470792, EPI_ISL_2470793, EPI_ISL_2470794, EPI_ISL_2470795, EPI_ISL_2470796, EPI_ISL_2470797, EPI_ISL_2470798, EPI_ISL_2470799, EPI_ISL_2470800, EPI_ISL_2470801, EPI_ISL_2470802, EPI_ISL_2470803, EPI_ISL_2470804, EPI_ISL_2470805, EPI_ISL_2470806, EPI_ISL_2632645, EPI_ISL_2632647, EPI_ISL_2632648, EPI_ISL_2632650, EPI_ISL_2632651, EPI_ISL_2632653, EPI_ISL_2632655, EPI_ISL_2632656, EPI_ISL_2632657, EPI_ISL_2632658, EPI_ISL_2632659, EPI_ISL_2632661, EPI_ISL_2632664, EPI_ISL_2632665, EPI_ISL_2632668, EPI_ISL_2632669, EPI_ISL_2632671, EPI_ISL_2632673, EPI_ISL_2632675, EPI_ISL_2632676, EPI_ISL_2632678, EPI_ISL_2632679, EPI_ISL_2632683, EPI_ISL_2632685, EPI_ISL_2632686, EPI_ISL_2632688, EPI_ISL_2632689, EPI_ISL_2632690, EPI_ISL_2632692, EPI_ISL_2632697, EPI_ISL_2632698, EPI_ISL_2632699, EPI_ISL_2632702, EPI_ISL_2632703, EPI_ISL_2632704, EPI_ISL_2632707, EPI_ISL_2632709, EPI_ISL_2632711, EPI_ISL_2632712, EPI_ISL_2632714, EPI_ISL_2632716, EPI_ISL_2632717, EPI_ISL_2632719, EPI_ISL_2632720, EPI_ISL_2632721, EPI_ISL_2632723, EPI_ISL_2632724, EPI_ISL_2632726, EPI_ISL_2632727, EPI_ISL_2632728, EPI_ISL_2632729, EPI_ISL_2632731, EPI_ISL_2632732, EPI_ISL_2632734, EPI_ISL_2632735, EPI_ISL_2632737, EPI_ISL_2632738, EPI_ISL_2632740, EPI_ISL_2632741, EPI_ISL_2632745, EPI_ISL_2632747, EPI_ISL_2632748, EPI_ISL_2632750, EPI_ISL_2632752, EPI_ISL_2632754, EPI_ISL_2632755, EPI_ISL_2632757, EPI_ISL_2632758, EPI_ISL_2632759, EPI_ISL_2632760, EPI_ISL_2632761, EPI_ISL_2632762, EPI_ISL_2632765, EPI_ISL_2632766, EPI_ISL_2632767, EPI_ISL_2632769, EPI_ISL_2632770, EPI_ISL_2632772, EPI_ISL_2632773, EPI_ISL_2632774, EPI_ISL_2632776, EPI_ISL_2632777, EPI_ISL_2632778, EPI_ISL_2632782, EPI_ISL_2632784, EPI_ISL_2632786, EPI_ISL_2632787, EPI_ISL_2632789, EPI_ISL_2632790

see above    Limbach - MVZ Humangenetik Ulm    Robert Koch Institute

EPI_ISL_2316612, EPI_ISL_2386212, EPI_ISL_2386214, EPI_ISL_2386219, EPI_ISL_2386221, EPI_ISL_2386226, EPI_ISL_2386227, EPI_ISL_2386232, EPI_ISL_2390093, EPI_ISL_2390098, EPI_ISL_2390100, EPI_ISL_2390104, EPI_ISL_2390106, EPI_ISL_2471467, EPI_ISL_2471474, EPI_ISL_2471479, EPI_ISL_2471480, EPI_ISL_2471483, EPI_ISL_2472542, EPI_ISL_2472706, EPI_ISL_2472710, EPI_ISL_2472716, EPI_ISL_2472721, EPI_ISL_2472722, EPI_ISL_2472726, EPI_ISL_2632843

see above    Limbach - MVZ Labor Eveld & Kollegen Essen    Robert Koch Institute

EPI_ISL_2472522, EPI_ISL_2632968, EPI_ISL_2632971, EPI_ISL_2632978    Limbach - MVZ Labor Ludwigsburg    Robert Koch Institute

EPI_ISL_2316493, EPI_ISL_2316497, EPI_ISL_2316499, EPI_ISL_2386166, EPI_ISL_2386168, EPI_ISL_2386176, EPI_ISL_2386178, EPI_ISL_2386196, EPI_ISL_2386197, EPI_ISL_2386198, EPI_ISL_2386199, EPI_ISL_2386200, EPI_ISL_2386201, EPI_ISL_2386202, EPI_ISL_2386203, EPI_ISL_2386204, EPI_ISL_2386205, EPI_ISL_2386206, EPI_ISL_2386207, EPI_ISL_2386208, EPI_ISL_2386209, EPI_ISL_2386210, EPI_ISL_2386211, EPI_ISL_2390094, EPI_ISL_2390103, EPI_ISL_2390198, EPI_ISL_2390199, EPI_ISL_2390200, EPI_ISL_2390201, EPI_ISL_2390202, EPI_ISL_2390203, EPI_ISL_2390204, EPI_ISL_2390205, EPI_ISL_2390206, EPI_ISL_2390207, EPI_ISL_2390208, EPI_ISL_2390209, EPI_ISL_2390210, EPI_ISL_2390211, EPI_ISL_2390212, EPI_ISL_2390213, EPI_ISL_2390214, EPI_ISL_2390216, EPI_ISL_2390220, EPI_ISL_2390260, EPI_ISL_2390263, EPI_ISL_2390266, EPI_ISL_2390268, EPI_ISL_2390269, EPI_ISL_2390270, EPI_ISL_2390271, EPI_ISL_2390273, EPI_ISL_2390276, EPI_ISL_2390279, EPI_ISL_2390280, EPI_ISL_2390281, EPI_ISL_2390283, EPI_ISL_2390285, EPI_ISL_2390287, EPI_ISL_2390288, EPI_ISL_2390290, EPI_ISL_2390291, EPI_ISL_2390292, EPI_ISL_2390293, EPI_ISL_2390294, EPI_ISL_2390295, EPI_ISL_2390296, EPI_ISL_2390298, EPI_ISL_2471484, EPI_ISL_2471485, EPI_ISL_2471486, EPI_ISL_2471487, EPI_ISL_2471490, EPI_ISL_2471491, EPI_ISL_2471492, EPI_ISL_2471494, EPI_ISL_2471496, EPI_ISL_2471497, EPI_ISL_2471498, EPI_ISL_2471500, EPI_ISL_2471501, EPI_ISL_2471502, EPI_ISL_2471503, EPI_ISL_2471504, EPI_ISL_2471506, EPI_ISL_2471507, EPI_ISL_2471509, EPI_ISL_2471511, EPI_ISL_2471514, EPI_ISL_2471516, EPI_ISL_2471517, EPI_ISL_2471519, EPI_ISL_2471521, EPI_ISL_2471523, EPI_ISL_2471526, EPI_ISL_2471527, EPI_ISL_2471530, EPI_ISL_2471551, EPI_ISL_2471553, EPI_ISL_2471555, EPI_ISL_2471568, EPI_ISL_2471573, EPI_ISL_2471579, EPI_ISL_2471581, EPI_ISL_2471584, EPI_ISL_2471589, EPI_ISL_2471590, EPI_ISL_2471591, EPI_ISL_2471592, EPI_ISL_2471594, EPI_ISL_2471733, EPI_ISL_2471734, EPI_ISL_2471735, EPI_ISL_2471736, EPI_ISL_2471737, EPI_ISL_2471739, EPI_ISL_2471740, EPI_ISL_2471741, EPI_ISL_2471743, EPI_ISL_2471744, EPI_ISL_2471746, EPI_ISL_2471747, EPI_ISL_2471748, EPI_ISL_2471895, EPI_ISL_2471902, EPI_ISL_2471903, EPI_ISL_2471906, EPI_ISL_2471907, EPI_ISL_2471909, EPI_ISL_2471911, EPI_ISL_2471912, EPI_ISL_2471913, EPI_ISL_2472484, EPI_ISL_2472518, EPI_ISL_2472520, EPI_ISL_2472523, EPI_ISL_2472524, EPI_ISL_2472525, EPI_ISL_2472527, EPI_ISL_2472528, EPI_ISL_2472529, EPI_ISL_2472532, EPI_ISL_2472534, EPI_ISL_2472535, EPI_ISL_2472536, EPI_ISL_2472537, EPI_ISL_2472700, EPI_ISL_2472701, EPI_ISL_2472703, EPI_ISL_2472707, EPI_ISL_2472715, EPI_ISL_2631764, EPI_ISL_2631767, EPI_ISL_2631768, EPI_ISL_2631769, EPI_ISL_2631771, EPI_ISL_2631772, EPI_ISL_2631773, EPI_ISL_2631776, EPI_ISL_2631777, EPI_ISL_2631778, EPI_ISL_2631780, EPI_ISL_2631781, EPI_ISL_2631782, EPI_ISL_2631783, EPI_ISL_2631786, EPI_ISL_2631789, EPI_ISL_2631791, EPI_ISL_2631796, EPI_ISL_2631797, EPI_ISL_2631799, EPI_ISL_2631800, EPI_ISL_2631801, EPI_ISL_2631803, EPI_ISL_2631806

see above    Limbach - MVZ Labor Ravensburg Labor Dr. Gärtner    Robert Koch Institute

EPI_ISL_2314774, EPI_ISL_2314776, EPI_ISL_2314777, EPI_ISL_2314779, EPI_ISL_2314780, EPI_ISL_2314781, EPI_ISL_2314782, EPI_ISL_2314785, EPI_ISL_2314786, EPI_ISL_2314789, EPI_ISL_2314790, EPI_ISL_2314795, EPI_ISL_2314796, EPI_ISL_2314798, EPI_ISL_2314799, EPI_ISL_2314800, EPI_ISL_2386164, EPI_ISL_2386167, EPI_ISL_2386169, EPI_ISL_2386170, EPI_ISL_2386174, EPI_ISL_2386175, EPI_ISL_2386179, EPI_ISL_2390250, EPI_ISL_2390253, EPI_ISL_2390255, EPI_ISL_2390256, EPI_ISL_2390262, EPI_ISL_2390264, EPI_ISL_2390265, EPI_ISL_2390267, EPI_ISL_2390272, EPI_ISL_2390274, EPI_ISL_2390277, EPI_ISL_2390278, EPI_ISL_2390284, EPI_ISL_2390289, EPI_ISL_2390299, EPI_ISL_2390300, EPI_ISL_2471513, EPI_ISL_2471518, EPI_ISL_2471520, EPI_ISL_2471524, EPI_ISL_2471525, EPI_ISL_2471691, EPI_ISL_2471692, EPI_ISL_2471742, EPI_ISL_2471922, EPI_ISL_2471923, EPI_ISL_2471924, EPI_ISL_2471925, EPI_ISL_2471926, EPI_ISL_2471927, EPI_ISL_2471928, EPI_ISL_2471929, EPI_ISL_2471930, EPI_ISL_2471931, EPI_ISL_2471932, EPI_ISL_2471933, EPI_ISL_2471934, EPI_ISL_2471935, EPI_ISL_2471936, EPI_ISL_2471937, EPI_ISL_2471938, EPI_ISL_2471939, EPI_ISL_2471940, EPI_ISL_2471941, EPI_ISL_2471942, EPI_ISL_2471943, EPI_ISL_2471944, EPI_ISL_2471945, EPI_ISL_2471946, EPI_ISL_2471947, EPI_ISL_2471948, EPI_ISL_2471949, EPI_ISL_2471950, EPI_ISL_2471951, EPI_ISL_2471952, EPI_ISL_2471953, EPI_ISL_2471954, EPI_ISL_2471955, EPI_ISL_2471956, EPI_ISL_2471957, EPI_ISL_2471958, EPI_ISL_2471959, EPI_ISL_2471960, EPI_ISL_2471961, EPI_ISL_2471962, EPI_ISL_2471963, EPI_ISL_2471964, EPI_ISL_2471965, EPI_ISL_2471966, EPI_ISL_2471967, EPI_ISL_2471968, EPI_ISL_2471969, EPI_ISL_2471970, EPI_ISL_2471971, EPI_ISL_2472366, EPI_ISL_2472367, EPI_ISL_2472368, EPI_ISL_2472369, EPI_ISL_2472370, EPI_ISL_2472371, EPI_ISL_2472372, EPI_ISL_2472373, EPI_ISL_2472374, EPI_ISL_2472375, EPI_ISL_2472376, EPI_ISL_2472377, EPI_ISL_2472378, EPI_ISL_2472379, EPI_ISL_2472380, EPI_ISL_2472381, EPI_ISL_2472382, EPI_ISL_2472383, EPI_ISL_2472384, EPI_ISL_2472385, EPI_ISL_2472386, EPI_ISL_2472387, EPI_ISL_2472388, EPI_ISL_2472389, EPI_ISL_2472390, EPI_ISL_2472391, EPI_ISL_2472392, EPI_ISL_2472393, EPI_ISL_2472394, EPI_ISL_2472395, EPI_ISL_2472396, EPI_ISL_2472397, EPI_ISL_2472398, EPI_ISL_2472399, EPI_ISL_2472400, EPI_ISL_2472401, EPI_ISL_2472402, EPI_ISL_2472412, EPI_ISL_2472483, EPI_ISL_2472702, EPI_ISL_2472704, EPI_ISL_2472708, EPI_ISL_2472718, EPI_ISL_2472727, EPI_ISL_2472728, EPI_ISL_2472729, EPI_ISL_2472730, EPI_ISL_2472732, EPI_ISL_2472733, EPI_ISL_2472734, EPI_ISL_2472739, EPI_ISL_2472740, EPI_ISL_2472742, EPI_ISL_2472745, EPI_ISL_2472747, EPI_ISL_2472748, EPI_ISL_2472749, EPI_ISL_2472751, EPI_ISL_2472752, EPI_ISL_2472753, EPI_ISL_2472754, EPI_ISL_2472755, EPI_ISL_2472756, EPI_ISL_2472757, EPI_ISL_2472758, EPI_ISL_2472760, EPI_ISL_2472763, EPI_ISL_2472765, EPI_ISL_2472766, EPI_ISL_2472768, EPI_ISL_2472769, EPI_ISL_2631787, EPI_ISL_2631790, EPI_ISL_2631794, EPI_ISL_2631795, EPI_ISL_2631798, EPI_ISL_2631804, EPI_ISL_2631851, EPI_ISL_2631855, EPI_ISL_2631858, EPI_ISL_2632920, EPI_ISL_2632931, EPI_ISL_2635824, EPI_ISL_2635831, EPI_ISL_2635834, EPI_ISL_2635842

see above    MDI Limbach Berlin GmbH; MVZ Labor Berlin    Robert Koch Institute

EPI_ISL_2689467    MEDILYS Laborgesellschaft mbH    Heinrich Pette Institute, Leibniz Institute for Experimental Virology    Adam Grundhoff; Alexis Robitaille; Johannes Knobloch; Martin Aepfelbacher; Nicole Fischer; Thomas Günther

EPI_ISL_2772711, EPI_ISL_2772712, EPI_ISL_2772713    MKK Mühlenkreiskliniken, Johannes Wesling Klinikum Minden    Hannover Medical School, Institute of Virology    Lars Steinbrück

EPI_ISL_2387347, EPI_ISL_2387356, EPI_ISL_2387357, EPI_ISL_2387358, EPI_ISL_2387359, EPI_ISL_2387361, EPI_ISL_2387363, EPI_ISL_2387371, EPI_ISL_2387374, EPI_ISL_2387381, EPI_ISL_2387386, EPI_ISL_2387388, EPI_ISL_2387389, EPI_ISL_2387392, EPI_ISL_2387396, EPI_ISL_2387398, EPI_ISL_2387399, EPI_ISL_2387400, EPI_ISL_2387409, EPI_ISL_2387411, EPI_ISL_2387414, EPI_ISL_2387417, EPI_ISL_2387420, EPI_ISL_2387421, EPI_ISL_2387423, EPI_ISL_2387426, EPI_ISL_2387427, EPI_ISL_2387429, EPI_ISL_2387432, EPI_ISL_2387433, EPI_ISL_2387436, EPI_ISL_2387437, EPI_ISL_2387441, EPI_ISL_2387444, EPI_ISL_2387445, EPI_ISL_2387447, EPI_ISL_2387448, EPI_ISL_2387452, EPI_ISL_2388184, EPI_ISL_2388185, EPI_ISL_2388186, EPI_ISL_2388187, EPI_ISL_2388188, EPI_ISL_2388189, EPI_ISL_2388190, EPI_ISL_2388192, EPI_ISL_2388193, EPI_ISL_2388194, EPI_ISL_2388195, EPI_ISL_2388201, EPI_ISL_2388203, EPI_ISL_2388204, EPI_ISL_2388205, EPI_ISL_2388208, EPI_ISL_2388209, EPI_ISL_2388212, EPI_ISL_2388213, EPI_ISL_2388214, EPI_ISL_2388215, EPI_ISL_2388216, EPI_ISL_2388224, EPI_ISL_2388226, EPI_ISL_2388229, EPI_ISL_2388230, EPI_ISL_2388232, EPI_ISL_2388233, EPI_ISL_2388235, EPI_ISL_2388237, EPI_ISL_2388240, EPI_ISL_2388244, EPI_ISL_2388265, EPI_ISL_2388269, EPI_ISL_2388273, EPI_ISL_2388275, EPI_ISL_2388279, EPI_ISL_2388287, EPI_ISL_2388299, EPI_ISL_2388306, EPI_ISL_2388313, EPI_ISL_2469836, EPI_ISL_2469837, EPI_ISL_2469838, EPI_ISL_2469839, EPI_ISL_2469840, EPI_ISL_2469841, EPI_ISL_2469842, EPI_ISL_2469843, EPI_ISL_2469844, EPI_ISL_2469845, EPI_ISL_2469846, EPI_ISL_2469847, EPI_ISL_2469848, EPI_ISL_2469849, EPI_ISL_2469851, EPI_ISL_2469852, EPI_ISL_2469853, EPI_ISL_2469854, EPI_ISL_2469855, EPI_ISL_2469856, EPI_ISL_2469857, EPI_ISL_2469862, EPI_ISL_2469864, EPI_ISL_2469865, EPI_ISL_2469866, EPI_ISL_2469869, EPI_ISL_2469870, EPI_ISL_2469871, EPI_ISL_2469873, EPI_ISL_2469874, EPI_ISL_2469875, EPI_ISL_2469876, EPI_ISL_2469877, EPI_ISL_2469879, EPI_ISL_2469880, EPI_ISL_2469881, EPI_ISL_2469882, EPI_ISL_2469883, EPI_ISL_2469884, EPI_ISL_2469887, EPI_ISL_2469888, EPI_ISL_2469889, EPI_ISL_2469890, EPI_ISL_2469893, EPI_ISL_2469894, EPI_ISL_2469895, EPI_ISL_2469896, EPI_ISL_2469898, EPI_ISL_2469899, EPI_ISL_2469900, EPI_ISL_2469901, EPI_ISL_2469902, EPI_ISL_2469903, EPI_ISL_2469905, EPI_ISL_2469907, EPI_ISL_2469908, EPI_ISL_2469909, EPI_ISL_2469910, EPI_ISL_2469911, EPI_ISL_2469912, EPI_ISL_2469913, EPI_ISL_2469915, EPI_ISL_2469916, EPI_ISL_2469917, EPI_ISL_2469918, EPI_ISL_2469919, EPI_ISL_2469920, EPI_ISL_2469921, EPI_ISL_2469922, EPI_ISL_2469923, EPI_ISL_2471694, EPI_ISL_2471695, EPI_ISL_2471696, EPI_ISL_2471697, EPI_ISL_2471698, EPI_ISL_2471700, EPI_ISL_2471701, EPI_ISL_2471703, EPI_ISL_2471704, EPI_ISL_2471705, EPI_ISL_2471706, EPI_ISL_2471707, EPI_ISL_2471708, EPI_ISL_2471709, EPI_ISL_2471710, EPI_ISL_2471712, EPI_ISL_2471713, EPI_ISL_2471714, EPI_ISL_2471715, EPI_ISL_2471716, EPI_ISL_2471717, EPI_ISL_2471718, EPI_ISL_2471719, EPI_ISL_2471720, EPI_ISL_2471721, EPI_ISL_2471723, EPI_ISL_2471724, EPI_ISL_2471725, EPI_ISL_2471726, EPI_ISL_2471727, EPI_ISL_2471728, EPI_ISL_2471729, EPI_ISL_2471730, EPI_ISL_2471731, EPI_ISL_2632024, EPI_ISL_2632025, EPI_ISL_2632027, EPI_ISL_2632028, EPI_ISL_2632031, EPI_ISL_2632032, EPI_ISL_2632033, EPI_ISL_2632034, EPI_ISL_2632035, EPI_ISL_2632036, EPI_ISL_2632040, EPI_ISL_2632042, EPI_ISL_2632045, EPI_ISL_2632046, EPI_ISL_2632047, EPI_ISL_2632048, EPI_ISL_2632050, EPI_ISL_2632053, EPI_ISL_2632055, EPI_ISL_2632057, EPI_ISL_2632061, EPI_ISL_2632062, EPI_ISL_2632066, EPI_ISL_2632068, EPI_ISL_2632069, EPI_ISL_2632070, EPI_ISL_2632087, EPI_ISL_2632088, EPI_ISL_2632098, EPI_ISL_2632105, EPI_ISL_2632106, EPI_ISL_2632107, EPI_ISL_2632112, EPI_ISL_2633403, EPI_ISL_2633409, EPI_ISL_2633410, EPI_ISL_2633415, EPI_ISL_2633419, EPI_ISL_2633424, EPI_ISL_2633427, EPI_ISL_2633431, EPI_ISL_2633432, EPI_ISL_2633439, EPI_ISL_2633445, EPI_ISL_2633446, EPI_ISL_2633453, EPI_ISL_2761941

see above    MVZ Dr. Eberhard & Partner Dortmund    Robert Koch Institute

EPI_ISL_2390117, EPI_ISL_2390118, EPI_ISL_2390119, EPI_ISL_2390120, EPI_ISL_2390121, EPI_ISL_2390122, EPI_ISL_2390123, EPI_ISL_2390124, EPI_ISL_2390125, EPI_ISL_2390126, EPI_ISL_2390127, EPI_ISL_2390128, EPI_ISL_2390129, EPI_ISL_2390130, EPI_ISL_2390131, EPI_ISL_2390132, EPI_ISL_2390133, EPI_ISL_2390134, EPI_ISL_2390135, EPI_ISL_2390136, EPI_ISL_2390137, EPI_ISL_2390138, EPI_ISL_2390139, EPI_ISL_2390140, EPI_ISL_2390141, EPI_ISL_2390142, EPI_ISL_2390143, EPI_ISL_2390144, EPI_ISL_2390145, EPI_ISL_2390146, EPI_ISL_2390147, EPI_ISL_2390148, EPI_ISL_2390149, EPI_ISL_2390150, EPI_ISL_2390151, EPI_ISL_2390152, EPI_ISL_2390153, EPI_ISL_2390154, EPI_ISL_2390155, EPI_ISL_2390157, EPI_ISL_2390159, EPI_ISL_2390160, EPI_ISL_2390161, EPI_ISL_2390162, EPI_ISL_2390163, EPI_ISL_2390164, EPI_ISL_2390165, EPI_ISL_2390166, EPI_ISL_2390168, EPI_ISL_2390169, EPI_ISL_2390170, EPI_ISL_2390171, EPI_ISL_2390172, EPI_ISL_2390173, EPI_ISL_2390174, EPI_ISL_2390176, EPI_ISL_2390177, EPI_ISL_2390178, EPI_ISL_2390179, EPI_ISL_2390180, EPI_ISL_2471531, EPI_ISL_2471532, EPI_ISL_2471533, EPI_ISL_2471534, EPI_ISL_2471535, EPI_ISL_2471536, EPI_ISL_2471537, EPI_ISL_2471538, EPI_ISL_2471539, EPI_ISL_2471540, EPI_ISL_2471541, EPI_ISL_2471542, EPI_ISL_2471543, EPI_ISL_2471544, EPI_ISL_2471545, EPI_ISL_2471546, EPI_ISL_2471547, EPI_ISL_2471548, EPI_ISL_2471549, EPI_ISL_2636747, EPI_ISL_2636755

see above    MVZ Labor Dr. Fenner und Kollegen (Standort Hamburg)    Robert Koch Institute

EPI_ISL_2316418, EPI_ISL_2316436, EPI_ISL_2316455, EPI_ISL_2388012, EPI_ISL_2388013, EPI_ISL_2388015, EPI_ISL_2388018, EPI_ISL_2388019, EPI_ISL_2388023, EPI_ISL_2388028, EPI_ISL_2388029, EPI_ISL_2388031, EPI_ISL_2388033, EPI_ISL_2388034, EPI_ISL_2388035, EPI_ISL_2388036, EPI_ISL_2388037, EPI_ISL_2388038, EPI_ISL_2388039, EPI_ISL_2388040, EPI_ISL_2388041, EPI_ISL_2388042, EPI_ISL_2388043, EPI_ISL_2388044, EPI_ISL_2388045, EPI_ISL_2388046, EPI_ISL_2388047, EPI_ISL_2388048, EPI_ISL_2388049, EPI_ISL_2388050, EPI_ISL_2388051, EPI_ISL_2388052, EPI_ISL_2388053, EPI_ISL_2388054, EPI_ISL_2388055, EPI_ISL_2388056, EPI_ISL_2388057, EPI_ISL_2388058, EPI_ISL_2388059, EPI_ISL_2388060, EPI_ISL_2388061, EPI_ISL_2388062, EPI_ISL_2388063, EPI_ISL_2388064, EPI_ISL_2388065, EPI_ISL_2388066, EPI_ISL_2388067, EPI_ISL_2388069, EPI_ISL_2388070, EPI_ISL_2388071, EPI_ISL_2388073, EPI_ISL_2388074, EPI_ISL_2388075, EPI_ISL_2388076, EPI_ISL_2388077, EPI_ISL_2388078, EPI_ISL_2388079, EPI_ISL_2388081, EPI_ISL_2388083, EPI_ISL_2388085, EPI_ISL_2388087, EPI_ISL_2388088, EPI_ISL_2388093, EPI_ISL_2388094, EPI_ISL_2388095, EPI_ISL_2388096, EPI_ISL_2388098, EPI_ISL_2388099, EPI_ISL_2388100, EPI_ISL_2388101, EPI_ISL_2388103, EPI_ISL_2388104, EPI_ISL_2388105, EPI_ISL_2388106, EPI_ISL_2388107, EPI_ISL_2388109, EPI_ISL_2388110, EPI_ISL_2388111, EPI_ISL_2388112, EPI_ISL_2388113, EPI_ISL_2388116, EPI_ISL_2388117, EPI_ISL_2388120, EPI_ISL_2388123, EPI_ISL_2388124, EPI_ISL_2388126, EPI_ISL_2388128, EPI_ISL_2388129, EPI_ISL_2388131, EPI_ISL_2388132, EPI_ISL_2388133, EPI_ISL_2388134, EPI_ISL_2388138, EPI_ISL_2388140, EPI_ISL_2388141, EPI_ISL_2388143, EPI_ISL_2388144, EPI_ISL_2388145, EPI_ISL_2388146, EPI_ISL_2388148, EPI_ISL_2388149, EPI_ISL_2388150, EPI_ISL_2388151, EPI_ISL_2388152, EPI_ISL_2388153, EPI_ISL_2388154, EPI_ISL_2388155, EPI_ISL_2388156, EPI_ISL_2388158, EPI_ISL_2388159, EPI_ISL_2388160, EPI_ISL_2388161, EPI_ISL_2388164, EPI_ISL_2388165, EPI_ISL_2388167, EPI_ISL_2388168, EPI_ISL_2388169, EPI_ISL_2388170, EPI_ISL_2388171, EPI_ISL_2388172, EPI_ISL_2388173, EPI_ISL_2388174, EPI_ISL_2388175, EPI_ISL_2388177, EPI_ISL_2388178, EPI_ISL_2388179, EPI_ISL_2388180, EPI_ISL_2390099, EPI_ISL_2390316, EPI_ISL_2390373, EPI_ISL_2390374, EPI_ISL_2471464, EPI_ISL_2471732, EPI_ISL_2471785, EPI_ISL_2471786, EPI_ISL_2471810, EPI_ISL_2471811, EPI_ISL_2471813, EPI_ISL_2471890, EPI_ISL_2471891, EPI_ISL_2471892, EPI_ISL_2471893, EPI_ISL_2471894, EPI_ISL_2471896, EPI_ISL_2471897, EPI_ISL_2471898, EPI_ISL_2471901, EPI_ISL_2471904, EPI_ISL_2471905, EPI_ISL_2471908, EPI_ISL_2471910, EPI_ISL_2471914, EPI_ISL_2472409, EPI_ISL_2472410, EPI_ISL_2472411, EPI_ISL_2472413, EPI_ISL_2472414, EPI_ISL_2472415, EPI_ISL_2472416, EPI_ISL_2472417, EPI_ISL_2472418, EPI_ISL_2472419, EPI_ISL_2472420, EPI_ISL_2472421, EPI_ISL_2472422, EPI_ISL_2472423, EPI_ISL_2472424, EPI_ISL_2472426, EPI_ISL_2472427, EPI_ISL_2472428, EPI_ISL_2472429, EPI_ISL_2472430, EPI_ISL_2472431, EPI_ISL_2472432, EPI_ISL_2472433, EPI_ISL_2472434, EPI_ISL_2472435, EPI_ISL_2472436, EPI_ISL_2472438, EPI_ISL_2472439, EPI_ISL_2472440, EPI_ISL_2472441, EPI_ISL_2472442, EPI_ISL_2472443, EPI_ISL_2472444, EPI_ISL_2472445, EPI_ISL_2472446, EPI_ISL_2472447, EPI_ISL_2472448, EPI_ISL_2472450, EPI_ISL_2472451, EPI_ISL_2472452, EPI_ISL_2472453, EPI_ISL_2472454, EPI_ISL_2472455, EPI_ISL_2472456, EPI_ISL_2472457, EPI_ISL_2472458, EPI_ISL_2472459, EPI_ISL_2472460, EPI_ISL_2472461, EPI_ISL_2472462, EPI_ISL_2472464, EPI_ISL_2472465, EPI_ISL_2472466, EPI_ISL_2472467, EPI_ISL_2472468, EPI_ISL_2472469, EPI_ISL_2472470, EPI_ISL_2472471, EPI_ISL_2472472, EPI_ISL_2472473, EPI_ISL_2472474, EPI_ISL_2472475, EPI_ISL_2472476, EPI_ISL_2472477, EPI_ISL_2472478, EPI_ISL_2472479, EPI_ISL_2472480, EPI_ISL_2472482, EPI_ISL_2472488, EPI_ISL_2472491, EPI_ISL_2472494, EPI_ISL_2472499, EPI_ISL_2472541, EPI_ISL_2632795, EPI_ISL_2632796, EPI_ISL_2632797, EPI_ISL_2632798, EPI_ISL_2632799, EPI_ISL_2632800, EPI_ISL_2632801, EPI_ISL_2632802, EPI_ISL_2632803, EPI_ISL_2632804, EPI_ISL_2632805, EPI_ISL_2632806, EPI_ISL_2632807, EPI_ISL_2632808, EPI_ISL_2632809, EPI_ISL_2632810, EPI_ISL_2632811, EPI_ISL_2632812, EPI_ISL_2632813, EPI_ISL_2632830, EPI_ISL_2632831, EPI_ISL_2632832, EPI_ISL_2632833, EPI_ISL_2632834, EPI_ISL_2632835, EPI_ISL_2632836, EPI_ISL_2632837, EPI_ISL_2632838, EPI_ISL_2632839, EPI_ISL_2632840, EPI_ISL_2632841, EPI_ISL_2632842, EPI_ISL_2632844, EPI_ISL_2632909, EPI_ISL_2632910, EPI_ISL_2632912, EPI_ISL_2632913, EPI_ISL_2632914, EPI_ISL_2632916, EPI_ISL_2632917, EPI_ISL_2632919, EPI_ISL_2632921, EPI_ISL_2632923, EPI_ISL_2632926, EPI_ISL_2632927, EPI_ISL_2632928, EPI_ISL_2632930, EPI_ISL_2632932, EPI_ISL_2632934, EPI_ISL_2632935, EPI_ISL_2632937, EPI_ISL_2632938, EPI_ISL_2632967, EPI_ISL_2632974, EPI_ISL_2632982, EPI_ISL_2632984, EPI_ISL_2635154, EPI_ISL_2635157, EPI_ISL_2635162, EPI_ISL_2635168, EPI_ISL_2635170, EPI_ISL_2635171, EPI_ISL_2635172, EPI_ISL_2635880, EPI_ISL_2635881, EPI_ISL_2635884, EPI_ISL_2760567, EPI_ISL_2760574, EPI_ISL_2760583, EPI_ISL_2760586, EPI_ISL_2760596, EPI_ISL_2760606

see above    MVZ Labor Dr. Limbach & Kollegen GbR    Robert Koch Institute

EPI_ISL_2390011, EPI_ISL_2390013, EPI_ISL_2390024, EPI_ISL_2390025, EPI_ISL_2390030, EPI_ISL_2390036, EPI_ISL_2390037, EPI_ISL_2390042, EPI_ISL_2390044, EPI_ISL_2390045, EPI_ISL_2390050, EPI_ISL_2390065, EPI_ISL_2390067, EPI_ISL_2390073, EPI_ISL_2678499, EPI_ISL_2678500, EPI_ISL_2678501, EPI_ISL_2678502, EPI_ISL_2678503, EPI_ISL_2678505, EPI_ISL_2678506, EPI_ISL_2678507, EPI_ISL_2678509, EPI_ISL_2678510, EPI_ISL_2678511, EPI_ISL_2678512, EPI_ISL_2678514, EPI_ISL_2678516, EPI_ISL_2678518, EPI_ISL_2678519, EPI_ISL_2678524, EPI_ISL_2678528, EPI_ISL_2678529, EPI_ISL_2678534, EPI_ISL_2678538, EPI_ISL_2678541, EPI_ISL_2678543, EPI_ISL_2678550, EPI_ISL_2678555, EPI_ISL_2678556, EPI_ISL_2678557, EPI_ISL_2678559, EPI_ISL_2678563, EPI_ISL_2678564, EPI_ISL_2678565, EPI_ISL_2678567, EPI_ISL_2678568, EPI_ISL_2678569, EPI_ISL_2678570, EPI_ISL_2678571, EPI_ISL_2678572, EPI_ISL_2678574, EPI_ISL_2678578, EPI_ISL_2678583, EPI_ISL_2678588, EPI_ISL_2678589, EPI_ISL_2678601, EPI_ISL_2678605, EPI_ISL_2678608, EPI_ISL_2678618, EPI_ISL_2678620, EPI_ISL_2678624, EPI_ISL_2678626, EPI_ISL_2678627, EPI_ISL_2678628, EPI_ISL_2678632,

| | | | |
|---|---|---|---|
| EPI_ISL_2678635, EPI_ISL_2678637, EPI_ISL_2678642, EPI_ISL_2678645, EPI_ISL_2678648, EPI_ISL_2678650, EPI_ISL_2678651 | | | |
| see above | MVZ Labor Dr. Quade & Kollegen GmbH | Robert Koch Institute | |
| EPI_ISL_2314659, EPI_ISL_2386224, EPI_ISL_2386230, EPI_ISL_2386240, EPI_ISL_2390091, EPI_ISL_2390096, EPI_ISL_2390101, EPI_ISL_2390105, EPI_ISL_2471465, EPI_ISL_2472437, EPI_ISL_2472449, EPI_ISL_2472713 | | | |
| see above | MVZ Labor Dr. Reising-Ackermann und Kollegen | Robert Koch Institute | |
| EPI_ISL_2316725, EPI_ISL_2316764, EPI_ISL_2316820, EPI_ISL_2389461, EPI_ISL_2389464, EPI_ISL_2389470, EPI_ISL_2389473, EPI_ISL_2389479, EPI_ISL_2389482, EPI_ISL_2389500, EPI_ISL_2389507, EPI_ISL_2390752, EPI_ISL_2390753, EPI_ISL_2390754, EPI_ISL_2390755, EPI_ISL_2390756, EPI_ISL_2390757, EPI_ISL_2390758, EPI_ISL_2390759, EPI_ISL_2390760, EPI_ISL_2390761, EPI_ISL_2390762, EPI_ISL_2390763, EPI_ISL_2390764, EPI_ISL_2390765, EPI_ISL_2390766, EPI_ISL_2390767, EPI_ISL_2390768, EPI_ISL_2390769, EPI_ISL_2390770, EPI_ISL_2390771, EPI_ISL_2390772, EPI_ISL_2390773, EPI_ISL_2390774, EPI_ISL_2390775, EPI_ISL_2390776, EPI_ISL_2390777, EPI_ISL_2390778, EPI_ISL_2390779, EPI_ISL_2390780, EPI_ISL_2390781, EPI_ISL_2390782, EPI_ISL_2390783, EPI_ISL_2390784, EPI_ISL_2390785, EPI_ISL_2390786, EPI_ISL_2390787, EPI_ISL_2390788, EPI_ISL_2390789, EPI_ISL_2390790, EPI_ISL_2390791, EPI_ISL_2390792, EPI_ISL_2390793, EPI_ISL_2390794, EPI_ISL_2471061, EPI_ISL_2471062, EPI_ISL_2471063, EPI_ISL_2471064, EPI_ISL_2471065, EPI_ISL_2471066, EPI_ISL_2471067, EPI_ISL_2471068, EPI_ISL_2471069, EPI_ISL_2471070, EPI_ISL_2471071, EPI_ISL_2471072, EPI_ISL_2471073, EPI_ISL_2471074, EPI_ISL_2471075, EPI_ISL_2471076, EPI_ISL_2471077, EPI_ISL_2471078, EPI_ISL_2471079, EPI_ISL_2471080, EPI_ISL_2471081, EPI_ISL_2471082, EPI_ISL_2471083, EPI_ISL_2471084, EPI_ISL_2471085, EPI_ISL_2471086, EPI_ISL_2471087, EPI_ISL_2471088, EPI_ISL_2471089, EPI_ISL_2471090, EPI_ISL_2471091, EPI_ISL_2471092, EPI_ISL_2471093, EPI_ISL_2471094, EPI_ISL_2471095, EPI_ISL_2471096, EPI_ISL_2471097, EPI_ISL_2471098, EPI_ISL_2471099, EPI_ISL_2471100, EPI_ISL_2471101, EPI_ISL_2471102, EPI_ISL_2471103, EPI_ISL_2471104, EPI_ISL_2471105, EPI_ISL_2471106, EPI_ISL_2471107, EPI_ISL_2471108, EPI_ISL_2471109, EPI_ISL_2471110, EPI_ISL_2471111, EPI_ISL_2471112, EPI_ISL_2471113, EPI_ISL_2471114, EPI_ISL_2471115, EPI_ISL_2471116, EPI_ISL_2471117, EPI_ISL_2471118, EPI_ISL_2471119, EPI_ISL_2471120, EPI_ISL_2471121, EPI_ISL_2471122, EPI_ISL_2471123, EPI_ISL_2471124, EPI_ISL_2471125, EPI_ISL_2471126, EPI_ISL_2471127, EPI_ISL_2471128, EPI_ISL_2471129, EPI_ISL_2471130, EPI_ISL_2471131, EPI_ISL_2631932, EPI_ISL_2631933, EPI_ISL_2631934, EPI_ISL_2631935, EPI_ISL_2631936, EPI_ISL_2631937, EPI_ISL_2631938, EPI_ISL_2631939, EPI_ISL_2631940, EPI_ISL_2631941, EPI_ISL_2631942, EPI_ISL_2631943, EPI_ISL_2631944, EPI_ISL_2631945, EPI_ISL_2631946, EPI_ISL_2631947, EPI_ISL_2631948, EPI_ISL_2631949, EPI_ISL_2631950, EPI_ISL_2631951, EPI_ISL_2631952, EPI_ISL_2631953, EPI_ISL_2631954, EPI_ISL_2631955, EPI_ISL_2631956, EPI_ISL_2631957, EPI_ISL_2631958, EPI_ISL_2631959, EPI_ISL_2631960, EPI_ISL_2631961, EPI_ISL_2631962, EPI_ISL_2631963, EPI_ISL_2631964, EPI_ISL_2631965, EPI_ISL_2631966, EPI_ISL_2631967, EPI_ISL_2631973, EPI_ISL_2631974, EPI_ISL_2631975, EPI_ISL_2631976, EPI_ISL_2631978, EPI_ISL_2631979, EPI_ISL_2631980, EPI_ISL_2631981, EPI_ISL_2631982, EPI_ISL_2631983, EPI_ISL_2631984, EPI_ISL_2631985, EPI_ISL_2631986, EPI_ISL_2631987, EPI_ISL_2631988, EPI_ISL_2631989, EPI_ISL_2631990, EPI_ISL_2631991, EPI_ISL_2631992, EPI_ISL_2631993, EPI_ISL_2631994, EPI_ISL_2631995, EPI_ISL_2631996, EPI_ISL_2631997, EPI_ISL_2631998, EPI_ISL_2631999, EPI_ISL_2632000, EPI_ISL_2632001, EPI_ISL_2632002, EPI_ISL_2632003, EPI_ISL_2632004, EPI_ISL_2632005, EPI_ISL_2632006, EPI_ISL_2632007, EPI_ISL_2632008, EPI_ISL_2632009, EPI_ISL_2632010, EPI_ISL_2632011, EPI_ISL_2632012, EPI_ISL_2632013, EPI_ISL_2632014, EPI_ISL_2632015, EPI_ISL_2632016, EPI_ISL_2632017, EPI_ISL_2632018, EPI_ISL_2632019, EPI_ISL_2632020, EPI_ISL_2678806, EPI_ISL_2678807, EPI_ISL_2678808, EPI_ISL_2678809, EPI_ISL_2678810, EPI_ISL_2678811, EPI_ISL_2678812, EPI_ISL_2678813, EPI_ISL_2678814, EPI_ISL_2678815, EPI_ISL_2678816, EPI_ISL_2678817, EPI_ISL_2678818, EPI_ISL_2678819, EPI_ISL_2678820, EPI_ISL_2678821, EPI_ISL_2678822, EPI_ISL_2678823, EPI_ISL_2678824, EPI_ISL_2678825, EPI_ISL_2678826, EPI_ISL_2678827, EPI_ISL_2678828, EPI_ISL_2678829, EPI_ISL_2678830, EPI_ISL_2678832, EPI_ISL_2678833, EPI_ISL_2678836, EPI_ISL_2678838, EPI_ISL_2678840, EPI_ISL_2678842, EPI_ISL_2678844, EPI_ISL_2678846, EPI_ISL_2678849, EPI_ISL_2678851, EPI_ISL_2678853, EPI_ISL_2678855, EPI_ISL_2678858, EPI_ISL_2678860, EPI_ISL_2678862, EPI_ISL_2678864, EPI_ISL_2678867, EPI_ISL_2678869, EPI_ISL_2678871, EPI_ISL_2678873, EPI_ISL_2678875, EPI_ISL_2678878, EPI_ISL_2678880, EPI_ISL_2678882, EPI_ISL_2678884, EPI_ISL_2678886, EPI_ISL_2678889, EPI_ISL_2678891, EPI_ISL_2678893 | | | |
| see above | MVZ Labor Krone GbR | Robert Koch Institute | |
| EPI_ISL_2316433, EPI_ISL_2316449, EPI_ISL_2390275, EPI_ISL_2390297, EPI_ISL_2471529, EPI_ISL_2471738, EPI_ISL_2471745, EPI_ISL_2472737, EPI_ISL_2472741 | | | |
| see above | MVZ Medizinisches Labor Hannover GmbH | Robert Koch Institute | |
| EPI_ISL_2314772, EPI_ISL_2314775, EPI_ISL_2314778, EPI_ISL_2314783, EPI_ISL_2314784, EPI_ISL_2314787, EPI_ISL_2314788, EPI_ISL_2314791, EPI_ISL_2314792, EPI_ISL_2314793, EPI_ISL_2314794, EPI_ISL_2314801, EPI_ISL_2314803, EPI_ISL_2314804, EPI_ISL_2316421, EPI_ISL_2316425, EPI_ISL_2316429, EPI_ISL_2316439, EPI_ISL_2316443, EPI_ISL_2316474, EPI_ISL_2316481, EPI_ISL_2316490, EPI_ISL_2389441, EPI_ISL_2390097, EPI_ISL_2390215, EPI_ISL_2390217, EPI_ISL_2390218, EPI_ISL_2390219, EPI_ISL_2390221, EPI_ISL_2390223, EPI_ISL_2390224, EPI_ISL_2390225, EPI_ISL_2390226, EPI_ISL_2390227, EPI_ISL_2390228, EPI_ISL_2390229, EPI_ISL_2390230, EPI_ISL_2390231, EPI_ISL_2390232, EPI_ISL_2390235, EPI_ISL_2390236, EPI_ISL_2390237, EPI_ISL_2390238, EPI_ISL_2390239, EPI_ISL_2390240, EPI_ISL_2390241, EPI_ISL_2390242, EPI_ISL_2390243, EPI_ISL_2390244, EPI_ISL_2390245, EPI_ISL_2390246, EPI_ISL_2390247, EPI_ISL_2390248, EPI_ISL_2390249, EPI_ISL_2390251, EPI_ISL_2390252, EPI_ISL_2390254, EPI_ISL_2390257, EPI_ISL_2390258, EPI_ISL_2390259, EPI_ISL_2471463, EPI_ISL_2471468, EPI_ISL_2471469, EPI_ISL_2471470, EPI_ISL_2471472, EPI_ISL_2471473, EPI_ISL_2471475, EPI_ISL_2471477, EPI_ISL_2471478, EPI_ISL_2471481, EPI_ISL_2471482, EPI_ISL_2471488, EPI_ISL_2471489, EPI_ISL_2471528, EPI_ISL_2471550, EPI_ISL_2471552, EPI_ISL_2471554, EPI_ISL_2471556, EPI_ISL_2471558, EPI_ISL_2471560, EPI_ISL_2471562, EPI_ISL_2471563, EPI_ISL_2471564, EPI_ISL_2471565, EPI_ISL_2471566, EPI_ISL_2471567, EPI_ISL_2471569, EPI_ISL_2471570, EPI_ISL_2471571, EPI_ISL_2471572, EPI_ISL_2471574, EPI_ISL_2471575, EPI_ISL_2471576, EPI_ISL_2471577, EPI_ISL_2471578, EPI_ISL_2471580, EPI_ISL_2471582, EPI_ISL_2471583, EPI_ISL_2471585, EPI_ISL_2471586, EPI_ISL_2471587, EPI_ISL_2471588, EPI_ISL_2471593, EPI_ISL_2471595, EPI_ISL_2471596, EPI_ISL_2471597, EPI_ISL_2471598, EPI_ISL_2471599, EPI_ISL_2471600, EPI_ISL_2471601, EPI_ISL_2471602, EPI_ISL_2471603, EPI_ISL_2471604, EPI_ISL_2471605, EPI_ISL_2471606, EPI_ISL_2471607, EPI_ISL_2471608, EPI_ISL_2472403, EPI_ISL_2472404, EPI_ISL_2472405, EPI_ISL_2472406, EPI_ISL_2472407, EPI_ISL_2472408, EPI_ISL_2472425, EPI_ISL_2472696, EPI_ISL_2472697, EPI_ISL_2472698, EPI_ISL_2472699, EPI_ISL_2472709, EPI_ISL_2472711, EPI_ISL_2472712, EPI_ISL_2472714, EPI_ISL_2472717, EPI_ISL_2472719, EPI_ISL_2472720, EPI_ISL_2472724, EPI_ISL_2472735, EPI_ISL_2472736, EPI_ISL_2472738, EPI_ISL_2472743, EPI_ISL_2472762, EPI_ISL_2472767, EPI_ISL_2631765, EPI_ISL_2631766, EPI_ISL_2631770, EPI_ISL_2631774, EPI_ISL_2631775, EPI_ISL_2631779, EPI_ISL_2631784, EPI_ISL_2631785, EPI_ISL_2631788, EPI_ISL_2631792, EPI_ISL_2631793, EPI_ISL_2631802, EPI_ISL_2631805, EPI_ISL_2631807, EPI_ISL_2631808, EPI_ISL_2631809, EPI_ISL_2631810, EPI_ISL_2631811, EPI_ISL_2631812, EPI_ISL_2631813, EPI_ISL_2631814, EPI_ISL_2631815, EPI_ISL_2631816, EPI_ISL_2631817, EPI_ISL_2631818, EPI_ISL_2631819, EPI_ISL_2631820, EPI_ISL_2631821, EPI_ISL_2631822, EPI_ISL_2631823, EPI_ISL_2631824, EPI_ISL_2631825, EPI_ISL_2631826, EPI_ISL_2631827, EPI_ISL_2631836, EPI_ISL_2631837, EPI_ISL_2631838, EPI_ISL_2631839, EPI_ISL_2631841, EPI_ISL_2631842, EPI_ISL_2631843, EPI_ISL_2631844, EPI_ISL_2631845, EPI_ISL_2631846, EPI_ISL_2631847, EPI_ISL_2631848, EPI_ISL_2631849, EPI_ISL_2631850, EPI_ISL_2631852, EPI_ISL_2631853, EPI_ISL_2631854, EPI_ISL_2631856, EPI_ISL_2631857, EPI_ISL_2631859, EPI_ISL_2632956, EPI_ISL_2635807, EPI_ISL_2635810, EPI_ISL_2635821, EPI_ISL_2635822, EPI_ISL_2635841, EPI_ISL_2636169, EPI_ISL_2636197, EPI_ISL_2636212 | | | |
| see above | MVZ für Laboratoriumsmedizin und Mikrobiologie Koblenz-Mittelrhein (Labor Koblenz) | Robert Koch Institute | |
| EPI_ISL_2471693, EPI_ISL_2633433 | MVZ für Laboratoriumsmedizin und Mikrobiologie Würzburg | Robert Koch Institute | |
| EPI_ISL_2450267, EPI_ISL_2450268, EPI_ISL_2450269, EPI_ISL_2450270, EPI_ISL_2450271, EPI_ISL_2450272, EPI_ISL_2450273, EPI_ISL_2450274, EPI_ISL_2450275, EPI_ISL_2450276, EPI_ISL_2450277, EPI_ISL_2450321, EPI_ISL_2450322, EPI_ISL_2450323, EPI_ISL_2450324, EPI_ISL_2450325, EPI_ISL_2450327, EPI_ISL_2772662, EPI_ISL_2772663, EPI_ISL_2772678, EPI_ISL_2772679, EPI_ISL_2772681, EPI_ISL_2772685, EPI_ISL_2772692, EPI_ISL_2772693, EPI_ISL_2772694 | | | |
| see above | Max von Pettenkofer Institute, Virology, National Reference Center for Retroviruses, LMU Munich | Laboratory for Functional Genome Analysis; Dept. Genomics; Gene Center of the LMU Munich | Alexander Graf; Helmut Blum; Max Muenchhoff; Oliver Keppler; Stefan Krebs |
| EPI_ISL_2389361, EPI_ISL_2389362, EPI_ISL_2389363, EPI_ISL_2389364, EPI_ISL_2389365, EPI_ISL_2389366, EPI_ISL_2389367, EPI_ISL_2389369, EPI_ISL_2469753, EPI_ISL_2469754, EPI_ISL_2469755, EPI_ISL_2469756, EPI_ISL_2469757, EPI_ISL_2469758, EPI_ISL_2469759, EPI_ISL_2469760, EPI_ISL_2469761, EPI_ISL_2635785 | | | |
| see above | Med. Labor Prof. Schenk Dr. Ansorge & Kollegen | Robert Koch Institute | |
| EPI_ISL_2638051, EPI_ISL_2638053, EPI_ISL_2638054, EPI_ISL_2638057, EPI_ISL_2638058, EPI_ISL_2638066, EPI_ISL_2638082, EPI_ISL_2638085, EPI_ISL_2638160 | | | |
| see above | Medical Laboratories Duesseldorf | Center of Medical Microbiology, Virology, and Hospital Hygiene, University of Duesseldorf | Alexander Dilthey; Andreas Walker; Angelika Helmer; Christian Lange; Daniel Strelow; Jessica Nicolai; Jörg Timm; Klaus Pfeffer; Lisanna Hülse; Malte Kohns Vasconcelos; Maximilian Damagnez; Nadine Lübke; Tobias Wienemann; Torsten Houwaart |
| EPI_ISL_2636079, EPI_ISL_2636080, EPI_ISL_2636081, EPI_ISL_2636083, EPI_ISL_2636084, EPI_ISL_2636086, EPI_ISL_2636088, EPI_ISL_2636090, EPI_ISL_2636091, EPI_ISL_2636093, EPI_ISL_2636094, EPI_ISL_2636095, EPI_ISL_2636098, EPI_ISL_2636099, EPI_ISL_2636101, EPI_ISL_2636104, EPI_ISL_2636105, EPI_ISL_2636106, EPI_ISL_2636107, EPI_ISL_2636108, EPI_ISL_2636109, EPI_ISL_2636110, EPI_ISL_2636111, EPI_ISL_2636114, EPI_ISL_2636115, EPI_ISL_2636117, EPI_ISL_2636118, EPI_ISL_2636119, EPI_ISL_2636120, EPI_ISL_2636122, EPI_ISL_2636123, EPI_ISL_2636124, EPI_ISL_2636794, EPI_ISL_2636799, EPI_ISL_2636801, EPI_ISL_2636808, EPI_ISL_2636813, EPI_ISL_2636814, EPI_ISL_2636816, EPI_ISL_2636818, EPI_ISL_2636820, EPI_ISL_2636821, EPI_ISL_2636822, EPI_ISL_2636824, EPI_ISL_2636825, EPI_ISL_2636827, EPI_ISL_2636833, EPI_ISL_2636834, EPI_ISL_2636839, EPI_ISL_2636841, EPI_ISL_2636842, EPI_ISL_2636848, EPI_ISL_2636849, EPI_ISL_2636850, EPI_ISL_2636858, EPI_ISL_2636859, EPI_ISL_2636867, EPI_ISL_2761913 | | | |
| see above | Medizinisch-Diagnostisches Labor Kempten allgäulab | Robert Koch Institute | |
| EPI_ISL_2389081, EPI_ISL_2389083, EPI_ISL_2389086, EPI_ISL_2389090, EPI_ISL_2389091, EPI_ISL_2389093, EPI_ISL_2389100, EPI_ISL_2389101, EPI_ISL_2389103, EPI_ISL_2389110, EPI_ISL_2389113, EPI_ISL_2389114, EPI_ISL_2389121, EPI_ISL_2389122, EPI_ISL_2389123, EPI_ISL_2389124, EPI_ISL_2389125, EPI_ISL_2389127, EPI_ISL_2470817, EPI_ISL_2470818, EPI_ISL_2470819, EPI_ISL_2470820, EPI_ISL_2470821, EPI_ISL_2470822, EPI_ISL_2470823, EPI_ISL_2470824, EPI_ISL_2470825, EPI_ISL_2470826, EPI_ISL_2470827, EPI_ISL_2470828, EPI_ISL_2470829, EPI_ISL_2470830, EPI_ISL_2470831, EPI_ISL_2470832, EPI_ISL_2470833, EPI_ISL_2470834, EPI_ISL_2470835, EPI_ISL_2470836, EPI_ISL_2470837, EPI_ISL_2470838, EPI_ISL_2470839, EPI_ISL_2470840, EPI_ISL_2470841, EPI_ISL_2470842, EPI_ISL_2470843, EPI_ISL_2470844, EPI_ISL_2470845, EPI_ISL_2470846, EPI_ISL_2470847, EPI_ISL_2470848, EPI_ISL_2470849, EPI_ISL_2470850, EPI_ISL_2470851, EPI_ISL_2470852, EPI_ISL_2470853, EPI_ISL_2470854, EPI_ISL_2470855, EPI_ISL_2470856, EPI_ISL_2470857, EPI_ISL_2470858, EPI_ISL_2470859, EPI_ISL_2470860, EPI_ISL_2470861, EPI_ISL_2470862, EPI_ISL_2632177, EPI_ISL_2632178, EPI_ISL_2632179, EPI_ISL_2632180, EPI_ISL_2632181, EPI_ISL_2632182, EPI_ISL_2632183, EPI_ISL_2632184, EPI_ISL_2632185, EPI_ISL_2632186, EPI_ISL_2632187, EPI_ISL_2632188, EPI_ISL_2632189, EPI_ISL_2632190, EPI_ISL_2632191, EPI_ISL_2632192, EPI_ISL_2632193, EPI_ISL_2632194, EPI_ISL_2632195, EPI_ISL_2632196, EPI_ISL_2632197, EPI_ISL_2632198, EPI_ISL_2632199, EPI_ISL_2632200, EPI_ISL_2632201, EPI_ISL_2632202, EPI_ISL_2632203, EPI_ISL_2632204, EPI_ISL_2632205, EPI_ISL_2632206, EPI_ISL_2632207, EPI_ISL_2632208, EPI_ISL_2632209, EPI_ISL_2632210, EPI_ISL_2632211, EPI_ISL_2632212, EPI_ISL_2632213, EPI_ISL_2632214, EPI_ISL_2632215, EPI_ISL_2632216, EPI_ISL_2632217, EPI_ISL_2632218, EPI_ISL_2632219, EPI_ISL_2632220, EPI_ISL_2632221, EPI_ISL_2632222, EPI_ISL_2632223, EPI_ISL_2632224, EPI_ISL_2632225, EPI_ISL_2632226, EPI_ISL_2632227, EPI_ISL_2632228, EPI_ISL_2632229, EPI_ISL_2632230, EPI_ISL_2632231, EPI_ISL_2632232, EPI_ISL_2632233, EPI_ISL_2632234, EPI_ISL_2632235, EPI_ISL_2632236, EPI_ISL_2632237, EPI_ISL_2632238, EPI_ISL_2632239 | | | |
| see above | Medizinische Laboratorien Düsseldorf | Robert Koch Institute | |
| EPI_ISL_2386181, EPI_ISL_2386182, EPI_ISL_2386183, EPI_ISL_2386184, EPI_ISL_2386185, EPI_ISL_2386186, EPI_ISL_2386187, EPI_ISL_2386190, EPI_ISL_2386191, EPI_ISL_2386192, EPI_ISL_2386193, EPI_ISL_2386194, EPI_ISL_2386195, EPI_ISL_2390108, EPI_ISL_2390303, EPI_ISL_2390305, EPI_ISL_2390308, EPI_ISL_2390309, EPI_ISL_2390310, EPI_ISL_2390311, EPI_ISL_2390313, EPI_ISL_2390319, EPI_ISL_2390326, EPI_ISL_2390332, EPI_ISL_2390333, EPI_ISL_2390337, EPI_ISL_2390338, EPI_ISL_2390341, EPI_ISL_2390343, EPI_ISL_2390344, EPI_ISL_2390345, EPI_ISL_2390346, EPI_ISL_2390347, EPI_ISL_2390352, EPI_ISL_2390353, EPI_ISL_2390354, EPI_ISL_2390357, EPI_ISL_2390362, EPI_ISL_2390370, EPI_ISL_2471749, EPI_ISL_2471843, EPI_ISL_2471845, EPI_ISL_2471854, EPI_ISL_2471856, EPI_ISL_2471857, EPI_ISL_2471859, EPI_ISL_2471862, EPI_ISL_2471873, EPI_ISL_2471879, EPI_ISL_2471881, EPI_ISL_2471888, EPI_ISL_2471889, EPI_ISL_2632939, EPI_ISL_2632943, EPI_ISL_2632946, EPI_ISL_2632947, EPI_ISL_2632948, EPI_ISL_2632950, EPI_ISL_2632951, EPI_ISL_2632954, EPI_ISL_2632958, EPI_ISL_2632962 | | | |
| see above | Medizinisches Labor Ostsachsen MVZ GbR Nebenbetriebsstätte Dresden | Robert Koch Institute | |
| EPI_ISL_2387348, EPI_ISL_2387351, EPI_ISL_2387353, EPI_ISL_2387382, EPI_ISL_2387418, EPI_ISL_2388223, EPI_ISL_2388225, EPI_ISL_2388308, EPI_ISL_2469850, EPI_ISL_2469860, EPI_ISL_2469878, EPI_ISL_2469886, EPI_ISL_2469914, EPI_ISL_2632026, EPI_ISL_2632089, EPI_ISL_2632102, EPI_ISL_2632103 | | | |
| see above | Medizinisches Labor Wahl Lüdenscheid | Robert Koch Institute | |
| EPI_ISL_2386426, EPI_ISL_2632021, EPI_ISL_2632022, EPI_ISL_2632114, EPI_ISL_2632115, EPI_ISL_2632116, EPI_ISL_2632117, EPI_ISL_2632118, EPI_ISL_2636732, EPI_ISL_2636733, EPI_ISL_2636734, EPI_ISL_2636735 | | | |
| see above | Medizinisches Zentrallabor Altenburg GmbH & Co.KG | Robert Koch Institute | |
| EPI_ISL_2471036, EPI_ISL_2471037, EPI_ISL_2471038, EPI_ISL_2471039, EPI_ISL_2471040, EPI_ISL_2632330, EPI_ISL_2632331, EPI_ISL_2632332, EPI_ISL_2632333, EPI_ISL_2632334, EPI_ISL_2632814, EPI_ISL_2632815, EPI_ISL_2632816, EPI_ISL_2632817, EPI_ISL_2632818, EPI_ISL_2632819, EPI_ISL_2632820, EPI_ISL_2632822, EPI_ISL_2632823, EPI_ISL_2635801, EPI_ISL_2635802, EPI_ISL_2636736 | | | |
| see above | Niedersächsisches Landesgesundheitsamt (NLGA) | Robert Koch Institute | |
| EPI_ISL_2387488, EPI_ISL_2389639, EPI_ISL_2389656, EPI_ISL_2389821, EPI_ISL_2389824, EPI_ISL_2469541, EPI_ISL_2469545, EPI_ISL_2472222, EPI_ISL_2631595 | | | |
| see above | Praxisgemeinschaft für Laboratoriumsmedizin Labor Blumenstraße; Praxis Dr. E. Wietschel & Dr. F. Wietschel | Robert Koch Institute | |
| EPI_ISL_2316187, EPI_ISL_2387463, EPI_ISL_2387469, EPI_ISL_2387476, EPI_ISL_2387591, EPI_ISL_2389754, EPI_ISL_2389760, EPI_ISL_2389765, EPI_ISL_2389841, EPI_ISL_2472236 | | | |
| see above | Praxisgemeinschaft für Laboratoriumsmedizin Labor Blumenstraße; Praxis Dr. Sel und Dr. Späte | Robert Koch Institute | |
| EPI_ISL_2471702, EPI_ISL_2471711, EPI_ISL_2471722 | Procomcure Biotech Germany GmbH | Robert Koch Institute | |
| EPI_ISL_2313339, EPI_ISL_2313340, EPI_ISL_2313341, EPI_ISL_2313342, EPI_ISL_2313343, EPI_ISL_2313344, EPI_ISL_2313345, EPI_ISL_2313346, EPI_ISL_2313347, EPI_ISL_2313348, EPI_ISL_2389371, EPI_ISL_2389372, EPI_ISL_2389373, EPI_ISL_2389374, EPI_ISL_2389375, EPI_ISL_2389376, EPI_ISL_2389377 | | | |
| see above | Robert Koch-Institut ZBS1 | Robert Koch Institute | Annika Brinkmann |
| EPI_ISL_2470046, EPI_ISL_2633988, EPI_ISL_2634239, EPI_ISL_2636915 | Robert-Bosch-Krankenhaus | Robert Koch Institute | |
| EPI_ISL_2390107, EPI_ISL_2390301, EPI_ISL_2390304, EPI_ISL_2390315, EPI_ISL_2471900 | SLK-Kliniken Heilbronn GmbH - Institut für Laboratoriumsmedizin | Robert Koch Institute | |
| EPI_ISL_2316071, EPI_ISL_2316084, EPI_ISL_2316204, EPI_ISL_2316254, EPI_ISL_2389552, EPI_ISL_2389674, EPI_ISL_2389785, EPI_ISL_2469634, EPI_ISL_2469637, EPI_ISL_2469669, EPI_ISL_2631548, EPI_ISL_2633095, EPI_ISL_2633111, EPI_ISL_2633117, EPI_ISL_2633180, EPI_ISL_2633182, EPI_ISL_2633185, EPI_ISL_2633192, EPI_ISL_2633213, EPI_ISL_2633273, EPI_ISL_2633313 | | | |
| see above | SYNLAB Jena Oncoscreen | Robert Koch Institute | |
| EPI_ISL_2386319, EPI_ISL_2389582, EPI_ISL_2389758, EPI_ISL_2469631, EPI_ISL_2469685, EPI_ISL_2469702, EPI_ISL_2469705, EPI_ISL_2469732, EPI_ISL_2472365, EPI_ISL_2631643 | | | |
| see above | SYNLAB MVZ Berlin | Robert Koch Institute | |
| EPI_ISL_2389787, EPI_ISL_2389791, EPI_ISL_2469703 | SYNLAB MVZ Hamburg | Robert Koch Institute | |
| EPI_ISL_2387473, EPI_ISL_2387499, EPI_ISL_2387504, EPI_ISL_2387516, EPI_ISL_2387520, EPI_ISL_2387534, EPI_ISL_2387535, EPI_ISL_2387570, EPI_ISL_2387578, EPI_ISL_2387593, EPI_ISL_2387595, EPI_ISL_2387602, EPI_ISL_2387629, EPI_ISL_2387732, EPI_ISL_2387776, EPI_ISL_2387787, EPI_ISL_2387816, EPI_ISL_2387822, EPI_ISL_2387829, EPI_ISL_2387830, EPI_ISL_2387840, EPI_ISL_2387843, EPI_ISL_2387848, EPI_ISL_2387851, EPI_ISL_2387864, EPI_ISL_2387907, EPI_ISL_2387929, EPI_ISL_2387937, EPI_ISL_2387945, EPI_ISL_2387965, EPI_ISL_2388000, EPI_ISL_2389514, EPI_ISL_2389540, EPI_ISL_2389554, EPI_ISL_2389566, EPI_ISL_2389587, EPI_ISL_2389668, EPI_ISL_2389697, EPI_ISL_2389721, EPI_ISL_2389733, EPI_ISL_2389757, EPI_ISL_2389771, EPI_ISL_2389772, EPI_ISL_2389779, EPI_ISL_2389783, EPI_ISL_2389784, EPI_ISL_2389794, EPI_ISL_2389799, EPI_ISL_2389805, EPI_ISL_2389846, EPI_ISL_2389848, EPI_ISL_2472283, EPI_ISL_2633301 | | | |
| see above | SYNLAB MVZ Heidelberg | Robert Koch Institute | |
| EPI_ISL_2314627, EPI_ISL_2316039, EPI_ISL_2316042, EPI_ISL_2316051, EPI_ISL_2316091, EPI_ISL_2316110, EPI_ISL_2316194, EPI_ISL_2316218, EPI_ISL_2316221, EPI_ISL_2316230, EPI_ISL_2316242, EPI_ISL_2316265, EPI_ISL_2316274, EPI_ISL_2316299, EPI_ISL_2316312, EPI_ISL_2316331, EPI_ISL_2386309, EPI_ISL_2386314, EPI_ISL_2386322, EPI_ISL_2386323, EPI_ISL_2386343, EPI_ISL_2386363, EPI_ISL_2386380, EPI_ISL_2386387, EPI_ISL_2386409, EPI_ISL_2386411, EPI_ISL_2386413, EPI_ISL_2386445, EPI_ISL_2386446, EPI_ISL_2386463, EPI_ISL_2386471, EPI_ISL_2386478, EPI_ISL_2386481, EPI_ISL_2386484, EPI_ISL_2386511, EPI_ISL_2386514, EPI_ISL_2386525, EPI_ISL_2386531, EPI_ISL_2386537, EPI_ISL_2387454, EPI_ISL_2387459, EPI_ISL_2387462, EPI_ISL_2387467, EPI_ISL_2387468, EPI_ISL_2387470, EPI_ISL_2387471, EPI_ISL_2387478, EPI_ISL_2387479, EPI_ISL_2387480, EPI_ISL_2387481, EPI_ISL_2387483, EPI_ISL_2387484, EPI_ISL_2387486, EPI_ISL_2387487, EPI_ISL_2387489, EPI_ISL_2387491, EPI_ISL_2387493, EPI_ISL_2387495, EPI_ISL_2387496, EPI_ISL_2387497, EPI_ISL_2387498, EPI_ISL_2387500, EPI_ISL_2387501, EPI_ISL_2387502, EPI_ISL_2387503, EPI_ISL_2387506, EPI_ISL_2387510, EPI_ISL_2387513, EPI_ISL_2387514, EPI_ISL_2387515, EPI_ISL_2387522, EPI_ISL_2387525, EPI_ISL_2387526, EPI_ISL_2387529, EPI_ISL_2387531, EPI_ISL_2387532, EPI_ISL_2387533, EPI_ISL_2387536, EPI_ISL_2387538, EPI_ISL_2387539, EPI_ISL_2387542, EPI_ISL_2387543, EPI_ISL_2387544, EPI_ISL_2387546, EPI_ISL_2387548, EPI_ISL_2387551, EPI_ISL_2387555, EPI_ISL_2387556, EPI_ISL_2387557, EPI_ISL_2387558, EPI_ISL_2387559, EPI_ISL_2387560, EPI_ISL_2387562, EPI_ISL_2387564, EPI_ISL_2387565, EPI_ISL_2387567, EPI_ISL_2387568, EPI_ISL_2387571, EPI_ISL_2387572, EPI_ISL_2387573, EPI_ISL_2387574, EPI_ISL_2387575, EPI_ISL_2387579, EPI_ISL_2387583, EPI_ISL_2387584, EPI_ISL_2387586, EPI_ISL_2387587, EPI_ISL_2387589, EPI_ISL_2387590, EPI_ISL_2387592, EPI_ISL_2387597, EPI_ISL_2387599, EPI_ISL_2387600, EPI_ISL_2387601, EPI_ISL_2387603, EPI_ISL_2387609, EPI_ISL_2387610, EPI_ISL_2387613, EPI_ISL_2387618, EPI_ISL_2387621, EPI_ISL_2387622, EPI_ISL_2387623, EPI_ISL_2387624, EPI_ISL_2387626, EPI_ISL_2387627, EPI_ISL_2387628, EPI_ISL_2387631, EPI_ISL_2387636, EPI_ISL_2387639, EPI_ISL_2387641, EPI_ISL_2387647, EPI_ISL_2387649, EPI_ISL_2387651, EPI_ISL_2387657, EPI_ISL_2387661, EPI_ISL_2387662, EPI_ISL_2387664, EPI_ISL_2387665, EPI_ISL_2387666, EPI_ISL_2387668, EPI_ISL_2387670, EPI_ISL_2387672, EPI_ISL_2387673, EPI_ISL_2387674, EPI_ISL_2387675, EPI_ISL_2387676, EPI_ISL_2387677, EPI_ISL_2387680, EPI_ISL_2387685, EPI_ISL_2387686, EPI_ISL_2387688, EPI_ISL_2387690, EPI_ISL_2387694, EPI_ISL_2387695, EPI_ISL_2387698, EPI_ISL_2387700, EPI_ISL_2387703, EPI_ISL_2387704, EPI_ISL_2387705, EPI_ISL_2387706, EPI_ISL_2387708, EPI_ISL_2387709, EPI_ISL_2387710, EPI_ISL_2387712, EPI_ISL_2387715, EPI_ISL_2387716, EPI_ISL_2387717, EPI_ISL_2387718, EPI_ISL_2387720, EPI_ISL_2387721, EPI_ISL_2387722, EPI_ISL_2387725, EPI_ISL_2387726, EPI_ISL_2387731, EPI_ISL_2387733, EPI_ISL_2387735, | | | |

EPI_ISL_2387737, EPI_ISL_2387738, EPI_ISL_2387741, EPI_ISL_2387744, EPI_ISL_2387745, EPI_ISL_2387746, EPI_ISL_2387747, EPI_ISL_2387748, EPI_ISL_2387751, EPI_ISL_2387754, EPI_ISL_2387756, EPI_ISL_2387758, EPI_ISL_2387761, EPI_ISL_2387763, EPI_ISL_2387764, EPI_ISL_2387765, EPI_ISL_2387766, EPI_ISL_2387769, EPI_ISL_2387774, EPI_ISL_2387775, EPI_ISL_2387778, EPI_ISL_2387780, EPI_ISL_2387782, EPI_ISL_2387783, EPI_ISL_2387784, EPI_ISL_2387785, EPI_ISL_2387786, EPI_ISL_2387788, EPI_ISL_2387789, EPI_ISL_2387790, EPI_ISL_2387791, EPI_ISL_2387794, EPI_ISL_2387796, EPI_ISL_2387797, EPI_ISL_2387800, EPI_ISL_2387802, EPI_ISL_2387804, EPI_ISL_2387807, EPI_ISL_2387809, EPI_ISL_2387810, EPI_ISL_2387813, EPI_ISL_2387815, EPI_ISL_2387817, EPI_ISL_2387818, EPI_ISL_2387820, EPI_ISL_2387821, EPI_ISL_2387824, EPI_ISL_2387827, EPI_ISL_2387832, EPI_ISL_2387834, EPI_ISL_2387835, EPI_ISL_2387836, EPI_ISL_2387837, EPI_ISL_2387838, EPI_ISL_2387842, EPI_ISL_2387844, EPI_ISL_2387845, EPI_ISL_2387846, EPI_ISL_2387849, EPI_ISL_2387852, EPI_ISL_2387855, EPI_ISL_2387856, EPI_ISL_2387858, EPI_ISL_2387859, EPI_ISL_2387862, EPI_ISL_2387868, EPI_ISL_2387869, EPI_ISL_2387874, EPI_ISL_2387875, EPI_ISL_2387877, EPI_ISL_2387879, EPI_ISL_2387880, EPI_ISL_2387883, EPI_ISL_2387884, EPI_ISL_2387886, EPI_ISL_2387887, EPI_ISL_2387888, EPI_ISL_2387889, EPI_ISL_2387890, EPI_ISL_2387891, EPI_ISL_2387892, EPI_ISL_2387895, EPI_ISL_2387896, EPI_ISL_2387899, EPI_ISL_2387900, EPI_ISL_2387905, EPI_ISL_2387906, EPI_ISL_2387908, EPI_ISL_2387910, EPI_ISL_2387911, EPI_ISL_2387912, EPI_ISL_2387913, EPI_ISL_2387916, EPI_ISL_2387917, EPI_ISL_2387918, EPI_ISL_2387919, EPI_ISL_2387921, EPI_ISL_2387922, EPI_ISL_2387925, EPI_ISL_2387926, EPI_ISL_2387927, EPI_ISL_2387928, EPI_ISL_2387930, EPI_ISL_2387931, EPI_ISL_2387932, EPI_ISL_2387933, EPI_ISL_2387934, EPI_ISL_2387938, EPI_ISL_2387939, EPI_ISL_2387940, EPI_ISL_2387941, EPI_ISL_2387943, EPI_ISL_2387944, EPI_ISL_2387946, EPI_ISL_2387947, EPI_ISL_2387950, EPI_ISL_2387951, EPI_ISL_2387952, EPI_ISL_2387953, EPI_ISL_2387955, EPI_ISL_2387956, EPI_ISL_2387957, EPI_ISL_2387958, EPI_ISL_2387959, EPI_ISL_2387960, EPI_ISL_2387964, EPI_ISL_2387966, EPI_ISL_2387969, EPI_ISL_2387971, EPI_ISL_2387973, EPI_ISL_2387974, EPI_ISL_2387975, EPI_ISL_2387977, EPI_ISL_2387979, EPI_ISL_2387983, EPI_ISL_2387984, EPI_ISL_2387986, EPI_ISL_2387988, EPI_ISL_2387989, EPI_ISL_2387996, EPI_ISL_2387997, EPI_ISL_2388001, EPI_ISL_2388002, EPI_ISL_2388003, EPI_ISL_2388004, EPI_ISL_2388005, EPI_ISL_2388006, EPI_ISL_2388008, EPI_ISL_2388010, EPI_ISL_2389513, EPI_ISL_2389516, EPI_ISL_2389519, EPI_ISL_2389524, EPI_ISL_2389525, EPI_ISL_2389528, EPI_ISL_2389530, EPI_ISL_2389531, EPI_ISL_2389532, EPI_ISL_2389533, EPI_ISL_2389534, EPI_ISL_2389535, EPI_ISL_2389538, EPI_ISL_2389539, EPI_ISL_2389541, EPI_ISL_2389542, EPI_ISL_2389543, EPI_ISL_2389544, EPI_ISL_2389546, EPI_ISL_2389547, EPI_ISL_2389548, EPI_ISL_2389549, EPI_ISL_2389550, EPI_ISL_2389551, EPI_ISL_2389553, EPI_ISL_2389555, EPI_ISL_2389558, EPI_ISL_2389559, EPI_ISL_2389561, EPI_ISL_2389567, EPI_ISL_2389568, EPI_ISL_2389569, EPI_ISL_2389574, EPI_ISL_2389577, EPI_ISL_2389585, EPI_ISL_2389586, EPI_ISL_2389590, EPI_ISL_2389591, EPI_ISL_2389595, EPI_ISL_2389599, EPI_ISL_2389601, EPI_ISL_2389602, EPI_ISL_2389604, EPI_ISL_2389607, EPI_ISL_2389608, EPI_ISL_2389612, EPI_ISL_2389614, EPI_ISL_2389615, EPI_ISL_2389616, EPI_ISL_2389633, EPI_ISL_2389635, EPI_ISL_2389637, EPI_ISL_2389642, EPI_ISL_2389644, EPI_ISL_2389646, EPI_ISL_2389652, EPI_ISL_2389658, EPI_ISL_2389660, EPI_ISL_2389663, EPI_ISL_2389665, EPI_ISL_2389666, EPI_ISL_2389667, EPI_ISL_2389671, EPI_ISL_2389677, EPI_ISL_2389680, EPI_ISL_2389682, EPI_ISL_2389684, EPI_ISL_2389687, EPI_ISL_2389691, EPI_ISL_2389692, EPI_ISL_2389693, EPI_ISL_2389696, EPI_ISL_2389701, EPI_ISL_2389705, EPI_ISL_2389706, EPI_ISL_2389707, EPI_ISL_2389709, EPI_ISL_2389710, EPI_ISL_2389711, EPI_ISL_2389716, EPI_ISL_2389727, EPI_ISL_2389728, EPI_ISL_2389731, EPI_ISL_2389732, EPI_ISL_2389737, EPI_ISL_2389741, EPI_ISL_2389745, EPI_ISL_2389747, EPI_ISL_2389749, EPI_ISL_2389752, EPI_ISL_2389761, EPI_ISL_2389764, EPI_ISL_2389766, EPI_ISL_2389767, EPI_ISL_2389770, EPI_ISL_2389775, EPI_ISL_2389776, EPI_ISL_2389777, EPI_ISL_2389780, EPI_ISL_2389781, EPI_ISL_2389782, EPI_ISL_2389788, EPI_ISL_2389789, EPI_ISL_2389796, EPI_ISL_2389797, EPI_ISL_2389798, EPI_ISL_2389800, EPI_ISL_2389802, EPI_ISL_2389803, EPI_ISL_2389804, EPI_ISL_2389806, EPI_ISL_2389815, EPI_ISL_2389816, EPI_ISL_2389819, EPI_ISL_2389822, EPI_ISL_2389825, EPI_ISL_2389826, EPI_ISL_2389830, EPI_ISL_2389832, EPI_ISL_2389833, EPI_ISL_2389834, EPI_ISL_2389835, EPI_ISL_2389840, EPI_ISL_2389842, EPI_ISL_2389843, EPI_ISL_2389844, EPI_ISL_2389850, EPI_ISL_2469472, EPI_ISL_2469473, EPI_ISL_2469475, EPI_ISL_2469478, EPI_ISL_2469479, EPI_ISL_2469480, EPI_ISL_2469485, EPI_ISL_2469486, EPI_ISL_2469487, EPI_ISL_2469489, EPI_ISL_2469491, EPI_ISL_2469493, EPI_ISL_2469494, EPI_ISL_2469497, EPI_ISL_2469501, EPI_ISL_2469505, EPI_ISL_2469506, EPI_ISL_2469507, EPI_ISL_2469511, EPI_ISL_2469512, EPI_ISL_2469515, EPI_ISL_2469516, EPI_ISL_2469519, EPI_ISL_2469521, EPI_ISL_2469527, EPI_ISL_2469528, EPI_ISL_2469529, EPI_ISL_2469530, EPI_ISL_2469532, EPI_ISL_2469533, EPI_ISL_2469534, EPI_ISL_2469539, EPI_ISL_2469540, EPI_ISL_2469542, EPI_ISL_2469543, EPI_ISL_2469544, EPI_ISL_2469549, EPI_ISL_2469550, EPI_ISL_2469554, EPI_ISL_2469556, EPI_ISL_2469557, EPI_ISL_2469559, EPI_ISL_2469561, EPI_ISL_2469562, EPI_ISL_2469563, EPI_ISL_2469564, EPI_ISL_2469565, EPI_ISL_2469566, EPI_ISL_2469567, EPI_ISL_2469568, EPI_ISL_2469569, EPI_ISL_2469570, EPI_ISL_2469571, EPI_ISL_2469572, EPI_ISL_2469573, EPI_ISL_2469576, EPI_ISL_2469579, EPI_ISL_2469583, EPI_ISL_2469584, EPI_ISL_2469588, EPI_ISL_2469589, EPI_ISL_2469590, EPI_ISL_2469591, EPI_ISL_2469592, EPI_ISL_2469593, EPI_ISL_2469595, EPI_ISL_2469596, EPI_ISL_2469600, EPI_ISL_2469602, EPI_ISL_2469604, EPI_ISL_2469605, EPI_ISL_2469609, EPI_ISL_2469610, EPI_ISL_2469613, EPI_ISL_2469616, EPI_ISL_2469617, EPI_ISL_2469618, EPI_ISL_2469622, EPI_ISL_2469623, EPI_ISL_2469625, EPI_ISL_2469626, EPI_ISL_2469627, EPI_ISL_2469635, EPI_ISL_2469636, EPI_ISL_2469641, EPI_ISL_2469643, EPI_ISL_2469648, EPI_ISL_2469655, EPI_ISL_2469658, EPI_ISL_2469659, EPI_ISL_2469663, EPI_ISL_2469664, EPI_ISL_2469668, EPI_ISL_2469670, EPI_ISL_2469672, EPI_ISL_2469674, EPI_ISL_2469676, EPI_ISL_2469677, EPI_ISL_2469681, EPI_ISL_2469691, EPI_ISL_2469692, EPI_ISL_2469694, EPI_ISL_2469695, EPI_ISL_2469696, EPI_ISL_2469697, EPI_ISL_2469699, EPI_ISL_2469701, EPI_ISL_2469714, EPI_ISL_2469715, EPI_ISL_2469717, EPI_ISL_2469719, EPI_ISL_2469721, EPI_ISL_2469724, EPI_ISL_2469728, EPI_ISL_2469734, EPI_ISL_2469735, EPI_ISL_2469737, EPI_ISL_2469741, EPI_ISL_2469744, EPI_ISL_2469747, EPI_ISL_2472042, EPI_ISL_2472047, EPI_ISL_2472052, EPI_ISL_2472055, EPI_ISL_2472065, EPI_ISL_2472069, EPI_ISL_2472106, EPI_ISL_2472113, EPI_ISL_2472121, EPI_ISL_2472150, EPI_ISL_2472154, EPI_ISL_2472161, EPI_ISL_2472165, EPI_ISL_2472167, EPI_ISL_2472170, EPI_ISL_2472181, EPI_ISL_2472183, EPI_ISL_2472187, EPI_ISL_2472189, EPI_ISL_2472194, EPI_ISL_2472196, EPI_ISL_2472197, EPI_ISL_2472202, EPI_ISL_2472206, EPI_ISL_2472209, EPI_ISL_2472210, EPI_ISL_2472213, EPI_ISL_2472216, EPI_ISL_2472218, EPI_ISL_2472221, EPI_ISL_2472228, EPI_ISL_2472229, EPI_ISL_2472232, EPI_ISL_2472241, EPI_ISL_2472242, EPI_ISL_2472244, EPI_ISL_2472246, EPI_ISL_2472251, EPI_ISL_2472256, EPI_ISL_2472257, EPI_ISL_2472260, EPI_ISL_2472261, EPI_ISL_2472263, EPI_ISL_2472270, EPI_ISL_2472272, EPI_ISL_2472279, EPI_ISL_2472281, EPI_ISL_2472287, EPI_ISL_2472289, EPI_ISL_2472294, EPI_ISL_2472295, EPI_ISL_2472296, EPI_ISL_2472299, EPI_ISL_2472304, EPI_ISL_2472310, EPI_ISL_2472311, EPI_ISL_2472314, EPI_ISL_2472318, EPI_ISL_2472320, EPI_ISL_2472322, EPI_ISL_2472323, EPI_ISL_2472324, EPI_ISL_2472326, EPI_ISL_2472328, EPI_ISL_2472329, EPI_ISL_2472331, EPI_ISL_2472332, EPI_ISL_2472337, EPI_ISL_2472341, EPI_ISL_2472342, EPI_ISL_2472343, EPI_ISL_2472344, EPI_ISL_2472347, EPI_ISL_2472348, EPI_ISL_2472352, EPI_ISL_2472353, EPI_ISL_2472360, EPI_ISL_2631545, EPI_ISL_2631549, EPI_ISL_2631553, EPI_ISL_2631554, EPI_ISL_2631555, EPI_ISL_2631557, EPI_ISL_2631558, EPI_ISL_2631559, EPI_ISL_2631566, EPI_ISL_2631568, EPI_ISL_2631572, EPI_ISL_2631578, EPI_ISL_2631582, EPI_ISL_2631585, EPI_ISL_2631586, EPI_ISL_2631588, EPI_ISL_2631592, EPI_ISL_2631594, EPI_ISL_2631604, EPI_ISL_2631605, EPI_ISL_2631616, EPI_ISL_2631617, EPI_ISL_2631620, EPI_ISL_2631624, EPI_ISL_2631626, EPI_ISL_2631631, EPI_ISL_2631632, EPI_ISL_2631635, EPI_ISL_2631637, EPI_ISL_2631638, EPI_ISL_2631655, EPI_ISL_2631669, EPI_ISL_2631672, EPI_ISL_2631682, EPI_ISL_2631684, EPI_ISL_2631686, EPI_ISL_2631687, EPI_ISL_2631693, EPI_ISL_2631694, EPI_ISL_2631695, EPI_ISL_2631702, EPI_ISL_2631709, EPI_ISL_2631712, EPI_ISL_2631715, EPI_ISL_2631718, EPI_ISL_2631720, EPI_ISL_2631723, EPI_ISL_2631724, EPI_ISL_2631727, EPI_ISL_2631729, EPI_ISL_2631730, EPI_ISL_2631732, EPI_ISL_2631737, EPI_ISL_2631741, EPI_ISL_2631744, EPI_ISL_2631749, EPI_ISL_2631753, EPI_ISL_2631757, EPI_ISL_2631759, EPI_ISL_2631761, EPI_ISL_2631763, EPI_ISL_2633040, EPI_ISL_2633058, EPI_ISL_2633205

see above SYNLAB MVZ Leinfelden-Echterdingen Robert Koch Institute

EPI_ISL_2267599, EPI_ISL_2267631, EPI_ISL_2267647, EPI_ISL_2267681, EPI_ISL_2267684, EPI_ISL_2267725, EPI_ISL_2267726, EPI_ISL_2267733, EPI_ISL_2267735, EPI_ISL_2267743, EPI_ISL_2267772, EPI_ISL_2267776, EPI_ISL_2267787, EPI_ISL_2267788, EPI_ISL_2267789, EPI_ISL_2267790, EPI_ISL_2267791, EPI_ISL_2267792, EPI_ISL_2267808, EPI_ISL_2267826, EPI_ISL_2267831, EPI_ISL_2267839, EPI_ISL_2267862, EPI_ISL_2267864, EPI_ISL_2267866, EPI_ISL_2267872, EPI_ISL_2267889, EPI_ISL_2267891, EPI_ISL_2267899, EPI_ISL_2315117, EPI_ISL_2315118, EPI_ISL_2315119, EPI_ISL_2315120, EPI_ISL_2315121, EPI_ISL_2315122, EPI_ISL_2315123, EPI_ISL_2315124, EPI_ISL_2315125, EPI_ISL_2315126, EPI_ISL_2315127, EPI_ISL_2315128, EPI_ISL_2315129, EPI_ISL_2315130, EPI_ISL_2315131, EPI_ISL_2315132, EPI_ISL_2315133, EPI_ISL_2315134, EPI_ISL_2315135, EPI_ISL_2315136, EPI_ISL_2315137, EPI_ISL_2315138, EPI_ISL_2315139, EPI_ISL_2315140, EPI_ISL_2315141, EPI_ISL_2315142, EPI_ISL_2315143, EPI_ISL_2315144, EPI_ISL_2315145, EPI_ISL_2315146, EPI_ISL_2315147, EPI_ISL_2315148, EPI_ISL_2315149, EPI_ISL_2315150, EPI_ISL_2315151, EPI_ISL_2315152, EPI_ISL_2315153, EPI_ISL_2315154, EPI_ISL_2315155, EPI_ISL_2315156, EPI_ISL_2315157, EPI_ISL_2315158, EPI_ISL_2315159, EPI_ISL_2315160, EPI_ISL_2315161, EPI_ISL_2315162, EPI_ISL_2315163, EPI_ISL_2315164, EPI_ISL_2315165, EPI_ISL_2315166, EPI_ISL_2315167, EPI_ISL_2315168, EPI_ISL_2315169, EPI_ISL_2315170, EPI_ISL_2315171, EPI_ISL_2315172, EPI_ISL_2315173, EPI_ISL_2315174, EPI_ISL_2315175, EPI_ISL_2315176, EPI_ISL_2315177, EPI_ISL_2315178, EPI_ISL_2315179, EPI_ISL_2315180, EPI_ISL_2315181, EPI_ISL_2315182, EPI_ISL_2315183, EPI_ISL_2315184, EPI_ISL_2315185, EPI_ISL_2315186, EPI_ISL_2315187, EPI_ISL_2315188, EPI_ISL_2315189, EPI_ISL_2315190, EPI_ISL_2315191, EPI_ISL_2315192, EPI_ISL_2315193, EPI_ISL_2315194, EPI_ISL_2315195, EPI_ISL_2315196, EPI_ISL_2315197, EPI_ISL_2315198, EPI_ISL_2315199, EPI_ISL_2315200, EPI_ISL_2315201, EPI_ISL_2315202, EPI_ISL_2315203, EPI_ISL_2315204, EPI_ISL_2315205, EPI_ISL_2315206, EPI_ISL_2315207, EPI_ISL_2315208, EPI_ISL_2315209, EPI_ISL_2315210, EPI_ISL_2315211, EPI_ISL_2315212, EPI_ISL_2315213, EPI_ISL_2315214, EPI_ISL_2315215, EPI_ISL_2315216, EPI_ISL_2315218, EPI_ISL_2315219, EPI_ISL_2315220, EPI_ISL_2315221, EPI_ISL_2315222, EPI_ISL_2315223, EPI_ISL_2315224, EPI_ISL_2315225, EPI_ISL_2315226, EPI_ISL_2315227, EPI_ISL_2315228, EPI_ISL_2315229, EPI_ISL_2315230, EPI_ISL_2315232, EPI_ISL_2315233, EPI_ISL_2315234, EPI_ISL_2315235, EPI_ISL_2315236, EPI_ISL_2315237, EPI_ISL_2315238, EPI_ISL_2315239, EPI_ISL_2315240, EPI_ISL_2315241, EPI_ISL_2315242, EPI_ISL_2315243, EPI_ISL_2315244, EPI_ISL_2315245, EPI_ISL_2315246, EPI_ISL_2315247, EPI_ISL_2315248, EPI_ISL_2315249, EPI_ISL_2315250, EPI_ISL_2315251, EPI_ISL_2315252, EPI_ISL_2315253, EPI_ISL_2315254, EPI_ISL_2315255, EPI_ISL_2315256, EPI_ISL_2315257, EPI_ISL_2315258, EPI_ISL_2315259, EPI_ISL_2315260, EPI_ISL_2315261, EPI_ISL_2315262, EPI_ISL_2315263, EPI_ISL_2315264, EPI_ISL_2315265, EPI_ISL_2315266, EPI_ISL_2315267, EPI_ISL_2315268, EPI_ISL_2315269, EPI_ISL_2315270, EPI_ISL_2315271, EPI_ISL_2315272, EPI_ISL_2315273, EPI_ISL_2315274, EPI_ISL_2315275, EPI_ISL_2315276, EPI_ISL_2315277, EPI_ISL_2315278, EPI_ISL_2315279, EPI_ISL_2315280, EPI_ISL_2315281, EPI_ISL_2315282, EPI_ISL_2315283, EPI_ISL_2315284, EPI_ISL_2315286, EPI_ISL_2315287, EPI_ISL_2315288, EPI_ISL_2315289, EPI_ISL_2315290, EPI_ISL_2315291, EPI_ISL_2315292, EPI_ISL_2315293, EPI_ISL_2315294, EPI_ISL_2315295, EPI_ISL_2315296, EPI_ISL_2315297, EPI_ISL_2315298, EPI_ISL_2315299, EPI_ISL_2315300, EPI_ISL_2315301, EPI_ISL_2315302, EPI_ISL_2315303, EPI_ISL_2315304, EPI_ISL_2315305, EPI_ISL_2315306, EPI_ISL_2315307, EPI_ISL_2315308, EPI_ISL_2315309, EPI_ISL_2315310, EPI_ISL_2315311, EPI_ISL_2315312, EPI_ISL_2315313, EPI_ISL_2315314, EPI_ISL_2315315, EPI_ISL_2315316, EPI_ISL_2315317, EPI_ISL_2315318, EPI_ISL_2315319, EPI_ISL_2315320, EPI_ISL_2315321, EPI_ISL_2315322, EPI_ISL_2315323, EPI_ISL_2315324, EPI_ISL_2315325, EPI_ISL_2315326, EPI_ISL_2315327, EPI_ISL_2315328, EPI_ISL_2315329, EPI_ISL_2315331, EPI_ISL_2315332, EPI_ISL_2315333, EPI_ISL_2315334, EPI_ISL_2315335, EPI_ISL_2315336, EPI_ISL_2315337, EPI_ISL_2315338, EPI_ISL_2315339, EPI_ISL_2315340, EPI_ISL_2315341, EPI_ISL_2315342, EPI_ISL_2315343, EPI_ISL_2315344, EPI_ISL_2315345, EPI_ISL_2315346, EPI_ISL_2315347, EPI_ISL_2315348, EPI_ISL_2315349, EPI_ISL_2315350, EPI_ISL_2315351, EPI_ISL_2315352, EPI_ISL_2315353, EPI_ISL_2315354, EPI_ISL_2315355, EPI_ISL_2315356, EPI_ISL_2315357, EPI_ISL_2315358, EPI_ISL_2315359, EPI_ISL_2315360, EPI_ISL_2315361, EPI_ISL_2315362, EPI_ISL_2315363, EPI_ISL_2315364, EPI_ISL_2315365, EPI_ISL_2315366, EPI_ISL_2315367, EPI_ISL_2315368, EPI_ISL_2315369, EPI_ISL_2315370, EPI_ISL_2315371, EPI_ISL_2315372, EPI_ISL_2315373, EPI_ISL_2315374, EPI_ISL_2315375, EPI_ISL_2315376, EPI_ISL_2315377, EPI_ISL_2315378, EPI_ISL_2315379, EPI_ISL_2315380, EPI_ISL_2315381, EPI_ISL_2315382, EPI_ISL_2315383, EPI_ISL_2315384, EPI_ISL_2315385, EPI_ISL_2315386, EPI_ISL_2315387, EPI_ISL_2315388, EPI_ISL_2315389, EPI_ISL_2315390, EPI_ISL_2315391, EPI_ISL_2315392, EPI_ISL_2315393, EPI_ISL_2315394, EPI_ISL_2315395, EPI_ISL_2315396, EPI_ISL_2315397, EPI_ISL_2315398, EPI_ISL_2315399, EPI_ISL_2315400, EPI_ISL_2315401, EPI_ISL_2315402, EPI_ISL_2315403, EPI_ISL_2315404, EPI_ISL_2315405, EPI_ISL_2315406, EPI_ISL_2315407, EPI_ISL_2315408, EPI_ISL_2315409, EPI_ISL_2315410, EPI_ISL_2315411, EPI_ISL_2315412, EPI_ISL_2315413, EPI_ISL_2315414, EPI_ISL_2315415, EPI_ISL_2315416, EPI_ISL_2315417, EPI_ISL_2315418, EPI_ISL_2315419, EPI_ISL_2315420, EPI_ISL_2315421, EPI_ISL_2315422, EPI_ISL_2315423, EPI_ISL_2315424, EPI_ISL_2315425, EPI_ISL_2315426, EPI_ISL_2315427, EPI_ISL_2315428, EPI_ISL_2315429, EPI_ISL_2315430, EPI_ISL_2315431, EPI_ISL_2315432, EPI_ISL_2315434, EPI_ISL_2315435, EPI_ISL_2315436, EPI_ISL_2315437, EPI_ISL_2315438, EPI_ISL_2315439, EPI_ISL_2315440, EPI_ISL_2315441, EPI_ISL_2315442, EPI_ISL_2315443, EPI_ISL_2315444, EPI_ISL_2315445, EPI_ISL_2315446, EPI_ISL_2315447, EPI_ISL_2315448, EPI_ISL_2315449, EPI_ISL_2315450, EPI_ISL_2315451, EPI_ISL_2315452, EPI_ISL_2315453, EPI_ISL_2315455, EPI_ISL_2386300, EPI_ISL_2386302, EPI_ISL_2386304, EPI_ISL_2386305, EPI_ISL_2386306, EPI_ISL_2386307, EPI_ISL_2386308, EPI_ISL_2386310, EPI_ISL_2386312, EPI_ISL_2386313, EPI_ISL_2386316, EPI_ISL_2386318, EPI_ISL_2386321, EPI_ISL_2386324, EPI_ISL_2386325, EPI_ISL_2386326, EPI_ISL_2386327, EPI_ISL_2386328, EPI_ISL_2386329, EPI_ISL_2386330, EPI_ISL_2386331, EPI_ISL_2386332, EPI_ISL_2386333, EPI_ISL_2386335, EPI_ISL_2386336, EPI_ISL_2386338, EPI_ISL_2386339, EPI_ISL_2386340, EPI_ISL_2386342, EPI_ISL_2386344, EPI_ISL_2386347, EPI_ISL_2386348, EPI_ISL_2386350, EPI_ISL_2386351, EPI_ISL_2386352, EPI_ISL_2386353, EPI_ISL_2386354, EPI_ISL_2386355, EPI_ISL_2386356, EPI_ISL_2386357, EPI_ISL_2386358, EPI_ISL_2386359, EPI_ISL_2386360, EPI_ISL_2386362, EPI_ISL_2386364, EPI_ISL_2386365, EPI_ISL_2386366, EPI_ISL_2386367, EPI_ISL_2386369, EPI_ISL_2386370, EPI_ISL_2386372, EPI_ISL_2386373, EPI_ISL_2386374, EPI_ISL_2386375, EPI_ISL_2386376, EPI_ISL_2386377, EPI_ISL_2386378, EPI_ISL_2386379, EPI_ISL_2386381, EPI_ISL_2386382, EPI_ISL_2386383, EPI_ISL_2386384, EPI_ISL_2386385, EPI_ISL_2386386, EPI_ISL_2386388, EPI_ISL_2386389, EPI_ISL_2386390, EPI_ISL_2386391, EPI_ISL_2386392, EPI_ISL_2386393, EPI_ISL_2386394, EPI_ISL_2386395, EPI_ISL_2386396, EPI_ISL_2386397, EPI_ISL_2386398, EPI_ISL_2386399, EPI_ISL_2386400, EPI_ISL_2386402, EPI_ISL_2386404, EPI_ISL_2386405, EPI_ISL_2386406, EPI_ISL_2386407, EPI_ISL_2386408, EPI_ISL_2386410, EPI_ISL_2386412, EPI_ISL_2386414, EPI_ISL_2386415, EPI_ISL_2386416, EPI_ISL_2386418, EPI_ISL_2386419, EPI_ISL_2386420, EPI_ISL_2386421, EPI_ISL_2386422, EPI_ISL_2386425, EPI_ISL_2386427, EPI_ISL_2386428, EPI_ISL_2386429, EPI_ISL_2386430, EPI_ISL_2386431, EPI_ISL_2386432, EPI_ISL_2386434, EPI_ISL_2386435, EPI_ISL_2386436, EPI_ISL_2386437, EPI_ISL_2386438, EPI_ISL_2386439, EPI_ISL_2386440, EPI_ISL_2386441, EPI_ISL_2386442, EPI_ISL_2386443, EPI_ISL_2386447, EPI_ISL_2386448, EPI_ISL_2386449, EPI_ISL_2386450, EPI_ISL_2386451, EPI_ISL_2386452, EPI_ISL_2386453, EPI_ISL_2386454, EPI_ISL_2386455, EPI_ISL_2386456, EPI_ISL_2386457, EPI_ISL_2386458, EPI_ISL_2386459, EPI_ISL_2386461, EPI_ISL_2386462, EPI_ISL_2386464, EPI_ISL_2386465, EPI_ISL_2386466, EPI_ISL_2386467, EPI_ISL_2386468, EPI_ISL_2386469, EPI_ISL_2386470, EPI_ISL_2386472, EPI_ISL_2386473, EPI_ISL_2386474, EPI_ISL_2386475, EPI_ISL_2386476, EPI_ISL_2386477, EPI_ISL_2386479, EPI_ISL_2386482, EPI_ISL_2386483, EPI_ISL_2386485, EPI_ISL_2386486, EPI_ISL_2386487, EPI_ISL_2386488, EPI_ISL_2386489, EPI_ISL_2386490, EPI_ISL_2386491, EPI_ISL_2386492, EPI_ISL_2386493, EPI_ISL_2386495, EPI_ISL_2386496, EPI_ISL_2386497, EPI_ISL_2386498, EPI_ISL_2386499, EPI_ISL_2386500, EPI_ISL_2386501, EPI_ISL_2386502, EPI_ISL_2386503, EPI_ISL_2386504, EPI_ISL_2386505, EPI_ISL_2386506, EPI_ISL_2386507, EPI_ISL_2386508, EPI_ISL_2386509, EPI_ISL_2386510, EPI_ISL_2386512, EPI_ISL_2386513, EPI_ISL_2386515, EPI_ISL_2386516, EPI_ISL_2386517, EPI_ISL_2386518, EPI_ISL_2386519, EPI_ISL_2386520, EPI_ISL_2386521, EPI_ISL_2386522, EPI_ISL_2386523, EPI_ISL_2386524, EPI_ISL_2386526, EPI_ISL_2386527, EPI_ISL_2386528, EPI_ISL_2386529, EPI_ISL_2386530, EPI_ISL_2386532, EPI_ISL_2386533, EPI_ISL_2386534, EPI_ISL_2386535, EPI_ISL_2386536, EPI_ISL_2386538, EPI_ISL_2386539, EPI_ISL_2387455, EPI_ISL_2387456, EPI_ISL_2387457, EPI_ISL_2387458, EPI_ISL_2387460, EPI_ISL_2387461, EPI_ISL_2387464, EPI_ISL_2387474, EPI_ISL_2387475, EPI_ISL_2387477, EPI_ISL_2387482, EPI_ISL_2387485, EPI_ISL_2387490, EPI_ISL_2387492, EPI_ISL_2387494, EPI_ISL_2387507, EPI_ISL_2387508, EPI_ISL_2387509, EPI_ISL_2387511, EPI_ISL_2387512, EPI_ISL_2387518, EPI_ISL_2387519, EPI_ISL_2387521, EPI_ISL_2387523, EPI_ISL_2387524, EPI_ISL_2387527, EPI_ISL_2387528, EPI_ISL_2387530, EPI_ISL_2387537, EPI_ISL_2387540, EPI_ISL_2387541, EPI_ISL_2387545, EPI_ISL_2387547, EPI_ISL_2387549, EPI_ISL_2387550, EPI_ISL_2387552, EPI_ISL_2387553, EPI_ISL_2387554, EPI_ISL_2387563, EPI_ISL_2387566, EPI_ISL_2387569, EPI_ISL_2387576, EPI_ISL_2387577, EPI_ISL_2387580, EPI_ISL_2387581, EPI_ISL_2387582, EPI_ISL_2387588, EPI_ISL_2387594, EPI_ISL_2387596, EPI_ISL_2387598, EPI_ISL_2387604, EPI_ISL_2387605, EPI_ISL_2387606, EPI_ISL_2387607, EPI_ISL_2387608, EPI_ISL_2387611, EPI_ISL_2387612, EPI_ISL_2387614, EPI_ISL_2387615, EPI_ISL_2387617, EPI_ISL_2387619, EPI_ISL_2387620, EPI_ISL_2387625, EPI_ISL_2387630, EPI_ISL_2387632, EPI_ISL_2387633, EPI_ISL_2387634, EPI_ISL_2387635, EPI_ISL_2387637, EPI_ISL_2387638, EPI_ISL_2387640, EPI_ISL_2387643, EPI_ISL_2387644, EPI_ISL_2387645, EPI_ISL_2387646, EPI_ISL_2387648, EPI_ISL_2387650, EPI_ISL_2387652, EPI_ISL_2387653, EPI_ISL_2387654, EPI_ISL_2387656, EPI_ISL_2387658, EPI_ISL_2387659, EPI_ISL_2387660, EPI_ISL_2387663, EPI_ISL_2387669, EPI_ISL_2387671, EPI_ISL_2387678, EPI_ISL_2387679, EPI_ISL_2387681, EPI_ISL_2387682, EPI_ISL_2387683, EPI_ISL_2387684, EPI_ISL_2387689, EPI_ISL_2387691, EPI_ISL_2387692, EPI_ISL_2387693, EPI_ISL_2387696, EPI_ISL_2387697, EPI_ISL_2387699, EPI_ISL_2387702, EPI_ISL_2387707, EPI_ISL_2387711, EPI_ISL_2387714, EPI_ISL_2387719, EPI_ISL_2387723, EPI_ISL_2387728, EPI_ISL_2387729, EPI_ISL_2387734, EPI_ISL_2387736, EPI_ISL_2387739, EPI_ISL_2387740, EPI_ISL_2387743, EPI_ISL_2387749, EPI_ISL_2387750, EPI_ISL_2387752, EPI_ISL_2387753, EPI_ISL_2387755, EPI_ISL_2387757, EPI_ISL_2387759, EPI_ISL_2387760, EPI_ISL_2387762, EPI_ISL_2387767, EPI_ISL_2387768, EPI_ISL_2387770, EPI_ISL_2387771, EPI_ISL_2387772, EPI_ISL_2387773, EPI_ISL_2387776, EPI_ISL_2387777, EPI_ISL_2387779, EPI_ISL_2387781, EPI_ISL_2387795, EPI_ISL_2387798, EPI_ISL_2387799, EPI_ISL_2387801, EPI_ISL_2387803, EPI_ISL_2387805, EPI_ISL_2387808, EPI_ISL_2387811, EPI_ISL_2387812, EPI_ISL_2387814, EPI_ISL_2387819, EPI_ISL_2387823, EPI_ISL_2387825, EPI_ISL_2387826, EPI_ISL_2387828, EPI_ISL_2387831, EPI_ISL_2387833, EPI_ISL_2387839, EPI_ISL_2387841, EPI_ISL_2387847, EPI_ISL_2387853, EPI_ISL_2387854, EPI_ISL_2387860, EPI_ISL_2387861, EPI_ISL_2387866, EPI_ISL_2387867, EPI_ISL_2387870, EPI_ISL_2387871, EPI_ISL_2387872, EPI_ISL_2387873, EPI_ISL_2387876, EPI_ISL_2387878, EPI_ISL_2387881, EPI_ISL_2387882, EPI_ISL_2387885, EPI_ISL_2387893, EPI_ISL_2387894, EPI_ISL_2387897, EPI_ISL_2387898, EPI_ISL_2387901, EPI_ISL_2387902, EPI_ISL_2387903, EPI_ISL_2387904, EPI_ISL_2387909, EPI_ISL_2387914, EPI_ISL_2387915, EPI_ISL_2387920, EPI_ISL_2387923, EPI_ISL_2387924, EPI_ISL_2387935, EPI_ISL_2387936, EPI_ISL_2387942, EPI_ISL_2387948, EPI_ISL_2387949, EPI_ISL_2387954, EPI_ISL_2387961, EPI_ISL_2387963, EPI_ISL_2387967, EPI_ISL_2387968, EPI_ISL_2387970, EPI_ISL_2387972, EPI_ISL_2387976, EPI_ISL_2387978, EPI_ISL_2387980, EPI_ISL_2387981, EPI_ISL_2387982, EPI_ISL_2387985, EPI_ISL_2387987, EPI_ISL_2387990, EPI_ISL_2387991, EPI_ISL_2387992, EPI_ISL_2387993, EPI_ISL_2387994, EPI_ISL_2387995, EPI_ISL_2387998, EPI_ISL_2387999, EPI_ISL_2388007, EPI_ISL_2388009, EPI_ISL_2389511, EPI_ISL_2389512, EPI_ISL_2389517, EPI_ISL_2389518, EPI_ISL_2389520, EPI_ISL_2389521, EPI_ISL_2389523, EPI_ISL_2389526, EPI_ISL_2389527, EPI_ISL_2389529, EPI_ISL_2389536, EPI_ISL_2389537, EPI_ISL_2389545, EPI_ISL_2389557, EPI_ISL_2389560, EPI_ISL_2389563, EPI_ISL_2389570, EPI_ISL_2389571, EPI_ISL_2389572, EPI_ISL_2389573, EPI_ISL_2389575, EPI_ISL_2389578, EPI_ISL_2389579, EPI_ISL_2389580, EPI_ISL_2389581, EPI_ISL_2389583, EPI_ISL_2389588, EPI_ISL_2389589, EPI_ISL_2389592, EPI_ISL_2389593, EPI_ISL_2389594, EPI_ISL_2389597, EPI_ISL_2389598, EPI_ISL_2389600, EPI_ISL_2389605, EPI_ISL_2389606, EPI_ISL_2389609, EPI_ISL_2389610, EPI_ISL_2389613, EPI_ISL_2389617, EPI_ISL_2389619, EPI_ISL_2389621, EPI_ISL_2389623, EPI_ISL_2389625, EPI_ISL_2389627, EPI_ISL_2389629, EPI_ISL_2389631, EPI_ISL_2389648, EPI_ISL_2389650, EPI_ISL_2389654, EPI_ISL_2389659, EPI_ISL_2389661, EPI_ISL_2389662, EPI_ISL_2389664, EPI_ISL_2389669, EPI_ISL_2389670, EPI_ISL_2389672, EPI_ISL_2389676, EPI_ISL_2389678, EPI_ISL_2389679, EPI_ISL_2389681, EPI_ISL_2389683, EPI_ISL_2389685, EPI_ISL_2389686, EPI_ISL_2389688, EPI_ISL_2389690, EPI_ISL_2389694, EPI_ISL_2389695, EPI_ISL_2389699, EPI_ISL_2389700, EPI_ISL_2389702, EPI_ISL_2389703, EPI_ISL_2389704, EPI_ISL_2389713, EPI_ISL_2389714, EPI_ISL_2389715, EPI_ISL_2389717, EPI_ISL_2389718, EPI_ISL_2389720, EPI_ISL_2389723, EPI_ISL_2389724, EPI_ISL_2389725, EPI_ISL_2389734, EPI_ISL_2389735, EPI_ISL_2389736, EPI_ISL_2389738, EPI_ISL_2389739, EPI_ISL_2389740, EPI_ISL_2389742, EPI_ISL_2389743, EPI_ISL_2389746, EPI_ISL_2389748, EPI_ISL_2389750, EPI_ISL_2389753, EPI_ISL_2389755, EPI_ISL_2389756, EPI_ISL_2389759, EPI_ISL_2389762, EPI_ISL_2389763, EPI_ISL_2389769, EPI_ISL_2389773, EPI_ISL_2389774, EPI_ISL_2389778, EPI_ISL_2389792, EPI_ISL_2389795, EPI_ISL_2389801, EPI_ISL_2389807, EPI_ISL_2389808, EPI_ISL_2389809, EPI_ISL_2389810, EPI_ISL_2389811, EPI_ISL_2389813, EPI_ISL_2389814, EPI_ISL_2389817, EPI_ISL_2389818, EPI_ISL_2389820, EPI_ISL_2389823, EPI_ISL_2389828, EPI_ISL_2389836, EPI_ISL_2389837, EPI_ISL_2389838, EPI_ISL_2389839, EPI_ISL_2389845, EPI_ISL_2389849, EPI_ISL_2469474, EPI_ISL_2469476, EPI_ISL_2469477, EPI_ISL_2469481, EPI_ISL_2469482, EPI_ISL_2469484, EPI_ISL_2469488, EPI_ISL_2469490, EPI_ISL_2469492, EPI_ISL_2469495, EPI_ISL_2469496, EPI_ISL_2469498, EPI_ISL_2469499, EPI_ISL_2469500, EPI_ISL_2469502, EPI_ISL_2469503, EPI_ISL_2469504, EPI_ISL_2469508, EPI_ISL_2469509, EPI_ISL_2469510, EPI_ISL_2469513, EPI_ISL_2469514, EPI_ISL_2469517, EPI_ISL_2469518, EPI_ISL_2469520, EPI_ISL_2469522, EPI_ISL_2469523, EPI_ISL_2469524, EPI_ISL_2469525, EPI_ISL_2469526, EPI_ISL_2469531, EPI_ISL_2469535, EPI_ISL_2469536, EPI_ISL_2469538, EPI_ISL_2469546, EPI_ISL_2469547, EPI_ISL_2469548, EPI_ISL_2469551, EPI_ISL_2469552, EPI_ISL_2469553, EPI_ISL_2469555, EPI_ISL_2469558, EPI_ISL_2469560, EPI_ISL_2469574, EPI_ISL_2469575, EPI_ISL_2469578, EPI_ISL_2469580, EPI_ISL_2469582, EPI_ISL_2469585, EPI_ISL_2469586, EPI_ISL_2469587, EPI_ISL_2469594, EPI_ISL_2469598, EPI_ISL_2469599, EPI_ISL_2469601, EPI_ISL_2469603, EPI_ISL_2469606, EPI_ISL_2469607, EPI_ISL_2469608, EPI_ISL_2469611, EPI_ISL_2469612, EPI_ISL_2469614, EPI_ISL_2469615, EPI_ISL_2469619, EPI_ISL_2469620, EPI_ISL_2469621, EPI_ISL_2469629, EPI_ISL_2469630, EPI_ISL_2469632, EPI_ISL_2469633, EPI_ISL_2469638, EPI_ISL_2469639, EPI_ISL_2469640, EPI_ISL_2469642, EPI_ISL_2469644, EPI_ISL_2469645, EPI_ISL_2469647, EPI_ISL_2469649, EPI_ISL_2469650, EPI_ISL_2469651, EPI_ISL_2469652, EPI_ISL_2469653, EPI_ISL_2469656, EPI_ISL_2469657, EPI_ISL_2469660, EPI_ISL_2469661, EPI_ISL_2469662, EPI_ISL_2469665, EPI_ISL_2469666, EPI_ISL_2469667, EPI_ISL_2469671, EPI_ISL_2469673, EPI_ISL_2469675, EPI_ISL_2469678, EPI_ISL_2469679, EPI_ISL_2469682, EPI_ISL_2469683, EPI_ISL_2469684, EPI_ISL_2469686, EPI_ISL_2469688, EPI_ISL_2469689, EPI_ISL_2469690, EPI_ISL_2469693, EPI_ISL_2469698, EPI_ISL_2469700, EPI_ISL_2469704, EPI_ISL_2469706, EPI_ISL_2469707, EPI_ISL_2469708, EPI_ISL_2469709, EPI_ISL_2469710, EPI_ISL_2469711, EPI_ISL_2469712, EPI_ISL_2469713, EPI_ISL_2469716, EPI_ISL_2469718, EPI_ISL_2469720, EPI_ISL_2469722, EPI_ISL_2469723, EPI_ISL_2469725, EPI_ISL_2469726, EPI_ISL_2469727, EPI_ISL_2469729, EPI_ISL_2469730, EPI_ISL_2469731, EPI_ISL_2469733, EPI_ISL_2469736, EPI_ISL_2469738, EPI_ISL_2469739, EPI_ISL_2469740, EPI_ISL_2469742, EPI_ISL_2469743, EPI_ISL_2469745, EPI_ISL_2469746, EPI_ISL_2469748, EPI_ISL_2469749, EPI_ISL_2469750, EPI_ISL_2469751, EPI_ISL_2472029, EPI_ISL_2472030, EPI_ISL_2472031, EPI_ISL_2472032, EPI_ISL_2472033, EPI_ISL_2472034, EPI_ISL_2472035, EPI_ISL_2472036, EPI_ISL_2472037, EPI_ISL_2472038, EPI_ISL_2472039, EPI_ISL_2472041, EPI_ISL_2472044, EPI_ISL_2472045, EPI_ISL_2472046, EPI_ISL_2472048, EPI_ISL_2472049, EPI_ISL_2472051, EPI_ISL_2472054, EPI_ISL_2472056, EPI_ISL_2472058, EPI_ISL_2472062, EPI_ISL_2472063, EPI_ISL_2472064, EPI_ISL_2472067, EPI_ISL_2472068, EPI_ISL_2472074, EPI_ISL_2472075, EPI_ISL_2472076, EPI_ISL_2472078, EPI_ISL_2472079, EPI_ISL_2472082, EPI_ISL_2472084, EPI_ISL_2472085, EPI_ISL_2472086, EPI_ISL_2472088, EPI_ISL_2472092, EPI_ISL_2472094, EPI_ISL_2472097, EPI_ISL_2472098, EPI_ISL_2472100, EPI_ISL_2472102, EPI_ISL_2472103, EPI_ISL_2472104, EPI_ISL_2472107, EPI_ISL_2472108, EPI_ISL_2472109, EPI_ISL_2472110, EPI_ISL_2472111, EPI_ISL_2472114, EPI_ISL_2472115, EPI_ISL_2472116, EPI_ISL_2472117, EPI_ISL_2472119, EPI_ISL_2472123, EPI_ISL_2472124, EPI_ISL_2472126, EPI_ISL_2472127, EPI_ISL_2472129, EPI_ISL_2472130, EPI_ISL_2472131, EPI_ISL_2472135, EPI_ISL_2472137, EPI_ISL_2472138, EPI_ISL_2472139, EPI_ISL_2472141, EPI_ISL_2472142, EPI_ISL_2472143, EPI_ISL_2472144, EPI_ISL_2472145, EPI_ISL_2472146, EPI_ISL_2472148, EPI_ISL_2472149, EPI_ISL_2472151, EPI_ISL_2472152, EPI_ISL_2472153, EPI_ISL_2472155, EPI_ISL_2472156, EPI_ISL_2472158, EPI_ISL_2472159, EPI_ISL_2472160, EPI_ISL_2472162, EPI_ISL_2472163, EPI_ISL_2472164, EPI_ISL_2472166, EPI_ISL_2472168, EPI_ISL_2472169, EPI_ISL_2472171, EPI_ISL_2472172, EPI_ISL_2472173, EPI_ISL_2472174, EPI_ISL_2472175, EPI_ISL_2472176, EPI_ISL_2472178, EPI_ISL_2472179, EPI_ISL_2472180, EPI_ISL_2472182, EPI_ISL_2472184, EPI_ISL_2472188, EPI_ISL_2472190, EPI_ISL_2472191, EPI_ISL_2472192, EPI_ISL_2472193, EPI_ISL_2472195, EPI_ISL_2472199, EPI_ISL_2472200, EPI_ISL_2472201, EPI_ISL_2472203, EPI_ISL_2472204, EPI_ISL_2472205, EPI_ISL_2472207, EPI_ISL_2472208, EPI_ISL_2472211, EPI_ISL_2472212, EPI_ISL_2472214, EPI_ISL_2472215, EPI_ISL_2472219, EPI_ISL_2472223, EPI_ISL_2472230, EPI_ISL_2472234, EPI_ISL_2472235, EPI_ISL_2472239, EPI_ISL_2472240, EPI_ISL_2472243, EPI_ISL_2472248, EPI_ISL_2472250, EPI_ISL_2472252, EPI_ISL_2472253, EPI_ISL_2472254, EPI_ISL_2472264, EPI_ISL_2472265, EPI_ISL_2472269, EPI_ISL_2472271, EPI_ISL_2472273, EPI_ISL_2472274, EPI_ISL_2472276, EPI_ISL_2472277, EPI_ISL_2472282, EPI_ISL_2472284, EPI_ISL_2472285, EPI_ISL_2472286, EPI_ISL_2472288, EPI_ISL_2472291, EPI_ISL_2472292, EPI_ISL_2472293, EPI_ISL_2472297, EPI_ISL_2472298, EPI_ISL_2472300, EPI_ISL_2472301, EPI_ISL_2472302, EPI_ISL_2472303, EPI_ISL_2472306, EPI_ISL_2472307, EPI_ISL_2472308, EPI_ISL_2472309, EPI_ISL_2472312, EPI_ISL_2472313, EPI_ISL_2472315, EPI_ISL_2472316, EPI_ISL_2472317, EPI_ISL_2472319, EPI_ISL_2472321, EPI_ISL_2472327, EPI_ISL_2472330, EPI_ISL_2472333, EPI_ISL_2472334, EPI_ISL_2472335, EPI_ISL_2472336, EPI_ISL_2472338, EPI_ISL_2472339, EPI_ISL_2472340, EPI_ISL_2472345, EPI_ISL_2472346, EPI_ISL_2472349, EPI_ISL_2472350, EPI_ISL_2472351, EPI_ISL_2472354, EPI_ISL_2472355, EPI_ISL_2472356, EPI_ISL_2472357, EPI_ISL_2472358, EPI_ISL_2472359, EPI_ISL_2472361, EPI_ISL_2472362, EPI_ISL_2472363, EPI_ISL_2472364, EPI_ISL_2631546, EPI_ISL_2631547, EPI_ISL_2631550, EPI_ISL_2631551, EPI_ISL_2631552, EPI_ISL_2631556, EPI_ISL_2631560, EPI_ISL_2631561, EPI_ISL_2631562, EPI_ISL_2631563, EPI_ISL_2631564, EPI_ISL_2631565, EPI_ISL_2631567, EPI_ISL_2631569, EPI_ISL_2631570, EPI_ISL_2631571, EPI_ISL_2631573, EPI_ISL_2631574, EPI_ISL_2631575, EPI_ISL_2631576, EPI_ISL_2631577, EPI_ISL_2631579, EPI_ISL_2631580, EPI_ISL_2631581, EPI_ISL_2631583, EPI_ISL_2631584, EPI_ISL_2631589, EPI_ISL_2631591, EPI_ISL_2631593, EPI_ISL_2631597, EPI_ISL_2631598, EPI_ISL_2631599, EPI_ISL_2631600, EPI_ISL_2631602, EPI_ISL_2631603, EPI_ISL_2631606, EPI_ISL_2631607, EPI_ISL_2631608, EPI_ISL_2631609, EPI_ISL_2631610, EPI_ISL_2631611, EPI_ISL_2631612, EPI_ISL_2631613, EPI_ISL_2631614, EPI_ISL_2631615, EPI_ISL_2631618, EPI_ISL_2631619, EPI_ISL_2631622, EPI_ISL_2631625, EPI_ISL_2631627, EPI_ISL_2631628, EPI_ISL_2631630, EPI_ISL_2631633, EPI_ISL_2631634, EPI_ISL_2631636, EPI_ISL_2631640, EPI_ISL_2631641, EPI_ISL_2631642, EPI_ISL_2631645, EPI_ISL_2631646, EPI_ISL_2631648, EPI_ISL_2631650, EPI_ISL_2631651, EPI_ISL_2631652, EPI_ISL_2631653, EPI_ISL_2631654, EPI_ISL_2631656, EPI_ISL_2631658, EPI_ISL_2631659, EPI_ISL_2631660, EPI_ISL_2631661, EPI_ISL_2631662, EPI_ISL_2631663, EPI_ISL_2631664, EPI_ISL_2631665, EPI_ISL_2631666, EPI_ISL_2631667, EPI_ISL_2631668, EPI_ISL_2631670, EPI_ISL_2631671, EPI_ISL_2631673, EPI_ISL_2631674, EPI_ISL_2631675, EPI_ISL_2631676, EPI_ISL_2631677, EPI_ISL_2631678, EPI_ISL_2631679, EPI_ISL_2631680, EPI_ISL_2631681, EPI_ISL_2631683, EPI_ISL_2631688, EPI_ISL_2631689, EPI_ISL_2631690, EPI_ISL_2631691, EPI_ISL_2631692, EPI_ISL_2631696, EPI_ISL_2631697, EPI_ISL_2631698, EPI_ISL_2631699, EPI_ISL_2631700, EPI_ISL_2631703, EPI_ISL_2631704, EPI_ISL_2631705, EPI_ISL_2631707, EPI_ISL_2631708, EPI_ISL_2631710, EPI_ISL_2631711, EPI_ISL_2631713, EPI_ISL_2631714, EPI_ISL_2631716, EPI_ISL_2631717, EPI_ISL_2631719, EPI_ISL_2631721, EPI_ISL_2631722, EPI_ISL_2631725, EPI_ISL_2631726, EPI_ISL_2631728, EPI_ISL_2631731, EPI_ISL_2631733, EPI_ISL_2631734, EPI_ISL_2631735, EPI_ISL_2631736, EPI_ISL_2631738, EPI_ISL_2631739, EPI_ISL_2631740, EPI_ISL_2631742, EPI_ISL_2631743, EPI_ISL_2631745, EPI_ISL_2631746, EPI_ISL_2631748, EPI_ISL_2631750, EPI_ISL_2631751, EPI_ISL_2631752, EPI_ISL_2631754, EPI_ISL_2631755, EPI_ISL_2631756, EPI_ISL_2631758, EPI_ISL_2631760, EPI_ISL_2631762, EPI_ISL_2633395

see above SYNLAB MVZ Leverkusen Robert Koch Institute

EPI_ISL_2387585, EPI_ISL_2387642, EPI_ISL_2387655, EPI_ISL_2387701, EPI_ISL_2387713, EPI_ISL_2387724, EPI_ISL_2387727, EPI_ISL_2387730, EPI_ISL_2387850, EPI_ISL_2389562, EPI_ISL_2389564, EPI_ISL_2389596, EPI_ISL_2389722, EPI_ISL_2389729, EPI_ISL_2389751, EPI_ISL_2389793, EPI_ISL_2389847, EPI_ISL_2389851, EPI_ISL_2472101

see above SYNLAB MVZ Neustadt a. d. Weinstraße Robert Koch Institute

EPI_ISL_2315231, EPI_ISL_2315433, EPI_ISL_2315454, EPI_ISL_2386301, EPI_ISL_2386303, EPI_ISL_2386334, EPI_ISL_2386337, EPI_ISL_2386341, EPI_ISL_2386349, EPI_ISL_2386368, EPI_ISL_2386371, EPI_ISL_2386401, EPI_ISL_2386423, EPI_ISL_2386444, EPI_ISL_2386480, EPI_ISL_2387465, EPI_ISL_2387505, EPI_ISL_2387561, EPI_ISL_2387806, EPI_ISL_2387865, EPI_ISL_2389515, EPI_ISL_2389565, EPI_ISL_2389584, EPI_ISL_2389603, EPI_ISL_2389698, EPI_ISL_2389744, EPI_ISL_2389786, EPI_ISL_2389831, EPI_ISL_2469537, EPI_ISL_2469577, EPI_ISL_2469624, EPI_ISL_2469646, EPI_ISL_2469654, EPI_ISL_2469687, EPI_ISL_2472061, EPI_ISL_2472073, EPI_ISL_2472080, EPI_ISL_2472089, EPI_ISL_2472096, EPI_ISL_2472217, EPI_ISL_2472220, EPI_ISL_2472224, EPI_ISL_2472227, EPI_ISL_2472231, EPI_ISL_2472249, EPI_ISL_2472290, EPI_ISL_2472305, EPI_ISL_2472325, EPI_ISL_2631590, EPI_ISL_2631596, EPI_ISL_2631649, EPI_ISL_2631657, EPI_ISL_2633041, EPI_ISL_2633355

see above SYNLAB MVZ Trier Robert Koch Institute

EPI_ISL_2386315, EPI_ISL_2386317, EPI_ISL_2386320, EPI_ISL_2386345, EPI_ISL_2386361, EPI_ISL_2386424, EPI_ISL_2386433, EPI_ISL_2387466, EPI_ISL_2387472, EPI_ISL_2387687, EPI_ISL_2387742, EPI_ISL_2387793, EPI_ISL_2387857, EPI_ISL_2387863, EPI_ISL_2387962, EPI_ISL_2389556, EPI_ISL_2389673, EPI_ISL_2389675, EPI_ISL_2389708, EPI_ISL_2389712, EPI_ISL_2389719, EPI_ISL_2389726, EPI_ISL_2389768, EPI_ISL_2389790, EPI_ISL_2389812, EPI_ISL_2389827, EPI_ISL_2389829, EPI_ISL_2469483, EPI_ISL_2469628, EPI_ISL_2472059, EPI_ISL_2472071, EPI_ISL_2472091, EPI_ISL_2472093, EPI_ISL_2472105, EPI_ISL_2472147, EPI_ISL_2472157, EPI_ISL_2472177, EPI_ISL_2472185, EPI_ISL_2472198, EPI_ISL_2472225, EPI_ISL_2472226, EPI_ISL_2472233, EPI_ISL_2472237, EPI_ISL_2472238, EPI_ISL_2472245, EPI_ISL_2472247, EPI_ISL_2472255, EPI_ISL_2472258, EPI_ISL_2472259, EPI_ISL_2472262, EPI_ISL_2472266, EPI_ISL_2472267, EPI_ISL_2472275, EPI_ISL_2472278, EPI_ISL_2472280, EPI_ISL_2631587, EPI_ISL_2631621, EPI_ISL_2631629, EPI_ISL_2631644, EPI_ISL_2631685, EPI_ISL_2631701, EPI_ISL_2631706, EPI_ISL_2631747, EPI_ISL_2633012, EPI_ISL_2633024, EPI_ISL_2633073, EPI_ISL_2633141, EPI_ISL_2633216, EPI_ISL_2633229, EPI_ISL_2633252, EPI_ISL_2633375

see above SYNLAB MVZ Weiden Robert Koch Institute

EPI_ISL_2469997, EPI_ISL_2469998, EPI_ISL_2470082, EPI_ISL_2470156, EPI_ISL_2470212, EPI_ISL_2470232, EPI_ISL_2470244, EPI_ISL_2470257, EPI_ISL_2470304, EPI_ISL_2470309, EPI_ISL_2470313, EPI_ISL_2470315, EPI_ISL_2470332, EPI_ISL_2470335, EPI_ISL_2470338, EPI_ISL_2470343, EPI_ISL_2470355, EPI_ISL_2470391, EPI_ISL_2470405, EPI_ISL_2470408, EPI_ISL_2470412, EPI_ISL_2470417, EPI_ISL_2470418, EPI_ISL_2470447, EPI_ISL_2470451, EPI_ISL_2632337, EPI_ISL_2632345, EPI_ISL_2632381, EPI_ISL_2632382, EPI_ISL_2632384, EPI_ISL_2632400, EPI_ISL_2632412, EPI_ISL_2632414, EPI_ISL_2632416, EPI_ISL_2632441, EPI_ISL_2632442, EPI_ISL_2632457, EPI_ISL_2632458, EPI_ISL_2632466, EPI_ISL_2632471, EPI_ISL_2632477, EPI_ISL_2632515, EPI_ISL_2632565, EPI_ISL_2632566, EPI_ISL_2632569, EPI_ISL_2632590, EPI_ISL_2632598, EPI_ISL_2632602, EPI_ISL_2632613, EPI_ISL_2632617, EPI_ISL_2632634, EPI_ISL_2632642, EPI_ISL_2633906, EPI_ISL_2633932, EPI_ISL_2633962, EPI_ISL_2633995, EPI_ISL_2634001, EPI_ISL_2634004, EPI_ISL_2634007, EPI_ISL_2634049, EPI_ISL_2634060, EPI_ISL_2634064, EPI_ISL_2634066, EPI_ISL_2634072, EPI_ISL_2634075, EPI_ISL_2634124, EPI_ISL_2634135, EPI_ISL_2634140, EPI_ISL_2634177, EPI_ISL_2634195, EPI_ISL_2634269, EPI_ISL_2634278, EPI_ISL_2634292, EPI_ISL_2634301, EPI_ISL_2634326, EPI_ISL_2634397, EPI_ISL_2634409, EPI_ISL_2634418, EPI_ISL_2634457, EPI_ISL_2636950, EPI_ISL_2637037, EPI_ISL_2637043, EPI_ISL_2637047, EPI_ISL_2637053, EPI_ISL_2637086, EPI_ISL_2637118, EPI_ISL_2637152, EPI_ISL_2761331, EPI_ISL_2761377, EPI_ISL_2761521

| | | | |
|---|---|---|---|
| see above | Sonic - Bioscientia - MVZ Labor Saar GmbH | Robert Koch Institute | |
| EPI_ISL_2388949, EPI_ISL_2389016, EPI_ISL_2469969, EPI_ISL_2469993, EPI_ISL_2470005, EPI_ISL_2470039, EPI_ISL_2470043, EPI_ISL_2470079, EPI_ISL_2470120, EPI_ISL_2470229, EPI_ISL_2470318, EPI_ISL_2470422, EPI_ISL_2470425, EPI_ISL_2470467, EPI_ISL_2633965, EPI_ISL_2634016, EPI_ISL_2634067, EPI_ISL_2634197, EPI_ISL_2634341, EPI_ISL_2634368, EPI_ISL_2634385, EPI_ISL_2636981, EPI_ISL_2637066 | | | |
| see above | Sonic - Dr. Staber & Kollegen GmbH München | Robert Koch Institute | |
| EPI_ISL_2469929, EPI_ISL_2469938, EPI_ISL_2469968, EPI_ISL_2469977, EPI_ISL_2469979, EPI_ISL_2470025, EPI_ISL_2470037, EPI_ISL_2470067, EPI_ISL_2470093, EPI_ISL_2470098, EPI_ISL_2470113, EPI_ISL_2470116, EPI_ISL_2470141, EPI_ISL_2470147, EPI_ISL_2470170, EPI_ISL_2470171, EPI_ISL_2470234, EPI_ISL_2470265, EPI_ISL_2470330, EPI_ISL_2470333, EPI_ISL_2470336, EPI_ISL_2470353, EPI_ISL_2470392, EPI_ISL_2470442, EPI_ISL_2470482, EPI_ISL_2633908, EPI_ISL_2633913, EPI_ISL_2633938, EPI_ISL_2633941, EPI_ISL_2633947, EPI_ISL_2633959, EPI_ISL_2633971, EPI_ISL_2634009, EPI_ISL_2634055, EPI_ISL_2634092, EPI_ISL_2634114, EPI_ISL_2634155, EPI_ISL_2634182, EPI_ISL_2634217, EPI_ISL_2634230, EPI_ISL_2634232, EPI_ISL_2634271, EPI_ISL_2634287, EPI_ISL_2634303, EPI_ISL_2634330, EPI_ISL_2634366, EPI_ISL_2634373, EPI_ISL_2634380, EPI_ISL_2634404, EPI_ISL_2634435, EPI_ISL_2634455, EPI_ISL_2636937 | | | |
| see above | Sonic - Labor Dr. von Froreich GmbH | Robert Koch Institute | |
| EPI_ISL_2388906, EPI_ISL_2388937, EPI_ISL_2388990, EPI_ISL_2469986, EPI_ISL_2470100, EPI_ISL_2470122, EPI_ISL_2470128, EPI_ISL_2470196, EPI_ISL_2634439, EPI_ISL_2637006 | | | |
| see above | Sonic - Labor Lademannbogen MVZ GmbH | Robert Koch Institute | |
| EPI_ISL_2470109, EPI_ISL_2470135, EPI_ISL_2470279, EPI_ISL_2633903, EPI_ISL_2633993, EPI_ISL_2634059, EPI_ISL_2634357, EPI_ISL_2636934, EPI_ISL_2636945, EPI_ISL_2636952, EPI_ISL_2636959, EPI_ISL_2637031, EPI_ISL_2637058, EPI_ISL_2637104 | | | |
| see above | Sonic - Labor Staber Bayreuth | Robert Koch Institute | |
| EPI_ISL_2389053, EPI_ISL_2469964, EPI_ISL_2470029, EPI_ISL_2470086, EPI_ISL_2470140, EPI_ISL_2470146, EPI_ISL_2470179, EPI_ISL_2470184, EPI_ISL_2470240, EPI_ISL_2470305, EPI_ISL_2470402, EPI_ISL_2470435, EPI_ISL_2634015, EPI_ISL_2634022, EPI_ISL_2634037, EPI_ISL_2634044, EPI_ISL_2634259, EPI_ISL_2634408, EPI_ISL_2636939, EPI_ISL_2637107, EPI_ISL_2637109, EPI_ISL_2637115 | | | |
| see above | Sonic - Labor Staber Dresden (Klipphausen) | Robert Koch Institute | |
| EPI_ISL_2388935, EPI_ISL_2469961, EPI_ISL_2470017, EPI_ISL_2470044, EPI_ISL_2470365, EPI_ISL_2470371, EPI_ISL_2633975, EPI_ISL_2634063, EPI_ISL_2634392, EPI_ISL_2634406 | | | |
| see above | Sonic - Labor Staber Heilbronn | Robert Koch Institute | |
| EPI_ISL_2389055, EPI_ISL_2469975, EPI_ISL_2470010, EPI_ISL_2470214, EPI_ISL_2470227, EPI_ISL_2633945, EPI_ISL_2633956, EPI_ISL_2634027, EPI_ISL_2634079, EPI_ISL_2634094, EPI_ISL_2634111, EPI_ISL_2634220, EPI_ISL_2634274, EPI_ISL_2634308, EPI_ISL_2634363, EPI_ISL_2634378, EPI_ISL_2634383, EPI_ISL_2634441, EPI_ISL_2637027 | | | |
| see above | Sonic - Labor Staber Nürnberg | Robert Koch Institute | |
| EPI_ISL_2388855, EPI_ISL_2388867, EPI_ISL_2388880, EPI_ISL_2388887, EPI_ISL_2389076, EPI_ISL_2469954, EPI_ISL_2470251, EPI_ISL_2470349, EPI_ISL_2633953, EPI_ISL_2633998, EPI_ISL_2634005, EPI_ISL_2634008, EPI_ISL_2634013, EPI_ISL_2634034, EPI_ISL_2634035, EPI_ISL_2634038, EPI_ISL_2634048, EPI_ISL_2634159, EPI_ISL_2634212, EPI_ISL_2634246, EPI_ISL_2634255, EPI_ISL_2634262, EPI_ISL_2634306, EPI_ISL_2634350, EPI_ISL_2634371, EPI_ISL_2634387, EPI_ISL_2634399, EPI_ISL_2634401, EPI_ISL_2634413, EPI_ISL_2634419 | | | |
| see above | Sonic - MVZ Medizinisches Labor Bremen GmbH | Robert Koch Institute | |
| EPI_ISL_2470180, EPI_ISL_2470253, EPI_ISL_2470307, EPI_ISL_2634209 | Sonic - MVZ Medizinisches Labor Oldenburg | Robert Koch Institute | |
| EPI_ISL_2632109 | St. Elisabeth-Gruppe GmbH - Katholische Kliniken Rhein-Ruhr | Robert Koch Institute | |
| EPI_ISL_2386543 | St. Franziskus-Hospital Münster | Robert Koch Institute | |
| EPI_ISL_2469891 | St. Marien-Hospital Hamm | Robert Koch Institute | |
| EPI_ISL_2390318, EPI_ISL_2390324, EPI_ISL_2390325, EPI_ISL_2390327, EPI_ISL_2390328, EPI_ISL_2390329, EPI_ISL_2390331, EPI_ISL_2390342, EPI_ISL_2390349, EPI_ISL_2390350, EPI_ISL_2390356, EPI_ISL_2390358, EPI_ISL_2390363, EPI_ISL_2390371, EPI_ISL_2471849, EPI_ISL_2471853, EPI_ISL_2471855, EPI_ISL_2471860, EPI_ISL_2471865, EPI_ISL_2471867, EPI_ISL_2471872, EPI_ISL_2471875, EPI_ISL_2471882 | | | |
| see above | Städtisches Klinikum Dresden Institut für Klinische Chemie und Laboratoriumsmedizin | Robert Koch Institute | |
| EPI_ISL_2386311, EPI_ISL_2386417, EPI_ISL_2387517, EPI_ISL_2389576, EPI_ISL_2389730, EPI_ISL_2469680, EPI_ISL_2472133, EPI_ISL_2631623, EPI_ISL_2631639, EPI_ISL_2631647 | | | |
| see above | Synlab MVZ Augsburg | Robert Koch Institute | |
| EPI_ISL_2471819, EPI_ISL_2471821, EPI_ISL_2471825, EPI_ISL_2471828, EPI_ISL_2471836, EPI_ISL_2471839, EPI_ISL_2634818, EPI_ISL_2634819, EPI_ISL_2634821, EPI_ISL_2634823, EPI_ISL_2634826, EPI_ISL_2634828, EPI_ISL_2634830, EPI_ISL_2634833, EPI_ISL_2634835, EPI_ISL_2634838, EPI_ISL_2634840, EPI_ISL_2634842, EPI_ISL_2634844, EPI_ISL_2634846, EPI_ISL_2634848, EPI_ISL_2634851, EPI_ISL_2634853, EPI_ISL_2678654, EPI_ISL_2678655 | | | |
| see above | Thüringer Landesamtes für Verbraucherschutz | Robert Koch Institute | |
| EPI_ISL_2715354, EPI_ISL_2715355, EPI_ISL_2715356, EPI_ISL_2715357, EPI_ISL_2715359, EPI_ISL_2715360, EPI_ISL_2715362, EPI_ISL_2715363 | | | |
| see above | US Air Force School of Aerospace Medicine | US Air Force School of Aerospace Medicine | Amanda Javorina; Anthony Fries; Carol Garrett; Clarise Starr; Cole Anderson; Elizabeth Macias; Fritz Castillo; Jennifer Meyer; Sarah Purves; William Gruner |
| EPI_ISL_2390321, EPI_ISL_2390330, EPI_ISL_2390335, EPI_ISL_2390336, EPI_ISL_2390339, EPI_ISL_2390340, EPI_ISL_2390360, EPI_ISL_2471846, EPI_ISL_2471863, EPI_ISL_2471864, EPI_ISL_2471870, EPI_ISL_2471871, EPI_ISL_2471876, EPI_ISL_2471880, EPI_ISL_2471884, EPI_ISL_2471885, EPI_ISL_2471886 | | | |
| see above | Uniklinikum Carl Gustav Carus an der TU Dresden; Institut für Virologie | Robert Koch Institute | |
| EPI_ISL_2389899, EPI_ISL_2389903, EPI_ISL_2389910, EPI_ISL_2471816, EPI_ISL_2471841, EPI_ISL_2634855 | Uniklinikum Jena | Robert Koch Institute | |
| EPI_ISL_2472021, EPI_ISL_2472023, EPI_ISL_2472025, EPI_ISL_2472028, EPI_ISL_2759820, EPI_ISL_2759821, EPI_ISL_2759822 | | | |
| see above | Uniklinikum Regensburg; Institut für klinische Mikrobiologie und Hygiene | Robert Koch Institute | |
| EPI_ISL_2689460, EPI_ISL_2689461, EPI_ISL_2689465, EPI_ISL_2689469, EPI_ISL_2689470, EPI_ISL_2689471, EPI_ISL_2689473, EPI_ISL_2689474, EPI_ISL_2689477, EPI_ISL_2689478, EPI_ISL_2689479 | | | |
| see above | University Medical Center Hamburg Eppendorf | Heinrich Pette Institute, Leibniz Institute for Experimental Virology | Adam Grundhoff; Alexis Robitaille; Johannes Knobloch; Martin Aepfelbacher; Nicole Fischer; Thomas Günther |
| EPI_ISL_2633751, EPI_ISL_2633769, EPI_ISL_2633772, EPI_ISL_2633773, EPI_ISL_2633776, EPI_ISL_2633823, EPI_ISL_2633839, EPI_ISL_2633854, EPI_ISL_2759816, EPI_ISL_2759817 | | | |
| see above | Universitätsklinikum Bonn - Institut für Virologie | Robert Koch Institute | |
| EPI_ISL_2470363, EPI_ISL_2470387, EPI_ISL_2470395, EPI_ISL_2470416, EPI_ISL_2470463, EPI_ISL_2470471, EPI_ISL_2634046, EPI_ISL_2634264, EPI_ISL_2634290, EPI_ISL_2634317 | | | |
| see above | Universitätsklinikum Frankfurt - Institut für Medizinische Virologie | Robert Koch Institute | |
| EPI_ISL_2678372 | Universitätsklinikum Gießen - Institut für Medizinische Virologie | Robert Koch Institute | |
| EPI_ISL_2760272, EPI_ISL_2760275, EPI_ISL_2760277, EPI_ISL_2845768, EPI_ISL_2845769, EPI_ISL_2845770, EPI_ISL_2845771 | | | |
| see above | Universitätsklinikum Heidelberg | Robert Koch Institute | |
| EPI_ISL_2386541, EPI_ISL_2470816 | Universitätsklinikum Münster Institut für Virologie | Robert Koch Institute | |
| EPI_ISL_2631970, EPI_ISL_2762205, EPI_ISL_2762206 | Universitätsklinikum Schleswig-Holstein (standortübergreifend inkl. Kiel) - Institut für Klinische Chemie und Laboratoriumsdiagnostik | Robert Koch Institute | |
| EPI_ISL_2389387, EPI_ISL_2389392, EPI_ISL_2389399, EPI_ISL_2389400, EPI_ISL_2389404, EPI_ISL_2389416, EPI_ISL_2389420, EPI_ISL_2389422, EPI_ISL_2389426, EPI_ISL_2389427, EPI_ISL_2389437, EPI_ISL_2389439, EPI_ISL_2389440, EPI_ISL_2633741, EPI_ISL_2633749, EPI_ISL_2633809, EPI_ISL_2633815, EPI_ISL_2633878, EPI_ISL_2633884, EPI_ISL_2633889, EPI_ISL_2633896, EPI_ISL_2633900 | | | |
| see above | Universitätsklinikum des Saarlandes - Institut für Virologie | Robert Koch Institute | |
| EPI_ISL_2678379, EPI_ISL_2678380, EPI_ISL_2678381, EPI_ISL_2678383, EPI_ISL_2678384 | Virologisches Institut des Universitätsklinikums Erlangen | Robert Koch Institute | |
| EPI_ISL_2529472, EPI_ISL_2529474, EPI_ISL_2529475, EPI_ISL_2529477, EPI_ISL_2529479 | Virology, Universitätsklinikum des Saarlandes | Epigenetics, Saarland University | Jörn Walter; Kathrin Kattler; Nastasia Seiwert; Sascha Tierling; Sigrun Smola; Stefan Lohse; Thorsten Pfuhl |
| EPI_ISL_2448376, EPI_ISL_2448377, EPI_ISL_2448378, EPI_ISL_2448379, EPI_ISL_2448380, EPI_ISL_2448381, EPI_ISL_2448382, EPI_ISL_2448383, EPI_ISL_2448384, EPI_ISL_2448385, EPI_ISL_2448386, EPI_ISL_2448387, EPI_ISL_2448388, EPI_ISL_2448389, EPI_ISL_2448390, EPI_ISL_2448391, EPI_ISL_2448392, EPI_ISL_2448393 | | | |
| see above | Virology, Universitätsklinikum des Saarlandes | Epigenetics, Saarland University | Jörn Walter; Kathrin Kattler; Sascha Tierling; Sigrun Smola; Stefan Lohse; Thorsten Pfuhl |
| EPI_ISL_2381678, EPI_ISL_2381680, EPI_ISL_2381681, EPI_ISL_2381682, EPI_ISL_2381683, EPI_ISL_2381686, EPI_ISL_2381690, EPI_ISL_2381691, EPI_ISL_2381692, EPI_ISL_2381693, EPI_ISL_2381694, EPI_ISL_2381695, EPI_ISL_2381696, EPI_ISL_2381697, EPI_ISL_2381698, EPI_ISL_2381699, EPI_ISL_2381700, EPI_ISL_2381701, EPI_ISL_2381702, EPI_ISL_2381704, EPI_ISL_2381706, EPI_ISL_2381707, EPI_ISL_2381708, EPI_ISL_2381710, EPI_ISL_2381713, EPI_ISL_2381714, EPI_ISL_2381715, EPI_ISL_2381717, EPI_ISL_2381718, EPI_ISL_2381719, EPI_ISL_2381720, EPI_ISL_2381721, EPI_ISL_2381722, EPI_ISL_2381723, EPI_ISL_2381725, EPI_ISL_2381727, EPI_ISL_2381728, EPI_ISL_2381729, EPI_ISL_2381730, EPI_ISL_2381731, EPI_ISL_2381732, EPI_ISL_2381734, EPI_ISL_2381735, EPI_ISL_2381736, EPI_ISL_2381739, EPI_ISL_2381740, EPI_ISL_2381741, EPI_ISL_2381743, EPI_ISL_2381744, EPI_ISL_2381745, EPI_ISL_2381746, EPI_ISL_2381747, EPI_ISL_2381748, EPI_ISL_2381750, EPI_ISL_2381752, EPI_ISL_2381753, EPI_ISL_2381754, EPI_ISL_2381755, EPI_ISL_2381779, EPI_ISL_2381780, EPI_ISL_2381781, EPI_ISL_2381782, EPI_ISL_2381784, EPI_ISL_2381785, EPI_ISL_2381786, EPI_ISL_2381787, EPI_ISL_2381788, EPI_ISL_2381789, EPI_ISL_2381790, EPI_ISL_2381791, EPI_ISL_2381792, EPI_ISL_2381793, EPI_ISL_2381794, EPI_ISL_2381795, EPI_ISL_2381796, EPI_ISL_2381797, EPI_ISL_2381798, EPI_ISL_2381799, EPI_ISL_2381802, EPI_ISL_2381803, EPI_ISL_2381804, EPI_ISL_2381805, EPI_ISL_2381806, EPI_ISL_2381807, EPI_ISL_2381808, EPI_ISL_2381809, EPI_ISL_2381810, EPI_ISL_2381811, EPI_ISL_2381816, EPI_ISL_2381817, EPI_ISL_2381818, EPI_ISL_2381819, EPI_ISL_2381820, EPI_ISL_2381821, EPI_ISL_2381822, EPI_ISL_2381823, EPI_ISL_2381824, EPI_ISL_2381825, EPI_ISL_2381826, EPI_ISL_2381827, EPI_ISL_2381828, EPI_ISL_2381829, EPI_ISL_2381831, EPI_ISL_2381832, EPI_ISL_2381833, EPI_ISL_2381834, EPI_ISL_2381836, EPI_ISL_2381837, EPI_ISL_2381838, EPI_ISL_2381839, EPI_ISL_2381840, EPI_ISL_2381841, EPI_ISL_2381843, EPI_ISL_2381844, EPI_ISL_2381845, EPI_ISL_2381846, EPI_ISL_2381847, EPI_ISL_2381848, EPI_ISL_2381849, EPI_ISL_2381850, EPI_ISL_2381851, EPI_ISL_2381852, EPI_ISL_2381853, EPI_ISL_2381854, EPI_ISL_2381855, EPI_ISL_2381856, EPI_ISL_2381859, EPI_ISL_2381863, EPI_ISL_2381865, EPI_ISL_2381867, EPI_ISL_2381868, EPI_ISL_2381869, EPI_ISL_2381870, EPI_ISL_2381871, EPI_ISL_2381873, EPI_ISL_2381874, EPI_ISL_2381875, EPI_ISL_2381876, EPI_ISL_2381878, EPI_ISL_2381879, EPI_ISL_2381880, EPI_ISL_2381881, EPI_ISL_2381882, EPI_ISL_2381883, EPI_ISL_2381884, EPI_ISL_2381885, EPI_ISL_2381886, EPI_ISL_2381887, EPI_ISL_2381888, EPI_ISL_2381889, EPI_ISL_2381890, EPI_ISL_2381891, EPI_ISL_2381892, EPI_ISL_2381894, EPI_ISL_2381895, EPI_ISL_2381896, EPI_ISL_2381897, EPI_ISL_2381898, EPI_ISL_2381899 | | | |
| see above | ZOTZ KLIMAS MVZ Düsseldorf-Centrum GbR ÜBAG für Labormedizin, Genetik, Zytologie, Pathologie | Center of Medical Microbiology, Virology, and Hospital Hygiene, University of Duesseldorf | Alexander Dilthey; Andreas Walker; Daniel Strelow; Jessica Nicolai; Jörg Timm; Katrin Hoffmann; Klaus Pfeffer; Lisanna Hülse; Malte Kohns Vasconcelos; Maximilian Damagnez; Nadine Lübke; Patrick Finzer; Rainer Zotz; Tobias Wienemann; Torsten Houwaart |
| EPI_ISL_2389316, EPI_ISL_2389317, EPI_ISL_2389318, EPI_ISL_2389319, EPI_ISL_2389320, EPI_ISL_2389321, EPI_ISL_2389323, EPI_ISL_2470808, EPI_ISL_2470809, EPI_ISL_2470810, EPI_ISL_2470811, EPI_ISL_2470813, EPI_ISL_2470814, EPI_ISL_2470815, EPI_ISL_2632898, EPI_ISL_2632899, EPI_ISL_2632902, EPI_ISL_2632903, EPI_ISL_2632905, EPI_ISL_2632906, EPI_ISL_2632908 | | | |
| see above | alphaomega Labor GbR (Medizinische Kooperationsgemeinschaft Dres. Ackermann/Tennert/Zaiß) | Robert Koch Institute | |
| EPI_ISL_2470878, EPI_ISL_2470882, EPI_ISL_2470883, EPI_ISL_2470892, EPI_ISL_2470910, EPI_ISL_2470919, EPI_ISL_2470924, EPI_ISL_2470971, EPI_ISL_2470975, EPI_ISL_2470982, EPI_ISL_2470985, EPI_ISL_2470989, EPI_ISL_2470993, EPI_ISL_2470997, EPI_ISL_2471002, EPI_ISL_2471007, EPI_ISL_2632247, EPI_ISL_2632289, EPI_ISL_2632297, EPI_ISL_2632298, EPI_ISL_2632314, EPI_ISL_2632317, EPI_ISL_2632321, EPI_ISL_2635623, EPI_ISL_2635628, EPI_ISL_2635634 | | | |
| see above | amedes Göttingen | Robert Koch Institute | |
| EPI_ISL_2387193, EPI_ISL_2387203, EPI_ISL_2387208, EPI_ISL_2387210, EPI_ISL_2387215, EPI_ISL_2387225, EPI_ISL_2387243, EPI_ISL_2387253, EPI_ISL_2387284, EPI_ISL_2387303, EPI_ISL_2387307, EPI_ISL_2387310, EPI_ISL_2387316, EPI_ISL_2470864, EPI_ISL_2470868, EPI_ISL_2470884, EPI_ISL_2470887, EPI_ISL_2470890, EPI_ISL_2470891, EPI_ISL_2470893, EPI_ISL_2470894, EPI_ISL_2470896, EPI_ISL_2470906, EPI_ISL_2470907, EPI_ISL_2470908, EPI_ISL_2470912, EPI_ISL_2470928, EPI_ISL_2470932, EPI_ISL_2470941, EPI_ISL_2470955, EPI_ISL_2470958, EPI_ISL_2632249, EPI_ISL_2632257, EPI_ISL_2632272, EPI_ISL_2632273, EPI_ISL_2632276, EPI_ISL_2632280, EPI_ISL_2632282, EPI_ISL_2632292, EPI_ISL_2632294, EPI_ISL_2632295, EPI_ISL_2632303, EPI_ISL_2632316 | | | |
| see above | amedes MVZ DIAMEDIS Sennestadt | Robert Koch Institute | |
| EPI_ISL_2470863, EPI_ISL_2470876, EPI_ISL_2470879, EPI_ISL_2470888, EPI_ISL_2470897, EPI_ISL_2470900, EPI_ISL_2470903, EPI_ISL_2470915, EPI_ISL_2470917, EPI_ISL_2470921, EPI_ISL_2470922, EPI_ISL_2470923, EPI_ISL_2470935, EPI_ISL_2470936, EPI_ISL_2470938, EPI_ISL_2470940, EPI_ISL_2470953, EPI_ISL_2470960, EPI_ISL_2470966, EPI_ISL_2470967, EPI_ISL_2470970, EPI_ISL_2470972, EPI_ISL_2470976, EPI_ISL_2470992, EPI_ISL_2471000, EPI_ISL_2471005, EPI_ISL_2471013, EPI_ISL_2632254, EPI_ISL_2632275, EPI_ISL_2632277, EPI_ISL_2632302, EPI_ISL_2632306, EPI_ISL_2632309, EPI_ISL_2632320 | | | |
| see above | amedes MVZ Halle/Leipzig | Robert Koch Institute | |

| | | |
|---|---|---|
| EPI_ISL_2387191, EPI_ISL_2387267, EPI_ISL_2387278, EPI_ISL_2387296, EPI_ISL_2387299, EPI_ISL_2387313, EPI_ISL_2387329, EPI_ISL_2387343, EPI_ISL_2470880, EPI_ISL_2470901, EPI_ISL_2470904, EPI_ISL_2470926, EPI_ISL_2470931, EPI_ISL_2470949, EPI_ISL_2470950, EPI_ISL_2470969, EPI_ISL_2470981, EPI_ISL_2470988, EPI_ISL_2470990, EPI_ISL_2470994, EPI_ISL_2471001, EPI_ISL_2471009, EPI_ISL_2471014, EPI_ISL_2632243, EPI_ISL_2632246, EPI_ISL_2632251, EPI_ISL_2632253, EPI_ISL_2632262, EPI_ISL_2632266, EPI_ISL_2632269, EPI_ISL_2632283, EPI_ISL_2632307, EPI_ISL_2632313, EPI_ISL_2632315 | | |
| see above | amedes MVZ Hannover | Robert Koch Institute |
| EPI_ISL_2387201, EPI_ISL_2387223, EPI_ISL_2387230, EPI_ISL_2387232, EPI_ISL_2387234, EPI_ISL_2387254, EPI_ISL_2387271, EPI_ISL_2387309, EPI_ISL_2387312, EPI_ISL_2470866, EPI_ISL_2470867, EPI_ISL_2470871, EPI_ISL_2470881, EPI_ISL_2470885, EPI_ISL_2470889, EPI_ISL_2470899, EPI_ISL_2470902, EPI_ISL_2470911, EPI_ISL_2470916, EPI_ISL_2470920, EPI_ISL_2470927, EPI_ISL_2470937, EPI_ISL_2470942, EPI_ISL_2470946, EPI_ISL_2470951, EPI_ISL_2470962, EPI_ISL_2470974, EPI_ISL_2470979, EPI_ISL_2470996, EPI_ISL_2632241, EPI_ISL_2632245, EPI_ISL_2632252, EPI_ISL_2632255, EPI_ISL_2632256, EPI_ISL_2632263, EPI_ISL_2632265, EPI_ISL_2632274, EPI_ISL_2632278, EPI_ISL_2632293, EPI_ISL_2632305, EPI_ISL_2632308, EPI_ISL_2632318 | | |
| see above | amedes MVZ Jena | Robert Koch Institute |
| EPI_ISL_2632250 | amedes MVZ Zentrum für Pathologie und Zytodiagnostik GmbH (Köln) | Robert Koch Institute |
| EPI_ISL_2387260, EPI_ISL_2470873, EPI_ISL_2470895, EPI_ISL_2470914, EPI_ISL_2470930, EPI_ISL_2470954, EPI_ISL_2470984, EPI_ISL_2470986, EPI_ISL_2471010, EPI_ISL_2632248, EPI_ISL_2632258, EPI_ISL_2632284, EPI_ISL_2632296, EPI_ISL_2632301, EPI_ISL_2632311 | | |
| see above | amedes MVZ für Laboratoriumsdiagnostik Raubling GmbH | Robert Koch Institute |
| EPI_ISL_2470909, EPI_ISL_2470918, EPI_ISL_2470959, EPI_ISL_2632267 | amedes MVZ für Laboratoriumsmedizin Fürstenfeldbruck | Robert Koch Institute |
| EPI_ISL_2387288, EPI_ISL_2387325 | amedes MVZ für Laboratoriumsmedizin München | Robert Koch Institute |
| EPI_ISL_2387220, EPI_ISL_2387240, EPI_ISL_2387244, EPI_ISL_2470925, EPI_ISL_2470952, EPI_ISL_2632260, EPI_ISL_2632312 | | |
| see above | amedes MVZ für Laboratoriumsmedizin Rhein/Ruhr | Robert Koch Institute |
| EPI_ISL_2470869, EPI_ISL_2470870, EPI_ISL_2470945, EPI_ISL_2470961, EPI_ISL_2470991, EPI_ISL_2635608, EPI_ISL_2635612, EPI_ISL_2635630, EPI_ISL_2635640 | | |
| see above | amedes MVZ für Labordiagnostik Rhein-Main | Robert Koch Institute |
| EPI_ISL_2470956, EPI_ISL_2470978 | amedes Medlab Arnold Analytik MVZ GmbH | Robert Koch Institute |
| EPI_ISL_2386550, EPI_ISL_2386551, EPI_ISL_2386552, EPI_ISL_2386553, EPI_ISL_2386554, EPI_ISL_2386555, EPI_ISL_2386556, EPI_ISL_2386557, EPI_ISL_2386558, EPI_ISL_2386559, EPI_ISL_2386560, EPI_ISL_2386561, EPI_ISL_2386562, EPI_ISL_2386563, EPI_ISL_2386564, EPI_ISL_2386565, EPI_ISL_2386566, EPI_ISL_2386567, EPI_ISL_2386568, EPI_ISL_2386569, EPI_ISL_2386570, EPI_ISL_2386571, EPI_ISL_2386572, EPI_ISL_2386573, EPI_ISL_2386574, EPI_ISL_2386575, EPI_ISL_2469807, EPI_ISL_2469808, EPI_ISL_2469809, EPI_ISL_2469810, EPI_ISL_2469811, EPI_ISL_2469812, EPI_ISL_2469813, EPI_ISL_2469814, EPI_ISL_2469815, EPI_ISL_2469816, EPI_ISL_2469817, EPI_ISL_2469818, EPI_ISL_2469819, EPI_ISL_2469820, EPI_ISL_2469821, EPI_ISL_2469822, EPI_ISL_2469823, EPI_ISL_2469824, EPI_ISL_2469825, EPI_ISL_2469826, EPI_ISL_2469827, EPI_ISL_2469828, EPI_ISL_2469829, EPI_ISL_2469830, EPI_ISL_2469831, EPI_ISL_2469832, EPI_ISL_2469833, EPI_ISL_2469834, EPI_ISL_2469835, EPI_ISL_2634459, EPI_ISL_2634461, EPI_ISL_2634463, EPI_ISL_2634466, EPI_ISL_2634468, EPI_ISL_2634470, EPI_ISL_2634474, EPI_ISL_2634479, EPI_ISL_2634481, EPI_ISL_2634486, EPI_ISL_2634490, EPI_ISL_2634492, EPI_ISL_2634495, EPI_ISL_2634497, EPI_ISL_2634499, EPI_ISL_2634500, EPI_ISL_2634501, EPI_ISL_2634504 | | |
| see above | labopart - Medizinische Laboratorien Dresden | Robert Koch Institute |
| EPI_ISL_2387190, EPI_ISL_2387192, EPI_ISL_2387194, EPI_ISL_2387195, EPI_ISL_2387211, EPI_ISL_2387218, EPI_ISL_2387221, EPI_ISL_2387226, EPI_ISL_2387227, EPI_ISL_2387246, EPI_ISL_2387248, EPI_ISL_2387251, EPI_ISL_2387256, EPI_ISL_2387259, EPI_ISL_2387261, EPI_ISL_2387263, EPI_ISL_2387266, EPI_ISL_2387268, EPI_ISL_2387272, EPI_ISL_2387279, EPI_ISL_2387281, EPI_ISL_2387291, EPI_ISL_2387292, EPI_ISL_2387298, EPI_ISL_2387308, EPI_ISL_2387311, EPI_ISL_2387314, EPI_ISL_2387315, EPI_ISL_2387322, EPI_ISL_2387326, EPI_ISL_2387328, EPI_ISL_2387330, EPI_ISL_2387331, EPI_ISL_2387332, EPI_ISL_2387335, EPI_ISL_2387336, EPI_ISL_2387337, EPI_ISL_2387338, EPI_ISL_2387341, EPI_ISL_2387342, EPI_ISL_2470874, EPI_ISL_2470877, EPI_ISL_2470886, EPI_ISL_2470905, EPI_ISL_2470913, EPI_ISL_2470929, EPI_ISL_2470939, EPI_ISL_2470944, EPI_ISL_2470948, EPI_ISL_2470957, EPI_ISL_2470964, EPI_ISL_2470965, EPI_ISL_2470973, EPI_ISL_2470977, EPI_ISL_2470980, EPI_ISL_2470987, EPI_ISL_2470995, EPI_ISL_2470999, EPI_ISL_2471003, EPI_ISL_2471004, EPI_ISL_2471006, EPI_ISL_2471011, EPI_ISL_2471012, EPI_ISL_2471015, EPI_ISL_2471016 | | |
| see above | nordlab - Partnerschaftspraxis für Laboratoriumsmedizin | Robert Koch Institute |

We gratefully acknowledge the following Authors from the Originating laboratories responsible for obtaining the specimens, as well as the Submitting laboratories where the genome data were generated and shared via GISAID, on which this research is based.

All Submitters of data may be contacted directly via www.gisaid.org

Authors are sorted alphabetically.

| Accession ID | Originating Laboratory | Submitting Laboratory | Authors |
|---|---|---|---|
| EPI_ISL_2833500, EPI_ISL_2833501, EPI_ISL_2833502, EPI_ISL_2833503, EPI_ISL_2833504, EPI_ISL_2833505, EPI_ISL_2833506, EPI_ISL_2833507, EPI_ISL_2833508, EPI_ISL_2833509, EPI_ISL_2833510, EPI_ISL_2833511, EPI_ISL_2833512, EPI_ISL_2833513, EPI_ISL_2833514 | | | |
| see above | A. Krumbholz, Labor Dr. Krause und Kollegen MVZ GmbH, Kiel | Charité Universitätsmedizin Berlin, Institut für Virologie | Andi Krumbholz; Barbara Muehlemann; Christian Drosten; Julia Schneider; Julia Tesch; Jörn Beheim-Schwarzbach; Talitha Veith; Terry Jones; Tobias Bleicker; Victor M Corman |
| EPI_ISL_2504058, EPI_ISL_2504059, EPI_ISL_2504060, EPI_ISL_2504061, EPI_ISL_2504063, EPI_ISL_2504066, EPI_ISL_2833515, EPI_ISL_2833516, EPI_ISL_2833517, EPI_ISL_2833518, EPI_ISL_2833519, EPI_ISL_2833520, EPI_ISL_2833521, EPI_ISL_2833522, EPI_ISL_2833523, EPI_ISL_2833525, EPI_ISL_2833529, EPI_ISL_2833530, EPI_ISL_2833533, EPI_ISL_2833535, EPI_ISL_2833537, EPI_ISL_2833539, EPI_ISL_2833540 | | | |
| see above | A. Krumbholz, Labor Dr. Krause und Kollegen MVZ GmbH, Kiel | Charité Universitätsmedizin Berlin, Institut für Virologie/Labor Berlin | Andi Krumbholz; Barbara Muehlemann; Barbara Mühlemann; Barbara Müehlemann; Christian Drosten; Christine Stephan; Joern Beheim-Schwarzbach; Julia Schneider; Julia Tesch; Peter Menzel; Rolf Schwarzer; Talitha Veith; Terry Jones; Tobias Bleicker; Victor M Corman |
| EPI_ISL_2693902, EPI_ISL_2693903, EPI_ISL_2693904, EPI_ISL_2894997, EPI_ISL_2894998, EPI_ISL_2894999, EPI_ISL_2895000 | | | |
| see above | A. Krumbholz, Labor Dr. Krause und Kollegen MVZ GmbH, Kiel | Charité Universitätsmedizin Berlin, Institute of Virology | Barbara Mühlemann; Christian Drosten; Julia Schneider; Julia Tesch; Jörn Beheim-Schwarzbach; Talitha Veith; Terry Jones; Tobias Bleicker; Victor M Corman |
| EPI_ISL_2637080, EPI_ISL_2637125, EPI_ISL_2759826, EPI_ISL_2761105, EPI_ISL_2761125, EPI_ISL_2761143, EPI_ISL_2845356 | | | |
| see above | ALB FILS KLINIKEN GmbH - Institut für Laboratoriumsmedizin Klinik am Eichert Göppingen | Robert Koch Institute | |
| EPI_ISL_2760022, EPI_ISL_2760023, EPI_ISL_2760025, EPI_ISL_2760054, EPI_ISL_2760055, EPI_ISL_2760056, EPI_ISL_2760058, EPI_ISL_2760067, EPI_ISL_2760084, EPI_ISL_2760098, EPI_ISL_2760119, EPI_ISL_2760131, EPI_ISL_2760141, EPI_ISL_2762207, EPI_ISL_2762208, EPI_ISL_2762209, EPI_ISL_2762210, EPI_ISL_2762212, EPI_ISL_2762214, EPI_ISL_2762215, EPI_ISL_2762217, EPI_ISL_2762218, EPI_ISL_2762219, EPI_ISL_2762220, EPI_ISL_2762221, EPI_ISL_2762222, EPI_ISL_2762223, EPI_ISL_2762224, EPI_ISL_2762225, EPI_ISL_2762226, EPI_ISL_2762228, EPI_ISL_2762229, EPI_ISL_2762230, EPI_ISL_2762231, EPI_ISL_2762232, EPI_ISL_2762233, EPI_ISL_2762234, EPI_ISL_2762236, EPI_ISL_2762237, EPI_ISL_2762239, EPI_ISL_2762240, EPI_ISL_2762242, EPI_ISL_2762244, EPI_ISL_2762245, EPI_ISL_2762246, EPI_ISL_2762247, EPI_ISL_2762248, EPI_ISL_2762250, EPI_ISL_2762251, EPI_ISL_2762252, EPI_ISL_2762253, EPI_ISL_2762254, EPI_ISL_2762255, EPI_ISL_2762256, EPI_ISL_2762257, EPI_ISL_2762258, EPI_ISL_2762259, EPI_ISL_2762260, EPI_ISL_2762261, EPI_ISL_2762262, EPI_ISL_2762263, EPI_ISL_2762264, EPI_ISL_2762265, EPI_ISL_2762267, EPI_ISL_2762268, EPI_ISL_2762269, EPI_ISL_2762272, EPI_ISL_2762273, EPI_ISL_2762274, EPI_ISL_2762275, EPI_ISL_2845886, EPI_ISL_2845887, EPI_ISL_2845888, EPI_ISL_2845889, EPI_ISL_2845890, EPI_ISL_2845891, EPI_ISL_2845892, EPI_ISL_2845893, EPI_ISL_2845894, EPI_ISL_2845895, EPI_ISL_2845896, EPI_ISL_2845897, EPI_ISL_2845898, EPI_ISL_2845899, EPI_ISL_2845900, EPI_ISL_2845901, EPI_ISL_2845902, EPI_ISL_2845903, EPI_ISL_2845904, EPI_ISL_2845905, EPI_ISL_2845906, EPI_ISL_2845907, EPI_ISL_2845908, EPI_ISL_2845909, EPI_ISL_2845910, EPI_ISL_2845911, EPI_ISL_2845912, EPI_ISL_2845913, EPI_ISL_2845914, EPI_ISL_2845915, EPI_ISL_2845916, EPI_ISL_2845917, EPI_ISL_2845918, EPI_ISL_2845919, EPI_ISL_2845920, EPI_ISL_2845921, EPI_ISL_2845922, EPI_ISL_2845923, EPI_ISL_2845924, EPI_ISL_2845925, EPI_ISL_2845926, EPI_ISL_2845927, EPI_ISL_2845928, EPI_ISL_2845929, EPI_ISL_2845930, EPI_ISL_2845931, EPI_ISL_2845932, EPI_ISL_2845933, EPI_ISL_2845934 | | | |
| see above | Bayerisches Landesamt für Gesundheit und Lebensmittelsicherheit (LGL) | Robert Koch Institute | |
| EPI_ISL_2636914, EPI_ISL_2636921, EPI_ISL_2636922, EPI_ISL_2636924, EPI_ISL_2636935, EPI_ISL_2636938, EPI_ISL_2636947, EPI_ISL_2636949, EPI_ISL_2636955, EPI_ISL_2636957, EPI_ISL_2636960, EPI_ISL_2636963, EPI_ISL_2636976, EPI_ISL_2636978, EPI_ISL_2636979, EPI_ISL_2636983, EPI_ISL_2636986, EPI_ISL_2636988, EPI_ISL_2636991, EPI_ISL_2636993, EPI_ISL_2636995, EPI_ISL_2636996, EPI_ISL_2637001, EPI_ISL_2637002, EPI_ISL_2637003, EPI_ISL_2637004, EPI_ISL_2637010, EPI_ISL_2637014, EPI_ISL_2637020, EPI_ISL_2637021, EPI_ISL_2637034, EPI_ISL_2637038, EPI_ISL_2637046, EPI_ISL_2637054, EPI_ISL_2637056, EPI_ISL_2637057, EPI_ISL_2637060, EPI_ISL_2637063, EPI_ISL_2637065, EPI_ISL_2637070, EPI_ISL_2637075, EPI_ISL_2637092, EPI_ISL_2637098, EPI_ISL_2637099, EPI_ISL_2637103, EPI_ISL_2637110, EPI_ISL_2637112, EPI_ISL_2637121, EPI_ISL_2637122, EPI_ISL_2637124, EPI_ISL_2637135, EPI_ISL_2637139, EPI_ISL_2637141, EPI_ISL_2637145, EPI_ISL_2637146, EPI_ISL_2637153, EPI_ISL_2637156, EPI_ISL_2637157, EPI_ISL_2637160, EPI_ISL_2637172, EPI_ISL_2637173, EPI_ISL_2637175, EPI_ISL_2637182, EPI_ISL_2637188, EPI_ISL_2759829, EPI_ISL_2759831, EPI_ISL_2759839, EPI_ISL_2759840, EPI_ISL_2759842, EPI_ISL_2759848, EPI_ISL_2761070, EPI_ISL_2761078, EPI_ISL_2761086, EPI_ISL_2761088, EPI_ISL_2761091, EPI_ISL_2761093, EPI_ISL_2761108, EPI_ISL_2761112, EPI_ISL_2761114, EPI_ISL_2761117, EPI_ISL_2761122, EPI_ISL_2761127, EPI_ISL_2761132, EPI_ISL_2761141, EPI_ISL_2761146, EPI_ISL_2761153, EPI_ISL_2761156, EPI_ISL_2761159, EPI_ISL_2761162, EPI_ISL_2761164, EPI_ISL_2761178, EPI_ISL_2761182, EPI_ISL_2761184, EPI_ISL_2761208, EPI_ISL_2761232, EPI_ISL_2761234, EPI_ISL_2761240, EPI_ISL_2761247, EPI_ISL_2761250, EPI_ISL_2761254, EPI_ISL_2761259, EPI_ISL_2761269, EPI_ISL_2761292, EPI_ISL_2761297, EPI_ISL_2761299, EPI_ISL_2761309, EPI_ISL_2761316, EPI_ISL_2761318, EPI_ISL_2761321, EPI_ISL_2761326, EPI_ISL_2761328, EPI_ISL_2761347, EPI_ISL_2761349, EPI_ISL_2761357, EPI_ISL_2761360, EPI_ISL_2761367, EPI_ISL_2761374, EPI_ISL_2761379, EPI_ISL_2761384, EPI_ISL_2761389, EPI_ISL_2761395, EPI_ISL_2761400, EPI_ISL_2761403, EPI_ISL_2761413, EPI_ISL_2761417, EPI_ISL_2761434, EPI_ISL_2761441, EPI_ISL_2761451, EPI_ISL_2761453, EPI_ISL_2761455, EPI_ISL_2761460, EPI_ISL_2761462, EPI_ISL_2761464, EPI_ISL_2761471, EPI_ISL_2761477, EPI_ISL_2761484, EPI_ISL_2761489, EPI_ISL_2761491, EPI_ISL_2761493, EPI_ISL_2761496, EPI_ISL_2761500, EPI_ISL_2761510, EPI_ISL_2761524, EPI_ISL_2761528, EPI_ISL_2761533, EPI_ISL_2761536, EPI_ISL_2761538, EPI_ISL_2761551, EPI_ISL_2761556, EPI_ISL_2761561, EPI_ISL_2761568, EPI_ISL_2761570, EPI_ISL_2761578, EPI_ISL_2761588, EPI_ISL_2761593, EPI_ISL_2761598, EPI_ISL_2761600, EPI_ISL_2761603, EPI_ISL_2761972, EPI_ISL_2761974, EPI_ISL_2761976, EPI_ISL_2761978, EPI_ISL_2761979, EPI_ISL_2761983, EPI_ISL_2761989, EPI_ISL_2761990, EPI_ISL_2761991, EPI_ISL_2761992, EPI_ISL_2761995, EPI_ISL_2761998, EPI_ISL_2762003, EPI_ISL_2762004, EPI_ISL_2762009, EPI_ISL_2762017, EPI_ISL_2762018, EPI_ISL_2762019, EPI_ISL_2762021, EPI_ISL_2762023, EPI_ISL_2762026, EPI_ISL_2762031, EPI_ISL_2762033, EPI_ISL_2762034, EPI_ISL_2762036, EPI_ISL_2762038, EPI_ISL_2762044, EPI_ISL_2762045, EPI_ISL_2762047, EPI_ISL_2762048, EPI_ISL_2762049, EPI_ISL_2762051, EPI_ISL_2762054, EPI_ISL_2762055, EPI_ISL_2762061, EPI_ISL_2762062, EPI_ISL_2762064, EPI_ISL_2762065, EPI_ISL_2762068, EPI_ISL_2762070, EPI_ISL_2762071, EPI_ISL_2762073, EPI_ISL_2762075, EPI_ISL_2762076, EPI_ISL_2762078, EPI_ISL_2762079, EPI_ISL_2762080, EPI_ISL_2762081, EPI_ISL_2762082, EPI_ISL_2762083, EPI_ISL_2762084, EPI_ISL_2762086, EPI_ISL_2762087, EPI_ISL_2762090, EPI_ISL_2762091, EPI_ISL_2762096, EPI_ISL_2762098, EPI_ISL_2762102, EPI_ISL_2762641, EPI_ISL_2762647, EPI_ISL_2762652, EPI_ISL_2762654, EPI_ISL_2762656, EPI_ISL_2762664, EPI_ISL_2762666, EPI_ISL_2762669, EPI_ISL_2762671, EPI_ISL_2762673, EPI_ISL_2762676, EPI_ISL_2762679, EPI_ISL_2762681, EPI_ISL_2762684, EPI_ISL_2762686, EPI_ISL_2762688, EPI_ISL_2762693, EPI_ISL_2762695, EPI_ISL_2762700, EPI_ISL_2762703, EPI_ISL_2762705, EPI_ISL_2762710, EPI_ISL_2762712, EPI_ISL_2762718, EPI_ISL_2762720, EPI_ISL_2762722, EPI_ISL_2762727, EPI_ISL_2762729, EPI_ISL_2762733, EPI_ISL_2762735, EPI_ISL_2762737, EPI_ISL_2762739, EPI_ISL_2762742, EPI_ISL_2762744, EPI_ISL_2762747, EPI_ISL_2762751, EPI_ISL_2762753, EPI_ISL_2762755, EPI_ISL_2762758, EPI_ISL_2762760, EPI_ISL_2762762, EPI_ISL_2762766, EPI_ISL_2762768, EPI_ISL_2762770, EPI_ISL_2762775, EPI_ISL_2762778, EPI_ISL_2762785, EPI_ISL_2762788, EPI_ISL_2762791, EPI_ISL_2762793, EPI_ISL_2762798, EPI_ISL_2762810, EPI_ISL_2762812, EPI_ISL_2762815, EPI_ISL_2762817, EPI_ISL_2762820, EPI_ISL_2762822, EPI_ISL_2845320, EPI_ISL_2845323, EPI_ISL_2845325, EPI_ISL_2845327, EPI_ISL_2845330, EPI_ISL_2845331, EPI_ISL_2845332, EPI_ISL_2845334, EPI_ISL_2845335, EPI_ISL_2845336, EPI_ISL_2845337, EPI_ISL_2845338, EPI_ISL_2845340, EPI_ISL_2845341, EPI_ISL_2845342, EPI_ISL_2845345, EPI_ISL_2845347, EPI_ISL_2845349, EPI_ISL_2845352, EPI_ISL_2845353, EPI_ISL_2845354, EPI_ISL_2845358, EPI_ISL_2845361, EPI_ISL_2845365, EPI_ISL_2845366, EPI_ISL_2845369, EPI_ISL_2845370, EPI_ISL_2845371, EPI_ISL_2845377, EPI_ISL_2845381, EPI_ISL_2845382, EPI_ISL_2845384, EPI_ISL_2845387, EPI_ISL_2845389, EPI_ISL_2845399, EPI_ISL_2845400, EPI_ISL_2845410, EPI_ISL_2845413, EPI_ISL_2845417, EPI_ISL_2845418, EPI_ISL_2845419, EPI_ISL_2845421, EPI_ISL_2845422, EPI_ISL_2845426, EPI_ISL_2845432, EPI_ISL_2845433, EPI_ISL_2845439, EPI_ISL_2845440, EPI_ISL_2845441 | | | |
| see above | Bioscientia Labor Wermsdorf | Robert Koch Institute | |
| EPI_ISL_2636913, EPI_ISL_2637000, EPI_ISL_2637111, EPI_ISL_2759837, EPI_ISL_2761130, EPI_ISL_2761180, EPI_ISL_2761261, EPI_ISL_2761277, EPI_ISL_2761285, EPI_ISL_2761314, EPI_ISL_2761387, EPI_ISL_2761405, EPI_ISL_2761436, EPI_ISL_2761469, EPI_ISL_2761486, EPI_ISL_2761508, EPI_ISL_2761602, EPI_ISL_2761985, EPI_ISL_2762000, EPI_ISL_2762007, EPI_ISL_2762013, EPI_ISL_2762030, EPI_ISL_2762041, EPI_ISL_2762094, EPI_ISL_2762662, EPI_ISL_2762725, EPI_ISL_2762796, EPI_ISL_2845317, EPI_ISL_2845339, EPI_ISL_2845344, EPI_ISL_2845374, EPI_ISL_2845375, EPI_ISL_2845406, EPI_ISL_2845423 | | | |
| see above | Bioscientia MVZ Labor Karlsruhe GmbH | Robert Koch Institute | |
| EPI_ISL_2727664, EPI_ISL_2727665 | Bundeswehrkrankenhaus Berlin | Bundeswehr Institute of Microbiology | Alexandra Rehn; Enrico Georgi; Malena Bestehorn-Willmann; Markus Antwerpen; Mathias Walter; Mike Pillukat; Roman Wölfel; Sabine Zange |
| EPI_ISL_2727667 | Bundeswehrkrankenhaus Ulm | Bundeswehr Institute of Microbiology | Alexandra Rehn; Enrico Georgi; Malena Bestehorn-Willmann; Markus Antwerpen; Mathias Walter; Mike Pillukat; Roman Wölfel; Sabine Zange |
| EPI_ISL_2633464, EPI_ISL_2633466, EPI_ISL_2633479, EPI_ISL_2633502, EPI_ISL_2633503, EPI_ISL_2633551, EPI_ISL_2633557, EPI_ISL_2633565, EPI_ISL_2633581, EPI_ISL_2633584, EPI_ISL_2633586, EPI_ISL_2633590, EPI_ISL_2633595, EPI_ISL_2633596, EPI_ISL_2633606, EPI_ISL_2633610, EPI_ISL_2633613, EPI_ISL_2633614, EPI_ISL_2633637, EPI_ISL_2633650, EPI_ISL_2633655, EPI_ISL_2633658, EPI_ISL_2633661, EPI_ISL_2633664, EPI_ISL_2633679, EPI_ISL_2633681, EPI_ISL_2633688, EPI_ISL_2761736, EPI_ISL_2761737, EPI_ISL_2761738, EPI_ISL_2761739, EPI_ISL_2761740, EPI_ISL_2761741, EPI_ISL_2761742, EPI_ISL_2761743, EPI_ISL_2761744, EPI_ISL_2761745, EPI_ISL_2761746, EPI_ISL_2761747, EPI_ISL_2761748, EPI_ISL_2761749, EPI_ISL_2761750, EPI_ISL_2761751, EPI_ISL_2761752, EPI_ISL_2761753, EPI_ISL_2761754, EPI_ISL_2761755, EPI_ISL_2761756, EPI_ISL_2761757, EPI_ISL_2761758, EPI_ISL_2761759, EPI_ISL_2761760, EPI_ISL_2761761, EPI_ISL_2761762, EPI_ISL_2761763, EPI_ISL_2761764, EPI_ISL_2761765, EPI_ISL_2761766, EPI_ISL_2761767, EPI_ISL_2761768, EPI_ISL_2761769, EPI_ISL_2761770, EPI_ISL_2761771, EPI_ISL_2761772, EPI_ISL_2761773, EPI_ISL_2761774, EPI_ISL_2761775, EPI_ISL_2761776, EPI_ISL_2761777, EPI_ISL_2761778, EPI_ISL_2761779, EPI_ISL_2761780, EPI_ISL_2761781, EPI_ISL_2761782, EPI_ISL_2761783, EPI_ISL_2761784, EPI_ISL_2761785, EPI_ISL_2761786, EPI_ISL_2761787, EPI_ISL_2761788, EPI_ISL_2761789, EPI_ISL_2761790, EPI_ISL_2761791, EPI_ISL_2761792, EPI_ISL_2761793, EPI_ISL_2761794, EPI_ISL_2761795, EPI_ISL_2761796, EPI_ISL_2761797, EPI_ISL_2761798, EPI_ISL_2761799, EPI_ISL_2761800, EPI_ISL_2761801, EPI_ISL_2761802, EPI_ISL_2761803, EPI_ISL_2761804, EPI_ISL_2761805, EPI_ISL_2761806, EPI_ISL_2761807, EPI_ISL_2761808, EPI_ISL_2761809, EPI_ISL_2761810, EPI_ISL_2761811, EPI_ISL_2761812, EPI_ISL_2761813, EPI_ISL_2761814, EPI_ISL_2761815, EPI_ISL_2761816, EPI_ISL_2761817, EPI_ISL_2761818, EPI_ISL_2761819, EPI_ISL_2761820, EPI_ISL_2761821, EPI_ISL_2761822, EPI_ISL_2761823, EPI_ISL_2761824, EPI_ISL_2761825, EPI_ISL_2761826, EPI_ISL_2761827, EPI_ISL_2761828, EPI_ISL_2761829, EPI_ISL_2761830, EPI_ISL_2761831, EPI_ISL_2761832, EPI_ISL_2761833, EPI_ISL_2761834, EPI_ISL_2761835, EPI_ISL_2761836, EPI_ISL_2761837, EPI_ISL_2761838, EPI_ISL_2761839, EPI_ISL_2761840, EPI_ISL_2761841, EPI_ISL_2761842, EPI_ISL_2761843, EPI_ISL_2761844, EPI_ISL_2761845, EPI_ISL_2761846, EPI_ISL_2761847, EPI_ISL_2761848, EPI_ISL_2761849, EPI_ISL_2761850, EPI_ISL_2761851, EPI_ISL_2761852, EPI_ISL_2761853, EPI_ISL_2761854, EPI_ISL_2761855, EPI_ISL_2761856, EPI_ISL_2761857, EPI_ISL_2761858, EPI_ISL_2761859, EPI_ISL_2761860, EPI_ISL_2761861, EPI_ISL_2761862, EPI_ISL_2761863, EPI_ISL_2761864, EPI_ISL_2761865, EPI_ISL_2761866, EPI_ISL_2761867, EPI_ISL_2845466, EPI_ISL_2845467, EPI_ISL_2845468, EPI_ISL_2845469, EPI_ISL_2845470, EPI_ISL_2845471, EPI_ISL_2845472, EPI_ISL_2845473, EPI_ISL_2845474, EPI_ISL_2845475, EPI_ISL_2845476, EPI_ISL_2845477, EPI_ISL_2845478, EPI_ISL_2845479, EPI_ISL_2845480, EPI_ISL_2845481, EPI_ISL_2845482, EPI_ISL_2845483, EPI_ISL_2845484, EPI_ISL_2845485, EPI_ISL_2845486, EPI_ISL_2845487, EPI_ISL_2845488, EPI_ISL_2845489, EPI_ISL_2845490, EPI_ISL_2845491, EPI_ISL_2845492, EPI_ISL_2845493, EPI_ISL_2845494, EPI_ISL_2845495, EPI_ISL_2845496, EPI_ISL_2845497, EPI_ISL_2845498, EPI_ISL_2845499, EPI_ISL_2845500, EPI_ISL_2845501, EPI_ISL_2845502, EPI_ISL_2845503, EPI_ISL_2845504, EPI_ISL_2845505, EPI_ISL_2845506, EPI_ISL_2845507, EPI_ISL_2845508, EPI_ISL_2845509, EPI_ISL_2845510, EPI_ISL_2845511, EPI_ISL_2845512, EPI_ISL_2845513, EPI_ISL_2845514, EPI_ISL_2845515, EPI_ISL_2845516, EPI_ISL_2845517, EPI_ISL_2845518, EPI_ISL_2845519, EPI_ISL_2845520, EPI_ISL_2845521, EPI_ISL_2845522, EPI_ISL_2845523, EPI_ISL_2845524, EPI_ISL_2845525, EPI_ISL_2845526, EPI_ISL_2845527, EPI_ISL_2845528, EPI_ISL_2845529, EPI_ISL_2845530, EPI_ISL_2845531, EPI_ISL_2845532, EPI_ISL_2845533, EPI_ISL_2845534, EPI_ISL_2845535, EPI_ISL_2845536, EPI_ISL_2845537, EPI_ISL_2845538, EPI_ISL_2845539, EPI_ISL_2845540, EPI_ISL_2845541, EPI_ISL_2845542 | | | |
| see above | CENTOGENE Frankfurt Laboratory: Niederlassung Industriepark Höchst | Robert Koch Institute | |
| EPI_ISL_2760265, EPI_ISL_2844846, EPI_ISL_2844847, EPI_ISL_2844848, EPI_ISL_2844849 | Chemische und Veterinäruntersuchungsamt Münsterland-Emscher-Lippe (CVUA-MEL) | Robert Koch Institute | |
| EPI_ISL_2634713, EPI_ISL_2634715, EPI_ISL_2760362, EPI_ISL_2760364, EPI_ISL_2760366, EPI_ISL_2760368, EPI_ISL_2760371, EPI_ISL_2760373, EPI_ISL_2760376, EPI_ISL_2760378, EPI_ISL_2760381, EPI_ISL_2760383, EPI_ISL_2760385, EPI_ISL_2760387, EPI_ISL_2760390, EPI_ISL_2760392, EPI_ISL_2845935, EPI_ISL_2845936, EPI_ISL_2845937, EPI_ISL_2845938, EPI_ISL_2845939, EPI_ISL_2845940, EPI_ISL_2845941, EPI_ISL_2845942, EPI_ISL_2845943, EPI_ISL_2845944, EPI_ISL_2845945, EPI_ISL_2845946, EPI_ISL_2845947, EPI_ISL_2845948, EPI_ISL_2845949, EPI_ISL_2845950, EPI_ISL_2845951 | | | |
| see above | Clemenshospital - Alexianer Clemenshospital Münster | Robert Koch Institute | |
| EPI_ISL_2636126, EPI_ISL_2636139 | Corona-Testzentrum ifp Institut für Produktqualität GmbH | Robert Koch Institute | |
| EPI_ISL_2762180, EPI_ISL_2762187, EPI_ISL_2762198, EPI_ISL_2845750, EPI_ISL_2845751, EPI_ISL_2845754, EPI_ISL_2845758 | | | |
| see above | DKMS Life Science Lab GmbH Dresden | Robert Koch Institute | |
| EPI_ISL_2759781, EPI_ISL_2759789, EPI_ISL_2759793, EPI_ISL_2759798, EPI_ISL_2759802, EPI_ISL_2759812, EPI_ISL_2759814, EPI_ISL_2759815 | | | |
| see above | Department für Labormedizin Abteilung III Bereich Molekulare Diagnostik Universitätsklinikum Halle(Saale) | Robert Koch Institute | |
| EPI_ISL_2762307, EPI_ISL_2762308, EPI_ISL_2762309, EPI_ISL_2762310, EPI_ISL_2762311, EPI_ISL_2762312, EPI_ISL_2762314, EPI_ISL_2762316, EPI_ISL_2762317, EPI_ISL_2762318, EPI_ISL_2762319, EPI_ISL_2762320, EPI_ISL_2762321, EPI_ISL_2762322, EPI_ISL_2762323, EPI_ISL_2762324, EPI_ISL_2762325, EPI_ISL_2762326, EPI_ISL_2762327, EPI_ISL_2762328, EPI_ISL_2762329, EPI_ISL_2762330, EPI_ISL_2762331, EPI_ISL_2762332, EPI_ISL_2762333, EPI_ISL_2762334, EPI_ISL_2762335, EPI_ISL_2762336, EPI_ISL_2762337, EPI_ISL_2762338, EPI_ISL_2762339, EPI_ISL_2762340, EPI_ISL_2762341, EPI_ISL_2762342, EPI_ISL_2762343, EPI_ISL_2762349, EPI_ISL_2762351, EPI_ISL_2762354, EPI_ISL_2762356, EPI_ISL_2762359, EPI_ISL_2762361, EPI_ISL_2762363, EPI_ISL_2762365, EPI_ISL_2762370, EPI_ISL_2762372, EPI_ISL_2762374, EPI_ISL_2762377, EPI_ISL_2762380, EPI_ISL_2762383, EPI_ISL_2762385, EPI_ISL_2762390, EPI_ISL_2762392, EPI_ISL_2762395, EPI_ISL_2762397, EPI_ISL_2762400, EPI_ISL_2762402, EPI_ISL_2762405, EPI_ISL_2762407, EPI_ISL_2762409, EPI_ISL_2762412, EPI_ISL_2762415, EPI_ISL_2762417, EPI_ISL_2762420, EPI_ISL_2762421, EPI_ISL_2762424, EPI_ISL_2762429, EPI_ISL_2762431, EPI_ISL_2762433, EPI_ISL_2762436, EPI_ISL_2762438, EPI_ISL_2762440, EPI_ISL_2762442, EPI_ISL_2762445, EPI_ISL_2762447, EPI_ISL_2762450 | | | |
| see above | Diagnosticum - Labor Neukirchen | Robert Koch Institute | |
| EPI_ISL_2635961, EPI_ISL_2635962, EPI_ISL_2635964, EPI_ISL_2635965, EPI_ISL_2635966, EPI_ISL_2636009, EPI_ISL_2636010, EPI_ISL_2636011, EPI_ISL_2636012, EPI_ISL_2636013, EPI_ISL_2636015, EPI_ISL_2636016, EPI_ISL_2678439, EPI_ISL_2678440, EPI_ISL_2678441, EPI_ISL_2678442, EPI_ISL_2678443, EPI_ISL_2678444, EPI_ISL_2678445, EPI_ISL_2845778, EPI_ISL_2845779, EPI_ISL_2845780, EPI_ISL_2845781, EPI_ISL_2845782, EPI_ISL_2845783, EPI_ISL_2845784, EPI_ISL_2845785, EPI_ISL_2845787, EPI_ISL_2845788, EPI_ISL_2846005, EPI_ISL_2846006, EPI_ISL_2846007 | | | |
| see above | Dianovis GmbH Greiz | Robert Koch Institute | |
| EPI_ISL_2634789, EPI_ISL_2634793 | ELBLAB GmbH Zentrum für LaborMedizin | Robert Koch Institute | |
| EPI_ISL_2762141, EPI_ISL_2762142, EPI_ISL_2762143, EPI_ISL_2762144, EPI_ISL_2762145, EPI_ISL_2762146, EPI_ISL_2762147, EPI_ISL_2762148, EPI_ISL_2762149, EPI_ISL_2762150, EPI_ISL_2762151, EPI_ISL_2762152, EPI_ISL_2762153, EPI_ISL_2762154, EPI_ISL_2762155, EPI_ISL_2762156, EPI_ISL_2762157, EPI_ISL_2762158, EPI_ISL_2762159, EPI_ISL_2762160, EPI_ISL_2762161, EPI_ISL_2762162, EPI_ISL_2762163, EPI_ISL_2762164, EPI_ISL_2762165, EPI_ISL_2845834, EPI_ISL_2845835, EPI_ISL_2845836, EPI_ISL_2845837, EPI_ISL_2845838, EPI_ISL_2845839, EPI_ISL_2845840, EPI_ISL_2845841, EPI_ISL_2845842, EPI_ISL_2845843, EPI_ISL_2845844, EPI_ISL_2845845, EPI_ISL_2845846, EPI_ISL_2845847, EPI_ISL_2845848, EPI_ISL_2845849, EPI_ISL_2845850, EPI_ISL_2845851, EPI_ISL_2845852, EPI_ISL_2845853, EPI_ISL_2845854, EPI_ISL_2845855, EPI_ISL_2845856, EPI_ISL_2845857, EPI_ISL_2845858, EPI_ISL_2845859, EPI_ISL_2845860, EPI_ISL_2845861, EPI_ISL_2845862, EPI_ISL_2845863, EPI_ISL_2845864, EPI_ISL_2845865, EPI_ISL_2845866, EPI_ISL_2845867, EPI_ISL_2845868, EPI_ISL_2845869, EPI_ISL_2845870, EPI_ISL_2845871, EPI_ISL_2845872, EPI_ISL_2845873, EPI_ISL_2845874, EPI_ISL_2845875, EPI_ISL_2845876, EPI_ISL_2845877, EPI_ISL_2845878, EPI_ISL_2845879, EPI_ISL_2845880, EPI_ISL_2845881, EPI_ISL_2845882, EPI_ISL_2845883, EPI_ISL_2845884, EPI_ISL_2845885 | | | |
| see above | Eurofins LifeCodexx GmbH | Robert Koch Institute | |
| EPI_ISL_2471915, EPI_ISL_2678367, EPI_ISL_2678368, EPI_ISL_2678369, EPI_ISL_2678370, EPI_ISL_2762133, EPI_ISL_2762134, EPI_ISL_2762135 | | | |
| see above | Eurofins MVZ Labor Gelsenkirchen | Robert Koch Institute | |
| EPI_ISL_2762166, EPI_ISL_2762167, EPI_ISL_2762168 | Fraunhofer-Institut für Zelltherapie und Immunologie IZI AG Next-Generation Diagnostics | Robert Koch Institute | |
| EPI_ISL_2635974, EPI_ISL_2635975 | Gemeinschaftspraxis für Pathologie Dr. Blume Prof. Dr. Dämmrich Dr. Heine Dr. Dämmrich | Robert Koch Institute | |
| EPI_ISL_2635178, EPI_ISL_2635179, EPI_ISL_2635180, EPI_ISL_2635181, EPI_ISL_2635182, EPI_ISL_2635183, EPI_ISL_2635184, EPI_ISL_2635185, EPI_ISL_2635186, EPI_ISL_2635187, EPI_ISL_2635188, EPI_ISL_2635189, EPI_ISL_2635190, EPI_ISL_2635191, EPI_ISL_2760228, EPI_ISL_2760230, EPI_ISL_2760233, EPI_ISL_2760235, EPI_ISL_2760238, EPI_ISL_2760241, EPI_ISL_2760243, EPI_ISL_2760246, | | | |

| | | | |
|---|---|---|---|
| EPI_ISL_2760249, EPI_ISL_2760251, EPI_ISL_2760254, EPI_ISL_2760258, EPI_ISL_2760260, EPI_ISL_2845822, EPI_ISL_2845823, EPI_ISL_2845824, EPI_ISL_2845825, EPI_ISL_2845826, EPI_ISL_2845827, EPI_ISL_2845828, EPI_ISL_2845829, EPI_ISL_2845830, EPI_ISL_2845831, EPI_ISL_2845832, EPI_ISL_2845833 | | | |
| see above | Helios Universitätsklinikum Wuppertal: Universität Witten/Herdecke: Institut für Medizinische Labordiagnostik | Robert Koch Institute | |
| EPI_ISL_2678656 | Hessisches Landesprüfungs- und Untersuchungsamt im Gesundheitswesen (HLPUG) | Robert Koch Institute | |
| EPI_ISL_2637205, EPI_ISL_2845680, EPI_ISL_2845681, EPI_ISL_2845682, EPI_ISL_2845684, EPI_ISL_2845685, EPI_ISL_2845686, EPI_ISL_2845688, EPI_ISL_2845690, EPI_ISL_2845692, EPI_ISL_2845693 | | | |
| see above | IMD - Labor Oderland | Robert Koch Institute | |
| EPI_ISL_2637198, EPI_ISL_2637201, EPI_ISL_2637206, EPI_ISL_2762136, EPI_ISL_2762138 | IMD - MVZ Labor Greifswald | Robert Koch Institute | |
| EPI_ISL_2637199, EPI_ISL_2637200, EPI_ISL_2762137, EPI_ISL_2762139, EPI_ISL_2762140, EPI_ISL_2845679, EPI_ISL_2845683, EPI_ISL_2845687, EPI_ISL_2845689, EPI_ISL_2845691 | | | |
| see above | IMD - MVZ Labor Martinsried | Robert Koch Institute | |
| EPI_ISL_2633780, EPI_ISL_2633786, EPI_ISL_2633849, EPI_ISL_2636908, EPI_ISL_2844855, EPI_ISL_2844856, EPI_ISL_2844857, EPI_ISL_2844861, EPI_ISL_2844867, EPI_ISL_2844875 | | | |
| see above | IMD Labor Frankfurt | Robert Koch Institute | |
| EPI_ISL_2762170, EPI_ISL_2762171, EPI_ISL_2762172, EPI_ISL_2762173 | Institut für Hygiene und Umwelt Hamburg | Robert Koch Institute | |
| EPI_ISL_2636909 | Institut für Immunologie und Genetik Kaiserslautern: Medizinisches Labor Dr. med. B. Thiele | Robert Koch Institute | |
| EPI_ISL_2761940, EPI_ISL_2845115 | Institut für Klinische Chemie Transfusions- und Laboratoriumsmedizin Berufsgenossenschaftliches Universitätsklinikum Bergmannsheil | Robert Koch Institute | |
| EPI_ISL_2761729, EPI_ISL_2761730, EPI_ISL_2761731, EPI_ISL_2761732, EPI_ISL_2761733, EPI_ISL_2761734 | Institut für Labormedizin Mikrobiologie und Hygiene | Robert Koch Institute | |
| EPI_ISL_2844882 | Institut für Medizinische Mikrobiologie - Universitätsmedizin Göttingen | Robert Koch Institute | |
| EPI_ISL_2845178 | Kerckhoff-Klinik GmbH | Robert Koch Institute | |
| EPI_ISL_2762638 | Klinikum Darmstadt GmbH | Robert Koch Institute | |
| EPI_ISL_2678739, EPI_ISL_2678741, EPI_ISL_2678742, EPI_ISL_2678743, EPI_ISL_2845767 | Klinikum Ernst von Bergmann gemeinnützige GmbH - stationärer Bereich | Robert Koch Institute | |
| EPI_ISL_2637162, EPI_ISL_2761256, EPI_ISL_2761513, EPI_ISL_2762035, EPI_ISL_2845411, EPI_ISL_2846004 | Klinikum Fulda gAG | Robert Koch Institute | |
| EPI_ISL_2761964, EPI_ISL_2761965, EPI_ISL_2761967 | Klinikum Stuttgart | Robert Koch Institute | |
| EPI_ISL_2634747, EPI_ISL_2634784, EPI_ISL_2678447, EPI_ISL_2678449, EPI_ISL_2678458 | LABOR CHEMNITZ – Zentrum für Diagnostik am Klinikum Chemnitz | Robert Koch Institute | |
| EPI_ISL_2636912, EPI_ISL_2636916, EPI_ISL_2636920, EPI_ISL_2636928, EPI_ISL_2636930, EPI_ISL_2636931, EPI_ISL_2636933, EPI_ISL_2636942, EPI_ISL_2636948, EPI_ISL_2636954, EPI_ISL_2636982, EPI_ISL_2636984, EPI_ISL_2636990, EPI_ISL_2636994, EPI_ISL_2636997, EPI_ISL_2637017, EPI_ISL_2637042, EPI_ISL_2637071, EPI_ISL_2637074, EPI_ISL_2637088, EPI_ISL_2637101, EPI_ISL_2637120, EPI_ISL_2637126, EPI_ISL_2637132, EPI_ISL_2637143, EPI_ISL_2637144, EPI_ISL_2637154, EPI_ISL_2637174, EPI_ISL_2637183, EPI_ISL_2637191, EPI_ISL_2637192, EPI_ISL_2759827, EPI_ISL_2759830, EPI_ISL_2759835, EPI_ISL_2759845, EPI_ISL_2761076, EPI_ISL_2761080, EPI_ISL_2761135, EPI_ISL_2761151, EPI_ISL_2761166, EPI_ISL_2761187, EPI_ISL_2761200, EPI_ISL_2761227, EPI_ISL_2761230, EPI_ISL_2761242, EPI_ISL_2761279, EPI_ISL_2761282, EPI_ISL_2761295, EPI_ISL_2761323, EPI_ISL_2761342, EPI_ISL_2761344, EPI_ISL_2761354, EPI_ISL_2761422, EPI_ISL_2761431, EPI_ISL_2761554, EPI_ISL_2761576, EPI_ISL_2845380, EPI_ISL_2845383, EPI_ISL_2845390, EPI_ISL_2845391, EPI_ISL_2845392, EPI_ISL_2845395, EPI_ISL_2845396, EPI_ISL_2845398, EPI_ISL_2845401, EPI_ISL_2845403, EPI_ISL_2845404, EPI_ISL_2845408, EPI_ISL_2845409, EPI_ISL_2845412, EPI_ISL_2845420, EPI_ISL_2845425, EPI_ISL_2845427, EPI_ISL_2845429, EPI_ISL_2845430, EPI_ISL_2845431, EPI_ISL_2845434, EPI_ISL_2845436, EPI_ISL_2845437 | | | |
| see above | LabKom - Labor Augsburg MVZ GmbH | Robert Koch Institute | |
| EPI_ISL_2761968, EPI_ISL_2762001 | LabKom - Labor Hamburg-Lübeck | Robert Koch Institute | |
| EPI_ISL_2759841, EPI_ISL_2761119, EPI_ISL_2761245, EPI_ISL_2761439, EPI_ISL_2761541, EPI_ISL_2762029, EPI_ISL_2845416 | | | |
| see above | LabKom - Labor Hannover MVZ GmbH | Robert Koch Institute | |
| EPI_ISL_2636987, EPI_ISL_2637024, EPI_ISL_2637025, EPI_ISL_2637030, EPI_ISL_2637062, EPI_ISL_2637108, EPI_ISL_2637185, EPI_ISL_2761971, EPI_ISL_2762043, EPI_ISL_2762052, EPI_ISL_2762105, EPI_ISL_2845386, EPI_ISL_2845407 | | | |
| see above | LabKom - Labor Mainz MVZ GmbH | Robert Koch Institute | |
| EPI_ISL_2636919, EPI_ISL_2636927, EPI_ISL_2636967, EPI_ISL_2636973, EPI_ISL_2637012, EPI_ISL_2637036, EPI_ISL_2637100, EPI_ISL_2637131, EPI_ISL_2637134, EPI_ISL_2637155, EPI_ISL_2637159, EPI_ISL_2637167, EPI_ISL_2637189, EPI_ISL_2637193, EPI_ISL_2761192, EPI_ISL_2761973, EPI_ISL_2761980, EPI_ISL_2761993, EPI_ISL_2762022, EPI_ISL_2762042, EPI_ISL_2845405, EPI_ISL_2845414 | | | |
| see above | LabKom - Labor an der Salzbrücke MVZ GmbH | Robert Koch Institute | |
| EPI_ISL_2636925, EPI_ISL_2636936, EPI_ISL_2637015, EPI_ISL_2637019, EPI_ISL_2637041, EPI_ISL_2637044, EPI_ISL_2637050, EPI_ISL_2637059, EPI_ISL_2637079, EPI_ISL_2637081, EPI_ISL_2637129, EPI_ISL_2637136, EPI_ISL_2637151, EPI_ISL_2637161, EPI_ISL_2637177, EPI_ISL_2637178, EPI_ISL_2759834, EPI_ISL_2761264, EPI_ISL_2761275, EPI_ISL_2761333, EPI_ISL_2761382, EPI_ISL_2761392, EPI_ISL_2761420, EPI_ISL_2761474, EPI_ISL_2761573, EPI_ISL_2762016, EPI_ISL_2762039, EPI_ISL_2762088, EPI_ISL_2762106, EPI_ISL_2762649, EPI_ISL_2845424 | | | |
| see above | LabKom - MVZ Labor Bochum MLB GmbH | Robert Koch Institute | |
| EPI_ISL_2637028, EPI_ISL_2637035, EPI_ISL_2637090, EPI_ISL_2637169, EPI_ISL_2637170, EPI_ISL_2637179, EPI_ISL_2761168, EPI_ISL_2761195, EPI_ISL_2761302, EPI_ISL_2761304, EPI_ISL_2761369, EPI_ISL_2761448, EPI_ISL_2761583, EPI_ISL_2762698, EPI_ISL_2845385, EPI_ISL_2845394, EPI_ISL_2845415, EPI_ISL_2845428, EPI_ISL_2845438 | | | |
| see above | Labor 28 MVZ GmbH | Robert Koch Institute | |
| EPI_ISL_2845721, EPI_ISL_2845722, EPI_ISL_2845723, EPI_ISL_2845724, EPI_ISL_2845725, EPI_ISL_2845726, EPI_ISL_2845727, EPI_ISL_2845728, EPI_ISL_2845729, EPI_ISL_2845730, EPI_ISL_2845731, EPI_ISL_2845732, EPI_ISL_2845733, EPI_ISL_2845734, EPI_ISL_2845735, EPI_ISL_2845736, EPI_ISL_2845737, EPI_ISL_2845738, EPI_ISL_2845739, EPI_ISL_2845740, EPI_ISL_2845741, EPI_ISL_2845789, EPI_ISL_2845791, EPI_ISL_2845797, EPI_ISL_2845798, EPI_ISL_2845799, EPI_ISL_2845801, EPI_ISL_2845804, EPI_ISL_2845807, EPI_ISL_2845810 | | | |
| see above | Labor Becker & Kollegen (Standort München) | Robert Koch Institute | |
| EPI_ISL_2504020, EPI_ISL_2504021, EPI_ISL_2504025, EPI_ISL_2504029, EPI_ISL_2694801, EPI_ISL_2694802, EPI_ISL_2694803, EPI_ISL_2694804, EPI_ISL_2694805, EPI_ISL_2694806, EPI_ISL_2694807, EPI_ISL_2694808, EPI_ISL_2694809, EPI_ISL_2694810, EPI_ISL_2694811, EPI_ISL_2694812, EPI_ISL_2694813, EPI_ISL_2694814, EPI_ISL_2694815, EPI_ISL_2694816, EPI_ISL_2694817, EPI_ISL_2694818, EPI_ISL_2694819, EPI_ISL_2694820, EPI_ISL_2694821, EPI_ISL_2694822, EPI_ISL_2694823, EPI_ISL_2694824, EPI_ISL_2694825, EPI_ISL_2694826, EPI_ISL_2694827, EPI_ISL_2694828, EPI_ISL_2694829, EPI_ISL_2694830, EPI_ISL_2694831, EPI_ISL_2694832, EPI_ISL_2694833, EPI_ISL_2694834, EPI_ISL_2694835, EPI_ISL_2694836, EPI_ISL_2694837, EPI_ISL_2694838, EPI_ISL_2694839, EPI_ISL_2694840, EPI_ISL_2694841, EPI_ISL_2694842, EPI_ISL_2694843, EPI_ISL_2694844, EPI_ISL_2694845, EPI_ISL_2694846, EPI_ISL_2694847, EPI_ISL_2694848, EPI_ISL_2694849, EPI_ISL_2694850, EPI_ISL_2694851, EPI_ISL_2694852, EPI_ISL_2694853, EPI_ISL_2694854, EPI_ISL_2694855, EPI_ISL_2694856, EPI_ISL_2694857, EPI_ISL_2694858, EPI_ISL_2694859, EPI_ISL_2694860, EPI_ISL_2833524, EPI_ISL_2833526, EPI_ISL_2833527, EPI_ISL_2833528, EPI_ISL_2833531, EPI_ISL_2833534, EPI_ISL_2833536, EPI_ISL_2833538 | | | |
| see above | Labor Berlin Charité Vivantes GmbH / Institut für Virologie | Charité Universitätsmedizin Berlin, Institut für Virologie/Labor Berlin | Barbara Mühlemann; Christian Drosten; Christine Stephan; Peter Menzel; Rolf Schwarzer; Terry Jones; Victor M Corman |
| EPI_ISL_2678533, EPI_ISL_2678542, EPI_ISL_2678551, EPI_ISL_2678575, EPI_ISL_2678579, EPI_ISL_2678580, EPI_ISL_2678581, EPI_ISL_2678582, EPI_ISL_2678584, EPI_ISL_2678585, EPI_ISL_2678587, EPI_ISL_2678590, EPI_ISL_2678591, EPI_ISL_2678592, EPI_ISL_2678593, EPI_ISL_2678594, EPI_ISL_2678595, EPI_ISL_2678598, EPI_ISL_2678599, EPI_ISL_2678600, EPI_ISL_2678602, EPI_ISL_2678603, EPI_ISL_2678604, EPI_ISL_2678607, EPI_ISL_2678609, EPI_ISL_2678610, EPI_ISL_2678611, EPI_ISL_2678614, EPI_ISL_2678617, EPI_ISL_2678630, EPI_ISL_2678634, EPI_ISL_2678636, EPI_ISL_2678639, EPI_ISL_2678640, EPI_ISL_2678643, EPI_ISL_2678644, EPI_ISL_2678647, EPI_ISL_2678649, EPI_ISL_2845129, EPI_ISL_2845130, EPI_ISL_2845131, EPI_ISL_2845132, EPI_ISL_2845134, EPI_ISL_2845135, EPI_ISL_2845136, EPI_ISL_2845137, EPI_ISL_2845139, EPI_ISL_2845141, EPI_ISL_2845143, EPI_ISL_2845144, EPI_ISL_2845145 | | | |
| see above | Labor Blackholm MVZ | Robert Koch Institute | |
| EPI_ISL_2635844, EPI_ISL_2635845, EPI_ISL_2635847, EPI_ISL_2635849, EPI_ISL_2635850, EPI_ISL_2635851, EPI_ISL_2635852, EPI_ISL_2635853, EPI_ISL_2635854, EPI_ISL_2635856, EPI_ISL_2635857, EPI_ISL_2635858, EPI_ISL_2635859, EPI_ISL_2635860, EPI_ISL_2761684, EPI_ISL_2761687, EPI_ISL_2761689, EPI_ISL_2761692, EPI_ISL_2761694, EPI_ISL_2761695, EPI_ISL_2761696, EPI_ISL_2761697, EPI_ISL_2761698, EPI_ISL_2761699, EPI_ISL_2761700, EPI_ISL_2761701, EPI_ISL_2761702, EPI_ISL_2761703, EPI_ISL_2761704, EPI_ISL_2761705, EPI_ISL_2761706, EPI_ISL_2761707, EPI_ISL_2845216, EPI_ISL_2845217, EPI_ISL_2845218, EPI_ISL_2845219, EPI_ISL_2845220, EPI_ISL_2845221, EPI_ISL_2845222, EPI_ISL_2845223, EPI_ISL_2845224, EPI_ISL_2845225, EPI_ISL_2845226, EPI_ISL_2845227, EPI_ISL_2845228, EPI_ISL_2845229, EPI_ISL_2845230, EPI_ISL_2845231, EPI_ISL_2845232, EPI_ISL_2845233, EPI_ISL_2845234, EPI_ISL_2845235, EPI_ISL_2845236 | | | |
| see above | Labor Dr. Heidrich & Kollegen MVZ GmbH Hamburg | Robert Koch Institute | |
| EPI_ISL_2636929, EPI_ISL_2636985, EPI_ISL_2637009, EPI_ISL_2637023, EPI_ISL_2637067, EPI_ISL_2637073, EPI_ISL_2637087, EPI_ISL_2637163, EPI_ISL_2637164, EPI_ISL_2637186, EPI_ISL_2761173, EPI_ISL_2761216, EPI_ISL_2761372, EPI_ISL_2761478, EPI_ISL_2761498, EPI_ISL_2761996, EPI_ISL_2762005, EPI_ISL_2762101, EPI_ISL_2762659, EPI_ISL_2762708, EPI_ISL_2762715 | | | |
| see above | Labor Dr. Schumacher MVZ | Robert Koch Institute | |
| EPI_ISL_2636901 | Labor Dr. Spranger | Robert Koch Institute | |
| EPI_ISL_2631830, EPI_ISL_2635757, EPI_ISL_2635758, EPI_ISL_2635759, EPI_ISL_2635760, EPI_ISL_2678377, EPI_ISL_2678652, EPI_ISL_2678653, EPI_ISL_2845244, EPI_ISL_2845245, EPI_ISL_2845246, EPI_ISL_2845247, EPI_ISL_2845248, EPI_ISL_2845249, EPI_ISL_2845250, EPI_ISL_2845251, EPI_ISL_2845252, EPI_ISL_2845253, EPI_ISL_2845254, EPI_ISL_2845255, EPI_ISL_2845256, EPI_ISL_2845257, EPI_ISL_2845258, EPI_ISL_2845259, EPI_ISL_2845260, EPI_ISL_2845261, EPI_ISL_2845262, EPI_ISL_2845263, EPI_ISL_2845264, EPI_ISL_2845265, EPI_ISL_2845266, EPI_ISL_2845267, EPI_ISL_2845268, EPI_ISL_2845269, EPI_ISL_2845270, EPI_ISL_2845271, EPI_ISL_2845272, EPI_ISL_2845273, EPI_ISL_2845274, EPI_ISL_2845275, EPI_ISL_2845276, EPI_ISL_2845277, EPI_ISL_2845278, EPI_ISL_2845279, EPI_ISL_2845280, EPI_ISL_2845281, EPI_ISL_2845282, EPI_ISL_2845283, EPI_ISL_2845284, EPI_ISL_2845285, EPI_ISL_2845286, EPI_ISL_2845287, EPI_ISL_2845288, EPI_ISL_2845289, EPI_ISL_2845290, EPI_ISL_2845291 | | | |
| see above | Labor Dr. Wisplinghoff - Köln | Robert Koch Institute | |
| EPI_ISL_2635957, EPI_ISL_2635958, EPI_ISL_2762204 | Labor Kneißler GmbH & Co. KG | Robert Koch Institute | |
| EPI_ISL_2635477, EPI_ISL_2635486, EPI_ISL_2760419, EPI_ISL_2760428, EPI_ISL_2760449, EPI_ISL_2845103, EPI_ISL_2845104 | | | |
| see above | Labor Lübeck bzw. Laborärztliche Gemeinschaftspraxis Lübeck | Robert Koch Institute | |
| EPI_ISL_2635159, EPI_ISL_2635160, EPI_ISL_2635165, EPI_ISL_2635166, EPI_ISL_2635169, EPI_ISL_2635733, EPI_ISL_2635734, EPI_ISL_2635735, EPI_ISL_2635736, EPI_ISL_2635737, EPI_ISL_2635738, EPI_ISL_2635739, EPI_ISL_2635741, EPI_ISL_2635742, EPI_ISL_2635744, EPI_ISL_2635745, EPI_ISL_2635746, EPI_ISL_2635747, EPI_ISL_2635748, EPI_ISL_2635749, EPI_ISL_2635750, EPI_ISL_2635751, EPI_ISL_2635752, EPI_ISL_2635753, EPI_ISL_2635754, EPI_ISL_2635755, EPI_ISL_2635756, EPI_ISL_2635804, EPI_ISL_2635806, EPI_ISL_2635825, EPI_ISL_2636165, EPI_ISL_2636167, EPI_ISL_2636172, EPI_ISL_2636175, EPI_ISL_2636177, EPI_ISL_2636179, EPI_ISL_2636181, EPI_ISL_2636184, EPI_ISL_2636193, EPI_ISL_2636200, EPI_ISL_2636202, EPI_ISL_2636205, EPI_ISL_2636207, EPI_ISL_2636210, EPI_ISL_2636214, EPI_ISL_2636218, EPI_ISL_2636222, EPI_ISL_2636227, EPI_ISL_2678721, EPI_ISL_2678722, EPI_ISL_2678725, EPI_ISL_2678726, EPI_ISL_2761604, EPI_ISL_2761605, EPI_ISL_2761606, EPI_ISL_2761607, EPI_ISL_2761609, EPI_ISL_2761610, EPI_ISL_2761611, EPI_ISL_2761612, EPI_ISL_2761613, EPI_ISL_2761614, EPI_ISL_2761616, EPI_ISL_2761617, EPI_ISL_2761618, EPI_ISL_2761619, EPI_ISL_2761620, EPI_ISL_2761621, EPI_ISL_2761622, EPI_ISL_2761623, EPI_ISL_2761626, EPI_ISL_2761629, EPI_ISL_2761630, EPI_ISL_2761633, EPI_ISL_2761634, EPI_ISL_2761637, EPI_ISL_2761640, EPI_ISL_2761708, EPI_ISL_2761710, EPI_ISL_2761711, EPI_ISL_2761712, EPI_ISL_2761713, EPI_ISL_2761714, EPI_ISL_2761716, EPI_ISL_2761718, EPI_ISL_2761719, EPI_ISL_2761720, EPI_ISL_2761721, EPI_ISL_2761722, EPI_ISL_2761723, EPI_ISL_2761725, EPI_ISL_2761726, EPI_ISL_2761727, EPI_ISL_2761728, EPI_ISL_2762522, EPI_ISL_2845118, EPI_ISL_2845120, EPI_ISL_2845121, EPI_ISL_2845123, EPI_ISL_2845124, EPI_ISL_2845146, EPI_ISL_2845147, EPI_ISL_2845148, EPI_ISL_2845149, EPI_ISL_2845150, EPI_ISL_2845162, EPI_ISL_2845164, EPI_ISL_2845166, EPI_ISL_2845168, EPI_ISL_2845170, EPI_ISL_2845174, EPI_ISL_2845177, EPI_ISL_2845180, EPI_ISL_2845187, EPI_ISL_2845188, EPI_ISL_2845189, EPI_ISL_2845190, EPI_ISL_2845191, EPI_ISL_2845192, EPI_ISL_2845193, EPI_ISL_2845194, EPI_ISL_2845195, EPI_ISL_2845196 | | | |
| see above | Labor Mönchengladbach MVZ Dr. Stein + Kollegen GbR | Robert Koch Institute | |
| EPI_ISL_2635148, EPI_ISL_2635829, EPI_ISL_2635837, EPI_ISL_2635840, EPI_ISL_2636478, EPI_ISL_2636479, EPI_ISL_2636484, EPI_ISL_2678661, EPI_ISL_2678669, EPI_ISL_2678670, EPI_ISL_2678675, EPI_ISL_2678676, EPI_ISL_2762539, EPI_ISL_2762548, EPI_ISL_2762552, EPI_ISL_2762559, EPI_ISL_2762561, EPI_ISL_2762564, EPI_ISL_2845960, EPI_ISL_2845972, EPI_ISL_2845973 | | | |
| see above | Labor Prof. Dr. G. Enders MVZ GbR | Robert Koch Institute | |
| EPI_ISL_2678749, EPI_ISL_2678750, EPI_ISL_2678752, EPI_ISL_2678753, EPI_ISL_2678754, EPI_ISL_2678755, EPI_ISL_2678756, EPI_ISL_2678757, EPI_ISL_2678758, EPI_ISL_2678759, EPI_ISL_2678760, EPI_ISL_2678762, EPI_ISL_2678763, EPI_ISL_2678765, EPI_ISL_2678766, EPI_ISL_2678767, EPI_ISL_2678768, EPI_ISL_2678769, EPI_ISL_2678770, EPI_ISL_2678771, EPI_ISL_2678772, EPI_ISL_2678773, EPI_ISL_2678774, EPI_ISL_2678775, EPI_ISL_2678776, EPI_ISL_2678777, EPI_ISL_2678778, EPI_ISL_2678779, EPI_ISL_2678780, EPI_ISL_2678781, EPI_ISL_2678782, EPI_ISL_2678783, EPI_ISL_2678784, EPI_ISL_2678785, EPI_ISL_2678786, EPI_ISL_2678787, EPI_ISL_2678788, EPI_ISL_2678789, EPI_ISL_2678790, EPI_ISL_2678791, EPI_ISL_2678793, EPI_ISL_2678794, EPI_ISL_2678795, EPI_ISL_2678797, EPI_ISL_2678798, EPI_ISL_2678799, EPI_ISL_2678800, EPI_ISL_2678801, EPI_ISL_2678802, EPI_ISL_2678803 | | | |
| see above | Labor ZOTZ\|KLIMAS; MVZ Düsseldorf-Centrum | Robert Koch Institute | |

| | | | |
|---|---|---|---|
| EPI_ISL_2762452, EPI_ISL_2762454, EPI_ISL_2762456 | Labor für medizinische Mikrobiologie Bakteriologie Mykologie Virologie & Infektionsserologie - Labor Mölbis | Robert Koch Institute | |
| EPI_ISL_2636002, EPI_ISL_2636003, EPI_ISL_2636004, EPI_ISL_2636005, EPI_ISL_2636006, EPI_ISL_2636008, EPI_ISL_2678744, EPI_ISL_2678745, EPI_ISL_2762277, EPI_ISL_2762278, EPI_ISL_2762279, EPI_ISL_2762280, EPI_ISL_2762281, EPI_ISL_2762282, EPI_ISL_2762283, EPI_ISL_2846008, EPI_ISL_2846009 | | | |
| see above | Laborarztpraxis Dres. med. Walther Weindel & Kollegen | Robert Koch Institute | |
| EPI_ISL_2678691, EPI_ISL_2678698, EPI_ISL_2761624, EPI_ISL_2761641 | Laborarztpraxis Jochen Hüter | Robert Koch Institute | |
| EPI_ISL_2636879, EPI_ISL_2636880, EPI_ISL_2636883, EPI_ISL_2636887, EPI_ISL_2636888, EPI_ISL_2636889, EPI_ISL_2636890, EPI_ISL_2636892, EPI_ISL_2636893, EPI_ISL_2636895, EPI_ISL_2636897, EPI_ISL_2636900, EPI_ISL_2761925, EPI_ISL_2761928, EPI_ISL_2761931, EPI_ISL_2761935, EPI_ISL_2761942 | | | |
| see above | Laborarztpraxis Osnabrück | Robert Koch Institute | |
| EPI_ISL_2636903, EPI_ISL_2636904, EPI_ISL_2636905, EPI_ISL_2636907, EPI_ISL_2760394, EPI_ISL_2760396, EPI_ISL_2844850, EPI_ISL_2844851, EPI_ISL_2844852, EPI_ISL_2844853, EPI_ISL_2844854, EPI_ISL_2844858, EPI_ISL_2844859, EPI_ISL_2844860, EPI_ISL_2844862, EPI_ISL_2844863, EPI_ISL_2844864, EPI_ISL_2844865, EPI_ISL_2844866, EPI_ISL_2844868, EPI_ISL_2844869, EPI_ISL_2844870, EPI_ISL_2844872, EPI_ISL_2844873, EPI_ISL_2844874, EPI_ISL_2844876, EPI_ISL_2844878, EPI_ISL_2844879 | | | |
| see above | Labormedizin Darmstadt | Robert Koch Institute | |
| EPI_ISL_2614388, EPI_ISL_2614389, EPI_ISL_2614390, EPI_ISL_2614391, EPI_ISL_2614392, EPI_ISL_2614393, EPI_ISL_2614394, EPI_ISL_2614395, EPI_ISL_2614396, EPI_ISL_2614397 | | | |
| see above | Landesamt für Verbraucherschutz Sachsen Anhalt, Magdeburg | Institute of Medical Microbiology and Hospital Hygiene | Aljoscha Tersteegen; Prof. Dr. Achim Kaasch |
| EPI_ISL_2635564, EPI_ISL_2635565, EPI_ISL_2635566, EPI_ISL_2845243 | Landesamt für Verbraucherschutz des Landes Sachsen-Anhalt | Robert Koch Institute | |
| EPI_ISL_2760153, EPI_ISL_2760157, EPI_ISL_2760159, EPI_ISL_2760161, EPI_ISL_2760166, EPI_ISL_2760181, EPI_ISL_2760184, EPI_ISL_2760186, EPI_ISL_2760191, EPI_ISL_2761642, EPI_ISL_2761643, EPI_ISL_2761644, EPI_ISL_2761645, EPI_ISL_2761648, EPI_ISL_2761649, EPI_ISL_2761650, EPI_ISL_2761652, EPI_ISL_2761653, EPI_ISL_2761654, EPI_ISL_2761655, EPI_ISL_2761657, EPI_ISL_2761658, EPI_ISL_2761659, EPI_ISL_2761661, EPI_ISL_2761662, EPI_ISL_2761663 | | | |
| see above | Landesgesundheitsamt Baden-Württemberg | Robert Koch Institute | |
| EPI_ISL_2634730, EPI_ISL_2634732, EPI_ISL_2634742, EPI_ISL_2634750, EPI_ISL_2634756, EPI_ISL_2634758, EPI_ISL_2634762, EPI_ISL_2634770, EPI_ISL_2634775, EPI_ISL_2634779, EPI_ISL_2634806, EPI_ISL_2678448, EPI_ISL_2678450, EPI_ISL_2678460, EPI_ISL_2678468, EPI_ISL_2762182, EPI_ISL_2762191, EPI_ISL_2845746, EPI_ISL_2845747, EPI_ISL_2845753, EPI_ISL_2845755, EPI_ISL_2845761, EPI_ISL_2845763, EPI_ISL_2845764, EPI_ISL_2845765, EPI_ISL_2845766 | | | |
| see above | Landesuntersuchungsanstalt für das Gesundheits- und Veterinärwesen (LUA) Sachsen | Robert Koch Institute | |
| EPI_ISL_2632853, EPI_ISL_2632879, EPI_ISL_2635146, EPI_ISL_2635761, EPI_ISL_2635763, EPI_ISL_2635767, EPI_ISL_2635769, EPI_ISL_2635809, EPI_ISL_2635830, EPI_ISL_2635832, EPI_ISL_2635839, EPI_ISL_2635890, EPI_ISL_2635893, EPI_ISL_2635894, EPI_ISL_2635895, EPI_ISL_2635899, EPI_ISL_2636474, EPI_ISL_2636476, EPI_ISL_2636487, EPI_ISL_2636509, EPI_ISL_2678664, EPI_ISL_2762599, EPI_ISL_2845967, EPI_ISL_2845971 | | | |
| see above | Limbach - MVZ Gemeinschaftslabor Suhl | Robert Koch Institute | |
| EPI_ISL_2632646, EPI_ISL_2632649, EPI_ISL_2632652, EPI_ISL_2632654, EPI_ISL_2632660, EPI_ISL_2632662, EPI_ISL_2632663, EPI_ISL_2632666, EPI_ISL_2632667, EPI_ISL_2632670, EPI_ISL_2632672, EPI_ISL_2632674, EPI_ISL_2632677, EPI_ISL_2632680, EPI_ISL_2632681, EPI_ISL_2632682, EPI_ISL_2632684, EPI_ISL_2632687, EPI_ISL_2632691, EPI_ISL_2632693, EPI_ISL_2632694, EPI_ISL_2632695, EPI_ISL_2632696, EPI_ISL_2632700, EPI_ISL_2632701, EPI_ISL_2632705, EPI_ISL_2632706, EPI_ISL_2632708, EPI_ISL_2632710, EPI_ISL_2632713, EPI_ISL_2632715, EPI_ISL_2632718, EPI_ISL_2632722, EPI_ISL_2632725, EPI_ISL_2632730, EPI_ISL_2632733, EPI_ISL_2632736, EPI_ISL_2632739, EPI_ISL_2632742, EPI_ISL_2632743, EPI_ISL_2632744, EPI_ISL_2632746, EPI_ISL_2632749, EPI_ISL_2632751, EPI_ISL_2632753, EPI_ISL_2632756, EPI_ISL_2632763, EPI_ISL_2632764, EPI_ISL_2632768, EPI_ISL_2632771, EPI_ISL_2632775, EPI_ISL_2632779, EPI_ISL_2632780, EPI_ISL_2632781, EPI_ISL_2632783, EPI_ISL_2632785, EPI_ISL_2632788, EPI_ISL_2632791, EPI_ISL_2632792, EPI_ISL_2632793, EPI_ISL_2632794, EPI_ISL_2635663, EPI_ISL_2635664, EPI_ISL_2635665, EPI_ISL_2635666, EPI_ISL_2635667, EPI_ISL_2635668, EPI_ISL_2635669, EPI_ISL_2635670, EPI_ISL_2635671, EPI_ISL_2635672, EPI_ISL_2635673, EPI_ISL_2635674, EPI_ISL_2635675, EPI_ISL_2635676, EPI_ISL_2635677, EPI_ISL_2635678, EPI_ISL_2635679, EPI_ISL_2635680, EPI_ISL_2635681, EPI_ISL_2635682, EPI_ISL_2635683, EPI_ISL_2635684, EPI_ISL_2635685, EPI_ISL_2635686, EPI_ISL_2635687, EPI_ISL_2635688, EPI_ISL_2635689, EPI_ISL_2635690, EPI_ISL_2635691, EPI_ISL_2635692, EPI_ISL_2635693, EPI_ISL_2635694, EPI_ISL_2635695, EPI_ISL_2635696, EPI_ISL_2635697, EPI_ISL_2635698, EPI_ISL_2635699, EPI_ISL_2635700, EPI_ISL_2635701, EPI_ISL_2635702, EPI_ISL_2635703, EPI_ISL_2635704, EPI_ISL_2635705, EPI_ISL_2635706, EPI_ISL_2635707, EPI_ISL_2635708, EPI_ISL_2635709, EPI_ISL_2635710, EPI_ISL_2635711, EPI_ISL_2635712, EPI_ISL_2635713, EPI_ISL_2635714, EPI_ISL_2635715, EPI_ISL_2635716, EPI_ISL_2635717, EPI_ISL_2635718, EPI_ISL_2635719, EPI_ISL_2635720, EPI_ISL_2635721, EPI_ISL_2635722, EPI_ISL_2635723, EPI_ISL_2635724, EPI_ISL_2635725, EPI_ISL_2635726, EPI_ISL_2635727, EPI_ISL_2635728, EPI_ISL_2635729, EPI_ISL_2635730, EPI_ISL_2635731, EPI_ISL_2635732, EPI_ISL_2845015, EPI_ISL_2845016, EPI_ISL_2845017, EPI_ISL_2845018, EPI_ISL_2845019, EPI_ISL_2845020, EPI_ISL_2845021, EPI_ISL_2845022, EPI_ISL_2845023, EPI_ISL_2845024, EPI_ISL_2845025, EPI_ISL_2845026, EPI_ISL_2845027, EPI_ISL_2845028, EPI_ISL_2845029, EPI_ISL_2845030, EPI_ISL_2845031, EPI_ISL_2845032, EPI_ISL_2845033, EPI_ISL_2845034, EPI_ISL_2845035, EPI_ISL_2845036, EPI_ISL_2845037, EPI_ISL_2845038, EPI_ISL_2845039, EPI_ISL_2845040, EPI_ISL_2845041, EPI_ISL_2845042, EPI_ISL_2845043, EPI_ISL_2845044, EPI_ISL_2845045, EPI_ISL_2845046, EPI_ISL_2845047, EPI_ISL_2845048, EPI_ISL_2845049, EPI_ISL_2845050, EPI_ISL_2845051, EPI_ISL_2845052, EPI_ISL_2845053, EPI_ISL_2845054, EPI_ISL_2845055, EPI_ISL_2845056, EPI_ISL_2845057, EPI_ISL_2845058, EPI_ISL_2845059, EPI_ISL_2845060 | | | |
| see above | Limbach - MVZ Humangenetik Ulm | Robert Koch Institute | |
| EPI_ISL_2632845, EPI_ISL_2635152, EPI_ISL_2635173, EPI_ISL_2635740, EPI_ISL_2635743, EPI_ISL_2635805, EPI_ISL_2635863, EPI_ISL_2635866, EPI_ISL_2635868, EPI_ISL_2635878, EPI_ISL_2636224, EPI_ISL_2678713, EPI_ISL_2678715, EPI_ISL_2678723, EPI_ISL_2678724, EPI_ISL_2678728, EPI_ISL_2678730, EPI_ISL_2762499, EPI_ISL_2762506, EPI_ISL_2845963 | | | |
| see above | Limbach - MVZ Labor Eveld & Kollegen Essen | Robert Koch Institute | |
| EPI_ISL_2632952, EPI_ISL_2632960, EPI_ISL_2635903, EPI_ISL_2635905, EPI_ISL_2678692, EPI_ISL_2678702, EPI_ISL_2761880, EPI_ISL_2762625, EPI_ISL_2762630, EPI_ISL_2762636 | | | |
| see above | Limbach - MVZ Labor Ludwigsburg | Robert Koch Institute | |
| EPI_ISL_2632846, EPI_ISL_2632848, EPI_ISL_2632849, EPI_ISL_2632851, EPI_ISL_2632854, EPI_ISL_2632855, EPI_ISL_2632856, EPI_ISL_2632857, EPI_ISL_2632859, EPI_ISL_2632860, EPI_ISL_2632861, EPI_ISL_2632863, EPI_ISL_2632865, EPI_ISL_2632869, EPI_ISL_2632870, EPI_ISL_2632871, EPI_ISL_2632874, EPI_ISL_2632875, EPI_ISL_2632885, EPI_ISL_2632886, EPI_ISL_2632891, EPI_ISL_2632963, EPI_ISL_2632964, EPI_ISL_2632970, EPI_ISL_2632975, EPI_ISL_2632977, EPI_ISL_2632980, EPI_ISL_2632981, EPI_ISL_2635132, EPI_ISL_2635134, EPI_ISL_2635139, EPI_ISL_2635141, EPI_ISL_2635150, EPI_ISL_2635762, EPI_ISL_2635765, EPI_ISL_2635768, EPI_ISL_2635772, EPI_ISL_2635773, EPI_ISL_2635774, EPI_ISL_2635775, EPI_ISL_2635776, EPI_ISL_2635777, EPI_ISL_2635778, EPI_ISL_2635779, EPI_ISL_2635780, EPI_ISL_2635781, EPI_ISL_2635782, EPI_ISL_2635783, EPI_ISL_2635784, EPI_ISL_2635808, EPI_ISL_2635811, EPI_ISL_2635812, EPI_ISL_2635826, EPI_ISL_2635827, EPI_ISL_2635828, EPI_ISL_2636220, EPI_ISL_2636230, EPI_ISL_2636473, EPI_ISL_2636477, EPI_ISL_2636482, EPI_ISL_2636483, EPI_ISL_2636488, EPI_ISL_2636489, EPI_ISL_2636490, EPI_ISL_2636496, EPI_ISL_2636500, EPI_ISL_2636501, EPI_ISL_2636505, EPI_ISL_2636506, EPI_ISL_2636508, EPI_ISL_2636515, EPI_ISL_2636516, EPI_ISL_2636517, EPI_ISL_2678657, EPI_ISL_2678658, EPI_ISL_2678659, EPI_ISL_2678660, EPI_ISL_2678662, EPI_ISL_2678663, EPI_ISL_2678665, EPI_ISL_2678666, EPI_ISL_2678667, EPI_ISL_2678668, EPI_ISL_2678671, EPI_ISL_2678672, EPI_ISL_2678673, EPI_ISL_2678674, EPI_ISL_2678677, EPI_ISL_2678678, EPI_ISL_2678679, EPI_ISL_2678680, EPI_ISL_2678687, EPI_ISL_2678688, EPI_ISL_2678689, EPI_ISL_2678690, EPI_ISL_2678693, EPI_ISL_2678694, EPI_ISL_2678696, EPI_ISL_2678697, EPI_ISL_2678699, EPI_ISL_2678700, EPI_ISL_2678701, EPI_ISL_2678703, EPI_ISL_2678704, EPI_ISL_2678705, EPI_ISL_2678706, EPI_ISL_2678707, EPI_ISL_2678719, EPI_ISL_2760581, EPI_ISL_2761627, EPI_ISL_2761869, EPI_ISL_2761870, EPI_ISL_2761871, EPI_ISL_2761873, EPI_ISL_2761877, EPI_ISL_2761881, EPI_ISL_2761882, EPI_ISL_2761883, EPI_ISL_2761886, EPI_ISL_2761888, EPI_ISL_2761892, EPI_ISL_2761894, EPI_ISL_2761897, EPI_ISL_2761898, EPI_ISL_2761900, EPI_ISL_2761903, EPI_ISL_2761905, EPI_ISL_2762569, EPI_ISL_2762582, EPI_ISL_2762584, EPI_ISL_2762601, EPI_ISL_2762605, EPI_ISL_2762609, EPI_ISL_2762611, EPI_ISL_2762613, EPI_ISL_2762616, EPI_ISL_2762620, EPI_ISL_2762623, EPI_ISL_2762627, EPI_ISL_2762633, EPI_ISL_2845155, EPI_ISL_2845157, EPI_ISL_2845159, EPI_ISL_2845167, EPI_ISL_2845176, EPI_ISL_2845198, EPI_ISL_2845199, EPI_ISL_2845200, EPI_ISL_2845201, EPI_ISL_2845202, EPI_ISL_2845204, EPI_ISL_2845206, EPI_ISL_2845207, EPI_ISL_2845209, EPI_ISL_2845211, EPI_ISL_2845212, EPI_ISL_2845214, EPI_ISL_2845962, EPI_ISL_2845977, EPI_ISL_2845980, EPI_ISL_2845986, EPI_ISL_2845993, EPI_ISL_2845998 | | | |
| see above | MDI Limbach Berlin GmbH; MVZ Labor Berlin | Robert Koch Institute | |
| EPI_ISL_2772709, EPI_ISL_2772715 | MKK Mühlenkreiskliniken, Johannes Wesling Klinikum Minden | Hannover Medical School, Institute of Virology | Lars Steinbrück |
| EPI_ISL_2633412, EPI_ISL_2633416, EPI_ISL_2633417, EPI_ISL_2633425, EPI_ISL_2635471, EPI_ISL_2635472, EPI_ISL_2635473, EPI_ISL_2635476, EPI_ISL_2635478, EPI_ISL_2635479, EPI_ISL_2635481, EPI_ISL_2635482, EPI_ISL_2635483, EPI_ISL_2635484, EPI_ISL_2635485, EPI_ISL_2635488, EPI_ISL_2635489, EPI_ISL_2636881, EPI_ISL_2636882, EPI_ISL_2636884, EPI_ISL_2636894, EPI_ISL_2636898, EPI_ISL_2636899, EPI_ISL_2760399, EPI_ISL_2760402, EPI_ISL_2760404, EPI_ISL_2760407, EPI_ISL_2760412, EPI_ISL_2760422, EPI_ISL_2760424, EPI_ISL_2760431, EPI_ISL_2760433, EPI_ISL_2760436, EPI_ISL_2760441, EPI_ISL_2760444, EPI_ISL_2760447, EPI_ISL_2760452, EPI_ISL_2761923, EPI_ISL_2761927, EPI_ISL_2761929, EPI_ISL_2761932, EPI_ISL_2761934, EPI_ISL_2761938, EPI_ISL_2761939, EPI_ISL_2761943, EPI_ISL_2845101, EPI_ISL_2845102, EPI_ISL_2845105, EPI_ISL_2845106, EPI_ISL_2845107, EPI_ISL_2845108, EPI_ISL_2845109, EPI_ISL_2845111, EPI_ISL_2845114, EPI_ISL_2845116, EPI_ISL_2845237, EPI_ISL_2845238, EPI_ISL_2845239, EPI_ISL_2845240, EPI_ISL_2845241 | | | |
| see above | MVZ Dr. Eberhard & Partner Dortmund | Robert Koch Institute | |
| EPI_ISL_2636737, EPI_ISL_2636738, EPI_ISL_2636739, EPI_ISL_2636740, EPI_ISL_2636741, EPI_ISL_2636742, EPI_ISL_2636743, EPI_ISL_2636744, EPI_ISL_2636745, EPI_ISL_2636746, EPI_ISL_2636748, EPI_ISL_2636749, EPI_ISL_2636750, EPI_ISL_2636751, EPI_ISL_2636752, EPI_ISL_2636753, EPI_ISL_2636754, EPI_ISL_2636756, EPI_ISL_2636757, EPI_ISL_2636758, EPI_ISL_2636759, EPI_ISL_2761664, EPI_ISL_2761665, EPI_ISL_2761666, EPI_ISL_2761667, EPI_ISL_2761668, EPI_ISL_2761669, EPI_ISL_2761670, EPI_ISL_2761671, EPI_ISL_2761673, EPI_ISL_2761677, EPI_ISL_2761679, EPI_ISL_2761682, EPI_ISL_2762176, EPI_ISL_2762284, EPI_ISL_2762285, EPI_ISL_2762286, EPI_ISL_2762287, EPI_ISL_2762288, EPI_ISL_2762289, EPI_ISL_2762290, EPI_ISL_2762291, EPI_ISL_2762292, EPI_ISL_2762293, EPI_ISL_2762294, EPI_ISL_2762295, EPI_ISL_2762296, EPI_ISL_2762297, EPI_ISL_2762298, EPI_ISL_2762299, EPI_ISL_2762300, EPI_ISL_2762301, EPI_ISL_2845813, EPI_ISL_2845814, EPI_ISL_2845815, EPI_ISL_2845816, EPI_ISL_2845817, EPI_ISL_2845818, EPI_ISL_2845819, EPI_ISL_2845820, EPI_ISL_2845821 | | | |
| see above | MVZ Labor Dr. Fenner und Kollegen (Standort Hamburg) | Robert Koch Institute | |
| EPI_ISL_2472509, EPI_ISL_2635161, EPI_ISL_2635163, EPI_ISL_2635164, EPI_ISL_2635167, EPI_ISL_2635174, EPI_ISL_2635175, EPI_ISL_2635176, EPI_ISL_2635177, EPI_ISL_2635770, EPI_ISL_2635771, EPI_ISL_2635861, EPI_ISL_2635862, EPI_ISL_2635864, EPI_ISL_2635865, EPI_ISL_2635867, EPI_ISL_2635869, EPI_ISL_2635870, EPI_ISL_2635871, EPI_ISL_2635872, EPI_ISL_2635873, EPI_ISL_2635874, EPI_ISL_2635875, EPI_ISL_2635876, EPI_ISL_2635877, EPI_ISL_2635879, EPI_ISL_2635882, EPI_ISL_2635883, EPI_ISL_2635885, EPI_ISL_2635886, EPI_ISL_2635887, EPI_ISL_2635888, EPI_ISL_2635889, EPI_ISL_2635891, EPI_ISL_2635892, EPI_ISL_2635897, EPI_ISL_2635898, EPI_ISL_2635900, EPI_ISL_2635901, EPI_ISL_2635902, EPI_ISL_2635904, EPI_ISL_2636493, EPI_ISL_2636498, EPI_ISL_2636876, EPI_ISL_2636877, EPI_ISL_2636878, EPI_ISL_2678695, EPI_ISL_2678709, EPI_ISL_2678714, EPI_ISL_2678727, EPI_ISL_2678746, EPI_ISL_2678747, EPI_ISL_2678748, EPI_ISL_2760558, EPI_ISL_2760560, EPI_ISL_2760562, EPI_ISL_2760565, EPI_ISL_2760570, EPI_ISL_2760576, EPI_ISL_2760578, EPI_ISL_2760589, EPI_ISL_2760592, EPI_ISL_2760594, EPI_ISL_2760599, EPI_ISL_2760601, EPI_ISL_2760603, EPI_ISL_2760609, EPI_ISL_2760611, EPI_ISL_2761615, EPI_ISL_2761631, EPI_ISL_2761709, EPI_ISL_2761715, EPI_ISL_2761717, EPI_ISL_2761724, EPI_ISL_2761875, EPI_ISL_2761893, EPI_ISL_2762465, EPI_ISL_2762468, EPI_ISL_2762470, EPI_ISL_2762472, EPI_ISL_2762474, EPI_ISL_2762477, EPI_ISL_2762479, EPI_ISL_2762481, EPI_ISL_2762483, EPI_ISL_2762485, EPI_ISL_2762488, EPI_ISL_2762490, EPI_ISL_2762492, EPI_ISL_2762494, EPI_ISL_2762497, EPI_ISL_2762501, EPI_ISL_2762503, EPI_ISL_2762508, EPI_ISL_2762510, EPI_ISL_2762513, EPI_ISL_2762515, EPI_ISL_2762520, EPI_ISL_2762525, EPI_ISL_2762529, EPI_ISL_2762532, EPI_ISL_2762536, EPI_ISL_2762543, EPI_ISL_2762545, EPI_ISL_2762566, EPI_ISL_2762618, EPI_ISL_2845092, EPI_ISL_2845093, EPI_ISL_2845094, EPI_ISL_2845095, EPI_ISL_2845097, EPI_ISL_2845098, EPI_ISL_2845100, EPI_ISL_2845119, EPI_ISL_2845122, EPI_ISL_2845125, EPI_ISL_2845126, EPI_ISL_2845127, EPI_ISL_2845203, EPI_ISL_2845205, EPI_ISL_2845208, EPI_ISL_2845210, EPI_ISL_2845213, EPI_ISL_2845292, EPI_ISL_2845952, EPI_ISL_2845953, EPI_ISL_2845954, EPI_ISL_2845955, EPI_ISL_2845957, EPI_ISL_2845958, EPI_ISL_2845959, EPI_ISL_2845961, EPI_ISL_2845995, EPI_ISL_2845997, EPI_ISL_2846003 | | | |
| see above | MVZ Labor Dr. Limbach & Kollegen GbR | Robert Koch Institute | |
| EPI_ISL_2678545, EPI_ISL_2678548, EPI_ISL_2678560, EPI_ISL_2678562, EPI_ISL_2678566, EPI_ISL_2678573, EPI_ISL_2678606, EPI_ISL_2678613, EPI_ISL_2678619, EPI_ISL_2678629, EPI_ISL_2678633, EPI_ISL_2678638, EPI_ISL_2678646, EPI_ISL_2845128, EPI_ISL_2845133, EPI_ISL_2845138, EPI_ISL_2845140, EPI_ISL_2845142 | | | |
| see above | MVZ Labor Dr. Quade & Kollegen GmbH | Robert Koch Institute | |
| EPI_ISL_2632924, EPI_ISL_2635764, EPI_ISL_2635823, EPI_ISL_2635833, EPI_ISL_2635843, EPI_ISL_2636190, EPI_ISL_2636195, EPI_ISL_2636216, EPI_ISL_2678716, EPI_ISL_2678717, EPI_ISL_2678718, EPI_ISL_2678720, EPI_ISL_2761625, EPI_ISL_2845096, EPI_ISL_2845099, EPI_ISL_2845964 | | | |
| see above | MVZ Labor Dr. Reising-Ackermann und Kollegen | Robert Koch Institute | |
| EPI_ISL_2631977, EPI_ISL_2635789, EPI_ISL_2635790, EPI_ISL_2635791, EPI_ISL_2635792, EPI_ISL_2635793, EPI_ISL_2635794, EPI_ISL_2636772, EPI_ISL_2636773, EPI_ISL_2636774, EPI_ISL_2636775, EPI_ISL_2636776, EPI_ISL_2636777, EPI_ISL_2636778, EPI_ISL_2636779, EPI_ISL_2636780, EPI_ISL_2636781, EPI_ISL_2636782, EPI_ISL_2636783, EPI_ISL_2636784, EPI_ISL_2636785, EPI_ISL_2636786, EPI_ISL_2636787, EPI_ISL_2760280, EPI_ISL_2760283, EPI_ISL_2760285, EPI_ISL_2760288, EPI_ISL_2760291, EPI_ISL_2760293, EPI_ISL_2760296, EPI_ISL_2760298, EPI_ISL_2760301, EPI_ISL_2760308, EPI_ISL_2760310, EPI_ISL_2760313, EPI_ISL_2760315, EPI_ISL_2760317, EPI_ISL_2760320, EPI_ISL_2760322, EPI_ISL_2760330, EPI_ISL_2760332, EPI_ISL_2760334, EPI_ISL_2760336, EPI_ISL_2760339, EPI_ISL_2760341, EPI_ISL_2760344, EPI_ISL_2760347, EPI_ISL_2760349, EPI_ISL_2760352, EPI_ISL_2760354, EPI_ISL_2760356, EPI_ISL_2760359, EPI_ISL_2762107, EPI_ISL_2762108, EPI_ISL_2762109, EPI_ISL_2762110, EPI_ISL_2762111, EPI_ISL_2762112, EPI_ISL_2762113, EPI_ISL_2762114, EPI_ISL_2762115, EPI_ISL_2762116, EPI_ISL_2762117, EPI_ISL_2762118, EPI_ISL_2762119, EPI_ISL_2762120, EPI_ISL_2762121, EPI_ISL_2762122, EPI_ISL_2762123, EPI_ISL_2762124, EPI_ISL_2762125, EPI_ISL_2762126, EPI_ISL_2762127, EPI_ISL_2762128, EPI_ISL_2762129, EPI_ISL_2762276, EPI_ISL_2845293, EPI_ISL_2845294, EPI_ISL_2845295, EPI_ISL_2845296, EPI_ISL_2845297, EPI_ISL_2845298, EPI_ISL_2845299, EPI_ISL_2845300, EPI_ISL_2845301, EPI_ISL_2845302, EPI_ISL_2845303, EPI_ISL_2845304, EPI_ISL_2845305, EPI_ISL_2845306, EPI_ISL_2845307, EPI_ISL_2845308, EPI_ISL_2845309, EPI_ISL_2845310, EPI_ISL_2845311, EPI_ISL_2845312, EPI_ISL_2845313, EPI_ISL_2845314, EPI_ISL_2845315, EPI_ISL_2845316 | | | |
| see above | MVZ Labor Krone GbR | Robert Koch Institute | |
| EPI_ISL_2761608, EPI_ISL_2761628, EPI_ISL_2761636, EPI_ISL_2761876, EPI_ISL_2761896 | MVZ Labor Passau GbR | Robert Koch Institute | |
| EPI_ISL_2632877, EPI_ISL_2632881, EPI_ISL_2632892, EPI_ISL_2635766, EPI_ISL_2635896, EPI_ISL_2678708, EPI_ISL_2678712, EPI_ISL_2678729, EPI_ISL_2761632, EPI_ISL_2761874, EPI_ISL_2762517, EPI_ISL_2762527, EPI_ISL_2762550, EPI_ISL_2762557, EPI_ISL_2762571, EPI_ISL_2762587, EPI_ISL_2762591, EPI_ISL_2762596, EPI_ISL_2762603, EPI_ISL_2845151, EPI_ISL_2845153, EPI_ISL_2845154, EPI_ISL_2845966, EPI_ISL_2845978, EPI_ISL_2845988, EPI_ISL_2845991 | | | |
| see above | MVZ Medizinisches Labor Hannover GmbH | Robert Koch Institute | |
| EPI_ISL_2632847, EPI_ISL_2632850, EPI_ISL_2632852, EPI_ISL_2632858, EPI_ISL_2632862, EPI_ISL_2632864, EPI_ISL_2632866, EPI_ISL_2632867, EPI_ISL_2632868, EPI_ISL_2632872, EPI_ISL_2632873, EPI_ISL_2632878, EPI_ISL_2632882, EPI_ISL_2632883, EPI_ISL_2632888, EPI_ISL_2632889, EPI_ISL_2632894, EPI_ISL_2632895, EPI_ISL_2632896, EPI_ISL_2635136, EPI_ISL_2635143, EPI_ISL_2635836, EPI_ISL_2635838, EPI_ISL_2636485, EPI_ISL_2636491, EPI_ISL_2636492, EPI_ISL_2636494, EPI_ISL_2636495, EPI_ISL_2636497, EPI_ISL_2636499, EPI_ISL_2636502, EPI_ISL_2636503, EPI_ISL_2636504, EPI_ISL_2636507, EPI_ISL_2636510, EPI_ISL_2636511, EPI_ISL_2636512, EPI_ISL_2636513, EPI_ISL_2636514, EPI_ISL_2761868, EPI_ISL_2761878, EPI_ISL_2761879, EPI_ISL_2761884, EPI_ISL_2761885, EPI_ISL_2761887, EPI_ISL_2761889, EPI_ISL_2761890, EPI_ISL_2761891, EPI_ISL_2761895, EPI_ISL_2761899, EPI_ISL_2761901, EPI_ISL_2761902, EPI_ISL_2761904, EPI_ISL_2761906, EPI_ISL_2762534, EPI_ISL_2762541, EPI_ISL_2762555, EPI_ISL_2762573, EPI_ISL_2762575, EPI_ISL_2762577, EPI_ISL_2762580, EPI_ISL_2762589, EPI_ISL_2762594, EPI_ISL_2845152, EPI_ISL_2845156, EPI_ISL_2845158, EPI_ISL_2845160, EPI_ISL_2845161, EPI_ISL_2845163, EPI_ISL_2845165, EPI_ISL_2845169, EPI_ISL_2845171, EPI_ISL_2845172, EPI_ISL_2845173, EPI_ISL_2845175, EPI_ISL_2845179, EPI_ISL_2845956, EPI_ISL_2845965, EPI_ISL_2845968, EPI_ISL_2845969, EPI_ISL_2845974, EPI_ISL_2845975, EPI_ISL_2845976, EPI_ISL_2845982, EPI_ISL_2845990 | | | |
| see above | MVZ für Laboratoriumsmedizin und Mikrobiologie Koblenz-Mittelrhein (Labor Koblenz) | Robert Koch Institute | |
| EPI_ISL_2633435, EPI_ISL_2635480, EPI_ISL_2635490, EPI_ISL_2760409, EPI_ISL_2760414, EPI_ISL_2760417, EPI_ISL_2760426, EPI_ISL_2760439, EPI_ISL_2761930, EPI_ISL_2761933, EPI_ISL_2845110, EPI_ISL_2845113, EPI_ISL_2845117, EPI_ISL_2845242 | | | |
| see above | MVZ für Laboratoriumsmedizin und Mikrobiologie Würzburg | Robert Koch Institute | |
| EPI_ISL_2772664, EPI_ISL_2772665, EPI_ISL_2772666, EPI_ISL_2772667, EPI_ISL_2772668, EPI_ISL_2772669, EPI_ISL_2772670, EPI_ISL_2772671, EPI_ISL_2772676, EPI_ISL_2772677, EPI_ISL_2772680, EPI_ISL_2772682, EPI_ISL_2772683, EPI_ISL_2772684, EPI_ISL_2772686, EPI_ISL_2772687, EPI_ISL_2772688, EPI_ISL_2772695, EPI_ISL_2772696, EPI_ISL_2772697, EPI_ISL_2772698, EPI_ISL_2772699, EPI_ISL_2772700, EPI_ISL_2772701, EPI_ISL_2772702, EPI_ISL_2772703, EPI_ISL_2772704, EPI_ISL_2772705 | | | |
| see above | Max von Pettenkofer Institute, Virology, National Reference Center for Retroviruses, LMU Munich | Laboratory for Functional Genome Analysis; Dept. Genomics; Gene Center of the LMU Munich | Alexander Graf; Helmut Blum; Marcel Stern; Max Muenchhoff; Oliver Keppler; Stefan Krebs |
| EPI_ISL_2635786, EPI_ISL_2635787, EPI_ISL_2635788 | Med. Labor Prof. Schenk Dr. Ansorge & Kollegen | Robert Koch Institute | |

| | | | |
|---|---|---|---|
| EPI_ISL_2928405, EPI_ISL_2928406, EPI_ISL_2928407, EPI_ISL_2928408, EPI_ISL_2928409, EPI_ISL_2928412, EPI_ISL_2928413, EPI_ISL_2928414, EPI_ISL_2928416, EPI_ISL_2928417, EPI_ISL_2928418 | | | |
| see above | Medical Laboratories Duesseldorf | Center of Medical Microbiology, Virology, and Hospital Hygiene, University of Duesseldorf | Alexander Dilthey; Andreas Walker; Angelika Helmer; Christian Lange; Daniel Strelow; Jessica Nicolai; Jörg Timm; Klaus Pfeffer; Lisanna Hülse; Malte Kohns Vasconcelos; Maximilian Damagnez; Nadine Lübke; Tobias Wienemann; Torsten Houwaart |
| EPI_ISL_2636791, EPI_ISL_2636792, EPI_ISL_2636793, EPI_ISL_2636795, EPI_ISL_2636796, EPI_ISL_2636798, EPI_ISL_2636800, EPI_ISL_2636809, EPI_ISL_2636811, EPI_ISL_2636815, EPI_ISL_2636817, EPI_ISL_2636828, EPI_ISL_2636830, EPI_ISL_2636832, EPI_ISL_2636835, EPI_ISL_2636837, EPI_ISL_2636843, EPI_ISL_2636847, EPI_ISL_2636853, EPI_ISL_2636855, EPI_ISL_2636857, EPI_ISL_2761907, EPI_ISL_2761908, EPI_ISL_2761909, EPI_ISL_2761910, EPI_ISL_2761911, EPI_ISL_2761912, EPI_ISL_2761914, EPI_ISL_2761915, EPI_ISL_2761917, EPI_ISL_2761918, EPI_ISL_2761919, EPI_ISL_2761920, EPI_ISL_2761921, EPI_ISL_2761922 | | | |
| see above | Medizinisch-Diagnostisches Labor Kempten allgäulab | Robert Koch Institute | |
| EPI_ISL_2635567, EPI_ISL_2635568, EPI_ISL_2635569, EPI_ISL_2635570, EPI_ISL_2635571, EPI_ISL_2635572, EPI_ISL_2635573, EPI_ISL_2635574, EPI_ISL_2635575, EPI_ISL_2635576, EPI_ISL_2635577, EPI_ISL_2635578, EPI_ISL_2635579, EPI_ISL_2635580, EPI_ISL_2635581, EPI_ISL_2635582, EPI_ISL_2635583, EPI_ISL_2635584, EPI_ISL_2635585, EPI_ISL_2635586, EPI_ISL_2635587, EPI_ISL_2635588, EPI_ISL_2635589, EPI_ISL_2635590, EPI_ISL_2635591, EPI_ISL_2635592, EPI_ISL_2635593, EPI_ISL_2635594, EPI_ISL_2635595, EPI_ISL_2635596, EPI_ISL_2635597, EPI_ISL_2635598, EPI_ISL_2760613, EPI_ISL_2760615, EPI_ISL_2760618, EPI_ISL_2760621, EPI_ISL_2760623, EPI_ISL_2760625, EPI_ISL_2760627, EPI_ISL_2760630, EPI_ISL_2760632, EPI_ISL_2760634, EPI_ISL_2760637, EPI_ISL_2760639, EPI_ISL_2760642, EPI_ISL_2760644, EPI_ISL_2760647, EPI_ISL_2760649, EPI_ISL_2760652, EPI_ISL_2760654, EPI_ISL_2760656, EPI_ISL_2760658, EPI_ISL_2760661, EPI_ISL_2760664, EPI_ISL_2845061, EPI_ISL_2845062, EPI_ISL_2845063, EPI_ISL_2845064, EPI_ISL_2845065, EPI_ISL_2845066, EPI_ISL_2845067, EPI_ISL_2845068, EPI_ISL_2845069, EPI_ISL_2845070, EPI_ISL_2845071, EPI_ISL_2845072, EPI_ISL_2845073, EPI_ISL_2845074, EPI_ISL_2845075, EPI_ISL_2845076, EPI_ISL_2845077 | | | |
| see above | Medizinische Laboratorien Düsseldorf | Robert Koch Institute | |
| EPI_ISL_2634744, EPI_ISL_2634772, EPI_ISL_2634777, EPI_ISL_2634795, EPI_ISL_2634797, EPI_ISL_2634809, EPI_ISL_2634815, EPI_ISL_2636475, EPI_ISL_2636481, EPI_ISL_2636486, EPI_ISL_2678454, EPI_ISL_2678457, EPI_ISL_2678462, EPI_ISL_2678463, EPI_ISL_2678471, EPI_ISL_2761635, EPI_ISL_2761638, EPI_ISL_2762177, EPI_ISL_2762179, EPI_ISL_2762184, EPI_ISL_2762185, EPI_ISL_2762186, EPI_ISL_2762193, EPI_ISL_2762194, EPI_ISL_2762195, EPI_ISL_2762196, EPI_ISL_2762197, EPI_ISL_2762202, EPI_ISL_2762203, EPI_ISL_2845744, EPI_ISL_2845745, EPI_ISL_2845748, EPI_ISL_2845749, EPI_ISL_2845756, EPI_ISL_2845757, EPI_ISL_2845762 | | | |
| see above | Medizinisches Labor Ostsachsen MVZ GbR Nebenbetriebsstätte Dresden | Robert Koch Institute | |
| EPI_ISL_2633436 | Medizinisches Labor Wahl Lüdenscheid | Robert Koch Institute | |
| EPI_ISL_2636896, EPI_ISL_2761924 | Medizinisches Versorgungszentrum für Labormedizin und Mikrobiologie Ruhr GmbH - mvzlm RUHR GmbH | Robert Koch Institute | |
| EPI_ISL_2635813, EPI_ISL_2635814, EPI_ISL_2635815, EPI_ISL_2635816, EPI_ISL_2635817, EPI_ISL_2635818, EPI_ISL_2635819, EPI_ISL_2635820, EPI_ISL_2760545, EPI_ISL_2760547, EPI_ISL_2760549, EPI_ISL_2760553, EPI_ISL_2760555, EPI_ISL_2845181, EPI_ISL_2845182, EPI_ISL_2845183, EPI_ISL_2845184, EPI_ISL_2845185, EPI_ISL_2845186 | | | |
| see above | Niedersächsisches Landesgesundheitsamt (NLGA) | Robert Koch Institute | |
| EPI_ISL_2678375, EPI_ISL_2678376 | Philipps-Universität Marburg Fachbereich Medizin Institut für Virologie | Robert Koch Institute | |
| EPI_ISL_2633134, EPI_ISL_2633149, EPI_ISL_2633314, EPI_ISL_2759921 | Praxisgemeinschaft für Laboratoriumsmedizin Labor Blumenstraße; Praxis Dr. E. Wietschel & Dr. F. Wietschel | Robert Koch Institute | |
| EPI_ISL_2633283, EPI_ISL_2635339, EPI_ISL_2760014, EPI_ISL_2844925, EPI_ISL_2844927 | Praxisgemeinschaft für Laboratoriumsmedizin Labor Blumenstraße; Praxis Dr. Sel und Dr. Späte | Robert Koch Institute | |
| EPI_ISL_2636885, EPI_ISL_2761926, EPI_ISL_2761937, EPI_ISL_2845112 | Procomcure Biotech Germany GmbH | Robert Koch Institute | |
| EPI_ISL_2761222, EPI_ISL_2761312, EPI_ISL_2762024, EPI_ISL_2762032, EPI_ISL_2762099 | Robert-Bosch-Krankenhaus | Robert Koch Institute | |
| EPI_ISL_2632966, EPI_ISL_2632972, EPI_ISL_2761872 | SLK-Kliniken Heilbronn GmbH - Institut für Laboratoriumsmedizin | Robert Koch Institute | |
| EPI_ISL_2759695, EPI_ISL_2759710, EPI_ISL_2759729, EPI_ISL_2759749, EPI_ISL_2759765, EPI_ISL_2759981, EPI_ISL_2760796, EPI_ISL_2760843, EPI_ISL_2844892, EPI_ISL_2844974, EPI_ISL_2845011 | | | |
| see above | SYNLAB Jena Oncoscreen | Robert Koch Institute | |
| EPI_ISL_2759689, EPI_ISL_2759735, EPI_ISL_2759738, EPI_ISL_2759987, EPI_ISL_2759989, EPI_ISL_2760700, EPI_ISL_2760722 | | | |
| see above | SYNLAB Labor München Zentrum LMZ | Robert Koch Institute | |
| EPI_ISL_2633199, EPI_ISL_2633233, EPI_ISL_2633342, EPI_ISL_2635428, EPI_ISL_2635431, EPI_ISL_2759931, EPI_ISL_2844978, EPI_ISL_2844995 | | | |
| see above | SYNLAB MVZ Berlin | Robert Koch Institute | |
| EPI_ISL_2633212, EPI_ISL_2635435, EPI_ISL_2635451, EPI_ISL_2845000 | SYNLAB MVZ Dachau | Robert Koch Institute | |
| EPI_ISL_2635455, EPI_ISL_2635456, EPI_ISL_2635467 | SYNLAB MVZ Hamburg | Robert Koch Institute | |
| EPI_ISL_2633059, EPI_ISL_2633308, EPI_ISL_2635423, EPI_ISL_2635424, EPI_ISL_2635427, EPI_ISL_2635429, EPI_ISL_2635432, EPI_ISL_2635437, EPI_ISL_2635440, EPI_ISL_2635441, EPI_ISL_2635444, EPI_ISL_2635445, EPI_ISL_2635447, EPI_ISL_2635454, EPI_ISL_2635461, EPI_ISL_2635469, EPI_ISL_2759666, EPI_ISL_2759669, EPI_ISL_2759701, EPI_ISL_2759712, EPI_ISL_2759717, EPI_ISL_2759913, EPI_ISL_2759927, EPI_ISL_2759935, EPI_ISL_2759947, EPI_ISL_2759949, EPI_ISL_2759951, EPI_ISL_2760009, EPI_ISL_2760012, EPI_ISL_2760019, EPI_ISL_2760691, EPI_ISL_2760698, EPI_ISL_2760717, EPI_ISL_2760756, EPI_ISL_2760780, EPI_ISL_2760789, EPI_ISL_2760804, EPI_ISL_2760838, EPI_ISL_2760875, EPI_ISL_2844917, EPI_ISL_2844919, EPI_ISL_2844921, EPI_ISL_2844929, EPI_ISL_2844937, EPI_ISL_2844938, EPI_ISL_2844945, EPI_ISL_2844947, EPI_ISL_2844950, EPI_ISL_2844986, EPI_ISL_2844989, EPI_ISL_2845003 | | | |
| see above | SYNLAB MVZ Heidelberg | Robert Koch Institute | |
| EPI_ISL_2632987, EPI_ISL_2632991, EPI_ISL_2632995, EPI_ISL_2632996, EPI_ISL_2632999, EPI_ISL_2633002, EPI_ISL_2633006, EPI_ISL_2633007, EPI_ISL_2633015, EPI_ISL_2633016, EPI_ISL_2633018, EPI_ISL_2633019, EPI_ISL_2633020, EPI_ISL_2633022, EPI_ISL_2633026, EPI_ISL_2633027, EPI_ISL_2633029, EPI_ISL_2633034, EPI_ISL_2633036, EPI_ISL_2633037, EPI_ISL_2633047, EPI_ISL_2633053, EPI_ISL_2633054, EPI_ISL_2633057, EPI_ISL_2633062, EPI_ISL_2633063, EPI_ISL_2633065, EPI_ISL_2633066, EPI_ISL_2633069, EPI_ISL_2633071, EPI_ISL_2633078, EPI_ISL_2633080, EPI_ISL_2633083, EPI_ISL_2633084, EPI_ISL_2633086, EPI_ISL_2633087, EPI_ISL_2633088, EPI_ISL_2633090, EPI_ISL_2633091, EPI_ISL_2633098, EPI_ISL_2633099, EPI_ISL_2633102, EPI_ISL_2633104, EPI_ISL_2633106, EPI_ISL_2633108, EPI_ISL_2633110, EPI_ISL_2633113, EPI_ISL_2633115, EPI_ISL_2633118, EPI_ISL_2633123, EPI_ISL_2633124, EPI_ISL_2633126, EPI_ISL_2633128, EPI_ISL_2633130, EPI_ISL_2633131, EPI_ISL_2633132, EPI_ISL_2633139, EPI_ISL_2633142, EPI_ISL_2633145, EPI_ISL_2633152, EPI_ISL_2633155, EPI_ISL_2633156, EPI_ISL_2633157, EPI_ISL_2633161, EPI_ISL_2633163, EPI_ISL_2633166, EPI_ISL_2633167, EPI_ISL_2633168, EPI_ISL_2633171, EPI_ISL_2633172, EPI_ISL_2633177, EPI_ISL_2633178, EPI_ISL_2633187, EPI_ISL_2633188, EPI_ISL_2633190, EPI_ISL_2633194, EPI_ISL_2633195, EPI_ISL_2633198, EPI_ISL_2633202, EPI_ISL_2633211, EPI_ISL_2633215, EPI_ISL_2633218, EPI_ISL_2633219, EPI_ISL_2633228, EPI_ISL_2633232, EPI_ISL_2633239, EPI_ISL_2633246, EPI_ISL_2633249, EPI_ISL_2633257, EPI_ISL_2633266, EPI_ISL_2633272, EPI_ISL_2633277, EPI_ISL_2633281, EPI_ISL_2633286, EPI_ISL_2633294, EPI_ISL_2633296, EPI_ISL_2633299, EPI_ISL_2633303, EPI_ISL_2633317, EPI_ISL_2633321, EPI_ISL_2633322, EPI_ISL_2633323, EPI_ISL_2633324, EPI_ISL_2633328, EPI_ISL_2633329, EPI_ISL_2633331, EPI_ISL_2633333, EPI_ISL_2633334, EPI_ISL_2633335, EPI_ISL_2633341, EPI_ISL_2633343, EPI_ISL_2633346, EPI_ISL_2633349, EPI_ISL_2633350, EPI_ISL_2633353, EPI_ISL_2633356, EPI_ISL_2633364, EPI_ISL_2633367, EPI_ISL_2633369, EPI_ISL_2633371, EPI_ISL_2633373, EPI_ISL_2633377, EPI_ISL_2633379, EPI_ISL_2633380, EPI_ISL_2633381, EPI_ISL_2633386, EPI_ISL_2633387, EPI_ISL_2633388, EPI_ISL_2633391, EPI_ISL_2633392, EPI_ISL_2633394, EPI_ISL_2635193, EPI_ISL_2635194, EPI_ISL_2635195, EPI_ISL_2635197, EPI_ISL_2635202, EPI_ISL_2635203, EPI_ISL_2635206, EPI_ISL_2635211, EPI_ISL_2635218, EPI_ISL_2635224, EPI_ISL_2635230, EPI_ISL_2635231, EPI_ISL_2635234, EPI_ISL_2635240, EPI_ISL_2635249, EPI_ISL_2635252, EPI_ISL_2635257, EPI_ISL_2635262, EPI_ISL_2635264, EPI_ISL_2635266, EPI_ISL_2635267, EPI_ISL_2635271, EPI_ISL_2635272, EPI_ISL_2635275, EPI_ISL_2635278, EPI_ISL_2635279, EPI_ISL_2635283, EPI_ISL_2635284, EPI_ISL_2635285, EPI_ISL_2635289, EPI_ISL_2635293, EPI_ISL_2635295, EPI_ISL_2635298, EPI_ISL_2635299, EPI_ISL_2635300, EPI_ISL_2635307, EPI_ISL_2635311, EPI_ISL_2635313, EPI_ISL_2635326, EPI_ISL_2635328, EPI_ISL_2635329, EPI_ISL_2635341, EPI_ISL_2635343, EPI_ISL_2635348, EPI_ISL_2635353, EPI_ISL_2635362, EPI_ISL_2635367, EPI_ISL_2635368, EPI_ISL_2635375, EPI_ISL_2635385, EPI_ISL_2635386, EPI_ISL_2635388, EPI_ISL_2635390, EPI_ISL_2635393, EPI_ISL_2635395, EPI_ISL_2635396, EPI_ISL_2635403, EPI_ISL_2635404, EPI_ISL_2635405, EPI_ISL_2635406, EPI_ISL_2635408, EPI_ISL_2635412, EPI_ISL_2635414, EPI_ISL_2759657, EPI_ISL_2759658, EPI_ISL_2759659, EPI_ISL_2759661, EPI_ISL_2759663, EPI_ISL_2759664, EPI_ISL_2759667, EPI_ISL_2759670, EPI_ISL_2759673, EPI_ISL_2759675, EPI_ISL_2759676, EPI_ISL_2759677, EPI_ISL_2759678, EPI_ISL_2759680, EPI_ISL_2759681, EPI_ISL_2759682, EPI_ISL_2759684, EPI_ISL_2759685, EPI_ISL_2759686, EPI_ISL_2759687, EPI_ISL_2759690, EPI_ISL_2759693, EPI_ISL_2759697, EPI_ISL_2759699, EPI_ISL_2759700, EPI_ISL_2759702, EPI_ISL_2759703, EPI_ISL_2759704, EPI_ISL_2759705, EPI_ISL_2759706, EPI_ISL_2759707, EPI_ISL_2759708, EPI_ISL_2759713, EPI_ISL_2759718, EPI_ISL_2759719, EPI_ISL_2759723, EPI_ISL_2759724, EPI_ISL_2759725, EPI_ISL_2759728, EPI_ISL_2759730, EPI_ISL_2759731, EPI_ISL_2759746, EPI_ISL_2759753, EPI_ISL_2759760, EPI_ISL_2759762, EPI_ISL_2759763, EPI_ISL_2759772, EPI_ISL_2759773, EPI_ISL_2759774, EPI_ISL_2759775, EPI_ISL_2759887, EPI_ISL_2759888, EPI_ISL_2759890, EPI_ISL_2759891, EPI_ISL_2759896, EPI_ISL_2759900, EPI_ISL_2759901, EPI_ISL_2759902, EPI_ISL_2759903, EPI_ISL_2759907, EPI_ISL_2759910, EPI_ISL_2759911, EPI_ISL_2759912, EPI_ISL_2759914, EPI_ISL_2759915, EPI_ISL_2759917, EPI_ISL_2759919, EPI_ISL_2759922, EPI_ISL_2759923, EPI_ISL_2759924, EPI_ISL_2759925, EPI_ISL_2759926, EPI_ISL_2759928, EPI_ISL_2759934, EPI_ISL_2759937, EPI_ISL_2759938, EPI_ISL_2759939, EPI_ISL_2759941, EPI_ISL_2759942, EPI_ISL_2759945, EPI_ISL_2759946, EPI_ISL_2759948, EPI_ISL_2759950, EPI_ISL_2759952, EPI_ISL_2759953, EPI_ISL_2759954, EPI_ISL_2759955, EPI_ISL_2759957, EPI_ISL_2759958, EPI_ISL_2759959, EPI_ISL_2759960, EPI_ISL_2759961, EPI_ISL_2759963, EPI_ISL_2759965, EPI_ISL_2759966, EPI_ISL_2759967, EPI_ISL_2759969, EPI_ISL_2759970, EPI_ISL_2759971, EPI_ISL_2759973, EPI_ISL_2759974, EPI_ISL_2759975, EPI_ISL_2759976, EPI_ISL_2759977, EPI_ISL_2759978, EPI_ISL_2759980, EPI_ISL_2759982, EPI_ISL_2759985, EPI_ISL_2759986, EPI_ISL_2759988, EPI_ISL_2759991, EPI_ISL_2759992, EPI_ISL_2759993, EPI_ISL_2759994, EPI_ISL_2759995, EPI_ISL_2759997, EPI_ISL_2759998, EPI_ISL_2760002, EPI_ISL_2760003, EPI_ISL_2760004, EPI_ISL_2760005, EPI_ISL_2760008, EPI_ISL_2760010, EPI_ISL_2760011, EPI_ISL_2760016, EPI_ISL_2760017, EPI_ISL_2760018, EPI_ISL_2760667, EPI_ISL_2760669, EPI_ISL_2760671, EPI_ISL_2760674, EPI_ISL_2760678, EPI_ISL_2760680, EPI_ISL_2760683, EPI_ISL_2760693, EPI_ISL_2760695, EPI_ISL_2760705, EPI_ISL_2760713, EPI_ISL_2760715, EPI_ISL_2760720, EPI_ISL_2760724, EPI_ISL_2760727, EPI_ISL_2760730, EPI_ISL_2760734, EPI_ISL_2760740, EPI_ISL_2760743, EPI_ISL_2760761, EPI_ISL_2760771, EPI_ISL_2760791, EPI_ISL_2760794, EPI_ISL_2760806, EPI_ISL_2760816, EPI_ISL_2760818, EPI_ISL_2760826, EPI_ISL_2760829, EPI_ISL_2760831, EPI_ISL_2760846, EPI_ISL_2760854, EPI_ISL_2760859, EPI_ISL_2760862, EPI_ISL_2760877, EPI_ISL_2844884, EPI_ISL_2844885, EPI_ISL_2844890, EPI_ISL_2844893, EPI_ISL_2844895, EPI_ISL_2844897, EPI_ISL_2844898, EPI_ISL_2844899, EPI_ISL_2844905, EPI_ISL_2844908, EPI_ISL_2844916, EPI_ISL_2844922, EPI_ISL_2844923, EPI_ISL_2844924, EPI_ISL_2844936, EPI_ISL_2844939, EPI_ISL_2844941, EPI_ISL_2844942, EPI_ISL_2844949, EPI_ISL_2844953, EPI_ISL_2844955, EPI_ISL_2844957, EPI_ISL_2844959, EPI_ISL_2844962, EPI_ISL_2844965, EPI_ISL_2844966, EPI_ISL_2844968, EPI_ISL_2844970, EPI_ISL_2844973, EPI_ISL_2844975, EPI_ISL_2844977, EPI_ISL_2844987, EPI_ISL_2844990, EPI_ISL_2845001, EPI_ISL_2845004, EPI_ISL_2845006, EPI_ISL_2845012, EPI_ISL_2845013, EPI_ISL_2845014 | | | |
| see above | SYNLAB MVZ Leinfelden-Echterdingen | Robert Koch Institute | |
| EPI_ISL_2632985, EPI_ISL_2632989, EPI_ISL_2632992, EPI_ISL_2632993, EPI_ISL_2633000, EPI_ISL_2633003, EPI_ISL_2633004, EPI_ISL_2633008, EPI_ISL_2633010, EPI_ISL_2633011, EPI_ISL_2633014, EPI_ISL_2633023, EPI_ISL_2633030, EPI_ISL_2633038, EPI_ISL_2633042, EPI_ISL_2633044, EPI_ISL_2633045, EPI_ISL_2633046, EPI_ISL_2633049, EPI_ISL_2633050, EPI_ISL_2633052, EPI_ISL_2633055, EPI_ISL_2633061, EPI_ISL_2633068, EPI_ISL_2633070, EPI_ISL_2633074, EPI_ISL_2633075, EPI_ISL_2633077, EPI_ISL_2633082, EPI_ISL_2633094, EPI_ISL_2633096, EPI_ISL_2633100, EPI_ISL_2633114, EPI_ISL_2633120, EPI_ISL_2633121, EPI_ISL_2633127, EPI_ISL_2633144, EPI_ISL_2633148, EPI_ISL_2633153, EPI_ISL_2633159, EPI_ISL_2633160, EPI_ISL_2633164, EPI_ISL_2633174, EPI_ISL_2633175, EPI_ISL_2633181, EPI_ISL_2633184, EPI_ISL_2633197, EPI_ISL_2633209, EPI_ISL_2633220, EPI_ISL_2633222, EPI_ISL_2633223, EPI_ISL_2633225, EPI_ISL_2633226, EPI_ISL_2633231, EPI_ISL_2633235, EPI_ISL_2633236, EPI_ISL_2633238, EPI_ISL_2633241, EPI_ISL_2633242, EPI_ISL_2633243, EPI_ISL_2633245, EPI_ISL_2633247, EPI_ISL_2633250, EPI_ISL_2633255, EPI_ISL_2633261, EPI_ISL_2633264, EPI_ISL_2633268, EPI_ISL_2633270, EPI_ISL_2633275, EPI_ISL_2633279, EPI_ISL_2633290, EPI_ISL_2633292, EPI_ISL_2633298, EPI_ISL_2633302, EPI_ISL_2633304, EPI_ISL_2633306, EPI_ISL_2633310, EPI_ISL_2633311, EPI_ISL_2633312, EPI_ISL_2633316, EPI_ISL_2633319, EPI_ISL_2633325, EPI_ISL_2633336, EPI_ISL_2633337, EPI_ISL_2633339, EPI_ISL_2633348, EPI_ISL_2633357, EPI_ISL_2633359, EPI_ISL_2633362, EPI_ISL_2633363, EPI_ISL_2633366, EPI_ISL_2633376, EPI_ISL_2633384, EPI_ISL_2633390, EPI_ISL_2635192, EPI_ISL_2635196, EPI_ISL_2635198, EPI_ISL_2635199, EPI_ISL_2635200, EPI_ISL_2635201, EPI_ISL_2635204, EPI_ISL_2635205, EPI_ISL_2635207, EPI_ISL_2635208, EPI_ISL_2635209, EPI_ISL_2635210, EPI_ISL_2635212, EPI_ISL_2635213, EPI_ISL_2635214, EPI_ISL_2635215, EPI_ISL_2635216, EPI_ISL_2635217, EPI_ISL_2635219, EPI_ISL_2635220, EPI_ISL_2635221, EPI_ISL_2635222, EPI_ISL_2635223, EPI_ISL_2635225, EPI_ISL_2635226, EPI_ISL_2635227, EPI_ISL_2635228, EPI_ISL_2635229, EPI_ISL_2635232, EPI_ISL_2635233, EPI_ISL_2635235, EPI_ISL_2635236, EPI_ISL_2635237, EPI_ISL_2635238, EPI_ISL_2635239, EPI_ISL_2635241, EPI_ISL_2635242, EPI_ISL_2635243, EPI_ISL_2635244, EPI_ISL_2635245, EPI_ISL_2635246, EPI_ISL_2635247, EPI_ISL_2635248, EPI_ISL_2635250, EPI_ISL_2635251, EPI_ISL_2635253, EPI_ISL_2635254, EPI_ISL_2635255, EPI_ISL_2635256, EPI_ISL_2635258, EPI_ISL_2635259, EPI_ISL_2635260, EPI_ISL_2635261, EPI_ISL_2635263, EPI_ISL_2635265, EPI_ISL_2635268, EPI_ISL_2635269, EPI_ISL_2635270, EPI_ISL_2635273, EPI_ISL_2635274, EPI_ISL_2635276, EPI_ISL_2635277, EPI_ISL_2635280, EPI_ISL_2635281, EPI_ISL_2635282, EPI_ISL_2635286, EPI_ISL_2635287, EPI_ISL_2635288, EPI_ISL_2635290, EPI_ISL_2635291, EPI_ISL_2635292, EPI_ISL_2635294, EPI_ISL_2635296, EPI_ISL_2635297, EPI_ISL_2635301, EPI_ISL_2635302, EPI_ISL_2635303, EPI_ISL_2635304, EPI_ISL_2635305, EPI_ISL_2635306, EPI_ISL_2635308, EPI_ISL_2635309, EPI_ISL_2635310, EPI_ISL_2635312, EPI_ISL_2635314, EPI_ISL_2635315, EPI_ISL_2635316, EPI_ISL_2635317, EPI_ISL_2635318, EPI_ISL_2635319, EPI_ISL_2635320, EPI_ISL_2635321, EPI_ISL_2635322, EPI_ISL_2635323, EPI_ISL_2635324, EPI_ISL_2635325, EPI_ISL_2635327, EPI_ISL_2635330, EPI_ISL_2635331, EPI_ISL_2635332, EPI_ISL_2635333, EPI_ISL_2635334, EPI_ISL_2635335, EPI_ISL_2635336, EPI_ISL_2635337, EPI_ISL_2635338, EPI_ISL_2635340, EPI_ISL_2635342, EPI_ISL_2635344, EPI_ISL_2635345, EPI_ISL_2635346, EPI_ISL_2635347, EPI_ISL_2635350, EPI_ISL_2635351, EPI_ISL_2635352, EPI_ISL_2635354, EPI_ISL_2635355, EPI_ISL_2635356, EPI_ISL_2635357, EPI_ISL_2635358, EPI_ISL_2635359, EPI_ISL_2635360, EPI_ISL_2635361, EPI_ISL_2635363, EPI_ISL_2635364, EPI_ISL_2635365, EPI_ISL_2635366, EPI_ISL_2635369, EPI_ISL_2635370, EPI_ISL_2635371, EPI_ISL_2635372, EPI_ISL_2635373, EPI_ISL_2635374, EPI_ISL_2635376, EPI_ISL_2635377, EPI_ISL_2635378, EPI_ISL_2635379, EPI_ISL_2635380, EPI_ISL_2635381, EPI_ISL_2635382, EPI_ISL_2635383, EPI_ISL_2635384, EPI_ISL_2635387, EPI_ISL_2635389, EPI_ISL_2635391, EPI_ISL_2635392, EPI_ISL_2635394, EPI_ISL_2635397, EPI_ISL_2635398, EPI_ISL_2635399, EPI_ISL_2635400, EPI_ISL_2635401, EPI_ISL_2635402, EPI_ISL_2635407, EPI_ISL_2635409, EPI_ISL_2635410, EPI_ISL_2635411, EPI_ISL_2635413, EPI_ISL_2635415, EPI_ISL_2635416, EPI_ISL_2635417, EPI_ISL_2635418, EPI_ISL_2635419, EPI_ISL_2759656, EPI_ISL_2759660, EPI_ISL_2759662, EPI_ISL_2759665, EPI_ISL_2759668, EPI_ISL_2759671, EPI_ISL_2759674, EPI_ISL_2759679, EPI_ISL_2759683, EPI_ISL_2759688, EPI_ISL_2759691, EPI_ISL_2759692, EPI_ISL_2759694, EPI_ISL_2759696, EPI_ISL_2759698, EPI_ISL_2759709, EPI_ISL_2759711, EPI_ISL_2759714, EPI_ISL_2759715, EPI_ISL_2759716, EPI_ISL_2759720, EPI_ISL_2759722, EPI_ISL_2759727, EPI_ISL_2759732, EPI_ISL_2759733, EPI_ISL_2759734, EPI_ISL_2759736, EPI_ISL_2759737, EPI_ISL_2759742, EPI_ISL_2759743, EPI_ISL_2759745, EPI_ISL_2759748, EPI_ISL_2759751, EPI_ISL_2759752, EPI_ISL_2759755, EPI_ISL_2759758, EPI_ISL_2759759, EPI_ISL_2759761, EPI_ISL_2759764, EPI_ISL_2759766, EPI_ISL_2759767, EPI_ISL_2759768, EPI_ISL_2759769, EPI_ISL_2759770, EPI_ISL_2759771, EPI_ISL_2759776, EPI_ISL_2759777, EPI_ISL_2759778, EPI_ISL_2759892, EPI_ISL_2759893, EPI_ISL_2759894, EPI_ISL_2759895, EPI_ISL_2759897, EPI_ISL_2759898, EPI_ISL_2759899, EPI_ISL_2759905, EPI_ISL_2759906, EPI_ISL_2759908, EPI_ISL_2759916, EPI_ISL_2759918, EPI_ISL_2759920, EPI_ISL_2759929, EPI_ISL_2759930, EPI_ISL_2759932, EPI_ISL_2759936, EPI_ISL_2759943, EPI_ISL_2759956, EPI_ISL_2759962, EPI_ISL_2759964, EPI_ISL_2759972, EPI_ISL_2759979, EPI_ISL_2759983, EPI_ISL_2759984, EPI_ISL_2759990, EPI_ISL_2759996, EPI_ISL_2760006, EPI_ISL_2760007, EPI_ISL_2760013, EPI_ISL_2760015, EPI_ISL_2760676, EPI_ISL_2760685, EPI_ISL_2760688, EPI_ISL_2760703, EPI_ISL_2760707, EPI_ISL_2760710, EPI_ISL_2760732, EPI_ISL_2760745, EPI_ISL_2760748, EPI_ISL_2760753, EPI_ISL_2760758, EPI_ISL_2760766, EPI_ISL_2760768, EPI_ISL_2760773, EPI_ISL_2760776, EPI_ISL_2760783, EPI_ISL_2760786, EPI_ISL_2760799, EPI_ISL_2760801, EPI_ISL_2760808, EPI_ISL_2760813, EPI_ISL_2760821, EPI_ISL_2760823, EPI_ISL_2760834, EPI_ISL_2760836, EPI_ISL_2760841, EPI_ISL_2760849, EPI_ISL_2760852, EPI_ISL_2760857, EPI_ISL_2760864, EPI_ISL_2760867, EPI_ISL_2760870, EPI_ISL_2844886, EPI_ISL_2844900, EPI_ISL_2844901, EPI_ISL_2844902, EPI_ISL_2844904, EPI_ISL_2844906, EPI_ISL_2844910, EPI_ISL_2844912, EPI_ISL_2844914, EPI_ISL_2844915, EPI_ISL_2844931, EPI_ISL_2844935, EPI_ISL_2844940, EPI_ISL_2844943, EPI_ISL_2844944, EPI_ISL_2844954, EPI_ISL_2844956, EPI_ISL_2844958, EPI_ISL_2844960, EPI_ISL_2844961, EPI_ISL_2844963, EPI_ISL_2844972, EPI_ISL_2844979, EPI_ISL_2844980, EPI_ISL_2844984, EPI_ISL_2844985, EPI_ISL_2844996, EPI_ISL_2844998, EPI_ISL_2844999, EPI_ISL_2845007, EPI_ISL_2845008 | | | |
| see above | SYNLAB MVZ Leverkusen | Robert Koch Institute | |
| EPI_ISL_2635422, EPI_ISL_2635458, EPI_ISL_2635459, EPI_ISL_2635460, EPI_ISL_2635463, EPI_ISL_2635464, EPI_ISL_2759741, EPI_ISL_2759889, EPI_ISL_2759904, EPI_ISL_2844920, EPI_ISL_2844933 | | | |
| see above | SYNLAB MVZ Neustadt a. d. Weinstraße | Robert Koch Institute | |
| EPI_ISL_2632997, EPI_ISL_2633092, EPI_ISL_2633201, EPI_ISL_2633330, EPI_ISL_2633340, EPI_ISL_2635425, EPI_ISL_2635434, EPI_ISL_2635436, EPI_ISL_2635443, EPI_ISL_2635452, EPI_ISL_2635457, EPI_ISL_2635468, EPI_ISL_2635470, EPI_ISL_2759739, EPI_ISL_2759740, EPI_ISL_2759933, EPI_ISL_2759940, EPI_ISL_2759944, EPI_ISL_2760763, EPI_ISL_2760872, EPI_ISL_2844909, EPI_ISL_2844934, EPI_ISL_2844948 | | | |
| see above | SYNLAB MVZ Trier | Robert Koch Institute | |
| EPI_ISL_2632988, EPI_ISL_2633103, EPI_ISL_2633146, EPI_ISL_2633150, EPI_ISL_2633170, EPI_ISL_2633191, EPI_ISL_2633206, EPI_ISL_2633208, EPI_ISL_2633253, EPI_ISL_2633259, EPI_ISL_2633276, EPI_ISL_2633288, EPI_ISL_2633307, EPI_ISL_2633318, EPI_ISL_2633327, EPI_ISL_2633352, EPI_ISL_2633360, EPI_ISL_2633383, EPI_ISL_2635349, EPI_ISL_2635421, EPI_ISL_2635426, EPI_ISL_2635438, EPI_ISL_2635439, EPI_ISL_2635442, EPI_ISL_2635446, EPI_ISL_2635448, EPI_ISL_2635449, EPI_ISL_2635450, EPI_ISL_2635453, EPI_ISL_2635465, EPI_ISL_2635466, EPI_ISL_2759672, EPI_ISL_2759721, EPI_ISL_2759726, EPI_ISL_2759757, EPI_ISL_2759909, EPI_ISL_2759968, EPI_ISL_2759999, EPI_ISL_2760000, EPI_ISL_2760001, EPI_ISL_2760751, EPI_ISL_2760811, EPI_ISL_2844883, EPI_ISL_2844888, EPI_ISL_2844889, EPI_ISL_2844891, EPI_ISL_2844894, EPI_ISL_2844907, EPI_ISL_2844911, EPI_ISL_2844918, EPI_ISL_2844926, EPI_ISL_2844928, EPI_ISL_2844930, EPI_ISL_2844932, EPI_ISL_2844946, EPI_ISL_2844952, EPI_ISL_2844964, EPI_ISL_2844967, EPI_ISL_2844969, EPI_ISL_2844971, EPI_ISL_2844976, EPI_ISL_2844981, EPI_ISL_2844982, EPI_ISL_2844983, EPI_ISL_2844988, EPI_ISL_2844991, EPI_ISL_2844992, EPI_ISL_2844994, EPI_ISL_2844997, EPI_ISL_2845002, EPI_ISL_2845005, EPI_ISL_2845009, EPI_ISL_2845010 | | | |
| see above | SYNLAB MVZ Weiden | Robert Koch Institute | |
| EPI_ISL_2636964, EPI_ISL_2636972, EPI_ISL_2637072, EPI_ISL_2637091, EPI_ISL_2637116, EPI_ISL_2637117, EPI_ISL_2637133, EPI_ISL_2759825, EPI_ISL_2759833, EPI_ISL_2761098, EPI_ISL_2761103, EPI_ISL_2761138, EPI_ISL_2761171, EPI_ISL_2761175, EPI_ISL_2761190, EPI_ISL_2761205, EPI_ISL_2761224, EPI_ISL_2761252, EPI_ISL_2761337, EPI_ISL_2761339, EPI_ISL_2761428, EPI_ISL_2761444, EPI_ISL_2761467, EPI_ISL_2761480, EPI_ISL_2761549, EPI_ISL_2761586, EPI_ISL_2761590, EPI_ISL_2761970, EPI_ISL_2761975, EPI_ISL_2761982, EPI_ISL_2761987, EPI_ISL_2762010, EPI_ISL_2762020, EPI_ISL_2762025, EPI_ISL_2762027, EPI_ISL_2762037, EPI_ISL_2762046, EPI_ISL_2762067, EPI_ISL_2762069, EPI_ISL_2762074, EPI_ISL_2762104, EPI_ISL_2762691, EPI_ISL_2762808, EPI_ISL_2845324, EPI_ISL_2845333, EPI_ISL_2845360, EPI_ISL_2845435 | | | |
| see above | Sonic - Bioscientia - MVZ Labor Saar GmbH | Robert Koch Institute | |

| | | | |
|---|---|---|---|
| EPI_ISL_2759838, EPI_ISL_2761110, EPI_ISL_2761408, EPI_ISL_2761977, EPI_ISL_2761986, EPI_ISL_2761997, EPI_ISL_2762012, EPI_ISL_2762056, EPI_ISL_2762100, EPI_ISL_2762103, EPI_ISL_2762645, EPI_ISL_2762801, EPI_ISL_2845319, EPI_ISL_2845326 | | | |
| see above | Sonic - Dr. Staber & Kollegen GmbH München | Robert Koch Institute | |
| EPI_ISL_2759824, EPI_ISL_2759836, EPI_ISL_2759844, EPI_ISL_2759846, EPI_ISL_2761096, EPI_ISL_2761197, EPI_ISL_2761210, EPI_ISL_2761237, EPI_ISL_2761272, EPI_ISL_2761290, EPI_ISL_2761351, EPI_ISL_2761364, EPI_ISL_2761415, EPI_ISL_2761457, EPI_ISL_2761526, EPI_ISL_2761558, EPI_ISL_2761601, EPI_ISL_2845318, EPI_ISL_2845321, EPI_ISL_2845343, EPI_ISL_2845348, EPI_ISL_2845350, EPI_ISL_2845359, EPI_ISL_2845363, EPI_ISL_2845378, EPI_ISL_2845379 | | | |
| see above | Sonic - Labor Dr. von Froreich GmbH | Robert Koch Institute | |
| EPI_ISL_2637083 | Sonic - Labor Lademannbogen MVZ GmbH | Robert Koch Institute | |
| EPI_ISL_2759850, EPI_ISL_2761213, EPI_ISL_2761515, EPI_ISL_2761969, EPI_ISL_2762060, EPI_ISL_2762093, EPI_ISL_2845402 | | | |
| see above | Sonic - Labor Staber Bayreuth | Robert Koch Institute | |
| EPI_ISL_2636962, EPI_ISL_2636965, EPI_ISL_2637089, EPI_ISL_2759828, EPI_ISL_2759832, EPI_ISL_2759843, EPI_ISL_2761083, EPI_ISL_2761148, EPI_ISL_2761202, EPI_ISL_2761307, EPI_ISL_2761335, EPI_ISL_2761362, EPI_ISL_2761410, EPI_ISL_2761425, EPI_ISL_2761505, EPI_ISL_2761518, EPI_ISL_2761566, EPI_ISL_2761595, EPI_ISL_2761988, EPI_ISL_2762058, EPI_ISL_2762072, EPI_ISL_2762092, EPI_ISL_2845376, EPI_ISL_2845388, EPI_ISL_2845393 | | | |
| see above | Sonic - Labor Staber Dresden (Klipphausen) | Robert Koch Institute | |
| EPI_ISL_2636968, EPI_ISL_2637032, EPI_ISL_2637078, EPI_ISL_2637084, EPI_ISL_2761073, EPI_ISL_2761267, EPI_ISL_2761398, EPI_ISL_2761446, EPI_ISL_2761502, EPI_ISL_2761544, EPI_ISL_2761546, EPI_ISL_2762059, EPI_ISL_2762773, EPI_ISL_2762780 | | | |
| see above | Sonic - Labor Staber Nürnberg | Robert Koch Institute | |
| EPI_ISL_2636926, EPI_ISL_2636941, EPI_ISL_2636977, EPI_ISL_2637013, EPI_ISL_2637040, EPI_ISL_2637052, EPI_ISL_2637093, EPI_ISL_2637113, EPI_ISL_2637130, EPI_ISL_2637138, EPI_ISL_2637140, EPI_ISL_2637165, EPI_ISL_2759847, EPI_ISL_2761100, EPI_ISL_2761219, EPI_ISL_2761288, EPI_ISL_2761531, EPI_ISL_2761563, EPI_ISL_2761581, EPI_ISL_2761981, EPI_ISL_2761984, EPI_ISL_2761999, EPI_ISL_2762002, EPI_ISL_2762006, EPI_ISL_2762008, EPI_ISL_2762011, EPI_ISL_2762014, EPI_ISL_2762015, EPI_ISL_2762028, EPI_ISL_2762040, EPI_ISL_2762050, EPI_ISL_2762053, EPI_ISL_2762057, EPI_ISL_2762063, EPI_ISL_2762066, EPI_ISL_2762085, EPI_ISL_2762097, EPI_ISL_2845322, EPI_ISL_2845328, EPI_ISL_2845329, EPI_ISL_2845346, EPI_ISL_2845351, EPI_ISL_2845355, EPI_ISL_2845357, EPI_ISL_2845362, EPI_ISL_2845367, EPI_ISL_2845368, EPI_ISL_2845372, EPI_ISL_2845373 | | | |
| see above | Sonic - MVZ Medizinisches Labor Bremen GmbH | Robert Koch Institute | |
| EPI_ISL_2762095 | Sonic - MVZ Medizinisches Labor Oldenburg | Robert Koch Institute | |
| EPI_ISL_2635662 | St. Franziskus-Hospital Münster | Robert Koch Institute | |
| EPI_ISL_2634726, EPI_ISL_2634734, EPI_ISL_2634740, EPI_ISL_2634791, EPI_ISL_2634802, EPI_ISL_2634804, EPI_ISL_2634811, EPI_ISL_2634813, EPI_ISL_2678446, EPI_ISL_2678453, EPI_ISL_2678455, EPI_ISL_2678456, EPI_ISL_2678464, EPI_ISL_2678466, EPI_ISL_2678467, EPI_ISL_2678469, EPI_ISL_2762181, EPI_ISL_2762190, EPI_ISL_2845742, EPI_ISL_2845760 | | | |
| see above | Städtisches Klinikum Dresden Institut für Klinische Chemie und Laboratoriumsmedizin | Robert Koch Institute | |
| EPI_ISL_2633032, EPI_ISL_2633033, EPI_ISL_2633203, EPI_ISL_2633345, EPI_ISL_2635420, EPI_ISL_2635430, EPI_ISL_2635433, EPI_ISL_2635462, EPI_ISL_2844887, EPI_ISL_2844896, EPI_ISL_2844903, EPI_ISL_2844913, EPI_ISL_2844951, EPI_ISL_2844993 | | | |
| see above | Synlab MVZ Augsburg | Robert Koch Institute | |
| EPI_ISL_2762130, EPI_ISL_2762131, EPI_ISL_2762132 | Thüringer Landesamtes für Verbraucherschutz | Robert Koch Institute | |
| EPI_ISL_2634728, EPI_ISL_2634736, EPI_ISL_2634738, EPI_ISL_2634760, EPI_ISL_2634765, EPI_ISL_2634781, EPI_ISL_2634799, EPI_ISL_2678451, EPI_ISL_2678452, EPI_ISL_2678459, EPI_ISL_2678461, EPI_ISL_2678465, EPI_ISL_2678470, EPI_ISL_2762178, EPI_ISL_2762183, EPI_ISL_2762188, EPI_ISL_2762189, EPI_ISL_2762192, EPI_ISL_2762199, EPI_ISL_2762200, EPI_ISL_2762201, EPI_ISL_2845743, EPI_ISL_2845752, EPI_ISL_2845759 | | | |
| see above | Uniklinikum Carl Gustav Carus an der TU Dresden; Institut für Virologie | Robert Koch Institute | |
| EPI_ISL_2759818, EPI_ISL_2759819, EPI_ISL_2759823 | Uniklinikum Regensburg; Institut für klinische Mikrobiologie und Hygiene | Robert Koch Institute | |
| EPI_ISL_2845786 | Universitätsklinikum Essen | Robert Koch Institute | |
| EPI_ISL_2636980, EPI_ISL_2637150, EPI_ISL_2759849, EPI_ISL_2761960, EPI_ISL_2761961, EPI_ISL_2761962, EPI_ISL_2761994, EPI_ISL_2762077, EPI_ISL_2845364, EPI_ISL_2845397, EPI_ISL_2845442 | | | |
| see above | Universitätsklinikum Frankfurt - Institut für Medizinische Virologie | Robert Koch Institute | |
| EPI_ISL_2678374 | Universitätsklinikum Gießen - Institut für Medizinische Virologie | Robert Koch Institute | |
| EPI_ISL_2845772, EPI_ISL_2845773, EPI_ISL_2845774, EPI_ISL_2845775, EPI_ISL_2845776, EPI_ISL_2845777 | Universitätsklinikum Heidelberg | Robert Koch Institute | |
| EPI_ISL_2761735 | Universitätsklinikum Münster Institut für Virologie | Robert Koch Institute | |
| EPI_ISL_2633825, EPI_ISL_2633845, EPI_ISL_2633856, EPI_ISL_2633859, EPI_ISL_2633864, EPI_ISL_2636902, EPI_ISL_2636906, EPI_ISL_2844871, EPI_ISL_2844877 | | | |
| see above | Universitätsklinikum des Saarlandes - Institut für Virologie | Robert Koch Institute | |
| EPI_ISL_2628195, EPI_ISL_2628196, EPI_ISL_2628197, EPI_ISL_2628198, EPI_ISL_2628199, EPI_ISL_2628200, EPI_ISL_2628201, EPI_ISL_2628202, EPI_ISL_2628203, EPI_ISL_2628204, EPI_ISL_2628205, EPI_ISL_2628206, EPI_ISL_2628207 | | | |
| see above | Virology, Universitätsklinikum des Saarlandes | Epigenetics, Saarland University | Jörn Walter; Kathrin Kattler; Nastasia Seiwert; Sascha Tierling; Sigrun Smola; Stefan Lohse; Thorsten Pfuhl |
| EPI_ISL_2762174, EPI_ISL_2762175 | alphaomega Labor GbR (Medizinische Kooperationsgemeinschaft Dres. Ackermann/Tennert/Zaiß) | Robert Koch Institute | |
| EPI_ISL_2635616, EPI_ISL_2635632, EPI_ISL_2635641, EPI_ISL_2635644, EPI_ISL_2635646, EPI_ISL_2635654, EPI_ISL_2635659, EPI_ISL_2760491, EPI_ISL_2845082 | | | |
| see above | amedes Göttingen | Robert Koch Institute | |
| EPI_ISL_2635599, EPI_ISL_2635600, EPI_ISL_2635606, EPI_ISL_2635607, EPI_ISL_2635617, EPI_ISL_2635627, EPI_ISL_2635643, EPI_ISL_2635658, EPI_ISL_2635661, EPI_ISL_2760460, EPI_ISL_2760466, EPI_ISL_2760493, EPI_ISL_2760504, EPI_ISL_2760518, EPI_ISL_2845083, EPI_ISL_2845090 | | | |
| see above | amedes MVZ DIAMEDIS Sennestadt | Robert Koch Institute | |
| EPI_ISL_2632281, EPI_ISL_2635618, EPI_ISL_2635626, EPI_ISL_2635638, EPI_ISL_2635649, EPI_ISL_2635651, EPI_ISL_2845078, EPI_ISL_2845086 | | | |
| see above | amedes MVZ Halle/Leipzig | Robert Koch Institute | |
| EPI_ISL_2635614, EPI_ISL_2635619, EPI_ISL_2635625, EPI_ISL_2635635, EPI_ISL_2635637, EPI_ISL_2635642, EPI_ISL_2635645, EPI_ISL_2760455, EPI_ISL_2760515, EPI_ISL_2760535, EPI_ISL_2845084 | | | |
| see above | amedes MVZ Hannover | Robert Koch Institute | |
| EPI_ISL_2635636, EPI_ISL_2635652, EPI_ISL_2760498, EPI_ISL_2760511 | amedes MVZ Jena | Robert Koch Institute | |
| EPI_ISL_2635609, EPI_ISL_2760483 | amedes MVZ Zentrum für Pathologie und Zytodiagnostik GmbH (Köln) | Robert Koch Institute | |
| EPI_ISL_2635605, EPI_ISL_2635620, EPI_ISL_2635631, EPI_ISL_2635655, EPI_ISL_2760463, EPI_ISL_2760475, EPI_ISL_2760508, EPI_ISL_2845085, EPI_ISL_2845091 | | | |
| see above | amedes MVZ für Laboratoriumsdiagnostik Raubling GmbH | Robert Koch Institute | |
| EPI_ISL_2760506 | amedes MVZ für Laboratoriumsmedizin Fürstenfeldbruck | Robert Koch Institute | |
| EPI_ISL_2635650 | amedes MVZ für Laboratoriumsmedizin Hamburg | Robert Koch Institute | |
| EPI_ISL_2760543 | amedes MVZ für Laboratoriumsmedizin München | Robert Koch Institute | |
| EPI_ISL_2635601, EPI_ISL_2635622, EPI_ISL_2635656, EPI_ISL_2760468, EPI_ISL_2760480 | amedes MVZ für Laboratoriumsmedizin Rhein/Ruhr | Robert Koch Institute | |
| EPI_ISL_2635604, EPI_ISL_2635610, EPI_ISL_2635621, EPI_ISL_2635653, EPI_ISL_2760525, EPI_ISL_2760530, EPI_ISL_2760538 | | | |
| see above | amedes MVZ für Labordiagnostik Rhein-Main | Robert Koch Institute | |
| EPI_ISL_2760473, EPI_ISL_2760521, EPI_ISL_2760523, EPI_ISL_2845079, EPI_ISL_2845088 | amedes Medlab Arnold Analytik MVZ GmbH | Robert Koch Institute | |
| EPI_ISL_2634472, EPI_ISL_2634477, EPI_ISL_2634483, EPI_ISL_2634488, EPI_ISL_2634503, EPI_ISL_2761944, EPI_ISL_2761945, EPI_ISL_2761946, EPI_ISL_2761947, EPI_ISL_2761948, EPI_ISL_2761949, EPI_ISL_2761950, EPI_ISL_2761951, EPI_ISL_2761952, EPI_ISL_2761953, EPI_ISL_2761954, EPI_ISL_2761955, EPI_ISL_2761956, EPI_ISL_2761957, EPI_ISL_2761958, EPI_ISL_2761959, EPI_ISL_2845197 | | | |
| see above | labopart - Medizinische Laboratorien Dresden | Robert Koch Institute | |
| EPI_ISL_2632242, EPI_ISL_2632244, EPI_ISL_2632259, EPI_ISL_2632264, EPI_ISL_2632268, EPI_ISL_2632270, EPI_ISL_2632271, EPI_ISL_2632279, EPI_ISL_2632285, EPI_ISL_2632286, EPI_ISL_2632287, EPI_ISL_2632288, EPI_ISL_2632290, EPI_ISL_2632291, EPI_ISL_2632300, EPI_ISL_2632304, EPI_ISL_2632310, EPI_ISL_2632319, EPI_ISL_2632322, EPI_ISL_2635602, EPI_ISL_2635603, EPI_ISL_2635611, EPI_ISL_2635613, EPI_ISL_2635615, EPI_ISL_2635624, EPI_ISL_2635629, EPI_ISL_2635633, EPI_ISL_2635639, EPI_ISL_2635647, EPI_ISL_2635648, EPI_ISL_2635657, EPI_ISL_2635660, EPI_ISL_2760457, EPI_ISL_2760471, EPI_ISL_2760478, EPI_ISL_2760485, EPI_ISL_2760488, EPI_ISL_2760496, EPI_ISL_2760501, EPI_ISL_2760513, EPI_ISL_2760528, EPI_ISL_2760533, EPI_ISL_2760540, EPI_ISL_2845080, EPI_ISL_2845081, EPI_ISL_2845087, EPI_ISL_2845089 | | | |
| see above | nordlab - Partnerschaftspraxis für Laboratoriumsmedizin | Robert Koch Institute | |

We gratefully acknowledge the following Authors from the Originating laboratories responsible for obtaining the specimens, as well as the Submitting laboratories where the genome data were generated and shared via GISAID, on which this research is based.

All Submitters of data may be contacted directly via www.gisaid.org

Authors are sorted alphabetically.

| Accession ID | Originating Laboratory | Submitting Laboratory | Authors |
|---|---|---|---|
| EPI_ISL_2047561, EPI_ISL_2047562, EPI_ISL_2047563, EPI_ISL_2047564, EPI_ISL_2047565, EPI_ISL_2047566, EPI_ISL_2047567, EPI_ISL_2047568, EPI_ISL_2047569 | | | |
| see above | Balai Litbangkes Tanah Bumbu | National Institute of Health Research and Development | Arie Ardiansyah Nugraha; Hana Apsari Pawestri; Hartanti Dian Ikawati; Juhairiyah; Kartika Dewi Puspa; Krisna Pangesti; Nelly Puspandari; Subangkit; Syarif Hidayat; Triyani Soekarso; Vivi Setiawaty |
| EPI_ISL_1915572, EPI_ISL_1915573, EPI_ISL_1915574, EPI_ISL_1915575 | Balitbang Tanah Bumbu | National Institute of Health Research and Development | Arie Ardiansyah Nugraha; Hana Apsari Pawestri; Hartanti Dian Ikawati; Kartika Dewi Puspa; Krisna Pangesti; Nelly Puspandari; Subangkit; Vivi Setiawaty |
| EPI_ISL_1252402, EPI_ISL_1252403, EPI_ISL_1252405, EPI_ISL_1252407, EPI_ISL_1252409, EPI_ISL_1252412, EPI_ISL_1252413, EPI_ISL_1252416, EPI_ISL_1252417, EPI_ISL_1252418, EPI_ISL_1252438, EPI_ISL_1315346, EPI_ISL_1315351, EPI_ISL_1315353, EPI_ISL_1315354, EPI_ISL_1315355, EPI_ISL_1315356, EPI_ISL_1315362, EPI_ISL_1315363, EPI_ISL_1315364, EPI_ISL_1315365, EPI_ISL_1315366, EPI_ISL_1315367, EPI_ISL_1315368, EPI_ISL_1315369, EPI_ISL_1315370, EPI_ISL_1315371, EPI_ISL_1315372, EPI_ISL_1315373, EPI_ISL_1315377, EPI_ISL_1315378, EPI_ISL_1315379, EPI_ISL_1315380, EPI_ISL_1315381, EPI_ISL_1315382, EPI_ISL_1318989, EPI_ISL_1318992, EPI_ISL_1318994, EPI_ISL_1318998, EPI_ISL_1318999, EPI_ISL_1319000, EPI_ISL_1319001, EPI_ISL_1319002, EPI_ISL_1319003, EPI_ISL_1319004, EPI_ISL_1319005, EPI_ISL_1319006, EPI_ISL_1319007, EPI_ISL_1319008, EPI_ISL_1319009, EPI_ISL_1319010, EPI_ISL_1319011, EPI_ISL_1319012, EPI_ISL_1319013, EPI_ISL_1319014, EPI_ISL_1319015, EPI_ISL_1319016, EPI_ISL_1319017, EPI_ISL_1319018, EPI_ISL_1319019, EPI_ISL_1319022, EPI_ISL_1319023, EPI_ISL_1319024, EPI_ISL_1319025, EPI_ISL_1319026, EPI_ISL_1319027, EPI_ISL_1319028, EPI_ISL_1319029, EPI_ISL_1319030, EPI_ISL_1319031, EPI_ISL_1319032, EPI_ISL_1319033, EPI_ISL_1319034, EPI_ISL_1319035, EPI_ISL_1319036, EPI_ISL_1319037, EPI_ISL_1319038, EPI_ISL_1319039, EPI_ISL_1319040, EPI_ISL_1319041, EPI_ISL_1489527, EPI_ISL_1489529, EPI_ISL_1489530, EPI_ISL_1489531, EPI_ISL_1489532, EPI_ISL_1489533, EPI_ISL_1489534, EPI_ISL_1489535, EPI_ISL_1489536, EPI_ISL_1489537, EPI_ISL_1489538, EPI_ISL_1489539, EPI_ISL_1489540, EPI_ISL_1489541, EPI_ISL_1489542, EPI_ISL_1489543, EPI_ISL_1489546, EPI_ISL_1489547, EPI_ISL_1489548, EPI_ISL_1489549, EPI_ISL_1489550, EPI_ISL_1489551, EPI_ISL_1489553, EPI_ISL_1489554, EPI_ISL_1489555, EPI_ISL_1489556, EPI_ISL_1489559, EPI_ISL_1489560, EPI_ISL_1489561, EPI_ISL_1489563, EPI_ISL_1489564, EPI_ISL_1489566, EPI_ISL_1489568, EPI_ISL_1489569, EPI_ISL_1489570, EPI_ISL_1489571, EPI_ISL_1489572, EPI_ISL_1489573, EPI_ISL_1489574, EPI_ISL_1489575, EPI_ISL_1489576, EPI_ISL_1489577, EPI_ISL_1489578, EPI_ISL_1489579, EPI_ISL_1489580, EPI_ISL_1489581, EPI_ISL_1489582, EPI_ISL_1489583, EPI_ISL_1489584, EPI_ISL_1489585, EPI_ISL_1489586, EPI_ISL_1489587, EPI_ISL_1489588, EPI_ISL_1489589, EPI_ISL_1489590, EPI_ISL_1489591, EPI_ISL_1489592, EPI_ISL_1489593, EPI_ISL_1489594, EPI_ISL_1489595, EPI_ISL_1489596, EPI_ISL_1489597, EPI_ISL_1489598, EPI_ISL_1489599, EPI_ISL_1489600, EPI_ISL_1489601, EPI_ISL_1489602, EPI_ISL_1489603, EPI_ISL_1489604, EPI_ISL_1489605, EPI_ISL_1489606, EPI_ISL_1489607, EPI_ISL_1489608, EPI_ISL_1489609, EPI_ISL_1489610, EPI_ISL_1489611, EPI_ISL_1489612, EPI_ISL_1489613, EPI_ISL_1489614, EPI_ISL_1489615, EPI_ISL_1489616, EPI_ISL_1489617, EPI_ISL_1489618, EPI_ISL_1489619, EPI_ISL_1489620, EPI_ISL_1489621, EPI_ISL_1489622, EPI_ISL_1489623, EPI_ISL_1489624, EPI_ISL_1489625, EPI_ISL_1489626, EPI_ISL_1489627, EPI_ISL_1489628, EPI_ISL_1489629, EPI_ISL_1489630, EPI_ISL_1489631, EPI_ISL_1489632, EPI_ISL_1489633, EPI_ISL_1489634, EPI_ISL_1489635, EPI_ISL_1489636, EPI_ISL_1489637, EPI_ISL_1489638, EPI_ISL_1489639, EPI_ISL_1489640, EPI_ISL_1489641, EPI_ISL_1489642, EPI_ISL_1489643, EPI_ISL_1489644, EPI_ISL_1489651, EPI_ISL_1489652, EPI_ISL_1489653, EPI_ISL_1489654, EPI_ISL_1489655, EPI_ISL_1489656, EPI_ISL_1489942, EPI_ISL_1489943, EPI_ISL_1489944, EPI_ISL_1489945, EPI_ISL_1489946, EPI_ISL_1489947, EPI_ISL_1489948, EPI_ISL_1489949, EPI_ISL_1489950, EPI_ISL_1489951, EPI_ISL_1489952, EPI_ISL_1489953, EPI_ISL_1489954, EPI_ISL_1489955, EPI_ISL_1489956, EPI_ISL_1489957, EPI_ISL_1489958, EPI_ISL_1489959, EPI_ISL_1489960, EPI_ISL_1489961, EPI_ISL_1489962, EPI_ISL_1489963, EPI_ISL_1489964, EPI_ISL_1489965, EPI_ISL_1489966, EPI_ISL_1489967, EPI_ISL_1489968, EPI_ISL_1489969, EPI_ISL_1489970, EPI_ISL_1489971, EPI_ISL_1489972, EPI_ISL_1489973, EPI_ISL_1489974, EPI_ISL_1489975, EPI_ISL_1489976, EPI_ISL_1489977, EPI_ISL_1489978, EPI_ISL_1489979, EPI_ISL_1489980, EPI_ISL_1489981, EPI_ISL_1489982, EPI_ISL_1489983, EPI_ISL_1489984, EPI_ISL_1489985, EPI_ISL_1489986, EPI_ISL_1489987, EPI_ISL_1489988, EPI_ISL_1489989, EPI_ISL_1489990, EPI_ISL_1489991, EPI_ISL_1489992, EPI_ISL_1489993, EPI_ISL_1489994, EPI_ISL_1489995, EPI_ISL_1489996, EPI_ISL_1489997, EPI_ISL_1489998, EPI_ISL_1489999, EPI_ISL_1490000, EPI_ISL_1490001, EPI_ISL_1490002, EPI_ISL_1490003, EPI_ISL_1490004, EPI_ISL_1490005, EPI_ISL_1490006, EPI_ISL_1490007, EPI_ISL_1490008, EPI_ISL_1490009, EPI_ISL_1490010, EPI_ISL_1490011, EPI_ISL_1490012, EPI_ISL_1490013, EPI_ISL_1490014, EPI_ISL_1490015, EPI_ISL_1490016, EPI_ISL_1490017, EPI_ISL_1490018, EPI_ISL_1490019, EPI_ISL_1490020, EPI_ISL_1490021, EPI_ISL_1490022, EPI_ISL_1490023, EPI_ISL_1490024, EPI_ISL_1490025, EPI_ISL_1490026, EPI_ISL_1490027, EPI_ISL_1490028, EPI_ISL_1490029, EPI_ISL_1490030, EPI_ISL_1490031, EPI_ISL_1490032, EPI_ISL_1490033, EPI_ISL_1490034, EPI_ISL_1490035, EPI_ISL_1490036, EPI_ISL_1490037, EPI_ISL_1490038, EPI_ISL_1490039, EPI_ISL_1490040, EPI_ISL_1490041, EPI_ISL_1490042, EPI_ISL_1490043, EPI_ISL_1490044, EPI_ISL_1490045, EPI_ISL_1490046, EPI_ISL_1490047, EPI_ISL_1490048, EPI_ISL_1490049, EPI_ISL_1490050, EPI_ISL_1490051, EPI_ISL_1490052, EPI_ISL_1490053, EPI_ISL_1490054, EPI_ISL_1490055, EPI_ISL_1490056, EPI_ISL_1490057, EPI_ISL_1490058, EPI_ISL_1490059, EPI_ISL_1490060, EPI_ISL_1490061, EPI_ISL_1490062, EPI_ISL_1490063, EPI_ISL_1490064, EPI_ISL_1490065, EPI_ISL_1490066, EPI_ISL_1490067, EPI_ISL_1490068, EPI_ISL_1490069, EPI_ISL_1490070, EPI_ISL_1490071, EPI_ISL_1490072, EPI_ISL_1490073, EPI_ISL_1490074, EPI_ISL_1490075, EPI_ISL_1490076, EPI_ISL_1490077, EPI_ISL_1490078, EPI_ISL_1490079, EPI_ISL_1490080, EPI_ISL_1490081, EPI_ISL_1490082, EPI_ISL_1490083, EPI_ISL_1490084, EPI_ISL_1490085, EPI_ISL_1490086, EPI_ISL_1490087, EPI_ISL_1490088, EPI_ISL_1490090, EPI_ISL_1490091, EPI_ISL_1490092, EPI_ISL_1490093, EPI_ISL_1490095, EPI_ISL_1490096, EPI_ISL_1490097, EPI_ISL_1490099, EPI_ISL_1490100, EPI_ISL_1490101, EPI_ISL_1490102, EPI_ISL_1490103, EPI_ISL_1490104, EPI_ISL_1490105, EPI_ISL_1490106, EPI_ISL_1490107, EPI_ISL_1490108, EPI_ISL_1490109, EPI_ISL_1490110, EPI_ISL_1490111, EPI_ISL_1490112, EPI_ISL_1490113, EPI_ISL_1490114, EPI_ISL_1490115, EPI_ISL_1490116, EPI_ISL_1490118, EPI_ISL_1490119, EPI_ISL_1490120, EPI_ISL_1490121, EPI_ISL_1490122, EPI_ISL_1490123, EPI_ISL_1490124, EPI_ISL_1490125, EPI_ISL_1490126, EPI_ISL_1490127, EPI_ISL_1490128, EPI_ISL_1490129, EPI_ISL_1490130, EPI_ISL_1490131, EPI_ISL_1490132, EPI_ISL_1490133, EPI_ISL_1490134, EPI_ISL_1490135, EPI_ISL_1490136, EPI_ISL_1490137, EPI_ISL_1490138, EPI_ISL_1490139, EPI_ISL_1490140, EPI_ISL_1490141, EPI_ISL_1490142, EPI_ISL_1490143, EPI_ISL_1490144, EPI_ISL_1490145, EPI_ISL_1490146, EPI_ISL_1490147, EPI_ISL_1490148, EPI_ISL_1490149, EPI_ISL_1490150, EPI_ISL_1490151, EPI_ISL_1490152, EPI_ISL_1490153, EPI_ISL_1490154, EPI_ISL_1490155, EPI_ISL_1490157, EPI_ISL_1490158, EPI_ISL_1490159, EPI_ISL_1490160, EPI_ISL_1490161, EPI_ISL_1490162, EPI_ISL_1490163, EPI_ISL_1490164, EPI_ISL_1587452, EPI_ISL_1587453, EPI_ISL_1587454, EPI_ISL_1587455, EPI_ISL_1587456, EPI_ISL_1587457, EPI_ISL_1587458, EPI_ISL_1587459, EPI_ISL_1587460, EPI_ISL_1587461, EPI_ISL_1587462, EPI_ISL_1587463, EPI_ISL_1587464, EPI_ISL_1587465, EPI_ISL_1587466, EPI_ISL_1587467, EPI_ISL_1587468, EPI_ISL_1587469, EPI_ISL_1587470, EPI_ISL_1587471, EPI_ISL_1587472, EPI_ISL_1587473, EPI_ISL_1587474, EPI_ISL_1587475, EPI_ISL_1587476, EPI_ISL_1587477, EPI_ISL_1587478, EPI_ISL_1587479, EPI_ISL_1587480, EPI_ISL_1587481, EPI_ISL_1587482, EPI_ISL_1587483, EPI_ISL_1587484, EPI_ISL_1587485, EPI_ISL_1587486, EPI_ISL_1587487, EPI_ISL_1587488, EPI_ISL_1587489, EPI_ISL_1587490, EPI_ISL_1587491, EPI_ISL_1587492, EPI_ISL_1587493, EPI_ISL_1587494, EPI_ISL_1587495, EPI_ISL_1587496, EPI_ISL_1587497, EPI_ISL_1587498, EPI_ISL_1587499, EPI_ISL_1587500, EPI_ISL_1587501, EPI_ISL_1587502, EPI_ISL_1587503, EPI_ISL_1587504, EPI_ISL_1587505, EPI_ISL_1587506, EPI_ISL_1587507, EPI_ISL_1587508, EPI_ISL_1587509, EPI_ISL_1587510, EPI_ISL_1587511, EPI_ISL_1587512, EPI_ISL_1587513, EPI_ISL_1587514, EPI_ISL_1587515, EPI_ISL_1587516, EPI_ISL_1587517, EPI_ISL_1587518, EPI_ISL_1587519, EPI_ISL_1587520, EPI_ISL_1587521, EPI_ISL_1587522, EPI_ISL_1587523, EPI_ISL_1587524, EPI_ISL_1587525, EPI_ISL_1587526, EPI_ISL_1587527, EPI_ISL_1587528, EPI_ISL_1587529, EPI_ISL_1587530, EPI_ISL_1587531, EPI_ISL_1587532, EPI_ISL_1587533, EPI_ISL_1587534, EPI_ISL_1587535, EPI_ISL_1587536, EPI_ISL_1587537, EPI_ISL_1587538, EPI_ISL_1587539, EPI_ISL_1587540, EPI_ISL_1587541, EPI_ISL_1587542, EPI_ISL_1587543, EPI_ISL_1587544, EPI_ISL_1587545, EPI_ISL_1587546, EPI_ISL_1587547, EPI_ISL_1587548, EPI_ISL_1587549, EPI_ISL_1587550, EPI_ISL_1622336, EPI_ISL_1622337, EPI_ISL_1622338, EPI_ISL_1622352, EPI_ISL_1622353, EPI_ISL_1622354, EPI_ISL_1622355, EPI_ISL_1622356, EPI_ISL_1622357, EPI_ISL_1622358, EPI_ISL_1622436, EPI_ISL_1622437, EPI_ISL_1622438, EPI_ISL_1622452, EPI_ISL_1622453, EPI_ISL_1622454, EPI_ISL_1622455, EPI_ISL_1622456, EPI_ISL_1622457, EPI_ISL_1622458, EPI_ISL_1622459, EPI_ISL_1622460, EPI_ISL_1622461, EPI_ISL_1622462, EPI_ISL_1622463, EPI_ISL_1622464, EPI_ISL_1622465, EPI_ISL_1622466, EPI_ISL_1622467, EPI_ISL_1622468, EPI_ISL_1622469, EPI_ISL_1622470, EPI_ISL_1622471, EPI_ISL_1622472, EPI_ISL_1622473, EPI_ISL_1622474, EPI_ISL_1622475, EPI_ISL_1622476, EPI_ISL_1622477, EPI_ISL_1622479, EPI_ISL_1622480, EPI_ISL_1622481, EPI_ISL_1622482, EPI_ISL_1622483, EPI_ISL_1622484, EPI_ISL_1622485, EPI_ISL_1622486, EPI_ISL_1622487, EPI_ISL_1622488, EPI_ISL_1622489, EPI_ISL_1622490, EPI_ISL_1622491, EPI_ISL_1622492, EPI_ISL_1622493, EPI_ISL_1622494, EPI_ISL_1622495, EPI_ISL_1622496, EPI_ISL_1622497, EPI_ISL_1622498, EPI_ISL_1622499, EPI_ISL_1647347, EPI_ISL_1647348, EPI_ISL_1647349, EPI_ISL_1647350, EPI_ISL_1647351, EPI_ISL_1647352, EPI_ISL_1675253, EPI_ISL_1675254, EPI_ISL_1675255, EPI_ISL_1675256, EPI_ISL_1675257, EPI_ISL_1675258, EPI_ISL_1675259, EPI_ISL_1675260, EPI_ISL_1675261, EPI_ISL_1675262, EPI_ISL_1675263, EPI_ISL_1675264, EPI_ISL_1675265, EPI_ISL_1675266, EPI_ISL_1675267, EPI_ISL_1675268, EPI_ISL_1675269, EPI_ISL_1675270, EPI_ISL_1675271, EPI_ISL_1675272, EPI_ISL_1675273, EPI_ISL_1675274, EPI_ISL_1675275, EPI_ISL_1675276, EPI_ISL_1675278, EPI_ISL_1675279, EPI_ISL_1675280, EPI_ISL_1675281, EPI_ISL_1675282, EPI_ISL_1675283, EPI_ISL_1675284, EPI_ISL_1675285, EPI_ISL_1675286, EPI_ISL_1675287, EPI_ISL_1675290, EPI_ISL_1675291, EPI_ISL_1675292, EPI_ISL_1934642, EPI_ISL_1934643, EPI_ISL_1934644, EPI_ISL_1934645, EPI_ISL_1934646, EPI_ISL_1934647, EPI_ISL_1934648, EPI_ISL_1934649, EPI_ISL_1934650, EPI_ISL_1934651, EPI_ISL_1934652, EPI_ISL_1934653, EPI_ISL_1934654, EPI_ISL_1934655, EPI_ISL_1934656, EPI_ISL_1934657, EPI_ISL_1934658, EPI_ISL_1934659, EPI_ISL_1934660, EPI_ISL_1934661, EPI_ISL_1934662, EPI_ISL_1934663, EPI_ISL_1934664, EPI_ISL_1934665, EPI_ISL_1934666, EPI_ISL_1934667, EPI_ISL_1934668, EPI_ISL_1934669, EPI_ISL_1934670, EPI_ISL_1934671, EPI_ISL_1934672, EPI_ISL_1934673, EPI_ISL_1934674, EPI_ISL_1934675, EPI_ISL_1934676, EPI_ISL_1934677, EPI_ISL_1934678, EPI_ISL_1934679, EPI_ISL_1934680, EPI_ISL_1934681, EPI_ISL_1934682, EPI_ISL_1934683, EPI_ISL_1934684, EPI_ISL_1934685, EPI_ISL_1934686, EPI_ISL_1934687, EPI_ISL_1934688, EPI_ISL_1934689, EPI_ISL_1934690, EPI_ISL_1934691, EPI_ISL_1934692, EPI_ISL_1934693, EPI_ISL_1934694, EPI_ISL_1934695, EPI_ISL_1934696, EPI_ISL_1934697, EPI_ISL_1934698, EPI_ISL_1934699, EPI_ISL_1934700, EPI_ISL_1934701, EPI_ISL_1934702, EPI_ISL_1934703, EPI_ISL_1934704, EPI_ISL_1934705, EPI_ISL_1934706, EPI_ISL_1934707, EPI_ISL_1934708, EPI_ISL_1934709, EPI_ISL_1934710, EPI_ISL_1934711, EPI_ISL_1934712, EPI_ISL_1934713, EPI_ISL_1934714, EPI_ISL_1934715, EPI_ISL_1934716, EPI_ISL_1934717, EPI_ISL_1934718, EPI_ISL_1934719, EPI_ISL_1934720, EPI_ISL_1934721, EPI_ISL_1934722, EPI_ISL_1934723, EPI_ISL_1934724, EPI_ISL_1934725, EPI_ISL_1934726, EPI_ISL_1934727, EPI_ISL_1934728, EPI_ISL_1934729, EPI_ISL_1934730, EPI_ISL_1934731, EPI_ISL_1934732, EPI_ISL_1934733, EPI_ISL_1934734, EPI_ISL_1934735, EPI_ISL_1934736, EPI_ISL_1934737, EPI_ISL_1934738, EPI_ISL_1934739, EPI_ISL_1934740, EPI_ISL_1934741, EPI_ISL_1934742, EPI_ISL_1934743, EPI_ISL_1934744, EPI_ISL_1934745, EPI_ISL_1934746, EPI_ISL_1934747, EPI_ISL_1934748, EPI_ISL_1934749, EPI_ISL_1934750, EPI_ISL_1934751, EPI_ISL_1934752, EPI_ISL_1934753, EPI_ISL_1934754, EPI_ISL_1934755, EPI_ISL_1934756, EPI_ISL_1934757, EPI_ISL_1934758, EPI_ISL_1934759, EPI_ISL_1934760, EPI_ISL_1934761, EPI_ISL_1934762, EPI_ISL_1934763, EPI_ISL_1934764, EPI_ISL_1934765, EPI_ISL_1934766, EPI_ISL_1934767, EPI_ISL_1934768, EPI_ISL_1934769, EPI_ISL_1934770, EPI_ISL_1934771, EPI_ISL_1934772, EPI_ISL_1934773, EPI_ISL_1934774, EPI_ISL_1934775, EPI_ISL_1934776, EPI_ISL_1934777, EPI_ISL_1934778, EPI_ISL_1934779, EPI_ISL_1934780, EPI_ISL_1934781, EPI_ISL_1934782, EPI_ISL_1934783, EPI_ISL_1934784, EPI_ISL_1934785, EPI_ISL_1934786, EPI_ISL_1934787, EPI_ISL_1934788, EPI_ISL_1934789, EPI_ISL_1934790, EPI_ISL_1934791, EPI_ISL_1934792, EPI_ISL_1934794, EPI_ISL_1934795, EPI_ISL_1934796, EPI_ISL_1934797, EPI_ISL_1934798, EPI_ISL_1934799, EPI_ISL_1934800, EPI_ISL_1934801, EPI_ISL_1934802, EPI_ISL_1934803, EPI_ISL_1934804, EPI_ISL_1934805, EPI_ISL_1934806, EPI_ISL_1934807, EPI_ISL_1934808, EPI_ISL_1934809, EPI_ISL_1934810, EPI_ISL_1934811, EPI_ISL_1934812, EPI_ISL_1934813, EPI_ISL_1934814, EPI_ISL_1934815, EPI_ISL_1934816, EPI_ISL_1934817, EPI_ISL_1934818, EPI_ISL_1934819, EPI_ISL_1934820, EPI_ISL_1934821, EPI_ISL_1934822, EPI_ISL_1934823, EPI_ISL_1934824, EPI_ISL_1934825, EPI_ISL_1934826, EPI_ISL_1934827, EPI_ISL_1934828, EPI_ISL_1934829, EPI_ISL_1934830, EPI_ISL_1934831, EPI_ISL_1934832, EPI_ISL_1934833, EPI_ISL_1934834, EPI_ISL_1934835, EPI_ISL_1934836, EPI_ISL_1934837, EPI_ISL_1934838, EPI_ISL_1934839, EPI_ISL_1934840, EPI_ISL_1934841, EPI_ISL_1934842, EPI_ISL_1934843, EPI_ISL_1934844, EPI_ISL_1934845, EPI_ISL_1934846, EPI_ISL_1934847, EPI_ISL_1934848, EPI_ISL_1934849, EPI_ISL_1934850, EPI_ISL_1934851, EPI_ISL_1934852, EPI_ISL_1934853, EPI_ISL_1936467, EPI_ISL_1936468, EPI_ISL_1936469, EPI_ISL_1936470, EPI_ISL_1936471, EPI_ISL_1936472, EPI_ISL_1936473, EPI_ISL_1936474, EPI_ISL_1936475, EPI_ISL_1936476, EPI_ISL_1936477, EPI_ISL_1936478, EPI_ISL_1936479, EPI_ISL_1936480, EPI_ISL_1936481, EPI_ISL_1936482, EPI_ISL_1936483, EPI_ISL_1936484, EPI_ISL_1936485, EPI_ISL_1936486, EPI_ISL_1936487, EPI_ISL_1936488, EPI_ISL_1936489, EPI_ISL_1936490, EPI_ISL_1936491, EPI_ISL_1936492, EPI_ISL_1936493, EPI_ISL_1936494, EPI_ISL_1936495, EPI_ISL_1936496, EPI_ISL_1936497, EPI_ISL_1936498, EPI_ISL_1936499, EPI_ISL_1936500, EPI_ISL_1936501, EPI_ISL_1936502, EPI_ISL_1936503, EPI_ISL_1936504, EPI_ISL_1936505, EPI_ISL_1936506, EPI_ISL_1936507, EPI_ISL_1936508, EPI_ISL_1936509, EPI_ISL_1936510, EPI_ISL_1936511, EPI_ISL_1936512, EPI_ISL_1936513, EPI_ISL_1936514, EPI_ISL_1936515, EPI_ISL_1936516, EPI_ISL_1936517, EPI_ISL_1936518, EPI_ISL_1936519, EPI_ISL_1936520, EPI_ISL_1936521, EPI_ISL_1936522, EPI_ISL_1936523, EPI_ISL_1936524, EPI_ISL_1936525, EPI_ISL_1936526, EPI_ISL_1936527, EPI_ISL_1936528, EPI_ISL_1936529, EPI_ISL_1936530, EPI_ISL_1936531, EPI_ISL_1936532, EPI_ISL_1936533, EPI_ISL_1936534, EPI_ISL_1936535, EPI_ISL_1936536, EPI_ISL_1936537, EPI_ISL_1936538, EPI_ISL_1936539, EPI_ISL_1936540, EPI_ISL_1936541, EPI_ISL_1936542, EPI_ISL_1936543, EPI_ISL_1936544, EPI_ISL_1936545, EPI_ISL_1936546, EPI_ISL_1936547, EPI_ISL_1936548, EPI_ISL_1936549, EPI_ISL_1936550, EPI_ISL_1936551, EPI_ISL_1936552, EPI_ISL_1936553, EPI_ISL_1936554, EPI_ISL_1936555, EPI_ISL_1936556, EPI_ISL_1936557, EPI_ISL_1936558, EPI_ISL_1936559, EPI_ISL_1936560, EPI_ISL_1936561, EPI_ISL_1936562, EPI_ISL_1936563, EPI_ISL_1936564, EPI_ISL_1936565, EPI_ISL_1936566, EPI_ISL_1936567, EPI_ISL_1936568, EPI_ISL_1936569, EPI_ISL_1936570, EPI_ISL_1936571, EPI_ISL_1936572, EPI_ISL_1936573, EPI_ISL_1936574, EPI_ISL_1936575, EPI_ISL_1936576, EPI_ISL_1936577, EPI_ISL_1936578, EPI_ISL_1936579, EPI_ISL_1936580, EPI_ISL_1936581, EPI_ISL_1936582, EPI_ISL_1936583, EPI_ISL_1936584, EPI_ISL_1936585, EPI_ISL_1936586, EPI_ISL_1936587, EPI_ISL_1936588, EPI_ISL_1936589, EPI_ISL_1936590, EPI_ISL_1936591, EPI_ISL_1936592, EPI_ISL_1936593, EPI_ISL_1936594, EPI_ISL_1936595, EPI_ISL_1936596, EPI_ISL_1936597, EPI_ISL_1936598, EPI_ISL_1936599, EPI_ISL_1936600, EPI_ISL_1936601, EPI_ISL_1936602, EPI_ISL_1936603, EPI_ISL_1936604, EPI_ISL_1936605, EPI_ISL_1936606, EPI_ISL_1936607, EPI_ISL_1936608, EPI_ISL_1936609, EPI_ISL_1936610, EPI_ISL_1936611, EPI_ISL_1936612, EPI_ISL_1936613, EPI_ISL_1936614, EPI_ISL_1936615, EPI_ISL_1936616, EPI_ISL_1936617, EPI_ISL_1936618, EPI_ISL_1936619, EPI_ISL_1936620, EPI_ISL_1936621, EPI_ISL_1936622, EPI_ISL_1936623, EPI_ISL_1936624, EPI_ISL_1936625, EPI_ISL_1936626, EPI_ISL_1936627, EPI_ISL_1936628, EPI_ISL_1936629, EPI_ISL_1936630, EPI_ISL_1936631, EPI_ISL_1936632, EPI_ISL_1936633, EPI_ISL_1936634, EPI_ISL_1936635, EPI_ISL_1936636, EPI_ISL_1936637, EPI_ISL_1936638, EPI_ISL_1936639, EPI_ISL_1936640, EPI_ISL_1936641, EPI_ISL_1936642, EPI_ISL_1936643, EPI_ISL_1936644, EPI_ISL_1936645, EPI_ISL_1936646, EPI_ISL_1936647, EPI_ISL_1936648, EPI_ISL_1936649, EPI_ISL_1936650, EPI_ISL_1936651, EPI_ISL_1936652, EPI_ISL_1936653, EPI_ISL_1936654, EPI_ISL_1936655, EPI_ISL_1936656, EPI_ISL_1936657, EPI_ISL_1936658, EPI_ISL_1936659, EPI_ISL_1936660, EPI_ISL_1936661, EPI_ISL_1936662, EPI_ISL_1936663, EPI_ISL_1936664, EPI_ISL_1936665, EPI_ISL_1936666, EPI_ISL_1936667, EPI_ISL_1936668, EPI_ISL_1936669, EPI_ISL_1936670, EPI_ISL_1936671, EPI_ISL_1936672, EPI_ISL_1936673, EPI_ISL_1936674, EPI_ISL_1936675, EPI_ISL_1936676, EPI_ISL_1936677, EPI_ISL_1936678, EPI_ISL_1936679 | | | |
| see above | Division of Emerging Infectious Diseases, Bureau of Infectious Diseases Diagnosis Control, Korea Disease Control and Prevention Agency | Division of Emerging Infectious Diseases, Bureau of Infectious Diseases Diagnosis Control, Korea Disease Control and Prevention Agency | Ae Kyung Park; Chae Young Lee; Eun-Jin Kim; Heui Man Kim; Il-Hwan Kim; Jeong-Ah Kim; Jeong-Ah Kimg; Jeong-Min Kim; Jin Sun No |
| EPI_ISL_1919436, EPI_ISL_1919471, EPI_ISL_1919486 | HELIX LLC | WHO National Influenza Centre Russian Federation | Andrey Komissarov; Artem Fadeev; Daria Danilenko; Dmitry Lioznov; Elena Nabieva; Georgii Bazykin; Kirill Varchenko; Ksenia Safina; Kseniya Komissarova; Maria Pisareva; Maria Timofeeva; Mikhail Bakaev; Nikita Yolshin; Oula Masour; Tamila Musaeva; Veronika Eder |
| EPI_ISL_1495762, EPI_ISL_1495766, EPI_ISL_1495789, EPI_ISL_1495794, EPI_ISL_1495798, EPI_ISL_1495799 | Klinikum Wels-Grieskirchen | Bergthaler laboratory, CeMM Research Center for Molecular Medicine of the Austrian Academy of Sciences | Andreas Bergthaler; Anna Schedl; Bekir Erguner; Benedikt Agerer; Christoph Bock; Fabian Amman; Jan Laine; Lukas Endler; Maelle Le Moing; Martin Senekowitsch; Michael Schuster; Petr Triska; Thomas Penz |
| EPI_ISL_1469276 | National Institute Health Research and Development | National Institute of Health Research and Development | Agustiningsih; Arie Ardiansyah Nugraha; Fauzul Muna; Hana Apsari Pawestri; Hartanti Dian Ikawati; Herna; Holy Arif Wibowo; Irene Lorinda Indalao; Kartika Dewi Puspa; Kindi Adam; Krisna Nur Andriana Pangesti; Natalie Laurencia Kipuw; Nelly Puspandari; Ni Ketut Susilarini; Nike Susanti; Nurika Hariastuti; Reni Herman; Ririn Ramadhany; Subangkit; Tati Febriyanti; Triyani Soekarso; Ully Alfi Nikmah; Vivi Setiawaty.; Yuni Rukminiati |

We gratefully acknowledge the following Authors from the Originating laboratories responsible for obtaining the specimens, as well as the Submitting laboratories where the genome data were generated and shared via GISAID, on which this research is based.

All Submitters of data may be contacted directly via www.gisaid.org

Authors are sorted alphabetically.

| Accession ID | Originating Laboratory | Submitting Laboratory | Authors |
|---|---|---|---|
| EPI_ISL_2617550, EPI_ISL_2617551, EPI_ISL_2617552, EPI_ISL_2617553, EPI_ISL_2617554 | DINKES Kab TAPIN KalSel | National Institute of Health Research and Development | Arie Ardiansyah Nugraha; Hana Apsari Pawestri; Hartanti Dian Ikawati; Kartika Dewi Puspa; Krisna Pangesti; Nelly Puspandari; Subangkit; Triyani Soekarso; Vivi Setiawaty |
| EPI_ISL_2620713 | Dinas Kesehatan Kabupaten Tapin Kalimantan Selatan | National Institute of Health Research and Development | Arie Ardiansyah Nugraha; Hana Apsari Pawestri; Hartanti Dian Ikawati; Kartika Dewi Puspa; Krisna Pangesti; Nelly Puspandari; Subangkit; Triyani Soekarso; Vivi Setiawaty |
| EPI_ISL_2284553, EPI_ISL_2284554, EPI_ISL_2284555, EPI_ISL_2284556, EPI_ISL_2284558, EPI_ISL_2284571, EPI_ISL_2284580, EPI_ISL_2284585, EPI_ISL_2284586, EPI_ISL_2284589, EPI_ISL_2284590, EPI_ISL_2284595, EPI_ISL_2284596, EPI_ISL_2284597, EPI_ISL_2284618, EPI_ISL_2284625, EPI_ISL_2284628, EPI_ISL_2284629, EPI_ISL_2284630, EPI_ISL_2284634, EPI_ISL_2284635, EPI_ISL_2284636, EPI_ISL_2284638, EPI_ISL_2284639, EPI_ISL_2284640, EPI_ISL_2284645, EPI_ISL_2284648, EPI_ISL_2284649, EPI_ISL_2284650, EPI_ISL_2284653, EPI_ISL_2284654, EPI_ISL_2284655, EPI_ISL_2284656, EPI_ISL_2284657, EPI_ISL_2284660, EPI_ISL_2284661, EPI_ISL_2284663, EPI_ISL_2284664, EPI_ISL_2284665, EPI_ISL_2284666, EPI_ISL_2284667, EPI_ISL_2284668, EPI_ISL_2284669, EPI_ISL_2284670, EPI_ISL_2284671, EPI_ISL_2332450, EPI_ISL_2332451, EPI_ISL_2332457, EPI_ISL_2332458, EPI_ISL_2332460, EPI_ISL_2332461, EPI_ISL_2332462, EPI_ISL_2332464, EPI_ISL_2332465, EPI_ISL_2332467, EPI_ISL_2332487, EPI_ISL_2332492, EPI_ISL_2332493, EPI_ISL_2332494, EPI_ISL_2332495, EPI_ISL_2332496, EPI_ISL_2332497, EPI_ISL_2332502, EPI_ISL_2332503, EPI_ISL_2332507, EPI_ISL_2332509, EPI_ISL_2332510, EPI_ISL_2332511, EPI_ISL_2332512, EPI_ISL_2332513, EPI_ISL_2332514, EPI_ISL_2332515, EPI_ISL_2332516, EPI_ISL_2332517, EPI_ISL_2332518, EPI_ISL_2332519, EPI_ISL_2332521, EPI_ISL_2332522, EPI_ISL_2332523, EPI_ISL_2332524, EPI_ISL_2332526, EPI_ISL_2332527, EPI_ISL_2332528, EPI_ISL_2332529, EPI_ISL_2332531, EPI_ISL_2332532, EPI_ISL_2332533, EPI_ISL_2332534, EPI_ISL_2332535, EPI_ISL_2332536, EPI_ISL_2332537, EPI_ISL_2332538, EPI_ISL_2332539, EPI_ISL_2332540, EPI_ISL_2332541, EPI_ISL_2332542, EPI_ISL_2332543, EPI_ISL_2332546, EPI_ISL_2332547, EPI_ISL_2332549, EPI_ISL_2332550, EPI_ISL_2332551, EPI_ISL_2332552, EPI_ISL_2332553, EPI_ISL_2332554, EPI_ISL_2332555, EPI_ISL_2332556, EPI_ISL_2332557, EPI_ISL_2332558, EPI_ISL_2332559, EPI_ISL_2332560, EPI_ISL_2332561, EPI_ISL_2332562, EPI_ISL_2332564, EPI_ISL_2332565, EPI_ISL_2332566, EPI_ISL_2332567, EPI_ISL_2332568, EPI_ISL_2332569, EPI_ISL_2332570, EPI_ISL_2332571, EPI_ISL_2332572, EPI_ISL_2332573, EPI_ISL_2332574, EPI_ISL_2332575, EPI_ISL_2332576, EPI_ISL_2332577, EPI_ISL_2332578, EPI_ISL_2332579, EPI_ISL_2361237, EPI_ISL_2361238, EPI_ISL_2361239, EPI_ISL_2361240, EPI_ISL_2361241, EPI_ISL_2361242, EPI_ISL_2361243, EPI_ISL_2361244, EPI_ISL_2361245, EPI_ISL_2361246, EPI_ISL_2361247, EPI_ISL_2361248, EPI_ISL_2361249, EPI_ISL_2361250, EPI_ISL_2361251, EPI_ISL_2361252, EPI_ISL_2361253, EPI_ISL_2361254, EPI_ISL_2361255, EPI_ISL_2361256, EPI_ISL_2361257, EPI_ISL_2361258, EPI_ISL_2361259, EPI_ISL_2361260, EPI_ISL_2361262, EPI_ISL_2361263, EPI_ISL_2361264, EPI_ISL_2361265, EPI_ISL_2464355, EPI_ISL_2464356, EPI_ISL_2464357, EPI_ISL_2464358, EPI_ISL_2464359, EPI_ISL_2464360, EPI_ISL_2464361, EPI_ISL_2464362, EPI_ISL_2464363, EPI_ISL_2464364, EPI_ISL_2464365, EPI_ISL_2464366, EPI_ISL_2464367, EPI_ISL_2464368, EPI_ISL_2464369, EPI_ISL_2464370, EPI_ISL_2464371, EPI_ISL_2464372, EPI_ISL_2464373, EPI_ISL_2464374, EPI_ISL_2464375, EPI_ISL_2710387, EPI_ISL_2710388, EPI_ISL_2710389, EPI_ISL_2710390, EPI_ISL_2710391, EPI_ISL_2710392, EPI_ISL_2710393, EPI_ISL_2710394, EPI_ISL_2710395, EPI_ISL_2710396, EPI_ISL_2710397, EPI_ISL_2710398, EPI_ISL_2710399, EPI_ISL_2710400, EPI_ISL_2710401, EPI_ISL_2710402, EPI_ISL_2710403, EPI_ISL_2710404, EPI_ISL_2710405, EPI_ISL_2710406, EPI_ISL_2710407, EPI_ISL_2710408, EPI_ISL_2710409, EPI_ISL_2710410, EPI_ISL_2710411, EPI_ISL_2710412, EPI_ISL_2710413, EPI_ISL_2710414, EPI_ISL_2710415, EPI_ISL_2710416, EPI_ISL_2710417, EPI_ISL_2710418, EPI_ISL_2710419, EPI_ISL_2710420, EPI_ISL_2710421, EPI_ISL_2710422, EPI_ISL_2710423, EPI_ISL_2710424, EPI_ISL_2710425, EPI_ISL_2710426, EPI_ISL_2710427, EPI_ISL_2710428, EPI_ISL_2710429, EPI_ISL_2710430, EPI_ISL_2710431, EPI_ISL_2710432, EPI_ISL_2710433, EPI_ISL_2710434, EPI_ISL_2710435, EPI_ISL_2710436, EPI_ISL_2710437, EPI_ISL_2710438, EPI_ISL_2710439, EPI_ISL_2710440, EPI_ISL_2710441, EPI_ISL_2710442, EPI_ISL_2710443, EPI_ISL_2710444, EPI_ISL_2710445, EPI_ISL_2710446, EPI_ISL_2710447, EPI_ISL_2710448, EPI_ISL_2710449, EPI_ISL_2710450, EPI_ISL_2710451, EPI_ISL_2710452, EPI_ISL_2710453, EPI_ISL_2710454, EPI_ISL_2710455, EPI_ISL_2710456, EPI_ISL_2710457, EPI_ISL_2710458, EPI_ISL_2710459, EPI_ISL_2710460, EPI_ISL_2710461, EPI_ISL_2710462, EPI_ISL_2710463, EPI_ISL_2710464, EPI_ISL_2710465, EPI_ISL_2710466, EPI_ISL_2710467, EPI_ISL_2710468, EPI_ISL_2710469, EPI_ISL_2710470, EPI_ISL_2710471, EPI_ISL_2710472, EPI_ISL_2710473, EPI_ISL_2710474, EPI_ISL_2710475, EPI_ISL_2710476, EPI_ISL_2710477, EPI_ISL_2710478, EPI_ISL_2710479, EPI_ISL_2710480, EPI_ISL_2710481, EPI_ISL_2710482, EPI_ISL_2710483, EPI_ISL_2710484, EPI_ISL_2710485, EPI_ISL_2710486, EPI_ISL_2710487, EPI_ISL_2710488, EPI_ISL_2710489, EPI_ISL_2710490, EPI_ISL_2710491, EPI_ISL_2710492, EPI_ISL_2710493, EPI_ISL_2710494, EPI_ISL_2710495, EPI_ISL_2710496, EPI_ISL_2710497, EPI_ISL_2710498, EPI_ISL_2710499, EPI_ISL_2710500, EPI_ISL_2710501, EPI_ISL_2710502, EPI_ISL_2710503, EPI_ISL_2710504, EPI_ISL_2710505, EPI_ISL_2710506, EPI_ISL_2710507, EPI_ISL_2710508, EPI_ISL_2710509, EPI_ISL_2710510, EPI_ISL_2710511, EPI_ISL_2710512, EPI_ISL_2710513, EPI_ISL_2710514, EPI_ISL_2710515, EPI_ISL_2710516, EPI_ISL_2710517, EPI_ISL_2710518, EPI_ISL_2710519, EPI_ISL_2710520, EPI_ISL_2710521, EPI_ISL_2710522, EPI_ISL_2710523, EPI_ISL_2710524, EPI_ISL_2710525, EPI_ISL_2710526, EPI_ISL_2710527, EPI_ISL_2710528, EPI_ISL_2710529, EPI_ISL_2710530, EPI_ISL_2710531, EPI_ISL_2710532, EPI_ISL_2710533, EPI_ISL_2710534, EPI_ISL_2710535, EPI_ISL_2710536, EPI_ISL_2710537, EPI_ISL_2710538, EPI_ISL_2710539, EPI_ISL_2710540, EPI_ISL_2710541, EPI_ISL_2710542, EPI_ISL_2710543, EPI_ISL_2710544, EPI_ISL_2710545, EPI_ISL_2710546, EPI_ISL_2710547, EPI_ISL_2710548, EPI_ISL_2710549, EPI_ISL_2754377, EPI_ISL_2754378, EPI_ISL_2754379, EPI_ISL_2754380, EPI_ISL_2754381, EPI_ISL_2754382, EPI_ISL_2754383, EPI_ISL_2754384, EPI_ISL_2754385, EPI_ISL_2754386, EPI_ISL_2754387, EPI_ISL_2754388, EPI_ISL_2754389, EPI_ISL_2754390, EPI_ISL_2754391, EPI_ISL_2754392, EPI_ISL_2754393, EPI_ISL_2754394, EPI_ISL_2754395, EPI_ISL_2754396, EPI_ISL_2754397, EPI_ISL_2754399, EPI_ISL_2754400, EPI_ISL_2754401, EPI_ISL_2754402, EPI_ISL_2754403, EPI_ISL_2754404, EPI_ISL_2754405, EPI_ISL_2754406, EPI_ISL_2754407, EPI_ISL_2754408, EPI_ISL_2754409, EPI_ISL_2754410, EPI_ISL_2754411, EPI_ISL_2754412, EPI_ISL_2754413, EPI_ISL_2754414, EPI_ISL_2754415, EPI_ISL_2754416, EPI_ISL_2754417, EPI_ISL_2754418, EPI_ISL_2754419, EPI_ISL_2754420, EPI_ISL_2754421, EPI_ISL_2754422, EPI_ISL_2754423, EPI_ISL_2754424, EPI_ISL_2754425, EPI_ISL_2754426, EPI_ISL_2754427, EPI_ISL_2754428, EPI_ISL_2754429, EPI_ISL_2754430, EPI_ISL_2754431, EPI_ISL_2754432, EPI_ISL_2754433, EPI_ISL_2754434, EPI_ISL_2754435, EPI_ISL_2754436, EPI_ISL_2754437, EPI_ISL_2754438, EPI_ISL_2754439, EPI_ISL_2754440, EPI_ISL_2754441, EPI_ISL_2754442, EPI_ISL_2754443, EPI_ISL_2754444, EPI_ISL_2754445, EPI_ISL_2754446, EPI_ISL_2754447, EPI_ISL_2754448, EPI_ISL_2754449, EPI_ISL_2754450, EPI_ISL_2754451, EPI_ISL_2754452, EPI_ISL_2754453, EPI_ISL_2754454, EPI_ISL_2754455, EPI_ISL_2754456, EPI_ISL_2754457, EPI_ISL_2754458, EPI_ISL_2754459, EPI_ISL_2754460, EPI_ISL_2754461, EPI_ISL_2754462, EPI_ISL_2754463, EPI_ISL_2754464, EPI_ISL_2754465, EPI_ISL_2754466, EPI_ISL_2754467, EPI_ISL_2754469, EPI_ISL_2754470, EPI_ISL_2754471, EPI_ISL_2754472, EPI_ISL_2754473, EPI_ISL_2754474, EPI_ISL_2754475, EPI_ISL_2754476, EPI_ISL_2754477, EPI_ISL_2754478, EPI_ISL_2754479, EPI_ISL_2754480, EPI_ISL_2754481, EPI_ISL_2754482, EPI_ISL_2754483, EPI_ISL_2754484, EPI_ISL_2754485, EPI_ISL_2754486, EPI_ISL_2754487, EPI_ISL_2754488, EPI_ISL_2754489, EPI_ISL_2754490, EPI_ISL_2754491, EPI_ISL_2754492, EPI_ISL_2754493, EPI_ISL_2754494, EPI_ISL_2754495, EPI_ISL_2754496, EPI_ISL_2754497, EPI_ISL_2754498, EPI_ISL_2754499, EPI_ISL_2754500, EPI_ISL_2754501, EPI_ISL_2754502, EPI_ISL_2754503, EPI_ISL_2754504, EPI_ISL_2754505, EPI_ISL_2754506, EPI_ISL_2754507, EPI_ISL_2754508, EPI_ISL_2754509, EPI_ISL_2754510, EPI_ISL_2754511, EPI_ISL_2754512, EPI_ISL_2754513, EPI_ISL_2754514, EPI_ISL_2754515, EPI_ISL_2754516, EPI_ISL_2754518, EPI_ISL_2754519, EPI_ISL_2754520, EPI_ISL_2754521, EPI_ISL_2754522, EPI_ISL_2754523, EPI_ISL_2754524, EPI_ISL_2754525, EPI_ISL_2754526, EPI_ISL_2754527, EPI_ISL_2754528, EPI_ISL_2754529, EPI_ISL_2754530, EPI_ISL_2754531, EPI_ISL_2754532, EPI_ISL_2754533, EPI_ISL_2754534, EPI_ISL_2754535, EPI_ISL_2754536, EPI_ISL_2754537, EPI_ISL_2754538, EPI_ISL_2754539, EPI_ISL_2754540, EPI_ISL_2754541, EPI_ISL_2754542, EPI_ISL_2754543, EPI_ISL_2754544, EPI_ISL_2754545, EPI_ISL_2754546, EPI_ISL_2754547, EPI_ISL_2754548, EPI_ISL_2754550, EPI_ISL_2754551, EPI_ISL_2754552, EPI_ISL_2754553, EPI_ISL_2754554, EPI_ISL_2754555, EPI_ISL_2754556, EPI_ISL_2754557, EPI_ISL_2754558, EPI_ISL_2754559, EPI_ISL_2754560, EPI_ISL_2754561, EPI_ISL_2754562, EPI_ISL_2754563, EPI_ISL_2754564, EPI_ISL_2754565, EPI_ISL_2754566, EPI_ISL_2754567, EPI_ISL_2754568, EPI_ISL_2754569, EPI_ISL_2754570, EPI_ISL_2754571, EPI_ISL_2754572, EPI_ISL_2754573, EPI_ISL_2754574, EPI_ISL_2754575, EPI_ISL_2754576, EPI_ISL_2754577, EPI_ISL_2754578, EPI_ISL_2754579, EPI_ISL_2754580, EPI_ISL_2754582, EPI_ISL_2754583, EPI_ISL_2754584, EPI_ISL_2754585, EPI_ISL_2754586, EPI_ISL_2754587, EPI_ISL_2754588, EPI_ISL_2754589, EPI_ISL_2754590, EPI_ISL_2754591, EPI_ISL_2754592, EPI_ISL_2754593, EPI_ISL_2754594, EPI_ISL_2754595, EPI_ISL_2754596, EPI_ISL_2754597, EPI_ISL_2754598, EPI_ISL_2754599, EPI_ISL_2754600, EPI_ISL_2754601, EPI_ISL_2754602, EPI_ISL_2754603, EPI_ISL_2754604, EPI_ISL_2754605, EPI_ISL_2754606, EPI_ISL_2754607, EPI_ISL_2754609, EPI_ISL_2754610, EPI_ISL_2754611, EPI_ISL_2754612, EPI_ISL_2754613, EPI_ISL_2754614, EPI_ISL_2754615, EPI_ISL_2754616, EPI_ISL_2754617, EPI_ISL_2754618, EPI_ISL_2754619, EPI_ISL_2754620, EPI_ISL_2754621, EPI_ISL_2754622, EPI_ISL_2754623, EPI_ISL_2754624, EPI_ISL_2754625, EPI_ISL_2754626, EPI_ISL_2754627, EPI_ISL_2754629, EPI_ISL_2754630, EPI_ISL_2754631, EPI_ISL_2754632, EPI_ISL_2754633, EPI_ISL_2754634, EPI_ISL_2754635, EPI_ISL_2754636, EPI_ISL_2754637, EPI_ISL_2754638, EPI_ISL_2754639, EPI_ISL_2754640, EPI_ISL_2754641, EPI_ISL_2754642, EPI_ISL_2754643, EPI_ISL_2754644, EPI_ISL_2754645, EPI_ISL_2754646, EPI_ISL_2754647, EPI_ISL_2754648, EPI_ISL_2754649, EPI_ISL_2754650, EPI_ISL_2754651, EPI_ISL_2754652, EPI_ISL_2754653, EPI_ISL_2754654, EPI_ISL_2754655, EPI_ISL_2754656, EPI_ISL_2754657, EPI_ISL_2754658, EPI_ISL_2754659, EPI_ISL_2754660, EPI_ISL_2754661, EPI_ISL_2754662, EPI_ISL_2754663, EPI_ISL_2754664, EPI_ISL_2754666, EPI_ISL_2754667, EPI_ISL_2754668, EPI_ISL_2754669, EPI_ISL_2754670, EPI_ISL_2754671, EPI_ISL_2754672, EPI_ISL_2754673, EPI_ISL_2754674, EPI_ISL_2754675, EPI_ISL_2754676, EPI_ISL_2754677, EPI_ISL_2754678, EPI_ISL_2754679, EPI_ISL_2754680, EPI_ISL_2754681, EPI_ISL_2754682, EPI_ISL_2754683, EPI_ISL_2754684, EPI_ISL_2754685, EPI_ISL_2754686, EPI_ISL_2754687, EPI_ISL_2754688, EPI_ISL_2754689, EPI_ISL_2754690, EPI_ISL_2754691, EPI_ISL_2754692, EPI_ISL_2754693, EPI_ISL_2754694, EPI_ISL_2754695, EPI_ISL_2754696, EPI_ISL_2754697, EPI_ISL_2754698, EPI_ISL_2754699, EPI_ISL_2754700, EPI_ISL_2754701, EPI_ISL_2754702, EPI_ISL_2754703, EPI_ISL_2754704, EPI_ISL_2754705, EPI_ISL_2754706, EPI_ISL_2754707, EPI_ISL_2754708, EPI_ISL_2754709, EPI_ISL_2754710, EPI_ISL_2754711, EPI_ISL_2754712, EPI_ISL_2754713, EPI_ISL_2754714, EPI_ISL_2754715, EPI_ISL_2754716, EPI_ISL_2754717, EPI_ISL_2754718, EPI_ISL_2754719, EPI_ISL_2754720, EPI_ISL_2754721, EPI_ISL_2754722, EPI_ISL_2754723, EPI_ISL_2754724, EPI_ISL_2754725, EPI_ISL_2754726, EPI_ISL_2754727, EPI_ISL_2754728, EPI_ISL_2754729, EPI_ISL_2754730, EPI_ISL_2754731, EPI_ISL_2754732, EPI_ISL_2754733, EPI_ISL_2754734, EPI_ISL_2754735, EPI_ISL_2754736, EPI_ISL_2754737, EPI_ISL_2754738, EPI_ISL_2754739, EPI_ISL_2754740, EPI_ISL_2754741, EPI_ISL_2754742, EPI_ISL_2754743, EPI_ISL_2754744, EPI_ISL_2754745, EPI_ISL_2754746, EPI_ISL_2754747, EPI_ISL_2754748, EPI_ISL_2754749, EPI_ISL_2754750, EPI_ISL_2754751, EPI_ISL_2754752, EPI_ISL_2754753, EPI_ISL_2754754, EPI_ISL_2754755, EPI_ISL_2754756, EPI_ISL_2754757, EPI_ISL_2754758, EPI_ISL_2754759, EPI_ISL_2754760, EPI_ISL_2754761, EPI_ISL_2754762, EPI_ISL_2754763, EPI_ISL_2754764, EPI_ISL_2754765, EPI_ISL_2754766, EPI_ISL_2754767, EPI_ISL_2754768, EPI_ISL_2754769, EPI_ISL_2754770, EPI_ISL_2754771, EPI_ISL_2754772, EPI_ISL_2754773, EPI_ISL_2754774, EPI_ISL_2754775, EPI_ISL_2754776, EPI_ISL_2754777, EPI_ISL_2754778, EPI_ISL_2754779, EPI_ISL_2754780, EPI_ISL_2754781, EPI_ISL_2754782, EPI_ISL_2754783, EPI_ISL_2754784, EPI_ISL_2754785, EPI_ISL_2754786, EPI_ISL_2754787, EPI_ISL_2754788, EPI_ISL_2754789, EPI_ISL_2754790, EPI_ISL_2754791, EPI_ISL_2754792, EPI_ISL_2754793, EPI_ISL_2754794, EPI_ISL_2754795, EPI_ISL_2754796, EPI_ISL_2754797, EPI_ISL_2754798, EPI_ISL_2754799, EPI_ISL_2754800, EPI_ISL_2754801, EPI_ISL_2754802, EPI_ISL_2754804, EPI_ISL_2754805, EPI_ISL_2754807, EPI_ISL_2754808, EPI_ISL_2754809, EPI_ISL_2754810, EPI_ISL_2754811, EPI_ISL_2754812, EPI_ISL_2754813, EPI_ISL_2754814, EPI_ISL_2754815, EPI_ISL_2754816, EPI_ISL_2754817, EPI_ISL_2754818, EPI_ISL_2754819, EPI_ISL_2754820, EPI_ISL_2754821, EPI_ISL_2754822, EPI_ISL_2754823, EPI_ISL_2754824, EPI_ISL_2754826, EPI_ISL_2754827, EPI_ISL_2754828, EPI_ISL_2754829, EPI_ISL_2754830, EPI_ISL_2754831, EPI_ISL_2754832, EPI_ISL_2754833, EPI_ISL_2754834, EPI_ISL_2754835, EPI_ISL_2754836, EPI_ISL_2754837, EPI_ISL_2754838, EPI_ISL_2754839, EPI_ISL_2754840, EPI_ISL_2754841, EPI_ISL_2754842, EPI_ISL_2754844, EPI_ISL_2754845, EPI_ISL_2754846, EPI_ISL_2754847, EPI_ISL_2754848, EPI_ISL_2754849, EPI_ISL_2754850, EPI_ISL_2754851, EPI_ISL_2754852, EPI_ISL_2754853, EPI_ISL_2887360, EPI_ISL_2887361, EPI_ISL_2887362, EPI_ISL_2887363, EPI_ISL_2887364, EPI_ISL_2887365, EPI_ISL_2887366, EPI_ISL_2887367, EPI_ISL_2887368, EPI_ISL_2887369, EPI_ISL_2887370, EPI_ISL_2887371, EPI_ISL_2887372, EPI_ISL_2887373, EPI_ISL_2887374, EPI_ISL_2887375, EPI_ISL_2887376, EPI_ISL_2887377, EPI_ISL_2887378, EPI_ISL_2887379, EPI_ISL_2887380, EPI_ISL_2887381, EPI_ISL_2887382, EPI_ISL_2887383, EPI_ISL_2887384, EPI_ISL_2887385, EPI_ISL_2887386, EPI_ISL_2887387, EPI_ISL_2887388, EPI_ISL_2887389, EPI_ISL_2887390, EPI_ISL_2887391, EPI_ISL_2887392, EPI_ISL_2887393, EPI_ISL_2887394, EPI_ISL_2887395, EPI_ISL_2887396, EPI_ISL_2887397, EPI_ISL_2887398, EPI_ISL_2887399, EPI_ISL_2887400, EPI_ISL_2887401, EPI_ISL_2887402, EPI_ISL_2887403, EPI_ISL_2887404, EPI_ISL_2887405, EPI_ISL_2887406, EPI_ISL_2887407, EPI_ISL_2887408, EPI_ISL_2887409, EPI_ISL_2887410, EPI_ISL_2887411, EPI_ISL_2887412, EPI_ISL_2887413, EPI_ISL_2887414, EPI_ISL_2887415, EPI_ISL_2887416 | | | |
| see above | Division of Emerging Infectious Diseases, Bureau of Infectious Diseases Diagnosis Control, Korea Disease Control and Prevention Agency | Division of Emerging Infectious Diseases, Bureau of Infectious Diseases Diagnosis Control, Korea Disease Control and Prevention Agency | Ae Kyung Park; Chae Young Lee; Eun-Jin Kim; Heui Man Kim; Il-Hwan Kim; Jeong-Ah Kim; Jeong-Min Kim; Jin Sun No |
| EPI_ISL_2881558, EPI_ISL_2881559, EPI_ISL_2881560, EPI_ISL_2881561, EPI_ISL_2881562, EPI_ISL_2881563, EPI_ISL_2881626, EPI_ISL_2881628, EPI_ISL_2881629, EPI_ISL_2881630, EPI_ISL_2881631 | | | |
| see above | INSACOG Surveillance | INSACOG at CSIR Institute of Genomics and Integrative Biology | INSACOG |

We gratefully acknowledge the following Authors from the Originating laboratories responsible for obtaining the specimens, as well as the Submitting laboratories where the genome data were generated and shared via GISAID, on which this research is based.

All Submitters of data may be contacted directly via www.gisaid.org

Authors are sorted alphabetically.

| Accession ID | Originating Laboratory | Submitting Laboratory | Authors |
|---|---|---|---|
| EPI_ISL_1940022, EPI_ISL_1940023, EPI_ISL_1940024, EPI_ISL_1940025, EPI_ISL_1940026, EPI_ISL_1940027, EPI_ISL_1940028, EPI_ISL_1940036, EPI_ISL_2015537, EPI_ISL_2015598 | | | |
| see above | A D HALLI | inStem NCBS - INSACOG | Uma Ramakrishnan Dasaradhi Palakodeti Aswin SaiNarain |
| EPI_ISL_1940091, EPI_ISL_1940092, EPI_ISL_1940130, EPI_ISL_1940131, EPI_ISL_1940132, EPI_ISL_2015553 | AD HALLI | inStem NCBS - INSACOG | Uma Ramakrishnan Dasaradhi Palakodeti Aswin SaiNarain |
| EPI_ISL_2015498 | AGAR PHC | inStem NCBS - INSACOG | Uma Ramakrishnan Dasaradhi Palakodeti Aswin SaiNarain |
| EPI_ISL_2015501 | AGARA UPHC | inStem NCBS - INSACOG | Uma Ramakrishnan Dasaradhi Palakodeti Aswin SaiNarain |
| EPI_ISL_2015473, EPI_ISL_2015474, EPI_ISL_2015476, EPI_ISL_2015477, EPI_ISL_2015484, EPI_ISL_2015488, EPI_ISL_2015491, EPI_ISL_2015496, EPI_ISL_2015535 | | | |
| see above | AGRAHARA PHC | inStem NCBS - INSACOG | Uma Ramakrishnan Dasaradhi Palakodeti Aswin SaiNarain |
| EPI_ISL_1663504, EPI_ISL_1663507, EPI_ISL_1663509, EPI_ISL_1663515, EPI_ISL_1663518, EPI_ISL_1663519, EPI_ISL_1663520, EPI_ISL_1663521, EPI_ISL_1663522, EPI_ISL_1663523 | | | |
| see above | AIIMS, Patna | Institute of Life Sciences - INSACOG | Ajay Parida; Amol M. Kanampalliwar; Arup Ghosh; Atimukta Jha; INSACOG Consortium; Punit Prasad; Rajeeb Swain; Rupesh Dash; Safal Walia; Shifu Aggarwal; Sunil K. Raghav |
| EPI_ISL_1372268, EPI_ISL_1372271, EPI_ISL_1372273, EPI_ISL_1372275, EPI_ISL_1372277, EPI_ISL_1547799, EPI_ISL_1547800 | | | |
| see above | AP SSO | CSIR-Centre for Cellular and Molecular Biology - INSACOG | Amareshwar Vodapalli; Ara Sreenivas; B Himasri; Divya Tej Sowpati; Karthik Bharadwaj Tallapaka; Lamuk Zaveri; Onkar Kulkarni; Rakesh K Mishra; Sharath Chandra Thota; Shreekant Verma; Sofia Banu; Viswagithe S L |
| EPI_ISL_1940093, EPI_ISL_1940094 | APOLLO | inStem NCBS - INSACOG | Uma Ramakrishnan Dasaradhi Palakodeti Aswin SaiNarain |
| EPI_ISL_2100679, EPI_ISL_2100680, EPI_ISL_2100681, EPI_ISL_2100682, EPI_ISL_2100683, EPI_ISL_2100684, EPI_ISL_2100685, EPI_ISL_2100686, EPI_ISL_2100687, EPI_ISL_2100688, EPI_ISL_2100689, EPI_ISL_2100690, EPI_ISL_2100691, EPI_ISL_2100692, EPI_ISL_2100693, EPI_ISL_2100694, EPI_ISL_2100695, EPI_ISL_2100696, EPI_ISL_2100697, EPI_ISL_2100698, EPI_ISL_2100699, EPI_ISL_2100700, EPI_ISL_2105583, EPI_ISL_2105584, EPI_ISL_2105585 | | | |
| see above | All India Institute of Medical Sciences, Ansari Nagar Delhi | CSIR-Institute of Genomics and Integrative Biology | Animesh Ray; Asangala Kamai; Ashwin Varadarajan; Ayush Goel; Bharathram Uppilli*; Devasish Desai; Manish Kumar; Mohammed Ahmed; Mohammed Faruq; Naveet Wig; Pooja Sharma*; Rajesh Pandey; Sarafaraz Alam; Saruchi Wadhwa; Satish Swain; Sheeba Saifi; Sushma Rajpoot; Umang Arora |
| EPI_ISL_1972130, EPI_ISL_1972131, EPI_ISL_1972132, EPI_ISL_1972133, EPI_ISL_1972134, EPI_ISL_1972135, EPI_ISL_1972136, EPI_ISL_1972141, EPI_ISL_1972142, EPI_ISL_1972147 | | | |
| see above | All India Institute of Medical Sciences, Bhopal | All India Institute of Medical Sciences, Bhopal | Anvita Gupta Malhotra; Arun Raghuvanshi; Ashvini Kumar Yadav; Debasis Biswas and Sarman Singh; Dipesh Kale; Jitendra Singh; Prem Shankar; Shashwati Nema |
| EPI_ISL_1544115, EPI_ISL_1544116, EPI_ISL_1544117 | B.J. Medical College and Civil hospital, Ahmedabad | Gujarat Biotechnology Research Centre | Chaitanya Joshi; Dinesh Kumar; Dipa Kinariwala; Janvi Raval; Kamlesh J Upadhyay; Madhvi Joshi; Nitesh Shah; Nitin Savaliya; Pranay Shah; Ramesh Pandit; Sanjay Kapadia; Sonal Sharma; Twinkle Soni; Umang Mishra; Zarna Patel; Zuber Saiyed |
| EPI_ISL_1595865, EPI_ISL_1595866 | BANGALORE MEDICAL COLLEGE AND RESEARCH INSTITUTE | INSACOG-KA, NIMHANS | Ananthapadmanabha Kotambail; Anita S Desai; Anson Kunjumon George; Chetan G K; Chitra Pattabiraman; Darshan Sreenivas; Gautham Arunachal Udupi; Pramada Prasad; V Ravi |
| EPI_ISL_1595855, EPI_ISL_1595860, EPI_ISL_1595861, EPI_ISL_1595862, EPI_ISL_1595863, EPI_ISL_1595894, EPI_ISL_1595901, EPI_ISL_1595902, EPI_ISL_1595903, EPI_ISL_1595904, EPI_ISL_1595905, EPI_ISL_1595906, EPI_ISL_1595907, EPI_ISL_1595908, EPI_ISL_1595909, EPI_ISL_1595910, EPI_ISL_1595911, EPI_ISL_1595912, EPI_ISL_1595915, EPI_ISL_1595916, EPI_ISL_1818595, EPI_ISL_1818603, EPI_ISL_1818604, EPI_ISL_1818605, EPI_ISL_1818609, EPI_ISL_1818610, EPI_ISL_1818611, EPI_ISL_1818612, EPI_ISL_1818613, EPI_ISL_1818624, EPI_ISL_1818625, EPI_ISL_1818626, EPI_ISL_1818627, EPI_ISL_1818628, EPI_ISL_1818631, EPI_ISL_1818632, EPI_ISL_1818633, EPI_ISL_1818634, EPI_ISL_1818635, EPI_ISL_1818636, EPI_ISL_1818637, EPI_ISL_1818638, EPI_ISL_1818651, EPI_ISL_1818652 | | | |
| see above | BBMP Urban PHC | INSACOG-KA, NIMHANS | Ananthapadmanabha Kotambail; Anita S Desai; Anson Kunjumon George; Chetan G K; Chitra Pattabiraman; Darshan Sreenivas; Gautham Arunachal Udupi; Pramada Prasad; V Ravi |
| EPI_ISL_2017765, EPI_ISL_2017766, EPI_ISL_2017767, EPI_ISL_2017768, EPI_ISL_2017769, EPI_ISL_2017770, EPI_ISL_2017771, EPI_ISL_2017772, EPI_ISL_2017773, EPI_ISL_2017774, EPI_ISL_2017775, EPI_ISL_2017776, EPI_ISL_2017777, EPI_ISL_2017778, EPI_ISL_2017779, EPI_ISL_2017780, EPI_ISL_2017781, EPI_ISL_2017782, EPI_ISL_2017783, EPI_ISL_2017784, EPI_ISL_2017785, EPI_ISL_2017786, EPI_ISL_2017787, EPI_ISL_2017788 | | | |
| see above | BJ Medical College and Civil Hospital, Ahmedabad | Gujarat Biotechnology Research Centre | Chaitanya Joshi; Dinesh Kumar; Janvi Raval; Madhvi Joshi; Nitesh Shah; Nitin Savaliya; Pranay Shah; Ramesh Pandit; Sonal Sharma; Twinkle Soni; Umang Mishra; Zarna Patel; Zuber Saiyed |
| EPI_ISL_1662387, EPI_ISL_1662401, EPI_ISL_1663239, EPI_ISL_1663240, EPI_ISL_1663241, EPI_ISL_1663242, EPI_ISL_1663243, EPI_ISL_1663244, EPI_ISL_1663245, EPI_ISL_1663246, EPI_ISL_1663247, EPI_ISL_1663248, EPI_ISL_1663249, EPI_ISL_1663250, EPI_ISL_1663251, EPI_ISL_1663255, EPI_ISL_1663260 | | | |
| see above | BRLSABVM Government Medical College, Rajnandgaon, Chhattisgarh | Institute of Life Sciences - INSACOG | Ajay Parida; Amol M. Kanampalliwar; Arup Ghosh; Atimukta Jha; INSACOG Consortium; Punit Prasad; Rajeeb Swain; Rupesh Dash; Safal Walia; Shifu Aggarwal; Sunil K. Raghav |
| EPI_ISL_1908685, EPI_ISL_1908686, EPI_ISL_1908687, EPI_ISL_1908688, EPI_ISL_1908689, EPI_ISL_1908690, EPI_ISL_1908691, EPI_ISL_1908692, EPI_ISL_1908693, EPI_ISL_1908694, EPI_ISL_1908695, EPI_ISL_1908696, EPI_ISL_1908697, EPI_ISL_1908698, EPI_ISL_1908699, EPI_ISL_1908700, EPI_ISL_1908701, EPI_ISL_1908702, EPI_ISL_1908703, EPI_ISL_1908704, EPI_ISL_1908705, EPI_ISL_1908706, EPI_ISL_1908707, EPI_ISL_1908708, EPI_ISL_1908709, EPI_ISL_1908710, EPI_ISL_1908711, EPI_ISL_1908712, EPI_ISL_1908713, EPI_ISL_1908714, EPI_ISL_1908715, EPI_ISL_1908716, EPI_ISL_1908717, EPI_ISL_1908718, EPI_ISL_1908722, EPI_ISL_1908723, EPI_ISL_1908724, EPI_ISL_1908725, EPI_ISL_1908726, EPI_ISL_1908727, EPI_ISL_1908728, EPI_ISL_1908729, EPI_ISL_1908742, EPI_ISL_1908743, EPI_ISL_1908750, EPI_ISL_1908751, EPI_ISL_1908752, EPI_ISL_1908753, EPI_ISL_1908754, EPI_ISL_1908755, EPI_ISL_1908756, EPI_ISL_1908765, EPI_ISL_1908766, EPI_ISL_1908767, EPI_ISL_1908768, EPI_ISL_1908769, EPI_ISL_1908770, EPI_ISL_1908771, EPI_ISL_1908777, EPI_ISL_1908778, EPI_ISL_1908779, EPI_ISL_1908780, EPI_ISL_1908781, EPI_ISL_1908782, EPI_ISL_1908783, EPI_ISL_1908784, EPI_ISL_1908785, EPI_ISL_1908786, EPI_ISL_1970088, EPI_ISL_1970089, EPI_ISL_1970090, EPI_ISL_1970091, EPI_ISL_1970092, EPI_ISL_1970093, EPI_ISL_1970094, EPI_ISL_1970095, EPI_ISL_1970096, EPI_ISL_1970097, EPI_ISL_1970098, EPI_ISL_1970099, EPI_ISL_1970100, EPI_ISL_1970101, EPI_ISL_1970102, EPI_ISL_1970103, EPI_ISL_1970104, EPI_ISL_1970105, EPI_ISL_1970106, EPI_ISL_1970107, EPI_ISL_1970108, EPI_ISL_1970109, EPI_ISL_1970110, EPI_ISL_1970111, EPI_ISL_1970112, EPI_ISL_1970113, EPI_ISL_1970114, EPI_ISL_1970115, EPI_ISL_1970116, EPI_ISL_1970117, EPI_ISL_1970118, EPI_ISL_1970119, EPI_ISL_1970120, EPI_ISL_1970121, EPI_ISL_1970122, EPI_ISL_1970123, EPI_ISL_1970124, EPI_ISL_1970125, EPI_ISL_1970126, EPI_ISL_1970127, EPI_ISL_1970128, EPI_ISL_1970129, EPI_ISL_1970130, EPI_ISL_1970131, EPI_ISL_1970132, EPI_ISL_1970133, EPI_ISL_1970134, EPI_ISL_1970135, EPI_ISL_1970136, EPI_ISL_1970138, EPI_ISL_1970139, EPI_ISL_1970140, EPI_ISL_1970141, EPI_ISL_1970142, EPI_ISL_1970143, EPI_ISL_1970144, EPI_ISL_1970145, EPI_ISL_1970146, EPI_ISL_1970147, EPI_ISL_1970148, EPI_ISL_1970149, EPI_ISL_1970150, EPI_ISL_1970151, EPI_ISL_1970152, EPI_ISL_1970153, EPI_ISL_1970154, EPI_ISL_1970155, EPI_ISL_1970156, EPI_ISL_1970157, EPI_ISL_1970158, EPI_ISL_1970159, EPI_ISL_1970160, EPI_ISL_1970161, EPI_ISL_1970162, EPI_ISL_1970163, EPI_ISL_1970164, EPI_ISL_1970165, EPI_ISL_1970166, EPI_ISL_1970167, EPI_ISL_1970168, EPI_ISL_1970169, EPI_ISL_1970170, EPI_ISL_1970171, EPI_ISL_1970172, EPI_ISL_1970173, EPI_ISL_1970174, EPI_ISL_1970175, EPI_ISL_1970176, EPI_ISL_1970177, EPI_ISL_1970178, EPI_ISL_1970179, EPI_ISL_1970207, EPI_ISL_1970208, EPI_ISL_1970209, EPI_ISL_1970210, EPI_ISL_1970211, EPI_ISL_1970212, EPI_ISL_1970213, EPI_ISL_1970214, EPI_ISL_1970215, EPI_ISL_1970216, EPI_ISL_1970217, EPI_ISL_1970218, EPI_ISL_1970219, EPI_ISL_1970220, EPI_ISL_1970221, EPI_ISL_1970222, EPI_ISL_1970223, EPI_ISL_1970224, EPI_ISL_1970225, EPI_ISL_1970234, EPI_ISL_1970235, EPI_ISL_1970236, EPI_ISL_1970237, EPI_ISL_1970238, EPI_ISL_1970239, EPI_ISL_1970240, EPI_ISL_1970241, EPI_ISL_1970242, EPI_ISL_1970243, EPI_ISL_1970244, EPI_ISL_1970245, EPI_ISL_1970246, EPI_ISL_1970247, EPI_ISL_1970248, EPI_ISL_1970249, EPI_ISL_1970250, EPI_ISL_1970251, EPI_ISL_1970252, EPI_ISL_1970253, EPI_ISL_1970254, EPI_ISL_1970255, EPI_ISL_1970256, EPI_ISL_1970257, EPI_ISL_1970258, EPI_ISL_1970259, EPI_ISL_1970260, EPI_ISL_1970261, EPI_ISL_1970262, EPI_ISL_1970263, EPI_ISL_1970264, EPI_ISL_1970265, EPI_ISL_1970266, EPI_ISL_1970267, EPI_ISL_1970268, EPI_ISL_1970269, EPI_ISL_1970270, EPI_ISL_1970271, EPI_ISL_1970272, EPI_ISL_1970273, EPI_ISL_1970274, EPI_ISL_1970275, EPI_ISL_1970276, EPI_ISL_1970277, EPI_ISL_1970278, EPI_ISL_1970279, EPI_ISL_1970280, EPI_ISL_1970281, EPI_ISL_1970282, EPI_ISL_1970283, EPI_ISL_1970284, EPI_ISL_1970285, EPI_ISL_1970286, EPI_ISL_1970287, EPI_ISL_1970288, EPI_ISL_1970289, EPI_ISL_1970290, EPI_ISL_1970291, EPI_ISL_1970292, EPI_ISL_1970293, EPI_ISL_1970294, EPI_ISL_1970295, EPI_ISL_1970296, EPI_ISL_1970297, EPI_ISL_1970298, EPI_ISL_1970299, EPI_ISL_1970300, EPI_ISL_1970301, EPI_ISL_1970302, EPI_ISL_1970303, EPI_ISL_1970304, EPI_ISL_1970305, EPI_ISL_1970306, EPI_ISL_1970307, EPI_ISL_1970308, EPI_ISL_1970309, EPI_ISL_1970310, EPI_ISL_1970311, EPI_ISL_1970312, EPI_ISL_1970313, EPI_ISL_1970314, EPI_ISL_1970315, EPI_ISL_1970316, EPI_ISL_1970317, EPI_ISL_1970318, EPI_ISL_1970319, EPI_ISL_1970320, EPI_ISL_1970321, EPI_ISL_1970322, EPI_ISL_1970323, EPI_ISL_1970324, EPI_ISL_1970325, EPI_ISL_1970326, EPI_ISL_1970327, EPI_ISL_1970328, EPI_ISL_1970329, EPI_ISL_1970330, EPI_ISL_1970331, EPI_ISL_1970332, EPI_ISL_1970333, EPI_ISL_1970334, EPI_ISL_1970335, EPI_ISL_1970336, EPI_ISL_1970337, EPI_ISL_1970338, EPI_ISL_1970339, EPI_ISL_1970340, EPI_ISL_1970341, EPI_ISL_1970342, EPI_ISL_1970343 | | | |
| see above | Biotechnology Division, NCDC Delhi | NCDC Delhi, Biotechnology Division | Hema Gogia; Hemlata Lall; Kalaiarasan Ponnusamy; Mahesh S Dhar; Manoj K Singh; Meena Datta; Partha Rakshit; Preeti Madan; Priyanka Singh; Radhakrishnan V. S; Robin Marwal; Sandhya Kabra; Sujeet K Singh; Uma Sharma |
| EPI_ISL_1663303, EPI_ISL_1663304, EPI_ISL_1663305, EPI_ISL_1663306, EPI_ISL_1663307, EPI_ISL_1663308, EPI_ISL_1663309, EPI_ISL_1663310, EPI_ISL_1663311, EPI_ISL_1663312, EPI_ISL_1663313, EPI_ISL_1663314, EPI_ISL_1663315, EPI_ISL_1663316, EPI_ISL_1663317, EPI_ISL_1663318, EPI_ISL_1663319, EPI_ISL_1663320, EPI_ISL_1663321, EPI_ISL_1663322 | | | |
| see above | CIIMS, Bilaspur, Chhattisgarh | Institute of Life Sciences - INSACOG | Ajay Parida; Amol M. Kanampalliwar; Arup Ghosh; Atimukta Jha; INSACOG Consortium; Punit Prasad; Rajeeb Swain; Rupesh Dash; Safal Walia; Shifu Aggarwal; Sunil K. Raghav |
| EPI_ISL_2015502, EPI_ISL_2015503, EPI_ISL_2015504, EPI_ISL_2015505, EPI_ISL_2015507, EPI_ISL_2015508, EPI_ISL_2015509, EPI_ISL_2015510, EPI_ISL_2015513, EPI_ISL_2015517, EPI_ISL_2015519, EPI_ISL_2015521, EPI_ISL_2015524, EPI_ISL_2015526, EPI_ISL_2015531, EPI_ISL_2015533, EPI_ISL_2015536, EPI_ISL_2015539, EPI_ISL_2015552, EPI_ISL_2015559, EPI_ISL_2015560, EPI_ISL_2015563, EPI_ISL_2015569, EPI_ISL_2015570, EPI_ISL_2015572, EPI_ISL_2015574, EPI_ISL_2015583, EPI_ISL_2015584, EPI_ISL_2015586, EPI_ISL_2015589, EPI_ISL_2015590, EPI_ISL_2015592, EPI_ISL_2015593, EPI_ISL_2015595, EPI_ISL_2015596, EPI_ISL_2015597, EPI_ISL_2015602, EPI_ISL_2015606, EPI_ISL_2015608, EPI_ISL_2015610, EPI_ISL_2015612, EPI_ISL_2015613, EPI_ISL_2015617, EPI_ISL_2015618 | | | |
| see above | CMC Vellore | inStem NCBS - INSACOG | Uma Ramakrishnan Dasaradhi Palakodeti Aswin SaiNarain |
| EPI_ISL_1818586 | COMMAND HOSPITAL | INSACOG-KA, NIMHANS | Ananthapadmanabha Kotambail; Anita S Desai; Anson Kunjumon George; Chetan G K; Chitra Pattabiraman; Darshan Sreenivas; Gautham Arunachal Udupi; Pramada Prasad; V Ravi |
| EPI_ISL_1838348, EPI_ISL_1838350, EPI_ISL_1838352, EPI_ISL_1838356, EPI_ISL_1838359, EPI_ISL_1838361, EPI_ISL_1838362, EPI_ISL_1838364, EPI_ISL_1838366, EPI_ISL_1838367, EPI_ISL_1838369, EPI_ISL_1838370, EPI_ISL_1838372, EPI_ISL_1838374, EPI_ISL_1838377, EPI_ISL_1838380, EPI_ISL_1838384, EPI_ISL_1838385, EPI_ISL_1838387, EPI_ISL_1838389, EPI_ISL_1838390, EPI_ISL_1838392, EPI_ISL_1838393, EPI_ISL_1838395, EPI_ISL_1838397, EPI_ISL_1838398, EPI_ISL_1838400, EPI_ISL_1838402, EPI_ISL_1838403, EPI_ISL_1838405, EPI_ISL_1838406, EPI_ISL_1838408, EPI_ISL_1838410, EPI_ISL_1838411, EPI_ISL_1838413, EPI_ISL_1838414, EPI_ISL_1838418, EPI_ISL_1838422, EPI_ISL_1838426, EPI_ISL_1838430, EPI_ISL_1838435, EPI_ISL_1838439, EPI_ISL_1838443, EPI_ISL_1838445, EPI_ISL_1838446, EPI_ISL_1838448, EPI_ISL_1838450, EPI_ISL_1838452, EPI_ISL_1838453, EPI_ISL_1838454, EPI_ISL_1838455, EPI_ISL_1838456, EPI_ISL_1838457, EPI_ISL_1838458, EPI_ISL_1838459, EPI_ISL_1838460, EPI_ISL_1838461, EPI_ISL_1838462, EPI_ISL_1838463, EPI_ISL_1838464, EPI_ISL_1838465, EPI_ISL_1838466, EPI_ISL_1838467, EPI_ISL_1838468, EPI_ISL_1838469, EPI_ISL_1838470, EPI_ISL_1838471, EPI_ISL_1838472, EPI_ISL_1838473, EPI_ISL_1838474, EPI_ISL_1838475, EPI_ISL_1838476, EPI_ISL_1838477, EPI_ISL_1838478, EPI_ISL_1838479, EPI_ISL_1838480, EPI_ISL_1838481, EPI_ISL_1838482, EPI_ISL_1838483, EPI_ISL_1838484, EPI_ISL_1838485, EPI_ISL_1838486, EPI_ISL_1838487, EPI_ISL_1838488, EPI_ISL_1838489, EPI_ISL_1838490, EPI_ISL_1838491, EPI_ISL_1838492, EPI_ISL_1838493, EPI_ISL_1838494, EPI_ISL_1838495, EPI_ISL_1838496, EPI_ISL_1838497, EPI_ISL_1838498, EPI_ISL_1838499, EPI_ISL_1838500, EPI_ISL_1838501, EPI_ISL_1838502, EPI_ISL_1838503, EPI_ISL_1838504, EPI_ISL_1838505, EPI_ISL_1838506, EPI_ISL_1838507, EPI_ISL_1838508, EPI_ISL_1838509, EPI_ISL_1838510, EPI_ISL_1838511, EPI_ISL_1838512, EPI_ISL_1838513, EPI_ISL_1838514, EPI_ISL_1838515, EPI_ISL_1838516, EPI_ISL_1838517, EPI_ISL_1838518, EPI_ISL_1838519, EPI_ISL_1838520, EPI_ISL_1838521, EPI_ISL_1838522, EPI_ISL_1838523, EPI_ISL_1838524, EPI_ISL_1838525, EPI_ISL_1838526, EPI_ISL_1838527, EPI_ISL_1838528, EPI_ISL_1838529, EPI_ISL_1838530, EPI_ISL_1838531, EPI_ISL_1838532, EPI_ISL_1838533, EPI_ISL_1838534, EPI_ISL_1838535, EPI_ISL_1838536, EPI_ISL_1838537, EPI_ISL_1838538, EPI_ISL_1838539, EPI_ISL_1838541, EPI_ISL_1838542, EPI_ISL_1838543, EPI_ISL_1838544, EPI_ISL_1838545, EPI_ISL_1838546, EPI_ISL_1838547, EPI_ISL_1838548, EPI_ISL_1838549, EPI_ISL_1838550, EPI_ISL_1838551, EPI_ISL_1838552, EPI_ISL_1838553, EPI_ISL_1838554, EPI_ISL_1838555, EPI_ISL_1838556, EPI_ISL_1838557, EPI_ISL_1838558, EPI_ISL_1838559, EPI_ISL_1838560, EPI_ISL_1838561, EPI_ISL_1838562, EPI_ISL_1838563, EPI_ISL_1838564, EPI_ISL_1838565, EPI_ISL_1838566, EPI_ISL_1838567, EPI_ISL_1838568, EPI_ISL_1838569, EPI_ISL_1838570, EPI_ISL_1838571, EPI_ISL_1838572, EPI_ISL_1838573, EPI_ISL_1838574, EPI_ISL_1838575, EPI_ISL_1838576, EPI_ISL_1838577, EPI_ISL_1838578, EPI_ISL_1838579, EPI_ISL_1838580, EPI_ISL_1838581, EPI_ISL_1838582, EPI_ISL_1838583, EPI_ISL_1838584, EPI_ISL_1838585, EPI_ISL_1838586, EPI_ISL_1838587, EPI_ISL_1838588, EPI_ISL_1838589, EPI_ISL_1838590, EPI_ISL_1838591, EPI_ISL_1838592, EPI_ISL_1838593, EPI_ISL_1838594, EPI_ISL_1838595, EPI_ISL_1838596, EPI_ISL_1838597, EPI_ISL_1838598, EPI_ISL_1838599, EPI_ISL_1838600, EPI_ISL_1838601, EPI_ISL_1838602, EPI_ISL_1838603, EPI_ISL_1838604, EPI_ISL_1838605, EPI_ISL_1838606, EPI_ISL_1838607, EPI_ISL_1838608, EPI_ISL_1838609, EPI_ISL_1838610, EPI_ISL_1838611, EPI_ISL_1838612, EPI_ISL_1838613, EPI_ISL_1838614, EPI_ISL_1838615, EPI_ISL_1838616, EPI_ISL_1838617, EPI_ISL_1838618, EPI_ISL_1838619, EPI_ISL_1838620, EPI_ISL_1838621, EPI_ISL_1838622, EPI_ISL_1838623, EPI_ISL_1838624, EPI_ISL_1838625, EPI_ISL_1838626, EPI_ISL_1838627, EPI_ISL_1838628, EPI_ISL_1838629, EPI_ISL_1838630, EPI_ISL_1838631, EPI_ISL_1838632, EPI_ISL_1838633, EPI_ISL_1838634, EPI_ISL_1838635, EPI_ISL_1838636, EPI_ISL_1838637, EPI_ISL_1838638, EPI_ISL_1838639, EPI_ISL_1838640, EPI_ISL_1838641, EPI_ISL_1838642, EPI_ISL_1838643, EPI_ISL_1838644, EPI_ISL_1838645, EPI_ISL_1838646, EPI_ISL_1838647, EPI_ISL_1838648, EPI_ISL_1838649, EPI_ISL_1838650, EPI_ISL_1838651, EPI_ISL_1838652, EPI_ISL_1838653, EPI_ISL_1838654, EPI_ISL_1838655, EPI_ISL_1838656, EPI_ISL_1838657, EPI_ISL_1838658, EPI_ISL_1838659, EPI_ISL_1838660, EPI_ISL_1838661, EPI_ISL_1838662, EPI_ISL_1838663, EPI_ISL_1838664, EPI_ISL_1838665, EPI_ISL_1838666, EPI_ISL_1838667, EPI_ISL_1838668, EPI_ISL_1838669, EPI_ISL_1838670, EPI_ISL_1838671, EPI_ISL_1838672, EPI_ISL_1838673, EPI_ISL_1838674, EPI_ISL_1838675, EPI_ISL_1838676, EPI_ISL_1838677, EPI_ISL_1838678, EPI_ISL_1838679, EPI_ISL_1838680, EPI_ISL_1838681, EPI_ISL_1838682, EPI_ISL_1838683, EPI_ISL_1838684, EPI_ISL_1838685, EPI_ISL_1838686, EPI_ISL_1838687, EPI_ISL_1838688, EPI_ISL_1838689, EPI_ISL_1838690, EPI_ISL_1838691, EPI_ISL_1838692, EPI_ISL_1838693, EPI_ISL_1838694, EPI_ISL_1838695, EPI_ISL_1838696, EPI_ISL_1838697, EPI_ISL_1838698, EPI_ISL_1838699, EPI_ISL_1838700, EPI_ISL_1838701, EPI_ISL_1838702, EPI_ISL_1838703, EPI_ISL_1838704, EPI_ISL_1838705, EPI_ISL_1838706, EPI_ISL_1838707, EPI_ISL_1838708, EPI_ISL_1838709, EPI_ISL_1838710, EPI_ISL_1838711, EPI_ISL_1838712, EPI_ISL_1838713, EPI_ISL_1838714, EPI_ISL_1838715, EPI_ISL_1838716, EPI_ISL_1838717, EPI_ISL_1838718, EPI_ISL_1838719, EPI_ISL_1838720, EPI_ISL_1838721, EPI_ISL_1838722, EPI_ISL_1838723, EPI_ISL_1838724, EPI_ISL_1838725, EPI_ISL_1838726, EPI_ISL_1838727, EPI_ISL_1838728, EPI_ISL_1838729, EPI_ISL_1838730, EPI_ISL_1838731, EPI_ISL_1838732, EPI_ISL_1838733, EPI_ISL_1838734, EPI_ISL_1838735, EPI_ISL_1838736, EPI_ISL_1838737, EPI_ISL_1838738, EPI_ISL_1838739, EPI_ISL_1838740, EPI_ISL_1838741, EPI_ISL_1838742, EPI_ISL_1838743, EPI_ISL_1838744, EPI_ISL_1838745, EPI_ISL_1838746, EPI_ISL_1838747, EPI_ISL_1838748, EPI_ISL_1838749, EPI_ISL_1838750, EPI_ISL_1838751, EPI_ISL_1838752, EPI_ISL_1838753, EPI_ISL_1838754, EPI_ISL_1838755, EPI_ISL_1838756, EPI_ISL_1838757, EPI_ISL_1838758, EPI_ISL_1838759, EPI_ISL_1838760, EPI_ISL_1838761, EPI_ISL_1838762, EPI_ISL_1838763, EPI_ISL_1838764, EPI_ISL_1838765, EPI_ISL_1838766, EPI_ISL_1838767, EPI_ISL_1838768, EPI_ISL_1838769, EPI_ISL_1838770, EPI_ISL_1838771, EPI_ISL_1838772, EPI_ISL_1838773, EPI_ISL_1838774, EPI_ISL_1838775, EPI_ISL_1838776, EPI_ISL_1838777, EPI_ISL_1838778, EPI_ISL_1838779, EPI_ISL_1838780, EPI_ISL_1838781, EPI_ISL_1838782, EPI_ISL_1838783, EPI_ISL_1838784, EPI_ISL_1838785, EPI_ISL_1838786, EPI_ISL_1838787, EPI_ISL_1838788, EPI_ISL_1838789, EPI_ISL_1838790, EPI_ISL_1838792, EPI_ISL_1838793, EPI_ISL_1838794, EPI_ISL_1838795, EPI_ISL_1838796, EPI_ISL_1838797, EPI_ISL_1838798, EPI_ISL_1838799, EPI_ISL_1838800, EPI_ISL_1838801, EPI_ISL_1838802, EPI_ISL_1838803, EPI_ISL_1838804, EPI_ISL_1838805, EPI_ISL_1838806, EPI_ISL_1838807, EPI_ISL_1838808, EPI_ISL_1838809, EPI_ISL_1838810, EPI_ISL_1838811, EPI_ISL_1838812, EPI_ISL_1838813, EPI_ISL_1838814, EPI_ISL_1838815, EPI_ISL_1838816, EPI_ISL_1838817, EPI_ISL_1838818, EPI_ISL_1838819, EPI_ISL_1838820, EPI_ISL_1838821, EPI_ISL_1838822, EPI_ISL_1838823, EPI_ISL_1838824, EPI_ISL_1838825, EPI_ISL_1838826, EPI_ISL_1838827, EPI_ISL_1838828, EPI_ISL_1838829, EPI_ISL_1838830, EPI_ISL_1838831, EPI_ISL_1838832, EPI_ISL_1838833, EPI_ISL_1838834, EPI_ISL_1838835, EPI_ISL_1838836, EPI_ISL_1838837, EPI_ISL_1838838, EPI_ISL_1838839, EPI_ISL_1838840, EPI_ISL_1838841, EPI_ISL_1838842, EPI_ISL_1838843, EPI_ISL_1838844, EPI_ISL_1838845, EPI_ISL_1838846, EPI_ISL_1838847, EPI_ISL_1838848, EPI_ISL_1838849, EPI_ISL_1838850, EPI_ISL_1838851, EPI_ISL_1838852, EPI_ISL_1838853, EPI_ISL_1838854, EPI_ISL_1838855, EPI_ISL_1838856, EPI_ISL_1838857, EPI_ISL_1838858, EPI_ISL_1838859, EPI_ISL_1838860, EPI_ISL_1838861, EPI_ISL_1838862, EPI_ISL_1838863, EPI_ISL_1838864, EPI_ISL_1838865, EPI_ISL_1838866, EPI_ISL_1838867, EPI_ISL_1838868, EPI_ISL_1838869, EPI_ISL_1838870, EPI_ISL_1838871, EPI_ISL_1838872, EPI_ISL_1838873, EPI_ISL_1838874, EPI_ISL_1838875, EPI_ISL_1838876, EPI_ISL_1838877, EPI_ISL_1838878, EPI_ISL_1838879, EPI_ISL_1838880, EPI_ISL_1838881, EPI_ISL_1838882, EPI_ISL_1838883, EPI_ISL_1838884, EPI_ISL_1838885, EPI_ISL_1838886, EPI_ISL_1838887, EPI_ISL_1838888, EPI_ISL_1838889, EPI_ISL_1838890, EPI_ISL_1838891, EPI_ISL_1838892, EPI_ISL_1838893, EPI_ISL_1838894, EPI_ISL_1838895, EPI_ISL_1838896, EPI_ISL_1838897, EPI_ISL_1838898, EPI_ISL_1838899, EPI_ISL_1838900, EPI_ISL_1838901, EPI_ISL_1838902, EPI_ISL_1838903, EPI_ISL_1838904, EPI_ISL_1838905, EPI_ISL_1838906, EPI_ISL_1838907, EPI_ISL_1838908, EPI_ISL_1838909, EPI_ISL_1838910, EPI_ISL_1838911, EPI_ISL_1838912, EPI_ISL_1838913, EPI_ISL_1838914, EPI_ISL_1838915, EPI_ISL_1838916, EPI_ISL_1838917, EPI_ISL_1838918, EPI_ISL_1838919, EPI_ISL_1972343, EPI_ISL_1972344, EPI_ISL_1972345, EPI_ISL_1972347, EPI_ISL_1972349, EPI_ISL_1972351, EPI_ISL_1972353, EPI_ISL_1972354, EPI_ISL_1972355, EPI_ISL_2156830, EPI_ISL_2156831, EPI_ISL_2156832, EPI_ISL_2156833, EPI_ISL_2156834, EPI_ISL_2156835, EPI_ISL_2156836, EPI_ISL_2156837, EPI_ISL_2156838, EPI_ISL_2156839, EPI_ISL_2156840, EPI_ISL_2156841, EPI_ISL_2156842, EPI_ISL_2156843, EPI_ISL_2156844, EPI_ISL_2156845, EPI_ISL_2156846, EPI_ISL_2156847, EPI_ISL_2156848, EPI_ISL_2156849, EPI_ISL_2156850, EPI_ISL_2156851, EPI_ISL_2156852, EPI_ISL_2156853, EPI_ISL_2156854, EPI_ISL_2156855, EPI_ISL_2156856, EPI_ISL_2156857, EPI_ISL_2156858, EPI_ISL_2156859, EPI_ISL_2156860, EPI_ISL_2156861, EPI_ISL_2156862, EPI_ISL_2156863, EPI_ISL_2156864, EPI_ISL_2156865, EPI_ISL_2156866, EPI_ISL_2156867, EPI_ISL_2156868, EPI_ISL_2156869, EPI_ISL_2156870, EPI_ISL_2156871, EPI_ISL_2156872, EPI_ISL_2156873, EPI_ISL_2156874, EPI_ISL_2156875, EPI_ISL_2156876, EPI_ISL_2156877, EPI_ISL_2156878, EPI_ISL_2156879, EPI_ISL_2156880, EPI_ISL_2156881, EPI_ISL_2156882, EPI_ISL_2156883, EPI_ISL_2156884, EPI_ISL_2156885, EPI_ISL_2156886, EPI_ISL_2156887, EPI_ISL_2156888, EPI_ISL_2156889, EPI_ISL_2156890, EPI_ISL_2156891, EPI_ISL_2156892, EPI_ISL_2156893, EPI_ISL_2156894, EPI_ISL_2156895, EPI_ISL_2156896, EPI_ISL_2156897, EPI_ISL_2156898, EPI_ISL_2156899, EPI_ISL_2156900, EPI_ISL_2156901, EPI_ISL_2156902, EPI_ISL_2156903, EPI_ISL_2156904, EPI_ISL_2156905, EPI_ISL_2156906, EPI_ISL_2156907, EPI_ISL_2156908, EPI_ISL_2156909, EPI_ISL_2156910, EPI_ISL_2156911, EPI_ISL_2156912, EPI_ISL_2156913, EPI_ISL_2156914, EPI_ISL_2156938, EPI_ISL_2156939, EPI_ISL_2156940, EPI_ISL_2156941, EPI_ISL_2156942, EPI_ISL_2156943, EPI_ISL_2156944, EPI_ISL_2156945, EPI_ISL_2156946, EPI_ISL_2156947, EPI_ISL_2156948, EPI_ISL_2156949, EPI_ISL_2156950, EPI_ISL_2156951, EPI_ISL_2156952, EPI_ISL_2156953, EPI_ISL_2156954, EPI_ISL_2156955, EPI_ISL_2156956, EPI_ISL_2156957, EPI_ISL_2156958, EPI_ISL_2156959, EPI_ISL_2156960, EPI_ISL_2156961, EPI_ISL_2156962, EPI_ISL_2156963, EPI_ISL_2156964, EPI_ISL_2156965, EPI_ISL_2156966, EPI_ISL_2156967, EPI_ISL_2156968, EPI_ISL_2156969, EPI_ISL_2156983, EPI_ISL_2156984, EPI_ISL_2156985, EPI_ISL_2156986, EPI_ISL_2156987, EPI_ISL_2156988, EPI_ISL_2156989, EPI_ISL_2156990, EPI_ISL_2156991, EPI_ISL_2156992, EPI_ISL_2156993, EPI_ISL_2156994, EPI_ISL_2156995, EPI_ISL_2156996, EPI_ISL_2156997, EPI_ISL_2156998, EPI_ISL_2156999, EPI_ISL_2157000, EPI_ISL_2157001, EPI_ISL_2157010, EPI_ISL_2157011, EPI_ISL_2157012, EPI_ISL_2157013, EPI_ISL_2157014, EPI_ISL_2157015, EPI_ISL_2157016, EPI_ISL_2157017, EPI_ISL_2157018, EPI_ISL_2157033, EPI_ISL_2157036, EPI_ISL_2157037, EPI_ISL_2157038, EPI_ISL_2157039, EPI_ISL_2157040, EPI_ISL_2157041, EPI_ISL_2157042, EPI_ISL_2157043, EPI_ISL_2157044, EPI_ISL_2157045, EPI_ISL_2157046, EPI_ISL_2157047, EPI_ISL_2157048, EPI_ISL_2157049, | | | |

| | | | |
|---|---|---|---|
| EPI_ISL_2157050, EPI_ISL_2157051, EPI_ISL_2157052, EPI_ISL_2157053, EPI_ISL_2157054, EPI_ISL_2157055, EPI_ISL_2157056, EPI_ISL_2157057, EPI_ISL_2157058, EPI_ISL_2157059, EPI_ISL_2157060, EPI_ISL_2157061, EPI_ISL_2157062, EPI_ISL_2157063, EPI_ISL_2157064, EPI_ISL_2157065, EPI_ISL_2157066, EPI_ISL_2157067, EPI_ISL_2157068, EPI_ISL_2157069, EPI_ISL_2157070, EPI_ISL_2157076, EPI_ISL_2157077, EPI_ISL_2157078, EPI_ISL_2157079, EPI_ISL_2157080, EPI_ISL_2157081, EPI_ISL_2157082, EPI_ISL_2157083, EPI_ISL_2157084, EPI_ISL_2157085, EPI_ISL_2157091, EPI_ISL_2157092, EPI_ISL_2157093, EPI_ISL_2157094, EPI_ISL_2157095, EPI_ISL_2157096, EPI_ISL_2157097, EPI_ISL_2157098, EPI_ISL_2157099, EPI_ISL_2157100, EPI_ISL_2157101, EPI_ISL_2157102, EPI_ISL_2157103, EPI_ISL_2157104, EPI_ISL_2157105, EPI_ISL_2157106, EPI_ISL_2157107, EPI_ISL_2157108, EPI_ISL_2157109, EPI_ISL_2157110, EPI_ISL_2157111, EPI_ISL_2157112, EPI_ISL_2157113, EPI_ISL_2157114, EPI_ISL_2157115, EPI_ISL_2157116, EPI_ISL_2157117, EPI_ISL_2157118, EPI_ISL_2157119, EPI_ISL_2157120, EPI_ISL_2157121, EPI_ISL_2157122, EPI_ISL_2157123, EPI_ISL_2157124, EPI_ISL_2157125, EPI_ISL_2157126, EPI_ISL_2157127, EPI_ISL_2157128, EPI_ISL_2157129, EPI_ISL_2157130, EPI_ISL_2157131, EPI_ISL_2157154, EPI_ISL_2157155, EPI_ISL_2157156 | | | |
| see above | CSIR-Centre for Cellular and Molecular Biology | CSIR-Centre for Cellular and Molecular Biology-INSACOG | Amareshwar Vodapalli; Ara Sreenivas; Archana Bharadwaj Siva; B Himasri; Blessy B John; Divya Tej Sowpati; Karthik Bharadwaj Tallapaka; Lamuk Zaveri; Onkar Kulkarni; Payel Mukherjee; Rakesh K Mishra; Sharath Chandra Thota; Shreekant Verma; Sofia Banu; Tulasi Nagabandi; Valli Nagalakshmi Undamatla; Viswagithe S L |
| EPI_ISL_1360380, EPI_ISL_1360381, EPI_ISL_1360382, EPI_ISL_1360383, EPI_ISL_1360384, EPI_ISL_1360385, EPI_ISL_1360386, EPI_ISL_1360387, EPI_ISL_1360388, EPI_ISL_1547802, EPI_ISL_1547803, EPI_ISL_1547804, EPI_ISL_1547805, EPI_ISL_1916408, EPI_ISL_1916409, EPI_ISL_1916410, EPI_ISL_1916411, EPI_ISL_1916412, EPI_ISL_1916413, EPI_ISL_1916414, EPI_ISL_1916418, EPI_ISL_1916419, EPI_ISL_1916420, EPI_ISL_1916421, EPI_ISL_1916422, EPI_ISL_1916423, EPI_ISL_1916424, EPI_ISL_1916430, EPI_ISL_1916431, EPI_ISL_1916432, EPI_ISL_1916433, EPI_ISL_1916434, EPI_ISL_1916440, EPI_ISL_1916441, EPI_ISL_1916442, EPI_ISL_1916443, EPI_ISL_1916444, EPI_ISL_1916445, EPI_ISL_1916446, EPI_ISL_1916451, EPI_ISL_1916452, EPI_ISL_1916453, EPI_ISL_1916454, EPI_ISL_1916455, EPI_ISL_1916456, EPI_ISL_1916457, EPI_ISL_1916463, EPI_ISL_1916464, EPI_ISL_1916465, EPI_ISL_1916466, EPI_ISL_1916467, EPI_ISL_1916468, EPI_ISL_1916472, EPI_ISL_1916473, EPI_ISL_1916474, EPI_ISL_1916475, EPI_ISL_1916476, EPI_ISL_1916477, EPI_ISL_1916479, EPI_ISL_1916483, EPI_ISL_1916484, EPI_ISL_1916485, EPI_ISL_1916486, EPI_ISL_1916487, EPI_ISL_1916488, EPI_ISL_1916489, EPI_ISL_1916490, EPI_ISL_1970628, EPI_ISL_1970629 | | | |
| see above | CSIR-National Environmental Engineering Research Institute | CSIR-Centre for Cellular and Molecular Biology - INSACOG | Amareshwar Vodapalli; Ara Sreenivas; B Himasri; Divya Tej Sowpati; Karthik Bharadwaj Tallapaka; Krishna Khairnar; Lamuk Zaveri; Onkar Kulkarni; Rakesh K Mishra; Sharath Chandra Thota; Shreekant Verma; Sofia Banu; Viswagithe S L |
| EPI_ISL_1662424, EPI_ISL_1662425, EPI_ISL_1662426, EPI_ISL_1662427, EPI_ISL_1662428, EPI_ISL_1662429, EPI_ISL_1662430, EPI_ISL_1662431, EPI_ISL_1662432, EPI_ISL_1662433, EPI_ISL_1662434, EPI_ISL_1662435, EPI_ISL_1662436, EPI_ISL_1662437, EPI_ISL_1662438, EPI_ISL_1662439, EPI_ISL_1662440, EPI_ISL_1662441, EPI_ISL_1662442, EPI_ISL_1662443, EPI_ISL_1662444, EPI_ISL_1663323, EPI_ISL_1663324, EPI_ISL_1663356, EPI_ISL_1663357, EPI_ISL_1663488, EPI_ISL_1663489, EPI_ISL_1663490, EPI_ISL_1663491, EPI_ISL_1663492, EPI_ISL_1663493, EPI_ISL_1663494, EPI_ISL_1663495, EPI_ISL_1663496 | | | |
| see above | Capital hospital, Bhubaneswar | Institute of Life Sciences - INSACOG | Ajay Parida; Amol M. Kanampalliwar; Arup Ghosh; Atimukta Jha; INSACOG Consortium; Punit Prasad; Rajeeb Swain; Rupesh Dash; Safal Walia; Shifu Aggarwal; Sunil K. Raghav |
| EPI_ISL_2106943, EPI_ISL_2106944, EPI_ISL_2106945, EPI_ISL_2106946, EPI_ISL_2106947, EPI_ISL_2106948, EPI_ISL_2106949, EPI_ISL_2106950, EPI_ISL_2106951, EPI_ISL_2106952, EPI_ISL_2106953, EPI_ISL_2106954, EPI_ISL_2106955, EPI_ISL_2106956, EPI_ISL_2106957, EPI_ISL_2106958, EPI_ISL_2106959, EPI_ISL_2106960, EPI_ISL_2106961, EPI_ISL_2106962, EPI_ISL_2106963, EPI_ISL_2106964, EPI_ISL_2106965, EPI_ISL_2106966, EPI_ISL_2106967, EPI_ISL_2106968, EPI_ISL_2106969, EPI_ISL_2106970, EPI_ISL_2106971, EPI_ISL_2106972, EPI_ISL_2106973, EPI_ISL_2106974 | | | |
| see above | Centre for DNA Fingerprinting and Diagnostics | CDFD -INSACOG | Ashwin Dalal; Asmita Gupta; Divya Vashisht; Murali Bashyam; Pratyusha Bala; Vinay Donipadi |
| EPI_ISL_2714694, EPI_ISL_2714695, EPI_ISL_2714696, EPI_ISL_2714697, EPI_ISL_2714698 | Communicable Diseases, Interactive Research School for Health Affairs (IRSHA) | Communicable Diseases, Interactive Research School for Health Affairs (IRSHA) | A.C.; Arankalle; Mishra; Modak, M.; Pisal; S.R.; Shrivastava, S.; V.A.; Virkar, R. |
| EPI_ISL_1710598 | Communicable Diseases, Interactive Research School for Health Affairs | Communicable Diseases, Interactive Research School for Health Affairs | Akhilesh Chandra Mishra; Meera Modak; Rashmi Virkar; Shamburaje S Pisal; Shubham Shrivastava; Vidya A Arankalle |
| EPI_ISL_1595867, EPI_ISL_1595868, EPI_ISL_1595869, EPI_ISL_1595870, EPI_ISL_1595871, EPI_ISL_1595872, EPI_ISL_1595873 | | | |
| see above | DSU KALABURAGI | INSACOG-KA, NIMHANS | Ananthapadmanabha Kotambail; Anita S Desai; Anson Kunjumon George; Chetan G K; Chitra Pattabiraman; Darshan Sreenivas; Gautham Arunachal Udupi; Pramada Prasad; V Ravi |
| EPI_ISL_2100635, EPI_ISL_2100636, EPI_ISL_2100637, EPI_ISL_2100638, EPI_ISL_2100639, EPI_ISL_2100640, EPI_ISL_2100641, EPI_ISL_2100642, EPI_ISL_2100643, EPI_ISL_2100644, EPI_ISL_2100645, EPI_ISL_2100646, EPI_ISL_2100647, EPI_ISL_2100648, EPI_ISL_2100649, EPI_ISL_2100650, EPI_ISL_2100651, EPI_ISL_2100652, EPI_ISL_2100653, EPI_ISL_2100654, EPI_ISL_2100655, EPI_ISL_2100656, EPI_ISL_2100657, EPI_ISL_2100658, EPI_ISL_2100659, EPI_ISL_2100660, EPI_ISL_2100661, EPI_ISL_2100662, EPI_ISL_2100663, EPI_ISL_2100664, EPI_ISL_2100665, EPI_ISL_2100666, EPI_ISL_2100667, EPI_ISL_2100668, EPI_ISL_2100669, EPI_ISL_2100670, EPI_ISL_2100671, EPI_ISL_2100672, EPI_ISL_2100673, EPI_ISL_2100674, EPI_ISL_2100675, EPI_ISL_2100676, EPI_ISL_2100677, EPI_ISL_2100678, EPI_ISL_2105576, EPI_ISL_2105577, EPI_ISL_2105578, EPI_ISL_2105579, EPI_ISL_2105580, EPI_ISL_2105581, EPI_ISL_2105582, EPI_ISL_2106198 | | | |
| see above | Delhi North District | CSIR-Institute of Genomics and Integrative Biology | Animesh Ray; Asangala Kamai; Ashwin Varadarajan; Ayush Goel; Bharathram Uppilli*; Devasish Desai; Manish Kumar; Mohammed Ahmed; Mohammed Faruq; Naveet Wig; Pooja Sharma*; Rajesh Pandey; Sarafaraz Alam; Saruchi Wadhwa; Satish Swain; Sheeba Saifi; Sushma Rajpoot; Umang Arora |
| EPI_ISL_1662287, EPI_ISL_1662288, EPI_ISL_1662289, EPI_ISL_1662290, EPI_ISL_1662291 | Dept. Of Microbiology, Lt.. Baliram Kashyap Memorial Govt. Medical college, Dimrapal, Jagdalpur | Institute of Life Sciences - INSACOG | Ajay Parida; Amol M. Kanampalliwar; Arup Ghosh; Atimukta Jha; INSACOG Consortium; Punit Prasad; Rajeeb Swain; Rupesh Dash; Safal Walia; Shifu Aggarwal; Sunil K. Raghav |
| EPI_ISL_1939849, EPI_ISL_1939850, EPI_ISL_1939851, EPI_ISL_1939852, EPI_ISL_1939853, EPI_ISL_1939854, EPI_ISL_1939855, EPI_ISL_1939856, EPI_ISL_1939857, EPI_ISL_1939858, EPI_ISL_1939859, EPI_ISL_1939860, EPI_ISL_1939861, EPI_ISL_1939862 | | | |
| see above | Dr S raju, Director of public heath and preventive medicine | inStem NCBS - INSACOG | Uma Ramakrishnan Dasaradhi Palakodeti Aswin SaiNarain |
| EPI_ISL_1940051, EPI_ISL_1940052 | GANGONAHALLI | inStem NCBS - INSACOG | Uma Ramakrishnan Dasaradhi Palakodeti Aswin SaiNarain |
| EPI_ISL_1940077 | GANGONDAHALLI | inStem NCBS - INSACOG | Uma Ramakrishnan Dasaradhi Palakodeti Aswin SaiNarain |
| EPI_ISL_2015532 | GANGONDNAHALLI | inStem NCBS - INSACOG | Uma Ramakrishnan Dasaradhi Palakodeti Aswin SaiNarain |
| EPI_ISL_2001178, EPI_ISL_2001179, EPI_ISL_2001180, EPI_ISL_2001181, EPI_ISL_2001182, EPI_ISL_2001183, EPI_ISL_2001184, EPI_ISL_2001185, EPI_ISL_2001186, EPI_ISL_2017752, EPI_ISL_2017753, EPI_ISL_2017754, EPI_ISL_2017755, EPI_ISL_2017756, EPI_ISL_2017757, EPI_ISL_2017758 | | | |
| see above | GMERS, Government Medical College (GMC), Gotri | Gujarat Biotechnology Research Centre | Bithika Duttaroy; Chaitanya Joshi; Dinesh Kumar; Janvi Raval; Madhvi Joshi; Nitesh Shah; Nitin Savaliya; Ramesh Pandit; Sonal Sharma; Twinkle Soni; Umang Mishra; Zarna Patel; Zuber Saiyed |
| EPI_ISL_2001121, EPI_ISL_2001122, EPI_ISL_2001123, EPI_ISL_2001124, EPI_ISL_2001125, EPI_ISL_2001126, EPI_ISL_2001127, EPI_ISL_2001128, EPI_ISL_2001129, EPI_ISL_2001130, EPI_ISL_2001131, EPI_ISL_2001132, EPI_ISL_2001133, EPI_ISL_2001134, EPI_ISL_2001135, EPI_ISL_2001136, EPI_ISL_2001137, EPI_ISL_2001138, EPI_ISL_2001139, EPI_ISL_2001140, EPI_ISL_2001141, EPI_ISL_2001142, EPI_ISL_2001143, EPI_ISL_2001144, EPI_ISL_2001145, EPI_ISL_2001146, EPI_ISL_2001147, EPI_ISL_2001148 | | | |
| see above | GMERS, Government Medical College, Gandhinagar | Gujarat Biotechnology Research Centre | Chaitanya Joshi; Dinesh Kumar; Gaurishankar Shrimali; Janvi Raval; Madhvi Joshi; Nitesh Shah; Nitin Savaliya; Ramesh Pandit; Sonal Sharma; Twinkle Soni; Umang Mishra; Zarna Patel; Zuber Saiyed |
| EPI_ISL_1544133, EPI_ISL_1544134 | GMERS, Medical College, Gandhinagar | Gujarat Biotechnology Research Centre | Chaitanya Joshi; Dinesh Kumar; Gaurishankar Shrimali; Janvi Raval; Madhvi Joshi; Nitesh Shah; Nitin Savaliya; Ramesh Pandit; Sonal Sharma; Twinkle Soni; Umang Mishra; Zarna Patel; Zuber Saiyed |
| EPI_ISL_2015549, EPI_ISL_2015639 | GOVIDRAJNAGAR | inStem NCBS - INSACOG | Uma Ramakrishnan Dasaradhi Palakodeti Aswin SaiNarain |
| EPI_ISL_1940017, EPI_ISL_1940018, EPI_ISL_1940019, EPI_ISL_1940020, EPI_ISL_1940021 | GOVIMDRAJ NAGAR | inStem NCBS - INSACOG | Uma Ramakrishnan Dasaradhi Palakodeti Aswin SaiNarain |
| EPI_ISL_1940071, EPI_ISL_1940072, EPI_ISL_1940073, EPI_ISL_1940074, EPI_ISL_1940075, EPI_ISL_1940076 | GOVINDRAJ NAGAR | inStem NCBS - INSACOG | Uma Ramakrishnan Dasaradhi Palakodeti Aswin SaiNarain |
| EPI_ISL_1940049, EPI_ISL_1940050, EPI_ISL_1940109, EPI_ISL_1940110, EPI_ISL_1940111, EPI_ISL_2015470, EPI_ISL_2015500, EPI_ISL_2015514, EPI_ISL_2015538, EPI_ISL_2015619 | | | |
| see above | GOVINDRAJNAGAR | inStem NCBS - INSACOG | Uma Ramakrishnan Dasaradhi Palakodeti Aswin SaiNarain |
| EPI_ISL_2000678, EPI_ISL_2000679 | Government Medical College Ernakulam | CSIR Institute of Genomics and Integrative Biology | Abhinav Jain; Afra Shamnath; Anjali Bajaj; Anoop Joseph; Arvinden V.R.; Bani Jolly; Beena Philomina J; Bindhu Vasudevan; Chandni Radhakrishnan; Fairoz C. P; Mercy Rophina; Mohamed Imran; Mohit Kumar Divakar; Neethu John; Nisha Majeed; P. Geetha; P. Jayesh Kumar; Prasanth Viswanathan; Priyanka R Nair; Rahul C. Bhoyar; Samatha Mathew; Srashti Jyoti Agrawal; Sridhar Sivasubbu; Vigneshwar Senthivel; Vinod Scaria; Vishu Gupta |
| EPI_ISL_2000674 | Government Medical College Idukki | CSIR Institute of Genomics and Integrative Biology | Abhinav Jain; Afra Shamnath; Anjali Bajaj; Anoop Joseph; Arvinden V.R.; Bani Jolly; Beena Philomina J; Bindhu Vasudevan; Chandni Radhakrishnan; Fairoz C. P; Mercy Rophina; Mohamed Imran; Mohit Kumar Divakar; Neethu John; Nisha Majeed; P. Geetha; P. Jayesh Kumar; Prasanth Viswanathan; Priyanka R Nair; Rahul C. Bhoyar; Samatha Mathew; Srashti Jyoti Agrawal; Sridhar Sivasubbu; Vigneshwar Senthivel; Vinod Scaria; Vishu Gupta |
| EPI_ISL_2000675, EPI_ISL_2000676, EPI_ISL_2000677 | Government Medical College Kozhikode | CSIR Institute of Genomics and Integrative Biology | Abhinav Jain; Afra Shamnath; Anjali Bajaj; Anoop Joseph; Arvinden V.R.; Bani Jolly; Beena Philomina J; Bindhu Vasudevan; Chandni Radhakrishnan; Fairoz C. P; Mercy Rophina; Mohamed Imran; Mohit Kumar Divakar; Neethu John; Nisha Majeed; P. Geetha; P. Jayesh Kumar; Prasanth Viswanathan; Priyanka R Nair; Rahul C. Bhoyar; Samatha Mathew; Srashti Jyoti Agrawal; Sridhar Sivasubbu; Vigneshwar Senthivel; Vinod Scaria; Vishu Gupta |
| EPI_ISL_2017789 | Gujarat Biotechnology Research Centre | Gujarat Biotechnology Research Centre | Chaitanya Joshi; Dinesh Kumar; Janvi Raval; Madhvi Joshi; Nitesh Shah; Nitin Savaliya; Ramesh Pandit; Sonal Sharma; Twinkle Soni; Umang Mishra; Zarna Patel; Zuber Saiyed |
| EPI_ISL_1677768, EPI_ISL_1677769 | Gujarat Biotechnology Research Centre (GBRC), Gandhinagar | Gujarat Biotechnology Research Centre | Chaitanya Joshi; Dinesh Kumar; Janvi Raval; Madhvi Joshi; Nitesh Shah; Nitin Savaliya; Ramesh Pandit; Sonal Sharma; Twinkle Soni; Umang Mishra; Zarna Patel; Zuber Saiyed |
| EPI_ISL_2015471, EPI_ISL_2015499, EPI_ISL_2015506, EPI_ISL_2015520, EPI_ISL_2015523, EPI_ISL_2015545, EPI_ISL_2015573, EPI_ISL_2015614, EPI_ISL_2015634 | | | |
| see above | HOSA HALLI | inStem NCBS - INSACOG | Uma Ramakrishnan Dasaradhi Palakodeti Aswin SaiNarain |
| EPI_ISL_1940000, EPI_ISL_1940001, EPI_ISL_1940002, EPI_ISL_1940003, EPI_ISL_1940004, EPI_ISL_1940005, EPI_ISL_1940006, EPI_ISL_1940007, EPI_ISL_1940008, EPI_ISL_1940009, EPI_ISL_1940010, EPI_ISL_1940011, EPI_ISL_1940012, EPI_ISL_1940013, EPI_ISL_1940014, EPI_ISL_1940015, EPI_ISL_1940016, EPI_ISL_1940037, EPI_ISL_1940038, EPI_ISL_1940039, EPI_ISL_1940040, EPI_ISL_1940041, EPI_ISL_1940042, EPI_ISL_1940043, EPI_ISL_1940044, EPI_ISL_1940045, EPI_ISL_1940046, EPI_ISL_1940047, EPI_ISL_1940048, EPI_ISL_1940062, EPI_ISL_1940063, EPI_ISL_1940064, EPI_ISL_1940065, EPI_ISL_1940066, EPI_ISL_1940067, EPI_ISL_1940068, EPI_ISL_1940078, EPI_ISL_1940079, EPI_ISL_1940080, EPI_ISL_1940081, EPI_ISL_1940082, EPI_ISL_1940083, EPI_ISL_1940084, EPI_ISL_1940085, EPI_ISL_1940086, EPI_ISL_1940087, EPI_ISL_1940088, EPI_ISL_1940089, EPI_ISL_1940090, EPI_ISL_1940112, EPI_ISL_1940113, EPI_ISL_1940114, EPI_ISL_1940115, EPI_ISL_1940116, EPI_ISL_1940117, EPI_ISL_1940118, EPI_ISL_1940119, EPI_ISL_1940120, EPI_ISL_1940121, EPI_ISL_1940122, EPI_ISL_1940123, EPI_ISL_1940124, EPI_ISL_1940125, EPI_ISL_1940126, EPI_ISL_1940127, EPI_ISL_1940128, EPI_ISL_1940129, EPI_ISL_1940142, EPI_ISL_2015493, EPI_ISL_2015516, EPI_ISL_2015527, EPI_ISL_2015547, EPI_ISL_2015554, EPI_ISL_2015565, EPI_ISL_2015567, EPI_ISL_2015575, EPI_ISL_2015616 | | | |
| see above | HOSAHALLI | inStem NCBS - INSACOG | Uma Ramakrishnan Dasaradhi Palakodeti Aswin SaiNarain |
| EPI_ISL_2100187, EPI_ISL_2100188, EPI_ISL_2100190, EPI_ISL_2100192, EPI_ISL_2100193, EPI_ISL_2100195, EPI_ISL_2100197, EPI_ISL_2100198, EPI_ISL_2100202, EPI_ISL_2100212, EPI_ISL_2100216 | | | |
| see above | ICMR-National Institute for Research In Tuberculosis | NIV Influenza | Dr Luke Elizabeth Hanna; Dr Padmapriyadarsini.C; Dr Radha Gopalaswamy; Dr S Siva Kumar; Mr Ramesh K |
| EPI_ISL_2189568, EPI_ISL_2189569, EPI_ISL_2189570, EPI_ISL_2189571, EPI_ISL_2189572, EPI_ISL_2189573, EPI_ISL_2189574, EPI_ISL_2189575, EPI_ISL_2189576, EPI_ISL_2189577, EPI_ISL_2189578, EPI_ISL_2189579, EPI_ISL_2189580, EPI_ISL_2189581, EPI_ISL_2189582, EPI_ISL_2189583, EPI_ISL_2189584, EPI_ISL_2189585, EPI_ISL_2189586, EPI_ISL_2189587, EPI_ISL_2189588, EPI_ISL_2189589, EPI_ISL_2189590, EPI_ISL_2189591, EPI_ISL_2189592, EPI_ISL_2189593, EPI_ISL_2189594, EPI_ISL_2189595, EPI_ISL_2189596, EPI_ISL_2189597, EPI_ISL_2189598, EPI_ISL_2189599, EPI_ISL_2189600, EPI_ISL_2189601, EPI_ISL_2189602, EPI_ISL_2189603, EPI_ISL_2189604, EPI_ISL_2189605, EPI_ISL_2189606, EPI_ISL_2189607, EPI_ISL_2189608, EPI_ISL_2189609, EPI_ISL_2189610, EPI_ISL_2189611, EPI_ISL_2189612, EPI_ISL_2189613, EPI_ISL_2189614, EPI_ISL_2189615, EPI_ISL_2189616, EPI_ISL_2189617, EPI_ISL_2189618, EPI_ISL_2189619, EPI_ISL_2189620, EPI_ISL_2189621, EPI_ISL_2189622, EPI_ISL_2189623, EPI_ISL_2189624, EPI_ISL_2189625, EPI_ISL_2189626, EPI_ISL_2189627, EPI_ISL_2189628, EPI_ISL_2189629, EPI_ISL_2189630, EPI_ISL_2189631, EPI_ISL_2189632, EPI_ISL_2189633, EPI_ISL_2189634, EPI_ISL_2189635, EPI_ISL_2189636, EPI_ISL_2189637, EPI_ISL_2189638, EPI_ISL_2189639, EPI_ISL_2189640, EPI_ISL_2189641, EPI_ISL_2189642, EPI_ISL_2189643, EPI_ISL_2189644, EPI_ISL_2189645, EPI_ISL_2189646, EPI_ISL_2189647, EPI_ISL_2189648, EPI_ISL_2189649, EPI_ISL_2189650, EPI_ISL_2189651, EPI_ISL_2189652, EPI_ISL_2189653, EPI_ISL_2189654, EPI_ISL_2189655, EPI_ISL_2189656, EPI_ISL_2189657, EPI_ISL_2189658, EPI_ISL_2189659, EPI_ISL_2189660, EPI_ISL_2189661, EPI_ISL_2189662, EPI_ISL_2189663, EPI_ISL_2189664, EPI_ISL_2189665, EPI_ISL_2189666, EPI_ISL_2189667, EPI_ISL_2189668, EPI_ISL_2189669, EPI_ISL_2189670, EPI_ISL_2189671, EPI_ISL_2189672, EPI_ISL_2189673, EPI_ISL_2189674, EPI_ISL_2189675, EPI_ISL_2189676, EPI_ISL_2189677, EPI_ISL_2189678, EPI_ISL_2189679, EPI_ISL_2189680, EPI_ISL_2189681, EPI_ISL_2189682, EPI_ISL_2189683, EPI_ISL_2189684, EPI_ISL_2189685, EPI_ISL_2189686, EPI_ISL_2189687, EPI_ISL_2189688, EPI_ISL_2189689, EPI_ISL_2189690, EPI_ISL_2189691, EPI_ISL_2189692, EPI_ISL_2189693, EPI_ISL_2189694, EPI_ISL_2189695, EPI_ISL_2189696, EPI_ISL_2189697, EPI_ISL_2189698, EPI_ISL_2189699, EPI_ISL_2189700, EPI_ISL_2189701, EPI_ISL_2189702, EPI_ISL_2189703, EPI_ISL_2189704, EPI_ISL_2189705, EPI_ISL_2189706, EPI_ISL_2189707, EPI_ISL_2189708, EPI_ISL_2189709, EPI_ISL_2189710, EPI_ISL_2189711, EPI_ISL_2189712, EPI_ISL_2189713, EPI_ISL_2189714, EPI_ISL_2189715, EPI_ISL_2189716, EPI_ISL_2189717, EPI_ISL_2189718, EPI_ISL_2189719, EPI_ISL_2189720, EPI_ISL_2189721, EPI_ISL_2189722, EPI_ISL_2189723, EPI_ISL_2189724, EPI_ISL_2189725, EPI_ISL_2189726, EPI_ISL_2189727, EPI_ISL_2189728, EPI_ISL_2189729, EPI_ISL_2189730, EPI_ISL_2189731, EPI_ISL_2189732, EPI_ISL_2189733, EPI_ISL_2189734, EPI_ISL_2189735, EPI_ISL_2189736, EPI_ISL_2189737, EPI_ISL_2189738, EPI_ISL_2189739, EPI_ISL_2189740, EPI_ISL_2189741, EPI_ISL_2189742, EPI_ISL_2189743, EPI_ISL_2189744, EPI_ISL_2189745, EPI_ISL_2189746, EPI_ISL_2189747, EPI_ISL_2189748, EPI_ISL_2189749, EPI_ISL_2189750, EPI_ISL_2189751, EPI_ISL_2189752, EPI_ISL_2189753, EPI_ISL_2189754, EPI_ISL_2189755, EPI_ISL_2189756, EPI_ISL_2189757, EPI_ISL_2189758, EPI_ISL_2189759, EPI_ISL_2189760, EPI_ISL_2189761, EPI_ISL_2189762, EPI_ISL_2189763, EPI_ISL_2189764, EPI_ISL_2189765, EPI_ISL_2189766, EPI_ISL_2189767, EPI_ISL_2189768, EPI_ISL_2189769, EPI_ISL_2189770, EPI_ISL_2189771, EPI_ISL_2189772, EPI_ISL_2189773, EPI_ISL_2189774, EPI_ISL_2189775, EPI_ISL_2189776, EPI_ISL_2189777, EPI_ISL_2189778, EPI_ISL_2189779, EPI_ISL_2189780, EPI_ISL_2189781, EPI_ISL_2189782, EPI_ISL_2189783, EPI_ISL_2189784, EPI_ISL_2189785, EPI_ISL_2189786, EPI_ISL_2189787, EPI_ISL_2189788, EPI_ISL_2189789, EPI_ISL_2189790, EPI_ISL_2189791, EPI_ISL_2189792, EPI_ISL_2189793, EPI_ISL_2189794, EPI_ISL_2189795, EPI_ISL_2189796, EPI_ISL_2189797, EPI_ISL_2189798, EPI_ISL_2189799, EPI_ISL_2189800, EPI_ISL_2189801, EPI_ISL_2189802, EPI_ISL_2189803, EPI_ISL_2189804, EPI_ISL_2189805, EPI_ISL_2189806, EPI_ISL_2189807, EPI_ISL_2189808, EPI_ISL_2189809, EPI_ISL_2189810, EPI_ISL_2189811, EPI_ISL_2189812, EPI_ISL_2189813, EPI_ISL_2189814, EPI_ISL_2189815, EPI_ISL_2189816, EPI_ISL_2189817, EPI_ISL_2189818, EPI_ISL_2189819, EPI_ISL_2189820, EPI_ISL_2189821, EPI_ISL_2189822, EPI_ISL_2189823, EPI_ISL_2189824, EPI_ISL_2189825, EPI_ISL_2189826, EPI_ISL_2189827, EPI_ISL_2189828, EPI_ISL_2189829, EPI_ISL_2189830, EPI_ISL_2189831, EPI_ISL_2189832, EPI_ISL_2189833, EPI_ISL_2189834, EPI_ISL_2189835, EPI_ISL_2189836, EPI_ISL_2189837, EPI_ISL_2189838, EPI_ISL_2189839, EPI_ISL_2189840, EPI_ISL_2189841, EPI_ISL_2189842, EPI_ISL_2189843, EPI_ISL_2189844, EPI_ISL_2189845, EPI_ISL_2189846, EPI_ISL_2189847, EPI_ISL_2189848, EPI_ISL_2189849, EPI_ISL_2189850, EPI_ISL_2189851, EPI_ISL_2189852, EPI_ISL_2189853, EPI_ISL_2189854, EPI_ISL_2189855, EPI_ISL_2189856, EPI_ISL_2189857, EPI_ISL_2189858, EPI_ISL_2189859, EPI_ISL_2189860, EPI_ISL_2189861, EPI_ISL_2189862, EPI_ISL_2189863 | | | |
| see above | ICMR-National Institute of Virology | NCDC Delhi, Biotechnology Division | A Walimbe; A.Awhale; A.Titkare; B.Apoorva; G.Divekar; H.Kengale; Hema Gogia; Hemlata Lall; K P.Shinde; K.Iyengar; K.Korabu; Kalaiarasan Ponnusamy; M L Choudhary; M.Das; Mahesh S Dhar; Manoj K Singh; Meena Datta; Partha Rakshit; Preeti Madan; Priyanka Singh; R.Verma; Radhakrishnan V. S; Robin Marwal; S Jadhav; S.Bhorekar; S.Jadhav; S.Shelkande; Sandhya Kabra; Sujeet K Singh; T.Sanjeevi; U.Saha; Uma Sharma; V Malik; V.Autude; V.Vipat; Z. Sayyed |
| EPI_ISL_1703640, EPI_ISL_1703645, EPI_ISL_1703648, EPI_ISL_1703662, EPI_ISL_1703664, EPI_ISL_1703668, EPI_ISL_1703673, EPI_ISL_1703684, EPI_ISL_1703701, EPI_ISL_1703717, EPI_ISL_1703727, EPI_ISL_1703730, EPI_ISL_1703731, EPI_ISL_1703735, EPI_ISL_1703739, EPI_ISL_1703740, EPI_ISL_1703768, EPI_ISL_1703769, EPI_ISL_1703770, EPI_ISL_1703772, EPI_ISL_1703777, EPI_ISL_1703789, EPI_ISL_1703798, EPI_ISL_1703800, EPI_ISL_1703822, EPI_ISL_1703831, EPI_ISL_1703860, EPI_ISL_1703870, EPI_ISL_1703873, EPI_ISL_1703878, EPI_ISL_1703879, EPI_ISL_1703880, EPI_ISL_1703884, EPI_ISL_1703889, EPI_ISL_1703904, EPI_ISL_1703918, EPI_ISL_1703922, EPI_ISL_1703923, EPI_ISL_1703924, EPI_ISL_1703927, EPI_ISL_1703943, EPI_ISL_1703945, EPI_ISL_1703960, EPI_ISL_1703961, EPI_ISL_1703964, EPI_ISL_1703969, EPI_ISL_1703983, EPI_ISL_1703984, EPI_ISL_1703994, EPI_ISL_1704009, EPI_ISL_1704028, EPI_ISL_1704030, EPI_ISL_1704032, EPI_ISL_1704033, EPI_ISL_1704034, EPI_ISL_1704042, EPI_ISL_1704043, EPI_ISL_1704064, EPI_ISL_1704076, EPI_ISL_1704080, EPI_ISL_1704093, EPI_ISL_1704094, EPI_ISL_1704196, EPI_ISL_1704197, EPI_ISL_1704198, EPI_ISL_1704199, EPI_ISL_1704201, EPI_ISL_1704202, EPI_ISL_1704227, EPI_ISL_1704229, EPI_ISL_1704230, EPI_ISL_1704231, EPI_ISL_1704232, EPI_ISL_1704233, EPI_ISL_1704234, EPI_ISL_1704235, EPI_ISL_1704236, EPI_ISL_1704341, EPI_ISL_1704342, EPI_ISL_1704343, EPI_ISL_1704344, EPI_ISL_1704345, EPI_ISL_1704346, EPI_ISL_1704347, EPI_ISL_1704348, EPI_ISL_1704349, EPI_ISL_1704470, EPI_ISL_1704471, | | | |

| | | | |
|---|---|---|---|
| EPI_ISL_1704472, EPI_ISL_1704473, EPI_ISL_1704474, EPI_ISL_1704475, EPI_ISL_1704476, EPI_ISL_1704494, EPI_ISL_1704495, EPI_ISL_1704496, EPI_ISL_1704497, EPI_ISL_1704498, EPI_ISL_1704499, EPI_ISL_1704500, EPI_ISL_1704501, EPI_ISL_1704502, EPI_ISL_1704503, EPI_ISL_1704504, EPI_ISL_1704505, EPI_ISL_1704506, EPI_ISL_1704507, EPI_ISL_1704508, EPI_ISL_1704509, EPI_ISL_1704510, EPI_ISL_1704511, EPI_ISL_1704512, EPI_ISL_1704513, EPI_ISL_1704514, EPI_ISL_1704516, EPI_ISL_1704518, EPI_ISL_1704519, EPI_ISL_1704520, EPI_ISL_1704521, EPI_ISL_1704522, EPI_ISL_1704523, EPI_ISL_1704524, EPI_ISL_1704525, EPI_ISL_1704526, EPI_ISL_1704527, EPI_ISL_1704528, EPI_ISL_1704529, EPI_ISL_1704530, EPI_ISL_1704531, EPI_ISL_1704533, EPI_ISL_1704534, EPI_ISL_1704535, EPI_ISL_1704536, EPI_ISL_1704537, EPI_ISL_1704538, EPI_ISL_1704539, EPI_ISL_1704540, EPI_ISL_1704541, EPI_ISL_1704542, EPI_ISL_1704543, EPI_ISL_1704544, EPI_ISL_1704545, EPI_ISL_1704546, EPI_ISL_1704547, EPI_ISL_1704548, EPI_ISL_1704549, EPI_ISL_1704550, EPI_ISL_1704551, EPI_ISL_1704552, EPI_ISL_1704553, EPI_ISL_1704554, EPI_ISL_1704555, EPI_ISL_1704556, EPI_ISL_1704557, EPI_ISL_1704558, EPI_ISL_1704559, EPI_ISL_1704560, EPI_ISL_1704561, EPI_ISL_1704577, EPI_ISL_1704578, EPI_ISL_1704579, EPI_ISL_1704580, EPI_ISL_1704581, EPI_ISL_1704588, EPI_ISL_1704589, EPI_ISL_1704590, EPI_ISL_1704591, EPI_ISL_1704592, EPI_ISL_1704593, EPI_ISL_1704594, EPI_ISL_1704595, EPI_ISL_1704596, EPI_ISL_1704597, EPI_ISL_1704598, EPI_ISL_1704599, EPI_ISL_1704600, EPI_ISL_1704601, EPI_ISL_1704602, EPI_ISL_1704603, EPI_ISL_1704604, EPI_ISL_1704605, EPI_ISL_1704606, EPI_ISL_1704607, EPI_ISL_1704608, EPI_ISL_1704609, EPI_ISL_1704610, EPI_ISL_1704611, EPI_ISL_1704612, EPI_ISL_1704613, EPI_ISL_1704614, EPI_ISL_1704615, EPI_ISL_1704617, EPI_ISL_1704618, EPI_ISL_1704619, EPI_ISL_1704620, EPI_ISL_1704621, EPI_ISL_1704622, EPI_ISL_1704623, EPI_ISL_1704624, EPI_ISL_1704625, EPI_ISL_1704626, EPI_ISL_1704627, EPI_ISL_1704628, EPI_ISL_1704629, EPI_ISL_1704630, EPI_ISL_1704631, EPI_ISL_1704632, EPI_ISL_1704633, EPI_ISL_1704634, EPI_ISL_1704635, EPI_ISL_1704636, EPI_ISL_1704637, EPI_ISL_1704638, EPI_ISL_1704639, EPI_ISL_1841236, EPI_ISL_1841237, EPI_ISL_1841238, EPI_ISL_1841239, EPI_ISL_1841240, EPI_ISL_1841241, EPI_ISL_1841242, EPI_ISL_1841243, EPI_ISL_1841244, EPI_ISL_1841245, EPI_ISL_1841246, EPI_ISL_1841247, EPI_ISL_1841248, EPI_ISL_1841249, EPI_ISL_1841250, EPI_ISL_1841251, EPI_ISL_1841252, EPI_ISL_1841253, EPI_ISL_1841254, EPI_ISL_1841255, EPI_ISL_1841256, EPI_ISL_1841257, EPI_ISL_1841258, EPI_ISL_1841259, EPI_ISL_1841260, EPI_ISL_1841261, EPI_ISL_1841262, EPI_ISL_1841263, EPI_ISL_1841264, EPI_ISL_1841265, EPI_ISL_1841266, EPI_ISL_1841267, EPI_ISL_1841268, EPI_ISL_1841269, EPI_ISL_1841270, EPI_ISL_1841271, EPI_ISL_1841272, EPI_ISL_1841273, EPI_ISL_1841274, EPI_ISL_1841275, EPI_ISL_1841276, EPI_ISL_1841277, EPI_ISL_1841278, EPI_ISL_1841279, EPI_ISL_1841280, EPI_ISL_1841281, EPI_ISL_1841282, EPI_ISL_1841283, EPI_ISL_1841284, EPI_ISL_1841285, EPI_ISL_1841286, EPI_ISL_1841287, EPI_ISL_1841288, EPI_ISL_1841289, EPI_ISL_1841290, EPI_ISL_1841291, EPI_ISL_1841292, EPI_ISL_1841293, EPI_ISL_1841294, EPI_ISL_1841295, EPI_ISL_1841296, EPI_ISL_1841297, EPI_ISL_1841298, EPI_ISL_1841299, EPI_ISL_1841305, EPI_ISL_1841306, EPI_ISL_1841307, EPI_ISL_1841308, EPI_ISL_1841309, EPI_ISL_1841310, EPI_ISL_1841311, EPI_ISL_1841312, EPI_ISL_1841313, EPI_ISL_1841314, EPI_ISL_1841315, EPI_ISL_1841316, EPI_ISL_1841317, EPI_ISL_1841318, EPI_ISL_1841319, EPI_ISL_1841320, EPI_ISL_1841321, EPI_ISL_1841324, EPI_ISL_1841325, EPI_ISL_1841326, EPI_ISL_1841327, EPI_ISL_1841328, EPI_ISL_1841329, EPI_ISL_1841330, EPI_ISL_1841331, EPI_ISL_1841332, EPI_ISL_1841333, EPI_ISL_1841334, EPI_ISL_1841335, EPI_ISL_1841336, EPI_ISL_1841337, EPI_ISL_1841338, EPI_ISL_1841339, EPI_ISL_1841340, EPI_ISL_1841341, EPI_ISL_1841342, EPI_ISL_1841343, EPI_ISL_1841345, EPI_ISL_1841346, EPI_ISL_1841347, EPI_ISL_1841348, EPI_ISL_1841349, EPI_ISL_1841350, EPI_ISL_1841351, EPI_ISL_1841352, EPI_ISL_1841353, EPI_ISL_1841354, EPI_ISL_1841355, EPI_ISL_1841356, EPI_ISL_1841357, EPI_ISL_1841358, EPI_ISL_1841359, EPI_ISL_1841360, EPI_ISL_1841361, EPI_ISL_1841362, EPI_ISL_1841363, EPI_ISL_1841364, EPI_ISL_1841365, EPI_ISL_1841366, EPI_ISL_1841367, EPI_ISL_1841368, EPI_ISL_1841369, EPI_ISL_1841370, EPI_ISL_1841371, EPI_ISL_1841372, EPI_ISL_1841373, EPI_ISL_1841374, EPI_ISL_1841375, EPI_ISL_1841376, EPI_ISL_1841377, EPI_ISL_1841378, EPI_ISL_1841379, EPI_ISL_1841380, EPI_ISL_1841381, EPI_ISL_1841382, EPI_ISL_1841383, EPI_ISL_1928396, EPI_ISL_1928398, EPI_ISL_1928399, EPI_ISL_1928401, EPI_ISL_1928402, EPI_ISL_1928404, EPI_ISL_1928405, EPI_ISL_1928407, EPI_ISL_1928408, EPI_ISL_1928410, EPI_ISL_1928411, EPI_ISL_1928413, EPI_ISL_1928414, EPI_ISL_1928416, EPI_ISL_1928417, EPI_ISL_1928419, EPI_ISL_1928420, EPI_ISL_1928422, EPI_ISL_1928423, EPI_ISL_1928425, EPI_ISL_1928426, EPI_ISL_1928427, EPI_ISL_1928429, EPI_ISL_1928430, EPI_ISL_1928432, EPI_ISL_1928433, EPI_ISL_1928435, EPI_ISL_1928436, EPI_ISL_1928438, EPI_ISL_1928439, EPI_ISL_1928441, EPI_ISL_1928442, EPI_ISL_1928444, EPI_ISL_1928445, EPI_ISL_1928447, EPI_ISL_1928448, EPI_ISL_1928450, EPI_ISL_1928451, EPI_ISL_1928453, EPI_ISL_1928454, EPI_ISL_1928456, EPI_ISL_1928457, EPI_ISL_1928459, EPI_ISL_1928460, EPI_ISL_1928462, EPI_ISL_1928463, EPI_ISL_1928464, EPI_ISL_1928465, EPI_ISL_1928466, EPI_ISL_1928467, EPI_ISL_1928468, EPI_ISL_1928469, EPI_ISL_1928471, EPI_ISL_1928472, EPI_ISL_1928474, EPI_ISL_1928475, EPI_ISL_1928477, EPI_ISL_1928478, EPI_ISL_1928480, EPI_ISL_1928481, EPI_ISL_1928483, EPI_ISL_1928484, EPI_ISL_1928485, EPI_ISL_1928487, EPI_ISL_1928488, EPI_ISL_1928490, EPI_ISL_1928491, EPI_ISL_1928493, EPI_ISL_1928494, EPI_ISL_1928496, EPI_ISL_1928497, EPI_ISL_1928499, EPI_ISL_1928500, EPI_ISL_1928502, EPI_ISL_1928503, EPI_ISL_1928505, EPI_ISL_1928506, EPI_ISL_1928507, EPI_ISL_1928509, EPI_ISL_1928510, EPI_ISL_1928512, EPI_ISL_1928513, EPI_ISL_1928515, EPI_ISL_1928517, EPI_ISL_1928518, EPI_ISL_1928520, EPI_ISL_1928521, EPI_ISL_1928522, EPI_ISL_1928524, EPI_ISL_1928525, EPI_ISL_1928527, EPI_ISL_1928528, EPI_ISL_1928530, EPI_ISL_1928531, EPI_ISL_1928533, EPI_ISL_1928534, EPI_ISL_1928536, EPI_ISL_1928537, EPI_ISL_1928539, EPI_ISL_1928540, EPI_ISL_1928541, EPI_ISL_1928543, EPI_ISL_1928544, EPI_ISL_1928546, EPI_ISL_1928547, EPI_ISL_1928549, EPI_ISL_1928550, EPI_ISL_1928552, EPI_ISL_1928553, EPI_ISL_1928555, EPI_ISL_1928556, EPI_ISL_1928558, EPI_ISL_1928559, EPI_ISL_1928561, EPI_ISL_1928562, EPI_ISL_1928563, EPI_ISL_1928565, EPI_ISL_1928566, EPI_ISL_1928568, EPI_ISL_1928569, EPI_ISL_1928571, EPI_ISL_1928572, EPI_ISL_1928573, EPI_ISL_1928575, EPI_ISL_1928576, EPI_ISL_1928578, EPI_ISL_1928579, EPI_ISL_1928581, EPI_ISL_1928583, EPI_ISL_1928584, EPI_ISL_1928586, EPI_ISL_1928587, EPI_ISL_1928588, EPI_ISL_1928590, EPI_ISL_1928592, EPI_ISL_1928593, EPI_ISL_1928595, EPI_ISL_1928596, EPI_ISL_1928598, EPI_ISL_1928599, EPI_ISL_1928601, EPI_ISL_1928602, EPI_ISL_1928603, EPI_ISL_1928605, EPI_ISL_1928607, EPI_ISL_1928608, EPI_ISL_1928610, EPI_ISL_1928611, EPI_ISL_1928613, EPI_ISL_1928614, EPI_ISL_1928615, EPI_ISL_1928617, EPI_ISL_1928618, EPI_ISL_1928620, EPI_ISL_1928622, EPI_ISL_1928623, EPI_ISL_1928624, EPI_ISL_1928626, EPI_ISL_1928627, EPI_ISL_1928629, EPI_ISL_1928631, EPI_ISL_1928632, EPI_ISL_1928633, EPI_ISL_1928635, EPI_ISL_1928636, EPI_ISL_1928638, EPI_ISL_1928639, EPI_ISL_1928641, EPI_ISL_1928642, EPI_ISL_1928644, EPI_ISL_1928645, EPI_ISL_1928647, EPI_ISL_1928648, EPI_ISL_1928650, EPI_ISL_1970419, EPI_ISL_1970420, EPI_ISL_1970421, EPI_ISL_1970422, EPI_ISL_1970423, EPI_ISL_1970424, EPI_ISL_1970425, EPI_ISL_1970426, EPI_ISL_1970427, EPI_ISL_1970428, EPI_ISL_1970429, EPI_ISL_1970430, EPI_ISL_1970431, EPI_ISL_1970432, EPI_ISL_1970433, EPI_ISL_1970434, EPI_ISL_1970435, EPI_ISL_1970436, EPI_ISL_1970437, EPI_ISL_1970438, EPI_ISL_1970439, EPI_ISL_1970440, EPI_ISL_1970441, EPI_ISL_1970442, EPI_ISL_1970443, EPI_ISL_1970444, EPI_ISL_1970445, EPI_ISL_1970446, EPI_ISL_1970447, EPI_ISL_1970448, EPI_ISL_1970449, EPI_ISL_1970450, EPI_ISL_1970451, EPI_ISL_1970452, EPI_ISL_1970453, EPI_ISL_1970454, EPI_ISL_1970455, EPI_ISL_1970456, EPI_ISL_1970457, EPI_ISL_1970458, EPI_ISL_1970459, EPI_ISL_1970460, EPI_ISL_1970461, EPI_ISL_1970462, EPI_ISL_1970463, EPI_ISL_1970464, EPI_ISL_1970465, EPI_ISL_1970466, EPI_ISL_1970467, EPI_ISL_1970468, EPI_ISL_1970469, EPI_ISL_1970470, EPI_ISL_1970471, EPI_ISL_1970472, EPI_ISL_1970473, EPI_ISL_1970474, EPI_ISL_1970475, EPI_ISL_1970476, EPI_ISL_1970477, EPI_ISL_1970478, EPI_ISL_1970479, EPI_ISL_1970480, EPI_ISL_1970481, EPI_ISL_1970482, EPI_ISL_1970483, EPI_ISL_1970484, EPI_ISL_1970485, EPI_ISL_1970486, EPI_ISL_1970487, EPI_ISL_1970488, EPI_ISL_1970489, EPI_ISL_1970490, EPI_ISL_1970491, EPI_ISL_1970492, EPI_ISL_1970493, EPI_ISL_1970494, EPI_ISL_1970495, EPI_ISL_1970496, EPI_ISL_1970497, EPI_ISL_1970498, EPI_ISL_1970499, EPI_ISL_1970500, EPI_ISL_1970501, EPI_ISL_1970502, EPI_ISL_1970503, EPI_ISL_1970504, EPI_ISL_1970505, EPI_ISL_1970506, EPI_ISL_1970507, EPI_ISL_1970508, EPI_ISL_1970509, EPI_ISL_1970510, EPI_ISL_1970511, EPI_ISL_1970512, EPI_ISL_1970513, EPI_ISL_1970514, EPI_ISL_1970515, EPI_ISL_1970516, EPI_ISL_1970517, EPI_ISL_1970518, EPI_ISL_1970519, EPI_ISL_1970520, EPI_ISL_1970521, EPI_ISL_1970522, EPI_ISL_1970523, EPI_ISL_1970524, EPI_ISL_1970525, EPI_ISL_1970526, EPI_ISL_1970527, EPI_ISL_1970528, EPI_ISL_1970529, EPI_ISL_1970530, EPI_ISL_1970531, EPI_ISL_1970532, EPI_ISL_1970533, EPI_ISL_1970534, EPI_ISL_1970535, EPI_ISL_1970536, EPI_ISL_1970537, EPI_ISL_1970538, EPI_ISL_1970539, EPI_ISL_1970540, EPI_ISL_1970541, EPI_ISL_1970542, EPI_ISL_1970543, EPI_ISL_1970544, EPI_ISL_1970545, EPI_ISL_1970546, EPI_ISL_2131510, EPI_ISL_2131511, EPI_ISL_2131512, EPI_ISL_2131513, EPI_ISL_2131514, EPI_ISL_2131535, EPI_ISL_2131536, EPI_ISL_2131537, EPI_ISL_2131538, EPI_ISL_2131539, EPI_ISL_2131540, EPI_ISL_2131541, EPI_ISL_2131542, EPI_ISL_2131543, EPI_ISL_2131544, EPI_ISL_2131545, EPI_ISL_2131546, EPI_ISL_2131555, EPI_ISL_2131556, EPI_ISL_2131557, EPI_ISL_2131558, EPI_ISL_2131559, EPI_ISL_2131560, EPI_ISL_2131561, EPI_ISL_2131562, EPI_ISL_2131564, EPI_ISL_2131565 | | | |
| see above | ICMR-National Institute of Virology - INSACOG | NIV Influenza | Dr. Varsha Potdar |
| EPI_ISL_2151084, EPI_ISL_2151085, EPI_ISL_2151086, EPI_ISL_2151087, EPI_ISL_2151088, EPI_ISL_2151089, EPI_ISL_2151090, EPI_ISL_2151091, EPI_ISL_2151092, EPI_ISL_2151093, EPI_ISL_2151094, EPI_ISL_2151095, EPI_ISL_2151096, EPI_ISL_2151097, EPI_ISL_2151098, EPI_ISL_2151099, EPI_ISL_2151100, EPI_ISL_2151101, EPI_ISL_2151102, EPI_ISL_2151103, EPI_ISL_2151104, EPI_ISL_2170802 | | | |
| see above | INSACOG-Sikkim | National Institute of Biomedical Genomics – INSACOG | Arindam Maitra; Kaden Zangmu Bhutia; Nidhan Kumar Biswas; Saumitra Das; Sreedhar Chinnaswamy Tshering Ongmu Bhutia; Tara D Sharma |
| EPI_ISL_1357693, EPI_ISL_1357695, EPI_ISL_1357696, EPI_ISL_1357697, EPI_ISL_1357698, EPI_ISL_1357699, EPI_ISL_1357700, EPI_ISL_1357701, EPI_ISL_1357702, EPI_ISL_1357703, EPI_ISL_1357705, EPI_ISL_1357706 | | | |
| see above | INSACOG-WB | National Institute of Biomedical Genomics | Ajay Chakraborti; Arindam Maitra; Bhaswati Bandyopadhyay; Nidhan Kumar Biswas; Saumitra Das; Sreedhar Chinnaswamy; Tamal Ghosh |
| EPI_ISL_1419048, EPI_ISL_1419049, EPI_ISL_1419050, EPI_ISL_1419051, EPI_ISL_1419152, EPI_ISL_1419163, EPI_ISL_1419164, EPI_ISL_1419165, EPI_ISL_1419166, EPI_ISL_1419170, EPI_ISL_1419404, EPI_ISL_1419405, EPI_ISL_1419406, EPI_ISL_1419407, EPI_ISL_1419408, EPI_ISL_1419411, EPI_ISL_1419412, EPI_ISL_1419413, EPI_ISL_1419414, EPI_ISL_1419415, EPI_ISL_1419416, EPI_ISL_1419417, EPI_ISL_1419418, EPI_ISL_1419426, EPI_ISL_1419427, EPI_ISL_1419582, EPI_ISL_1419583, EPI_ISL_1419584, EPI_ISL_1419585, EPI_ISL_1419596, EPI_ISL_1419597, EPI_ISL_1419598, EPI_ISL_1419599, EPI_ISL_1419612, EPI_ISL_1419613, EPI_ISL_1419614, EPI_ISL_1419615, EPI_ISL_1419616, EPI_ISL_1419617, EPI_ISL_1419618, EPI_ISL_1419619, EPI_ISL_1419620, EPI_ISL_1419621, EPI_ISL_1419625, EPI_ISL_1419626, EPI_ISL_1419627, EPI_ISL_1419628, EPI_ISL_1419629, EPI_ISL_1419630, EPI_ISL_1419631, EPI_ISL_1419632, EPI_ISL_1419633, EPI_ISL_1419634, EPI_ISL_1419635, EPI_ISL_1419636, EPI_ISL_1419637, EPI_ISL_1419638, EPI_ISL_1419639, EPI_ISL_1419640, EPI_ISL_1419641, EPI_ISL_1419642, EPI_ISL_1419643, EPI_ISL_1419644, EPI_ISL_1419645, EPI_ISL_1419647, EPI_ISL_1419648, EPI_ISL_1419684, EPI_ISL_1419685, EPI_ISL_1419686, EPI_ISL_1419689, EPI_ISL_1419690, EPI_ISL_1419780, EPI_ISL_1419781, EPI_ISL_1419782, EPI_ISL_1419797, EPI_ISL_1419798, EPI_ISL_1419799, EPI_ISL_1419800, EPI_ISL_1419804, EPI_ISL_1419805, EPI_ISL_1419806, EPI_ISL_1419807, EPI_ISL_1419808, EPI_ISL_1419809, EPI_ISL_1419862, EPI_ISL_1419887, EPI_ISL_1419890, EPI_ISL_1419891, EPI_ISL_1419892, EPI_ISL_1419893, EPI_ISL_1419894, EPI_ISL_1419921, EPI_ISL_1419923, EPI_ISL_1589655, EPI_ISL_1589656, EPI_ISL_1589657, EPI_ISL_1589658, EPI_ISL_1589659, EPI_ISL_1589660, EPI_ISL_1589661, EPI_ISL_1589662, EPI_ISL_1589663, EPI_ISL_1589664, EPI_ISL_1589665, EPI_ISL_1589666, EPI_ISL_1589667, EPI_ISL_1589670, EPI_ISL_1589671, EPI_ISL_1589673, EPI_ISL_1589674, EPI_ISL_1589675, EPI_ISL_1589676, EPI_ISL_1589677, EPI_ISL_1589678, EPI_ISL_1589679, EPI_ISL_1589680, EPI_ISL_1589681, EPI_ISL_1589683, EPI_ISL_1589684, EPI_ISL_1589685, EPI_ISL_1589686, EPI_ISL_1589687, EPI_ISL_1589688, EPI_ISL_1589689, EPI_ISL_1589690, EPI_ISL_1589691, EPI_ISL_1589692, EPI_ISL_1589693, EPI_ISL_1589695, EPI_ISL_1589696, EPI_ISL_1589697, EPI_ISL_1589698, EPI_ISL_1589699, EPI_ISL_1589700, EPI_ISL_1589701, EPI_ISL_1589702, EPI_ISL_1589703, EPI_ISL_1589704, EPI_ISL_1589705, EPI_ISL_1589706, EPI_ISL_1589708, EPI_ISL_1589709, EPI_ISL_1589710, EPI_ISL_1589711, EPI_ISL_1589712, EPI_ISL_1589713, EPI_ISL_1589714, EPI_ISL_1589715, EPI_ISL_1589716, EPI_ISL_1589717, EPI_ISL_1589718, EPI_ISL_1589719, EPI_ISL_1589720, EPI_ISL_1589721, EPI_ISL_1589722, EPI_ISL_1589724, EPI_ISL_1589725, EPI_ISL_1589726, EPI_ISL_1589727, EPI_ISL_1589728, EPI_ISL_1589729, EPI_ISL_1589730, EPI_ISL_1589731, EPI_ISL_1589732, EPI_ISL_1589733, EPI_ISL_1589734, EPI_ISL_1589735, EPI_ISL_1589736, EPI_ISL_1589737, EPI_ISL_1589738, EPI_ISL_1589739, EPI_ISL_1589740, EPI_ISL_1589741, EPI_ISL_1589742, EPI_ISL_1589743, EPI_ISL_1589745, EPI_ISL_1589746, EPI_ISL_1589747, EPI_ISL_1589748, EPI_ISL_1589750, EPI_ISL_1589751, EPI_ISL_1589753, EPI_ISL_1589754, EPI_ISL_1589755, EPI_ISL_1589756, EPI_ISL_1589757, EPI_ISL_1589758, EPI_ISL_1589759, EPI_ISL_1589760, EPI_ISL_1589761, EPI_ISL_1589762, EPI_ISL_1589763, EPI_ISL_1589764, EPI_ISL_1589765, EPI_ISL_1589766, EPI_ISL_1589767, EPI_ISL_1589768, EPI_ISL_1589770, EPI_ISL_1589771, EPI_ISL_1589772, EPI_ISL_1589773, EPI_ISL_1589774, EPI_ISL_1589775, EPI_ISL_1589776, EPI_ISL_1589777, EPI_ISL_1589779, EPI_ISL_1589780, EPI_ISL_1589781, EPI_ISL_1589782, EPI_ISL_1589783, EPI_ISL_1589784, EPI_ISL_1589785, EPI_ISL_1589787, EPI_ISL_1589788, EPI_ISL_1589789, EPI_ISL_1589790, EPI_ISL_1589793, EPI_ISL_1589794, EPI_ISL_1589795, EPI_ISL_1589796, EPI_ISL_1589797, EPI_ISL_1589800, EPI_ISL_1589801, EPI_ISL_1589802, EPI_ISL_1589803, EPI_ISL_1589804, EPI_ISL_1589805, EPI_ISL_1589806, EPI_ISL_1589807, EPI_ISL_1589808, EPI_ISL_1589809, EPI_ISL_1589810, EPI_ISL_1589811, EPI_ISL_1589812, EPI_ISL_1589813, EPI_ISL_1589814, EPI_ISL_1589815, EPI_ISL_1589816, EPI_ISL_1589817, EPI_ISL_1589818, EPI_ISL_1589819, EPI_ISL_1589820, EPI_ISL_1589821, EPI_ISL_1589823, EPI_ISL_1589824, EPI_ISL_1589825, EPI_ISL_1589826, EPI_ISL_1589827, EPI_ISL_1589828, EPI_ISL_1589829, EPI_ISL_1589830, EPI_ISL_1589831, EPI_ISL_1589832, EPI_ISL_1589833, EPI_ISL_1589834, EPI_ISL_1589835, EPI_ISL_1589836, EPI_ISL_1589837, EPI_ISL_1589838, EPI_ISL_1589839, EPI_ISL_1589840, EPI_ISL_1589841, EPI_ISL_1589842, EPI_ISL_1589843, EPI_ISL_1589844, EPI_ISL_1589845, EPI_ISL_1589846, EPI_ISL_1589847, EPI_ISL_1589849, EPI_ISL_1589850, EPI_ISL_1589852, EPI_ISL_1589853, EPI_ISL_1589854, EPI_ISL_1589855, EPI_ISL_1589856, EPI_ISL_1589857, EPI_ISL_1589859, EPI_ISL_1589860, EPI_ISL_1589861, EPI_ISL_1589862, EPI_ISL_1589863, EPI_ISL_1589864, EPI_ISL_1589865, EPI_ISL_1589866, EPI_ISL_1589867, EPI_ISL_1589868, EPI_ISL_1589869, EPI_ISL_1589870, EPI_ISL_1589871, EPI_ISL_1589872, EPI_ISL_1589873, EPI_ISL_1589874, EPI_ISL_1589875, EPI_ISL_1589876, EPI_ISL_1589877, EPI_ISL_1589878, EPI_ISL_1589879, EPI_ISL_1589880, EPI_ISL_1589881, EPI_ISL_1589882, EPI_ISL_1589883, EPI_ISL_1589884, EPI_ISL_1589885, EPI_ISL_1589886, EPI_ISL_1589887, EPI_ISL_1589888, EPI_ISL_1589889, EPI_ISL_1589890, EPI_ISL_1589891, EPI_ISL_1589892, EPI_ISL_1589893, EPI_ISL_1589894, EPI_ISL_1589895, EPI_ISL_1589896, EPI_ISL_1589897, EPI_ISL_1589898, EPI_ISL_1589899, EPI_ISL_1589900, EPI_ISL_1589901, EPI_ISL_1589902, EPI_ISL_1589903, EPI_ISL_1589905, EPI_ISL_1589906, EPI_ISL_1589907, EPI_ISL_1589908, EPI_ISL_1589909, EPI_ISL_1589910, EPI_ISL_1589911, EPI_ISL_1589912, EPI_ISL_1589913, EPI_ISL_1589914, EPI_ISL_1589915, EPI_ISL_1589916, EPI_ISL_1589917, EPI_ISL_1589918, EPI_ISL_1589919, EPI_ISL_1589920, EPI_ISL_1589921, EPI_ISL_1589922, EPI_ISL_1589923, EPI_ISL_1589924, EPI_ISL_1589925, EPI_ISL_1589926, EPI_ISL_1589927, EPI_ISL_1589928, EPI_ISL_1589929, EPI_ISL_1589930, EPI_ISL_1589931, EPI_ISL_1589932, EPI_ISL_1589933, EPI_ISL_1589934, EPI_ISL_1589935, EPI_ISL_1589936, EPI_ISL_1589937, EPI_ISL_1589938, EPI_ISL_1589939, EPI_ISL_1589940, EPI_ISL_1589941, EPI_ISL_1589942, EPI_ISL_1589943, EPI_ISL_1589944, EPI_ISL_1589945, EPI_ISL_1589946, EPI_ISL_1589947, EPI_ISL_1589948, EPI_ISL_1589949, EPI_ISL_1589950, EPI_ISL_1589951, EPI_ISL_1589955, EPI_ISL_1589956, EPI_ISL_1589957, EPI_ISL_1589958, EPI_ISL_1589959, EPI_ISL_1589960, EPI_ISL_1589961, EPI_ISL_1589962, EPI_ISL_1589963, EPI_ISL_1589964, EPI_ISL_1589965, EPI_ISL_1589966, EPI_ISL_1589968, EPI_ISL_1589969, EPI_ISL_1589970, EPI_ISL_1589971, EPI_ISL_1589972, EPI_ISL_1589973, EPI_ISL_2132706, EPI_ISL_2132707, EPI_ISL_2132708, EPI_ISL_2132713, EPI_ISL_2132714, EPI_ISL_2132715, EPI_ISL_2132716, EPI_ISL_2132717, EPI_ISL_2132718, EPI_ISL_2132719, EPI_ISL_2132720, EPI_ISL_2132721 | | | |
| see above | INSACOG-WB | National Institute of Biomedical Genomics – INSACOG | Ajay Chakraborti; Arindam Maitra; Bhaswati Bandyopadhyay; Nidhan Kumar Biswas; Saumitra Das; Sreedhar Chinnaswamy; Tamal Ghosh |
| EPI_ISL_1663354, EPI_ISL_1663355 | ITKI Arogyashala | Institute of Life Sciences - INSACOG | Ajay Parida; Amol M. Kanampalliwar; Arup Ghosh; Atimukta Jha; INSACOG Consortium; Punit Prasad; Rajeeb Swain; Rupesh Dash; Safal Walia; Shifu Aggarwal; Sunil K. Raghav |
| EPI_ISL_1662445, EPI_ISL_1662446, EPI_ISL_1662447, EPI_ISL_1662448, EPI_ISL_1662449, EPI_ISL_1662450, EPI_ISL_1662451, EPI_ISL_1662452, EPI_ISL_1662453, EPI_ISL_1662454, EPI_ISL_1662455, EPI_ISL_1662456, EPI_ISL_1662457, EPI_ISL_1662458, EPI_ISL_1662459, EPI_ISL_1662460, EPI_ISL_1662461, EPI_ISL_1662462, EPI_ISL_1662463, EPI_ISL_1663552, EPI_ISL_1663553, EPI_ISL_1663554, EPI_ISL_1663555, EPI_ISL_1663557, EPI_ISL_1663558, EPI_ISL_1663559, EPI_ISL_1663560, EPI_ISL_1663561, EPI_ISL_1663562, EPI_ISL_1663563, EPI_ISL_1663564 | | | |
| see above | Immunogenomics lab, Institute of Life Sciences, Bhubaneswar | Institute of Life Sciences - INSACOG | Ajay Parida; Amol M. Kanampalliwar; Arup Ghosh; Atimukta Jha; INSACOG Consortium; Punit Prasad; Rajeeb Swain; Rupesh Dash; Safal Walia; Shifu Aggarwal; Sunil K. Raghav |
| EPI_ISL_2036281, EPI_ISL_2036282, EPI_ISL_2036283, EPI_ISL_2036284, EPI_ISL_2036285, EPI_ISL_2036286 | Indian Council of Medical Research-National Institute of Virology, Microbial Containment Complex | Indian Council of Medical Research-National Institute of Virology, Microbial Containment Complex | Pragya D Yadav |
| EPI_ISL_1372094 | Ispat General Hospital (IGH), Rourkela | Institute of Life Sciences - INSACOG | Ajay Parida; Amol M. Kanampalliwar; Arup Ghosh; Atimukta Jha; INSACOG Consortium; Punit Prasad; Rajeeb Swain; Rupesh Dash; Safal Walia; Shifu Aggarwal; Sunil K. Raghav |
| EPI_ISL_1939845, EPI_ISL_1939846, EPI_ISL_1939847, EPI_ISL_1939848, EPI_ISL_1939863, EPI_ISL_1939864, EPI_ISL_1939865, EPI_ISL_1939866, EPI_ISL_1939867, EPI_ISL_1939868, EPI_ISL_1939869, EPI_ISL_1939870, EPI_ISL_1939871, EPI_ISL_1939872, EPI_ISL_1939873, EPI_ISL_1939874, EPI_ISL_1939929, EPI_ISL_1939930, EPI_ISL_1939931, EPI_ISL_1939932, EPI_ISL_1939933, EPI_ISL_1939934, EPI_ISL_1939935, EPI_ISL_1939936, EPI_ISL_1939937, EPI_ISL_1939938, EPI_ISL_1939939, EPI_ISL_1939940, EPI_ISL_1939941, EPI_ISL_1939942, EPI_ISL_1939943, EPI_ISL_1939944, EPI_ISL_1939945, EPI_ISL_1939946, EPI_ISL_1939947, EPI_ISL_1939948, EPI_ISL_1939949, EPI_ISL_1939950, EPI_ISL_1939951, EPI_ISL_1939952, EPI_ISL_1939953, EPI_ISL_1939954, EPI_ISL_1939955, EPI_ISL_1939956, EPI_ISL_1939957, EPI_ISL_1939958, EPI_ISL_1939959, EPI_ISL_1939960, EPI_ISL_1939961, EPI_ISL_1939962, EPI_ISL_1939963, EPI_ISL_1939964, EPI_ISL_1939965, EPI_ISL_1939966, EPI_ISL_1939967, EPI_ISL_1939968 | | | |
| see above | JIPMER Puducherry | inStem NCBS - INSACOG | Uma Ramakrishnan Dasaradhi Palakodeti Aswin SaiNarain |
| EPI_ISL_1818630 | Jayanagar General Hospital | INSACOG-KA, NIMHANS | Ananthapadmanabha Kotambail; Anita S Desai; Anson Kunjumon George; Chetan G K; Chitra Pattabiraman; Darshan Sreenivas; Gautham Arunachal Udupi; Pramada Prasad; V Ravi |
| EPI_ISL_1595857, EPI_ISL_1595858, EPI_ISL_1595859, EPI_ISL_1595891, EPI_ISL_1595892, EPI_ISL_1595893, EPI_ISL_1595895, EPI_ISL_1595896, EPI_ISL_1595897, EPI_ISL_1595898, EPI_ISL_1595899, EPI_ISL_1595900, EPI_ISL_1818583, EPI_ISL_1818584, EPI_ISL_1818585, EPI_ISL_1818587, EPI_ISL_1818588, EPI_ISL_1818589, EPI_ISL_1818614, EPI_ISL_1818615, EPI_ISL_1818616, EPI_ISL_1818617, EPI_ISL_1818618, EPI_ISL_1818622 | | | |
| see above | KEMPEGOWDA INTERNATIONAL AIRPORT | INSACOG-KA, NIMHANS | Ananthapadmanabha Kotambail; Anita S Desai; Anson Kunjumon George; Chetan G K; Chitra Pattabiraman; Darshan Sreenivas; Gautham Arunachal Udupi; Pramada Prasad; V Ravi |
| EPI_ISL_1940100, EPI_ISL_2015555, EPI_ISL_2015633 | KEMPEGOWDA INTERNATIONAL AIRPORT | inStem NCBS - INSACOG | Uma Ramakrishnan Dasaradhi Palakodeti Aswin SaiNarain |
| EPI_ISL_1595914 | KIMS, KARWAR | INSACOG-KA, NIMHANS | Ananthapadmanabha Kotambail; Anita S Desai; Anson Kunjumon George; Chetan G K; Chitra Pattabiraman; Darshan Sreenivas; Gautham Arunachal Udupi; Pramada Prasad; V Ravi |
| EPI_ISL_1663539, EPI_ISL_1663540, EPI_ISL_1663541, EPI_ISL_1663542, EPI_ISL_1663543, EPI_ISL_1663544, EPI_ISL_1663545, EPI_ISL_1663546, EPI_ISL_1663547 | | | |
| see above | MGM Medical College, Jamshedpur | Institute of Life Sciences - INSACOG | Ajay Parida; Amol M. Kanampalliwar; Arup Ghosh; Atimukta Jha; INSACOG Consortium; Punit Prasad; Rajeeb Swain; Rupesh Dash; Safal Walia; Shifu Aggarwal; Sunil K. Raghav |
| EPI_ISL_1372269, EPI_ISL_1372270, EPI_ISL_1372272, EPI_ISL_1372274, EPI_ISL_1372276, EPI_ISL_1372278, EPI_ISL_1547801, EPI_ISL_2157167 | | | |
| see above | Mapmygenome | CSIR-Centre for Cellular and Molecular Biology - INSACOG | Amareshwar Vodapalli; Ara Sreenivas; B Himasri; Divya Tej Sowpati; Karthik Bharadwaj Tallapaka; Lamuk Zaveri; Onkar Kulkarni; Rakesh K Mishra; Sharath Chandra Thota; Shreekant Verma; Sofia Banu; Viswagithe S L |
| EPI_ISL_2156828, EPI_ISL_2156829, EPI_ISL_2156915, EPI_ISL_2156916, EPI_ISL_2156917, EPI_ISL_2156918, EPI_ISL_2156919, EPI_ISL_2156920, EPI_ISL_2156921, EPI_ISL_2156922, EPI_ISL_2156923, EPI_ISL_2156924, EPI_ISL_2156925, EPI_ISL_2156926, EPI_ISL_2156927, EPI_ISL_2156928, EPI_ISL_2156929, EPI_ISL_2156930, EPI_ISL_2156931, EPI_ISL_2156932, EPI_ISL_2156933, EPI_ISL_2156934, EPI_ISL_2156935, EPI_ISL_2156936, EPI_ISL_2156937, EPI_ISL_2156970, EPI_ISL_2156971, EPI_ISL_2156972, EPI_ISL_2156973, EPI_ISL_2156974, EPI_ISL_2156975, EPI_ISL_2156976, EPI_ISL_2156977, EPI_ISL_2156978, EPI_ISL_2156979, EPI_ISL_2156980, EPI_ISL_2156981, EPI_ISL_2156982, EPI_ISL_2157002, EPI_ISL_2157003, EPI_ISL_2157004, EPI_ISL_2157005, EPI_ISL_2157006, EPI_ISL_2157007, EPI_ISL_2157008, EPI_ISL_2157009, EPI_ISL_2157019, EPI_ISL_2157020, EPI_ISL_2157021, EPI_ISL_2157022, EPI_ISL_2157023, EPI_ISL_2157024, EPI_ISL_2157025, EPI_ISL_2157026, EPI_ISL_2157027, EPI_ISL_2157028, EPI_ISL_2157029, EPI_ISL_2157030, EPI_ISL_2157031, EPI_ISL_2157032, EPI_ISL_2157034, EPI_ISL_2157035, EPI_ISL_2157071, EPI_ISL_2157072, EPI_ISL_2157073, EPI_ISL_2157074, EPI_ISL_2157075, EPI_ISL_2157086, EPI_ISL_2157087, EPI_ISL_2157088, EPI_ISL_2157089, EPI_ISL_2157090, EPI_ISL_2157132, EPI_ISL_2157133, EPI_ISL_2157134, EPI_ISL_2157135, EPI_ISL_2157136, EPI_ISL_2157137, EPI_ISL_2157138, EPI_ISL_2157139, EPI_ISL_2157140, EPI_ISL_2157141, EPI_ISL_2157142, EPI_ISL_2157143, EPI_ISL_2157144, EPI_ISL_2157145, EPI_ISL_2157146, EPI_ISL_2157147, EPI_ISL_2157148, EPI_ISL_2157149, EPI_ISL_2157150, EPI_ISL_2157151, EPI_ISL_2157152, EPI_ISL_2157153, EPI_ISL_2157157, EPI_ISL_2157158, EPI_ISL_2157159, EPI_ISL_2157160, EPI_ISL_2157161, EPI_ISL_2157162, EPI_ISL_2157163, EPI_ISL_2157164, EPI_ISL_2157165 | | | |
| see above | Mapmygenome | CSIR-Centre for Cellular and Molecular Biology- INSACOG | Amareshwar Vodapalli; Ara Sreenivas; Archana Bharadwaj Siva; B Himasri; Blessy B John; Divya Tej Sowpati; Karthik Bharadwaj Tallapaka; Lamuk Zaveri; Payel Mukherjee; Rakesh K Mishra; Sharath Chandra Thota; Shreekant Verma; Sofia Banu; Tulasi Nagabandi; Valli Nagalakshmi Undamatla; Viswagithe S L |
| EPI_ISL_2017759, EPI_ISL_2017760, EPI_ISL_2017761, EPI_ISL_2017762 | N.H.L Municipal Medical College, Ahmedabad | Gujarat Biotechnology Research Centre | Chaitanya Joshi; Dinesh Kumar; Janvi Raval; Jayshri Pethani; Madhvi Joshi; Nitesh Shah; Nitin Savaliya; Ramesh Pandit; Sonal Sharma; Twinkle Soni; Umang Mishra; Zarna Patel; Zuber Saiyed |
| EPI_ISL_1939875, EPI_ISL_1939876, EPI_ISL_1939877, EPI_ISL_1939878, EPI_ISL_1939879, EPI_ISL_1939880, EPI_ISL_1939881, EPI_ISL_1939882, EPI_ISL_1939883, EPI_ISL_1939884, EPI_ISL_1939885, EPI_ISL_1939886, EPI_ISL_1939887, EPI_ISL_1939888, EPI_ISL_1939889, EPI_ISL_1939890, EPI_ISL_1939891, EPI_ISL_1939892, EPI_ISL_1939893, EPI_ISL_1939894, EPI_ISL_1939895, EPI_ISL_1939896, EPI_ISL_1939897, EPI_ISL_1939898, EPI_ISL_1939899, EPI_ISL_1939900, EPI_ISL_1939901, EPI_ISL_1939902, EPI_ISL_1939903, EPI_ISL_1939904, EPI_ISL_1939905, EPI_ISL_1939906, EPI_ISL_1939907, EPI_ISL_1939908, EPI_ISL_1939909, EPI_ISL_1939910, EPI_ISL_1939911, EPI_ISL_1939912, EPI_ISL_1939913, EPI_ISL_1939914, EPI_ISL_1939915, EPI_ISL_1939916, EPI_ISL_1939917, EPI_ISL_1939918, EPI_ISL_1939919, EPI_ISL_1939920, EPI_ISL_1939921, EPI_ISL_1939922, EPI_ISL_1939923, EPI_ISL_1939924, EPI_ISL_1939925, EPI_ISL_1939926, EPI_ISL_1939927, EPI_ISL_1939928 | | | |
| see above | NCCS | inStem NCBS - INSACOG | Uma Ramakrishnan Dasaradhi Palakodeti Aswin SaiNarain |
| EPI_ISL_1663363, EPI_ISL_1663364, EPI_ISL_1663365, EPI_ISL_1663366, EPI_ISL_1663367, EPI_ISL_1663368, EPI_ISL_1663369, EPI_ISL_1663370, EPI_ISL_1663371, EPI_ISL_1663372, EPI_ISL_1663373, EPI_ISL_1663374, EPI_ISL_1663375, EPI_ISL_1663376, EPI_ISL_1663377, EPI_ISL_1663378, EPI_ISL_1663379, EPI_ISL_1663380, EPI_ISL_1663381, EPI_ISL_1663382, EPI_ISL_1663383, EPI_ISL_1663384, EPI_ISL_1663385, EPI_ISL_1663386, EPI_ISL_1663387, EPI_ISL_1663388, EPI_ISL_1663389, EPI_ISL_1663390, EPI_ISL_1663391, EPI_ISL_1663392, EPI_ISL_1663393, EPI_ISL_1663394, EPI_ISL_1663395, EPI_ISL_1663396, EPI_ISL_1663397, EPI_ISL_1663398, EPI_ISL_1663399, EPI_ISL_1663400, EPI_ISL_1663401, EPI_ISL_1663402, EPI_ISL_1663403, EPI_ISL_1663405, EPI_ISL_1663406, EPI_ISL_1663407, EPI_ISL_1663408, EPI_ISL_1663409, EPI_ISL_1663410, EPI_ISL_1663411, EPI_ISL_1663412, EPI_ISL_1663413, EPI_ISL_1663414, EPI_ISL_1663415, EPI_ISL_1663416, EPI_ISL_1663417, EPI_ISL_1663418, EPI_ISL_1663419, EPI_ISL_1663420, EPI_ISL_1663421, EPI_ISL_1663422, EPI_ISL_1663423, EPI_ISL_1663424, EPI_ISL_1663425, EPI_ISL_1663426, EPI_ISL_1663427, EPI_ISL_1663428, EPI_ISL_1663429, EPI_ISL_1663430, EPI_ISL_1663431, EPI_ISL_1663432, EPI_ISL_1663433, EPI_ISL_1663434, EPI_ISL_1663435, EPI_ISL_1663436, EPI_ISL_1663437, EPI_ISL_1663438, EPI_ISL_1663439, EPI_ISL_1663440, EPI_ISL_1663441, EPI_ISL_1663442, EPI_ISL_1663443, EPI_ISL_1663444, EPI_ISL_1663445, EPI_ISL_1663446, EPI_ISL_1663447, EPI_ISL_1663448, EPI_ISL_1663449, EPI_ISL_1663450, EPI_ISL_1663451, EPI_ISL_1663452, EPI_ISL_1663453, EPI_ISL_1663454, EPI_ISL_1663455, EPI_ISL_1663456, EPI_ISL_1663457, EPI_ISL_1663458, EPI_ISL_1663459, EPI_ISL_1663460, EPI_ISL_1663461, EPI_ISL_1663462, EPI_ISL_1663463, EPI_ISL_1663464, EPI_ISL_1663465, EPI_ISL_1663466, EPI_ISL_1663467, EPI_ISL_1663468, EPI_ISL_1663469, EPI_ISL_1663470, EPI_ISL_1663471, EPI_ISL_1663472, EPI_ISL_1663473, | | | |

| | | | |
|---|---|---|---|
| EPI_ISL_1663474, EPI_ISL_1663475 | | | |
| see above | NCCS, Pune | Institute of Life Sciences - INSACOG | Ajay Parida; Amol M. Kanampalliwar; Arup Ghosh; Atimukta Jha; INSACOG Consortium; Punit Prasad; Rajeeb Swain; Rupesh Dash; Safal Walia; Shifu Aggarwal; Sunil K. Raghav |
| EPI_ISL_1939985, EPI_ISL_1939987, EPI_ISL_1939988, EPI_ISL_1939989, EPI_ISL_1939999, EPI_ISL_1940029, EPI_ISL_1940035, EPI_ISL_1940053, EPI_ISL_1940054, EPI_ISL_1940055, EPI_ISL_1940056, EPI_ISL_1940057, EPI_ISL_1940058, EPI_ISL_1940069, EPI_ISL_1940070, EPI_ISL_1940095, EPI_ISL_1940096, EPI_ISL_1940097, EPI_ISL_1940098, EPI_ISL_1940099, EPI_ISL_1940107, EPI_ISL_1940108, EPI_ISL_1940133, EPI_ISL_1940134, EPI_ISL_1940135, EPI_ISL_1940136, EPI_ISL_1940137, EPI_ISL_1940138, EPI_ISL_1940139, EPI_ISL_1940140, EPI_ISL_1940141, EPI_ISL_2015475, EPI_ISL_2015478, EPI_ISL_2015486, EPI_ISL_2015492, EPI_ISL_2015497, EPI_ISL_2015542, EPI_ISL_2015546, EPI_ISL_2015564, EPI_ISL_2015576, EPI_ISL_2015578, EPI_ISL_2015580, EPI_ISL_2015588, EPI_ISL_2015594, EPI_ISL_2015603, EPI_ISL_2015604, EPI_ISL_2015621, EPI_ISL_2015628, EPI_ISL_2015631 | | | |
| see above | NIMHANS | inStem NCBS - INSACOG | Uma Ramakrishnan Dasaradhi Palakodeti Aswin SaiNarain |
| EPI_ISL_1662277, EPI_ISL_1662278, EPI_ISL_1662279, EPI_ISL_1662280, EPI_ISL_1662281 | Nalanda Medical College & Hospital, Patna | Institute of Life Sciences - INSACOG | Ajay Parida; Amol M. Kanampalliwar; Arup Ghosh; Atimukta Jha; INSACOG Consortium; Punit Prasad; Rajeeb Swain; Rupesh Dash; Safal Walia; Shifu Aggarwal; Sunil K. Raghav |
| EPI_ISL_1372095 | Nalanda Medical College Hospital, Patna | Institute of Life Sciences - INSACOG | Ajay Parida; Amol M. Kanampalliwar; Arup Ghosh; Atimukta Jha; INSACOG Consortium; Punit Prasad; Rajeeb Swain; Rupesh Dash; Safal Walia; Shifu Aggarwal; Sunil K. Raghav |
| EPI_ISL_1415094, EPI_ISL_1415186, EPI_ISL_1540456, EPI_ISL_1543988, EPI_ISL_1543989, EPI_ISL_1544057, EPI_ISL_1544058, EPI_ISL_1544059, EPI_ISL_1544060, EPI_ISL_1544061, EPI_ISL_1544062, EPI_ISL_1544063, EPI_ISL_1544070, EPI_ISL_1544071, EPI_ISL_1544072, EPI_ISL_1544073, EPI_ISL_1544074, EPI_ISL_2162386, EPI_ISL_2162387, EPI_ISL_2162388, EPI_ISL_2162389, EPI_ISL_2162390, EPI_ISL_2162391, EPI_ISL_2162392, EPI_ISL_2162393, EPI_ISL_2162394, EPI_ISL_2162395, EPI_ISL_2162396, EPI_ISL_2162397, EPI_ISL_2162398, EPI_ISL_2162399, EPI_ISL_2162406, EPI_ISL_2162407, EPI_ISL_2162411, EPI_ISL_2162412, EPI_ISL_2162413, EPI_ISL_2162414, EPI_ISL_2162415, EPI_ISL_2162416, EPI_ISL_2162417, EPI_ISL_2162418, EPI_ISL_2162419, EPI_ISL_2162420, EPI_ISL_2162421, EPI_ISL_2162422, EPI_ISL_2162423, EPI_ISL_2162424, EPI_ISL_2162425, EPI_ISL_2162426, EPI_ISL_2162427, EPI_ISL_2162428, EPI_ISL_2162429, EPI_ISL_2162430, EPI_ISL_2162431, EPI_ISL_2162432, EPI_ISL_2162433, EPI_ISL_2162434, EPI_ISL_2162435, EPI_ISL_2162436, EPI_ISL_2162437, EPI_ISL_2162438, EPI_ISL_2162439, EPI_ISL_2162440, EPI_ISL_2162441, EPI_ISL_2162442, EPI_ISL_2162443, EPI_ISL_2162444, EPI_ISL_2162445, EPI_ISL_2162446, EPI_ISL_2162447, EPI_ISL_2162448, EPI_ISL_2162449, EPI_ISL_2162450, EPI_ISL_2162451, EPI_ISL_2162452, EPI_ISL_2162453, EPI_ISL_2162454, EPI_ISL_2162455, EPI_ISL_2162456, EPI_ISL_2162457, EPI_ISL_2162458, EPI_ISL_2162459, EPI_ISL_2162460, EPI_ISL_2162461, EPI_ISL_2162462, EPI_ISL_2162463, EPI_ISL_2162464, EPI_ISL_2162465 | | | |
| see above | National Centre For Cell Science | National Centre For Cell Science – INSACOG | Ajay Pillai; Dhiraj Paul; INSACOG Consortium team; Manoj Kumar Bhat; Mitali Inamdar; Mohak P Gujare; Shivang P. Bhanushali; Sonal Manik Chavan; Yogesh Shouche; Yogesh Shouche. |
| EPI_ISL_2107008, EPI_ISL_2107009, EPI_ISL_2107010, EPI_ISL_2107011, EPI_ISL_2107012, EPI_ISL_2107013, EPI_ISL_2107014, EPI_ISL_2107015, EPI_ISL_2107016, EPI_ISL_2107017, EPI_ISL_2107018, EPI_ISL_2107019, EPI_ISL_2107020, EPI_ISL_2107021, EPI_ISL_2107022, EPI_ISL_2107023, EPI_ISL_2107024, EPI_ISL_2107025, EPI_ISL_2107026, EPI_ISL_2107027, EPI_ISL_2107028, EPI_ISL_2107029, EPI_ISL_2107030, EPI_ISL_2107031, EPI_ISL_2107032, EPI_ISL_2107033, EPI_ISL_2107034, EPI_ISL_2107035, EPI_ISL_2107037, EPI_ISL_2107043, EPI_ISL_2107044, EPI_ISL_2107046, EPI_ISL_2107047, EPI_ISL_2107048, EPI_ISL_2107049, EPI_ISL_2107051, EPI_ISL_2107052, EPI_ISL_2107053, EPI_ISL_2107054, EPI_ISL_2107055, EPI_ISL_2107056, EPI_ISL_2107057, EPI_ISL_2107058, EPI_ISL_2107059, EPI_ISL_2107060, EPI_ISL_2107061, EPI_ISL_2107062, EPI_ISL_2107063, EPI_ISL_2107064, EPI_ISL_2107065, EPI_ISL_2107066, EPI_ISL_2107067, EPI_ISL_2107068, EPI_ISL_2107069, EPI_ISL_2107070, EPI_ISL_2107071, EPI_ISL_2107072, EPI_ISL_2107073, EPI_ISL_2107074, EPI_ISL_2107075, EPI_ISL_2107076, EPI_ISL_2107077, EPI_ISL_2107078, EPI_ISL_2107079, EPI_ISL_2107080, EPI_ISL_2107081, EPI_ISL_2107082, EPI_ISL_2107083, EPI_ISL_2107084, EPI_ISL_2107085, EPI_ISL_2107086, EPI_ISL_2107087, EPI_ISL_2107088, EPI_ISL_2107089, EPI_ISL_2107090 | | | |
| see above | National Centre For Cell Science – INSACOG | National Centre For Cell Science | Ajay Pillai; Dhiraj Paul; INSACOG Consortium team; Manoj Kumar Bhat; Mitali Inamdar; Mohak P Gujare; Shivang P. Bhanushali; Yogesh Shouche |
| EPI_ISL_2099674, EPI_ISL_2099675, EPI_ISL_2099676, EPI_ISL_2099677 | National Centre for Disease Control | CDFD -INSACOG | To be addded later |
| EPI_ISL_1595856, EPI_ISL_1595889, EPI_ISL_1595917, EPI_ISL_1818590, EPI_ISL_1818596, EPI_ISL_1818597, EPI_ISL_1818598, EPI_ISL_1818599, EPI_ISL_1818600, EPI_ISL_1818601, EPI_ISL_1818602, EPI_ISL_1818607, EPI_ISL_1818608, EPI_ISL_1818629, EPI_ISL_1818639, EPI_ISL_1818640, EPI_ISL_1818641, EPI_ISL_1818642, EPI_ISL_1818643, EPI_ISL_1818644 | | | |
| see above | National Institute of Mental Health and Neurosciences (NIMHANS) | INSACOG-KA, NIMHANS | Ananthapadmanabha Kotambail; Anita S Desai; Anson Kunjumon George; Chetan G K; Chitra Pattabiraman; Darshan Sreenivas; Gautham Arunachal Udupi; Pramada Prasad; V Ravi |
| EPI_ISL_1663333, EPI_ISL_1663334, EPI_ISL_1663335, EPI_ISL_1663336, EPI_ISL_1663337 | PRMMCCH, Baripada | Institute of Life Sciences - INSACOG | Ajay Parida; Amol M. Kanampalliwar; Arup Ghosh; Atimukta Jha; INSACOG Consortium; Punit Prasad; Rajeeb Swain; Rupesh Dash; Safal Walia; Shifu Aggarwal; Sunil K. Raghav |
| EPI_ISL_1939992, EPI_ISL_1939993, EPI_ISL_1939994, EPI_ISL_1939995, EPI_ISL_1939996, EPI_ISL_1939997, EPI_ISL_1939998, EPI_ISL_1940030, EPI_ISL_1940031, EPI_ISL_1940032, EPI_ISL_1940033, EPI_ISL_1940034, EPI_ISL_1940102, EPI_ISL_1940103, EPI_ISL_1940104, EPI_ISL_1940105, EPI_ISL_1940106, EPI_ISL_2015511, EPI_ISL_2015548, EPI_ISL_2015558, EPI_ISL_2015605, EPI_ISL_2015620, EPI_ISL_2015622, EPI_ISL_2015623, EPI_ISL_2015624, EPI_ISL_2015625, EPI_ISL_2015626, EPI_ISL_2015629, EPI_ISL_2015630, EPI_ISL_2015632, EPI_ISL_2015635, EPI_ISL_2015638 | | | |
| see above | RAILWAY | inStem NCBS - INSACOG | Uma Ramakrishnan Dasaradhi Palakodeti Aswin SaiNarain |
| EPI_ISL_1818591, EPI_ISL_1818592, EPI_ISL_1818593, EPI_ISL_1818594, EPI_ISL_1818606, EPI_ISL_1818619, EPI_ISL_1818620, EPI_ISL_1818621, EPI_ISL_1818623, EPI_ISL_1818645, EPI_ISL_1818646, EPI_ISL_1818647, EPI_ISL_1818648, EPI_ISL_1818649 | | | |
| see above | RAILWAY H | INSACOG-KA, NIMHANS | Ananthapadmanabha Kotambail; Anita S Desai; Anson Kunjumon George; Chetan G K; Chitra Pattabiraman; Darshan Sreenivas; Gautham Arunachal Udupi; Pramada Prasad; V Ravi |
| EPI_ISL_1940059, EPI_ISL_1940060, EPI_ISL_1940061, EPI_ISL_2015490, EPI_ISL_2015512, EPI_ISL_2015522, EPI_ISL_2015529, EPI_ISL_2015530, EPI_ISL_2015550, EPI_ISL_2015571, EPI_ISL_2015587, EPI_ISL_2015591, EPI_ISL_2015599, EPI_ISL_2015609 | | | |
| see above | RAILWAY H | inStem NCBS - INSACOG | Uma Ramakrishnan Dasaradhi Palakodeti Aswin SaiNarain |
| EPI_ISL_2015515, EPI_ISL_2015518, EPI_ISL_2015615 | RAILWAY HOSPITAL | inStem NCBS - INSACOG | Uma Ramakrishnan Dasaradhi Palakodeti Aswin SaiNarain |
| EPI_ISL_1663524, EPI_ISL_1663525, EPI_ISL_1663526, EPI_ISL_1663527, EPI_ISL_1663528, EPI_ISL_1663529, EPI_ISL_1663530, EPI_ISL_1663531, EPI_ISL_1663532, EPI_ISL_1663533, EPI_ISL_1663534, EPI_ISL_1663535 | | | |
| see above | REGIONAL VRDL, ICMR-RMRC BBSR | Institute of Life Sciences - INSACOG | Ajay Parida; Amol M. Kanampalliwar; Arup Ghosh; Atimukta Jha; INSACOG Consortium; Punit Prasad; Rajeeb Swain; Rupesh Dash; Safal Walia; Shifu Aggarwal; Sunil K. Raghav |
| EPI_ISL_1663339, EPI_ISL_1663340, EPI_ISL_1663343, EPI_ISL_1663344, EPI_ISL_1663346, EPI_ISL_1663347, EPI_ISL_1663350, EPI_ISL_1663351, EPI_ISL_1663352 | | | |
| see above | RIMS, Ranchi | Institute of Life Sciences - INSACOG | Ajay Parida; Amol M. Kanampalliwar; Arup Ghosh; Atimukta Jha; INSACOG Consortium; Punit Prasad; Rajeeb Swain; Rupesh Dash; Safal Walia; Shifu Aggarwal; Sunil K. Raghav |
| EPI_ISL_1663358, EPI_ISL_1663359, EPI_ISL_1663360, EPI_ISL_1663361, EPI_ISL_1663362 | RMRI, Patna | Institute of Life Sciences - INSACOG | Ajay Parida; Amol M. Kanampalliwar; Arup Ghosh; Atimukta Jha; INSACOG Consortium; Punit Prasad; Rajeeb Swain; Rupesh Dash; Safal Walia; Shifu Aggarwal; Sunil K. Raghav |
| EPI_ISL_2015627 | Railway | inStem NCBS - INSACOG | Uma Ramakrishnan Dasaradhi Palakodeti Aswin SaiNarain |
| EPI_ISL_1662422, EPI_ISL_1662423 | S.C.B. Medical College and Hospital, Cuttack | Institute of Life Sciences - INSACOG | Ajay Parida; Amol M. Kanampalliwar; Arup Ghosh; Atimukta Jha; INSACOG Consortium; Punit Prasad; Rajeeb Swain; Rupesh Dash; Safal Walia; Shifu Aggarwal; Sunil K. Raghav |
| EPI_ISL_1939990, EPI_ISL_1939991, EPI_ISL_1940101, EPI_ISL_2015640 | SANJAY GANDHI | inStem NCBS - INSACOG | Uma Ramakrishnan Dasaradhi Palakodeti Aswin SaiNarain |
| EPI_ISL_1580503 | SMS Medical College, Jaipur | S.M.S. Medical College | ; Bharti Malhotra; Himanshu Sharma; Pratibha Mishra; Swati Gautam |
| EPI_ISL_1595864, EPI_ISL_1595913, EPI_ISL_1818650 | Sri Jayadeva Institute of Cardiovascular Sciences and Research | INSACOG-KA, NIMHANS | Ananthapadmanabha Kotambail; Anita S Desai; Anson Kunjumon George; Chetan G K; Chitra Pattabiraman; Darshan Sreenivas; Gautham Arunachal Udupi; Pramada Prasad; V Ravi |
| EPI_ISL_1731751, EPI_ISL_1731752, EPI_ISL_1731753, EPI_ISL_1731754, EPI_ISL_1731755, EPI_ISL_1731756, EPI_ISL_1731757, EPI_ISL_1914579, EPI_ISL_1914580, EPI_ISL_1914581, EPI_ISL_1914582, EPI_ISL_1914583, EPI_ISL_1914584, EPI_ISL_1914585, EPI_ISL_1914586, EPI_ISL_1914587, EPI_ISL_1914588, EPI_ISL_1914589, EPI_ISL_1914590, EPI_ISL_1914592, EPI_ISL_1914593, EPI_ISL_1914594, EPI_ISL_1914595, EPI_ISL_1914596, EPI_ISL_1914597, EPI_ISL_1914598, EPI_ISL_1969752, EPI_ISL_1969753, EPI_ISL_2098710, EPI_ISL_2098711, EPI_ISL_2098712, EPI_ISL_2098713, EPI_ISL_2098714, EPI_ISL_2098715, EPI_ISL_2098716, EPI_ISL_2098717, EPI_ISL_2098718, EPI_ISL_2098719, EPI_ISL_2098720, EPI_ISL_2098721, EPI_ISL_2098722 | | | |
| see above | State Virus Research and Diagnostic Laboratory (VRDL), AIIMS Raipur | State Virus Research and Diagnostic Laboratory (VRDL), AIIMS Raipur | Anudita Bhargava; Jiyan Chandrawanshi; Kuldeep Sharma; MD Rafiiqullah Khan; Priyanka Singh; Pushpendra Singh; Sanjay Singh Negi; Somya Sharma |
| EPI_ISL_2023383 | The Foundation for Medical Research | The Foundation for Medical Research | Ambreen Shaikh; Ayan Mandal; Grishma Patel; Kayzad Nilgiriwala; Nerges Mistry; Smriti Vaswani; Swapneil Parikh |
| EPI_ISL_1837988, EPI_ISL_1837991, EPI_ISL_1837993, EPI_ISL_1837996, EPI_ISL_1837998, EPI_ISL_1838001, EPI_ISL_1838004, EPI_ISL_1838007, EPI_ISL_1838010, EPI_ISL_1838012, EPI_ISL_1838015, EPI_ISL_1838018, EPI_ISL_1838021, EPI_ISL_1838023, EPI_ISL_1838026, EPI_ISL_1838029, EPI_ISL_1838032, EPI_ISL_1838035, EPI_ISL_1838037, EPI_ISL_1838040, EPI_ISL_1838042, EPI_ISL_1838045, EPI_ISL_1838047, EPI_ISL_1838050, EPI_ISL_1838052, EPI_ISL_1838055, EPI_ISL_1838057, EPI_ISL_1838060, EPI_ISL_1838063, EPI_ISL_1838065, EPI_ISL_1838068, EPI_ISL_1838070, EPI_ISL_1838073, EPI_ISL_1838075, EPI_ISL_1838078, EPI_ISL_1838080, EPI_ISL_1838083, EPI_ISL_1838086, EPI_ISL_1838088, EPI_ISL_1838091, EPI_ISL_1838094, EPI_ISL_1838097, EPI_ISL_1838100, EPI_ISL_1838102, EPI_ISL_1838106, EPI_ISL_1838109, EPI_ISL_1838111, EPI_ISL_1838114, EPI_ISL_1838117, EPI_ISL_1838119, EPI_ISL_1838122, EPI_ISL_1838123, EPI_ISL_1838125, EPI_ISL_1838126, EPI_ISL_1838128, EPI_ISL_1838129, EPI_ISL_1838131, EPI_ISL_1838132, EPI_ISL_1838134, EPI_ISL_1838136, EPI_ISL_1838139, EPI_ISL_1838140, EPI_ISL_1838142, EPI_ISL_1838143, EPI_ISL_1838145, EPI_ISL_1838148, EPI_ISL_1838149, EPI_ISL_1838151, EPI_ISL_1838152, EPI_ISL_1838153, EPI_ISL_1838155, EPI_ISL_1838156, EPI_ISL_1838158, EPI_ISL_1838159, EPI_ISL_1838161, EPI_ISL_1838162, EPI_ISL_1838164, EPI_ISL_1838165, EPI_ISL_1838166, EPI_ISL_1838168, EPI_ISL_1838169, EPI_ISL_1838171, EPI_ISL_1838172, EPI_ISL_1838173, EPI_ISL_1838175, EPI_ISL_1838176, EPI_ISL_1838178, EPI_ISL_1838179, EPI_ISL_1838182, EPI_ISL_1838185, EPI_ISL_1838186, EPI_ISL_1838188, EPI_ISL_1838189, EPI_ISL_1838191, EPI_ISL_1838194, EPI_ISL_1838195, EPI_ISL_1838196, EPI_ISL_1838198, EPI_ISL_1838200, EPI_ISL_1838201, EPI_ISL_1838203, EPI_ISL_1838205, EPI_ISL_1838207, EPI_ISL_1838209, EPI_ISL_1838210, EPI_ISL_1838212, EPI_ISL_1838214, EPI_ISL_1838215, EPI_ISL_1838217, EPI_ISL_1838219, EPI_ISL_1838220, EPI_ISL_1838222, EPI_ISL_1838224, EPI_ISL_1838225, EPI_ISL_1838227, EPI_ISL_1838229, EPI_ISL_1838230, EPI_ISL_1838232, EPI_ISL_1838234, EPI_ISL_1838235, EPI_ISL_1838237, EPI_ISL_1838239, EPI_ISL_1838241, EPI_ISL_1838242, EPI_ISL_1838244, EPI_ISL_1838246, EPI_ISL_1838248, EPI_ISL_1838250, EPI_ISL_1838251, EPI_ISL_1838253, EPI_ISL_1838254, EPI_ISL_1838256, EPI_ISL_1838257, EPI_ISL_1838259, EPI_ISL_1838260, EPI_ISL_1838262, EPI_ISL_1838264, EPI_ISL_1838265, EPI_ISL_1838267, EPI_ISL_1838269, EPI_ISL_1838271, EPI_ISL_1838272, EPI_ISL_1838274, EPI_ISL_1838275, EPI_ISL_1838277, EPI_ISL_1838278, EPI_ISL_1838280, EPI_ISL_1838282, EPI_ISL_1838284, EPI_ISL_1838285, EPI_ISL_1838287, EPI_ISL_1838288, EPI_ISL_1838290, EPI_ISL_1838292, EPI_ISL_1838293, EPI_ISL_1838295, EPI_ISL_1838296, EPI_ISL_1838298, EPI_ISL_1838299, EPI_ISL_1838301, EPI_ISL_1838303, EPI_ISL_1838304, EPI_ISL_1838306, EPI_ISL_1838307, EPI_ISL_1838309, EPI_ISL_1838311, EPI_ISL_1838312, EPI_ISL_1838314, EPI_ISL_1838315, EPI_ISL_1838317, EPI_ISL_1838318, EPI_ISL_1838320, EPI_ISL_1838322, EPI_ISL_1838323, EPI_ISL_1838325, EPI_ISL_1838327, EPI_ISL_1838328, EPI_ISL_1838330, EPI_ISL_1838332, EPI_ISL_1838333, EPI_ISL_1838335, EPI_ISL_1838337, EPI_ISL_1838338, EPI_ISL_1838340, EPI_ISL_1838342, EPI_ISL_1838343, EPI_ISL_1838345, EPI_ISL_1838346, EPI_ISL_1972346, EPI_ISL_1972348, EPI_ISL_1972350 | | | |
| see above | The National Centre for Cell Science | CSIR-Centre for Cellular and Molecular Biology-INSACOG | Ajay Pillai; Amareshwar Vodapalli; Ara Sreenivas; Archana Bharadwaj Siva; B Himasri; Blessy B John; Dhiraj Paul; Divya Tej Sowpati; INSACOG Consortium team; Karthik Bharadwaj Tallapaka; Lamuk Zaveri; Manoj Kumar Bhat; Mitali Inamdar; Mohak P Gujare; Onkar Kulkarni; Payel Mukherjee; Rakesh K Mishra; Sharath Chandra Thota; Shivang P. Bhanushali; Shreekant Verma; Sofia Banu; Sonal Manik Chavan; Tulasi Nagabandi; Valli Nagalakshmi Undamatla; Viswagithe S L; Yogesh Shouche |
| EPI_ISL_2033743, EPI_ISL_2033745, EPI_ISL_2033747 | Translational Health Science and Technology Institute -ESIC medical college and hospital, Faridabad | THSTI Bioassay laboratory | Akshay Kanakan; Anil Pandey; Guruprasad R Medigeshi; Heena Shaman; Janani Srinivasa Vasudevan; Jigme Wangchuk; Naseem Ahmed Khan; Neha Jha; Priyanka Mehta; Rajesh Pandey; Ranjeet Maurya |
| EPI_ISL_2020930, EPI_ISL_2020931, EPI_ISL_2020932, EPI_ISL_2020933, EPI_ISL_2020934, EPI_ISL_2020935, EPI_ISL_2020936, EPI_ISL_2020937, EPI_ISL_2020938, EPI_ISL_2020939, EPI_ISL_2020940, EPI_ISL_2020941, EPI_ISL_2020942, EPI_ISL_2020943, EPI_ISL_2020944, EPI_ISL_2020945, EPI_ISL_2020946, EPI_ISL_2020947, EPI_ISL_2020948, EPI_ISL_2020949, EPI_ISL_2020950, EPI_ISL_2020951, EPI_ISL_2020952, EPI_ISL_2020953, EPI_ISL_2020954, EPI_ISL_2020955, EPI_ISL_2020956, EPI_ISL_2020957, EPI_ISL_2020958, EPI_ISL_2020959, EPI_ISL_2020960 | | | |
| see above | Urban Public Health Centre (UPHC), SECTOR-2, Gandhinagar | Gujarat Biotechnology Research Centre | Amit Kanani; Amrutlal Patel; Bhargav Waghela; Bhumika Prajapati; Chaitanya Joshi; Dinesh Kumar; Janvi Raval; Kalpesh Goswami; Madhvi Joshi; Nilen Patel; Nitesh Shah; Nitin Savaliya; Ramesh Pandit; Sonal Sharma; Tejendra Purohit; Twinkle Soni; Umang Mishra; Zarna Patel; Zuber Saiyed |
| EPI_ISL_2017746, EPI_ISL_2017747, EPI_ISL_2017748, EPI_ISL_2017749, EPI_ISL_2017750, EPI_ISL_2017751 | VRDL, Goverment Medical College (GMC) and Hospital, Valsad | Gujarat Biotechnology Research Centre | Chaitanya Joshi; Dinesh Kumar; Janvi Raval; Madhvi Joshi; Nitesh Shah; Nitin Savaliya; Ramesh Pandit; Sonal Sharma; Twinkle Soni; Umang Mishra; Vicky Gandhi; Zarna Patel; Zuber Saiyed |
| EPI_ISL_2001187, EPI_ISL_2001188, EPI_ISL_2001189, EPI_ISL_2001190, EPI_ISL_2001191, EPI_ISL_2001192, EPI_ISL_2001193, EPI_ISL_2001194, EPI_ISL_2001195, EPI_ISL_2001196, EPI_ISL_2001197, EPI_ISL_2001198, EPI_ISL_2001199, EPI_ISL_2001200, EPI_ISL_2001201, EPI_ISL_2001202, EPI_ISL_2001203, EPI_ISL_2001204, EPI_ISL_2001205, EPI_ISL_2001206, EPI_ISL_2001207, EPI_ISL_2001208, EPI_ISL_2001209, EPI_ISL_2001210, EPI_ISL_2001211, EPI_ISL_2001212, EPI_ISL_2017728, EPI_ISL_2017729, EPI_ISL_2017730, EPI_ISL_2017731, EPI_ISL_2017732, EPI_ISL_2017733, EPI_ISL_2017734, EPI_ISL_2017735, EPI_ISL_2017736, EPI_ISL_2017737, EPI_ISL_2017738, EPI_ISL_2017739, EPI_ISL_2017740, EPI_ISL_2017741, EPI_ISL_2017742, EPI_ISL_2017743, EPI_ISL_2017744, EPI_ISL_2017745, EPI_ISL_2017763, EPI_ISL_2017764 | | | |
| see above | VRDL, Goverment Medical College (GMC), Surat | Gujarat Biotechnology Research Centre | Chaitanya Joshi; Dinesh Kumar; Janvi Raval; Madhvi Joshi; Neeta Khandelwal; Nitesh Shah; Nitin Savaliya; Ramesh Pandit; Sonal Sharma; Twinkle Soni; Umang Mishra; Zarna Patel; Zuber Saiyed |
| EPI_ISL_1662283, EPI_ISL_1662284, EPI_ISL_1662285, EPI_ISL_1662286, EPI_ISL_1663325, EPI_ISL_1663326, EPI_ISL_1663327, EPI_ISL_1663328, EPI_ISL_1663329, EPI_ISL_1663330, EPI_ISL_1663331, EPI_ISL_1663332, EPI_ISL_1663497, EPI_ISL_1663498, EPI_ISL_1663499, EPI_ISL_1663500, EPI_ISL_1663501, EPI_ISL_1663502, EPI_ISL_1663503 | | | |
| see above | Veer Surendra Sai Institute of Medical Sciences and Research, Burla, Sambalpur | Institute of Life Sciences - INSACOG | Ajay Parida; Amol M. Kanampalliwar; Arup Ghosh; Atimukta Jha; INSACOG Consortium; Punit Prasad; Rajeeb Swain; Rupesh Dash; Safal Walia; Shifu Aggarwal; Sunil K. Raghav |
| EPI_ISL_1595876, EPI_ISL_1595877, EPI_ISL_1595878, EPI_ISL_1595879, EPI_ISL_1595880, EPI_ISL_1595881, EPI_ISL_1595882, EPI_ISL_1595883, EPI_ISL_1595884, EPI_ISL_1595885, EPI_ISL_1595886, EPI_ISL_1595887, EPI_ISL_1595888 | | | |
| see above | WENLOCK HOSPITAL, MANGALORE | INSACOG-KA, NIMHANS | Ananthapadmanabha Kotambail; Anita S Desai; Anson Kunjumon George; Chetan G K; Chitra Pattabiraman; Darshan Sreenivas; Gautham Arunachal Udupi; Pramada Prasad; V Ravi |

We gratefully acknowledge the following Authors from the Originating laboratories responsible for obtaining the specimens, as well as the Submitting laboratories where the genome data were generated and shared via GISAID, on which this research is based.

All Submitters of data may be contacted directly via www.gisaid.org

Authors are sorted alphabetically.

| Accession ID | Originating Laboratory | Submitting Laboratory | Authors |
|---|---|---|---|
| EPI_ISL_2379358, EPI_ISL_2379359, EPI_ISL_2379361, EPI_ISL_2379362, EPI_ISL_2379378, EPI_ISL_2379390 | AD HALLI | INSACOG-KA, NIMHANS | Ananthapadmanabha Kotambail; Anita S Desai; Anson Kunjumon George; Chetan G K; Chitra Pattabiraman; Darshan Sreenivas; Gautham Arunachal Udupi; Pramada Prasad; V Ravi |
| EPI_ISL_2373137, EPI_ISL_2373138, EPI_ISL_2373139, EPI_ISL_2373140, EPI_ISL_2373141, EPI_ISL_2373142, EPI_ISL_2373143, EPI_ISL_2373144, EPI_ISL_2373145, EPI_ISL_2373146, EPI_ISL_2373147, EPI_ISL_2373148, EPI_ISL_2373149, EPI_ISL_2373150, EPI_ISL_2373151, EPI_ISL_2373152, EPI_ISL_2373153, EPI_ISL_2373154, EPI_ISL_2373155, EPI_ISL_2373156, EPI_ISL_2373157, EPI_ISL_2373158, EPI_ISL_2373159, EPI_ISL_2373160, EPI_ISL_2373161, EPI_ISL_2373162, EPI_ISL_2373163, EPI_ISL_2373164, EPI_ISL_2373165, EPI_ISL_2373166, EPI_ISL_2373167, EPI_ISL_2373168, EPI_ISL_2373169, EPI_ISL_2373170, EPI_ISL_2373171, EPI_ISL_2373172, EPI_ISL_2373173, EPI_ISL_2373174, EPI_ISL_2373175, EPI_ISL_2373176, EPI_ISL_2373177, EPI_ISL_2373178, EPI_ISL_2373179, EPI_ISL_2373180, EPI_ISL_2373181, EPI_ISL_2373182, EPI_ISL_2373183, EPI_ISL_2373184, EPI_ISL_2373185, EPI_ISL_2373186, EPI_ISL_2373187, EPI_ISL_2373188, EPI_ISL_2373189, EPI_ISL_2373190, EPI_ISL_2373191, EPI_ISL_2373192, EPI_ISL_2373193, EPI_ISL_2373194, EPI_ISL_2373195, EPI_ISL_2373196, EPI_ISL_2373197, EPI_ISL_2373198, EPI_ISL_2373199, EPI_ISL_2373200, EPI_ISL_2373201, EPI_ISL_2373202, EPI_ISL_2373203, EPI_ISL_2373204, EPI_ISL_2373205, EPI_ISL_2373206, EPI_ISL_2373207, EPI_ISL_2373208, EPI_ISL_2373209, EPI_ISL_2373210, EPI_ISL_2373211, EPI_ISL_2373212, EPI_ISL_2373213, EPI_ISL_2373214, EPI_ISL_2373215, EPI_ISL_2373216, EPI_ISL_2373217, EPI_ISL_2373218, EPI_ISL_2373219, EPI_ISL_2373220, EPI_ISL_2373221, EPI_ISL_2373222, EPI_ISL_2373223, EPI_ISL_2373224, EPI_ISL_2373225, EPI_ISL_2373226, EPI_ISL_2373227, EPI_ISL_2373228, EPI_ISL_2373229, EPI_ISL_2373230, EPI_ISL_2373231, EPI_ISL_2373232, EPI_ISL_2373233, EPI_ISL_2373234, EPI_ISL_2373235, EPI_ISL_2373236, EPI_ISL_2373237, EPI_ISL_2373238, EPI_ISL_2373239, EPI_ISL_2373240, EPI_ISL_2373241, EPI_ISL_2373242, EPI_ISL_2441103, EPI_ISL_2441104, EPI_ISL_2441105, EPI_ISL_2441106, EPI_ISL_2441107, EPI_ISL_2441108, EPI_ISL_2441109, EPI_ISL_2441110, EPI_ISL_2441111, EPI_ISL_2441112, EPI_ISL_2441113, EPI_ISL_2441114, EPI_ISL_2441115, EPI_ISL_2441116, EPI_ISL_2441117, EPI_ISL_2441118, EPI_ISL_2441119, EPI_ISL_2441120, EPI_ISL_2441121, EPI_ISL_2441122, EPI_ISL_2441123, EPI_ISL_2441124, EPI_ISL_2441125, EPI_ISL_2441126, EPI_ISL_2441127, EPI_ISL_2441128, EPI_ISL_2441129, EPI_ISL_2441130, EPI_ISL_2441131, EPI_ISL_2441132, EPI_ISL_2441133, EPI_ISL_2441134, EPI_ISL_2441135, EPI_ISL_2441136, EPI_ISL_2441137, EPI_ISL_2441138, EPI_ISL_2441139, EPI_ISL_2441140, EPI_ISL_2441141, EPI_ISL_2441142, EPI_ISL_2441143, EPI_ISL_2441144, EPI_ISL_2441145, EPI_ISL_2441146, EPI_ISL_2441147, EPI_ISL_2441148, EPI_ISL_2441149, EPI_ISL_2441150, EPI_ISL_2441151, EPI_ISL_2441152, EPI_ISL_2441153, EPI_ISL_2441154, EPI_ISL_2441155, EPI_ISL_2441156, EPI_ISL_2441157, EPI_ISL_2441158, EPI_ISL_2441159, EPI_ISL_2441160, EPI_ISL_2441161, EPI_ISL_2441162, EPI_ISL_2441163, EPI_ISL_2441164, EPI_ISL_2441165, EPI_ISL_2441166, EPI_ISL_2441167, EPI_ISL_2441168, EPI_ISL_2441169, EPI_ISL_2441170, EPI_ISL_2441171, EPI_ISL_2441172, EPI_ISL_2441173, EPI_ISL_2441174, EPI_ISL_2441175, EPI_ISL_2441176, EPI_ISL_2441177, EPI_ISL_2441178, EPI_ISL_2441179, EPI_ISL_2441180, EPI_ISL_2441181, EPI_ISL_2441182, EPI_ISL_2441183, EPI_ISL_2441184, EPI_ISL_2441185, EPI_ISL_2441186, EPI_ISL_2441187, EPI_ISL_2441188, EPI_ISL_2441189, EPI_ISL_2441190, EPI_ISL_2441191, EPI_ISL_2441192, EPI_ISL_2441193, EPI_ISL_2441194, EPI_ISL_2441195, EPI_ISL_2441196, EPI_ISL_2441197, EPI_ISL_2441198, EPI_ISL_2441199, EPI_ISL_2441200, EPI_ISL_2441201, EPI_ISL_2441202, EPI_ISL_2441203, EPI_ISL_2441204, EPI_ISL_2441205, EPI_ISL_2441206, EPI_ISL_2441207, EPI_ISL_2441208, EPI_ISL_2441209, EPI_ISL_2441210, EPI_ISL_2441211, EPI_ISL_2441212, EPI_ISL_2441213, EPI_ISL_2441214, EPI_ISL_2441215, EPI_ISL_2441216, EPI_ISL_2441217, EPI_ISL_2441218, EPI_ISL_2441219, EPI_ISL_2441220, EPI_ISL_2441221, EPI_ISL_2441222, EPI_ISL_2441223, EPI_ISL_2441224, EPI_ISL_2441225, EPI_ISL_2441226, EPI_ISL_2441227, EPI_ISL_2441228, EPI_ISL_2441229, EPI_ISL_2441230, EPI_ISL_2441231, EPI_ISL_2441232, EPI_ISL_2441233, EPI_ISL_2441234, EPI_ISL_2441235, EPI_ISL_2441236, EPI_ISL_2441237, EPI_ISL_2441238, EPI_ISL_2441239, EPI_ISL_2441240, EPI_ISL_2441241, EPI_ISL_2441242, EPI_ISL_2441243, EPI_ISL_2441244, EPI_ISL_2441245, EPI_ISL_2441246, EPI_ISL_2441247, EPI_ISL_2441248, EPI_ISL_2441249, EPI_ISL_2441250, EPI_ISL_2441251, EPI_ISL_2441252, EPI_ISL_2441253, EPI_ISL_2441254, EPI_ISL_2441255, EPI_ISL_2441256, EPI_ISL_2441257, EPI_ISL_2441738, EPI_ISL_2441739, EPI_ISL_2441740, EPI_ISL_2441741, EPI_ISL_2441742, EPI_ISL_2441743, EPI_ISL_2441744, EPI_ISL_2441745, EPI_ISL_2441746, EPI_ISL_2441747, EPI_ISL_2441748, EPI_ISL_2441749, EPI_ISL_2441750, EPI_ISL_2441751, EPI_ISL_2441752, EPI_ISL_2441753, EPI_ISL_2441754, EPI_ISL_2441755, EPI_ISL_2441756, EPI_ISL_2441757, EPI_ISL_2441758, EPI_ISL_2441759, EPI_ISL_2441760, EPI_ISL_2441761, EPI_ISL_2441762, EPI_ISL_2441763, EPI_ISL_2441764, EPI_ISL_2441765, EPI_ISL_2441766, EPI_ISL_2441767, EPI_ISL_2441768, EPI_ISL_2441769, EPI_ISL_2441770, EPI_ISL_2441771, EPI_ISL_2660698, EPI_ISL_2660699, EPI_ISL_2660700, EPI_ISL_2660701, EPI_ISL_2660702, EPI_ISL_2660703, EPI_ISL_2660704, EPI_ISL_2660705, EPI_ISL_2660706, EPI_ISL_2660707, EPI_ISL_2660708, EPI_ISL_2660709, EPI_ISL_2660710, EPI_ISL_2660711, EPI_ISL_2660712, EPI_ISL_2660713, EPI_ISL_2660714, EPI_ISL_2660715, EPI_ISL_2660716, EPI_ISL_2660717, EPI_ISL_2660718, EPI_ISL_2660719, EPI_ISL_2660720, EPI_ISL_2660721, EPI_ISL_2660722, EPI_ISL_2660723, EPI_ISL_2660724, EPI_ISL_2660725, EPI_ISL_2660726, EPI_ISL_2660727, EPI_ISL_2660728, EPI_ISL_2660729, EPI_ISL_2660730, EPI_ISL_2660731, EPI_ISL_2660732, EPI_ISL_2660733, EPI_ISL_2660734, EPI_ISL_2660735, EPI_ISL_2660736, EPI_ISL_2660737, EPI_ISL_2660738, EPI_ISL_2660739, EPI_ISL_2660740, EPI_ISL_2660741, EPI_ISL_2660742, EPI_ISL_2660743, EPI_ISL_2660744, EPI_ISL_2660745, EPI_ISL_2660746, EPI_ISL_2660747, EPI_ISL_2660748, EPI_ISL_2660749, EPI_ISL_2660750, EPI_ISL_2660751, EPI_ISL_2660752, EPI_ISL_2660753, EPI_ISL_2660754, EPI_ISL_2660755, EPI_ISL_2660756, EPI_ISL_2660757, EPI_ISL_2660758, EPI_ISL_2660759, EPI_ISL_2660760, EPI_ISL_2660761, EPI_ISL_2660762, EPI_ISL_2660763, EPI_ISL_2660764, EPI_ISL_2660765, EPI_ISL_2660766, EPI_ISL_2660767, EPI_ISL_2660768, EPI_ISL_2660769, EPI_ISL_2660770, EPI_ISL_2660771, EPI_ISL_2660772, EPI_ISL_2660773, EPI_ISL_2660774, EPI_ISL_2660775, EPI_ISL_2660776, EPI_ISL_2660777, EPI_ISL_2660778, EPI_ISL_2660779, EPI_ISL_2660780, EPI_ISL_2660781, EPI_ISL_2660782, EPI_ISL_2660783, EPI_ISL_2660784, EPI_ISL_2660785, EPI_ISL_2660786, EPI_ISL_2660787, EPI_ISL_2660788, EPI_ISL_2660789, EPI_ISL_2660790, EPI_ISL_2660791, EPI_ISL_2660792, EPI_ISL_2660793, EPI_ISL_2660794, EPI_ISL_2660795, EPI_ISL_2660796, EPI_ISL_2660797, EPI_ISL_2660798, EPI_ISL_2660799, EPI_ISL_2660800, EPI_ISL_2660801, EPI_ISL_2660802, EPI_ISL_2660803, EPI_ISL_2660804, EPI_ISL_2660805, EPI_ISL_2660806, EPI_ISL_2660807, EPI_ISL_2660808, EPI_ISL_2660809, EPI_ISL_2660810, EPI_ISL_2660811, EPI_ISL_2660812, EPI_ISL_2660813, EPI_ISL_2660814, EPI_ISL_2660815, EPI_ISL_2688250, EPI_ISL_2688251, EPI_ISL_2688252, EPI_ISL_2688253, EPI_ISL_2774730, EPI_ISL_2774731, EPI_ISL_2774732, EPI_ISL_2774733, EPI_ISL_2774734, EPI_ISL_2774735, EPI_ISL_2774736, EPI_ISL_2774737, EPI_ISL_2774738, EPI_ISL_2774739, EPI_ISL_2774740, EPI_ISL_2774741, EPI_ISL_2774742, EPI_ISL_2774743, EPI_ISL_2774744, EPI_ISL_2774745, EPI_ISL_2774746, EPI_ISL_2774747, EPI_ISL_2774748, EPI_ISL_2774749, EPI_ISL_2774750, EPI_ISL_2774751, EPI_ISL_2774752, EPI_ISL_2774753, EPI_ISL_2774754, EPI_ISL_2774755, EPI_ISL_2774756, EPI_ISL_2774757, EPI_ISL_2774758, EPI_ISL_2774759, EPI_ISL_2774760, EPI_ISL_2774761, EPI_ISL_2774762, EPI_ISL_2774763, EPI_ISL_2774764, EPI_ISL_2774765, EPI_ISL_2774766, EPI_ISL_2774767, EPI_ISL_2774768, EPI_ISL_2774769, EPI_ISL_2774770, EPI_ISL_2774771, EPI_ISL_2774772, EPI_ISL_2774773, EPI_ISL_2774774, EPI_ISL_2774775, EPI_ISL_2774776, EPI_ISL_2774777, EPI_ISL_2774778, EPI_ISL_2774779, EPI_ISL_2774780, EPI_ISL_2774781, EPI_ISL_2774782, EPI_ISL_2774783, EPI_ISL_2774784, EPI_ISL_2774785, EPI_ISL_2774786, EPI_ISL_2774787, EPI_ISL_2774788, EPI_ISL_2774789, EPI_ISL_2774790, EPI_ISL_2774791, EPI_ISL_2774792, EPI_ISL_2774793, EPI_ISL_2774794, EPI_ISL_2774795, EPI_ISL_2774796, EPI_ISL_2774797, EPI_ISL_2774798, EPI_ISL_2774799, EPI_ISL_2774800, EPI_ISL_2774801, EPI_ISL_2774802, EPI_ISL_2774803, EPI_ISL_2774804, EPI_ISL_2774805, EPI_ISL_2774806, EPI_ISL_2774807, EPI_ISL_2774808, EPI_ISL_2774809, EPI_ISL_2774810, EPI_ISL_2774811, EPI_ISL_2774812, EPI_ISL_2774813, EPI_ISL_2774814, EPI_ISL_2774815, EPI_ISL_2774816, EPI_ISL_2774817, EPI_ISL_2774818, EPI_ISL_2774819, EPI_ISL_2774820, EPI_ISL_2774821, EPI_ISL_2774822, EPI_ISL_2774823, EPI_ISL_2774824, EPI_ISL_2774825, EPI_ISL_2774826, EPI_ISL_2774827, EPI_ISL_2774828, EPI_ISL_2774829, EPI_ISL_2774830, EPI_ISL_2774831, EPI_ISL_2774832, EPI_ISL_2774833, EPI_ISL_2775288, EPI_ISL_2775289, EPI_ISL_2775290, EPI_ISL_2775291, EPI_ISL_2775292, EPI_ISL_2775293, EPI_ISL_2775294, EPI_ISL_2775295, EPI_ISL_2775296, EPI_ISL_2775297, EPI_ISL_2775298, EPI_ISL_2775299, EPI_ISL_2775300, EPI_ISL_2775301, EPI_ISL_2775302, EPI_ISL_2775303, EPI_ISL_2775304, EPI_ISL_2775305, EPI_ISL_2775306, EPI_ISL_2775307, EPI_ISL_2775308, EPI_ISL_2775309, EPI_ISL_2775310, EPI_ISL_2775311, EPI_ISL_2775312, EPI_ISL_2775313, EPI_ISL_2775314, EPI_ISL_2775315, EPI_ISL_2775316, EPI_ISL_2775317, EPI_ISL_2775318, EPI_ISL_2775319, EPI_ISL_2775320, EPI_ISL_2775321, EPI_ISL_2775322, EPI_ISL_2775323, EPI_ISL_2775324, EPI_ISL_2775325, EPI_ISL_2775326, EPI_ISL_2775327, EPI_ISL_2775328, EPI_ISL_2775329, EPI_ISL_2775330, EPI_ISL_2775331, EPI_ISL_2775332, EPI_ISL_2775333, EPI_ISL_2775334, EPI_ISL_2775335, EPI_ISL_2775336, EPI_ISL_2775337, EPI_ISL_2775338, EPI_ISL_2775339, EPI_ISL_2775340, EPI_ISL_2775341, EPI_ISL_2775342, EPI_ISL_2775343, EPI_ISL_2775344, EPI_ISL_2775345, EPI_ISL_2775346, EPI_ISL_2775347, EPI_ISL_2775348, EPI_ISL_2775349, EPI_ISL_2775350, EPI_ISL_2775351, EPI_ISL_2775352, EPI_ISL_2775353, EPI_ISL_2775354, EPI_ISL_2775355, EPI_ISL_2775356, EPI_ISL_2775357, EPI_ISL_2775358, EPI_ISL_2775359, EPI_ISL_2775360, EPI_ISL_2775361, EPI_ISL_2775362, EPI_ISL_2775363, EPI_ISL_2775364, EPI_ISL_2775365, EPI_ISL_2775366, EPI_ISL_2775367, EPI_ISL_2775368, EPI_ISL_2775369, EPI_ISL_2775370, EPI_ISL_2775371, EPI_ISL_2775372, EPI_ISL_2775373, EPI_ISL_2775374, EPI_ISL_2775375, EPI_ISL_2775376, EPI_ISL_2775377, EPI_ISL_2775378, EPI_ISL_2775379, EPI_ISL_2775380, EPI_ISL_2789349, EPI_ISL_2789350 | | | |
| see above | AP SSO | CSIR-Centre for Cellular and Molecular Biology-INSACOG | ; Amareshwar Vodapalli; Ara Sreenivas; Archana Bharadwaj Siva; B Himasri; Blessy B John; Divya Tej Sowpati; Jandhyala Sai Krishna; Karthik Bharadwaj Tallapaka; Lamuk Zaveri; Onkar Kulkarni; Payel Mukherjee; Priya Nurkurthy; Rakesh K Mishra; Sharath Chandra Thota; Shreekant Verma; Sofia Banu; Sumedha Avadhanula; Tulasi Nagabandi; Valli Nagalakshmi Undamatla; Vidhyadhari Methuku; Viswagithe S L |
| EPI_ISL_2379394, EPI_ISL_2379395, EPI_ISL_2379401 | ASHOKPURAM | INSACOG-KA, NIMHANS | Ananthapadmanabha Kotambail; Anita S Desai; Anson Kunjumon George; Chetan G K; Chitra Pattabiraman; Darshan Sreenivas; Gautham Arunachal Udupi; Pramada Prasad; V Ravi |
| EPI_ISL_2426193, EPI_ISL_2426195, EPI_ISL_2426197, EPI_ISL_2426198, EPI_ISL_2426199, EPI_ISL_2426200 | All India Institute of Medical Sciences, Delhi | CSIR Institute of Genomics and Integrative Biology | Abhinav Jain; Afra Shamnath; Anjali Bajaj; Arvinden VR; Bani Jolly; COVID CBNAAT CORE GROUP; Kiran Bala; Mercy Rophina; Mohamed Imran; Mohit Kumar Divakar; Nayer Jamshed; Praveen Aggarwal; Rahul C. Bhoyar; Rama Chaudhry; Randeep Guleria; Ritu Gupta; Sridhar Sivasubbu; Subrata Sinha; Urvashi B Singh; Vigneshwar Senthivel; Vinod Scaria |
| EPI_ISL_2651296, EPI_ISL_2651297, EPI_ISL_2651299, EPI_ISL_2651300, EPI_ISL_2651301, EPI_ISL_2651302, EPI_ISL_2651304, EPI_ISL_2651305, EPI_ISL_2651306, EPI_ISL_2651310, EPI_ISL_2651311, EPI_ISL_2651315, EPI_ISL_2651320, EPI_ISL_2651326, EPI_ISL_2651327, EPI_ISL_2651328, EPI_ISL_2651329, EPI_ISL_2651332, EPI_ISL_2651333, EPI_ISL_2651334, EPI_ISL_2651335, EPI_ISL_2651336, EPI_ISL_2651337, EPI_ISL_2651338, EPI_ISL_2651339, EPI_ISL_2651340, EPI_ISL_2651344, EPI_ISL_2651345, EPI_ISL_2651346, EPI_ISL_2651347, EPI_ISL_2651349, EPI_ISL_2651351, EPI_ISL_2651352, EPI_ISL_2651353, EPI_ISL_2651356, EPI_ISL_2651357, EPI_ISL_2651358, EPI_ISL_2651360, EPI_ISL_2651361, EPI_ISL_2725525, EPI_ISL_2725526, EPI_ISL_2725527, EPI_ISL_2725528, EPI_ISL_2725530, EPI_ISL_2725531, EPI_ISL_2725532, EPI_ISL_2725533, EPI_ISL_2725534, EPI_ISL_2725535, EPI_ISL_2725536, EPI_ISL_2728230, EPI_ISL_2728231, EPI_ISL_2728233, EPI_ISL_2728234, EPI_ISL_2728235, EPI_ISL_2728237, EPI_ISL_2728238, EPI_ISL_2728239, EPI_ISL_2728240, EPI_ISL_2728241, EPI_ISL_2728242, EPI_ISL_2728243, EPI_ISL_2728244, EPI_ISL_2728245, EPI_ISL_2728246, EPI_ISL_2728247, EPI_ISL_2728248, EPI_ISL_2728250, EPI_ISL_2728251, EPI_ISL_2728252, EPI_ISL_2728253, EPI_ISL_2728254, EPI_ISL_2728255, EPI_ISL_2728256, EPI_ISL_2728257, EPI_ISL_2728258, EPI_ISL_2728259, EPI_ISL_2728260, EPI_ISL_2728261, EPI_ISL_2728262, EPI_ISL_2728263, EPI_ISL_2728264, EPI_ISL_2728266, EPI_ISL_2728267, EPI_ISL_2728268, EPI_ISL_2728269, EPI_ISL_2728272, EPI_ISL_2728273, EPI_ISL_2728274, EPI_ISL_2728275, EPI_ISL_2728276, EPI_ISL_2728277, EPI_ISL_2728278, EPI_ISL_2728279, EPI_ISL_2728280, EPI_ISL_2728281, EPI_ISL_2728282, EPI_ISL_2728283, EPI_ISL_2728284, EPI_ISL_2728285, EPI_ISL_2728286, EPI_ISL_2728287, EPI_ISL_2728288, EPI_ISL_2728289, EPI_ISL_2728290, EPI_ISL_2728294, EPI_ISL_2728295, EPI_ISL_2728296, EPI_ISL_2728309, EPI_ISL_2728310, EPI_ISL_2728311, EPI_ISL_2728313, EPI_ISL_2728315, EPI_ISL_2728342, EPI_ISL_2728430, EPI_ISL_2728431, EPI_ISL_2728432, EPI_ISL_2728433, EPI_ISL_2728434, EPI_ISL_2728435, EPI_ISL_2728436, EPI_ISL_2728437, EPI_ISL_2728438, EPI_ISL_2842103, EPI_ISL_2842119, EPI_ISL_2842121, EPI_ISL_2842122, EPI_ISL_2842123, EPI_ISL_2842124, EPI_ISL_2842125, EPI_ISL_2842126, EPI_ISL_2842130, EPI_ISL_2842131, EPI_ISL_2842145, EPI_ISL_2842146, EPI_ISL_2842147, EPI_ISL_2842148, EPI_ISL_2842150, EPI_ISL_2842151, EPI_ISL_2842152, EPI_ISL_2842153, EPI_ISL_2842154, EPI_ISL_2842157, EPI_ISL_2842158, EPI_ISL_2842159, EPI_ISL_2842160, EPI_ISL_2842161, EPI_ISL_2842162, EPI_ISL_2842163, EPI_ISL_2842164, EPI_ISL_2842172, EPI_ISL_2842173, EPI_ISL_2842174, EPI_ISL_2842175, EPI_ISL_2842176, EPI_ISL_2842179, EPI_ISL_2842180, EPI_ISL_2842183, EPI_ISL_2842184, EPI_ISL_2842185, EPI_ISL_2842291, EPI_ISL_2842292, EPI_ISL_2842293, EPI_ISL_2842298, EPI_ISL_2842299, EPI_ISL_2842300, EPI_ISL_2842301, EPI_ISL_2842302, EPI_ISL_2842303, EPI_ISL_2842305, EPI_ISL_2842307, EPI_ISL_2842308, EPI_ISL_2842309, EPI_ISL_2842310, EPI_ISL_2842311, EPI_ISL_2842312, EPI_ISL_2842317, EPI_ISL_2842319, EPI_ISL_2842320, EPI_ISL_2842323, EPI_ISL_2842324, EPI_ISL_2842325, EPI_ISL_2842326, EPI_ISL_2842327, EPI_ISL_2842329, EPI_ISL_2842330, EPI_ISL_2842331, EPI_ISL_2842332, EPI_ISL_2842333, EPI_ISL_2842334, EPI_ISL_2842335, EPI_ISL_2842336, EPI_ISL_2842337, EPI_ISL_2842338, EPI_ISL_2842339, EPI_ISL_2842340, EPI_ISL_2842341, EPI_ISL_2842342, EPI_ISL_2842343, EPI_ISL_2842344, EPI_ISL_2842345, EPI_ISL_2842346, EPI_ISL_2842348, EPI_ISL_2842349, EPI_ISL_2842350, EPI_ISL_2842351, EPI_ISL_2842352, EPI_ISL_2842354, EPI_ISL_2842356, EPI_ISL_2842357, EPI_ISL_2842358, EPI_ISL_2842359, EPI_ISL_2842360, EPI_ISL_2842361, EPI_ISL_2842362, EPI_ISL_2842363, EPI_ISL_2842364, EPI_ISL_2842365, EPI_ISL_2842366, EPI_ISL_2842367, EPI_ISL_2842368, EPI_ISL_2842369, EPI_ISL_2842370, EPI_ISL_2842371, EPI_ISL_2842372, EPI_ISL_2842375, EPI_ISL_2842376, EPI_ISL_2842377, EPI_ISL_2842378, EPI_ISL_2842381, EPI_ISL_2842382, EPI_ISL_2842383, EPI_ISL_2842384, EPI_ISL_2842385, EPI_ISL_2842386, EPI_ISL_2842387, EPI_ISL_2842388, EPI_ISL_2842389, EPI_ISL_2842390, EPI_ISL_2842391, EPI_ISL_2842393, EPI_ISL_2842394, EPI_ISL_2842395, EPI_ISL_2842398, EPI_ISL_2842399, EPI_ISL_2842400, EPI_ISL_2842409, EPI_ISL_2842410, EPI_ISL_2842411, EPI_ISL_2842438, EPI_ISL_2842439, EPI_ISL_2842442, EPI_ISL_2842445, EPI_ISL_2842446, EPI_ISL_2842447, EPI_ISL_2842449, EPI_ISL_2842450, EPI_ISL_2842451, EPI_ISL_2842453, EPI_ISL_2842454, EPI_ISL_2842456, EPI_ISL_2842457 | | | |
| see above | BBMP Urban | INSACOG-KA, NIMHANS | Ananthapadmanabha Kotambail; Anita S Desai; Anson Kunjumon George; Chetan G K; Chitra Pattabiraman; Darshan Sreenivas; Ellango Ramasamy; Gautham Arunachal Udupi; Mahesh Kumar.C.S; Pramada Prasad; Sony Sharma; V Ravi |
| EPI_ISL_2405143 | BJ Medical College and Civil Hospital, Ahmedabad | Gujarat Biotechnology Research Centre | Chaitanya Joshi; Dinesh Kumar; Janvi Raval; Madhvi Joshi; Nitesh Shah; Nitin Savaliya; Pranay Shah; Ramesh Pandit; Sonal Sharma; Twinkle Soni; Umang Mishra; Zarna Patel; Zuber Saiyed |
| EPI_ISL_2918941, EPI_ISL_2918942, EPI_ISL_2918943, EPI_ISL_2918944, EPI_ISL_2918945, EPI_ISL_2918946, EPI_ISL_2918947, EPI_ISL_2918948, EPI_ISL_2918949, EPI_ISL_2918950, EPI_ISL_2918951, EPI_ISL_2918952, EPI_ISL_2918953, EPI_ISL_2918954, EPI_ISL_2918955, EPI_ISL_2918956, EPI_ISL_2918957, EPI_ISL_2918958, EPI_ISL_2918959 | | | |
| see above | Babina Diagnostics | CDFD | Ashwin Dalal; Asmita Gupta; Divya Vashisht; Murali Bashyam; Reelina Basu; Vinay Donipadi |
| EPI_ISL_2724304, EPI_ISL_2724305, EPI_ISL_2724306, EPI_ISL_2724307, EPI_ISL_2724308, EPI_ISL_2724309, EPI_ISL_2724310, EPI_ISL_2724311, EPI_ISL_2724314, EPI_ISL_2724315, EPI_ISL_2724316, EPI_ISL_2724317 | | | |
| see above | Bangalore Baptist Hospital | inStem NCBS – INSACOG | Uma Ramakrishnan Dasaradhi Palakodeti Aswin SaiNarain |
| EPI_ISL_2897840, EPI_ISL_2897841, EPI_ISL_2897842, EPI_ISL_2897843, EPI_ISL_2897844, EPI_ISL_2897845, EPI_ISL_2897846, EPI_ISL_2897847, EPI_ISL_2897848, EPI_ISL_2897849, EPI_ISL_2897850, EPI_ISL_2897876, EPI_ISL_2897877, EPI_ISL_2897878, EPI_ISL_2897879, EPI_ISL_2897880, EPI_ISL_2897881, EPI_ISL_2897882, EPI_ISL_2897883, EPI_ISL_2897884, EPI_ISL_2897885, EPI_ISL_2897886, EPI_ISL_2897887, EPI_ISL_2897888, EPI_ISL_2897889, EPI_ISL_2897890, EPI_ISL_2897891, EPI_ISL_2897892, EPI_ISL_2897907, EPI_ISL_2897908, EPI_ISL_2897909 | | | |
| see above | CDFD | CDFD | Ashwin Dalal; Asmita Gupta; Divya Vashisht; Murali Bashyam; Nagamani Kammili; Reelina Basu; Vinay Donipadi |
| EPI_ISL_2373377, EPI_ISL_2373378, EPI_ISL_2373379, EPI_ISL_2373380, EPI_ISL_2373381, EPI_ISL_2373382, EPI_ISL_2373383, EPI_ISL_2373384, EPI_ISL_2373385, EPI_ISL_2373386, EPI_ISL_2373387, EPI_ISL_2373388, EPI_ISL_2373389, EPI_ISL_2373390, EPI_ISL_2373391, EPI_ISL_2373392, EPI_ISL_2373393, EPI_ISL_2373394, EPI_ISL_2373395, EPI_ISL_2373396, EPI_ISL_2373397, EPI_ISL_2373398, EPI_ISL_2373399, EPI_ISL_2373400, EPI_ISL_2373401, EPI_ISL_2373402, EPI_ISL_2373403, EPI_ISL_2373404, EPI_ISL_2373405, EPI_ISL_2373406, EPI_ISL_2373407, EPI_ISL_2373408, EPI_ISL_2373409, EPI_ISL_2373410, EPI_ISL_2373411, EPI_ISL_2373412, EPI_ISL_2373413, EPI_ISL_2373414, EPI_ISL_2373415, EPI_ISL_2373416, EPI_ISL_2373417, EPI_ISL_2373418, EPI_ISL_2373419, EPI_ISL_2373420, EPI_ISL_2373421, EPI_ISL_2373422, EPI_ISL_2373423, EPI_ISL_2373424, EPI_ISL_2373425, EPI_ISL_2373426, EPI_ISL_2373427, EPI_ISL_2373428, EPI_ISL_2373429, EPI_ISL_2373430, EPI_ISL_2373431, EPI_ISL_2373432, EPI_ISL_2373433, EPI_ISL_2373434, EPI_ISL_2373435, EPI_ISL_2373436, EPI_ISL_2373437, EPI_ISL_2373438, EPI_ISL_2373439, EPI_ISL_2373440, EPI_ISL_2373441, EPI_ISL_2373442, EPI_ISL_2373443, EPI_ISL_2373444, EPI_ISL_2373445, EPI_ISL_2373446, EPI_ISL_2373447, EPI_ISL_2373448, EPI_ISL_2373449, EPI_ISL_2373450, EPI_ISL_2373451, EPI_ISL_2373452, EPI_ISL_2373453, EPI_ISL_2373454, EPI_ISL_2373455, EPI_ISL_2373456, EPI_ISL_2373457, EPI_ISL_2373458, EPI_ISL_2373459, EPI_ISL_2373460, EPI_ISL_2373461, EPI_ISL_2373462, EPI_ISL_2373463, EPI_ISL_2373464, EPI_ISL_2373465, EPI_ISL_2373466, EPI_ISL_2373467, EPI_ISL_2373468, EPI_ISL_2373469, EPI_ISL_2373470, EPI_ISL_2373471, EPI_ISL_2373472, EPI_ISL_2373473, EPI_ISL_2373474, EPI_ISL_2373475, EPI_ISL_2373476, EPI_ISL_2373477, EPI_ISL_2373478, EPI_ISL_2373479, EPI_ISL_2373480, EPI_ISL_2373481, EPI_ISL_2373482, EPI_ISL_2373483, EPI_ISL_2373484, EPI_ISL_2373485, EPI_ISL_2373486, EPI_ISL_2373487, EPI_ISL_2373488, EPI_ISL_2373489, EPI_ISL_2373490, EPI_ISL_2373491, EPI_ISL_2373492, EPI_ISL_2373493, EPI_ISL_2373494, EPI_ISL_2373495, EPI_ISL_2373496, EPI_ISL_2373497, EPI_ISL_2373498, EPI_ISL_2373499, EPI_ISL_2373500, EPI_ISL_2400317, EPI_ISL_2441367, EPI_ISL_2441368, EPI_ISL_2441369, EPI_ISL_2441370, EPI_ISL_2441371, EPI_ISL_2441478, EPI_ISL_2441479, EPI_ISL_2441480, EPI_ISL_2441481, EPI_ISL_2441482, EPI_ISL_2441483, EPI_ISL_2441484, EPI_ISL_2441485, EPI_ISL_2441486, EPI_ISL_2441487, EPI_ISL_2441488, EPI_ISL_2441489, EPI_ISL_2441490, EPI_ISL_2441491, EPI_ISL_2441492, EPI_ISL_2441493, EPI_ISL_2441494, EPI_ISL_2441495, EPI_ISL_2441496, EPI_ISL_2441497, EPI_ISL_2441498, EPI_ISL_2441499, EPI_ISL_2441500, EPI_ISL_2441501, EPI_ISL_2441502, EPI_ISL_2441503, EPI_ISL_2441504, EPI_ISL_2441505, EPI_ISL_2441506, EPI_ISL_2441507, EPI_ISL_2441508, EPI_ISL_2441509, EPI_ISL_2441510, EPI_ISL_2441511, EPI_ISL_2441512, EPI_ISL_2441513, EPI_ISL_2441514, EPI_ISL_2441515, EPI_ISL_2441516, EPI_ISL_2441517, EPI_ISL_2441518, EPI_ISL_2441519, EPI_ISL_2441520, EPI_ISL_2441521, EPI_ISL_2441522, EPI_ISL_2441523, EPI_ISL_2441524, EPI_ISL_2441525, EPI_ISL_2441526, EPI_ISL_2441527, EPI_ISL_2441528, EPI_ISL_2441529, EPI_ISL_2441530, EPI_ISL_2441531, EPI_ISL_2441532, EPI_ISL_2441533, EPI_ISL_2441534, EPI_ISL_2441535, EPI_ISL_2441536, EPI_ISL_2441537, EPI_ISL_2441538, EPI_ISL_2441539, EPI_ISL_2441540, EPI_ISL_2441541, EPI_ISL_2441542, EPI_ISL_2441543, EPI_ISL_2441544, EPI_ISL_2441545, EPI_ISL_2441546, EPI_ISL_2441547, EPI_ISL_2441548, EPI_ISL_2441549, EPI_ISL_2441550, EPI_ISL_2441551, EPI_ISL_2441552, EPI_ISL_2441553, EPI_ISL_2441554, EPI_ISL_2441555, EPI_ISL_2441556, EPI_ISL_2441557, EPI_ISL_2441558, EPI_ISL_2441559, EPI_ISL_2441560, EPI_ISL_2441561, EPI_ISL_2441562, EPI_ISL_2441563, EPI_ISL_2441564, EPI_ISL_2441565, EPI_ISL_2441566, EPI_ISL_2441567, EPI_ISL_2441568, EPI_ISL_2441569, EPI_ISL_2441570, EPI_ISL_2441571, EPI_ISL_2441572, EPI_ISL_2441573, EPI_ISL_2441574, EPI_ISL_2441575, EPI_ISL_2441576, EPI_ISL_2441577, EPI_ISL_2441578, EPI_ISL_2441579, EPI_ISL_2441580, EPI_ISL_2441581, EPI_ISL_2441582, EPI_ISL_2441583, EPI_ISL_2441584, EPI_ISL_2441585, EPI_ISL_2441586, EPI_ISL_2441587, EPI_ISL_2441588, EPI_ISL_2441589, EPI_ISL_2441590, EPI_ISL_2441591, EPI_ISL_2441592, EPI_ISL_2441593, EPI_ISL_2441594, EPI_ISL_2441595, EPI_ISL_2441596, EPI_ISL_2441597, EPI_ISL_2441598, EPI_ISL_2441599, EPI_ISL_2441600, EPI_ISL_2441601, EPI_ISL_2441602, EPI_ISL_2441603, EPI_ISL_2441604, EPI_ISL_2441605, EPI_ISL_2441606, EPI_ISL_2441607, EPI_ISL_2441608, EPI_ISL_2441609, EPI_ISL_2441610, EPI_ISL_2441611, EPI_ISL_2441612, EPI_ISL_2441613, EPI_ISL_2441614, EPI_ISL_2441615, EPI_ISL_2441616, EPI_ISL_2441617, EPI_ISL_2441618, EPI_ISL_2441619, EPI_ISL_2441620, EPI_ISL_2441621, EPI_ISL_2441622, EPI_ISL_2441623, EPI_ISL_2441624, EPI_ISL_2441625, EPI_ISL_2441626, EPI_ISL_2441627, EPI_ISL_2441628, EPI_ISL_2441629, EPI_ISL_2441630, EPI_ISL_2441631, EPI_ISL_2441632, EPI_ISL_2441633, EPI_ISL_2441634, EPI_ISL_2441635, EPI_ISL_2441636, EPI_ISL_2441637, EPI_ISL_2441638, EPI_ISL_2441639, EPI_ISL_2441640, EPI_ISL_2441641, EPI_ISL_2441642, EPI_ISL_2441643, EPI_ISL_2441644, EPI_ISL_2441645, EPI_ISL_2441646, EPI_ISL_2441647, EPI_ISL_2441648, EPI_ISL_2441649, EPI_ISL_2441650, EPI_ISL_2441651, EPI_ISL_2441652, EPI_ISL_2441653, EPI_ISL_2441654, EPI_ISL_2441655, EPI_ISL_2441656, EPI_ISL_2441657, EPI_ISL_2441658, EPI_ISL_2441659, EPI_ISL_2441660, EPI_ISL_2441661, EPI_ISL_2441662, EPI_ISL_2441663, EPI_ISL_2441664, EPI_ISL_2441665, EPI_ISL_2441666, EPI_ISL_2441667, EPI_ISL_2441668, EPI_ISL_2441669, EPI_ISL_2441670, EPI_ISL_2441671, EPI_ISL_2441672, EPI_ISL_2441673, EPI_ISL_2441674, EPI_ISL_2441675, EPI_ISL_2441676, EPI_ISL_2441677, EPI_ISL_2441678, EPI_ISL_2441679, EPI_ISL_2441680, EPI_ISL_2441681, EPI_ISL_2441682, EPI_ISL_2441683, EPI_ISL_2441684, EPI_ISL_2441685, EPI_ISL_2441686, EPI_ISL_2441687, EPI_ISL_2441688, EPI_ISL_2441689, EPI_ISL_2441690, EPI_ISL_2441691, EPI_ISL_2441692, EPI_ISL_2441693, EPI_ISL_2441694, EPI_ISL_2441695, EPI_ISL_2441696, EPI_ISL_2441697, EPI_ISL_2441698, EPI_ISL_2441699, EPI_ISL_2441700, EPI_ISL_2441701, EPI_ISL_2441702, EPI_ISL_2441703, EPI_ISL_2441704, EPI_ISL_2441705, EPI_ISL_2441706, EPI_ISL_2441707, EPI_ISL_2441708, EPI_ISL_2441709, EPI_ISL_2441710, EPI_ISL_2441711, EPI_ISL_2441712, EPI_ISL_2441713, EPI_ISL_2441714, EPI_ISL_2441715, EPI_ISL_2441716, EPI_ISL_2441717, EPI_ISL_2441718, EPI_ISL_2441719, EPI_ISL_2441720, EPI_ISL_2441721, EPI_ISL_2441722, EPI_ISL_2441723, EPI_ISL_2441724, EPI_ISL_2441725, EPI_ISL_2441726, EPI_ISL_2441727, EPI_ISL_2441728, EPI_ISL_2441729, EPI_ISL_2441730, EPI_ISL_2441731, EPI_ISL_2441732, EPI_ISL_2441733, EPI_ISL_2441734, EPI_ISL_2441735, EPI_ISL_2441736, EPI_ISL_2441737, EPI_ISL_2449862, EPI_ISL_2661097, EPI_ISL_2661098, EPI_ISL_2661099, EPI_ISL_2661134, EPI_ISL_2661155, EPI_ISL_2661156, EPI_ISL_2661157, EPI_ISL_2661158, EPI_ISL_2661159, EPI_ISL_2661160, EPI_ISL_2661161, EPI_ISL_2661162, EPI_ISL_2661163, EPI_ISL_2661164, EPI_ISL_2661165, EPI_ISL_2661166, EPI_ISL_2661167, EPI_ISL_2661168, EPI_ISL_2661169, EPI_ISL_2661170, EPI_ISL_2661171, EPI_ISL_2661172, EPI_ISL_2661173, EPI_ISL_2661174, EPI_ISL_2661175, EPI_ISL_2661176, EPI_ISL_2661177, EPI_ISL_2661178, EPI_ISL_2661179, EPI_ISL_2661180, EPI_ISL_2661181, EPI_ISL_2661182, EPI_ISL_2661183, EPI_ISL_2661184, EPI_ISL_2661185, EPI_ISL_2661186, EPI_ISL_2661187, EPI_ISL_2661188, EPI_ISL_2661189, EPI_ISL_2661190, EPI_ISL_2661191, EPI_ISL_2661192, EPI_ISL_2661193, EPI_ISL_2661194, EPI_ISL_2661195, EPI_ISL_2661196, EPI_ISL_2661197, EPI_ISL_2661198, EPI_ISL_2661199, EPI_ISL_2661200, EPI_ISL_2661201, EPI_ISL_2661202, EPI_ISL_2661203, EPI_ISL_2661204, EPI_ISL_2661205, EPI_ISL_2661206, EPI_ISL_2661207, EPI_ISL_2661208, EPI_ISL_2661209, EPI_ISL_2661210, EPI_ISL_2661211, EPI_ISL_2661212, EPI_ISL_2661213, EPI_ISL_2661214, EPI_ISL_2661215, EPI_ISL_2661216, EPI_ISL_2661217, EPI_ISL_2661218, EPI_ISL_2661219, EPI_ISL_2661220, EPI_ISL_2661221, EPI_ISL_2661222, EPI_ISL_2661223, EPI_ISL_2661224, EPI_ISL_2661225, EPI_ISL_2661226, EPI_ISL_2661227, EPI_ISL_2661228, EPI_ISL_2661229, EPI_ISL_2661230, EPI_ISL_2661231, EPI_ISL_2661232, EPI_ISL_2661233, EPI_ISL_2661234, EPI_ISL_2661235, EPI_ISL_2661236, EPI_ISL_2661237, EPI_ISL_2661238, EPI_ISL_2661239, EPI_ISL_2661240, EPI_ISL_2661241, EPI_ISL_2661242, EPI_ISL_2661243, EPI_ISL_2661244, EPI_ISL_2661245, EPI_ISL_2661246, EPI_ISL_2661247, EPI_ISL_2661248, EPI_ISL_2661249, EPI_ISL_2661250, EPI_ISL_2661251, EPI_ISL_2661252, EPI_ISL_2661253, EPI_ISL_2661254, EPI_ISL_2661255, EPI_ISL_2661256, EPI_ISL_2661257, EPI_ISL_2661258, EPI_ISL_2661259, EPI_ISL_2661260, EPI_ISL_2661261, EPI_ISL_2661262, EPI_ISL_2661263, EPI_ISL_2661264, EPI_ISL_2661265, EPI_ISL_2661266, EPI_ISL_2661267, EPI_ISL_2661268, EPI_ISL_2661269, EPI_ISL_2661270, EPI_ISL_2661271, EPI_ISL_2661272, EPI_ISL_2661273, EPI_ISL_2661274, EPI_ISL_2661275, EPI_ISL_2661276, EPI_ISL_2661277, EPI_ISL_2661278, EPI_ISL_2661279, EPI_ISL_2661280, EPI_ISL_2661281, EPI_ISL_2661282, EPI_ISL_2661283, EPI_ISL_2661284, EPI_ISL_2661285, EPI_ISL_2661286, EPI_ISL_2661287, EPI_ISL_2661288, EPI_ISL_2661289, EPI_ISL_2661290, EPI_ISL_2661291, EPI_ISL_2661292, EPI_ISL_2661293, EPI_ISL_2661294, EPI_ISL_2661295, EPI_ISL_2661296, EPI_ISL_2661297, EPI_ISL_2661298, EPI_ISL_2661299, EPI_ISL_2661300, EPI_ISL_2661301, EPI_ISL_2661302, EPI_ISL_2661303, EPI_ISL_2661304, EPI_ISL_2661305, EPI_ISL_2661306, EPI_ISL_2661307, EPI_ISL_2661308, EPI_ISL_2661309, EPI_ISL_2661310, EPI_ISL_2661311, EPI_ISL_2661312, EPI_ISL_2661313, EPI_ISL_2661314, EPI_ISL_2661315, EPI_ISL_2661316, EPI_ISL_2661317, EPI_ISL_2661318, EPI_ISL_2661319, EPI_ISL_2661320, EPI_ISL_2661321, EPI_ISL_2661322, EPI_ISL_2661323, EPI_ISL_2661324, EPI_ISL_2661325, EPI_ISL_2661326, EPI_ISL_2661327, EPI_ISL_2661328, EPI_ISL_2661329, EPI_ISL_2661330, EPI_ISL_2661331, EPI_ISL_2661332, EPI_ISL_2661333, EPI_ISL_2661334, EPI_ISL_2661335, EPI_ISL_2661336, EPI_ISL_2661337, EPI_ISL_2661338, EPI_ISL_2661339, EPI_ISL_2661340, EPI_ISL_2661341, EPI_ISL_2661342, EPI_ISL_2661343, EPI_ISL_2661344, EPI_ISL_2661345, EPI_ISL_2661346, EPI_ISL_2661347, EPI_ISL_2661348, EPI_ISL_2661349, EPI_ISL_2661350, EPI_ISL_2661351, EPI_ISL_2661352, EPI_ISL_2661353, EPI_ISL_2661354, EPI_ISL_2661355, EPI_ISL_2661356, EPI_ISL_2661357, EPI_ISL_2661358, EPI_ISL_2661359, EPI_ISL_2661360, EPI_ISL_2661361, EPI_ISL_2661362, EPI_ISL_2661363, EPI_ISL_2661364, EPI_ISL_2661365, EPI_ISL_2661366, EPI_ISL_2661367, EPI_ISL_2661368, EPI_ISL_2661369, EPI_ISL_2661370, EPI_ISL_2661371, EPI_ISL_2661372, EPI_ISL_2661373, EPI_ISL_2661374, EPI_ISL_2661375, EPI_ISL_2661376, EPI_ISL_2661377, EPI_ISL_2661378, EPI_ISL_2688257, EPI_ISL_2688258, EPI_ISL_2774835, EPI_ISL_2774916, EPI_ISL_2774917, EPI_ISL_2774918, EPI_ISL_2774919, EPI_ISL_2774920, EPI_ISL_2774921, EPI_ISL_2774922, EPI_ISL_2774923, EPI_ISL_2774924, EPI_ISL_2774925, EPI_ISL_2774926, EPI_ISL_2774927, EPI_ISL_2774928, EPI_ISL_2774929, EPI_ISL_2774930, EPI_ISL_2774931, EPI_ISL_2774932, EPI_ISL_2774933, EPI_ISL_2774934, EPI_ISL_2774935, EPI_ISL_2774936, EPI_ISL_2774937, EPI_ISL_2774938, EPI_ISL_2774939, EPI_ISL_2774940, EPI_ISL_2774941, EPI_ISL_2774942, EPI_ISL_2774943, EPI_ISL_2774944, EPI_ISL_2774945, EPI_ISL_2774946, EPI_ISL_2774947, EPI_ISL_2774948, EPI_ISL_2774949, EPI_ISL_2774950, EPI_ISL_2774951, EPI_ISL_2774952, EPI_ISL_2774953, EPI_ISL_2774954, EPI_ISL_2774955, EPI_ISL_2774956, EPI_ISL_2774957, EPI_ISL_2774958, EPI_ISL_2774959, EPI_ISL_2774960, EPI_ISL_2774961, EPI_ISL_2774962, EPI_ISL_2774963, EPI_ISL_2774964, EPI_ISL_2774965, EPI_ISL_2774966, EPI_ISL_2774967, EPI_ISL_2774968, EPI_ISL_2774969, EPI_ISL_2774970, EPI_ISL_2774971, EPI_ISL_2774972, EPI_ISL_2774973, EPI_ISL_2774974, EPI_ISL_2774975, EPI_ISL_2774976, EPI_ISL_2774977, EPI_ISL_2774978, EPI_ISL_2774979, EPI_ISL_2774980, EPI_ISL_2774981, EPI_ISL_2774982, EPI_ISL_2774983, EPI_ISL_2774984, EPI_ISL_2774985, EPI_ISL_2774986, EPI_ISL_2774987, EPI_ISL_2774988, EPI_ISL_2774989, EPI_ISL_2774990, EPI_ISL_2774991, EPI_ISL_2774992, EPI_ISL_2774993, EPI_ISL_2774994, EPI_ISL_2774995, EPI_ISL_2774996, EPI_ISL_2774997, EPI_ISL_2774998, EPI_ISL_2774999, EPI_ISL_2775000, EPI_ISL_2775001, EPI_ISL_2775002, EPI_ISL_2775003, EPI_ISL_2775004, EPI_ISL_2775005, EPI_ISL_2775006, EPI_ISL_2775007, EPI_ISL_2775008, EPI_ISL_2775009, EPI_ISL_2775010, EPI_ISL_2775011, EPI_ISL_2775012, EPI_ISL_2775013, EPI_ISL_2775014, EPI_ISL_2775015, EPI_ISL_2775016, EPI_ISL_2775017, EPI_ISL_2775018, EPI_ISL_2775019, | | | |

| | | | |
|---|---|---|---|
| EPI_ISL_2775020, EPI_ISL_2775021, EPI_ISL_2775022, EPI_ISL_2775023, EPI_ISL_2775024, EPI_ISL_2775025, EPI_ISL_2775026, EPI_ISL_2775027, EPI_ISL_2775028, EPI_ISL_2775029, EPI_ISL_2775030, EPI_ISL_2775031, EPI_ISL_2775032, EPI_ISL_2775033, EPI_ISL_2775034, EPI_ISL_2775035, EPI_ISL_2775036, EPI_ISL_2775037, EPI_ISL_2775038, EPI_ISL_2775039, EPI_ISL_2775040, EPI_ISL_2775041, EPI_ISL_2775042, EPI_ISL_2775043, EPI_ISL_2775044, EPI_ISL_2775045, EPI_ISL_2775046, EPI_ISL_2775047, EPI_ISL_2775048, EPI_ISL_2775049, EPI_ISL_2775050, EPI_ISL_2775051, EPI_ISL_2775052, EPI_ISL_2775053, EPI_ISL_2775054, EPI_ISL_2775055, EPI_ISL_2775056, EPI_ISL_2775057, EPI_ISL_2775058, EPI_ISL_2775059, EPI_ISL_2775060, EPI_ISL_2775061, EPI_ISL_2775062, EPI_ISL_2775063, EPI_ISL_2775064, EPI_ISL_2775065, EPI_ISL_2775066, EPI_ISL_2775067, EPI_ISL_2775068, EPI_ISL_2775069, EPI_ISL_2775070, EPI_ISL_2775071, EPI_ISL_2775072, EPI_ISL_2775073, EPI_ISL_2775074, EPI_ISL_2775075, EPI_ISL_2775076, EPI_ISL_2775077, EPI_ISL_2775078, EPI_ISL_2775079, EPI_ISL_2775080, EPI_ISL_2775081, EPI_ISL_2775082, EPI_ISL_2775083, EPI_ISL_2775084, EPI_ISL_2775085, EPI_ISL_2775086, EPI_ISL_2775087, EPI_ISL_2775088, EPI_ISL_2775089, EPI_ISL_2775090, EPI_ISL_2775091, EPI_ISL_2775092, EPI_ISL_2775093, EPI_ISL_2775094, EPI_ISL_2775095, EPI_ISL_2775096, EPI_ISL_2775097, EPI_ISL_2775098, EPI_ISL_2775099, EPI_ISL_2775100, EPI_ISL_2775101, EPI_ISL_2775102, EPI_ISL_2775103, EPI_ISL_2775104, EPI_ISL_2775105, EPI_ISL_2775106, EPI_ISL_2775107, EPI_ISL_2775108, EPI_ISL_2775109, EPI_ISL_2775148, EPI_ISL_2775149, EPI_ISL_2775150, EPI_ISL_2775151, EPI_ISL_2775152, EPI_ISL_2775153, EPI_ISL_2775154, EPI_ISL_2775155, EPI_ISL_2775156, EPI_ISL_2775157, EPI_ISL_2775158, EPI_ISL_2775159, EPI_ISL_2775160, EPI_ISL_2775161, EPI_ISL_2775162, EPI_ISL_2775163, EPI_ISL_2775164, EPI_ISL_2775165, EPI_ISL_2775166, EPI_ISL_2775167, EPI_ISL_2775168, EPI_ISL_2775169, EPI_ISL_2775170, EPI_ISL_2775171, EPI_ISL_2775172, EPI_ISL_2775173, EPI_ISL_2775174, EPI_ISL_2775175, EPI_ISL_2775176, EPI_ISL_2775177, EPI_ISL_2775178, EPI_ISL_2775179, EPI_ISL_2775180, EPI_ISL_2775181, EPI_ISL_2775182, EPI_ISL_2775183, EPI_ISL_2775184, EPI_ISL_2775185, EPI_ISL_2775186, EPI_ISL_2775187, EPI_ISL_2775188, EPI_ISL_2775189, EPI_ISL_2775190, EPI_ISL_2775191, EPI_ISL_2775192, EPI_ISL_2775193, EPI_ISL_2775194, EPI_ISL_2775195, EPI_ISL_2775196, EPI_ISL_2775197, EPI_ISL_2775198, EPI_ISL_2775199, EPI_ISL_2775200, EPI_ISL_2775201, EPI_ISL_2775202, EPI_ISL_2775203, EPI_ISL_2775204, EPI_ISL_2775205, EPI_ISL_2775206, EPI_ISL_2775207, EPI_ISL_2775208, EPI_ISL_2775209, EPI_ISL_2775210, EPI_ISL_2775211, EPI_ISL_2775212, EPI_ISL_2775213, EPI_ISL_2775214, EPI_ISL_2775215, EPI_ISL_2775216, EPI_ISL_2775217, EPI_ISL_2775218, EPI_ISL_2775219, EPI_ISL_2775220, EPI_ISL_2775221, EPI_ISL_2775222, EPI_ISL_2775223, EPI_ISL_2775224, EPI_ISL_2775225, EPI_ISL_2775226, EPI_ISL_2775227, EPI_ISL_2775228, EPI_ISL_2775229, EPI_ISL_2775230, EPI_ISL_2775231, EPI_ISL_2775232, EPI_ISL_2775233, EPI_ISL_2775234, EPI_ISL_2775235, EPI_ISL_2775236, EPI_ISL_2775237, EPI_ISL_2775238, EPI_ISL_2775239, EPI_ISL_2775240, EPI_ISL_2775241, EPI_ISL_2775242, EPI_ISL_2775243, EPI_ISL_2775244, EPI_ISL_2775245, EPI_ISL_2775246, EPI_ISL_2775247, EPI_ISL_2775248, EPI_ISL_2775249, EPI_ISL_2775250, EPI_ISL_2775251, EPI_ISL_2775252, EPI_ISL_2775253, EPI_ISL_2775254, EPI_ISL_2775255, EPI_ISL_2775256, EPI_ISL_2775257, EPI_ISL_2775258, EPI_ISL_2775259, EPI_ISL_2775260, EPI_ISL_2775261, EPI_ISL_2775262, EPI_ISL_2775263, EPI_ISL_2775264, EPI_ISL_2775265, EPI_ISL_2775266, EPI_ISL_2775267, EPI_ISL_2775268, EPI_ISL_2775269, EPI_ISL_2775270, EPI_ISL_2775271, EPI_ISL_2775272, EPI_ISL_2775273, EPI_ISL_2775274, EPI_ISL_2775275, EPI_ISL_2775276, EPI_ISL_2775277, EPI_ISL_2775278, EPI_ISL_2775279, EPI_ISL_2775280, EPI_ISL_2775281, EPI_ISL_2775282, EPI_ISL_2775283, EPI_ISL_2775284, EPI_ISL_2775285, EPI_ISL_2775286, EPI_ISL_2775287, EPI_ISL_2789347 | | | |
| see above | CSIR-Centre for Cellular and Molecular Biology | CSIR-Centre for Cellular and Molecular Biology-INSACOG | ; Amareshwar Vodapalli; Ara Sreenivas; Archana Bharadwaj Siva; B Himasri; Blessy B John; Divya Tej Sowpati; Jandhyala Sai Krishna; Karthik Bharadwaj Tallapaka; Lamuk Zaveri; Onkar Kulkarni; Payel Mukherjee; Priya Nurkurthy; Rakesh K Mishra; Sharath Chandra Thota; Shreekant Verma; Sofia Banu; Sumedha Avadhanula; Tulasi Nagabandi; Valli Nagalakshmi Undamatla; Vidhyadhari Methuku; Viswagithe S L |
| EPI_ISL_2441442, EPI_ISL_2441443, EPI_ISL_2441444, EPI_ISL_2441445, EPI_ISL_2441446, EPI_ISL_2441447, EPI_ISL_2441448, EPI_ISL_2441449, EPI_ISL_2441450, EPI_ISL_2441451, EPI_ISL_2441452, EPI_ISL_2441453, EPI_ISL_2441454, EPI_ISL_2441455, EPI_ISL_2441456, EPI_ISL_2441457, EPI_ISL_2441458, EPI_ISL_2441459, EPI_ISL_2441460, EPI_ISL_2441461, EPI_ISL_2441462, EPI_ISL_2441463, EPI_ISL_2441464, EPI_ISL_2441465, EPI_ISL_2441466, EPI_ISL_2441467, EPI_ISL_2441468, EPI_ISL_2441469, EPI_ISL_2441470, EPI_ISL_2441471 | | | |
| see above | CSIR-National Environmental Engineering Research Institute | CSIR-Centre for Cellular and Molecular Biology-INSACOG | Amareshwar Vodapalli; Ara Sreenivas; Archana Bharadwaj Siva; B Himasri; Divya Tej Sowpati; Karthik Bharadwaj Tallapaka; Krishna Khairnar; Lamuk Zaveri; Onkar Kulkarni; Payel Mukherjee; Priya Nurkurthy; Rakesh K Mishra; Shreekant Verma; Sofia Banu; Sumedha Avadhanula; Tulasi Nagabandi; Valli Nagalakshmi Undamatla; Vidhyadhari Methuku |
| EPI_ISL_2424135, EPI_ISL_2426145 | Clinical Virology Laboratory, Institute of Liver and Biliary Sciences | ILBS - IGIB | Amit Pandey; Anurag Agrawal; Aparna S; Bharathram Uppili; Chhagan Bihari Sharma; Ekta Gupta; Manish Kumar; Mohammed Faruq; Neha Jha; Partha Chattopadhyay; Pooja Sharma; Pratibha Kale; Priti Devi; Priyanka Mehta; Rajesh Pandey; Ranjeet Maurya; Sarfaraz Alam; Sheeba Saifi; Sheetalnath Rooge; Shiv Kumar Sarin |
| EPI_ISL_2379418, EPI_ISL_2379419, EPI_ISL_2379420, EPI_ISL_2379421, EPI_ISL_2379422, EPI_ISL_2842475, EPI_ISL_2842476, EPI_ISL_2842477 | | | |
| see above | DSU CHAMRAJANAGARA | INSACOG-KA, NIMHANS | Ananthapadmanabha Kotambail; Anita S Desai; Anson Kunjumon George; Chetan G K; Chitra Pattabiraman; Darshan Sreenivas; Ellango Ramasamy; Gautham Arunachal Udupi; Mahesh Kumar.C.S; Pramada Prasad; Sony Sharma; V Ravi |
| EPI_ISL_2728343, EPI_ISL_2728344, EPI_ISL_2728345, EPI_ISL_2728346, EPI_ISL_2728347 | DSU DAVANAGERE | INSACOG-KA, NIMHANS | Ananthapadmanabha Kotambail; Anita S Desai; Anson Kunjumon George; Chetan G K; Chitra Pattabiraman; Darshan Sreenivas; Ellango Ramasamy; Gautham Arunachal Udupi; Mahesh Kumar.C.S; Sony Sharma; V Ravi |
| EPI_ISL_2728308, EPI_ISL_2728317, EPI_ISL_2728318, EPI_ISL_2728319, EPI_ISL_2728320, EPI_ISL_2728321, EPI_ISL_2728322, EPI_ISL_2728323, EPI_ISL_2728325, EPI_ISL_2728326, EPI_ISL_2728327, EPI_ISL_2728328, EPI_ISL_2728329, EPI_ISL_2842491, EPI_ISL_2842493, EPI_ISL_2842494, EPI_ISL_2842495, EPI_ISL_2842496, EPI_ISL_2842497, EPI_ISL_2842498, EPI_ISL_2842499 | | | |
| see above | DSU HASSAN | INSACOG-KA, NIMHANS | Ananthapadmanabha Kotambail; Anita S Desai; Anson Kunjumon George; Chetan G K; Chitra Pattabiraman; Darshan Sreenivas; Ellango Ramasamy; Gautham Arunachal Udupi; Mahesh Kumar.C.S; Sony Sharma; V Ravi |
| EPI_ISL_2728330, EPI_ISL_2728331, EPI_ISL_2728332, EPI_ISL_2728333, EPI_ISL_2728334, EPI_ISL_2728335, EPI_ISL_2728336, EPI_ISL_2728337, EPI_ISL_2728338, EPI_ISL_2728339, EPI_ISL_2728340, EPI_ISL_2728341, EPI_ISL_2842481, EPI_ISL_2842482, EPI_ISL_2842483, EPI_ISL_2842484, EPI_ISL_2842485, EPI_ISL_2842486, EPI_ISL_2842487, EPI_ISL_2842488, EPI_ISL_2842489, EPI_ISL_2842490 | | | |
| see above | DSU KALABURAGI | INSACOG-KA, NIMHANS | Ananthapadmanabha Kotambail; Anita S Desai; Anson Kunjumon George; Chetan G K; Chitra Pattabiraman; Darshan Sreenivas; Ellango Ramasamy; Gautham Arunachal Udupi; Mahesh Kumar.C.S; Sony Sharma; V Ravi |
| EPI_ISL_2379423, EPI_ISL_2379424, EPI_ISL_2379425, EPI_ISL_2379429, EPI_ISL_2842478, EPI_ISL_2842479, EPI_ISL_2842480 | | | |
| see above | DSU KODAGU | INSACOG-KA, NIMHANS | Ananthapadmanabha Kotambail; Anita S Desai; Anson Kunjumon George; Chetan G K; Chitra Pattabiraman; Darshan Sreenivas; Ellango Ramasamy; Gautham Arunachal Udupi; Mahesh Kumar.C.S; Pramada Prasad; Sony Sharma; V Ravi |
| EPI_ISL_2379412, EPI_ISL_2379413, EPI_ISL_2379416, EPI_ISL_2379417, EPI_ISL_2842473, EPI_ISL_2842474 | DSU MANDYA | INSACOG-KA, NIMHANS | Ananthapadmanabha Kotambail; Anita S Desai; Anson Kunjumon George; Chetan G K; Chitra Pattabiraman; Darshan Sreenivas; Ellango Ramasamy; Gautham Arunachal Udupi; Mahesh Kumar.C.S; Pramada Prasad; Sony Sharma; V Ravi |
| EPI_ISL_2379404, EPI_ISL_2379405, EPI_ISL_2379406, EPI_ISL_2379407, EPI_ISL_2379408, EPI_ISL_2379409, EPI_ISL_2379410, EPI_ISL_2379411, EPI_ISL_2842467, EPI_ISL_2842468, EPI_ISL_2842469, EPI_ISL_2842470, EPI_ISL_2842471, EPI_ISL_2842472 | | | |
| see above | DSU MYSURU | INSACOG-KA, NIMHANS | Ananthapadmanabha Kotambail; Anita S Desai; Anson Kunjumon George; Chetan G K; Chitra Pattabiraman; Darshan Sreenivas; Ellango Ramasamy; Gautham Arunachal Udupi; Mahesh Kumar.C.S; Pramada Prasad; Sony Sharma; V Ravi |
| EPI_ISL_2728353, EPI_ISL_2728355, EPI_ISL_2728356, EPI_ISL_2728357, EPI_ISL_2728365, EPI_ISL_2728367, EPI_ISL_2728368, EPI_ISL_2728369, EPI_ISL_2728371, EPI_ISL_2728372, EPI_ISL_2728373, EPI_ISL_2728374, EPI_ISL_2728376, EPI_ISL_2728377, EPI_ISL_2728378, EPI_ISL_2728379, EPI_ISL_2728383, EPI_ISL_2728393, EPI_ISL_2728394, EPI_ISL_2728397, EPI_ISL_2728402, EPI_ISL_2728403, EPI_ISL_2728405, EPI_ISL_2728410, EPI_ISL_2728413, EPI_ISL_2728416, EPI_ISL_2728422, EPI_ISL_2728423, EPI_ISL_2728426, EPI_ISL_2728427, EPI_ISL_2842431, EPI_ISL_2842432, EPI_ISL_2842433, EPI_ISL_2842434, EPI_ISL_2842435, EPI_ISL_2842436 | | | |
| see above | DSU VIJAYAPURA | INSACOG-KA, NIMHANS | Ananthapadmanabha Kotambail; Anita S Desai; Anson Kunjumon George; Chetan G K; Chitra Pattabiraman; Darshan Sreenivas; Ellango Ramasamy; Gautham Arunachal Udupi; Mahesh Kumar.C.S; Sony Sharma; V Ravi |
| EPI_ISL_2897960, EPI_ISL_2897961, EPI_ISL_2897962, EPI_ISL_2897963, EPI_ISL_2897964, EPI_ISL_2897965, EPI_ISL_2897966, EPI_ISL_2897967, EPI_ISL_2897968, EPI_ISL_2897969, EPI_ISL_2897970, EPI_ISL_2897971, EPI_ISL_2897972, EPI_ISL_2897973, EPI_ISL_2897974, EPI_ISL_2897975, EPI_ISL_2897976, EPI_ISL_2897977, EPI_ISL_2897978, EPI_ISL_2897979, EPI_ISL_2897980, EPI_ISL_2897981, EPI_ISL_2897982, EPI_ISL_2897983, EPI_ISL_2897984, EPI_ISL_2897985, EPI_ISL_2897986, EPI_ISL_2897987, EPI_ISL_2897988, EPI_ISL_2897989, EPI_ISL_2897990, EPI_ISL_2897991 | | | |
| see above | District Hospital | CDFD | Ashwin Dalal; Asmita Gupta; Divya Vashisht; Murali Bashyam; Nagamani Kammili; Reelina Basu; Vinay Donipadi |
| EPI_ISL_2379463, EPI_ISL_2379466, EPI_ISL_2379474, EPI_ISL_2379482, EPI_ISL_2379483, EPI_ISL_2379484, EPI_ISL_2379485, EPI_ISL_2379489, EPI_ISL_2379490, EPI_ISL_2379510, EPI_ISL_2379511, EPI_ISL_2379521, EPI_ISL_2379531, EPI_ISL_2379535, EPI_ISL_2379542, EPI_ISL_2379546, EPI_ISL_2379547, EPI_ISL_2379548, EPI_ISL_2379549, EPI_ISL_2379551, EPI_ISL_2379552, EPI_ISL_2379553, EPI_ISL_2379554, EPI_ISL_2379555, EPI_ISL_2379556, EPI_ISL_2379557, EPI_ISL_2379563, EPI_ISL_2379567, EPI_ISL_2379579, EPI_ISL_2379588, EPI_ISL_2379589, EPI_ISL_2379590, EPI_ISL_2379591, EPI_ISL_2379592, EPI_ISL_2379593, EPI_ISL_2379614, EPI_ISL_2379630, EPI_ISL_2379631, EPI_ISL_2379636, EPI_ISL_2379637, EPI_ISL_2379643, EPI_ISL_2379644 | | | |
| see above | Dr S Raju, Director of Public Heath and Preventive Medicine | inStem NCBS - INSACOG | Uma Ramakrishnan Dasaradhi Palakodeti Aswin SaiNarain |
| EPI_ISL_2521778, EPI_ISL_2521779, EPI_ISL_2521780, EPI_ISL_2521781, EPI_ISL_2521782, EPI_ISL_2521783, EPI_ISL_2521784, EPI_ISL_2521785, EPI_ISL_2521786, EPI_ISL_2521787, EPI_ISL_2521788, EPI_ISL_2521789, EPI_ISL_2521790, EPI_ISL_2521799, EPI_ISL_2521800, EPI_ISL_2521811, EPI_ISL_2521822, EPI_ISL_2521843, EPI_ISL_2521846, EPI_ISL_2521847, EPI_ISL_2521848, EPI_ISL_2521857, EPI_ISL_2521868, EPI_ISL_2521870, EPI_ISL_2521871, EPI_ISL_2521874, EPI_ISL_2521875, EPI_ISL_2521879, EPI_ISL_2521880, EPI_ISL_2521881, EPI_ISL_2521882, EPI_ISL_2521883, EPI_ISL_2521884, EPI_ISL_2521885, EPI_ISL_2521886, EPI_ISL_2521887, EPI_ISL_2521888, EPI_ISL_2521889, EPI_ISL_2521890, EPI_ISL_2521891, EPI_ISL_2521892, EPI_ISL_2521893, EPI_ISL_2521903, EPI_ISL_2521904, EPI_ISL_2521905, EPI_ISL_2521906, EPI_ISL_2521908, EPI_ISL_2521909, EPI_ISL_2521910, EPI_ISL_2521911, EPI_ISL_2521912, EPI_ISL_2521913, EPI_ISL_2521914, EPI_ISL_2521915, EPI_ISL_2521916, EPI_ISL_2521917, EPI_ISL_2521918, EPI_ISL_2521919, EPI_ISL_2521920, EPI_ISL_2521921, EPI_ISL_2521922, EPI_ISL_2521923, EPI_ISL_2521924, EPI_ISL_2521925, EPI_ISL_2521926, EPI_ISL_2521927, EPI_ISL_2521947, EPI_ISL_2521948, EPI_ISL_2521949, EPI_ISL_2521950, EPI_ISL_2521959 | | | |
| see above | Dr S raju, Director of public heath and preventive medicine | inStem NCBS - INSACOG | Uma Ramakrishnan Dasaradhi Palakodeti Aswin SaiNarain |
| EPI_ISL_2723773, EPI_ISL_2723774, EPI_ISL_2723775, EPI_ISL_2723776, EPI_ISL_2723781, EPI_ISL_2723783, EPI_ISL_2723784, EPI_ISL_2723785, EPI_ISL_2723786, EPI_ISL_2723794, EPI_ISL_2723795, EPI_ISL_2723796, EPI_ISL_2723797, EPI_ISL_2723798, EPI_ISL_2723799, EPI_ISL_2723907, EPI_ISL_2723908, EPI_ISL_2723909, EPI_ISL_2723910, EPI_ISL_2723913, EPI_ISL_2723914, EPI_ISL_2723915, EPI_ISL_2723935, EPI_ISL_2723936, EPI_ISL_2723937, EPI_ISL_2723947, EPI_ISL_2723949, EPI_ISL_2723952, EPI_ISL_2723953, EPI_ISL_2723954, EPI_ISL_2723955, EPI_ISL_2723956, EPI_ISL_2723957, EPI_ISL_2723958, EPI_ISL_2723959, EPI_ISL_2723960, EPI_ISL_2724010, EPI_ISL_2724025, EPI_ISL_2724026, EPI_ISL_2724027, EPI_ISL_2724028, EPI_ISL_2724029, EPI_ISL_2724030, EPI_ISL_2724031, EPI_ISL_2724032, EPI_ISL_2724033, EPI_ISL_2724034, EPI_ISL_2724035, EPI_ISL_2724036, EPI_ISL_2724037, EPI_ISL_2724038, EPI_ISL_2724039, EPI_ISL_2724040, EPI_ISL_2724055, EPI_ISL_2724056, EPI_ISL_2724058, EPI_ISL_2724059, EPI_ISL_2724060, EPI_ISL_2724061, EPI_ISL_2724064, EPI_ISL_2724065, EPI_ISL_2724066, EPI_ISL_2724067, EPI_ISL_2724068, EPI_ISL_2724069, EPI_ISL_2724070, EPI_ISL_2724071, EPI_ISL_2724072, EPI_ISL_2724073, EPI_ISL_2724074, EPI_ISL_2724075, EPI_ISL_2724076, EPI_ISL_2724077, EPI_ISL_2724078, EPI_ISL_2724079, EPI_ISL_2724086, EPI_ISL_2724091, EPI_ISL_2724092, EPI_ISL_2724093, EPI_ISL_2724139, EPI_ISL_2724140, EPI_ISL_2724141, EPI_ISL_2724142, EPI_ISL_2724143, EPI_ISL_2724168, EPI_ISL_2724169, EPI_ISL_2724170, EPI_ISL_2724171, EPI_ISL_2724172, EPI_ISL_2724173, EPI_ISL_2724174, EPI_ISL_2724175, EPI_ISL_2724176, EPI_ISL_2724177, EPI_ISL_2724178, EPI_ISL_2724179, EPI_ISL_2724180, EPI_ISL_2724181, EPI_ISL_2724182, EPI_ISL_2724183, EPI_ISL_2724184, EPI_ISL_2724185, EPI_ISL_2724186, EPI_ISL_2724187, EPI_ISL_2724188, EPI_ISL_2724189, EPI_ISL_2724190, EPI_ISL_2724191, EPI_ISL_2724192, EPI_ISL_2724193, EPI_ISL_2724194, EPI_ISL_2724196, EPI_ISL_2724199, EPI_ISL_2724200, EPI_ISL_2724201, EPI_ISL_2724202, EPI_ISL_2724204, EPI_ISL_2724208, EPI_ISL_2724209, EPI_ISL_2724217, EPI_ISL_2724218, EPI_ISL_2724219, EPI_ISL_2724220, EPI_ISL_2724221, EPI_ISL_2724222, EPI_ISL_2724223, EPI_ISL_2724224, EPI_ISL_2724225, EPI_ISL_2724226, EPI_ISL_2724227, EPI_ISL_2724228, EPI_ISL_2724229, EPI_ISL_2724230, EPI_ISL_2724231, EPI_ISL_2724232, EPI_ISL_2724233, EPI_ISL_2724234, EPI_ISL_2724235, EPI_ISL_2724236, EPI_ISL_2724237, EPI_ISL_2724238, EPI_ISL_2724239, EPI_ISL_2724261, EPI_ISL_2724262, EPI_ISL_2724263, EPI_ISL_2724264, EPI_ISL_2724275, EPI_ISL_2724276, EPI_ISL_2724279, EPI_ISL_2724284, EPI_ISL_2724285, EPI_ISL_2724318, EPI_ISL_2724319, EPI_ISL_2724320, EPI_ISL_2724321, EPI_ISL_2724322, EPI_ISL_2724323, EPI_ISL_2724324, EPI_ISL_2724325, EPI_ISL_2724326, EPI_ISL_2724332, EPI_ISL_2724333, EPI_ISL_2724334, EPI_ISL_2724335, EPI_ISL_2724342, EPI_ISL_2724345, EPI_ISL_2724346, EPI_ISL_2724347, EPI_ISL_2724349, EPI_ISL_2724350, EPI_ISL_2724351 | | | |
| see above | Dr S raju, Director of public heath and preventive medicine | inStem NCBS – INSACOG | Uma Ramakrishnan Dasaradhi Palakodeti Aswin SaiNarain |
| EPI_ISL_2723771, EPI_ISL_2723814, EPI_ISL_2723815, EPI_ISL_2723823, EPI_ISL_2723824, EPI_ISL_2723825, EPI_ISL_2723826, EPI_ISL_2723827, EPI_ISL_2723828, EPI_ISL_2723829, EPI_ISL_2723980, EPI_ISL_2723981, EPI_ISL_2723982, EPI_ISL_2723983, EPI_ISL_2723984, EPI_ISL_2723985, EPI_ISL_2723986, EPI_ISL_2723987, EPI_ISL_2723988, EPI_ISL_2723989, EPI_ISL_2723990, EPI_ISL_2723991, EPI_ISL_2723992, EPI_ISL_2723993, EPI_ISL_2723994, EPI_ISL_2723995, EPI_ISL_2723996, EPI_ISL_2723997, EPI_ISL_2723998, EPI_ISL_2723999, EPI_ISL_2724000, EPI_ISL_2724001, EPI_ISL_2724002, EPI_ISL_2724352, EPI_ISL_2724353, EPI_ISL_2724354, EPI_ISL_2724355, EPI_ISL_2724356, EPI_ISL_2724357 | | | |
| see above | Dr. Madhusudhan, MBBD, MD, ICMR VRDL | inStem NCBS – INSACOG | Uma Ramakrishnan Dasaradhi Palakodeti Aswin SaiNarain |
| EPI_ISL_2379384, EPI_ISL_2379385, EPI_ISL_2379386 | GANDHIGRAMA | INSACOG-KA, NIMHANS | Ananthapadmanabha Kotambail; Anita S Desai; Anson Kunjumon George; Chetan G K; Chitra Pattabiraman; Darshan Sreenivas; Gautham Arunachal Udupi; Pramada Prasad; V Ravi |
| EPI_ISL_2544812, EPI_ISL_2544816, EPI_ISL_2544817, EPI_ISL_2544818, EPI_ISL_2544819, EPI_ISL_2544820, EPI_ISL_2544821 | | | |
| see above | GMERS Government Medical College (GMC), Gandhinagar | Gujarat Biotechnology Research Centre | Chaitanya Joshi; Dinesh Kumar; Gaurishankar Shrimali; Janvi Raval; Madhvi Joshi; Nitesh Shah; Nitin Savaliya; Ramesh Pandit; Sonal Sharma; Twinkle Soni; Umang Mishra; Zarna Patel; Zuber Saiyed |
| EPI_ISL_2544823, EPI_ISL_2544824, EPI_ISL_2544825, EPI_ISL_2544826, EPI_ISL_2544827, EPI_ISL_2544828, EPI_ISL_2544829, EPI_ISL_2544830, EPI_ISL_2544831, EPI_ISL_2544832, EPI_ISL_2544833, EPI_ISL_2544834, EPI_ISL_2544835, EPI_ISL_2562085, EPI_ISL_2562086, EPI_ISL_2562087, EPI_ISL_2562088, EPI_ISL_2562089, EPI_ISL_2562090, EPI_ISL_2562091, EPI_ISL_2562092, EPI_ISL_2562093, EPI_ISL_2562094, EPI_ISL_2562095, EPI_ISL_2562096, EPI_ISL_2562097, EPI_ISL_2562098, EPI_ISL_2562099, EPI_ISL_2562100, EPI_ISL_2562101, EPI_ISL_2562102, EPI_ISL_2562103, EPI_ISL_2562104, EPI_ISL_2562105, EPI_ISL_2562106, EPI_ISL_2562107, EPI_ISL_2562108, EPI_ISL_2562109, EPI_ISL_2562110, EPI_ISL_2562111 | | | |
| see above | GMERS Government Medical College (GMC), Gotri | Gujarat Biotechnology Research Centre | Bithika Duttaroy; Chaitanya Joshi; Dinesh Kumar; Janvi Raval; Madhvi Joshi; Nitesh Shah; Nitin Savaliya; Ramesh Pandit; Sonal Sharma; Twinkle Soni; Umang Mishra; Zarna Patel; Zuber Saiyed |
| EPI_ISL_2379372, EPI_ISL_2379373, EPI_ISL_2379374 | GOVIDRAJNAGAR | INSACOG-KA, NIMHANS | Ananthapadmanabha Kotambail; Anita S Desai; Anson Kunjumon George; Chetan G K; Chitra Pattabiraman; Darshan Sreenivas; Gautham Arunachal Udupi; Pramada Prasad; V Ravi |
| EPI_ISL_2897906, EPI_ISL_2897910, EPI_ISL_2897911, EPI_ISL_2897912, EPI_ISL_2897913, EPI_ISL_2897914, EPI_ISL_2897915, EPI_ISL_2897916, EPI_ISL_2897917, EPI_ISL_2897918, EPI_ISL_2897919, EPI_ISL_2897920, EPI_ISL_2897921, EPI_ISL_2897922, EPI_ISL_2897923, EPI_ISL_2897924, EPI_ISL_2897925, EPI_ISL_2897926, EPI_ISL_2897927, EPI_ISL_2897928, EPI_ISL_2897929, EPI_ISL_2897930, EPI_ISL_2897931, EPI_ISL_2897932, EPI_ISL_2897933, EPI_ISL_2897934, EPI_ISL_2897935, EPI_ISL_2924152, EPI_ISL_2924153, EPI_ISL_2924154, EPI_ISL_2924155, EPI_ISL_2924156, EPI_ISL_2924157, EPI_ISL_2924158, EPI_ISL_2924159, EPI_ISL_2924160, EPI_ISL_2924161, EPI_ISL_2924162, EPI_ISL_2924163, EPI_ISL_2924164, EPI_ISL_2924165, EPI_ISL_2924166, EPI_ISL_2924167, EPI_ISL_2924168, EPI_ISL_2924169, EPI_ISL_2924170, EPI_ISL_2924217, EPI_ISL_2924218, EPI_ISL_2924219, EPI_ISL_2924220, EPI_ISL_2924221, EPI_ISL_2924222, EPI_ISL_2924223, EPI_ISL_2924224, EPI_ISL_2924225, EPI_ISL_2924226, EPI_ISL_2924227, EPI_ISL_2924228, EPI_ISL_2924229, EPI_ISL_2924230, EPI_ISL_2924231, EPI_ISL_2924232, EPI_ISL_2924234, EPI_ISL_2924235, EPI_ISL_2924264, EPI_ISL_2924265, EPI_ISL_2924266, EPI_ISL_2924267 | | | |
| see above | Gandhi Hospital | CDFD | Ashwin Dalal; Asmita Gupta; Divya Vashisht; Murali Bashyam; Nagamani Kammili; Reelina Basu; Vinay Donipadi |
| EPI_ISL_2897851, EPI_ISL_2897852, EPI_ISL_2897853, EPI_ISL_2897854, EPI_ISL_2897855, EPI_ISL_2897856, EPI_ISL_2897857, EPI_ISL_2897858, EPI_ISL_2897859, EPI_ISL_2897860, EPI_ISL_2897861, EPI_ISL_2897862, EPI_ISL_2897863, EPI_ISL_2897864, EPI_ISL_2897865, EPI_ISL_2897866, EPI_ISL_2897867, EPI_ISL_2897868, EPI_ISL_2897869, EPI_ISL_2897870, EPI_ISL_2897871, EPI_ISL_2897872, EPI_ISL_2897873, EPI_ISL_2897874, EPI_ISL_2897875, EPI_ISL_2924203, EPI_ISL_2924204, EPI_ISL_2924205, EPI_ISL_2924206, EPI_ISL_2924207, EPI_ISL_2924208, EPI_ISL_2924209, EPI_ISL_2924210, EPI_ISL_2924211, EPI_ISL_2924212, EPI_ISL_2924213, EPI_ISL_2924214, EPI_ISL_2924215 | | | |
| see above | Government Fever Hospital | CDFD | Ashwin Dalal; Asmita Gupta; Divya Vashisht; Murali Bashyam; Nagamani Kammili; Reelina Basu; Vinay Donipadi |
| EPI_ISL_2897943, EPI_ISL_2897944, EPI_ISL_2897945, EPI_ISL_2897946, EPI_ISL_2897947, EPI_ISL_2897948, EPI_ISL_2897949 | | | |
| see above | Government General Hospital | CDFD | Ashwin Dalal; Asmita Gupta; Divya Vashisht; Murali Bashyam; Nagamani Kammili; Reelina Basu; Vinay Donipadi |
| EPI_ISL_2897950, EPI_ISL_2897951, EPI_ISL_2897952, EPI_ISL_2897953, EPI_ISL_2897954, EPI_ISL_2897955, EPI_ISL_2897956, EPI_ISL_2897957, EPI_ISL_2897958, EPI_ISL_2897959 | | | |
| see above | Government Maternity Hospital | CDFD | Ashwin Dalal; Asmita Gupta; Divya Vashisht; Murali Bashyam; Nagamani Kammili; Reelina Basu; Vinay Donipadi |
| EPI_ISL_2897936, EPI_ISL_2897937, EPI_ISL_2897938, EPI_ISL_2897939, EPI_ISL_2897940, EPI_ISL_2897941, EPI_ISL_2897942 | | | |
| see above | Government Medical College | CDFD | Ashwin Dalal; Asmita Gupta; Divya Vashisht; Murali Bashyam; Nagamani Kammili; Reelina Basu; Vinay Donipadi |
| EPI_ISL_2379363, EPI_ISL_2379364, | HOSAHALLI | INSACOG-KA, NIMHANS | Ananthapadmanabha Kotambail; Anita S Desai; Anson Kunjumon George; Chetan G K; Chitra Pattabiraman; Darshan Sreenivas; Gautham Arunachal Udupi; Pramada Prasad; V Ravi |

EPI_ISL_2379365, EPI_ISL_2379366, EPI_ISL_2379380

EPI_ISL_2530250, EPI_ISL_2530252, EPI_ISL_2530253, EPI_ISL_2530255, EPI_ISL_2530256, EPI_ISL_2530258, EPI_ISL_2530259, EPI_ISL_2530261, EPI_ISL_2530262, EPI_ISL_2530264, EPI_ISL_2530268, EPI_ISL_2530270, EPI_ISL_2530286, EPI_ISL_2530290, EPI_ISL_2530384, EPI_ISL_2530388, EPI_ISL_2530390, EPI_ISL_2530391, EPI_ISL_2530392, EPI_ISL_2530394

see above
Hematopathology Laboratory, ACTREC, TMC
Hematopathology Laboratory, ACTREC, TMC
ACTREC; Hematopathology Laboratory

EPI_ISL_2361830, EPI_ISL_2361831, EPI_ISL_2361832, EPI_ISL_2361833, EPI_ISL_2361834, EPI_ISL_2361835, EPI_ISL_2361836, EPI_ISL_2361837, EPI_ISL_2361838, EPI_ISL_2361839, EPI_ISL_2361840, EPI_ISL_2361841, EPI_ISL_2361842, EPI_ISL_2361843, EPI_ISL_2361844, EPI_ISL_2361845, EPI_ISL_2361846, EPI_ISL_2361847, EPI_ISL_2361848, EPI_ISL_2361849, EPI_ISL_2361850, EPI_ISL_2361851, EPI_ISL_2361852, EPI_ISL_2361853, EPI_ISL_2361854, EPI_ISL_2361855, EPI_ISL_2361856, EPI_ISL_2361857, EPI_ISL_2361858, EPI_ISL_2361859, EPI_ISL_2361860, EPI_ISL_2361861, EPI_ISL_2361862, EPI_ISL_2361863, EPI_ISL_2361864, EPI_ISL_2361865, EPI_ISL_2361866, EPI_ISL_2361867, EPI_ISL_2361868, EPI_ISL_2361869, EPI_ISL_2361870, EPI_ISL_2361871, EPI_ISL_2361872, EPI_ISL_2361873, EPI_ISL_2361874, EPI_ISL_2361875, EPI_ISL_2361876, EPI_ISL_2361877, EPI_ISL_2361878

see above
ICMR-National Institute for Research In Tuberculosis
NIV Influenza
Dr Luke Elizabeth Hanna; Dr Nandhini Palani; Dr Padmapriyadarsini.C; Dr S Siva Kumar; Mr Anbazagan Selvaraj

EPI_ISL_2131515, EPI_ISL_2131516, EPI_ISL_2131517, EPI_ISL_2131518, EPI_ISL_2131519, EPI_ISL_2131520, EPI_ISL_2131521, EPI_ISL_2131522, EPI_ISL_2131523, EPI_ISL_2131524, EPI_ISL_2131525, EPI_ISL_2131526, EPI_ISL_2131527, EPI_ISL_2131528, EPI_ISL_2131529, EPI_ISL_2131530, EPI_ISL_2131531, EPI_ISL_2131532, EPI_ISL_2131533, EPI_ISL_2131534, EPI_ISL_2131547, EPI_ISL_2131548, EPI_ISL_2131549, EPI_ISL_2131550, EPI_ISL_2131551, EPI_ISL_2131552, EPI_ISL_2131553, EPI_ISL_2131554, EPI_ISL_2131563, EPI_ISL_2131566, EPI_ISL_2131567, EPI_ISL_2131568, EPI_ISL_2131569, EPI_ISL_2131570, EPI_ISL_2131571, EPI_ISL_2131572, EPI_ISL_2546689, EPI_ISL_2546690, EPI_ISL_2546691, EPI_ISL_2546692, EPI_ISL_2546693, EPI_ISL_2546694, EPI_ISL_2546695, EPI_ISL_2546696, EPI_ISL_2546697, EPI_ISL_2546710, EPI_ISL_2546711, EPI_ISL_2546712, EPI_ISL_2546713, EPI_ISL_2546714, EPI_ISL_2546715, EPI_ISL_2546716, EPI_ISL_2546717, EPI_ISL_2546718, EPI_ISL_2546719, EPI_ISL_2546720, EPI_ISL_2546721, EPI_ISL_2546722, EPI_ISL_2546723, EPI_ISL_2546724, EPI_ISL_2546725, EPI_ISL_2546726, EPI_ISL_2546727, EPI_ISL_2546728, EPI_ISL_2546729, EPI_ISL_2546730, EPI_ISL_2546731, EPI_ISL_2546732, EPI_ISL_2546733, EPI_ISL_2546734, EPI_ISL_2546735, EPI_ISL_2546736, EPI_ISL_2546737, EPI_ISL_2546738, EPI_ISL_2546739, EPI_ISL_2546740, EPI_ISL_2546741, EPI_ISL_2546742, EPI_ISL_2546743, EPI_ISL_2546744, EPI_ISL_2546745, EPI_ISL_2546746, EPI_ISL_2546747, EPI_ISL_2546748, EPI_ISL_2546749, EPI_ISL_2546750, EPI_ISL_2546751, EPI_ISL_2546752, EPI_ISL_2546753, EPI_ISL_2546754, EPI_ISL_2546755, EPI_ISL_2546756, EPI_ISL_2546757, EPI_ISL_2546758, EPI_ISL_2546759, EPI_ISL_2546760, EPI_ISL_2546761, EPI_ISL_2546762, EPI_ISL_2546763, EPI_ISL_2546764, EPI_ISL_2546765, EPI_ISL_2546766, EPI_ISL_2546767, EPI_ISL_2546768, EPI_ISL_2546769, EPI_ISL_2546770, EPI_ISL_2546771, EPI_ISL_2546772, EPI_ISL_2546773, EPI_ISL_2546774, EPI_ISL_2546775, EPI_ISL_2546776, EPI_ISL_2546777, EPI_ISL_2546778, EPI_ISL_2546779, EPI_ISL_2546780, EPI_ISL_2546781, EPI_ISL_2546782, EPI_ISL_2546783, EPI_ISL_2546784, EPI_ISL_2546785, EPI_ISL_2546786, EPI_ISL_2546787, EPI_ISL_2546788, EPI_ISL_2546789, EPI_ISL_2546790, EPI_ISL_2546791, EPI_ISL_2546792, EPI_ISL_2546793, EPI_ISL_2546794, EPI_ISL_2546795, EPI_ISL_2546796, EPI_ISL_2546797, EPI_ISL_2546798, EPI_ISL_2546799, EPI_ISL_2546800, EPI_ISL_2546801, EPI_ISL_2546802, EPI_ISL_2546803, EPI_ISL_2546804, EPI_ISL_2546805, EPI_ISL_2546806, EPI_ISL_2546807, EPI_ISL_2546808, EPI_ISL_2546809, EPI_ISL_2546810, EPI_ISL_2546811, EPI_ISL_2546812, EPI_ISL_2546813, EPI_ISL_2546814, EPI_ISL_2546815, EPI_ISL_2546816, EPI_ISL_2546817, EPI_ISL_2546818, EPI_ISL_2546819, EPI_ISL_2546820, EPI_ISL_2546821, EPI_ISL_2546822, EPI_ISL_2546823, EPI_ISL_2546824, EPI_ISL_2546825, EPI_ISL_2546826, EPI_ISL_2546827, EPI_ISL_2546828, EPI_ISL_2546830, EPI_ISL_2546831, EPI_ISL_2546832, EPI_ISL_2546833, EPI_ISL_2546834, EPI_ISL_2546835, EPI_ISL_2546836, EPI_ISL_2546837, EPI_ISL_2546838, EPI_ISL_2546839, EPI_ISL_2546840, EPI_ISL_2546841, EPI_ISL_2546842, EPI_ISL_2546843, EPI_ISL_2546844, EPI_ISL_2546845, EPI_ISL_2546846, EPI_ISL_2546847, EPI_ISL_2546848, EPI_ISL_2546849, EPI_ISL_2546850, EPI_ISL_2546851, EPI_ISL_2546852, EPI_ISL_2546853, EPI_ISL_2546854, EPI_ISL_2546855, EPI_ISL_2546856, EPI_ISL_2546857, EPI_ISL_2546858, EPI_ISL_2546859, EPI_ISL_2546860, EPI_ISL_2546861, EPI_ISL_2546862, EPI_ISL_2546863, EPI_ISL_2546864, EPI_ISL_2546865, EPI_ISL_2546866, EPI_ISL_2546867, EPI_ISL_2546868, EPI_ISL_2546869, EPI_ISL_2546870, EPI_ISL_2546871, EPI_ISL_2546872, EPI_ISL_2546873, EPI_ISL_2546874, EPI_ISL_2546875, EPI_ISL_2546876, EPI_ISL_2546877, EPI_ISL_2546878, EPI_ISL_2546879, EPI_ISL_2546880, EPI_ISL_2546881, EPI_ISL_2546882, EPI_ISL_2546883, EPI_ISL_2546884, EPI_ISL_2546885, EPI_ISL_2546886, EPI_ISL_2546887, EPI_ISL_2546888, EPI_ISL_2546889, EPI_ISL_2546890, EPI_ISL_2546891, EPI_ISL_2546892, EPI_ISL_2546893, EPI_ISL_2546894, EPI_ISL_2546895, EPI_ISL_2546896, EPI_ISL_2546897, EPI_ISL_2546898, EPI_ISL_2546899, EPI_ISL_2546900, EPI_ISL_2547069, EPI_ISL_2547070, EPI_ISL_2547071, EPI_ISL_2547072, EPI_ISL_2547073, EPI_ISL_2547074, EPI_ISL_2547075, EPI_ISL_2547076, EPI_ISL_2547077, EPI_ISL_2547078, EPI_ISL_2547079, EPI_ISL_2547080, EPI_ISL_2547081, EPI_ISL_2547082, EPI_ISL_2547083, EPI_ISL_2547084, EPI_ISL_2547085, EPI_ISL_2547086, EPI_ISL_2547087, EPI_ISL_2547088, EPI_ISL_2547089, EPI_ISL_2547090, EPI_ISL_2547091, EPI_ISL_2547092, EPI_ISL_2547093, EPI_ISL_2547094, EPI_ISL_2547095, EPI_ISL_2547096, EPI_ISL_2547097, EPI_ISL_2547098, EPI_ISL_2547099, EPI_ISL_2547100, EPI_ISL_2547101, EPI_ISL_2547102, EPI_ISL_2547103, EPI_ISL_2547104, EPI_ISL_2547105, EPI_ISL_2547106, EPI_ISL_2547107, EPI_ISL_2547108, EPI_ISL_2547109, EPI_ISL_2547110, EPI_ISL_2547111, EPI_ISL_2547112, EPI_ISL_2547113, EPI_ISL_2547114, EPI_ISL_2547115, EPI_ISL_2547116, EPI_ISL_2547117, EPI_ISL_2547118, EPI_ISL_2547119, EPI_ISL_2547120, EPI_ISL_2547121, EPI_ISL_2547122, EPI_ISL_2547123, EPI_ISL_2547124, EPI_ISL_2547125, EPI_ISL_2547126, EPI_ISL_2547127, EPI_ISL_2547128, EPI_ISL_2547129, EPI_ISL_2547130, EPI_ISL_2547131, EPI_ISL_2547132, EPI_ISL_2547133, EPI_ISL_2547134, EPI_ISL_2547135, EPI_ISL_2547136, EPI_ISL_2547137, EPI_ISL_2547138, EPI_ISL_2547139, EPI_ISL_2547140, EPI_ISL_2547141, EPI_ISL_2547142, EPI_ISL_2547143, EPI_ISL_2547144, EPI_ISL_2547145, EPI_ISL_2547146, EPI_ISL_2547147, EPI_ISL_2547148, EPI_ISL_2547149, EPI_ISL_2547150, EPI_ISL_2547151, EPI_ISL_2547152, EPI_ISL_2547153, EPI_ISL_2547154, EPI_ISL_2547155, EPI_ISL_2547156, EPI_ISL_2547157, EPI_ISL_2547158, EPI_ISL_2547159, EPI_ISL_2547160, EPI_ISL_2547161, EPI_ISL_2547162, EPI_ISL_2547163, EPI_ISL_2547164, EPI_ISL_2547165, EPI_ISL_2547168, EPI_ISL_2547169, EPI_ISL_2547170, EPI_ISL_2547171, EPI_ISL_2547172, EPI_ISL_2547173, EPI_ISL_2547174, EPI_ISL_2547175, EPI_ISL_2547176, EPI_ISL_2547177, EPI_ISL_2547178, EPI_ISL_2547179, EPI_ISL_2547180, EPI_ISL_2547181, EPI_ISL_2547182, EPI_ISL_2547183, EPI_ISL_2547184, EPI_ISL_2547185, EPI_ISL_2547186, EPI_ISL_2547187, EPI_ISL_2547188, EPI_ISL_2547189, EPI_ISL_2547190, EPI_ISL_2547191, EPI_ISL_2547192, EPI_ISL_2547193, EPI_ISL_2547194, EPI_ISL_2547195, EPI_ISL_2547196, EPI_ISL_2547197, EPI_ISL_2547198, EPI_ISL_2547199, EPI_ISL_2547200, EPI_ISL_2547201, EPI_ISL_2547202, EPI_ISL_2547203, EPI_ISL_2547204, EPI_ISL_2547205, EPI_ISL_2547206, EPI_ISL_2547207, EPI_ISL_2547208, EPI_ISL_2547209, EPI_ISL_2547210, EPI_ISL_2547211, EPI_ISL_2547212, EPI_ISL_2547213, EPI_ISL_2547214, EPI_ISL_2547215, EPI_ISL_2547216, EPI_ISL_2547217, EPI_ISL_2547218, EPI_ISL_2547219, EPI_ISL_2547220, EPI_ISL_2547221, EPI_ISL_2547222, EPI_ISL_2547223, EPI_ISL_2547224, EPI_ISL_2547225, EPI_ISL_2547226, EPI_ISL_2547227, EPI_ISL_2547228, EPI_ISL_2547229, EPI_ISL_2547230, EPI_ISL_2547231, EPI_ISL_2547232, EPI_ISL_2547233, EPI_ISL_2547234, EPI_ISL_2547235, EPI_ISL_2547236, EPI_ISL_2547237, EPI_ISL_2547238, EPI_ISL_2547239, EPI_ISL_2547240, EPI_ISL_2547241, EPI_ISL_2547242, EPI_ISL_2547243, EPI_ISL_2547244, EPI_ISL_2547245, EPI_ISL_2547246, EPI_ISL_2547247, EPI_ISL_2547248, EPI_ISL_2547249, EPI_ISL_2547250, EPI_ISL_2547251, EPI_ISL_2547252, EPI_ISL_2547253, EPI_ISL_2547254, EPI_ISL_2547255, EPI_ISL_2547256, EPI_ISL_2547257, EPI_ISL_2547258, EPI_ISL_2547259, EPI_ISL_2547260, EPI_ISL_2547261, EPI_ISL_2547262, EPI_ISL_2547263, EPI_ISL_2547264, EPI_ISL_2547265, EPI_ISL_2547266, EPI_ISL_2547267, EPI_ISL_2547268, EPI_ISL_2547269, EPI_ISL_2547270, EPI_ISL_2547271, EPI_ISL_2547272, EPI_ISL_2547273, EPI_ISL_2547274, EPI_ISL_2547275, EPI_ISL_2547276, EPI_ISL_2547277, EPI_ISL_2547278, EPI_ISL_2547279, EPI_ISL_2547280, EPI_ISL_2547281, EPI_ISL_2547282, EPI_ISL_2547283, EPI_ISL_2547284

see above
ICMR-National Institute of Virology - INSACOG
NIV Influenza
Dr. Varsha Potdar; Dr. Varsha Potdar and NIC Team

EPI_ISL_2880356, EPI_ISL_2880357, EPI_ISL_2880358, EPI_ISL_2880359, EPI_ISL_2880360, EPI_ISL_2880361, EPI_ISL_2880362, EPI_ISL_2880363, EPI_ISL_2880364, EPI_ISL_2880365, EPI_ISL_2880366, EPI_ISL_2880367, EPI_ISL_2880368, EPI_ISL_2880369, EPI_ISL_2880370, EPI_ISL_2880371, EPI_ISL_2880372, EPI_ISL_2880373, EPI_ISL_2880374, EPI_ISL_2880375, EPI_ISL_2880376, EPI_ISL_2880377, EPI_ISL_2880378, EPI_ISL_2880379, EPI_ISL_2880380, EPI_ISL_2880381, EPI_ISL_2880382, EPI_ISL_2880383, EPI_ISL_2880384, EPI_ISL_2880385, EPI_ISL_2880386, EPI_ISL_2880387, EPI_ISL_2880388, EPI_ISL_2880389, EPI_ISL_2880390, EPI_ISL_2880391, EPI_ISL_2880392, EPI_ISL_2880393, EPI_ISL_2880394, EPI_ISL_2880395, EPI_ISL_2880396, EPI_ISL_2880397, EPI_ISL_2880398, EPI_ISL_2880399, EPI_ISL_2880400, EPI_ISL_2880401, EPI_ISL_2880402, EPI_ISL_2880403, EPI_ISL_2880404, EPI_ISL_2880405, EPI_ISL_2880406, EPI_ISL_2880407, EPI_ISL_2880408, EPI_ISL_2880409, EPI_ISL_2880410, EPI_ISL_2880411, EPI_ISL_2880412, EPI_ISL_2880413, EPI_ISL_2880414, EPI_ISL_2880415, EPI_ISL_2880416, EPI_ISL_2880417, EPI_ISL_2880418, EPI_ISL_2880419, EPI_ISL_2880420, EPI_ISL_2880421, EPI_ISL_2880422, EPI_ISL_2880423, EPI_ISL_2880424, EPI_ISL_2880425, EPI_ISL_2880426, EPI_ISL_2880427, EPI_ISL_2880428, EPI_ISL_2880429, EPI_ISL_2880430, EPI_ISL_2880431, EPI_ISL_2880432, EPI_ISL_2880433, EPI_ISL_2880434, EPI_ISL_2880435, EPI_ISL_2880436, EPI_ISL_2880437, EPI_ISL_2880438, EPI_ISL_2880439, EPI_ISL_2880440, EPI_ISL_2880441, EPI_ISL_2880442, EPI_ISL_2880443, EPI_ISL_2880444, EPI_ISL_2880445, EPI_ISL_2880446, EPI_ISL_2880447, EPI_ISL_2880448, EPI_ISL_2880449, EPI_ISL_2880450, EPI_ISL_2880451, EPI_ISL_2880452, EPI_ISL_2880453, EPI_ISL_2880454, EPI_ISL_2880455, EPI_ISL_2880456, EPI_ISL_2880457, EPI_ISL_2880458, EPI_ISL_2880459, EPI_ISL_2880460, EPI_ISL_2880461, EPI_ISL_2880462, EPI_ISL_2880463, EPI_ISL_2880464, EPI_ISL_2880465, EPI_ISL_2880466, EPI_ISL_2880467, EPI_ISL_2880468, EPI_ISL_2880469, EPI_ISL_2880470, EPI_ISL_2880471, EPI_ISL_2880472, EPI_ISL_2880473, EPI_ISL_2880474, EPI_ISL_2880475, EPI_ISL_2880476, EPI_ISL_2880477, EPI_ISL_2880478, EPI_ISL_2880479, EPI_ISL_2880480, EPI_ISL_2880481, EPI_ISL_2880482, EPI_ISL_2880483, EPI_ISL_2880484, EPI_ISL_2880485, EPI_ISL_2880486, EPI_ISL_2880487, EPI_ISL_2880488, EPI_ISL_2880489, EPI_ISL_2880490, EPI_ISL_2880491, EPI_ISL_2880492, EPI_ISL_2880493, EPI_ISL_2880494, EPI_ISL_2880495, EPI_ISL_2880496, EPI_ISL_2880497, EPI_ISL_2880498, EPI_ISL_2880499, EPI_ISL_2880500, EPI_ISL_2880501, EPI_ISL_2880502, EPI_ISL_2880503, EPI_ISL_2880504, EPI_ISL_2880505, EPI_ISL_2880506, EPI_ISL_2880507, EPI_ISL_2880508, EPI_ISL_2880509, EPI_ISL_2880510, EPI_ISL_2880511, EPI_ISL_2880512, EPI_ISL_2880513, EPI_ISL_2880514, EPI_ISL_2880515, EPI_ISL_2880516, EPI_ISL_2880517, EPI_ISL_2880518, EPI_ISL_2880519, EPI_ISL_2880520, EPI_ISL_2880521, EPI_ISL_2880522, EPI_ISL_2880523, EPI_ISL_2880524, EPI_ISL_2880525, EPI_ISL_2880526, EPI_ISL_2880527, EPI_ISL_2880528, EPI_ISL_2880529, EPI_ISL_2880530, EPI_ISL_2880531, EPI_ISL_2880532, EPI_ISL_2880533, EPI_ISL_2880534, EPI_ISL_2880535, EPI_ISL_2880536, EPI_ISL_2880537, EPI_ISL_2880538, EPI_ISL_2880539, EPI_ISL_2880540, EPI_ISL_2880541, EPI_ISL_2880542, EPI_ISL_2880543, EPI_ISL_2880544, EPI_ISL_2880545, EPI_ISL_2880546, EPI_ISL_2880547, EPI_ISL_2880548, EPI_ISL_2880549, EPI_ISL_2880550, EPI_ISL_2880551, EPI_ISL_2880552, EPI_ISL_2880553, EPI_ISL_2880554, EPI_ISL_2880555, EPI_ISL_2880556, EPI_ISL_2880557, EPI_ISL_2880558, EPI_ISL_2880559, EPI_ISL_2880560, EPI_ISL_2880561, EPI_ISL_2880562, EPI_ISL_2880563, EPI_ISL_2880564, EPI_ISL_2880565, EPI_ISL_2880566, EPI_ISL_2880567, EPI_ISL_2880568, EPI_ISL_2880569, EPI_ISL_2880570, EPI_ISL_2880571, EPI_ISL_2880572, EPI_ISL_2880573, EPI_ISL_2880574, EPI_ISL_2880575, EPI_ISL_2880576, EPI_ISL_2880577, EPI_ISL_2880578, EPI_ISL_2880579, EPI_ISL_2880580, EPI_ISL_2880581, EPI_ISL_2880582, EPI_ISL_2880583, EPI_ISL_2880584, EPI_ISL_2880585, EPI_ISL_2880586, EPI_ISL_2880587, EPI_ISL_2880588, EPI_ISL_2880589, EPI_ISL_2880590, EPI_ISL_2880591, EPI_ISL_2880592, EPI_ISL_2880593, EPI_ISL_2880594, EPI_ISL_2880595, EPI_ISL_2880596, EPI_ISL_2880597, EPI_ISL_2880598, EPI_ISL_2880599, EPI_ISL_2880600, EPI_ISL_2880601, EPI_ISL_2880602, EPI_ISL_2880603, EPI_ISL_2880604, EPI_ISL_2880605, EPI_ISL_2880606, EPI_ISL_2880607, EPI_ISL_2880608, EPI_ISL_2880609, EPI_ISL_2880610, EPI_ISL_2880611, EPI_ISL_2880612, EPI_ISL_2880613, EPI_ISL_2880614, EPI_ISL_2880615, EPI_ISL_2880616, EPI_ISL_2880617, EPI_ISL_2880618, EPI_ISL_2880619, EPI_ISL_2880620, EPI_ISL_2880621, EPI_ISL_2880622, EPI_ISL_2880623, EPI_ISL_2880624, EPI_ISL_2880625, EPI_ISL_2880626, EPI_ISL_2880627, EPI_ISL_2880628, EPI_ISL_2880629, EPI_ISL_2880630, EPI_ISL_2880631, EPI_ISL_2880632, EPI_ISL_2880633, EPI_ISL_2880634, EPI_ISL_2880635, EPI_ISL_2880636, EPI_ISL_2880637, EPI_ISL_2880638, EPI_ISL_2880639, EPI_ISL_2880640, EPI_ISL_2880641, EPI_ISL_2880642, EPI_ISL_2880643, EPI_ISL_2880644, EPI_ISL_2880645, EPI_ISL_2880646, EPI_ISL_2880647, EPI_ISL_2880648, EPI_ISL_2880649, EPI_ISL_2880650, EPI_ISL_2880651, EPI_ISL_2880652, EPI_ISL_2880653, EPI_ISL_2880654, EPI_ISL_2880655, EPI_ISL_2880656, EPI_ISL_2880657, EPI_ISL_2880658, EPI_ISL_2880659, EPI_ISL_2880660, EPI_ISL_2880661, EPI_ISL_2880662, EPI_ISL_2880663, EPI_ISL_2880664, EPI_ISL_2880665, EPI_ISL_2880666, EPI_ISL_2880667, EPI_ISL_2880668, EPI_ISL_2880669, EPI_ISL_2880670, EPI_ISL_2880671, EPI_ISL_2880672, EPI_ISL_2880673, EPI_ISL_2880674, EPI_ISL_2880675, EPI_ISL_2880676, EPI_ISL_2880677, EPI_ISL_2880678, EPI_ISL_2880679, EPI_ISL_2880680, EPI_ISL_2880681, EPI_ISL_2880682, EPI_ISL_2880683, EPI_ISL_2880684, EPI_ISL_2880685, EPI_ISL_2880686, EPI_ISL_2880687, EPI_ISL_2880688, EPI_ISL_2880689, EPI_ISL_2880690, EPI_ISL_2880691, EPI_ISL_2880692, EPI_ISL_2880693, EPI_ISL_2880694, EPI_ISL_2880695, EPI_ISL_2880696, EPI_ISL_2880697, EPI_ISL_2880698, EPI_ISL_2880699, EPI_ISL_2880935, EPI_ISL_2880936, EPI_ISL_2880937, EPI_ISL_2880938, EPI_ISL_2880939, EPI_ISL_2880940, EPI_ISL_2880941, EPI_ISL_2880942, EPI_ISL_2880943, EPI_ISL_2880944, EPI_ISL_2880945, EPI_ISL_2880946, EPI_ISL_2880947, EPI_ISL_2880948, EPI_ISL_2880949, EPI_ISL_2880950, EPI_ISL_2880951, EPI_ISL_2880952, EPI_ISL_2880953, EPI_ISL_2880954, EPI_ISL_2880955, EPI_ISL_2880956, EPI_ISL_2880957, EPI_ISL_2880958, EPI_ISL_2880959, EPI_ISL_2880960, EPI_ISL_2880961, EPI_ISL_2880962, EPI_ISL_2880963, EPI_ISL_2880964, EPI_ISL_2880965, EPI_ISL_2880966, EPI_ISL_2880967, EPI_ISL_2880968, EPI_ISL_2880969, EPI_ISL_2880970, EPI_ISL_2880971, EPI_ISL_2880972, EPI_ISL_2880973, EPI_ISL_2880974, EPI_ISL_2880975, EPI_ISL_2880976, EPI_ISL_2880977, EPI_ISL_2880978, EPI_ISL_2880980, EPI_ISL_2880981, EPI_ISL_2880982, EPI_ISL_2880983, EPI_ISL_2880984, EPI_ISL_2880985, EPI_ISL_2880986, EPI_ISL_2880987, EPI_ISL_2880988, EPI_ISL_2880989, EPI_ISL_2880990, EPI_ISL_2880991, EPI_ISL_2880992, EPI_ISL_2880993, EPI_ISL_2880994, EPI_ISL_2880995, EPI_ISL_2880996, EPI_ISL_2880997, EPI_ISL_2880998, EPI_ISL_2880999, EPI_ISL_2881000, EPI_ISL_2881001, EPI_ISL_2881002, EPI_ISL_2881003, EPI_ISL_2881004, EPI_ISL_2881005, EPI_ISL_2881006, EPI_ISL_2881007, EPI_ISL_2881008, EPI_ISL_2881009, EPI_ISL_2881010, EPI_ISL_2881011, EPI_ISL_2881012, EPI_ISL_2881013, EPI_ISL_2881014, EPI_ISL_2881015, EPI_ISL_2881016, EPI_ISL_2881017, EPI_ISL_2881018, EPI_ISL_2881019, EPI_ISL_2881020, EPI_ISL_2881021, EPI_ISL_2881022, EPI_ISL_2881023, EPI_ISL_2881024, EPI_ISL_2881025, EPI_ISL_2881026, EPI_ISL_2881027, EPI_ISL_2881028, EPI_ISL_2881029, EPI_ISL_2881030, EPI_ISL_2881031, EPI_ISL_2881032, EPI_ISL_2881033, EPI_ISL_2881034, EPI_ISL_2881035, EPI_ISL_2881036, EPI_ISL_2881037, EPI_ISL_2881038, EPI_ISL_2881039, EPI_ISL_2881040, EPI_ISL_2881041, EPI_ISL_2881042, EPI_ISL_2881043, EPI_ISL_2881044, EPI_ISL_2881045, EPI_ISL_2881046, EPI_ISL_2881047, EPI_ISL_2881048, EPI_ISL_2881049, EPI_ISL_2881050, EPI_ISL_2881051, EPI_ISL_2881052, EPI_ISL_2881053, EPI_ISL_2881054, EPI_ISL_2881055, EPI_ISL_2881056, EPI_ISL_2881057, EPI_ISL_2881058, EPI_ISL_2881059, EPI_ISL_2881071, EPI_ISL_2881072, EPI_ISL_2881073, EPI_ISL_2881074, EPI_ISL_2881075, EPI_ISL_2881076, EPI_ISL_2881077, EPI_ISL_2881078, EPI_ISL_2881079, EPI_ISL_2881080, EPI_ISL_2881106, EPI_ISL_2881107, EPI_ISL_2881108, EPI_ISL_2881109, EPI_ISL_2881110, EPI_ISL_2881111, EPI_ISL_2881112, EPI_ISL_2881113, EPI_ISL_2881114, EPI_ISL_2881115, EPI_ISL_2881116, EPI_ISL_2881117, EPI_ISL_2881118, EPI_ISL_2881119, EPI_ISL_2881120, EPI_ISL_2881121, EPI_ISL_2881122, EPI_ISL_2881123, EPI_ISL_2881124, EPI_ISL_2881125, EPI_ISL_2881126, EPI_ISL_2881127, EPI_ISL_2881128, EPI_ISL_2881129, EPI_ISL_2881130, EPI_ISL_2881131, EPI_ISL_2881132, EPI_ISL_2881133, EPI_ISL_2881134, EPI_ISL_2881145, EPI_ISL_2881146, EPI_ISL_2881147, EPI_ISL_2881148, EPI_ISL_2881149, EPI_ISL_2881150, EPI_ISL_2881151, EPI_ISL_2881152, EPI_ISL_2881153, EPI_ISL_2881154, EPI_ISL_2881155, EPI_ISL_2881156, EPI_ISL_2881157, EPI_ISL_2881158, EPI_ISL_2881159, EPI_ISL_2881160, EPI_ISL_2881161, EPI_ISL_2881162, EPI_ISL_2881163, EPI_ISL_2881164, EPI_ISL_2881165, EPI_ISL_2881166, EPI_ISL_2881167, EPI_ISL_2881168, EPI_ISL_2881169, EPI_ISL_2881170, EPI_ISL_2881171, EPI_ISL_2881172, EPI_ISL_2881173, EPI_ISL_2881174, EPI_ISL_2881175, EPI_ISL_2881176, EPI_ISL_2881177, EPI_ISL_2881178, EPI_ISL_2881179, EPI_ISL_2881187, EPI_ISL_2881188, EPI_ISL_2881190, EPI_ISL_2881191, EPI_ISL_2881192, EPI_ISL_2881195, EPI_ISL_2881196, EPI_ISL_2881197, EPI_ISL_2881198, EPI_ISL_2881199, EPI_ISL_2881200, EPI_ISL_2881201, EPI_ISL_2881202, EPI_ISL_2881203, EPI_ISL_2881204, EPI_ISL_2881205, EPI_ISL_2881216, EPI_ISL_2881217, EPI_ISL_2881218, EPI_ISL_2881219, EPI_ISL_2881220, EPI_ISL_2881221, EPI_ISL_2881222, EPI_ISL_2881251, EPI_ISL_2881263, EPI_ISL_2881264, EPI_ISL_2881265, EPI_ISL_2881266, EPI_ISL_2881267, EPI_ISL_2881268, EPI_ISL_2881269, EPI_ISL_2881270, EPI_ISL_2881271, EPI_ISL_2881272, EPI_ISL_2881274, EPI_ISL_2881275, EPI_ISL_2881276, EPI_ISL_2881277, EPI_ISL_2881278, EPI_ISL_2881279, EPI_ISL_2881280, EPI_ISL_2881281, EPI_ISL_2881282, EPI_ISL_2881283, EPI_ISL_2881284, EPI_ISL_2881285, EPI_ISL_2881286, EPI_ISL_2881287, EPI_ISL_2881288, EPI_ISL_2881289, EPI_ISL_2881290, EPI_ISL_2881291, EPI_ISL_2881292, EPI_ISL_2881293, EPI_ISL_2881294, EPI_ISL_2881295, EPI_ISL_2881296, EPI_ISL_2881297, EPI_ISL_2881298, EPI_ISL_2881299, EPI_ISL_2881300, EPI_ISL_2881301, EPI_ISL_2881302, EPI_ISL_2881303, EPI_ISL_2881304, EPI_ISL_2881305, EPI_ISL_2881306, EPI_ISL_2881307, EPI_ISL_2881308, EPI_ISL_2881309, EPI_ISL_2881310, EPI_ISL_2881311, EPI_ISL_2881312, EPI_ISL_2881313, EPI_ISL_2881314, EPI_ISL_2881315, EPI_ISL_2881316, EPI_ISL_2881317, EPI_ISL_2881318, EPI_ISL_2881319, EPI_ISL_2881320, EPI_ISL_2881321, EPI_ISL_2881322, EPI_ISL_2881323, EPI_ISL_2881376, EPI_ISL_2881377, EPI_ISL_2881378, EPI_ISL_2881379, EPI_ISL_2881380, EPI_ISL_2881381, EPI_ISL_2881382, EPI_ISL_2881383, EPI_ISL_2881384, EPI_ISL_2881385, EPI_ISL_2881386, EPI_ISL_2881387, EPI_ISL_2881388, EPI_ISL_2881389, EPI_ISL_2881390, EPI_ISL_2881391, EPI_ISL_2881392, EPI_ISL_2881393, EPI_ISL_2881394, EPI_ISL_2881395, EPI_ISL_2881396, EPI_ISL_2881397, EPI_ISL_2881398, EPI_ISL_2881399, EPI_ISL_2881400, EPI_ISL_2881401, EPI_ISL_2881402, EPI_ISL_2881403, EPI_ISL_2881404, EPI_ISL_2881405, EPI_ISL_2881406, EPI_ISL_2881407, EPI_ISL_2881408, EPI_ISL_2881409, EPI_ISL_2881410, EPI_ISL_2881411, EPI_ISL_2881412, EPI_ISL_2881413, EPI_ISL_2881414, EPI_ISL_2881415, EPI_ISL_2881416, EPI_ISL_2881417, EPI_ISL_2881418, EPI_ISL_2881419, EPI_ISL_2881420, EPI_ISL_2881421, EPI_ISL_2881422, EPI_ISL_2881423, EPI_ISL_2881424, EPI_ISL_2881425, EPI_ISL_2881426, EPI_ISL_2881427, EPI_ISL_2881428, EPI_ISL_2881429, EPI_ISL_2881430, EPI_ISL_2881431, EPI_ISL_2881432, EPI_ISL_2881435, EPI_ISL_2881436, EPI_ISL_2881437, EPI_ISL_2881438, EPI_ISL_2881439, EPI_ISL_2881440, EPI_ISL_2881441, EPI_ISL_2881443, EPI_ISL_2881444, EPI_ISL_2881445, EPI_ISL_2881446, EPI_ISL_2881448, EPI_ISL_2881455, EPI_ISL_2881456, EPI_ISL_2881457, EPI_ISL_2881458, EPI_ISL_2881459, EPI_ISL_2881460, EPI_ISL_2881461, EPI_ISL_2881462, EPI_ISL_2881463, EPI_ISL_2881464, EPI_ISL_2881465, EPI_ISL_2881466, EPI_ISL_2881468, EPI_ISL_2881469, EPI_ISL_2881470, EPI_ISL_2881471, EPI_ISL_2881472, EPI_ISL_2881473, EPI_ISL_2881474, EPI_ISL_2881475, EPI_ISL_2881476, EPI_ISL_2881477, EPI_ISL_2881478, EPI_ISL_2881479, EPI_ISL_2881480, EPI_ISL_2881481, EPI_ISL_2881500, EPI_ISL_2881501, EPI_ISL_2881502, EPI_ISL_2881503, EPI_ISL_2881504, EPI_ISL_2881505, EPI_ISL_2881506, EPI_ISL_2881507, EPI_ISL_2881508, EPI_ISL_2881509, EPI_ISL_2881510, EPI_ISL_2881511, EPI_ISL_2881512, EPI_ISL_2881517, EPI_ISL_2881519, EPI_ISL_2881520, EPI_ISL_2881521, EPI_ISL_2881523, EPI_ISL_2881524, EPI_ISL_2881525, EPI_ISL_2881526, EPI_ISL_2881527, EPI_ISL_2881528, EPI_ISL_2881529, EPI_ISL_2881530, EPI_ISL_2881531, EPI_ISL_2881532, EPI_ISL_2881533, EPI_ISL_2881534, EPI_ISL_2881535, EPI_ISL_2881536, EPI_ISL_2881537, EPI_ISL_2881538, EPI_ISL_2881539, EPI_ISL_2881543, EPI_ISL_2881544, EPI_ISL_2881545, EPI_ISL_2881546, EPI_ISL_2881547, EPI_ISL_2881548, EPI_ISL_2881549, EPI_ISL_2881550, EPI_ISL_2881551, EPI_ISL_2881552, EPI_ISL_2881553, EPI_ISL_2881554, EPI_ISL_2881555, EPI_ISL_2881556, EPI_ISL_2881558, EPI_ISL_2881559, EPI_ISL_2881560, EPI_ISL_2881561, EPI_ISL_2881562, EPI_ISL_2881563, EPI_ISL_2881564, EPI_ISL_2881565, EPI_ISL_2881566, EPI_ISL_2881567, EPI_ISL_2881568, EPI_ISL_2881569, EPI_ISL_2881570, EPI_ISL_2881571, EPI_ISL_2881572, EPI_ISL_2881573, EPI_ISL_2881574, EPI_ISL_2881575, EPI_ISL_2881576, EPI_ISL_2881577, EPI_ISL_2881578, EPI_ISL_2881579, EPI_ISL_2881580, EPI_ISL_2881581, EPI_ISL_2881582, EPI_ISL_2881583, EPI_ISL_2881584, EPI_ISL_2881585, EPI_ISL_2881586, EPI_ISL_2881587, EPI_ISL_2881588, EPI_ISL_2881589, EPI_ISL_2881590, EPI_ISL_2881591, EPI_ISL_2881592, EPI_ISL_2881593, EPI_ISL_2881594, EPI_ISL_2881595, EPI_ISL_2881596, EPI_ISL_2881597, EPI_ISL_2881598, EPI_ISL_2881599, EPI_ISL_2881600, EPI_ISL_2881601, EPI_ISL_2881602, EPI_ISL_2881603, EPI_ISL_2881604, EPI_ISL_2881605, EPI_ISL_2881606, EPI_ISL_2881607, EPI_ISL_2881608, EPI_ISL_2881609, EPI_ISL_2881610, EPI_ISL_2881611, EPI_ISL_2881612, EPI_ISL_2881614, EPI_ISL_2881615, EPI_ISL_2881616, EPI_ISL_2881617, EPI_ISL_2881618, EPI_ISL_2881619, EPI_ISL_2881620, EPI_ISL_2881621, EPI_ISL_2881622, EPI_ISL_2881623, EPI_ISL_2881624, EPI_ISL_2881625, EPI_ISL_2881626, EPI_ISL_2881627, EPI_ISL_2881628, EPI_ISL_2881629, EPI_ISL_2881630, EPI_ISL_2881631, EPI_ISL_2881632, EPI_ISL_2881633, EPI_ISL_2881634, EPI_ISL_2881635, EPI_ISL_2881636, EPI_ISL_2881637, EPI_ISL_2881638, EPI_ISL_2881639, EPI_ISL_2881640, EPI_ISL_2881641, EPI_ISL_2881642, EPI_ISL_2881643, EPI_ISL_2881644, EPI_ISL_2881645, EPI_ISL_2881646, EPI_ISL_2881647, EPI_ISL_2881648, EPI_ISL_2881649, EPI_ISL_2881650, EPI_ISL_2881651, EPI_ISL_2881652, EPI_ISL_2881653, EPI_ISL_2881654, EPI_ISL_2881655, EPI_ISL_2881656, EPI_ISL_2881657, EPI_ISL_2881658, EPI_ISL_2881659, EPI_ISL_2881672, EPI_ISL_2881673, EPI_ISL_2881674, EPI_ISL_2881675, EPI_ISL_2881676, EPI_ISL_2881677, EPI_ISL_2881678, EPI_ISL_2881679, EPI_ISL_2881680, EPI_ISL_2881681, EPI_ISL_2881682, EPI_ISL_2881683, EPI_ISL_2881684, EPI_ISL_2881685, EPI_ISL_2881686, EPI_ISL_2881687, EPI_ISL_2881688, EPI_ISL_2881689, EPI_ISL_2881690, EPI_ISL_2881691, EPI_ISL_2881692, EPI_ISL_2881693, EPI_ISL_2881694, EPI_ISL_2881695, EPI_ISL_2881696, EPI_ISL_2881697, EPI_ISL_2881698, EPI_ISL_2881699, EPI_ISL_2881700, EPI_ISL_2881701, EPI_ISL_2881702, EPI_ISL_2881703, EPI_ISL_2881704, EPI_ISL_2881705, EPI_ISL_2881706, EPI_ISL_2881707, EPI_ISL_2881708, EPI_ISL_2881709, EPI_ISL_2881710, EPI_ISL_2881711, EPI_ISL_2881712, EPI_ISL_2881713, EPI_ISL_2881714, EPI_ISL_2881715, EPI_ISL_2881716, EPI_ISL_2881717, EPI_ISL_2881718, EPI_ISL_2881719, EPI_ISL_2881720

see above
INSACOG Surveillance
INSACOG at CSIR Institute of Genomics and Integrative Biology
INSACOG

EPI_ISL_2502353, EPI_ISL_2502366, EPI_ISL_2502368, EPI_ISL_2502369, EPI_ISL_2502370, EPI_ISL_2502371, EPI_ISL_2502372, EPI_ISL_2502373, EPI_ISL_2502374, EPI_ISL_2877746, EPI_ISL_2877747, EPI_ISL_2877748, EPI_ISL_2877749, EPI_ISL_2877750, EPI_ISL_2877751, EPI_ISL_2877752, EPI_ISL_2877753, EPI_ISL_2877754, EPI_ISL_2877755, EPI_ISL_2877756, EPI_ISL_2877757, EPI_ISL_2877758, EPI_ISL_2877759, EPI_ISL_2877760, EPI_ISL_2877761, EPI_ISL_2877762, EPI_ISL_2877763, EPI_ISL_2877764, EPI_ISL_2877765, EPI_ISL_2877766, EPI_ISL_2877767, EPI_ISL_2877768, EPI_ISL_2877769, EPI_ISL_2877770, EPI_ISL_2877771, EPI_ISL_2877772, EPI_ISL_2877773, EPI_ISL_2877774, EPI_ISL_2877775, EPI_ISL_2877776, EPI_ISL_2877777, EPI_ISL_2877778, EPI_ISL_2877779, EPI_ISL_2877780, EPI_ISL_2877781, EPI_ISL_2877782, EPI_ISL_2877783, EPI_ISL_2877784, EPI_ISL_2877785, EPI_ISL_2877786, EPI_ISL_2877787, EPI_ISL_2877788, EPI_ISL_2877789, EPI_ISL_2877790, EPI_ISL_2877791, EPI_ISL_2877792, EPI_ISL_2877793, EPI_ISL_2877794, EPI_ISL_2877795, EPI_ISL_2877796, EPI_ISL_2877797, EPI_ISL_2877798, EPI_ISL_2877799, EPI_ISL_2877800, EPI_ISL_2877801, EPI_ISL_2877802, EPI_ISL_2877803, EPI_ISL_2877804, EPI_ISL_2877805, EPI_ISL_2877806, EPI_ISL_2877807, EPI_ISL_2877808, EPI_ISL_2877809, EPI_ISL_2877810, EPI_ISL_2877811, EPI_ISL_2877812, EPI_ISL_2877813, EPI_ISL_2877814, EPI_ISL_2877815, EPI_ISL_2877816, EPI_ISL_2877831, EPI_ISL_2877832, EPI_ISL_2877833, EPI_ISL_2877834, EPI_ISL_2877835, EPI_ISL_2877836, EPI_ISL_2877837, EPI_ISL_2877838, EPI_ISL_2877839, EPI_ISL_2877840, EPI_ISL_2877841, EPI_ISL_2877842, EPI_ISL_2877843, EPI_ISL_2877844, EPI_ISL_2877845, EPI_ISL_2877846, EPI_ISL_2877847, EPI_ISL_2877848, EPI_ISL_2877852, EPI_ISL_2877855, EPI_ISL_2877856, EPI_ISL_2877857, EPI_ISL_2877858, EPI_ISL_2877859, EPI_ISL_2877860, EPI_ISL_2877861, EPI_ISL_2877862, EPI_ISL_2877863, EPI_ISL_2877864, EPI_ISL_2877865, EPI_ISL_2877866, EPI_ISL_2877867, EPI_ISL_2877868, EPI_ISL_2877869, EPI_ISL_2877870, EPI_ISL_2877871, EPI_ISL_2877872, EPI_ISL_2877873, EPI_ISL_2877874, EPI_ISL_2877875, EPI_ISL_2877876, EPI_ISL_2877877, EPI_ISL_2877878, EPI_ISL_2877879, EPI_ISL_2877880, EPI_ISL_2877881, EPI_ISL_2877882, EPI_ISL_2877883, EPI_ISL_2877884, EPI_ISL_2877885, EPI_ISL_2877886, EPI_ISL_2877887, EPI_ISL_2877888, EPI_ISL_2877889

see above
INSACOG-Assam
National Institute of Biomedical Genomics – INSACOG
Ajanta Sharma; Arindam Maitra; Kailash Chamuah; Lahari Saikia; Nidhan Kumar Biswas; Saumitra Das; Sreedhar Chinnaswamy

EPI_ISL_2812579, EPI_ISL_2812580, EPI_ISL_2812581, EPI_ISL_2812582, EPI_ISL_2812583, EPI_ISL_2812584, EPI_ISL_2812585, EPI_ISL_2812586, EPI_ISL_2812587, EPI_ISL_2812588, EPI_ISL_2812589, EPI_ISL_2812590, EPI_ISL_2812591, EPI_ISL_2812592, EPI_ISL_2812593, EPI_ISL_2812594, EPI_ISL_2812595, EPI_ISL_2812596, EPI_ISL_2812597, EPI_ISL_2812598, EPI_ISL_2812599, EPI_ISL_2812600, EPI_ISL_2812601, EPI_ISL_2812602, EPI_ISL_2812603, EPI_ISL_2812604, EPI_ISL_2812605, EPI_ISL_2812606, EPI_ISL_2812607, EPI_ISL_2812608, EPI_ISL_2812609, EPI_ISL_2812610, EPI_ISL_2812611, EPI_ISL_2812612, EPI_ISL_2812613, EPI_ISL_2812614, EPI_ISL_2812615, EPI_ISL_2812616, EPI_ISL_2812617, EPI_ISL_2812618, EPI_ISL_2812619, EPI_ISL_2812620, EPI_ISL_2812621, EPI_ISL_2812622, EPI_ISL_2812623, EPI_ISL_2812624, EPI_ISL_2812625, EPI_ISL_2812626, EPI_ISL_2812627, EPI_ISL_2812628, EPI_ISL_2812629, EPI_ISL_2812630, EPI_ISL_2812631, EPI_ISL_2812632, EPI_ISL_2812633, EPI_ISL_2812634, EPI_ISL_2812635, EPI_ISL_2812636, EPI_ISL_2812637, EPI_ISL_2812638, EPI_ISL_2812639, EPI_ISL_2812640, EPI_ISL_2812641, EPI_ISL_2812642, EPI_ISL_2812643, EPI_ISL_2812644, EPI_ISL_2812645, EPI_ISL_2812646

see above
INSACOG-MANIPUR
National Institute of Biomedical Genomics – INSACOG
Arindam Maitra; Khumanthem Ranjana; Nanaocha Sharma; Nidhan Kumar Biswas; R K Manglem Singh; Saumitra Das; Sreedhar Chinnaswamy

EPI_ISL_2675307, EPI_ISL_2675308, EPI_ISL_2675309, EPI_ISL_2675310, EPI_ISL_2675311, EPI_ISL_2675312, EPI_ISL_2675313, EPI_ISL_2675314, EPI_ISL_2675315, EPI_ISL_2675316, EPI_ISL_2675317, EPI_ISL_2675318, EPI_ISL_2675319, EPI_ISL_2675320, EPI_ISL_2675321, EPI_ISL_2675322, EPI_ISL_2675323, EPI_ISL_2675324, EPI_ISL_2675325, EPI_ISL_2675326, EPI_ISL_2675327, EPI_ISL_2675328, EPI_ISL_2675329, EPI_ISL_2675330, EPI_ISL_2675331, EPI_ISL_2675332, EPI_ISL_2675333, EPI_ISL_2675334, EPI_ISL_2675335, EPI_ISL_2675336, EPI_ISL_2675337, EPI_ISL_2675338, EPI_ISL_2675339, EPI_ISL_2675340, EPI_ISL_2675341, EPI_ISL_2675342, EPI_ISL_2675343, EPI_ISL_2675344, EPI_ISL_2675345, EPI_ISL_2675346, EPI_ISL_2675347, EPI_ISL_2675348, EPI_ISL_2675349, EPI_ISL_2675350, EPI_ISL_2675351, EPI_ISL_2675352, EPI_ISL_2675353, EPI_ISL_2675354, EPI_ISL_2675355, EPI_ISL_2675356, EPI_ISL_2675357, EPI_ISL_2675360, EPI_ISL_2675361, EPI_ISL_2675362, EPI_ISL_2675363, EPI_ISL_2675364, EPI_ISL_2675365, EPI_ISL_2675366, EPI_ISL_2675367, EPI_ISL_2675368, EPI_ISL_2675369, EPI_ISL_2675370, EPI_ISL_2675371, EPI_ISL_2675373, EPI_ISL_2675374, EPI_ISL_2675380, EPI_ISL_2675382, EPI_ISL_2675383, EPI_ISL_2675384, EPI_ISL_2675386, EPI_ISL_2675388, EPI_ISL_2675390, EPI_ISL_2675391

see above
INSACOG-NAGALAND
National Institute of Biomedical Genomics – INSACOG
Arindam Maitra; Chenole Keppen; Nidhan Kumar Biswas; Saumitra Das; Sreedhar Chinnaswamy; Vinotsole Khamo; Wetetsho Kapfo

EPI_ISL_2877955, EPI_ISL_2877956, EPI_ISL_2877957, EPI_ISL_2877958, EPI_ISL_2877959, EPI_ISL_2877960, EPI_ISL_2877961, EPI_ISL_2877962, EPI_ISL_2877963, EPI_ISL_2877964, EPI_ISL_2877965, EPI_ISL_2877966, EPI_ISL_2877967, EPI_ISL_2877968, EPI_ISL_2877969, EPI_ISL_2877970, EPI_ISL_2877971, EPI_ISL_2877972, EPI_ISL_2877973, EPI_ISL_2877974, EPI_ISL_2877975, EPI_ISL_2877976, EPI_ISL_2877977, EPI_ISL_2877978, EPI_ISL_2877979, EPI_ISL_2877980, EPI_ISL_2877981, EPI_ISL_2877982, EPI_ISL_2877983, EPI_ISL_2877984, EPI_ISL_2877985, EPI_ISL_2877986, EPI_ISL_2877987, EPI_ISL_2877988, EPI_ISL_2877989, EPI_ISL_2877990, EPI_ISL_2877991, EPI_ISL_2877992, EPI_ISL_2877993, EPI_ISL_2877994, EPI_ISL_2877995, EPI_ISL_2877996, EPI_ISL_2877997, EPI_ISL_2877998, EPI_ISL_2877999, EPI_ISL_2878000, EPI_ISL_2878001, EPI_ISL_2878002, EPI_ISL_2878003, EPI_ISL_2878004, EPI_ISL_2878005, EPI_ISL_2878006, EPI_ISL_2878007, EPI_ISL_2878008, EPI_ISL_2878009, EPI_ISL_2878010, EPI_ISL_2878011, EPI_ISL_2878012, EPI_ISL_2878013, EPI_ISL_2878014, EPI_ISL_2878015, EPI_ISL_2878016, EPI_ISL_2878017, EPI_ISL_2878018, EPI_ISL_2878019, EPI_ISL_2878020, EPI_ISL_2878021, EPI_ISL_2878022, EPI_ISL_2878023, EPI_ISL_2878024, EPI_ISL_2878025, EPI_ISL_2878026, EPI_ISL_2878027, EPI_ISL_2878028, EPI_ISL_2878029, EPI_ISL_2878030, EPI_ISL_2878031, EPI_ISL_2878032, EPI_ISL_2878033, EPI_ISL_2878034, EPI_ISL_2878035, EPI_ISL_2878036, EPI_ISL_2878037, EPI_ISL_2878038, EPI_ISL_2878039

see above
INSACOG-Tripura
National Institute of Biomedical Genomics – INSACOG
Apurba Sarkar; Arindam Maitra; Nidhan Kumar Biswas; Saikat Majumder; Saumitra Das; Sreedhar Chinnaswamy; Tapan Majumdar

| | | | |
|---|---|---|---|
| EPI_ISL_2548379, EPI_ISL_2548380, EPI_ISL_2548381, EPI_ISL_2548382, EPI_ISL_2548383, EPI_ISL_2548384, EPI_ISL_2548385, EPI_ISL_2548387, EPI_ISL_2548388, EPI_ISL_2548389, EPI_ISL_2548390, EPI_ISL_2548391, EPI_ISL_2548392, EPI_ISL_2548393, EPI_ISL_2548394, EPI_ISL_2548395, EPI_ISL_2548396, EPI_ISL_2548397, EPI_ISL_2548398, EPI_ISL_2548400, EPI_ISL_2548401, EPI_ISL_2548402, EPI_ISL_2548403, EPI_ISL_2548404, EPI_ISL_2548405, EPI_ISL_2548406, EPI_ISL_2548407, EPI_ISL_2548408, EPI_ISL_2548409, EPI_ISL_2548410, EPI_ISL_2548411, EPI_ISL_2548412, EPI_ISL_2548413, EPI_ISL_2548414, EPI_ISL_2548415, EPI_ISL_2548416, EPI_ISL_2548417, EPI_ISL_2548418, EPI_ISL_2548419, EPI_ISL_2548420, EPI_ISL_2548421, EPI_ISL_2548422, EPI_ISL_2548423, EPI_ISL_2548424, EPI_ISL_2548425, EPI_ISL_2548426, EPI_ISL_2548427, EPI_ISL_2548428, EPI_ISL_2548429, EPI_ISL_2548430, EPI_ISL_2548431, EPI_ISL_2548432, EPI_ISL_2548433, EPI_ISL_2548434, EPI_ISL_2548435, EPI_ISL_2548436, EPI_ISL_2548437, EPI_ISL_2548438, EPI_ISL_2548439, EPI_ISL_2548440, EPI_ISL_2548441, EPI_ISL_2548442, EPI_ISL_2548443, EPI_ISL_2548444, EPI_ISL_2548445, EPI_ISL_2548446, EPI_ISL_2548447, EPI_ISL_2548448, EPI_ISL_2548449, EPI_ISL_2548450, EPI_ISL_2548451, EPI_ISL_2548452, EPI_ISL_2548453, EPI_ISL_2548454, EPI_ISL_2548455, EPI_ISL_2548456, EPI_ISL_2548457, EPI_ISL_2548458, EPI_ISL_2548459, EPI_ISL_2548460, EPI_ISL_2548461, EPI_ISL_2548462, EPI_ISL_2548463, EPI_ISL_2548464, EPI_ISL_2548465, EPI_ISL_2548466, EPI_ISL_2548467, EPI_ISL_2548468, EPI_ISL_2548469, EPI_ISL_2548470, EPI_ISL_2548471, EPI_ISL_2548472, EPI_ISL_2548473, EPI_ISL_2548474, EPI_ISL_2548475, EPI_ISL_2548476, EPI_ISL_2548477, EPI_ISL_2548478, EPI_ISL_2548479, EPI_ISL_2548480, EPI_ISL_2548481, EPI_ISL_2548482, EPI_ISL_2548483, EPI_ISL_2548484, EPI_ISL_2548485, EPI_ISL_2548486, EPI_ISL_2548487, EPI_ISL_2548488, EPI_ISL_2548489, EPI_ISL_2548490, EPI_ISL_2548491, EPI_ISL_2548492, EPI_ISL_2548493, EPI_ISL_2548494, EPI_ISL_2548495, EPI_ISL_2548496, EPI_ISL_2548497, EPI_ISL_2548498, EPI_ISL_2548499, EPI_ISL_2548500, EPI_ISL_2548501, EPI_ISL_2548502, EPI_ISL_2548503, EPI_ISL_2548504, EPI_ISL_2548505, EPI_ISL_2548506, EPI_ISL_2548507, EPI_ISL_2548508, EPI_ISL_2548509, EPI_ISL_2548510, EPI_ISL_2548511, EPI_ISL_2548512, EPI_ISL_2548513, EPI_ISL_2548514, EPI_ISL_2548515, EPI_ISL_2548516, EPI_ISL_2548517, EPI_ISL_2548518, EPI_ISL_2548519, EPI_ISL_2548520, EPI_ISL_2548521, EPI_ISL_2548522, EPI_ISL_2548523, EPI_ISL_2548524, EPI_ISL_2548525, EPI_ISL_2548526, EPI_ISL_2548527, EPI_ISL_2548528, EPI_ISL_2548529, EPI_ISL_2548530, EPI_ISL_2548531, EPI_ISL_2548532, EPI_ISL_2548533, EPI_ISL_2548534, EPI_ISL_2548535, EPI_ISL_2548536, EPI_ISL_2548537, EPI_ISL_2548538, EPI_ISL_2548540, EPI_ISL_2548541, EPI_ISL_2548542, EPI_ISL_2548543, EPI_ISL_2548544, EPI_ISL_2548545, EPI_ISL_2548546, EPI_ISL_2548547, EPI_ISL_2548548, EPI_ISL_2548549, EPI_ISL_2548550, EPI_ISL_2548551, EPI_ISL_2548552, EPI_ISL_2548553, EPI_ISL_2548554, EPI_ISL_2548555, EPI_ISL_2548556, EPI_ISL_2548557, EPI_ISL_2548558, EPI_ISL_2548559, EPI_ISL_2548560, EPI_ISL_2548561, EPI_ISL_2548562, EPI_ISL_2548563, EPI_ISL_2548564, EPI_ISL_2548565, EPI_ISL_2548566, EPI_ISL_2548567, EPI_ISL_2548568, EPI_ISL_2548569, EPI_ISL_2548570, EPI_ISL_2548571, EPI_ISL_2548572, EPI_ISL_2548582, EPI_ISL_2548583, EPI_ISL_2548584, EPI_ISL_2548585, EPI_ISL_2548586, EPI_ISL_2548587, EPI_ISL_2548588, EPI_ISL_2548589, EPI_ISL_2548590, EPI_ISL_2548591, EPI_ISL_2548592, EPI_ISL_2548608, EPI_ISL_2548609, EPI_ISL_2548610, EPI_ISL_2548822, EPI_ISL_2548823, EPI_ISL_2548824, EPI_ISL_2548844, EPI_ISL_2548845, EPI_ISL_2548846, EPI_ISL_2548848, EPI_ISL_2548849, EPI_ISL_2548850, EPI_ISL_2548851, EPI_ISL_2548852, EPI_ISL_2548853, EPI_ISL_2548854, EPI_ISL_2548855, EPI_ISL_2548856, EPI_ISL_2548858, EPI_ISL_2548859, EPI_ISL_2548860, EPI_ISL_2548861, EPI_ISL_2548862, EPI_ISL_2548863, EPI_ISL_2548864, EPI_ISL_2548865, EPI_ISL_2548866, EPI_ISL_2548867, EPI_ISL_2548868, EPI_ISL_2548869, EPI_ISL_2548870, EPI_ISL_2548871, EPI_ISL_2548872, EPI_ISL_2548873, EPI_ISL_2548874, EPI_ISL_2548875, EPI_ISL_2548876, EPI_ISL_2548877, EPI_ISL_2548878, EPI_ISL_2548879, EPI_ISL_2548880, EPI_ISL_2548881, EPI_ISL_2548882, EPI_ISL_2548883, EPI_ISL_2548884, EPI_ISL_2548885, EPI_ISL_2548886, EPI_ISL_2548887, EPI_ISL_2548888, EPI_ISL_2548889, EPI_ISL_2548890, EPI_ISL_2548891, EPI_ISL_2548892, EPI_ISL_2548893, EPI_ISL_2548894, EPI_ISL_2548895, EPI_ISL_2548896, EPI_ISL_2548897, EPI_ISL_2548898, EPI_ISL_2548899, EPI_ISL_2548900, EPI_ISL_2548901, EPI_ISL_2548902, EPI_ISL_2548903, EPI_ISL_2548904, EPI_ISL_2548906, EPI_ISL_2548907, EPI_ISL_2548908, EPI_ISL_2549009, EPI_ISL_2549010, EPI_ISL_2549055, EPI_ISL_2549056, EPI_ISL_2549057, EPI_ISL_2549058, EPI_ISL_2549059, EPI_ISL_2549060, EPI_ISL_2549061, EPI_ISL_2549062, EPI_ISL_2549063, EPI_ISL_2549064, EPI_ISL_2549065, EPI_ISL_2549066, EPI_ISL_2549067, EPI_ISL_2549068, EPI_ISL_2549069, EPI_ISL_2549070, EPI_ISL_2549071, EPI_ISL_2549072, EPI_ISL_2675890, EPI_ISL_2675891, EPI_ISL_2675892, EPI_ISL_2675893, EPI_ISL_2675894, EPI_ISL_2675895, EPI_ISL_2675896, EPI_ISL_2675897, EPI_ISL_2675898, EPI_ISL_2675899, EPI_ISL_2675900, EPI_ISL_2675901, EPI_ISL_2675902, EPI_ISL_2675903, EPI_ISL_2675904, EPI_ISL_2675905, EPI_ISL_2675906, EPI_ISL_2675907, EPI_ISL_2675908, EPI_ISL_2675909, EPI_ISL_2675910, EPI_ISL_2675911, EPI_ISL_2675912, EPI_ISL_2675913, EPI_ISL_2675914, EPI_ISL_2675915, EPI_ISL_2675916, EPI_ISL_2675917, EPI_ISL_2675918, EPI_ISL_2675919, EPI_ISL_2675920, EPI_ISL_2675921, EPI_ISL_2675922, EPI_ISL_2675923, EPI_ISL_2675924, EPI_ISL_2675925, EPI_ISL_2675926, EPI_ISL_2675927, EPI_ISL_2675928, EPI_ISL_2675929, EPI_ISL_2675930, EPI_ISL_2675931, EPI_ISL_2675932, EPI_ISL_2675933, EPI_ISL_2675934, EPI_ISL_2675935, EPI_ISL_2675973, EPI_ISL_2675974, EPI_ISL_2675975, EPI_ISL_2675976, EPI_ISL_2675977, EPI_ISL_2675978, EPI_ISL_2675979, EPI_ISL_2676077, EPI_ISL_2676085, EPI_ISL_2676086, EPI_ISL_2676089, EPI_ISL_2676090, EPI_ISL_2676091, EPI_ISL_2676092, EPI_ISL_2676093, EPI_ISL_2676094, EPI_ISL_2676095, EPI_ISL_2676096, EPI_ISL_2676097, EPI_ISL_2676098, EPI_ISL_2676099, EPI_ISL_2676100, EPI_ISL_2676101, EPI_ISL_2676102, EPI_ISL_2676103, EPI_ISL_2676104, EPI_ISL_2676105, EPI_ISL_2676106, EPI_ISL_2676107, EPI_ISL_2676108, EPI_ISL_2676109, EPI_ISL_2676110, EPI_ISL_2676111, EPI_ISL_2676112, EPI_ISL_2676113, EPI_ISL_2676114, EPI_ISL_2676115, EPI_ISL_2676116, EPI_ISL_2676117, EPI_ISL_2676118, EPI_ISL_2676119, EPI_ISL_2676120, EPI_ISL_2676121, EPI_ISL_2676122, EPI_ISL_2676123, EPI_ISL_2676124, EPI_ISL_2676125, EPI_ISL_2676126, EPI_ISL_2676127, EPI_ISL_2676128, EPI_ISL_2676129, EPI_ISL_2676130, EPI_ISL_2676131, EPI_ISL_2676132, EPI_ISL_2676133, EPI_ISL_2676134, EPI_ISL_2676135, EPI_ISL_2676136, EPI_ISL_2676137, EPI_ISL_2676138, EPI_ISL_2676139, EPI_ISL_2676140, EPI_ISL_2676141, EPI_ISL_2676142, EPI_ISL_2676143, EPI_ISL_2676144, EPI_ISL_2676145, EPI_ISL_2676146, EPI_ISL_2676147, EPI_ISL_2676148, EPI_ISL_2676149, EPI_ISL_2676150, EPI_ISL_2676151, EPI_ISL_2676152, EPI_ISL_2676153, EPI_ISL_2676154, EPI_ISL_2676155, EPI_ISL_2676156, EPI_ISL_2676157, EPI_ISL_2676158, EPI_ISL_2676159, EPI_ISL_2676160, EPI_ISL_2676161, EPI_ISL_2676162, EPI_ISL_2676163, EPI_ISL_2676164, EPI_ISL_2676165, EPI_ISL_2676166, EPI_ISL_2676167, EPI_ISL_2676168, EPI_ISL_2676169, EPI_ISL_2676170, EPI_ISL_2676171, EPI_ISL_2676172, EPI_ISL_2676173, EPI_ISL_2676174, EPI_ISL_2676175, EPI_ISL_2676176, EPI_ISL_2676177, EPI_ISL_2676178, EPI_ISL_2676179, EPI_ISL_2676180, EPI_ISL_2676181, EPI_ISL_2676182, EPI_ISL_2676183, EPI_ISL_2676184, EPI_ISL_2676185, EPI_ISL_2676186, EPI_ISL_2676187, EPI_ISL_2676188, EPI_ISL_2676189, EPI_ISL_2676190, EPI_ISL_2676191, EPI_ISL_2676192, EPI_ISL_2676193, EPI_ISL_2676194, EPI_ISL_2676195, EPI_ISL_2676196, EPI_ISL_2676197, EPI_ISL_2676198, EPI_ISL_2676199, EPI_ISL_2676200, EPI_ISL_2676201, EPI_ISL_2676202, EPI_ISL_2676203, EPI_ISL_2676204, EPI_ISL_2676205, EPI_ISL_2676206, EPI_ISL_2676207, EPI_ISL_2676208, EPI_ISL_2676209, EPI_ISL_2676210, EPI_ISL_2676211, EPI_ISL_2676212, EPI_ISL_2676213, EPI_ISL_2676214, EPI_ISL_2676215, EPI_ISL_2676216, EPI_ISL_2676217, EPI_ISL_2676218, EPI_ISL_2676219, EPI_ISL_2676220, EPI_ISL_2676221, EPI_ISL_2676222, EPI_ISL_2676224, EPI_ISL_2676225, EPI_ISL_2676226, EPI_ISL_2676227, EPI_ISL_2676228, EPI_ISL_2676229, EPI_ISL_2676230, EPI_ISL_2676231, EPI_ISL_2676232, EPI_ISL_2676233, EPI_ISL_2676234, EPI_ISL_2676235, EPI_ISL_2676236, EPI_ISL_2676237, EPI_ISL_2676238, EPI_ISL_2676239, EPI_ISL_2676240, EPI_ISL_2676241, EPI_ISL_2676242, EPI_ISL_2676243, EPI_ISL_2676244, EPI_ISL_2676245, EPI_ISL_2676246, EPI_ISL_2676247, EPI_ISL_2676248, EPI_ISL_2676249, EPI_ISL_2676250, EPI_ISL_2676251, EPI_ISL_2676252, EPI_ISL_2676253, EPI_ISL_2676254, EPI_ISL_2676255, EPI_ISL_2676256, EPI_ISL_2676257, EPI_ISL_2676258, EPI_ISL_2676259, EPI_ISL_2676260, EPI_ISL_2676261, EPI_ISL_2676262, EPI_ISL_2676263, EPI_ISL_2676264, EPI_ISL_2676265, EPI_ISL_2676266, EPI_ISL_2676267, EPI_ISL_2676268, EPI_ISL_2676269, EPI_ISL_2676270, EPI_ISL_2676271, EPI_ISL_2676272, EPI_ISL_2676273, EPI_ISL_2676274, EPI_ISL_2676275, EPI_ISL_2676276, EPI_ISL_2676277, EPI_ISL_2676278, EPI_ISL_2676279, EPI_ISL_2676280, EPI_ISL_2676281, EPI_ISL_2676282, EPI_ISL_2676283, EPI_ISL_2676284, EPI_ISL_2676285, EPI_ISL_2676286, EPI_ISL_2676287, EPI_ISL_2676288, EPI_ISL_2676289, EPI_ISL_2676290, EPI_ISL_2676291, EPI_ISL_2676292, EPI_ISL_2676293, EPI_ISL_2676294, EPI_ISL_2676295, EPI_ISL_2676296, EPI_ISL_2676297, EPI_ISL_2676298, EPI_ISL_2676299, EPI_ISL_2676300, EPI_ISL_2676301, EPI_ISL_2676302, EPI_ISL_2676303, EPI_ISL_2676304, EPI_ISL_2676305, EPI_ISL_2676306, EPI_ISL_2676307, EPI_ISL_2676308, EPI_ISL_2676309, EPI_ISL_2676310, EPI_ISL_2676311, EPI_ISL_2676312, EPI_ISL_2676313, EPI_ISL_2676314, EPI_ISL_2676315, EPI_ISL_2676316, EPI_ISL_2676317, EPI_ISL_2676318, EPI_ISL_2676319, EPI_ISL_2676320, EPI_ISL_2676321, EPI_ISL_2676322, EPI_ISL_2676323, EPI_ISL_2676324, EPI_ISL_2676325, EPI_ISL_2676326, EPI_ISL_2676327, EPI_ISL_2676328, EPI_ISL_2676329, EPI_ISL_2676330, EPI_ISL_2676331, EPI_ISL_2676332, EPI_ISL_2676333, EPI_ISL_2676334, EPI_ISL_2676335, EPI_ISL_2676336, EPI_ISL_2676337, EPI_ISL_2676338, EPI_ISL_2676339, EPI_ISL_2676340, EPI_ISL_2676341, EPI_ISL_2676342, EPI_ISL_2676343, EPI_ISL_2676344, EPI_ISL_2676345, EPI_ISL_2676346, EPI_ISL_2676347, EPI_ISL_2676348, EPI_ISL_2676349, EPI_ISL_2676350, EPI_ISL_2676351, EPI_ISL_2676352, EPI_ISL_2676353, EPI_ISL_2676354, EPI_ISL_2676355, EPI_ISL_2676356, EPI_ISL_2676357, EPI_ISL_2676358, EPI_ISL_2676359, EPI_ISL_2676360, EPI_ISL_2676361, EPI_ISL_2676362, EPI_ISL_2676363, EPI_ISL_2676364, EPI_ISL_2676365, EPI_ISL_2676366, EPI_ISL_2676367, EPI_ISL_2676368, EPI_ISL_2676369, EPI_ISL_2676370, EPI_ISL_2676371, EPI_ISL_2676372, EPI_ISL_2676373, EPI_ISL_2676374, EPI_ISL_2676375, EPI_ISL_2676376, EPI_ISL_2676378, EPI_ISL_2676379, EPI_ISL_2676380, EPI_ISL_2676381, EPI_ISL_2676382, EPI_ISL_2676383, EPI_ISL_2676384, EPI_ISL_2676385, EPI_ISL_2676386, EPI_ISL_2676387, EPI_ISL_2676388, EPI_ISL_2676389, EPI_ISL_2676390, EPI_ISL_2676391, EPI_ISL_2676392, EPI_ISL_2676393, EPI_ISL_2676394, EPI_ISL_2676395, EPI_ISL_2676396, EPI_ISL_2676397, EPI_ISL_2676398, EPI_ISL_2676399, EPI_ISL_2676400, EPI_ISL_2676401, EPI_ISL_2676402, EPI_ISL_2860930, EPI_ISL_2860931, EPI_ISL_2860932, EPI_ISL_2860933, EPI_ISL_2860934, EPI_ISL_2860935, EPI_ISL_2860936, EPI_ISL_2860937, EPI_ISL_2860938, EPI_ISL_2860939, EPI_ISL_2860940, EPI_ISL_2860941, EPI_ISL_2860942, EPI_ISL_2860943, EPI_ISL_2860944, EPI_ISL_2860945, EPI_ISL_2860946, EPI_ISL_2860947, EPI_ISL_2860978, EPI_ISL_2860979, EPI_ISL_2860980, EPI_ISL_2860981, EPI_ISL_2860982, EPI_ISL_2860983, EPI_ISL_2860984, EPI_ISL_2860985, EPI_ISL_2860986, EPI_ISL_2860987, EPI_ISL_2860988, EPI_ISL_2860989, EPI_ISL_2860990, EPI_ISL_2860991, EPI_ISL_2860992, EPI_ISL_2860993, EPI_ISL_2860994, EPI_ISL_2860995, EPI_ISL_2860996, EPI_ISL_2860997, EPI_ISL_2860998, EPI_ISL_2860999, EPI_ISL_2861000, EPI_ISL_2861001, EPI_ISL_2861002, EPI_ISL_2861003, EPI_ISL_2861004, EPI_ISL_2861005, EPI_ISL_2861006, EPI_ISL_2861007, EPI_ISL_2861008, EPI_ISL_2861009 | | | |
| see above | INSACOG-WB | National Institute of Biomedical Genomics – INSACOG | Ajay Chakraborti; Arindam Maitra; Bhaswati Bandyopadhyay; Nidhan Kumar Biswas; Saumitra Das; Sreedhar Chinnaswamy; Tamal Ghosh |
| EPI_ISL_2897893, EPI_ISL_2897894, EPI_ISL_2897895, EPI_ISL_2897896, EPI_ISL_2897897, EPI_ISL_2897898, EPI_ISL_2897899, EPI_ISL_2897900, EPI_ISL_2897901, EPI_ISL_2897902, EPI_ISL_2897903, EPI_ISL_2897904, EPI_ISL_2897905, EPI_ISL_2924188, EPI_ISL_2924189, EPI_ISL_2924190, EPI_ISL_2924191, EPI_ISL_2924192, EPI_ISL_2924193, EPI_ISL_2924194, EPI_ISL_2924195, EPI_ISL_2924196, EPI_ISL_2924197, EPI_ISL_2924198, EPI_ISL_2924199, EPI_ISL_2924200, EPI_ISL_2924201, EPI_ISL_2924202, EPI_ISL_2924238, EPI_ISL_2924239, EPI_ISL_2924240, EPI_ISL_2924241, EPI_ISL_2924242, EPI_ISL_2924243, EPI_ISL_2924244, EPI_ISL_2924245, EPI_ISL_2924246, EPI_ISL_2924247, EPI_ISL_2924248, EPI_ISL_2924249 | | | |
| see above | Institute of Preventive Medicine | CDFD | Ashwin Dalal; Asmita Gupta; Divya Vashisht; Murali Bashyam; Nagamani Kammili; Reelina Basu; Vinay Donipadi |
| EPI_ISL_2842169, EPI_ISL_2842170, EPI_ISL_2842306, EPI_ISL_2842379, EPI_ISL_2842380 | JAYADEVA H | INSACOG-KA, NIMHANS | Ananthapadmanabha Kotambail; Anita S Desai; Anson Kunjumon George; Chetan G K; Chitra Pattabiraman; Darshan Sreenivas; Ellango Ramasamy; Gautham Arunachal Udupi; Mahesh Kumar.C.S; Sony Sharma; V Ravi |
| EPI_ISL_2379376 | JAYDEVA | INSACOG-KA, NIMHANS | Ananthapadmanabha Kotambail; Anita S Desai; Anson Kunjumon George; Chetan G K; Chitra Pattabiraman; Darshan Sreenivas; Gautham Arunachal Udupi; Pramada Prasad; V Ravi |
| EPI_ISL_2723821, EPI_ISL_2723822 | JIPMER Asia / India / Puducherry / Puducherry | inStem NCBS – INSACOG | Uma Ramakrishnan Dasaradhi Palakodeti Aswin SaiNarain |
| EPI_ISL_2272804, EPI_ISL_2272805, EPI_ISL_2272806, EPI_ISL_2272807, EPI_ISL_2272808, EPI_ISL_2272809, EPI_ISL_2272810, EPI_ISL_2272811, EPI_ISL_2272855, EPI_ISL_2272873, EPI_ISL_2272883, EPI_ISL_2272884, EPI_ISL_2272885, EPI_ISL_2272889, EPI_ISL_2272890 | | | |
| see above | JIPMER Puducherry | inStem NCBS - INSACOG | Uma Ramakrishnan Dasaradhi Palakodeti Aswin SaiNarain |
| EPI_ISL_2379392, EPI_ISL_2379402, EPI_ISL_2379403, EPI_ISL_2379427, EPI_ISL_2379428 | MAHALAKSHMI LAYOUT | INSACOG-KA, NIMHANS | Ananthapadmanabha Kotambail; Anita S Desai; Anson Kunjumon George; Chetan G K; Chitra Pattabiraman; Darshan Sreenivas; Gautham Arunachal Udupi; Pramada Prasad; V Ravi |
| EPI_ISL_2379367, EPI_ISL_2379368, EPI_ISL_2379369, EPI_ISL_2379370, EPI_ISL_2379371, EPI_ISL_2379381, EPI_ISL_2379382 | | | |
| see above | MOODALAPALYA | INSACOG-KA, NIMHANS | Ananthapadmanabha Kotambail; Anita S Desai; Anson Kunjumon George; Chetan G K; Chitra Pattabiraman; Darshan Sreenivas; Gautham Arunachal Udupi; Pramada Prasad; V Ravi |
| EPI_ISL_2157166, EPI_ISL_2157168, EPI_ISL_2157169, EPI_ISL_2157170, EPI_ISL_2157172, EPI_ISL_2157173, EPI_ISL_2157174, EPI_ISL_2157175, EPI_ISL_2157176, EPI_ISL_2157177, EPI_ISL_2157178, EPI_ISL_2157180, EPI_ISL_2157181, EPI_ISL_2157182, EPI_ISL_2157186, EPI_ISL_2157187, EPI_ISL_2157188, EPI_ISL_2157189, EPI_ISL_2157191, EPI_ISL_2157193, EPI_ISL_2323023, EPI_ISL_2373615, EPI_ISL_2373617, EPI_ISL_2373621, EPI_ISL_2373624, EPI_ISL_2373626, EPI_ISL_2373632, EPI_ISL_2373633, EPI_ISL_2373635, EPI_ISL_2373636, EPI_ISL_2373638, EPI_ISL_2373639, EPI_ISL_2373640, EPI_ISL_2373641, EPI_ISL_2373644, EPI_ISL_2400300, EPI_ISL_2400301, EPI_ISL_2400302, EPI_ISL_2400303, EPI_ISL_2400304, EPI_ISL_2400305, EPI_ISL_2400306, EPI_ISL_2400307, EPI_ISL_2400308, EPI_ISL_2400309, EPI_ISL_2400310, EPI_ISL_2400311 | | | |
| see above | Mapmygenome | CSIR-Centre for Cellular and Molecular Biology - INSACOG | Amareshwar Vodapalli; Ara Sreenivas; B Himasri; Divya Tej Sowpati; Karthik Bharadwaj Tallapaka; Lamuk Zaveri; Onkar Kulkarni; Rakesh K Mishra; Sharath Chandra Thota; Shreekant Verma; Sofia Banu; Viswagithe S L |
| EPI_ISL_2226261, EPI_ISL_2226262, EPI_ISL_2226263, EPI_ISL_2373350, EPI_ISL_2373351, EPI_ISL_2373352, EPI_ISL_2373353, EPI_ISL_2373354, EPI_ISL_2373355, EPI_ISL_2373356, EPI_ISL_2373357, EPI_ISL_2373358, EPI_ISL_2373359, EPI_ISL_2373360, EPI_ISL_2373361, EPI_ISL_2373362, EPI_ISL_2373363, EPI_ISL_2373364, EPI_ISL_2373365, EPI_ISL_2373366, EPI_ISL_2373367, EPI_ISL_2373368, EPI_ISL_2373369, EPI_ISL_2373370, EPI_ISL_2373371, EPI_ISL_2373372, EPI_ISL_2373373, EPI_ISL_2373374, EPI_ISL_2373375, EPI_ISL_2373376, EPI_ISL_2661100, EPI_ISL_2661101, EPI_ISL_2661102, EPI_ISL_2661103, EPI_ISL_2661104, EPI_ISL_2661105, EPI_ISL_2661106, EPI_ISL_2661107, EPI_ISL_2661108, EPI_ISL_2661109, EPI_ISL_2661110, EPI_ISL_2661111, EPI_ISL_2661112, EPI_ISL_2661113, EPI_ISL_2661114, EPI_ISL_2661115, EPI_ISL_2661116, EPI_ISL_2661117, EPI_ISL_2661118, EPI_ISL_2661119, EPI_ISL_2661120, EPI_ISL_2661121, EPI_ISL_2661122, EPI_ISL_2661123, EPI_ISL_2661124, EPI_ISL_2661125, EPI_ISL_2661126, EPI_ISL_2661127, EPI_ISL_2661128, EPI_ISL_2661129, EPI_ISL_2661130, EPI_ISL_2661131, EPI_ISL_2661132, EPI_ISL_2661133, EPI_ISL_2661135, EPI_ISL_2661136, EPI_ISL_2661137, EPI_ISL_2661138, EPI_ISL_2661139, EPI_ISL_2661140, EPI_ISL_2661141, EPI_ISL_2661142, EPI_ISL_2661143, EPI_ISL_2661144, EPI_ISL_2661145, EPI_ISL_2774678, EPI_ISL_2774679, EPI_ISL_2774680, EPI_ISL_2774681, EPI_ISL_2774682, EPI_ISL_2774683, EPI_ISL_2774684, EPI_ISL_2774685, EPI_ISL_2774686, EPI_ISL_2774687, EPI_ISL_2774688, EPI_ISL_2774689, EPI_ISL_2774690, EPI_ISL_2774691, EPI_ISL_2774692, EPI_ISL_2774693, EPI_ISL_2774694, EPI_ISL_2774695, EPI_ISL_2774696, EPI_ISL_2774697, EPI_ISL_2774698, EPI_ISL_2774699, EPI_ISL_2774700, EPI_ISL_2774701, EPI_ISL_2774702, EPI_ISL_2774703, EPI_ISL_2774704, EPI_ISL_2774705, EPI_ISL_2774706, EPI_ISL_2774707, EPI_ISL_2774708, EPI_ISL_2774709, EPI_ISL_2774710, EPI_ISL_2774711, EPI_ISL_2774712, EPI_ISL_2774713, EPI_ISL_2774714, EPI_ISL_2774715, EPI_ISL_2774716, EPI_ISL_2774717, EPI_ISL_2774718, EPI_ISL_2774719, EPI_ISL_2774720, EPI_ISL_2774721, EPI_ISL_2774722, EPI_ISL_2774723, EPI_ISL_2775128, EPI_ISL_2775130, EPI_ISL_2775131, EPI_ISL_2775136, EPI_ISL_2775137, EPI_ISL_2775138, EPI_ISL_2775139, EPI_ISL_2775140, EPI_ISL_2775141, EPI_ISL_2775142, EPI_ISL_2775143, EPI_ISL_2775144, EPI_ISL_2775145, EPI_ISL_2775146, EPI_ISL_2775147 | | | |
| see above | Mapmygenome | CSIR-Centre for Cellular and Molecular Biology- INSACOG | ; Amareshwar Vodapalli; Ara Sreenivas; Archana Bharadwaj Siva; B Himasri; Blessy B John; Divya Tej Sowpati; Jandhyala Sai Krishna; Karthik Bharadwaj Tallapaka; Lamuk Zaveri; Onkar Kulkarni; Payel Mukherjee; Priya Nurkurthy; Rakesh K Mishra; Sharath Chandra Thota; Shreekant Verma; Sofia Banu; Sumedha Avadhanula; Tulasi Nagabandi; Valli Nagalakshmi Undamatla; Vidhyadhari Methuku; Viswagithe S L |
| EPI_ISL_2350828 | Medical Bioinformatics, Sri Ramachandra institute of higher education and research | Medical Bioinformatics, Sri Ramachandra institute of higher education and research | Velu, P. |
| EPI_ISL_2348743 | Msc Stem cell, Ramachandra | Msc Stem cell, Ramachandra | Kumar, N. |
| EPI_ISL_2227163 | Multidisiciplinary Research Unit, Institute of Medical Sciences, Banaras Hindu University, Varanasi - 221005 | Multidisiciplinary Research Unit, Institute of Medical Sciences, Banaras Hindu University, Varanasi - 221005 | Abhay Kumar Yadav; Ashish; Chetan Sahni; Deepa Devadas; Manpreet Kaur; Nitish Kumar Singh; Priyoneel Basu; Royana Singh; Saumya Singh; Shani Vishwakarma; Shivani Mishra; Surendra Pratap Mishra; Tribhuwan Mohan Mohapatra |
| EPI_ISL_2543900, EPI_ISL_2543901, EPI_ISL_2543902, EPI_ISL_2543903, EPI_ISL_2543904, EPI_ISL_2543905, EPI_ISL_2543906, EPI_ISL_2543907, EPI_ISL_2543908, EPI_ISL_2543909, EPI_ISL_2543910, EPI_ISL_2543911, EPI_ISL_2543912, EPI_ISL_2543913, EPI_ISL_2543914, EPI_ISL_2543915, EPI_ISL_2544822 | | | |
| see above | N.H.L Municipal Medical College, Ahmedabad | Gujarat Biotechnology Research Centre | Chaitanya Joshi; Dinesh Kumar; Janvi Raval; Jayshri Pethani; Madhvi Joshi; Nitesh Shah; Nitin Savaliya; Ramesh Pandit; Sonal Sharma; Twinkle Soni; Umang Mishra; Zarna Patel; Zuber Saiyed |
| EPI_ISL_2379387 | NAGAPPA BLOCK | INSACOG-KA, NIMHANS | Ananthapadmanabha Kotambail; Anita S Desai; Anson Kunjumon George; Chetan G K; Chitra Pattabiraman; Darshan Sreenivas; Gautham Arunachal Udupi; Pramada Prasad; V Ravi |
| EPI_ISL_2379383 | NETHAJI CIRCLE | INSACOG-KA, NIMHANS | Ananthapadmanabha Kotambail; Anita S Desai; Anson Kunjumon George; Chetan G K; Chitra Pattabiraman; Darshan Sreenivas; Gautham Arunachal Udupi; Pramada Prasad; V Ravi |
| EPI_ISL_2379360, EPI_ISL_2379388, EPI_ISL_2379398, EPI_ISL_2450354, EPI_ISL_2450355, EPI_ISL_2450356, EPI_ISL_2651298, EPI_ISL_2651312, EPI_ISL_2651313, EPI_ISL_2651314, EPI_ISL_2651321, EPI_ISL_2651330, EPI_ISL_2651331, EPI_ISL_2651350, EPI_ISL_2651355, EPI_ISL_2651359, EPI_ISL_2725537, EPI_ISL_2728232, EPI_ISL_2728249, EPI_ISL_2728291, EPI_ISL_2728292, EPI_ISL_2728312, EPI_ISL_2728316, EPI_ISL_2728428, EPI_ISL_2728501, EPI_ISL_2728502, EPI_ISL_2728529, EPI_ISL_2842101, EPI_ISL_2842132, EPI_ISL_2842133, EPI_ISL_2842149, EPI_ISL_2842165, EPI_ISL_2842166, EPI_ISL_2842171, EPI_ISL_2842177, EPI_ISL_2842181, EPI_ISL_2842182, EPI_ISL_2842290, EPI_ISL_2842313, EPI_ISL_2842316, EPI_ISL_2842328, EPI_ISL_2842347, EPI_ISL_2842355, EPI_ISL_2842396, EPI_ISL_2842443 | | | |
| see above | NIMHANS | INSACOG-KA, NIMHANS | Ananthapadmanabha Kotambail; Anita S Desai; Anson Kunjumon George; Chetan G K; Chitra Pattabiraman; Darshan Sreenivas; Ellango Ramasamy; Gautham Arunachal Udupi; Mahesh Kumar.C.S; Pramada Prasad; Sony Sharma; V Ravi |
| EPI_ISL_2811888 | National Centre For Cell Science | National Centre For Cell Science – INSACOG | Ajay Pillai; Dhiraj Paul; Girish R Nair; Harshada Ghode; INSACOG Consortium team; Kalpesh Chandrakant Bavkar; Kartik Jayant Deopujari; Manoj Kumar Bhat; Mitali Inamdar; Shivang P. Bhanushali; Yogesh Shouche |
| EPI_ISL_2382915, EPI_ISL_2382916, EPI_ISL_2382917, EPI_ISL_2382918, EPI_ISL_2382919, EPI_ISL_2382920, EPI_ISL_2382921, EPI_ISL_2382922, EPI_ISL_2382923, EPI_ISL_2382924, EPI_ISL_2382925, EPI_ISL_2382926, EPI_ISL_2382927, EPI_ISL_2382928, EPI_ISL_2382929, EPI_ISL_2382930, EPI_ISL_2382931, EPI_ISL_2382932, EPI_ISL_2382933, EPI_ISL_2382936, EPI_ISL_2382937, EPI_ISL_2382938, EPI_ISL_2382939, EPI_ISL_2382940, EPI_ISL_2382941, EPI_ISL_2382942, EPI_ISL_2382943, EPI_ISL_2382944, EPI_ISL_2382945, EPI_ISL_2382946, EPI_ISL_2382947, EPI_ISL_2382948, EPI_ISL_2382949, EPI_ISL_2382950, EPI_ISL_2382951, EPI_ISL_2382952, EPI_ISL_2382953, EPI_ISL_2382954, EPI_ISL_2382955, EPI_ISL_2382956, EPI_ISL_2382957, EPI_ISL_2382958, EPI_ISL_2382959, EPI_ISL_2382960, EPI_ISL_2382961, EPI_ISL_2382962, EPI_ISL_2382963, EPI_ISL_2382964, EPI_ISL_2382965, EPI_ISL_2382966, EPI_ISL_2382967, EPI_ISL_2382968, EPI_ISL_2382969, EPI_ISL_2382973, EPI_ISL_2382976, EPI_ISL_2382978, EPI_ISL_2382979, EPI_ISL_2382980, EPI_ISL_2382981, EPI_ISL_2382982, EPI_ISL_2382983 | | | |
| see above | National Centre For Cell Science | National Centre For Cell Science-INSACOG | Ajay Pillai; Dhiraj Paul; INSACOG Consortium team; Manoj Kumar Bhat; Mitali Inamdar; Mohak P Gujare; Shivang P. Bhanushali; Yogesh Shouche |
| EPI_ISL_2897666, EPI_ISL_2897667, EPI_ISL_2897668, EPI_ISL_2897669, EPI_ISL_2897670, EPI_ISL_2897672, EPI_ISL_2897673, EPI_ISL_2897674, EPI_ISL_2897675, EPI_ISL_2897676, EPI_ISL_2897677, EPI_ISL_2897692, EPI_ISL_2897693, EPI_ISL_2897694, EPI_ISL_2897819, EPI_ISL_2897820, EPI_ISL_2897821, EPI_ISL_2897822, EPI_ISL_2897823 | | | |
| see above | National Centre for Cell Science | CDFD | Ashwin Dalal; Asmita Gupta; Divya Vashisht; Murali Bashyam; Reelina Basu; Vinay Donipadi |
| EPI_ISL_2461008, EPI_ISL_2461009, EPI_ISL_2461100, EPI_ISL_2461101, EPI_ISL_2461102, EPI_ISL_2461103, EPI_ISL_2461104, EPI_ISL_2461105, EPI_ISL_2461106, EPI_ISL_2461107, EPI_ISL_2461108, EPI_ISL_2461110, EPI_ISL_2461121, EPI_ISL_2461129, EPI_ISL_2461130, EPI_ISL_2555968, EPI_ISL_2556059, EPI_ISL_2556091, EPI_ISL_2556092, EPI_ISL_2556093, EPI_ISL_2556094, EPI_ISL_2556095, EPI_ISL_2556096, EPI_ISL_2556097, EPI_ISL_2556098, EPI_ISL_2556099 | | | |
| see above | National Centre for Disease Control (NCDC) Biotechnology Division, Delhi | NCDC Delhi, Biotechnology Division INSACOG | Hema Gogia; Hemlata Lall; Kalaiarasan Ponnusamy; Mahesh S Dhar; Manoj K Singh; Meena Datta; Partha Rakshit; Preeti Madan; Priyanka Singh; Radhakrishnan V. S; Robin Marwal; Sandhya Kabra; Sujeet K Singh; Uma Sharma |
| EPI_ISL_2924250, EPI_ISL_2924251, EPI_ISL_2924252, EPI_ISL_2924253, EPI_ISL_2924254, EPI_ISL_2924255, EPI_ISL_2924256, EPI_ISL_2924257, EPI_ISL_2924258, EPI_ISL_2924259, EPI_ISL_2924260, EPI_ISL_2924261, EPI_ISL_2924262, EPI_ISL_2924263 | | | |
| see above | Osmania Medical College | CDFD | Ashwin Dalal; Asmita Gupta; Divya Vashisht; Murali Bashyam; Nagamani Kammili; Reelina Basu; Vinay Donipadi |
| EPI_ISL_2379379 | PANTHRPALYA | INSACOG-KA, NIMHANS | Ananthapadmanabha Kotambail; Anita S Desai; Anson Kunjumon George; Chetan G K; Chitra Pattabiraman; Darshan Sreenivas; Gautham Arunachal Udupi; Pramada Prasad; V Ravi |
| EPI_ISL_2660816, EPI_ISL_2660817, EPI_ISL_2660818, EPI_ISL_2660819, EPI_ISL_2660820, EPI_ISL_2660821, EPI_ISL_2660822, EPI_ISL_2660823, EPI_ISL_2660824, EPI_ISL_2660825, EPI_ISL_2660826, EPI_ISL_2660827, EPI_ISL_2660828, EPI_ISL_2660830, EPI_ISL_2660831, EPI_ISL_2660832, EPI_ISL_2660833, EPI_ISL_2660834, EPI_ISL_2660835, EPI_ISL_2660836, EPI_ISL_2660837, EPI_ISL_2660838, EPI_ISL_2660839, EPI_ISL_2660840, EPI_ISL_2660841, EPI_ISL_2660842, EPI_ISL_2660843, EPI_ISL_2660844, EPI_ISL_2660845, EPI_ISL_2660846, EPI_ISL_2660847, EPI_ISL_2660848, EPI_ISL_2660849, EPI_ISL_2660850, EPI_ISL_2660851, EPI_ISL_2660852, EPI_ISL_2660853, EPI_ISL_2660854, EPI_ISL_2660855, EPI_ISL_2660856, EPI_ISL_2660857, EPI_ISL_2660858, EPI_ISL_2660859, EPI_ISL_2660860, | | | |

| | | | |
|---|---|---|---|
| EPI_ISL_2660861, EPI_ISL_2660862, EPI_ISL_2660863, EPI_ISL_2660864, EPI_ISL_2660865, EPI_ISL_2660866, EPI_ISL_2660867, EPI_ISL_2660868, EPI_ISL_2660869, EPI_ISL_2660870, EPI_ISL_2660871, EPI_ISL_2660872, EPI_ISL_2660873, EPI_ISL_2660874, EPI_ISL_2660875, EPI_ISL_2660876, EPI_ISL_2660877, EPI_ISL_2660878, EPI_ISL_2660879, EPI_ISL_2660880, EPI_ISL_2660881, EPI_ISL_2660882, EPI_ISL_2660883, EPI_ISL_2660884, EPI_ISL_2660885, EPI_ISL_2660886, EPI_ISL_2660888, EPI_ISL_2660889, EPI_ISL_2660890, EPI_ISL_2660892, EPI_ISL_2660893, EPI_ISL_2660894, EPI_ISL_2660895, EPI_ISL_2660896, EPI_ISL_2660897, EPI_ISL_2660898, EPI_ISL_2660899, EPI_ISL_2660900, EPI_ISL_2660901, EPI_ISL_2660902, EPI_ISL_2660903, EPI_ISL_2660904, EPI_ISL_2660905, EPI_ISL_2660906, EPI_ISL_2660907, EPI_ISL_2660908, EPI_ISL_2660909, EPI_ISL_2688255 | | | |
| see above | PASTEUR MOLECULAR LABORATORY - INSACOG MEGHALAYA | CSIR-Centre for Cellular and Molecular Biology-INSACOG | Amareshwar Vodapalli; Ara Sreenivas; Archana Bharadwaj Siva; B Himasri; Divya Tej Sowpati; E Marak; E Shadap; Karthik Bharadwaj Tallapaka; Lamuk Zaveri; Onkar Kulkarni; Payel Mukherjee; Priya Nurkurthy; R. Lyngdoh; Rakesh K Mishra; Shreekant Verma; Sofia Banu; Sumedha Avadhanula; Tulasi Nagabandi; Valli Nagalakshmi Undamatla; Vidhyadhari Methuku |
| EPI_ISL_2379399, EPI_ISL_2379400 | PEENYA KAMALA NAGAR | INSACOG-KA, NIMHANS | Ananthapadmanabha Kotambail; Anita S Desai; Anson Kunjumon George; Chetan G K; Chitra Pattabiraman; Darshan Sreenivas; Gautham Arunachal Udupi; Pramada Prasad; V Ravi |
| EPI_ISL_2379393, EPI_ISL_2379415 | PEENYA KAMALANAGAR | INSACOG-KA, NIMHANS | Ananthapadmanabha Kotambail; Anita S Desai; Anson Kunjumon George; Chetan G K; Chitra Pattabiraman; Darshan Sreenivas; Gautham Arunachal Udupi; Pramada Prasad; V Ravi |
| EPI_ISL_2379396, EPI_ISL_2379397 | RAILWAY | INSACOG-KA, NIMHANS | Ananthapadmanabha Kotambail; Anita S Desai; Anson Kunjumon George; Chetan G K; Chitra Pattabiraman; Darshan Sreenivas; Gautham Arunachal Udupi; Pramada Prasad; V Ravi |
| EPI_ISL_2379375, EPI_ISL_2379377, EPI_ISL_2379389, EPI_ISL_2379430, EPI_ISL_2379431, EPI_ISL_2651307, EPI_ISL_2651308, EPI_ISL_2651309, EPI_ISL_2651316, EPI_ISL_2651317, EPI_ISL_2651318, EPI_ISL_2651319, EPI_ISL_2651322, EPI_ISL_2651323, EPI_ISL_2651324, EPI_ISL_2651325, EPI_ISL_2651341, EPI_ISL_2651342, EPI_ISL_2651343, EPI_ISL_2651348 | | | |
| see above | RAILWAY H | INSACOG-KA, NIMHANS | Ananthapadmanabha Kotambail; Anita S Desai; Anson Kunjumon George; Chetan G K; Chitra Pattabiraman; Darshan Sreenivas; Ellango Ramasamy; Gautham Arunachal Udupi; Mahesh Kumar.C.S; Pramada Prasad; Sony Sharma; V Ravi |
| EPI_ISL_2450359, EPI_ISL_2450360, EPI_ISL_2450361, EPI_ISL_2450362, EPI_ISL_2450363, EPI_ISL_2450364, EPI_ISL_2725529, EPI_ISL_2728265, EPI_ISL_2728348, EPI_ISL_2728503, EPI_ISL_2842102, EPI_ISL_2842118, EPI_ISL_2842120, EPI_ISL_2842167, EPI_ISL_2842168, EPI_ISL_2842178, EPI_ISL_2842294, EPI_ISL_2842295, EPI_ISL_2842296, EPI_ISL_2842297, EPI_ISL_2842304, EPI_ISL_2842314, EPI_ISL_2842315, EPI_ISL_2842373, EPI_ISL_2842374, EPI_ISL_2842397, EPI_ISL_2842437, EPI_ISL_2842440, EPI_ISL_2842441, EPI_ISL_2842444, EPI_ISL_2842448, EPI_ISL_2842452, EPI_ISL_2842455 | | | |
| see above | RAILWAY HOSPITAL | INSACOG-KA, NIMHANS | Ananthapadmanabha Kotambail; Anita S Desai; Anson Kunjumon George; Chetan G K; Chitra Pattabiraman; Darshan Sreenivas; Ellango Ramasamy; Gautham Arunachal Udupi; Mahesh Kumar.C.S; Sony Sharma; V Ravi |
| EPI_ISL_2661385, EPI_ISL_2661386, EPI_ISL_2661387, EPI_ISL_2661388, EPI_ISL_2661389, EPI_ISL_2661390, EPI_ISL_2661391, EPI_ISL_2661392, EPI_ISL_2661393, EPI_ISL_2661394, EPI_ISL_2661395, EPI_ISL_2661396, EPI_ISL_2661397, EPI_ISL_2661398, EPI_ISL_2661399, EPI_ISL_2661400, EPI_ISL_2661401, EPI_ISL_2661402, EPI_ISL_2661403, EPI_ISL_2661404, EPI_ISL_2661405, EPI_ISL_2661406, EPI_ISL_2661407, EPI_ISL_2661408, EPI_ISL_2661409, EPI_ISL_2661410, EPI_ISL_2661411, EPI_ISL_2661415, EPI_ISL_2661416, EPI_ISL_2661421, EPI_ISL_2661422, EPI_ISL_2661423, EPI_ISL_2661424, EPI_ISL_2661425, EPI_ISL_2661426, EPI_ISL_2661427, EPI_ISL_2661428, EPI_ISL_2661429, EPI_ISL_2661430, EPI_ISL_2661431, EPI_ISL_2661432, EPI_ISL_2661433, EPI_ISL_2661434, EPI_ISL_2661435, EPI_ISL_2661436, EPI_ISL_2661437, EPI_ISL_2661438, EPI_ISL_2661439, EPI_ISL_2661440, EPI_ISL_2661441, EPI_ISL_2661442, EPI_ISL_2661443, EPI_ISL_2661444, EPI_ISL_2661445, EPI_ISL_2661446, EPI_ISL_2661447, EPI_ISL_2661448, EPI_ISL_2661449, EPI_ISL_2661450, EPI_ISL_2661451, EPI_ISL_2661452, EPI_ISL_2661453, EPI_ISL_2661454, EPI_ISL_2661455, EPI_ISL_2661456, EPI_ISL_2661457, EPI_ISL_2661458, EPI_ISL_2661459, EPI_ISL_2661460, EPI_ISL_2661461, EPI_ISL_2661462, EPI_ISL_2661463, EPI_ISL_2661464 | | | |
| see above | RUHS College of Medical Sciences | CSIR-Centre for Cellular and Molecular Biology - INSACOG | Amareshwar Vodapalli; Ara Sreenivas; Archana Bharadwaj Siva; B Himasri; Divya Tej Sowpati; Dr. Gaurav Dalela; Dr. Jitendra Panda; Dr. Nilofer Khayyam; Dr. Raja Babu Panwar; Dr. Rajeev Gupta; Dr. Ramesh Sharma; Dr. Sonali Sharma; Dr. Sudhanshu Kacker; Dr. Vaseem Naheed Baig; Jandhyala Sai Krishna; Karthik Bharadwaj Tallapaka; Lamuk Zaveri; Onkar Kulkarni; Payel Mukherjee; Priya Nurkurthy; Rakesh K Mishra; Shreekant Verma; Sofia Banu; Sumedha Avadhanula; Tulasi Nagabandi; Valli Nagalakshmi Undamatla; Vidhyadhari Methuku |
| EPI_ISL_2774834 | RUHS College of Medical Sciences | CSIR-Centre for Cellular and Molecular Biology-INSACOG | Amareshwar Vodapalli; Ara Sreenivas; Archana Bharadwaj Siva; B Himasri; Divya Tej Sowpati; Dr. Gaurav Dalela; Dr. Jitendra Panda; Dr. Nilofer Khayyam; Dr. Raja Babu Panwar; Dr. Rajeev Gupta; Dr. Ramesh Sharma; Dr. Sonali Sharma; Dr. Sudhanshu Kacker; Dr. Vaseem Naheed Baig; Jandhyala Sai Krishna; Karthik Bharadwaj Tallapaka; Lamuk Zaveri; Onkar Kulkarni; Payel Mukherjee; Priya Nurkurthy; Rakesh K Mishra; Shreekant Verma; Sofia Banu; Sumedha Avadhanula; Tulasi Nagabandi; Valli Nagalakshmi Undamatla; Vidhyadhari Methuku |
| EPI_ISL_2710005, EPI_ISL_2710006, EPI_ISL_2710010, EPI_ISL_2710011 | Regional VRDL,Dibrugarh | Regional VRDL,Dibrugarh | A Jakharia; B Borkakoty; C Bhattacharya; D Biswas; K Narain; Mandakini Das Sarma; R Hazarika |
| EPI_ISL_2728293, EPI_ISL_2728314 | SANJAY GANDHI INSTITUTE OF TRAUMA AND ORTHOPAEDICS | INSACOG-KA, NIMHANS | Ananthapadmanabha Kotambail; Anita S Desai; Anson Kunjumon George; Chetan G K; Chitra Pattabiraman; Darshan Sreenivas; Ellango Ramasamy; Gautham Arunachal Udupi; Mahesh Kumar.C.S; Sony Sharma; V Ravi |
| EPI_ISL_2379391 | SHANKARNAGAR | INSACOG-KA, NIMHANS | Ananthapadmanabha Kotambail; Anita S Desai; Anson Kunjumon George; Chetan G K; Chitra Pattabiraman; Darshan Sreenivas; Gautham Arunachal Udupi; Pramada Prasad; V Ravi |
| EPI_ISL_2379414 | SHANKARNAGR | INSACOG-KA, NIMHANS | Ananthapadmanabha Kotambail; Anita S Desai; Anson Kunjumon George; Chetan G K; Chitra Pattabiraman; Darshan Sreenivas; Gautham Arunachal Udupi; Pramada Prasad; V Ravi |
| EPI_ISL_2922000 | SMS Medical College | SMS Medical College Jaipur | Bharti Malhotra; Dinesh Parsoya; Himanshu Sharma; Neha Bhomia; Pratibha Sharma; Sudheer Bhandari; Swati Gautam |
| EPI_ISL_2922001 | SMS Medical College, Jaipur | SMS Medical College | Bharti Malhotra; Dinesh Parsoya; Himanshu Sharma; Neha Bhomia; Pratibha Mishra; Sudheer Bhandari; Swati Gautam |
| EPI_ISL_2918762 | SMS, Medical College, Jaipur | SMS, Medical College, Jaipur | Bharti Malhotra; Dinesh parsoya; Himanshu Sharma; Neha Bhomia; Pratibha Mishra; Sudhher Bhandari; Swati Gatuam |
| EPI_ISL_2921999 | SMS, Medical college | SMS, Medical college | Bharti Malhotra; Dinesh Parsoya; Himanshu Sharma; Neha Bhomia; Pratibha Mishra; Sudheer Bhandari; Swati Gautam |
| EPI_ISL_2450365, EPI_ISL_2728236, EPI_ISL_2728270, EPI_ISL_2728271, EPI_ISL_2842392 | SRI JAYADEVA INSTITUTE OF CARDIOLOGY | INSACOG-KA, NIMHANS | Ananthapadmanabha Kotambail; Anita S Desai; Anson Kunjumon George; Chetan G K; Chitra Pattabiraman; Darshan Sreenivas; Ellango Ramasamy; Gautham Arunachal Udupi; Mahesh Kumar.C.S; Sony Sharma; V Ravi |
| EPI_ISL_2651303, EPI_ISL_2651354 | Sanjay Gandhi Institute Of Trauma & Orthopaedics Centre | INSACOG-KA, NIMHANS | Ananthapadmanabha Kotambail; Anita S Desai; Anson Kunjumon George; Chetan G K; Chitra Pattabiraman; Darshan Sreenivas; Ellango Ramasamy; Gautham Arunachal Udupi; Mahesh Kumar.C.S; Pramada Prasad; Sony Sharma; V Ravi |
| EPI_ISL_2307111, EPI_ISL_2307112, EPI_ISL_2307113, EPI_ISL_2307114, EPI_ISL_2307115, EPI_ISL_2307117, EPI_ISL_2307118, EPI_ISL_2307119, EPI_ISL_2307121, EPI_ISL_2307122, EPI_ISL_2307124, EPI_ISL_2307125, EPI_ISL_2307126, EPI_ISL_2307127, EPI_ISL_2307128, EPI_ISL_2307129, EPI_ISL_2307130, EPI_ISL_2307134, EPI_ISL_2307135, EPI_ISL_2307136, EPI_ISL_2620743 | | | |
| see above | State Virus Research and Diagnostic Laboratory (VRDL), AIIMS Raipur | State Virus Research and Diagnostic Laboratory (VRDL), AIIMS Raipur | Anudita Bhargava; Jiyan Chandrawanshi; Kuldeep Sharma; MD Rafiiqullah Khan; Priyanka Singh; Pushpendra Singh; Sanjay Singh Negi; Somya Sharma |
| EPI_ISL_2543899 | Sterling Hospital, Memnagar, Ahmedabad | Gujarat Biotechnology Research Centre | Atulbhai Patel; Chaitanya Joshi; Dinesh Kumar; Janvi Raval; Madhvi Joshi; Nitesh Shah; Nitin Savaliya; Ramesh Pandit; Sonal Sharma; Twinkle Soni; Umang Mishra; Zarna Patel; Zuber Saiyed |
| EPI_ISL_2450357, EPI_ISL_2450358 | UPHC A D HALLI | INSACOG-KA, NIMHANS | Ananthapadmanabha Kotambail; Anita S Desai; Anson Kunjumon George; Chetan G K; Chitra Pattabiraman; Darshan Sreenivas; Gautham Arunachal Udupi; V Ravi |
| EPI_ISL_2842458, EPI_ISL_2842459, EPI_ISL_2842460, EPI_ISL_2842461, EPI_ISL_2842462, EPI_ISL_2842463, EPI_ISL_2842464, EPI_ISL_2842465, EPI_ISL_2842466 | | | |
| see above | VICTORIA HOSPITAL | INSACOG-KA, NIMHANS | Ananthapadmanabha Kotambail; Anita S Desai; Anson Kunjumon George; Chetan G K; Chitra Pattabiraman; Darshan Sreenivas; Ellango Ramasamy; Gautham Arunachal Udupi; Mahesh Kumar.C.S; Sony Sharma; V Ravi |
| EPI_ISL_2842134, EPI_ISL_2842135, EPI_ISL_2842136, EPI_ISL_2842137, EPI_ISL_2842138, EPI_ISL_2842139, EPI_ISL_2842140, EPI_ISL_2842141, EPI_ISL_2842142, EPI_ISL_2842143, EPI_ISL_2842144 | | | |
| see above | VRDL VIJAYAPURA | INSACOG-KA, NIMHANS | Ananthapadmanabha Kotambail; Anita S Desai; Anson Kunjumon George; Chetan G K; Chitra Pattabiraman; Darshan Sreenivas; Ellango Ramasamy; Gautham Arunachal Udupi; Mahesh Kumar.C.S; Sony Sharma; V Ravi |

We gratefully acknowledge the following Authors from the Originating laboratories responsible for obtaining the specimens, as well as the Submitting laboratories where the genome data were generated and shared via GISAID, on which this research is based.

All Submitters of data may be contacted directly via www.gisaid.org

Authors are sorted alphabetically.

| Accession ID | Originating Laboratory | Submitting Laboratory | Authors |
|---|---|---|---|
| EPI_ISL_1431615, EPI_ISL_1431669, EPI_ISL_1431670, EPI_ISL_1431676, EPI_ISL_1431677, EPI_ISL_1431678, EPI_ISL_1431679, EPI_ISL_1431680, EPI_ISL_1431685, EPI_ISL_1431686, EPI_ISL_1431687, EPI_ISL_1431688, EPI_ISL_1431689, EPI_ISL_1431690, EPI_ISL_1431691, EPI_ISL_1431692, EPI_ISL_1431693, EPI_ISL_1431694, EPI_ISL_1431695, EPI_ISL_1431696, EPI_ISL_1431697, EPI_ISL_1431700, EPI_ISL_1431701, EPI_ISL_1431707, EPI_ISL_1431708, EPI_ISL_1432054, EPI_ISL_1432056, EPI_ISL_1432058, EPI_ISL_1432060, EPI_ISL_1432063, EPI_ISL_1432073, EPI_ISL_1432076, EPI_ISL_1432077, EPI_ISL_1432079, EPI_ISL_1432081, EPI_ISL_1432083, EPI_ISL_1927241, EPI_ISL_1927242, EPI_ISL_1927244, EPI_ISL_1927245, EPI_ISL_1927247, EPI_ISL_1927248, EPI_ISL_1927249, EPI_ISL_1927251, EPI_ISL_1927252, EPI_ISL_1927254, EPI_ISL_1927255, EPI_ISL_1927257, EPI_ISL_1927258, EPI_ISL_1927260, EPI_ISL_1927261, EPI_ISL_1927263, EPI_ISL_1927264, EPI_ISL_1927266, EPI_ISL_1927267, EPI_ISL_1927269, EPI_ISL_1927270, EPI_ISL_1927271, EPI_ISL_1927273, EPI_ISL_1927275, EPI_ISL_1927460, EPI_ISL_1927462, EPI_ISL_1927463, EPI_ISL_1927849, EPI_ISL_1929239, EPI_ISL_1929399, EPI_ISL_1929401, EPI_ISL_1929402, EPI_ISL_1929404, EPI_ISL_1929405, EPI_ISL_1929424, EPI_ISL_1929426, EPI_ISL_1929427, EPI_ISL_1929435, EPI_ISL_1929545, EPI_ISL_1929546, EPI_ISL_1929548, EPI_ISL_1929549, EPI_ISL_1929551, EPI_ISL_1929552, EPI_ISL_1929554, EPI_ISL_1929555, EPI_ISL_1929557, EPI_ISL_1929558, EPI_ISL_1929560, EPI_ISL_1929561, EPI_ISL_1929563, EPI_ISL_1929564, EPI_ISL_1929566, EPI_ISL_1929567, EPI_ISL_1929569, EPI_ISL_1929570, EPI_ISL_1929572, EPI_ISL_1929573, EPI_ISL_1929575, EPI_ISL_1929584, EPI_ISL_1929585, EPI_ISL_1929587, EPI_ISL_1929588, EPI_ISL_1929590, EPI_ISL_1929591, EPI_ISL_1929593, EPI_ISL_1929594, EPI_ISL_1929596, EPI_ISL_1929597, EPI_ISL_1929599, EPI_ISL_1929600, EPI_ISL_1929602, EPI_ISL_1929603, EPI_ISL_1929605, EPI_ISL_1929606, EPI_ISL_1929608, EPI_ISL_1929609, EPI_ISL_1929611, EPI_ISL_1929612, EPI_ISL_1929614, EPI_ISL_1929615, EPI_ISL_1929617, EPI_ISL_1929618, EPI_ISL_1929620, EPI_ISL_1929621, EPI_ISL_1929623, EPI_ISL_1929624, EPI_ISL_1929626, EPI_ISL_1929627, EPI_ISL_1929629, EPI_ISL_1929630, EPI_ISL_1929632, EPI_ISL_1929633, EPI_ISL_1929635, EPI_ISL_1929636, EPI_ISL_1929638, EPI_ISL_1929639, EPI_ISL_1929641, EPI_ISL_1929642, EPI_ISL_1929643, EPI_ISL_1929645, EPI_ISL_1929646, EPI_ISL_1929648, EPI_ISL_1929649, EPI_ISL_1929651, EPI_ISL_1929652, EPI_ISL_1929654, EPI_ISL_1929655, EPI_ISL_1929656, EPI_ISL_1929658, EPI_ISL_1929659, EPI_ISL_1929661, EPI_ISL_1929662, EPI_ISL_1929664, EPI_ISL_1929665, EPI_ISL_1929667, EPI_ISL_1929668, EPI_ISL_1929670, EPI_ISL_1929671, EPI_ISL_1929673, EPI_ISL_1929674, EPI_ISL_1929675, EPI_ISL_1929677, EPI_ISL_1929678, EPI_ISL_1929680, EPI_ISL_1929681, EPI_ISL_1929683, EPI_ISL_1929684, EPI_ISL_1929686, EPI_ISL_1929687, EPI_ISL_1929689, EPI_ISL_1929690, EPI_ISL_1929692, EPI_ISL_1929693, EPI_ISL_1929695, EPI_ISL_1929696, EPI_ISL_1929698, EPI_ISL_1929699, EPI_ISL_1929701, EPI_ISL_1929702, EPI_ISL_1929704, EPI_ISL_1929705, EPI_ISL_1929707, EPI_ISL_1929708, EPI_ISL_1929710, EPI_ISL_1929711, EPI_ISL_1929713, EPI_ISL_1929714, EPI_ISL_1929716, EPI_ISL_1929717, EPI_ISL_1929719, EPI_ISL_1929720, EPI_ISL_1929722, EPI_ISL_1929723, EPI_ISL_1929725, EPI_ISL_1929727, EPI_ISL_1929728, EPI_ISL_1929730, EPI_ISL_1929731, EPI_ISL_1929732, EPI_ISL_1929734, EPI_ISL_1929735, EPI_ISL_1929737, EPI_ISL_1929739, EPI_ISL_1929740, EPI_ISL_1929742, EPI_ISL_1929743, EPI_ISL_1929745, EPI_ISL_1929746, EPI_ISL_1929748, EPI_ISL_1930243, EPI_ISL_1930245, EPI_ISL_1930246, EPI_ISL_1930248, EPI_ISL_1930249, EPI_ISL_1930251, EPI_ISL_1930252, EPI_ISL_1930254, EPI_ISL_1930256, EPI_ISL_1930257, EPI_ISL_1930259, EPI_ISL_1930260, EPI_ISL_1930262, EPI_ISL_1930263, EPI_ISL_1930265, EPI_ISL_1930266, EPI_ISL_1930268, EPI_ISL_1930269, EPI_ISL_1930271, EPI_ISL_1930272, EPI_ISL_1930274, EPI_ISL_1930275, EPI_ISL_1930277, EPI_ISL_1930279, EPI_ISL_1930280, EPI_ISL_1930282, EPI_ISL_1930283, EPI_ISL_1930285, EPI_ISL_1930286, EPI_ISL_1930288, EPI_ISL_1931618, EPI_ISL_1931970, EPI_ISL_1931971, EPI_ISL_1931972, EPI_ISL_1931974, EPI_ISL_1931976, EPI_ISL_1931977, EPI_ISL_1931980, EPI_ISL_1931984, EPI_ISL_1931988, EPI_ISL_1931992, EPI_ISL_1931995, EPI_ISL_1931998, EPI_ISL_1932002, EPI_ISL_1932004, EPI_ISL_1932005, EPI_ISL_1932007, EPI_ISL_1932008, EPI_ISL_1932010, EPI_ISL_1932011, EPI_ISL_1932013, EPI_ISL_1932014, EPI_ISL_1932016, EPI_ISL_1932017, EPI_ISL_1932019, EPI_ISL_1932020, EPI_ISL_1932022, EPI_ISL_1932023, EPI_ISL_1932025, EPI_ISL_1932027, EPI_ISL_1932028, EPI_ISL_1932030, EPI_ISL_1932032, EPI_ISL_1932033, EPI_ISL_1932034, EPI_ISL_1932036, EPI_ISL_1932038, EPI_ISL_1932039, EPI_ISL_1932041, EPI_ISL_1932308, EPI_ISL_1932309, EPI_ISL_1932310, EPI_ISL_1932311, EPI_ISL_1932312, EPI_ISL_1932313, EPI_ISL_1932725, EPI_ISL_1933148, EPI_ISL_1933149, EPI_ISL_1933150, EPI_ISL_1933151, EPI_ISL_1933152, EPI_ISL_1933153, EPI_ISL_1933154, EPI_ISL_1933155, EPI_ISL_1933156, EPI_ISL_1933157, EPI_ISL_1933158, EPI_ISL_1933159, EPI_ISL_1933160, EPI_ISL_1933161, EPI_ISL_1933162, EPI_ISL_1933163, EPI_ISL_1933164, EPI_ISL_1933165, EPI_ISL_1933166, EPI_ISL_1933167, EPI_ISL_1933168, EPI_ISL_1933169, EPI_ISL_1933170, EPI_ISL_1933171, EPI_ISL_1933172, EPI_ISL_1933173, EPI_ISL_1933174, EPI_ISL_1933175, EPI_ISL_1933176, EPI_ISL_1933177, EPI_ISL_1933178, EPI_ISL_1933179, EPI_ISL_1933180, EPI_ISL_1933181, EPI_ISL_1933182, EPI_ISL_1933183, EPI_ISL_1933184, EPI_ISL_1933185, EPI_ISL_1933186, EPI_ISL_1933187, EPI_ISL_1933188, EPI_ISL_1933189, EPI_ISL_1933190, EPI_ISL_1933191, EPI_ISL_1933192, EPI_ISL_1933193, EPI_ISL_1933194, EPI_ISL_1933195, EPI_ISL_1933196, EPI_ISL_1933197, EPI_ISL_1933198, EPI_ISL_1933199, EPI_ISL_1933200, EPI_ISL_1933201, EPI_ISL_1933202, EPI_ISL_1933203, EPI_ISL_1933204, EPI_ISL_1933205, EPI_ISL_1933206, EPI_ISL_1933207, EPI_ISL_1933208, EPI_ISL_1933209, EPI_ISL_1933210, EPI_ISL_1933211, EPI_ISL_1933212, EPI_ISL_1933213, EPI_ISL_1933214, EPI_ISL_1933215, EPI_ISL_1933382, EPI_ISL_1933383, EPI_ISL_1933384, EPI_ISL_1933385, EPI_ISL_1933437, EPI_ISL_1933438, EPI_ISL_1933439, EPI_ISL_1933440, EPI_ISL_1933441, EPI_ISL_1933442, EPI_ISL_1933443, EPI_ISL_1933444, EPI_ISL_1933445, EPI_ISL_1933446, EPI_ISL_1933638, EPI_ISL_1933725, EPI_ISL_1933726, EPI_ISL_1933727, EPI_ISL_1933728, EPI_ISL_1933729, EPI_ISL_1933730, EPI_ISL_1933731, EPI_ISL_1933732, EPI_ISL_1933733, EPI_ISL_1933734, EPI_ISL_1933735, EPI_ISL_1933749, EPI_ISL_1933933, EPI_ISL_1933934, EPI_ISL_1933935, EPI_ISL_1933936, EPI_ISL_1933937 | | | |
| see above | Chiba Prefectural Institute of Public Health | Pathogen Genomics Center, National Institute of Infectious Diseases | Ai Kawana-Tachikawa; Chang-Kweng Lim; Eri Nakayama; Hazuka Y Furihata; Hidemasa Izumiya; Ken-ichi Lee; Kentaro Itokawa; Koichi Ishikawa; Makoto Kuroda; Masanori Hashino; Masatomo Morita; Masumichi Saito; Midori Nakamura-Hoshi; Motohiko Ogawa; Naomi Nojiri; Nobuo Koizumi; Nozomu Hanaoka; Rina Tanaka; Sana Uchikoba; Sayuri Seki; Shigeru Kusagawa; Shigeru Tajima; Shouji Yamamoto; Sunao Iyoda; Takahiro Maeki; Tsuguto Fujimoto; Tsuyoshi Sekizuka |
| EPI_ISL_1425487, EPI_ISL_1425488, EPI_ISL_1425489, EPI_ISL_1425495, EPI_ISL_1425496, EPI_ISL_1425497, EPI_ISL_1425498, EPI_ISL_1425499, EPI_ISL_1425500, EPI_ISL_1425501, EPI_ISL_1425502, EPI_ISL_1425503, EPI_ISL_1425504, EPI_ISL_1425505, EPI_ISL_1425506, EPI_ISL_1425507, EPI_ISL_1425508, EPI_ISL_1425509, EPI_ISL_1425510, EPI_ISL_1425511, EPI_ISL_1425512, EPI_ISL_1425513, EPI_ISL_1425514, EPI_ISL_1425515, EPI_ISL_1425516, EPI_ISL_1425517, EPI_ISL_1425518, EPI_ISL_1425519, EPI_ISL_1425520, EPI_ISL_1425521, EPI_ISL_1425522, EPI_ISL_1425523, EPI_ISL_1425524, EPI_ISL_1425525, EPI_ISL_1425526, EPI_ISL_1425527, EPI_ISL_1425528, EPI_ISL_1425529, EPI_ISL_1425530, EPI_ISL_1425531, EPI_ISL_1425532, EPI_ISL_1425533, EPI_ISL_1425534, EPI_ISL_1425535, EPI_ISL_1425536, EPI_ISL_1425537, EPI_ISL_1425538, EPI_ISL_1425539, EPI_ISL_1425540, EPI_ISL_1941590, EPI_ISL_1941592, EPI_ISL_1941593, EPI_ISL_1941596, EPI_ISL_1941598, EPI_ISL_1941600, EPI_ISL_1941601, EPI_ISL_1941603, EPI_ISL_1941606, EPI_ISL_1941607, EPI_ISL_1941610, EPI_ISL_1941611, EPI_ISL_1941614, EPI_ISL_1941615, EPI_ISL_1941616, EPI_ISL_1941617, EPI_ISL_1941621, EPI_ISL_1941623, EPI_ISL_1941627, EPI_ISL_1941630, EPI_ISL_1941632, EPI_ISL_1941634, EPI_ISL_1941635, EPI_ISL_1941638, EPI_ISL_1941640, EPI_ISL_1941641, EPI_ISL_1941644, EPI_ISL_1941645, EPI_ISL_1941647, EPI_ISL_1941649, EPI_ISL_1941650, EPI_ISL_1941651, EPI_ISL_1941652, EPI_ISL_1941653, EPI_ISL_1941654, EPI_ISL_1941655, EPI_ISL_1941656, EPI_ISL_1941657, EPI_ISL_1941658, EPI_ISL_1941659, EPI_ISL_1941662, EPI_ISL_1941663, EPI_ISL_1941665, EPI_ISL_1941667, EPI_ISL_1941669, EPI_ISL_1941671, EPI_ISL_1941673, EPI_ISL_1941676, EPI_ISL_1941678, EPI_ISL_1941680, EPI_ISL_1941682, EPI_ISL_1941683, EPI_ISL_1941686, EPI_ISL_1941687, EPI_ISL_1941689, EPI_ISL_1941692, EPI_ISL_1941694, EPI_ISL_1941696, EPI_ISL_1941697, EPI_ISL_1941699, EPI_ISL_1941701, EPI_ISL_1941703, EPI_ISL_1941705, EPI_ISL_1941707, EPI_ISL_1941709, EPI_ISL_1941711, EPI_ISL_1941713, EPI_ISL_1941716, EPI_ISL_1941718, EPI_ISL_1941721, EPI_ISL_1941725, EPI_ISL_1941728, EPI_ISL_1941730, EPI_ISL_1941733, EPI_ISL_1941735, EPI_ISL_1941737, EPI_ISL_1941740, EPI_ISL_1941741, EPI_ISL_1941743, EPI_ISL_1941745, EPI_ISL_1941746, EPI_ISL_1941748, EPI_ISL_1941750, EPI_ISL_1941752, EPI_ISL_1941753, EPI_ISL_1941756, EPI_ISL_1941758, EPI_ISL_1941760, EPI_ISL_1941762, EPI_ISL_1941764, EPI_ISL_1941766, EPI_ISL_1941767, EPI_ISL_1941769, EPI_ISL_1941771, EPI_ISL_1941772, EPI_ISL_1941774, EPI_ISL_1941775, EPI_ISL_1941777, EPI_ISL_1941780, EPI_ISL_1941781, EPI_ISL_1941783, EPI_ISL_1941784, EPI_ISL_1941785, EPI_ISL_1941786, EPI_ISL_1941788, EPI_ISL_1941789, EPI_ISL_1941791, EPI_ISL_1941794, EPI_ISL_1941795, EPI_ISL_1941797, EPI_ISL_1941799, EPI_ISL_1941802, EPI_ISL_1941803, EPI_ISL_1941806, EPI_ISL_1941807, EPI_ISL_1941808, EPI_ISL_1941810, EPI_ISL_1941811, EPI_ISL_1941814, EPI_ISL_1941815, EPI_ISL_1941817, EPI_ISL_1941819, EPI_ISL_1941822, EPI_ISL_1941823, EPI_ISL_1941826, EPI_ISL_1941827, EPI_ISL_1941830, EPI_ISL_1941832, EPI_ISL_1941833, EPI_ISL_1941835, EPI_ISL_1941838, EPI_ISL_1941839, EPI_ISL_1941842, EPI_ISL_1941843, EPI_ISL_1941846, EPI_ISL_1941848, EPI_ISL_1941849, EPI_ISL_1941851, EPI_ISL_1941852, EPI_ISL_1941853, EPI_ISL_1941854, EPI_ISL_1941855, EPI_ISL_1941856, EPI_ISL_1941857, EPI_ISL_1941858, EPI_ISL_1941859, EPI_ISL_1941860, EPI_ISL_1941861, EPI_ISL_1941862, EPI_ISL_1941863, EPI_ISL_1941864, EPI_ISL_1941865, EPI_ISL_1941866, EPI_ISL_1941867, EPI_ISL_1941868, EPI_ISL_1941869, EPI_ISL_1941870, EPI_ISL_1941871, EPI_ISL_1941872, EPI_ISL_1941873, EPI_ISL_1941874, EPI_ISL_1941876, EPI_ISL_1941877, EPI_ISL_1941878, EPI_ISL_1941879, EPI_ISL_1941880, EPI_ISL_1941881, EPI_ISL_1941882, EPI_ISL_1941883, EPI_ISL_1941884, EPI_ISL_1941885, EPI_ISL_1941886, EPI_ISL_1941887, EPI_ISL_1941888, EPI_ISL_1941889, EPI_ISL_1941890, EPI_ISL_1941891, EPI_ISL_1941892, EPI_ISL_1941893, EPI_ISL_1941894, EPI_ISL_1941895, EPI_ISL_1941896, EPI_ISL_1941897, EPI_ISL_1941898, EPI_ISL_1941899, EPI_ISL_1941900, EPI_ISL_1941901, EPI_ISL_1941902, EPI_ISL_1941903, EPI_ISL_1941904, EPI_ISL_1941905, EPI_ISL_1941906, EPI_ISL_1941907, EPI_ISL_1941908, EPI_ISL_1941909, EPI_ISL_1941910, EPI_ISL_1941911, EPI_ISL_1941912, EPI_ISL_1941913, EPI_ISL_1941914, EPI_ISL_1941915, EPI_ISL_1941916, EPI_ISL_1941917, EPI_ISL_1941918, EPI_ISL_1941919, EPI_ISL_1941920, EPI_ISL_1941922, EPI_ISL_1941923, EPI_ISL_1941924, EPI_ISL_1941925, EPI_ISL_1941926, EPI_ISL_1941927, EPI_ISL_1941928, EPI_ISL_1941929, EPI_ISL_1941930, EPI_ISL_1941931, EPI_ISL_1941932, EPI_ISL_1941933, EPI_ISL_1941934, EPI_ISL_1941935, EPI_ISL_1941936, EPI_ISL_1941937, EPI_ISL_1941938, EPI_ISL_1941939, EPI_ISL_1941940, EPI_ISL_1941941, EPI_ISL_1941942, EPI_ISL_1941943, EPI_ISL_1941944, EPI_ISL_1941945, EPI_ISL_1941946, EPI_ISL_1941947, EPI_ISL_1941948, EPI_ISL_1941949, EPI_ISL_1941950, EPI_ISL_1941951, EPI_ISL_1941952, EPI_ISL_1941953, EPI_ISL_1941954, EPI_ISL_1941955, EPI_ISL_1941956, EPI_ISL_1941957, EPI_ISL_1941958, EPI_ISL_1941959, EPI_ISL_1941960, EPI_ISL_1941961, EPI_ISL_1941962, EPI_ISL_1941963, EPI_ISL_1941964, EPI_ISL_1941965, EPI_ISL_1941966, EPI_ISL_1941967, EPI_ISL_1941968, EPI_ISL_1941969, EPI_ISL_1941970, EPI_ISL_1941971, EPI_ISL_1941972, EPI_ISL_1941973, EPI_ISL_1941974, EPI_ISL_1941975, EPI_ISL_1941976, EPI_ISL_1941977, EPI_ISL_1941978, EPI_ISL_1941979, EPI_ISL_1941980, EPI_ISL_1941981, EPI_ISL_1941982, EPI_ISL_1941983, EPI_ISL_1941984, EPI_ISL_1941985, EPI_ISL_1941986, EPI_ISL_1941987, EPI_ISL_1941988, EPI_ISL_1941989, EPI_ISL_1941990, EPI_ISL_1941991, EPI_ISL_1941992, EPI_ISL_1941993, EPI_ISL_1941994, EPI_ISL_1941995, EPI_ISL_1941996, EPI_ISL_1941997, EPI_ISL_1941998, EPI_ISL_1941999, EPI_ISL_1942000, EPI_ISL_1942001, EPI_ISL_1942002, EPI_ISL_1942003, EPI_ISL_1942004, EPI_ISL_1942005, EPI_ISL_1942006, EPI_ISL_1942007, EPI_ISL_1942008, EPI_ISL_1942009, EPI_ISL_1942010, EPI_ISL_1942011, EPI_ISL_1942012, EPI_ISL_1942013, EPI_ISL_1942014, EPI_ISL_1942015, EPI_ISL_1942016, EPI_ISL_1942017, EPI_ISL_1942018, EPI_ISL_1942019, EPI_ISL_1942020, EPI_ISL_1942021, EPI_ISL_1942022, EPI_ISL_1942023, EPI_ISL_1942024, EPI_ISL_1942025, EPI_ISL_1942026, EPI_ISL_1942027, EPI_ISL_1942028, EPI_ISL_1942029, EPI_ISL_1942030, EPI_ISL_1942031, EPI_ISL_1942032, EPI_ISL_1942033, EPI_ISL_1942034, EPI_ISL_1942035, EPI_ISL_1942036, EPI_ISL_1942037, EPI_ISL_1942038, EPI_ISL_1942039, EPI_ISL_1942040, EPI_ISL_1942041, EPI_ISL_1942042, EPI_ISL_1942043, EPI_ISL_1942044, EPI_ISL_1942045, EPI_ISL_1942046, EPI_ISL_1942047, EPI_ISL_1942048, EPI_ISL_1942049, EPI_ISL_1942050, EPI_ISL_1942051, EPI_ISL_1942052, EPI_ISL_1942053, EPI_ISL_1942054, EPI_ISL_1942055, EPI_ISL_1942056, EPI_ISL_1942057, EPI_ISL_1942058, EPI_ISL_1942059, EPI_ISL_1942060, EPI_ISL_1942061, EPI_ISL_1942062, EPI_ISL_1942063, EPI_ISL_1942064, EPI_ISL_1942065, EPI_ISL_1942066, EPI_ISL_1942067, EPI_ISL_1942068, EPI_ISL_1942069, EPI_ISL_1942070, EPI_ISL_1942071, EPI_ISL_1942072, EPI_ISL_1942073, EPI_ISL_1942074, EPI_ISL_1942075, EPI_ISL_1942076, EPI_ISL_1942077, EPI_ISL_1942078, EPI_ISL_1942079, EPI_ISL_1942080, EPI_ISL_1942081, EPI_ISL_1942082, EPI_ISL_1942083, EPI_ISL_1942084, EPI_ISL_1942085, EPI_ISL_1942086, EPI_ISL_1942087, EPI_ISL_1942088, EPI_ISL_1942089, EPI_ISL_1942090, EPI_ISL_1942091, EPI_ISL_1942092, EPI_ISL_1942093, EPI_ISL_1942094, EPI_ISL_1942095, EPI_ISL_1942096, EPI_ISL_1942097, EPI_ISL_1942098, EPI_ISL_1942099, EPI_ISL_1942100, EPI_ISL_1942101, EPI_ISL_1942102, EPI_ISL_1942103, EPI_ISL_1942104, EPI_ISL_1942105, EPI_ISL_1942106, EPI_ISL_1942107, EPI_ISL_1942108, EPI_ISL_1942109, EPI_ISL_1942110, EPI_ISL_1942111, EPI_ISL_1942112, EPI_ISL_1942113, EPI_ISL_1942114, EPI_ISL_1942115, EPI_ISL_1942116, EPI_ISL_1942117, EPI_ISL_1942118, EPI_ISL_1942119, EPI_ISL_1942120, EPI_ISL_1942121, EPI_ISL_1942122, EPI_ISL_1942123, EPI_ISL_1942124, EPI_ISL_1942125, EPI_ISL_1942126, EPI_ISL_1942127, EPI_ISL_1942128, EPI_ISL_1942129, EPI_ISL_1942130, EPI_ISL_1942131, EPI_ISL_1942132, EPI_ISL_1942133, EPI_ISL_1942134, EPI_ISL_1942135, EPI_ISL_1968981, EPI_ISL_1968982, EPI_ISL_1968983, EPI_ISL_1968984, EPI_ISL_1968985, EPI_ISL_1968986, EPI_ISL_1968987, EPI_ISL_1968988, EPI_ISL_1968989, EPI_ISL_1968990, EPI_ISL_1968991, EPI_ISL_1968992, EPI_ISL_1968993, EPI_ISL_1968994, EPI_ISL_1968995, EPI_ISL_1968996, EPI_ISL_1968997, EPI_ISL_1968998, EPI_ISL_1968999 | | | |
| see above | Department of Infectious Diseases, Kobe Institute of Health | Department of Infectious Diseases, Kobe Institute of Health | Kentaro Itokawa; Makoto Kuroda; Masanori Hashino; Noriko Nakanishi; Rina Tanaka; Ryohei Nomoto; Tomotada Iwamoto; Tsuyoshi Sekizuka |
| EPI_ISL_1708321 | Department of Infectious, Respiratory and Digestive Medicine, Graduate School of Medicine, University of the Ryukyus | Genome Information Research Center, Research Institute for Microbial Diseases, Osaka University | Daisuke Motooka; Hiroya Oki; Shota Nakamura; Takeshi Kinjo; Wakaki Kami; Wakako Arakaki; and Jiro Fujita |
| EPI_ISL_2107432, EPI_ISL_2107433, EPI_ISL_2107434 | Department of Intensive Care Medicine, and clinical lab., Osaka Women's and Children's Hospital | Department of Developmental Medicine, Research Institute, Osaka Women's and Children's Hospital | Itaru Yanagihara; Muneyuki Takeuchi; Shinobu Ida; Yukiko Nakura |
| EPI_ISL_2080355, EPI_ISL_2080356 | Department of Microbiology, Tochigi Prefectural Institute of Public Health and Enviromental Science | Genome Analysis Center, Kamma Memorial Hospital | Fuminori Mizukoshi; Hanako Yazawa; Hiroshi Kamma; Jun Ishii; Kaori Watanabe; Michiaki Masuda; Satoko Soma; Takuya Yazawa; Yoshiko Hitomi |
| EPI_ISL_2080354 | Department of Microbiology, Tochigi Prefectural Institute of Public Health and Environmental Science | Genome Analysis Center, Kamma Memorial Hospital | Fuminori Mizukoshi; Hanako Yazawa; Hiroshi Kamma; Jun Ishii; Kaori Watanabe; Michiaki Masuda; Satoko Soma; Takuya Yazawa; Yoshiko Hitomi |
| EPI_ISL_2158613, EPI_ISL_2158614, EPI_ISL_2158615, EPI_ISL_2158616, EPI_ISL_2158617 | Department of Veterinary Science, National Institute of Infectious Diseases | Department of Veterinary Science, National Institute of Infectious Diseases | Keita Ishijima; Ken Maeda; Shuetsu Fukushi; Souichi Yamada; Tsukasa Yamamoto; Yudai Kuroda |
| EPI_ISL_2107438 | Dr. Yoshitaka Tamura Department of Clinical Laboratory, Osaka Habikino Medical | Department of Developmental Medicine, Research Institute, Osaka Women's and Children's Hospital | Itaru Yanagihara; Yukiko Nakura |
| EPI_ISL_2107437, EPI_ISL_2107439, EPI_ISL_2107440, EPI_ISL_2107441, EPI_ISL_2107460, EPI_ISL_2107461, EPI_ISL_2107462, EPI_ISL_2107463, EPI_ISL_2107465, EPI_ISL_2107466, EPI_ISL_2107467, EPI_ISL_2107468 | | | |
| see above | Dr. Yoshitaka Tamura Department of Clinical Laboratory, Osaka Habikino Medical Center | Department of Developmental Medicine, Research Institute, Osaka Women's and Children's Hospital | Itaru Yanagihara; Yukiko Nakura |
| EPI_ISL_2107464 | Dr. Yoshitaka Tamura Department of Clinical Laboratory, Osaka Habikino Medical Center Dr. Yoshitaka Tamura Department of Clinical Laboratory, Osaka Habikino Medical Center | Department of Developmental Medicine, Research Institute, Osaka Women's and Children's Hospital | Itaru Yanagihara; Yukiko Nakura |
| EPI_ISL_1472258, EPI_ISL_1472259, EPI_ISL_1472273, EPI_ISL_2014961, EPI_ISL_2014962, EPI_ISL_2014963, EPI_ISL_2014964, EPI_ISL_2014965, EPI_ISL_2014966, EPI_ISL_2014967, EPI_ISL_2014968, EPI_ISL_2014969, EPI_ISL_2014970, EPI_ISL_2014971, EPI_ISL_2014972, EPI_ISL_2014973, EPI_ISL_2014974, EPI_ISL_2014975, EPI_ISL_2014976, EPI_ISL_2014977, EPI_ISL_2014978, EPI_ISL_2014979, EPI_ISL_2014980, EPI_ISL_2014981, EPI_ISL_2014982, EPI_ISL_2014983, EPI_ISL_2014984, EPI_ISL_2014985, EPI_ISL_2014988, EPI_ISL_2014989, EPI_ISL_2014990, EPI_ISL_2014991, EPI_ISL_2014992, EPI_ISL_2014993, EPI_ISL_2014994, EPI_ISL_2014995, EPI_ISL_2014996, EPI_ISL_2014997, EPI_ISL_2014998, EPI_ISL_2014999, EPI_ISL_2015000, EPI_ISL_2015001, EPI_ISL_2015002, EPI_ISL_2015003, EPI_ISL_2015004, EPI_ISL_2015005, EPI_ISL_2015006, EPI_ISL_2015007, EPI_ISL_2015008, EPI_ISL_2015009, EPI_ISL_2015010, EPI_ISL_2015011, EPI_ISL_2015012, EPI_ISL_2015013, EPI_ISL_2015014, EPI_ISL_2015019, EPI_ISL_2015020, EPI_ISL_2015021, EPI_ISL_2015022, EPI_ISL_2015023, EPI_ISL_2015024, EPI_ISL_2015025, EPI_ISL_2015026, EPI_ISL_2015027, EPI_ISL_2015028, EPI_ISL_2015029, EPI_ISL_2015030, EPI_ISL_2015031, EPI_ISL_2015032, EPI_ISL_2015033, EPI_ISL_2015034, EPI_ISL_2015035, EPI_ISL_2015036, EPI_ISL_2015037, EPI_ISL_2015038, EPI_ISL_2015039, EPI_ISL_2015040, EPI_ISL_2015041, EPI_ISL_2015042, EPI_ISL_2015043, EPI_ISL_2015044, EPI_ISL_2015045, EPI_ISL_2015046, EPI_ISL_2015047, EPI_ISL_2015048, EPI_ISL_2015049, EPI_ISL_2015050, EPI_ISL_2015051, EPI_ISL_2015052, EPI_ISL_2015053, EPI_ISL_2015054, EPI_ISL_2015055, EPI_ISL_2015056, EPI_ISL_2015057, EPI_ISL_2015058, EPI_ISL_2015059, EPI_ISL_2015060, EPI_ISL_2015061, EPI_ISL_2015062, EPI_ISL_2015063, EPI_ISL_2015064, EPI_ISL_2015065, EPI_ISL_2015066, EPI_ISL_2015067, EPI_ISL_2015068, EPI_ISL_2015069, EPI_ISL_2015070, EPI_ISL_2015071, EPI_ISL_2015072, EPI_ISL_2015073, EPI_ISL_2015074, EPI_ISL_2015075, EPI_ISL_2015076, EPI_ISL_2015077, EPI_ISL_2015078, EPI_ISL_2015079, EPI_ISL_2015080, EPI_ISL_2015081, EPI_ISL_2015082, EPI_ISL_2015083, EPI_ISL_2015084, EPI_ISL_2015085, EPI_ISL_2015086, EPI_ISL_2015087, EPI_ISL_2015088, EPI_ISL_2015089, EPI_ISL_2015090, EPI_ISL_2015091, EPI_ISL_2015092, EPI_ISL_2015093, EPI_ISL_2015094, EPI_ISL_2015095 | | | |
| see above | Fukuoka Institute of Health and Environmental Sciences | Fukuoka Institute of Health and Environmental Sciences | Akira Ohishi; Asako Nakamura; Chiharu Katamune; Hiroaki Shigemura; Kentaro Itokawa; Makoto Kuroda; Masanori Hashino; Mitsuhiro Hamasaki; Rina Tanaka; Saori Ueda; Takayuki Kobayashi; Tsuyoshi Sekizuka; Yoshiki Etoh; Yuki Ashizuka |
| EPI_ISL_1929141, EPI_ISL_1929143 | Fukuoka Institute of Health and Environmental Sciences | Pathogen Genomics Center, National Institute of Infectious Diseases | Hidemasa Izumiya; Ken Shimuta; Kentaro Itokawa; Makoto Kuroda; Masanori Hashino; Nobuo Koizumi; Rina Tanaka; Sunao Iyoda; Tsuyoshi Sekizuka |
| EPI_ISL_1324384, EPI_ISL_1324466, EPI_ISL_1530875, EPI_ISL_1530876 | Genome Analysis Center, Kamma Memorial Hospital | Genome Analysis Center, Kamma Memorial Hospital | Hanako Yazawa; Hiroshi Kamma; Jun Ishii; Kaori Watanabe; Michiaki Masuda; Satoko Soma; Takuya Yazawa |
| EPI_ISL_1578114, EPI_ISL_1578115, EPI_ISL_1578116, EPI_ISL_1587580, EPI_ISL_1587581, EPI_ISL_1587582, EPI_ISL_1595846, EPI_ISL_1595847, EPI_ISL_1595848, EPI_ISL_1595849, EPI_ISL_1595850, EPI_ISL_1595851, EPI_ISL_1669760, EPI_ISL_1669761, EPI_ISL_1669762, EPI_ISL_1669763, EPI_ISL_1669764, EPI_ISL_1669765, EPI_ISL_1793646, EPI_ISL_1793647, EPI_ISL_1793648, EPI_ISL_1793649, EPI_ISL_1793650, EPI_ISL_1793651, EPI_ISL_1793652, EPI_ISL_1793653 | | | |
| see above | Genome Analysis Center, Yamanashi Central Hospital | Genome Analysis Center, Yamanashi Central Hospital | Yosuke Hirotsu |
| EPI_ISL_1442959, EPI_ISL_1442960, EPI_ISL_1442961, EPI_ISL_1442962, EPI_ISL_1442963, EPI_ISL_1442992, EPI_ISL_1922003, EPI_ISL_1922004, EPI_ISL_1922005, EPI_ISL_1922006, EPI_ISL_1922007, EPI_ISL_1922008, EPI_ISL_1922009, EPI_ISL_1922010, EPI_ISL_1922011, EPI_ISL_1922012, EPI_ISL_1922013, EPI_ISL_1922014, EPI_ISL_1922015, EPI_ISL_1922016, EPI_ISL_1922017, EPI_ISL_1922018, EPI_ISL_1922019, EPI_ISL_1922020, EPI_ISL_1922021, EPI_ISL_1922022, EPI_ISL_1922023, EPI_ISL_1922024, EPI_ISL_1922025, EPI_ISL_1922026, EPI_ISL_1922027, EPI_ISL_1922028, EPI_ISL_1922029, EPI_ISL_1922030, EPI_ISL_1922031, EPI_ISL_1922032, EPI_ISL_1922033, EPI_ISL_1922034, EPI_ISL_1922035, EPI_ISL_1922036, EPI_ISL_1922037, EPI_ISL_1922038, EPI_ISL_1922039, EPI_ISL_1922040, EPI_ISL_1922041 | | | |

| | | | |
|---|---|---|---|
| see above | Gunma Prefectural Institute of Public Health and Environmental Sciences | Gunma Prefectural Institute of Public Health and Environmental Sciences | Daisuke Shinoda; Hiroyuki Tsukagoshi; Kentaro Itokawa; Makoto Kuroda; Mariko Saito; Masanori Hashino; Rina Tanaka; Tsuyoshi Sekizuka |
| EPI_ISL_1932420, EPI_ISL_1933570, EPI_ISL_1933571, EPI_ISL_1933572 | Gunma Prefectural Institute of Public Health and Environmental Sciences | Pathogen Genomics Center, National Institute of Infectious Diseases | Hazuka Y Furihata; Kentaro Itokawa; Makoto Kuroda; Masanori Hashino; Masumichi Saito; Naomi Nojiri; Nozomu Hanaoka; Rina Tanaka; Sana Uchikoba; Tsuguto Fujimoto; Tsuyoshi Sekizuka |
| EPI_ISL_1472316, EPI_ISL_1472317, EPI_ISL_1472318, EPI_ISL_1472319, EPI_ISL_1472320, EPI_ISL_1472321, EPI_ISL_1472322, EPI_ISL_1472323, EPI_ISL_1472324, EPI_ISL_1472325, EPI_ISL_1472331, EPI_ISL_1472332, EPI_ISL_1472333, EPI_ISL_1472334, EPI_ISL_1472335, EPI_ISL_1472336, EPI_ISL_1472337, EPI_ISL_1967393, EPI_ISL_1967394, EPI_ISL_1967395, EPI_ISL_1967396, EPI_ISL_1967397, EPI_ISL_1967398, EPI_ISL_1967399, EPI_ISL_1967400, EPI_ISL_1967401, EPI_ISL_1967402, EPI_ISL_1967403, EPI_ISL_1967404, EPI_ISL_1967405, EPI_ISL_1967406, EPI_ISL_1967407, EPI_ISL_1967408, EPI_ISL_1967409, EPI_ISL_1967410, EPI_ISL_1967411, EPI_ISL_1967412, EPI_ISL_1967413, EPI_ISL_1967414, EPI_ISL_1967415, EPI_ISL_1967416, EPI_ISL_1967417, EPI_ISL_1967418, EPI_ISL_1967419, EPI_ISL_1967420, EPI_ISL_1967421, EPI_ISL_1967422, EPI_ISL_1967423, EPI_ISL_1967424, EPI_ISL_1967425, EPI_ISL_1967426, EPI_ISL_1967427, EPI_ISL_1967428, EPI_ISL_1967429, EPI_ISL_1967430, EPI_ISL_1967431, EPI_ISL_1967432, EPI_ISL_1967433, EPI_ISL_1967434, EPI_ISL_1967435, EPI_ISL_1967436, EPI_ISL_1967437, EPI_ISL_1967438, EPI_ISL_1967439, EPI_ISL_1967440, EPI_ISL_1967441, EPI_ISL_1967442, EPI_ISL_1967443, EPI_ISL_1967444, EPI_ISL_1967445, EPI_ISL_1967446, EPI_ISL_1967447, EPI_ISL_1967448, EPI_ISL_1967449, EPI_ISL_1967450, EPI_ISL_1967451, EPI_ISL_1967452, EPI_ISL_1967453, EPI_ISL_1967454, EPI_ISL_1967455, EPI_ISL_1967456, EPI_ISL_1967457, EPI_ISL_1967458, EPI_ISL_1967459, EPI_ISL_1967460, EPI_ISL_1967461, EPI_ISL_1967462, EPI_ISL_1967463, EPI_ISL_1967464, EPI_ISL_1967465, EPI_ISL_1967466, EPI_ISL_1967467, EPI_ISL_1967468, EPI_ISL_1967469, EPI_ISL_1967470, EPI_ISL_1967471, EPI_ISL_1967472, EPI_ISL_1967473 | | | |
| see above | Hokkaido Institute of Public Health | Hokkaido Institute of Public Health | Hiroki Yamaguchi; Kazuhiro Okubo; Kentaro Itokawa; Makoto Kuroda; Masanori Hashino; Rina Tanaka; Tetsuya Ikeda; Tsuyoshi Sekizuka; Yuta Ohno |
| EPI_ISL_2007394, EPI_ISL_2007395, EPI_ISL_2007396, EPI_ISL_2007397, EPI_ISL_2007398, EPI_ISL_2007399, EPI_ISL_2007400, EPI_ISL_2007401, EPI_ISL_2007402, EPI_ISL_2007403, EPI_ISL_2007404, EPI_ISL_2007405, EPI_ISL_2007406, EPI_ISL_2007407, EPI_ISL_2007408, EPI_ISL_2007409, EPI_ISL_2007410, EPI_ISL_2007411, EPI_ISL_2007412, EPI_ISL_2007413, EPI_ISL_2007414, EPI_ISL_2007415, EPI_ISL_2007416, EPI_ISL_2007417, EPI_ISL_2007418, EPI_ISL_2007419, EPI_ISL_2007420, EPI_ISL_2007421, EPI_ISL_2007422, EPI_ISL_2007423, EPI_ISL_2007424, EPI_ISL_2007425, EPI_ISL_2007426, EPI_ISL_2007427, EPI_ISL_2007428, EPI_ISL_2007429, EPI_ISL_2007430, EPI_ISL_2007431, EPI_ISL_2007432, EPI_ISL_2007433, EPI_ISL_2007434, EPI_ISL_2007435, EPI_ISL_2007436, EPI_ISL_2007437, EPI_ISL_2007438, EPI_ISL_2007439, EPI_ISL_2007440, EPI_ISL_2007441, EPI_ISL_2007442, EPI_ISL_2007443, EPI_ISL_2007444, EPI_ISL_2007445, EPI_ISL_2007446, EPI_ISL_2007447, EPI_ISL_2007448, EPI_ISL_2007449, EPI_ISL_2007450, EPI_ISL_2007451, EPI_ISL_2007452, EPI_ISL_2007453, EPI_ISL_2007454, EPI_ISL_2007455, EPI_ISL_2007456, EPI_ISL_2007457, EPI_ISL_2007458 | | | |
| see above | Hyogo Prefectural Institute of Public Health science | Pathogen Genomics Center, National Institute of Infectious Diseases | Denshi Takai; Etsuko Saito; Kenichi Ogita; Kentaro Itokawa; Kyoko Suzuki; Makoto Kuroda; Masanori Hashino; Masatsugu Chikahira; Miki Ogi; Rina Tanaka; Tomohiro Oshibe; Tsuyoshi Sekizuka; Yumi Akiyama |
| EPI_ISL_1434464, EPI_ISL_1434527, EPI_ISL_1435187, EPI_ISL_1435260, EPI_ISL_1972918, EPI_ISL_1972923, EPI_ISL_1972924, EPI_ISL_1972925, EPI_ISL_1972926, EPI_ISL_1972927, EPI_ISL_1972928, EPI_ISL_1972929, EPI_ISL_1972930, EPI_ISL_1972931, EPI_ISL_1972932, EPI_ISL_1972933, EPI_ISL_1972934, EPI_ISL_1972935, EPI_ISL_1972936, EPI_ISL_1972937, EPI_ISL_1972938, EPI_ISL_1972939, EPI_ISL_1972940, EPI_ISL_1972941, EPI_ISL_1972942 | | | |
| see above | Ibaraki Prefectural Institute of Public Health | Ibaraki Prefectural Institute of Public Health | Keiko Goto; Kentaro Itokawa; Makoto Kuroda; Masanori Hashino; Rina Tanaka; Tsuyoshi Sekizuka |
| EPI_ISL_1930966, EPI_ISL_1930968, EPI_ISL_1931499, EPI_ISL_1933479, EPI_ISL_1933480, EPI_ISL_1933537, EPI_ISL_1933538, EPI_ISL_1933539 | | | |
| see above | Ibaraki Prefectural Institute of Public Health | Pathogen Genomics Center, National Institute of Infectious Diseases | Hazuka Y Furihata; Kentaro Itokawa; Makoto Kuroda; Masanori Hashino; Masumichi Saito; Naomi Nojiri; Nozomu Hanaoka; Rina Tanaka; Sana Uchikoba; Tsuguto Fujimoto; Tsuyoshi Sekizuka |
| EPI_ISL_1809482 | Illinois Department of Public Health | Illinois Department of Public Health - Chicago Lab | Ira Heimler; Joel Price; Vineet K. Dhiman |
| EPI_ISL_1754897, EPI_ISL_1754898, EPI_ISL_1754899, EPI_ISL_1754902, EPI_ISL_1754903, EPI_ISL_1754904, EPI_ISL_1754905, EPI_ISL_1754906, EPI_ISL_1754907, EPI_ISL_1754908, EPI_ISL_1754909, EPI_ISL_1754910, EPI_ISL_1754911, EPI_ISL_1754912, EPI_ISL_1754913, EPI_ISL_1754914, EPI_ISL_1754915, EPI_ISL_1754916, EPI_ISL_1754917, EPI_ISL_1754918, EPI_ISL_1754919, EPI_ISL_1755002, EPI_ISL_1755003, EPI_ISL_1755004, EPI_ISL_1755005, EPI_ISL_1755006, EPI_ISL_1755007, EPI_ISL_1755908, EPI_ISL_1755909, EPI_ISL_1755910, EPI_ISL_1755911, EPI_ISL_1755912, EPI_ISL_1755913, EPI_ISL_1755914, EPI_ISL_1755915, EPI_ISL_1755916, EPI_ISL_1972571, EPI_ISL_1972572 | | | |
| see above | Kanagawa Prefectural Institute of Public Health | Kanagawa Prefectural Institute of Public Health | Ichiro Furukawa; Junichi Sakuragi; Kentaro Itokawa; Makiko Kondo; Makoto Kuroda; Masanori Hashino; Rieko Suzuki; Rina Tanaka; Sumi Watanabe; Takako Sano; Takayuki Hishiki; Tomohiko Takasaki; Tsuyoshi Sekizuka |
| EPI_ISL_1430582, EPI_ISL_1930941, EPI_ISL_1930942, EPI_ISL_1931336 | Kanagawa Prefectural Institute of Public Health | Pathogen Genomics Center, National Institute of Infectious Diseases | Hazuka Y Furihata; Hiromizu Takahashi; Ken Miyazawa; Kentaro Itokawa; Makoto Kuroda; Masanori Hashino; Masumichi Saito; Minoru Nagi; Naomi Nojiri; Nozomu Hanaoka; Rina Tanaka; Takashi Sakudoh; Tsuguto Fujimoto; Tsuyoshi Sekizuka |
| EPI_ISL_2107805, EPI_ISL_2107806, EPI_ISL_2107808, EPI_ISL_2107810, EPI_ISL_2107821 | Kawakita General Hospital | Tokyo Medical University Department of Microbiology | Hirofumi Miyake; Jun Suzuki; Kenji Sadamasu; Mami Nagashima; Masaki Hayashi; Shigeki Nakamura; Takahiro Okai; Takako Yamazaki; Takushi Fujiwara; Tatsuya Inukai |
| EPI_ISL_1973283, EPI_ISL_1973284, EPI_ISL_1973285, EPI_ISL_1973286, EPI_ISL_1973287 | Keio University School of Medicine | Keio University School of Medicine | Haruhiko Siomi; Hirotsugu Ishizu; Kenjiro Kosaki; Kodai Abe; Yuka Iwasaki |
| EPI_ISL_2020237, EPI_ISL_2020242 | Kumamoto Prefectural Institute of Public-Health and Environmental Science | Kumamoto Prefectural Institute of Public-Health and Environmental Science | Kentaro Itokawa; Makoto Kuroda; Mami Sato; Masanori Hashino; Rina Tanaka; Shunsuke Yahiro; Tsuyoshi Sekizuka |
| EPI_ISL_1430709, EPI_ISL_1430710, EPI_ISL_1430711, EPI_ISL_1430712, EPI_ISL_1430713, EPI_ISL_1430714, EPI_ISL_1430854, EPI_ISL_1431522, EPI_ISL_1431523 | | | |
| see above | Niigata Prefectural Institute of Public Health and Environmental Sciences | Pathogen Genomics Center, National Institute of Infectious Diseases | Kentaro Itokawa; Makoto Kuroda; Masanori Hashino; Rina Tanaka; Tsuyoshi Sekizuka |
| EPI_ISL_1430617, EPI_ISL_1430618, EPI_ISL_1430619, EPI_ISL_1430620, EPI_ISL_1430621, EPI_ISL_1430622, EPI_ISL_1430623, EPI_ISL_1430624, EPI_ISL_1430626, EPI_ISL_1430638, EPI_ISL_1430653, EPI_ISL_1430654, EPI_ISL_1430655, EPI_ISL_1430668, EPI_ISL_1430669, EPI_ISL_1430681, EPI_ISL_1432340, EPI_ISL_1926970, EPI_ISL_1926972, EPI_ISL_1926973, EPI_ISL_1926975, EPI_ISL_1926979, EPI_ISL_1926981, EPI_ISL_1926982, EPI_ISL_1926984, EPI_ISL_1926987, EPI_ISL_1926988, EPI_ISL_1927000, EPI_ISL_1927002, EPI_ISL_1927004, EPI_ISL_1927023, EPI_ISL_1927024, EPI_ISL_1927026, EPI_ISL_1927027, EPI_ISL_1927032, EPI_ISL_1927034, EPI_ISL_1927035, EPI_ISL_1927069, EPI_ISL_1927071, EPI_ISL_1927073, EPI_ISL_1927074, EPI_ISL_1927076, EPI_ISL_1927203, EPI_ISL_1927318, EPI_ISL_1927681, EPI_ISL_1928054, EPI_ISL_1928056, EPI_ISL_1928057, EPI_ISL_1928059, EPI_ISL_1928060, EPI_ISL_1928062, EPI_ISL_1928069, EPI_ISL_1928071, EPI_ISL_1928080, EPI_ISL_1928112, EPI_ISL_1929077, EPI_ISL_1929079, EPI_ISL_1929080, EPI_ISL_1929151, EPI_ISL_1929155, EPI_ISL_1929156, EPI_ISL_1929158, EPI_ISL_1929159, EPI_ISL_1929160, EPI_ISL_1929162, EPI_ISL_1929164, EPI_ISL_1929165, EPI_ISL_1929167, EPI_ISL_1929168, EPI_ISL_1929170, EPI_ISL_1929171, EPI_ISL_1929173, EPI_ISL_1929174, EPI_ISL_1929176, EPI_ISL_1929177, EPI_ISL_1929179, EPI_ISL_1929180, EPI_ISL_1929182, EPI_ISL_1929183, EPI_ISL_1929185, EPI_ISL_1929189, EPI_ISL_1929191, EPI_ISL_1929193, EPI_ISL_1929194, EPI_ISL_1929196, EPI_ISL_1929250, EPI_ISL_1929252, EPI_ISL_1929256, EPI_ISL_1929286, EPI_ISL_1929287, EPI_ISL_1929288, EPI_ISL_1929290, EPI_ISL_1929291, EPI_ISL_1929293, EPI_ISL_1929294, EPI_ISL_1929296, EPI_ISL_1929302, EPI_ISL_1929303, EPI_ISL_1929305, EPI_ISL_1929306, EPI_ISL_1929307, EPI_ISL_1929310, EPI_ISL_1929312, EPI_ISL_1929313, EPI_ISL_1929315, EPI_ISL_1929316, EPI_ISL_1929329, EPI_ISL_1929330, EPI_ISL_1929331, EPI_ISL_1929333, EPI_ISL_1929334, EPI_ISL_1929336, EPI_ISL_1929337, EPI_ISL_1929339, EPI_ISL_1929342, EPI_ISL_1929381, EPI_ISL_1929383, EPI_ISL_1929384, EPI_ISL_1929386, EPI_ISL_1929387, EPI_ISL_1929389, EPI_ISL_1930894, EPI_ISL_1930896, EPI_ISL_1930897, EPI_ISL_1930899, EPI_ISL_1930900, EPI_ISL_1930902, EPI_ISL_1930903, EPI_ISL_1930905, EPI_ISL_1930906, EPI_ISL_1930908, EPI_ISL_1930910, EPI_ISL_1930911, EPI_ISL_1930913, EPI_ISL_1930914, EPI_ISL_1930915, EPI_ISL_1930917, EPI_ISL_1930918, EPI_ISL_1930920, EPI_ISL_1930921, EPI_ISL_1930923, EPI_ISL_1930974, EPI_ISL_1930975, EPI_ISL_1930976, EPI_ISL_1930978, EPI_ISL_1930979, EPI_ISL_1930981, EPI_ISL_1930983, EPI_ISL_1930984, EPI_ISL_1930985, EPI_ISL_1930987, EPI_ISL_1930988, EPI_ISL_1930990, EPI_ISL_1930991, EPI_ISL_1930993, EPI_ISL_1930994, EPI_ISL_1930996, EPI_ISL_1930997, EPI_ISL_1930999, EPI_ISL_1931000, EPI_ISL_1931002, EPI_ISL_1931004, EPI_ISL_1931005, EPI_ISL_1931006, EPI_ISL_1931008, EPI_ISL_1931009, EPI_ISL_1931011, EPI_ISL_1931012, EPI_ISL_1931014, EPI_ISL_1931015, EPI_ISL_1931017, EPI_ISL_1931018, EPI_ISL_1931020, EPI_ISL_1931132, EPI_ISL_1931135, EPI_ISL_1931338, EPI_ISL_1931339, EPI_ISL_1931341, EPI_ISL_1931342, EPI_ISL_1931344, EPI_ISL_1931345, EPI_ISL_1931347, EPI_ISL_1931362, EPI_ISL_1931363, EPI_ISL_1931365, EPI_ISL_1931366, EPI_ISL_1931368, EPI_ISL_1931370, EPI_ISL_1931371, EPI_ISL_1931372, EPI_ISL_1931374, EPI_ISL_1931375, EPI_ISL_1931377, EPI_ISL_1931378, EPI_ISL_1931380, EPI_ISL_1931382, EPI_ISL_1931383, EPI_ISL_1931385, EPI_ISL_1931386, EPI_ISL_1931388, EPI_ISL_1931389, EPI_ISL_1931393, EPI_ISL_1931395, EPI_ISL_1931396, EPI_ISL_1931398, EPI_ISL_1931399, EPI_ISL_1931401, EPI_ISL_1931402, EPI_ISL_1931404, EPI_ISL_1931405, EPI_ISL_1931407, EPI_ISL_1931408, EPI_ISL_1931410, EPI_ISL_1931411, EPI_ISL_1931413, EPI_ISL_1931414, EPI_ISL_1931416, EPI_ISL_1931417, EPI_ISL_1931419, EPI_ISL_1931421, EPI_ISL_1931422, EPI_ISL_1931424, EPI_ISL_1931426, EPI_ISL_1931427, EPI_ISL_1931429, EPI_ISL_1931430, EPI_ISL_1931432, EPI_ISL_1931433, EPI_ISL_1931435, EPI_ISL_1931436, EPI_ISL_1931438, EPI_ISL_1931439, EPI_ISL_1931441, EPI_ISL_1931621, EPI_ISL_1931660, EPI_ISL_1931944, EPI_ISL_1931945, EPI_ISL_1931947, EPI_ISL_1931948, EPI_ISL_1932090, EPI_ISL_1932091, EPI_ISL_1932092, EPI_ISL_1932370, EPI_ISL_1932371, EPI_ISL_1932372, EPI_ISL_1932373, EPI_ISL_1932374, EPI_ISL_1932445, EPI_ISL_1932446, EPI_ISL_1932447, EPI_ISL_1932448, EPI_ISL_1932449, EPI_ISL_1932450, EPI_ISL_1932451, EPI_ISL_1932452, EPI_ISL_1932453, EPI_ISL_1932454, EPI_ISL_1932455, EPI_ISL_1932456, EPI_ISL_1932457, EPI_ISL_1932458, EPI_ISL_1932459, EPI_ISL_1932460, EPI_ISL_1932581, EPI_ISL_1932582, EPI_ISL_1932584, EPI_ISL_1932585, EPI_ISL_1932586, EPI_ISL_1932587, EPI_ISL_1932588, EPI_ISL_1932589, EPI_ISL_1932590, EPI_ISL_1932591, EPI_ISL_1932592, EPI_ISL_1932593, EPI_ISL_1932594, EPI_ISL_1932595, EPI_ISL_1932596, EPI_ISL_1932597, EPI_ISL_1932598, EPI_ISL_1932599, EPI_ISL_1932600, EPI_ISL_1932601, EPI_ISL_1932602, EPI_ISL_1932603, EPI_ISL_1932604, EPI_ISL_1932605, EPI_ISL_1932606, EPI_ISL_1932607, EPI_ISL_1932608, EPI_ISL_1932609, EPI_ISL_1932610, EPI_ISL_1932611, EPI_ISL_1932612, EPI_ISL_1932613, EPI_ISL_1932614, EPI_ISL_1932615, EPI_ISL_1932616, EPI_ISL_1932617, EPI_ISL_1932618, EPI_ISL_1932619, EPI_ISL_1932620, EPI_ISL_1932621, EPI_ISL_1932622, EPI_ISL_1932698, EPI_ISL_1932699, EPI_ISL_1932714, EPI_ISL_1932759, EPI_ISL_1932760, EPI_ISL_1932906, EPI_ISL_1932907, EPI_ISL_1932908, EPI_ISL_1932909, EPI_ISL_1933423, EPI_ISL_1933424, EPI_ISL_1933425, EPI_ISL_1933426, EPI_ISL_1933427, EPI_ISL_1933487, EPI_ISL_1933488, EPI_ISL_1933489, EPI_ISL_1933490, EPI_ISL_1933491, EPI_ISL_1933492, EPI_ISL_1933493, EPI_ISL_1933494, EPI_ISL_1933495, EPI_ISL_1933496, EPI_ISL_1933497, EPI_ISL_1933498, EPI_ISL_1933499, EPI_ISL_1933500, EPI_ISL_1933501, EPI_ISL_1933502, EPI_ISL_1933503, EPI_ISL_1933544, EPI_ISL_1933573, EPI_ISL_1933574, EPI_ISL_1933575, EPI_ISL_1933576, EPI_ISL_1933577, EPI_ISL_1933578, EPI_ISL_1933579, EPI_ISL_1933645, EPI_ISL_1933646, EPI_ISL_1933647, EPI_ISL_1933648, EPI_ISL_1933649, EPI_ISL_1933650, EPI_ISL_1933701, EPI_ISL_1933711, EPI_ISL_1933712, EPI_ISL_1933713, EPI_ISL_1933752, EPI_ISL_1933786, EPI_ISL_1933787, EPI_ISL_1933788, EPI_ISL_1933789, EPI_ISL_1933790, EPI_ISL_1933791, EPI_ISL_1933792, EPI_ISL_1933793, EPI_ISL_1933794, EPI_ISL_1933795, EPI_ISL_1933796, EPI_ISL_1933797, EPI_ISL_1933798, EPI_ISL_1933799, EPI_ISL_1933800, EPI_ISL_1933801, EPI_ISL_1933802, EPI_ISL_1933803, EPI_ISL_1933804, EPI_ISL_1933805, EPI_ISL_1933806, EPI_ISL_1933807, EPI_ISL_1933808, EPI_ISL_1933809, EPI_ISL_1933810, EPI_ISL_1933811, EPI_ISL_1933812, EPI_ISL_1933813, EPI_ISL_1933814, EPI_ISL_1933815, EPI_ISL_1933816, EPI_ISL_1933817, EPI_ISL_1933818, EPI_ISL_1933819, EPI_ISL_1933820, EPI_ISL_1933821, EPI_ISL_1933822, EPI_ISL_1933823, EPI_ISL_1933824, EPI_ISL_1933825, EPI_ISL_1933826, EPI_ISL_1933827, EPI_ISL_1933828, EPI_ISL_1933829, EPI_ISL_1933830, EPI_ISL_1933831, EPI_ISL_1933832, EPI_ISL_1933833, EPI_ISL_1933834, EPI_ISL_1933835, EPI_ISL_1933836, EPI_ISL_1934105, EPI_ISL_1934106, EPI_ISL_1934107, EPI_ISL_1934108, EPI_ISL_1934206, EPI_ISL_1934207, EPI_ISL_1934208, EPI_ISL_1934209, EPI_ISL_1934210, EPI_ISL_1934211, EPI_ISL_1934212, EPI_ISL_1934213, EPI_ISL_1934214, EPI_ISL_1934215, EPI_ISL_1934216, EPI_ISL_1934217, EPI_ISL_1934218, EPI_ISL_1934219, EPI_ISL_1934220, EPI_ISL_1934221, EPI_ISL_1934222, EPI_ISL_1934223, EPI_ISL_1934224, EPI_ISL_1934225, EPI_ISL_1934226, EPI_ISL_1934227, EPI_ISL_1934228, EPI_ISL_1934229, EPI_ISL_1934230, EPI_ISL_1934231, EPI_ISL_1934408, EPI_ISL_1934409 | | | |
| see above | Osaka Institute of Public Health, Morinomiya Center | Pathogen Genomics Center, National Institute of Infectious Diseases | Ai Kawana-Tachikawa; Chang-Kweng Lim; Chikako Shimokawa; Eri Nakayama; Eunsil Park; Hazuka Y Furihata; Hidemasa Izumiya; Hiromizu Takahashi; Ken Shimuta; Ken-ichi Lee; Kentaro Itokawa; Koichi Ishikawa; Makoto Kuroda; Masanori Hashino; Masatomo Morita; Masumichi Saito; Midori Nakamura-Hoshi; Motohiko Ogawa; Naomi Nojiri; Nobuo Koizumi; Nozomu Hanaoka; Rina Tanaka; Sana Uchikoba; Sayuri Seki; Shigeru Kusagawa; Shigeru Tajima; Shigeru Tanjima; Shouji Yamamoto; Sunao Iyoda; Takahiro Maeki; Tsuguto Fujimoto; Tsuyoshi Sekizuka; Yasutaka Hoshino; Yoshihiro Kaku |
| EPI_ISL_1430775, EPI_ISL_1430776, EPI_ISL_1430777, EPI_ISL_1430778, EPI_ISL_1430843, EPI_ISL_1430844, EPI_ISL_1430845, EPI_ISL_1430846, EPI_ISL_1430847, EPI_ISL_1430848, EPI_ISL_1430867, EPI_ISL_1431220, EPI_ISL_1431221, EPI_ISL_1431494, EPI_ISL_1431495, EPI_ISL_1431512, EPI_ISL_1431513, EPI_ISL_1431514, EPI_ISL_1431515, EPI_ISL_1431516, EPI_ISL_1431517, EPI_ISL_1431518, EPI_ISL_1431519, EPI_ISL_1431520, EPI_ISL_1431579, EPI_ISL_1431585, EPI_ISL_1431590, EPI_ISL_1431591, EPI_ISL_1431592, EPI_ISL_1431616, EPI_ISL_1431623, EPI_ISL_1431624, EPI_ISL_1431628, EPI_ISL_1431857, EPI_ISL_1431858, EPI_ISL_1431859, EPI_ISL_1431860, EPI_ISL_1431861, EPI_ISL_1431862, EPI_ISL_1431863, EPI_ISL_1431864, EPI_ISL_1431865, EPI_ISL_1431866, EPI_ISL_1431867, EPI_ISL_1431868, EPI_ISL_1431869, EPI_ISL_1431870, EPI_ISL_1431871, EPI_ISL_1431872, EPI_ISL_1431873, EPI_ISL_1431874, EPI_ISL_1431875, EPI_ISL_1431876, EPI_ISL_1431877, EPI_ISL_1431878, EPI_ISL_1431879, EPI_ISL_1431880, EPI_ISL_1431881, EPI_ISL_1431884, EPI_ISL_1431885, EPI_ISL_1431886, EPI_ISL_1431887, EPI_ISL_1431889, EPI_ISL_1431890, EPI_ISL_1431892, EPI_ISL_1431893, EPI_ISL_1431894, EPI_ISL_1431895, EPI_ISL_1431897, EPI_ISL_1431898, EPI_ISL_1431899, EPI_ISL_1431900, EPI_ISL_1431901, EPI_ISL_1431902, EPI_ISL_1431903, EPI_ISL_1431904, EPI_ISL_1431905, EPI_ISL_1431906, EPI_ISL_1431907, EPI_ISL_1431908, EPI_ISL_1431909, EPI_ISL_1431910, EPI_ISL_1431911, EPI_ISL_1431912, EPI_ISL_1431913, EPI_ISL_1431914, EPI_ISL_1431915, EPI_ISL_1431916, EPI_ISL_1431917, EPI_ISL_1431918, EPI_ISL_1431919, EPI_ISL_1431920, EPI_ISL_1431921, EPI_ISL_1431922, EPI_ISL_1431923, EPI_ISL_1431924, EPI_ISL_1431925, EPI_ISL_1431926, EPI_ISL_1431927, EPI_ISL_1431928, EPI_ISL_1431929, EPI_ISL_1431930, EPI_ISL_1431931, EPI_ISL_1431932, EPI_ISL_1431933, EPI_ISL_1431934, EPI_ISL_1431935, EPI_ISL_1431936, EPI_ISL_1431937, EPI_ISL_1431938, EPI_ISL_1432137, EPI_ISL_1432139, EPI_ISL_1432606, EPI_ISL_1432608, EPI_ISL_1432610, EPI_ISL_1432732, EPI_ISL_1432734, EPI_ISL_1432736, EPI_ISL_1432738, EPI_ISL_1432739, EPI_ISL_1432767, EPI_ISL_1432789, EPI_ISL_1432819, EPI_ISL_1432821, EPI_ISL_1432823, EPI_ISL_1432825, EPI_ISL_1432826, EPI_ISL_1432830, EPI_ISL_1432832, EPI_ISL_1432834, EPI_ISL_1432835, EPI_ISL_1432837, EPI_ISL_1432839, EPI_ISL_1432841, EPI_ISL_1432842, EPI_ISL_1432845, EPI_ISL_1432846, EPI_ISL_1432848, EPI_ISL_1432850, EPI_ISL_1432852, EPI_ISL_1432854, EPI_ISL_1432856, EPI_ISL_1432858, EPI_ISL_1432860, EPI_ISL_1432862, EPI_ISL_1432863, EPI_ISL_1432865, EPI_ISL_1432867, EPI_ISL_1432868, EPI_ISL_1432870, EPI_ISL_1432872, EPI_ISL_1432874, EPI_ISL_1432876, EPI_ISL_1432877, EPI_ISL_1926969, EPI_ISL_1926976, EPI_ISL_1926978, EPI_ISL_1926985, EPI_ISL_1926990, EPI_ISL_1926992, EPI_ISL_1926993, EPI_ISL_1926994, EPI_ISL_1926996, EPI_ISL_1926997, EPI_ISL_1926999, EPI_ISL_1927005, EPI_ISL_1927007, EPI_ISL_1927008, EPI_ISL_1927010, EPI_ISL_1927011, EPI_ISL_1927013, EPI_ISL_1927014, EPI_ISL_1927016, EPI_ISL_1927017, EPI_ISL_1927019, EPI_ISL_1927020, EPI_ISL_1927022, EPI_ISL_1927029, EPI_ISL_1927030, EPI_ISL_1927044, EPI_ISL_1927045, EPI_ISL_1927047, EPI_ISL_1927048, EPI_ISL_1927050, EPI_ISL_1927051, EPI_ISL_1927053, EPI_ISL_1927054, EPI_ISL_1927056, EPI_ISL_1927058, EPI_ISL_1927059, EPI_ISL_1927061, EPI_ISL_1927062, EPI_ISL_1927064, EPI_ISL_1927065, EPI_ISL_1927067, EPI_ISL_1927068, EPI_ISL_1927077, EPI_ISL_1927079, EPI_ISL_1927080, EPI_ISL_1927082, EPI_ISL_1927083, EPI_ISL_1927084, EPI_ISL_1927086, EPI_ISL_1927087, EPI_ISL_1927089, EPI_ISL_1927090, EPI_ISL_1927092, EPI_ISL_1927093, EPI_ISL_1927095, EPI_ISL_1927096, EPI_ISL_1927099, EPI_ISL_1927101, EPI_ISL_1927102, EPI_ISL_1927190, EPI_ISL_1927192, EPI_ISL_1927193, EPI_ISL_1927195, EPI_ISL_1927196, EPI_ISL_1927197, EPI_ISL_1927199, EPI_ISL_1927200, EPI_ISL_1927202, EPI_ISL_1927207, EPI_ISL_1927208, EPI_ISL_1927209, EPI_ISL_1927211, EPI_ISL_1927212, EPI_ISL_1927214, EPI_ISL_1927215, EPI_ISL_1927217, EPI_ISL_1927236, EPI_ISL_1927294, EPI_ISL_1927309, EPI_ISL_1927311, EPI_ISL_1927319, EPI_ISL_1927321, EPI_ISL_1927322, EPI_ISL_1927328, EPI_ISL_1927341, EPI_ISL_1927342, EPI_ISL_1927395, EPI_ISL_1927398, EPI_ISL_1927465, EPI_ISL_1927466, EPI_ISL_1927468, EPI_ISL_1927469, EPI_ISL_1927471, EPI_ISL_1927472, EPI_ISL_1927473, EPI_ISL_1927475, EPI_ISL_1927476, EPI_ISL_1927478, EPI_ISL_1927508, EPI_ISL_1927517, EPI_ISL_1927518, EPI_ISL_1927520, EPI_ISL_1927521, EPI_ISL_1927523, EPI_ISL_1927524, EPI_ISL_1927525, EPI_ISL_1927527, EPI_ISL_1927528, EPI_ISL_1927530, EPI_ISL_1927531, EPI_ISL_1927533, EPI_ISL_1927536, EPI_ISL_1927537, EPI_ISL_1927539, EPI_ISL_1927540, EPI_ISL_1927542, EPI_ISL_1927543, EPI_ISL_1927545, EPI_ISL_1927546, EPI_ISL_1927547, EPI_ISL_1927549, EPI_ISL_1927550, EPI_ISL_1927552, EPI_ISL_1927553, EPI_ISL_1927555, EPI_ISL_1927556, EPI_ISL_1927558, EPI_ISL_1927559, EPI_ISL_1927561, EPI_ISL_1927563, EPI_ISL_1927564, EPI_ISL_1927566, EPI_ISL_1927567, EPI_ISL_1927570, EPI_ISL_1927571, EPI_ISL_1927573, EPI_ISL_1927575, EPI_ISL_1927576, EPI_ISL_1927578, EPI_ISL_1927579, EPI_ISL_1927581, EPI_ISL_1927582, EPI_ISL_1927583, EPI_ISL_1927585, EPI_ISL_1927586, EPI_ISL_1927588, EPI_ISL_1927589, EPI_ISL_1927591, EPI_ISL_1927592, EPI_ISL_1927594, EPI_ISL_1927595, EPI_ISL_1927596, EPI_ISL_1927598, EPI_ISL_1927599, EPI_ISL_1927601, EPI_ISL_1927602, EPI_ISL_1927604, EPI_ISL_1927605, EPI_ISL_1927606, EPI_ISL_1927608, EPI_ISL_1927609, EPI_ISL_1927611, EPI_ISL_1927612, EPI_ISL_1927614, EPI_ISL_1927615, EPI_ISL_1927617, EPI_ISL_1927618, EPI_ISL_1927620, EPI_ISL_1927621, EPI_ISL_1927623, EPI_ISL_1927624, EPI_ISL_1927626, EPI_ISL_1927627, EPI_ISL_1927629, EPI_ISL_1927630, EPI_ISL_1927632, EPI_ISL_1927633, EPI_ISL_1927635, EPI_ISL_1927637, EPI_ISL_1927638, EPI_ISL_1927640, EPI_ISL_1927641, EPI_ISL_1927643, EPI_ISL_1927644, EPI_ISL_1927645, EPI_ISL_1927647, EPI_ISL_1927649, EPI_ISL_1927650, EPI_ISL_1927652, EPI_ISL_1927653, EPI_ISL_1927655, EPI_ISL_1927656, EPI_ISL_1927658, EPI_ISL_1927659, EPI_ISL_1927660, EPI_ISL_1927662, EPI_ISL_1927663, EPI_ISL_1927665, EPI_ISL_1927667, EPI_ISL_1927668, EPI_ISL_1927670, EPI_ISL_1927671, EPI_ISL_1927673, EPI_ISL_1927674, EPI_ISL_1927683, EPI_ISL_1927684, EPI_ISL_1927686, EPI_ISL_1927687, EPI_ISL_1927689, EPI_ISL_1927690, EPI_ISL_1927695, EPI_ISL_1927696, EPI_ISL_1927698, EPI_ISL_1927699, EPI_ISL_1927701, EPI_ISL_1927702, EPI_ISL_1927704, EPI_ISL_1927705, EPI_ISL_1927707, EPI_ISL_1927708, EPI_ISL_1927710, EPI_ISL_1927714, EPI_ISL_1927716, EPI_ISL_1927717, EPI_ISL_1927719, EPI_ISL_1927720, EPI_ISL_1927722, EPI_ISL_1927725, EPI_ISL_1927726, EPI_ISL_1927727, EPI_ISL_1927785, EPI_ISL_1927786, EPI_ISL_1927788, EPI_ISL_1927789, EPI_ISL_1927791, EPI_ISL_1927792, EPI_ISL_1927794, EPI_ISL_1927795, EPI_ISL_1927797, EPI_ISL_1927798, EPI_ISL_1927800, EPI_ISL_1927801, EPI_ISL_1927803, EPI_ISL_1927805, EPI_ISL_1927806, EPI_ISL_1927808, EPI_ISL_1927809, EPI_ISL_1927813, EPI_ISL_1927816, EPI_ISL_1927818, EPI_ISL_1927819, EPI_ISL_1927821, EPI_ISL_1927822, EPI_ISL_1927824, EPI_ISL_1927825, EPI_ISL_1927851, EPI_ISL_1927931, EPI_ISL_1927954, EPI_ISL_1927955, EPI_ISL_1927996, EPI_ISL_1927997, EPI_ISL_1927999, EPI_ISL_1928000, EPI_ISL_1928002, EPI_ISL_1928003, EPI_ISL_1928005, EPI_ISL_1928006, EPI_ISL_1928008, EPI_ISL_1928009, EPI_ISL_1928011, EPI_ISL_1928012, EPI_ISL_1928014, EPI_ISL_1928015, EPI_ISL_1928017, EPI_ISL_1928018, EPI_ISL_1928020, EPI_ISL_1928021, EPI_ISL_1928023, EPI_ISL_1928024, EPI_ISL_1928042, EPI_ISL_1928044, EPI_ISL_1928045, EPI_ISL_1928047, EPI_ISL_1928048, EPI_ISL_1928050, EPI_ISL_1928051, EPI_ISL_1928053, EPI_ISL_1928113, EPI_ISL_1928114, EPI_ISL_1928116, EPI_ISL_1928117, EPI_ISL_1928119, EPI_ISL_1928120, EPI_ISL_1928122, EPI_ISL_1928123, EPI_ISL_1928125, EPI_ISL_1928126, EPI_ISL_1928128, EPI_ISL_1928129, EPI_ISL_1928131, EPI_ISL_1928132, EPI_ISL_1928134, EPI_ISL_1928136, EPI_ISL_1928137, EPI_ISL_1928138, EPI_ISL_1928140, EPI_ISL_1928141, EPI_ISL_1928143, EPI_ISL_1928144, EPI_ISL_1928146, EPI_ISL_1928147, EPI_ISL_1928150, EPI_ISL_1928156, EPI_ISL_1928346, EPI_ISL_1928348, EPI_ISL_1928349, EPI_ISL_1928350, EPI_ISL_1928352, EPI_ISL_1928353, EPI_ISL_1928355, EPI_ISL_1928356, EPI_ISL_1928358, EPI_ISL_1928359, EPI_ISL_1928361, EPI_ISL_1928362, EPI_ISL_1928364, EPI_ISL_1928365, EPI_ISL_1928367, EPI_ISL_1928368, EPI_ISL_1928370, EPI_ISL_1929073, EPI_ISL_1929074, EPI_ISL_1929076, EPI_ISL_1929082, EPI_ISL_1929083, EPI_ISL_1929085, EPI_ISL_1929086, EPI_ISL_1929088, EPI_ISL_1929089, EPI_ISL_1929091, EPI_ISL_1929092, EPI_ISL_1929094, EPI_ISL_1929095, EPI_ISL_1929096, EPI_ISL_1929098, EPI_ISL_1929099, EPI_ISL_1929101, EPI_ISL_1929102, EPI_ISL_1929104, EPI_ISL_1929106, EPI_ISL_1929107, EPI_ISL_1929109, EPI_ISL_1929110, EPI_ISL_1929112, EPI_ISL_1929113, EPI_ISL_1929115, EPI_ISL_1929116, EPI_ISL_1929117, EPI_ISL_1929119, EPI_ISL_1929121, EPI_ISL_1929122, EPI_ISL_1929124, EPI_ISL_1929125, EPI_ISL_1929127, EPI_ISL_1929128, EPI_ISL_1929130, EPI_ISL_1929131, EPI_ISL_1929132, EPI_ISL_1929133, EPI_ISL_1929134, EPI_ISL_1929135, EPI_ISL_1929136, EPI_ISL_1929138, EPI_ISL_1929139, EPI_ISL_1929144, EPI_ISL_1929145, EPI_ISL_1929147, EPI_ISL_1929148, EPI_ISL_1929150, EPI_ISL_1929197, EPI_ISL_1929211, EPI_ISL_1929212, EPI_ISL_1929214, EPI_ISL_1929216, EPI_ISL_1929217, EPI_ISL_1929219, EPI_ISL_1929220, EPI_ISL_1929221, EPI_ISL_1929223, EPI_ISL_1929225, EPI_ISL_1929226, EPI_ISL_1929228, EPI_ISL_1929229, EPI_ISL_1929231, EPI_ISL_1929233, EPI_ISL_1929234, EPI_ISL_1929236, EPI_ISL_1929237, EPI_ISL_1929240, EPI_ISL_1929241, EPI_ISL_1929243, EPI_ISL_1929244, EPI_ISL_1929246, EPI_ISL_1929247, EPI_ISL_1929249, EPI_ISL_1929258, EPI_ISL_1929259, EPI_ISL_1929261, EPI_ISL_1929262, EPI_ISL_1929264, EPI_ISL_1929265, EPI_ISL_1929266, EPI_ISL_1929268, EPI_ISL_1929269, EPI_ISL_1929271, EPI_ISL_1929272, EPI_ISL_1929274, EPI_ISL_1929275, EPI_ISL_1929277, EPI_ISL_1929278, EPI_ISL_1929280, EPI_ISL_1929281, EPI_ISL_1929283, EPI_ISL_1929284, EPI_ISL_1929297, EPI_ISL_1929299, EPI_ISL_1929300, EPI_ISL_1929309, EPI_ISL_1929318, EPI_ISL_1929319, EPI_ISL_1929321, EPI_ISL_1929322, EPI_ISL_1929324, EPI_ISL_1929326, EPI_ISL_1929327, EPI_ISL_1929343, EPI_ISL_1929345, EPI_ISL_1929346, EPI_ISL_1929348, EPI_ISL_1929349, EPI_ISL_1929351, EPI_ISL_1929352, EPI_ISL_1929354, EPI_ISL_1929355, EPI_ISL_1929357, EPI_ISL_1929358, EPI_ISL_1929360, EPI_ISL_1929361, EPI_ISL_1929363, EPI_ISL_1929364, EPI_ISL_1929366, EPI_ISL_1929368, EPI_ISL_1929369, EPI_ISL_1929371, EPI_ISL_1929372, EPI_ISL_1929373, EPI_ISL_1929375, EPI_ISL_1929390, EPI_ISL_1929391, EPI_ISL_1929393, EPI_ISL_1929395, EPI_ISL_1929396, EPI_ISL_1929407, EPI_ISL_1929408, EPI_ISL_1929413, EPI_ISL_1929414, EPI_ISL_1929417, EPI_ISL_1929419, EPI_ISL_1929432, EPI_ISL_1929433, EPI_ISL_1929436, EPI_ISL_1929440, EPI_ISL_1929441, EPI_ISL_1929446, EPI_ISL_1929447, EPI_ISL_1929449, EPI_ISL_1929450, EPI_ISL_1929452, EPI_ISL_1929453, EPI_ISL_1929455, EPI_ISL_1929456, EPI_ISL_1929458, EPI_ISL_1929459, EPI_ISL_1929461, EPI_ISL_1929462, EPI_ISL_1929463, EPI_ISL_1929465, EPI_ISL_1929467, EPI_ISL_1929468, EPI_ISL_1929470, EPI_ISL_1929471, EPI_ISL_1929473, EPI_ISL_1929474, EPI_ISL_1929476, EPI_ISL_1929477, EPI_ISL_1929479, EPI_ISL_1929480, EPI_ISL_1929482, EPI_ISL_1929483, EPI_ISL_1929485, EPI_ISL_1929486, EPI_ISL_1929488, EPI_ISL_1929489, EPI_ISL_1929491, EPI_ISL_1929492, EPI_ISL_1929494, EPI_ISL_1929495, EPI_ISL_1929497, EPI_ISL_1929500, EPI_ISL_1929501, EPI_ISL_1929503, EPI_ISL_1929504, EPI_ISL_1929506, EPI_ISL_1929507, EPI_ISL_1929509, EPI_ISL_1929510, EPI_ISL_1929512, EPI_ISL_1929525, EPI_ISL_1929527, EPI_ISL_1929749, EPI_ISL_1929755, EPI_ISL_1929757, EPI_ISL_1929758, EPI_ISL_1929760, EPI_ISL_1929761, EPI_ISL_1929763, EPI_ISL_1929764, EPI_ISL_1929766, EPI_ISL_1929768, EPI_ISL_1929769, EPI_ISL_1929771, EPI_ISL_1929772, EPI_ISL_1929774, EPI_ISL_1929783, EPI_ISL_1929795, EPI_ISL_1929796, EPI_ISL_1929797, EPI_ISL_1929799, EPI_ISL_1929800, EPI_ISL_1929802, EPI_ISL_1929803, EPI_ISL_1929805, EPI_ISL_1929806, EPI_ISL_1929808, EPI_ISL_1929809, EPI_ISL_1929811, EPI_ISL_1929812, EPI_ISL_1929814, EPI_ISL_1929815, EPI_ISL_1929817, | | | |

EPI_ISL_1929818, EPI_ISL_1929820, EPI_ISL_1929821, EPI_ISL_1929823, EPI_ISL_1929824, EPI_ISL_1929826, EPI_ISL_1929827, EPI_ISL_1929829, EPI_ISL_1929831, EPI_ISL_1929832, EPI_ISL_1929834, EPI_ISL_1929835, EPI_ISL_1929837, EPI_ISL_1929838, EPI_ISL_1929840, EPI_ISL_1929841, EPI_ISL_1929843, EPI_ISL_1929844, EPI_ISL_1929845, EPI_ISL_1929847, EPI_ISL_1929848, EPI_ISL_1929849,
EPI_ISL_1929851, EPI_ISL_1929852, EPI_ISL_1929854, EPI_ISL_1929855, EPI_ISL_1929857, EPI_ISL_1929858, EPI_ISL_1929860, EPI_ISL_1929862, EPI_ISL_1929863, EPI_ISL_1929865, EPI_ISL_1929866, EPI_ISL_1929868, EPI_ISL_1929869, EPI_ISL_1929871, EPI_ISL_1929872, EPI_ISL_1929874, EPI_ISL_1929875, EPI_ISL_1929877, EPI_ISL_1929878, EPI_ISL_1929880, EPI_ISL_1929881, EPI_ISL_1929883,
EPI_ISL_1929884, EPI_ISL_1929886, EPI_ISL_1929887, EPI_ISL_1929888, EPI_ISL_1929895, EPI_ISL_1929898, EPI_ISL_1929900, EPI_ISL_1929906, EPI_ISL_1929907, EPI_ISL_1929912, EPI_ISL_1929913, EPI_ISL_1929915, EPI_ISL_1929916, EPI_ISL_1929918, EPI_ISL_1929919, EPI_ISL_1929921, EPI_ISL_1929922, EPI_ISL_1929924, EPI_ISL_1929925, EPI_ISL_1929927, EPI_ISL_1929928, EPI_ISL_1929930,
EPI_ISL_1929931, EPI_ISL_1929933, EPI_ISL_1929934, EPI_ISL_1929936, EPI_ISL_1929939, EPI_ISL_1929940, EPI_ISL_1929942, EPI_ISL_1929943, EPI_ISL_1929945, EPI_ISL_1929946, EPI_ISL_1929948, EPI_ISL_1929949, EPI_ISL_1929951, EPI_ISL_1929952, EPI_ISL_1929954, EPI_ISL_1929955, EPI_ISL_1929957, EPI_ISL_1929958, EPI_ISL_1929960, EPI_ISL_1929962, EPI_ISL_1929963, EPI_ISL_1929964,
EPI_ISL_1929966, EPI_ISL_1929968, EPI_ISL_1929969, EPI_ISL_1929970, EPI_ISL_1929972, EPI_ISL_1929973, EPI_ISL_1929975, EPI_ISL_1929976, EPI_ISL_1929978, EPI_ISL_1929979, EPI_ISL_1929981, EPI_ISL_1929982, EPI_ISL_1929983, EPI_ISL_1929985, EPI_ISL_1929988, EPI_ISL_1929989, EPI_ISL_1929991, EPI_ISL_1929992, EPI_ISL_1929993, EPI_ISL_1929995, EPI_ISL_1929997, EPI_ISL_1929998,
EPI_ISL_1930000, EPI_ISL_1930001, EPI_ISL_1930018, EPI_ISL_1930036, EPI_ISL_1930038, EPI_ISL_1930039, EPI_ISL_1930041, EPI_ISL_1930042, EPI_ISL_1930044, EPI_ISL_1930046, EPI_ISL_1930047, EPI_ISL_1930049, EPI_ISL_1930055, EPI_ISL_1930056, EPI_ISL_1930058, EPI_ISL_1930059, EPI_ISL_1930061, EPI_ISL_1930064, EPI_ISL_1930065, EPI_ISL_1930067, EPI_ISL_1930069, EPI_ISL_1930070,
EPI_ISL_1930072, EPI_ISL_1930073, EPI_ISL_1930075, EPI_ISL_1930076, EPI_ISL_1930078, EPI_ISL_1930080, EPI_ISL_1930082, EPI_ISL_1930083, EPI_ISL_1930085, EPI_ISL_1930086, EPI_ISL_1930088, EPI_ISL_1930090, EPI_ISL_1930091, EPI_ISL_1930093, EPI_ISL_1930094, EPI_ISL_1930096, EPI_ISL_1930098, EPI_ISL_1930099, EPI_ISL_1930101, EPI_ISL_1930102, EPI_ISL_1930104, EPI_ISL_1930105,
EPI_ISL_1930107, EPI_ISL_1930108, EPI_ISL_1930110, EPI_ISL_1930111, EPI_ISL_1930113, EPI_ISL_1930223, EPI_ISL_1930224, EPI_ISL_1930226, EPI_ISL_1930227, EPI_ISL_1930229, EPI_ISL_1930232, EPI_ISL_1930233, EPI_ISL_1930235, EPI_ISL_1930236, EPI_ISL_1930238, EPI_ISL_1930240, EPI_ISL_1930306, EPI_ISL_1930308, EPI_ISL_1930309, EPI_ISL_1930311, EPI_ISL_1930312, EPI_ISL_1930314,
EPI_ISL_1930316, EPI_ISL_1930317, EPI_ISL_1930319, EPI_ISL_1930341, EPI_ISL_1930342, EPI_ISL_1930344, EPI_ISL_1930366, EPI_ISL_1930367, EPI_ISL_1930369, EPI_ISL_1930370, EPI_ISL_1930372, EPI_ISL_1930373, EPI_ISL_1930375, EPI_ISL_1930376, EPI_ISL_1930378, EPI_ISL_1930380, EPI_ISL_1930381, EPI_ISL_1930382, EPI_ISL_1930384, EPI_ISL_1930385, EPI_ISL_1930387, EPI_ISL_1930395,
EPI_ISL_1930396, EPI_ISL_1930397, EPI_ISL_1930399, EPI_ISL_1930401, EPI_ISL_1930402, EPI_ISL_1930417, EPI_ISL_1930419, EPI_ISL_1930420, EPI_ISL_1930422, EPI_ISL_1930423, EPI_ISL_1930425, EPI_ISL_1930426, EPI_ISL_1930428, EPI_ISL_1930486, EPI_ISL_1930489, EPI_ISL_1930490, EPI_ISL_1930492, EPI_ISL_1930503, EPI_ISL_1930506, EPI_ISL_1930507, EPI_ISL_1930509, EPI_ISL_1930510,
EPI_ISL_1930512, EPI_ISL_1930514, EPI_ISL_1930515, EPI_ISL_1930516, EPI_ISL_1930518, EPI_ISL_1930520, EPI_ISL_1930521, EPI_ISL_1930522, EPI_ISL_1930524, EPI_ISL_1930526, EPI_ISL_1930527, EPI_ISL_1930529, EPI_ISL_1930530, EPI_ISL_1930532, EPI_ISL_1930534, EPI_ISL_1930535, EPI_ISL_1930537, EPI_ISL_1930538, EPI_ISL_1930540, EPI_ISL_1930541, EPI_ISL_1930543, EPI_ISL_1930544,
EPI_ISL_1930546, EPI_ISL_1930547, EPI_ISL_1930549, EPI_ISL_1930550, EPI_ISL_1930552, EPI_ISL_1930553, EPI_ISL_1930558, EPI_ISL_1930863, EPI_ISL_1930864, EPI_ISL_1930866, EPI_ISL_1930875, EPI_ISL_1930876, EPI_ISL_1930878, EPI_ISL_1930879, EPI_ISL_1930881, EPI_ISL_1930882, EPI_ISL_1930884, EPI_ISL_1930885, EPI_ISL_1930887, EPI_ISL_1930889, EPI_ISL_1930890, EPI_ISL_1930924,
EPI_ISL_1930926, EPI_ISL_1930927, EPI_ISL_1930929, EPI_ISL_1930930, EPI_ISL_1930932, EPI_ISL_1930933, EPI_ISL_1930935, EPI_ISL_1930936, EPI_ISL_1930938, EPI_ISL_1930939, EPI_ISL_1930944, EPI_ISL_1930945, EPI_ISL_1930947, EPI_ISL_1930948, EPI_ISL_1930957, EPI_ISL_1930959, EPI_ISL_1930960, EPI_ISL_1930962, EPI_ISL_1930963, EPI_ISL_1930965, EPI_ISL_1930969, EPI_ISL_1930971,
EPI_ISL_1930972, EPI_ISL_1931021, EPI_ISL_1931023, EPI_ISL_1931025, EPI_ISL_1931026, EPI_ISL_1931027, EPI_ISL_1931029, EPI_ISL_1931030, EPI_ISL_1931032, EPI_ISL_1931033, EPI_ISL_1931035, EPI_ISL_1931036, EPI_ISL_1931038, EPI_ISL_1931039, EPI_ISL_1931041, EPI_ISL_1931043, EPI_ISL_1931044, EPI_ISL_1931045, EPI_ISL_1931047, EPI_ISL_1931048, EPI_ISL_1931050, EPI_ISL_1931051,
EPI_ISL_1931053, EPI_ISL_1931054, EPI_ISL_1931056, EPI_ISL_1931057, EPI_ISL_1931059, EPI_ISL_1931060, EPI_ISL_1931062, EPI_ISL_1931063, EPI_ISL_1931065, EPI_ISL_1931066, EPI_ISL_1931068, EPI_ISL_1931069, EPI_ISL_1931071, EPI_ISL_1931072, EPI_ISL_1931074, EPI_ISL_1931088, EPI_ISL_1931090, EPI_ISL_1931091, EPI_ISL_1931093, EPI_ISL_1931094, EPI_ISL_1931096, EPI_ISL_1931097,
EPI_ISL_1931099, EPI_ISL_1931100, EPI_ISL_1931102, EPI_ISL_1931103, EPI_ISL_1931104, EPI_ISL_1931105, EPI_ISL_1931107, EPI_ISL_1931108, EPI_ISL_1931110, EPI_ISL_1931111, EPI_ISL_1931113, EPI_ISL_1931114, EPI_ISL_1931118, EPI_ISL_1931121, EPI_ISL_1931125, EPI_ISL_1931128, EPI_ISL_1931138, EPI_ISL_1931141, EPI_ISL_1931144, EPI_ISL_1931147, EPI_ISL_1931150, EPI_ISL_1931153,
EPI_ISL_1931156, EPI_ISL_1931159, EPI_ISL_1931162, EPI_ISL_1931166, EPI_ISL_1931169, EPI_ISL_1931172, EPI_ISL_1931175, EPI_ISL_1931178, EPI_ISL_1931181, EPI_ISL_1931184, EPI_ISL_1931187, EPI_ISL_1931191, EPI_ISL_1931194, EPI_ISL_1931197, EPI_ISL_1931200, EPI_ISL_1931204, EPI_ISL_1931207, EPI_ISL_1931210, EPI_ISL_1931212, EPI_ISL_1931228, EPI_ISL_1931231, EPI_ISL_1931233,
EPI_ISL_1931234, EPI_ISL_1931236, EPI_ISL_1931237, EPI_ISL_1931239, EPI_ISL_1931240, EPI_ISL_1931242, EPI_ISL_1931243, EPI_ISL_1931245, EPI_ISL_1931249, EPI_ISL_1931252, EPI_ISL_1931255, EPI_ISL_1931259, EPI_ISL_1931262, EPI_ISL_1931265, EPI_ISL_1931269, EPI_ISL_1931271, EPI_ISL_1931274, EPI_ISL_1931277, EPI_ISL_1931280, EPI_ISL_1931283, EPI_ISL_1931286, EPI_ISL_1931289,
EPI_ISL_1931292, EPI_ISL_1931295, EPI_ISL_1931298, EPI_ISL_1931301, EPI_ISL_1931305, EPI_ISL_1931308, EPI_ISL_1931310, EPI_ISL_1931313, EPI_ISL_1931317, EPI_ISL_1931320, EPI_ISL_1931323, EPI_ISL_1931326, EPI_ISL_1931327, EPI_ISL_1931329, EPI_ISL_1931330, EPI_ISL_1931332, EPI_ISL_1931333, EPI_ISL_1931335, EPI_ISL_1931349, EPI_ISL_1931350, EPI_ISL_1931352, EPI_ISL_1931353,
EPI_ISL_1931355, EPI_ISL_1931356, EPI_ISL_1931358, EPI_ISL_1931359, EPI_ISL_1931442, EPI_ISL_1931444, EPI_ISL_1931446, EPI_ISL_1931447, EPI_ISL_1931456, EPI_ISL_1931458, EPI_ISL_1931459, EPI_ISL_1931461, EPI_ISL_1931462, EPI_ISL_1931464, EPI_ISL_1931465, EPI_ISL_1931466, EPI_ISL_1931467, EPI_ISL_1931468, EPI_ISL_1931469, EPI_ISL_1931470, EPI_ISL_1931472, EPI_ISL_1931474,
EPI_ISL_1931475, EPI_ISL_1931477, EPI_ISL_1931478, EPI_ISL_1931480, EPI_ISL_1931482, EPI_ISL_1931483, EPI_ISL_1931485, EPI_ISL_1931486, EPI_ISL_1931488, EPI_ISL_1931489, EPI_ISL_1931491, EPI_ISL_1931493, EPI_ISL_1931494, EPI_ISL_1931496, EPI_ISL_1931497, EPI_ISL_1931502, EPI_ISL_1931503, EPI_ISL_1931505, EPI_ISL_1931506, EPI_ISL_1931508, EPI_ISL_1931509, EPI_ISL_1931511,
EPI_ISL_1931512, EPI_ISL_1931536, EPI_ISL_1931538, EPI_ISL_1931539, EPI_ISL_1931541, EPI_ISL_1931542, EPI_ISL_1931544, EPI_ISL_1931546, EPI_ISL_1931547, EPI_ISL_1931549, EPI_ISL_1931550, EPI_ISL_1931552, EPI_ISL_1931553, EPI_ISL_1931558, EPI_ISL_1931561, EPI_ISL_1931564, EPI_ISL_1931568, EPI_ISL_1931571, EPI_ISL_1931575, EPI_ISL_1931578, EPI_ISL_1931579, EPI_ISL_1931581,
EPI_ISL_1931582, EPI_ISL_1931590, EPI_ISL_1931592, EPI_ISL_1931597, EPI_ISL_1931599, EPI_ISL_1931600, EPI_ISL_1931602, EPI_ISL_1931603, EPI_ISL_1931605, EPI_ISL_1931607, EPI_ISL_1931608, EPI_ISL_1931610, EPI_ISL_1931611, EPI_ISL_1931613, EPI_ISL_1931614, EPI_ISL_1931625, EPI_ISL_1931628, EPI_ISL_1931631, EPI_ISL_1931641, EPI_ISL_1931643, EPI_ISL_1931644, EPI_ISL_1931646,
EPI_ISL_1931648, EPI_ISL_1931649, EPI_ISL_1931652, EPI_ISL_1931653, EPI_ISL_1931655, EPI_ISL_1931657, EPI_ISL_1931664, EPI_ISL_1931677, EPI_ISL_1931681, EPI_ISL_1931682, EPI_ISL_1931684, EPI_ISL_1931686, EPI_ISL_1931687, EPI_ISL_1931689, EPI_ISL_1931690, EPI_ISL_1931692, EPI_ISL_1931693, EPI_ISL_1931695, EPI_ISL_1931696, EPI_ISL_1931698, EPI_ISL_1931699, EPI_ISL_1931701,
EPI_ISL_1931702, EPI_ISL_1931704, EPI_ISL_1931705, EPI_ISL_1931707, EPI_ISL_1931709, EPI_ISL_1931712, EPI_ISL_1931715, EPI_ISL_1931719, EPI_ISL_1931722, EPI_ISL_1931725, EPI_ISL_1931727, EPI_ISL_1931729, EPI_ISL_1931730, EPI_ISL_1931732, EPI_ISL_1931733, EPI_ISL_1931735, EPI_ISL_1931736, EPI_ISL_1931738, EPI_ISL_1931739, EPI_ISL_1931741, EPI_ISL_1931743, EPI_ISL_1931744,
EPI_ISL_1931746, EPI_ISL_1931747, EPI_ISL_1931749, EPI_ISL_1931750, EPI_ISL_1931752, EPI_ISL_1931757, EPI_ISL_1931761, EPI_ISL_1931764, EPI_ISL_1931765, EPI_ISL_1931767, EPI_ISL_1931768, EPI_ISL_1931770, EPI_ISL_1931772, EPI_ISL_1931773, EPI_ISL_1931775, EPI_ISL_1931776, EPI_ISL_1931778, EPI_ISL_1931779, EPI_ISL_1931781, EPI_ISL_1931786, EPI_ISL_1931787, EPI_ISL_1931789,
EPI_ISL_1931790, EPI_ISL_1931792, EPI_ISL_1931794, EPI_ISL_1931795, EPI_ISL_1931797, EPI_ISL_1931798, EPI_ISL_1931800, EPI_ISL_1931801, EPI_ISL_1931803, EPI_ISL_1931804, EPI_ISL_1931806, EPI_ISL_1931810, EPI_ISL_1931813, EPI_ISL_1931817, EPI_ISL_1931820, EPI_ISL_1931823, EPI_ISL_1931827, EPI_ISL_1931830, EPI_ISL_1931833, EPI_ISL_1931835, EPI_ISL_1931836, EPI_ISL_1931838,
EPI_ISL_1931840, EPI_ISL_1931841, EPI_ISL_1931843, EPI_ISL_1931844, EPI_ISL_1931846, EPI_ISL_1931847, EPI_ISL_1931849, EPI_ISL_1931850, EPI_ISL_1931852, EPI_ISL_1931853, EPI_ISL_1931855, EPI_ISL_1931856, EPI_ISL_1931858, EPI_ISL_1931859, EPI_ISL_1931861, EPI_ISL_1931862, EPI_ISL_1931864, EPI_ISL_1931865, EPI_ISL_1931867, EPI_ISL_1931868, EPI_ISL_1931872, EPI_ISL_1931876,
EPI_ISL_1931878, EPI_ISL_1931879, EPI_ISL_1931881, EPI_ISL_1931882, EPI_ISL_1931884, EPI_ISL_1931887, EPI_ISL_1931891, EPI_ISL_1931892, EPI_ISL_1931894, EPI_ISL_1931895, EPI_ISL_1931897, EPI_ISL_1931898, EPI_ISL_1931902, EPI_ISL_1931904, EPI_ISL_1931905, EPI_ISL_1931907, EPI_ISL_1931909, EPI_ISL_1931910, EPI_ISL_1931912, EPI_ISL_1931913, EPI_ISL_1931915, EPI_ISL_1931916,
EPI_ISL_1931918, EPI_ISL_1931919, EPI_ISL_1931921, EPI_ISL_1931922, EPI_ISL_1931924, EPI_ISL_1931926, EPI_ISL_1931929, EPI_ISL_1931931, EPI_ISL_1931933, EPI_ISL_1931934, EPI_ISL_1931936, EPI_ISL_1931937, EPI_ISL_1931939, EPI_ISL_1931941, EPI_ISL_1931942, EPI_ISL_1931949, EPI_ISL_1931951, EPI_ISL_1931955, EPI_ISL_1931959, EPI_ISL_1931963, EPI_ISL_1931965, EPI_ISL_1931966,
EPI_ISL_1931968, EPI_ISL_1932043, EPI_ISL_1932044, EPI_ISL_1932046, EPI_ISL_1932047, EPI_ISL_1932049, EPI_ISL_1932052, EPI_ISL_1932055, EPI_ISL_1932056, EPI_ISL_1932057, EPI_ISL_1932058, EPI_ISL_1932059, EPI_ISL_1932060, EPI_ISL_1932061, EPI_ISL_1932062, EPI_ISL_1932063, EPI_ISL_1932064, EPI_ISL_1932065, EPI_ISL_1932066, EPI_ISL_1932067, EPI_ISL_1932068, EPI_ISL_1932069,
EPI_ISL_1932070, EPI_ISL_1932071, EPI_ISL_1932072, EPI_ISL_1932073, EPI_ISL_1932074, EPI_ISL_1932075, EPI_ISL_1932076, EPI_ISL_1932077, EPI_ISL_1932078, EPI_ISL_1932079, EPI_ISL_1932080, EPI_ISL_1932081, EPI_ISL_1932082, EPI_ISL_1932083, EPI_ISL_1932084, EPI_ISL_1932085, EPI_ISL_1932086, EPI_ISL_1932087, EPI_ISL_1932088, EPI_ISL_1932089, EPI_ISL_1932093, EPI_ISL_1932094,
EPI_ISL_1932095, EPI_ISL_1932096, EPI_ISL_1932097, EPI_ISL_1932098, EPI_ISL_1932099, EPI_ISL_1932100, EPI_ISL_1932101, EPI_ISL_1932102, EPI_ISL_1932103, EPI_ISL_1932104, EPI_ISL_1932105, EPI_ISL_1932106, EPI_ISL_1932107, EPI_ISL_1932108, EPI_ISL_1932109, EPI_ISL_1932110, EPI_ISL_1932111, EPI_ISL_1932112, EPI_ISL_1932113, EPI_ISL_1932114, EPI_ISL_1932115, EPI_ISL_1932116,
EPI_ISL_1932117, EPI_ISL_1932118, EPI_ISL_1932119, EPI_ISL_1932120, EPI_ISL_1932121, EPI_ISL_1932122, EPI_ISL_1932123, EPI_ISL_1932124, EPI_ISL_1932125, EPI_ISL_1932126, EPI_ISL_1932127, EPI_ISL_1932128, EPI_ISL_1932129, EPI_ISL_1932130, EPI_ISL_1932131, EPI_ISL_1932132, EPI_ISL_1932133, EPI_ISL_1932134, EPI_ISL_1932135, EPI_ISL_1932136, EPI_ISL_1932137, EPI_ISL_1932138,
EPI_ISL_1932139, EPI_ISL_1932140, EPI_ISL_1932141, EPI_ISL_1932142, EPI_ISL_1932143, EPI_ISL_1932144, EPI_ISL_1932145, EPI_ISL_1932146, EPI_ISL_1932147, EPI_ISL_1932148, EPI_ISL_1932149, EPI_ISL_1932150, EPI_ISL_1932151, EPI_ISL_1932152, EPI_ISL_1932153, EPI_ISL_1932265, EPI_ISL_1932266, EPI_ISL_1932267, EPI_ISL_1932268, EPI_ISL_1932269, EPI_ISL_1932270, EPI_ISL_1932271,
EPI_ISL_1932272, EPI_ISL_1932281, EPI_ISL_1932282, EPI_ISL_1932283, EPI_ISL_1932284, EPI_ISL_1932285, EPI_ISL_1932286, EPI_ISL_1932287, EPI_ISL_1932288, EPI_ISL_1932289, EPI_ISL_1932290, EPI_ISL_1932291, EPI_ISL_1932292, EPI_ISL_1932293, EPI_ISL_1932294, EPI_ISL_1932295, EPI_ISL_1932297, EPI_ISL_1932298, EPI_ISL_1932299, EPI_ISL_1932300, EPI_ISL_1932301, EPI_ISL_1932302,
EPI_ISL_1932303, EPI_ISL_1932304, EPI_ISL_1932305, EPI_ISL_1932306, EPI_ISL_1932307, EPI_ISL_1932314, EPI_ISL_1932315, EPI_ISL_1932316, EPI_ISL_1932317, EPI_ISL_1932318, EPI_ISL_1932319, EPI_ISL_1932320, EPI_ISL_1932321, EPI_ISL_1932322, EPI_ISL_1932323, EPI_ISL_1932377, EPI_ISL_1932378, EPI_ISL_1932379, EPI_ISL_1932380, EPI_ISL_1932381, EPI_ISL_1932382, EPI_ISL_1932383,
EPI_ISL_1932385, EPI_ISL_1932386, EPI_ISL_1932387, EPI_ISL_1932388, EPI_ISL_1932389, EPI_ISL_1932390, EPI_ISL_1932391, EPI_ISL_1932392, EPI_ISL_1932393, EPI_ISL_1932394, EPI_ISL_1932395, EPI_ISL_1932396, EPI_ISL_1932397, EPI_ISL_1932398, EPI_ISL_1932399, EPI_ISL_1932400, EPI_ISL_1932401, EPI_ISL_1932402, EPI_ISL_1932403, EPI_ISL_1932404, EPI_ISL_1932405, EPI_ISL_1932406,
EPI_ISL_1932407, EPI_ISL_1932408, EPI_ISL_1932409, EPI_ISL_1932410, EPI_ISL_1932411, EPI_ISL_1932418, EPI_ISL_1932421, EPI_ISL_1932422, EPI_ISL_1932423, EPI_ISL_1932424, EPI_ISL_1932425, EPI_ISL_1932426, EPI_ISL_1932427, EPI_ISL_1932428, EPI_ISL_1932429, EPI_ISL_1932430, EPI_ISL_1932431, EPI_ISL_1932432, EPI_ISL_1932433, EPI_ISL_1932434, EPI_ISL_1932435, EPI_ISL_1932436,
EPI_ISL_1932437, EPI_ISL_1932438, EPI_ISL_1932439, EPI_ISL_1932440, EPI_ISL_1932441, EPI_ISL_1932442, EPI_ISL_1932443, EPI_ISL_1932444, EPI_ISL_1932461, EPI_ISL_1932462, EPI_ISL_1932463, EPI_ISL_1932464, EPI_ISL_1932465, EPI_ISL_1932466, EPI_ISL_1932467, EPI_ISL_1932468, EPI_ISL_1932469, EPI_ISL_1932470, EPI_ISL_1932471, EPI_ISL_1932472, EPI_ISL_1932473, EPI_ISL_1932474,
EPI_ISL_1932475, EPI_ISL_1932476, EPI_ISL_1932477, EPI_ISL_1932478, EPI_ISL_1932479, EPI_ISL_1932480, EPI_ISL_1932481, EPI_ISL_1932482, EPI_ISL_1932483, EPI_ISL_1932484, EPI_ISL_1932485, EPI_ISL_1932487, EPI_ISL_1932488, EPI_ISL_1932489, EPI_ISL_1932490, EPI_ISL_1932491, EPI_ISL_1932492, EPI_ISL_1932493, EPI_ISL_1932494, EPI_ISL_1932495, EPI_ISL_1932496, EPI_ISL_1932497,
EPI_ISL_1932498, EPI_ISL_1932499, EPI_ISL_1932500, EPI_ISL_1932501, EPI_ISL_1932502, EPI_ISL_1932503, EPI_ISL_1932504, EPI_ISL_1932505, EPI_ISL_1932506, EPI_ISL_1932507, EPI_ISL_1932508, EPI_ISL_1932509, EPI_ISL_1932510, EPI_ISL_1932511, EPI_ISL_1932512, EPI_ISL_1932513, EPI_ISL_1932514, EPI_ISL_1932515, EPI_ISL_1932516, EPI_ISL_1932517, EPI_ISL_1932518, EPI_ISL_1932519,
EPI_ISL_1932520, EPI_ISL_1932521, EPI_ISL_1932522, EPI_ISL_1932523, EPI_ISL_1932524, EPI_ISL_1932525, EPI_ISL_1932526, EPI_ISL_1932527, EPI_ISL_1932528, EPI_ISL_1932529, EPI_ISL_1932530, EPI_ISL_1932541, EPI_ISL_1932542, EPI_ISL_1932543, EPI_ISL_1932544, EPI_ISL_1932545, EPI_ISL_1932546, EPI_ISL_1932547, EPI_ISL_1932548, EPI_ISL_1932549, EPI_ISL_1932550, EPI_ISL_1932551,
EPI_ISL_1932552, EPI_ISL_1932553, EPI_ISL_1932554, EPI_ISL_1932555, EPI_ISL_1932556, EPI_ISL_1932557, EPI_ISL_1932558, EPI_ISL_1932559, EPI_ISL_1932560, EPI_ISL_1932561, EPI_ISL_1932562, EPI_ISL_1932563, EPI_ISL_1932564, EPI_ISL_1932565, EPI_ISL_1932566, EPI_ISL_1932567, EPI_ISL_1932568, EPI_ISL_1932569, EPI_ISL_1932570, EPI_ISL_1932571, EPI_ISL_1932572, EPI_ISL_1932573,
EPI_ISL_1932574, EPI_ISL_1932575, EPI_ISL_1932576, EPI_ISL_1932577, EPI_ISL_1932578, EPI_ISL_1932579, EPI_ISL_1932580, EPI_ISL_1932583, EPI_ISL_1932623, EPI_ISL_1932624, EPI_ISL_1932625, EPI_ISL_1932626, EPI_ISL_1932627, EPI_ISL_1932628, EPI_ISL_1932629, EPI_ISL_1932630, EPI_ISL_1932631, EPI_ISL_1932632, EPI_ISL_1932633, EPI_ISL_1932634, EPI_ISL_1932635, EPI_ISL_1932636,
EPI_ISL_1932637, EPI_ISL_1932638, EPI_ISL_1932639, EPI_ISL_1932640, EPI_ISL_1932641, EPI_ISL_1932642, EPI_ISL_1932643, EPI_ISL_1932644, EPI_ISL_1932645, EPI_ISL_1932646, EPI_ISL_1932647, EPI_ISL_1932648, EPI_ISL_1932649, EPI_ISL_1932650, EPI_ISL_1932651, EPI_ISL_1932652, EPI_ISL_1932653, EPI_ISL_1932654, EPI_ISL_1932655, EPI_ISL_1932656, EPI_ISL_1932657, EPI_ISL_1932658,
EPI_ISL_1932659, EPI_ISL_1932660, EPI_ISL_1932661, EPI_ISL_1932662, EPI_ISL_1932663, EPI_ISL_1932664, EPI_ISL_1932665, EPI_ISL_1932666, EPI_ISL_1932667, EPI_ISL_1932668, EPI_ISL_1932669, EPI_ISL_1932670, EPI_ISL_1932671, EPI_ISL_1932672, EPI_ISL_1932673, EPI_ISL_1932674, EPI_ISL_1932675, EPI_ISL_1932676, EPI_ISL_1932677, EPI_ISL_1932678, EPI_ISL_1932679, EPI_ISL_1932680,
EPI_ISL_1932681, EPI_ISL_1932682, EPI_ISL_1932683, EPI_ISL_1932684, EPI_ISL_1932685, EPI_ISL_1932686, EPI_ISL_1932687, EPI_ISL_1932688, EPI_ISL_1932689, EPI_ISL_1932690, EPI_ISL_1932691, EPI_ISL_1932697, EPI_ISL_1932700, EPI_ISL_1932702, EPI_ISL_1932703, EPI_ISL_1932704, EPI_ISL_1932705, EPI_ISL_1932711, EPI_ISL_1932712, EPI_ISL_1932713, EPI_ISL_1932715, EPI_ISL_1932716,
EPI_ISL_1932717, EPI_ISL_1932719, EPI_ISL_1932720, EPI_ISL_1932721, EPI_ISL_1932722, EPI_ISL_1932723, EPI_ISL_1932724, EPI_ISL_1932726, EPI_ISL_1932727, EPI_ISL_1932728, EPI_ISL_1932730, EPI_ISL_1932731, EPI_ISL_1932732, EPI_ISL_1932733, EPI_ISL_1932737, EPI_ISL_1932738, EPI_ISL_1932739, EPI_ISL_1932740, EPI_ISL_1932741, EPI_ISL_1932742, EPI_ISL_1932743, EPI_ISL_1932744,
EPI_ISL_1932745, EPI_ISL_1932746, EPI_ISL_1932747, EPI_ISL_1932748, EPI_ISL_1932749, EPI_ISL_1932750, EPI_ISL_1932751, EPI_ISL_1932752, EPI_ISL_1932753, EPI_ISL_1932754, EPI_ISL_1932755, EPI_ISL_1932756, EPI_ISL_1932757, EPI_ISL_1932758, EPI_ISL_1932761, EPI_ISL_1932762, EPI_ISL_1932763, EPI_ISL_1932764, EPI_ISL_1932765, EPI_ISL_1932766, EPI_ISL_1932767, EPI_ISL_1932768,
EPI_ISL_1932769, EPI_ISL_1932770, EPI_ISL_1932771, EPI_ISL_1932772, EPI_ISL_1932773, EPI_ISL_1932774, EPI_ISL_1932775, EPI_ISL_1932776, EPI_ISL_1932777, EPI_ISL_1932778, EPI_ISL_1932779, EPI_ISL_1932780, EPI_ISL_1932781, EPI_ISL_1932782, EPI_ISL_1932783, EPI_ISL_1932784, EPI_ISL_1932785, EPI_ISL_1932786, EPI_ISL_1932787, EPI_ISL_1932788, EPI_ISL_1932789, EPI_ISL_1932790,
EPI_ISL_1932791, EPI_ISL_1932792, EPI_ISL_1932793, EPI_ISL_1932794, EPI_ISL_1932795, EPI_ISL_1932796, EPI_ISL_1932797, EPI_ISL_1932798, EPI_ISL_1932799, EPI_ISL_1932800, EPI_ISL_1932801, EPI_ISL_1932802, EPI_ISL_1932803, EPI_ISL_1932804, EPI_ISL_1932805, EPI_ISL_1932806, EPI_ISL_1932807, EPI_ISL_1932808, EPI_ISL_1932809, EPI_ISL_1932810, EPI_ISL_1932811, EPI_ISL_1932812,
EPI_ISL_1932813, EPI_ISL_1932814, EPI_ISL_1932815, EPI_ISL_1932816, EPI_ISL_1932817, EPI_ISL_1932818, EPI_ISL_1932819, EPI_ISL_1932820, EPI_ISL_1932821, EPI_ISL_1932822, EPI_ISL_1932823, EPI_ISL_1932824, EPI_ISL_1932825, EPI_ISL_1932826, EPI_ISL_1932827, EPI_ISL_1932828, EPI_ISL_1932829, EPI_ISL_1932830, EPI_ISL_1932831, EPI_ISL_1932849, EPI_ISL_1932850, EPI_ISL_1932851,
EPI_ISL_1932852, EPI_ISL_1932853, EPI_ISL_1932854, EPI_ISL_1932855, EPI_ISL_1932856, EPI_ISL_1932857, EPI_ISL_1932858, EPI_ISL_1932859, EPI_ISL_1932860, EPI_ISL_1932861, EPI_ISL_1932862, EPI_ISL_1932863, EPI_ISL_1932864, EPI_ISL_1932865, EPI_ISL_1932866, EPI_ISL_1932867, EPI_ISL_1932868, EPI_ISL_1932869, EPI_ISL_1932870, EPI_ISL_1932871, EPI_ISL_1932872, EPI_ISL_1932873,
EPI_ISL_1932874, EPI_ISL_1932875, EPI_ISL_1932876, EPI_ISL_1932877, EPI_ISL_1932878, EPI_ISL_1932879, EPI_ISL_1932880, EPI_ISL_1932881, EPI_ISL_1932882, EPI_ISL_1932883, EPI_ISL_1932884, EPI_ISL_1932885, EPI_ISL_1932886, EPI_ISL_1932887, EPI_ISL_1932888, EPI_ISL_1932889, EPI_ISL_1932890, EPI_ISL_1932891, EPI_ISL_1932892, EPI_ISL_1932893, EPI_ISL_1932894, EPI_ISL_1932895,
EPI_ISL_1932897, EPI_ISL_1932898, EPI_ISL_1932899, EPI_ISL_1932900, EPI_ISL_1932901, EPI_ISL_1932902, EPI_ISL_1932903, EPI_ISL_1932904, EPI_ISL_1932905, EPI_ISL_1932910, EPI_ISL_1932911, EPI_ISL_1932912, EPI_ISL_1932913, EPI_ISL_1932914, EPI_ISL_1932915, EPI_ISL_1932916, EPI_ISL_1932917, EPI_ISL_1932918, EPI_ISL_1932919, EPI_ISL_1932920, EPI_ISL_1932921, EPI_ISL_1932922,
EPI_ISL_1932923, EPI_ISL_1932924, EPI_ISL_1932925, EPI_ISL_1932926, EPI_ISL_1932927, EPI_ISL_1932928, EPI_ISL_1932929, EPI_ISL_1932930, EPI_ISL_1932931, EPI_ISL_1932932, EPI_ISL_1932933, EPI_ISL_1932934, EPI_ISL_1932935, EPI_ISL_1932936, EPI_ISL_1932937, EPI_ISL_1932938, EPI_ISL_1932939, EPI_ISL_1932940, EPI_ISL_1932941, EPI_ISL_1932942, EPI_ISL_1932943, EPI_ISL_1932944,
EPI_ISL_1932945, EPI_ISL_1932946, EPI_ISL_1932947, EPI_ISL_1932948, EPI_ISL_1932949, EPI_ISL_1932950, EPI_ISL_1932951, EPI_ISL_1932952, EPI_ISL_1932953, EPI_ISL_1932954, EPI_ISL_1932955, EPI_ISL_1932956, EPI_ISL_1932957, EPI_ISL_1932958, EPI_ISL_1932959, EPI_ISL_1932960, EPI_ISL_1932961, EPI_ISL_1932962, EPI_ISL_1932963, EPI_ISL_1932964, EPI_ISL_1932965, EPI_ISL_1932966,
EPI_ISL_1932967, EPI_ISL_1932968, EPI_ISL_1932969, EPI_ISL_1932970, EPI_ISL_1932971, EPI_ISL_1932972, EPI_ISL_1932973, EPI_ISL_1932974, EPI_ISL_1932975, EPI_ISL_1932976, EPI_ISL_1932977, EPI_ISL_1932978, EPI_ISL_1932979, EPI_ISL_1932980, EPI_ISL_1932981, EPI_ISL_1932982, EPI_ISL_1932983, EPI_ISL_1932984, EPI_ISL_1932985, EPI_ISL_1932986, EPI_ISL_1932987, EPI_ISL_1932988,
EPI_ISL_1932989, EPI_ISL_1932990, EPI_ISL_1932991, EPI_ISL_1932992, EPI_ISL_1932993, EPI_ISL_1932994, EPI_ISL_1932995, EPI_ISL_1932996, EPI_ISL_1932997, EPI_ISL_1932998, EPI_ISL_1932999, EPI_ISL_1933000, EPI_ISL_1933001, EPI_ISL_1933002, EPI_ISL_1933003, EPI_ISL_1933005, EPI_ISL_1933006, EPI_ISL_1933007, EPI_ISL_1933008, EPI_ISL_1933009, EPI_ISL_1933010, EPI_ISL_1933011,
EPI_ISL_1933012, EPI_ISL_1933013, EPI_ISL_1933014, EPI_ISL_1933015, EPI_ISL_1933016, EPI_ISL_1933017, EPI_ISL_1933018, EPI_ISL_1933019, EPI_ISL_1933020, EPI_ISL_1933021, EPI_ISL_1933022, EPI_ISL_1933023, EPI_ISL_1933024, EPI_ISL_1933025, EPI_ISL_1933026, EPI_ISL_1933027, EPI_ISL_1933028, EPI_ISL_1933029, EPI_ISL_1933030, EPI_ISL_1933031, EPI_ISL_1933032, EPI_ISL_1933033,
EPI_ISL_1933034, EPI_ISL_1933035, EPI_ISL_1933036, EPI_ISL_1933037, EPI_ISL_1933038, EPI_ISL_1933039, EPI_ISL_1933040, EPI_ISL_1933041, EPI_ISL_1933042, EPI_ISL_1933043, EPI_ISL_1933044, EPI_ISL_1933045, EPI_ISL_1933046, EPI_ISL_1933047, EPI_ISL_1933048, EPI_ISL_1933049, EPI_ISL_1933050, EPI_ISL_1933051, EPI_ISL_1933052, EPI_ISL_1933053, EPI_ISL_1933054, EPI_ISL_1933055,
EPI_ISL_1933056, EPI_ISL_1933057, EPI_ISL_1933058, EPI_ISL_1933059, EPI_ISL_1933060, EPI_ISL_1933061, EPI_ISL_1933062, EPI_ISL_1933063, EPI_ISL_1933064, EPI_ISL_1933065, EPI_ISL_1933066, EPI_ISL_1933067, EPI_ISL_1933068, EPI_ISL_1933069, EPI_ISL_1933070, EPI_ISL_1933071, EPI_ISL_1933072, EPI_ISL_1933073, EPI_ISL_1933074, EPI_ISL_1933075, EPI_ISL_1933076, EPI_ISL_1933077,
EPI_ISL_1933078, EPI_ISL_1933079, EPI_ISL_1933080, EPI_ISL_1933081, EPI_ISL_1933082, EPI_ISL_1933083, EPI_ISL_1933084, EPI_ISL_1933085, EPI_ISL_1933086, EPI_ISL_1933087, EPI_ISL_1933088, EPI_ISL_1933089, EPI_ISL_1933090, EPI_ISL_1933091, EPI_ISL_1933092, EPI_ISL_1933093, EPI_ISL_1933094, EPI_ISL_1933095, EPI_ISL_1933096, EPI_ISL_1933097, EPI_ISL_1933098, EPI_ISL_1933099,
EPI_ISL_1933101, EPI_ISL_1933102, EPI_ISL_1933103, EPI_ISL_1933104, EPI_ISL_1933105, EPI_ISL_1933106, EPI_ISL_1933107, EPI_ISL_1933108, EPI_ISL_1933109, EPI_ISL_1933110, EPI_ISL_1933111, EPI_ISL_1933112, EPI_ISL_1933113, EPI_ISL_1933114, EPI_ISL_1933115, EPI_ISL_1933116, EPI_ISL_1933117, EPI_ISL_1933118, EPI_ISL_1933119, EPI_ISL_1933120, EPI_ISL_1933121, EPI_ISL_1933122,
EPI_ISL_1933123, EPI_ISL_1933124, EPI_ISL_1933125, EPI_ISL_1933126, EPI_ISL_1933127, EPI_ISL_1933128, EPI_ISL_1933131, EPI_ISL_1933132, EPI_ISL_1933133, EPI_ISL_1933134, EPI_ISL_1933135, EPI_ISL_1933136, EPI_ISL_1933137, EPI_ISL_1933138, EPI_ISL_1933140, EPI_ISL_1933141, EPI_ISL_1933142, EPI_ISL_1933143, EPI_ISL_1933144, EPI_ISL_1933145, EPI_ISL_1933146, EPI_ISL_1933147,
EPI_ISL_1933216, EPI_ISL_1933217, EPI_ISL_1933218, EPI_ISL_1933219, EPI_ISL_1933220, EPI_ISL_1933221, EPI_ISL_1933222, EPI_ISL_1933223, EPI_ISL_1933224, EPI_ISL_1933225, EPI_ISL_1933226, EPI_ISL_1933227, EPI_ISL_1933228, EPI_ISL_1933229, EPI_ISL_1933230, EPI_ISL_1933231, EPI_ISL_1933232, EPI_ISL_1933233, EPI_ISL_1933234, EPI_ISL_1933235, EPI_ISL_1933236, EPI_ISL_1933237,
EPI_ISL_1933238, EPI_ISL_1933239, EPI_ISL_1933240, EPI_ISL_1933241, EPI_ISL_1933242, EPI_ISL_1933243, EPI_ISL_1933244, EPI_ISL_1933245, EPI_ISL_1933246, EPI_ISL_1933247, EPI_ISL_1933248, EPI_ISL_1933249, EPI_ISL_1933255, EPI_ISL_1933256, EPI_ISL_1933257, EPI_ISL_1933258, EPI_ISL_1933260, EPI_ISL_1933261, EPI_ISL_1933262, EPI_ISL_1933263, EPI_ISL_1933264, EPI_ISL_1933265,
EPI_ISL_1933272, EPI_ISL_1933280, EPI_ISL_1933281, EPI_ISL_1933282, EPI_ISL_1933283, EPI_ISL_1933284, EPI_ISL_1933285, EPI_ISL_1933286, EPI_ISL_1933287, EPI_ISL_1933288, EPI_ISL_1933289, EPI_ISL_1933290, EPI_ISL_1933291, EPI_ISL_1933292, EPI_ISL_1933293, EPI_ISL_1933294, EPI_ISL_1933295, EPI_ISL_1933296, EPI_ISL_1933297, EPI_ISL_1933298, EPI_ISL_1933299, EPI_ISL_1933300,
EPI_ISL_1933301, EPI_ISL_1933302, EPI_ISL_1933303, EPI_ISL_1933304, EPI_ISL_1933305, EPI_ISL_1933306, EPI_ISL_1933307, EPI_ISL_1933308, EPI_ISL_1933309, EPI_ISL_1933310, EPI_ISL_1933311, EPI_ISL_1933312, EPI_ISL_1933313, EPI_ISL_1933314, EPI_ISL_1933315, EPI_ISL_1933316, EPI_ISL_1933317, EPI_ISL_1933318, EPI_ISL_1933319, EPI_ISL_1933320, EPI_ISL_1933321, EPI_ISL_1933322,
EPI_ISL_1933323, EPI_ISL_1933324, EPI_ISL_1933325, EPI_ISL_1933326, EPI_ISL_1933327, EPI_ISL_1933328, EPI_ISL_1933329, EPI_ISL_1933330, EPI_ISL_1933331, EPI_ISL_1933332, EPI_ISL_1933333, EPI_ISL_1933334, EPI_ISL_1933335, EPI_ISL_1933336, EPI_ISL_1933337, EPI_ISL_1933338, EPI_ISL_1933339, EPI_ISL_1933340, EPI_ISL_1933341, EPI_ISL_1933342, EPI_ISL_1933343, EPI_ISL_1933344,
EPI_ISL_1933345, EPI_ISL_1933346, EPI_ISL_1933347, EPI_ISL_1933348, EPI_ISL_1933349, EPI_ISL_1933350, EPI_ISL_1933351, EPI_ISL_1933352, EPI_ISL_1933353, EPI_ISL_1933354, EPI_ISL_1933355, EPI_ISL_1933356, EPI_ISL_1933357, EPI_ISL_1933358, EPI_ISL_1933359, EPI_ISL_1933360, EPI_ISL_1933361, EPI_ISL_1933362, EPI_ISL_1933363, EPI_ISL_1933364, EPI_ISL_1933365, EPI_ISL_1933366,
EPI_ISL_1933367, EPI_ISL_1933368, EPI_ISL_1933369, EPI_ISL_1933370, EPI_ISL_1933371, EPI_ISL_1933372, EPI_ISL_1933373, EPI_ISL_1933374, EPI_ISL_1933375, EPI_ISL_1933376, EPI_ISL_1933377, EPI_ISL_1933378, EPI_ISL_1933379, EPI_ISL_1933380, EPI_ISL_1933381, EPI_ISL_1933388, EPI_ISL_1933389, EPI_ISL_1933390, EPI_ISL_1933391, EPI_ISL_1933392, EPI_ISL_1933393, EPI_ISL_1933394,
EPI_ISL_1933395, EPI_ISL_1933396, EPI_ISL_1933397, EPI_ISL_1933398, EPI_ISL_1933399, EPI_ISL_1933400, EPI_ISL_1933401, EPI_ISL_1933402, EPI_ISL_1933403, EPI_ISL_1933404, EPI_ISL_1933405, EPI_ISL_1933406, EPI_ISL_1933407, EPI_ISL_1933408, EPI_ISL_1933409, EPI_ISL_1933410, EPI_ISL_1933411, EPI_ISL_1933412, EPI_ISL_1933413, EPI_ISL_1933415, EPI_ISL_1933428, EPI_ISL_1933429,
EPI_ISL_1933430, EPI_ISL_1933431, EPI_ISL_1933432, EPI_ISL_1933433, EPI_ISL_1933434, EPI_ISL_1933435, EPI_ISL_1933436, EPI_ISL_1933447, EPI_ISL_1933448, EPI_ISL_1933449, EPI_ISL_1933450, EPI_ISL_1933451, EPI_ISL_1933452, EPI_ISL_1933453, EPI_ISL_1933454, EPI_ISL_1933455, EPI_ISL_1933456, EPI_ISL_1933457, EPI_ISL_1933458, EPI_ISL_1933459, EPI_ISL_1933460, EPI_ISL_1933461,
EPI_ISL_1933462, EPI_ISL_1933463, EPI_ISL_1933465, EPI_ISL_1933466, EPI_ISL_1933467, EPI_ISL_1933468, EPI_ISL_1933469, EPI_ISL_1933470, EPI_ISL_1933471, EPI_ISL_1933472, EPI_ISL_1933473, EPI_ISL_1933474, EPI_ISL_1933475, EPI_ISL_1933476, EPI_ISL_1933477, EPI_ISL_1933478, EPI_ISL_1933504, EPI_ISL_1933505, EPI_ISL_1933506, EPI_ISL_1933507, EPI_ISL_1933508,
EPI_ISL_1933509, EPI_ISL_1933510, EPI_ISL_1933511, EPI_ISL_1933512, EPI_ISL_1933513, EPI_ISL_1933514, EPI_ISL_1933515, EPI_ISL_1933516, EPI_ISL_1933517, EPI_ISL_1933518, EPI_ISL_1933519, EPI_ISL_1933520, EPI_ISL_1933521, EPI_ISL_1933522, EPI_ISL_1933523, EPI_ISL_1933524, EPI_ISL_1933525, EPI_ISL_1933526, EPI_ISL_1933527, EPI_ISL_1933528, EPI_ISL_1933529, EPI_ISL_1933530,
EPI_ISL_1933531, EPI_ISL_1933532, EPI_ISL_1933534, EPI_ISL_1933535, EPI_ISL_1933536, EPI_ISL_1933540, EPI_ISL_1933541, EPI_ISL_1933542, EPI_ISL_1933543, EPI_ISL_1933553, EPI_ISL_1933554, EPI_ISL_1933555, EPI_ISL_1933556, EPI_ISL_1933557, EPI_ISL_1933558, EPI_ISL_1933559, EPI_ISL_1933560, EPI_ISL_1933561, EPI_ISL_1933562, EPI_ISL_1933563, EPI_ISL_1933564,
EPI_ISL_1933565, EPI_ISL_1933566, EPI_ISL_1933567, EPI_ISL_1933568, EPI_ISL_1933569, EPI_ISL_1933580, EPI_ISL_1933581, EPI_ISL_1933582, EPI_ISL_1933583, EPI_ISL_1933584, EPI_ISL_1933585, EPI_ISL_1933586, EPI_ISL_1933587, EPI_ISL_1933588, EPI_ISL_1933589, EPI_ISL_1933590, EPI_ISL_1933591, EPI_ISL_1933592, EPI_ISL_1933593, EPI_ISL_1933594, EPI_ISL_1933595, EPI_ISL_1933596,
EPI_ISL_1933597, EPI_ISL_1933598, EPI_ISL_1933599, EPI_ISL_1933600, EPI_ISL_1933601, EPI_ISL_1933602, EPI_ISL_1933603, EPI_ISL_1933604, EPI_ISL_1933605, EPI_ISL_1933606, EPI_ISL_1933607, EPI_ISL_1933608, EPI_ISL_1933609, EPI_ISL_1933610, EPI_ISL_1933611, EPI_ISL_1933612, EPI_ISL_1933613, EPI_ISL_1933614, EPI_ISL_1933615, EPI_ISL_1933616, EPI_ISL_1933617, EPI_ISL_1933618,
EPI_ISL_1933619, EPI_ISL_1933620, EPI_ISL_1933621, EPI_ISL_1933622, EPI_ISL_1933623, EPI_ISL_1933624, EPI_ISL_1933625, EPI_ISL_1933626, EPI_ISL_1933627, EPI_ISL_1933628, EPI_ISL_1933629, EPI_ISL_1933630, EPI_ISL_1933631, EPI_ISL_1933632, EPI_ISL_1933633, EPI_ISL_1933634, EPI_ISL_1933635, EPI_ISL_1933636, EPI_ISL_1933637, EPI_ISL_1933639, EPI_ISL_1933640, EPI_ISL_1933641,
EPI_ISL_1933642, EPI_ISL_1933643, EPI_ISL_1933644, EPI_ISL_1933651, EPI_ISL_1933652, EPI_ISL_1933653, EPI_ISL_1933654, EPI_ISL_1933655, EPI_ISL_1933656, EPI_ISL_1933657, EPI_ISL_1933658, EPI_ISL_1933659, EPI_ISL_1933660, EPI_ISL_1933661, EPI_ISL_1933662, EPI_ISL_1933663, EPI_ISL_1933664, EPI_ISL_1933665, EPI_ISL_1933666, EPI_ISL_1933667, EPI_ISL_1933668, EPI_ISL_1933669,
EPI_ISL_1933670, EPI_ISL_1933671, EPI_ISL_1933672, EPI_ISL_1933673, EPI_ISL_1933674, EPI_ISL_1933675, EPI_ISL_1933676, EPI_ISL_1933677, EPI_ISL_1933678, EPI_ISL_1933679, EPI_ISL_1933680, EPI_ISL_1933681, EPI_ISL_1933682, EPI_ISL_1933683, EPI_ISL_1933684, EPI_ISL_1933685, EPI_ISL_1933686, EPI_ISL_1933687, EPI_ISL_1933688, EPI_ISL_1933689, EPI_ISL_1933690, EPI_ISL_1933691,
EPI_ISL_1933692, EPI_ISL_1933693, EPI_ISL_1933694, EPI_ISL_1933695, EPI_ISL_1933696, EPI_ISL_1933697, EPI_ISL_1933698, EPI_ISL_1933699, EPI_ISL_1933702, EPI_ISL_1933703, EPI_ISL_1933707, EPI_ISL_1933708, EPI_ISL_1933709, EPI_ISL_1933710, EPI_ISL_1933714, EPI_ISL_1933715, EPI_ISL_1933716, EPI_ISL_1933717, EPI_ISL_1933718, EPI_ISL_1933719, EPI_ISL_1933720,
EPI_ISL_1933721, EPI_ISL_1933722, EPI_ISL_1933723, EPI_ISL_1933724, EPI_ISL_1933736, EPI_ISL_1933737, EPI_ISL_1933738, EPI_ISL_1933739, EPI_ISL_1933740, EPI_ISL_1933741, EPI_ISL_1933742, EPI_ISL_1933743, EPI_ISL_1933744, EPI_ISL_1933745, EPI_ISL_1933746, EPI_ISL_1933747, EPI_ISL_1933748, EPI_ISL_1933753, EPI_ISL_1933754, EPI_ISL_1933755, EPI_ISL_1933756, EPI_ISL_1933757,
EPI_ISL_1933758, EPI_ISL_1933759, EPI_ISL_1933760, EPI_ISL_1933761, EPI_ISL_1933762, EPI_ISL_1933763, EPI_ISL_1933764, EPI_ISL_1933765, EPI_ISL_1933766, EPI_ISL_1933767, EPI_ISL_1933768, EPI_ISL_1933769, EPI_ISL_1933770, EPI_ISL_1933771, EPI_ISL_1933772, EPI_ISL_1933773, EPI_ISL_1933774, EPI_ISL_1933775, EPI_ISL_1933776, EPI_ISL_1933777, EPI_ISL_1933778, EPI_ISL_1933779,
EPI_ISL_1933780, EPI_ISL_1933781, EPI_ISL_1933782, EPI_ISL_1933783, EPI_ISL_1933784, EPI_ISL_1933785, EPI_ISL_1933837, EPI_ISL_1933838, EPI_ISL_1933839, EPI_ISL_1933840, EPI_ISL_1933841, EPI_ISL_1933842, EPI_ISL_1933843, EPI_ISL_1933844, EPI_ISL_1933845, EPI_ISL_1933846, EPI_ISL_1933847, EPI_ISL_1933848, EPI_ISL_1933849, EPI_ISL_1933850, EPI_ISL_1933851, EPI_ISL_1933852,
EPI_ISL_1933853, EPI_ISL_1933854, EPI_ISL_1933855, EPI_ISL_1933856, EPI_ISL_1933857, EPI_ISL_1933858, EPI_ISL_1933859, EPI_ISL_1933860, EPI_ISL_1933861, EPI_ISL_1933862, EPI_ISL_1933863, EPI_ISL_1933864, EPI_ISL_1933865, EPI_ISL_1933866, EPI_ISL_1933867, EPI_ISL_1933868, EPI_ISL_1933869, EPI_ISL_1933870, EPI_ISL_1933871, EPI_ISL_1933872, EPI_ISL_1933873, EPI_ISL_1933874,
EPI_ISL_1933875, EPI_ISL_1933876, EPI_ISL_1933877, EPI_ISL_1933878, EPI_ISL_1933879, EPI_ISL_1933880, EPI_ISL_1933881, EPI_ISL_1933882, EPI_ISL_1933883, EPI_ISL_1933884, EPI_ISL_1933885, EPI_ISL_1933886, EPI_ISL_1933887, EPI_ISL_1933888, EPI_ISL_1933889, EPI_ISL_1933890, EPI_ISL_1933891, EPI_ISL_1933892, EPI_ISL_1933893, EPI_ISL_1933894, EPI_ISL_1933895, EPI_ISL_1933896,
EPI_ISL_1933897, EPI_ISL_1933898, EPI_ISL_1933899, EPI_ISL_1933900, EPI_ISL_1933901, EPI_ISL_1933902, EPI_ISL_1933903, EPI_ISL_1933904, EPI_ISL_1933905, EPI_ISL_1933906, EPI_ISL_1933907, EPI_ISL_1933908, EPI_ISL_1933909, EPI_ISL_1933910, EPI_ISL_1933911, EPI_ISL_1933912, EPI_ISL_1933913, EPI_ISL_1933914, EPI_ISL_1933915, EPI_ISL_1933916, EPI_ISL_1933917, EPI_ISL_1933918,
EPI_ISL_1933919, EPI_ISL_1933920, EPI_ISL_1933921, EPI_ISL_1933922, EPI_ISL_1933923, EPI_ISL_1933924, EPI_ISL_1933925, EPI_ISL_1933926, EPI_ISL_1933927, EPI_ISL_1933928, EPI_ISL_1933929, EPI_ISL_1933930, EPI_ISL_1933931, EPI_ISL_1933932, EPI_ISL_1933938, EPI_ISL_1933939, EPI_ISL_1933940, EPI_ISL_1933941, EPI_ISL_1933942, EPI_ISL_1933943, EPI_ISL_1933944, EPI_ISL_1933945,
EPI_ISL_1933946, EPI_ISL_1933947, EPI_ISL_1933948, EPI_ISL_1933949, EPI_ISL_1933950, EPI_ISL_1933951, EPI_ISL_1933952, EPI_ISL_1933953, EPI_ISL_1933954, EPI_ISL_1933955, EPI_ISL_1933956, EPI_ISL_1933957, EPI_ISL_1933959, EPI_ISL_1933960, EPI_ISL_1933961, EPI_ISL_1933962, EPI_ISL_1933963, EPI_ISL_1933964, EPI_ISL_1933965, EPI_ISL_1933966, EPI_ISL_1933967,
EPI_ISL_1933968, EPI_ISL_1933969, EPI_ISL_1933970, EPI_ISL_1933971, EPI_ISL_1933972, EPI_ISL_1933973, EPI_ISL_1933974, EPI_ISL_1933975, EPI_ISL_1933976, EPI_ISL_1933977, EPI_ISL_1933978, EPI_ISL_1933979, EPI_ISL_1933980, EPI_ISL_1933981, EPI_ISL_1933982, EPI_ISL_1933983, EPI_ISL_1933984, EPI_ISL_1933985, EPI_ISL_1933986, EPI_ISL_1933987, EPI_ISL_1933988, EPI_ISL_1933989,
EPI_ISL_1933990, EPI_ISL_1933991, EPI_ISL_1933992, EPI_ISL_1933993, EPI_ISL_1933994, EPI_ISL_1933995, EPI_ISL_1933996, EPI_ISL_1933997, EPI_ISL_1933998, EPI_ISL_1933999, EPI_ISL_1934000, EPI_ISL_1934001, EPI_ISL_1934002, EPI_ISL_1934003, EPI_ISL_1934004, EPI_ISL_1934005, EPI_ISL_1934006, EPI_ISL_1934007, EPI_ISL_1934008, EPI_ISL_1934009, EPI_ISL_1934010, EPI_ISL_1934011,
EPI_ISL_1934012, EPI_ISL_1934013, EPI_ISL_1934014, EPI_ISL_1934015, EPI_ISL_1934016, EPI_ISL_1934017, EPI_ISL_1934018, EPI_ISL_1934019, EPI_ISL_1934020, EPI_ISL_1934021, EPI_ISL_1934022, EPI_ISL_1934023, EPI_ISL_1934024, EPI_ISL_1934025, EPI_ISL_1934026, EPI_ISL_1934027, EPI_ISL_1934028, EPI_ISL_1934029, EPI_ISL_1934030, EPI_ISL_1934031, EPI_ISL_1934032, EPI_ISL_1934033,
EPI_ISL_1934034, EPI_ISL_1934035, EPI_ISL_1934036, EPI_ISL_1934037, EPI_ISL_1934038, EPI_ISL_1934039, EPI_ISL_1934040, EPI_ISL_1934041, EPI_ISL_1934042, EPI_ISL_1934043, EPI_ISL_1934044, EPI_ISL_1934045, EPI_ISL_1934046, EPI_ISL_1934047, EPI_ISL_1934048, EPI_ISL_1934049, EPI_ISL_1934050, EPI_ISL_1934051, EPI_ISL_1934052, EPI_ISL_1934053, EPI_ISL_1934054, EPI_ISL_1934055,
EPI_ISL_1934056, EPI_ISL_1934057, EPI_ISL_1934058, EPI_ISL_1934059, EPI_ISL_1934060, EPI_ISL_1934061, EPI_ISL_1934062, EPI_ISL_1934063, EPI_ISL_1934064, EPI_ISL_1934065, EPI_ISL_1934066, EPI_ISL_1934067, EPI_ISL_1934068, EPI_ISL_1934069, EPI_ISL_1934070, EPI_ISL_1934071, EPI_ISL_1934072, EPI_ISL_1934073, EPI_ISL_1934074, EPI_ISL_1934075, EPI_ISL_1934076, EPI_ISL_1934077,
EPI_ISL_1934078, EPI_ISL_1934079, EPI_ISL_1934080, EPI_ISL_1934081, EPI_ISL_1934082, EPI_ISL_1934083, EPI_ISL_1934084, EPI_ISL_1934085, EPI_ISL_1934086, EPI_ISL_1934087, EPI_ISL_1934088, EPI_ISL_1934089, EPI_ISL_1934091, EPI_ISL_1934092, EPI_ISL_1934093, EPI_ISL_1934094, EPI_ISL_1934095, EPI_ISL_1934096, EPI_ISL_1934097, EPI_ISL_1934098, EPI_ISL_1934099, EPI_ISL_1934100,
EPI_ISL_1934101, EPI_ISL_1934102, EPI_ISL_1934103, EPI_ISL_1934104, EPI_ISL_1934109, EPI_ISL_1934110, EPI_ISL_1934111, EPI_ISL_1934112, EPI_ISL_1934113, EPI_ISL_1934114, EPI_ISL_1934115, EPI_ISL_1934116, EPI_ISL_1934117, EPI_ISL_1934118, EPI_ISL_1934119, EPI_ISL_1934120, EPI_ISL_1934121, EPI_ISL_1934122, EPI_ISL_1934123, EPI_ISL_1934124, EPI_ISL_1934125, EPI_ISL_1934126,
EPI_ISL_1934127, EPI_ISL_1934128, EPI_ISL_1934129, EPI_ISL_1934130, EPI_ISL_1934131, EPI_ISL_1934132, EPI_ISL_1934133, EPI_ISL_1934134, EPI_ISL_1934135, EPI_ISL_1934136, EPI_ISL_1934137, EPI_ISL_1934138, EPI_ISL_1934139, EPI_ISL_1934140, EPI_ISL_1934141, EPI_ISL_1934142, EPI_ISL_1934143, EPI_ISL_1934144, EPI_ISL_1934145, EPI_ISL_1934146, EPI_ISL_1934147, EPI_ISL_1934148,
EPI_ISL_1934149, EPI_ISL_1934150, EPI_ISL_1934151, EPI_ISL_1934152, EPI_ISL_1934153, EPI_ISL_1934154, EPI_ISL_1934155, EPI_ISL_1934156, EPI_ISL_1934157, EPI_ISL_1934158, EPI_ISL_1934159, EPI_ISL_1934160, EPI_ISL_1934161, EPI_ISL_1934162, EPI_ISL_1934163, EPI_ISL_1934164, EPI_ISL_1934165, EPI_ISL_1934166, EPI_ISL_1934167, EPI_ISL_1934168, EPI_ISL_1934169, EPI_ISL_1934170,
EPI_ISL_1934171, EPI_ISL_1934172, EPI_ISL_1934173, EPI_ISL_1934174, EPI_ISL_1934175, EPI_ISL_1934176, EPI_ISL_1934177, EPI_ISL_1934178, EPI_ISL_1934179, EPI_ISL_1934180, EPI_ISL_1934181, EPI_ISL_1934182, EPI_ISL_1934183, EPI_ISL_1934184, EPI_ISL_1934185, EPI_ISL_1934186, EPI_ISL_1934187, EPI_ISL_1934188, EPI_ISL_1934189, EPI_ISL_1934190, EPI_ISL_1934191, EPI_ISL_1934192,
EPI_ISL_1934193, EPI_ISL_1934194, EPI_ISL_1934195, EPI_ISL_1934196, EPI_ISL_1934197, EPI_ISL_1934198, EPI_ISL_1934199, EPI_ISL_1934200, EPI_ISL_1934201, EPI_ISL_1934202, EPI_ISL_1934203, EPI_ISL_1934204, EPI_ISL_1934205, EPI_ISL_1934238, EPI_ISL_1934239, EPI_ISL_1934240, EPI_ISL_1934241, EPI_ISL_1934242, EPI_ISL_1934243, EPI_ISL_1934244, EPI_ISL_1934245, EPI_ISL_1934246,
EPI_ISL_1934247, EPI_ISL_1934248, EPI_ISL_1934249, EPI_ISL_1934250, EPI_ISL_1934251, EPI_ISL_1934252, EPI_ISL_1934253, EPI_ISL_1934254, EPI_ISL_1934255, EPI_ISL_1934256, EPI_ISL_1934257, EPI_ISL_1934258, EPI_ISL_1934259, EPI_ISL_1934260, EPI_ISL_1934261, EPI_ISL_1934262, EPI_ISL_1934263, EPI_ISL_1934264, EPI_ISL_1934265, EPI_ISL_1934266, EPI_ISL_1934267, EPI_ISL_1934268,
EPI_ISL_1934269, EPI_ISL_1934270, EPI_ISL_1934271, EPI_ISL_1934272, EPI_ISL_1934273, EPI_ISL_1934274, EPI_ISL_1934275, EPI_ISL_1934276, EPI_ISL_1934277, EPI_ISL_1934278, EPI_ISL_1934279, EPI_ISL_1934280, EPI_ISL_1934281, EPI_ISL_1934282, EPI_ISL_1934283, EPI_ISL_1934302, EPI_ISL_1934303, EPI_ISL_1934304, EPI_ISL_1934305, EPI_ISL_1934306, EPI_ISL_1934307, EPI_ISL_1934308,
EPI_ISL_1934309, EPI_ISL_1934310, EPI_ISL_1934311, EPI_ISL_1934312, EPI_ISL_1934313, EPI_ISL_1934314, EPI_ISL_1934315, EPI_ISL_1934316, EPI_ISL_1934317, EPI_ISL_1934318, EPI_ISL_1934319, EPI_ISL_1934320, EPI_ISL_1934321, EPI_ISL_1934322, EPI_ISL_1934323, EPI_ISL_1934324, EPI_ISL_1934325, EPI_ISL_1934326, EPI_ISL_1934327, EPI_ISL_1934328, EPI_ISL_1934329, EPI_ISL_1934330,
EPI_ISL_1934331, EPI_ISL_1934332, EPI_ISL_1934333, EPI_ISL_1934334, EPI_ISL_1934335, EPI_ISL_1934336, EPI_ISL_1934337, EPI_ISL_1934338, EPI_ISL_1934339, EPI_ISL_1934340, EPI_ISL_1934341, EPI_ISL_1934342, EPI_ISL_1934343, EPI_ISL_1934344, EPI_ISL_1934345, EPI_ISL_1934346, EPI_ISL_1934347, EPI_ISL_1934348, EPI_ISL_1934349, EPI_ISL_1934350, EPI_ISL_1934351, EPI_ISL_1934352,
EPI_ISL_1934353, EPI_ISL_1934354, EPI_ISL_1934355, EPI_ISL_1934356, EPI_ISL_1934357, EPI_ISL_1934358, EPI_ISL_1934359, EPI_ISL_1934360, EPI_ISL_1934361, EPI_ISL_1934362, EPI_ISL_1934363, EPI_ISL_1934364, EPI_ISL_1934365, EPI_ISL_1934366, EPI_ISL_1934367, EPI_ISL_1934368, EPI_ISL_1934369, EPI_ISL_1934370, EPI_ISL_1934371, EPI_ISL_1934372, EPI_ISL_1934373, EPI_ISL_1934374,
EPI_ISL_1934375, EPI_ISL_1934376, EPI_ISL_1934377, EPI_ISL_1934378, EPI_ISL_1934379, EPI_ISL_1934380, EPI_ISL_1934381, EPI_ISL_1934382, EPI_ISL_1934383, EPI_ISL_1934384, EPI_ISL_1934385, EPI_ISL_1934386, EPI_ISL_1934387, EPI_ISL_1934388, EPI_ISL_1934389, EPI_ISL_1934390, EPI_ISL_1934391, EPI_ISL_1934392, EPI_ISL_1934393, EPI_ISL_1934394, EPI_ISL_1934395, EPI_ISL_1934396,
EPI_ISL_1934397, EPI_ISL_1934398, EPI_ISL_1934399, EPI_ISL_1934400, EPI_ISL_1934401, EPI_ISL_1934402, EPI_ISL_1934403, EPI_ISL_1934404, EPI_ISL_1934405, EPI_ISL_1934406, EPI_ISL_1934407, EPI_ISL_1934410, EPI_ISL_1934412, EPI_ISL_1934413, EPI_ISL_1934414, EPI_ISL_1934415, EPI_ISL_1934416, EPI_ISL_1934417, EPI_ISL_1934418, EPI_ISL_1934419, EPI_ISL_1934420, EPI_ISL_1934421,

| | | | |
|---|---|---|---|
| EPI_ISL_1934422, EPI_ISL_1934423, EPI_ISL_1934424, EPI_ISL_1934425, EPI_ISL_1934427, EPI_ISL_1934428, EPI_ISL_1934430, EPI_ISL_1934431, EPI_ISL_1934432, EPI_ISL_1934433, EPI_ISL_1934434, EPI_ISL_1934435, EPI_ISL_1934436, EPI_ISL_1934437, EPI_ISL_1934438, EPI_ISL_1934439, EPI_ISL_1934440, EPI_ISL_1934441, EPI_ISL_1934442, EPI_ISL_1934443, EPI_ISL_1934444, EPI_ISL_1934445, EPI_ISL_1934456, EPI_ISL_1934457, EPI_ISL_1934458, EPI_ISL_1934459, EPI_ISL_1934460, EPI_ISL_1934461, EPI_ISL_1934462, EPI_ISL_1934463, EPI_ISL_1934464, EPI_ISL_1934465, EPI_ISL_1934466, EPI_ISL_1934467, EPI_ISL_1934468, EPI_ISL_1934469, EPI_ISL_1934470, EPI_ISL_1934471, EPI_ISL_1934472, EPI_ISL_1934473, EPI_ISL_1934474, EPI_ISL_1934475, EPI_ISL_1934476, EPI_ISL_1934477, EPI_ISL_1934478, EPI_ISL_1934479, EPI_ISL_1934480, EPI_ISL_1934481, EPI_ISL_1934482, EPI_ISL_1934484, EPI_ISL_1934485, EPI_ISL_1934486, EPI_ISL_1934487, EPI_ISL_1934488, EPI_ISL_1934489, EPI_ISL_1934490, EPI_ISL_1934491, EPI_ISL_1934492, EPI_ISL_1934493, EPI_ISL_1934494, EPI_ISL_1934495, EPI_ISL_1934496, EPI_ISL_1934497, EPI_ISL_1934498, EPI_ISL_1934499, EPI_ISL_1934500, EPI_ISL_1934501, EPI_ISL_1934502, EPI_ISL_1934503, EPI_ISL_1934504, EPI_ISL_1934505, EPI_ISL_1934506, EPI_ISL_1934507, EPI_ISL_1934508, EPI_ISL_1934509, EPI_ISL_1934510, EPI_ISL_1934511, EPI_ISL_1934523, EPI_ISL_1934524, EPI_ISL_1934525, EPI_ISL_1934526, EPI_ISL_1934527, EPI_ISL_1934528, EPI_ISL_1934529, EPI_ISL_1934530, EPI_ISL_1934531, EPI_ISL_1934532, EPI_ISL_1934533, EPI_ISL_1934534, EPI_ISL_1934535, EPI_ISL_1934536, EPI_ISL_1934537, EPI_ISL_1934538, EPI_ISL_1934539, EPI_ISL_1934540, EPI_ISL_1934541, EPI_ISL_1934542, EPI_ISL_1934543, EPI_ISL_1934544, EPI_ISL_1934545, EPI_ISL_1934546, EPI_ISL_1934547, EPI_ISL_1934548, EPI_ISL_1934549, EPI_ISL_1934550, EPI_ISL_1934551, EPI_ISL_1934552, EPI_ISL_1934553, EPI_ISL_1934554, EPI_ISL_1934555, EPI_ISL_1934556, EPI_ISL_1934557, EPI_ISL_1934558, EPI_ISL_1934559, EPI_ISL_1934560, EPI_ISL_1934561, EPI_ISL_1934562, EPI_ISL_1934563, EPI_ISL_1934564, EPI_ISL_1934565, EPI_ISL_1934566, EPI_ISL_1934567, EPI_ISL_1934568, EPI_ISL_1934569, EPI_ISL_1934570, EPI_ISL_1934571, EPI_ISL_1934572, EPI_ISL_1934573, EPI_ISL_1934574, EPI_ISL_1934575, EPI_ISL_1934576, EPI_ISL_1934577, EPI_ISL_1934578, EPI_ISL_1934579, EPI_ISL_1934580, EPI_ISL_1934581, EPI_ISL_1934582, EPI_ISL_1934583, EPI_ISL_1934584, EPI_ISL_1934585, EPI_ISL_1934586, EPI_ISL_1934587, EPI_ISL_1934588, EPI_ISL_1934589, EPI_ISL_1934590, EPI_ISL_1934591, EPI_ISL_1934592, EPI_ISL_1934593, EPI_ISL_1934632, EPI_ISL_1934633, EPI_ISL_1934634, EPI_ISL_1934635, EPI_ISL_1934636, EPI_ISL_1934637, EPI_ISL_1934638, EPI_ISL_1934639, EPI_ISL_1934640, EPI_ISL_1934641, EPI_ISL_1963571, EPI_ISL_1963572, EPI_ISL_1963573 | | | |
| see above | Pathogen Genomics Center, National Institute of Infectious Diseases | Pathogen Genomics Center, National Institute of Infectious Diseases | Ai Kawana-Tachikawa; Chang-Kweng Lim; Eri Nakayama; Hazuka Y Furihata; Hidemasa Izumiya; Hiromizu Takahashi; Ken Shimuta; Ken-ichi Lee; Kentaro Itokawa; Koichi Ishikawa; Makoto Kuroda; Masanori Hashino; Masatomo Morita; Masumichi Saito; Midori Nakamura-Hoshi; Motohiko Ogawa; Naomi Nojiri; Nobuo Koizumi; Nozomu Hanaoka; Rina Tanaka; Sana Uchikoba; Sayuri Seki; Shigeru Kusagawa; Shigeru Tajima; Shouji Yamamoto; Sunao Iyoda; Takahiro Maeki; Tsuguto Fujimoto; Tsuyoshi Sekizuka |
| EPI_ISL_1198843, EPI_ISL_1198847, EPI_ISL_1425573, EPI_ISL_1425574, EPI_ISL_1425575, EPI_ISL_1425576, EPI_ISL_1425605, EPI_ISL_1425606, EPI_ISL_1425607, EPI_ISL_1425608, EPI_ISL_1425609, EPI_ISL_1425610, EPI_ISL_1425611, EPI_ISL_1425612, EPI_ISL_1425613, EPI_ISL_1425617, EPI_ISL_1425618, EPI_ISL_1425619, EPI_ISL_1425699, EPI_ISL_1425712, EPI_ISL_1425714, EPI_ISL_1425715, EPI_ISL_1425716, EPI_ISL_1425720, EPI_ISL_1425749, EPI_ISL_1425750, EPI_ISL_1425751, EPI_ISL_1430584, EPI_ISL_1430631, EPI_ISL_1430634, EPI_ISL_1430640, EPI_ISL_1430641, EPI_ISL_1430642, EPI_ISL_1430643, EPI_ISL_1430644, EPI_ISL_1430645, EPI_ISL_1430646, EPI_ISL_1430647, EPI_ISL_1430648, EPI_ISL_1430649, EPI_ISL_1430650, EPI_ISL_1430651, EPI_ISL_1430652, EPI_ISL_1430658, EPI_ISL_1430659, EPI_ISL_1430665, EPI_ISL_1430666, EPI_ISL_1430737, EPI_ISL_1430825, EPI_ISL_1430826, EPI_ISL_1430827, EPI_ISL_1430828, EPI_ISL_1430868, EPI_ISL_1430873, EPI_ISL_1430874, EPI_ISL_1431578, EPI_ISL_1433306, EPI_ISL_1926885, EPI_ISL_1926887, EPI_ISL_1926888, EPI_ISL_1926890, EPI_ISL_1926891, EPI_ISL_1926893, EPI_ISL_1926894, EPI_ISL_1926896, EPI_ISL_1926898, EPI_ISL_1926899, EPI_ISL_1926901, EPI_ISL_1926902, EPI_ISL_1926903, EPI_ISL_1926905, EPI_ISL_1926906, EPI_ISL_1926908, EPI_ISL_1926909, EPI_ISL_1926911, EPI_ISL_1926912, EPI_ISL_1926914, EPI_ISL_1926915, EPI_ISL_1926917, EPI_ISL_1926918, EPI_ISL_1926919, EPI_ISL_1926921, EPI_ISL_1926922, EPI_ISL_1926924, EPI_ISL_1926925, EPI_ISL_1926927, EPI_ISL_1926929, EPI_ISL_1926930, EPI_ISL_1926931, EPI_ISL_1926933, EPI_ISL_1926934, EPI_ISL_1926936, EPI_ISL_1926937, EPI_ISL_1926939, EPI_ISL_1926941, EPI_ISL_1926942, EPI_ISL_1926944, EPI_ISL_1926945, EPI_ISL_1926946, EPI_ISL_1926948, EPI_ISL_1926949, EPI_ISL_1926951, EPI_ISL_1926952, EPI_ISL_1926954, EPI_ISL_1926955, EPI_ISL_1926957, EPI_ISL_1926958, EPI_ISL_1926960, EPI_ISL_1926961, EPI_ISL_1926962, EPI_ISL_1926963, EPI_ISL_1926964, EPI_ISL_1926965, EPI_ISL_1926966, EPI_ISL_1926967, EPI_ISL_1927104, EPI_ISL_1927105, EPI_ISL_1927107, EPI_ISL_1927108, EPI_ISL_1927110, EPI_ISL_1927111, EPI_ISL_1927113, EPI_ISL_1927114, EPI_ISL_1927116, EPI_ISL_1927117, EPI_ISL_1927119, EPI_ISL_1927120, EPI_ISL_1927122, EPI_ISL_1927123, EPI_ISL_1927125, EPI_ISL_1927126, EPI_ISL_1927127, EPI_ISL_1927129, EPI_ISL_1927130, EPI_ISL_1927132, EPI_ISL_1927133, EPI_ISL_1927135, EPI_ISL_1927136, EPI_ISL_1927138, EPI_ISL_1927139, EPI_ISL_1927141, EPI_ISL_1927143, EPI_ISL_1927144, EPI_ISL_1927145, EPI_ISL_1927147, EPI_ISL_1927148, EPI_ISL_1927150, EPI_ISL_1927151, EPI_ISL_1927153, EPI_ISL_1927154, EPI_ISL_1927156, EPI_ISL_1927157, EPI_ISL_1927159, EPI_ISL_1927160, EPI_ISL_1927162, EPI_ISL_1927163, EPI_ISL_1927165, EPI_ISL_1927166, EPI_ISL_1927168, EPI_ISL_1927169, EPI_ISL_1927171, EPI_ISL_1927172, EPI_ISL_1927174, EPI_ISL_1927175, EPI_ISL_1927177, EPI_ISL_1927178, EPI_ISL_1927180, EPI_ISL_1927181, EPI_ISL_1927183, EPI_ISL_1927184, EPI_ISL_1927218, EPI_ISL_1927220, EPI_ISL_1927224, EPI_ISL_1927226, EPI_ISL_1927227, EPI_ISL_1927230, EPI_ISL_1927232, EPI_ISL_1927233, EPI_ISL_1927235, EPI_ISL_1927238, EPI_ISL_1927239, EPI_ISL_1927276, EPI_ISL_1927278, EPI_ISL_1927279, EPI_ISL_1927280, EPI_ISL_1927282, EPI_ISL_1927283, EPI_ISL_1927285, EPI_ISL_1927289, EPI_ISL_1927291, EPI_ISL_1927297, EPI_ISL_1927299, EPI_ISL_1927300, EPI_ISL_1927302, EPI_ISL_1927303, EPI_ISL_1927305, EPI_ISL_1927306, EPI_ISL_1927308, EPI_ISL_1927325, EPI_ISL_1927345, EPI_ISL_1927346, EPI_ISL_1927348, EPI_ISL_1927358, EPI_ISL_1927360, EPI_ISL_1927361, EPI_ISL_1927362, EPI_ISL_1927364, EPI_ISL_1927365, EPI_ISL_1927367, EPI_ISL_1927368, EPI_ISL_1927370, EPI_ISL_1927371, EPI_ISL_1927373, EPI_ISL_1927374, EPI_ISL_1927376, EPI_ISL_1927377, EPI_ISL_1927378, EPI_ISL_1927380, EPI_ISL_1927381, EPI_ISL_1927383, EPI_ISL_1927384, EPI_ISL_1927386, EPI_ISL_1927388, EPI_ISL_1927389, EPI_ISL_1927391, EPI_ISL_1927392, EPI_ISL_1927394, EPI_ISL_1927397, EPI_ISL_1927400, EPI_ISL_1927401, EPI_ISL_1927403, EPI_ISL_1927404, EPI_ISL_1927405, EPI_ISL_1927407, EPI_ISL_1927408, EPI_ISL_1927410, EPI_ISL_1927411, EPI_ISL_1927413, EPI_ISL_1927414, EPI_ISL_1927416, EPI_ISL_1927417, EPI_ISL_1927419, EPI_ISL_1927420, EPI_ISL_1927422, EPI_ISL_1927423, EPI_ISL_1927424, EPI_ISL_1927426, EPI_ISL_1927427, EPI_ISL_1927429, EPI_ISL_1927432, EPI_ISL_1927433, EPI_ISL_1927435, EPI_ISL_1927436, EPI_ISL_1927438, EPI_ISL_1927439, EPI_ISL_1927441, EPI_ISL_1927442, EPI_ISL_1927444, EPI_ISL_1927445, EPI_ISL_1927811, EPI_ISL_1927812, EPI_ISL_1927815, EPI_ISL_1927828, EPI_ISL_1927830, EPI_ISL_1927831, EPI_ISL_1927833, EPI_ISL_1927834, EPI_ISL_1927836, EPI_ISL_1927837, EPI_ISL_1927839, EPI_ISL_1927840, EPI_ISL_1927842, EPI_ISL_1927843, EPI_ISL_1927845, EPI_ISL_1927846, EPI_ISL_1927848, EPI_ISL_1929787, EPI_ISL_1929789, EPI_ISL_1929790, EPI_ISL_1929792, EPI_ISL_1929793, EPI_ISL_1931782, EPI_ISL_2131621, EPI_ISL_2131622, EPI_ISL_2131623, EPI_ISL_2131624, EPI_ISL_2131625, EPI_ISL_2131626, EPI_ISL_2131627, EPI_ISL_2131628, EPI_ISL_2131629, EPI_ISL_2131631, EPI_ISL_2131635, EPI_ISL_2131636, EPI_ISL_2131637, EPI_ISL_2131638, EPI_ISL_2131639, EPI_ISL_2131640, EPI_ISL_2131641, EPI_ISL_2131642, EPI_ISL_2131643, EPI_ISL_2131644, EPI_ISL_2131645, EPI_ISL_2131646, EPI_ISL_2131647, EPI_ISL_2131648, EPI_ISL_2131649, EPI_ISL_2131656, EPI_ISL_2131658, EPI_ISL_2131659, EPI_ISL_2131660, EPI_ISL_2131662, EPI_ISL_2131667, EPI_ISL_2131669, EPI_ISL_2131670, EPI_ISL_2131671, EPI_ISL_2131674, EPI_ISL_2131676, EPI_ISL_2131677, EPI_ISL_2131678, EPI_ISL_2131679, EPI_ISL_2131680, EPI_ISL_2131681, EPI_ISL_2131682, EPI_ISL_2131683, EPI_ISL_2131685, EPI_ISL_2131686, EPI_ISL_2131691, EPI_ISL_2131692, EPI_ISL_2131693, EPI_ISL_2131694, EPI_ISL_2131695, EPI_ISL_2131696, EPI_ISL_2131697, EPI_ISL_2131698, EPI_ISL_2131699, EPI_ISL_2131700, EPI_ISL_2131701, EPI_ISL_2131702, EPI_ISL_2131703, EPI_ISL_2131704, EPI_ISL_2131705, EPI_ISL_2131706, EPI_ISL_2131707, EPI_ISL_2131708, EPI_ISL_2131709, EPI_ISL_2131710, EPI_ISL_2131711, EPI_ISL_2131712, EPI_ISL_2131713, EPI_ISL_2131714, EPI_ISL_2131715, EPI_ISL_2131716, EPI_ISL_2131717, EPI_ISL_2131718, EPI_ISL_2131719, EPI_ISL_2131730, EPI_ISL_2131734, EPI_ISL_2131735, EPI_ISL_2131790, EPI_ISL_2131791, EPI_ISL_2131792, EPI_ISL_2131797, EPI_ISL_2131798, EPI_ISL_2131799, EPI_ISL_2131800, EPI_ISL_2131801, EPI_ISL_2131802 | | | |
| see above | SARS-CoV-2 testing team, National Institute of Infectious Diseases | Pathogen Genomics Center, National Institute of Infectious Diseases | Ai Kawana-Tachikawa; Chang-Kweng Lim; Chikako Shimokawa; Eri Nakayama; Eunsil Park; Hazuka Y Furihata; Hazuka Yoshida; Hidemasa Izumiya; Hiromizu Takahashi; Ken Miyazawa; Ken Shimuta; Ken-ichi Lee; Kentaro Itokawa; Koichi Ishikawa; Makoto Kuroda; Masanori Hashino; Masatomo Morita; Masumichi Saito; Midori Nakamura-Hoshi; Minoru Nagi; Motohiko Ogawa; Naomi Nojiri; Nobuo Koizumi; Nozomu Hanaoka; Rina Tanaka; Sana Uchikoba; Sayuri Seki; Shigeru Kusagawa; Shigeru Tanjima; Shouji Yamamoto; Sunao Iyoda; Takahiro Maeki; Takashi Sakudoh; Tsuguto Fujimoto; Tsuyoshi Sekizuka; Yasutaka Hoshino; Yoshihiro Kaku |
| EPI_ISL_1430698, EPI_ISL_1437618, EPI_ISL_2029031, EPI_ISL_2029032, EPI_ISL_2029033, EPI_ISL_2029034, EPI_ISL_2029035, EPI_ISL_2029036, EPI_ISL_2029037, EPI_ISL_2029038, EPI_ISL_2029039, EPI_ISL_2029040, EPI_ISL_2029041, EPI_ISL_2029042, EPI_ISL_2029043, EPI_ISL_2029044, EPI_ISL_2029045, EPI_ISL_2029046, EPI_ISL_2029047, EPI_ISL_2029048, EPI_ISL_2029049, EPI_ISL_2029050, EPI_ISL_2029051, EPI_ISL_2029052, EPI_ISL_2029053, EPI_ISL_2029054, EPI_ISL_2029055, EPI_ISL_2029056, EPI_ISL_2029057, EPI_ISL_2029058, EPI_ISL_2029059, EPI_ISL_2029060, EPI_ISL_2029061, EPI_ISL_2029062, EPI_ISL_2029063, EPI_ISL_2029064, EPI_ISL_2029065, EPI_ISL_2029069, EPI_ISL_2029070, EPI_ISL_2029071, EPI_ISL_2029072, EPI_ISL_2029073, EPI_ISL_2029074 | | | |
| see above | Saitama Prefectural Institute of Public Health | Pathogen Genomics Center, National Institute of Infectious Diseases | Chang-Kweng Lim; Eri Nakayama; Kentaro Itokawa; Makoto Kuroda; Masanori Hashino; Motohiko Ogawa; Rina Tanaka; Shigeru Tanjima; Takahiro Maeki; Tsuyoshi Sekizuka |
| EPI_ISL_1929253, EPI_ISL_1929254, EPI_ISL_1929420, EPI_ISL_1929422, EPI_ISL_1929891, EPI_ISL_1929893, EPI_ISL_1929894, EPI_ISL_1930389, EPI_ISL_1930390, EPI_ISL_1930392, EPI_ISL_1930891, EPI_ISL_1930893, EPI_ISL_1931360, EPI_ISL_1931531, EPI_ISL_1931533, EPI_ISL_1931535, EPI_ISL_1933481, EPI_ISL_1933482, EPI_ISL_1933483, EPI_ISL_1933484, EPI_ISL_1933485, EPI_ISL_1933486, EPI_ISL_1933705, EPI_ISL_1933706 | | | |
| see above | Sakai City Institute of Public Health | Pathogen Genomics Center, National Institute of Infectious Diseases | Hazuka Y Furihata; Hidemasa Izumiya; Hiromizu Takahashi; Ken-ichi Lee; Kentaro Itokawa; Makoto Kuroda; Masanori Hashino; Masatomo Morita; Masumichi Saito; Naomi Nojiri; Nobuo Koizumi; Nozomu Hanaoka; Rina Tanaka; Sana Uchikoba; Shouji Yamamoto; Sunao Iyoda; Tsuguto Fujimoto; Tsuyoshi Sekizuka |
| EPI_ISL_1971002, EPI_ISL_1971003, EPI_ISL_1971004, EPI_ISL_1971005, EPI_ISL_1971006, EPI_ISL_1971007, EPI_ISL_1971008, EPI_ISL_1971009, EPI_ISL_1971010, EPI_ISL_1971011, EPI_ISL_1971012, EPI_ISL_1971013, EPI_ISL_1971014, EPI_ISL_1971015, EPI_ISL_1971016, EPI_ISL_1971017, EPI_ISL_1971018, EPI_ISL_1971019, EPI_ISL_1971020, EPI_ISL_1971021, EPI_ISL_1971022, EPI_ISL_1971023, EPI_ISL_1971024, EPI_ISL_1971025, EPI_ISL_1971026, EPI_ISL_1971027, EPI_ISL_1971028, EPI_ISL_1971029, EPI_ISL_1971030, EPI_ISL_1971031, EPI_ISL_1971032, EPI_ISL_1971033 | | | |
| see above | Sapporo City Institute of Public Health | Sapporo City Institute of Public Health | Asami Ohnishi; Kentaro Itokawa; Makoto Kuroda; Masanori Hashino; Masayuki Kikuchi; Rie Shimazaki; Rina Tanaka; Tsuyoshi Sekizuka; Yusuke Oguchi |
| EPI_ISL_1761362, EPI_ISL_1761364, EPI_ISL_1761365, EPI_ISL_1761366, EPI_ISL_1761367, EPI_ISL_1761368, EPI_ISL_1761369 | | | |
| see above | Tokyo Medical and Dental University | Tokyo Medical and Dental University | Akinori Kimura; Hiroaki Takeuchi; Kousuke Tanimoto; Yoko Nukui; Yukie Tanaka |
| EPI_ISL_1430682, EPI_ISL_1927037, EPI_ISL_1927038, EPI_ISL_1927039, EPI_ISL_1927041, EPI_ISL_1927042, EPI_ISL_1927186, EPI_ISL_1927187, EPI_ISL_1927189, EPI_ISL_1927205, EPI_ISL_1929377, EPI_ISL_1929378, EPI_ISL_1929380, EPI_ISL_1929398, EPI_ISL_1929410, EPI_ISL_1929411, EPI_ISL_1930867, EPI_ISL_1930869, EPI_ISL_1930950, EPI_ISL_1930951, EPI_ISL_1930953, EPI_ISL_1930954, EPI_ISL_1930956, EPI_ISL_1931075, EPI_ISL_1931077, EPI_ISL_1931078, EPI_ISL_1931080, EPI_ISL_1931081, EPI_ISL_1931083, EPI_ISL_1931084, EPI_ISL_1931085, EPI_ISL_1931087, EPI_ISL_1931216, EPI_ISL_1931219, EPI_ISL_1931222, EPI_ISL_1931225, EPI_ISL_1931449, EPI_ISL_1931450, EPI_ISL_1931452, EPI_ISL_1931453, EPI_ISL_1931455, EPI_ISL_1931514, EPI_ISL_1931515, EPI_ISL_1931517, EPI_ISL_1931520, EPI_ISL_1931524, EPI_ISL_1931527, EPI_ISL_1931528, EPI_ISL_1931530, EPI_ISL_1931555, EPI_ISL_1931584, EPI_ISL_1931586, EPI_ISL_1931587, EPI_ISL_1931589, EPI_ISL_1931593, EPI_ISL_1931595, EPI_ISL_1931596, EPI_ISL_1933414, EPI_ISL_1933416, EPI_ISL_1933417, EPI_ISL_1933418, EPI_ISL_1933419, EPI_ISL_1933420, EPI_ISL_1933421, EPI_ISL_1933422, EPI_ISL_1933545, EPI_ISL_1933546, EPI_ISL_1933547, EPI_ISL_1933548, EPI_ISL_1933549, EPI_ISL_1933550, EPI_ISL_1933551, EPI_ISL_1933552, EPI_ISL_1933700, EPI_ISL_1934411, EPI_ISL_1934426 | | | |
| see above | Tokyo Metropolitan Institute of Public Health | Pathogen Genomics Center, National Institute of Infectious Diseases | Ai Kawana-Tachikawa; Chang-Kweng Lim; Chikako Shimokawa; Eri Nakayama; Eunsil Park; Hazuka Y Furihata; Hidemasa Izumiya; Hiromizu Takahashi; Ken Miyazawa; Ken-ichi Lee; Kentaro Itokawa; Koichi Ishikawa; Makoto Kuroda; Masanori Hashino; Masatomo Morita; Masumichi Saito; Midori Nakamura-Hoshi; Minoru Nagi; Motohiko Ogawa; Naomi Nojiri; Nobuo Koizumi; Nozomu Hanaoka; Rina Tanaka; Sana Uchikoba; Sayuri Seki; Shigeru Kusagawa; Shigeru Tajima; Shouji Yamamoto; Sunao Iyoda; Takahiro Maeki; Takashi Sakudoh; Tsuguto Fujimoto; Tsuyoshi Sekizuka; Yasutaka Hoshino; Yoshihiro Kaku |
| EPI_ISL_2080576, EPI_ISL_2080577, EPI_ISL_2080578, EPI_ISL_2080579, EPI_ISL_2080580, EPI_ISL_2080581, EPI_ISL_2080582, EPI_ISL_2080583, EPI_ISL_2080586, EPI_ISL_2080587, EPI_ISL_2080591, EPI_ISL_2080592, EPI_ISL_2080593, EPI_ISL_2080594, EPI_ISL_2080609, EPI_ISL_2080610, EPI_ISL_2080611, EPI_ISL_2080612, EPI_ISL_2080613, EPI_ISL_2080614, EPI_ISL_2080615 | | | |
| see above | Tokyo Metropolitan Institute of Public Health | Tokyo Metropolitan Institute of Public Health | Akane Negishi; Hirofumi Miyake; Jun Suzuki; Kayo Hagino; Kenji Sadamasu; Mami Nagashima; Masaki Hayashi; Rio Suzuki; Takako Yamazaki; Takeo Sasamoto; Takushi Fujiwara |
| EPI_ISL_1431611, EPI_ISL_1432005, EPI_ISL_1929186, EPI_ISL_1929188, EPI_ISL_1930870, EPI_ISL_1930872, EPI_ISL_1930873, EPI_ISL_1932531, EPI_ISL_1932532, EPI_ISL_1932533, EPI_ISL_1932534, EPI_ISL_1932535, EPI_ISL_1932536, EPI_ISL_1932537, EPI_ISL_1932538, EPI_ISL_1932539, EPI_ISL_1932540 | | | |
| see above | Toyama Institute of Health | Pathogen Genomics Center, National Institute of Infectious Diseases | Hazuka Y Furihata; Hidemasa Izumiya; Hiromizu Takahashi; Ken Shimuta; Kentaro Itokawa; Makoto Kuroda; Masanori Hashino; Masumichi Saito; Naomi Nojiri; Nobuo Koizumi; Nozomu Hanaoka; Rina Tanaka; Sana Uchikoba; Sunao Iyoda; Tsuguto Fujimoto; Tsuyoshi Sekizuka |
| EPI_ISL_1929901, EPI_ISL_1929903, EPI_ISL_1929904, EPI_ISL_1930488 | Yamagata Prefectural Institute of Public Health | Pathogen Genomics Center, National Institute of Infectious Diseases | Kentaro Itokawa; Makoto Kuroda; Masanori Hashino; Rina Tanaka; Tsuyoshi Sekizuka |

We gratefully acknowledge the following Authors from the Originating laboratories responsible for obtaining the specimens, as well as the Submitting laboratories where the genome data were generated and shared via GISAID, on which this research is based.

All Submitters of data may be contacted directly via www.gisaid.org

Authors are sorted alphabetically.

| Accession ID | Originating Laboratory | Submitting Laboratory | Authors |
|---|---|---|---|
| EPI_ISL_2329678, EPI_ISL_2329679, EPI_ISL_2337258, EPI_ISL_2337259, EPI_ISL_2337260, EPI_ISL_2337263, EPI_ISL_2337264, EPI_ISL_2337265, EPI_ISL_2337266, EPI_ISL_2337267, EPI_ISL_2337270, EPI_ISL_2337271, EPI_ISL_2337272, EPI_ISL_2337273, EPI_ISL_2337274, EPI_ISL_2337275, EPI_ISL_2337276, EPI_ISL_2337277, EPI_ISL_2337278, EPI_ISL_2337279, EPI_ISL_2337280, EPI_ISL_2337281, EPI_ISL_2337282, EPI_ISL_2337285, EPI_ISL_2760079, EPI_ISL_2760812, EPI_ISL_2760814, EPI_ISL_2760815, EPI_ISL_2760817, EPI_ISL_2760819, EPI_ISL_2760820, EPI_ISL_2760822, EPI_ISL_2760824, EPI_ISL_2760825, EPI_ISL_2760827, EPI_ISL_2760828, EPI_ISL_2760830, EPI_ISL_2760832, EPI_ISL_2760833, EPI_ISL_2760835, EPI_ISL_2760837, EPI_ISL_2760839, EPI_ISL_2764039, EPI_ISL_2764040, EPI_ISL_2764041, EPI_ISL_2764042, EPI_ISL_2764043, EPI_ISL_2764044, EPI_ISL_2764045, EPI_ISL_2764046, EPI_ISL_2764047, EPI_ISL_2764048, EPI_ISL_2764049, EPI_ISL_2768844 | | | |
| see above | Chiba Prefectural Institute of Public Health | Pathogen Genomics Center, National Institute of Infectious Diseases | Kentaro Itokawa; Makoto Kuroda; Masanori Hashino; Rina Tanaka; Tsuyoshi Sekizuka |
| EPI_ISL_2292088, EPI_ISL_2292089, EPI_ISL_2292090, EPI_ISL_2292091, EPI_ISL_2292092, EPI_ISL_2292093, EPI_ISL_2292094, EPI_ISL_2292095 | | | |
| see above | Department of Infectious Diseases and Immunology, National Hospital Organization Nagoya Medical Center | Clinical Research Center, National Hospital Organization Nagoya Medical Center | Hirotaka Ode; Kazuhiro Matsuoka; Mai Kubota; Masakazu Matsuda; Mayumi Imahashi; Miho Nakasuji; Mikiko Mori; Yasumasa Iwatani; Yoshihiro Nakata; Yoshiyuki Yokomaku |
| EPI_ISL_2359483, EPI_ISL_2359485, EPI_ISL_2359487, EPI_ISL_2359489, EPI_ISL_2359493, EPI_ISL_2359495, EPI_ISL_2359497, EPI_ISL_2359499, EPI_ISL_2359501, EPI_ISL_2359503, EPI_ISL_2359504, EPI_ISL_2359507, EPI_ISL_2359508, EPI_ISL_2359510, EPI_ISL_2359512, EPI_ISL_2359514, EPI_ISL_2359516, EPI_ISL_2359518, EPI_ISL_2359569, EPI_ISL_2359571, EPI_ISL_2359573, EPI_ISL_2359575, EPI_ISL_2359577, EPI_ISL_2359579, EPI_ISL_2359581, EPI_ISL_2359583, EPI_ISL_2359585, EPI_ISL_2359587, EPI_ISL_2359589, EPI_ISL_2359590, EPI_ISL_2359592, EPI_ISL_2359594, EPI_ISL_2359596, EPI_ISL_2359598, EPI_ISL_2359600, EPI_ISL_2359602, EPI_ISL_2359604, EPI_ISL_2359605, EPI_ISL_2359608, EPI_ISL_2359610, EPI_ISL_2359611, EPI_ISL_2359612, EPI_ISL_2359613, EPI_ISL_2359615, EPI_ISL_2359617, EPI_ISL_2359619, EPI_ISL_2359621, EPI_ISL_2359623, EPI_ISL_2359625, EPI_ISL_2359627, EPI_ISL_2359629, EPI_ISL_2359631, EPI_ISL_2359633, EPI_ISL_2359635, EPI_ISL_2359637, EPI_ISL_2359639, EPI_ISL_2359641, EPI_ISL_2359643, EPI_ISL_2359645, EPI_ISL_2359647, EPI_ISL_2359648, EPI_ISL_2359650, EPI_ISL_2359661, EPI_ISL_2359664, EPI_ISL_2359666, EPI_ISL_2359667, EPI_ISL_2359670, EPI_ISL_2359746, EPI_ISL_2359748, EPI_ISL_2359750, EPI_ISL_2359752, EPI_ISL_2359754, EPI_ISL_2359755, EPI_ISL_2359757, EPI_ISL_2359759, EPI_ISL_2359760, EPI_ISL_2359822, EPI_ISL_2359824, EPI_ISL_2359826, EPI_ISL_2359828, EPI_ISL_2359830, EPI_ISL_2359832, EPI_ISL_2359834, EPI_ISL_2359836, EPI_ISL_2359838, EPI_ISL_2359840, EPI_ISL_2359842, EPI_ISL_2359844, EPI_ISL_2359847, EPI_ISL_2359849, EPI_ISL_2359850, EPI_ISL_2359852, EPI_ISL_2359854, EPI_ISL_2359855, EPI_ISL_2359857, EPI_ISL_2359859, EPI_ISL_2359892, EPI_ISL_2359893, EPI_ISL_2359895, EPI_ISL_2359897, EPI_ISL_2359900, EPI_ISL_2359901, EPI_ISL_2359903, EPI_ISL_2359905, EPI_ISL_2359942, EPI_ISL_2359947, EPI_ISL_2359949, EPI_ISL_2359951, EPI_ISL_2359953, EPI_ISL_2359954, EPI_ISL_2359955, EPI_ISL_2359956, EPI_ISL_2359957, EPI_ISL_2359958, EPI_ISL_2359959, EPI_ISL_2359960, EPI_ISL_2359961, EPI_ISL_2359962, EPI_ISL_2359963, EPI_ISL_2359964, EPI_ISL_2359965, EPI_ISL_2359966, EPI_ISL_2359967, EPI_ISL_2359968, EPI_ISL_2359969, EPI_ISL_2359970, EPI_ISL_2359971, EPI_ISL_2359972, EPI_ISL_2359973, EPI_ISL_2359974, EPI_ISL_2359975, EPI_ISL_2359976, EPI_ISL_2359977, EPI_ISL_2359978, EPI_ISL_2359979, EPI_ISL_2359980, EPI_ISL_2359981, EPI_ISL_2359982, EPI_ISL_2359983, EPI_ISL_2359984, EPI_ISL_2359985, EPI_ISL_2359986, EPI_ISL_2359987, EPI_ISL_2359988, EPI_ISL_2359989, EPI_ISL_2359990, EPI_ISL_2359991, EPI_ISL_2359992, EPI_ISL_2359993, EPI_ISL_2359994, EPI_ISL_2359995, EPI_ISL_2359996, EPI_ISL_2359997, EPI_ISL_2359998, EPI_ISL_2359999, EPI_ISL_2360000, EPI_ISL_2360001, EPI_ISL_2360002, EPI_ISL_2360003, EPI_ISL_2360004, EPI_ISL_2360005, EPI_ISL_2360006, EPI_ISL_2360007, EPI_ISL_2360008, EPI_ISL_2360009, EPI_ISL_2360010, EPI_ISL_2360011, EPI_ISL_2360012, EPI_ISL_2360013, EPI_ISL_2360014, EPI_ISL_2360015, EPI_ISL_2360016, EPI_ISL_2360017, EPI_ISL_2360018, EPI_ISL_2360019, EPI_ISL_2360020, EPI_ISL_2360021, EPI_ISL_2360022, EPI_ISL_2360023, EPI_ISL_2360024, EPI_ISL_2360025, EPI_ISL_2360026, EPI_ISL_2360027, EPI_ISL_2360028, EPI_ISL_2360029, EPI_ISL_2360030, EPI_ISL_2360031, EPI_ISL_2360032, EPI_ISL_2360033, EPI_ISL_2360034, EPI_ISL_2360035, EPI_ISL_2360036, EPI_ISL_2360037, EPI_ISL_2360038, EPI_ISL_2360039, EPI_ISL_2360040, EPI_ISL_2360057, EPI_ISL_2360058, EPI_ISL_2360059, EPI_ISL_2360060, EPI_ISL_2360061, EPI_ISL_2360062, EPI_ISL_2360064, EPI_ISL_2360065, EPI_ISL_2360081, EPI_ISL_2360082, EPI_ISL_2360083, EPI_ISL_2360084, EPI_ISL_2360085, EPI_ISL_2360086, EPI_ISL_2360087, EPI_ISL_2360088, EPI_ISL_2360089, EPI_ISL_2360090, EPI_ISL_2360091, EPI_ISL_2360092, EPI_ISL_2360093, EPI_ISL_2360094, EPI_ISL_2360095, EPI_ISL_2360096, EPI_ISL_2360097, EPI_ISL_2360098, EPI_ISL_2360099, EPI_ISL_2360100, EPI_ISL_2360101, EPI_ISL_2360102, EPI_ISL_2360103, EPI_ISL_2360104, EPI_ISL_2360105, EPI_ISL_2360106, EPI_ISL_2360107, EPI_ISL_2360108, EPI_ISL_2360109, EPI_ISL_2360110, EPI_ISL_2360127, EPI_ISL_2360128, EPI_ISL_2360129, EPI_ISL_2360130, EPI_ISL_2360131, EPI_ISL_2360132, EPI_ISL_2360133, EPI_ISL_2360134, EPI_ISL_2360135, EPI_ISL_2360136, EPI_ISL_2360137, EPI_ISL_2360138, EPI_ISL_2360139, EPI_ISL_2360140, EPI_ISL_2360141, EPI_ISL_2360142, EPI_ISL_2360143, EPI_ISL_2360144, EPI_ISL_2360145, EPI_ISL_2360146, EPI_ISL_2360147, EPI_ISL_2360148, EPI_ISL_2360149, EPI_ISL_2360150, EPI_ISL_2360151, EPI_ISL_2360152, EPI_ISL_2360153, EPI_ISL_2360154, EPI_ISL_2360155, EPI_ISL_2360156, EPI_ISL_2360157, EPI_ISL_2360158, EPI_ISL_2360159, EPI_ISL_2360160, EPI_ISL_2360161, EPI_ISL_2360162, EPI_ISL_2360163, EPI_ISL_2360164, EPI_ISL_2360165, EPI_ISL_2360166, EPI_ISL_2360167, EPI_ISL_2360168, EPI_ISL_2360169, EPI_ISL_2360170, EPI_ISL_2360171, EPI_ISL_2360172, EPI_ISL_2360173, EPI_ISL_2360174, EPI_ISL_2360175, EPI_ISL_2360176, EPI_ISL_2360177, EPI_ISL_2360178, EPI_ISL_2360179, EPI_ISL_2360183, EPI_ISL_2360184, EPI_ISL_2360185, EPI_ISL_2360186, EPI_ISL_2360187, EPI_ISL_2360188, EPI_ISL_2360189, EPI_ISL_2360190, EPI_ISL_2360191, EPI_ISL_2360192, EPI_ISL_2360193, EPI_ISL_2360194, EPI_ISL_2360195, EPI_ISL_2360196, EPI_ISL_2360197, EPI_ISL_2360198, EPI_ISL_2360199, EPI_ISL_2360200, EPI_ISL_2360201, EPI_ISL_2360202, EPI_ISL_2360203, EPI_ISL_2360204, EPI_ISL_2360215, EPI_ISL_2360216, EPI_ISL_2810786, EPI_ISL_2810787, EPI_ISL_2810788, EPI_ISL_2810789, EPI_ISL_2810790, EPI_ISL_2810791, EPI_ISL_2810792, EPI_ISL_2810793, EPI_ISL_2810794, EPI_ISL_2810795, EPI_ISL_2810796, EPI_ISL_2810797, EPI_ISL_2810798, EPI_ISL_2810799, EPI_ISL_2810800, EPI_ISL_2810801, EPI_ISL_2810802, EPI_ISL_2810803, EPI_ISL_2810804, EPI_ISL_2810805, EPI_ISL_2810806, EPI_ISL_2810807, EPI_ISL_2810808, EPI_ISL_2810809, EPI_ISL_2810810, EPI_ISL_2810811, EPI_ISL_2810812, EPI_ISL_2810813, EPI_ISL_2810814, EPI_ISL_2810815, EPI_ISL_2810816, EPI_ISL_2810817, EPI_ISL_2810818, EPI_ISL_2810819, EPI_ISL_2810820, EPI_ISL_2810821, EPI_ISL_2810822, EPI_ISL_2810823, EPI_ISL_2810824, EPI_ISL_2810825, EPI_ISL_2810826, EPI_ISL_2810827, EPI_ISL_2810828, EPI_ISL_2810829, EPI_ISL_2810830, EPI_ISL_2810831, EPI_ISL_2810832, EPI_ISL_2810833, EPI_ISL_2810834, EPI_ISL_2810835, EPI_ISL_2810836, EPI_ISL_2810837, EPI_ISL_2810838, EPI_ISL_2810839, EPI_ISL_2810840, EPI_ISL_2810841, EPI_ISL_2810842, EPI_ISL_2810843, EPI_ISL_2810844, EPI_ISL_2810845, EPI_ISL_2810846, EPI_ISL_2810847, EPI_ISL_2810848, EPI_ISL_2810849, EPI_ISL_2810850, EPI_ISL_2810851, EPI_ISL_2810852, EPI_ISL_2810853, EPI_ISL_2810854, EPI_ISL_2810855, EPI_ISL_2810856, EPI_ISL_2810857, EPI_ISL_2810858, EPI_ISL_2810859, EPI_ISL_2810860, EPI_ISL_2810861, EPI_ISL_2810862, EPI_ISL_2810863, EPI_ISL_2810864, EPI_ISL_2810865, EPI_ISL_2810866, EPI_ISL_2810867, EPI_ISL_2810868, EPI_ISL_2810869, EPI_ISL_2810870, EPI_ISL_2810871, EPI_ISL_2810872, EPI_ISL_2810873, EPI_ISL_2810874, EPI_ISL_2810875, EPI_ISL_2810876, EPI_ISL_2810877, EPI_ISL_2810878, EPI_ISL_2810879, EPI_ISL_2810880, EPI_ISL_2810881, EPI_ISL_2810882, EPI_ISL_2810883, EPI_ISL_2810884, EPI_ISL_2810885, EPI_ISL_2810886, EPI_ISL_2810887, EPI_ISL_2810888, EPI_ISL_2810889, EPI_ISL_2810890, EPI_ISL_2810891, EPI_ISL_2810892, EPI_ISL_2810893, EPI_ISL_2810894, EPI_ISL_2810895, EPI_ISL_2810896, EPI_ISL_2810897, EPI_ISL_2810898, EPI_ISL_2810899, EPI_ISL_2810900, EPI_ISL_2810901, EPI_ISL_2810902, EPI_ISL_2810903, EPI_ISL_2810904, EPI_ISL_2810905, EPI_ISL_2810906, EPI_ISL_2810907, EPI_ISL_2810908, EPI_ISL_2810909, EPI_ISL_2810910, EPI_ISL_2810911, EPI_ISL_2810912, EPI_ISL_2810913, EPI_ISL_2810914, EPI_ISL_2810915, EPI_ISL_2810916, EPI_ISL_2810917, EPI_ISL_2810918, EPI_ISL_2810919, EPI_ISL_2810920, EPI_ISL_2810921, EPI_ISL_2810922, EPI_ISL_2810923, EPI_ISL_2810924, EPI_ISL_2810925, EPI_ISL_2810926, EPI_ISL_2810927, EPI_ISL_2810928, EPI_ISL_2810929, EPI_ISL_2810930, EPI_ISL_2810931, EPI_ISL_2810932, EPI_ISL_2810933, EPI_ISL_2810934, EPI_ISL_2810935, EPI_ISL_2810936, EPI_ISL_2810937, EPI_ISL_2810938, EPI_ISL_2810939, EPI_ISL_2810940, EPI_ISL_2810941, EPI_ISL_2810942, EPI_ISL_2810943, EPI_ISL_2810944, EPI_ISL_2810945, EPI_ISL_2810946, EPI_ISL_2810947, EPI_ISL_2810948, EPI_ISL_2810949, EPI_ISL_2810950, EPI_ISL_2810951, EPI_ISL_2810952, EPI_ISL_2810953, EPI_ISL_2810954, EPI_ISL_2810955, EPI_ISL_2810956, EPI_ISL_2810957, EPI_ISL_2810958, EPI_ISL_2810959, EPI_ISL_2810960, EPI_ISL_2810961, EPI_ISL_2810962, EPI_ISL_2810963, EPI_ISL_2810964, EPI_ISL_2810965, EPI_ISL_2810966, EPI_ISL_2810967, EPI_ISL_2810968, EPI_ISL_2810969, EPI_ISL_2810970, EPI_ISL_2810971, EPI_ISL_2810972, EPI_ISL_2810973, EPI_ISL_2810974, EPI_ISL_2810975, EPI_ISL_2810976, EPI_ISL_2810977, EPI_ISL_2810978, EPI_ISL_2810979, EPI_ISL_2810980, EPI_ISL_2810981, EPI_ISL_2810982, EPI_ISL_2810983, EPI_ISL_2810984, EPI_ISL_2810985, EPI_ISL_2810986, EPI_ISL_2810987, EPI_ISL_2810988, EPI_ISL_2810989, EPI_ISL_2810990, EPI_ISL_2810991, EPI_ISL_2810993, EPI_ISL_2810994, EPI_ISL_2810995, EPI_ISL_2810996, EPI_ISL_2810997, EPI_ISL_2810998, EPI_ISL_2810999, EPI_ISL_2811000, EPI_ISL_2811001, EPI_ISL_2811002, EPI_ISL_2811003, EPI_ISL_2811004, EPI_ISL_2811005, EPI_ISL_2811006, EPI_ISL_2811007, EPI_ISL_2811008, EPI_ISL_2811009, EPI_ISL_2811010, EPI_ISL_2811011, EPI_ISL_2811012, EPI_ISL_2811013, EPI_ISL_2811014, EPI_ISL_2811015, EPI_ISL_2811016, EPI_ISL_2811017, EPI_ISL_2811018, EPI_ISL_2811019, EPI_ISL_2811020, EPI_ISL_2811021, EPI_ISL_2811022, EPI_ISL_2811023, EPI_ISL_2811024, EPI_ISL_2811025, EPI_ISL_2811026, EPI_ISL_2811027, EPI_ISL_2811028, EPI_ISL_2811029, EPI_ISL_2811030, EPI_ISL_2811031, EPI_ISL_2811032, EPI_ISL_2811033, EPI_ISL_2811034, EPI_ISL_2811035, EPI_ISL_2811036, EPI_ISL_2811037, EPI_ISL_2811038, EPI_ISL_2811039, EPI_ISL_2811040, EPI_ISL_2811041, EPI_ISL_2811042, EPI_ISL_2811043, EPI_ISL_2811044, EPI_ISL_2811045, EPI_ISL_2811049, EPI_ISL_2811050, EPI_ISL_2811051, EPI_ISL_2811052, EPI_ISL_2811053, EPI_ISL_2811054, EPI_ISL_2811055, EPI_ISL_2811056, EPI_ISL_2811057, EPI_ISL_2811058, EPI_ISL_2811059, EPI_ISL_2811060, EPI_ISL_2811061, EPI_ISL_2811062, EPI_ISL_2811063, EPI_ISL_2811064, EPI_ISL_2811065, EPI_ISL_2811066, EPI_ISL_2811067, EPI_ISL_2811068, EPI_ISL_2811069, EPI_ISL_2811070, EPI_ISL_2811071, EPI_ISL_2811072, EPI_ISL_2811073, EPI_ISL_2811074, EPI_ISL_2811075, EPI_ISL_2811076, EPI_ISL_2811077, EPI_ISL_2811078, EPI_ISL_2811079, EPI_ISL_2811080, EPI_ISL_2811081, EPI_ISL_2811082, EPI_ISL_2811083, EPI_ISL_2811084, EPI_ISL_2811085, EPI_ISL_2811089, EPI_ISL_2811092, EPI_ISL_2811094, EPI_ISL_2811096, EPI_ISL_2811097, EPI_ISL_2811098, EPI_ISL_2811099, EPI_ISL_2811100, EPI_ISL_2811101, EPI_ISL_2811102, EPI_ISL_2811103, EPI_ISL_2811104, EPI_ISL_2811106, EPI_ISL_2811107, EPI_ISL_2811108, EPI_ISL_2811109, EPI_ISL_2811111, EPI_ISL_2811112, EPI_ISL_2811113, EPI_ISL_2811114, EPI_ISL_2811115, EPI_ISL_2811116, EPI_ISL_2811140, EPI_ISL_2811146, EPI_ISL_2811160, EPI_ISL_2811174, EPI_ISL_2811175, EPI_ISL_2811186, EPI_ISL_2811187, EPI_ISL_2811194, EPI_ISL_2811254, EPI_ISL_2811255, EPI_ISL_2811256, EPI_ISL_2811257, EPI_ISL_2811302, EPI_ISL_2811303, EPI_ISL_2811304, EPI_ISL_2811305, EPI_ISL_2811306, EPI_ISL_2811307, EPI_ISL_2811308, EPI_ISL_2811309, EPI_ISL_2811310, EPI_ISL_2811311, EPI_ISL_2811312, EPI_ISL_2811313, EPI_ISL_2811314, EPI_ISL_2811315, EPI_ISL_2811316, EPI_ISL_2811317, EPI_ISL_2811318, EPI_ISL_2811319, EPI_ISL_2811320, EPI_ISL_2811321, EPI_ISL_2811322, EPI_ISL_2811323, EPI_ISL_2811324, EPI_ISL_2811325, EPI_ISL_2811326, EPI_ISL_2811327, EPI_ISL_2811328, EPI_ISL_2811329, EPI_ISL_2811330, EPI_ISL_2811331, EPI_ISL_2811332, EPI_ISL_2811333 | | | |
| see above | Department of Infectious Diseases, Kobe Institute of Health | Department of Infectious Diseases, Kobe Institute of Health | Kentaro Itokawa; Makoto Kuroda; Masanori Hashino; Noriko Nakanishi; Rina Tanaka; Ryohei Nomoto; Tomotada Iwamoto; Tsuyoshi Sekizuka |
| EPI_ISL_2080479, EPI_ISL_2101018, EPI_ISL_2101020 | Department of Microbiology, Tochigi Prefectural Institute of Public Health and Enviromental Science | Genome Analysis Center, Kamma Memorial Hospital | Fuminori Mizukoshi; Hanako Yazawa; Hiroshi Kamma; Jun Ishii; Kaori Watanabe; Michiaki Masuda; Satoko Soma; Takuya Yazawa; Yoshiko Hitomi |
| EPI_ISL_2080357, EPI_ISL_2080358, EPI_ISL_2080359, EPI_ISL_2101019, EPI_ISL_2551183, EPI_ISL_2551519, EPI_ISL_2551520 | | | |
| see above | Department of Microbiology, Tochigi Prefectural Institute of Public Health and Environmental Science | Genome Analysis Center, Kamma Memorial Hospital | Fuminori Mizukoshi; Hanako Yazawa; Hiroshi Kamma; Jun Ishii; Kaori Watanabe; Michiaki Masuda; Satoko Soma; Takuya Yazawa; Yoshiko Hitomi |
| EPI_ISL_2476406, EPI_ISL_2476407 | Department of Pathology, National Institute of Infectious Diseases | Department of Pathology, National Institute of Infectious Diseases | Harutaka Katano; Kentaro Itokawa; Makoto Kuroda; Tadaki Suzuki; Tsuyoshi Sekizuka; Yuichiro Hirata |
| EPI_ISL_2509397, EPI_ISL_2509806, EPI_ISL_2509933, EPI_ISL_2509961, EPI_ISL_2510089, EPI_ISL_2510297, EPI_ISL_2510298, EPI_ISL_2510299 | | | |
| see above | Dr. Yoshitaka Tamura Department of Clinical Laboratory, Osaka Habikino Medical Center | Department of Developmental Medicine, Research Institute, Osaka Women's and Children's Hospital | Itaru Yanagihara; Yukiko Nakura |
| EPI_ISL_2772734, EPI_ISL_2772735, EPI_ISL_2772736, EPI_ISL_2772737, EPI_ISL_2772738, EPI_ISL_2772739, EPI_ISL_2772740, EPI_ISL_2772741, EPI_ISL_2772742, EPI_ISL_2772743, EPI_ISL_2772744, EPI_ISL_2772745, EPI_ISL_2772746, EPI_ISL_2772747, EPI_ISL_2772748, EPI_ISL_2772749, EPI_ISL_2772750, EPI_ISL_2772751, EPI_ISL_2772752, EPI_ISL_2772753, EPI_ISL_2772754, EPI_ISL_2772755, EPI_ISL_2772756, EPI_ISL_2772757, EPI_ISL_2772758, EPI_ISL_2772759, EPI_ISL_2772760, EPI_ISL_2772761, EPI_ISL_2772762, EPI_ISL_2772763, EPI_ISL_2772764, EPI_ISL_2772765, EPI_ISL_2772766, EPI_ISL_2772767, EPI_ISL_2772768, EPI_ISL_2772769, EPI_ISL_2772770, EPI_ISL_2772789, EPI_ISL_2772790, EPI_ISL_2772791, EPI_ISL_2772792, EPI_ISL_2772793, EPI_ISL_2772797, EPI_ISL_2772798, EPI_ISL_2772799, EPI_ISL_2772800, EPI_ISL_2772801, EPI_ISL_2772802, EPI_ISL_2772803, EPI_ISL_2772804, EPI_ISL_2772805, EPI_ISL_2772806, EPI_ISL_2772807, EPI_ISL_2772808, EPI_ISL_2772809, EPI_ISL_2772810, EPI_ISL_2772811, EPI_ISL_2772812, EPI_ISL_2772813, EPI_ISL_2772814, EPI_ISL_2772815, EPI_ISL_2772816, EPI_ISL_2772817, EPI_ISL_2772818, EPI_ISL_2772819, EPI_ISL_2772820, EPI_ISL_2772821, EPI_ISL_2772822, EPI_ISL_2772823, EPI_ISL_2772824, EPI_ISL_2772825, EPI_ISL_2772826, EPI_ISL_2772827, EPI_ISL_2772828, EPI_ISL_2772829, EPI_ISL_2772830, EPI_ISL_2772831, EPI_ISL_2772832, EPI_ISL_2772833, EPI_ISL_2772834, EPI_ISL_2772835, EPI_ISL_2772836, EPI_ISL_2772837, EPI_ISL_2772838, EPI_ISL_2772839, EPI_ISL_2772840, EPI_ISL_2772841, EPI_ISL_2772842, EPI_ISL_2772843, EPI_ISL_2772844, EPI_ISL_2772845, EPI_ISL_2772846, EPI_ISL_2772847, EPI_ISL_2772848, EPI_ISL_2772849, EPI_ISL_2772850, EPI_ISL_2772851, EPI_ISL_2772852, EPI_ISL_2772853, EPI_ISL_2772854, EPI_ISL_2772855, EPI_ISL_2772856, EPI_ISL_2772857, EPI_ISL_2772858, EPI_ISL_2772859, EPI_ISL_2772860, EPI_ISL_2772861, EPI_ISL_2772862, EPI_ISL_2772863, EPI_ISL_2772864, EPI_ISL_2772865, EPI_ISL_2772866, EPI_ISL_2772867, EPI_ISL_2772868, EPI_ISL_2772869, EPI_ISL_2772870, EPI_ISL_2772871, EPI_ISL_2772872, EPI_ISL_2772873, EPI_ISL_2772874, EPI_ISL_2772875, EPI_ISL_2772876, EPI_ISL_2772877, EPI_ISL_2772878, EPI_ISL_2772879, EPI_ISL_2772880, EPI_ISL_2772881, EPI_ISL_2772882, EPI_ISL_2772883, EPI_ISL_2772884, EPI_ISL_2772885, EPI_ISL_2772886, EPI_ISL_2772887, EPI_ISL_2772888, EPI_ISL_2772889, EPI_ISL_2772890, EPI_ISL_2772891, EPI_ISL_2772892, EPI_ISL_2772893, EPI_ISL_2772894, EPI_ISL_2772895, EPI_ISL_2772896, EPI_ISL_2772897, EPI_ISL_2772898, EPI_ISL_2772899, EPI_ISL_2772900, EPI_ISL_2772901, EPI_ISL_2772902, EPI_ISL_2772903, EPI_ISL_2772904, EPI_ISL_2772905, EPI_ISL_2772906, EPI_ISL_2772907, EPI_ISL_2772908, EPI_ISL_2772909, EPI_ISL_2772936, EPI_ISL_2772937, EPI_ISL_2772938, EPI_ISL_2772939, EPI_ISL_2772940, EPI_ISL_2772941, EPI_ISL_2772942, EPI_ISL_2772943, EPI_ISL_2772944, EPI_ISL_2772945, EPI_ISL_2772946, EPI_ISL_2772947, EPI_ISL_2772948, EPI_ISL_2772949, EPI_ISL_2772950, EPI_ISL_2772951, EPI_ISL_2772952, EPI_ISL_2772953, EPI_ISL_2772954, EPI_ISL_2772955, EPI_ISL_2772956, EPI_ISL_2772957, EPI_ISL_2772958, EPI_ISL_2772959, EPI_ISL_2772960, EPI_ISL_2772961, EPI_ISL_2772962, EPI_ISL_2772963, EPI_ISL_2772964, EPI_ISL_2772965, EPI_ISL_2772966, EPI_ISL_2772967, EPI_ISL_2772968, EPI_ISL_2772969, EPI_ISL_2772970, EPI_ISL_2772971, EPI_ISL_2772972, EPI_ISL_2772973, EPI_ISL_2772974, EPI_ISL_2772975, EPI_ISL_2772976, EPI_ISL_2772977, EPI_ISL_2772978, EPI_ISL_2772979, EPI_ISL_2772980, EPI_ISL_2772981, EPI_ISL_2772982, EPI_ISL_2772983, EPI_ISL_2772984, EPI_ISL_2772985, EPI_ISL_2772986, EPI_ISL_2772987, EPI_ISL_2772988, EPI_ISL_2772989, EPI_ISL_2772990, EPI_ISL_2772991, EPI_ISL_2772992, EPI_ISL_2772993, EPI_ISL_2772994, EPI_ISL_2772995, EPI_ISL_2772996, EPI_ISL_2772997, EPI_ISL_2772998, EPI_ISL_2772999, EPI_ISL_2773000, EPI_ISL_2773001, EPI_ISL_2773002, EPI_ISL_2773003, EPI_ISL_2773004, EPI_ISL_2773005, EPI_ISL_2773006, EPI_ISL_2773007, EPI_ISL_2773008, EPI_ISL_2773009, EPI_ISL_2773010, EPI_ISL_2773011, EPI_ISL_2773012, EPI_ISL_2773013, EPI_ISL_2773014, EPI_ISL_2773015, EPI_ISL_2773016, EPI_ISL_2773017, EPI_ISL_2773018, EPI_ISL_2773019, EPI_ISL_2773020, EPI_ISL_2773021, EPI_ISL_2773022, EPI_ISL_2773023, EPI_ISL_2773024, EPI_ISL_2773025, EPI_ISL_2773026, EPI_ISL_2773027, EPI_ISL_2773028, EPI_ISL_2773029, EPI_ISL_2773030, EPI_ISL_2773031, EPI_ISL_2773032, EPI_ISL_2773033, EPI_ISL_2773034, EPI_ISL_2773035, EPI_ISL_2773036, EPI_ISL_2773037, EPI_ISL_2773038, EPI_ISL_2773039, EPI_ISL_2773040, EPI_ISL_2773041, EPI_ISL_2773042, EPI_ISL_2773043, EPI_ISL_2773044, EPI_ISL_2773045, EPI_ISL_2773046, EPI_ISL_2773047, EPI_ISL_2773048, EPI_ISL_2773049, EPI_ISL_2773050, EPI_ISL_2773051, EPI_ISL_2773052, EPI_ISL_2773053, EPI_ISL_2773054, EPI_ISL_2773055, EPI_ISL_2773056, EPI_ISL_2773057, EPI_ISL_2773058, EPI_ISL_2773059, EPI_ISL_2773060, EPI_ISL_2773061, EPI_ISL_2773062, EPI_ISL_2773063, EPI_ISL_2773064, EPI_ISL_2773065, EPI_ISL_2773066, EPI_ISL_2773067, EPI_ISL_2773068, EPI_ISL_2773069, EPI_ISL_2773070, EPI_ISL_2773071, EPI_ISL_2773072, EPI_ISL_2773073, EPI_ISL_2773074, EPI_ISL_2773075, EPI_ISL_2773076, EPI_ISL_2773077, EPI_ISL_2773078, EPI_ISL_2773079, EPI_ISL_2773080, EPI_ISL_2773081, EPI_ISL_2773095, EPI_ISL_2773096, EPI_ISL_2773097, EPI_ISL_2773098, EPI_ISL_2773099, EPI_ISL_2773100, EPI_ISL_2773101, EPI_ISL_2773102, EPI_ISL_2773103, EPI_ISL_2773104, EPI_ISL_2773105, EPI_ISL_2773106, EPI_ISL_2773107, EPI_ISL_2773108, EPI_ISL_2773109, EPI_ISL_2773110, EPI_ISL_2773111, EPI_ISL_2773112, EPI_ISL_2773113, EPI_ISL_2773114, EPI_ISL_2773115, EPI_ISL_2773116, EPI_ISL_2773117, EPI_ISL_2773118, EPI_ISL_2773119, EPI_ISL_2773120, EPI_ISL_2773121, EPI_ISL_2773122, EPI_ISL_2773123, EPI_ISL_2773124, EPI_ISL_2773125, EPI_ISL_2773126, EPI_ISL_2773127, EPI_ISL_2773128, EPI_ISL_2773129, EPI_ISL_2773130, EPI_ISL_2773131, EPI_ISL_2773132, EPI_ISL_2773133, EPI_ISL_2773134, EPI_ISL_2773135, EPI_ISL_2773136, EPI_ISL_2773137, EPI_ISL_2773138, EPI_ISL_2773139, EPI_ISL_2773140, EPI_ISL_2773141, EPI_ISL_2773142, EPI_ISL_2773143, EPI_ISL_2773144, EPI_ISL_2773145, EPI_ISL_2773146, EPI_ISL_2773147, EPI_ISL_2773148, EPI_ISL_2773149, EPI_ISL_2773150, EPI_ISL_2773151, EPI_ISL_2773152, EPI_ISL_2773153, EPI_ISL_2773154, EPI_ISL_2773155, EPI_ISL_2773156, EPI_ISL_2773157, EPI_ISL_2773158, EPI_ISL_2773159, EPI_ISL_2773160, EPI_ISL_2773161, EPI_ISL_2773162, EPI_ISL_2773163, EPI_ISL_2773164, EPI_ISL_2773165, EPI_ISL_2773166, EPI_ISL_2773167, EPI_ISL_2773168, EPI_ISL_2773169, EPI_ISL_2773170, EPI_ISL_2773171, EPI_ISL_2773172, EPI_ISL_2773173, EPI_ISL_2773174, EPI_ISL_2773175, EPI_ISL_2773176, EPI_ISL_2773177, EPI_ISL_2773178, EPI_ISL_2773179, EPI_ISL_2773180, EPI_ISL_2773181, EPI_ISL_2773182, EPI_ISL_2773183, EPI_ISL_2773184, EPI_ISL_2773185, EPI_ISL_2773186, EPI_ISL_2773187, EPI_ISL_2773188, EPI_ISL_2773189, EPI_ISL_2773190, EPI_ISL_2773191, EPI_ISL_2773192, EPI_ISL_2773193, EPI_ISL_2773194, EPI_ISL_2773195, EPI_ISL_2773196, EPI_ISL_2773197, EPI_ISL_2773198, EPI_ISL_2773199, EPI_ISL_2773200, EPI_ISL_2773201, EPI_ISL_2773202, EPI_ISL_2773203, EPI_ISL_2773204, EPI_ISL_2773205, EPI_ISL_2773206, EPI_ISL_2773207, EPI_ISL_2773208, EPI_ISL_2773209, EPI_ISL_2773210, EPI_ISL_2773211, EPI_ISL_2773212, EPI_ISL_2773213, EPI_ISL_2773214, EPI_ISL_2773215, EPI_ISL_2773216, EPI_ISL_2773217, EPI_ISL_2773218, EPI_ISL_2773219, EPI_ISL_2773220, EPI_ISL_2773221, EPI_ISL_2773222, EPI_ISL_2773223, EPI_ISL_2773224, EPI_ISL_2773225, EPI_ISL_2773226, EPI_ISL_2773227, EPI_ISL_2773228, EPI_ISL_2773229, EPI_ISL_2773230, EPI_ISL_2773231, EPI_ISL_2773232, EPI_ISL_2773233, EPI_ISL_2773234, EPI_ISL_2773235, EPI_ISL_2773236, EPI_ISL_2773237, EPI_ISL_2773238, EPI_ISL_2773239, EPI_ISL_2773240, EPI_ISL_2773241, EPI_ISL_2773242, EPI_ISL_2773243, EPI_ISL_2773244, EPI_ISL_2773245, EPI_ISL_2773246, EPI_ISL_2773247, EPI_ISL_2773248, EPI_ISL_2773249, EPI_ISL_2773250, EPI_ISL_2773251, EPI_ISL_2773252, EPI_ISL_2773253, EPI_ISL_2773254, EPI_ISL_2773255, EPI_ISL_2773256, EPI_ISL_2773257, EPI_ISL_2773258, EPI_ISL_2773259, EPI_ISL_2773260, EPI_ISL_2773261, EPI_ISL_2773262, EPI_ISL_2773263, EPI_ISL_2773264, EPI_ISL_2773265, EPI_ISL_2773266, EPI_ISL_2773267, EPI_ISL_2773268, EPI_ISL_2773269, EPI_ISL_2773270, EPI_ISL_2773271, EPI_ISL_2773272, EPI_ISL_2773273, EPI_ISL_2773274, EPI_ISL_2773275, EPI_ISL_2773276, EPI_ISL_2773277, EPI_ISL_2773278, EPI_ISL_2773279, EPI_ISL_2773280, EPI_ISL_2773281, EPI_ISL_2773282, EPI_ISL_2773283, EPI_ISL_2773284, EPI_ISL_2773285, EPI_ISL_2773286, EPI_ISL_2773287, EPI_ISL_2773288, EPI_ISL_2773289, EPI_ISL_2773290, EPI_ISL_2773291, EPI_ISL_2773292, EPI_ISL_2773293, EPI_ISL_2773294, EPI_ISL_2773295, EPI_ISL_2773296, EPI_ISL_2773297, EPI_ISL_2773298, EPI_ISL_2773299, EPI_ISL_2773300, EPI_ISL_2773301, EPI_ISL_2773302, EPI_ISL_2773303, EPI_ISL_2773304, EPI_ISL_2773305, EPI_ISL_2773306, EPI_ISL_2773307, EPI_ISL_2773308, EPI_ISL_2773309, EPI_ISL_2773310, EPI_ISL_2773311, EPI_ISL_2773312, EPI_ISL_2773313, EPI_ISL_2773314, EPI_ISL_2773315, EPI_ISL_2773316, EPI_ISL_2773317, EPI_ISL_2773318, EPI_ISL_2773319, EPI_ISL_2773320, EPI_ISL_2773321, EPI_ISL_2773322, EPI_ISL_2773323, EPI_ISL_2773324, EPI_ISL_2773325, EPI_ISL_2773326, EPI_ISL_2773327, EPI_ISL_2773328, EPI_ISL_2773329, EPI_ISL_2773330, EPI_ISL_2773331, EPI_ISL_2773332, EPI_ISL_2773333, EPI_ISL_2773334, EPI_ISL_2773335, EPI_ISL_2773336, EPI_ISL_2773337, EPI_ISL_2773338, EPI_ISL_2773339, EPI_ISL_2773340, EPI_ISL_2773366, EPI_ISL_2773367, EPI_ISL_2773368, EPI_ISL_2773369, EPI_ISL_2773370, EPI_ISL_2773371, EPI_ISL_2773372, EPI_ISL_2773373, EPI_ISL_2773374, EPI_ISL_2773375, EPI_ISL_2773376, EPI_ISL_2773377, EPI_ISL_2773378, EPI_ISL_2773379, EPI_ISL_2773380, EPI_ISL_2773381, EPI_ISL_2773382, EPI_ISL_2773383, EPI_ISL_2773384, EPI_ISL_2773385, EPI_ISL_2773386, EPI_ISL_2773387, EPI_ISL_2773388, EPI_ISL_2773389, EPI_ISL_2773390, EPI_ISL_2773391, EPI_ISL_2773392, EPI_ISL_2773393, EPI_ISL_2773394, EPI_ISL_2773395, EPI_ISL_2773396, EPI_ISL_2773397, EPI_ISL_2773398, EPI_ISL_2773399, EPI_ISL_2773400, EPI_ISL_2773401, EPI_ISL_2773402, EPI_ISL_2773403, EPI_ISL_2773404, EPI_ISL_2773405, EPI_ISL_2773406, EPI_ISL_2773407, EPI_ISL_2773408, EPI_ISL_2773409, EPI_ISL_2773410, EPI_ISL_2773411, EPI_ISL_2773412, EPI_ISL_2773413, EPI_ISL_2773414, EPI_ISL_2773415, EPI_ISL_2773416, EPI_ISL_2773417, EPI_ISL_2773418, EPI_ISL_2773419, EPI_ISL_2773420, EPI_ISL_2773421, EPI_ISL_2773422, EPI_ISL_2773423, EPI_ISL_2773424, EPI_ISL_2773425, EPI_ISL_2773426, EPI_ISL_2773427, EPI_ISL_2773428, EPI_ISL_2773429, EPI_ISL_2773430, EPI_ISL_2773431, EPI_ISL_2773432, EPI_ISL_2773433, EPI_ISL_2773434, EPI_ISL_2773435, EPI_ISL_2773436, EPI_ISL_2773437, EPI_ISL_2773438, EPI_ISL_2773439, EPI_ISL_2773440, EPI_ISL_2773441, EPI_ISL_2773442, EPI_ISL_2773443, EPI_ISL_2773444, EPI_ISL_2773445, EPI_ISL_2773446, EPI_ISL_2773447, EPI_ISL_2773448, EPI_ISL_2773449, EPI_ISL_2773450, EPI_ISL_2773451, EPI_ISL_2773452, EPI_ISL_2773453, EPI_ISL_2773454, EPI_ISL_2773455, EPI_ISL_2773456, EPI_ISL_2773457, EPI_ISL_2773458, EPI_ISL_2773459, EPI_ISL_2773460, EPI_ISL_2773461, EPI_ISL_2773462, EPI_ISL_2773463, EPI_ISL_2773464, EPI_ISL_2773465, EPI_ISL_2773466, EPI_ISL_2773467, EPI_ISL_2773468, EPI_ISL_2773469, EPI_ISL_2773470, EPI_ISL_2773471, EPI_ISL_2773472, EPI_ISL_2773473, EPI_ISL_2773474, EPI_ISL_2773475, EPI_ISL_2773476, EPI_ISL_2773477, EPI_ISL_2773478, EPI_ISL_2773479, EPI_ISL_2773480, EPI_ISL_2773481, EPI_ISL_2773482, EPI_ISL_2773483, EPI_ISL_2773484, EPI_ISL_2773485, EPI_ISL_2773486, EPI_ISL_2773487, EPI_ISL_2773488, EPI_ISL_2773532, EPI_ISL_2773533, EPI_ISL_2773534, EPI_ISL_2773539, EPI_ISL_2773543, EPI_ISL_2773544, EPI_ISL_2773545, EPI_ISL_2773546, EPI_ISL_2773547, EPI_ISL_2773548, EPI_ISL_2773549, EPI_ISL_2773550, EPI_ISL_2773551, EPI_ISL_2773552, EPI_ISL_2773553, EPI_ISL_2773554, EPI_ISL_2773555, EPI_ISL_2773556, EPI_ISL_2773557, EPI_ISL_2773558, EPI_ISL_2773559, EPI_ISL_2773560, EPI_ISL_2773561, EPI_ISL_2773562, EPI_ISL_2773563 | | | |
| see above | Fukuoka Institute of Health and Environmental Sciences | Fukuoka Institute of Health and Environmental Sciences | Akira Ohishi; Carl Yuki; Chiharu Katamune; Hiroaki Shigemura; Kentaro Itokawa; Makoto Kuroda; Masanori Hashino; Mitsuhiro Hamasaki; Rina Tanaka; Saori Ueda; Takayuki Kobayashi; Tsuyoshi Sekizuka; Yoshiki Etoh; Yuki Ashizuka |
| EPI_ISL_2391285, EPI_ISL_2391286, EPI_ISL_2391287, EPI_ISL_2812186, EPI_ISL_2812203, EPI_ISL_2812204, EPI_ISL_2812205, EPI_ISL_2812206, EPI_ISL_2812207, EPI_ISL_2812208, EPI_ISL_2812209, EPI_ISL_2812210, EPI_ISL_2812211, EPI_ISL_2812212, EPI_ISL_2812213, EPI_ISL_2812214, EPI_ISL_2812215, EPI_ISL_2812216, EPI_ISL_2812217, EPI_ISL_2812218, EPI_ISL_2812219, EPI_ISL_2812220, EPI_ISL_2812221, EPI_ISL_2812222, EPI_ISL_2812226, EPI_ISL_2812227 | | | |
| see above | Fukushima Prefectural Institute of Public Health | Fukushima Prefectural Institute of Public Health | Kadzuhiro Kitakawa; Kazunori Suzuki; Kentaro Itokawa; Makoto Kuroda; Masanori Hashino; Naoko Kashiwabara; Rie Suzuki; Rina Tanaka; Shohei Fujita; Tsuyoshi Sekizuka |
| EPI_ISL_2080250, EPI_ISL_2080251, EPI_ISL_2080252, EPI_ISL_2080253, EPI_ISL_2080254, EPI_ISL_2080269, EPI_ISL_2080351 | | | |
| see above | Genome Analysis Center, Kamma Memorial Hospital | Genome Analysis Center, Kamma Memorial Hospital | Hanako Yazawa; Hiroshi Kamma; Jun Ishii; Kaori Watanabe; Masaki Sato; Michiaki Masuda; Naruhisa Takano; Noriyuki Tannai; Satoko Soma; Takuya Yazawa |
| EPI_ISL_2235516, EPI_ISL_2235517, EPI_ISL_2235518, EPI_ISL_2235519, EPI_ISL_2235520, EPI_ISL_2349693, EPI_ISL_2349694, EPI_ISL_2349695, EPI_ISL_2349696, EPI_ISL_2349697, EPI_ISL_2349698, EPI_ISL_2349699, EPI_ISL_2349700, EPI_ISL_2349701, EPI_ISL_2349702, EPI_ISL_2349703, EPI_ISL_2349704, EPI_ISL_2349705, EPI_ISL_2349706, EPI_ISL_2349707, EPI_ISL_2349708, EPI_ISL_2462044, EPI_ISL_2462045, EPI_ISL_2462046, EPI_ISL_2462047, EPI_ISL_2462048, EPI_ISL_2462049, EPI_ISL_2462050, EPI_ISL_2462051, EPI_ISL_2462052, EPI_ISL_2462053, EPI_ISL_2462054, EPI_ISL_2462055, EPI_ISL_2462056, EPI_ISL_2497479, EPI_ISL_2497480, EPI_ISL_2497481, EPI_ISL_2497482, EPI_ISL_2497483, EPI_ISL_2497484, EPI_ISL_2497485, EPI_ISL_2497486, EPI_ISL_2497487, EPI_ISL_2497488, EPI_ISL_2497489, EPI_ISL_2497490, EPI_ISL_2497491, EPI_ISL_2497492, EPI_ISL_2497493, EPI_ISL_2497494, EPI_ISL_2497495, EPI_ISL_2621731, EPI_ISL_2644656, EPI_ISL_2644657, EPI_ISL_2657371 | | | |

| | | | |
|---|---|---|---|
| see above | Genome Analysis Center, Yamanashi Central Hospital | Genome Analysis Center, Yamanashi Central Hospital | Yosuke Hirotsu |
| EPI_ISL_2349250, EPI_ISL_2349251, EPI_ISL_2349252, EPI_ISL_2349253, EPI_ISL_2349254, EPI_ISL_2349255, EPI_ISL_2349256, EPI_ISL_2349257, EPI_ISL_2349258, EPI_ISL_2349259, EPI_ISL_2349260, EPI_ISL_2349261, EPI_ISL_2349262, EPI_ISL_2349263, EPI_ISL_2349264, EPI_ISL_2349265, EPI_ISL_2349266, EPI_ISL_2349267, EPI_ISL_2349268, EPI_ISL_2788645, EPI_ISL_2788646, EPI_ISL_2788647, EPI_ISL_2788648, EPI_ISL_2788649, EPI_ISL_2788650, EPI_ISL_2788651, EPI_ISL_2788652, EPI_ISL_2788653, EPI_ISL_2788654, EPI_ISL_2788655, EPI_ISL_2788656, EPI_ISL_2788657, EPI_ISL_2788658, EPI_ISL_2788659, EPI_ISL_2788660, EPI_ISL_2788661, EPI_ISL_2788662, EPI_ISL_2788663, EPI_ISL_2788664, EPI_ISL_2788665, EPI_ISL_2788666, EPI_ISL_2788667, EPI_ISL_2788668, EPI_ISL_2788669, EPI_ISL_2788670, EPI_ISL_2788671, EPI_ISL_2788672, EPI_ISL_2788673, EPI_ISL_2788674, EPI_ISL_2788675, EPI_ISL_2788676, EPI_ISL_2788677, EPI_ISL_2788678, EPI_ISL_2788679, EPI_ISL_2788680, EPI_ISL_2788681, EPI_ISL_2788682, EPI_ISL_2788683, EPI_ISL_2788684, EPI_ISL_2788685, EPI_ISL_2788686, EPI_ISL_2788687, EPI_ISL_2788688, EPI_ISL_2788692, EPI_ISL_2788703, EPI_ISL_2788704, EPI_ISL_2788705, EPI_ISL_2788711 | | | |
| see above | Gunma Prefectural Institute of Public Health and Environmental Sciences | Gunma Prefectural Institute of Public Health and Environmental Sciences | Daisuke Shinoda; Hiroyuki Tsukagoshi; Kentaro Itokawa; Makoto Kuroda; Mariko Saito; Masanori Hashino; Rina Tanaka; Tsuyoshi Sekizuka |
| EPI_ISL_2773701, EPI_ISL_2773702, EPI_ISL_2773703, EPI_ISL_2773704, EPI_ISL_2773705, EPI_ISL_2773706, EPI_ISL_2773707, EPI_ISL_2773708, EPI_ISL_2773709, EPI_ISL_2773710, EPI_ISL_2773711, EPI_ISL_2773712, EPI_ISL_2773713, EPI_ISL_2773714, EPI_ISL_2773715, EPI_ISL_2773716, EPI_ISL_2773717, EPI_ISL_2773718, EPI_ISL_2773719, EPI_ISL_2773720, EPI_ISL_2773721, EPI_ISL_2773722, EPI_ISL_2773723, EPI_ISL_2773724, EPI_ISL_2773725, EPI_ISL_2773726, EPI_ISL_2773727, EPI_ISL_2773728, EPI_ISL_2773729, EPI_ISL_2773730, EPI_ISL_2773731, EPI_ISL_2773732, EPI_ISL_2773733, EPI_ISL_2773734, EPI_ISL_2773735, EPI_ISL_2773736, EPI_ISL_2773737, EPI_ISL_2773738, EPI_ISL_2773739, EPI_ISL_2773740, EPI_ISL_2773742, EPI_ISL_2773743, EPI_ISL_2773744, EPI_ISL_2773745, EPI_ISL_2773746, EPI_ISL_2773747, EPI_ISL_2773748, EPI_ISL_2773749, EPI_ISL_2773750, EPI_ISL_2773751, EPI_ISL_2773752, EPI_ISL_2773753, EPI_ISL_2773754, EPI_ISL_2773755, EPI_ISL_2773756, EPI_ISL_2773757, EPI_ISL_2773758, EPI_ISL_2773759, EPI_ISL_2773760, EPI_ISL_2773761, EPI_ISL_2773762, EPI_ISL_2773763, EPI_ISL_2773764, EPI_ISL_2773765, EPI_ISL_2773766, EPI_ISL_2773768, EPI_ISL_2773769, EPI_ISL_2773770, EPI_ISL_2773771, EPI_ISL_2773774, EPI_ISL_2773798, EPI_ISL_2773807, EPI_ISL_2773813, EPI_ISL_2773836 | | | |
| see above | Hiroshima Prefectural Technology Research Institute, Public Health and Environment Center | Hiroshima Prefectural Technology Research Institute, Public Health and Environment Center | Akie Nakashima; Ayano Ito; Hiroko Akita; Kanako Masuda; Kentaro Itokawa; Makoto Kuroda; Mana Suei; Masanori Hashino; Naoki Shigemoto; Rina Tanaka; Shinichi Takao; Syuhei Ikeda; Takahiro Hiratsuka; Tsuyoshi Sekizuka; Yukie Shimazu |
| EPI_ISL_2358164, EPI_ISL_2358165, EPI_ISL_2358166, EPI_ISL_2358167, EPI_ISL_2358168, EPI_ISL_2358169, EPI_ISL_2358173, EPI_ISL_2358174, EPI_ISL_2358175, EPI_ISL_2358176, EPI_ISL_2358177, EPI_ISL_2358179, EPI_ISL_2358184, EPI_ISL_2358185, EPI_ISL_2358186, EPI_ISL_2358187, EPI_ISL_2358188, EPI_ISL_2358189, EPI_ISL_2358190, EPI_ISL_2358191, EPI_ISL_2358192, EPI_ISL_2358193, EPI_ISL_2358194, EPI_ISL_2358195, EPI_ISL_2358196, EPI_ISL_2358197, EPI_ISL_2358198, EPI_ISL_2358199, EPI_ISL_2358200, EPI_ISL_2358201, EPI_ISL_2358202, EPI_ISL_2358205, EPI_ISL_2358206, EPI_ISL_2358207, EPI_ISL_2358209, EPI_ISL_2358210, EPI_ISL_2358211, EPI_ISL_2358212, EPI_ISL_2358213, EPI_ISL_2358214, EPI_ISL_2358215, EPI_ISL_2358216, EPI_ISL_2358217, EPI_ISL_2358218, EPI_ISL_2358219, EPI_ISL_2358220, EPI_ISL_2358221, EPI_ISL_2358222, EPI_ISL_2358223, EPI_ISL_2358224, EPI_ISL_2358225, EPI_ISL_2884639, EPI_ISL_2884640, EPI_ISL_2884641, EPI_ISL_2884642, EPI_ISL_2884643, EPI_ISL_2884644, EPI_ISL_2884645, EPI_ISL_2884646, EPI_ISL_2884647, EPI_ISL_2884648, EPI_ISL_2884649, EPI_ISL_2884650, EPI_ISL_2884651, EPI_ISL_2884652, EPI_ISL_2884653, EPI_ISL_2884654, EPI_ISL_2884655, EPI_ISL_2884656, EPI_ISL_2884657, EPI_ISL_2884658, EPI_ISL_2884659, EPI_ISL_2884660, EPI_ISL_2884661, EPI_ISL_2884662, EPI_ISL_2884663, EPI_ISL_2884664, EPI_ISL_2884665, EPI_ISL_2884666, EPI_ISL_2884667, EPI_ISL_2884668, EPI_ISL_2884669, EPI_ISL_2884670, EPI_ISL_2884671, EPI_ISL_2884672, EPI_ISL_2884673, EPI_ISL_2884674, EPI_ISL_2884676, EPI_ISL_2884677, EPI_ISL_2884678, EPI_ISL_2884679, EPI_ISL_2884680, EPI_ISL_2884681, EPI_ISL_2884682, EPI_ISL_2884683, EPI_ISL_2884684, EPI_ISL_2884685, EPI_ISL_2884686, EPI_ISL_2884687, EPI_ISL_2884688, EPI_ISL_2884689, EPI_ISL_2884690, EPI_ISL_2884691, EPI_ISL_2884692, EPI_ISL_2884694, EPI_ISL_2884695, EPI_ISL_2884696, EPI_ISL_2884697, EPI_ISL_2884698, EPI_ISL_2884699, EPI_ISL_2884700, EPI_ISL_2884701, EPI_ISL_2884702, EPI_ISL_2884703, EPI_ISL_2884704, EPI_ISL_2884705, EPI_ISL_2884706, EPI_ISL_2884707, EPI_ISL_2884708, EPI_ISL_2884709, EPI_ISL_2884710, EPI_ISL_2884711, EPI_ISL_2884712, EPI_ISL_2884713, EPI_ISL_2884714, EPI_ISL_2884715, EPI_ISL_2884716, EPI_ISL_2884717, EPI_ISL_2884718, EPI_ISL_2884720, EPI_ISL_2884721, EPI_ISL_2884723, EPI_ISL_2884724, EPI_ISL_2884725, EPI_ISL_2884726, EPI_ISL_2884727, EPI_ISL_2884728, EPI_ISL_2884729, EPI_ISL_2884730, EPI_ISL_2884731, EPI_ISL_2884732, EPI_ISL_2884736, EPI_ISL_2884737, EPI_ISL_2884738, EPI_ISL_2884742, EPI_ISL_2884743, EPI_ISL_2884747, EPI_ISL_2884748, EPI_ISL_2884749, EPI_ISL_2884754, EPI_ISL_2884787, EPI_ISL_2884816, EPI_ISL_2884817, EPI_ISL_2884818, EPI_ISL_2884823, EPI_ISL_2884824, EPI_ISL_2884825, EPI_ISL_2884832, EPI_ISL_2884833, EPI_ISL_2884838, EPI_ISL_2884839, EPI_ISL_2884840, EPI_ISL_2884841, EPI_ISL_2884842, EPI_ISL_2884843, EPI_ISL_2884844, EPI_ISL_2884845, EPI_ISL_2884846, EPI_ISL_2884847, EPI_ISL_2884848, EPI_ISL_2884849, EPI_ISL_2884850, EPI_ISL_2884851, EPI_ISL_2884853, EPI_ISL_2884854, EPI_ISL_2884855, EPI_ISL_2884856, EPI_ISL_2884857, EPI_ISL_2884858, EPI_ISL_2884859, EPI_ISL_2884863, EPI_ISL_2884864, EPI_ISL_2884865, EPI_ISL_2884866, EPI_ISL_2884867, EPI_ISL_2884868, EPI_ISL_2884869, EPI_ISL_2884870, EPI_ISL_2884871, EPI_ISL_2884872, EPI_ISL_2884873, EPI_ISL_2884874, EPI_ISL_2884875, EPI_ISL_2884876, EPI_ISL_2884877, EPI_ISL_2884878 | | | |
| see above | Hokkaido Institute of Public Health | Hokkaido Institute of Public Health | Kazuhiro Okubo; Kentaro Itokawa; Makoto Kuroda; Masanori Hashino; Rina Tanaka; Tsuyoshi Sekizuka; Yuta Ohno |
| EPI_ISL_2364544, EPI_ISL_2364545, EPI_ISL_2364546, EPI_ISL_2364547, EPI_ISL_2364548, EPI_ISL_2364549, EPI_ISL_2364550, EPI_ISL_2364551, EPI_ISL_2364552, EPI_ISL_2364553, EPI_ISL_2364554, EPI_ISL_2364555, EPI_ISL_2364556, EPI_ISL_2364557, EPI_ISL_2364558, EPI_ISL_2364559, EPI_ISL_2364560, EPI_ISL_2364561, EPI_ISL_2364562, EPI_ISL_2364563, EPI_ISL_2364564, EPI_ISL_2364565, EPI_ISL_2364566, EPI_ISL_2860648, EPI_ISL_2860649, EPI_ISL_2860650, EPI_ISL_2860651, EPI_ISL_2860652, EPI_ISL_2860653, EPI_ISL_2860654, EPI_ISL_2860655, EPI_ISL_2860656, EPI_ISL_2860657, EPI_ISL_2860658, EPI_ISL_2860659, EPI_ISL_2860660, EPI_ISL_2860661, EPI_ISL_2860662, EPI_ISL_2860663, EPI_ISL_2860664, EPI_ISL_2860665, EPI_ISL_2860666, EPI_ISL_2860667, EPI_ISL_2860668, EPI_ISL_2860669, EPI_ISL_2860670, EPI_ISL_2860671, EPI_ISL_2860672, EPI_ISL_2860673, EPI_ISL_2860674, EPI_ISL_2860675, EPI_ISL_2860676, EPI_ISL_2860677, EPI_ISL_2860678, EPI_ISL_2860679, EPI_ISL_2860680, EPI_ISL_2860701, EPI_ISL_2860702, EPI_ISL_2860703, EPI_ISL_2860704, EPI_ISL_2860705, EPI_ISL_2860724, EPI_ISL_2860725, EPI_ISL_2860726, EPI_ISL_2860728, EPI_ISL_2860729, EPI_ISL_2860730, EPI_ISL_2860731, EPI_ISL_2860732, EPI_ISL_2860754, EPI_ISL_2860755, EPI_ISL_2860763, EPI_ISL_2860764 | | | |
| see above | Hyogo Prefectural Institute of Public Health science | Pathogen Genomics Center, National Institute of Infectious Diseases | Denshi Takai; Etsuko Saito; Kenichi Ogita; Kentaro Itokawa; Kyoko Suzuki; Makoto Kuroda; Masanori Hashino; Masatsugu Chikahira; Miki Ogi; Rina Tanaka; Tomohiro Oshibe; Tsuyoshi Sekizuka; Yumi Akiyama |
| EPI_ISL_2350621, EPI_ISL_2350628, EPI_ISL_2350630, EPI_ISL_2350631, EPI_ISL_2350632, EPI_ISL_2350633, EPI_ISL_2350634, EPI_ISL_2350635, EPI_ISL_2350636, EPI_ISL_2350638, EPI_ISL_2350639, EPI_ISL_2350640, EPI_ISL_2350641, EPI_ISL_2350642, EPI_ISL_2350643, EPI_ISL_2350644, EPI_ISL_2350645, EPI_ISL_2350646, EPI_ISL_2350647, EPI_ISL_2350648, EPI_ISL_2350649, EPI_ISL_2350650, EPI_ISL_2350651, EPI_ISL_2350652, EPI_ISL_2350653, EPI_ISL_2350654, EPI_ISL_2350655, EPI_ISL_2350656, EPI_ISL_2350694, EPI_ISL_2786706, EPI_ISL_2786707, EPI_ISL_2786708, EPI_ISL_2786709, EPI_ISL_2786710, EPI_ISL_2786711, EPI_ISL_2786712, EPI_ISL_2786713, EPI_ISL_2786714, EPI_ISL_2786715, EPI_ISL_2786716, EPI_ISL_2786717, EPI_ISL_2786718, EPI_ISL_2786719, EPI_ISL_2786720, EPI_ISL_2786721, EPI_ISL_2786722, EPI_ISL_2786723, EPI_ISL_2786734, EPI_ISL_2786735, EPI_ISL_2786736, EPI_ISL_2786737, EPI_ISL_2786738, EPI_ISL_2786794, EPI_ISL_2786795, EPI_ISL_2786796, EPI_ISL_2786797, EPI_ISL_2786798, EPI_ISL_2786799, EPI_ISL_2786820, EPI_ISL_2786821, EPI_ISL_2786822, EPI_ISL_2786823, EPI_ISL_2786824, EPI_ISL_2786845, EPI_ISL_2786846, EPI_ISL_2786847, EPI_ISL_2786848, EPI_ISL_2786859, EPI_ISL_2786870, EPI_ISL_2786871, EPI_ISL_2786872, EPI_ISL_2786873, EPI_ISL_2786874, EPI_ISL_2786875, EPI_ISL_2786876, EPI_ISL_2786886 | | | |
| see above | Ibaraki Prefectural Institute of Public Health | Ibaraki Prefectural Institute of Public Health | Keiko Goto; Kentaro Itokawa; Makoto Kuroda; Masanori Hashino; Rina Tanaka; Tsuyoshi Sekizuka |
| EPI_ISL_2787099, EPI_ISL_2787100, EPI_ISL_2787101, EPI_ISL_2787103, EPI_ISL_2787104, EPI_ISL_2787105, EPI_ISL_2787106, EPI_ISL_2787107, EPI_ISL_2787108, EPI_ISL_2787110 | | | |
| see above | Kagawa Prefectural Research Institute for Environmental Sciences and Public Health | Pathogen Genomics Center, National Institute of Infectious Diseases | Kenji Kuwahara; Kentaro Itokawa; Makoto Kuroda; Masanori Hashino; Mayumi Arizuka; Rina Tanaka; Tsuyoshi Sekizuka; Yukari Terajima |
| EPI_ISL_2842531, EPI_ISL_2842532, EPI_ISL_2842533, EPI_ISL_2842534, EPI_ISL_2842535, EPI_ISL_2842536, EPI_ISL_2842539, EPI_ISL_2842540, EPI_ISL_2842541, EPI_ISL_2842545, EPI_ISL_2842547, EPI_ISL_2842555, EPI_ISL_2842557, EPI_ISL_2842562 | | | |
| see above | Kagoshima Prefectural Institute for Environmental Research and Public Health | Kagoshima Prefectural Institute for Environmental Research and Public Health | Kentaro Itokawa; Makoto Kuroda; Masanori Hashino; Rina Tanaka; Tsuyoshi Sekizuka; Yuka Hamada |
| EPI_ISL_2361601, EPI_ISL_2361602, EPI_ISL_2361603, EPI_ISL_2361613, EPI_ISL_2361614, EPI_ISL_2361615, EPI_ISL_2361616, EPI_ISL_2544681, EPI_ISL_2544682, EPI_ISL_2544683, EPI_ISL_2544684, EPI_ISL_2544685, EPI_ISL_2544691, EPI_ISL_2544692, EPI_ISL_2544694 | | | |
| see above | Kanagawa Prefectural Institute of Public Health | Kanagawa Prefectural Institute of Public Health | Itsumi Toyokura; Junichi Sakuragi; Kentaro Itokawa; Makiko Kondo; Makoto Kuroda; Masanori Hashino; Rieko Suzuki; Rina Tanaka; Sumi Watanabe; Takako Sano; Takayuki Hishiki; Tomohiko Takasaki; Tsuyoshi Sekizuka |
| EPI_ISL_2758646, EPI_ISL_2758807, EPI_ISL_2758808, EPI_ISL_2758828 | Kawakita General Hospital | Tokyo Medical University Department of Microbiology | Hirofumi Miyake; Jun Suzuki; Kenji Sadamas; Kenshirou Kroki; Mami Nagashima; Maya Isogai; Shigeki Nakamura; Takahiro Okai; Takako Yamazaki; Takushi Fujiwara; Tatsuya Inukai |
| EPI_ISL_2828096, EPI_ISL_2828097, EPI_ISL_2828098 | Kyoto Prefectural Institute Of Public Health And Environment | Kyoto Prefectural Institute Of Public Health And Environment | Akira Kotera; Kenji Konaka; Kentaro Itokawa; Makoto Kuroda; Masanori Hashino; Miki Tsuda; Nana Horikawa; Norio Asai; Rina Tanaka; Tsuyoshi Sekizuka; Yasushi Fujimoto; Yoshihiro Tanabe; Yoshiyuki Kawahara; Yuri Sakai |
| EPI_ISL_2841491, EPI_ISL_2841492, EPI_ISL_2841493, EPI_ISL_2841494, EPI_ISL_2841496, EPI_ISL_2841504 | Medical Microbiology and Zoology Section, Okinawa Prefectural Institute of Health and Environment | Pathogen Genomics Center, National Institute of Infectious Diseases | Hisako Kyan; Kentaro Itokawa; Makoto Kuroda; Masanori Hashino; Minoru Nidaira; Noriyuki Maeshiro; Rina Tanaka; Taketoshi Takara; Tetsuya Kakita; Tsuyoshi Kudeken; Tsuyoshi Sekizuka; Yumani Kuba |
| EPI_ISL_2768437, EPI_ISL_2768438, EPI_ISL_2768439, EPI_ISL_2768440, EPI_ISL_2768441, EPI_ISL_2768442, EPI_ISL_2768443, EPI_ISL_2768444, EPI_ISL_2768445, EPI_ISL_2768446, EPI_ISL_2768447, EPI_ISL_2768448, EPI_ISL_2768449, EPI_ISL_2768450, EPI_ISL_2768451, EPI_ISL_2768452, EPI_ISL_2768453, EPI_ISL_2768454, EPI_ISL_2768455, EPI_ISL_2768456, EPI_ISL_2768457, EPI_ISL_2768458, EPI_ISL_2768459, EPI_ISL_2768460, EPI_ISL_2768461, EPI_ISL_2768462, EPI_ISL_2768463, EPI_ISL_2768464 | | | |
| see above | Niigata Prefectural Institute of Public Health and Environmental Sciences | Pathogen Genomics Center, National Institute of Infectious Diseases | Kentaro Itokawa; Makoto Kuroda; Masanori Hashino; Rina Tanaka; Tsuyoshi Sekizuka |
| EPI_ISL_2330520, EPI_ISL_2338398, EPI_ISL_2338399, EPI_ISL_2338400, EPI_ISL_2760212, EPI_ISL_2760213, EPI_ISL_2760215, EPI_ISL_2760216, EPI_ISL_2760840, EPI_ISL_2760842, EPI_ISL_2760844 | | | |
| see above | Osaka Institute of Public Health, Morinomiya Center | Pathogen Genomics Center, National Institute of Infectious Diseases | Kentaro Itokawa; Makoto Kuroda; Masanori Hashino; Rina Tanaka; Tsuyoshi Sekizuka |
| EPI_ISL_2663786, EPI_ISL_2663788, EPI_ISL_2663789, EPI_ISL_2663790, EPI_ISL_2663792, EPI_ISL_2663794, EPI_ISL_2663795, EPI_ISL_2663796 | | | |
| see above | Osaka University Hospital | Genome Information Research Center, Research Institute for Microbial Diseases, Osaka University | Daisuke Motooka; Haruhiko Hirata; Hiroshi Ogura; Hiroya Oki; Hisatake Matsumoto; Kentaro Tanaka; Shota Nakamura; Takeshi Ebihara; Yasuhiro Kato; and Atsushi Kumanogoh |
| EPI_ISL_2329642, EPI_ISL_2329643, EPI_ISL_2329644, EPI_ISL_2329645, EPI_ISL_2329646, EPI_ISL_2329647, EPI_ISL_2329648, EPI_ISL_2329649, EPI_ISL_2329650, EPI_ISL_2329651, EPI_ISL_2329652, EPI_ISL_2329653, EPI_ISL_2329654, EPI_ISL_2329655, EPI_ISL_2329656, EPI_ISL_2329657, EPI_ISL_2329658, EPI_ISL_2329659, EPI_ISL_2329660, EPI_ISL_2329661, EPI_ISL_2329662, EPI_ISL_2329663, EPI_ISL_2329664, EPI_ISL_2329665, EPI_ISL_2329666, EPI_ISL_2329667, EPI_ISL_2329668, EPI_ISL_2329669, EPI_ISL_2329670, EPI_ISL_2329671, EPI_ISL_2329672, EPI_ISL_2329673, EPI_ISL_2329677, EPI_ISL_2329736, EPI_ISL_2329807, EPI_ISL_2329808, EPI_ISL_2329809, EPI_ISL_2329810, EPI_ISL_2330338, EPI_ISL_2330339, EPI_ISL_2330340, EPI_ISL_2330341, EPI_ISL_2330342, EPI_ISL_2330343, EPI_ISL_2330350, EPI_ISL_2330375, EPI_ISL_2330379, EPI_ISL_2330380, EPI_ISL_2330381, EPI_ISL_2330382, EPI_ISL_2330383, EPI_ISL_2330384, EPI_ISL_2330385, EPI_ISL_2330386, EPI_ISL_2330387, EPI_ISL_2330407, EPI_ISL_2330408, EPI_ISL_2330409, EPI_ISL_2330410, EPI_ISL_2330411, EPI_ISL_2330412, EPI_ISL_2330413, EPI_ISL_2330414, EPI_ISL_2330415, EPI_ISL_2330416, EPI_ISL_2330420, EPI_ISL_2330421, EPI_ISL_2330422, EPI_ISL_2330425, EPI_ISL_2330426, EPI_ISL_2330432, EPI_ISL_2330433, EPI_ISL_2330434, EPI_ISL_2330435, EPI_ISL_2330436, EPI_ISL_2330437, EPI_ISL_2330438, EPI_ISL_2330439, EPI_ISL_2330440, EPI_ISL_2330441, EPI_ISL_2330442, EPI_ISL_2330443, EPI_ISL_2330444, EPI_ISL_2330445, EPI_ISL_2330446, EPI_ISL_2330447, EPI_ISL_2330448, EPI_ISL_2330449, EPI_ISL_2330450, EPI_ISL_2330451, EPI_ISL_2330452, EPI_ISL_2330453, EPI_ISL_2330454, EPI_ISL_2330455, EPI_ISL_2330456, EPI_ISL_2330457, EPI_ISL_2330458, EPI_ISL_2330459, EPI_ISL_2330460, EPI_ISL_2330461, EPI_ISL_2330462, EPI_ISL_2330463, EPI_ISL_2330464, EPI_ISL_2330465, EPI_ISL_2330466, EPI_ISL_2330467, EPI_ISL_2330468, EPI_ISL_2330469, EPI_ISL_2330470, EPI_ISL_2330471, EPI_ISL_2330472, EPI_ISL_2330473, EPI_ISL_2330474, EPI_ISL_2330475, EPI_ISL_2330476, EPI_ISL_2330477, EPI_ISL_2330478, EPI_ISL_2330479, EPI_ISL_2330480, EPI_ISL_2330481, EPI_ISL_2330482, EPI_ISL_2330483, EPI_ISL_2330484, EPI_ISL_2330485, EPI_ISL_2330486, EPI_ISL_2330487, EPI_ISL_2330488, EPI_ISL_2330489, EPI_ISL_2330490, EPI_ISL_2330491, EPI_ISL_2330492, EPI_ISL_2330493, EPI_ISL_2330494, EPI_ISL_2330495, EPI_ISL_2330496, EPI_ISL_2330497, EPI_ISL_2330498, EPI_ISL_2330499, EPI_ISL_2330500, EPI_ISL_2330501, EPI_ISL_2330502, EPI_ISL_2330503, EPI_ISL_2330504, EPI_ISL_2330505, EPI_ISL_2330506, EPI_ISL_2330507, EPI_ISL_2330623, EPI_ISL_2330624, EPI_ISL_2330625, EPI_ISL_2330626, EPI_ISL_2330627, EPI_ISL_2330628, EPI_ISL_2336371, EPI_ISL_2336372, EPI_ISL_2336373, EPI_ISL_2336374, EPI_ISL_2336375, EPI_ISL_2336834, EPI_ISL_2336835, EPI_ISL_2336836, EPI_ISL_2336837, EPI_ISL_2336838, EPI_ISL_2336839, EPI_ISL_2336840, EPI_ISL_2336883, EPI_ISL_2336896, EPI_ISL_2337397, EPI_ISL_2337400, EPI_ISL_2337401, EPI_ISL_2337402, EPI_ISL_2337403, EPI_ISL_2337405, EPI_ISL_2337406, EPI_ISL_2337407, EPI_ISL_2337410, EPI_ISL_2337411, EPI_ISL_2337413, EPI_ISL_2337414, EPI_ISL_2337415, EPI_ISL_2337417, EPI_ISL_2337418, EPI_ISL_2337419, EPI_ISL_2337420, EPI_ISL_2337422, EPI_ISL_2337424, EPI_ISL_2337429, EPI_ISL_2337431, EPI_ISL_2337434, EPI_ISL_2337442, EPI_ISL_2337445, EPI_ISL_2337446, EPI_ISL_2337468, EPI_ISL_2337469, EPI_ISL_2337470, EPI_ISL_2337471, EPI_ISL_2337472, EPI_ISL_2337473, EPI_ISL_2337474, EPI_ISL_2337475, EPI_ISL_2337476, EPI_ISL_2337477, EPI_ISL_2337478, EPI_ISL_2337479, EPI_ISL_2337480, EPI_ISL_2337481, EPI_ISL_2337482, EPI_ISL_2337483, EPI_ISL_2337484, EPI_ISL_2337485, EPI_ISL_2337486, EPI_ISL_2337487, EPI_ISL_2337488, EPI_ISL_2337489, EPI_ISL_2337490, EPI_ISL_2337491, EPI_ISL_2337492, EPI_ISL_2337493, EPI_ISL_2337513, EPI_ISL_2337514, EPI_ISL_2337515, EPI_ISL_2337516, EPI_ISL_2337517, EPI_ISL_2337518, EPI_ISL_2337519, EPI_ISL_2337520, EPI_ISL_2337521, EPI_ISL_2337522, EPI_ISL_2337523, EPI_ISL_2337524, EPI_ISL_2337560, EPI_ISL_2337561, EPI_ISL_2337562, EPI_ISL_2337563, EPI_ISL_2337564, EPI_ISL_2337565, EPI_ISL_2337566, EPI_ISL_2337567, EPI_ISL_2337578, EPI_ISL_2337579, EPI_ISL_2337580, EPI_ISL_2337581, EPI_ISL_2337582, EPI_ISL_2337583, EPI_ISL_2337584, EPI_ISL_2337590, EPI_ISL_2337592, EPI_ISL_2337597, EPI_ISL_2338346, EPI_ISL_2338347, EPI_ISL_2338348, EPI_ISL_2338349, EPI_ISL_2338350, EPI_ISL_2338356, EPI_ISL_2338357, EPI_ISL_2338358, EPI_ISL_2338359, EPI_ISL_2338360, EPI_ISL_2338361, EPI_ISL_2338362, EPI_ISL_2338363, EPI_ISL_2338364, EPI_ISL_2338365, EPI_ISL_2338391, EPI_ISL_2338392, EPI_ISL_2338393, EPI_ISL_2338402, EPI_ISL_2338403, EPI_ISL_2338404, EPI_ISL_2338405, EPI_ISL_2339239, EPI_ISL_2339240, EPI_ISL_2339243, EPI_ISL_2339245, EPI_ISL_2339246, EPI_ISL_2339247, EPI_ISL_2339248, EPI_ISL_2339249, EPI_ISL_2339254, EPI_ISL_2339255, EPI_ISL_2339256, EPI_ISL_2392159, EPI_ISL_2491348, EPI_ISL_2491350, EPI_ISL_2491351, EPI_ISL_2491354, EPI_ISL_2491356, EPI_ISL_2491357, EPI_ISL_2491358, EPI_ISL_2491360, EPI_ISL_2491361, EPI_ISL_2491362, EPI_ISL_2491363, EPI_ISL_2760080, EPI_ISL_2760082, EPI_ISL_2760205, EPI_ISL_2760207, EPI_ISL_2760210, EPI_ISL_2760355, EPI_ISL_2760357, EPI_ISL_2760358, EPI_ISL_2760360, EPI_ISL_2760361, EPI_ISL_2760363, EPI_ISL_2760365, EPI_ISL_2760367, EPI_ISL_2760369, EPI_ISL_2760370, EPI_ISL_2760374, EPI_ISL_2760375, EPI_ISL_2760377, EPI_ISL_2760379, EPI_ISL_2760380, EPI_ISL_2760382, EPI_ISL_2760384, EPI_ISL_2760386, EPI_ISL_2760388, EPI_ISL_2760389, EPI_ISL_2760391, EPI_ISL_2760393, EPI_ISL_2760395, EPI_ISL_2760397, EPI_ISL_2760398, EPI_ISL_2760400, EPI_ISL_2760401, EPI_ISL_2760403, EPI_ISL_2760405, EPI_ISL_2760406, EPI_ISL_2760408, EPI_ISL_2760410, EPI_ISL_2760411, EPI_ISL_2760413, EPI_ISL_2760415, EPI_ISL_2760416, EPI_ISL_2760418, EPI_ISL_2760421, EPI_ISL_2760423, EPI_ISL_2760425, EPI_ISL_2760427, EPI_ISL_2760429, EPI_ISL_2760430, EPI_ISL_2760432, EPI_ISL_2760434, EPI_ISL_2760435, EPI_ISL_2760437, EPI_ISL_2760438, EPI_ISL_2760440, EPI_ISL_2760442, EPI_ISL_2760443, EPI_ISL_2760445, EPI_ISL_2760446, EPI_ISL_2760448, EPI_ISL_2760450, EPI_ISL_2760451, EPI_ISL_2760453, EPI_ISL_2760454, EPI_ISL_2760456, EPI_ISL_2760458, EPI_ISL_2760459, EPI_ISL_2760461, EPI_ISL_2760465, EPI_ISL_2760805, EPI_ISL_2760807, EPI_ISL_2760809, EPI_ISL_2760810, EPI_ISL_2760845, EPI_ISL_2760847, EPI_ISL_2760848, EPI_ISL_2760850, EPI_ISL_2760851, EPI_ISL_2760853, EPI_ISL_2760855, EPI_ISL_2760856, EPI_ISL_2760858, EPI_ISL_2760860, EPI_ISL_2760861, EPI_ISL_2760863, EPI_ISL_2760865, EPI_ISL_2760866, EPI_ISL_2760868, EPI_ISL_2760869, EPI_ISL_2762957, EPI_ISL_2762958, EPI_ISL_2762959, EPI_ISL_2762960, EPI_ISL_2762961, EPI_ISL_2762962, EPI_ISL_2762976, EPI_ISL_2762977, EPI_ISL_2762978, EPI_ISL_2762979, EPI_ISL_2762980, EPI_ISL_2762981, EPI_ISL_2762982, EPI_ISL_2762983, EPI_ISL_2762984, EPI_ISL_2762985, EPI_ISL_2762986, EPI_ISL_2762987, EPI_ISL_2762988, EPI_ISL_2762989, EPI_ISL_2762990, EPI_ISL_2762991, EPI_ISL_2762993, EPI_ISL_2762994, EPI_ISL_2762995, EPI_ISL_2762996, EPI_ISL_2762997, EPI_ISL_2762998, EPI_ISL_2762999, EPI_ISL_2763000, EPI_ISL_2763001, EPI_ISL_2763002, EPI_ISL_2763003, EPI_ISL_2763011, EPI_ISL_2763133, EPI_ISL_2763134, EPI_ISL_2763135, EPI_ISL_2763136, EPI_ISL_2763137, EPI_ISL_2763138, EPI_ISL_2763139, EPI_ISL_2763140, EPI_ISL_2763141, EPI_ISL_2763142, EPI_ISL_2763143, EPI_ISL_2763144, EPI_ISL_2763145, EPI_ISL_2763146, EPI_ISL_2763147, EPI_ISL_2763148, EPI_ISL_2763149, EPI_ISL_2763150, EPI_ISL_2763151, EPI_ISL_2763962, EPI_ISL_2763963, EPI_ISL_2763964, EPI_ISL_2763965, EPI_ISL_2763966, EPI_ISL_2763967, EPI_ISL_2763968, EPI_ISL_2763969, EPI_ISL_2763970, EPI_ISL_2763971, EPI_ISL_2763972, EPI_ISL_2763973, EPI_ISL_2763974, EPI_ISL_2763975, EPI_ISL_2763977, EPI_ISL_2763978, EPI_ISL_2763979, EPI_ISL_2763980, EPI_ISL_2763981, EPI_ISL_2763982, EPI_ISL_2763983, EPI_ISL_2763984, EPI_ISL_2763986, EPI_ISL_2763987, EPI_ISL_2763988, EPI_ISL_2763989, EPI_ISL_2763990, EPI_ISL_2763991, EPI_ISL_2763992, EPI_ISL_2763993, EPI_ISL_2763994, EPI_ISL_2763995, EPI_ISL_2763996, EPI_ISL_2763997, EPI_ISL_2763998, EPI_ISL_2763999, EPI_ISL_2764000, EPI_ISL_2764001, EPI_ISL_2764002, EPI_ISL_2764003, EPI_ISL_2764004, EPI_ISL_2764005, EPI_ISL_2764006, EPI_ISL_2764007, EPI_ISL_2764008, EPI_ISL_2764009, EPI_ISL_2764010, EPI_ISL_2764011, EPI_ISL_2764012, EPI_ISL_2764013, EPI_ISL_2764014, EPI_ISL_2764015, EPI_ISL_2764016, EPI_ISL_2764017, EPI_ISL_2764018, EPI_ISL_2764019, EPI_ISL_2764020, EPI_ISL_2764021, EPI_ISL_2764023, EPI_ISL_2764024, EPI_ISL_2764025, EPI_ISL_2764026, EPI_ISL_2764027, EPI_ISL_2764028, EPI_ISL_2764038, EPI_ISL_2764091, EPI_ISL_2764092, EPI_ISL_2764093, EPI_ISL_2764094, EPI_ISL_2764095, EPI_ISL_2764097, EPI_ISL_2764098, EPI_ISL_2764099, EPI_ISL_2764100, EPI_ISL_2764101, EPI_ISL_2764103, EPI_ISL_2768312, EPI_ISL_2768313, EPI_ISL_2768314, EPI_ISL_2768316, EPI_ISL_2768317, EPI_ISL_2768318, EPI_ISL_2768320, EPI_ISL_2768321, EPI_ISL_2768323, EPI_ISL_2768376, EPI_ISL_2768377, EPI_ISL_2768378, EPI_ISL_2768379, EPI_ISL_2768380, EPI_ISL_2768381, EPI_ISL_2768382, EPI_ISL_2768383, EPI_ISL_2768384, EPI_ISL_2768392, EPI_ISL_2768393, EPI_ISL_2768394, EPI_ISL_2768416, EPI_ISL_2768417, EPI_ISL_2768418, EPI_ISL_2768419, EPI_ISL_2768420, EPI_ISL_2768421, EPI_ISL_2768422, EPI_ISL_2768423, EPI_ISL_2768424, EPI_ISL_2768425, EPI_ISL_2768426, EPI_ISL_2768848, EPI_ISL_2768872, EPI_ISL_2768873, EPI_ISL_2768874, EPI_ISL_2768875, EPI_ISL_2768876, EPI_ISL_2768877, EPI_ISL_2768878, EPI_ISL_2768879, EPI_ISL_2768880, EPI_ISL_2768881, EPI_ISL_2768882, EPI_ISL_2768883, EPI_ISL_2768884, EPI_ISL_2768885, EPI_ISL_2768886, EPI_ISL_2768887, EPI_ISL_2768888, EPI_ISL_2768889, EPI_ISL_2768890, EPI_ISL_2768891, EPI_ISL_2768892, EPI_ISL_2768893, EPI_ISL_2768894, EPI_ISL_2768895, EPI_ISL_2768896, EPI_ISL_2768897, EPI_ISL_2768898, EPI_ISL_2768899, EPI_ISL_2768900, EPI_ISL_2768901, EPI_ISL_2768902, EPI_ISL_2768903, EPI_ISL_2768904, EPI_ISL_2768905, EPI_ISL_2768906, EPI_ISL_2768907, EPI_ISL_2768908, EPI_ISL_2768909, EPI_ISL_2768910, EPI_ISL_2768911, EPI_ISL_2768912, EPI_ISL_2768913, EPI_ISL_2768914, EPI_ISL_2768915, EPI_ISL_2768916, EPI_ISL_2768917, EPI_ISL_2768918, EPI_ISL_2768919, EPI_ISL_2768920, EPI_ISL_2768921, EPI_ISL_2768922, EPI_ISL_2768923, EPI_ISL_2768924, EPI_ISL_2768925, EPI_ISL_2768926, EPI_ISL_2768927, EPI_ISL_2768928, EPI_ISL_2768929, EPI_ISL_2768930, EPI_ISL_2768931, EPI_ISL_2768932, EPI_ISL_2768959, EPI_ISL_2768960, EPI_ISL_2768961, EPI_ISL_2768962, EPI_ISL_2768963, EPI_ISL_2768964, EPI_ISL_2768965, EPI_ISL_2768966, EPI_ISL_2768967, EPI_ISL_2768968, EPI_ISL_2768969, EPI_ISL_2768970, EPI_ISL_2768971, EPI_ISL_2768972, EPI_ISL_2768973, EPI_ISL_2768974, EPI_ISL_2768975, EPI_ISL_2768976, EPI_ISL_2768977, EPI_ISL_2768978, EPI_ISL_2768979, EPI_ISL_2768980, EPI_ISL_2768981, EPI_ISL_2768982, EPI_ISL_2768983, EPI_ISL_2768984, EPI_ISL_2768985, EPI_ISL_2768986, EPI_ISL_2768987, EPI_ISL_2768988, EPI_ISL_2769004, EPI_ISL_2769005, EPI_ISL_2769006, EPI_ISL_2769007, EPI_ISL_2769008, EPI_ISL_2769009, EPI_ISL_2769010, EPI_ISL_2769011, EPI_ISL_2769012, EPI_ISL_2769013, EPI_ISL_2769014, EPI_ISL_2769015, EPI_ISL_2769016, EPI_ISL_2769017, EPI_ISL_2770256, EPI_ISL_2770257, EPI_ISL_2770258, EPI_ISL_2770259, EPI_ISL_2770260, EPI_ISL_2770261, EPI_ISL_2770262, EPI_ISL_2770263, EPI_ISL_2770264, EPI_ISL_2770265, EPI_ISL_2770266, EPI_ISL_2770267, EPI_ISL_2770268, EPI_ISL_2770269, EPI_ISL_2770270, EPI_ISL_2770271, EPI_ISL_2770272, EPI_ISL_2770273, EPI_ISL_2770274, EPI_ISL_2770275, EPI_ISL_2770276, EPI_ISL_2770277, EPI_ISL_2770278, EPI_ISL_2770279, EPI_ISL_2770280, EPI_ISL_2770281, EPI_ISL_2770282, EPI_ISL_2770283, EPI_ISL_2770284, EPI_ISL_2770285, EPI_ISL_2770286, EPI_ISL_2770287, EPI_ISL_2770288, EPI_ISL_2770289, EPI_ISL_2770290, EPI_ISL_2770291, EPI_ISL_2770292, EPI_ISL_2770293, EPI_ISL_2770294, EPI_ISL_2770295, EPI_ISL_2770296, EPI_ISL_2770297, EPI_ISL_2770298, EPI_ISL_2770299, EPI_ISL_2770300, EPI_ISL_2770301, EPI_ISL_2770302, EPI_ISL_2770303, EPI_ISL_2770304, EPI_ISL_2770305, EPI_ISL_2770306, EPI_ISL_2770307, EPI_ISL_2770308, EPI_ISL_2770309, EPI_ISL_2770310, EPI_ISL_2770311, EPI_ISL_2770312, EPI_ISL_2770313, EPI_ISL_2770314, EPI_ISL_2770315, EPI_ISL_2770316, EPI_ISL_2770317, EPI_ISL_2770318, EPI_ISL_2770319, EPI_ISL_2770320, EPI_ISL_2770321, EPI_ISL_2770322, EPI_ISL_2770323, EPI_ISL_2770324, EPI_ISL_2770325, EPI_ISL_2770326, EPI_ISL_2770327, EPI_ISL_2770328, EPI_ISL_2770329, EPI_ISL_2770330, EPI_ISL_2770331, EPI_ISL_2770332, EPI_ISL_2770333, EPI_ISL_2770334, EPI_ISL_2770335, EPI_ISL_2770336, EPI_ISL_2770337, EPI_ISL_2770338, EPI_ISL_2770339, EPI_ISL_2770340, EPI_ISL_2770341, EPI_ISL_2770342, EPI_ISL_2770343, EPI_ISL_2770344, EPI_ISL_2770345, EPI_ISL_2770346, EPI_ISL_2770347, EPI_ISL_2770348, EPI_ISL_2770718, EPI_ISL_2770719, EPI_ISL_2770720, EPI_ISL_2770721, EPI_ISL_2770722, EPI_ISL_2770723, EPI_ISL_2770724, EPI_ISL_2770725, EPI_ISL_2770726, EPI_ISL_2770727, EPI_ISL_2770728, EPI_ISL_2770729, EPI_ISL_2770730, EPI_ISL_2770731, EPI_ISL_2770732, EPI_ISL_2770733, EPI_ISL_2770734, EPI_ISL_2770735, EPI_ISL_2770736, EPI_ISL_2770737, EPI_ISL_2770738, EPI_ISL_2770739, EPI_ISL_2770740, EPI_ISL_2770741, EPI_ISL_2770742, EPI_ISL_2770755, EPI_ISL_2771520, EPI_ISL_2771521, EPI_ISL_2771522, EPI_ISL_2771523, EPI_ISL_2771524, EPI_ISL_2771525, EPI_ISL_2771526, EPI_ISL_2771527, EPI_ISL_2771528, EPI_ISL_2771529, EPI_ISL_2771530, EPI_ISL_2771531, EPI_ISL_2771532, EPI_ISL_2771533, EPI_ISL_2771534, EPI_ISL_2771535, EPI_ISL_2771536, EPI_ISL_2771537, EPI_ISL_2771538, EPI_ISL_2771539, EPI_ISL_2771540, EPI_ISL_2771541, EPI_ISL_2771550, EPI_ISL_2771551, EPI_ISL_2771552, EPI_ISL_2771553, EPI_ISL_2772130, EPI_ISL_2772131, EPI_ISL_2772132, EPI_ISL_2772133, EPI_ISL_2772134, EPI_ISL_2772135, EPI_ISL_2772136, EPI_ISL_2772137, EPI_ISL_2772138, EPI_ISL_2772139, EPI_ISL_2772140, EPI_ISL_2772141, EPI_ISL_2772142, EPI_ISL_2772143, EPI_ISL_2772144, EPI_ISL_2772145, EPI_ISL_2772146, EPI_ISL_2772147, EPI_ISL_2772148, EPI_ISL_2772149, EPI_ISL_2772150, EPI_ISL_2772151, EPI_ISL_2772152, EPI_ISL_2772153, EPI_ISL_2772154, EPI_ISL_2772155, EPI_ISL_2772156, EPI_ISL_2772157, EPI_ISL_2772158, EPI_ISL_2772159, EPI_ISL_2772160, EPI_ISL_2772161, EPI_ISL_2772162, EPI_ISL_2772163, EPI_ISL_2772164, EPI_ISL_2772165, EPI_ISL_2772166, EPI_ISL_2772167, EPI_ISL_2772168, EPI_ISL_2772169, EPI_ISL_2772170, EPI_ISL_2772171, EPI_ISL_2772172, EPI_ISL_2772173, EPI_ISL_2772174, EPI_ISL_2772175, EPI_ISL_2772176, EPI_ISL_2772177, EPI_ISL_2772178, EPI_ISL_2772179, EPI_ISL_2772180, EPI_ISL_2772181, EPI_ISL_2772182, EPI_ISL_2772183, EPI_ISL_2772184, EPI_ISL_2772185, EPI_ISL_2772186, EPI_ISL_2772187, EPI_ISL_2772188, EPI_ISL_2772189, EPI_ISL_2772190, EPI_ISL_2772191, EPI_ISL_2772192, EPI_ISL_2772193, EPI_ISL_2772194, EPI_ISL_2772195, EPI_ISL_2772196, EPI_ISL_2772197, EPI_ISL_2772198, EPI_ISL_2772199, EPI_ISL_2772200, EPI_ISL_2772201, EPI_ISL_2772202, EPI_ISL_2772203, EPI_ISL_2772204, EPI_ISL_2772205, EPI_ISL_2772206, EPI_ISL_2772207, EPI_ISL_2772208, EPI_ISL_2772209, EPI_ISL_2772210, EPI_ISL_2772211, EPI_ISL_2772212, EPI_ISL_2772213, EPI_ISL_2772214, EPI_ISL_2772215, EPI_ISL_2772216, EPI_ISL_2773981, EPI_ISL_2773982, EPI_ISL_2773983, EPI_ISL_2773984, EPI_ISL_2773985, EPI_ISL_2775676, EPI_ISL_2777137, EPI_ISL_2777138, EPI_ISL_2777139, EPI_ISL_2777140, EPI_ISL_2777141, EPI_ISL_2777142, EPI_ISL_2777143, EPI_ISL_2777144, EPI_ISL_2777145, EPI_ISL_2777146, EPI_ISL_2777147, EPI_ISL_2777148, EPI_ISL_2777149, EPI_ISL_2777150, EPI_ISL_2777151, EPI_ISL_2777152, EPI_ISL_2777153, EPI_ISL_2777154, EPI_ISL_2777155, EPI_ISL_2777156, EPI_ISL_2777157, EPI_ISL_2777158, EPI_ISL_2777159, EPI_ISL_2777160, EPI_ISL_2777161, EPI_ISL_2777162, EPI_ISL_2777163, EPI_ISL_2777164, EPI_ISL_2777165, EPI_ISL_2777166, EPI_ISL_2777185, EPI_ISL_2777186, EPI_ISL_2777187, EPI_ISL_2777188, EPI_ISL_2777189, EPI_ISL_2777190, EPI_ISL_2777191, EPI_ISL_2932266, EPI_ISL_2932267, EPI_ISL_2932268, EPI_ISL_2932269, EPI_ISL_2932270, EPI_ISL_2932271, EPI_ISL_2932272, EPI_ISL_2932273, EPI_ISL_2932274, EPI_ISL_2932276, EPI_ISL_2932293, EPI_ISL_2932294, EPI_ISL_2932295, EPI_ISL_2932296, EPI_ISL_2932297, EPI_ISL_2932298, EPI_ISL_2932299, EPI_ISL_2932300, EPI_ISL_2932301, EPI_ISL_2932302, EPI_ISL_2932303, EPI_ISL_2932304, EPI_ISL_2932309, EPI_ISL_2932310, | | | |

EPI_ISL_2932312, EPI_ISL_2932313, EPI_ISL_2932314

| | | | |
|---|---|---|---|
| see above | Pathogen Genomics Center, National Institute of Infectious Diseases | Pathogen Genomics Center, National Institute of Infectious Diseases | Kentaro Itokawa; Makoto Kuroda; Masanori Hashino; Rina Tanaka; Tsuyoshi Sekizuka |

EPI_ISL_2131620, EPI_ISL_2131630, EPI_ISL_2131632, EPI_ISL_2131633, EPI_ISL_2131634, EPI_ISL_2131650, EPI_ISL_2131651, EPI_ISL_2131652, EPI_ISL_2131653, EPI_ISL_2131654, EPI_ISL_2131655, EPI_ISL_2131657, EPI_ISL_2131661, EPI_ISL_2131663, EPI_ISL_2131664, EPI_ISL_2131665, EPI_ISL_2131666, EPI_ISL_2131668, EPI_ISL_2131672, EPI_ISL_2131673, EPI_ISL_2131675, EPI_ISL_2131684,
EPI_ISL_2131687, EPI_ISL_2131688, EPI_ISL_2131689, EPI_ISL_2131690, EPI_ISL_2131720, EPI_ISL_2131721, EPI_ISL_2131723, EPI_ISL_2131724, EPI_ISL_2131725, EPI_ISL_2131726, EPI_ISL_2131727, EPI_ISL_2131728, EPI_ISL_2131729, EPI_ISL_2131731, EPI_ISL_2131732, EPI_ISL_2131733, EPI_ISL_2131736, EPI_ISL_2131737, EPI_ISL_2131738, EPI_ISL_2131739, EPI_ISL_2131740,
EPI_ISL_2131741, EPI_ISL_2131742, EPI_ISL_2131743, EPI_ISL_2131745, EPI_ISL_2131746, EPI_ISL_2131747, EPI_ISL_2131748, EPI_ISL_2131749, EPI_ISL_2131750, EPI_ISL_2131751, EPI_ISL_2131752, EPI_ISL_2131753, EPI_ISL_2131754, EPI_ISL_2131755, EPI_ISL_2131756, EPI_ISL_2131757, EPI_ISL_2131758, EPI_ISL_2131759, EPI_ISL_2131760, EPI_ISL_2131761, EPI_ISL_2131762,
EPI_ISL_2131763, EPI_ISL_2131764, EPI_ISL_2131765, EPI_ISL_2131766, EPI_ISL_2131767, EPI_ISL_2131768, EPI_ISL_2131769, EPI_ISL_2131770, EPI_ISL_2131771, EPI_ISL_2131772, EPI_ISL_2131773, EPI_ISL_2131774, EPI_ISL_2131775, EPI_ISL_2131776, EPI_ISL_2131777, EPI_ISL_2131778, EPI_ISL_2131779, EPI_ISL_2131780, EPI_ISL_2131781, EPI_ISL_2131783, EPI_ISL_2131784,
EPI_ISL_2131785, EPI_ISL_2131786, EPI_ISL_2131787, EPI_ISL_2131788, EPI_ISL_2131789, EPI_ISL_2131793, EPI_ISL_2131794, EPI_ISL_2131795, EPI_ISL_2131796, EPI_ISL_2253110, EPI_ISL_2253111, EPI_ISL_2253112, EPI_ISL_2253113, EPI_ISL_2253114, EPI_ISL_2253115, EPI_ISL_2253116, EPI_ISL_2253117, EPI_ISL_2253118, EPI_ISL_2253119, EPI_ISL_2253120, EPI_ISL_2253121, EPI_ISL_2253122,
EPI_ISL_2253123, EPI_ISL_2253124, EPI_ISL_2329811, EPI_ISL_2329812, EPI_ISL_2329813, EPI_ISL_2329814, EPI_ISL_2329860, EPI_ISL_2329861, EPI_ISL_2329862, EPI_ISL_2329863, EPI_ISL_2329864, EPI_ISL_2329865, EPI_ISL_2329866, EPI_ISL_2329883, EPI_ISL_2329884, EPI_ISL_2329885, EPI_ISL_2329886, EPI_ISL_2329887, EPI_ISL_2329888, EPI_ISL_2329889, EPI_ISL_2329899, EPI_ISL_2329900,
EPI_ISL_2329901, EPI_ISL_2329902, EPI_ISL_2329903, EPI_ISL_2329904, EPI_ISL_2329905, EPI_ISL_2329906, EPI_ISL_2329907, EPI_ISL_2329908, EPI_ISL_2329909, EPI_ISL_2329910, EPI_ISL_2329911, EPI_ISL_2329912, EPI_ISL_2329913, EPI_ISL_2329914, EPI_ISL_2329915, EPI_ISL_2329916, EPI_ISL_2329918, EPI_ISL_2329919, EPI_ISL_2329920, EPI_ISL_2329921, EPI_ISL_2329922, EPI_ISL_2329923,
EPI_ISL_2329924, EPI_ISL_2329926, EPI_ISL_2329927, EPI_ISL_2329928, EPI_ISL_2329929, EPI_ISL_2329930, EPI_ISL_2329931, EPI_ISL_2329932, EPI_ISL_2329933, EPI_ISL_2329934, EPI_ISL_2329935, EPI_ISL_2329936, EPI_ISL_2329937, EPI_ISL_2329938, EPI_ISL_2329939, EPI_ISL_2329940, EPI_ISL_2329974, EPI_ISL_2329975, EPI_ISL_2329976, EPI_ISL_2329977, EPI_ISL_2329978, EPI_ISL_2329979,
EPI_ISL_2329980, EPI_ISL_2329981, EPI_ISL_2329982, EPI_ISL_2329983, EPI_ISL_2329984, EPI_ISL_2329985, EPI_ISL_2329986, EPI_ISL_2329987, EPI_ISL_2329988, EPI_ISL_2329989, EPI_ISL_2329990, EPI_ISL_2329991, EPI_ISL_2329992, EPI_ISL_2329993, EPI_ISL_2330028, EPI_ISL_2330029, EPI_ISL_2330044, EPI_ISL_2330045, EPI_ISL_2330046, EPI_ISL_2330048, EPI_ISL_2330049, EPI_ISL_2330050,
EPI_ISL_2330051, EPI_ISL_2330052, EPI_ISL_2330053, EPI_ISL_2330054, EPI_ISL_2330065, EPI_ISL_2330066, EPI_ISL_2330067, EPI_ISL_2330068, EPI_ISL_2330069, EPI_ISL_2330070, EPI_ISL_2330071, EPI_ISL_2330072, EPI_ISL_2330073, EPI_ISL_2330074, EPI_ISL_2330082, EPI_ISL_2330083, EPI_ISL_2330084, EPI_ISL_2330085, EPI_ISL_2330086, EPI_ISL_2330087, EPI_ISL_2330088, EPI_ISL_2330113,
EPI_ISL_2330115, EPI_ISL_2330116, EPI_ISL_2330117, EPI_ISL_2330118, EPI_ISL_2330119, EPI_ISL_2330120, EPI_ISL_2330121, EPI_ISL_2330122, EPI_ISL_2330123, EPI_ISL_2330124, EPI_ISL_2330125, EPI_ISL_2330126, EPI_ISL_2330129, EPI_ISL_2330130, EPI_ISL_2330131, EPI_ISL_2330132, EPI_ISL_2330133, EPI_ISL_2330134, EPI_ISL_2330164, EPI_ISL_2330165, EPI_ISL_2330166, EPI_ISL_2330217,
EPI_ISL_2330218, EPI_ISL_2330220, EPI_ISL_2330221, EPI_ISL_2330222, EPI_ISL_2330223, EPI_ISL_2330226, EPI_ISL_2330227, EPI_ISL_2330228, EPI_ISL_2330229, EPI_ISL_2330230, EPI_ISL_2330231, EPI_ISL_2330232, EPI_ISL_2330233, EPI_ISL_2330234, EPI_ISL_2330235, EPI_ISL_2330236, EPI_ISL_2330237, EPI_ISL_2330255, EPI_ISL_2330256, EPI_ISL_2330257, EPI_ISL_2330258, EPI_ISL_2330259,
EPI_ISL_2330260, EPI_ISL_2330261, EPI_ISL_2330262, EPI_ISL_2330263, EPI_ISL_2330264, EPI_ISL_2330265, EPI_ISL_2330266, EPI_ISL_2330267, EPI_ISL_2330268, EPI_ISL_2330269, EPI_ISL_2330270, EPI_ISL_2330271, EPI_ISL_2330272, EPI_ISL_2330273, EPI_ISL_2330274, EPI_ISL_2330275, EPI_ISL_2330276, EPI_ISL_2330277, EPI_ISL_2330278, EPI_ISL_2330279, EPI_ISL_2330280, EPI_ISL_2330281,
EPI_ISL_2330282, EPI_ISL_2330283, EPI_ISL_2330284, EPI_ISL_2330285, EPI_ISL_2330286, EPI_ISL_2330287, EPI_ISL_2330288, EPI_ISL_2330289, EPI_ISL_2330290, EPI_ISL_2330291, EPI_ISL_2330292, EPI_ISL_2330293, EPI_ISL_2330294, EPI_ISL_2330295, EPI_ISL_2330296, EPI_ISL_2330297, EPI_ISL_2330298, EPI_ISL_2330299, EPI_ISL_2330300, EPI_ISL_2330301, EPI_ISL_2330302, EPI_ISL_2330303,
EPI_ISL_2330304, EPI_ISL_2330305, EPI_ISL_2330306, EPI_ISL_2330307, EPI_ISL_2330308, EPI_ISL_2330309, EPI_ISL_2330310, EPI_ISL_2330311, EPI_ISL_2330312, EPI_ISL_2330313, EPI_ISL_2330314, EPI_ISL_2330315, EPI_ISL_2330316, EPI_ISL_2330317, EPI_ISL_2330318, EPI_ISL_2330319, EPI_ISL_2330320, EPI_ISL_2330321, EPI_ISL_2330322, EPI_ISL_2330323, EPI_ISL_2330324, EPI_ISL_2330325,
EPI_ISL_2330326, EPI_ISL_2330327, EPI_ISL_2330640, EPI_ISL_2330641, EPI_ISL_2330642, EPI_ISL_2330643, EPI_ISL_2330644, EPI_ISL_2330645, EPI_ISL_2330646, EPI_ISL_2330647, EPI_ISL_2330648, EPI_ISL_2330649, EPI_ISL_2330650, EPI_ISL_2330651, EPI_ISL_2330654, EPI_ISL_2330655, EPI_ISL_2330656, EPI_ISL_2330657, EPI_ISL_2330658, EPI_ISL_2330666, EPI_ISL_2330667, EPI_ISL_2330668,
EPI_ISL_2330669, EPI_ISL_2330677, EPI_ISL_2330678, EPI_ISL_2330679, EPI_ISL_2330680, EPI_ISL_2330681, EPI_ISL_2330682, EPI_ISL_2330683, EPI_ISL_2330684, EPI_ISL_2330685, EPI_ISL_2330686, EPI_ISL_2330687, EPI_ISL_2330688, EPI_ISL_2330689, EPI_ISL_2330690, EPI_ISL_2330716, EPI_ISL_2330717, EPI_ISL_2330718, EPI_ISL_2330719, EPI_ISL_2330720, EPI_ISL_2330721, EPI_ISL_2330722,
EPI_ISL_2330723, EPI_ISL_2330724, EPI_ISL_2330725, EPI_ISL_2330726, EPI_ISL_2330727, EPI_ISL_2330805, EPI_ISL_2330806, EPI_ISL_2330813, EPI_ISL_2330814, EPI_ISL_2330815, EPI_ISL_2330816, EPI_ISL_2330817, EPI_ISL_2330818, EPI_ISL_2330819, EPI_ISL_2330820, EPI_ISL_2330821, EPI_ISL_2330822, EPI_ISL_2330823, EPI_ISL_2330824, EPI_ISL_2330825, EPI_ISL_2330826, EPI_ISL_2330827,
EPI_ISL_2330828, EPI_ISL_2330829, EPI_ISL_2330830, EPI_ISL_2330831, EPI_ISL_2330832, EPI_ISL_2330833, EPI_ISL_2330840, EPI_ISL_2330841, EPI_ISL_2330842, EPI_ISL_2330843, EPI_ISL_2330844, EPI_ISL_2330845, EPI_ISL_2330846, EPI_ISL_2330847, EPI_ISL_2330848, EPI_ISL_2330849, EPI_ISL_2330850, EPI_ISL_2330851, EPI_ISL_2330852, EPI_ISL_2330853, EPI_ISL_2330854, EPI_ISL_2330885,
EPI_ISL_2330886, EPI_ISL_2330887, EPI_ISL_2330888, EPI_ISL_2330889, EPI_ISL_2330890, EPI_ISL_2330891, EPI_ISL_2330892, EPI_ISL_2330893, EPI_ISL_2330894, EPI_ISL_2330895, EPI_ISL_2330896, EPI_ISL_2330897, EPI_ISL_2330898, EPI_ISL_2330899, EPI_ISL_2330900, EPI_ISL_2330901, EPI_ISL_2330902, EPI_ISL_2330903, EPI_ISL_2330904, EPI_ISL_2330905, EPI_ISL_2330906, EPI_ISL_2330907,
EPI_ISL_2330908, EPI_ISL_2330909, EPI_ISL_2330910, EPI_ISL_2330911, EPI_ISL_2330912, EPI_ISL_2330913, EPI_ISL_2330914, EPI_ISL_2330915, EPI_ISL_2330916, EPI_ISL_2330917, EPI_ISL_2330918, EPI_ISL_2330919, EPI_ISL_2330920, EPI_ISL_2330921, EPI_ISL_2330922, EPI_ISL_2330923, EPI_ISL_2330924, EPI_ISL_2330925, EPI_ISL_2330926, EPI_ISL_2330927, EPI_ISL_2330930, EPI_ISL_2330934,
EPI_ISL_2330935, EPI_ISL_2330936, EPI_ISL_2330937, EPI_ISL_2330938, EPI_ISL_2330939, EPI_ISL_2330940, EPI_ISL_2330941, EPI_ISL_2330942, EPI_ISL_2330946, EPI_ISL_2330947, EPI_ISL_2330948, EPI_ISL_2330949, EPI_ISL_2330950, EPI_ISL_2330951, EPI_ISL_2330952, EPI_ISL_2330953, EPI_ISL_2330954, EPI_ISL_2330955, EPI_ISL_2330956, EPI_ISL_2330957, EPI_ISL_2330958, EPI_ISL_2330959,
EPI_ISL_2330960, EPI_ISL_2330961, EPI_ISL_2330962, EPI_ISL_2330963, EPI_ISL_2330964, EPI_ISL_2330965, EPI_ISL_2330966, EPI_ISL_2330967, EPI_ISL_2330968, EPI_ISL_2330969, EPI_ISL_2330970, EPI_ISL_2330971, EPI_ISL_2330972, EPI_ISL_2330973, EPI_ISL_2330974, EPI_ISL_2330975, EPI_ISL_2330976, EPI_ISL_2330977, EPI_ISL_2330978, EPI_ISL_2330979, EPI_ISL_2330980, EPI_ISL_2330981,
EPI_ISL_2330982, EPI_ISL_2330983, EPI_ISL_2330984, EPI_ISL_2330985, EPI_ISL_2330987, EPI_ISL_2330988, EPI_ISL_2330989, EPI_ISL_2330990, EPI_ISL_2331004, EPI_ISL_2331005, EPI_ISL_2331006, EPI_ISL_2331007, EPI_ISL_2331008, EPI_ISL_2331009, EPI_ISL_2331010, EPI_ISL_2331011, EPI_ISL_2331012, EPI_ISL_2331013, EPI_ISL_2331014, EPI_ISL_2331015, EPI_ISL_2331016, EPI_ISL_2331017,
EPI_ISL_2331018, EPI_ISL_2331019, EPI_ISL_2331020, EPI_ISL_2331021, EPI_ISL_2331022, EPI_ISL_2331023, EPI_ISL_2331024, EPI_ISL_2331058, EPI_ISL_2331059, EPI_ISL_2331061, EPI_ISL_2331062, EPI_ISL_2331064, EPI_ISL_2331065, EPI_ISL_2331066, EPI_ISL_2331067, EPI_ISL_2331068, EPI_ISL_2331069, EPI_ISL_2331070, EPI_ISL_2331071, EPI_ISL_2331072, EPI_ISL_2331073, EPI_ISL_2331074,
EPI_ISL_2331075, EPI_ISL_2331076, EPI_ISL_2331077, EPI_ISL_2331078, EPI_ISL_2331079, EPI_ISL_2331080, EPI_ISL_2331081, EPI_ISL_2331082, EPI_ISL_2331083, EPI_ISL_2331084, EPI_ISL_2331085, EPI_ISL_2331086, EPI_ISL_2331087, EPI_ISL_2331088, EPI_ISL_2331089, EPI_ISL_2331090, EPI_ISL_2331091, EPI_ISL_2331092, EPI_ISL_2331093, EPI_ISL_2331094, EPI_ISL_2331095, EPI_ISL_2331096,
EPI_ISL_2331097, EPI_ISL_2331098, EPI_ISL_2331099, EPI_ISL_2331100, EPI_ISL_2331101, EPI_ISL_2331102, EPI_ISL_2331103, EPI_ISL_2331104, EPI_ISL_2331105, EPI_ISL_2331106, EPI_ISL_2331108, EPI_ISL_2331109, EPI_ISL_2331110, EPI_ISL_2331111, EPI_ISL_2331112, EPI_ISL_2331113, EPI_ISL_2331114, EPI_ISL_2331115, EPI_ISL_2331116, EPI_ISL_2331117, EPI_ISL_2331118, EPI_ISL_2331119,
EPI_ISL_2331120, EPI_ISL_2331121, EPI_ISL_2331122, EPI_ISL_2331123, EPI_ISL_2331124, EPI_ISL_2331125, EPI_ISL_2331126, EPI_ISL_2331127, EPI_ISL_2331128, EPI_ISL_2331129, EPI_ISL_2331130, EPI_ISL_2331131, EPI_ISL_2331132, EPI_ISL_2331133, EPI_ISL_2331134, EPI_ISL_2331135, EPI_ISL_2331137, EPI_ISL_2331138, EPI_ISL_2331139, EPI_ISL_2331140, EPI_ISL_2331141, EPI_ISL_2331142,
EPI_ISL_2331143, EPI_ISL_2331144, EPI_ISL_2331147, EPI_ISL_2331148, EPI_ISL_2331149, EPI_ISL_2331150, EPI_ISL_2331151, EPI_ISL_2331152, EPI_ISL_2331153, EPI_ISL_2331154, EPI_ISL_2331158, EPI_ISL_2331159, EPI_ISL_2331160, EPI_ISL_2331161, EPI_ISL_2331162, EPI_ISL_2331163, EPI_ISL_2331164, EPI_ISL_2331165, EPI_ISL_2331166, EPI_ISL_2331167, EPI_ISL_2331168, EPI_ISL_2331169,
EPI_ISL_2331170, EPI_ISL_2331171, EPI_ISL_2331172, EPI_ISL_2331182, EPI_ISL_2331183, EPI_ISL_2331184, EPI_ISL_2331185, EPI_ISL_2331186, EPI_ISL_2331187, EPI_ISL_2331188, EPI_ISL_2331193, EPI_ISL_2331194, EPI_ISL_2331195, EPI_ISL_2331196, EPI_ISL_2331206, EPI_ISL_2331207, EPI_ISL_2331208, EPI_ISL_2331209, EPI_ISL_2331210, EPI_ISL_2331211, EPI_ISL_2331212, EPI_ISL_2331217,
EPI_ISL_2331218, EPI_ISL_2331219, EPI_ISL_2331220, EPI_ISL_2331221, EPI_ISL_2331222, EPI_ISL_2331223, EPI_ISL_2331232, EPI_ISL_2331247, EPI_ISL_2331248, EPI_ISL_2331249, EPI_ISL_2331250, EPI_ISL_2331251, EPI_ISL_2331252, EPI_ISL_2331253, EPI_ISL_2331254, EPI_ISL_2331255, EPI_ISL_2331256, EPI_ISL_2331257, EPI_ISL_2331258, EPI_ISL_2331259, EPI_ISL_2331260, EPI_ISL_2331261,
EPI_ISL_2331262, EPI_ISL_2331263, EPI_ISL_2331264, EPI_ISL_2331265, EPI_ISL_2331266, EPI_ISL_2331267, EPI_ISL_2331268, EPI_ISL_2331269, EPI_ISL_2331270, EPI_ISL_2331271, EPI_ISL_2331272, EPI_ISL_2331273, EPI_ISL_2331274, EPI_ISL_2331275, EPI_ISL_2331276, EPI_ISL_2331277, EPI_ISL_2331278, EPI_ISL_2331279, EPI_ISL_2331280, EPI_ISL_2331281, EPI_ISL_2331282, EPI_ISL_2331283,
EPI_ISL_2331284, EPI_ISL_2331285, EPI_ISL_2331286, EPI_ISL_2331287, EPI_ISL_2331302, EPI_ISL_2331303, EPI_ISL_2331304, EPI_ISL_2331305, EPI_ISL_2331306, EPI_ISL_2336077, EPI_ISL_2336078, EPI_ISL_2336079, EPI_ISL_2336080, EPI_ISL_2336105, EPI_ISL_2336106, EPI_ISL_2336107, EPI_ISL_2336108, EPI_ISL_2336109, EPI_ISL_2336119, EPI_ISL_2336141, EPI_ISL_2336142, EPI_ISL_2336143,
EPI_ISL_2336144, EPI_ISL_2336145, EPI_ISL_2336146, EPI_ISL_2336147, EPI_ISL_2336254, EPI_ISL_2336255, EPI_ISL_2336256, EPI_ISL_2336258, EPI_ISL_2336259, EPI_ISL_2336260, EPI_ISL_2336261, EPI_ISL_2336555, EPI_ISL_2336556, EPI_ISL_2336563, EPI_ISL_2336571, EPI_ISL_2336574, EPI_ISL_2336576, EPI_ISL_2336577, EPI_ISL_2336578, EPI_ISL_2336579, EPI_ISL_2336580, EPI_ISL_2336581,
EPI_ISL_2336583, EPI_ISL_2336584, EPI_ISL_2336585, EPI_ISL_2336586, EPI_ISL_2336590, EPI_ISL_2336591, EPI_ISL_2336610, EPI_ISL_2336611, EPI_ISL_2336614, EPI_ISL_2336615, EPI_ISL_2336616, EPI_ISL_2336621, EPI_ISL_2336622, EPI_ISL_2336623, EPI_ISL_2336624, EPI_ISL_2336628, EPI_ISL_2336629, EPI_ISL_2336630, EPI_ISL_2336631, EPI_ISL_2336632, EPI_ISL_2336633, EPI_ISL_2336634,
EPI_ISL_2336635, EPI_ISL_2336637, EPI_ISL_2336736, EPI_ISL_2336737, EPI_ISL_2336738, EPI_ISL_2336739, EPI_ISL_2336740, EPI_ISL_2336741, EPI_ISL_2336742, EPI_ISL_2336743, EPI_ISL_2336744, EPI_ISL_2336748, EPI_ISL_2336897, EPI_ISL_2336898, EPI_ISL_2336899, EPI_ISL_2336900, EPI_ISL_2336901, EPI_ISL_2336902, EPI_ISL_2336903, EPI_ISL_2336904, EPI_ISL_2336905, EPI_ISL_2336906,
EPI_ISL_2336907, EPI_ISL_2336908, EPI_ISL_2336909, EPI_ISL_2336910, EPI_ISL_2336911, EPI_ISL_2336912, EPI_ISL_2336913, EPI_ISL_2336914, EPI_ISL_2336915, EPI_ISL_2336916, EPI_ISL_2336917, EPI_ISL_2336918, EPI_ISL_2336919, EPI_ISL_2336920, EPI_ISL_2336921, EPI_ISL_2336922, EPI_ISL_2336923, EPI_ISL_2336924, EPI_ISL_2336925, EPI_ISL_2336926, EPI_ISL_2336927, EPI_ISL_2336928,
EPI_ISL_2336929, EPI_ISL_2336930, EPI_ISL_2336931, EPI_ISL_2336932, EPI_ISL_2336933, EPI_ISL_2336941, EPI_ISL_2336942, EPI_ISL_2336943, EPI_ISL_2336944, EPI_ISL_2336945, EPI_ISL_2336946, EPI_ISL_2336947, EPI_ISL_2336948, EPI_ISL_2336949, EPI_ISL_2336950, EPI_ISL_2336951, EPI_ISL_2336952, EPI_ISL_2336953, EPI_ISL_2336954, EPI_ISL_2336955, EPI_ISL_2336956, EPI_ISL_2336957,
EPI_ISL_2336958, EPI_ISL_2336959, EPI_ISL_2336960, EPI_ISL_2336961, EPI_ISL_2336962, EPI_ISL_2336974, EPI_ISL_2336975, EPI_ISL_2336976, EPI_ISL_2336977, EPI_ISL_2336978, EPI_ISL_2336991, EPI_ISL_2336992, EPI_ISL_2336994, EPI_ISL_2336995, EPI_ISL_2336996, EPI_ISL_2336997, EPI_ISL_2336998, EPI_ISL_2336999, EPI_ISL_2337000, EPI_ISL_2337001, EPI_ISL_2337002, EPI_ISL_2337003,
EPI_ISL_2337004, EPI_ISL_2337005, EPI_ISL_2337006, EPI_ISL_2337007, EPI_ISL_2337008, EPI_ISL_2337009, EPI_ISL_2337010, EPI_ISL_2337011, EPI_ISL_2337012, EPI_ISL_2337013, EPI_ISL_2337014, EPI_ISL_2337015, EPI_ISL_2337016, EPI_ISL_2337017, EPI_ISL_2337018, EPI_ISL_2337019, EPI_ISL_2337020, EPI_ISL_2337043, EPI_ISL_2337127, EPI_ISL_2337128, EPI_ISL_2337137, EPI_ISL_2337138,
EPI_ISL_2337139, EPI_ISL_2337140, EPI_ISL_2337141, EPI_ISL_2337142, EPI_ISL_2337143, EPI_ISL_2337144, EPI_ISL_2337145, EPI_ISL_2337147, EPI_ISL_2337148, EPI_ISL_2337149, EPI_ISL_2337150, EPI_ISL_2337151, EPI_ISL_2337152, EPI_ISL_2337169, EPI_ISL_2337170, EPI_ISL_2337172, EPI_ISL_2337173, EPI_ISL_2337174, EPI_ISL_2337175, EPI_ISL_2337176, EPI_ISL_2337177, EPI_ISL_2337178,
EPI_ISL_2337179, EPI_ISL_2337181, EPI_ISL_2337182, EPI_ISL_2337186, EPI_ISL_2337187, EPI_ISL_2337188, EPI_ISL_2337189, EPI_ISL_2337190, EPI_ISL_2337191, EPI_ISL_2337192, EPI_ISL_2337193, EPI_ISL_2337194, EPI_ISL_2337196, EPI_ISL_2337197, EPI_ISL_2337198, EPI_ISL_2337199, EPI_ISL_2337200, EPI_ISL_2337201, EPI_ISL_2337202, EPI_ISL_2337203, EPI_ISL_2337204, EPI_ISL_2337205,
EPI_ISL_2337206, EPI_ISL_2337208, EPI_ISL_2337209, EPI_ISL_2337210, EPI_ISL_2337211, EPI_ISL_2337212, EPI_ISL_2337213, EPI_ISL_2337214, EPI_ISL_2337215, EPI_ISL_2337216, EPI_ISL_2337217, EPI_ISL_2337218, EPI_ISL_2337227, EPI_ISL_2337228, EPI_ISL_2337229, EPI_ISL_2337230, EPI_ISL_2337231, EPI_ISL_2337232, EPI_ISL_2337233, EPI_ISL_2337234, EPI_ISL_2337235, EPI_ISL_2337236,
EPI_ISL_2337237, EPI_ISL_2337238, EPI_ISL_2337634, EPI_ISL_2337635, EPI_ISL_2337636, EPI_ISL_2337637, EPI_ISL_2337638, EPI_ISL_2337639, EPI_ISL_2337640, EPI_ISL_2337641, EPI_ISL_2337642, EPI_ISL_2337643, EPI_ISL_2337644, EPI_ISL_2337645, EPI_ISL_2337646, EPI_ISL_2337647, EPI_ISL_2337648, EPI_ISL_2337649, EPI_ISL_2337650, EPI_ISL_2337651, EPI_ISL_2337652, EPI_ISL_2337653,
EPI_ISL_2337654, EPI_ISL_2337655, EPI_ISL_2337656, EPI_ISL_2337657, EPI_ISL_2337658, EPI_ISL_2337659, EPI_ISL_2337660, EPI_ISL_2337661, EPI_ISL_2337662, EPI_ISL_2337663, EPI_ISL_2337664, EPI_ISL_2337665, EPI_ISL_2337666, EPI_ISL_2337667, EPI_ISL_2337668, EPI_ISL_2337749, EPI_ISL_2337750, EPI_ISL_2337751, EPI_ISL_2337752, EPI_ISL_2337763, EPI_ISL_2337764, EPI_ISL_2337765,
EPI_ISL_2337767, EPI_ISL_2337768, EPI_ISL_2337769, EPI_ISL_2337770, EPI_ISL_2337771, EPI_ISL_2337772, EPI_ISL_2337773, EPI_ISL_2337774, EPI_ISL_2337775, EPI_ISL_2337811, EPI_ISL_2337812, EPI_ISL_2337813, EPI_ISL_2337814, EPI_ISL_2337815, EPI_ISL_2337816, EPI_ISL_2337817, EPI_ISL_2337818, EPI_ISL_2337819, EPI_ISL_2337820, EPI_ISL_2337821, EPI_ISL_2337822, EPI_ISL_2337823,
EPI_ISL_2337824, EPI_ISL_2337825, EPI_ISL_2337826, EPI_ISL_2337827, EPI_ISL_2337828, EPI_ISL_2337829, EPI_ISL_2337830, EPI_ISL_2337831, EPI_ISL_2337873, EPI_ISL_2337874, EPI_ISL_2337875, EPI_ISL_2337876, EPI_ISL_2337877, EPI_ISL_2337878, EPI_ISL_2337879, EPI_ISL_2337880, EPI_ISL_2337881, EPI_ISL_2337882, EPI_ISL_2337883, EPI_ISL_2337884, EPI_ISL_2337885, EPI_ISL_2337886,
EPI_ISL_2337887, EPI_ISL_2337888, EPI_ISL_2337890, EPI_ISL_2337891, EPI_ISL_2337892, EPI_ISL_2337965, EPI_ISL_2337982, EPI_ISL_2337983, EPI_ISL_2337984, EPI_ISL_2337985, EPI_ISL_2337986, EPI_ISL_2337987, EPI_ISL_2337988, EPI_ISL_2337989, EPI_ISL_2337990, EPI_ISL_2337991, EPI_ISL_2337992, EPI_ISL_2337993, EPI_ISL_2337994, EPI_ISL_2337995, EPI_ISL_2337996,
EPI_ISL_2337997, EPI_ISL_2337998, EPI_ISL_2337999, EPI_ISL_2338000, EPI_ISL_2338001, EPI_ISL_2338002, EPI_ISL_2338003, EPI_ISL_2338004, EPI_ISL_2338005, EPI_ISL_2338006, EPI_ISL_2338007, EPI_ISL_2338008, EPI_ISL_2338009, EPI_ISL_2338010, EPI_ISL_2338011, EPI_ISL_2338012, EPI_ISL_2338015, EPI_ISL_2338016, EPI_ISL_2338017, EPI_ISL_2338018, EPI_ISL_2338019, EPI_ISL_2338020,
EPI_ISL_2338021, EPI_ISL_2338022, EPI_ISL_2338023, EPI_ISL_2338024, EPI_ISL_2338025, EPI_ISL_2338026, EPI_ISL_2338027, EPI_ISL_2338028, EPI_ISL_2338029, EPI_ISL_2338030, EPI_ISL_2338031, EPI_ISL_2338032, EPI_ISL_2338033, EPI_ISL_2338034, EPI_ISL_2338035, EPI_ISL_2338056, EPI_ISL_2338057, EPI_ISL_2338058, EPI_ISL_2338059, EPI_ISL_2338060, EPI_ISL_2338061, EPI_ISL_2338062,
EPI_ISL_2338063, EPI_ISL_2338064, EPI_ISL_2338065, EPI_ISL_2338066, EPI_ISL_2338067, EPI_ISL_2338068, EPI_ISL_2338069, EPI_ISL_2338070, EPI_ISL_2338071, EPI_ISL_2338072, EPI_ISL_2338073, EPI_ISL_2338074, EPI_ISL_2338075, EPI_ISL_2338076, EPI_ISL_2338077, EPI_ISL_2338089, EPI_ISL_2338090, EPI_ISL_2338091, EPI_ISL_2338092, EPI_ISL_2338093, EPI_ISL_2338094, EPI_ISL_2338095,
EPI_ISL_2338096, EPI_ISL_2338097, EPI_ISL_2338098, EPI_ISL_2338099, EPI_ISL_2338100, EPI_ISL_2338101, EPI_ISL_2338102, EPI_ISL_2338103, EPI_ISL_2338104, EPI_ISL_2338105, EPI_ISL_2338106, EPI_ISL_2338107, EPI_ISL_2338108, EPI_ISL_2338109, EPI_ISL_2338110, EPI_ISL_2338111, EPI_ISL_2338113, EPI_ISL_2338114, EPI_ISL_2338115, EPI_ISL_2338116, EPI_ISL_2338117, EPI_ISL_2338118,
EPI_ISL_2338119, EPI_ISL_2338120, EPI_ISL_2338121, EPI_ISL_2338122, EPI_ISL_2338123, EPI_ISL_2338124, EPI_ISL_2338125, EPI_ISL_2338126, EPI_ISL_2338127, EPI_ISL_2338128, EPI_ISL_2338129, EPI_ISL_2338130, EPI_ISL_2338131, EPI_ISL_2338132, EPI_ISL_2338133, EPI_ISL_2338134, EPI_ISL_2338135, EPI_ISL_2338136, EPI_ISL_2338137, EPI_ISL_2338138, EPI_ISL_2338139, EPI_ISL_2338140,
EPI_ISL_2338141, EPI_ISL_2338142, EPI_ISL_2338174, EPI_ISL_2338175, EPI_ISL_2338176, EPI_ISL_2338177, EPI_ISL_2338178, EPI_ISL_2338179, EPI_ISL_2338180, EPI_ISL_2338181, EPI_ISL_2338186, EPI_ISL_2338187, EPI_ISL_2338189, EPI_ISL_2338190, EPI_ISL_2338205, EPI_ISL_2338206, EPI_ISL_2338207, EPI_ISL_2338208, EPI_ISL_2338209, EPI_ISL_2338210, EPI_ISL_2338211, EPI_ISL_2338212,
EPI_ISL_2338213, EPI_ISL_2338214, EPI_ISL_2338215, EPI_ISL_2338216, EPI_ISL_2338217, EPI_ISL_2338218, EPI_ISL_2338219, EPI_ISL_2338220, EPI_ISL_2338221, EPI_ISL_2338222, EPI_ISL_2338223, EPI_ISL_2338224, EPI_ISL_2338225, EPI_ISL_2338226, EPI_ISL_2338227, EPI_ISL_2338228, EPI_ISL_2338229, EPI_ISL_2338230, EPI_ISL_2338231, EPI_ISL_2338232, EPI_ISL_2338233, EPI_ISL_2338234,
EPI_ISL_2338235, EPI_ISL_2338236, EPI_ISL_2338237, EPI_ISL_2338238, EPI_ISL_2338239, EPI_ISL_2338240, EPI_ISL_2338241, EPI_ISL_2338242, EPI_ISL_2338243, EPI_ISL_2338245, EPI_ISL_2338246, EPI_ISL_2338247, EPI_ISL_2338248, EPI_ISL_2338249, EPI_ISL_2338250, EPI_ISL_2338251, EPI_ISL_2338252, EPI_ISL_2338253, EPI_ISL_2338254, EPI_ISL_2338255, EPI_ISL_2338256, EPI_ISL_2338257,
EPI_ISL_2338258, EPI_ISL_2338259, EPI_ISL_2338303, EPI_ISL_2338304, EPI_ISL_2338305, EPI_ISL_2338306, EPI_ISL_2338307, EPI_ISL_2338308, EPI_ISL_2338309, EPI_ISL_2338310, EPI_ISL_2338311, EPI_ISL_2338321, EPI_ISL_2338923, EPI_ISL_2338924, EPI_ISL_2338927, EPI_ISL_2338928, EPI_ISL_2338929, EPI_ISL_2338930, EPI_ISL_2338931, EPI_ISL_2338932, EPI_ISL_2338933, EPI_ISL_2338934,
EPI_ISL_2338941, EPI_ISL_2338942, EPI_ISL_2338943, EPI_ISL_2338944, EPI_ISL_2338945, EPI_ISL_2338946, EPI_ISL_2338947, EPI_ISL_2338948, EPI_ISL_2338949, EPI_ISL_2338950, EPI_ISL_2338951, EPI_ISL_2338952, EPI_ISL_2338953, EPI_ISL_2338954, EPI_ISL_2338955, EPI_ISL_2338956, EPI_ISL_2338957, EPI_ISL_2338958, EPI_ISL_2338959, EPI_ISL_2338960, EPI_ISL_2338961, EPI_ISL_2338962,
EPI_ISL_2338963, EPI_ISL_2338964, EPI_ISL_2447604, EPI_ISL_2447605, EPI_ISL_2447606, EPI_ISL_2447607, EPI_ISL_2447610, EPI_ISL_2447612, EPI_ISL_2447617, EPI_ISL_2760068, EPI_ISL_2760070, EPI_ISL_2760071, EPI_ISL_2760074, EPI_ISL_2760075, EPI_ISL_2760077, EPI_ISL_2760078, EPI_ISL_2760087, EPI_ISL_2760088, EPI_ISL_2760090, EPI_ISL_2760092, EPI_ISL_2760094, EPI_ISL_2760095,
EPI_ISL_2760097, EPI_ISL_2760099, EPI_ISL_2760100, EPI_ISL_2760110, EPI_ISL_2760112, EPI_ISL_2760113, EPI_ISL_2760115, EPI_ISL_2760116, EPI_ISL_2760118, EPI_ISL_2760120, EPI_ISL_2760121, EPI_ISL_2760123, EPI_ISL_2760125, EPI_ISL_2760127, EPI_ISL_2760128, EPI_ISL_2760130, EPI_ISL_2760140, EPI_ISL_2760142, EPI_ISL_2760144, EPI_ISL_2760145, EPI_ISL_2760147, EPI_ISL_2760148,
EPI_ISL_2760150, EPI_ISL_2760152, EPI_ISL_2760154, EPI_ISL_2760156, EPI_ISL_2760158, EPI_ISL_2760160, EPI_ISL_2760162, EPI_ISL_2760163, EPI_ISL_2760165, EPI_ISL_2760167, EPI_ISL_2760169, EPI_ISL_2760170, EPI_ISL_2760172, EPI_ISL_2760173, EPI_ISL_2760175, EPI_ISL_2760177, EPI_ISL_2760179, EPI_ISL_2760187, EPI_ISL_2760188, EPI_ISL_2760192, EPI_ISL_2760194, EPI_ISL_2760195,
EPI_ISL_2760197, EPI_ISL_2760199, EPI_ISL_2760200, EPI_ISL_2760467, EPI_ISL_2760472, EPI_ISL_2760474, EPI_ISL_2760476, EPI_ISL_2760477, EPI_ISL_2760479, EPI_ISL_2760481, EPI_ISL_2760482, EPI_ISL_2760484, EPI_ISL_2760486, EPI_ISL_2760487, EPI_ISL_2760489, EPI_ISL_2760490, EPI_ISL_2760492, EPI_ISL_2760494, EPI_ISL_2760495, EPI_ISL_2760497, EPI_ISL_2760499, EPI_ISL_2760500,
EPI_ISL_2760502, EPI_ISL_2760503, EPI_ISL_2760505, EPI_ISL_2760507, EPI_ISL_2760509, EPI_ISL_2760510, EPI_ISL_2760512, EPI_ISL_2760514, EPI_ISL_2760516, EPI_ISL_2760517, EPI_ISL_2760519, EPI_ISL_2760520, EPI_ISL_2760522, EPI_ISL_2760524, EPI_ISL_2760526, EPI_ISL_2760527, EPI_ISL_2760529, EPI_ISL_2760531, EPI_ISL_2760532, EPI_ISL_2760534, EPI_ISL_2760537, EPI_ISL_2760539,
EPI_ISL_2760541, EPI_ISL_2760542, EPI_ISL_2760544, EPI_ISL_2760546, EPI_ISL_2760548, EPI_ISL_2760550, EPI_ISL_2760551, EPI_ISL_2760552, EPI_ISL_2760554, EPI_ISL_2760556, EPI_ISL_2760557, EPI_ISL_2760559, EPI_ISL_2760561, EPI_ISL_2760563, EPI_ISL_2760564, EPI_ISL_2760566, EPI_ISL_2760568, EPI_ISL_2760569, EPI_ISL_2760571, EPI_ISL_2760573, EPI_ISL_2760575, EPI_ISL_2760577,
EPI_ISL_2760579, EPI_ISL_2760580, EPI_ISL_2760582, EPI_ISL_2760584, EPI_ISL_2760590, EPI_ISL_2760591, EPI_ISL_2760593, EPI_ISL_2760595, EPI_ISL_2760597, EPI_ISL_2760598, EPI_ISL_2760600, EPI_ISL_2760602, EPI_ISL_2760604, EPI_ISL_2760605, EPI_ISL_2760607, EPI_ISL_2760608, EPI_ISL_2760610, EPI_ISL_2760612, EPI_ISL_2760614, EPI_ISL_2760616, EPI_ISL_2760617, EPI_ISL_2760619,
EPI_ISL_2760620, EPI_ISL_2760622, EPI_ISL_2760624, EPI_ISL_2760626, EPI_ISL_2760628, EPI_ISL_2760629, EPI_ISL_2760631, EPI_ISL_2760633, EPI_ISL_2760635, EPI_ISL_2760636, EPI_ISL_2760638, EPI_ISL_2760640, EPI_ISL_2760641, EPI_ISL_2760643, EPI_ISL_2760645, EPI_ISL_2760646, EPI_ISL_2760648, EPI_ISL_2760650, EPI_ISL_2760651, EPI_ISL_2760653, EPI_ISL_2760655, EPI_ISL_2760657,
EPI_ISL_2760659, EPI_ISL_2760660, EPI_ISL_2760663, EPI_ISL_2760665, EPI_ISL_2760666, EPI_ISL_2760668, EPI_ISL_2760670, EPI_ISL_2760672, EPI_ISL_2760673, EPI_ISL_2760675, EPI_ISL_2760677, EPI_ISL_2760679, EPI_ISL_2760681, EPI_ISL_2760682, EPI_ISL_2760684, EPI_ISL_2760686, EPI_ISL_2760687, EPI_ISL_2760690, EPI_ISL_2760692, EPI_ISL_2760694, EPI_ISL_2760696,
EPI_ISL_2760697, EPI_ISL_2760699, EPI_ISL_2760701, EPI_ISL_2760702, EPI_ISL_2760704, EPI_ISL_2760706, EPI_ISL_2760708, EPI_ISL_2760709, EPI_ISL_2760711, EPI_ISL_2760712, EPI_ISL_2760714, EPI_ISL_2760716, EPI_ISL_2760718, EPI_ISL_2760719, EPI_ISL_2760721, EPI_ISL_2760723, EPI_ISL_2760725, EPI_ISL_2760726, EPI_ISL_2760729, EPI_ISL_2760731, EPI_ISL_2760733, EPI_ISL_2760735,
EPI_ISL_2760736, EPI_ISL_2760737, EPI_ISL_2760738, EPI_ISL_2760739, EPI_ISL_2760741, EPI_ISL_2760742, EPI_ISL_2760744, EPI_ISL_2760746, EPI_ISL_2760747, EPI_ISL_2760749, EPI_ISL_2760750, EPI_ISL_2760752, EPI_ISL_2760755, EPI_ISL_2760757, EPI_ISL_2760759, EPI_ISL_2760760, EPI_ISL_2760762, EPI_ISL_2760764, EPI_ISL_2760765, EPI_ISL_2760767, EPI_ISL_2760769,
EPI_ISL_2760770, EPI_ISL_2760772, EPI_ISL_2760774, EPI_ISL_2760775, EPI_ISL_2760777, EPI_ISL_2760778, EPI_ISL_2760779, EPI_ISL_2760781, EPI_ISL_2760782, EPI_ISL_2760784, EPI_ISL_2760785, EPI_ISL_2760787, EPI_ISL_2760788, EPI_ISL_2760790, EPI_ISL_2760792, EPI_ISL_2760793, EPI_ISL_2760795, EPI_ISL_2760797, EPI_ISL_2760798, EPI_ISL_2760800, EPI_ISL_2760802, EPI_ISL_2760803,
EPI_ISL_2760871, EPI_ISL_2760873, EPI_ISL_2760874, EPI_ISL_2760876, EPI_ISL_2760878, EPI_ISL_2760879, EPI_ISL_2760880, EPI_ISL_2760881, EPI_ISL_2760882, EPI_ISL_2760883, EPI_ISL_2760884, EPI_ISL_2760885, EPI_ISL_2760886, EPI_ISL_2760887, EPI_ISL_2760888, EPI_ISL_2760889, EPI_ISL_2760891, EPI_ISL_2760892, EPI_ISL_2760893, EPI_ISL_2760894, EPI_ISL_2760895, EPI_ISL_2760896,
EPI_ISL_2760897, EPI_ISL_2760898, EPI_ISL_2760899, EPI_ISL_2760900, EPI_ISL_2760901, EPI_ISL_2760902, EPI_ISL_2760903, EPI_ISL_2760904, EPI_ISL_2760905, EPI_ISL_2760906, EPI_ISL_2760907, EPI_ISL_2760908, EPI_ISL_2760910, EPI_ISL_2760911, EPI_ISL_2760912, EPI_ISL_2760913, EPI_ISL_2760914, EPI_ISL_2760915, EPI_ISL_2760916, EPI_ISL_2760917, EPI_ISL_2760918, EPI_ISL_2760919,
EPI_ISL_2760920, EPI_ISL_2760921, EPI_ISL_2760922, EPI_ISL_2760923, EPI_ISL_2760924, EPI_ISL_2760925, EPI_ISL_2760926, EPI_ISL_2760927, EPI_ISL_2760928, EPI_ISL_2760929, EPI_ISL_2760930, EPI_ISL_2760931, EPI_ISL_2760932, EPI_ISL_2760933, EPI_ISL_2760934, EPI_ISL_2760935, EPI_ISL_2760936, EPI_ISL_2760937, EPI_ISL_2760938, EPI_ISL_2760939, EPI_ISL_2760942, EPI_ISL_2760943,
EPI_ISL_2760944, EPI_ISL_2760945, EPI_ISL_2760946, EPI_ISL_2760947, EPI_ISL_2760948, EPI_ISL_2760949, EPI_ISL_2760950, EPI_ISL_2760951, EPI_ISL_2760952, EPI_ISL_2760953, EPI_ISL_2760954, EPI_ISL_2760955, EPI_ISL_2760956, EPI_ISL_2760957, EPI_ISL_2760958, EPI_ISL_2760959, EPI_ISL_2760960, EPI_ISL_2760961, EPI_ISL_2760962, EPI_ISL_2760963, EPI_ISL_2760964, EPI_ISL_2760965,
EPI_ISL_2760966, EPI_ISL_2760967, EPI_ISL_2760968, EPI_ISL_2760969, EPI_ISL_2760970, EPI_ISL_2760971, EPI_ISL_2760972, EPI_ISL_2760973, EPI_ISL_2760974, EPI_ISL_2760975, EPI_ISL_2760976, EPI_ISL_2760977, EPI_ISL_2760978, EPI_ISL_2760979, EPI_ISL_2760980, EPI_ISL_2760981, EPI_ISL_2760982, EPI_ISL_2760983, EPI_ISL_2760984, EPI_ISL_2760986, EPI_ISL_2760987, EPI_ISL_2760988,
EPI_ISL_2760989, EPI_ISL_2760990, EPI_ISL_2760991, EPI_ISL_2760992, EPI_ISL_2760993, EPI_ISL_2760994, EPI_ISL_2760995, EPI_ISL_2760996, EPI_ISL_2760997, EPI_ISL_2760998, EPI_ISL_2760999, EPI_ISL_2761000, EPI_ISL_2761001, EPI_ISL_2761002, EPI_ISL_2761003, EPI_ISL_2761004, EPI_ISL_2761005, EPI_ISL_2761006, EPI_ISL_2761007, EPI_ISL_2761008, EPI_ISL_2761009, EPI_ISL_2761010,
EPI_ISL_2761011, EPI_ISL_2761012, EPI_ISL_2761013, EPI_ISL_2761014, EPI_ISL_2761015, EPI_ISL_2761016, EPI_ISL_2761017, EPI_ISL_2761018, EPI_ISL_2761019, EPI_ISL_2761020, EPI_ISL_2761021, EPI_ISL_2761022, EPI_ISL_2761023, EPI_ISL_2761024, EPI_ISL_2761025, EPI_ISL_2761026, EPI_ISL_2761027, EPI_ISL_2761028, EPI_ISL_2761030, EPI_ISL_2761031, EPI_ISL_2761032,
EPI_ISL_2761033, EPI_ISL_2761034, EPI_ISL_2761035, EPI_ISL_2761036, EPI_ISL_2761037, EPI_ISL_2761038, EPI_ISL_2761039, EPI_ISL_2761040, EPI_ISL_2761041, EPI_ISL_2761042, EPI_ISL_2761043, EPI_ISL_2761054, EPI_ISL_2761055, EPI_ISL_2761056, EPI_ISL_2761057, EPI_ISL_2761058, EPI_ISL_2761059, EPI_ISL_2761060, EPI_ISL_2761061, EPI_ISL_2761062, EPI_ISL_2761063, EPI_ISL_2761064,
EPI_ISL_2761065, EPI_ISL_2761066, EPI_ISL_2761067, EPI_ISL_2761068, EPI_ISL_2761069, EPI_ISL_2761071, EPI_ISL_2761072, EPI_ISL_2761074, EPI_ISL_2761075, EPI_ISL_2761077, EPI_ISL_2761079, EPI_ISL_2761081, EPI_ISL_2761084, EPI_ISL_2761085, EPI_ISL_2761087, EPI_ISL_2761089, EPI_ISL_2761090, EPI_ISL_2761092, EPI_ISL_2761094, EPI_ISL_2761097, EPI_ISL_2761099,
EPI_ISL_2761101, EPI_ISL_2761102, EPI_ISL_2761104, EPI_ISL_2761106, EPI_ISL_2761107, EPI_ISL_2761109, EPI_ISL_2761111, EPI_ISL_2761113, EPI_ISL_2761115, EPI_ISL_2761116, EPI_ISL_2761118, EPI_ISL_2761120, EPI_ISL_2761121, EPI_ISL_2761123, EPI_ISL_2761124, EPI_ISL_2761126, EPI_ISL_2761128, EPI_ISL_2761129, EPI_ISL_2761131, EPI_ISL_2761133, EPI_ISL_2761134, EPI_ISL_2761136,
EPI_ISL_2761137, EPI_ISL_2761139, EPI_ISL_2761140, EPI_ISL_2761142, EPI_ISL_2761144, EPI_ISL_2761145, EPI_ISL_2761147, EPI_ISL_2761149, EPI_ISL_2761150, EPI_ISL_2761152, EPI_ISL_2761154, EPI_ISL_2761155, EPI_ISL_2761157, EPI_ISL_2761158, EPI_ISL_2761160, EPI_ISL_2761161, EPI_ISL_2761163, EPI_ISL_2761165, EPI_ISL_2761167, EPI_ISL_2761169, EPI_ISL_2761170, EPI_ISL_2761172,
EPI_ISL_2761174, EPI_ISL_2761176, EPI_ISL_2761177, EPI_ISL_2761179, EPI_ISL_2761181, EPI_ISL_2761183, EPI_ISL_2761185, EPI_ISL_2761186, EPI_ISL_2761188, EPI_ISL_2761189, EPI_ISL_2761191, EPI_ISL_2761193, EPI_ISL_2761194, EPI_ISL_2761196, EPI_ISL_2761198, EPI_ISL_2761199, EPI_ISL_2761201, EPI_ISL_2761203, EPI_ISL_2761204, EPI_ISL_2761206, EPI_ISL_2761207, EPI_ISL_2761209,
EPI_ISL_2761211, EPI_ISL_2761212, EPI_ISL_2761214, EPI_ISL_2761215, EPI_ISL_2761217, EPI_ISL_2761218, EPI_ISL_2761220, EPI_ISL_2761221, EPI_ISL_2761223, EPI_ISL_2761225, EPI_ISL_2761226, EPI_ISL_2761228, EPI_ISL_2761229, EPI_ISL_2761231, EPI_ISL_2761233, EPI_ISL_2761235, EPI_ISL_2761236, EPI_ISL_2761238, EPI_ISL_2761239, EPI_ISL_2761241, EPI_ISL_2761243, EPI_ISL_2761244,
EPI_ISL_2761246, EPI_ISL_2761248, EPI_ISL_2761249, EPI_ISL_2761251, EPI_ISL_2761253, EPI_ISL_2761255, EPI_ISL_2761257, EPI_ISL_2761258, EPI_ISL_2761260, EPI_ISL_2761262, EPI_ISL_2761263, EPI_ISL_2761265, EPI_ISL_2761266, EPI_ISL_2761268, EPI_ISL_2761270, EPI_ISL_2761271, EPI_ISL_2761273, EPI_ISL_2761274, EPI_ISL_2761276, EPI_ISL_2761278, EPI_ISL_2761280, EPI_ISL_2761281,
EPI_ISL_2761283, EPI_ISL_2761284, EPI_ISL_2761286, EPI_ISL_2761287, EPI_ISL_2761289, EPI_ISL_2761293, EPI_ISL_2761294, EPI_ISL_2761296, EPI_ISL_2761298, EPI_ISL_2761300, EPI_ISL_2761301, EPI_ISL_2761303, EPI_ISL_2761305, EPI_ISL_2761306, EPI_ISL_2761308, EPI_ISL_2761310, EPI_ISL_2761311, EPI_ISL_2761313, EPI_ISL_2761315, EPI_ISL_2761317, EPI_ISL_2761319,
EPI_ISL_2761320, EPI_ISL_2761322, EPI_ISL_2761324, EPI_ISL_2761325, EPI_ISL_2761327, EPI_ISL_2761329, EPI_ISL_2761330, EPI_ISL_2761332, EPI_ISL_2761334, EPI_ISL_2761336, EPI_ISL_2761338, EPI_ISL_2761340, EPI_ISL_2761341, EPI_ISL_2761343, EPI_ISL_2761345, EPI_ISL_2761346, EPI_ISL_2761348, EPI_ISL_2761350, EPI_ISL_2761352, EPI_ISL_2761353, EPI_ISL_2761355, EPI_ISL_2761356,
EPI_ISL_2761358, EPI_ISL_2761359, EPI_ISL_2761361, EPI_ISL_2761363, EPI_ISL_2761365, EPI_ISL_2761366, EPI_ISL_2761368, EPI_ISL_2761370, EPI_ISL_2761371, EPI_ISL_2761373, EPI_ISL_2761375, EPI_ISL_2761376, EPI_ISL_2761378, EPI_ISL_2761380, EPI_ISL_2761381, EPI_ISL_2761383, EPI_ISL_2761385, EPI_ISL_2761386, EPI_ISL_2761388, EPI_ISL_2761390, EPI_ISL_2761391, EPI_ISL_2761393,
EPI_ISL_2761394, EPI_ISL_2761396, EPI_ISL_2761397, EPI_ISL_2761399, EPI_ISL_2761401, EPI_ISL_2761402, EPI_ISL_2761404, EPI_ISL_2761473, EPI_ISL_2761475, EPI_ISL_2761476, EPI_ISL_2761479, EPI_ISL_2761481, EPI_ISL_2761547, EPI_ISL_2761548, EPI_ISL_2761550, EPI_ISL_2761552, EPI_ISL_2761553, EPI_ISL_2761555, EPI_ISL_2761557, EPI_ISL_2761559, EPI_ISL_2761560, EPI_ISL_2761562,
EPI_ISL_2761564, EPI_ISL_2761565, EPI_ISL_2761567, EPI_ISL_2761569, EPI_ISL_2761571, EPI_ISL_2761572, EPI_ISL_2761574, EPI_ISL_2761575, EPI_ISL_2761577, EPI_ISL_2761579, EPI_ISL_2761580, EPI_ISL_2761582, EPI_ISL_2761584, EPI_ISL_2761585, EPI_ISL_2761587, EPI_ISL_2761589, EPI_ISL_2761591, EPI_ISL_2761592, EPI_ISL_2761594, EPI_ISL_2761596, EPI_ISL_2761597, EPI_ISL_2761599,
EPI_ISL_2762344, EPI_ISL_2762345, EPI_ISL_2762346, EPI_ISL_2762348, EPI_ISL_2762357, EPI_ISL_2762358, EPI_ISL_2762360, EPI_ISL_2762362, EPI_ISL_2762364, EPI_ISL_2762366, EPI_ISL_2762367, EPI_ISL_2762369, EPI_ISL_2762371, EPI_ISL_2762373, EPI_ISL_2762375, EPI_ISL_2762376, EPI_ISL_2762378, EPI_ISL_2762379, EPI_ISL_2762381, EPI_ISL_2762382, EPI_ISL_2762384, EPI_ISL_2762386,
EPI_ISL_2762387, EPI_ISL_2762389, EPI_ISL_2762391, EPI_ISL_2762393, EPI_ISL_2762394, EPI_ISL_2762396, EPI_ISL_2762398, EPI_ISL_2762399, EPI_ISL_2762401, EPI_ISL_2762403, EPI_ISL_2762404, EPI_ISL_2762406, EPI_ISL_2762408, EPI_ISL_2762410, EPI_ISL_2762411, EPI_ISL_2762413, EPI_ISL_2762414, EPI_ISL_2762416, EPI_ISL_2762418, EPI_ISL_2762419, EPI_ISL_2762422, EPI_ISL_2762423,
EPI_ISL_2762425, EPI_ISL_2762427, EPI_ISL_2762428, EPI_ISL_2762430, EPI_ISL_2762432, EPI_ISL_2762434, EPI_ISL_2762435, EPI_ISL_2762437, EPI_ISL_2762439, EPI_ISL_2762441, EPI_ISL_2762443, EPI_ISL_2762444, EPI_ISL_2762446, EPI_ISL_2762448, EPI_ISL_2762449, EPI_ISL_2762451, EPI_ISL_2762453, EPI_ISL_2762455, EPI_ISL_2762457, EPI_ISL_2762458, EPI_ISL_2762460, EPI_ISL_2762462,
EPI_ISL_2762464, EPI_ISL_2762466, EPI_ISL_2762467, EPI_ISL_2762469, EPI_ISL_2762471, EPI_ISL_2762473, EPI_ISL_2762475, EPI_ISL_2762476, EPI_ISL_2762478, EPI_ISL_2762480, EPI_ISL_2762482, EPI_ISL_2762484, EPI_ISL_2762486, EPI_ISL_2762487, EPI_ISL_2762489, EPI_ISL_2762491, EPI_ISL_2762493, EPI_ISL_2762495, EPI_ISL_2762496, EPI_ISL_2762498, EPI_ISL_2762500, EPI_ISL_2762502,
EPI_ISL_2762504, EPI_ISL_2762505, EPI_ISL_2762507, EPI_ISL_2762509, EPI_ISL_2762511, EPI_ISL_2762512, EPI_ISL_2762514, EPI_ISL_2762516, EPI_ISL_2762518, EPI_ISL_2762519, EPI_ISL_2762521, EPI_ISL_2762523, EPI_ISL_2762524, EPI_ISL_2762526, EPI_ISL_2762528, EPI_ISL_2762530, EPI_ISL_2762531, EPI_ISL_2762533, EPI_ISL_2762535, EPI_ISL_2762537, EPI_ISL_2762538, EPI_ISL_2762540,
EPI_ISL_2762542, EPI_ISL_2762544, EPI_ISL_2762546, EPI_ISL_2762547, EPI_ISL_2762549, EPI_ISL_2762551, EPI_ISL_2762553, EPI_ISL_2762554, EPI_ISL_2762556, EPI_ISL_2762558, EPI_ISL_2762560, EPI_ISL_2762562, EPI_ISL_2762565, EPI_ISL_2762567, EPI_ISL_2762568, EPI_ISL_2762570, EPI_ISL_2762572, EPI_ISL_2762574, EPI_ISL_2762576, EPI_ISL_2762578, EPI_ISL_2762579,
EPI_ISL_2762581, EPI_ISL_2762583, EPI_ISL_2762585, EPI_ISL_2762586, EPI_ISL_2762588, EPI_ISL_2762590, EPI_ISL_2762592, EPI_ISL_2762595, EPI_ISL_2762597, EPI_ISL_2762598, EPI_ISL_2762600, EPI_ISL_2762602, EPI_ISL_2762604, EPI_ISL_2762606, EPI_ISL_2762607, EPI_ISL_2762608, EPI_ISL_2762610, EPI_ISL_2762612, EPI_ISL_2762614, EPI_ISL_2762615, EPI_ISL_2762617, EPI_ISL_2762619,
EPI_ISL_2762621, EPI_ISL_2762622, EPI_ISL_2762624, EPI_ISL_2762626, EPI_ISL_2762628, EPI_ISL_2762629, EPI_ISL_2762631, EPI_ISL_2762632, EPI_ISL_2762634, EPI_ISL_2762635, EPI_ISL_2762637, EPI_ISL_2762639, EPI_ISL_2762640, EPI_ISL_2762642, EPI_ISL_2762643, EPI_ISL_2762644, EPI_ISL_2762646, EPI_ISL_2762648, EPI_ISL_2762650, EPI_ISL_2762651, EPI_ISL_2762653, EPI_ISL_2762655,
EPI_ISL_2762657, EPI_ISL_2762658, EPI_ISL_2762660, EPI_ISL_2762661, EPI_ISL_2762663, EPI_ISL_2762665, EPI_ISL_2762667, EPI_ISL_2762668, EPI_ISL_2762670, EPI_ISL_2762672, EPI_ISL_2762674, EPI_ISL_2762675, EPI_ISL_2762677, EPI_ISL_2762678, EPI_ISL_2762680, EPI_ISL_2762682, EPI_ISL_2762683, EPI_ISL_2762685, EPI_ISL_2762687, EPI_ISL_2762689, EPI_ISL_2762690, EPI_ISL_2762692,
EPI_ISL_2762694, EPI_ISL_2762696, EPI_ISL_2762697, EPI_ISL_2762699, EPI_ISL_2762701, EPI_ISL_2762702, EPI_ISL_2762704, EPI_ISL_2762706, EPI_ISL_2762707, EPI_ISL_2762709, EPI_ISL_2762711, EPI_ISL_2762713, EPI_ISL_2762714, EPI_ISL_2762716, EPI_ISL_2762717, EPI_ISL_2762719, EPI_ISL_2762721, EPI_ISL_2762723, EPI_ISL_2762724, EPI_ISL_2762726, EPI_ISL_2762728, EPI_ISL_2762730,
EPI_ISL_2762731, EPI_ISL_2762732, EPI_ISL_2762734, EPI_ISL_2762736, EPI_ISL_2762738, EPI_ISL_2762740, EPI_ISL_2762741, EPI_ISL_2762743, EPI_ISL_2762745, EPI_ISL_2762746, EPI_ISL_2762748, EPI_ISL_2762749, EPI_ISL_2762750, EPI_ISL_2762752, EPI_ISL_2762754, EPI_ISL_2762756, EPI_ISL_2762757, EPI_ISL_2762759, EPI_ISL_2762761, EPI_ISL_2762763, EPI_ISL_2762764, EPI_ISL_2762765,
EPI_ISL_2762767, EPI_ISL_2762769, EPI_ISL_2762771, EPI_ISL_2762772, EPI_ISL_2762774, EPI_ISL_2762776, EPI_ISL_2762777, EPI_ISL_2762779, EPI_ISL_2762782, EPI_ISL_2762783, EPI_ISL_2762784, EPI_ISL_2762786, EPI_ISL_2762787, EPI_ISL_2762789, EPI_ISL_2762790, EPI_ISL_2762792, EPI_ISL_2762794, EPI_ISL_2762795, EPI_ISL_2762797, EPI_ISL_2762799, EPI_ISL_2762800, EPI_ISL_2762802,
EPI_ISL_2762803, EPI_ISL_2762804, EPI_ISL_2762805, EPI_ISL_2762806, EPI_ISL_2762807, EPI_ISL_2762809, EPI_ISL_2762811, EPI_ISL_2762813, EPI_ISL_2762814, EPI_ISL_2762816, EPI_ISL_2762818, EPI_ISL_2762819, EPI_ISL_2762821, EPI_ISL_2763152, EPI_ISL_2763153, EPI_ISL_2763154, EPI_ISL_2763155, EPI_ISL_2763156, EPI_ISL_2763157, EPI_ISL_2763158, EPI_ISL_2763159, EPI_ISL_2763174,
EPI_ISL_2763175, EPI_ISL_2763183, EPI_ISL_2763184, EPI_ISL_2763185, EPI_ISL_2763186, EPI_ISL_2763187, EPI_ISL_2763188, EPI_ISL_2763189, EPI_ISL_2763190, EPI_ISL_2763191, EPI_ISL_2763192, EPI_ISL_2763193, EPI_ISL_2763194, EPI_ISL_2763199, EPI_ISL_2763200, EPI_ISL_2763201, EPI_ISL_2763202, EPI_ISL_2763203, EPI_ISL_2763204, EPI_ISL_2763205, EPI_ISL_2763206, EPI_ISL_2763207,
EPI_ISL_2763208, EPI_ISL_2763209, EPI_ISL_2763210, EPI_ISL_2763211, EPI_ISL_2763212, EPI_ISL_2763213, EPI_ISL_2763214, EPI_ISL_2763215, EPI_ISL_2763216, EPI_ISL_2763217, EPI_ISL_2763218, EPI_ISL_2763219, EPI_ISL_2763220, EPI_ISL_2763221, EPI_ISL_2763222, EPI_ISL_2763223, EPI_ISL_2763224, EPI_ISL_2763225, EPI_ISL_2763226, EPI_ISL_2763227, EPI_ISL_2763228, EPI_ISL_2763229,
EPI_ISL_2763230, EPI_ISL_2763231, EPI_ISL_2763232, EPI_ISL_2763233, EPI_ISL_2763234, EPI_ISL_2763235, EPI_ISL_2763236, EPI_ISL_2763237, EPI_ISL_2763238, EPI_ISL_2763239, EPI_ISL_2763240, EPI_ISL_2763241, EPI_ISL_2763242, EPI_ISL_2763243, EPI_ISL_2763244, EPI_ISL_2763245, EPI_ISL_2763246, EPI_ISL_2763247, EPI_ISL_2763248, EPI_ISL_2763249, EPI_ISL_2763250, EPI_ISL_2763251,
EPI_ISL_2763252, EPI_ISL_2763253, EPI_ISL_2763254, EPI_ISL_2763255, EPI_ISL_2763279, EPI_ISL_2763280, EPI_ISL_2763281, EPI_ISL_2763282, EPI_ISL_2763283, EPI_ISL_2763284, EPI_ISL_2763285, EPI_ISL_2763286, EPI_ISL_2763287, EPI_ISL_2763288, EPI_ISL_2763289, EPI_ISL_2763290, EPI_ISL_2763291, EPI_ISL_2763292, EPI_ISL_2763293, EPI_ISL_2763294, EPI_ISL_2763295, EPI_ISL_2763296,
EPI_ISL_2763297, EPI_ISL_2763298, EPI_ISL_2763299, EPI_ISL_2763311, EPI_ISL_2763312, EPI_ISL_2763313, EPI_ISL_2763314, EPI_ISL_2763315, EPI_ISL_2763316, EPI_ISL_2763317, EPI_ISL_2763318, EPI_ISL_2763319, EPI_ISL_2763320, EPI_ISL_2763321, EPI_ISL_2763322, EPI_ISL_2763323, EPI_ISL_2763345, EPI_ISL_2763346, EPI_ISL_2763347, EPI_ISL_2763348, EPI_ISL_2763349, EPI_ISL_2763350,

EPI_ISL_2763351, EPI_ISL_2763352, EPI_ISL_2763353, EPI_ISL_2763354, EPI_ISL_2763355, EPI_ISL_2763356, EPI_ISL_2763357, EPI_ISL_2763358, EPI_ISL_2763359, EPI_ISL_2763360, EPI_ISL_2763361, EPI_ISL_2763362, EPI_ISL_2763363, EPI_ISL_2763364, EPI_ISL_2763365, EPI_ISL_2763366, EPI_ISL_2763367, EPI_ISL_2763368, EPI_ISL_2763369, EPI_ISL_2763370, EPI_ISL_2763371, EPI_ISL_2763372,
EPI_ISL_2763373, EPI_ISL_2763374, EPI_ISL_2763375, EPI_ISL_2763376, EPI_ISL_2763377, EPI_ISL_2763378, EPI_ISL_2763379, EPI_ISL_2763380, EPI_ISL_2763381, EPI_ISL_2763382, EPI_ISL_2763383, EPI_ISL_2763384, EPI_ISL_2763385, EPI_ISL_2763386, EPI_ISL_2763387, EPI_ISL_2763388, EPI_ISL_2763389, EPI_ISL_2763390, EPI_ISL_2763391, EPI_ISL_2763392, EPI_ISL_2763393, EPI_ISL_2763394,
EPI_ISL_2763395, EPI_ISL_2763396, EPI_ISL_2763397, EPI_ISL_2763398, EPI_ISL_2763399, EPI_ISL_2763400, EPI_ISL_2763401, EPI_ISL_2763402, EPI_ISL_2763403, EPI_ISL_2763404, EPI_ISL_2763405, EPI_ISL_2763406, EPI_ISL_2763407, EPI_ISL_2763408, EPI_ISL_2763409, EPI_ISL_2763410, EPI_ISL_2763411, EPI_ISL_2763412, EPI_ISL_2763413, EPI_ISL_2763414, EPI_ISL_2763415, EPI_ISL_2763416,
EPI_ISL_2763417, EPI_ISL_2763418, EPI_ISL_2763419, EPI_ISL_2763420, EPI_ISL_2763421, EPI_ISL_2763422, EPI_ISL_2763423, EPI_ISL_2763424, EPI_ISL_2763425, EPI_ISL_2763426, EPI_ISL_2763427, EPI_ISL_2763428, EPI_ISL_2763429, EPI_ISL_2763430, EPI_ISL_2763431, EPI_ISL_2763432, EPI_ISL_2763433, EPI_ISL_2763434, EPI_ISL_2763435, EPI_ISL_2763436, EPI_ISL_2763437, EPI_ISL_2763438,
EPI_ISL_2763439, EPI_ISL_2763440, EPI_ISL_2763441, EPI_ISL_2763442, EPI_ISL_2763443, EPI_ISL_2763444, EPI_ISL_2763445, EPI_ISL_2763446, EPI_ISL_2763447, EPI_ISL_2763448, EPI_ISL_2763449, EPI_ISL_2763450, EPI_ISL_2763451, EPI_ISL_2763452, EPI_ISL_2763453, EPI_ISL_2763454, EPI_ISL_2763455, EPI_ISL_2763456, EPI_ISL_2763457, EPI_ISL_2763458, EPI_ISL_2763459, EPI_ISL_2763463,
EPI_ISL_2763464, EPI_ISL_2763465, EPI_ISL_2763466, EPI_ISL_2763467, EPI_ISL_2763468, EPI_ISL_2763469, EPI_ISL_2763470, EPI_ISL_2763471, EPI_ISL_2763472, EPI_ISL_2763473, EPI_ISL_2763474, EPI_ISL_2763475, EPI_ISL_2763476, EPI_ISL_2763477, EPI_ISL_2763478, EPI_ISL_2763479, EPI_ISL_2763480, EPI_ISL_2763481, EPI_ISL_2763482, EPI_ISL_2763483, EPI_ISL_2763484, EPI_ISL_2763485,
EPI_ISL_2763486, EPI_ISL_2763487, EPI_ISL_2763488, EPI_ISL_2763489, EPI_ISL_2763490, EPI_ISL_2763491, EPI_ISL_2763492, EPI_ISL_2763493, EPI_ISL_2763494, EPI_ISL_2763495, EPI_ISL_2763496, EPI_ISL_2763497, EPI_ISL_2763498, EPI_ISL_2763499, EPI_ISL_2763500, EPI_ISL_2763501, EPI_ISL_2763502, EPI_ISL_2763503, EPI_ISL_2763504, EPI_ISL_2763505, EPI_ISL_2763506, EPI_ISL_2763507,
EPI_ISL_2763508, EPI_ISL_2763509, EPI_ISL_2763510, EPI_ISL_2763511, EPI_ISL_2763512, EPI_ISL_2763513, EPI_ISL_2763514, EPI_ISL_2763515, EPI_ISL_2763516, EPI_ISL_2763517, EPI_ISL_2763518, EPI_ISL_2763519, EPI_ISL_2763520, EPI_ISL_2763521, EPI_ISL_2763522, EPI_ISL_2763523, EPI_ISL_2763524, EPI_ISL_2763525, EPI_ISL_2763526, EPI_ISL_2763527, EPI_ISL_2763528, EPI_ISL_2763529,
EPI_ISL_2763530, EPI_ISL_2763531, EPI_ISL_2763532, EPI_ISL_2763533, EPI_ISL_2763534, EPI_ISL_2763535, EPI_ISL_2763536, EPI_ISL_2763537, EPI_ISL_2763538, EPI_ISL_2763539, EPI_ISL_2763540, EPI_ISL_2763541, EPI_ISL_2763542, EPI_ISL_2763543, EPI_ISL_2763544, EPI_ISL_2763545, EPI_ISL_2763546, EPI_ISL_2763547, EPI_ISL_2763548, EPI_ISL_2763549, EPI_ISL_2763550, EPI_ISL_2763551,
EPI_ISL_2763552, EPI_ISL_2763553, EPI_ISL_2763554, EPI_ISL_2763555, EPI_ISL_2763556, EPI_ISL_2763557, EPI_ISL_2763558, EPI_ISL_2763559, EPI_ISL_2763560, EPI_ISL_2763561, EPI_ISL_2763562, EPI_ISL_2763563, EPI_ISL_2763564, EPI_ISL_2763565, EPI_ISL_2763566, EPI_ISL_2763567, EPI_ISL_2763569, EPI_ISL_2763570, EPI_ISL_2763571, EPI_ISL_2763572, EPI_ISL_2763573, EPI_ISL_2763574,
EPI_ISL_2763575, EPI_ISL_2763576, EPI_ISL_2763577, EPI_ISL_2763578, EPI_ISL_2763579, EPI_ISL_2763580, EPI_ISL_2763581, EPI_ISL_2763582, EPI_ISL_2763583, EPI_ISL_2763584, EPI_ISL_2763585, EPI_ISL_2763586, EPI_ISL_2763587, EPI_ISL_2763588, EPI_ISL_2763589, EPI_ISL_2763590, EPI_ISL_2763591, EPI_ISL_2763592, EPI_ISL_2763593, EPI_ISL_2763600, EPI_ISL_2763601, EPI_ISL_2763602,
EPI_ISL_2763603, EPI_ISL_2763604, EPI_ISL_2763605, EPI_ISL_2763606, EPI_ISL_2763607, EPI_ISL_2763608, EPI_ISL_2763609, EPI_ISL_2763610, EPI_ISL_2763655, EPI_ISL_2763656, EPI_ISL_2763657, EPI_ISL_2763658, EPI_ISL_2763659, EPI_ISL_2763660, EPI_ISL_2763661, EPI_ISL_2763662, EPI_ISL_2763663, EPI_ISL_2763664, EPI_ISL_2763665, EPI_ISL_2763666, EPI_ISL_2763667, EPI_ISL_2763668,
EPI_ISL_2763669, EPI_ISL_2763670, EPI_ISL_2763671, EPI_ISL_2763672, EPI_ISL_2763673, EPI_ISL_2763674, EPI_ISL_2763676, EPI_ISL_2763677, EPI_ISL_2763678, EPI_ISL_2763679, EPI_ISL_2763680, EPI_ISL_2763681, EPI_ISL_2763682, EPI_ISL_2763683, EPI_ISL_2763684, EPI_ISL_2763685, EPI_ISL_2763686, EPI_ISL_2763687, EPI_ISL_2763688, EPI_ISL_2763689, EPI_ISL_2763690, EPI_ISL_2763691,
EPI_ISL_2763692, EPI_ISL_2763693, EPI_ISL_2763694, EPI_ISL_2763695, EPI_ISL_2763696, EPI_ISL_2763697, EPI_ISL_2763698, EPI_ISL_2763699, EPI_ISL_2763700, EPI_ISL_2763701, EPI_ISL_2763702, EPI_ISL_2763703, EPI_ISL_2763704, EPI_ISL_2763705, EPI_ISL_2763706, EPI_ISL_2763707, EPI_ISL_2763708, EPI_ISL_2763709, EPI_ISL_2763710, EPI_ISL_2763711, EPI_ISL_2763712, EPI_ISL_2763713,
EPI_ISL_2763714, EPI_ISL_2763715, EPI_ISL_2763716, EPI_ISL_2763717, EPI_ISL_2763718, EPI_ISL_2763719, EPI_ISL_2763720, EPI_ISL_2763721, EPI_ISL_2763722, EPI_ISL_2763723, EPI_ISL_2763724, EPI_ISL_2763725, EPI_ISL_2763726, EPI_ISL_2763727, EPI_ISL_2763728, EPI_ISL_2763729, EPI_ISL_2763730, EPI_ISL_2763731, EPI_ISL_2763732, EPI_ISL_2763733, EPI_ISL_2763734, EPI_ISL_2763735,
EPI_ISL_2763736, EPI_ISL_2763737, EPI_ISL_2763738, EPI_ISL_2763739, EPI_ISL_2763740, EPI_ISL_2763741, EPI_ISL_2763742, EPI_ISL_2763743, EPI_ISL_2763744, EPI_ISL_2763745, EPI_ISL_2763746, EPI_ISL_2763747, EPI_ISL_2763748, EPI_ISL_2763749, EPI_ISL_2763750, EPI_ISL_2763751, EPI_ISL_2763752, EPI_ISL_2763753, EPI_ISL_2763754, EPI_ISL_2763755, EPI_ISL_2763756, EPI_ISL_2763757,
EPI_ISL_2763758, EPI_ISL_2763759, EPI_ISL_2763760, EPI_ISL_2763761, EPI_ISL_2763762, EPI_ISL_2763763, EPI_ISL_2763764, EPI_ISL_2763765, EPI_ISL_2763766, EPI_ISL_2763767, EPI_ISL_2763768, EPI_ISL_2763769, EPI_ISL_2763770, EPI_ISL_2763771, EPI_ISL_2763772, EPI_ISL_2763773, EPI_ISL_2763774, EPI_ISL_2763775, EPI_ISL_2763776, EPI_ISL_2763777, EPI_ISL_2763778, EPI_ISL_2763779,
EPI_ISL_2763780, EPI_ISL_2763781, EPI_ISL_2763782, EPI_ISL_2763783, EPI_ISL_2763794, EPI_ISL_2763795, EPI_ISL_2763796, EPI_ISL_2763797, EPI_ISL_2763798, EPI_ISL_2763799, EPI_ISL_2763800, EPI_ISL_2763801, EPI_ISL_2763802, EPI_ISL_2763803, EPI_ISL_2763804, EPI_ISL_2763805, EPI_ISL_2763806, EPI_ISL_2763807, EPI_ISL_2763808, EPI_ISL_2763809, EPI_ISL_2763810, EPI_ISL_2763811,
EPI_ISL_2763812, EPI_ISL_2763813, EPI_ISL_2763814, EPI_ISL_2763815, EPI_ISL_2763816, EPI_ISL_2763817, EPI_ISL_2763818, EPI_ISL_2763819, EPI_ISL_2763820, EPI_ISL_2763821, EPI_ISL_2763822, EPI_ISL_2763824, EPI_ISL_2763825, EPI_ISL_2763826, EPI_ISL_2763827, EPI_ISL_2763828, EPI_ISL_2763829, EPI_ISL_2763830, EPI_ISL_2763831, EPI_ISL_2763832, EPI_ISL_2763833, EPI_ISL_2763834,
EPI_ISL_2763835, EPI_ISL_2763836, EPI_ISL_2763837, EPI_ISL_2763838, EPI_ISL_2763839, EPI_ISL_2763840, EPI_ISL_2763841, EPI_ISL_2763842, EPI_ISL_2763843, EPI_ISL_2763844, EPI_ISL_2763845, EPI_ISL_2763846, EPI_ISL_2763847, EPI_ISL_2763848, EPI_ISL_2763849, EPI_ISL_2763850, EPI_ISL_2763851, EPI_ISL_2763852, EPI_ISL_2763853, EPI_ISL_2763854, EPI_ISL_2763855, EPI_ISL_2763856,
EPI_ISL_2763857, EPI_ISL_2763858, EPI_ISL_2763859, EPI_ISL_2763860, EPI_ISL_2763861, EPI_ISL_2763864, EPI_ISL_2763865, EPI_ISL_2763901, EPI_ISL_2763902, EPI_ISL_2763903, EPI_ISL_2763904, EPI_ISL_2763905, EPI_ISL_2763906, EPI_ISL_2763907, EPI_ISL_2763908, EPI_ISL_2763909, EPI_ISL_2763910, EPI_ISL_2763911, EPI_ISL_2763912, EPI_ISL_2763913, EPI_ISL_2763914, EPI_ISL_2763915,
EPI_ISL_2763916, EPI_ISL_2763917, EPI_ISL_2763918, EPI_ISL_2763919, EPI_ISL_2763920, EPI_ISL_2763921, EPI_ISL_2763922, EPI_ISL_2763923, EPI_ISL_2763924, EPI_ISL_2763925, EPI_ISL_2763939, EPI_ISL_2763940, EPI_ISL_2763941, EPI_ISL_2763942, EPI_ISL_2763943, EPI_ISL_2763944, EPI_ISL_2763945, EPI_ISL_2763946, EPI_ISL_2763947, EPI_ISL_2763948, EPI_ISL_2763949, EPI_ISL_2763950,
EPI_ISL_2763951, EPI_ISL_2763952, EPI_ISL_2763953, EPI_ISL_2763954, EPI_ISL_2763955, EPI_ISL_2763956, EPI_ISL_2763957, EPI_ISL_2763958, EPI_ISL_2763959, EPI_ISL_2763960, EPI_ISL_2763961, EPI_ISL_2764111, EPI_ISL_2764112, EPI_ISL_2764113, EPI_ISL_2764114, EPI_ISL_2764115, EPI_ISL_2764116, EPI_ISL_2764117, EPI_ISL_2764118, EPI_ISL_2764119, EPI_ISL_2764120, EPI_ISL_2764121,
EPI_ISL_2764122, EPI_ISL_2764123, EPI_ISL_2764124, EPI_ISL_2764125, EPI_ISL_2764126, EPI_ISL_2764127, EPI_ISL_2764128, EPI_ISL_2764129, EPI_ISL_2764130, EPI_ISL_2764131, EPI_ISL_2764132, EPI_ISL_2764133, EPI_ISL_2764134, EPI_ISL_2764135, EPI_ISL_2764136, EPI_ISL_2764137, EPI_ISL_2764138, EPI_ISL_2764139, EPI_ISL_2764140, EPI_ISL_2764141, EPI_ISL_2764142, EPI_ISL_2764143,
EPI_ISL_2764144, EPI_ISL_2764145, EPI_ISL_2764146, EPI_ISL_2764147, EPI_ISL_2764148, EPI_ISL_2764149, EPI_ISL_2764150, EPI_ISL_2764151, EPI_ISL_2764152, EPI_ISL_2764153, EPI_ISL_2764154, EPI_ISL_2764155, EPI_ISL_2764156, EPI_ISL_2764157, EPI_ISL_2764158, EPI_ISL_2764159, EPI_ISL_2764160, EPI_ISL_2764161, EPI_ISL_2764162, EPI_ISL_2764163, EPI_ISL_2764164, EPI_ISL_2764165,
EPI_ISL_2764166, EPI_ISL_2764167, EPI_ISL_2764168, EPI_ISL_2764169, EPI_ISL_2764170, EPI_ISL_2764171, EPI_ISL_2764172, EPI_ISL_2764173, EPI_ISL_2764174, EPI_ISL_2764175, EPI_ISL_2764177, EPI_ISL_2764178, EPI_ISL_2764179, EPI_ISL_2764190, EPI_ISL_2764191, EPI_ISL_2764192, EPI_ISL_2764193, EPI_ISL_2764194, EPI_ISL_2764195, EPI_ISL_2764196, EPI_ISL_2764197, EPI_ISL_2764198,
EPI_ISL_2764199, EPI_ISL_2764200, EPI_ISL_2764201, EPI_ISL_2764202, EPI_ISL_2764203, EPI_ISL_2764204, EPI_ISL_2764205, EPI_ISL_2764206, EPI_ISL_2764207, EPI_ISL_2764208, EPI_ISL_2764209, EPI_ISL_2764210, EPI_ISL_2764211, EPI_ISL_2764212, EPI_ISL_2764213, EPI_ISL_2764214, EPI_ISL_2764215, EPI_ISL_2764216, EPI_ISL_2764217, EPI_ISL_2764218, EPI_ISL_2764219, EPI_ISL_2764220,
EPI_ISL_2764221, EPI_ISL_2764222, EPI_ISL_2764223, EPI_ISL_2764224, EPI_ISL_2764225, EPI_ISL_2764226, EPI_ISL_2764227, EPI_ISL_2764228, EPI_ISL_2764229, EPI_ISL_2764230, EPI_ISL_2764231, EPI_ISL_2764232, EPI_ISL_2764233, EPI_ISL_2764234, EPI_ISL_2764235, EPI_ISL_2764236, EPI_ISL_2764237, EPI_ISL_2764238, EPI_ISL_2764239, EPI_ISL_2764240, EPI_ISL_2764241, EPI_ISL_2764242,
EPI_ISL_2764243, EPI_ISL_2764244, EPI_ISL_2764245, EPI_ISL_2764246, EPI_ISL_2764247, EPI_ISL_2764248, EPI_ISL_2764249, EPI_ISL_2764250, EPI_ISL_2764251, EPI_ISL_2764252, EPI_ISL_2764253, EPI_ISL_2764254, EPI_ISL_2764255, EPI_ISL_2764256, EPI_ISL_2764257, EPI_ISL_2764263, EPI_ISL_2764264, EPI_ISL_2764265, EPI_ISL_2764266, EPI_ISL_2764267, EPI_ISL_2764268, EPI_ISL_2764269,
EPI_ISL_2764270, EPI_ISL_2764271, EPI_ISL_2764272, EPI_ISL_2764273, EPI_ISL_2764274, EPI_ISL_2764286, EPI_ISL_2764287, EPI_ISL_2764288, EPI_ISL_2764289, EPI_ISL_2764290, EPI_ISL_2764291, EPI_ISL_2764292, EPI_ISL_2764293, EPI_ISL_2764306, EPI_ISL_2764307, EPI_ISL_2764308, EPI_ISL_2764309, EPI_ISL_2764310, EPI_ISL_2764311, EPI_ISL_2764312, EPI_ISL_2764313, EPI_ISL_2764314,
EPI_ISL_2764315, EPI_ISL_2764316, EPI_ISL_2764317, EPI_ISL_2764319, EPI_ISL_2764320, EPI_ISL_2764321, EPI_ISL_2764322, EPI_ISL_2764323, EPI_ISL_2764324, EPI_ISL_2764325, EPI_ISL_2764326, EPI_ISL_2764327, EPI_ISL_2764328, EPI_ISL_2764329, EPI_ISL_2764330, EPI_ISL_2764331, EPI_ISL_2764332, EPI_ISL_2764333, EPI_ISL_2764334, EPI_ISL_2764335, EPI_ISL_2764336, EPI_ISL_2764337,
EPI_ISL_2764338, EPI_ISL_2764339, EPI_ISL_2764340, EPI_ISL_2764341, EPI_ISL_2764342, EPI_ISL_2764343, EPI_ISL_2764344, EPI_ISL_2764345, EPI_ISL_2764346, EPI_ISL_2764347, EPI_ISL_2764348, EPI_ISL_2764349, EPI_ISL_2764350, EPI_ISL_2764351, EPI_ISL_2764352, EPI_ISL_2764353, EPI_ISL_2764354, EPI_ISL_2764355, EPI_ISL_2764356, EPI_ISL_2764357, EPI_ISL_2764358, EPI_ISL_2764359,
EPI_ISL_2764360, EPI_ISL_2764361, EPI_ISL_2764362, EPI_ISL_2764363, EPI_ISL_2764364, EPI_ISL_2764365, EPI_ISL_2764366, EPI_ISL_2764367, EPI_ISL_2764371, EPI_ISL_2764372, EPI_ISL_2764377, EPI_ISL_2764378, EPI_ISL_2764379, EPI_ISL_2764380, EPI_ISL_2764381, EPI_ISL_2764382, EPI_ISL_2764383, EPI_ISL_2764384, EPI_ISL_2764385, EPI_ISL_2764386, EPI_ISL_2764387, EPI_ISL_2764388,
EPI_ISL_2764389, EPI_ISL_2764390, EPI_ISL_2764391, EPI_ISL_2764392, EPI_ISL_2764393, EPI_ISL_2764394, EPI_ISL_2764395, EPI_ISL_2764396, EPI_ISL_2764397, EPI_ISL_2764398, EPI_ISL_2764399, EPI_ISL_2764400, EPI_ISL_2764401, EPI_ISL_2764402, EPI_ISL_2764412, EPI_ISL_2764413, EPI_ISL_2764414, EPI_ISL_2764415, EPI_ISL_2764416, EPI_ISL_2764417, EPI_ISL_2764418, EPI_ISL_2764419,
EPI_ISL_2764420, EPI_ISL_2764422, EPI_ISL_2764423, EPI_ISL_2764427, EPI_ISL_2764428, EPI_ISL_2764429, EPI_ISL_2764430, EPI_ISL_2764431, EPI_ISL_2764432, EPI_ISL_2764433, EPI_ISL_2764434, EPI_ISL_2764435, EPI_ISL_2764436, EPI_ISL_2764437, EPI_ISL_2764438, EPI_ISL_2764439, EPI_ISL_2764440, EPI_ISL_2764441, EPI_ISL_2764442, EPI_ISL_2764443, EPI_ISL_2764444, EPI_ISL_2764445,
EPI_ISL_2764446, EPI_ISL_2764447, EPI_ISL_2764448, EPI_ISL_2764449, EPI_ISL_2764450, EPI_ISL_2764451, EPI_ISL_2764452, EPI_ISL_2764453, EPI_ISL_2764454, EPI_ISL_2764455, EPI_ISL_2764456, EPI_ISL_2764457, EPI_ISL_2764458, EPI_ISL_2764459, EPI_ISL_2764460, EPI_ISL_2764461, EPI_ISL_2764462, EPI_ISL_2764463, EPI_ISL_2764464, EPI_ISL_2764465, EPI_ISL_2764466, EPI_ISL_2764467,
EPI_ISL_2764468, EPI_ISL_2764469, EPI_ISL_2764470, EPI_ISL_2764471, EPI_ISL_2764472, EPI_ISL_2764473, EPI_ISL_2764474, EPI_ISL_2764475, EPI_ISL_2764476, EPI_ISL_2764477, EPI_ISL_2764478, EPI_ISL_2764479, EPI_ISL_2764480, EPI_ISL_2764481, EPI_ISL_2764483, EPI_ISL_2764484, EPI_ISL_2764485, EPI_ISL_2764486, EPI_ISL_2764487, EPI_ISL_2764488, EPI_ISL_2764489, EPI_ISL_2764490,
EPI_ISL_2764491, EPI_ISL_2764492, EPI_ISL_2764493, EPI_ISL_2764494, EPI_ISL_2764495, EPI_ISL_2764496, EPI_ISL_2764497, EPI_ISL_2764498, EPI_ISL_2764499, EPI_ISL_2764500, EPI_ISL_2764501, EPI_ISL_2764502, EPI_ISL_2764503, EPI_ISL_2764504, EPI_ISL_2764505, EPI_ISL_2764506, EPI_ISL_2764507, EPI_ISL_2764508, EPI_ISL_2764509, EPI_ISL_2764510, EPI_ISL_2764511, EPI_ISL_2764512,
EPI_ISL_2764513, EPI_ISL_2764514, EPI_ISL_2764515, EPI_ISL_2764516, EPI_ISL_2764517, EPI_ISL_2764518, EPI_ISL_2764519, EPI_ISL_2764520, EPI_ISL_2764521, EPI_ISL_2764525, EPI_ISL_2764526, EPI_ISL_2764527, EPI_ISL_2764528, EPI_ISL_2764533, EPI_ISL_2764534, EPI_ISL_2764535, EPI_ISL_2764536, EPI_ISL_2764537, EPI_ISL_2764538, EPI_ISL_2764539, EPI_ISL_2764540, EPI_ISL_2764541,
EPI_ISL_2764542, EPI_ISL_2764545, EPI_ISL_2764546, EPI_ISL_2764547, EPI_ISL_2764548, EPI_ISL_2764549, EPI_ISL_2764550, EPI_ISL_2764551, EPI_ISL_2764552, EPI_ISL_2764553, EPI_ISL_2764554, EPI_ISL_2764555, EPI_ISL_2764563, EPI_ISL_2764564, EPI_ISL_2764565, EPI_ISL_2764566, EPI_ISL_2764568, EPI_ISL_2764569, EPI_ISL_2764570, EPI_ISL_2764576, EPI_ISL_2764577, EPI_ISL_2764578,
EPI_ISL_2764579, EPI_ISL_2764580, EPI_ISL_2764581, EPI_ISL_2764582, EPI_ISL_2764583, EPI_ISL_2764584, EPI_ISL_2764585, EPI_ISL_2764586, EPI_ISL_2764587, EPI_ISL_2764588, EPI_ISL_2764589, EPI_ISL_2764590, EPI_ISL_2764591, EPI_ISL_2764592, EPI_ISL_2764593, EPI_ISL_2764594, EPI_ISL_2764595, EPI_ISL_2764596, EPI_ISL_2764597, EPI_ISL_2764598, EPI_ISL_2764599, EPI_ISL_2764600,
EPI_ISL_2764601, EPI_ISL_2764602, EPI_ISL_2764603, EPI_ISL_2764604, EPI_ISL_2764605, EPI_ISL_2764606, EPI_ISL_2764607, EPI_ISL_2764608, EPI_ISL_2764609, EPI_ISL_2764610, EPI_ISL_2764611, EPI_ISL_2764612, EPI_ISL_2764613, EPI_ISL_2764614, EPI_ISL_2764615, EPI_ISL_2764618, EPI_ISL_2764619, EPI_ISL_2764620, EPI_ISL_2764621, EPI_ISL_2764625, EPI_ISL_2764626, EPI_ISL_2764627,
EPI_ISL_2764628, EPI_ISL_2764629, EPI_ISL_2764630, EPI_ISL_2764631, EPI_ISL_2764632, EPI_ISL_2764637, EPI_ISL_2764640, EPI_ISL_2764641, EPI_ISL_2764642, EPI_ISL_2764643, EPI_ISL_2764644, EPI_ISL_2764645, EPI_ISL_2764646, EPI_ISL_2764647, EPI_ISL_2764648, EPI_ISL_2764649, EPI_ISL_2764650, EPI_ISL_2764651, EPI_ISL_2764652, EPI_ISL_2764653, EPI_ISL_2764658, EPI_ISL_2764659,
EPI_ISL_2764660, EPI_ISL_2764661, EPI_ISL_2764662, EPI_ISL_2764663, EPI_ISL_2764664, EPI_ISL_2764665, EPI_ISL_2764678, EPI_ISL_2764680, EPI_ISL_2764681, EPI_ISL_2764682, EPI_ISL_2764683, EPI_ISL_2764684, EPI_ISL_2764685, EPI_ISL_2764686, EPI_ISL_2764687, EPI_ISL_2764688, EPI_ISL_2764689, EPI_ISL_2764690, EPI_ISL_2764691, EPI_ISL_2764692, EPI_ISL_2764693, EPI_ISL_2764694,
EPI_ISL_2764695, EPI_ISL_2764696, EPI_ISL_2764697, EPI_ISL_2764698, EPI_ISL_2764699, EPI_ISL_2764700, EPI_ISL_2764701, EPI_ISL_2764702, EPI_ISL_2764703, EPI_ISL_2764704, EPI_ISL_2764705, EPI_ISL_2764706, EPI_ISL_2764707, EPI_ISL_2764708, EPI_ISL_2764709, EPI_ISL_2764710, EPI_ISL_2764711, EPI_ISL_2764712, EPI_ISL_2764713, EPI_ISL_2764714, EPI_ISL_2764715, EPI_ISL_2764716,
EPI_ISL_2764717, EPI_ISL_2764718, EPI_ISL_2764719, EPI_ISL_2764720, EPI_ISL_2764721, EPI_ISL_2764722, EPI_ISL_2764723, EPI_ISL_2764724, EPI_ISL_2764725, EPI_ISL_2764726, EPI_ISL_2764727, EPI_ISL_2764728, EPI_ISL_2764729, EPI_ISL_2764730, EPI_ISL_2764731, EPI_ISL_2764732, EPI_ISL_2764733, EPI_ISL_2764734, EPI_ISL_2764735, EPI_ISL_2764736, EPI_ISL_2764737, EPI_ISL_2764738,
EPI_ISL_2764739, EPI_ISL_2764740, EPI_ISL_2764741, EPI_ISL_2764742, EPI_ISL_2764743, EPI_ISL_2764744, EPI_ISL_2764745, EPI_ISL_2764746, EPI_ISL_2764747, EPI_ISL_2764748, EPI_ISL_2764749, EPI_ISL_2764752, EPI_ISL_2764753, EPI_ISL_2764754, EPI_ISL_2764755, EPI_ISL_2764756, EPI_ISL_2764757, EPI_ISL_2764758, EPI_ISL_2764759, EPI_ISL_2764760, EPI_ISL_2764761, EPI_ISL_2764764,
EPI_ISL_2764765, EPI_ISL_2764766, EPI_ISL_2764767, EPI_ISL_2764768, EPI_ISL_2764769, EPI_ISL_2764770, EPI_ISL_2764771, EPI_ISL_2764772, EPI_ISL_2764773, EPI_ISL_2764774, EPI_ISL_2764775, EPI_ISL_2764776, EPI_ISL_2764777, EPI_ISL_2764778, EPI_ISL_2764779, EPI_ISL_2764781, EPI_ISL_2764782, EPI_ISL_2764783, EPI_ISL_2764786, EPI_ISL_2764787, EPI_ISL_2764790, EPI_ISL_2764791,
EPI_ISL_2764793, EPI_ISL_2764794, EPI_ISL_2764795, EPI_ISL_2764796, EPI_ISL_2764797, EPI_ISL_2764798, EPI_ISL_2764799, EPI_ISL_2764800, EPI_ISL_2764801, EPI_ISL_2764802, EPI_ISL_2764803, EPI_ISL_2764804, EPI_ISL_2764805, EPI_ISL_2764806, EPI_ISL_2764807, EPI_ISL_2764808, EPI_ISL_2764809, EPI_ISL_2764810, EPI_ISL_2764811, EPI_ISL_2764812, EPI_ISL_2764813, EPI_ISL_2764814,
EPI_ISL_2764815, EPI_ISL_2764816, EPI_ISL_2764817, EPI_ISL_2764818, EPI_ISL_2764819, EPI_ISL_2764820, EPI_ISL_2764821, EPI_ISL_2764822, EPI_ISL_2764824, EPI_ISL_2764825, EPI_ISL_2764826, EPI_ISL_2764827, EPI_ISL_2764828, EPI_ISL_2764829, EPI_ISL_2764830, EPI_ISL_2764831, EPI_ISL_2764832, EPI_ISL_2764833, EPI_ISL_2764834, EPI_ISL_2764835, EPI_ISL_2764836, EPI_ISL_2764837,
EPI_ISL_2764842, EPI_ISL_2764843, EPI_ISL_2764844, EPI_ISL_2764845, EPI_ISL_2764846, EPI_ISL_2764854, EPI_ISL_2764855, EPI_ISL_2764856, EPI_ISL_2764857, EPI_ISL_2764858, EPI_ISL_2764859, EPI_ISL_2764860, EPI_ISL_2764861, EPI_ISL_2764862, EPI_ISL_2764863, EPI_ISL_2764864, EPI_ISL_2764865, EPI_ISL_2764866, EPI_ISL_2764867, EPI_ISL_2764868, EPI_ISL_2764869, EPI_ISL_2764870,
EPI_ISL_2764871, EPI_ISL_2764872, EPI_ISL_2764873, EPI_ISL_2764874, EPI_ISL_2764875, EPI_ISL_2764876, EPI_ISL_2764877, EPI_ISL_2764878, EPI_ISL_2764879, EPI_ISL_2764880, EPI_ISL_2764881, EPI_ISL_2764882, EPI_ISL_2764883, EPI_ISL_2764884, EPI_ISL_2764885, EPI_ISL_2764886, EPI_ISL_2764895, EPI_ISL_2764904, EPI_ISL_2764907, EPI_ISL_2764908, EPI_ISL_2764910, EPI_ISL_2764911,
EPI_ISL_2764912, EPI_ISL_2764920, EPI_ISL_2764959, EPI_ISL_2764965, EPI_ISL_2764966, EPI_ISL_2764967, EPI_ISL_2764968, EPI_ISL_2764969, EPI_ISL_2764970, EPI_ISL_2764971, EPI_ISL_2764972, EPI_ISL_2764973, EPI_ISL_2764974, EPI_ISL_2764975, EPI_ISL_2764976, EPI_ISL_2764977, EPI_ISL_2764978, EPI_ISL_2764979, EPI_ISL_2764980, EPI_ISL_2764981, EPI_ISL_2764982, EPI_ISL_2764983,
EPI_ISL_2764984, EPI_ISL_2764985, EPI_ISL_2764986, EPI_ISL_2765004, EPI_ISL_2765005, EPI_ISL_2765006, EPI_ISL_2765007, EPI_ISL_2765008, EPI_ISL_2765009, EPI_ISL_2765010, EPI_ISL_2765011, EPI_ISL_2765012, EPI_ISL_2765013, EPI_ISL_2765014, EPI_ISL_2765015, EPI_ISL_2765016, EPI_ISL_2765017, EPI_ISL_2765018, EPI_ISL_2765019, EPI_ISL_2765104, EPI_ISL_2765105, EPI_ISL_2765106,
EPI_ISL_2765107, EPI_ISL_2765108, EPI_ISL_2765109, EPI_ISL_2765110, EPI_ISL_2765111, EPI_ISL_2765112, EPI_ISL_2765113, EPI_ISL_2765114, EPI_ISL_2765115, EPI_ISL_2765116, EPI_ISL_2765117, EPI_ISL_2765118, EPI_ISL_2765119, EPI_ISL_2765120, EPI_ISL_2765121, EPI_ISL_2765122, EPI_ISL_2765123, EPI_ISL_2765124, EPI_ISL_2765125, EPI_ISL_2765126, EPI_ISL_2765127, EPI_ISL_2765128,
EPI_ISL_2765129, EPI_ISL_2765130, EPI_ISL_2765131, EPI_ISL_2765132, EPI_ISL_2765133, EPI_ISL_2765134, EPI_ISL_2765135, EPI_ISL_2765136, EPI_ISL_2765137, EPI_ISL_2765138, EPI_ISL_2765139, EPI_ISL_2765140, EPI_ISL_2765141, EPI_ISL_2765142, EPI_ISL_2765143, EPI_ISL_2765144, EPI_ISL_2765145, EPI_ISL_2765146, EPI_ISL_2765147, EPI_ISL_2765148, EPI_ISL_2765149, EPI_ISL_2765150,
EPI_ISL_2765151, EPI_ISL_2765152, EPI_ISL_2765153, EPI_ISL_2765154, EPI_ISL_2765155, EPI_ISL_2765156, EPI_ISL_2765157, EPI_ISL_2765158, EPI_ISL_2765159, EPI_ISL_2765167, EPI_ISL_2765183, EPI_ISL_2765184, EPI_ISL_2765185, EPI_ISL_2765186, EPI_ISL_2765187, EPI_ISL_2765188, EPI_ISL_2765189, EPI_ISL_2765190, EPI_ISL_2765191, EPI_ISL_2765192, EPI_ISL_2765193, EPI_ISL_2765194,
EPI_ISL_2765195, EPI_ISL_2765196, EPI_ISL_2765197, EPI_ISL_2765198, EPI_ISL_2765199, EPI_ISL_2765200, EPI_ISL_2765201, EPI_ISL_2765202, EPI_ISL_2765203, EPI_ISL_2765204, EPI_ISL_2765205, EPI_ISL_2765206, EPI_ISL_2765207, EPI_ISL_2765208, EPI_ISL_2765209, EPI_ISL_2765210, EPI_ISL_2765211, EPI_ISL_2765212, EPI_ISL_2765213, EPI_ISL_2765214, EPI_ISL_2765215, EPI_ISL_2765216,
EPI_ISL_2765217, EPI_ISL_2765218, EPI_ISL_2765219, EPI_ISL_2765220, EPI_ISL_2765221, EPI_ISL_2765222, EPI_ISL_2765223, EPI_ISL_2765224, EPI_ISL_2765225, EPI_ISL_2765226, EPI_ISL_2765227, EPI_ISL_2765228, EPI_ISL_2765229, EPI_ISL_2765230, EPI_ISL_2765231, EPI_ISL_2765232, EPI_ISL_2765233, EPI_ISL_2765234, EPI_ISL_2765235, EPI_ISL_2765236, EPI_ISL_2765237, EPI_ISL_2765238,
EPI_ISL_2765239, EPI_ISL_2765240, EPI_ISL_2765241, EPI_ISL_2765242, EPI_ISL_2765243, EPI_ISL_2765244, EPI_ISL_2765245, EPI_ISL_2765246, EPI_ISL_2765247, EPI_ISL_2765248, EPI_ISL_2765249, EPI_ISL_2765250, EPI_ISL_2765251, EPI_ISL_2765252, EPI_ISL_2765253, EPI_ISL_2765254, EPI_ISL_2765255, EPI_ISL_2765256, EPI_ISL_2765267, EPI_ISL_2765268, EPI_ISL_2765269, EPI_ISL_2765270,
EPI_ISL_2765271, EPI_ISL_2765280, EPI_ISL_2765281, EPI_ISL_2765282, EPI_ISL_2765283, EPI_ISL_2765284, EPI_ISL_2765289, EPI_ISL_2765299, EPI_ISL_2765300, EPI_ISL_2765301, EPI_ISL_2765302, EPI_ISL_2765329, EPI_ISL_2765330, EPI_ISL_2765331, EPI_ISL_2765332, EPI_ISL_2765333, EPI_ISL_2765364, EPI_ISL_2765365, EPI_ISL_2765366, EPI_ISL_2765367, EPI_ISL_2765368, EPI_ISL_2765369,
EPI_ISL_2765370, EPI_ISL_2765371, EPI_ISL_2765372, EPI_ISL_2765373, EPI_ISL_2765374, EPI_ISL_2765375, EPI_ISL_2765376, EPI_ISL_2765377, EPI_ISL_2765378, EPI_ISL_2765379, EPI_ISL_2765380, EPI_ISL_2765381, EPI_ISL_2765382, EPI_ISL_2765383, EPI_ISL_2765384, EPI_ISL_2765385, EPI_ISL_2765386, EPI_ISL_2765387, EPI_ISL_2765388, EPI_ISL_2765389, EPI_ISL_2765390, EPI_ISL_2765391,
EPI_ISL_2765392, EPI_ISL_2765393, EPI_ISL_2765394, EPI_ISL_2765395, EPI_ISL_2765396, EPI_ISL_2765397, EPI_ISL_2765398, EPI_ISL_2765399, EPI_ISL_2765400, EPI_ISL_2765401, EPI_ISL_2765402, EPI_ISL_2765403, EPI_ISL_2765404, EPI_ISL_2765405, EPI_ISL_2765406, EPI_ISL_2765407, EPI_ISL_2765408, EPI_ISL_2765409, EPI_ISL_2765410, EPI_ISL_2765411, EPI_ISL_2765412, EPI_ISL_2765413,
EPI_ISL_2765414, EPI_ISL_2765415, EPI_ISL_2765416, EPI_ISL_2765417, EPI_ISL_2765418, EPI_ISL_2765419, EPI_ISL_2765420, EPI_ISL_2765421, EPI_ISL_2765422, EPI_ISL_2765423, EPI_ISL_2765424, EPI_ISL_2765425, EPI_ISL_2765426, EPI_ISL_2765427, EPI_ISL_2765428, EPI_ISL_2765429, EPI_ISL_2765430, EPI_ISL_2765444, EPI_ISL_2765445, EPI_ISL_2765446, EPI_ISL_2765447, EPI_ISL_2765448,
EPI_ISL_2765449, EPI_ISL_2765450, EPI_ISL_2765451, EPI_ISL_2765452, EPI_ISL_2765453, EPI_ISL_2765454, EPI_ISL_2765455, EPI_ISL_2765456, EPI_ISL_2765457, EPI_ISL_2765458, EPI_ISL_2765459, EPI_ISL_2765460, EPI_ISL_2765461, EPI_ISL_2765462, EPI_ISL_2765463, EPI_ISL_2765464, EPI_ISL_2765465, EPI_ISL_2765466, EPI_ISL_2765467, EPI_ISL_2765468, EPI_ISL_2765469, EPI_ISL_2765470,
EPI_ISL_2765471, EPI_ISL_2765472, EPI_ISL_2765473, EPI_ISL_2765474, EPI_ISL_2765475, EPI_ISL_2765476, EPI_ISL_2765477, EPI_ISL_2765478, EPI_ISL_2765479, EPI_ISL_2765480, EPI_ISL_2765481, EPI_ISL_2765482, EPI_ISL_2765483, EPI_ISL_2765484, EPI_ISL_2765485, EPI_ISL_2765486, EPI_ISL_2765487, EPI_ISL_2765488, EPI_ISL_2765489, EPI_ISL_2765490, EPI_ISL_2765491, EPI_ISL_2765492,
EPI_ISL_2765493, EPI_ISL_2765494, EPI_ISL_2765495, EPI_ISL_2765496, EPI_ISL_2765497, EPI_ISL_2765498, EPI_ISL_2765499, EPI_ISL_2765500, EPI_ISL_2765501, EPI_ISL_2765505, EPI_ISL_2765506, EPI_ISL_2765507, EPI_ISL_2765508, EPI_ISL_2765509, EPI_ISL_2765510, EPI_ISL_2765511, EPI_ISL_2765512, EPI_ISL_2765513, EPI_ISL_2765514, EPI_ISL_2765515, EPI_ISL_2765516, EPI_ISL_2765517,
EPI_ISL_2765518, EPI_ISL_2765519, EPI_ISL_2765520, EPI_ISL_2765521, EPI_ISL_2765522, EPI_ISL_2765523, EPI_ISL_2765524, EPI_ISL_2765525, EPI_ISL_2765526, EPI_ISL_2765527, EPI_ISL_2765528, EPI_ISL_2765529, EPI_ISL_2765530, EPI_ISL_2765531, EPI_ISL_2765532, EPI_ISL_2765533, EPI_ISL_2765534, EPI_ISL_2765535, EPI_ISL_2765536, EPI_ISL_2765537, EPI_ISL_2765538, EPI_ISL_2765539,
EPI_ISL_2765540, EPI_ISL_2765541, EPI_ISL_2765542, EPI_ISL_2768107, EPI_ISL_2768112, EPI_ISL_2768115, EPI_ISL_2768116, EPI_ISL_2768117, EPI_ISL_2768118, EPI_ISL_2768119, EPI_ISL_2768128, EPI_ISL_2768129, EPI_ISL_2768130, EPI_ISL_2768131, EPI_ISL_2768132, EPI_ISL_2768133, EPI_ISL_2768134, EPI_ISL_2768135, EPI_ISL_2768136, EPI_ISL_2768137, EPI_ISL_2768138, EPI_ISL_2768139,
EPI_ISL_2768140, EPI_ISL_2768141, EPI_ISL_2768142, EPI_ISL_2768143, EPI_ISL_2768144, EPI_ISL_2768145, EPI_ISL_2768146, EPI_ISL_2768147, EPI_ISL_2768148, EPI_ISL_2768149, EPI_ISL_2768150, EPI_ISL_2768151, EPI_ISL_2768152, EPI_ISL_2768153, EPI_ISL_2768154, EPI_ISL_2768155, EPI_ISL_2768156, EPI_ISL_2768157, EPI_ISL_2768158, EPI_ISL_2768159, EPI_ISL_2768160, EPI_ISL_2768161,
EPI_ISL_2768162, EPI_ISL_2768185, EPI_ISL_2768186, EPI_ISL_2768187, EPI_ISL_2768188, EPI_ISL_2768189, EPI_ISL_2768190, EPI_ISL_2768191, EPI_ISL_2768193, EPI_ISL_2768194, EPI_ISL_2768195, EPI_ISL_2768196, EPI_ISL_2768197, EPI_ISL_2768198, EPI_ISL_2768221, EPI_ISL_2768465, EPI_ISL_2768466, EPI_ISL_2768467, EPI_ISL_2768468, EPI_ISL_2768469, EPI_ISL_2768470, EPI_ISL_2768471,
EPI_ISL_2768472, EPI_ISL_2768473, EPI_ISL_2768474, EPI_ISL_2768475, EPI_ISL_2768476, EPI_ISL_2768477, EPI_ISL_2768478, EPI_ISL_2768479, EPI_ISL_2768480, EPI_ISL_2768481, EPI_ISL_2768482, EPI_ISL_2768483, EPI_ISL_2768484, EPI_ISL_2768485, EPI_ISL_2768486, EPI_ISL_2768487, EPI_ISL_2768488, EPI_ISL_2768489, EPI_ISL_2768490, EPI_ISL_2768491, EPI_ISL_2768492, EPI_ISL_2768493,
EPI_ISL_2768494, EPI_ISL_2768495, EPI_ISL_2768496, EPI_ISL_2768497, EPI_ISL_2768498, EPI_ISL_2768499, EPI_ISL_2768500, EPI_ISL_2768501, EPI_ISL_2768502, EPI_ISL_2768503, EPI_ISL_2768504, EPI_ISL_2768505, EPI_ISL_2768506, EPI_ISL_2768507, EPI_ISL_2768508, EPI_ISL_2768509, EPI_ISL_2768510, EPI_ISL_2768511, EPI_ISL_2768512, EPI_ISL_2768513, EPI_ISL_2768514, EPI_ISL_2768515,
EPI_ISL_2768516, EPI_ISL_2768528, EPI_ISL_2768529, EPI_ISL_2768530, EPI_ISL_2768531, EPI_ISL_2768532, EPI_ISL_2768533, EPI_ISL_2768534, EPI_ISL_2768537, EPI_ISL_2768626, EPI_ISL_2768627, EPI_ISL_2768630, EPI_ISL_2768631, EPI_ISL_2768632, EPI_ISL_2768633, EPI_ISL_2768634, EPI_ISL_2768635, EPI_ISL_2768636, EPI_ISL_2768647, EPI_ISL_2768648, EPI_ISL_2768649, EPI_ISL_2768650,
EPI_ISL_2768651, EPI_ISL_2768652, EPI_ISL_2768653, EPI_ISL_2768654, EPI_ISL_2768655, EPI_ISL_2768656, EPI_ISL_2768657, EPI_ISL_2768658, EPI_ISL_2768659, EPI_ISL_2768660, EPI_ISL_2768661, EPI_ISL_2768662, EPI_ISL_2768663, EPI_ISL_2768664, EPI_ISL_2768665, EPI_ISL_2768666, EPI_ISL_2768667, EPI_ISL_2768668, EPI_ISL_2768669, EPI_ISL_2768670, EPI_ISL_2768671, EPI_ISL_2768672,
EPI_ISL_2768673, EPI_ISL_2768674, EPI_ISL_2768675, EPI_ISL_2768676, EPI_ISL_2768677, EPI_ISL_2768678, EPI_ISL_2768679, EPI_ISL_2768680, EPI_ISL_2768681, EPI_ISL_2768682, EPI_ISL_2768683, EPI_ISL_2768684, EPI_ISL_2768685, EPI_ISL_2768686, EPI_ISL_2768687, EPI_ISL_2768688, EPI_ISL_2768689, EPI_ISL_2768690, EPI_ISL_2768691, EPI_ISL_2768692, EPI_ISL_2768693, EPI_ISL_2768694,
EPI_ISL_2768695, EPI_ISL_2768696, EPI_ISL_2768697, EPI_ISL_2768698, EPI_ISL_2768699, EPI_ISL_2768700, EPI_ISL_2768701, EPI_ISL_2768702, EPI_ISL_2768703, EPI_ISL_2768704, EPI_ISL_2768705, EPI_ISL_2768706, EPI_ISL_2768707, EPI_ISL_2768708, EPI_ISL_2768709, EPI_ISL_2768710, EPI_ISL_2768711, EPI_ISL_2768712, EPI_ISL_2768725, EPI_ISL_2768726, EPI_ISL_2768727, EPI_ISL_2768728,
EPI_ISL_2768729, EPI_ISL_2768730, EPI_ISL_2768731, EPI_ISL_2768732, EPI_ISL_2768733, EPI_ISL_2768734, EPI_ISL_2768735, EPI_ISL_2768736, EPI_ISL_2768737, EPI_ISL_2768738, EPI_ISL_2768739, EPI_ISL_2768741, EPI_ISL_2768742, EPI_ISL_2768743, EPI_ISL_2768744, EPI_ISL_2768745, EPI_ISL_2768746, EPI_ISL_2768747, EPI_ISL_2768748, EPI_ISL_2768749, EPI_ISL_2768750, EPI_ISL_2768751,
EPI_ISL_2768752, EPI_ISL_2768753, EPI_ISL_2768754, EPI_ISL_2768755, EPI_ISL_2768756, EPI_ISL_2768757, EPI_ISL_2768758, EPI_ISL_2768759, EPI_ISL_2768760, EPI_ISL_2768761, EPI_ISL_2768762, EPI_ISL_2768763, EPI_ISL_2768764, EPI_ISL_2768765, EPI_ISL_2768766, EPI_ISL_2768767, EPI_ISL_2768768, EPI_ISL_2768769, EPI_ISL_2768770, EPI_ISL_2768781, EPI_ISL_2768782, EPI_ISL_2768783,
EPI_ISL_2768784, EPI_ISL_2768785, EPI_ISL_2768786, EPI_ISL_2768787, EPI_ISL_2768788, EPI_ISL_2768789, EPI_ISL_2768790, EPI_ISL_2768791, EPI_ISL_2768792, EPI_ISL_2768793, EPI_ISL_2768794, EPI_ISL_2768795, EPI_ISL_2768796, EPI_ISL_2768797, EPI_ISL_2768798, EPI_ISL_2768799, EPI_ISL_2768800, EPI_ISL_2768801, EPI_ISL_2768802, EPI_ISL_2768803, EPI_ISL_2768804, EPI_ISL_2768805,
EPI_ISL_2768806, EPI_ISL_2768807, EPI_ISL_2769030, EPI_ISL_2769031, EPI_ISL_2769032, EPI_ISL_2769033, EPI_ISL_2769034, EPI_ISL_2769035, EPI_ISL_2769036, EPI_ISL_2769037, EPI_ISL_2769038, EPI_ISL_2769039, EPI_ISL_2769040, EPI_ISL_2769041, EPI_ISL_2769042, EPI_ISL_2769043, EPI_ISL_2769044, EPI_ISL_2769045, EPI_ISL_2769046, EPI_ISL_2769047, EPI_ISL_2769048, EPI_ISL_2769049,
EPI_ISL_2769050, EPI_ISL_2769051, EPI_ISL_2769052, EPI_ISL_2769053, EPI_ISL_2769054, EPI_ISL_2769055, EPI_ISL_2769056, EPI_ISL_2769057, EPI_ISL_2769058, EPI_ISL_2769059, EPI_ISL_2769060, EPI_ISL_2769061, EPI_ISL_2769062, EPI_ISL_2769063, EPI_ISL_2769064, EPI_ISL_2769065, EPI_ISL_2769066, EPI_ISL_2769095, EPI_ISL_2769476, EPI_ISL_2769477, EPI_ISL_2769478, EPI_ISL_2769481,
EPI_ISL_2769482, EPI_ISL_2769483, EPI_ISL_2769484, EPI_ISL_2769485, EPI_ISL_2769486, EPI_ISL_2769487, EPI_ISL_2769488, EPI_ISL_2769489, EPI_ISL_2769490, EPI_ISL_2769491, EPI_ISL_2769492, EPI_ISL_2769493, EPI_ISL_2769494, EPI_ISL_2769495, EPI_ISL_2769496, EPI_ISL_2769497, EPI_ISL_2769498, EPI_ISL_2769499, EPI_ISL_2769500, EPI_ISL_2769501, EPI_ISL_2769502, EPI_ISL_2769503,
EPI_ISL_2769504, EPI_ISL_2769505, EPI_ISL_2769506, EPI_ISL_2769507, EPI_ISL_2769508, EPI_ISL_2769509, EPI_ISL_2769510, EPI_ISL_2769511, EPI_ISL_2769512, EPI_ISL_2769513, EPI_ISL_2769514, EPI_ISL_2769515, EPI_ISL_2769516, EPI_ISL_2769517, EPI_ISL_2769518, EPI_ISL_2769519, EPI_ISL_2769520, EPI_ISL_2769521, EPI_ISL_2769522, EPI_ISL_2769523, EPI_ISL_2769524, EPI_ISL_2769525,
EPI_ISL_2769526, EPI_ISL_2769527, EPI_ISL_2769528, EPI_ISL_2769529, EPI_ISL_2769530, EPI_ISL_2769531, EPI_ISL_2769532, EPI_ISL_2769533, EPI_ISL_2769534, EPI_ISL_2769535, EPI_ISL_2769536, EPI_ISL_2769537, EPI_ISL_2769538, EPI_ISL_2769539, EPI_ISL_2769540, EPI_ISL_2769541, EPI_ISL_2769542, EPI_ISL_2769543, EPI_ISL_2769544, EPI_ISL_2769545, EPI_ISL_2769546, EPI_ISL_2769547,
EPI_ISL_2769548, EPI_ISL_2769549, EPI_ISL_2769550, EPI_ISL_2769551, EPI_ISL_2769552, EPI_ISL_2769553, EPI_ISL_2769554, EPI_ISL_2769555, EPI_ISL_2769556, EPI_ISL_2769557, EPI_ISL_2769573, EPI_ISL_2769574, EPI_ISL_2769575, EPI_ISL_2769576, EPI_ISL_2769577, EPI_ISL_2769578, EPI_ISL_2769579, EPI_ISL_2769580, EPI_ISL_2769581, EPI_ISL_2769582, EPI_ISL_2769583, EPI_ISL_2769584,
EPI_ISL_2769585, EPI_ISL_2769586, EPI_ISL_2769587, EPI_ISL_2769588, EPI_ISL_2769589, EPI_ISL_2769590, EPI_ISL_2769591, EPI_ISL_2769592, EPI_ISL_2769593, EPI_ISL_2769594, EPI_ISL_2769595, EPI_ISL_2769596, EPI_ISL_2769597, EPI_ISL_2769599, EPI_ISL_2769600, EPI_ISL_2769630, EPI_ISL_2769659, EPI_ISL_2769660, EPI_ISL_2769661, EPI_ISL_2769662, EPI_ISL_2769663, EPI_ISL_2769664,
EPI_ISL_2769665, EPI_ISL_2769666, EPI_ISL_2769668, EPI_ISL_2769669, EPI_ISL_2769670, EPI_ISL_2769671, EPI_ISL_2769672, EPI_ISL_2769673, EPI_ISL_2769674, EPI_ISL_2769675, EPI_ISL_2769683, EPI_ISL_2769684, EPI_ISL_2769685, EPI_ISL_2769686, EPI_ISL_2769687, EPI_ISL_2769697, EPI_ISL_2769698, EPI_ISL_2769699, EPI_ISL_2769700, EPI_ISL_2769701, EPI_ISL_2769717, EPI_ISL_2769718,
EPI_ISL_2769719, EPI_ISL_2769720, EPI_ISL_2769721, EPI_ISL_2769722, EPI_ISL_2769729, EPI_ISL_2769730, EPI_ISL_2769731, EPI_ISL_2769732, EPI_ISL_2769733, EPI_ISL_2769734, EPI_ISL_2769735, EPI_ISL_2769736, EPI_ISL_2769737, EPI_ISL_2769738, EPI_ISL_2769739, EPI_ISL_2769740, EPI_ISL_2769741, EPI_ISL_2769742, EPI_ISL_2769743, EPI_ISL_2769744, EPI_ISL_2769745, EPI_ISL_2769746,
EPI_ISL_2769747, EPI_ISL_2769748, EPI_ISL_2769749, EPI_ISL_2769750, EPI_ISL_2769751, EPI_ISL_2769752, EPI_ISL_2769753, EPI_ISL_2769754, EPI_ISL_2769755, EPI_ISL_2769756, EPI_ISL_2769757, EPI_ISL_2769758, EPI_ISL_2769759, EPI_ISL_2769760, EPI_ISL_2769761, EPI_ISL_2769762, EPI_ISL_2769763, EPI_ISL_2769764, EPI_ISL_2769765, EPI_ISL_2769766, EPI_ISL_2769767, EPI_ISL_2769768,
EPI_ISL_2769769, EPI_ISL_2769770, EPI_ISL_2769771, EPI_ISL_2769772, EPI_ISL_2769773, EPI_ISL_2769774, EPI_ISL_2769775, EPI_ISL_2769776, EPI_ISL_2769777, EPI_ISL_2769778, EPI_ISL_2769779, EPI_ISL_2769780, EPI_ISL_2769781, EPI_ISL_2769782, EPI_ISL_2769783, EPI_ISL_2769784, EPI_ISL_2769785, EPI_ISL_2769786, EPI_ISL_2769787, EPI_ISL_2769788, EPI_ISL_2769789, EPI_ISL_2769790,
EPI_ISL_2769791, EPI_ISL_2769792, EPI_ISL_2769793, EPI_ISL_2769794, EPI_ISL_2769795, EPI_ISL_2769796, EPI_ISL_2769807, EPI_ISL_2769828, EPI_ISL_2769829, EPI_ISL_2769830, EPI_ISL_2769831, EPI_ISL_2769832, EPI_ISL_2769836, EPI_ISL_2769837, EPI_ISL_2769850, EPI_ISL_2769851, EPI_ISL_2769852, EPI_ISL_2769853, EPI_ISL_2769854, EPI_ISL_2769855, EPI_ISL_2769856, EPI_ISL_2769857,
EPI_ISL_2769858, EPI_ISL_2769859, EPI_ISL_2769860, EPI_ISL_2769861, EPI_ISL_2769897, EPI_ISL_2769909, EPI_ISL_2769914, EPI_ISL_2769926, EPI_ISL_2769927, EPI_ISL_2769932, EPI_ISL_2769933, EPI_ISL_2769934, EPI_ISL_2769935, EPI_ISL_2769938, EPI_ISL_2769940, EPI_ISL_2769941, EPI_ISL_2769949, EPI_ISL_2769950, EPI_ISL_2769951, EPI_ISL_2769952, EPI_ISL_2769953, EPI_ISL_2769970,
EPI_ISL_2769971, EPI_ISL_2769972, EPI_ISL_2769973, EPI_ISL_2769974, EPI_ISL_2769975, EPI_ISL_2769976, EPI_ISL_2769977, EPI_ISL_2769978, EPI_ISL_2769979, EPI_ISL_2769980, EPI_ISL_2769981, EPI_ISL_2769982, EPI_ISL_2769983, EPI_ISL_2769984, EPI_ISL_2769985, EPI_ISL_2769986, EPI_ISL_2769987, EPI_ISL_2769988, EPI_ISL_2769989, EPI_ISL_2769990, EPI_ISL_2769991, EPI_ISL_2769992,
EPI_ISL_2769993, EPI_ISL_2769994, EPI_ISL_2769995, EPI_ISL_2769996, EPI_ISL_2769997, EPI_ISL_2769998, EPI_ISL_2769999, EPI_ISL_2770000, EPI_ISL_2770001, EPI_ISL_2770002, EPI_ISL_2770003, EPI_ISL_2770004, EPI_ISL_2770005, EPI_ISL_2770006, EPI_ISL_2770007, EPI_ISL_2770008, EPI_ISL_2770009, EPI_ISL_2770010, EPI_ISL_2770011, EPI_ISL_2770012, EPI_ISL_2770013, EPI_ISL_2770014,
EPI_ISL_2770015, EPI_ISL_2770016, EPI_ISL_2770017, EPI_ISL_2770018, EPI_ISL_2770019, EPI_ISL_2770020, EPI_ISL_2770021, EPI_ISL_2770146, EPI_ISL_2770204, EPI_ISL_2770205, EPI_ISL_2770206, EPI_ISL_2770207, EPI_ISL_2770208, EPI_ISL_2770209, EPI_ISL_2770210, EPI_ISL_2770211, EPI_ISL_2770220, EPI_ISL_2770221, EPI_ISL_2770222, EPI_ISL_2770223, EPI_ISL_2770224, EPI_ISL_2770225,
EPI_ISL_2770226, EPI_ISL_2770227, EPI_ISL_2770228, EPI_ISL_2770229, EPI_ISL_2770230, EPI_ISL_2770231, EPI_ISL_2770232, EPI_ISL_2770233, EPI_ISL_2770234, EPI_ISL_2770235, EPI_ISL_2770236, EPI_ISL_2770237, EPI_ISL_2770238, EPI_ISL_2770239, EPI_ISL_2770240, EPI_ISL_2770241, EPI_ISL_2770247, EPI_ISL_2770253, EPI_ISL_2770255, EPI_ISL_2770422, EPI_ISL_2770423, EPI_ISL_2770424,
EPI_ISL_2770426, EPI_ISL_2770427, EPI_ISL_2770429, EPI_ISL_2770430, EPI_ISL_2770431, EPI_ISL_2770432, EPI_ISL_2770433, EPI_ISL_2770434, EPI_ISL_2770435, EPI_ISL_2770436, EPI_ISL_2770437, EPI_ISL_2770438, EPI_ISL_2770491, EPI_ISL_2770492, EPI_ISL_2770493, EPI_ISL_2770494, EPI_ISL_2770495, EPI_ISL_2770496, EPI_ISL_2770497, EPI_ISL_2770498, EPI_ISL_2770499, EPI_ISL_2770500,
EPI_ISL_2770501, EPI_ISL_2770502, EPI_ISL_2770503, EPI_ISL_2770504, EPI_ISL_2770505, EPI_ISL_2770506, EPI_ISL_2770507, EPI_ISL_2770508, EPI_ISL_2770509, EPI_ISL_2770510, EPI_ISL_2770511, EPI_ISL_2770512, EPI_ISL_2770513, EPI_ISL_2770514, EPI_ISL_2770515, EPI_ISL_2770516, EPI_ISL_2770517, EPI_ISL_2770518, EPI_ISL_2770519, EPI_ISL_2770520, EPI_ISL_2770521, EPI_ISL_2770522,
EPI_ISL_2770523, EPI_ISL_2770524, EPI_ISL_2770525, EPI_ISL_2770526, EPI_ISL_2770527, EPI_ISL_2770528, EPI_ISL_2770529, EPI_ISL_2770530, EPI_ISL_2770531, EPI_ISL_2770532, EPI_ISL_2770533, EPI_ISL_2770534, EPI_ISL_2770535, EPI_ISL_2770536, EPI_ISL_2770537, EPI_ISL_2770538, EPI_ISL_2770539, EPI_ISL_2770540, EPI_ISL_2770541, EPI_ISL_2770542, EPI_ISL_2770543, EPI_ISL_2770544,
EPI_ISL_2770545, EPI_ISL_2770546, EPI_ISL_2770547, EPI_ISL_2770548, EPI_ISL_2770549, EPI_ISL_2770550, EPI_ISL_2770551, EPI_ISL_2770552, EPI_ISL_2770553, EPI_ISL_2770554, EPI_ISL_2770555, EPI_ISL_2770556, EPI_ISL_2770557, EPI_ISL_2770558, EPI_ISL_2770559, EPI_ISL_2770560, EPI_ISL_2770561, EPI_ISL_2770562, EPI_ISL_2770563, EPI_ISL_2770564, EPI_ISL_2770565, EPI_ISL_2770566,
EPI_ISL_2770567, EPI_ISL_2770568, EPI_ISL_2770569, EPI_ISL_2770570, EPI_ISL_2770571, EPI_ISL_2770572, EPI_ISL_2770573, EPI_ISL_2770574, EPI_ISL_2770575, EPI_ISL_2770576, EPI_ISL_2770577, EPI_ISL_2770578, EPI_ISL_2770579, EPI_ISL_2770580, EPI_ISL_2770581, EPI_ISL_2770582, EPI_ISL_2770583, EPI_ISL_2770584, EPI_ISL_2770585, EPI_ISL_2770586, EPI_ISL_2770587, EPI_ISL_2770588,
EPI_ISL_2770589, EPI_ISL_2770590, EPI_ISL_2770591, EPI_ISL_2770592, EPI_ISL_2770593, EPI_ISL_2770594, EPI_ISL_2770595, EPI_ISL_2770596, EPI_ISL_2770597, EPI_ISL_2770598, EPI_ISL_2770599, EPI_ISL_2770600, EPI_ISL_2770601, EPI_ISL_2770602, EPI_ISL_2770603, EPI_ISL_2770604, EPI_ISL_2770605, EPI_ISL_2770606, EPI_ISL_2770607, EPI_ISL_2770608, EPI_ISL_2770609, EPI_ISL_2770610,
EPI_ISL_2770611, EPI_ISL_2770612, EPI_ISL_2770613, EPI_ISL_2770614, EPI_ISL_2770615, EPI_ISL_2770616, EPI_ISL_2770617, EPI_ISL_2770618, EPI_ISL_2770619, EPI_ISL_2770620, EPI_ISL_2770621, EPI_ISL_2770622, EPI_ISL_2770623, EPI_ISL_2770624, EPI_ISL_2770625, EPI_ISL_2770626, EPI_ISL_2770627, EPI_ISL_2770628, EPI_ISL_2770629, EPI_ISL_2770630, EPI_ISL_2770631, EPI_ISL_2770632,
EPI_ISL_2770633, EPI_ISL_2770634, EPI_ISL_2770635, EPI_ISL_2770636, EPI_ISL_2770637, EPI_ISL_2770638, EPI_ISL_2770639, EPI_ISL_2770640, EPI_ISL_2770641, EPI_ISL_2770642, EPI_ISL_2770643, EPI_ISL_2770644, EPI_ISL_2770645, EPI_ISL_2770646, EPI_ISL_2770647, EPI_ISL_2770648, EPI_ISL_2770649, EPI_ISL_2770650, EPI_ISL_2770651, EPI_ISL_2770652, EPI_ISL_2770653, EPI_ISL_2770654,
EPI_ISL_2770655, EPI_ISL_2770656, EPI_ISL_2770657, EPI_ISL_2770658, EPI_ISL_2770659, EPI_ISL_2770660, EPI_ISL_2770661, EPI_ISL_2770662, EPI_ISL_2770663, EPI_ISL_2770664, EPI_ISL_2770665, EPI_ISL_2770666, EPI_ISL_2770667, EPI_ISL_2770668, EPI_ISL_2770669, EPI_ISL_2770670, EPI_ISL_2770671, EPI_ISL_2770672, EPI_ISL_2770673, EPI_ISL_2770674, EPI_ISL_2770675, EPI_ISL_2770676,
EPI_ISL_2770677, EPI_ISL_2770678, EPI_ISL_2770679, EPI_ISL_2770680, EPI_ISL_2770681, EPI_ISL_2770682, EPI_ISL_2770885, EPI_ISL_2770886, EPI_ISL_2770887, EPI_ISL_2771015, EPI_ISL_2771016, EPI_ISL_2771017, EPI_ISL_2771018, EPI_ISL_2771019, EPI_ISL_2771020, EPI_ISL_2771021, EPI_ISL_2771022, EPI_ISL_2771023, EPI_ISL_2771024, EPI_ISL_2771025, EPI_ISL_2771026, EPI_ISL_2771027,
EPI_ISL_2771028, EPI_ISL_2771029, EPI_ISL_2771030, EPI_ISL_2771031, EPI_ISL_2771033, EPI_ISL_2771034, EPI_ISL_2771035, EPI_ISL_2771036, EPI_ISL_2771037, EPI_ISL_2771038, EPI_ISL_2771039, EPI_ISL_2771040, EPI_ISL_2771041, EPI_ISL_2771042, EPI_ISL_2771043, EPI_ISL_2771044, EPI_ISL_2771045, EPI_ISL_2771046, EPI_ISL_2771047, EPI_ISL_2771048, EPI_ISL_2771049, EPI_ISL_2771050,
EPI_ISL_2771051, EPI_ISL_2771052, EPI_ISL_2771053, EPI_ISL_2771054, EPI_ISL_2771055, EPI_ISL_2771056, EPI_ISL_2771057, EPI_ISL_2771058, EPI_ISL_2771059, EPI_ISL_2771060, EPI_ISL_2771061, EPI_ISL_2771062, EPI_ISL_2771063, EPI_ISL_2771064, EPI_ISL_2771065, EPI_ISL_2771066, EPI_ISL_2771067, EPI_ISL_2771068, EPI_ISL_2771069, EPI_ISL_2771070, EPI_ISL_2771071, EPI_ISL_2771072,
EPI_ISL_2771073, EPI_ISL_2771074, EPI_ISL_2771075, EPI_ISL_2771076, EPI_ISL_2771077, EPI_ISL_2771078, EPI_ISL_2771079, EPI_ISL_2771080, EPI_ISL_2771081, EPI_ISL_2771082, EPI_ISL_2771083, EPI_ISL_2771084, EPI_ISL_2771085, EPI_ISL_2771086, EPI_ISL_2771087, EPI_ISL_2771088, EPI_ISL_2771089, EPI_ISL_2771090, EPI_ISL_2771091, EPI_ISL_2771092, EPI_ISL_2771093, EPI_ISL_2771094,

| | | | |
|---|---|---|---|
| EPI_ISL_2771095, EPI_ISL_2771096, EPI_ISL_2771097, EPI_ISL_2771098, EPI_ISL_2771099, EPI_ISL_2771100, EPI_ISL_2771101, EPI_ISL_2771102, EPI_ISL_2771103, EPI_ISL_2771104, EPI_ISL_2771105, EPI_ISL_2771106, EPI_ISL_2771107, EPI_ISL_2771108, EPI_ISL_2771109, EPI_ISL_2771110, EPI_ISL_2771111, EPI_ISL_2771112, EPI_ISL_2771113, EPI_ISL_2771114, EPI_ISL_2771116, EPI_ISL_2771117, EPI_ISL_2771118, EPI_ISL_2771119, EPI_ISL_2771120, EPI_ISL_2771121, EPI_ISL_2771122, EPI_ISL_2771123, EPI_ISL_2771124, EPI_ISL_2771125, EPI_ISL_2771126, EPI_ISL_2771127, EPI_ISL_2771130, EPI_ISL_2771131, EPI_ISL_2771132, EPI_ISL_2771133, EPI_ISL_2771134, EPI_ISL_2771135, EPI_ISL_2771136, EPI_ISL_2771137, EPI_ISL_2771138, EPI_ISL_2771139, EPI_ISL_2771140, EPI_ISL_2771141, EPI_ISL_2771142, EPI_ISL_2771143, EPI_ISL_2771144, EPI_ISL_2771145, EPI_ISL_2771146, EPI_ISL_2771147, EPI_ISL_2771148, EPI_ISL_2771149, EPI_ISL_2771154, EPI_ISL_2771185, EPI_ISL_2771211, EPI_ISL_2771212, EPI_ISL_2771213, EPI_ISL_2771214, EPI_ISL_2771215, EPI_ISL_2771216, EPI_ISL_2771217, EPI_ISL_2771218, EPI_ISL_2771219, EPI_ISL_2771220, EPI_ISL_2771221, EPI_ISL_2771222, EPI_ISL_2771223, EPI_ISL_2771224, EPI_ISL_2771225, EPI_ISL_2771226, EPI_ISL_2771227, EPI_ISL_2771228, EPI_ISL_2771229, EPI_ISL_2771230, EPI_ISL_2771231, EPI_ISL_2771232, EPI_ISL_2771233, EPI_ISL_2771234, EPI_ISL_2771235, EPI_ISL_2771236, EPI_ISL_2771237, EPI_ISL_2771238, EPI_ISL_2771239, EPI_ISL_2771240, EPI_ISL_2771241, EPI_ISL_2771242, EPI_ISL_2771243, EPI_ISL_2771244, EPI_ISL_2771245, EPI_ISL_2771246, EPI_ISL_2771247, EPI_ISL_2771248, EPI_ISL_2771249, EPI_ISL_2771250, EPI_ISL_2771251, EPI_ISL_2771252, EPI_ISL_2771253, EPI_ISL_2771254, EPI_ISL_2771255, EPI_ISL_2771256, EPI_ISL_2771257, EPI_ISL_2771258, EPI_ISL_2771259, EPI_ISL_2771260, EPI_ISL_2771261, EPI_ISL_2771262, EPI_ISL_2771263, EPI_ISL_2771264, EPI_ISL_2771265, EPI_ISL_2771266, EPI_ISL_2771267, EPI_ISL_2771268, EPI_ISL_2771269, EPI_ISL_2771270, EPI_ISL_2771271, EPI_ISL_2771272, EPI_ISL_2771273, EPI_ISL_2771274, EPI_ISL_2771275, EPI_ISL_2771276, EPI_ISL_2771277, EPI_ISL_2771278, EPI_ISL_2771279, EPI_ISL_2771280, EPI_ISL_2771281, EPI_ISL_2771282, EPI_ISL_2771283, EPI_ISL_2771284, EPI_ISL_2771285, EPI_ISL_2771286, EPI_ISL_2771287, EPI_ISL_2771288, EPI_ISL_2771289, EPI_ISL_2771290, EPI_ISL_2771291, EPI_ISL_2771292, EPI_ISL_2771293, EPI_ISL_2771294, EPI_ISL_2771295, EPI_ISL_2771296, EPI_ISL_2771297, EPI_ISL_2771298, EPI_ISL_2771299, EPI_ISL_2771300, EPI_ISL_2771301, EPI_ISL_2771302, EPI_ISL_2771303, EPI_ISL_2771304, EPI_ISL_2771305, EPI_ISL_2771306, EPI_ISL_2771307, EPI_ISL_2771308, EPI_ISL_2771309, EPI_ISL_2771310, EPI_ISL_2771311, EPI_ISL_2771312, EPI_ISL_2771313, EPI_ISL_2771314, EPI_ISL_2771315, EPI_ISL_2771316, EPI_ISL_2771317, EPI_ISL_2771318, EPI_ISL_2771319, EPI_ISL_2771320, EPI_ISL_2771321, EPI_ISL_2771322, EPI_ISL_2771323, EPI_ISL_2771324, EPI_ISL_2771325, EPI_ISL_2771326, EPI_ISL_2771327, EPI_ISL_2771328, EPI_ISL_2771329, EPI_ISL_2771330, EPI_ISL_2771331, EPI_ISL_2771332, EPI_ISL_2771333, EPI_ISL_2771334, EPI_ISL_2771335, EPI_ISL_2771336, EPI_ISL_2771337, EPI_ISL_2771338, EPI_ISL_2771339, EPI_ISL_2771340, EPI_ISL_2771341, EPI_ISL_2771342, EPI_ISL_2771343, EPI_ISL_2771344, EPI_ISL_2771345, EPI_ISL_2771346, EPI_ISL_2771347, EPI_ISL_2771348, EPI_ISL_2771349, EPI_ISL_2771350, EPI_ISL_2771351, EPI_ISL_2771352, EPI_ISL_2771353, EPI_ISL_2771354, EPI_ISL_2771355, EPI_ISL_2771356, EPI_ISL_2771357, EPI_ISL_2771358, EPI_ISL_2771359, EPI_ISL_2771360, EPI_ISL_2771361, EPI_ISL_2771362, EPI_ISL_2771363, EPI_ISL_2771364, EPI_ISL_2771365, EPI_ISL_2771366, EPI_ISL_2771367, EPI_ISL_2771368, EPI_ISL_2771369, EPI_ISL_2771370, EPI_ISL_2771371, EPI_ISL_2771372, EPI_ISL_2771373, EPI_ISL_2771374, EPI_ISL_2771375, EPI_ISL_2771376, EPI_ISL_2771377, EPI_ISL_2771378, EPI_ISL_2771379, EPI_ISL_2771380, EPI_ISL_2771381, EPI_ISL_2771382, EPI_ISL_2771383, EPI_ISL_2771384, EPI_ISL_2771385, EPI_ISL_2771386, EPI_ISL_2771387, EPI_ISL_2771388, EPI_ISL_2771395, EPI_ISL_2771647, EPI_ISL_2771648, EPI_ISL_2771649, EPI_ISL_2771650, EPI_ISL_2771651, EPI_ISL_2771652, EPI_ISL_2771653, EPI_ISL_2771654, EPI_ISL_2771655, EPI_ISL_2771656, EPI_ISL_2771657, EPI_ISL_2771658, EPI_ISL_2771659, EPI_ISL_2771660, EPI_ISL_2771661, EPI_ISL_2771662, EPI_ISL_2771663, EPI_ISL_2771664, EPI_ISL_2771665, EPI_ISL_2771666, EPI_ISL_2771667, EPI_ISL_2771668, EPI_ISL_2771669, EPI_ISL_2771670, EPI_ISL_2771671, EPI_ISL_2771672, EPI_ISL_2771673, EPI_ISL_2771674, EPI_ISL_2771675, EPI_ISL_2771676, EPI_ISL_2771677, EPI_ISL_2771678, EPI_ISL_2771679, EPI_ISL_2771680, EPI_ISL_2771681, EPI_ISL_2771682, EPI_ISL_2771683, EPI_ISL_2771684, EPI_ISL_2771685, EPI_ISL_2771686, EPI_ISL_2771687, EPI_ISL_2771688, EPI_ISL_2771689, EPI_ISL_2771690, EPI_ISL_2771691, EPI_ISL_2771692, EPI_ISL_2771693, EPI_ISL_2771694, EPI_ISL_2771695, EPI_ISL_2771696, EPI_ISL_2771697, EPI_ISL_2771698, EPI_ISL_2771699, EPI_ISL_2771700, EPI_ISL_2771701, EPI_ISL_2771702, EPI_ISL_2771703, EPI_ISL_2771704, EPI_ISL_2771705, EPI_ISL_2771706, EPI_ISL_2771707, EPI_ISL_2771708, EPI_ISL_2771709, EPI_ISL_2771710, EPI_ISL_2771711, EPI_ISL_2771712, EPI_ISL_2771713, EPI_ISL_2771714, EPI_ISL_2771715, EPI_ISL_2771716, EPI_ISL_2771717, EPI_ISL_2771718, EPI_ISL_2771719, EPI_ISL_2771720, EPI_ISL_2771721, EPI_ISL_2771722, EPI_ISL_2771723, EPI_ISL_2771724, EPI_ISL_2771725, EPI_ISL_2771726, EPI_ISL_2771727, EPI_ISL_2771728, EPI_ISL_2771729, EPI_ISL_2771730, EPI_ISL_2771731, EPI_ISL_2771732, EPI_ISL_2771733, EPI_ISL_2771734, EPI_ISL_2771735, EPI_ISL_2771736, EPI_ISL_2771737, EPI_ISL_2771738, EPI_ISL_2771739, EPI_ISL_2771740, EPI_ISL_2771741, EPI_ISL_2771742, EPI_ISL_2771743, EPI_ISL_2771744, EPI_ISL_2771745, EPI_ISL_2771746, EPI_ISL_2771747, EPI_ISL_2771748, EPI_ISL_2771749, EPI_ISL_2771750, EPI_ISL_2771751, EPI_ISL_2771752, EPI_ISL_2771753, EPI_ISL_2771754, EPI_ISL_2771755, EPI_ISL_2771756, EPI_ISL_2771757, EPI_ISL_2771758, EPI_ISL_2771759, EPI_ISL_2771760, EPI_ISL_2771761, EPI_ISL_2771762, EPI_ISL_2771763, EPI_ISL_2771764, EPI_ISL_2771765, EPI_ISL_2771766, EPI_ISL_2771767, EPI_ISL_2771768, EPI_ISL_2771769, EPI_ISL_2771770, EPI_ISL_2771771, EPI_ISL_2771772, EPI_ISL_2771773, EPI_ISL_2771774, EPI_ISL_2771775, EPI_ISL_2771776, EPI_ISL_2771777, EPI_ISL_2771778, EPI_ISL_2771779, EPI_ISL_2771780, EPI_ISL_2771781, EPI_ISL_2771782, EPI_ISL_2771783, EPI_ISL_2771784, EPI_ISL_2771785, EPI_ISL_2771786, EPI_ISL_2771787, EPI_ISL_2771788, EPI_ISL_2771789, EPI_ISL_2771790, EPI_ISL_2771791, EPI_ISL_2771792, EPI_ISL_2771793, EPI_ISL_2771794, EPI_ISL_2771795, EPI_ISL_2771796, EPI_ISL_2771797, EPI_ISL_2771798, EPI_ISL_2771799, EPI_ISL_2771800, EPI_ISL_2771801, EPI_ISL_2771802, EPI_ISL_2771803, EPI_ISL_2771804, EPI_ISL_2771805, EPI_ISL_2771806, EPI_ISL_2771807, EPI_ISL_2771808, EPI_ISL_2771809, EPI_ISL_2771810, EPI_ISL_2771811, EPI_ISL_2771812, EPI_ISL_2771813, EPI_ISL_2771814, EPI_ISL_2771815, EPI_ISL_2771816, EPI_ISL_2771817, EPI_ISL_2771818, EPI_ISL_2771819, EPI_ISL_2771820, EPI_ISL_2771821, EPI_ISL_2771822, EPI_ISL_2771823, EPI_ISL_2771824, EPI_ISL_2771825, EPI_ISL_2771826, EPI_ISL_2771827, EPI_ISL_2771828, EPI_ISL_2771829, EPI_ISL_2771830, EPI_ISL_2771831, EPI_ISL_2771832, EPI_ISL_2771833, EPI_ISL_2771834, EPI_ISL_2771835, EPI_ISL_2771836, EPI_ISL_2771837, EPI_ISL_2771838, EPI_ISL_2771839, EPI_ISL_2771840, EPI_ISL_2771841, EPI_ISL_2771842, EPI_ISL_2771843, EPI_ISL_2771844, EPI_ISL_2771845, EPI_ISL_2771846, EPI_ISL_2771847, EPI_ISL_2771848, EPI_ISL_2771849, EPI_ISL_2771850, EPI_ISL_2771851, EPI_ISL_2771852, EPI_ISL_2771853, EPI_ISL_2771854, EPI_ISL_2771855, EPI_ISL_2771856, EPI_ISL_2771857, EPI_ISL_2771858, EPI_ISL_2771859, EPI_ISL_2771860, EPI_ISL_2771861, EPI_ISL_2771862, EPI_ISL_2771863, EPI_ISL_2771864, EPI_ISL_2771865, EPI_ISL_2771866, EPI_ISL_2771867, EPI_ISL_2771868, EPI_ISL_2771869, EPI_ISL_2771870, EPI_ISL_2771871, EPI_ISL_2771872, EPI_ISL_2771873, EPI_ISL_2771874, EPI_ISL_2771875, EPI_ISL_2771876, EPI_ISL_2771877, EPI_ISL_2771878, EPI_ISL_2771879, EPI_ISL_2771880, EPI_ISL_2771881, EPI_ISL_2771882, EPI_ISL_2771883, EPI_ISL_2771884, EPI_ISL_2771885, EPI_ISL_2771886, EPI_ISL_2771887, EPI_ISL_2771888, EPI_ISL_2771889, EPI_ISL_2771890, EPI_ISL_2771891, EPI_ISL_2771892, EPI_ISL_2771893, EPI_ISL_2771894, EPI_ISL_2771895, EPI_ISL_2771896, EPI_ISL_2771897, EPI_ISL_2771898, EPI_ISL_2771899, EPI_ISL_2771900, EPI_ISL_2771901, EPI_ISL_2771902, EPI_ISL_2771903, EPI_ISL_2771904, EPI_ISL_2771905, EPI_ISL_2771906, EPI_ISL_2771907, EPI_ISL_2771908, EPI_ISL_2771909, EPI_ISL_2771910, EPI_ISL_2771911, EPI_ISL_2771912, EPI_ISL_2771913, EPI_ISL_2771914, EPI_ISL_2771915, EPI_ISL_2771916, EPI_ISL_2771917, EPI_ISL_2771918, EPI_ISL_2771919, EPI_ISL_2771920, EPI_ISL_2771921, EPI_ISL_2771922, EPI_ISL_2771925, EPI_ISL_2771926, EPI_ISL_2771927, EPI_ISL_2771928, EPI_ISL_2771929, EPI_ISL_2771930, EPI_ISL_2771931, EPI_ISL_2771932, EPI_ISL_2771933, EPI_ISL_2771934, EPI_ISL_2771955, EPI_ISL_2771956, EPI_ISL_2771957, EPI_ISL_2771958, EPI_ISL_2771959, EPI_ISL_2771960, EPI_ISL_2771961, EPI_ISL_2771962, EPI_ISL_2771963, EPI_ISL_2771964, EPI_ISL_2771965, EPI_ISL_2771966, EPI_ISL_2771967, EPI_ISL_2771968, EPI_ISL_2771969, EPI_ISL_2771970, EPI_ISL_2771971, EPI_ISL_2771972, EPI_ISL_2771973, EPI_ISL_2771974, EPI_ISL_2771975, EPI_ISL_2771976, EPI_ISL_2771977, EPI_ISL_2771978, EPI_ISL_2771979, EPI_ISL_2771987, EPI_ISL_2771992, EPI_ISL_2771993, EPI_ISL_2772007, EPI_ISL_2772017, EPI_ISL_2772018, EPI_ISL_2772019, EPI_ISL_2772054, EPI_ISL_2772088, EPI_ISL_2772089, EPI_ISL_2772095, EPI_ISL_2772096, EPI_ISL_2772097, EPI_ISL_2772098, EPI_ISL_2772217, EPI_ISL_2772218, EPI_ISL_2772219, EPI_ISL_2772220, EPI_ISL_2772221, EPI_ISL_2772222, EPI_ISL_2772223, EPI_ISL_2772224, EPI_ISL_2772225, EPI_ISL_2772226, EPI_ISL_2772227, EPI_ISL_2772228, EPI_ISL_2772229, EPI_ISL_2772230, EPI_ISL_2772231, EPI_ISL_2772232, EPI_ISL_2772233, EPI_ISL_2772234, EPI_ISL_2772235, EPI_ISL_2772236, EPI_ISL_2772237, EPI_ISL_2772238, EPI_ISL_2772239, EPI_ISL_2772240, EPI_ISL_2772241, EPI_ISL_2772242, EPI_ISL_2772243, EPI_ISL_2772244, EPI_ISL_2772245, EPI_ISL_2772246, EPI_ISL_2772247, EPI_ISL_2772248, EPI_ISL_2772249, EPI_ISL_2772250, EPI_ISL_2772251, EPI_ISL_2772252, EPI_ISL_2772253, EPI_ISL_2772254, EPI_ISL_2772255, EPI_ISL_2772256, EPI_ISL_2772257, EPI_ISL_2772258, EPI_ISL_2772259, EPI_ISL_2772260, EPI_ISL_2772261, EPI_ISL_2772262, EPI_ISL_2772263, EPI_ISL_2772264, EPI_ISL_2772265, EPI_ISL_2772266, EPI_ISL_2772267, EPI_ISL_2772268, EPI_ISL_2772269, EPI_ISL_2772270, EPI_ISL_2772271, EPI_ISL_2772272, EPI_ISL_2772273, EPI_ISL_2772274, EPI_ISL_2772275, EPI_ISL_2772276, EPI_ISL_2772277, EPI_ISL_2772278, EPI_ISL_2772279, EPI_ISL_2772280, EPI_ISL_2772281, EPI_ISL_2772282, EPI_ISL_2772283, EPI_ISL_2772284, EPI_ISL_2772285, EPI_ISL_2772286, EPI_ISL_2772287, EPI_ISL_2772288, EPI_ISL_2772289, EPI_ISL_2772290, EPI_ISL_2772291, EPI_ISL_2772292, EPI_ISL_2772293, EPI_ISL_2772294, EPI_ISL_2772295, EPI_ISL_2772296, EPI_ISL_2772297, EPI_ISL_2772298, EPI_ISL_2772299, EPI_ISL_2772300, EPI_ISL_2772301, EPI_ISL_2772302, EPI_ISL_2772303, EPI_ISL_2772304, EPI_ISL_2772305, EPI_ISL_2772306, EPI_ISL_2772307, EPI_ISL_2772308, EPI_ISL_2772309, EPI_ISL_2772310, EPI_ISL_2772311, EPI_ISL_2772312, EPI_ISL_2772313, EPI_ISL_2772314, EPI_ISL_2772315, EPI_ISL_2772316, EPI_ISL_2772317, EPI_ISL_2772318, EPI_ISL_2772319, EPI_ISL_2772320, EPI_ISL_2772321, EPI_ISL_2772322, EPI_ISL_2772323, EPI_ISL_2772324, EPI_ISL_2772325, EPI_ISL_2772326, EPI_ISL_2772327, EPI_ISL_2772328, EPI_ISL_2772329, EPI_ISL_2772330, EPI_ISL_2772331, EPI_ISL_2772332, EPI_ISL_2772333, EPI_ISL_2772334, EPI_ISL_2772335, EPI_ISL_2772336, EPI_ISL_2772337, EPI_ISL_2772338, EPI_ISL_2772339, EPI_ISL_2772340, EPI_ISL_2772341, EPI_ISL_2772342, EPI_ISL_2772343, EPI_ISL_2772344, EPI_ISL_2772345, EPI_ISL_2772346, EPI_ISL_2772347, EPI_ISL_2772348, EPI_ISL_2772349, EPI_ISL_2772350, EPI_ISL_2772351, EPI_ISL_2772352, EPI_ISL_2772353, EPI_ISL_2772354, EPI_ISL_2772355, EPI_ISL_2772356, EPI_ISL_2772357, EPI_ISL_2772358, EPI_ISL_2772398, EPI_ISL_2772399, EPI_ISL_2772400, EPI_ISL_2772401, EPI_ISL_2772402, EPI_ISL_2772403, EPI_ISL_2772404, EPI_ISL_2772405, EPI_ISL_2772406, EPI_ISL_2772407, EPI_ISL_2772408, EPI_ISL_2772409, EPI_ISL_2772410, EPI_ISL_2772411, EPI_ISL_2772412, EPI_ISL_2772413, EPI_ISL_2772414, EPI_ISL_2772423, EPI_ISL_2772424, EPI_ISL_2772425, EPI_ISL_2772426, EPI_ISL_2772427, EPI_ISL_2772428, EPI_ISL_2772441, EPI_ISL_2772442, EPI_ISL_2772443, EPI_ISL_2772444, EPI_ISL_2772445, EPI_ISL_2772446, EPI_ISL_2772447, EPI_ISL_2772448, EPI_ISL_2772449, EPI_ISL_2772450, EPI_ISL_2772451, EPI_ISL_2772452, EPI_ISL_2772453, EPI_ISL_2772454, EPI_ISL_2772455, EPI_ISL_2772456, EPI_ISL_2772457, EPI_ISL_2772458, EPI_ISL_2772459, EPI_ISL_2772460, EPI_ISL_2772461, EPI_ISL_2772477, EPI_ISL_2772485, EPI_ISL_2772486, EPI_ISL_2772488, EPI_ISL_2772491, EPI_ISL_2772495, EPI_ISL_2772497, EPI_ISL_2772498, EPI_ISL_2776433, EPI_ISL_2776436, EPI_ISL_2776437, EPI_ISL_2776438, EPI_ISL_2776439, EPI_ISL_2776440, EPI_ISL_2776441, EPI_ISL_2776442, EPI_ISL_2776443, EPI_ISL_2776444, EPI_ISL_2776445, EPI_ISL_2776446, EPI_ISL_2776447, EPI_ISL_2776449, EPI_ISL_2776493, EPI_ISL_2776494, EPI_ISL_2776495, EPI_ISL_2776496, EPI_ISL_2776497, EPI_ISL_2776498, EPI_ISL_2776499, EPI_ISL_2776500, EPI_ISL_2776505, EPI_ISL_2776506, EPI_ISL_2776507, EPI_ISL_2776508, EPI_ISL_2776509, EPI_ISL_2776510, EPI_ISL_2776511, EPI_ISL_2776512, EPI_ISL_2776513, EPI_ISL_2776514, EPI_ISL_2776515, EPI_ISL_2776516, EPI_ISL_2776517, EPI_ISL_2776518, EPI_ISL_2776519, EPI_ISL_2776520, EPI_ISL_2776521, EPI_ISL_2776522, EPI_ISL_2776523, EPI_ISL_2776524, EPI_ISL_2776525, EPI_ISL_2776526, EPI_ISL_2776527, EPI_ISL_2776528, EPI_ISL_2776539, EPI_ISL_2776540, EPI_ISL_2776541, EPI_ISL_2776542, EPI_ISL_2776543, EPI_ISL_2776544, EPI_ISL_2776545, EPI_ISL_2776546, EPI_ISL_2776547, EPI_ISL_2776548, EPI_ISL_2776549, EPI_ISL_2776550, EPI_ISL_2776551, EPI_ISL_2776552, EPI_ISL_2776553, EPI_ISL_2776554, EPI_ISL_2776555, EPI_ISL_2776556, EPI_ISL_2776557, EPI_ISL_2776558, EPI_ISL_2776559, EPI_ISL_2776560, EPI_ISL_2776562, EPI_ISL_2776563, EPI_ISL_2776564, EPI_ISL_2776565, EPI_ISL_2776623, EPI_ISL_2776624, EPI_ISL_2776625, EPI_ISL_2776626, EPI_ISL_2776627, EPI_ISL_2776628, EPI_ISL_2776629, EPI_ISL_2776681, EPI_ISL_2776682, EPI_ISL_2776683, EPI_ISL_2776684, EPI_ISL_2776685, EPI_ISL_2776686, EPI_ISL_2776687, EPI_ISL_2776688, EPI_ISL_2776689, EPI_ISL_2776690, EPI_ISL_2776691, EPI_ISL_2776692, EPI_ISL_2776693, EPI_ISL_2776694, EPI_ISL_2776695, EPI_ISL_2776696, EPI_ISL_2776697, EPI_ISL_2776698, EPI_ISL_2776699, EPI_ISL_2776700, EPI_ISL_2776701, EPI_ISL_2776702, EPI_ISL_2776703, EPI_ISL_2776704, EPI_ISL_2776705, EPI_ISL_2776706, EPI_ISL_2776707, EPI_ISL_2776708, EPI_ISL_2776709, EPI_ISL_2776710, EPI_ISL_2776711, EPI_ISL_2776712, EPI_ISL_2776713, EPI_ISL_2776714, EPI_ISL_2776715, EPI_ISL_2776716, EPI_ISL_2776717, EPI_ISL_2776718, EPI_ISL_2776719, EPI_ISL_2776720, EPI_ISL_2776721, EPI_ISL_2776722, EPI_ISL_2776723, EPI_ISL_2776724, EPI_ISL_2776725, EPI_ISL_2776726, EPI_ISL_2776727, EPI_ISL_2776728, EPI_ISL_2776729, EPI_ISL_2776730, EPI_ISL_2776731, EPI_ISL_2776732, EPI_ISL_2776733, EPI_ISL_2776734, EPI_ISL_2776735, EPI_ISL_2776736, EPI_ISL_2776737, EPI_ISL_2776738, EPI_ISL_2776739, EPI_ISL_2776740, EPI_ISL_2776741, EPI_ISL_2776742, EPI_ISL_2776743, EPI_ISL_2776744, EPI_ISL_2776745, EPI_ISL_2776746, EPI_ISL_2776809, EPI_ISL_2776810, EPI_ISL_2776811, EPI_ISL_2776812, EPI_ISL_2776813, EPI_ISL_2776814, EPI_ISL_2776815, EPI_ISL_2776816, EPI_ISL_2776817, EPI_ISL_2776818, EPI_ISL_2776819, EPI_ISL_2776820, EPI_ISL_2776821, EPI_ISL_2776822, EPI_ISL_2776823, EPI_ISL_2776839, EPI_ISL_2776840, EPI_ISL_2776841, EPI_ISL_2776842, EPI_ISL_2776843, EPI_ISL_2776844, EPI_ISL_2776845, EPI_ISL_2776972, EPI_ISL_2776973, EPI_ISL_2776975, EPI_ISL_2776976, EPI_ISL_2776977, EPI_ISL_2776978, EPI_ISL_2776979, EPI_ISL_2776980, EPI_ISL_2776981, EPI_ISL_2776982, EPI_ISL_2776983, EPI_ISL_2776984, EPI_ISL_2776985, EPI_ISL_2776986, EPI_ISL_2776987, EPI_ISL_2776988, EPI_ISL_2776989, EPI_ISL_2776990, EPI_ISL_2776991, EPI_ISL_2776992, EPI_ISL_2776993, EPI_ISL_2776994, EPI_ISL_2776995, EPI_ISL_2776996, EPI_ISL_2776997, EPI_ISL_2776998, EPI_ISL_2776999, EPI_ISL_2777000, EPI_ISL_2777001, EPI_ISL_2777002, EPI_ISL_2777003, EPI_ISL_2777004, EPI_ISL_2777005, EPI_ISL_2777006, EPI_ISL_2777007, EPI_ISL_2777008, EPI_ISL_2777009, EPI_ISL_2777010, EPI_ISL_2777011, EPI_ISL_2777012, EPI_ISL_2777013, EPI_ISL_2777014, EPI_ISL_2777015, EPI_ISL_2777016, EPI_ISL_2777017, EPI_ISL_2777018, EPI_ISL_2777019, EPI_ISL_2777020, EPI_ISL_2777021, EPI_ISL_2777022, EPI_ISL_2777023, EPI_ISL_2777024, EPI_ISL_2777025, EPI_ISL_2777026, EPI_ISL_2777027, EPI_ISL_2777028, EPI_ISL_2777029, EPI_ISL_2777030, EPI_ISL_2777031, EPI_ISL_2777032, EPI_ISL_2777033, EPI_ISL_2777034, EPI_ISL_2777035, EPI_ISL_2777036, EPI_ISL_2777037, EPI_ISL_2777038, EPI_ISL_2777039, EPI_ISL_2777040, EPI_ISL_2777041, EPI_ISL_2777042, EPI_ISL_2777043, EPI_ISL_2777044, EPI_ISL_2777045, EPI_ISL_2777046, EPI_ISL_2777047, EPI_ISL_2777048, EPI_ISL_2777049, EPI_ISL_2777050, EPI_ISL_2777051, EPI_ISL_2777052, EPI_ISL_2777053, EPI_ISL_2777054, EPI_ISL_2777055, EPI_ISL_2777056, EPI_ISL_2777057, EPI_ISL_2777058, EPI_ISL_2777059, EPI_ISL_2777060, EPI_ISL_2777061, EPI_ISL_2777062, EPI_ISL_2777063, EPI_ISL_2777064, EPI_ISL_2777065, EPI_ISL_2777066, EPI_ISL_2777067, EPI_ISL_2777068, EPI_ISL_2777069, EPI_ISL_2777070, EPI_ISL_2777071, EPI_ISL_2777072, EPI_ISL_2777073, EPI_ISL_2777075, EPI_ISL_2777105, EPI_ISL_2777106, EPI_ISL_2777107, EPI_ISL_2777108, EPI_ISL_2777109, EPI_ISL_2777110, EPI_ISL_2777111, EPI_ISL_2777112, EPI_ISL_2777126 | | | |
| see above | SARS-CoV-2 testing team, National Institute of Infectious Diseases | Pathogen Genomics Center, National Institute of Infectious Diseases | Hazuka Y Furihata; Hiromizu Takahashi; Kentaro Itokawa; Makoto Kuroda; Masanori Hashino; Masumichi Saito; Naomi Nojiri; Nozomu Hanaoka; Rina Tanaka; Sana Uchikoba; Tsuguto Fujimoto; Tsuyoshi Sekizuka |
| EPI_ISL_2350459, EPI_ISL_2350460, EPI_ISL_2350461, EPI_ISL_2350462, EPI_ISL_2350463, EPI_ISL_2350464, EPI_ISL_2350465, EPI_ISL_2350466, EPI_ISL_2350468, EPI_ISL_2350469, EPI_ISL_2350470, EPI_ISL_2350471, EPI_ISL_2350472, EPI_ISL_2350473, EPI_ISL_2350474, EPI_ISL_2350475, EPI_ISL_2350476, EPI_ISL_2350477, EPI_ISL_2350478, EPI_ISL_2350479, EPI_ISL_2350480, EPI_ISL_2350481, EPI_ISL_2350482, EPI_ISL_2350483, EPI_ISL_2350484, EPI_ISL_2812030, EPI_ISL_2812031, EPI_ISL_2812032, EPI_ISL_2812033, EPI_ISL_2812034, EPI_ISL_2812035, EPI_ISL_2812036, EPI_ISL_2812037, EPI_ISL_2812038, EPI_ISL_2812039, EPI_ISL_2812040, EPI_ISL_2812041, EPI_ISL_2812042, EPI_ISL_2812043, EPI_ISL_2812044, EPI_ISL_2812045, EPI_ISL_2812046, EPI_ISL_2812047, EPI_ISL_2812048, EPI_ISL_2812049, EPI_ISL_2812050, EPI_ISL_2812051, EPI_ISL_2812052, EPI_ISL_2812053, EPI_ISL_2812054, EPI_ISL_2812058, EPI_ISL_2812059, EPI_ISL_2812060, EPI_ISL_2812061, EPI_ISL_2812062, EPI_ISL_2812063, EPI_ISL_2812064, EPI_ISL_2812065, EPI_ISL_2812066, EPI_ISL_2812067, EPI_ISL_2812068, EPI_ISL_2812069, EPI_ISL_2812070, EPI_ISL_2812071, EPI_ISL_2812072, EPI_ISL_2812073 | | | |
| see above | Saitama Prefectural Institute of Public Health | Saitama Prefectural Institute of Public Health | Eri Aonuma; Hayato Ehara; Kentaro Itokawa; Makoto Kuroda; Masanori Hashino; Rina Tanaka; Satoshi Daisaki; Tsuyoshi Sekizuka |
| EPI_ISL_2357938, EPI_ISL_2357955, EPI_ISL_2357956, EPI_ISL_2357957, EPI_ISL_2357958 | Sapporo City Institute of Public Health | Sapporo City Institute of Public Health | Asami Ohnishi; Kentaro Itokawa; Makoto Kuroda; Masanori Hashino; Masayuki Kikuchi; Rie Shimazaki; Rina Tanaka; Tsuyoshi Sekizuka; Yusuke Oguchi |
| EPI_ISL_2405225, EPI_ISL_2405226, EPI_ISL_2405227, EPI_ISL_2405228, EPI_ISL_2405229, EPI_ISL_2405230, EPI_ISL_2405231, EPI_ISL_2885192, EPI_ISL_2885193, EPI_ISL_2885194, EPI_ISL_2885195, EPI_ISL_2885196, EPI_ISL_2885197, EPI_ISL_2885220 | | | |
| see above | Shimane Prefectural Institute of Public Health and Environmental Science | Shimane Prefectural Institute of Public Health and Environmental Science | Aiko Fukuma; Kentaro Itokawa; Makoto Kuroda; Masanori Hashino; Naoki Fujisawa; Rie Onishi; Rina Tanaka; Tsuyoshi Sekizuka; Yurie Kanba |
| EPI_ISL_2860257, EPI_ISL_2860258, EPI_ISL_2860259, EPI_ISL_2860260, EPI_ISL_2860261, EPI_ISL_2860262, EPI_ISL_2860263, EPI_ISL_2860264, EPI_ISL_2860265, EPI_ISL_2860266, EPI_ISL_2860267, EPI_ISL_2860268, EPI_ISL_2860269, EPI_ISL_2860270, EPI_ISL_2860271, EPI_ISL_2860272, EPI_ISL_2860273, EPI_ISL_2860274, EPI_ISL_2860275, EPI_ISL_2860276, EPI_ISL_2860277 | | | |
| see above | Tokushima Prefectural Public Health, Pharmaceutical and Environmental Sciences Center | Tokushima Prefectural Public Health, Pharmaceutical and Environmental Sciences Center | Go Sato; Kentaro Itokawa; Makoto Kuroda; Manami Iwaya; Masanori Hashino; Rina Tanaka; Tsuyoshi Sekizuka; Yumiko Kawakami |
| EPI_ISL_2406026, EPI_ISL_2786936, EPI_ISL_2786937, EPI_ISL_2786938, EPI_ISL_2786939, EPI_ISL_2786940, EPI_ISL_2786941, EPI_ISL_2786942, EPI_ISL_2786943, EPI_ISL_2786944, EPI_ISL_2786945, EPI_ISL_2786946, EPI_ISL_2786947, EPI_ISL_2786948, EPI_ISL_2786949, EPI_ISL_2786950, EPI_ISL_2786952, EPI_ISL_2786953, EPI_ISL_2786965 | | | |
| see above | Toyama Institute of Health | Toyama Institute of Health | Emiko Igarashi; Hideki Tani; Kazunori Oishi; Kentaro Itokawa; Makoto Kuroda; Masae Itamochi; Masanori Hashino; Noriko Inasaki; Rina Tanaka; Shunsuke Yazawa; Sumiyo Hasegawa; Tsuyoshi Sekizuka; Yumiko Saga |
| EPI_ISL_2825137, EPI_ISL_2827756, EPI_ISL_2827757, EPI_ISL_2827758 | Virology section, Okayama Prefectural Institute for Environmental Science and Public Health | Virology section, Okayama Prefectural Institute for Environmental Science and Public Health | Gou Tsurumi; Kazuhiko Nagao; Keisuke Funahashi; Kouji Kida; Manabu Ishii; Mayumi Hirata; Naoko Okamoto; Sachie Tsuchimoto |
| EPI_ISL_2841528, EPI_ISL_2841529, EPI_ISL_2841530, EPI_ISL_2841531 | Yamagata Prefectural Institute of Public Health | Pathogen Genomics Center, National Institute of Infectious Diseases | Akinobu Wada; Hideyuki Shinohara; Junji Seto; Katsumi Mizuta; Kenichi Komabayashi; Kentaro Itokawa; Makoto Kuroda; Masanori Hashino; Midori Taruishi; Mika Sampei; Naomi Ogawa; Rina Tanaka; Tsuyoshi Sekizuka; Waka Tanaka; Yoko Aoki; Yoko Ikeda; Yoshitaka Ishida |
| EPI_ISL_2859484, EPI_ISL_2859486, EPI_ISL_2859487, EPI_ISL_2859488, EPI_ISL_2859489 | Yamaguchi Prefectural Institute of Public Health and Environment | Yamaguchi Prefectural Institute of Public Health and Environment | Kanako Kawasaki; Kentaro Itokawa; Makoto Kuroda; Masanori Hashino; Mitushiro Kameyama; Rina Tanaka; Tsuyoshi Sekizuka |

We gratefully acknowledge the following Authors from the Originating laboratories responsible for obtaining the specimens, as well as the Submitting laboratories where the genome data were generated and shared via GISAID, on which this research is based.

All Submitters of data may be contacted directly via www.gisaid.org

Authors are sorted alphabetically.

| Accession ID | Originating Laboratory | Submitting Laboratory | Authors |
|---|---|---|---|
| EPI_ISL_2768845, EPI_ISL_2768846, EPI_ISL_2773953, EPI_ISL_2773954, EPI_ISL_2773955, EPI_ISL_2773956, EPI_ISL_2773957, EPI_ISL_2773958, EPI_ISL_2773959, EPI_ISL_2773960, EPI_ISL_2773961, EPI_ISL_2773962, EPI_ISL_2773963, EPI_ISL_2773964, EPI_ISL_2773965, EPI_ISL_2773966, EPI_ISL_2773967, EPI_ISL_2773968, EPI_ISL_2773969, EPI_ISL_2773970, EPI_ISL_2773971, EPI_ISL_2773972, EPI_ISL_2773973, EPI_ISL_2773974, EPI_ISL_2773975, EPI_ISL_2773976, EPI_ISL_2773977, EPI_ISL_2773978, EPI_ISL_2773979, EPI_ISL_2773980 | | | |
| see above | Chiba Prefectural Institute of Public Health | Pathogen Genomics Center, National Institute of Infectious Diseases | Akihiro Takemura; Haruna Nishijima; Kentaro Itokawa; Makoto Kuroda; Mari Ohta; Masakatsu Taira; Masanori Hashino; Mika Takeuchi; Rina Tanaka; Shigenori Sato; Tsuyoshi Sekizuka; Yuji Hanada; Yuki Fujinuma |
| EPI_ISL_2695962 | Department of Developmental Medicine, Research Institute, Osaka Women's and Children's Hospital | Department of Developmental Medicine, Research Institute, Osaka Women's and Children's Hospital | Itaru Yanagihara; Yukiko Nakura |
| EPI_ISL_2693952, EPI_ISL_2693953, EPI_ISL_2693954, EPI_ISL_2693955, EPI_ISL_2693956, EPI_ISL_2693957 | Department of Infectious Diseases and Immunology, National Hospital Organization Nagoya Medical Center | Clinical Research Center, National Hospital Organization Nagoya Medical Center | Hirotaka Ode; Kazuhiro Matsuoka; Mai Kubota; Masakazu Matsuda; Mayumi Imahashi; Miho Nakasuji; Mikiko Mori; Yasumasa Iwatani; Yoshihiro Nakata; Yoshiyuki Yokomaku |
| EPI_ISL_2810992, EPI_ISL_2811086, EPI_ISL_2811087, EPI_ISL_2811088, EPI_ISL_2811090, EPI_ISL_2811091, EPI_ISL_2811093, EPI_ISL_2811095, EPI_ISL_2811105, EPI_ISL_2811110, EPI_ISL_2811117, EPI_ISL_2811118, EPI_ISL_2811119, EPI_ISL_2811120, EPI_ISL_2811121, EPI_ISL_2811122, EPI_ISL_2811123, EPI_ISL_2811124, EPI_ISL_2811125, EPI_ISL_2811126, EPI_ISL_2811127, EPI_ISL_2811128, EPI_ISL_2811129, EPI_ISL_2811130, EPI_ISL_2811131, EPI_ISL_2811132, EPI_ISL_2811133, EPI_ISL_2811134, EPI_ISL_2811135, EPI_ISL_2811136, EPI_ISL_2811137, EPI_ISL_2811138, EPI_ISL_2811139, EPI_ISL_2811141, EPI_ISL_2811142, EPI_ISL_2811143, EPI_ISL_2811144, EPI_ISL_2811145, EPI_ISL_2811147, EPI_ISL_2811148, EPI_ISL_2811149, EPI_ISL_2811150, EPI_ISL_2811151, EPI_ISL_2811152, EPI_ISL_2811153, EPI_ISL_2811154, EPI_ISL_2811155, EPI_ISL_2811156, EPI_ISL_2811157, EPI_ISL_2811158, EPI_ISL_2811159, EPI_ISL_2811161, EPI_ISL_2811162, EPI_ISL_2811163, EPI_ISL_2811164, EPI_ISL_2811165, EPI_ISL_2811166, EPI_ISL_2811167, EPI_ISL_2811168, EPI_ISL_2811169, EPI_ISL_2811170, EPI_ISL_2811171, EPI_ISL_2811172, EPI_ISL_2811173, EPI_ISL_2811176, EPI_ISL_2811177, EPI_ISL_2811178, EPI_ISL_2811179, EPI_ISL_2811180, EPI_ISL_2811181, EPI_ISL_2811182, EPI_ISL_2811183, EPI_ISL_2811184, EPI_ISL_2811185, EPI_ISL_2811188, EPI_ISL_2811189, EPI_ISL_2811190, EPI_ISL_2811191, EPI_ISL_2811192, EPI_ISL_2811193, EPI_ISL_2811195, EPI_ISL_2811196, EPI_ISL_2811197, EPI_ISL_2811198, EPI_ISL_2811199, EPI_ISL_2811200, EPI_ISL_2811201, EPI_ISL_2811202, EPI_ISL_2811203, EPI_ISL_2811204, EPI_ISL_2811205, EPI_ISL_2811206, EPI_ISL_2811207, EPI_ISL_2811208, EPI_ISL_2811209, EPI_ISL_2811210, EPI_ISL_2811211, EPI_ISL_2811212, EPI_ISL_2811213, EPI_ISL_2811214, EPI_ISL_2811215, EPI_ISL_2811216, EPI_ISL_2811217, EPI_ISL_2811218, EPI_ISL_2811219, EPI_ISL_2811220, EPI_ISL_2811221, EPI_ISL_2811222, EPI_ISL_2811223, EPI_ISL_2811224, EPI_ISL_2811225, EPI_ISL_2811226, EPI_ISL_2811227, EPI_ISL_2811228, EPI_ISL_2811229, EPI_ISL_2811230, EPI_ISL_2811231, EPI_ISL_2811232, EPI_ISL_2811233, EPI_ISL_2811234, EPI_ISL_2811235, EPI_ISL_2811236, EPI_ISL_2811237, EPI_ISL_2811238, EPI_ISL_2811239, EPI_ISL_2811240, EPI_ISL_2811241, EPI_ISL_2811242, EPI_ISL_2811243, EPI_ISL_2811244, EPI_ISL_2811245, EPI_ISL_2811246, EPI_ISL_2811247, EPI_ISL_2811248, EPI_ISL_2811249, EPI_ISL_2811250, EPI_ISL_2811251, EPI_ISL_2811252, EPI_ISL_2811253, EPI_ISL_2811258, EPI_ISL_2811259, EPI_ISL_2811260, EPI_ISL_2811261, EPI_ISL_2811262, EPI_ISL_2811263, EPI_ISL_2811264, EPI_ISL_2811265, EPI_ISL_2811266, EPI_ISL_2811267, EPI_ISL_2811268, EPI_ISL_2811269, EPI_ISL_2811270, EPI_ISL_2811271, EPI_ISL_2811272, EPI_ISL_2811273, EPI_ISL_2811274, EPI_ISL_2811275, EPI_ISL_2811276, EPI_ISL_2811277, EPI_ISL_2811278, EPI_ISL_2811279, EPI_ISL_2811280, EPI_ISL_2811281, EPI_ISL_2811282, EPI_ISL_2811283, EPI_ISL_2811284, EPI_ISL_2811285, EPI_ISL_2811286, EPI_ISL_2811287, EPI_ISL_2811288, EPI_ISL_2811289, EPI_ISL_2811290, EPI_ISL_2811291, EPI_ISL_2811292, EPI_ISL_2811293, EPI_ISL_2811294, EPI_ISL_2811295, EPI_ISL_2811296, EPI_ISL_2811297, EPI_ISL_2811298, EPI_ISL_2811299, EPI_ISL_2811300, EPI_ISL_2811301, EPI_ISL_2811334, EPI_ISL_2811335, EPI_ISL_2811336, EPI_ISL_2811337, EPI_ISL_2811338, EPI_ISL_2811339, EPI_ISL_2811340, EPI_ISL_2811341, EPI_ISL_2811342, EPI_ISL_2811343, EPI_ISL_2811344, EPI_ISL_2811345, EPI_ISL_2811346, EPI_ISL_2811347, EPI_ISL_2811348, EPI_ISL_2811349, EPI_ISL_2811350, EPI_ISL_2811351, EPI_ISL_2811352, EPI_ISL_2811353, EPI_ISL_2811354, EPI_ISL_2811355, EPI_ISL_2811356, EPI_ISL_2811357, EPI_ISL_2811358, EPI_ISL_2811359, EPI_ISL_2811360, EPI_ISL_2811361, EPI_ISL_2811362, EPI_ISL_2811363, EPI_ISL_2811364, EPI_ISL_2811365, EPI_ISL_2811366, EPI_ISL_2811367, EPI_ISL_2811368, EPI_ISL_2811369, EPI_ISL_2811370, EPI_ISL_2811371, EPI_ISL_2811372, EPI_ISL_2811373, EPI_ISL_2811374, EPI_ISL_2811375, EPI_ISL_2811376, EPI_ISL_2811377, EPI_ISL_2811378, EPI_ISL_2811379, EPI_ISL_2811380, EPI_ISL_2811381, EPI_ISL_2811382, EPI_ISL_2811383, EPI_ISL_2811384, EPI_ISL_2811385, EPI_ISL_2811386, EPI_ISL_2811387, EPI_ISL_2811388, EPI_ISL_2811389, EPI_ISL_2811390, EPI_ISL_2811391, EPI_ISL_2811392, EPI_ISL_2811393 | | | |
| see above | Department of Infectious Diseases, Kobe Institute of Health | Department of Infectious Diseases, Kobe Institute of Health | Kentaro Itokawa; Makoto Kuroda; Masanori Hashino; Noriko Nakanishi; Rina Tanaka; Ryohei Nomoto; Tomotada Iwamoto; Tsuyoshi Sekizuka |
| EPI_ISL_2637224, EPI_ISL_2811397, EPI_ISL_2811398, EPI_ISL_2811399, EPI_ISL_2811400, EPI_ISL_2811401, EPI_ISL_2811402, EPI_ISL_2811403, EPI_ISL_2811404, EPI_ISL_2811405, EPI_ISL_2811406, EPI_ISL_2811407, EPI_ISL_2811408, EPI_ISL_2811409 | | | |
| see above | Department of Microbiology, Yamanashi Institute for Public Health and Environment | Genome Analysis Center, Yamanashi Central Hospital | Yosuke Hirotsu |
| EPI_ISL_2861261, EPI_ISL_2861262, EPI_ISL_2861263, EPI_ISL_2861264, EPI_ISL_2861265, EPI_ISL_2861266 | Dr. Yoshitaka Tamura Department of Clinical Laboratory, Osaka Habikino Medical Center | Department of Developmental Medicine, Research Institute, Osaka Women's and Children's Hospital | Itaru Yanagihara; Yukiko Nakura |
| EPI_ISL_2614634, EPI_ISL_2614635, EPI_ISL_2614636, EPI_ISL_2614637, EPI_ISL_2614638, EPI_ISL_2614641, EPI_ISL_2614642 | | | |
| see above | Dr. Yuichiro Oba Department of General Medicine, Osaka General Medical Center | Department of Developmental Medicine, Research Institute, Osaka Women's and Children's Hospital | Itaru Yanagihara; Yukiko Nakura |
| EPI_ISL_2772910, EPI_ISL_2772911, EPI_ISL_2772912, EPI_ISL_2772913, EPI_ISL_2772914, EPI_ISL_2772915, EPI_ISL_2772916, EPI_ISL_2772917, EPI_ISL_2772918, EPI_ISL_2772919, EPI_ISL_2772920, EPI_ISL_2772921, EPI_ISL_2772922, EPI_ISL_2772923, EPI_ISL_2772924, EPI_ISL_2772925, EPI_ISL_2772926, EPI_ISL_2772927, EPI_ISL_2772928, EPI_ISL_2772929, EPI_ISL_2772930, EPI_ISL_2772931, EPI_ISL_2772932, EPI_ISL_2772933, EPI_ISL_2772934, EPI_ISL_2772935, EPI_ISL_2773082, EPI_ISL_2773083, EPI_ISL_2773084, EPI_ISL_2773085, EPI_ISL_2773086, EPI_ISL_2773087, EPI_ISL_2773088, EPI_ISL_2773089, EPI_ISL_2773090, EPI_ISL_2773091, EPI_ISL_2773092, EPI_ISL_2773093, EPI_ISL_2773094, EPI_ISL_2773341, EPI_ISL_2773342, EPI_ISL_2773343, EPI_ISL_2773344, EPI_ISL_2773345, EPI_ISL_2773346, EPI_ISL_2773347, EPI_ISL_2773348, EPI_ISL_2773349, EPI_ISL_2773350, EPI_ISL_2773351, EPI_ISL_2773352, EPI_ISL_2773353, EPI_ISL_2773354, EPI_ISL_2773355, EPI_ISL_2773356, EPI_ISL_2773357, EPI_ISL_2773358, EPI_ISL_2773359, EPI_ISL_2773360, EPI_ISL_2773361, EPI_ISL_2773362, EPI_ISL_2773363, EPI_ISL_2773364, EPI_ISL_2773365, EPI_ISL_2773489, EPI_ISL_2773490, EPI_ISL_2773491, EPI_ISL_2773492, EPI_ISL_2773493, EPI_ISL_2773494, EPI_ISL_2773495, EPI_ISL_2773496, EPI_ISL_2773497, EPI_ISL_2773498, EPI_ISL_2773499, EPI_ISL_2773500, EPI_ISL_2773541, EPI_ISL_2773542, EPI_ISL_2773564, EPI_ISL_2773565, EPI_ISL_2773566, EPI_ISL_2773567, EPI_ISL_2773568, EPI_ISL_2773569, EPI_ISL_2773570, EPI_ISL_2773571, EPI_ISL_2773572, EPI_ISL_2773573, EPI_ISL_2773574, EPI_ISL_2773575, EPI_ISL_2773576, EPI_ISL_2773577, EPI_ISL_2773578, EPI_ISL_2773579, EPI_ISL_2773580, EPI_ISL_2773581, EPI_ISL_2773582, EPI_ISL_2773583, EPI_ISL_2773584, EPI_ISL_2773585, EPI_ISL_2773586, EPI_ISL_2773587, EPI_ISL_2773588, EPI_ISL_2773589, EPI_ISL_2773590, EPI_ISL_2773591, EPI_ISL_2773592, EPI_ISL_2773593, EPI_ISL_2773594, EPI_ISL_2773595, EPI_ISL_2773596, EPI_ISL_2773597, EPI_ISL_2773598, EPI_ISL_2773599, EPI_ISL_2773600, EPI_ISL_2773601, EPI_ISL_2773602, EPI_ISL_2773603, EPI_ISL_2773604, EPI_ISL_2773605, EPI_ISL_2773606, EPI_ISL_2773607, EPI_ISL_2773608, EPI_ISL_2773609, EPI_ISL_2773610, EPI_ISL_2773611, EPI_ISL_2773612, EPI_ISL_2773613, EPI_ISL_2773614, EPI_ISL_2773615, EPI_ISL_2773616, EPI_ISL_2773617, EPI_ISL_2773618, EPI_ISL_2773619, EPI_ISL_2773620, EPI_ISL_2773621, EPI_ISL_2773622, EPI_ISL_2773623, EPI_ISL_2773624, EPI_ISL_2773625, EPI_ISL_2773626, EPI_ISL_2773627, EPI_ISL_2773628, EPI_ISL_2773629, EPI_ISL_2773630, EPI_ISL_2773631, EPI_ISL_2773632, EPI_ISL_2773633, EPI_ISL_2773634, EPI_ISL_2773635, EPI_ISL_2773636, EPI_ISL_2773637, EPI_ISL_2773638, EPI_ISL_2773639, EPI_ISL_2773640, EPI_ISL_2773641, EPI_ISL_2773642, EPI_ISL_2773643, EPI_ISL_2773644, EPI_ISL_2773645, EPI_ISL_2773646, EPI_ISL_2773647, EPI_ISL_2773648, EPI_ISL_2773649, EPI_ISL_2773650, EPI_ISL_2773651, EPI_ISL_2773652, EPI_ISL_2773653, EPI_ISL_2773654, EPI_ISL_2773655, EPI_ISL_2773656, EPI_ISL_2773657, EPI_ISL_2773658, EPI_ISL_2773659, EPI_ISL_2773660, EPI_ISL_2773661, EPI_ISL_2773662 | | | |
| see above | Fukuoka Institute of Health and Environmental Sciences | Fukuoka Institute of Health and Environmental Sciences | Akira Ohishi; Carl Yuki; Chiharu Katamune; Hiroaki Shigemura; Kentaro Itokawa; Makoto Kuroda; Masanori Hashino; Mitsuhiro Hamasaki; Rina Tanaka; Saori Ueda; Takayuki Kobayashi; Tsuyoshi Sekizuka; Yoshiki Etoh; Yuki Ashizuka |
| EPI_ISL_2812223, EPI_ISL_2812224, EPI_ISL_2812225, EPI_ISL_2812228, EPI_ISL_2812229, EPI_ISL_2812230, EPI_ISL_2812231, EPI_ISL_2812232 | | | |
| see above | Fukushima Prefectural Institute of Public Health | Fukushima Prefectural Institute of Public Health | Kadzuhiro Kitakawa; Kazunori Suzuki; Kentaro Itokawa; Makoto Kuroda; Masanori Hashino; Naoko Kashiwabara; Rie Suzuki; Rina Tanaka; Shohei Fujita; Tsuyoshi Sekizuka |
| EPI_ISL_2566681 | Genome Analysis Center, Kamma Memorial Hospital | Genome Analysis Center, Kamma Memorial Hospital | Hanako Yazawa; Hiroshi Kamma; Jun Ishii; Kaori Watanabe; Masaki Sato; Michiaki Masuda; Naruhisa Takano; Noriyuki Tannai; Satoko Soma; Takuya Yazawa |
| EPI_ISL_2462057, EPI_ISL_2462058, EPI_ISL_2565984, EPI_ISL_2565985, EPI_ISL_2565986, EPI_ISL_2565987, EPI_ISL_2565988, EPI_ISL_2565989, EPI_ISL_2565990, EPI_ISL_2565991, EPI_ISL_2565992, EPI_ISL_2565993, EPI_ISL_2565994, EPI_ISL_2565995, EPI_ISL_2565996, EPI_ISL_2565997, EPI_ISL_2621732, EPI_ISL_2621733, EPI_ISL_2621734, EPI_ISL_2621735, EPI_ISL_2621736, EPI_ISL_2621737, EPI_ISL_2637225, EPI_ISL_2637226, EPI_ISL_2637227, EPI_ISL_2637228, EPI_ISL_2637229, EPI_ISL_2637230, EPI_ISL_2637231, EPI_ISL_2644658, EPI_ISL_2644659, EPI_ISL_2644660, EPI_ISL_2644661, EPI_ISL_2644662, EPI_ISL_2644663, EPI_ISL_2644664, EPI_ISL_2644665, EPI_ISL_2644666, EPI_ISL_2644667, EPI_ISL_2644668, EPI_ISL_2644669, EPI_ISL_2644670, EPI_ISL_2644671, EPI_ISL_2657372, EPI_ISL_2657373, EPI_ISL_2657374, EPI_ISL_2696282, EPI_ISL_2696283, EPI_ISL_2696284, EPI_ISL_2696285, EPI_ISL_2696286, EPI_ISL_2696287, EPI_ISL_2696288, EPI_ISL_2696289, EPI_ISL_2696290, EPI_ISL_2696291, EPI_ISL_2696292, EPI_ISL_2696293, EPI_ISL_2696294, EPI_ISL_2696295, EPI_ISL_2696296, EPI_ISL_2696297, EPI_ISL_2754133, EPI_ISL_2754134, EPI_ISL_2754135, EPI_ISL_2754136, EPI_ISL_2754137, EPI_ISL_2754138, EPI_ISL_2790309, EPI_ISL_2790310, EPI_ISL_2790311, EPI_ISL_2790312, EPI_ISL_2790313, EPI_ISL_2790314, EPI_ISL_2790315, EPI_ISL_2876291, EPI_ISL_2876292, EPI_ISL_2876293, EPI_ISL_2876294, EPI_ISL_2876295, EPI_ISL_2876296, EPI_ISL_2876297, EPI_ISL_2876298, EPI_ISL_2876299, EPI_ISL_2876300, EPI_ISL_2876301, EPI_ISL_2907483 | | | |
| see above | Genome Analysis Center, Yamanashi Central Hospital | Genome Analysis Center, Yamanashi Central Hospital | Yosuke Hirotsu |
| EPI_ISL_2565983, EPI_ISL_2727654, EPI_ISL_2758192, EPI_ISL_2759101, EPI_ISL_2759102, EPI_ISL_2759103, EPI_ISL_2759104, EPI_ISL_2759105, EPI_ISL_2759106, EPI_ISL_2759107, EPI_ISL_2759108, EPI_ISL_2759109, EPI_ISL_2759110, EPI_ISL_2759111, EPI_ISL_2759112, EPI_ISL_2759113, EPI_ISL_2759114, EPI_ISL_2759115, EPI_ISL_2759116, EPI_ISL_2759117, EPI_ISL_2759118, EPI_ISL_2759119, EPI_ISL_2759120, EPI_ISL_2759121, EPI_ISL_2759122, EPI_ISL_2759123, EPI_ISL_2759124, EPI_ISL_2759125, EPI_ISL_2759126, EPI_ISL_2759127, EPI_ISL_2759128, EPI_ISL_2759129, EPI_ISL_2759130, EPI_ISL_2759131, EPI_ISL_2759132, EPI_ISL_2759133, EPI_ISL_2759134, EPI_ISL_2759135, EPI_ISL_2759136, EPI_ISL_2759137, EPI_ISL_2759138, EPI_ISL_2759139, EPI_ISL_2759140, EPI_ISL_2759141, EPI_ISL_2759645, EPI_ISL_2759646, EPI_ISL_2759647, EPI_ISL_2759648, EPI_ISL_2759649, EPI_ISL_2759650, EPI_ISL_2759651, EPI_ISL_2759652, EPI_ISL_2759653, EPI_ISL_2759654, EPI_ISL_2759655, EPI_ISL_2759744, EPI_ISL_2759747, EPI_ISL_2759750, EPI_ISL_2759754, EPI_ISL_2759756, EPI_ISL_2759851, EPI_ISL_2759852, EPI_ISL_2759853, EPI_ISL_2759854, EPI_ISL_2759855, EPI_ISL_2759856, EPI_ISL_2759857, EPI_ISL_2759858, EPI_ISL_2759859, EPI_ISL_2759860, EPI_ISL_2759861, EPI_ISL_2759862, EPI_ISL_2759863, EPI_ISL_2759864, EPI_ISL_2759865, EPI_ISL_2759866, EPI_ISL_2759867, EPI_ISL_2759868, EPI_ISL_2759869, EPI_ISL_2759870, EPI_ISL_2759871, EPI_ISL_2759872, EPI_ISL_2759873, EPI_ISL_2759874, EPI_ISL_2759875, EPI_ISL_2773863, EPI_ISL_2773864, EPI_ISL_2773865, EPI_ISL_2773866, EPI_ISL_2773867, EPI_ISL_2773868, EPI_ISL_2773869, EPI_ISL_2773870, EPI_ISL_2773871, EPI_ISL_2773945, EPI_ISL_2773946, EPI_ISL_2854210, EPI_ISL_2861561, EPI_ISL_2861562, EPI_ISL_2861718, EPI_ISL_2861720, EPI_ISL_2861721, EPI_ISL_2861722, EPI_ISL_2861723, EPI_ISL_2861724, EPI_ISL_2861799, EPI_ISL_2861800, EPI_ISL_2861801, EPI_ISL_2861802, EPI_ISL_2861803, EPI_ISL_2877625, EPI_ISL_2877626, EPI_ISL_2877627, EPI_ISL_2877628, EPI_ISL_2877629, EPI_ISL_2877630, EPI_ISL_2877631, EPI_ISL_2877632, EPI_ISL_2877633, EPI_ISL_2877634, EPI_ISL_2877635, EPI_ISL_2877636, EPI_ISL_2877637, EPI_ISL_2877638, EPI_ISL_2877639, EPI_ISL_2877640, EPI_ISL_2877641, EPI_ISL_2877642, EPI_ISL_2877643, EPI_ISL_2877644, EPI_ISL_2877645, EPI_ISL_2877646, EPI_ISL_2877647, EPI_ISL_2877648, EPI_ISL_2877649, EPI_ISL_2877650, EPI_ISL_2877651, EPI_ISL_2877652, EPI_ISL_2877653, EPI_ISL_2877654 | | | |
| see above | Genome Information Research Center, Research Institute for Microbial Diseases, Osaka University | Genome Information Research Center, Research Institute for Microbial Diseases, Osaka University | Daisuke Motooka; Hiroya Oki; Kentaro Tanaka; and Shota Nakamura |
| EPI_ISL_2788689, EPI_ISL_2788690, EPI_ISL_2788691, EPI_ISL_2788693, EPI_ISL_2788694, EPI_ISL_2788695, EPI_ISL_2788696, EPI_ISL_2788697, EPI_ISL_2788698, EPI_ISL_2788699, EPI_ISL_2788700, EPI_ISL_2788701, EPI_ISL_2788702, EPI_ISL_2788706, EPI_ISL_2788707, EPI_ISL_2788708, EPI_ISL_2788709, EPI_ISL_2788710, EPI_ISL_2788712, EPI_ISL_2788713, EPI_ISL_2788714, EPI_ISL_2788715, EPI_ISL_2788716, EPI_ISL_2788717, EPI_ISL_2788718, EPI_ISL_2788719 | | | |
| see above | Gunma Prefectural Institute of Public Health and Environmental Sciences | Gunma Prefectural Institute of Public Health and Environmental Sciences | Daisuke Shinoda; Hiroyuki Tsukagoshi; Kentaro Itokawa; Makoto Kuroda; Mariko Saito; Masanori Hashino; Rina Tanaka; Tsuyoshi Sekizuka |
| EPI_ISL_2773741, EPI_ISL_2773767, EPI_ISL_2773772, EPI_ISL_2773773, EPI_ISL_2773775, EPI_ISL_2773776, EPI_ISL_2773777, EPI_ISL_2773778, EPI_ISL_2773779, EPI_ISL_2773780, EPI_ISL_2773781, EPI_ISL_2773782, EPI_ISL_2773783, EPI_ISL_2773784, EPI_ISL_2773785, EPI_ISL_2773787, EPI_ISL_2773788, EPI_ISL_2773789, EPI_ISL_2773790, EPI_ISL_2773791, EPI_ISL_2773792, EPI_ISL_2773793, EPI_ISL_2773794, EPI_ISL_2773795, EPI_ISL_2773796, EPI_ISL_2773797, EPI_ISL_2773799, EPI_ISL_2773800, EPI_ISL_2773801, EPI_ISL_2773802, EPI_ISL_2773803, EPI_ISL_2773804, EPI_ISL_2773805, EPI_ISL_2773806, EPI_ISL_2773808, EPI_ISL_2773809, EPI_ISL_2773810, EPI_ISL_2773811, EPI_ISL_2773812, EPI_ISL_2773814, EPI_ISL_2773815, EPI_ISL_2773816, EPI_ISL_2773817, EPI_ISL_2773818, EPI_ISL_2773819, EPI_ISL_2773820, EPI_ISL_2773821, EPI_ISL_2773822, EPI_ISL_2773823, EPI_ISL_2773824, EPI_ISL_2773825, EPI_ISL_2773826, EPI_ISL_2773827, EPI_ISL_2773828, EPI_ISL_2773829, EPI_ISL_2773830, EPI_ISL_2773831, EPI_ISL_2773832, EPI_ISL_2773833, EPI_ISL_2773834, EPI_ISL_2773835, EPI_ISL_2773837, EPI_ISL_2773838, EPI_ISL_2773839, EPI_ISL_2773840 | | | |
| see above | Hiroshima Prefectural Technology Research Institute, Public Health and Environment Center | Hiroshima Prefectural Technology Research Institute, Public Health and Environment Center | Akie Nakashima; Ayano Ito; Hiroko Akita; Kanako Masuda; Kentaro Itokawa; Makoto Kuroda; Mana Suei; Masanori Hashino; Naoki Shigemoto; Rina Tanaka; Shinichi Takao; Syuhei Ikeda; Takahiro Hiratsuka; Tsuyoshi Sekizuka; Yukie Shimazu |
| EPI_ISL_2884693, EPI_ISL_2884719, EPI_ISL_2884722, EPI_ISL_2884733, EPI_ISL_2884734, EPI_ISL_2884735, EPI_ISL_2884739, EPI_ISL_2884740, EPI_ISL_2884741, EPI_ISL_2884744, EPI_ISL_2884745, EPI_ISL_2884746, EPI_ISL_2884750, EPI_ISL_2884751, EPI_ISL_2884752, EPI_ISL_2884753, EPI_ISL_2884755, EPI_ISL_2884756, EPI_ISL_2884757, EPI_ISL_2884758, EPI_ISL_2884759, EPI_ISL_2884760, EPI_ISL_2884761, EPI_ISL_2884762, EPI_ISL_2884763, EPI_ISL_2884764, EPI_ISL_2884765, EPI_ISL_2884766, EPI_ISL_2884767, EPI_ISL_2884768, EPI_ISL_2884769, EPI_ISL_2884770, EPI_ISL_2884771, EPI_ISL_2884772, EPI_ISL_2884773, EPI_ISL_2884774, EPI_ISL_2884775, EPI_ISL_2884776, EPI_ISL_2884777, EPI_ISL_2884778, EPI_ISL_2884779, EPI_ISL_2884780, EPI_ISL_2884781, EPI_ISL_2884782, EPI_ISL_2884783, EPI_ISL_2884784, EPI_ISL_2884785, EPI_ISL_2884786, EPI_ISL_2884788, EPI_ISL_2884789, EPI_ISL_2884790, EPI_ISL_2884791, EPI_ISL_2884792, EPI_ISL_2884793, EPI_ISL_2884794, EPI_ISL_2884795, EPI_ISL_2884796, EPI_ISL_2884797, EPI_ISL_2884798, EPI_ISL_2884799, EPI_ISL_2884800, EPI_ISL_2884801, EPI_ISL_2884802, EPI_ISL_2884803, EPI_ISL_2884804, EPI_ISL_2884805, EPI_ISL_2884806, EPI_ISL_2884807, EPI_ISL_2884808, EPI_ISL_2884809, EPI_ISL_2884810, EPI_ISL_2884811, EPI_ISL_2884812, EPI_ISL_2884813, EPI_ISL_2884814, EPI_ISL_2884815, EPI_ISL_2884819, EPI_ISL_2884820, EPI_ISL_2884821, EPI_ISL_2884822, EPI_ISL_2884826, EPI_ISL_2884827, EPI_ISL_2884828, EPI_ISL_2884829, EPI_ISL_2884830, EPI_ISL_2884831, EPI_ISL_2884834, EPI_ISL_2884835, EPI_ISL_2884836, EPI_ISL_2884837, EPI_ISL_2884852, EPI_ISL_2884860, EPI_ISL_2884861, EPI_ISL_2884862 | | | |
| see above | Hokkaido Institute of Public Health | Hokkaido Institute of Public Health | Kazuhiro Okubo; Kentaro Itokawa; Makoto Kuroda; Masanori Hashino; Rina Tanaka; Tsuyoshi Sekizuka; Yuta Ohno |
| EPI_ISL_2860706, EPI_ISL_2860707, EPI_ISL_2860708, EPI_ISL_2860709, EPI_ISL_2860710, EPI_ISL_2860711, EPI_ISL_2860712, EPI_ISL_2860713, EPI_ISL_2860714, EPI_ISL_2860715, EPI_ISL_2860716, EPI_ISL_2860717, EPI_ISL_2860718, EPI_ISL_2860719, EPI_ISL_2860720, EPI_ISL_2860721, EPI_ISL_2860722, EPI_ISL_2860723, EPI_ISL_2860727, EPI_ISL_2860733, EPI_ISL_2860734, EPI_ISL_2860735, EPI_ISL_2860736, EPI_ISL_2860737, EPI_ISL_2860738, EPI_ISL_2860739, EPI_ISL_2860740, EPI_ISL_2860741, EPI_ISL_2860742, EPI_ISL_2860743, EPI_ISL_2860744, EPI_ISL_2860745, EPI_ISL_2860746, EPI_ISL_2860747, EPI_ISL_2860748, EPI_ISL_2860749, EPI_ISL_2860750, EPI_ISL_2860756, EPI_ISL_2860757, EPI_ISL_2860758, EPI_ISL_2860759, EPI_ISL_2860760, EPI_ISL_2860761, EPI_ISL_2860765, EPI_ISL_2860766, EPI_ISL_2860767, EPI_ISL_2860768, EPI_ISL_2860769, EPI_ISL_2860770, EPI_ISL_2860771, EPI_ISL_2860772, EPI_ISL_2860773 | | | |
| see above | Hyogo Prefectural Institute of Public Health science | Pathogen Genomics Center, National Institute of Infectious Diseases | Denshi Takai; Etsuko Saito; Kenichi Ogita; Kentaro Itokawa; Kyoko Suzuki; Makoto Kuroda; Masanori Hashino; Masatsugu Chikahira; Miki Ogi; Rina Tanaka; Tomohiro Oshibe; Tsuyoshi Sekizuka; Yumi Akiyama |
| EPI_ISL_2786701, EPI_ISL_2786724, EPI_ISL_2786725, EPI_ISL_2786726, EPI_ISL_2786727, EPI_ISL_2786728, EPI_ISL_2786729, EPI_ISL_2786761, EPI_ISL_2786762, EPI_ISL_2786763, EPI_ISL_2786764, EPI_ISL_2786765, EPI_ISL_2786766, EPI_ISL_2786767, EPI_ISL_2786772, EPI_ISL_2786773, EPI_ISL_2786774, EPI_ISL_2786775, EPI_ISL_2786776, EPI_ISL_2786777, EPI_ISL_2786778, EPI_ISL_2786779, EPI_ISL_2786780, EPI_ISL_2786781, EPI_ISL_2786782, EPI_ISL_2786783, EPI_ISL_2786784, EPI_ISL_2786800, EPI_ISL_2786801, EPI_ISL_2786849, EPI_ISL_2786850, EPI_ISL_2786851, EPI_ISL_2786852, EPI_ISL_2786877, EPI_ISL_2786878 | | | |
| see above | Ibaraki Prefectural Institute of Public Health | Ibaraki Prefectural Institute of Public Health | Keiko Goto; Kentaro Itokawa; Makoto Kuroda; Masanori Hashino; Rina Tanaka; Tsuyoshi Sekizuka |
| EPI_ISL_2787102, EPI_ISL_2787109 | Kagawa Prefectural Research Institute for Environmental Sciences and Public Health | Pathogen Genomics Center, National Institute of Infectious Diseases | Kenji Kuwahara; Kentaro Itokawa; Makoto Kuroda; Masanori Hashino; Mayumi Arizuka; Rina Tanaka; Tsuyoshi Sekizuka; Yukari Terajima |
| EPI_ISL_2842537, EPI_ISL_2842538, EPI_ISL_2842542, EPI_ISL_2842543, EPI_ISL_2842544, EPI_ISL_2842546, EPI_ISL_2842548, EPI_ISL_2842549, EPI_ISL_2842550, EPI_ISL_2842551, EPI_ISL_2842552, EPI_ISL_2842553, EPI_ISL_2842554, EPI_ISL_2842556, EPI_ISL_2842558, EPI_ISL_2842559, EPI_ISL_2842560, EPI_ISL_2842561, EPI_ISL_2842563, EPI_ISL_2842564, EPI_ISL_2842565, EPI_ISL_2842566, EPI_ISL_2842567, EPI_ISL_2842568, EPI_ISL_2842569, EPI_ISL_2842570, EPI_ISL_2842571, EPI_ISL_2842572, EPI_ISL_2842573, EPI_ISL_2842574, EPI_ISL_2842575, EPI_ISL_2842576, EPI_ISL_2842577, EPI_ISL_2842578 | | | |
| see above | Kagoshima Prefectural Institute for Environmental Research and | Kagoshima Prefectural Institute for Environmental Research and Public Health | Kentaro Itokawa; Makoto Kuroda; Masanori Hashino; Rina Tanaka; Tsuyoshi Sekizuka; Yuka Hamada |

| | | | |
|---|---|---|---|
| | Public Health | | |
| EPI_ISL_2544686, EPI_ISL_2544688, EPI_ISL_2544690, EPI_ISL_2544693 | Kanagawa Prefectural Institute of Public Health | Kanagawa Prefectural Institute of Public Health | Itsumi Toyokura; Junichi Sakuragi; Kentaro Itokawa; Makiko Kondo; Makoto Kuroda; Masanori Hashino; Rieko Suzuki; Rina Tanaka; Sumi Watanabe; Takako Sano; Takayuki Hishiki; Tomohiko Takasaki; Tsuyoshi Sekizuka |
| EPI_ISL_2839671, EPI_ISL_2839672, EPI_ISL_2839673 | Kumamoto City Medical Association PCR Center | Sato Laboratory, Division of Genomics and Transcriptomics, The Joint Research Center for Human Retrovirus Infection, Kumamoto University | Fumiya Ishikawa; Ryota Irie; Samiul Alam Rajib; Yorifumi Sato |
| EPI_ISL_2828099, EPI_ISL_2828100, EPI_ISL_2828101, EPI_ISL_2828102, EPI_ISL_2828103, EPI_ISL_2828104, EPI_ISL_2828105 | | | |
| see above | Kyoto Prefectural Institute Of Public Health And Environment | Kyoto Prefectural Institute Of Public Health And Environment | Akira Kotera; Kenji Konaka; Kentaro Itokawa; Makoto Kuroda; Masanori Hashino; Miki Tsuda; Nana Horikawa; Norio Asai; Rina Tanaka; Tsuyoshi Sekizuka; Yasushi Fujimoto; Yoshihiro Tanabe; Yoshiyuki Kawahara; Yuri Sakai |
| EPI_ISL_2841495, EPI_ISL_2841497, EPI_ISL_2841498, EPI_ISL_2841499, EPI_ISL_2841500, EPI_ISL_2841501, EPI_ISL_2841502, EPI_ISL_2841503, EPI_ISL_2841505, EPI_ISL_2841506, EPI_ISL_2841507, EPI_ISL_2841508, EPI_ISL_2841509, EPI_ISL_2841510, EPI_ISL_2841511, EPI_ISL_2841512 | | | |
| see above | Medical Microbiology and Zoology Section, Okinawa Prefectural Institute of Health and Environment | Pathogen Genomics Center, National Institute of Infectious Diseases | Hisako Kyan; Kentaro Itokawa; Makoto Kuroda; Masanori Hashino; Minoru Nidaira; Noriyuki Maeshiro; Rina Tanaka; Taketoshi Takara; Tetsuya Kakita; Tsuyoshi Kudeken; Tsuyoshi Sekizuka; Yumani Kuba |
| EPI_ISL_2695878, EPI_ISL_2695879, EPI_ISL_2695880, EPI_ISL_2695881, EPI_ISL_2695883, EPI_ISL_2861267, EPI_ISL_2861268, EPI_ISL_2896258 | | | |
| see above | Osaka General Medical Center | Department of Developmental Medicine, Research Institute, Osaka Women's and Children's Hospital | Itaru Yanagihara; Yukiko Nakura |
| EPI_ISL_2768839, EPI_ISL_2768840, EPI_ISL_2768841, EPI_ISL_2768842, EPI_ISL_2768843 | Osaka Institute of Public Health, Morinomiya Center | Pathogen Genomics Center, National Institute of Infectious Diseases | Kentaro Itokawa; Makoto Kuroda; Masanori Hashino; Rina Tanaka; Tsuyoshi Sekizuka |
| EPI_ISL_2876423 | Osaka Prefecture | Clinical Laboratory Medicine, Kyoto University Graduate School of Medicine | Masaki Yamamoto; Miki Nagao; Yasufumi Matsumura |
| EPI_ISL_2764096, EPI_ISL_2764102, EPI_ISL_2764949, EPI_ISL_2764950, EPI_ISL_2768322, EPI_ISL_2768324, EPI_ISL_2768325, EPI_ISL_2768326, EPI_ISL_2768327, EPI_ISL_2768328, EPI_ISL_2768329, EPI_ISL_2768330, EPI_ISL_2768331, EPI_ISL_2768332, EPI_ISL_2768333, EPI_ISL_2768334, EPI_ISL_2768335, EPI_ISL_2768336, EPI_ISL_2768337, EPI_ISL_2768338, EPI_ISL_2768339, EPI_ISL_2768340, EPI_ISL_2768341, EPI_ISL_2768342, EPI_ISL_2768343, EPI_ISL_2768344, EPI_ISL_2768345, EPI_ISL_2768346, EPI_ISL_2768347, EPI_ISL_2768348, EPI_ISL_2768349, EPI_ISL_2768350, EPI_ISL_2768351, EPI_ISL_2768352, EPI_ISL_2768353, EPI_ISL_2768354, EPI_ISL_2768355, EPI_ISL_2768356, EPI_ISL_2768357, EPI_ISL_2768358, EPI_ISL_2768359, EPI_ISL_2768360, EPI_ISL_2768361, EPI_ISL_2768362, EPI_ISL_2768363, EPI_ISL_2768364, EPI_ISL_2768366, EPI_ISL_2768367, EPI_ISL_2768368, EPI_ISL_2768370, EPI_ISL_2768371, EPI_ISL_2768372, EPI_ISL_2768373, EPI_ISL_2768374, EPI_ISL_2768375, EPI_ISL_2768385, EPI_ISL_2768386, EPI_ISL_2768387, EPI_ISL_2768388, EPI_ISL_2768847, EPI_ISL_2768849, EPI_ISL_2768850, EPI_ISL_2768851, EPI_ISL_2768852, EPI_ISL_2768853, EPI_ISL_2768854, EPI_ISL_2768855, EPI_ISL_2768856, EPI_ISL_2768857, EPI_ISL_2768858, EPI_ISL_2768859, EPI_ISL_2768860, EPI_ISL_2768861, EPI_ISL_2768862, EPI_ISL_2768863, EPI_ISL_2768864, EPI_ISL_2768865, EPI_ISL_2768866, EPI_ISL_2768867, EPI_ISL_2768868, EPI_ISL_2768869, EPI_ISL_2768870, EPI_ISL_2768871, EPI_ISL_2770683, EPI_ISL_2770684, EPI_ISL_2770685, EPI_ISL_2770686, EPI_ISL_2770687, EPI_ISL_2770688, EPI_ISL_2770689, EPI_ISL_2770690, EPI_ISL_2770691, EPI_ISL_2770692, EPI_ISL_2770693, EPI_ISL_2770694, EPI_ISL_2770695, EPI_ISL_2770696, EPI_ISL_2770697, EPI_ISL_2770698, EPI_ISL_2770700, EPI_ISL_2770701, EPI_ISL_2770702, EPI_ISL_2770703, EPI_ISL_2770704, EPI_ISL_2770705, EPI_ISL_2770706, EPI_ISL_2770743, EPI_ISL_2770744, EPI_ISL_2770745, EPI_ISL_2770746, EPI_ISL_2770747, EPI_ISL_2770748, EPI_ISL_2770749, EPI_ISL_2770750, EPI_ISL_2770751, EPI_ISL_2770752, EPI_ISL_2770753, EPI_ISL_2770754, EPI_ISL_2770756, EPI_ISL_2770757, EPI_ISL_2771542, EPI_ISL_2771543, EPI_ISL_2771544, EPI_ISL_2771545, EPI_ISL_2771546, EPI_ISL_2771547, EPI_ISL_2771548, EPI_ISL_2771549, EPI_ISL_2771554, EPI_ISL_2771555, EPI_ISL_2771556, EPI_ISL_2771557, EPI_ISL_2771558, EPI_ISL_2771559, EPI_ISL_2771560, EPI_ISL_2771561, EPI_ISL_2771562, EPI_ISL_2771563, EPI_ISL_2771564, EPI_ISL_2771565, EPI_ISL_2771566, EPI_ISL_2771567, EPI_ISL_2771568, EPI_ISL_2771569, EPI_ISL_2771570, EPI_ISL_2771571, EPI_ISL_2771572, EPI_ISL_2771573, EPI_ISL_2771574, EPI_ISL_2772109, EPI_ISL_2772110, EPI_ISL_2772111, EPI_ISL_2772112, EPI_ISL_2772113, EPI_ISL_2772114, EPI_ISL_2772115, EPI_ISL_2772116, EPI_ISL_2772117, EPI_ISL_2772118, EPI_ISL_2772119, EPI_ISL_2772120, EPI_ISL_2772121, EPI_ISL_2772122, EPI_ISL_2772123, EPI_ISL_2772124, EPI_ISL_2772125, EPI_ISL_2772126, EPI_ISL_2772127, EPI_ISL_2772128, EPI_ISL_2772129, EPI_ISL_2777167, EPI_ISL_2777168, EPI_ISL_2777169, EPI_ISL_2777170, EPI_ISL_2777171, EPI_ISL_2777172, EPI_ISL_2777173, EPI_ISL_2777174, EPI_ISL_2777175, EPI_ISL_2777176, EPI_ISL_2777177, EPI_ISL_2777178, EPI_ISL_2777179, EPI_ISL_2777180, EPI_ISL_2777181, EPI_ISL_2777182, EPI_ISL_2777183, EPI_ISL_2777192, EPI_ISL_2777193, EPI_ISL_2777194, EPI_ISL_2777195, EPI_ISL_2777196, EPI_ISL_2777197, EPI_ISL_2777198, EPI_ISL_2777199, EPI_ISL_2777200, EPI_ISL_2777201, EPI_ISL_2777202, EPI_ISL_2777203, EPI_ISL_2777204, EPI_ISL_2777205, EPI_ISL_2777206, EPI_ISL_2777207, EPI_ISL_2777208, EPI_ISL_2777209, EPI_ISL_2777210, EPI_ISL_2932277, EPI_ISL_2932278, EPI_ISL_2932279, EPI_ISL_2932305, EPI_ISL_2932306, EPI_ISL_2932307, EPI_ISL_2932308, EPI_ISL_2932315 | | | |
| see above | Pathogen Genomics Center, National Institute of Infectious Diseases | Pathogen Genomics Center, National Institute of Infectious Diseases | Kentaro Itokawa; Makoto Kuroda; Masanori Hashino; Rina Tanaka; Tsuyoshi Sekizuka |
| EPI_ISL_2447608, EPI_ISL_2447609, EPI_ISL_2447611, EPI_ISL_2447613, EPI_ISL_2447615, EPI_ISL_2447620, EPI_ISL_2447623, EPI_ISL_2447625, EPI_ISL_2447628, EPI_ISL_2447631, EPI_ISL_2447634, EPI_ISL_2447636, EPI_ISL_2447639, EPI_ISL_2447642, EPI_ISL_2447644, EPI_ISL_2447648, EPI_ISL_2447650, EPI_ISL_2447653, EPI_ISL_2447656, EPI_ISL_2447659, EPI_ISL_2447662, EPI_ISL_2447664, EPI_ISL_2447667, EPI_ISL_2447670, EPI_ISL_2447673, EPI_ISL_2447676, EPI_ISL_2447678, EPI_ISL_2447680, EPI_ISL_2447683, EPI_ISL_2447686, EPI_ISL_2447689, EPI_ISL_2447692, EPI_ISL_2447694, EPI_ISL_2447697, EPI_ISL_2447700, EPI_ISL_2447988, EPI_ISL_2621738, EPI_ISL_2621739, EPI_ISL_2621740, EPI_ISL_2621741, EPI_ISL_2621742, EPI_ISL_2621743, EPI_ISL_2621744, EPI_ISL_2621745, EPI_ISL_2621746, EPI_ISL_2621747, EPI_ISL_2621748, EPI_ISL_2621749, EPI_ISL_2621750, EPI_ISL_2621751, EPI_ISL_2621752, EPI_ISL_2621753, EPI_ISL_2621754, EPI_ISL_2621755, EPI_ISL_2621756, EPI_ISL_2621757, EPI_ISL_2621758, EPI_ISL_2621759, EPI_ISL_2621760, EPI_ISL_2621761, EPI_ISL_2621762, EPI_ISL_2621763, EPI_ISL_2621764, EPI_ISL_2621765, EPI_ISL_2621766, EPI_ISL_2621767, EPI_ISL_2621768, EPI_ISL_2621769, EPI_ISL_2621770, EPI_ISL_2621771, EPI_ISL_2621772, EPI_ISL_2621773, EPI_ISL_2621774, EPI_ISL_2621775, EPI_ISL_2621776, EPI_ISL_2621777, EPI_ISL_2621778, EPI_ISL_2621779, EPI_ISL_2621780, EPI_ISL_2621781, EPI_ISL_2621782, EPI_ISL_2674806, EPI_ISL_2674807, EPI_ISL_2674808, EPI_ISL_2674809, EPI_ISL_2674810, EPI_ISL_2674811, EPI_ISL_2760890, EPI_ISL_2762823, EPI_ISL_2762824, EPI_ISL_2762826, EPI_ISL_2763460, EPI_ISL_2763461, EPI_ISL_2763462, EPI_ISL_2763788, EPI_ISL_2763789, EPI_ISL_2763790, EPI_ISL_2763791, EPI_ISL_2763792, EPI_ISL_2763793, EPI_ISL_2763823, EPI_ISL_2763862, EPI_ISL_2763866, EPI_ISL_2763867, EPI_ISL_2763926, EPI_ISL_2763927, EPI_ISL_2763928, EPI_ISL_2763929, EPI_ISL_2763930, EPI_ISL_2763931, EPI_ISL_2763932, EPI_ISL_2763933, EPI_ISL_2763934, EPI_ISL_2763935, EPI_ISL_2763936, EPI_ISL_2763937, EPI_ISL_2763938, EPI_ISL_2764104, EPI_ISL_2764105, EPI_ISL_2764106, EPI_ISL_2764180, EPI_ISL_2764181, EPI_ISL_2764182, EPI_ISL_2764183, EPI_ISL_2764184, EPI_ISL_2764185, EPI_ISL_2764186, EPI_ISL_2764187, EPI_ISL_2764188, EPI_ISL_2764189, EPI_ISL_2764616, EPI_ISL_2764617, EPI_ISL_2764622, EPI_ISL_2764623, EPI_ISL_2764624, EPI_ISL_2764633, EPI_ISL_2764634, EPI_ISL_2764635, EPI_ISL_2764636, EPI_ISL_2764638, EPI_ISL_2764639, EPI_ISL_2764654, EPI_ISL_2764655, EPI_ISL_2764656, EPI_ISL_2764657, EPI_ISL_2764666, EPI_ISL_2764667, EPI_ISL_2764668, EPI_ISL_2764669, EPI_ISL_2764670, EPI_ISL_2764671, EPI_ISL_2764672, EPI_ISL_2764673, EPI_ISL_2764674, EPI_ISL_2764675, EPI_ISL_2764676, EPI_ISL_2764677, EPI_ISL_2764750, EPI_ISL_2764751, EPI_ISL_2764762, EPI_ISL_2764763, EPI_ISL_2764780, EPI_ISL_2764784, EPI_ISL_2764785, EPI_ISL_2764788, EPI_ISL_2764789, EPI_ISL_2764792, EPI_ISL_2764839, EPI_ISL_2764840, EPI_ISL_2764841, EPI_ISL_2764847, EPI_ISL_2764848, EPI_ISL_2764849, EPI_ISL_2764850, EPI_ISL_2764851, EPI_ISL_2764852, EPI_ISL_2764853, EPI_ISL_2764887, EPI_ISL_2764888, EPI_ISL_2764889, EPI_ISL_2764890, EPI_ISL_2764891, EPI_ISL_2764892, EPI_ISL_2764893, EPI_ISL_2764894, EPI_ISL_2764896, EPI_ISL_2764897, EPI_ISL_2764898, EPI_ISL_2764899, EPI_ISL_2764900, EPI_ISL_2764901, EPI_ISL_2764902, EPI_ISL_2764903, EPI_ISL_2764905, EPI_ISL_2764906, EPI_ISL_2764909, EPI_ISL_2764913, EPI_ISL_2764914, EPI_ISL_2764915, EPI_ISL_2764916, EPI_ISL_2764917, EPI_ISL_2764918, EPI_ISL_2764919, EPI_ISL_2764921, EPI_ISL_2764922, EPI_ISL_2764923, EPI_ISL_2764924, EPI_ISL_2764925, EPI_ISL_2764926, EPI_ISL_2764927, EPI_ISL_2764928, EPI_ISL_2764929, EPI_ISL_2764930, EPI_ISL_2764931, EPI_ISL_2764932, EPI_ISL_2764933, EPI_ISL_2764934, EPI_ISL_2764935, EPI_ISL_2764936, EPI_ISL_2764937, EPI_ISL_2764938, EPI_ISL_2764939, EPI_ISL_2764940, EPI_ISL_2764941, EPI_ISL_2764942, EPI_ISL_2764943, EPI_ISL_2764944, EPI_ISL_2764945, EPI_ISL_2764946, EPI_ISL_2764947, EPI_ISL_2764948, EPI_ISL_2764960, EPI_ISL_2764961, EPI_ISL_2764962, EPI_ISL_2764963, EPI_ISL_2764964, EPI_ISL_2764987, EPI_ISL_2764988, EPI_ISL_2764989, EPI_ISL_2764990, EPI_ISL_2764991, EPI_ISL_2764992, EPI_ISL_2764993, EPI_ISL_2764994, EPI_ISL_2764995, EPI_ISL_2764996, EPI_ISL_2765160, EPI_ISL_2765161, EPI_ISL_2765162, EPI_ISL_2765163, EPI_ISL_2765164, EPI_ISL_2765165, EPI_ISL_2765166, EPI_ISL_2765257, EPI_ISL_2765258, EPI_ISL_2765259, EPI_ISL_2765260, EPI_ISL_2765261, EPI_ISL_2765262, EPI_ISL_2765263, EPI_ISL_2765264, EPI_ISL_2765265, EPI_ISL_2765266, EPI_ISL_2765272, EPI_ISL_2765273, EPI_ISL_2765274, EPI_ISL_2765275, EPI_ISL_2765276, EPI_ISL_2765277, EPI_ISL_2765278, EPI_ISL_2765279, EPI_ISL_2765285, EPI_ISL_2765286, EPI_ISL_2765287, EPI_ISL_2765288, EPI_ISL_2765290, EPI_ISL_2765291, EPI_ISL_2765292, EPI_ISL_2765293, EPI_ISL_2765294, EPI_ISL_2765295, EPI_ISL_2765296, EPI_ISL_2765297, EPI_ISL_2765298, EPI_ISL_2765303, EPI_ISL_2765304, EPI_ISL_2765305, EPI_ISL_2765306, EPI_ISL_2765307, EPI_ISL_2765308, EPI_ISL_2765309, EPI_ISL_2765310, EPI_ISL_2765311, EPI_ISL_2765312, EPI_ISL_2765313, EPI_ISL_2765314, EPI_ISL_2765315, EPI_ISL_2765316, EPI_ISL_2765317, EPI_ISL_2765318, EPI_ISL_2765319, EPI_ISL_2765320, EPI_ISL_2765321, EPI_ISL_2765322, EPI_ISL_2765323, EPI_ISL_2765324, EPI_ISL_2765325, EPI_ISL_2765326, EPI_ISL_2765327, EPI_ISL_2765328, EPI_ISL_2765334, EPI_ISL_2765335, EPI_ISL_2765336, EPI_ISL_2765431, EPI_ISL_2765432, EPI_ISL_2765433, EPI_ISL_2765434, EPI_ISL_2765435, EPI_ISL_2765436, EPI_ISL_2765437, EPI_ISL_2765438, EPI_ISL_2765439, EPI_ISL_2765440, EPI_ISL_2765441, EPI_ISL_2765442, EPI_ISL_2765443, EPI_ISL_2765502, EPI_ISL_2765503, EPI_ISL_2765504, EPI_ISL_2765543, EPI_ISL_2765544, EPI_ISL_2768105, EPI_ISL_2768106, EPI_ISL_2768108, EPI_ISL_2768109, EPI_ISL_2768110, EPI_ISL_2768111, EPI_ISL_2768113, EPI_ISL_2768114, EPI_ISL_2768120, EPI_ISL_2768121, EPI_ISL_2768122, EPI_ISL_2768123, EPI_ISL_2768124, EPI_ISL_2768125, EPI_ISL_2768126, EPI_ISL_2768127, EPI_ISL_2768163, EPI_ISL_2768164, EPI_ISL_2768165, EPI_ISL_2768166, EPI_ISL_2768167, EPI_ISL_2768168, EPI_ISL_2768169, EPI_ISL_2768170, EPI_ISL_2768171, EPI_ISL_2768172, EPI_ISL_2768173, EPI_ISL_2768174, EPI_ISL_2768175, EPI_ISL_2768176, EPI_ISL_2768177, EPI_ISL_2768178, EPI_ISL_2768179, EPI_ISL_2768180, EPI_ISL_2768181, EPI_ISL_2768182, EPI_ISL_2768183, EPI_ISL_2768184, EPI_ISL_2768192, EPI_ISL_2768199, EPI_ISL_2768200, EPI_ISL_2768201, EPI_ISL_2768202, EPI_ISL_2768203, EPI_ISL_2768204, EPI_ISL_2768205, EPI_ISL_2768206, EPI_ISL_2768207, EPI_ISL_2768208, EPI_ISL_2768209, EPI_ISL_2768210, EPI_ISL_2768211, EPI_ISL_2768212, EPI_ISL_2768213, EPI_ISL_2768214, EPI_ISL_2768215, EPI_ISL_2768216, EPI_ISL_2768217, EPI_ISL_2768218, EPI_ISL_2768219, EPI_ISL_2768220, EPI_ISL_2768222, EPI_ISL_2768223, EPI_ISL_2768224, EPI_ISL_2768225, EPI_ISL_2768226, EPI_ISL_2768227, EPI_ISL_2768228, EPI_ISL_2768517, EPI_ISL_2768518, EPI_ISL_2768519, EPI_ISL_2768520, EPI_ISL_2768521, EPI_ISL_2768522, EPI_ISL_2768523, EPI_ISL_2768524, EPI_ISL_2768525, EPI_ISL_2768526, EPI_ISL_2768527, EPI_ISL_2768535, EPI_ISL_2768536, EPI_ISL_2768538, EPI_ISL_2768540, EPI_ISL_2768541, EPI_ISL_2768542, EPI_ISL_2768543, EPI_ISL_2768544, EPI_ISL_2768545, EPI_ISL_2768546, EPI_ISL_2768547, EPI_ISL_2768548, EPI_ISL_2768549, EPI_ISL_2768550, EPI_ISL_2768551, EPI_ISL_2768552, EPI_ISL_2768553, EPI_ISL_2768554, EPI_ISL_2768555, EPI_ISL_2768556, EPI_ISL_2768558, EPI_ISL_2768559, EPI_ISL_2768601, EPI_ISL_2768602, EPI_ISL_2768603, EPI_ISL_2768604, EPI_ISL_2768605, EPI_ISL_2768606, EPI_ISL_2768607, EPI_ISL_2768608, EPI_ISL_2768609, EPI_ISL_2768610, EPI_ISL_2768611, EPI_ISL_2768612, EPI_ISL_2768613, EPI_ISL_2768614, EPI_ISL_2768615, EPI_ISL_2768616, EPI_ISL_2768617, EPI_ISL_2768618, EPI_ISL_2768619, EPI_ISL_2768620, EPI_ISL_2768621, EPI_ISL_2768622, EPI_ISL_2768623, EPI_ISL_2768624, EPI_ISL_2768625, EPI_ISL_2768628, EPI_ISL_2768629, EPI_ISL_2768637, EPI_ISL_2768638, EPI_ISL_2768771, EPI_ISL_2768772, EPI_ISL_2768773, EPI_ISL_2768774, EPI_ISL_2768775, EPI_ISL_2768776, EPI_ISL_2768777, EPI_ISL_2768778, EPI_ISL_2768779, EPI_ISL_2768780, EPI_ISL_2768808, EPI_ISL_2768809, EPI_ISL_2768810, EPI_ISL_2768811, EPI_ISL_2768812, EPI_ISL_2768813, EPI_ISL_2768814, EPI_ISL_2768815, EPI_ISL_2768816, EPI_ISL_2768817, EPI_ISL_2768818, EPI_ISL_2768819, EPI_ISL_2768820, EPI_ISL_2768821, EPI_ISL_2768822, EPI_ISL_2768823, EPI_ISL_2768824, EPI_ISL_2768825, EPI_ISL_2768826, EPI_ISL_2768827, EPI_ISL_2768828, EPI_ISL_2768829, EPI_ISL_2768830, EPI_ISL_2768831, EPI_ISL_2768832, EPI_ISL_2768833, EPI_ISL_2768834, EPI_ISL_2768835, EPI_ISL_2768836, EPI_ISL_2768837, EPI_ISL_2768838, EPI_ISL_2769018, EPI_ISL_2769019, EPI_ISL_2769020, EPI_ISL_2769021, EPI_ISL_2769022, EPI_ISL_2769023, EPI_ISL_2769024, EPI_ISL_2769025, EPI_ISL_2769026, EPI_ISL_2769027, EPI_ISL_2769028, EPI_ISL_2769029, EPI_ISL_2769067, EPI_ISL_2769068, EPI_ISL_2769069, EPI_ISL_2769070, EPI_ISL_2769071, EPI_ISL_2769072, EPI_ISL_2769073, EPI_ISL_2769074, EPI_ISL_2769075, EPI_ISL_2769076, EPI_ISL_2769077, EPI_ISL_2769078, EPI_ISL_2769079, EPI_ISL_2769080, EPI_ISL_2769081, EPI_ISL_2769082, EPI_ISL_2769083, EPI_ISL_2769084, EPI_ISL_2769085, EPI_ISL_2769086, EPI_ISL_2769087, EPI_ISL_2769088, EPI_ISL_2769089, EPI_ISL_2769090, EPI_ISL_2769091, EPI_ISL_2769092, EPI_ISL_2769093, EPI_ISL_2769094, EPI_ISL_2769096, EPI_ISL_2769097, EPI_ISL_2769098, EPI_ISL_2769099, EPI_ISL_2769100, EPI_ISL_2769101, EPI_ISL_2769102, EPI_ISL_2769103, EPI_ISL_2769104, EPI_ISL_2769439, EPI_ISL_2769440, EPI_ISL_2769441, EPI_ISL_2769442, EPI_ISL_2769443, EPI_ISL_2769444, EPI_ISL_2769445, EPI_ISL_2769446, EPI_ISL_2769447, EPI_ISL_2769448, EPI_ISL_2769449, EPI_ISL_2769450, EPI_ISL_2769451, EPI_ISL_2769452, EPI_ISL_2769453, EPI_ISL_2769454, EPI_ISL_2769455, EPI_ISL_2769456, EPI_ISL_2769457, EPI_ISL_2769458, EPI_ISL_2769459, EPI_ISL_2769460, EPI_ISL_2769461, EPI_ISL_2769462, EPI_ISL_2769463, EPI_ISL_2769464, EPI_ISL_2769465, EPI_ISL_2769466, EPI_ISL_2769467, EPI_ISL_2769468, EPI_ISL_2769469, EPI_ISL_2769470, EPI_ISL_2769471, EPI_ISL_2769472, EPI_ISL_2769473, EPI_ISL_2769474, EPI_ISL_2769475, EPI_ISL_2769601, EPI_ISL_2769602, EPI_ISL_2769603, EPI_ISL_2769604, EPI_ISL_2769605, EPI_ISL_2769606, EPI_ISL_2769607, EPI_ISL_2769608, EPI_ISL_2769609, EPI_ISL_2769610, EPI_ISL_2769611, EPI_ISL_2769612, EPI_ISL_2769613, EPI_ISL_2769614, EPI_ISL_2769615, EPI_ISL_2769616, EPI_ISL_2769617, EPI_ISL_2769618, EPI_ISL_2769619, EPI_ISL_2769620, EPI_ISL_2769621, EPI_ISL_2769622, EPI_ISL_2769623, EPI_ISL_2769624, EPI_ISL_2769625, EPI_ISL_2769626, EPI_ISL_2769627, EPI_ISL_2769628, EPI_ISL_2769629, EPI_ISL_2769631, EPI_ISL_2769632, EPI_ISL_2769633, EPI_ISL_2769634, EPI_ISL_2769635, EPI_ISL_2769636, EPI_ISL_2769637, EPI_ISL_2769638, EPI_ISL_2769639, EPI_ISL_2769640, EPI_ISL_2769641, EPI_ISL_2769642, EPI_ISL_2769643, EPI_ISL_2769644, EPI_ISL_2769645, EPI_ISL_2769646, EPI_ISL_2769647, EPI_ISL_2769648, EPI_ISL_2769649, EPI_ISL_2769650, EPI_ISL_2769651, EPI_ISL_2769652, EPI_ISL_2769653, EPI_ISL_2769654, EPI_ISL_2769655, EPI_ISL_2769656, EPI_ISL_2769657, EPI_ISL_2769658, EPI_ISL_2769667, EPI_ISL_2769676, EPI_ISL_2769677, EPI_ISL_2769678, EPI_ISL_2769679, EPI_ISL_2769680, EPI_ISL_2769681, EPI_ISL_2769682, EPI_ISL_2769688, EPI_ISL_2769689, EPI_ISL_2769690, EPI_ISL_2769691, EPI_ISL_2769692, EPI_ISL_2769693, EPI_ISL_2769694, EPI_ISL_2769695, EPI_ISL_2769696, EPI_ISL_2769702, EPI_ISL_2769703, EPI_ISL_2769704, EPI_ISL_2769705, EPI_ISL_2769706, EPI_ISL_2769707, EPI_ISL_2769708, EPI_ISL_2769709, EPI_ISL_2769710, EPI_ISL_2769711, EPI_ISL_2769712, EPI_ISL_2769713, EPI_ISL_2769714, EPI_ISL_2769715, EPI_ISL_2769716, EPI_ISL_2769723, EPI_ISL_2769724, EPI_ISL_2769725, EPI_ISL_2769726, EPI_ISL_2769727, EPI_ISL_2769728, EPI_ISL_2769797, EPI_ISL_2769798, EPI_ISL_2769799, EPI_ISL_2769800, EPI_ISL_2769801, EPI_ISL_2769802, EPI_ISL_2769803, EPI_ISL_2769804, EPI_ISL_2769805, EPI_ISL_2769806, EPI_ISL_2769808, EPI_ISL_2769809, EPI_ISL_2769810, EPI_ISL_2769811, EPI_ISL_2769812, EPI_ISL_2769813, EPI_ISL_2769814, EPI_ISL_2769815, EPI_ISL_2769816, EPI_ISL_2769817, EPI_ISL_2769818, EPI_ISL_2769819, EPI_ISL_2769820, EPI_ISL_2769821, EPI_ISL_2769822, EPI_ISL_2769823, EPI_ISL_2769824, EPI_ISL_2769825, EPI_ISL_2769826, EPI_ISL_2769827, EPI_ISL_2769833, EPI_ISL_2769834, EPI_ISL_2769835, EPI_ISL_2769838, EPI_ISL_2769839, EPI_ISL_2769840, EPI_ISL_2769841, EPI_ISL_2769842, EPI_ISL_2769843, EPI_ISL_2769844, EPI_ISL_2769845, EPI_ISL_2769846, EPI_ISL_2769847, EPI_ISL_2769848, EPI_ISL_2769849, EPI_ISL_2769862, EPI_ISL_2769863, EPI_ISL_2769864, EPI_ISL_2769865, EPI_ISL_2769866, EPI_ISL_2769867, EPI_ISL_2769868, EPI_ISL_2769869, EPI_ISL_2769870, EPI_ISL_2769871, EPI_ISL_2769872, EPI_ISL_2769873, EPI_ISL_2769874, EPI_ISL_2769875, EPI_ISL_2769876, EPI_ISL_2769877, EPI_ISL_2769878, EPI_ISL_2769879, EPI_ISL_2769880, EPI_ISL_2769881, EPI_ISL_2769882, EPI_ISL_2769883, EPI_ISL_2769884, EPI_ISL_2769885, EPI_ISL_2769886, EPI_ISL_2769887, EPI_ISL_2769888, EPI_ISL_2769889, EPI_ISL_2769890, EPI_ISL_2769891, EPI_ISL_2769892, EPI_ISL_2769893, EPI_ISL_2769894, EPI_ISL_2769895, EPI_ISL_2769896, EPI_ISL_2769898, EPI_ISL_2769899, EPI_ISL_2769900, EPI_ISL_2769901, EPI_ISL_2769902, EPI_ISL_2769903, EPI_ISL_2769904, EPI_ISL_2769905, EPI_ISL_2769906, EPI_ISL_2769907, EPI_ISL_2769908, EPI_ISL_2769910, EPI_ISL_2769911, EPI_ISL_2769912, EPI_ISL_2769913, EPI_ISL_2769915, EPI_ISL_2769916, EPI_ISL_2769917, EPI_ISL_2769918, EPI_ISL_2769919, EPI_ISL_2769920, EPI_ISL_2769921, EPI_ISL_2769922, EPI_ISL_2769923, EPI_ISL_2769924, EPI_ISL_2769925, EPI_ISL_2769928, EPI_ISL_2769929, EPI_ISL_2769930, EPI_ISL_2769931, EPI_ISL_2769936, EPI_ISL_2769937, EPI_ISL_2769939, EPI_ISL_2769942, EPI_ISL_2769943, EPI_ISL_2769944, EPI_ISL_2769945, EPI_ISL_2769946, EPI_ISL_2769947, EPI_ISL_2769948, EPI_ISL_2769954, EPI_ISL_2769955, EPI_ISL_2769956, EPI_ISL_2769957, EPI_ISL_2769958, EPI_ISL_2769959, EPI_ISL_2769960, EPI_ISL_2769961, EPI_ISL_2770022, EPI_ISL_2770023, EPI_ISL_2770024, EPI_ISL_2770025, EPI_ISL_2770026, EPI_ISL_2770027, EPI_ISL_2770028, EPI_ISL_2770029, EPI_ISL_2770030, EPI_ISL_2770031, EPI_ISL_2770032, EPI_ISL_2770033, EPI_ISL_2770034, EPI_ISL_2770035, EPI_ISL_2770036, EPI_ISL_2770037, EPI_ISL_2770038, EPI_ISL_2770039, EPI_ISL_2770040, EPI_ISL_2770041, EPI_ISL_2770042, EPI_ISL_2770043, EPI_ISL_2770044, EPI_ISL_2770045, EPI_ISL_2770046, EPI_ISL_2770047, EPI_ISL_2770048, EPI_ISL_2770049, EPI_ISL_2770050, EPI_ISL_2770051, EPI_ISL_2770052, EPI_ISL_2770053, EPI_ISL_2770054, EPI_ISL_2770055, EPI_ISL_2770056, EPI_ISL_2770057, EPI_ISL_2770058, EPI_ISL_2770059, EPI_ISL_2770060, EPI_ISL_2770061, EPI_ISL_2770062, EPI_ISL_2770063, EPI_ISL_2770064, EPI_ISL_2770065, EPI_ISL_2770066, EPI_ISL_2770067, EPI_ISL_2770068, EPI_ISL_2770069, EPI_ISL_2770070, EPI_ISL_2770071, EPI_ISL_2770072, EPI_ISL_2770073, EPI_ISL_2770074, EPI_ISL_2770075, EPI_ISL_2770076, EPI_ISL_2770077, EPI_ISL_2770078, EPI_ISL_2770079, EPI_ISL_2770080, EPI_ISL_2770081, EPI_ISL_2770082, EPI_ISL_2770083, EPI_ISL_2770084, EPI_ISL_2770085, EPI_ISL_2770086, EPI_ISL_2770087, EPI_ISL_2770088, EPI_ISL_2770089, EPI_ISL_2770090, EPI_ISL_2770091, EPI_ISL_2770092, EPI_ISL_2770093, EPI_ISL_2770094, EPI_ISL_2770095, EPI_ISL_2770096, EPI_ISL_2770097, EPI_ISL_2770098, EPI_ISL_2770099, EPI_ISL_2770100, EPI_ISL_2770101, EPI_ISL_2770102, EPI_ISL_2770103, EPI_ISL_2770104, EPI_ISL_2770105, EPI_ISL_2770106, EPI_ISL_2770107, EPI_ISL_2770108, EPI_ISL_2770109, EPI_ISL_2770110, EPI_ISL_2770111, EPI_ISL_2770112, EPI_ISL_2770113, EPI_ISL_2770114, EPI_ISL_2770115, EPI_ISL_2770116, EPI_ISL_2770117, EPI_ISL_2770118, EPI_ISL_2770119, EPI_ISL_2770120, EPI_ISL_2770121, EPI_ISL_2770122, EPI_ISL_2770123, EPI_ISL_2770124, EPI_ISL_2770125, EPI_ISL_2770126, EPI_ISL_2770127, EPI_ISL_2770128, EPI_ISL_2770129, EPI_ISL_2770130, EPI_ISL_2770131, EPI_ISL_2770132, EPI_ISL_2770133, EPI_ISL_2770134, EPI_ISL_2770135, EPI_ISL_2770136, EPI_ISL_2770137, EPI_ISL_2770138, EPI_ISL_2770139, EPI_ISL_2770140, EPI_ISL_2770141, EPI_ISL_2770142, EPI_ISL_2770143, EPI_ISL_2770144, EPI_ISL_2770145, EPI_ISL_2770147, EPI_ISL_2770148, EPI_ISL_2770149, EPI_ISL_2770150, EPI_ISL_2770151, EPI_ISL_2770152, EPI_ISL_2770153, EPI_ISL_2770154, EPI_ISL_2770155, EPI_ISL_2770156, EPI_ISL_2770157, EPI_ISL_2770158, EPI_ISL_2770159, EPI_ISL_2770160, EPI_ISL_2770161, EPI_ISL_2770162, EPI_ISL_2770163, EPI_ISL_2770164, EPI_ISL_2770165, EPI_ISL_2770166, EPI_ISL_2770167, EPI_ISL_2770168, EPI_ISL_2770169, EPI_ISL_2770170, EPI_ISL_2770171, EPI_ISL_2770172, EPI_ISL_2770173, EPI_ISL_2770174, EPI_ISL_2770175, EPI_ISL_2770176, EPI_ISL_2770177, EPI_ISL_2770178, EPI_ISL_2770179, EPI_ISL_2770180, EPI_ISL_2770181, EPI_ISL_2770182, EPI_ISL_2770183, EPI_ISL_2770184, EPI_ISL_2770185, EPI_ISL_2770186, EPI_ISL_2770187, EPI_ISL_2770188, EPI_ISL_2770189, EPI_ISL_2770190, EPI_ISL_2770191, EPI_ISL_2770192, EPI_ISL_2770193, EPI_ISL_2770194, EPI_ISL_2770195, EPI_ISL_2770196, EPI_ISL_2770197, EPI_ISL_2770198, EPI_ISL_2770199, EPI_ISL_2770200, EPI_ISL_2770201, EPI_ISL_2770202, EPI_ISL_2770203, EPI_ISL_2770212, EPI_ISL_2770213, EPI_ISL_2770214, EPI_ISL_2770215, EPI_ISL_2770216, EPI_ISL_2770217, EPI_ISL_2770218, EPI_ISL_2770219, EPI_ISL_2770242, EPI_ISL_2770243, EPI_ISL_2770244, EPI_ISL_2770245, EPI_ISL_2770246, EPI_ISL_2770248, EPI_ISL_2770249, EPI_ISL_2770250, EPI_ISL_2770251, EPI_ISL_2770252, EPI_ISL_2770254, EPI_ISL_2770402, EPI_ISL_2770758, EPI_ISL_2770759, EPI_ISL_2770760, EPI_ISL_2770761, EPI_ISL_2770762, EPI_ISL_2770763, EPI_ISL_2770764, EPI_ISL_2770765, EPI_ISL_2770766, EPI_ISL_2770767, EPI_ISL_2770768, EPI_ISL_2770769, EPI_ISL_2770770, EPI_ISL_2770771, EPI_ISL_2770772, EPI_ISL_2770773, EPI_ISL_2770774, EPI_ISL_2770775, EPI_ISL_2770776, EPI_ISL_2770777, EPI_ISL_2770778, EPI_ISL_2770779, EPI_ISL_2770780, EPI_ISL_2770781, EPI_ISL_2770782, EPI_ISL_2770783, EPI_ISL_2770784, EPI_ISL_2770785, EPI_ISL_2770786, EPI_ISL_2770787, EPI_ISL_2770788, EPI_ISL_2770789, EPI_ISL_2770790, EPI_ISL_2770791, EPI_ISL_2770792, EPI_ISL_2770793, EPI_ISL_2770794, EPI_ISL_2770795, EPI_ISL_2770796, EPI_ISL_2770797, EPI_ISL_2770798, EPI_ISL_2770799, EPI_ISL_2770800, EPI_ISL_2770801, EPI_ISL_2770802, EPI_ISL_2770803, EPI_ISL_2770804, EPI_ISL_2770805, EPI_ISL_2770806, EPI_ISL_2770807, EPI_ISL_2770808, EPI_ISL_2770809, EPI_ISL_2770810, EPI_ISL_2770811, EPI_ISL_2770812, EPI_ISL_2770813, EPI_ISL_2770814, EPI_ISL_2770815, EPI_ISL_2770816, EPI_ISL_2770817, EPI_ISL_2770818, EPI_ISL_2770819, EPI_ISL_2770820, EPI_ISL_2770821, EPI_ISL_2770822, EPI_ISL_2770823, EPI_ISL_2770824, EPI_ISL_2770825, EPI_ISL_2770826, EPI_ISL_2770827, EPI_ISL_2770828, EPI_ISL_2770829, EPI_ISL_2770830, EPI_ISL_2770831, EPI_ISL_2770832, EPI_ISL_2770888, EPI_ISL_2770889, EPI_ISL_2770890, EPI_ISL_2770891, EPI_ISL_2770892, EPI_ISL_2770893, EPI_ISL_2770894, EPI_ISL_2770895, EPI_ISL_2770896, EPI_ISL_2770897, EPI_ISL_2770898, EPI_ISL_2770899, EPI_ISL_2770900, EPI_ISL_2770901, EPI_ISL_2770902, EPI_ISL_2770903, EPI_ISL_2770904, EPI_ISL_2770905, EPI_ISL_2770906, EPI_ISL_2770907, EPI_ISL_2770908, EPI_ISL_2770909, EPI_ISL_2770910, EPI_ISL_2770911, EPI_ISL_2770912, EPI_ISL_2770913, EPI_ISL_2770914, EPI_ISL_2770915, EPI_ISL_2770916, EPI_ISL_2770917, EPI_ISL_2770918, EPI_ISL_2770919, EPI_ISL_2770920, EPI_ISL_2770921, EPI_ISL_2770922, EPI_ISL_2770923, EPI_ISL_2770924, EPI_ISL_2770925, EPI_ISL_2770926, EPI_ISL_2770927, EPI_ISL_2770928, EPI_ISL_2770929, EPI_ISL_2770930, EPI_ISL_2770931, EPI_ISL_2770932, EPI_ISL_2770933, EPI_ISL_2770934, EPI_ISL_2770935, EPI_ISL_2770936, EPI_ISL_2770937, EPI_ISL_2770938, EPI_ISL_2770939, EPI_ISL_2770940, EPI_ISL_2770941, EPI_ISL_2770942, EPI_ISL_2770943, EPI_ISL_2770944, EPI_ISL_2770945, EPI_ISL_2770946, EPI_ISL_2770947, EPI_ISL_2770948, EPI_ISL_2770949, EPI_ISL_2770950, EPI_ISL_2770951, EPI_ISL_2770952, EPI_ISL_2770953, EPI_ISL_2770954, EPI_ISL_2770955, EPI_ISL_2770956, EPI_ISL_2770957, EPI_ISL_2770958, EPI_ISL_2770959, EPI_ISL_2770960, EPI_ISL_2770961, EPI_ISL_2770962, EPI_ISL_2770963, EPI_ISL_2770964, EPI_ISL_2770965, EPI_ISL_2770966, EPI_ISL_2770967, EPI_ISL_2770968, EPI_ISL_2770969, EPI_ISL_2770970, EPI_ISL_2770971, EPI_ISL_2770972, EPI_ISL_2770973, EPI_ISL_2770974, EPI_ISL_2770975, EPI_ISL_2770976, EPI_ISL_2770977, EPI_ISL_2770978, EPI_ISL_2770979, EPI_ISL_2770980, EPI_ISL_2770981, EPI_ISL_2770982, EPI_ISL_2770983, EPI_ISL_2770984, EPI_ISL_2770985, EPI_ISL_2770986, EPI_ISL_2770987, EPI_ISL_2770988, EPI_ISL_2770989, EPI_ISL_2770990, EPI_ISL_2770991, EPI_ISL_2770992, EPI_ISL_2770993, EPI_ISL_2770994, EPI_ISL_2770995, EPI_ISL_2770996, EPI_ISL_2770997, EPI_ISL_2770998, EPI_ISL_2770999, EPI_ISL_2771000, EPI_ISL_2771001, EPI_ISL_2771002, EPI_ISL_2771003, EPI_ISL_2771004, EPI_ISL_2771005, EPI_ISL_2771006, EPI_ISL_2771007, EPI_ISL_2771008, EPI_ISL_2771009, EPI_ISL_2771010, EPI_ISL_2771011, EPI_ISL_2771012, EPI_ISL_2771013, EPI_ISL_2771014, EPI_ISL_2771115, EPI_ISL_2771128, EPI_ISL_2771129, EPI_ISL_2771150, EPI_ISL_2771151, EPI_ISL_2771152, EPI_ISL_2771153, EPI_ISL_2771155, EPI_ISL_2771156, EPI_ISL_2771157, EPI_ISL_2771158, EPI_ISL_2771159, EPI_ISL_2771160, EPI_ISL_2771161, EPI_ISL_2771162, EPI_ISL_2771163, EPI_ISL_2771164, EPI_ISL_2771165, EPI_ISL_2771166, EPI_ISL_2771167, EPI_ISL_2771168, EPI_ISL_2771169, EPI_ISL_2771170, EPI_ISL_2771171, EPI_ISL_2771172, EPI_ISL_2771173, EPI_ISL_2771174, EPI_ISL_2771175, EPI_ISL_2771176, EPI_ISL_2771177, EPI_ISL_2771178, EPI_ISL_2771179, EPI_ISL_2771180, EPI_ISL_2771181, EPI_ISL_2771182, EPI_ISL_2771183, EPI_ISL_2771184, EPI_ISL_2771186, EPI_ISL_2771187, EPI_ISL_2771188, EPI_ISL_2771189, EPI_ISL_2771190, EPI_ISL_2771191, EPI_ISL_2771192, EPI_ISL_2771193, EPI_ISL_2771194, EPI_ISL_2771195, EPI_ISL_2771196, EPI_ISL_2771197, EPI_ISL_2771198, EPI_ISL_2771199, EPI_ISL_2771200, EPI_ISL_2771201, EPI_ISL_2771202, EPI_ISL_2771203, EPI_ISL_2771204, EPI_ISL_2771205, EPI_ISL_2771206, EPI_ISL_2771207, EPI_ISL_2771208, EPI_ISL_2771209, EPI_ISL_2771210, EPI_ISL_2771389, EPI_ISL_2771390, EPI_ISL_2771391, EPI_ISL_2771392, EPI_ISL_2771393, EPI_ISL_2771394, EPI_ISL_2771396, EPI_ISL_2771397, EPI_ISL_2771398, EPI_ISL_2771399, EPI_ISL_2771400, EPI_ISL_2771401, EPI_ISL_2771402, EPI_ISL_2771403, EPI_ISL_2771404, EPI_ISL_2771405, EPI_ISL_2771406, EPI_ISL_2771407, EPI_ISL_2771408, EPI_ISL_2771409, EPI_ISL_2771410, EPI_ISL_2771411, EPI_ISL_2771412, EPI_ISL_2771413, EPI_ISL_2771414, EPI_ISL_2771415, EPI_ISL_2771416, EPI_ISL_2771417, EPI_ISL_2771418, EPI_ISL_2771419, EPI_ISL_2771420, EPI_ISL_2771421, EPI_ISL_2771422, EPI_ISL_2771423, EPI_ISL_2771424, EPI_ISL_2771425, EPI_ISL_2771426, EPI_ISL_2771427, EPI_ISL_2771428, EPI_ISL_2771429, EPI_ISL_2771430, EPI_ISL_2771431, EPI_ISL_2771432, EPI_ISL_2771433, EPI_ISL_2771434, EPI_ISL_2771435, EPI_ISL_2771436, EPI_ISL_2771437, EPI_ISL_2771438, EPI_ISL_2771439, EPI_ISL_2771440, EPI_ISL_2771441, EPI_ISL_2771442, EPI_ISL_2771443, EPI_ISL_2771444, EPI_ISL_2771445, EPI_ISL_2771446, EPI_ISL_2771447, EPI_ISL_2771448, EPI_ISL_2771449, EPI_ISL_2771450, EPI_ISL_2771451, EPI_ISL_2771452, EPI_ISL_2771453, EPI_ISL_2771454, EPI_ISL_2771455, EPI_ISL_2771456, EPI_ISL_2771457, EPI_ISL_2771458, EPI_ISL_2771459, EPI_ISL_2771460, EPI_ISL_2771461, EPI_ISL_2771462, EPI_ISL_2771463, EPI_ISL_2771464, EPI_ISL_2771465, EPI_ISL_2771466, EPI_ISL_2771467, EPI_ISL_2771468, EPI_ISL_2771469, EPI_ISL_2771470, EPI_ISL_2771471, EPI_ISL_2771472, EPI_ISL_2771473, EPI_ISL_2771474, EPI_ISL_2771475, EPI_ISL_2771476, EPI_ISL_2771477, EPI_ISL_2771478, EPI_ISL_2771479, EPI_ISL_2771480, EPI_ISL_2771481, EPI_ISL_2771482, EPI_ISL_2771483, EPI_ISL_2771484, EPI_ISL_2771485, EPI_ISL_2771486, EPI_ISL_2771487, EPI_ISL_2771488, EPI_ISL_2771489, EPI_ISL_2771490, EPI_ISL_2771491, EPI_ISL_2771500, EPI_ISL_2771501, EPI_ISL_2771502, EPI_ISL_2771503, EPI_ISL_2771504, EPI_ISL_2771505, EPI_ISL_2771506, EPI_ISL_2771507, EPI_ISL_2771508, EPI_ISL_2771509, EPI_ISL_2771510, EPI_ISL_2771511, EPI_ISL_2771512, EPI_ISL_2771513, EPI_ISL_2771514, EPI_ISL_2771515, EPI_ISL_2771516, EPI_ISL_2771517, EPI_ISL_2771518, EPI_ISL_2771519, EPI_ISL_2771575, EPI_ISL_2771576, EPI_ISL_2771577, EPI_ISL_2771578, EPI_ISL_2771579, EPI_ISL_2771580, EPI_ISL_2771581, EPI_ISL_2771582, EPI_ISL_2771583, EPI_ISL_2771584, EPI_ISL_2771585, EPI_ISL_2771586, EPI_ISL_2771587, EPI_ISL_2771588, EPI_ISL_2771589, EPI_ISL_2771590, EPI_ISL_2771591, EPI_ISL_2771592, EPI_ISL_2771593, EPI_ISL_2771594, EPI_ISL_2771595, EPI_ISL_2771596, EPI_ISL_2771597, EPI_ISL_2771598, EPI_ISL_2771599, EPI_ISL_2771600, EPI_ISL_2771601, EPI_ISL_2771602, EPI_ISL_2771603, EPI_ISL_2771604, EPI_ISL_2771605, EPI_ISL_2771606, EPI_ISL_2771607, EPI_ISL_2771608, EPI_ISL_2771609, EPI_ISL_2771610, EPI_ISL_2771611, EPI_ISL_2771612, EPI_ISL_2771613, EPI_ISL_2771614, EPI_ISL_2771615, EPI_ISL_2771616, EPI_ISL_2771617, EPI_ISL_2771618, EPI_ISL_2771619, EPI_ISL_2771620, EPI_ISL_2771621, EPI_ISL_2771622, EPI_ISL_2771623, EPI_ISL_2771624, EPI_ISL_2771625, EPI_ISL_2771626, EPI_ISL_2771627, EPI_ISL_2771628, EPI_ISL_2771629, EPI_ISL_2771630, EPI_ISL_2771631, EPI_ISL_2771632, EPI_ISL_2771633, EPI_ISL_2771634, EPI_ISL_2771635, EPI_ISL_2771636, EPI_ISL_2771637, EPI_ISL_2771638, EPI_ISL_2771639, EPI_ISL_2771641, EPI_ISL_2771642, EPI_ISL_2771643, EPI_ISL_2771644, EPI_ISL_2771645, EPI_ISL_2771646, EPI_ISL_2771923, EPI_ISL_2771924, EPI_ISL_2771935, EPI_ISL_2771936, EPI_ISL_2771937, EPI_ISL_2771938, EPI_ISL_2771939, EPI_ISL_2771940, EPI_ISL_2771941, EPI_ISL_2771942, EPI_ISL_2771943, EPI_ISL_2771944, EPI_ISL_2771945, EPI_ISL_2771946, EPI_ISL_2771947, EPI_ISL_2771948, EPI_ISL_2771949, EPI_ISL_2771950, EPI_ISL_2771951, EPI_ISL_2771952, EPI_ISL_2771953, EPI_ISL_2771954, EPI_ISL_2771980, EPI_ISL_2771981, EPI_ISL_2771982, EPI_ISL_2771983, EPI_ISL_2771984, EPI_ISL_2771985, EPI_ISL_2771986, EPI_ISL_2771988, EPI_ISL_2771989, EPI_ISL_2771990, EPI_ISL_2771991, EPI_ISL_2771994, EPI_ISL_2771995, EPI_ISL_2771996, EPI_ISL_2771997, EPI_ISL_2771998, EPI_ISL_2771999, EPI_ISL_2772000, EPI_ISL_2772001, EPI_ISL_2772002, EPI_ISL_2772003, EPI_ISL_2772004, EPI_ISL_2772005, EPI_ISL_2772006, EPI_ISL_2772008, EPI_ISL_2772009, EPI_ISL_2772010, EPI_ISL_2772011, EPI_ISL_2772012, EPI_ISL_2772013, EPI_ISL_2772014, EPI_ISL_2772015, EPI_ISL_2772016, EPI_ISL_2772020, EPI_ISL_2772021, EPI_ISL_2772022, EPI_ISL_2772023, EPI_ISL_2772024, EPI_ISL_2772025, EPI_ISL_2772026, EPI_ISL_2772027, EPI_ISL_2772028, EPI_ISL_2772029, EPI_ISL_2772030, EPI_ISL_2772031, EPI_ISL_2772032, EPI_ISL_2772033, EPI_ISL_2772034, EPI_ISL_2772035, EPI_ISL_2772036, EPI_ISL_2772037, EPI_ISL_2772038, EPI_ISL_2772039, EPI_ISL_2772040, EPI_ISL_2772041, EPI_ISL_2772042, EPI_ISL_2772043, EPI_ISL_2772044, EPI_ISL_2772045, EPI_ISL_2772046, EPI_ISL_2772047, EPI_ISL_2772048, EPI_ISL_2772049, EPI_ISL_2772050, EPI_ISL_2772051, EPI_ISL_2772052, EPI_ISL_2772053, EPI_ISL_2772055, EPI_ISL_2772056, EPI_ISL_2772057, EPI_ISL_2772058, EPI_ISL_2772059, EPI_ISL_2772060, EPI_ISL_2772061, EPI_ISL_2772062, EPI_ISL_2772063, EPI_ISL_2772064, EPI_ISL_2772065, EPI_ISL_2772066, EPI_ISL_2772067, EPI_ISL_2772068, EPI_ISL_2772069, EPI_ISL_2772070, EPI_ISL_2772071, EPI_ISL_2772072, EPI_ISL_2772073, EPI_ISL_2772074, EPI_ISL_2772075, EPI_ISL_2772076, EPI_ISL_2772077, EPI_ISL_2772078, EPI_ISL_2772079, EPI_ISL_2772080, EPI_ISL_2772081, EPI_ISL_2772082, EPI_ISL_2772083, EPI_ISL_2772084, EPI_ISL_2772085, EPI_ISL_2772086, EPI_ISL_2772087, EPI_ISL_2772090, EPI_ISL_2772091, EPI_ISL_2772092, EPI_ISL_2772093, EPI_ISL_2772094, EPI_ISL_2772099, EPI_ISL_2772100, EPI_ISL_2772101, EPI_ISL_2772102, EPI_ISL_2772103, EPI_ISL_2772104, EPI_ISL_2772105, EPI_ISL_2772106, EPI_ISL_2772107, EPI_ISL_2772108, EPI_ISL_2772359, EPI_ISL_2772360, EPI_ISL_2772361, EPI_ISL_2772362, EPI_ISL_2772363, EPI_ISL_2772364, EPI_ISL_2772365, EPI_ISL_2772366, EPI_ISL_2772367, EPI_ISL_2772368, EPI_ISL_2772369, EPI_ISL_2772370, EPI_ISL_2772371, EPI_ISL_2772372, EPI_ISL_2772373, EPI_ISL_2772374, EPI_ISL_2772375, EPI_ISL_2772376, EPI_ISL_2772377, EPI_ISL_2772378, EPI_ISL_2772379, EPI_ISL_2772380, EPI_ISL_2772381, EPI_ISL_2772382, EPI_ISL_2772383, EPI_ISL_2772384, EPI_ISL_2772385, EPI_ISL_2772386, EPI_ISL_2772387, EPI_ISL_2772388, EPI_ISL_2772389, EPI_ISL_2772390, EPI_ISL_2772391, EPI_ISL_2772392, EPI_ISL_2772393, EPI_ISL_2772394, EPI_ISL_2772395, EPI_ISL_2772396, EPI_ISL_2772397, EPI_ISL_2772415, EPI_ISL_2772416, EPI_ISL_2772417, EPI_ISL_2772418, EPI_ISL_2772419, EPI_ISL_2772429, EPI_ISL_2772430, EPI_ISL_2772431, EPI_ISL_2772432, EPI_ISL_2772433, EPI_ISL_2772434, EPI_ISL_2772435, EPI_ISL_2772436, EPI_ISL_2772437, EPI_ISL_2772438, EPI_ISL_2772439, EPI_ISL_2772440, EPI_ISL_2772462, EPI_ISL_2772463, EPI_ISL_2772464, EPI_ISL_2772465, EPI_ISL_2772466, EPI_ISL_2772467, EPI_ISL_2772468, EPI_ISL_2772469, EPI_ISL_2772470, EPI_ISL_2772471, EPI_ISL_2772472, EPI_ISL_2772473, EPI_ISL_2772474, EPI_ISL_2776448, EPI_ISL_2776450, EPI_ISL_2776451, EPI_ISL_2776452, EPI_ISL_2776453, EPI_ISL_2776454, EPI_ISL_2776455, EPI_ISL_2776456, EPI_ISL_2776457, EPI_ISL_2776458, EPI_ISL_2776459, EPI_ISL_2776460, EPI_ISL_2776461, EPI_ISL_2776462, EPI_ISL_2776463, EPI_ISL_2776464, EPI_ISL_2776465, EPI_ISL_2776466, EPI_ISL_2776467, | | | |

| | | | |
|---|---|---|---|
| EPI_ISL_2776468, EPI_ISL_2776469, EPI_ISL_2776470, EPI_ISL_2776471, EPI_ISL_2776472, EPI_ISL_2776473, EPI_ISL_2776474, EPI_ISL_2776475, EPI_ISL_2776476, EPI_ISL_2776477, EPI_ISL_2776478, EPI_ISL_2776479, EPI_ISL_2776480, EPI_ISL_2776481, EPI_ISL_2776482, EPI_ISL_2776483, EPI_ISL_2776484, EPI_ISL_2776485, EPI_ISL_2776486, EPI_ISL_2776487, EPI_ISL_2776488, EPI_ISL_2776489, EPI_ISL_2776490, EPI_ISL_2776491, EPI_ISL_2776492, EPI_ISL_2776501, EPI_ISL_2776502, EPI_ISL_2776503, EPI_ISL_2776504, EPI_ISL_2776529, EPI_ISL_2776530, EPI_ISL_2776531, EPI_ISL_2776532, EPI_ISL_2776533, EPI_ISL_2776534, EPI_ISL_2776535, EPI_ISL_2776536, EPI_ISL_2776537, EPI_ISL_2776538, EPI_ISL_2776561, EPI_ISL_2776566, EPI_ISL_2776567, EPI_ISL_2776568, EPI_ISL_2776569, EPI_ISL_2776570, EPI_ISL_2776571, EPI_ISL_2776572, EPI_ISL_2776573, EPI_ISL_2776574, EPI_ISL_2776575, EPI_ISL_2776576, EPI_ISL_2776577, EPI_ISL_2776578, EPI_ISL_2776579, EPI_ISL_2776580, EPI_ISL_2776581, EPI_ISL_2776582, EPI_ISL_2776583, EPI_ISL_2776584, EPI_ISL_2776585, EPI_ISL_2776586, EPI_ISL_2776587, EPI_ISL_2776588, EPI_ISL_2776589, EPI_ISL_2776590, EPI_ISL_2776591, EPI_ISL_2776592, EPI_ISL_2776593, EPI_ISL_2776594, EPI_ISL_2776595, EPI_ISL_2776596, EPI_ISL_2776597, EPI_ISL_2776598, EPI_ISL_2776599, EPI_ISL_2776600, EPI_ISL_2776601, EPI_ISL_2776602, EPI_ISL_2776603, EPI_ISL_2776604, EPI_ISL_2776605, EPI_ISL_2776606, EPI_ISL_2776607, EPI_ISL_2776608, EPI_ISL_2776609, EPI_ISL_2776610, EPI_ISL_2776611, EPI_ISL_2776612, EPI_ISL_2776613, EPI_ISL_2776614, EPI_ISL_2776615, EPI_ISL_2776616, EPI_ISL_2776617, EPI_ISL_2776618, EPI_ISL_2776619, EPI_ISL_2776620, EPI_ISL_2776621, EPI_ISL_2776622, EPI_ISL_2776630, EPI_ISL_2776631, EPI_ISL_2776632, EPI_ISL_2776633, EPI_ISL_2776634, EPI_ISL_2776635, EPI_ISL_2776636, EPI_ISL_2776637, EPI_ISL_2776638, EPI_ISL_2776639, EPI_ISL_2776640, EPI_ISL_2776641, EPI_ISL_2776642, EPI_ISL_2776643, EPI_ISL_2776644, EPI_ISL_2776645, EPI_ISL_2776646, EPI_ISL_2776647, EPI_ISL_2776648, EPI_ISL_2776649, EPI_ISL_2776650, EPI_ISL_2776651, EPI_ISL_2776652, EPI_ISL_2776653, EPI_ISL_2776654, EPI_ISL_2776655, EPI_ISL_2776656, EPI_ISL_2776657, EPI_ISL_2776658, EPI_ISL_2776659, EPI_ISL_2776660, EPI_ISL_2776661, EPI_ISL_2776662, EPI_ISL_2776663, EPI_ISL_2776664, EPI_ISL_2776747, EPI_ISL_2776748, EPI_ISL_2776749, EPI_ISL_2776750, EPI_ISL_2776751, EPI_ISL_2776752, EPI_ISL_2776753, EPI_ISL_2776754, EPI_ISL_2776755, EPI_ISL_2776756, EPI_ISL_2776757, EPI_ISL_2776758, EPI_ISL_2776759, EPI_ISL_2776760, EPI_ISL_2776761, EPI_ISL_2776762, EPI_ISL_2776763, EPI_ISL_2776764, EPI_ISL_2776765, EPI_ISL_2776766, EPI_ISL_2776767, EPI_ISL_2776768, EPI_ISL_2776769, EPI_ISL_2776770, EPI_ISL_2776771, EPI_ISL_2776772, EPI_ISL_2776773, EPI_ISL_2776774, EPI_ISL_2776775, EPI_ISL_2776776, EPI_ISL_2776777, EPI_ISL_2776778, EPI_ISL_2776779, EPI_ISL_2776780, EPI_ISL_2776781, EPI_ISL_2776782, EPI_ISL_2776783, EPI_ISL_2776824, EPI_ISL_2776825, EPI_ISL_2776826, EPI_ISL_2776827, EPI_ISL_2776828, EPI_ISL_2776829, EPI_ISL_2776830, EPI_ISL_2776831, EPI_ISL_2776832, EPI_ISL_2776846, EPI_ISL_2776847, EPI_ISL_2776848, EPI_ISL_2776849, EPI_ISL_2776850, EPI_ISL_2776851, EPI_ISL_2776852, EPI_ISL_2776853, EPI_ISL_2776854, EPI_ISL_2776855, EPI_ISL_2776856, EPI_ISL_2776857, EPI_ISL_2776858, EPI_ISL_2776859, EPI_ISL_2776860, EPI_ISL_2776861, EPI_ISL_2776862, EPI_ISL_2776863, EPI_ISL_2776864, EPI_ISL_2776865, EPI_ISL_2776866, EPI_ISL_2776867, EPI_ISL_2776868, EPI_ISL_2776869, EPI_ISL_2776870, EPI_ISL_2776871, EPI_ISL_2776872, EPI_ISL_2776873, EPI_ISL_2776874, EPI_ISL_2776875, EPI_ISL_2776876, EPI_ISL_2776877, EPI_ISL_2776878, EPI_ISL_2776879, EPI_ISL_2776880, EPI_ISL_2776881, EPI_ISL_2776882, EPI_ISL_2776883, EPI_ISL_2776884, EPI_ISL_2776885, EPI_ISL_2776886, EPI_ISL_2776887, EPI_ISL_2776888, EPI_ISL_2776889, EPI_ISL_2776890, EPI_ISL_2776891, EPI_ISL_2776892, EPI_ISL_2776893, EPI_ISL_2776894, EPI_ISL_2776895, EPI_ISL_2776896, EPI_ISL_2776897, EPI_ISL_2776898, EPI_ISL_2776899, EPI_ISL_2776900, EPI_ISL_2776901, EPI_ISL_2776902, EPI_ISL_2776903, EPI_ISL_2776904, EPI_ISL_2776905, EPI_ISL_2776906, EPI_ISL_2776907, EPI_ISL_2776908, EPI_ISL_2776909, EPI_ISL_2776910, EPI_ISL_2776911, EPI_ISL_2776912, EPI_ISL_2776913, EPI_ISL_2776914, EPI_ISL_2776915, EPI_ISL_2776916, EPI_ISL_2776917, EPI_ISL_2776918, EPI_ISL_2776919, EPI_ISL_2776920, EPI_ISL_2776921, EPI_ISL_2776922, EPI_ISL_2777074, EPI_ISL_2777076, EPI_ISL_2777077, EPI_ISL_2777078, EPI_ISL_2777079, EPI_ISL_2777080, EPI_ISL_2777081, EPI_ISL_2777082, EPI_ISL_2777083, EPI_ISL_2777084, EPI_ISL_2777085, EPI_ISL_2777086, EPI_ISL_2777087, EPI_ISL_2777088, EPI_ISL_2777089, EPI_ISL_2777090, EPI_ISL_2777091, EPI_ISL_2777092, EPI_ISL_2777093, EPI_ISL_2777094, EPI_ISL_2777095, EPI_ISL_2777096, EPI_ISL_2777097, EPI_ISL_2777098, EPI_ISL_2777099, EPI_ISL_2777100, EPI_ISL_2777101, EPI_ISL_2777102, EPI_ISL_2777103, EPI_ISL_2777104, EPI_ISL_2777113, EPI_ISL_2777114, EPI_ISL_2777115, EPI_ISL_2777116, EPI_ISL_2777117, EPI_ISL_2777118, EPI_ISL_2777119, EPI_ISL_2777120, EPI_ISL_2777121, EPI_ISL_2777122, EPI_ISL_2777123, EPI_ISL_2777124, EPI_ISL_2777125, EPI_ISL_2777127, EPI_ISL_2777128, EPI_ISL_2777129, EPI_ISL_2777130, EPI_ISL_2777131, EPI_ISL_2777132, EPI_ISL_2777133, EPI_ISL_2777134, EPI_ISL_2777135, EPI_ISL_2777136, EPI_ISL_2828254, EPI_ISL_2828255, EPI_ISL_2828256, EPI_ISL_2828257, EPI_ISL_2828258, EPI_ISL_2828259, EPI_ISL_2828260, EPI_ISL_2828261, EPI_ISL_2828262, EPI_ISL_2828263, EPI_ISL_2828264, EPI_ISL_2828265, EPI_ISL_2828266, EPI_ISL_2828267, EPI_ISL_2828268, EPI_ISL_2828269, EPI_ISL_2828270, EPI_ISL_2828271, EPI_ISL_2828272, EPI_ISL_2828273, EPI_ISL_2828274, EPI_ISL_2828275, EPI_ISL_2828276, EPI_ISL_2828277, EPI_ISL_2828278, EPI_ISL_2828279, EPI_ISL_2828280, EPI_ISL_2828281, EPI_ISL_2828282, EPI_ISL_2828283, EPI_ISL_2828284, EPI_ISL_2828285, EPI_ISL_2828286, EPI_ISL_2828287, EPI_ISL_2828288, EPI_ISL_2828289, EPI_ISL_2828290, EPI_ISL_2828291, EPI_ISL_2828292, EPI_ISL_2828293, EPI_ISL_2828294, EPI_ISL_2828295, EPI_ISL_2828296, EPI_ISL_2828297, EPI_ISL_2828298, EPI_ISL_2828299, EPI_ISL_2828300, EPI_ISL_2828301, EPI_ISL_2828302, EPI_ISL_2828303, EPI_ISL_2828304, EPI_ISL_2828305, EPI_ISL_2828306, EPI_ISL_2828307, EPI_ISL_2828308, EPI_ISL_2828309, EPI_ISL_2828310, EPI_ISL_2828311, EPI_ISL_2828312, EPI_ISL_2828313, EPI_ISL_2828314, EPI_ISL_2828315, EPI_ISL_2828316, EPI_ISL_2828317, EPI_ISL_2828318, EPI_ISL_2828319, EPI_ISL_2828320, EPI_ISL_2828321, EPI_ISL_2828322, EPI_ISL_2828323, EPI_ISL_2828324, EPI_ISL_2828325, EPI_ISL_2930562, EPI_ISL_2930563, EPI_ISL_2930567, EPI_ISL_2933813 | | | |
| see above | SARS-CoV-2 testing team, National Institute of Infectious Diseases | Pathogen Genomics Center, National Institute of Infectious Diseases | Hazuka Y Furihata; Hiromizu Takahashi; Kentaro Itokawa; Makoto Kuroda; Masanori Hashino; Masumichi Saito; Naomi Nojiri; Nozomu Hanaoka; Rina Tanaka; Sana Uchikoba; Tsuguto Fujimoto; Tsuyoshi Sekizuka |
| EPI_ISL_2812074, EPI_ISL_2812094, EPI_ISL_2812095, EPI_ISL_2812096, EPI_ISL_2812097, EPI_ISL_2812098, EPI_ISL_2812099, EPI_ISL_2812113, EPI_ISL_2812114, EPI_ISL_2812115, EPI_ISL_2812116, EPI_ISL_2812117, EPI_ISL_2812118, EPI_ISL_2812145 | | | |
| see above | Saitama Prefectural Institute of Public Health | Saitama Prefectural Institute of Public Health | Eri Aonuma; Hayato Ehara; Kentaro Itokawa; Makoto Kuroda; Masanori Hashino; Rina Tanaka; Satoshi Daisaki; Tsuyoshi Sekizuka |
| EPI_ISL_2885221, EPI_ISL_2885222, EPI_ISL_2885223, EPI_ISL_2885224, EPI_ISL_2885225, EPI_ISL_2885226, EPI_ISL_2885227, EPI_ISL_2885228, EPI_ISL_2885229 | | | |
| see above | Shimane Prefectural Institute of Public Health and Environmental Science | Shimane Prefectural Institute of Public Health and Environmental Science | Aiko Fukuma; Kentaro Itokawa; Makoto Kuroda; Masanori Hashino; Naoki Fujisawa; Rie Onishi; Rina Tanaka; Tsuyoshi Sekizuka; Yurie Kanba |
| EPI_ISL_2551582 | Tochigi Prefectural Institute of Environment and Health | Genome Analysis Center, Kamma Memorial Hospital | Fuminori Mizukoshi; Hanako Yazawa; Hiroshi Kamma; Jun Ishii; Kaori Watanabe; Michiaki Masuda; Satoko Soma; Takuya Yazawa; Yoshiko Hitomi |
| EPI_ISL_2562055, EPI_ISL_2562056, EPI_ISL_2562057, EPI_ISL_2562058, EPI_ISL_2651240, EPI_ISL_2651241, EPI_ISL_2651242, EPI_ISL_2651243, EPI_ISL_2651244, EPI_ISL_2651245, EPI_ISL_2651246, EPI_ISL_2651247, EPI_ISL_2651248, EPI_ISL_2651249, EPI_ISL_2709594, EPI_ISL_2709595, EPI_ISL_2709596, EPI_ISL_2709597, EPI_ISL_2709598, EPI_ISL_2709599, EPI_ISL_2709600, EPI_ISL_2709601, EPI_ISL_2709602, EPI_ISL_2784335, EPI_ISL_2784336, EPI_ISL_2784337, EPI_ISL_2784338, EPI_ISL_2784339, EPI_ISL_2784340, EPI_ISL_2784341, EPI_ISL_2784342, EPI_ISL_2784343, EPI_ISL_2786970, EPI_ISL_2786971, EPI_ISL_2786972, EPI_ISL_2786973, EPI_ISL_2786974, EPI_ISL_2786975, EPI_ISL_2786976, EPI_ISL_2786977, EPI_ISL_2786978, EPI_ISL_2786979, EPI_ISL_2786980, EPI_ISL_2786981, EPI_ISL_2786982, EPI_ISL_2786983, EPI_ISL_2786984, EPI_ISL_2786985, EPI_ISL_2786986, EPI_ISL_2786987, EPI_ISL_2786988, EPI_ISL_2786989, EPI_ISL_2786990 | | | |
| see above | Tokyo Metropolitan Institute of Public Health | Tokyo Metropolitan Institute of Public Health | Akane Negishi; Hirofumi Miyake; Jun Suzuki; Kenji Sadamasu; Kenshirou Kuroki; Mami Nagashima; Masaki Hayashi; Maya Isogai; Takako Yamazaki; Takushi Fujiwara |
| EPI_ISL_2786951, EPI_ISL_2786954, EPI_ISL_2786955, EPI_ISL_2786956, EPI_ISL_2786957, EPI_ISL_2786958, EPI_ISL_2786959, EPI_ISL_2786960, EPI_ISL_2786961, EPI_ISL_2786962, EPI_ISL_2786963, EPI_ISL_2786964, EPI_ISL_2786966, EPI_ISL_2786967 | | | |
| see above | Toyama Institute of Health | Toyama Institute of Health | Emiko Igarashi; Hideki Tani; Kazunori Oishi; Kentaro Itokawa; Makoto Kuroda; Masae Itamochi; Masanori Hashino; Noriko Inasaki; Rina Tanaka; Shunsuke Yazawa; Sumiyo Hasegawa; Tsuyoshi Sekizuka; Yumiko Saga |
| EPI_ISL_2827759 | Virology section, Okayama Prefectural Institute for Environmental Science and Public Health | Virology section, Okayama Prefectural Institute for Environmental Science and Public Health | Gou Tsurumi; Kazuhiko Nagao; Keisuke Funahashi; Kouji Kida; Manabu Ishii; Mayumi Hirata; Naoko Okamoto; Sachie Tsuchimoto |
| EPI_ISL_2841532, EPI_ISL_2841533 | Yamagata Prefectural Institute of Public Health | Pathogen Genomics Center, National Institute of Infectious Diseases | Akinobu Wada; Hideyuki Shinohara; Junji Seto; Katsumi Mizuta; Kenichi Komabayashi; Kentaro Itokawa; Makoto Kuroda; Masanori Hashino; Midori Taruishi; Mika Sampei; Naomi Ogawa; Rina Tanaka; Tsuyoshi Sekizuka; Waka Tanaka; Yoko Aoki; Yoko Ikeda; Yoshitaka Ishida |
| EPI_ISL_2859490 | Yamaguchi Prefectural Institute of Public Health and Environment | Yamaguchi Prefectural Institute of Public Health and Environment | Kanako Kawasaki; Kentaro Itokawa; Makoto Kuroda; Masanori Hashino; Mitushiro Kameyama; Rina Tanaka; Tsuyoshi Sekizuka |

We gratefully acknowledge the following Authors from the Originating laboratories responsible for obtaining the specimens, as well as the Submitting laboratories where the genome data were generated and shared via GISAID, on which this research is based.

All Submitters of data may be contacted directly via www.gisaid.org

Authors are sorted alphabetically.

| Accession ID | Originating Laboratory | Submitting Laboratory | Authors |
|---|---|---|---|
| EPI_ISL_1973325, EPI_ISL_1973326, EPI_ISL_1973327, EPI_ISL_1973328, EPI_ISL_2020238, EPI_ISL_2020239, EPI_ISL_2020240 | | | |
| see above | Bintulu Hospital | Institute for Medical Research, Infectious Disease Research Centre, National Institutes of Health, Ministry of Health Malaysia | Azizan MA; Kamel K; Mohd Zawawi Z; Ramly N; Robert F; Suppiah J; Thayan R |
| EPI_ISL_1919861 | Duchess of Kent Hospital, Sandakan | Institute for Medical Research, Infectious Disease Research Centre, National Institutes of Health, Ministry of Health Malaysia | Kamel K; Mohd Zawawi Z; Suppiah J; Thayan R |
| EPI_ISL_1673257 | Hospital Lahad Datu | Institute for Medical Research, Infectious Disease Research Centre, National Institutes of Health, Ministry of Health Malaysia Jalan Setia Murni U13/52, Bandar Setia Alam, 40170 Selangor, Malaysia | Kamel K; Mohd Zawawi Z; Suppiah J; Thayan R |
| EPI_ISL_1673288 | Hospital Segamat Johor | Institute for Medical Research, Infectious Disease Research Centre, National Institutes of Health, Ministry of Health Malaysia | Kamel K; Mohd Zawawi Z; Suppiah J; Thayan R |
| EPI_ISL_1673280 | Hospital Sultan Ismail | Institute for Medical Research, Infectious Disease Research Centre, National Institutes of Health, Ministry of Health Malaysia | Kamel K; Mohd Zawawi Z; Suppiah J; Thayan R |
| EPI_ISL_1673283 | Hospital Sultanah Aminah | Institute for Medical Research, Infectious Disease Research Centre, National Institutes of Health, Ministry of Health Malaysia | Kamel K; Mohd Zawawi Z; Suppiah J; Thayan R |
| EPI_ISL_1673284 | Hospital Sultanah Nora Ismail | Institute for Medical Research, Infectious Disease Research Centre, National Institutes of Health, Ministry of Health Malaysia | Kamel K; Mohd Zawawi Z; Suppiah J; Thayan R |
| EPI_ISL_1673309 | Hospital Temenggong Seri Maharaja Tun Ibrahim | Institute for Medical Research, Infectious Disease Research Centre, National Institutes of Health, Ministry of Health Malaysia | Kamel K; Mohd Zawawi Z; Suppiah J; Thayan R |
| EPI_ISL_1672633 | Hospital Tuanku Jaafar Seremban | Institute for Medical Research, Infectious Disease Research Centre, National Institutes of Health, Ministry of Health Malaysia | Kamel K; Mohd Zawawi Z; Suppiah J; Thayan R |
| EPI_ISL_1424467, EPI_ISL_1424468, EPI_ISL_1424470, EPI_ISL_1424471, EPI_ISL_1424510, EPI_ISL_1424513, EPI_ISL_1424518, EPI_ISL_1583006, EPI_ISL_1583007, EPI_ISL_1585242, EPI_ISL_1673287, EPI_ISL_1673293, EPI_ISL_1673296, EPI_ISL_1673299, EPI_ISL_1673304, EPI_ISL_1673307, EPI_ISL_1673332, EPI_ISL_1673336, EPI_ISL_1673338, EPI_ISL_1673340, EPI_ISL_1673341, EPI_ISL_1673342, EPI_ISL_1673344, EPI_ISL_1673345, EPI_ISL_1673420, EPI_ISL_1673683, EPI_ISL_1673735, EPI_ISL_1696746, EPI_ISL_1696747, EPI_ISL_1696748, EPI_ISL_1696749, EPI_ISL_1696750, EPI_ISL_1696751, EPI_ISL_1696752, EPI_ISL_1696753, EPI_ISL_1696754, EPI_ISL_1696755, EPI_ISL_1696756, EPI_ISL_1696757, EPI_ISL_1696758, EPI_ISL_1714796, EPI_ISL_1714797, EPI_ISL_1787254, EPI_ISL_1787317, EPI_ISL_1787318, EPI_ISL_1787319, EPI_ISL_1787320, EPI_ISL_1787321, EPI_ISL_1787322, EPI_ISL_1787323, EPI_ISL_1787324, EPI_ISL_1787689, EPI_ISL_1787983, EPI_ISL_1788079, EPI_ISL_1788080, EPI_ISL_1788081, EPI_ISL_1788082, EPI_ISL_1789203, EPI_ISL_1789204, EPI_ISL_1805505, EPI_ISL_1805508, EPI_ISL_1805509, EPI_ISL_1807185, EPI_ISL_1807186, EPI_ISL_1807188, EPI_ISL_1807206, EPI_ISL_1807207, EPI_ISL_1919650, EPI_ISL_1919651, EPI_ISL_1919652, EPI_ISL_1919653, EPI_ISL_1919654, EPI_ISL_1919860, EPI_ISL_1972534, EPI_ISL_1972535, EPI_ISL_1972536, EPI_ISL_1972537, EPI_ISL_2090891, EPI_ISL_2091000 | | | |
| see above | Institute for Medical Research, Infectious Disease Research Centre, National Institutes of Health, Ministry of Health Malaysia | Institute for Medical Research, Infectious Disease Research Centre, National Institutes of Health, Ministry of Health Malaysia | Azizan MA; Kamel K; Mohd Zawawi Z; Ramly N; Robert F; Suppiah J; Thayan R |
| EPI_ISL_1919422, EPI_ISL_1919645, EPI_ISL_1919646 | Ipoh Public Health Laboratory (MKAI), Ministry of Health Malaysia | Institute for Medical Research, Infectious Disease Research Centre, National Institutes of Health, Ministry | Kamel K; Mohd Zawawi Z; Suppiah J; Thayan R |
| EPI_ISL_1821883, EPI_ISL_1821884, EPI_ISL_1919272, EPI_ISL_1919273, EPI_ISL_1919647, EPI_ISL_1919648, EPI_ISL_1919649, EPI_ISL_1919858, EPI_ISL_1919859, EPI_ISL_1919944, EPI_ISL_1919945, EPI_ISL_1919946, EPI_ISL_1919947, EPI_ISL_1919949, EPI_ISL_1919950, EPI_ISL_1919951, EPI_ISL_1919952, EPI_ISL_1919986, EPI_ISL_1919987, EPI_ISL_1921767, EPI_ISL_1921769, EPI_ISL_1921770 | | | |
| see above | Ipoh Public Health Laboratory (MKAI), Ministry of Health Malaysia | Institute for Medical Research, Infectious Disease Research Centre, National Institutes of Health, Ministry of Health Malaysia | Kamel K; Mohd Zawawi Z; Suppiah J; Thayan R |
| EPI_ISL_1972549, EPI_ISL_1972550 | Klinik Kesihatan Bentong | Institute for Medical Research, Infectious Disease Research Centre, National Institutes of Health, Ministry of Health Malaysia | Azizan MA; Kamel K; Mohd Zawawi Z; Ramly N; Robert F; Suppiah J; Thayan R |
| EPI_ISL_1972553 | Klinik Kesihatan Karak | Institute for Medical Research, Infectious Disease Research Centre, National Institutes of Health, Ministry of Health Malaysia | Azizan MA; Kamel K; Mohd Zawawi Z; Ramly N; Robert F; Suppiah J; Thayan R |
| EPI_ISL_1972548, EPI_ISL_1972551, EPI_ISL_1972552 | Klinik Kesihatan Lurah Bilut (Felda) | Institute for Medical Research, Infectious Disease Research Centre, National Institutes of Health, Ministry of Health Malaysia | Azizan MA; Kamel K; Mohd Zawawi Z; Ramly N; Robert F; Suppiah J; Thayan R |
| EPI_ISL_1973324, EPI_ISL_2020075, EPI_ISL_2020076, EPI_ISL_2020077, EPI_ISL_2020078, EPI_ISL_2020165, EPI_ISL_2020166, EPI_ISL_2020167 | | | |
| see above | Kota Bharu Public Health Laboratory | Institute for Medical Research, Infectious Disease Research Centre, National Institutes of Health, Ministry of Health Malaysia | Azizan MA; Kamel K; Mohd Zawawi Z; Ramly N; Robert F; Suppiah J; Thayan R; uppiah J |
| EPI_ISL_1585270, EPI_ISL_1585271 | Labuan Hospital | Institute for Medical Research, Infectious Disease Research Centre, National Institutes of Health, Ministry of Health Malaysia | Kamel K; Mohd Zawawi Z; Ramly N; Robert F; Suppiah J; Thayan R |
| EPI_ISL_1972365, EPI_ISL_1972366, EPI_ISL_1972533 | Sultanah Bahiyah Hospital, Alor Setar | Institute for Medical Research, Infectious Disease Research Centre, National Institutes of Health, Ministry of Health Malaysia | Azizan MA; Kamel K; Mohd Zawawi Z; Ramly N; Robert F; Suppiah J; Thayan R |
| EPI_ISL_1972356, EPI_ISL_2091020 | Sungai Buloh Hospital | Institute for Medical Research, Infectious Disease Research Centre, National Institutes of Health, Ministry of Health Malaysia | Azizan MA; Kamel K; Mohd Zawawi Z; Ramly N; Robert F; Suppiah J; Thayan R |
| EPI_ISL_1972538, EPI_ISL_1972539, EPI_ISL_1972540, EPI_ISL_1972547 | Tampin Hospital | Institute for Medical Research, Infectious Disease Research Centre, National Institutes of Health, Ministry of Health Malaysia | Azizan MA; Kamel K; Mohd Zawawi Z; Ramly N; Robert F; Suppiah J; Thayan R |
| EPI_ISL_1919862, EPI_ISL_1919863 | Tawau Hospital | Institute for Medical Research, Infectious Disease Research Centre, National Institutes of Health, Ministry of Health Malaysia | Kamel K; Mohd Zawawi Z; Suppiah J; Thayan R |

We gratefully acknowledge the following Authors from the Originating laboratories responsible for obtaining the specimens, as well as the Submitting laboratories where the genome data were generated and shared via GISAID, on which this research is based.

All Submitters of data may be contacted directly via www.gisaid.org

Authors are sorted alphabetically.

| Accession ID | Originating Laboratory | Submitting Laboratory | Authors |
|---|---|---|---|
| EPI_ISL_2854052 | Ampang Hospital | Institute for Medical Research, Infectious Disease Research Centre, National Institutes of Health, Ministry of Health Malaysia | Anasir MI; Azizan MA; Kamel K; Mohd Zawawi Z; Ramly N; Robert F; Suppiah J; Thayan R |
| EPI_ISL_2811998 | Bandar Kuantan Health Clinic | Institute for Medical Research, Infectious Disease Research Centre, National Institutes of Health, Ministry of Health Malaysia | Azizan MA; Kamel K; Mohd Zawawi Z; Ramly N; Robert F; Suppiah J; Thayan R |
| EPI_ISL_2854069 | Bayan Lepas Health Clinic | Institute for Medical Research, Infectious Disease Research Centre, National Institutes of Health, Ministry of Health Malaysia | Anasir MI; Azizan MA; Kamel K; Mohd Zawawi Z; Ramly N; Robert F; Suppiah J; Thayan R |
| EPI_ISL_2535780, EPI_ISL_2608379, EPI_ISL_2608380, EPI_ISL_2608381, EPI_ISL_2608382, EPI_ISL_2608383, EPI_ISL_2608384, EPI_ISL_2608385, EPI_ISL_2608386, EPI_ISL_2608387, EPI_ISL_2608388, EPI_ISL_2608389, EPI_ISL_2608390, EPI_ISL_2608391, EPI_ISL_2608392, EPI_ISL_2608393, EPI_ISL_2608394, EPI_ISL_2608395, EPI_ISL_2608396, EPI_ISL_2608397, EPI_ISL_2608398, EPI_ISL_2608399, EPI_ISL_2608400, EPI_ISL_2608401, EPI_ISL_2608402, EPI_ISL_2608403, EPI_ISL_2608404, EPI_ISL_2608405 | | | |
| see above | Borneo Medical Centre | Institute of Health and Community Medicine | Chan Chia Jui; Chua Hock Hin; David Perera; Ooi Mong How; Tonnii Sia Loong Loong; Wong Jyn Shan; Wong Kiing Aik |
| EPI_ISL_2608408, EPI_ISL_2608409, EPI_ISL_2608410, EPI_ISL_2608411, EPI_ISL_2608412, EPI_ISL_2608413, EPI_ISL_2608414 | | | |
| see above | Clinical Research Centre, Sarawak General Hospital | Institute of Health and Community Medicine | Chan Chia Jui; Chua Hock Hin; David Perera; Ooi Mong How; Tonnii Sia Loong Loong; Wong Jyn Shan; Wong Kiing Aik |
| EPI_ISL_2622089 | Department of Pathology | Centre for Research in Advanced Tropical Bioscience Universiti Malaysia Pahang, 26300 Gambang, Pahang, Malaysia | Hajar Fauzan Bin Ahmad; Norhidayah Binti Kamarudin; Ummu Afeera Zainulabid |
| EPI_ISL_2549562 | Gribbles Pathology | Institute for Medical Research, Infectious Disease Research Centre, National Institutes of Health, Ministry of Health Malaysia | Azizan MA; Kamel K; Mohd Zawawi Z; Ramly N; Robert F; Suppiah J; Thayan R |
| EPI_ISL_2233383 | Gribbles Pathology Malaysia | Institute for Medical Research, Infectious Disease Research Centre, National Institutes of Health, Ministry of Health Malaysia | Azizan MA; Kamel K; Mohd Zawawi Z; Ramly N; Robert F; Suppiah J; Thayan R |
| EPI_ISL_2727836 | Hospital Angkatan Tentera Kota Kinabalu | Institute for Medical Research, Infectious Disease Research Centre, National Institutes of Health, Ministry of Health Malaysia | Azizan MA; Kamel K; Mohd Zawawi Z; Ramly N; Robert F; Suppiah J; Thayan R |
| EPI_ISL_2727837 | Hospital Angkatan Tentera Wilayah Kota Kinabalu | Institute for Medical Research, Infectious Disease Research Centre, National Institutes of Health, Ministry of Health Malaysia | Azizan MA; Kamel K; Mohd Zawawi Z; Ramly N; Robert F; Suppiah J; Thayan R |
| EPI_ISL_2233382, EPI_ISL_2550733 | Hospital Labuan | Institute for Medical Research, Infectious Disease Research Centre, National Institutes of Health, Ministry of Health Malaysia | Azizan MA; Kamel K; Mohd Zawawi Z; Ramly N; Robert F; Suppiah J; Thayan R |
| EPI_ISL_2379461 | Hospital Melaka | Institute for Medical Research, Infectious Disease Research Centre, National Inst | Azizan MA; Kamel K; Mohd Zawawi Z; Ramly N; Robert F; Suppiah J; Thayan R |
| EPI_ISL_2650001, EPI_ISL_2650002 | Hospital Sultanah Nur Zahirah Kuala Terengganu | Institute for Medical Research, Infectious Disease Research Centre, National Institutes of Health, Ministry of Health Malaysia | Azizan MA; Kamel K; Mohd Zawawi Z; Ramly N; Robert F; Suppiah J; Thayan R |
| EPI_ISL_2549576 | Hospital Tuanku Fauziah | Institute for Medical Research, Infectious Disease Research Centre, National Institutes of Health, Ministry of Health Malaysia | Azizan MA; Kamel K; Mohd Zawawi Z; Ramly N; Robert F; Suppiah J; Thayan R |
| EPI_ISL_2620716 | Hospital Tuanku Fauziah, Kangar | Institute for Medical Research, Infectious Disease Research Centre, National Institutes of Health, Ministry of Health | Azizan MA; Kamel K; Mohd Zawawi Z; Ramly N; Robert F; Suppiah J; Thayan R |
| EPI_ISL_2620715 | Hospital Tuanku Fauziah, Kangar | Institute for Medical Research, Infectious Disease Research Centre, National Institutes of Health, Ministry of Health Malaysia | Azizan MA; Kamel K; Mohd Zawawi Z; Ramly N; Robert F; Suppiah J; Thayan R |
| EPI_ISL_2650003 | Hospital Wanita Dan Kanak-Kanak Sabah | Institute for Medical Research, Infectious Disease Research Centre, National Institutes of Health, Ministry of Health Malaysia | 40170 Selangor; Bandar Setia Alam; Jalan Setia Murni U13/52; Malaysia |
| EPI_ISL_2090886, EPI_ISL_2090887, EPI_ISL_2090889, EPI_ISL_2090890, EPI_ISL_2379433, EPI_ISL_2379443, EPI_ISL_2379650, EPI_ISL_2429134, EPI_ISL_2429136, EPI_ISL_2429137, EPI_ISL_2429138, EPI_ISL_2429141, EPI_ISL_2429142, EPI_ISL_2429143, EPI_ISL_2429145, EPI_ISL_2625633, EPI_ISL_2625635, EPI_ISL_2684231, EPI_ISL_2684232, EPI_ISL_2684233, EPI_ISL_2684545, EPI_ISL_2685359, EPI_ISL_2685766, EPI_ISL_2685835, EPI_ISL_2685836, EPI_ISL_2685837, EPI_ISL_2685838, EPI_ISL_2685839, EPI_ISL_2685840, EPI_ISL_2685841, EPI_ISL_2685843, EPI_ISL_2685844, EPI_ISL_2685845, EPI_ISL_2811887, EPI_ISL_2815375, EPI_ISL_2819024, EPI_ISL_2819571, EPI_ISL_2819919 | | | |
| see above | Institute for Medical Research, Infectious Disease Research Centre, National Institutes of Health, Ministry of Health Malaysia | Institute for Medical Research, Infectious Disease Research Centre, National Institutes of Health, Ministry of Health Malaysia | Azizan MA; Kamel K; Mohd Zawawi Z; Ramly N; Robert F; Suppiah J; Thayan R |
| EPI_ISL_2650007, EPI_ISL_2683498, EPI_ISL_2815329, EPI_ISL_2815330, EPI_ISL_2921400 | Ipoh Public Health Laboratory (MKAI), Ministry of Health Malaysia | Institute for Medical Research, Infectious Disease Research Centre, National Institutes of Health, Ministry of Health Malaysia | Anasir MI; Azizan MA; Kamel K; Mohd Zawawi Z; Ramly N; Robert F; Suppiah J; Thayan R |
| EPI_ISL_2091022, EPI_ISL_2233384, EPI_ISL_2550714, EPI_ISL_2550731, EPI_ISL_2625621, EPI_ISL_2625624, EPI_ISL_2625625, EPI_ISL_2625626, EPI_ISL_2625627, EPI_ISL_2625628, EPI_ISL_2625629, EPI_ISL_2625630, EPI_ISL_2625631, EPI_ISL_2649996, EPI_ISL_2650256 | | | |
| see above | Johor Bahru Public Health Laboratory | Institute for Medical Research, Infectious Disease Research Centre, National Institutes of Health, Ministry of Health Malaysia | Azizan MA; Kamel K; Mohd Zawawi Z; Ramly N; Robert F; Suppiah J; Thayan R |
| EPI_ISL_2757930, EPI_ISL_2757951, EPI_ISL_2812981 | Johor State Health Department | Institute for Medical Research, Infectious Disease Research Centre, National Institutes of Health, Ministry of Health Malaysia | Azizan MA; Kamel K; Mohd Zawawi Z; Ramly N; Robert F; Suppiah J; Thayan R |
| EPI_ISL_2812576, EPI_ISL_2854053 | Kemaman Hospital | Institute for Medical Research, Infectious Disease Research Centre, National Institutes of Health, Ministry of Health Malaysia | Anasir MI; Azizan MA; Kamel K; Mohd Zawawi Z; Ramly N; Robert F; Suppiah J; Thayan R |
| EPI_ISL_2815331 | Kuala Lumpur General Hospital | Institute for Medical Research, Infectious Disease Research Centre, National Institutes of Health, Ministry of Health Malaysia | Azizan MA; Kamel K; Mohd Zawawi Z; Ramly N; Robert F; Suppiah J; Thayan R |
| EPI_ISL_2649993 | Kuala Lumpur Hospital | Institute for Medical Research, Infectious Disease Research Centre, National Institutes of Health, Ministry of Health Malaysia | Azizan MA; Kamel K; Mohd Zawawi Z; Ramly N; Robert F; Suppiah J; Thayan R |
| EPI_ISL_2811865, EPI_ISL_2811866, EPI_ISL_2811867, EPI_ISL_2811868, EPI_ISL_2811869, EPI_ISL_2811870 | Kuala Lumpur International Healthcare Centre (KLIHC) | Institute for Medical Research, Infectious Disease Research Centre, National Institutes of Health, Ministry of Health Malaysia | Azizan MA; Kamel K; Mohd Zawawi Z; Ramly N; Robert F; Suppiah J; Thaya; Thayan R |
| EPI_ISL_2854187 | Kuala Terengganu District Health Office | Institute for Medical Research, Infectious Disease Research Centre, National Institutes of Health, Ministry of Health Malaysia | Anasir MI; Azizan MA; Kamel K; Mohd Zawawi Z; Ramly N; Robert F; Suppiah J; Thayan R |
| EPI_ISL_2649991 | Kudat Hospital | Institute for Medical Research, Infectious Disease Research Centre, National Institutes of Health, Ministry of Health Malaysia | Azizan MA; Kamel K; Mohd Zawawi Z; Ramly N; Robert F; Suppiah J; Thayan R |
| EPI_ISL_2549741, EPI_ISL_2757931, EPI_ISL_2812651, EPI_ISL_2812971, EPI_ISL_2854048, EPI_ISL_2921611, EPI_ISL_2924057, EPI_ISL_2931921 | | | |
| see above | Labuan Hospital | Institute for Medical Research, Infectious Disease Research Centre, National Institutes of Health, Ministry of Health Malaysia | Anasir MI; Azizan MA; Kamel K; Mohd Zawawi Z; Ramly N; Robert F; Suppiah J; Thayan R |
| EPI_ISL_2854068 | Lahad Datu Hospital | Institute for Medical Research, Infectious Disease Research Centre, National Institutes of Health, Ministry of Health Malaysia | Anasir MI; Azizan MA; Kamel K; Mohd Zawawi Z; Ramly N; Robert F; Suppiah J; Thayan R |
| EPI_ISL_2854188 | Marang District Health Office | Institute for Medical Research, Infectious Disease Research Centre, National Institutes of Health, Ministry of Health Malaysia | Anasir MI; Azizan MA; Kamel K; Mohd Zawawi Z; Ramly N; Robert F; Suppiah J; Thayan R |
| EPI_ISL_2684234, EPI_ISL_2684235, EPI_ISL_2814916, EPI_ISL_2815327, EPI_ISL_2815328, EPI_ISL_2854049, EPI_ISL_2854050, EPI_ISL_2854051, EPI_ISL_2921996 | | | |
| see above | Melaka Hospital | Institute for Medical Research, Infectious Disease Research Centre, National Institutes of Health, Ministry of Health Malaysia | Anasir MI; Azizan MA; Kamel K; Mohd Zawawi Z; Ramly N; Robert F; Suppiah J; Thayan R |
| EPI_ISL_2535778, EPI_ISL_2535779 | Ministry of Health Hospitals | Institute of Health and Community Medicine | Chan Chia Jui; Chua Hock Hin; David Perera; Ooi Mong How; Tonnii Sia Loong Loong; Wong Jyn Shan; Wong Kiing Aik |
| EPI_ISL_2614740, EPI_ISL_2614742 | Miri Hospital | Institute for Medical Research, Infectious Disease Research Centre, National Institutes of Health, Ministry of Health Malaysia | Azizan MA; Kamel K; Mohd Zawawi Z; Ramly N; Robert F; Suppiah J; Thayan R |
| EPI_ISL_2379748 | National Heart Institute | Institute for Medical Research, Infectious Disease Research Centre, National Institutes of Health, Ministry of Health Malaysia | Azizan MA; Kamel K; Mohd Zawawi Z; Ramly N; Robert F; Suppiah J; Thayan R |
| EPI_ISL_2921263, EPI_ISL_2921264, EPI_ISL_2921265, EPI_ISL_2921266 | Pahang State Health Department | Institute for Medical Research, Infectious Disease Research Centre, National Institutes of Health, Ministry of Health Malaysia | Anasir MI; Azizan MA; Kamel K; Mohd Zawawi Z; Ramly N; Robert F; Suppiah J; Thayan R |
| EPI_ISL_2921262 | Pantai Hospital Kuala Lumpur | Institute for Medical Research, Infectious Disease Research Centre, National Institutes of Health, Ministry of Health Malaysia | Anasir MI; Azizan MA; Kamel K; Mohd Zawawi Z; Ramly N; Robert F; Suppiah J; Thayan R |
| EPI_ISL_2811871, EPI_ISL_2811873 | Pasir Mas Town Health Clinic | Institute for Medical Research, Infectious Disease Research Centre, National Institutes of Health, Ministry of Health Malaysia | Azizan MA; Kamel K; Mohd Zawawi Z; Ramly N; Robert F; Suppiah J; Thayan R |
| EPI_ISL_2811984 | Pekan Tajau Health Clinic | Institute for Medical Research, Infectious Disease Research Centre, National Institutes of Health, Ministry of Health Malaysia | Azizan MA; Kamel K; Mohd Zawawi Z; Ramly N; Robert F; Suppiah J; Thayan R |

| | | | |
|---|---|---|---|
| EPI_ISL_2649998, EPI_ISL_2649999 | Penang General Hospital | Institute for Medical Research, Infectious Disease Research Centre, National Institutes of Health, Ministry of Health Malaysia | Azizan MA; Kamel K; Mohd Zawawi Z; Ramly N; Robert F; Suppiah J; Thayan R |
| EPI_ISL_2550758, EPI_ISL_2550759 | Port Dickson Hospital | Institute for Medical Research, Infectious Disease Research Centre, National Institutes of Health, Ministry of Health Malaysia | Azizan MA; Kamel K; Mohd Zawawi Z; Ramly N; Robert F; Suppiah J; Thayan R |
| EPI_ISL_2650027, EPI_ISL_2650430, EPI_ISL_2650432, EPI_ISL_2650499, EPI_ISL_2650505, EPI_ISL_2854106 | Queen Elizabeth Hospital | Institute for Medical Research, Infectious Disease Research Centre, National Institutes of Health, Ministry of Health Malaysia | Anasir MI; Azizan MA; Kamel K; Mohd Zawawi Z; Ramly N; Robert F; Suppiah J; Thayan R |
| EPI_ISL_2757941, EPI_ISL_2757942, EPI_ISL_2931924 | Queen Elizabeth Hospital, Kota Kinabalu | Institute for Medical Research, Infectious Disease Research Centre, National Institutes of Health, Ministry of Health Malaysia | Anasir MI; Azizan MA; Kamel K; Mohd Zawawi Z; Ramly N; Robert F; Suppiah J; Thayan R |
| EPI_ISL_2811886 | Raja Perempuan Zainab II Hospital, Kota Bharu | Institute for Medical Research, Infectious Disease Research Centre, National Institutes of Health, Ministry of Health Malaysia | Azizan MA; Kamel K; Mohd Zawawi Z; Ramly N; Robert F; Suppiah J; Thayan R |
| EPI_ISL_2812026, EPI_ISL_2812027, EPI_ISL_2812028 | Rembau Health Clinic | Institute for Medical Research, Infectious Disease Research Centre, National Institutes of Health, Ministry of Health Malaysia | Azizan MA; Kamel K; Mohd Zawawi Z; Ramly N; Robert F; Suppiah J; Thayan R |
| EPI_ISL_2614745 | Sarawak General Hospital | Institute for Medical Research, Infectious Disease Research Centre, National Institutes of Health, Ministry of Health Malaysia | Azizan MA; Kamel K; Mohd Zawawi Z; Ramly N; Robert F; Suppiah J; Thayan R |
| EPI_ISL_2649995 | Segamat Hospital | Institute for Medical Research, Infectious Disease Research Centre, National Institutes of Health, Ministry of Health Malaysia | Azizan MA; Kamel K; Mohd Zawawi Z; Ramly N; Robert F; Suppiah J; Thayan R |
| EPI_ISL_2812706, EPI_ISL_2921612, EPI_ISL_2924137, EPI_ISL_2924139 | Sultan Haji Ahmad Shah Hospital | Institute for Medical Research, Infectious Disease Research Centre, National Institutes of Health, Ministry of Health Malaysia | Anasir MI; Azizan MA; Kamel K; Mohd Zawawi Z; Ramly N; Robert F; Suppiah J; Thayan R |
| EPI_ISL_2649997, EPI_ISL_2854107 | Sultan Ismail Hospital | Institute for Medical Research, Infectious Disease Research Centre, National Institutes of Health, Ministry of Health Malaysia | Anasir MI; Azizan MA; Kamel K; Mohd Zawawi Z; Ramly N; Robert F; Suppiah J; Thayan R |
| EPI_ISL_2650028 | Sultanah Aminah Hospital | Institute for Medical Research, Infectious Disease Research Centre, National Institutes of Health, Ministry of Health Malaysia | Azizan MA; Kamel K; Mohd Zawawi Z; Ramly N; Robert F; Suppiah J; Thayan R |
| EPI_ISL_2091024, EPI_ISL_2625637 | Sultanah Aminah Hospital, Johor Bahru | Institute for Medical Research, Infectious Disease Research Centre, National Institutes of Health, Ministry of Health Malaysia | Azizan MA; Kamel K; Mohd Zawawi Z; Ramly N; Robert F; Suppiah J; Thayan R |
| EPI_ISL_2650014, EPI_ISL_2650015 | Sultanah Bahiyah Hospital | Institute for Medical Research, Infectious Disease Research Centre, National Institutes of Health, Ministry of Health Malaysia | Azizan MA; Kamel K; Mohd Zawawi Z; Ramly N; Robert F; Suppiah J; Thayan R |
| EPI_ISL_2614739, EPI_ISL_2683247, EPI_ISL_2683496, EPI_ISL_2684236, EPI_ISL_2811885, EPI_ISL_2812654, EPI_ISL_2812704, EPI_ISL_2815656, EPI_ISL_2815971 | | | |
| see above | Sultanah Bahiyah Hospital, Alor Setar | Institute for Medical Research, Infectious Disease Research Centre, National Institutes of Health, Ministry of Health Malaysia | Azizan MA; Kamel K; Mohd Zawawi Z; Ramly N; Robert F; Suppiah J; Thayan R |
| EPI_ISL_2931920, EPI_ISL_2931923 | Sultanah Fatimah Specialist Hospital, Muar | Institute for Medical Research, Infectious Disease Research Centre, National Institutes of Health, Ministry of Health Malaysia | Anasir MI; Azizan MA; Kamel K; Mohd Zawawi Z; Ramly N; Robert F; Suppiah J; Thayan R |
| EPI_ISL_2811879, EPI_ISL_2811880, EPI_ISL_2811881, EPI_ISL_2811882, EPI_ISL_2811883 | Sultanah Nora Ismail Hospital, Batu Pahat | Institute for Medical Research, Infectious Disease Research Centre, National Institutes of Health, Ministry of Health Malaysia | Azizan MA; Kamel K; Mohd Zawawi Z; Ramly N; Robert F; Suppiah J; Thayan R |
| EPI_ISL_2650000 | Sultanah Nur Zahirah Hospital | Institute for Medical Research, Infectious Disease Research Centre, National Institutes of Health, Ministry of Health Malaysia | Azizan MA; Kamel K; Mohd Zawawi Z; Ramly N; Robert F; Suppiah J; Thayan R |
| EPI_ISL_2614743, EPI_ISL_2650004, EPI_ISL_2650005, EPI_ISL_2650010, EPI_ISL_2683956 | Sungai Buloh Hospital | Institute for Medical Research, Infectious Disease Research Centre, National Institutes of Health, Ministry of Health Malaysia | Azizan MA; Kamel K; Mohd Zawawi Z; Ramly N; Robert F; Suppiah J; Thayan R |
| EPI_ISL_2379749, EPI_ISL_2650506, EPI_ISL_2854070, EPI_ISL_2854103, EPI_ISL_2854104, EPI_ISL_2854105, EPI_ISL_2854108, EPI_ISL_2854109, EPI_ISL_2854174, EPI_ISL_2854186 | | | |
| see above | Tawau Hospital | Institute for Medical Research, Infectious Disease Research Centre, National Institutes of Health, Ministry of Health Malaysia | 40170 Selangor; Anasir MI; Azizan MA; Bandar Setia Alam; Jalan Setia Murni U13/52; Kamel K; Malaysia; Mohd Zawawi Z; Ramly N; Robert F; Suppiah J; Thayan R |
| EPI_ISL_2811959 | Tengku Ampuan Afzan Hospital (HTAA) | Institute for Medical Research, Infectious Disease Research Centre, National Institutes of Health, Ministry of Health Malaysia | Azizan MA; Kamel K; Mohd Zawawi Z; Ramly N; Robert F; Suppiah J; Thayan R |
| EPI_ISL_2921997 | Tengku Ampuan Rahimah Hospital | Institute for Medical Research, Infectious Disease Research Centre, National Institutes of Health, Ministry of Health Malaysia | Anasir MI; Azizan MA; Kamel K; Mohd Zawawi Z; Ramly N; Robert F; Suppiah J; Thayan R |
| EPI_ISL_2921260, EPI_ISL_2921261 | Terengganu State Health Department | Institute for Medical Research, Infectious Disease Research Centre, National Institutes of Health, Ministry of Health Malaysia | Anasir MI; Azizan MA; Kamel K; Mohd Zawawi Z; Ramly N; Robert F; Suppiah J; Thayan R |
| EPI_ISL_2614744, EPI_ISL_2649992 | Tuanku Ampuan Najihah Hospital, Kuala Pilah | Institute for Medical Research, Infectious Disease Research Centre, National Institutes of Health, Ministry of Health Malaysia | Azizan MA; Kamel K; Mohd Zawawi Z; Ramly N; Robert F; Suppiah J; Thayan R |
| EPI_ISL_2649994 | Tuanku Fauziah Hopsital | Institute for Medical Research, Infectious Disease Research Centre, National Institutes of Health, Ministry of Health Malaysia | Azizan MA; Kamel K; Mohd Zawawi Z; Ramly N; Robert F; Suppiah J; Thayan R |
| EPI_ISL_2931922 | Tuanku Fauziah Hospital | Institute for Medical Research, Infectious Disease Research Centre, National Institutes of Health, Ministry of Health Malaysia | Anasir MI; Azizan MA; Kamel K; Mohd Zawawi Z; Ramly N; Robert F; Suppiah J; Thayan R |
| EPI_ISL_2650006, EPI_ISL_2811872 | Tumpat Hospital | Institute for Medical Research, Infectious Disease Research Centre, National Institutes of Health, Ministry of Health Malaysia | Azizan MA; Kamel K; Mohd Zawawi Z; Ramly N; Robert F; Suppiah J; Thayan R |
| EPI_ISL_2811875 | Universiti Sains Malaysia Hospital | Institute for Medical Research, Infectious Disease Research Centre, National Institutes of Health, Ministry of Health Malaysia | Azizan MA; Kamel K; Mohd Zawawi Z; Ramly N; Robert F; Suppiah J; Thayan R |

We gratefully acknowledge the following Authors from the Originating laboratories responsible for obtaining the specimens, as well as the Submitting laboratories where the genome data were generated and shared via GISAID, on which this research is based.

All Submitters of data may be contacted directly via www.gisaid.org

Authors are sorted alphabetically.

| Accession ID | Originating Laboratory | Submitting Laboratory | Authors |
| --- | --- | --- | --- |
| EPI_ISL_1446386, EPI_ISL_1446389, EPI_ISL_1446392, EPI_ISL_1446395, EPI_ISL_1446398, EPI_ISL_1446401, EPI_ISL_1446404, EPI_ISL_1446406, EPI_ISL_1446409, EPI_ISL_1446632, EPI_ISL_1446633, EPI_ISL_1446635, EPI_ISL_1446636, EPI_ISL_1446637, EPI_ISL_1446638, EPI_ISL_1446639, EPI_ISL_1528378, EPI_ISL_1528379, EPI_ISL_1528380, EPI_ISL_1528381, EPI_ISL_1528382, EPI_ISL_1528383, EPI_ISL_1528384, EPI_ISL_1528385, EPI_ISL_1678081, EPI_ISL_1678082, EPI_ISL_1678266, EPI_ISL_1678267, EPI_ISL_1678268, EPI_ISL_1678269, EPI_ISL_1678270, EPI_ISL_1711719, EPI_ISL_1711723, EPI_ISL_1711729, EPI_ISL_1711737, EPI_ISL_1711741, EPI_ISL_1711743, EPI_ISL_1711746, EPI_ISL_1711748, EPI_ISL_1711749, EPI_ISL_1823774, EPI_ISL_1823775, EPI_ISL_1823776, EPI_ISL_1823777, EPI_ISL_1823778, EPI_ISL_1823779, EPI_ISL_1823780 | | | |
| see above | "NM Dept. Health, Scientific Laboratory Division " | Centers for Disease Control and Prevention Division of Viral Diseases, Pathogen Discovery | Alison Laufer Halpin; Ben L. Rambo-Martin; Clinton R. Paden; Dakota Howard; Darlene Wagner; Dave Wentworth; Dhwani Batra; Jasmine Padilla; Justin Lee; Katie Dillon; Krista Queen; Kristen Knipe; Kristine Lacek; Mark Burroughs; Matthew Schmerer; Mili Sheth; Peter Cook; Sam Shepard; Sarah Nobles; Shoshona Le; Suxiang Tong; Vivien Dugan; Yvette Unoarumhi |
| EPI_ISL_1445423, EPI_ISL_1479554, EPI_ISL_1479819, EPI_ISL_1479820, EPI_ISL_1491845, EPI_ISL_1491871, EPI_ISL_1513357, EPI_ISL_1513445, EPI_ISL_1550703, EPI_ISL_1550765, EPI_ISL_1551297, EPI_ISL_1560356, EPI_ISL_1562315, EPI_ISL_1562504, EPI_ISL_1562505, EPI_ISL_1649330, EPI_ISL_1685919, EPI_ISL_1685921, EPI_ISL_1685923, EPI_ISL_1685926, EPI_ISL_1685935, EPI_ISL_1686200, EPI_ISL_1686394, EPI_ISL_1686401, EPI_ISL_1687013, EPI_ISL_1687548, EPI_ISL_1687573, EPI_ISL_1687574, EPI_ISL_1687865, EPI_ISL_1688781, EPI_ISL_1690089, EPI_ISL_1690219, EPI_ISL_1735929, EPI_ISL_1736462, EPI_ISL_1736466, EPI_ISL_1736765, EPI_ISL_1736770, EPI_ISL_1736960, EPI_ISL_1753541, EPI_ISL_1834633, EPI_ISL_1835447, EPI_ISL_1835731, EPI_ISL_1836201, EPI_ISL_1836508, EPI_ISL_1838373, EPI_ISL_1925192, EPI_ISL_1925196, EPI_ISL_1942219, EPI_ISL_1995315, EPI_ISL_1997772, EPI_ISL_1997921, EPI_ISL_1998413, EPI_ISL_1998562, EPI_ISL_1999081, EPI_ISL_1999278, EPI_ISL_1999300, EPI_ISL_1999918, EPI_ISL_2000405, EPI_ISL_2039050, EPI_ISL_2039633, EPI_ISL_2039775, EPI_ISL_2039801, EPI_ISL_2039854, EPI_ISL_2040114, EPI_ISL_2040380, EPI_ISL_2040426, EPI_ISL_2040728, EPI_ISL_2040806, EPI_ISL_2041644, EPI_ISL_2041696, EPI_ISL_2041726, EPI_ISL_2041928, EPI_ISL_2041967, EPI_ISL_2043407, EPI_ISL_2043428, EPI_ISL_2089650, EPI_ISL_2089653, EPI_ISL_2089654, EPI_ISL_2089655, EPI_ISL_2089861, EPI_ISL_2089983, EPI_ISL_2090085, EPI_ISL_2132573, EPI_ISL_2148022, EPI_ISL_2148698, EPI_ISL_2150184, EPI_ISL_2150742, EPI_ISL_2185937, EPI_ISL_2186001 | | | |
| see above | Aegis Sciences Corporation | Centers for Disease Control and Prevention Division of Viral Diseases, Pathogen Discovery | Adrian Paskey; Alec Vest; Benjamin Rambo-Martin; Christopher Gulvick; Clinton R. Paden; Cyndi Clark; Dakota Howard; Darlene Wagner; Dhwani Batra; Dillon Nall; Duncan MacCannell; Ethan Sanders; Holly Houdeshell; Jason Caravas; Kara Moser; Matthew Hardison; Matthew Schmerer; Ola Kvalvaag; Patrick Campbell; Peter W. Cook; Rob Case; Scott Sammons; Shatavia Morrison; Shaun Westlund; Vikramsinha Ghorpade; Yvette Unoarumhi |
| EPI_ISL_2105724, EPI_ISL_2105725, EPI_ISL_2105726, EPI_ISL_2105727, EPI_ISL_2105728, EPI_ISL_2105729, EPI_ISL_2105730, EPI_ISL_2105732, EPI_ISL_2105733, EPI_ISL_2105734, EPI_ISL_2105735, EPI_ISL_2105736, EPI_ISL_2105737, EPI_ISL_2105740 | | | |
| see above | Arión Genética | Instituto Nacional de Medicina Genomica | Cedro-Tanda A; Escobar-Arrazola M; Gonzalez-Barrera D; Herrera-Montalvo LA.; Hidalgo-Miranda A; Mendoza-Vargas A; Munguia-Garza P; Ramirez-Vega O; Rangel-DeLeon D; Reyes-Grajeda JP; Roldan-Castillo Magaly; Uribe-Figueroa Laura; Verea Jazmin |
| EPI_ISL_1661979, EPI_ISL_1662062, EPI_ISL_1662063, EPI_ISL_1662064, EPI_ISL_1662065, EPI_ISL_1662066, EPI_ISL_1662067, EPI_ISL_1662068, EPI_ISL_1662069, EPI_ISL_1662075, EPI_ISL_1662076, EPI_ISL_1662077, EPI_ISL_1662078, EPI_ISL_1662079, EPI_ISL_1662080, EPI_ISL_1662081, EPI_ISL_1662082, EPI_ISL_1662083, EPI_ISL_1662084, EPI_ISL_1662085, EPI_ISL_1662086, EPI_ISL_1662087, EPI_ISL_1662088, EPI_ISL_1662089, EPI_ISL_1662090, EPI_ISL_1662091, EPI_ISL_1662092, EPI_ISL_1662093, EPI_ISL_1662094, EPI_ISL_1662095, EPI_ISL_1662096, EPI_ISL_1662097, EPI_ISL_1662098, EPI_ISL_1662099, EPI_ISL_1662100, EPI_ISL_1662101, EPI_ISL_1662102, EPI_ISL_1662103, EPI_ISL_1662123, EPI_ISL_1662124, EPI_ISL_1662125, EPI_ISL_1662126, EPI_ISL_1662127, EPI_ISL_1662131, EPI_ISL_1662132, EPI_ISL_1662186, EPI_ISL_1662187, EPI_ISL_1662188, EPI_ISL_1662189, EPI_ISL_1662190, EPI_ISL_1662191, EPI_ISL_1662192, EPI_ISL_1662193, EPI_ISL_1662197, EPI_ISL_1662198 | | | |
| see above | Centro de Investigacion Biomedica de Occidente (CIBO) | Unidad de Genomica Avanzada | Alejandro Sanchez-Flores; Alfredo Herrera-Estrella; Alicia Ocana-Mondragon; Angel Gustavo Salas-Lais; Bernardo Martinez-Miguel; Blanca Taboada; Brenda Irasema Maldonado-Meza; Carla Ivon Herrera-Najera; Carlos F. Arias; Celia Boukadida; Celida Duque Molina; Clara Esperanza Santacruz-Tinoco; Concepcion Grajales-Muniz; Consorcio Mexicano de Vigilancia Genomica (CoViGen-Mex). Authors (in alphabetical order): Julio Elias Alvarado-Yaah; Fernando Fontove-Herrera; Francisco Pulido; Gloria Elena Espinosa-Ayala; Gloria Maria Molina-Salinas; Gloria Vazquez; Hector Esteban Paz-Juarez; Hector Montoya-Fuentes; Helen Haydee Fernanda Ramirez-Plascencia; Jose Antonio Enciso-Moreno; Jose Esteban Munoz-Medina; Jose de Jesus Nunez-Contreras; Juan Bautista Chale-Dzul; Luis Alberto Ochoa-Carrera; Margarita Matias-Florentino; Maria Guadalupe Santiago-Mauricio; Maria Guadalupe de Jesus Mireles-Rivera; Nelly Selem-Mojica; Pavel Isa; Ricardo Grande; Santiago Ávila-Rios; Victor Eduardo Garcia-Arias; Victor Hugo Borja-Aburto |
| EPI_ISL_1381234, EPI_ISL_1381235, EPI_ISL_1381236, EPI_ISL_1381237, EPI_ISL_1381238, EPI_ISL_1381239, EPI_ISL_1381240, EPI_ISL_1381241, EPI_ISL_1381242, EPI_ISL_1381243, EPI_ISL_1381244, EPI_ISL_1381245, EPI_ISL_1381246, EPI_ISL_1381247, EPI_ISL_1381248, EPI_ISL_1381249, EPI_ISL_1381250, EPI_ISL_1381251, EPI_ISL_1381252, EPI_ISL_1381253, EPI_ISL_1381254, EPI_ISL_1381255, EPI_ISL_1381256, EPI_ISL_1381258, EPI_ISL_1661982, EPI_ISL_1661983, EPI_ISL_1661984, EPI_ISL_1661985, EPI_ISL_1661986, EPI_ISL_1661987, EPI_ISL_1661988, EPI_ISL_1661989, EPI_ISL_1661990, EPI_ISL_1662035, EPI_ISL_1662036, EPI_ISL_1662037, EPI_ISL_1662038, EPI_ISL_1662039, EPI_ISL_1662040, EPI_ISL_1662041, EPI_ISL_1662042, EPI_ISL_1662043, EPI_ISL_1662044, EPI_ISL_1662045, EPI_ISL_1662046, EPI_ISL_1662047, EPI_ISL_1662048, EPI_ISL_1662049, EPI_ISL_1662050, EPI_ISL_1662051, EPI_ISL_1662052, EPI_ISL_1662053, EPI_ISL_1662054, EPI_ISL_1662055, EPI_ISL_1662056, EPI_ISL_1662057, EPI_ISL_1662133, EPI_ISL_1662134, EPI_ISL_1662135, EPI_ISL_1662136, EPI_ISL_1662137, EPI_ISL_1662138, EPI_ISL_1662139, EPI_ISL_1662140, EPI_ISL_1662141, EPI_ISL_1662142, EPI_ISL_1662143, EPI_ISL_1662144, EPI_ISL_1662199, EPI_ISL_1662200 | | | |
| see above | Centro de Investigacion Biomedica del Noreste (CIBIN) | Unidad de Genomica Avanzada | Alejandro Sanchez-Flores; Alfredo Herrera-Estrella; Alicia Ocana-Mondragon; Angel Gustavo Salas-Lais; Bernardo Martinez-Miguel; Blanca Taboada; Brenda Irasema Maldonado-Meza; Carla Ivon Herrera-Najera; Carlos F. Arias; Celia Boukadida; Celida Duque Molina; Clara Esperanza Santacruz-Tinoco; Concepcion Grajales-Muniz; Consorcio Mexicano de Vigilancia Genomica (CoViGen-Mex). Authors (in alphabetical order): Julio Elias Alvarado-Yaah; Fernando Fontove-Herrera; Francisco Pulido; Gloria Elena Espinosa-Ayala; Gloria Elena Espinoza-Ayala; Gloria Maria Molina-Salinas; Gloria Vazquez; Hector Esteban Paz-Juarez; Hector Montoya-Fuentes; Helen Haydee Fernanda Ramirez-Plascencia; Jorge Ivan Salinal-Nevarez; Jose Antonio Enciso-Moreno; Jose Esteban Munoz-Medina; Jose de Jesus Nunez-Contreras; Juan Bautista Chale-Dzul; Luis Alberto Ochoa-Carrera; Margarita Matias-Florentino; Maria Guadalupe Santiago-Mauricio; Maria Guadalupe de Jesus Mireles-Rivera; Nelly Selem-Mojica; Pavel Isa; Ricardo Grande; Santiago Avila-Rios; Santiago Ávila-Rios; Victor Eduardo Garcia-Arias; Victor Hugo Borja-Aburto |
| EPI_ISL_1351511, EPI_ISL_1351517, EPI_ISL_1351520, EPI_ISL_1351526, EPI_ISL_1351529, EPI_ISL_1351532, EPI_ISL_1351535, EPI_ISL_1351538, EPI_ISL_1351540, EPI_ISL_1351543, EPI_ISL_1351555, EPI_ISL_1351558, EPI_ISL_1351561, EPI_ISL_1351564, EPI_ISL_1351567, EPI_ISL_1351571, EPI_ISL_1351574, EPI_ISL_1351577, EPI_ISL_1351580, EPI_ISL_1351583, EPI_ISL_1351585, EPI_ISL_1351589, EPI_ISL_1351592, EPI_ISL_1351595, EPI_ISL_1351598, EPI_ISL_1351601, EPI_ISL_1351605, EPI_ISL_1351608, EPI_ISL_1351611, EPI_ISL_1351614, EPI_ISL_1351617, EPI_ISL_1351620, EPI_ISL_1351623, EPI_ISL_1351626, EPI_ISL_1351629, EPI_ISL_1351632, EPI_ISL_1351635, EPI_ISL_1351638, EPI_ISL_1351642, EPI_ISL_1351645, EPI_ISL_1351648, EPI_ISL_1351651, EPI_ISL_1351654, EPI_ISL_1351656, EPI_ISL_1351659, EPI_ISL_1351662, EPI_ISL_1351665, EPI_ISL_1351668, EPI_ISL_1351670, EPI_ISL_1351672, EPI_ISL_1351675, EPI_ISL_1351679, EPI_ISL_1351682, EPI_ISL_1351685, EPI_ISL_1351688, EPI_ISL_1351691, EPI_ISL_1351694, EPI_ISL_1351697, EPI_ISL_1351700, EPI_ISL_1351703, EPI_ISL_1351706, EPI_ISL_1351709 | | | |
| see above | Centro de InvestigaciÃ³n BiomÃ©dica de Occidente (CIBO) | Unidad de Genomica Avanzada | Alejandro Sanchez-Flores; Alfredo Herrera-Estrella; Alicia Ocana-Mondragon; Angel Gustavo Salas-Lais; Bernardo Martinez-Miguel; Blanca Taboada; Brenda Irasema Maldonado-Meza; Carla Ivon Herrera-Najera; Carlos F. Arias; Celia Boukadida; Clara Esperanza Santacruz-Tinoco; Concepcion Grajales-Muniz; Consorcio Mexicano de Vigilancia Genomica (CoViGen-Mex). Authors (in alphabetical order): Julio Elias Alvarado-Yaah; Fernando Fontove-Herrera; Francisco Pulido; Gloria Elena Espinoza-Ayala; Gloria Maria Molina-Salinas; Gloria Vazquez; Hector Esteban Paz-Juarez; Hector Montoya-Fuentes; Helen Haydee Fernanda Ramirez-Plascencia; Jorge Ivan Salinal-Nevarez; Jose Antonio Enciso-Moreno; Jose Esteban Munoz-Medina; Jose de Jesus Nunez-Contreras; Juan Bautista Chale-Dzul; Luis Alberto Ochoa-Carrera; Margarita Matias-Florentino; Maria Guadalupe Santiago-Mauricio; Maria Guadalupe de Jesus Mireles-Rivera; Nelly Selem-Mojica; Pavel Isa; Ricardo Grande; Santiago Avila-Rios; Victor Hugo Borja-Aburto |
| EPI_ISL_1351441, EPI_ISL_1351443, EPI_ISL_1351445, EPI_ISL_1351448, EPI_ISL_1351451, EPI_ISL_1351454, EPI_ISL_1351457, EPI_ISL_1351460, EPI_ISL_1351462 | | | |
| see above | Centro de InvestigaciÃ³n BiomÃ©dica del Noreste (CIBIN) | Unidad de Genomica Avanzada | Alejandro Sanchez-Flores; Alfredo Herrera-Estrella; Alicia Ocana-Mondragon; Angel Gustavo Salas-Lais; Bernardo Martinez-Miguel; Blanca Taboada; Brenda Irasema Maldonado-Meza; Carla Ivon Herrera-Najera; Carlos F. Arias; Celia Boukadida; Clara Esperanza Santacruz-Tinoco; Concepcion Grajales-Muniz; Consorcio Mexicano de Vigilancia Genomica (CoViGen-Mex). Authors (in alphabetical order): Julio Elias Alvarado-Yaah; Fernando Fontove-Herrera; Francisco Pulido; Gloria Elena Espinoza-Ayala; Gloria Maria Molina-Salinas; Gloria Vazquez; Hector Esteban Paz-Juarez; Hector Montoya-Fuentes; Helen Haydee Fernanda Ramirez-Plascencia; Jorge Ivan Salinal-Nevarez; Jose Antonio Enciso-Moreno; Jose Esteban Munoz-Medina; Jose de Jesus Nunez-Contreras; Juan Bautista Chale-Dzul; Luis Alberto Ochoa-Carrera; Margarita Matias-Florentino; Maria Guadalupe Santiago-Mauricio; Maria Guadalupe de Jesus Mireles-Rivera; Nelly Selem-Mojica; Pavel Isa; Ricardo Grande; Santiago Avila-Rios; Victor Hugo Borja-Aburto |
| EPI_ISL_2091140, EPI_ISL_2091141, EPI_ISL_2091142, EPI_ISL_2091145, EPI_ISL_2091258, EPI_ISL_2091259, EPI_ISL_2091260, EPI_ISL_2091261, EPI_ISL_2091262, EPI_ISL_2091263, EPI_ISL_2091264, EPI_ISL_2091265, EPI_ISL_2091266, EPI_ISL_2091267, EPI_ISL_2091268, EPI_ISL_2091269, EPI_ISL_2091270, EPI_ISL_2091271, EPI_ISL_2091272, EPI_ISL_2091273, EPI_ISL_2091274, EPI_ISL_2091275, EPI_ISL_2091276, EPI_ISL_2091277, EPI_ISL_2091278, EPI_ISL_2091279, EPI_ISL_2091280, EPI_ISL_2091281, EPI_ISL_2091282, EPI_ISL_2091283, EPI_ISL_2091284, EPI_ISL_2091285, EPI_ISL_2091286, EPI_ISL_2091287, EPI_ISL_2091288, EPI_ISL_2091289, EPI_ISL_2091290, EPI_ISL_2091291, EPI_ISL_2091292, EPI_ISL_2091293, EPI_ISL_2091294, EPI_ISL_2091295, EPI_ISL_2091296, EPI_ISL_2091297, EPI_ISL_2091298, EPI_ISL_2091299, EPI_ISL_2091300, EPI_ISL_2091301, EPI_ISL_2091302, EPI_ISL_2091303, EPI_ISL_2091304, EPI_ISL_2091305, EPI_ISL_2091306, EPI_ISL_2091307, EPI_ISL_2091308, EPI_ISL_2091309, EPI_ISL_2091310, EPI_ISL_2091311, EPI_ISL_2091312, EPI_ISL_2091313, EPI_ISL_2091314, EPI_ISL_2091315, EPI_ISL_2091316, EPI_ISL_2091317, EPI_ISL_2091318, EPI_ISL_2091319, EPI_ISL_2178729 | | | |
| see above | Centro de Investigación Biomédica de Occidente (CIBO) | Centro de Investigación en Enfermedades Infecciosas (CIENI), Instituto Nacional de Enfermedades Respiratorias (INER) | Alejandro Sanchez-Flores; Alfredo Herrera-Estrella; Alicia Ocaña-Mondragón; Angel Gustavo Salas-Lais; Bernardo Martínez-Miguel; Blanca Taboada; Brenda Irasema Maldonado-Meza; Carla Ivón Herrera-Najera; Carlos F. Arias; Celia Boukadida; Clara Esperanza Santacruz-Tinoco; Concepción Grajales-Muñiz; Consorcio Mexicano de Vigilancia Genómica (CoViGen-Mex). Authors (in alphabetical order): Julio Elias Alvarado-Yaah; Célida Duque Molina; Fernando Fontove-Herrera; Francisco Pulido; Gloria Elena Espinosa-Ayala; Gloria María Molina-Salinas; Gloria Vazquez; Hector Esteban Paz-Juárez; Hector Montoya-Fuentes; Helen Haydee Fernanda Ramírez-Plascencia; José Antonio Enciso-Moreno; José Esteban Muñoz-Medina; José de Jesús Nuñez-Contreras; Juan Bautista Chale-Dzul; Luis Alberto Ochoa-Carrera; Margarita Matías-Florentino; María Guadalupe Santiago-Mauricio; María Guadalupe de Jesús Mireles-Rivera; Nelly Sélem-Mojica; Pavel Isa; Ricardo Grande; Santiago Ávila-Ríos; Victor Eduardo Garcia-Arias; Víctor Hugo Borja-Aburto |
| EPI_ISL_1585480, EPI_ISL_1585481, EPI_ISL_1585482, EPI_ISL_1585483, EPI_ISL_1585484, EPI_ISL_1585485, EPI_ISL_1585486, EPI_ISL_1585487, EPI_ISL_1585488, EPI_ISL_1585489, EPI_ISL_1585490, EPI_ISL_1585491, EPI_ISL_1585492, EPI_ISL_1585493, EPI_ISL_1585494, EPI_ISL_1585495, EPI_ISL_1585496, EPI_ISL_1585497, EPI_ISL_1585498, EPI_ISL_1585499, EPI_ISL_1585500, EPI_ISL_1585501, EPI_ISL_1585502, EPI_ISL_1585503, EPI_ISL_1585504, EPI_ISL_1585505, EPI_ISL_1585506, EPI_ISL_1585507, EPI_ISL_1585508, EPI_ISL_1585509, EPI_ISL_1585510, EPI_ISL_1585511, EPI_ISL_1585512, EPI_ISL_1585513, EPI_ISL_1585514, EPI_ISL_1585515, EPI_ISL_1585516, EPI_ISL_1585517, EPI_ISL_1585518, EPI_ISL_1585519, EPI_ISL_1585520, EPI_ISL_1585521, EPI_ISL_1585522, EPI_ISL_1585523, EPI_ISL_1585524, EPI_ISL_1585525, EPI_ISL_1585526, EPI_ISL_1585527, EPI_ISL_1585528, EPI_ISL_1585529, EPI_ISL_1585530, EPI_ISL_1585531, EPI_ISL_1585532, EPI_ISL_1585533, EPI_ISL_1585534, EPI_ISL_1585535, EPI_ISL_1585536, EPI_ISL_1585537, EPI_ISL_1585538, EPI_ISL_1585539, EPI_ISL_1585540, EPI_ISL_1585541, EPI_ISL_1585542, EPI_ISL_1585543, EPI_ISL_1585544, EPI_ISL_1585545, EPI_ISL_1585546, EPI_ISL_1585547, EPI_ISL_1585548, EPI_ISL_1585549, EPI_ISL_1585550, EPI_ISL_1585551, EPI_ISL_1585552, EPI_ISL_1585553, EPI_ISL_1585554, EPI_ISL_1585555, EPI_ISL_1585556, EPI_ISL_1585557, EPI_ISL_1585558, EPI_ISL_1585559, EPI_ISL_1585560, EPI_ISL_1585561, EPI_ISL_1585562, EPI_ISL_1585563, EPI_ISL_1595601, EPI_ISL_1595602, EPI_ISL_1595604, EPI_ISL_1595605, EPI_ISL_1595606, EPI_ISL_1595607 | | | |
| see above | Centro de Investigación Biomédica de Occidente (CIBO) | Instituto Nacional de Enfermedades Respiratorias (INER): Centro de Investigación en Enfermedades Infecciosas (CIENI) | Alejandro Sanchez-Flores; Alfredo Herrera-Estrella; Alicia Ocaña-Mondragón; Angel Gustavo Salas-Lais; Bernardo Martínez-Miguel; Blanca Taboada; Brenda Irasema Maldonado-Meza; Carla Ivón Herrera-Najera; Carlos F. Arias; Celia Boukadida; Clara Esperanza Santacruz-Tinoco; Concepción Grajales-Muñiz; Consorcio Mexicano de Vigilancia Genómica (CoViGen-Mex). Authors (in alphabetical order): Julio Elias Alvarado-Yaah; Célida Duque Molina; Fernando Fontove-Herrera; Francisco Pulido; Gloria Elena Espinosa-Ayala; Gloria María Molina-Salinas; Gloria Vazquez; Hector Esteban Paz-Juárez; Hector Montoya-Fuentes; Helen Haydee Fernanda Ramírez-Plascencia; José Antonio Enciso-Moreno; José Esteban Muñoz-Medina; José de Jesús Nuñez-Contreras; Juan Bautista Chale-Dzul; Luis Alberto Ochoa-Carrera; Margarita Matías-Florentino; María Guadalupe Santiago-Mauricio; María Guadalupe de Jesús Mireles-Rivera; Nelly Sélem-Mojica; Pavel Isa; Ricardo Grande; Santiago Ávila-Ríos; Victor Eduardo Garcia-Arias; Víctor Hugo Borja-Aburto |
| EPI_ISL_2091138, EPI_ISL_2091139, EPI_ISL_2091160, EPI_ISL_2091161, EPI_ISL_2091162, EPI_ISL_2091163, EPI_ISL_2091164, EPI_ISL_2091165, EPI_ISL_2091166, EPI_ISL_2091167, EPI_ISL_2091168, EPI_ISL_2091169, EPI_ISL_2091170, EPI_ISL_2091171, EPI_ISL_2091172, EPI_ISL_2091173, EPI_ISL_2091174, EPI_ISL_2091175, EPI_ISL_2091176, EPI_ISL_2091177, EPI_ISL_2091178, EPI_ISL_2091179, EPI_ISL_2091180, EPI_ISL_2091182, EPI_ISL_2091183, EPI_ISL_2091184, EPI_ISL_2091185, EPI_ISL_2091186, EPI_ISL_2091187, EPI_ISL_2091188, EPI_ISL_2091189, EPI_ISL_2091190, EPI_ISL_2091191, EPI_ISL_2091192, EPI_ISL_2091193, EPI_ISL_2091194, EPI_ISL_2091195, EPI_ISL_2091196, EPI_ISL_2091197, EPI_ISL_2091198, EPI_ISL_2091199, EPI_ISL_2091200, EPI_ISL_2091201, EPI_ISL_2091202, EPI_ISL_2091203, EPI_ISL_2091204, EPI_ISL_2091205, EPI_ISL_2091206, EPI_ISL_2091207, EPI_ISL_2091208, EPI_ISL_2091209, EPI_ISL_2091210, EPI_ISL_2091211, EPI_ISL_2091212, EPI_ISL_2091213, EPI_ISL_2091214, EPI_ISL_2091215, EPI_ISL_2091216, EPI_ISL_2091217, EPI_ISL_2091218, EPI_ISL_2091219, EPI_ISL_2091220, EPI_ISL_2091221, EPI_ISL_2091222, EPI_ISL_2091223, EPI_ISL_2091224, EPI_ISL_2178725, EPI_ISL_2178726 | | | |
| see above | Centro de Investigación Biomédica del Noreste (CIBIN) | Centro de Investigación en Enfermedades Infecciosas (CIENI), Instituto Nacional de Enfermedades Respiratorias (INER) | Alejandro Sanchez-Flores; Alfredo Herrera-Estrella; Alicia Ocaña-Mondragón; Angel Gustavo Salas-Lais; Bernardo Martínez-Miguel; Blanca Taboada; Brenda Irasema Maldonado-Meza; Carla Ivón Herrera-Najera; Carlos F. Arias; Celia Boukadida; Clara Esperanza Santacruz-Tinoco; Concepción Grajales-Muñiz; Consorcio Mexicano de Vigilancia Genómica (CoViGen-Mex). Authors (in alphabetical order): Julio Elias Alvarado-Yaah; Célida Duque Molina; Fernando Fontove-Herrera; Francisco Pulido; Gloria Elena Espinosa-Ayala; Gloria María Molina-Salinas; Gloria Vazquez; Hector Esteban Paz-Juárez; Hector Montoya-Fuentes; Helen Haydee Fernanda Ramírez-Plascencia; José Antonio Enciso-Moreno; José Esteban Muñoz-Medina; José de Jesús Nuñez-Contreras; Juan Bautista Chale-Dzul; Luis Alberto Ochoa-Carrera; Margarita Matías-Florentino; María Guadalupe Santiago-Mauricio; María Guadalupe de Jesús Mireles-Rivera; Nelly Sélem-Mojica; Pavel Isa; Ricardo Grande; Santiago Ávila-Ríos; Victor Eduardo Garcia-Arias; Víctor Hugo Borja-Aburto |
| EPI_ISL_1585384, EPI_ISL_1585385, EPI_ISL_1585386, EPI_ISL_1585388, EPI_ISL_1585389, EPI_ISL_1585390, EPI_ISL_1585391, EPI_ISL_1585392, EPI_ISL_1585393, EPI_ISL_1585394, EPI_ISL_1585395, EPI_ISL_1585396, EPI_ISL_1585397, EPI_ISL_1585398, EPI_ISL_1585399, EPI_ISL_1585400, EPI_ISL_1585401, EPI_ISL_1585402, EPI_ISL_1585403, EPI_ISL_1585404, EPI_ISL_1585405, EPI_ISL_1585406, EPI_ISL_1585407, EPI_ISL_1585408, EPI_ISL_1585409, EPI_ISL_1585410, EPI_ISL_1585411, EPI_ISL_1585412, EPI_ISL_1585413, EPI_ISL_1585414, EPI_ISL_1585415, EPI_ISL_1585416, EPI_ISL_1585417, EPI_ISL_1585418, EPI_ISL_1585419, EPI_ISL_1585420, EPI_ISL_1585421, EPI_ISL_1585422, EPI_ISL_1585423, EPI_ISL_1585424, EPI_ISL_1585425, EPI_ISL_1585426, EPI_ISL_1585427, EPI_ISL_1595596, EPI_ISL_1595597, EPI_ISL_1595598 | | | |
| see above | Centro de Investigación Biomédica del Noreste (CIBIN) | Instituto Nacional de Enfermedades Respiratorias (INER): Centro de Investigación en Enfermedades Infecciosas (CIENI) | Alejandro Sanchez-Flores; Alfredo Herrera-Estrella; Alicia Ocaña-Mondragón; Angel Gustavo Salas-Lais; Bernardo Martínez-Miguel; Blanca Taboada; Brenda Irasema Maldonado-Meza; Carla Ivón Herrera-Najera; Carlos F. Arias; Celia Boukadida; Clara Esperanza Santacruz-Tinoco; Concepción Grajales-Muñiz; Consorcio Mexicano de Vigilancia Genómica (CoViGen-Mex). Authors (in alphabetical order): Julio Elias Alvarado-Yaah; Célida Duque Molina; Fernando Fontove-Herrera; Francisco Pulido; Gloria Elena Espinosa-Ayala; Gloria María Molina-Salinas; Gloria Vazquez; Hector Esteban Paz-Juárez; Hector Montoya-Fuentes; Helen Haydee Fernanda Ramírez-Plascencia; José Antonio Enciso-Moreno; José Esteban Muñoz-Medina; José de Jesús Nuñez-Contreras; Juan Bautista Chale-Dzul; Luis Alberto Ochoa-Carrera; Margarita Matías-Florentino; María Guadalupe Santiago-Mauricio; María Guadalupe de Jesús Mireles-Rivera; Nelly Sélem-Mojica; Pavel Isa; Ricardo Grande; Santiago Ávila-Ríos; Victor Eduardo Garcia-Arias; Víctor Hugo Borja-Aburto |
| EPI_ISL_1970836, EPI_ISL_1970837 | Centro de Investigación en Enfermedades Infecciosas (CIENI), Instituto Nacional de Enfermedades Respiratorias (INER) | Centro de Investigación en Enfermedades Infecciosas (CIENI), Instituto Nacional de Enfermedades Respiratorias (INER) | Alejandra Hernández-Terán; Alejandro Sanchez-Flores; Blanca Taboada; Carlos F. Arias; Celia Boukadida; Eduardo Becerril-Vargas; Fidencio Mejía-Nepomuceno; Francisco Pulido; Hector Esteban Paz-Juárez; Joel Armando Vázquez-Pérez and Consorcio Mexicano de Vigilancia Genómica (CoViGen-Mex); Jorge Salas-Hernández; José Arturo Martínez-Orozco; Margarita Matías-Florentino; Mario Mújica-Sánchez; Pavel Isa; Ricardo Grande; Santiago Ávila-Ríos |
| EPI_ISL_1582426, EPI_ISL_1582427, EPI_ISL_1582428, EPI_ISL_1582429, EPI_ISL_1582430, EPI_ISL_1582431, EPI_ISL_1582432, EPI_ISL_1582433, EPI_ISL_1582434, EPI_ISL_1582435, EPI_ISL_1582436, EPI_ISL_1582437, EPI_ISL_1582438, EPI_ISL_1582439, EPI_ISL_1582441, EPI_ISL_1582442, EPI_ISL_1582443, EPI_ISL_1805328, EPI_ISL_1805329, EPI_ISL_1805331, EPI_ISL_1805334, EPI_ISL_1805336, EPI_ISL_1805338, EPI_ISL_1805339, EPI_ISL_1805340, EPI_ISL_1805341, EPI_ISL_1805342, EPI_ISL_1805343, EPI_ISL_1805344, EPI_ISL_1805345, EPI_ISL_1805346, EPI_ISL_1805350, EPI_ISL_1805351, EPI_ISL_1805353, EPI_ISL_1805354, EPI_ISL_1805355, EPI_ISL_1805356, EPI_ISL_1805357, EPI_ISL_1805358, EPI_ISL_1805359, EPI_ISL_1805360, EPI_ISL_1805362, EPI_ISL_1805363, EPI_ISL_1805364, EPI_ISL_1805365, EPI_ISL_1805367, EPI_ISL_1825118, EPI_ISL_1970846, EPI_ISL_1970847, EPI_ISL_1970848, EPI_ISL_1970849, EPI_ISL_1970855, EPI_ISL_1970857, EPI_ISL_1970859, EPI_ISL_1970860, EPI_ISL_1970861, EPI_ISL_1970863, EPI_ISL_1970864, EPI_ISL_1970865, EPI_ISL_1970867, EPI_ISL_1970868, EPI_ISL_1970869, EPI_ISL_1970870, EPI_ISL_1970871, EPI_ISL_1970872, EPI_ISL_1970873, EPI_ISL_1970875, EPI_ISL_1970876, EPI_ISL_1970878, EPI_ISL_1970879, EPI_ISL_1970880, EPI_ISL_1970881, EPI_ISL_1970883, EPI_ISL_1970887, EPI_ISL_1970888, EPI_ISL_1970889, EPI_ISL_1970891, EPI_ISL_1970892, EPI_ISL_1970893, EPI_ISL_2185537, EPI_ISL_2185538, EPI_ISL_2185539, EPI_ISL_2185540, EPI_ISL_2185541, EPI_ISL_2185542, EPI_ISL_2185543, EPI_ISL_2185544, EPI_ISL_2185545, EPI_ISL_2185546, EPI_ISL_2185549, EPI_ISL_2185550, EPI_ISL_2185552, EPI_ISL_2185553, EPI_ISL_2185554, EPI_ISL_2185555, EPI_ISL_2185556, EPI_ISL_2185557, EPI_ISL_2185558, EPI_ISL_2185559, EPI_ISL_2185560, EPI_ISL_2185561, EPI_ISL_2185562, EPI_ISL_2185564, EPI_ISL_2185565, EPI_ISL_2185566, EPI_ISL_2185567, EPI_ISL_2185568, EPI_ISL_2185569, EPI_ISL_2185570, EPI_ISL_2185571, EPI_ISL_2185572, EPI_ISL_2185573, EPI_ISL_2185574 | | | |
| see above | Curative | New Mexico Department of Health | Anastacia Griego-Fisher; D'eldra Malone; Ellie Johnson; Jennifer Benoit; Linda Salazar; Mark Willmon; Ratheesh Rajan |

| | | | |
|---|---|---|---|
| | | Scientific Laboratory | |
| EPI_ISL_1459019, EPI_ISL_1459197, EPI_ISL_1459198, EPI_ISL_1460234, EPI_ISL_1460244, EPI_ISL_1481709, EPI_ISL_1481712, EPI_ISL_1481821, EPI_ISL_1525403, EPI_ISL_1525458, EPI_ISL_1556575, EPI_ISL_1614335, EPI_ISL_1665924, EPI_ISL_1666477, EPI_ISL_1733977, EPI_ISL_1734020, EPI_ISL_1734225, EPI_ISL_1734293, EPI_ISL_1839086, EPI_ISL_1840141, EPI_ISL_1992215, EPI_ISL_1992216, EPI_ISL_1992462, EPI_ISL_1992512, EPI_ISL_1992557, EPI_ISL_1992558, EPI_ISL_1992559, EPI_ISL_1992636, EPI_ISL_1992698, EPI_ISL_1992766, EPI_ISL_2037935, EPI_ISL_2038108, EPI_ISL_2038109, EPI_ISL_2038110, EPI_ISL_2043830, EPI_ISL_2043831 | | | |
| see above | Fulgent Genetics | Centers for Disease Control and Prevention Division of Viral Diseases, Pathogen Discovery | Adrian Paskey; Becky Tsai; Benafsh Sapra; Benjamin Rambo-Martin; Christopher Gulvick; Clinton R. Paden; Dakota Howard; Darlene Wagner; Dhwani Batra; Doreen Ng; Duncan MacCannell; Harry Gao; James Xie; Jason Caravas; John Gao; Joseph Fierro; Kara Moser; Matthew Schmerer; Mickey Li; Peter W. Cook; Scott Sammons; Shatavia Morrison; Yan Meng; Yvette Unoarumhi |
| EPI_ISL_1340922 | Gallup Indian Medical Center | New Mexico Department of Health Scientific Laboratory | Anastacia Griego-Fisher; D'eldra Malone; Ellie Johnson; Jennifer Benoit |
| EPI_ISL_1421747, EPI_ISL_1512261, EPI_ISL_1512715, EPI_ISL_1576590, EPI_ISL_1679317, EPI_ISL_1803976, EPI_ISL_1991070 | | | |
| see above | Helix/Illumina | Centers for Disease Control and Prevention Division of Viral Diseases, Pathogen Discovery | Adrian Paskey; Alexandre Bolze; Ary Ascencio; Ben L. Rambo-Martin; Benjamin Rambo-Martin; Brad Sickler; Charlotte Rivera-Garcia; Christine Tran; Christopher Gulvick; Clinton R. Paden; Dakota Howard; Darlene Wagner; David Becker; Dhwani Batra; Duncan MacCannell; Efren Sandoval; Eileen de Feo; Elizabeth Cirulli; Eric Allen; Geraint Levan; James Lu; Jan Antico; Jason Caravas; Jason Nguyen; Jimmy Ramirez; Jingtao Liu; Kara Moser; Kelly Schiabor Barrett; Kim Gietzen; Magnus Isaksson; Marc Laurent; Matthew Schmerer; Matthew Tolentino; Nicole L. Washington; Peter W. Cook; Phil Febbo; Ryan Cho; Scott Sammons; Shannon Wickline; Shatavia Morrison; Sherry Wang; Simon White; Summer Galloway; Suxiang Tong; Tyler Cassens; William Lee; Yvette Unoarumhi |
| EPI_ISL_1805395 | Holy Cross Hospital | New Mexico Department of Health Scientific Laboratory | Anastacia Griego-Fisher; D'eldra Malone; Ellie Johnson; Jennifer Benoit; Linda Salazar; Ratheesh Rajan |
| EPI_ISL_1627083, EPI_ISL_1627084, EPI_ISL_1660617, EPI_ISL_1660620, EPI_ISL_1660621, EPI_ISL_1660622, EPI_ISL_1660624, EPI_ISL_1660628, EPI_ISL_1972328, EPI_ISL_1972329, EPI_ISL_1972330, EPI_ISL_1972331, EPI_ISL_1972332, EPI_ISL_1972333, EPI_ISL_1972334, EPI_ISL_2026394, EPI_ISL_2026400, EPI_ISL_2026402, EPI_ISL_2026404, EPI_ISL_2026409, EPI_ISL_2026411, EPI_ISL_2026413, EPI_ISL_2026415, EPI_ISL_2026417, EPI_ISL_2026426, EPI_ISL_2026436, EPI_ISL_2026437, EPI_ISL_2026440, EPI_ISL_2026446, EPI_ISL_2026448, EPI_ISL_2026450, EPI_ISL_2026452, EPI_ISL_2026454, EPI_ISL_2026458 | | | |
| see above | Hospital Margarita Maza de Juárez | Microbial Genomics Laboratory | Alejandra Garcia-Gasca; Bruno Gomez-Gil; Daniel Fregoso-Rueda; Julissa Enciso-Ibarra |
| EPI_ISL_1482629, EPI_ISL_1482631, EPI_ISL_1627080, EPI_ISL_1627082, EPI_ISL_1627085, EPI_ISL_1660618, EPI_ISL_1660623, EPI_ISL_1660625, EPI_ISL_1660626, EPI_ISL_1972338, EPI_ISL_1972339, EPI_ISL_1972340, EPI_ISL_1972341, EPI_ISL_1972342, EPI_ISL_2026396, EPI_ISL_2026398, EPI_ISL_2026407, EPI_ISL_2026418, EPI_ISL_2026420, EPI_ISL_2026422, EPI_ISL_2026424, EPI_ISL_2026428, EPI_ISL_2026429, EPI_ISL_2026432, EPI_ISL_2026434, EPI_ISL_2026442, EPI_ISL_2026444, EPI_ISL_2026456, EPI_ISL_2026460, EPI_ISL_2026462 | | | |
| see above | Hospital Sharp | Microbial Genomics Laboratory | Alejandra Garcia-Gasca; Bruno Gomez-Gil; Daniel Fregoso-Rueda; Julissa Enciso-Ibarra |
| EPI_ISL_1491362, EPI_ISL_1627081, EPI_ISL_1660619, EPI_ISL_1660627, EPI_ISL_1972335, EPI_ISL_1972336, EPI_ISL_1972337, EPI_ISL_2026406 | | | |
| see above | Imágenes Diagnósticas de Mazatlán | Microbial Genomics Laboratory | Alejandra Garcia-Gasca; Bruno Gomez-Gil; Daniel Fregoso-Rueda; Julissa Enciso-Ibarra |
| EPI_ISL_1628596, EPI_ISL_1628597, EPI_ISL_1628598, EPI_ISL_1628599, EPI_ISL_1628600, EPI_ISL_1628601, EPI_ISL_1628602, EPI_ISL_1628603, EPI_ISL_1628604, EPI_ISL_1628605, EPI_ISL_1628606, EPI_ISL_1628607, EPI_ISL_1628608, EPI_ISL_1628609, EPI_ISL_1628610, EPI_ISL_1628611, EPI_ISL_1628612, EPI_ISL_1628613, EPI_ISL_1628614, EPI_ISL_1628615, EPI_ISL_1628616, EPI_ISL_1628617, EPI_ISL_1628618, EPI_ISL_1628619, EPI_ISL_1628620, EPI_ISL_1628621, EPI_ISL_1628622, EPI_ISL_1628623, EPI_ISL_1628624, EPI_ISL_1628625, EPI_ISL_1628626, EPI_ISL_1628627, EPI_ISL_1628628, EPI_ISL_1628629, EPI_ISL_1628630, EPI_ISL_1628631, EPI_ISL_1628632, EPI_ISL_1628633, EPI_ISL_1628634, EPI_ISL_1628635, EPI_ISL_1628636, EPI_ISL_1628637, EPI_ISL_1628638, EPI_ISL_1628639, EPI_ISL_1628640, EPI_ISL_1628641, EPI_ISL_1628642, EPI_ISL_1628643, EPI_ISL_1628644, EPI_ISL_1628645, EPI_ISL_1628646, EPI_ISL_1628647, EPI_ISL_1628648, EPI_ISL_1628649, EPI_ISL_1628650, EPI_ISL_1628651, EPI_ISL_1628652, EPI_ISL_1628653, EPI_ISL_1628654, EPI_ISL_1628655, EPI_ISL_1628656, EPI_ISL_1628657, EPI_ISL_1628658, EPI_ISL_1628659, EPI_ISL_1628660, EPI_ISL_1628661, EPI_ISL_1628662, EPI_ISL_1628663, EPI_ISL_1628664, EPI_ISL_1628665, EPI_ISL_1628666, EPI_ISL_1628667, EPI_ISL_1628668, EPI_ISL_1628669, EPI_ISL_1628670, EPI_ISL_1628671, EPI_ISL_1628672, EPI_ISL_1628673, EPI_ISL_1628674, EPI_ISL_1628675, EPI_ISL_1628676, EPI_ISL_1628677, EPI_ISL_1628678, EPI_ISL_1628679, EPI_ISL_1628680, EPI_ISL_1628681, EPI_ISL_1628682, EPI_ISL_1628683, EPI_ISL_1628684, EPI_ISL_1810831, EPI_ISL_1810832, EPI_ISL_1810833, EPI_ISL_1810834, EPI_ISL_1810835, EPI_ISL_1810836, EPI_ISL_1810837, EPI_ISL_1810838, EPI_ISL_1810839, EPI_ISL_1810840, EPI_ISL_1810841, EPI_ISL_1810842, EPI_ISL_1810843, EPI_ISL_1810844, EPI_ISL_1810845, EPI_ISL_1810846, EPI_ISL_1810847, EPI_ISL_1810848, EPI_ISL_1810849, EPI_ISL_1810850, EPI_ISL_1810851, EPI_ISL_1810852, EPI_ISL_1810853, EPI_ISL_1810854, EPI_ISL_1810855, EPI_ISL_1810856, EPI_ISL_1810857, EPI_ISL_1810858, EPI_ISL_1810859, EPI_ISL_1810860, EPI_ISL_1810861, EPI_ISL_1810862, EPI_ISL_1810863, EPI_ISL_1810864, EPI_ISL_1810865, EPI_ISL_1810866, EPI_ISL_1810867, EPI_ISL_1810868, EPI_ISL_1810869, EPI_ISL_1810870, EPI_ISL_1810871, EPI_ISL_1810872, EPI_ISL_1810873, EPI_ISL_1810874, EPI_ISL_1810875, EPI_ISL_1810876, EPI_ISL_1810877, EPI_ISL_1810878, EPI_ISL_1810879, EPI_ISL_1810880, EPI_ISL_1810881, EPI_ISL_1810882, EPI_ISL_1810883, EPI_ISL_1810884, EPI_ISL_1810885, EPI_ISL_1810886, EPI_ISL_1810887, EPI_ISL_1810888, EPI_ISL_1810889, EPI_ISL_1810890, EPI_ISL_1810891, EPI_ISL_1810892, EPI_ISL_1810893, EPI_ISL_1810894, EPI_ISL_1810895, EPI_ISL_1810896, EPI_ISL_1810897, EPI_ISL_1810898, EPI_ISL_1810899, EPI_ISL_1810900, EPI_ISL_1810901, EPI_ISL_1810902, EPI_ISL_1810903, EPI_ISL_1810904, EPI_ISL_1810905, EPI_ISL_1810906, EPI_ISL_1810907, EPI_ISL_1810908, EPI_ISL_1810909, EPI_ISL_1810910, EPI_ISL_1810911, EPI_ISL_1810912, EPI_ISL_1810913, EPI_ISL_1810914, EPI_ISL_1810915, EPI_ISL_1810916, EPI_ISL_1810917, EPI_ISL_1810918, EPI_ISL_1810919, EPI_ISL_1810920, EPI_ISL_1810921, EPI_ISL_1810922, EPI_ISL_1810923, EPI_ISL_2090648, EPI_ISL_2090649 | | | |
| see above | InDRE | Instituto Nacional de Medicina Genomica | Alcaraz N; Canseco Mendez JC; Cedro-Tanda A; Cisneros-Villanueva M; Garcia-Cardenas FJ; Gisela Barrera-Badillo; Gonzalez-Barrera D; Gonzalez-Woge MA; Herrera-Montalvo LA.; Hidalgo-Miranda A; Irma Lopez-Martinez; Jose Ernesto Ramirez González; Mendoza-Vargas A; Miranda-Ortiz H; Munguia-Garza P; Orjuela-Rodríguez M; Ramirez-Vega O; Rangel-DeLeon D; Reyes-Grajeda JP; Rosas-Escobar P |
| EPI_ISL_1512851, EPI_ISL_1513033, EPI_ISL_1587780, EPI_ISL_1587792, EPI_ISL_1587914, EPI_ISL_1587997, EPI_ISL_1588056, EPI_ISL_1588187, EPI_ISL_1588274, EPI_ISL_1588374, EPI_ISL_1691550, EPI_ISL_1692461, EPI_ISL_1693892, EPI_ISL_1694062, EPI_ISL_1694319, EPI_ISL_1694381, EPI_ISL_1702220, EPI_ISL_1795461, EPI_ISL_1795468, EPI_ISL_1818085, EPI_ISL_2013832, EPI_ISL_2014264, EPI_ISL_2087301, EPI_ISL_2087303, EPI_ISL_2087763, EPI_ISL_2087882, EPI_ISL_2088639, EPI_ISL_2088646, EPI_ISL_2088812, EPI_ISL_2089224, EPI_ISL_2089229, EPI_ISL_2089289, EPI_ISL_2089292, EPI_ISL_2089313, EPI_ISL_2089332, EPI_ISL_2089342, EPI_ISL_2089372, EPI_ISL_2089414, EPI_ISL_2089417, EPI_ISL_2089451, EPI_ISL_2132526, EPI_ISL_2186323 | | | |
| see above | Infinity Biologix | Centers for Disease Control and Prevention Division of Viral Diseases, Pathogen Discovery | Adrian Paskey; Benjamin Rambo-Martin; Chirayu Goswami; Christian Bixby; Christopher Gulvick; Clinton R. Paden; Dakota Howard; Darlene Wagner; Dhwani Batra; Duncan MacCannell; Jason Caravas; Jonathan Schultz; Kara Moser; Matthew Schmerer; Peter W. Cook; Robin Grimwood; Russ Hager; Scott Sammons; Shatavia Morrison; Yihe Wang; Yvette Unoarumhi |
| EPI_ISL_1824430, EPI_ISL_1824431, EPI_ISL_1824432, EPI_ISL_1824433, EPI_ISL_1824434, EPI_ISL_1824435, EPI_ISL_1824436, EPI_ISL_1824437, EPI_ISL_1824438, EPI_ISL_1824439, EPI_ISL_1824440, EPI_ISL_1824441, EPI_ISL_1824451, EPI_ISL_1824452, EPI_ISL_1824453, EPI_ISL_1824454, EPI_ISL_1824455, EPI_ISL_1824456, EPI_ISL_1824457, EPI_ISL_1824458, EPI_ISL_1824459, EPI_ISL_1824460, EPI_ISL_1824461, EPI_ISL_1824462, EPI_ISL_1824463, EPI_ISL_1824464, EPI_ISL_1824465, EPI_ISL_1824466, EPI_ISL_1824467, EPI_ISL_1824468, EPI_ISL_1824469, EPI_ISL_1824470, EPI_ISL_1824471, EPI_ISL_1824472, EPI_ISL_1824473, EPI_ISL_1824474, EPI_ISL_1824475, EPI_ISL_1824476, EPI_ISL_1824477, EPI_ISL_1824478, EPI_ISL_1824479, EPI_ISL_1824480, EPI_ISL_1824481, EPI_ISL_1824482, EPI_ISL_1824483, EPI_ISL_1824484, EPI_ISL_1824485, EPI_ISL_1824486, EPI_ISL_1824487, EPI_ISL_1824488, EPI_ISL_1824489, EPI_ISL_1824490, EPI_ISL_1824491, EPI_ISL_1824492, EPI_ISL_1824493, EPI_ISL_1824494, EPI_ISL_1824495, EPI_ISL_1824496, EPI_ISL_1824497, EPI_ISL_1824498, EPI_ISL_1970839, EPI_ISL_1970840, EPI_ISL_1970841, EPI_ISL_2160642, EPI_ISL_2160644, EPI_ISL_2160646, EPI_ISL_2160647, EPI_ISL_2160649, EPI_ISL_2160650, EPI_ISL_2160651, EPI_ISL_2160653, EPI_ISL_2160655, EPI_ISL_2160656, EPI_ISL_2160658, EPI_ISL_2160659, EPI_ISL_2160661, EPI_ISL_2160662, EPI_ISL_2160665, EPI_ISL_2160667, EPI_ISL_2160668, EPI_ISL_2160670, EPI_ISL_2160671, EPI_ISL_2160673, EPI_ISL_2160674, EPI_ISL_2160676, EPI_ISL_2160678, EPI_ISL_2160679, EPI_ISL_2160681, EPI_ISL_2160682, EPI_ISL_2160684, EPI_ISL_2160685, EPI_ISL_2160687, EPI_ISL_2160689, EPI_ISL_2160690, EPI_ISL_2160692, EPI_ISL_2160693, EPI_ISL_2160695, EPI_ISL_2178742, EPI_ISL_2178743 | | | |
| see above | Instituto Nacional de Medicina Genomica | Centro de Investigación en Enfermedades Infecciosas (CIENI), Instituto Nacional de Enfermedades Respiratorias (INER) | Arriaga-Canon C; Avila-Rios S; Boukadida C; Cedro-Tanda A; Herrera-Montalvo LA; Hidalgo-Miranda A; Matias-Florentino M; Mendoza-Vargas A; Perez-Garcia M; Reyes-Grajeda JP; Reyes-Teran G |
| EPI_ISL_1406733, EPI_ISL_1406734, EPI_ISL_1406735, EPI_ISL_1406736, EPI_ISL_1406737, EPI_ISL_1406738, EPI_ISL_1406739, EPI_ISL_1406740, EPI_ISL_1406741, EPI_ISL_1406742, EPI_ISL_1406743, EPI_ISL_1406744, EPI_ISL_1406745, EPI_ISL_1406746, EPI_ISL_1406747, EPI_ISL_1406748, EPI_ISL_1406749, EPI_ISL_1406750, EPI_ISL_1406777, EPI_ISL_1423996, EPI_ISL_1423997, EPI_ISL_1423998, EPI_ISL_1423999 | | | |
| see above | Instituto Nacional de Medicina Genomica | Instituto Nacional de Enfermedades Respiratorias (INER), Centro de Investigación en Enfermedades Infecciosas (CIENI) | Arriaga-Canon C; Avila-Rios S; Boukadida C; Cedro-Tanda A; Herrera-Montalvo LA; Hidalgo-Miranda A; Matias-Florentino M; Mendoza-Vargas A; Reyes-Grajeda JP; Reyes-Teran G |
| EPI_ISL_1262630, EPI_ISL_1262632, EPI_ISL_1262633, EPI_ISL_1262634, EPI_ISL_1262635, EPI_ISL_1262636, EPI_ISL_1262637, EPI_ISL_1262639, EPI_ISL_1262640, EPI_ISL_1262641, EPI_ISL_1262643, EPI_ISL_1262644, EPI_ISL_1262645, EPI_ISL_1262646, EPI_ISL_1262647, EPI_ISL_1262648, EPI_ISL_1262649, EPI_ISL_1262650, EPI_ISL_1262651, EPI_ISL_1262652, EPI_ISL_1262653, EPI_ISL_1262654, EPI_ISL_1262655, EPI_ISL_1262656, EPI_ISL_1262657, EPI_ISL_1262658, EPI_ISL_1262659, EPI_ISL_1262660, EPI_ISL_1262661, EPI_ISL_1262662, EPI_ISL_1262663, EPI_ISL_1262664, EPI_ISL_1262665, EPI_ISL_1262666, EPI_ISL_1262667, EPI_ISL_1262668, EPI_ISL_1262669, EPI_ISL_1262670, EPI_ISL_1262671, EPI_ISL_1262672, EPI_ISL_1262673, EPI_ISL_1262674, EPI_ISL_1262675, EPI_ISL_1262676, EPI_ISL_1262677, EPI_ISL_1262678, EPI_ISL_1262679, EPI_ISL_1310808, EPI_ISL_1315480, EPI_ISL_1315481, EPI_ISL_1315482, EPI_ISL_1315483, EPI_ISL_1315484, EPI_ISL_1315485, EPI_ISL_1315486, EPI_ISL_1315488, EPI_ISL_1315489, EPI_ISL_1315490, EPI_ISL_1315491, EPI_ISL_1315494, EPI_ISL_1315495, EPI_ISL_1315496, EPI_ISL_1315497, EPI_ISL_1315498, EPI_ISL_1315499, EPI_ISL_1315500, EPI_ISL_1315501, EPI_ISL_1315502, EPI_ISL_1315503, EPI_ISL_1315504, EPI_ISL_1315505, EPI_ISL_1315506, EPI_ISL_1315507, EPI_ISL_1315508, EPI_ISL_1315509, EPI_ISL_1315510, EPI_ISL_1315511, EPI_ISL_1315512, EPI_ISL_1315513, EPI_ISL_1315514, EPI_ISL_1315515, EPI_ISL_1315516, EPI_ISL_1315517, EPI_ISL_1315518, EPI_ISL_1315519, EPI_ISL_1315520, EPI_ISL_1315521, EPI_ISL_1315523, EPI_ISL_1315524, EPI_ISL_1315525, EPI_ISL_1315526, EPI_ISL_1315527, EPI_ISL_1315528, EPI_ISL_1315529, EPI_ISL_1315530, EPI_ISL_1315531, EPI_ISL_1315532, EPI_ISL_1315533, EPI_ISL_1315534, EPI_ISL_1315535, EPI_ISL_1315536, EPI_ISL_1315537, EPI_ISL_1315538, EPI_ISL_1315539, EPI_ISL_1315540, EPI_ISL_1315541, EPI_ISL_1315542, EPI_ISL_1315543, EPI_ISL_1315544, EPI_ISL_1315545, EPI_ISL_1315546, EPI_ISL_1315547, EPI_ISL_1315548, EPI_ISL_1315549, EPI_ISL_1315550, EPI_ISL_1315551, EPI_ISL_1315552, EPI_ISL_1315553, EPI_ISL_1315554, EPI_ISL_1315555, EPI_ISL_1315556, EPI_ISL_1315557, EPI_ISL_1315558, EPI_ISL_1315559, EPI_ISL_1315560, EPI_ISL_1315561, EPI_ISL_1315562, EPI_ISL_1315563, EPI_ISL_1315564, EPI_ISL_1315565, EPI_ISL_1315566, EPI_ISL_1315567, EPI_ISL_1315568, EPI_ISL_1315569, EPI_ISL_1315570, EPI_ISL_1315571, EPI_ISL_1315572, EPI_ISL_1392922, EPI_ISL_1400795, EPI_ISL_1400796, EPI_ISL_1400797, EPI_ISL_1400798, EPI_ISL_1400799, EPI_ISL_1406083, EPI_ISL_1406084, EPI_ISL_1406085, EPI_ISL_1406086, EPI_ISL_1406087, EPI_ISL_1406088, EPI_ISL_1406089, EPI_ISL_1406090, EPI_ISL_1406091, EPI_ISL_1406092, EPI_ISL_1406093, EPI_ISL_1406094, EPI_ISL_1406095, EPI_ISL_1406096, EPI_ISL_1406097, EPI_ISL_1406098, EPI_ISL_1406099, EPI_ISL_1406100, EPI_ISL_1406101, EPI_ISL_1406102, EPI_ISL_1406103, EPI_ISL_1406104, EPI_ISL_1406105, EPI_ISL_1406106, EPI_ISL_1406107, EPI_ISL_1406108, EPI_ISL_1406109, EPI_ISL_1406110, EPI_ISL_1406111, EPI_ISL_1406112, EPI_ISL_1406113, EPI_ISL_1406114, EPI_ISL_1406115, EPI_ISL_1406116, EPI_ISL_1406117, EPI_ISL_1406118, EPI_ISL_1406119, EPI_ISL_1406120, EPI_ISL_1406121, EPI_ISL_1406122, EPI_ISL_1406123, EPI_ISL_1406124, EPI_ISL_1406125, EPI_ISL_1406135, EPI_ISL_1406136, EPI_ISL_1406137, EPI_ISL_1406138, EPI_ISL_1406139, EPI_ISL_1406140, EPI_ISL_1406141, EPI_ISL_1406142, EPI_ISL_1416873, EPI_ISL_1417442, EPI_ISL_1558802, EPI_ISL_1558803, EPI_ISL_1558804, EPI_ISL_1558805, EPI_ISL_1558806, EPI_ISL_1558807, EPI_ISL_1558808, EPI_ISL_1558809, EPI_ISL_1558810, EPI_ISL_1558811, EPI_ISL_1558812, EPI_ISL_1558813, EPI_ISL_1558814, EPI_ISL_1558815, EPI_ISL_1558816, EPI_ISL_1558817, EPI_ISL_1558818, EPI_ISL_1558819, EPI_ISL_1558820, EPI_ISL_1558821, EPI_ISL_1558822, EPI_ISL_1558823, EPI_ISL_1558824, EPI_ISL_1558825, EPI_ISL_1558826, EPI_ISL_1558827, EPI_ISL_1585916, EPI_ISL_1585917, EPI_ISL_1585918, EPI_ISL_1585919, EPI_ISL_1591464, EPI_ISL_1591465, EPI_ISL_1591466, EPI_ISL_1591472, EPI_ISL_1591473, EPI_ISL_1591474, EPI_ISL_1591480, EPI_ISL_1591481, EPI_ISL_1591486, EPI_ISL_1591487, EPI_ISL_1591488, EPI_ISL_1591492, EPI_ISL_1591493, EPI_ISL_1591494, EPI_ISL_1591498, EPI_ISL_1591499, EPI_ISL_1591500, EPI_ISL_1591501, EPI_ISL_1591502, EPI_ISL_1591506, EPI_ISL_1591507, EPI_ISL_1591508, EPI_ISL_1591509, EPI_ISL_1591510, EPI_ISL_1591514, EPI_ISL_1591515, EPI_ISL_1591516, EPI_ISL_1591517, EPI_ISL_1591521, EPI_ISL_1591522, EPI_ISL_1591523, EPI_ISL_1591526, EPI_ISL_1591527, EPI_ISL_1591528, EPI_ISL_1591529, EPI_ISL_1591533, EPI_ISL_1591534, EPI_ISL_1591535, EPI_ISL_1591540, EPI_ISL_1591541, EPI_ISL_1591542, EPI_ISL_1591543, EPI_ISL_1591544, EPI_ISL_1591545, EPI_ISL_1591546, EPI_ISL_1591547, EPI_ISL_1591548, EPI_ISL_1591549, EPI_ISL_1591550, EPI_ISL_1591551, EPI_ISL_1591552, EPI_ISL_1591553, EPI_ISL_1591554, EPI_ISL_1591555, EPI_ISL_1591556, EPI_ISL_1591557, EPI_ISL_1591558, EPI_ISL_1591559, EPI_ISL_1591560, EPI_ISL_1591561, EPI_ISL_1591562, EPI_ISL_1591563, EPI_ISL_1591564, EPI_ISL_1591565, EPI_ISL_1591566, EPI_ISL_1591567, EPI_ISL_1591568, EPI_ISL_1591569, EPI_ISL_1591570, EPI_ISL_1591571, EPI_ISL_1591572, EPI_ISL_1591573, EPI_ISL_1591574, EPI_ISL_1591575, EPI_ISL_1591576, EPI_ISL_1591577, EPI_ISL_1591578, EPI_ISL_1591579, EPI_ISL_1591580, EPI_ISL_1591581, EPI_ISL_1591582, EPI_ISL_1591583, EPI_ISL_1591584, EPI_ISL_1591585, EPI_ISL_1591586, EPI_ISL_1591587, EPI_ISL_1591588, EPI_ISL_1591589, EPI_ISL_1591590, EPI_ISL_1591591, EPI_ISL_1591592, EPI_ISL_1591593, EPI_ISL_1591594, EPI_ISL_1591595, EPI_ISL_1591596, EPI_ISL_1591597, EPI_ISL_1591598, EPI_ISL_1591599, EPI_ISL_1591600, EPI_ISL_1591601, EPI_ISL_1591602, EPI_ISL_1591603, EPI_ISL_1591604, EPI_ISL_1591605, EPI_ISL_1591606, EPI_ISL_1591607, EPI_ISL_1591608, EPI_ISL_1591609, EPI_ISL_1591610, EPI_ISL_1591611, EPI_ISL_1591612, EPI_ISL_1591613, EPI_ISL_1591614, EPI_ISL_1591615, EPI_ISL_1591616, EPI_ISL_1591617, EPI_ISL_1591618, EPI_ISL_1591619, EPI_ISL_1591620, EPI_ISL_1591621, EPI_ISL_1591622, EPI_ISL_1591623, EPI_ISL_1591624, EPI_ISL_1591625, EPI_ISL_1591626, EPI_ISL_1591627, EPI_ISL_1591628, EPI_ISL_1591629, EPI_ISL_1591630, EPI_ISL_1591631, EPI_ISL_1591632, EPI_ISL_1591633, EPI_ISL_1591634, EPI_ISL_1591635, EPI_ISL_1591636, EPI_ISL_1591637, EPI_ISL_1591638, EPI_ISL_1591639, EPI_ISL_1591640, EPI_ISL_1591641, EPI_ISL_1591642, EPI_ISL_1591643, EPI_ISL_1591644, EPI_ISL_1591645, EPI_ISL_1591646, EPI_ISL_1591647, EPI_ISL_1591648, EPI_ISL_1591649, EPI_ISL_1591650, EPI_ISL_1591651, EPI_ISL_1591652, EPI_ISL_1591653, EPI_ISL_1591654, EPI_ISL_1591655, EPI_ISL_1591656, EPI_ISL_1591657, EPI_ISL_1591658, EPI_ISL_1591659, EPI_ISL_1591660, EPI_ISL_1591661, EPI_ISL_1591662, EPI_ISL_1591663, EPI_ISL_1591664, EPI_ISL_1591665, EPI_ISL_1591666, EPI_ISL_1591667, EPI_ISL_1591668, EPI_ISL_1591669, EPI_ISL_1591670, EPI_ISL_1591671, EPI_ISL_1591672, EPI_ISL_1591673, EPI_ISL_1591674, EPI_ISL_1591675, EPI_ISL_1591676, EPI_ISL_1591677, EPI_ISL_1591678, EPI_ISL_1591679, EPI_ISL_1591680, EPI_ISL_1591681, EPI_ISL_1591682, EPI_ISL_1591683, EPI_ISL_1591684, EPI_ISL_1591685, EPI_ISL_1591686, EPI_ISL_1591687, EPI_ISL_1591688, EPI_ISL_1591689, EPI_ISL_1591690, EPI_ISL_1591691, EPI_ISL_1591692, EPI_ISL_1591693, EPI_ISL_1591694, EPI_ISL_1591695, EPI_ISL_1591696, EPI_ISL_1591697, EPI_ISL_1591698, EPI_ISL_1591699, EPI_ISL_1591700, EPI_ISL_1591701, EPI_ISL_1591702, EPI_ISL_1591703, EPI_ISL_1591704, EPI_ISL_1591705, EPI_ISL_1591706, EPI_ISL_1591707, EPI_ISL_1591708, EPI_ISL_1591709, EPI_ISL_1591710, EPI_ISL_1591711, EPI_ISL_1591712, EPI_ISL_1591713, EPI_ISL_1591714, EPI_ISL_1591715, EPI_ISL_1591716, EPI_ISL_1591717, EPI_ISL_1594356, EPI_ISL_1594357, EPI_ISL_1594358, EPI_ISL_1594359, EPI_ISL_1594360, EPI_ISL_1594361, EPI_ISL_1594362, EPI_ISL_1594363, EPI_ISL_1594364, EPI_ISL_1594365, EPI_ISL_1594366, EPI_ISL_1594367, EPI_ISL_1594368, EPI_ISL_1594369, EPI_ISL_1594370, EPI_ISL_1594371, EPI_ISL_1594372, EPI_ISL_1594373, EPI_ISL_1594374, EPI_ISL_1594375, EPI_ISL_1594376, EPI_ISL_1594377, EPI_ISL_1594378, EPI_ISL_1614744, EPI_ISL_1615069, EPI_ISL_1626614, EPI_ISL_1628523, EPI_ISL_1628524, EPI_ISL_1628525, EPI_ISL_1628526, EPI_ISL_1628527, EPI_ISL_1628529, EPI_ISL_1628530, EPI_ISL_1628531, EPI_ISL_1628532, EPI_ISL_1628533, EPI_ISL_1628535, EPI_ISL_1628536, EPI_ISL_1628537, EPI_ISL_1628538, EPI_ISL_1628539, EPI_ISL_1628540, EPI_ISL_1628542, EPI_ISL_1628543, EPI_ISL_1628544, EPI_ISL_1628545, EPI_ISL_1628546, EPI_ISL_1628547, EPI_ISL_1628550, EPI_ISL_1628551, EPI_ISL_1628552, EPI_ISL_1628553, EPI_ISL_1628554, EPI_ISL_1628557, EPI_ISL_1628558, EPI_ISL_1628559, EPI_ISL_1628562, EPI_ISL_1628564, EPI_ISL_1628565, EPI_ISL_1628568, EPI_ISL_1628569, EPI_ISL_1628570, EPI_ISL_1628571, EPI_ISL_1628572, EPI_ISL_1628574, EPI_ISL_1628575, EPI_ISL_1628576, EPI_ISL_1628577, EPI_ISL_1628578, EPI_ISL_1628581, EPI_ISL_1628582, EPI_ISL_1628583, EPI_ISL_1628584, EPI_ISL_1628585, EPI_ISL_1628586, EPI_ISL_1628587, EPI_ISL_1628588, EPI_ISL_1628589, EPI_ISL_1628592, EPI_ISL_1628593, EPI_ISL_1628594, EPI_ISL_1628595, EPI_ISL_1661253, EPI_ISL_1661254, EPI_ISL_1711276, EPI_ISL_1711277, EPI_ISL_1711278, EPI_ISL_1711279, EPI_ISL_1711280, EPI_ISL_1711281, EPI_ISL_1711282, EPI_ISL_1711283, EPI_ISL_1711284, EPI_ISL_1711285, EPI_ISL_1711286, EPI_ISL_1711287, EPI_ISL_1711288, EPI_ISL_1711289, EPI_ISL_1711290, EPI_ISL_1711291, EPI_ISL_1711292, EPI_ISL_1711293, EPI_ISL_1711294, EPI_ISL_1711295, EPI_ISL_1711296, EPI_ISL_1711297, EPI_ISL_1711298, EPI_ISL_1711299, EPI_ISL_1711300, EPI_ISL_1711301, EPI_ISL_1711302, EPI_ISL_1711303, EPI_ISL_1711304, EPI_ISL_1711305, EPI_ISL_1711306, EPI_ISL_1711308, EPI_ISL_1711310, EPI_ISL_1711311, EPI_ISL_1711312, EPI_ISL_1711313, EPI_ISL_1711314, EPI_ISL_1711315, EPI_ISL_1711316, EPI_ISL_1711317, EPI_ISL_1711318, EPI_ISL_1711319, EPI_ISL_1711320, EPI_ISL_1711321, EPI_ISL_1711322, EPI_ISL_1711323, EPI_ISL_1711324, EPI_ISL_1711325, EPI_ISL_1711326, EPI_ISL_1711327, EPI_ISL_1711328, EPI_ISL_1711329, EPI_ISL_1711330, EPI_ISL_1711331, EPI_ISL_1711332, EPI_ISL_1711333, EPI_ISL_1711334, EPI_ISL_1711335, EPI_ISL_1711336, EPI_ISL_1711337, EPI_ISL_1711338, EPI_ISL_1711339, EPI_ISL_1711340, EPI_ISL_1711341, EPI_ISL_1711342, EPI_ISL_1711343, EPI_ISL_1711344, EPI_ISL_1711345, EPI_ISL_1711346, EPI_ISL_1711347, EPI_ISL_1758186, EPI_ISL_1807319, EPI_ISL_1807320, EPI_ISL_1807321, EPI_ISL_1807322, EPI_ISL_1807323, EPI_ISL_1807324, EPI_ISL_1807325, EPI_ISL_1807326, EPI_ISL_1807327, EPI_ISL_1807328, EPI_ISL_1807329, EPI_ISL_1807330, EPI_ISL_1807331, EPI_ISL_1807332, EPI_ISL_1807333, EPI_ISL_1807334, EPI_ISL_1807335, EPI_ISL_1807336, EPI_ISL_1807337, EPI_ISL_1807338, EPI_ISL_1807339, EPI_ISL_1807340, EPI_ISL_1807341, EPI_ISL_1807342, EPI_ISL_1807343, EPI_ISL_1807344, EPI_ISL_1807345, EPI_ISL_1807346, EPI_ISL_1807347, EPI_ISL_1807348, EPI_ISL_1807349, EPI_ISL_1807350, EPI_ISL_1807351, EPI_ISL_1807352, EPI_ISL_1807353, EPI_ISL_1807354, EPI_ISL_1807355, EPI_ISL_1807356, EPI_ISL_1807357, EPI_ISL_1807358, EPI_ISL_1807359, EPI_ISL_1807360, EPI_ISL_1807361, EPI_ISL_1807362, EPI_ISL_1807363, EPI_ISL_1807364, EPI_ISL_1807365, EPI_ISL_1807366, EPI_ISL_1807367, EPI_ISL_1807368, EPI_ISL_1807369, EPI_ISL_1807370, EPI_ISL_1807371, EPI_ISL_1807372, EPI_ISL_1807373, EPI_ISL_1807374, EPI_ISL_2080616, EPI_ISL_2080617, EPI_ISL_2080624, EPI_ISL_2080625, EPI_ISL_2080626, EPI_ISL_2080627, EPI_ISL_2080628, EPI_ISL_2080629, EPI_ISL_2080630, EPI_ISL_2080631, EPI_ISL_2080632, EPI_ISL_2080633, EPI_ISL_2080634, EPI_ISL_2080635, EPI_ISL_2080636, EPI_ISL_2080637, EPI_ISL_2080638, EPI_ISL_2080639, EPI_ISL_2080640, EPI_ISL_2080641, EPI_ISL_2080642, EPI_ISL_2080643, EPI_ISL_2080644, EPI_ISL_2080645, EPI_ISL_2080646, EPI_ISL_2080647, EPI_ISL_2080648, EPI_ISL_2080649, EPI_ISL_2080650, EPI_ISL_2080651, EPI_ISL_2080652, EPI_ISL_2080653, EPI_ISL_2080654, EPI_ISL_2080655, EPI_ISL_2080656, EPI_ISL_2080657, EPI_ISL_2080658, EPI_ISL_2080659, EPI_ISL_2080660, EPI_ISL_2080661, EPI_ISL_2080662, EPI_ISL_2080663, EPI_ISL_2080664, EPI_ISL_2080665, EPI_ISL_2080666, EPI_ISL_2080667, EPI_ISL_2080668, EPI_ISL_2080669, EPI_ISL_2080670, EPI_ISL_2080671, EPI_ISL_2080672, EPI_ISL_2080673, EPI_ISL_2080674, EPI_ISL_2080675, EPI_ISL_2080676, EPI_ISL_2080677, EPI_ISL_2080678, EPI_ISL_2080679, EPI_ISL_2080680, EPI_ISL_2080681, EPI_ISL_2080682, EPI_ISL_2080683, EPI_ISL_2080684, EPI_ISL_2080685, EPI_ISL_2080686, EPI_ISL_2080687, EPI_ISL_2080688, EPI_ISL_2080689, EPI_ISL_2080690, EPI_ISL_2080691, EPI_ISL_2080692, EPI_ISL_2080693, EPI_ISL_2080694, EPI_ISL_2096054, EPI_ISL_2096055, EPI_ISL_2096056, EPI_ISL_2103199 | | | |
| see above | Instituto Nacional de Medicina Genomica | Instituto Nacional de Medicina Genomica | Alcaraz N; Arriaga-Canon C; Canseco Mendez JC; Canseco-Mendez JC; Cedro-Tanda A; Cisneros-Villanueva M; Escobar-Arrazola; Frías-Jimenez E; Garcia-Cardenas FJ; Garcia-Cárdenas FJ; Gonzalez-Barrera D; Gonzalez-Woge MA; González-Woge MA; Herrera-Montalvo LA; Herrera-Montalvo LA.; Hidalgo-Miranda A; M; Mendoza-Vargas A; Miranda-Ortiz H; Miranda-Ortíz H; Munguia-Garza; Munguia-Garza P; Orjuela-Rodríguez M; Ramirez-Vega O; Rangel-DeLeon D; Reyes-Grajeda JP; Rosas-Escobar P |
| EPI_ISL_1337390, EPI_ISL_1399280, EPI_ISL_1504108, EPI_ISL_1532264, EPI_ISL_1558798, EPI_ISL_1661550, EPI_ISL_1700837, EPI_ISL_1700838, EPI_ISL_1710971, EPI_ISL_1710972, EPI_ISL_1754141, EPI_ISL_1805493, EPI_ISL_1821201, EPI_ISL_1821202, EPI_ISL_1821203, EPI_ISL_1857271, EPI_ISL_2101880, EPI_ISL_2105688, EPI_ISL_2139908, EPI_ISL_2139909, EPI_ISL_2139914, EPI_ISL_2157328, EPI_ISL_2158108 | | | |
| see above | LESP Baja California | Instituto de Diagnostico y Referencia Epidemiologicos (INDRE) | Abril Rodriguez-Maldonado; Ariadna Medina-Benitez; Claudia Wong-Arambula; Ernesto Ramirez-Gonzalez.; Gisela Barrera-Badillo; Irma Lopez-Martinez; Joaquin Quiroz-Mercado; Lucia Hernandez-Rivas; Natividad Cruz-Ortiz; Sergio Rangel-Guerrero; Tatiana Nunez-Garcia; Vanessa Rivero-Arredondo |
| EPI_ISL_1504105, EPI_ISL_1532243, EPI_ISL_1532244, EPI_ISL_1532245, EPI_ISL_1558801, EPI_ISL_1626811, EPI_ISL_1626812, EPI_ISL_1626813, EPI_ISL_1626814, EPI_ISL_1626815, EPI_ISL_1651890, EPI_ISL_1651891, EPI_ISL_1651892, EPI_ISL_1651893, EPI_ISL_1651894, EPI_ISL_1651895, EPI_ISL_1651900, EPI_ISL_1700819, EPI_ISL_1700822, EPI_ISL_1805487, EPI_ISL_1821104, EPI_ISL_1821105, EPI_ISL_1821180, EPI_ISL_1821181, EPI_ISL_1821182, EPI_ISL_1821183, EPI_ISL_1821184, EPI_ISL_1857224, EPI_ISL_1857225, EPI_ISL_1857226, EPI_ISL_1857227, EPI_ISL_1857235, EPI_ISL_1857266, EPI_ISL_1857267, EPI_ISL_2139913, EPI_ISL_2158231 | | | |
| see above | LESP Baja California Sur | Instituto de Diagnostico y Referencia Epidemiologicos (INDRE) | Abril Rodriguez-Maldonado; Ariadna Medina-Benitez; Claudia Wong-Arambula; Ernesto Ramirez-Gonzalez.; Gisela Barrera-Badillo; Irma Lopez-Martinez; Joaquin Quiroz-Mercado; Lucia Hernandez-Rivas; Natividad Cruz-Ortiz; Sergio Rangel-Guerrero; Tatiana Nunez-Garcia; Vanessa Rivero-Arredondo |

| | | | |
|---|---|---|---|
| EPI_ISL_1483066, EPI_ISL_1483067, EPI_ISL_1700816, EPI_ISL_1754139, EPI_ISL_1754140, EPI_ISL_2094509, EPI_ISL_2094510, EPI_ISL_2094511, EPI_ISL_2094512, EPI_ISL_2139921, EPI_ISL_2157327 | | | |
| see above | LESP Campeche | Instituto de Diagnostico y Referencia Epidemiologicos (INDRE) | Abril Rodriguez-Maldonado; Ariadna Medina-Benitez; Claudia Wong-Arambula; Ernesto Ramirez-Gonzalez.; Gisela Barrera-Badillo; Irma Lopez-Martinez; Joaquin Quiroz-Mercado; Lucia Hernandez-Rivas; Natividad Cruz-Ortiz; Sergio Rangel-Guerrero; Tatiana Nunez-Garcia; Vanessa Rivero-Arredondo |
| EPI_ISL_1532246, EPI_ISL_1532247, EPI_ISL_1626810, EPI_ISL_1651229, EPI_ISL_1651230, EPI_ISL_1700795, EPI_ISL_1857207, EPI_ISL_2158230 | | | |
| see above | LESP Chiapas | Instituto de Diagnostico y Referencia Epidemiologicos (INDRE) | Abril Rodriguez-Maldonado; Ariadna Medina-Benitez; Claudia Wong-Arambula; Ernesto Ramirez-Gonzalez.; Gisela Barrera-Badillo; Irma Lopez-Martinez; Joaquin Quiroz-Mercado; Lucia Hernandez-Rivas; Natividad Cruz-Ortiz; Sergio Rangel-Guerrero; Tatiana Nunez-Garcia; Vanessa Rivero-Arredondo |
| EPI_ISL_1423506, EPI_ISL_1423507, EPI_ISL_1423508, EPI_ISL_1423509, EPI_ISL_1423512, EPI_ISL_1483074, EPI_ISL_1494626, EPI_ISL_1494627, EPI_ISL_1494628, EPI_ISL_1494629, EPI_ISL_1494630, EPI_ISL_1504084, EPI_ISL_1504085, EPI_ISL_1504086, EPI_ISL_1504087, EPI_ISL_1504088, EPI_ISL_1516766, EPI_ISL_1516767, EPI_ISL_1516768, EPI_ISL_1516816, EPI_ISL_1516818, EPI_ISL_1516819, EPI_ISL_1516820, EPI_ISL_1516821, EPI_ISL_1516822, EPI_ISL_1516823, EPI_ISL_1558791, EPI_ISL_1558792, EPI_ISL_1558793, EPI_ISL_1626794, EPI_ISL_1626795, EPI_ISL_1626796, EPI_ISL_1626797, EPI_ISL_1626798, EPI_ISL_1626799, EPI_ISL_1626800, EPI_ISL_1626801, EPI_ISL_1626803, EPI_ISL_1626804, EPI_ISL_1626805, EPI_ISL_1651877, EPI_ISL_1651878, EPI_ISL_1651879, EPI_ISL_1651880, EPI_ISL_1661481, EPI_ISL_1661482, EPI_ISL_1661483, EPI_ISL_1661484, EPI_ISL_1661485, EPI_ISL_1661486, EPI_ISL_1661487, EPI_ISL_1661488, EPI_ISL_1661489, EPI_ISL_1673267, EPI_ISL_1673268, EPI_ISL_1700807, EPI_ISL_1700808, EPI_ISL_1700809, EPI_ISL_1805466, EPI_ISL_1805467, EPI_ISL_1805468, EPI_ISL_1805469, EPI_ISL_1805474, EPI_ISL_1805475, EPI_ISL_1805476, EPI_ISL_1805477, EPI_ISL_1805478, EPI_ISL_1805479, EPI_ISL_1821103, EPI_ISL_1821125, EPI_ISL_1821127, EPI_ISL_1821167, EPI_ISL_1821168, EPI_ISL_1857219, EPI_ISL_1857220, EPI_ISL_1857221, EPI_ISL_1857222, EPI_ISL_1857223, EPI_ISL_1857258, EPI_ISL_1857259, EPI_ISL_1857260, EPI_ISL_1857261, EPI_ISL_1857262, EPI_ISL_2101871, EPI_ISL_2101872, EPI_ISL_2101873, EPI_ISL_2101874, EPI_ISL_2101875, EPI_ISL_2101876, EPI_ISL_2101877, EPI_ISL_2101878, EPI_ISL_2101881, EPI_ISL_2101882, EPI_ISL_2101883, EPI_ISL_2101884, EPI_ISL_2101885, EPI_ISL_2101886, EPI_ISL_2101887, EPI_ISL_2101888, EPI_ISL_2101889, EPI_ISL_2101890, EPI_ISL_2101903, EPI_ISL_2101904, EPI_ISL_2139916, EPI_ISL_2157316, EPI_ISL_2157317, EPI_ISL_2157318, EPI_ISL_2157322, EPI_ISL_2158136, EPI_ISL_2158162, EPI_ISL_2158163, EPI_ISL_2158164, EPI_ISL_2158165, EPI_ISL_2158167, EPI_ISL_2158168, EPI_ISL_2158186, EPI_ISL_2158187, EPI_ISL_2158188, EPI_ISL_2158189, EPI_ISL_2158190, EPI_ISL_2158191, EPI_ISL_2158192, EPI_ISL_2158193, EPI_ISL_2158194, EPI_ISL_2158195, EPI_ISL_2158196, EPI_ISL_2158198, EPI_ISL_2158309, EPI_ISL_2158311, EPI_ISL_2158320 | | | |
| see above | LESP Chihuahua | Instituto de Diagnostico y Referencia Epidemiologicos (INDRE) | Abril Rodriguez-Maldonado; Ariadna Medina-Benitez; Claudia Wong-Arambula; Ernesto Ramirez-Gonzalez.; Gisela Barrera-Badillo; Irma Lopez-Martinez; Joaquin Quiroz-Mercado; Lucia Hernandez-Rivas; Natividad Cruz-Ortiz; Sergio Rangel-Guerrero; Tatiana Nunez-Garcia; Vanessa Rivero-Arredondo |
| EPI_ISL_1423510, EPI_ISL_1469941, EPI_ISL_1494636, EPI_ISL_1494637, EPI_ISL_1494638, EPI_ISL_1494639, EPI_ISL_1494640, EPI_ISL_1494641, EPI_ISL_1494642, EPI_ISL_1494643, EPI_ISL_1494644, EPI_ISL_1494645, EPI_ISL_1494646, EPI_ISL_1494647, EPI_ISL_1504083, EPI_ISL_1516787, EPI_ISL_1516788, EPI_ISL_1516817, EPI_ISL_1532265, EPI_ISL_1532279, EPI_ISL_1626807, EPI_ISL_1651888, EPI_ISL_1651899, EPI_ISL_1651901, EPI_ISL_1651902, EPI_ISL_1661490, EPI_ISL_1661493, EPI_ISL_1661494, EPI_ISL_1700824, EPI_ISL_1710964, EPI_ISL_1710965, EPI_ISL_1710966, EPI_ISL_1805480, EPI_ISL_1805486, EPI_ISL_1821106, EPI_ISL_1821169, EPI_ISL_1821170, EPI_ISL_1821187, EPI_ISL_1821188, EPI_ISL_1821189, EPI_ISL_1821190, EPI_ISL_1857102, EPI_ISL_1857103, EPI_ISL_1857104, EPI_ISL_1857105, EPI_ISL_1857217, EPI_ISL_1857268, EPI_ISL_1857269, EPI_ISL_1857272, EPI_ISL_1857273, EPI_ISL_1857274, EPI_ISL_1857275, EPI_ISL_2101906, EPI_ISL_2101907, EPI_ISL_2101908, EPI_ISL_2101909, EPI_ISL_2101910, EPI_ISL_2101911, EPI_ISL_2101912, EPI_ISL_2105694, EPI_ISL_2105716, EPI_ISL_2105717, EPI_ISL_2105718, EPI_ISL_2139920, EPI_ISL_2139922, EPI_ISL_2157333, EPI_ISL_2157334, EPI_ISL_2158166, EPI_ISL_2158238, EPI_ISL_2158239, EPI_ISL_2158240, EPI_ISL_2158241, EPI_ISL_2158250, EPI_ISL_2158251, EPI_ISL_2158257, EPI_ISL_2158258, EPI_ISL_2158259, EPI_ISL_2158260, EPI_ISL_2158261, EPI_ISL_2158262, EPI_ISL_2158263, EPI_ISL_2158315, EPI_ISL_2158316, EPI_ISL_2158317, EPI_ISL_2158318 | | | |
| see above | LESP Ciudad de Mexico | Instituto de Diagnostico y Referencia Epidemiologicos (INDRE) | Abril Rodriguez-Maldonado; Ariadna Medina-Benitez; Claudia Wong-Arambula; Ernesto Ramirez-Gonzalez.; Gisela Barrera-Badillo; Irma Lopez-Martinez; Joaquin Quiroz-Mercado; Lucia Hernandez-Rivas; Natividad Cruz-Ortiz; Sergio Rangel-Guerrero; Tatiana Nunez-Garcia; Vanessa Rivero-Arredondo |
| EPI_ISL_2158252, EPI_ISL_2158253 | LESP Ciudad de Mexico / Hospital Angeles | Instituto de Diagnostico y Referencia Epidemiologicos (INDRE) | Abril Rodriguez-Maldonado; Ariadna Medina-Benitez; Claudia Wong-Arambula; Ernesto Ramirez-Gonzalez.; Gisela Barrera-Badillo; Irma Lopez-Martinez; Joaquin Quiroz-Mercado; Lucia Hernandez-Rivas; Natividad Cruz-Ortiz; Sergio Rangel-Guerrero; Tatiana Nunez-Garcia; Vanessa Rivero-Arredondo |
| EPI_ISL_1516791, EPI_ISL_1516792, EPI_ISL_1516793, EPI_ISL_1532280, EPI_ISL_1651889, EPI_ISL_1700818, EPI_ISL_1708605, EPI_ISL_1805471, EPI_ISL_1857232, EPI_ISL_1857263, EPI_ISL_2101901, EPI_ISL_2157319, EPI_ISL_2158169, EPI_ISL_2158234 | | | |
| see above | LESP Coahuila | Instituto de Diagnostico y Referencia Epidemiologicos (INDRE) | Abril Rodriguez-Maldonado; Ariadna Medina-Benitez; Claudia Wong-Arambula; Ernesto Ramirez-Gonzalez.; Gisela Barrera-Badillo; Irma Lopez-Martinez; Joaquin Quiroz-Mercado; Lucia Hernandez-Rivas; Natividad Cruz-Ortiz; Sergio Rangel-Guerrero; Tatiana Nunez-Garcia; Vanessa Rivero-Arredondo |
| EPI_ISL_1821107, EPI_ISL_1857276, EPI_ISL_2139912, EPI_ISL_2158138, EPI_ISL_2158139, EPI_ISL_2158140 | LESP Colima | Instituto de Diagnostico y Referencia Epidemiologicos (INDRE) | Abril Rodriguez-Maldonado; Ariadna Medina-Benitez; Claudia Wong-Arambula; Ernesto Ramirez-Gonzalez.; Gisela Barrera-Badillo; Irma Lopez-Martinez; Joaquin Quiroz-Mercado; Lucia Hernandez-Rivas; Natividad Cruz-Ortiz; Sergio Rangel-Guerrero; Tatiana Nunez-Garcia; Vanessa Rivero-Arredondo |
| EPI_ISL_1532249, EPI_ISL_1651903, EPI_ISL_1661491, EPI_ISL_1661492, EPI_ISL_1661521, EPI_ISL_1700825, EPI_ISL_1857270, EPI_ISL_2158255, EPI_ISL_2158256 | | | |
| see above | LESP Estado de Mexico | Instituto de Diagnostico y Referencia Epidemiologicos (INDRE) | Abril Rodriguez-Maldonado; Ariadna Medina-Benitez; Claudia Wong-Arambula; Ernesto Ramirez-Gonzalez.; Gisela Barrera-Badillo; Irma Lopez-Martinez; Joaquin Quiroz-Mercado; Lucia Hernandez-Rivas; Natividad Cruz-Ortiz; Sergio Rangel-Guerrero; Tatiana Nunez-Garcia; Vanessa Rivero-Arredondo |
| EPI_ISL_1424034, EPI_ISL_1469942, EPI_ISL_1469943, EPI_ISL_1469944, EPI_ISL_1483048, EPI_ISL_1494657, EPI_ISL_1494658, EPI_ISL_1494659, EPI_ISL_1494660, EPI_ISL_1494661, EPI_ISL_1494662, EPI_ISL_1494663, EPI_ISL_1494664, EPI_ISL_1494665, EPI_ISL_1494666, EPI_ISL_1494667, EPI_ISL_1494668, EPI_ISL_1494669, EPI_ISL_1516794, EPI_ISL_1516796, EPI_ISL_1532238, EPI_ISL_1626809, EPI_ISL_1626816, EPI_ISL_1626819, EPI_ISL_1626820, EPI_ISL_1661495, EPI_ISL_1661496, EPI_ISL_1661497, EPI_ISL_1661498, EPI_ISL_1661499, EPI_ISL_1661500, EPI_ISL_1661501, EPI_ISL_1661502, EPI_ISL_1661503, EPI_ISL_1661504, EPI_ISL_1661505, EPI_ISL_1661506, EPI_ISL_1661507, EPI_ISL_1661508, EPI_ISL_1661509, EPI_ISL_1661510, EPI_ISL_1661511, EPI_ISL_1661512, EPI_ISL_1661513, EPI_ISL_1661514, EPI_ISL_1661515, EPI_ISL_1661516, EPI_ISL_1661517, EPI_ISL_1661518, EPI_ISL_1661519, EPI_ISL_1661520, EPI_ISL_1661565, EPI_ISL_1661566, EPI_ISL_1661567, EPI_ISL_1661568, EPI_ISL_1661569, EPI_ISL_1673269, EPI_ISL_1673270, EPI_ISL_1673271, EPI_ISL_1673272, EPI_ISL_1673273, EPI_ISL_1700828, EPI_ISL_1700829, EPI_ISL_1700830, EPI_ISL_1708611, EPI_ISL_1708612, EPI_ISL_1710968, EPI_ISL_1710969, EPI_ISL_1805498, EPI_ISL_1821108, EPI_ISL_1821109, EPI_ISL_1821110, EPI_ISL_1821111, EPI_ISL_1857101, EPI_ISL_1857236, EPI_ISL_1857237, EPI_ISL_1857238, EPI_ISL_1857239, EPI_ISL_1857240, EPI_ISL_2101893, EPI_ISL_2101894, EPI_ISL_2105681, EPI_ISL_2105682, EPI_ISL_2105683, EPI_ISL_2105684, EPI_ISL_2105685, EPI_ISL_2105686, EPI_ISL_2105687, EPI_ISL_2139911, EPI_ISL_2157324, EPI_ISL_2157325, EPI_ISL_2157326, EPI_ISL_2158132, EPI_ISL_2158133, EPI_ISL_2158134, EPI_ISL_2158207, EPI_ISL_2158208, EPI_ISL_2158209, EPI_ISL_2158210, EPI_ISL_2158211, EPI_ISL_2158212, EPI_ISL_2158213, EPI_ISL_2158214, EPI_ISL_2158215, EPI_ISL_2158216, EPI_ISL_2158217, EPI_ISL_2158218, EPI_ISL_2158219, EPI_ISL_2158220, EPI_ISL_2158221, EPI_ISL_2158222, EPI_ISL_2158223, EPI_ISL_2158224, EPI_ISL_2158225, EPI_ISL_2158249, EPI_ISL_2158308 | | | |
| see above | LESP Guanajuato | Instituto de Diagnostico y Referencia Epidemiologicos (INDRE) | Abril Rodriguez-Maldonado; Ariadna Medina-Benitez; Claudia Wong-Arambula; Ernesto Ramirez-Gonzalez.; Gisela Barrera-Badillo; Irma Lopez-Martinez; Joaquin Quiroz-Mercado; Lucia Hernandez-Rivas; Natividad Cruz-Ortiz; Sergio Rangel-Guerrero; Tatiana Nunez-Garcia; Vanessa Rivero-Arredondo |
| EPI_ISL_1483073, EPI_ISL_1504104, EPI_ISL_1532240, EPI_ISL_1532241, EPI_ISL_1532242, EPI_ISL_1661570, EPI_ISL_1673265, EPI_ISL_1700810, EPI_ISL_1821123, EPI_ISL_1821149, EPI_ISL_1821150, EPI_ISL_1821151, EPI_ISL_1857201, EPI_ISL_1857252, EPI_ISL_2157329, EPI_ISL_2157330, EPI_ISL_2158170, EPI_ISL_2158232 | | | |
| see above | LESP Guerrero | Instituto de Diagnostico y Referencia Epidemiologicos (INDRE) | Abril Rodriguez-Maldonado; Ariadna Medina-Benitez; Claudia Wong-Arambula; Ernesto Ramirez-Gonzalez.; Gisela Barrera-Badillo; Irma Lopez-Martinez; Joaquin Quiroz-Mercado; Lucia Hernandez-Rivas; Natividad Cruz-Ortiz; Sergio Rangel-Guerrero; Tatiana Nunez-Garcia; Vanessa Rivero-Arredondo |
| EPI_ISL_1483058, EPI_ISL_1494655, EPI_ISL_1494656, EPI_ISL_1516763, EPI_ISL_1626808, EPI_ISL_1661534, EPI_ISL_1661539, EPI_ISL_1661540, EPI_ISL_1661541, EPI_ISL_1661551, EPI_ISL_1661552, EPI_ISL_1805483, EPI_ISL_1821194, EPI_ISL_1821195, EPI_ISL_1821196, EPI_ISL_1857233, EPI_ISL_1857234, EPI_ISL_2158121, EPI_ISL_2158122, EPI_ISL_2158123, EPI_ISL_2158124, EPI_ISL_2158125, EPI_ISL_2158126, EPI_ISL_2158127, EPI_ISL_2158203, EPI_ISL_2158227 | | | |
| see above | LESP Hidalgo | Instituto de Diagnostico y Referencia Epidemiologicos (INDRE) | Abril Rodriguez-Maldonado; Ariadna Medina-Benitez; Claudia Wong-Arambula; Ernesto Ramirez-Gonzalez.; Gisela Barrera-Badillo; Irma Lopez-Martinez; Joaquin Quiroz-Mercado; Lucia Hernandez-Rivas; Natividad Cruz-Ortiz; Sergio Rangel-Guerrero; Tatiana Nunez-Garcia; Vanessa Rivero-Arredondo |
| EPI_ISL_1469937, EPI_ISL_1483068, EPI_ISL_1483069, EPI_ISL_1483070, EPI_ISL_1483071, EPI_ISL_1483072, EPI_ISL_1504109, EPI_ISL_1516765, EPI_ISL_1516795, EPI_ISL_1532266, EPI_ISL_1532267, EPI_ISL_1532268, EPI_ISL_1532269, EPI_ISL_1532270, EPI_ISL_1558799, EPI_ISL_1558800, EPI_ISL_1626817, EPI_ISL_1651250, EPI_ISL_1651251, EPI_ISL_1673276, EPI_ISL_1700800, EPI_ISL_1700820, EPI_ISL_1700821, EPI_ISL_1700839, EPI_ISL_1708606, EPI_ISL_1805488, EPI_ISL_1805489, EPI_ISL_1805491, EPI_ISL_1805497, EPI_ISL_1821129, EPI_ISL_1821130, EPI_ISL_1821131, EPI_ISL_1821185, EPI_ISL_1821186, EPI_ISL_1821197, EPI_ISL_1821198, EPI_ISL_1821200, EPI_ISL_1857230, EPI_ISL_1857231, EPI_ISL_1857244, EPI_ISL_2094505, EPI_ISL_2101869, EPI_ISL_2101891, EPI_ISL_2101895, EPI_ISL_2101900, EPI_ISL_2101905, EPI_ISL_2105715, EPI_ISL_2139915, EPI_ISL_2139919, EPI_ISL_2158135, EPI_ISL_2158171, EPI_ISL_2158172, EPI_ISL_2158201, EPI_ISL_2158202, EPI_ISL_2158233, EPI_ISL_2158310 | | | |
| see above | LESP Jalisco | Instituto de Diagnostico y Referencia Epidemiologicos (INDRE) | Abril Rodriguez-Maldonado; Ariadna Medina-Benitez; Claudia Wong-Arambula; Ernesto Ramirez-Gonzalez.; Gisela Barrera-Badillo; Irma Lopez-Martinez; Joaquin Quiroz-Mercado; Lucia Hernandez-Rivas; Natividad Cruz-Ortiz; Sergio Rangel-Guerrero; Tatiana Nunez-Garcia; Vanessa Rivero-Arredondo |
| EPI_ISL_1492456 | LESP Jalisco | Instituto de diagnóstico y Referencia Epidemiologicos (INDRE) | Abril Rodriguez-Maldonado; Ariadna Medina-Benitez; Claudia Wong-Arambula; Ernesto Ramirez-Gonzalez.; Gisela Barrera-Badillo; Irma Lopez-Martinez; Joaquin Quiroz-Mercado; Lucia Hernandez-Rivas; Natividad Cruz-Ortiz; Sergio Rangel-Guerrero; Tatiana Nunez-Garcia; Vanessa Rivero-Arredondo |
| EPI_ISL_2101902 | LESP Jalisco /Unidad de Patologia | Instituto de Diagnostico y Referencia Epidemiologicos (INDRE) | Abril Rodriguez-Maldonado; Ariadna Medina-Benitez; Claudia Wong-Arambula; Ernesto Ramirez-Gonzalez.; Gisela Barrera-Badillo; Irma Lopez-Martinez; Joaquin Quiroz-Mercado; Lucia Hernandez-Rivas; Natividad Cruz-Ortiz; Sergio Rangel-Guerrero; Tatiana Nunez-Garcia; Vanessa Rivero-Arredondo |
| EPI_ISL_1516762 | LESP Jalisco/UdeG | Instituto de Diagnostico y Referencia Epidemiologicos (INDRE) | Abril Rodriguez-Maldonado; Ariadna Medina-Benitez; Claudia Wong-Arambula; Ernesto Ramirez-Gonzalez.; Gisela Barrera-Badillo; Irma Lopez-Martinez; Joaquin Quiroz-Mercado; Lucia Hernandez-Rivas; Natividad Cruz-Ortiz; Sergio Rangel-Guerrero; Tatiana Nunez-Garcia; Vanessa Rivero-Arredondo |
| EPI_ISL_1805495, EPI_ISL_1805496 | LESP Jalisco/Unidad de Patologia | Instituto de Diagnostico y Referencia Epidemiologicos (INDRE) | Abril Rodriguez-Maldonado; Ariadna Medina-Benitez; Claudia Wong-Arambula; Ernesto Ramirez-Gonzalez.; Gisela Barrera-Badillo; Irma Lopez-Martinez; Joaquin Quiroz-Mercado; Lucia Hernandez-Rivas; Natividad Cruz-Ortiz; Sergio Rangel-Guerrero; Tatiana Nunez-Garcia; Vanessa Rivero-Arredondo |
| EPI_ISL_1661548, EPI_ISL_2139917 | LESP Michoacan | Instituto de Diagnostico y Referencia Epidemiologicos (INDRE) | Abril Rodriguez-Maldonado; Ariadna Medina-Benitez; Claudia Wong-Arambula; Ernesto Ramirez-Gonzalez.; Gisela Barrera-Badillo; Irma Lopez-Martinez; Joaquin Quiroz-Mercado; Lucia Hernandez-Rivas; Natividad Cruz-Ortiz; Sergio Rangel-Guerrero; Tatiana Nunez-Garcia; Vanessa Rivero-Arredondo |
| EPI_ISL_1504068, EPI_ISL_1504069, EPI_ISL_1504070, EPI_ISL_1504071, EPI_ISL_1504072, EPI_ISL_1504073, EPI_ISL_1504074, EPI_ISL_1504075, EPI_ISL_1504076, EPI_ISL_1504077, EPI_ISL_1504078, EPI_ISL_1504079, EPI_ISL_1504080, EPI_ISL_1504081, EPI_ISL_1504082, EPI_ISL_1516808, EPI_ISL_1516809, EPI_ISL_1516810, EPI_ISL_1516811, EPI_ISL_1516812, EPI_ISL_1516813, EPI_ISL_1516814, EPI_ISL_1516815, EPI_ISL_1651255, EPI_ISL_1651256, EPI_ISL_1651257, EPI_ISL_1651258, EPI_ISL_1651259, EPI_ISL_1651260, EPI_ISL_1651261, EPI_ISL_1651262, EPI_ISL_1651263, EPI_ISL_1651264, EPI_ISL_1651265, EPI_ISL_1651266, EPI_ISL_1651267, EPI_ISL_1651268, EPI_ISL_1651269, EPI_ISL_1651270, EPI_ISL_1651271, EPI_ISL_1651272, EPI_ISL_1651273, EPI_ISL_1651274, EPI_ISL_1651275, EPI_ISL_1651276, EPI_ISL_1651904, EPI_ISL_1651905, EPI_ISL_1651906, EPI_ISL_1651907, EPI_ISL_1651908, EPI_ISL_1661474, EPI_ISL_1661475, EPI_ISL_1661476, EPI_ISL_1661477, EPI_ISL_1661479, EPI_ISL_1661480, EPI_ISL_1661542, EPI_ISL_1661543, EPI_ISL_1673266, EPI_ISL_1700802, EPI_ISL_1700826, EPI_ISL_1700827, EPI_ISL_1700834, EPI_ISL_1700835, EPI_ISL_1700840, EPI_ISL_1710967, EPI_ISL_1821137, EPI_ISL_1821138, EPI_ISL_1821139, EPI_ISL_1821140, EPI_ISL_1821141, EPI_ISL_1821142, EPI_ISL_1821143, EPI_ISL_1821144, EPI_ISL_1821145, EPI_ISL_1821146, EPI_ISL_1821147, EPI_ISL_1821148, EPI_ISL_1857247, EPI_ISL_1857248, EPI_ISL_1857249, EPI_ISL_1857250, EPI_ISL_1857251, EPI_ISL_2101870, EPI_ISL_2157315, EPI_ISL_2158152, EPI_ISL_2158153, EPI_ISL_2158154, EPI_ISL_2158155, EPI_ISL_2158156, EPI_ISL_2158157, EPI_ISL_2158158, EPI_ISL_2158159, EPI_ISL_2158160, EPI_ISL_2158161, EPI_ISL_2158174, EPI_ISL_2158175, EPI_ISL_2158176, EPI_ISL_2158177, EPI_ISL_2158178, EPI_ISL_2158179, EPI_ISL_2158180, EPI_ISL_2158181, EPI_ISL_2158182, EPI_ISL_2158183, EPI_ISL_2158184, EPI_ISL_2158314 | | | |
| see above | LESP Morelos | Instituto de Diagnostico y Referencia Epidemiologicos (INDRE) | Abril Rodriguez-Maldonado; Ariadna Medina-Benitez; Claudia Wong-Arambula; Ernesto Ramirez-Gonzalez.; Gisela Barrera-Badillo; Irma Lopez-Martinez; Joaquin Quiroz-Mercado; Lucia Hernandez-Rivas; Natividad Cruz-Ortiz; Sergio Rangel-Guerrero; Tatiana Nunez-Garcia; Vanessa Rivero-Arredondo |
| EPI_ISL_1483040, EPI_ISL_1483041, EPI_ISL_1700823, EPI_ISL_1821161, EPI_ISL_2158204 | LESP Nayarit | Instituto de Diagnostico y Referencia Epidemiologicos (INDRE) | Abril Rodriguez-Maldonado; Ariadna Medina-Benitez; Claudia Wong-Arambula; Ernesto Ramirez-Gonzalez.; Gisela Barrera-Badillo; Irma Lopez-Martinez; Joaquin Quiroz-Mercado; Lucia Hernandez-Rivas; Natividad Cruz-Ortiz; Sergio Rangel-Guerrero; Tatiana Nunez-Garcia; Vanessa Rivero-Arredondo |
| EPI_ISL_1469938, EPI_ISL_1469939, EPI_ISL_1483032, EPI_ISL_1483033, EPI_ISL_1483034, EPI_ISL_1483035, EPI_ISL_1483036, EPI_ISL_1483037, EPI_ISL_1494631, EPI_ISL_1494632, EPI_ISL_1494633, EPI_ISL_1494634, EPI_ISL_1494635, EPI_ISL_1494648, EPI_ISL_1494649, EPI_ISL_1494673, EPI_ISL_1504058, EPI_ISL_1504059, EPI_ISL_1504060, EPI_ISL_1504061, EPI_ISL_1504062, EPI_ISL_1504063, EPI_ISL_1504064, EPI_ISL_1504065, EPI_ISL_1504066, EPI_ISL_1504067, EPI_ISL_1516769, EPI_ISL_1516770, EPI_ISL_1516771, EPI_ISL_1516772, EPI_ISL_1516773, EPI_ISL_1516797, EPI_ISL_1516798, EPI_ISL_1516799, EPI_ISL_1516800, EPI_ISL_1516801, EPI_ISL_1516802, EPI_ISL_1516803, EPI_ISL_1516804, EPI_ISL_1516805, EPI_ISL_1516806, EPI_ISL_1516807, EPI_ISL_1532271, EPI_ISL_1626806, EPI_ISL_1651235, EPI_ISL_1651236, EPI_ISL_1651237, EPI_ISL_1651238, EPI_ISL_1651239, EPI_ISL_1651240, EPI_ISL_1651241, EPI_ISL_1651242, EPI_ISL_1651243, EPI_ISL_1651244, EPI_ISL_1651245, EPI_ISL_1651246, EPI_ISL_1651247, EPI_ISL_1651248, EPI_ISL_1651249, EPI_ISL_1661553, EPI_ISL_1661554, EPI_ISL_1661555, EPI_ISL_1661556, EPI_ISL_1661558, EPI_ISL_1661559, EPI_ISL_1661560, EPI_ISL_1661561, EPI_ISL_1661562, EPI_ISL_1661563, EPI_ISL_1661564, EPI_ISL_1700796, EPI_ISL_1700797, EPI_ISL_1700798, EPI_ISL_1700799, EPI_ISL_1805473, EPI_ISL_1805484, EPI_ISL_1805490, EPI_ISL_1805499, EPI_ISL_1805500, EPI_ISL_1805503, EPI_ISL_1805504, EPI_ISL_1821112, EPI_ISL_1821113, EPI_ISL_1821114, EPI_ISL_1821115, EPI_ISL_1821116, EPI_ISL_1821117, EPI_ISL_1821118, EPI_ISL_1857099, EPI_ISL_1912939, EPI_ISL_2105680, EPI_ISL_2139910, EPI_ISL_2157310, EPI_ISL_2157323, EPI_ISL_2158109, EPI_ISL_2158110, EPI_ISL_2158111, EPI_ISL_2158112, EPI_ISL_2158113, EPI_ISL_2158115, EPI_ISL_2158116, EPI_ISL_2158117, EPI_ISL_2158118, EPI_ISL_2158119, EPI_ISL_2158120, EPI_ISL_2158137, EPI_ISL_2158197, EPI_ISL_2158312, EPI_ISL_2158313, EPI_ISL_2158319 | | | |
| see above | LESP Nuevo Leon | Instituto de Diagnostico y Referencia Epidemiologicos (INDRE) | Abril Rodriguez-Maldonado; Ariadna Medina-Benitez; Claudia Wong-Arambula; Ernesto Ramirez-Gonzalez.; Gisela Barrera-Badillo; Irma Lopez-Martinez; Joaquin Quiroz-Mercado; Lucia Hernandez-Rivas; Natividad Cruz-Ortiz; Sergio Rangel-Guerrero; Tatiana Nunez-Garcia; Vanessa Rivero-Arredondo |
| EPI_ISL_1651252, EPI_ISL_1805482, EPI_ISL_1805485, EPI_ISL_1821162, EPI_ISL_1857255, EPI_ISL_1857256, EPI_ISL_2157331 | | | |
| see above | LESP Oaxaca | Instituto de Diagnostico y Referencia Epidemiologicos (INDRE) | Abril Rodriguez-Maldonado; Ariadna Medina-Benitez; Claudia Wong-Arambula; Ernesto Ramirez-Gonzalez.; Gisela Barrera-Badillo; Irma Lopez-Martinez; Joaquin Quiroz-Mercado; Lucia Hernandez-Rivas; Natividad Cruz-Ortiz; Sergio Rangel-Guerrero; Tatiana Nunez-Garcia; Vanessa Rivero-Arredondo |
| EPI_ISL_1483059, EPI_ISL_1483060, EPI_ISL_1483061, EPI_ISL_1483062, EPI_ISL_1483063, EPI_ISL_1483064, EPI_ISL_1483065, EPI_ISL_1516764, EPI_ISL_1651231, EPI_ISL_1651232, EPI_ISL_1651233, EPI_ISL_1651234, EPI_ISL_1661478, EPI_ISL_1661535, EPI_ISL_1708607, EPI_ISL_1708608, EPI_ISL_1708609, EPI_ISL_1821119, EPI_ISL_1821120, EPI_ISL_1821121, EPI_ISL_1821122, EPI_ISL_1821124, EPI_ISL_1857100, EPI_ISL_1857241, EPI_ISL_1857242, EPI_ISL_1857243, EPI_ISL_2157332, EPI_ISL_2158128, EPI_ISL_2158129, EPI_ISL_2158130, EPI_ISL_2158151, EPI_ISL_2158199, EPI_ISL_2158200, EPI_ISL_2158226, EPI_ISL_2158243, EPI_ISL_2158244, EPI_ISL_2158245, EPI_ISL_2158246, EPI_ISL_2158247, EPI_ISL_2158248 | | | |
| see above | LESP Puebla | Instituto de Diagnostico y Referencia Epidemiologicos (INDRE) | Abril Rodriguez-Maldonado; Ariadna Medina-Benitez; Claudia Wong-Arambula; Ernesto Ramirez-Gonzalez.; Gisela Barrera-Badillo; Irma Lopez-Martinez; Joaquin Quiroz-Mercado; Lucia Hernandez-Rivas; Natividad Cruz-Ortiz; Sergio Rangel-Guerrero; Tatiana Nunez-Garcia; Vanessa Rivero-Arredondo |
| EPI_ISL_1399272, EPI_ISL_1423513, EPI_ISL_1483044, EPI_ISL_1483045, EPI_ISL_1483046, EPI_ISL_1483047, EPI_ISL_1483049, EPI_ISL_1483050, EPI_ISL_1504089, EPI_ISL_1504090, EPI_ISL_1504091, EPI_ISL_1504092, EPI_ISL_1504093, EPI_ISL_1504094, EPI_ISL_1504095, EPI_ISL_1504096, EPI_ISL_1504097, EPI_ISL_1504098, EPI_ISL_1504099, EPI_ISL_1504100, EPI_ISL_1504101, EPI_ISL_1504102, EPI_ISL_1504103, EPI_ISL_1504110, EPI_ISL_1504111, EPI_ISL_1516775, EPI_ISL_1516824, EPI_ISL_1516825, EPI_ISL_1516826, EPI_ISL_1532236, EPI_ISL_1532237, EPI_ISL_1532239, EPI_ISL_1532272, EPI_ISL_1532273, EPI_ISL_1532274, EPI_ISL_1532275, EPI_ISL_1532276, EPI_ISL_1532277, EPI_ISL_1532278, EPI_ISL_1558794, EPI_ISL_1558795, EPI_ISL_1651277, EPI_ISL_1651278, EPI_ISL_1651279, EPI_ISL_1651280, EPI_ISL_1651281, EPI_ISL_1651282, EPI_ISL_1651283, EPI_ISL_1651284, EPI_ISL_1651285, EPI_ISL_1651286, EPI_ISL_1651287, EPI_ISL_1651876, EPI_ISL_1661522, EPI_ISL_1661523, EPI_ISL_1661524, EPI_ISL_1661525, EPI_ISL_1661526, EPI_ISL_1661527, EPI_ISL_1661528, EPI_ISL_1661529, EPI_ISL_1661530, EPI_ISL_1661531, EPI_ISL_1661532, EPI_ISL_1661533, EPI_ISL_1673264, EPI_ISL_1673274, EPI_ISL_1700803, EPI_ISL_1700804, EPI_ISL_1700805, EPI_ISL_1700806, EPI_ISL_1700831, EPI_ISL_1700832, EPI_ISL_1805465, EPI_ISL_1821102, EPI_ISL_1821132, EPI_ISL_1821133, EPI_ISL_1821134, EPI_ISL_1821135, EPI_ISL_1821136, EPI_ISL_1821191, EPI_ISL_1821192, EPI_ISL_1857208, EPI_ISL_1857209, EPI_ISL_1857210, EPI_ISL_1857211, EPI_ISL_1857212, EPI_ISL_1857213, | | | |

| | | | |
|---|---|---|---|
| EPI_ISL_1857214, EPI_ISL_1857215, EPI_ISL_1857216, EPI_ISL_1857218, EPI_ISL_1857245, EPI_ISL_1857246, EPI_ISL_1912938, EPI_ISL_1912940, EPI_ISL_1912941, EPI_ISL_2157311, EPI_ISL_2157312, EPI_ISL_2157313, EPI_ISL_2157314, EPI_ISL_2158141, EPI_ISL_2158143, EPI_ISL_2158144, EPI_ISL_2158145, EPI_ISL_2158146, EPI_ISL_2158147, EPI_ISL_2158148, EPI_ISL_2158149, EPI_ISL_2158150, EPI_ISL_2158235, EPI_ISL_2158236, EPI_ISL_2158321 | | | |
| see above | LESP Queretaro | Instituto de Diagnostico y Referencia Epidemiologicos (INDRE) | Abril Rodriguez-Maldonado; Ariadna Medina-Benitez; Claudia Wong-Arambula; Ernesto Ramirez-Gonzalez.; Gisela Barrera-Badillo; Irma Lopez-Martinez; Joaquin Quiroz-Mercado; Lucia Hernandez-Rivas; Natividad Cruz-Ortiz; Sergio Rangel-Guerrero; Tatiana Nunez-Garcia; Vanessa Rivero-Arredondo |
| EPI_ISL_2158242 | LESP Queretaro /BIMODI | Instituto de Diagnostico y Referencia Epidemiologicos (INDRE) | Abril Rodriguez-Maldonado; Ariadna Medina-Benitez; Claudia Wong-Arambula; Ernesto Ramirez-Gonzalez.; Gisela Barrera-Badillo; Irma Lopez-Martinez; Joaquin Quiroz-Mercado; Lucia Hernandez-Rivas; Natividad Cruz-Ortiz; Sergio Rangel-Guerrero; Tatiana Nunez-Garcia; Vanessa Rivero-Arredondo |
| EPI_ISL_1532281, EPI_ISL_1626821, EPI_ISL_1651896, EPI_ISL_1651897, EPI_ISL_1651898, EPI_ISL_1710963, EPI_ISL_1805470, EPI_ISL_1805481, EPI_ISL_1821171, EPI_ISL_1821172, EPI_ISL_1857228, EPI_ISL_1857229, EPI_ISL_2101879, EPI_ISL_2101898, EPI_ISL_2101899, EPI_ISL_2105689, EPI_ISL_2105690, EPI_ISL_2105691, EPI_ISL_2105692, EPI_ISL_2105693, EPI_ISL_2157320, EPI_ISL_2158237 | | | |
| see above | LESP Quintana Roo | Instituto de Diagnostico y Referencia Epidemiologicos (INDRE) | Abril Rodriguez-Maldonado; Ariadna Medina-Benitez; Claudia Wong-Arambula; Ernesto Ramirez-Gonzalez.; Gisela Barrera-Badillo; Irma Lopez-Martinez; Joaquin Quiroz-Mercado; Lucia Hernandez-Rivas; Natividad Cruz-Ortiz; Sergio Rangel-Guerrero; Tatiana Nunez-Garcia; Vanessa Rivero-Arredondo |
| EPI_ISL_1661557, EPI_ISL_2094506, EPI_ISL_2094507, EPI_ISL_2094508 | LESP San Luis Potosi | Instituto de Diagnostico y Referencia Epidemiologicos (INDRE) | Abril Rodriguez-Maldonado; Ariadna Medina-Benitez; Claudia Wong-Arambula; Ernesto Ramirez-Gonzalez.; Gisela Barrera-Badillo; Irma Lopez-Martinez; Joaquin Quiroz-Mercado; Lucia Hernandez-Rivas; Natividad Cruz-Ortiz; Sergio Rangel-Guerrero; Tatiana Nunez-Garcia; Vanessa Rivero-Arredondo |
| EPI_ISL_1469940, EPI_ISL_1483042, EPI_ISL_1483043, EPI_ISL_1494670, EPI_ISL_1494671, EPI_ISL_1494672, EPI_ISL_1651253, EPI_ISL_1673263, EPI_ISL_1708610, EPI_ISL_1821156, EPI_ISL_1821157, EPI_ISL_1821199, EPI_ISL_2101896, EPI_ISL_2139918, EPI_ISL_2158173 | | | |
| see above | LESP Sinaloa | Instituto de Diagnostico y Referencia Epidemiologicos (INDRE) | Abril Rodriguez-Maldonado; Ariadna Medina-Benitez; Claudia Wong-Arambula; Ernesto Ramirez-Gonzalez.; Gisela Barrera-Badillo; Irma Lopez-Martinez; Joaquin Quiroz-Mercado; Lucia Hernandez-Rivas; Natividad Cruz-Ortiz; Sergio Rangel-Guerrero; Tatiana Nunez-Garcia; Vanessa Rivero-Arredondo |
| EPI_ISL_1651254, EPI_ISL_1700801, EPI_ISL_1805472, EPI_ISL_1805492, EPI_ISL_1821126, EPI_ISL_1821128, EPI_ISL_2101897, EPI_ISL_2157321, EPI_ISL_2158254 | | | |
| see above | LESP Sonora | Instituto de Diagnostico y Referencia Epidemiologicos (INDRE) | Abril Rodriguez-Maldonado; Ariadna Medina-Benitez; Claudia Wong-Arambula; Ernesto Ramirez-Gonzalez.; Gisela Barrera-Badillo; Irma Lopez-Martinez; Joaquin Quiroz-Mercado; Lucia Hernandez-Rivas; Natividad Cruz-Ortiz; Sergio Rangel-Guerrero; Tatiana Nunez-Garcia; Vanessa Rivero-Arredondo |
| EPI_ISL_1821193 | LESP Tabasco | Instituto de Diagnostico y Referencia Epidemiologicos (INDRE) | Abril Rodriguez-Maldonado; Ariadna Medina-Benitez; Claudia Wong-Arambula; Ernesto Ramirez-Gonzalez.; Gisela Barrera-Badillo; Irma Lopez-Martinez; Joaquin Quiroz-Mercado; Lucia Hernandez-Rivas; Natividad Cruz-Ortiz; Sergio Rangel-Guerrero; Tatiana Nunez-Garcia; Vanessa Rivero-Arredondo |
| EPI_ISL_1423511, EPI_ISL_1494650, EPI_ISL_1494651, EPI_ISL_1494652, EPI_ISL_1494653, EPI_ISL_1494654, EPI_ISL_1516776, EPI_ISL_1516789, EPI_ISL_1516790, EPI_ISL_1651887, EPI_ISL_1700817, EPI_ISL_1821158, EPI_ISL_1821159, EPI_ISL_1821160, EPI_ISL_1857200, EPI_ISL_1857254, EPI_ISL_1912942, EPI_ISL_1912943, EPI_ISL_1912944, EPI_ISL_1912945, EPI_ISL_2158114, EPI_ISL_2158206, EPI_ISL_2158229 | | | |
| see above | LESP Tamaulipas | Instituto de Diagnostico y Referencia Epidemiologicos (INDRE) | Abril Rodriguez-Maldonado; Ariadna Medina-Benitez; Claudia Wong-Arambula; Ernesto Ramirez-Gonzalez.; Gisela Barrera-Badillo; Irma Lopez-Martinez; Joaquin Quiroz-Mercado; Lucia Hernandez-Rivas; Natividad Cruz-Ortiz; Sergio Rangel-Guerrero; Tatiana Nunez-Garcia; Vanessa Rivero-Arredondo |
| EPI_ISL_1821166, EPI_ISL_1857257, EPI_ISL_2158131 | LESP Tlaxcala | Instituto de Diagnostico y Referencia Epidemiologicos (INDRE) | Abril Rodriguez-Maldonado; Ariadna Medina-Benitez; Claudia Wong-Arambula; Ernesto Ramirez-Gonzalez.; Gisela Barrera-Badillo; Irma Lopez-Martinez; Joaquin Quiroz-Mercado; Lucia Hernandez-Rivas; Natividad Cruz-Ortiz; Sergio Rangel-Guerrero; Tatiana Nunez-Garcia; Vanessa Rivero-Arredondo |
| EPI_ISL_1483055, EPI_ISL_1483056, EPI_ISL_1483057, EPI_ISL_1651881, EPI_ISL_1651882, EPI_ISL_1700811, EPI_ISL_1700812, EPI_ISL_1700813, EPI_ISL_1700814, EPI_ISL_1700815, EPI_ISL_1821163, EPI_ISL_1821164, EPI_ISL_1821165, EPI_ISL_1912946, EPI_ISL_2101892, EPI_ISL_2158142, EPI_ISL_2158205, EPI_ISL_2158228 | | | |
| see above | LESP Veracruz | Instituto de Diagnostico y Referencia Epidemiologicos (INDRE) | Abril Rodriguez-Maldonado; Ariadna Medina-Benitez; Claudia Wong-Arambula; Ernesto Ramirez-Gonzalez.; Gisela Barrera-Badillo; Irma Lopez-Martinez; Joaquin Quiroz-Mercado; Lucia Hernandez-Rivas; Natividad Cruz-Ortiz; Sergio Rangel-Guerrero; Tatiana Nunez-Garcia; Vanessa Rivero-Arredondo |
| EPI_ISL_1423514, EPI_ISL_1424035, EPI_ISL_1424036, EPI_ISL_1424037, EPI_ISL_1424038, EPI_ISL_1483038, EPI_ISL_1483039, EPI_ISL_1516774, EPI_ISL_1532248, EPI_ISL_1626802, EPI_ISL_1651883, EPI_ISL_1651884, EPI_ISL_1651885, EPI_ISL_1651886, EPI_ISL_1805501, EPI_ISL_1805502, EPI_ISL_1821152, EPI_ISL_1821153, EPI_ISL_1821154, EPI_ISL_1821155, EPI_ISL_1857202, EPI_ISL_1857203, EPI_ISL_1857204, EPI_ISL_1857205, EPI_ISL_1857206, EPI_ISL_1857253 | | | |
| see above | LESP Yucatan | Instituto de Diagnostico y Referencia Epidemiologicos (INDRE) | Abril Rodriguez-Maldonado; Ariadna Medina-Benitez; Claudia Wong-Arambula; Ernesto Ramirez-Gonzalez.; Gisela Barrera-Badillo; Irma Lopez-Martinez; Joaquin Quiroz-Mercado; Lucia Hernandez-Rivas; Natividad Cruz-Ortiz; Sergio Rangel-Guerrero; Tatiana Nunez-Garcia; Vanessa Rivero-Arredondo |
| EPI_ISL_1821173, EPI_ISL_1821174, EPI_ISL_1821175, EPI_ISL_1821176, EPI_ISL_1821177, EPI_ISL_1821178, EPI_ISL_1821179, EPI_ISL_1857264, EPI_ISL_1857265 | | | |
| see above | LESP Zacatecas | Instituto de Diagnostico y Referencia Epidemiologicos (INDRE) | Abril Rodriguez-Maldonado; Ariadna Medina-Benitez; Claudia Wong-Arambula; Ernesto Ramirez-Gonzalez.; Gisela Barrera-Badillo; Irma Lopez-Martinez; Joaquin Quiroz-Mercado; Lucia Hernandez-Rivas; Natividad Cruz-Ortiz; Sergio Rangel-Guerrero; Tatiana Nunez-Garcia; Vanessa Rivero-Arredondo |
| EPI_ISL_1911253, EPI_ISL_1911254, EPI_ISL_1911255, EPI_ISL_1911256, EPI_ISL_1911257, EPI_ISL_1911258, EPI_ISL_1911259, EPI_ISL_1911260, EPI_ISL_1911261, EPI_ISL_1911262, EPI_ISL_1911263, EPI_ISL_1911264, EPI_ISL_1911265, EPI_ISL_1911266, EPI_ISL_1911267, EPI_ISL_1911268, EPI_ISL_1911269, EPI_ISL_1911270, EPI_ISL_1911272, EPI_ISL_1911273, EPI_ISL_1911274, EPI_ISL_1911276, EPI_ISL_1911277, EPI_ISL_1911278, EPI_ISL_1911279, EPI_ISL_1911280, EPI_ISL_1911281, EPI_ISL_1911282, EPI_ISL_1911285, EPI_ISL_1911286, EPI_ISL_1911287, EPI_ISL_1911288, EPI_ISL_1911289, EPI_ISL_1911290, EPI_ISL_1911291, EPI_ISL_1911292, EPI_ISL_1911293, EPI_ISL_1911294, EPI_ISL_1911295, EPI_ISL_1911296, EPI_ISL_1911297, EPI_ISL_1911298, EPI_ISL_1911299, EPI_ISL_1911300, EPI_ISL_1911301, EPI_ISL_1911302, EPI_ISL_1911303, EPI_ISL_1911304, EPI_ISL_1911305, EPI_ISL_1911306, EPI_ISL_1911307, EPI_ISL_1911309, EPI_ISL_1911310, EPI_ISL_1911311 | | | |
| see above | LESPNL | LESPNL | (in alphabetical order) Consuelo Treviño-Garza; Eduardo Isaac de la Rosa-Moreno; Else del Carmen Garcia-Garcia; Gloria Alejandra Jasso-de la Peña; Manuel Enrique de la O-Cavazos; Olín Medina-Chávez; Yulianna Mayre Cordero-Cruz |
| EPI_ISL_1504106, EPI_ISL_1504107, EPI_ISL_1532250, EPI_ISL_1532251, EPI_ISL_1532252, EPI_ISL_1532253, EPI_ISL_1532254, EPI_ISL_1532255, EPI_ISL_1532256, EPI_ISL_1532257, EPI_ISL_1532258, EPI_ISL_1532259, EPI_ISL_1532260, EPI_ISL_1532261, EPI_ISL_1532262, EPI_ISL_1532263, EPI_ISL_1558796, EPI_ISL_1558797, EPI_ISL_1661536, EPI_ISL_1661537, EPI_ISL_1661538, EPI_ISL_1661544, EPI_ISL_1661545, EPI_ISL_1661546, EPI_ISL_1661547, EPI_ISL_1700833, EPI_ISL_1700836, EPI_ISL_1710970, EPI_ISL_1805494 | | | |
| see above | LESPZacatecas | Instituto de Diagnostico y Referencia Epidemiologicos (INDRE) | Abril Rodriguez-Maldonado; Ariadna Medina-Benitez; Claudia Wong-Arambula; Ernesto Ramirez-Gonzalez.; Gisela Barrera-Badillo; Irma Lopez-Martinez; Joaquin Quiroz-Mercado; Lucia Hernandez-Rivas; Natividad Cruz-Ortiz; Sergio Rangel-Guerrero; Tatiana Nunez-Garcia; Vanessa Rivero-Arredondo |
| EPI_ISL_1661977, EPI_ISL_1661978, EPI_ISL_1661980, EPI_ISL_1661981, EPI_ISL_1661991, EPI_ISL_1661992, EPI_ISL_1661993, EPI_ISL_1661994, EPI_ISL_1661995, EPI_ISL_1661996, EPI_ISL_1661997, EPI_ISL_1661998, EPI_ISL_1661999, EPI_ISL_1662000, EPI_ISL_1662001, EPI_ISL_1662002, EPI_ISL_1662003, EPI_ISL_1662004, EPI_ISL_1662005, EPI_ISL_1662006, EPI_ISL_1662007, EPI_ISL_1662008, EPI_ISL_1662009, EPI_ISL_1662010, EPI_ISL_1662011, EPI_ISL_1662012, EPI_ISL_1662013, EPI_ISL_1662014, EPI_ISL_1662015, EPI_ISL_1662016, EPI_ISL_1662017, EPI_ISL_1662018, EPI_ISL_1662019, EPI_ISL_1662020, EPI_ISL_1662021, EPI_ISL_1662022, EPI_ISL_1662023, EPI_ISL_1662024, EPI_ISL_1662025, EPI_ISL_1662026, EPI_ISL_1662027, EPI_ISL_1662028, EPI_ISL_1662029, EPI_ISL_1662030, EPI_ISL_1662031, EPI_ISL_1662032, EPI_ISL_1662033, EPI_ISL_1662034, EPI_ISL_1662058, EPI_ISL_1662059, EPI_ISL_1662060, EPI_ISL_1662061, EPI_ISL_1662070, EPI_ISL_1662071, EPI_ISL_1662072, EPI_ISL_1662073, EPI_ISL_1662074, EPI_ISL_1662104, EPI_ISL_1662105, EPI_ISL_1662106, EPI_ISL_1662107, EPI_ISL_1662108, EPI_ISL_1662109, EPI_ISL_1662110, EPI_ISL_1662111, EPI_ISL_1662112, EPI_ISL_1662113, EPI_ISL_1662114, EPI_ISL_1662115, EPI_ISL_1662116, EPI_ISL_1662117, EPI_ISL_1662118, EPI_ISL_1662119, EPI_ISL_1662120, EPI_ISL_1662121, EPI_ISL_1662122, EPI_ISL_1662128, EPI_ISL_1662129, EPI_ISL_1662130, EPI_ISL_1662145, EPI_ISL_1662146, EPI_ISL_1662147, EPI_ISL_1662148, EPI_ISL_1662149, EPI_ISL_1662150, EPI_ISL_1662151, EPI_ISL_1662152, EPI_ISL_1662153, EPI_ISL_1662154, EPI_ISL_1662155, EPI_ISL_1662156, EPI_ISL_1662157, EPI_ISL_1662158, EPI_ISL_1662159, EPI_ISL_1662160, EPI_ISL_1662161, EPI_ISL_1662162, EPI_ISL_1662163, EPI_ISL_1662164, EPI_ISL_1662165, EPI_ISL_1662166, EPI_ISL_1662167, EPI_ISL_1662168, EPI_ISL_1662169, EPI_ISL_1662170, EPI_ISL_1662171, EPI_ISL_1662172, EPI_ISL_1662173, EPI_ISL_1662174, EPI_ISL_1662175, EPI_ISL_1662176, EPI_ISL_1662177, EPI_ISL_1662178, EPI_ISL_1662179, EPI_ISL_1662180, EPI_ISL_1662181, EPI_ISL_1662182, EPI_ISL_1662183, EPI_ISL_1662194, EPI_ISL_1662195, EPI_ISL_1662196, EPI_ISL_1662201, EPI_ISL_1662202, EPI_ISL_1662203, EPI_ISL_1662204, EPI_ISL_1662205, EPI_ISL_1662206, EPI_ISL_1662207, EPI_ISL_1662208, EPI_ISL_1662209, EPI_ISL_1662210, EPI_ISL_1662211, EPI_ISL_1662212 | | | |
| see above | Laboratorio Central de Epidemiologia (LCE) | Unidad de Genomica Avanzada | Alejandro Sanchez-Flores; Alfredo Herrera-Estrella; Alicia Ocana-Mondragon; Angel Gustavo Salas-Lais; Bernardo Martinez-Miguel; Blanca Taboada; Brenda Irasema Maldonado-Meza; Carla Ivon Herrera-Najera; Carlos F. Arias; Celia Boukadida; Celida Duque Molina; Clara Esperanza Santacruz-Tinoco; Concepcion Grajales-Muniz; Consorcio Mexicano de Vigilancia Genomica (CoViGen-Mex). Authors (in alphabetical order): Julio Elias Alvarado-Yaah; Fernando Fontove-Herrera; Francisco Pulido; Gloria Elena Espinosa-Ayala; Gloria Maria Molina-Salinas; Gloria Vazquez; Hector Esteban Paz-Juarez; Hector Montoya-Fuentes; Helen Haydee Fernanda Ramirez-Plascencia; Jose Antonio Enciso-Moreno; Jose Esteban Munoz-Medina; Jose de Jesus Nunez-Contreras; Juan Bautista Chale-Dzul; Luis Alberto Ochoa-Carrera; Margarita Matias-Florentino; Maria Guadalupe Santiago-Mauricio; Maria Guadalupe de Jesus Mireles-Rivera; Nelly Selem-Mojica; Pavel Isa; Ricardo Grande; Santiago Ávila-Rios; Victor Eduardo Garcia-Arias; Victor Hugo Borja-Aburto |
| EPI_ISL_1351713, EPI_ISL_1351716, EPI_ISL_1351719, EPI_ISL_1351725, EPI_ISL_1351728, EPI_ISL_1351731, EPI_ISL_1351734, EPI_ISL_1351737, EPI_ISL_1351739, EPI_ISL_1351743, EPI_ISL_1351746, EPI_ISL_1351749, EPI_ISL_1351752, EPI_ISL_1351754, EPI_ISL_1351757, EPI_ISL_1351760, EPI_ISL_1351764, EPI_ISL_1351767, EPI_ISL_1351770, EPI_ISL_1351774, EPI_ISL_1351777, EPI_ISL_1351780, EPI_ISL_1351783, EPI_ISL_1351787, EPI_ISL_1351790, EPI_ISL_1351796, EPI_ISL_1351799, EPI_ISL_1351802, EPI_ISL_1351805, EPI_ISL_1351809, EPI_ISL_1351810, EPI_ISL_1351811, EPI_ISL_1351815, EPI_ISL_1351818, EPI_ISL_1351821, EPI_ISL_1351824, EPI_ISL_1351827, EPI_ISL_1351830, EPI_ISL_1351833, EPI_ISL_1351836, EPI_ISL_1351839, EPI_ISL_1351842, EPI_ISL_1351845, EPI_ISL_1351848, EPI_ISL_1351851, EPI_ISL_1351854, EPI_ISL_1351857, EPI_ISL_1351860, EPI_ISL_1351863, EPI_ISL_1351866, EPI_ISL_1351869, EPI_ISL_1351872, EPI_ISL_1351875, EPI_ISL_1351878, EPI_ISL_1351880, EPI_ISL_1351883, EPI_ISL_1351886, EPI_ISL_1351890, EPI_ISL_1351893, EPI_ISL_1351896, EPI_ISL_1351899, EPI_ISL_1351902, EPI_ISL_1351905, EPI_ISL_1351908, EPI_ISL_1351911, EPI_ISL_1351914, EPI_ISL_1351917, EPI_ISL_1351921, EPI_ISL_1351924, EPI_ISL_1351927, EPI_ISL_1351930, EPI_ISL_1351931, EPI_ISL_1351933, EPI_ISL_1351935, EPI_ISL_1351937, EPI_ISL_1351941, EPI_ISL_1351944, EPI_ISL_1351947, EPI_ISL_1351950, EPI_ISL_1351952, EPI_ISL_1351954, EPI_ISL_1351956, EPI_ISL_1351958, EPI_ISL_1351960, EPI_ISL_1351963, EPI_ISL_1351966, EPI_ISL_1351969, EPI_ISL_1351972, EPI_ISL_1351975, EPI_ISL_1351978, EPI_ISL_1351981, EPI_ISL_1351984, EPI_ISL_1351987, EPI_ISL_1351990, EPI_ISL_1351993, EPI_ISL_1351997, EPI_ISL_1352000, EPI_ISL_1352002, EPI_ISL_1352006, EPI_ISL_1352009, EPI_ISL_1352012, EPI_ISL_1352015, EPI_ISL_1352018, EPI_ISL_1352020, EPI_ISL_1352023, EPI_ISL_1352026, EPI_ISL_1352029, EPI_ISL_1352032, EPI_ISL_1352035, EPI_ISL_1352038, EPI_ISL_1352041, EPI_ISL_1352043, EPI_ISL_1352047, EPI_ISL_1352048, EPI_ISL_1352050, EPI_ISL_1352051, EPI_ISL_1352053, EPI_ISL_1352055, EPI_ISL_1352057, EPI_ISL_1352059, EPI_ISL_1352060, EPI_ISL_1352062, EPI_ISL_1352066, EPI_ISL_1352068, EPI_ISL_1352070, EPI_ISL_1352072 | | | |
| see above | Laboratorio Central de EpidemiologÃa (LCE) | Unidad de Genomica Avanzada | Alejandro Sanchez-Flores; Alfredo Herrera-Estrella; Alicia Ocana-Mondragon; Angel Gustavo Salas-Lais; Bernardo Martinez-Miguel; Blanca Taboada; Brenda Irasema Maldonado-Meza; Carla Ivon Herrera-Najera; Carlos F. Arias; Celia Boukadida; Clara Esperanza Santacruz-Tinoco; Concepcion Grajales-Muniz; Consorcio Mexicano de Vigilancia Genomica (CoViGen-Mex). Authors (in alphabetical order): Julio Elias Alvarado-Yaah; Fernando Fontove-Herrera; Francisco Pulido; Gloria Elena Espinoza-Ayala; Gloria Maria Molina-Salinas; Gloria Vazquez; Hector Esteban Paz-Juarez; Hector Montoya-Fuentes; Helen Haydee Fernanda Ramirez-Plascencia; Jorge Ivan Salinal-Nevarez; Jose Antonio Enciso-Moreno; Jose Esteban Munoz-Medina; Jose de Jesus Nunez-Contreras; Juan Bautista Chale-Dzul; Luis Alberto Ochoa-Carrera; Margarita Matias-Florentino; Maria Guadalupe Santiago-Mauricio; Maria Guadalupe de Jesus Mireles-Rivera; Nelly Selem-Mojica; Pavel Isa; Ricardo Grande; Santiago Avila-Rios; Victor Hugo Borja-Aburto |
| EPI_ISL_2091143, EPI_ISL_2091144, EPI_ISL_2091146, EPI_ISL_2091147, EPI_ISL_2091148, EPI_ISL_2091149, EPI_ISL_2091150, EPI_ISL_2091151, EPI_ISL_2091152, EPI_ISL_2091153, EPI_ISL_2091154, EPI_ISL_2091155, EPI_ISL_2091156, EPI_ISL_2091157, EPI_ISL_2091158, EPI_ISL_2091159, EPI_ISL_2091320, EPI_ISL_2091321, EPI_ISL_2091322, EPI_ISL_2091323, EPI_ISL_2091324, EPI_ISL_2091325, EPI_ISL_2091326, EPI_ISL_2091327, EPI_ISL_2091328, EPI_ISL_2091329, EPI_ISL_2091330, EPI_ISL_2091331, EPI_ISL_2091332, EPI_ISL_2091333, EPI_ISL_2091334, EPI_ISL_2091335, EPI_ISL_2091336, EPI_ISL_2091337, EPI_ISL_2091338, EPI_ISL_2091339, EPI_ISL_2091340, EPI_ISL_2091341, EPI_ISL_2091342, EPI_ISL_2091343, EPI_ISL_2091344, EPI_ISL_2091345, EPI_ISL_2091346, EPI_ISL_2091347, EPI_ISL_2091348, EPI_ISL_2091349, EPI_ISL_2091350, EPI_ISL_2091351, EPI_ISL_2091352, EPI_ISL_2091353, EPI_ISL_2091354, EPI_ISL_2091355, EPI_ISL_2091356, EPI_ISL_2091357, EPI_ISL_2091358, EPI_ISL_2091359, EPI_ISL_2091360, EPI_ISL_2091361, EPI_ISL_2091362, EPI_ISL_2091363, EPI_ISL_2091364, EPI_ISL_2091365, EPI_ISL_2091366, EPI_ISL_2091367, EPI_ISL_2091368, EPI_ISL_2091369, EPI_ISL_2091370, EPI_ISL_2091371, EPI_ISL_2091372, EPI_ISL_2091373, EPI_ISL_2091374, EPI_ISL_2091375, EPI_ISL_2091376, EPI_ISL_2091377, EPI_ISL_2091378, EPI_ISL_2091379, EPI_ISL_2091380, EPI_ISL_2091381, EPI_ISL_2091382, EPI_ISL_2091383, EPI_ISL_2091384, EPI_ISL_2091385, EPI_ISL_2091386, EPI_ISL_2091387, EPI_ISL_2091388, EPI_ISL_2091389, EPI_ISL_2091390, EPI_ISL_2091391, EPI_ISL_2091392, EPI_ISL_2091393, EPI_ISL_2091394, EPI_ISL_2091395, EPI_ISL_2091396, EPI_ISL_2091397, EPI_ISL_2091398, EPI_ISL_2091399, EPI_ISL_2091400, EPI_ISL_2091401, EPI_ISL_2091402, EPI_ISL_2091403, EPI_ISL_2091404, EPI_ISL_2091405, EPI_ISL_2091406, EPI_ISL_2091407, EPI_ISL_2091408, EPI_ISL_2091409, EPI_ISL_2091410, EPI_ISL_2091411, EPI_ISL_2091412, EPI_ISL_2091413, EPI_ISL_2091414, EPI_ISL_2091415, EPI_ISL_2091416, EPI_ISL_2091417, EPI_ISL_2091418, EPI_ISL_2091419, EPI_ISL_2091420, EPI_ISL_2091421, EPI_ISL_2091422, EPI_ISL_2091423, EPI_ISL_2091424, EPI_ISL_2091425, EPI_ISL_2091426, EPI_ISL_2091427, EPI_ISL_2091428, EPI_ISL_2091429, EPI_ISL_2091430, EPI_ISL_2091431, EPI_ISL_2091432, EPI_ISL_2091433, EPI_ISL_2091434, EPI_ISL_2091435, EPI_ISL_2091436, EPI_ISL_2091437, EPI_ISL_2091438, EPI_ISL_2091439, EPI_ISL_2091440, EPI_ISL_2091441, EPI_ISL_2091442, EPI_ISL_2091443, EPI_ISL_2091444, EPI_ISL_2091445, EPI_ISL_2091446, EPI_ISL_2091447, EPI_ISL_2091448, EPI_ISL_2091449, EPI_ISL_2091450, EPI_ISL_2091451, EPI_ISL_2091452, EPI_ISL_2091453, EPI_ISL_2091454, EPI_ISL_2091455, EPI_ISL_2091456, EPI_ISL_2178730, EPI_ISL_2178731, EPI_ISL_2178732, EPI_ISL_2178733, EPI_ISL_2178734, EPI_ISL_2178735, EPI_ISL_2178736, EPI_ISL_2178737, EPI_ISL_2178738, EPI_ISL_2178739, EPI_ISL_2178740, EPI_ISL_2178741 | | | |
| see above | Laboratorio Central de Epidemiología (LCE) | Centro de Investigación en Enfermedades Infecciosas (CIENI), Instituto Nacional de Enfermedades Respiratorias (INER) | Alejandro Sanchez-Flores; Alfredo Herrera-Estrella; Alicia Ocaña-Mondragón; Angel Gustavo Salas-Lais; Bernardo Martínez-Miguel; Blanca Taboada; Brenda Irasema Maldonado-Meza; Carla Ivón Herrera-Najera; Carlos F. Arias; Celia Boukadida; Clara Esperanza Santacruz-Tinoco; Concepción Grajales-Muñiz; Consorcio Mexicano de Vigilancia Genómica (CoViGen-Mex). Authors (in alphabetical order): Julio Elias Alvarado-Yaah; Célida Duque Molina; Fernando Fontove-Herrera; Francisco Pulido; Gloria Elena Espinosa-Ayala; Gloria María Molina-Salinas; Gloria Vazquez; Hector Esteban Paz-Juárez; Hector Montoya-Fuentes; Helen Haydee Fernanda Ramírez-Plascencia; José Antonio Enciso-Moreno; José Esteban Muñoz-Medina; José de Jesús Nuñez-Contreras; Juan Bautista Chale-Dzul; Luis Alberto Ochoa-Carrera; Margarita Matías-Florentino; María Guadalupe Santiago-Mauricio; María Guadalupe de Jesús Mireles-Rivera; Nelly Sélem-Mojica; Pavel Isa; Ricardo Grande; Santiago Ávila-Ríos; Victor Eduardo Garcia-Arias; Víctor Hugo Borja-Aburto |
| EPI_ISL_1585564, EPI_ISL_1585565, EPI_ISL_1585566, EPI_ISL_1585567, EPI_ISL_1585568, EPI_ISL_1585569, EPI_ISL_1585570, EPI_ISL_1585571, EPI_ISL_1585572, EPI_ISL_1585573, EPI_ISL_1585574, EPI_ISL_1585575, EPI_ISL_1585576, EPI_ISL_1585577, EPI_ISL_1585578, EPI_ISL_1585579, EPI_ISL_1585580, EPI_ISL_1585581, EPI_ISL_1585582, EPI_ISL_1585583, EPI_ISL_1585584, EPI_ISL_1585585, EPI_ISL_1585586, EPI_ISL_1585587, EPI_ISL_1585588, EPI_ISL_1585589, EPI_ISL_1585590, EPI_ISL_1585591, EPI_ISL_1585592, EPI_ISL_1585593, EPI_ISL_1585594, EPI_ISL_1585595, EPI_ISL_1585596, EPI_ISL_1585597, EPI_ISL_1585598, EPI_ISL_1585599, EPI_ISL_1585600, EPI_ISL_1585601, EPI_ISL_1585602, EPI_ISL_1585603, EPI_ISL_1585604, EPI_ISL_1585605, EPI_ISL_1585606, EPI_ISL_1585607, EPI_ISL_1585608, EPI_ISL_1585609, EPI_ISL_1585610, EPI_ISL_1585611, EPI_ISL_1585612, EPI_ISL_1585613, EPI_ISL_1585614, EPI_ISL_1585615, EPI_ISL_1585616, EPI_ISL_1585617, EPI_ISL_1585618, EPI_ISL_1585619, EPI_ISL_1585620, EPI_ISL_1585621, EPI_ISL_1585622, EPI_ISL_1585623, EPI_ISL_1585624, EPI_ISL_1585625, EPI_ISL_1585626, EPI_ISL_1585627, EPI_ISL_1585628, EPI_ISL_1585629, EPI_ISL_1585630, EPI_ISL_1585631, EPI_ISL_1585632, EPI_ISL_1585633, EPI_ISL_1585634, EPI_ISL_1585635, EPI_ISL_1585636, EPI_ISL_1585637, EPI_ISL_1585638, EPI_ISL_1585639, EPI_ISL_1585640, EPI_ISL_1585641, EPI_ISL_1585642, EPI_ISL_1585643, EPI_ISL_1585644, EPI_ISL_1585645, EPI_ISL_1585646, EPI_ISL_1585647, EPI_ISL_1585648, EPI_ISL_1585649, EPI_ISL_1585650, EPI_ISL_1585651, EPI_ISL_1585652, EPI_ISL_1585653, EPI_ISL_1585654, EPI_ISL_1585655, EPI_ISL_1585656, EPI_ISL_1585657, EPI_ISL_1585658, EPI_ISL_1585659, EPI_ISL_1585660, EPI_ISL_1585661, EPI_ISL_1585662, EPI_ISL_1585663, EPI_ISL_1585664, EPI_ISL_1585665, EPI_ISL_1585666, EPI_ISL_1585667, EPI_ISL_1585668, EPI_ISL_1585669, EPI_ISL_1585670, EPI_ISL_1595608, EPI_ISL_1595609, EPI_ISL_1595610, EPI_ISL_1595611 | | | |
| see above | Laboratorio Central de Epidemiología (LCE) | Instituto Nacional de Enfermedades Respiratorias (INER): Centro de Investigación en Enfermedades Infecciosas (CIENI) | Alejandro Sanchez-Flores; Alfredo Herrera-Estrella; Alicia Ocaña-Mondragón; Angel Gustavo Salas-Lais; Bernardo Martínez-Miguel; Blanca Taboada; Brenda Irasema Maldonado-Meza; Carla Ivón Herrera-Najera; Carlos F. Arias; Celia Boukadida; Clara Esperanza Santacruz-Tinoco; Concepción Grajales-Muñiz; Consorcio Mexicano de Vigilancia Genómica (CoViGen-Mex). Authors (in alphabetical order): Julio Elias Alvarado-Yaah; Célida Duque Molina; Fernando Fontove-Herrera; Francisco Pulido; Gloria Elena Espinosa-Ayala; Gloria María Molina-Salinas; Gloria Vazquez; Hector Esteban Paz-Juárez; Hector Montoya-Fuentes; Helen Haydee Fernanda Ramírez-Plascencia; José Antonio Enciso-Moreno; José Esteban Muñoz-Medina; José de Jesús Nuñez-Contreras; Juan Bautista Chale-Dzul; Luis Alberto Ochoa-Carrera; Margarita Matías-Florentino; María Guadalupe Santiago-Mauricio; María Guadalupe de Jesús Mireles-Rivera; Nelly Sélem-Mojica; Pavel Isa; Ricardo Grande; Santiago Ávila-Ríos; Victor Eduardo Garcia-Arias; Víctor Hugo Borja-Aburto |
| EPI_ISL_1416332, EPI_ISL_1416333, EPI_ISL_1416334, EPI_ISL_1416335, EPI_ISL_1416336, EPI_ISL_1416337, EPI_ISL_1416338, EPI_ISL_1416339, EPI_ISL_1416340, EPI_ISL_1416341, EPI_ISL_1416342, EPI_ISL_1416343, EPI_ISL_1416344, EPI_ISL_1416345, EPI_ISL_1416346, EPI_ISL_1416347, EPI_ISL_1416348, EPI_ISL_1416349, EPI_ISL_1416350, EPI_ISL_1416351, EPI_ISL_1416352, EPI_ISL_1416353, EPI_ISL_1416354, EPI_ISL_1416355, EPI_ISL_1416356, EPI_ISL_1416357, EPI_ISL_1416358, EPI_ISL_1416359, EPI_ISL_1416360, EPI_ISL_1416361, EPI_ISL_1416362, EPI_ISL_1416363, EPI_ISL_1416364, EPI_ISL_1416365, EPI_ISL_1416366, EPI_ISL_1416367, EPI_ISL_1416368, EPI_ISL_1416369, EPI_ISL_1416370, EPI_ISL_1416371, EPI_ISL_1416372, EPI_ISL_1416373, EPI_ISL_1416374, EPI_ISL_1416375, EPI_ISL_1416376, EPI_ISL_1416377, EPI_ISL_1416378, EPI_ISL_1416379, EPI_ISL_1416380, EPI_ISL_1416381, EPI_ISL_1416382, EPI_ISL_1416383, EPI_ISL_1416384, EPI_ISL_1416385, EPI_ISL_1416386, EPI_ISL_1416387, EPI_ISL_1416388, EPI_ISL_1416389, EPI_ISL_1416390, EPI_ISL_1416391, EPI_ISL_1416392, EPI_ISL_1416393, EPI_ISL_1416394, EPI_ISL_1416395, EPI_ISL_1416396, EPI_ISL_1416397, EPI_ISL_1416398, EPI_ISL_1416399, EPI_ISL_1416400, EPI_ISL_1416401, EPI_ISL_1416402, EPI_ISL_1416403, EPI_ISL_1416404, EPI_ISL_1416405, EPI_ISL_1416406, EPI_ISL_1416407, EPI_ISL_1416408, EPI_ISL_1416409, EPI_ISL_1416410, EPI_ISL_1416411, EPI_ISL_1416412, EPI_ISL_1416413, EPI_ISL_1416414, EPI_ISL_1416415, EPI_ISL_1416416, EPI_ISL_1416417, EPI_ISL_1416418, EPI_ISL_1416419, EPI_ISL_1416420, EPI_ISL_1416421, EPI_ISL_1416422, EPI_ISL_1416423, EPI_ISL_1416424, EPI_ISL_1416425, EPI_ISL_1416426, EPI_ISL_1416427, EPI_ISL_1416428, EPI_ISL_1416429, EPI_ISL_1416430, EPI_ISL_1416431, EPI_ISL_1416432, EPI_ISL_1416433, EPI_ISL_1416434, EPI_ISL_1416435, EPI_ISL_1416436, EPI_ISL_1416437, EPI_ISL_1416438, EPI_ISL_1416439, EPI_ISL_1416440, EPI_ISL_1416441, EPI_ISL_1416442, EPI_ISL_1416443, EPI_ISL_1416444, EPI_ISL_1416445, EPI_ISL_1416446, EPI_ISL_1416447, EPI_ISL_1416448, EPI_ISL_1416449, EPI_ISL_1416450, EPI_ISL_1416451, EPI_ISL_1416452, EPI_ISL_1416453, EPI_ISL_1416454, EPI_ISL_1416455, EPI_ISL_1416456, EPI_ISL_1416457, EPI_ISL_1416458, EPI_ISL_1416459, EPI_ISL_1416460, EPI_ISL_1416461, EPI_ISL_1416462, EPI_ISL_1416463, | | | |

EPI_ISL_1416464, EPI_ISL_1416465, EPI_ISL_1416466, EPI_ISL_1416467, EPI_ISL_1416468, EPI_ISL_1416469, EPI_ISL_1416470, EPI_ISL_1416471, EPI_ISL_1416472, EPI_ISL_1416473, EPI_ISL_1416474, EPI_ISL_1416475, EPI_ISL_1416476, EPI_ISL_1416477, EPI_ISL_1416478, EPI_ISL_1416479, EPI_ISL_1416480, EPI_ISL_1416481, EPI_ISL_1416482, EPI_ISL_1416483, EPI_ISL_1416484, EPI_ISL_1416485, EPI_ISL_1416486, EPI_ISL_1416487, EPI_ISL_1416488, EPI_ISL_1416489, EPI_ISL_1416490, EPI_ISL_1416491, EPI_ISL_1416492, EPI_ISL_1416493, EPI_ISL_1416494, EPI_ISL_1416495, EPI_ISL_1416496, EPI_ISL_1416497, EPI_ISL_1416498, EPI_ISL_1416499, EPI_ISL_1416500, EPI_ISL_1416501, EPI_ISL_1416502, EPI_ISL_1416503, EPI_ISL_1416504, EPI_ISL_1416505, EPI_ISL_1416506, EPI_ISL_1416507, EPI_ISL_1416508, EPI_ISL_1416509, EPI_ISL_1416510, EPI_ISL_1416511, EPI_ISL_1416512, EPI_ISL_1416513, EPI_ISL_1416514, EPI_ISL_1416515, EPI_ISL_1416516, EPI_ISL_1416517, EPI_ISL_1416518, EPI_ISL_1416519, EPI_ISL_1416520, EPI_ISL_1416521, EPI_ISL_1416522, EPI_ISL_1416523, EPI_ISL_1416524, EPI_ISL_1416525, EPI_ISL_1416526, EPI_ISL_1416527, EPI_ISL_1416528, EPI_ISL_1416529, EPI_ISL_1416530, EPI_ISL_1416531, EPI_ISL_1416532, EPI_ISL_1416533, EPI_ISL_1416534, EPI_ISL_1416535, EPI_ISL_1416536, EPI_ISL_1416537, EPI_ISL_1416538, EPI_ISL_1416539, EPI_ISL_1416540, EPI_ISL_1416541, EPI_ISL_1416542, EPI_ISL_1416543, EPI_ISL_1416544, EPI_ISL_1416545, EPI_ISL_1416546, EPI_ISL_1416547, EPI_ISL_1416548, EPI_ISL_1416549, EPI_ISL_1416550, EPI_ISL_1416551, EPI_ISL_1416552, EPI_ISL_1416553, EPI_ISL_1416554, EPI_ISL_1416555, EPI_ISL_1416556, EPI_ISL_1416557, EPI_ISL_1416558, EPI_ISL_1416559, EPI_ISL_1416560, EPI_ISL_1416561, EPI_ISL_1416562, EPI_ISL_1416563, EPI_ISL_1416564, EPI_ISL_1416565, EPI_ISL_1416566, EPI_ISL_1416567, EPI_ISL_1416568, EPI_ISL_1416569, EPI_ISL_1416570, EPI_ISL_1416571, EPI_ISL_1416572, EPI_ISL_1416573, EPI_ISL_1416574, EPI_ISL_1416575, EPI_ISL_1416576, EPI_ISL_1416577, EPI_ISL_1416578, EPI_ISL_1416579, EPI_ISL_1416580, EPI_ISL_1416581, EPI_ISL_1416582, EPI_ISL_1416583, EPI_ISL_1416584, EPI_ISL_1416585, EPI_ISL_1416586, EPI_ISL_1416587, EPI_ISL_1416588, EPI_ISL_1416589, EPI_ISL_1416590, EPI_ISL_1416591, EPI_ISL_1416592, EPI_ISL_1416593, EPI_ISL_1416594, EPI_ISL_1416595, EPI_ISL_1416596, EPI_ISL_1416597, EPI_ISL_1416598, EPI_ISL_1416599, EPI_ISL_1416600, EPI_ISL_1416601, EPI_ISL_1416602, EPI_ISL_1416603, EPI_ISL_1416604, EPI_ISL_1416605, EPI_ISL_1416606, EPI_ISL_1416607, EPI_ISL_1416608, EPI_ISL_1416609, EPI_ISL_1416610, EPI_ISL_1416611, EPI_ISL_1416612, EPI_ISL_1416613, EPI_ISL_1416614, EPI_ISL_1416615, EPI_ISL_1416616, EPI_ISL_1416617, EPI_ISL_1416618, EPI_ISL_1416619, EPI_ISL_1416620, EPI_ISL_1416621, EPI_ISL_1416622, EPI_ISL_1416623, EPI_ISL_1416624, EPI_ISL_1416625, EPI_ISL_1416626, EPI_ISL_1416627, EPI_ISL_1416628, EPI_ISL_1416629, EPI_ISL_1416630, EPI_ISL_1416631, EPI_ISL_1416632, EPI_ISL_1416633, EPI_ISL_1416634, EPI_ISL_1416635, EPI_ISL_1416636, EPI_ISL_1416637, EPI_ISL_1416638, EPI_ISL_1416639, EPI_ISL_1416640, EPI_ISL_1416641, EPI_ISL_1416642, EPI_ISL_1416643, EPI_ISL_1416644, EPI_ISL_1416645, EPI_ISL_1416646, EPI_ISL_1416647, EPI_ISL_1416648, EPI_ISL_1416649, EPI_ISL_1416650, EPI_ISL_1416651, EPI_ISL_1416652, EPI_ISL_1416653, EPI_ISL_1416654, EPI_ISL_1416655, EPI_ISL_1416656, EPI_ISL_1416657, EPI_ISL_1416658, EPI_ISL_1416659, EPI_ISL_1416660, EPI_ISL_1416661, EPI_ISL_1416662, EPI_ISL_1416663, EPI_ISL_1416664, EPI_ISL_1416665, EPI_ISL_1416666, EPI_ISL_1416667, EPI_ISL_1416668, EPI_ISL_1416669, EPI_ISL_1416670, EPI_ISL_1416671, EPI_ISL_1416672, EPI_ISL_1416673, EPI_ISL_1416674, EPI_ISL_1416675, EPI_ISL_1416676, EPI_ISL_1416677, EPI_ISL_1416678, EPI_ISL_1416679, EPI_ISL_1416680, EPI_ISL_1416681, EPI_ISL_1811379, EPI_ISL_1811380, EPI_ISL_1811381, EPI_ISL_1811382, EPI_ISL_1811383, EPI_ISL_1811384, EPI_ISL_1811385, EPI_ISL_1811386, EPI_ISL_1811387, EPI_ISL_1811388, EPI_ISL_1811389, EPI_ISL_1811390, EPI_ISL_1811391, EPI_ISL_1811392, EPI_ISL_1811393, EPI_ISL_1811394, EPI_ISL_1811395, EPI_ISL_1811396, EPI_ISL_1811397, EPI_ISL_1811398, EPI_ISL_1811399, EPI_ISL_1811400, EPI_ISL_1811401, EPI_ISL_1811402, EPI_ISL_1811403, EPI_ISL_1811404, EPI_ISL_1811405, EPI_ISL_1811406, EPI_ISL_1811407, EPI_ISL_1811408, EPI_ISL_1811409, EPI_ISL_1811410, EPI_ISL_1811411, EPI_ISL_1811412, EPI_ISL_1811413, EPI_ISL_1811414, EPI_ISL_1811415, EPI_ISL_1811416, EPI_ISL_1811417, EPI_ISL_1811418, EPI_ISL_1811419, EPI_ISL_1811420, EPI_ISL_1811421, EPI_ISL_1811422, EPI_ISL_1811423, EPI_ISL_1811424, EPI_ISL_1811425, EPI_ISL_1811426, EPI_ISL_1811427, EPI_ISL_1811428, EPI_ISL_1811429, EPI_ISL_1811430, EPI_ISL_1811431, EPI_ISL_1811432, EPI_ISL_1811433, EPI_ISL_1811434, EPI_ISL_1811435, EPI_ISL_1811436, EPI_ISL_1811437, EPI_ISL_1811438, EPI_ISL_1811439, EPI_ISL_1811440, EPI_ISL_1811441, EPI_ISL_1811442, EPI_ISL_1811443, EPI_ISL_1811444, EPI_ISL_1811445, EPI_ISL_1811446, EPI_ISL_1811447, EPI_ISL_1811448, EPI_ISL_1811449, EPI_ISL_1811450, EPI_ISL_1811451, EPI_ISL_1811452, EPI_ISL_1811453, EPI_ISL_1811454, EPI_ISL_1811455, EPI_ISL_1811456, EPI_ISL_1811457, EPI_ISL_1811458, EPI_ISL_1811459, EPI_ISL_1811460, EPI_ISL_1811461, EPI_ISL_1811462, EPI_ISL_1811463, EPI_ISL_1811464, EPI_ISL_1811465, EPI_ISL_1811466, EPI_ISL_1811467, EPI_ISL_1811468, EPI_ISL_1811469, EPI_ISL_1811470, EPI_ISL_1811471, EPI_ISL_1811472, EPI_ISL_1811473, EPI_ISL_1811474, EPI_ISL_1811475, EPI_ISL_1811476, EPI_ISL_1811477, EPI_ISL_1811478, EPI_ISL_1811479, EPI_ISL_1811480, EPI_ISL_1811481, EPI_ISL_1811482, EPI_ISL_1811483, EPI_ISL_1811484, EPI_ISL_1811485, EPI_ISL_1811486, EPI_ISL_1811487, EPI_ISL_1811488, EPI_ISL_1811489, EPI_ISL_1811490, EPI_ISL_1811491, EPI_ISL_1811492, EPI_ISL_1811493, EPI_ISL_1811494, EPI_ISL_1811495, EPI_ISL_1811496, EPI_ISL_1811497, EPI_ISL_1811498, EPI_ISL_1811499, EPI_ISL_1811500, EPI_ISL_1811501, EPI_ISL_1811502, EPI_ISL_1811503, EPI_ISL_1811504, EPI_ISL_1811505, EPI_ISL_1811506, EPI_ISL_1811507, EPI_ISL_1811508, EPI_ISL_1811509, EPI_ISL_1811510, EPI_ISL_1811511, EPI_ISL_1811512, EPI_ISL_1811513, EPI_ISL_1811514, EPI_ISL_1811515, EPI_ISL_1811516, EPI_ISL_1811517, EPI_ISL_1811518, EPI_ISL_1811519, EPI_ISL_1811520, EPI_ISL_1811521, EPI_ISL_1811522, EPI_ISL_1811523, EPI_ISL_1811524, EPI_ISL_1811525, EPI_ISL_1811526, EPI_ISL_1811527, EPI_ISL_1811528, EPI_ISL_1811529, EPI_ISL_1811530, EPI_ISL_1811531, EPI_ISL_1811532, EPI_ISL_1811533, EPI_ISL_1811534, EPI_ISL_1811535, EPI_ISL_1811536, EPI_ISL_1811537, EPI_ISL_1811538, EPI_ISL_1811539, EPI_ISL_1811540, EPI_ISL_1811541, EPI_ISL_1811542, EPI_ISL_1811543, EPI_ISL_1811544, EPI_ISL_1811545

| | | | |
|---|---|---|---|
| see above | Laboratorio Central de Epidemiología (LCE) | Instituto de Biotecnología de la UNAM | Alejandro Sanchez-Flores; Alfredo Herrera-Estrella; Alicia Ocaña-Mondragón; Angel Gustavo Salas-Lais; Bernardo Martínez-Miguel; Blanca Taboada; Brenda Irasema Maldonado-Meza; Carla Ivón Herrera-Najera; Carlos F. Arias; Celia Boukadida; Clara Esperanza Santacruz-Tinoco; Concepción Grajales-Muñiz; Consorcio Mexicano de Vigilancia Genómica (CoViGen-Mex). Authors (in alphabetical order): Julio Elias Alvarado-Yaah; Célida Duque-Molina; Fernando Fontove-Herrera; Francisco Pulido; Gloria Elena Espinosa-Ayala; Gloria Elena Espinoza-Ayala; Gloria María Molina-Salinas; Gloria Vazquez; Hector Esteban Paz-Juárez; Hector Montoya-Fuentes; Helen Haydee Fernanda Ramírez-Plascencia; Jorge Ivan Salinal-Nevarez; José Antonio Enciso-Moreno; José Esteban Muñoz-Medina; José de Jesús Nuñez-Contreras; Juan Bautista Chale-Dzul; Julio Elias Alvarado-Yaah; Luis Alberto Ochoa-Carrera; Margarita Matías-Florentino; María Guadalupe Santiago-Mauricio; María Guadalupe de Jesús Mireles-Rivera; Nelly Sélem-Mojica; Pavel Isa; Ricardo Grande; Santiago Ávila-Ríos; Victor Eduardo Garcia-Arias; Víctor Hugo Borja-Aburto |

EPI_ISL_2102599, EPI_ISL_2102600, EPI_ISL_2102602, EPI_ISL_2102606, EPI_ISL_2102614, EPI_ISL_2102615, EPI_ISL_2102616, EPI_ISL_2102617, EPI_ISL_2102618, EPI_ISL_2102619, EPI_ISL_2102623, EPI_ISL_2102624, EPI_ISL_2102626, EPI_ISL_2102637, EPI_ISL_2102638, EPI_ISL_2102639, EPI_ISL_2102640, EPI_ISL_2102641, EPI_ISL_2102642, EPI_ISL_2102660, EPI_ISL_2102663, EPI_ISL_2102664, EPI_ISL_2102665, EPI_ISL_2102666, EPI_ISL_2102667, EPI_ISL_2102668, EPI_ISL_2102675, EPI_ISL_2102676, EPI_ISL_2102677, EPI_ISL_2102680

| | | | |
|---|---|---|---|
| see above | Laboratorio Estatal de Salud Publica de Nuevo Leon | Laboratorio Estatal de Salud Publica de Nuevo Leon | (in alphabetical order) Consuelo Treviño-Garza; Eduardo Isaac de la Rosa-Moreno; Else del Carmen Garcia-Garcia; Gloria Alejandra Jasso-de la Peña; Manuel Enrique de la O-Cavazos; Olín Medina-Chávez; Yulianna Mayre Cordero-Cruz |
| EPI_ISL_1789696, EPI_ISL_1789697, EPI_ISL_1789698, EPI_ISL_1789699, EPI_ISL_1794455 | Laboratorio PGM | Laboratorio de Infectología Molecular, Departamento de Bioquímica y Medicina Molecular,Facultad de Medicina - Universidad Autónoma de Nuevo León | Ana M. Rivas-Estilla; Daniel Arellanos-Soto; Eduardo Garza-de-la-Peña; Gabriela Elizondo; Javier Ramos-Jimenez; Kame A. Galán-Huerta; María F. Herrera-Saldivar; Natalia Martínez-Acuña; Sonia A. Lozano-Sepúlveda |
| EPI_ISL_1334383, EPI_ISL_1334384 | Laboratorios Lister | Instituto de Diagnostico y Referencia Epidemiologicos (INDRE) | Abril Rodriguez-Maldonado; Ariadna Medina-Benitez; Claudia Wong-Arambula; Ernesto Ramirez-Gonzalez.; Gisela Barrera-Badillo; Irma Lopez-Martinez; Joaquin Quiroz-Mercado; Lucia Hernandez-Rivas; Natividad Cruz-Ortiz; Sergio Rangel-Guerrero; Tatiana Nunez-Garcia; Vanessa Rivero-Arredondo |

EPI_ISL_1321310, EPI_ISL_1321311, EPI_ISL_1321339, EPI_ISL_1321348, EPI_ISL_1321349, EPI_ISL_1321350, EPI_ISL_1338916, EPI_ISL_1338920, EPI_ISL_1338960, EPI_ISL_1338961, EPI_ISL_1338962, EPI_ISL_1338965, EPI_ISL_1338976, EPI_ISL_1338984, EPI_ISL_1338985, EPI_ISL_1338987, EPI_ISL_1338990, EPI_ISL_1338996, EPI_ISL_1339035, EPI_ISL_1339047, EPI_ISL_1339048, EPI_ISL_1339049, EPI_ISL_1339206, EPI_ISL_1422210, EPI_ISL_1461775, EPI_ISL_1461834, EPI_ISL_1461858, EPI_ISL_1461861, EPI_ISL_1461862, EPI_ISL_1461863, EPI_ISL_1461864, EPI_ISL_1461865, EPI_ISL_1461880, EPI_ISL_1513754, EPI_ISL_1513782, EPI_ISL_1513813, EPI_ISL_1513815, EPI_ISL_1513816, EPI_ISL_1514801, EPI_ISL_1514802, EPI_ISL_1514803, EPI_ISL_1514835, EPI_ISL_1514836, EPI_ISL_1514873, EPI_ISL_1514874, EPI_ISL_1514875, EPI_ISL_1514888, EPI_ISL_1514889, EPI_ISL_1514890, EPI_ISL_1514891, EPI_ISL_1514892, EPI_ISL_1514894, EPI_ISL_1514919, EPI_ISL_1514921, EPI_ISL_1514950, EPI_ISL_1514951, EPI_ISL_1514953, EPI_ISL_1514955, EPI_ISL_1514965, EPI_ISL_1515243, EPI_ISL_1515249, EPI_ISL_1515272, EPI_ISL_1515273, EPI_ISL_1515274, EPI_ISL_1515275, EPI_ISL_1515276, EPI_ISL_1515301, EPI_ISL_1515351, EPI_ISL_1515384, EPI_ISL_1515385, EPI_ISL_1515386, EPI_ISL_1515387, EPI_ISL_1515532, EPI_ISL_1515533, EPI_ISL_1515534, EPI_ISL_1515535, EPI_ISL_1515564, EPI_ISL_1515579, EPI_ISL_1515617, EPI_ISL_1515657, EPI_ISL_1515658, EPI_ISL_1515659, EPI_ISL_1515753, EPI_ISL_1549201, EPI_ISL_1549208, EPI_ISL_1549872, EPI_ISL_1549873, EPI_ISL_1549982, EPI_ISL_1550001, EPI_ISL_1550002, EPI_ISL_1550003, EPI_ISL_1550063, EPI_ISL_1550267, EPI_ISL_1550307, EPI_ISL_1550308, EPI_ISL_1550310, EPI_ISL_1550362, EPI_ISL_1550363, EPI_ISL_1612019, EPI_ISL_1612022, EPI_ISL_1612023, EPI_ISL_1612024, EPI_ISL_1612025, EPI_ISL_1612028, EPI_ISL_1612094, EPI_ISL_1612095, EPI_ISL_1612097, EPI_ISL_1612218, EPI_ISL_1612219, EPI_ISL_1612220, EPI_ISL_1612221, EPI_ISL_1612235, EPI_ISL_1612236, EPI_ISL_1612237, EPI_ISL_1612470, EPI_ISL_1612471, EPI_ISL_1612473, EPI_ISL_1612663, EPI_ISL_1612674, EPI_ISL_1612675, EPI_ISL_1684395, EPI_ISL_1684396, EPI_ISL_1684481, EPI_ISL_1684482, EPI_ISL_1684489, EPI_ISL_1684576, EPI_ISL_1684578, EPI_ISL_1684579, EPI_ISL_1684633, EPI_ISL_1684751, EPI_ISL_1684752, EPI_ISL_1684753, EPI_ISL_1684762, EPI_ISL_1684763, EPI_ISL_1684764, EPI_ISL_1684804, EPI_ISL_1684806, EPI_ISL_1684813, EPI_ISL_1684816, EPI_ISL_1684840, EPI_ISL_1684893, EPI_ISL_1684946, EPI_ISL_1684976, EPI_ISL_1701874, EPI_ISL_1798197, EPI_ISL_1798198, EPI_ISL_1798199, EPI_ISL_1798201, EPI_ISL_1798203, EPI_ISL_1798245, EPI_ISL_1798246, EPI_ISL_1798267, EPI_ISL_1798287, EPI_ISL_1798288, EPI_ISL_1798289, EPI_ISL_1798290, EPI_ISL_1798291, EPI_ISL_1798315, EPI_ISL_1798362, EPI_ISL_1798931, EPI_ISL_1798932, EPI_ISL_1798956, EPI_ISL_1799026, EPI_ISL_1799027, EPI_ISL_1799045, EPI_ISL_1799081, EPI_ISL_1799082, EPI_ISL_1799211, EPI_ISL_1799212, EPI_ISL_1799316, EPI_ISL_1799317, EPI_ISL_1799319, EPI_ISL_1799320, EPI_ISL_1799384, EPI_ISL_1799385, EPI_ISL_1799386, EPI_ISL_1799434, EPI_ISL_1799439, EPI_ISL_1799440, EPI_ISL_1799442, EPI_ISL_1799514, EPI_ISL_1799515, EPI_ISL_1799529, EPI_ISL_1799568, EPI_ISL_1799569, EPI_ISL_1799607, EPI_ISL_1930678, EPI_ISL_1930712, EPI_ISL_1930714, EPI_ISL_1930737, EPI_ISL_1930738, EPI_ISL_1930748, EPI_ISL_1930800, EPI_ISL_1930801, EPI_ISL_1930814, EPI_ISL_1930815, EPI_ISL_1930816, EPI_ISL_1930817, EPI_ISL_1930818, EPI_ISL_1930822, EPI_ISL_1930823, EPI_ISL_1931092, EPI_ISL_1931168, EPI_ISL_1931276, EPI_ISL_1931278, EPI_ISL_1931279, EPI_ISL_1931281, EPI_ISL_1931282, EPI_ISL_1931337, EPI_ISL_1931397, EPI_ISL_1931400, EPI_ISL_1931569, EPI_ISL_1931594, EPI_ISL_1931598, EPI_ISL_1931669, EPI_ISL_1931697, EPI_ISL_1931700, EPI_ISL_1931932, EPI_ISL_1931935, EPI_ISL_1931938, EPI_ISL_1931940, EPI_ISL_1931943, EPI_ISL_1931978, EPI_ISL_1931987, EPI_ISL_1931989, EPI_ISL_1931990, EPI_ISL_2046016, EPI_ISL_2046017, EPI_ISL_2046018, EPI_ISL_2046019, EPI_ISL_2046053, EPI_ISL_2046056, EPI_ISL_2046057, EPI_ISL_2046060, EPI_ISL_2046199, EPI_ISL_2046200, EPI_ISL_2046201, EPI_ISL_2046203, EPI_ISL_2046205, EPI_ISL_2046206, EPI_ISL_2046207, EPI_ISL_2046208, EPI_ISL_2046275, EPI_ISL_2046304, EPI_ISL_2046305, EPI_ISL_2046306, EPI_ISL_2046309, EPI_ISL_2046310, EPI_ISL_2046318, EPI_ISL_2046531, EPI_ISL_2046532, EPI_ISL_2046534, EPI_ISL_2046601, EPI_ISL_2046639, EPI_ISL_2046674

| | | | |
|---|---|---|---|
| see above | Laboratory Corporation of America | Centers for Disease Control and Prevention Division of Viral Diseases, Pathogen Discovery | Adrian Paskey; Amanda Douglas; Amanda Suchanek; Andrea Throop; Ayla Burns; Ben L. Rambo-Martin; Benjamin Rambo-Martin; Bobbi Croy; Brian Krueger; Brian Norvell; Christopher Gulvick; Christos Petropoulos; Clinton R. Paden; Craig Lukasik; Dakota Howard; Darlene Wagner; Debbie Boles; Dhwani Batra; Duncan MacCannell; Eyad Almasri; Goran Stevovic; Howard Engler; Hrushikesh Deshmukh; Jake Humphrey; Jana Schroth; Jason Caravas; Joe Voshell; John Pruitt; Jonathan Meltzer; Jonathan Williams; Kara Moser; Kimberly Wagner; Lax Iyer; Lyndon Tilson; Manoj Jain; Marcia Eisenberg; Mary Ann Cristobal; Mary Williamson; Matthew Schmerer; Michael Levandoski; Mike Sapeta; Mindy Nye; Minoo Agarwal; Mohan Kolli; Nuthawin Charoensri; Oren Cohen; Peter W. Cook; Prashant Gupta; Qian Zeng; Rama Ghatti; Scott Parker; Scott Ryan; Scott Sammons; Shatavia Morrison; Stanley Letovsky; Steven Ragan; Suresh Babu Selvaraju; Susan Countryman; Susan Hicks; Suxiang Tong; Suzanne Dale; Thomas Urban; Tim Kuphal; Tricia Zwiefelhofer; Vincent Drouillon; Yvette Unoarumhi |

EPI_ISL_1516236, EPI_ISL_1516237, EPI_ISL_1516238, EPI_ISL_1516239, EPI_ISL_1516240, EPI_ISL_1516241, EPI_ISL_1516242, EPI_ISL_1516243, EPI_ISL_1516244, EPI_ISL_1695781, EPI_ISL_1695887, EPI_ISL_1695888, EPI_ISL_2018173, EPI_ISL_2018174, EPI_ISL_2018175, EPI_ISL_2018176, EPI_ISL_2018177, EPI_ISL_2018178, EPI_ISL_2018179, EPI_ISL_2018180, EPI_ISL_2018181

| | | | |
|---|---|---|---|
| see above | NM Dept. Health, Scientific Laboratory Division | Centers for Disease Control and Prevention Division of Viral Diseases, Pathogen Discovery | Alison Laufer Halpin; Ben L. Rambo-Martin; Clinton R. Paden; Dakota Howard; Darlene Wagner; Dave Wentworth; Dhwani Batra; Jasmine Padilla; Justin Lee; Katie Dillon; Krista Queen; Kristen Knipe; Kristine Lacek; Mark Burroughs; Matthew Schmerer; Mili Sheth; Peter Cook; Sam Shepard; Sarah Nobles; Shoshona Le; Suxiang Tong; Vivien Dugan; Yvette Unoarumhi |
| EPI_ISL_1759376 | NM Dept. Health, Scientific Laboratory Division | Pathogen Discovery, Respiratory Viruses Branch, Division of Viral Diseases, Centers for Disease Control and Prevention | Adam Retchless; Anna Kelleher; Anna Montmayeur; Anna Uehara; Brian Lynch; Clinton R. Paden; Haibin Wang; Han Jia Justin Ng; Jing Zhang; Justin Lee; Krista Queen; Mark Burroughs; Peter Cook; Rachel Marine; Suxiang Tong; Yan Li; Ying Tao |

EPI_ISL_1255982, EPI_ISL_1255985, EPI_ISL_1255988, EPI_ISL_1255991, EPI_ISL_1255994, EPI_ISL_1255997, EPI_ISL_1255999, EPI_ISL_1256002, EPI_ISL_1256005, EPI_ISL_1256008, EPI_ISL_1256011, EPI_ISL_1256014, EPI_ISL_1256016, EPI_ISL_1256019, EPI_ISL_1256021, EPI_ISL_1256024, EPI_ISL_1256027, EPI_ISL_1256029, EPI_ISL_1256032, EPI_ISL_1256035, EPI_ISL_1256039, EPI_ISL_1256042, EPI_ISL_1256045, EPI_ISL_1256048, EPI_ISL_1256051, EPI_ISL_1256053, EPI_ISL_1261545, EPI_ISL_1261584, EPI_ISL_1261585, EPI_ISL_1340880, EPI_ISL_1340881, EPI_ISL_1340882, EPI_ISL_1340883, EPI_ISL_1340884, EPI_ISL_1340885, EPI_ISL_1340886, EPI_ISL_1340887, EPI_ISL_1340888, EPI_ISL_1340889, EPI_ISL_1340890, EPI_ISL_1340891, EPI_ISL_1340892, EPI_ISL_1340893, EPI_ISL_1340898, EPI_ISL_1340899, EPI_ISL_1340901, EPI_ISL_1340902, EPI_ISL_1340903, EPI_ISL_1340904, EPI_ISL_1340905, EPI_ISL_1340906, EPI_ISL_1340907, EPI_ISL_1340908, EPI_ISL_1340909, EPI_ISL_1340910, EPI_ISL_1340911, EPI_ISL_1340917, EPI_ISL_1340918, EPI_ISL_1340919, EPI_ISL_1340920, EPI_ISL_1340921, EPI_ISL_1340923, EPI_ISL_1340924, EPI_ISL_1340925, EPI_ISL_1340926, EPI_ISL_1400942, EPI_ISL_1400943, EPI_ISL_1400944, EPI_ISL_1400945, EPI_ISL_1400946, EPI_ISL_1400947, EPI_ISL_1400948, EPI_ISL_1400949, EPI_ISL_1400950, EPI_ISL_1400951, EPI_ISL_1400952, EPI_ISL_1400953, EPI_ISL_1400954, EPI_ISL_1400955, EPI_ISL_1400956, EPI_ISL_1400957, EPI_ISL_1400958, EPI_ISL_1400959, EPI_ISL_1400960, EPI_ISL_1400961, EPI_ISL_1400962, EPI_ISL_1400963, EPI_ISL_1400967, EPI_ISL_1400968, EPI_ISL_1400969, EPI_ISL_1400970, EPI_ISL_1400971, EPI_ISL_1400972, EPI_ISL_1400973, EPI_ISL_1400974, EPI_ISL_1400975, EPI_ISL_1400976, EPI_ISL_1400977, EPI_ISL_1400978, EPI_ISL_1400979, EPI_ISL_1400980, EPI_ISL_1400981, EPI_ISL_1400984, EPI_ISL_1530948, EPI_ISL_1530949, EPI_ISL_1530950, EPI_ISL_1530951, EPI_ISL_1530952, EPI_ISL_1530953, EPI_ISL_1530954, EPI_ISL_1530955, EPI_ISL_1530956, EPI_ISL_1530957, EPI_ISL_1530958, EPI_ISL_1530959, EPI_ISL_1530960, EPI_ISL_1530961, EPI_ISL_1530962, EPI_ISL_1530963, EPI_ISL_1530964, EPI_ISL_1530965, EPI_ISL_1530966, EPI_ISL_1530967, EPI_ISL_1530968, EPI_ISL_1530969, EPI_ISL_1530970, EPI_ISL_1530971, EPI_ISL_1530972, EPI_ISL_1530973, EPI_ISL_1530974, EPI_ISL_1530975, EPI_ISL_1530976, EPI_ISL_1530977, EPI_ISL_1530978, EPI_ISL_1530979, EPI_ISL_1530980, EPI_ISL_1530981, EPI_ISL_1530982, EPI_ISL_1530983, EPI_ISL_1530984, EPI_ISL_1530985, EPI_ISL_1530986, EPI_ISL_1530987, EPI_ISL_1530988, EPI_ISL_1530989, EPI_ISL_1582408, EPI_ISL_1582409, EPI_ISL_1582424, EPI_ISL_1582425, EPI_ISL_1582444, EPI_ISL_1582445, EPI_ISL_1582446, EPI_ISL_1582447, EPI_ISL_1582448, EPI_ISL_1582449, EPI_ISL_1582450, EPI_ISL_1582451, EPI_ISL_1582452, EPI_ISL_1582453, EPI_ISL_1582454, EPI_ISL_1582455, EPI_ISL_1582456, EPI_ISL_1582457, EPI_ISL_1582458, EPI_ISL_1582459, EPI_ISL_1582460, EPI_ISL_1582461, EPI_ISL_1582462, EPI_ISL_1582463, EPI_ISL_1582468, EPI_ISL_1582469, EPI_ISL_1710561, EPI_ISL_1710562, EPI_ISL_1710563, EPI_ISL_1710564, EPI_ISL_1710565, EPI_ISL_1710566, EPI_ISL_1710567, EPI_ISL_1710568, EPI_ISL_1710569, EPI_ISL_1710570, EPI_ISL_1710571, EPI_ISL_1710572, EPI_ISL_1710573, EPI_ISL_1710574, EPI_ISL_1710575, EPI_ISL_1710576, EPI_ISL_1710577, EPI_ISL_1710578, EPI_ISL_1710579, EPI_ISL_1710584, EPI_ISL_1710585, EPI_ISL_1710586, EPI_ISL_1710587, EPI_ISL_1710588, EPI_ISL_1710589, EPI_ISL_1710590, EPI_ISL_1710591, EPI_ISL_1710592, EPI_ISL_1710593, EPI_ISL_1710594, EPI_ISL_1710595, EPI_ISL_1710596, EPI_ISL_1710597, EPI_ISL_1805368, EPI_ISL_1805369, EPI_ISL_1805370, EPI_ISL_1805371, EPI_ISL_1805372, EPI_ISL_1805373, EPI_ISL_1805374, EPI_ISL_1805375, EPI_ISL_1805376, EPI_ISL_1805377, EPI_ISL_1805378, EPI_ISL_1805379, EPI_ISL_1805380, EPI_ISL_1805381, EPI_ISL_1805382, EPI_ISL_1805383, EPI_ISL_1805384, EPI_ISL_1805385, EPI_ISL_1805386, EPI_ISL_1805387, EPI_ISL_1805388, EPI_ISL_1805389, EPI_ISL_1805390, EPI_ISL_1805391, EPI_ISL_1805393, EPI_ISL_1805394, EPI_ISL_1805396, EPI_ISL_1805397, EPI_ISL_1805398, EPI_ISL_1970894, EPI_ISL_1970895, EPI_ISL_1970896, EPI_ISL_1970897, EPI_ISL_1970898, EPI_ISL_1970899, EPI_ISL_1970900, EPI_ISL_1970901, EPI_ISL_1970902, EPI_ISL_1970903, EPI_ISL_1970904, EPI_ISL_1970905, EPI_ISL_1970906, EPI_ISL_1970907, EPI_ISL_1970908, EPI_ISL_1970909, EPI_ISL_1970910, EPI_ISL_1970911, EPI_ISL_1970912, EPI_ISL_1970913, EPI_ISL_1970914, EPI_ISL_1970915, EPI_ISL_1970916, EPI_ISL_1970917, EPI_ISL_1970918, EPI_ISL_1970919, EPI_ISL_1970934, EPI_ISL_1970935, EPI_ISL_1970936, EPI_ISL_1970938, EPI_ISL_1970939, EPI_ISL_1970941, EPI_ISL_1970942, EPI_ISL_1970943, EPI_ISL_1970944, EPI_ISL_1970945, EPI_ISL_1970946, EPI_ISL_1970947, EPI_ISL_1970957, EPI_ISL_1970958, EPI_ISL_1970959, EPI_ISL_1970960, EPI_ISL_1970961, EPI_ISL_1970962, EPI_ISL_1970964, EPI_ISL_1970965, EPI_ISL_1970966, EPI_ISL_1970967, EPI_ISL_1970968, EPI_ISL_1970969, EPI_ISL_1970970, EPI_ISL_1970971, EPI_ISL_1970972, EPI_ISL_1970973, EPI_ISL_1970974

| | | | |
|---|---|---|---|
| see above | New Mexico Department of Health Scientific Laboratory | New Mexico Department of Health Scientific Laboratory | Anastacia Griego-Fisher; D'eldra Malone; Ellie Johnson; Jennifer Benoit; Linda Salazar; Mark Willmon; Ratheesh Rajan |
| EPI_ISL_1661549 | PEMEX | Instituto de Diagnostico y Referencia Epidemiologicos (INDRE) | Abril Rodriguez-Maldonado; Ariadna Medina-Benitez; Claudia Wong-Arambula; Ernesto Ramirez-Gonzalez.; Gisela Barrera-Badillo; Irma Lopez-Martinez; Joaquin Quiroz-Mercado; Lucia Hernandez-Rivas; Natividad Cruz-Ortiz; Sergio Rangel-Guerrero; Tatiana Nunez-Garcia; Vanessa Rivero-Arredondo |
| EPI_ISL_1340914, EPI_ISL_1340915, EPI_ISL_1340916, EPI_ISL_1805392, EPI_ISL_1805409, EPI_ISL_1970940 | PHS/IHS Hospital-Jicarilla Service Unit | New Mexico Department of Health Scientific Laboratory | Anastacia Griego-Fisher; D'eldra Malone; Ellie Johnson; Jennifer Benoit; Linda Salazar; Mark Willmon; Ratheesh Rajan |

EPI_ISL_1267027, EPI_ISL_1267093, EPI_ISL_1267168, EPI_ISL_1267173, EPI_ISL_1267197, EPI_ISL_1267461, EPI_ISL_1297502, EPI_ISL_1297516, EPI_ISL_1297529, EPI_ISL_1297536, EPI_ISL_1297545, EPI_ISL_1297546, EPI_ISL_1297549, EPI_ISL_1297553, EPI_ISL_1297748, EPI_ISL_1314823, EPI_ISL_1314895, EPI_ISL_1314960, EPI_ISL_1339859, EPI_ISL_1339868, EPI_ISL_1339869, EPI_ISL_1367271, EPI_ISL_1367274, EPI_ISL_1367303, EPI_ISL_1391430, EPI_ISL_1391519, EPI_ISL_1391683, EPI_ISL_1392278, EPI_ISL_1392368, EPI_ISL_1392376, EPI_ISL_1392378, EPI_ISL_1392434, EPI_ISL_1422723, EPI_ISL_1422816, EPI_ISL_1422821, EPI_ISL_1444423, EPI_ISL_1444591, EPI_ISL_1444610, EPI_ISL_1460745, EPI_ISL_1479712, EPI_ISL_1479719, EPI_ISL_1480763, EPI_ISL_1492059, EPI_ISL_1551732, EPI_ISL_1552036, EPI_ISL_1552048, EPI_ISL_1552097, EPI_ISL_1552110, EPI_ISL_1552128, EPI_ISL_1559415, EPI_ISL_1559567, EPI_ISL_1559584, EPI_ISL_1668467, EPI_ISL_1668479, EPI_ISL_1668692, EPI_ISL_1668859, EPI_ISL_1668973, EPI_ISL_1669005, EPI_ISL_1669076, EPI_ISL_1669186, EPI_ISL_1738133, EPI_ISL_1818282, EPI_ISL_1906170, EPI_ISL_1924673, EPI_ISL_2010966, EPI_ISL_2011347, EPI_ISL_2011426, EPI_ISL_2046890, EPI_ISL_2133165, EPI_ISL_2133278, EPI_ISL_2133554, EPI_ISL_2133562, EPI_ISL_2133826, EPI_ISL_2143686, EPI_ISL_2143758, EPI_ISL_2143785, EPI_ISL_2143861, EPI_ISL_2144000, EPI_ISL_2179953, EPI_ISL_2179967, EPI_ISL_2180473

| | | | |
|---|---|---|---|
| see above | Quest Diagnostics Incorporated | Centers for Disease Control and Prevention Division of Viral Diseases, Pathogen Discovery | A. Gerasimova; A. Perez; Adrian Paskey; B. Anderson; Ben L. Rambo-Martin; Benjamin Rambo-Martin; Christopher Gulvick; Clinton R. Paden; Dakota Howard; Darlene Wagner; Dhwani Batra; Duncan MacCannell; F. Lacbawan; I. A. Shlyakhter; Jason Caravas; K.E. Livingston; Kara Moser; L.E. Bernstein; M. Hua; Matthew Schmerer; P. Tanpaiboon; Peter W. Cook; R. M. Kagan; R. Owen; R. V. Rolando; S. H. Rosenthal; Scott Sammons; Shatavia Morrison; Suxiang Tong; Y. Liu; Yvette Unoarumhi |
| EPI_ISL_2313597, EPI_ISL_2313675, EPI_ISL_2313852 | Respiratory Viruses Branch, Centers for Disease Control and Prevention | Respiratory Viruses Branch, Centers for Disease Control and Prevention | Batra, D.; Bixby, C.; C.R.; Caravas, J.; Cook; Goswami, C.; Grimwood, R.; Gulvick, C.; Hager, R.; Howard, D.; MacCannell, D.; Morrison, S.; Moser, K.; P.W.; Paden; Paskey, A.; Rambo-Martin, B.; Sammons, S.; Schmerer, M.; Schultz, J.; Unoarumhi, Y.; Wagner, D.; Wang, Y. |

EPI_ISL_2105742, EPI_ISL_2105743, EPI_ISL_2105744, EPI_ISL_2105745, EPI_ISL_2105746, EPI_ISL_2105747, EPI_ISL_2105748, EPI_ISL_2105749, EPI_ISL_2105750, EPI_ISL_2105751, EPI_ISL_2105752, EPI_ISL_2105753, EPI_ISL_2105754, EPI_ISL_2105755, EPI_ISL_2105756, EPI_ISL_2105757, EPI_ISL_2105758, EPI_ISL_2105759, EPI_ISL_2105760, EPI_ISL_2105761, EPI_ISL_2105762, EPI_ISL_2105763, EPI_ISL_2105764, EPI_ISL_2105765, EPI_ISL_2105766, EPI_ISL_2105767, EPI_ISL_2105768, EPI_ISL_2105769, EPI_ISL_2105770, EPI_ISL_2105771, EPI_ISL_2105772, EPI_ISL_2105773, EPI_ISL_2105774, EPI_ISL_2105775, EPI_ISL_2105776, EPI_ISL_2105777, EPI_ISL_2105778, EPI_ISL_2105779, EPI_ISL_2105780, EPI_ISL_2105781, EPI_ISL_2105782, EPI_ISL_2105783, EPI_ISL_2105784, EPI_ISL_2105785, EPI_ISL_2105786, EPI_ISL_2105787, EPI_ISL_2105788, EPI_ISL_2105789, EPI_ISL_2105790, EPI_ISL_2105791, EPI_ISL_2105792, EPI_ISL_2105793, EPI_ISL_2105794, EPI_ISL_2105795, EPI_ISL_2105796, EPI_ISL_2105797, EPI_ISL_2105798, EPI_ISL_2105799, EPI_ISL_2105800, EPI_ISL_2105801, EPI_ISL_2105802, EPI_ISL_2105803, EPI_ISL_2105804, EPI_ISL_2105805, EPI_ISL_2105806, EPI_ISL_2105807, EPI_ISL_2105808, EPI_ISL_2105809, EPI_ISL_2105810, EPI_ISL_2105811, EPI_ISL_2105812, EPI_ISL_2105813, EPI_ISL_2105814, EPI_ISL_2105815, EPI_ISL_2105816, EPI_ISL_2105817, EPI_ISL_2105818, EPI_ISL_2105819, EPI_ISL_2105820, EPI_ISL_2178749, EPI_ISL_2178750, EPI_ISL_2178754, EPI_ISL_2178755, EPI_ISL_2178756, EPI_ISL_2178757

| | | | |
|---|---|---|---|
| see above | Salud Digna | Instituto Nacional de Medicina Genomica | Abraham Campos-Romero; Cedro-Tanda A; Cisneros-Villanueva M; Escobar-Arrazola; Gonzalez-Barrera D; Herrera-Montalvo LA.; Hidalgo-Miranda A; Luna-Ruiz Marco; M; Mendoza-Vargas A; Moreno-Camacho José Luis; Munguia-Garza P; Orjuela-Rodríguez M; Ramirez-Vega O; Rangel-DeLeon D; Reyes-Grajeda JP; Rodriguez-Gallegos Jorge |

EPI_ISL_1340912, EPI_ISL_1340913, EPI_ISL_1400982, EPI_ISL_1400983, EPI_ISL_1805401, EPI_ISL_1805402, EPI_ISL_1805403, EPI_ISL_1805404, EPI_ISL_1805405, EPI_ISL_1805406, EPI_ISL_1805407, EPI_ISL_1805408, EPI_ISL_1970920, EPI_ISL_1970921, EPI_ISL_1970922, EPI_ISL_1970923, EPI_ISL_1970924, EPI_ISL_1970925, EPI_ISL_1970926, EPI_ISL_1970927, EPI_ISL_1970950, EPI_ISL_1970951, EPI_ISL_1970952, EPI_ISL_1970953, EPI_ISL_1970954, EPI_ISL_1970955, EPI_ISL_1970956, EPI_ISL_1970963, EPI_ISL_1970975, EPI_ISL_1970976, EPI_ISL_1970977, EPI_ISL_1970978, EPI_ISL_1970979, EPI_ISL_1970980, EPI_ISL_1970981, EPI_ISL_1970982, EPI_ISL_1970983, EPI_ISL_1970984

| | | | |
|---|---|---|---|
| see above | San Juan Regional Medical Center | New Mexico Department of Health Scientific Laboratory | Anastacia Griego-Fisher; D'eldra Malone; Ellie Johnson; Jennifer Benoit; Linda Salazar; Mark Willmon; Ratheesh Rajan |

| | | | |
|---|---|---|---|
| EPI_ISL_1468835, EPI_ISL_1468846, EPI_ISL_1468919, EPI_ISL_1468958, EPI_ISL_1531778, EPI_ISL_1531787, EPI_ISL_1794698, EPI_ISL_1794744, EPI_ISL_1794747, EPI_ISL_1794890, EPI_ISL_1969312 | | | |
| see above | Sharp HealthCare Laboratory | Andersen lab at Scripps Research | Art Mendoza; Cathy Woerle; Jacquelyn Berumen; Liam McGinnis; Omid Bakhtar; SEARCH Alliance San Diego with Aaron Harding |
| EPI_ISL_1805399, EPI_ISL_1805400 | Sierra Vista Hospital | New Mexico Department of Health Scientific Laboratory | Anastacia Griego-Fisher; D'eldra Malone; Ellie Johnson; Jennifer Benoit; Linda Salazar; Ratheesh Rajan |
| EPI_ISL_1582464, EPI_ISL_1582465, EPI_ISL_1582466, EPI_ISL_1582467, EPI_ISL_1970928, EPI_ISL_1970929, EPI_ISL_1970930, EPI_ISL_1970931, EPI_ISL_1970932, EPI_ISL_1970933, EPI_ISL_1970937 | | | |
| see above | TriCore Reference Laboratories | New Mexico Department of Health Scientific Laboratory | Anastacia Griego-Fisher; D'eldra Malone; Ellie Johnson; Jennifer Benoit; Linda Salazar; Mark Willmon; Ratheesh Rajan |
| EPI_ISL_1970948, EPI_ISL_1970949 | TriCore Reference Laboratory | New Mexico Department of Health Scientific Laboratory | Anastacia Griego-Fisher; D'eldra Malone; Ellie Johnson; Jennifer Benoit; Linda Salazar; Mark Willmon; Ratheesh Rajan |
| EPI_ISL_1340894, EPI_ISL_1340895, EPI_ISL_1340896, EPI_ISL_1340897, EPI_ISL_1400964, EPI_ISL_1400965, EPI_ISL_1400966, EPI_ISL_1710580, EPI_ISL_1710581, EPI_ISL_1710582, EPI_ISL_1710583, EPI_ISL_1739339 | | | |
| see above | TriCore Reference Laboratory Inc. | New Mexico Department of Health Scientific Laboratory | Anastacia Griego-Fisher; D'eldra Malone; Ellie Johnson; Jennifer Benoit; Linda Salazar; Ratheesh Rajan |
| EPI_ISL_1711089 | US Air Force School of Aerospace Medicine | US Air Force School of Aerospace Medicine | Amanda Javorina; Anthony Fries; Clarise Starr; Elizabeth Macias; Jennifer Meyer; Sarah Purves; William Buggele; William Gruner |
| EPI_ISL_1662221, EPI_ISL_1662222, EPI_ISL_1662223, EPI_ISL_1662224, EPI_ISL_1662225, EPI_ISL_1662226, EPI_ISL_1662227, EPI_ISL_1662228, EPI_ISL_1662229, EPI_ISL_1662230, EPI_ISL_1662231, EPI_ISL_1662232 | | | |
| see above | Unidad de Investigacion Biomedica de Zacatecas (UIBZ) | Unidad de Genomica Avanzada | Alejandro Sanchez-Flores; Alfredo Herrera-Estrella; Alicia Ocana-Mondragon; Angel Gustavo Salas-Lais; Bernardo Martinez-Miguel; Blanca Taboada; Brenda Irasema Maldonado-Meza; Carla Ivon Herrera-Najera; Carlos F. Arias; Celia Boukadida; Celida Duque Molina; Clara Esperanza Santacruz-Tinoco; Concepcion Grajales-Muniz; Consorcio Mexicano de Vigilancia Genomica (CoViGen-Mex). Authors (in alphabetical order): Julio Elias Alvarado-Yaah; Fernando Fontove-Herrera; Francisco Pulido; Gloria Elena Espinosa-Ayala; Gloria Maria Molina-Salinas; Gloria Vazquez; Hector Esteban Paz-Juarez; Hector Montoya-Fuentes; Helen Haydee Fernanda Ramirez-Plascencia; Jose Antonio Enciso-Moreno; Jose Esteban Munoz-Medina; Jose de Jesus Nunez-Contreras; Juan Bautista Chale-Dzul; Luis Alberto Ochoa-Carrera; Margarita Matias-Florentino; Maria Guadalupe Santiago-Mauricio; Maria Guadalupe de Jesus Mireles-Rivera; Nelly Selem-Mojica; Pavel Isa; Ricardo Grande; Santiago Ávila-Rios; Victor Eduardo Garcia-Arias; Victor Hugo Borja-Aburto |
| EPI_ISL_1662184, EPI_ISL_1662185, EPI_ISL_1662213, EPI_ISL_1662214, EPI_ISL_1662215, EPI_ISL_1662216, EPI_ISL_1662217, EPI_ISL_1662218, EPI_ISL_1662219, EPI_ISL_1662220 | | | |
| see above | Unidad de Investigacion Medica de Yucatan (UIMY) | Unidad de Genomica Avanzada | Alejandro Sanchez-Flores; Alfredo Herrera-Estrella; Alicia Ocana-Mondragon; Angel Gustavo Salas-Lais; Bernardo Martinez-Miguel; Blanca Taboada; Brenda Irasema Maldonado-Meza; Carla Ivon Herrera-Najera; Carlos F. Arias; Celia Boukadida; Celida Duque Molina; Clara Esperanza Santacruz-Tinoco; Concepcion Grajales-Muniz; Consorcio Mexicano de Vigilancia Genomica (CoViGen-Mex). Authors (in alphabetical order): Julio Elias Alvarado-Yaah; Fernando Fontove-Herrera; Francisco Pulido; Gloria Elena Espinosa-Ayala; Gloria Maria Molina-Salinas; Gloria Vazquez; Hector Esteban Paz-Juarez; Hector Montoya-Fuentes; Helen Haydee Fernanda Ramirez-Plascencia; Jose Antonio Enciso-Moreno; Jose Esteban Munoz-Medina; Jose de Jesus Nunez-Contreras; Juan Bautista Chale-Dzul; Luis Alberto Ochoa-Carrera; Margarita Matias-Florentino; Maria Guadalupe Santiago-Mauricio; Maria Guadalupe de Jesus Mireles-Rivera; Nelly Selem-Mojica; Pavel Isa; Ricardo Grande; Santiago Ávila-Rios; Victor Eduardo Garcia-Arias; Victor Hugo Borja-Aburto |
| EPI_ISL_1352076, EPI_ISL_1352079, EPI_ISL_1352083, EPI_ISL_1352085 | Unidad de InvestigaciÃ³n BiomÃ©dica de Zacatecas (UIBZ) | Unidad de Genomica Avanzada | Alejandro Sanchez-Flores; Alfredo Herrera-Estrella; Alicia Ocana-Mondragon; Angel Gustavo Salas-Lais; Bernardo Martinez-Miguel; Blanca Taboada; Brenda Irasema Maldonado-Meza; Carla Ivon Herrera-Najera; Carlos F. Arias; Celia Boukadida; Clara Esperanza Santacruz-Tinoco; Concepcion Grajales-Muniz; Consorcio Mexicano de Vigilancia Genomica (CoViGen-Mex). Authors (in alphabetical order): Julio Elias Alvarado-Yaah; Fernando Fontove-Herrera; Francisco Pulido; Gloria Elena Espinoza-Ayala; Gloria Maria Molina-Salinas; Gloria Vazquez; Hector Esteban Paz-Juarez; Hector Montoya-Fuentes; Helen Haydee Fernanda Ramirez-Plascencia; Jorge Ivan Salinal-Nevarez; Jose Antonio Enciso-Moreno; Jose Esteban Munoz-Medina; Jose de Jesus Nunez-Contreras; Juan Bautista Chale-Dzul; Luis Alberto Ochoa-Carrera; Margarita Matias-Florentino; Maria Guadalupe Santiago-Mauricio; Maria Guadalupe de Jesus Mireles-Rivera; Nelly Selem-Mojica; Pavel Isa; Ricardo Grande; Santiago Avila-Rios; Victor Hugo Borja-Aburto |
| EPI_ISL_1351500, EPI_ISL_1351506, EPI_ISL_1351509 | Unidad de InvestigaciÃ³n MÃ©dica de YucatÃ¡n (UIMY) | Unidad de Genomica Avanzada | Alejandro Sanchez-Flores; Alfredo Herrera-Estrella; Alicia Ocana-Mondragon; Angel Gustavo Salas-Lais; Bernardo Martinez-Miguel; Blanca Taboada; Brenda Irasema Maldonado-Meza; Carla Ivon Herrera-Najera; Carlos F. Arias; Celia Boukadida; Clara Esperanza Santacruz-Tinoco; Concepcion Grajales-Muniz; Consorcio Mexicano de Vigilancia Genomica (CoViGen-Mex). Authors (in alphabetical order): Julio Elias Alvarado-Yaah; Fernando Fontove-Herrera; Francisco Pulido; Gloria Elena Espinoza-Ayala; Gloria Maria Molina-Salinas; Gloria Vazquez; Hector Esteban Paz-Juarez; Hector Montoya-Fuentes; Helen Haydee Fernanda Ramirez-Plascencia; Jorge Ivan Salinal-Nevarez; Jose Antonio Enciso-Moreno; Jose Esteban Munoz-Medina; Jose de Jesus Nunez-Contreras; Juan Bautista Chale-Dzul; Luis Alberto Ochoa-Carrera; Margarita Matias-Florentino; Maria Guadalupe Santiago-Mauricio; Maria Guadalupe de Jesus Mireles-Rivera; Nelly Selem-Mojica; Pavel Isa; Ricardo Grande; Santiago Avila-Rios; Victor Hugo Borja-Aburto |
| EPI_ISL_1585455, EPI_ISL_1585456, EPI_ISL_1585457, EPI_ISL_1585458, EPI_ISL_1585459, EPI_ISL_1585460, EPI_ISL_1585461, EPI_ISL_1585462, EPI_ISL_1585463, EPI_ISL_1585464, EPI_ISL_1585465, EPI_ISL_1585466, EPI_ISL_1585467, EPI_ISL_1585468, EPI_ISL_1585469, EPI_ISL_1585470, EPI_ISL_1585471, EPI_ISL_1585472, EPI_ISL_1585473, EPI_ISL_1585474, EPI_ISL_1585475, EPI_ISL_1585476, EPI_ISL_1585477, EPI_ISL_1585478, EPI_ISL_1585479, EPI_ISL_1595600 | | | |
| see above | Unidad de Investigación Biomédica de Zacatecas (UIBZ) | Instituto Nacional de Enfermedades Respiratorias (INER): Centro de Investigación en Enfermedades Infecciosas (CIENI) | Alejandro Sanchez-Flores; Alfredo Herrera-Estrella; Alicia Ocaña-Mondragón; Angel Gustavo Salas-Lais; Bernardo Martínez-Miguel; Blanca Taboada; Brenda Irasema Maldonado-Meza; Carla Ivón Herrera-Najera; Carlos F. Arias; Celia Boukadida; Clara Esperanza Santacruz-Tinoco; Concepción Grajales-Muñiz; Consorcio Mexicano de Vigilancia Genómica (CoViGen-Mex). Authors (in alphabetical order): Julio Elias Alvarado-Yaah; Célida Duque Molina; Fernando Fontove-Herrera; Francisco Pulido; Gloria Elena Espinosa-Ayala; Gloria María Molina-Salinas; Gloria Vazquez; Hector Esteban Paz-Juárez; Hector Montoya-Fuentes; Helen Haydee Fernanda Ramírez-Plascencia; José Antonio Enciso-Moreno; José Esteban Muñoz-Medina; José de Jesús Nuñez-Contreras; Juan Bautista Chale-Dzul; Luis Alberto Ochoa-Carrera; Margarita Matías-Florentino; María Guadalupe Santiago-Mauricio; María Guadalupe de Jesús Mireles-Rivera; Nelly Sélem-Mojica; Pavel Isa; Ricardo Grande; Santiago Ávila-Ríos; Victor Eduardo Garcia-Arias; Víctor Hugo Borja-Aburto |
| EPI_ISL_2091225, EPI_ISL_2091226, EPI_ISL_2091227, EPI_ISL_2091228, EPI_ISL_2091229, EPI_ISL_2091230, EPI_ISL_2091231, EPI_ISL_2091232, EPI_ISL_2091233, EPI_ISL_2091234, EPI_ISL_2091235, EPI_ISL_2091236, EPI_ISL_2091237, EPI_ISL_2091238, EPI_ISL_2091239, EPI_ISL_2091240, EPI_ISL_2091241, EPI_ISL_2091242, EPI_ISL_2091243, EPI_ISL_2091244, EPI_ISL_2091245, EPI_ISL_2091246, EPI_ISL_2091247, EPI_ISL_2091248, EPI_ISL_2091249, EPI_ISL_2091250, EPI_ISL_2091251, EPI_ISL_2091252, EPI_ISL_2091253, EPI_ISL_2091254, EPI_ISL_2091255, EPI_ISL_2091256, EPI_ISL_2091257, EPI_ISL_2178727, EPI_ISL_2178728 | | | |
| see above | Unidad de Investigación Médica de Yucatán (UIMY) | Centro de Investigación en Enfermedades Infecciosas (CIENI), Instituto Nacional de Enfermedades Respiratorias (INER) | Alejandro Sanchez-Flores; Alfredo Herrera-Estrella; Alicia Ocaña-Mondragón; Angel Gustavo Salas-Lais; Bernardo Martínez-Miguel; Blanca Taboada; Brenda Irasema Maldonado-Meza; Carla Ivón Herrera-Najera; Carlos F. Arias; Celia Boukadida; Clara Esperanza Santacruz-Tinoco; Concepción Grajales-Muñiz; Consorcio Mexicano de Vigilancia Genómica (CoViGen-Mex). Authors (in alphabetical order): Julio Elias Alvarado-Yaah; Célida Duque Molina; Fernando Fontove-Herrera; Francisco Pulido; Gloria Elena Espinosa-Ayala; Gloria María Molina-Salinas; Gloria Vazquez; Hector Esteban Paz-Juárez; Hector Montoya-Fuentes; Helen Haydee Fernanda Ramírez-Plascencia; José Antonio Enciso-Moreno; José Esteban Muñoz-Medina; José de Jesús Nuñez-Contreras; Juan Bautista Chale-Dzul; Luis Alberto Ochoa-Carrera; Margarita Matías-Florentino; María Guadalupe Santiago-Mauricio; María Guadalupe de Jesús Mireles-Rivera; Nelly Sélem-Mojica; Pavel Isa; Ricardo Grande; Santiago Ávila-Ríos; Victor Eduardo Garcia-Arias; Víctor Hugo Borja-Aburto |
| EPI_ISL_1585428, EPI_ISL_1585429, EPI_ISL_1585430, EPI_ISL_1585431, EPI_ISL_1585432, EPI_ISL_1585433, EPI_ISL_1585434, EPI_ISL_1585435, EPI_ISL_1585436, EPI_ISL_1585437, EPI_ISL_1585438, EPI_ISL_1585439, EPI_ISL_1585440, EPI_ISL_1585441, EPI_ISL_1585442, EPI_ISL_1585443, EPI_ISL_1585444, EPI_ISL_1585445, EPI_ISL_1585446, EPI_ISL_1585447, EPI_ISL_1585448, EPI_ISL_1585449, EPI_ISL_1585450, EPI_ISL_1585451, EPI_ISL_1585452, EPI_ISL_1585453, EPI_ISL_1585454, EPI_ISL_1595599 | | | |
| see above | Unidad de Investigación Médica de Yucatán (UIMY) | Instituto Nacional de Enfermedades Respiratorias (INER): Centro de Investigación en Enfermedades Infecciosas (CIENI) | Alejandro Sanchez-Flores; Alfredo Herrera-Estrella; Alicia Ocaña-Mondragón; Angel Gustavo Salas-Lais; Bernardo Martínez-Miguel; Blanca Taboada; Brenda Irasema Maldonado-Meza; Carla Ivón Herrera-Najera; Carlos F. Arias; Celia Boukadida; Clara Esperanza Santacruz-Tinoco; Concepción Grajales-Muñiz; Consorcio Mexicano de Vigilancia Genómica (CoViGen-Mex). Authors (in alphabetical order): Julio Elias Alvarado-Yaah; Célida Duque Molina; Fernando Fontove-Herrera; Francisco Pulido; Gloria Elena Espinosa-Ayala; Gloria María Molina-Salinas; Gloria Vazquez; Hector Esteban Paz-Juárez; Hector Montoya-Fuentes; Helen Haydee Fernanda Ramírez-Plascencia; José Antonio Enciso-Moreno; José Esteban Muñoz-Medina; José de Jesús Nuñez-Contreras; Juan Bautista Chale-Dzul; Luis Alberto Ochoa-Carrera; Margarita Matías-Florentino; María Guadalupe Santiago-Mauricio; María Guadalupe de Jesús Mireles-Rivera; Nelly Sélem-Mojica; Pavel Isa; Ricardo Grande; Santiago Ávila-Ríos; Victor Eduardo Garcia-Arias; Víctor Hugo Borja-Aburto |
| EPI_ISL_1423505 | Unidad de Patologia Clinica | Instituto de Diagnostico y Referencia Epidemiologicos (INDRE) | Abril Rodriguez-Maldonado; Ariadna Medina-Benitez; Claudia Wong-Arambula; Ernesto Ramirez-Gonzalez.; Gisela Barrera-Badillo; Irma Lopez-Martinez; Joaquin Quiroz-Mercado; Lucia Hernandez-Rivas; Natividad Cruz-Ortiz; Sergio Rangel-Guerrero; Tatiana Nunez-Garcia; Vanessa Rivero-Arredondo |
| EPI_ISL_2101994, EPI_ISL_2101996, EPI_ISL_2101998 | Univeristy of New Mexico Hospital | Center for Global Health, University of New Mexico Health Sciences Center | Darrell Dinwiddie; Daryl Domman; Jon Femling; Justin Bacca; Kurt Schwalm; Valerie Morley |
| EPI_ISL_1340900 | VA Hospital Path & Lab | New Mexico Department of Health Scientific Laboratory | Anastacia Griego-Fisher; D'eldra Malone; Ellie Johnson; Jennifer Benoit |

We gratefully acknowledge the following Authors from the Originating laboratories responsible for obtaining the specimens, as well as the Submitting laboratories where the genome data were generated and shared via GISAID, on which this research is based.

All Submitters of data may be contacted directly via www.gisaid.org

Authors are sorted alphabetically.

| Accession ID | Originating Laboratory | Submitting Laboratory | Authors |
|---|---|---|---|
| EPI_ISL_2229283, EPI_ISL_2229284, EPI_ISL_2229285, EPI_ISL_2229286, EPI_ISL_2229287, EPI_ISL_2229288, EPI_ISL_2229289, EPI_ISL_2229290, EPI_ISL_2248733, EPI_ISL_2690043, EPI_ISL_2690044, EPI_ISL_2690045, EPI_ISL_2690046, EPI_ISL_2709562, EPI_ISL_2787464, EPI_ISL_2787467, EPI_ISL_2787471, EPI_ISL_2787474, EPI_ISL_2787477, EPI_ISL_2787480, EPI_ISL_2787483, EPI_ISL_2787485, EPI_ISL_2787488, EPI_ISL_2787491, EPI_ISL_2787494, EPI_ISL_2787497, EPI_ISL_2787500, EPI_ISL_2787503, EPI_ISL_2787506, EPI_ISL_2787508, EPI_ISL_2840264, EPI_ISL_2840265, EPI_ISL_2840266, EPI_ISL_2840267 | | | |
| see above | "NM Dept. Health, Scientific Laboratory Division " | Centers for Disease Control and Prevention Division of Viral Diseases, Pathogen Discovery | Alison Laufer Halpin; Ben L. Rambo-Martin; Clinton R. Paden; Dakota Howard; Darlene Wagner; Dave Wentworth; Dhwani Batra; Jasmine Padilla; Justin Lee; Katie Dillon; Krista Queen; Kristen Knipe; Kristine Lacek; Mark Burroughs; Matthew Schmerer; Mili Sheth; Peter Cook; Sam Shepard; Sarah Nobles; Shoshona Le; Suxiang Tong; Vivien Dugan; Yvette Unoarumhi |
| EPI_ISL_2146116, EPI_ISL_2146308, EPI_ISL_2147199, EPI_ISL_2147699, EPI_ISL_2147701, EPI_ISL_2147745, EPI_ISL_2147864, EPI_ISL_2148185, EPI_ISL_2148186, EPI_ISL_2158859, EPI_ISL_2158878, EPI_ISL_2159068, EPI_ISL_2159071, EPI_ISL_2159293, EPI_ISL_2181473, EPI_ISL_2181655, EPI_ISL_2181656, EPI_ISL_2181657, EPI_ISL_2187385, EPI_ISL_2204675, EPI_ISL_2205016, EPI_ISL_2205020, EPI_ISL_2241660, EPI_ISL_2280509, EPI_ISL_2280754, EPI_ISL_2280920, EPI_ISL_2283872, EPI_ISL_2321325, EPI_ISL_2321326, EPI_ISL_2368275, EPI_ISL_2489105, EPI_ISL_2489253, EPI_ISL_2528125, EPI_ISL_2528135, EPI_ISL_2528295, EPI_ISL_2528727, EPI_ISL_2528819, EPI_ISL_2687196, EPI_ISL_2687490, EPI_ISL_2687517, EPI_ISL_2687525, EPI_ISL_2687579, EPI_ISL_2785250, EPI_ISL_2875588, EPI_ISL_2875751, EPI_ISL_2875753, EPI_ISL_2875768 | | | |
| see above | Aegis Sciences Corporation | Centers for Disease Control and Prevention Division of Viral Diseases, Pathogen Discovery | Adrian Paskey; Alec Vest; Benjamin Rambo-Martin; Christopher Gulvick; Clinton R. Paden; Cyndi Clark; Dakota Howard; Darlene Wagner; Dhwani Batra; Dillon Nall; Duncan MacCannell; Ethan Sanders; Holly Houdeshell; Jason Caravas; Kara Moser; Matthew Hardison; Matthew Schmerer; Ola Kvalvaag; Patrick Campbell; Peter W. Cook; Rob Case; Scott Sammons; Shatavia Morrison; Shaun Westlund; Vikramsinha Ghorpade; Yvette Unoarumhi |
| EPI_ISL_2105739, EPI_ISL_2105741, EPI_ISL_2810100, EPI_ISL_2810101, EPI_ISL_2810107, EPI_ISL_2810109, EPI_ISL_2810110, EPI_ISL_2810112, EPI_ISL_2810113, EPI_ISL_2810117, EPI_ISL_2810120, EPI_ISL_2810132, EPI_ISL_2810140, EPI_ISL_2810149, EPI_ISL_2810150, EPI_ISL_2810151, EPI_ISL_2810152, EPI_ISL_2810154, EPI_ISL_2810159, EPI_ISL_2810163, EPI_ISL_2810165, EPI_ISL_2928489, EPI_ISL_2928490 | | | |
| see above | Arión Genética | Instituto Nacional de Medicina Genomica | Blancas S; Cedro-Tanda A; Cisneros-Villanueva M; Díaz-Quiñonez José Alberto; Escobar-Arrazola M; Gonzalez-Barrera D; Herrera-Montalvo LA.; Hidalgo-Miranda A; Mendoza-Vargas A; Munguia-Garza P; Orjuela-Rodríguez M; Ramirez-Vega O; Rangel-DeLeon D; Reyes-Grajeda JP; Roldan-Castillo Magaly; Uribe-Figueroa Laura; Verea Jazmin |
| EPI_ISL_2918940, EPI_ISL_2928138, EPI_ISL_2928140, EPI_ISL_2928142, EPI_ISL_2928143, EPI_ISL_2928144, EPI_ISL_2928146, EPI_ISL_2928148, EPI_ISL_2928221, EPI_ISL_2928224, EPI_ISL_2928225, EPI_ISL_2928297, EPI_ISL_2928298, EPI_ISL_2928299, EPI_ISL_2928303, EPI_ISL_2928304, EPI_ISL_2928305, EPI_ISL_2928339, EPI_ISL_2928340, EPI_ISL_2928342, EPI_ISL_2928343 | | | |
| see above | Biomédicos de Mérida | Biomédicos de Mérida | CANCHE-PECH JOSE REYES; CAUICH-VERA KRISTIAN; CEBALLOS-LOPEZ ADRIAN; CHIN-HAU LETICIA; GARCIA-GONZALEZ IGRID; GARCIA-GONZALEZ IGRID; LOPEZ-NOVELO MARIA E; MENDOZA-OLIVEROS TAHALI; SAURI-MEDINA EMILY |
| EPI_ISL_2943172, EPI_ISL_2943231 | CENTRO MEDICO NAVAL SEMAR LAB BIOLOGIA MOLECULAR | CENTRO MEDICO NAVAL SEMAR LAB PATOLOGIA MOLECULAR | MEIXUEIRO CALDERON CLAUDIA. MORALES RAMIREZ OMAR. BALTIERRA URIBE SHANTAL. ZUNO CORONADO TEUTLI. GARCIA ALEGRIA IRMA.ALVAREZ JIMENEZ VIOLETA DEJANIRA. LOPEZ BUSTOS ABRAHAM. CABRERA GONZALEZ MOISES. ROMERO ROMAN EDGAR. LIEVANO ESQUINCA JAZMIN. FIGUEROA ANTONIO VICTOR. DIAZ PONCE MEDRANO JUAN ALBERTO. |
| EPI_ISL_2943127, EPI_ISL_2943148, EPI_ISL_2943233, EPI_ISL_2943236 | CENTRO MEDICO NAVAL SEMAR LAB BIOLOGIA MOLECULAR | CENTRO MEDICO NAVAL SEMAR LABORATORIO DE PATOLOGIA MOLECULAR | MEIXUEIRO CALDERON CLAUDIA. MORALES RAMIREZ OMAR. BALTIERRA URIBE SHANTAL. ZUNO CORONADO TEUTLI. GARCIA ALEGRIA IRMA.ALVAREZ JIMENEZ VIOLETA DEJANIRA. LOPEZ BUSTOS ABRAHAM. CABRERA GONZALEZ MOISES. ROMERO ROMAN EDGAR. LIEVANO ESQUINCA JAZMIN. FIGUEROA ANTONIO VICTOR. DIAZ PONCE MEDRANO JUAN ALBERTO.; MEIXUEIRO CALDERON CLAUDIA. MORALES RAMIREZ OMAR. BALTIERRA URIBE SHANTAL. ZUNO CORONADO TEUTLI. GARCIA ALEGRIA IRMA.ALVAREZ JIMENEZ VIOLETA DEJANIRA. LOPEZ BUSTOS ABRAHAM. CABRERA GONZALEZ MOISES. ROMERO ROMAN EDGAR.RIVERA ROSAS PATRICIA. LIEVANO ESQUINCA JAZMIN. FIGUEROA ANTONIO VICTOR. DIAZ PONCE MEDRANO JUAN ALBERTO. |
| EPI_ISL_2894566, EPI_ISL_2894567, EPI_ISL_2894568, EPI_ISL_2894569, EPI_ISL_2927979, EPI_ISL_2927980 | CIAD Hermosillo | CIAD Hermosillo | ; Alejandra García-Gasca; Alejandra Hernández-Terán; Alejandro Sánchez-Flores; Alfredo Herrera-Estrella; Alicia Ocaña-Mondragón; Andreu Comas-García; Angel Gustavo Salas-Lais; Antonio Loza Román; Bernardo Martínez-Miguel; Blanca Taboada; Brenda Irasema Maldonado-Meza; Bruno Gómez-Gil; Carla Ivón Herrera-Najera; Carlos F. Arias; Celia Boukadida; Clara Esperanza Santacruz-Tinoco; Concepción Grajales-Muñiz; Consorcio Mexicano de Vigilancia Genómica (CoViGen-Mex). Authors (in alphabetical order): Julio Elias Alvarado-Yaah; Cristóbal Cháidez-Quiróz; Célida Duque Molina; Célida Martínez- Rodríguez; Daniel Fregoso-Rueda; Daniel Lira Morales; Eduardo Becerril-Vargas; Fernando Fontove-Herrera; Fidencio Mejía-Nepomuceno; Francisco Pulido; Gloria Elena Espinosa-Ayala; Gloria María Molina-Salinas; Gloria Vazquez; Hector Esteban Paz-Juárez; Hector Montoya-Fuentes; Helen Haydee Fernanda Ramírez-Plascencia; Irvin González-López; Jean Pierre González; Jesús Hernández; Joel Armando Vázquez-Pérez.; Jorge Salas-Hernández; José Antonio Enciso-Moreno; José Arturo Martínez-Orozco; José Esteban Muñoz-Medina; José de Jesús Nuñez-Contreras; Juan Bautista Chale-Dzul; Julissa Enciso-Ibarra; Luis Alberto Ochoa-Carrera; Margarita Matías-Florentino; Mario Mújica-Sánchez; Marissa Perez-Garcia; María Guadalupe Santiago-Mauricio; María Guadalupe de Jesús Mireles-Rivera; Nelly Sélem-Mojica; Pavel Isa; Ricardo Ciria Merce; Ricardo Grande; Rosa María Gutiérrez Rios; Santiago Ávila-Ríos; Selene Zárate; Susana Lopez; Verónica Mata-Haro; Victor Eduardo García-Arias; Víctor Hugo Borja-Aburto |
| EPI_ISL_2533931, EPI_ISL_2533932 | CIAD-Hermosillo | Immunology Laboratory | ; Alejandra García-Gasca; Alejandra Hernández-Terán; Alejandro Sánchez-Flores; Alfredo Herrera-Estrella; Alicia Ocaña-Mondragón; Andreu Comas-García; Angel Gustavo Salas-Lais; Antonio Loza Román; Bernardo Martínez-Miguel; Blanca Taboada; Brenda Irasema Maldonado-Meza; Bruno Gómez-Gil; Carla Ivón Herrera-Najera; Carlos F. Arias; Celia Boukadida; Clara Esperanza Santacruz-Tinoco; Concepción Grajales-Muñiz; Consorcio Mexicano de Vigilancia Genómica (CoViGen-Mex). Authors (in alphabetical order): Julio Elias Alvarado-Yaah; Cristóbal Cháidez-Quiróz; Célida Duque Molina; Célida Martínez- Rodríguez; Daniel Fregoso-Rueda; Daniel Lira Morales; Eduardo Becerril-Vargas; Fernando Fontove-Herrera; Fidencio Mejía-Nepomuceno; Francisco Pulido; Gloria Elena Espinosa-Ayala; Gloria María Molina-Salinas; Gloria Vazquez; Hector Esteban Paz-Juárez; Hector Montoya-Fuentes; Helen Haydee Fernanda Ramírez-Plascencia; Irvin González-López; Jean Pierre González; Jesús Hernández; Joel Armando Vázquez-Pérez.; Jorge Salas-Hernández; José Antonio Enciso-Moreno; José Arturo Martínez-Orozco; José Esteban Muñoz-Medina; José de Jesús Nuñez-Contreras; Juan Bautista Chale-Dzul; Julissa Enciso-Ibarra; Luis Alberto Ochoa-Carrera; Margarita Matías-Florentino; Mario Mújica-Sánchez; Marissa Perez-Garcia; María Guadalupe Santiago-Mauricio; María Guadalupe de Jesús Mireles-Rivera; Nelly Sélem-Mojica; Pavel Isa; Ricardo Ciria Merce; Ricardo Grande; Rosa María Gutiérrez Rios; Santiago Ávila-Ríos; Selene Zárate; Susana Lopez; Verónica Mata-Haro; Victor Eduardo García-Arias; Víctor Hugo Borja-Aburto |
| EPI_ISL_2842780, EPI_ISL_2842781, EPI_ISL_2842782, EPI_ISL_2842783, EPI_ISL_2842784, EPI_ISL_2842785, EPI_ISL_2842803, EPI_ISL_2842804 | | | |
| see above | Centro Medico ABC | Instituto Nacional de Medicina Genomica | Cedro-Tanda A; Cesar Lara; Daniel Aguirre; Escobar-Arrazola; Gonzalez-Barrera D; Herrera-Montalvo LA.; Hidalgo-Miranda A; Jacomine Reyes-Carrasco; M; Mendoza-Vargas A; Munguia-Garza P; Ramirez-Vega O; Rangel-DeLeon D; Reyes-Grajeda JP; Roxana Trejo-González |
| EPI_ISL_2671484, EPI_ISL_2671548, EPI_ISL_2671549, EPI_ISL_2671550, EPI_ISL_2671551, EPI_ISL_2671552, EPI_ISL_2671553, EPI_ISL_2671554, EPI_ISL_2671555, EPI_ISL_2671556, EPI_ISL_2671557, EPI_ISL_2671564, EPI_ISL_2671565, EPI_ISL_2671566, EPI_ISL_2671567, EPI_ISL_2671568, EPI_ISL_2671570, EPI_ISL_2671571, EPI_ISL_2671572, EPI_ISL_2671573, EPI_ISL_2671574, EPI_ISL_2671575, EPI_ISL_2671576, EPI_ISL_2671577, EPI_ISL_2671578, EPI_ISL_2671579, EPI_ISL_2671580, EPI_ISL_2671581, EPI_ISL_2671582, EPI_ISL_2671583, EPI_ISL_2671584, EPI_ISL_2671585, EPI_ISL_2671586, EPI_ISL_2671608, EPI_ISL_2671609, EPI_ISL_2671610, EPI_ISL_2671611, EPI_ISL_2671612, EPI_ISL_2671616, EPI_ISL_2671617, EPI_ISL_2671618, EPI_ISL_2671698, EPI_ISL_2671699, EPI_ISL_2671703, EPI_ISL_2942408, EPI_ISL_2942409, EPI_ISL_2942410, EPI_ISL_2942411, EPI_ISL_2942412, EPI_ISL_2942413, EPI_ISL_2942414, EPI_ISL_2942415, EPI_ISL_2942416, EPI_ISL_2942417, EPI_ISL_2942418, EPI_ISL_2942419, EPI_ISL_2942420, EPI_ISL_2942421, EPI_ISL_2942422, EPI_ISL_2942423, EPI_ISL_2942424, EPI_ISL_2942425, EPI_ISL_2942426, EPI_ISL_2942427, EPI_ISL_2942428, EPI_ISL_2942429, EPI_ISL_2942430, EPI_ISL_2942431, EPI_ISL_2942432, EPI_ISL_2942433, EPI_ISL_2942434, EPI_ISL_2942435, EPI_ISL_2942436, EPI_ISL_2942437, EPI_ISL_2942438, EPI_ISL_2942439, EPI_ISL_2942440, EPI_ISL_2942441, EPI_ISL_2942442, EPI_ISL_2942443, EPI_ISL_2942444, EPI_ISL_2942445, EPI_ISL_2942446, EPI_ISL_2942447, EPI_ISL_2942448, EPI_ISL_2942449, EPI_ISL_2942450, EPI_ISL_2942451, EPI_ISL_2942452, EPI_ISL_2942453, EPI_ISL_2942454, EPI_ISL_2942455, EPI_ISL_2942456, EPI_ISL_2942457, EPI_ISL_2942458, EPI_ISL_2942459, EPI_ISL_2942460, EPI_ISL_2942461, EPI_ISL_2942462, EPI_ISL_2942463, EPI_ISL_2942464, EPI_ISL_2942465, EPI_ISL_2942466, EPI_ISL_2942720, EPI_ISL_2942721, EPI_ISL_2942722, EPI_ISL_2942723, EPI_ISL_2942724, EPI_ISL_2942725, EPI_ISL_2942726, EPI_ISL_2942727, EPI_ISL_2942728, EPI_ISL_2942729, EPI_ISL_2942733, EPI_ISL_2942734, EPI_ISL_2942735, EPI_ISL_2942736, EPI_ISL_2942737 | | | |
| see above | Centro de Investigacion Biomedica de Occidente (CIBO) | Unidad de Genomica Avanzada | ; Alejandra Garcia-Gasca; Alejandra Hernandez-Teran; Alejandro Sanchez-Flores; Alfredo Herrera-Estrella; Alicia Ocaña-Mondragon; Andreu Comas-Garcia; Angel Gustavo Salas-Lais; Antonio Loza Roman; Bernardo Martinez-Miguel; Blanca Taboada; Brenda Irasema Maldonado-Meza; Bruno Gomez-Gil; Carla Ivon Herrera-Najera; Carlos F. Arias; Celia Boukadida; Celida Duque Molina; Celida Martinez- Rodriguez; Clara Esperanza Santacruz-Tinoco; Concepcion Grajales-Muñiz; Consorcio Mexicano de Vigilancia Genomica (CoViGen-Mex). Authors (in alphabetical order): Julio Elias Alvarado-Yaah; Cristobal Chaidez-Quiroz; Daniel Fregoso-Rueda; Daniel Lira Morales; Eduardo Becerril-Vargas; Fernando Fontove-Herrera; Fidencio Mejia-Nepomuceno; Francisco Pulido; Gloria Elena Espinosa-Ayala; Gloria Maria Molina-Salinas; Gloria Vazquez; Hector Esteban Paz-Juarez; Hector Montoya-Fuentes; Helen Haydee Fernanda Ramirez-Plascencia; Irvin Gonzalez-Lopez; Jean Pierre Gonzalez; Jesus Hernandez; Joel Armando Vazquez-Perez.; Jorge Salas-Hernandez; Jose Antonio Enciso-Moreno; Jose Arturo Martinez-Orozco; Jose Esteban Muñoz-Medina; Jose de Jesus Nuñez-Contreras; Juan Bautista Chale-Dzul; Julissa Enciso-Ibarra; Luis Alberto Ochoa-Carrera; Margarita Matias-Florentino; Maria Guadalupe Santiago-Mauricio; Maria Guadalupe de Jesus Mireles-Rivera; Mario Mujica-Sanchez; Marissa Perez-Garcia; Nelly Selem-Mojica; Pavel Isa; Ricardo Ciria Merce; Ricardo Grande; Rosa Maria Gutierrez Rios; Santiago avila-Rios; Selene Zarate; Susana Lopez; Veronica Mata-Haro; Victor Eduardo Garcia-Arias; Victor Hugo Borja-Aburto |
| EPI_ISL_2671505, EPI_ISL_2671506, EPI_ISL_2671507, EPI_ISL_2671508, EPI_ISL_2671509, EPI_ISL_2671510, EPI_ISL_2671511, EPI_ISL_2671512, EPI_ISL_2671513, EPI_ISL_2671514, EPI_ISL_2671515, EPI_ISL_2671516, EPI_ISL_2671517, EPI_ISL_2671518, EPI_ISL_2671519, EPI_ISL_2671520, EPI_ISL_2671521, EPI_ISL_2671522, EPI_ISL_2671523, EPI_ISL_2671524, EPI_ISL_2671525, EPI_ISL_2671543, EPI_ISL_2671544, EPI_ISL_2671545, EPI_ISL_2671546, EPI_ISL_2671547, EPI_ISL_2671558, EPI_ISL_2671559, EPI_ISL_2671560, EPI_ISL_2671619, EPI_ISL_2671620, EPI_ISL_2671621, EPI_ISL_2671622, EPI_ISL_2671623, EPI_ISL_2671624, EPI_ISL_2671625, EPI_ISL_2671626, EPI_ISL_2671627, EPI_ISL_2671628, EPI_ISL_2671629, EPI_ISL_2671630, EPI_ISL_2671631, EPI_ISL_2671632, EPI_ISL_2671633, EPI_ISL_2671634, EPI_ISL_2671635, EPI_ISL_2671636, EPI_ISL_2671716, EPI_ISL_2671717, EPI_ISL_2671718, EPI_ISL_2671719, EPI_ISL_2671720, EPI_ISL_2671721, EPI_ISL_2671722, EPI_ISL_2942481, EPI_ISL_2942482, EPI_ISL_2942483, EPI_ISL_2942484, EPI_ISL_2942485, EPI_ISL_2942486, EPI_ISL_2942487, EPI_ISL_2942488, EPI_ISL_2942489, EPI_ISL_2942490, EPI_ISL_2942491, EPI_ISL_2942492, EPI_ISL_2942493, EPI_ISL_2942494, EPI_ISL_2942495, EPI_ISL_2942496, EPI_ISL_2942497, EPI_ISL_2942498, EPI_ISL_2942499, EPI_ISL_2942500, EPI_ISL_2942501, EPI_ISL_2942502, EPI_ISL_2942503, EPI_ISL_2942504, EPI_ISL_2942505, EPI_ISL_2942506, EPI_ISL_2942507, EPI_ISL_2942508, EPI_ISL_2942509, EPI_ISL_2942510, EPI_ISL_2942511, EPI_ISL_2942512, EPI_ISL_2942754, EPI_ISL_2942755, EPI_ISL_2942756, EPI_ISL_2942757, EPI_ISL_2942758, EPI_ISL_2942759, EPI_ISL_2942760, EPI_ISL_2942761, EPI_ISL_2942762, EPI_ISL_2942763, EPI_ISL_2942764, EPI_ISL_2942765, EPI_ISL_2942766, EPI_ISL_2942767, EPI_ISL_2942768, EPI_ISL_2942769, EPI_ISL_2942770, EPI_ISL_2942772, EPI_ISL_2942773, EPI_ISL_2942774, EPI_ISL_2942775, EPI_ISL_2942776, EPI_ISL_2942777, EPI_ISL_2942781, EPI_ISL_2942782, EPI_ISL_2942783, EPI_ISL_2942784, EPI_ISL_2942785, EPI_ISL_2942786, EPI_ISL_2942787 | | | |
| see above | Centro de Investigacion Biomedica del Noreste (CIBIN) | Unidad de Genomica Avanzada | ; Alejandra Garcia-Gasca; Alejandra Hernandez-Teran; Alejandro Sanchez-Flores; Alfredo Herrera-Estrella; Alicia Ocaña-Mondragon; Andreu Comas-Garcia; Angel Gustavo Salas-Lais; Antonio Loza Roman; Bernardo Martinez-Miguel; Blanca Taboada; Brenda Irasema Maldonado-Meza; Bruno Gomez-Gil; Carla Ivon Herrera-Najera; Carlos F. Arias; Celia Boukadida; Celida Duque Molina; Celida Martinez- Rodriguez; Clara Esperanza Santacruz-Tinoco; Concepcion Grajales-Muñiz; Consorcio Mexicano de Vigilancia Genomica (CoViGen-Mex). Authors (in alphabetical order): Julio Elias Alvarado-Yaah; Cristobal Chaidez-Quiroz; Daniel Fregoso-Rueda; Daniel Lira Morales; Eduardo Becerril-Vargas; Fernando Fontove-Herrera; Fidencio Mejia-Nepomuceno; Francisco Pulido; Gloria Elena Espinosa-Ayala; Gloria Maria Molina-Salinas; Gloria Vazquez; Hector Esteban Paz-Juarez; Hector Montoya-Fuentes; Helen Haydee Fernanda Ramirez-Plascencia; Irvin Gonzalez-Lopez; Jean Pierre Gonzalez; Jesus Hernandez; Joel Armando Vazquez-Perez.; Jorge Salas-Hernandez; Jose Antonio Enciso-Moreno; Jose Arturo Martinez-Orozco; Jose Esteban Muñoz-Medina; Jose de Jesus Nuñez-Contreras; Juan Bautista Chale-Dzul; Julissa Enciso-Ibarra; Luis Alberto Ochoa-Carrera; Margarita Matias-Florentino; Maria Guadalupe Santiago-Mauricio; Maria Guadalupe de Jesus Mireles-Rivera; Mario Mujica-Sanchez; Marissa Perez-Garcia; Nelly Selem-Mojica; Pavel Isa; Ricardo Ciria Merce; Ricardo Grande; Rosa Maria Gutierrez Rios; Santiago avila-Rios; Selene Zarate; Susana Lopez; Veronica Mata-Haro; Victor Eduardo Garcia-Arias; Victor Hugo Borja-Aburto |
| EPI_ISL_2490331, EPI_ISL_2490332, EPI_ISL_2490382, EPI_ISL_2490383, EPI_ISL_2490384, EPI_ISL_2490395, EPI_ISL_2490397, EPI_ISL_2490398, EPI_ISL_2490399, EPI_ISL_2490400, EPI_ISL_2490404, EPI_ISL_2490493, EPI_ISL_2490494, EPI_ISL_2490495, EPI_ISL_2490496, EPI_ISL_2490497, EPI_ISL_2490498, EPI_ISL_2490499, EPI_ISL_2835934, EPI_ISL_2835935, EPI_ISL_2835936 | | | |
| see above | Centro de Investigación Biomédica de Occidente (CIBO) | Centro de Investigación en Enfermedades Infecciosas (CIENI), Instituto Nacional de Enfermedades Respiratorias (INER) | ; Alejandra Garcia-Gasca; Alejandra Hernández-Terán; Alejandro Sanchez-Flores; Alfredo Herrera-Estrella; Alicia Ocaña-Mondragón; Andreu Comas-García; Angel Gustavo Salas-Lais; Antonio Loza Román; Bernardo Martínez-Miguel; Blanca Taboada; Brenda Irasema Maldonado-Meza; Bruno Gomez-Gil; Carla Ivón Herrera-Najera; Carlos F. Arias; Celia Boukadida; Clara Esperanza Santacruz-Tinoco; Concepción Grajales-Muñiz; Consorcio Mexicano de Vigilancia Genómica (CoViGen-Mex). Authors (in alphabetical order): Julio Elias Alvarado-Yaah; Cristóbal Cháidez-Quiróz; Célida Duque Molina; Célida Martínez- Rodríguez; Daniel Fregoso-Rueda; Daniel Lira Morales; Eduardo Becerril-Vargas; Fernando Fontove-Herrera; Fidencio Mejía-Nepomuceno; Francisco Pulido; Gloria Elena Espinosa-Ayala; Gloria María Molina-Salinas; Gloria Vazquez; Hector Esteban Paz-Juárez; Hector Montoya-Fuentes; Helen Haydee Fernanda Ramírez-Plascencia; Irvin González-López; Jean Pierre González; Joel Armando Vázquez-Pérez.; Jorge Salas-Hernández; José Antonio Enciso-Moreno; José Arturo Martínez-Orozco; José Esteban Muñoz-Medina; José de Jesús Nuñez-Contreras; Juan Bautista Chale-Dzul; Julissa Enciso-Ibarra; Luis Alberto Ochoa-Carrera; Margarita Matías-Florentino; Mario Mújica-Sánchez; Marissa Perez-Garcia; María Guadalupe Santiago-Mauricio; María Guadalupe de Jesús Mireles-Rivera; Nelly Sélem-Mojica; Pavel Isa; Ricardo Ciria Merce; Ricardo Grande; Rosa María Gutierrez Rios; Santiago Ávila-Ríos; Selene Zárate; Susana Lopez; Victor Eduardo Garcia-Arias; Víctor Hugo Borja-Aburto |
| EPI_ISL_2801707, EPI_ISL_2801708, EPI_ISL_2801709, EPI_ISL_2801710, EPI_ISL_2801711, EPI_ISL_2801712, EPI_ISL_2801713, EPI_ISL_2801714, EPI_ISL_2801715, EPI_ISL_2801716, EPI_ISL_2801717, EPI_ISL_2801718, EPI_ISL_2801719, EPI_ISL_2801720, EPI_ISL_2801721, EPI_ISL_2801722, EPI_ISL_2801723, EPI_ISL_2801724, EPI_ISL_2801725, EPI_ISL_2801726, EPI_ISL_2801727, EPI_ISL_2801728, EPI_ISL_2801729, EPI_ISL_2801730, EPI_ISL_2801731, EPI_ISL_2801732, EPI_ISL_2801733, EPI_ISL_2801734, EPI_ISL_2801735, EPI_ISL_2801736, EPI_ISL_2801737, EPI_ISL_2801738, EPI_ISL_2801739, EPI_ISL_2801740, EPI_ISL_2801741, EPI_ISL_2801742, EPI_ISL_2801743, EPI_ISL_2801744, EPI_ISL_2801745, EPI_ISL_2801746, EPI_ISL_2801747, EPI_ISL_2801748, EPI_ISL_2801749, EPI_ISL_2801750, EPI_ISL_2801751, EPI_ISL_2801752 | | | |
| see above | Centro de Investigación Biomédica de Occidente (CIBO) | Instituto de Biotecnología de la UNAM | ; Alejandra García-Gasca; Alejandra Hernández-Terán; Alejandro Sánchez-Flores; Alfredo Herrera-Estrella; Alicia Ocaña-Mondragón; Andreu Comas-García; Angel Gustavo Salas-Lais; Antonio Loza Román; Bernardo Martínez-Miguel; Blanca Taboada; Brenda Irasema Maldonado-Meza; Bruno Gómez-Gil; Carla Ivón Herrera-Najera; Carlos F. Arias; Celia Boukadida; Clara Esperanza Santacruz-Tinoco; Concepción Grajales-Muñiz; Consorcio Mexicano de Vigilancia Genómica (CoViGen-Mex). Authors (in alphabetical order): Julio Elias Alvarado-Yaah; Cristóbal Cháidez-Quiróz; Célida Duque Molina; Célida Martínez- Rodríguez; Daniel Fregoso-Rueda; Daniel Lira Morales; Eduardo Becerril-Vargas; Fernando Fontove-Herrera; Fidencio Mejía-Nepomuceno; Francisco Pulido; Gloria Elena Espinosa-Ayala; Gloria María Molina-Salinas; Gloria Vazquez; Hector Esteban Paz-Juárez; Hector Montoya-Fuentes; Helen Haydee Fernanda Ramírez-Plascencia; Irvin González-López; Jean Pierre González; Jesús Hernández; Joel Armando Vázquez-Pérez.; Jorge Salas-Hernández; José Antonio Enciso-Moreno; José Arturo Martínez-Orozco; José Esteban Muñoz-Medina; José de Jesús Nuñez-Contreras; Juan Bautista Chale-Dzul; Julissa Enciso-Ibarra; Luis Alberto Ochoa-Carrera; Margarita Matías-Florentino; Mario Mújica-Sánchez; Marissa Perez-Garcia; María Guadalupe Santiago-Mauricio; María Guadalupe de Jesús Mireles-Rivera; Nelly Sélem-Mojica; Pavel Isa; Ricardo Ciria Merce; Ricardo Grande; Rosa María Gutiérrez Rios; Santiago Ávila-Ríos; Selene Zárate; Susana Lopez; Verónica Mata-Haro; Victor Eduardo García-Arias; Víctor Hugo Borja-Aburto |
| EPI_ISL_2402175, EPI_ISL_2402176 | Centro de Investigación Biomédica de Occidente (CIBO) | Unidad de Genomica Avanzada | Alejandro Sanchez-Flores; Alfredo Herrera-Estrella; Alicia Ocana-Mondragon; Angel Gustavo Salas-Lais; Bernardo Martinez-Miguel; Blanca Taboada; Brenda Irasema Maldonado-Meza; Carla Ivon Herrera-Najera; Carlos F. Arias; Celia Boukadida; Clara Esperanza Santacruz-Tinoco; Concepcion Grajales-Muniz; Consorcio Mexicano de Vigilancia Genomica (CoViGen-Mex). Authors (in alphabetical order): Julio Elias Alvarado-Yaah; Fernando Fontove-Herrera; Francisco Pulido; Gloria Elena Espinoza-Ayala; Gloria Maria Molina-Salinas; Gloria Vazquez; Hector Esteban Paz-Juarez; Hector Montoya-Fuentes; Helen Haydee Fernanda Ramirez-Plascencia; Jorge Ivan Salinal-Nevarez; Jose Antonio Enciso-Moreno; Jose Esteban Munoz-Medina; Jose de Jesus Nunez-Contreras; Juan Bautista Chale-Dzul; Luis Alberto Ochoa-Carrera; Margarita Matias-Florentino; Maria Guadalupe Santiago-Mauricio; Maria Guadalupe de Jesus Mireles-Rivera; Nelly Selem-Mojica; Pavel Isa; Ricardo Grande; Santiago Avila-Rios; Victor Hugo Borja-Aburto |
| EPI_ISL_2490312, EPI_ISL_2490313, EPI_ISL_2490314, EPI_ISL_2490315, EPI_ISL_2490316, EPI_ISL_2490317, EPI_ISL_2490318, EPI_ISL_2490319, EPI_ISL_2490320, EPI_ISL_2490321, EPI_ISL_2490322, EPI_ISL_2490323, EPI_ISL_2490324, EPI_ISL_2490325, EPI_ISL_2490326, EPI_ISL_2490334, EPI_ISL_2490337, EPI_ISL_2490338, EPI_ISL_2490339, EPI_ISL_2490340, EPI_ISL_2490341, EPI_ISL_2490342, EPI_ISL_2490356, EPI_ISL_2490359, EPI_ISL_2490360, EPI_ISL_2490361, EPI_ISL_2490362, EPI_ISL_2490363, EPI_ISL_2490364, EPI_ISL_2490365, EPI_ISL_2490366, EPI_ISL_2490394, EPI_ISL_2490405, EPI_ISL_2490406, EPI_ISL_2490409, EPI_ISL_2490410, EPI_ISL_2490411, EPI_ISL_2490426, EPI_ISL_2490427, EPI_ISL_2490428, EPI_ISL_2490429, EPI_ISL_2490430, EPI_ISL_2490431, EPI_ISL_2490432, | | | |

| | | | |
|---|---|---|---|
| EPI_ISL_2490439, EPI_ISL_2490440, EPI_ISL_2490441, EPI_ISL_2490442, EPI_ISL_2490443, EPI_ISL_2490444, EPI_ISL_2490445, EPI_ISL_2490446, EPI_ISL_2490447, EPI_ISL_2490448, EPI_ISL_2490449, EPI_ISL_2490450, EPI_ISL_2490451, EPI_ISL_2490452, EPI_ISL_2490453, EPI_ISL_2490454, EPI_ISL_2490455, EPI_ISL_2490456, EPI_ISL_2490457, EPI_ISL_2490458, EPI_ISL_2490459, EPI_ISL_2490460, EPI_ISL_2490461, EPI_ISL_2490462, EPI_ISL_2490463, EPI_ISL_2490464, EPI_ISL_2490465, EPI_ISL_2835915, EPI_ISL_2835916, EPI_ISL_2835917, EPI_ISL_2835918, EPI_ISL_2835919, EPI_ISL_2835920, EPI_ISL_2835921, EPI_ISL_2835922, EPI_ISL_2835923, EPI_ISL_2835924, EPI_ISL_2835925, EPI_ISL_2835926, EPI_ISL_2835927, EPI_ISL_2835928, EPI_ISL_2835929 | | | |
| see above | Centro de Investigación Biomédica del Noreste (CIBIN) | Centro de Investigación en Enfermedades Infecciosas (CIENI), Instituto Nacional de Enfermedades Respiratorias (INER) | ; Alejandra Garcia-Gasca; Alejandra Hernández-Terán; Alejandro Sánchez-Flores; Alfredo Herrera-Estrella; Alicia Ocaña-Mondragón; Andreu Comas-García; Angel Gustavo Salas-Lais; Antonio Loza Román; Bernardo Martínez-Miguel; Blanca Taboada; Brenda Irasema Maldonado-Meza; Bruno Gómez-Gil; Carla Ivón Herrera-Najera; Carlos F. Arias; Celia Boukadida; Clara Esperanza Santacruz-Tinoco; Concepción Grajales-Muñiz; Consorcio Mexicano de Vigilancia Genómica (CoViGen-Mex). Authors (in alphabetical order): Julio Elias Alvarado-Yaah; Cristóbal Cháidez-Quiróz; Célida Duque Molina; Célida Martínez- Rodríguez; Daniel Fregoso-Rueda; Daniel Lira Morales; Eduardo Becerril-Vargas; Fernando Fontove-Herrera; Fidencio Mejía-Nepomuceno; Francisco Pulido; Gloria Elena Espinosa-Ayala; Gloria María Molina-Salinas; Gloria Vazquez; Hector Esteban Paz-Juárez; Hector Montoya-Fuentes; Helen Haydee Fernanda Ramírez-Plascencia; Irvin González-López; Jean Pierre González; Joel Armando Vázquez-Pérez.; Jorge Salas-Hernández; José Antonio Enciso-Moreno; José Arturo Martínez-Orozco; José Esteban Muñoz-Medina; José de Jesús Nuñez-Contreras; Juan Bautista Chale-Dzul; Julissa Enciso-Ibarra; Luis Alberto Ochoa-Carrera; Margarita Matías-Florentino; Mario Mújica-Sánchez; Marissa Perez-Garcia; María Guadalupe Santiago-Mauricio; María Guadalupe de Jesús Mireles-Rivera; Nelly Sélem-Mojica; Pavel Isa; Ricardo Ciria Merce; Ricardo Grande; Rosa María Gutierrez Rios; Santiago Ávila-Ríos; Selene Zárate; Susana Lopez; Victor Eduardo Garcia-Arias; Víctor Hugo Borja-Aburto |
| EPI_ISL_2681086, EPI_ISL_2681095, EPI_ISL_2681096, EPI_ISL_2681102, EPI_ISL_2681103, EPI_ISL_2681104, EPI_ISL_2681110, EPI_ISL_2681127, EPI_ISL_2681137, EPI_ISL_2681197, EPI_ISL_2681201, EPI_ISL_2681219, EPI_ISL_2681227, EPI_ISL_2681228, EPI_ISL_2681248, EPI_ISL_2681260, EPI_ISL_2681265, EPI_ISL_2681271, EPI_ISL_2681277, EPI_ISL_2681285, EPI_ISL_2681289, EPI_ISL_2681296, EPI_ISL_2681303, EPI_ISL_2681308, EPI_ISL_2681316, EPI_ISL_2681324, EPI_ISL_2681331, EPI_ISL_2681336, EPI_ISL_2681348, EPI_ISL_2681357, EPI_ISL_2681358, EPI_ISL_2681359, EPI_ISL_2681380, EPI_ISL_2801658, EPI_ISL_2801659, EPI_ISL_2801660, EPI_ISL_2801661, EPI_ISL_2801662, EPI_ISL_2801663, EPI_ISL_2801664, EPI_ISL_2801665, EPI_ISL_2801666, EPI_ISL_2801667, EPI_ISL_2801668, EPI_ISL_2801669, EPI_ISL_2801670, EPI_ISL_2801671, EPI_ISL_2801672, EPI_ISL_2801673, EPI_ISL_2801674, EPI_ISL_2801675, EPI_ISL_2801676, EPI_ISL_2801677, EPI_ISL_2801678, EPI_ISL_2801679, EPI_ISL_2801680, EPI_ISL_2801681, EPI_ISL_2801682, EPI_ISL_2801683, EPI_ISL_2801684, EPI_ISL_2801685, EPI_ISL_2801686, EPI_ISL_2801687, EPI_ISL_2801688, EPI_ISL_2801689, EPI_ISL_2801690, EPI_ISL_2801691, EPI_ISL_2801692, EPI_ISL_2801693, EPI_ISL_2801694, EPI_ISL_2801695, EPI_ISL_2801696 | | | |
| see above | Centro de Investigación Biomédica del Noreste (CIBIN) | Instituto de Biotecnología de la UNAM | ; Alejandra García-Gasca; Alejandra Hernández-Terán; Alejandro Sánchez-Flores; Alfredo Herrera-Estrella; Alicia Ocaña-Mondragón; Andreu Comas-García; Angel Gustavo Salas-Lais; Antonio Loza Román; Bernardo Martínez-Miguel; Blanca Taboada; Brenda Irasema Maldonado-Meza; Bruno Gómez-Gil; Carla Ivón Herrera-Najera; Carlos F. Arias; Celia Boukadida; Clara Esperanza Santacruz-Tinoco; Concepción Grajales-Muñiz; Consorcio Mexicano de Vigilancia Genómica (CoViGen-Mex). Authors (in alphabetical order): Julio Elias Alvarado-Yaah; Cristóbal Cháidez-Quiróz; Célida Duque Molina; Célida Martínez- Rodríguez; Daniel Fregoso-Rueda; Daniel Lira Morales; Eduardo Becerril-Vargas; Fernando Fontove-Herrera; Fidencio Mejía-Nepomuceno; Francisco Pulido; Gloria Elena Espinosa-Ayala; Gloria María Molina-Salinas; Gloria Vazquez; Hector Esteban Paz-Juárez; Hector Montoya-Fuentes; Helen Haydee Fernanda Ramírez-Plascencia; Irvin González-López; Jean Pierre González; Jesús Hernández; Joel Armando Vázquez-Pérez.; Jorge Salas-Hernández; José Antonio Enciso-Moreno; José Arturo Martínez-Orozco; José Esteban Muñoz-Medina; José de Jesús Nuñez-Contreras; Juan Bautista Chale-Dzul; Julissa Enciso-Ibarra; Luis Alberto Ochoa-Carrera; Margarita Matías-Florentino; Mario Mújica-Sánchez; Marissa Perez-Garcia; María Guadalupe Santiago-Mauricio; María Guadalupe de Jesús Mireles-Rivera; Nelly Sélem-Mojica; Pavel Isa; Ricardo Ciria Merce; Ricardo Grande; Rosa María Gutiérrez Rios; Santiago Ávila-Ríos; Selene Zárate; Susana Lopez; Verónica Mata-Haro; Victor Eduardo García-Arias; Víctor Hugo Borja-Aburto |
| EPI_ISL_2402006, EPI_ISL_2402007 | Centro de Investigación Biomédica del Noreste (CIBIN) | Unidad de Genomica Avanzada | Alejandro Sanchez-Flores; Alfredo Herrera-Estrella; Alicia Ocana-Mondragon; Angel Gustavo Salas-Lais; Bernardo Martinez-Miguel; Blanca Taboada; Brenda Irasema Maldonado-Meza; Carla Ivon Herrera-Najera; Carlos F. Arias; Celia Boukadida; Clara Esperanza Santacruz-Tinoco; Concepcion Grajales-Muniz; Consorcio Mexicano de Vigilancia Genomica (CoViGen-Mex). Authors (in alphabetical order): Julio Elias Alvarado-Yaah; Fernando Fontove-Herrera; Francisco Pulido; Gloria Elena Espinoza-Ayala; Gloria Maria Molina-Salinas; Gloria Vazquez; Hector Esteban Paz-Juarez; Hector Montoya-Fuentes; Helen Haydee Fernanda Ramirez-Plascencia; Jorge Ivan Salinal-Nevarez; Jose Antonio Enciso-Moreno; Jose Esteban Munoz-Medina; Jose de Jesus Nunez-Contreras; Juan Bautista Chale-Dzul; Luis Alberto Ochoa-Carrera; Margarita Matias-Florentino; Maria Guadalupe Santiago-Mauricio; Maria Guadalupe de Jesus Mireles-Rivera; Nelly Selem-Mojica; Pavel Isa; Ricardo Grande; Santiago Avila-Rios; Victor Hugo Borja-Aburto |
| EPI_ISL_2755740, EPI_ISL_2755741, EPI_ISL_2783609, EPI_ISL_2783610, EPI_ISL_2783735, EPI_ISL_2783756, EPI_ISL_2783757, EPI_ISL_2783758, EPI_ISL_2784046, EPI_ISL_2784140, EPI_ISL_2784223, EPI_ISL_2784224, EPI_ISL_2784232, EPI_ISL_2784261, EPI_ISL_2784262, EPI_ISL_2784263, EPI_ISL_2784300, EPI_ISL_2839695, EPI_ISL_2839696, EPI_ISL_2839713, EPI_ISL_2839762, EPI_ISL_2839810, EPI_ISL_2839830, EPI_ISL_2941881, EPI_ISL_2941893 | | | |
| see above | Curative Labs | Curative Labs | Elias L. Salfati; Eugenia Khorosheva; George Way; J.Cesar Ignacio-Espinoza; Janet Chen; Mikhail Hanewich-Hollatz; Nabjot Sandhu; Sophia Quasem; Vladimir Slepnev; Zhiyi Xie |
| EPI_ISL_2832464, EPI_ISL_2832465, EPI_ISL_2832466, EPI_ISL_2832467, EPI_ISL_2832468, EPI_ISL_2832469, EPI_ISL_2832470, EPI_ISL_2832471, EPI_ISL_2832472, EPI_ISL_2832473, EPI_ISL_2832474, EPI_ISL_2832475, EPI_ISL_2832476, EPI_ISL_2832477, EPI_ISL_2832478, EPI_ISL_2832479, EPI_ISL_2832480, EPI_ISL_2832481, EPI_ISL_2832482, EPI_ISL_2832483, EPI_ISL_2832484, EPI_ISL_2832485, EPI_ISL_2832486, EPI_ISL_2832487, EPI_ISL_2832488, EPI_ISL_2832489, EPI_ISL_2832490, EPI_ISL_2858901, EPI_ISL_2858902, EPI_ISL_2858903, EPI_ISL_2858930, EPI_ISL_2858931, EPI_ISL_2858932, EPI_ISL_2858933, EPI_ISL_2928093, EPI_ISL_2928094, EPI_ISL_2928095, EPI_ISL_2928096, EPI_ISL_2928097, EPI_ISL_2928098, EPI_ISL_2928099, EPI_ISL_2928100, EPI_ISL_2928101, EPI_ISL_2928102, EPI_ISL_2928103, EPI_ISL_2928104, EPI_ISL_2928105, EPI_ISL_2928106, EPI_ISL_2928107, EPI_ISL_2928108, EPI_ISL_2928109, EPI_ISL_2928110, EPI_ISL_2928111, EPI_ISL_2928112, EPI_ISL_2928113, EPI_ISL_2928114, EPI_ISL_2928115, EPI_ISL_2941654, EPI_ISL_2941655, EPI_ISL_2941656, EPI_ISL_2941657, EPI_ISL_2941658, EPI_ISL_2941659, EPI_ISL_2941660, EPI_ISL_2941661, EPI_ISL_2941662, EPI_ISL_2941663, EPI_ISL_2941664, EPI_ISL_2941665, EPI_ISL_2941666, EPI_ISL_2941667, EPI_ISL_2941668, EPI_ISL_2941669, EPI_ISL_2941670, EPI_ISL_2941671, EPI_ISL_2941672, EPI_ISL_2941673, EPI_ISL_2941674, EPI_ISL_2941675, EPI_ISL_2941676, EPI_ISL_2941677, EPI_ISL_2941679, EPI_ISL_2941680, EPI_ISL_2941681, EPI_ISL_2941682, EPI_ISL_2941683, EPI_ISL_2941684, EPI_ISL_2941685, EPI_ISL_2941686, EPI_ISL_2941687 | | | |
| see above | Curative Labs | New Mexico Department of Health Scientific Laboratory | ; Anastacia Griego-Fisher; D'eldra Malone; Ellie Johnson; Jennifer Benoit; Keila Gutierrez; Linda Salazar; Ratheesh Rajan |
| EPI_ISL_2134130, EPI_ISL_2160788, EPI_ISL_2269263, EPI_ISL_2269425, EPI_ISL_2371615, EPI_ISL_2686549 | Fulgent Genetics | Centers for Disease Control and Prevention Division of Viral Diseases, Pathogen Discovery | Adrian Paskey; Becky Tsai; Benafsh Sapra; Benjamin Rambo-Martin; Christopher Gulvick; Clinton R. Paden; Dakota Howard; Darlene Wagner; Dhwani Batra; Doreen Ng; Duncan MacCannell; Harry Gao; James Xie; Jason Caravas; John Gao; Joseph Fierro; Kara Moser; Matthew Schmerer; Mickey Li; Peter W. Cook; Scott Sammons; Shatavia Morrison; Yan Meng; Yvette Unoarumhi |
| EPI_ISL_2894595, EPI_ISL_2894596, EPI_ISL_2894597, EPI_ISL_2894598, EPI_ISL_2894599, EPI_ISL_2894600, EPI_ISL_2894601, EPI_ISL_2894602, EPI_ISL_2894603 | | | |
| see above | Genolife | Genolife | Yenelli Cedano-Thomas |
| EPI_ISL_2537082, EPI_ISL_2602131, EPI_ISL_2854946 | Gravity Diagnostics, LLC | Gravity Diagnostics, LLC | Gravity Diagnostics |
| EPI_ISL_2160408 | Helix/Illumina | Centers for Disease Control and Prevention Division of Viral Diseases, Pathogen Discovery | Adrian Paskey; Alexandre Bolze; Ary Ascencio; Benjamin Rambo-Martin; Brad Sickler; Charlotte Rivera-Garcia; Christine Tran; Christopher Gulvick; Clinton R. Paden; Dakota Howard; Darlene Wagner; David Becker; Dhwani Batra; Duncan MacCannell; Efren Sandoval; Eileen de Feo; Elizabeth Cirulli; Eric Allen; Geraint Levan; James Lu; Jan Antico; Jason Caravas; Jason Nguyen; Jimmy Ramirez; Jingtao Liu; Kara Moser; Kelly Schiabor Barrett; Kim Gietzen; Magnus Isaksson; Marc Laurent; Matthew Schmerer; Matthew Tolentino; Nicole L. Washington; Peter W. Cook; Phil Febbo; Ryan Cho; Scott Sammons; Shannon Wickline; Shatavia Morrison; Sherry Wang; Simon White; Tyler Cassens; William Lee; Yvette Unoarumhi |
| EPI_ISL_2533936 | Hospital Fátima S.A. de C.V. | Laboratorio Nacional-LANIIA-CIAD | ; Alejandra García-Gasca; Alejandra Hernández-Terán; Alejandro Sánchez-Flores; Alfredo Herrera-Estrella; Alicia Ocaña-Mondragón; Andreu Comas-García; Angel Gustavo Salas-Lais; Antonio Loza Román; Bernardo Martínez-Miguel; Blanca Taboada; Brenda Irasema Maldonado-Meza; Bruno Gómez-Gil; Carla Ivón Herrera-Najera; Carlos F. Arias; Celia Boukadida; Clara Esperanza Santacruz-Tinoco; Concepción Grajales-Muñiz; Consorcio Mexicano de Vigilancia Genómica (CoViGen-Mex). Authors (in alphabetical order): Julio Elias Alvarado-Yaah; Cristóbal Cháidez-Quiróz; Célida Duque Molina; Célida Martínez- Rodríguez; Daniel Fregoso-Rueda; Daniel Lira Morales; Eduardo Becerril-Vargas; Fernando Fontove-Herrera; Fidencio Mejía-Nepomuceno; Francisco Pulido; Gloria Elena Espinosa-Ayala; Gloria María Molina-Salinas; Gloria Vazquez; Hector Esteban Paz-Juárez; Hector Montoya-Fuentes; Helen Haydee Fernanda Ramírez-Plascencia; Irvin González-López; Jean Pierre González; Jesús Hernández; Joel Armando Vázquez-Pérez.; Jorge Salas-Hernández; José Antonio Enciso-Moreno; José Arturo Martínez-Orozco; José Esteban Muñoz-Medina; José de Jesús Nuñez-Contreras; Juan Bautista Chale-Dzul; Julissa Enciso-Ibarra; Luis Alberto Ochoa-Carrera; Margarita Matías-Florentino; Mario Mújica-Sánchez; Marissa Perez-Garcia; María Guadalupe Santiago-Mauricio; María Guadalupe de Jesús Mireles-Rivera; Nelly Sélem-Mojica; Pavel Isa; Ricardo Ciria Merce; Ricardo Grande; Rosa María Gutiérrez Rios; Santiago Ávila-Ríos; Selene Zárate; Susana Lopez; Verónica Mata-Haro; Victor Eduardo García-Arias; Víctor Hugo Borja-Aburto |
| EPI_ISL_2680928, EPI_ISL_2680929 | Hospital Fátima S.A. de C.V. | Laboratorio nacional-LANIIA-CIAD | ; Alejandra García-Gasca; Alejandra Hernández-Terán; Alejandro Sánchez-Flores; Alfredo Herrera-Estrella; Alicia Ocaña-Mondragón; Andreu Comas-García; Angel Gustavo Salas-Lais; Antonio Loza Román; Bernardo Martínez-Miguel; Blanca Taboada; Brenda Irasema Maldonado-Meza; Bruno Gómez-Gil; Carla Ivón Herrera-Najera; Carlos F. Arias; Celia Boukadida; Clara Esperanza Santacruz-Tinoco; Concepción Grajales-Muñiz; Consorcio Mexicano de Vigilancia Genómica (CoViGen-Mex). Authors (in alphabetical order): Julio Elias Alvarado-Yaah; Cristóbal Cháidez-Quiróz; Célida Duque Molina; Célida Martínez- Rodríguez; Daniel Fregoso-Rueda; Daniel Lira Morales; Eduardo Becerril-Vargas; Fernando Fontove-Herrera; Fidencio Mejía-Nepomuceno; Francisco Pulido; Gloria Elena Espinosa-Ayala; Gloria María Molina-Salinas; Gloria Vazquez; Hector Esteban Paz-Juárez; Hector Montoya-Fuentes; Helen Haydee Fernanda Ramírez-Plascencia; Irvin González-López; Jean Pierre González; Jesús Hernández; Joel Armando Vázquez-Pérez.; Jorge Salas-Hernández; José Antonio Enciso-Moreno; José Arturo Martínez-Orozco; José Esteban Muñoz-Medina; José de Jesús Nuñez-Contreras; Juan Bautista Chale-Dzul; Julissa Enciso-Ibarra; Luis Alberto Ochoa-Carrera; Margarita Matías-Florentino; Mario Mújica-Sánchez; Marissa Perez-Garcia; María Guadalupe Santiago-Mauricio; María Guadalupe de Jesús Mireles-Rivera; Nelly Sélem-Mojica; Pavel Isa; Ricardo Ciria Merce; Ricardo Grande; Rosa María Gutiérrez Rios; Santiago Ávila-Ríos; Selene Zárate; Susana Lopez; Verónica Mata-Haro; Victor Eduardo García-Arias; Víctor Hugo Borja-Aburto |
| EPI_ISL_2249235, EPI_ISL_2249239, EPI_ISL_2249241, EPI_ISL_2249242, EPI_ISL_2249243, EPI_ISL_2249244, EPI_ISL_2249245, EPI_ISL_2249250, EPI_ISL_2249254, EPI_ISL_2533905, EPI_ISL_2533907, EPI_ISL_2533917, EPI_ISL_2680931, EPI_ISL_2680932, EPI_ISL_2680933, EPI_ISL_2680936, EPI_ISL_2680937, EPI_ISL_2894553, EPI_ISL_2894554, EPI_ISL_2894555, EPI_ISL_2894556, EPI_ISL_2894557, EPI_ISL_2894558, EPI_ISL_2894559, EPI_ISL_2894560, EPI_ISL_2894561, EPI_ISL_2927976, EPI_ISL_2927977, EPI_ISL_2927978 | | | |
| see above | Hospital Margarita Maza de Juárez | Microbial Genomics Laboratory | ; Alejandra Garcia-Gasca; Alejandra García-Gasca; Alejandra Hernández-Terán; Alejandro Sánchez-Flores; Alfredo Herrera-Estrella; Alicia Ocaña-Mondragón; Andreu Comas-García; Angel Gustavo Salas-Lais; Antonio Loza Román; Bernardo Martínez-Miguel; Blanca Taboada; Brenda Irasema Maldonado-Meza; Bruno Gomez-Gil; Bruno Gómez-Gil; Carla Ivón Herrera-Najera; Carlos F. Arias; Celia Boukadida; Clara Esperanza Santacruz-Tinoco; Concepción Grajales-Muñiz; Consorcio Mexicano de Vigilancia Genómica (CoViGen-Mex). Authors (in alphabetical order): Julio Elias Alvarado-Yaah; Cristóbal Cháidez-Quiróz; Célida Duque Molina; Célida Martínez- Rodríguez; Daniel Fregoso-Rueda; Daniel Lira Morales; Eduardo Becerril-Vargas; Fernando Fontove-Herrera; Fidencio Mejía-Nepomuceno; Francisco Pulido; Gloria Elena Espinosa-Ayala; Gloria María Molina-Salinas; Gloria Vazquez; Hector Esteban Paz-Juárez; Hector Montoya-Fuentes; Helen Haydee Fernanda Ramírez-Plascencia; Irvin González-López; Jean Pierre González; Jesús Hernández; Joel Armando Vázquez-Pérez.; Jorge Salas-Hernández; José Antonio Enciso-Moreno; José Arturo Martínez-Orozco; José Esteban Muñoz-Medina; José de Jesús Nuñez-Contreras; Juan Bautista Chale-Dzul; Julissa Enciso-Ibarra; Luis Alberto Ochoa-Carrera; Margarita Matías-Florentino; Mario Mújica-Sánchez; Marissa Perez-Garcia; María Guadalupe Santiago-Mauricio; María Guadalupe de Jesús Mireles-Rivera; Nelly Sélem-Mojica; Pavel Isa; Ricardo Ciria Merce; Ricardo Grande; Rosa María Gutiérrez Rios; Santiago Ávila-Ríos; Selene Zárate; Susana Lopez; Verónica Mata-Haro; Victor Eduardo García-Arias; Víctor Hugo Borja-Aburto |
| EPI_ISL_1972570, EPI_ISL_2001050, EPI_ISL_2249236, EPI_ISL_2249237, EPI_ISL_2249238, EPI_ISL_2249240, EPI_ISL_2249246, EPI_ISL_2249247, EPI_ISL_2249248, EPI_ISL_2249249, EPI_ISL_2249251, EPI_ISL_2249252, EPI_ISL_2249253, EPI_ISL_2249255, EPI_ISL_2277642, EPI_ISL_2533904, EPI_ISL_2533908, EPI_ISL_2533909, EPI_ISL_2533910, EPI_ISL_2533911, EPI_ISL_2533912, EPI_ISL_2533914, EPI_ISL_2533915, EPI_ISL_2533916, EPI_ISL_2533919, EPI_ISL_2533920, EPI_ISL_2533921, EPI_ISL_2533922, EPI_ISL_2533923, EPI_ISL_2533924, EPI_ISL_2533926, EPI_ISL_2533927, EPI_ISL_2680935, EPI_ISL_2680938, EPI_ISL_2680939, EPI_ISL_2680940, EPI_ISL_2680941, EPI_ISL_2680942, EPI_ISL_2894562, EPI_ISL_2894563, EPI_ISL_2894564, EPI_ISL_2894565, EPI_ISL_2927974, EPI_ISL_2927975 | | | |
| see above | Hospital Sharp | Microbial Genomics Laboratory | ; Alejandra Garcia-Gasca; Alejandra García-Gasca; Alejandra Hernández-Terán; Alejandro Sánchez-Flores; Alfredo Herrera-Estrella; Alicia Ocaña-Mondragón; Andreu Comas-García; Angel Gustavo Salas-Lais; Antonio Loza Román; Bernardo Martínez-Miguel; Blanca Taboada; Brenda Irasema Maldonado-Meza; Bruno Gomez-Gil; Bruno Gómez-Gil; Carla Ivón Herrera-Najera; Carlos F. Arias; Celia Boukadida; Clara Esperanza Santacruz-Tinoco; Concepción Grajales-Muñiz; Consorcio Mexicano de Vigilancia Genómica (CoViGen-Mex). Authors (in alphabetical order): Julio Elias Alvarado-Yaah; Cristóbal Cháidez-Quiróz; Célida Duque Molina; Célida Martínez- Rodríguez; Daniel Fregoso-Rueda; Daniel Lira Morales; Eduardo Becerril-Vargas; Fernando Fontove-Herrera; Fidencio Mejía-Nepomuceno; Francisco Pulido; Gloria Elena Espinosa-Ayala; Gloria María Molina-Salinas; Gloria Vazquez; Hector Esteban Paz-Juárez; Hector Montoya-Fuentes; Helen Haydee Fernanda Ramírez-Plascencia; Irvin González-López; Jean Pierre González; Jesús Hernández; Joel Armando Vázquez-Pérez.; Jorge Salas-Hernández; José Antonio Enciso-Moreno; José Arturo Martínez-Orozco; José Esteban Muñoz-Medina; José de Jesús Nuñez-Contreras; Juan Bautista Chale-Dzul; Julissa Enciso-Ibarra; Luis Alberto Ochoa-Carrera; Margarita Matías-Florentino; Mario Mújica-Sánchez; Marissa Perez-Garcia; María Guadalupe Santiago-Mauricio; María Guadalupe de Jesús Mireles-Rivera; Nelly Sélem-Mojica; Pavel Isa; Ricardo Ciria Merce; Ricardo Grande; Rosa María Gutiérrez Rios; Santiago Ávila-Ríos; Selene Zárate; Susana Lopez; Verónica Mata-Haro; Victor Eduardo García-Arias; Víctor Hugo Borja-Aburto |
| EPI_ISL_2319313, EPI_ISL_2319314, EPI_ISL_2319315, EPI_ISL_2319316, EPI_ISL_2319317, EPI_ISL_2319318, EPI_ISL_2319319, EPI_ISL_2319320, EPI_ISL_2319321, EPI_ISL_2319322, EPI_ISL_2319323, EPI_ISL_2319324, EPI_ISL_2319325, EPI_ISL_2319326, EPI_ISL_2319327, EPI_ISL_2319328, EPI_ISL_2319329, EPI_ISL_2319330, EPI_ISL_2319331, EPI_ISL_2319332, EPI_ISL_2319333, EPI_ISL_2319334, EPI_ISL_2319335, EPI_ISL_2319336, EPI_ISL_2319337, EPI_ISL_2319338, EPI_ISL_2319339, EPI_ISL_2319340, EPI_ISL_2319341, EPI_ISL_2319342, EPI_ISL_2319343, EPI_ISL_2319344, EPI_ISL_2319345, EPI_ISL_2319346, EPI_ISL_2319347, EPI_ISL_2319348, EPI_ISL_2319349, EPI_ISL_2319350, EPI_ISL_2319351, EPI_ISL_2319352, EPI_ISL_2319353, EPI_ISL_2319354, EPI_ISL_2319355, EPI_ISL_2319356, EPI_ISL_2319357, EPI_ISL_2319358, EPI_ISL_2319359, EPI_ISL_2319360, EPI_ISL_2319361, EPI_ISL_2319362, EPI_ISL_2319363, EPI_ISL_2319364, EPI_ISL_2319365, EPI_ISL_2319366, EPI_ISL_2319367, EPI_ISL_2319368, EPI_ISL_2319369, EPI_ISL_2319370, EPI_ISL_2319371, EPI_ISL_2319372, EPI_ISL_2319373, EPI_ISL_2319374, EPI_ISL_2319375, EPI_ISL_2319376, EPI_ISL_2319377, EPI_ISL_2319378, EPI_ISL_2319379, EPI_ISL_2319380, EPI_ISL_2319381, EPI_ISL_2341029, EPI_ISL_2341031, EPI_ISL_2341040, EPI_ISL_2341041 | | | |
| see above | INDRE | Instituto Nacional de Medicina Genomica | Cedro-Tanda A; Cisneros-Villanueva M; Garcia-Cardenas FJ; Gisela Barrera-Badillo; Gonzalez-Barrera D; Gonzalez-Woge MA; Herrera-Montalvo LA.; Hidalgo-Miranda A; Irma Lopez-Martinez; Jose Ernesto Ramirez González; Mendoza-Vargas A; Munguia-Garza P; Orjuela-Rodríguez M; Rangel-DeLeon D; Reyes-Grajeda JP |
| EPI_ISL_2249234, EPI_ISL_2533903, EPI_ISL_2533906, EPI_ISL_2533913, EPI_ISL_2533918, EPI_ISL_2533925, EPI_ISL_2533928, EPI_ISL_2533929, EPI_ISL_2680934, EPI_ISL_2927972, EPI_ISL_2927973 | | | |
| see above | Imágenes Diagnósticas de Mazatlán | Microbial Genomics Laboratory | ; Alejandra Garcia-Gasca; Alejandra García-Gasca; Alejandra Hernández-Terán; Alejandro Sánchez-Flores; Alfredo Herrera-Estrella; Alicia Ocaña-Mondragón; Andreu Comas-García; Angel Gustavo Salas-Lais; Antonio Loza Román; Bernardo Martínez-Miguel; Blanca Taboada; Brenda Irasema Maldonado-Meza; Bruno Gomez-Gil; Bruno Gómez-Gil; Carla Ivón Herrera-Najera; Carlos F. Arias; Celia Boukadida; Clara Esperanza Santacruz-Tinoco; Concepción Grajales-Muñiz; Consorcio Mexicano de Vigilancia Genómica (CoViGen-Mex). Authors (in alphabetical order): Julio Elias Alvarado-Yaah; Cristóbal Cháidez-Quiróz; Célida Duque Molina; Célida Martínez- Rodríguez; Daniel Fregoso-Rueda; Daniel Lira Morales; Eduardo Becerril-Vargas; Fernando Fontove-Herrera; Fidencio Mejía-Nepomuceno; Francisco Pulido; Gloria Elena Espinosa-Ayala; Gloria María Molina-Salinas; Gloria Vazquez; Hector Esteban Paz-Juárez; Hector Montoya-Fuentes; Helen Haydee Fernanda Ramírez-Plascencia; Irvin González-López; Jean Pierre González; Jesús Hernández; Joel Armando Vázquez-Pérez.; Jorge Salas-Hernández; José Antonio Enciso-Moreno; José Arturo Martínez-Orozco; José Esteban Muñoz-Medina; José de Jesús Nuñez-Contreras; Juan Bautista Chale-Dzul; Julissa Enciso-Ibarra; Luis Alberto Ochoa-Carrera; Margarita Matías-Florentino; Mario Mújica-Sánchez; Marissa Perez-Garcia; María Guadalupe Santiago-Mauricio; María Guadalupe de Jesús Mireles-Rivera; Nelly Sélem-Mojica; Pavel Isa; Ricardo Ciria Merce; Ricardo Grande; Rosa María Gutiérrez Rios; Santiago Ávila-Ríos; Selene Zárate; Susana Lopez; Verónica Mata-Haro; Victor Eduardo García-Arias; Víctor Hugo Borja-Aburto |
| EPI_ISL_2230880, EPI_ISL_2603595, EPI_ISL_2603596, EPI_ISL_2603597, EPI_ISL_2603598, EPI_ISL_2603599, EPI_ISL_2603600, EPI_ISL_2603601, EPI_ISL_2603602, EPI_ISL_2603603, EPI_ISL_2603604, EPI_ISL_2603605, EPI_ISL_2603606, EPI_ISL_2603607, EPI_ISL_2603608, EPI_ISL_2603609, EPI_ISL_2603610, EPI_ISL_2603611, EPI_ISL_2603612, EPI_ISL_2603613, EPI_ISL_2603614, EPI_ISL_2603615, EPI_ISL_2603616, EPI_ISL_2603617, EPI_ISL_2603618, EPI_ISL_2603619, EPI_ISL_2603620, EPI_ISL_2603621, EPI_ISL_2603622, EPI_ISL_2603623, EPI_ISL_2603624, EPI_ISL_2603625, EPI_ISL_2603626, EPI_ISL_2603627, EPI_ISL_2603628, EPI_ISL_2603629, EPI_ISL_2603630, EPI_ISL_2603631, EPI_ISL_2603632, EPI_ISL_2603633, EPI_ISL_2603634, EPI_ISL_2603635, EPI_ISL_2603636, EPI_ISL_2603637, EPI_ISL_2603638, EPI_ISL_2603639, EPI_ISL_2603640, EPI_ISL_2603641, EPI_ISL_2603642, EPI_ISL_2603643, EPI_ISL_2603644, EPI_ISL_2603645, EPI_ISL_2603646, EPI_ISL_2603647, EPI_ISL_2603648, EPI_ISL_2603649, EPI_ISL_2603650, EPI_ISL_2603651, EPI_ISL_2603652, EPI_ISL_2603653, EPI_ISL_2603654, EPI_ISL_2603655, EPI_ISL_2603656, EPI_ISL_2603657, EPI_ISL_2603658, EPI_ISL_2790209, EPI_ISL_2790210, EPI_ISL_2790211, EPI_ISL_2790212, EPI_ISL_2790213, EPI_ISL_2790214, EPI_ISL_2790215, EPI_ISL_2790216, EPI_ISL_2790217, EPI_ISL_2881767, EPI_ISL_2881768, EPI_ISL_2881769, EPI_ISL_2881770, EPI_ISL_2881771, EPI_ISL_2881772, EPI_ISL_2881773, EPI_ISL_2881774, EPI_ISL_2881812, EPI_ISL_2881813, EPI_ISL_2881814, EPI_ISL_2881815, EPI_ISL_2881816 | | | |
| see above | InDRE | Instituto Nacional de Medicina Genomica | Cedro-Tanda A; Cisneros-Villanueva M; Garcia-Cardenas FJ; Gisela Barrera-Badillo; Gonzalez-Barrera D; Gonzalez-Woge MA; Herrera-Montalvo LA.; Hidalgo-Miranda A; Irma Lopez-Martinez; Jose Ernesto Ramirez González; Mendoza-Vargas A; Munguia-Garza P; Orjuela-Rodríguez M; Rangel-DeLeon D; Reyes-Grajeda JP |
| EPI_ISL_2088675, EPI_ISL_2088764, EPI_ISL_2089040, EPI_ISL_2186828, EPI_ISL_2186840, EPI_ISL_2186888, EPI_ISL_2282293, EPI_ISL_2282314, EPI_ISL_2282333, EPI_ISL_2282434, EPI_ISL_2282477, EPI_ISL_2282489, EPI_ISL_2282580, EPI_ISL_2282805, EPI_ISL_2323967, EPI_ISL_2323969, EPI_ISL_2323977, EPI_ISL_2324004, EPI_ISL_2324009, EPI_ISL_2324048, EPI_ISL_2372795, EPI_ISL_2375611, EPI_ISL_2557558, EPI_ISL_2652519, EPI_ISL_2652536, EPI_ISL_2652540, EPI_ISL_2652608, EPI_ISL_2652654, EPI_ISL_2652723, EPI_ISL_2652824, EPI_ISL_2652859, EPI_ISL_2652865, EPI_ISL_2652972, EPI_ISL_2686859, EPI_ISL_2686874 | | | |
| see above | Infinity Biologix | Centers for Disease Control and Prevention | Adrian Paskey; Benjamin Rambo-Martin; Chirayu Goswami; Christian Bixby; Christopher Gulvick; Clinton R. Paden; Dakota Howard; Darlene Wagner; Dhwani Batra; Duncan MacCannell; Jason Caravas; Jonathan Schultz; Kara Moser; Matthew Schmerer; Peter W. Cook; Robin Grimwood; Russ Hager; Scott Sammons; Shatavia Morrison; Yihe Wang; Yvette Unoarumhi |

| | | | |
|---|---|---|---|
| | | Division of Viral Diseases, Pathogen Discovery | |
| EPI_ISL_2648114, EPI_ISL_2777215, EPI_ISL_2777221, EPI_ISL_2777222, EPI_ISL_2777223, EPI_ISL_2777224, EPI_ISL_2777225, EPI_ISL_2777226, EPI_ISL_2777227, EPI_ISL_2777228, EPI_ISL_2777229, EPI_ISL_2777230, EPI_ISL_2777231, EPI_ISL_2777232, EPI_ISL_2777233, EPI_ISL_2777234, EPI_ISL_2790132, EPI_ISL_2790133, EPI_ISL_2790134, EPI_ISL_2790135, EPI_ISL_2790136 | | | |
| see above | Instituto Nacional de Cancerología | Centro de Investigación en Enfermedades Infecciosas (CIENI), Instituto Nacional de Enfermedades Respiratorias (INER) | ; Alejandra Garcia-Gasca; Alejandra Hernández-Terán; Alejandro Sanchez-Flores; Alfredo Herrera-Estrella; Alicia Ocaña-Mondragón; Andreu Comas-García; Angel Gustavo Salas-Lais; Antonio Loza Román; Bernardo Martínez-Miguel; Blanca Taboada; Brenda Irasema Maldonado-Meza; Bruno Gomez-Gil; Carla Ivón Herrera-Najera; Carlos F. Arias; Celia Boukadida; Clara Esperanza Santacruz-Tinoco; Concepción Grajales-Muñiz; Consorcio Mexicano de Vigilancia Genómica (CoViGen-Mex). Authors (in alphabetical order): Julio Elias Alvarado-Yaah; Cristóbal Cháidez-Quiróz; Célida Duque Molina; Célida Martínez- Rodríguez; Daniel Fregoso-Rueda; Daniel Lira Morales; Eduardo Becerril-Vargas; Fernando Fontove-Herrera; Fidencio Mejía-Nepomuceno; Francisco Pulido; Gloria Elena Espinosa-Ayala; Gloria María Molina-Salinas; Gloria Vazquez; Hector Esteban Paz-Juárez; Hector Montoya-Fuentes; Helen Haydee Fernanda Ramírez-Plascencia; Irvin González-López; Jean Pierre González; Joel Armando Vázquez-Pérez.; Jorge Salas-Hernández; José Antonio Enciso-Moreno; José Arturo Martínez-Orozco; José Esteban Muñoz-Medina; José de Jesús Nuñez-Contreras; Juan Bautista Chale-Dzul; Julissa Enciso-Ibarra; Kathia Elizabeth Tapia-Diaz; Luis Alberto Ochoa-Carrera; Margarita Matías-Florentino; Mario Mújica-Sánchez; Marissa Pérez-García; María Guadalupe Santiago-Mauricio; María Guadalupe de Jesús Mireles-Rivera; Nelly Sélem-Mojica; Pavel Isa; Ricardo Ciria Merce; Ricardo Grande; Rosa María Gutierrez Rios; Santiago Ávila-Ríos; Selene Zárate; Susana Lopez; Victor Eduardo Garcia-Arias; Víctor Hugo Borja-Aburto |
| EPI_ISL_2490585, EPI_ISL_2490586, EPI_ISL_2490587, EPI_ISL_2490588, EPI_ISL_2490589, EPI_ISL_2490590, EPI_ISL_2490591, EPI_ISL_2490593, EPI_ISL_2490594, EPI_ISL_2490595, EPI_ISL_2490596, EPI_ISL_2490597, EPI_ISL_2490598, EPI_ISL_2490599, EPI_ISL_2490600, EPI_ISL_2490601, EPI_ISL_2490602, EPI_ISL_2490603, EPI_ISL_2490604, EPI_ISL_2490605, EPI_ISL_2490606, EPI_ISL_2490607, EPI_ISL_2490608, EPI_ISL_2648115, EPI_ISL_2648116, EPI_ISL_2648117, EPI_ISL_2648118, EPI_ISL_2673696, EPI_ISL_2777211, EPI_ISL_2777212, EPI_ISL_2777213, EPI_ISL_2777214, EPI_ISL_2777216, EPI_ISL_2777217, EPI_ISL_2777218, EPI_ISL_2777219, EPI_ISL_2777220, EPI_ISL_2790175, EPI_ISL_2790176 | | | |
| see above | Instituto Nacional de Enfermedades Respiratorias (INER) | Centro de Investigación en Enfermedades Infecciosas (CIENI), Instituto Nacional de Enfermedades Respiratorias (INER) | ; Alejandra Garcia-Gasca; Alejandra Hernández-Terán; Alejandro Sanchez-Flores; Alfredo Herrera-Estrella; Alicia Ocaña-Mondragón; Andreu Comas-García; Angel Gustavo Salas-Lais; Antonio Loza Román; Bernardo Martínez-Miguel; Blanca Taboada; Brenda Irasema Maldonado-Meza; Bruno Gomez-Gil; Carla Ivón Herrera-Najera; Carlos F. Arias; Celia Boukadida; Clara Esperanza Santacruz-Tinoco; Concepción Grajales-Muñiz; Consorcio Mexicano de Vigilancia Genómica (CoViGen-Mex). Authors (in alphabetical order): Julio Elias Alvarado-Yaah; Cristóbal Cháidez-Quiróz; Célida Duque Molina; Célida Martínez- Rodríguez; Daniel Fregoso-Rueda; Daniel Lira Morales; Eduardo Becerril-Vargas; Fernando Fontove-Herrera; Fidencio Mejía-Nepomuceno; Francisco Pulido; Gloria Elena Espinosa-Ayala; Gloria María Molina-Salinas; Gloria Vazquez; Hector Esteban Paz-Juárez; Hector Montoya-Fuentes; Helen Haydee Fernanda Ramírez-Plascencia; Irvin González-López; Jean Pierre González; Joel Armando Vázquez-Pérez.; Jorge Salas-Hernández; José Antonio Enciso-Moreno; José Arturo Martínez-Orozco; José Esteban Muñoz-Medina; José de Jesús Nuñez-Contreras; Juan Bautista Chale-Dzul; Julissa Enciso-Ibarra; Kathia Elizabeth Tapia-Diaz; Luis Alberto Ochoa-Carrera; Margarita Matías-Florentino; Mario Mújica-Sánchez; Marissa Perez-Garcia; Marissa Pérez-García; María Guadalupe Santiago-Mauricio; María Guadalupe de Jesús Mireles-Rivera; Nelly Sélem-Mojica; Pavel Isa; Ricardo Ciria Merce; Ricardo Grande; Rosa María Gutierrez Rios; Santiago Ávila-Ríos; Selene Zárate; Susana Lopez; Victor Eduardo Garcia-Arias; Víctor Hugo Borja-Aburto |
| EPI_ISL_2646123, EPI_ISL_2646124, EPI_ISL_2646125, EPI_ISL_2646126, EPI_ISL_2646127, EPI_ISL_2646128, EPI_ISL_2646129, EPI_ISL_2646130, EPI_ISL_2646131, EPI_ISL_2646132, EPI_ISL_2646133, EPI_ISL_2646134, EPI_ISL_2646135, EPI_ISL_2646136, EPI_ISL_2646137, EPI_ISL_2646138, EPI_ISL_2646139, EPI_ISL_2778010, EPI_ISL_2778011, EPI_ISL_2778012, EPI_ISL_2778017, EPI_ISL_2790177, EPI_ISL_2790178, EPI_ISL_2790179 | | | |
| see above | Instituto Nacional de Medicina Genomica | Centro de Investigación en Enfermedades Infecciosas (CIENI), Instituto Nacional de Enfermedades Respiratorias (INER) | Avila-Rios S; Boukadida C; Cedro-Tanda A; Herrera-Montalvo LA; Hidalgo-Miranda A; Matias-Florentino M; Mendoza-Vargas A; Perez-Garcia M; Reyes-Grajeda JP; Reyes-Teran G |
| EPI_ISL_2105721, EPI_ISL_2105722, EPI_ISL_2105723, EPI_ISL_2178748, EPI_ISL_2230802, EPI_ISL_2230807, EPI_ISL_2230809, EPI_ISL_2230818, EPI_ISL_2230819, EPI_ISL_2230820, EPI_ISL_2230826, EPI_ISL_2230829, EPI_ISL_2230839, EPI_ISL_2230847, EPI_ISL_2230861, EPI_ISL_2230892, EPI_ISL_2230913, EPI_ISL_2230920, EPI_ISL_2230921, EPI_ISL_2230936, EPI_ISL_2230937, EPI_ISL_2230938, EPI_ISL_2230939, EPI_ISL_2230940, EPI_ISL_2230941, EPI_ISL_2230942, EPI_ISL_2230992, EPI_ISL_2230993, EPI_ISL_2230994, EPI_ISL_2276104, EPI_ISL_2276105, EPI_ISL_2276108, EPI_ISL_2277643, EPI_ISL_2277644, EPI_ISL_2319387, EPI_ISL_2319388, EPI_ISL_2319389, EPI_ISL_2319390, EPI_ISL_2350098, EPI_ISL_2350099, EPI_ISL_2350100, EPI_ISL_2350101, EPI_ISL_2350102, EPI_ISL_2350103, EPI_ISL_2350104, EPI_ISL_2350105, EPI_ISL_2350106, EPI_ISL_2350107, EPI_ISL_2350108, EPI_ISL_2350109, EPI_ISL_2350110, EPI_ISL_2350111, EPI_ISL_2350112, EPI_ISL_2350113, EPI_ISL_2443082, EPI_ISL_2443083, EPI_ISL_2692539, EPI_ISL_2692546, EPI_ISL_2692825, EPI_ISL_2692833, EPI_ISL_2692834, EPI_ISL_2692835, EPI_ISL_2692836, EPI_ISL_2692837, EPI_ISL_2692838, EPI_ISL_2692839, EPI_ISL_2692840, EPI_ISL_2692841, EPI_ISL_2692842, EPI_ISL_2692843, EPI_ISL_2692844, EPI_ISL_2692845, EPI_ISL_2692846, EPI_ISL_2692847, EPI_ISL_2778967, EPI_ISL_2778968, EPI_ISL_2810031, EPI_ISL_2810040, EPI_ISL_2810041, EPI_ISL_2810046, EPI_ISL_2810048, EPI_ISL_2810053, EPI_ISL_2810057, EPI_ISL_2810063, EPI_ISL_2810068, EPI_ISL_2881724, EPI_ISL_2881725, EPI_ISL_2881726, EPI_ISL_2881727, EPI_ISL_2881728, EPI_ISL_2881729, EPI_ISL_2881730, EPI_ISL_2881731, EPI_ISL_2881732, EPI_ISL_2881733, EPI_ISL_2881748, EPI_ISL_2881749, EPI_ISL_2881750, EPI_ISL_2881780, EPI_ISL_2881781, EPI_ISL_2881833, EPI_ISL_2881855, EPI_ISL_2894236, EPI_ISL_2894237, EPI_ISL_2894238, EPI_ISL_2894239, EPI_ISL_2894240, EPI_ISL_2894241, EPI_ISL_2894242, EPI_ISL_2894243, EPI_ISL_2894244, EPI_ISL_2894245, EPI_ISL_2894246, EPI_ISL_2894247, EPI_ISL_2894248, EPI_ISL_2894249, EPI_ISL_2894250, EPI_ISL_2894251, EPI_ISL_2894252, EPI_ISL_2894253, EPI_ISL_2894254, EPI_ISL_2894255, EPI_ISL_2894256, EPI_ISL_2894257, EPI_ISL_2894258, EPI_ISL_2894259, EPI_ISL_2894260, EPI_ISL_2894261, EPI_ISL_2894262, EPI_ISL_2894263, EPI_ISL_2894266, EPI_ISL_2894267, EPI_ISL_2894268, EPI_ISL_2894269, EPI_ISL_2894270, EPI_ISL_2894271, EPI_ISL_2894272, EPI_ISL_2894273, EPI_ISL_2894274, EPI_ISL_2894275, EPI_ISL_2894276, EPI_ISL_2894277, EPI_ISL_2894278, EPI_ISL_2894279, EPI_ISL_2894280, EPI_ISL_2894281, EPI_ISL_2894282, EPI_ISL_2894283, EPI_ISL_2894284, EPI_ISL_2894285, EPI_ISL_2894286, EPI_ISL_2894287, EPI_ISL_2894288, EPI_ISL_2894289, EPI_ISL_2894290, EPI_ISL_2894291, EPI_ISL_2894292, EPI_ISL_2894293, EPI_ISL_2894294, EPI_ISL_2894295, EPI_ISL_2894296, EPI_ISL_2894297, EPI_ISL_2894298, EPI_ISL_2894299, EPI_ISL_2894300, EPI_ISL_2894301, EPI_ISL_2894302, EPI_ISL_2894303, EPI_ISL_2894304, EPI_ISL_2894305, EPI_ISL_2894306, EPI_ISL_2894307, EPI_ISL_2894308, EPI_ISL_2894309, EPI_ISL_2894310, EPI_ISL_2894311, EPI_ISL_2894312, EPI_ISL_2894313, EPI_ISL_2894314, EPI_ISL_2894315, EPI_ISL_2894316, EPI_ISL_2894317, EPI_ISL_2894318, EPI_ISL_2894319, EPI_ISL_2894320, EPI_ISL_2894321, EPI_ISL_2894322, EPI_ISL_2894323, EPI_ISL_2894324, EPI_ISL_2894325, EPI_ISL_2894326, EPI_ISL_2894327, EPI_ISL_2894328, EPI_ISL_2894329, EPI_ISL_2894330 | | | |
| see above | Instituto Nacional de Medicina Genomica | Instituto Nacional de Medicina Genomica | Abraham Campos-Romero; Cedro-Tanda A; Cisneros- Villanueva M; Cisneros-Villanueva M; Escobar-Arrazola; Gonzalez-Barrera D; Herrera-Montalvo LA.; Hidalgo-Miranda A; Luna-Ruiz Marco; M; Mendoza-Vargas A; Moreno-Camacho José Luis; Munguia-Garza P; Orjuela-Rodríguez M; PENDIENTE; Pendiente; Ramirez-Vega O; Rangel-DeLeon D; Reyes-Grajeda JP; Rodriguez-Gallegos Jorge |
| EPI_ISL_2272498, EPI_ISL_2272499, EPI_ISL_2272500, EPI_ISL_2293023, EPI_ISL_2293024 | LANIIA | LANIIA | Bruno Gomez-Gil; Cristobal Chaidez; Célida Isabel Martínez-Rodríguez; Irvin González-López; Jean Pierre González-Gómez; Juan Daniel Lira-Morales; Julissa Enciso-Ibarra |
| EPI_ISL_2455934, EPI_ISL_2495924, EPI_ISL_2835022, EPI_ISL_2835071, EPI_ISL_2861186, EPI_ISL_2861187, EPI_ISL_2920747, EPI_ISL_2920748, EPI_ISL_2920791, EPI_ISL_2937750, EPI_ISL_2937900, EPI_ISL_2937901, EPI_ISL_2937902, EPI_ISL_2937903, EPI_ISL_2937905 | | | |
| see above | LESP Aguascalientes | Instituto de Diagnostico y Referencia Epidemiologicos (INDRE) | Abril Rodriguez-Maldonado; Ariadna Medina-Benitez; Claudia Wong-Arambula; Ernesto Ramirez-Gonzalez.; Gisela Barrera-Badillo; Irma Lopez-Martinez; Joaquin Quiroz-Mercado; Lucia Hernandez-Rivas; Maribel Gonzalez-Villa; Natividad Cruz-Ortiz; Sergio Rangel-Guerrero; Tatiana Nunez-Garcia; Vanessa Rivero-Arredondo |
| EPI_ISL_2296064, EPI_ISL_2296104, EPI_ISL_2455998, EPI_ISL_2455999, EPI_ISL_2456000, EPI_ISL_2476439, EPI_ISL_2476443, EPI_ISL_2479884, EPI_ISL_2479888, EPI_ISL_2545705, EPI_ISL_2545807, EPI_ISL_2709687, EPI_ISL_2736832, EPI_ISL_2835023, EPI_ISL_2858980, EPI_ISL_2920685 | | | |
| see above | LESP Baja California | Instituto de Diagnostico y Referencia Epidemiologicos (INDRE) | Abril Rodriguez-Maldonado; Ariadna Medina-Benitez; Claudia Wong-Arambula; Ernesto Ramirez-Gonzalez.; Gisela Barrera-Badillo; Irma Lopez-Martinez; Joaquin Quiroz-Mercado; Lucia Hernandez-Rivas; Maribel Gonzalez-Villa; Natividad Cruz-Ortiz; Sergio Rangel-Guerrero; Tatiana Nunez-Garcia; Vanessa Rivero-Arredondo |
| EPI_ISL_2341013, EPI_ISL_2533818, EPI_ISL_2545729, EPI_ISL_2545730, EPI_ISL_2663353, EPI_ISL_2779197, EPI_ISL_2937836, EPI_ISL_2937847, EPI_ISL_2937848 | | | |
| see above | LESP Baja California Sur | Instituto de Diagnostico y Referencia Epidemiologicos (INDRE) | Abril Rodriguez-Maldonado; Ariadna Medina-Benitez; Claudia Wong-Arambula; Ernesto Ramirez-Gonzalez.; Gisela Barrera-Badillo; Irma Lopez-Martinez; Joaquin Quiroz-Mercado; Lucia Hernandez-Rivas; Maribel Gonzalez-Villa; Natividad Cruz-Ortiz; Sergio Rangel-Guerrero; Tatiana Nunez-Garcia; Vanessa Rivero-Arredondo |
| EPI_ISL_2283689, EPI_ISL_2340887, EPI_ISL_2341023, EPI_ISL_2455913, EPI_ISL_2476442, EPI_ISL_2495993, EPI_ISL_2496005, EPI_ISL_2533816, EPI_ISL_2545699, EPI_ISL_2545724, EPI_ISL_2545725, EPI_ISL_2545726, EPI_ISL_2545727, EPI_ISL_2545806, EPI_ISL_2559361, EPI_ISL_2736797, EPI_ISL_2736857, EPI_ISL_2779001, EPI_ISL_2779059, EPI_ISL_2779195, EPI_ISL_2835024, EPI_ISL_2835086, EPI_ISL_2835087, EPI_ISL_2858938, EPI_ISL_2859035, EPI_ISL_2859036, EPI_ISL_2859037, EPI_ISL_2859038, EPI_ISL_2937784, EPI_ISL_2937789, EPI_ISL_2937790, EPI_ISL_2937791, EPI_ISL_2937792 | | | |
| see above | LESP Campeche | Instituto de Diagnostico y Referencia Epidemiologicos (INDRE) | Abril Rodriguez-Maldonado; Ariadna Medina-Benitez; Claudia Wong-Arambula; Ernesto Ramirez-Gonzalez.; Gisela Barrera-Badillo; Irma Lopez-Martinez; Joaquin Quiroz-Mercado; Lucia Hernandez-Rivas; Maribel Gonzalez-Villa; Natividad Cruz-Ortiz; Sergio Rangel-Guerrero; Tatiana Nunez-Garcia; Vanessa Rivero-Arredondo |
| EPI_ISL_2479885, EPI_ISL_2492489, EPI_ISL_2492494, EPI_ISL_2689795, EPI_ISL_2709732, EPI_ISL_2778997, EPI_ISL_2859008, EPI_ISL_2859009, EPI_ISL_2859010, EPI_ISL_2859011, EPI_ISL_2861171, EPI_ISL_2937807 | | | |
| see above | LESP Chiapas | Instituto de Diagnostico y Referencia Epidemiologicos (INDRE) | Abril Rodriguez-Maldonado; Ariadna Medina-Benitez; Claudia Wong-Arambula; Ernesto Ramirez-Gonzalez.; Gisela Barrera-Badillo; Irma Lopez-Martinez; Joaquin Quiroz-Mercado; Lucia Hernandez-Rivas; Maribel Gonzalez-Villa; Natividad Cruz-Ortiz; Sergio Rangel-Guerrero; Tatiana Nunez-Garcia; Vanessa Rivero-Arredondo |
| EPI_ISL_2283691, EPI_ISL_2283692, EPI_ISL_2296092, EPI_ISL_2340889, EPI_ISL_2455914, EPI_ISL_2455915, EPI_ISL_2455917, EPI_ISL_2455919, EPI_ISL_2455920, EPI_ISL_2455926, EPI_ISL_2455927, EPI_ISL_2455928, EPI_ISL_2455929, EPI_ISL_2476441, EPI_ISL_2484164, EPI_ISL_2496003, EPI_ISL_2533759, EPI_ISL_2533760, EPI_ISL_2533761, EPI_ISL_2533762, EPI_ISL_2545707, EPI_ISL_2545708, EPI_ISL_2545709, EPI_ISL_2559263, EPI_ISL_2559265, EPI_ISL_2559267, EPI_ISL_2559269, EPI_ISL_2559271, EPI_ISL_2559273, EPI_ISL_2559275, EPI_ISL_2559356, EPI_ISL_2559358, EPI_ISL_2663394, EPI_ISL_2663395, EPI_ISL_2663396, EPI_ISL_2663397, EPI_ISL_2663398, EPI_ISL_2674699, EPI_ISL_2688957, EPI_ISL_2688980, EPI_ISL_2688981, EPI_ISL_2688982, EPI_ISL_2688983, EPI_ISL_2688984, EPI_ISL_2689827, EPI_ISL_2689828, EPI_ISL_2835045, EPI_ISL_2835055, EPI_ISL_2835056, EPI_ISL_2858939, EPI_ISL_2858940, EPI_ISL_2858954, EPI_ISL_2858955, EPI_ISL_2858956, EPI_ISL_2858979, EPI_ISL_2859076, EPI_ISL_2920702, EPI_ISL_2920703, EPI_ISL_2920704 | | | |
| see above | LESP Chihuahua | Instituto de Diagnostico y Referencia Epidemiologicos (INDRE) | Abril Rodriguez-Maldonado; Ariadna Medina-Benitez; Claudia Wong-Arambula; Ernesto Ramirez-Gonzalez.; Gisela Barrera-Badillo; Irma Lopez-Martinez; Joaquin Quiroz-Mercado; Lucia Hernandez-Rivas; Maribel Gonzalez-Villa; Natividad Cruz-Ortiz; Sergio Rangel-Guerrero; Tatiana Nunez-Garcia; Vanessa Rivero-Arredondo |
| EPI_ISL_2246838, EPI_ISL_2283705, EPI_ISL_2283706, EPI_ISL_2283707, EPI_ISL_2283708, EPI_ISL_2295588, EPI_ISL_2296072, EPI_ISL_2296074, EPI_ISL_2296075, EPI_ISL_2296076, EPI_ISL_2296077, EPI_ISL_2296078, EPI_ISL_2296083, EPI_ISL_2296110, EPI_ISL_2341006, EPI_ISL_2342996, EPI_ISL_2362694, EPI_ISL_2443048, EPI_ISL_2455959, EPI_ISL_2476409, EPI_ISL_2476410, EPI_ISL_2476411, EPI_ISL_2476412, EPI_ISL_2476423, EPI_ISL_2476427, EPI_ISL_2476428, EPI_ISL_2476429, EPI_ISL_2479882, EPI_ISL_2479883, EPI_ISL_2492466, EPI_ISL_2492484, EPI_ISL_2492490, EPI_ISL_2492491, EPI_ISL_2496015, EPI_ISL_2496016, EPI_ISL_2496017, EPI_ISL_2496018, EPI_ISL_2496019, EPI_ISL_2533807, EPI_ISL_2533808, EPI_ISL_2533810, EPI_ISL_2533811, EPI_ISL_2533812, EPI_ISL_2533813, EPI_ISL_2533814, EPI_ISL_2533815, EPI_ISL_2533817, EPI_ISL_2533824, EPI_ISL_2545695, EPI_ISL_2545696, EPI_ISL_2545697, EPI_ISL_2545722, EPI_ISL_2545815, EPI_ISL_2545816, EPI_ISL_2545818, EPI_ISL_2559456, EPI_ISL_2559458, EPI_ISL_2559463, EPI_ISL_2559465, EPI_ISL_2559467, EPI_ISL_2559471, EPI_ISL_2559472, EPI_ISL_2559473, EPI_ISL_2559475, EPI_ISL_2559477, EPI_ISL_2559479, EPI_ISL_2597995, EPI_ISL_2598004, EPI_ISL_2598005, EPI_ISL_2598006, EPI_ISL_2598007, EPI_ISL_2598008, EPI_ISL_2598009, EPI_ISL_2598010, EPI_ISL_2598011, EPI_ISL_2598012, EPI_ISL_2598013, EPI_ISL_2598014, EPI_ISL_2598015, EPI_ISL_2663328, EPI_ISL_2663329, EPI_ISL_2663330, EPI_ISL_2663342, EPI_ISL_2663343, EPI_ISL_2663344, EPI_ISL_2663358, EPI_ISL_2663393, EPI_ISL_2674632, EPI_ISL_2674698, EPI_ISL_2674900, EPI_ISL_2674901, EPI_ISL_2674902, EPI_ISL_2674904, EPI_ISL_2688911, EPI_ISL_2688919, EPI_ISL_2688920, EPI_ISL_2688921, EPI_ISL_2688922, EPI_ISL_2688923, EPI_ISL_2688926, EPI_ISL_2688960, EPI_ISL_2688961, EPI_ISL_2688962, EPI_ISL_2688963, EPI_ISL_2688964, EPI_ISL_2688966, EPI_ISL_2688967, EPI_ISL_2688968, EPI_ISL_2689796, EPI_ISL_2689797, EPI_ISL_2689798, EPI_ISL_2689800, EPI_ISL_2689801, EPI_ISL_2689802, EPI_ISL_2689803, EPI_ISL_2689804, EPI_ISL_2689814, EPI_ISL_2689815, EPI_ISL_2689816, EPI_ISL_2689817, EPI_ISL_2689818, EPI_ISL_2689819, EPI_ISL_2689821, EPI_ISL_2689822, EPI_ISL_2689823, EPI_ISL_2709703, EPI_ISL_2709712, EPI_ISL_2709713, EPI_ISL_2709714, EPI_ISL_2709715, EPI_ISL_2709716, EPI_ISL_2709720, EPI_ISL_2709730, EPI_ISL_2779173, EPI_ISL_2835005, EPI_ISL_2835006, EPI_ISL_2835009, EPI_ISL_2835010, EPI_ISL_2835011, EPI_ISL_2835012, EPI_ISL_2835013, EPI_ISL_2835026, EPI_ISL_2835027, EPI_ISL_2835028, EPI_ISL_2835029, EPI_ISL_2835030, EPI_ISL_2835031, EPI_ISL_2835032, EPI_ISL_2835033, EPI_ISL_2835034, EPI_ISL_2835035, EPI_ISL_2835036, EPI_ISL_2835037, EPI_ISL_2835038, EPI_ISL_2835039, EPI_ISL_2835041, EPI_ISL_2835042, EPI_ISL_2835043, EPI_ISL_2835050, EPI_ISL_2835051, EPI_ISL_2835052, EPI_ISL_2835053, EPI_ISL_2835091, EPI_ISL_2858948, EPI_ISL_2858949, EPI_ISL_2858950, EPI_ISL_2858951, EPI_ISL_2859061, EPI_ISL_2859062, EPI_ISL_2859066, EPI_ISL_2859067, EPI_ISL_2859068, EPI_ISL_2859069, EPI_ISL_2859070, EPI_ISL_2859071, EPI_ISL_2859072, EPI_ISL_2859073, EPI_ISL_2859074, EPI_ISL_2859075, EPI_ISL_2861158, EPI_ISL_2861192, EPI_ISL_2861193, EPI_ISL_2861194, EPI_ISL_2861195, EPI_ISL_2861196, EPI_ISL_2920707, EPI_ISL_2920708, EPI_ISL_2920709, EPI_ISL_2920710, EPI_ISL_2920711, EPI_ISL_2920712, EPI_ISL_2920743, EPI_ISL_2920744, EPI_ISL_2920745, EPI_ISL_2920752, EPI_ISL_2937752, EPI_ISL_2937769, EPI_ISL_2937770, EPI_ISL_2937802, EPI_ISL_2937803, EPI_ISL_2937804, EPI_ISL_2937805, EPI_ISL_2937832, EPI_ISL_2937833, EPI_ISL_2937834, EPI_ISL_2937838, EPI_ISL_2937839, EPI_ISL_2937840, EPI_ISL_2937841, EPI_ISL_2937842, EPI_ISL_2937843, EPI_ISL_2937857, EPI_ISL_2937858, EPI_ISL_2937859, EPI_ISL_2937860, EPI_ISL_2937862, EPI_ISL_2937863, EPI_ISL_2937864, EPI_ISL_2937865, EPI_ISL_2937866, EPI_ISL_2937867, EPI_ISL_2937868, EPI_ISL_2937869, EPI_ISL_2937870, EPI_ISL_2937871, EPI_ISL_2937872, EPI_ISL_2937873, EPI_ISL_2937874, EPI_ISL_2937875, EPI_ISL_2937876, EPI_ISL_2937877, EPI_ISL_2937878, EPI_ISL_2937879, EPI_ISL_2937880, EPI_ISL_2937881, EPI_ISL_2937882, EPI_ISL_2937883, EPI_ISL_2937884, EPI_ISL_2937885, EPI_ISL_2937886, EPI_ISL_2937887, EPI_ISL_2937888, EPI_ISL_2937889, EPI_ISL_2937890, EPI_ISL_2937891, EPI_ISL_2937892, EPI_ISL_2937893, EPI_ISL_2937894, EPI_ISL_2937895, EPI_ISL_2937906, EPI_ISL_2937908, EPI_ISL_2937909, EPI_ISL_2937910, EPI_ISL_2937911, EPI_ISL_2937912, EPI_ISL_2937913, EPI_ISL_2937914, EPI_ISL_2937915, EPI_ISL_2937916, EPI_ISL_2937917, EPI_ISL_2937918, EPI_ISL_2937919, EPI_ISL_2937920, EPI_ISL_2937921, EPI_ISL_2937922, EPI_ISL_2937923, EPI_ISL_2937924, EPI_ISL_2937925, EPI_ISL_2937928 | | | |
| see above | LESP Ciudad de Mexico | Instituto de Diagnostico y | Abril Rodriguez-Maldonado; Ariadna Medina-Benitez; Claudia Wong-Arambula; Ernesto Ramirez-Gonzalez.; Gisela Barrera-Badillo; Irma Lopez-Martinez; Joaquin Quiroz-Mercado; Lucia Hernandez-Rivas; Maribel Gonzalez-Villa; Natividad Cruz-Ortiz; Sergio Rangel-Guerrero; Tatiana Nunez-Garcia; Vanessa Rivero-Arredondo |

| | | | |
|---|---|---|---|
| | | Referencia Epidemiologicos (INDRE) | |
| EPI_ISL_2343020 | LESP Ciudad de Mexico /Hospital Angeles | Instituto de Diagnostico y Referencia Epidemiologicos (INDRE) | Abril Rodriguez-Maldonado; Ariadna Medina-Benitez; Claudia Wong-Arambula; Ernesto Ramirez-Gonzalez.; Gisela Barrera-Badillo; Irma Lopez-Martinez; Joaquin Quiroz-Mercado; Lucia Hernandez-Rivas; Natividad Cruz-Ortiz; Sergio Rangel-Guerrero; Tatiana Nunez-Garcia; Vanessa Rivero-Arredondo |
| EPI_ISL_2246827, EPI_ISL_2246828 | LESP Ciudad de Mexico /LANS | Instituto de Diagnostico y Referencia Epidemiologicos (INDRE) | Abril Rodriguez-Maldonado; Ariadna Medina-Benitez; Claudia Wong-Arambula; Ernesto Ramirez-Gonzalez.; Gisela Barrera-Badillo; Irma Lopez-Martinez; Joaquin Quiroz-Mercado; Lucia Hernandez-Rivas; Natividad Cruz-Ortiz; Sergio Rangel-Guerrero; Tatiana Nunez-Garcia; Vanessa Rivero-Arredondo |
| EPI_ISL_2533821 | LESP Ciudad de México /DIAGNOMOL | Instituto de Diagnostico y Referencia Epidemiologicos (INDRE) | Abril Rodriguez-Maldonado; Ariadna Medina-Benitez; Claudia Wong-Arambula; Ernesto Ramirez-Gonzalez.; Gisela Barrera-Badillo; Irma Lopez-Martinez; Joaquin Quiroz-Mercado; Lucia Hernandez-Rivas; Natividad Cruz-Ortiz; Sergio Rangel-Guerrero; Tatiana Nunez-Garcia; Vanessa Rivero-Arredondo |
| EPI_ISL_2533763, EPI_ISL_2533803, EPI_ISL_2545728, EPI_ISL_2598003, EPI_ISL_2674689, EPI_ISL_2688985, EPI_ISL_2709731, EPI_ISL_2736829, EPI_ISL_2779034, EPI_ISL_2861180, EPI_ISL_2920683, EPI_ISL_2920705, EPI_ISL_2937907 | | | |
| see above | LESP Coahuila | Instituto de Diagnostico y Referencia Epidemiologicos (INDRE) | Abril Rodriguez-Maldonado; Ariadna Medina-Benitez; Claudia Wong-Arambula; Ernesto Ramirez-Gonzalez.; Gisela Barrera-Badillo; Irma Lopez-Martinez; Joaquin Quiroz-Mercado; Lucia Hernandez-Rivas; Maribel Gonzalez-Villa; Natividad Cruz-Ortiz; Sergio Rangel-Guerrero; Tatiana Nunez-Garcia; Vanessa Rivero-Arredondo |
| EPI_ISL_2283697, EPI_ISL_2296067, EPI_ISL_2343017, EPI_ISL_2455980, EPI_ISL_2456001, EPI_ISL_2476438, EPI_ISL_2479892, EPI_ISL_2479909, EPI_ISL_2479910, EPI_ISL_2559452, EPI_ISL_2663345, EPI_ISL_2920686 | | | |
| see above | LESP Colima | Instituto de Diagnostico y Referencia Epidemiologicos (INDRE) | Abril Rodriguez-Maldonado; Ariadna Medina-Benitez; Claudia Wong-Arambula; Ernesto Ramirez-Gonzalez.; Gisela Barrera-Badillo; Irma Lopez-Martinez; Joaquin Quiroz-Mercado; Lucia Hernandez-Rivas; Natividad Cruz-Ortiz; Sergio Rangel-Guerrero; Tatiana Nunez-Garcia; Vanessa Rivero-Arredondo |
| EPI_ISL_2545805, EPI_ISL_2559448, EPI_ISL_2674642, EPI_ISL_2778993, EPI_ISL_2778994, EPI_ISL_2778996, EPI_ISL_2779000, EPI_ISL_2800933, EPI_ISL_2800934, EPI_ISL_2835049, EPI_ISL_2861168, EPI_ISL_2861169, EPI_ISL_2861170 | | | |
| see above | LESP Durango | Instituto de Diagnostico y Referencia Epidemiologicos (INDRE) | Abril Rodriguez-Maldonado; Ariadna Medina-Benitez; Claudia Wong-Arambula; Ernesto Ramirez-Gonzalez.; Gisela Barrera-Badillo; Irma Lopez-Martinez; Joaquin Quiroz-Mercado; Lucia Hernandez-Rivas; Maribel Gonzalez-Villa; Natividad Cruz-Ortiz; Sergio Rangel-Guerrero; Tatiana Nunez-Garcia; Vanessa Rivero-Arredondo |
| EPI_ISL_2296073, EPI_ISL_2455992, EPI_ISL_2533809, EPI_ISL_2545698, EPI_ISL_2545723, EPI_ISL_2545817, EPI_ISL_2559460, EPI_ISL_2559469, EPI_ISL_2674664, EPI_ISL_2674668, EPI_ISL_2674669, EPI_ISL_2674670, EPI_ISL_2688959, EPI_ISL_2688965, EPI_ISL_2709704, EPI_ISL_2709705, EPI_ISL_2709719, EPI_ISL_2709721, EPI_ISL_2778999, EPI_ISL_2835040, EPI_ISL_2920742, EPI_ISL_2920751, EPI_ISL_2937771, EPI_ISL_2937861 | | | |
| see above | LESP Estado de Mexico | Instituto de Diagnostico y Referencia Epidemiologicos (INDRE) | Abril Rodriguez-Maldonado; Ariadna Medina-Benitez; Claudia Wong-Arambula; Ernesto Ramirez-Gonzalez.; Gisela Barrera-Badillo; Irma Lopez-Martinez; Joaquin Quiroz-Mercado; Lucia Hernandez-Rivas; Maribel Gonzalez-Villa; Natividad Cruz-Ortiz; Sergio Rangel-Guerrero; Tatiana Nunez-Garcia; Vanessa Rivero-Arredondo |
| EPI_ISL_2283721, EPI_ISL_2283722, EPI_ISL_2296095, EPI_ISL_2296096, EPI_ISL_2341016, EPI_ISL_2341017, EPI_ISL_2341018, EPI_ISL_2341019, EPI_ISL_2341020, EPI_ISL_2455986, EPI_ISL_2455987, EPI_ISL_2455988, EPI_ISL_2455990, EPI_ISL_2455991, EPI_ISL_2476413, EPI_ISL_2476415, EPI_ISL_2476418, EPI_ISL_2476419, EPI_ISL_2476420, EPI_ISL_2476421, EPI_ISL_2476422, EPI_ISL_2476424, EPI_ISL_2476426, EPI_ISL_2479899, EPI_ISL_2492465, EPI_ISL_2492471, EPI_ISL_2492478, EPI_ISL_2492479, EPI_ISL_2492480, EPI_ISL_2492481, EPI_ISL_2492482, EPI_ISL_2492483, EPI_ISL_2492485, EPI_ISL_2495964, EPI_ISL_2495982, EPI_ISL_2495983, EPI_ISL_2495984, EPI_ISL_2495985, EPI_ISL_2495986, EPI_ISL_2495987, EPI_ISL_2495989, EPI_ISL_2495990, EPI_ISL_2495991, EPI_ISL_2495997, EPI_ISL_2495999, EPI_ISL_2496000, EPI_ISL_2496002, EPI_ISL_2663326, EPI_ISL_2663383, EPI_ISL_2663384, EPI_ISL_2663385, EPI_ISL_2663386, EPI_ISL_2663387, EPI_ISL_2663388, EPI_ISL_2674628, EPI_ISL_2674656, EPI_ISL_2674657, EPI_ISL_2674691, EPI_ISL_2674692, EPI_ISL_2674693, EPI_ISL_2674694, EPI_ISL_2674695, EPI_ISL_2674696, EPI_ISL_2688974, EPI_ISL_2688976, EPI_ISL_2688977, EPI_ISL_2688978, EPI_ISL_2688979, EPI_ISL_2709724, EPI_ISL_2709725, EPI_ISL_2736787, EPI_ISL_2736827, EPI_ISL_2736859, EPI_ISL_2736861, EPI_ISL_2736864, EPI_ISL_2778990, EPI_ISL_2778991, EPI_ISL_2778992, EPI_ISL_2779019, EPI_ISL_2779047, EPI_ISL_2779048, EPI_ISL_2779056, EPI_ISL_2779057, EPI_ISL_2779058, EPI_ISL_2779120, EPI_ISL_2779186, EPI_ISL_2779187, EPI_ISL_2779188, EPI_ISL_2779189, EPI_ISL_2779190, EPI_ISL_2779191, EPI_ISL_2779192, EPI_ISL_2779193, EPI_ISL_2779215, EPI_ISL_2779216, EPI_ISL_2779217, EPI_ISL_2779218, EPI_ISL_2779276, EPI_ISL_2779281, EPI_ISL_2800913, EPI_ISL_2800914, EPI_ISL_2800915, EPI_ISL_2800917, EPI_ISL_2800924, EPI_ISL_2800926, EPI_ISL_2800932, EPI_ISL_2800941, EPI_ISL_2800943, EPI_ISL_2858991, EPI_ISL_2858992, EPI_ISL_2858993, EPI_ISL_2861167, EPI_ISL_2937772 | | | |
| see above | LESP Guanajuato | Instituto de Diagnostico y Referencia Epidemiologicos (INDRE) | Abril Rodriguez-Maldonado; Ariadna Medina-Benitez; Claudia Wong-Arambula; Ernesto Ramirez-Gonzalez.; Gisela Barrera-Badillo; Irma Lopez-Martinez; Joaquin Quiroz-Mercado; Lucia Hernandez-Rivas; Maribel Gonzalez-Villa; Natividad Cruz-Ortiz; Sergio Rangel-Guerrero; Tatiana Nunez-Garcia; Vanessa Rivero-Arredondo |
| EPI_ISL_2361362 | LESP Guanajuato | Instituto de diagnóstico y Referencia Epidemiologicos (INDRE) Departamento de Virología | Abril Rodriguez-Maldonado; Ariadna Medina-Benitez; Claudia Wong-Arambula; Ernesto Ramirez-Gonzalez.; Gisela Barrera-Badillo; Irma Lopez-Martinez; Joaquin Quiroz-Mercado; Lucia Hernandez-Rivas; Natividad Cruz-Ortiz; Sergio Rangel-Guerrero; Tatiana Nunez-Garcia; Vanessa Rivero-Arredondo |
| EPI_ISL_2495988, EPI_ISL_2736841, EPI_ISL_2779050, EPI_ISL_2779196, EPI_ISL_2779214, EPI_ISL_2779227, EPI_ISL_2920749, EPI_ISL_2920750 | | | |
| see above | LESP Guerrero | Instituto de Diagnostico y Referencia Epidemiologicos (INDRE) | Abril Rodriguez-Maldonado; Ariadna Medina-Benitez; Claudia Wong-Arambula; Ernesto Ramirez-Gonzalez.; Gisela Barrera-Badillo; Irma Lopez-Martinez; Joaquin Quiroz-Mercado; Lucia Hernandez-Rivas; Maribel Gonzalez-Villa; Natividad Cruz-Ortiz; Sergio Rangel-Guerrero; Tatiana Nunez-Garcia; Vanessa Rivero-Arredondo |
| EPI_ISL_2644834, EPI_ISL_2644835, EPI_ISL_2644836 | LESP HIDALGO | LESP HIDALGO GENE2LIFE | CASTILLO-ORTEGA LAURA SOFIA; GARCIA-GONZALEZ OCTAVIO; GARCIA-REYES MELITO; HERREGA-GOMEZ CONNIE ALEJANDRA; HERRERA-GOMEZ CONNIE ALEJANDRA; VIGUERAS-MORALES YAJAIRA SULIM; VILLAFUENTES-TELLEZ HECTOR ALBERTO |
| EPI_ISL_2283688, EPI_ISL_2479901, EPI_ISL_2479911, EPI_ISL_2479912, EPI_ISL_2495976, EPI_ISL_2663360, EPI_ISL_2663361, EPI_ISL_2663362, EPI_ISL_2663363, EPI_ISL_2663364, EPI_ISL_2663365, EPI_ISL_2663366, EPI_ISL_2663367, EPI_ISL_2663368, EPI_ISL_2663369, EPI_ISL_2663370, EPI_ISL_2663371, EPI_ISL_2663372, EPI_ISL_2663373, EPI_ISL_2663374, EPI_ISL_2663375, EPI_ISL_2674658, EPI_ISL_2674659, EPI_ISL_2674660, EPI_ISL_2674661, EPI_ISL_2674662, EPI_ISL_2674663, EPI_ISL_2674665, EPI_ISL_2674666, EPI_ISL_2674667, EPI_ISL_2674671, EPI_ISL_2674672, EPI_ISL_2674674, EPI_ISL_2674675, EPI_ISL_2674676, EPI_ISL_2674677, EPI_ISL_2674678, EPI_ISL_2674679, EPI_ISL_2674680, EPI_ISL_2674681, EPI_ISL_2674682, EPI_ISL_2674690, EPI_ISL_2688972, EPI_ISL_2709722, EPI_ISL_2835090, EPI_ISL_2858947, EPI_ISL_2859012, EPI_ISL_2861173, EPI_ISL_2937768 | | | |
| see above | LESP Hidalgo | Instituto de Diagnostico y Referencia Epidemiologicos (INDRE) | Abril Rodriguez-Maldonado; Ariadna Medina-Benitez; Claudia Wong-Arambula; Ernesto Ramirez-Gonzalez.; Gisela Barrera-Badillo; Irma Lopez-Martinez; Joaquin Quiroz-Mercado; Lucia Hernandez-Rivas; Maribel Gonzalez-Villa; Natividad Cruz-Ortiz; Sergio Rangel-Guerrero; Tatiana Nunez-Garcia; Vanessa Rivero-Arredondo |
| EPI_ISL_2795657, EPI_ISL_2800345 | LESP Hidalgo | LESP Hidalgo GENE2LIFE | CASTILLO-ORTEGA LAURA SOFIA; GARCIA-GONZALEZ OCTAVIO; GARCIA-REYES MELITO; HERRERA-GOMEZ CONNIE ALEJANDRA; VIGUERAS-MORALES YAJAIRA SULIM; VILLAFUENTES-TELLEZ HECTOR ALBERTO |
| EPI_ISL_2341007, EPI_ISL_2455958, EPI_ISL_2479916, EPI_ISL_2495930, EPI_ISL_2496023, EPI_ISL_2496024, EPI_ISL_2533767, EPI_ISL_2545683, EPI_ISL_2545706, EPI_ISL_2545721, EPI_ISL_2559261, EPI_ISL_2674650, EPI_ISL_2674903, EPI_ISL_2688913, EPI_ISL_2688914, EPI_ISL_2688975, EPI_ISL_2709699, EPI_ISL_2709717, EPI_ISL_2779175, EPI_ISL_2835016, EPI_ISL_2858972, EPI_ISL_2858994, EPI_ISL_2858995, EPI_ISL_2859028, EPI_ISL_2859107, EPI_ISL_2937751, EPI_ISL_2937757, EPI_ISL_2937801, EPI_ISL_2937815, EPI_ISL_2937896, EPI_ISL_2937897, EPI_ISL_2937898 | | | |
| see above | LESP Jalisco | Instituto de Diagnostico y Referencia Epidemiologicos (INDRE) | Abril Rodriguez-Maldonado; Ariadna Medina-Benitez; Claudia Wong-Arambula; Ernesto Ramirez-Gonzalez.; Gisela Barrera-Badillo; Irma Lopez-Martinez; Joaquin Quiroz-Mercado; Lucia Hernandez-Rivas; Maribel Gonzalez-Villa; Natividad Cruz-Ortiz; Sergio Rangel-Guerrero; Tatiana Nunez-Garcia; Vanessa Rivero-Arredondo |
| EPI_ISL_2246829, EPI_ISL_2246830, EPI_ISL_2246831, EPI_ISL_2246832, EPI_ISL_2246833, EPI_ISL_2340870, EPI_ISL_2443050, EPI_ISL_2495936, EPI_ISL_2495970, EPI_ISL_2496006, EPI_ISL_2496007, EPI_ISL_2496008, EPI_ISL_2496009, EPI_ISL_2496010, EPI_ISL_2496011, EPI_ISL_2496012, EPI_ISL_2499967, EPI_ISL_2545702, EPI_ISL_2545804, EPI_ISL_2559425, EPI_ISL_2559427, EPI_ISL_2559429, EPI_ISL_2559431, EPI_ISL_2559433, EPI_ISL_2559435, EPI_ISL_2559437, EPI_ISL_2559439, EPI_ISL_2559442, EPI_ISL_2559444, EPI_ISL_2559446, EPI_ISL_2674641, EPI_ISL_2688943, EPI_ISL_2688944, EPI_ISL_2688945, EPI_ISL_2688946, EPI_ISL_2688947, EPI_ISL_2688970, EPI_ISL_2689811, EPI_ISL_2689812, EPI_ISL_2689825, EPI_ISL_2709688, EPI_ISL_2736784, EPI_ISL_2736825, EPI_ISL_2736834, EPI_ISL_2736848, EPI_ISL_2736849, EPI_ISL_2736851, EPI_ISL_2779226, EPI_ISL_2779232, EPI_ISL_2779233, EPI_ISL_2800945, EPI_ISL_2858960, EPI_ISL_2859064, EPI_ISL_2859065, EPI_ISL_2861172, EPI_ISL_2920753, EPI_ISL_2937846 | | | |
| see above | LESP Michoacan | Instituto de Diagnostico y Referencia Epidemiologicos (INDRE) | Abril Rodriguez-Maldonado; Ariadna Medina-Benitez; Claudia Wong-Arambula; Ernesto Ramirez-Gonzalez.; Gisela Barrera-Badillo; Irma Lopez-Martinez; Joaquin Quiroz-Mercado; Lucia Hernandez-Rivas; Maribel Gonzalez-Villa; Natividad Cruz-Ortiz; Sergio Rangel-Guerrero; Tatiana Nunez-Garcia; Vanessa Rivero-Arredondo |
| EPI_ISL_2342999 | LESP Michoacan/LANS | Instituto de Diagnostico y Referencia Epidemiologicos (INDRE) | Abril Rodriguez-Maldonado; Ariadna Medina-Benitez; Claudia Wong-Arambula; Ernesto Ramirez-Gonzalez.; Gisela Barrera-Badillo; Irma Lopez-Martinez; Joaquin Quiroz-Mercado; Lucia Hernandez-Rivas; Natividad Cruz-Ortiz; Sergio Rangel-Guerrero; Tatiana Nunez-Garcia; Vanessa Rivero-Arredondo |

| | | | |
|---|---|---|---|
| EPI_ISL_2296094, EPI_ISL_2341014, EPI_ISL_2341015, EPI_ISL_2476440, EPI_ISL_2479900, EPI_ISL_2495933, EPI_ISL_2495935, EPI_ISL_2495992, EPI_ISL_2495994, EPI_ISL_2495995, EPI_ISL_2495996, EPI_ISL_2559354, EPI_ISL_2663382, EPI_ISL_2688915, EPI_ISL_2688916, EPI_ISL_2688917, EPI_ISL_2688918, EPI_ISL_2688973, EPI_ISL_2709700, EPI_ISL_2709701, EPI_ISL_2709702, EPI_ISL_2835008 | | | |
| see above | LESP Morelos | Instituto de Diagnostico y Referencia Epidemiologicos (INDRE) | Abril Rodriguez-Maldonado; Ariadna Medina-Benitez; Claudia Wong-Arambula; Ernesto Ramirez-Gonzalez.; Gisela Barrera-Badillo; Irma Lopez-Martinez; Joaquin Quiroz-Mercado; Lucia Hernandez-Rivas; Natividad Cruz-Ortiz; Sergio Rangel-Guerrero; Tatiana Nunez-Garcia; Vanessa Rivero-Arredondo |
| EPI_ISL_2778995, EPI_ISL_2937774 | LESP Nayarit | Instituto de Diagnostico y Referencia Epidemiologicos (INDRE) | Abril Rodriguez-Maldonado; Ariadna Medina-Benitez; Claudia Wong-Arambula; Ernesto Ramirez-Gonzalez.; Gisela Barrera-Badillo; Irma Lopez-Martinez; Joaquin Quiroz-Mercado; Lucia Hernandez-Rivas; Maribel Gonzalez-Villa; Natividad Cruz-Ortiz; Sergio Rangel-Guerrero; Tatiana Nunez-Garcia; Vanessa Rivero-Arredondo |
| EPI_ISL_2246839, EPI_ISL_2341021, EPI_ISL_2341022, EPI_ISL_2455982, EPI_ISL_2455983, EPI_ISL_2455984, EPI_ISL_2455985, EPI_ISL_2479894, EPI_ISL_2479895, EPI_ISL_2479896, EPI_ISL_2479897, EPI_ISL_2479898, EPI_ISL_2479902, EPI_ISL_2492464, EPI_ISL_2492467, EPI_ISL_2492475, EPI_ISL_2492476, EPI_ISL_2492477, EPI_ISL_2492486, EPI_ISL_2495934, EPI_ISL_2495963, EPI_ISL_2495980, EPI_ISL_2495981, EPI_ISL_2495998, EPI_ISL_2658953, EPI_ISL_2663327, EPI_ISL_2663389, EPI_ISL_2663390, EPI_ISL_2663391, EPI_ISL_2663392, EPI_ISL_2674629, EPI_ISL_2674630, EPI_ISL_2674631, EPI_ISL_2674697, EPI_ISL_2688912, EPI_ISL_2709689, EPI_ISL_2709690, EPI_ISL_2709691, EPI_ISL_2709692, EPI_ISL_2709693, EPI_ISL_2709694, EPI_ISL_2709695, EPI_ISL_2709696, EPI_ISL_2709697, EPI_ISL_2709698, EPI_ISL_2709726, EPI_ISL_2709727, EPI_ISL_2709728, EPI_ISL_2709729, EPI_ISL_2835044, EPI_ISL_2835074, EPI_ISL_2835075, EPI_ISL_2835076, EPI_ISL_2858935, EPI_ISL_2858936, EPI_ISL_2858937, EPI_ISL_2858983, EPI_ISL_2859054, EPI_ISL_2859121, EPI_ISL_2861159, EPI_ISL_2861160, EPI_ISL_2861161, EPI_ISL_2861163, EPI_ISL_2920687, EPI_ISL_2920688, EPI_ISL_2920689, EPI_ISL_2920690, EPI_ISL_2920691, EPI_ISL_2920692, EPI_ISL_2920739, EPI_ISL_2937844 | | | |
| see above | LESP Nuevo Leon | Instituto de Diagnostico y Referencia Epidemiologicos (INDRE) | Abril Rodriguez-Maldonado; Ariadna Medina-Benitez; Claudia Wong-Arambula; Ernesto Ramirez-Gonzalez.; Gisela Barrera-Badillo; Irma Lopez-Martinez; Joaquin Quiroz-Mercado; Lucia Hernandez-Rivas; Maribel Gonzalez-Villa; Natividad Cruz-Ortiz; Sergio Rangel-Guerrero; Tatiana Nunez-Garcia; Vanessa Rivero-Arredondo |
| EPI_ISL_2283694, EPI_ISL_2455922, EPI_ISL_2476430, EPI_ISL_2492472, EPI_ISL_2663352, EPI_ISL_2920724 | LESP Oaxaca | Instituto de Diagnostico y Referencia Epidemiologicos (INDRE) | Abril Rodriguez-Maldonado; Ariadna Medina-Benitez; Claudia Wong-Arambula; Ernesto Ramirez-Gonzalez.; Gisela Barrera-Badillo; Irma Lopez-Martinez; Joaquin Quiroz-Mercado; Lucia Hernandez-Rivas; Maribel Gonzalez-Villa; Natividad Cruz-Ortiz; Sergio Rangel-Guerrero; Tatiana Nunez-Garcia; Vanessa Rivero-Arredondo |
| EPI_ISL_2455994, EPI_ISL_2455995, EPI_ISL_2479903, EPI_ISL_2479904, EPI_ISL_2479905, EPI_ISL_2495929, EPI_ISL_2559363, EPI_ISL_2559365, EPI_ISL_2597996, EPI_ISL_2663399, EPI_ISL_2674673, EPI_ISL_2674700, EPI_ISL_2688986, EPI_ISL_2688987, EPI_ISL_2736785, EPI_ISL_2779035, EPI_ISL_2779036, EPI_ISL_2779037, EPI_ISL_2779038, EPI_ISL_2779039, EPI_ISL_2779040, EPI_ISL_2779041, EPI_ISL_2779042, EPI_ISL_2779174, EPI_ISL_2779211, EPI_ISL_2779212, EPI_ISL_2779257, EPI_ISL_2779258, EPI_ISL_2779259, EPI_ISL_2779260, EPI_ISL_2779261, EPI_ISL_2779262, EPI_ISL_2779263, EPI_ISL_2779264, EPI_ISL_2779265, EPI_ISL_2779266, EPI_ISL_2779267, EPI_ISL_2779268, EPI_ISL_2779269, EPI_ISL_2779280, EPI_ISL_2800936, EPI_ISL_2937756 | | | |
| see above | LESP Puebla | Instituto de Diagnostico y Referencia Epidemiologicos (INDRE) | Abril Rodriguez-Maldonado; Ariadna Medina-Benitez; Claudia Wong-Arambula; Ernesto Ramirez-Gonzalez.; Gisela Barrera-Badillo; Irma Lopez-Martinez; Joaquin Quiroz-Mercado; Lucia Hernandez-Rivas; Maribel Gonzalez-Villa; Natividad Cruz-Ortiz; Sergio Rangel-Guerrero; Tatiana Nunez-Garcia; Vanessa Rivero-Arredondo |
| EPI_ISL_2283720, EPI_ISL_2340881, EPI_ISL_2341008, EPI_ISL_2455904, EPI_ISL_2455905, EPI_ISL_2455906, EPI_ISL_2455907, EPI_ISL_2455908, EPI_ISL_2455909, EPI_ISL_2455910, EPI_ISL_2455911, EPI_ISL_2455912, EPI_ISL_2455960, EPI_ISL_2455961, EPI_ISL_2455962, EPI_ISL_2455963, EPI_ISL_2455981, EPI_ISL_2455989, EPI_ISL_2456002, EPI_ISL_2479877, EPI_ISL_2479878, EPI_ISL_2479879, EPI_ISL_2479880, EPI_ISL_2479881, EPI_ISL_2479893, EPI_ISL_2479913, EPI_ISL_2492458, EPI_ISL_2492459, EPI_ISL_2492460, EPI_ISL_2492461, EPI_ISL_2492462, EPI_ISL_2492463, EPI_ISL_2492468, EPI_ISL_2492469, EPI_ISL_2492470, EPI_ISL_2492474, EPI_ISL_2492497, EPI_ISL_2492498, EPI_ISL_2495922, EPI_ISL_2495923, EPI_ISL_2495925, EPI_ISL_2495927, EPI_ISL_2495928, EPI_ISL_2495938, EPI_ISL_2495939, EPI_ISL_2495940, EPI_ISL_2495941, EPI_ISL_2495942, EPI_ISL_2495943, EPI_ISL_2495944, EPI_ISL_2495945, EPI_ISL_2495946, EPI_ISL_2495947, EPI_ISL_2495948, EPI_ISL_2495949, EPI_ISL_2495950, EPI_ISL_2495951, EPI_ISL_2495952, EPI_ISL_2495953, EPI_ISL_2495954, EPI_ISL_2495955, EPI_ISL_2495956, EPI_ISL_2495957, EPI_ISL_2495958, EPI_ISL_2495959, EPI_ISL_2495960, EPI_ISL_2495961, EPI_ISL_2495962, EPI_ISL_2495967, EPI_ISL_2495968, EPI_ISL_2495969, EPI_ISL_2495977, EPI_ISL_2495978, EPI_ISL_2496020, EPI_ISL_2496021, EPI_ISL_2496022, EPI_ISL_2533764, EPI_ISL_2533765, EPI_ISL_2533766, EPI_ISL_2533822, EPI_ISL_2545682, EPI_ISL_2545700, EPI_ISL_2545701, EPI_ISL_2545713, EPI_ISL_2545714, EPI_ISL_2545715, EPI_ISL_2545716, EPI_ISL_2545717, EPI_ISL_2545718, EPI_ISL_2545719, EPI_ISL_2545720, EPI_ISL_2545731, EPI_ISL_2545732, EPI_ISL_2559283, EPI_ISL_2559285, EPI_ISL_2559287, EPI_ISL_2559289, EPI_ISL_2559291, EPI_ISL_2559293, EPI_ISL_2559295, EPI_ISL_2559297, EPI_ISL_2559299, EPI_ISL_2559302, EPI_ISL_2559303, EPI_ISL_2559305, EPI_ISL_2559307, EPI_ISL_2559309, EPI_ISL_2559311, EPI_ISL_2559314, EPI_ISL_2559316, EPI_ISL_2559317, EPI_ISL_2559319, EPI_ISL_2559321, EPI_ISL_2559323, EPI_ISL_2559325, EPI_ISL_2559327, EPI_ISL_2559329, EPI_ISL_2559331, EPI_ISL_2559333, EPI_ISL_2559336, EPI_ISL_2559338, EPI_ISL_2559340, EPI_ISL_2559342, EPI_ISL_2559343, EPI_ISL_2559346, EPI_ISL_2559347, EPI_ISL_2559350, EPI_ISL_2559352, EPI_ISL_2559367, EPI_ISL_2559369, EPI_ISL_2559371, EPI_ISL_2559373, EPI_ISL_2597997, EPI_ISL_2597998, EPI_ISL_2597999, EPI_ISL_2598000, EPI_ISL_2598001, EPI_ISL_2663331, EPI_ISL_2663332, EPI_ISL_2663377, EPI_ISL_2663380, EPI_ISL_2663381, EPI_ISL_2674683, EPI_ISL_2688829, EPI_ISL_2688830, EPI_ISL_2688924, EPI_ISL_2688925, EPI_ISL_2688969, EPI_ISL_2689824, EPI_ISL_2709686, EPI_ISL_2709706, EPI_ISL_2709707, EPI_ISL_2709708, EPI_ISL_2779049, EPI_ISL_2835014, EPI_ISL_2835089, EPI_ISL_2861174, EPI_ISL_2861175, EPI_ISL_2920713, EPI_ISL_2920754, EPI_ISL_2937926, EPI_ISL_2937927 | | | |
| see above | LESP Queretaro | Instituto de Diagnostico y Referencia Epidemiologicos (INDRE) | Abril Rodriguez-Maldonado; Ariadna Medina-Benitez; Claudia Wong-Arambula; Ernesto Ramirez-Gonzalez.; Gisela Barrera-Badillo; Irma Lopez-Martinez; Joaquin Quiroz-Mercado; Lucia Hernandez-Rivas; Maribel Gonzalez-Villa; Natividad Cruz-Ortiz; Sergio Rangel-Guerrero; Tatiana Nunez-Garcia; Vanessa Rivero-Arredondo |
| EPI_ISL_2443049, EPI_ISL_2455997, EPI_ISL_2479889, EPI_ISL_2479890, EPI_ISL_2479891, EPI_ISL_2479914, EPI_ISL_2479915, EPI_ISL_2492495, EPI_ISL_2492496, EPI_ISL_2495966, EPI_ISL_2496025, EPI_ISL_2533804, EPI_ISL_2533805, EPI_ISL_2533820, EPI_ISL_2545693, EPI_ISL_2545694, EPI_ISL_2545808, EPI_ISL_2545809, EPI_ISL_2545810, EPI_ISL_2545811, EPI_ISL_2663356, EPI_ISL_2663357, EPI_ISL_2674651, EPI_ISL_2674652, EPI_ISL_2674653, EPI_ISL_2674654, EPI_ISL_2674655, EPI_ISL_2674714, EPI_ISL_2779051, EPI_ISL_2779052, EPI_ISL_2779176, EPI_ISL_2779177, EPI_ISL_2779178, EPI_ISL_2779179, EPI_ISL_2779180, EPI_ISL_2779181, EPI_ISL_2800938, EPI_ISL_2859052, EPI_ISL_2859053, EPI_ISL_2937758, EPI_ISL_2937759, EPI_ISL_2937760, EPI_ISL_2937761, EPI_ISL_2937762, EPI_ISL_2937763, EPI_ISL_2937764, EPI_ISL_2937765, EPI_ISL_2937766, EPI_ISL_2937798, EPI_ISL_2937799, EPI_ISL_2937800, EPI_ISL_2937856 | | | |
| see above | LESP Quintana Roo | Instituto de Diagnostico y Referencia Epidemiologicos (INDRE) | Abril Rodriguez-Maldonado; Ariadna Medina-Benitez; Claudia Wong-Arambula; Ernesto Ramirez-Gonzalez.; Gisela Barrera-Badillo; Irma Lopez-Martinez; Joaquin Quiroz-Mercado; Lucia Hernandez-Rivas; Maribel Gonzalez-Villa; Natividad Cruz-Ortiz; Sergio Rangel-Guerrero; Tatiana Nunez-Garcia; Vanessa Rivero-Arredondo |
| EPI_ISL_2455953, EPI_ISL_2492473, EPI_ISL_2533806, EPI_ISL_2545813, EPI_ISL_2545814, EPI_ISL_2545819, EPI_ISL_2559454, EPI_ISL_2658951, EPI_ISL_2658952, EPI_ISL_2674716, EPI_ISL_2779159, EPI_ISL_2858987, EPI_ISL_2858988, EPI_ISL_2858989, EPI_ISL_2858990, EPI_ISL_2859058, EPI_ISL_2859059, EPI_ISL_2859060, EPI_ISL_2859108, EPI_ISL_2937773 | | | |
| see above | LESP San Luis Potosi | Instituto de Diagnostico y Referencia Epidemiologicos (INDRE) | Abril Rodriguez-Maldonado; Ariadna Medina-Benitez; Claudia Wong-Arambula; Ernesto Ramirez-Gonzalez.; Gisela Barrera-Badillo; Irma Lopez-Martinez; Joaquin Quiroz-Mercado; Lucia Hernandez-Rivas; Maribel Gonzalez-Villa; Natividad Cruz-Ortiz; Sergio Rangel-Guerrero; Tatiana Nunez-Garcia; Vanessa Rivero-Arredondo |
| EPI_ISL_2246834, EPI_ISL_2246835, EPI_ISL_2246836, EPI_ISL_2246837, EPI_ISL_2283700, EPI_ISL_2283701, EPI_ISL_2283703, EPI_ISL_2296071, EPI_ISL_2296102, EPI_ISL_2296103, EPI_ISL_2340960, EPI_ISL_2340961, EPI_ISL_2342993, EPI_ISL_2343005, EPI_ISL_2476437, EPI_ISL_2476444, EPI_ISL_2479908, EPI_ISL_2496013, EPI_ISL_2496014, EPI_ISL_2499966, EPI_ISL_2533823, EPI_ISL_2545692, EPI_ISL_2545703, EPI_ISL_2545704, EPI_ISL_2559450, EPI_ISL_2663333, EPI_ISL_2663334, EPI_ISL_2674633, EPI_ISL_2674634, EPI_ISL_2689805, EPI_ISL_2689806, EPI_ISL_2736780, EPI_ISL_2736781, EPI_ISL_2736783, EPI_ISL_2778998, EPI_ISL_2779005, EPI_ISL_2779006, EPI_ISL_2779007, EPI_ISL_2779220, EPI_ISL_2835017, EPI_ISL_2835018, EPI_ISL_2835019, EPI_ISL_2835020, EPI_ISL_2835021, EPI_ISL_2835054, EPI_ISL_2835077, EPI_ISL_2858973, EPI_ISL_2859013, EPI_ISL_2859014, EPI_ISL_2859015, EPI_ISL_2859034 | | | |
| see above | LESP Sinaloa | Instituto de Diagnostico y Referencia Epidemiologicos (INDRE) | Abril Rodriguez-Maldonado; Ariadna Medina-Benitez; Claudia Wong-Arambula; Ernesto Ramirez-Gonzalez.; Gisela Barrera-Badillo; Irma Lopez-Martinez; Joaquin Quiroz-Mercado; Lucia Hernandez-Rivas; Maribel Gonzalez-Villa; Natividad Cruz-Ortiz; Sergio Rangel-Guerrero; Tatiana Nunez-Garcia; Vanessa Rivero-Arredondo |
| EPI_ISL_2479907, EPI_ISL_2545812, EPI_ISL_2663354, EPI_ISL_2663355, EPI_ISL_2663359, EPI_ISL_2663403, EPI_ISL_2674643, EPI_ISL_2674644, EPI_ISL_2674645, EPI_ISL_2674646, EPI_ISL_2674647, EPI_ISL_2674648, EPI_ISL_2674649, EPI_ISL_2674718, EPI_ISL_2674719, EPI_ISL_2689794, EPI_ISL_2689829, EPI_ISL_2709718, EPI_ISL_2736856, EPI_ISL_2778989, EPI_ISL_2779055, EPI_ISL_2779184, EPI_ISL_2779185, EPI_ISL_2779213, EPI_ISL_2800930, EPI_ISL_2800931, EPI_ISL_2800937, EPI_ISL_2835025, EPI_ISL_2835072, EPI_ISL_2835073, EPI_ISL_2858961, EPI_ISL_2858981, EPI_ISL_2858982, EPI_ISL_2859051, EPI_ISL_2920706, EPI_ISL_2937767, EPI_ISL_2937795, EPI_ISL_2937796, EPI_ISL_2937797, EPI_ISL_2937831, EPI_ISL_2937851, EPI_ISL_2937852, EPI_ISL_2937853, EPI_ISL_2937854, EPI_ISL_2937855 | | | |
| see above | LESP Sonora | Instituto de Diagnostico y Referencia Epidemiologicos (INDRE) | Abril Rodriguez-Maldonado; Ariadna Medina-Benitez; Claudia Wong-Arambula; Ernesto Ramirez-Gonzalez.; Gisela Barrera-Badillo; Irma Lopez-Martinez; Joaquin Quiroz-Mercado; Lucia Hernandez-Rivas; Maribel Gonzalez-Villa; Natividad Cruz-Ortiz; Sergio Rangel-Guerrero; Tatiana Nunez-Garcia; Vanessa Rivero-Arredondo |
| EPI_ISL_2495979, EPI_ISL_2533768, EPI_ISL_2533769, EPI_ISL_2559375, EPI_ISL_2559377, EPI_ISL_2663336, EPI_ISL_2663337, EPI_ISL_2663338, EPI_ISL_2663339, EPI_ISL_2663340, EPI_ISL_2663341, EPI_ISL_2663346, EPI_ISL_2663347, EPI_ISL_2663348, EPI_ISL_2663349, EPI_ISL_2663351, EPI_ISL_2674635, EPI_ISL_2674636, EPI_ISL_2674637, EPI_ISL_2674638, EPI_ISL_2674639, EPI_ISL_2674640, EPI_ISL_2688824, EPI_ISL_2688825, EPI_ISL_2688826, EPI_ISL_2688827, EPI_ISL_2688828, EPI_ISL_2688927, EPI_ISL_2688928, EPI_ISL_2688929, EPI_ISL_2688930, EPI_ISL_2688931, EPI_ISL_2688932, EPI_ISL_2688933, EPI_ISL_2688934, EPI_ISL_2688935, EPI_ISL_2688936, EPI_ISL_2688937, EPI_ISL_2688938, EPI_ISL_2688939, EPI_ISL_2688940, EPI_ISL_2688941, EPI_ISL_2688942, EPI_ISL_2688948, EPI_ISL_2688949, EPI_ISL_2688950, EPI_ISL_2688951, EPI_ISL_2688952, EPI_ISL_2688953, EPI_ISL_2688954, EPI_ISL_2688955, EPI_ISL_2688956, EPI_ISL_2688958, EPI_ISL_2689808, EPI_ISL_2689809, EPI_ISL_2689810, EPI_ISL_2689813, EPI_ISL_2709709, EPI_ISL_2709710, EPI_ISL_2709711, EPI_ISL_2736790, EPI_ISL_2736792, EPI_ISL_2736795, EPI_ISL_2736800, EPI_ISL_2736802, EPI_ISL_2736804, EPI_ISL_2736807, EPI_ISL_2736810, EPI_ISL_2736811, EPI_ISL_2736813, EPI_ISL_2736815, EPI_ISL_2736817, EPI_ISL_2736819, EPI_ISL_2736821, EPI_ISL_2736823, EPI_ISL_2736837, EPI_ISL_2736839, EPI_ISL_2736853, EPI_ISL_2778984, EPI_ISL_2778985, EPI_ISL_2778986, EPI_ISL_2778987, EPI_ISL_2778988, EPI_ISL_2779024, EPI_ISL_2779025, EPI_ISL_2779026, EPI_ISL_2779027, EPI_ISL_2779028, EPI_ISL_2779029, EPI_ISL_2779030, EPI_ISL_2779031, EPI_ISL_2779032, EPI_ISL_2779033, EPI_ISL_2779043, EPI_ISL_2779044, EPI_ISL_2779045, EPI_ISL_2779046, EPI_ISL_2779053, EPI_ISL_2779054, EPI_ISL_2779160, EPI_ISL_2779161, EPI_ISL_2779162, EPI_ISL_2779163, EPI_ISL_2779164, EPI_ISL_2779165, EPI_ISL_2779166, EPI_ISL_2779167, EPI_ISL_2779168, EPI_ISL_2779169, EPI_ISL_2779170, EPI_ISL_2779171, EPI_ISL_2779172, EPI_ISL_2779182, EPI_ISL_2779183, EPI_ISL_2779194, EPI_ISL_2779198, EPI_ISL_2779199, EPI_ISL_2779200, EPI_ISL_2779201, EPI_ISL_2779202, EPI_ISL_2779203, EPI_ISL_2779204, EPI_ISL_2779205, EPI_ISL_2779206, EPI_ISL_2779207, EPI_ISL_2779208, EPI_ISL_2779209, EPI_ISL_2779210, EPI_ISL_2779234, EPI_ISL_2779235, EPI_ISL_2779236, EPI_ISL_2779237, EPI_ISL_2779238, EPI_ISL_2779239, EPI_ISL_2779240, EPI_ISL_2779241, EPI_ISL_2779242, EPI_ISL_2779243, EPI_ISL_2779244, EPI_ISL_2779245, EPI_ISL_2779246, EPI_ISL_2779247, EPI_ISL_2779248, EPI_ISL_2779249, EPI_ISL_2779250, EPI_ISL_2779251, EPI_ISL_2779252, EPI_ISL_2779253, EPI_ISL_2779254, EPI_ISL_2779255, EPI_ISL_2779256, EPI_ISL_2779271, EPI_ISL_2779272, EPI_ISL_2779273, EPI_ISL_2779274, EPI_ISL_2779275, EPI_ISL_2779277, EPI_ISL_2779278, EPI_ISL_2779279, EPI_ISL_2800925, EPI_ISL_2800927, EPI_ISL_2800928, EPI_ISL_2800929, EPI_ISL_2800939, EPI_ISL_2800940, EPI_ISL_2800942, EPI_ISL_2835015, EPI_ISL_2835057, EPI_ISL_2835058, EPI_ISL_2835059, EPI_ISL_2835060, EPI_ISL_2835061, EPI_ISL_2835062, EPI_ISL_2835063, EPI_ISL_2835064, EPI_ISL_2835065, EPI_ISL_2835066, EPI_ISL_2835067, EPI_ISL_2835068, EPI_ISL_2835069, EPI_ISL_2835070, EPI_ISL_2835078, EPI_ISL_2835079, EPI_ISL_2835080, EPI_ISL_2835081, EPI_ISL_2835082, EPI_ISL_2835083, EPI_ISL_2835084, EPI_ISL_2835088, EPI_ISL_2835092, EPI_ISL_2835093, EPI_ISL_2835094, EPI_ISL_2835095, EPI_ISL_2835096, EPI_ISL_2835097, EPI_ISL_2835098, EPI_ISL_2835099, EPI_ISL_2835100, EPI_ISL_2835101, EPI_ISL_2835102, EPI_ISL_2835103, EPI_ISL_2835104, EPI_ISL_2835105, EPI_ISL_2835106, EPI_ISL_2858957, EPI_ISL_2858958, EPI_ISL_2858959, EPI_ISL_2858962, EPI_ISL_2858963, EPI_ISL_2858964, EPI_ISL_2858974, EPI_ISL_2858975, EPI_ISL_2858976, EPI_ISL_2858977, EPI_ISL_2858978, EPI_ISL_2858984, EPI_ISL_2858985, EPI_ISL_2858986, EPI_ISL_2859016, EPI_ISL_2859017, EPI_ISL_2859018, EPI_ISL_2859019, EPI_ISL_2859020, EPI_ISL_2859021, EPI_ISL_2859022, EPI_ISL_2859023, EPI_ISL_2859024, EPI_ISL_2859025, EPI_ISL_2859026, EPI_ISL_2859027, EPI_ISL_2859039, EPI_ISL_2859040, EPI_ISL_2859041, EPI_ISL_2859042, EPI_ISL_2859043, EPI_ISL_2859044, EPI_ISL_2859045, EPI_ISL_2859046, EPI_ISL_2859047, EPI_ISL_2859048, EPI_ISL_2859055, EPI_ISL_2859056, EPI_ISL_2859057, EPI_ISL_2859077, EPI_ISL_2859078, EPI_ISL_2859079, EPI_ISL_2859080, EPI_ISL_2859081, EPI_ISL_2859082, EPI_ISL_2859083, EPI_ISL_2859084, EPI_ISL_2859085, EPI_ISL_2859086, EPI_ISL_2859087, EPI_ISL_2859088, EPI_ISL_2859089, EPI_ISL_2859090, EPI_ISL_2859091, EPI_ISL_2859092, EPI_ISL_2859093, EPI_ISL_2859094, EPI_ISL_2859095, EPI_ISL_2859096, EPI_ISL_2859097, EPI_ISL_2859098, EPI_ISL_2859099, EPI_ISL_2859100, EPI_ISL_2859101, EPI_ISL_2859102, EPI_ISL_2859103, EPI_ISL_2859104, EPI_ISL_2859105, EPI_ISL_2859106, EPI_ISL_2859109, EPI_ISL_2859110, EPI_ISL_2859111, EPI_ISL_2859112, EPI_ISL_2859113, EPI_ISL_2859114, EPI_ISL_2859115, EPI_ISL_2859116, EPI_ISL_2859117, EPI_ISL_2859118, EPI_ISL_2859119, EPI_ISL_2859120, EPI_ISL_2859122, EPI_ISL_2859123, EPI_ISL_2859124, EPI_ISL_2859125, EPI_ISL_2861176, EPI_ISL_2861177, EPI_ISL_2861178, EPI_ISL_2861179, EPI_ISL_2861183, EPI_ISL_2861184, EPI_ISL_2861185, EPI_ISL_2861188, EPI_ISL_2861189, EPI_ISL_2861190, EPI_ISL_2861191, EPI_ISL_2920714, EPI_ISL_2920715, EPI_ISL_2920716, EPI_ISL_2920717, EPI_ISL_2920718, EPI_ISL_2920719, EPI_ISL_2920720, EPI_ISL_2920721, EPI_ISL_2920722, EPI_ISL_2920723, EPI_ISL_2920732, EPI_ISL_2920733, EPI_ISL_2920734, EPI_ISL_2920735, EPI_ISL_2920736, EPI_ISL_2920737, EPI_ISL_2920738, EPI_ISL_2920741, EPI_ISL_2920746, EPI_ISL_2920755, EPI_ISL_2920756, EPI_ISL_2920757, EPI_ISL_2920758, EPI_ISL_2920759, EPI_ISL_2920760, EPI_ISL_2920761, EPI_ISL_2920762, EPI_ISL_2920763, EPI_ISL_2920764, EPI_ISL_2920765, EPI_ISL_2920766, EPI_ISL_2920767, EPI_ISL_2920768, EPI_ISL_2920769, EPI_ISL_2920770, EPI_ISL_2920771, EPI_ISL_2920772, EPI_ISL_2920773, EPI_ISL_2920774, EPI_ISL_2920775, EPI_ISL_2920776, EPI_ISL_2920777, EPI_ISL_2920778, EPI_ISL_2920779, EPI_ISL_2920780, EPI_ISL_2920781, EPI_ISL_2920782, EPI_ISL_2920783, EPI_ISL_2920784, EPI_ISL_2920785, EPI_ISL_2920786, EPI_ISL_2920787, EPI_ISL_2920788, EPI_ISL_2920789, EPI_ISL_2920790, EPI_ISL_2937747, EPI_ISL_2937749, EPI_ISL_2937753, EPI_ISL_2937754, EPI_ISL_2937755, EPI_ISL_2937806, EPI_ISL_2937808, EPI_ISL_2937809, EPI_ISL_2937810, EPI_ISL_2937811, EPI_ISL_2937812, EPI_ISL_2937813, EPI_ISL_2937814, EPI_ISL_2937816, EPI_ISL_2937817, EPI_ISL_2937818, EPI_ISL_2937819, EPI_ISL_2937820, EPI_ISL_2937821, EPI_ISL_2937822, EPI_ISL_2937823, EPI_ISL_2937824, EPI_ISL_2937825, EPI_ISL_2937826, EPI_ISL_2937827, EPI_ISL_2937899 | | | |
| see above | LESP Tabasco | Instituto de Diagnostico y Referencia Epidemiologicos (INDRE) | Abril Rodriguez-Maldonado; Ariadna Medina-Benitez; Claudia Wong-Arambula; Ernesto Ramirez-Gonzalez.; Gisela Barrera-Badillo; Irma Lopez-Martinez; Joaquin Quiroz-Mercado; Lucia Hernandez-Rivas; Maribel Gonzalez-Villa; Natividad Cruz-Ortiz; Sergio Rangel-Guerrero; Tatiana Nunez-Garcia; Vanessa Rivero-Arredondo |
| EPI_ISL_2283693, EPI_ISL_2455916, EPI_ISL_2455918, EPI_ISL_2455923, EPI_ISL_2455924, EPI_ISL_2455925, EPI_ISL_2455930, EPI_ISL_2455931, EPI_ISL_2455932, EPI_ISL_2455933, EPI_ISL_2455935, EPI_ISL_2455936, EPI_ISL_2455937, EPI_ISL_2455938, EPI_ISL_2455939, EPI_ISL_2455940, EPI_ISL_2455941, EPI_ISL_2455942, EPI_ISL_2455943, EPI_ISL_2455944, EPI_ISL_2455945, EPI_ISL_2455946, EPI_ISL_2455947, EPI_ISL_2455948, EPI_ISL_2455949, EPI_ISL_2455950, EPI_ISL_2455951, EPI_ISL_2455952, EPI_ISL_2455954, EPI_ISL_2455955, EPI_ISL_2455956, EPI_ISL_2455957, EPI_ISL_2476416, EPI_ISL_2476425, EPI_ISL_2476431, EPI_ISL_2476432, EPI_ISL_2476433, EPI_ISL_2476434, EPI_ISL_2476435, EPI_ISL_2476436, EPI_ISL_2479906, EPI_ISL_2492487, EPI_ISL_2492492, EPI_ISL_2492493, | | | |

EPI_ISL_2496004, EPI_ISL_2533797, EPI_ISL_2533798, EPI_ISL_2533799, EPI_ISL_2533800, EPI_ISL_2533801, EPI_ISL_2545680, EPI_ISL_2545681, EPI_ISL_2545689, EPI_ISL_2545690, EPI_ISL_2545691, EPI_ISL_2598002, EPI_ISL_2663350, EPI_ISL_2663400, EPI_ISL_2663401, EPI_ISL_2779002, EPI_ISL_2779003, EPI_ISL_2779004, EPI_ISL_2779060, EPI_ISL_2779061, EPI_ISL_2779062, EPI_ISL_2779063, EPI_ISL_2779064, EPI_ISL_2779065, EPI_ISL_2779219, EPI_ISL_2800935, EPI_ISL_2835046, EPI_ISL_2835048, EPI_ISL_2835085, EPI_ISL_2858941, EPI_ISL_2858942, EPI_ISL_2858943, EPI_ISL_2858952, EPI_ISL_2858953, EPI_ISL_2858965, EPI_ISL_2858966, EPI_ISL_2858967, EPI_ISL_2858968, EPI_ISL_2858969, EPI_ISL_2858970, EPI_ISL_2858971, EPI_ISL_2858996, EPI_ISL_2858997, EPI_ISL_2858998, EPI_ISL_2858999, EPI_ISL_2859029, EPI_ISL_2859030, EPI_ISL_2859031, EPI_ISL_2859032, EPI_ISL_2859033, EPI_ISL_2861162, EPI_ISL_2861164, EPI_ISL_2861165, EPI_ISL_2861181, EPI_ISL_2861182, EPI_ISL_2920693, EPI_ISL_2920694, EPI_ISL_2920695, EPI_ISL_2920696, EPI_ISL_2920697, EPI_ISL_2920698, EPI_ISL_2920699, EPI_ISL_2920700, EPI_ISL_2920701, EPI_ISL_2920725, EPI_ISL_2920726, EPI_ISL_2920727, EPI_ISL_2920728, EPI_ISL_2920729, EPI_ISL_2920730, EPI_ISL_2937775, EPI_ISL_2937776, EPI_ISL_2937777, EPI_ISL_2937828, EPI_ISL_2937829, EPI_ISL_2937830, EPI_ISL_2937929

| | | | |
|---|---|---|---|
| see above | LESP Tamaulipas | Instituto de Diagnostico y Referencia Epidemiologicos (INDRE) | Abril Rodriguez-Maldonado; Ariadna Medina-Benitez; Claudia Wong-Arambula; Ernesto Ramirez-Gonzalez.; Gisela Barrera-Badillo; Irma Lopez-Martinez; Joaquin Quiroz-Mercado; Lucia Hernandez-Rivas; Maribel Gonzalez-Villa; Natividad Cruz-Ortiz; Sergio Rangel-Guerrero; Tatiana Nunez-Garcia; Vanessa Rivero-Arredondo |
| EPI_ISL_2937845 | LESP Tlaxcala | Instituto de Diagnostico y Referencia Epidemiologicos (INDRE) | Abril Rodriguez-Maldonado; Ariadna Medina-Benitez; Claudia Wong-Arambula; Ernesto Ramirez-Gonzalez.; Gisela Barrera-Badillo; Irma Lopez-Martinez; Joaquin Quiroz-Mercado; Lucia Hernandez-Rivas; Maribel Gonzalez-Villa; Natividad Cruz-Ortiz; Sergio Rangel-Guerrero; Tatiana Nunez-Garcia; Vanessa Rivero-Arredondo |

EPI_ISL_2545710, EPI_ISL_2545711, EPI_ISL_2559277, EPI_ISL_2559379, EPI_ISL_2688831, EPI_ISL_2688988, EPI_ISL_2688989, EPI_ISL_2689826, EPI_ISL_2709723, EPI_ISL_2709733, EPI_ISL_2835007, EPI_ISL_2835047, EPI_ISL_2858944, EPI_ISL_2858945, EPI_ISL_2859000, EPI_ISL_2859049, EPI_ISL_2859050, EPI_ISL_2861166, EPI_ISL_2920740, EPI_ISL_2937793, EPI_ISL_2937794, EPI_ISL_2937849, EPI_ISL_2937850, EPI_ISL_2937930

| | | | |
|---|---|---|---|
| see above | LESP Veracruz | Instituto de Diagnostico y Referencia Epidemiologicos (INDRE) | Abril Rodriguez-Maldonado; Ariadna Medina-Benitez; Claudia Wong-Arambula; Ernesto Ramirez-Gonzalez.; Gisela Barrera-Badillo; Irma Lopez-Martinez; Joaquin Quiroz-Mercado; Lucia Hernandez-Rivas; Maribel Gonzalez-Villa; Natividad Cruz-Ortiz; Sergio Rangel-Guerrero; Tatiana Nunez-Garcia; Vanessa Rivero-Arredondo |

EPI_ISL_2476414, EPI_ISL_2479886, EPI_ISL_2479887, EPI_ISL_2495965, EPI_ISL_2533770, EPI_ISL_2533771, EPI_ISL_2533772, EPI_ISL_2533773, EPI_ISL_2533774, EPI_ISL_2533775, EPI_ISL_2533776, EPI_ISL_2533777, EPI_ISL_2533778, EPI_ISL_2533779, EPI_ISL_2533780, EPI_ISL_2533781, EPI_ISL_2533782, EPI_ISL_2533783, EPI_ISL_2533784, EPI_ISL_2533785, EPI_ISL_2533786, EPI_ISL_2533787, EPI_ISL_2533788, EPI_ISL_2533789, EPI_ISL_2533790, EPI_ISL_2533791, EPI_ISL_2533792, EPI_ISL_2533793, EPI_ISL_2533794, EPI_ISL_2533795, EPI_ISL_2533796, EPI_ISL_2533819, EPI_ISL_2545684, EPI_ISL_2545685, EPI_ISL_2545686, EPI_ISL_2545687, EPI_ISL_2545688, EPI_ISL_2545733, EPI_ISL_2545734, EPI_ISL_2545735, EPI_ISL_2545736, EPI_ISL_2545737, EPI_ISL_2545738, EPI_ISL_2545739, EPI_ISL_2545740, EPI_ISL_2545741, EPI_ISL_2545742, EPI_ISL_2545743, EPI_ISL_2545744, EPI_ISL_2545745, EPI_ISL_2545746, EPI_ISL_2545747, EPI_ISL_2545748, EPI_ISL_2545749, EPI_ISL_2545750, EPI_ISL_2545751, EPI_ISL_2545752, EPI_ISL_2545753, EPI_ISL_2545754, EPI_ISL_2545755, EPI_ISL_2545756, EPI_ISL_2545757, EPI_ISL_2545758, EPI_ISL_2545759, EPI_ISL_2545760, EPI_ISL_2545761, EPI_ISL_2545762, EPI_ISL_2545763, EPI_ISL_2545764, EPI_ISL_2545765, EPI_ISL_2545766, EPI_ISL_2545767, EPI_ISL_2545768, EPI_ISL_2545769, EPI_ISL_2545770, EPI_ISL_2545771, EPI_ISL_2545772, EPI_ISL_2545773, EPI_ISL_2545774, EPI_ISL_2545775, EPI_ISL_2545776, EPI_ISL_2545777, EPI_ISL_2545778, EPI_ISL_2545779, EPI_ISL_2545780, EPI_ISL_2545781, EPI_ISL_2545782, EPI_ISL_2545783, EPI_ISL_2545784, EPI_ISL_2545785, EPI_ISL_2545786, EPI_ISL_2545787, EPI_ISL_2545788, EPI_ISL_2545789, EPI_ISL_2545790, EPI_ISL_2545791, EPI_ISL_2545792, EPI_ISL_2545793, EPI_ISL_2545794, EPI_ISL_2545795, EPI_ISL_2545796, EPI_ISL_2545797, EPI_ISL_2545798, EPI_ISL_2545799, EPI_ISL_2545800, EPI_ISL_2545801, EPI_ISL_2545802, EPI_ISL_2545803, EPI_ISL_2559381, EPI_ISL_2559384, EPI_ISL_2559386, EPI_ISL_2559388, EPI_ISL_2559389, EPI_ISL_2559391, EPI_ISL_2559393, EPI_ISL_2559395, EPI_ISL_2559397, EPI_ISL_2559399, EPI_ISL_2559402, EPI_ISL_2559404, EPI_ISL_2559406, EPI_ISL_2559408, EPI_ISL_2559410, EPI_ISL_2559412, EPI_ISL_2559414, EPI_ISL_2559416, EPI_ISL_2559419, EPI_ISL_2559421, EPI_ISL_2559423, EPI_ISL_2663378, EPI_ISL_2663379, EPI_ISL_2663402, EPI_ISL_2674684, EPI_ISL_2674685, EPI_ISL_2674686, EPI_ISL_2674687, EPI_ISL_2674688, EPI_ISL_2674701, EPI_ISL_2674703, EPI_ISL_2674704, EPI_ISL_2674705, EPI_ISL_2674706, EPI_ISL_2674707, EPI_ISL_2674708, EPI_ISL_2674709, EPI_ISL_2674710, EPI_ISL_2674711, EPI_ISL_2674712, EPI_ISL_2674713, EPI_ISL_2674715, EPI_ISL_2674905, EPI_ISL_2688990, EPI_ISL_2736845, EPI_ISL_2779008, EPI_ISL_2779009, EPI_ISL_2779010, EPI_ISL_2779011, EPI_ISL_2779012, EPI_ISL_2779013, EPI_ISL_2779014, EPI_ISL_2779015, EPI_ISL_2779016, EPI_ISL_2779017, EPI_ISL_2779018, EPI_ISL_2779020, EPI_ISL_2779021, EPI_ISL_2779022, EPI_ISL_2779023, EPI_ISL_2779066, EPI_ISL_2779067, EPI_ISL_2779068, EPI_ISL_2779069, EPI_ISL_2779070, EPI_ISL_2779071, EPI_ISL_2779072, EPI_ISL_2779073, EPI_ISL_2779074, EPI_ISL_2779075, EPI_ISL_2779076, EPI_ISL_2779077, EPI_ISL_2779078, EPI_ISL_2779079, EPI_ISL_2779080, EPI_ISL_2779081, EPI_ISL_2779082, EPI_ISL_2779083, EPI_ISL_2779084, EPI_ISL_2779085, EPI_ISL_2779086, EPI_ISL_2779087, EPI_ISL_2779088, EPI_ISL_2779089, EPI_ISL_2779090, EPI_ISL_2779091, EPI_ISL_2779092, EPI_ISL_2779093, EPI_ISL_2779094, EPI_ISL_2779095, EPI_ISL_2779096, EPI_ISL_2779097, EPI_ISL_2779098, EPI_ISL_2779099, EPI_ISL_2779100, EPI_ISL_2779101, EPI_ISL_2779102, EPI_ISL_2779103, EPI_ISL_2779104, EPI_ISL_2779105, EPI_ISL_2779106, EPI_ISL_2779107, EPI_ISL_2779108, EPI_ISL_2779109, EPI_ISL_2779110, EPI_ISL_2779111, EPI_ISL_2779112, EPI_ISL_2779113, EPI_ISL_2779114, EPI_ISL_2779115, EPI_ISL_2779116, EPI_ISL_2779117, EPI_ISL_2779118, EPI_ISL_2779119, EPI_ISL_2779121, EPI_ISL_2779122, EPI_ISL_2779123, EPI_ISL_2779124, EPI_ISL_2779125, EPI_ISL_2779126, EPI_ISL_2779127, EPI_ISL_2779128, EPI_ISL_2779129, EPI_ISL_2779130, EPI_ISL_2779131, EPI_ISL_2779132, EPI_ISL_2779133, EPI_ISL_2779134, EPI_ISL_2779135, EPI_ISL_2779136, EPI_ISL_2779137, EPI_ISL_2779138, EPI_ISL_2779139, EPI_ISL_2779140, EPI_ISL_2779141, EPI_ISL_2779142, EPI_ISL_2779143, EPI_ISL_2779144, EPI_ISL_2779145, EPI_ISL_2779146, EPI_ISL_2779147, EPI_ISL_2779148, EPI_ISL_2779149, EPI_ISL_2779150, EPI_ISL_2779151, EPI_ISL_2779152, EPI_ISL_2779153, EPI_ISL_2779154, EPI_ISL_2779155, EPI_ISL_2779156, EPI_ISL_2779157, EPI_ISL_2779158, EPI_ISL_2779221, EPI_ISL_2779222, EPI_ISL_2779223, EPI_ISL_2779224, EPI_ISL_2779225, EPI_ISL_2779228, EPI_ISL_2779229, EPI_ISL_2779230, EPI_ISL_2779231, EPI_ISL_2800916, EPI_ISL_2800918, EPI_ISL_2800919, EPI_ISL_2800920, EPI_ISL_2800921, EPI_ISL_2800922, EPI_ISL_2800923, EPI_ISL_2800944, EPI_ISL_2858946, EPI_ISL_2859001, EPI_ISL_2859002, EPI_ISL_2859003, EPI_ISL_2859004, EPI_ISL_2859005, EPI_ISL_2859006, EPI_ISL_2859007, EPI_ISL_2859063, EPI_ISL_2920731, EPI_ISL_2937778, EPI_ISL_2937779, EPI_ISL_2937780, EPI_ISL_2937781, EPI_ISL_2937782, EPI_ISL_2937783, EPI_ISL_2937785, EPI_ISL_2937786, EPI_ISL_2937787, EPI_ISL_2937788

| | | | |
|---|---|---|---|
| see above | LESP Yucatan | Instituto de Diagnostico y Referencia Epidemiologicos (INDRE) | Abril Rodriguez-Maldonado; Ariadna Medina-Benitez; Claudia Wong-Arambula; Ernesto Ramirez-Gonzalez.; Gisela Barrera-Badillo; Irma Lopez-Martinez; Joaquin Quiroz-Mercado; Lucia Hernandez-Rivas; Maribel Gonzalez-Villa; Natividad Cruz-Ortiz; Sergio Rangel-Guerrero; Tatiana Nunez-Garcia; Vanessa Rivero-Arredondo |

EPI_ISL_2476417, EPI_ISL_2484166, EPI_ISL_2495971, EPI_ISL_2495972, EPI_ISL_2495973, EPI_ISL_2495974, EPI_ISL_2495975, EPI_ISL_2688971, EPI_ISL_2689807, EPI_ISL_2920684, EPI_ISL_2937837

| | | | |
|---|---|---|---|
| see above | LESP Zacatecas | Instituto de Diagnostico y Referencia Epidemiologicos (INDRE) | Abril Rodriguez-Maldonado; Ariadna Medina-Benitez; Claudia Wong-Arambula; Ernesto Ramirez-Gonzalez.; Gisela Barrera-Badillo; Irma Lopez-Martinez; Joaquin Quiroz-Mercado; Lucia Hernandez-Rivas; Maribel Gonzalez-Villa; Natividad Cruz-Ortiz; Sergio Rangel-Guerrero; Tatiana Nunez-Garcia; Vanessa Rivero-Arredondo |

EPI_ISL_2942513, EPI_ISL_2942514, EPI_ISL_2942515, EPI_ISL_2942516, EPI_ISL_2942517, EPI_ISL_2942518, EPI_ISL_2942519, EPI_ISL_2942520, EPI_ISL_2942521, EPI_ISL_2942522, EPI_ISL_2942523, EPI_ISL_2942524, EPI_ISL_2942525, EPI_ISL_2942526, EPI_ISL_2942527, EPI_ISL_2942528, EPI_ISL_2942529, EPI_ISL_2942530, EPI_ISL_2942531, EPI_ISL_2942532, EPI_ISL_2942533, EPI_ISL_2942534, EPI_ISL_2942535, EPI_ISL_2942536, EPI_ISL_2942537, EPI_ISL_2942538, EPI_ISL_2942539, EPI_ISL_2942540, EPI_ISL_2942541, EPI_ISL_2942542, EPI_ISL_2942543, EPI_ISL_2942544, EPI_ISL_2942545, EPI_ISL_2942546, EPI_ISL_2942547, EPI_ISL_2942548, EPI_ISL_2942549, EPI_ISL_2942550, EPI_ISL_2942551, EPI_ISL_2942552, EPI_ISL_2942553, EPI_ISL_2942554, EPI_ISL_2942555, EPI_ISL_2942556, EPI_ISL_2942557, EPI_ISL_2942558, EPI_ISL_2942559, EPI_ISL_2942560, EPI_ISL_2942561, EPI_ISL_2942562, EPI_ISL_2942563, EPI_ISL_2942564, EPI_ISL_2942565, EPI_ISL_2942566, EPI_ISL_2942567, EPI_ISL_2942568, EPI_ISL_2942569, EPI_ISL_2942570, EPI_ISL_2942571, EPI_ISL_2942572, EPI_ISL_2942573, EPI_ISL_2942574, EPI_ISL_2942575, EPI_ISL_2942576, EPI_ISL_2942577, EPI_ISL_2942578, EPI_ISL_2942579, EPI_ISL_2942580, EPI_ISL_2942581, EPI_ISL_2942582, EPI_ISL_2942583, EPI_ISL_2942584, EPI_ISL_2942585, EPI_ISL_2942586, EPI_ISL_2942587, EPI_ISL_2942588, EPI_ISL_2942589, EPI_ISL_2942590, EPI_ISL_2942591, EPI_ISL_2942592, EPI_ISL_2942593, EPI_ISL_2942594, EPI_ISL_2942595, EPI_ISL_2942596, EPI_ISL_2942597, EPI_ISL_2942598, EPI_ISL_2942599, EPI_ISL_2942600, EPI_ISL_2942601, EPI_ISL_2942602, EPI_ISL_2942603, EPI_ISL_2942604, EPI_ISL_2942605, EPI_ISL_2942606, EPI_ISL_2942607, EPI_ISL_2942608, EPI_ISL_2942609, EPI_ISL_2942610, EPI_ISL_2942611, EPI_ISL_2942612, EPI_ISL_2942613, EPI_ISL_2942614, EPI_ISL_2942615, EPI_ISL_2942616, EPI_ISL_2942617, EPI_ISL_2942618, EPI_ISL_2942619, EPI_ISL_2942620, EPI_ISL_2942621, EPI_ISL_2942622, EPI_ISL_2942623, EPI_ISL_2942624, EPI_ISL_2942793, EPI_ISL_2942794, EPI_ISL_2942795, EPI_ISL_2942796, EPI_ISL_2942797, EPI_ISL_2942798, EPI_ISL_2942799, EPI_ISL_2942800, EPI_ISL_2942801, EPI_ISL_2942802, EPI_ISL_2942803, EPI_ISL_2942804, EPI_ISL_2942805, EPI_ISL_2942806, EPI_ISL_2942807, EPI_ISL_2942808, EPI_ISL_2942809, EPI_ISL_2942810, EPI_ISL_2942811, EPI_ISL_2942812, EPI_ISL_2942813, EPI_ISL_2942814, EPI_ISL_2942815, EPI_ISL_2942816, EPI_ISL_2942817, EPI_ISL_2942818, EPI_ISL_2942819, EPI_ISL_2942820, EPI_ISL_2942821, EPI_ISL_2942822, EPI_ISL_2942823, EPI_ISL_2942824, EPI_ISL_2942825, EPI_ISL_2942826, EPI_ISL_2942827, EPI_ISL_2942828, EPI_ISL_2942829, EPI_ISL_2942830, EPI_ISL_2942831, EPI_ISL_2942832, EPI_ISL_2942833, EPI_ISL_2942834, EPI_ISL_2942835, EPI_ISL_2942836, EPI_ISL_2942837, EPI_ISL_2942838, EPI_ISL_2942839, EPI_ISL_2942840, EPI_ISL_2942841, EPI_ISL_2942842, EPI_ISL_2942843, EPI_ISL_2942844, EPI_ISL_2942845, EPI_ISL_2942846, EPI_ISL_2942847, EPI_ISL_2942848, EPI_ISL_2942849, EPI_ISL_2942850, EPI_ISL_2942851, EPI_ISL_2942852, EPI_ISL_2942853, EPI_ISL_2942854, EPI_ISL_2942855, EPI_ISL_2942856, EPI_ISL_2942857, EPI_ISL_2942858, EPI_ISL_2942859, EPI_ISL_2942860, EPI_ISL_2942861, EPI_ISL_2942862, EPI_ISL_2942863, EPI_ISL_2942864, EPI_ISL_2942865, EPI_ISL_2942866, EPI_ISL_2942867, EPI_ISL_2942868, EPI_ISL_2942869, EPI_ISL_2942870, EPI_ISL_2942871, EPI_ISL_2942872, EPI_ISL_2942873, EPI_ISL_2942874, EPI_ISL_2942875, EPI_ISL_2942876, EPI_ISL_2942877, EPI_ISL_2942878, EPI_ISL_2942879, EPI_ISL_2942880, EPI_ISL_2942881, EPI_ISL_2942882, EPI_ISL_2942883, EPI_ISL_2942884, EPI_ISL_2942885, EPI_ISL_2942887, EPI_ISL_2942888, EPI_ISL_2942889, EPI_ISL_2942890, EPI_ISL_2942893, EPI_ISL_2942894, EPI_ISL_2942897, EPI_ISL_2942898, EPI_ISL_2942899, EPI_ISL_2942900, EPI_ISL_2942901, EPI_ISL_2942902, EPI_ISL_2942903, EPI_ISL_2942904, EPI_ISL_2942905, EPI_ISL_2942906, EPI_ISL_2942907, EPI_ISL_2942908, EPI_ISL_2942909, EPI_ISL_2942914, EPI_ISL_2942916, EPI_ISL_2942917, EPI_ISL_2942918, EPI_ISL_2942919, EPI_ISL_2942921, EPI_ISL_2942922, EPI_ISL_2942923, EPI_ISL_2942924, EPI_ISL_2942925, EPI_ISL_2942926, EPI_ISL_2942927, EPI_ISL_2942928, EPI_ISL_2942930, EPI_ISL_2942931, EPI_ISL_2942935

| | | | |
|---|---|---|---|
| see above | Laboratorio Central de Epidemiologia (LCE) | Unidad de Genomica Avanzada | ; Alejandra Garcia-Gasca; Alejandra Hernandez-Teran; Alejandro Sanchez-Flores; Alfredo Herrera-Estrella; Alicia Ocaña-Mondragon; Andreu Comas-Garcia; Angel Gustavo Salas-Lais; Antonio Loza Roman; Bernardo Martinez-Miguel; Blanca Taboada; Brenda Irasema Maldonado-Meza; Bruno Gomez-Gil; Carla Ivon Herrera-Najera; Carlos F. Arias; Celia Boukadida; Celida Duque Molina; Celida Martinez- Rodriguez; Clara Esperanza Santacruz-Tinoco; Concepcion Grajales-Muñiz; Consorcio Mexicano de Vigilancia Genomica (CoViGen-Mex). Authors (in alphabetical order): Julio Elias Alvarado-Yaah; Cristobal Chaidez-Quiroz; Daniel Fregoso-Rueda; Daniel Lira Morales; Eduardo Becerril-Vargas; Fernando Fontove-Herrera; Fidencio Mejia-Nepomuceno; Francisco Pulido; Gloria Elena Espinosa-Ayala; Gloria Maria Molina-Salinas; Gloria Vazquez; Hector Esteban Paz-Juarez; Hector Montoya-Fuentes; Helen Haydee Fernanda Ramirez-Plascencia; Irvin Gonzalez-Lopez; Jean Pierre Gonzalez; Jesus Hernandez; Joel Armando Vazquez-Perez.; Jorge Salas-Hernandez; Jose Antonio Enciso-Moreno; Jose Arturo Martinez-Orozco; Jose Esteban Muñoz-Medina; Jose de Jesus Nuñez-Contreras; Juan Bautista Chale-Dzul; Julissa Enciso-Ibarra; Luis Alberto Ochoa-Carrera; Margarita Matias-Florentino; Maria Guadalupe Santiago-Mauricio; Maria Guadalupe de Jesus Mireles-Rivera; Mario Mujica-Sanchez; Marissa Perez-Garcia; Nelly Selem-Mojica; Pavel Isa; Ricardo Ciria Merce; Ricardo Grande; Rosa Maria Gutierrez Rios; Santiago avila-Rios; Selene Zarate; Susana Lopez; Veronica Mata-Haro; Victor Eduardo Garcia-Arias; Victor Hugo Borja-Aburto |

EPI_ISL_2671485, EPI_ISL_2671486, EPI_ISL_2671487, EPI_ISL_2671488, EPI_ISL_2671489, EPI_ISL_2671490, EPI_ISL_2671491, EPI_ISL_2671492, EPI_ISL_2671526, EPI_ISL_2671527, EPI_ISL_2671528, EPI_ISL_2671529, EPI_ISL_2671530, EPI_ISL_2671531, EPI_ISL_2671532, EPI_ISL_2671533, EPI_ISL_2671534, EPI_ISL_2671535, EPI_ISL_2671536, EPI_ISL_2671537, EPI_ISL_2671538, EPI_ISL_2671539, EPI_ISL_2671540, EPI_ISL_2671541, EPI_ISL_2671542, EPI_ISL_2671561, EPI_ISL_2671562, EPI_ISL_2671563, EPI_ISL_2671569, EPI_ISL_2671587, EPI_ISL_2671588, EPI_ISL_2671589, EPI_ISL_2671590, EPI_ISL_2671591, EPI_ISL_2671592, EPI_ISL_2671593, EPI_ISL_2671594, EPI_ISL_2671595, EPI_ISL_2671596, EPI_ISL_2671597, EPI_ISL_2671598, EPI_ISL_2671599, EPI_ISL_2671600, EPI_ISL_2671601, EPI_ISL_2671602, EPI_ISL_2671603, EPI_ISL_2671605, EPI_ISL_2671606, EPI_ISL_2671607, EPI_ISL_2671613, EPI_ISL_2671615, EPI_ISL_2671637, EPI_ISL_2671638, EPI_ISL_2671639, EPI_ISL_2671640, EPI_ISL_2671641, EPI_ISL_2671642, EPI_ISL_2671643, EPI_ISL_2671644, EPI_ISL_2671645, EPI_ISL_2671646, EPI_ISL_2671647, EPI_ISL_2671648, EPI_ISL_2671649, EPI_ISL_2671650, EPI_ISL_2671651, EPI_ISL_2671652, EPI_ISL_2671723, EPI_ISL_2671724, EPI_ISL_2671725, EPI_ISL_2671726, EPI_ISL_2671727, EPI_ISL_2671728, EPI_ISL_2671729, EPI_ISL_2671730, EPI_ISL_2671731, EPI_ISL_2671732, EPI_ISL_2671733, EPI_ISL_2671734, EPI_ISL_2671735, EPI_ISL_2671736, EPI_ISL_2671737, EPI_ISL_2671738, EPI_ISL_2671739, EPI_ISL_2671740, EPI_ISL_2671741, EPI_ISL_2671742, EPI_ISL_2671743, EPI_ISL_2671744, EPI_ISL_2671745

| | | | |
|---|---|---|---|
| see above | Laboratorio Central de Epidemiologia (LCE) | Unidad de Genomica Avanzada | ; Alejandra Garcia-Gasca; Alejandra Hernandez-Teran; Alejandro Sanchez-Flores; Alfredo Herrera-Estrella; Alicia Ocaña-Mondragon; Andreu Comas-Garcia; Angel Gustavo Salas-Lais; Antonio Loza Roman; Bernardo Martinez-Miguel; Blanca Taboada; Brenda Irasema Maldonado-Meza; Bruno Gomez-Gil; Carla Ivon Herrera-Najera; Carlos F. Arias; Celia Boukadida; Celida Duque Molina; Celida Martinez- Rodriguez; Clara Esperanza Santacruz-Tinoco; Concepcion Grajales-Muñiz; Consorcio Mexicano de Vigilancia Genomica (CoViGen-Mex). Authors (in alphabetical order): Julio Elias Alvarado-Yaah; Cristobal Chaidez-Quiroz; Daniel Fregoso-Rueda; Daniel Lira Morales; Eduardo Becerril-Vargas; Fernando Fontove-Herrera; Fidencio Mejia-Nepomuceno; Francisco Pulido; Gloria Elena Espinosa-Ayala; Gloria Maria Molina-Salinas; Gloria Vazquez; Hector Esteban Paz-Juarez; Hector Montoya-Fuentes; Helen Haydee Fernanda Ramirez-Plascencia; Irvin Gonzalez-Lopez; Jean Pierre Gonzalez; Jesus Hernandez; Joel Armando Vazquez-Perez.; Jorge Salas-Hernandez; Jose Antonio Enciso-Moreno; Jose Arturo Martinez-Orozco; Jose Esteban Muñoz-Medina; Jose de Jesus Nuñez-Contreras; Juan Bautista Chale-Dzul; Julissa Enciso-Ibarra; Luis Alberto Ochoa-Carrera; Margarita Matias-Florentino; Maria Guadalupe Santiago-Mauricio; Maria Guadalupe de Jesus Mireles-Rivera; Mario Mujica-Sanchez; Marissa Perez-Garcia; Nelly Selem-Mojica; Pavel Isa; Ricardo Ciria Merce; Ricardo Grande; Rosa Maria Gutierrez Rios; Santiago avila-Rios; Selene Zarate; Susana Lopez; Veronica Mata-Haro; Victor Eduardo Garcia-Arias; Victor Hugo Borja-Aburto |

EPI_ISL_2490336, EPI_ISL_2490349, EPI_ISL_2490350, EPI_ISL_2490351, EPI_ISL_2490352, EPI_ISL_2490353, EPI_ISL_2490354, EPI_ISL_2490355, EPI_ISL_2490387, EPI_ISL_2490388, EPI_ISL_2490389, EPI_ISL_2490390, EPI_ISL_2490391, EPI_ISL_2490392, EPI_ISL_2490393, EPI_ISL_2490396, EPI_ISL_2490402, EPI_ISL_2490403, EPI_ISL_2490407, EPI_ISL_2490408, EPI_ISL_2490412, EPI_ISL_2490413, EPI_ISL_2490414, EPI_ISL_2490415, EPI_ISL_2490416, EPI_ISL_2490417, EPI_ISL_2490423, EPI_ISL_2490425, EPI_ISL_2490433, EPI_ISL_2490434, EPI_ISL_2490435, EPI_ISL_2490436, EPI_ISL_2490437, EPI_ISL_2490438, EPI_ISL_2490542, EPI_ISL_2490543, EPI_ISL_2490544, EPI_ISL_2490545, EPI_ISL_2490546, EPI_ISL_2490547, EPI_ISL_2490548, EPI_ISL_2490549, EPI_ISL_2490550, EPI_ISL_2490551, EPI_ISL_2490552, EPI_ISL_2490553, EPI_ISL_2490554, EPI_ISL_2490555, EPI_ISL_2490556, EPI_ISL_2490557, EPI_ISL_2490558, EPI_ISL_2490559, EPI_ISL_2490560, EPI_ISL_2490561, EPI_ISL_2490562, EPI_ISL_2490563, EPI_ISL_2490564, EPI_ISL_2490565, EPI_ISL_2490566, EPI_ISL_2490567, EPI_ISL_2490568, EPI_ISL_2490569, EPI_ISL_2490570, EPI_ISL_2490571, EPI_ISL_2490572, EPI_ISL_2490573, EPI_ISL_2490574, EPI_ISL_2490575, EPI_ISL_2490576, EPI_ISL_2490577, EPI_ISL_2490578, EPI_ISL_2490579, EPI_ISL_2490580, EPI_ISL_2490581, EPI_ISL_2490582, EPI_ISL_2490583, EPI_ISL_2490584, EPI_ISL_2835946, EPI_ISL_2835947, EPI_ISL_2835948, EPI_ISL_2835949, EPI_ISL_2835950, EPI_ISL_2835951

| | | | |
|---|---|---|---|
| see above | Laboratorio Central de Epidemiología (LCE) | Centro de Investigación en Enfermedades Infecciosas (CIENI), Instituto Nacional de Enfermedades Respiratorias (INER) | ; Alejandra Garcia-Gasca; Alejandra Hernández-Terán; Alejandro Sanchez-Flores; Alfredo Herrera-Estrella; Alicia Ocaña-Mondragón; Andreu Comas-García; Angel Gustavo Salas-Lais; Antonio Loza Román; Bernardo Martínez-Miguel; Blanca Taboada; Brenda Irasema Maldonado-Meza; Bruno Gomez-Gil; Carla Ivón Herrera-Najera; Carlos F. Arias; Celia Boukadida; Clara Esperanza Santacruz-Tinoco; Concepción Grajales-Muñiz; Consorcio Mexicano de Vigilancia Genómica (CoViGen-Mex). Authors (in alphabetical order): Julio Elías Alvarado-Yaah; Cristóbal Cháidez-Quiróz; Célida Duque-Molina; Célida Martínez- Rodríguez; Daniel Fregoso-Rueda; Daniel Lira Morales; Eduardo Becerril-Vargas; Fernando Fontove-Herrera; Fidencio Mejía-Nepomuceno; Francisco Pulido; Gloria Elena Espinosa-Ayala; Gloria María Molina-Salinas; Gloria Vazquez; Hector Esteban Paz-Juárez; Hector Montoya-Fuentes; Helen Haydee Fernanda Ramírez-Plascencia; Irvin González-López; Jean Pierre González; Joel Armando Vázquez-Pérez.; Jorge Salas-Hernández; José Antonio Enciso-Moreno; José Arturo Martínez-Orozco; José Esteban Muñoz-Medina; José de Jesús Nuñez-Contreras; Juan Bautista Chale-Dzul; Julissa Enciso-Ibarra; Luis Alberto Ochoa-Carrera; Margarita Matías-Florentino; Mario Mújica-Sánchez; Marissa Perez-Garcia; María Guadalupe Santiago-Mauricio; María Guadalupe de Jesús Mireles-Rivera; Nelly Sélem-Mojica; Pavel Isa; Ricardo Ciria Merce; Ricardo Grande; Rosa María Gutierrez Rios; Santiago Ávila-Ríos; Selene Zárate; Susana Lopez; Victor Eduardo Garcia-Arias; Víctor Hugo Borja-Aburto |

EPI_ISL_2391508, EPI_ISL_2391515, EPI_ISL_2391516, EPI_ISL_2391517, EPI_ISL_2391519, EPI_ISL_2391520, EPI_ISL_2391521, EPI_ISL_2391522, EPI_ISL_2391530, EPI_ISL_2391541, EPI_ISL_2391543, EPI_ISL_2391545, EPI_ISL_2391549, EPI_ISL_2391556, EPI_ISL_2391557, EPI_ISL_2391558, EPI_ISL_2391559, EPI_ISL_2391560, EPI_ISL_2391563, EPI_ISL_2391564, EPI_ISL_2391567, EPI_ISL_2391579, EPI_ISL_2391580, EPI_ISL_2391582, EPI_ISL_2391586, EPI_ISL_2391587, EPI_ISL_2391594, EPI_ISL_2391595, EPI_ISL_2391600, EPI_ISL_2391609, EPI_ISL_2391611, EPI_ISL_2391617, EPI_ISL_2391618, EPI_ISL_2391620, EPI_ISL_2391621, EPI_ISL_2391622, EPI_ISL_2391623, EPI_ISL_2391628, EPI_ISL_2391629, EPI_ISL_2391630, EPI_ISL_2391634, EPI_ISL_2391635, EPI_ISL_2391636, EPI_ISL_2391637, EPI_ISL_2391638, EPI_ISL_2391640, EPI_ISL_2391641, EPI_ISL_2391643, EPI_ISL_2391644, EPI_ISL_2391646, EPI_ISL_2391647, EPI_ISL_2391652, EPI_ISL_2391653, EPI_ISL_2391654, EPI_ISL_2391658, EPI_ISL_2391659, EPI_ISL_2391662, EPI_ISL_2391670, EPI_ISL_2391676, EPI_ISL_2391677, EPI_ISL_2391678, EPI_ISL_2391681, EPI_ISL_2391682, EPI_ISL_2391687, EPI_ISL_2391692, EPI_ISL_2391693, EPI_ISL_2391699, EPI_ISL_2391700, EPI_ISL_2391701, EPI_ISL_2391703, EPI_ISL_2391708, EPI_ISL_2391716, EPI_ISL_2391722, EPI_ISL_2391724, EPI_ISL_2391725, EPI_ISL_2391729, EPI_ISL_2391731, EPI_ISL_2391734, EPI_ISL_2391737, EPI_ISL_2391740, EPI_ISL_2391741, EPI_ISL_2391742, EPI_ISL_2391743, EPI_ISL_2391748, EPI_ISL_2681046, EPI_ISL_2681051, EPI_ISL_2681054, EPI_ISL_2681056, EPI_ISL_2681058, EPI_ISL_2681059, EPI_ISL_2681060, EPI_ISL_2681061, EPI_ISL_2681062, EPI_ISL_2681063, EPI_ISL_2681064, EPI_ISL_2681065, EPI_ISL_2681066, EPI_ISL_2681069, EPI_ISL_2681070, EPI_ISL_2681071, EPI_ISL_2681072, EPI_ISL_2681073, EPI_ISL_2681076, EPI_ISL_2681081, EPI_ISL_2681087, EPI_ISL_2681093, EPI_ISL_2681105, EPI_ISL_2681111, EPI_ISL_2681112, EPI_ISL_2681113, EPI_ISL_2681124, EPI_ISL_2681125, EPI_ISL_2681128, EPI_ISL_2681129, EPI_ISL_2681131, EPI_ISL_2681132, EPI_ISL_2681134, EPI_ISL_2681135, EPI_ISL_2681138, EPI_ISL_2681139, EPI_ISL_2681140, EPI_ISL_2681143, EPI_ISL_2681144, EPI_ISL_2681145, EPI_ISL_2681146, EPI_ISL_2681147, EPI_ISL_2681148, EPI_ISL_2681149, EPI_ISL_2681150, EPI_ISL_2681151, EPI_ISL_2681152, EPI_ISL_2681153, EPI_ISL_2681154, EPI_ISL_2681155, EPI_ISL_2681156, EPI_ISL_2681157, EPI_ISL_2681158, EPI_ISL_2681159, EPI_ISL_2681161, EPI_ISL_2681162, EPI_ISL_2681163, EPI_ISL_2681164, EPI_ISL_2681165, EPI_ISL_2681166, EPI_ISL_2681167, EPI_ISL_2681168, EPI_ISL_2681169, EPI_ISL_2681170, EPI_ISL_2681171, EPI_ISL_2681172, EPI_ISL_2681173, EPI_ISL_2681174, EPI_ISL_2681175, EPI_ISL_2681177, EPI_ISL_2681178, EPI_ISL_2681179, EPI_ISL_2681180, EPI_ISL_2681181, EPI_ISL_2681182, EPI_ISL_2681183, EPI_ISL_2681184, EPI_ISL_2681185, EPI_ISL_2681186, EPI_ISL_2681187, EPI_ISL_2681188, EPI_ISL_2681189, EPI_ISL_2681191, EPI_ISL_2681192, EPI_ISL_2681193, EPI_ISL_2681194, EPI_ISL_2681195, EPI_ISL_2681196, EPI_ISL_2681198, EPI_ISL_2681199, EPI_ISL_2681200, EPI_ISL_2681202, EPI_ISL_2681203, EPI_ISL_2681204, EPI_ISL_2681205, EPI_ISL_2681206, EPI_ISL_2681208, EPI_ISL_2681210, EPI_ISL_2681213, EPI_ISL_2681214, EPI_ISL_2681215, EPI_ISL_2681216, EPI_ISL_2681217, EPI_ISL_2681220, EPI_ISL_2681222, EPI_ISL_2681224, EPI_ISL_2681225, EPI_ISL_2681226, EPI_ISL_2681229, EPI_ISL_2681230, EPI_ISL_2681231, EPI_ISL_2681237, EPI_ISL_2681238, EPI_ISL_2681240, EPI_ISL_2681243, EPI_ISL_2681245, EPI_ISL_2681247, EPI_ISL_2681249, EPI_ISL_2681250, EPI_ISL_2681251, EPI_ISL_2681252, EPI_ISL_2681253, EPI_ISL_2681254, EPI_ISL_2681255, EPI_ISL_2681256, EPI_ISL_2681257, EPI_ISL_2681258, EPI_ISL_2681259, EPI_ISL_2681262, EPI_ISL_2681263, EPI_ISL_2681264, EPI_ISL_2681266, EPI_ISL_2681267, EPI_ISL_2681268, EPI_ISL_2681269, EPI_ISL_2681270, EPI_ISL_2681273, EPI_ISL_2681276, EPI_ISL_2681278, EPI_ISL_2681279, EPI_ISL_2681280, EPI_ISL_2681281, EPI_ISL_2681282, EPI_ISL_2681283, EPI_ISL_2681284, EPI_ISL_2681287, EPI_ISL_2681288, EPI_ISL_2681290, EPI_ISL_2681294, EPI_ISL_2681297, EPI_ISL_2681299, EPI_ISL_2681301, EPI_ISL_2681302, EPI_ISL_2681305, EPI_ISL_2681306, EPI_ISL_2681307, EPI_ISL_2681310, EPI_ISL_2681312, EPI_ISL_2681313, EPI_ISL_2681314, EPI_ISL_2681318, EPI_ISL_2681321, EPI_ISL_2681323, EPI_ISL_2681329, EPI_ISL_2681330, EPI_ISL_2681332, EPI_ISL_2681334, EPI_ISL_2681335, EPI_ISL_2681337, EPI_ISL_2681343, EPI_ISL_2681344, EPI_ISL_2681347, EPI_ISL_2681350, EPI_ISL_2681352, EPI_ISL_2681369, EPI_ISL_2681371, EPI_ISL_2681373, EPI_ISL_2681386, EPI_ISL_2681387, EPI_ISL_2681388, EPI_ISL_2801753, EPI_ISL_2801754, EPI_ISL_2801755, EPI_ISL_2801756, EPI_ISL_2801757, EPI_ISL_2801758, EPI_ISL_2801759, EPI_ISL_2801760, EPI_ISL_2801761, EPI_ISL_2801762, EPI_ISL_2801763, EPI_ISL_2801764, EPI_ISL_2801765, EPI_ISL_2801766, EPI_ISL_2801767, EPI_ISL_2801768, EPI_ISL_2801769, EPI_ISL_2801770, EPI_ISL_2801771, EPI_ISL_2801772, EPI_ISL_2801773, EPI_ISL_2801774, EPI_ISL_2801775, EPI_ISL_2801776, EPI_ISL_2801777, EPI_ISL_2801778, EPI_ISL_2801779, EPI_ISL_2801780, EPI_ISL_2801781, EPI_ISL_2801782, EPI_ISL_2801783, EPI_ISL_2801784, EPI_ISL_2801785, EPI_ISL_2801786, EPI_ISL_2801787, EPI_ISL_2801788, EPI_ISL_2801789, EPI_ISL_2801790, EPI_ISL_2801791, EPI_ISL_2801792, EPI_ISL_2801793, EPI_ISL_2801794, EPI_ISL_2801795, EPI_ISL_2801796, EPI_ISL_2801797, EPI_ISL_2801798, EPI_ISL_2801799, EPI_ISL_2801800, EPI_ISL_2801801, EPI_ISL_2801802, EPI_ISL_2801803, EPI_ISL_2801804, EPI_ISL_2801805, EPI_ISL_2801806, EPI_ISL_2801807, EPI_ISL_2801808, EPI_ISL_2801809, EPI_ISL_2801810, EPI_ISL_2801811, EPI_ISL_2801812, EPI_ISL_2801813, EPI_ISL_2801814, EPI_ISL_2801815, EPI_ISL_2801816, EPI_ISL_2801817, EPI_ISL_2801818, EPI_ISL_2801819, EPI_ISL_2801820, EPI_ISL_2801821, EPI_ISL_2801822, EPI_ISL_2801823, EPI_ISL_2801824, EPI_ISL_2801825, EPI_ISL_2801826, EPI_ISL_2801827, EPI_ISL_2801828, EPI_ISL_2801829, EPI_ISL_2801830, EPI_ISL_2801831, EPI_ISL_2801832, EPI_ISL_2801833, EPI_ISL_2801834, EPI_ISL_2801835, EPI_ISL_2801836, EPI_ISL_2801837, EPI_ISL_2801838, EPI_ISL_2801839, EPI_ISL_2801840, EPI_ISL_2801841, EPI_ISL_2801842, EPI_ISL_2801843, EPI_ISL_2801844, EPI_ISL_2801845, EPI_ISL_2801846, EPI_ISL_2801847, EPI_ISL_2801848, EPI_ISL_2801849, EPI_ISL_2801850, EPI_ISL_2801851, EPI_ISL_2801852, EPI_ISL_2801853, EPI_ISL_2801854, EPI_ISL_2801855, EPI_ISL_2801856, EPI_ISL_2801857, EPI_ISL_2801858, EPI_ISL_2801859, EPI_ISL_2801860, EPI_ISL_2801861, EPI_ISL_2801862, EPI_ISL_2801863, EPI_ISL_2801864, EPI_ISL_2801865, EPI_ISL_2801866, EPI_ISL_2801867, EPI_ISL_2801868, EPI_ISL_2801869, EPI_ISL_2801870, EPI_ISL_2801871, EPI_ISL_2801872, EPI_ISL_2801873, EPI_ISL_2801874, EPI_ISL_2801875, EPI_ISL_2801876, EPI_ISL_2801877, EPI_ISL_2801878, EPI_ISL_2801879, EPI_ISL_2801880, EPI_ISL_2801881, EPI_ISL_2801882, EPI_ISL_2801883, EPI_ISL_2801884, EPI_ISL_2801885, EPI_ISL_2801886, EPI_ISL_2801887, EPI_ISL_2801888, EPI_ISL_2801889, EPI_ISL_2801890, EPI_ISL_2801891

| | | | |
|---|---|---|---|
| see above | Laboratorio Central de | Instituto de | ; Alejandra Garcia-Gasca; Alejandra García-Gasca; Alejandra Hernández-Terán; Alejandro Sanchez-Flores; Alejandro Sánchez-Flores; Alfredo Herrera-Estrella; Alicia Ocaña-Mondragón; Andreu Comas-García; Angel Gustavo Salas-Lais; Antonio Loza Román; Bernardo Martínez-Miguel; Blanca Taboada; Brenda Irasema Maldonado-Meza; |

| | | | |
|---|---|---|---|
| | Epidemiología (LCE) | Biotecnología de la UNAM | Bruno Gomez-Gil; Bruno Gómez-Gil; Carla Ivón Herrera-Najera; Carlos F. Arias; Celia Boukadida; Clara Esperanza Santacruz-Tinoco; Concepción Grajales-Muñiz; Consorcio Mexicano de Vigilancia Genómica (CoViGen-Mex). Authors (in alphabetical order): Julio Elias Alvarado-Yaah; Cristóbal Cháidez-Quiróz; Célida Duque Molina; Célida Martínez- Rodríguez; Daniel Fregoso-Rueda; Daniel Lira Morales; Eduardo Becerril-Vargas; Fernando Fontove-Herrera; Fidencio Mejía-Nepomuceno; Francisco Pulido; Gloria Elena Espinosa-Ayala; Gloria María Molina-Salinas; Gloria Vazquez; Hector Esteban Paz-Juárez; Hector Montoya-Fuentes; Helen Haydee Fernanda Ramírez-Plascencia; Irvin González-López; Jean Pierre González; Jesús Hernández; Joel Armando Vázquez-Pérez.; Jorge Salas-Hernández; José Antonio Enciso-Moreno; José Arturo Martínez-Orozco; José Esteban Muñoz-Medina; José de Jesús Nuñez-Contreras; Juan Bautista Chale-Dzul; Julissa Enciso-Ibarra; Luis Alberto Ochoa-Carrera; Margarita Matías-Florentino; Mario Mújica-Sánchez; Marissa Perez-Garcia; María Guadalupe Santiago-Mauricio; María Guadalupe de Jesús Mireles-Rivera; Nelly Sélem-Mojica; Pavel Isa; Ricardo Ciria Merce; Ricardo Grande; Rosa María Gutierrez Rios; Rosa María Gutiérrez Rios; Santiago Ávila-Ríos; Selene Zárate; Susana Lopez; Verónica Mata-Haro; Victor Eduardo Garcia-Arias; Victor Eduardo García-Arias; Víctor Hugo Borja-Aburto |
| EPI_ISL_2102601, EPI_ISL_2102603, EPI_ISL_2102604, EPI_ISL_2102605, EPI_ISL_2102607, EPI_ISL_2102608, EPI_ISL_2102609, EPI_ISL_2102610, EPI_ISL_2102611, EPI_ISL_2102612, EPI_ISL_2102613, EPI_ISL_2102620, EPI_ISL_2102621, EPI_ISL_2102622, EPI_ISL_2102625, EPI_ISL_2102627, EPI_ISL_2102628, EPI_ISL_2102629, EPI_ISL_2102630, EPI_ISL_2102631, EPI_ISL_2102632, EPI_ISL_2102633, EPI_ISL_2102634, EPI_ISL_2102635, EPI_ISL_2102636, EPI_ISL_2102643, EPI_ISL_2102644, EPI_ISL_2102645, EPI_ISL_2102646, EPI_ISL_2102647, EPI_ISL_2102648, EPI_ISL_2102649, EPI_ISL_2102650, EPI_ISL_2102651, EPI_ISL_2102652, EPI_ISL_2102653, EPI_ISL_2102654, EPI_ISL_2102655, EPI_ISL_2102656, EPI_ISL_2102657, EPI_ISL_2102658, EPI_ISL_2102659, EPI_ISL_2102661, EPI_ISL_2102662, EPI_ISL_2102669, EPI_ISL_2102670, EPI_ISL_2102671, EPI_ISL_2102672, EPI_ISL_2102673, EPI_ISL_2102674, EPI_ISL_2102678, EPI_ISL_2102679 | | | |
| see above | Laboratorio Estatal de Salud Publica de Nuevo Leon | Laboratorio Estatal de Salud Publica de Nuevo Leon | (in alphabetical order) Consuelo Treviño-Garza; Eduardo Isaac de la Rosa-Moreno; Else del Carmen Garcia-Garcia; Gloria Alejandra Jasso-de la Peña; Manuel Enrique de la O-Cavazos; Olín Medina-Chávez; Yulianna Mayre Cordero-Cruz |
| EPI_ISL_2680916, EPI_ISL_2680917, EPI_ISL_2680918, EPI_ISL_2680919, EPI_ISL_2680920, EPI_ISL_2680921, EPI_ISL_2680922, EPI_ISL_2680923, EPI_ISL_2680924, EPI_ISL_2680925, EPI_ISL_2680926, EPI_ISL_2680927, EPI_ISL_2894549, EPI_ISL_2894550, EPI_ISL_2894551, EPI_ISL_2894552, EPI_ISL_2927967, EPI_ISL_2927968, EPI_ISL_2927969 | | | |
| see above | Laboratorios Delia Barraza | Laboratorio nacional-LANIIA-CIAD | ; Alejandra García-Gasca; Alejandra Hernández-Terán; Alejandro Sánchez-Flores; Alfredo Herrera-Estrella; Alicia Ocaña-Mondragón; Andreu Comas-García; Angel Gustavo Salas-Lais; Antonio Loza Román; Bernardo Martínez-Miguel; Blanca Taboada; Brenda Irasema Maldonado-Meza; Bruno Gómez-Gil; Carla Ivón Herrera-Najera; Carlos F. Arias; Celia Boukadida; Clara Esperanza Santacruz-Tinoco; Claudia Soto Félix; Concepción Grajales-Muñiz; Consorcio Mexicano de Vigilancia Genómica (CoViGen-Mex). Authors (in alphabetical order): Julio Elias Alvarado-Yaah; Cristóbal Cháidez-Quiróz; Célida Duque Molina; Célida Martínez- Rodríguez; Daniel Fregoso-Rueda; Daniel Lira Morales; Eduardo Becerril-Vargas; Fernando Fontove-Herrera; Fidencio Mejía-Nepomuceno; Francisco Pulido; Gloria Elena Espinosa-Ayala; Gloria María Molina-Salinas; Gloria Vazquez; Hector Esteban Paz-Juárez; Hector Montoya-Fuentes; Helen Haydee Fernanda Ramírez-Plascencia; Irvin González-López; Jean Pierre González; Jesús Hernández; Joel Armando Vázquez-Pérez. Laboratorios Delia Barraza: María Delia Barraza Sámano; Jorge Salas-Hernández; José Antonio Enciso-Moreno; José Arturo Martínez-Orozco; José Esteban Muñoz-Medina; José de Jesús Nuñez-Contreras; Juan Bautista Chale-Dzul; Julissa Enciso-Ibarra; Luis Alberto Ochoa-Carrera; Margarita Matías-Florentino; Mario Mújica-Sánchez; Marissa Perez-Garcia; María Guadalupe Santiago-Mauricio; María Guadalupe de Jesús Mireles-Rivera; Melissa García Angulo.; Nelly Sélem-Mojica; Pavel Isa; Ricardo Ciria Merce; Ricardo Grande; Rosa María Gutiérrez Rios; Santiago Ávila-Ríos; Selene Zárate; Susana Lopez; Verónica Mata-Haro; Victor Eduardo García-Arias; Víctor Hugo Borja-Aburto |
| EPI_ISL_2046706, EPI_ISL_2046730, EPI_ISL_2046738, EPI_ISL_2046739, EPI_ISL_2046740, EPI_ISL_2046741, EPI_ISL_2046751, EPI_ISL_2046759, EPI_ISL_2046760, EPI_ISL_2046816, EPI_ISL_2046817, EPI_ISL_2046818, EPI_ISL_2046819, EPI_ISL_2184886, EPI_ISL_2184887, EPI_ISL_2184905, EPI_ISL_2184928, EPI_ISL_2184929, EPI_ISL_2184930, EPI_ISL_2184932, EPI_ISL_2185069, EPI_ISL_2185070, EPI_ISL_2185114, EPI_ISL_2185116, EPI_ISL_2185120, EPI_ISL_2185121, EPI_ISL_2185167, EPI_ISL_2185172, EPI_ISL_2185208, EPI_ISL_2185210, EPI_ISL_2185221, EPI_ISL_2185222, EPI_ISL_2185271, EPI_ISL_2185272, EPI_ISL_2185273, EPI_ISL_2185397, EPI_ISL_2185435, EPI_ISL_2185438, EPI_ISL_2185439, EPI_ISL_2185441, EPI_ISL_2185460, EPI_ISL_2185475, EPI_ISL_2185476, EPI_ISL_2185478, EPI_ISL_2185479, EPI_ISL_2185480, EPI_ISL_2185481, EPI_ISL_2185482, EPI_ISL_2185483, EPI_ISL_2185489, EPI_ISL_2185490, EPI_ISL_2185491, EPI_ISL_2304897, EPI_ISL_2304898, EPI_ISL_2304899, EPI_ISL_2304900, EPI_ISL_2304901, EPI_ISL_2304949, EPI_ISL_2304950, EPI_ISL_2304952, EPI_ISL_2304953, EPI_ISL_2304954, EPI_ISL_2304955, EPI_ISL_2304956, EPI_ISL_2304957, EPI_ISL_2304958, EPI_ISL_2305044, EPI_ISL_2305077, EPI_ISL_2305078, EPI_ISL_2305079, EPI_ISL_2305080, EPI_ISL_2305174, EPI_ISL_2305175, EPI_ISL_2305191, EPI_ISL_2305193, EPI_ISL_2305194, EPI_ISL_2305276, EPI_ISL_2305279, EPI_ISL_2305338, EPI_ISL_2305339, EPI_ISL_2305382, EPI_ISL_2305385, EPI_ISL_2305387, EPI_ISL_2305416, EPI_ISL_2305450, EPI_ISL_2305470, EPI_ISL_2305471, EPI_ISL_2305472, EPI_ISL_2305483, EPI_ISL_2305521, EPI_ISL_2305564, EPI_ISL_2305565, EPI_ISL_2305580, EPI_ISL_2397849, EPI_ISL_2397850, EPI_ISL_2397864, EPI_ISL_2397866, EPI_ISL_2397867, EPI_ISL_2397868, EPI_ISL_2397869, EPI_ISL_2397870, EPI_ISL_2397965, EPI_ISL_2398014, EPI_ISL_2398024, EPI_ISL_2481566, EPI_ISL_2481567, EPI_ISL_2481571, EPI_ISL_2481606, EPI_ISL_2481607, EPI_ISL_2481612, EPI_ISL_2481626, EPI_ISL_2481628, EPI_ISL_2481630, EPI_ISL_2481631, EPI_ISL_2481695, EPI_ISL_2481697, EPI_ISL_2481698, EPI_ISL_2481740, EPI_ISL_2481746, EPI_ISL_2481751, EPI_ISL_2481752, EPI_ISL_2481824, EPI_ISL_2481826, EPI_ISL_2481832, EPI_ISL_2481833, EPI_ISL_2481834, EPI_ISL_2481835, EPI_ISL_2481926, EPI_ISL_2481984, EPI_ISL_2481985, EPI_ISL_2482020, EPI_ISL_2482115, EPI_ISL_2482116, EPI_ISL_2482148, EPI_ISL_2482219, EPI_ISL_2873152, EPI_ISL_2873199, EPI_ISL_2873200, EPI_ISL_2873201, EPI_ISL_2873202, EPI_ISL_2873287, EPI_ISL_2873288, EPI_ISL_2926674, EPI_ISL_2926761, EPI_ISL_2926780, EPI_ISL_2926781, EPI_ISL_2926794, EPI_ISL_2926795, EPI_ISL_2926873, EPI_ISL_2926874, EPI_ISL_2927045, EPI_ISL_2927046, EPI_ISL_2927119, EPI_ISL_2927120, EPI_ISL_2927228, EPI_ISL_2927229, EPI_ISL_2927230, EPI_ISL_2927304, EPI_ISL_2927305, EPI_ISL_2927306 | | | |
| see above | Laboratory Corporation of America | Centers for Disease Control and Prevention Division of Viral Diseases, Pathogen Discovery | Adrian Paskey; Amanda Douglas; Amanda Suchanek; Andrea Throop; Ayla Burns; Benjamin Rambo-Martin; Bobbi Croy; Brian Krueger; Brian Norvell; Christopher Gulvick; Christos Petropoulos; Clinton R. Paden; Craig Lukasik; Dakota Howard; Darlene Wagner; Debbie Boles; Dhwani Batra; Duncan MacCannell; Eyad Almasri; Goran Stevovic; Howard Engler; Hrushikesh Deshmukh; Jake Humphrey; Jana Schroth; Jason Caravas; Joe Voshell; John Pruitt; Jonathan Meltzer; Jonathan Williams; Kara Moser; Kimberly Wagner; Lax Iyer; Lisa Pfefferle; Lyndon Tilson; Manoj Jain; Marcia Eisenberg; Mary Ann Cristobal; Mary Williamson; Matthew Robinson; Matthew Schmerer; Michael Levandoski; Mike Sapeta; Mindy Nye; Minoo Agarwal; Mohan Kolli; Nuthawin Charoensri; Oren Cohen; Peter W. Cook; Prashant Gupta; Qian Zeng; Rama Ghatti; Scott Parker; Scott Ryan; Scott Sammons; Shatavia Morrison; Stanley Letovsky; Steven Ragan; Suresh Babu Selvaraju; Susan Countryman; Susan Hicks; Suzanne Dale; Thomas Urban; Tim Kuphal; Tricia Zwiefelhofer; Vincent Drouillon; Yvette Unoarumhi |
| EPI_ISL_2696448, EPI_ISL_3025611 | Laboratory, Gravity Diagnostics | Laboratory, Gravity Diagnostics | Diagnostics, G. |
| EPI_ISL_2533802 | Lesp Jalisco /Unidad de Patologia | Instituto de Diagnostico y Referencia Epidemiologicos (INDRE) | Abril Rodriguez-Maldonado; Ariadna Medina-Benitez; Claudia Wong-Arambula; Ernesto Ramirez-Gonzalez.; Gisela Barrera-Badillo; Irma Lopez-Martinez; Joaquin Quiroz-Mercado; Lucia Hernandez-Rivas; Natividad Cruz-Ortiz; Sergio Rangel-Guerrero; Tatiana Nunez-Garcia; Vanessa Rivero-Arredondo |
| EPI_ISL_2724009 | Molecular Biology Laboratory, Infectious Diseases Department, Instituto Nacional de Ciencias Medicas y Nutricion Salvador Zubiran | Instituto Nacional de Ciencias Medicas y Nutricion Infectious Diseases | Arturo Galindo; Eric Ochoa; Fernando Ledesma Barrientos; Guillermo Ruiz-Palacios y Santos; Patricia Leal; Pilar Ramos Cervantes; Violeta Ibarra Gonzalez |
| EPI_ISL_2384037, EPI_ISL_2384038, EPI_ISL_2384039, EPI_ISL_2384040, EPI_ISL_2384041, EPI_ISL_2384042, EPI_ISL_2384043, EPI_ISL_2384044, EPI_ISL_2384045, EPI_ISL_2384046, EPI_ISL_2452099, EPI_ISL_2452100, EPI_ISL_2452102, EPI_ISL_2452103, EPI_ISL_2452105, EPI_ISL_2452107, EPI_ISL_2452108 | | | |
| see above | NM Dept. Health, Scientific Laboratory Division | Centers for Disease Control and Prevention Division of Viral Diseases, Pathogen Discovery | Alison Laufer Halpin; Ben L. Rambo-Martin; Clinton R. Paden; Dakota Howard; Darlene Wagner; Dave Wentworth; Dhwani Batra; Jasmine Padilla; Justin Lee; Katie Dillon; Krista Queen; Kristen Knipe; Kristine Lacek; Mark Burroughs; Matthew Schmerer; Mili Sheth; Peter Cook; Sam Shepard; Sarah Nobles; Shoshona Le; Suxiang Tong; Vivien Dugan; Yvette Unoarumhi |
| EPI_ISL_2562732, EPI_ISL_2562733, EPI_ISL_2689602 | NM Dept. Health, Scientific Laboratory Division | Genomics and Discovery, Respiratory Viruses Branch, Division of Viral Diseases, Centers for Disease Control and Prevention | Adam Retchless; Anna Kelleher; Anna Uehara; Brian Lynch; Clinton R. Paden; Dhwani Batra; Haibin Wang; Han Jia Justin Ng; Jasmine Padilla; Jing Zhang; Justin Lee; Krista Queen; Mark Burroughs; Mili Sheth; Morgan Davis; Peter Cook; Rachel Marine; Sarah Nobles; Suxiang Tong; Tara Coalter; Yan Li; Ying Tao |
| EPI_ISL_2598135, EPI_ISL_2598138 | New Mexico Department of Health Scientific Laboratory | Center for Global Health, University of New Mexico Health Sciences Center | Anastacia Griego; Darrell Dinwiddie; Daryl Domman; Joseph Hicks; Kurt Schwalm; Michael Edwards; Twila Kunde; Valerie Morley |
| EPI_ISL_2211365, EPI_ISL_2211370, EPI_ISL_2211376, EPI_ISL_2211380, EPI_ISL_2211386, EPI_ISL_2287816, EPI_ISL_2287820, EPI_ISL_2287822, EPI_ISL_2287824, EPI_ISL_2287827, EPI_ISL_2287830, EPI_ISL_2287853, EPI_ISL_2287856, EPI_ISL_2287859, EPI_ISL_2287862, EPI_ISL_2287865, EPI_ISL_2287869, EPI_ISL_2287874, EPI_ISL_2287875, EPI_ISL_2287886, EPI_ISL_2287899, EPI_ISL_2287906, EPI_ISL_2287908, EPI_ISL_2287912, EPI_ISL_2287915, EPI_ISL_2287918, EPI_ISL_2287921, EPI_ISL_2287933, EPI_ISL_2287943, EPI_ISL_2322800, EPI_ISL_2322801, EPI_ISL_2322802, EPI_ISL_2322803, EPI_ISL_2322804, EPI_ISL_2322805, EPI_ISL_2322806, EPI_ISL_2322807, EPI_ISL_2322808, EPI_ISL_2322809, EPI_ISL_2322810, EPI_ISL_2322812, EPI_ISL_2322813, EPI_ISL_2322814, EPI_ISL_2322815, EPI_ISL_2322816, EPI_ISL_2322817, EPI_ISL_2322818, EPI_ISL_2322819, EPI_ISL_2322820, EPI_ISL_2322821, EPI_ISL_2322822, EPI_ISL_2322823, EPI_ISL_2322824, EPI_ISL_2322825, EPI_ISL_2322826, EPI_ISL_2322827, EPI_ISL_2322828, EPI_ISL_2322829, EPI_ISL_2322830, EPI_ISL_2322831, EPI_ISL_2322832, EPI_ISL_2322833, EPI_ISL_2322834, EPI_ISL_2322835, EPI_ISL_2322836, EPI_ISL_2322837, EPI_ISL_2322838, EPI_ISL_2322839, EPI_ISL_2322840, EPI_ISL_2322843, EPI_ISL_2322845, EPI_ISL_2322846, EPI_ISL_2322847, EPI_ISL_2322848, EPI_ISL_2322849, EPI_ISL_2322850, EPI_ISL_2322851, EPI_ISL_2322852, EPI_ISL_2322853, EPI_ISL_2322854, EPI_ISL_2322855, EPI_ISL_2322856, EPI_ISL_2322857, EPI_ISL_2322858, EPI_ISL_2381902, EPI_ISL_2381903, EPI_ISL_2381904, EPI_ISL_2381905, EPI_ISL_2381906, EPI_ISL_2381907, EPI_ISL_2381908, EPI_ISL_2381909, EPI_ISL_2381910, EPI_ISL_2381911, EPI_ISL_2381912, EPI_ISL_2381913, EPI_ISL_2381914, EPI_ISL_2381915, EPI_ISL_2381916, EPI_ISL_2381917, EPI_ISL_2381918, EPI_ISL_2381919, EPI_ISL_2381920, EPI_ISL_2381921, EPI_ISL_2381922, EPI_ISL_2381923, EPI_ISL_2381924, EPI_ISL_2381925, EPI_ISL_2381926, EPI_ISL_2381927, EPI_ISL_2381928, EPI_ISL_2381929, EPI_ISL_2381930, EPI_ISL_2381931, EPI_ISL_2381932, EPI_ISL_2381933, EPI_ISL_2381934, EPI_ISL_2381935, EPI_ISL_2381936, EPI_ISL_2381937, EPI_ISL_2381938, EPI_ISL_2381939, EPI_ISL_2381940, EPI_ISL_2381941, EPI_ISL_2381942, EPI_ISL_2381943, EPI_ISL_2381944, EPI_ISL_2381945, EPI_ISL_2381946, EPI_ISL_2381947, EPI_ISL_2381948, EPI_ISL_2381949, EPI_ISL_2381950, EPI_ISL_2381951, EPI_ISL_2381952, EPI_ISL_2381953, EPI_ISL_2381954, EPI_ISL_2381955, EPI_ISL_2381956, EPI_ISL_2381957, EPI_ISL_2381958, EPI_ISL_2381959, EPI_ISL_2381960, EPI_ISL_2381961, EPI_ISL_2381962, EPI_ISL_2381963, EPI_ISL_2381964, EPI_ISL_2381965, EPI_ISL_2381966, EPI_ISL_2381967, EPI_ISL_2381968, EPI_ISL_2381969, EPI_ISL_2381970, EPI_ISL_2381971, EPI_ISL_2381972, EPI_ISL_2381973, EPI_ISL_2381974, EPI_ISL_2381975, EPI_ISL_2381976, EPI_ISL_2381977, EPI_ISL_2381978, EPI_ISL_2381979, EPI_ISL_2381980, EPI_ISL_2381981, EPI_ISL_2381982, EPI_ISL_2381983, EPI_ISL_2381984, EPI_ISL_2381985, EPI_ISL_2381986, EPI_ISL_2381987, EPI_ISL_2381988, EPI_ISL_2381989, EPI_ISL_2381990, EPI_ISL_2381991, EPI_ISL_2652009, EPI_ISL_2652010, EPI_ISL_2652011, EPI_ISL_2652012, EPI_ISL_2652013, EPI_ISL_2652014, EPI_ISL_2652015, EPI_ISL_2652016, EPI_ISL_2652017, EPI_ISL_2652018, EPI_ISL_2652019, EPI_ISL_2652020, EPI_ISL_2652021, EPI_ISL_2652022, EPI_ISL_2652023, EPI_ISL_2652024, EPI_ISL_2652025, EPI_ISL_2652026, EPI_ISL_2652027, EPI_ISL_2652028, EPI_ISL_2652029, EPI_ISL_2652030, EPI_ISL_2652031, EPI_ISL_2652032, EPI_ISL_2652033, EPI_ISL_2652034, EPI_ISL_2652035, EPI_ISL_2652036, EPI_ISL_2652037, EPI_ISL_2652038, EPI_ISL_2652039, EPI_ISL_2652040, EPI_ISL_2652041, EPI_ISL_2652042, EPI_ISL_2652043, EPI_ISL_2652044, EPI_ISL_2652045, EPI_ISL_2652046, EPI_ISL_2652047, EPI_ISL_2652048, EPI_ISL_2652050, EPI_ISL_2652053, EPI_ISL_2652056, EPI_ISL_2652060, EPI_ISL_2652062, EPI_ISL_2652065, EPI_ISL_2652067, EPI_ISL_2652068, EPI_ISL_2652069, EPI_ISL_2652070, EPI_ISL_2652071, EPI_ISL_2652072, EPI_ISL_2652073, EPI_ISL_2652074, EPI_ISL_2652075, EPI_ISL_2652076, EPI_ISL_2652077, EPI_ISL_2652078, EPI_ISL_2652079, EPI_ISL_2652080, EPI_ISL_2652081, EPI_ISL_2652082, EPI_ISL_2652083, EPI_ISL_2652084, EPI_ISL_2652085, EPI_ISL_2652086, EPI_ISL_2652087, EPI_ISL_2652088, EPI_ISL_2652089, EPI_ISL_2652090, EPI_ISL_2652091, EPI_ISL_2652092, EPI_ISL_2652093, EPI_ISL_2652094, EPI_ISL_2652095, EPI_ISL_2652096, EPI_ISL_2652097, EPI_ISL_2652098, EPI_ISL_2652099, EPI_ISL_2652100, EPI_ISL_2652101, EPI_ISL_2652102, EPI_ISL_2652103, EPI_ISL_2652104, EPI_ISL_2652105, EPI_ISL_2652106, EPI_ISL_2652107, EPI_ISL_2652108, EPI_ISL_2652109, EPI_ISL_2832491, EPI_ISL_2832492, EPI_ISL_2832493, EPI_ISL_2832494, EPI_ISL_2832495, EPI_ISL_2832496, EPI_ISL_2832497, EPI_ISL_2832498, EPI_ISL_2832499, EPI_ISL_2832500, EPI_ISL_2832501, EPI_ISL_2832502, EPI_ISL_2832503, EPI_ISL_2832504, EPI_ISL_2832506, EPI_ISL_2832508, EPI_ISL_2832509, EPI_ISL_2832510, EPI_ISL_2858897, EPI_ISL_2858898, EPI_ISL_2858899, EPI_ISL_2858900, EPI_ISL_2858904, EPI_ISL_2858905, EPI_ISL_2858906, EPI_ISL_2858907, EPI_ISL_2858908, EPI_ISL_2858909, EPI_ISL_2858910, EPI_ISL_2858911, EPI_ISL_2858912, EPI_ISL_2858913, EPI_ISL_2858914, EPI_ISL_2858915, EPI_ISL_2858916, EPI_ISL_2858918, EPI_ISL_2858919, EPI_ISL_2858920, EPI_ISL_2858921, EPI_ISL_2858922, EPI_ISL_2858923, EPI_ISL_2858924, EPI_ISL_2858925, EPI_ISL_2858926, EPI_ISL_2858927, EPI_ISL_2858928, EPI_ISL_2858929, EPI_ISL_2858934, EPI_ISL_2918751, EPI_ISL_2918752, EPI_ISL_2918753, EPI_ISL_2918754, EPI_ISL_2928040, EPI_ISL_2928041, EPI_ISL_2928042, EPI_ISL_2928043, EPI_ISL_2928044, EPI_ISL_2928088, EPI_ISL_2928089, EPI_ISL_2928090, EPI_ISL_2928091, EPI_ISL_2928092, EPI_ISL_2928116, EPI_ISL_2928117, EPI_ISL_2928118, EPI_ISL_2928119, EPI_ISL_2928120, EPI_ISL_2928121, EPI_ISL_2928122, EPI_ISL_2928123, EPI_ISL_2928124, EPI_ISL_2928125, EPI_ISL_2928126, EPI_ISL_2928127, EPI_ISL_2928128, EPI_ISL_2928129, EPI_ISL_2928130, EPI_ISL_2928131, EPI_ISL_2928132, EPI_ISL_2928133, EPI_ISL_2928134, EPI_ISL_2941641, EPI_ISL_2941642, EPI_ISL_2941643, EPI_ISL_2941644, EPI_ISL_2941645, EPI_ISL_2941646, EPI_ISL_2941647, EPI_ISL_2941648, EPI_ISL_2941649, EPI_ISL_2941650, EPI_ISL_2941651, EPI_ISL_2941652, EPI_ISL_2941653 | | | |
| see above | New Mexico Department of Health Scientific Laboratory | New Mexico Department of Health Scientific Laboratory | ; Anastacia Griego-Fisher; D'eldra Malone; Ellie Johnson; Jennifer Benoit; Keila Gutierrez; Linda Salazar; Ratheesh Rajan |
| EPI_ISL_2566354, EPI_ISL_2566355, EPI_ISL_2566356, EPI_ISL_2566357, EPI_ISL_2566358 | Operation Infectious Diseases, Naval Health Research Center | Operation Infectious Diseases, Naval Health Research Center | A.E.; Balansay-Ames; C.A.; Fierro, M.; Iniguez-Stevens, E.; Kriner, P.; M.S.; Moser, K.; Myers; Pan; Phippard; R.S.; R.W.; Underwood |
| EPI_ISL_2247095, EPI_ISL_2247102, EPI_ISL_2247103, EPI_ISL_2247141, EPI_ISL_2247182, EPI_ISL_2248588, EPI_ISL_2248680, EPI_ISL_2268854, EPI_ISL_2280103, EPI_ISL_2304577, EPI_ISL_2304585, EPI_ISL_2304592, EPI_ISL_2304601, EPI_ISL_2304793, EPI_ISL_2307149, EPI_ISL_2367773, EPI_ISL_2368040, EPI_ISL_2371751, EPI_ISL_2371771, EPI_ISL_2371809, EPI_ISL_2371874, EPI_ISL_2371962, EPI_ISL_2371983, EPI_ISL_2439336, EPI_ISL_2439340, EPI_ISL_2439394, EPI_ISL_2439617, EPI_ISL_2439658, EPI_ISL_2439740, EPI_ISL_2439779, EPI_ISL_2439798, EPI_ISL_2440571, EPI_ISL_2440611, EPI_ISL_2440880, EPI_ISL_2451272, EPI_ISL_2463144, EPI_ISL_2463150, EPI_ISL_2599842, EPI_ISL_2599851, EPI_ISL_2599894, EPI_ISL_2599921, EPI_ISL_2652383, EPI_ISL_2652464, EPI_ISL_2652470, EPI_ISL_2869226, EPI_ISL_2869244, EPI_ISL_2869379, EPI_ISL_2871358, EPI_ISL_2871395, EPI_ISL_2871652, EPI_ISL_2874001, EPI_ISL_2929328 | | | |
| see above | Quest Diagnostics Incorporated | Centers for Disease Control and Prevention Division of Viral Diseases, Pathogen Discovery | A. Gerasimova; A. Perez; Adrian Paskey; B. Anderson; Benjamin Rambo-Martin; Christopher Gulvick; Clinton R. Paden; Dakota Howard; Darlene Wagner; Dhwani Batra; Duncan MacCannell; F. Lacbawan; I. A. Shlyakhter; Jason Caravas; K.E. Livingston; Kara Moser; L.E. Bernstein; M. Hua; Matthew Schmerer; P. Tanpaiboon; Peter W. Cook; R. M. Kagan; R. Owen; R. V. Rolando; S. H. Rosenthal; Scott Sammons; Shatavia Morrison; Y. Liu; Yvette Unoarumhi |
| EPI_ISL_3091581, EPI_ISL_3091631, EPI_ISL_3091692, EPI_ISL_3091697, EPI_ISL_3091748, EPI_ISL_3091829, EPI_ISL_3091873 | | | |
| see above | Respiratory Viruses Branch, Centers for Disease Control and Prevention | Respiratory Viruses Branch, Centers for Disease Control and Prevention | Anderson, B.; Batra, D.; Bernstein, L.; C.R.; Caravas, J.; Cook; Gerasimova, A.; Gulvick, C.; Howard, D.; Hua, M.; I.A.; Kagan; Lacbawan, F.; Liu, Y.; Livingston, K.; MacCannell, D.; Morrison, S.; Moser, K.; Owen, R.; P.W.; Paden; Paskey, A.; Perez, A.; R.M.; R.V.; Rambo-Martin, B.; Rolando; Rosenthal; S.H.; Sammons, S.; Schmerer, M.; Shlyakhter; Tanpaiboon, P.; Unoarumhi, Y.; Wagner, D. |
| EPI_ISL_2105731, EPI_ISL_2105738, EPI_ISL_2105821, EPI_ISL_2105822, EPI_ISL_2105823, EPI_ISL_2105824, EPI_ISL_2105825, EPI_ISL_2105826, EPI_ISL_2105827, EPI_ISL_2105828, EPI_ISL_2105829, EPI_ISL_2105830, EPI_ISL_2105831, EPI_ISL_2105832, EPI_ISL_2105833, EPI_ISL_2105834, EPI_ISL_2105835, EPI_ISL_2105836, EPI_ISL_2105837, EPI_ISL_2105838, EPI_ISL_2105839, EPI_ISL_2105840, | | | |

| | | | |
|---|---|---|---|
| EPI_ISL_2105841, EPI_ISL_2105842, EPI_ISL_2105843, EPI_ISL_2105844, EPI_ISL_2105845, EPI_ISL_2105846, EPI_ISL_2105847, EPI_ISL_2105848, EPI_ISL_2105849, EPI_ISL_2105850, EPI_ISL_2105851, EPI_ISL_2105852, EPI_ISL_2105853, EPI_ISL_2105854, EPI_ISL_2105855, EPI_ISL_2105856, EPI_ISL_2105857, EPI_ISL_2105858, EPI_ISL_2105859, EPI_ISL_2105860, EPI_ISL_2105861, EPI_ISL_2105862, EPI_ISL_2105863, EPI_ISL_2105864, EPI_ISL_2105865, EPI_ISL_2105866, EPI_ISL_2105867, EPI_ISL_2105868, EPI_ISL_2105869, EPI_ISL_2105870, EPI_ISL_2105871, EPI_ISL_2105872, EPI_ISL_2105873, EPI_ISL_2105874, EPI_ISL_2178751, EPI_ISL_2178752, EPI_ISL_2178753, EPI_ISL_2178758, EPI_ISL_2178759, EPI_ISL_2230806, EPI_ISL_2230811, EPI_ISL_2230813, EPI_ISL_2230816, EPI_ISL_2230817, EPI_ISL_2230825, EPI_ISL_2230831, EPI_ISL_2230834, EPI_ISL_2230835, EPI_ISL_2230838, EPI_ISL_2230841, EPI_ISL_2230844, EPI_ISL_2230851, EPI_ISL_2230854, EPI_ISL_2230857, EPI_ISL_2230859, EPI_ISL_2230862, EPI_ISL_2230865, EPI_ISL_2230866, EPI_ISL_2230867, EPI_ISL_2230868, EPI_ISL_2230869, EPI_ISL_2230877, EPI_ISL_2230878, EPI_ISL_2230879, EPI_ISL_2230881, EPI_ISL_2230882, EPI_ISL_2230883, EPI_ISL_2230884, EPI_ISL_2230885, EPI_ISL_2230886, EPI_ISL_2230893, EPI_ISL_2230900, EPI_ISL_2230906, EPI_ISL_2230911, EPI_ISL_2230912, EPI_ISL_2230914, EPI_ISL_2230915, EPI_ISL_2230917, EPI_ISL_2230918, EPI_ISL_2230919, EPI_ISL_2230922, EPI_ISL_2230923, EPI_ISL_2230975, EPI_ISL_2230976, EPI_ISL_2230977, EPI_ISL_2230978, EPI_ISL_2230979, EPI_ISL_2230980, EPI_ISL_2230981, EPI_ISL_2230982, EPI_ISL_2230983, EPI_ISL_2230984, EPI_ISL_2230985, EPI_ISL_2230986, EPI_ISL_2230987, EPI_ISL_2230988, EPI_ISL_2230989, EPI_ISL_2230990, EPI_ISL_2230991, EPI_ISL_2231000, EPI_ISL_2231010, EPI_ISL_2231011, EPI_ISL_2231027, EPI_ISL_2231028, EPI_ISL_2231029, EPI_ISL_2231030, EPI_ISL_2231031, EPI_ISL_2231032, EPI_ISL_2231033, EPI_ISL_2231034, EPI_ISL_2231035, EPI_ISL_2231036, EPI_ISL_2231037, EPI_ISL_2276107, EPI_ISL_2319171, EPI_ISL_2319172, EPI_ISL_2319173, EPI_ISL_2319174, EPI_ISL_2319175, EPI_ISL_2319176, EPI_ISL_2319177, EPI_ISL_2319178, EPI_ISL_2319179, EPI_ISL_2319180, EPI_ISL_2319181, EPI_ISL_2319182, EPI_ISL_2319183, EPI_ISL_2319184, EPI_ISL_2319185, EPI_ISL_2319186, EPI_ISL_2319187, EPI_ISL_2319188, EPI_ISL_2319189, EPI_ISL_2319190, EPI_ISL_2319191, EPI_ISL_2319192, EPI_ISL_2319193, EPI_ISL_2319194, EPI_ISL_2319195, EPI_ISL_2319196, EPI_ISL_2319197, EPI_ISL_2319198, EPI_ISL_2319199, EPI_ISL_2319200, EPI_ISL_2319201, EPI_ISL_2319202, EPI_ISL_2319203, EPI_ISL_2319204, EPI_ISL_2319205, EPI_ISL_2319206, EPI_ISL_2319207, EPI_ISL_2319208, EPI_ISL_2319209, EPI_ISL_2319210, EPI_ISL_2319211, EPI_ISL_2319212, EPI_ISL_2319213, EPI_ISL_2319214, EPI_ISL_2319215, EPI_ISL_2319216, EPI_ISL_2319217, EPI_ISL_2319218, EPI_ISL_2319219, EPI_ISL_2319220, EPI_ISL_2319221, EPI_ISL_2319222, EPI_ISL_2319223, EPI_ISL_2319224, EPI_ISL_2319225, EPI_ISL_2319226, EPI_ISL_2319227, EPI_ISL_2319228, EPI_ISL_2319229, EPI_ISL_2319230, EPI_ISL_2319231, EPI_ISL_2319232, EPI_ISL_2319233, EPI_ISL_2319234, EPI_ISL_2319235, EPI_ISL_2319236, EPI_ISL_2319237, EPI_ISL_2319238, EPI_ISL_2319239, EPI_ISL_2319240, EPI_ISL_2319241, EPI_ISL_2319242, EPI_ISL_2319243, EPI_ISL_2319244, EPI_ISL_2319245, EPI_ISL_2319246, EPI_ISL_2319247, EPI_ISL_2319248, EPI_ISL_2319249, EPI_ISL_2319250, EPI_ISL_2319251, EPI_ISL_2319252, EPI_ISL_2319253, EPI_ISL_2319254, EPI_ISL_2319255, EPI_ISL_2319256, EPI_ISL_2319257, EPI_ISL_2319258, EPI_ISL_2319259, EPI_ISL_2319260, EPI_ISL_2319261, EPI_ISL_2319262, EPI_ISL_2319263, EPI_ISL_2319264, EPI_ISL_2319265, EPI_ISL_2319266, EPI_ISL_2319267, EPI_ISL_2319268, EPI_ISL_2319269, EPI_ISL_2319270, EPI_ISL_2319271, EPI_ISL_2319272, EPI_ISL_2319273, EPI_ISL_2319274, EPI_ISL_2319275, EPI_ISL_2319276, EPI_ISL_2319277, EPI_ISL_2319278, EPI_ISL_2319279, EPI_ISL_2319280, EPI_ISL_2319281, EPI_ISL_2319282, EPI_ISL_2319283, EPI_ISL_2319284, EPI_ISL_2319285, EPI_ISL_2319286, EPI_ISL_2319287, EPI_ISL_2319288, EPI_ISL_2319289, EPI_ISL_2319290, EPI_ISL_2319291, EPI_ISL_2319292, EPI_ISL_2319293, EPI_ISL_2319294, EPI_ISL_2319295, EPI_ISL_2319296, EPI_ISL_2319297, EPI_ISL_2319298, EPI_ISL_2319299, EPI_ISL_2319300, EPI_ISL_2319301, EPI_ISL_2319302, EPI_ISL_2319303, EPI_ISL_2319304, EPI_ISL_2319305, EPI_ISL_2319306, EPI_ISL_2319307, EPI_ISL_2319308, EPI_ISL_2319309, EPI_ISL_2319310, EPI_ISL_2319311, EPI_ISL_2319312, EPI_ISL_2319382, EPI_ISL_2319383, EPI_ISL_2319384, EPI_ISL_2319385, EPI_ISL_2319386, EPI_ISL_2319391, EPI_ISL_2319392, EPI_ISL_2319393, EPI_ISL_2319394, EPI_ISL_2319395, EPI_ISL_2319396, EPI_ISL_2319397, EPI_ISL_2319398, EPI_ISL_2319399, EPI_ISL_2319400, EPI_ISL_2319401, EPI_ISL_2319402, EPI_ISL_2319403, EPI_ISL_2319404, EPI_ISL_2319405, EPI_ISL_2319406, EPI_ISL_2319407, EPI_ISL_2319408, EPI_ISL_2319409, EPI_ISL_2319410, EPI_ISL_2319411, EPI_ISL_2319412, EPI_ISL_2341028, EPI_ISL_2341030, EPI_ISL_2341032, EPI_ISL_2341033, EPI_ISL_2341034, EPI_ISL_2341035, EPI_ISL_2341036, EPI_ISL_2341037, EPI_ISL_2341038, EPI_ISL_2341039, EPI_ISL_2341042, EPI_ISL_2341043, EPI_ISL_2341044, EPI_ISL_2341045, EPI_ISL_2348837, EPI_ISL_2349881, EPI_ISL_2349882, EPI_ISL_2349883, EPI_ISL_2349884, EPI_ISL_2349885, EPI_ISL_2349886, EPI_ISL_2349887, EPI_ISL_2349888, EPI_ISL_2349889, EPI_ISL_2349890, EPI_ISL_2349891, EPI_ISL_2349892, EPI_ISL_2349893, EPI_ISL_2349894, EPI_ISL_2349895, EPI_ISL_2349896, EPI_ISL_2349897, EPI_ISL_2349898, EPI_ISL_2349899, EPI_ISL_2349900, EPI_ISL_2349901, EPI_ISL_2349902, EPI_ISL_2349903, EPI_ISL_2349904, EPI_ISL_2349905, EPI_ISL_2349906, EPI_ISL_2349907, EPI_ISL_2349908, EPI_ISL_2349909, EPI_ISL_2349910, EPI_ISL_2349911, EPI_ISL_2349912, EPI_ISL_2349913, EPI_ISL_2349914, EPI_ISL_2349915, EPI_ISL_2349916, EPI_ISL_2349917, EPI_ISL_2349918, EPI_ISL_2349919, EPI_ISL_2349920, EPI_ISL_2349921, EPI_ISL_2349922, EPI_ISL_2349923, EPI_ISL_2349924, EPI_ISL_2349925, EPI_ISL_2349926, EPI_ISL_2349927, EPI_ISL_2349928, EPI_ISL_2349929, EPI_ISL_2349930, EPI_ISL_2349931, EPI_ISL_2349932, EPI_ISL_2349933, EPI_ISL_2349934, EPI_ISL_2349935, EPI_ISL_2349936, EPI_ISL_2349937, EPI_ISL_2349938, EPI_ISL_2349939, EPI_ISL_2349940, EPI_ISL_2349941, EPI_ISL_2349942, EPI_ISL_2349943, EPI_ISL_2349944, EPI_ISL_2349945, EPI_ISL_2349946, EPI_ISL_2349947, EPI_ISL_2349948, EPI_ISL_2349949, EPI_ISL_2349950, EPI_ISL_2349951, EPI_ISL_2349952, EPI_ISL_2349953, EPI_ISL_2349954, EPI_ISL_2349955, EPI_ISL_2349956, EPI_ISL_2349957, EPI_ISL_2349958, EPI_ISL_2349959, EPI_ISL_2349960, EPI_ISL_2349961, EPI_ISL_2349962, EPI_ISL_2349963, EPI_ISL_2349964, EPI_ISL_2349965, EPI_ISL_2349966, EPI_ISL_2349967, EPI_ISL_2349968, EPI_ISL_2349969, EPI_ISL_2349970, EPI_ISL_2349971, EPI_ISL_2349972, EPI_ISL_2349973, EPI_ISL_2349974, EPI_ISL_2349975, EPI_ISL_2349976, EPI_ISL_2349977, EPI_ISL_2349978, EPI_ISL_2349979, EPI_ISL_2349980, EPI_ISL_2349981, EPI_ISL_2349982, EPI_ISL_2349983, EPI_ISL_2349984, EPI_ISL_2349985, EPI_ISL_2349986, EPI_ISL_2349987, EPI_ISL_2349988, EPI_ISL_2349989, EPI_ISL_2349990, EPI_ISL_2349991, EPI_ISL_2349992, EPI_ISL_2349993, EPI_ISL_2349994, EPI_ISL_2349995, EPI_ISL_2349996, EPI_ISL_2349997, EPI_ISL_2349998, EPI_ISL_2349999, EPI_ISL_2350000, EPI_ISL_2350001, EPI_ISL_2350002, EPI_ISL_2350003, EPI_ISL_2350004, EPI_ISL_2350005, EPI_ISL_2350006, EPI_ISL_2350007, EPI_ISL_2350008, EPI_ISL_2350009, EPI_ISL_2350010, EPI_ISL_2350011, EPI_ISL_2350012, EPI_ISL_2350013, EPI_ISL_2350014, EPI_ISL_2350015, EPI_ISL_2350016, EPI_ISL_2350017, EPI_ISL_2350018, EPI_ISL_2350019, EPI_ISL_2350020, EPI_ISL_2350021, EPI_ISL_2350022, EPI_ISL_2350023, EPI_ISL_2350024, EPI_ISL_2350025, EPI_ISL_2350026, EPI_ISL_2350027, EPI_ISL_2350028, EPI_ISL_2350029, EPI_ISL_2350030, EPI_ISL_2350031, EPI_ISL_2350032, EPI_ISL_2350033, EPI_ISL_2350034, EPI_ISL_2350035, EPI_ISL_2350036, EPI_ISL_2350037, EPI_ISL_2350038, EPI_ISL_2350039, EPI_ISL_2350040, EPI_ISL_2350041, EPI_ISL_2350042, EPI_ISL_2350043, EPI_ISL_2350044, EPI_ISL_2350045, EPI_ISL_2350046, EPI_ISL_2350047, EPI_ISL_2350048, EPI_ISL_2350049, EPI_ISL_2350050, EPI_ISL_2350051, EPI_ISL_2350052, EPI_ISL_2350053, EPI_ISL_2350054, EPI_ISL_2350055, EPI_ISL_2350056, EPI_ISL_2350057, EPI_ISL_2350058, EPI_ISL_2350059, EPI_ISL_2350060, EPI_ISL_2350061, EPI_ISL_2350062, EPI_ISL_2350063, EPI_ISL_2350064, EPI_ISL_2350065, EPI_ISL_2350066, EPI_ISL_2350067, EPI_ISL_2350068, EPI_ISL_2350069, EPI_ISL_2350070, EPI_ISL_2350071, EPI_ISL_2350072, EPI_ISL_2350073, EPI_ISL_2350074, EPI_ISL_2350075, EPI_ISL_2350076, EPI_ISL_2350077, EPI_ISL_2350078, EPI_ISL_2350079, EPI_ISL_2350080, EPI_ISL_2350081, EPI_ISL_2350082, EPI_ISL_2350083, EPI_ISL_2350084, EPI_ISL_2350085, EPI_ISL_2350086, EPI_ISL_2350087, EPI_ISL_2350088, EPI_ISL_2350089, EPI_ISL_2350090, EPI_ISL_2350091, EPI_ISL_2350092, EPI_ISL_2350093, EPI_ISL_2350094, EPI_ISL_2350114, EPI_ISL_2350115, EPI_ISL_2350116, EPI_ISL_2350117, EPI_ISL_2350118, EPI_ISL_2350119, EPI_ISL_2350120, EPI_ISL_2350121, EPI_ISL_2350122, EPI_ISL_2350123, EPI_ISL_2350124, EPI_ISL_2350125, EPI_ISL_2350126, EPI_ISL_2350127, EPI_ISL_2350128, EPI_ISL_2350129, EPI_ISL_2350130, EPI_ISL_2350131, EPI_ISL_2350132, EPI_ISL_2350133, EPI_ISL_2350134, EPI_ISL_2350135, EPI_ISL_2350136, EPI_ISL_2350137, EPI_ISL_2350138, EPI_ISL_2350139, EPI_ISL_2350140, EPI_ISL_2350141, EPI_ISL_2443071, EPI_ISL_2443072, EPI_ISL_2443073, EPI_ISL_2443074, EPI_ISL_2443075, EPI_ISL_2443076, EPI_ISL_2443077, EPI_ISL_2443078, EPI_ISL_2443079, EPI_ISL_2443080, EPI_ISL_2443081, EPI_ISL_2443084, EPI_ISL_2443085, EPI_ISL_2444397, EPI_ISL_2444398, EPI_ISL_2444399, EPI_ISL_2444400, EPI_ISL_2444401, EPI_ISL_2444402, EPI_ISL_2444403, EPI_ISL_2444404, EPI_ISL_2444405, EPI_ISL_2444406, EPI_ISL_2444407, EPI_ISL_2444408, EPI_ISL_2444409, EPI_ISL_2444410, EPI_ISL_2444411, EPI_ISL_2444412, EPI_ISL_2444413, EPI_ISL_2444414, EPI_ISL_2444415, EPI_ISL_2444416, EPI_ISL_2444417, EPI_ISL_2444418, EPI_ISL_2444419, EPI_ISL_2444420, EPI_ISL_2444421, EPI_ISL_2444422, EPI_ISL_2444423, EPI_ISL_2444424, EPI_ISL_2444425, EPI_ISL_2444426, EPI_ISL_2444427, EPI_ISL_2444428, EPI_ISL_2444429, EPI_ISL_2444430, EPI_ISL_2444431, EPI_ISL_2444432, EPI_ISL_2444433, EPI_ISL_2444434, EPI_ISL_2444435, EPI_ISL_2444436, EPI_ISL_2444437, EPI_ISL_2444438, EPI_ISL_2444439, EPI_ISL_2444440, EPI_ISL_2444441, EPI_ISL_2444442, EPI_ISL_2444443, EPI_ISL_2444444, EPI_ISL_2444445, EPI_ISL_2444446, EPI_ISL_2444447, EPI_ISL_2444448, EPI_ISL_2444449, EPI_ISL_2444450, EPI_ISL_2444451, EPI_ISL_2444452, EPI_ISL_2444453, EPI_ISL_2444454, EPI_ISL_2444455, EPI_ISL_2444456, EPI_ISL_2444457, EPI_ISL_2444458, EPI_ISL_2444459, EPI_ISL_2444460, EPI_ISL_2444461, EPI_ISL_2444462, EPI_ISL_2444463, EPI_ISL_2444464, EPI_ISL_2444465, EPI_ISL_2444466, EPI_ISL_2444467, EPI_ISL_2444468, EPI_ISL_2444469, EPI_ISL_2444470, EPI_ISL_2444471, EPI_ISL_2444472, EPI_ISL_2444473, EPI_ISL_2444474, EPI_ISL_2444475, EPI_ISL_2444476, EPI_ISL_2444477, EPI_ISL_2444478, EPI_ISL_2444479, EPI_ISL_2444480, EPI_ISL_2444481, EPI_ISL_2444482, EPI_ISL_2444483, EPI_ISL_2444484, EPI_ISL_2444485, EPI_ISL_2444486, EPI_ISL_2444487, EPI_ISL_2444488, EPI_ISL_2444489, EPI_ISL_2444490, EPI_ISL_2444491, EPI_ISL_2444492, EPI_ISL_2444493, EPI_ISL_2444494, EPI_ISL_2444495, EPI_ISL_2444496, EPI_ISL_2444497, EPI_ISL_2444498, EPI_ISL_2444499, EPI_ISL_2444500, EPI_ISL_2444501, EPI_ISL_2444502, EPI_ISL_2444503, EPI_ISL_2444504, EPI_ISL_2444505, EPI_ISL_2444506, EPI_ISL_2444507, EPI_ISL_2444508, EPI_ISL_2444509, EPI_ISL_2444510, EPI_ISL_2444511, EPI_ISL_2444512, EPI_ISL_2444513, EPI_ISL_2444514, EPI_ISL_2444515, EPI_ISL_2444516, EPI_ISL_2444517, EPI_ISL_2444518, EPI_ISL_2444519, EPI_ISL_2444520, EPI_ISL_2444521, EPI_ISL_2444522, EPI_ISL_2444523, EPI_ISL_2444524, EPI_ISL_2444525, EPI_ISL_2444526, EPI_ISL_2444527, EPI_ISL_2444528, EPI_ISL_2444529, EPI_ISL_2444530, EPI_ISL_2444531, EPI_ISL_2444532, EPI_ISL_2444533, EPI_ISL_2444534, EPI_ISL_2444535, EPI_ISL_2444536, EPI_ISL_2444537, EPI_ISL_2444538, EPI_ISL_2444539, EPI_ISL_2444540, EPI_ISL_2444541, EPI_ISL_2444542, EPI_ISL_2444543, EPI_ISL_2444544, EPI_ISL_2444545, EPI_ISL_2444546, EPI_ISL_2444547, EPI_ISL_2444548, EPI_ISL_2444549, EPI_ISL_2444550, EPI_ISL_2444551, EPI_ISL_2444552, EPI_ISL_2444553, EPI_ISL_2444554, EPI_ISL_2444555, EPI_ISL_2444556, EPI_ISL_2444557, EPI_ISL_2444558, EPI_ISL_2444559, EPI_ISL_2444560, EPI_ISL_2550734, EPI_ISL_2550735, EPI_ISL_2550736, EPI_ISL_2550737, EPI_ISL_2550738, EPI_ISL_2550739, EPI_ISL_2550740, EPI_ISL_2550741, EPI_ISL_2550742, EPI_ISL_2550743, EPI_ISL_2550744, EPI_ISL_2550745, EPI_ISL_2550746, EPI_ISL_2550747, EPI_ISL_2550748, EPI_ISL_2550749, EPI_ISL_2550750, EPI_ISL_2550751, EPI_ISL_2550752, EPI_ISL_2550753, EPI_ISL_2550754, EPI_ISL_2550755, EPI_ISL_2550756, EPI_ISL_2550757, EPI_ISL_2603659, EPI_ISL_2603660, EPI_ISL_2603661, EPI_ISL_2603662, EPI_ISL_2603663, EPI_ISL_2603664, EPI_ISL_2603665, EPI_ISL_2603666, EPI_ISL_2603667, EPI_ISL_2603668, EPI_ISL_2603669, EPI_ISL_2603670, EPI_ISL_2603671, EPI_ISL_2603672, EPI_ISL_2603673, EPI_ISL_2603674, EPI_ISL_2603675, EPI_ISL_2603676, EPI_ISL_2603677, EPI_ISL_2603678, EPI_ISL_2603679, EPI_ISL_2603680, EPI_ISL_2603681, EPI_ISL_2603682, EPI_ISL_2603683, EPI_ISL_2603684, EPI_ISL_2603685, EPI_ISL_2603686, EPI_ISL_2603687, EPI_ISL_2603688, EPI_ISL_2603689, EPI_ISL_2603690, EPI_ISL_2603691, EPI_ISL_2603692, EPI_ISL_2603693, EPI_ISL_2603694, EPI_ISL_2603695, EPI_ISL_2603696, EPI_ISL_2603697, EPI_ISL_2603698, EPI_ISL_2603699, EPI_ISL_2603700, EPI_ISL_2603701, EPI_ISL_2603702, EPI_ISL_2603703, EPI_ISL_2603704, EPI_ISL_2603705, EPI_ISL_2603706, EPI_ISL_2603707, EPI_ISL_2603708, EPI_ISL_2603709, EPI_ISL_2603710, EPI_ISL_2603711, EPI_ISL_2603712, EPI_ISL_2603713, EPI_ISL_2603714, EPI_ISL_2603715, EPI_ISL_2603716, EPI_ISL_2603717, EPI_ISL_2603718, EPI_ISL_2603719, EPI_ISL_2603720, EPI_ISL_2603721, EPI_ISL_2603722, EPI_ISL_2603723, EPI_ISL_2603724, EPI_ISL_2603725, EPI_ISL_2603726, EPI_ISL_2603727, EPI_ISL_2603728, EPI_ISL_2603729, EPI_ISL_2603730, EPI_ISL_2603731, EPI_ISL_2603732, EPI_ISL_2603733, EPI_ISL_2603734, EPI_ISL_2603735, EPI_ISL_2603736, EPI_ISL_2603737, EPI_ISL_2603738, EPI_ISL_2603739, EPI_ISL_2603740, EPI_ISL_2603741, EPI_ISL_2603742, EPI_ISL_2603743, EPI_ISL_2603744, EPI_ISL_2603745, EPI_ISL_2603746, EPI_ISL_2603747, EPI_ISL_2603748, EPI_ISL_2603749, EPI_ISL_2603750, EPI_ISL_2603751, EPI_ISL_2603752, EPI_ISL_2603753, EPI_ISL_2603754, EPI_ISL_2603755, EPI_ISL_2603756, EPI_ISL_2603757, EPI_ISL_2603758, EPI_ISL_2603759, EPI_ISL_2603760, EPI_ISL_2603761, EPI_ISL_2603762, EPI_ISL_2603763, EPI_ISL_2603764, EPI_ISL_2603765, EPI_ISL_2603766, EPI_ISL_2603767, EPI_ISL_2603768, EPI_ISL_2603769, EPI_ISL_2603770, EPI_ISL_2603771, EPI_ISL_2603772, EPI_ISL_2603773, EPI_ISL_2603774, EPI_ISL_2603775, EPI_ISL_2603776, EPI_ISL_2603777, EPI_ISL_2603778, EPI_ISL_2603779, EPI_ISL_2603780, EPI_ISL_2603781, EPI_ISL_2603782, EPI_ISL_2603783, EPI_ISL_2603784, EPI_ISL_2603785, EPI_ISL_2603786, EPI_ISL_2603787, EPI_ISL_2603788, EPI_ISL_2603789, EPI_ISL_2603790, EPI_ISL_2603791, EPI_ISL_2603792, EPI_ISL_2603793, EPI_ISL_2603794, EPI_ISL_2603795, EPI_ISL_2603796, EPI_ISL_2603797, EPI_ISL_2603798, EPI_ISL_2603799, EPI_ISL_2603800, EPI_ISL_2603801, EPI_ISL_2603802, EPI_ISL_2603803, EPI_ISL_2603804, EPI_ISL_2603805, EPI_ISL_2603806, EPI_ISL_2603807, EPI_ISL_2603808, EPI_ISL_2603809, EPI_ISL_2603810, EPI_ISL_2603811, EPI_ISL_2603812, EPI_ISL_2603813, EPI_ISL_2603814, EPI_ISL_2603815, EPI_ISL_2616784, EPI_ISL_2616785, EPI_ISL_2616786, EPI_ISL_2616787, EPI_ISL_2616788, EPI_ISL_2616789, EPI_ISL_2616790, EPI_ISL_2616791, EPI_ISL_2616792, EPI_ISL_2616793, EPI_ISL_2616794, EPI_ISL_2616795, EPI_ISL_2616796, EPI_ISL_2616797, EPI_ISL_2616798, EPI_ISL_2616799, EPI_ISL_2616800, EPI_ISL_2616801, EPI_ISL_2616802, EPI_ISL_2616803, EPI_ISL_2616804, EPI_ISL_2616805, EPI_ISL_2616806, EPI_ISL_2616807, EPI_ISL_2616808, EPI_ISL_2616809, EPI_ISL_2616810, EPI_ISL_2616811, EPI_ISL_2616812, EPI_ISL_2616813, EPI_ISL_2616814, EPI_ISL_2616815, EPI_ISL_2616816, EPI_ISL_2616817, EPI_ISL_2616818, EPI_ISL_2616819, EPI_ISL_2616820, EPI_ISL_2616821, EPI_ISL_2616822, EPI_ISL_2616823, EPI_ISL_2616824, EPI_ISL_2616825, EPI_ISL_2616826, EPI_ISL_2616827, EPI_ISL_2616828, EPI_ISL_2616829, EPI_ISL_2616830, EPI_ISL_2616831, EPI_ISL_2616832, EPI_ISL_2616833, EPI_ISL_2616834, EPI_ISL_2616835, EPI_ISL_2616836, EPI_ISL_2616837, EPI_ISL_2616838, EPI_ISL_2616839, EPI_ISL_2616840, EPI_ISL_2616841, EPI_ISL_2616842, EPI_ISL_2616843, EPI_ISL_2616844, EPI_ISL_2616845, EPI_ISL_2616846, EPI_ISL_2616847, EPI_ISL_2616848, EPI_ISL_2616849, EPI_ISL_2616850, EPI_ISL_2616851, EPI_ISL_2616852, EPI_ISL_2616853, EPI_ISL_2616854, EPI_ISL_2616855, EPI_ISL_2616856, EPI_ISL_2616857, EPI_ISL_2616858, EPI_ISL_2616859, EPI_ISL_2616860, EPI_ISL_2616861, EPI_ISL_2616862, EPI_ISL_2616863, EPI_ISL_2616864, EPI_ISL_2616865, EPI_ISL_2616866, EPI_ISL_2616867, EPI_ISL_2616868, EPI_ISL_2616869, EPI_ISL_2616870, EPI_ISL_2616871, EPI_ISL_2616872, EPI_ISL_2616873, EPI_ISL_2616874, EPI_ISL_2616875, EPI_ISL_2616876, EPI_ISL_2616877, EPI_ISL_2616878, EPI_ISL_2616879, EPI_ISL_2616880, EPI_ISL_2616881, EPI_ISL_2616882, EPI_ISL_2616883, EPI_ISL_2616884, EPI_ISL_2616885, EPI_ISL_2616886, EPI_ISL_2616887, EPI_ISL_2616888, EPI_ISL_2616889, EPI_ISL_2616890, EPI_ISL_2616891, EPI_ISL_2616892, EPI_ISL_2616893, EPI_ISL_2616894, EPI_ISL_2616895, EPI_ISL_2616896, EPI_ISL_2616897, EPI_ISL_2616898, EPI_ISL_2616899, EPI_ISL_2616900, EPI_ISL_2616901, EPI_ISL_2616902, EPI_ISL_2616903, EPI_ISL_2616904, EPI_ISL_2616905, EPI_ISL_2616906, EPI_ISL_2616907, EPI_ISL_2616908, EPI_ISL_2616909, EPI_ISL_2616910, EPI_ISL_2616911, EPI_ISL_2616912, EPI_ISL_2616913, EPI_ISL_2616914, EPI_ISL_2616915, EPI_ISL_2616916, EPI_ISL_2616917, EPI_ISL_2616918, EPI_ISL_2616919, EPI_ISL_2616920, EPI_ISL_2616921, EPI_ISL_2616922, EPI_ISL_2616923, EPI_ISL_2616924, EPI_ISL_2616925, EPI_ISL_2616926, EPI_ISL_2616927, EPI_ISL_2616928, EPI_ISL_2616929, EPI_ISL_2616930, EPI_ISL_2616931, EPI_ISL_2616932, EPI_ISL_2616933, EPI_ISL_2616934, EPI_ISL_2616935, EPI_ISL_2616936, EPI_ISL_2616937, EPI_ISL_2616938, EPI_ISL_2616939, EPI_ISL_2616940, EPI_ISL_2616941, EPI_ISL_2616942, EPI_ISL_2616943, EPI_ISL_2616944, EPI_ISL_2616945, EPI_ISL_2616946, EPI_ISL_2616947, EPI_ISL_2616948, EPI_ISL_2616949, EPI_ISL_2616950, EPI_ISL_2616951, EPI_ISL_2616952, EPI_ISL_2616953, EPI_ISL_2616954, EPI_ISL_2616955, EPI_ISL_2616956, EPI_ISL_2616957, EPI_ISL_2616958, EPI_ISL_2616959, EPI_ISL_2616960, EPI_ISL_2616961, EPI_ISL_2616962, EPI_ISL_2616963, EPI_ISL_2616964, EPI_ISL_2616965, EPI_ISL_2616966, EPI_ISL_2616967, EPI_ISL_2616968, EPI_ISL_2616969, EPI_ISL_2616970, EPI_ISL_2616971, EPI_ISL_2616972, EPI_ISL_2616973, EPI_ISL_2616974, EPI_ISL_2616975, EPI_ISL_2616976, EPI_ISL_2616977, EPI_ISL_2616978, EPI_ISL_2616979, EPI_ISL_2616980, EPI_ISL_2616981, EPI_ISL_2616982, EPI_ISL_2616983, EPI_ISL_2616984, EPI_ISL_2616985, EPI_ISL_2616986, EPI_ISL_2616987, EPI_ISL_2616988, EPI_ISL_2616989, EPI_ISL_2616990, EPI_ISL_2616991, EPI_ISL_2616992, EPI_ISL_2616993, EPI_ISL_2616994, EPI_ISL_2616995, EPI_ISL_2616996, EPI_ISL_2616997, EPI_ISL_2616998, EPI_ISL_2616999, EPI_ISL_2617000, EPI_ISL_2617001, EPI_ISL_2617002, EPI_ISL_2617003, EPI_ISL_2617004, EPI_ISL_2617005, EPI_ISL_2617006, EPI_ISL_2617007, EPI_ISL_2617008, EPI_ISL_2617009, EPI_ISL_2617010, EPI_ISL_2617011, EPI_ISL_2617012, EPI_ISL_2617013, EPI_ISL_2617014, EPI_ISL_2617015, EPI_ISL_2617016, EPI_ISL_2617017, EPI_ISL_2617018, EPI_ISL_2617019, EPI_ISL_2617020, EPI_ISL_2617021, EPI_ISL_2617022, EPI_ISL_2617023, EPI_ISL_2617024, EPI_ISL_2617025, EPI_ISL_2617026, EPI_ISL_2617027, EPI_ISL_2617028, EPI_ISL_2617029, EPI_ISL_2617030, EPI_ISL_2617031, EPI_ISL_2617032, EPI_ISL_2617033, EPI_ISL_2617034, EPI_ISL_2692540, EPI_ISL_2692541, EPI_ISL_2692542, EPI_ISL_2692543, EPI_ISL_2692544, EPI_ISL_2692545, EPI_ISL_2692547, EPI_ISL_2692548, EPI_ISL_2692549, EPI_ISL_2692550, EPI_ISL_2692551, EPI_ISL_2692552, EPI_ISL_2692553, EPI_ISL_2692554, EPI_ISL_2692555, EPI_ISL_2692556, EPI_ISL_2692557, EPI_ISL_2692558, EPI_ISL_2692559, EPI_ISL_2692560, EPI_ISL_2692561, EPI_ISL_2692562, EPI_ISL_2692563, EPI_ISL_2692564, EPI_ISL_2692565, EPI_ISL_2692566, EPI_ISL_2692567, EPI_ISL_2692568, EPI_ISL_2692569, EPI_ISL_2692570, EPI_ISL_2692571, EPI_ISL_2692572, EPI_ISL_2692573, EPI_ISL_2692574, EPI_ISL_2692575, EPI_ISL_2692576, EPI_ISL_2692577, EPI_ISL_2692578, EPI_ISL_2692579, EPI_ISL_2692580, EPI_ISL_2692581, EPI_ISL_2692582, EPI_ISL_2692583, EPI_ISL_2692584, EPI_ISL_2692585, EPI_ISL_2692586, EPI_ISL_2692587, EPI_ISL_2692588, EPI_ISL_2692589, EPI_ISL_2692590, EPI_ISL_2692591, EPI_ISL_2692592, EPI_ISL_2692593, EPI_ISL_2692594, EPI_ISL_2692595, EPI_ISL_2692596, EPI_ISL_2692597, EPI_ISL_2692598, EPI_ISL_2692599, EPI_ISL_2692600, EPI_ISL_2692601, EPI_ISL_2692602, EPI_ISL_2692603, EPI_ISL_2692604, EPI_ISL_2692605, EPI_ISL_2692606, EPI_ISL_2692607, EPI_ISL_2692608, EPI_ISL_2692609, EPI_ISL_2692610, EPI_ISL_2692611, EPI_ISL_2692612, EPI_ISL_2692613, EPI_ISL_2692614, EPI_ISL_2692615, EPI_ISL_2692616, EPI_ISL_2692617, EPI_ISL_2692618, EPI_ISL_2692619, EPI_ISL_2692620, EPI_ISL_2692621, EPI_ISL_2692622, EPI_ISL_2692623, EPI_ISL_2692624, EPI_ISL_2692625, EPI_ISL_2692626, EPI_ISL_2692627, EPI_ISL_2692628, EPI_ISL_2692629, EPI_ISL_2692630, EPI_ISL_2692631, EPI_ISL_2692632, EPI_ISL_2692633, EPI_ISL_2692634, EPI_ISL_2692635, EPI_ISL_2692636, EPI_ISL_2692637, EPI_ISL_2692638, EPI_ISL_2692639, EPI_ISL_2692640, EPI_ISL_2692641, EPI_ISL_2692642, EPI_ISL_2692643, EPI_ISL_2692644, EPI_ISL_2692645, EPI_ISL_2692646, EPI_ISL_2692647, EPI_ISL_2692648, EPI_ISL_2692649, EPI_ISL_2692650, EPI_ISL_2692651, EPI_ISL_2692652, EPI_ISL_2692653, EPI_ISL_2692654, EPI_ISL_2692655, EPI_ISL_2692656, EPI_ISL_2692657, EPI_ISL_2692658, EPI_ISL_2692659, EPI_ISL_2692660, EPI_ISL_2692661, EPI_ISL_2692662, EPI_ISL_2692663, EPI_ISL_2692664, EPI_ISL_2692665, EPI_ISL_2692666, EPI_ISL_2692667, EPI_ISL_2692668, EPI_ISL_2692669, EPI_ISL_2692670, EPI_ISL_2692671, EPI_ISL_2692672, EPI_ISL_2692673, EPI_ISL_2692674, EPI_ISL_2692675, EPI_ISL_2692676, EPI_ISL_2692677, EPI_ISL_2692678, EPI_ISL_2692679, EPI_ISL_2692680, EPI_ISL_2692681, EPI_ISL_2692682, EPI_ISL_2692683, EPI_ISL_2692684, EPI_ISL_2692685, EPI_ISL_2692686, EPI_ISL_2692687, EPI_ISL_2692688, EPI_ISL_2692689, EPI_ISL_2692690, EPI_ISL_2692691, EPI_ISL_2692692, EPI_ISL_2692693, EPI_ISL_2692694, EPI_ISL_2692695, EPI_ISL_2692696, EPI_ISL_2692697, EPI_ISL_2692698, EPI_ISL_2692699, EPI_ISL_2692700, EPI_ISL_2692701, EPI_ISL_2692702, EPI_ISL_2692703, EPI_ISL_2692704, EPI_ISL_2692705, EPI_ISL_2692707, EPI_ISL_2692708, EPI_ISL_2692709, EPI_ISL_2692710, EPI_ISL_2692711, EPI_ISL_2692712, EPI_ISL_2692713, EPI_ISL_2692714, EPI_ISL_2692715, EPI_ISL_2692716, EPI_ISL_2692717, EPI_ISL_2692718, EPI_ISL_2692719, EPI_ISL_2692720, EPI_ISL_2692721, EPI_ISL_2692722, EPI_ISL_2692723, EPI_ISL_2692724, EPI_ISL_2692725, EPI_ISL_2692726, EPI_ISL_2692727, EPI_ISL_2692728, EPI_ISL_2692729, EPI_ISL_2692730, EPI_ISL_2692731, EPI_ISL_2692732, EPI_ISL_2692733, EPI_ISL_2692734, EPI_ISL_2692735, EPI_ISL_2692736, EPI_ISL_2692737, EPI_ISL_2692738, EPI_ISL_2692739, EPI_ISL_2692740, EPI_ISL_2692741, EPI_ISL_2692742, EPI_ISL_2692743, EPI_ISL_2692744, EPI_ISL_2692745, EPI_ISL_2692746, EPI_ISL_2692747, EPI_ISL_2692748, EPI_ISL_2692749, EPI_ISL_2692750, EPI_ISL_2692751, EPI_ISL_2692752, EPI_ISL_2692753, EPI_ISL_2692754, EPI_ISL_2692755, EPI_ISL_2692756, EPI_ISL_2692757, EPI_ISL_2692758, EPI_ISL_2692759, EPI_ISL_2692760, EPI_ISL_2692761, EPI_ISL_2692762, EPI_ISL_2692763, EPI_ISL_2692764, EPI_ISL_2692765, EPI_ISL_2692766, EPI_ISL_2692767, EPI_ISL_2692768, EPI_ISL_2692769, EPI_ISL_2692770, EPI_ISL_2692771, EPI_ISL_2692772, EPI_ISL_2692773, EPI_ISL_2692774, EPI_ISL_2692775, EPI_ISL_2692776, EPI_ISL_2692777, EPI_ISL_2692778, EPI_ISL_2692779, EPI_ISL_2692780, EPI_ISL_2692781, EPI_ISL_2692782, EPI_ISL_2692783, EPI_ISL_2692784, EPI_ISL_2692785, EPI_ISL_2692786, EPI_ISL_2692787, EPI_ISL_2692788, EPI_ISL_2692789, EPI_ISL_2692790, EPI_ISL_2692791, EPI_ISL_2692792, EPI_ISL_2692793, EPI_ISL_2692794, EPI_ISL_2692795, EPI_ISL_2692796, EPI_ISL_2692797, EPI_ISL_2692798, EPI_ISL_2692799, EPI_ISL_2692800, EPI_ISL_2692801, EPI_ISL_2692802, EPI_ISL_2692803, EPI_ISL_2692804, EPI_ISL_2692805, EPI_ISL_2692806, EPI_ISL_2692807, EPI_ISL_2692808, EPI_ISL_2692809, EPI_ISL_2692810, EPI_ISL_2692811, EPI_ISL_2692812, EPI_ISL_2692813, EPI_ISL_2692814, EPI_ISL_2692815, EPI_ISL_2692816, EPI_ISL_2692817, EPI_ISL_2692818, EPI_ISL_2692819, EPI_ISL_2692820, EPI_ISL_2692821, EPI_ISL_2692822, EPI_ISL_2692823, EPI_ISL_2692824, EPI_ISL_2692826, EPI_ISL_2692827, EPI_ISL_2692828, EPI_ISL_2692829, EPI_ISL_2692830, EPI_ISL_2692831, EPI_ISL_2692832, EPI_ISL_2692848, EPI_ISL_2692849, EPI_ISL_2692850, EPI_ISL_2692851, EPI_ISL_2692852, EPI_ISL_2774493, EPI_ISL_2778955, EPI_ISL_2778956, EPI_ISL_2778957, EPI_ISL_2778958, EPI_ISL_2778959, EPI_ISL_2778960, EPI_ISL_2778961, EPI_ISL_2778962, EPI_ISL_2778963, EPI_ISL_2778964, EPI_ISL_2778965, EPI_ISL_2778966, EPI_ISL_2779614, EPI_ISL_2779615, EPI_ISL_2779616, EPI_ISL_2779617, EPI_ISL_2779618, EPI_ISL_2779619, EPI_ISL_2779620, EPI_ISL_2779621, EPI_ISL_2779622, EPI_ISL_2779623, EPI_ISL_2779624, EPI_ISL_2779625, EPI_ISL_2779626, EPI_ISL_2779627, EPI_ISL_2779628, EPI_ISL_2779629, EPI_ISL_2779630, EPI_ISL_2779631, EPI_ISL_2779632, EPI_ISL_2779633, EPI_ISL_2779634, EPI_ISL_2779635, EPI_ISL_2779636, EPI_ISL_2779637, EPI_ISL_2790218, EPI_ISL_2790219, EPI_ISL_2790220, EPI_ISL_2790221, EPI_ISL_2790222, EPI_ISL_2790223, EPI_ISL_2790224, EPI_ISL_2790225, EPI_ISL_2790226, EPI_ISL_2790227, EPI_ISL_2790228, EPI_ISL_2790229, EPI_ISL_2790230, EPI_ISL_2790231, EPI_ISL_2810032, EPI_ISL_2810033, EPI_ISL_2810034, EPI_ISL_2810035, EPI_ISL_2810036, EPI_ISL_2810037, EPI_ISL_2810038, EPI_ISL_2810039, EPI_ISL_2810042, EPI_ISL_2810043, EPI_ISL_2810044, EPI_ISL_2810045, EPI_ISL_2810047, EPI_ISL_2810049, EPI_ISL_2810050, EPI_ISL_2810051, EPI_ISL_2810052, EPI_ISL_2810054, EPI_ISL_2810055, EPI_ISL_2810056, EPI_ISL_2810058, EPI_ISL_2810059, EPI_ISL_2810060, EPI_ISL_2810061, EPI_ISL_2810062, EPI_ISL_2810064, EPI_ISL_2810065, EPI_ISL_2810066, EPI_ISL_2810067, EPI_ISL_2810069, EPI_ISL_2810070, EPI_ISL_2810071, EPI_ISL_2810072, EPI_ISL_2810073, EPI_ISL_2810074, EPI_ISL_2810075, EPI_ISL_2810076, EPI_ISL_2810077, EPI_ISL_2810078, EPI_ISL_2810079, EPI_ISL_2810080, EPI_ISL_2810081, EPI_ISL_2810082, EPI_ISL_2810083, EPI_ISL_2810084, EPI_ISL_2810085, EPI_ISL_2810086, EPI_ISL_2810087, EPI_ISL_2810088, EPI_ISL_2810089, EPI_ISL_2810090, EPI_ISL_2810091, EPI_ISL_2810092, EPI_ISL_2810093, EPI_ISL_2810094, EPI_ISL_2810095, EPI_ISL_2810096, EPI_ISL_2810097, EPI_ISL_2810098, EPI_ISL_2810099, EPI_ISL_2810102, EPI_ISL_2810103, EPI_ISL_2810104, EPI_ISL_2810105, EPI_ISL_2810106, EPI_ISL_2810108, EPI_ISL_2810111, EPI_ISL_2810114, EPI_ISL_2810115, EPI_ISL_2810116, EPI_ISL_2810118, EPI_ISL_2810119, EPI_ISL_2810121, EPI_ISL_2810122, EPI_ISL_2810123, EPI_ISL_2810124, EPI_ISL_2810125, EPI_ISL_2810126, EPI_ISL_2810127, EPI_ISL_2810128, EPI_ISL_2810129, EPI_ISL_2810130, EPI_ISL_2810131, EPI_ISL_2810133, EPI_ISL_2810134, EPI_ISL_2810135, EPI_ISL_2810136, EPI_ISL_2810137, EPI_ISL_2810138, EPI_ISL_2810139, EPI_ISL_2810141, EPI_ISL_2810142, EPI_ISL_2810143, EPI_ISL_2810144, EPI_ISL_2810145, EPI_ISL_2810146, EPI_ISL_2810147, EPI_ISL_2810148, EPI_ISL_2810153, EPI_ISL_2810155, EPI_ISL_2810156, EPI_ISL_2810157, EPI_ISL_2810158, EPI_ISL_2810160, EPI_ISL_2810161, EPI_ISL_2810162, EPI_ISL_2810164, EPI_ISL_2810166, EPI_ISL_2810167, EPI_ISL_2810168, EPI_ISL_2810169, EPI_ISL_2810170, EPI_ISL_2810171, EPI_ISL_2810172, EPI_ISL_2810173, EPI_ISL_2810174, EPI_ISL_2810175, EPI_ISL_2810176, EPI_ISL_2810177, EPI_ISL_2810178, EPI_ISL_2810179, EPI_ISL_2810180, EPI_ISL_2810181, EPI_ISL_2810182, EPI_ISL_2810183, EPI_ISL_2810184, EPI_ISL_2810185, EPI_ISL_2810186, EPI_ISL_2810187, EPI_ISL_2810188, EPI_ISL_2810189, EPI_ISL_2810190, EPI_ISL_2810191, EPI_ISL_2810192, EPI_ISL_2810193, EPI_ISL_2810194, EPI_ISL_2810195, EPI_ISL_2810196, EPI_ISL_2810197, EPI_ISL_2810198, EPI_ISL_2810199, EPI_ISL_2810200, EPI_ISL_2810201, EPI_ISL_2810202, EPI_ISL_2810203, EPI_ISL_2810204, EPI_ISL_2810205, EPI_ISL_2810206, EPI_ISL_2810207, EPI_ISL_2810208, EPI_ISL_2810209, EPI_ISL_2810210, EPI_ISL_2810211, EPI_ISL_2810212, EPI_ISL_2810213, EPI_ISL_2810214, EPI_ISL_2810215, EPI_ISL_2810216, EPI_ISL_2810217, EPI_ISL_2810218, EPI_ISL_2810219, EPI_ISL_2810220, EPI_ISL_2810221, EPI_ISL_2810222, EPI_ISL_2810223, EPI_ISL_2810224, EPI_ISL_2810225, EPI_ISL_2810226, EPI_ISL_2810227, EPI_ISL_2810228, EPI_ISL_2810229, EPI_ISL_2810230, EPI_ISL_2810231, EPI_ISL_2810232, EPI_ISL_2810233, EPI_ISL_2810234, EPI_ISL_2810235, EPI_ISL_2810236, EPI_ISL_2810237, EPI_ISL_2810238, EPI_ISL_2810239, EPI_ISL_2810240, EPI_ISL_2810241, EPI_ISL_2810242, EPI_ISL_2810243, EPI_ISL_2810244, EPI_ISL_2810245, EPI_ISL_2810246, EPI_ISL_2810247, EPI_ISL_2810248, EPI_ISL_2810249, EPI_ISL_2810250, EPI_ISL_2810251, EPI_ISL_2810252, EPI_ISL_2810253, EPI_ISL_2810254, EPI_ISL_2810255, EPI_ISL_2810256, EPI_ISL_2810257, EPI_ISL_2810258, EPI_ISL_2810259, EPI_ISL_2810260, EPI_ISL_2810261, EPI_ISL_2810262, EPI_ISL_2810263, EPI_ISL_2810264, EPI_ISL_2810265, EPI_ISL_2810266, EPI_ISL_2810267, EPI_ISL_2810268, EPI_ISL_2810269, EPI_ISL_2810270, EPI_ISL_2810271, EPI_ISL_2810272, EPI_ISL_2810273, EPI_ISL_2810274, EPI_ISL_2810275, EPI_ISL_2810276, EPI_ISL_2810277, EPI_ISL_2810278, EPI_ISL_2810279, EPI_ISL_2810280, EPI_ISL_2810281, EPI_ISL_2810282, EPI_ISL_2810283, EPI_ISL_2810284, EPI_ISL_2810285, EPI_ISL_2810286, EPI_ISL_2810287, EPI_ISL_2810288, EPI_ISL_2810289, EPI_ISL_2810290, EPI_ISL_2810291, EPI_ISL_2810292, EPI_ISL_2810293, EPI_ISL_2810294, EPI_ISL_2810295, EPI_ISL_2810296, EPI_ISL_2810297, EPI_ISL_2810298, EPI_ISL_2810299, EPI_ISL_2810300, EPI_ISL_2810301, EPI_ISL_2810302, EPI_ISL_2810303, EPI_ISL_2810304, EPI_ISL_2810305, EPI_ISL_2810306, EPI_ISL_2810307, EPI_ISL_2810308, EPI_ISL_2810309, EPI_ISL_2810310, EPI_ISL_2810311, EPI_ISL_2810312, EPI_ISL_2810313, EPI_ISL_2810314, EPI_ISL_2810315, EPI_ISL_2810316, EPI_ISL_2810317, EPI_ISL_2810318, EPI_ISL_2810319, EPI_ISL_2810320, EPI_ISL_2810321, EPI_ISL_2810322, EPI_ISL_2810323, EPI_ISL_2810324, EPI_ISL_2810325, EPI_ISL_2881746, EPI_ISL_2881747, EPI_ISL_2881758, EPI_ISL_2881759, EPI_ISL_2881760, EPI_ISL_2881761, EPI_ISL_2881762, EPI_ISL_2881763, EPI_ISL_2881764, EPI_ISL_2881765, EPI_ISL_2881766, EPI_ISL_2881775, EPI_ISL_2881776, EPI_ISL_2881777, EPI_ISL_2881778, EPI_ISL_2881779, EPI_ISL_2881782, EPI_ISL_2881783, EPI_ISL_2881784, EPI_ISL_2881785, EPI_ISL_2881786, EPI_ISL_2881787, EPI_ISL_2881788, EPI_ISL_2881789, EPI_ISL_2881790, EPI_ISL_2881791, EPI_ISL_2881792, EPI_ISL_2881793, EPI_ISL_2881794, EPI_ISL_2881795, EPI_ISL_2881796, EPI_ISL_2881797, EPI_ISL_2881798, EPI_ISL_2881799, EPI_ISL_2881800, EPI_ISL_2881801, EPI_ISL_2881802, EPI_ISL_2881803, EPI_ISL_2881804, EPI_ISL_2881805, EPI_ISL_2881806, EPI_ISL_2881807, EPI_ISL_2881808, EPI_ISL_2881809, EPI_ISL_2881810, EPI_ISL_2881811, EPI_ISL_2881817, EPI_ISL_2881818, EPI_ISL_2881819, EPI_ISL_2881820, EPI_ISL_2881821, EPI_ISL_2881822, EPI_ISL_2881823, EPI_ISL_2881824, EPI_ISL_2881825, EPI_ISL_2881826, EPI_ISL_2881827, EPI_ISL_2881828, EPI_ISL_2881829, EPI_ISL_2881830, EPI_ISL_2881831, EPI_ISL_2881832, EPI_ISL_2881834, EPI_ISL_2881835, EPI_ISL_2881836, EPI_ISL_2881837, EPI_ISL_2881838, EPI_ISL_2881839, EPI_ISL_2881840, EPI_ISL_2881841, EPI_ISL_2881842, EPI_ISL_2881843, EPI_ISL_2881844, EPI_ISL_2881845, EPI_ISL_2881846, EPI_ISL_2881847, EPI_ISL_2881848, EPI_ISL_2881849, EPI_ISL_2881850, EPI_ISL_2881851, EPI_ISL_2881852, EPI_ISL_2881853, EPI_ISL_2881854, EPI_ISL_2881856, EPI_ISL_2881857, EPI_ISL_2894152, EPI_ISL_2894153, EPI_ISL_2894154, EPI_ISL_2894155, EPI_ISL_2894156, EPI_ISL_2894157, EPI_ISL_2894158, EPI_ISL_2894159, EPI_ISL_2894160, EPI_ISL_2894161, EPI_ISL_2894162, EPI_ISL_2894163, EPI_ISL_2894164, EPI_ISL_2894165, EPI_ISL_2894166, EPI_ISL_2894167, EPI_ISL_2894168, EPI_ISL_2894169, EPI_ISL_2894170, EPI_ISL_2894171, EPI_ISL_2894172, EPI_ISL_2894173, EPI_ISL_2894174, EPI_ISL_2894175, EPI_ISL_2894176, EPI_ISL_2894177, EPI_ISL_2894178, EPI_ISL_2894179, EPI_ISL_2894180, EPI_ISL_2894181, EPI_ISL_2894182, EPI_ISL_2894183, EPI_ISL_2894184, EPI_ISL_2894185, EPI_ISL_2894186, EPI_ISL_2894187, EPI_ISL_2894188, EPI_ISL_2894189, EPI_ISL_2894190, EPI_ISL_2894191, EPI_ISL_2894192, EPI_ISL_2894193, EPI_ISL_2894194, EPI_ISL_2894195, EPI_ISL_2894196, EPI_ISL_2894197, EPI_ISL_2894198, EPI_ISL_2894199, EPI_ISL_2894200, EPI_ISL_2894201, EPI_ISL_2894202, EPI_ISL_2894203, EPI_ISL_2894204, EPI_ISL_2894205, EPI_ISL_2894206, EPI_ISL_2894207, EPI_ISL_2894208, EPI_ISL_2894209, EPI_ISL_2894210, EPI_ISL_2894211, EPI_ISL_2894212, EPI_ISL_2894213, EPI_ISL_2894214, EPI_ISL_2894215, EPI_ISL_2894216, EPI_ISL_2894217, EPI_ISL_2894218, EPI_ISL_2894219, EPI_ISL_2894220, EPI_ISL_2894221, EPI_ISL_2894222, EPI_ISL_2894223, EPI_ISL_2894224, EPI_ISL_2894225, EPI_ISL_2894226, EPI_ISL_2894227, EPI_ISL_2894228, EPI_ISL_2894229, EPI_ISL_2894230, EPI_ISL_2894231, EPI_ISL_2894232, EPI_ISL_2894233, EPI_ISL_2894234, EPI_ISL_2894235, EPI_ISL_2894264, EPI_ISL_2894265, EPI_ISL_2894331, EPI_ISL_2894332, EPI_ISL_2894333, EPI_ISL_2894334, EPI_ISL_2894335, EPI_ISL_2894336, EPI_ISL_2894337, EPI_ISL_2894338, EPI_ISL_2894339, EPI_ISL_2894340, EPI_ISL_2894341, EPI_ISL_2894342, EPI_ISL_2894343, EPI_ISL_2894344, EPI_ISL_2894345, EPI_ISL_2894346, EPI_ISL_2894347, EPI_ISL_2894348, EPI_ISL_2894349, EPI_ISL_2894350, EPI_ISL_2894351, EPI_ISL_2894352, EPI_ISL_2894353, EPI_ISL_2894354, EPI_ISL_2894355, EPI_ISL_2894356, EPI_ISL_2894357, EPI_ISL_2894358, EPI_ISL_2894359, EPI_ISL_2894360, EPI_ISL_2894361, EPI_ISL_2894362, EPI_ISL_2894363, EPI_ISL_2894364, EPI_ISL_2894365, EPI_ISL_2894366, EPI_ISL_2894367, EPI_ISL_2894368, EPI_ISL_2894369, EPI_ISL_2894370, EPI_ISL_2894371, EPI_ISL_2894372, EPI_ISL_2894373, EPI_ISL_2894374, EPI_ISL_2894375, EPI_ISL_2894376, EPI_ISL_2894377, EPI_ISL_2894378, EPI_ISL_2894379, EPI_ISL_2894380, EPI_ISL_2894381, EPI_ISL_2894382, EPI_ISL_2894383, EPI_ISL_2894384, EPI_ISL_2894385, EPI_ISL_2894386, EPI_ISL_2894387, EPI_ISL_2894388, EPI_ISL_2894389, EPI_ISL_2894390, EPI_ISL_2894391, EPI_ISL_2894392, EPI_ISL_2894393, EPI_ISL_2894406, EPI_ISL_2894407, EPI_ISL_2894408, EPI_ISL_2894409, EPI_ISL_2894410, EPI_ISL_2894411, EPI_ISL_2894412, EPI_ISL_2894413, EPI_ISL_2894414, EPI_ISL_2894415, EPI_ISL_2928479, EPI_ISL_2928480, EPI_ISL_2928481, EPI_ISL_2928482, EPI_ISL_2928483, EPI_ISL_2928484, EPI_ISL_2928485, EPI_ISL_2928486, EPI_ISL_2928487, EPI_ISL_2928488, EPI_ISL_2928491, EPI_ISL_2928492, EPI_ISL_2928493, EPI_ISL_2928494, EPI_ISL_2928495, EPI_ISL_2928496, EPI_ISL_2928497, EPI_ISL_2928498, EPI_ISL_2928499, EPI_ISL_2928500, EPI_ISL_2928501, EPI_ISL_2928502, EPI_ISL_2928503, EPI_ISL_2928504, EPI_ISL_2928505, EPI_ISL_2928506 | | | |
| see above | Salud Digna | Instituto Nacional de Medicina Genomica | Abraham Campos-Romero; Blancas S; Cedro-Tanda A; Cisneros-Villanueva M; Escobar-Arrazola; Gonzalez-Barrera D; Herrera-Montalvo LA.; Hidalgo-Miranda A; Luna-Ruiz Marco; M; Mendoza-Vargas A; Moreno-Camacho José Luis; Munguia-Garza P; Orjuela-Rodríguez M; Ramirez-Vega O; Rangel-DeLeon D; Reyes-Grajeda JP; Rodriguez-Gallegos Jorge |
| EPI_ISL_2287804, EPI_ISL_2287807, EPI_ISL_2287810, EPI_ISL_2287813, EPI_ISL_2287834, EPI_ISL_2287837, EPI_ISL_2287840, EPI_ISL_2287843, EPI_ISL_2287846, EPI_ISL_2287849, EPI_ISL_2287876, EPI_ISL_2287880, EPI_ISL_2287883, EPI_ISL_2287889, EPI_ISL_2287893, EPI_ISL_2287896, EPI_ISL_2287924, EPI_ISL_2287928, EPI_ISL_2287931 | | | |
| see above | San Juan Regional Medical Center | New Mexico Department of Health Scientific Laboratory | Anastacia Griego-Fisher; D'eldra Malone; Ellie Johnson; Jennifer Benoit |
| EPI_ISL_2272520, EPI_ISL_2272521 | Servicio Diagnóstico Clínico | LANIIA | Bruno Gomez-Gil; Cristobal Chaidez; Célida Isabel Martínez-Rodríguez; Irvin González-López; Jean Pierre González-Gómez; Juan Daniel Lira-Morales; Julissa Enciso-Ibarra |
| EPI_ISL_2533933, EPI_ISL_2533934, EPI_ISL_2533935 | Servicio Diagnóstico Clínico | Laboratorio Nacional-LANIIA-CIAD | ; Alejandra García-Gasca; Alejandra Hernández-Terán; Alejandro Sánchez-Flores; Alfredo Herrera-Estrella; Alicia Ocaña-Mondragón; Andreu Comas-García; Angel Gustavo Salas-Lais; Antonio Loza Román; Bernardo Martínez-Miguel; Blanca Taboada; Brenda Irasema Maldonado-Meza; Bruno Gómez-Gil; Carla Ivón Herrera-Najera; Carlos F. Arias; Celia Boukadida; Clara Esperanza Santacruz-Tinoco; Concepción Grajales-Muñiz; Consorcio Mexicano de Vigilancia Genómica (CoViGen-Mex). Authors (in alphabetical order): Julio Elias Alvarado-Yaah; Cristóbal Cháidez-Quiróz; Célida Duque Molina; Célida Martínez- Rodríguez; Daniel Fregoso-Rueda; Daniel Lira Morales; Eduardo Becerril-Vargas; Fernando Fontove-Herrera; Fidencio Mejía-Nepomuceno; Francisco Pulido; Gloria Elena Espinosa-Ayala; Gloria María Molina-Salinas; Gloria Vazquez; Hector Esteban Paz-Juárez; Hector Montoya-Fuentes; Helen Haydee Fernanda Ramírez-Plascencia; Irvin González-López; Jean Pierre González; Jesús Hernández; Joel Armando Vázquez-Pérez.; Jorge Salas-Hernández; José Antonio Enciso-Moreno; José Arturo Martínez-Orozco; José Esteban Muñoz-Medina; José de Jesús Nuñez-Contreras; Juan Bautista Chale-Dzul; Julissa Enciso-Ibarra; Luis Alberto Ochoa-Carrera; Margarita Matías-Florentino; Mario Mújica-Sánchez; Marissa Perez-Garcia; María Guadalupe Santiago-Mauricio; María Guadalupe de Jesús Mireles-Rivera; Nelly Sélem-Mojica; Pavel Isa; Ricardo Ciria Merce; Ricardo Grande; Rosa María Gutiérrez Rios; Santiago Ávila-Ríos; Selene Zárate; Susana Lopez; Verónica Mata-Haro; Victor Eduardo García-Arias; Víctor Hugo Borja-Aburto |
| EPI_ISL_2680930, | Servicio Diagnóstico | Laboratorio nacional- | ; Alejandra García-Gasca; Alejandra Hernández-Terán; Alejandro Sánchez-Flores; Alfredo Herrera-Estrella; Alicia Ocaña-Mondragón; Andreu Comas-García; Angel Gustavo Salas-Lais; Antonio Loza Román; Bernardo Martínez-Miguel; Blanca Taboada; Brenda Irasema Maldonado-Meza; Bruno Gómez-Gil; Carla Ivón Herrera-Najera; Carlos |

| | | | |
|---|---|---|---|
| EPI_ISL_2927970, EPI_ISL_2927971 | Clínico | LANIIA-CIAD | F. Arias; Celia Boukadida; Clara Esperanza Santacruz-Tinoco; Concepción Grajales-Muñiz; Consorcio Mexicano de Vigilancia Genómica (CoViGen-Mex). Authors (in alphabetical order): Julio Elias Alvarado-Yaah; Cristóbal Cháidez-Quiróz; Célida Duque Molina; Célida Martínez- Rodríguez; Daniel Fregoso-Rueda; Daniel Lira Morales; Eduardo Becerril-Vargas; Fernando Fontove-Herrera; Fidencio Mejía-Nepomuceno; Francisco Pulido; Gloria Elena Espinosa-Ayala; Gloria María Molina-Salinas; Gloria Vazquez; Hector Esteban Paz-Juárez; Hector Montoya-Fuentes; Helen Haydee Fernanda Ramírez-Plascencia; Irvin González-López; Jean Pierre González; Jesús Hernández; Joel Armando Vázquez-Pérez.; Jorge Salas-Hernández; José Antonio Enciso-Moreno; José Arturo Martínez-Orozco; José Esteban Muñoz-Medina; José de Jesús Nuñez-Contreras; Juan Bautista Chale-Dzul; Julissa Enciso-Ibarra; Luis Alberto Ochoa-Carrera; Margarita Matías-Florentino; Mario Mújica-Sánchez; Marissa Perez-Garcia; María Guadalupe Santiago-Mauricio; María Guadalupe de Jesús Mireles-Rivera; Nelly Sélem-Mojica; Pavel Isa; Ricardo Ciria Merce; Ricardo Grande; Rosa María Gutiérrez Rios; Santiago Ávila-Ríos; Selene Zárate; Susana Lopez; Verónica Mata-Haro; Victor Eduardo García-Arias; Víctor Hugo Borja-Aburto |
| EPI_ISL_2329447, EPI_ISL_2329458, EPI_ISL_2422818, EPI_ISL_2422832, EPI_ISL_2628540 | Sharp HealthCare Laboratory | Andersen lab at Scripps Research | Art Mendoza; Cathy Woerle; Jacquelyn Berumen; Liam McGinnis; Omid Bakhtar; SEARCH Alliance San Diego with Aaron Harding |
| EPI_ISL_2287902 | Sierra Vista Hospital | New Mexico Department of Health Scientific Laboratory | Anastacia Griego-Fisher; D'eldra Malone; Ellie Johnson; Jennifer Benoit |
| EPI_ISL_2454309, EPI_ISL_2454319, EPI_ISL_2454321, EPI_ISL_2454322, EPI_ISL_2454323, EPI_ISL_2454325, EPI_ISL_2454331, EPI_ISL_2454332, EPI_ISL_2454334, EPI_ISL_2454335, EPI_ISL_2454347, EPI_ISL_2454348, EPI_ISL_2454357, EPI_ISL_2454358, EPI_ISL_2454359 | | | |
| see above | TriCore Reference Laboratories | Center for Global Health, University of New Mexico Health Sciences Center | Cecilia Thompson; Darrell Dinwiddie; Daryl Domman; Karissa Culbreath; Kendra Pesko; Kurt Schwalm; Valerie Morley |
| EPI_ISL_2287937, EPI_ISL_2287940 | TriCore Reference Laboratory Inc. | New Mexico Department of Health Scientific Laboratory | Anastacia Griego-Fisher; D'eldra Malone; Ellie Johnson; Jennifer Benoit |
| EPI_ISL_2562753, EPI_ISL_2562761, EPI_ISL_2562763, EPI_ISL_2562765, EPI_ISL_2562766, EPI_ISL_2562767, EPI_ISL_2562768, EPI_ISL_2562769, EPI_ISL_2562770, EPI_ISL_2562771, EPI_ISL_2562772, EPI_ISL_2562773, EPI_ISL_2562774, EPI_ISL_2562775, EPI_ISL_2562776, EPI_ISL_2562777, EPI_ISL_2562778, EPI_ISL_2562779, EPI_ISL_2562780, EPI_ISL_2562781, EPI_ISL_2562782, EPI_ISL_2562783, EPI_ISL_2562784, EPI_ISL_2562785, EPI_ISL_2562786, EPI_ISL_2562787, EPI_ISL_2562788, EPI_ISL_2562789, EPI_ISL_2562790, EPI_ISL_2562791, EPI_ISL_2562792, EPI_ISL_2562793, EPI_ISL_2562794, EPI_ISL_2562795, EPI_ISL_2598087, EPI_ISL_2598088, EPI_ISL_2598089, EPI_ISL_2598090, EPI_ISL_2598091, EPI_ISL_2598092, EPI_ISL_2598093, EPI_ISL_2598094, EPI_ISL_2598095, EPI_ISL_2598096, EPI_ISL_2598097, EPI_ISL_2598098, EPI_ISL_2598099, EPI_ISL_2598100, EPI_ISL_2598101, EPI_ISL_2598102, EPI_ISL_2598103, EPI_ISL_2598104, EPI_ISL_2598105, EPI_ISL_2598106, EPI_ISL_2598107, EPI_ISL_2598108, EPI_ISL_2598109, EPI_ISL_2598110, EPI_ISL_2598111, EPI_ISL_2598112, EPI_ISL_2598113, EPI_ISL_2598114, EPI_ISL_2598115, EPI_ISL_2598116, EPI_ISL_2598117, EPI_ISL_2598118, EPI_ISL_2598119 | | | |
| see above | TriCore Reference Laboratories | Center for Global Health, University of New Mexico Health Sciences Center | Cecilia Thompson; Darrell Dinwiddie; Daryl Domman; Karissa Culbreath; Kendra Pesko; Kurt Schwalm; Valerie Morley |
| EPI_ISL_2671483, EPI_ISL_2671700, EPI_ISL_2671701, EPI_ISL_2671702, EPI_ISL_2671758, EPI_ISL_2671759, EPI_ISL_2671760, EPI_ISL_2671761, EPI_ISL_2671762, EPI_ISL_2671763, EPI_ISL_2671764, EPI_ISL_2671765, EPI_ISL_2671766, EPI_ISL_2671767, EPI_ISL_2671768, EPI_ISL_2671769, EPI_ISL_2671770, EPI_ISL_2671771, EPI_ISL_2671772, EPI_ISL_2671773, EPI_ISL_2671774, EPI_ISL_2671775, EPI_ISL_2671776, EPI_ISL_2671777, EPI_ISL_2671778, EPI_ISL_2671779, EPI_ISL_2671780, EPI_ISL_2942467, EPI_ISL_2942468, EPI_ISL_2942469, EPI_ISL_2942470, EPI_ISL_2942471, EPI_ISL_2942472, EPI_ISL_2942473, EPI_ISL_2942474, EPI_ISL_2942475, EPI_ISL_2942476, EPI_ISL_2942477, EPI_ISL_2942478, EPI_ISL_2942479, EPI_ISL_2942480, EPI_ISL_2942750 | | | |
| see above | Unidad de Investigacion Biomedica de Zacatecas (UIBZ) | Unidad de Genomica Avanzada | ; Alejandra Garcia-Gasca; Alejandra Hernandez-Teran; Alejandro Sanchez-Flores; Alfredo Herrera-Estrella; Alicia Ocaña-Mondragon; Andreu Comas-Garcia; Angel Gustavo Salas-Lais; Antonio Loza Roman; Bernardo Martinez-Miguel; Blanca Taboada; Brenda Irasema Maldonado-Meza; Bruno Gomez-Gil; Carla Ivon Herrera-Najera; Carlos F. Arias; Celia Boukadida; Celida Duque Molina; Celida Martinez- Rodriguez; Clara Esperanza Santacruz-Tinoco; Concepcion Grajales-Muñiz; Consorcio Mexicano de Vigilancia Genomica (CoViGen-Mex). Authors (in alphabetical order): Julio Elias Alvarado-Yaah; Cristobal Chaidez-Quiroz; Daniel Fregoso-Rueda; Daniel Lira Morales; Eduardo Becerril-Vargas; Fernando Fontove-Herrera; Fidencio Mejia-Nepomuceno; Francisco Pulido; Gloria Elena Espinosa-Ayala; Gloria Maria Molina-Salinas; Gloria Vazquez; Hector Esteban Paz-Juarez; Hector Montoya-Fuentes; Helen Haydee Fernanda Ramirez-Plascencia; Irvin Gonzalez-Lopez; Jean Pierre Gonzalez; Jesus Hernandez; Joel Armando Vazquez-Perez.; Jorge Salas-Hernandez; Jose Antonio Enciso-Moreno; Jose Arturo Martinez-Orozco; Jose Esteban Muñoz-Medina; Jose de Jesus Nuñez-Contreras; Juan Bautista Chale-Dzul; Julissa Enciso-Ibarra; Luis Alberto Ochoa-Carrera; Margarita Matias-Florentino; Maria Guadalupe Santiago-Mauricio; Maria Guadalupe de Jesus Mireles-Rivera; Mario Mujica-Sanchez; Marissa Perez-Garcia; Nelly Selem-Mojica; Pavel Isa; Ricardo Ciria Merce; Ricardo Grande; Rosa Maria Gutierrez Rios; Santiago avila-Rios; Selene Zarate; Susana Lopez; Veronica Mata-Haro; Victor Eduardo Garcia-Arias; Victor Hugo Borja-Aburto |
| EPI_ISL_2671493, EPI_ISL_2671494, EPI_ISL_2671495, EPI_ISL_2671496, EPI_ISL_2671497, EPI_ISL_2671498, EPI_ISL_2671499, EPI_ISL_2671500, EPI_ISL_2671501, EPI_ISL_2671502, EPI_ISL_2671503, EPI_ISL_2671504, EPI_ISL_2671653, EPI_ISL_2671654, EPI_ISL_2671655, EPI_ISL_2671656, EPI_ISL_2671657, EPI_ISL_2671658, EPI_ISL_2671659, EPI_ISL_2671660, EPI_ISL_2671661, EPI_ISL_2671662, EPI_ISL_2671663, EPI_ISL_2671664, EPI_ISL_2671665, EPI_ISL_2671666, EPI_ISL_2671667, EPI_ISL_2671668, EPI_ISL_2671669, EPI_ISL_2671670, EPI_ISL_2671671, EPI_ISL_2671672, EPI_ISL_2671673, EPI_ISL_2671674, EPI_ISL_2671675, EPI_ISL_2671676, EPI_ISL_2671677, EPI_ISL_2671678, EPI_ISL_2671679, EPI_ISL_2671680, EPI_ISL_2671681, EPI_ISL_2671682, EPI_ISL_2671683, EPI_ISL_2671684, EPI_ISL_2671685, EPI_ISL_2671686, EPI_ISL_2671687, EPI_ISL_2671688, EPI_ISL_2671689, EPI_ISL_2671690, EPI_ISL_2671691, EPI_ISL_2671692, EPI_ISL_2671693, EPI_ISL_2671694, EPI_ISL_2671695, EPI_ISL_2671696, EPI_ISL_2671697, EPI_ISL_2671704, EPI_ISL_2671705, EPI_ISL_2671706, EPI_ISL_2671707, EPI_ISL_2671708, EPI_ISL_2671709, EPI_ISL_2671710, EPI_ISL_2671711, EPI_ISL_2671712, EPI_ISL_2671713, EPI_ISL_2671714, EPI_ISL_2671715, EPI_ISL_2671746, EPI_ISL_2671747, EPI_ISL_2671748, EPI_ISL_2671749, EPI_ISL_2671750, EPI_ISL_2671751, EPI_ISL_2671752, EPI_ISL_2671753, EPI_ISL_2671754, EPI_ISL_2671755, EPI_ISL_2671756, EPI_ISL_2671757, EPI_ISL_2671781, EPI_ISL_2671782, EPI_ISL_2671783, EPI_ISL_2671784, EPI_ISL_2671785, EPI_ISL_2942326, EPI_ISL_2942327, EPI_ISL_2942328, EPI_ISL_2942329, EPI_ISL_2942330, EPI_ISL_2942331, EPI_ISL_2942332, EPI_ISL_2942333, EPI_ISL_2942334, EPI_ISL_2942335, EPI_ISL_2942336, EPI_ISL_2942337, EPI_ISL_2942338, EPI_ISL_2942339, EPI_ISL_2942340, EPI_ISL_2942341, EPI_ISL_2942342, EPI_ISL_2942343, EPI_ISL_2942344, EPI_ISL_2942345, EPI_ISL_2942346, EPI_ISL_2942347, EPI_ISL_2942348, EPI_ISL_2942349, EPI_ISL_2942350, EPI_ISL_2942351, EPI_ISL_2942352, EPI_ISL_2942353, EPI_ISL_2942354, EPI_ISL_2942355, EPI_ISL_2942356, EPI_ISL_2942357, EPI_ISL_2942358, EPI_ISL_2942359, EPI_ISL_2942360, EPI_ISL_2942361, EPI_ISL_2942362, EPI_ISL_2942363, EPI_ISL_2942364, EPI_ISL_2942365, EPI_ISL_2942366, EPI_ISL_2942367, EPI_ISL_2942368, EPI_ISL_2942369, EPI_ISL_2942370, EPI_ISL_2942371, EPI_ISL_2942372, EPI_ISL_2942373, EPI_ISL_2942374, EPI_ISL_2942375, EPI_ISL_2942376, EPI_ISL_2942377, EPI_ISL_2942378, EPI_ISL_2942379, EPI_ISL_2942380, EPI_ISL_2942381, EPI_ISL_2942382, EPI_ISL_2942383, EPI_ISL_2942384, EPI_ISL_2942385, EPI_ISL_2942386, EPI_ISL_2942387, EPI_ISL_2942388, EPI_ISL_2942389, EPI_ISL_2942390, EPI_ISL_2942391, EPI_ISL_2942392, EPI_ISL_2942393, EPI_ISL_2942394, EPI_ISL_2942395, EPI_ISL_2942396, EPI_ISL_2942397, EPI_ISL_2942398, EPI_ISL_2942399, EPI_ISL_2942400, EPI_ISL_2942401, EPI_ISL_2942402, EPI_ISL_2942403, EPI_ISL_2942404, EPI_ISL_2942405, EPI_ISL_2942406, EPI_ISL_2942407, EPI_ISL_2942626, EPI_ISL_2942627, EPI_ISL_2942628, EPI_ISL_2942629, EPI_ISL_2942630, EPI_ISL_2942631, EPI_ISL_2942632, EPI_ISL_2942633, EPI_ISL_2942634, EPI_ISL_2942635, EPI_ISL_2942636, EPI_ISL_2942637, EPI_ISL_2942638, EPI_ISL_2942639, EPI_ISL_2942640, EPI_ISL_2942641, EPI_ISL_2942642, EPI_ISL_2942643, EPI_ISL_2942644, EPI_ISL_2942645, EPI_ISL_2942646, EPI_ISL_2942647, EPI_ISL_2942648, EPI_ISL_2942649, EPI_ISL_2942650, EPI_ISL_2942651, EPI_ISL_2942652, EPI_ISL_2942653, EPI_ISL_2942654, EPI_ISL_2942655, EPI_ISL_2942656, EPI_ISL_2942657, EPI_ISL_2942658, EPI_ISL_2942659, EPI_ISL_2942660, EPI_ISL_2942661, EPI_ISL_2942662, EPI_ISL_2942663, EPI_ISL_2942664, EPI_ISL_2942665, EPI_ISL_2942666, EPI_ISL_2942667, EPI_ISL_2942668, EPI_ISL_2942669, EPI_ISL_2942670, EPI_ISL_2942671, EPI_ISL_2942672, EPI_ISL_2942673, EPI_ISL_2942674, EPI_ISL_2942675, EPI_ISL_2942676, EPI_ISL_2942677, EPI_ISL_2942678, EPI_ISL_2942679, EPI_ISL_2942680, EPI_ISL_2942681, EPI_ISL_2942682, EPI_ISL_2942683, EPI_ISL_2942684, EPI_ISL_2942685, EPI_ISL_2942686, EPI_ISL_2942687, EPI_ISL_2942688, EPI_ISL_2942689, EPI_ISL_2942690, EPI_ISL_2942691, EPI_ISL_2942692, EPI_ISL_2942693, EPI_ISL_2942694, EPI_ISL_2942695, EPI_ISL_2942696, EPI_ISL_2942697, EPI_ISL_2942698, EPI_ISL_2942699, EPI_ISL_2942700, EPI_ISL_2942701, EPI_ISL_2942702, EPI_ISL_2942703, EPI_ISL_2942704, EPI_ISL_2942705, EPI_ISL_2942706, EPI_ISL_2942707, EPI_ISL_2942708, EPI_ISL_2942709, EPI_ISL_2942710, EPI_ISL_2942711, EPI_ISL_2942712, EPI_ISL_2942713, EPI_ISL_2942714, EPI_ISL_2942715, EPI_ISL_2942716, EPI_ISL_2942717, EPI_ISL_2942718, EPI_ISL_2942719 | | | |
| see above | Unidad de Investigacion Medica de Yucatan (UIMY) | Unidad de Genomica Avanzada | ; Alejandra Garcia-Gasca; Alejandra Hernandez-Teran; Alejandro Sanchez-Flores; Alfredo Herrera-Estrella; Alicia Ocaña-Mondragon; Andreu Comas-Garcia; Angel Gustavo Salas-Lais; Antonio Loza Roman; Bernardo Martinez-Miguel; Blanca Taboada; Brenda Irasema Maldonado-Meza; Bruno Gomez-Gil; Carla Ivon Herrera-Najera; Carlos F. Arias; Celia Boukadida; Celida Duque Molina; Celida Martinez- Rodriguez; Clara Esperanza Santacruz-Tinoco; Concepcion Grajales-Muñiz; Consorcio Mexicano de Vigilancia Genomica (CoViGen-Mex). Authors (in alphabetical order): Julio Elias Alvarado-Yaah; Cristobal Chaidez-Quiroz; Daniel Fregoso-Rueda; Daniel Lira Morales; Eduardo Becerril-Vargas; Fernando Fontove-Herrera; Fidencio Mejia-Nepomuceno; Francisco Pulido; Gloria Elena Espinosa-Ayala; Gloria Maria Molina-Salinas; Gloria Vazquez; Hector Esteban Paz-Juarez; Hector Montoya-Fuentes; Helen Haydee Fernanda Ramirez-Plascencia; Irvin Gonzalez-Lopez; Jean Pierre Gonzalez; Jesus Hernandez; Joel Armando Vazquez-Perez.; Jorge Salas-Hernandez; Jose Antonio Enciso-Moreno; Jose Arturo Martinez-Orozco; Jose Esteban Muñoz-Medina; Jose de Jesus Nuñez-Contreras; Juan Bautista Chale-Dzul; Julissa Enciso-Ibarra; Luis Alberto Ochoa-Carrera; Margarita Matias-Florentino; Maria Guadalupe Santiago-Mauricio; Maria Guadalupe de Jesus Mireles-Rivera; Mario Mujica-Sanchez; Marissa Perez-Garcia; Nelly Selem-Mojica; Pavel Isa; Ricardo Ciria Merce; Ricardo Grande; Rosa Maria Gutierrez Rios; Santiago avila-Rios; Selene Zarate; Susana Lopez; Veronica Mata-Haro; Victor Eduardo Garcia-Arias; Victor Hugo Borja-Aburto |
| EPI_ISL_2490310, EPI_ISL_2490333, EPI_ISL_2490335, EPI_ISL_2490348, EPI_ISL_2490358, EPI_ISL_2490385, EPI_ISL_2490386, EPI_ISL_2490401, EPI_ISL_2490424, EPI_ISL_2490500, EPI_ISL_2490501, EPI_ISL_2490502, EPI_ISL_2490503, EPI_ISL_2490504, EPI_ISL_2490505, EPI_ISL_2490506, EPI_ISL_2490507, EPI_ISL_2490508, EPI_ISL_2490509, EPI_ISL_2490510, EPI_ISL_2490511, EPI_ISL_2490512, EPI_ISL_2490513, EPI_ISL_2490514, EPI_ISL_2490515, EPI_ISL_2490516, EPI_ISL_2490517, EPI_ISL_2490518, EPI_ISL_2490519, EPI_ISL_2490520, EPI_ISL_2490521, EPI_ISL_2490522, EPI_ISL_2490523, EPI_ISL_2490524, EPI_ISL_2490525, EPI_ISL_2490526, EPI_ISL_2490527, EPI_ISL_2490528, EPI_ISL_2490529, EPI_ISL_2490530, EPI_ISL_2490531, EPI_ISL_2490532, EPI_ISL_2490533, EPI_ISL_2490535, EPI_ISL_2490536, EPI_ISL_2490537, EPI_ISL_2490538, EPI_ISL_2490539, EPI_ISL_2490540, EPI_ISL_2490541, EPI_ISL_2835937, EPI_ISL_2835938, EPI_ISL_2835939, EPI_ISL_2835940, EPI_ISL_2835941, EPI_ISL_2835942, EPI_ISL_2835943, EPI_ISL_2835944, EPI_ISL_2835945 | | | |
| see above | Unidad de Investigación Biomédica de Zacatecas (UIBZ) | Centro de Investigación en Enfermedades Infecciosas (CIENI), Instituto Nacional de Enfermedades Respiratorias (INER) | ; Alejandra Garcia-Gasca; Alejandra Hernández-Terán; Alejandro Sanchez-Flores; Alfredo Herrera-Estrella; Alicia Ocaña-Mondragón; Andreu Comas-García; Angel Gustavo Salas-Lais; Antonio Loza Román; Bernardo Martínez-Miguel; Blanca Taboada; Brenda Irasema Maldonado-Meza; Bruno Gomez-Gil; Carla Ivón Herrera-Najera; Carlos F. Arias; Celia Boukadida; Clara Esperanza Santacruz-Tinoco; Concepción Grajales-Muñiz; Consorcio Mexicano de Vigilancia Genómica (CoViGen-Mex). Authors (in alphabetical order): Julio Elias Alvarado-Yaah; Cristóbal Cháidez-Quiróz; Célida Duque Molina; Célida Martínez- Rodríguez; Daniel Fregoso-Rueda; Daniel Lira Morales; Eduardo Becerril-Vargas; Fernando Fontove-Herrera; Fidencio Mejía-Nepomuceno; Francisco Pulido; Gloria Elena Espinosa-Ayala; Gloria María Molina-Salinas; Gloria Vazquez; Hector Esteban Paz-Juárez; Hector Montoya-Fuentes; Helen Haydee Fernanda Ramírez-Plascencia; Irvin González-López; Jean Pierre González; Joel Armando Vázquez-Pérez.; Jorge Salas-Hernández; José Antonio Enciso-Moreno; José Arturo Martínez-Orozco; José Esteban Muñoz-Medina; José de Jesús Nuñez-Contreras; Juan Bautista Chale-Dzul; Julissa Enciso-Ibarra; Luis Alberto Ochoa-Carrera; Margarita Matías-Florentino; Mario Mújica-Sánchez; Marissa Perez-Garcia; María Guadalupe Santiago-Mauricio; María Guadalupe de Jesús Mireles-Rivera; Nelly Sélem-Mojica; Pavel Isa; Ricardo Ciria Merce; Ricardo Grande; Rosa María Gutierrez Rios; Santiago Ávila-Ríos; Selene Zárate; Susana Lopez; Victor Eduardo Garcia-Arias; Víctor Hugo Borja-Aburto |
| EPI_ISL_2681047, EPI_ISL_2681048, EPI_ISL_2681049, EPI_ISL_2681052, EPI_ISL_2681057, EPI_ISL_2681068, EPI_ISL_2681092, EPI_ISL_2681097, EPI_ISL_2681119, EPI_ISL_2681120, EPI_ISL_2681121, EPI_ISL_2681122, EPI_ISL_2681123, EPI_ISL_2681142, EPI_ISL_2681160, EPI_ISL_2681190, EPI_ISL_2681207, EPI_ISL_2681209, EPI_ISL_2681212, EPI_ISL_2681221, EPI_ISL_2681223, EPI_ISL_2681234, EPI_ISL_2681235, EPI_ISL_2681239, EPI_ISL_2681242, EPI_ISL_2681246, EPI_ISL_2681272, EPI_ISL_2681286, EPI_ISL_2681291, EPI_ISL_2681293, EPI_ISL_2681309, EPI_ISL_2681315, EPI_ISL_2681317, EPI_ISL_2681339, EPI_ISL_2681340, EPI_ISL_2681346, EPI_ISL_2681349, EPI_ISL_2681354, EPI_ISL_2681355, EPI_ISL_2681362, EPI_ISL_2681367, EPI_ISL_2681376, EPI_ISL_2681381, EPI_ISL_2801697, EPI_ISL_2801698, EPI_ISL_2801699, EPI_ISL_2801700, EPI_ISL_2801701, EPI_ISL_2801702, EPI_ISL_2801703, EPI_ISL_2801704, EPI_ISL_2801705, EPI_ISL_2801706 | | | |
| see above | Unidad de Investigación Biomédica de Zacatecas (UIBZ) | Instituto de Biotecnología de la UNAM | ; Alejandra García-Gasca; Alejandra Hernández-Terán; Alejandro Sánchez-Flores; Alfredo Herrera-Estrella; Alicia Ocaña-Mondragón; Andreu Comas-García; Angel Gustavo Salas-Lais; Antonio Loza Román; Bernardo Martínez-Miguel; Blanca Taboada; Brenda Irasema Maldonado-Meza; Bruno Gómez-Gil; Carla Ivón Herrera-Najera; Carlos F. Arias; Celia Boukadida; Clara Esperanza Santacruz-Tinoco; Concepción Grajales-Muñiz; Consorcio Mexicano de Vigilancia Genómica (CoViGen-Mex). Authors (in alphabetical order): Julio Elias Alvarado-Yaah; Cristóbal Cháidez-Quiróz; Célida Duque Molina; Célida Martínez- Rodríguez; Daniel Fregoso-Rueda; Daniel Lira Morales; Eduardo Becerril-Vargas; Fernando Fontove-Herrera; Fidencio Mejía-Nepomuceno; Francisco Pulido; Gloria Elena Espinosa-Ayala; Gloria María Molina-Salinas; Gloria Vazquez; Hector Esteban Paz-Juárez; Hector Montoya-Fuentes; Helen Haydee Fernanda Ramírez-Plascencia; Irvin González-López; Jean Pierre González; Jesús Hernández; Joel Armando Vázquez-Pérez.; Jorge Salas-Hernández; José Antonio Enciso-Moreno; José Arturo Martínez-Orozco; José Esteban Muñoz-Medina; José de Jesús Nuñez-Contreras; Juan Bautista Chale-Dzul; Julissa Enciso-Ibarra; Luis Alberto Ochoa-Carrera; Margarita Matías-Florentino; Mario Mújica-Sánchez; Marissa Perez-Garcia; María Guadalupe Santiago-Mauricio; María Guadalupe de Jesús Mireles-Rivera; Nelly Sélem-Mojica; Pavel Isa; Ricardo Ciria Merce; Ricardo Grande; Rosa María Gutiérrez Rios; Santiago Ávila-Ríos; Selene Zárate; Susana Lopez; Verónica Mata-Haro; Victor Eduardo García-Arias; Víctor Hugo Borja-Aburto |
| EPI_ISL_2402126, EPI_ISL_2402127, EPI_ISL_2402129, EPI_ISL_2402130, EPI_ISL_2402131 | Unidad de Investigación Biomédica de Zacatecas (UIBZ) | Unidad de Genomica Avanzada | Alejandro Sanchez-Flores; Alfredo Herrera-Estrella; Alicia Ocana-Mondragon; Angel Gustavo Salas-Lais; Bernardo Martinez-Miguel; Blanca Taboada; Brenda Irasema Maldonado-Meza; Carla Ivon Herrera-Najera; Carlos F. Arias; Celia Boukadida; Clara Esperanza Santacruz-Tinoco; Concepcion Grajales-Muniz; Consorcio Mexicano de Vigilancia Genomica (CoViGen-Mex). Authors (in alphabetical order): Julio Elias Alvarado-Yaah; Fernando Fontove-Herrera; Francisco Pulido; Gloria Elena Espinoza-Ayala; Gloria Maria Molina-Salinas; Gloria Vazquez; Hector Esteban Paz-Juarez; Hector Montoya-Fuentes; Helen Haydee Fernanda Ramirez-Plascencia; Jorge Ivan Salinal-Nevarez; Jose Antonio Enciso-Moreno; Jose Esteban Munoz-Medina; Jose de Jesus Nunez-Contreras; Juan Bautista Chale-Dzul; Luis Alberto Ochoa-Carrera; Margarita Matias-Florentino; Maria Guadalupe Santiago-Mauricio; Maria Guadalupe de Jesus Mireles-Rivera; Nelly Selem-Mojica; Pavel Isa; Ricardo Grande; Santiago Avila-Rios; Victor Hugo Borja-Aburto |
| EPI_ISL_2490311, EPI_ISL_2490328, EPI_ISL_2490329, EPI_ISL_2490330, EPI_ISL_2490346, EPI_ISL_2490347, EPI_ISL_2490357, EPI_ISL_2490369, EPI_ISL_2490370, EPI_ISL_2490371, EPI_ISL_2490372, EPI_ISL_2490373, EPI_ISL_2490374, EPI_ISL_2490375, EPI_ISL_2490376, EPI_ISL_2490377, EPI_ISL_2490378, EPI_ISL_2490379, EPI_ISL_2490380, EPI_ISL_2490381, EPI_ISL_2490420, EPI_ISL_2490421, EPI_ISL_2490422, EPI_ISL_2490475, EPI_ISL_2490476, EPI_ISL_2490477, EPI_ISL_2490478, EPI_ISL_2490479, EPI_ISL_2490480, EPI_ISL_2490481, EPI_ISL_2490482, EPI_ISL_2490483, EPI_ISL_2490484, EPI_ISL_2490485, EPI_ISL_2490486, EPI_ISL_2490487, EPI_ISL_2490488, EPI_ISL_2490489, EPI_ISL_2490490, EPI_ISL_2490491, EPI_ISL_2490492, EPI_ISL_2835930, EPI_ISL_2835931, EPI_ISL_2835932, EPI_ISL_2835933 | | | |
| see above | Unidad de Investigación Médica de Yucatán (UIMY) | Centro de Investigación en Enfermedades Infecciosas (CIENI), Instituto Nacional de Enfermedades Respiratorias (INER) | ; Alejandra Garcia-Gasca; Alejandra Hernández-Terán; Alejandro Sanchez-Flores; Alfredo Herrera-Estrella; Alicia Ocaña-Mondragón; Andreu Comas-García; Angel Gustavo Salas-Lais; Antonio Loza Román; Bernardo Martínez-Miguel; Blanca Taboada; Brenda Irasema Maldonado-Meza; Bruno Gomez-Gil; Carla Ivón Herrera-Najera; Carlos F. Arias; Celia Boukadida; Clara Esperanza Santacruz-Tinoco; Concepción Grajales-Muñiz; Consorcio Mexicano de Vigilancia Genómica (CoViGen-Mex). Authors (in alphabetical order): Julio Elias Alvarado-Yaah; Cristóbal Cháidez-Quiróz; Célida Duque Molina; Célida Martínez- Rodríguez; Daniel Fregoso-Rueda; Daniel Lira Morales; Eduardo Becerril-Vargas; Fernando Fontove-Herrera; Fidencio Mejía-Nepomuceno; Francisco Pulido; Gloria Elena Espinosa-Ayala; Gloria María Molina-Salinas; Gloria Vazquez; Hector Esteban Paz-Juárez; Hector Montoya-Fuentes; Helen Haydee Fernanda Ramírez-Plascencia; Irvin González-López; Jean Pierre González; Joel Armando Vázquez-Pérez.; Jorge Salas-Hernández; José Antonio Enciso-Moreno; José Arturo Martínez-Orozco; José Esteban Muñoz-Medina; José de Jesús Nuñez-Contreras; Juan Bautista Chale-Dzul; Julissa Enciso-Ibarra; Luis Alberto Ochoa-Carrera; Margarita Matías-Florentino; Mario Mújica-Sánchez; Marissa Perez-Garcia; María Guadalupe Santiago-Mauricio; María Guadalupe de Jesús Mireles-Rivera; Nelly Sélem-Mojica; Pavel Isa; Ricardo Ciria Merce; Ricardo Grande; Rosa María Gutierrez Rios; Santiago Ávila-Ríos; Selene Zárate; Susana Lopez; Victor Eduardo Garcia-Arias; Víctor Hugo Borja-Aburto |
| EPI_ISL_2681050, EPI_ISL_2681055, EPI_ISL_2681067, EPI_ISL_2681074, EPI_ISL_2681075, EPI_ISL_2681077, EPI_ISL_2681078, EPI_ISL_2681079, EPI_ISL_2681080, EPI_ISL_2681082, EPI_ISL_2681083, EPI_ISL_2681084, EPI_ISL_2681085, EPI_ISL_2681088, EPI_ISL_2681089, EPI_ISL_2681090, EPI_ISL_2681091, EPI_ISL_2681094, EPI_ISL_2681098, EPI_ISL_2681099, EPI_ISL_2681100, EPI_ISL_2681101, EPI_ISL_2681106, EPI_ISL_2681107, EPI_ISL_2681108, EPI_ISL_2681109, EPI_ISL_2681114, EPI_ISL_2681115, EPI_ISL_2681116, EPI_ISL_2681117, EPI_ISL_2681118, EPI_ISL_2681126, EPI_ISL_2681130, EPI_ISL_2681133, EPI_ISL_2681136, EPI_ISL_2681141, EPI_ISL_2681176, EPI_ISL_2681211, EPI_ISL_2681218, EPI_ISL_2681232, EPI_ISL_2681233, EPI_ISL_2681236, EPI_ISL_2681241, EPI_ISL_2681244, EPI_ISL_2681261, EPI_ISL_2681274, EPI_ISL_2681275, EPI_ISL_2681292, EPI_ISL_2681295, EPI_ISL_2681298, EPI_ISL_2681300, EPI_ISL_2681304, EPI_ISL_2681311, EPI_ISL_2681319, EPI_ISL_2681320, EPI_ISL_2681322, EPI_ISL_2681325, EPI_ISL_2681326, EPI_ISL_2681327, EPI_ISL_2681328, EPI_ISL_2681333, EPI_ISL_2681338, EPI_ISL_2681341, EPI_ISL_2681342, EPI_ISL_2681345, EPI_ISL_2681351, EPI_ISL_2681353, EPI_ISL_2681356, EPI_ISL_2681360, EPI_ISL_2681361, EPI_ISL_2681363, EPI_ISL_2681364, EPI_ISL_2681365, EPI_ISL_2681366, EPI_ISL_2681368, EPI_ISL_2681370, EPI_ISL_2681372, EPI_ISL_2681374, EPI_ISL_2681375, EPI_ISL_2681377, EPI_ISL_2681378, EPI_ISL_2681379, EPI_ISL_2681382, EPI_ISL_2681383, EPI_ISL_2681384, EPI_ISL_2681385, EPI_ISL_2801564, EPI_ISL_2801565, EPI_ISL_2801566, EPI_ISL_2801567, EPI_ISL_2801568, EPI_ISL_2801569, EPI_ISL_2801570, EPI_ISL_2801571, EPI_ISL_2801572, EPI_ISL_2801573, EPI_ISL_2801574, EPI_ISL_2801575, EPI_ISL_2801576, EPI_ISL_2801577, EPI_ISL_2801578, EPI_ISL_2801579, EPI_ISL_2801580, EPI_ISL_2801581, EPI_ISL_2801582, EPI_ISL_2801583, EPI_ISL_2801584, EPI_ISL_2801585, EPI_ISL_2801586, EPI_ISL_2801587, EPI_ISL_2801588, EPI_ISL_2801589, EPI_ISL_2801590, EPI_ISL_2801591, EPI_ISL_2801592, EPI_ISL_2801593, EPI_ISL_2801594, EPI_ISL_2801595, EPI_ISL_2801596, EPI_ISL_2801597, EPI_ISL_2801598, EPI_ISL_2801599, EPI_ISL_2801600, EPI_ISL_2801601, EPI_ISL_2801602, EPI_ISL_2801603, EPI_ISL_2801604, EPI_ISL_2801605, EPI_ISL_2801606, EPI_ISL_2801607, EPI_ISL_2801608, EPI_ISL_2801609, EPI_ISL_2801610, EPI_ISL_2801611, EPI_ISL_2801612, EPI_ISL_2801613, EPI_ISL_2801614, EPI_ISL_2801615, EPI_ISL_2801616, EPI_ISL_2801617, EPI_ISL_2801618, EPI_ISL_2801619, EPI_ISL_2801620, EPI_ISL_2801621, EPI_ISL_2801622, EPI_ISL_2801623, EPI_ISL_2801624, EPI_ISL_2801625, EPI_ISL_2801626, EPI_ISL_2801627, EPI_ISL_2801628, EPI_ISL_2801629, EPI_ISL_2801630, EPI_ISL_2801631, EPI_ISL_2801632, EPI_ISL_2801633, EPI_ISL_2801634, EPI_ISL_2801635, EPI_ISL_2801636, EPI_ISL_2801637, EPI_ISL_2801638, EPI_ISL_2801639, EPI_ISL_2801640, EPI_ISL_2801641, EPI_ISL_2801642, EPI_ISL_2801643, EPI_ISL_2801644, EPI_ISL_2801645, EPI_ISL_2801646, EPI_ISL_2801647, EPI_ISL_2801648, EPI_ISL_2801649, EPI_ISL_2801650, EPI_ISL_2801651, EPI_ISL_2801652, EPI_ISL_2801653, EPI_ISL_2801654, EPI_ISL_2801655, EPI_ISL_2801656, EPI_ISL_2801657 | | | |
| see above | Unidad de Investigación Médica de Yucatán (UIMY) | Instituto de Biotecnología de la UNAM | ; Alejandra García-Gasca; Alejandra Hernández-Terán; Alejandro Sánchez-Flores; Alfredo Herrera-Estrella; Alicia Ocaña-Mondragón; Andreu Comas-García; Angel Gustavo Salas-Lais; Antonio Loza Román; Bernardo Martínez-Miguel; Blanca Taboada; Brenda Irasema Maldonado-Meza; Bruno Gómez-Gil; Carla Ivón Herrera-Najera; Carlos F. Arias; Celia Boukadida; Clara Esperanza Santacruz-Tinoco; Concepción Grajales-Muñiz; Consorcio Mexicano de Vigilancia Genómica (CoViGen-Mex). Authors (in alphabetical order): Julio Elias Alvarado-Yaah; Cristóbal Cháidez-Quiróz; Célida Duque Molina; Célida Martínez- Rodríguez; Daniel Fregoso-Rueda; Daniel Lira Morales; Eduardo Becerril-Vargas; Fernando Fontove-Herrera; Fidencio Mejía-Nepomuceno; Francisco Pulido; Gloria Elena Espinosa-Ayala; Gloria María Molina-Salinas; Gloria Vazquez; Hector Esteban Paz-Juárez; Hector Montoya-Fuentes; Helen Haydee Fernanda Ramírez-Plascencia; Irvin González-López; Jean Pierre González; Jesús Hernández; Joel Armando Vázquez-Pérez.; Jorge Salas-Hernández; José Antonio Enciso-Moreno; José Arturo Martínez-Orozco; José Esteban Muñoz-Medina; José de Jesús Nuñez-Contreras; Juan Bautista Chale-Dzul; Julissa Enciso-Ibarra; Luis Alberto Ochoa-Carrera; Margarita Matías-Florentino; Mario Mújica-Sánchez; Marissa Perez-Garcia; María Guadalupe Santiago-Mauricio; María Guadalupe de Jesús Mireles-Rivera; Nelly Sélem-Mojica; Pavel Isa; Ricardo Ciria Merce; Ricardo Grande; Rosa María Gutiérrez Rios; Santiago Ávila-Ríos; Selene Zárate; Susana Lopez; Verónica Mata-Haro; Victor Eduardo García-Arias; Víctor Hugo Borja-Aburto |
| EPI_ISL_2207266, EPI_ISL_2248704, EPI_ISL_2248705, EPI_ISL_2248706, EPI_ISL_2248707, EPI_ISL_2248708, EPI_ISL_2248709, EPI_ISL_2248710, EPI_ISL_2248711, EPI_ISL_2248714, EPI_ISL_2248717, EPI_ISL_2445972, EPI_ISL_2445974 | | | |

| | | | |
|---|---|---|---|
| see above | Univeristy of New Mexico Hospital | Center for Global Health, University of New Mexico Health Sciences Center | Darrell Dinwiddie; Daryl Domman; Jon Femling; Justin Bacca; Kurt Schwalm; Valerie Morley |
| EPI_ISL_2648122 | Willis-Knighton Medical Center Hospital Laboratory | LSUHS Emerging Viral Threat Laboratory | Alexander Mijalis; Andrew D. Yurochko; April N. Johnson; Christopher G. Kevil; Gregory L. Ware; Jennifer L. Carroll; Jeremy P. Kamil; John A. Vanchiere; Joseph A. Bocchini; Lorie M. Atkins; Maarten Van Diest; Rona S. Scott |

We gratefully acknowledge the following Authors from the Originating laboratories responsible for obtaining the specimens, as well as the Submitting laboratories where the genome data were generated and shared via GISAID, on which this research is based.

All Submitters of data may be contacted directly via www.gisaid.org

Authors are sorted alphabetically.

| Accession ID | Originating Laboratory | Submitting Laboratory | Authors |
|---|---|---|---|
| EPI_ISL_1446676, EPI_ISL_1446677 | "NYSDOH Wadsworth Center, Virology Lab" | Centers for Disease Control and Prevention Division of Viral Diseases, Pathogen Discovery | Alison Laufer Halpin; Ben L. Rambo-Martin; Clinton R. Paden; Dakota Howard; Darlene Wagner; Dave Wentworth; Dhwani Batra; Jasmine Padilla; Justin Lee; Katie Dillon; Krista Queen; Kristen Knipe; Kristine Lacek; Mark Burroughs; Matthew Schmerer; Mili Sheth; Peter Cook; Sam Shepard; Sarah Nobles; Shoshona Le; Suxiang Tong; Vivien Dugan; Yvette Unoarumhi |
| EPI_ISL_1397853, EPI_ISL_1397854, EPI_ISL_1397855, EPI_ISL_1397856, EPI_ISL_1397858, EPI_ISL_1397861, EPI_ISL_1397866, EPI_ISL_1397867, EPI_ISL_1397868, EPI_ISL_1397869, EPI_ISL_1398027, EPI_ISL_1398153, EPI_ISL_1398276, EPI_ISL_1398277, EPI_ISL_1398278, EPI_ISL_1398279, EPI_ISL_1398280, EPI_ISL_1398281, EPI_ISL_1398282, EPI_ISL_1398283, EPI_ISL_1461295, EPI_ISL_1461296, EPI_ISL_1461297, EPI_ISL_1461298, EPI_ISL_1461299, EPI_ISL_1462506, EPI_ISL_1462508, EPI_ISL_1465639, EPI_ISL_1482455, EPI_ISL_1482456, EPI_ISL_1482457, EPI_ISL_1482458, EPI_ISL_1482459, EPI_ISL_1482460, EPI_ISL_1482461, EPI_ISL_1482462, EPI_ISL_1482463, EPI_ISL_1482464, EPI_ISL_1482465, EPI_ISL_1482466, EPI_ISL_1482467, EPI_ISL_1482468, EPI_ISL_1482469, EPI_ISL_1576824, EPI_ISL_1580700, EPI_ISL_1580741, EPI_ISL_1582154, EPI_ISL_1582218, EPI_ISL_1582260, EPI_ISL_1582407, EPI_ISL_1671315, EPI_ISL_1671317, EPI_ISL_1671318, EPI_ISL_1671323, EPI_ISL_1671324, EPI_ISL_1671325, EPI_ISL_1671337, EPI_ISL_1671357, EPI_ISL_1671358, EPI_ISL_1755838, EPI_ISL_1755839, EPI_ISL_1756329 | | | |
| see above | ALBANY MEDICAL CENTER | Wadsworth Center, New York State Department of Health | Alexis Russel; Alexis Russell; Catharine Prussing; Daryl M. Lamson; Erasmus Schneider; Erica Lasek-Nesselquist; John Kelly; Jonathan Plitnick; Kirsten St. George; Matthew Shudt; Melissa A Leisner; Navjot Singh |
| EPI_ISL_1445307, EPI_ISL_1445308, EPI_ISL_1445380, EPI_ISL_1445424, EPI_ISL_1445443, EPI_ISL_1445702, EPI_ISL_1445705, EPI_ISL_1445878, EPI_ISL_1445881, EPI_ISL_1445889, EPI_ISL_1445906, EPI_ISL_1445928, EPI_ISL_1445943, EPI_ISL_1446075, EPI_ISL_1446114, EPI_ISL_1446139, EPI_ISL_1446140, EPI_ISL_1446144, EPI_ISL_1446148, EPI_ISL_1446152, EPI_ISL_1446190, EPI_ISL_1479449, EPI_ISL_1479456, EPI_ISL_1479460, EPI_ISL_1479461, EPI_ISL_1479477, EPI_ISL_1479549, EPI_ISL_1479604, EPI_ISL_1479823, EPI_ISL_1491743, EPI_ISL_1491783, EPI_ISL_1491784, EPI_ISL_1491815, EPI_ISL_1491849, EPI_ISL_1491887, EPI_ISL_1491937, EPI_ISL_1493507, EPI_ISL_1493519, EPI_ISL_1493520, EPI_ISL_1493534, EPI_ISL_1493540, EPI_ISL_1493559, EPI_ISL_1513202, EPI_ISL_1513204, EPI_ISL_1513205, EPI_ISL_1513229, EPI_ISL_1513231, EPI_ISL_1513233, EPI_ISL_1513257, EPI_ISL_1513258, EPI_ISL_1513259, EPI_ISL_1513261, EPI_ISL_1513337, EPI_ISL_1513339, EPI_ISL_1513368, EPI_ISL_1513473, EPI_ISL_1513482, EPI_ISL_1513562, EPI_ISL_1513564, EPI_ISL_1513567, EPI_ISL_1513590, EPI_ISL_1513592, EPI_ISL_1525019, EPI_ISL_1525020, EPI_ISL_1525037, EPI_ISL_1525038, EPI_ISL_1525043, EPI_ISL_1525050, EPI_ISL_1525057, EPI_ISL_1525058, EPI_ISL_1525108, EPI_ISL_1525137, EPI_ISL_1525181, EPI_ISL_1525183, EPI_ISL_1525218, EPI_ISL_1525239, EPI_ISL_1525243, EPI_ISL_1550397, EPI_ISL_1550503, EPI_ISL_1550549, EPI_ISL_1550668, EPI_ISL_1550684, EPI_ISL_1550694, EPI_ISL_1550820, EPI_ISL_1550871, EPI_ISL_1550989, EPI_ISL_1551037, EPI_ISL_1551052, EPI_ISL_1551082, EPI_ISL_1551091, EPI_ISL_1551103, EPI_ISL_1551116, EPI_ISL_1551213, EPI_ISL_1551330, EPI_ISL_1551343, EPI_ISL_1551344, EPI_ISL_1551390, EPI_ISL_1551457, EPI_ISL_1551499, EPI_ISL_1551502, EPI_ISL_1551504, EPI_ISL_1551511, EPI_ISL_1551512, EPI_ISL_1551612, EPI_ISL_1551626, EPI_ISL_1559638, EPI_ISL_1559639, EPI_ISL_1559646, EPI_ISL_1559688, EPI_ISL_1559691, EPI_ISL_1559698, EPI_ISL_1559704, EPI_ISL_1559705, EPI_ISL_1559715, EPI_ISL_1559717, EPI_ISL_1559751, EPI_ISL_1559791, EPI_ISL_1559800, EPI_ISL_1559817, EPI_ISL_1559826, EPI_ISL_1559884, EPI_ISL_1559888, EPI_ISL_1559890, EPI_ISL_1559892, EPI_ISL_1559893, EPI_ISL_1559920, EPI_ISL_1559929, EPI_ISL_1559946, EPI_ISL_1559947, EPI_ISL_1559948, EPI_ISL_1559949, EPI_ISL_1560017, EPI_ISL_1560022, EPI_ISL_1560024, EPI_ISL_1560048, EPI_ISL_1560049, EPI_ISL_1560071, EPI_ISL_1560186, EPI_ISL_1560187, EPI_ISL_1560199, EPI_ISL_1560244, EPI_ISL_1560265, EPI_ISL_1560269, EPI_ISL_1560287, EPI_ISL_1560291, EPI_ISL_1560358, EPI_ISL_1560402, EPI_ISL_1560403, EPI_ISL_1560462, EPI_ISL_1560493, EPI_ISL_1560494, EPI_ISL_1560667, EPI_ISL_1560668, EPI_ISL_1560698, EPI_ISL_1560855, EPI_ISL_1560859, EPI_ISL_1560871, EPI_ISL_1560875, EPI_ISL_1560901, EPI_ISL_1560902, EPI_ISL_1560907, EPI_ISL_1560968, EPI_ISL_1560971, EPI_ISL_1561025, EPI_ISL_1561031, EPI_ISL_1561124, EPI_ISL_1561127, EPI_ISL_1561228, EPI_ISL_1561230, EPI_ISL_1561257, EPI_ISL_1561259, EPI_ISL_1561270, EPI_ISL_1561273, EPI_ISL_1561274, EPI_ISL_1561275, EPI_ISL_1561276, EPI_ISL_1561292, EPI_ISL_1561293, EPI_ISL_1561388, EPI_ISL_1561408, EPI_ISL_1561415, EPI_ISL_1561424, EPI_ISL_1561503, EPI_ISL_1561512, EPI_ISL_1561513, EPI_ISL_1561532, EPI_ISL_1561653, EPI_ISL_1561654, EPI_ISL_1561655, EPI_ISL_1561670, EPI_ISL_1561671, EPI_ISL_1561672, EPI_ISL_1561691, EPI_ISL_1561720, EPI_ISL_1561732, EPI_ISL_1561743, EPI_ISL_1561766, EPI_ISL_1561771, EPI_ISL_1561775, EPI_ISL_1561776, EPI_ISL_1561777, EPI_ISL_1561778, EPI_ISL_1561835, EPI_ISL_1561837, EPI_ISL_1561839, EPI_ISL_1561840, EPI_ISL_1561843, EPI_ISL_1561854, EPI_ISL_1561865, EPI_ISL_1561890, EPI_ISL_1561905, EPI_ISL_1561909, EPI_ISL_1561913, EPI_ISL_1562066, EPI_ISL_1562068, EPI_ISL_1562150, EPI_ISL_1562154, EPI_ISL_1562220, EPI_ISL_1562234, EPI_ISL_1562245, EPI_ISL_1562246, EPI_ISL_1562251, EPI_ISL_1562275, EPI_ISL_1562276, EPI_ISL_1562293, EPI_ISL_1562303, EPI_ISL_1562316, EPI_ISL_1562369, EPI_ISL_1562370, EPI_ISL_1562372, EPI_ISL_1562417, EPI_ISL_1562452, EPI_ISL_1562496, EPI_ISL_1562507, EPI_ISL_1562510, EPI_ISL_1562545, EPI_ISL_1562555, EPI_ISL_1562569, EPI_ISL_1562609, EPI_ISL_1562614, EPI_ISL_1562615, EPI_ISL_1562622, EPI_ISL_1562623, EPI_ISL_1562625, EPI_ISL_1562752, EPI_ISL_1562784, EPI_ISL_1562785, EPI_ISL_1562786, EPI_ISL_1562787, EPI_ISL_1562788, EPI_ISL_1562789, EPI_ISL_1562814, EPI_ISL_1562937, EPI_ISL_1562949, EPI_ISL_1562952, EPI_ISL_1562956, EPI_ISL_1562959, EPI_ISL_1562968, EPI_ISL_1562969, EPI_ISL_1562994, EPI_ISL_1563070, EPI_ISL_1563156, EPI_ISL_1563184, EPI_ISL_1563241, EPI_ISL_1563249, EPI_ISL_1563295, EPI_ISL_1563299, EPI_ISL_1563330, EPI_ISL_1563332, EPI_ISL_1563355, EPI_ISL_1563356, EPI_ISL_1563377, EPI_ISL_1563381, EPI_ISL_1563578, EPI_ISL_1563587, EPI_ISL_1563593, EPI_ISL_1563606, EPI_ISL_1563617, EPI_ISL_1563618, EPI_ISL_1649006, EPI_ISL_1649008, EPI_ISL_1649024, EPI_ISL_1649040, EPI_ISL_1649113, EPI_ISL_1649116, EPI_ISL_1649154, EPI_ISL_1649155, EPI_ISL_1649185, EPI_ISL_1649209, EPI_ISL_1649223, EPI_ISL_1649228, EPI_ISL_1649230, EPI_ISL_1649234, EPI_ISL_1649235, EPI_ISL_1649236, EPI_ISL_1649266, EPI_ISL_1649277, EPI_ISL_1649278, EPI_ISL_1649293, EPI_ISL_1649295, EPI_ISL_1649385, EPI_ISL_1649392, EPI_ISL_1649393, EPI_ISL_1649394, EPI_ISL_1649459, EPI_ISL_1649462, EPI_ISL_1649463, EPI_ISL_1649482, EPI_ISL_1649483, EPI_ISL_1649523, EPI_ISL_1649589, EPI_ISL_1649646, EPI_ISL_1649685, EPI_ISL_1649687, EPI_ISL_1649729, EPI_ISL_1649753, EPI_ISL_1649759, EPI_ISL_1649763, EPI_ISL_1649820, EPI_ISL_1649840, EPI_ISL_1649845, EPI_ISL_1650018, EPI_ISL_1650049, EPI_ISL_1650053, EPI_ISL_1650054, EPI_ISL_1650055, EPI_ISL_1650056, EPI_ISL_1650061, EPI_ISL_1650132, EPI_ISL_1650143, EPI_ISL_1650144, EPI_ISL_1650158, EPI_ISL_1650159, EPI_ISL_1650233, EPI_ISL_1650237, EPI_ISL_1650240, EPI_ISL_1650241, EPI_ISL_1650242, EPI_ISL_1650243, EPI_ISL_1650259, EPI_ISL_1650287, EPI_ISL_1650293, EPI_ISL_1650313, EPI_ISL_1650326, EPI_ISL_1650352, EPI_ISL_1650383, EPI_ISL_1650384, EPI_ISL_1650411, EPI_ISL_1650436, EPI_ISL_1650555, EPI_ISL_1650657, EPI_ISL_1650664, EPI_ISL_1650678, EPI_ISL_1650686, EPI_ISL_1650687, EPI_ISL_1650711, EPI_ISL_1650839, EPI_ISL_1650997, EPI_ISL_1651147, EPI_ISL_1651148, EPI_ISL_1667754, EPI_ISL_1667773, EPI_ISL_1667838, EPI_ISL_1667839, EPI_ISL_1667840, EPI_ISL_1667951, EPI_ISL_1668130, EPI_ISL_1668180, EPI_ISL_1668181, EPI_ISL_1668185, EPI_ISL_1668190, EPI_ISL_1668373, EPI_ISL_1668374, EPI_ISL_1668375, EPI_ISL_1685483, EPI_ISL_1685561, EPI_ISL_1685578, EPI_ISL_1685586, EPI_ISL_1685611, EPI_ISL_1685612, EPI_ISL_1685689, EPI_ISL_1685703, EPI_ISL_1685713, EPI_ISL_1685824, EPI_ISL_1685834, EPI_ISL_1685835, EPI_ISL_1685842, EPI_ISL_1686008, EPI_ISL_1686294, EPI_ISL_1686373, EPI_ISL_1686480, EPI_ISL_1686515, EPI_ISL_1686522, EPI_ISL_1686525, EPI_ISL_1686527, EPI_ISL_1686535, EPI_ISL_1686536, EPI_ISL_1686538, EPI_ISL_1686691, EPI_ISL_1686760, EPI_ISL_1686769, EPI_ISL_1686771, EPI_ISL_1686772, EPI_ISL_1686773, EPI_ISL_1686774, EPI_ISL_1686776, EPI_ISL_1686778, EPI_ISL_1686782, EPI_ISL_1686783, EPI_ISL_1686786, EPI_ISL_1686792, EPI_ISL_1686793, EPI_ISL_1686795, EPI_ISL_1686817, EPI_ISL_1686863, EPI_ISL_1686865, EPI_ISL_1686867, EPI_ISL_1686869, EPI_ISL_1686879, EPI_ISL_1686958, EPI_ISL_1686972, EPI_ISL_1686973, EPI_ISL_1686975, EPI_ISL_1687061, EPI_ISL_1687120, EPI_ISL_1687230, EPI_ISL_1687262, EPI_ISL_1687579, EPI_ISL_1687736, EPI_ISL_1687737, EPI_ISL_1687738, EPI_ISL_1687748, EPI_ISL_1687781, EPI_ISL_1687782, EPI_ISL_1687783, EPI_ISL_1687809, EPI_ISL_1687824, EPI_ISL_1687832, EPI_ISL_1687833, EPI_ISL_1687834, EPI_ISL_1687835, EPI_ISL_1687838, EPI_ISL_1687842, EPI_ISL_1687848, EPI_ISL_1687856, EPI_ISL_1687858, EPI_ISL_1687871, EPI_ISL_1688064, EPI_ISL_1688065, EPI_ISL_1688071, EPI_ISL_1688132, EPI_ISL_1688159, EPI_ISL_1688306, EPI_ISL_1688421, EPI_ISL_1688548, EPI_ISL_1688551, EPI_ISL_1688609, EPI_ISL_1688616, EPI_ISL_1688620, EPI_ISL_1695135, EPI_ISL_1695305, EPI_ISL_1695382, EPI_ISL_1695469, EPI_ISL_1695545, EPI_ISL_1695578, EPI_ISL_1701900, EPI_ISL_1701973, EPI_ISL_1701980, EPI_ISL_1702014, EPI_ISL_1702030, EPI_ISL_1702035, EPI_ISL_1702076, EPI_ISL_1702107, EPI_ISL_1737512, EPI_ISL_1834830, EPI_ISL_1925228, EPI_ISL_1925248, EPI_ISL_1925249, EPI_ISL_1925250, EPI_ISL_1925251, EPI_ISL_1925252, EPI_ISL_1925275, EPI_ISL_1925303, EPI_ISL_1925304, EPI_ISL_1925311, EPI_ISL_1925329, EPI_ISL_1925358, EPI_ISL_1925360, EPI_ISL_1925380, EPI_ISL_1925382, EPI_ISL_1925421, EPI_ISL_1925454, EPI_ISL_1925459, EPI_ISL_1925464, EPI_ISL_1925477, EPI_ISL_1925498, EPI_ISL_1925502, EPI_ISL_1925541, EPI_ISL_1925545, EPI_ISL_1925550, EPI_ISL_1925551, EPI_ISL_1925584, EPI_ISL_1925604, EPI_ISL_1925617, EPI_ISL_2185809, EPI_ISL_2185920, EPI_ISL_2186066, EPI_ISL_2186121, EPI_ISL_2186197, EPI_ISL_2186211, EPI_ISL_2186248, EPI_ISL_2186249 | | | |
| see above | Aegis Sciences Corporation | Centers for Disease Control and Prevention Division of Viral Diseases, Pathogen Discovery | Adrian Paskey; Alec Vest; Benjamin Rambo-Martin; Christopher Gulvick; Clinton R. Paden; Cyndi Clark; Dakota Howard; Darlene Wagner; Dhwani Batra; Dillon Nall; Duncan MacCannell; Ethan Sanders; Holly Houdeshell; Jason Caravas; Kara Moser; Matthew Hardison; Matthew Schmerer; Ola Kvalvaag; Patrick Campbell; Peter W. Cook; Rob Case; Scott Sammons; Shatavia Morrison; Shaun Westlund; Vikramsinha Ghorpade; Yvette Unoarumhi |
| EPI_ISL_1397695, EPI_ISL_1397696, EPI_ISL_1397697, EPI_ISL_1397698, EPI_ISL_1397699, EPI_ISL_1397912, EPI_ISL_1397913 | | | |
| see above | BIO-REFERENCE LABORATORIES | Wadsworth Center, New York State Department of Health | Alexis Russel; Daryl M. Lamson; Erasmus Schneider; Erica Lasek-Nesselquist; John Kelly; Jonathan Plitnick; Kirsten St. George; Matthew Shudt; Melissa A Leisner; Navjot Singh |
| EPI_ISL_1482408, EPI_ISL_1482409, EPI_ISL_1482446, EPI_ISL_1482447, EPI_ISL_1482448, EPI_ISL_1482449, EPI_ISL_1482450, EPI_ISL_1482451, EPI_ISL_1482452, EPI_ISL_1482453, EPI_ISL_1575477, EPI_ISL_1580667, EPI_ISL_1580668, EPI_ISL_1580669, EPI_ISL_1580670, EPI_ISL_1624409, EPI_ISL_1671446, EPI_ISL_1671454, EPI_ISL_1671476, EPI_ISL_1671495, EPI_ISL_1740510, EPI_ISL_1755849, EPI_ISL_2098126 | | | |
| see above | BOSTON HEART DIAGNOSTICS CORP | Wadsworth Center, New York State Department of Health | Alexis Russell; Catharine Prussing; Daryl M. Lamson; Erasmus Schneider; Erica Lasek-Nesselquist; John Kelly; Jonathan Plitnick; Kirsten St. George; Matthew Shudt; Melissa A Leisner; Navjot Singh |
| EPI_ISL_1413241, EPI_ISL_1413243, EPI_ISL_1413246, EPI_ISL_1413263, EPI_ISL_1413350, EPI_ISL_1516525, EPI_ISL_1516526, EPI_ISL_1516527, EPI_ISL_1516528, EPI_ISL_1516530, EPI_ISL_1516531, EPI_ISL_1516532, EPI_ISL_1516533, EPI_ISL_1516536, EPI_ISL_1516537, EPI_ISL_1516538, EPI_ISL_1516539, EPI_ISL_1516541, EPI_ISL_1516542, EPI_ISL_1516543, EPI_ISL_1516544, EPI_ISL_1516563, EPI_ISL_1516571, EPI_ISL_1578133, EPI_ISL_1578152, EPI_ISL_1578170, EPI_ISL_1578181, EPI_ISL_1578201, EPI_ISL_1578202, EPI_ISL_1578280, EPI_ISL_1578316, EPI_ISL_1578318, EPI_ISL_1578319, EPI_ISL_1578320, EPI_ISL_1578321, EPI_ISL_1578325, EPI_ISL_1578334, EPI_ISL_1578374, EPI_ISL_1578386, EPI_ISL_1578396, EPI_ISL_1578398, EPI_ISL_1578401, EPI_ISL_1578405, EPI_ISL_1578406, EPI_ISL_1578432 | | | |
| see above | Broad Institute Clinical Research Sequencing Platform | Infectious Disease Program, Broad Institute of Harvard and MIT | Adams, G.; B.L.; B.W.; Bauer, M.; Birren; Blumenstiel, B.; Brown, C.; Carter, A.; Chaluvadi, S.; D.J.; DeFelice, M.; DeRuff, K.; Dodge, S.; Gabriel, S.; Gallagher, G.; Gladden-Young, A.; Granger, B.; J.E.; K.J.; Lagerborg, K.; Larkin, K.; Lee, M.; Lemieux; Lennon, N.; Loreth, C.; Madoff, L.; McGovern, S.; Meldrim, J.; Normandin, E.; P.C.; Park; Pearlman, L.; Reilly, S.; Rudy, M.; Sabeti; Siddle; Smole, S.; Tomkins-Tinch, C.; Vicente, G.; and MacInnis |
| EPI_ISL_2101788, EPI_ISL_2101800 | Bronx Veterans Medical Research Foundation | Pathogen Discovery, Respiratory Viruses Branch, Division of Viral Diseases, Centers for Disease Control and Prevention | Adam Retchless; Anna Kelleher; Anna Uehara; Brian Lynch; Clinton R. Paden; Dhwani Batra; Haibin Wang; Han Jia Justin Ng; Jasmine Padilla; Jing Zhang; Justin Lee; Krista Queen; Mark Burroughs; Mili Sheth; Morgan Davis; Peter Cook; Rachel Marine; Sarah Nobles; Suxiang Tong; Tara Coalter; Yan Li; Ying Tao |
| EPI_ISL_1965759, EPI_ISL_1965760, EPI_ISL_1965761, EPI_ISL_1965762, EPI_ISL_1965763, EPI_ISL_1965764, EPI_ISL_1965765, EPI_ISL_1965766, EPI_ISL_1965767, EPI_ISL_1965768, EPI_ISL_1965769, EPI_ISL_1965770, EPI_ISL_1965771, EPI_ISL_1965772, EPI_ISL_1965773, EPI_ISL_1965774, EPI_ISL_1965775, EPI_ISL_1965776, EPI_ISL_1965777, EPI_ISL_1965778, EPI_ISL_1965779, EPI_ISL_1965780, EPI_ISL_1965781, EPI_ISL_1965782, EPI_ISL_1965783, EPI_ISL_1965784, EPI_ISL_1965785, EPI_ISL_1965786, EPI_ISL_1965787, EPI_ISL_1965788, EPI_ISL_1965789, EPI_ISL_1965790, EPI_ISL_1965791, EPI_ISL_1965792, EPI_ISL_1965793, EPI_ISL_1965794, EPI_ISL_1965795, EPI_ISL_1965796, EPI_ISL_1965797, EPI_ISL_1965798, EPI_ISL_1965799, EPI_ISL_1965800, EPI_ISL_1965801, EPI_ISL_1965802, EPI_ISL_1965803, EPI_ISL_1965804, EPI_ISL_1965805, EPI_ISL_1965806, EPI_ISL_1965807, EPI_ISL_1965808, EPI_ISL_1965809, EPI_ISL_1965810, EPI_ISL_1965811, EPI_ISL_1965812, EPI_ISL_1965813, EPI_ISL_1965814, EPI_ISL_1965815, EPI_ISL_1965816, EPI_ISL_1965817, EPI_ISL_1965818, EPI_ISL_1965819, EPI_ISL_1965820, EPI_ISL_1965821, EPI_ISL_1965822, EPI_ISL_1965823, EPI_ISL_1965824, EPI_ISL_1965825, EPI_ISL_1965826, EPI_ISL_1965827, EPI_ISL_1965828, EPI_ISL_1965829, EPI_ISL_1965830, EPI_ISL_1965831, EPI_ISL_1965832, EPI_ISL_1965833, EPI_ISL_1965834, EPI_ISL_1965835, EPI_ISL_1965836, EPI_ISL_1965837, EPI_ISL_1965838, EPI_ISL_1965839, EPI_ISL_1965840, EPI_ISL_1965841, EPI_ISL_1965842, EPI_ISL_1965843, EPI_ISL_1965844, EPI_ISL_1965845, EPI_ISL_1965846, EPI_ISL_1965847, EPI_ISL_1965848, EPI_ISL_1965849, EPI_ISL_1965850, EPI_ISL_1965851, EPI_ISL_1965852, EPI_ISL_1965853, EPI_ISL_1965854, EPI_ISL_1965855, EPI_ISL_1965856, EPI_ISL_1965857, EPI_ISL_1965858, EPI_ISL_1965859, EPI_ISL_1965860, EPI_ISL_1965861, EPI_ISL_1965862, EPI_ISL_1965863, EPI_ISL_1965864, EPI_ISL_1965865, EPI_ISL_1965866, EPI_ISL_1965867, EPI_ISL_1965868, EPI_ISL_1965869, EPI_ISL_1965870, EPI_ISL_1965871, EPI_ISL_1965872, EPI_ISL_1965873, EPI_ISL_1965874, EPI_ISL_1965875, EPI_ISL_1965876, EPI_ISL_1965877, EPI_ISL_1965878, EPI_ISL_1965879 | | | |
| see above | Clinical Microbiology Laboratory, NewYork Presbyterian Hospital/Columbia University Irving Medical Center | Uhlemann Laboratory, Columbia University Irving Medical Center | Anne Kelley; Anne-Catrin Uhlemann; Maya Tagliavia; Medini K. Annavajhala |
| EPI_ISL_2031642, EPI_ISL_2031643, EPI_ISL_2031647, EPI_ISL_2151440 | Columbia University | Pathogen Discovery, Respiratory Viruses Branch, Division of Viral Diseases, Centers for Disease Control and Prevention | Adam Retchless; Anna Kelleher; Anna Uehara; Brian Lynch; Clinton R. Paden; Dhwani Batra; Haibin Wang; Han Jia Justin Ng; Jasmine Padilla; Jing Zhang; Justin Lee; Krista Queen; Mark Burroughs; Mili Sheth; Morgan Davis; Peter Cook; Rachel Marine; Sarah Nobles; Suxiang Tong; Tara Coalter; Yan Li; Ying Tao |
| EPI_ISL_1397824, EPI_ISL_1398163, EPI_ISL_1398164, EPI_ISL_1398165, EPI_ISL_1398166, EPI_ISL_1398167, EPI_ISL_1398168, EPI_ISL_1398169, EPI_ISL_1398170, EPI_ISL_1398171, EPI_ISL_1398172, EPI_ISL_1398173, EPI_ISL_1398174, EPI_ISL_1398175, EPI_ISL_1398176, EPI_ISL_1398177, EPI_ISL_1398178, EPI_ISL_1398179, EPI_ISL_1398180, EPI_ISL_1398181, EPI_ISL_1398182, EPI_ISL_1398183, EPI_ISL_1398184, EPI_ISL_1398185, EPI_ISL_1398186, EPI_ISL_1398187, EPI_ISL_1398188, EPI_ISL_1398189, EPI_ISL_1398190, EPI_ISL_1398191, EPI_ISL_1398192, EPI_ISL_1398193, EPI_ISL_1398194, EPI_ISL_1398195, EPI_ISL_1398196, EPI_ISL_1398197, EPI_ISL_1398198, EPI_ISL_1398199, EPI_ISL_1398200, EPI_ISL_1398201, EPI_ISL_1398202, EPI_ISL_1398203, EPI_ISL_1465693, EPI_ISL_1465694, EPI_ISL_1465695, EPI_ISL_1465804, EPI_ISL_1482277, EPI_ISL_1482410, EPI_ISL_1575472, EPI_ISL_1580687, EPI_ISL_1580688, EPI_ISL_1580689, EPI_ISL_1580690, EPI_ISL_1580691, EPI_ISL_1580692, EPI_ISL_1580693, EPI_ISL_1580694, EPI_ISL_1580695, EPI_ISL_1580696, EPI_ISL_1580697, EPI_ISL_1580698, EPI_ISL_1582214, EPI_ISL_1582215, EPI_ISL_1582216, EPI_ISL_1582217, EPI_ISL_1582261, EPI_ISL_1582262, EPI_ISL_1582263, EPI_ISL_1582264, EPI_ISL_1582265, EPI_ISL_1582266, EPI_ISL_1582267, EPI_ISL_1582268, EPI_ISL_1582269, EPI_ISL_1582270, EPI_ISL_1582271, EPI_ISL_1582337, EPI_ISL_1582368, EPI_ISL_1756328, EPI_ISL_1920139, EPI_ISL_1920148 | | | |
| see above | Columbia University Irving Medical Center | Wadsworth Center, New York State Department of Health | Alexis Russel; Alexis Russell; Catharine Prussing; Daryl M. Lamson; Erasmus Schneider; Erica Lasek-Nesselquist; John Kelly; Jonathan Plitnick; Kirsten St. George; Matthew Shudt; Melissa A Leisner; Navjot Singh |
| EPI_ISL_1200522, EPI_ISL_1200625, EPI_ISL_1261801, EPI_ISL_1261804, EPI_ISL_1261807, EPI_ISL_1261808, EPI_ISL_1261824, EPI_ISL_1261843, EPI_ISL_1261856, EPI_ISL_1261858, EPI_ISL_1261864, EPI_ISL_1337465, EPI_ISL_1337476, EPI_ISL_1337489, EPI_ISL_1337517, EPI_ISL_1337520, EPI_ISL_1337536, EPI_ISL_1337548, EPI_ISL_1337549, EPI_ISL_1447101, EPI_ISL_1447109, EPI_ISL_1447116, EPI_ISL_1447129, EPI_ISL_1447130, EPI_ISL_1447134, EPI_ISL_1447167, EPI_ISL_1447168, EPI_ISL_1447186, EPI_ISL_1593720, EPI_ISL_1593722, EPI_ISL_1593742, EPI_ISL_1593743, EPI_ISL_1593744, EPI_ISL_1593745, EPI_ISL_1593751, EPI_ISL_1593752, EPI_ISL_1593763, EPI_ISL_1593764, EPI_ISL_1593765, EPI_ISL_1593766, EPI_ISL_1593767 | | | |
| see above | DOHMH Central Harlem | New York City Public Health Laboratory | Jade Wang; et al. |
| EPI_ISL_1200534, EPI_ISL_1200574, EPI_ISL_1261817, EPI_ISL_1261852, EPI_ISL_1261855, EPI_ISL_1261862, EPI_ISL_1261863, EPI_ISL_1337464, EPI_ISL_1337479, EPI_ISL_1337500, EPI_ISL_1337542, EPI_ISL_1447093, EPI_ISL_1447096, EPI_ISL_1447118, EPI_ISL_1447123, EPI_ISL_1447133, EPI_ISL_1447156, EPI_ISL_1447157, EPI_ISL_1447169, EPI_ISL_1447193, EPI_ISL_1593733, EPI_ISL_1593754, EPI_ISL_1593755 | | | |
| see above | DOHMH Chelsea | New York City Public Health Laboratory | Jade Wang; et al. |
| EPI_ISL_1200540, EPI_ISL_1200546, EPI_ISL_1200603, EPI_ISL_1200626, EPI_ISL_1261806, EPI_ISL_1261810, EPI_ISL_1261812, EPI_ISL_1261813, EPI_ISL_1261832, EPI_ISL_1261833, EPI_ISL_1261835, EPI_ISL_1261836, EPI_ISL_1261837, EPI_ISL_1261844, EPI_ISL_1261859, EPI_ISL_1337467, EPI_ISL_1337475, EPI_ISL_1337477, EPI_ISL_1337484, EPI_ISL_1337491, EPI_ISL_1337507, EPI_ISL_1337511, EPI_ISL_1337512, EPI_ISL_1337514, EPI_ISL_1337515, EPI_ISL_1337531, EPI_ISL_1337551, EPI_ISL_1447088, EPI_ISL_1447095, EPI_ISL_1447126, EPI_ISL_1447136, EPI_ISL_1447137, EPI_ISL_1447139, EPI_ISL_1447140, EPI_ISL_1447141, EPI_ISL_1447142, EPI_ISL_1447144, EPI_ISL_1447147, EPI_ISL_1447155, EPI_ISL_1447160, EPI_ISL_1447165, EPI_ISL_1447171, EPI_ISL_1447172, EPI_ISL_1447185, EPI_ISL_1593721, EPI_ISL_1593737, EPI_ISL_1593746, EPI_ISL_1593759, EPI_ISL_1593760, EPI_ISL_1593761 | | | |
| see above | DOHMH Corona | New York City Public Health Laboratory | Jade Wang; et al. |
| EPI_ISL_1200524, EPI_ISL_1200543, EPI_ISL_1200554, EPI_ISL_1200571, EPI_ISL_1200605, EPI_ISL_1261803, EPI_ISL_1261821, EPI_ISL_1261825, EPI_ISL_1261834, EPI_ISL_1261861, EPI_ISL_1337474, EPI_ISL_1337488, EPI_ISL_1337490, EPI_ISL_1337492, EPI_ISL_1337493, EPI_ISL_1337494, EPI_ISL_1337499, EPI_ISL_1337528, EPI_ISL_1337534, EPI_ISL_1337535, EPI_ISL_1337544, EPI_ISL_1447089, EPI_ISL_1447113, EPI_ISL_1447128, EPI_ISL_1447148, EPI_ISL_1447161, EPI_ISL_1447162, EPI_ISL_1447166, EPI_ISL_1447170, EPI_ISL_1447191, EPI_ISL_1593741, EPI_ISL_1593749 | | | |

| | | | |
|---|---|---|---|
| see above | DOHMH Crown Heights | New York City Public Health Laboratory | Jade Wang; et al. |
| EPI_ISL_1261809, EPI_ISL_1261818, EPI_ISL_1261847, EPI_ISL_1261850, EPI_ISL_1337468, EPI_ISL_1337471, EPI_ISL_1337510, EPI_ISL_1447098, EPI_ISL_1447112, EPI_ISL_1447115, EPI_ISL_1447127, EPI_ISL_1447135, EPI_ISL_1447176, EPI_ISL_1447187, EPI_ISL_1593740, EPI_ISL_1593750, EPI_ISL_1593758 | | | |
| see above | DOHMH Fort Greene | New York City Public Health Laboratory | Jade Wang; et al. |
| EPI_ISL_1200553, EPI_ISL_1200572, EPI_ISL_1200573, EPI_ISL_1200581, EPI_ISL_1261790, EPI_ISL_1261791, EPI_ISL_1261792, EPI_ISL_1261793, EPI_ISL_1261794, EPI_ISL_1261802, EPI_ISL_1261815, EPI_ISL_1261820, EPI_ISL_1261822, EPI_ISL_1261831, EPI_ISL_1261857, EPI_ISL_1261860, EPI_ISL_1261866, EPI_ISL_1337469, EPI_ISL_1337486, EPI_ISL_1337487, EPI_ISL_1337495, EPI_ISL_1337496, EPI_ISL_1337497, EPI_ISL_1337498, EPI_ISL_1337501, EPI_ISL_1337504, EPI_ISL_1337505, EPI_ISL_1337513, EPI_ISL_1337521, EPI_ISL_1337522, EPI_ISL_1337525, EPI_ISL_1337532, EPI_ISL_1337533, EPI_ISL_1337537, EPI_ISL_1337538, EPI_ISL_1337550, EPI_ISL_1447092, EPI_ISL_1447094, EPI_ISL_1447103, EPI_ISL_1447106, EPI_ISL_1447117, EPI_ISL_1447120, EPI_ISL_1447121, EPI_ISL_1447122, EPI_ISL_1447124, EPI_ISL_1447125, EPI_ISL_1447131, EPI_ISL_1447132, EPI_ISL_1447146, EPI_ISL_1447151, EPI_ISL_1447152, EPI_ISL_1447153, EPI_ISL_1447158, EPI_ISL_1447159, EPI_ISL_1447163, EPI_ISL_1447164, EPI_ISL_1447175, EPI_ISL_1447179, EPI_ISL_1447180, EPI_ISL_1447188, EPI_ISL_1447189, EPI_ISL_1447190, EPI_ISL_1447192, EPI_ISL_1593730, EPI_ISL_1593732, EPI_ISL_1593735, EPI_ISL_1593738, EPI_ISL_1593739, EPI_ISL_1593747, EPI_ISL_1593748, EPI_ISL_1593762 | | | |
| see above | DOHMH Jamaica | New York City Public Health Laboratory | Jade Wang; et al. |
| EPI_ISL_1200557, EPI_ISL_1200558, EPI_ISL_1200559, EPI_ISL_1200575, EPI_ISL_1200579, EPI_ISL_1200580, EPI_ISL_1200600, EPI_ISL_1200604, EPI_ISL_1261814, EPI_ISL_1261826, EPI_ISL_1261838, EPI_ISL_1261839, EPI_ISL_1261848, EPI_ISL_1261851, EPI_ISL_1337463, EPI_ISL_1337470, EPI_ISL_1337473, EPI_ISL_1337478, EPI_ISL_1337481, EPI_ISL_1337485, EPI_ISL_1337502, EPI_ISL_1337506, EPI_ISL_1337523, EPI_ISL_1337540, EPI_ISL_1337545, EPI_ISL_1447087, EPI_ISL_1447090, EPI_ISL_1447097, EPI_ISL_1447108, EPI_ISL_1447110, EPI_ISL_1447181, EPI_ISL_1593753 | | | |
| see above | DOHMH Morrisania | New York City Public Health Laboratory | Jade Wang; et al. |
| EPI_ISL_1200538, EPI_ISL_1200601, EPI_ISL_1200602, EPI_ISL_1261798, EPI_ISL_1261805, EPI_ISL_1261823, EPI_ISL_1337508, EPI_ISL_1337509, EPI_ISL_1337516, EPI_ISL_1337530, EPI_ISL_1447099, EPI_ISL_1447107, EPI_ISL_1447145, EPI_ISL_1447149, EPI_ISL_1447150, EPI_ISL_1447184, EPI_ISL_1593731, EPI_ISL_1593734 | | | |
| see above | DOHMH PHL | New York City Public Health Laboratory | Jade Wang; et al. |
| EPI_ISL_1200525, EPI_ISL_1200537, EPI_ISL_1200606, EPI_ISL_1200611, EPI_ISL_1261795, EPI_ISL_1261799, EPI_ISL_1261816, EPI_ISL_1261840, EPI_ISL_1261841, EPI_ISL_1261842, EPI_ISL_1261845, EPI_ISL_1261849, EPI_ISL_1337480, EPI_ISL_1337483, EPI_ISL_1337503, EPI_ISL_1337526, EPI_ISL_1337543, EPI_ISL_1337547, EPI_ISL_1447105, EPI_ISL_1447154, EPI_ISL_1447173, EPI_ISL_1593736, EPI_ISL_1694696 | | | |
| see above | DOHMH Riverside | New York City Public Health Laboratory | Jade Wang; et al. |
| EPI_ISL_1261800, EPI_ISL_1261830, EPI_ISL_1261853, EPI_ISL_1337466, EPI_ISL_1337472, EPI_ISL_1447102, EPI_ISL_1447174, EPI_ISL_1447182, EPI_ISL_1593771 | | | |
| see above | Department of Homeless Services | New York City Public Health Laboratory | Jade Wang; et al. |
| EPI_ISL_1444911, EPI_ISL_1444912, EPI_ISL_1444915, EPI_ISL_1444916, EPI_ISL_1444917, EPI_ISL_1444918, EPI_ISL_1444919, EPI_ISL_1444920, EPI_ISL_1444921, EPI_ISL_1444922, EPI_ISL_1444949, EPI_ISL_1444966, EPI_ISL_1444967, EPI_ISL_1444982, EPI_ISL_1444998, EPI_ISL_1444999, EPI_ISL_1445000, EPI_ISL_1445001, EPI_ISL_1445002, EPI_ISL_1445003, EPI_ISL_1445004, EPI_ISL_1445005, EPI_ISL_1445006, EPI_ISL_1445007, EPI_ISL_1445009, EPI_ISL_1445010 | | | |
| see above | Erie County Public Health (ECPHL) | University at Buffalo Genomics and Bioinformatics Core | Alyssa Pohlman; Amanda Boccolucci; Brandon Marzullo; Donald Yergeau; Jennifer Surtees; Jonathan Bard; Natalie Lamb; Norma Nowak |
| EPI_ISL_1481835 | Fulgent Genetics | Centers for Disease Control and Prevention Division of Viral Diseases, Pathogen Discovery | Adrian Paskey; Becky Tsai; Benafsh Sapra; Benjamin Rambo-Martin; Christopher Gulvick; Clinton R. Paden; Dakota Howard; Darlene Wagner; Dhwani Batra; Doreen Ng; Duncan MacCannell; Harry Gao; James Xie; Jason Caravas; John Gao; Joseph Fierro; Kara Moser; Matthew Schmerer; Mickey Li; Peter W. Cook; Scott Sammons; Shatavia Morrison; Yan Meng; Yvette Unoarumhi |
| EPI_ISL_1465853, EPI_ISL_1465854, EPI_ISL_1465855, EPI_ISL_1465856, EPI_ISL_1482333, EPI_ISL_1482334, EPI_ISL_1482335, EPI_ISL_1582406, EPI_ISL_1755488 | | | |
| see above | GLENS FALLS HOSPITAL LABORATORY | Wadsworth Center, New York State Department of Health | Alexis Russell; Catharine Prussing; Daryl M. Lamson; Erasmus Schneider; Erica Lasek-Nesselquist; John Kelly; Jonathan Plitnick; Kirsten St. George; Matthew Shudt; Melissa A Leisner; Navjot Singh |
| EPI_ISL_1465848, EPI_ISL_1465858, EPI_ISL_1582255 | GOOD SAMARITAN HOSPITAL LABORATORY | Wadsworth Center, New York State Department of Health | Alexis Russell; Catharine Prussing; Daryl M. Lamson; Erasmus Schneider; Erica Lasek-Nesselquist; John Kelly; Jonathan Plitnick; Kirsten St. George; Matthew Shudt; Melissa A Leisner; Navjot Singh |
| EPI_ISL_1920119, EPI_ISL_1920120 | Glens Falls Hospital Laboratory | Wadsworth Center, New York State Department of Health | Alexis Russell; Catharine Prussing; Daryl M. Lamson; Erasmus Schneider; Erica Lasek-Nesselquist; John Kelly; Jonathan Plitnick; Kirsten St. George; Matthew Shudt; Melissa A Leisner; Navjot Singh |
| EPI_ISL_1482470 | Good Samaritan Hospital Laboratory | Wadsworth Center, New York State Department of Health | Alexis Russell; Catharine Prussing; Daryl M. Lamson; Erasmus Schneider; Erica Lasek-Nesselquist; John Kelly; Jonathan Plitnick; Kirsten St. George; Matthew Shudt; Melissa A Leisner; Navjot Singh |
| EPI_ISL_2151397, EPI_ISL_2151401, EPI_ISL_2179166 | HealthQuest Esoterics | New York City Public Health Laboratory | Jade Wang; et al. |
| EPI_ISL_1338149, EPI_ISL_1338229, EPI_ISL_1338263, EPI_ISL_1340378, EPI_ISL_1340381, EPI_ISL_1340392, EPI_ISL_1340441, EPI_ISL_1443911, EPI_ISL_1479386, EPI_ISL_1480025, EPI_ISL_1480043, EPI_ISL_1480059, EPI_ISL_1480155, EPI_ISL_1480290, EPI_ISL_1480429, EPI_ISL_1493799, EPI_ISL_1511926, EPI_ISL_1512010, EPI_ISL_1512135, EPI_ISL_1512243, EPI_ISL_1512415, EPI_ISL_1512449, EPI_ISL_1512483, EPI_ISL_1553421, EPI_ISL_1553555, EPI_ISL_1554114, EPI_ISL_1554306, EPI_ISL_1575510, EPI_ISL_1575846, EPI_ISL_1575913, EPI_ISL_1576513, EPI_ISL_1576617, EPI_ISL_1576668, EPI_ISL_1580918, EPI_ISL_1581703, EPI_ISL_1592417, EPI_ISL_1592426, EPI_ISL_1615131, EPI_ISL_1679009, EPI_ISL_1690309, EPI_ISL_1690352, EPI_ISL_1690546, EPI_ISL_1691214 | | | |
| see above | Helix/Illumina | Centers for Disease Control and Prevention Division of Viral Diseases, Pathogen Discovery | Adrian Paskey; Alexandre Bolze; Ary Ascencio; Ben L. Rambo-Martin; Benjamin Rambo-Martin; Brad Sickler; Charlotte Rivera-Garcia; Christine Tran; Christopher Gulvick; Clinton R. Paden; Dakota Howard; Darlene Wagner; David Becker; Dhwani Batra; Duncan MacCannell; Efren Sandoval; Eileen de Feo; Elizabeth Cirulli; Eric Allen; Geraint Levan; James Lu; Jan Antico; Jason Caravas; Jason Nguyen; Jimmy Ramirez; Jingtao Liu; Kara Moser; Kelly Schiabor Barrett; Kim Gietzen; Magnus Isaksson; Marc Laurent; Matthew Schmerer; Matthew Tolentino; Nicole L. Washington; Peter W. Cook; Phil Febbo; Ryan Cho; Scott Sammons; Shannon Wickline; Shatavia Morrison; Sherry Wang; Simon White; Summer Galloway; Suxiang Tong; Tyler Cassens; William Lee; Yvette Unoarumhi |
| EPI_ISL_1465857 | ITHACA COLLEGE LABORATORY HAMMOND HEALTH CENTER | Wadsworth Center, New York State Department of Health | Alexis Russell; Catharine Prussing; Daryl M. Lamson; Erasmus Schneider; Erica Lasek-Nesselquist; John Kelly; Jonathan Plitnick; Kirsten St. George; Matthew Shudt; Melissa A Leisner; Navjot Singh |
| EPI_ISL_1512832, EPI_ISL_1512837, EPI_ISL_1512853, EPI_ISL_1512854, EPI_ISL_1512855, EPI_ISL_1512857, EPI_ISL_1512862, EPI_ISL_1512896, EPI_ISL_1512917, EPI_ISL_1512935, EPI_ISL_1512948, EPI_ISL_1512949, EPI_ISL_1512950, EPI_ISL_1512952, EPI_ISL_1513036, EPI_ISL_1513039, EPI_ISL_1513044, EPI_ISL_1513074, EPI_ISL_1513092, EPI_ISL_1513094, EPI_ISL_1513096, EPI_ISL_1513101, EPI_ISL_1513102, EPI_ISL_1513131, EPI_ISL_1587942, EPI_ISL_1587954, EPI_ISL_1588003, EPI_ISL_1588190, EPI_ISL_1588288, EPI_ISL_1588418, EPI_ISL_1691557, EPI_ISL_1691562, EPI_ISL_1691674, EPI_ISL_1691686, EPI_ISL_1691716, EPI_ISL_1691720, EPI_ISL_1691721, EPI_ISL_1691756, EPI_ISL_1691959, EPI_ISL_1691975, EPI_ISL_1691986, EPI_ISL_1691995, EPI_ISL_1692005, EPI_ISL_1692010, EPI_ISL_1692028, EPI_ISL_1692030, EPI_ISL_1692084, EPI_ISL_1692129, EPI_ISL_1692160, EPI_ISL_1692198, EPI_ISL_1692224, EPI_ISL_1692266, EPI_ISL_1692284, EPI_ISL_1692285, EPI_ISL_1692292, EPI_ISL_1692310, EPI_ISL_1692327, EPI_ISL_1692341, EPI_ISL_1692371, EPI_ISL_1692408, EPI_ISL_1692409, EPI_ISL_1692448, EPI_ISL_1692460, EPI_ISL_1692516, EPI_ISL_1692524, EPI_ISL_1692575, EPI_ISL_1692578, EPI_ISL_1692581, EPI_ISL_1692587, EPI_ISL_1692592, EPI_ISL_1692600, EPI_ISL_1692606, EPI_ISL_1692634, EPI_ISL_1692655, EPI_ISL_1692658, EPI_ISL_1692669, EPI_ISL_1692687, EPI_ISL_1692708, EPI_ISL_1693102, EPI_ISL_1693220, EPI_ISL_1693274, EPI_ISL_1693317, EPI_ISL_1693319, EPI_ISL_1693320, EPI_ISL_1693321, EPI_ISL_1693868, EPI_ISL_1694388, EPI_ISL_1702185, EPI_ISL_1702186, EPI_ISL_1702187, EPI_ISL_1702255, EPI_ISL_1702267, EPI_ISL_1702281, EPI_ISL_2013321 | | | |
| see above | Infinity Biologix | Centers for Disease Control and Prevention Division of Viral Diseases, Pathogen Discovery | Adrian Paskey; Benjamin Rambo-Martin; Chirayu Goswami; Christian Bixby; Christopher Gulvick; Clinton R. Paden; Dakota Howard; Darlene Wagner; Dhwani Batra; Duncan MacCannell; Jason Caravas; Jonathan Schultz; Kara Moser; Matthew Schmerer; Peter W. Cook; Robin Grimwood; Russ Hager; Scott Sammons; Shatavia Morrison; Yihe Wang; Yvette Unoarumhi |
| EPI_ISL_1319176, EPI_ISL_1319180, EPI_ISL_1319189, EPI_ISL_1319190, EPI_ISL_1319646, EPI_ISL_1319647, EPI_ISL_1319648, EPI_ISL_1319653, EPI_ISL_1319654, EPI_ISL_1319679, EPI_ISL_1319683, EPI_ISL_1319686, EPI_ISL_1319687, EPI_ISL_1319704, EPI_ISL_1319720, EPI_ISL_1319740, EPI_ISL_1319779, EPI_ISL_1319951, EPI_ISL_1319972, EPI_ISL_1320000, EPI_ISL_1320007, EPI_ISL_1320531, EPI_ISL_1320544, EPI_ISL_1320550, EPI_ISL_1320551, EPI_ISL_1320553, EPI_ISL_1320554, EPI_ISL_1320564, EPI_ISL_1320586, EPI_ISL_1320610, EPI_ISL_1320611, EPI_ISL_1320618, EPI_ISL_1320622, EPI_ISL_1320632, EPI_ISL_1320633, EPI_ISL_1320644, EPI_ISL_1320890, EPI_ISL_1320891, EPI_ISL_1320892, EPI_ISL_1320893, EPI_ISL_1320894, EPI_ISL_1320895, EPI_ISL_1320896, EPI_ISL_1320897, EPI_ISL_1320898, EPI_ISL_1320899, EPI_ISL_1320900, EPI_ISL_1320901, EPI_ISL_1320902, EPI_ISL_1320903, EPI_ISL_1320906, EPI_ISL_1320907, EPI_ISL_1320919, EPI_ISL_1320952, EPI_ISL_1320953, EPI_ISL_1320954, EPI_ISL_1320955, EPI_ISL_1320956, EPI_ISL_1320957, EPI_ISL_1320958, EPI_ISL_1320959, EPI_ISL_1320960, EPI_ISL_1320961, EPI_ISL_1320962, EPI_ISL_1320966, EPI_ISL_1320984, EPI_ISL_1321226, EPI_ISL_1338400, EPI_ISL_1338401, EPI_ISL_1338521, EPI_ISL_1338525, EPI_ISL_1338526, EPI_ISL_1338527, EPI_ISL_1338530, EPI_ISL_1338532, EPI_ISL_1338533, EPI_ISL_1338534, EPI_ISL_1338535, EPI_ISL_1338536, EPI_ISL_1338541, EPI_ISL_1338625, EPI_ISL_1338627, EPI_ISL_1338629, EPI_ISL_1338644, EPI_ISL_1338649, EPI_ISL_1338653, EPI_ISL_1338658, EPI_ISL_1338660, EPI_ISL_1338680, EPI_ISL_1338681, EPI_ISL_1338682, EPI_ISL_1338683, EPI_ISL_1338687, EPI_ISL_1338705, EPI_ISL_1338720, EPI_ISL_1338721, EPI_ISL_1338730, EPI_ISL_1338737, EPI_ISL_1338846, EPI_ISL_1338847, EPI_ISL_1339180, EPI_ISL_1339237, EPI_ISL_1339238, EPI_ISL_1339241, EPI_ISL_1339242, EPI_ISL_1339247, EPI_ISL_1339498, EPI_ISL_1339505, EPI_ISL_1339522, EPI_ISL_1339526, EPI_ISL_1339529, EPI_ISL_1339535, EPI_ISL_1339536, EPI_ISL_1339543, EPI_ISL_1339544, EPI_ISL_1339573, EPI_ISL_1339577, EPI_ISL_1339584, EPI_ISL_1339585, EPI_ISL_1339591, EPI_ISL_1339628, EPI_ISL_1339638, EPI_ISL_1339639, EPI_ISL_1339642, EPI_ISL_1421894, EPI_ISL_1421895, EPI_ISL_1421897, EPI_ISL_1421961, EPI_ISL_1421965, EPI_ISL_1421967, EPI_ISL_1421976, EPI_ISL_1421977, EPI_ISL_1422025, EPI_ISL_1422044, EPI_ISL_1422046, EPI_ISL_1422094, EPI_ISL_1422095, EPI_ISL_1422106, EPI_ISL_1422107, EPI_ISL_1422108, EPI_ISL_1422109, EPI_ISL_1422110, EPI_ISL_1422111, EPI_ISL_1422112, EPI_ISL_1422113, EPI_ISL_1422114, EPI_ISL_1422115, EPI_ISL_1422116, EPI_ISL_1422146, EPI_ISL_1422147, EPI_ISL_1422148, EPI_ISL_1422182, EPI_ISL_1422186, EPI_ISL_1422187, EPI_ISL_1461826, EPI_ISL_1461842, EPI_ISL_1461983, EPI_ISL_1461996, EPI_ISL_1461997, EPI_ISL_1461998, EPI_ISL_1462005, EPI_ISL_1462239, EPI_ISL_1462242, EPI_ISL_1462357, EPI_ISL_1462358, EPI_ISL_1462359, EPI_ISL_1462360, EPI_ISL_1462361, EPI_ISL_1462362, EPI_ISL_1462363, EPI_ISL_1462371, EPI_ISL_1462372, EPI_ISL_1462375, EPI_ISL_1462376, EPI_ISL_1462377, EPI_ISL_1462425, EPI_ISL_1462426, EPI_ISL_1462427, EPI_ISL_1462428, EPI_ISL_1462429, EPI_ISL_1462430, EPI_ISL_1462431, EPI_ISL_1462433, EPI_ISL_1462434, EPI_ISL_1462440, EPI_ISL_1462441, EPI_ISL_1462473, EPI_ISL_1462474, EPI_ISL_1462475, EPI_ISL_1462478, EPI_ISL_1462479, EPI_ISL_1462480, EPI_ISL_1462483, EPI_ISL_1462484, EPI_ISL_1462485, EPI_ISL_1462497, EPI_ISL_1462498, EPI_ISL_1462499, EPI_ISL_1462501, EPI_ISL_1462504, EPI_ISL_1462507, EPI_ISL_1462517, EPI_ISL_1462520, EPI_ISL_1462571, EPI_ISL_1462838, EPI_ISL_1463082, EPI_ISL_1463106, EPI_ISL_1463109, EPI_ISL_1463110, EPI_ISL_1463112, EPI_ISL_1463121, EPI_ISL_1463122, EPI_ISL_1463125, EPI_ISL_1463126, EPI_ISL_1463139, EPI_ISL_1463140, EPI_ISL_1463141, EPI_ISL_1463142, EPI_ISL_1463173, EPI_ISL_1463187, EPI_ISL_1463188, EPI_ISL_1463189, EPI_ISL_1463191, EPI_ISL_1463194, EPI_ISL_1463207, EPI_ISL_1463208, EPI_ISL_1463209, EPI_ISL_1463210, EPI_ISL_1463211, EPI_ISL_1463212, EPI_ISL_1463213, EPI_ISL_1463214, EPI_ISL_1463215, EPI_ISL_1463216, EPI_ISL_1463217, EPI_ISL_1463218, EPI_ISL_1463222, EPI_ISL_1463229, EPI_ISL_1463260, EPI_ISL_1463261, EPI_ISL_1463262, EPI_ISL_1463263, EPI_ISL_1463264, EPI_ISL_1463265, EPI_ISL_1463266, EPI_ISL_1463267, EPI_ISL_1463268, EPI_ISL_1463269, EPI_ISL_1463270, EPI_ISL_1463271, EPI_ISL_1463272, EPI_ISL_1463278, EPI_ISL_1463281, EPI_ISL_1463282, EPI_ISL_1463307, EPI_ISL_1463309, EPI_ISL_1463310, EPI_ISL_1463311, EPI_ISL_1463313, EPI_ISL_1463314, EPI_ISL_1463316, EPI_ISL_1463317, EPI_ISL_1463318, EPI_ISL_1463319, EPI_ISL_1463320, EPI_ISL_1463321, EPI_ISL_1463335, EPI_ISL_1463355, EPI_ISL_1463356, EPI_ISL_1463357, EPI_ISL_1463358, EPI_ISL_1463359, EPI_ISL_1463360, EPI_ISL_1463362, EPI_ISL_1463393, EPI_ISL_1463394, EPI_ISL_1463395, EPI_ISL_1463399, EPI_ISL_1463400, EPI_ISL_1480977, EPI_ISL_1481057, EPI_ISL_1481058, EPI_ISL_1481061, EPI_ISL_1481067, EPI_ISL_1481105, EPI_ISL_1481110, EPI_ISL_1481111, EPI_ISL_1481132, EPI_ISL_1481136, EPI_ISL_1481137, EPI_ISL_1481138, EPI_ISL_1481154, EPI_ISL_1481182, EPI_ISL_1481187, EPI_ISL_1481373, EPI_ISL_1481388, EPI_ISL_1481392, EPI_ISL_1481396, EPI_ISL_1481398, EPI_ISL_1481400, EPI_ISL_1481404, EPI_ISL_1481405, EPI_ISL_1481409, EPI_ISL_1481429, EPI_ISL_1481450, EPI_ISL_1481451, EPI_ISL_1481475, EPI_ISL_1481480, EPI_ISL_1481481, EPI_ISL_1513637, EPI_ISL_1513640, EPI_ISL_1513641, EPI_ISL_1513642, EPI_ISL_1513647, EPI_ISL_1513652, EPI_ISL_1513653, EPI_ISL_1513989, EPI_ISL_1513990, EPI_ISL_1513992, EPI_ISL_1513993, EPI_ISL_1513994, EPI_ISL_1513996, EPI_ISL_1513997, EPI_ISL_1514220, EPI_ISL_1514224, EPI_ISL_1514284, EPI_ISL_1514329, EPI_ISL_1514364, EPI_ISL_1514575, EPI_ISL_1514582, EPI_ISL_1514583, EPI_ISL_1514585, EPI_ISL_1514587, EPI_ISL_1514588, EPI_ISL_1514590, EPI_ISL_1514591, EPI_ISL_1514592, EPI_ISL_1514604, EPI_ISL_1514605, EPI_ISL_1514606, EPI_ISL_1514621, EPI_ISL_1514624, EPI_ISL_1514625, EPI_ISL_1514627, EPI_ISL_1514628, EPI_ISL_1514632, EPI_ISL_1514633, EPI_ISL_1514634, EPI_ISL_1514635, EPI_ISL_1514636, EPI_ISL_1514637, EPI_ISL_1514638, EPI_ISL_1514647, EPI_ISL_1514648, EPI_ISL_1514649, EPI_ISL_1514653, EPI_ISL_1514657, EPI_ISL_1514659, EPI_ISL_1514660, EPI_ISL_1514661, EPI_ISL_1514663, EPI_ISL_1514692, EPI_ISL_1514693, EPI_ISL_1514702, EPI_ISL_1514706, EPI_ISL_1514707, EPI_ISL_1514708, EPI_ISL_1514709, EPI_ISL_1514710, EPI_ISL_1514711, EPI_ISL_1514716, EPI_ISL_1514718, EPI_ISL_1514719, EPI_ISL_1514721, EPI_ISL_1515063, EPI_ISL_1515076, EPI_ISL_1515077, EPI_ISL_1515079, EPI_ISL_1515083, EPI_ISL_1515084, EPI_ISL_1515085, EPI_ISL_1515086, EPI_ISL_1515087, EPI_ISL_1515088, EPI_ISL_1515089, EPI_ISL_1515125, EPI_ISL_1515128, EPI_ISL_1515129, EPI_ISL_1515132, EPI_ISL_1515137, EPI_ISL_1515138, EPI_ISL_1515154, EPI_ISL_1515323, EPI_ISL_1515326, EPI_ISL_1515327, EPI_ISL_1515329, EPI_ISL_1515520, EPI_ISL_1548152, EPI_ISL_1548166, EPI_ISL_1548207, EPI_ISL_1548208, EPI_ISL_1548210, EPI_ISL_1548211, EPI_ISL_1548213, EPI_ISL_1548235, EPI_ISL_1548237, EPI_ISL_1548241, EPI_ISL_1548242, EPI_ISL_1548248, EPI_ISL_1548249, EPI_ISL_1548250, EPI_ISL_1548260, EPI_ISL_1548262, EPI_ISL_1548375, EPI_ISL_1548382, EPI_ISL_1548384, EPI_ISL_1548438, EPI_ISL_1548439, EPI_ISL_1548440, EPI_ISL_1548441, EPI_ISL_1548442, EPI_ISL_1548445, EPI_ISL_1548446, EPI_ISL_1548447, EPI_ISL_1548474, EPI_ISL_1548475, EPI_ISL_1548476, EPI_ISL_1548477, EPI_ISL_1548478, EPI_ISL_1548479, EPI_ISL_1548506, EPI_ISL_1548507, EPI_ISL_1548508, EPI_ISL_1548509, EPI_ISL_1548952, EPI_ISL_1548979, EPI_ISL_1548986, EPI_ISL_1548993, EPI_ISL_1549000, EPI_ISL_1549017, EPI_ISL_1549019, EPI_ISL_1549024, EPI_ISL_1549032, EPI_ISL_1549033, EPI_ISL_1549034, EPI_ISL_1549038, EPI_ISL_1549044, EPI_ISL_1549045, EPI_ISL_1549054, EPI_ISL_1549063, EPI_ISL_1549064, EPI_ISL_1549065, EPI_ISL_1549066, EPI_ISL_1549067, EPI_ISL_1549068, EPI_ISL_1549080, EPI_ISL_1549081, EPI_ISL_1549082, EPI_ISL_1549102, EPI_ISL_1549106, EPI_ISL_1549173, EPI_ISL_1549376, EPI_ISL_1549377, EPI_ISL_1549378, EPI_ISL_1549379, EPI_ISL_1549380, EPI_ISL_1549558, EPI_ISL_1549564, EPI_ISL_1549632, EPI_ISL_1549635, EPI_ISL_1549645, EPI_ISL_1549650, EPI_ISL_1549674, EPI_ISL_1549758, EPI_ISL_1549760, EPI_ISL_1549768, EPI_ISL_1549769, EPI_ISL_1549771, EPI_ISL_1549776, EPI_ISL_1549777, EPI_ISL_1549779, EPI_ISL_1549790, EPI_ISL_1549791, EPI_ISL_1549919, EPI_ISL_1549940, EPI_ISL_1549942, EPI_ISL_1549943, EPI_ISL_1550039, EPI_ISL_1550040, EPI_ISL_1609076, EPI_ISL_1609079, EPI_ISL_1609080, EPI_ISL_1609139, EPI_ISL_1609147, EPI_ISL_1609151, EPI_ISL_1609174, EPI_ISL_1609178, EPI_ISL_1609179, EPI_ISL_1609180, EPI_ISL_1609216, EPI_ISL_1609238, EPI_ISL_1609239, EPI_ISL_1609240, EPI_ISL_1609241, EPI_ISL_1609249, EPI_ISL_1609250, EPI_ISL_1609251, EPI_ISL_1609314, EPI_ISL_1609315, EPI_ISL_1609316, EPI_ISL_1609319, EPI_ISL_1609328, EPI_ISL_1609332, EPI_ISL_1609333, EPI_ISL_1609345, EPI_ISL_1609346, EPI_ISL_1609348, EPI_ISL_1609351, EPI_ISL_1609352, EPI_ISL_1609353, EPI_ISL_1609354, EPI_ISL_1609355, EPI_ISL_1609356, EPI_ISL_1609357, EPI_ISL_1609368, EPI_ISL_1609369, EPI_ISL_1609406, EPI_ISL_1609430, EPI_ISL_1609432, EPI_ISL_1609433, EPI_ISL_1609434, EPI_ISL_1609437, EPI_ISL_1609438, EPI_ISL_1609439, EPI_ISL_1609444, EPI_ISL_1609446, EPI_ISL_1609454, EPI_ISL_1609457, EPI_ISL_1609464, EPI_ISL_1609465, EPI_ISL_1609471, EPI_ISL_1609472, EPI_ISL_1609475, EPI_ISL_1609476, EPI_ISL_1609477, EPI_ISL_1609495, EPI_ISL_1609499, EPI_ISL_1609504, EPI_ISL_1609513, EPI_ISL_1609527, EPI_ISL_1609535, EPI_ISL_1609537, EPI_ISL_1609538, EPI_ISL_1609539, EPI_ISL_1609554, EPI_ISL_1609556, EPI_ISL_1609557, EPI_ISL_1609558, EPI_ISL_1609559, EPI_ISL_1609583, EPI_ISL_1609584, EPI_ISL_1609585, EPI_ISL_1609586, EPI_ISL_1609604, EPI_ISL_1609608, EPI_ISL_1609609, EPI_ISL_1609610, EPI_ISL_1609611, EPI_ISL_1609612, EPI_ISL_1609613, EPI_ISL_1609614, EPI_ISL_1609619, EPI_ISL_1609620, EPI_ISL_1609623, EPI_ISL_1609624, EPI_ISL_1609633, EPI_ISL_1609635, EPI_ISL_1609636, EPI_ISL_1609640, EPI_ISL_1609641, EPI_ISL_1609666, EPI_ISL_1609667, EPI_ISL_1609668, EPI_ISL_1609669, EPI_ISL_1609670, EPI_ISL_1609683, EPI_ISL_1609687, EPI_ISL_1609689, EPI_ISL_1609690, EPI_ISL_1609698, EPI_ISL_1609706, EPI_ISL_1609714, EPI_ISL_1609716, EPI_ISL_1609719, EPI_ISL_1609720, EPI_ISL_1609731, EPI_ISL_1609733, EPI_ISL_1609741, EPI_ISL_1609751, EPI_ISL_1609754, EPI_ISL_1609757, EPI_ISL_1609770, EPI_ISL_1609778, EPI_ISL_1609779, EPI_ISL_1609785, EPI_ISL_1609786, EPI_ISL_1609789, EPI_ISL_1609792, EPI_ISL_1609799, EPI_ISL_1609800, EPI_ISL_1609801, EPI_ISL_1609802, EPI_ISL_1609803, EPI_ISL_1609804, EPI_ISL_1609805, EPI_ISL_1609806, EPI_ISL_1609807, EPI_ISL_1609808, EPI_ISL_1609809, EPI_ISL_1609810, EPI_ISL_1609811, EPI_ISL_1609812, EPI_ISL_1609813, EPI_ISL_1609814, EPI_ISL_1609815, EPI_ISL_1609874, EPI_ISL_1609880, EPI_ISL_1609883, EPI_ISL_1609887, EPI_ISL_1609889, EPI_ISL_1609890, EPI_ISL_1609891, EPI_ISL_1609893, EPI_ISL_1609894, EPI_ISL_1609900, EPI_ISL_1609904, EPI_ISL_1609909, EPI_ISL_1609910, EPI_ISL_1609912, EPI_ISL_1609917, EPI_ISL_1609921, EPI_ISL_1609922, EPI_ISL_1609923, EPI_ISL_1609924, EPI_ISL_1609926, EPI_ISL_1609927, EPI_ISL_1609938, EPI_ISL_1609946, EPI_ISL_1609949, EPI_ISL_1609952, EPI_ISL_1609954, EPI_ISL_1609955, EPI_ISL_1609957, EPI_ISL_1609964, EPI_ISL_1609967, EPI_ISL_1610041, EPI_ISL_1610047, EPI_ISL_1610236, EPI_ISL_1610237, EPI_ISL_1610238, EPI_ISL_1610239, EPI_ISL_1610240, EPI_ISL_1610241, EPI_ISL_1610242, EPI_ISL_1610243, EPI_ISL_1610245, EPI_ISL_1610246, EPI_ISL_1610247, EPI_ISL_1610248, EPI_ISL_1610249, EPI_ISL_1610293, EPI_ISL_1610294, EPI_ISL_1610295, EPI_ISL_1610296, EPI_ISL_1610297, EPI_ISL_1610298, EPI_ISL_1610299, EPI_ISL_1610300, EPI_ISL_1610302, EPI_ISL_1610303, EPI_ISL_1610321, EPI_ISL_1610322, EPI_ISL_1610323, EPI_ISL_1610401, EPI_ISL_1610404, EPI_ISL_1610406, EPI_ISL_1610408, EPI_ISL_1610410, EPI_ISL_1610411, EPI_ISL_1610432, EPI_ISL_1610441, EPI_ISL_1610467, EPI_ISL_1610494, EPI_ISL_1610568, EPI_ISL_1610569, EPI_ISL_1610571, EPI_ISL_1610573, EPI_ISL_1610595, EPI_ISL_1610597, EPI_ISL_1610599, EPI_ISL_1610601, EPI_ISL_1610602, EPI_ISL_1610604, EPI_ISL_1610623, EPI_ISL_1610625, EPI_ISL_1610900, EPI_ISL_1610901, EPI_ISL_1610902, EPI_ISL_1610903, EPI_ISL_1610904, EPI_ISL_1610905, EPI_ISL_1610906, EPI_ISL_1610907, EPI_ISL_1610908, EPI_ISL_1610920, EPI_ISL_1610949, EPI_ISL_1610951, EPI_ISL_1610953, EPI_ISL_1611111, EPI_ISL_1611173, EPI_ISL_1611206, EPI_ISL_1611213, EPI_ISL_1611219, EPI_ISL_1611220, EPI_ISL_1611221, EPI_ISL_1611242, EPI_ISL_1611243, EPI_ISL_1611244, EPI_ISL_1611268, EPI_ISL_1611282, EPI_ISL_1611297, EPI_ISL_1611433, EPI_ISL_1611436, EPI_ISL_1611464, EPI_ISL_1611472, EPI_ISL_1611474, EPI_ISL_1611475, EPI_ISL_1611476, EPI_ISL_1611477, EPI_ISL_1611479, EPI_ISL_1611480, EPI_ISL_1611482, EPI_ISL_1611526, EPI_ISL_1611527, EPI_ISL_1611528, EPI_ISL_1611534, EPI_ISL_1611542, EPI_ISL_1611543, EPI_ISL_1611544, EPI_ISL_1611719, EPI_ISL_1611721, EPI_ISL_1611722, EPI_ISL_1611738, EPI_ISL_1611739, EPI_ISL_1611741, EPI_ISL_1611743, EPI_ISL_1611744, EPI_ISL_1611768, EPI_ISL_1611769, EPI_ISL_1611776, EPI_ISL_1611784, EPI_ISL_1611785, EPI_ISL_1611792, EPI_ISL_1611795, EPI_ISL_1611796, EPI_ISL_1611813, EPI_ISL_1611814, EPI_ISL_1611825, EPI_ISL_1680473, EPI_ISL_1680474, EPI_ISL_1680477, EPI_ISL_1680483, EPI_ISL_1695014, EPI_ISL_1695015 | | | |
| see above | Laboratory Corporation of | Centers for Disease Control and | Adrian Paskey; Amanda Douglas; Amanda Suchanek; Andrea Throop; Ayla Burns; Ben L. Rambo-Martin; Benjamin Rambo-Martin; Bobbi Croy; Brian Krueger; Brian Norvell; Christopher Gulvick; Christos Petropoulos; Clinton R. Paden; Craig Lukasik; Dakota Howard; Darlene Wagner; Debbie Boles; Dhwani |

America — Prevention Division of Viral Diseases, Pathogen Discovery — Batra; Duncan MacCannell; Eyad Almasri; Goran Stevovic; Howard Engler; Hrushikesh Deshmukh; Jake Humphrey; Jana Schroth; Jason Caravas; Joe Voshell; John Pruitt; Jonathan Meltzer; Jonathan Williams; Kara Moser; Kimberly Wagner; Lax Iyer; Lyndon Tilson; Manoj Jain; Marcia Eisenberg; Mary Ann Cristobal; Mary Williamson; Matthew Schmerer; Michael Levandoski; Mike Sapeta; Mindy Nye; Minoo Agarwal; Mohan Kolli; Nuthawin Charoensri; Oren Cohen; Peter W. Cook; Prashant Gupta; Qian Zeng; Rama Ghatti; Scott Parker; Scott Ryan; Scott Sammons; Shatavia Morrison; Stanley Letovsky; Steven Ragan; Suresh Babu Selvaraju; Susan Countryman; Susan Hicks; Suxiang Tong; Suzanne Dale; Thomas Urban; Tim Kuphal; Tricia Zwiefelhofer; Vincent Drouillon; Yvette Unoarumhi

EPI_ISL_1397622, EPI_ISL_1397791, EPI_ISL_1397792, EPI_ISL_1397794, EPI_ISL_1397795, EPI_ISL_1397796, EPI_ISL_1397798, EPI_ISL_1397799, EPI_ISL_1397800, EPI_ISL_1397803, EPI_ISL_1397804, EPI_ISL_1397805, EPI_ISL_1397806, EPI_ISL_1397807, EPI_ISL_1397808, EPI_ISL_1397809, EPI_ISL_1398034, EPI_ISL_1398035, EPI_ISL_1398036, EPI_ISL_1398037, EPI_ISL_1398038, EPI_ISL_1398048, EPI_ISL_1398049, EPI_ISL_1398050, EPI_ISL_1398051, EPI_ISL_1398052, EPI_ISL_1398053, EPI_ISL_1398054, EPI_ISL_1398055, EPI_ISL_1398056, EPI_ISL_1398057, EPI_ISL_1398058, EPI_ISL_1398059, EPI_ISL_1398060, EPI_ISL_1398061, EPI_ISL_1398062, EPI_ISL_1398063, EPI_ISL_1398064, EPI_ISL_1398065, EPI_ISL_1398066, EPI_ISL_1398067, EPI_ISL_1398136, EPI_ISL_1398331, EPI_ISL_1398333, EPI_ISL_1398334, EPI_ISL_1398335, EPI_ISL_1398336, EPI_ISL_1398337, EPI_ISL_1398338, EPI_ISL_1398339, EPI_ISL_1462551, EPI_ISL_1465668, EPI_ISL_1465669, EPI_ISL_1465670, EPI_ISL_1465671, EPI_ISL_1465672, EPI_ISL_1465701, EPI_ISL_1465702, EPI_ISL_1465703, EPI_ISL_1465704, EPI_ISL_1465705, EPI_ISL_1465706, EPI_ISL_1465707, EPI_ISL_1465708, EPI_ISL_1465709, EPI_ISL_1465710, EPI_ISL_1465711, EPI_ISL_1465712, EPI_ISL_1465713, EPI_ISL_1465794, EPI_ISL_1465795, EPI_ISL_1465796, EPI_ISL_1465797, EPI_ISL_1465798, EPI_ISL_1465799, EPI_ISL_1465800, EPI_ISL_1465801, EPI_ISL_1575470, EPI_ISL_1576816, EPI_ISL_1576819, EPI_ISL_1580686, EPI_ISL_1582159, EPI_ISL_1582162, EPI_ISL_1582165, EPI_ISL_1582168, EPI_ISL_1582170, EPI_ISL_1582173, EPI_ISL_1582174, EPI_ISL_1582175, EPI_ISL_1582176, EPI_ISL_1582177, EPI_ISL_1582178, EPI_ISL_1582319, EPI_ISL_1582320, EPI_ISL_1582321, EPI_ISL_1593868, EPI_ISL_1593869, EPI_ISL_1593881, EPI_ISL_1671499, EPI_ISL_1671642, EPI_ISL_1671654, EPI_ISL_1671664, EPI_ISL_1755820, EPI_ISL_1755821, EPI_ISL_1755822, EPI_ISL_1755823, EPI_ISL_1755824, EPI_ISL_1755825, EPI_ISL_1755826, EPI_ISL_1755827, EPI_ISL_1755828, EPI_ISL_1755829, EPI_ISL_1755830, EPI_ISL_1755831, EPI_ISL_1755832, EPI_ISL_1755833, EPI_ISL_1755834, EPI_ISL_1755836, EPI_ISL_1755837, EPI_ISL_1755948, EPI_ISL_1755949, EPI_ISL_1755950, EPI_ISL_1755951, EPI_ISL_1755952, EPI_ISL_1755953, EPI_ISL_1755954, EPI_ISL_1755955, EPI_ISL_1755956, EPI_ISL_1755957, EPI_ISL_1755958, EPI_ISL_1755959, EPI_ISL_1755960, EPI_ISL_1755961, EPI_ISL_1755962, EPI_ISL_1755963, EPI_ISL_1755964, EPI_ISL_1789710, EPI_ISL_1789711, EPI_ISL_1789715, EPI_ISL_1855103, EPI_ISL_1855215, EPI_ISL_1855216, EPI_ISL_1855217, EPI_ISL_1855218, EPI_ISL_1855219, EPI_ISL_2134791

see above — MONTEFIORE MEDICAL CENTER LABORATORIES — Wadsworth Center, New York State Department of Health — Alexis Russel; Alexis Russell; Catharine Prussing; Daryl M. Lamson; Erasmus Schneider; Erica Lasek-Nesselquist; John Kelly; Jonathan Plitnick; Kirsten St. George; Matthew Shudt; Melissa A Leisner; Navjot Singh

EPI_ISL_1465802, EPI_ISL_1465849, EPI_ISL_1465850, EPI_ISL_1465851, EPI_ISL_1465852, EPI_ISL_1482336, EPI_ISL_1482337, EPI_ISL_1482338, EPI_ISL_1482339, EPI_ISL_1482340, EPI_ISL_1482341, EPI_ISL_1482342, EPI_ISL_1482343, EPI_ISL_1482344, EPI_ISL_1482345

see above — MONTEFIORE NEW ROCHELLE HOSPITAL LABORATORY — Wadsworth Center, New York State Department of Health — Alexis Russell; Catharine Prussing; Daryl M. Lamson; Erasmus Schneider; Erica Lasek-Nesselquist; John Kelly; Jonathan Plitnick; Kirsten St. George; Matthew Shudt; Melissa A Leisner; Navjot Singh

EPI_ISL_1300819, EPI_ISL_1709108, EPI_ISL_1709109, EPI_ISL_1709110, EPI_ISL_1709111, EPI_ISL_1709112, EPI_ISL_1709113, EPI_ISL_1709114, EPI_ISL_1709115, EPI_ISL_1709116, EPI_ISL_1709117, EPI_ISL_1709118, EPI_ISL_1709119, EPI_ISL_1709120, EPI_ISL_1709121, EPI_ISL_1709122, EPI_ISL_1709123, EPI_ISL_1709124, EPI_ISL_1709125, EPI_ISL_1709126, EPI_ISL_1709127, EPI_ISL_1709128, EPI_ISL_1709129, EPI_ISL_1709130, EPI_ISL_1709131, EPI_ISL_1709132, EPI_ISL_1709133, EPI_ISL_1709134, EPI_ISL_1709135, EPI_ISL_1709136, EPI_ISL_1709137, EPI_ISL_1709138, EPI_ISL_1709139, EPI_ISL_1709140, EPI_ISL_1709141, EPI_ISL_1709142, EPI_ISL_1709143, EPI_ISL_1709144, EPI_ISL_1709145, EPI_ISL_1709146, EPI_ISL_1709147, EPI_ISL_1709148, EPI_ISL_1709149, EPI_ISL_1709150, EPI_ISL_1709151, EPI_ISL_1709152, EPI_ISL_1709153, EPI_ISL_1709154, EPI_ISL_1709155, EPI_ISL_1709156, EPI_ISL_1709157, EPI_ISL_1709158, EPI_ISL_1709159, EPI_ISL_1709160, EPI_ISL_1709161, EPI_ISL_1709162, EPI_ISL_1709163, EPI_ISL_1709164, EPI_ISL_1709165, EPI_ISL_1709166, EPI_ISL_1709167, EPI_ISL_1709168, EPI_ISL_1709169, EPI_ISL_1709170, EPI_ISL_1709171, EPI_ISL_1709172, EPI_ISL_1709173, EPI_ISL_1709174, EPI_ISL_1709175, EPI_ISL_1709176, EPI_ISL_1709177, EPI_ISL_1709178, EPI_ISL_1709179, EPI_ISL_1709180, EPI_ISL_1709181, EPI_ISL_1709182, EPI_ISL_1709183, EPI_ISL_1709184, EPI_ISL_1709185, EPI_ISL_1709186, EPI_ISL_1709187, EPI_ISL_1709188, EPI_ISL_1709189, EPI_ISL_1709190, EPI_ISL_1709191, EPI_ISL_1709192, EPI_ISL_1709193, EPI_ISL_1709194, EPI_ISL_1709195, EPI_ISL_1709196, EPI_ISL_1709197, EPI_ISL_1709198, EPI_ISL_1709199, EPI_ISL_1709200, EPI_ISL_1709201, EPI_ISL_1709202, EPI_ISL_1709203, EPI_ISL_1709204, EPI_ISL_1709205, EPI_ISL_1709206, EPI_ISL_1709207, EPI_ISL_1709208, EPI_ISL_1709209, EPI_ISL_1709210, EPI_ISL_1709211, EPI_ISL_1709212, EPI_ISL_1709213, EPI_ISL_1709214, EPI_ISL_1709215, EPI_ISL_1709216, EPI_ISL_1709217, EPI_ISL_1709218, EPI_ISL_1709219, EPI_ISL_1709220, EPI_ISL_1709221, EPI_ISL_1709222, EPI_ISL_1709223, EPI_ISL_1709224, EPI_ISL_1709225, EPI_ISL_1709226, EPI_ISL_1709227, EPI_ISL_1709228, EPI_ISL_1709229, EPI_ISL_1709230, EPI_ISL_1709231, EPI_ISL_1709232, EPI_ISL_1709233, EPI_ISL_1709234, EPI_ISL_1709235, EPI_ISL_1709236, EPI_ISL_1709237, EPI_ISL_1709238, EPI_ISL_1709239, EPI_ISL_1709240, EPI_ISL_1709241, EPI_ISL_1709242, EPI_ISL_1709243, EPI_ISL_1709244, EPI_ISL_1709245, EPI_ISL_1709246, EPI_ISL_1709247, EPI_ISL_1709248, EPI_ISL_1709249, EPI_ISL_1709250, EPI_ISL_1709251, EPI_ISL_1709252, EPI_ISL_1709253, EPI_ISL_1709254, EPI_ISL_1709255, EPI_ISL_1709256, EPI_ISL_1709257, EPI_ISL_1709258, EPI_ISL_1709259, EPI_ISL_1709260, EPI_ISL_1709261, EPI_ISL_1709262, EPI_ISL_1709263, EPI_ISL_1709264, EPI_ISL_1709265, EPI_ISL_1709266, EPI_ISL_1709267, EPI_ISL_1709268, EPI_ISL_1709269, EPI_ISL_1709270, EPI_ISL_1709271, EPI_ISL_1709272, EPI_ISL_1709273, EPI_ISL_1709274, EPI_ISL_1709275, EPI_ISL_1709276, EPI_ISL_1709277, EPI_ISL_1709278, EPI_ISL_1709280, EPI_ISL_1709281, EPI_ISL_1709282, EPI_ISL_1709283, EPI_ISL_1709284, EPI_ISL_1709285, EPI_ISL_1709286, EPI_ISL_1709287, EPI_ISL_1709288, EPI_ISL_1709289, EPI_ISL_1709290, EPI_ISL_1709291, EPI_ISL_1709292, EPI_ISL_1709293, EPI_ISL_1709294, EPI_ISL_1709295, EPI_ISL_1709296, EPI_ISL_1709297, EPI_ISL_1709298, EPI_ISL_1709299, EPI_ISL_1709301, EPI_ISL_1709302, EPI_ISL_1709303, EPI_ISL_1709304, EPI_ISL_1709305, EPI_ISL_1709306, EPI_ISL_1709307, EPI_ISL_1709308, EPI_ISL_1709309, EPI_ISL_1709310, EPI_ISL_1709311, EPI_ISL_1709312, EPI_ISL_1709313, EPI_ISL_1709314, EPI_ISL_1709315, EPI_ISL_1709316, EPI_ISL_1709317, EPI_ISL_1709318, EPI_ISL_1709319, EPI_ISL_1709320, EPI_ISL_1709321, EPI_ISL_1709322, EPI_ISL_1709323, EPI_ISL_1709324, EPI_ISL_1709325, EPI_ISL_1709326, EPI_ISL_1709327, EPI_ISL_1709328, EPI_ISL_1709329, EPI_ISL_1709330, EPI_ISL_1709331, EPI_ISL_1709332, EPI_ISL_1709333, EPI_ISL_1709334, EPI_ISL_1709335, EPI_ISL_1709336, EPI_ISL_1709337, EPI_ISL_1709338, EPI_ISL_1709339, EPI_ISL_1709340, EPI_ISL_1709341, EPI_ISL_1709342, EPI_ISL_1709343, EPI_ISL_1709344, EPI_ISL_1709345, EPI_ISL_1709346, EPI_ISL_1709347, EPI_ISL_1709348, EPI_ISL_1709349, EPI_ISL_1709350, EPI_ISL_1709351, EPI_ISL_1709352, EPI_ISL_1709353, EPI_ISL_1709354, EPI_ISL_1709355, EPI_ISL_1709356, EPI_ISL_1709357, EPI_ISL_1709358, EPI_ISL_1709359, EPI_ISL_1709360, EPI_ISL_1709361, EPI_ISL_1709362, EPI_ISL_1709363, EPI_ISL_1709364, EPI_ISL_1709365, EPI_ISL_1709366, EPI_ISL_1709367, EPI_ISL_1709368, EPI_ISL_1709369, EPI_ISL_1709370, EPI_ISL_1709371, EPI_ISL_1709372, EPI_ISL_1709373, EPI_ISL_1709374, EPI_ISL_1709375, EPI_ISL_1709376, EPI_ISL_1709377, EPI_ISL_1709378, EPI_ISL_1709379, EPI_ISL_1709380, EPI_ISL_1709381, EPI_ISL_1709382, EPI_ISL_1709383, EPI_ISL_1709384, EPI_ISL_1709385, EPI_ISL_1709386, EPI_ISL_1709387, EPI_ISL_1709388, EPI_ISL_1709389, EPI_ISL_1709390, EPI_ISL_1709391, EPI_ISL_1709392, EPI_ISL_1709393, EPI_ISL_1709394, EPI_ISL_1709395, EPI_ISL_1709396, EPI_ISL_1709397, EPI_ISL_1709398, EPI_ISL_1709399, EPI_ISL_1709400, EPI_ISL_1709401, EPI_ISL_1709402, EPI_ISL_1709403, EPI_ISL_1709404, EPI_ISL_1709405, EPI_ISL_1709406, EPI_ISL_1709407, EPI_ISL_1709408, EPI_ISL_1709409, EPI_ISL_1709410, EPI_ISL_1709411, EPI_ISL_1709412, EPI_ISL_1709413, EPI_ISL_1709414, EPI_ISL_1709415, EPI_ISL_1709416, EPI_ISL_1709417, EPI_ISL_1709418, EPI_ISL_1709419, EPI_ISL_1709420, EPI_ISL_1709422, EPI_ISL_1709423, EPI_ISL_1709424, EPI_ISL_1709425, EPI_ISL_1709426, EPI_ISL_1709427, EPI_ISL_1709428, EPI_ISL_1709429, EPI_ISL_1709430, EPI_ISL_1709431, EPI_ISL_1709432, EPI_ISL_1709434, EPI_ISL_1709435, EPI_ISL_1709436, EPI_ISL_1709437, EPI_ISL_1709438, EPI_ISL_1709439, EPI_ISL_1709440, EPI_ISL_1709441, EPI_ISL_1709442, EPI_ISL_1709443, EPI_ISL_1709444, EPI_ISL_1709445, EPI_ISL_1709446, EPI_ISL_1709447, EPI_ISL_1709448, EPI_ISL_1709449, EPI_ISL_1709450, EPI_ISL_1709451, EPI_ISL_1709452, EPI_ISL_1709453, EPI_ISL_1709454, EPI_ISL_1709455, EPI_ISL_1709456, EPI_ISL_1709457, EPI_ISL_1709458, EPI_ISL_1709459, EPI_ISL_1709460, EPI_ISL_1709461, EPI_ISL_1709462, EPI_ISL_1709463, EPI_ISL_1709464, EPI_ISL_1709465, EPI_ISL_1709466, EPI_ISL_1709467, EPI_ISL_1709468, EPI_ISL_1709469, EPI_ISL_1709470, EPI_ISL_1709472, EPI_ISL_1709473, EPI_ISL_1709474, EPI_ISL_1709475, EPI_ISL_1709476, EPI_ISL_1709477, EPI_ISL_1709478, EPI_ISL_1709479, EPI_ISL_1709480, EPI_ISL_1709481, EPI_ISL_1709482, EPI_ISL_1709483, EPI_ISL_1709484, EPI_ISL_1709485, EPI_ISL_1709486, EPI_ISL_1709487, EPI_ISL_1709488, EPI_ISL_1709489, EPI_ISL_1709490, EPI_ISL_1709492, EPI_ISL_1709493, EPI_ISL_1709494, EPI_ISL_1709495, EPI_ISL_1709496, EPI_ISL_1709497, EPI_ISL_1709498, EPI_ISL_1709499, EPI_ISL_1709500, EPI_ISL_1709501, EPI_ISL_1709502, EPI_ISL_1709503, EPI_ISL_1709504, EPI_ISL_1709505, EPI_ISL_1709506, EPI_ISL_1709507, EPI_ISL_1709508, EPI_ISL_1709509, EPI_ISL_1709510, EPI_ISL_1709511, EPI_ISL_1709512, EPI_ISL_1709513, EPI_ISL_1709514, EPI_ISL_1709515, EPI_ISL_1709516, EPI_ISL_1709517, EPI_ISL_1709518, EPI_ISL_1709519, EPI_ISL_1709520, EPI_ISL_1709521, EPI_ISL_1709522, EPI_ISL_1709523, EPI_ISL_1709524, EPI_ISL_1709525, EPI_ISL_1709526, EPI_ISL_1709527, EPI_ISL_1709528, EPI_ISL_1709529, EPI_ISL_1709530, EPI_ISL_1709531, EPI_ISL_1709532, EPI_ISL_1709533, EPI_ISL_1709534, EPI_ISL_1709535, EPI_ISL_1709536, EPI_ISL_1709537, EPI_ISL_1709538, EPI_ISL_1709539, EPI_ISL_1709540, EPI_ISL_1709541, EPI_ISL_1709542, EPI_ISL_1709543, EPI_ISL_1709544, EPI_ISL_1709545, EPI_ISL_1709546, EPI_ISL_1709547, EPI_ISL_1709548, EPI_ISL_1709549, EPI_ISL_1709550, EPI_ISL_1709551, EPI_ISL_1709552, EPI_ISL_1709553, EPI_ISL_1709554, EPI_ISL_1709555, EPI_ISL_1709556, EPI_ISL_1709558, EPI_ISL_1709559, EPI_ISL_1709560, EPI_ISL_1709561, EPI_ISL_1709562, EPI_ISL_1709563, EPI_ISL_1709564, EPI_ISL_1709565, EPI_ISL_1709566, EPI_ISL_1709567, EPI_ISL_1709568, EPI_ISL_1709569, EPI_ISL_1709570, EPI_ISL_1709571, EPI_ISL_1709572, EPI_ISL_1709573, EPI_ISL_1709574, EPI_ISL_1709575, EPI_ISL_1709576, EPI_ISL_1709577, EPI_ISL_1709578, EPI_ISL_1709579, EPI_ISL_1709580, EPI_ISL_1709581, EPI_ISL_1709582, EPI_ISL_1709583, EPI_ISL_1709584, EPI_ISL_1709585, EPI_ISL_1709586, EPI_ISL_1709587, EPI_ISL_1709588, EPI_ISL_1709589, EPI_ISL_1709590, EPI_ISL_1709591, EPI_ISL_1709592, EPI_ISL_1709593, EPI_ISL_1709594, EPI_ISL_1709595, EPI_ISL_1709596, EPI_ISL_1709597, EPI_ISL_1709598, EPI_ISL_1709599, EPI_ISL_1709600, EPI_ISL_1709601, EPI_ISL_1709602, EPI_ISL_1709603, EPI_ISL_1709604, EPI_ISL_1709605, EPI_ISL_1709606, EPI_ISL_1709607, EPI_ISL_1709608, EPI_ISL_1709609, EPI_ISL_1709610, EPI_ISL_1709611, EPI_ISL_1709612, EPI_ISL_1709613, EPI_ISL_1709614, EPI_ISL_1709615, EPI_ISL_1709616, EPI_ISL_1709617, EPI_ISL_1709618, EPI_ISL_1709619, EPI_ISL_1709620, EPI_ISL_1709621, EPI_ISL_1709622, EPI_ISL_1709623, EPI_ISL_1709624, EPI_ISL_1709625, EPI_ISL_1709626, EPI_ISL_1709627, EPI_ISL_1709628, EPI_ISL_1709629, EPI_ISL_1709630, EPI_ISL_1709631, EPI_ISL_1709632, EPI_ISL_1709633, EPI_ISL_1709634, EPI_ISL_1709635, EPI_ISL_1709636, EPI_ISL_1709637, EPI_ISL_1709638, EPI_ISL_1709639, EPI_ISL_1709640, EPI_ISL_1709641, EPI_ISL_1709642, EPI_ISL_1709643, EPI_ISL_1709644, EPI_ISL_1709645, EPI_ISL_1709646, EPI_ISL_1709647, EPI_ISL_1709648, EPI_ISL_1709649, EPI_ISL_1709650, EPI_ISL_1709651, EPI_ISL_1709652, EPI_ISL_1709653, EPI_ISL_1709654, EPI_ISL_1709655, EPI_ISL_1709656, EPI_ISL_1709657, EPI_ISL_1709658, EPI_ISL_1709659, EPI_ISL_1709660, EPI_ISL_1709661, EPI_ISL_1709662, EPI_ISL_1709663, EPI_ISL_1709664, EPI_ISL_1709665, EPI_ISL_1709666, EPI_ISL_1709667, EPI_ISL_1709668, EPI_ISL_1709669, EPI_ISL_1709670, EPI_ISL_1709671, EPI_ISL_1709672, EPI_ISL_1709673, EPI_ISL_1709674, EPI_ISL_1709675, EPI_ISL_1709676, EPI_ISL_1709677, EPI_ISL_1709678, EPI_ISL_1709679, EPI_ISL_1709680, EPI_ISL_1709681, EPI_ISL_1709682, EPI_ISL_1709683, EPI_ISL_1709684, EPI_ISL_1709685, EPI_ISL_1709686, EPI_ISL_1709687, EPI_ISL_1709688, EPI_ISL_1709689, EPI_ISL_1709690, EPI_ISL_1709691, EPI_ISL_1709692, EPI_ISL_1709693, EPI_ISL_1709694, EPI_ISL_1709695, EPI_ISL_1709696, EPI_ISL_1709697, EPI_ISL_1709698, EPI_ISL_1709699, EPI_ISL_1709700, EPI_ISL_1709701, EPI_ISL_1709702, EPI_ISL_1709703, EPI_ISL_1709704, EPI_ISL_1709705, EPI_ISL_1709706, EPI_ISL_1709707, EPI_ISL_1709708, EPI_ISL_1709709, EPI_ISL_1709710, EPI_ISL_1709711, EPI_ISL_1709712, EPI_ISL_1709713, EPI_ISL_1709714, EPI_ISL_1709715, EPI_ISL_1709716, EPI_ISL_1709717, EPI_ISL_1709718, EPI_ISL_1709719, EPI_ISL_1709720, EPI_ISL_1709721, EPI_ISL_1709722, EPI_ISL_1709723, EPI_ISL_1709724, EPI_ISL_1709725, EPI_ISL_1709726, EPI_ISL_1709727, EPI_ISL_1709728, EPI_ISL_1709729, EPI_ISL_1709731, EPI_ISL_1709732, EPI_ISL_1709733, EPI_ISL_1709734, EPI_ISL_1709735, EPI_ISL_1709736, EPI_ISL_1709737, EPI_ISL_1709738, EPI_ISL_1709739, EPI_ISL_1709740, EPI_ISL_1709741, EPI_ISL_1709742, EPI_ISL_1709743, EPI_ISL_1709744, EPI_ISL_1709745, EPI_ISL_1709746, EPI_ISL_1709747, EPI_ISL_1709748, EPI_ISL_1709749, EPI_ISL_1709750, EPI_ISL_1709751, EPI_ISL_1709752, EPI_ISL_1709753, EPI_ISL_1709754, EPI_ISL_1709755, EPI_ISL_1709756, EPI_ISL_1709757, EPI_ISL_1709758, EPI_ISL_1709759, EPI_ISL_1709760, EPI_ISL_1709761, EPI_ISL_1709762, EPI_ISL_1709763, EPI_ISL_1709764, EPI_ISL_1709765, EPI_ISL_1709766, EPI_ISL_1709767, EPI_ISL_1709768, EPI_ISL_1709769, EPI_ISL_1709770, EPI_ISL_1709771, EPI_ISL_1709772, EPI_ISL_1709773, EPI_ISL_1709774, EPI_ISL_1709775, EPI_ISL_1709776, EPI_ISL_1709777, EPI_ISL_1709778, EPI_ISL_1709779, EPI_ISL_1709780, EPI_ISL_1709781, EPI_ISL_1709782, EPI_ISL_1709783, EPI_ISL_1709784, EPI_ISL_1709785, EPI_ISL_1709786, EPI_ISL_1709787, EPI_ISL_1709788, EPI_ISL_1709789, EPI_ISL_1709790, EPI_ISL_1709791, EPI_ISL_1709792, EPI_ISL_1709793, EPI_ISL_1709794, EPI_ISL_1709795, EPI_ISL_1709796, EPI_ISL_1709797, EPI_ISL_1709798, EPI_ISL_1709799, EPI_ISL_1709800, EPI_ISL_1709801, EPI_ISL_1709802, EPI_ISL_1709803, EPI_ISL_1709804, EPI_ISL_1709805, EPI_ISL_1709806, EPI_ISL_1709807, EPI_ISL_1709808, EPI_ISL_1709809, EPI_ISL_1709810, EPI_ISL_1709811, EPI_ISL_1709812, EPI_ISL_1709813, EPI_ISL_1709814, EPI_ISL_1709815, EPI_ISL_1709816

see above — MSHS Clinical Microbiology Laboratories — MSHS Pathogen Surveillance Program — Adolfo García-Sastre; Adriana van de Guchte; Ajay Obla; Alberto Paniz-Mondolfi; Ana S. Gonzalez-Reiche; Angela Amoako; Ashley Salimbangon; Betsaida Salom Melo; Bremy Alburquerque; Brianne Ciferri; Charles Gleason; Daniel Floda; Deena R. Altman; Denise Jurczyszak; Emilia Mia Sordillo; Gintaras Deikus; Giulio Kleiner; Gopi Patel; Hala Alshammary; Harm van Bakel; Irina Oussenko; Jayeeta Dutta; Juan Soto; Julia Matthews; Katherine Beach; Kathryn Twyman; Kayla Russo; Komal Srivastava; Levy Sominsky; Mahmoud Awawda; Marta Luksza; Matthew M. Hernandez; Melissa Gitman; Michael D. Nowak; Mitchell J. Sullivan; Nancy Francoeur; Robert Sebra; Sarah Schaefer; Shelcie Fabre; Shwetha Hara Sridhar; Viviana Simon; Ying-Chih Wang; Zenab Khan

EPI_ISL_2009144, EPI_ISL_2009176 — MSK Microbiology Lab — MSK Microbiology Lab — Esther Babady; Krupa Jani; Tracy McMillen

EPI_ISL_2011717 — Moderna Inc. — Moderna Inc. — Groves Dixon; Rolando Pajon; Yamuna Paila

EPI_ISL_1919998, EPI_ISL_1920001 — Montefiore Medical Center Laboratories — Wadsworth Center, New York State Department of Health — Alexis Russell; Catharine Prussing; Daryl M. Lamson; Erasmus Schneider; Erica Lasek-Nesselquist; John Kelly; Jonathan Plitnick; Kirsten St. George; Matthew Shudt; Melissa A Leisner; Navjot Singh

EPI_ISL_1378824, EPI_ISL_1378825, EPI_ISL_1378826 — Murphy Medical Association — Grubaugh Lab - Yale School of Public Health — Anderson Brito; Annie Watkins; Caleb Neal; Chaney Kalinich; Chantal Vogels; Eva Laszlo; Isabell Ott; Jessica Rothman; Joseph Fauver; Mallery Breban; Mary Petrone; Nathan Grubaugh; Steven Murphy; Tara Alpert

EPI_ISL_1395756, EPI_ISL_1395757, EPI_ISL_1395758, EPI_ISL_1395760, EPI_ISL_1395763, EPI_ISL_1397655, EPI_ISL_1397656, EPI_ISL_1397658, EPI_ISL_1397659, EPI_ISL_1397660, EPI_ISL_1397661, EPI_ISL_1397662, EPI_ISL_1397663, EPI_ISL_1397664, EPI_ISL_1397665, EPI_ISL_1397666, EPI_ISL_1397667, EPI_ISL_1397668, EPI_ISL_1397669, EPI_ISL_1397670, EPI_ISL_1397671, EPI_ISL_1397672, EPI_ISL_1397673, EPI_ISL_1397674, EPI_ISL_1397675, EPI_ISL_1397676, EPI_ISL_1397677, EPI_ISL_1397678, EPI_ISL_1397679, EPI_ISL_1397680, EPI_ISL_1397681, EPI_ISL_1397682, EPI_ISL_1397683, EPI_ISL_1397684, EPI_ISL_1397685, EPI_ISL_1397686, EPI_ISL_1397687, EPI_ISL_1397688, EPI_ISL_1397689, EPI_ISL_1482346, EPI_ISL_1482347, EPI_ISL_1482348, EPI_ISL_1482349, EPI_ISL_1482350, EPI_ISL_1482351, EPI_ISL_1482352, EPI_ISL_1482353, EPI_ISL_1582256, EPI_ISL_1582257, EPI_ISL_1582258, EPI_ISL_1582259

see above — NORTH SHORE UNIVERSITY HOSPITAL — Wadsworth Center, New York State Department of Health — Alexis Russel; Alexis Russell; Catharine Prussing; Daryl M. Lamson; Erasmus Schneider; Erica Lasek-Nesselquist; John Kelly; Jonathan Plitnick; Kirsten St. George; Matthew Shudt; Melissa A Leisner; Navjot Singh

EPI_ISL_1397830, EPI_ISL_1397909, EPI_ISL_1397911, EPI_ISL_1398068, EPI_ISL_1398069, EPI_ISL_1398070, EPI_ISL_1398071, EPI_ISL_1398072, EPI_ISL_1398073, EPI_ISL_1398074, EPI_ISL_1398075, EPI_ISL_1398076, EPI_ISL_1398077, EPI_ISL_1398078, EPI_ISL_1398079, EPI_ISL_1398080, EPI_ISL_1398081, EPI_ISL_1398082, EPI_ISL_1398083, EPI_ISL_1398084, EPI_ISL_1398085, EPI_ISL_1398086, EPI_ISL_1398087, EPI_ISL_1398088, EPI_ISL_1398089, EPI_ISL_1398090, EPI_ISL_1398091, EPI_ISL_1398092, EPI_ISL_1398093, EPI_ISL_1398094, EPI_ISL_1398095, EPI_ISL_1398096, EPI_ISL_1398097, EPI_ISL_1398098, EPI_ISL_1398099, EPI_ISL_1398100, EPI_ISL_1398101, EPI_ISL_1398102, EPI_ISL_1398103, EPI_ISL_1398104, EPI_ISL_1398105, EPI_ISL_1398106, EPI_ISL_1398107, EPI_ISL_1398108, EPI_ISL_1398109, EPI_ISL_1398110, EPI_ISL_1398111, EPI_ISL_1398112, EPI_ISL_1398113, EPI_ISL_1398114, EPI_ISL_1398115, EPI_ISL_1398116, EPI_ISL_1398117, EPI_ISL_1398118, EPI_ISL_1398119, EPI_ISL_1462556, EPI_ISL_1462558, EPI_ISL_1462559, EPI_ISL_1462561, EPI_ISL_1462563, EPI_ISL_1462564, EPI_ISL_1462566, EPI_ISL_1462567, EPI_ISL_1462569, EPI_ISL_1462570, EPI_ISL_1462576, EPI_ISL_1462578, EPI_ISL_1462579, EPI_ISL_1462581, EPI_ISL_1462582, EPI_ISL_1462584, EPI_ISL_1462585, EPI_ISL_1462587, EPI_ISL_1462588, EPI_ISL_1462589, EPI_ISL_1462591, EPI_ISL_1462593, EPI_ISL_1462594, EPI_ISL_1462596, EPI_ISL_1462597, EPI_ISL_1462599, EPI_ISL_1462600, EPI_ISL_1462601, EPI_ISL_1462603, EPI_ISL_1462604, EPI_ISL_1462606, EPI_ISL_1462607, EPI_ISL_1462609, EPI_ISL_1462610, EPI_ISL_1462612, EPI_ISL_1462613, EPI_ISL_1462615, EPI_ISL_1462617, EPI_ISL_1462618, EPI_ISL_1462620, EPI_ISL_1462621, EPI_ISL_1462623, EPI_ISL_1462624, EPI_ISL_1465678, EPI_ISL_1465679, EPI_ISL_1465680, EPI_ISL_1465681, EPI_ISL_1465682, EPI_ISL_1465755, EPI_ISL_1465756, EPI_ISL_1465757, EPI_ISL_1465758, EPI_ISL_1465759, EPI_ISL_1465761, EPI_ISL_1465762, EPI_ISL_1465763, EPI_ISL_1465764, EPI_ISL_1465765, EPI_ISL_1465766, EPI_ISL_1465835, EPI_ISL_1465836, EPI_ISL_1465837, EPI_ISL_1482556, EPI_ISL_1482557, EPI_ISL_1482558, EPI_ISL_1482559, EPI_ISL_1482560, EPI_ISL_1482561, EPI_ISL_1482562, EPI_ISL_1482563, EPI_ISL_1482564, EPI_ISL_1482565, EPI_ISL_1482566, EPI_ISL_1482567, EPI_ISL_1482568, EPI_ISL_1482569, EPI_ISL_1482570, EPI_ISL_1482571, EPI_ISL_1482572, EPI_ISL_1482580, EPI_ISL_1482581, EPI_ISL_1482582, EPI_ISL_1482583, EPI_ISL_1482584, EPI_ISL_1482585, EPI_ISL_1482586, EPI_ISL_1482587, EPI_ISL_1482588, EPI_ISL_1482589, EPI_ISL_1482590, EPI_ISL_1482591, EPI_ISL_1482592, EPI_ISL_1482593, EPI_ISL_1482594, EPI_ISL_1482595, EPI_ISL_1482596, EPI_ISL_1482597, EPI_ISL_1482598, EPI_ISL_1482599, EPI_ISL_1482600, EPI_ISL_1482601, EPI_ISL_1482602, EPI_ISL_1482603, EPI_ISL_1482604, EPI_ISL_1482605, EPI_ISL_1482606, EPI_ISL_1482607, EPI_ISL_1482608, EPI_ISL_1482609, EPI_ISL_1482610, EPI_ISL_1482611, EPI_ISL_1482612, EPI_ISL_1482613, EPI_ISL_1482614, EPI_ISL_1482615, EPI_ISL_1482616, EPI_ISL_1575449, EPI_ISL_1575469, EPI_ISL_1575478, EPI_ISL_1576815, EPI_ISL_1576825, EPI_ISL_1576826, EPI_ISL_1580723, EPI_ISL_1580724, EPI_ISL_1580725, EPI_ISL_1580726, EPI_ISL_1580727, EPI_ISL_1580728, EPI_ISL_1580729, EPI_ISL_1580730, EPI_ISL_1580731, EPI_ISL_1580732, EPI_ISL_1580733, EPI_ISL_1580734, EPI_ISL_1580735, EPI_ISL_1580736, EPI_ISL_1580737, EPI_ISL_1580738, EPI_ISL_1582156, EPI_ISL_1582225, EPI_ISL_1582226, EPI_ISL_1582283, EPI_ISL_1582286, EPI_ISL_1582287, EPI_ISL_1582288, EPI_ISL_1582291, EPI_ISL_1582293, EPI_ISL_1582294, EPI_ISL_1582295, EPI_ISL_1582296, EPI_ISL_1582297, EPI_ISL_1582298, EPI_ISL_1582299, EPI_ISL_1582300, EPI_ISL_1582301, EPI_ISL_1582302, EPI_ISL_1582303, EPI_ISL_1582304, EPI_ISL_1582305, EPI_ISL_1582306, EPI_ISL_1582307, EPI_ISL_1582317, EPI_ISL_1582354, EPI_ISL_1582355, EPI_ISL_1582356, EPI_ISL_1582370, EPI_ISL_1582371, EPI_ISL_1582373, EPI_ISL_1582374, EPI_ISL_1582375, EPI_ISL_1582376, EPI_ISL_1582377, EPI_ISL_1582378, EPI_ISL_1582379, EPI_ISL_1582380, EPI_ISL_1582381, EPI_ISL_1582382, EPI_ISL_1582383, EPI_ISL_1582384, EPI_ISL_1582385, EPI_ISL_1582386, EPI_ISL_1582387, EPI_ISL_1582388, EPI_ISL_1582389, EPI_ISL_1582405, EPI_ISL_1593876, EPI_ISL_1593877, EPI_ISL_1593887, EPI_ISL_1756327, EPI_ISL_1935609

see above — NORTHWELL HEALTH LABORATORIES — Wadsworth Center, New York State Department of Health — Alexis Russel; Alexis Russell; Catharine Prussing; Daryl M. Lamson; Erasmus Schneider; Erica Lasek-Nesselquist; John Kelly; Jonathan Plitnick; Kirsten St. George; Matthew Shudt; Melissa A Leisner; Navjot Singh

EPI_ISL_1318347, EPI_ISL_1318348, EPI_ISL_1318349, EPI_ISL_1446974, EPI_ISL_1446975, EPI_ISL_1516205, EPI_ISL_1516206, EPI_ISL_1516207, EPI_ISL_1516208, EPI_ISL_1516209, EPI_ISL_1516210, EPI_ISL_1516211, EPI_ISL_1516212, EPI_ISL_1678056, EPI_ISL_1678057, EPI_ISL_1678058, EPI_ISL_1678059, EPI_ISL_1678060, EPI_ISL_1678061, EPI_ISL_1678062, EPI_ISL_1678063, EPI_ISL_1678064, EPI_ISL_1678065, EPI_ISL_1678066, EPI_ISL_1678067, EPI_ISL_1678068, EPI_ISL_1678069, EPI_ISL_1678070, EPI_ISL_1678071, EPI_ISL_1678072, EPI_ISL_1678073, EPI_ISL_1678074, EPI_ISL_1678075, EPI_ISL_1678076, EPI_ISL_1678077, EPI_ISL_1678078, EPI_ISL_1678079, EPI_ISL_1678080, EPI_ISL_1678257, EPI_ISL_1678258, EPI_ISL_1678259, EPI_ISL_1678260, EPI_ISL_1678261, EPI_ISL_1678262, EPI_ISL_1678263, EPI_ISL_1678264, EPI_ISL_1678265, EPI_ISL_1695970, EPI_ISL_1823946, EPI_ISL_1823958, EPI_ISL_1823968, EPI_ISL_1823969

see above — NYC Department of Health and Mental Hygiene — Centers for Disease Control and Prevention Division of Viral Diseases, Pathogen Discovery — Alison Laufer Halpin; Anna Montmayeur; Anna Uehara; Ben L. Rambo-Martin; Clinton R. Paden; Dakota Howard; Darlene Wagner; Dave Wentworth; Dhwani Batra; Haibin Wang; Jasmine Padilla; Jing Zhang; Justin Lee; Katie Dillon; Krista Queen; Kristen Knipe; Kristine Lacek; Lori Rowe; Mark Burroughs; Matthew Schmerer; Mili Sheth; Peter Cook; Peter W. Cook; Rachel Marine; Sam Shepard; Sarah Nobles; Shoshona Le; Suxiang Tong; Vivien Dugan; Yan Li; Ying Tao; Yvette Unoarumhi

EPI_ISL_1700669, EPI_ISL_1700670, EPI_ISL_1700671, EPI_ISL_1700672, EPI_ISL_1700673 — NYC Department of Health and Mental Hygiene — Pathogen Discovery, Respiratory Viruses Branch, Division of Viral Diseases, Centers for Disease Control and Prevention — Adam Retchless; Anna Kelleher; Anna Montmayeur; Anna Uehara; Brian Lynch; Clinton R. Paden; Haibin Wang; Han Jia Justin Ng; Jing Zhang; Justin Lee; Krista Queen; Mark Burroughs; Peter Cook; Rachel Marine; Suxiang Tong; Yan Li; Ying Tao

EPI_ISL_1398320, EPI_ISL_1398321, EPI_ISL_1398322, EPI_ISL_1398323, EPI_ISL_1398324, EPI_ISL_1398325, EPI_ISL_1398326, EPI_ISL_1398327, EPI_ISL_1398328, EPI_ISL_1398329, EPI_ISL_1398330, EPI_ISL_1461350, EPI_ISL_1461351, EPI_ISL_1465714, EPI_ISL_1465715, EPI_ISL_1465716, EPI_ISL_1465717, EPI_ISL_1465718, EPI_ISL_1465719, EPI_ISL_1465720, EPI_ISL_1465721, EPI_ISL_1465722, EPI_ISL_1465834, EPI_ISL_1582179, EPI_ISL_1582180, EPI_ISL_1582181, EPI_ISL_1582182, EPI_ISL_1582183, EPI_ISL_1582184, EPI_ISL_1582185, EPI_ISL_1582186, EPI_ISL_1582187, EPI_ISL_1582188, EPI_ISL_1582189, EPI_ISL_1582190, EPI_ISL_1582191, EPI_ISL_1582192, EPI_ISL_1582193, EPI_ISL_1582194, EPI_ISL_1582195, EPI_ISL_1582196, EPI_ISL_1582197, EPI_ISL_1582198, EPI_ISL_1582199, EPI_ISL_1582200, EPI_ISL_1582201, EPI_ISL_1582202, EPI_ISL_1582203, EPI_ISL_1582204, EPI_ISL_1582205, EPI_ISL_1582206, EPI_ISL_1582207, EPI_ISL_1582208, EPI_ISL_1582209, EPI_ISL_1582210, EPI_ISL_1582211, EPI_ISL_1582212, EPI_ISL_1582213, EPI_ISL_1582230, EPI_ISL_1582322, EPI_ISL_1582323, EPI_ISL_1582324, EPI_ISL_1582325, EPI_ISL_1582326, EPI_ISL_1582327, EPI_ISL_1582328, EPI_ISL_1582329, EPI_ISL_1582330, EPI_ISL_1582331, EPI_ISL_1582332, EPI_ISL_1582333, EPI_ISL_1582334, EPI_ISL_1582335, EPI_ISL_1582336, EPI_ISL_1593870, EPI_ISL_1593871, EPI_ISL_1593878, EPI_ISL_1593883, EPI_ISL_1605704, EPI_ISL_1755495, EPI_ISL_1755496, EPI_ISL_1755498, EPI_ISL_1755499, EPI_ISL_1755501, EPI_ISL_1854696, EPI_ISL_1854697, EPI_ISL_1854698, EPI_ISL_1854699, EPI_ISL_1854700, EPI_ISL_1854701, EPI_ISL_1854702, EPI_ISL_1854704, EPI_ISL_1854705, EPI_ISL_1854706, EPI_ISL_1854707, EPI_ISL_1854708, EPI_ISL_1854709, EPI_ISL_1854710, EPI_ISL_1854711, EPI_ISL_1854712, EPI_ISL_1854713, EPI_ISL_1854714, EPI_ISL_1854715, EPI_ISL_1854716, EPI_ISL_1854717, EPI_ISL_1854718, EPI_ISL_1854719, EPI_ISL_1854720, EPI_ISL_1854721, EPI_ISL_1854722, EPI_ISL_1854723, EPI_ISL_1854724, EPI_ISL_1854725, EPI_ISL_1854726, EPI_ISL_1854727, EPI_ISL_1854728, EPI_ISL_1854729, EPI_ISL_1854730, EPI_ISL_1854731, EPI_ISL_1854732, EPI_ISL_1854733, EPI_ISL_1854734, EPI_ISL_1854735, EPI_ISL_1935619, EPI_ISL_1935628

see above — NYC Pandemic Response Lab — Wadsworth Center, New York State Department of Health — Alexis Russel; Alexis Russell; Catharine Prussing; Daryl M. Lamson; Erasmus Schneider; Erica Lasek-Nesselquist; John Kelly; Jonathan Plitnick; Kirsten St. George; Matthew Shudt; Melissa A Leisner; Navjot Singh

EPI_ISL_1379345, EPI_ISL_1379346, EPI_ISL_1379347, EPI_ISL_1379348, EPI_ISL_1379351, EPI_ISL_1379352, EPI_ISL_1379353, EPI_ISL_1379354, EPI_ISL_1379362, EPI_ISL_1379363, EPI_ISL_1379364, EPI_ISL_1379365, EPI_ISL_1379366, EPI_ISL_1379367, EPI_ISL_1379368, EPI_ISL_1379369, EPI_ISL_1379370, EPI_ISL_1379371, EPI_ISL_1379372, EPI_ISL_1379373, EPI_ISL_1379374, EPI_ISL_1379375, EPI_ISL_1379376, EPI_ISL_1379382, EPI_ISL_1379383, EPI_ISL_1379384, EPI_ISL_1379387, EPI_ISL_1379390

see above — NYP-WCM — New York Genome Center — Andre Corvelo; Arryn Craney; Chris Mason; Dayna M. Oschwald; Hanna Rennert; Lars F Westblade; Margaret Elizabeth Ross; Melissa Cushing; Michael Zody; Olivier Elemento; Priya Velu; Samantha Fennessey; Tom Maniatis

EPI_ISL_1695889, EPI_ISL_1695890, — NYSDOH Wadsworth Center, Virology Lab — Centers for Disease Control and Prevention Division of Viral — Alison Laufer Halpin; Ben L. Rambo-Martin; Clinton R. Paden; Dakota Howard; Darlene Wagner; Dave Wentworth; Dhwani Batra; Jasmine Padilla; Justin Lee; Katie Dillon; Krista Queen; Kristen Knipe; Kristine Lacek; Mark Burroughs; Matthew Schmerer; Mili Sheth; Peter Cook; Sam Shepard; Sarah Nobles; Shoshona Le; Suxiang Tong; Vivien Dugan; Yvette Unoarumhi

| | | |
|---|---|---|
| EPI_ISL_1695891, EPI_ISL_1695892 | | Diseases, Pathogen Discovery |

EPI_ISL_1262501, EPI_ISL_1262502, EPI_ISL_1262503, EPI_ISL_1262505, EPI_ISL_1262506, EPI_ISL_1262507, EPI_ISL_1262508, EPI_ISL_1262510, EPI_ISL_1262511, EPI_ISL_1262512, EPI_ISL_1262513, EPI_ISL_1262515, EPI_ISL_1262516, EPI_ISL_1262517, EPI_ISL_1262518, EPI_ISL_1262519, EPI_ISL_1262520, EPI_ISL_1262521, EPI_ISL_1262523, EPI_ISL_1262524, EPI_ISL_1262525, EPI_ISL_1262526, EPI_ISL_1262527, EPI_ISL_1262528, EPI_ISL_1262529, EPI_ISL_1262530, EPI_ISL_1262531, EPI_ISL_1262532, EPI_ISL_1262533, EPI_ISL_1262534, EPI_ISL_1262535, EPI_ISL_1262536, EPI_ISL_1262537, EPI_ISL_1262538, EPI_ISL_1262539, EPI_ISL_1262540, EPI_ISL_1262541, EPI_ISL_1262542, EPI_ISL_1262543, EPI_ISL_1262544, EPI_ISL_1262545, EPI_ISL_1262546, EPI_ISL_1262547, EPI_ISL_1262548, EPI_ISL_1262550, EPI_ISL_1262551, EPI_ISL_1262552, EPI_ISL_1262553, EPI_ISL_1262554, EPI_ISL_1262555, EPI_ISL_1262556, EPI_ISL_1262557, EPI_ISL_1262558, EPI_ISL_1262559, EPI_ISL_1262560, EPI_ISL_1262561, EPI_ISL_1262562, EPI_ISL_1262563, EPI_ISL_1262564, EPI_ISL_1262565, EPI_ISL_1262567, EPI_ISL_1262568, EPI_ISL_1262569, EPI_ISL_1262573, EPI_ISL_1262574, EPI_ISL_1262575, EPI_ISL_1262576, EPI_ISL_1262577, EPI_ISL_1262578, EPI_ISL_1262579, EPI_ISL_1262580, EPI_ISL_1262581, EPI_ISL_1262582, EPI_ISL_1262583, EPI_ISL_1262584, EPI_ISL_1300175, EPI_ISL_1300176, EPI_ISL_1300177, EPI_ISL_1300178, EPI_ISL_1300179, EPI_ISL_1300180, EPI_ISL_1300181, EPI_ISL_1300183, EPI_ISL_1300184, EPI_ISL_1300185, EPI_ISL_1300186, EPI_ISL_1300187, EPI_ISL_1300188, EPI_ISL_1300189, EPI_ISL_1300190, EPI_ISL_1300191, EPI_ISL_1300192, EPI_ISL_1300193, EPI_ISL_1300194, EPI_ISL_1300195, EPI_ISL_1300196, EPI_ISL_1300197, EPI_ISL_1300198, EPI_ISL_1300199, EPI_ISL_1300200, EPI_ISL_1300201, EPI_ISL_1300202, EPI_ISL_1300203, EPI_ISL_1300204, EPI_ISL_1300205, EPI_ISL_1300206, EPI_ISL_1300207, EPI_ISL_1300208, EPI_ISL_1300209, EPI_ISL_1300210, EPI_ISL_1300211, EPI_ISL_1300212, EPI_ISL_1300213, EPI_ISL_1300214, EPI_ISL_1300215, EPI_ISL_1300216, EPI_ISL_1300217, EPI_ISL_1300218, EPI_ISL_1300219, EPI_ISL_1300220, EPI_ISL_1300221, EPI_ISL_1300222, EPI_ISL_1300223, EPI_ISL_1300224, EPI_ISL_1300225, EPI_ISL_1300226, EPI_ISL_1300227, EPI_ISL_1300228, EPI_ISL_1300229, EPI_ISL_1300230, EPI_ISL_1300231, EPI_ISL_1300232, EPI_ISL_1300233, EPI_ISL_1300234, EPI_ISL_1300235, EPI_ISL_1300236, EPI_ISL_1300237, EPI_ISL_1300238, EPI_ISL_1300239, EPI_ISL_1300240, EPI_ISL_1300241, EPI_ISL_1300242, EPI_ISL_1300243, EPI_ISL_1300244, EPI_ISL_1300245, EPI_ISL_1300246, EPI_ISL_1300247, EPI_ISL_1300248, EPI_ISL_1300249, EPI_ISL_1300250, EPI_ISL_1300251, EPI_ISL_1300252, EPI_ISL_1300253, EPI_ISL_1300254, EPI_ISL_1300255, EPI_ISL_1300256, EPI_ISL_1300257, EPI_ISL_1300258, EPI_ISL_1300259, EPI_ISL_1300261, EPI_ISL_1300262, EPI_ISL_1300263, EPI_ISL_1300264, EPI_ISL_1300265, EPI_ISL_1300266, EPI_ISL_1300267, EPI_ISL_1300268, EPI_ISL_1300269, EPI_ISL_1300270, EPI_ISL_1300271, EPI_ISL_1300272, EPI_ISL_1300273, EPI_ISL_1300274, EPI_ISL_1322745, EPI_ISL_1322746, EPI_ISL_1322747, EPI_ISL_1322748, EPI_ISL_1322749, EPI_ISL_1322750, EPI_ISL_1322751, EPI_ISL_1322752, EPI_ISL_1322753, EPI_ISL_1322754, EPI_ISL_1322755, EPI_ISL_1322756, EPI_ISL_1322757, EPI_ISL_1322758, EPI_ISL_1322759, EPI_ISL_1322760, EPI_ISL_1322761, EPI_ISL_1322762, EPI_ISL_1322763, EPI_ISL_1322764, EPI_ISL_1322765, EPI_ISL_1322766, EPI_ISL_1322767, EPI_ISL_1322768, EPI_ISL_1322769, EPI_ISL_1322770, EPI_ISL_1322771, EPI_ISL_1322772, EPI_ISL_1322773, EPI_ISL_1322774, EPI_ISL_1322775, EPI_ISL_1322776, EPI_ISL_1322777, EPI_ISL_1322778, EPI_ISL_1322779, EPI_ISL_1322780, EPI_ISL_1322781, EPI_ISL_1322782, EPI_ISL_1322783, EPI_ISL_1322784, EPI_ISL_1322785, EPI_ISL_1322786, EPI_ISL_1322787, EPI_ISL_1322788, EPI_ISL_1322789, EPI_ISL_1322790, EPI_ISL_1322791, EPI_ISL_1322792, EPI_ISL_1322793, EPI_ISL_1322794, EPI_ISL_1322795, EPI_ISL_1322796, EPI_ISL_1322797, EPI_ISL_1322798, EPI_ISL_1322799, EPI_ISL_1322800, EPI_ISL_1322801, EPI_ISL_1322802, EPI_ISL_1322803, EPI_ISL_1322804, EPI_ISL_1322806, EPI_ISL_1322807, EPI_ISL_1322808, EPI_ISL_1322809, EPI_ISL_1322810, EPI_ISL_1322811, EPI_ISL_1322812, EPI_ISL_1322813, EPI_ISL_1322814, EPI_ISL_1322815, EPI_ISL_1322816, EPI_ISL_1322817, EPI_ISL_1322818, EPI_ISL_1322819, EPI_ISL_1322821, EPI_ISL_1322822, EPI_ISL_1322823, EPI_ISL_1322824, EPI_ISL_1322825, EPI_ISL_1322826, EPI_ISL_1322827, EPI_ISL_1322828, EPI_ISL_1322829, EPI_ISL_1322830, EPI_ISL_1322831, EPI_ISL_1322832, EPI_ISL_1322833, EPI_ISL_1322834, EPI_ISL_1423394, EPI_ISL_1423397, EPI_ISL_1423398, EPI_ISL_1423399, EPI_ISL_1423401, EPI_ISL_1423402, EPI_ISL_1423403, EPI_ISL_1423404, EPI_ISL_1423405, EPI_ISL_1423406, EPI_ISL_1423407, EPI_ISL_1423408, EPI_ISL_1423409, EPI_ISL_1423410, EPI_ISL_1423411, EPI_ISL_1423412, EPI_ISL_1423413, EPI_ISL_1423414, EPI_ISL_1423415, EPI_ISL_1423416, EPI_ISL_1423417, EPI_ISL_1423418, EPI_ISL_1423419, EPI_ISL_1423420, EPI_ISL_1423421, EPI_ISL_1423422, EPI_ISL_1423423, EPI_ISL_1423424, EPI_ISL_1423425, EPI_ISL_1423426, EPI_ISL_1423427, EPI_ISL_1423428, EPI_ISL_1423429, EPI_ISL_1423430, EPI_ISL_1423431, EPI_ISL_1423432, EPI_ISL_1423433, EPI_ISL_1423434, EPI_ISL_1423435, EPI_ISL_1423436, EPI_ISL_1423437, EPI_ISL_1423438, EPI_ISL_1423439, EPI_ISL_1423440, EPI_ISL_1423441, EPI_ISL_1423442, EPI_ISL_1423443, EPI_ISL_1423444, EPI_ISL_1423447, EPI_ISL_1423448, EPI_ISL_1423449, EPI_ISL_1423450, EPI_ISL_1423451, EPI_ISL_1423452, EPI_ISL_1423453, EPI_ISL_1423454, EPI_ISL_1423455, EPI_ISL_1423456, EPI_ISL_1423457, EPI_ISL_1423458, EPI_ISL_1423459, EPI_ISL_1423460, EPI_ISL_1423461, EPI_ISL_1423462, EPI_ISL_1423463, EPI_ISL_1423464, EPI_ISL_1423465, EPI_ISL_1423466, EPI_ISL_1423467, EPI_ISL_1423468, EPI_ISL_1423469, EPI_ISL_1423470, EPI_ISL_1423471, EPI_ISL_1423472, EPI_ISL_1423473, EPI_ISL_1423474, EPI_ISL_1423475, EPI_ISL_1423476, EPI_ISL_1423477, EPI_ISL_1423478, EPI_ISL_1423479, EPI_ISL_1423480, EPI_ISL_1423481, EPI_ISL_1423483, EPI_ISL_1423484, EPI_ISL_1423485, EPI_ISL_1423486, EPI_ISL_1423487, EPI_ISL_1423488, EPI_ISL_1423489, EPI_ISL_1423490, EPI_ISL_1423492, EPI_ISL_1423493, EPI_ISL_1423494, EPI_ISL_1423495, EPI_ISL_1423496, EPI_ISL_1423497, EPI_ISL_1423498, EPI_ISL_1423499, EPI_ISL_1423500, EPI_ISL_1423501, EPI_ISL_1423502, EPI_ISL_1423503, EPI_ISL_1532731, EPI_ISL_1532732, EPI_ISL_1532733, EPI_ISL_1532734, EPI_ISL_1532735, EPI_ISL_1532736, EPI_ISL_1532737, EPI_ISL_1532738, EPI_ISL_1532739, EPI_ISL_1532740, EPI_ISL_1532741, EPI_ISL_1532742, EPI_ISL_1532743, EPI_ISL_1532744, EPI_ISL_1532746, EPI_ISL_1532747, EPI_ISL_1532748, EPI_ISL_1532749, EPI_ISL_1532750, EPI_ISL_1532751, EPI_ISL_1532752, EPI_ISL_1532753, EPI_ISL_1532754, EPI_ISL_1532755, EPI_ISL_1532756, EPI_ISL_1532757, EPI_ISL_1532758, EPI_ISL_1532759, EPI_ISL_1532760, EPI_ISL_1532761, EPI_ISL_1532762, EPI_ISL_1532763, EPI_ISL_1532765, EPI_ISL_1532766, EPI_ISL_1532767, EPI_ISL_1532769, EPI_ISL_1532770, EPI_ISL_1532771, EPI_ISL_1532772, EPI_ISL_1532773, EPI_ISL_1532774, EPI_ISL_1532775, EPI_ISL_1532776, EPI_ISL_1532777, EPI_ISL_1532778, EPI_ISL_1532779, EPI_ISL_1532780, EPI_ISL_1532781, EPI_ISL_1532783, EPI_ISL_1532784, EPI_ISL_1532785, EPI_ISL_1576872, EPI_ISL_1576873, EPI_ISL_1576874, EPI_ISL_1576875, EPI_ISL_1576880, EPI_ISL_1576881, EPI_ISL_1576882, EPI_ISL_1576883, EPI_ISL_1576906, EPI_ISL_1576907, EPI_ISL_1576908, EPI_ISL_1576909, EPI_ISL_1576910, EPI_ISL_1576911, EPI_ISL_1576945, EPI_ISL_1576946, EPI_ISL_1654574, EPI_ISL_1654575, EPI_ISL_1654576, EPI_ISL_1654577, EPI_ISL_1654578, EPI_ISL_1654579, EPI_ISL_1654580, EPI_ISL_1654581, EPI_ISL_1654582, EPI_ISL_1654583, EPI_ISL_1654584, EPI_ISL_1654585, EPI_ISL_1654587, EPI_ISL_1654588, EPI_ISL_1654589, EPI_ISL_1654590, EPI_ISL_1654591, EPI_ISL_1654592, EPI_ISL_1654593, EPI_ISL_1654594, EPI_ISL_1654595, EPI_ISL_1654596, EPI_ISL_1654597, EPI_ISL_1654599, EPI_ISL_1654600, EPI_ISL_1654601, EPI_ISL_1654602, EPI_ISL_1654603, EPI_ISL_1654604, EPI_ISL_1654605, EPI_ISL_1654606, EPI_ISL_1654607, EPI_ISL_1654608, EPI_ISL_1654610, EPI_ISL_1922854, EPI_ISL_1922855, EPI_ISL_1922856, EPI_ISL_1922860, EPI_ISL_1922864, EPI_ISL_1922865, EPI_ISL_1922868

| | | | |
|---|---|---|---|
| see above | NYU Langone Health | Departments of Pathology and Medicine, New York University School of Medicine | Adriana Heguy; Christian Marier; Dacia Dimartino; Emily Guzman; Gael Westby; Guiqing Wang; Paolo Cotzia; Paul Zappile; Peter Meyn; Sitharam Ramaswami; Yutong Zhang |
| EPI_ISL_1920132, EPI_ISL_1920151, EPI_ISL_1920166, EPI_ISL_1920168, EPI_ISL_1920169, EPI_ISL_1920170 | Northwell Health Laboratories | Wadsworth Center, New York State Department of Health | Alexis Russell; Catharine Prussing; Daryl M. Lamson; Erasmus Schneider; Erica Lasek-Nesselquist; John Kelly; Jonathan Plitnick; Kirsten St. George; Matthew Shudt; Melissa A Leisner; Navjot Singh |

EPI_ISL_1261796, EPI_ISL_1261797, EPI_ISL_1261811, EPI_ISL_1261819, EPI_ISL_1261827, EPI_ISL_1261828, EPI_ISL_1261829, EPI_ISL_1261846, EPI_ISL_1261854, EPI_ISL_1261865, EPI_ISL_1337482, EPI_ISL_1337518, EPI_ISL_1337519, EPI_ISL_1337524, EPI_ISL_1337527, EPI_ISL_1337529, EPI_ISL_1337539, EPI_ISL_1337541, EPI_ISL_1337546, EPI_ISL_1447091, EPI_ISL_1447100, EPI_ISL_1447104, EPI_ISL_1447111, EPI_ISL_1447114, EPI_ISL_1447119, EPI_ISL_1447138, EPI_ISL_1447177, EPI_ISL_1447178, EPI_ISL_1447183, EPI_ISL_1593756, EPI_ISL_1593757

| | | | |
|---|---|---|---|
| see above | OCME Office Of Chief Medical Examiner | New York City Public Health Laboratory | Jade Wang; et al. |

EPI_ISL_1172898, EPI_ISL_1172899, EPI_ISL_1172900, EPI_ISL_1172901, EPI_ISL_1172902, EPI_ISL_1172903, EPI_ISL_1172904, EPI_ISL_1172905, EPI_ISL_1172906, EPI_ISL_1172907, EPI_ISL_1172908, EPI_ISL_1172909, EPI_ISL_1172910, EPI_ISL_1172911, EPI_ISL_1172912, EPI_ISL_1172914, EPI_ISL_1172915, EPI_ISL_1172916, EPI_ISL_1172922, EPI_ISL_1172927, EPI_ISL_1172928, EPI_ISL_1172929, EPI_ISL_1172930, EPI_ISL_1172931, EPI_ISL_1172932, EPI_ISL_1172933, EPI_ISL_1172934, EPI_ISL_1172935, EPI_ISL_1172937, EPI_ISL_1172938, EPI_ISL_1172939, EPI_ISL_1172940, EPI_ISL_1172941, EPI_ISL_1172942, EPI_ISL_1172944, EPI_ISL_1172945, EPI_ISL_1172946, EPI_ISL_1172947, EPI_ISL_1172948, EPI_ISL_1172949, EPI_ISL_1172950, EPI_ISL_1172951, EPI_ISL_1172952, EPI_ISL_1172953, EPI_ISL_1172954, EPI_ISL_1172955, EPI_ISL_1172956, EPI_ISL_1172957, EPI_ISL_1172959, EPI_ISL_1172960, EPI_ISL_1172961, EPI_ISL_1172962, EPI_ISL_1172963, EPI_ISL_1172964, EPI_ISL_1172965, EPI_ISL_1172966, EPI_ISL_1172967, EPI_ISL_1172968, EPI_ISL_1172969, EPI_ISL_1172970, EPI_ISL_1172971, EPI_ISL_1172972, EPI_ISL_1172973, EPI_ISL_1172974, EPI_ISL_1172975, EPI_ISL_1172976, EPI_ISL_1172978, EPI_ISL_1172982, EPI_ISL_1172983, EPI_ISL_1172984, EPI_ISL_1172985, EPI_ISL_1172986, EPI_ISL_1172987, EPI_ISL_1172988, EPI_ISL_1172989, EPI_ISL_1172990, EPI_ISL_1172991, EPI_ISL_1172992, EPI_ISL_1172993, EPI_ISL_1172995, EPI_ISL_1172996, EPI_ISL_1172997, EPI_ISL_1172998, EPI_ISL_1172999, EPI_ISL_1173000, EPI_ISL_1173001, EPI_ISL_1173002, EPI_ISL_1173003, EPI_ISL_1173004, EPI_ISL_1173005, EPI_ISL_1173006, EPI_ISL_1173007, EPI_ISL_1173008, EPI_ISL_1173009, EPI_ISL_1173010, EPI_ISL_1173011, EPI_ISL_1173012, EPI_ISL_1173013, EPI_ISL_1173014, EPI_ISL_1173015, EPI_ISL_1173016, EPI_ISL_1173017, EPI_ISL_1173018, EPI_ISL_1173019, EPI_ISL_1173020, EPI_ISL_1173021, EPI_ISL_1173022, EPI_ISL_1173023, EPI_ISL_1173024, EPI_ISL_1173025, EPI_ISL_1173026, EPI_ISL_1173027, EPI_ISL_1173029, EPI_ISL_1173030, EPI_ISL_1173031, EPI_ISL_1173032, EPI_ISL_1173033, EPI_ISL_1173034, EPI_ISL_1173035, EPI_ISL_1173036, EPI_ISL_1173037, EPI_ISL_1173038, EPI_ISL_1173039, EPI_ISL_1173040, EPI_ISL_1173041, EPI_ISL_1173042, EPI_ISL_1173043, EPI_ISL_1173044, EPI_ISL_1173045, EPI_ISL_1173046, EPI_ISL_1173047, EPI_ISL_1173048, EPI_ISL_1173049, EPI_ISL_1173050, EPI_ISL_1173051, EPI_ISL_1173052, EPI_ISL_1173053, EPI_ISL_1173054, EPI_ISL_1173055, EPI_ISL_1173056, EPI_ISL_1173057, EPI_ISL_1173058, EPI_ISL_1173059, EPI_ISL_1173060, EPI_ISL_1173061, EPI_ISL_1173062, EPI_ISL_1173063, EPI_ISL_1173064, EPI_ISL_1173065, EPI_ISL_1173066, EPI_ISL_1173067, EPI_ISL_1173068, EPI_ISL_1173069, EPI_ISL_1173070, EPI_ISL_1173071, EPI_ISL_1173072, EPI_ISL_1173073, EPI_ISL_1173074, EPI_ISL_1173075, EPI_ISL_1173076, EPI_ISL_1173077, EPI_ISL_1173078, EPI_ISL_1173079, EPI_ISL_1173080, EPI_ISL_1173081, EPI_ISL_1173082, EPI_ISL_1173083, EPI_ISL_1173084, EPI_ISL_1173085, EPI_ISL_1173086, EPI_ISL_1173087, EPI_ISL_1173088, EPI_ISL_1173089, EPI_ISL_1173090, EPI_ISL_1173091, EPI_ISL_1173092, EPI_ISL_1173093, EPI_ISL_1173094, EPI_ISL_1173095, EPI_ISL_1173096, EPI_ISL_1173097, EPI_ISL_1173098, EPI_ISL_1173099, EPI_ISL_1173100, EPI_ISL_1173101, EPI_ISL_1173102, EPI_ISL_1173103, EPI_ISL_1173104, EPI_ISL_1173105, EPI_ISL_1173106, EPI_ISL_1173107, EPI_ISL_1173108, EPI_ISL_1173109, EPI_ISL_1173110, EPI_ISL_1173111, EPI_ISL_1173112, EPI_ISL_1173113, EPI_ISL_1173114, EPI_ISL_1173115, EPI_ISL_1173116, EPI_ISL_1173117, EPI_ISL_1173118, EPI_ISL_1173119, EPI_ISL_1173120, EPI_ISL_1173121, EPI_ISL_1173122, EPI_ISL_1173123, EPI_ISL_1173124, EPI_ISL_1173125, EPI_ISL_1173126, EPI_ISL_1173127, EPI_ISL_1173128, EPI_ISL_1173129, EPI_ISL_1173130, EPI_ISL_1173131, EPI_ISL_1173132, EPI_ISL_1173133, EPI_ISL_1173134, EPI_ISL_1173135, EPI_ISL_1173136, EPI_ISL_1173137, EPI_ISL_1173138, EPI_ISL_1173139, EPI_ISL_1173140, EPI_ISL_1173141, EPI_ISL_1173142, EPI_ISL_1173143, EPI_ISL_1173144, EPI_ISL_1173145, EPI_ISL_1173146, EPI_ISL_1173147, EPI_ISL_1173148, EPI_ISL_1173149, EPI_ISL_1173150, EPI_ISL_1173151, EPI_ISL_1173152, EPI_ISL_1173153, EPI_ISL_1173154, EPI_ISL_1173155, EPI_ISL_1173156, EPI_ISL_1173157, EPI_ISL_1173158, EPI_ISL_1258466, EPI_ISL_1258467, EPI_ISL_1258468, EPI_ISL_1258469, EPI_ISL_1258470, EPI_ISL_1258471, EPI_ISL_1258472, EPI_ISL_1258473, EPI_ISL_1258474, EPI_ISL_1258475, EPI_ISL_1258476, EPI_ISL_1258477, EPI_ISL_1258478, EPI_ISL_1258479, EPI_ISL_1258480, EPI_ISL_1258481, EPI_ISL_1258482, EPI_ISL_1258483, EPI_ISL_1258484, EPI_ISL_1258485, EPI_ISL_1258486, EPI_ISL_1258487, EPI_ISL_1258488, EPI_ISL_1258489, EPI_ISL_1258490, EPI_ISL_1258491, EPI_ISL_1258492, EPI_ISL_1258493, EPI_ISL_1258494, EPI_ISL_1258495, EPI_ISL_1258496, EPI_ISL_1258497, EPI_ISL_1258498, EPI_ISL_1258499, EPI_ISL_1258500, EPI_ISL_1258501, EPI_ISL_1258502, EPI_ISL_1258503, EPI_ISL_1258504, EPI_ISL_1258505, EPI_ISL_1258506, EPI_ISL_1258507, EPI_ISL_1258508, EPI_ISL_1258509, EPI_ISL_1258510, EPI_ISL_1258511, EPI_ISL_1258512, EPI_ISL_1258513, EPI_ISL_1258514, EPI_ISL_1258515, EPI_ISL_1258516, EPI_ISL_1258517, EPI_ISL_1258518, EPI_ISL_1258519, EPI_ISL_1258520, EPI_ISL_1258521, EPI_ISL_1258522, EPI_ISL_1258523, EPI_ISL_1258524, EPI_ISL_1258525, EPI_ISL_1258526, EPI_ISL_1258527, EPI_ISL_1258528, EPI_ISL_1258529, EPI_ISL_1258530, EPI_ISL_1258531, EPI_ISL_1258532, EPI_ISL_1258533, EPI_ISL_1258534, EPI_ISL_1258535, EPI_ISL_1258536, EPI_ISL_1258537, EPI_ISL_1258538, EPI_ISL_1258539, EPI_ISL_1258540, EPI_ISL_1258541, EPI_ISL_1258542, EPI_ISL_1258543, EPI_ISL_1258544, EPI_ISL_1258545, EPI_ISL_1258546, EPI_ISL_1258547, EPI_ISL_1258548, EPI_ISL_1258549, EPI_ISL_1258550, EPI_ISL_1258551, EPI_ISL_1258552, EPI_ISL_1258553, EPI_ISL_1258554, EPI_ISL_1258555, EPI_ISL_1258556, EPI_ISL_1258557, EPI_ISL_1258558, EPI_ISL_1258559, EPI_ISL_1258560, EPI_ISL_1258561, EPI_ISL_1258562, EPI_ISL_1258563, EPI_ISL_1258564, EPI_ISL_1258565, EPI_ISL_1258566, EPI_ISL_1258567, EPI_ISL_1258568, EPI_ISL_1258569, EPI_ISL_1258570, EPI_ISL_1258571, EPI_ISL_1258572, EPI_ISL_1258573, EPI_ISL_1258574, EPI_ISL_1258575, EPI_ISL_1258576, EPI_ISL_1258577, EPI_ISL_1258578, EPI_ISL_1258579, EPI_ISL_1258580, EPI_ISL_1258581, EPI_ISL_1258582, EPI_ISL_1258583, EPI_ISL_1258584, EPI_ISL_1258585, EPI_ISL_1258586, EPI_ISL_1258587, EPI_ISL_1258588, EPI_ISL_1258589, EPI_ISL_1258590, EPI_ISL_1258591, EPI_ISL_1258592, EPI_ISL_1258593, EPI_ISL_1258594, EPI_ISL_1258595, EPI_ISL_1258596, EPI_ISL_1258597, EPI_ISL_1258598, EPI_ISL_1258599, EPI_ISL_1258600, EPI_ISL_1258601, EPI_ISL_1258602, EPI_ISL_1258603, EPI_ISL_1258604, EPI_ISL_1258605, EPI_ISL_1258606, EPI_ISL_1258607, EPI_ISL_1258608, EPI_ISL_1258609, EPI_ISL_1258610, EPI_ISL_1258611, EPI_ISL_1258612, EPI_ISL_1258613, EPI_ISL_1258614, EPI_ISL_1258615, EPI_ISL_1258616, EPI_ISL_1258617, EPI_ISL_1258618, EPI_ISL_1258619, EPI_ISL_1258620, EPI_ISL_1258621, EPI_ISL_1258622, EPI_ISL_1258623, EPI_ISL_1258624, EPI_ISL_1258625, EPI_ISL_1258626, EPI_ISL_1258627, EPI_ISL_1258628, EPI_ISL_1258629, EPI_ISL_1258630, EPI_ISL_1258631, EPI_ISL_1258632, EPI_ISL_1258633, EPI_ISL_1258634, EPI_ISL_1258635, EPI_ISL_1258636, EPI_ISL_1258637, EPI_ISL_1258638, EPI_ISL_1258639, EPI_ISL_1258640, EPI_ISL_1258641, EPI_ISL_1258642, EPI_ISL_1258643, EPI_ISL_1258644, EPI_ISL_1258645, EPI_ISL_1258646, EPI_ISL_1258647, EPI_ISL_1258649, EPI_ISL_1258650, EPI_ISL_1258651, EPI_ISL_1258652, EPI_ISL_1258653, EPI_ISL_1258654, EPI_ISL_1258655, EPI_ISL_1258656, EPI_ISL_1258657, EPI_ISL_1258658, EPI_ISL_1258659, EPI_ISL_1258660, EPI_ISL_1258661, EPI_ISL_1258662, EPI_ISL_1258663, EPI_ISL_1258664, EPI_ISL_1258665, EPI_ISL_1258666, EPI_ISL_1258667, EPI_ISL_1258668, EPI_ISL_1258669, EPI_ISL_1258670, EPI_ISL_1258671, EPI_ISL_1258672, EPI_ISL_1258673, EPI_ISL_1258674, EPI_ISL_1258675, EPI_ISL_1258676, EPI_ISL_1258677, EPI_ISL_1258678, EPI_ISL_1258679, EPI_ISL_1258680, EPI_ISL_1258681, EPI_ISL_1258682, EPI_ISL_1258683, EPI_ISL_1258684, EPI_ISL_1258685, EPI_ISL_1258686, EPI_ISL_1258687, EPI_ISL_1258688, EPI_ISL_1258689, EPI_ISL_1258690, EPI_ISL_1258691, EPI_ISL_1258692, EPI_ISL_1258693, EPI_ISL_1258694, EPI_ISL_1258695, EPI_ISL_1258696, EPI_ISL_1258697, EPI_ISL_1258698, EPI_ISL_1258699, EPI_ISL_1258700, EPI_ISL_1258701, EPI_ISL_1258702, EPI_ISL_1258703, EPI_ISL_1258704, EPI_ISL_1258705, EPI_ISL_1258706, EPI_ISL_1258707, EPI_ISL_1258708, EPI_ISL_1258709, EPI_ISL_1258710, EPI_ISL_1258711, EPI_ISL_1258712, EPI_ISL_1258713, EPI_ISL_1258714, EPI_ISL_1258715, EPI_ISL_1258716, EPI_ISL_1258717, EPI_ISL_1258718, EPI_ISL_1258719, EPI_ISL_1258720, EPI_ISL_1258721, EPI_ISL_1258722, EPI_ISL_1258723, EPI_ISL_1258724, EPI_ISL_1258725, EPI_ISL_1258726, EPI_ISL_1258727, EPI_ISL_1258728, EPI_ISL_1258729, EPI_ISL_1258730, EPI_ISL_1258731, EPI_ISL_1258732, EPI_ISL_1258733, EPI_ISL_1258734, EPI_ISL_1258735, EPI_ISL_1258736, EPI_ISL_1258737, EPI_ISL_1258738, EPI_ISL_1258739, EPI_ISL_1258740, EPI_ISL_1258741, EPI_ISL_1258742, EPI_ISL_1258743, EPI_ISL_1258744, EPI_ISL_1258745, EPI_ISL_1258746, EPI_ISL_1258747, EPI_ISL_1258748, EPI_ISL_1258749, EPI_ISL_1258750, EPI_ISL_1258751, EPI_ISL_1258752, EPI_ISL_1258753, EPI_ISL_1258754, EPI_ISL_1258755, EPI_ISL_1258756, EPI_ISL_1258757, EPI_ISL_1258758, EPI_ISL_1258759, EPI_ISL_1258760, EPI_ISL_1258761, EPI_ISL_1258762, EPI_ISL_1258763, EPI_ISL_1258764, EPI_ISL_1258765, EPI_ISL_1258766, EPI_ISL_1258767, EPI_ISL_1258768, EPI_ISL_1258769, EPI_ISL_1258770, EPI_ISL_1258771, EPI_ISL_1258772, EPI_ISL_1258773, EPI_ISL_1258774, EPI_ISL_1258775, EPI_ISL_1258776, EPI_ISL_1258777, EPI_ISL_1258778, EPI_ISL_1258779, EPI_ISL_1258780, EPI_ISL_1258781, EPI_ISL_1258782, EPI_ISL_1258783, EPI_ISL_1258784, EPI_ISL_1258785, EPI_ISL_1258786, EPI_ISL_1258787, EPI_ISL_1258788, EPI_ISL_1258789, EPI_ISL_1258790, EPI_ISL_1258791, EPI_ISL_1258792, EPI_ISL_1258793, EPI_ISL_1258794, EPI_ISL_1258795, EPI_ISL_1258796, EPI_ISL_1258797, EPI_ISL_1258798, EPI_ISL_1258799, EPI_ISL_1258800, EPI_ISL_1258801, EPI_ISL_1258802, EPI_ISL_1258803, EPI_ISL_1258804, EPI_ISL_1258805, EPI_ISL_1258806, EPI_ISL_1258807, EPI_ISL_1258808, EPI_ISL_1258809, EPI_ISL_1258810, EPI_ISL_1258811, EPI_ISL_1258812, EPI_ISL_1258813, EPI_ISL_1258814, EPI_ISL_1258815, EPI_ISL_1258816, EPI_ISL_1258817, EPI_ISL_1258818, EPI_ISL_1258819, EPI_ISL_1258820, EPI_ISL_1258821, EPI_ISL_1258822, EPI_ISL_1258823, EPI_ISL_1258824, EPI_ISL_1258825, EPI_ISL_1258826, EPI_ISL_1258827, EPI_ISL_1258828, EPI_ISL_1258829, EPI_ISL_1258830, EPI_ISL_1258831, EPI_ISL_1258832, EPI_ISL_1258833, EPI_ISL_1258834, EPI_ISL_1258835, EPI_ISL_1258836, EPI_ISL_1258837, EPI_ISL_1258838, EPI_ISL_1258839, EPI_ISL_1258840, EPI_ISL_1258841, EPI_ISL_1258842, EPI_ISL_1258843, EPI_ISL_1258844, EPI_ISL_1258845, EPI_ISL_1258846, EPI_ISL_1258847, EPI_ISL_1258848, EPI_ISL_1258849, EPI_ISL_1258850, EPI_ISL_1258851, EPI_ISL_1258852, EPI_ISL_1258853, EPI_ISL_1258854, EPI_ISL_1258855, EPI_ISL_1258856, EPI_ISL_1258857, EPI_ISL_1258858, EPI_ISL_1258859, EPI_ISL_1258860, EPI_ISL_1258861, EPI_ISL_1258862, EPI_ISL_1258863, EPI_ISL_1258864, EPI_ISL_1258865, EPI_ISL_1258866, EPI_ISL_1258867, EPI_ISL_1258868, EPI_ISL_1258869, EPI_ISL_1258870, EPI_ISL_1258871, EPI_ISL_1258872, EPI_ISL_1258873, EPI_ISL_1258874, EPI_ISL_1258875, EPI_ISL_1258876, EPI_ISL_1258877, EPI_ISL_1258878, EPI_ISL_1258879, EPI_ISL_1258880, EPI_ISL_1258881, EPI_ISL_1258882, EPI_ISL_1258883, EPI_ISL_1258884, EPI_ISL_1258885, EPI_ISL_1258886, EPI_ISL_1258887, EPI_ISL_1258888, EPI_ISL_1258889, EPI_ISL_1258890, EPI_ISL_1258891, EPI_ISL_1258892, EPI_ISL_1258893, EPI_ISL_1258894, EPI_ISL_1258895, EPI_ISL_1258896, EPI_ISL_1258897, EPI_ISL_1258898, EPI_ISL_1258899, EPI_ISL_1258900, EPI_ISL_1258901, EPI_ISL_1258902, EPI_ISL_1258903, EPI_ISL_1258904, EPI_ISL_1258905, EPI_ISL_1258906, EPI_ISL_1258907, EPI_ISL_1258908, EPI_ISL_1258909, EPI_ISL_1258910, EPI_ISL_1258911, EPI_ISL_1258912, EPI_ISL_1258913, EPI_ISL_1258914, EPI_ISL_1258915, EPI_ISL_1258916, EPI_ISL_1258917, EPI_ISL_1258918, EPI_ISL_1258919, EPI_ISL_1258920, EPI_ISL_1258921, EPI_ISL_1258922, EPI_ISL_1258923, EPI_ISL_1258924, EPI_ISL_1258925, EPI_ISL_1258926, EPI_ISL_1258927, EPI_ISL_1258928, EPI_ISL_1258929, EPI_ISL_1258930, EPI_ISL_1258931, EPI_ISL_1258932, EPI_ISL_1258933, EPI_ISL_1258934, EPI_ISL_1258935, EPI_ISL_1258936, EPI_ISL_1258937, EPI_ISL_1258938, EPI_ISL_1258939, EPI_ISL_1258940, EPI_ISL_1258941, EPI_ISL_1258942, EPI_ISL_1258943, EPI_ISL_1258944, EPI_ISL_1258945, EPI_ISL_1258946, EPI_ISL_1258947, EPI_ISL_1258948, EPI_ISL_1258949, EPI_ISL_1258950, EPI_ISL_1258951, EPI_ISL_1258952, EPI_ISL_1258953, EPI_ISL_1258954, EPI_ISL_1258955, EPI_ISL_1258956, EPI_ISL_1258957, EPI_ISL_1258958, EPI_ISL_1258959, EPI_ISL_1258960, EPI_ISL_1258961, EPI_ISL_1258962, EPI_ISL_1258963, EPI_ISL_1258964, EPI_ISL_1258965, EPI_ISL_1258966, EPI_ISL_1258967, EPI_ISL_1258968, EPI_ISL_1258969, EPI_ISL_1258970, EPI_ISL_1258971, EPI_ISL_1258972, EPI_ISL_1258973, EPI_ISL_1258974, EPI_ISL_1258975, EPI_ISL_1258976, EPI_ISL_1258977, EPI_ISL_1258978, EPI_ISL_1258979, EPI_ISL_1258980, EPI_ISL_1258981, EPI_ISL_1258982, EPI_ISL_1258983, EPI_ISL_1258984, EPI_ISL_1258985, EPI_ISL_1258986, EPI_ISL_1258987, EPI_ISL_1306253, EPI_ISL_1306254, EPI_ISL_1306255, EPI_ISL_1306256, EPI_ISL_1306257, EPI_ISL_1306258, EPI_ISL_1306259, EPI_ISL_1306260, EPI_ISL_1306261, EPI_ISL_1306262, EPI_ISL_1306263, EPI_ISL_1306264, EPI_ISL_1306265, EPI_ISL_1306266, EPI_ISL_1306267, EPI_ISL_1306268, EPI_ISL_1306269, EPI_ISL_1306270, EPI_ISL_1306271, EPI_ISL_1306273, EPI_ISL_1306274, EPI_ISL_1306275, EPI_ISL_1306276, EPI_ISL_1306277, EPI_ISL_1306278, EPI_ISL_1306279, EPI_ISL_1306280, EPI_ISL_1306281, EPI_ISL_1306282, EPI_ISL_1306283, EPI_ISL_1306284, EPI_ISL_1306285, EPI_ISL_1306286, EPI_ISL_1306287, EPI_ISL_1306288, EPI_ISL_1306289, EPI_ISL_1306290, EPI_ISL_1306291, EPI_ISL_1306292, EPI_ISL_1306293, EPI_ISL_1306294, EPI_ISL_1306295, EPI_ISL_1306296, EPI_ISL_1306297, EPI_ISL_1306298, EPI_ISL_1306299, EPI_ISL_1306300, EPI_ISL_1306301, EPI_ISL_1306302, EPI_ISL_1306303, EPI_ISL_1306304, EPI_ISL_1306305, EPI_ISL_1306306, EPI_ISL_1306307, EPI_ISL_1306308, EPI_ISL_1306309, EPI_ISL_1306310, EPI_ISL_1306311, EPI_ISL_1306312, EPI_ISL_1306313, EPI_ISL_1306314, EPI_ISL_1306315, EPI_ISL_1306316, EPI_ISL_1306317, EPI_ISL_1306318, EPI_ISL_1306319, EPI_ISL_1306320, EPI_ISL_1306321, EPI_ISL_1306322, EPI_ISL_1306323, EPI_ISL_1306324, EPI_ISL_1306325, EPI_ISL_1306326, EPI_ISL_1306327, EPI_ISL_1306328, EPI_ISL_1306329, EPI_ISL_1306330, EPI_ISL_1306331, EPI_ISL_1306332, EPI_ISL_1306333, EPI_ISL_1306334, EPI_ISL_1306335, EPI_ISL_1306336, EPI_ISL_1306337, EPI_ISL_1306338, EPI_ISL_1306339, EPI_ISL_1306340, EPI_ISL_1306341, EPI_ISL_1306342, EPI_ISL_1306343, EPI_ISL_1306344, EPI_ISL_1306345, EPI_ISL_1306346, EPI_ISL_1306347, EPI_ISL_1306348, EPI_ISL_1306349, EPI_ISL_1306350, EPI_ISL_1306351, EPI_ISL_1306352, EPI_ISL_1306353, EPI_ISL_1306354, EPI_ISL_1306355, EPI_ISL_1306356, EPI_ISL_1306357, EPI_ISL_1306358, EPI_ISL_1306359, EPI_ISL_1306360, EPI_ISL_1306361, EPI_ISL_1306362, EPI_ISL_1306363, EPI_ISL_1306364, EPI_ISL_1306365, EPI_ISL_1306366, EPI_ISL_1306367, EPI_ISL_1306368, EPI_ISL_1306369, EPI_ISL_1306370, EPI_ISL_1306371, EPI_ISL_1306372, EPI_ISL_1306373, EPI_ISL_1306374, EPI_ISL_1306375, EPI_ISL_1306376, EPI_ISL_1306377, EPI_ISL_1306378, EPI_ISL_1306379, EPI_ISL_1306380, EPI_ISL_1306381, EPI_ISL_1306382, EPI_ISL_1306383, EPI_ISL_1306384, EPI_ISL_1306385, EPI_ISL_1306386, EPI_ISL_1306387, EPI_ISL_1306388, EPI_ISL_1306389, EPI_ISL_1306390, EPI_ISL_1306391, EPI_ISL_1306392, EPI_ISL_1306393, EPI_ISL_1306394, EPI_ISL_1306395, EPI_ISL_1306396, EPI_ISL_1306397, EPI_ISL_1306398, EPI_ISL_1306399, EPI_ISL_1306400, EPI_ISL_1306401, EPI_ISL_1306402, EPI_ISL_1306403, EPI_ISL_1306404, EPI_ISL_1306405, EPI_ISL_1306406, EPI_ISL_1306407, EPI_ISL_1306408, EPI_ISL_1306409, EPI_ISL_1306410, EPI_ISL_1306411, EPI_ISL_1306412, EPI_ISL_1306413, EPI_ISL_1306414, EPI_ISL_1306415, EPI_ISL_1306416, EPI_ISL_1306417, EPI_ISL_1306418, EPI_ISL_1306419, EPI_ISL_1306420, EPI_ISL_1306421, EPI_ISL_1306422, EPI_ISL_1306423, EPI_ISL_1306424, EPI_ISL_1306425, EPI_ISL_1306426, EPI_ISL_1306427, EPI_ISL_1306428, EPI_ISL_1306429, EPI_ISL_1306430, EPI_ISL_1306431, EPI_ISL_1306432, EPI_ISL_1306433, EPI_ISL_1306434, EPI_ISL_1306435, EPI_ISL_1306436, EPI_ISL_1306437, EPI_ISL_1306438, EPI_ISL_1306439, EPI_ISL_1306440, EPI_ISL_1306441, EPI_ISL_1306442, EPI_ISL_1306443, EPI_ISL_1306444, EPI_ISL_1306445, EPI_ISL_1306446, EPI_ISL_1306447, EPI_ISL_1306448, EPI_ISL_1306449, EPI_ISL_1306450, EPI_ISL_1306451, EPI_ISL_1306452, EPI_ISL_1306453, EPI_ISL_1306454, EPI_ISL_1306455, EPI_ISL_1306456, EPI_ISL_1306457, EPI_ISL_1306458, EPI_ISL_1306459, EPI_ISL_1306460, EPI_ISL_1306461, EPI_ISL_1306462, EPI_ISL_1306463, EPI_ISL_1306464, EPI_ISL_1306465, EPI_ISL_1306466, EPI_ISL_1306467, EPI_ISL_1306468, EPI_ISL_1306469, EPI_ISL_1306470, EPI_ISL_1306471, EPI_ISL_1306472, EPI_ISL_1306473, EPI_ISL_1306474, EPI_ISL_1306475, EPI_ISL_1306476, EPI_ISL_1306477, EPI_ISL_1306478, EPI_ISL_1306479, EPI_ISL_1306480, EPI_ISL_1306481, EPI_ISL_1306482, EPI_ISL_1306483, EPI_ISL_1306484, EPI_ISL_1306485, EPI_ISL_1306486, EPI_ISL_1306487, EPI_ISL_1306488, EPI_ISL_1306489, EPI_ISL_1306490, EPI_ISL_1306491, EPI_ISL_1306492, EPI_ISL_1306493, EPI_ISL_1306494, EPI_ISL_1306495, EPI_ISL_1306496, EPI_ISL_1306497, EPI_ISL_1306498, EPI_ISL_1306499, EPI_ISL_1306500, EPI_ISL_1306501, EPI_ISL_1306502, EPI_ISL_1306503, EPI_ISL_1306504, EPI_ISL_1306505, EPI_ISL_1306506, EPI_ISL_1306507, EPI_ISL_1306508, EPI_ISL_1306509, EPI_ISL_1306510, EPI_ISL_1306511, EPI_ISL_1306512, EPI_ISL_1306513, EPI_ISL_1306514, EPI_ISL_1306515, EPI_ISL_1306516, EPI_ISL_1306517, EPI_ISL_1306518, EPI_ISL_1306519, EPI_ISL_1306520, EPI_ISL_1306521, EPI_ISL_1306522, EPI_ISL_1306523, EPI_ISL_1306524, EPI_ISL_1306525, EPI_ISL_1306526, EPI_ISL_1306527, EPI_ISL_1306528, EPI_ISL_1306529, EPI_ISL_1306530, EPI_ISL_1306531, EPI_ISL_1306532, EPI_ISL_1306533, EPI_ISL_1306534, EPI_ISL_1306535, EPI_ISL_1306536, EPI_ISL_1306537, EPI_ISL_1306538, EPI_ISL_1306539, EPI_ISL_1306540, EPI_ISL_1306541, EPI_ISL_1306542, EPI_ISL_1306543, EPI_ISL_1306544, EPI_ISL_1306545, EPI_ISL_1306546, EPI_ISL_1306547, EPI_ISL_1306548, EPI_ISL_1306549, EPI_ISL_1306550, EPI_ISL_1306551, EPI_ISL_1306552, EPI_ISL_1306553, EPI_ISL_1306554, EPI_ISL_1306555, EPI_ISL_1306556, EPI_ISL_1306557, EPI_ISL_1306558, EPI_ISL_1306559, EPI_ISL_1306560, EPI_ISL_1306561, EPI_ISL_1306562, EPI_ISL_1306563, EPI_ISL_1306564, EPI_ISL_1306565, EPI_ISL_1306566, EPI_ISL_1306567, EPI_ISL_1306568, EPI_ISL_1306569, EPI_ISL_1306570, EPI_ISL_1306571, EPI_ISL_1306572, EPI_ISL_1306573, EPI_ISL_1306574, EPI_ISL_1306575, EPI_ISL_1306576, EPI_ISL_1306577, EPI_ISL_1306578, EPI_ISL_1306579, EPI_ISL_1306580, EPI_ISL_1306581, EPI_ISL_1306582, EPI_ISL_1306583, EPI_ISL_1306584, EPI_ISL_1306585, EPI_ISL_1306586, EPI_ISL_1306587, EPI_ISL_1306588, EPI_ISL_1306589, EPI_ISL_1306590, EPI_ISL_1306591, EPI_ISL_1306592, EPI_ISL_1306593, EPI_ISL_1306594, EPI_ISL_1306595, EPI_ISL_1306596, EPI_ISL_1306597, EPI_ISL_1306598, EPI_ISL_1306599, EPI_ISL_1306600, EPI_ISL_1306601, EPI_ISL_1306602, EPI_ISL_1306603, EPI_ISL_1306604, EPI_ISL_1306605, EPI_ISL_1306606, EPI_ISL_1306607, EPI_ISL_1306608, EPI_ISL_1306609, EPI_ISL_1306610, EPI_ISL_1306611, EPI_ISL_1306612, EPI_ISL_1306613, EPI_ISL_1306614, EPI_ISL_1306615, EPI_ISL_1306616, EPI_ISL_1306617, EPI_ISL_1306618, EPI_ISL_1306619, EPI_ISL_1306620, EPI_ISL_1306621, EPI_ISL_1306622, EPI_ISL_1306623, EPI_ISL_1306624, EPI_ISL_1306625, EPI_ISL_1306626, EPI_ISL_1306627, EPI_ISL_1306628, EPI_ISL_1306629, EPI_ISL_1306630, EPI_ISL_1306631, EPI_ISL_1306632, EPI_ISL_1306633, EPI_ISL_1306634, EPI_ISL_1306635, EPI_ISL_1306636, EPI_ISL_1306637, EPI_ISL_1306638, EPI_ISL_1306639, EPI_ISL_1306640, EPI_ISL_1306641, EPI_ISL_1306642, EPI_ISL_1306643, EPI_ISL_1306644, EPI_ISL_1306645, EPI_ISL_1306646, EPI_ISL_1306647, EPI_ISL_1306648, EPI_ISL_1306649, EPI_ISL_1306650, EPI_ISL_1306651, EPI_ISL_1306652, EPI_ISL_1306653, EPI_ISL_1306654, EPI_ISL_1306655, EPI_ISL_1306656, EPI_ISL_1306657, EPI_ISL_1306658, EPI_ISL_1306659, EPI_ISL_1306660, EPI_ISL_1306661, EPI_ISL_1306662, EPI_ISL_1306663, EPI_ISL_1306664, EPI_ISL_1306665, EPI_ISL_1306666, EPI_ISL_1306667, EPI_ISL_1306668, EPI_ISL_1306669, EPI_ISL_1306670, EPI_ISL_1306671, EPI_ISL_1306672, EPI_ISL_1306673, EPI_ISL_1306674, EPI_ISL_1306675, EPI_ISL_1306676, EPI_ISL_1306677, EPI_ISL_1306678, EPI_ISL_1306679, EPI_ISL_1306680, EPI_ISL_1306681, EPI_ISL_1306682, EPI_ISL_1306683, EPI_ISL_1306684, EPI_ISL_1306685, EPI_ISL_1306686, EPI_ISL_1306687, EPI_ISL_1306688, EPI_ISL_1306689, EPI_ISL_1306690, EPI_ISL_1306691, EPI_ISL_1306692, EPI_ISL_1306693, EPI_ISL_1306694, EPI_ISL_1306695, EPI_ISL_1306696, EPI_ISL_1306697, EPI_ISL_1306698, EPI_ISL_1306699, EPI_ISL_1306700, EPI_ISL_1306701, EPI_ISL_1306702, EPI_ISL_1306703, EPI_ISL_1306704, EPI_ISL_1306705, EPI_ISL_1306706, EPI_ISL_1306707, EPI_ISL_1306708, EPI_ISL_1306709, EPI_ISL_1306710, EPI_ISL_1306711, EPI_ISL_1306712, EPI_ISL_1306713, EPI_ISL_1306714, EPI_ISL_1306715, EPI_ISL_1306716, EPI_ISL_1306717, EPI_ISL_1306719, EPI_ISL_1306720, EPI_ISL_1306721, EPI_ISL_1306722, EPI_ISL_1306723, EPI_ISL_1306724, EPI_ISL_1306725, EPI_ISL_1306726, EPI_ISL_1306727, EPI_ISL_1306728, EPI_ISL_1306729, EPI_ISL_1306730, EPI_ISL_1306731, EPI_ISL_1306732, EPI_ISL_1306733, EPI_ISL_1306734, EPI_ISL_1306735, EPI_ISL_1306736, EPI_ISL_1306737, EPI_ISL_1306738, EPI_ISL_1306739, EPI_ISL_1306740, EPI_ISL_1306741, EPI_ISL_1306742, EPI_ISL_1306743, EPI_ISL_1306744, EPI_ISL_1306745, EPI_ISL_1306746, EPI_ISL_1306747, EPI_ISL_1306748, EPI_ISL_1306749, EPI_ISL_1306750, EPI_ISL_1306751, EPI_ISL_1306752, EPI_ISL_1306753, EPI_ISL_1306754, EPI_ISL_1306755, EPI_ISL_1306756, EPI_ISL_1306757, EPI_ISL_1306758, EPI_ISL_1306759, EPI_ISL_1306760, EPI_ISL_1306761, EPI_ISL_1306762, EPI_ISL_1306763, EPI_ISL_1306764, EPI_ISL_1306765, EPI_ISL_1306766, EPI_ISL_1306767, EPI_ISL_1306768, EPI_ISL_1306769, EPI_ISL_1306770, EPI_ISL_1306771, EPI_ISL_1306772, EPI_ISL_1306773, EPI_ISL_1306774, EPI_ISL_1306775, EPI_ISL_1306776, EPI_ISL_1306777, EPI_ISL_1306778, EPI_ISL_1306779, EPI_ISL_1306780, EPI_ISL_1306781, EPI_ISL_1306782, EPI_ISL_1306783, EPI_ISL_1306784, EPI_ISL_1306785, EPI_ISL_1306786, EPI_ISL_1306787, EPI_ISL_1306788, EPI_ISL_1306789, EPI_ISL_1306790, EPI_ISL_1306791, EPI_ISL_1306792, EPI_ISL_1306793, EPI_ISL_1306794, EPI_ISL_1306795, EPI_ISL_1306796, EPI_ISL_1306797, EPI_ISL_1306798, EPI_ISL_1306799, EPI_ISL_1306800, EPI_ISL_1306801, EPI_ISL_1306802, EPI_ISL_1306803, EPI_ISL_1306804, EPI_ISL_1306805, EPI_ISL_1306806, EPI_ISL_1306807, EPI_ISL_1306808, EPI_ISL_1306809, EPI_ISL_1306810, EPI_ISL_1306811, EPI_ISL_1306812, EPI_ISL_1306813, EPI_ISL_1306814, EPI_ISL_1306815, EPI_ISL_1306816, EPI_ISL_1306817, EPI_ISL_1306818, EPI_ISL_1306819, EPI_ISL_1306820, EPI_ISL_1306821, EPI_ISL_1306822, EPI_ISL_1306823, EPI_ISL_1306824, EPI_ISL_1306825, EPI_ISL_1306826, EPI_ISL_1306827, EPI_ISL_1306828, EPI_ISL_1306829, EPI_ISL_1306830, EPI_ISL_1306831, EPI_ISL_1306832, EPI_ISL_1306833, EPI_ISL_1306834, EPI_ISL_1306835, EPI_ISL_1306836, EPI_ISL_1306837, EPI_ISL_1306838, EPI_ISL_1306839, EPI_ISL_1306840, EPI_ISL_1306841, EPI_ISL_1306842, EPI_ISL_1306843, EPI_ISL_1306844, EPI_ISL_1306845, EPI_ISL_1306846, EPI_ISL_1306847, EPI_ISL_1306848, EPI_ISL_1306849, EPI_ISL_1306850, EPI_ISL_1306851, EPI_ISL_1306852, EPI_ISL_1306853, EPI_ISL_1306854, EPI_ISL_1306855, EPI_ISL_1306856, EPI_ISL_1306857, EPI_ISL_1306858, EPI_ISL_1306859, EPI_ISL_1306860, EPI_ISL_1306861, EPI_ISL_1306862, EPI_ISL_1306863, EPI_ISL_1306864, EPI_ISL_1306865, EPI_ISL_1306866, EPI_ISL_1306867, EPI_ISL_1306868, EPI_ISL_1306869, EPI_ISL_1306870, EPI_ISL_1306871, EPI_ISL_1306872, EPI_ISL_1306873, EPI_ISL_1306874, EPI_ISL_1306875, EPI_ISL_1306876, EPI_ISL_1306877, EPI_ISL_1306878, EPI_ISL_1306879, EPI_ISL_1306880, EPI_ISL_1306881, EPI_ISL_1306882, EPI_ISL_1306883, EPI_ISL_1306884, EPI_ISL_1306885, EPI_ISL_1306886, EPI_ISL_1306887, EPI_ISL_1306888, EPI_ISL_1306889, EPI_ISL_1306890, EPI_ISL_1306891, EPI_ISL_1306892, EPI_ISL_1306893, EPI_ISL_1306894, EPI_ISL_1306895, EPI_ISL_1306896, EPI_ISL_1306897, EPI_ISL_1306898, EPI_ISL_1306899, EPI_ISL_1306900, EPI_ISL_1306901, EPI_ISL_1306902, EPI_ISL_1306903, EPI_ISL_1306904, EPI_ISL_1306905, EPI_ISL_1306906, EPI_ISL_1306907, EPI_ISL_1306908, EPI_ISL_1306909, EPI_ISL_1306910, EPI_ISL_1306911, EPI_ISL_1306912, EPI_ISL_1306913, EPI_ISL_1306914, EPI_ISL_1306915, EPI_ISL_1306916, EPI_ISL_1306917, EPI_ISL_1306918, EPI_ISL_1306919, EPI_ISL_1306920, EPI_ISL_1306921, EPI_ISL_1306922, EPI_ISL_1306923, EPI_ISL_1306924, EPI_ISL_1306925, EPI_ISL_1306926, EPI_ISL_1306927, EPI_ISL_1306928, EPI_ISL_1306929, EPI_ISL_1306930, EPI_ISL_1306931, EPI_ISL_1306932, EPI_ISL_1306933, EPI_ISL_1306934, EPI_ISL_1306935, EPI_ISL_1306936, EPI_ISL_1306937, EPI_ISL_1306938, EPI_ISL_1306939, EPI_ISL_1306940, EPI_ISL_1306941, EPI_ISL_1306942, EPI_ISL_1306943, EPI_ISL_1306944, EPI_ISL_1306945, EPI_ISL_1306946, EPI_ISL_1306947, EPI_ISL_1306948, EPI_ISL_1306949, EPI_ISL_1306950, EPI_ISL_1306951, EPI_ISL_1306952, EPI_ISL_1306953, EPI_ISL_1306954, EPI_ISL_1306955, EPI_ISL_1306956, EPI_ISL_1306957, EPI_ISL_1306958, EPI_ISL_1306959, EPI_ISL_1306960, EPI_ISL_1306961, EPI_ISL_1306962, EPI_ISL_1306963, EPI_ISL_1306964, EPI_ISL_1306965, EPI_ISL_1306966, EPI_ISL_1306967, EPI_ISL_1306968, EPI_ISL_1306969, EPI_ISL_1306970, EPI_ISL_1306971, EPI_ISL_1306972, EPI_ISL_1306973, EPI_ISL_1306974, EPI_ISL_1306975, EPI_ISL_1306976, EPI_ISL_1306977, EPI_ISL_1306978, EPI_ISL_1306979, EPI_ISL_1306980, EPI_ISL_1306981, EPI_ISL_1306982, EPI_ISL_1306983, EPI_ISL_1306984, EPI_ISL_1306985, EPI_ISL_1306986, EPI_ISL_1306987, EPI_ISL_1306988, EPI_ISL_1306989, EPI_ISL_1306990, EPI_ISL_1306991, EPI_ISL_1306992, EPI_ISL_1306993, EPI_ISL_1306994, EPI_ISL_1306995, EPI_ISL_1306996, EPI_ISL_1306997, EPI_ISL_1306998, EPI_ISL_1306999, EPI_ISL_1307000, EPI_ISL_1307001, EPI_ISL_1307002, EPI_ISL_1307003, EPI_ISL_1307004, EPI_ISL_1307005, EPI_ISL_1307006, EPI_ISL_1307007, EPI_ISL_1307008, EPI_ISL_1307009, EPI_ISL_1307010, EPI_ISL_1307011, EPI_ISL_1307012, EPI_ISL_1307013, EPI_ISL_1307014, EPI_ISL_1307015, EPI_ISL_1307016, EPI_ISL_1307017, EPI_ISL_1307018, EPI_ISL_1307019, EPI_ISL_1307020, EPI_ISL_1307021, EPI_ISL_1307022, EPI_ISL_1307023, EPI_ISL_1307024, EPI_ISL_1307025, EPI_ISL_1307026, EPI_ISL_1307027, EPI_ISL_1307028, EPI_ISL_1307029,

EPI_ISL_1307030, EPI_ISL_1307031, EPI_ISL_1307032, EPI_ISL_1307033, EPI_ISL_1307034, EPI_ISL_1307035, EPI_ISL_1307036, EPI_ISL_1307037, EPI_ISL_1307038, EPI_ISL_1307039, EPI_ISL_1307040, EPI_ISL_1307041, EPI_ISL_1307042, EPI_ISL_1307043, EPI_ISL_1307044, EPI_ISL_1307045, EPI_ISL_1307046, EPI_ISL_1307047, EPI_ISL_1307048, EPI_ISL_1307049, EPI_ISL_1307050, EPI_ISL_1307051,
EPI_ISL_1307052, EPI_ISL_1307053, EPI_ISL_1307054, EPI_ISL_1307055, EPI_ISL_1307056, EPI_ISL_1307057, EPI_ISL_1307058, EPI_ISL_1307059, EPI_ISL_1307060, EPI_ISL_1307061, EPI_ISL_1307062, EPI_ISL_1307063, EPI_ISL_1307064, EPI_ISL_1307065, EPI_ISL_1307066, EPI_ISL_1307067, EPI_ISL_1307068, EPI_ISL_1307069, EPI_ISL_1307070, EPI_ISL_1307071, EPI_ISL_1307072, EPI_ISL_1307073,
EPI_ISL_1307074, EPI_ISL_1307075, EPI_ISL_1307076, EPI_ISL_1307077, EPI_ISL_1307078, EPI_ISL_1307079, EPI_ISL_1307080, EPI_ISL_1307081, EPI_ISL_1307082, EPI_ISL_1307083, EPI_ISL_1307084, EPI_ISL_1307085, EPI_ISL_1307086, EPI_ISL_1307087, EPI_ISL_1307088, EPI_ISL_1307089, EPI_ISL_1307090, EPI_ISL_1307091, EPI_ISL_1307092, EPI_ISL_1307093, EPI_ISL_1307094, EPI_ISL_1307095,
EPI_ISL_1307096, EPI_ISL_1307097, EPI_ISL_1307098, EPI_ISL_1307099, EPI_ISL_1307100, EPI_ISL_1307101, EPI_ISL_1307102, EPI_ISL_1307103, EPI_ISL_1307104, EPI_ISL_1307105, EPI_ISL_1307106, EPI_ISL_1307107, EPI_ISL_1307108, EPI_ISL_1307109, EPI_ISL_1307110, EPI_ISL_1307111, EPI_ISL_1307112, EPI_ISL_1307113, EPI_ISL_1307114, EPI_ISL_1307115, EPI_ISL_1307116, EPI_ISL_1307117,
EPI_ISL_1307118, EPI_ISL_1307119, EPI_ISL_1307120, EPI_ISL_1307121, EPI_ISL_1307122, EPI_ISL_1307123, EPI_ISL_1307124, EPI_ISL_1307125, EPI_ISL_1307126, EPI_ISL_1307127, EPI_ISL_1307128, EPI_ISL_1307129, EPI_ISL_1307130, EPI_ISL_1307131, EPI_ISL_1307132, EPI_ISL_1307133, EPI_ISL_1307134, EPI_ISL_1307135, EPI_ISL_1307136, EPI_ISL_1307137, EPI_ISL_1307138, EPI_ISL_1307139,
EPI_ISL_1307140, EPI_ISL_1307141, EPI_ISL_1307142, EPI_ISL_1307143, EPI_ISL_1307144, EPI_ISL_1307145, EPI_ISL_1307146, EPI_ISL_1307147, EPI_ISL_1307148, EPI_ISL_1307149, EPI_ISL_1307150, EPI_ISL_1307151, EPI_ISL_1307152, EPI_ISL_1307153, EPI_ISL_1307154, EPI_ISL_1307155, EPI_ISL_1307156, EPI_ISL_1307157, EPI_ISL_1307158, EPI_ISL_1307159, EPI_ISL_1307160, EPI_ISL_1307161,
EPI_ISL_1307162, EPI_ISL_1307163, EPI_ISL_1307164, EPI_ISL_1307165, EPI_ISL_1307166, EPI_ISL_1307167, EPI_ISL_1307168, EPI_ISL_1307169, EPI_ISL_1307170, EPI_ISL_1307171, EPI_ISL_1307172, EPI_ISL_1307173, EPI_ISL_1307174, EPI_ISL_1307175, EPI_ISL_1307176, EPI_ISL_1307178, EPI_ISL_1307179, EPI_ISL_1307180, EPI_ISL_1307181, EPI_ISL_1307182, EPI_ISL_1307183, EPI_ISL_1307184,
EPI_ISL_1307185, EPI_ISL_1307186, EPI_ISL_1307187, EPI_ISL_1307188, EPI_ISL_1307189, EPI_ISL_1307190, EPI_ISL_1307191, EPI_ISL_1307192, EPI_ISL_1307193, EPI_ISL_1307194, EPI_ISL_1307195, EPI_ISL_1307196, EPI_ISL_1307197, EPI_ISL_1307198, EPI_ISL_1307199, EPI_ISL_1307200, EPI_ISL_1307201, EPI_ISL_1307202, EPI_ISL_1307203, EPI_ISL_1307204, EPI_ISL_1307205, EPI_ISL_1307206,
EPI_ISL_1307207, EPI_ISL_1307208, EPI_ISL_1307209, EPI_ISL_1307210, EPI_ISL_1307211, EPI_ISL_1307212, EPI_ISL_1307213, EPI_ISL_1307214, EPI_ISL_1307215, EPI_ISL_1307216, EPI_ISL_1307217, EPI_ISL_1307218, EPI_ISL_1307219, EPI_ISL_1307220, EPI_ISL_1307221, EPI_ISL_1307222, EPI_ISL_1307223, EPI_ISL_1307224, EPI_ISL_1307225, EPI_ISL_1307226, EPI_ISL_1307227, EPI_ISL_1307228,
EPI_ISL_1307229, EPI_ISL_1307230, EPI_ISL_1307231, EPI_ISL_1307232, EPI_ISL_1307233, EPI_ISL_1307234, EPI_ISL_1307235, EPI_ISL_1307236, EPI_ISL_1307237, EPI_ISL_1307238, EPI_ISL_1307239, EPI_ISL_1307240, EPI_ISL_1307241, EPI_ISL_1307242, EPI_ISL_1307243, EPI_ISL_1307244, EPI_ISL_1307245, EPI_ISL_1307246, EPI_ISL_1307247, EPI_ISL_1307248, EPI_ISL_1307249, EPI_ISL_1307250,
EPI_ISL_1307251, EPI_ISL_1307252, EPI_ISL_1307253, EPI_ISL_1307254, EPI_ISL_1307255, EPI_ISL_1307256, EPI_ISL_1307257, EPI_ISL_1307258, EPI_ISL_1307259, EPI_ISL_1307260, EPI_ISL_1307261, EPI_ISL_1307262, EPI_ISL_1307263, EPI_ISL_1307264, EPI_ISL_1307265, EPI_ISL_1307266, EPI_ISL_1307267, EPI_ISL_1307268, EPI_ISL_1307269, EPI_ISL_1307270, EPI_ISL_1307271, EPI_ISL_1307272,
EPI_ISL_1307273, EPI_ISL_1307274, EPI_ISL_1307275, EPI_ISL_1307276, EPI_ISL_1307277, EPI_ISL_1307278, EPI_ISL_1307279, EPI_ISL_1307280, EPI_ISL_1307281, EPI_ISL_1307282, EPI_ISL_1307283, EPI_ISL_1307284, EPI_ISL_1307285, EPI_ISL_1307286, EPI_ISL_1307287, EPI_ISL_1307288, EPI_ISL_1307289, EPI_ISL_1307290, EPI_ISL_1307291, EPI_ISL_1307292, EPI_ISL_1307293, EPI_ISL_1307294,
EPI_ISL_1307295, EPI_ISL_1307296, EPI_ISL_1307297, EPI_ISL_1307298, EPI_ISL_1307299, EPI_ISL_1307300, EPI_ISL_1307301, EPI_ISL_1307302, EPI_ISL_1307303, EPI_ISL_1307304, EPI_ISL_1307305, EPI_ISL_1307306, EPI_ISL_1307307, EPI_ISL_1307308, EPI_ISL_1307309, EPI_ISL_1307310, EPI_ISL_1307311, EPI_ISL_1307312, EPI_ISL_1307313, EPI_ISL_1307314, EPI_ISL_1307315, EPI_ISL_1307316,
EPI_ISL_1307317, EPI_ISL_1307318, EPI_ISL_1307319, EPI_ISL_1307320, EPI_ISL_1307321, EPI_ISL_1307322, EPI_ISL_1307323, EPI_ISL_1307324, EPI_ISL_1307325, EPI_ISL_1307326, EPI_ISL_1307327, EPI_ISL_1307328, EPI_ISL_1307329, EPI_ISL_1307330, EPI_ISL_1307331, EPI_ISL_1307332, EPI_ISL_1307333, EPI_ISL_1307334, EPI_ISL_1307335, EPI_ISL_1307336, EPI_ISL_1307337, EPI_ISL_1307338,
EPI_ISL_1307339, EPI_ISL_1307340, EPI_ISL_1307341, EPI_ISL_1307342, EPI_ISL_1307343, EPI_ISL_1307344, EPI_ISL_1307345, EPI_ISL_1307346, EPI_ISL_1307347, EPI_ISL_1307348, EPI_ISL_1307349, EPI_ISL_1307350, EPI_ISL_1307351, EPI_ISL_1307352, EPI_ISL_1307353, EPI_ISL_1307354, EPI_ISL_1307355, EPI_ISL_1307356, EPI_ISL_1307357, EPI_ISL_1307358, EPI_ISL_1307359, EPI_ISL_1307360,
EPI_ISL_1307361, EPI_ISL_1307362, EPI_ISL_1307363, EPI_ISL_1307364, EPI_ISL_1307365, EPI_ISL_1307366, EPI_ISL_1307367, EPI_ISL_1307368, EPI_ISL_1307369, EPI_ISL_1307370, EPI_ISL_1307371, EPI_ISL_1307372, EPI_ISL_1307373, EPI_ISL_1307374, EPI_ISL_1307375, EPI_ISL_1307376, EPI_ISL_1307377, EPI_ISL_1307378, EPI_ISL_1307379, EPI_ISL_1307380, EPI_ISL_1307381, EPI_ISL_1307382,
EPI_ISL_1307383, EPI_ISL_1307384, EPI_ISL_1307385, EPI_ISL_1307386, EPI_ISL_1307387, EPI_ISL_1307388, EPI_ISL_1307389, EPI_ISL_1307390, EPI_ISL_1307391, EPI_ISL_1307392, EPI_ISL_1307393, EPI_ISL_1307394, EPI_ISL_1307395, EPI_ISL_1307396, EPI_ISL_1307397, EPI_ISL_1307398, EPI_ISL_1307399, EPI_ISL_1307400, EPI_ISL_1307401, EPI_ISL_1307402, EPI_ISL_1307403, EPI_ISL_1307404,
EPI_ISL_1307405, EPI_ISL_1307406, EPI_ISL_1307407, EPI_ISL_1307408, EPI_ISL_1307409, EPI_ISL_1307410, EPI_ISL_1307411, EPI_ISL_1307412, EPI_ISL_1307413, EPI_ISL_1307414, EPI_ISL_1307415, EPI_ISL_1307416, EPI_ISL_1307417, EPI_ISL_1307418, EPI_ISL_1307419, EPI_ISL_1307420, EPI_ISL_1307421, EPI_ISL_1307422, EPI_ISL_1307423, EPI_ISL_1307424, EPI_ISL_1307425, EPI_ISL_1307426,
EPI_ISL_1307427, EPI_ISL_1307428, EPI_ISL_1307429, EPI_ISL_1307430, EPI_ISL_1307431, EPI_ISL_1307432, EPI_ISL_1307433, EPI_ISL_1307434, EPI_ISL_1307435, EPI_ISL_1307436, EPI_ISL_1307437, EPI_ISL_1307438, EPI_ISL_1307439, EPI_ISL_1307440, EPI_ISL_1307441, EPI_ISL_1307442, EPI_ISL_1307443, EPI_ISL_1307444, EPI_ISL_1307445, EPI_ISL_1307446, EPI_ISL_1307447, EPI_ISL_1307448,
EPI_ISL_1307449, EPI_ISL_1307450, EPI_ISL_1307451, EPI_ISL_1307452, EPI_ISL_1307453, EPI_ISL_1307454, EPI_ISL_1307455, EPI_ISL_1307456, EPI_ISL_1307457, EPI_ISL_1307458, EPI_ISL_1307459, EPI_ISL_1307460, EPI_ISL_1307461, EPI_ISL_1307462, EPI_ISL_1307463, EPI_ISL_1307464, EPI_ISL_1307465, EPI_ISL_1307466, EPI_ISL_1307467, EPI_ISL_1307468, EPI_ISL_1307469, EPI_ISL_1307470,
EPI_ISL_1307471, EPI_ISL_1307472, EPI_ISL_1307473, EPI_ISL_1307474, EPI_ISL_1307475, EPI_ISL_1307476, EPI_ISL_1307477, EPI_ISL_1307478, EPI_ISL_1307479, EPI_ISL_1307480, EPI_ISL_1307481, EPI_ISL_1307482, EPI_ISL_1307483, EPI_ISL_1307484, EPI_ISL_1307485, EPI_ISL_1307486, EPI_ISL_1307487, EPI_ISL_1307488, EPI_ISL_1307489, EPI_ISL_1307490, EPI_ISL_1307491, EPI_ISL_1307492,
EPI_ISL_1307493, EPI_ISL_1307494, EPI_ISL_1307495, EPI_ISL_1307496, EPI_ISL_1307497, EPI_ISL_1307498, EPI_ISL_1307499, EPI_ISL_1307500, EPI_ISL_1307501, EPI_ISL_1307502, EPI_ISL_1307503, EPI_ISL_1307504, EPI_ISL_1307505, EPI_ISL_1307506, EPI_ISL_1307507, EPI_ISL_1307508, EPI_ISL_1307509, EPI_ISL_1307510, EPI_ISL_1307511, EPI_ISL_1307512, EPI_ISL_1307513, EPI_ISL_1307514,
EPI_ISL_1307515, EPI_ISL_1307516, EPI_ISL_1307517, EPI_ISL_1307518, EPI_ISL_1307519, EPI_ISL_1307520, EPI_ISL_1307521, EPI_ISL_1307522, EPI_ISL_1307523, EPI_ISL_1307524, EPI_ISL_1307525, EPI_ISL_1307526, EPI_ISL_1307527, EPI_ISL_1307528, EPI_ISL_1307529, EPI_ISL_1307530, EPI_ISL_1307531, EPI_ISL_1307532, EPI_ISL_1307533, EPI_ISL_1307534, EPI_ISL_1307535, EPI_ISL_1307536,
EPI_ISL_1307537, EPI_ISL_1307538, EPI_ISL_1307539, EPI_ISL_1307540, EPI_ISL_1307541, EPI_ISL_1307542, EPI_ISL_1307543, EPI_ISL_1307544, EPI_ISL_1307545, EPI_ISL_1307546, EPI_ISL_1307547, EPI_ISL_1307548, EPI_ISL_1307549, EPI_ISL_1307550, EPI_ISL_1307551, EPI_ISL_1307552, EPI_ISL_1307553, EPI_ISL_1307554, EPI_ISL_1307555, EPI_ISL_1307556, EPI_ISL_1307557, EPI_ISL_1307558,
EPI_ISL_1307559, EPI_ISL_1307560, EPI_ISL_1307561, EPI_ISL_1307562, EPI_ISL_1307563, EPI_ISL_1307564, EPI_ISL_1307565, EPI_ISL_1307566, EPI_ISL_1307567, EPI_ISL_1307568, EPI_ISL_1307569, EPI_ISL_1307570, EPI_ISL_1307571, EPI_ISL_1307572, EPI_ISL_1307573, EPI_ISL_1307574, EPI_ISL_1307575, EPI_ISL_1307576, EPI_ISL_1307577, EPI_ISL_1307578, EPI_ISL_1307579, EPI_ISL_1307580,
EPI_ISL_1307581, EPI_ISL_1307582, EPI_ISL_1307583, EPI_ISL_1307584, EPI_ISL_1307585, EPI_ISL_1307586, EPI_ISL_1307587, EPI_ISL_1307588, EPI_ISL_1307589, EPI_ISL_1307590, EPI_ISL_1307591, EPI_ISL_1307592, EPI_ISL_1307593, EPI_ISL_1307594, EPI_ISL_1307595, EPI_ISL_1307596, EPI_ISL_1307597, EPI_ISL_1307598, EPI_ISL_1307599, EPI_ISL_1307600, EPI_ISL_1307601, EPI_ISL_1307602,
EPI_ISL_1307603, EPI_ISL_1307604, EPI_ISL_1307605, EPI_ISL_1307606, EPI_ISL_1307607, EPI_ISL_1307608, EPI_ISL_1307609, EPI_ISL_1307610, EPI_ISL_1307611, EPI_ISL_1307612, EPI_ISL_1307613, EPI_ISL_1307614, EPI_ISL_1307615, EPI_ISL_1307616, EPI_ISL_1307617, EPI_ISL_1307618, EPI_ISL_1307619, EPI_ISL_1307620, EPI_ISL_1307621, EPI_ISL_1307622, EPI_ISL_1307623, EPI_ISL_1307624,
EPI_ISL_1307625, EPI_ISL_1307626, EPI_ISL_1307627, EPI_ISL_1307628, EPI_ISL_1307629, EPI_ISL_1307630, EPI_ISL_1307631, EPI_ISL_1307632, EPI_ISL_1307633, EPI_ISL_1307634, EPI_ISL_1307635, EPI_ISL_1384885, EPI_ISL_1384886, EPI_ISL_1384887, EPI_ISL_1384888, EPI_ISL_1384889, EPI_ISL_1384890, EPI_ISL_1384891, EPI_ISL_1384892, EPI_ISL_1384893, EPI_ISL_1384894, EPI_ISL_1384895,
EPI_ISL_1384896, EPI_ISL_1384897, EPI_ISL_1384898, EPI_ISL_1384899, EPI_ISL_1384900, EPI_ISL_1384901, EPI_ISL_1384902, EPI_ISL_1384903, EPI_ISL_1384904, EPI_ISL_1384905, EPI_ISL_1384906, EPI_ISL_1384907, EPI_ISL_1384908, EPI_ISL_1384909, EPI_ISL_1384910, EPI_ISL_1384911, EPI_ISL_1384912, EPI_ISL_1384913, EPI_ISL_1384914, EPI_ISL_1384915, EPI_ISL_1384916, EPI_ISL_1384917,
EPI_ISL_1384918, EPI_ISL_1384919, EPI_ISL_1384920, EPI_ISL_1384921, EPI_ISL_1384922, EPI_ISL_1384923, EPI_ISL_1384924, EPI_ISL_1384925, EPI_ISL_1384926, EPI_ISL_1384927, EPI_ISL_1384928, EPI_ISL_1384929, EPI_ISL_1384930, EPI_ISL_1384931, EPI_ISL_1384932, EPI_ISL_1384933, EPI_ISL_1384934, EPI_ISL_1384935, EPI_ISL_1384936, EPI_ISL_1384937, EPI_ISL_1384938, EPI_ISL_1384939,
EPI_ISL_1384940, EPI_ISL_1384941, EPI_ISL_1384942, EPI_ISL_1384943, EPI_ISL_1384944, EPI_ISL_1384945, EPI_ISL_1384946, EPI_ISL_1384947, EPI_ISL_1384948, EPI_ISL_1384949, EPI_ISL_1384950, EPI_ISL_1384951, EPI_ISL_1384952, EPI_ISL_1384953, EPI_ISL_1384954, EPI_ISL_1384955, EPI_ISL_1384956, EPI_ISL_1384957, EPI_ISL_1384958, EPI_ISL_1384959, EPI_ISL_1384960, EPI_ISL_1384961,
EPI_ISL_1384962, EPI_ISL_1384963, EPI_ISL_1384964, EPI_ISL_1384965, EPI_ISL_1384966, EPI_ISL_1384967, EPI_ISL_1384968, EPI_ISL_1384969, EPI_ISL_1384970, EPI_ISL_1384971, EPI_ISL_1384972, EPI_ISL_1384973, EPI_ISL_1384974, EPI_ISL_1384975, EPI_ISL_1384976, EPI_ISL_1384977, EPI_ISL_1384978, EPI_ISL_1384979, EPI_ISL_1384980, EPI_ISL_1384981, EPI_ISL_1384982, EPI_ISL_1384983,
EPI_ISL_1384984, EPI_ISL_1384985, EPI_ISL_1384986, EPI_ISL_1384987, EPI_ISL_1384988, EPI_ISL_1384989, EPI_ISL_1384990, EPI_ISL_1384991, EPI_ISL_1384992, EPI_ISL_1384993, EPI_ISL_1384994, EPI_ISL_1384995, EPI_ISL_1384996, EPI_ISL_1384997, EPI_ISL_1384998, EPI_ISL_1384999, EPI_ISL_1385000, EPI_ISL_1385001, EPI_ISL_1385002, EPI_ISL_1385003, EPI_ISL_1385004, EPI_ISL_1385005,
EPI_ISL_1385006, EPI_ISL_1385007, EPI_ISL_1385008, EPI_ISL_1385009, EPI_ISL_1385010, EPI_ISL_1385011, EPI_ISL_1385012, EPI_ISL_1385013, EPI_ISL_1385014, EPI_ISL_1385015, EPI_ISL_1385016, EPI_ISL_1385017, EPI_ISL_1385018, EPI_ISL_1385019, EPI_ISL_1385020, EPI_ISL_1385021, EPI_ISL_1385022, EPI_ISL_1385023, EPI_ISL_1385024, EPI_ISL_1385025, EPI_ISL_1385026, EPI_ISL_1385027,
EPI_ISL_1385028, EPI_ISL_1385029, EPI_ISL_1385030, EPI_ISL_1385031, EPI_ISL_1385032, EPI_ISL_1385033, EPI_ISL_1385034, EPI_ISL_1385035, EPI_ISL_1385036, EPI_ISL_1385037, EPI_ISL_1385038, EPI_ISL_1385039, EPI_ISL_1385040, EPI_ISL_1385041, EPI_ISL_1385042, EPI_ISL_1385043, EPI_ISL_1385044, EPI_ISL_1385045, EPI_ISL_1385046, EPI_ISL_1385047, EPI_ISL_1385048, EPI_ISL_1385049,
EPI_ISL_1385050, EPI_ISL_1385051, EPI_ISL_1385052, EPI_ISL_1385053, EPI_ISL_1385054, EPI_ISL_1385055, EPI_ISL_1385056, EPI_ISL_1385057, EPI_ISL_1385058, EPI_ISL_1385059, EPI_ISL_1385060, EPI_ISL_1385061, EPI_ISL_1385062, EPI_ISL_1385063, EPI_ISL_1385064, EPI_ISL_1385065, EPI_ISL_1385066, EPI_ISL_1385067, EPI_ISL_1385068, EPI_ISL_1385069, EPI_ISL_1385070, EPI_ISL_1385071,
EPI_ISL_1385072, EPI_ISL_1385073, EPI_ISL_1385074, EPI_ISL_1385075, EPI_ISL_1385076, EPI_ISL_1385077, EPI_ISL_1385078, EPI_ISL_1385079, EPI_ISL_1385080, EPI_ISL_1385081, EPI_ISL_1385082, EPI_ISL_1385083, EPI_ISL_1385084, EPI_ISL_1385085, EPI_ISL_1385086, EPI_ISL_1385087, EPI_ISL_1385088, EPI_ISL_1385089, EPI_ISL_1385090, EPI_ISL_1385091, EPI_ISL_1385092, EPI_ISL_1385093,
EPI_ISL_1385094, EPI_ISL_1385095, EPI_ISL_1385096, EPI_ISL_1385097, EPI_ISL_1385098, EPI_ISL_1385099, EPI_ISL_1385100, EPI_ISL_1385101, EPI_ISL_1385102, EPI_ISL_1385103, EPI_ISL_1385104, EPI_ISL_1385105, EPI_ISL_1385106, EPI_ISL_1385107, EPI_ISL_1385108, EPI_ISL_1385109, EPI_ISL_1385110, EPI_ISL_1385111, EPI_ISL_1385112, EPI_ISL_1385113, EPI_ISL_1385114, EPI_ISL_1385115,
EPI_ISL_1385116, EPI_ISL_1385117, EPI_ISL_1385118, EPI_ISL_1385119, EPI_ISL_1385120, EPI_ISL_1385121, EPI_ISL_1385122, EPI_ISL_1385123, EPI_ISL_1385124, EPI_ISL_1385125, EPI_ISL_1385126, EPI_ISL_1385127, EPI_ISL_1385128, EPI_ISL_1385129, EPI_ISL_1385130, EPI_ISL_1385131, EPI_ISL_1385132, EPI_ISL_1385133, EPI_ISL_1385134, EPI_ISL_1385135, EPI_ISL_1385136, EPI_ISL_1385137,
EPI_ISL_1385138, EPI_ISL_1385139, EPI_ISL_1385140, EPI_ISL_1385141, EPI_ISL_1385142, EPI_ISL_1385143, EPI_ISL_1385144, EPI_ISL_1385145, EPI_ISL_1385146, EPI_ISL_1385147, EPI_ISL_1385148, EPI_ISL_1385149, EPI_ISL_1385150, EPI_ISL_1385151, EPI_ISL_1385152, EPI_ISL_1385153, EPI_ISL_1385154, EPI_ISL_1385155, EPI_ISL_1385156, EPI_ISL_1385157, EPI_ISL_1385158, EPI_ISL_1385159,
EPI_ISL_1385160, EPI_ISL_1385161, EPI_ISL_1385162, EPI_ISL_1385163, EPI_ISL_1385164, EPI_ISL_1385165, EPI_ISL_1385166, EPI_ISL_1385167, EPI_ISL_1385168, EPI_ISL_1385169, EPI_ISL_1385170, EPI_ISL_1385171, EPI_ISL_1385172, EPI_ISL_1385173, EPI_ISL_1385174, EPI_ISL_1385175, EPI_ISL_1385176, EPI_ISL_1385177, EPI_ISL_1385178, EPI_ISL_1385179, EPI_ISL_1385180, EPI_ISL_1385181,
EPI_ISL_1385182, EPI_ISL_1385183, EPI_ISL_1385184, EPI_ISL_1385185, EPI_ISL_1385186, EPI_ISL_1385187, EPI_ISL_1385188, EPI_ISL_1385189, EPI_ISL_1385190, EPI_ISL_1385191, EPI_ISL_1385192, EPI_ISL_1385193, EPI_ISL_1385194, EPI_ISL_1385195, EPI_ISL_1385196, EPI_ISL_1385197, EPI_ISL_1385198, EPI_ISL_1385199, EPI_ISL_1385200, EPI_ISL_1385201, EPI_ISL_1385202, EPI_ISL_1385203,
EPI_ISL_1385204, EPI_ISL_1385205, EPI_ISL_1385206, EPI_ISL_1385207, EPI_ISL_1385208, EPI_ISL_1385209, EPI_ISL_1385210, EPI_ISL_1385211, EPI_ISL_1385212, EPI_ISL_1385213, EPI_ISL_1385214, EPI_ISL_1385215, EPI_ISL_1385216, EPI_ISL_1385217, EPI_ISL_1385218, EPI_ISL_1385219, EPI_ISL_1385220, EPI_ISL_1385221, EPI_ISL_1385222, EPI_ISL_1385223, EPI_ISL_1385224, EPI_ISL_1385225,
EPI_ISL_1385226, EPI_ISL_1385227, EPI_ISL_1385228, EPI_ISL_1385229, EPI_ISL_1385230, EPI_ISL_1385231, EPI_ISL_1385232, EPI_ISL_1385233, EPI_ISL_1385234, EPI_ISL_1385235, EPI_ISL_1385236, EPI_ISL_1385237, EPI_ISL_1385238, EPI_ISL_1385239, EPI_ISL_1385240, EPI_ISL_1385241, EPI_ISL_1385242, EPI_ISL_1385243, EPI_ISL_1385244, EPI_ISL_1385245, EPI_ISL_1385246, EPI_ISL_1385247,
EPI_ISL_1385248, EPI_ISL_1385249, EPI_ISL_1385250, EPI_ISL_1385251, EPI_ISL_1385252, EPI_ISL_1385253, EPI_ISL_1385254, EPI_ISL_1385255, EPI_ISL_1385256, EPI_ISL_1385257, EPI_ISL_1385258, EPI_ISL_1385259, EPI_ISL_1385260, EPI_ISL_1385261, EPI_ISL_1385262, EPI_ISL_1385263, EPI_ISL_1385264, EPI_ISL_1385265, EPI_ISL_1385266, EPI_ISL_1385267, EPI_ISL_1385268, EPI_ISL_1385269,
EPI_ISL_1385270, EPI_ISL_1385271, EPI_ISL_1385272, EPI_ISL_1385273, EPI_ISL_1385274, EPI_ISL_1385275, EPI_ISL_1385276, EPI_ISL_1385277, EPI_ISL_1385278, EPI_ISL_1385279, EPI_ISL_1385280, EPI_ISL_1385281, EPI_ISL_1385282, EPI_ISL_1385283, EPI_ISL_1385284, EPI_ISL_1385285, EPI_ISL_1385286, EPI_ISL_1385287, EPI_ISL_1385288, EPI_ISL_1385289, EPI_ISL_1385290, EPI_ISL_1385291,
EPI_ISL_1385292, EPI_ISL_1385293, EPI_ISL_1385294, EPI_ISL_1385295, EPI_ISL_1385296, EPI_ISL_1385297, EPI_ISL_1385298, EPI_ISL_1385299, EPI_ISL_1385300, EPI_ISL_1385301, EPI_ISL_1385302, EPI_ISL_1385303, EPI_ISL_1385304, EPI_ISL_1385305, EPI_ISL_1385306, EPI_ISL_1385307, EPI_ISL_1385308, EPI_ISL_1385309, EPI_ISL_1385310, EPI_ISL_1385311, EPI_ISL_1385312, EPI_ISL_1385313,
EPI_ISL_1385314, EPI_ISL_1385315, EPI_ISL_1385316, EPI_ISL_1385317, EPI_ISL_1385318, EPI_ISL_1385319, EPI_ISL_1385320, EPI_ISL_1385321, EPI_ISL_1385322, EPI_ISL_1385323, EPI_ISL_1385324, EPI_ISL_1385325, EPI_ISL_1385326, EPI_ISL_1385327, EPI_ISL_1385328, EPI_ISL_1385329, EPI_ISL_1385330, EPI_ISL_1385331, EPI_ISL_1385332, EPI_ISL_1385333, EPI_ISL_1385334, EPI_ISL_1385335,
EPI_ISL_1385336, EPI_ISL_1385337, EPI_ISL_1385338, EPI_ISL_1385339, EPI_ISL_1385340, EPI_ISL_1385341, EPI_ISL_1385342, EPI_ISL_1385343, EPI_ISL_1385344, EPI_ISL_1385345, EPI_ISL_1385346, EPI_ISL_1385347, EPI_ISL_1385348, EPI_ISL_1385350, EPI_ISL_1385351, EPI_ISL_1385352, EPI_ISL_1385353, EPI_ISL_1385354, EPI_ISL_1385355, EPI_ISL_1385356, EPI_ISL_1385357, EPI_ISL_1385358,
EPI_ISL_1385359, EPI_ISL_1385360, EPI_ISL_1385361, EPI_ISL_1385362, EPI_ISL_1385363, EPI_ISL_1385364, EPI_ISL_1385365, EPI_ISL_1385366, EPI_ISL_1385367, EPI_ISL_1385368, EPI_ISL_1385369, EPI_ISL_1385370, EPI_ISL_1385371, EPI_ISL_1385372, EPI_ISL_1385373, EPI_ISL_1385374, EPI_ISL_1385375, EPI_ISL_1385376, EPI_ISL_1385377, EPI_ISL_1385378, EPI_ISL_1385379, EPI_ISL_1385380,
EPI_ISL_1385381, EPI_ISL_1385382, EPI_ISL_1385383, EPI_ISL_1385384, EPI_ISL_1385385, EPI_ISL_1385386, EPI_ISL_1385387, EPI_ISL_1385388, EPI_ISL_1385389, EPI_ISL_1385390, EPI_ISL_1385391, EPI_ISL_1385392, EPI_ISL_1385393, EPI_ISL_1385394, EPI_ISL_1385395, EPI_ISL_1385396, EPI_ISL_1385397, EPI_ISL_1385398, EPI_ISL_1385399, EPI_ISL_1385400, EPI_ISL_1385401, EPI_ISL_1385402,
EPI_ISL_1385403, EPI_ISL_1385404, EPI_ISL_1385406, EPI_ISL_1385407, EPI_ISL_1385408, EPI_ISL_1385409, EPI_ISL_1385410, EPI_ISL_1385411, EPI_ISL_1385412, EPI_ISL_1385413, EPI_ISL_1385414, EPI_ISL_1385415, EPI_ISL_1385416, EPI_ISL_1385417, EPI_ISL_1385418, EPI_ISL_1385419, EPI_ISL_1385420, EPI_ISL_1385421, EPI_ISL_1385422, EPI_ISL_1385423, EPI_ISL_1385424, EPI_ISL_1385425,
EPI_ISL_1385426, EPI_ISL_1385427, EPI_ISL_1385428, EPI_ISL_1385429, EPI_ISL_1385430, EPI_ISL_1385431, EPI_ISL_1385432, EPI_ISL_1385433, EPI_ISL_1385434, EPI_ISL_1385435, EPI_ISL_1385436, EPI_ISL_1385437, EPI_ISL_1385438, EPI_ISL_1385439, EPI_ISL_1385440, EPI_ISL_1385441, EPI_ISL_1385442, EPI_ISL_1385443, EPI_ISL_1385444, EPI_ISL_1385445, EPI_ISL_1385446, EPI_ISL_1385447,
EPI_ISL_1385448, EPI_ISL_1385449, EPI_ISL_1385450, EPI_ISL_1385451, EPI_ISL_1385452, EPI_ISL_1385453, EPI_ISL_1385454, EPI_ISL_1385455, EPI_ISL_1385456, EPI_ISL_1385457, EPI_ISL_1385458, EPI_ISL_1385459, EPI_ISL_1385460, EPI_ISL_1385461, EPI_ISL_1385462, EPI_ISL_1385463, EPI_ISL_1385464, EPI_ISL_1385465, EPI_ISL_1385466, EPI_ISL_1385467, EPI_ISL_1385468, EPI_ISL_1385469,
EPI_ISL_1385470, EPI_ISL_1385471, EPI_ISL_1385472, EPI_ISL_1385473, EPI_ISL_1385474, EPI_ISL_1385475, EPI_ISL_1385476, EPI_ISL_1385477, EPI_ISL_1385478, EPI_ISL_1385479, EPI_ISL_1385480, EPI_ISL_1385481, EPI_ISL_1385482, EPI_ISL_1385483, EPI_ISL_1385484, EPI_ISL_1385485, EPI_ISL_1385486, EPI_ISL_1385487, EPI_ISL_1385488, EPI_ISL_1385489, EPI_ISL_1385490, EPI_ISL_1385491,
EPI_ISL_1385492, EPI_ISL_1385493, EPI_ISL_1385494, EPI_ISL_1385495, EPI_ISL_1385496, EPI_ISL_1385497, EPI_ISL_1385498, EPI_ISL_1385499, EPI_ISL_1385500, EPI_ISL_1385501, EPI_ISL_1385502, EPI_ISL_1385503, EPI_ISL_1385504, EPI_ISL_1385505, EPI_ISL_1385506, EPI_ISL_1385507, EPI_ISL_1385508, EPI_ISL_1385509, EPI_ISL_1385510, EPI_ISL_1385511, EPI_ISL_1385512, EPI_ISL_1385513,
EPI_ISL_1385514, EPI_ISL_1385515, EPI_ISL_1385516, EPI_ISL_1385517, EPI_ISL_1385518, EPI_ISL_1385519, EPI_ISL_1385520, EPI_ISL_1385521, EPI_ISL_1385522, EPI_ISL_1385523, EPI_ISL_1385524, EPI_ISL_1385525, EPI_ISL_1385526, EPI_ISL_1385527, EPI_ISL_1385528, EPI_ISL_1385529, EPI_ISL_1385530, EPI_ISL_1385531, EPI_ISL_1385532, EPI_ISL_1385533, EPI_ISL_1385534, EPI_ISL_1385535,
EPI_ISL_1385536, EPI_ISL_1385537, EPI_ISL_1385538, EPI_ISL_1385539, EPI_ISL_1385540, EPI_ISL_1385541, EPI_ISL_1385542, EPI_ISL_1385543, EPI_ISL_1385544, EPI_ISL_1385545, EPI_ISL_1385546, EPI_ISL_1385547, EPI_ISL_1385548, EPI_ISL_1385549, EPI_ISL_1385550, EPI_ISL_1385551, EPI_ISL_1385552, EPI_ISL_1385553, EPI_ISL_1385554, EPI_ISL_1385555, EPI_ISL_1385556, EPI_ISL_1385557,
EPI_ISL_1385558, EPI_ISL_1385559, EPI_ISL_1385560, EPI_ISL_1385561, EPI_ISL_1385562, EPI_ISL_1385563, EPI_ISL_1385564, EPI_ISL_1385565, EPI_ISL_1385566, EPI_ISL_1385567, EPI_ISL_1385568, EPI_ISL_1385569, EPI_ISL_1385570, EPI_ISL_1385571, EPI_ISL_1385572, EPI_ISL_1385573, EPI_ISL_1385574, EPI_ISL_1385575, EPI_ISL_1385576, EPI_ISL_1385577, EPI_ISL_1385578, EPI_ISL_1385579,
EPI_ISL_1385580, EPI_ISL_1385581, EPI_ISL_1385582, EPI_ISL_1385583, EPI_ISL_1385584, EPI_ISL_1385585, EPI_ISL_1385586, EPI_ISL_1385587, EPI_ISL_1385588, EPI_ISL_1385589, EPI_ISL_1385590, EPI_ISL_1385591, EPI_ISL_1385592, EPI_ISL_1385593, EPI_ISL_1385594, EPI_ISL_1385595, EPI_ISL_1385596, EPI_ISL_1385597, EPI_ISL_1385598, EPI_ISL_1385599, EPI_ISL_1385600, EPI_ISL_1385601,
EPI_ISL_1385602, EPI_ISL_1385603, EPI_ISL_1385604, EPI_ISL_1385605, EPI_ISL_1385606, EPI_ISL_1385607, EPI_ISL_1385608, EPI_ISL_1385609, EPI_ISL_1385610, EPI_ISL_1385611, EPI_ISL_1385612, EPI_ISL_1385613, EPI_ISL_1385614, EPI_ISL_1385615, EPI_ISL_1385616, EPI_ISL_1385617, EPI_ISL_1385618, EPI_ISL_1385619, EPI_ISL_1385620, EPI_ISL_1385621, EPI_ISL_1385622, EPI_ISL_1385623,
EPI_ISL_1385624, EPI_ISL_1385625, EPI_ISL_1385626, EPI_ISL_1385627, EPI_ISL_1385628, EPI_ISL_1385629, EPI_ISL_1385630, EPI_ISL_1385631, EPI_ISL_1385632, EPI_ISL_1385633, EPI_ISL_1385634, EPI_ISL_1385635, EPI_ISL_1385636, EPI_ISL_1385637, EPI_ISL_1385638, EPI_ISL_1385639, EPI_ISL_1385640, EPI_ISL_1385641, EPI_ISL_1385642, EPI_ISL_1385643, EPI_ISL_1385644, EPI_ISL_1385645,
EPI_ISL_1385646, EPI_ISL_1385647, EPI_ISL_1385648, EPI_ISL_1385649, EPI_ISL_1385650, EPI_ISL_1385651, EPI_ISL_1385652, EPI_ISL_1385653, EPI_ISL_1385654, EPI_ISL_1385655, EPI_ISL_1385656, EPI_ISL_1385657, EPI_ISL_1385658, EPI_ISL_1385659, EPI_ISL_1385660, EPI_ISL_1385661, EPI_ISL_1385662, EPI_ISL_1385663, EPI_ISL_1385664, EPI_ISL_1385665, EPI_ISL_1385666, EPI_ISL_1385667,
EPI_ISL_1385668, EPI_ISL_1385669, EPI_ISL_1385670, EPI_ISL_1385671, EPI_ISL_1385672, EPI_ISL_1385673, EPI_ISL_1385674, EPI_ISL_1385675, EPI_ISL_1385676, EPI_ISL_1385677, EPI_ISL_1385678, EPI_ISL_1385679, EPI_ISL_1385680, EPI_ISL_1385681, EPI_ISL_1385682, EPI_ISL_1385683, EPI_ISL_1385684, EPI_ISL_1385685, EPI_ISL_1385686, EPI_ISL_1385687, EPI_ISL_1385688, EPI_ISL_1385689,
EPI_ISL_1385690, EPI_ISL_1385691, EPI_ISL_1385692, EPI_ISL_1385693, EPI_ISL_1385694, EPI_ISL_1385695, EPI_ISL_1385696, EPI_ISL_1385697, EPI_ISL_1385698, EPI_ISL_1385699, EPI_ISL_1385700, EPI_ISL_1385701, EPI_ISL_1385702, EPI_ISL_1385703, EPI_ISL_1385704, EPI_ISL_1385705, EPI_ISL_1385706, EPI_ISL_1385707, EPI_ISL_1385708, EPI_ISL_1385709, EPI_ISL_1385710, EPI_ISL_1385711,
EPI_ISL_1385712, EPI_ISL_1385713, EPI_ISL_1385714, EPI_ISL_1385715, EPI_ISL_1385716, EPI_ISL_1385717, EPI_ISL_1385718, EPI_ISL_1385719, EPI_ISL_1385720, EPI_ISL_1385721, EPI_ISL_1385722, EPI_ISL_1385723, EPI_ISL_1385724, EPI_ISL_1385725, EPI_ISL_1385726, EPI_ISL_1385727, EPI_ISL_1385728, EPI_ISL_1385729, EPI_ISL_1385730, EPI_ISL_1385731, EPI_ISL_1385732, EPI_ISL_1385733,
EPI_ISL_1385734, EPI_ISL_1385735, EPI_ISL_1385736, EPI_ISL_1385737, EPI_ISL_1385738, EPI_ISL_1385739, EPI_ISL_1385740, EPI_ISL_1385741, EPI_ISL_1385742, EPI_ISL_1385743, EPI_ISL_1385744, EPI_ISL_1385745, EPI_ISL_1385746, EPI_ISL_1385747, EPI_ISL_1385748, EPI_ISL_1385749, EPI_ISL_1385750, EPI_ISL_1385751, EPI_ISL_1385752, EPI_ISL_1385753, EPI_ISL_1385754, EPI_ISL_1385755,
EPI_ISL_1385756, EPI_ISL_1385757, EPI_ISL_1385758, EPI_ISL_1385759, EPI_ISL_1385760, EPI_ISL_1385761, EPI_ISL_1385762, EPI_ISL_1385763, EPI_ISL_1385764, EPI_ISL_1385765, EPI_ISL_1385766, EPI_ISL_1385767, EPI_ISL_1385768, EPI_ISL_1385769, EPI_ISL_1385770, EPI_ISL_1385771, EPI_ISL_1385772, EPI_ISL_1385773, EPI_ISL_1385774, EPI_ISL_1385775, EPI_ISL_1385776, EPI_ISL_1385777,
EPI_ISL_1385778, EPI_ISL_1385779, EPI_ISL_1385780, EPI_ISL_1385781, EPI_ISL_1385782, EPI_ISL_1385783, EPI_ISL_1470741, EPI_ISL_1470742, EPI_ISL_1470743, EPI_ISL_1470744, EPI_ISL_1470745, EPI_ISL_1470746, EPI_ISL_1470747, EPI_ISL_1470748, EPI_ISL_1470749, EPI_ISL_1470750, EPI_ISL_1470751, EPI_ISL_1470752, EPI_ISL_1470753, EPI_ISL_1470754, EPI_ISL_1470755, EPI_ISL_1470756,
EPI_ISL_1470757, EPI_ISL_1470758, EPI_ISL_1470759, EPI_ISL_1470760, EPI_ISL_1470761, EPI_ISL_1470762, EPI_ISL_1470763, EPI_ISL_1470764, EPI_ISL_1470765, EPI_ISL_1470766, EPI_ISL_1470767, EPI_ISL_1470768, EPI_ISL_1470769, EPI_ISL_1470770, EPI_ISL_1470771, EPI_ISL_1470772, EPI_ISL_1470773, EPI_ISL_1470774, EPI_ISL_1470775, EPI_ISL_1470776, EPI_ISL_1470777, EPI_ISL_1470778,
EPI_ISL_1470779, EPI_ISL_1470780, EPI_ISL_1470781, EPI_ISL_1470782, EPI_ISL_1470783, EPI_ISL_1470784, EPI_ISL_1470785, EPI_ISL_1470786, EPI_ISL_1470787, EPI_ISL_1470788, EPI_ISL_1470789, EPI_ISL_1470790, EPI_ISL_1470791, EPI_ISL_1470792, EPI_ISL_1470793, EPI_ISL_1470794, EPI_ISL_1470795, EPI_ISL_1470796, EPI_ISL_1470797, EPI_ISL_1470798, EPI_ISL_1470799, EPI_ISL_1470800,
EPI_ISL_1470801, EPI_ISL_1470802, EPI_ISL_1470803, EPI_ISL_1470804, EPI_ISL_1470805, EPI_ISL_1470806, EPI_ISL_1470807, EPI_ISL_1470808, EPI_ISL_1470809, EPI_ISL_1470810, EPI_ISL_1470811, EPI_ISL_1470812, EPI_ISL_1470813, EPI_ISL_1470814, EPI_ISL_1470815, EPI_ISL_1470816, EPI_ISL_1470817, EPI_ISL_1470818, EPI_ISL_1470819, EPI_ISL_1470820, EPI_ISL_1470821, EPI_ISL_1470822,
EPI_ISL_1470823, EPI_ISL_1470824, EPI_ISL_1470825, EPI_ISL_1470826, EPI_ISL_1470827, EPI_ISL_1470828, EPI_ISL_1470829, EPI_ISL_1470830, EPI_ISL_1470831, EPI_ISL_1470832, EPI_ISL_1470833, EPI_ISL_1470834, EPI_ISL_1470835, EPI_ISL_1470836, EPI_ISL_1470837, EPI_ISL_1470838, EPI_ISL_1470839, EPI_ISL_1470840, EPI_ISL_1470841, EPI_ISL_1470842, EPI_ISL_1470843, EPI_ISL_1470844,
EPI_ISL_1470845, EPI_ISL_1470846, EPI_ISL_1470847, EPI_ISL_1470848, EPI_ISL_1470849, EPI_ISL_1470850, EPI_ISL_1470851, EPI_ISL_1470852, EPI_ISL_1470853, EPI_ISL_1470854, EPI_ISL_1470855, EPI_ISL_1470856, EPI_ISL_1470857, EPI_ISL_1470858, EPI_ISL_1470859, EPI_ISL_1470860, EPI_ISL_1470861, EPI_ISL_1470862, EPI_ISL_1470863, EPI_ISL_1470864, EPI_ISL_1470865, EPI_ISL_1470866,
EPI_ISL_1470867, EPI_ISL_1470868, EPI_ISL_1470869, EPI_ISL_1470870, EPI_ISL_1470871, EPI_ISL_1470872, EPI_ISL_1470873, EPI_ISL_1470874, EPI_ISL_1470875, EPI_ISL_1470876, EPI_ISL_1470877, EPI_ISL_1470878, EPI_ISL_1470879, EPI_ISL_1470880, EPI_ISL_1470881, EPI_ISL_1470882, EPI_ISL_1470883, EPI_ISL_1470884, EPI_ISL_1470885, EPI_ISL_1470886, EPI_ISL_1470887, EPI_ISL_1470888,
EPI_ISL_1470889, EPI_ISL_1470890, EPI_ISL_1470891, EPI_ISL_1470892, EPI_ISL_1470893, EPI_ISL_1470894, EPI_ISL_1470895, EPI_ISL_1470896, EPI_ISL_1470897, EPI_ISL_1470898, EPI_ISL_1470899, EPI_ISL_1470900, EPI_ISL_1470901, EPI_ISL_1470902, EPI_ISL_1470903, EPI_ISL_1470904, EPI_ISL_1470905, EPI_ISL_1470906, EPI_ISL_1470907, EPI_ISL_1470908, EPI_ISL_1470909, EPI_ISL_1470910,
EPI_ISL_1470911, EPI_ISL_1470912, EPI_ISL_1470913, EPI_ISL_1470914, EPI_ISL_1470915, EPI_ISL_1470916, EPI_ISL_1470917, EPI_ISL_1470918, EPI_ISL_1470919, EPI_ISL_1470920, EPI_ISL_1470921, EPI_ISL_1470922, EPI_ISL_1470923, EPI_ISL_1470924, EPI_ISL_1470925, EPI_ISL_1470926, EPI_ISL_1470927, EPI_ISL_1470928, EPI_ISL_1470929, EPI_ISL_1470930, EPI_ISL_1470931, EPI_ISL_1470932,
EPI_ISL_1470933, EPI_ISL_1470934, EPI_ISL_1470935, EPI_ISL_1470936, EPI_ISL_1470937, EPI_ISL_1470938, EPI_ISL_1470939, EPI_ISL_1470940, EPI_ISL_1470941, EPI_ISL_1470942, EPI_ISL_1470943, EPI_ISL_1470944, EPI_ISL_1470945, EPI_ISL_1470946, EPI_ISL_1470947, EPI_ISL_1470948, EPI_ISL_1470949, EPI_ISL_1470950, EPI_ISL_1470951, EPI_ISL_1470952, EPI_ISL_1470953, EPI_ISL_1470954,
EPI_ISL_1470955, EPI_ISL_1470956, EPI_ISL_1470957, EPI_ISL_1470958, EPI_ISL_1470959, EPI_ISL_1470960, EPI_ISL_1470961, EPI_ISL_1470962, EPI_ISL_1470963, EPI_ISL_1470964, EPI_ISL_1470965, EPI_ISL_1470966, EPI_ISL_1470967, EPI_ISL_1470968, EPI_ISL_1470969, EPI_ISL_1470970, EPI_ISL_1470971, EPI_ISL_1470972, EPI_ISL_1470973, EPI_ISL_1470974, EPI_ISL_1470975, EPI_ISL_1470976,
EPI_ISL_1470977, EPI_ISL_1470978, EPI_ISL_1470979, EPI_ISL_1470980, EPI_ISL_1470981, EPI_ISL_1470982, EPI_ISL_1470983, EPI_ISL_1470984, EPI_ISL_1470985, EPI_ISL_1470986, EPI_ISL_1470987, EPI_ISL_1470988, EPI_ISL_1470989, EPI_ISL_1470990, EPI_ISL_1470991, EPI_ISL_1470992, EPI_ISL_1470993, EPI_ISL_1470994, EPI_ISL_1470995, EPI_ISL_1470996, EPI_ISL_1470997, EPI_ISL_1470998,
EPI_ISL_1470999, EPI_ISL_1471000, EPI_ISL_1471001, EPI_ISL_1471002, EPI_ISL_1471003, EPI_ISL_1471004, EPI_ISL_1471005, EPI_ISL_1471006, EPI_ISL_1471007, EPI_ISL_1471008, EPI_ISL_1471009, EPI_ISL_1471010, EPI_ISL_1471011, EPI_ISL_1471012, EPI_ISL_1471013, EPI_ISL_1471014, EPI_ISL_1471015, EPI_ISL_1471016, EPI_ISL_1471017, EPI_ISL_1471018, EPI_ISL_1471019, EPI_ISL_1471020,
EPI_ISL_1471021, EPI_ISL_1471022, EPI_ISL_1471023, EPI_ISL_1471024, EPI_ISL_1471025, EPI_ISL_1471026, EPI_ISL_1471027, EPI_ISL_1471028, EPI_ISL_1471029, EPI_ISL_1471030, EPI_ISL_1471031, EPI_ISL_1471032, EPI_ISL_1471033, EPI_ISL_1471034, EPI_ISL_1471035, EPI_ISL_1471036, EPI_ISL_1471037, EPI_ISL_1471038, EPI_ISL_1471039, EPI_ISL_1471040, EPI_ISL_1471041, EPI_ISL_1471042,
EPI_ISL_1471043, EPI_ISL_1471044, EPI_ISL_1471045, EPI_ISL_1471046, EPI_ISL_1471047, EPI_ISL_1471048, EPI_ISL_1471049, EPI_ISL_1471050, EPI_ISL_1471051, EPI_ISL_1471052, EPI_ISL_1471053, EPI_ISL_1471054, EPI_ISL_1471055, EPI_ISL_1471056, EPI_ISL_1471057, EPI_ISL_1471058, EPI_ISL_1471059, EPI_ISL_1471060, EPI_ISL_1471061, EPI_ISL_1471062, EPI_ISL_1471063, EPI_ISL_1471064,
EPI_ISL_1471065, EPI_ISL_1471066, EPI_ISL_1471067, EPI_ISL_1471068, EPI_ISL_1471069, EPI_ISL_1471070, EPI_ISL_1471071, EPI_ISL_1471072, EPI_ISL_1471073, EPI_ISL_1471074, EPI_ISL_1471075, EPI_ISL_1471076, EPI_ISL_1471077, EPI_ISL_1471078, EPI_ISL_1471079, EPI_ISL_1471080, EPI_ISL_1471081, EPI_ISL_1471082, EPI_ISL_1471083, EPI_ISL_1471084, EPI_ISL_1471085, EPI_ISL_1471086,
EPI_ISL_1471087, EPI_ISL_1471088, EPI_ISL_1471089, EPI_ISL_1471090, EPI_ISL_1471091, EPI_ISL_1471092, EPI_ISL_1471093, EPI_ISL_1471094, EPI_ISL_1471095, EPI_ISL_1471096, EPI_ISL_1471097, EPI_ISL_1471098, EPI_ISL_1471099, EPI_ISL_1471100, EPI_ISL_1471101, EPI_ISL_1471102, EPI_ISL_1471103, EPI_ISL_1471104, EPI_ISL_1471105, EPI_ISL_1471106, EPI_ISL_1471107, EPI_ISL_1471108,
EPI_ISL_1471109, EPI_ISL_1471110, EPI_ISL_1471111, EPI_ISL_1471112, EPI_ISL_1471113, EPI_ISL_1471114, EPI_ISL_1471115, EPI_ISL_1471116, EPI_ISL_1471117, EPI_ISL_1471118, EPI_ISL_1471119, EPI_ISL_1471120, EPI_ISL_1471121, EPI_ISL_1471122, EPI_ISL_1471123, EPI_ISL_1471124, EPI_ISL_1471125, EPI_ISL_1471126, EPI_ISL_1471127, EPI_ISL_1471128, EPI_ISL_1471129, EPI_ISL_1471130,
EPI_ISL_1471131, EPI_ISL_1471132, EPI_ISL_1471133, EPI_ISL_1471134, EPI_ISL_1471135, EPI_ISL_1471136, EPI_ISL_1471137, EPI_ISL_1471138, EPI_ISL_1471139, EPI_ISL_1471140, EPI_ISL_1471141, EPI_ISL_1471142, EPI_ISL_1471143, EPI_ISL_1471144, EPI_ISL_1471145, EPI_ISL_1471146, EPI_ISL_1471147, EPI_ISL_1471148, EPI_ISL_1471149, EPI_ISL_1471150, EPI_ISL_1471151, EPI_ISL_1471152,
EPI_ISL_1471153, EPI_ISL_1471154, EPI_ISL_1471155, EPI_ISL_1471156, EPI_ISL_1471157, EPI_ISL_1471158, EPI_ISL_1471159, EPI_ISL_1471160, EPI_ISL_1471161, EPI_ISL_1471162, EPI_ISL_1471163, EPI_ISL_1471164, EPI_ISL_1471165, EPI_ISL_1471166, EPI_ISL_1471167, EPI_ISL_1471168, EPI_ISL_1471169, EPI_ISL_1471170, EPI_ISL_1471171, EPI_ISL_1471172, EPI_ISL_1471173, EPI_ISL_1471174,
EPI_ISL_1471175, EPI_ISL_1471176, EPI_ISL_1471177, EPI_ISL_1471178, EPI_ISL_1471179, EPI_ISL_1471180, EPI_ISL_1471181, EPI_ISL_1471182, EPI_ISL_1471183, EPI_ISL_1471184, EPI_ISL_1471185, EPI_ISL_1471186, EPI_ISL_1471187, EPI_ISL_1471188, EPI_ISL_1471189, EPI_ISL_1471190, EPI_ISL_1471191, EPI_ISL_1471192, EPI_ISL_1471193, EPI_ISL_1471194, EPI_ISL_1471195, EPI_ISL_1471196,
EPI_ISL_1471197, EPI_ISL_1471198, EPI_ISL_1471199, EPI_ISL_1471200, EPI_ISL_1471201, EPI_ISL_1471202, EPI_ISL_1471203, EPI_ISL_1471204, EPI_ISL_1471205, EPI_ISL_1471206, EPI_ISL_1471207, EPI_ISL_1471208, EPI_ISL_1471209, EPI_ISL_1471210, EPI_ISL_1471211, EPI_ISL_1471212, EPI_ISL_1471213, EPI_ISL_1471214, EPI_ISL_1471215, EPI_ISL_1471216, EPI_ISL_1471217, EPI_ISL_1471218,
EPI_ISL_1471219, EPI_ISL_1471220, EPI_ISL_1471221, EPI_ISL_1471222, EPI_ISL_1471223, EPI_ISL_1471224, EPI_ISL_1471225, EPI_ISL_1471226, EPI_ISL_1471227, EPI_ISL_1471228, EPI_ISL_1471229, EPI_ISL_1471230, EPI_ISL_1471231, EPI_ISL_1471232, EPI_ISL_1471233, EPI_ISL_1471234, EPI_ISL_1471235, EPI_ISL_1471236, EPI_ISL_1471237, EPI_ISL_1471238, EPI_ISL_1471239, EPI_ISL_1471240,
EPI_ISL_1471241, EPI_ISL_1471242, EPI_ISL_1471243, EPI_ISL_1471244, EPI_ISL_1471245, EPI_ISL_1471246, EPI_ISL_1471247, EPI_ISL_1471248, EPI_ISL_1471249, EPI_ISL_1471250, EPI_ISL_1471251, EPI_ISL_1471252, EPI_ISL_1471253, EPI_ISL_1471254, EPI_ISL_1471255, EPI_ISL_1471256, EPI_ISL_1471257, EPI_ISL_1471258, EPI_ISL_1471259, EPI_ISL_1471260, EPI_ISL_1471261, EPI_ISL_1471262,
EPI_ISL_1471263, EPI_ISL_1471264, EPI_ISL_1471265, EPI_ISL_1471266, EPI_ISL_1471267, EPI_ISL_1471268, EPI_ISL_1471269, EPI_ISL_1471270, EPI_ISL_1471271, EPI_ISL_1471272, EPI_ISL_1471273, EPI_ISL_1471274, EPI_ISL_1471275, EPI_ISL_1471276, EPI_ISL_1471277, EPI_ISL_1471278, EPI_ISL_1471279, EPI_ISL_1471280, EPI_ISL_1471281, EPI_ISL_1471282, EPI_ISL_1471283, EPI_ISL_1471284,
EPI_ISL_1471285, EPI_ISL_1471286, EPI_ISL_1471287, EPI_ISL_1471288, EPI_ISL_1471289, EPI_ISL_1471290, EPI_ISL_1471291, EPI_ISL_1471292, EPI_ISL_1471293, EPI_ISL_1471294, EPI_ISL_1471295, EPI_ISL_1471296, EPI_ISL_1471297, EPI_ISL_1471298, EPI_ISL_1471299, EPI_ISL_1471300, EPI_ISL_1471301, EPI_ISL_1471302, EPI_ISL_1471303, EPI_ISL_1471304, EPI_ISL_1471305, EPI_ISL_1471306,
EPI_ISL_1471307, EPI_ISL_1471308, EPI_ISL_1471309, EPI_ISL_1471310, EPI_ISL_1471311, EPI_ISL_1471312, EPI_ISL_1471313, EPI_ISL_1471314, EPI_ISL_1471315, EPI_ISL_1471316, EPI_ISL_1471317, EPI_ISL_1471318, EPI_ISL_1471319, EPI_ISL_1471320, EPI_ISL_1471321, EPI_ISL_1471322, EPI_ISL_1471323, EPI_ISL_1471324, EPI_ISL_1471325, EPI_ISL_1471326, EPI_ISL_1471327, EPI_ISL_1471328,
EPI_ISL_1471329, EPI_ISL_1471330, EPI_ISL_1471331, EPI_ISL_1471332, EPI_ISL_1471333, EPI_ISL_1471334, EPI_ISL_1471335, EPI_ISL_1471336, EPI_ISL_1471337, EPI_ISL_1471338, EPI_ISL_1471339, EPI_ISL_1471340, EPI_ISL_1471341, EPI_ISL_1471342, EPI_ISL_1471343, EPI_ISL_1471344, EPI_ISL_1471345, EPI_ISL_1471346, EPI_ISL_1471347, EPI_ISL_1471348, EPI_ISL_1471349, EPI_ISL_1471350,
EPI_ISL_1471351, EPI_ISL_1471352, EPI_ISL_1471353, EPI_ISL_1471354, EPI_ISL_1471355, EPI_ISL_1471356, EPI_ISL_1471357, EPI_ISL_1471358, EPI_ISL_1471359, EPI_ISL_1471360, EPI_ISL_1471361, EPI_ISL_1471362, EPI_ISL_1471363, EPI_ISL_1471364, EPI_ISL_1471365, EPI_ISL_1471366, EPI_ISL_1471367, EPI_ISL_1471368, EPI_ISL_1471369, EPI_ISL_1471370, EPI_ISL_1471371, EPI_ISL_1471372,
EPI_ISL_1471373, EPI_ISL_1471374, EPI_ISL_1471375, EPI_ISL_1471376, EPI_ISL_1471377, EPI_ISL_1471378, EPI_ISL_1471379, EPI_ISL_1471380, EPI_ISL_1471381, EPI_ISL_1471382, EPI_ISL_1471383, EPI_ISL_1471384, EPI_ISL_1471385, EPI_ISL_1471386, EPI_ISL_1471387, EPI_ISL_1471388, EPI_ISL_1471389, EPI_ISL_1471390, EPI_ISL_1471391, EPI_ISL_1471392, EPI_ISL_1471393, EPI_ISL_1471394,
EPI_ISL_1471395, EPI_ISL_1471396, EPI_ISL_1471397, EPI_ISL_1471398, EPI_ISL_1471399, EPI_ISL_1471400, EPI_ISL_1471401, EPI_ISL_1471402, EPI_ISL_1471403, EPI_ISL_1471404, EPI_ISL_1471405, EPI_ISL_1471406, EPI_ISL_1471407, EPI_ISL_1471408, EPI_ISL_1471409, EPI_ISL_1471410, EPI_ISL_1471411, EPI_ISL_1471412, EPI_ISL_1471413, EPI_ISL_1471414, EPI_ISL_1471415, EPI_ISL_1471416,
EPI_ISL_1471417, EPI_ISL_1471418, EPI_ISL_1471419, EPI_ISL_1471420, EPI_ISL_1471421, EPI_ISL_1471422, EPI_ISL_1471423, EPI_ISL_1471424, EPI_ISL_1471425, EPI_ISL_1471426, EPI_ISL_1471427, EPI_ISL_1471428, EPI_ISL_1471429, EPI_ISL_1471430, EPI_ISL_1471431, EPI_ISL_1471432, EPI_ISL_1471433, EPI_ISL_1471434, EPI_ISL_1471435, EPI_ISL_1471436, EPI_ISL_1471437, EPI_ISL_1471438,
EPI_ISL_1471439, EPI_ISL_1471440, EPI_ISL_1471441, EPI_ISL_1471442, EPI_ISL_1471443, EPI_ISL_1471444, EPI_ISL_1471445, EPI_ISL_1471446, EPI_ISL_1471447, EPI_ISL_1471448, EPI_ISL_1471449, EPI_ISL_1471450, EPI_ISL_1471451, EPI_ISL_1471452, EPI_ISL_1471453, EPI_ISL_1471454, EPI_ISL_1471455, EPI_ISL_1471456, EPI_ISL_1471457, EPI_ISL_1471458, EPI_ISL_1471459, EPI_ISL_1471460,
EPI_ISL_1471461, EPI_ISL_1471462, EPI_ISL_1471463, EPI_ISL_1471464, EPI_ISL_1471465, EPI_ISL_1471466, EPI_ISL_1471467, EPI_ISL_1471468, EPI_ISL_1471469, EPI_ISL_1471470, EPI_ISL_1471471, EPI_ISL_1471472, EPI_ISL_1471473, EPI_ISL_1471474, EPI_ISL_1471475, EPI_ISL_1471476, EPI_ISL_1471477, EPI_ISL_1471478, EPI_ISL_1471479, EPI_ISL_1471480, EPI_ISL_1471481, EPI_ISL_1471482,
EPI_ISL_1471483, EPI_ISL_1471484, EPI_ISL_1471485, EPI_ISL_1471486, EPI_ISL_1471487, EPI_ISL_1471488, EPI_ISL_1471489, EPI_ISL_1471490, EPI_ISL_1471491, EPI_ISL_1471492, EPI_ISL_1471493, EPI_ISL_1471494, EPI_ISL_1471495, EPI_ISL_1471496, EPI_ISL_1471497, EPI_ISL_1471498, EPI_ISL_1471499, EPI_ISL_1471500, EPI_ISL_1471501, EPI_ISL_1471502, EPI_ISL_1471503, EPI_ISL_1471504,
EPI_ISL_1471505, EPI_ISL_1471506, EPI_ISL_1471507, EPI_ISL_1471508, EPI_ISL_1471509, EPI_ISL_1471510, EPI_ISL_1471511, EPI_ISL_1471512, EPI_ISL_1471513, EPI_ISL_1471514, EPI_ISL_1471515, EPI_ISL_1471516, EPI_ISL_1471517, EPI_ISL_1471518, EPI_ISL_1471519, EPI_ISL_1471520, EPI_ISL_1471521, EPI_ISL_1471522, EPI_ISL_1471523, EPI_ISL_1471524, EPI_ISL_1471525, EPI_ISL_1471526,
EPI_ISL_1471527, EPI_ISL_1471528, EPI_ISL_1471529, EPI_ISL_1471530, EPI_ISL_1471531, EPI_ISL_1471532, EPI_ISL_1471533, EPI_ISL_1471534, EPI_ISL_1471535, EPI_ISL_1471536, EPI_ISL_1471537, EPI_ISL_1471538, EPI_ISL_1471539, EPI_ISL_1471540, EPI_ISL_1471541, EPI_ISL_1471542, EPI_ISL_1471543, EPI_ISL_1471544, EPI_ISL_1471545, EPI_ISL_1471546, EPI_ISL_1471547, EPI_ISL_1471548,
EPI_ISL_1471549, EPI_ISL_1471550, EPI_ISL_1471551, EPI_ISL_1471552, EPI_ISL_1471553, EPI_ISL_1471554, EPI_ISL_1471555, EPI_ISL_1471556, EPI_ISL_1471557, EPI_ISL_1471558, EPI_ISL_1471559, EPI_ISL_1471560, EPI_ISL_1471561, EPI_ISL_1471562, EPI_ISL_1471563, EPI_ISL_1471564, EPI_ISL_1471565, EPI_ISL_1471566, EPI_ISL_1471567, EPI_ISL_1471568, EPI_ISL_1471569, EPI_ISL_1471570,
EPI_ISL_1471571, EPI_ISL_1471572, EPI_ISL_1471573, EPI_ISL_1471574, EPI_ISL_1471575, EPI_ISL_1471576, EPI_ISL_1471577, EPI_ISL_1471578, EPI_ISL_1471579, EPI_ISL_1471580, EPI_ISL_1471581, EPI_ISL_1471582, EPI_ISL_1471583, EPI_ISL_1471584, EPI_ISL_1471585, EPI_ISL_1471586, EPI_ISL_1471587, EPI_ISL_1471588, EPI_ISL_1471589, EPI_ISL_1471590, EPI_ISL_1471591, EPI_ISL_1471592,
EPI_ISL_1471593, EPI_ISL_1471594, EPI_ISL_1471595, EPI_ISL_1471596, EPI_ISL_1471597, EPI_ISL_1471598, EPI_ISL_1471599, EPI_ISL_1471600, EPI_ISL_1471601, EPI_ISL_1471602, EPI_ISL_1471603, EPI_ISL_1471604, EPI_ISL_1471605, EPI_ISL_1471606, EPI_ISL_1471607, EPI_ISL_1471608, EPI_ISL_1471609, EPI_ISL_1471610, EPI_ISL_1471611, EPI_ISL_1471612, EPI_ISL_1471613, EPI_ISL_1471614,
EPI_ISL_1471615, EPI_ISL_1471616, EPI_ISL_1471617, EPI_ISL_1471618, EPI_ISL_1471619, EPI_ISL_1471620, EPI_ISL_1471621, EPI_ISL_1471622, EPI_ISL_1471623, EPI_ISL_1471624, EPI_ISL_1471625, EPI_ISL_1471626, EPI_ISL_1471627, EPI_ISL_1471628, EPI_ISL_1471629, EPI_ISL_1471630, EPI_ISL_1471631, EPI_ISL_1471632, EPI_ISL_1471633, EPI_ISL_1471634, EPI_ISL_1471635, EPI_ISL_1471636,
EPI_ISL_1471637, EPI_ISL_1471638, EPI_ISL_1471639, EPI_ISL_1471640, EPI_ISL_1471641, EPI_ISL_1471642, EPI_ISL_1471643, EPI_ISL_1471644, EPI_ISL_1471645, EPI_ISL_1471646, EPI_ISL_1471647, EPI_ISL_1471648, EPI_ISL_1471649, EPI_ISL_1471650, EPI_ISL_1471651, EPI_ISL_1471652, EPI_ISL_1471653, EPI_ISL_1471654, EPI_ISL_1471655, EPI_ISL_1471656, EPI_ISL_1471657, EPI_ISL_1471658,

EPI_ISL_1471659, EPI_ISL_1471660, EPI_ISL_1471661, EPI_ISL_1471662, EPI_ISL_1471663, EPI_ISL_1471664, EPI_ISL_1471665, EPI_ISL_1471666, EPI_ISL_1471667, EPI_ISL_1471668, EPI_ISL_1471669, EPI_ISL_1471670, EPI_ISL_1471671, EPI_ISL_1471672, EPI_ISL_1471673, EPI_ISL_1471674, EPI_ISL_1471675, EPI_ISL_1471676, EPI_ISL_1471677, EPI_ISL_1471678, EPI_ISL_1471679, EPI_ISL_1471680, EPI_ISL_1471681, EPI_ISL_1471682, EPI_ISL_1471683, EPI_ISL_1471684, EPI_ISL_1471685, EPI_ISL_1471686, EPI_ISL_1471687, EPI_ISL_1471688, EPI_ISL_1471689, EPI_ISL_1471690, EPI_ISL_1471691, EPI_ISL_1471692, EPI_ISL_1471693, EPI_ISL_1471694, EPI_ISL_1471695, EPI_ISL_1471696, EPI_ISL_1471697, EPI_ISL_1471698, EPI_ISL_1471699, EPI_ISL_1471700, EPI_ISL_1471701, EPI_ISL_1471702, EPI_ISL_1471703, EPI_ISL_1471704, EPI_ISL_1471705, EPI_ISL_1471706, EPI_ISL_1471707, EPI_ISL_1471708, EPI_ISL_1471709, EPI_ISL_1471710, EPI_ISL_1471711, EPI_ISL_1471712, EPI_ISL_1471713, EPI_ISL_1471714, EPI_ISL_1471715, EPI_ISL_1471716, EPI_ISL_1471717, EPI_ISL_1471718, EPI_ISL_1471719, EPI_ISL_1471720, EPI_ISL_1471721, EPI_ISL_1471722, EPI_ISL_1471723, EPI_ISL_1471724, EPI_ISL_1471725, EPI_ISL_1471726, EPI_ISL_1471727, EPI_ISL_1471728, EPI_ISL_1471729, EPI_ISL_1471730, EPI_ISL_1471731, EPI_ISL_1471732, EPI_ISL_1471733, EPI_ISL_1471734, EPI_ISL_1471735, EPI_ISL_1471736, EPI_ISL_1471737, EPI_ISL_1471738, EPI_ISL_1471739, EPI_ISL_1471740, EPI_ISL_1471741, EPI_ISL_1471742, EPI_ISL_1471743, EPI_ISL_1471744, EPI_ISL_1471745, EPI_ISL_1471746, EPI_ISL_1471747, EPI_ISL_1471748, EPI_ISL_1471749, EPI_ISL_1471750, EPI_ISL_1471751, EPI_ISL_1471752, EPI_ISL_1471753, EPI_ISL_1471754, EPI_ISL_1471755, EPI_ISL_1471756, EPI_ISL_1471757, EPI_ISL_1471758, EPI_ISL_1471759, EPI_ISL_1471760, EPI_ISL_1471761, EPI_ISL_1471762, EPI_ISL_1471763, EPI_ISL_1471764, EPI_ISL_1471765, EPI_ISL_1471766, EPI_ISL_1471767, EPI_ISL_1471768, EPI_ISL_1471769, EPI_ISL_1471770, EPI_ISL_1471771, EPI_ISL_1471772, EPI_ISL_1471773, EPI_ISL_1471774, EPI_ISL_1471775, EPI_ISL_1471776, EPI_ISL_1471777, EPI_ISL_1471778, EPI_ISL_1471779, EPI_ISL_1471780, EPI_ISL_1471781, EPI_ISL_1471782, EPI_ISL_1471783, EPI_ISL_1471784, EPI_ISL_1471785, EPI_ISL_1471786, EPI_ISL_1471787, EPI_ISL_1471788, EPI_ISL_1471789, EPI_ISL_1471790, EPI_ISL_1471791, EPI_ISL_1471792, EPI_ISL_1471793, EPI_ISL_1471794, EPI_ISL_1471795, EPI_ISL_1471796, EPI_ISL_1471797, EPI_ISL_1471798, EPI_ISL_1471799, EPI_ISL_1471800, EPI_ISL_1471801, EPI_ISL_1471802, EPI_ISL_1471803, EPI_ISL_1471804, EPI_ISL_1471805, EPI_ISL_1471806, EPI_ISL_1471807, EPI_ISL_1471808, EPI_ISL_1471809, EPI_ISL_1471810, EPI_ISL_1471811, EPI_ISL_1471812, EPI_ISL_1471813, EPI_ISL_1471814, EPI_ISL_1471815, EPI_ISL_1471816, EPI_ISL_1471817, EPI_ISL_1471818, EPI_ISL_1471819, EPI_ISL_1471820, EPI_ISL_1471821, EPI_ISL_1471822, EPI_ISL_1471823, EPI_ISL_1471824, EPI_ISL_1471825, EPI_ISL_1471826, EPI_ISL_1471827, EPI_ISL_1471828, EPI_ISL_1471829, EPI_ISL_1471830, EPI_ISL_1471831, EPI_ISL_1471832, EPI_ISL_1471833, EPI_ISL_1471834, EPI_ISL_1471835, EPI_ISL_1471836, EPI_ISL_1471837, EPI_ISL_1471838, EPI_ISL_1471839, EPI_ISL_1471840, EPI_ISL_1471841, EPI_ISL_1471842, EPI_ISL_1471843, EPI_ISL_1471844, EPI_ISL_1471845, EPI_ISL_1471846, EPI_ISL_1471847, EPI_ISL_1471848, EPI_ISL_1471849, EPI_ISL_1471850, EPI_ISL_1471851, EPI_ISL_1471852, EPI_ISL_1471853, EPI_ISL_1471854, EPI_ISL_1471855, EPI_ISL_1471856, EPI_ISL_1471857, EPI_ISL_1471858, EPI_ISL_1471860, EPI_ISL_1471861, EPI_ISL_1471862, EPI_ISL_1471863, EPI_ISL_1471864, EPI_ISL_1471865, EPI_ISL_1471866, EPI_ISL_1471867, EPI_ISL_1471868, EPI_ISL_1471871, EPI_ISL_1471872, EPI_ISL_1471873, EPI_ISL_1471874, EPI_ISL_1471875, EPI_ISL_1471876, EPI_ISL_1471877, EPI_ISL_1471878, EPI_ISL_1471879, EPI_ISL_1471881, EPI_ISL_1471885, EPI_ISL_1471886, EPI_ISL_1471887, EPI_ISL_1471888, EPI_ISL_1471889, EPI_ISL_1471890, EPI_ISL_1471891, EPI_ISL_1471893, EPI_ISL_1471894, EPI_ISL_1471896, EPI_ISL_1471897, EPI_ISL_1471899, EPI_ISL_1471906, EPI_ISL_1471907, EPI_ISL_1471908, EPI_ISL_1471912, EPI_ISL_1471913, EPI_ISL_1471914, EPI_ISL_1471915, EPI_ISL_1471916, EPI_ISL_1471918, EPI_ISL_1471919, EPI_ISL_1471920, EPI_ISL_1471921, EPI_ISL_1471922, EPI_ISL_1471923, EPI_ISL_1471924, EPI_ISL_1471926, EPI_ISL_1471927, EPI_ISL_1471928, EPI_ISL_1471930, EPI_ISL_1471931, EPI_ISL_1471933, EPI_ISL_1471935, EPI_ISL_1471936, EPI_ISL_1471937, EPI_ISL_1471939, EPI_ISL_1471941, EPI_ISL_1471942, EPI_ISL_1471944, EPI_ISL_1471945, EPI_ISL_1471946, EPI_ISL_1471948, EPI_ISL_1471951, EPI_ISL_1471952, EPI_ISL_1471954, EPI_ISL_1471956, EPI_ISL_1471957, EPI_ISL_1471959, EPI_ISL_1471960, EPI_ISL_1471961, EPI_ISL_1471963, EPI_ISL_1471964, EPI_ISL_1471965, EPI_ISL_1471966, EPI_ISL_1471967, EPI_ISL_1471968, EPI_ISL_1471969, EPI_ISL_1471972, EPI_ISL_1471977, EPI_ISL_1471979, EPI_ISL_1471980, EPI_ISL_1471981, EPI_ISL_1471982, EPI_ISL_1471983, EPI_ISL_1471985, EPI_ISL_1471987, EPI_ISL_1471988, EPI_ISL_1471990, EPI_ISL_1471991, EPI_ISL_1471992, EPI_ISL_1471996, EPI_ISL_1471999, EPI_ISL_1472000, EPI_ISL_1472003, EPI_ISL_1472004, EPI_ISL_1472005, EPI_ISL_1472006, EPI_ISL_1472007, EPI_ISL_1472008, EPI_ISL_1472010, EPI_ISL_1472011, EPI_ISL_1472012, EPI_ISL_1472013, EPI_ISL_1472014, EPI_ISL_1472015, EPI_ISL_1472016, EPI_ISL_1472018, EPI_ISL_1472020, EPI_ISL_1472021, EPI_ISL_1472022, EPI_ISL_1472024, EPI_ISL_1472025, EPI_ISL_1472026, EPI_ISL_1472028, EPI_ISL_1472030, EPI_ISL_1472031, EPI_ISL_1472033, EPI_ISL_1472035, EPI_ISL_1472036, EPI_ISL_1472037, EPI_ISL_1472038, EPI_ISL_1472039, EPI_ISL_1472041, EPI_ISL_1472042, EPI_ISL_1472043, EPI_ISL_1472046, EPI_ISL_1472047, EPI_ISL_1472048, EPI_ISL_1472049, EPI_ISL_1472052, EPI_ISL_1472053, EPI_ISL_1472054, EPI_ISL_1472055, EPI_ISL_1472056, EPI_ISL_1472058, EPI_ISL_1472060, EPI_ISL_1472064, EPI_ISL_1472065, EPI_ISL_1472066, EPI_ISL_1472067, EPI_ISL_1472069, EPI_ISL_1472070, EPI_ISL_1472071, EPI_ISL_1472073, EPI_ISL_1472074, EPI_ISL_1472076, EPI_ISL_1542199, EPI_ISL_1542902, EPI_ISL_1542903, EPI_ISL_1542906, EPI_ISL_1542910, EPI_ISL_1542915, EPI_ISL_1542916, EPI_ISL_1542919, EPI_ISL_1542920, EPI_ISL_1542923, EPI_ISL_1542925, EPI_ISL_1542927, EPI_ISL_1542928, EPI_ISL_1542929, EPI_ISL_1542931, EPI_ISL_1542936, EPI_ISL_1542938, EPI_ISL_1542940, EPI_ISL_1542943, EPI_ISL_1542945, EPI_ISL_1542946, EPI_ISL_1542948, EPI_ISL_1542949, EPI_ISL_1542953, EPI_ISL_1542956, EPI_ISL_1542958, EPI_ISL_1542959, EPI_ISL_1542961, EPI_ISL_1542962, EPI_ISL_1542963, EPI_ISL_1542964, EPI_ISL_1542965, EPI_ISL_1542967, EPI_ISL_1542968, EPI_ISL_1542969, EPI_ISL_1542974, EPI_ISL_1542975, EPI_ISL_1542978, EPI_ISL_1542979, EPI_ISL_1542981, EPI_ISL_1542985, EPI_ISL_1542987, EPI_ISL_1542988, EPI_ISL_1542989, EPI_ISL_1542990, EPI_ISL_1542995, EPI_ISL_1542996, EPI_ISL_1543002, EPI_ISL_1543004, EPI_ISL_1543006, EPI_ISL_1543009, EPI_ISL_1543010, EPI_ISL_1543013, EPI_ISL_1543016, EPI_ISL_1543022, EPI_ISL_1543023, EPI_ISL_1543025, EPI_ISL_1543030, EPI_ISL_1543031, EPI_ISL_1543032, EPI_ISL_1543034, EPI_ISL_1543036, EPI_ISL_1543037, EPI_ISL_1543041, EPI_ISL_1543042, EPI_ISL_1543043, EPI_ISL_1543044, EPI_ISL_1543046, EPI_ISL_1543048, EPI_ISL_1543051, EPI_ISL_1543053, EPI_ISL_1543058, EPI_ISL_1543063, EPI_ISL_1543067, EPI_ISL_1543069, EPI_ISL_1543072, EPI_ISL_1543076, EPI_ISL_1543081, EPI_ISL_1543083, EPI_ISL_1543085, EPI_ISL_1543090, EPI_ISL_1543092, EPI_ISL_1543093, EPI_ISL_1543094, EPI_ISL_1543099, EPI_ISL_1543100, EPI_ISL_1543101, EPI_ISL_1543105, EPI_ISL_1543106, EPI_ISL_1543107, EPI_ISL_1543111, EPI_ISL_1543115, EPI_ISL_1543118

| | | | |
|---|---|---|---|
| see above | Pandemic Response Lab - NYC | Pandemic Response Lab, R&D | Cybill del Castillo; Dylan Law; Haiping Hao; Henry Lee; Jon Laurent; Katharine Nelson; Melissa Hopkins; Michael Hammerling; Pradeep Bugga; Shinyoung Clair Kang; Sol Rey; William Ward |
| EPI_ISL_1694720 | Poplar Healthcare | New York City Public Health Laboratory | Jade Wang; et al. |

EPI_ISL_1267456, EPI_ISL_1297766, EPI_ISL_1297769, EPI_ISL_1314552, EPI_ISL_1314553, EPI_ISL_1314556, EPI_ISL_1314568, EPI_ISL_1314573, EPI_ISL_1314589, EPI_ISL_1314590, EPI_ISL_1314608, EPI_ISL_1314613, EPI_ISL_1314624, EPI_ISL_1314630, EPI_ISL_1314643, EPI_ISL_1314646, EPI_ISL_1314659, EPI_ISL_1314664, EPI_ISL_1314677, EPI_ISL_1314700, EPI_ISL_1314708, EPI_ISL_1314709, EPI_ISL_1314739, EPI_ISL_1314760, EPI_ISL_1314782, EPI_ISL_1314809, EPI_ISL_1314812, EPI_ISL_1314813, EPI_ISL_1314829, EPI_ISL_1314830, EPI_ISL_1314914, EPI_ISL_1314922, EPI_ISL_1339842, EPI_ISL_1340452, EPI_ISL_1340472, EPI_ISL_1340475, EPI_ISL_1340478, EPI_ISL_1340511, EPI_ISL_1340521, EPI_ISL_1340735, EPI_ISL_1367423, EPI_ISL_1367441, EPI_ISL_1367453, EPI_ISL_1367467, EPI_ISL_1367479, EPI_ISL_1422804, EPI_ISL_1422813, EPI_ISL_1422814, EPI_ISL_1422823, EPI_ISL_1422859, EPI_ISL_1422864, EPI_ISL_1422886, EPI_ISL_1422888, EPI_ISL_1422902, EPI_ISL_1422905, EPI_ISL_1422953, EPI_ISL_1422954, EPI_ISL_1422959, EPI_ISL_1422974, EPI_ISL_1479876, EPI_ISL_1479881, EPI_ISL_1479891, EPI_ISL_1479911, EPI_ISL_1480604, EPI_ISL_1480608, EPI_ISL_1480617, EPI_ISL_1480632, EPI_ISL_1480654, EPI_ISL_1480859, EPI_ISL_1480943, EPI_ISL_1480971, EPI_ISL_1480976, EPI_ISL_1491965, EPI_ISL_1491986, EPI_ISL_1491994, EPI_ISL_1492112, EPI_ISL_1493627, EPI_ISL_1511714, EPI_ISL_1511768, EPI_ISL_1511769, EPI_ISL_1511770, EPI_ISL_1511774, EPI_ISL_1511776, EPI_ISL_1511779, EPI_ISL_1511781, EPI_ISL_1511782, EPI_ISL_1511783, EPI_ISL_1511789, EPI_ISL_1511791, EPI_ISL_1511793, EPI_ISL_1511794, EPI_ISL_1511797, EPI_ISL_1511802, EPI_ISL_1511805, EPI_ISL_1511809, EPI_ISL_1511813, EPI_ISL_1511814, EPI_ISL_1511815, EPI_ISL_1511816, EPI_ISL_1511817, EPI_ISL_1511820, EPI_ISL_1511821, EPI_ISL_1511822, EPI_ISL_1511824, EPI_ISL_1511829, EPI_ISL_1511830, EPI_ISL_1511831, EPI_ISL_1511832, EPI_ISL_1511834, EPI_ISL_1511836, EPI_ISL_1511838, EPI_ISL_1511842, EPI_ISL_1511846, EPI_ISL_1511848, EPI_ISL_1511850, EPI_ISL_1511852, EPI_ISL_1511856, EPI_ISL_1511858, EPI_ISL_1551720, EPI_ISL_1551723, EPI_ISL_1551725, EPI_ISL_1551741, EPI_ISL_1551746, EPI_ISL_1551766, EPI_ISL_1551772, EPI_ISL_1552235, EPI_ISL_1552259, EPI_ISL_1552275, EPI_ISL_1552278, EPI_ISL_1552301, EPI_ISL_1552713, EPI_ISL_1552918, EPI_ISL_1553024, EPI_ISL_1553038, EPI_ISL_1553044, EPI_ISL_1553049, EPI_ISL_1553053, EPI_ISL_1553056, EPI_ISL_1553061, EPI_ISL_1553064, EPI_ISL_1553069, EPI_ISL_1553070, EPI_ISL_1553071, EPI_ISL_1553075, EPI_ISL_1553076, EPI_ISL_1553080, EPI_ISL_1553082, EPI_ISL_1553085, EPI_ISL_1553086, EPI_ISL_1553087, EPI_ISL_1553092, EPI_ISL_1553093, EPI_ISL_1559420, EPI_ISL_1559430, EPI_ISL_1559442, EPI_ISL_1559557, EPI_ISL_1581871, EPI_ISL_1581898, EPI_ISL_1581931, EPI_ISL_1581934, EPI_ISL_1581962, EPI_ISL_1582034, EPI_ISL_1582039, EPI_ISL_1582051, EPI_ISL_1582058, EPI_ISL_1582066, EPI_ISL_1582100, EPI_ISL_1582109, EPI_ISL_1582116, EPI_ISL_1582119, EPI_ISL_1582133, EPI_ISL_1582142, EPI_ISL_1582148, EPI_ISL_1614918, EPI_ISL_1614935, EPI_ISL_1614940, EPI_ISL_1614948, EPI_ISL_1614957, EPI_ISL_1614977, EPI_ISL_1614981, EPI_ISL_1615021, EPI_ISL_1615032, EPI_ISL_1615037, EPI_ISL_1615039, EPI_ISL_1615047, EPI_ISL_1615061, EPI_ISL_1621744, EPI_ISL_1621807, EPI_ISL_1621850, EPI_ISL_1621854, EPI_ISL_1621859, EPI_ISL_1621866, EPI_ISL_1621873, EPI_ISL_1621876, EPI_ISL_1621884, EPI_ISL_1621906, EPI_ISL_1621909, EPI_ISL_1621925, EPI_ISL_1621928, EPI_ISL_1621945, EPI_ISL_1621947, EPI_ISL_1621956, EPI_ISL_1621958, EPI_ISL_1622003, EPI_ISL_1622005, EPI_ISL_1622008, EPI_ISL_1622038, EPI_ISL_1622055, EPI_ISL_1622059, EPI_ISL_1622079, EPI_ISL_1622103, EPI_ISL_1648211, EPI_ISL_1648236, EPI_ISL_1648386, EPI_ISL_1648392, EPI_ISL_1648401, EPI_ISL_1648417, EPI_ISL_1648437, EPI_ISL_1648461, EPI_ISL_1648538, EPI_ISL_1648757, EPI_ISL_1648829, EPI_ISL_1648842, EPI_ISL_1648885, EPI_ISL_1694671, EPI_ISL_1694672, EPI_ISL_1694673, EPI_ISL_1695037, EPI_ISL_1695051, EPI_ISL_1695067

| | | | |
|---|---|---|---|
| see above | Quest Diagnostics Incorporated | Centers for Disease Control and Prevention Division of Viral Diseases, Pathogen Discovery | A. Gerasimova; A. Perez; Adrian Paskey; B. Anderson; Ben L. Rambo-Martin; Benjamin Rambo-Martin; Christopher Gulvick; Clinton R. Paden; Dakota Howard; Darlene Wagner; Dhwani Batra; Duncan MacCannell; F. Lacbawan; I. A. Shlyakhter; Jason Caravas; K.E. Livingston; Kara Moser; L.E. Bernstein; M. Hua; Matthew Schmerer; P. Tanpaiboon; Peter W. Cook; R. M. Kagan; R. Owen; R. V. Rolando; S. H. Rosenthal; Scott Sammons; Shatavia Morrison; Suxiang Tong; Y. Liu; Yvette Unoarumhi |

EPI_ISL_2313517, EPI_ISL_2313606, EPI_ISL_2313617, EPI_ISL_2313633, EPI_ISL_2313635, EPI_ISL_2313637, EPI_ISL_2313667, EPI_ISL_2313705, EPI_ISL_2313707, EPI_ISL_2313797, EPI_ISL_2313844

| | | | |
|---|---|---|---|
| see above | Respiratory Viruses Branch, Centers for Disease Control and Prevention | Respiratory Viruses Branch, Centers for Disease Control and Prevention | Batra, D.; Bixby, C.; C.R.; Caravas, J.; Cook; Goswami, C.; Grimwood, R.; Gulvick, C.; Hager, R.; Howard, D.; MacCannell, D.; Morrison, S.; Moser, K.; P.W.; Paden; Paskey, A.; Rambo-Martin, B.; Sammons, S.; Schmerer, M.; Schultz, J.; Unoarumhi, Y.; Wagner, D.; Wang, Y. |
| EPI_ISL_1447143 | Rockefeller University Hospital | New York City Public Health Laboratory | Jade Wang; et al. |

EPI_ISL_1580739, EPI_ISL_1580740, EPI_ISL_1582227, EPI_ISL_1582228, EPI_ISL_1582229, EPI_ISL_1624484, EPI_ISL_1624486, EPI_ISL_1651224, EPI_ISL_2098627

| | | | |
|---|---|---|---|
| see above | SARATOGA HOSPITAL LABORATORY | Wadsworth Center, New York State Department of Health | Alexis Russell; Catharine Prussing; Daryl M. Lamson; Erasmus Schneider; Erica Lasek-Nesselquist; John Kelly; Jonathan Plitnick; Kirsten St. George; Matthew Shudt; Melissa A Leisner; Navjot Singh |
| EPI_ISL_1482454, EPI_ISL_1582272, EPI_ISL_1671468 | ST. FRANCIS HOSPITAL LABORATORY | Wadsworth Center, New York State Department of Health | Alexis Russell; Catharine Prussing; Daryl M. Lamson; Erasmus Schneider; Erica Lasek-Nesselquist; John Kelly; Jonathan Plitnick; Kirsten St. George; Matthew Shudt; Melissa A Leisner; Navjot Singh |

EPI_ISL_1397831, EPI_ISL_1397832, EPI_ISL_1465838, EPI_ISL_1465839, EPI_ISL_1465840, EPI_ISL_1465841, EPI_ISL_1465842, EPI_ISL_1465843, EPI_ISL_1465844, EPI_ISL_1465845, EPI_ISL_1465846, EPI_ISL_1465847, EPI_ISL_1582245, EPI_ISL_1582246, EPI_ISL_1582247, EPI_ISL_1582248, EPI_ISL_1582249, EPI_ISL_1582250, EPI_ISL_1582251, EPI_ISL_1582252, EPI_ISL_1582253, EPI_ISL_1582254, EPI_ISL_1671322, EPI_ISL_1671328, EPI_ISL_1671356, EPI_ISL_1740508

| | | | |
|---|---|---|---|
| see above | STONY BROOK UNIVERSITY HOSPITAL | Wadsworth Center, New York State Department of Health | Alexis Russel; Alexis Russell; Catharine Prussing; Daryl M. Lamson; Erasmus Schneider; Erica Lasek-Nesselquist; John Kelly; Jonathan Plitnick; Kirsten St. George; Matthew Shudt; Melissa A Leisner; Navjot Singh |

EPI_ISL_1395751, EPI_ISL_1395752, EPI_ISL_1395755, EPI_ISL_1395759, EPI_ISL_1395761, EPI_ISL_1395762, EPI_ISL_1395764, EPI_ISL_1395765, EPI_ISL_1395767, EPI_ISL_1395770, EPI_ISL_1395772, EPI_ISL_1395774, EPI_ISL_1395775, EPI_ISL_1395776, EPI_ISL_1397594, EPI_ISL_1397595, EPI_ISL_1397596, EPI_ISL_1397597, EPI_ISL_1397598, EPI_ISL_1397599, EPI_ISL_1397600, EPI_ISL_1397602, EPI_ISL_1397603, EPI_ISL_1397608, EPI_ISL_1397784, EPI_ISL_1397785, EPI_ISL_1398156, EPI_ISL_1398157, EPI_ISL_1398158, EPI_ISL_1398159, EPI_ISL_1398160, EPI_ISL_1398161, EPI_ISL_1398162, EPI_ISL_1461294, EPI_ISL_1461310, EPI_ISL_1461311, EPI_ISL_1461312, EPI_ISL_1461313, EPI_ISL_1461314, EPI_ISL_1461315, EPI_ISL_1461316, EPI_ISL_1461317, EPI_ISL_1462500, EPI_ISL_1462502, EPI_ISL_1462503, EPI_ISL_1462505, EPI_ISL_1462536, EPI_ISL_1462537, EPI_ISL_1462539, EPI_ISL_1462540, EPI_ISL_1462542, EPI_ISL_1462543, EPI_ISL_1462545, EPI_ISL_1462546, EPI_ISL_1462548, EPI_ISL_1462549, EPI_ISL_1465638, EPI_ISL_1465665, EPI_ISL_1465666, EPI_ISL_1465689, EPI_ISL_1465690, EPI_ISL_1465691, EPI_ISL_1465692, EPI_ISL_1465697, EPI_ISL_1465698, EPI_ISL_1465699, EPI_ISL_1465700, EPI_ISL_1482274, EPI_ISL_1482275, EPI_ISL_1482276, EPI_ISL_1482406, EPI_ISL_1482407, EPI_ISL_1575445, EPI_ISL_1576812, EPI_ISL_1576813, EPI_ISL_1580678, EPI_ISL_1580679, EPI_ISL_1580680, EPI_ISL_1580681, EPI_ISL_1580682, EPI_ISL_1580683, EPI_ISL_1580684, EPI_ISL_1580685, EPI_ISL_1582318, EPI_ISL_1582360, EPI_ISL_1582361, EPI_ISL_1582362, EPI_ISL_1582363, EPI_ISL_1582364, EPI_ISL_1582365, EPI_ISL_1582366, EPI_ISL_1582367, EPI_ISL_1582399, EPI_ISL_1624406, EPI_ISL_1624407, EPI_ISL_1624408, EPI_ISL_1624420, EPI_ISL_1671463, EPI_ISL_1755489, EPI_ISL_1755490, EPI_ISL_1755842, EPI_ISL_1789709

| | | | |
|---|---|---|---|
| see above | SUNY UPSTATE MEDICAL UNIVERSITY | Wadsworth Center, New York State Department of Health | Alexis Russel; Alexis Russell; Catharine Prussing; Daryl M. Lamson; Erasmus Schneider; Erica Lasek-Nesselquist; John Kelly; Jonathan Plitnick; Kirsten St. George; Matthew Shudt; Melissa A Leisner; Navjot Singh |
| EPI_ISL_1967685, EPI_ISL_1967687 | Stony Brook Medicine | New York Genome Center | Alaa B. Salaheldin; Andre Corvelo; Dayna M. Oschwald; Dimitri Gnatenko; Eric D. Spitzer; Jiang Chen; Josue M. Flores Acuna; Kenneth R. Shroyer; Margaret M. McGovern; Michael Zody; Richard R. Kew; Samantha Fennessey; Sarah A. Halbohn; Sarah A. Malik; Silvia G. Spitzer; Tom Maniatis |

EPI_ISL_1397581, EPI_ISL_1397582, EPI_ISL_1397583, EPI_ISL_1397584, EPI_ISL_1397700, EPI_ISL_1397701, EPI_ISL_1397702, EPI_ISL_1397703, EPI_ISL_1397704, EPI_ISL_1397836, EPI_ISL_1397838, EPI_ISL_1397839, EPI_ISL_1397840, EPI_ISL_1397841, EPI_ISL_1397842, EPI_ISL_1397844, EPI_ISL_1397845, EPI_ISL_1397846, EPI_ISL_1397847, EPI_ISL_1397848, EPI_ISL_1397849, EPI_ISL_1397850, EPI_ISL_1397851, EPI_ISL_1397852, EPI_ISL_1397857, EPI_ISL_1397859, EPI_ISL_1397860, EPI_ISL_1397862, EPI_ISL_1397863, EPI_ISL_1397864, EPI_ISL_1397865, EPI_ISL_1461356, EPI_ISL_1461357, EPI_ISL_1461358, EPI_ISL_1461359, EPI_ISL_1461360, EPI_ISL_1461361, EPI_ISL_1461362, EPI_ISL_1461363, EPI_ISL_1461364, EPI_ISL_1461365, EPI_ISL_1461366, EPI_ISL_1461367, EPI_ISL_1461368, EPI_ISL_1461369, EPI_ISL_1461370, EPI_ISL_1461371, EPI_ISL_1461372, EPI_ISL_1461373, EPI_ISL_1461374, EPI_ISL_1461375, EPI_ISL_1461376, EPI_ISL_1461377, EPI_ISL_1462552, EPI_ISL_1465859, EPI_ISL_1465860, EPI_ISL_1465861, EPI_ISL_1465862, EPI_ISL_1465863, EPI_ISL_1465864, EPI_ISL_1465865, EPI_ISL_1465866, EPI_ISL_1465867, EPI_ISL_1465868, EPI_ISL_1465869, EPI_ISL_1465870, EPI_ISL_1465871, EPI_ISL_1465872, EPI_ISL_1465873, EPI_ISL_1465874, EPI_ISL_1465875, EPI_ISL_1465876, EPI_ISL_1465877, EPI_ISL_1465878, EPI_ISL_1575471, EPI_ISL_1576814, EPI_ISL_1576821, EPI_ISL_1576822, EPI_ISL_1582232, EPI_ISL_1582233, EPI_ISL_1582234, EPI_ISL_1582235, EPI_ISL_1582236, EPI_ISL_1582237, EPI_ISL_1582238, EPI_ISL_1582239, EPI_ISL_1582240, EPI_ISL_1582241, EPI_ISL_1582242, EPI_ISL_1582243, EPI_ISL_1582244, EPI_ISL_1582357, EPI_ISL_1582358, EPI_ISL_1582359, EPI_ISL_1582403, EPI_ISL_1593873, EPI_ISL_1593874, EPI_ISL_1593875, EPI_ISL_1593884, EPI_ISL_1593885, EPI_ISL_1605699

| | | | |
|---|---|---|---|
| see above | TEMPUS LABS INC | Wadsworth Center, New York State Department of Health | Alexis Russel; Alexis Russell; Catharine Prussing; Daryl M. Lamson; Erasmus Schneider; Erica Lasek-Nesselquist; John Kelly; Jonathan Plitnick; Kirsten St. George; Matthew Shudt; Melissa A Leisner; Navjot Singh |

EPI_ISL_1395766, EPI_ISL_1395768, EPI_ISL_1395769, EPI_ISL_1395771, EPI_ISL_1395773, EPI_ISL_1397613, EPI_ISL_1397617, EPI_ISL_1462572, EPI_ISL_1462573, EPI_ISL_1462575, EPI_ISL_1465673, EPI_ISL_1465674, EPI_ISL_1465675, EPI_ISL_1465676, EPI_ISL_1465677, EPI_ISL_1465760

| | | | |
|---|---|---|---|
| see above | THE MARY IMOGENE BASSETT HOSPITAL | Wadsworth Center, New York State Department of Health | Alexis Russel; Alexis Russell; Catharine Prussing; Daryl M. Lamson; Erasmus Schneider; Erica Lasek-Nesselquist; John Kelly; Jonathan Plitnick; Kirsten St. George; Matthew Shudt; Melissa A Leisner; Navjot Singh |

EPI_ISL_1378790, EPI_ISL_1378791, EPI_ISL_1378792, EPI_ISL_1378793, EPI_ISL_1378794, EPI_ISL_1378795, EPI_ISL_1378796

| | | | |
|---|---|---|---|
| see above | Tempus | Grubaugh Lab - Yale School of Public Health | Anderson Brito; Annie Watkins; Chaney Kalinich; Chantal Vogels; Christopher E. Mason; Gaurav Khullar; Isabell Ott; Jessica Metti; Jessica Rothman; Joel T. Dudley; Joseph Fauver; Mallery Breban; Mary Petrone; Matthew J. MacKay; Megan Nash; Nathan Grubaugh; Nike Beaubier; Tara Alpert |
| EPI_ISL_1470626 | The Jackson Laboratory | The Jackson Laboratory | Adams M; Bergeron D; Kelly K; Li L; Omerza G; Renzette N |

EPI_ISL_1397630, EPI_ISL_1397631, EPI_ISL_1397632, EPI_ISL_1397633, EPI_ISL_1397634, EPI_ISL_1397810, EPI_ISL_1397811, EPI_ISL_1397812, EPI_ISL_1397813, EPI_ISL_1397814, EPI_ISL_1397815, EPI_ISL_1397816, EPI_ISL_1397817, EPI_ISL_1397818, EPI_ISL_1397819, EPI_ISL_1397820, EPI_ISL_1397821, EPI_ISL_1397823, EPI_ISL_1398005, EPI_ISL_1398006, EPI_ISL_1398007, EPI_ISL_1398008, EPI_ISL_1398009, EPI_ISL_1398012, EPI_ISL_1398013, EPI_ISL_1398014, EPI_ISL_1398015, EPI_ISL_1398016, EPI_ISL_1398017, EPI_ISL_1398018, EPI_ISL_1398019, EPI_ISL_1398020, EPI_ISL_1398137, EPI_ISL_1398142, EPI_ISL_1398143, EPI_ISL_1398312, EPI_ISL_1398313, EPI_ISL_1398314, EPI_ISL_1398315, EPI_ISL_1398316, EPI_ISL_1398317, EPI_ISL_1461300, EPI_ISL_1461301, EPI_ISL_1461302, EPI_ISL_1461303, EPI_ISL_1461304, EPI_ISL_1461305, EPI_ISL_1461306, EPI_ISL_1461307, EPI_ISL_1461308, EPI_ISL_1461309, EPI_ISL_1462509, EPI_ISL_1462510, EPI_ISL_1462512, EPI_ISL_1462513, EPI_ISL_1462515, EPI_ISL_1462516, EPI_ISL_1462518, EPI_ISL_1462519, EPI_ISL_1462521, EPI_ISL_1462523, EPI_ISL_1462524, EPI_ISL_1462525, EPI_ISL_1462527, EPI_ISL_1462528, EPI_ISL_1462530, EPI_ISL_1462531, EPI_ISL_1462533, EPI_ISL_1462534, EPI_ISL_1465632, EPI_ISL_1465641, EPI_ISL_1465642, EPI_ISL_1465643, EPI_ISL_1465644, EPI_ISL_1465645, EPI_ISL_1465646, EPI_ISL_1465647, EPI_ISL_1465648, EPI_ISL_1465649, EPI_ISL_1465650, EPI_ISL_1465651, EPI_ISL_1465652, EPI_ISL_1465654, EPI_ISL_1465655, EPI_ISL_1465656, EPI_ISL_1465657, EPI_ISL_1465658, EPI_ISL_1465659, EPI_ISL_1465660, EPI_ISL_1465661, EPI_ISL_1465662, EPI_ISL_1465663, EPI_ISL_1465696, EPI_ISL_1465723, EPI_ISL_1465724, EPI_ISL_1465725, EPI_ISL_1465726, EPI_ISL_1465727, EPI_ISL_1465728, EPI_ISL_1465729, EPI_ISL_1465730, EPI_ISL_1465731, EPI_ISL_1465732, EPI_ISL_1465733, EPI_ISL_1465734, EPI_ISL_1465735, EPI_ISL_1465736, EPI_ISL_1465737, EPI_ISL_1465738, EPI_ISL_1465739, EPI_ISL_1465740, EPI_ISL_1465741, EPI_ISL_1465742, EPI_ISL_1465743, EPI_ISL_1465744, EPI_ISL_1465745, EPI_ISL_1465746, EPI_ISL_1465747, EPI_ISL_1465748, EPI_ISL_1465749, EPI_ISL_1465750, EPI_ISL_1465751, EPI_ISL_1465752, EPI_ISL_1465753, EPI_ISL_1465754, EPI_ISL_1482278, EPI_ISL_1482279, EPI_ISL_1482280, EPI_ISL_1482281, EPI_ISL_1482282, EPI_ISL_1482283, EPI_ISL_1482284, EPI_ISL_1482285, EPI_ISL_1482286, EPI_ISL_1482296, EPI_ISL_1482297, EPI_ISL_1482298, EPI_ISL_1482299, EPI_ISL_1482300, EPI_ISL_1482301, EPI_ISL_1482302, EPI_ISL_1482303, EPI_ISL_1482304, EPI_ISL_1482305, EPI_ISL_1482306, EPI_ISL_1482307, EPI_ISL_1482308, EPI_ISL_1482309, EPI_ISL_1482310, EPI_ISL_1482311, EPI_ISL_1482312, EPI_ISL_1482313, EPI_ISL_1482314, EPI_ISL_1482315, EPI_ISL_1482316, EPI_ISL_1482317, EPI_ISL_1482318, EPI_ISL_1482319, EPI_ISL_1482320, EPI_ISL_1482321, EPI_ISL_1482322, EPI_ISL_1482323, EPI_ISL_1482324, EPI_ISL_1482325, EPI_ISL_1482326, EPI_ISL_1482327, EPI_ISL_1482328, EPI_ISL_1482329, EPI_ISL_1482330, EPI_ISL_1482331, EPI_ISL_1482332, EPI_ISL_1482625, EPI_ISL_1482626, EPI_ISL_1575446, EPI_ISL_1575447, EPI_ISL_1575448, EPI_ISL_1575455, EPI_ISL_1575456, EPI_ISL_1575457, EPI_ISL_1575458, EPI_ISL_1575460, EPI_ISL_1575461, EPI_ISL_1575462, EPI_ISL_1575463, EPI_ISL_1575464, EPI_ISL_1575465, EPI_ISL_1575466, EPI_ISL_1575467, EPI_ISL_1575468, EPI_ISL_1575473, EPI_ISL_1575474, EPI_ISL_1575475, EPI_ISL_1575476, EPI_ISL_1576817, EPI_ISL_1576818, EPI_ISL_1576823, EPI_ISL_1580701, EPI_ISL_1580702, EPI_ISL_1580703, EPI_ISL_1580704, EPI_ISL_1580705, EPI_ISL_1580706, EPI_ISL_1580707, EPI_ISL_1580708, EPI_ISL_1580709, EPI_ISL_1580710, EPI_ISL_1580711, EPI_ISL_1580712, EPI_ISL_1580713, EPI_ISL_1580714, EPI_ISL_1580715, EPI_ISL_1580716, EPI_ISL_1580717, EPI_ISL_1580718, EPI_ISL_1580719, EPI_ISL_1580720, EPI_ISL_1580721, EPI_ISL_1580722, EPI_ISL_1582151, EPI_ISL_1582219, EPI_ISL_1582220, EPI_ISL_1582221, EPI_ISL_1582222, EPI_ISL_1582223, EPI_ISL_1582224, EPI_ISL_1582339, EPI_ISL_1582340, EPI_ISL_1582341, EPI_ISL_1582342, EPI_ISL_1582343, EPI_ISL_1582344, EPI_ISL_1582345, EPI_ISL_1582346, EPI_ISL_1582347, EPI_ISL_1582348, EPI_ISL_1582349, EPI_ISL_1582350, EPI_ISL_1582351, EPI_ISL_1582352, EPI_ISL_1582353, EPI_ISL_1582404, EPI_ISL_1593866, EPI_ISL_1593867, EPI_ISL_1593882, EPI_ISL_1624410, EPI_ISL_1624411, EPI_ISL_1624412, EPI_ISL_1624413, EPI_ISL_1624414, EPI_ISL_1624415, EPI_ISL_1624416, EPI_ISL_1624417, EPI_ISL_1624418, EPI_ISL_1624419, EPI_ISL_1624421, EPI_ISL_1624422, EPI_ISL_1624423, EPI_ISL_1624424, EPI_ISL_1624425, EPI_ISL_1624426, EPI_ISL_1624427, EPI_ISL_1624428, EPI_ISL_1624429, EPI_ISL_1624430, EPI_ISL_1624431, EPI_ISL_1624432, EPI_ISL_1624433, EPI_ISL_1624434, EPI_ISL_1624435, EPI_ISL_1624436, EPI_ISL_1624437, EPI_ISL_1624438, EPI_ISL_1624439, EPI_ISL_1624440, EPI_ISL_1624441, EPI_ISL_1624442, EPI_ISL_1624443, EPI_ISL_1624444, EPI_ISL_1624445, EPI_ISL_1624446, EPI_ISL_1624447, EPI_ISL_1624448, EPI_ISL_1624449, EPI_ISL_1624450, EPI_ISL_1624451, EPI_ISL_1624452, EPI_ISL_1624453, EPI_ISL_1624454, EPI_ISL_1624455, EPI_ISL_1624456, EPI_ISL_1624457, EPI_ISL_1624458, EPI_ISL_1624459, EPI_ISL_1624460, EPI_ISL_1624461, EPI_ISL_1624462, EPI_ISL_1624463, EPI_ISL_1624464, EPI_ISL_1624465, EPI_ISL_1624466, EPI_ISL_1624467, EPI_ISL_1624468, EPI_ISL_1624469, EPI_ISL_1624470, EPI_ISL_1624471, EPI_ISL_1624472, EPI_ISL_1624473, EPI_ISL_1624474, EPI_ISL_1624475, EPI_ISL_1624476, EPI_ISL_1624477, EPI_ISL_1624478, EPI_ISL_1624479, EPI_ISL_1624480, EPI_ISL_1624481, EPI_ISL_1624482, EPI_ISL_1651222, EPI_ISL_1651223, EPI_ISL_1651227, EPI_ISL_1651228, EPI_ISL_1671613, EPI_ISL_1755835, EPI_ISL_1755876, EPI_ISL_1755877, EPI_ISL_1755878, EPI_ISL_1755879, EPI_ISL_1755880, EPI_ISL_1755881, EPI_ISL_1755882, EPI_ISL_1755883, EPI_ISL_1755884, EPI_ISL_1755896, EPI_ISL_1755904, EPI_ISL_1920066, EPI_ISL_1920067, EPI_ISL_1920068, EPI_ISL_1920129, EPI_ISL_1920131, EPI_ISL_1935618, EPI_ISL_2098161, EPI_ISL_2098162, EPI_ISL_2098164, EPI_ISL_2098166, EPI_ISL_2098167, EPI_ISL_2098169, EPI_ISL_2098170, EPI_ISL_2098172, EPI_ISL_2098173, EPI_ISL_2098175, EPI_ISL_2098176, EPI_ISL_2098178, EPI_ISL_2098180, EPI_ISL_2098181, EPI_ISL_2098183, EPI_ISL_2098184, EPI_ISL_2098186, EPI_ISL_2098187, EPI_ISL_2098189, EPI_ISL_2098191, EPI_ISL_2098192, EPI_ISL_2098193, EPI_ISL_2098195, EPI_ISL_2098196, EPI_ISL_2098198, EPI_ISL_2098199, EPI_ISL_2098201, EPI_ISL_2098202, EPI_ISL_2098204, EPI_ISL_2098206, EPI_ISL_2098208, EPI_ISL_2098209, EPI_ISL_2098211, EPI_ISL_2098212, EPI_ISL_2098214, EPI_ISL_2098215, EPI_ISL_2098217, EPI_ISL_2098222, EPI_ISL_2098227, EPI_ISL_2098228, EPI_ISL_2134793, EPI_ISL_2134794

| | | | |
|---|---|---|---|
| see above | URMC LABS | Wadsworth Center, New York State Department of Health | Alexis Russel; Alexis Russell; Catharine Prussing; Daryl M. Lamson; Erasmus Schneider; Erica Lasek-Nesselquist; John Kelly; Jonathan Plitnick; Kirsten St. George; Matthew Shudt; Melissa A Leisner; Navjot Singh |
| EPI_ISL_1531475 | US Air Force School of Aerospace Medicine | US Air Force School of Aerospace Medicine | Amanda Javorina; Anthony Fries; Clarise Starr; Elizabeth Macias; Jennifer Meyer; Sarah Purves; William Buggele; William Gruner |

| | | | |
|---|---|---|---|
| EPI_ISL_1397935, EPI_ISL_1397936, EPI_ISL_1397937, EPI_ISL_1397938, EPI_ISL_1397939, EPI_ISL_1397940, EPI_ISL_1397941, EPI_ISL_1397942, EPI_ISL_1397943, EPI_ISL_1397944, EPI_ISL_1397945, EPI_ISL_1397946, EPI_ISL_1397947, EPI_ISL_1397948, EPI_ISL_1397949, EPI_ISL_1397950, EPI_ISL_1397951, EPI_ISL_1397952, EPI_ISL_1398033, EPI_ISL_1462555, EPI_ISL_1465637, EPI_ISL_1482394, EPI_ISL_1482395, EPI_ISL_1482396, EPI_ISL_1482397, EPI_ISL_1482398, EPI_ISL_1482399, EPI_ISL_1482400, EPI_ISL_1482401, EPI_ISL_1482402, EPI_ISL_1482403, EPI_ISL_1482404, EPI_ISL_1482405, EPI_ISL_1482411, EPI_ISL_1482412, EPI_ISL_1482413, EPI_ISL_1482414, EPI_ISL_1482415, EPI_ISL_1482416, EPI_ISL_1482417, EPI_ISL_1482418, EPI_ISL_1482419, EPI_ISL_1482420, EPI_ISL_1482421, EPI_ISL_1482422, EPI_ISL_1482423, EPI_ISL_1482424, EPI_ISL_1482425, EPI_ISL_1482426, EPI_ISL_1482427, EPI_ISL_1482428, EPI_ISL_1482429, EPI_ISL_1482430, EPI_ISL_1482431, EPI_ISL_1482432, EPI_ISL_1482433, EPI_ISL_1482434, EPI_ISL_1482435, EPI_ISL_1482436, EPI_ISL_1482437, EPI_ISL_1482438, EPI_ISL_1482439, EPI_ISL_1482440, EPI_ISL_1482441, EPI_ISL_1482442, EPI_ISL_1482443, EPI_ISL_1482444, EPI_ISL_1482445, EPI_ISL_1482539, EPI_ISL_1482540, EPI_ISL_1482541, EPI_ISL_1482542, EPI_ISL_1482543, EPI_ISL_1482544, EPI_ISL_1482545, EPI_ISL_1482546, EPI_ISL_1482547, EPI_ISL_1482548, EPI_ISL_1482549, EPI_ISL_1482550, EPI_ISL_1482551, EPI_ISL_1482552, EPI_ISL_1482553, EPI_ISL_1482554, EPI_ISL_1482555, EPI_ISL_1482617, EPI_ISL_1482618, EPI_ISL_1482619, EPI_ISL_1482620, EPI_ISL_1482621, EPI_ISL_1482622, EPI_ISL_1482623, EPI_ISL_1482624, EPI_ISL_1580673, EPI_ISL_1580674, EPI_ISL_1580675, EPI_ISL_1580676, EPI_ISL_1580677, EPI_ISL_1580699, EPI_ISL_1582231, EPI_ISL_1582308, EPI_ISL_1582309, EPI_ISL_1582310, EPI_ISL_1582311, EPI_ISL_1582312, EPI_ISL_1582313, EPI_ISL_1582314, EPI_ISL_1582315, EPI_ISL_1582316, EPI_ISL_1582338, EPI_ISL_1593864, EPI_ISL_1593865, EPI_ISL_1593872, EPI_ISL_1671459, EPI_ISL_1671460, EPI_ISL_1671467, EPI_ISL_1671481, EPI_ISL_1671482, EPI_ISL_1671483, EPI_ISL_1671492, EPI_ISL_1671508, EPI_ISL_1671509, EPI_ISL_1671510, EPI_ISL_1671619, EPI_ISL_1671624, EPI_ISL_1671625, EPI_ISL_1671637, EPI_ISL_1671639, EPI_ISL_1671646, EPI_ISL_1671647, EPI_ISL_1671657, EPI_ISL_1671658, EPI_ISL_1671659, EPI_ISL_1671660, EPI_ISL_1671666, EPI_ISL_1755851, EPI_ISL_1755852, EPI_ISL_1755853, EPI_ISL_1755867, EPI_ISL_1755868, EPI_ISL_1755966, EPI_ISL_1755967, EPI_ISL_1935614 | | | |
| see above | WESTCHESTER MEDICAL CENTER | Wadsworth Center, New York State Department of Health | Alexis Russel; Alexis Russell; Catharine Prussing; Daryl M. Lamson; Erasmus Schneider; Erica Lasek-Nesselquist; John Kelly; Jonathan Plitnick; Kirsten St. George; Matthew Shudt; Melissa A Leisner; Navjot Singh |
| EPI_ISL_1398268, EPI_ISL_1482573, EPI_ISL_1482574, EPI_ISL_1482575, EPI_ISL_1482576, EPI_ISL_1482577, EPI_ISL_1482578, EPI_ISL_1482579, EPI_ISL_1582273, EPI_ISL_1582274, EPI_ISL_1582275, EPI_ISL_1582276, EPI_ISL_1582277, EPI_ISL_1582278, EPI_ISL_1582279, EPI_ISL_1582280, EPI_ISL_1582281, EPI_ISL_1582282, EPI_ISL_1593886 | | | |
| see above | WHITE PLAINS HOSPITAL CENTER LABORATORY | Wadsworth Center, New York State Department of Health | Alexis Russel; Alexis Russell; Catharine Prussing; Daryl M. Lamson; Erasmus Schneider; Erica Lasek-Nesselquist; John Kelly; Jonathan Plitnick; Kirsten St. George; Matthew Shudt; Melissa A Leisner; Navjot Singh |
| EPI_ISL_1967664, EPI_ISL_1967677, EPI_ISL_1967679, EPI_ISL_1968623, EPI_ISL_1971088, EPI_ISL_1971089, EPI_ISL_1971090, EPI_ISL_1971091, EPI_ISL_1971092, EPI_ISL_1971093, EPI_ISL_1971094, EPI_ISL_1971095, EPI_ISL_1971096, EPI_ISL_1971097, EPI_ISL_1971098, EPI_ISL_1971099, EPI_ISL_1971100, EPI_ISL_1971101, EPI_ISL_1971102, EPI_ISL_1971103, EPI_ISL_1971104, EPI_ISL_1971105, EPI_ISL_1971106, EPI_ISL_1971107, EPI_ISL_1971108, EPI_ISL_1971109, EPI_ISL_1971110, EPI_ISL_1971111, EPI_ISL_1971112, EPI_ISL_1971113, EPI_ISL_1971114, EPI_ISL_1971115, EPI_ISL_1971116, EPI_ISL_1971117, EPI_ISL_1971118, EPI_ISL_1971119, EPI_ISL_1971120, EPI_ISL_1971121, EPI_ISL_1971122, EPI_ISL_1971123, EPI_ISL_1971124, EPI_ISL_1971125, EPI_ISL_1971126, EPI_ISL_1971127, EPI_ISL_1971128, EPI_ISL_1971129, EPI_ISL_1971130, EPI_ISL_1971131, EPI_ISL_1971132, EPI_ISL_1971133, EPI_ISL_1971134, EPI_ISL_1971135, EPI_ISL_1971136, EPI_ISL_1971137, EPI_ISL_1971138, EPI_ISL_1971139, EPI_ISL_1971140, EPI_ISL_1971141, EPI_ISL_1971142, EPI_ISL_1971143, EPI_ISL_1971144, EPI_ISL_1971145, EPI_ISL_1971146, EPI_ISL_1971147, EPI_ISL_1971148, EPI_ISL_1971149, EPI_ISL_1971150, EPI_ISL_1971151, EPI_ISL_1971152, EPI_ISL_1971153, EPI_ISL_1971154, EPI_ISL_1971157, EPI_ISL_1971159, EPI_ISL_1971160, EPI_ISL_1971161, EPI_ISL_1971163, EPI_ISL_1971164, EPI_ISL_1971165, EPI_ISL_1971166, EPI_ISL_1971167, EPI_ISL_1971168, EPI_ISL_1971169, EPI_ISL_1971170, EPI_ISL_1971171, EPI_ISL_1971172, EPI_ISL_1971173, EPI_ISL_1971174, EPI_ISL_1971175, EPI_ISL_1971176, EPI_ISL_1971177, EPI_ISL_1971179, EPI_ISL_1971180, EPI_ISL_1971181, EPI_ISL_1971182, EPI_ISL_1971183, EPI_ISL_1971184, EPI_ISL_1971187, EPI_ISL_1971188, EPI_ISL_1971189, EPI_ISL_1971191, EPI_ISL_1971192, EPI_ISL_1971193, EPI_ISL_1971194, EPI_ISL_1971195, EPI_ISL_1971197, EPI_ISL_1971198, EPI_ISL_1971199, EPI_ISL_1971200, EPI_ISL_1971201, EPI_ISL_1971202, EPI_ISL_1971203, EPI_ISL_1971204, EPI_ISL_1971205, EPI_ISL_1971206, EPI_ISL_1971209, EPI_ISL_1971212, EPI_ISL_1971215, EPI_ISL_1971216, EPI_ISL_1971217, EPI_ISL_1971218, EPI_ISL_1971220, EPI_ISL_1971222, EPI_ISL_1971224, EPI_ISL_1971225, EPI_ISL_1971226, EPI_ISL_1971229, EPI_ISL_1971230, EPI_ISL_1971231, EPI_ISL_2031764, EPI_ISL_2031765, EPI_ISL_2031766, EPI_ISL_2031767, EPI_ISL_2086300, EPI_ISL_2086301, EPI_ISL_2086302, EPI_ISL_2086303 | | | |
| see above | Weill Cornell Medicine | New York Genome Center | Andre Corvelo; Arryn Craney; Chris Mason; Dayna M. Oschwald; Hanna Rennert; Lars F Westblade; Margaret Elizabeth Ross; Melissa Cushing; Michael Zody; Olivier Elemento; Priya Velu; Samantha Fennessey; Tom Maniatis |
| EPI_ISL_1920153, EPI_ISL_1920173 | Westchester Medical Center | Wadsworth Center, New York State Department of Health | Alexis Russell; Catharine Prussing; Daryl M. Lamson; Erasmus Schneider; Erica Lasek-Nesselquist; John Kelly; Jonathan Plitnick; Kirsten St. George; Matthew Shudt; Melissa A Leisner; Navjot Singh |
| EPI_ISL_1378687, EPI_ISL_1447572, EPI_ISL_1447577, EPI_ISL_1509180, EPI_ISL_1509238, EPI_ISL_1587343, EPI_ISL_1587390, EPI_ISL_1963661, EPI_ISL_1963667 | | | |
| see above | Yale Clinical Virology Lab | Grubaugh Lab - Yale School of Public Health | Anderson Brito; Annie Watkins; Chaney Kalinich; Chantal Vogels; Isabel Ott; Isabell Ott; Jessica Rothman; Joseph Fauver; Mallery Breban; Marie L. Landry; Mary Petrone; Nathan Grubaugh; Tara Alpert |
| EPI_ISL_2034813, EPI_ISL_2034818 | Yale Clinical Virology Lab | Yale Center for Genomic Analysis | Brooke Sullivan; Curt Scharfe; Irina Tikhonova; Kaya Bilguvar; Shrikant Mane |

We gratefully acknowledge the following Authors from the Originating laboratories responsible for obtaining the specimens, as well as the Submitting laboratories where the genome data were generated and shared via GISAID, on which this research is based.

All Submitters of data may be contacted directly via www.gisaid.org

Authors are sorted alphabetically.

| Accession ID | Originating Laboratory | Submitting Laboratory | Authors |
| --- | --- | --- | --- |
| EPI_ISL_1824033, EPI_ISL_1824034, EPI_ISL_1824035 | "NYSDOH Wadsworth Center, Virology Lab" | Centers for Disease Control and Prevention Division of Viral Diseases, Pathogen Discovery | Alison Laufer Halpin; Ben L. Rambo-Martin; Clinton R. Paden; Dakota Howard; Darlene Wagner; Dave Wentworth; Dhwani Batra; Jasmine Padilla; Justin Lee; Katie Dillon; Krista Queen; Kristen Knipe; Kristine Lacek; Mark Burroughs; Matthew Schmerer; Mili Sheth; Peter Cook; Sam Shepard; Sarah Nobles; Shoshona Le; Suxiang Tong; Vivien Dugan; Yvette Unoarumhi |
| EPI_ISL_2098499 | ADIRONDACK MEDICAL CENTER | Wadsworth Center, New York State Department of Health | Alexis Russell; Catharine Prussing; Daryl M. Lamson; Erasmus Schneider; Erica Lasek-Nesselquist; John Kelly; Jonathan Plitnick; Kirsten St. George; Matthew Shudt; Melissa A Leisner; Navjot Singh |
| EPI_ISL_1671313, EPI_ISL_1671319, EPI_ISL_1671320, EPI_ISL_1671321, EPI_ISL_1671326, EPI_ISL_1671327, EPI_ISL_1671341, EPI_ISL_1671350, EPI_ISL_1671351, EPI_ISL_1671354, EPI_ISL_1671359, EPI_ISL_1671450, EPI_ISL_1671451, EPI_ISL_1671462, EPI_ISL_1671599, EPI_ISL_1671623, EPI_ISL_1671645, EPI_ISL_1740509, EPI_ISL_1755840, EPI_ISL_1755841, EPI_ISL_1756001, EPI_ISL_1756002, EPI_ISL_1756003, EPI_ISL_1756004, EPI_ISL_1756005, EPI_ISL_1756006, EPI_ISL_1756007, EPI_ISL_1756008, EPI_ISL_1756009, EPI_ISL_1756010, EPI_ISL_1756011, EPI_ISL_1756012, EPI_ISL_1756013, EPI_ISL_1756014, EPI_ISL_1756015, EPI_ISL_1756016, EPI_ISL_1756017, EPI_ISL_1756018, EPI_ISL_1756019, EPI_ISL_1756020, EPI_ISL_1789716, EPI_ISL_1789717, EPI_ISL_1789718, EPI_ISL_1854692, EPI_ISL_1855116, EPI_ISL_1855117, EPI_ISL_1855118, EPI_ISL_1935607, EPI_ISL_1935638, EPI_ISL_1935644, EPI_ISL_2097232, EPI_ISL_2097233, EPI_ISL_2097234, EPI_ISL_2097235, EPI_ISL_2097236, EPI_ISL_2097237, EPI_ISL_2097238, EPI_ISL_2098115, EPI_ISL_2098117, EPI_ISL_2098119, EPI_ISL_2098120, EPI_ISL_2098122, EPI_ISL_2098123, EPI_ISL_2098125, EPI_ISL_2098500 | | | |
| see above | ALBANY MEDICAL CENTER | Wadsworth Center, New York State Department of Health | Alexis Russell; Catharine Prussing; Daryl M. Lamson; Erasmus Schneider; Erica Lasek-Nesselquist; John Kelly; Jonathan Plitnick; Kirsten St. George; Matthew Shudt; Melissa A Leisner; Navjot Singh |
| EPI_ISL_1667463, EPI_ISL_1667465, EPI_ISL_1667473, EPI_ISL_1667476, EPI_ISL_1667543, EPI_ISL_1667544, EPI_ISL_1667549, EPI_ISL_1667569, EPI_ISL_1667572, EPI_ISL_1667618, EPI_ISL_1667660, EPI_ISL_1667704, EPI_ISL_1667740, EPI_ISL_1667744, EPI_ISL_1667746, EPI_ISL_1667747, EPI_ISL_1668015, EPI_ISL_1668022, EPI_ISL_1668025, EPI_ISL_1668038, EPI_ISL_1668214, EPI_ISL_1668304, EPI_ISL_1668328, EPI_ISL_1668354, EPI_ISL_1668364, EPI_ISL_1685759, EPI_ISL_1685928, EPI_ISL_1685938, EPI_ISL_1685948, EPI_ISL_1685956, EPI_ISL_1685966, EPI_ISL_1685981, EPI_ISL_1685983, EPI_ISL_1685992, EPI_ISL_1686100, EPI_ISL_1686169, EPI_ISL_1686170, EPI_ISL_1686173, EPI_ISL_1686207, EPI_ISL_1686213, EPI_ISL_1686217, EPI_ISL_1686220, EPI_ISL_1686224, EPI_ISL_1686227, EPI_ISL_1686264, EPI_ISL_1686281, EPI_ISL_1686282, EPI_ISL_1687299, EPI_ISL_1687316, EPI_ISL_1687343, EPI_ISL_1687358, EPI_ISL_1687387, EPI_ISL_1687426, EPI_ISL_1687491, EPI_ISL_1687502, EPI_ISL_1687506, EPI_ISL_1687507, EPI_ISL_1687508, EPI_ISL_1687587, EPI_ISL_1687615, EPI_ISL_1687650, EPI_ISL_1687651, EPI_ISL_1687674, EPI_ISL_1687680, EPI_ISL_1687681, EPI_ISL_1688634, EPI_ISL_1688638, EPI_ISL_1688689, EPI_ISL_1688937, EPI_ISL_1688963, EPI_ISL_1689021, EPI_ISL_1689086, EPI_ISL_1689186, EPI_ISL_1689197, EPI_ISL_1689200, EPI_ISL_1689202, EPI_ISL_1689207, EPI_ISL_1689214, EPI_ISL_1689389, EPI_ISL_1689391, EPI_ISL_1689393, EPI_ISL_1689396, EPI_ISL_1689397, EPI_ISL_1689596, EPI_ISL_1689599, EPI_ISL_1689603, EPI_ISL_1689608, EPI_ISL_1689612, EPI_ISL_1689629, EPI_ISL_1689631, EPI_ISL_1689732, EPI_ISL_1689750, EPI_ISL_1689816, EPI_ISL_1689843, EPI_ISL_1689847, EPI_ISL_1689848, EPI_ISL_1689853, EPI_ISL_1689862, EPI_ISL_1689864, EPI_ISL_1689871, EPI_ISL_1689883, EPI_ISL_1689977, EPI_ISL_1690008, EPI_ISL_1690013, EPI_ISL_1690021, EPI_ISL_1690039, EPI_ISL_1690040, EPI_ISL_1690052, EPI_ISL_1690079, EPI_ISL_1690081, EPI_ISL_1690087, EPI_ISL_1690099, EPI_ISL_1690100, EPI_ISL_1690101, EPI_ISL_1690115, EPI_ISL_1690121, EPI_ISL_1690189, EPI_ISL_1690197, EPI_ISL_1690203, EPI_ISL_1690204, EPI_ISL_1690250, EPI_ISL_1690267, EPI_ISL_1690268, EPI_ISL_1695487, EPI_ISL_1695554, EPI_ISL_1701890, EPI_ISL_1701941, EPI_ISL_1701968, EPI_ISL_1701969, EPI_ISL_1701996, EPI_ISL_1701997, EPI_ISL_1702127, EPI_ISL_1702138, EPI_ISL_1702728, EPI_ISL_1702778, EPI_ISL_1702788, EPI_ISL_1702789, EPI_ISL_1702816, EPI_ISL_1702829, EPI_ISL_1702838, EPI_ISL_1702865, EPI_ISL_1702867, EPI_ISL_1702884, EPI_ISL_1702954, EPI_ISL_1702970, EPI_ISL_1735719, EPI_ISL_1735723, EPI_ISL_1735724, EPI_ISL_1735772, EPI_ISL_1735773, EPI_ISL_1735774, EPI_ISL_1735775, EPI_ISL_1735783, EPI_ISL_1735855, EPI_ISL_1735914, EPI_ISL_1735940, EPI_ISL_1736007, EPI_ISL_1736116, EPI_ISL_1736118, EPI_ISL_1736119, EPI_ISL_1736120, EPI_ISL_1736146, EPI_ISL_1736163, EPI_ISL_1736218, EPI_ISL_1736219, EPI_ISL_1736235, EPI_ISL_1736375, EPI_ISL_1736384, EPI_ISL_1736495, EPI_ISL_1736516, EPI_ISL_1736525, EPI_ISL_1736550, EPI_ISL_1736626, EPI_ISL_1736656, EPI_ISL_1736685, EPI_ISL_1736695, EPI_ISL_1736702, EPI_ISL_1736720, EPI_ISL_1736763, EPI_ISL_1736796, EPI_ISL_1736798, EPI_ISL_1736799, EPI_ISL_1736806, EPI_ISL_1736809, EPI_ISL_1736842, EPI_ISL_1736847, EPI_ISL_1736852, EPI_ISL_1736862, EPI_ISL_1736882, EPI_ISL_1736890, EPI_ISL_1736899, EPI_ISL_1736927, EPI_ISL_1736930, EPI_ISL_1737063, EPI_ISL_1737198, EPI_ISL_1737202, EPI_ISL_1737223, EPI_ISL_1737238, EPI_ISL_1737239, EPI_ISL_1737284, EPI_ISL_1737289, EPI_ISL_1737290, EPI_ISL_1737291, EPI_ISL_1737292, EPI_ISL_1737296, EPI_ISL_1737399, EPI_ISL_1737478, EPI_ISL_1737517, EPI_ISL_1737589, EPI_ISL_1737590, EPI_ISL_1737597, EPI_ISL_1737689, EPI_ISL_1737780, EPI_ISL_1737813, EPI_ISL_1737847, EPI_ISL_1738769, EPI_ISL_1753533, EPI_ISL_1753552, EPI_ISL_1753557, EPI_ISL_1753572, EPI_ISL_1753574, EPI_ISL_1753582, EPI_ISL_1834544, EPI_ISL_1834546, EPI_ISL_1834549, EPI_ISL_1834558, EPI_ISL_1834564, EPI_ISL_1834732, EPI_ISL_1834785, EPI_ISL_1834786, EPI_ISL_1834791, EPI_ISL_1834792, EPI_ISL_1834793, EPI_ISL_1834794, EPI_ISL_1834796, EPI_ISL_1834807, EPI_ISL_1834837, EPI_ISL_1834876, EPI_ISL_1834887, EPI_ISL_1834899, EPI_ISL_1834915, EPI_ISL_1834928, EPI_ISL_1834929, EPI_ISL_1834976, EPI_ISL_1835090, EPI_ISL_1835091, EPI_ISL_1835093, EPI_ISL_1835113, EPI_ISL_1835160, EPI_ISL_1835194, EPI_ISL_1835207, EPI_ISL_1835246, EPI_ISL_1835297, EPI_ISL_1835336, EPI_ISL_1835337, EPI_ISL_1835353, EPI_ISL_1835355, EPI_ISL_1835362, EPI_ISL_1835476, EPI_ISL_1835525, EPI_ISL_1835566, EPI_ISL_1835665, EPI_ISL_1835701, EPI_ISL_1835727, EPI_ISL_1835780, EPI_ISL_1835801, EPI_ISL_1835884, EPI_ISL_1835940, EPI_ISL_1835989, EPI_ISL_1836019, EPI_ISL_1836037, EPI_ISL_1836170, EPI_ISL_1836175, EPI_ISL_1836256, EPI_ISL_1836280, EPI_ISL_1836303, EPI_ISL_1836305, EPI_ISL_1836306, EPI_ISL_1836309, EPI_ISL_1836313, EPI_ISL_1836348, EPI_ISL_1836381, EPI_ISL_1836384, EPI_ISL_1836386, EPI_ISL_1836479, EPI_ISL_1836620, EPI_ISL_1836689, EPI_ISL_1836696, EPI_ISL_1836698, EPI_ISL_1836785, EPI_ISL_1836814, EPI_ISL_1836876, EPI_ISL_1836965, EPI_ISL_1837051, EPI_ISL_1837154, EPI_ISL_1837168, EPI_ISL_1837252, EPI_ISL_1837254, EPI_ISL_1837281, EPI_ISL_1837287, EPI_ISL_1837295, EPI_ISL_1837340, EPI_ISL_1837400, EPI_ISL_1837517, EPI_ISL_1837518, EPI_ISL_1837519, EPI_ISL_1837540, EPI_ISL_1837591, EPI_ISL_1837637, EPI_ISL_1837693, EPI_ISL_1837694, EPI_ISL_1837797, EPI_ISL_1837904, EPI_ISL_1837936, EPI_ISL_1837941, EPI_ISL_1837947, EPI_ISL_1838017, EPI_ISL_1838027, EPI_ISL_1838034, EPI_ISL_1838334, EPI_ISL_1838438, EPI_ISL_1900231, EPI_ISL_1900249, EPI_ISL_1900283, EPI_ISL_1900354, EPI_ISL_1900419, EPI_ISL_1922972, EPI_ISL_1922974, EPI_ISL_1922995, EPI_ISL_1923020, EPI_ISL_1923029, EPI_ISL_1923040, EPI_ISL_1923080, EPI_ISL_1923092, EPI_ISL_1923116, EPI_ISL_1923149, EPI_ISL_1942142, EPI_ISL_1994977, EPI_ISL_1994994, EPI_ISL_1995028, EPI_ISL_1995029, EPI_ISL_1995039, EPI_ISL_1995083, EPI_ISL_1995095, EPI_ISL_1995114, EPI_ISL_1995124, EPI_ISL_1995125, EPI_ISL_1995126, EPI_ISL_1995130, EPI_ISL_1995132, EPI_ISL_1995133, EPI_ISL_1995142, EPI_ISL_1995164, EPI_ISL_1995289, EPI_ISL_1995299, EPI_ISL_1995300, EPI_ISL_1995302, EPI_ISL_1995381, EPI_ISL_1995406, EPI_ISL_1995412, EPI_ISL_1995518, EPI_ISL_1995549, EPI_ISL_1995553, EPI_ISL_1995554, EPI_ISL_1995682, EPI_ISL_1995683, EPI_ISL_1995684, EPI_ISL_1995731, EPI_ISL_1995733, EPI_ISL_1995734, EPI_ISL_1995761, EPI_ISL_1995887, EPI_ISL_1995915, EPI_ISL_1995916, EPI_ISL_1995938, EPI_ISL_1995949, EPI_ISL_1995951, EPI_ISL_1995957, EPI_ISL_1996018, EPI_ISL_1996068, EPI_ISL_1996073, EPI_ISL_1996074, EPI_ISL_1996113, EPI_ISL_1996114, EPI_ISL_1996115, EPI_ISL_1996132, EPI_ISL_1996137, EPI_ISL_1996208, EPI_ISL_1996277, EPI_ISL_1996297, EPI_ISL_1996475, EPI_ISL_1996503, EPI_ISL_1996510, EPI_ISL_1996602, EPI_ISL_1996612, EPI_ISL_1996844, EPI_ISL_1996845, EPI_ISL_1996846, EPI_ISL_1996854, EPI_ISL_1996938, EPI_ISL_1996945, EPI_ISL_1996946, EPI_ISL_1996947, EPI_ISL_1996983, EPI_ISL_1996997, EPI_ISL_1997025, EPI_ISL_1997067, EPI_ISL_1997069, EPI_ISL_1997120, EPI_ISL_1997129, EPI_ISL_1997147, EPI_ISL_1997148, EPI_ISL_1997149, EPI_ISL_1997172, EPI_ISL_1997251, EPI_ISL_1997283, EPI_ISL_1997298, EPI_ISL_1997329, EPI_ISL_1997330, EPI_ISL_1997381, EPI_ISL_1997383, EPI_ISL_1997398, EPI_ISL_1997449, EPI_ISL_1997450, EPI_ISL_1997452, EPI_ISL_1997453, EPI_ISL_1997463, EPI_ISL_1997465, EPI_ISL_1997475, EPI_ISL_1997476, EPI_ISL_1997499, EPI_ISL_1997502, EPI_ISL_1997518, EPI_ISL_1997519, EPI_ISL_1997545, EPI_ISL_1997550, EPI_ISL_1997576, EPI_ISL_1997625, EPI_ISL_1997627, EPI_ISL_1997681, EPI_ISL_1997683, EPI_ISL_1997736, EPI_ISL_1997737, EPI_ISL_1997742, EPI_ISL_1997752, EPI_ISL_1997753, EPI_ISL_1997755, EPI_ISL_1997827, EPI_ISL_1997939, EPI_ISL_1997940, EPI_ISL_1997998, EPI_ISL_1998084, EPI_ISL_1998189, EPI_ISL_1998251, EPI_ISL_1998286, EPI_ISL_1998288, EPI_ISL_1998305, EPI_ISL_1998338, EPI_ISL_1998606, EPI_ISL_1998608, EPI_ISL_1998680, EPI_ISL_1998688, EPI_ISL_1998715, EPI_ISL_1998733, EPI_ISL_1998736, EPI_ISL_1998770, EPI_ISL_1999110, EPI_ISL_1999124, EPI_ISL_1999232, EPI_ISL_1999246, EPI_ISL_1999250, EPI_ISL_1999262, EPI_ISL_1999353, EPI_ISL_1999476, EPI_ISL_1999477, EPI_ISL_1999572, EPI_ISL_1999684, EPI_ISL_1999715, EPI_ISL_1999807, EPI_ISL_1999808, EPI_ISL_1999816, EPI_ISL_1999868, EPI_ISL_1999870, EPI_ISL_1999892, EPI_ISL_1999941, EPI_ISL_1999974, EPI_ISL_1999982, EPI_ISL_2000182, EPI_ISL_2000186, EPI_ISL_2000187, EPI_ISL_2000190, EPI_ISL_2000260, EPI_ISL_2000278, EPI_ISL_2000286, EPI_ISL_2000386, EPI_ISL_2000389, EPI_ISL_2000390, EPI_ISL_2000391, EPI_ISL_2000438, EPI_ISL_2000507, EPI_ISL_2000542, EPI_ISL_2035107, EPI_ISL_2035130, EPI_ISL_2035243, EPI_ISL_2035253, EPI_ISL_2035263, EPI_ISL_2038970, EPI_ISL_2038975, EPI_ISL_2038976, EPI_ISL_2038977, EPI_ISL_2039013, EPI_ISL_2039137, EPI_ISL_2039138, EPI_ISL_2039255, EPI_ISL_2039259, EPI_ISL_2039263, EPI_ISL_2039276, EPI_ISL_2039278, EPI_ISL_2039279, EPI_ISL_2039280, EPI_ISL_2039281, EPI_ISL_2039283, EPI_ISL_2039380, EPI_ISL_2039403, EPI_ISL_2039447, EPI_ISL_2039454, EPI_ISL_2039456, EPI_ISL_2039547, EPI_ISL_2039631, EPI_ISL_2039635, EPI_ISL_2039746, EPI_ISL_2039808, EPI_ISL_2039867, EPI_ISL_2039872, EPI_ISL_2039979, EPI_ISL_2039981, EPI_ISL_2039999, EPI_ISL_2040092, EPI_ISL_2040240, EPI_ISL_2040241, EPI_ISL_2040242, EPI_ISL_2040249, EPI_ISL_2040256, EPI_ISL_2040262, EPI_ISL_2040286, EPI_ISL_2040299, EPI_ISL_2040310, EPI_ISL_2040316, EPI_ISL_2040466, EPI_ISL_2040467, EPI_ISL_2040505, EPI_ISL_2040517, EPI_ISL_2040546, EPI_ISL_2040567, EPI_ISL_2040593, EPI_ISL_2040626, EPI_ISL_2040687, EPI_ISL_2040795, EPI_ISL_2040807, EPI_ISL_2040810, EPI_ISL_2040823, EPI_ISL_2040847, EPI_ISL_2040869, EPI_ISL_2040870, EPI_ISL_2040891, EPI_ISL_2040935, EPI_ISL_2040940, EPI_ISL_2040997, EPI_ISL_2041057, EPI_ISL_2041060, EPI_ISL_2041163, EPI_ISL_2041167, EPI_ISL_2041170, EPI_ISL_2041187, EPI_ISL_2041204, EPI_ISL_2041335, EPI_ISL_2041354, EPI_ISL_2041356, EPI_ISL_2041364, EPI_ISL_2041383, EPI_ISL_2041488, EPI_ISL_2041583, EPI_ISL_2041700, EPI_ISL_2041703, EPI_ISL_2041707, EPI_ISL_2041708, EPI_ISL_2041709, EPI_ISL_2041736, EPI_ISL_2041742, EPI_ISL_2041747, EPI_ISL_2041749, EPI_ISL_2041755, EPI_ISL_2041787, EPI_ISL_2041956, EPI_ISL_2041958, EPI_ISL_2042096, EPI_ISL_2042115, EPI_ISL_2042123, EPI_ISL_2042164, EPI_ISL_2042237, EPI_ISL_2042325, EPI_ISL_2042337, EPI_ISL_2042347, EPI_ISL_2042508, EPI_ISL_2042510, EPI_ISL_2042533, EPI_ISL_2042540, EPI_ISL_2042543, EPI_ISL_2042606, EPI_ISL_2042611, EPI_ISL_2042621, EPI_ISL_2042622, EPI_ISL_2042629, EPI_ISL_2042630, EPI_ISL_2042678, EPI_ISL_2042679, EPI_ISL_2042693, EPI_ISL_2042697, EPI_ISL_2042698, EPI_ISL_2042706, EPI_ISL_2042744, EPI_ISL_2042745, EPI_ISL_2042767, EPI_ISL_2042769, EPI_ISL_2042775, EPI_ISL_2043139, EPI_ISL_2043151, EPI_ISL_2043166, EPI_ISL_2043188, EPI_ISL_2043207, EPI_ISL_2043213, EPI_ISL_2043216, EPI_ISL_2043230, EPI_ISL_2043239, EPI_ISL_2043255, EPI_ISL_2043327, EPI_ISL_2043367, EPI_ISL_2043425, EPI_ISL_2089530, EPI_ISL_2089599, EPI_ISL_2089607, EPI_ISL_2089673, EPI_ISL_2089704, EPI_ISL_2089736, EPI_ISL_2089737, EPI_ISL_2089765, EPI_ISL_2089768, EPI_ISL_2089769, EPI_ISL_2089770, EPI_ISL_2089810, EPI_ISL_2089952, EPI_ISL_2089982, EPI_ISL_2089986, EPI_ISL_2090007, EPI_ISL_2090133, EPI_ISL_2090151, EPI_ISL_2090170, EPI_ISL_2090181, EPI_ISL_2090182, EPI_ISL_2090210, EPI_ISL_2090211, EPI_ISL_2090219, EPI_ISL_2090220, EPI_ISL_2090235, EPI_ISL_2090239, EPI_ISL_2090240, EPI_ISL_2090241, EPI_ISL_2090408, EPI_ISL_2090426, EPI_ISL_2090458, EPI_ISL_2090468, EPI_ISL_2132561, EPI_ISL_2145807, EPI_ISL_2145831, EPI_ISL_2145933, EPI_ISL_2145936, EPI_ISL_2145998, EPI_ISL_2146005, EPI_ISL_2146007, EPI_ISL_2148021, EPI_ISL_2148055, EPI_ISL_2148637, EPI_ISL_2148690, EPI_ISL_2148718, EPI_ISL_2149297, EPI_ISL_2149312, EPI_ISL_2149341, EPI_ISL_2149342, EPI_ISL_2149386, EPI_ISL_2149388, EPI_ISL_2149402, EPI_ISL_2149443, EPI_ISL_2149510, EPI_ISL_2149518, EPI_ISL_2149521, EPI_ISL_2149532, EPI_ISL_2149535, EPI_ISL_2149586, EPI_ISL_2149812, EPI_ISL_2149816, EPI_ISL_2149848, EPI_ISL_2149876, EPI_ISL_2149877, EPI_ISL_2149879, EPI_ISL_2149967, EPI_ISL_2150008, EPI_ISL_2150026, EPI_ISL_2150028, EPI_ISL_2150029, EPI_ISL_2150066, EPI_ISL_2150070, EPI_ISL_2150096, EPI_ISL_2150097, EPI_ISL_2150120, EPI_ISL_2150156, EPI_ISL_2150157, EPI_ISL_2150161, EPI_ISL_2150177, EPI_ISL_2150272, EPI_ISL_2150398, EPI_ISL_2150476, EPI_ISL_2150515, EPI_ISL_2150595, EPI_ISL_2150598, EPI_ISL_2150658, EPI_ISL_2150694, EPI_ISL_2150793, EPI_ISL_2150838, EPI_ISL_2150840, EPI_ISL_2150841, EPI_ISL_2159213, EPI_ISL_2159242, EPI_ISL_2159253, EPI_ISL_2159429, EPI_ISL_2187508, EPI_ISL_2187591, EPI_ISL_2187607 | | | |
| see above | Aegis Sciences Corporation | Centers for Disease Control and Prevention Division of Viral Diseases, Pathogen Discovery | Adrian Paskey; Alec Vest; Benjamin Rambo-Martin; Christopher Gulvick; Clinton R. Paden; Cyndi Clark; Dakota Howard; Darlene Wagner; Dhwani Batra; Dillon Nall; Duncan MacCannell; Ethan Sanders; Holly Houdeshell; Jason Caravas; Kara Moser; Matthew Hardison; Matthew Schmerer; Ola Kvalvaag; Patrick Campbell; Peter W. Cook; Rob Case; Scott Sammons; Shatavia Morrison; Shaun Westlund; Vikramsinha Ghorpade; Yvette Unoarumhi |
| EPI_ISL_1920050, EPI_ISL_1920051, EPI_ISL_1920052, EPI_ISL_1920053, EPI_ISL_1920054, EPI_ISL_1920055, EPI_ISL_1920056, EPI_ISL_1920057, EPI_ISL_1920058, EPI_ISL_1920059, EPI_ISL_1920060, EPI_ISL_1921018, EPI_ISL_1921022, EPI_ISL_1921028, EPI_ISL_1921029, EPI_ISL_1921030, EPI_ISL_1921031, EPI_ISL_1921032, EPI_ISL_1921033, EPI_ISL_1921034, EPI_ISL_1921035, EPI_ISL_1921036 | | | |
| see above | Albany Medical Center | Wadsworth Center, New York State Department of Health | Alexis Russell; Catharine Prussing; Daryl M. Lamson; Erasmus Schneider; Erica Lasek-Nesselquist; John Kelly; Jonathan Plitnick; Kirsten St. George; Matthew Shudt; Melissa A Leisner; Navjot Singh |
| EPI_ISL_1756356 | BIO-REFERENCE LABORATORIES | Wadsworth Center, New York State Department of Health | Alexis Russell; Catharine Prussing; Daryl M. Lamson; Erasmus Schneider; Erica Lasek-Nesselquist; John Kelly; Jonathan Plitnick; Kirsten St. George; Matthew Shudt; Melissa A Leisner; Navjot Singh |
| EPI_ISL_1671444, EPI_ISL_1671445, EPI_ISL_1671447, EPI_ISL_1671455, EPI_ISL_1671456, EPI_ISL_1671457, EPI_ISL_1671496, EPI_ISL_1671596, EPI_ISL_1671618, EPI_ISL_1740511, EPI_ISL_1755850, EPI_ISL_1789712, EPI_ISL_1855310, EPI_ISL_1855311, EPI_ISL_1855312, EPI_ISL_1855313, EPI_ISL_1855314, EPI_ISL_1855315, EPI_ISL_1855316, EPI_ISL_1855317, EPI_ISL_1855318, EPI_ISL_1855319, EPI_ISL_1855320, EPI_ISL_1855321, EPI_ISL_1855322, EPI_ISL_1855323, EPI_ISL_1935617, EPI_ISL_1935620, EPI_ISL_2097239, EPI_ISL_2097240, EPI_ISL_2097241, EPI_ISL_2097242, EPI_ISL_2097243, EPI_ISL_2097244, EPI_ISL_2097245, EPI_ISL_2097246, EPI_ISL_2097247, EPI_ISL_2097248, EPI_ISL_2097249, EPI_ISL_2097250, EPI_ISL_2097251, EPI_ISL_2098128, EPI_ISL_2098589, EPI_ISL_2098590, EPI_ISL_2098591, EPI_ISL_2098592, EPI_ISL_2098593, EPI_ISL_2098594, EPI_ISL_2098595, EPI_ISL_2098596, EPI_ISL_2098597, EPI_ISL_2098598, EPI_ISL_2098599, EPI_ISL_2134784, EPI_ISL_2134790, EPI_ISL_2134796, EPI_ISL_2134797 | | | |
| see above | BOSTON HEART DIAGNOSTICS CORP | Wadsworth Center, New York State Department of Health | Alexis Russell; Catharine Prussing; Daryl M. Lamson; Erasmus Schneider; Erica Lasek-Nesselquist; John Kelly; Jonathan Plitnick; Kirsten St. George; Matthew Shudt; Melissa A Leisner; Navjot Singh |
| EPI_ISL_1710048, EPI_ISL_1710050, EPI_ISL_1710060, EPI_ISL_1710064, EPI_ISL_1710067, EPI_ISL_1710075, EPI_ISL_1710081, EPI_ISL_1710085, EPI_ISL_1710086, EPI_ISL_1710095, EPI_ISL_1710100, EPI_ISL_1710102, EPI_ISL_1710103, EPI_ISL_1710105, EPI_ISL_1710136, EPI_ISL_1710138, EPI_ISL_1757072, EPI_ISL_1757073, EPI_ISL_1757074, EPI_ISL_1757076, EPI_ISL_1757077, EPI_ISL_1757089, EPI_ISL_1757091, EPI_ISL_1757092, EPI_ISL_1757279, EPI_ISL_1757284, EPI_ISL_1757286, EPI_ISL_1787259, EPI_ISL_1787260, EPI_ISL_1787261, EPI_ISL_1825233, EPI_ISL_1825481, EPI_ISL_1825483, EPI_ISL_1825484, EPI_ISL_1825487, EPI_ISL_1825488, EPI_ISL_1825492, EPI_ISL_1825494, EPI_ISL_1918967, EPI_ISL_1918968, EPI_ISL_1919089, EPI_ISL_1919090, EPI_ISL_1919091, EPI_ISL_1919092, EPI_ISL_1919093, EPI_ISL_1919094, EPI_ISL_1919095, EPI_ISL_1919096, EPI_ISL_1919097, EPI_ISL_1919098, EPI_ISL_1919099, EPI_ISL_1919100, EPI_ISL_1919101, EPI_ISL_1919102, EPI_ISL_1919103, EPI_ISL_1919104, EPI_ISL_1919105, EPI_ISL_1968508, EPI_ISL_1968509, EPI_ISL_1968510, EPI_ISL_1972014, EPI_ISL_1972015, EPI_ISL_1972016, EPI_ISL_1972017, EPI_ISL_1972018, EPI_ISL_1972019, EPI_ISL_1972020, EPI_ISL_1972021, EPI_ISL_1972022, EPI_ISL_1972023, EPI_ISL_1972024, EPI_ISL_1972025, EPI_ISL_1972026, EPI_ISL_1972028, EPI_ISL_1972029, EPI_ISL_1972030, EPI_ISL_1972031, EPI_ISL_2096715 | | | |
| see above | Broad Institute Clinical Research Sequencing Platform | Infectious Disease Program, Broad Institute of Harvard and MIT | Adams, G.; B.L.; B.W.; Bauer, M.; Birren; Blumenstiel, B.; Brown, C.; Carter, A.; Chaluvadi, S.; D.J.; DeFelice, M.; DeRuff, K.; Dodge, S.; Gabriel, S.; Gallagher, G.; Gladden-Young, A.; Granger, B.; J.E.; K.J.; Lagerborg, K.; Larkin, K.; Lee, M.; Lemieux; Lennon, N.; Loreth, C.; Madoff, L.; McGovern, S.; Meldrim, J.; Normandin, E.; P.C.; Park; Pearlman, L.; Reilly, S.; Rudy, M.; Sabeti; Siddle; Smole, S.; Tomkins-Tinch, C.; Vicente, G.; and MacInnis |
| EPI_ISL_2101789, EPI_ISL_2101790 | Bronx Veterans Medical Research Foundation | Pathogen Discovery, Respiratory Viruses Branch, Division of Viral Diseases, Centers for Disease Control and Prevention | Adam Retchless; Anna Kelleher; Anna Uehara; Brian Lynch; Clinton R. Paden; Dhwani Batra; Haibin Wang; Han Jia Justin Ng; Jasmine Padilla; Jing Zhang; Justin Lee; Krista Queen; Mark Burroughs; Mili Sheth; Morgan Davis; Peter Cook; Rachel Marine; Sarah Nobles; Suxiang Tong; Tara Coalter; Yan Li; Ying Tao |
| EPI_ISL_2097252 | CENTERS LAB NJ, LLC (DBA MEDLABS DIAGNOSTICS) | Wadsworth Center, New York State Department of Health | Alexis Russell; Catharine Prussing; Daryl M. Lamson; Erasmus Schneider; Erica Lasek-Nesselquist; John Kelly; Jonathan Plitnick; Kirsten St. George; Matthew Shudt; Melissa A Leisner; Navjot Singh |
| EPI_ISL_1624487 | CHAMPLAIN VALLEY PHYSICIANS HOSPITAL MEDICAL CENTER | Wadsworth Center, New York State Department of Health | Alexis Russell; Catharine Prussing; Daryl M. Lamson; Erasmus Schneider; Erica Lasek-Nesselquist; John Kelly; Jonathan Plitnick; Kirsten St. George; Matthew Shudt; Melissa A Leisner; Navjot Singh |
| EPI_ISL_2036759 | CT Department of Public Health | CT Department of Public Health | Claire_Pearson; Tu_N_Nguyen |
| EPI_ISL_1965723, EPI_ISL_1965724, EPI_ISL_1965725, EPI_ISL_1965726, EPI_ISL_1965727, EPI_ISL_1965728, EPI_ISL_1965729, EPI_ISL_1965730, EPI_ISL_1965731, EPI_ISL_1965732, EPI_ISL_1965733, EPI_ISL_1965734, EPI_ISL_1965735, EPI_ISL_1965736, EPI_ISL_1965737, EPI_ISL_1965738, EPI_ISL_1965739, EPI_ISL_1965740, EPI_ISL_1965741, EPI_ISL_1965742, EPI_ISL_1965743, EPI_ISL_1965744, EPI_ISL_1965745, EPI_ISL_1965746, EPI_ISL_1965747, EPI_ISL_1965748, EPI_ISL_1965749, EPI_ISL_1965750, EPI_ISL_1965751, EPI_ISL_1965752, EPI_ISL_1965753, EPI_ISL_1965754, EPI_ISL_1965755, EPI_ISL_1965756, EPI_ISL_1965757, EPI_ISL_1965758 | | | |
| see above | Clinical Microbiology Laboratory, NewYork Presbyterian Hospital/Columbia University Irving Medical Center | Uhlemann Laboratory, Columbia University Irving Medical Center | Anne Kelley; Anne-Catrin Uhlemann; Maya Tagliavia; Medini K. Annavajhala |
| EPI_ISL_2151436 | Columbia University | Pathogen Discovery, Respiratory Viruses Branch, Division of Viral Diseases, Centers for Disease Control and Prevention | Adam Retchless; Anna Kelleher; Anna Uehara; Brian Lynch; Clinton R. Paden; Dhwani Batra; Haibin Wang; Han Jia Justin Ng; Jasmine Padilla; Jing Zhang; Justin Lee; Krista Queen; Mark Burroughs; Mili Sheth; Morgan Davis; Peter Cook; Rachel Marine; Sarah Nobles; Suxiang Tong; Tara Coalter; Yan Li; Ying Tao |
| EPI_ISL_1755854, EPI_ISL_1755855, EPI_ISL_1755856, EPI_ISL_1755857, EPI_ISL_1755858, EPI_ISL_1755859, EPI_ISL_1755860, EPI_ISL_1755861, EPI_ISL_1755862, EPI_ISL_1755863, EPI_ISL_1755864, EPI_ISL_1755865, EPI_ISL_1755866, EPI_ISL_1789713, EPI_ISL_1855286, EPI_ISL_1855287, EPI_ISL_1855288, EPI_ISL_1855289, EPI_ISL_1855290, EPI_ISL_1855291, EPI_ISL_1855292, EPI_ISL_1855293 | | | |
| see above | Columbia University Irving Medical Center | Wadsworth Center, New York State Department of Health | Alexis Russell; Catharine Prussing; Daryl M. Lamson; Erasmus Schneider; Erica Lasek-Nesselquist; John Kelly; Jonathan Plitnick; Kirsten St. George; Matthew Shudt; Melissa A Leisner; Navjot Singh |

| | | | |
|---|---|---|---|
| EPI_ISL_2098600 | DELHI REHABILITATION AND NURSING CENTER | Wadsworth Center, New York State Department of Health | Alexis Russell; Catharine Prussing; Daryl M. Lamson; Erasmus Schneider; Erica Lasek-Nesselquist; John Kelly; Jonathan Plitnick; Kirsten St. George; Matthew Shudt; Melissa A Leisner; Navjot Singh |
| EPI_ISL_1694702, EPI_ISL_1694703, EPI_ISL_1694709, EPI_ISL_1694718, EPI_ISL_1694759, EPI_ISL_1694773, EPI_ISL_1694774, EPI_ISL_1694775, EPI_ISL_1804945, EPI_ISL_1804954, EPI_ISL_1804955, EPI_ISL_2151368, EPI_ISL_2151382, EPI_ISL_2151387, EPI_ISL_2151391 | | | |
| see above | DOHMH Central Harlem | New York City Public Health Laboratory | Jade Wang; et al. |
| EPI_ISL_1694697, EPI_ISL_1694698, EPI_ISL_1694706, EPI_ISL_1694730, EPI_ISL_1804946, EPI_ISL_1804950, EPI_ISL_2151383, EPI_ISL_2151390 | | | |
| see above | DOHMH Chelsea | New York City Public Health Laboratory | Jade Wang; et al. |
| EPI_ISL_1593768, EPI_ISL_1694707, EPI_ISL_1694710, EPI_ISL_1694713, EPI_ISL_1694714, EPI_ISL_1694734, EPI_ISL_1694735, EPI_ISL_1694750, EPI_ISL_1694751, EPI_ISL_1694752, EPI_ISL_1694753, EPI_ISL_1804943, EPI_ISL_1804957, EPI_ISL_1804960 | | | |
| see above | DOHMH Corona | New York City Public Health Laboratory | Jade Wang; et al. |
| EPI_ISL_1593769, EPI_ISL_1694711, EPI_ISL_1694728, EPI_ISL_1804952, EPI_ISL_1804953, EPI_ISL_2151367, EPI_ISL_2151375, EPI_ISL_2151376, EPI_ISL_2151377, EPI_ISL_2151379, EPI_ISL_2151380, EPI_ISL_2151385 | | | |
| see above | DOHMH Crown Heights | New York City Public Health Laboratory | Jade Wang; et al. |
| EPI_ISL_1593770, EPI_ISL_1694719, EPI_ISL_1694733, EPI_ISL_1694746, EPI_ISL_1694747, EPI_ISL_2151384 | DOHMH Fort Greene | New York City Public Health Laboratory | Jade Wang; et al. |
| EPI_ISL_1694699, EPI_ISL_1694708, EPI_ISL_1694712, EPI_ISL_1694736, EPI_ISL_1694737, EPI_ISL_1694738, EPI_ISL_1694739, EPI_ISL_1694740, EPI_ISL_1694754, EPI_ISL_1694755, EPI_ISL_1694756, EPI_ISL_1694757, EPI_ISL_1694758, EPI_ISL_1804942, EPI_ISL_1804958, EPI_ISL_1804959, EPI_ISL_2151369, EPI_ISL_2151370, EPI_ISL_2151389, EPI_ISL_2151392 | | | |
| see above | DOHMH Jamaica | New York City Public Health Laboratory | Jade Wang; et al. |
| EPI_ISL_1694704, EPI_ISL_1694705, EPI_ISL_1694717, EPI_ISL_1694729, EPI_ISL_1694776, EPI_ISL_1804956, EPI_ISL_2151381 | | | |
| see above | DOHMH Morrisania | New York City Public Health Laboratory | Jade Wang; et al. |
| EPI_ISL_1694715, EPI_ISL_1694716, EPI_ISL_1694741, EPI_ISL_1694742, EPI_ISL_1694743, EPI_ISL_1694744, EPI_ISL_1694745, EPI_ISL_1694748, EPI_ISL_1694749, EPI_ISL_1804944, EPI_ISL_1804951, EPI_ISL_2151386, EPI_ISL_2151393, EPI_ISL_2151394 | | | |
| see above | DOHMH PHL | New York City Public Health Laboratory | Jade Wang; et al. |
| EPI_ISL_1694700, EPI_ISL_1694701, EPI_ISL_2151371, EPI_ISL_2151372, EPI_ISL_2151373, EPI_ISL_2151374, EPI_ISL_2151388 | | | |
| see above | DOHMH Riverside | New York City Public Health Laboratory | Jade Wang; et al. |
| EPI_ISL_1694760, EPI_ISL_1694761, EPI_ISL_1694762, EPI_ISL_1804941, EPI_ISL_1804947, EPI_ISL_2151378, EPI_ISL_2151402, EPI_ISL_2151403 | | | |
| see above | Department of Homeless Services | New York City Public Health Laboratory | Jade Wang; et al. |
| EPI_ISL_1937166, EPI_ISL_1937357 | Fulgent Genetics | Fulgent Genetics | Becky Tsai; Benafsh Sapra; Doreen Ng; Harry Gao; James Xie; John Gao; Joseph Fierro; Mickey Li; Yan Meng |
| EPI_ISL_1855144, EPI_ISL_1855145, EPI_ISL_1855146, EPI_ISL_1855147, EPI_ISL_1855148, EPI_ISL_1855149, EPI_ISL_1855241, EPI_ISL_1855242, EPI_ISL_1855243 | | | |
| see above | GLENS FALLS HOSPITAL LABORATORY | Wadsworth Center, New York State Department of Health | Alexis Russell; Catharine Prussing; Daryl M. Lamson; Erasmus Schneider; Erica Lasek-Nesselquist; John Kelly; Jonathan Plitnick; Kirsten St. George; Matthew Shudt; Melissa A Leisner; Navjot Singh |
| EPI_ISL_1624089, EPI_ISL_1624090 | GOOD SAMARITAN HOSPITAL LABORATORY | Wadsworth Center, New York State Department of Health | Alexis Russell; Catharine Prussing; Daryl M. Lamson; Erasmus Schneider; Erica Lasek-Nesselquist; John Kelly; Jonathan Plitnick; Kirsten St. George; Matthew Shudt; Melissa A Leisner; Navjot Singh |
| EPI_ISL_1921037 | Glens Falls Hospital Laboratory | Wadsworth Center, New York State Department of Health | Alexis Russell; Catharine Prussing; Daryl M. Lamson; Erasmus Schneider; Erica Lasek-Nesselquist; John Kelly; Jonathan Plitnick; Kirsten St. George; Matthew Shudt; Melissa A Leisner; Navjot Singh |
| EPI_ISL_2151399 | HealthQuest Esoterics | New York City Public Health Laboratory | Jade Wang; et al. |
| EPI_ISL_1679008, EPI_ISL_1679029, EPI_ISL_1679426, EPI_ISL_1679621, EPI_ISL_1679640, EPI_ISL_1679734, EPI_ISL_1690890, EPI_ISL_1691266, EPI_ISL_1691275, EPI_ISL_1734943, EPI_ISL_1735142, EPI_ISL_1735643, EPI_ISL_1796130, EPI_ISL_1796436, EPI_ISL_1796534, EPI_ISL_1796840, EPI_ISL_1797051, EPI_ISL_1797367, EPI_ISL_1803619, EPI_ISL_1803731, EPI_ISL_1803819, EPI_ISL_1906926, EPI_ISL_1907154, EPI_ISL_1907210, EPI_ISL_1907414, EPI_ISL_1907867, EPI_ISL_1907911, EPI_ISL_1923513, EPI_ISL_1923597, EPI_ISL_1923605, EPI_ISL_1923719, EPI_ISL_1924033, EPI_ISL_1924207, EPI_ISL_1990722, EPI_ISL_1990941, EPI_ISL_1991481, EPI_ISL_1991836, EPI_ISL_2009862, EPI_ISL_2010081, EPI_ISL_2010284, EPI_ISL_2010619, EPI_ISL_2010810, EPI_ISL_2010860, EPI_ISL_2144378, EPI_ISL_2144388, EPI_ISL_2144820, EPI_ISL_2159854, EPI_ISL_2160675 | | | |
| see above | Helix/Illumina | Centers for Disease Control and Prevention Division of Viral Diseases, Pathogen Discovery | Adrian Paskey; Alexandre Bolze; Ary Ascencio; Benjamin Rambo-Martin; Brad Sickler; Charlotte Rivera-Garcia; Christine Tran; Christopher Gulvick; Clinton R. Paden; Dakota Howard; Darlene Wagner; David Becker; Dhwani Batra; Duncan MacCannell; Efren Sandoval; Eileen de Feo; Elizabeth Cirulli; Eric Allen; Geraint Levan; James Lu; Jan Antico; Jason Caravas; Jason Nguyen; Jimmy Ramirez; Jingtao Liu; Kara Moser; Kelly Schiabor Barrett; Kim Gietzen; Magnus Isaksson; Marc Laurent; Matthew Schmerer; Matthew Tolentino; Nicole L. Washington; Peter W. Cook; Phil Febbo; Ryan Cho; Scott Sammons; Shannon Wickline; Shatavia Morrison; Sherry Wang; Simon White; Tyler Cassens; William Lee; Yvette Unoarumhi |
| EPI_ISL_1587589, EPI_ISL_1587712, EPI_ISL_1587757, EPI_ISL_1587786, EPI_ISL_1587941, EPI_ISL_1587944, EPI_ISL_1587951, EPI_ISL_1587952, EPI_ISL_1587968, EPI_ISL_1587975, EPI_ISL_1588112, EPI_ISL_1588322, EPI_ISL_1588324, EPI_ISL_1588405, EPI_ISL_1588406, EPI_ISL_1588412, EPI_ISL_1588415, EPI_ISL_1588419, EPI_ISL_1588427, EPI_ISL_1588433, EPI_ISL_1694015, EPI_ISL_1694028, EPI_ISL_1694242, EPI_ISL_1694519, EPI_ISL_1733367, EPI_ISL_1733640, EPI_ISL_1733721, EPI_ISL_1795506, EPI_ISL_1795589, EPI_ISL_1795597, EPI_ISL_1795605, EPI_ISL_1795615, EPI_ISL_1795620, EPI_ISL_1795621, EPI_ISL_1795622, EPI_ISL_1802835, EPI_ISL_1817992, EPI_ISL_2014189, EPI_ISL_2087865, EPI_ISL_2088170, EPI_ISL_2088271, EPI_ISL_2089216, EPI_ISL_2089245, EPI_ISL_2089442 | | | |
| see above | Infinity Biologix | Centers for Disease Control and Prevention Division of Viral Diseases, Pathogen Discovery | Adrian Paskey; Benjamin Rambo-Martin; Chirayu Goswami; Christian Bixby; Christopher Gulvick; Clinton R. Paden; Dakota Howard; Darlene Wagner; Dhwani Batra; Duncan MacCannell; Jason Caravas; Jonathan Schultz; Kara Moser; Matthew Schmerer; Peter W. Cook; Robin Grimwood; Russ Hager; Scott Sammons; Shatavia Morrison; Yihe Wang; Yvette Unoarumhi |
| EPI_ISL_1611300, EPI_ISL_1611308, EPI_ISL_1611311, EPI_ISL_1611314, EPI_ISL_1611315, EPI_ISL_1611316, EPI_ISL_1611363, EPI_ISL_1611366, EPI_ISL_1611368, EPI_ISL_1611371, EPI_ISL_1611374, EPI_ISL_1611375, EPI_ISL_1611376, EPI_ISL_1611385, EPI_ISL_1611398, EPI_ISL_1611399, EPI_ISL_1611417, EPI_ISL_1611420, EPI_ISL_1611547, EPI_ISL_1611549, EPI_ISL_1611564, EPI_ISL_1611565, EPI_ISL_1611566, EPI_ISL_1611567, EPI_ISL_1611568, EPI_ISL_1611570, EPI_ISL_1611572, EPI_ISL_1611624, EPI_ISL_1611625, EPI_ISL_1611628, EPI_ISL_1611632, EPI_ISL_1611640, EPI_ISL_1611644, EPI_ISL_1611647, EPI_ISL_1611650, EPI_ISL_1611651, EPI_ISL_1611653, EPI_ISL_1611654, EPI_ISL_1611659, EPI_ISL_1611660, EPI_ISL_1611667, EPI_ISL_1611668, EPI_ISL_1611690, EPI_ISL_1611693, EPI_ISL_1611695, EPI_ISL_1611839, EPI_ISL_1611911, EPI_ISL_1611913, EPI_ISL_1611917, EPI_ISL_1611919, EPI_ISL_1611948, EPI_ISL_1611952, EPI_ISL_1611954, EPI_ISL_1611955, EPI_ISL_1611957, EPI_ISL_1611959, EPI_ISL_1680475, EPI_ISL_1680479, EPI_ISL_1680480, EPI_ISL_1680482, EPI_ISL_1680484, EPI_ISL_1680485, EPI_ISL_1680486, EPI_ISL_1680487, EPI_ISL_1680488, EPI_ISL_1680489, EPI_ISL_1680490, EPI_ISL_1680536, EPI_ISL_1680540, EPI_ISL_1680541, EPI_ISL_1680542, EPI_ISL_1680543, EPI_ISL_1680544, EPI_ISL_1680545, EPI_ISL_1680549, EPI_ISL_1680550, EPI_ISL_1680551, EPI_ISL_1680552, EPI_ISL_1680553, EPI_ISL_1680554, EPI_ISL_1680566, EPI_ISL_1680585, EPI_ISL_1680586, EPI_ISL_1680591, EPI_ISL_1680592, EPI_ISL_1680593, EPI_ISL_1680594, EPI_ISL_1680595, EPI_ISL_1680596, EPI_ISL_1680597, EPI_ISL_1680598, EPI_ISL_1680599, EPI_ISL_1680603, EPI_ISL_1680638, EPI_ISL_1680639, EPI_ISL_1680640, EPI_ISL_1680641, EPI_ISL_1680642, EPI_ISL_1680643, EPI_ISL_1680644, EPI_ISL_1680672, EPI_ISL_1680674, EPI_ISL_1680747, EPI_ISL_1680748, EPI_ISL_1680765, EPI_ISL_1680767, EPI_ISL_1680768, EPI_ISL_1680769, EPI_ISL_1680802, EPI_ISL_1680847, EPI_ISL_1680848, EPI_ISL_1681168, EPI_ISL_1681169, EPI_ISL_1681170, EPI_ISL_1681171, EPI_ISL_1681172, EPI_ISL_1681173, EPI_ISL_1681187, EPI_ISL_1681188, EPI_ISL_1681190, EPI_ISL_1681193, EPI_ISL_1681195, EPI_ISL_1681249, EPI_ISL_1681250, EPI_ISL_1681251, EPI_ISL_1681309, EPI_ISL_1681310, EPI_ISL_1681311, EPI_ISL_1681314, EPI_ISL_1681315, EPI_ISL_1681316, EPI_ISL_1681318, EPI_ISL_1681321, EPI_ISL_1681370, EPI_ISL_1681371, EPI_ISL_1681372, EPI_ISL_1681409, EPI_ISL_1681415, EPI_ISL_1681416, EPI_ISL_1681417, EPI_ISL_1681418, EPI_ISL_1681419, EPI_ISL_1681420, EPI_ISL_1681444, EPI_ISL_1681445, EPI_ISL_1681446, EPI_ISL_1681447, EPI_ISL_1681448, EPI_ISL_1681451, EPI_ISL_1681452, EPI_ISL_1681453, EPI_ISL_1681461, EPI_ISL_1681495, EPI_ISL_1681496, EPI_ISL_1681497, EPI_ISL_1681498, EPI_ISL_1681499, EPI_ISL_1681500, EPI_ISL_1681542, EPI_ISL_1681543, EPI_ISL_1681544, EPI_ISL_1681545, EPI_ISL_1681546, EPI_ISL_1681547, EPI_ISL_1681548, EPI_ISL_1681549, EPI_ISL_1681550, EPI_ISL_1681551, EPI_ISL_1681552, EPI_ISL_1681644, EPI_ISL_1681645, EPI_ISL_1681646, EPI_ISL_1681647, EPI_ISL_1681648, EPI_ISL_1681649, EPI_ISL_1682025, EPI_ISL_1682026, EPI_ISL_1682061, EPI_ISL_1682062, EPI_ISL_1682071, EPI_ISL_1682085, EPI_ISL_1682109, EPI_ISL_1682111, EPI_ISL_1682137, EPI_ISL_1682161, EPI_ISL_1682162, EPI_ISL_1682522, EPI_ISL_1682525, EPI_ISL_1682596, EPI_ISL_1682608, EPI_ISL_1682787, EPI_ISL_1682808, EPI_ISL_1682811, EPI_ISL_1682929, EPI_ISL_1682961, EPI_ISL_1683040, EPI_ISL_1683058, EPI_ISL_1683061, EPI_ISL_1683064, EPI_ISL_1683088, EPI_ISL_1683104, EPI_ISL_1683107, EPI_ISL_1683126, EPI_ISL_1683132, EPI_ISL_1683136, EPI_ISL_1683180, EPI_ISL_1683181, EPI_ISL_1683185, EPI_ISL_1683187, EPI_ISL_1683197, EPI_ISL_1683202, EPI_ISL_1683203, EPI_ISL_1683210, EPI_ISL_1683212, EPI_ISL_1683214, EPI_ISL_1683215, EPI_ISL_1683216, EPI_ISL_1683248, EPI_ISL_1683249, EPI_ISL_1683254, EPI_ISL_1683258, EPI_ISL_1683260, EPI_ISL_1683262, EPI_ISL_1683265, EPI_ISL_1683266, EPI_ISL_1683267, EPI_ISL_1683277, EPI_ISL_1683281, EPI_ISL_1683289, EPI_ISL_1683290, EPI_ISL_1683292, EPI_ISL_1683293, EPI_ISL_1683294, EPI_ISL_1683295, EPI_ISL_1683299, EPI_ISL_1683321, EPI_ISL_1683322, EPI_ISL_1683323, EPI_ISL_1683325, EPI_ISL_1683326, EPI_ISL_1683330, EPI_ISL_1683331, EPI_ISL_1683332, EPI_ISL_1683333, EPI_ISL_1683336, EPI_ISL_1683351, EPI_ISL_1683358, EPI_ISL_1683361, EPI_ISL_1683381, EPI_ISL_1683409, EPI_ISL_1683410, EPI_ISL_1683413, EPI_ISL_1683458, EPI_ISL_1683461, EPI_ISL_1683464, EPI_ISL_1683485, EPI_ISL_1683494, EPI_ISL_1683497, EPI_ISL_1683499, EPI_ISL_1683501, EPI_ISL_1683503, EPI_ISL_1683504, EPI_ISL_1683531, EPI_ISL_1683532, EPI_ISL_1683536, EPI_ISL_1683537, EPI_ISL_1683542, EPI_ISL_1683543, EPI_ISL_1683558, EPI_ISL_1683566, EPI_ISL_1683599, EPI_ISL_1683617, EPI_ISL_1683634, EPI_ISL_1683635, EPI_ISL_1683661, EPI_ISL_1683662, EPI_ISL_1683663, EPI_ISL_1683664, EPI_ISL_1683666, EPI_ISL_1683667, EPI_ISL_1683668, EPI_ISL_1683669, EPI_ISL_1683670, EPI_ISL_1683672, EPI_ISL_1683675, EPI_ISL_1683701, EPI_ISL_1683749, EPI_ISL_1683756, EPI_ISL_1683760, EPI_ISL_1683768, EPI_ISL_1683769, EPI_ISL_1683772, EPI_ISL_1683799, EPI_ISL_1683800, EPI_ISL_1683808, EPI_ISL_1683811, EPI_ISL_1683833, EPI_ISL_1683835, EPI_ISL_1683836, EPI_ISL_1683837, EPI_ISL_1683839, EPI_ISL_1683910, EPI_ISL_1684063, EPI_ISL_1684065, EPI_ISL_1684071, EPI_ISL_1684084, EPI_ISL_1684085, EPI_ISL_1684153, EPI_ISL_1684162, EPI_ISL_1684164, EPI_ISL_1684165, EPI_ISL_1684167, EPI_ISL_1684168, EPI_ISL_1684173, EPI_ISL_1684174, EPI_ISL_1684177, EPI_ISL_1684211, EPI_ISL_1684212, EPI_ISL_1684213, EPI_ISL_1684214, EPI_ISL_1684253, EPI_ISL_1684254, EPI_ISL_1684255, EPI_ISL_1684256, EPI_ISL_1684261, EPI_ISL_1684263, EPI_ISL_1684265, EPI_ISL_1684266, EPI_ISL_1684267, EPI_ISL_1684271, EPI_ISL_1684276, EPI_ISL_1684277, EPI_ISL_1684279, EPI_ISL_1684287, EPI_ISL_1684289, EPI_ISL_1684303, EPI_ISL_1684305, EPI_ISL_1684307, EPI_ISL_1684541, EPI_ISL_1685066, EPI_ISL_1799429, EPI_ISL_1799495, EPI_ISL_1799693, EPI_ISL_1799740, EPI_ISL_1799741, EPI_ISL_1799744, EPI_ISL_1799746, EPI_ISL_1799747, EPI_ISL_1799769, EPI_ISL_1799781, EPI_ISL_1799784, EPI_ISL_1799790, EPI_ISL_1799841, EPI_ISL_1799842, EPI_ISL_1799877, EPI_ISL_1799938, EPI_ISL_1799950, EPI_ISL_1799970, EPI_ISL_1799974, EPI_ISL_1799975, EPI_ISL_1799980, EPI_ISL_1799983, EPI_ISL_1799984, EPI_ISL_1799985, EPI_ISL_1799986, EPI_ISL_1799992, EPI_ISL_1800344, EPI_ISL_1800349, EPI_ISL_1800355, EPI_ISL_1800357, EPI_ISL_1800359, EPI_ISL_1800402, EPI_ISL_1800404, EPI_ISL_1800405, EPI_ISL_1800406, EPI_ISL_1800407, EPI_ISL_1800409, EPI_ISL_1800413, EPI_ISL_1800455, EPI_ISL_1800457, EPI_ISL_1800460, EPI_ISL_1800461, EPI_ISL_1800470, EPI_ISL_1800471, EPI_ISL_1800472, EPI_ISL_1800473, EPI_ISL_1800475, EPI_ISL_1800479, EPI_ISL_1800488, EPI_ISL_1800489, EPI_ISL_1800490, EPI_ISL_1800491, EPI_ISL_1800492, EPI_ISL_1800513, EPI_ISL_1800515, EPI_ISL_1800516, EPI_ISL_1800517, EPI_ISL_1800527, EPI_ISL_1800533, EPI_ISL_1800541, EPI_ISL_1800542, EPI_ISL_1800666, EPI_ISL_1800695, EPI_ISL_1800701, EPI_ISL_1800747, EPI_ISL_1800753, EPI_ISL_1800765, EPI_ISL_1800766, EPI_ISL_1800773, EPI_ISL_1800774, EPI_ISL_1800778, EPI_ISL_1800780, EPI_ISL_1800781, EPI_ISL_1800799, EPI_ISL_1800816, EPI_ISL_1800817, EPI_ISL_1800822, EPI_ISL_1800824, EPI_ISL_1800825, EPI_ISL_1800828, EPI_ISL_1800829, EPI_ISL_1800859, EPI_ISL_1800860, EPI_ISL_1800866, EPI_ISL_1800889, EPI_ISL_1800891, EPI_ISL_1800892, EPI_ISL_1801293, EPI_ISL_1801320, EPI_ISL_1801321, EPI_ISL_1801323, EPI_ISL_1801324, EPI_ISL_1801325, EPI_ISL_1801327, EPI_ISL_1801403, EPI_ISL_1801437, EPI_ISL_1801487, EPI_ISL_1801488, EPI_ISL_1801507, EPI_ISL_1801508, EPI_ISL_1801509, EPI_ISL_1801510, EPI_ISL_1801513, EPI_ISL_1801514, EPI_ISL_1801516, EPI_ISL_1801517, EPI_ISL_1801518, EPI_ISL_1801519, EPI_ISL_1801520, EPI_ISL_1801521, EPI_ISL_1801522, EPI_ISL_1801523, EPI_ISL_1801524, EPI_ISL_1801525, EPI_ISL_1801526, EPI_ISL_1801527, EPI_ISL_1801528, EPI_ISL_1801574, EPI_ISL_1801575, EPI_ISL_1801576, EPI_ISL_1801577, EPI_ISL_1801579, EPI_ISL_1801580, EPI_ISL_1801609, EPI_ISL_1801610, EPI_ISL_1801611, EPI_ISL_1801612, EPI_ISL_1801634, EPI_ISL_1801663, EPI_ISL_1801664, EPI_ISL_1801666, EPI_ISL_1801667, EPI_ISL_1801668, EPI_ISL_1801669, EPI_ISL_1801670, EPI_ISL_1801671, EPI_ISL_1801672, EPI_ISL_1801673, EPI_ISL_1801762, EPI_ISL_1801846, EPI_ISL_1801852, EPI_ISL_1801860, EPI_ISL_1801866, EPI_ISL_1802295, EPI_ISL_1802304, EPI_ISL_1802307, EPI_ISL_1802310, EPI_ISL_1802312, EPI_ISL_1802324, EPI_ISL_1802512, EPI_ISL_1802513, EPI_ISL_1802574, EPI_ISL_1802576, EPI_ISL_1802663, EPI_ISL_1802664, EPI_ISL_1925748, EPI_ISL_1925789, EPI_ISL_1925924, EPI_ISL_1925980, EPI_ISL_1925983, EPI_ISL_1925986, EPI_ISL_1926045, EPI_ISL_1926108, EPI_ISL_1926229, EPI_ISL_1926230, EPI_ISL_1926231, EPI_ISL_1926253, EPI_ISL_1926262, EPI_ISL_1926292, EPI_ISL_1926296, EPI_ISL_1926297, EPI_ISL_1926300, EPI_ISL_1926301, EPI_ISL_1926302, EPI_ISL_1926303, EPI_ISL_1926312, EPI_ISL_1926323, EPI_ISL_1926340, EPI_ISL_1926372, EPI_ISL_1926412, EPI_ISL_1926413, EPI_ISL_1926414, EPI_ISL_1926415, EPI_ISL_1926424, EPI_ISL_1926425, EPI_ISL_1926426, EPI_ISL_1926427, EPI_ISL_1926428, EPI_ISL_1926548, EPI_ISL_1926569, EPI_ISL_1926571, EPI_ISL_1926572, EPI_ISL_1926573, EPI_ISL_1926574, EPI_ISL_1926575, EPI_ISL_1926576, EPI_ISL_1926577, EPI_ISL_1926578, EPI_ISL_1927480, EPI_ISL_1927554, EPI_ISL_1927587, EPI_ISL_1927610, EPI_ISL_1927613, EPI_ISL_1927616, EPI_ISL_1927619, EPI_ISL_1927622, EPI_ISL_1927625, EPI_ISL_1927639, EPI_ISL_1927868, EPI_ISL_1927871, EPI_ISL_1927899, EPI_ISL_1927947, EPI_ISL_1927977, EPI_ISL_1927989, EPI_ISL_1927992, EPI_ISL_1927998, EPI_ISL_1928001, EPI_ISL_1928004, EPI_ISL_1928007, EPI_ISL_1928010, EPI_ISL_1928013, EPI_ISL_1928034, EPI_ISL_1928040, EPI_ISL_1928179, EPI_ISL_1928182, EPI_ISL_1928210, EPI_ISL_1928231, EPI_ISL_1928268, EPI_ISL_1928282, EPI_ISL_1928292, EPI_ISL_1928295, EPI_ISL_1928314, EPI_ISL_1928371, EPI_ISL_1928372, EPI_ISL_1928373, EPI_ISL_1928374, EPI_ISL_1928384, EPI_ISL_1928390, EPI_ISL_1928393, EPI_ISL_1928589, EPI_ISL_1928606, EPI_ISL_1928609, EPI_ISL_1928646, EPI_ISL_1928651, EPI_ISL_1928652, EPI_ISL_1928653, EPI_ISL_1928663, EPI_ISL_1928664, EPI_ISL_1928665, EPI_ISL_1928706, EPI_ISL_1928712, EPI_ISL_1928835, EPI_ISL_1928867, EPI_ISL_1928931, EPI_ISL_1928938, EPI_ISL_1928945, EPI_ISL_1929024, EPI_ISL_1929041, EPI_ISL_1929056, EPI_ISL_1929057, EPI_ISL_1929285, EPI_ISL_1929301, EPI_ISL_1929325, EPI_ISL_1929328, EPI_ISL_1929338, EPI_ISL_1929392, EPI_ISL_1929657, EPI_ISL_1929870, EPI_ISL_1929902, EPI_ISL_1930006, EPI_ISL_1930009, EPI_ISL_1930022, EPI_ISL_1930060, EPI_ISL_1930062, EPI_ISL_1930066, EPI_ISL_1930158, EPI_ISL_1930162, EPI_ISL_1930184, EPI_ISL_1930244, EPI_ISL_1930247, EPI_ISL_1930250, EPI_ISL_1930261, EPI_ISL_1930264, EPI_ISL_1930292, EPI_ISL_1930295, EPI_ISL_1930356, EPI_ISL_1930358, EPI_ISL_1942239, EPI_ISL_2044008, EPI_ISL_2044009, EPI_ISL_2044074, EPI_ISL_2044076, EPI_ISL_2044077, EPI_ISL_2044079, EPI_ISL_2044080, EPI_ISL_2044155, EPI_ISL_2044165, EPI_ISL_2044174, EPI_ISL_2044181, EPI_ISL_2044208, EPI_ISL_2044210, EPI_ISL_2044211, EPI_ISL_2044216, EPI_ISL_2044217, EPI_ISL_2044218, EPI_ISL_2044219, EPI_ISL_2044222, EPI_ISL_2044254, EPI_ISL_2044512, EPI_ISL_2044521, EPI_ISL_2044522, EPI_ISL_2044555, EPI_ISL_2044576, EPI_ISL_2044580, EPI_ISL_2044581, EPI_ISL_2044638, EPI_ISL_2044644, EPI_ISL_2044647, EPI_ISL_2044663, EPI_ISL_2044671, EPI_ISL_2044713, EPI_ISL_2044761, EPI_ISL_2044934, EPI_ISL_2044935, EPI_ISL_2044936, EPI_ISL_2044937, EPI_ISL_2044981, EPI_ISL_2044993, EPI_ISL_2044995, EPI_ISL_2044998, EPI_ISL_2045017, EPI_ISL_2045036, EPI_ISL_2045037, EPI_ISL_2045039, EPI_ISL_2045087, EPI_ISL_2045089, EPI_ISL_2045090, EPI_ISL_2045095, EPI_ISL_2045469, EPI_ISL_2045470, EPI_ISL_2045471, EPI_ISL_2045472, EPI_ISL_2045473, EPI_ISL_2045474, EPI_ISL_2045475, EPI_ISL_2045476, EPI_ISL_2045559, EPI_ISL_2045571, EPI_ISL_2045572, EPI_ISL_2045720, EPI_ISL_2045725, EPI_ISL_2045726, EPI_ISL_2045727, EPI_ISL_2045728, EPI_ISL_2045729, EPI_ISL_2045750, EPI_ISL_2045751, EPI_ISL_2045754, EPI_ISL_2045756, EPI_ISL_2045785, EPI_ISL_2045786, EPI_ISL_2045793, EPI_ISL_2045794, EPI_ISL_2045795, EPI_ISL_2045796, EPI_ISL_2045797, EPI_ISL_2045798, EPI_ISL_2045799, EPI_ISL_2045800, EPI_ISL_2045815, EPI_ISL_2045816, EPI_ISL_2045817, EPI_ISL_2045818, EPI_ISL_2045820, EPI_ISL_2045821, EPI_ISL_2045822, EPI_ISL_2045823, EPI_ISL_2045853, EPI_ISL_2045859 | | | |
| see above | Laboratory Corporation of America | Centers for Disease Control and Prevention Division of Viral Diseases, Pathogen Discovery | Adrian Paskey; Amanda Douglas; Amanda Suchanek; Andrea Throop; Ayla Burns; Benjamin Rambo-Martin; Bobbi Croy; Brian Krueger; Brian Norvell; Christopher Gulvick; Christos Petropoulos; Clinton R. Paden; Craig Lukasik; Dakota Howard; Darlene Wagner; Debbie Boles; Dhwani Batra; Duncan MacCannell; Eyad Almasri; Goran Stevovic; Howard Engler; Hrushikesh Deshmukh; Jake Humphrey; Jana Schroth; Jason Caravas; Joe Voshell; John Pruitt; Jonathan Meltzer; Jonathan Williams; Kara Moser; Kimberly Wagner; Lax Iyer; Lyndon Tilson; Manoj Jain; Marcia Eisenberg; Mary Ann Cristobal; Mary Williamson; Matthew Schmerer; Michael Levandoski; Mike Sapeta; Mindy Nye; Minoo Agarwal; Mohan Kolli; Nuthawin Charoensri; Oren Cohen; Peter W. Cook; Prashant Gupta; Qian Zeng; Rama Ghatti; Scott Parker; Scott Ryan; Scott Sammons; Shatavia Morrison; Stanley Letovsky; Steven Ragan; Suresh Babu Selvaraju; Susan Countryman; Susan Hicks; Suzanne Dale; Thomas Urban; Tim Kuphal; Tricia Zwiefelhofer; Vincent Drouillon; Yvette Unoarumhi |
| EPI_ISL_1671466, EPI_ISL_1671477, EPI_ISL_1671478, EPI_ISL_1671479, EPI_ISL_1671497, EPI_ISL_1671498, EPI_ISL_1740512, EPI_ISL_1740513, EPI_ISL_1756342, EPI_ISL_1756343, EPI_ISL_1756344, EPI_ISL_1756345, EPI_ISL_1756346, EPI_ISL_1756347, EPI_ISL_1756348, EPI_ISL_1756349, EPI_ISL_1756350, EPI_ISL_1756351, EPI_ISL_1756352, EPI_ISL_1756353, EPI_ISL_1756354, EPI_ISL_1756355, | | | |

| | | | |
|---|---|---|---|
| EPI_ISL_1789720, EPI_ISL_1789721, EPI_ISL_1855324, EPI_ISL_1855325, EPI_ISL_1855326, EPI_ISL_1855327, EPI_ISL_1855328, EPI_ISL_1855329, EPI_ISL_1855330, EPI_ISL_1855331, EPI_ISL_1855332, EPI_ISL_1855333, EPI_ISL_2097253, EPI_ISL_2097254, EPI_ISL_2097255, EPI_ISL_2097256, EPI_ISL_2097257, EPI_ISL_2097258, EPI_ISL_2097259, EPI_ISL_2097260, EPI_ISL_2097261, EPI_ISL_2097262, EPI_ISL_2097263, EPI_ISL_2097264, EPI_ISL_2097265, EPI_ISL_2134785 | | | |
| see above | MONTEFIORE MEDICAL CENTER LABORATORIES | Wadsworth Center, New York State Department of Health | Alexis Russell; Catharine Prussing; Daryl M. Lamson; Erasmus Schneider; Erica Lasek-Nesselquist; John Kelly; Jonathan Plitnick; Kirsten St. George; Matthew Shudt; Melissa A Leisner; Navjot Singh |
| EPI_ISL_1709817, EPI_ISL_1709818, EPI_ISL_1709819, EPI_ISL_1709820, EPI_ISL_1709821, EPI_ISL_1709822, EPI_ISL_1709823, EPI_ISL_1709824, EPI_ISL_1709825, EPI_ISL_1709826, EPI_ISL_1709827, EPI_ISL_1709828, EPI_ISL_1709829, EPI_ISL_1709830, EPI_ISL_1709831, EPI_ISL_1709832, EPI_ISL_1709833, EPI_ISL_1709834, EPI_ISL_1709835, EPI_ISL_1709836, EPI_ISL_1709837, EPI_ISL_1709838, EPI_ISL_1709839, EPI_ISL_1709840, EPI_ISL_1709841, EPI_ISL_1709842, EPI_ISL_1709843, EPI_ISL_1709844, EPI_ISL_1709845, EPI_ISL_1709846, EPI_ISL_1709847, EPI_ISL_1709848, EPI_ISL_1709849, EPI_ISL_1709850, EPI_ISL_1709851, EPI_ISL_1709852, EPI_ISL_1709853, EPI_ISL_1709854, EPI_ISL_1709855, EPI_ISL_1709856, EPI_ISL_1709857, EPI_ISL_1709858, EPI_ISL_1709859, EPI_ISL_1709860, EPI_ISL_1709861, EPI_ISL_1709862, EPI_ISL_1709863, EPI_ISL_1709864, EPI_ISL_1709865, EPI_ISL_1709866, EPI_ISL_1709867, EPI_ISL_1709868, EPI_ISL_1709869, EPI_ISL_1709870, EPI_ISL_1709871, EPI_ISL_1709872, EPI_ISL_1709873, EPI_ISL_1709874, EPI_ISL_1709875, EPI_ISL_1709876, EPI_ISL_1709877, EPI_ISL_1709878, EPI_ISL_1709879, EPI_ISL_1709880, EPI_ISL_1709881, EPI_ISL_1709882, EPI_ISL_1709883, EPI_ISL_1709884, EPI_ISL_1709885, EPI_ISL_1709886, EPI_ISL_1709887, EPI_ISL_1709888, EPI_ISL_1709890, EPI_ISL_1709891, EPI_ISL_1709892, EPI_ISL_1709893, EPI_ISL_1709894, EPI_ISL_1709895, EPI_ISL_1709896, EPI_ISL_1709897, EPI_ISL_1709898, EPI_ISL_1709899, EPI_ISL_1709900, EPI_ISL_1709901, EPI_ISL_1709902, EPI_ISL_1709903, EPI_ISL_1709904, EPI_ISL_1709905, EPI_ISL_1709906, EPI_ISL_1709907, EPI_ISL_1709908, EPI_ISL_1709909, EPI_ISL_1709910, EPI_ISL_1709911, EPI_ISL_1709912, EPI_ISL_1709913, EPI_ISL_1709914, EPI_ISL_1709915, EPI_ISL_1709916, EPI_ISL_1709917, EPI_ISL_1709918, EPI_ISL_1709919, EPI_ISL_1709920, EPI_ISL_1709921, EPI_ISL_1709922, EPI_ISL_1709923, EPI_ISL_1709924, EPI_ISL_1709925, EPI_ISL_1709926, EPI_ISL_1709927, EPI_ISL_1709928, EPI_ISL_1709929, EPI_ISL_1709930, EPI_ISL_1709931, EPI_ISL_1709932, EPI_ISL_1709933, EPI_ISL_1709934, EPI_ISL_1709935, EPI_ISL_1709936, EPI_ISL_1709937, EPI_ISL_1709938, EPI_ISL_1709939, EPI_ISL_1709940, EPI_ISL_1709941, EPI_ISL_1709942, EPI_ISL_1709943, EPI_ISL_1709944, EPI_ISL_1709945, EPI_ISL_1709947, EPI_ISL_1709948, EPI_ISL_1709950, EPI_ISL_1709951, EPI_ISL_1709952, EPI_ISL_1709953, EPI_ISL_1709954, EPI_ISL_1709955, EPI_ISL_1709956, EPI_ISL_1709957, EPI_ISL_1709958, EPI_ISL_1709959, EPI_ISL_1709960, EPI_ISL_1709961, EPI_ISL_1709962, EPI_ISL_1709963, EPI_ISL_1709964, EPI_ISL_1709965, EPI_ISL_1709966, EPI_ISL_1709967, EPI_ISL_1709968, EPI_ISL_1709969, EPI_ISL_1709970, EPI_ISL_1709971, EPI_ISL_1709972, EPI_ISL_1709974, EPI_ISL_1709975, EPI_ISL_1709976, EPI_ISL_1709977, EPI_ISL_1709978, EPI_ISL_1709979, EPI_ISL_1709980, EPI_ISL_1709981, EPI_ISL_1709982, EPI_ISL_1709983, EPI_ISL_1709984, EPI_ISL_1709985, EPI_ISL_1709986, EPI_ISL_1709987, EPI_ISL_1709988, EPI_ISL_1709989, EPI_ISL_1709991, EPI_ISL_1709992, EPI_ISL_1709993, EPI_ISL_1709994, EPI_ISL_1709995, EPI_ISL_1709996, EPI_ISL_1709997, EPI_ISL_1709998, EPI_ISL_1709999, EPI_ISL_1710000, EPI_ISL_1710001, EPI_ISL_1710002, EPI_ISL_1710003, EPI_ISL_1710004, EPI_ISL_1710005, EPI_ISL_1710006, EPI_ISL_1710007, EPI_ISL_1710008, EPI_ISL_1710009, EPI_ISL_1710010, EPI_ISL_1710011, EPI_ISL_1710012, EPI_ISL_1710013, EPI_ISL_1710014 | | | |
| see above | MSHS Clinical Microbiology Laboratories | MSHS Pathogen Surveillance Program | Adolfo García-Sastre; Adriana van de Guchte; Ajay Obla; Alberto Paniz-Mondolfi; Ana S. Gonzalez-Reiche; Angela Amoako; Ashley Salimbangon; Betsaida Salom Melo; Bremy Alburquerque; Brianne Ciferri; Charles Gleason; Daniel Floda; Deena R. Altman; Denise Jurczyszak; Emilia Mia Sordillo; Gintaras Deikus; Giulio Kleiner; Gopi Patel; Hala Alshammary; Harm van Bakel; Irina Oussenko; Jayeeta Dutta; Juan Soto; Julia Matthews; Katherine Beach; Kathryn Twyman; Kayla Russo; Komal Srivastava; Levy Sominsky; Mahmoud Awawda; Marta Luksza; Matthew M. Hernandez; Melissa Gitman; Michael D. Nowak; Mitchell J. Sullivan; Nancy Francoeur; Robert Sebra; Sarah Schaefer; Shelcie Fabre; Shwetha Hara Sridhar; Viviana Simon; Ying-Chih Wang; Zenab Khan |
| EPI_ISL_2009117, EPI_ISL_2009122, EPI_ISL_2009123, EPI_ISL_2009125, EPI_ISL_2009127, EPI_ISL_2009131, EPI_ISL_2009132, EPI_ISL_2009133, EPI_ISL_2009134, EPI_ISL_2009137, EPI_ISL_2009138, EPI_ISL_2009139, EPI_ISL_2009140, EPI_ISL_2009141, EPI_ISL_2009142, EPI_ISL_2009143, EPI_ISL_2009150, EPI_ISL_2009152, EPI_ISL_2009158, EPI_ISL_2009160, EPI_ISL_2009165, EPI_ISL_2009166, EPI_ISL_2009167, EPI_ISL_2009168, EPI_ISL_2009169, EPI_ISL_2009177 | | | |
| see above | MSK Microbiology Lab | MSK Microbiology Lab | Esther Babady; Krupa Jani; Tracy McMillen |
| EPI_ISL_1919997, EPI_ISL_1919999, EPI_ISL_1920000, EPI_ISL_1920002, EPI_ISL_1920003, EPI_ISL_1920004, EPI_ISL_1920005, EPI_ISL_1920006 | | | |
| see above | Montefiore Medical Center Laboratories | Wadsworth Center, New York State Department of Health | Alexis Russell; Catharine Prussing; Daryl M. Lamson; Erasmus Schneider; Erica Lasek-Nesselquist; John Kelly; Jonathan Plitnick; Kirsten St. George; Matthew Shudt; Melissa A Leisner; Navjot Singh |
| EPI_ISL_1674703 | Murphy Medical Association | Grubaugh Lab - Yale School of Public Health | Anderson Brito; Annie Watkins; Caleb Neal; Chaney Kalinich; Chantal Vogels; Eva Laszlo; Isabel Ott; Jessica Rothman; Joseph Fauver; Mallery Breban; Mary Petrone; Nathan Grubaugh; Steven Murphy; Tara Alpert |
| EPI_ISL_1671448, EPI_ISL_1671452, EPI_ISL_1671453, EPI_ISL_1671493, EPI_ISL_1855348, EPI_ISL_1855349, EPI_ISL_1855350, EPI_ISL_1855351, EPI_ISL_1855352, EPI_ISL_1855353 | | | |
| see above | NORTH SHORE UNIVERSITY HOSPITAL | Wadsworth Center, New York State Department of Health | Alexis Russell; Catharine Prussing; Daryl M. Lamson; Erasmus Schneider; Erica Lasek-Nesselquist; John Kelly; Jonathan Plitnick; Kirsten St. George; Matthew Shudt; Melissa A Leisner; Navjot Singh |
| EPI_ISL_1582284, EPI_ISL_1582285, EPI_ISL_1582289, EPI_ISL_1582290, EPI_ISL_1582292, EPI_ISL_1582369, EPI_ISL_1582372, EPI_ISL_1671601, EPI_ISL_1671602, EPI_ISL_1671603, EPI_ISL_1671604, EPI_ISL_1671605, EPI_ISL_1671606, EPI_ISL_1671608, EPI_ISL_1671609, EPI_ISL_1671614, EPI_ISL_1671615, EPI_ISL_1671616, EPI_ISL_1671617, EPI_ISL_1671620, EPI_ISL_1671621, EPI_ISL_1671628, EPI_ISL_1671629, EPI_ISL_1671630, EPI_ISL_1671631, EPI_ISL_1671632, EPI_ISL_1671633, EPI_ISL_1671638, EPI_ISL_1671640, EPI_ISL_1671641, EPI_ISL_1671648, EPI_ISL_1671649, EPI_ISL_1671650, EPI_ISL_1671651, EPI_ISL_1671652, EPI_ISL_1671653, EPI_ISL_1671661, EPI_ISL_1671662, EPI_ISL_1671663, EPI_ISL_1671668, EPI_ISL_1671669, EPI_ISL_1671670, EPI_ISL_1671671, EPI_ISL_1671672, EPI_ISL_1671673, EPI_ISL_1671674, EPI_ISL_1671675, EPI_ISL_1671676, EPI_ISL_1740518, EPI_ISL_1740519, EPI_ISL_1740520, EPI_ISL_1740521, EPI_ISL_1740522, EPI_ISL_1756330, EPI_ISL_1935608, EPI_ISL_1935621, EPI_ISL_1935624, EPI_ISL_1935626, EPI_ISL_1935629, EPI_ISL_1935637, EPI_ISL_2097266, EPI_ISL_2097267, EPI_ISL_2097268, EPI_ISL_2097269, EPI_ISL_2097270, EPI_ISL_2097271, EPI_ISL_2097272, EPI_ISL_2097273, EPI_ISL_2097274, EPI_ISL_2097275, EPI_ISL_2097276, EPI_ISL_2097277, EPI_ISL_2097278, EPI_ISL_2097279, EPI_ISL_2097280, EPI_ISL_2097281, EPI_ISL_2097282, EPI_ISL_2097283, EPI_ISL_2097284, EPI_ISL_2097285, EPI_ISL_2097286, EPI_ISL_2097287, EPI_ISL_2097288, EPI_ISL_2097289, EPI_ISL_2097290, EPI_ISL_2097291, EPI_ISL_2097292, EPI_ISL_2097293, EPI_ISL_2097294, EPI_ISL_2098132, EPI_ISL_2134786, EPI_ISL_2134787 | | | |
| see above | NORTHWELL HEALTH LABORATORIES | Wadsworth Center, New York State Department of Health | Alexis Russell; Catharine Prussing; Daryl M. Lamson; Erasmus Schneider; Erica Lasek-Nesselquist; John Kelly; Jonathan Plitnick; Kirsten St. George; Matthew Shudt; Melissa A Leisner; Navjot Singh |
| EPI_ISL_1823944, EPI_ISL_1823945, EPI_ISL_1823947, EPI_ISL_1823948, EPI_ISL_1823949, EPI_ISL_1823950, EPI_ISL_1823951, EPI_ISL_1823952, EPI_ISL_1823953, EPI_ISL_1823954, EPI_ISL_1823955, EPI_ISL_1823956, EPI_ISL_1823957, EPI_ISL_1823959, EPI_ISL_1823960, EPI_ISL_1823961, EPI_ISL_1823962, EPI_ISL_1823963, EPI_ISL_1823964, EPI_ISL_1823965, EPI_ISL_1823966, EPI_ISL_1823967, EPI_ISL_1823970, EPI_ISL_1823971, EPI_ISL_1823972, EPI_ISL_1823973, EPI_ISL_1937596, EPI_ISL_1937598, EPI_ISL_1937600, EPI_ISL_1937601, EPI_ISL_1937603, EPI_ISL_1937605, EPI_ISL_1937606, EPI_ISL_1937608, EPI_ISL_1937610, EPI_ISL_1937611, EPI_ISL_1937613, EPI_ISL_1937615, EPI_ISL_1937616, EPI_ISL_1937618, EPI_ISL_1937620, EPI_ISL_1937622, EPI_ISL_1937623, EPI_ISL_2018418, EPI_ISL_2018419, EPI_ISL_2018420, EPI_ISL_2018421, EPI_ISL_2018422, EPI_ISL_2018423, EPI_ISL_2018424, EPI_ISL_2018425, EPI_ISL_2018426, EPI_ISL_2018427, EPI_ISL_2018428, EPI_ISL_2018429, EPI_ISL_2018430, EPI_ISL_2018431, EPI_ISL_2018432, EPI_ISL_2018433, EPI_ISL_2047506 | | | |
| see above | NYC Department of Health and Mental Hygiene | Centers for Disease Control and Prevention Division of Viral Diseases, Pathogen Discovery | Alison Laufer Halpin; Ben L. Rambo-Martin; Clinton R. Paden; Dakota Howard; Darlene Wagner; Dave Wentworth; Dhwani Batra; Jasmine Padilla; Justin Lee; Katie Dillon; Krista Queen; Kristen Knipe; Kristine Lacek; Mark Burroughs; Matthew Schmerer; Mili Sheth; Peter Cook; Sam Shepard; Sarah Nobles; Shoshona Le; Suxiang Tong; Vivien Dugan; Yvette Unoarumhi |
| EPI_ISL_1854703, EPI_ISL_1854736, EPI_ISL_1854737, EPI_ISL_1854738, EPI_ISL_1854739, EPI_ISL_1935631, EPI_ISL_1935636, EPI_ISL_2098601, EPI_ISL_2098602, EPI_ISL_2098603, EPI_ISL_2098604, EPI_ISL_2098605, EPI_ISL_2098606, EPI_ISL_2098607, EPI_ISL_2098608, EPI_ISL_2098609, EPI_ISL_2098610, EPI_ISL_2098611, EPI_ISL_2098612, EPI_ISL_2098613, EPI_ISL_2098614, EPI_ISL_2098615, EPI_ISL_2098616, EPI_ISL_2098617, EPI_ISL_2098618, EPI_ISL_2098619, EPI_ISL_2098620, EPI_ISL_2098621 | | | |
| see above | NYC Pandemic Response Lab | Wadsworth Center, New York State Department of Health | Alexis Russell; Catharine Prussing; Daryl M. Lamson; Erasmus Schneider; Erica Lasek-Nesselquist; John Kelly; Jonathan Plitnick; Kirsten St. George; Matthew Shudt; Melissa A Leisner; Navjot Singh |
| EPI_ISL_2018243, EPI_ISL_2018244, EPI_ISL_2018245, EPI_ISL_2018246 | NYSDOH Wadsworth Center, Virology Lab | Centers for Disease Control and Prevention Division of Viral Diseases, Pathogen Discovery | Alison Laufer Halpin; Ben L. Rambo-Martin; Clinton R. Paden; Dakota Howard; Darlene Wagner; Dave Wentworth; Dhwani Batra; Jasmine Padilla; Justin Lee; Katie Dillon; Krista Queen; Kristen Knipe; Kristine Lacek; Mark Burroughs; Matthew Schmerer; Mili Sheth; Peter Cook; Sam Shepard; Sarah Nobles; Shoshona Le; Suxiang Tong; Vivien Dugan; Yvette Unoarumhi |
| EPI_ISL_1576862, EPI_ISL_1576863, EPI_ISL_1576864, EPI_ISL_1576865, EPI_ISL_1576866, EPI_ISL_1576867, EPI_ISL_1576868, EPI_ISL_1576869, EPI_ISL_1576870, EPI_ISL_1576871, EPI_ISL_1576876, EPI_ISL_1576877, EPI_ISL_1576878, EPI_ISL_1576879, EPI_ISL_1576884, EPI_ISL_1576885, EPI_ISL_1576886, EPI_ISL_1576887, EPI_ISL_1576888, EPI_ISL_1576889, EPI_ISL_1576890, EPI_ISL_1576891, EPI_ISL_1576892, EPI_ISL_1576893, EPI_ISL_1576894, EPI_ISL_1576895, EPI_ISL_1576896, EPI_ISL_1576897, EPI_ISL_1576898, EPI_ISL_1576899, EPI_ISL_1576900, EPI_ISL_1576901, EPI_ISL_1576902, EPI_ISL_1576904, EPI_ISL_1576912, EPI_ISL_1576913, EPI_ISL_1576914, EPI_ISL_1576915, EPI_ISL_1576916, EPI_ISL_1576917, EPI_ISL_1576918, EPI_ISL_1576919, EPI_ISL_1576920, EPI_ISL_1576921, EPI_ISL_1576922, EPI_ISL_1576923, EPI_ISL_1576924, EPI_ISL_1576925, EPI_ISL_1576926, EPI_ISL_1576927, EPI_ISL_1576928, EPI_ISL_1576929, EPI_ISL_1576930, EPI_ISL_1576931, EPI_ISL_1576932, EPI_ISL_1576934, EPI_ISL_1576935, EPI_ISL_1576936, EPI_ISL_1576937, EPI_ISL_1576938, EPI_ISL_1576939, EPI_ISL_1576940, EPI_ISL_1576941, EPI_ISL_1576942, EPI_ISL_1576944, EPI_ISL_1576948, EPI_ISL_1654611, EPI_ISL_1654613, EPI_ISL_1654614, EPI_ISL_1654615, EPI_ISL_1654616, EPI_ISL_1654617, EPI_ISL_1654618, EPI_ISL_1654619, EPI_ISL_1654620, EPI_ISL_1654621, EPI_ISL_1654622, EPI_ISL_1654623, EPI_ISL_1654624, EPI_ISL_1654625, EPI_ISL_1654626, EPI_ISL_1654627, EPI_ISL_1654628, EPI_ISL_1654629, EPI_ISL_1654630, EPI_ISL_1654631, EPI_ISL_1654632, EPI_ISL_1654633, EPI_ISL_1654634, EPI_ISL_1654635, EPI_ISL_1654636, EPI_ISL_1654637, EPI_ISL_1654638, EPI_ISL_1654639, EPI_ISL_1654640, EPI_ISL_1654641, EPI_ISL_1654644, EPI_ISL_1654645, EPI_ISL_1654646, EPI_ISL_1654647, EPI_ISL_1669801, EPI_ISL_1669802, EPI_ISL_1669803, EPI_ISL_1669821, EPI_ISL_1789903, EPI_ISL_1789904, EPI_ISL_1789905, EPI_ISL_1789906, EPI_ISL_1789907, EPI_ISL_1789908, EPI_ISL_1789909, EPI_ISL_1789910, EPI_ISL_1789911, EPI_ISL_1789912, EPI_ISL_1789913, EPI_ISL_1789914, EPI_ISL_1789915, EPI_ISL_1789916, EPI_ISL_1789917, EPI_ISL_1789918, EPI_ISL_1789919, EPI_ISL_1789920, EPI_ISL_1789921, EPI_ISL_1789922, EPI_ISL_1789923, EPI_ISL_1789924, EPI_ISL_1789925, EPI_ISL_1789926, EPI_ISL_1789928, EPI_ISL_1789929, EPI_ISL_1789930, EPI_ISL_1789931, EPI_ISL_1789932, EPI_ISL_1789933, EPI_ISL_1789934, EPI_ISL_1789935, EPI_ISL_1789936, EPI_ISL_1789938, EPI_ISL_1789939, EPI_ISL_1789940, EPI_ISL_1789941, EPI_ISL_1789942, EPI_ISL_1789943, EPI_ISL_1789944, EPI_ISL_1789945, EPI_ISL_1789946, EPI_ISL_1789947, EPI_ISL_1789948, EPI_ISL_1789949, EPI_ISL_1789950, EPI_ISL_1789951, EPI_ISL_1789952, EPI_ISL_1789953, EPI_ISL_1789954, EPI_ISL_1789955, EPI_ISL_1789956, EPI_ISL_1789957, EPI_ISL_1789958, EPI_ISL_1789959, EPI_ISL_1789960, EPI_ISL_1789961, EPI_ISL_1789962, EPI_ISL_1789963, EPI_ISL_1789964, EPI_ISL_1789966, EPI_ISL_1789967, EPI_ISL_1789968, EPI_ISL_1789969, EPI_ISL_1789970, EPI_ISL_1789971, EPI_ISL_1789972, EPI_ISL_1789973, EPI_ISL_1789974, EPI_ISL_1789975, EPI_ISL_1789976, EPI_ISL_1789977, EPI_ISL_1789978, EPI_ISL_1789979, EPI_ISL_1789980, EPI_ISL_1789981, EPI_ISL_1789982, EPI_ISL_1789983, EPI_ISL_1789984, EPI_ISL_1789985, EPI_ISL_1789986, EPI_ISL_1789987, EPI_ISL_1789988, EPI_ISL_1789989, EPI_ISL_1789990, EPI_ISL_1789991, EPI_ISL_1789992, EPI_ISL_1789993, EPI_ISL_1789994, EPI_ISL_1789995, EPI_ISL_1789996, EPI_ISL_1789997, EPI_ISL_1789998, EPI_ISL_1789999, EPI_ISL_1790000, EPI_ISL_1790001, EPI_ISL_1790003, EPI_ISL_1790004, EPI_ISL_1790005, EPI_ISL_1790006, EPI_ISL_1790007, EPI_ISL_1790009, EPI_ISL_1790010, EPI_ISL_1790011, EPI_ISL_1790012, EPI_ISL_1790013, EPI_ISL_1790014, EPI_ISL_1790015, EPI_ISL_1790016, EPI_ISL_1790017, EPI_ISL_1790019, EPI_ISL_1790020, EPI_ISL_1790021, EPI_ISL_1790022, EPI_ISL_1790023, EPI_ISL_1790024, EPI_ISL_1790025, EPI_ISL_1790027, EPI_ISL_1790028, EPI_ISL_1790029, EPI_ISL_1790030, EPI_ISL_1790031, EPI_ISL_1790032, EPI_ISL_1790033, EPI_ISL_1790034, EPI_ISL_1790035, EPI_ISL_1790036, EPI_ISL_1790037, EPI_ISL_1790038, EPI_ISL_1790039, EPI_ISL_1790040, EPI_ISL_1790041, EPI_ISL_1790042, EPI_ISL_1790043, EPI_ISL_1790044, EPI_ISL_1790045, EPI_ISL_1790046, EPI_ISL_1790047, EPI_ISL_1922800, EPI_ISL_1922801, EPI_ISL_1922802, EPI_ISL_1922803, EPI_ISL_1922804, EPI_ISL_1922805, EPI_ISL_1922806, EPI_ISL_1922807, EPI_ISL_1922808, EPI_ISL_1922809, EPI_ISL_1922810, EPI_ISL_1922811, EPI_ISL_1922812, EPI_ISL_1922813, EPI_ISL_1922814, EPI_ISL_1922815, EPI_ISL_1922816, EPI_ISL_1922817, EPI_ISL_1922818, EPI_ISL_1922819, EPI_ISL_1922820, EPI_ISL_1922821, EPI_ISL_1922822, EPI_ISL_1922823, EPI_ISL_1922824, EPI_ISL_1922825, EPI_ISL_1922826, EPI_ISL_1922827, EPI_ISL_1922828, EPI_ISL_1922829, EPI_ISL_1922830, EPI_ISL_1922831, EPI_ISL_1922832, EPI_ISL_1922833, EPI_ISL_1922834, EPI_ISL_1922835, EPI_ISL_1922836, EPI_ISL_1922837, EPI_ISL_1922838, EPI_ISL_1922840, EPI_ISL_1922842, EPI_ISL_1922844, EPI_ISL_1922845, EPI_ISL_1922846, EPI_ISL_1922847, EPI_ISL_1922848, EPI_ISL_1922849, EPI_ISL_1922851, EPI_ISL_1922852, EPI_ISL_1922853, EPI_ISL_1922857, EPI_ISL_1922858, EPI_ISL_1922861, EPI_ISL_1922863, EPI_ISL_1922866, EPI_ISL_1922867, EPI_ISL_1922869, EPI_ISL_1922870, EPI_ISL_1922871, EPI_ISL_2161702, EPI_ISL_2161703, EPI_ISL_2161704, EPI_ISL_2161705, EPI_ISL_2161706, EPI_ISL_2161707, EPI_ISL_2161708, EPI_ISL_2161709, EPI_ISL_2161710, EPI_ISL_2161711, EPI_ISL_2161712, EPI_ISL_2161713, EPI_ISL_2161714, EPI_ISL_2161715, EPI_ISL_2161716, EPI_ISL_2161717, EPI_ISL_2161718, EPI_ISL_2161719, EPI_ISL_2161720, EPI_ISL_2161721, EPI_ISL_2161722, EPI_ISL_2161724, EPI_ISL_2161725, EPI_ISL_2161726, EPI_ISL_2161727, EPI_ISL_2161728, EPI_ISL_2161729, EPI_ISL_2161730, EPI_ISL_2161731, EPI_ISL_2161751, EPI_ISL_2161752, EPI_ISL_2161753, EPI_ISL_2161754, EPI_ISL_2161755, EPI_ISL_2161756, EPI_ISL_2161757, EPI_ISL_2161758, EPI_ISL_2161759, EPI_ISL_2161760, EPI_ISL_2161761, EPI_ISL_2161762, EPI_ISL_2161763, EPI_ISL_2161764, EPI_ISL_2161765, EPI_ISL_2161767, EPI_ISL_2161768, EPI_ISL_2161769, EPI_ISL_2161770, EPI_ISL_2161771, EPI_ISL_2161772, EPI_ISL_2161773, EPI_ISL_2161774, EPI_ISL_2161775, EPI_ISL_2161781, EPI_ISL_2161782 | | | |
| see above | NYU Langone Health | Departments of Pathology and Medicine, New York University School of Medicine | Adriana Heguy; Christian Marier; Dacia Dimartino; Emily Guzman; Gael Westby; Guiqing Wang; Paolo Cotzia; Paul Zappile; Peter Meyn; Sitharam Ramaswami; Yutong Zhang |
| EPI_ISL_1919988, EPI_ISL_1919989, EPI_ISL_1919993, EPI_ISL_1919994, EPI_ISL_1919995, EPI_ISL_1919996, EPI_ISL_1920008, EPI_ISL_1920010, EPI_ISL_1920011, EPI_ISL_1920012, EPI_ISL_1920013, EPI_ISL_1920014, EPI_ISL_1920015, EPI_ISL_1920016, EPI_ISL_1920017, EPI_ISL_1920018, EPI_ISL_1920019, EPI_ISL_1920020, EPI_ISL_1920021, EPI_ISL_1920022, EPI_ISL_1920023, EPI_ISL_1920024, EPI_ISL_1920025, EPI_ISL_1920026, EPI_ISL_1920027, EPI_ISL_1920028, EPI_ISL_1920031, EPI_ISL_1920032, EPI_ISL_1920033, EPI_ISL_1920034, EPI_ISL_1920036, EPI_ISL_1920037, EPI_ISL_1920038, EPI_ISL_1920039, EPI_ISL_1920040, EPI_ISL_1920042, EPI_ISL_1920043, EPI_ISL_1920045, EPI_ISL_1920101 | | | |
| see above | Northwell Health Laboratories | Wadsworth Center, New York State Department of Health | Alexis Russell; Catharine Prussing; Daryl M. Lamson; Erasmus Schneider; Erica Lasek-Nesselquist; John Kelly; Jonathan Plitnick; Kirsten St. George; Matthew Shudt; Melissa A Leisner; Navjot Singh |
| EPI_ISL_1704714 | Northwestern Memorial Hospital | Northwestern University - Ozer Lab | Chad J. Achenbach; Chao Qi; Egon A. Ozer; Judd F. Hultquist; Lacy M. Simons; Lawrence J. Jennings; Michael G. Ison; Ramon Lorenzo-Redondo; Taylor J. Dean |
| EPI_ISL_1593772, EPI_ISL_1694721, EPI_ISL_1694722, EPI_ISL_1694723, EPI_ISL_1694724, EPI_ISL_1694725, EPI_ISL_1694726, EPI_ISL_1694727, EPI_ISL_1694731, EPI_ISL_1694732, EPI_ISL_1694763, EPI_ISL_1694764, EPI_ISL_1694765, EPI_ISL_1694766, EPI_ISL_1694767, EPI_ISL_1694768, EPI_ISL_1694769, EPI_ISL_1694770, EPI_ISL_1694771, EPI_ISL_1694772, EPI_ISL_1804948, EPI_ISL_1804949, EPI_ISL_1804961, EPI_ISL_2151404, EPI_ISL_2151405, EPI_ISL_2151406, EPI_ISL_2151407, EPI_ISL_2151408, EPI_ISL_2151409, EPI_ISL_2151410 | | | |
| see above | OCME Office Of Chief Medical Examiner | New York City Public Health Laboratory | Jade Wang; et al. |
| EPI_ISL_1471859, EPI_ISL_1471869, EPI_ISL_1471870, EPI_ISL_1471880, EPI_ISL_1471882, EPI_ISL_1471883, EPI_ISL_1471884, EPI_ISL_1471892, EPI_ISL_1471895, EPI_ISL_1471898, EPI_ISL_1471900, EPI_ISL_1471901, EPI_ISL_1471903, EPI_ISL_1471904, EPI_ISL_1471905, EPI_ISL_1471909, EPI_ISL_1471910, EPI_ISL_1471911, EPI_ISL_1471917, EPI_ISL_1471925, EPI_ISL_1471929, EPI_ISL_1471932, EPI_ISL_1471934, EPI_ISL_1471938, EPI_ISL_1471940, EPI_ISL_1471943, EPI_ISL_1471947, EPI_ISL_1471949, EPI_ISL_1471950, EPI_ISL_1471953, EPI_ISL_1471955, EPI_ISL_1471958, EPI_ISL_1471962, EPI_ISL_1471970, EPI_ISL_1471971, EPI_ISL_1471973, EPI_ISL_1471974, EPI_ISL_1471975, EPI_ISL_1471976, EPI_ISL_1471978, EPI_ISL_1471984, EPI_ISL_1471986, EPI_ISL_1471989, EPI_ISL_1471993, EPI_ISL_1471994, EPI_ISL_1471995, EPI_ISL_1471997, EPI_ISL_1471998, EPI_ISL_1472001, EPI_ISL_1472002, EPI_ISL_1472009, EPI_ISL_1472017, EPI_ISL_1472019, EPI_ISL_1472023, EPI_ISL_1472027, EPI_ISL_1472029, EPI_ISL_1472032, EPI_ISL_1472034, EPI_ISL_1472040, EPI_ISL_1472044, EPI_ISL_1472045, EPI_ISL_1472050, EPI_ISL_1472051, EPI_ISL_1472057, EPI_ISL_1472059, EPI_ISL_1472061, EPI_ISL_1472062, EPI_ISL_1472063, EPI_ISL_1472068, EPI_ISL_1472072, EPI_ISL_1472075, EPI_ISL_1542074, EPI_ISL_1542075, EPI_ISL_1542076, EPI_ISL_1542077, EPI_ISL_1542078, EPI_ISL_1542079, EPI_ISL_1542080, EPI_ISL_1542081, EPI_ISL_1542082, EPI_ISL_1542083, EPI_ISL_1542084, EPI_ISL_1542085, EPI_ISL_1542086, EPI_ISL_1542087, EPI_ISL_1542088, EPI_ISL_1542089, EPI_ISL_1542090, EPI_ISL_1542091, EPI_ISL_1542092, EPI_ISL_1542093, EPI_ISL_1542094, EPI_ISL_1542095, EPI_ISL_1542096, EPI_ISL_1542097, EPI_ISL_1542098, EPI_ISL_1542099, EPI_ISL_1542100, EPI_ISL_1542101, EPI_ISL_1542102, EPI_ISL_1542103, EPI_ISL_1542104, EPI_ISL_1542105, EPI_ISL_1542106, EPI_ISL_1542107, EPI_ISL_1542108, EPI_ISL_1542109, EPI_ISL_1542110, EPI_ISL_1542111, EPI_ISL_1542112, EPI_ISL_1542113, EPI_ISL_1542114, EPI_ISL_1542115, EPI_ISL_1542116, EPI_ISL_1542117, EPI_ISL_1542118, EPI_ISL_1542119, EPI_ISL_1542120, EPI_ISL_1542121, EPI_ISL_1542122, EPI_ISL_1542123, EPI_ISL_1542124, EPI_ISL_1542125, EPI_ISL_1542126, EPI_ISL_1542127, EPI_ISL_1542128, EPI_ISL_1542129, EPI_ISL_1542130, EPI_ISL_1542131, EPI_ISL_1542132, EPI_ISL_1542133, EPI_ISL_1542134, EPI_ISL_1542135, EPI_ISL_1542136, EPI_ISL_1542137, EPI_ISL_1542138, EPI_ISL_1542139, EPI_ISL_1542140, EPI_ISL_1542141, EPI_ISL_1542142, EPI_ISL_1542143, EPI_ISL_1542144, EPI_ISL_1542145, EPI_ISL_1542146, EPI_ISL_1542147, EPI_ISL_1542148, EPI_ISL_1542149, EPI_ISL_1542150, EPI_ISL_1542151, EPI_ISL_1542152, EPI_ISL_1542153, EPI_ISL_1542154, EPI_ISL_1542155, EPI_ISL_1542156, EPI_ISL_1542157, EPI_ISL_1542158, EPI_ISL_1542159, EPI_ISL_1542160, EPI_ISL_1542161, EPI_ISL_1542162, EPI_ISL_1542163, EPI_ISL_1542164, EPI_ISL_1542165, EPI_ISL_1542166, EPI_ISL_1542167, EPI_ISL_1542168, EPI_ISL_1542169, EPI_ISL_1542170, EPI_ISL_1542171, EPI_ISL_1542172, EPI_ISL_1542173, EPI_ISL_1542174, EPI_ISL_1542175, EPI_ISL_1542176, EPI_ISL_1542177, EPI_ISL_1542178, EPI_ISL_1542179, EPI_ISL_1542180, EPI_ISL_1542181, EPI_ISL_1542182, EPI_ISL_1542183, EPI_ISL_1542184, EPI_ISL_1542185, EPI_ISL_1542186, EPI_ISL_1542187, EPI_ISL_1542188, EPI_ISL_1542189, EPI_ISL_1542190, EPI_ISL_1542191, EPI_ISL_1542192, EPI_ISL_1542193, EPI_ISL_1542194, EPI_ISL_1542195, EPI_ISL_1542196, EPI_ISL_1542197, EPI_ISL_1542198, EPI_ISL_1542200, EPI_ISL_1542201, EPI_ISL_1542202, EPI_ISL_1542203, EPI_ISL_1542204, EPI_ISL_1542205, EPI_ISL_1542206, EPI_ISL_1542207, EPI_ISL_1542208, EPI_ISL_1542209, EPI_ISL_1542210, EPI_ISL_1542211, EPI_ISL_1542212, EPI_ISL_1542213, EPI_ISL_1542214, EPI_ISL_1542215, EPI_ISL_1542216, EPI_ISL_1542217, EPI_ISL_1542218, EPI_ISL_1542219, EPI_ISL_1542220, EPI_ISL_1542221, EPI_ISL_1542222, EPI_ISL_1542223, EPI_ISL_1542224, EPI_ISL_1542225, EPI_ISL_1542226, EPI_ISL_1542227, EPI_ISL_1542228, EPI_ISL_1542229, EPI_ISL_1542230, EPI_ISL_1542231, EPI_ISL_1542232, EPI_ISL_1542233, EPI_ISL_1542234, EPI_ISL_1542235, EPI_ISL_1542236, EPI_ISL_1542237, EPI_ISL_1542238, EPI_ISL_1542239, EPI_ISL_1542240, EPI_ISL_1542241, EPI_ISL_1542242, EPI_ISL_1542243, EPI_ISL_1542244, EPI_ISL_1542245, EPI_ISL_1542246, EPI_ISL_1542247, EPI_ISL_1542248, EPI_ISL_1542249, EPI_ISL_1542250, EPI_ISL_1542251, EPI_ISL_1542252, EPI_ISL_1542253, EPI_ISL_1542254, EPI_ISL_1542255, EPI_ISL_1542256, EPI_ISL_1542257, EPI_ISL_1542258, EPI_ISL_1542259, EPI_ISL_1542260, EPI_ISL_1542261, EPI_ISL_1542262, EPI_ISL_1542263, EPI_ISL_1542264, EPI_ISL_1542265, EPI_ISL_1542266, EPI_ISL_1542267, EPI_ISL_1542268, EPI_ISL_1542269, EPI_ISL_1542270, EPI_ISL_1542271, EPI_ISL_1542272, EPI_ISL_1542273, EPI_ISL_1542274, EPI_ISL_1542275, EPI_ISL_1542276, EPI_ISL_1542277, EPI_ISL_1542278, EPI_ISL_1542279, EPI_ISL_1542280, EPI_ISL_1542281, EPI_ISL_1542282, EPI_ISL_1542283, EPI_ISL_1542284, EPI_ISL_1542285, EPI_ISL_1542286, EPI_ISL_1542287, EPI_ISL_1542288, EPI_ISL_1542289, EPI_ISL_1542290, EPI_ISL_1542291, EPI_ISL_1542292, EPI_ISL_1542293, EPI_ISL_1542294, EPI_ISL_1542295, EPI_ISL_1542296, EPI_ISL_1542297, EPI_ISL_1542298, EPI_ISL_1542299, EPI_ISL_1542300, EPI_ISL_1542301, EPI_ISL_1542302, EPI_ISL_1542303, EPI_ISL_1542304, EPI_ISL_1542305, EPI_ISL_1542306, EPI_ISL_1542307, EPI_ISL_1542308, EPI_ISL_1542309, EPI_ISL_1542310, EPI_ISL_1542311, EPI_ISL_1542312, EPI_ISL_1542313, EPI_ISL_1542314, EPI_ISL_1542315, EPI_ISL_1542316, EPI_ISL_1542317, EPI_ISL_1542318, EPI_ISL_1542319, EPI_ISL_1542320, EPI_ISL_1542321, EPI_ISL_1542322, EPI_ISL_1542323, EPI_ISL_1542324, EPI_ISL_1542325, EPI_ISL_1542326, EPI_ISL_1542327, EPI_ISL_1542328, EPI_ISL_1542329, EPI_ISL_1542330, EPI_ISL_1542331, EPI_ISL_1542332, EPI_ISL_1542333, EPI_ISL_1542334, EPI_ISL_1542335, EPI_ISL_1542336, EPI_ISL_1542337, EPI_ISL_1542338, EPI_ISL_1542339, EPI_ISL_1542340, EPI_ISL_1542341, EPI_ISL_1542342, EPI_ISL_1542343, EPI_ISL_1542344, EPI_ISL_1542345, EPI_ISL_1542346, EPI_ISL_1542347, EPI_ISL_1542348, EPI_ISL_1542349, EPI_ISL_1542350, EPI_ISL_1542351, EPI_ISL_1542352, EPI_ISL_1542353, EPI_ISL_1542354, EPI_ISL_1542355, EPI_ISL_1542356, EPI_ISL_1542357, EPI_ISL_1542358, EPI_ISL_1542359, EPI_ISL_1542360, EPI_ISL_1542361, EPI_ISL_1542362, EPI_ISL_1542363, EPI_ISL_1542364, EPI_ISL_1542365, EPI_ISL_1542366, EPI_ISL_1542367, EPI_ISL_1542368, EPI_ISL_1542369, EPI_ISL_1542370, EPI_ISL_1542371, EPI_ISL_1542372, EPI_ISL_1542373, EPI_ISL_1542374, EPI_ISL_1542375, EPI_ISL_1542376, EPI_ISL_1542377, EPI_ISL_1542378, EPI_ISL_1542379, EPI_ISL_1542380, EPI_ISL_1542381, EPI_ISL_1542382, EPI_ISL_1542383, EPI_ISL_1542384, EPI_ISL_1542385, EPI_ISL_1542386, EPI_ISL_1542387, EPI_ISL_1542388, EPI_ISL_1542389, EPI_ISL_1542390, EPI_ISL_1542391, EPI_ISL_1542392, EPI_ISL_1542393, EPI_ISL_1542394, EPI_ISL_1542395, EPI_ISL_1542396, EPI_ISL_1542397, EPI_ISL_1542398, EPI_ISL_1542399, EPI_ISL_1542400, EPI_ISL_1542401, EPI_ISL_1542402, EPI_ISL_1542403, EPI_ISL_1542404, EPI_ISL_1542405, EPI_ISL_1542406, EPI_ISL_1542407, EPI_ISL_1542408, EPI_ISL_1542409, EPI_ISL_1542410, EPI_ISL_1542411, EPI_ISL_1542412, EPI_ISL_1542413, EPI_ISL_1542414, EPI_ISL_1542415, EPI_ISL_1542416, EPI_ISL_1542417, EPI_ISL_1542418, EPI_ISL_1542419, EPI_ISL_1542420, EPI_ISL_1542421, EPI_ISL_1542422, EPI_ISL_1542423, EPI_ISL_1542424, EPI_ISL_1542425, EPI_ISL_1542426, EPI_ISL_1542427, EPI_ISL_1542428, EPI_ISL_1542429, EPI_ISL_1542430, EPI_ISL_1542431, EPI_ISL_1542432, EPI_ISL_1542433, EPI_ISL_1542434, EPI_ISL_1542435, EPI_ISL_1542436, EPI_ISL_1542437, EPI_ISL_1542438, EPI_ISL_1542439, EPI_ISL_1542440, EPI_ISL_1542441, EPI_ISL_1542442, EPI_ISL_1542443, EPI_ISL_1542444, EPI_ISL_1542445, EPI_ISL_1542446, EPI_ISL_1542447, EPI_ISL_1542448, EPI_ISL_1542449, EPI_ISL_1542450, EPI_ISL_1542451, EPI_ISL_1542452, EPI_ISL_1542453, EPI_ISL_1542454, EPI_ISL_1542455, EPI_ISL_1542456, EPI_ISL_1542457, EPI_ISL_1542458, EPI_ISL_1542459, EPI_ISL_1542460, EPI_ISL_1542461, EPI_ISL_1542462, EPI_ISL_1542463, EPI_ISL_1542464, EPI_ISL_1542465, | | | |

EPI_ISL_1542466, EPI_ISL_1542467, EPI_ISL_1542468, EPI_ISL_1542469, EPI_ISL_1542470, EPI_ISL_1542471, EPI_ISL_1542472, EPI_ISL_1542473, EPI_ISL_1542474, EPI_ISL_1542475, EPI_ISL_1542476, EPI_ISL_1542477, EPI_ISL_1542478, EPI_ISL_1542479, EPI_ISL_1542480, EPI_ISL_1542481, EPI_ISL_1542482, EPI_ISL_1542483, EPI_ISL_1542484, EPI_ISL_1542485, EPI_ISL_1542486, EPI_ISL_1542487,
EPI_ISL_1542488, EPI_ISL_1542489, EPI_ISL_1542490, EPI_ISL_1542491, EPI_ISL_1542492, EPI_ISL_1542493, EPI_ISL_1542494, EPI_ISL_1542495, EPI_ISL_1542496, EPI_ISL_1542497, EPI_ISL_1542498, EPI_ISL_1542499, EPI_ISL_1542500, EPI_ISL_1542501, EPI_ISL_1542502, EPI_ISL_1542503, EPI_ISL_1542504, EPI_ISL_1542505, EPI_ISL_1542506, EPI_ISL_1542507, EPI_ISL_1542508, EPI_ISL_1542509,
EPI_ISL_1542510, EPI_ISL_1542511, EPI_ISL_1542512, EPI_ISL_1542513, EPI_ISL_1542514, EPI_ISL_1542515, EPI_ISL_1542516, EPI_ISL_1542517, EPI_ISL_1542518, EPI_ISL_1542519, EPI_ISL_1542520, EPI_ISL_1542521, EPI_ISL_1542522, EPI_ISL_1542523, EPI_ISL_1542524, EPI_ISL_1542525, EPI_ISL_1542526, EPI_ISL_1542527, EPI_ISL_1542528, EPI_ISL_1542529, EPI_ISL_1542530, EPI_ISL_1542531,
EPI_ISL_1542532, EPI_ISL_1542533, EPI_ISL_1542534, EPI_ISL_1542535, EPI_ISL_1542536, EPI_ISL_1542537, EPI_ISL_1542538, EPI_ISL_1542539, EPI_ISL_1542540, EPI_ISL_1542541, EPI_ISL_1542542, EPI_ISL_1542543, EPI_ISL_1542544, EPI_ISL_1542545, EPI_ISL_1542546, EPI_ISL_1542547, EPI_ISL_1542548, EPI_ISL_1542549, EPI_ISL_1542550, EPI_ISL_1542551, EPI_ISL_1542552, EPI_ISL_1542553,
EPI_ISL_1542554, EPI_ISL_1542555, EPI_ISL_1542556, EPI_ISL_1542557, EPI_ISL_1542558, EPI_ISL_1542559, EPI_ISL_1542560, EPI_ISL_1542561, EPI_ISL_1542562, EPI_ISL_1542563, EPI_ISL_1542564, EPI_ISL_1542565, EPI_ISL_1542566, EPI_ISL_1542567, EPI_ISL_1542568, EPI_ISL_1542569, EPI_ISL_1542570, EPI_ISL_1542571, EPI_ISL_1542572, EPI_ISL_1542573, EPI_ISL_1542574, EPI_ISL_1542575,
EPI_ISL_1542576, EPI_ISL_1542577, EPI_ISL_1542578, EPI_ISL_1542579, EPI_ISL_1542580, EPI_ISL_1542581, EPI_ISL_1542582, EPI_ISL_1542583, EPI_ISL_1542584, EPI_ISL_1542585, EPI_ISL_1542586, EPI_ISL_1542587, EPI_ISL_1542588, EPI_ISL_1542589, EPI_ISL_1542590, EPI_ISL_1542591, EPI_ISL_1542592, EPI_ISL_1542593, EPI_ISL_1542594, EPI_ISL_1542595, EPI_ISL_1542596, EPI_ISL_1542597,
EPI_ISL_1542598, EPI_ISL_1542599, EPI_ISL_1542600, EPI_ISL_1542601, EPI_ISL_1542602, EPI_ISL_1542603, EPI_ISL_1542604, EPI_ISL_1542605, EPI_ISL_1542606, EPI_ISL_1542607, EPI_ISL_1542608, EPI_ISL_1542609, EPI_ISL_1542610, EPI_ISL_1542611, EPI_ISL_1542612, EPI_ISL_1542613, EPI_ISL_1542614, EPI_ISL_1542615, EPI_ISL_1542616, EPI_ISL_1542617, EPI_ISL_1542618, EPI_ISL_1542619,
EPI_ISL_1542620, EPI_ISL_1542621, EPI_ISL_1542622, EPI_ISL_1542623, EPI_ISL_1542624, EPI_ISL_1542625, EPI_ISL_1542626, EPI_ISL_1542627, EPI_ISL_1542628, EPI_ISL_1542629, EPI_ISL_1542630, EPI_ISL_1542631, EPI_ISL_1542632, EPI_ISL_1542633, EPI_ISL_1542634, EPI_ISL_1542635, EPI_ISL_1542636, EPI_ISL_1542637, EPI_ISL_1542638, EPI_ISL_1542639, EPI_ISL_1542640, EPI_ISL_1542641,
EPI_ISL_1542642, EPI_ISL_1542643, EPI_ISL_1542644, EPI_ISL_1542645, EPI_ISL_1542646, EPI_ISL_1542647, EPI_ISL_1542648, EPI_ISL_1542649, EPI_ISL_1542650, EPI_ISL_1542651, EPI_ISL_1542652, EPI_ISL_1542653, EPI_ISL_1542654, EPI_ISL_1542655, EPI_ISL_1542656, EPI_ISL_1542657, EPI_ISL_1542658, EPI_ISL_1542659, EPI_ISL_1542660, EPI_ISL_1542661, EPI_ISL_1542662, EPI_ISL_1542663,
EPI_ISL_1542664, EPI_ISL_1542665, EPI_ISL_1542666, EPI_ISL_1542667, EPI_ISL_1542668, EPI_ISL_1542669, EPI_ISL_1542670, EPI_ISL_1542671, EPI_ISL_1542672, EPI_ISL_1542673, EPI_ISL_1542674, EPI_ISL_1542675, EPI_ISL_1542676, EPI_ISL_1542677, EPI_ISL_1542678, EPI_ISL_1542679, EPI_ISL_1542680, EPI_ISL_1542681, EPI_ISL_1542682, EPI_ISL_1542683, EPI_ISL_1542684, EPI_ISL_1542685,
EPI_ISL_1542686, EPI_ISL_1542687, EPI_ISL_1542688, EPI_ISL_1542689, EPI_ISL_1542690, EPI_ISL_1542691, EPI_ISL_1542692, EPI_ISL_1542693, EPI_ISL_1542694, EPI_ISL_1542695, EPI_ISL_1542696, EPI_ISL_1542697, EPI_ISL_1542698, EPI_ISL_1542699, EPI_ISL_1542700, EPI_ISL_1542701, EPI_ISL_1542702, EPI_ISL_1542703, EPI_ISL_1542704, EPI_ISL_1542705, EPI_ISL_1542706, EPI_ISL_1542707,
EPI_ISL_1542708, EPI_ISL_1542709, EPI_ISL_1542710, EPI_ISL_1542711, EPI_ISL_1542712, EPI_ISL_1542713, EPI_ISL_1542714, EPI_ISL_1542715, EPI_ISL_1542716, EPI_ISL_1542717, EPI_ISL_1542718, EPI_ISL_1542719, EPI_ISL_1542720, EPI_ISL_1542721, EPI_ISL_1542722, EPI_ISL_1542723, EPI_ISL_1542724, EPI_ISL_1542725, EPI_ISL_1542726, EPI_ISL_1542727, EPI_ISL_1542728, EPI_ISL_1542729,
EPI_ISL_1542730, EPI_ISL_1542731, EPI_ISL_1542732, EPI_ISL_1542733, EPI_ISL_1542734, EPI_ISL_1542735, EPI_ISL_1542736, EPI_ISL_1542737, EPI_ISL_1542738, EPI_ISL_1542739, EPI_ISL_1542740, EPI_ISL_1542741, EPI_ISL_1542742, EPI_ISL_1542743, EPI_ISL_1542744, EPI_ISL_1542745, EPI_ISL_1542746, EPI_ISL_1542747, EPI_ISL_1542748, EPI_ISL_1542749, EPI_ISL_1542750, EPI_ISL_1542751,
EPI_ISL_1542752, EPI_ISL_1542753, EPI_ISL_1542754, EPI_ISL_1542755, EPI_ISL_1542756, EPI_ISL_1542757, EPI_ISL_1542758, EPI_ISL_1542759, EPI_ISL_1542760, EPI_ISL_1542761, EPI_ISL_1542762, EPI_ISL_1542763, EPI_ISL_1542764, EPI_ISL_1542765, EPI_ISL_1542766, EPI_ISL_1542767, EPI_ISL_1542768, EPI_ISL_1542769, EPI_ISL_1542770, EPI_ISL_1542771, EPI_ISL_1542772, EPI_ISL_1542773,
EPI_ISL_1542774, EPI_ISL_1542775, EPI_ISL_1542776, EPI_ISL_1542777, EPI_ISL_1542778, EPI_ISL_1542779, EPI_ISL_1542780, EPI_ISL_1542781, EPI_ISL_1542782, EPI_ISL_1542783, EPI_ISL_1542784, EPI_ISL_1542785, EPI_ISL_1542786, EPI_ISL_1542787, EPI_ISL_1542788, EPI_ISL_1542789, EPI_ISL_1542790, EPI_ISL_1542791, EPI_ISL_1542792, EPI_ISL_1542793, EPI_ISL_1542794, EPI_ISL_1542795,
EPI_ISL_1542796, EPI_ISL_1542797, EPI_ISL_1542798, EPI_ISL_1542799, EPI_ISL_1542800, EPI_ISL_1542801, EPI_ISL_1542802, EPI_ISL_1542803, EPI_ISL_1542804, EPI_ISL_1542805, EPI_ISL_1542806, EPI_ISL_1542807, EPI_ISL_1542808, EPI_ISL_1542809, EPI_ISL_1542810, EPI_ISL_1542811, EPI_ISL_1542812, EPI_ISL_1542813, EPI_ISL_1542814, EPI_ISL_1542815, EPI_ISL_1542816, EPI_ISL_1542817,
EPI_ISL_1542818, EPI_ISL_1542819, EPI_ISL_1542820, EPI_ISL_1542821, EPI_ISL_1542822, EPI_ISL_1542823, EPI_ISL_1542824, EPI_ISL_1542825, EPI_ISL_1542826, EPI_ISL_1542827, EPI_ISL_1542828, EPI_ISL_1542829, EPI_ISL_1542830, EPI_ISL_1542831, EPI_ISL_1542832, EPI_ISL_1542833, EPI_ISL_1542834, EPI_ISL_1542835, EPI_ISL_1542836, EPI_ISL_1542837, EPI_ISL_1542838, EPI_ISL_1542839,
EPI_ISL_1542840, EPI_ISL_1542841, EPI_ISL_1542842, EPI_ISL_1542843, EPI_ISL_1542844, EPI_ISL_1542845, EPI_ISL_1542846, EPI_ISL_1542847, EPI_ISL_1542848, EPI_ISL_1542849, EPI_ISL_1542850, EPI_ISL_1542851, EPI_ISL_1542852, EPI_ISL_1542853, EPI_ISL_1542854, EPI_ISL_1542855, EPI_ISL_1542856, EPI_ISL_1542857, EPI_ISL_1542858, EPI_ISL_1542859, EPI_ISL_1542860, EPI_ISL_1542861,
EPI_ISL_1542862, EPI_ISL_1542863, EPI_ISL_1542864, EPI_ISL_1542865, EPI_ISL_1542866, EPI_ISL_1542867, EPI_ISL_1542868, EPI_ISL_1542869, EPI_ISL_1542870, EPI_ISL_1542871, EPI_ISL_1542872, EPI_ISL_1542873, EPI_ISL_1542874, EPI_ISL_1542875, EPI_ISL_1542876, EPI_ISL_1542877, EPI_ISL_1542878, EPI_ISL_1542879, EPI_ISL_1542880, EPI_ISL_1542881, EPI_ISL_1542882, EPI_ISL_1542883,
EPI_ISL_1542884, EPI_ISL_1542885, EPI_ISL_1542886, EPI_ISL_1542887, EPI_ISL_1542888, EPI_ISL_1542889, EPI_ISL_1542890, EPI_ISL_1542891, EPI_ISL_1542892, EPI_ISL_1542893, EPI_ISL_1542894, EPI_ISL_1542895, EPI_ISL_1542896, EPI_ISL_1542897, EPI_ISL_1542898, EPI_ISL_1542899, EPI_ISL_1542900, EPI_ISL_1542901, EPI_ISL_1542904, EPI_ISL_1542905, EPI_ISL_1542907, EPI_ISL_1542908,
EPI_ISL_1542909, EPI_ISL_1542911, EPI_ISL_1542912, EPI_ISL_1542913, EPI_ISL_1542914, EPI_ISL_1542917, EPI_ISL_1542921, EPI_ISL_1542922, EPI_ISL_1542924, EPI_ISL_1542926, EPI_ISL_1542930, EPI_ISL_1542932, EPI_ISL_1542933, EPI_ISL_1542934, EPI_ISL_1542935, EPI_ISL_1542937, EPI_ISL_1542939, EPI_ISL_1542941, EPI_ISL_1542942, EPI_ISL_1542944, EPI_ISL_1542947, EPI_ISL_1542950,
EPI_ISL_1542951, EPI_ISL_1542952, EPI_ISL_1542954, EPI_ISL_1542955, EPI_ISL_1542957, EPI_ISL_1542960, EPI_ISL_1542966, EPI_ISL_1542970, EPI_ISL_1542971, EPI_ISL_1542972, EPI_ISL_1542973, EPI_ISL_1542976, EPI_ISL_1542977, EPI_ISL_1542980, EPI_ISL_1542982, EPI_ISL_1542983, EPI_ISL_1542984, EPI_ISL_1542986, EPI_ISL_1542991, EPI_ISL_1542992, EPI_ISL_1542993, EPI_ISL_1542994,
EPI_ISL_1542997, EPI_ISL_1542998, EPI_ISL_1542999, EPI_ISL_1543000, EPI_ISL_1543001, EPI_ISL_1543003, EPI_ISL_1543005, EPI_ISL_1543007, EPI_ISL_1543008, EPI_ISL_1543011, EPI_ISL_1543012, EPI_ISL_1543014, EPI_ISL_1543015, EPI_ISL_1543017, EPI_ISL_1543018, EPI_ISL_1543019, EPI_ISL_1543020, EPI_ISL_1543021, EPI_ISL_1543024, EPI_ISL_1543026, EPI_ISL_1543027, EPI_ISL_1543028,
EPI_ISL_1543029, EPI_ISL_1543033, EPI_ISL_1543035, EPI_ISL_1543038, EPI_ISL_1543039, EPI_ISL_1543040, EPI_ISL_1543045, EPI_ISL_1543047, EPI_ISL_1543049, EPI_ISL_1543050, EPI_ISL_1543052, EPI_ISL_1543054, EPI_ISL_1543055, EPI_ISL_1543056, EPI_ISL_1543057, EPI_ISL_1543059, EPI_ISL_1543060, EPI_ISL_1543061, EPI_ISL_1543062, EPI_ISL_1543064, EPI_ISL_1543065, EPI_ISL_1543066,
EPI_ISL_1543068, EPI_ISL_1543070, EPI_ISL_1543071, EPI_ISL_1543073, EPI_ISL_1543074, EPI_ISL_1543075, EPI_ISL_1543077, EPI_ISL_1543078, EPI_ISL_1543079, EPI_ISL_1543080, EPI_ISL_1543082, EPI_ISL_1543084, EPI_ISL_1543086, EPI_ISL_1543087, EPI_ISL_1543088, EPI_ISL_1543089, EPI_ISL_1543091, EPI_ISL_1543095, EPI_ISL_1543096, EPI_ISL_1543097, EPI_ISL_1543098, EPI_ISL_1543102,
EPI_ISL_1543103, EPI_ISL_1543104, EPI_ISL_1543108, EPI_ISL_1543109, EPI_ISL_1543110, EPI_ISL_1543112, EPI_ISL_1543113, EPI_ISL_1543114, EPI_ISL_1543116, EPI_ISL_1543117, EPI_ISL_1543119, EPI_ISL_1543120, EPI_ISL_1543121, EPI_ISL_1543122, EPI_ISL_1543123, EPI_ISL_1543124, EPI_ISL_1543125, EPI_ISL_1543126, EPI_ISL_1543127, EPI_ISL_1543128, EPI_ISL_1543129, EPI_ISL_1543130,
EPI_ISL_1543131, EPI_ISL_1543132, EPI_ISL_1543133, EPI_ISL_1543134, EPI_ISL_1543135, EPI_ISL_1543136, EPI_ISL_1543137, EPI_ISL_1543138, EPI_ISL_1543139, EPI_ISL_1543140, EPI_ISL_1543141, EPI_ISL_1543142, EPI_ISL_1543143, EPI_ISL_1543144, EPI_ISL_1543145, EPI_ISL_1543146, EPI_ISL_1543147, EPI_ISL_1543148, EPI_ISL_1543149, EPI_ISL_1543150, EPI_ISL_1543151, EPI_ISL_1543152,
EPI_ISL_1543153, EPI_ISL_1543154, EPI_ISL_1543155, EPI_ISL_1543156, EPI_ISL_1543157, EPI_ISL_1543158, EPI_ISL_1543159, EPI_ISL_1543160, EPI_ISL_1543161, EPI_ISL_1543162, EPI_ISL_1543163, EPI_ISL_1543164, EPI_ISL_1543165, EPI_ISL_1543166, EPI_ISL_1543167, EPI_ISL_1543168, EPI_ISL_1543169, EPI_ISL_1543170, EPI_ISL_1543171, EPI_ISL_1543172, EPI_ISL_1543173, EPI_ISL_1543174,
EPI_ISL_1543175, EPI_ISL_1543176, EPI_ISL_1543177, EPI_ISL_1543178, EPI_ISL_1543179, EPI_ISL_1543180, EPI_ISL_1543181, EPI_ISL_1543182, EPI_ISL_1543183, EPI_ISL_1543184, EPI_ISL_1543185, EPI_ISL_1543186, EPI_ISL_1543187, EPI_ISL_1543188, EPI_ISL_1543189, EPI_ISL_1543190, EPI_ISL_1543191, EPI_ISL_1543192, EPI_ISL_1543193, EPI_ISL_1543194, EPI_ISL_1543195, EPI_ISL_1543196,
EPI_ISL_1543197, EPI_ISL_1543198, EPI_ISL_1543199, EPI_ISL_1543200, EPI_ISL_1543201, EPI_ISL_1543202, EPI_ISL_1543203, EPI_ISL_1543204, EPI_ISL_1543205, EPI_ISL_1543206, EPI_ISL_1543207, EPI_ISL_1543208, EPI_ISL_1543209, EPI_ISL_1543210, EPI_ISL_1543211, EPI_ISL_1543212, EPI_ISL_1543213, EPI_ISL_1543214, EPI_ISL_1543215, EPI_ISL_1543216, EPI_ISL_1543217, EPI_ISL_1543218,
EPI_ISL_1543219, EPI_ISL_1543220, EPI_ISL_1543221, EPI_ISL_1543222, EPI_ISL_1543223, EPI_ISL_1543224, EPI_ISL_1543225, EPI_ISL_1543226, EPI_ISL_1543227, EPI_ISL_1543228, EPI_ISL_1543229, EPI_ISL_1543230, EPI_ISL_1543231, EPI_ISL_1543232, EPI_ISL_1543233, EPI_ISL_1543234, EPI_ISL_1543235, EPI_ISL_1543236, EPI_ISL_1543237, EPI_ISL_1543238, EPI_ISL_1543239, EPI_ISL_1543240,
EPI_ISL_1543241, EPI_ISL_1543242, EPI_ISL_1543243, EPI_ISL_1543244, EPI_ISL_1543245, EPI_ISL_1543246, EPI_ISL_1543247, EPI_ISL_1543248, EPI_ISL_1543249, EPI_ISL_1543250, EPI_ISL_1543251, EPI_ISL_1543252, EPI_ISL_1543253, EPI_ISL_1543254, EPI_ISL_1543255, EPI_ISL_1543256, EPI_ISL_1543257, EPI_ISL_1543258, EPI_ISL_1543259, EPI_ISL_1543260, EPI_ISL_1543261, EPI_ISL_1543262,
EPI_ISL_1543263, EPI_ISL_1543264, EPI_ISL_1543265, EPI_ISL_1543266, EPI_ISL_1543267, EPI_ISL_1543268, EPI_ISL_1543269, EPI_ISL_1543270, EPI_ISL_1543271, EPI_ISL_1543272, EPI_ISL_1543273, EPI_ISL_1543274, EPI_ISL_1543275, EPI_ISL_1543276, EPI_ISL_1543277, EPI_ISL_1543278, EPI_ISL_1543279, EPI_ISL_1543280, EPI_ISL_1543281, EPI_ISL_1543282, EPI_ISL_1543283, EPI_ISL_1543284,
EPI_ISL_1543285, EPI_ISL_1543286, EPI_ISL_1543287, EPI_ISL_1543288, EPI_ISL_1543289, EPI_ISL_1543290, EPI_ISL_1543291, EPI_ISL_1543292, EPI_ISL_1543293, EPI_ISL_1543294, EPI_ISL_1543295, EPI_ISL_1543296, EPI_ISL_1543297, EPI_ISL_1543298, EPI_ISL_1543299, EPI_ISL_1543300, EPI_ISL_1543301, EPI_ISL_1543302, EPI_ISL_1543303, EPI_ISL_1543304, EPI_ISL_1543305, EPI_ISL_1543306,
EPI_ISL_1543307, EPI_ISL_1543308, EPI_ISL_1543309, EPI_ISL_1543310, EPI_ISL_1543311, EPI_ISL_1543312, EPI_ISL_1543313, EPI_ISL_1543314, EPI_ISL_1543315, EPI_ISL_1543316, EPI_ISL_1543317, EPI_ISL_1543318, EPI_ISL_1543319, EPI_ISL_1543320, EPI_ISL_1543321, EPI_ISL_1543322, EPI_ISL_1543323, EPI_ISL_1543324, EPI_ISL_1543325, EPI_ISL_1543326, EPI_ISL_1543327, EPI_ISL_1543328,
EPI_ISL_1543329, EPI_ISL_1543330, EPI_ISL_1543331, EPI_ISL_1543332, EPI_ISL_1543333, EPI_ISL_1543334, EPI_ISL_1543335, EPI_ISL_1543336, EPI_ISL_1543337, EPI_ISL_1543338, EPI_ISL_1543339, EPI_ISL_1543340, EPI_ISL_1543341, EPI_ISL_1543342, EPI_ISL_1543343, EPI_ISL_1543344, EPI_ISL_1543345, EPI_ISL_1543346, EPI_ISL_1543347, EPI_ISL_1543348, EPI_ISL_1543349, EPI_ISL_1543350,
EPI_ISL_1543351, EPI_ISL_1543352, EPI_ISL_1543353, EPI_ISL_1543354, EPI_ISL_1543355, EPI_ISL_1543356, EPI_ISL_1543357, EPI_ISL_1543358, EPI_ISL_1543359, EPI_ISL_1543360, EPI_ISL_1543361, EPI_ISL_1543362, EPI_ISL_1543363, EPI_ISL_1543364, EPI_ISL_1543365, EPI_ISL_1543366, EPI_ISL_1543367, EPI_ISL_1543368, EPI_ISL_1543369, EPI_ISL_1543370, EPI_ISL_1543371, EPI_ISL_1543372,
EPI_ISL_1543373, EPI_ISL_1543374, EPI_ISL_1543375, EPI_ISL_1543376, EPI_ISL_1543377, EPI_ISL_1543378, EPI_ISL_1543379, EPI_ISL_1543380, EPI_ISL_1543381, EPI_ISL_1543382, EPI_ISL_1543383, EPI_ISL_1543384, EPI_ISL_1543385, EPI_ISL_1543386, EPI_ISL_1543387, EPI_ISL_1543388, EPI_ISL_1543389, EPI_ISL_1543390, EPI_ISL_1543391, EPI_ISL_1543392, EPI_ISL_1543393, EPI_ISL_1543394,
EPI_ISL_1543395, EPI_ISL_1543396, EPI_ISL_1543397, EPI_ISL_1543398, EPI_ISL_1543399, EPI_ISL_1543400, EPI_ISL_1543401, EPI_ISL_1543402, EPI_ISL_1543403, EPI_ISL_1543404, EPI_ISL_1543405, EPI_ISL_1543406, EPI_ISL_1543407, EPI_ISL_1543408, EPI_ISL_1543409, EPI_ISL_1543410, EPI_ISL_1543411, EPI_ISL_1543412, EPI_ISL_1543413, EPI_ISL_1543414, EPI_ISL_1543415, EPI_ISL_1543416,
EPI_ISL_1543417, EPI_ISL_1543418, EPI_ISL_1543419, EPI_ISL_1543420, EPI_ISL_1543421, EPI_ISL_1543422, EPI_ISL_1543423, EPI_ISL_1543424, EPI_ISL_1543425, EPI_ISL_1543426, EPI_ISL_1543427, EPI_ISL_1543428, EPI_ISL_1543429, EPI_ISL_1543430, EPI_ISL_1543431, EPI_ISL_1543433, EPI_ISL_1543434, EPI_ISL_1543435, EPI_ISL_1543436, EPI_ISL_1543437, EPI_ISL_1543438, EPI_ISL_1543439,
EPI_ISL_1543440, EPI_ISL_1543441, EPI_ISL_1543442, EPI_ISL_1543443, EPI_ISL_1543444, EPI_ISL_1543445, EPI_ISL_1543446, EPI_ISL_1543447, EPI_ISL_1543448, EPI_ISL_1543449, EPI_ISL_1543450, EPI_ISL_1543451, EPI_ISL_1543452, EPI_ISL_1543453, EPI_ISL_1543454, EPI_ISL_1543455, EPI_ISL_1543456, EPI_ISL_1543457, EPI_ISL_1543458, EPI_ISL_1543459, EPI_ISL_1543460, EPI_ISL_1543461,
EPI_ISL_1543462, EPI_ISL_1543463, EPI_ISL_1543464, EPI_ISL_1543465, EPI_ISL_1543466, EPI_ISL_1543467, EPI_ISL_1543468, EPI_ISL_1543469, EPI_ISL_1543470, EPI_ISL_1543471, EPI_ISL_1543472, EPI_ISL_1543473, EPI_ISL_1543474, EPI_ISL_1543475, EPI_ISL_1543476, EPI_ISL_1543477, EPI_ISL_1543478, EPI_ISL_1543479, EPI_ISL_1543480, EPI_ISL_1543481, EPI_ISL_1543482, EPI_ISL_1543483,
EPI_ISL_1543484, EPI_ISL_1543485, EPI_ISL_1543486, EPI_ISL_1543487, EPI_ISL_1543488, EPI_ISL_1543489, EPI_ISL_1543490, EPI_ISL_1543491, EPI_ISL_1543492, EPI_ISL_1543493, EPI_ISL_1543494, EPI_ISL_1543495, EPI_ISL_1543496, EPI_ISL_1543497, EPI_ISL_1543498, EPI_ISL_1543499, EPI_ISL_1543500, EPI_ISL_1543501, EPI_ISL_1543502, EPI_ISL_1543503, EPI_ISL_1543504, EPI_ISL_1543505,
EPI_ISL_1543506, EPI_ISL_1543507, EPI_ISL_1543508, EPI_ISL_1543509, EPI_ISL_1543510, EPI_ISL_1543511, EPI_ISL_1543512, EPI_ISL_1543513, EPI_ISL_1543514, EPI_ISL_1543515, EPI_ISL_1543516, EPI_ISL_1543517, EPI_ISL_1543518, EPI_ISL_1543519, EPI_ISL_1543520, EPI_ISL_1543521, EPI_ISL_1543522, EPI_ISL_1543523, EPI_ISL_1543524, EPI_ISL_1543525, EPI_ISL_1543526, EPI_ISL_1543527,
EPI_ISL_1543528, EPI_ISL_1543529, EPI_ISL_1543530, EPI_ISL_1543531, EPI_ISL_1543532, EPI_ISL_1543533, EPI_ISL_1543534, EPI_ISL_1543535, EPI_ISL_1543536, EPI_ISL_1543537, EPI_ISL_1543538, EPI_ISL_1543539, EPI_ISL_1543540, EPI_ISL_1543541, EPI_ISL_1543542, EPI_ISL_1543543, EPI_ISL_1543544, EPI_ISL_1543545, EPI_ISL_1543546, EPI_ISL_1543547, EPI_ISL_1543548, EPI_ISL_1543549,
EPI_ISL_1543550, EPI_ISL_1543551, EPI_ISL_1543552, EPI_ISL_1543553, EPI_ISL_1543554, EPI_ISL_1543555, EPI_ISL_1543556, EPI_ISL_1543557, EPI_ISL_1543558, EPI_ISL_1543559, EPI_ISL_1543560, EPI_ISL_1543561, EPI_ISL_1543562, EPI_ISL_1543563, EPI_ISL_1543564, EPI_ISL_1543565, EPI_ISL_1543566, EPI_ISL_1543567, EPI_ISL_1543568, EPI_ISL_1543569, EPI_ISL_1543570, EPI_ISL_1543571,
EPI_ISL_1543572, EPI_ISL_1543573, EPI_ISL_1543574, EPI_ISL_1543575, EPI_ISL_1543576, EPI_ISL_1543577, EPI_ISL_1543578, EPI_ISL_1543579, EPI_ISL_1543580, EPI_ISL_1543581, EPI_ISL_1543582, EPI_ISL_1543583, EPI_ISL_1543584, EPI_ISL_1543585, EPI_ISL_1543586, EPI_ISL_1543587, EPI_ISL_1543588, EPI_ISL_1543589, EPI_ISL_1543590, EPI_ISL_1543591, EPI_ISL_1543592, EPI_ISL_1543593,
EPI_ISL_1543594, EPI_ISL_1543595, EPI_ISL_1543596, EPI_ISL_1543597, EPI_ISL_1543598, EPI_ISL_1543599, EPI_ISL_1543600, EPI_ISL_1543601, EPI_ISL_1543602, EPI_ISL_1543603, EPI_ISL_1543604, EPI_ISL_1543605, EPI_ISL_1543606, EPI_ISL_1543607, EPI_ISL_1543608, EPI_ISL_1543609, EPI_ISL_1543610, EPI_ISL_1543611, EPI_ISL_1543612, EPI_ISL_1543613, EPI_ISL_1543614, EPI_ISL_1543615,
EPI_ISL_1543616, EPI_ISL_1543617, EPI_ISL_1543618, EPI_ISL_1543619, EPI_ISL_1543620, EPI_ISL_1543621, EPI_ISL_1543622, EPI_ISL_1543623, EPI_ISL_1543624, EPI_ISL_1543625, EPI_ISL_1543626, EPI_ISL_1543627, EPI_ISL_1543628, EPI_ISL_1543629, EPI_ISL_1543630, EPI_ISL_1543631, EPI_ISL_1543632, EPI_ISL_1543633, EPI_ISL_1543634, EPI_ISL_1543635, EPI_ISL_1543636, EPI_ISL_1543637,
EPI_ISL_1543638, EPI_ISL_1543639, EPI_ISL_1543640, EPI_ISL_1543641, EPI_ISL_1543642, EPI_ISL_1543643, EPI_ISL_1543644, EPI_ISL_1543645, EPI_ISL_1543646, EPI_ISL_1543647, EPI_ISL_1543648, EPI_ISL_1543649, EPI_ISL_1543650, EPI_ISL_1543651, EPI_ISL_1543652, EPI_ISL_1543653, EPI_ISL_1543654, EPI_ISL_1543655, EPI_ISL_1543656, EPI_ISL_1543657, EPI_ISL_1543658, EPI_ISL_1543659,
EPI_ISL_1543660, EPI_ISL_1543661, EPI_ISL_1543662, EPI_ISL_1543663, EPI_ISL_1543664, EPI_ISL_1543665, EPI_ISL_1543666, EPI_ISL_1543667, EPI_ISL_1543668, EPI_ISL_1543669, EPI_ISL_1543670, EPI_ISL_1543671, EPI_ISL_1543672, EPI_ISL_1543673, EPI_ISL_1543674, EPI_ISL_1543675, EPI_ISL_1543676, EPI_ISL_1543677, EPI_ISL_1543678, EPI_ISL_1543679, EPI_ISL_1543680, EPI_ISL_1543681,
EPI_ISL_1543682, EPI_ISL_1543683, EPI_ISL_1543684, EPI_ISL_1543685, EPI_ISL_1543686, EPI_ISL_1543687, EPI_ISL_1543688, EPI_ISL_1543689, EPI_ISL_1543690, EPI_ISL_1543691, EPI_ISL_1543692, EPI_ISL_1543693, EPI_ISL_1543694, EPI_ISL_1543695, EPI_ISL_1543696, EPI_ISL_1543697, EPI_ISL_1543698, EPI_ISL_1543699, EPI_ISL_1543700, EPI_ISL_1543701, EPI_ISL_1543702, EPI_ISL_1543703,
EPI_ISL_1543704, EPI_ISL_1543705, EPI_ISL_1543706, EPI_ISL_1543707, EPI_ISL_1543708, EPI_ISL_1543709, EPI_ISL_1543710, EPI_ISL_1543711, EPI_ISL_1543712, EPI_ISL_1543713, EPI_ISL_1543714, EPI_ISL_1543715, EPI_ISL_1543716, EPI_ISL_1543717, EPI_ISL_1543718, EPI_ISL_1543719, EPI_ISL_1543720, EPI_ISL_1543721, EPI_ISL_1543722, EPI_ISL_1543723, EPI_ISL_1543724, EPI_ISL_1543725,
EPI_ISL_1543726, EPI_ISL_1543727, EPI_ISL_1543728, EPI_ISL_1543729, EPI_ISL_1543730, EPI_ISL_1543731, EPI_ISL_1543732, EPI_ISL_1543733, EPI_ISL_1543734, EPI_ISL_1543735, EPI_ISL_1543736, EPI_ISL_1543737, EPI_ISL_1543738, EPI_ISL_1543739, EPI_ISL_1543740, EPI_ISL_1543741, EPI_ISL_1543742, EPI_ISL_1543743, EPI_ISL_1543744, EPI_ISL_1543745, EPI_ISL_1543746, EPI_ISL_1543747,
EPI_ISL_1543748, EPI_ISL_1543749, EPI_ISL_1543750, EPI_ISL_1543751, EPI_ISL_1543752, EPI_ISL_1543753, EPI_ISL_1543754, EPI_ISL_1543755, EPI_ISL_1543756, EPI_ISL_1543757, EPI_ISL_1543758, EPI_ISL_1543759, EPI_ISL_1543760, EPI_ISL_1543761, EPI_ISL_1543762, EPI_ISL_1543763, EPI_ISL_1543764, EPI_ISL_1543765, EPI_ISL_1543766, EPI_ISL_1543767, EPI_ISL_1543768, EPI_ISL_1543769,
EPI_ISL_1543770, EPI_ISL_1543771, EPI_ISL_1543772, EPI_ISL_1543773, EPI_ISL_1543774, EPI_ISL_1543775, EPI_ISL_1543776, EPI_ISL_1543777, EPI_ISL_1543778, EPI_ISL_1543779, EPI_ISL_1543780, EPI_ISL_1543781, EPI_ISL_1543782, EPI_ISL_1543783, EPI_ISL_1543784, EPI_ISL_1543785, EPI_ISL_1543786, EPI_ISL_1543787, EPI_ISL_1543788, EPI_ISL_1543789, EPI_ISL_1543790, EPI_ISL_1543791,
EPI_ISL_1543792, EPI_ISL_1543793, EPI_ISL_1543794, EPI_ISL_1543795, EPI_ISL_1543796, EPI_ISL_1543797, EPI_ISL_1543798, EPI_ISL_1543799, EPI_ISL_1543800, EPI_ISL_1543801, EPI_ISL_1543802, EPI_ISL_1543803, EPI_ISL_1543804, EPI_ISL_1543805, EPI_ISL_1543806, EPI_ISL_1543807, EPI_ISL_1543808, EPI_ISL_1543809, EPI_ISL_1543810, EPI_ISL_1543811, EPI_ISL_1543812, EPI_ISL_1543813,
EPI_ISL_1543814, EPI_ISL_1543815, EPI_ISL_1543816, EPI_ISL_1543817, EPI_ISL_1543818, EPI_ISL_1543819, EPI_ISL_1543820, EPI_ISL_1543821, EPI_ISL_1543822, EPI_ISL_1543823, EPI_ISL_1543824, EPI_ISL_1543825, EPI_ISL_1543826, EPI_ISL_1543827, EPI_ISL_1543828, EPI_ISL_1543829, EPI_ISL_1543830, EPI_ISL_1543831, EPI_ISL_1543832, EPI_ISL_1543833, EPI_ISL_1543834, EPI_ISL_1543835,
EPI_ISL_1543836, EPI_ISL_1543837, EPI_ISL_1543838, EPI_ISL_1543839, EPI_ISL_1543840, EPI_ISL_1543841, EPI_ISL_1543842, EPI_ISL_1543843, EPI_ISL_1543844, EPI_ISL_1543845, EPI_ISL_1543846, EPI_ISL_1543847, EPI_ISL_1543848, EPI_ISL_1543849, EPI_ISL_1543850, EPI_ISL_1543851, EPI_ISL_1543852, EPI_ISL_1543853, EPI_ISL_1543854, EPI_ISL_1543855, EPI_ISL_1543856, EPI_ISL_1543857,
EPI_ISL_1543858, EPI_ISL_1543859, EPI_ISL_1543860, EPI_ISL_1543861, EPI_ISL_1543862, EPI_ISL_1543863, EPI_ISL_1543864, EPI_ISL_1543865, EPI_ISL_1543866, EPI_ISL_1543867, EPI_ISL_1543868, EPI_ISL_1543869, EPI_ISL_1543870, EPI_ISL_1543871, EPI_ISL_1543872, EPI_ISL_1543873, EPI_ISL_1543874, EPI_ISL_1543875, EPI_ISL_1543876, EPI_ISL_1543877, EPI_ISL_1543878, EPI_ISL_1543879,
EPI_ISL_1543880, EPI_ISL_1543881, EPI_ISL_1543882, EPI_ISL_1543883, EPI_ISL_1543884, EPI_ISL_1543885, EPI_ISL_1543886, EPI_ISL_1543887, EPI_ISL_1543888, EPI_ISL_1543889, EPI_ISL_1543890, EPI_ISL_1543891, EPI_ISL_1543892, EPI_ISL_1543893, EPI_ISL_1543894, EPI_ISL_1543895, EPI_ISL_1543896, EPI_ISL_1543897, EPI_ISL_1543898, EPI_ISL_1543899, EPI_ISL_1543900, EPI_ISL_1543901,
EPI_ISL_1543902, EPI_ISL_1543903, EPI_ISL_1543904, EPI_ISL_1543905, EPI_ISL_1543906, EPI_ISL_1543907, EPI_ISL_1543908, EPI_ISL_1543909, EPI_ISL_1543910, EPI_ISL_1543911, EPI_ISL_1543912, EPI_ISL_1543913, EPI_ISL_1543914, EPI_ISL_1543915, EPI_ISL_1543916, EPI_ISL_1543917, EPI_ISL_1543918, EPI_ISL_1543919, EPI_ISL_1543920, EPI_ISL_1633685, EPI_ISL_1633686, EPI_ISL_1633687,
EPI_ISL_1633688, EPI_ISL_1633689, EPI_ISL_1633690, EPI_ISL_1633691, EPI_ISL_1633692, EPI_ISL_1633693, EPI_ISL_1633694, EPI_ISL_1633695, EPI_ISL_1633696, EPI_ISL_1633697, EPI_ISL_1633698, EPI_ISL_1633699, EPI_ISL_1633700, EPI_ISL_1633701, EPI_ISL_1633702, EPI_ISL_1633703, EPI_ISL_1633704, EPI_ISL_1633705, EPI_ISL_1633706, EPI_ISL_1633707, EPI_ISL_1633708, EPI_ISL_1633709,
EPI_ISL_1633710, EPI_ISL_1633711, EPI_ISL_1633712, EPI_ISL_1633713, EPI_ISL_1633714, EPI_ISL_1633715, EPI_ISL_1633716, EPI_ISL_1633717, EPI_ISL_1633718, EPI_ISL_1633719, EPI_ISL_1633720, EPI_ISL_1633721, EPI_ISL_1633722, EPI_ISL_1633723, EPI_ISL_1633724, EPI_ISL_1633725, EPI_ISL_1633726, EPI_ISL_1633727, EPI_ISL_1633728, EPI_ISL_1633729, EPI_ISL_1633730, EPI_ISL_1633731,
EPI_ISL_1633732, EPI_ISL_1633733, EPI_ISL_1633734, EPI_ISL_1633735, EPI_ISL_1633736, EPI_ISL_1633737, EPI_ISL_1633738, EPI_ISL_1633739, EPI_ISL_1633740, EPI_ISL_1633741, EPI_ISL_1633742, EPI_ISL_1633743, EPI_ISL_1633744, EPI_ISL_1633745, EPI_ISL_1633746, EPI_ISL_1633747, EPI_ISL_1633748, EPI_ISL_1633749, EPI_ISL_1633750, EPI_ISL_1633751, EPI_ISL_1633752, EPI_ISL_1633753,
EPI_ISL_1633754, EPI_ISL_1633755, EPI_ISL_1633756, EPI_ISL_1633757, EPI_ISL_1633758, EPI_ISL_1633759, EPI_ISL_1633760, EPI_ISL_1633761, EPI_ISL_1633762, EPI_ISL_1633763, EPI_ISL_1633764, EPI_ISL_1633765, EPI_ISL_1633766, EPI_ISL_1633767, EPI_ISL_1633768, EPI_ISL_1633769, EPI_ISL_1633770, EPI_ISL_1633771, EPI_ISL_1633772, EPI_ISL_1633773, EPI_ISL_1633774, EPI_ISL_1633775,
EPI_ISL_1633776, EPI_ISL_1633777, EPI_ISL_1633778, EPI_ISL_1633779, EPI_ISL_1633780, EPI_ISL_1633781, EPI_ISL_1633782, EPI_ISL_1633783, EPI_ISL_1633784, EPI_ISL_1633785, EPI_ISL_1633786, EPI_ISL_1633787, EPI_ISL_1633788, EPI_ISL_1633789, EPI_ISL_1633790, EPI_ISL_1633791, EPI_ISL_1633792, EPI_ISL_1633793, EPI_ISL_1633794, EPI_ISL_1633795, EPI_ISL_1633796, EPI_ISL_1633797,
EPI_ISL_1633798, EPI_ISL_1633799, EPI_ISL_1633800, EPI_ISL_1633801, EPI_ISL_1633802, EPI_ISL_1633803, EPI_ISL_1633804, EPI_ISL_1633805, EPI_ISL_1633806, EPI_ISL_1633807, EPI_ISL_1633808, EPI_ISL_1633809, EPI_ISL_1633810, EPI_ISL_1633811, EPI_ISL_1633812, EPI_ISL_1633813, EPI_ISL_1633814, EPI_ISL_1633815, EPI_ISL_1633816, EPI_ISL_1633817, EPI_ISL_1633818, EPI_ISL_1633819,
EPI_ISL_1633820, EPI_ISL_1633821, EPI_ISL_1633822, EPI_ISL_1633823, EPI_ISL_1633824, EPI_ISL_1633825, EPI_ISL_1633826, EPI_ISL_1633827, EPI_ISL_1633828, EPI_ISL_1633829, EPI_ISL_1633830, EPI_ISL_1633831, EPI_ISL_1633832, EPI_ISL_1633833, EPI_ISL_1633834, EPI_ISL_1633835, EPI_ISL_1633836, EPI_ISL_1633837, EPI_ISL_1633838, EPI_ISL_1633839, EPI_ISL_1633840, EPI_ISL_1633841,
EPI_ISL_1633842, EPI_ISL_1633843, EPI_ISL_1633844, EPI_ISL_1633845, EPI_ISL_1633846, EPI_ISL_1633847, EPI_ISL_1633848, EPI_ISL_1633849, EPI_ISL_1633850, EPI_ISL_1633851, EPI_ISL_1633852, EPI_ISL_1633853, EPI_ISL_1633854, EPI_ISL_1633855, EPI_ISL_1633856, EPI_ISL_1633857, EPI_ISL_1633858, EPI_ISL_1633859, EPI_ISL_1633860, EPI_ISL_1633861, EPI_ISL_1633862, EPI_ISL_1633863,
EPI_ISL_1633864, EPI_ISL_1633865, EPI_ISL_1633866, EPI_ISL_1633867, EPI_ISL_1633868, EPI_ISL_1633869, EPI_ISL_1633870, EPI_ISL_1633871, EPI_ISL_1633872, EPI_ISL_1633873, EPI_ISL_1633874, EPI_ISL_1633875, EPI_ISL_1633876, EPI_ISL_1633877, EPI_ISL_1633878, EPI_ISL_1633879, EPI_ISL_1633880, EPI_ISL_1633881, EPI_ISL_1633882, EPI_ISL_1633883, EPI_ISL_1633884, EPI_ISL_1633885,
EPI_ISL_1633886, EPI_ISL_1633887, EPI_ISL_1633888, EPI_ISL_1633889, EPI_ISL_1633890, EPI_ISL_1633891, EPI_ISL_1633892, EPI_ISL_1633893, EPI_ISL_1633894, EPI_ISL_1633895, EPI_ISL_1633896, EPI_ISL_1633897, EPI_ISL_1633899, EPI_ISL_1633901, EPI_ISL_1633902, EPI_ISL_1633903, EPI_ISL_1633905, EPI_ISL_1633906, EPI_ISL_1633907, EPI_ISL_1633909, EPI_ISL_1633910, EPI_ISL_1633911,
EPI_ISL_1633913, EPI_ISL_1633914, EPI_ISL_1633916, EPI_ISL_1633917, EPI_ISL_1633918, EPI_ISL_1633920, EPI_ISL_1633921, EPI_ISL_1633922, EPI_ISL_1633924, EPI_ISL_1633925, EPI_ISL_1633927, EPI_ISL_1633929, EPI_ISL_1633930, EPI_ISL_1633931, EPI_ISL_1633932, EPI_ISL_1633934, EPI_ISL_1633935, EPI_ISL_1633937, EPI_ISL_1633938, EPI_ISL_1633939, EPI_ISL_1633940, EPI_ISL_1633942,
EPI_ISL_1633943, EPI_ISL_1633944, EPI_ISL_1633946, EPI_ISL_1633947, EPI_ISL_1633949, EPI_ISL_1633951, EPI_ISL_1633953, EPI_ISL_1633954, EPI_ISL_1633956, EPI_ISL_1633958, EPI_ISL_1633959, EPI_ISL_1633960, EPI_ISL_1633962, EPI_ISL_1633963, EPI_ISL_1633964, EPI_ISL_1633966, EPI_ISL_1633967, EPI_ISL_1633968, EPI_ISL_1633970, EPI_ISL_1633971, EPI_ISL_1633972, EPI_ISL_1633973,
EPI_ISL_1633975, EPI_ISL_1633976, EPI_ISL_1633977, EPI_ISL_1633979, EPI_ISL_1633980, EPI_ISL_1633981, EPI_ISL_1633983, EPI_ISL_1633984, EPI_ISL_1633985, EPI_ISL_1633986, EPI_ISL_1633988, EPI_ISL_1633989, EPI_ISL_1633990, EPI_ISL_1633992, EPI_ISL_1633993, EPI_ISL_1633994, EPI_ISL_1633996, EPI_ISL_1633997, EPI_ISL_1633998, EPI_ISL_1634000, EPI_ISL_1634001, EPI_ISL_1634002,
EPI_ISL_1634004, EPI_ISL_1634005, EPI_ISL_1634007, EPI_ISL_1634008, EPI_ISL_1634009, EPI_ISL_1634011, EPI_ISL_1634012, EPI_ISL_1634014, EPI_ISL_1634016, EPI_ISL_1634018, EPI_ISL_1634019, EPI_ISL_1634021, EPI_ISL_1634022, EPI_ISL_1634023, EPI_ISL_1634025, EPI_ISL_1634026, EPI_ISL_1634028, EPI_ISL_1634029, EPI_ISL_1634030, EPI_ISL_1634032, EPI_ISL_1634033, EPI_ISL_1634034,
EPI_ISL_1634036, EPI_ISL_1634037, EPI_ISL_1634038, EPI_ISL_1634040, EPI_ISL_1634041, EPI_ISL_1634042, EPI_ISL_1634044, EPI_ISL_1634045, EPI_ISL_1634046, EPI_ISL_1634047, EPI_ISL_1634049, EPI_ISL_1634051, EPI_ISL_1634052, EPI_ISL_1634053, EPI_ISL_1634054, EPI_ISL_1634056, EPI_ISL_1634058, EPI_ISL_1634060, EPI_ISL_1634061, EPI_ISL_1634062, EPI_ISL_1634064, EPI_ISL_1634065,
EPI_ISL_1634066, EPI_ISL_1634068, EPI_ISL_1634069, EPI_ISL_1634071, EPI_ISL_1634072, EPI_ISL_1634074, EPI_ISL_1634076, EPI_ISL_1634077, EPI_ISL_1634079, EPI_ISL_1634080, EPI_ISL_1634082, EPI_ISL_1634083, EPI_ISL_1634084, EPI_ISL_1634086, EPI_ISL_1634087, EPI_ISL_1634088, EPI_ISL_1634089, EPI_ISL_1634091, EPI_ISL_1634092, EPI_ISL_1634093, EPI_ISL_1634095, EPI_ISL_1634096,
EPI_ISL_1634097, EPI_ISL_1634099, EPI_ISL_1634100, EPI_ISL_1634101, EPI_ISL_1634103, EPI_ISL_1634104, EPI_ISL_1634105, EPI_ISL_1634107, EPI_ISL_1634108, EPI_ISL_1634110, EPI_ISL_1634112, EPI_ISL_1634113, EPI_ISL_1634114, EPI_ISL_1634116, EPI_ISL_1634117, EPI_ISL_1634118, EPI_ISL_1634120, EPI_ISL_1634121, EPI_ISL_1634123, EPI_ISL_1634124, EPI_ISL_1634125, EPI_ISL_1634126,
EPI_ISL_1634128, EPI_ISL_1634129, EPI_ISL_1634130, EPI_ISL_1634132, EPI_ISL_1634133, EPI_ISL_1634135, EPI_ISL_1634136, EPI_ISL_1634137, EPI_ISL_1634139, EPI_ISL_1634140, EPI_ISL_1634141, EPI_ISL_1634143, EPI_ISL_1634150, EPI_ISL_1634152, EPI_ISL_1634153, EPI_ISL_1634155, EPI_ISL_1634157, EPI_ISL_1634158, EPI_ISL_1634159, EPI_ISL_1634161, EPI_ISL_1634162, EPI_ISL_1634163,
EPI_ISL_1634165, EPI_ISL_1634166, EPI_ISL_1634167, EPI_ISL_1634169, EPI_ISL_1634170, EPI_ISL_1634171, EPI_ISL_1634173, EPI_ISL_1634174, EPI_ISL_1634175, EPI_ISL_1634176, EPI_ISL_1634177, EPI_ISL_1634178, EPI_ISL_1634179, EPI_ISL_1634180, EPI_ISL_1634181, EPI_ISL_1634182, EPI_ISL_1634183, EPI_ISL_1634184, EPI_ISL_1634185, EPI_ISL_1634186, EPI_ISL_1634187, EPI_ISL_1634188,
EPI_ISL_1634189, EPI_ISL_1634190, EPI_ISL_1634191, EPI_ISL_1634192, EPI_ISL_1634193, EPI_ISL_1634194, EPI_ISL_1634195, EPI_ISL_1634196, EPI_ISL_1634197, EPI_ISL_1634198, EPI_ISL_1634199, EPI_ISL_1634200, EPI_ISL_1634201, EPI_ISL_1634202, EPI_ISL_1634203, EPI_ISL_1634204, EPI_ISL_1634205, EPI_ISL_1634206, EPI_ISL_1634207, EPI_ISL_1634208, EPI_ISL_1634209, EPI_ISL_1634210,
EPI_ISL_1634211, EPI_ISL_1634212, EPI_ISL_1634213, EPI_ISL_1634214, EPI_ISL_1634215, EPI_ISL_1634216, EPI_ISL_1634217, EPI_ISL_1634218, EPI_ISL_1634219, EPI_ISL_1634220, EPI_ISL_1634221, EPI_ISL_1634222, EPI_ISL_1634223, EPI_ISL_1634224, EPI_ISL_1634225, EPI_ISL_1634226, EPI_ISL_1634227, EPI_ISL_1634228, EPI_ISL_1634229, EPI_ISL_1634230, EPI_ISL_1634231, EPI_ISL_1634232,
EPI_ISL_1634233, EPI_ISL_1634234, EPI_ISL_1634235, EPI_ISL_1634236, EPI_ISL_1634237, EPI_ISL_1634238, EPI_ISL_1634239, EPI_ISL_1634240, EPI_ISL_1634241, EPI_ISL_1634242, EPI_ISL_1634243, EPI_ISL_1634244, EPI_ISL_1634245, EPI_ISL_1634246, EPI_ISL_1634247, EPI_ISL_1634248, EPI_ISL_1634249, EPI_ISL_1634250, EPI_ISL_1634251, EPI_ISL_1634252, EPI_ISL_1634253, EPI_ISL_1634254,
EPI_ISL_1634255, EPI_ISL_1634256, EPI_ISL_1634257, EPI_ISL_1634258, EPI_ISL_1634259, EPI_ISL_1634260, EPI_ISL_1634261, EPI_ISL_1634262, EPI_ISL_1634263, EPI_ISL_1634264, EPI_ISL_1634265, EPI_ISL_1634266, EPI_ISL_1634267, EPI_ISL_1634268, EPI_ISL_1634269, EPI_ISL_1634270, EPI_ISL_1634271, EPI_ISL_1634272, EPI_ISL_1634273, EPI_ISL_1634274, EPI_ISL_1634275, EPI_ISL_1634276,
EPI_ISL_1634277, EPI_ISL_1634278, EPI_ISL_1634279, EPI_ISL_1634280, EPI_ISL_1634281, EPI_ISL_1634282, EPI_ISL_1634283, EPI_ISL_1634284, EPI_ISL_1634285, EPI_ISL_1634286, EPI_ISL_1634287, EPI_ISL_1634288, EPI_ISL_1634289, EPI_ISL_1634290, EPI_ISL_1634291, EPI_ISL_1634292, EPI_ISL_1634293, EPI_ISL_1634294, EPI_ISL_1634295, EPI_ISL_1634296, EPI_ISL_1634297, EPI_ISL_1634298,
EPI_ISL_1634299, EPI_ISL_1634300, EPI_ISL_1634301, EPI_ISL_1634302, EPI_ISL_1634303, EPI_ISL_1634304, EPI_ISL_1634305, EPI_ISL_1634306, EPI_ISL_1634307, EPI_ISL_1634308, EPI_ISL_1634309, EPI_ISL_1634310, EPI_ISL_1634311, EPI_ISL_1634312, EPI_ISL_1634313, EPI_ISL_1634314, EPI_ISL_1634315, EPI_ISL_1634316, EPI_ISL_1634317, EPI_ISL_1634318, EPI_ISL_1634319, EPI_ISL_1634320,
EPI_ISL_1634321, EPI_ISL_1634322, EPI_ISL_1634323, EPI_ISL_1634324, EPI_ISL_1634325, EPI_ISL_1634326, EPI_ISL_1634327, EPI_ISL_1634328, EPI_ISL_1634329, EPI_ISL_1634330, EPI_ISL_1634331, EPI_ISL_1634332, EPI_ISL_1634333, EPI_ISL_1634334, EPI_ISL_1634335, EPI_ISL_1634336, EPI_ISL_1634337, EPI_ISL_1634338, EPI_ISL_1634339, EPI_ISL_1635437, EPI_ISL_1635438, EPI_ISL_1635439,
EPI_ISL_1635440, EPI_ISL_1635441, EPI_ISL_1635442, EPI_ISL_1635443, EPI_ISL_1635444, EPI_ISL_1635445, EPI_ISL_1635446, EPI_ISL_1635447, EPI_ISL_1635448, EPI_ISL_1635449, EPI_ISL_1635450, EPI_ISL_1635451, EPI_ISL_1635452, EPI_ISL_1635453, EPI_ISL_1635454, EPI_ISL_1635455, EPI_ISL_1635456, EPI_ISL_1635457, EPI_ISL_1635458, EPI_ISL_1635459, EPI_ISL_1635460, EPI_ISL_1635461,
EPI_ISL_1635462, EPI_ISL_1635463, EPI_ISL_1635464, EPI_ISL_1635465, EPI_ISL_1635466, EPI_ISL_1635467, EPI_ISL_1635468, EPI_ISL_1635469, EPI_ISL_1635470, EPI_ISL_1635471, EPI_ISL_1635472, EPI_ISL_1635473, EPI_ISL_1635474, EPI_ISL_1635475, EPI_ISL_1635476, EPI_ISL_1635477, EPI_ISL_1635478, EPI_ISL_1635479, EPI_ISL_1635480, EPI_ISL_1635481, EPI_ISL_1635482, EPI_ISL_1635483,
EPI_ISL_1635484, EPI_ISL_1635485, EPI_ISL_1635486, EPI_ISL_1635487, EPI_ISL_1635488, EPI_ISL_1635489, EPI_ISL_1635490, EPI_ISL_1635491, EPI_ISL_1635492, EPI_ISL_1635493, EPI_ISL_1635494, EPI_ISL_1635495, EPI_ISL_1635496, EPI_ISL_1635497, EPI_ISL_1635498, EPI_ISL_1635499, EPI_ISL_1635500, EPI_ISL_1635501, EPI_ISL_1635502, EPI_ISL_1635503, EPI_ISL_1635504, EPI_ISL_1635505,
EPI_ISL_1635506, EPI_ISL_1635507, EPI_ISL_1635508, EPI_ISL_1635509, EPI_ISL_1635510, EPI_ISL_1635511, EPI_ISL_1635512, EPI_ISL_1635513, EPI_ISL_1635514, EPI_ISL_1635515, EPI_ISL_1635516, EPI_ISL_1635517, EPI_ISL_1635518, EPI_ISL_1635519, EPI_ISL_1635520, EPI_ISL_1635521, EPI_ISL_1635522, EPI_ISL_1635523, EPI_ISL_1635524, EPI_ISL_1635525, EPI_ISL_1635526, EPI_ISL_1635527,
EPI_ISL_1635528, EPI_ISL_1635529, EPI_ISL_1635530, EPI_ISL_1635531, EPI_ISL_1635532, EPI_ISL_1635533, EPI_ISL_1635534, EPI_ISL_1635535, EPI_ISL_1635536, EPI_ISL_1635537, EPI_ISL_1635538, EPI_ISL_1635539, EPI_ISL_1635540, EPI_ISL_1635541, EPI_ISL_1635542, EPI_ISL_1635543, EPI_ISL_1635544, EPI_ISL_1635545, EPI_ISL_1635546, EPI_ISL_1635547, EPI_ISL_1635548, EPI_ISL_1635549,
EPI_ISL_1635550, EPI_ISL_1635551, EPI_ISL_1635552, EPI_ISL_1635553, EPI_ISL_1635554, EPI_ISL_1635555, EPI_ISL_1635556, EPI_ISL_1635557, EPI_ISL_1635558, EPI_ISL_1635559, EPI_ISL_1635560, EPI_ISL_1635561, EPI_ISL_1635562, EPI_ISL_1635563, EPI_ISL_1635564, EPI_ISL_1635565, EPI_ISL_1635566, EPI_ISL_1635567, EPI_ISL_1635568, EPI_ISL_1635569, EPI_ISL_1635570, EPI_ISL_1635571,
EPI_ISL_1635572, EPI_ISL_1635573, EPI_ISL_1635574, EPI_ISL_1635575, EPI_ISL_1635576, EPI_ISL_1635577, EPI_ISL_1635578, EPI_ISL_1635579, EPI_ISL_1635580, EPI_ISL_1635581, EPI_ISL_1635582, EPI_ISL_1635583, EPI_ISL_1635584, EPI_ISL_1635585, EPI_ISL_1635586, EPI_ISL_1635587, EPI_ISL_1635588, EPI_ISL_1635589, EPI_ISL_1635590, EPI_ISL_1635591, EPI_ISL_1635592, EPI_ISL_1635593,
EPI_ISL_1635594, EPI_ISL_1635595, EPI_ISL_1635596, EPI_ISL_1635597, EPI_ISL_1635598, EPI_ISL_1635599, EPI_ISL_1635600, EPI_ISL_1635601, EPI_ISL_1635602, EPI_ISL_1635603, EPI_ISL_1635604, EPI_ISL_1635605, EPI_ISL_1635606, EPI_ISL_1635607, EPI_ISL_1635608, EPI_ISL_1635609, EPI_ISL_1635610, EPI_ISL_1635611, EPI_ISL_1635612, EPI_ISL_1635613, EPI_ISL_1635614, EPI_ISL_1635615,
EPI_ISL_1635616, EPI_ISL_1635617, EPI_ISL_1635618, EPI_ISL_1635619, EPI_ISL_1635620, EPI_ISL_1635621, EPI_ISL_1635622, EPI_ISL_1635623, EPI_ISL_1635624, EPI_ISL_1635625, EPI_ISL_1635626, EPI_ISL_1635627, EPI_ISL_1635628, EPI_ISL_1635629, EPI_ISL_1635630, EPI_ISL_1635631, EPI_ISL_1635632, EPI_ISL_1635633, EPI_ISL_1635634, EPI_ISL_1635635, EPI_ISL_1635636, EPI_ISL_1635637,
EPI_ISL_1635638, EPI_ISL_1635639, EPI_ISL_1635640, EPI_ISL_1635641, EPI_ISL_1635642, EPI_ISL_1635643, EPI_ISL_1635644, EPI_ISL_1635645, EPI_ISL_1635646, EPI_ISL_1635647, EPI_ISL_1635648, EPI_ISL_1635649, EPI_ISL_1635650, EPI_ISL_1635651, EPI_ISL_1635652, EPI_ISL_1635653, EPI_ISL_1635654, EPI_ISL_1635655, EPI_ISL_1635656, EPI_ISL_1635657, EPI_ISL_1635658, EPI_ISL_1635659,
EPI_ISL_1635660, EPI_ISL_1635661, EPI_ISL_1635662, EPI_ISL_1635663, EPI_ISL_1635664, EPI_ISL_1635665, EPI_ISL_1635667, EPI_ISL_1635668, EPI_ISL_1635669, EPI_ISL_1635670, EPI_ISL_1635671, EPI_ISL_1635672, EPI_ISL_1635673, EPI_ISL_1635674, EPI_ISL_1635675, EPI_ISL_1635676, EPI_ISL_1635677, EPI_ISL_1635678, EPI_ISL_1635679, EPI_ISL_1635680, EPI_ISL_1635681, EPI_ISL_1635682,
EPI_ISL_1635683, EPI_ISL_1635684, EPI_ISL_1635685, EPI_ISL_1635686, EPI_ISL_1635687, EPI_ISL_1635688, EPI_ISL_1635689, EPI_ISL_1635690, EPI_ISL_1635691, EPI_ISL_1635692, EPI_ISL_1635693, EPI_ISL_1635694, EPI_ISL_1635695, EPI_ISL_1635696, EPI_ISL_1635697, EPI_ISL_1635698, EPI_ISL_1635699, EPI_ISL_1635700, EPI_ISL_1635701, EPI_ISL_1635702, EPI_ISL_1635703, EPI_ISL_1635704,
EPI_ISL_1635705, EPI_ISL_1635706, EPI_ISL_1635707, EPI_ISL_1635708, EPI_ISL_1635709, EPI_ISL_1635710, EPI_ISL_1635711, EPI_ISL_1635712, EPI_ISL_1635713, EPI_ISL_1635714, EPI_ISL_1635715, EPI_ISL_1635716, EPI_ISL_1635717, EPI_ISL_1635718, EPI_ISL_1635719, EPI_ISL_1635720, EPI_ISL_1635721, EPI_ISL_1635722, EPI_ISL_1635723, EPI_ISL_1635724, EPI_ISL_1635725, EPI_ISL_1635726,
EPI_ISL_1635727, EPI_ISL_1635728, EPI_ISL_1635729, EPI_ISL_1635730, EPI_ISL_1635731, EPI_ISL_1635732, EPI_ISL_1635733, EPI_ISL_1635734, EPI_ISL_1635735, EPI_ISL_1635736, EPI_ISL_1635737, EPI_ISL_1635738, EPI_ISL_1635739, EPI_ISL_1635740, EPI_ISL_1635741, EPI_ISL_1635742, EPI_ISL_1635743, EPI_ISL_1635744, EPI_ISL_1635745, EPI_ISL_1635746, EPI_ISL_1635747, EPI_ISL_1635748,
EPI_ISL_1635749, EPI_ISL_1635750, EPI_ISL_1635751, EPI_ISL_1635752, EPI_ISL_1635753, EPI_ISL_1635755, EPI_ISL_1635756, EPI_ISL_1635757, EPI_ISL_1635758, EPI_ISL_1635759, EPI_ISL_1635760, EPI_ISL_1635761, EPI_ISL_1635762, EPI_ISL_1635763, EPI_ISL_1635764, EPI_ISL_1635765, EPI_ISL_1635766, EPI_ISL_1635767, EPI_ISL_1635768, EPI_ISL_1635769, EPI_ISL_1635770, EPI_ISL_1635771,
EPI_ISL_1635772, EPI_ISL_1635773, EPI_ISL_1635774, EPI_ISL_1635775, EPI_ISL_1635776, EPI_ISL_1635777, EPI_ISL_1635778, EPI_ISL_1635779, EPI_ISL_1635780, EPI_ISL_1635781, EPI_ISL_1635782, EPI_ISL_1635783, EPI_ISL_1635784, EPI_ISL_1635785, EPI_ISL_1635786, EPI_ISL_1635787, EPI_ISL_1635788, EPI_ISL_1635789, EPI_ISL_1635790, EPI_ISL_1635791, EPI_ISL_1635792, EPI_ISL_1635793,
EPI_ISL_1635794, EPI_ISL_1635795, EPI_ISL_1635796, EPI_ISL_1635797, EPI_ISL_1635798, EPI_ISL_1635799, EPI_ISL_1635800, EPI_ISL_1635801, EPI_ISL_1635802, EPI_ISL_1635803, EPI_ISL_1635804, EPI_ISL_1635805, EPI_ISL_1635806, EPI_ISL_1635807, EPI_ISL_1635808, EPI_ISL_1635809, EPI_ISL_1635810, EPI_ISL_1635811, EPI_ISL_1635812, EPI_ISL_1635813, EPI_ISL_1635814, EPI_ISL_1635815,
EPI_ISL_1635816, EPI_ISL_1635817, EPI_ISL_1635818, EPI_ISL_1635819, EPI_ISL_1635820, EPI_ISL_1635821, EPI_ISL_1635822, EPI_ISL_1635823, EPI_ISL_1635824, EPI_ISL_1635825, EPI_ISL_1635826, EPI_ISL_1635827, EPI_ISL_1635828, EPI_ISL_1635829, EPI_ISL_1635830, EPI_ISL_1635831, EPI_ISL_1635832, EPI_ISL_1635833, EPI_ISL_1635834, EPI_ISL_1635835, EPI_ISL_1635836, EPI_ISL_1635837,
EPI_ISL_1635838, EPI_ISL_1635839, EPI_ISL_1635840, EPI_ISL_1635841, EPI_ISL_1635842, EPI_ISL_1635843, EPI_ISL_1635844, EPI_ISL_1635845, EPI_ISL_1635846, EPI_ISL_1635847, EPI_ISL_1635848, EPI_ISL_1635849, EPI_ISL_1635850, EPI_ISL_1635851, EPI_ISL_1635852, EPI_ISL_1635853, EPI_ISL_1635854, EPI_ISL_1635855, EPI_ISL_1635856, EPI_ISL_1635857, EPI_ISL_1635858, EPI_ISL_1635859,
EPI_ISL_1635860, EPI_ISL_1635861, EPI_ISL_1635862, EPI_ISL_1635863, EPI_ISL_1635864, EPI_ISL_1635865, EPI_ISL_1635866, EPI_ISL_1635867, EPI_ISL_1635868, EPI_ISL_1635869, EPI_ISL_1635870, EPI_ISL_1635871, EPI_ISL_1635872, EPI_ISL_1635873, EPI_ISL_1635874, EPI_ISL_1635875, EPI_ISL_1635876, EPI_ISL_1635877, EPI_ISL_1635878, EPI_ISL_1635879, EPI_ISL_1635880, EPI_ISL_1635881,
EPI_ISL_1635882, EPI_ISL_1635883, EPI_ISL_1635884, EPI_ISL_1635885, EPI_ISL_1635886, EPI_ISL_1635887, EPI_ISL_1635888, EPI_ISL_1635889, EPI_ISL_1635890, EPI_ISL_1635891, EPI_ISL_1635892, EPI_ISL_1635893, EPI_ISL_1635894, EPI_ISL_1635895, EPI_ISL_1635896, EPI_ISL_1635897, EPI_ISL_1635898, EPI_ISL_1635899, EPI_ISL_1635900, EPI_ISL_1635901, EPI_ISL_1635902, EPI_ISL_1635903,
EPI_ISL_1635904, EPI_ISL_1635905, EPI_ISL_1635906, EPI_ISL_1635907, EPI_ISL_1635908, EPI_ISL_1635909, EPI_ISL_1635910, EPI_ISL_1635911, EPI_ISL_1635912, EPI_ISL_1635913, EPI_ISL_1635914, EPI_ISL_1635915, EPI_ISL_1635916, EPI_ISL_1635917, EPI_ISL_1635918, EPI_ISL_1635919, EPI_ISL_1635920, EPI_ISL_1635921, EPI_ISL_1635922, EPI_ISL_1635923, EPI_ISL_1635924, EPI_ISL_1635925,

EPI_ISL_1635926, EPI_ISL_1635927, EPI_ISL_1635928, EPI_ISL_1635929, EPI_ISL_1635930, EPI_ISL_1635931, EPI_ISL_1635932, EPI_ISL_1635933, EPI_ISL_1635934, EPI_ISL_1635935, EPI_ISL_1635936, EPI_ISL_1635937, EPI_ISL_1635938, EPI_ISL_1635939, EPI_ISL_1635940, EPI_ISL_1635941, EPI_ISL_1635942, EPI_ISL_1635943, EPI_ISL_1635944, EPI_ISL_1635945, EPI_ISL_1635946, EPI_ISL_1635947,
EPI_ISL_1635948, EPI_ISL_1635949, EPI_ISL_1635950, EPI_ISL_1635951, EPI_ISL_1635952, EPI_ISL_1635953, EPI_ISL_1635954, EPI_ISL_1635955, EPI_ISL_1635956, EPI_ISL_1635957, EPI_ISL_1635958, EPI_ISL_1635959, EPI_ISL_1635960, EPI_ISL_1635961, EPI_ISL_1635962, EPI_ISL_1635963, EPI_ISL_1635965, EPI_ISL_1635966, EPI_ISL_1635967, EPI_ISL_1635968, EPI_ISL_1635969, EPI_ISL_1635970,
EPI_ISL_1635971, EPI_ISL_1635972, EPI_ISL_1635973, EPI_ISL_1635974, EPI_ISL_1635975, EPI_ISL_1635976, EPI_ISL_1635977, EPI_ISL_1635978, EPI_ISL_1635979, EPI_ISL_1635980, EPI_ISL_1635981, EPI_ISL_1635982, EPI_ISL_1635983, EPI_ISL_1635984, EPI_ISL_1635985, EPI_ISL_1635986, EPI_ISL_1635987, EPI_ISL_1635988, EPI_ISL_1635989, EPI_ISL_1635990, EPI_ISL_1635991, EPI_ISL_1635992,
EPI_ISL_1635993, EPI_ISL_1635994, EPI_ISL_1635995, EPI_ISL_1635996, EPI_ISL_1635997, EPI_ISL_1635998, EPI_ISL_1635999, EPI_ISL_1636000, EPI_ISL_1636001, EPI_ISL_1636002, EPI_ISL_1636003, EPI_ISL_1636004, EPI_ISL_1636005, EPI_ISL_1636006, EPI_ISL_1636007, EPI_ISL_1636008, EPI_ISL_1636009, EPI_ISL_1636010, EPI_ISL_1636011, EPI_ISL_1636012, EPI_ISL_1636013, EPI_ISL_1636014,
EPI_ISL_1636015, EPI_ISL_1636016, EPI_ISL_1636017, EPI_ISL_1636018, EPI_ISL_1636019, EPI_ISL_1636020, EPI_ISL_1636021, EPI_ISL_1636022, EPI_ISL_1636023, EPI_ISL_1636024, EPI_ISL_1636025, EPI_ISL_1636026, EPI_ISL_1636027, EPI_ISL_1636028, EPI_ISL_1636029, EPI_ISL_1636030, EPI_ISL_1636031, EPI_ISL_1636032, EPI_ISL_1636033, EPI_ISL_1636034, EPI_ISL_1636035, EPI_ISL_1636036,
EPI_ISL_1636037, EPI_ISL_1636038, EPI_ISL_1636039, EPI_ISL_1636040, EPI_ISL_1636041, EPI_ISL_1636042, EPI_ISL_1636043, EPI_ISL_1636044, EPI_ISL_1636045, EPI_ISL_1636046, EPI_ISL_1636047, EPI_ISL_1636048, EPI_ISL_1636049, EPI_ISL_1636050, EPI_ISL_1636051, EPI_ISL_1636052, EPI_ISL_1636053, EPI_ISL_1636054, EPI_ISL_1636055, EPI_ISL_1636056, EPI_ISL_1636057, EPI_ISL_1636058,
EPI_ISL_1636059, EPI_ISL_1636060, EPI_ISL_1636061, EPI_ISL_1636062, EPI_ISL_1636063, EPI_ISL_1636064, EPI_ISL_1636065, EPI_ISL_1636066, EPI_ISL_1636067, EPI_ISL_1636068, EPI_ISL_1636069, EPI_ISL_1636070, EPI_ISL_1636071, EPI_ISL_1636072, EPI_ISL_1636073, EPI_ISL_1636074, EPI_ISL_1636075, EPI_ISL_1636076, EPI_ISL_1636077, EPI_ISL_1636078, EPI_ISL_1636079, EPI_ISL_1636080,
EPI_ISL_1636081, EPI_ISL_1636082, EPI_ISL_1636083, EPI_ISL_1636084, EPI_ISL_1636085, EPI_ISL_1636086, EPI_ISL_1636087, EPI_ISL_1636088, EPI_ISL_1636089, EPI_ISL_1636090, EPI_ISL_1636091, EPI_ISL_1636092, EPI_ISL_1636093, EPI_ISL_1636094, EPI_ISL_1636095, EPI_ISL_1636096, EPI_ISL_1636097, EPI_ISL_1636098, EPI_ISL_1636099, EPI_ISL_1636100, EPI_ISL_1636101, EPI_ISL_1636102,
EPI_ISL_1636103, EPI_ISL_1636104, EPI_ISL_1636105, EPI_ISL_1636106, EPI_ISL_1636107, EPI_ISL_1636108, EPI_ISL_1636109, EPI_ISL_1636111, EPI_ISL_1636112, EPI_ISL_1636113, EPI_ISL_1636114, EPI_ISL_1636115, EPI_ISL_1636116, EPI_ISL_1636117, EPI_ISL_1636118, EPI_ISL_1636119, EPI_ISL_1636120, EPI_ISL_1636121, EPI_ISL_1636122, EPI_ISL_1636123, EPI_ISL_1636124, EPI_ISL_1636125,
EPI_ISL_1636126, EPI_ISL_1636127, EPI_ISL_1636128, EPI_ISL_1636129, EPI_ISL_1636130, EPI_ISL_1636131, EPI_ISL_1636132, EPI_ISL_1636133, EPI_ISL_1636134, EPI_ISL_1636135, EPI_ISL_1636136, EPI_ISL_1636137, EPI_ISL_1636138, EPI_ISL_1636139, EPI_ISL_1636140, EPI_ISL_1636141, EPI_ISL_1636142, EPI_ISL_1636143, EPI_ISL_1636144, EPI_ISL_1636145, EPI_ISL_1636146, EPI_ISL_1636147,
EPI_ISL_1636148, EPI_ISL_1636149, EPI_ISL_1636150, EPI_ISL_1636151, EPI_ISL_1636152, EPI_ISL_1636153, EPI_ISL_1636154, EPI_ISL_1636155, EPI_ISL_1636156, EPI_ISL_1636157, EPI_ISL_1636158, EPI_ISL_1636159, EPI_ISL_1636160, EPI_ISL_1636161, EPI_ISL_1636162, EPI_ISL_1636163, EPI_ISL_1636164, EPI_ISL_1636165, EPI_ISL_1636166, EPI_ISL_1636167, EPI_ISL_1636168, EPI_ISL_1636169,
EPI_ISL_1636170, EPI_ISL_1636171, EPI_ISL_1636172, EPI_ISL_1636173, EPI_ISL_1636174, EPI_ISL_1636175, EPI_ISL_1636176, EPI_ISL_1636177, EPI_ISL_1636178, EPI_ISL_1636179, EPI_ISL_1636180, EPI_ISL_1636181, EPI_ISL_1636182, EPI_ISL_1636183, EPI_ISL_1636184, EPI_ISL_1636185, EPI_ISL_1636186, EPI_ISL_1636187, EPI_ISL_1636188, EPI_ISL_1636189, EPI_ISL_1636190, EPI_ISL_1636191,
EPI_ISL_1636192, EPI_ISL_1636193, EPI_ISL_1636194, EPI_ISL_1636195, EPI_ISL_1636196, EPI_ISL_1636197, EPI_ISL_1636198, EPI_ISL_1636199, EPI_ISL_1636200, EPI_ISL_1636201, EPI_ISL_1636202, EPI_ISL_1636203, EPI_ISL_1636204, EPI_ISL_1636205, EPI_ISL_1636206, EPI_ISL_1636207, EPI_ISL_1636208, EPI_ISL_1636209, EPI_ISL_1636210, EPI_ISL_1636211, EPI_ISL_1636212, EPI_ISL_1636213,
EPI_ISL_1636214, EPI_ISL_1636215, EPI_ISL_1636217, EPI_ISL_1636218, EPI_ISL_1636219, EPI_ISL_1636220, EPI_ISL_1636221, EPI_ISL_1636222, EPI_ISL_1636223, EPI_ISL_1636224, EPI_ISL_1636225, EPI_ISL_1636226, EPI_ISL_1636227, EPI_ISL_1636228, EPI_ISL_1636229, EPI_ISL_1636230, EPI_ISL_1636231, EPI_ISL_1636232, EPI_ISL_1636233, EPI_ISL_1636234, EPI_ISL_1636235, EPI_ISL_1636236,
EPI_ISL_1636237, EPI_ISL_1636238, EPI_ISL_1636239, EPI_ISL_1636240, EPI_ISL_1636241, EPI_ISL_1636242, EPI_ISL_1636243, EPI_ISL_1636244, EPI_ISL_1636245, EPI_ISL_1636246, EPI_ISL_1636247, EPI_ISL_1636248, EPI_ISL_1636249, EPI_ISL_1636250, EPI_ISL_1636251, EPI_ISL_1636252, EPI_ISL_1636253, EPI_ISL_1636254, EPI_ISL_1636255, EPI_ISL_1636256, EPI_ISL_1636257, EPI_ISL_1636258,
EPI_ISL_1636259, EPI_ISL_1636260, EPI_ISL_1636261, EPI_ISL_1636262, EPI_ISL_1636263, EPI_ISL_1636264, EPI_ISL_1636265, EPI_ISL_1636266, EPI_ISL_1636267, EPI_ISL_1636268, EPI_ISL_1636269, EPI_ISL_1636270, EPI_ISL_1636271, EPI_ISL_1636272, EPI_ISL_1636273, EPI_ISL_1636274, EPI_ISL_1636275, EPI_ISL_1636276, EPI_ISL_1636277, EPI_ISL_1636278, EPI_ISL_1636279, EPI_ISL_1636280,
EPI_ISL_1636281, EPI_ISL_1636282, EPI_ISL_1636283, EPI_ISL_1636284, EPI_ISL_1636285, EPI_ISL_1636286, EPI_ISL_1636287, EPI_ISL_1636288, EPI_ISL_1636289, EPI_ISL_1636290, EPI_ISL_1636291, EPI_ISL_1636292, EPI_ISL_1636293, EPI_ISL_1636294, EPI_ISL_1636295, EPI_ISL_1636296, EPI_ISL_1636297, EPI_ISL_1636298, EPI_ISL_1636299, EPI_ISL_1636300, EPI_ISL_1636301, EPI_ISL_1636302,
EPI_ISL_1636303, EPI_ISL_1636304, EPI_ISL_1636305, EPI_ISL_1636306, EPI_ISL_1636307, EPI_ISL_1636308, EPI_ISL_1636309, EPI_ISL_1636310, EPI_ISL_1636311, EPI_ISL_1636312, EPI_ISL_1636313, EPI_ISL_1636314, EPI_ISL_1636315, EPI_ISL_1636316, EPI_ISL_1636317, EPI_ISL_1636318, EPI_ISL_1636319, EPI_ISL_1636320, EPI_ISL_1636321, EPI_ISL_1636322, EPI_ISL_1636323, EPI_ISL_1636324,
EPI_ISL_1636325, EPI_ISL_1636326, EPI_ISL_1636327, EPI_ISL_1636328, EPI_ISL_1636329, EPI_ISL_1636330, EPI_ISL_1636331, EPI_ISL_1636332, EPI_ISL_1636333, EPI_ISL_1636334, EPI_ISL_1636335, EPI_ISL_1636336, EPI_ISL_1636337, EPI_ISL_1636338, EPI_ISL_1636339, EPI_ISL_1636340, EPI_ISL_1636341, EPI_ISL_1636342, EPI_ISL_1636343, EPI_ISL_1636344, EPI_ISL_1636345, EPI_ISL_1636346,
EPI_ISL_1636347, EPI_ISL_1636348, EPI_ISL_1636349, EPI_ISL_1636350, EPI_ISL_1636351, EPI_ISL_1636352, EPI_ISL_1636353, EPI_ISL_1636354, EPI_ISL_1636355, EPI_ISL_1636356, EPI_ISL_1636357, EPI_ISL_1636358, EPI_ISL_1636359, EPI_ISL_1636360, EPI_ISL_1636361, EPI_ISL_1636362, EPI_ISL_1636363, EPI_ISL_1636364, EPI_ISL_1636366, EPI_ISL_1636367, EPI_ISL_1636368, EPI_ISL_1636369,
EPI_ISL_1636370, EPI_ISL_1636371, EPI_ISL_1636372, EPI_ISL_1636373, EPI_ISL_1636374, EPI_ISL_1636375, EPI_ISL_1636376, EPI_ISL_1636377, EPI_ISL_1636378, EPI_ISL_1636379, EPI_ISL_1636380, EPI_ISL_1636381, EPI_ISL_1636382, EPI_ISL_1636383, EPI_ISL_1636384, EPI_ISL_1636385, EPI_ISL_1636386, EPI_ISL_1636387, EPI_ISL_1636388, EPI_ISL_1636389, EPI_ISL_1636390, EPI_ISL_1636391,
EPI_ISL_1636392, EPI_ISL_1636393, EPI_ISL_1636394, EPI_ISL_1636395, EPI_ISL_1636396, EPI_ISL_1636397, EPI_ISL_1717107, EPI_ISL_1717108, EPI_ISL_1717109, EPI_ISL_1717110, EPI_ISL_1717111, EPI_ISL_1717112, EPI_ISL_1717113, EPI_ISL_1717114, EPI_ISL_1717115, EPI_ISL_1717116, EPI_ISL_1717117, EPI_ISL_1717118, EPI_ISL_1717119, EPI_ISL_1717120, EPI_ISL_1717121, EPI_ISL_1717122,
EPI_ISL_1717123, EPI_ISL_1717124, EPI_ISL_1717125, EPI_ISL_1717126, EPI_ISL_1717127, EPI_ISL_1717128, EPI_ISL_1717129, EPI_ISL_1717130, EPI_ISL_1717131, EPI_ISL_1717132, EPI_ISL_1717133, EPI_ISL_1717134, EPI_ISL_1717135, EPI_ISL_1717136, EPI_ISL_1717137, EPI_ISL_1717138, EPI_ISL_1717139, EPI_ISL_1717140, EPI_ISL_1717141, EPI_ISL_1717142, EPI_ISL_1717143, EPI_ISL_1717144,
EPI_ISL_1717145, EPI_ISL_1717146, EPI_ISL_1717147, EPI_ISL_1717148, EPI_ISL_1717149, EPI_ISL_1717150, EPI_ISL_1717151, EPI_ISL_1717152, EPI_ISL_1717153, EPI_ISL_1717154, EPI_ISL_1717155, EPI_ISL_1717156, EPI_ISL_1717157, EPI_ISL_1717158, EPI_ISL_1717159, EPI_ISL_1717160, EPI_ISL_1717161, EPI_ISL_1717162, EPI_ISL_1717163, EPI_ISL_1717164, EPI_ISL_1717165, EPI_ISL_1717166,
EPI_ISL_1717167, EPI_ISL_1717168, EPI_ISL_1717170, EPI_ISL_1717171, EPI_ISL_1717172, EPI_ISL_1717173, EPI_ISL_1717174, EPI_ISL_1717175, EPI_ISL_1717176, EPI_ISL_1717177, EPI_ISL_1717178, EPI_ISL_1717179, EPI_ISL_1717180, EPI_ISL_1717181, EPI_ISL_1717182, EPI_ISL_1717183, EPI_ISL_1717184, EPI_ISL_1717185, EPI_ISL_1717186, EPI_ISL_1717187, EPI_ISL_1717188, EPI_ISL_1717189,
EPI_ISL_1717190, EPI_ISL_1717191, EPI_ISL_1717192, EPI_ISL_1717193, EPI_ISL_1717194, EPI_ISL_1717195, EPI_ISL_1717196, EPI_ISL_1717197, EPI_ISL_1717198, EPI_ISL_1717199, EPI_ISL_1717200, EPI_ISL_1717201, EPI_ISL_1717202, EPI_ISL_1717203, EPI_ISL_1717204, EPI_ISL_1717205, EPI_ISL_1717206, EPI_ISL_1717207, EPI_ISL_1717208, EPI_ISL_1717209, EPI_ISL_1717210, EPI_ISL_1717211,
EPI_ISL_1717212, EPI_ISL_1717213, EPI_ISL_1717214, EPI_ISL_1717215, EPI_ISL_1717216, EPI_ISL_1717217, EPI_ISL_1717218, EPI_ISL_1717219, EPI_ISL_1717220, EPI_ISL_1717221, EPI_ISL_1717222, EPI_ISL_1717223, EPI_ISL_1717224, EPI_ISL_1717225, EPI_ISL_1717226, EPI_ISL_1717227, EPI_ISL_1717228, EPI_ISL_1717229, EPI_ISL_1717230, EPI_ISL_1717231, EPI_ISL_1717232, EPI_ISL_1717233,
EPI_ISL_1717234, EPI_ISL_1717235, EPI_ISL_1717236, EPI_ISL_1717237, EPI_ISL_1717238, EPI_ISL_1717239, EPI_ISL_1717240, EPI_ISL_1717241, EPI_ISL_1717242, EPI_ISL_1717243, EPI_ISL_1717244, EPI_ISL_1717245, EPI_ISL_1717246, EPI_ISL_1717247, EPI_ISL_1717248, EPI_ISL_1717249, EPI_ISL_1717250, EPI_ISL_1717251, EPI_ISL_1717252, EPI_ISL_1717253, EPI_ISL_1717254, EPI_ISL_1717255,
EPI_ISL_1717256, EPI_ISL_1717257, EPI_ISL_1717258, EPI_ISL_1717259, EPI_ISL_1717260, EPI_ISL_1717261, EPI_ISL_1717262, EPI_ISL_1717263, EPI_ISL_1717264, EPI_ISL_1717265, EPI_ISL_1717266, EPI_ISL_1717267, EPI_ISL_1717268, EPI_ISL_1717269, EPI_ISL_1717270, EPI_ISL_1717271, EPI_ISL_1717272, EPI_ISL_1717273, EPI_ISL_1717274, EPI_ISL_1717275, EPI_ISL_1717276, EPI_ISL_1717277,
EPI_ISL_1717278, EPI_ISL_1717279, EPI_ISL_1717280, EPI_ISL_1717281, EPI_ISL_1717282, EPI_ISL_1717283, EPI_ISL_1717284, EPI_ISL_1717285, EPI_ISL_1717286, EPI_ISL_1717287, EPI_ISL_1717288, EPI_ISL_1717289, EPI_ISL_1717290, EPI_ISL_1717291, EPI_ISL_1717292, EPI_ISL_1717293, EPI_ISL_1717294, EPI_ISL_1717295, EPI_ISL_1717296, EPI_ISL_1717297, EPI_ISL_1717298, EPI_ISL_1717299,
EPI_ISL_1717300, EPI_ISL_1717301, EPI_ISL_1717302, EPI_ISL_1717303, EPI_ISL_1717304, EPI_ISL_1717305, EPI_ISL_1717306, EPI_ISL_1717307, EPI_ISL_1717308, EPI_ISL_1717309, EPI_ISL_1717310, EPI_ISL_1717311, EPI_ISL_1717312, EPI_ISL_1717313, EPI_ISL_1717314, EPI_ISL_1717315, EPI_ISL_1717316, EPI_ISL_1717317, EPI_ISL_1717318, EPI_ISL_1717319, EPI_ISL_1717320, EPI_ISL_1717321,
EPI_ISL_1717322, EPI_ISL_1717323, EPI_ISL_1717324, EPI_ISL_1717325, EPI_ISL_1717326, EPI_ISL_1717327, EPI_ISL_1717328, EPI_ISL_1717329, EPI_ISL_1717330, EPI_ISL_1717331, EPI_ISL_1717332, EPI_ISL_1717333, EPI_ISL_1717334, EPI_ISL_1717335, EPI_ISL_1717336, EPI_ISL_1717337, EPI_ISL_1717338, EPI_ISL_1717339, EPI_ISL_1717340, EPI_ISL_1717341, EPI_ISL_1717342, EPI_ISL_1717343,
EPI_ISL_1717344, EPI_ISL_1717345, EPI_ISL_1717346, EPI_ISL_1717347, EPI_ISL_1717348, EPI_ISL_1717349, EPI_ISL_1717350, EPI_ISL_1717351, EPI_ISL_1717352, EPI_ISL_1717353, EPI_ISL_1717354, EPI_ISL_1717355, EPI_ISL_1717356, EPI_ISL_1717357, EPI_ISL_1717358, EPI_ISL_1717359, EPI_ISL_1717360, EPI_ISL_1717361, EPI_ISL_1717362, EPI_ISL_1717363, EPI_ISL_1717364, EPI_ISL_1717365,
EPI_ISL_1717366, EPI_ISL_1717367, EPI_ISL_1717368, EPI_ISL_1717369, EPI_ISL_1717370, EPI_ISL_1717371, EPI_ISL_1717372, EPI_ISL_1717373, EPI_ISL_1717374, EPI_ISL_1717375, EPI_ISL_1717376, EPI_ISL_1717377, EPI_ISL_1717378, EPI_ISL_1717379, EPI_ISL_1717380, EPI_ISL_1717381, EPI_ISL_1717382, EPI_ISL_1717383, EPI_ISL_1717384, EPI_ISL_1717385, EPI_ISL_1717386, EPI_ISL_1717387,
EPI_ISL_1717388, EPI_ISL_1717389, EPI_ISL_1717390, EPI_ISL_1717391, EPI_ISL_1717392, EPI_ISL_1717393, EPI_ISL_1717394, EPI_ISL_1717395, EPI_ISL_1717396, EPI_ISL_1717397, EPI_ISL_1717398, EPI_ISL_1717399, EPI_ISL_1717400, EPI_ISL_1717401, EPI_ISL_1717402, EPI_ISL_1717403, EPI_ISL_1717404, EPI_ISL_1717405, EPI_ISL_1717406, EPI_ISL_1717407, EPI_ISL_1717408, EPI_ISL_1717409,
EPI_ISL_1717410, EPI_ISL_1717411, EPI_ISL_1717412, EPI_ISL_1717413, EPI_ISL_1717414, EPI_ISL_1717415, EPI_ISL_1717416, EPI_ISL_1717417, EPI_ISL_1717418, EPI_ISL_1717419, EPI_ISL_1717420, EPI_ISL_1717421, EPI_ISL_1717422, EPI_ISL_1717423, EPI_ISL_1717424, EPI_ISL_1717425, EPI_ISL_1717426, EPI_ISL_1717427, EPI_ISL_1717428, EPI_ISL_1717429, EPI_ISL_1717430, EPI_ISL_1717431,
EPI_ISL_1717432, EPI_ISL_1717433, EPI_ISL_1717434, EPI_ISL_1717435, EPI_ISL_1717436, EPI_ISL_1717437, EPI_ISL_1717438, EPI_ISL_1717439, EPI_ISL_1717440, EPI_ISL_1717441, EPI_ISL_1717442, EPI_ISL_1717443, EPI_ISL_1717444, EPI_ISL_1717445, EPI_ISL_1717446, EPI_ISL_1717447, EPI_ISL_1717448, EPI_ISL_1717449, EPI_ISL_1717450, EPI_ISL_1717451, EPI_ISL_1717452, EPI_ISL_1717453,
EPI_ISL_1717454, EPI_ISL_1717455, EPI_ISL_1717456, EPI_ISL_1717457, EPI_ISL_1717458, EPI_ISL_1717459, EPI_ISL_1717460, EPI_ISL_1717461, EPI_ISL_1717462, EPI_ISL_1717463, EPI_ISL_1717464, EPI_ISL_1717465, EPI_ISL_1717466, EPI_ISL_1717467, EPI_ISL_1717468, EPI_ISL_1717469, EPI_ISL_1717470, EPI_ISL_1717471, EPI_ISL_1717472, EPI_ISL_1717473, EPI_ISL_1717474, EPI_ISL_1717475,
EPI_ISL_1717476, EPI_ISL_1717477, EPI_ISL_1717478, EPI_ISL_1717479, EPI_ISL_1717480, EPI_ISL_1717481, EPI_ISL_1717482, EPI_ISL_1717483, EPI_ISL_1717484, EPI_ISL_1717485, EPI_ISL_1717486, EPI_ISL_1717487, EPI_ISL_1717488, EPI_ISL_1717489, EPI_ISL_1717490, EPI_ISL_1717491, EPI_ISL_1717492, EPI_ISL_1717493, EPI_ISL_1717494, EPI_ISL_1717495, EPI_ISL_1717496, EPI_ISL_1717497,
EPI_ISL_1717498, EPI_ISL_1717499, EPI_ISL_1717500, EPI_ISL_1717501, EPI_ISL_1717502, EPI_ISL_1717503, EPI_ISL_1717504, EPI_ISL_1717505, EPI_ISL_1717506, EPI_ISL_1717507, EPI_ISL_1717508, EPI_ISL_1717509, EPI_ISL_1717510, EPI_ISL_1717511, EPI_ISL_1717512, EPI_ISL_1717513, EPI_ISL_1717514, EPI_ISL_1717515, EPI_ISL_1717516, EPI_ISL_1717517, EPI_ISL_1717518, EPI_ISL_1717519,
EPI_ISL_1717520, EPI_ISL_1717521, EPI_ISL_1717522, EPI_ISL_1717523, EPI_ISL_1717524, EPI_ISL_1717525, EPI_ISL_1717526, EPI_ISL_1717527, EPI_ISL_1717528, EPI_ISL_1717529, EPI_ISL_1717530, EPI_ISL_1717531, EPI_ISL_1717532, EPI_ISL_1717533, EPI_ISL_1717534, EPI_ISL_1717535, EPI_ISL_1717536, EPI_ISL_1717537, EPI_ISL_1717538, EPI_ISL_1717539, EPI_ISL_1717540, EPI_ISL_1717541,
EPI_ISL_1717542, EPI_ISL_1717543, EPI_ISL_1717544, EPI_ISL_1717545, EPI_ISL_1717546, EPI_ISL_1717547, EPI_ISL_1717548, EPI_ISL_1717549, EPI_ISL_1717550, EPI_ISL_1717551, EPI_ISL_1717552, EPI_ISL_1717553, EPI_ISL_1717554, EPI_ISL_1717555, EPI_ISL_1717556, EPI_ISL_1717557, EPI_ISL_1717558, EPI_ISL_1717559, EPI_ISL_1717560, EPI_ISL_1717561, EPI_ISL_1717562, EPI_ISL_1717563,
EPI_ISL_1717564, EPI_ISL_1717565, EPI_ISL_1717566, EPI_ISL_1717567, EPI_ISL_1717568, EPI_ISL_1717569, EPI_ISL_1717570, EPI_ISL_1717571, EPI_ISL_1717572, EPI_ISL_1717573, EPI_ISL_1717574, EPI_ISL_1717575, EPI_ISL_1717576, EPI_ISL_1717577, EPI_ISL_1717578, EPI_ISL_1717579, EPI_ISL_1717580, EPI_ISL_1717581, EPI_ISL_1717582, EPI_ISL_1717583, EPI_ISL_1717584, EPI_ISL_1717585,
EPI_ISL_1717586, EPI_ISL_1717587, EPI_ISL_1717588, EPI_ISL_1717589, EPI_ISL_1717590, EPI_ISL_1717591, EPI_ISL_1717592, EPI_ISL_1717593, EPI_ISL_1717594, EPI_ISL_1717595, EPI_ISL_1717596, EPI_ISL_1717597, EPI_ISL_1717598, EPI_ISL_1717599, EPI_ISL_1717600, EPI_ISL_1717601, EPI_ISL_1717602, EPI_ISL_1717603, EPI_ISL_1717604, EPI_ISL_1717605, EPI_ISL_1717606, EPI_ISL_1717607,
EPI_ISL_1717608, EPI_ISL_1717609, EPI_ISL_1717610, EPI_ISL_1717611, EPI_ISL_1717612, EPI_ISL_1717613, EPI_ISL_1717614, EPI_ISL_1717615, EPI_ISL_1717616, EPI_ISL_1717617, EPI_ISL_1717618, EPI_ISL_1717619, EPI_ISL_1717620, EPI_ISL_1717621, EPI_ISL_1717622, EPI_ISL_1717623, EPI_ISL_1717624, EPI_ISL_1717625, EPI_ISL_1717626, EPI_ISL_1717627, EPI_ISL_1717628, EPI_ISL_1717629,
EPI_ISL_1717630, EPI_ISL_1717631, EPI_ISL_1717632, EPI_ISL_1717633, EPI_ISL_1717634, EPI_ISL_1717635, EPI_ISL_1717636, EPI_ISL_1717637, EPI_ISL_1717638, EPI_ISL_1717639, EPI_ISL_1717640, EPI_ISL_1717641, EPI_ISL_1717642, EPI_ISL_1717643, EPI_ISL_1717644, EPI_ISL_1717645, EPI_ISL_1717646, EPI_ISL_1717647, EPI_ISL_1717648, EPI_ISL_1717649, EPI_ISL_1717650, EPI_ISL_1717651,
EPI_ISL_1717652, EPI_ISL_1717653, EPI_ISL_1717654, EPI_ISL_1717655, EPI_ISL_1717656, EPI_ISL_1717657, EPI_ISL_1717658, EPI_ISL_1717659, EPI_ISL_1717660, EPI_ISL_1717661, EPI_ISL_1717662, EPI_ISL_1717663, EPI_ISL_1717664, EPI_ISL_1717665, EPI_ISL_1717666, EPI_ISL_1717667, EPI_ISL_1717668, EPI_ISL_1717669, EPI_ISL_1717670, EPI_ISL_1717671, EPI_ISL_1717672, EPI_ISL_1717673,
EPI_ISL_1717674, EPI_ISL_1717675, EPI_ISL_1717676, EPI_ISL_1717677, EPI_ISL_1717678, EPI_ISL_1717679, EPI_ISL_1717680, EPI_ISL_1717681, EPI_ISL_1717682, EPI_ISL_1717683, EPI_ISL_1717684, EPI_ISL_1717685, EPI_ISL_1717686, EPI_ISL_1717687, EPI_ISL_1717688, EPI_ISL_1717689, EPI_ISL_1717690, EPI_ISL_1717691, EPI_ISL_1717692, EPI_ISL_1717693, EPI_ISL_1717694, EPI_ISL_1717695,
EPI_ISL_1717696, EPI_ISL_1717697, EPI_ISL_1717698, EPI_ISL_1717699, EPI_ISL_1717700, EPI_ISL_1717701, EPI_ISL_1717702, EPI_ISL_1717703, EPI_ISL_1717704, EPI_ISL_1717705, EPI_ISL_1717706, EPI_ISL_1717707, EPI_ISL_1717708, EPI_ISL_1717709, EPI_ISL_1717710, EPI_ISL_1717711, EPI_ISL_1717712, EPI_ISL_1717713, EPI_ISL_1717714, EPI_ISL_1717715, EPI_ISL_1717716, EPI_ISL_1717717,
EPI_ISL_1717718, EPI_ISL_1717719, EPI_ISL_1717720, EPI_ISL_1717721, EPI_ISL_1717722, EPI_ISL_1717723, EPI_ISL_1717724, EPI_ISL_1717725, EPI_ISL_1717726, EPI_ISL_1717727, EPI_ISL_1717728, EPI_ISL_1717729, EPI_ISL_1717730, EPI_ISL_1717731, EPI_ISL_1717732, EPI_ISL_1717733, EPI_ISL_1717734, EPI_ISL_1717735, EPI_ISL_1717736, EPI_ISL_1717737, EPI_ISL_1717738, EPI_ISL_1717739,
EPI_ISL_1717740, EPI_ISL_1717741, EPI_ISL_1717742, EPI_ISL_1717743, EPI_ISL_1717744, EPI_ISL_1717745, EPI_ISL_1717746, EPI_ISL_1717747, EPI_ISL_1717748, EPI_ISL_1717749, EPI_ISL_1717750, EPI_ISL_1717751, EPI_ISL_1717752, EPI_ISL_1717753, EPI_ISL_1717754, EPI_ISL_1717755, EPI_ISL_1717756, EPI_ISL_1717757, EPI_ISL_1717758, EPI_ISL_1717759, EPI_ISL_1717760, EPI_ISL_1717761,
EPI_ISL_1717762, EPI_ISL_1717763, EPI_ISL_1717764, EPI_ISL_1717765, EPI_ISL_1717766, EPI_ISL_1717767, EPI_ISL_1717768, EPI_ISL_1717769, EPI_ISL_1717770, EPI_ISL_1717771, EPI_ISL_1717772, EPI_ISL_1717773, EPI_ISL_1717774, EPI_ISL_1717775, EPI_ISL_1717776, EPI_ISL_1717777, EPI_ISL_1717778, EPI_ISL_1717779, EPI_ISL_1717780, EPI_ISL_1717781, EPI_ISL_1717782, EPI_ISL_1717783,
EPI_ISL_1717784, EPI_ISL_1717785, EPI_ISL_1717786, EPI_ISL_1717787, EPI_ISL_1717788, EPI_ISL_1717789, EPI_ISL_1717790, EPI_ISL_1717791, EPI_ISL_1717792, EPI_ISL_1717793, EPI_ISL_1717794, EPI_ISL_1717795, EPI_ISL_1717796, EPI_ISL_1717797, EPI_ISL_1717798, EPI_ISL_1717799, EPI_ISL_1717800, EPI_ISL_1717801, EPI_ISL_1717802, EPI_ISL_1717803, EPI_ISL_1717804, EPI_ISL_1717805,
EPI_ISL_1717806, EPI_ISL_1717807, EPI_ISL_1717808, EPI_ISL_1717809, EPI_ISL_1717810, EPI_ISL_1717811, EPI_ISL_1717812, EPI_ISL_1717813, EPI_ISL_1717814, EPI_ISL_1717815, EPI_ISL_1717816, EPI_ISL_1717817, EPI_ISL_1717818, EPI_ISL_1717819, EPI_ISL_1717820, EPI_ISL_1717821, EPI_ISL_1717822, EPI_ISL_1717823, EPI_ISL_1717824, EPI_ISL_1717825, EPI_ISL_1717826, EPI_ISL_1717827,
EPI_ISL_1717828, EPI_ISL_1717829, EPI_ISL_1717830, EPI_ISL_1717831, EPI_ISL_1717832, EPI_ISL_1717833, EPI_ISL_1717834, EPI_ISL_1717835, EPI_ISL_1717836, EPI_ISL_1717837, EPI_ISL_1717838, EPI_ISL_1717839, EPI_ISL_1717840, EPI_ISL_1717841, EPI_ISL_1717842, EPI_ISL_1717843, EPI_ISL_1717844, EPI_ISL_1717845, EPI_ISL_1717846, EPI_ISL_1717847, EPI_ISL_1717848, EPI_ISL_1717849,
EPI_ISL_1717850, EPI_ISL_1717851, EPI_ISL_1717852, EPI_ISL_1717853, EPI_ISL_1717854, EPI_ISL_1717855, EPI_ISL_1717856, EPI_ISL_1717857, EPI_ISL_1717858, EPI_ISL_1717859, EPI_ISL_1717860, EPI_ISL_1717861, EPI_ISL_1717862, EPI_ISL_1717863, EPI_ISL_1717864, EPI_ISL_1717865, EPI_ISL_1717866, EPI_ISL_1717867, EPI_ISL_1717868, EPI_ISL_1717869, EPI_ISL_1717870, EPI_ISL_1717871,
EPI_ISL_1717872, EPI_ISL_1717873, EPI_ISL_1717874, EPI_ISL_1717875, EPI_ISL_1717876, EPI_ISL_1717877, EPI_ISL_1717878, EPI_ISL_1717879, EPI_ISL_1717880, EPI_ISL_1717881, EPI_ISL_1717882, EPI_ISL_1717883, EPI_ISL_1717884, EPI_ISL_1717885, EPI_ISL_1717886, EPI_ISL_1717887, EPI_ISL_1717888, EPI_ISL_1717889, EPI_ISL_1717890, EPI_ISL_1717891, EPI_ISL_1717892, EPI_ISL_1717893,
EPI_ISL_1717894, EPI_ISL_1717895, EPI_ISL_1717896, EPI_ISL_1717897, EPI_ISL_1717898, EPI_ISL_1717899, EPI_ISL_1717900, EPI_ISL_1717901, EPI_ISL_1717902, EPI_ISL_1717903, EPI_ISL_1717904, EPI_ISL_1717905, EPI_ISL_1717906, EPI_ISL_1717907, EPI_ISL_1717908, EPI_ISL_1717909, EPI_ISL_1717910, EPI_ISL_1717911, EPI_ISL_1717912, EPI_ISL_1717913, EPI_ISL_1717914, EPI_ISL_1717915,
EPI_ISL_1717916, EPI_ISL_1717917, EPI_ISL_1717918, EPI_ISL_1717919, EPI_ISL_1717920, EPI_ISL_1717921, EPI_ISL_1717922, EPI_ISL_1717923, EPI_ISL_1717924, EPI_ISL_1717925, EPI_ISL_1717926, EPI_ISL_1717927, EPI_ISL_1717928, EPI_ISL_1717929, EPI_ISL_1717930, EPI_ISL_1717931, EPI_ISL_1717932, EPI_ISL_1717933, EPI_ISL_1717934, EPI_ISL_1717935, EPI_ISL_1717936, EPI_ISL_1717937,
EPI_ISL_1717938, EPI_ISL_1717939, EPI_ISL_1717940, EPI_ISL_1717941, EPI_ISL_1717942, EPI_ISL_1717943, EPI_ISL_1717944, EPI_ISL_1717945, EPI_ISL_1717946, EPI_ISL_1717947, EPI_ISL_1717948, EPI_ISL_1717949, EPI_ISL_1717950, EPI_ISL_1717951, EPI_ISL_1717952, EPI_ISL_1717953, EPI_ISL_1717954, EPI_ISL_1717955, EPI_ISL_1717956, EPI_ISL_1717957, EPI_ISL_1717958, EPI_ISL_1717959,
EPI_ISL_1717960, EPI_ISL_1717961, EPI_ISL_1717962, EPI_ISL_1717963, EPI_ISL_1717964, EPI_ISL_1717965, EPI_ISL_1717966, EPI_ISL_1717967, EPI_ISL_1717968, EPI_ISL_1717969, EPI_ISL_1717970, EPI_ISL_1717971, EPI_ISL_1717972, EPI_ISL_1717973, EPI_ISL_1717974, EPI_ISL_1717975, EPI_ISL_1717976, EPI_ISL_1717977, EPI_ISL_1717978, EPI_ISL_1717979, EPI_ISL_1717980, EPI_ISL_1717981,
EPI_ISL_1717982, EPI_ISL_1717983, EPI_ISL_1717984, EPI_ISL_1717985, EPI_ISL_1717986, EPI_ISL_1717987, EPI_ISL_1717988, EPI_ISL_1717989, EPI_ISL_1717990, EPI_ISL_1717991, EPI_ISL_1717992, EPI_ISL_1717993, EPI_ISL_1717994, EPI_ISL_1717995, EPI_ISL_1717996, EPI_ISL_1717997, EPI_ISL_1717998, EPI_ISL_1717999, EPI_ISL_1718000, EPI_ISL_1718001, EPI_ISL_1718002, EPI_ISL_1718003,
EPI_ISL_1718004, EPI_ISL_1718005, EPI_ISL_1718006, EPI_ISL_1718007, EPI_ISL_1718008, EPI_ISL_1718009, EPI_ISL_1718010, EPI_ISL_1718011, EPI_ISL_1718012, EPI_ISL_1718013, EPI_ISL_1718014, EPI_ISL_1718015, EPI_ISL_1718016, EPI_ISL_1718017, EPI_ISL_1718018, EPI_ISL_1718019, EPI_ISL_1718020, EPI_ISL_1718021, EPI_ISL_1718022, EPI_ISL_1718023, EPI_ISL_1718024, EPI_ISL_1718025,
EPI_ISL_1718026, EPI_ISL_1718027, EPI_ISL_1718028, EPI_ISL_1718029, EPI_ISL_1718030, EPI_ISL_1718031, EPI_ISL_1718032, EPI_ISL_1718033, EPI_ISL_1718034, EPI_ISL_1718035, EPI_ISL_1718036, EPI_ISL_1718037, EPI_ISL_1718038, EPI_ISL_1718039, EPI_ISL_1718040, EPI_ISL_1718041, EPI_ISL_1718042, EPI_ISL_1718043, EPI_ISL_1718044, EPI_ISL_1718045, EPI_ISL_1718046, EPI_ISL_1718047,
EPI_ISL_1718048, EPI_ISL_1718060, EPI_ISL_1718061, EPI_ISL_1718062, EPI_ISL_1718063, EPI_ISL_1718064, EPI_ISL_1718065, EPI_ISL_1718066, EPI_ISL_1718067, EPI_ISL_1718068, EPI_ISL_1718069, EPI_ISL_1718070, EPI_ISL_1718071, EPI_ISL_1718072, EPI_ISL_1718073, EPI_ISL_1718074, EPI_ISL_1718075, EPI_ISL_1718076, EPI_ISL_1718077, EPI_ISL_1718078, EPI_ISL_1718079, EPI_ISL_1718080,
EPI_ISL_1718081, EPI_ISL_1718082, EPI_ISL_1718083, EPI_ISL_1718084, EPI_ISL_1718085, EPI_ISL_1718086, EPI_ISL_1718087, EPI_ISL_1718088, EPI_ISL_1718089, EPI_ISL_1718090, EPI_ISL_1718091, EPI_ISL_1718092, EPI_ISL_1718093, EPI_ISL_1718094, EPI_ISL_1718095, EPI_ISL_1718096, EPI_ISL_1718097, EPI_ISL_1718098, EPI_ISL_1718099, EPI_ISL_1718100, EPI_ISL_1718101, EPI_ISL_1718102,
EPI_ISL_1718103, EPI_ISL_1718104, EPI_ISL_1718105, EPI_ISL_1718106, EPI_ISL_1718107, EPI_ISL_1718108, EPI_ISL_1718109, EPI_ISL_1718110, EPI_ISL_1718111, EPI_ISL_1718112, EPI_ISL_1718113, EPI_ISL_1718114, EPI_ISL_1718115, EPI_ISL_1718116, EPI_ISL_1718117, EPI_ISL_1718118, EPI_ISL_1718119, EPI_ISL_1718120, EPI_ISL_1718121, EPI_ISL_1718122, EPI_ISL_1718123, EPI_ISL_1718124,
EPI_ISL_1718125, EPI_ISL_1718126, EPI_ISL_1718127, EPI_ISL_1718128, EPI_ISL_1718129, EPI_ISL_1718130, EPI_ISL_1718131, EPI_ISL_1718132, EPI_ISL_1718133, EPI_ISL_1718134, EPI_ISL_1718135, EPI_ISL_1718136, EPI_ISL_1718137, EPI_ISL_1718138, EPI_ISL_1718139, EPI_ISL_1718140, EPI_ISL_1718141, EPI_ISL_1718142, EPI_ISL_1718143, EPI_ISL_1718144, EPI_ISL_1718145, EPI_ISL_1718146,
EPI_ISL_1718147, EPI_ISL_1718148, EPI_ISL_1718149, EPI_ISL_1718150, EPI_ISL_1718151, EPI_ISL_1718152, EPI_ISL_1718153, EPI_ISL_1718154, EPI_ISL_1718155, EPI_ISL_1718156, EPI_ISL_1718157, EPI_ISL_1718158, EPI_ISL_1718159, EPI_ISL_1718160, EPI_ISL_1718161, EPI_ISL_1718162, EPI_ISL_1718163, EPI_ISL_1718164, EPI_ISL_1718165, EPI_ISL_1718166, EPI_ISL_1718167, EPI_ISL_1718168,
EPI_ISL_1718169, EPI_ISL_1718170, EPI_ISL_1718171, EPI_ISL_1718172, EPI_ISL_1718173, EPI_ISL_1718174, EPI_ISL_1718175, EPI_ISL_1718176, EPI_ISL_1718177, EPI_ISL_1718178, EPI_ISL_1718179, EPI_ISL_1718180, EPI_ISL_1718181, EPI_ISL_1718182, EPI_ISL_1718183, EPI_ISL_1718184, EPI_ISL_1718185, EPI_ISL_1718186, EPI_ISL_1718187, EPI_ISL_1718188, EPI_ISL_1718189, EPI_ISL_1718190,
EPI_ISL_1718191, EPI_ISL_1718192, EPI_ISL_1718193, EPI_ISL_1718194, EPI_ISL_1718195, EPI_ISL_1718196, EPI_ISL_1718197, EPI_ISL_1718198, EPI_ISL_1718199, EPI_ISL_1718200, EPI_ISL_1718201, EPI_ISL_1718202, EPI_ISL_1718203, EPI_ISL_1718204, EPI_ISL_1718205, EPI_ISL_1718206, EPI_ISL_1718207, EPI_ISL_1718208, EPI_ISL_1718209, EPI_ISL_1718210, EPI_ISL_1718211, EPI_ISL_1718212,
EPI_ISL_1718213, EPI_ISL_1718214, EPI_ISL_1718215, EPI_ISL_1718216, EPI_ISL_1718217, EPI_ISL_1718218, EPI_ISL_1718219, EPI_ISL_1718220, EPI_ISL_1718221, EPI_ISL_1718222, EPI_ISL_1718223, EPI_ISL_1718224, EPI_ISL_1718225, EPI_ISL_1718226, EPI_ISL_1718227, EPI_ISL_1718228, EPI_ISL_1718229, EPI_ISL_1718230, EPI_ISL_1718231, EPI_ISL_1718232, EPI_ISL_1718233, EPI_ISL_1718234,
EPI_ISL_1718235, EPI_ISL_1718236, EPI_ISL_1718237, EPI_ISL_1718238, EPI_ISL_1718239, EPI_ISL_1718240, EPI_ISL_1718241, EPI_ISL_1718242, EPI_ISL_1718243, EPI_ISL_1718244, EPI_ISL_1718245, EPI_ISL_1718246, EPI_ISL_1718247, EPI_ISL_1827954, EPI_ISL_1827955, EPI_ISL_1827956, EPI_ISL_1827957, EPI_ISL_1827958, EPI_ISL_1827959, EPI_ISL_1827960, EPI_ISL_1827961, EPI_ISL_1827962,
EPI_ISL_1827963, EPI_ISL_1827964, EPI_ISL_1827965, EPI_ISL_1827966, EPI_ISL_1827967, EPI_ISL_1827968, EPI_ISL_1827969, EPI_ISL_1827970, EPI_ISL_1827971, EPI_ISL_1827972, EPI_ISL_1827973, EPI_ISL_1827974, EPI_ISL_1827975, EPI_ISL_1827976, EPI_ISL_1827977, EPI_ISL_1827978, EPI_ISL_1827979, EPI_ISL_1827980, EPI_ISL_1827981, EPI_ISL_1827982, EPI_ISL_1827983, EPI_ISL_1827984,
EPI_ISL_1827985, EPI_ISL_1827986, EPI_ISL_1827987, EPI_ISL_1827988, EPI_ISL_1827989, EPI_ISL_1827990, EPI_ISL_1827991, EPI_ISL_1827992, EPI_ISL_1827993, EPI_ISL_1827994, EPI_ISL_1827995, EPI_ISL_1827996, EPI_ISL_1827997, EPI_ISL_1827998, EPI_ISL_1827999, EPI_ISL_1828000, EPI_ISL_1828001, EPI_ISL_1828002, EPI_ISL_1828003, EPI_ISL_1828004, EPI_ISL_1828005, EPI_ISL_1828006,
EPI_ISL_1828007, EPI_ISL_1828008, EPI_ISL_1828009, EPI_ISL_1828010, EPI_ISL_1828011, EPI_ISL_1828012, EPI_ISL_1828013, EPI_ISL_1828014, EPI_ISL_1828015, EPI_ISL_1828016, EPI_ISL_1828017, EPI_ISL_1828018, EPI_ISL_1828019, EPI_ISL_1828020, EPI_ISL_1828021, EPI_ISL_1828022, EPI_ISL_1828023, EPI_ISL_1828024, EPI_ISL_1828025, EPI_ISL_1828026, EPI_ISL_1828027, EPI_ISL_1828028,
EPI_ISL_1828029, EPI_ISL_1828030, EPI_ISL_1828031, EPI_ISL_1828032, EPI_ISL_1828033, EPI_ISL_1828034, EPI_ISL_1828035, EPI_ISL_1828036, EPI_ISL_1828037, EPI_ISL_1828038, EPI_ISL_1828039, EPI_ISL_1828040, EPI_ISL_1828041, EPI_ISL_1828042, EPI_ISL_1828043, EPI_ISL_1828044, EPI_ISL_1828045, EPI_ISL_1828046, EPI_ISL_1828047, EPI_ISL_1828048, EPI_ISL_1828049, EPI_ISL_1828050,
EPI_ISL_1828051, EPI_ISL_1828052, EPI_ISL_1828053, EPI_ISL_1828054, EPI_ISL_1828055, EPI_ISL_1828056, EPI_ISL_1828057, EPI_ISL_1828058, EPI_ISL_1828059, EPI_ISL_1828060, EPI_ISL_1828061, EPI_ISL_1828062, EPI_ISL_1828063, EPI_ISL_1828064, EPI_ISL_1828065, EPI_ISL_1828066, EPI_ISL_1828067, EPI_ISL_1828068, EPI_ISL_1828069, EPI_ISL_1828070, EPI_ISL_1828071, EPI_ISL_1828072,
EPI_ISL_1828073, EPI_ISL_1828074, EPI_ISL_1828075, EPI_ISL_1828076, EPI_ISL_1828077, EPI_ISL_1828078, EPI_ISL_1828079, EPI_ISL_1828080, EPI_ISL_1828081, EPI_ISL_1828082, EPI_ISL_1828083, EPI_ISL_1828084, EPI_ISL_1828085, EPI_ISL_1828086, EPI_ISL_1828087, EPI_ISL_1828088, EPI_ISL_1828089, EPI_ISL_1828090, EPI_ISL_1828091, EPI_ISL_1828092, EPI_ISL_1828093, EPI_ISL_1828094,
EPI_ISL_1828095, EPI_ISL_1828096, EPI_ISL_1828098, EPI_ISL_1828099, EPI_ISL_1828100, EPI_ISL_1828101, EPI_ISL_1828102, EPI_ISL_1828103, EPI_ISL_1828104, EPI_ISL_1828105, EPI_ISL_1828106, EPI_ISL_1828107, EPI_ISL_1828108, EPI_ISL_1828109, EPI_ISL_1828110, EPI_ISL_1828111, EPI_ISL_1828112, EPI_ISL_1828113, EPI_ISL_1828114, EPI_ISL_1828115, EPI_ISL_1828116, EPI_ISL_1828117,
EPI_ISL_1828118, EPI_ISL_1828119, EPI_ISL_1828120, EPI_ISL_1828121, EPI_ISL_1828122, EPI_ISL_1828123, EPI_ISL_1828124, EPI_ISL_1828125, EPI_ISL_1828126, EPI_ISL_1828127, EPI_ISL_1828128, EPI_ISL_1828129, EPI_ISL_1828130, EPI_ISL_1828131, EPI_ISL_1828132, EPI_ISL_1828133, EPI_ISL_1828134, EPI_ISL_1828135, EPI_ISL_1828136, EPI_ISL_1828137, EPI_ISL_1828138, EPI_ISL_1828139,
EPI_ISL_1828140, EPI_ISL_1828141, EPI_ISL_1828142, EPI_ISL_1828143, EPI_ISL_1828144, EPI_ISL_1828145, EPI_ISL_1828146, EPI_ISL_1828147, EPI_ISL_1828148, EPI_ISL_1828149, EPI_ISL_1828150, EPI_ISL_1828151, EPI_ISL_1828152, EPI_ISL_1828153, EPI_ISL_1828154, EPI_ISL_1828155, EPI_ISL_1828156, EPI_ISL_1828157, EPI_ISL_1828158, EPI_ISL_1828159, EPI_ISL_1828160, EPI_ISL_1828161,
EPI_ISL_1828162, EPI_ISL_1828163, EPI_ISL_1828164, EPI_ISL_1828165, EPI_ISL_1828166, EPI_ISL_1828167, EPI_ISL_1828168, EPI_ISL_1828169, EPI_ISL_1828170, EPI_ISL_1828171, EPI_ISL_1828172, EPI_ISL_1828173, EPI_ISL_1828174, EPI_ISL_1828175, EPI_ISL_1828176, EPI_ISL_1828177, EPI_ISL_1828178, EPI_ISL_1828179, EPI_ISL_1828180, EPI_ISL_1828181, EPI_ISL_1828182, EPI_ISL_1828183,
EPI_ISL_1828184, EPI_ISL_1828185, EPI_ISL_1828186, EPI_ISL_1828187, EPI_ISL_1828188, EPI_ISL_1828189, EPI_ISL_1828190, EPI_ISL_1828191, EPI_ISL_1828192, EPI_ISL_1828193, EPI_ISL_1828194, EPI_ISL_1828195, EPI_ISL_1828196, EPI_ISL_1828197, EPI_ISL_1828198, EPI_ISL_1828199, EPI_ISL_1828200, EPI_ISL_1828201, EPI_ISL_1828202, EPI_ISL_1828203, EPI_ISL_1828204, EPI_ISL_1828205,
EPI_ISL_1828206, EPI_ISL_1828207, EPI_ISL_1828209, EPI_ISL_1828210, EPI_ISL_1828211, EPI_ISL_1828212, EPI_ISL_1828213, EPI_ISL_1828214, EPI_ISL_1828215, EPI_ISL_1828216, EPI_ISL_1828217, EPI_ISL_1828218, EPI_ISL_1828219, EPI_ISL_1828220, EPI_ISL_1828221, EPI_ISL_1828222, EPI_ISL_1828223, EPI_ISL_1828224, EPI_ISL_1828225, EPI_ISL_1828226, EPI_ISL_1828227, EPI_ISL_1828228,
EPI_ISL_1828229, EPI_ISL_1828230, EPI_ISL_1828231, EPI_ISL_1828232, EPI_ISL_1828233, EPI_ISL_1828234, EPI_ISL_1828235, EPI_ISL_1828236, EPI_ISL_1828237, EPI_ISL_1828238, EPI_ISL_1828239, EPI_ISL_1828240, EPI_ISL_1828241, EPI_ISL_1828242, EPI_ISL_1828243, EPI_ISL_1828244, EPI_ISL_1828245, EPI_ISL_1828246, EPI_ISL_1828247, EPI_ISL_1828248, EPI_ISL_1828249, EPI_ISL_1828250,
EPI_ISL_1828251, EPI_ISL_1828252, EPI_ISL_1828253, EPI_ISL_1828254, EPI_ISL_1828255, EPI_ISL_1828256, EPI_ISL_1828257, EPI_ISL_1828258, EPI_ISL_1828259, EPI_ISL_1828260, EPI_ISL_1828261, EPI_ISL_1828262, EPI_ISL_1828263, EPI_ISL_1828264, EPI_ISL_1828265, EPI_ISL_1828266, EPI_ISL_1828267, EPI_ISL_1828268, EPI_ISL_1828269, EPI_ISL_1828270, EPI_ISL_1828271, EPI_ISL_1828272,
EPI_ISL_1828273, EPI_ISL_1828274, EPI_ISL_1828275, EPI_ISL_1828276, EPI_ISL_1828277, EPI_ISL_1828278, EPI_ISL_1828279, EPI_ISL_1828280, EPI_ISL_1828281, EPI_ISL_1828282, EPI_ISL_1828283, EPI_ISL_1828284, EPI_ISL_1828285, EPI_ISL_1828286, EPI_ISL_1828287, EPI_ISL_1828288, EPI_ISL_1828289, EPI_ISL_1828290, EPI_ISL_1828291, EPI_ISL_1828292, EPI_ISL_1828293, EPI_ISL_1828294,
EPI_ISL_1828295, EPI_ISL_1828296, EPI_ISL_1828297, EPI_ISL_1828298, EPI_ISL_1828299, EPI_ISL_1828300, EPI_ISL_1828301, EPI_ISL_1828302, EPI_ISL_1828303, EPI_ISL_1828304, EPI_ISL_1828305, EPI_ISL_1828306, EPI_ISL_1828307, EPI_ISL_1828308, EPI_ISL_1828309, EPI_ISL_1828310, EPI_ISL_1828311, EPI_ISL_1828312, EPI_ISL_1828313, EPI_ISL_1828314, EPI_ISL_1828315, EPI_ISL_1828316,
EPI_ISL_1828317, EPI_ISL_1828318, EPI_ISL_1828319, EPI_ISL_1828320, EPI_ISL_1828321, EPI_ISL_1828322, EPI_ISL_1828323, EPI_ISL_1828324, EPI_ISL_1828325, EPI_ISL_1828326, EPI_ISL_1828327, EPI_ISL_1828328, EPI_ISL_1828329, EPI_ISL_1828330, EPI_ISL_1828331, EPI_ISL_1828332, EPI_ISL_1828333, EPI_ISL_1828334, EPI_ISL_1828335, EPI_ISL_1828336, EPI_ISL_1828337, EPI_ISL_1828338,
EPI_ISL_1828339, EPI_ISL_1828340, EPI_ISL_1828341, EPI_ISL_1828342, EPI_ISL_1828343, EPI_ISL_1828344, EPI_ISL_1828345, EPI_ISL_1828346, EPI_ISL_1828347, EPI_ISL_1828348, EPI_ISL_1828349, EPI_ISL_1828350, EPI_ISL_1828351, EPI_ISL_1828352, EPI_ISL_1828353, EPI_ISL_1828354, EPI_ISL_1828355, EPI_ISL_1828356, EPI_ISL_1828357, EPI_ISL_1828358, EPI_ISL_1828359, EPI_ISL_1828360,
EPI_ISL_1828361, EPI_ISL_1828362, EPI_ISL_1828363, EPI_ISL_1828364, EPI_ISL_1828365, EPI_ISL_1828366, EPI_ISL_1828367, EPI_ISL_1828368, EPI_ISL_1828369, EPI_ISL_1828370, EPI_ISL_1828371, EPI_ISL_1828372, EPI_ISL_1828373, EPI_ISL_1828374, EPI_ISL_1828375, EPI_ISL_1828376, EPI_ISL_1828377, EPI_ISL_1828378, EPI_ISL_1828379, EPI_ISL_1828380, EPI_ISL_1828381, EPI_ISL_1828382,
EPI_ISL_1828383, EPI_ISL_1828384, EPI_ISL_1828385, EPI_ISL_1828386, EPI_ISL_1828387, EPI_ISL_1828388, EPI_ISL_1828389, EPI_ISL_1828390, EPI_ISL_1828391, EPI_ISL_1828392, EPI_ISL_1828393, EPI_ISL_1828394, EPI_ISL_1828395, EPI_ISL_1828396, EPI_ISL_1828397, EPI_ISL_1828398, EPI_ISL_1828399, EPI_ISL_1828400, EPI_ISL_1828401, EPI_ISL_1828402, EPI_ISL_1828403, EPI_ISL_1828404,
EPI_ISL_1828405, EPI_ISL_1828406, EPI_ISL_1828407, EPI_ISL_1828408, EPI_ISL_1828409, EPI_ISL_1828410, EPI_ISL_1828411, EPI_ISL_1828412, EPI_ISL_1828413, EPI_ISL_1828414, EPI_ISL_1828415, EPI_ISL_1828416, EPI_ISL_1828417, EPI_ISL_1828418, EPI_ISL_1828419, EPI_ISL_1828420, EPI_ISL_1828421, EPI_ISL_1828422, EPI_ISL_1828423, EPI_ISL_1828424, EPI_ISL_1828425, EPI_ISL_1828426,
EPI_ISL_1828427, EPI_ISL_1828428, EPI_ISL_1828429, EPI_ISL_1828430, EPI_ISL_1828431, EPI_ISL_1828432, EPI_ISL_1828433, EPI_ISL_1828434, EPI_ISL_1828435, EPI_ISL_1828436, EPI_ISL_1828437, EPI_ISL_1828438, EPI_ISL_1828439, EPI_ISL_1828440, EPI_ISL_1828441, EPI_ISL_1828442, EPI_ISL_1828443, EPI_ISL_1828444, EPI_ISL_1828445, EPI_ISL_1828446, EPI_ISL_1828447, EPI_ISL_1828448,
EPI_ISL_1828449, EPI_ISL_1828450, EPI_ISL_1828451, EPI_ISL_1828452, EPI_ISL_1828453, EPI_ISL_1828454, EPI_ISL_1828455, EPI_ISL_1828456, EPI_ISL_1828457, EPI_ISL_1828458, EPI_ISL_1828459, EPI_ISL_1828460, EPI_ISL_1828461, EPI_ISL_1828462, EPI_ISL_1828463, EPI_ISL_1828464, EPI_ISL_1828465, EPI_ISL_1828466, EPI_ISL_1828467, EPI_ISL_1828468, EPI_ISL_1828469, EPI_ISL_1828470,
EPI_ISL_1828471, EPI_ISL_1828472, EPI_ISL_1828473, EPI_ISL_1828474, EPI_ISL_1828475, EPI_ISL_1828476, EPI_ISL_1828477, EPI_ISL_1828478, EPI_ISL_1828479, EPI_ISL_1828480, EPI_ISL_1828481, EPI_ISL_1828482, EPI_ISL_1828483, EPI_ISL_1828484, EPI_ISL_1828485, EPI_ISL_1828486, EPI_ISL_1828487, EPI_ISL_1828488, EPI_ISL_1828489, EPI_ISL_1828490, EPI_ISL_1828491, EPI_ISL_1828492,
EPI_ISL_1828493, EPI_ISL_1828494, EPI_ISL_1828495, EPI_ISL_1828496, EPI_ISL_1828497, EPI_ISL_1828498, EPI_ISL_1828499, EPI_ISL_1828500, EPI_ISL_1828501, EPI_ISL_1828502, EPI_ISL_1828503, EPI_ISL_1828504, EPI_ISL_1828505, EPI_ISL_1828506, EPI_ISL_1828507, EPI_ISL_1828508, EPI_ISL_1828509, EPI_ISL_1828510, EPI_ISL_1828511, EPI_ISL_1828512, EPI_ISL_1828513, EPI_ISL_1828514,
EPI_ISL_1828515, EPI_ISL_1828516, EPI_ISL_1828517, EPI_ISL_1828519, EPI_ISL_1828520, EPI_ISL_1828521, EPI_ISL_1828522, EPI_ISL_1828523, EPI_ISL_1828524, EPI_ISL_1828525, EPI_ISL_1828526, EPI_ISL_1828527, EPI_ISL_1828528, EPI_ISL_1828529, EPI_ISL_1828530, EPI_ISL_1828531, EPI_ISL_1828532, EPI_ISL_1828533, EPI_ISL_1828534, EPI_ISL_1828535, EPI_ISL_1828536, EPI_ISL_1828537,
EPI_ISL_1828538, EPI_ISL_1828539, EPI_ISL_1828540, EPI_ISL_1828541, EPI_ISL_1828542, EPI_ISL_1828543, EPI_ISL_1828544, EPI_ISL_1828545, EPI_ISL_1828546, EPI_ISL_1828547, EPI_ISL_1828548, EPI_ISL_1828549, EPI_ISL_1828550, EPI_ISL_1828551, EPI_ISL_1828552, EPI_ISL_1828553, EPI_ISL_1828554, EPI_ISL_1828555, EPI_ISL_1828556, EPI_ISL_1828557, EPI_ISL_1828558, EPI_ISL_1828559,
EPI_ISL_1828560, EPI_ISL_1828561, EPI_ISL_1828562, EPI_ISL_1828563, EPI_ISL_1828564, EPI_ISL_1828565, EPI_ISL_1828566, EPI_ISL_1828567, EPI_ISL_1828568, EPI_ISL_1828569, EPI_ISL_1828570, EPI_ISL_1828571, EPI_ISL_1828572, EPI_ISL_1828573, EPI_ISL_1828574, EPI_ISL_1828575, EPI_ISL_1828576, EPI_ISL_1828577, EPI_ISL_1828578, EPI_ISL_1828579, EPI_ISL_1828580, EPI_ISL_1828581,
EPI_ISL_1828582, EPI_ISL_1828583, EPI_ISL_1828584, EPI_ISL_1828585, EPI_ISL_1828586, EPI_ISL_1828587, EPI_ISL_1828588, EPI_ISL_1828589, EPI_ISL_1828590, EPI_ISL_1828591, EPI_ISL_1828592, EPI_ISL_1828593, EPI_ISL_1828594, EPI_ISL_1828595, EPI_ISL_1828596, EPI_ISL_1828597, EPI_ISL_1828598, EPI_ISL_1828599, EPI_ISL_1828600, EPI_ISL_1828601, EPI_ISL_1828602, EPI_ISL_1828603,
EPI_ISL_1828604, EPI_ISL_1828605, EPI_ISL_1828606, EPI_ISL_1828607, EPI_ISL_1828608, EPI_ISL_1828609, EPI_ISL_1828610, EPI_ISL_1828611, EPI_ISL_1828612, EPI_ISL_1828613, EPI_ISL_1828614, EPI_ISL_1828615, EPI_ISL_1828616, EPI_ISL_1828617, EPI_ISL_1828618, EPI_ISL_1828619, EPI_ISL_1828620, EPI_ISL_1828621, EPI_ISL_1828622, EPI_ISL_1828623, EPI_ISL_1828624, EPI_ISL_1828625,
EPI_ISL_1828626, EPI_ISL_1828627, EPI_ISL_1828628, EPI_ISL_1828629, EPI_ISL_1828630, EPI_ISL_1828631, EPI_ISL_1828632, EPI_ISL_1828633, EPI_ISL_1828634, EPI_ISL_1828635, EPI_ISL_1828636, EPI_ISL_1828637, EPI_ISL_1828638, EPI_ISL_1828639, EPI_ISL_1828640, EPI_ISL_1828641, EPI_ISL_1828642, EPI_ISL_1828643, EPI_ISL_1828644, EPI_ISL_1828645, EPI_ISL_1828646, EPI_ISL_1828647,
EPI_ISL_1828648, EPI_ISL_1828649, EPI_ISL_1828650, EPI_ISL_1828651, EPI_ISL_1828652, EPI_ISL_1828653, EPI_ISL_1828654, EPI_ISL_1828655, EPI_ISL_1828656, EPI_ISL_1828657, EPI_ISL_1828658, EPI_ISL_1828659, EPI_ISL_1828660, EPI_ISL_1828661, EPI_ISL_1828662, EPI_ISL_1828663, EPI_ISL_1828664, EPI_ISL_1828665, EPI_ISL_1828666, EPI_ISL_1828667, EPI_ISL_1828668, EPI_ISL_1828669,
EPI_ISL_1828670, EPI_ISL_1828671, EPI_ISL_1828672, EPI_ISL_1828673, EPI_ISL_1828674, EPI_ISL_1828675, EPI_ISL_1828676, EPI_ISL_1828677, EPI_ISL_1828678, EPI_ISL_1828679, EPI_ISL_1828680, EPI_ISL_1828681, EPI_ISL_1828682, EPI_ISL_1828683, EPI_ISL_1828684, EPI_ISL_1828685, EPI_ISL_1828686, EPI_ISL_1828687, EPI_ISL_1828688, EPI_ISL_1828689, EPI_ISL_1828690, EPI_ISL_1828691,
EPI_ISL_1828692, EPI_ISL_1828693, EPI_ISL_1828694, EPI_ISL_1828695, EPI_ISL_1828696, EPI_ISL_1828697, EPI_ISL_2036964, EPI_ISL_2036968, EPI_ISL_2036969, EPI_ISL_2036970, EPI_ISL_2036971, EPI_ISL_2036972, EPI_ISL_2036973, EPI_ISL_2036974, EPI_ISL_2036976, EPI_ISL_2036977, EPI_ISL_2036978, EPI_ISL_2036979, EPI_ISL_2036981, EPI_ISL_2036983, EPI_ISL_2036984, EPI_ISL_2036987,
EPI_ISL_2036988, EPI_ISL_2036991, EPI_ISL_2036992, EPI_ISL_2036993, EPI_ISL_2036999, EPI_ISL_2037000, EPI_ISL_2037001, EPI_ISL_2037004, EPI_ISL_2037005, EPI_ISL_2037006, EPI_ISL_2037008, EPI_ISL_2037010, EPI_ISL_2037011, EPI_ISL_2037012, EPI_ISL_2037013, EPI_ISL_2037014, EPI_ISL_2037016, EPI_ISL_2037017, EPI_ISL_2037020, EPI_ISL_2037021, EPI_ISL_2037023, EPI_ISL_2037024,
EPI_ISL_2037025, EPI_ISL_2037026, EPI_ISL_2037028, EPI_ISL_2037029, EPI_ISL_2037030, EPI_ISL_2037031, EPI_ISL_2037032, EPI_ISL_2037033, EPI_ISL_2037034, EPI_ISL_2037035, EPI_ISL_2037038, EPI_ISL_2037039, EPI_ISL_2037040, EPI_ISL_2037042, EPI_ISL_2037047, EPI_ISL_2037048, EPI_ISL_2037049, EPI_ISL_2037050, EPI_ISL_2037053, EPI_ISL_2037054, EPI_ISL_2037056, EPI_ISL_2037058,
EPI_ISL_2037059, EPI_ISL_2037061, EPI_ISL_2037062, EPI_ISL_2037063, EPI_ISL_2037064, EPI_ISL_2037065, EPI_ISL_2037070, EPI_ISL_2037071, EPI_ISL_2037073, EPI_ISL_2037074, EPI_ISL_2037076, EPI_ISL_2037078, EPI_ISL_2037079, EPI_ISL_2037080, EPI_ISL_2037082, EPI_ISL_2037083, EPI_ISL_2037084, EPI_ISL_2037086, EPI_ISL_2037087, EPI_ISL_2037088, EPI_ISL_2037089, EPI_ISL_2037092,

| | | | |
|---|---|---|---|
| EPI_ISL_2037093, EPI_ISL_2037094, EPI_ISL_2037095, EPI_ISL_2037096, EPI_ISL_2037097, EPI_ISL_2037098, EPI_ISL_2037099, EPI_ISL_2037100, EPI_ISL_2037101, EPI_ISL_2037102, EPI_ISL_2037103, EPI_ISL_2037104, EPI_ISL_2037106, EPI_ISL_2037108, EPI_ISL_2037110, EPI_ISL_2037115, EPI_ISL_2037118, EPI_ISL_2037120, EPI_ISL_2037121, EPI_ISL_2037123, EPI_ISL_2037124, EPI_ISL_2037129, EPI_ISL_2037131, EPI_ISL_2037133, EPI_ISL_2037134, EPI_ISL_2037135, EPI_ISL_2037137, EPI_ISL_2037138, EPI_ISL_2037140, EPI_ISL_2037141, EPI_ISL_2037145, EPI_ISL_2037146, EPI_ISL_2037149, EPI_ISL_2037152, EPI_ISL_2037153, EPI_ISL_2037154, EPI_ISL_2037155, EPI_ISL_2037156, EPI_ISL_2037157, EPI_ISL_2037159, EPI_ISL_2037162, EPI_ISL_2037163, EPI_ISL_2037164, EPI_ISL_2037166, EPI_ISL_2037167, EPI_ISL_2037168, EPI_ISL_2037169, EPI_ISL_2037172, EPI_ISL_2037174, EPI_ISL_2037175, EPI_ISL_2037176, EPI_ISL_2037177, EPI_ISL_2037180, EPI_ISL_2037181, EPI_ISL_2037182, EPI_ISL_2037183, EPI_ISL_2037187, EPI_ISL_2037188, EPI_ISL_2037190, EPI_ISL_2037191, EPI_ISL_2037192, EPI_ISL_2037193, EPI_ISL_2037199, EPI_ISL_2037201, EPI_ISL_2106476 | | | |
| see above | Pandemic Response Lab - NYC | Pandemic Response Lab, R&D | Cybill del Castillo; Dylan Law; Haiping Hao; Henry Lee; Jon Laurent; Katharine Nelson; Melissa Hopkins; Michael Hammerling; Pradeep Bugga; Shinyoung Clair Kang; Sol Rey; William Ward |
| EPI_ISL_1625770 | Pathology and Laboratory Medicine Institute, Cleveland Clinic, Ohio, USA | Pathology and Laboratory Medicine Institute, Cleveland Clinic, Ohio, USA | Daniel H. Farkas; Daniel Rhoads; David Bosler; David Plunkett; Gary Procop; Jay Brock; Jennifer Starbuck; Joy Nakitandwe; Kristen McDonnell; Yu-Wei Cheng; Zheng Jin Tu |
| EPI_ISL_2134820 | PrimBio Research institute | PrimBio Research institute | Yang Liu |
| EPI_ISL_1920122 | Pulaski Urgent Care | Wadsworth Center, New York State Department of Health | Alexis Russell; Catharine Prussing; Daryl M. Lamson; Erasmus Schneider; Erica Lasek-Nesselquist; John Kelly; Jonathan Plitnick; Kirsten St. George; Matthew Shudt; Melissa A Leisner; Navjot Singh |
| EPI_ISL_1582161, EPI_ISL_1614913, EPI_ISL_1614967, EPI_ISL_1614988, EPI_ISL_1621729, EPI_ISL_1621747, EPI_ISL_1621749, EPI_ISL_1621786, EPI_ISL_1621789, EPI_ISL_1621812, EPI_ISL_1621815, EPI_ISL_1621818, EPI_ISL_1621855, EPI_ISL_1621856, EPI_ISL_1621857, EPI_ISL_1621860, EPI_ISL_1621865, EPI_ISL_1621871, EPI_ISL_1621872, EPI_ISL_1621875, EPI_ISL_1621879, EPI_ISL_1621880, EPI_ISL_1621881, EPI_ISL_1621890, EPI_ISL_1621896, EPI_ISL_1621903, EPI_ISL_1621913, EPI_ISL_1621915, EPI_ISL_1621921, EPI_ISL_1621924, EPI_ISL_1621941, EPI_ISL_1621942, EPI_ISL_1621946, EPI_ISL_1621962, EPI_ISL_1621988, EPI_ISL_1621991, EPI_ISL_1621992, EPI_ISL_1622001, EPI_ISL_1622007, EPI_ISL_1622018, EPI_ISL_1622026, EPI_ISL_1622030, EPI_ISL_1622031, EPI_ISL_1622042, EPI_ISL_1622047, EPI_ISL_1622072, EPI_ISL_1622073, EPI_ISL_1622075, EPI_ISL_1622087, EPI_ISL_1622095, EPI_ISL_1622105, EPI_ISL_1622109, EPI_ISL_1648303, EPI_ISL_1648304, EPI_ISL_1648314, EPI_ISL_1648317, EPI_ISL_1648320, EPI_ISL_1648327, EPI_ISL_1648341, EPI_ISL_1648358, EPI_ISL_1648362, EPI_ISL_1648365, EPI_ISL_1648384, EPI_ISL_1648397, EPI_ISL_1648402, EPI_ISL_1648499, EPI_ISL_1648558, EPI_ISL_1648561, EPI_ISL_1648586, EPI_ISL_1648778, EPI_ISL_1648780, EPI_ISL_1648784, EPI_ISL_1648916, EPI_ISL_1668648, EPI_ISL_1668707, EPI_ISL_1668768, EPI_ISL_1669089, EPI_ISL_1691410, EPI_ISL_1691436, EPI_ISL_1691492, EPI_ISL_1695681, EPI_ISL_1702639, EPI_ISL_1738119, EPI_ISL_1738187, EPI_ISL_1738235, EPI_ISL_1738591, EPI_ISL_1738671, EPI_ISL_1738750, EPI_ISL_1753345, EPI_ISL_1753413, EPI_ISL_1753625, EPI_ISL_1753644, EPI_ISL_1761906, EPI_ISL_1761913, EPI_ISL_1761936, EPI_ISL_1761939, EPI_ISL_1761949, EPI_ISL_1761951, EPI_ISL_1761953, EPI_ISL_1761963, EPI_ISL_1761971, EPI_ISL_1761977, EPI_ISL_1761985, EPI_ISL_1762034, EPI_ISL_1762038, EPI_ISL_1798049, EPI_ISL_1798098, EPI_ISL_1798112, EPI_ISL_1798113, EPI_ISL_1798146, EPI_ISL_1798164, EPI_ISL_1804820, EPI_ISL_1818070, EPI_ISL_1818295, EPI_ISL_1818327, EPI_ISL_1818331, EPI_ISL_1818346, EPI_ISL_1818349, EPI_ISL_1818401, EPI_ISL_1818422, EPI_ISL_1818519, EPI_ISL_1834218, EPI_ISL_1834241, EPI_ISL_1834304, EPI_ISL_1834326, EPI_ISL_1834346, EPI_ISL_1834359, EPI_ISL_1834371, EPI_ISL_1834375, EPI_ISL_1834408, EPI_ISL_1834421, EPI_ISL_1834440, EPI_ISL_1838933, EPI_ISL_1838960, EPI_ISL_1838965, EPI_ISL_1838975, EPI_ISL_1838980, EPI_ISL_1838993, EPI_ISL_1838996, EPI_ISL_1839003, EPI_ISL_1839004, EPI_ISL_1839010, EPI_ISL_1839013, EPI_ISL_1839014, EPI_ISL_1839020, EPI_ISL_1839021, EPI_ISL_1839024, EPI_ISL_1839042, EPI_ISL_1839054, EPI_ISL_1839055, EPI_ISL_1840412, EPI_ISL_1840423, EPI_ISL_1840431, EPI_ISL_1840435, EPI_ISL_1840458, EPI_ISL_1840526, EPI_ISL_1840528, EPI_ISL_1840531, EPI_ISL_1840546, EPI_ISL_1840559, EPI_ISL_1840561, EPI_ISL_1840567, EPI_ISL_1840570, EPI_ISL_1840602, EPI_ISL_1840604, EPI_ISL_1840619, EPI_ISL_1840623, EPI_ISL_1840625, EPI_ISL_1906183, EPI_ISL_1906197, EPI_ISL_1906206, EPI_ISL_1906210, EPI_ISL_1906215, EPI_ISL_1906217, EPI_ISL_1906223, EPI_ISL_1906244, EPI_ISL_1906253, EPI_ISL_1906258, EPI_ISL_1906268, EPI_ISL_1906270, EPI_ISL_1906272, EPI_ISL_1906274, EPI_ISL_1906276, EPI_ISL_1906286, EPI_ISL_1906290, EPI_ISL_1906308, EPI_ISL_1906318, EPI_ISL_1906329, EPI_ISL_1906345, EPI_ISL_1906347, EPI_ISL_1906349, EPI_ISL_1906353, EPI_ISL_1906361, EPI_ISL_1906372, EPI_ISL_1906400, EPI_ISL_1906402, EPI_ISL_1906407, EPI_ISL_1906420, EPI_ISL_1906422, EPI_ISL_1906437, EPI_ISL_1906440, EPI_ISL_1906452, EPI_ISL_1906455, EPI_ISL_1906461, EPI_ISL_1906474, EPI_ISL_1906480, EPI_ISL_1906482, EPI_ISL_1906486, EPI_ISL_1906491, EPI_ISL_1906504, EPI_ISL_1906511, EPI_ISL_1906516, EPI_ISL_1906523, EPI_ISL_1906526, EPI_ISL_1906530, EPI_ISL_1906532, EPI_ISL_1906538, EPI_ISL_1906542, EPI_ISL_1906546, EPI_ISL_1906549, EPI_ISL_1906555, EPI_ISL_1906557, EPI_ISL_1906562, EPI_ISL_1906564, EPI_ISL_1906567, EPI_ISL_1906573, EPI_ISL_1906593, EPI_ISL_1906595, EPI_ISL_1906600, EPI_ISL_1906619, EPI_ISL_1906623, EPI_ISL_1906634, EPI_ISL_1906639, EPI_ISL_1906657, EPI_ISL_1906659, EPI_ISL_1906665, EPI_ISL_1906714, EPI_ISL_1906721, EPI_ISL_1906747, EPI_ISL_1906761, EPI_ISL_1906774, EPI_ISL_1906781, EPI_ISL_1906788, EPI_ISL_1906797, EPI_ISL_1906806, EPI_ISL_1906814, EPI_ISL_1906823, EPI_ISL_1906828, EPI_ISL_1906832, EPI_ISL_1906844, EPI_ISL_1906850, EPI_ISL_1906851, EPI_ISL_1906852, EPI_ISL_1906855, EPI_ISL_1906857, EPI_ISL_1906858, EPI_ISL_1906861, EPI_ISL_1906866, EPI_ISL_1906868, EPI_ISL_1906872, EPI_ISL_1906879, EPI_ISL_1924286, EPI_ISL_1924290, EPI_ISL_1924295, EPI_ISL_1924297, EPI_ISL_1924298, EPI_ISL_1924300, EPI_ISL_1924301, EPI_ISL_1924308, EPI_ISL_1924311, EPI_ISL_1924316, EPI_ISL_1924318, EPI_ISL_1924321, EPI_ISL_1924322, EPI_ISL_1924323, EPI_ISL_1924328, EPI_ISL_1924330, EPI_ISL_1924332, EPI_ISL_1924340, EPI_ISL_1924350, EPI_ISL_1924353, EPI_ISL_1924358, EPI_ISL_1924361, EPI_ISL_1924362, EPI_ISL_1924364, EPI_ISL_1924365, EPI_ISL_1924371, EPI_ISL_1924374, EPI_ISL_1924376, EPI_ISL_1924377, EPI_ISL_1924378, EPI_ISL_1924379, EPI_ISL_1924381, EPI_ISL_1924382, EPI_ISL_1924385, EPI_ISL_1924400, EPI_ISL_1924407, EPI_ISL_1924409, EPI_ISL_1924410, EPI_ISL_1924417, EPI_ISL_1924420, EPI_ISL_1924422, EPI_ISL_1924423, EPI_ISL_1924424, EPI_ISL_1924427, EPI_ISL_1924429, EPI_ISL_1924430, EPI_ISL_1924514, EPI_ISL_1935569, EPI_ISL_1935574, EPI_ISL_1989680, EPI_ISL_1989687, EPI_ISL_1989695, EPI_ISL_1989714, EPI_ISL_1989718, EPI_ISL_1989735, EPI_ISL_1989741, EPI_ISL_1989742, EPI_ISL_1989764, EPI_ISL_1989771, EPI_ISL_1989778, EPI_ISL_1989781, EPI_ISL_1989789, EPI_ISL_1989809, EPI_ISL_1989817, EPI_ISL_1989989, EPI_ISL_1990140, EPI_ISL_1990151, EPI_ISL_1990158, EPI_ISL_1990162, EPI_ISL_1990165, EPI_ISL_1990173, EPI_ISL_1990176, EPI_ISL_1990177, EPI_ISL_1990191, EPI_ISL_1990196, EPI_ISL_1990205, EPI_ISL_1990219, EPI_ISL_1990229, EPI_ISL_1990234, EPI_ISL_1990257, EPI_ISL_1990259, EPI_ISL_1990279, EPI_ISL_1990288, EPI_ISL_1990296, EPI_ISL_1990302, EPI_ISL_1990304, EPI_ISL_1990310, EPI_ISL_1990312, EPI_ISL_1990314, EPI_ISL_1990316, EPI_ISL_1990324, EPI_ISL_1990335, EPI_ISL_1990342, EPI_ISL_1990345, EPI_ISL_1990349, EPI_ISL_1990352, EPI_ISL_1990355, EPI_ISL_1990370, EPI_ISL_1990372, EPI_ISL_1990374, EPI_ISL_1990381, EPI_ISL_1990388, EPI_ISL_1990396, EPI_ISL_1990400, EPI_ISL_1990405, EPI_ISL_2010902, EPI_ISL_2010904, EPI_ISL_2010972, EPI_ISL_2010977, EPI_ISL_2010991, EPI_ISL_2034749, EPI_ISL_2047034, EPI_ISL_2047130, EPI_ISL_2047131, EPI_ISL_2047140, EPI_ISL_2047157, EPI_ISL_2047159, EPI_ISL_2047188, EPI_ISL_2047193, EPI_ISL_2047226, EPI_ISL_2047239, EPI_ISL_2047241, EPI_ISL_2047248, EPI_ISL_2047256, EPI_ISL_2047262, EPI_ISL_2047273, EPI_ISL_2047274, EPI_ISL_2047277, EPI_ISL_2098274, EPI_ISL_2098277, EPI_ISL_2098279, EPI_ISL_2098287, EPI_ISL_2098291, EPI_ISL_2098304, EPI_ISL_2098309, EPI_ISL_2098328, EPI_ISL_2098361, EPI_ISL_2098362, EPI_ISL_2098367, EPI_ISL_2098372, EPI_ISL_2098383, EPI_ISL_2098402, EPI_ISL_2098406, EPI_ISL_2132626, EPI_ISL_2133297, EPI_ISL_2133301, EPI_ISL_2133580, EPI_ISL_2133608, EPI_ISL_2133615, EPI_ISL_2133638, EPI_ISL_2133693, EPI_ISL_2133749, EPI_ISL_2133770, EPI_ISL_2133820, EPI_ISL_2133842, EPI_ISL_2140791, EPI_ISL_2142952, EPI_ISL_2143061, EPI_ISL_2143149, EPI_ISL_2143251, EPI_ISL_2143257, EPI_ISL_2187244 | | | |
| see above | Quest Diagnostics Incorporated | Centers for Disease Control and Prevention Division of Viral Diseases, Pathogen Discovery | A. Gerasimova; A. Perez; Adrian Paskey; B. Anderson; Benjamin Rambo-Martin; Christopher Gulvick; Clinton R. Paden; Dakota Howard; Darlene Wagner; Dhwani Batra; Duncan MacCannell; F. Lacbawan; I. A. Shlyakhter; Jason Caravas; K.E. Livingston; Kara Moser; L.E. Bernstein; M. Hua; Matthew Schmerer; P. Tanpaiboon; Peter W. Cook; R. M. Kagan; R. Owen; R. V. Rolando; S. H. Rosenthal; Scott Sammons; Shatavia Morrison; Y. Liu; Yvette Unoarumhi |
| EPI_ISL_2254333, EPI_ISL_2254348, EPI_ISL_2254384, EPI_ISL_2254417, EPI_ISL_2254419, EPI_ISL_2254479 | Respiratory Viruses Branch, Centers for Disease Control and Prevention | Respiratory Viruses Branch, Centers for Disease Control and Prevention | Batra, D.; C.R.; Campbell, P.; Caravas, J.; Case, R.; Clark, C.; Cook; Ghorpade, V.; Gulvick, C.; Hardison, M.; Houdeshell, H.; Howard, D.; Kvalvaag, O.; MacCannell, D.; Morrison, S.; Moser, K.; Nall, D.; P.W.; Paden; Paskey, A.; Rambo-Martin, B.; Sammons, S.; Sanders, E.; Schmerer, M.; Unoarumhi, Y.; Vest, A.; Wagner, D.; Westlund, S. |
| EPI_ISL_1624485, EPI_ISL_1755992, EPI_ISL_1755993, EPI_ISL_1755994, EPI_ISL_1755995, EPI_ISL_1755996, EPI_ISL_1755997, EPI_ISL_1755998, EPI_ISL_1755999, EPI_ISL_1855220, EPI_ISL_1855273, EPI_ISL_1855274, EPI_ISL_1855275, EPI_ISL_2098622, EPI_ISL_2098623, EPI_ISL_2098624, EPI_ISL_2098625, EPI_ISL_2098626, EPI_ISL_2098628 | | | |
| see above | SARATOGA HOSPITAL LABORATORY | Wadsworth Center, New York State Department of Health | Alexis Russell; Catharine Prussing; Daryl M. Lamson; Erasmus Schneider; Erica Lasek-Nesselquist; John Kelly; Jonathan Plitnick; Kirsten St. George; Matthew Shudt; Melissa A Leisner; Navjot Singh |
| EPI_ISL_2097295 | ST. CATHERINE OF SIENA HOSPITAL LABORATORY | Wadsworth Center, New York State Department of Health | Alexis Russell; Catharine Prussing; Daryl M. Lamson; Erasmus Schneider; Erica Lasek-Nesselquist; John Kelly; Jonathan Plitnick; Kirsten St. George; Matthew Shudt; Melissa A Leisner; Navjot Singh |
| EPI_ISL_1671484, EPI_ISL_1756368, EPI_ISL_2098629, EPI_ISL_2098630 | ST. FRANCIS HOSPITAL LABORATORY | Wadsworth Center, New York State Department of Health | Alexis Russell; Catharine Prussing; Daryl M. Lamson; Erasmus Schneider; Erica Lasek-Nesselquist; John Kelly; Jonathan Plitnick; Kirsten St. George; Matthew Shudt; Melissa A Leisner; Navjot Singh |
| EPI_ISL_1671316, EPI_ISL_1671329, EPI_ISL_1671330, EPI_ISL_1671336, EPI_ISL_1671349, EPI_ISL_1756000, EPI_ISL_1756331, EPI_ISL_1756332, EPI_ISL_1756333, EPI_ISL_1756334, EPI_ISL_1756335, EPI_ISL_1756336, EPI_ISL_1756337, EPI_ISL_1789719, EPI_ISL_2097296, EPI_ISL_2097297, EPI_ISL_2097298, EPI_ISL_2097299, EPI_ISL_2097300, EPI_ISL_2097301, EPI_ISL_2097302, EPI_ISL_2097303, EPI_ISL_2134788, EPI_ISL_2134789 | | | |
| see above | STONY BROOK UNIVERSITY HOSPITAL | Wadsworth Center, New York State Department of Health | Alexis Russell; Catharine Prussing; Daryl M. Lamson; Erasmus Schneider; Erica Lasek-Nesselquist; John Kelly; Jonathan Plitnick; Kirsten St. George; Matthew Shudt; Melissa A Leisner; Navjot Singh |
| EPI_ISL_2151420 | SUNY Downstate Medical Center | New York City Public Health Laboratory | Jade Wang; et al. |
| EPI_ISL_1671464, EPI_ISL_1671465, EPI_ISL_1671494, EPI_ISL_1671512, EPI_ISL_1671513, EPI_ISL_1671514, EPI_ISL_1671612, EPI_ISL_1755843, EPI_ISL_1755844, EPI_ISL_1755845, EPI_ISL_1755846, EPI_ISL_1755847, EPI_ISL_1755848, EPI_ISL_1756338, EPI_ISL_1756339, EPI_ISL_1756340, EPI_ISL_1756341, EPI_ISL_1854691, EPI_ISL_1855105, EPI_ISL_1855106, EPI_ISL_1855107, EPI_ISL_1855108, EPI_ISL_1855109, EPI_ISL_1855110, EPI_ISL_1855111, EPI_ISL_1855112, EPI_ISL_1855113, EPI_ISL_1855114, EPI_ISL_1855115, EPI_ISL_1855221, EPI_ISL_1855222, EPI_ISL_1855223, EPI_ISL_1855224, EPI_ISL_1855225, EPI_ISL_1855354, EPI_ISL_1855355, EPI_ISL_1855356, EPI_ISL_1855357, EPI_ISL_1855358, EPI_ISL_1855359, EPI_ISL_1855360, EPI_ISL_1935639, EPI_ISL_1935640, EPI_ISL_1935641, EPI_ISL_1935642, EPI_ISL_1935646, EPI_ISL_1935647, EPI_ISL_1935649, EPI_ISL_1935650, EPI_ISL_1935651, EPI_ISL_2097304, EPI_ISL_2097305, EPI_ISL_2098135, EPI_ISL_2098136, EPI_ISL_2098138, EPI_ISL_2098140, EPI_ISL_2098141, EPI_ISL_2098143, EPI_ISL_2098144, EPI_ISL_2098146, EPI_ISL_2098147, EPI_ISL_2098149, EPI_ISL_2098150, EPI_ISL_2098151, EPI_ISL_2098153, EPI_ISL_2098155, EPI_ISL_2098156, EPI_ISL_2098158, EPI_ISL_2098159, EPI_ISL_2098637, EPI_ISL_2098638, EPI_ISL_2098639, EPI_ISL_2098640, EPI_ISL_2098641, EPI_ISL_2098642, EPI_ISL_2098643, EPI_ISL_2098644, EPI_ISL_2098645, EPI_ISL_2098646, EPI_ISL_2098647, EPI_ISL_2098648, EPI_ISL_2098649, EPI_ISL_2098650, EPI_ISL_2098651, EPI_ISL_2098652, EPI_ISL_2098653, EPI_ISL_2098654, EPI_ISL_2098655, EPI_ISL_2098656, EPI_ISL_2098657, EPI_ISL_2098658 | | | |
| see above | SUNY UPSTATE MEDICAL UNIVERSITY | Wadsworth Center, New York State Department of Health | Alexis Russell; Catharine Prussing; Daryl M. Lamson; Erasmus Schneider; Erica Lasek-Nesselquist; John Kelly; Jonathan Plitnick; Kirsten St. George; Matthew Shudt; Melissa A Leisner; Navjot Singh |
| EPI_ISL_1921011, EPI_ISL_1921012, EPI_ISL_1921013, EPI_ISL_1921014, EPI_ISL_1921015, EPI_ISL_1921016, EPI_ISL_1921017, EPI_ISL_1921019, EPI_ISL_1921020, EPI_ISL_1921021, EPI_ISL_1921023, EPI_ISL_1921024, EPI_ISL_1921025, EPI_ISL_1921026, EPI_ISL_1921027, EPI_ISL_1921038, EPI_ISL_1921039 | | | |
| see above | SUNY Upstate Medical University | Wadsworth Center, New York State Department of Health | Alexis Russell; Catharine Prussing; Daryl M. Lamson; Erasmus Schneider; Erica Lasek-Nesselquist; John Kelly; Jonathan Plitnick; Kirsten St. George; Matthew Shudt; Melissa A Leisner; Navjot Singh |
| EPI_ISL_1920029 | St. Francis Hospital Laboratory | Wadsworth Center, New York State Department of Health | Alexis Russell; Catharine Prussing; Daryl M. Lamson; Erasmus Schneider; Erica Lasek-Nesselquist; John Kelly; Jonathan Plitnick; Kirsten St. George; Matthew Shudt; Melissa A Leisner; Navjot Singh |
| EPI_ISL_1967689, EPI_ISL_1967690, EPI_ISL_1967691, EPI_ISL_1967692, EPI_ISL_1967693, EPI_ISL_1967694, EPI_ISL_1967695 | | | |
| see above | Stony Brook Medicine | New York Genome Center | Alaa B. Salaheldin; Andre Corvelo; Dayna M. Oschwald; Dimitri Gnatenko; Eric D. Spitzer; Jiang Chen; Josue M. Flores Acuna; Kenneth R. Shroyer; Margaret M. McGovern; Michael Zody; Richard R. Kew; Samantha Fennessey; Sarah A. Halbohn; Sarah A. Malik; Silvia G. Spitzer; Susan V. Donelan; Tom Maniatis |
| EPI_ISL_1919991, EPI_ISL_1919992, EPI_ISL_1920030, EPI_ISL_1920035, EPI_ISL_1920041, EPI_ISL_1920044, EPI_ISL_1920046, EPI_ISL_1920047, EPI_ISL_1920048, EPI_ISL_1920049 | | | |
| see above | Stony Brook University Hospital | Wadsworth Center, New York State Department of Health | Alexis Russell; Catharine Prussing; Daryl M. Lamson; Erasmus Schneider; Erica Lasek-Nesselquist; John Kelly; Jonathan Plitnick; Kirsten St. George; Matthew Shudt; Melissa A Leisner; Navjot Singh |
| EPI_ISL_1919990 | Suny Upstate Medical University | Wadsworth Center, New York State Department of Health | Alexis Russell; Catharine Prussing; Daryl M. Lamson; Erasmus Schneider; Erica Lasek-Nesselquist; John Kelly; Jonathan Plitnick; Kirsten St. George; Matthew Shudt; Melissa A Leisner; Navjot Singh |
| EPI_ISL_1671312, EPI_ISL_1671314, EPI_ISL_1671331, EPI_ISL_1671332, EPI_ISL_1671333, EPI_ISL_1671334, EPI_ISL_1671335, EPI_ISL_1671338, EPI_ISL_1671339, EPI_ISL_1671340, EPI_ISL_1671342, EPI_ISL_1671343, EPI_ISL_1671344, EPI_ISL_1671345, EPI_ISL_1671346, EPI_ISL_1671347, EPI_ISL_1671348, EPI_ISL_1671352, EPI_ISL_1671353, EPI_ISL_1671355, EPI_ISL_1671643, EPI_ISL_1740506, EPI_ISL_1740507, EPI_ISL_1755968, EPI_ISL_1755969, EPI_ISL_1755970, EPI_ISL_1755971, EPI_ISL_1755972, EPI_ISL_1755973, EPI_ISL_1755974, EPI_ISL_1755975, EPI_ISL_1755976, EPI_ISL_1755977, EPI_ISL_1755978, EPI_ISL_1755979, EPI_ISL_1755980, EPI_ISL_1755981, EPI_ISL_1755982, EPI_ISL_1755983, EPI_ISL_1755984, EPI_ISL_1755985, EPI_ISL_1755986, EPI_ISL_1755987, EPI_ISL_1755988, EPI_ISL_1755989, EPI_ISL_1755990, EPI_ISL_1755991, EPI_ISL_1855104, EPI_ISL_1855150, EPI_ISL_1855151, EPI_ISL_1855152, EPI_ISL_1855153, EPI_ISL_1855154, EPI_ISL_1855155, EPI_ISL_1855156, EPI_ISL_1855157, EPI_ISL_1855158, EPI_ISL_1855159, EPI_ISL_1855160, EPI_ISL_1855161, EPI_ISL_1855162, EPI_ISL_1855163, EPI_ISL_1855164, EPI_ISL_1855165, EPI_ISL_1855166, EPI_ISL_1855167, EPI_ISL_1855244, EPI_ISL_1855245, EPI_ISL_1855246, EPI_ISL_1855247, EPI_ISL_1855248, EPI_ISL_1935613, EPI_ISL_1935627, EPI_ISL_1935632, EPI_ISL_1935633 | | | |
| see above | TEMPUS LABS INC | Wadsworth Center, New York State Department of Health | Alexis Russell; Catharine Prussing; Daryl M. Lamson; Erasmus Schneider; Erica Lasek-Nesselquist; John Kelly; Jonathan Plitnick; Kirsten St. George; Matthew Shudt; Melissa A Leisner; Navjot Singh |
| EPI_ISL_1920098, EPI_ISL_1920099, EPI_ISL_1920100, EPI_ISL_1920102, EPI_ISL_1920103, EPI_ISL_1920106, EPI_ISL_1920107, EPI_ISL_1920115, EPI_ISL_1920116, EPI_ISL_1920117, EPI_ISL_1920118, EPI_ISL_1920123 | | | |
| see above | Tempus Labs Inc | Wadsworth Center, New York State Department of Health | Alexis Russell; Catharine Prussing; Daryl M. Lamson; Erasmus Schneider; Erica Lasek-Nesselquist; John Kelly; Jonathan Plitnick; Kirsten St. George; Matthew Shudt; Melissa A Leisner; Navjot Singh |
| EPI_ISL_1540718, EPI_ISL_1625677, EPI_ISL_1739440 | The Jackson Laboratory | The Jackson Laboratory | Adams M; Bergeron D; Kelly K; Li L; Long J; Omerza G; Renzette N |
| EPI_ISL_1671626, EPI_ISL_1671627, EPI_ISL_1671634, EPI_ISL_1671667, EPI_ISL_1740516, EPI_ISL_1740517, EPI_ISL_1755869, EPI_ISL_1755870, EPI_ISL_1755871, EPI_ISL_1755872, EPI_ISL_1755873, EPI_ISL_1755874, EPI_ISL_1755875, EPI_ISL_1755885, EPI_ISL_1755886, EPI_ISL_1755887, EPI_ISL_1755888, EPI_ISL_1755889, EPI_ISL_1755890, EPI_ISL_1755891, EPI_ISL_1755892, EPI_ISL_1755893, EPI_ISL_1755894, EPI_ISL_1755895, EPI_ISL_1755897, EPI_ISL_1755898, EPI_ISL_1755899, EPI_ISL_1755900, EPI_ISL_1755901, EPI_ISL_1755902, EPI_ISL_1755903, EPI_ISL_1755905, EPI_ISL_1755906, EPI_ISL_1755907, EPI_ISL_1756357, EPI_ISL_1756358, EPI_ISL_1756359, EPI_ISL_1756360, EPI_ISL_1756361, EPI_ISL_1756362, EPI_ISL_1756363, EPI_ISL_1756364, EPI_ISL_1756365, EPI_ISL_1756366, EPI_ISL_1756367, EPI_ISL_1854693, EPI_ISL_1854694, EPI_ISL_1854695, EPI_ISL_1855119, EPI_ISL_1855120, EPI_ISL_1855121, EPI_ISL_1855122, EPI_ISL_1855123, EPI_ISL_1855124, EPI_ISL_1855125, EPI_ISL_1855126, EPI_ISL_1855127, EPI_ISL_1855128, EPI_ISL_1855129, EPI_ISL_1855130, EPI_ISL_1855131, EPI_ISL_1855132, EPI_ISL_1855133, EPI_ISL_1855134, EPI_ISL_1855135, EPI_ISL_1855136, EPI_ISL_1855137, EPI_ISL_1855138, EPI_ISL_1855139, EPI_ISL_1855140, EPI_ISL_1855141, EPI_ISL_1855142, EPI_ISL_1855143, EPI_ISL_1855168, EPI_ISL_1855169, EPI_ISL_1855170, EPI_ISL_1855171, EPI_ISL_1855172, EPI_ISL_1855173, EPI_ISL_1855174, EPI_ISL_1855175, EPI_ISL_1855176, EPI_ISL_1855177, EPI_ISL_1855178, EPI_ISL_1855179, EPI_ISL_1855180, EPI_ISL_1855181, EPI_ISL_1855182, EPI_ISL_1855183, EPI_ISL_1855184, EPI_ISL_1855185, EPI_ISL_1855226, EPI_ISL_1855227, EPI_ISL_1855228, EPI_ISL_1855229, EPI_ISL_1855230, EPI_ISL_1855231, EPI_ISL_1855232, EPI_ISL_1855233, EPI_ISL_1855234, EPI_ISL_1855235, EPI_ISL_1855236, EPI_ISL_1855237, EPI_ISL_1855238, EPI_ISL_1855239, EPI_ISL_1855240, EPI_ISL_1855249, EPI_ISL_1855250, EPI_ISL_1855251, EPI_ISL_1855252, EPI_ISL_1855253, EPI_ISL_1855254, EPI_ISL_1855255, EPI_ISL_1855256, EPI_ISL_1855257, EPI_ISL_1855258, EPI_ISL_1855259, EPI_ISL_1855260, EPI_ISL_1855261, EPI_ISL_1855262, EPI_ISL_1855263, EPI_ISL_1855264, EPI_ISL_1855265, EPI_ISL_1855266, EPI_ISL_1855267, EPI_ISL_1855268, EPI_ISL_1855269, EPI_ISL_1855270, EPI_ISL_1855271, EPI_ISL_1855272, EPI_ISL_1855276, EPI_ISL_1855277, EPI_ISL_1855278, EPI_ISL_1855279, EPI_ISL_1855280, EPI_ISL_1855281, EPI_ISL_1855282, EPI_ISL_1855283, EPI_ISL_1855284, EPI_ISL_1855285, EPI_ISL_1855294, EPI_ISL_1855295, EPI_ISL_1855296, EPI_ISL_1855297, EPI_ISL_1855298, EPI_ISL_1855299, EPI_ISL_1855300, EPI_ISL_1855301, EPI_ISL_1855302, EPI_ISL_1855303, EPI_ISL_1855304, EPI_ISL_1855305, EPI_ISL_1855306, EPI_ISL_1855307, EPI_ISL_1855308, EPI_ISL_1855309, EPI_ISL_1920069, EPI_ISL_1920124, EPI_ISL_1920125, EPI_ISL_1920126, EPI_ISL_1920128, EPI_ISL_1920130, EPI_ISL_1920997, EPI_ISL_1920998, EPI_ISL_1920999, EPI_ISL_1921000, EPI_ISL_1921001, EPI_ISL_1921002, EPI_ISL_1921003, EPI_ISL_1921004, EPI_ISL_1921005, EPI_ISL_1921006, EPI_ISL_1921007, EPI_ISL_1921008, EPI_ISL_1921009, EPI_ISL_1921010, EPI_ISL_1921040, EPI_ISL_1921041, EPI_ISL_1921042, EPI_ISL_1921043, EPI_ISL_1921044, EPI_ISL_1921045, EPI_ISL_1921046, EPI_ISL_1921047, EPI_ISL_1921048, EPI_ISL_1921049, EPI_ISL_1921050, EPI_ISL_1921051, EPI_ISL_1921052, EPI_ISL_1921053, EPI_ISL_1921054, EPI_ISL_1921055, EPI_ISL_1921056, EPI_ISL_1921057, EPI_ISL_1921058, EPI_ISL_1921059, EPI_ISL_1921060, EPI_ISL_1921061, EPI_ISL_1921062, EPI_ISL_1921063, EPI_ISL_1921064, EPI_ISL_1921065, EPI_ISL_1921066, EPI_ISL_1921067, EPI_ISL_1921068, EPI_ISL_1921069, EPI_ISL_1921070, EPI_ISL_1921071, EPI_ISL_1921072, EPI_ISL_1921073, EPI_ISL_1921074, EPI_ISL_1921075, EPI_ISL_1921076, EPI_ISL_1921077, EPI_ISL_1921078, EPI_ISL_1921079, EPI_ISL_1935611, EPI_ISL_1935616, EPI_ISL_1935622, EPI_ISL_1935623, EPI_ISL_1935634, EPI_ISL_1935635, EPI_ISL_1935645, EPI_ISL_1935652, EPI_ISL_2098218, EPI_ISL_2098220, EPI_ISL_2098223, EPI_ISL_2098224, EPI_ISL_2098225, EPI_ISL_2098230, EPI_ISL_2098231, EPI_ISL_2098502, EPI_ISL_2098503, EPI_ISL_2098504, EPI_ISL_2098505, EPI_ISL_2098506, EPI_ISL_2098507, EPI_ISL_2098508, EPI_ISL_2098509, EPI_ISL_2098510, EPI_ISL_2098511, EPI_ISL_2098512, | | | |

| | | | |
|---|---|---|---|
| EPI_ISL_2098513, EPI_ISL_2098514, EPI_ISL_2098515, EPI_ISL_2098516, EPI_ISL_2098517, EPI_ISL_2098518, EPI_ISL_2098519, EPI_ISL_2098520, EPI_ISL_2098521, EPI_ISL_2098522, EPI_ISL_2098523, EPI_ISL_2098524, EPI_ISL_2098525, EPI_ISL_2098526, EPI_ISL_2098527, EPI_ISL_2098528, EPI_ISL_2098529, EPI_ISL_2098530, EPI_ISL_2098531, EPI_ISL_2098532, EPI_ISL_2098533, EPI_ISL_2098534, EPI_ISL_2098535, EPI_ISL_2098536, EPI_ISL_2098537, EPI_ISL_2098538, EPI_ISL_2098539, EPI_ISL_2098540, EPI_ISL_2098541, EPI_ISL_2098542, EPI_ISL_2098543, EPI_ISL_2098544, EPI_ISL_2098545, EPI_ISL_2098546, EPI_ISL_2098547, EPI_ISL_2098548, EPI_ISL_2098549, EPI_ISL_2098550, EPI_ISL_2098551, EPI_ISL_2098552, EPI_ISL_2098553, EPI_ISL_2098554, EPI_ISL_2098555, EPI_ISL_2098556, EPI_ISL_2098557, EPI_ISL_2098558, EPI_ISL_2098559, EPI_ISL_2098560, EPI_ISL_2098561, EPI_ISL_2098562, EPI_ISL_2098563, EPI_ISL_2098564, EPI_ISL_2098565, EPI_ISL_2098566, EPI_ISL_2098567, EPI_ISL_2098568, EPI_ISL_2098569, EPI_ISL_2098570, EPI_ISL_2098571, EPI_ISL_2098572, EPI_ISL_2098573, EPI_ISL_2098574, EPI_ISL_2098575, EPI_ISL_2098576, EPI_ISL_2098577, EPI_ISL_2098578, EPI_ISL_2098579, EPI_ISL_2098580, EPI_ISL_2098581, EPI_ISL_2098582, EPI_ISL_2098583, EPI_ISL_2098584, EPI_ISL_2098585, EPI_ISL_2098586, EPI_ISL_2098587, EPI_ISL_2134795 | | | |
| see above | URMC LABS | Wadsworth Center, New York State Department of Health | Alexis Russell; Catharine Prussing; Daryl M. Lamson; Erasmus Schneider; Erica Lasek-Nesselquist; John Kelly; Jonathan Plitnick; Kirsten St. George; Matthew Shudt; Melissa A Leisner; Navjot Singh |
| EPI_ISL_1671449, EPI_ISL_1671458, EPI_ISL_1671461, EPI_ISL_1671469, EPI_ISL_1671470, EPI_ISL_1671471, EPI_ISL_1671472, EPI_ISL_1671473, EPI_ISL_1671474, EPI_ISL_1671475, EPI_ISL_1671480, EPI_ISL_1671485, EPI_ISL_1671486, EPI_ISL_1671487, EPI_ISL_1671488, EPI_ISL_1671489, EPI_ISL_1671490, EPI_ISL_1671491, EPI_ISL_1671500, EPI_ISL_1671501, EPI_ISL_1671502, EPI_ISL_1671503, EPI_ISL_1671504, EPI_ISL_1671505, EPI_ISL_1671506, EPI_ISL_1671507, EPI_ISL_1671511, EPI_ISL_1671597, EPI_ISL_1671635, EPI_ISL_1671636, EPI_ISL_1740514, EPI_ISL_1740515, EPI_ISL_1755965, EPI_ISL_1756369, EPI_ISL_1756370, EPI_ISL_1756371, EPI_ISL_1756372, EPI_ISL_1756373, EPI_ISL_1756374, EPI_ISL_1756375, EPI_ISL_1756376, EPI_ISL_1756377, EPI_ISL_1756378, EPI_ISL_1756379, EPI_ISL_1756380, EPI_ISL_1756381, EPI_ISL_1756382, EPI_ISL_1756383, EPI_ISL_1756384, EPI_ISL_1756385, EPI_ISL_1756386, EPI_ISL_1756387, EPI_ISL_1756388, EPI_ISL_1756389, EPI_ISL_1756390, EPI_ISL_1756391, EPI_ISL_1756392, EPI_ISL_1756393, EPI_ISL_1756394, EPI_ISL_1756395, EPI_ISL_1756396, EPI_ISL_1756397, EPI_ISL_1756398, EPI_ISL_1756399, EPI_ISL_1756400, EPI_ISL_1756401, EPI_ISL_1756402, EPI_ISL_1756403, EPI_ISL_1756404, EPI_ISL_1756405, EPI_ISL_1756406, EPI_ISL_1756407, EPI_ISL_1756408, EPI_ISL_1789722, EPI_ISL_1789723, EPI_ISL_1855334, EPI_ISL_1855335, EPI_ISL_1855336, EPI_ISL_1855337, EPI_ISL_1855338, EPI_ISL_1855339, EPI_ISL_1855340, EPI_ISL_1855341, EPI_ISL_1855342, EPI_ISL_1855343, EPI_ISL_1855344, EPI_ISL_1855345, EPI_ISL_1855346, EPI_ISL_1855347, EPI_ISL_2097306, EPI_ISL_2097307, EPI_ISL_2097308, EPI_ISL_2097309, EPI_ISL_2097310, EPI_ISL_2097311, EPI_ISL_2097312, EPI_ISL_2097313, EPI_ISL_2097314, EPI_ISL_2098233 | | | |
| see above | WESTCHESTER MEDICAL CENTER | Wadsworth Center, New York State Department of Health | Alexis Russell; Catharine Prussing; Daryl M. Lamson; Erasmus Schneider; Erica Lasek-Nesselquist; John Kelly; Jonathan Plitnick; Kirsten St. George; Matthew Shudt; Melissa A Leisner; Navjot Singh |
| EPI_ISL_1671598, EPI_ISL_1671600, EPI_ISL_1671607, EPI_ISL_1671610, EPI_ISL_1671611, EPI_ISL_1671622, EPI_ISL_1671644, EPI_ISL_1671655, EPI_ISL_1671656, EPI_ISL_1671665 | | | |
| see above | WHITE PLAINS HOSPITAL CENTER LABORATORY | Wadsworth Center, New York State Department of Health | Alexis Russell; Catharine Prussing; Daryl M. Lamson; Erasmus Schneider; Erica Lasek-Nesselquist; John Kelly; Jonathan Plitnick; Kirsten St. George; Matthew Shudt; Melissa A Leisner; Navjot Singh |
| EPI_ISL_1920113, EPI_ISL_1920114, EPI_ISL_1920121 | Wadsworth Center, New York State Department of Health | Wadsworth Center, New York State Department of Health | Alexis Russell; Catharine Prussing; Daryl M. Lamson; Erasmus Schneider; Erica Lasek-Nesselquist; John Kelly; Jonathan Plitnick; Kirsten St. George; Matthew Shudt; Melissa A Leisner; Navjot Singh |
| EPI_ISL_1967662, EPI_ISL_1967663, EPI_ISL_1967665, EPI_ISL_1967666, EPI_ISL_1967667, EPI_ISL_1967668, EPI_ISL_1967669, EPI_ISL_1967670, EPI_ISL_1967671, EPI_ISL_1967672, EPI_ISL_1967673, EPI_ISL_1967674, EPI_ISL_1967675, EPI_ISL_1967676, EPI_ISL_1967678, EPI_ISL_1967680, EPI_ISL_1967681, EPI_ISL_1967682, EPI_ISL_1967683, EPI_ISL_1967684, EPI_ISL_1971369, EPI_ISL_1971370, EPI_ISL_1971371, EPI_ISL_1971372, EPI_ISL_1971373, EPI_ISL_1971374, EPI_ISL_1971375, EPI_ISL_1971376, EPI_ISL_1971377, EPI_ISL_1971378, EPI_ISL_1971379, EPI_ISL_1971380, EPI_ISL_1971381, EPI_ISL_1971382, EPI_ISL_1971383, EPI_ISL_1971384, EPI_ISL_1971385, EPI_ISL_1971386, EPI_ISL_1971387, EPI_ISL_1971388, EPI_ISL_1971389, EPI_ISL_1971390, EPI_ISL_1971391, EPI_ISL_1971392, EPI_ISL_1971393, EPI_ISL_1971394, EPI_ISL_1971395, EPI_ISL_1971396, EPI_ISL_1971397, EPI_ISL_1971398, EPI_ISL_1971399, EPI_ISL_1971400, EPI_ISL_1971401, EPI_ISL_1971402, EPI_ISL_1971403, EPI_ISL_1971404, EPI_ISL_1971405, EPI_ISL_1971406, EPI_ISL_2081128, EPI_ISL_2081129, EPI_ISL_2081130, EPI_ISL_2081131, EPI_ISL_2081132, EPI_ISL_2081133, EPI_ISL_2081134, EPI_ISL_2081135, EPI_ISL_2081136, EPI_ISL_2081137, EPI_ISL_2081138, EPI_ISL_2081139, EPI_ISL_2081140, EPI_ISL_2081141, EPI_ISL_2081142, EPI_ISL_2081143, EPI_ISL_2081144, EPI_ISL_2081145, EPI_ISL_2081146, EPI_ISL_2081147, EPI_ISL_2081148, EPI_ISL_2081149, EPI_ISL_2081150, EPI_ISL_2081151, EPI_ISL_2081152, EPI_ISL_2081153, EPI_ISL_2081154, EPI_ISL_2081155, EPI_ISL_2081156, EPI_ISL_2081157, EPI_ISL_2081158, EPI_ISL_2081159, EPI_ISL_2081160, EPI_ISL_2081161, EPI_ISL_2081162, EPI_ISL_2081163, EPI_ISL_2081164, EPI_ISL_2081165, EPI_ISL_2081166, EPI_ISL_2081167, EPI_ISL_2081168, EPI_ISL_2082729, EPI_ISL_2086266 | | | |
| see above | Weill Cornell Medicine | New York Genome Center | Andre Corvelo; Arryn Craney; Chris Mason; Dayna M. Oschwald; Hanna Rennert; Lars F Westblade; Margaret Elizabeth Ross; Melissa Cushing; Michael Zody; Olivier Elemento; Priya Velu; Samantha Fennessey; Tom Maniatis |
| EPI_ISL_1793502, EPI_ISL_2020795, EPI_ISL_2020840, EPI_ISL_2034886, EPI_ISL_2034934, EPI_ISL_2158984 | Yale Clinical Virology Lab | Yale Center for Genomic Analysis | Brooke Sullivan; Christopher Castaldi; Curt Scharfe; Irina Tikhonova; Kaya Bilguvar; Shrikant Mane |

We gratefully acknowledge the following Authors from the Originating laboratories responsible for obtaining the specimens, as well as the Submitting laboratories where the genome data were generated and shared via GISAID, on which this research is based.

All Submitters of data may be contacted directly via www.gisaid.org

Authors are sorted alphabetically.

| Accession ID | Originating Laboratory | Submitting Laboratory | Authors |
|---|---|---|---|
| EPI_ISL_2274625, EPI_ISL_2274626, EPI_ISL_2274627, EPI_ISL_2274628, EPI_ISL_2274629, EPI_ISL_2274630, EPI_ISL_2274664, EPI_ISL_2278742, EPI_ISL_2279503, EPI_ISL_2279504, EPI_ISL_2279505, EPI_ISL_2308802, EPI_ISL_2421140, EPI_ISL_2421141, EPI_ISL_2421142, EPI_ISL_2421143, EPI_ISL_2422313, EPI_ISL_2448312, EPI_ISL_2626999, EPI_ISL_2627000, EPI_ISL_2627001, EPI_ISL_2627002, EPI_ISL_2627157, EPI_ISL_2627158, EPI_ISL_2627159, EPI_ISL_2627160, EPI_ISL_2627161, EPI_ISL_2627162, EPI_ISL_2627163, EPI_ISL_2727104, EPI_ISL_2727111, EPI_ISL_2727113, EPI_ISL_2727114, EPI_ISL_2868552 | | | |
| see above | ALBANY MEDICAL CENTER | Wadsworth Center, New York State Department of Health | Alexis Russell; Catharine Prussing; Daryl M. Lamson; Erasmus Schneider; Erica Lasek-Nesselquist; John Kelly; Jonathan Plitnick; Kirsten St. George; Matthew Shudt; Melissa A Leisner; Navjot Singh |
| EPI_ISL_2146058, EPI_ISL_2146067, EPI_ISL_2146071, EPI_ISL_2146093, EPI_ISL_2146141, EPI_ISL_2146178, EPI_ISL_2146203, EPI_ISL_2146249, EPI_ISL_2146251, EPI_ISL_2146270, EPI_ISL_2146343, EPI_ISL_2146406, EPI_ISL_2146473, EPI_ISL_2146474, EPI_ISL_2146483, EPI_ISL_2146525, EPI_ISL_2146663, EPI_ISL_2146703, EPI_ISL_2146721, EPI_ISL_2146825, EPI_ISL_2146826, EPI_ISL_2146839, EPI_ISL_2146914, EPI_ISL_2146964, EPI_ISL_2146979, EPI_ISL_2146992, EPI_ISL_2147007, EPI_ISL_2147034, EPI_ISL_2147035, EPI_ISL_2147150, EPI_ISL_2147155, EPI_ISL_2147191, EPI_ISL_2147230, EPI_ISL_2147240, EPI_ISL_2147246, EPI_ISL_2147356, EPI_ISL_2147439, EPI_ISL_2147441, EPI_ISL_2147442, EPI_ISL_2147476, EPI_ISL_2147480, EPI_ISL_2147499, EPI_ISL_2147500, EPI_ISL_2147533, EPI_ISL_2147536, EPI_ISL_2147571, EPI_ISL_2147591, EPI_ISL_2147614, EPI_ISL_2147615, EPI_ISL_2147741, EPI_ISL_2147840, EPI_ISL_2147844, EPI_ISL_2147920, EPI_ISL_2147969, EPI_ISL_2147992, EPI_ISL_2148173, EPI_ISL_2148191, EPI_ISL_2148216, EPI_ISL_2148217, EPI_ISL_2148288, EPI_ISL_2148352, EPI_ISL_2148481, EPI_ISL_2148576, EPI_ISL_2148587, EPI_ISL_2148589, EPI_ISL_2148784, EPI_ISL_2148785, EPI_ISL_2148806, EPI_ISL_2148856, EPI_ISL_2148887, EPI_ISL_2148902, EPI_ISL_2148905, EPI_ISL_2148906, EPI_ISL_2148937, EPI_ISL_2149182, EPI_ISL_2158971, EPI_ISL_2159089, EPI_ISL_2159170, EPI_ISL_2159187, EPI_ISL_2159198, EPI_ISL_2159201, EPI_ISL_2159457, EPI_ISL_2180576, EPI_ISL_2180579, EPI_ISL_2180665, EPI_ISL_2180742, EPI_ISL_2180764, EPI_ISL_2180772, EPI_ISL_2180797, EPI_ISL_2180824, EPI_ISL_2180907, EPI_ISL_2181043, EPI_ISL_2181085, EPI_ISL_2181120, EPI_ISL_2181137, EPI_ISL_2181201, EPI_ISL_2181216, EPI_ISL_2181295, EPI_ISL_2181325, EPI_ISL_2181326, EPI_ISL_2181327, EPI_ISL_2181329, EPI_ISL_2181334, EPI_ISL_2181340, EPI_ISL_2181360, EPI_ISL_2181365, EPI_ISL_2181390, EPI_ISL_2181391, EPI_ISL_2181392, EPI_ISL_2181502, EPI_ISL_2181514, EPI_ISL_2181574, EPI_ISL_2181575, EPI_ISL_2181601, EPI_ISL_2181640, EPI_ISL_2181652, EPI_ISL_2181653, EPI_ISL_2187370, EPI_ISL_2187445, EPI_ISL_2187452, EPI_ISL_2187491, EPI_ISL_2187596, EPI_ISL_2204599, EPI_ISL_2241646, EPI_ISL_2280327, EPI_ISL_2280340, EPI_ISL_2280341, EPI_ISL_2280385, EPI_ISL_2280423, EPI_ISL_2280424, EPI_ISL_2280425, EPI_ISL_2280460, EPI_ISL_2280468, EPI_ISL_2280499, EPI_ISL_2280504, EPI_ISL_2280505, EPI_ISL_2280530, EPI_ISL_2280537, EPI_ISL_2280756, EPI_ISL_2280760, EPI_ISL_2280771, EPI_ISL_2280774, EPI_ISL_2280782, EPI_ISL_2280906, EPI_ISL_2280940, EPI_ISL_2280942, EPI_ISL_2280960, EPI_ISL_2280961, EPI_ISL_2280996, EPI_ISL_2281008, EPI_ISL_2281031, EPI_ISL_2281076, EPI_ISL_2281100, EPI_ISL_2281172, EPI_ISL_2281182, EPI_ISL_2283819, EPI_ISL_2283853, EPI_ISL_2283871, EPI_ISL_2321336, EPI_ISL_2321345, EPI_ISL_2368228, EPI_ISL_2368236, EPI_ISL_2368304, EPI_ISL_2368318, EPI_ISL_2368343, EPI_ISL_2368344, EPI_ISL_2368351, EPI_ISL_2368363, EPI_ISL_2368364, EPI_ISL_2368485, EPI_ISL_2368507, EPI_ISL_2368531, EPI_ISL_2368569, EPI_ISL_2368727, EPI_ISL_2368942, EPI_ISL_2368954, EPI_ISL_2369952, EPI_ISL_2369953, EPI_ISL_2369979, EPI_ISL_2370004, EPI_ISL_2370026, EPI_ISL_2370211, EPI_ISL_2370609, EPI_ISL_2370630, EPI_ISL_2370664, EPI_ISL_2370675, EPI_ISL_2370678, EPI_ISL_2370716, EPI_ISL_2372335, EPI_ISL_2372364, EPI_ISL_2372366, EPI_ISL_2372393, EPI_ISL_2372425, EPI_ISL_2372436, EPI_ISL_2372456, EPI_ISL_2372459, EPI_ISL_2421982, EPI_ISL_2421983, EPI_ISL_2421985, EPI_ISL_2422105, EPI_ISL_2442328, EPI_ISL_2489124, EPI_ISL_2489210, EPI_ISL_2489211, EPI_ISL_2489288, EPI_ISL_2489322, EPI_ISL_2489333, EPI_ISL_2489351, EPI_ISL_2489352, EPI_ISL_2489384, EPI_ISL_2489387, EPI_ISL_2489451, EPI_ISL_2489510, EPI_ISL_2489593, EPI_ISL_2489627, EPI_ISL_2489628, EPI_ISL_2489640, EPI_ISL_2489677, EPI_ISL_2489704, EPI_ISL_2489719, EPI_ISL_2489772, EPI_ISL_2489775, EPI_ISL_2489778, EPI_ISL_2489780, EPI_ISL_2489788, EPI_ISL_2489794, EPI_ISL_2489796, EPI_ISL_2489799, EPI_ISL_2489812, EPI_ISL_2527874, EPI_ISL_2527876, EPI_ISL_2527878, EPI_ISL_2528010, EPI_ISL_2528470, EPI_ISL_2528494, EPI_ISL_2528889, EPI_ISL_2529020, EPI_ISL_2530983, EPI_ISL_2687051, EPI_ISL_2687092, EPI_ISL_2687121, EPI_ISL_2687160, EPI_ISL_2687186, EPI_ISL_2687260, EPI_ISL_2687293, EPI_ISL_2687329, EPI_ISL_2687572, EPI_ISL_2785286, EPI_ISL_2785333, EPI_ISL_2785337, EPI_ISL_2785366, EPI_ISL_2785421, EPI_ISL_2785515, EPI_ISL_2875636, EPI_ISL_2875657, EPI_ISL_2875762 | | | |
| see above | Aegis Sciences Corporation | Centers for Disease Control and Prevention Division of Viral Diseases, Pathogen Discovery | Adrian Paskey; Alec Vest; Benjamin Rambo-Martin; Christopher Gulvick; Clinton R. Paden; Cyndi Clark; Dakota Howard; Darlene Wagner; Dhwani Batra; Dillon Nall; Duncan MacCannell; Ethan Sanders; Holly Houdeshell; Jason Caravas; Kara Moser; Matthew Hardison; Matthew Schmerer; Ola Kvalvaag; Patrick Campbell; Peter W. Cook; Rob Case; Scott Sammons; Shatavia Morrison; Shaun Westlund; Vikramsinha Ghorpade; Yvette Unoarumhi |
| EPI_ISL_2279585 | BIO-REFERENCE LABORATORIES | Wadsworth Center, New York State Department of Health | Alexis Russell; Catharine Prussing; Daryl M. Lamson; Erasmus Schneider; Erica Lasek-Nesselquist; John Kelly; Jonathan Plitnick; Kirsten St. George; Matthew Shudt; Melissa A Leisner; Navjot Singh |
| EPI_ISL_2421082, EPI_ISL_2421083, EPI_ISL_2421084, EPI_ISL_2421085, EPI_ISL_2421086, EPI_ISL_2421087, EPI_ISL_2421088, EPI_ISL_2421089, EPI_ISL_2421090, EPI_ISL_2421092, EPI_ISL_2421093, EPI_ISL_2421094, EPI_ISL_2421095, EPI_ISL_2421189, EPI_ISL_2421190, EPI_ISL_2421191, EPI_ISL_2421192, EPI_ISL_2421193, EPI_ISL_2421194, EPI_ISL_2421195, EPI_ISL_2421196, EPI_ISL_2421197, EPI_ISL_2421198, EPI_ISL_2421199, EPI_ISL_2421200, EPI_ISL_2421201, EPI_ISL_2421202, EPI_ISL_2421203, EPI_ISL_2421204, EPI_ISL_2421205, EPI_ISL_2421206, EPI_ISL_2421207, EPI_ISL_2421208, EPI_ISL_2421209, EPI_ISL_2421210, EPI_ISL_2421211, EPI_ISL_2421230, EPI_ISL_2421231, EPI_ISL_2421232, EPI_ISL_2421233, EPI_ISL_2421234, EPI_ISL_2421235, EPI_ISL_2421236, EPI_ISL_2421237, EPI_ISL_2421238, EPI_ISL_2421239, EPI_ISL_2421240, EPI_ISL_2421241, EPI_ISL_2421242, EPI_ISL_2421243, EPI_ISL_2421244, EPI_ISL_2421245, EPI_ISL_2421246, EPI_ISL_2421247, EPI_ISL_2421248, EPI_ISL_2448304, EPI_ISL_2448308, EPI_ISL_2448309, EPI_ISL_2448311, EPI_ISL_2448313, EPI_ISL_2448314, EPI_ISL_2448315, EPI_ISL_2448316, EPI_ISL_2448317, EPI_ISL_2448319, EPI_ISL_2448320, EPI_ISL_2627135, EPI_ISL_2627136, EPI_ISL_2627137, EPI_ISL_2868461, EPI_ISL_2868462, EPI_ISL_2868464, EPI_ISL_2868465, EPI_ISL_2868468, EPI_ISL_2868475, EPI_ISL_2868478, EPI_ISL_2868484, EPI_ISL_2868521, EPI_ISL_2868543, EPI_ISL_2868544, EPI_ISL_2868554, EPI_ISL_2868609, EPI_ISL_2919191, EPI_ISL_2919192, EPI_ISL_2919193 | | | |
| see above | BOSTON HEART DIAGNOSTICS CORP | Wadsworth Center, New York State Department of Health | Alexis Russell; Catharine Prussing; Daryl M. Lamson; Erasmus Schneider; Erica Lasek-Nesselquist; John Kelly; Jonathan Plitnick; Kirsten St. George; Matthew Shudt; Melissa A Leisner; Navjot Singh |
| EPI_ISL_2991962, EPI_ISL_2991963, EPI_ISL_2991964, EPI_ISL_2991965 | Broad Institute Clinical Research Sequencing Platform | Infectious Disease Program, Broad Institute of Harvard and MIT | A.E.; Adams, G.; Anahtar, M.; B.L.; B.W.; Bauer, M.; Birren; Branda, J.; Carter, A.; Cerrato, F.; Chaluvadi, S.; Chapman; Cusick, C.; D.J.; DeRuff, K.; Flowers, K.; Gallagher, G.; Gladden-Young, A.; Gnirke, A.; Harris, J.; J.E.; K.J.; LaRocque, R.; Lagerborg, K.; Lemieux; Lin; Loreth, C.; MacInnis; Neumann, A.; Normandin, E.; P.C.; Park; Pierce, V.; Reilly, S.; Rosenberg, E.; Rudy, M.; Ryan, E.; S.B.; Sabeti; Shaw, B.; Siddle; Slater, D.; Smole, S.; Tomkins-Tinch, C.; Turbett, S. |
| EPI_ISL_1972027, EPI_ISL_2096714, EPI_ISL_2096716, EPI_ISL_2096717, EPI_ISL_2096718, EPI_ISL_2096719, EPI_ISL_2096720, EPI_ISL_2096721, EPI_ISL_2104331, EPI_ISL_2104332, EPI_ISL_2104333, EPI_ISL_2104334, EPI_ISL_2104335, EPI_ISL_2193151, EPI_ISL_2193152, EPI_ISL_2193153, EPI_ISL_2312443 | | | |
| see above | Broad Institute Clinical Research Sequencing Platform | Infectious Disease Program, Broad Institute of Harvard and MIT | Adams, G.; B.L.; B.W.; Bauer, M.; Birren; Blumenstiel, B.; Brown, C.; Carter, A.; Chaluvadi, S.; D.J.; DeFelice, M.; DeRuff, K.; Dodge, S.; Gabriel, S.; Gallagher, G.; Gladden-Young, A.; Granger, B.; J.E.; K.J.; Lagerborg, K.; Larkin, K.; Lee, M.; Lemieux; Lennon, N.; Loreth, C.; Madoff, L.; McGovern, S.; Meldrim, J.; Normandin, E.; P.C.; Park; Pearlman, L.; Reilly, S.; Rudy, M.; Sabeti; Siddle; Smole, S.; Tomkins-Tinch, C.; Vicente, G.; and MacInnis |
| EPI_ISL_2497184 | Cayuga Medical Center | Cornell Covid-19 Testing Laboratory | Brittany Cronk; Diego Diel; Elizabeth Plocharczyk; Leonardo Caserta; Melissa Laverack; Patrick Mitchell; Renee Anderson; Roopa Venugopalan |
| EPI_ISL_2500843, EPI_ISL_2500844, EPI_ISL_2500845, EPI_ISL_2500846, EPI_ISL_2500847, EPI_ISL_2500848 | Clinical Microbiology Laboratory, NewYork Presbyterian Hospital/Columbia University Irving Medical Center | Uhlemann Laboratory, Columbia University Irving Medical Center | Anne Kelley; Anne-Catrin Uhlemann; Maya Tagliavia; Medini K. Annavajhala |
| EPI_ISL_2151412, EPI_ISL_2151413, EPI_ISL_2179177, EPI_ISL_2179178, EPI_ISL_2322912, EPI_ISL_2322913, EPI_ISL_2400432, EPI_ISL_2466684, EPI_ISL_2466685 | | | |
| see above | DOHMH Central Harlem | New York City Public Health Laboratory | Jade Wang; et al. |
| EPI_ISL_2400426, EPI_ISL_2400436, EPI_ISL_2466687, EPI_ISL_2758638 | DOHMH Chelsea | New York City Public Health Laboratory | Jade Wang; et al. |
| EPI_ISL_2179169, EPI_ISL_2179181, EPI_ISL_2179182, EPI_ISL_2322903, EPI_ISL_2322904, EPI_ISL_2322905, EPI_ISL_2400434, EPI_ISL_2400435, EPI_ISL_2466682, EPI_ISL_2466683, EPI_ISL_2466688, EPI_ISL_2466690, EPI_ISL_2612553, EPI_ISL_2612554, EPI_ISL_2612555, EPI_ISL_2612556 | | | |
| see above | DOHMH Corona | New York City Public Health Laboratory | Jade Wang; et al. |
| EPI_ISL_2151414, EPI_ISL_2179174, EPI_ISL_2179185, EPI_ISL_2322906, EPI_ISL_2322907, EPI_ISL_2400430 | DOHMH Crown Heights | New York City Public Health Laboratory | Jade Wang; et al. |
| EPI_ISL_2151416, EPI_ISL_2151417, EPI_ISL_2151418, EPI_ISL_2179172, EPI_ISL_2179173, EPI_ISL_2322902, EPI_ISL_2322911, EPI_ISL_2758636 | | | |
| see above | DOHMH Fort Greene | New York City Public Health Laboratory | Jade Wang; et al. |
| EPI_ISL_2151415, EPI_ISL_2151419, EPI_ISL_2179180, EPI_ISL_2179186, EPI_ISL_2179187, EPI_ISL_2322914, EPI_ISL_2322915, EPI_ISL_2322916, EPI_ISL_2322917, EPI_ISL_2322918, EPI_ISL_2466686, EPI_ISL_2466689, EPI_ISL_2612557, EPI_ISL_2612558, EPI_ISL_2612559, EPI_ISL_2612562 | | | |
| see above | DOHMH Jamaica | New York City Public Health Laboratory | Jade Wang; et al. |
| EPI_ISL_2179167, EPI_ISL_2179168, EPI_ISL_2179176, EPI_ISL_2179183, EPI_ISL_2179184, EPI_ISL_2322900, EPI_ISL_2322901, EPI_ISL_2400425, EPI_ISL_2400433, EPI_ISL_2758635 | | | |
| see above | DOHMH Morrisania | New York City Public Health Laboratory | Jade Wang; et al. |
| EPI_ISL_2179170, EPI_ISL_2179171, EPI_ISL_2179179, EPI_ISL_2322908, EPI_ISL_2322909, EPI_ISL_2322910, EPI_ISL_2400431, EPI_ISL_2466681 | | | |
| see above | DOHMH PHL | New York City Public Health Laboratory | Jade Wang; et al. |
| EPI_ISL_2179175, EPI_ISL_2322919, EPI_ISL_2612560, EPI_ISL_2612561, EPI_ISL_2758637, EPI_ISL_2758639 | DOHMH Riverside | New York City Public Health Laboratory | Jade Wang; et al. |
| EPI_ISL_2179188, EPI_ISL_2179189, EPI_ISL_2400427, EPI_ISL_2466697 | Department of Homeless Services | New York City Public Health Laboratory | Jade Wang; et al. |
| EPI_ISL_2868259 | ELLIS HOSPITAL LABORATORY | Wadsworth Center, New | Alexis Russell; Catharine Prussing; Daryl M. Lamson; Erasmus Schneider; Erica Lasek-Nesselquist; John Kelly; Jonathan Plitnick; Kirsten St. George; Matthew Shudt; Melissa A Leisner; Navjot Singh |

| | | | |
|---|---|---|---|
| | | York State Department of Health | |
| EPI_ISL_2662829, EPI_ISL_2662830, EPI_ISL_2662831 | Erie County Medical Center (ECMC) | University at Buffalo Genomics and Bioinformatics Core | Alyssa Pohlman; Amanda Boccolucci; Brandon Marzullo; Donald Yergeau; Jennifer Surtees; Jonathan Bard; Natalie Lamb; Norma Nowak |
| EPI_ISL_2662917, EPI_ISL_2662918, EPI_ISL_2662919, EPI_ISL_2662920, EPI_ISL_2662921, EPI_ISL_2662922, EPI_ISL_2662923, EPI_ISL_2662924 | | | |
| see above | Erie County Public Health (ECPHL) | University at Buffalo Genomics and Bioinformatics Core | Alyssa Pohlman; Amanda Boccolucci; Brandon Marzullo; Donald Yergeau; Jennifer Surtees; Jonathan Bard; Natalie Lamb; Norma Nowak |
| EPI_ISL_2362354 | Flow Health | Infectious Disease Program, Broad Institute of Harvard and MIT | Adams, G.; B.L.; B.W.; Bauer, M.; Birren; Carter, A.; Chaluvadi, S.; D.J.; DeRuff, K.; Gallagher, G.; Gladden-Young, A.; J.E.; K.J.; Lagerborg, K.; Lemieux; Loreth, C.; MacInnis; Normandin, E.; P.C.; Park; Reilly, S.; Rudy, M.; Siddle; Smole, S.; Tomkins-Tinch, C.; and Sabeti |
| EPI_ISL_2280230, EPI_ISL_2280244, EPI_ISL_2280246, EPI_ISL_2480150, EPI_ISL_2480157, EPI_ISL_2480158, EPI_ISL_2480159, EPI_ISL_2480160, EPI_ISL_2480161, EPI_ISL_2480162, EPI_ISL_2480163, EPI_ISL_2480164, EPI_ISL_2480165, EPI_ISL_2480166, EPI_ISL_2480167, EPI_ISL_2480168, EPI_ISL_2525982, EPI_ISL_2525987, EPI_ISL_2525991, EPI_ISL_2526002, EPI_ISL_2526015, EPI_ISL_2526017, EPI_ISL_2526024, EPI_ISL_2526028, EPI_ISL_2598768, EPI_ISL_2598801, EPI_ISL_2598802, EPI_ISL_2598805, EPI_ISL_2598806, EPI_ISL_2686446, EPI_ISL_2686447, EPI_ISL_2686448, EPI_ISL_2686449, EPI_ISL_2686450, EPI_ISL_2686452, EPI_ISL_2686474, EPI_ISL_2686476, EPI_ISL_2686477, EPI_ISL_2686479, EPI_ISL_2686482, EPI_ISL_2686485, EPI_ISL_2686486, EPI_ISL_2686489, EPI_ISL_2686498, EPI_ISL_2686499, EPI_ISL_2686502, EPI_ISL_2686503, EPI_ISL_2686504, EPI_ISL_2686505, EPI_ISL_2686506, EPI_ISL_2686507, EPI_ISL_2686508, EPI_ISL_2686509, EPI_ISL_2686696, EPI_ISL_2686699, EPI_ISL_2686700, EPI_ISL_2686701, EPI_ISL_2686702, EPI_ISL_2686703, EPI_ISL_2686705, EPI_ISL_2686707, EPI_ISL_2686749, EPI_ISL_2686751, EPI_ISL_2686752, EPI_ISL_2686753, EPI_ISL_2686755, EPI_ISL_2686756, EPI_ISL_2686759, EPI_ISL_2686760, EPI_ISL_2686761, EPI_ISL_2785862, EPI_ISL_2785863, EPI_ISL_2785864, EPI_ISL_2785865, EPI_ISL_2785867, EPI_ISL_2785868, EPI_ISL_2785869, EPI_ISL_2785870, EPI_ISL_2785872, EPI_ISL_2873714, EPI_ISL_2873794, EPI_ISL_2873796, EPI_ISL_2873797, EPI_ISL_2873798, EPI_ISL_2873799, EPI_ISL_2873801, EPI_ISL_2873802, EPI_ISL_2873803, EPI_ISL_2873804, EPI_ISL_2873805, EPI_ISL_2873806, EPI_ISL_2873807, EPI_ISL_2873808, EPI_ISL_2873809, EPI_ISL_2873810, EPI_ISL_2873812, EPI_ISL_2873813, EPI_ISL_2873814, EPI_ISL_2873815, EPI_ISL_2929133, EPI_ISL_2929134, EPI_ISL_2929135, EPI_ISL_2929136, EPI_ISL_2929138, EPI_ISL_2929139, EPI_ISL_2929140, EPI_ISL_2929141, EPI_ISL_2929142, EPI_ISL_2929144, EPI_ISL_2929145, EPI_ISL_2929146, EPI_ISL_2929147, EPI_ISL_2929148, EPI_ISL_2929149, EPI_ISL_2929151, EPI_ISL_2929154, EPI_ISL_2929155, EPI_ISL_2929157, EPI_ISL_2929159, EPI_ISL_2929160, EPI_ISL_2929161 | | | |
| see above | Fulgent Genetics | Centers for Disease Control and Prevention Division of Viral Diseases, Pathogen Discovery | Adrian Paskey; Becky Tsai; Benafsh Sapra; Benjamin Rambo-Martin; Christopher Gulvick; Clinton R. Paden; Dakota Howard; Darlene Wagner; Dhwani Batra; Doreen Ng; Duncan MacCannell; Harry Gao; James Xie; Jason Caravas; John Gao; Joseph Fierro; Kara Moser; Matthew Schmerer; Mickey Li; Peter W. Cook; Scott Sammons; Shatavia Morrison; Yan Meng; Yvette Unoarumhi |
| EPI_ISL_2421229 | GLENS FALLS HOSPITAL LABORATORY | Wadsworth Center, New York State Department of Health | Alexis Russell; Catharine Prussing; Daryl M. Lamson; Erasmus Schneider; Erica Lasek-Nesselquist; John Kelly; Jonathan Plitnick; Kirsten St. George; Matthew Shudt; Melissa A Leisner; Navjot Singh |
| EPI_ISL_2421228 | Garnet Health Medical Center Laboratory | Wadsworth Center, New York State Department of Health | Alexis Russell; Catharine Prussing; Daryl M. Lamson; Erasmus Schneider; Erica Lasek-Nesselquist; John Kelly; Jonathan Plitnick; Kirsten St. George; Matthew Shudt; Melissa A Leisner; Navjot Singh |
| EPI_ISL_2537215, EPI_ISL_2602157, EPI_ISL_2614010 | Gravity Diagnostics, LLC | Gravity Diagnostics, LLC | Gravity Diagnostics |
| EPI_ISL_2248022, EPI_ISL_2269696, EPI_ISL_2269767, EPI_ISL_2270003, EPI_ISL_2320386, EPI_ISL_2320727, EPI_ISL_2321100, EPI_ISL_2440136, EPI_ISL_2440146, EPI_ISL_2599069 | | | |
| see above | Helix/Illumina | Centers for Disease Control and Prevention Division of Viral Diseases, Pathogen Discovery | Adrian Paskey; Alexandre Bolze; Ary Ascencio; Benjamin Rambo-Martin; Brad Sickler; Charlotte Rivera-Garcia; Christine Tran; Christopher Gulvick; Clinton R. Paden; Dakota Howard; Darlene Wagner; David Becker; Dhwani Batra; Duncan MacCannell; Efren Sandoval; Eileen de Feo; Elizabeth Cirulli; Eric Allen; Geraint Levan; James Lu; Jan Antico; Jason Caravas; Jason Nguyen; Jimmy Ramirez; Jingtao Liu; Kara Moser; Kelly Schiabor Barrett; Kim Gietzen; Magnus Isaksson; Marc Laurent; Matthew Schmerer; Matthew Tolentino; Nicole L. Washington; Peter W. Cook; Phil Febbo; Ryan Cho; Scott Sammons; Shannon Wickline; Shatavia Morrison; Sherry Wang; Simon White; Tyler Cassens; William Lee; Yvette Unoarumhi |
| EPI_ISL_2598286, EPI_ISL_2612299, EPI_ISL_2801459, EPI_ISL_2801471, EPI_ISL_2801472 | IVY3 Central Lab, Vanderbilt University Medical Center | Lauring Lab, University of Michigan, Department of Microbiology and Immunology | Valesano |
| EPI_ISL_2088913, EPI_ISL_2089185, EPI_ISL_2089237, EPI_ISL_2089329, EPI_ISL_2282329, EPI_ISL_2282900, EPI_ISL_2282902, EPI_ISL_2324063, EPI_ISL_2324139, EPI_ISL_2324144, EPI_ISL_2324145, EPI_ISL_2324151, EPI_ISL_2324153, EPI_ISL_2324168, EPI_ISL_2372802, EPI_ISL_2375782, EPI_ISL_2652711, EPI_ISL_2652793, EPI_ISL_2652852, EPI_ISL_2652967, EPI_ISL_2652969, EPI_ISL_2686896, EPI_ISL_2686897, EPI_ISL_2876049, EPI_ISL_2876068, EPI_ISL_2876069 | | | |
| see above | Infinity Biologix | Centers for Disease Control and Prevention Division of Viral Diseases, Pathogen Discovery | Adrian Paskey; Benjamin Rambo-Martin; Chirayu Goswami; Christian Bixby; Christopher Gulvick; Clinton R. Paden; Dakota Howard; Darlene Wagner; Dhwani Batra; Duncan MacCannell; Jason Caravas; Jonathan Schultz; Kara Moser; Matthew Schmerer; Peter W. Cook; Robin Grimwood; Russ Hager; Scott Sammons; Shatavia Morrison; Yihe Wang; Yvette Unoarumhi |
| EPI_ISL_2662969, EPI_ISL_2662970, EPI_ISL_2662971, EPI_ISL_2662972, EPI_ISL_2662973, EPI_ISL_2662974, EPI_ISL_2662975, EPI_ISL_2662976, EPI_ISL_2662977, EPI_ISL_2662978, EPI_ISL_2662979, EPI_ISL_2662980, EPI_ISL_2662981, EPI_ISL_2662982, EPI_ISL_2662983, EPI_ISL_2662984, EPI_ISL_2662985, EPI_ISL_2662986, EPI_ISL_2662987, EPI_ISL_2662988, EPI_ISL_2662989, EPI_ISL_2662990, EPI_ISL_2662991, EPI_ISL_2662992, EPI_ISL_2662993, EPI_ISL_2662994 | | | |
| see above | Kaleida Health Laboratories | University at Buffalo Genomics and Bioinformatics Core | Alyssa Pohlman; Amanda Boccolucci; Brandon Marzullo; Donald Yergeau; Jennifer Surtees; Jonathan Bard; Natalie Lamb; Norma Nowak |
| EPI_ISL_2045854, EPI_ISL_2045860, EPI_ISL_2181681, EPI_ISL_2181682, EPI_ISL_2181685, EPI_ISL_2181686, EPI_ISL_2181687, EPI_ISL_2181749, EPI_ISL_2181750, EPI_ISL_2181814, EPI_ISL_2181819, EPI_ISL_2181822, EPI_ISL_2181825, EPI_ISL_2181828, EPI_ISL_2181831, EPI_ISL_2181865, EPI_ISL_2181917, EPI_ISL_2182281, EPI_ISL_2182284, EPI_ISL_2182286, EPI_ISL_2182289, EPI_ISL_2182292, EPI_ISL_2182294, EPI_ISL_2182297, EPI_ISL_2182302, EPI_ISL_2182308, EPI_ISL_2182310, EPI_ISL_2182357, EPI_ISL_2182392, EPI_ISL_2182496, EPI_ISL_2182509, EPI_ISL_2182511, EPI_ISL_2182517, EPI_ISL_2182520, EPI_ISL_2182522, EPI_ISL_2182525, EPI_ISL_2182535, EPI_ISL_2182541, EPI_ISL_2182544, EPI_ISL_2182546, EPI_ISL_2182549, EPI_ISL_2182552, EPI_ISL_2182555, EPI_ISL_2182557, EPI_ISL_2182574, EPI_ISL_2182705, EPI_ISL_2182725, EPI_ISL_2182728, EPI_ISL_2182731, EPI_ISL_2182736, EPI_ISL_2182739, EPI_ISL_2182741, EPI_ISL_2182744, EPI_ISL_2182745, EPI_ISL_2182747, EPI_ISL_2182749, EPI_ISL_2182752, EPI_ISL_2182941, EPI_ISL_2182944, EPI_ISL_2182946, EPI_ISL_2182965, EPI_ISL_2182968, EPI_ISL_2183091, EPI_ISL_2183105, EPI_ISL_2183108, EPI_ISL_2183110, EPI_ISL_2183693, EPI_ISL_2183695, EPI_ISL_2183698, EPI_ISL_2183700, EPI_ISL_2183705, EPI_ISL_2183707, EPI_ISL_2183710, EPI_ISL_2183731, EPI_ISL_2183734, EPI_ISL_2183735, EPI_ISL_2183888, EPI_ISL_2183938, EPI_ISL_2183955, EPI_ISL_2184200, EPI_ISL_2184201, EPI_ISL_2184224, EPI_ISL_2184232, EPI_ISL_2184234, EPI_ISL_2184244, EPI_ISL_2184247, EPI_ISL_2184248, EPI_ISL_2184290, EPI_ISL_2184291, EPI_ISL_2184296, EPI_ISL_2184297, EPI_ISL_2184301, EPI_ISL_2184302, EPI_ISL_2184310, EPI_ISL_2184311, EPI_ISL_2184329, EPI_ISL_2184385, EPI_ISL_2184480, EPI_ISL_2184500, EPI_ISL_2184609, EPI_ISL_2184674, EPI_ISL_2184681, EPI_ISL_2184794, EPI_ISL_2184835, EPI_ISL_2184836, EPI_ISL_2184850, EPI_ISL_2305855, EPI_ISL_2305910, EPI_ISL_2305954, EPI_ISL_2305957, EPI_ISL_2306120, EPI_ISL_2306195, EPI_ISL_2306218, EPI_ISL_2306252, EPI_ISL_2306264, EPI_ISL_2306265, EPI_ISL_2306266, EPI_ISL_2306267, EPI_ISL_2306324, EPI_ISL_2306333, EPI_ISL_2306346, EPI_ISL_2306375, EPI_ISL_2306499, EPI_ISL_2306508, EPI_ISL_2306509, EPI_ISL_2306524, EPI_ISL_2306603, EPI_ISL_2306604, EPI_ISL_2306629, EPI_ISL_2306640, EPI_ISL_2306641, EPI_ISL_2306644, EPI_ISL_2306645, EPI_ISL_2306678, EPI_ISL_2306679, EPI_ISL_2306680, EPI_ISL_2306681, EPI_ISL_2306682, EPI_ISL_2306685, EPI_ISL_2306686, EPI_ISL_2306687, EPI_ISL_2306728, EPI_ISL_2306729, EPI_ISL_2306730, EPI_ISL_2306731, EPI_ISL_2306740, EPI_ISL_2306741, EPI_ISL_2306818, EPI_ISL_2306834, EPI_ISL_2306836, EPI_ISL_2306838, EPI_ISL_2306861, EPI_ISL_2306887, EPI_ISL_2306888, EPI_ISL_2306889, EPI_ISL_2306890, EPI_ISL_2306905, EPI_ISL_2306906, EPI_ISL_2306907, EPI_ISL_2306956, EPI_ISL_2306957, EPI_ISL_2306958, EPI_ISL_2306967, EPI_ISL_2306997, EPI_ISL_2307003, EPI_ISL_2307011, EPI_ISL_2324479, EPI_ISL_2324520, EPI_ISL_2324556, EPI_ISL_2324582, EPI_ISL_2324589, EPI_ISL_2398174, EPI_ISL_2398178, EPI_ISL_2398186, EPI_ISL_2398188, EPI_ISL_2398193, EPI_ISL_2398265, EPI_ISL_2398267, EPI_ISL_2398269, EPI_ISL_2398328, EPI_ISL_2398336, EPI_ISL_2398346, EPI_ISL_2398347, EPI_ISL_2398358, EPI_ISL_2398359, EPI_ISL_2398404, EPI_ISL_2398405, EPI_ISL_2398408, EPI_ISL_2398450, EPI_ISL_2398451, EPI_ISL_2398452, EPI_ISL_2398471, EPI_ISL_2398536, EPI_ISL_2398545, EPI_ISL_2398550, EPI_ISL_2398569, EPI_ISL_2398572, EPI_ISL_2398787, EPI_ISL_2398878, EPI_ISL_2398903, EPI_ISL_2398909, EPI_ISL_2399022, EPI_ISL_2399024, EPI_ISL_2399026, EPI_ISL_2399028, EPI_ISL_2399030, EPI_ISL_2399035, EPI_ISL_2399036, EPI_ISL_2399038, EPI_ISL_2399073, EPI_ISL_2399074, EPI_ISL_2399085, EPI_ISL_2399087, EPI_ISL_2399088, EPI_ISL_2399089, EPI_ISL_2399090, EPI_ISL_2399091, EPI_ISL_2399092, EPI_ISL_2399093, EPI_ISL_2399094, EPI_ISL_2399095, EPI_ISL_2399248, EPI_ISL_2399264, EPI_ISL_2399271, EPI_ISL_2399273, EPI_ISL_2399274, EPI_ISL_2399275, EPI_ISL_2399276, EPI_ISL_2399277, EPI_ISL_2399278, EPI_ISL_2399279, EPI_ISL_2399280, EPI_ISL_2399290, EPI_ISL_2399295, EPI_ISL_2399297, EPI_ISL_2399327, EPI_ISL_2399338, EPI_ISL_2399339, EPI_ISL_2399340, EPI_ISL_2480230, EPI_ISL_2480231, EPI_ISL_2480232, EPI_ISL_2480485, EPI_ISL_2480486, EPI_ISL_2480487, EPI_ISL_2480488, EPI_ISL_2480489, EPI_ISL_2480491, EPI_ISL_2480499, EPI_ISL_2480500, EPI_ISL_2480507, EPI_ISL_2480508, EPI_ISL_2480516, EPI_ISL_2480974, EPI_ISL_2481063, EPI_ISL_2481084, EPI_ISL_2481113, EPI_ISL_2481114, EPI_ISL_2481141, EPI_ISL_2481143, EPI_ISL_2481144, EPI_ISL_2481145, EPI_ISL_2481154, EPI_ISL_2481178, EPI_ISL_2481220, EPI_ISL_2481301, EPI_ISL_2481456, EPI_ISL_2611155, EPI_ISL_2611156, EPI_ISL_2611157, EPI_ISL_2611301, EPI_ISL_2611302, EPI_ISL_2611303, EPI_ISL_2611308, EPI_ISL_2611335, EPI_ISL_2611336, EPI_ISL_2611359, EPI_ISL_2611360, EPI_ISL_2611361, EPI_ISL_2611436, EPI_ISL_2611467, EPI_ISL_2611502, EPI_ISL_2611506, EPI_ISL_2611516, EPI_ISL_2611550, EPI_ISL_2611593, EPI_ISL_2611648, EPI_ISL_2611663, EPI_ISL_2611720, EPI_ISL_2611754, EPI_ISL_2611755, EPI_ISL_2872144, EPI_ISL_2872153, EPI_ISL_2872156, EPI_ISL_2872159, EPI_ISL_2872202, EPI_ISL_2872229, EPI_ISL_2872516, EPI_ISL_2872517, EPI_ISL_2872788, EPI_ISL_2872789, EPI_ISL_2872793, EPI_ISL_2872799, EPI_ISL_2872812, EPI_ISL_2872813, EPI_ISL_2872831, EPI_ISL_2872855, EPI_ISL_2872871, EPI_ISL_2872874, EPI_ISL_2872877, EPI_ISL_2872895, EPI_ISL_2872972, EPI_ISL_2872975, EPI_ISL_2873043, EPI_ISL_2873076, EPI_ISL_2926208, EPI_ISL_2926213, EPI_ISL_2926214, EPI_ISL_2926215, EPI_ISL_2926216, EPI_ISL_2926219, EPI_ISL_2926220, EPI_ISL_2926250, EPI_ISL_2926267, EPI_ISL_2926485, EPI_ISL_2926486, EPI_ISL_2926488, EPI_ISL_2926494, EPI_ISL_2927453, EPI_ISL_2927568, EPI_ISL_2927570, EPI_ISL_2927571, EPI_ISL_2927572, EPI_ISL_2927573, EPI_ISL_2927588, EPI_ISL_2927593, EPI_ISL_2927605, EPI_ISL_2927622, EPI_ISL_2927626, EPI_ISL_2927652, EPI_ISL_2927656, EPI_ISL_2927662, EPI_ISL_2927663, EPI_ISL_2927681, EPI_ISL_2927691, EPI_ISL_2927695 | | | |
| see above | Laboratory Corporation of America | Centers for Disease Control and Prevention Division of Viral Diseases, Pathogen Discovery | Adrian Paskey; Amanda Douglas; Amanda Suchanek; Andrea Throop; Ayla Burns; Benjamin Rambo-Martin; Bobbi Croy; Brian Krueger; Brian Norvell; Christopher Gulvick; Christos Petropoulos; Clinton R. Paden; Craig Lukasik; Dakota Howard; Darlene Wagner; Debbie Boles; Dhwani Batra; Duncan MacCannell; Eyad Almasri; Goran Stevovic; Howard Engler; Hrushikesh Deshmukh; Jake Humphrey; Jana Schroth; Jason Caravas; Joe Voshell; John Pruitt; Jonathan Meltzer; Jonathan Williams; Kara Moser; Kimberly Wagner; Lax Iyer; Lisa Pfefferle; Lyndon Tilson; Manoj Jain; Marcia Eisenberg; Mary Ann Cristobal; Mary Williamson; Matthew Robinson; Matthew Schmerer; Michael Levandoski; Mike Sapeta; Mindy Nye; Minoo Agarwal; Mohan Kolli; Nuthawin Charoensri; Oren Cohen; Peter W. Cook; Prashant Gupta; Qian Zeng; Rama Ghatti; Scott Parker; Scott Ryan; Scott Sammons; Shatavia Morrison; Stanley Letovsky; Steven Ragan; Suresh Babu Selvaraju; Susan Countryman; Susan Hicks; Suzanne Dale; Thomas Urban; Tim Kuphal; Tricia Zwiefelhofer; Vincent Drouillon; Yvette Unoarumhi |
| EPI_ISL_2696473 | Laboratory, Gravity Diagnostics | Laboratory, Gravity Diagnostics | Diagnostics, G. |
| EPI_ISL_2274614, EPI_ISL_2274615, EPI_ISL_2274616, EPI_ISL_2274617, EPI_ISL_2308788, EPI_ISL_2308789, EPI_ISL_2421096, EPI_ISL_2421097, EPI_ISL_2421098, EPI_ISL_2421099, EPI_ISL_2421100, EPI_ISL_2421101, EPI_ISL_2421102, EPI_ISL_2421103, EPI_ISL_2421104, EPI_ISL_2422323, EPI_ISL_2422324, EPI_ISL_2422325, EPI_ISL_2422326, EPI_ISL_2448298, EPI_ISL_2448302, EPI_ISL_2626993, EPI_ISL_2626994, EPI_ISL_2626995, EPI_ISL_2626996, EPI_ISL_2626997, EPI_ISL_2626998, EPI_ISL_2627143, EPI_ISL_2627144, EPI_ISL_2627145, EPI_ISL_2627146, EPI_ISL_2627147 | | | |
| see above | MONTEFIORE MEDICAL CENTER LABORATORIES | Wadsworth Center, New York State Department of Health | Alexis Russell; Catharine Prussing; Daryl M. Lamson; Erasmus Schneider; Erica Lasek-Nesselquist; John Kelly; Jonathan Plitnick; Kirsten St. George; Matthew Shudt; Melissa A Leisner; Navjot Singh |
| EPI_ISL_2689149, EPI_ISL_2690330, EPI_ISL_2690695, EPI_ISL_2690697, EPI_ISL_2690698 | MSK Microbiology Lab | MSK Microbiology Lab | Esther Babady; Krupa Jani; Tracy McMillen |
| EPI_ISL_2278797 | Maimonides Medical Center | Wadsworth Center, New York State Department of Health | Alexis Russell; Catharine Prussing; Daryl M. Lamson; Erasmus Schneider; Erica Lasek-Nesselquist; John Kelly; Jonathan Plitnick; Kirsten St. George; Matthew Shudt; Melissa A Leisner; Navjot Singh |
| EPI_ISL_2646943, EPI_ISL_2875488 | Mako Medical | Centers for Disease Control and Prevention Division of Viral Diseases, Pathogen Discovery | Adrian Paskey; Benjamin Rambo-Martin; Christopher Gulvick; Clinton R. Paden; Dakota Howard; Darlene Wagner; Dhwani Batra; Duncan MacCannell; Jason Caravas; Kara Moser; Lauren Moon; Matthew Schmerer; Matthew Tugwell; Peter W. Cook; Scott Sammons; Shatavia Morrison; Yvette Unoarumhi |
| EPI_ISL_2400443, EPI_ISL_2400444, EPI_ISL_2400445, EPI_ISL_2400446, EPI_ISL_2400447, EPI_ISL_2400448, EPI_ISL_2405094, EPI_ISL_2405095, EPI_ISL_2405096, EPI_ISL_2405097, EPI_ISL_2405107, EPI_ISL_2405108 | | | |
| see above | Molecular Biology Laboratory, Westchester Medical Center | Genomics Core Laboratory, New York Medical College | Changhong Yin; Jian Zhuge; Utsav Pandey; Weihua Huang |
| EPI_ISL_2627153, EPI_ISL_2627154, EPI_ISL_2627155, EPI_ISL_2627156, EPI_ISL_2627164, EPI_ISL_2627173, EPI_ISL_2627177, EPI_ISL_2627178, EPI_ISL_2627179, EPI_ISL_2727105, EPI_ISL_2868611, EPI_ISL_2868613, EPI_ISL_2868621, EPI_ISL_2868622 | | | |
| see above | NORTHWELL HEALTH LABORATORIES | Wadsworth Center, New York State Department of | Alexis Russell; Catharine Prussing; Daryl M. Lamson; Erasmus Schneider; Erica Lasek-Nesselquist; John Kelly; Jonathan Plitnick; Kirsten St. George; Matthew Shudt; Melissa A Leisner; Navjot Singh |

| | | | |
|---|---|---|---|
| | | Health | |
| EPI_ISL_2452047, EPI_ISL_2452048, EPI_ISL_2452050, EPI_ISL_2452052, EPI_ISL_2452064, EPI_ISL_2452066, EPI_ISL_2452067, EPI_ISL_2452071, EPI_ISL_2452073, EPI_ISL_2690057, EPI_ISL_2690058, EPI_ISL_2787777 | | | |
| see above | NYC Department of Health and Mental Hygiene | Centers for Disease Control and Prevention Division of Viral Diseases, Pathogen Discovery | Alison Laufer Halpin; Ben L. Rambo-Martin; Clinton R. Paden; Dakota Howard; Darlene Wagner; Dave Wentworth; Dhwani Batra; Jasmine Padilla; Justin Lee; Katie Dillon; Krista Queen; Kristen Knipe; Kristine Lacek; Mark Burroughs; Matthew Schmerer; Mili Sheth; Peter Cook; Sam Shepard; Sarah Nobles; Shoshona Le; Suxiang Tong; Vivien Dugan; Yvette Unoarumhi |
| EPI_ISL_2274678, EPI_ISL_2274680, EPI_ISL_2274681, EPI_ISL_2274682, EPI_ISL_2274683, EPI_ISL_2274684, EPI_ISL_2274689, EPI_ISL_2274690, EPI_ISL_2274693, EPI_ISL_2274694, EPI_ISL_2274697, EPI_ISL_2274698, EPI_ISL_2274703, EPI_ISL_2274704, EPI_ISL_2274705, EPI_ISL_2274706, EPI_ISL_2274707, EPI_ISL_2274709, EPI_ISL_2274712, EPI_ISL_2274713, EPI_ISL_2274714, EPI_ISL_2308797 | | | |
| see above | NYC Pandemic Response Lab | Wadsworth Center, New York State Department of Health | Alexis Russell; Catharine Prussing; Daryl M. Lamson; Erasmus Schneider; Erica Lasek-Nesselquist; John Kelly; Jonathan Plitnick; Kirsten St. George; Matthew Shudt; Melissa A Leisner; Navjot Singh |
| EPI_ISL_2161723, EPI_ISL_2161732, EPI_ISL_2161733, EPI_ISL_2161734, EPI_ISL_2161735, EPI_ISL_2161736, EPI_ISL_2161737, EPI_ISL_2161738, EPI_ISL_2161739, EPI_ISL_2161740, EPI_ISL_2161741, EPI_ISL_2161742, EPI_ISL_2161743, EPI_ISL_2161744, EPI_ISL_2161745, EPI_ISL_2161746, EPI_ISL_2161747, EPI_ISL_2161748, EPI_ISL_2161749, EPI_ISL_2161750, EPI_ISL_2161776, EPI_ISL_2161779, EPI_ISL_2161780, EPI_ISL_2202768, EPI_ISL_2202777, EPI_ISL_2202790, EPI_ISL_2202791, EPI_ISL_2202807, EPI_ISL_2427403, EPI_ISL_2427404, EPI_ISL_2427405, EPI_ISL_2427406, EPI_ISL_2427407, EPI_ISL_2427408, EPI_ISL_2427409, EPI_ISL_2427410, EPI_ISL_2427411, EPI_ISL_2427412, EPI_ISL_2427413, EPI_ISL_2427414, EPI_ISL_2427415, EPI_ISL_2427416, EPI_ISL_2427417, EPI_ISL_2427418, EPI_ISL_2427419, EPI_ISL_2427420, EPI_ISL_2427421, EPI_ISL_2427422, EPI_ISL_2427423, EPI_ISL_2427424, EPI_ISL_2427425, EPI_ISL_2427426, EPI_ISL_2427427, EPI_ISL_2427428, EPI_ISL_2427429, EPI_ISL_2427431, EPI_ISL_2427432, EPI_ISL_2427433, EPI_ISL_2427434, EPI_ISL_2427435, EPI_ISL_2427436, EPI_ISL_2427437, EPI_ISL_2427438, EPI_ISL_2427439, EPI_ISL_2427440, EPI_ISL_2427441, EPI_ISL_2427442, EPI_ISL_2427443, EPI_ISL_2427444, EPI_ISL_2427445, EPI_ISL_2427446, EPI_ISL_2427447, EPI_ISL_2427448, EPI_ISL_2427449, EPI_ISL_2427450, EPI_ISL_2427451, EPI_ISL_2427452, EPI_ISL_2427453, EPI_ISL_2427454, EPI_ISL_2427455, EPI_ISL_2427456, EPI_ISL_2427457, EPI_ISL_2427458, EPI_ISL_2427459, EPI_ISL_2427460, EPI_ISL_2427461, EPI_ISL_2427462, EPI_ISL_2427463, EPI_ISL_2427464, EPI_ISL_2427465, EPI_ISL_2427466, EPI_ISL_2427467, EPI_ISL_2427468, EPI_ISL_2427469, EPI_ISL_2427470, EPI_ISL_2427471, EPI_ISL_2427472, EPI_ISL_2427473, EPI_ISL_2427474, EPI_ISL_2427475, EPI_ISL_2427476, EPI_ISL_2427477, EPI_ISL_2427478, EPI_ISL_2427479, EPI_ISL_2427480, EPI_ISL_2427481, EPI_ISL_2427482, EPI_ISL_2427483, EPI_ISL_2443066, EPI_ISL_2443067, EPI_ISL_2458161, EPI_ISL_2458164, EPI_ISL_2458170, EPI_ISL_2458177, EPI_ISL_2458178, EPI_ISL_2458185, EPI_ISL_2458186, EPI_ISL_2458190, EPI_ISL_2458208, EPI_ISL_2663512, EPI_ISL_2663513, EPI_ISL_2663514, EPI_ISL_2663515, EPI_ISL_2663516, EPI_ISL_2663517, EPI_ISL_2663518, EPI_ISL_2663519, EPI_ISL_2663520, EPI_ISL_2663522, EPI_ISL_2663523, EPI_ISL_2663524, EPI_ISL_2663525, EPI_ISL_2663526, EPI_ISL_2663527, EPI_ISL_2663529, EPI_ISL_2663530, EPI_ISL_2663531, EPI_ISL_2663532, EPI_ISL_2663533, EPI_ISL_2663534, EPI_ISL_2663535, EPI_ISL_2663537, EPI_ISL_2663538, EPI_ISL_2663539, EPI_ISL_2663540, EPI_ISL_2663541, EPI_ISL_2663542, EPI_ISL_2663543, EPI_ISL_2663544, EPI_ISL_2663545, EPI_ISL_2663546, EPI_ISL_2663547, EPI_ISL_2663548, EPI_ISL_2663549, EPI_ISL_2663550, EPI_ISL_2663551, EPI_ISL_2663552, EPI_ISL_2753940, EPI_ISL_2753941, EPI_ISL_2753942, EPI_ISL_2753943, EPI_ISL_2753944, EPI_ISL_2753945, EPI_ISL_2753946, EPI_ISL_2753947, EPI_ISL_2753948, EPI_ISL_2753949, EPI_ISL_2753951, EPI_ISL_2753952 | | | |
| see above | NYU Langone Health | Departments of Pathology and Medicine, New York University School of Medicine | Adriana Heguy; Christian Marier; Dacia Dimartino; Emily Guzman; Gael Westby; Guiqing Wang; Paolo Cotzia; Paul Zappile; Peter Meyn; Sitharam Ramaswami; Yutong Zhang |
| EPI_ISL_2151411, EPI_ISL_2179190, EPI_ISL_2322920, EPI_ISL_2400428, EPI_ISL_2400429, EPI_ISL_2400442, EPI_ISL_2466700, EPI_ISL_2466701, EPI_ISL_2466702, EPI_ISL_2612563, EPI_ISL_2758645 | | | |
| see above | OCME Office Of Chief Medical Examiner | New York City Public Health Laboratory | Jade Wang; et al. |
| EPI_ISL_2274586 | OSWEGO HOSPITAL LABORATORY | Wadsworth Center, New York State Department of Health | Alexis Russell; Catharine Prussing; Daryl M. Lamson; Erasmus Schneider; Erica Lasek-Nesselquist; John Kelly; Jonathan Plitnick; Kirsten St. George; Matthew Shudt; Melissa A Leisner; Navjot Singh |
| EPI_ISL_2626972 | PECONIC BAY MEDICAL CENTER LABORATORY | Wadsworth Center, New York State Department of Health | Alexis Russell; Catharine Prussing; Daryl M. Lamson; Erasmus Schneider; Erica Lasek-Nesselquist; John Kelly; Jonathan Plitnick; Kirsten St. George; Matthew Shudt; Melissa A Leisner; Navjot Singh |
| EPI_ISL_2036965, EPI_ISL_2036966, EPI_ISL_2036967, EPI_ISL_2036975, EPI_ISL_2036980, EPI_ISL_2036982, EPI_ISL_2036985, EPI_ISL_2036986, EPI_ISL_2036989, EPI_ISL_2036990, EPI_ISL_2036994, EPI_ISL_2036995, EPI_ISL_2036996, EPI_ISL_2036997, EPI_ISL_2036998, EPI_ISL_2037002, EPI_ISL_2037003, EPI_ISL_2037007, EPI_ISL_2037009, EPI_ISL_2037015, EPI_ISL_2037018, EPI_ISL_2037019, EPI_ISL_2037022, EPI_ISL_2037027, EPI_ISL_2037036, EPI_ISL_2037037, EPI_ISL_2037041, EPI_ISL_2037043, EPI_ISL_2037044, EPI_ISL_2037045, EPI_ISL_2037046, EPI_ISL_2037051, EPI_ISL_2037052, EPI_ISL_2037055, EPI_ISL_2037057, EPI_ISL_2037060, EPI_ISL_2037066, EPI_ISL_2037067, EPI_ISL_2037068, EPI_ISL_2037069, EPI_ISL_2037072, EPI_ISL_2037075, EPI_ISL_2037077, EPI_ISL_2037081, EPI_ISL_2037085, EPI_ISL_2037090, EPI_ISL_2037091, EPI_ISL_2037105, EPI_ISL_2037107, EPI_ISL_2037109, EPI_ISL_2037111, EPI_ISL_2037112, EPI_ISL_2037113, EPI_ISL_2037114, EPI_ISL_2037116, EPI_ISL_2037117, EPI_ISL_2037119, EPI_ISL_2037122, EPI_ISL_2037125, EPI_ISL_2037126, EPI_ISL_2037127, EPI_ISL_2037130, EPI_ISL_2037132, EPI_ISL_2037136, EPI_ISL_2037139, EPI_ISL_2037142, EPI_ISL_2037143, EPI_ISL_2037144, EPI_ISL_2037147, EPI_ISL_2037148, EPI_ISL_2037150, EPI_ISL_2037151, EPI_ISL_2037158, EPI_ISL_2037160, EPI_ISL_2037161, EPI_ISL_2037165, EPI_ISL_2037170, EPI_ISL_2037171, EPI_ISL_2037173, EPI_ISL_2037178, EPI_ISL_2037179, EPI_ISL_2037184, EPI_ISL_2037185, EPI_ISL_2037186, EPI_ISL_2037189, EPI_ISL_2037194, EPI_ISL_2037195, EPI_ISL_2037196, EPI_ISL_2037197, EPI_ISL_2037198, EPI_ISL_2037200, EPI_ISL_2037202, EPI_ISL_2037203, EPI_ISL_2037204, EPI_ISL_2037205, EPI_ISL_2037206, EPI_ISL_2037207, EPI_ISL_2037208, EPI_ISL_2037209, EPI_ISL_2037210, EPI_ISL_2037211, EPI_ISL_2037212, EPI_ISL_2037213, EPI_ISL_2037214, EPI_ISL_2037215, EPI_ISL_2037216, EPI_ISL_2037217, EPI_ISL_2037218, EPI_ISL_2037219, EPI_ISL_2037220, EPI_ISL_2037221, EPI_ISL_2037222, EPI_ISL_2037223, EPI_ISL_2037224, EPI_ISL_2037225, EPI_ISL_2037226, EPI_ISL_2037227, EPI_ISL_2037228, EPI_ISL_2037229, EPI_ISL_2037230, EPI_ISL_2037231, EPI_ISL_2037232, EPI_ISL_2037233, EPI_ISL_2037234, EPI_ISL_2037235, EPI_ISL_2037236, EPI_ISL_2037237, EPI_ISL_2037238, EPI_ISL_2037239, EPI_ISL_2037240, EPI_ISL_2037241, EPI_ISL_2037242, EPI_ISL_2037243, EPI_ISL_2037244, EPI_ISL_2037245, EPI_ISL_2037246, EPI_ISL_2037247, EPI_ISL_2037248, EPI_ISL_2037249, EPI_ISL_2037250, EPI_ISL_2037251, EPI_ISL_2037252, EPI_ISL_2037253, EPI_ISL_2037254, EPI_ISL_2037255, EPI_ISL_2037256, EPI_ISL_2037257, EPI_ISL_2037258, EPI_ISL_2037259, EPI_ISL_2037260, EPI_ISL_2037261, EPI_ISL_2037262, EPI_ISL_2037263, EPI_ISL_2037264, EPI_ISL_2037265, EPI_ISL_2037266, EPI_ISL_2037267, EPI_ISL_2037268, EPI_ISL_2037269, EPI_ISL_2037270, EPI_ISL_2037271, EPI_ISL_2037272, EPI_ISL_2037274, EPI_ISL_2037275, EPI_ISL_2037276, EPI_ISL_2037277, EPI_ISL_2037278, EPI_ISL_2037279, EPI_ISL_2037280, EPI_ISL_2037281, EPI_ISL_2037282, EPI_ISL_2037283, EPI_ISL_2037284, EPI_ISL_2037285, EPI_ISL_2037286, EPI_ISL_2037287, EPI_ISL_2037288, EPI_ISL_2037289, EPI_ISL_2037290, EPI_ISL_2037291, EPI_ISL_2037292, EPI_ISL_2037293, EPI_ISL_2037294, EPI_ISL_2037295, EPI_ISL_2037296, EPI_ISL_2037297, EPI_ISL_2037298, EPI_ISL_2037299, EPI_ISL_2037300, EPI_ISL_2037301, EPI_ISL_2037302, EPI_ISL_2037303, EPI_ISL_2037304, EPI_ISL_2037305, EPI_ISL_2037306, EPI_ISL_2037307, EPI_ISL_2037308, EPI_ISL_2037309, EPI_ISL_2037310, EPI_ISL_2037311, EPI_ISL_2037312, EPI_ISL_2037313, EPI_ISL_2037314, EPI_ISL_2037315, EPI_ISL_2037316, EPI_ISL_2037317, EPI_ISL_2037318, EPI_ISL_2037319, EPI_ISL_2037320, EPI_ISL_2037321, EPI_ISL_2037322, EPI_ISL_2037323, EPI_ISL_2037324, EPI_ISL_2037325, EPI_ISL_2037326, EPI_ISL_2037327, EPI_ISL_2037328, EPI_ISL_2037329, EPI_ISL_2037330, EPI_ISL_2037331, EPI_ISL_2037332, EPI_ISL_2037333, EPI_ISL_2037334, EPI_ISL_2037335, EPI_ISL_2037336, EPI_ISL_2037337, EPI_ISL_2037338, EPI_ISL_2037339, EPI_ISL_2037340, EPI_ISL_2037341, EPI_ISL_2037342, EPI_ISL_2037343, EPI_ISL_2037344, EPI_ISL_2037345, EPI_ISL_2037346, EPI_ISL_2037347, EPI_ISL_2037348, EPI_ISL_2037349, EPI_ISL_2037350, EPI_ISL_2037351, EPI_ISL_2037352, EPI_ISL_2037353, EPI_ISL_2037354, EPI_ISL_2037355, EPI_ISL_2037356, EPI_ISL_2037357, EPI_ISL_2037358, EPI_ISL_2037359, EPI_ISL_2037360, EPI_ISL_2037361, EPI_ISL_2037362, EPI_ISL_2037363, EPI_ISL_2037364, EPI_ISL_2037365, EPI_ISL_2037366, EPI_ISL_2037367, EPI_ISL_2037368, EPI_ISL_2037369, EPI_ISL_2037370, EPI_ISL_2037371, EPI_ISL_2037372, EPI_ISL_2037373, EPI_ISL_2037374, EPI_ISL_2037375, EPI_ISL_2037376, EPI_ISL_2037377, EPI_ISL_2037378, EPI_ISL_2037379, EPI_ISL_2037380, EPI_ISL_2037381, EPI_ISL_2037382, EPI_ISL_2037383, EPI_ISL_2037384, EPI_ISL_2037385, EPI_ISL_2037386, EPI_ISL_2037387, EPI_ISL_2037388, EPI_ISL_2106283, EPI_ISL_2106284, EPI_ISL_2106285, EPI_ISL_2106286, EPI_ISL_2106287, EPI_ISL_2106288, EPI_ISL_2106289, EPI_ISL_2106290, EPI_ISL_2106291, EPI_ISL_2106292, EPI_ISL_2106293, EPI_ISL_2106294, EPI_ISL_2106295, EPI_ISL_2106296, EPI_ISL_2106297, EPI_ISL_2106298, EPI_ISL_2106299, EPI_ISL_2106300, EPI_ISL_2106301, EPI_ISL_2106302, EPI_ISL_2106303, EPI_ISL_2106304, EPI_ISL_2106305, EPI_ISL_2106306, EPI_ISL_2106307, EPI_ISL_2106308, EPI_ISL_2106309, EPI_ISL_2106310, EPI_ISL_2106311, EPI_ISL_2106312, EPI_ISL_2106313, EPI_ISL_2106314, EPI_ISL_2106315, EPI_ISL_2106316, EPI_ISL_2106317, EPI_ISL_2106318, EPI_ISL_2106319, EPI_ISL_2106320, EPI_ISL_2106321, EPI_ISL_2106322, EPI_ISL_2106323, EPI_ISL_2106324, EPI_ISL_2106325, EPI_ISL_2106326, EPI_ISL_2106327, EPI_ISL_2106328, EPI_ISL_2106329, EPI_ISL_2106330, EPI_ISL_2106331, EPI_ISL_2106332, EPI_ISL_2106333, EPI_ISL_2106334, EPI_ISL_2106335, EPI_ISL_2106336, EPI_ISL_2106337, EPI_ISL_2106338, EPI_ISL_2106339, EPI_ISL_2106340, EPI_ISL_2106341, EPI_ISL_2106342, EPI_ISL_2106343, EPI_ISL_2106344, EPI_ISL_2106345, EPI_ISL_2106346, EPI_ISL_2106347, EPI_ISL_2106348, EPI_ISL_2106349, EPI_ISL_2106350, EPI_ISL_2106351, EPI_ISL_2106352, EPI_ISL_2106353, EPI_ISL_2106354, EPI_ISL_2106355, EPI_ISL_2106356, EPI_ISL_2106357, EPI_ISL_2106358, EPI_ISL_2106359, EPI_ISL_2106360, EPI_ISL_2106361, EPI_ISL_2106362, EPI_ISL_2106363, EPI_ISL_2106364, EPI_ISL_2106365, EPI_ISL_2106366, EPI_ISL_2106367, EPI_ISL_2106368, EPI_ISL_2106369, EPI_ISL_2106370, EPI_ISL_2106371, EPI_ISL_2106372, EPI_ISL_2106373, EPI_ISL_2106374, EPI_ISL_2106375, EPI_ISL_2106376, EPI_ISL_2106377, EPI_ISL_2106378, EPI_ISL_2106379, EPI_ISL_2106380, EPI_ISL_2106381, EPI_ISL_2106382, EPI_ISL_2106383, EPI_ISL_2106384, EPI_ISL_2106385, EPI_ISL_2106386, EPI_ISL_2106387, EPI_ISL_2106388, EPI_ISL_2106389, EPI_ISL_2106390, EPI_ISL_2106391, EPI_ISL_2106392, EPI_ISL_2106393, EPI_ISL_2106394, EPI_ISL_2106395, EPI_ISL_2106396, EPI_ISL_2106397, EPI_ISL_2106398, EPI_ISL_2106399, EPI_ISL_2106400, EPI_ISL_2106401, EPI_ISL_2106402, EPI_ISL_2106403, EPI_ISL_2106404, EPI_ISL_2106405, EPI_ISL_2106406, EPI_ISL_2106407, EPI_ISL_2106408, EPI_ISL_2106409, EPI_ISL_2106410, EPI_ISL_2106411, EPI_ISL_2106412, EPI_ISL_2106413, EPI_ISL_2106414, EPI_ISL_2106415, EPI_ISL_2106416, EPI_ISL_2106417, EPI_ISL_2106418, EPI_ISL_2106419, EPI_ISL_2106420, EPI_ISL_2106421, EPI_ISL_2106422, EPI_ISL_2106423, EPI_ISL_2106424, EPI_ISL_2106425, EPI_ISL_2106426, EPI_ISL_2106427, EPI_ISL_2106428, EPI_ISL_2106429, EPI_ISL_2106430, EPI_ISL_2106431, EPI_ISL_2106432, EPI_ISL_2106433, EPI_ISL_2106434, EPI_ISL_2106435, EPI_ISL_2106436, EPI_ISL_2106437, EPI_ISL_2106438, EPI_ISL_2106439, EPI_ISL_2106440, EPI_ISL_2106441, EPI_ISL_2106442, EPI_ISL_2106443, EPI_ISL_2106444, EPI_ISL_2106445, EPI_ISL_2106446, EPI_ISL_2106447, EPI_ISL_2106448, EPI_ISL_2106449, EPI_ISL_2106450, EPI_ISL_2106451, EPI_ISL_2106452, EPI_ISL_2106453, EPI_ISL_2106454, EPI_ISL_2106455, EPI_ISL_2106456, EPI_ISL_2106457, EPI_ISL_2106458, EPI_ISL_2106459, EPI_ISL_2106460, EPI_ISL_2106461, EPI_ISL_2106462, EPI_ISL_2106463, EPI_ISL_2106464, EPI_ISL_2106465, EPI_ISL_2106466, EPI_ISL_2106467, EPI_ISL_2106468, EPI_ISL_2106469, EPI_ISL_2106470, EPI_ISL_2106471, EPI_ISL_2106472, EPI_ISL_2106473, EPI_ISL_2106474, EPI_ISL_2106475, EPI_ISL_2106477, EPI_ISL_2106478, EPI_ISL_2106479, EPI_ISL_2106480, EPI_ISL_2106481, EPI_ISL_2106482, EPI_ISL_2106483, EPI_ISL_2106484, EPI_ISL_2106485, EPI_ISL_2106486, EPI_ISL_2106487, EPI_ISL_2106488, EPI_ISL_2106489, EPI_ISL_2106490, EPI_ISL_2106491, EPI_ISL_2106492, EPI_ISL_2106493, EPI_ISL_2106494, EPI_ISL_2106495, EPI_ISL_2106496, EPI_ISL_2106497, EPI_ISL_2106498, EPI_ISL_2106499, EPI_ISL_2106500, EPI_ISL_2106501, EPI_ISL_2106502, EPI_ISL_2106503, EPI_ISL_2106504, EPI_ISL_2106505, EPI_ISL_2106506, EPI_ISL_2106507, EPI_ISL_2106508, EPI_ISL_2106509, EPI_ISL_2106510, EPI_ISL_2106512, EPI_ISL_2106513, EPI_ISL_2106514, EPI_ISL_2106515, EPI_ISL_2106516, EPI_ISL_2106517, EPI_ISL_2106518, EPI_ISL_2106519, EPI_ISL_2106520, EPI_ISL_2106521, EPI_ISL_2106522, EPI_ISL_2106523, EPI_ISL_2106524, EPI_ISL_2106525, EPI_ISL_2106526, EPI_ISL_2106527, EPI_ISL_2106528, EPI_ISL_2106529, EPI_ISL_2106530, EPI_ISL_2106531, EPI_ISL_2106532, EPI_ISL_2106533, EPI_ISL_2106534, EPI_ISL_2106535, EPI_ISL_2106536, EPI_ISL_2106537, EPI_ISL_2106538, EPI_ISL_2106539, EPI_ISL_2106540, EPI_ISL_2106541, EPI_ISL_2106542, EPI_ISL_2106543, EPI_ISL_2106544, EPI_ISL_2106545, EPI_ISL_2106546, EPI_ISL_2106547, EPI_ISL_2106548, EPI_ISL_2106549, EPI_ISL_2106550, EPI_ISL_2106551, EPI_ISL_2106552, EPI_ISL_2106553, EPI_ISL_2106554, EPI_ISL_2106555, EPI_ISL_2106556, EPI_ISL_2106557, EPI_ISL_2106558, EPI_ISL_2106559, EPI_ISL_2106560, EPI_ISL_2106561, EPI_ISL_2106562, EPI_ISL_2106563, EPI_ISL_2106564, EPI_ISL_2106565, EPI_ISL_2106566, EPI_ISL_2106567, EPI_ISL_2106568, EPI_ISL_2106569, EPI_ISL_2106570, EPI_ISL_2106571, EPI_ISL_2106572, EPI_ISL_2106573, EPI_ISL_2106574, EPI_ISL_2106575, EPI_ISL_2106576, EPI_ISL_2106577, EPI_ISL_2106578, EPI_ISL_2106579, EPI_ISL_2106580, EPI_ISL_2106581, EPI_ISL_2106582, EPI_ISL_2106583, EPI_ISL_2106584, EPI_ISL_2106585, EPI_ISL_2106586, EPI_ISL_2106587, EPI_ISL_2106588, EPI_ISL_2106589, EPI_ISL_2106590, EPI_ISL_2106591, EPI_ISL_2106592, EPI_ISL_2106593, EPI_ISL_2106594, EPI_ISL_2234908, EPI_ISL_2234909, EPI_ISL_2234910, EPI_ISL_2234911, EPI_ISL_2234912, EPI_ISL_2234913, EPI_ISL_2234914, EPI_ISL_2234915, EPI_ISL_2234916, EPI_ISL_2234917, EPI_ISL_2234918, EPI_ISL_2234919, EPI_ISL_2234920, EPI_ISL_2234921, EPI_ISL_2234922, EPI_ISL_2234923, EPI_ISL_2234924, EPI_ISL_2234925, EPI_ISL_2234926, EPI_ISL_2234927, EPI_ISL_2234928, EPI_ISL_2234929, EPI_ISL_2234930, EPI_ISL_2234931, EPI_ISL_2234932, EPI_ISL_2234933, EPI_ISL_2234934, EPI_ISL_2234935, EPI_ISL_2234936, EPI_ISL_2234937, EPI_ISL_2234938, EPI_ISL_2234939, EPI_ISL_2234940, EPI_ISL_2234941, EPI_ISL_2234942, EPI_ISL_2234943, EPI_ISL_2234944, EPI_ISL_2234945, EPI_ISL_2234946, EPI_ISL_2234947, EPI_ISL_2234948, EPI_ISL_2234949, EPI_ISL_2234950, EPI_ISL_2234951, EPI_ISL_2234952, EPI_ISL_2234953, EPI_ISL_2234954, EPI_ISL_2234955, EPI_ISL_2234956, EPI_ISL_2234957, EPI_ISL_2234958, EPI_ISL_2234959, EPI_ISL_2234960, EPI_ISL_2234961, EPI_ISL_2234962, EPI_ISL_2234963, EPI_ISL_2234964, EPI_ISL_2234965, EPI_ISL_2234966, EPI_ISL_2234967, EPI_ISL_2234968, EPI_ISL_2234969, EPI_ISL_2234970, EPI_ISL_2234971, EPI_ISL_2234972, EPI_ISL_2234973, EPI_ISL_2234974, EPI_ISL_2234975, EPI_ISL_2234976, EPI_ISL_2234977, EPI_ISL_2234978, EPI_ISL_2234979, EPI_ISL_2234980, EPI_ISL_2234981, EPI_ISL_2234982, EPI_ISL_2234983, EPI_ISL_2234984, EPI_ISL_2234985, EPI_ISL_2234986, EPI_ISL_2234987, EPI_ISL_2234988, EPI_ISL_2234989, EPI_ISL_2234990, EPI_ISL_2234991, EPI_ISL_2234992, EPI_ISL_2234993, EPI_ISL_2234994, EPI_ISL_2234995, EPI_ISL_2234996, EPI_ISL_2234997, EPI_ISL_2234998, EPI_ISL_2234999, EPI_ISL_2235000, EPI_ISL_2235001, EPI_ISL_2235002, EPI_ISL_2235003, EPI_ISL_2235004, EPI_ISL_2235005, EPI_ISL_2235006, EPI_ISL_2235007, EPI_ISL_2235008, EPI_ISL_2235009, EPI_ISL_2235010, EPI_ISL_2235011, EPI_ISL_2235012, EPI_ISL_2235013, EPI_ISL_2235014, EPI_ISL_2235015, EPI_ISL_2235016, EPI_ISL_2235017, EPI_ISL_2235018, EPI_ISL_2235019, EPI_ISL_2235020, EPI_ISL_2235021, EPI_ISL_2235022, EPI_ISL_2235023, EPI_ISL_2235024, EPI_ISL_2235025, EPI_ISL_2235026, EPI_ISL_2235027, EPI_ISL_2235028, EPI_ISL_2235029, EPI_ISL_2235030, EPI_ISL_2235031, EPI_ISL_2235032, EPI_ISL_2235033, EPI_ISL_2235034, EPI_ISL_2235035, EPI_ISL_2235036, EPI_ISL_2235037, EPI_ISL_2235038, EPI_ISL_2235039, EPI_ISL_2235040, EPI_ISL_2235041, EPI_ISL_2235042, EPI_ISL_2235043, EPI_ISL_2235044, EPI_ISL_2235045, EPI_ISL_2235046, EPI_ISL_2235047, EPI_ISL_2235048, EPI_ISL_2235049, EPI_ISL_2235050, EPI_ISL_2235051, EPI_ISL_2235052, EPI_ISL_2235053, EPI_ISL_2235054, EPI_ISL_2235055, EPI_ISL_2235056, EPI_ISL_2235057, EPI_ISL_2235058, EPI_ISL_2235059, EPI_ISL_2235060, EPI_ISL_2235061, EPI_ISL_2235062, EPI_ISL_2235063, EPI_ISL_2235064, EPI_ISL_2235065, EPI_ISL_2235066, EPI_ISL_2235067, EPI_ISL_2235068, EPI_ISL_2235069, EPI_ISL_2235070, EPI_ISL_2235071, EPI_ISL_2235072, EPI_ISL_2235073, EPI_ISL_2235074, EPI_ISL_2235075, EPI_ISL_2235076, EPI_ISL_2235077, EPI_ISL_2235078, EPI_ISL_2235079, EPI_ISL_2235080, EPI_ISL_2235081, EPI_ISL_2235082, EPI_ISL_2235083, EPI_ISL_2235084, EPI_ISL_2235085, EPI_ISL_2235086, EPI_ISL_2235087, EPI_ISL_2235088, EPI_ISL_2235089, EPI_ISL_2235090, EPI_ISL_2235091, EPI_ISL_2235092, EPI_ISL_2235093, EPI_ISL_2235094, EPI_ISL_2235095, EPI_ISL_2235096, EPI_ISL_2235097, EPI_ISL_2235098, EPI_ISL_2235099, EPI_ISL_2235100, EPI_ISL_2235101, EPI_ISL_2235102, EPI_ISL_2235103, EPI_ISL_2235104, EPI_ISL_2235105, EPI_ISL_2235106, EPI_ISL_2235107, EPI_ISL_2235108, EPI_ISL_2235109, EPI_ISL_2235110, EPI_ISL_2235111, EPI_ISL_2235112, EPI_ISL_2235113, EPI_ISL_2235114, EPI_ISL_2235115, EPI_ISL_2235116, EPI_ISL_2235117, EPI_ISL_2235118, EPI_ISL_2235119, EPI_ISL_2235120, EPI_ISL_2235121, EPI_ISL_2235123, EPI_ISL_2235124, EPI_ISL_2235125, EPI_ISL_2235126, EPI_ISL_2235127, EPI_ISL_2235128, EPI_ISL_2235129, EPI_ISL_2235130, EPI_ISL_2235131, EPI_ISL_2235132, EPI_ISL_2235133, EPI_ISL_2235134, EPI_ISL_2235135, EPI_ISL_2235136, EPI_ISL_2235137, EPI_ISL_2235138, EPI_ISL_2235139, EPI_ISL_2235140, EPI_ISL_2235141, EPI_ISL_2235142, EPI_ISL_2235143, EPI_ISL_2235144, EPI_ISL_2235145, EPI_ISL_2235146, EPI_ISL_2235147, EPI_ISL_2235148, EPI_ISL_2235149, EPI_ISL_2235150, EPI_ISL_2235151, EPI_ISL_2235152, EPI_ISL_2235153, EPI_ISL_2235154, EPI_ISL_2235155, EPI_ISL_2235156, EPI_ISL_2235157, EPI_ISL_2235158, EPI_ISL_2235159, EPI_ISL_2235160, EPI_ISL_2235161, EPI_ISL_2235162, EPI_ISL_2235163, EPI_ISL_2235164, EPI_ISL_2235165, EPI_ISL_2235166, EPI_ISL_2235167, EPI_ISL_2235168, EPI_ISL_2235169, EPI_ISL_2235170, EPI_ISL_2235171, EPI_ISL_2235172, EPI_ISL_2235173, EPI_ISL_2235174, EPI_ISL_2235175, EPI_ISL_2235176, EPI_ISL_2235177, EPI_ISL_2235178, EPI_ISL_2235179, EPI_ISL_2235180, EPI_ISL_2235181, EPI_ISL_2235182, EPI_ISL_2235183, EPI_ISL_2235184, EPI_ISL_2235185, EPI_ISL_2235186, EPI_ISL_2235187, EPI_ISL_2235188, EPI_ISL_2235189, EPI_ISL_2235190, EPI_ISL_2235191, EPI_ISL_2235192, EPI_ISL_2235193, EPI_ISL_2235194, EPI_ISL_2235195, EPI_ISL_2235196, EPI_ISL_2235197, EPI_ISL_2235198, EPI_ISL_2235199, EPI_ISL_2235200, EPI_ISL_2235201, EPI_ISL_2235202, EPI_ISL_2235203, EPI_ISL_2235204, EPI_ISL_2235205, EPI_ISL_2235206, EPI_ISL_2235207, EPI_ISL_2235208, EPI_ISL_2235209, EPI_ISL_2235210, EPI_ISL_2235211, EPI_ISL_2235212, EPI_ISL_2235213, EPI_ISL_2235214, EPI_ISL_2235215, EPI_ISL_2235216, EPI_ISL_2235217, EPI_ISL_2235218, EPI_ISL_2349615, EPI_ISL_2349616, EPI_ISL_2349617, EPI_ISL_2349618, EPI_ISL_2349619, EPI_ISL_2349620, EPI_ISL_2349621, EPI_ISL_2349623, EPI_ISL_2349624, EPI_ISL_2349625, EPI_ISL_2349626, EPI_ISL_2349627, EPI_ISL_2349628, EPI_ISL_2349629, EPI_ISL_2349630, EPI_ISL_2349631, EPI_ISL_2349632, EPI_ISL_2349633, EPI_ISL_2349634, EPI_ISL_2349635, EPI_ISL_2349636, EPI_ISL_2349637, EPI_ISL_2349638, EPI_ISL_2349639, EPI_ISL_2349640, EPI_ISL_2349641, EPI_ISL_2349642, EPI_ISL_2349643, EPI_ISL_2349644, EPI_ISL_2349645, EPI_ISL_2349646, EPI_ISL_2349647, EPI_ISL_2349648, EPI_ISL_2349649, EPI_ISL_2349650, EPI_ISL_2349651, EPI_ISL_2349652, EPI_ISL_2349653, EPI_ISL_2349654, EPI_ISL_2349655, EPI_ISL_2349656, EPI_ISL_2349657, EPI_ISL_2349658, EPI_ISL_2349659, EPI_ISL_2349660, EPI_ISL_2349661, EPI_ISL_2349662, EPI_ISL_2349663, EPI_ISL_2349664, EPI_ISL_2349665, EPI_ISL_2349666, EPI_ISL_2349667, EPI_ISL_2349668, EPI_ISL_2349669, EPI_ISL_2349670, EPI_ISL_2349671, EPI_ISL_2349672, EPI_ISL_2349673, EPI_ISL_2349674, EPI_ISL_2349675, EPI_ISL_2349676, EPI_ISL_2349677, EPI_ISL_2349678, EPI_ISL_2349679, EPI_ISL_2349680, EPI_ISL_2349681, EPI_ISL_2349682, EPI_ISL_2349683, EPI_ISL_2349684, EPI_ISL_2349685, EPI_ISL_2349686, EPI_ISL_2349687, EPI_ISL_2349688, EPI_ISL_2349689, EPI_ISL_2349690, EPI_ISL_2349691, EPI_ISL_2446035, EPI_ISL_2446036, EPI_ISL_2446037, EPI_ISL_2446038, EPI_ISL_2446039, EPI_ISL_2446040, EPI_ISL_2446041, EPI_ISL_2446042, EPI_ISL_2446043, EPI_ISL_2446044, EPI_ISL_2446045, EPI_ISL_2446046, EPI_ISL_2446047, EPI_ISL_2446048, EPI_ISL_2446049, EPI_ISL_2446050, EPI_ISL_2446051, EPI_ISL_2446052, EPI_ISL_2446053, EPI_ISL_2446054, EPI_ISL_2446055, EPI_ISL_2446056, EPI_ISL_2446057, EPI_ISL_2446058, EPI_ISL_2446059, EPI_ISL_2446060, EPI_ISL_2446061, EPI_ISL_2446062, EPI_ISL_2446063, EPI_ISL_2446064, EPI_ISL_2446065, EPI_ISL_2446066, EPI_ISL_2446067, EPI_ISL_2446068, EPI_ISL_2446069, EPI_ISL_2446070, EPI_ISL_2446071, EPI_ISL_2446072, EPI_ISL_2446073, EPI_ISL_2446074, EPI_ISL_2446075, EPI_ISL_2446076, EPI_ISL_2446077, EPI_ISL_2446078, EPI_ISL_2446079, EPI_ISL_2446080, EPI_ISL_2446081, EPI_ISL_2446082, EPI_ISL_2446083, EPI_ISL_2446084, EPI_ISL_2446085, EPI_ISL_2446086, EPI_ISL_2446087, EPI_ISL_2446088, EPI_ISL_2446089, EPI_ISL_2446090, EPI_ISL_2446091, EPI_ISL_2446092, EPI_ISL_2446093, EPI_ISL_2446094, EPI_ISL_2446096, EPI_ISL_2446099, EPI_ISL_2446102, EPI_ISL_2446105, EPI_ISL_2446107, EPI_ISL_2446111, EPI_ISL_2446114, EPI_ISL_2446117, EPI_ISL_2446120, EPI_ISL_2631338, EPI_ISL_2631339, EPI_ISL_2631340, EPI_ISL_2631341, EPI_ISL_2631342, EPI_ISL_2631343, EPI_ISL_2631344, EPI_ISL_2631345, EPI_ISL_2631346, EPI_ISL_2631347, EPI_ISL_2631348, EPI_ISL_2631349, EPI_ISL_2631350, EPI_ISL_2631351, EPI_ISL_2631352, EPI_ISL_2631353, EPI_ISL_2631354, EPI_ISL_2631355, EPI_ISL_2631356, EPI_ISL_2631357, EPI_ISL_2631358, EPI_ISL_2631359, EPI_ISL_2631360, EPI_ISL_2631361, EPI_ISL_2631362, EPI_ISL_2631363, EPI_ISL_2631364, EPI_ISL_2631365, EPI_ISL_2631366, EPI_ISL_2631367, EPI_ISL_2631368, EPI_ISL_2631369, EPI_ISL_2631370, EPI_ISL_2631371, EPI_ISL_2631372, EPI_ISL_2631373, EPI_ISL_2631374, EPI_ISL_2631375, EPI_ISL_2631376, EPI_ISL_2631377, EPI_ISL_2631378, EPI_ISL_2631379, EPI_ISL_2631380, EPI_ISL_2631381, EPI_ISL_2631382, EPI_ISL_2631383, EPI_ISL_2631384, EPI_ISL_2631385, EPI_ISL_2631386, EPI_ISL_2631387, EPI_ISL_2631388, EPI_ISL_2631389, EPI_ISL_2631390, EPI_ISL_2631391, EPI_ISL_2631392, EPI_ISL_2631393, EPI_ISL_2631394, EPI_ISL_2631395, EPI_ISL_2631396, EPI_ISL_2631397, EPI_ISL_2631398, EPI_ISL_2631399, EPI_ISL_2631400, EPI_ISL_2631401, EPI_ISL_2631402, EPI_ISL_2631403, EPI_ISL_2631404, EPI_ISL_2631405, EPI_ISL_2631406, EPI_ISL_2631407, EPI_ISL_2634916, EPI_ISL_2634918, EPI_ISL_2634920, EPI_ISL_2634922, EPI_ISL_2634923, EPI_ISL_2634925, EPI_ISL_2634927, EPI_ISL_2634929, EPI_ISL_2634931, EPI_ISL_2634933, EPI_ISL_2634934, EPI_ISL_2634936, EPI_ISL_2634938, EPI_ISL_2634940, EPI_ISL_2634942, EPI_ISL_2634943, EPI_ISL_2634945, EPI_ISL_2634947, EPI_ISL_2634949, EPI_ISL_2634951, EPI_ISL_2634952, EPI_ISL_2634954, EPI_ISL_2634956, EPI_ISL_2634958, EPI_ISL_2634959, EPI_ISL_2634961, EPI_ISL_2634963, EPI_ISL_2634965, EPI_ISL_2634967, EPI_ISL_2634969, EPI_ISL_2634971, EPI_ISL_2634973, EPI_ISL_2634974, EPI_ISL_2715652, EPI_ISL_2715653, EPI_ISL_2715654, EPI_ISL_2715655, EPI_ISL_2715656, EPI_ISL_2715657, EPI_ISL_2715658, EPI_ISL_2715659, EPI_ISL_2715660, EPI_ISL_2715661, EPI_ISL_2715662, EPI_ISL_2715663, EPI_ISL_2715664, EPI_ISL_2715665, EPI_ISL_2715666, EPI_ISL_2715667, EPI_ISL_2715668, EPI_ISL_2715669, EPI_ISL_2715670, EPI_ISL_2715671, EPI_ISL_2715672, EPI_ISL_2715673, EPI_ISL_2715674, EPI_ISL_2715675, EPI_ISL_2715676, EPI_ISL_2715677, EPI_ISL_2715678, EPI_ISL_2715679, EPI_ISL_2827469, EPI_ISL_2827470, EPI_ISL_2827471, EPI_ISL_2827472, EPI_ISL_2827473, EPI_ISL_2827474, EPI_ISL_2827475, EPI_ISL_2827476, EPI_ISL_2827477, EPI_ISL_2827478, EPI_ISL_2827479, EPI_ISL_2827480, EPI_ISL_2827481, EPI_ISL_2827482, EPI_ISL_2827483, EPI_ISL_2827484, EPI_ISL_2827485, EPI_ISL_2827486, EPI_ISL_2827487, EPI_ISL_2827488, EPI_ISL_2827489, EPI_ISL_2827490, EPI_ISL_2827491, EPI_ISL_2827492, EPI_ISL_2827493, EPI_ISL_2827494, EPI_ISL_2827495, EPI_ISL_2827496, EPI_ISL_2827497, EPI_ISL_2827498, EPI_ISL_2827499, EPI_ISL_2827500, EPI_ISL_2827501, EPI_ISL_2827502, EPI_ISL_2827503, EPI_ISL_2827504, EPI_ISL_2827505, EPI_ISL_2827506, EPI_ISL_2827507, EPI_ISL_2827508, EPI_ISL_2827509, EPI_ISL_2827510, EPI_ISL_2827511, EPI_ISL_2827512, EPI_ISL_2827513, EPI_ISL_2827514, EPI_ISL_2827515, EPI_ISL_2827516, EPI_ISL_2827517, EPI_ISL_2827518, EPI_ISL_2827519, EPI_ISL_2827520, EPI_ISL_2827521, EPI_ISL_2827522, EPI_ISL_2923907, EPI_ISL_2923909, EPI_ISL_2923914, EPI_ISL_2923915, EPI_ISL_2923917, EPI_ISL_2923918, EPI_ISL_2923920, EPI_ISL_2923925, EPI_ISL_2923928, EPI_ISL_2923929, EPI_ISL_2923930, EPI_ISL_2923932, EPI_ISL_2923934, EPI_ISL_2923936, EPI_ISL_2923939, EPI_ISL_2923942, EPI_ISL_2923943, EPI_ISL_2923946 | | | |
| see above | Pandemic Response Lab - NYC | Pandemic Response Lab, R&D | Cybill del Castillo; Dylan Law; Haiping Hao; Henry Lee; Isabel Fernandez Escapa; Jon Laurent; Katharine Nelson; Melissa Hopkins; Michael Hammerling; Pradeep Bugga; Shinyoung Clair Kang; Sol Rey; William Ward |
| EPI_ISL_2759146, EPI_ISL_2759147, EPI_ISL_2759148, EPI_ISL_2759149, EPI_ISL_2759150, EPI_ISL_2759151, EPI_ISL_2759191, EPI_ISL_2759193, EPI_ISL_2759194, EPI_ISL_2759195, EPI_ISL_2759196, EPI_ISL_2759197, EPI_ISL_2759221, EPI_ISL_2759222, EPI_ISL_2759223, EPI_ISL_2759224, EPI_ISL_2759225, EPI_ISL_2759226, EPI_ISL_2759227, EPI_ISL_2759228, EPI_ISL_2759229, EPI_ISL_2759230, EPI_ISL_2759231, EPI_ISL_2759232, EPI_ISL_2759233, EPI_ISL_2759234, EPI_ISL_2759235, EPI_ISL_2759236, EPI_ISL_2759237, EPI_ISL_2759238, EPI_ISL_2759239, EPI_ISL_2759240, EPI_ISL_2759241, EPI_ISL_2759242, EPI_ISL_2759243, EPI_ISL_2759244, EPI_ISL_2759245, EPI_ISL_2759246, EPI_ISL_2759247, EPI_ISL_2759248, EPI_ISL_2759249, EPI_ISL_2759250, EPI_ISL_2759251, EPI_ISL_2759252, EPI_ISL_2759253, EPI_ISL_2759254, EPI_ISL_2759255, EPI_ISL_2759256, EPI_ISL_2759257, EPI_ISL_2759258, EPI_ISL_2759259, EPI_ISL_2759260, EPI_ISL_2759261, EPI_ISL_2759262, EPI_ISL_2759263, EPI_ISL_2759264, EPI_ISL_2759265, EPI_ISL_2759266, EPI_ISL_2759267, EPI_ISL_2759268, EPI_ISL_2759269, EPI_ISL_2759270, EPI_ISL_2759328, EPI_ISL_2759329, EPI_ISL_2759330, EPI_ISL_2759331, EPI_ISL_2759332, EPI_ISL_2759333, EPI_ISL_2759334, EPI_ISL_2759335, EPI_ISL_2759336, EPI_ISL_2759337, EPI_ISL_2759338, EPI_ISL_2759339, EPI_ISL_2759340, EPI_ISL_2759341, EPI_ISL_2759342, EPI_ISL_2759343, EPI_ISL_2759344 | | | |
| see above | Population Medicine and Diagnostic Sciences, Cornell University | Population Medicine and Diagnostic Sciences, Cornell University | Anderson; B.D.; Caserta; Cronk; D.G.; Diel; L.C.; Laverack, M.; Mitchell; P.K.; Plocharczyk, E.; R.R.; Venugopalan, R. |
| EPI_ISL_2143046, EPI_ISL_2143068, EPI_ISL_2143164, EPI_ISL_2203953, EPI_ISL_2203980, EPI_ISL_2204038, EPI_ISL_2204054, EPI_ISL_2204139, EPI_ISL_2247107, EPI_ISL_2248467, EPI_ISL_2268297, EPI_ISL_2268675, EPI_ISL_2268737, EPI_ISL_2304502, EPI_ISL_2304509, EPI_ISL_2304536, EPI_ISL_2304551, EPI_ISL_2304567, EPI_ISL_2304583, EPI_ISL_2304584, EPI_ISL_2304594, EPI_ISL_2304610, EPI_ISL_2304647, EPI_ISL_2304649, EPI_ISL_2304653, EPI_ISL_2304656, EPI_ISL_2304659, EPI_ISL_2304663, EPI_ISL_2304667, EPI_ISL_2304669, EPI_ISL_2304672, EPI_ISL_2304673, EPI_ISL_2304675, EPI_ISL_2304678, EPI_ISL_2304682, EPI_ISL_2304685, EPI_ISL_2304687, EPI_ISL_2304697, EPI_ISL_2304701, EPI_ISL_2304702, EPI_ISL_2304708, EPI_ISL_2304709, EPI_ISL_2304715, EPI_ISL_2304718, EPI_ISL_2304733, EPI_ISL_2304735, EPI_ISL_2304746, EPI_ISL_2304750, EPI_ISL_2304753, EPI_ISL_2304754, EPI_ISL_2304759, EPI_ISL_2304760, EPI_ISL_2304764, EPI_ISL_2304768, EPI_ISL_2304769, EPI_ISL_2304794, EPI_ISL_2307143, EPI_ISL_2307160, EPI_ISL_2307161, EPI_ISL_2367700, EPI_ISL_2367705, EPI_ISL_2367724, EPI_ISL_2367756, EPI_ISL_2367764, EPI_ISL_2367801, EPI_ISL_2367829, EPI_ISL_2367864, EPI_ISL_2367896, EPI_ISL_2367922, EPI_ISL_2367931, EPI_ISL_2369028, EPI_ISL_2369070, EPI_ISL_2369106, EPI_ISL_2369116, EPI_ISL_2369158, EPI_ISL_2369161, EPI_ISL_2369170, EPI_ISL_2369186, EPI_ISL_2369200, EPI_ISL_2369203, EPI_ISL_2369228, EPI_ISL_2369231, EPI_ISL_2369255, EPI_ISL_2369258, EPI_ISL_2369276, EPI_ISL_2369296, EPI_ISL_2369360, EPI_ISL_2369384, EPI_ISL_2371692, EPI_ISL_2371698, EPI_ISL_2371883, EPI_ISL_2371902, EPI_ISL_2371913, EPI_ISL_2371914, EPI_ISL_2371921, EPI_ISL_2371964, EPI_ISL_2371985, EPI_ISL_2371995, EPI_ISL_2371996, EPI_ISL_2371997, EPI_ISL_2371998, EPI_ISL_2372030, EPI_ISL_2372040, EPI_ISL_2372042, EPI_ISL_2372043, EPI_ISL_2372056, EPI_ISL_2372060, EPI_ISL_2372062, EPI_ISL_2372077, EPI_ISL_2372078, EPI_ISL_2372096, EPI_ISL_2372100, EPI_ISL_2372102, EPI_ISL_2372103, EPI_ISL_2372108, EPI_ISL_2372109, EPI_ISL_2372110, EPI_ISL_2372124, EPI_ISL_2372125, EPI_ISL_2372133, EPI_ISL_2372134, EPI_ISL_2372138, EPI_ISL_2372422, EPI_ISL_2372929, EPI_ISL_2372932, EPI_ISL_2397416, EPI_ISL_2397433, EPI_ISL_2397439, EPI_ISL_2397459, EPI_ISL_2397464, EPI_ISL_2397475, EPI_ISL_2397493, EPI_ISL_2397509, EPI_ISL_2397546, EPI_ISL_2397563, EPI_ISL_2397572, EPI_ISL_2397577, EPI_ISL_2397605, EPI_ISL_2397606, EPI_ISL_2397630, EPI_ISL_2397635, EPI_ISL_2397640, EPI_ISL_2397648, EPI_ISL_2397668, EPI_ISL_2397696, EPI_ISL_2399475, EPI_ISL_2439300, EPI_ISL_2439427, EPI_ISL_2439478, EPI_ISL_2439494, EPI_ISL_2439506, EPI_ISL_2439518, EPI_ISL_2439536, EPI_ISL_2439538, | | | |

| | | | |
|---|---|---|---|
| EPI_ISL_2439832, EPI_ISL_2439834, EPI_ISL_2439852, EPI_ISL_2439855, EPI_ISL_2439856, EPI_ISL_2439867, EPI_ISL_2439868, EPI_ISL_2439887, EPI_ISL_2439889, EPI_ISL_2439896, EPI_ISL_2439900, EPI_ISL_2439903, EPI_ISL_2439904, EPI_ISL_2439906, EPI_ISL_2439912, EPI_ISL_2439918, EPI_ISL_2439919, EPI_ISL_2439921, EPI_ISL_2439922, EPI_ISL_2439925, EPI_ISL_2439933, EPI_ISL_2439935, EPI_ISL_2439942, EPI_ISL_2439944, EPI_ISL_2439959, EPI_ISL_2439961, EPI_ISL_2439962, EPI_ISL_2439971, EPI_ISL_2440648, EPI_ISL_2440800, EPI_ISL_2440829, EPI_ISL_2451376, EPI_ISL_2451388, EPI_ISL_2451404, EPI_ISL_2451424, EPI_ISL_2451433, EPI_ISL_2451478, EPI_ISL_2451488, EPI_ISL_2463182, EPI_ISL_2526777, EPI_ISL_2526792, EPI_ISL_2526811, EPI_ISL_2526993, EPI_ISL_2529340, EPI_ISL_2599757, EPI_ISL_2599846, EPI_ISL_2599860, EPI_ISL_2611828, EPI_ISL_2611830, EPI_ISL_2611834, EPI_ISL_2611841, EPI_ISL_2649967, EPI_ISL_2869040, EPI_ISL_2869125, EPI_ISL_2869161, EPI_ISL_2869252, EPI_ISL_2869309, EPI_ISL_2869310, EPI_ISL_2869478, EPI_ISL_2869559, EPI_ISL_2869584, EPI_ISL_2869614, EPI_ISL_2870243, EPI_ISL_2871078, EPI_ISL_2871131, EPI_ISL_2871161, EPI_ISL_2871202, EPI_ISL_2871208, EPI_ISL_2871337, EPI_ISL_2871352, EPI_ISL_2871376, EPI_ISL_2871465, EPI_ISL_2871515, EPI_ISL_2871569, EPI_ISL_2871589, EPI_ISL_2871595, EPI_ISL_2871846, EPI_ISL_2871849, EPI_ISL_2871931, EPI_ISL_2871957, EPI_ISL_2872024, EPI_ISL_2872071, EPI_ISL_2872074, EPI_ISL_2872109, EPI_ISL_2874026, EPI_ISL_2874713, EPI_ISL_2874793, EPI_ISL_2874803, EPI_ISL_2874830, EPI_ISL_2874853, EPI_ISL_2874893, EPI_ISL_2874934, EPI_ISL_2874963, EPI_ISL_2875160, EPI_ISL_2876668, EPI_ISL_2929220, EPI_ISL_2929286, EPI_ISL_2929350, EPI_ISL_2929361, EPI_ISL_2929405, EPI_ISL_2929426, EPI_ISL_2929429, EPI_ISL_2929528, EPI_ISL_2929571, EPI_ISL_2929573, EPI_ISL_2929594, EPI_ISL_2929724, EPI_ISL_2929746, EPI_ISL_2930082, EPI_ISL_2930245, EPI_ISL_2930283, EPI_ISL_2930318 | | | |
| see above | Quest Diagnostics Incorporated | Centers for Disease Control and Prevention Division of Viral Diseases, Pathogen Discovery | A. Gerasimova; A. Perez; Adrian Paskey; B. Anderson; Benjamin Rambo-Martin; Christopher Gulvick; Clinton R. Paden; Dakota Howard; Darlene Wagner; Dhwani Batra; Duncan MacCannell; F. Lacbawan; I. A. Shlyakhter; Jason Caravas; K.E. Livingston; Kara Moser; L.E. Bernstein; M. Hua; Matthew Schmerer; P. Tanpaiboon; Peter W. Cook; R. M. Kagan; R. Owen; R. V. Rolando; S. H. Rosenthal; Scott Sammons; Shatavia Morrison; Y. Liu; Yvette Unoarumhi |
| EPI_ISL_3091820, EPI_ISL_3091990, EPI_ISL_3092001 | Respiratory Viruses Branch, Centers for Disease Control and Prevention | Respiratory Viruses Branch, Centers for Disease Control and Prevention | Anderson, B.; Batra, D.; Bernstein, L.; C.R.; Caravas, J.; Cook; Gerasimova, A.; Gulvick, C.; Howard, D.; Hua, M.; I.A.; Kagan; Lacbawan, F.; Liu, Y.; Livingston, K.; MacCannell, D.; Morrison, S.; Moser, K.; Owen, R.; P.W.; Paden; Paskey, A.; Perez, A.; R.M.; R.V.; Rambo-Martin, B.; Rolando; Rosenthal; S.H.; Sammons, S.; Schmerer, M.; Shlyakhter; Tanpaiboon, P.; Unoarumhi, Y.; Wagner, D. |
| EPI_ISL_2627142 | SBH Health System | Wadsworth Center, New York State Department of Health | Alexis Russell; Catharine Prussing; Daryl M. Lamson; Erasmus Schneider; Erica Lasek-Nesselquist; John Kelly; Jonathan Plitnick; Kirsten St. George; Matthew Shudt; Melissa A Leisner; Navjot Singh |
| EPI_ISL_2274576, EPI_ISL_2274579, EPI_ISL_2274580, EPI_ISL_2274581, EPI_ISL_2274585, EPI_ISL_2421045, EPI_ISL_2421046, EPI_ISL_2421047, EPI_ISL_2421048, EPI_ISL_2421049, EPI_ISL_2421050, EPI_ISL_2421051, EPI_ISL_2421052, EPI_ISL_2421053, EPI_ISL_2422314, EPI_ISL_2422315, EPI_ISL_2422316, EPI_ISL_2422317, EPI_ISL_2422318, EPI_ISL_2422319, EPI_ISL_2422320, EPI_ISL_2422321, EPI_ISL_2422322, EPI_ISL_2448310, EPI_ISL_2627148, EPI_ISL_2627149, EPI_ISL_2627150, EPI_ISL_2627151, EPI_ISL_2627152, EPI_ISL_2868517, EPI_ISL_2868518, EPI_ISL_2868519, EPI_ISL_2868520, EPI_ISL_2868524, EPI_ISL_2868525, EPI_ISL_2868526, EPI_ISL_2868528, EPI_ISL_2868537 | | | |
| see above | STONY BROOK UNIVERSITY HOSPITAL | Wadsworth Center, New York State Department of Health | Alexis Russell; Catharine Prussing; Daryl M. Lamson; Erasmus Schneider; Erica Lasek-Nesselquist; John Kelly; Jonathan Plitnick; Kirsten St. George; Matthew Shudt; Melissa A Leisner; Navjot Singh |
| EPI_ISL_2098659, EPI_ISL_2274575, EPI_ISL_2274657, EPI_ISL_2274658, EPI_ISL_2274659, EPI_ISL_2274660, EPI_ISL_2274661, EPI_ISL_2274662, EPI_ISL_2274663, EPI_ISL_2278740, EPI_ISL_2278741, EPI_ISL_2279499, EPI_ISL_2279500, EPI_ISL_2279501, EPI_ISL_2279502, EPI_ISL_2421054, EPI_ISL_2421055, EPI_ISL_2421056, EPI_ISL_2421057, EPI_ISL_2421058, EPI_ISL_2421059, EPI_ISL_2421060, EPI_ISL_2421061, EPI_ISL_2421062, EPI_ISL_2421063, EPI_ISL_2421064, EPI_ISL_2421065, EPI_ISL_2421066, EPI_ISL_2421067, EPI_ISL_2421068, EPI_ISL_2421069, EPI_ISL_2421070, EPI_ISL_2421071, EPI_ISL_2421072, EPI_ISL_2421073, EPI_ISL_2421074, EPI_ISL_2421075, EPI_ISL_2421076, EPI_ISL_2421077, EPI_ISL_2421078, EPI_ISL_2421079, EPI_ISL_2421080, EPI_ISL_2421081, EPI_ISL_2422283, EPI_ISL_2422284, EPI_ISL_2422285, EPI_ISL_2422286, EPI_ISL_2422287, EPI_ISL_2422288, EPI_ISL_2422289, EPI_ISL_2422290, EPI_ISL_2422291, EPI_ISL_2422292, EPI_ISL_2422293, EPI_ISL_2422294, EPI_ISL_2422295, EPI_ISL_2422296, EPI_ISL_2422297, EPI_ISL_2422298, EPI_ISL_2422299, EPI_ISL_2422300, EPI_ISL_2422301, EPI_ISL_2422302, EPI_ISL_2422303, EPI_ISL_2422304, EPI_ISL_2422305, EPI_ISL_2422306, EPI_ISL_2422307, EPI_ISL_2422308, EPI_ISL_2422309, EPI_ISL_2422310, EPI_ISL_2422311, EPI_ISL_2422312, EPI_ISL_2448299, EPI_ISL_2868260, EPI_ISL_2868261, EPI_ISL_2868262, EPI_ISL_2868263, EPI_ISL_2868476, EPI_ISL_2868493, EPI_ISL_2868494, EPI_ISL_2868495, EPI_ISL_2868542, EPI_ISL_2868595, EPI_ISL_2919201 | | | |
| see above | SUNY UPSTATE MEDICAL UNIVERSITY | Wadsworth Center, New York State Department of Health | Alexis Russell; Catharine Prussing; Daryl M. Lamson; Erasmus Schneider; Erica Lasek-Nesselquist; John Kelly; Jonathan Plitnick; Kirsten St. George; Matthew Shudt; Melissa A Leisner; Navjot Singh |
| EPI_ISL_2421249, EPI_ISL_2421250, EPI_ISL_2421251, EPI_ISL_2421252, EPI_ISL_2627003, EPI_ISL_2627004, EPI_ISL_2627005, EPI_ISL_2627006 | | | |
| see above | Saratoga Hospital Laboratory | Wadsworth Center, New York State Department of Health | Alexis Russell; Catharine Prussing; Daryl M. Lamson; Erasmus Schneider; Erica Lasek-Nesselquist; John Kelly; Jonathan Plitnick; Kirsten St. George; Matthew Shudt; Melissa A Leisner; Navjot Singh |
| EPI_ISL_2466691, EPI_ISL_2466692, EPI_ISL_2466693, EPI_ISL_2466694, EPI_ISL_2466695, EPI_ISL_2466696 | Sherman Abrams Laboratory | New York City Public Health Laboratory | Jade Wang; et al. |
| EPI_ISL_2576919, EPI_ISL_2576922, EPI_ISL_2576923, EPI_ISL_2576924 | Sonic Reference Laboratory | Sonic Reference Laboratory | "Robert Moller; Bijaya Dhakal; Cora Lahey; Ellen Pizarek"; John Woody; Krishna Maddula |
| EPI_ISL_2466668, EPI_ISL_2466669, EPI_ISL_2466670 | Stony Brook Medicine | New York Genome Center | Alaa B. Salaheldin; Andre Corvelo; Dayna M. Oschwald; Dimitri Gnatenko; Eric D. Spitzer; Jiang Chen; Josue M. Flores Acuna; Kenneth R. Shroyer; Margaret M. McGovern; Michael Zody; Richard R. Kew; Samantha Fennessey; Sarah A. Halbohn; Sarah A. Malik; Silvia G. Spitzer; Susan V. Donelan; Tom Maniatis |
| EPI_ISL_2421161, EPI_ISL_2421162, EPI_ISL_2421163, EPI_ISL_2421164, EPI_ISL_2421165, EPI_ISL_2421166, EPI_ISL_2421167, EPI_ISL_2421168, EPI_ISL_2421169, EPI_ISL_2421170, EPI_ISL_2421171, EPI_ISL_2421172, EPI_ISL_2421173, EPI_ISL_2421174, EPI_ISL_2421175, EPI_ISL_2421176, EPI_ISL_2421177, EPI_ISL_2421178, EPI_ISL_2421179, EPI_ISL_2421180, EPI_ISL_2421181, EPI_ISL_2421182, EPI_ISL_2421183, EPI_ISL_2421184, EPI_ISL_2421185, EPI_ISL_2421186, EPI_ISL_2421187, EPI_ISL_2421188, EPI_ISL_2448300, EPI_ISL_2448305, EPI_ISL_2626925, EPI_ISL_2626926, EPI_ISL_2626927, EPI_ISL_2626928, EPI_ISL_2626929, EPI_ISL_2626930, EPI_ISL_2626931, EPI_ISL_2626932, EPI_ISL_2626933, EPI_ISL_2626934, EPI_ISL_2626935, EPI_ISL_2626936, EPI_ISL_2626937, EPI_ISL_2626938, EPI_ISL_2626939, EPI_ISL_2626940, EPI_ISL_2626941, EPI_ISL_2626942, EPI_ISL_2626943, EPI_ISL_2626944, EPI_ISL_2626945, EPI_ISL_2626946, EPI_ISL_2626947, EPI_ISL_2626948, EPI_ISL_2626949, EPI_ISL_2626950, EPI_ISL_2626951, EPI_ISL_2626952, EPI_ISL_2626953, EPI_ISL_2626954, EPI_ISL_2626955, EPI_ISL_2626956, EPI_ISL_2626957, EPI_ISL_2626958, EPI_ISL_2626959, EPI_ISL_2626960, EPI_ISL_2626961, EPI_ISL_2626962, EPI_ISL_2626963, EPI_ISL_2626964, EPI_ISL_2626965, EPI_ISL_2626966, EPI_ISL_2626967, EPI_ISL_2626968, EPI_ISL_2626969, EPI_ISL_2626970, EPI_ISL_2626971, EPI_ISL_2627115, EPI_ISL_2627116, EPI_ISL_2627117, EPI_ISL_2627118, EPI_ISL_2627119, EPI_ISL_2627120, EPI_ISL_2627121, EPI_ISL_2627122, EPI_ISL_2627123, EPI_ISL_2627124, EPI_ISL_2627125, EPI_ISL_2627126, EPI_ISL_2627127, EPI_ISL_2627128, EPI_ISL_2627129, EPI_ISL_2627130, EPI_ISL_2627131, EPI_ISL_2627132, EPI_ISL_2627133, EPI_ISL_2627134, EPI_ISL_2727112, EPI_ISL_2868548, EPI_ISL_2868550, EPI_ISL_2868560, EPI_ISL_2868564, EPI_ISL_2868565, EPI_ISL_2868566, EPI_ISL_2868567, EPI_ISL_2868569, EPI_ISL_2868573, EPI_ISL_2868577, EPI_ISL_2868579, EPI_ISL_2868580, EPI_ISL_2868582, EPI_ISL_2868584, EPI_ISL_2868586, EPI_ISL_2868588, EPI_ISL_2868598, EPI_ISL_2868602, EPI_ISL_2868604, EPI_ISL_2868605, EPI_ISL_2868607, EPI_ISL_2868615, EPI_ISL_2868617, EPI_ISL_2868619, EPI_ISL_2868620, EPI_ISL_2868623, EPI_ISL_2919203 | | | |
| see above | TEMPUS LABS INC | Wadsworth Center, New York State Department of Health | Alexis Russell; Catharine Prussing; Daryl M. Lamson; Erasmus Schneider; Erica Lasek-Nesselquist; John Kelly; Jonathan Plitnick; Kirsten St. George; Matthew Shudt; Melissa A Leisner; Navjot Singh |
| EPI_ISL_2230036 | The Jackson Laboratory | The Jackson Laboratory | Adams M; Bergeron D; Kelly K; Li L; Long J; Omerza G; Renzette N |
| EPI_ISL_2885699, EPI_ISL_2885700, EPI_ISL_2885701, EPI_ISL_2885702, EPI_ISL_2885703, EPI_ISL_2885704, EPI_ISL_2885705, EPI_ISL_2885706, EPI_ISL_2885707, EPI_ISL_2885708, EPI_ISL_2885709, EPI_ISL_2885710, EPI_ISL_2885711, EPI_ISL_2885712, EPI_ISL_2885713, EPI_ISL_2885714, EPI_ISL_2885715, EPI_ISL_2885716, EPI_ISL_2885717, EPI_ISL_2885718, EPI_ISL_2885719, EPI_ISL_2885720, EPI_ISL_2885721, EPI_ISL_2885722, EPI_ISL_2885723, EPI_ISL_2885724, EPI_ISL_2885725, EPI_ISL_2885726, EPI_ISL_2885727, EPI_ISL_2885728, EPI_ISL_2885729, EPI_ISL_2885730, EPI_ISL_2885731, EPI_ISL_2885732, EPI_ISL_2885733, EPI_ISL_2885734, EPI_ISL_2885735, EPI_ISL_2885736, EPI_ISL_2885737, EPI_ISL_2885738, EPI_ISL_2920207, EPI_ISL_2920208, EPI_ISL_2920209, EPI_ISL_2920210, EPI_ISL_2920211, EPI_ISL_2920212, EPI_ISL_2920213, EPI_ISL_2920214, EPI_ISL_2920215, EPI_ISL_2920216, EPI_ISL_2920217, EPI_ISL_2920218, EPI_ISL_2920219, EPI_ISL_2920220, EPI_ISL_2920221, EPI_ISL_2920222, EPI_ISL_2920223, EPI_ISL_2920224, EPI_ISL_2920225, EPI_ISL_2920226, EPI_ISL_2920227, EPI_ISL_2920228, EPI_ISL_2920229, EPI_ISL_2920230, EPI_ISL_2920231, EPI_ISL_2920232, EPI_ISL_2920233, EPI_ISL_2920234, EPI_ISL_2920235, EPI_ISL_2920236, EPI_ISL_2920237, EPI_ISL_2920238, EPI_ISL_2920239, EPI_ISL_2920240, EPI_ISL_2920241, EPI_ISL_2920242, EPI_ISL_2920243, EPI_ISL_2920244, EPI_ISL_2920245, EPI_ISL_2920246, EPI_ISL_2920247, EPI_ISL_2920248, EPI_ISL_2920249 | | | |
| see above | URMC LABS | URMC LABS | Andrew Cameron; Dwight Hardy; Ghinwa Dumyati; Kelly DeLary; Stepan Voljehn |
| EPI_ISL_2274631, EPI_ISL_2274635, EPI_ISL_2274636, EPI_ISL_2274637, EPI_ISL_2274638, EPI_ISL_2274639, EPI_ISL_2274640, EPI_ISL_2274641, EPI_ISL_2274642, EPI_ISL_2274643, EPI_ISL_2274644, EPI_ISL_2274645, EPI_ISL_2274646, EPI_ISL_2274665, EPI_ISL_2274666, EPI_ISL_2274667, EPI_ISL_2274668, EPI_ISL_2274669, EPI_ISL_2274670, EPI_ISL_2274671, EPI_ISL_2274672, EPI_ISL_2274673, EPI_ISL_2274674, EPI_ISL_2274675, EPI_ISL_2278762, EPI_ISL_2278764, EPI_ISL_2278766, EPI_ISL_2278768, EPI_ISL_2278769, EPI_ISL_2278773, EPI_ISL_2278774, EPI_ISL_2278775, EPI_ISL_2278776, EPI_ISL_2278777, EPI_ISL_2278778, EPI_ISL_2278779, EPI_ISL_2278780, EPI_ISL_2278781, EPI_ISL_2278782, EPI_ISL_2278783, EPI_ISL_2278784, EPI_ISL_2278785, EPI_ISL_2278786, EPI_ISL_2278787, EPI_ISL_2278788, EPI_ISL_2278789, EPI_ISL_2278790, EPI_ISL_2278791, EPI_ISL_2278792, EPI_ISL_2278793, EPI_ISL_2278794, EPI_ISL_2278795, EPI_ISL_2278796, EPI_ISL_2279506, EPI_ISL_2279507, EPI_ISL_2279508, EPI_ISL_2279509, EPI_ISL_2279510, EPI_ISL_2279511, EPI_ISL_2279512, EPI_ISL_2279513, EPI_ISL_2279514, EPI_ISL_2279515, EPI_ISL_2279516, EPI_ISL_2279517, EPI_ISL_2279518, EPI_ISL_2279519, EPI_ISL_2279520, EPI_ISL_2279521, EPI_ISL_2279522, EPI_ISL_2279523, EPI_ISL_2279524, EPI_ISL_2279525, EPI_ISL_2279526, EPI_ISL_2279527, EPI_ISL_2279528, EPI_ISL_2279529, EPI_ISL_2279530, EPI_ISL_2279531, EPI_ISL_2279532, EPI_ISL_2279533, EPI_ISL_2279534, EPI_ISL_2279535, EPI_ISL_2279536, EPI_ISL_2279537, EPI_ISL_2279538, EPI_ISL_2279539, EPI_ISL_2279540, EPI_ISL_2279541, EPI_ISL_2279542, EPI_ISL_2279543, EPI_ISL_2279544, EPI_ISL_2279545, EPI_ISL_2279546, EPI_ISL_2279547, EPI_ISL_2279548, EPI_ISL_2279549, EPI_ISL_2279550, EPI_ISL_2279551, EPI_ISL_2279552, EPI_ISL_2279553, EPI_ISL_2279554, EPI_ISL_2279555, EPI_ISL_2279556, EPI_ISL_2279557, EPI_ISL_2279558, EPI_ISL_2279559, EPI_ISL_2279560, EPI_ISL_2279561, EPI_ISL_2279562, EPI_ISL_2279563, EPI_ISL_2279564, EPI_ISL_2279565, EPI_ISL_2279566, EPI_ISL_2279567, EPI_ISL_2279568, EPI_ISL_2279569, EPI_ISL_2279570, EPI_ISL_2279571, EPI_ISL_2279572, EPI_ISL_2279573, EPI_ISL_2279574, EPI_ISL_2279575, EPI_ISL_2279576, EPI_ISL_2279577, EPI_ISL_2279578, EPI_ISL_2279579, EPI_ISL_2279580, EPI_ISL_2279581, EPI_ISL_2279582, EPI_ISL_2279583, EPI_ISL_2279584, EPI_ISL_2308791, EPI_ISL_2308793, EPI_ISL_2308794, EPI_ISL_2308800, EPI_ISL_2308801, EPI_ISL_2308803, EPI_ISL_2308804, EPI_ISL_2308805, EPI_ISL_2308806, EPI_ISL_2421144, EPI_ISL_2421145, EPI_ISL_2421146, EPI_ISL_2421147, EPI_ISL_2421148, EPI_ISL_2421149, EPI_ISL_2421150, EPI_ISL_2421151, EPI_ISL_2421152, EPI_ISL_2421153, EPI_ISL_2421154, EPI_ISL_2421155, EPI_ISL_2421156, EPI_ISL_2421223, EPI_ISL_2421224, EPI_ISL_2421225, EPI_ISL_2421226, EPI_ISL_2421227, EPI_ISL_2422327, EPI_ISL_2422328, EPI_ISL_2422329, EPI_ISL_2422330, EPI_ISL_2422331, EPI_ISL_2422332, EPI_ISL_2422333, EPI_ISL_2422334, EPI_ISL_2422335, EPI_ISL_2422336, EPI_ISL_2422337, EPI_ISL_2422338, EPI_ISL_2422339, EPI_ISL_2422340, EPI_ISL_2422341, EPI_ISL_2422342, EPI_ISL_2422343, EPI_ISL_2422344, EPI_ISL_2422345, EPI_ISL_2422346, EPI_ISL_2422347, EPI_ISL_2422348, EPI_ISL_2422349, EPI_ISL_2422350, EPI_ISL_2422351, EPI_ISL_2422352, EPI_ISL_2422353, EPI_ISL_2422354, EPI_ISL_2422355, EPI_ISL_2422356, EPI_ISL_2422357, EPI_ISL_2422358, EPI_ISL_2422359, EPI_ISL_2422360, EPI_ISL_2448303, EPI_ISL_2448306, EPI_ISL_2448307, EPI_ISL_2448318, EPI_ISL_2626973, EPI_ISL_2626974, EPI_ISL_2626975, EPI_ISL_2626976, EPI_ISL_2626977, EPI_ISL_2626978, EPI_ISL_2626979, EPI_ISL_2626980, EPI_ISL_2626981, EPI_ISL_2626982, EPI_ISL_2626983, EPI_ISL_2626984, EPI_ISL_2626985, EPI_ISL_2626986, EPI_ISL_2626987, EPI_ISL_2626988, EPI_ISL_2626989, EPI_ISL_2626990, EPI_ISL_2626991, EPI_ISL_2626992, EPI_ISL_2627007, EPI_ISL_2627008, EPI_ISL_2627009, EPI_ISL_2627010, EPI_ISL_2627012, EPI_ISL_2627013, EPI_ISL_2627014, EPI_ISL_2627015, EPI_ISL_2627016, EPI_ISL_2627017, EPI_ISL_2627018, EPI_ISL_2627019, EPI_ISL_2627020, EPI_ISL_2627021, EPI_ISL_2627022, EPI_ISL_2627023, EPI_ISL_2627024, EPI_ISL_2627025, EPI_ISL_2627026, EPI_ISL_2627027, EPI_ISL_2627028, EPI_ISL_2627029, EPI_ISL_2627030, EPI_ISL_2627031, EPI_ISL_2627032, EPI_ISL_2627033, EPI_ISL_2627034, EPI_ISL_2627035, EPI_ISL_2627036, EPI_ISL_2627037, EPI_ISL_2627038, EPI_ISL_2627039, EPI_ISL_2627040, EPI_ISL_2627041, EPI_ISL_2627042, EPI_ISL_2627043, EPI_ISL_2627044, EPI_ISL_2627045, EPI_ISL_2627046, EPI_ISL_2627047, EPI_ISL_2627048, EPI_ISL_2627049, EPI_ISL_2627050, EPI_ISL_2627051, EPI_ISL_2627052, EPI_ISL_2627053, EPI_ISL_2627054, EPI_ISL_2627055, EPI_ISL_2627056, EPI_ISL_2627057, EPI_ISL_2627058, EPI_ISL_2627059, EPI_ISL_2627060, EPI_ISL_2627061, EPI_ISL_2627062, EPI_ISL_2627063, EPI_ISL_2627064, EPI_ISL_2627065, EPI_ISL_2627066, EPI_ISL_2627067, EPI_ISL_2627068, EPI_ISL_2627069, EPI_ISL_2627070, EPI_ISL_2627071, EPI_ISL_2627072, EPI_ISL_2627073, EPI_ISL_2627074, EPI_ISL_2627075, EPI_ISL_2627076, EPI_ISL_2627077, EPI_ISL_2627078, EPI_ISL_2627079, EPI_ISL_2627080, EPI_ISL_2627081, EPI_ISL_2627082, EPI_ISL_2627083, EPI_ISL_2627084, EPI_ISL_2627085, EPI_ISL_2627086, EPI_ISL_2627087, EPI_ISL_2627088, EPI_ISL_2627089, EPI_ISL_2627090, EPI_ISL_2627091, EPI_ISL_2627092, EPI_ISL_2627093, EPI_ISL_2627094, EPI_ISL_2627095, EPI_ISL_2627096, EPI_ISL_2627097, EPI_ISL_2627098, EPI_ISL_2627099, EPI_ISL_2627100, EPI_ISL_2627101, EPI_ISL_2627102, EPI_ISL_2627103, EPI_ISL_2627104, EPI_ISL_2627105, EPI_ISL_2627106, EPI_ISL_2627107, EPI_ISL_2627108, EPI_ISL_2627109, EPI_ISL_2627110, EPI_ISL_2627111, EPI_ISL_2627112, EPI_ISL_2627113, EPI_ISL_2627114, EPI_ISL_2727102, EPI_ISL_2727106, EPI_ISL_2727107, EPI_ISL_2727108, EPI_ISL_2868238, EPI_ISL_2868239, EPI_ISL_2868240, EPI_ISL_2868241, EPI_ISL_2868242, EPI_ISL_2868243, EPI_ISL_2868244, EPI_ISL_2868245, EPI_ISL_2868246, EPI_ISL_2868247, EPI_ISL_2868248, EPI_ISL_2868249, EPI_ISL_2868250, EPI_ISL_2868251, EPI_ISL_2868252, EPI_ISL_2868253, EPI_ISL_2868254, EPI_ISL_2868255, EPI_ISL_2868256, EPI_ISL_2868257, EPI_ISL_2868258, EPI_ISL_2868264, EPI_ISL_2868265, EPI_ISL_2868266, EPI_ISL_2868267, EPI_ISL_2868268, EPI_ISL_2868269, EPI_ISL_2868270, EPI_ISL_2868271, EPI_ISL_2868272, EPI_ISL_2868273, EPI_ISL_2868274, EPI_ISL_2868275, EPI_ISL_2868276, EPI_ISL_2868277, EPI_ISL_2868278, EPI_ISL_2868279, EPI_ISL_2868280, EPI_ISL_2868281, EPI_ISL_2868282, EPI_ISL_2868283, EPI_ISL_2868284, EPI_ISL_2868285, EPI_ISL_2868286, EPI_ISL_2868287, EPI_ISL_2868288, EPI_ISL_2868289, EPI_ISL_2868290, EPI_ISL_2868291, EPI_ISL_2868292, EPI_ISL_2868293, EPI_ISL_2868294, EPI_ISL_2868295, EPI_ISL_2868296, EPI_ISL_2868297, EPI_ISL_2868298, EPI_ISL_2868299, EPI_ISL_2868300, EPI_ISL_2868301, EPI_ISL_2868302, EPI_ISL_2868303, EPI_ISL_2868304, EPI_ISL_2868305, EPI_ISL_2868306, EPI_ISL_2868307, EPI_ISL_2868308, EPI_ISL_2868309, EPI_ISL_2868310, EPI_ISL_2868311, EPI_ISL_2868312, EPI_ISL_2868313, EPI_ISL_2868314, EPI_ISL_2868315, EPI_ISL_2868316, EPI_ISL_2868317, EPI_ISL_2868318, EPI_ISL_2868319, EPI_ISL_2868320, EPI_ISL_2868321, EPI_ISL_2868322, EPI_ISL_2868323, EPI_ISL_2868324, EPI_ISL_2868325, EPI_ISL_2868326, EPI_ISL_2868460, EPI_ISL_2868463, EPI_ISL_2868466, EPI_ISL_2868467, EPI_ISL_2868469, EPI_ISL_2868470, EPI_ISL_2868471, EPI_ISL_2868472, EPI_ISL_2868473, EPI_ISL_2868474, EPI_ISL_2868477, EPI_ISL_2868479, EPI_ISL_2868480, EPI_ISL_2868481, EPI_ISL_2868482, EPI_ISL_2868483, EPI_ISL_2868485, EPI_ISL_2868486, EPI_ISL_2868487, EPI_ISL_2868488, EPI_ISL_2868489, EPI_ISL_2868490, EPI_ISL_2868491, EPI_ISL_2868492, EPI_ISL_2868496, EPI_ISL_2868497, EPI_ISL_2868498, EPI_ISL_2868499, EPI_ISL_2868500, EPI_ISL_2868501, EPI_ISL_2868502, EPI_ISL_2868503, EPI_ISL_2868504, EPI_ISL_2868505, EPI_ISL_2868506, EPI_ISL_2868507, EPI_ISL_2868508, EPI_ISL_2868509, EPI_ISL_2868510, EPI_ISL_2868511, EPI_ISL_2868512, EPI_ISL_2868513, EPI_ISL_2868514, EPI_ISL_2868515, EPI_ISL_2868516, EPI_ISL_2868522, EPI_ISL_2868523, EPI_ISL_2868527, EPI_ISL_2868529, EPI_ISL_2868530, EPI_ISL_2868531, EPI_ISL_2868532, EPI_ISL_2868533, EPI_ISL_2868534, EPI_ISL_2868535, EPI_ISL_2868536, EPI_ISL_2868538, EPI_ISL_2868539, EPI_ISL_2868540, EPI_ISL_2868541, EPI_ISL_2868545, EPI_ISL_2868546, EPI_ISL_2868549, EPI_ISL_2868551, EPI_ISL_2868556, EPI_ISL_2868557, EPI_ISL_2868558, EPI_ISL_2868559, EPI_ISL_2868562, EPI_ISL_2868571, EPI_ISL_2868575, EPI_ISL_2868590, EPI_ISL_2868592, EPI_ISL_2868594, EPI_ISL_2919190, EPI_ISL_2919194, EPI_ISL_2919195, EPI_ISL_2919196, EPI_ISL_2919197, EPI_ISL_2919198, EPI_ISL_2919199 | | | |
| see above | URMC LABS | Wadsworth Center, New York State Department of Health | Alexis Russell; Catharine Prussing; Daryl M. Lamson; Erasmus Schneider; Erica Lasek-Nesselquist; John Kelly; Jonathan Plitnick; Kirsten St. George; Matthew Shudt; Melissa A Leisner; Navjot Singh |
| EPI_ISL_2421253 | Vermont DoH | Wadsworth Center, New York State Department of Health | Alexis Russell; Catharine Prussing; Daryl M. Lamson; Erasmus Schneider; Erica Lasek-Nesselquist; John Kelly; Jonathan Plitnick; Kirsten St. George; Matthew Shudt; Melissa A Leisner; Navjot Singh |
| EPI_ISL_2274649, EPI_ISL_2274650, EPI_ISL_2274651, EPI_ISL_2274652, EPI_ISL_2274653, EPI_ISL_2274654, EPI_ISL_2308792, EPI_ISL_2421105, EPI_ISL_2421106, EPI_ISL_2421107, EPI_ISL_2448301, EPI_ISL_2626924, EPI_ISL_2627138, EPI_ISL_2627139, EPI_ISL_2627140, EPI_ISL_2627141, EPI_ISL_2727103 | | | |
| see above | WESTCHESTER MEDICAL CENTER | Wadsworth Center, New York State Department of Health | Alexis Russell; Catharine Prussing; Daryl M. Lamson; Erasmus Schneider; Erica Lasek-Nesselquist; John Kelly; Jonathan Plitnick; Kirsten St. George; Matthew Shudt; Melissa A Leisner; Navjot Singh |
| EPI_ISL_2273899, EPI_ISL_2273926, EPI_ISL_2273934, EPI_ISL_2273949, EPI_ISL_2273950, EPI_ISL_2273974, EPI_ISL_2273979, EPI_ISL_2273981, EPI_ISL_2402818, EPI_ISL_2402819, EPI_ISL_2402825, EPI_ISL_2402845, EPI_ISL_2402856, EPI_ISL_2402859, EPI_ISL_2402862, EPI_ISL_2402866, EPI_ISL_2402874, EPI_ISL_2402894, EPI_ISL_2402902, EPI_ISL_2402908, EPI_ISL_2450657, EPI_ISL_2450674, EPI_ISL_2450678, EPI_ISL_2450681, EPI_ISL_2450685, EPI_ISL_2450688, EPI_ISL_2455392, EPI_ISL_2758225, EPI_ISL_2758226, EPI_ISL_2758227, EPI_ISL_2758228, EPI_ISL_2758229, EPI_ISL_2758230, EPI_ISL_2758231, EPI_ISL_2758232, EPI_ISL_2758233, EPI_ISL_2758234, EPI_ISL_2758235, EPI_ISL_2774495, EPI_ISL_2774496, EPI_ISL_2774524, EPI_ISL_2774525, EPI_ISL_2774526 | | | |
| see above | Weill Cornell Medicine | New York Genome Center | Andre Corvelo; Arryn Craney; Chris Mason; Dayna M. Oschwald; Hanna Rennert; Lars F Westblade; Margaret Elizabeth Ross; Melissa Cushing; Michael Zody; Olivier Elemento; Priya Velu; Samantha Fennessey; Tom Maniatis |
| EPI_ISL_2716237, | Yale Clinical Virology Lab | Grubaugh Lab - Yale | Anderson Brito; Annie Watkins; Chaney Kalinich; Chantal Vogels; Isabel Ott; Jessica Rothman; Joseph Fauver; Mallery Breban; Marie L. Landry; Mary Petrone; Nathan Grubaugh; Tara Alpert |

| | |
|---|---|
| EPI_ISL_2716240, EPI_ISL_2716245 | School of Public Health |

We gratefully acknowledge the following Authors from the Originating laboratories responsible for obtaining the specimens, as well as the Submitting laboratories where the genome data were generated and shared via GISAID, on which this research is based.

All Submitters of data may be contacted directly via www.gisaid.org

Authors are sorted alphabetically.

| Accession ID | Originating Laboratory | Submitting Laboratory | Authors |
|---|---|---|---|
| EPI_ISL_2791488, EPI_ISL_2791489 | Gatuario Veterinary Clinic | Faculty of Veterinary Medicine, Universidad Peruana Cayetano Heredia | Francesca Schiaffino |
| EPI_ISL_1629795, EPI_ISL_1629798, EPI_ISL_1629799, EPI_ISL_1629802, EPI_ISL_1629803, EPI_ISL_1629804, EPI_ISL_1629805, EPI_ISL_1629806, EPI_ISL_1629807 | | | |
| see above | Instituto de Medicina Tropical Alexander Von Humboldt, Universidad Peruana Cayetano Heredia | Laboratorio de Genómica Microbiana, Universidad Peruana Cayetano Heredia | Alejandra Dávila-Barclay; Diego Cuicapuza; Guillermo Salvatierra; Janet Huancachoque; Lenin Maturrano; Luis González; Pablo Tsukayama; Pedro E. Romero; Pool Marcos |
| EPI_ISL_1629808 | Laboratorio de Referencia Nacional de Virus Respiratorios, Instituto Nacional de Salud Peru | Laboratorio de Genómica Microbiana, Universidad Peruana Cayetano Heredia | Alejandra Dávila-Barclay; Diego Cuicapuza; Guillermo Salvatierra; Janet Huancachoque; Lenin Maturrano; Luis González; Pablo Tsukayama; Pedro E. Romero; Pool Marcos |
| EPI_ISL_2536701, EPI_ISL_2536702, EPI_ISL_2536703, EPI_ISL_2536704, EPI_ISL_2536705, EPI_ISL_2536706, EPI_ISL_2536707, EPI_ISL_2536708, EPI_ISL_2536709, EPI_ISL_2536710, EPI_ISL_2536711, EPI_ISL_2536712, EPI_ISL_2536713, EPI_ISL_2536714, EPI_ISL_2536715, EPI_ISL_2536716, EPI_ISL_2536717, EPI_ISL_2536718, EPI_ISL_2536719, EPI_ISL_2536720, EPI_ISL_2536721, EPI_ISL_2536722, EPI_ISL_2536723, EPI_ISL_2536724, EPI_ISL_2536725, EPI_ISL_2536726, EPI_ISL_2536727, EPI_ISL_2536728, EPI_ISL_2536729, EPI_ISL_2536730, EPI_ISL_2536731, EPI_ISL_2536732, EPI_ISL_2536733, EPI_ISL_2536734, EPI_ISL_2536735, EPI_ISL_2536736, EPI_ISL_2536737, EPI_ISL_2536738, EPI_ISL_2536739, EPI_ISL_2536740, EPI_ISL_2536741, EPI_ISL_2536742, EPI_ISL_2536743, EPI_ISL_2536744, EPI_ISL_2536745, EPI_ISL_2536746, EPI_ISL_2536747, EPI_ISL_2536748, EPI_ISL_2536749, EPI_ISL_2536750, EPI_ISL_2536751, EPI_ISL_2536752, EPI_ISL_2536753, EPI_ISL_2536754, EPI_ISL_2536755, EPI_ISL_2536756, EPI_ISL_2536757, EPI_ISL_2536758, EPI_ISL_2536759, EPI_ISL_2536760, EPI_ISL_2536761, EPI_ISL_2536762, EPI_ISL_2536763, EPI_ISL_2536764, EPI_ISL_2536765, EPI_ISL_2536766, EPI_ISL_2536767, EPI_ISL_2536768, EPI_ISL_2536769, EPI_ISL_2536770, EPI_ISL_2536771, EPI_ISL_2536772, EPI_ISL_2536773, EPI_ISL_2536774, EPI_ISL_2536775, EPI_ISL_2536778, EPI_ISL_2536779, EPI_ISL_2536780, EPI_ISL_2536781, EPI_ISL_2536782, EPI_ISL_2536783, EPI_ISL_2536797, EPI_ISL_2536798, EPI_ISL_2536799, EPI_ISL_2536800, EPI_ISL_2536801, EPI_ISL_2536802, EPI_ISL_2536803, EPI_ISL_2536804, EPI_ISL_2536805, EPI_ISL_2536806, EPI_ISL_2536807, EPI_ISL_2536808, EPI_ISL_2536809, EPI_ISL_2536810, EPI_ISL_2536811, EPI_ISL_2536812, EPI_ISL_2536813, EPI_ISL_2536814, EPI_ISL_2536815, EPI_ISL_2536816, EPI_ISL_2536817, EPI_ISL_2536818, EPI_ISL_2536819, EPI_ISL_2536820, EPI_ISL_2536821, EPI_ISL_2536822, EPI_ISL_2536823, EPI_ISL_2536824, EPI_ISL_2536825, EPI_ISL_2536826, EPI_ISL_2536827, EPI_ISL_2536828, EPI_ISL_2536829, EPI_ISL_2536830, EPI_ISL_2536831, EPI_ISL_2536832, EPI_ISL_2536833, EPI_ISL_2536834, EPI_ISL_2536835, EPI_ISL_2536836, EPI_ISL_2536837, EPI_ISL_2536838, EPI_ISL_2536839, EPI_ISL_2536840, EPI_ISL_2536841, EPI_ISL_2536842, EPI_ISL_2536843, EPI_ISL_2536844, EPI_ISL_2536845, EPI_ISL_2536846, EPI_ISL_2536847, EPI_ISL_2536848, EPI_ISL_2536849, EPI_ISL_2536850, EPI_ISL_2536851, EPI_ISL_2536852, EPI_ISL_2536853, EPI_ISL_2536854, EPI_ISL_2536855, EPI_ISL_2536856, EPI_ISL_2536857, EPI_ISL_2536858, EPI_ISL_2536859, EPI_ISL_2536860, EPI_ISL_2536861, EPI_ISL_2536862, EPI_ISL_2536863, EPI_ISL_2536864, EPI_ISL_2536865, EPI_ISL_2536866, EPI_ISL_2536867, EPI_ISL_2536868, EPI_ISL_2536869, EPI_ISL_2536870, EPI_ISL_2536871, EPI_ISL_2536872, EPI_ISL_2536873, EPI_ISL_2536874, EPI_ISL_2536875, EPI_ISL_2536876, EPI_ISL_2536877, EPI_ISL_2536878, EPI_ISL_2536879, EPI_ISL_2536880, EPI_ISL_2536881, EPI_ISL_2536882, EPI_ISL_2536883, EPI_ISL_2536884, EPI_ISL_2536885, EPI_ISL_2536886, EPI_ISL_2536887, EPI_ISL_2536888, EPI_ISL_2536889, EPI_ISL_2536890, EPI_ISL_2536891, EPI_ISL_2536892, EPI_ISL_2562266, EPI_ISL_2844694, EPI_ISL_2844695, EPI_ISL_2844696, EPI_ISL_2844697, EPI_ISL_2844698, EPI_ISL_2844699, EPI_ISL_2844700, EPI_ISL_2844701, EPI_ISL_2844702, EPI_ISL_2844703, EPI_ISL_2844704, EPI_ISL_2844705, EPI_ISL_2844706, EPI_ISL_2844707, EPI_ISL_2844708, EPI_ISL_2844709, EPI_ISL_2844710, EPI_ISL_2844711, EPI_ISL_2844712, EPI_ISL_2844713, EPI_ISL_2844714, EPI_ISL_2844715, EPI_ISL_2844716, EPI_ISL_2844717, EPI_ISL_2844718, EPI_ISL_2844719, EPI_ISL_2844720, EPI_ISL_2844721, EPI_ISL_2844722, EPI_ISL_2844723, EPI_ISL_2844724, EPI_ISL_2844725, EPI_ISL_2844726, EPI_ISL_2844727, EPI_ISL_2844728, EPI_ISL_2844729, EPI_ISL_2844730, EPI_ISL_2844731, EPI_ISL_2844732, EPI_ISL_2844733, EPI_ISL_2844734, EPI_ISL_2844735, EPI_ISL_2844736, EPI_ISL_2844737, EPI_ISL_2844738, EPI_ISL_2844739, EPI_ISL_2844740, EPI_ISL_2844741, EPI_ISL_2844742, EPI_ISL_2844743, EPI_ISL_2844744, EPI_ISL_2844745, EPI_ISL_2844746, EPI_ISL_2844747, EPI_ISL_2844748, EPI_ISL_2844749, EPI_ISL_2844750, EPI_ISL_2844751, EPI_ISL_2844752, EPI_ISL_2844753, EPI_ISL_2844754, EPI_ISL_2844756, EPI_ISL_2844757, EPI_ISL_2844758, EPI_ISL_2844759, EPI_ISL_2844760, EPI_ISL_2844761, EPI_ISL_2844762, EPI_ISL_2844763, EPI_ISL_2844764, EPI_ISL_2844765, EPI_ISL_2844766, EPI_ISL_2844767, EPI_ISL_2844768, EPI_ISL_2844769, EPI_ISL_2844770, EPI_ISL_2844771, EPI_ISL_2844772, EPI_ISL_2844773, EPI_ISL_2844774, EPI_ISL_2844775, EPI_ISL_2844776, EPI_ISL_2844777, EPI_ISL_2844778, EPI_ISL_2844779, EPI_ISL_2844780, EPI_ISL_2844781, EPI_ISL_2844782, EPI_ISL_2844783, EPI_ISL_2844784, EPI_ISL_2844785, EPI_ISL_2844786, EPI_ISL_2844787, EPI_ISL_2844788, EPI_ISL_2844789, EPI_ISL_2844790, EPI_ISL_2844791, EPI_ISL_2921267, EPI_ISL_2921268, EPI_ISL_2921269, EPI_ISL_2921270, EPI_ISL_2921271, EPI_ISL_2921273, EPI_ISL_2921276, EPI_ISL_2921277, EPI_ISL_2921278, EPI_ISL_2921279, EPI_ISL_2921280, EPI_ISL_2921281, EPI_ISL_2921282, EPI_ISL_2921283, EPI_ISL_2921284, EPI_ISL_2921285, EPI_ISL_2921286, EPI_ISL_2921287, EPI_ISL_2921288, EPI_ISL_2921289, EPI_ISL_2921290, EPI_ISL_2921291, EPI_ISL_2921292, EPI_ISL_2921293, EPI_ISL_2921294, EPI_ISL_2921295, EPI_ISL_2921296, EPI_ISL_2921297, EPI_ISL_2921298, EPI_ISL_2921299, EPI_ISL_2921300, EPI_ISL_2921301, EPI_ISL_2921302, EPI_ISL_2921303, EPI_ISL_2921304, EPI_ISL_2921305, EPI_ISL_2921306, EPI_ISL_2921307, EPI_ISL_2921308, EPI_ISL_2921309, EPI_ISL_2921310, EPI_ISL_2921311, EPI_ISL_2921312, EPI_ISL_2921313, EPI_ISL_2921314, EPI_ISL_2921315, EPI_ISL_2921316, EPI_ISL_2921317, EPI_ISL_2921318, EPI_ISL_2921319, EPI_ISL_2921320, EPI_ISL_2921321, EPI_ISL_2921322, EPI_ISL_2921323, EPI_ISL_2921324, EPI_ISL_2921325, EPI_ISL_2921326, EPI_ISL_2921327, EPI_ISL_2921328, EPI_ISL_2921329, EPI_ISL_2921330, EPI_ISL_2921331, EPI_ISL_2921332, EPI_ISL_2921333, EPI_ISL_2921334, EPI_ISL_2921335, EPI_ISL_2921336, EPI_ISL_2921337, EPI_ISL_2921338, EPI_ISL_2921339, EPI_ISL_2921340, EPI_ISL_2921341, EPI_ISL_2921342, EPI_ISL_2921343, EPI_ISL_2921344, EPI_ISL_2921345, EPI_ISL_2921346, EPI_ISL_2921347, EPI_ISL_2921348, EPI_ISL_2921349, EPI_ISL_2921350, EPI_ISL_2921351, EPI_ISL_2921352, EPI_ISL_2921353, EPI_ISL_2921354, EPI_ISL_2921355, EPI_ISL_2921370, EPI_ISL_2921371, EPI_ISL_2921375, EPI_ISL_2921376, EPI_ISL_2921377, EPI_ISL_2921378, EPI_ISL_2921379, EPI_ISL_2921380, EPI_ISL_2921381, EPI_ISL_2921382, EPI_ISL_2921383, EPI_ISL_2921384, EPI_ISL_2921385, EPI_ISL_2921386, EPI_ISL_2921387, EPI_ISL_2921388, EPI_ISL_2921389, EPI_ISL_2921390, EPI_ISL_2921391, EPI_ISL_2921392, EPI_ISL_2921393, EPI_ISL_2921394, EPI_ISL_2921395, EPI_ISL_2921396, EPI_ISL_2921397, EPI_ISL_2921398, EPI_ISL_2921399, EPI_ISL_2921401, EPI_ISL_2921402, EPI_ISL_2921403, EPI_ISL_2921404, EPI_ISL_2921405, EPI_ISL_2921406, EPI_ISL_2921407, EPI_ISL_2921408, EPI_ISL_2921409, EPI_ISL_2921410, EPI_ISL_2921411, EPI_ISL_2921412, EPI_ISL_2921413, EPI_ISL_2921414, EPI_ISL_2921415, EPI_ISL_2921416, EPI_ISL_2921417, EPI_ISL_2921418, EPI_ISL_2921419, EPI_ISL_2921420, EPI_ISL_2921421, EPI_ISL_2921422, EPI_ISL_2921423, EPI_ISL_2921424, EPI_ISL_2921425, EPI_ISL_2921426, EPI_ISL_2921427, EPI_ISL_2921428, EPI_ISL_2921429, EPI_ISL_2921430, EPI_ISL_2921431, EPI_ISL_2921432, EPI_ISL_2921433, EPI_ISL_2921434, EPI_ISL_2921435, EPI_ISL_2921436, EPI_ISL_2921437, EPI_ISL_2921438, EPI_ISL_2921439, EPI_ISL_2921440, EPI_ISL_2921441, EPI_ISL_2921442, EPI_ISL_2921443, EPI_ISL_2921444, EPI_ISL_2921445, EPI_ISL_2921446, EPI_ISL_2921447, EPI_ISL_2921448, EPI_ISL_2921449, EPI_ISL_2921450, EPI_ISL_2921451, EPI_ISL_2921452, EPI_ISL_2921454, EPI_ISL_2921455, EPI_ISL_2921456, EPI_ISL_2921458, EPI_ISL_2921459, EPI_ISL_2921460, EPI_ISL_2921461, EPI_ISL_2921462, EPI_ISL_2921463, EPI_ISL_2921467, EPI_ISL_2921468, EPI_ISL_2921475, EPI_ISL_2921476, EPI_ISL_2921477, EPI_ISL_2921478, EPI_ISL_2921479, EPI_ISL_2921480, EPI_ISL_2921481, EPI_ISL_2921482, EPI_ISL_2921483, EPI_ISL_2921484, EPI_ISL_2921485, EPI_ISL_2921486, EPI_ISL_2921487, EPI_ISL_2921488, EPI_ISL_2921489, EPI_ISL_2921490, EPI_ISL_2921491, EPI_ISL_2921492, EPI_ISL_2921493, EPI_ISL_2921494, EPI_ISL_2921495, EPI_ISL_2921496, EPI_ISL_2921497, EPI_ISL_2921498, EPI_ISL_2921499, EPI_ISL_2921500, EPI_ISL_2921501, EPI_ISL_2921502, EPI_ISL_2921503, EPI_ISL_2921504, EPI_ISL_2921505, EPI_ISL_2921506, EPI_ISL_2921507, EPI_ISL_2921508, EPI_ISL_2921509, EPI_ISL_2921510, EPI_ISL_2921511, EPI_ISL_2921512, EPI_ISL_2921513, EPI_ISL_2921514, EPI_ISL_2921515, EPI_ISL_2921516, EPI_ISL_2921517, EPI_ISL_2921518, EPI_ISL_2921519, EPI_ISL_2921520, EPI_ISL_2921521, EPI_ISL_2921522, EPI_ISL_2921523, EPI_ISL_2921524, EPI_ISL_2921525, EPI_ISL_2921526, EPI_ISL_2921527, EPI_ISL_2921528, EPI_ISL_2921529, EPI_ISL_2921530, EPI_ISL_2921531, EPI_ISL_2921532 | | | |
| see above | Laboratorio de Referencial Nacional de Virus Respiratorios | Laboratorio de Referencial Nacional de Virus Respiratorios | Carlos Padilla Rojas; Henri Bailon Calderon; Iris Silva Molina; Joseph Huayra Niquen; Lely Solari Zerpa; Luis Barcena Flores; Marco Galarza Perez; Nancy Rojas Serrano; Omar Caceres Rey; Orson Mestanza Millones; Priscila Lope Pari; Sandra Morales Ruiz; Steve Acedo Lazo; Veronica Hurtado Vela |

We gratefully acknowledge the following Authors from the Originating laboratories responsible for obtaining the specimens, as well as the Submitting laboratories where the genome data were generated and shared via GISAID, on which this research is based.

All Submitters of data may be contacted directly via www.gisaid.org

Authors are sorted alphabetically.

| Accession ID | Originating Laboratory | Submitting Laboratory | Authors |
|---|---|---|---|
| EPI_ISL_2385508, EPI_ISL_2534912, EPI_ISL_2534913, EPI_ISL_2535035, EPI_ISL_2691315, EPI_ISL_2691316, EPI_ISL_2691317, EPI_ISL_2691318, EPI_ISL_2836982, EPI_ISL_2836983, EPI_ISL_2836984 | | | |
| see above | Laboratorio Central Noel Nutels | Bioinformatics Laboratory / LNCC | Alessandra P Lamarca; Alexandra L Gerber; Amilcar Tanuri; Ana Paula de C Guimaraes; Ana Tereza R Vasconcelos; Andrea Cony Cavalcanti; Caio Luiz Pereira Ribeiro; Cassia Alves; Cintia Policarpo; Claudia Maria Braga de Mello; Cristiane Gomes da Silva; Diana Mariani; Douglas Terra Machado; Flavio Dias da Silva; Gleidson da Silva de Oliveira; Leandro Magalhaes de Souza; Liliane Cavalcante; Luiz G P de Almeida; Marcio Henrique de Oliveira Garcia; Mario Sergio Ribeiro; Ronaldo da Silva F Jr; Silvia Carvalho |
| EPI_ISL_2536776, EPI_ISL_2536777, EPI_ISL_2536784, EPI_ISL_2536785, EPI_ISL_2536786, EPI_ISL_2536787, EPI_ISL_2536788, EPI_ISL_2536789, EPI_ISL_2536790, EPI_ISL_2536791, EPI_ISL_2536792, EPI_ISL_2536793, EPI_ISL_2536794, EPI_ISL_2536795, EPI_ISL_2536796, EPI_ISL_2562268, EPI_ISL_2652249, EPI_ISL_2844755, EPI_ISL_2921272, EPI_ISL_2921274, EPI_ISL_2921275, EPI_ISL_2921356, EPI_ISL_2921357, EPI_ISL_2921358, EPI_ISL_2921359, EPI_ISL_2921360, EPI_ISL_2921361, EPI_ISL_2921362, EPI_ISL_2921363, EPI_ISL_2921364, EPI_ISL_2921365, EPI_ISL_2921366, EPI_ISL_2921367, EPI_ISL_2921368, EPI_ISL_2921369, EPI_ISL_2921372, EPI_ISL_2921373, EPI_ISL_2921374, EPI_ISL_2921453, EPI_ISL_2921457, EPI_ISL_2921464, EPI_ISL_2921465, EPI_ISL_2921466, EPI_ISL_2921469, EPI_ISL_2921470, EPI_ISL_2921471, EPI_ISL_2921472, EPI_ISL_2921473, EPI_ISL_2921474 | | | |
| see above | Laboratorio de Referencial Nacional de Virus Respiratorios | Laboratorio de Referencial Nacional de Virus Respiratorios | Carlos Padilla Rojas; Henri Bailon Calderon; Iris Silva Molina; Joseph Huayra Niquen; Lely Solari Zerpa; Luis Barcena Flores; Marco Galarza Perez; Nancy Rojas Serrano; Omar Caceres Rey; Orson Mestanza Millones; Priscila Lope Pari; Sandra Morales Ruiz; Steve Acedo Lazo; Veronica Hurtado Vela |

We gratefully acknowledge the following Authors from the Originating laboratories responsible for obtaining the specimens, as well as the Submitting laboratories where the genome data were generated and shared via GISAID, on which this research is based.

All Submitters of data may be contacted directly via www.gisaid.org

Authors are sorted alphabetically.

| Accession ID | Originating Laboratory | Submitting Laboratory | Authors |
|---|---|---|---|
| EPI_ISL_2086667, EPI_ISL_2086676 | FEDERAL STATE BUDGETARY INSTITUTION OF HEALTHCARE HOSPITAL OF THE RUSSIAN ACADEMY OF SCIENCES (TROITSK), Academic Consortium | FEDERAL RESEARCH AND CLINICAL CENTER OF PHYSICAL-CHEMICAL MEDICINE OF FEDERAL MEDICAL BIOLOGICAL AGENCY, Academic Consortium | Alexander Manolov; Alexander Pavlenko; Alexander Rumyantsev; Alexander Samoilov; Alexander Troitsky; Alexander Tulichev; Alexander Vasiliev; Alexey Volnukhin; Alla Petrova; Andrey Koritsky; Anna Blagonravova; Anna Varizhuk; Daria Boldyreva; Daria Kharlampieva; Daria Matyushkina; Dmitry Fedorov; Dmitry Pushkar; Ekaterina Moskaleva; Elena Galova; Elena Ilina; Elizaveta Orlova; Galina Trifonova; Gaukhar Yusubalieva; Ilshat Gafurov; Irina Shirokova; Konstantin Ivanov; Konstantin Kolontarev; Ksenia Klimina; Larisa Balykova; Lyubov Rychkova; Nadezhda Kolyshkina; Natalia Pozhenko; Oleg Ogarkov; Pavel Bashkirov; Polina Khromova; Polina Tikhonova; Ramiz Bakhtyev; Raushaniya Gaifullina; Sergey Rumyantsev; Svetlana Solovieva; Vadim Govorun; Valentin Manuvera; Varvara Musarova; Vasily Lazarev; Vladimir Baklaushev; Vladimir Rumyantsev; Vladimir Veselovsky; Vladislav Babenko; Vyacheslav Kuropatkin; Vyacheslav Sinkov; Yulia Galeeva; Yuri Udalov |
| EPI_ISL_2038922, EPI_ISL_2038923, EPI_ISL_2038924, EPI_ISL_2038925 | Federal Budget Health Care Institution "Center of Hygiene and Epidemiology in Ivanovo region" | Group of Genomics and Postgenomic Technologies of Central Research Institute of Epidemiology | Akimkin VG; Bunina IV; Kaptelova VV; Kondrasheva LY; Korneenko EV; Nadtoka MI; Saenko SS; Samoilov AE; Shorohova EV; Smirnova YS; Speranskaya AS; Tivanova EV |
| EPI_ISL_2038895, EPI_ISL_2038896, EPI_ISL_2038897, EPI_ISL_2038898 | Federal Budget Health Care Institution "Center of Hygiene and Epidemiology in Ryazan region" | Group of Genomics and Postgenomic Technologies of Central Research Institute of Epidemiology | Akimkin VG; Baranova SV; Kaptelova VV; Kondrasheva LY; Korneenko EV; Nadtoka MI; Saenko SS; Samoilov AE; Smirnova YS; Speranskaya AS; Tivanova EV |
| EPI_ISL_2038866, EPI_ISL_2038867, EPI_ISL_2038868, EPI_ISL_2080489 | Federal Budget Health Care Institution "Center of Hygiene and Epidemiology in Tambov region" | Group of Genomics and Postgenomic Technologies of Central Research Institute of Epidemiology | Akimkin VG; Belova MV; Kaptelova VV; Kondrasheva LY; Korneenko EV; Nadtoka MI; Saenko SS; Samoilov AE; Smirnova YS; Speranskaya AS; Tivanova EV |
| EPI_ISL_2032644, EPI_ISL_2038893, EPI_ISL_2038894 | Federal Budget Health Care Institution "Center of Hygiene and Epidemiology in Tver region" | Group of Genomics and Postgenomic Technologies of Central Research Institute of Epidemiology | Akimkin VG; Kaptelova VV; Kondrasheva LY; Korneenko EV; Melushenkova TG; Nadtoka MI; Saenko SS; Samoilov AE; Smirnova YS; Speranskaya AS; Tivanova EV |
| EPI_ISL_2086647 | Federal State Public Scientific Institution «Scientific Centre for Family Health and Human Reproduction Problems», Academic Consortium | FEDERAL RESEARCH AND CLINICAL CENTER OF PHYSICAL-CHEMICAL MEDICINE OF FEDERAL MEDICAL BIOLOGICAL AGENCY, Academic Consortium | Alexander Manolov; Alexander Pavlenko; Alexander Rumyantsev; Alexander Samoilov; Alexander Troitsky; Alexander Tulichev; Alexander Vasiliev; Alexey Volnukhin; Alla Petrova; Andrey Koritsky; Anna Blagonravova; Anna Varizhuk; Daria Boldyreva; Daria Kharlampieva; Daria Matyushkina; Dmitry Fedorov; Dmitry Pushkar; Ekaterina Moskaleva; Elena Galova; Elena Ilina; Elizaveta Orlova; Galina Trifonova; Gaukhar Yusubalieva; Ilshat Gafurov; Irina Shirokova; Konstantin Ivanov; Konstantin Kolontarev; Ksenia Klimina; Larisa Balykova; Lyubov Rychkova; Nadezhda Kolyshkina; Natalia Pozhenko; Oleg Ogarkov; Pavel Bashkirov; Polina Khromova; Polina Tikhonova; Ramiz Bakhtyev; Raushaniya Gaifullina; Sergey Rumyantsev; Svetlana Solovieva; Vadim Govorun; Valentin Manuvera; Varvara Musarova; Vasily Lazarev; Vladimir Baklaushev; Vladimir Rumyantsev; Vladimir Veselovsky; Vladislav Babenko; Vyacheslav Kuropatkin; Vyacheslav Sinkov; Yulia Galeeva; Yuri Udalov |
| EPI_ISL_1652537, EPI_ISL_1652538, EPI_ISL_1652539, EPI_ISL_1652540, EPI_ISL_1652542, EPI_ISL_1652543, EPI_ISL_1652544, EPI_ISL_1652545, EPI_ISL_1652546, EPI_ISL_1652547, EPI_ISL_1652548, EPI_ISL_1652549, EPI_ISL_1652550, EPI_ISL_1652551, EPI_ISL_1652552, EPI_ISL_1652553, EPI_ISL_1652554, EPI_ISL_1652555, EPI_ISL_1652556, EPI_ISL_1652557, EPI_ISL_1652558, EPI_ISL_1652559, EPI_ISL_1652560, EPI_ISL_1652561, EPI_ISL_1652562, EPI_ISL_1652563, EPI_ISL_1652564, EPI_ISL_1652565, EPI_ISL_1652566, EPI_ISL_1652567, EPI_ISL_1652568, EPI_ISL_1652569, EPI_ISL_1652570, EPI_ISL_1652571, EPI_ISL_1652572, EPI_ISL_1652573, EPI_ISL_1652574, EPI_ISL_1652575, EPI_ISL_1652577, EPI_ISL_1652578, EPI_ISL_1652579, EPI_ISL_1652580, EPI_ISL_1652581, EPI_ISL_1652582, EPI_ISL_1652583, EPI_ISL_1652585, EPI_ISL_1652586, EPI_ISL_1652587, EPI_ISL_1652588, EPI_ISL_1652589, EPI_ISL_1652590, EPI_ISL_1652591, EPI_ISL_1652592, EPI_ISL_1652593, EPI_ISL_1652594, EPI_ISL_1652595, EPI_ISL_1652596, EPI_ISL_1652597, EPI_ISL_1652598, EPI_ISL_1652599, EPI_ISL_1652600, EPI_ISL_1652602, EPI_ISL_1652603, EPI_ISL_1652605, EPI_ISL_1652606, EPI_ISL_1652608, EPI_ISL_1652609, EPI_ISL_1652611, EPI_ISL_1652612, EPI_ISL_1652613, EPI_ISL_1652614, EPI_ISL_1652615, EPI_ISL_1652616, EPI_ISL_1652617, EPI_ISL_1652618, EPI_ISL_1652619, EPI_ISL_1652620, EPI_ISL_1652621, EPI_ISL_1652622, EPI_ISL_1652623, EPI_ISL_1652624, EPI_ISL_1652625, EPI_ISL_1652626, EPI_ISL_1652627, EPI_ISL_1652629, EPI_ISL_1652630, EPI_ISL_1652631, EPI_ISL_1652632, EPI_ISL_1652633, EPI_ISL_1652634, EPI_ISL_1652635, EPI_ISL_1652636, EPI_ISL_1652637, EPI_ISL_1652638, EPI_ISL_1652640, EPI_ISL_1652641, EPI_ISL_1652644, EPI_ISL_1652645, EPI_ISL_1652646, EPI_ISL_1652647, EPI_ISL_1652648, EPI_ISL_1652649, EPI_ISL_1652650, EPI_ISL_1652652, EPI_ISL_1652653, EPI_ISL_1652654, EPI_ISL_1652655, EPI_ISL_1652656, EPI_ISL_1652658, EPI_ISL_1652659, EPI_ISL_1652660, EPI_ISL_1652661, EPI_ISL_1652662, EPI_ISL_1652663, EPI_ISL_1652664, EPI_ISL_1652665, EPI_ISL_1652666, EPI_ISL_1652667, EPI_ISL_1652669, EPI_ISL_1652670, EPI_ISL_1652671, EPI_ISL_1652672, EPI_ISL_1652673, EPI_ISL_1652674, EPI_ISL_1652675, EPI_ISL_1652676, EPI_ISL_1652677, EPI_ISL_1652678, EPI_ISL_1652679, EPI_ISL_1652680, EPI_ISL_1652681, EPI_ISL_1652682, EPI_ISL_1652683, EPI_ISL_1652684, EPI_ISL_1919424, EPI_ISL_1919425, EPI_ISL_1919426, EPI_ISL_1919427, EPI_ISL_1919428, EPI_ISL_1919429, EPI_ISL_1919430, EPI_ISL_1919431, EPI_ISL_1919432, EPI_ISL_1919433, EPI_ISL_1919434, EPI_ISL_1919435, EPI_ISL_1919436, EPI_ISL_1919437, EPI_ISL_1919438, EPI_ISL_1919439, EPI_ISL_1919440, EPI_ISL_1919441, EPI_ISL_1919442, EPI_ISL_1919443, EPI_ISL_1919445, EPI_ISL_1919446, EPI_ISL_1919447, EPI_ISL_1919448, EPI_ISL_1919449, EPI_ISL_1919450, EPI_ISL_1919451, EPI_ISL_1919452, EPI_ISL_1919453, EPI_ISL_1919454, EPI_ISL_1919455, EPI_ISL_1919456, EPI_ISL_1919457, EPI_ISL_1919458, EPI_ISL_1919459, EPI_ISL_1919460, EPI_ISL_1919461, EPI_ISL_1919462, EPI_ISL_1919463, EPI_ISL_1919464, EPI_ISL_1919466, EPI_ISL_1919467, EPI_ISL_1919468, EPI_ISL_1919469, EPI_ISL_1919470, EPI_ISL_1919471, EPI_ISL_1919473, EPI_ISL_1919474, EPI_ISL_1919475, EPI_ISL_1919479, EPI_ISL_1919481, EPI_ISL_1919484, EPI_ISL_1919485, EPI_ISL_1919486, EPI_ISL_1919487, EPI_ISL_1919488, EPI_ISL_1919489, EPI_ISL_1919492, EPI_ISL_1919493, EPI_ISL_1919497, EPI_ISL_1919500, EPI_ISL_1919502, EPI_ISL_1919504, EPI_ISL_1919505, EPI_ISL_1919507, EPI_ISL_1919509, EPI_ISL_1919510, EPI_ISL_1919512, EPI_ISL_1919514, EPI_ISL_1919515, EPI_ISL_1919516, EPI_ISL_1919519, EPI_ISL_1919520, EPI_ISL_1919521, EPI_ISL_1919523, EPI_ISL_1919524, EPI_ISL_1919525, EPI_ISL_1919528, EPI_ISL_1919529, EPI_ISL_1919530, EPI_ISL_1919531, EPI_ISL_1919535, EPI_ISL_1919537, EPI_ISL_1919538, EPI_ISL_1919543, EPI_ISL_1919545, EPI_ISL_1919547, EPI_ISL_1919548, EPI_ISL_1919552, EPI_ISL_1919553, EPI_ISL_1919556, EPI_ISL_1919557, EPI_ISL_1919559, EPI_ISL_1919560, EPI_ISL_1919562, EPI_ISL_1919565, EPI_ISL_1919568, EPI_ISL_1919572, EPI_ISL_2007302, EPI_ISL_2007303, EPI_ISL_2007304, EPI_ISL_2007305, EPI_ISL_2007306 | | | |
| see above | HELIX LLC | WHO National Influenza Centre Russian Federation | Andrey Komissarov; Anna Ivanova; Artem Fadeev; Daria Danilenko; Dmitry Lioznov; Elena Nabieva; Georgii Bazykin; Kirill Varchenko; Ksenia Safina; Kseniya Komissarova; Maria Pisareva; Maria Timofeeva; Mikhail Bakaev; Nikita Yolshin; Oula Masour; Tamila Musaeva; Veronika Eder |
| EPI_ISL_2032642, EPI_ISL_2032645, EPI_ISL_2032648 | Infectious Diseases Hospital № 2 | Group of Genomics and Postgenomic Technologies of Central Research Institute of Epidemiology | Akimkin VG; Gordeeva TG; Kaptelova VV; Kondrasheva LY; Korneenko EV; Nadtoka MI; Saenko SS; Samoilov AE; Sergacheva IP; Smirnova YS; Speranskaya AS; Tivanova EV |
| EPI_ISL_1823191, EPI_ISL_1823192, EPI_ISL_1823193, EPI_ISL_1823194, EPI_ISL_1823195, EPI_ISL_2035729, EPI_ISL_2035731, EPI_ISL_2035732, EPI_ISL_2038899, EPI_ISL_2038900, EPI_ISL_2038901, EPI_ISL_2038902, EPI_ISL_2038903, EPI_ISL_2038904, EPI_ISL_2038905, EPI_ISL_2038906, EPI_ISL_2038907, EPI_ISL_2038908, EPI_ISL_2038909, EPI_ISL_2038910, EPI_ISL_2038911, EPI_ISL_2038912, EPI_ISL_2038913, EPI_ISL_2038914, EPI_ISL_2038915, EPI_ISL_2038916, EPI_ISL_2038917, EPI_ISL_2038918, EPI_ISL_2038919, EPI_ISL_2038920, EPI_ISL_2038921, EPI_ISL_2080490, EPI_ISL_2080491 | | | |
| see above | Laboratory of Polymerase Chain Reaction of Federal Budget Institution of Science "Central Research Institute of Epidemiology" of The Federal Service on Customers' Rights Protection and Human Well-being Surveillance | Group of Genomics and Postgenomic Technologies of Central Research Institute of Epidemiology | Akimkin VG; Berestkovskaya AY; Kaptelova VV; Kondrasheva LY; Korneenko EV; Nadtoka MI; Saenko SS; Samoilov AE; Smirnova YS; Speranskaya AS; Tivanova EV |
| EPI_ISL_2031862, EPI_ISL_2031863, EPI_ISL_2031864, EPI_ISL_2031865, EPI_ISL_2031866, EPI_ISL_2031867, EPI_ISL_2031868, EPI_ISL_2031869, EPI_ISL_2031870, EPI_ISL_2031871, EPI_ISL_2031872, EPI_ISL_2031873, EPI_ISL_2031874, EPI_ISL_2031875, EPI_ISL_2031876, EPI_ISL_2031877, EPI_ISL_2031878, EPI_ISL_2031879, EPI_ISL_2031880, EPI_ISL_2031881, EPI_ISL_2031882, EPI_ISL_2031883, EPI_ISL_2031884, EPI_ISL_2031885, EPI_ISL_2031886, EPI_ISL_2031887, EPI_ISL_2031888, EPI_ISL_2031889, EPI_ISL_2031890, EPI_ISL_2031891, EPI_ISL_2031892, EPI_ISL_2031893, EPI_ISL_2031894, EPI_ISL_2031895, EPI_ISL_2031896, EPI_ISL_2031897, EPI_ISL_2031898, EPI_ISL_2031899, EPI_ISL_2031900, EPI_ISL_2031901, EPI_ISL_2031902, EPI_ISL_2031903, EPI_ISL_2031904, EPI_ISL_2031905, EPI_ISL_2031906, EPI_ISL_2031907, EPI_ISL_2031908, EPI_ISL_2031909, EPI_ISL_2031910, EPI_ISL_2031911, EPI_ISL_2031912, EPI_ISL_2031913, EPI_ISL_2031914, EPI_ISL_2031915, EPI_ISL_2031916, EPI_ISL_2031917, EPI_ISL_2031918, EPI_ISL_2031919, EPI_ISL_2031920, EPI_ISL_2031921, EPI_ISL_2031922, EPI_ISL_2031923, EPI_ISL_2031924, EPI_ISL_2031925, EPI_ISL_2031926, EPI_ISL_2031927, EPI_ISL_2031928, EPI_ISL_2031929, EPI_ISL_2031930, EPI_ISL_2031931, EPI_ISL_2031932, EPI_ISL_2031933, EPI_ISL_2031934, EPI_ISL_2031935, EPI_ISL_2031936, EPI_ISL_2031937, EPI_ISL_2031938, EPI_ISL_2031939, EPI_ISL_2031940, EPI_ISL_2031941, EPI_ISL_2031942, EPI_ISL_2031943, EPI_ISL_2031944, EPI_ISL_2031945, EPI_ISL_2031947, EPI_ISL_2031948, EPI_ISL_2031949, EPI_ISL_2031950, EPI_ISL_2031951, EPI_ISL_2031952, EPI_ISL_2031953, EPI_ISL_2031955, EPI_ISL_2031956, EPI_ISL_2031958, EPI_ISL_2031959, EPI_ISL_2031960, EPI_ISL_2031961, EPI_ISL_2031962, EPI_ISL_2031963, EPI_ISL_2031964, EPI_ISL_2031965, EPI_ISL_2031966, EPI_ISL_2031967, EPI_ISL_2031968, EPI_ISL_2031969, EPI_ISL_2031970, EPI_ISL_2031971, EPI_ISL_2031972, EPI_ISL_2031973, EPI_ISL_2031974, EPI_ISL_2031975, EPI_ISL_2031977, EPI_ISL_2031978 | | | |
| see above | Molecular diagnostic laboratory of Federal Budget Institution of Science "Central Research Institute of Epidemiology" of The Federal Service on Customers' Rights Protection and Human Well-being Surveillance | Group of Genomics and Postgenomic Technologies of Central Research Institute of Epidemilology | Akimkin V.G.; Kaptelova V.V.; Kondrasheva L.Y.; Korneenko E.V.; Nadtoka M.I.; Saenko S.S.; Samojlov A.E.; Shipulina O.Y.; Sinicyn S.O.; Smirnova Y.S.; Speranskaya A.S.; Tivanova E.V. |
| EPI_ISL_1324653, EPI_ISL_1392640, EPI_ISL_1392641 | National Medical Research Center for Obstetrics, Gynecology and Perinatology named after Academician V.I.Kulakov of the Ministry of Healthcare of Russian Federation | National Medical Research Center for Obstetrics, Gynecology and Perinatology named after Academician V.I.Kulakov of the Ministry of Healthcare of Russian Federation | Andrey Donnikov; Andrey Goltsov; Denis Rebrikov; Dmitry Abramov; Dmitry Korostin; Dmitry Trofimov; Gennady Sukhikh; Jekaterina Shubina; Tatiana Priputnevich; Yegor Botsmanov |
| EPI_ISL_1627560, EPI_ISL_1627564, EPI_ISL_1627565, EPI_ISL_1627575, EPI_ISL_1627576, EPI_ISL_1627577 | Pathogenic Microorganisms Variability Laboratory | WHO National Influenza Centre Russian Federation | Alexander Gintsburg; Alexey Shchetinin; Alina Odintsova; Andrei Siniavin; Andrey Komissarov; Andrey Pochtovyy; Artem Fadeev; Artem Tkachuk; Daria Danilenko; Denis Logunov; Dmitry Lioznov; Elena Nabieva; Elena Shidlovskaya; Elizaveta Divisenko; Evgeny Usachev; Georgii Bazykin; Ksenia Safina; Kseniya Komissarova; Ludmila Kolobukhina; Lyudmila Vasilchenko; Maria Nikiforova; Nadezhda Kuznetsova; Olga Burgasova; Svetlana Smetanina; Vladimir Gushchin |
| EPI_ISL_1823178, EPI_ISL_1823179, EPI_ISL_1823180, EPI_ISL_1823181, EPI_ISL_1823182, EPI_ISL_1823183, EPI_ISL_1823184, EPI_ISL_1823185, EPI_ISL_1823186, EPI_ISL_1823187, EPI_ISL_1823188, EPI_ISL_1823189, EPI_ISL_1823190, EPI_ISL_2035722, EPI_ISL_2035723, EPI_ISL_2035724, EPI_ISL_2035725, EPI_ISL_2035726, EPI_ISL_2035727, EPI_ISL_2035728, EPI_ISL_2038869, EPI_ISL_2038870, EPI_ISL_2038871, EPI_ISL_2038872, EPI_ISL_2038873, EPI_ISL_2038874, EPI_ISL_2038875, EPI_ISL_2038876, EPI_ISL_2038877, EPI_ISL_2038878, EPI_ISL_2038879, EPI_ISL_2038880, EPI_ISL_2038881, EPI_ISL_2038882, EPI_ISL_2038883, EPI_ISL_2038884, EPI_ISL_2038885, EPI_ISL_2038886, EPI_ISL_2038887, EPI_ISL_2038888, EPI_ISL_2038889, EPI_ISL_2038890, EPI_ISL_2038891, EPI_ISL_2038892 | | | |
| see above | Public Institution of Health Care "Diagnostic Center ( Laboratory Testing Center) of Moscow Health Department " | Group of Genomics and Postgenomic Technologies of Central Research Institute of Epidemiology | Akimkin VG; Kaptelova VV; Kondrasheva LY; Korneenko EV; Nadtoka MI; Saenko SS; Samoilov AE; Smirnova YS; Spakova OG; Speranskaya AS; Tivanova EV |
| EPI_ISL_1824598 | Public Institution of Health Care "Diagnostic Center (Laboratory Testing Center) of Moscow Health Department " | Group of Genomics and Postgenomic Technologies of Central Research Institute of Epidemiology | Akimkin VG; Kaptelova VV; Kondrasheva LY; Korneenko EV; Nadtoka MI; Saenko SS; Samoilov AE; Smirnova YS; Spakova OG; Speranskaya AS; Tivanova EV |
| EPI_ISL_1652686, EPI_ISL_1789541, EPI_ISL_1789542, EPI_ISL_1797436, EPI_ISL_1797437, EPI_ISL_1919423, EPI_ISL_1919574, EPI_ISL_1919575 | | | |
| see above | WHO National Influenza Centre Russian Federation | WHO National Influenza Centre Russian Federation | Andrey Komissarov; Anna Ivanova; Artem Fadeev; Daria Danilenko; Dmitry Lioznov; Elena Nabieva; Georgii Bazykin; Kirill Varchenko; Ksenia Safina; Kseniya Komissarova; Maria Pisareva; Maria Timofeeva; Mikhail Bakaev; Nikita Yolshin; Oula Mansour; Oula Masour; Tamila Musaeva; Veronika Eder |

We gratefully acknowledge the following Authors from the Originating laboratories responsible for obtaining the specimens, as well as the Submitting laboratories where the genome data were generated and shared via GISAID, on which this research is based.

All Submitters of data may be contacted directly via www.gisaid.org

Authors are sorted alphabetically.

| Accession ID | Originating Laboratory | Submitting Laboratory | Authors |
|---|---|---|---|
| EPI_ISL_2385407, EPI_ISL_2385408, EPI_ISL_2385409, EPI_ISL_2385410, EPI_ISL_2385411, EPI_ISL_2385412, EPI_ISL_2385413, EPI_ISL_2385414, EPI_ISL_2385415, EPI_ISL_2385416, EPI_ISL_2385417, EPI_ISL_2385418, EPI_ISL_2385419, EPI_ISL_2385420, EPI_ISL_2385421, EPI_ISL_2385422, EPI_ISL_2385423, EPI_ISL_2385424, EPI_ISL_2385425 | | | |
| see above | Center of Hygiene and Epidemiology in Belgorod Region | WHO National Influenza Centre Russian Federation | Alexey Masharsky; Andrey Komissarov; Artem Fadeev; Daria Danilenko; Dmitry Lioznov; Elena Nabieva; Georgii Bazykin; Kirill Varchenko; Ksenia Safina; Kseniya Komissarova; Maria Baturova; Maria Pisareva; Maria Timofeeva; Mikhail Bakaev; Nikita Yolshin; Oula Masour; Tamila Musaeva; Veronika Eder |
| EPI_ISL_2450543, EPI_ISL_2450544, EPI_ISL_2450545, EPI_ISL_2450546, EPI_ISL_2450547, EPI_ISL_2450548, EPI_ISL_2450549, EPI_ISL_2450550, EPI_ISL_2450551, EPI_ISL_2450552, EPI_ISL_2450553, EPI_ISL_2450554, EPI_ISL_2450555, EPI_ISL_2450556, EPI_ISL_2450557, EPI_ISL_2450558, EPI_ISL_2450559, EPI_ISL_2450560, EPI_ISL_2450561, EPI_ISL_2450562, EPI_ISL_2450563, EPI_ISL_2450564, EPI_ISL_2450565, EPI_ISL_2462881, EPI_ISL_2523806, EPI_ISL_2523808, EPI_ISL_2523811, EPI_ISL_2523814, EPI_ISL_2523817, EPI_ISL_2523821, EPI_ISL_2523824, EPI_ISL_2523827, EPI_ISL_2523830, EPI_ISL_2523833, EPI_ISL_2523836, EPI_ISL_2523839, EPI_ISL_2523843, EPI_ISL_2523846, EPI_ISL_2523849, EPI_ISL_2523853, EPI_ISL_2523856, EPI_ISL_2523859, EPI_ISL_2899791, EPI_ISL_2899792 | | | |
| see above | City Hospital No 40 | WHO National Influenza Centre Russian Federation | Andrey Komissarov; Artem Fadeev; Daria Danilenko; Dmitry Lioznov; Elena Nabieva; Georgii Bazykin; Kirill Varchenko; Ksenia Safina; Kseniya Komissarova; Maria Pisareva; Maria Timofeeva; Mikhail Bakaev; Nikita Yolshin; Olga Shneider; Oula Mansour; Sergey Scherbak; Tamila Musaeva; Veronika Eder |
| EPI_ISL_2816195, EPI_ISL_2899793, EPI_ISL_2899794, EPI_ISL_2899795, EPI_ISL_2899796, EPI_ISL_2899797, EPI_ISL_2899798, EPI_ISL_2899799 | | | |
| see above | D. I. Ivanovskiy Institute of Virology | WHO National Influenza Centre Russian Federation | Andrey Komissarov; Artem Fadeev; Daria Danilenko; Dmitry Lioznov; Elena Burtseva; Elena Nabieva; Georgii Bazykin; Kirill Krasnoslobodtsev; Kirill Varchenko; Ksenia Safina; Kseniya Komissarova; Maria Pisareva; Mikhail Bakaev; Nikita Yolshin; Oula Mansour; Tamila Musaeva; Veronika Eder |
| EPI_ISL_2385214, EPI_ISL_2385215, EPI_ISL_2385217, EPI_ISL_2385219, EPI_ISL_2385220, EPI_ISL_2385233, EPI_ISL_2385242, EPI_ISL_2385246, EPI_ISL_2385249, EPI_ISL_2385259, EPI_ISL_2385264, EPI_ISL_2385267, EPI_ISL_2385272, EPI_ISL_2385273, EPI_ISL_2385275, EPI_ISL_2385278, EPI_ISL_2385279, EPI_ISL_2385282, EPI_ISL_2385283, EPI_ISL_2385284, EPI_ISL_2385285, EPI_ISL_2385286, EPI_ISL_2385288, EPI_ISL_2385289, EPI_ISL_2385290, EPI_ISL_2385292, EPI_ISL_2385294, EPI_ISL_2385295, EPI_ISL_2385296, EPI_ISL_2385297, EPI_ISL_2385299, EPI_ISL_2385300, EPI_ISL_2385301, EPI_ISL_2385302, EPI_ISL_2385304, EPI_ISL_2385305, EPI_ISL_2450589, EPI_ISL_2450593, EPI_ISL_2450595, EPI_ISL_2450596, EPI_ISL_2450605, EPI_ISL_2450606, EPI_ISL_2450607, EPI_ISL_2523386, EPI_ISL_2523398, EPI_ISL_2523608, EPI_ISL_2523611, EPI_ISL_2523614, EPI_ISL_2523617, EPI_ISL_2523620, EPI_ISL_2523623, EPI_ISL_2523627, EPI_ISL_2523629, EPI_ISL_2523633, EPI_ISL_2523636, EPI_ISL_2523639, EPI_ISL_2523641, EPI_ISL_2523644, EPI_ISL_2523648, EPI_ISL_2523651, EPI_ISL_2523654, EPI_ISL_2523657, EPI_ISL_2523661, EPI_ISL_2523664, EPI_ISL_2523862, EPI_ISL_2899759, EPI_ISL_2899760, EPI_ISL_2899761, EPI_ISL_2899762, EPI_ISL_2899763, EPI_ISL_2899764, EPI_ISL_2899765, EPI_ISL_2899766, EPI_ISL_2899767, EPI_ISL_2899768, EPI_ISL_2899769, EPI_ISL_2899770, EPI_ISL_2899771, EPI_ISL_2899772, EPI_ISL_2899773, EPI_ISL_2899774, EPI_ISL_2899775, EPI_ISL_2899776, EPI_ISL_2899777, EPI_ISL_2899778, EPI_ISL_2899779, EPI_ISL_2899780, EPI_ISL_2899781, EPI_ISL_2899782, EPI_ISL_2899783, EPI_ISL_2899784, EPI_ISL_2899785, EPI_ISL_2899786, EPI_ISL_2899787, EPI_ISL_2899788, EPI_ISL_2899789, EPI_ISL_2899790 | | | |
| see above | HELIX LLC | WHO National Influenza Centre Russian Federation | Andrey Komissarov; Artem Fadeev; Daria Danilenko; Dmitry Lioznov; Elena Nabieva; Georgii Bazykin; Kirill Varchenko; Ksenia Safina; Kseniya Komissarova; Maria Pisareva; Maria Timofeeva; Mikhail Bakaev; Nikita Yolshin; Oula Mansour; Oula Masour; Tamila Musaeva; Veronika Eder |
| EPI_ISL_2816224, EPI_ISL_2816225, EPI_ISL_2816226, EPI_ISL_2816227, EPI_ISL_2816228, EPI_ISL_2816229, EPI_ISL_2816230, EPI_ISL_2816231, EPI_ISL_2816232, EPI_ISL_2816233, EPI_ISL_2816234, EPI_ISL_2816235, EPI_ISL_2816236, EPI_ISL_2816237, EPI_ISL_2816238, EPI_ISL_2816239, EPI_ISL_2816240, EPI_ISL_2816241, EPI_ISL_2816242 | | | |
| see above | Infectious Diseases Hospital No 2 | WHO National Influenza Centre Russian Federation | Andrey Komissarov; Artem Fadeev; Daria Danilenko; Dmitry Lioznov; Elena Nabieva; Georgii Bazykin; Kirill Varchenko; Ksenia Safina; Kseniya Komissarova; Maria Pisareva; Mikhail Bakaev; Nikita Yolshin; Oula Mansour; Tamila Musaeva; Veronika Eder |
| EPI_ISL_2031976 | Molecular diagnostic laboratory of Federal Budget Institution of Science "Central Research Institute of Epidemiology" of The Federal Service on Customers' Rights Protection and Human Well-being Surveillance | Group of Genomics and Postgenomic Technologies of Central Research Institute of Epidemilology | Akimkin V.G.; Kaptelova V.V.; Kondrasheva L.Y.; Korneenko E.V.; Nadtoka M.I.; Saenko S.S.; Samojlov A.E.; Shipulina O.Y.; Sinicyn S.O.; Smirnova Y.S.; Speranskaya A.S.; Tivanova E.V. |
| EPI_ISL_2626223, EPI_ISL_2626225, EPI_ISL_2626230, EPI_ISL_2626234, EPI_ISL_2626235, EPI_ISL_2626244, EPI_ISL_2626350, EPI_ISL_2626351, EPI_ISL_2626352, EPI_ISL_2626353, EPI_ISL_2626354, EPI_ISL_2626355, EPI_ISL_2626375, EPI_ISL_2626392, EPI_ISL_2626393, EPI_ISL_2626394, EPI_ISL_2626395, EPI_ISL_2626396, EPI_ISL_2626401, EPI_ISL_2626402, EPI_ISL_2626403, EPI_ISL_2626404, EPI_ISL_2626405, EPI_ISL_2626408, EPI_ISL_2626409, EPI_ISL_2626410, EPI_ISL_2626411, EPI_ISL_2626412, EPI_ISL_2626413, EPI_ISL_2626414, EPI_ISL_2626415, EPI_ISL_2626416, EPI_ISL_2626417, EPI_ISL_2626418, EPI_ISL_2626419, EPI_ISL_2626420, EPI_ISL_2626421, EPI_ISL_2626422, EPI_ISL_2626423, EPI_ISL_2626424, EPI_ISL_2626425, EPI_ISL_2626426, EPI_ISL_2626427, EPI_ISL_2626428, EPI_ISL_2626430 | | | |
| see above | Molecular diagnostic laboratory of Federal Budget Institution of Science "Central Research Institute of Epidemiology" of The Federal Service on Customers' Rights Protection and Human Well-being Surveillance | Group of Genomics and Postgenomic Technologies of Central Research Institute of Epidemiology | Akimkin V.G.; Cherkashina A.S.; Kaptelova V.V.; Kondrasheva L.Y.; Korneenko E.V.; Nadtoka M.I.; Saenko S.S.; Samoilov A.E.; Sinitsyn S.O.; Smirnova Y.S.; Speranskaya A.S.; Tivanova E.V. |
| EPI_ISL_2662999, EPI_ISL_2663000, EPI_ISL_2663001, EPI_ISL_2663003, EPI_ISL_2663004, EPI_ISL_2663005, EPI_ISL_2663006, EPI_ISL_2663007, EPI_ISL_2663008, EPI_ISL_2663009, EPI_ISL_2663010, EPI_ISL_2663011, EPI_ISL_2663012, EPI_ISL_2663013, EPI_ISL_2663014, EPI_ISL_2663015, EPI_ISL_2663016, EPI_ISL_2663017, EPI_ISL_2663018, EPI_ISL_2663019, EPI_ISL_2663020, EPI_ISL_2663022, EPI_ISL_2663023, EPI_ISL_2663024, EPI_ISL_2663025, EPI_ISL_2663026, EPI_ISL_2663027, EPI_ISL_2663028, EPI_ISL_2663029, EPI_ISL_2663030, EPI_ISL_2663031, EPI_ISL_2663032, EPI_ISL_2663033, EPI_ISL_2663034, EPI_ISL_2663035, EPI_ISL_2663036, EPI_ISL_2663038, EPI_ISL_2663039, EPI_ISL_2663040, EPI_ISL_2663041, EPI_ISL_2663042, EPI_ISL_2663044, EPI_ISL_2663045, EPI_ISL_2663046, EPI_ISL_2663047, EPI_ISL_2663048, EPI_ISL_2663049, EPI_ISL_2663050, EPI_ISL_2663051, EPI_ISL_2663052, EPI_ISL_2663053, EPI_ISL_2663054, EPI_ISL_2663055, EPI_ISL_2663056, EPI_ISL_2663057, EPI_ISL_2663058, EPI_ISL_2663059, EPI_ISL_2663060, EPI_ISL_2663061, EPI_ISL_2663062, EPI_ISL_2663063, EPI_ISL_2663064, EPI_ISL_2663065, EPI_ISL_2663066, EPI_ISL_2663067, EPI_ISL_2663068, EPI_ISL_2663069, EPI_ISL_2663070, EPI_ISL_2663071, EPI_ISL_2663072, EPI_ISL_2663073, EPI_ISL_2663074, EPI_ISL_2663075, EPI_ISL_2663076, EPI_ISL_2663077, EPI_ISL_2663078, EPI_ISL_2663079, EPI_ISL_2663080, EPI_ISL_2663081, EPI_ISL_2663082, EPI_ISL_2663083, EPI_ISL_2663084, EPI_ISL_2663085, EPI_ISL_2663086, EPI_ISL_2663087, EPI_ISL_2663088, EPI_ISL_2663089, EPI_ISL_2663090, EPI_ISL_2663091, EPI_ISL_2663092, EPI_ISL_2663093, EPI_ISL_2663094, EPI_ISL_2663095, EPI_ISL_2663096, EPI_ISL_2663097, EPI_ISL_2663098, EPI_ISL_2663099, EPI_ISL_2663100, EPI_ISL_2663101, EPI_ISL_2663102, EPI_ISL_2663103, EPI_ISL_2663104, EPI_ISL_2663105, EPI_ISL_2663106, EPI_ISL_2663107, EPI_ISL_2663108, EPI_ISL_2663109, EPI_ISL_2663110, EPI_ISL_2663111, EPI_ISL_2663112, EPI_ISL_2663113, EPI_ISL_2663114, EPI_ISL_2663116, EPI_ISL_2663119, EPI_ISL_2663122, EPI_ISL_2663125, EPI_ISL_2663127, EPI_ISL_2663130, EPI_ISL_2663133, EPI_ISL_2663136, EPI_ISL_2663138, EPI_ISL_2663141, EPI_ISL_2663144, EPI_ISL_2663147, EPI_ISL_2663150, EPI_ISL_2663153, EPI_ISL_2663156, EPI_ISL_2663159, EPI_ISL_2663161, EPI_ISL_2663164, EPI_ISL_2663167, EPI_ISL_2663170, EPI_ISL_2663172, EPI_ISL_2663175, EPI_ISL_2663178, EPI_ISL_2663180, EPI_ISL_2663183, EPI_ISL_2663186, EPI_ISL_2663189, EPI_ISL_2663191, EPI_ISL_2816243, EPI_ISL_2816244, EPI_ISL_2816245, EPI_ISL_2816246, EPI_ISL_2816247, EPI_ISL_2816248, EPI_ISL_2816249, EPI_ISL_2816250, EPI_ISL_2816251, EPI_ISL_2816252, EPI_ISL_2816253, EPI_ISL_2816254, EPI_ISL_2816255, EPI_ISL_2816256, EPI_ISL_2816257, EPI_ISL_2816258, EPI_ISL_2816259, EPI_ISL_2816260, EPI_ISL_2816261, EPI_ISL_2816262, EPI_ISL_2816263, EPI_ISL_2816264, EPI_ISL_2816265, EPI_ISL_2816266, EPI_ISL_2816267, EPI_ISL_2816268, EPI_ISL_2816269, EPI_ISL_2816270, EPI_ISL_2816271, EPI_ISL_2816272, EPI_ISL_2816273, EPI_ISL_2816274, EPI_ISL_2816275, EPI_ISL_2816276, EPI_ISL_2816277, EPI_ISL_2816278, EPI_ISL_2816279, EPI_ISL_2816280, EPI_ISL_2816281, EPI_ISL_2816282, EPI_ISL_2816283, EPI_ISL_2816285, EPI_ISL_2816288, EPI_ISL_2816290, EPI_ISL_2816292, EPI_ISL_2816294, EPI_ISL_2816298, EPI_ISL_2816301, EPI_ISL_2816304, EPI_ISL_2816306, EPI_ISL_2816309, EPI_ISL_2816311, EPI_ISL_2816314, EPI_ISL_2816316, EPI_ISL_2816318, EPI_ISL_2816321, EPI_ISL_2816323, EPI_ISL_2816326, EPI_ISL_2816328, EPI_ISL_2816331, EPI_ISL_2816334, EPI_ISL_2816336, EPI_ISL_2816339, EPI_ISL_2816341, EPI_ISL_2816344, EPI_ISL_2816346, EPI_ISL_2816349, EPI_ISL_2816351, EPI_ISL_2816354, EPI_ISL_2816356, EPI_ISL_2816359, EPI_ISL_2816361, EPI_ISL_2816363, EPI_ISL_2816366, EPI_ISL_2816368, EPI_ISL_2816371, EPI_ISL_2816373, EPI_ISL_2816376, EPI_ISL_2816378, EPI_ISL_2816380, EPI_ISL_2816383, EPI_ISL_2816385, EPI_ISL_2816388, EPI_ISL_2816390, EPI_ISL_2816393, EPI_ISL_2816395, EPI_ISL_2816398, EPI_ISL_2816401, EPI_ISL_2816403, EPI_ISL_2816406, EPI_ISL_2816408, EPI_ISL_2816410, EPI_ISL_2899800, EPI_ISL_2899801, EPI_ISL_2899802, EPI_ISL_2899803, EPI_ISL_2899804, EPI_ISL_2899805, EPI_ISL_2899806, EPI_ISL_2899807, EPI_ISL_2899808, EPI_ISL_2899809, EPI_ISL_2899810, EPI_ISL_2899811, EPI_ISL_2899812, EPI_ISL_2899814, EPI_ISL_2899815, EPI_ISL_2899816, EPI_ISL_2899817, EPI_ISL_2899818, EPI_ISL_2899819, EPI_ISL_2899820, EPI_ISL_2899821, EPI_ISL_2899822, EPI_ISL_2899823, EPI_ISL_2899824, EPI_ISL_2899825, EPI_ISL_2899826, EPI_ISL_2899827, EPI_ISL_2899828, EPI_ISL_2899829, EPI_ISL_2899830, EPI_ISL_2899831, EPI_ISL_2899832, EPI_ISL_2899833, EPI_ISL_2899834, EPI_ISL_2899835, EPI_ISL_2899836, EPI_ISL_2899837, EPI_ISL_2899838, EPI_ISL_2899839, EPI_ISL_2899840, EPI_ISL_2899841, EPI_ISL_2899842, EPI_ISL_2899843, EPI_ISL_2899844, EPI_ISL_2899845, EPI_ISL_2899846, EPI_ISL_2899847, EPI_ISL_2899848, EPI_ISL_2899849, EPI_ISL_2899850, EPI_ISL_2899851, EPI_ISL_2899852, EPI_ISL_2899853, EPI_ISL_2899854, EPI_ISL_2899855, EPI_ISL_2899856, EPI_ISL_2899857, EPI_ISL_2899858, EPI_ISL_2899859, EPI_ISL_2899860, EPI_ISL_2899861, EPI_ISL_2899862, EPI_ISL_2899863, EPI_ISL_2899864, EPI_ISL_2899865, EPI_ISL_2899866, EPI_ISL_2899867, EPI_ISL_2899868, EPI_ISL_2899869, EPI_ISL_2899870, EPI_ISL_2899871, EPI_ISL_2899872, EPI_ISL_2899873, EPI_ISL_2899874, EPI_ISL_2899875, EPI_ISL_2899876, EPI_ISL_2899877, EPI_ISL_2899878, EPI_ISL_2899879, EPI_ISL_2899880, EPI_ISL_2899881, EPI_ISL_2899883, EPI_ISL_2899884, EPI_ISL_2899885, EPI_ISL_2899886, EPI_ISL_2899887, EPI_ISL_2899889, EPI_ISL_2899891, EPI_ISL_2899892, EPI_ISL_2899893, EPI_ISL_2899894, EPI_ISL_2899895, EPI_ISL_2899896, EPI_ISL_2899897, EPI_ISL_2899898, EPI_ISL_2899899, EPI_ISL_2899900, EPI_ISL_2899901, EPI_ISL_2899902, EPI_ISL_2899903, EPI_ISL_2899905, EPI_ISL_2899906, EPI_ISL_2899908, EPI_ISL_2899909, EPI_ISL_2899910, EPI_ISL_2899911, EPI_ISL_2899912, EPI_ISL_2899913, EPI_ISL_2899914, EPI_ISL_2899915, EPI_ISL_2899916, EPI_ISL_2899917, EPI_ISL_2899918, EPI_ISL_2899919, EPI_ISL_2899920, EPI_ISL_2899921, EPI_ISL_2899923, EPI_ISL_2899924, EPI_ISL_2899925 | | | |
| see above | N.F. Gamaleya Research Center for Epidemiology and Microbiology | WHO National Influenza Centre Russian Federation | Alexander Gintsburg; Alexey Shchetinin; Alina Odintsova; Andrei Botikov; Andrei Pochtovyi; Andrei Siniavin; Andrey Komissarov; Anna Kovyrshina; Artem Fadeev; Artem Tkachuk; Daria Danilenko; Denis Kleymenov; Denis Logunov; Dmitry Lioznov; Dmitry Shcheblyakov; Elena Mazunina; Elena Nabieva; Elena Shidlovskaya; Elizaveta Divisenko; Evgeniia Bykonia; Georgii Bazykin; Inna Dolzhikova; Kirill Varchenko; Ksenia Safina; Kseniya Komissarova; Liubov Popova; Ludmila Vasilchenko; Maria Nikiforova; Maria Pisareva; Maria Timofeeva; Mikhail Bakaev; Nadezhda Kuznetsova; Nikita Yolshin; Oula Mansour; Tamila Musaeva; Veronika Eder; Vladimir Gushchin |
| EPI_ISL_2450588 | SOGAZ PROFMEDICINA LLC | WHO National Influenza Centre Russian Federation | Andrey Komissarov; Artem Fadeev; Daria Danilenko; Dmitry Lioznov; Elena Nabieva; Galina Razbitskova; Georgii Bazykin; Kirill Varchenko; Ksenia Safina; Kseniya Komissarova; Larisa Gritsay; Maria Pisareva; Maria Timofeeva; Mikhail Bakaev; Nikita Yolshin; Oleg Lezhnev; Oula Mansour; Tamila Musaeva; Veronika Eder |
| EPI_ISL_2816196, EPI_ISL_2816197, EPI_ISL_2816198, EPI_ISL_2816199, EPI_ISL_2816200, EPI_ISL_2816201, EPI_ISL_2816202, EPI_ISL_2816203, EPI_ISL_2816204, EPI_ISL_2816205, EPI_ISL_2816206, EPI_ISL_2816207, EPI_ISL_2816208, EPI_ISL_2816209, EPI_ISL_2816210, EPI_ISL_2816211, EPI_ISL_2816212, EPI_ISL_2816213, EPI_ISL_2816214, EPI_ISL_2816215, EPI_ISL_2816216, EPI_ISL_2816217, EPI_ISL_2816218, EPI_ISL_2816219, EPI_ISL_2816220, EPI_ISL_2816221, EPI_ISL_2816222, EPI_ISL_2816223 | | | |
| see above | Sklifosovsky Research Institute of Emergency Medicine | WHO National Influenza Centre Russian Federation | Andrey Komissarov; Artem Fadeev; Daria Danilenko; Dmitry Lioznov; Elena Nabieva; Georgii Bazykin; Kirill Varchenko; Ksenia Safina; Kseniya Komissarova; Maria Pisareva; Mikhail Bakaev; Nikita Yolshin; Oula Mansour; Tamila Musaeva; Veronika Eder |
| EPI_ISL_2385074, EPI_ISL_2385075, EPI_ISL_2385076, EPI_ISL_2385077, EPI_ISL_2385078, EPI_ISL_2385079, EPI_ISL_2385080, EPI_ISL_2385081, EPI_ISL_2385082, EPI_ISL_2385083, EPI_ISL_2385084, EPI_ISL_2385085, EPI_ISL_2385086, EPI_ISL_2385087, EPI_ISL_2385088, EPI_ISL_2385089, EPI_ISL_2385090, EPI_ISL_2385091, EPI_ISL_2385092, EPI_ISL_2385093, EPI_ISL_2385094, EPI_ISL_2385095, EPI_ISL_2385096, EPI_ISL_2385097, EPI_ISL_2385098, EPI_ISL_2385099, EPI_ISL_2385100, EPI_ISL_2385101, EPI_ISL_2385102, EPI_ISL_2385103, EPI_ISL_2385104, EPI_ISL_2385105, EPI_ISL_2385106, EPI_ISL_2385107, EPI_ISL_2385108, EPI_ISL_2385109, EPI_ISL_2385110, EPI_ISL_2385111, EPI_ISL_2385112, EPI_ISL_2385113, EPI_ISL_2385114, EPI_ISL_2385115, EPI_ISL_2385116, EPI_ISL_2385117, EPI_ISL_2385118, EPI_ISL_2385119, EPI_ISL_2385120, EPI_ISL_2385121, EPI_ISL_2385122, EPI_ISL_2385123, EPI_ISL_2385124, EPI_ISL_2385125, EPI_ISL_2385126, EPI_ISL_2385127, EPI_ISL_2385128, EPI_ISL_2385129, EPI_ISL_2385130, EPI_ISL_2385131, EPI_ISL_2385132, EPI_ISL_2385133, EPI_ISL_2385134, EPI_ISL_2385135, EPI_ISL_2385136, EPI_ISL_2385137, EPI_ISL_2385138, EPI_ISL_2385139, EPI_ISL_2385140, EPI_ISL_2385141, EPI_ISL_2385142, EPI_ISL_2385143, EPI_ISL_2385144, EPI_ISL_2385145, EPI_ISL_2385146, EPI_ISL_2385147, EPI_ISL_2385148, EPI_ISL_2385149, EPI_ISL_2385150, EPI_ISL_2385151, EPI_ISL_2385152, EPI_ISL_2385153, EPI_ISL_2385154, EPI_ISL_2385155, EPI_ISL_2385156, EPI_ISL_2385157, EPI_ISL_2385158, EPI_ISL_2385159, EPI_ISL_2385160, EPI_ISL_2385161, EPI_ISL_2385162, EPI_ISL_2385163, EPI_ISL_2385164, EPI_ISL_2385165, EPI_ISL_2385166, EPI_ISL_2385167, EPI_ISL_2385168, EPI_ISL_2385169, EPI_ISL_2385170, EPI_ISL_2385171, EPI_ISL_2385172, EPI_ISL_2385173, EPI_ISL_2385174, EPI_ISL_2385175, EPI_ISL_2385176, EPI_ISL_2385177, EPI_ISL_2385178, EPI_ISL_2385179, EPI_ISL_2385180, EPI_ISL_2385181, EPI_ISL_2385182, EPI_ISL_2385183, EPI_ISL_2385184, EPI_ISL_2385185, EPI_ISL_2385186, EPI_ISL_2385187, EPI_ISL_2385188, EPI_ISL_2385189, EPI_ISL_2385190, EPI_ISL_2385191, EPI_ISL_2385192, EPI_ISL_2385193, EPI_ISL_2385194, EPI_ISL_2385195, EPI_ISL_2385196, EPI_ISL_2385197, EPI_ISL_2385198, EPI_ISL_2385199, EPI_ISL_2385200, EPI_ISL_2385201, EPI_ISL_2385202, EPI_ISL_2385203, EPI_ISL_2385204, EPI_ISL_2385205, EPI_ISL_2385206, EPI_ISL_2385207, EPI_ISL_2385208, EPI_ISL_2385209, EPI_ISL_2385210, EPI_ISL_2385211, EPI_ISL_2385309, EPI_ISL_2385310, EPI_ISL_2385311, EPI_ISL_2385312, EPI_ISL_2385313, EPI_ISL_2385314, EPI_ISL_2385315, EPI_ISL_2385316, EPI_ISL_2385317, EPI_ISL_2385318, EPI_ISL_2385319, EPI_ISL_2385320, EPI_ISL_2385321, EPI_ISL_2385322, EPI_ISL_2385323, EPI_ISL_2385324, EPI_ISL_2385325, EPI_ISL_2385326, EPI_ISL_2385327, EPI_ISL_2385328, EPI_ISL_2385329, EPI_ISL_2385330, EPI_ISL_2385331, EPI_ISL_2385332, EPI_ISL_2385333, EPI_ISL_2385334, EPI_ISL_2385335, EPI_ISL_2385336, EPI_ISL_2385426, EPI_ISL_2385427, EPI_ISL_2385428, EPI_ISL_2385429, EPI_ISL_2385430, EPI_ISL_2385431, EPI_ISL_2450454, EPI_ISL_2450455, EPI_ISL_2450456, EPI_ISL_2450457, EPI_ISL_2450458, EPI_ISL_2450459, EPI_ISL_2450460, EPI_ISL_2450461, EPI_ISL_2450462, EPI_ISL_2450463, EPI_ISL_2450464, EPI_ISL_2450465, EPI_ISL_2450466, EPI_ISL_2450467, EPI_ISL_2450468, EPI_ISL_2450469, EPI_ISL_2450470, EPI_ISL_2450471, EPI_ISL_2450472, EPI_ISL_2450473, EPI_ISL_2450474, EPI_ISL_2450475, EPI_ISL_2450476, EPI_ISL_2450477, EPI_ISL_2450478, EPI_ISL_2450479, EPI_ISL_2450480, EPI_ISL_2450481, EPI_ISL_2450482, EPI_ISL_2450483, EPI_ISL_2450484, EPI_ISL_2450485, EPI_ISL_2450486, EPI_ISL_2450487, EPI_ISL_2450488, EPI_ISL_2450489, EPI_ISL_2450490, EPI_ISL_2450491, EPI_ISL_2450492, EPI_ISL_2450493, EPI_ISL_2450494, EPI_ISL_2450495, EPI_ISL_2450496, EPI_ISL_2450497, EPI_ISL_2450498, EPI_ISL_2450499, EPI_ISL_2450500, EPI_ISL_2450501, EPI_ISL_2450502, EPI_ISL_2450503, EPI_ISL_2450504, EPI_ISL_2450505, EPI_ISL_2450506, EPI_ISL_2450507, EPI_ISL_2450508, EPI_ISL_2450509, EPI_ISL_2450510, EPI_ISL_2450511, EPI_ISL_2450512, EPI_ISL_2450513, EPI_ISL_2450514, EPI_ISL_2450515, EPI_ISL_2450516, EPI_ISL_2450517, EPI_ISL_2450518, EPI_ISL_2450519, EPI_ISL_2450520, EPI_ISL_2450521, EPI_ISL_2450522, EPI_ISL_2450523, EPI_ISL_2450524, EPI_ISL_2450525, EPI_ISL_2450526, EPI_ISL_2450527, EPI_ISL_2450528, EPI_ISL_2450529, EPI_ISL_2450530, EPI_ISL_2450531, EPI_ISL_2450532, EPI_ISL_2450533, EPI_ISL_2450534, EPI_ISL_2450535, EPI_ISL_2450536, EPI_ISL_2450537, EPI_ISL_2450538, EPI_ISL_2450539, EPI_ISL_2450540, EPI_ISL_2450541, EPI_ISL_2450542, EPI_ISL_2450597, EPI_ISL_2450598, EPI_ISL_2450599, EPI_ISL_2450600, EPI_ISL_2450601, EPI_ISL_2450602, EPI_ISL_2450603, EPI_ISL_2450604, EPI_ISL_2462879, EPI_ISL_2462880, EPI_ISL_2523389, EPI_ISL_2523392, EPI_ISL_2523395, EPI_ISL_2523400, EPI_ISL_2523403, EPI_ISL_2523406, EPI_ISL_2523409, EPI_ISL_2523420, EPI_ISL_2523422, EPI_ISL_2523425, EPI_ISL_2523428, EPI_ISL_2523430, EPI_ISL_2523433, EPI_ISL_2523436, EPI_ISL_2523438, EPI_ISL_2523442, EPI_ISL_2523444, EPI_ISL_2523448, EPI_ISL_2523451, EPI_ISL_2523454, EPI_ISL_2523457, EPI_ISL_2523460, EPI_ISL_2523463, EPI_ISL_2523466, EPI_ISL_2523469, EPI_ISL_2523473, EPI_ISL_2523476, EPI_ISL_2523480, EPI_ISL_2523482, EPI_ISL_2523485, EPI_ISL_2523488, EPI_ISL_2523491, EPI_ISL_2523494, EPI_ISL_2523497, EPI_ISL_2523500, EPI_ISL_2523503, EPI_ISL_2523507, EPI_ISL_2523511, EPI_ISL_2523513, EPI_ISL_2523516, EPI_ISL_2523518, EPI_ISL_2523522, EPI_ISL_2523525, EPI_ISL_2523528, EPI_ISL_2523531, EPI_ISL_2523534, EPI_ISL_2523536, EPI_ISL_2523538, EPI_ISL_2523540, EPI_ISL_2523543, EPI_ISL_2523546, EPI_ISL_2523549, EPI_ISL_2523552, EPI_ISL_2523555, EPI_ISL_2523559, EPI_ISL_2523562, EPI_ISL_2523565, EPI_ISL_2523568, EPI_ISL_2523571, EPI_ISL_2523575, EPI_ISL_2523578, EPI_ISL_2523581, EPI_ISL_2523584, EPI_ISL_2523587, EPI_ISL_2523590, EPI_ISL_2523594, EPI_ISL_2523596, EPI_ISL_2523599, EPI_ISL_2523602, EPI_ISL_2523605, EPI_ISL_2523667, EPI_ISL_2523669, EPI_ISL_2523670, EPI_ISL_2523672, EPI_ISL_2523675, EPI_ISL_2523678, EPI_ISL_2523682, EPI_ISL_2523684, EPI_ISL_2523687, EPI_ISL_2523690, EPI_ISL_2523694, EPI_ISL_2523696, EPI_ISL_2523700, EPI_ISL_2523702, EPI_ISL_2523705, EPI_ISL_2523708, EPI_ISL_2523711, EPI_ISL_2523714, EPI_ISL_2523718, EPI_ISL_2523721, EPI_ISL_2523722, EPI_ISL_2523724, EPI_ISL_2523726, EPI_ISL_2523729, EPI_ISL_2523732, EPI_ISL_2523736, EPI_ISL_2523738, EPI_ISL_2523741, EPI_ISL_2523744, EPI_ISL_2523747, EPI_ISL_2523752, EPI_ISL_2523755, EPI_ISL_2523759, EPI_ISL_2523762, EPI_ISL_2523765, EPI_ISL_2523768, EPI_ISL_2523771, EPI_ISL_2523774, EPI_ISL_2523777, EPI_ISL_2523780, EPI_ISL_2523783, EPI_ISL_2523786, EPI_ISL_2523789, EPI_ISL_2523793, EPI_ISL_2523796, EPI_ISL_2523799, EPI_ISL_2523803, EPI_ISL_2698425, EPI_ISL_2698433, EPI_ISL_2698439, EPI_ISL_2698442, EPI_ISL_2698444, EPI_ISL_2698447, EPI_ISL_2698450, EPI_ISL_2698455, EPI_ISL_2698458, EPI_ISL_2698461, EPI_ISL_2698467, EPI_ISL_2698470, EPI_ISL_2698473, EPI_ISL_2698476, EPI_ISL_2698478, EPI_ISL_2698481, EPI_ISL_2698484, EPI_ISL_2698487, EPI_ISL_2698490, EPI_ISL_2698493, EPI_ISL_2698496, EPI_ISL_2698499, EPI_ISL_2698502, EPI_ISL_2698504, EPI_ISL_2698507, EPI_ISL_2698509, EPI_ISL_2698512, EPI_ISL_2698515, EPI_ISL_2698517, EPI_ISL_2698520, EPI_ISL_2698522, EPI_ISL_2698527, EPI_ISL_2698530, EPI_ISL_2698532, EPI_ISL_2698535, EPI_ISL_2698537, EPI_ISL_2698539, EPI_ISL_2698542, EPI_ISL_2698545, EPI_ISL_2698547, EPI_ISL_2698549, EPI_ISL_2698552, EPI_ISL_2698554, EPI_ISL_2698557, EPI_ISL_2698559, EPI_ISL_2698562, EPI_ISL_2698564, EPI_ISL_2698567, EPI_ISL_2698570, EPI_ISL_2698572, EPI_ISL_2698575, EPI_ISL_2698577, EPI_ISL_2698579, EPI_ISL_2698582, EPI_ISL_2698585, EPI_ISL_2698586, EPI_ISL_2698588, EPI_ISL_2698590, EPI_ISL_2698591, EPI_ISL_2698593, EPI_ISL_2698595, EPI_ISL_2698598, EPI_ISL_2698601, EPI_ISL_2698604, EPI_ISL_2698607, EPI_ISL_2698610, EPI_ISL_2698612, EPI_ISL_2698615, EPI_ISL_2698617, EPI_ISL_2698620, EPI_ISL_2698622, EPI_ISL_2698625, EPI_ISL_2698628, EPI_ISL_2698631, EPI_ISL_2698634, EPI_ISL_2698636, EPI_ISL_2698639, EPI_ISL_2698641, EPI_ISL_2698644, EPI_ISL_2698646, EPI_ISL_2698648, EPI_ISL_2698651, EPI_ISL_2698653, EPI_ISL_2698655, EPI_ISL_2698658, EPI_ISL_2698661, EPI_ISL_2698664, EPI_ISL_2698666, EPI_ISL_2698669, EPI_ISL_2698672, EPI_ISL_2698675, EPI_ISL_2698678, EPI_ISL_2698680, EPI_ISL_2816144, EPI_ISL_2816146, EPI_ISL_2816149, EPI_ISL_2816150, EPI_ISL_2816151, EPI_ISL_2816152, EPI_ISL_2816153, EPI_ISL_2816154, EPI_ISL_2816155, EPI_ISL_2816156, EPI_ISL_2816157, EPI_ISL_2816158, EPI_ISL_2816159, EPI_ISL_2816160, EPI_ISL_2816161, EPI_ISL_2816162, EPI_ISL_2816163, EPI_ISL_2816164, EPI_ISL_2816165, EPI_ISL_2816166, EPI_ISL_2816167, EPI_ISL_2816168, EPI_ISL_2816169, EPI_ISL_2816170, EPI_ISL_2816171, EPI_ISL_2816172, EPI_ISL_2816173, EPI_ISL_2816174, EPI_ISL_2816175, EPI_ISL_2816176, EPI_ISL_2816177, EPI_ISL_2816178, EPI_ISL_2816179, EPI_ISL_2816180, EPI_ISL_2816181, EPI_ISL_2816182, EPI_ISL_2816183, EPI_ISL_2816184, EPI_ISL_2816185, EPI_ISL_2816186, EPI_ISL_2816187, EPI_ISL_2816188, EPI_ISL_2816189, EPI_ISL_2816190, EPI_ISL_2816191, EPI_ISL_2816192, EPI_ISL_2816193, EPI_ISL_2816194 | | | |
| see above | WHO National Influenza Centre Russian Federation | WHO National Influenza Centre Russian Federation | Alexey Masharsky; Andrey Komissarov; Artem Fadeev; Daria Danilenko; Dmitry Lioznov; Elena Nabieva; Georgii Bazykin; Kirill Varchenko; Ksenia Safina; Kseniya Komissarova; Maria Baturova; Maria Pisareva; Maria Timofeeva; Mikhail Bakaev; Nikita Yolshin; Oula Mansour; Oula Masour; Tamila Musaeva; Veronika Eder |

We gratefully acknowledge the following Authors from the Originating laboratories responsible for obtaining the specimens, as well as the Submitting laboratories where the genome data were generated and shared via GISAID, on which this research is based.

All Submitters of data may be contacted directly via www.gisaid.org

Authors are sorted alphabetically.

| Accession ID | Originating Laboratory | Submitting Laboratory | Authors |
|---|---|---|---|
| EPI_ISL_1534286, EPI_ISL_1817693, EPI_ISL_1817737, EPI_ISL_1817739, EPI_ISL_2140694, EPI_ISL_2140715, EPI_ISL_2140717 | | | |
| see above | 2 Military Hospital wc MAA | NHLS/UCT | Arash Iranzadeh; Bruna Galvao; Carolyn Williamson; Deelan Doolabh; Diana Hardie; Innocent Mudau; Kruger Marais; Lynn Tyers; Marvin Hsiao; Stephen Korsman |
| EPI_ISL_2087002, EPI_ISL_2087003, EPI_ISL_2087004, EPI_ISL_2087005, EPI_ISL_2087006, EPI_ISL_2087007, EPI_ISL_2087008, EPI_ISL_2087009, EPI_ISL_2087010, EPI_ISL_2087011, EPI_ISL_2087012, EPI_ISL_2087013, EPI_ISL_2087014, EPI_ISL_2087015, EPI_ISL_2087016, EPI_ISL_2087017, EPI_ISL_2087018, EPI_ISL_2087019, EPI_ISL_2087020, EPI_ISL_2087021, EPI_ISL_2087022, EPI_ISL_2087023, EPI_ISL_2087024, EPI_ISL_2087025, EPI_ISL_2087026, EPI_ISL_2087027, EPI_ISL_2087028, EPI_ISL_2087029 | | | |
| see above | AMPATH LABORATORIES | National Institute for Communicable Diseases of the National Health Laboratory Service | Amoako DG; Bhiman JN; Ismail A; Mahlangu B; Mohale T; Ntuli N; Scheepers C |
| EPI_ISL_1534281 | Albertinia Clinic wc AAP | NHLS/UCT | Arash Iranzadeh; Bruna Galvao; Carolyn Williamson; Deelan Doolabh; Diana Hardie; Innocent Mudau; Kruger Marais; Lynn Tyers; Marvin Hsiao; Stephen Korsman |
| EPI_ISL_1994608 | Ampath - Kingsway | KRISP, KZN Research Innovation and Sequencing Platform | Emmanuel SJ; Giandhari J; Khan S; Lessells R; Mdlalose K; Naidoo Y; Pillay S; Ramphal U; Tegally H; Wilkinson E; York D; de Oliveira T |
| EPI_ISL_2178779, EPI_ISL_2178780, EPI_ISL_2178781, EPI_ISL_2178782, EPI_ISL_2178783, EPI_ISL_2178784, EPI_ISL_2178785, EPI_ISL_2178786, EPI_ISL_2178787, EPI_ISL_2178788, EPI_ISL_2178789, EPI_ISL_2178790, EPI_ISL_2178791, EPI_ISL_2178792, EPI_ISL_2178793, EPI_ISL_2178794 | | | |
| see above | Ampath Laboratories | National Institute for Communicable Diseases of the National Health Laboratory Service | Amoako DG; Bhiman JN; Glass A; Gottberg A; Mahlangu B; Marshall T; Mohale T; Ntuli N; Scheepers C; Viana R |
| EPI_ISL_2087030, EPI_ISL_2087031, EPI_ISL_2087032, EPI_ISL_2087034, EPI_ISL_2087035, EPI_ISL_2087036, EPI_ISL_2087037, EPI_ISL_2087038, EPI_ISL_2087039, EPI_ISL_2087040, EPI_ISL_2087041, EPI_ISL_2087042, EPI_ISL_2087043, EPI_ISL_2087044, EPI_ISL_2087045, EPI_ISL_2087046, EPI_ISL_2153523, EPI_ISL_2153525, EPI_ISL_2153526, EPI_ISL_2153527, EPI_ISL_2153528, EPI_ISL_2153529, EPI_ISL_2153530, EPI_ISL_2153531, EPI_ISL_2153532, EPI_ISL_2153533, EPI_ISL_2153534, EPI_ISL_2153535, EPI_ISL_2153536, EPI_ISL_2153537 | | | |
| see above | CHRIS HANI BARAGWANATH LABORATORY | National Institute for Communicable Diseases of the National Health Laboratory Service | Amoako DG; Bhiman JN; Ismail A; Mahlangu B; Mohale T; Ntuli N; Scheepers C |
| EPI_ISL_1817660, EPI_ISL_1817661, EPI_ISL_1817672, EPI_ISL_1817678, EPI_ISL_1817685, EPI_ISL_1817705, EPI_ISL_1817708, EPI_ISL_1817717, EPI_ISL_1817724 | | | |
| see above | Calvinia Corr Centre | NHLS/UCT | Arash Iranzadeh; Bruna Galvao; Carolyn Williamson; Deelan Doolabh; Diana Hardie; Innocent Mudau; Kruger Marais; Lynn Tyers; Marvin Hsiao; Stephen Korsman |
| EPI_ISL_1817719 | Calvinia Hospital | NHLS/UCT | Arash Iranzadeh; Bruna Galvao; Carolyn Williamson; Deelan Doolabh; Diana Hardie; Innocent Mudau; Kruger Marais; Lynn Tyers; Marvin Hsiao; Stephen Korsman |
| EPI_ISL_1817666, EPI_ISL_1817677, EPI_ISL_1817695, EPI_ISL_1817718, EPI_ISL_1817721, EPI_ISL_1817722, EPI_ISL_1817726, EPI_ISL_1817727, EPI_ISL_1817729, EPI_ISL_1817730, EPI_ISL_1817731 | | | |
| see above | Calvinia RSC Clinic | NHLS/UCT | Arash Iranzadeh; Bruna Galvao; Carolyn Williamson; Deelan Doolabh; Diana Hardie; Innocent Mudau; Kruger Marais; Lynn Tyers; Marvin Hsiao; Stephen Korsman |
| EPI_ISL_1817733 | Conville CDC wc CVC | NHLS/UCT | Arash Iranzadeh; Bruna Galvao; Carolyn Williamson; Deelan Doolabh; Diana Hardie; Innocent Mudau; Kruger Marais; Lynn Tyers; Marvin Hsiao; Stephen Korsman |
| EPI_ISL_2140706, EPI_ISL_2140707 | D'Almeida Clinic wc DAL | NHLS/UCT | Arash Iranzadeh; Bruna Galvao; Carolyn Williamson; Deelan Doolabh; Diana Hardie; Innocent Mudau; Kruger Marais; Lynn Tyers; Marvin Hsiao; Stephen Korsman |
| EPI_ISL_2140727 | De Rust (Blommenek) Clinic wc DRC | NHLS/UCT | Arash Iranzadeh; Bruna Galvao; Carolyn Williamson; Deelan Doolabh; Diana Hardie; Innocent Mudau; Kruger Marais; Lynn Tyers; Marvin Hsiao; Stephen Korsman |
| EPI_ISL_1502146, EPI_ISL_1502147, EPI_ISL_1502148, EPI_ISL_1502149, EPI_ISL_1502150, EPI_ISL_1502151, EPI_ISL_1502152, EPI_ISL_1502153, EPI_ISL_1502154, EPI_ISL_1502155, EPI_ISL_1502156, EPI_ISL_1502157, EPI_ISL_1502158, EPI_ISL_1502159, EPI_ISL_1502160, EPI_ISL_1502161, EPI_ISL_1502162, EPI_ISL_1502163, EPI_ISL_1502164, EPI_ISL_1502165, EPI_ISL_1502166, EPI_ISL_1502167, EPI_ISL_1502168, EPI_ISL_1502169, EPI_ISL_1502170, EPI_ISL_1502171, EPI_ISL_1502172, EPI_ISL_1502173, EPI_ISL_1502174, EPI_ISL_1502175, EPI_ISL_1502176, EPI_ISL_1502177, EPI_ISL_1502178, EPI_ISL_1502179, EPI_ISL_1502180, EPI_ISL_1502181, EPI_ISL_1502182, EPI_ISL_1502183, EPI_ISL_1502184, EPI_ISL_1502185, EPI_ISL_1502186, EPI_ISL_1502187, EPI_ISL_1502188, EPI_ISL_1502189, EPI_ISL_1502190, EPI_ISL_1502191, EPI_ISL_1502192, EPI_ISL_1502193, EPI_ISL_1502194, EPI_ISL_1502195, EPI_ISL_1502196, EPI_ISL_1502197, EPI_ISL_1502198 | | | |
| see above | Division of Medical Virology, National Health Laboratory Service (NHLS), Tygerberg Hospital / Stellenbosch University | Division of Medical Virology, Stellenbosch University and NHLS Tygerberg Hospital | Bronwyn Kleinhans; Gert van Zyl; Susan Engelbrecht; Wolfgang Preiser |
| EPI_ISL_1534299, EPI_ISL_1817689, EPI_ISL_2140725, EPI_ISL_2140728 | Dysselsdorp Clinic wc DDC | NHLS/UCT | Arash Iranzadeh; Bruna Galvao; Carolyn Williamson; Deelan Doolabh; Diana Hardie; Innocent Mudau; Kruger Marais; Lynn Tyers; Marvin Hsiao; Stephen Korsman |
| EPI_ISL_1817670, EPI_ISL_1817734 | Fraserburg CHC | NHLS/UCT | Arash Iranzadeh; Bruna Galvao; Carolyn Williamson; Deelan Doolabh; Diana Hardie; Innocent Mudau; Kruger Marais; Lynn Tyers; Marvin Hsiao; Stephen Korsman |
| EPI_ISL_1817710, EPI_ISL_1817711, EPI_ISL_1817712 | George Central CDC wc GCC | NHLS/UCT | Arash Iranzadeh; Bruna Galvao; Carolyn Williamson; Deelan Doolabh; Diana Hardie; Innocent Mudau; Kruger Marais; Lynn Tyers; Marvin Hsiao; Stephen Korsman |
| EPI_ISL_2140696 | George Hospital wc GRH | NHLS/UCT | Arash Iranzadeh; Bruna Galvao; Carolyn Williamson; Deelan Doolabh; Diana Hardie; Innocent Mudau; Kruger Marais; Lynn Tyers; Marvin Hsiao; Stephen Korsman |
| EPI_ISL_1534308, EPI_ISL_1534309, EPI_ISL_1817663, EPI_ISL_1817667, EPI_ISL_1817681, EPI_ISL_1817702, EPI_ISL_1817703, EPI_ISL_1817704, EPI_ISL_1817744, EPI_ISL_1817745, EPI_ISL_1817746, EPI_ISL_1817747, EPI_ISL_1817748, EPI_ISL_1817749, EPI_ISL_2140691, EPI_ISL_2140700, EPI_ISL_2140701, EPI_ISL_2140702, EPI_ISL_2140703, EPI_ISL_2140704, EPI_ISL_2140712, EPI_ISL_2140713 | | | |
| see above | Groote Schuur Hospital wc GSH | NHLS/UCT | Arash Iranzadeh; Bruna Galvao; Carolyn Williamson; Deelan Doolabh; Diana Hardie; Innocent Mudau; Kruger Marais; Lynn Tyers; Marvin Hsiao; Stephen Korsman |
| EPI_ISL_1817673 | Guguletu CHC wc GDH | NHLS/UCT | Arash Iranzadeh; Bruna Galvao; Carolyn Williamson; Deelan Doolabh; Diana Hardie; Innocent Mudau; Kruger Marais; Lynn Tyers; Marvin Hsiao; Stephen Korsman |
| EPI_ISL_1817735 | Heideveld Emergency Centre | NHLS/UCT | Arash Iranzadeh; Bruna Galvao; Carolyn Williamson; Deelan Doolabh; Diana Hardie; Innocent Mudau; Kruger Marais; Lynn Tyers; Marvin Hsiao; Stephen Korsman |
| EPI_ISL_2140721 | Hillside Clinic wc BFW | NHLS/UCT | Arash Iranzadeh; Bruna Galvao; Carolyn Williamson; Deelan Doolabh; Diana Hardie; Innocent Mudau; Kruger Marais; Lynn Tyers; Marvin Hsiao; Stephen Korsman |
| EPI_ISL_2140722 | Knysna Hospital wc KNY | NHLS/UCT | Arash Iranzadeh; Bruna Galvao; Carolyn Williamson; Deelan Doolabh; Diana Hardie; Innocent Mudau; Kruger Marais; Lynn Tyers; Marvin Hsiao; Stephen Korsman |
| EPI_ISL_2013048, EPI_ISL_2013049, EPI_ISL_2013050, EPI_ISL_2013051, EPI_ISL_2013052 | Lancet Laboratories | National Institute for Communicable Diseases of the National Health Laboratory Service | Amoako DG; Bhiman JN; Glass A; Mahlangu B; Mohale T; Ntuli N; Scheepers C; Viana R |
| EPI_ISL_1817662, EPI_ISL_1817664, EPI_ISL_1817679, EPI_ISL_1817682, EPI_ISL_1817709, EPI_ISL_2140698, EPI_ISL_2140710 | | | |
| see above | Mitchells Plain Hospital wc MPH | NHLS/UCT | Arash Iranzadeh; Bruna Galvao; Carolyn Williamson; Deelan Doolabh; Diana Hardie; Innocent Mudau; Kruger Marais; Lynn Tyers; Marvin Hsiao; Stephen Korsman |
| EPI_ISL_2162339, EPI_ISL_2162340, EPI_ISL_2162341, EPI_ISL_2162342, EPI_ISL_2162343, EPI_ISL_2162344, EPI_ISL_2162345, EPI_ISL_2162346, EPI_ISL_2162347, EPI_ISL_2162348, EPI_ISL_2162349, EPI_ISL_2162350, EPI_ISL_2162351, EPI_ISL_2162352, EPI_ISL_2162353, EPI_ISL_2162354, EPI_ISL_2162355, EPI_ISL_2162356, EPI_ISL_2162357, EPI_ISL_2162358, EPI_ISL_2162359, EPI_ISL_2162360, EPI_ISL_2162361, EPI_ISL_2162362, EPI_ISL_2162363, EPI_ISL_2162364, EPI_ISL_2162365, EPI_ISL_2162366, EPI_ISL_2162367, EPI_ISL_2162368, EPI_ISL_2162369, EPI_ISL_2162370, EPI_ISL_2162371, EPI_ISL_2162372, EPI_ISL_2162373, EPI_ISL_2162374, EPI_ISL_2162375, EPI_ISL_2162376, EPI_ISL_2162377, EPI_ISL_2162378, EPI_ISL_2162379, EPI_ISL_2162380, EPI_ISL_2162381, EPI_ISL_2162382 | | | |
| see above | NHLS Universitas Academic | UFS Virology | D Goedhals; Emmanuel Ogunbayo; MM Nyaga; MT Mogotsi; P Nthiga; PA Bester; T de Oliveira |
| EPI_ISL_2140645, EPI_ISL_2140646, EPI_ISL_2140647, EPI_ISL_2140648, EPI_ISL_2140649, EPI_ISL_2140650, EPI_ISL_2140651, EPI_ISL_2140729 | | | |
| see above | NHLS/UCT | NHLS/UCT | Arash Iranzadeh; Bruna Galvao; Carolyn Williamson; Deelan Doolabh; Diana Hardie; Innocent Mudau; Kruger Marais; Lynn Tyers; Marvin Hsiao; Stephen Korsman |
| EPI_ISL_1550912, EPI_ISL_1550914, EPI_ISL_1550915, EPI_ISL_1550916, EPI_ISL_1550917, EPI_ISL_1550918, EPI_ISL_1550919, EPI_ISL_1550920, EPI_ISL_1550922, EPI_ISL_1550923, EPI_ISL_1550925, EPI_ISL_1550926, EPI_ISL_1550927, EPI_ISL_1550928, EPI_ISL_1550929, EPI_ISL_1550931, EPI_ISL_1550933, EPI_ISL_1550934, EPI_ISL_1550935, EPI_ISL_1550936, EPI_ISL_1550937, EPI_ISL_1550940, EPI_ISL_1550943, EPI_ISL_1550944, EPI_ISL_1550945, EPI_ISL_1550946, EPI_ISL_1550947, EPI_ISL_1550948, EPI_ISL_1550949, EPI_ISL_1550950, EPI_ISL_1550951, EPI_ISL_1550952, EPI_ISL_1550953, EPI_ISL_1550954, EPI_ISL_1550955, EPI_ISL_1550956, EPI_ISL_1550957, EPI_ISL_1550958, EPI_ISL_1550959, EPI_ISL_1550960, EPI_ISL_1550961, EPI_ISL_1550962, EPI_ISL_1817755, EPI_ISL_1817756, EPI_ISL_1817757, EPI_ISL_1817758, EPI_ISL_1817759, EPI_ISL_1817760, EPI_ISL_1817761, EPI_ISL_1817762, EPI_ISL_1817763, EPI_ISL_1817764, EPI_ISL_1817765, EPI_ISL_1817766, EPI_ISL_1817767, EPI_ISL_1817768, EPI_ISL_1817769, EPI_ISL_1817770, EPI_ISL_1817771, EPI_ISL_1817772, EPI_ISL_1817773, EPI_ISL_1817774, EPI_ISL_1817775, EPI_ISL_1817776, EPI_ISL_1817777, EPI_ISL_1817778, EPI_ISL_1817779, EPI_ISL_1817780, EPI_ISL_1817781, EPI_ISL_1817784, EPI_ISL_1817785, EPI_ISL_1817786, EPI_ISL_1817787, EPI_ISL_1817788, EPI_ISL_1817789, EPI_ISL_1817790, EPI_ISL_1817791, EPI_ISL_1817792, EPI_ISL_1817793, EPI_ISL_1817794, EPI_ISL_1817795, EPI_ISL_1817796, EPI_ISL_1817797, EPI_ISL_1817798, EPI_ISL_1817799, EPI_ISL_1817800, EPI_ISL_1817801, EPI_ISL_1817802, EPI_ISL_1817803, EPI_ISL_1817804, EPI_ISL_1817805, EPI_ISL_1817806, EPI_ISL_1817807, EPI_ISL_1817808, EPI_ISL_1817809, EPI_ISL_1817810, EPI_ISL_1817811, EPI_ISL_1817812, EPI_ISL_1817813, EPI_ISL_1817814, EPI_ISL_1817815, EPI_ISL_1817816, EPI_ISL_1817817, EPI_ISL_1817818, EPI_ISL_1817819, EPI_ISL_1817820, EPI_ISL_1817821, EPI_ISL_1817822, EPI_ISL_1817823, EPI_ISL_1817824, EPI_ISL_1817825, EPI_ISL_1817826, EPI_ISL_1817827, EPI_ISL_1817828, EPI_ISL_1817829, EPI_ISL_1817830, EPI_ISL_1817831, EPI_ISL_1817832, EPI_ISL_1817833, EPI_ISL_1817834, EPI_ISL_1817835, EPI_ISL_1817836, EPI_ISL_1817837, EPI_ISL_1817838, EPI_ISL_1817839, EPI_ISL_1817840, EPI_ISL_1817841, EPI_ISL_1817842, EPI_ISL_1817843, EPI_ISL_1817844, EPI_ISL_1817845, EPI_ISL_1817846, EPI_ISL_1817848, EPI_ISL_1817849, EPI_ISL_1817850, EPI_ISL_1817851, EPI_ISL_1817852, EPI_ISL_1817853, EPI_ISL_1817854, EPI_ISL_1817858, EPI_ISL_1817859, EPI_ISL_1817860, EPI_ISL_1817861, EPI_ISL_1817862, EPI_ISL_1817863, EPI_ISL_2086228, EPI_ISL_2086229, EPI_ISL_2086230, EPI_ISL_2086231, EPI_ISL_2086232, EPI_ISL_2086233, EPI_ISL_2086234, EPI_ISL_2086235, EPI_ISL_2086236, EPI_ISL_2086237, EPI_ISL_2086238, EPI_ISL_2086239, EPI_ISL_2086240, EPI_ISL_2086241, EPI_ISL_2086242, EPI_ISL_2086243, EPI_ISL_2086244, EPI_ISL_2086245, EPI_ISL_2086246, EPI_ISL_2086247 | | | |
| see above | National Health Laboratory Service, South Africa | KRISP, KZN Research Innovation and Sequencing Platform | Emmanuel SJ; Giandhari J; Khan S; Lessells R; Mdlalose K; Naidoo Y; Pillay S; Ramphal U; Tegally H; Wilkinson E; York D; de Oliveira T |
| EPI_ISL_1914928, EPI_ISL_1914929, EPI_ISL_1914930, EPI_ISL_1914931, EPI_ISL_1914932, EPI_ISL_1914933, EPI_ISL_1914934, EPI_ISL_1914935, EPI_ISL_1914936 | | | |
| see above | National Institute for Communicable Diseases,National Health Laboratory Services, Gauteng, South Africa | National Institute for Communicable Diseases of the National Health Laboratory Service | Amoako DG; Bhiman JN; Ismail A; Mahlangu B; Mohale T; Ntuli N; Scheepers C |
| EPI_ISL_2140697 | Nelspoort Clinic wc NPP | NHLS/UCT | Arash Iranzadeh; Bruna Galvao; Carolyn Williamson; Deelan Doolabh; Diana Hardie; Innocent Mudau; Kruger Marais; Lynn Tyers; Marvin Hsiao; Stephen Korsman |

| | | | |
|---|---|---|---|
| EPI_ISL_2140711 | New Horizon Clinic wc NZC | NHLS/UCT | Arash Iranzadeh; Bruna Galvao; Carolyn Williamson; Deelan Doolabh; Diana Hardie; Innocent Mudau; Kruger Marais; Lynn Tyers; Marvin Hsiao; Stephen Korsman |
| EPI_ISL_1534279, EPI_ISL_1817671 | Ocean View CDC wc OCE | NHLS/UCT | Arash Iranzadeh; Bruna Galvao; Carolyn Williamson; Deelan Doolabh; Diana Hardie; Innocent Mudau; Kruger Marais; Lynn Tyers; Marvin Hsiao; Stephen Korsman |
| EPI_ISL_1817669, EPI_ISL_2140699, EPI_ISL_2140723, EPI_ISL_2140724, EPI_ISL_2140726 | Oudtshoorn Hospital wc OUD | NHLS/UCT | Arash Iranzadeh; Bruna Galvao; Carolyn Williamson; Deelan Doolabh; Diana Hardie; Innocent Mudau; Kruger Marais; Lynn Tyers; Marvin Hsiao; Stephen Korsman |
| EPI_ISL_2086971, EPI_ISL_2086973, EPI_ISL_2086974, EPI_ISL_2086975, EPI_ISL_2086976, EPI_ISL_2086977, EPI_ISL_2086978, EPI_ISL_2086980, EPI_ISL_2086982 | | | |
| see above | PATH CARE | National Institute for Communicable Diseases of the National Health Laboratory Service | Amoako DG; Bhiman JN; Ismail A; Mahlangu B; Mohale T; Ntuli N; Scheepers C |
| EPI_ISL_2013036, EPI_ISL_2013039, EPI_ISL_2013041 | Pathcare-Vermaak Centurion | National Institute for Communicable Diseases of the National Health Laboratory Service | Amoako DG; Bhiman JN; Glass A; Gottberg A; Mahlangu B; Mohale T; Ntuli N; Oliveira TD; Scheepers C; Tegally H; Viana R |
| EPI_ISL_2140716 | Still Bay Sat Clinic wc SFV | NHLS/UCT | Arash Iranzadeh; Bruna Galvao; Carolyn Williamson; Deelan Doolabh; Diana Hardie; Innocent Mudau; Kruger Marais; Lynn Tyers; Marvin Hsiao; Stephen Korsman |
| EPI_ISL_1817675 | Thembalethu CDC wc THC | NHLS/UCT | Arash Iranzadeh; Bruna Galvao; Carolyn Williamson; Deelan Doolabh; Diana Hardie; Innocent Mudau; Kruger Marais; Lynn Tyers; Marvin Hsiao; Stephen Korsman |
| EPI_ISL_2140718 | Touwsranten Clinic wc TST | NHLS/UCT | Arash Iranzadeh; Bruna Galvao; Carolyn Williamson; Deelan Doolabh; Diana Hardie; Innocent Mudau; Kruger Marais; Lynn Tyers; Marvin Hsiao; Stephen Korsman |
| EPI_ISL_2140714 | Vanguard CHC wc VGC | NHLS/UCT | Arash Iranzadeh; Bruna Galvao; Carolyn Williamson; Deelan Doolabh; Diana Hardie; Innocent Mudau; Kruger Marais; Lynn Tyers; Marvin Hsiao; Stephen Korsman |
| EPI_ISL_2086968, EPI_ISL_2086972, EPI_ISL_2086979, EPI_ISL_2086981, EPI_ISL_2086983 | Vermaak & Partners Laboratory | National Institute for Communicable Diseases of the National Health Laboratory Service | Amoako DG; Bhiman JN; Ismail A; Mahlangu B; Mohale T; Ntuli N; Scheepers C |
| EPI_ISL_1534282, EPI_ISL_1534287, EPI_ISL_1534289, EPI_ISL_1534292, EPI_ISL_1534303, EPI_ISL_1534312, EPI_ISL_1817691, EPI_ISL_1817701, EPI_ISL_1817715, EPI_ISL_1817738, EPI_ISL_1817740, EPI_ISL_1817741, EPI_ISL_1817742, EPI_ISL_1817743, EPI_ISL_2140692, EPI_ISL_2140693, EPI_ISL_2140695, EPI_ISL_2140705, EPI_ISL_2140708, EPI_ISL_2140709, EPI_ISL_2140719, EPI_ISL_2140720 | | | |
| see above | Victoria Hospital wc VHW | NHLS/UCT | Arash Iranzadeh; Bruna Galvao; Carolyn Williamson; Deelan Doolabh; Diana Hardie; Innocent Mudau; Kruger Marais; Lynn Tyers; Marvin Hsiao; Stephen Korsman |
| EPI_ISL_1817696, EPI_ISL_1817697, EPI_ISL_1817713 | unknown | NHLS/UCT | Arash Iranzadeh; Bruna Galvao; Carolyn Williamson; Deelan Doolabh; Diana Hardie; Innocent Mudau; Kruger Marais; Lynn Tyers; Marvin Hsiao; Stephen Korsman |

We gratefully acknowledge the following Authors from the Originating laboratories responsible for obtaining the specimens, as well as the Submitting laboratories where the genome data were generated and shared via GISAID, on which this research is based.

All Submitters of data may be contacted directly via www.gisaid.org

Authors are sorted alphabetically.

| Accession ID | Originating Laboratory | Submitting Laboratory | Authors |
|---|---|---|---|
| EPI_ISL_2547412 | 2 Military Hospital VPA wc MAA | National Health Laboratory Service/University of Cape Town (NHLS/UCT) | Arash Iranzadeh; Bruna Galvao; Carolyn Williamson; Deelan Doolabh; Diana Hardie; Gert Marais; Innocent Mudau; Lynn Tyers; Marvin Hsiao; Stephen Korsman |
| EPI_ISL_2375994, EPI_ISL_2621134, EPI_ISL_2621136, EPI_ISL_2621144, EPI_ISL_2876944, EPI_ISL_2876945, EPI_ISL_2876957, EPI_ISL_2876958, EPI_ISL_2876982, EPI_ISL_2876993, EPI_ISL_2876997 | | | |
| see above | 2 Military Hospital wc MAA | NHLS/UCT | Arash Iranzadeh; Bruna Galvao; Carolyn Williamson; Deelan Doolabh; Diana Hardie; Gert Marais; Innocent Mudau; Kruger Marais; Lynn Tyers; Marvin Hsiao; Stephen Korsman |
| EPI_ISL_2547411, EPI_ISL_2547413, EPI_ISL_2547421, EPI_ISL_2547422, EPI_ISL_2547423 | 2 Military Hospital wc MAA | National Health Laboratory Service/University of Cape Town (NHLS/UCT) | Arash Iranzadeh; Bruna Galvao; Carolyn Williamson; Deelan Doolabh; Diana Hardie; Gert Marais; Innocent Mudau; Lynn Tyers; Marvin Hsiao; Stephen Korsman |
| EPI_ISL_2493041 | AMPATH | CERI, Centre for Epidemic Response and Innvoation, Stellenbosch University and KRISP, KZN Research Innovation and Sequencing Platform, UKZN. | Emmanuel SJ; Giandhari J; Khan S; Lessells R; Mdlalose K; Naidoo Y; Pillay S; Ramphal U; Tegally H; Wilkinson E; York D; de Oliveira T |
| EPI_ISL_2501090 | AMPATH | KRISP, KZN Research Innovation and Sequencing Platform | Emmanuel SJ; Giandhari J; Khan S; Lessells R; Mdlalose K; Naidoo Y; Pillay S; Ramphal U; Sisonke Team; Tegally H; Wilkinson E; York D; de Oliveira T |
| EPI_ISL_2621060, EPI_ISL_2876965 | Alma CDC wc AHC | NHLS/UCT | Arash Iranzadeh; Bruna Galvao; Carolyn Williamson; Deelan Doolabh; Diana Hardie; Gert Marais; Innocent Mudau; Lynn Tyers; Marvin Hsiao; Stephen Korsman |
| EPI_ISL_2547409 | Alma CDC wc AHC | National Health Laboratory Service/University of Cape Town (NHLS/UCT) | Arash Iranzadeh; Bruna Galvao; Carolyn Williamson; Deelan Doolabh; Diana Hardie; Gert Marais; Innocent Mudau; Lynn Tyers; Marvin Hsiao; Stephen Korsman |
| EPI_ISL_2494961 | Ampath | KRISP, KZN Research Innovation and Sequencing Platform | Emmanuel SJ; Giandhari J; Khan S; Lessells R; Mdlalose K; Naidoo Y; Pillay S; Ramphal U; Tegally H; Wilkinson E; York D; de Oliveira T |
| EPI_ISL_2727233, EPI_ISL_2727234, EPI_ISL_2727235, EPI_ISL_2727236, EPI_ISL_2727237, EPI_ISL_2727238 | Ampath Greenacres Hospital | KRISP, KZn Research Innovation and Sequencing Platform | Giandhari Jennifer; Naidoo Yeshnee; Pillay Sureshnee; San James; Tegally Houriiyah; Tshabuila Derek; Wilkinson Eduan; Yajna Ramphal; de Oliveira Tulio |
| EPI_ISL_2193354, EPI_ISL_2193355, EPI_ISL_2193356, EPI_ISL_2193357, EPI_ISL_2193358, EPI_ISL_2193359, EPI_ISL_2193360, EPI_ISL_2193361 | | | |
| see above | Ampath Laboratories | National Institute for Communicable Diseases of the National Health Laboratory Service | Amoako DG; Bhiman JN; Glass A; Gottberg A; Mahlangu B; Marshall T; Mohale T; Ntuli N; Scheepers C; Viana R |
| EPI_ISL_2693088 | Ampath laboratories- Sharks | KRISP, KZN Research Innovation and Sequencing Platform | Giandhari Jennifer; Naidoo Yeshnee; Pillay Sureshnee; San James; Tegally Houriiyah; Tshabuila Derek; Wilkinson Eduan; Yajna Ramphal; de Oliveira Tulio |
| EPI_ISL_2893242, EPI_ISL_2893300, EPI_ISL_2893301, EPI_ISL_2893302, EPI_ISL_2893303, EPI_ISL_2893304, EPI_ISL_2893305, EPI_ISL_2893306, EPI_ISL_2893307, EPI_ISL_2893308, EPI_ISL_2893309, EPI_ISL_2893310, EPI_ISL_2893311, EPI_ISL_2893312, EPI_ISL_2893313, EPI_ISL_2893314, EPI_ISL_2893315, EPI_ISL_2893316, EPI_ISL_2893317, EPI_ISL_2893318 | | | |
| see above | BARC - Sisonke | KRISP, KZN Research Innovation and Sequencing Platform | Giandhari Jennifer; Naidoo Yeshnee; Pillay Sureshnee; San James; Sisonke; Tegally Houriiyah; Tshabuila Derek; Wilkinson Eduan; Yajna Ramphal; de Oliveira Tulio |
| EPI_ISL_2710306, EPI_ISL_2710307, EPI_ISL_2710308, EPI_ISL_2710309, EPI_ISL_2710310, EPI_ISL_2710311, EPI_ISL_2710312, EPI_ISL_2710313, EPI_ISL_2710314, EPI_ISL_2710315, EPI_ISL_2710316, EPI_ISL_2710317, EPI_ISL_2710318, EPI_ISL_2710319, EPI_ISL_2710320, EPI_ISL_2710321, EPI_ISL_2710322 | | | |
| see above | BARC / Lancet | KRISP, KZN Research Innovation and Sequencing Platform | Giandhari Jennifer; Naidoo Yeshnee; Pillay Sureshnee; San James; Sisonke; Tegally Houriiyah; Tshabuila Derek; Wilkinson Eduan; Yajna Ramphal; de Oliveira Tulio |
| EPI_ISL_2802108 | Beaconsfield Clinic [Kimberley] | NHLS/UCT | Arash Iranzadeh; Bruna Galvao; Carolyn Williamson; Deelan Doolabh; Diana Hardie; Gert Marais; Innocent Mudau; Lynn Tyers; Marvin Hsiao; Stephen Korsman |
| EPI_ISL_2695506 | Bertha gxowa hospital | National Institute for Communicable Diseases of the National Health Laboratory Service | Amoako DG; Bhiman JN; Everatt J; Ismail A; Mahlangu B; Mnguni A; Mohale T; Ntuli N; Scheepers C |
| EPI_ISL_2693318, EPI_ISL_2802186 | Bothasig | NHLS/UCT | Arash Iranzadeh; Bruna Galvao; Carolyn Williamson; Deelan Doolabh; Diana Hardie; Gert Marais; Innocent Mudau; Lynn Tyers; Marvin Hsiao; Stephen Korsman |
| EPI_ISL_2547410 | Brandwacht Sat Clinic wc BBC | National Health Laboratory Service/University of Cape Town (NHLS/UCT) | Arash Iranzadeh; Bruna Galvao; Carolyn Williamson; Deelan Doolabh; Diana Hardie; Gert Marais; Innocent Mudau; Lynn Tyers; Marvin Hsiao; Stephen Korsman |
| EPI_ISL_2694627, EPI_ISL_2694628, EPI_ISL_2694629, EPI_ISL_2694630, EPI_ISL_2694631, EPI_ISL_2694633, EPI_ISL_2694634, EPI_ISL_2694635, EPI_ISL_2694636, EPI_ISL_2694637, EPI_ISL_2694639, EPI_ISL_2694640, EPI_ISL_2694641, EPI_ISL_2694642, EPI_ISL_2694643, EPI_ISL_2694644, EPI_ISL_2694645, EPI_ISL_2694646, EPI_ISL_2694647, EPI_ISL_2694648, EPI_ISL_2694649, EPI_ISL_2694650, EPI_ISL_2694651, EPI_ISL_2694652, EPI_ISL_2694653, EPI_ISL_2694654, EPI_ISL_2694655, EPI_ISL_2694656, EPI_ISL_2694657, EPI_ISL_2694658, EPI_ISL_2694659, EPI_ISL_2694660, EPI_ISL_2694661, EPI_ISL_2694662, EPI_ISL_2694663, EPI_ISL_2694664, EPI_ISL_2694665, EPI_ISL_2694666, EPI_ISL_2694667, EPI_ISL_2694668, EPI_ISL_2694669, EPI_ISL_2694670, EPI_ISL_2694671, EPI_ISL_2694672, EPI_ISL_2694673, EPI_ISL_2694674, EPI_ISL_2694675, EPI_ISL_2694676, EPI_ISL_2694677, EPI_ISL_2709879, EPI_ISL_2709880, EPI_ISL_2709881, EPI_ISL_2709882, EPI_ISL_2709883, EPI_ISL_2709884, EPI_ISL_2709887, EPI_ISL_2709888, EPI_ISL_2709889, EPI_ISL_2709890, EPI_ISL_2709891, EPI_ISL_2709892, EPI_ISL_2709893, EPI_ISL_2709894, EPI_ISL_2709895, EPI_ISL_2709896, EPI_ISL_2709897, EPI_ISL_2709898, EPI_ISL_2709899, EPI_ISL_2709900, EPI_ISL_2709901, EPI_ISL_2709902, EPI_ISL_2709903, EPI_ISL_2709904, EPI_ISL_2709905, EPI_ISL_2709906, EPI_ISL_2709907, EPI_ISL_2709908, EPI_ISL_2709909, EPI_ISL_2709910, EPI_ISL_2709911, EPI_ISL_2709912, EPI_ISL_2709913, EPI_ISL_2709914, EPI_ISL_2709915, EPI_ISL_2709917, EPI_ISL_2709918, EPI_ISL_2709919, EPI_ISL_2709920, EPI_ISL_2709921, EPI_ISL_2709922, EPI_ISL_2709923, EPI_ISL_2709924, EPI_ISL_2709925, EPI_ISL_2709926, EPI_ISL_2709927, EPI_ISL_2709928, EPI_ISL_2709930, EPI_ISL_2709931, EPI_ISL_2709932, EPI_ISL_2709933, EPI_ISL_2709934, EPI_ISL_2709935, EPI_ISL_2709936, EPI_ISL_2709937, EPI_ISL_2709938, EPI_ISL_2709939, EPI_ISL_2709940, EPI_ISL_2709941, EPI_ISL_2709942 | | | |
| see above | CHRIS HANI BARAGWANATH | National Institute for Communicable Diseases of the National Health Laboratory Service | Amoako DG; Bhiman JN; Everatt J; Ismail A; Mahlangu B; Mnguni A; Mohale T; Ntuli N; Scheepers C |
| EPI_ISL_2382225, EPI_ISL_2382226, EPI_ISL_2382227, EPI_ISL_2382229, EPI_ISL_2382230, EPI_ISL_2382232, EPI_ISL_2382233, EPI_ISL_2382234, EPI_ISL_2382235, EPI_ISL_2382238, EPI_ISL_2382239, EPI_ISL_2382240, EPI_ISL_2382242, EPI_ISL_2382243, EPI_ISL_2382244, EPI_ISL_2382246, EPI_ISL_2382248, EPI_ISL_2382249, EPI_ISL_2382251, EPI_ISL_2382252, EPI_ISL_2382253, EPI_ISL_2382254, EPI_ISL_2382255, EPI_ISL_2382256, EPI_ISL_2382257, EPI_ISL_2382259, EPI_ISL_2662618, EPI_ISL_2662619, EPI_ISL_2662620, EPI_ISL_2662622, EPI_ISL_2662623, EPI_ISL_2662628, EPI_ISL_2662629, EPI_ISL_2662630, EPI_ISL_2662632, EPI_ISL_2662633, EPI_ISL_2662635, EPI_ISL_2662636, EPI_ISL_2662637, EPI_ISL_2662638, EPI_ISL_2662639, EPI_ISL_2662640, EPI_ISL_2662641, EPI_ISL_2662642, EPI_ISL_2662644, EPI_ISL_2662645, EPI_ISL_2662651, EPI_ISL_2662652, EPI_ISL_2662654, EPI_ISL_2662655, EPI_ISL_2662656, EPI_ISL_2662658, EPI_ISL_2662659, EPI_ISL_2827156, EPI_ISL_2827157 | | | |
| see above | CHRIS HANI BARAGWANATH LABORATORY | National Institute for Communicable Diseases of the National Health Laboratory Service | Amoako DG; Bhiman JN; Everatt J; Ismail A; Mahlangu B; Mnguni A; Mohale T; Ntuli N; Scheepers C |
| EPI_ISL_2375928 | Caledon Hospital wc CLD | NHLS/UCT | Arash Iranzadeh; Bruna Galvao; Carolyn Williamson; Deelan Doolabh; Diana Hardie; Innocent Mudau; Kruger Marais; Lynn Tyers; Marvin Hsiao; Stephen Korsman |
| EPI_ISL_2547365 | Calvinia Hospital | National Health Laboratory Service/University of Cape Town (NHLS/UCT) | Arash Iranzadeh; Bruna Galvao; Carolyn Williamson; Deelan Doolabh; Diana Hardie; Gert Marais; Innocent Mudau; Lynn Tyers; Marvin Hsiao; Stephen Korsman |
| EPI_ISL_2621053 | Calvinia RSC Clinic | NHLS/UCT | Arash Iranzadeh; Bruna Galvao; Carolyn Williamson; Deelan Doolabh; Diana Hardie; Gert Marais; Innocent Mudau; Lynn Tyers; Marvin Hsiao; Stephen Korsman |
| EPI_ISL_2547355, EPI_ISL_2547356, EPI_ISL_2547357, EPI_ISL_2547358, EPI_ISL_2547359, EPI_ISL_2547360, EPI_ISL_2547361, EPI_ISL_2547362 | | | |
| see above | Calvinia RSC Clinic | National Health Laboratory Service/University of Cape Town (NHLS/UCT) | Arash Iranzadeh; Bruna Galvao; Carolyn Williamson; Deelan Doolabh; Diana Hardie; Gert Marais; Innocent Mudau; Lynn Tyers; Marvin Hsiao; Stephen Korsman |
| EPI_ISL_2375926, EPI_ISL_2375929 | Clanwilliam Hospital wc CLH | NHLS/UCT | Arash Iranzadeh; Bruna Galvao; Carolyn Williamson; Deelan Doolabh; Diana Hardie; Innocent Mudau; Kruger Marais; Lynn Tyers; Marvin Hsiao; Stephen Korsman |
| EPI_ISL_2876980 | Claremont Clinic wc CLA | NHLS/UCT | Arash Iranzadeh; Bruna Galvao; Carolyn Williamson; Deelan Doolabh; Diana Hardie; Gert Marais; Innocent Mudau; Lynn Tyers; Marvin Hsiao; Stephen Korsman |
| EPI_ISL_2876942, EPI_ISL_2876949, EPI_ISL_2876988 | Clinic-in-Asla | NHLS/UCT | Arash Iranzadeh; Bruna Galvao; Carolyn Williamson; Deelan Doolabh; Diana Hardie; Gert Marais; Innocent Mudau; Lynn Tyers; Marvin Hsiao; Stephen Korsman |
| EPI_ISL_2876947, EPI_ISL_2876954, EPI_ISL_2876987 | Conville CDC wc CVC | NHLS/UCT | Arash Iranzadeh; Bruna Galvao; Carolyn Williamson; Deelan Doolabh; Diana Hardie; Gert Marais; Innocent Mudau; Lynn Tyers; Marvin Hsiao; Stephen Korsman |
| EPI_ISL_2621141, EPI_ISL_2621143, EPI_ISL_2876952, EPI_ISL_2876953, EPI_ISL_2876990, EPI_ISL_2876992 | D'Almeida Clinic wc DAL | NHLS/UCT | Arash Iranzadeh; Bruna Galvao; Carolyn Williamson; Deelan Doolabh; Diana Hardie; Gert Marais; Innocent Mudau; Lynn Tyers; Marvin Hsiao; Stephen Korsman |
| EPI_ISL_2375930 | Delft CHC wc DFP | NHLS/UCT | Arash Iranzadeh; Bruna Galvao; Carolyn Williamson; Deelan Doolabh; Diana Hardie; Innocent Mudau; Kruger Marais; Lynn Tyers; Marvin Hsiao; Stephen Korsman |
| EPI_ISL_2695849, EPI_ISL_2695850, EPI_ISL_2695851, EPI_ISL_2695852, EPI_ISL_2695853, EPI_ISL_2695854, EPI_ISL_2695855, EPI_ISL_2695856, EPI_ISL_2695857, EPI_ISL_2695858, EPI_ISL_2695859, EPI_ISL_2695860, EPI_ISL_2695861, EPI_ISL_2695862, EPI_ISL_2695863, EPI_ISL_2695864, EPI_ISL_2695865, EPI_ISL_2695866, EPI_ISL_2695867, EPI_ISL_2695868, EPI_ISL_2695869, EPI_ISL_2695873, EPI_ISL_2876306, EPI_ISL_2876307, EPI_ISL_2876308, EPI_ISL_2876309, EPI_ISL_2876310, EPI_ISL_2876311, EPI_ISL_2876312, EPI_ISL_2876313, EPI_ISL_2876314, EPI_ISL_2876317, EPI_ISL_2876318, EPI_ISL_2876319, EPI_ISL_2876320, EPI_ISL_2876321, EPI_ISL_2876322, EPI_ISL_2876323, EPI_ISL_2876324, EPI_ISL_2876325, EPI_ISL_2876326, EPI_ISL_2876329, EPI_ISL_2876330, EPI_ISL_2876331, EPI_ISL_2876332, EPI_ISL_2876333, EPI_ISL_2876334, EPI_ISL_2876335, EPI_ISL_2876336, EPI_ISL_2876337, EPI_ISL_2876340, EPI_ISL_2876341, EPI_ISL_2876342, EPI_ISL_2876343, EPI_ISL_2876344, EPI_ISL_2876345, EPI_ISL_2876346, EPI_ISL_2876347, EPI_ISL_2876348, EPI_ISL_2876354, EPI_ISL_2876355, EPI_ISL_2876356, EPI_ISL_2876357, EPI_ISL_2876358, EPI_ISL_2876359, EPI_ISL_2876360, EPI_ISL_2876361, EPI_ISL_2876362, EPI_ISL_2876365, EPI_ISL_2876366, EPI_ISL_2876367, EPI_ISL_2876368, EPI_ISL_2876369, EPI_ISL_2876370, EPI_ISL_2876371, EPI_ISL_2876372, EPI_ISL_2876373, EPI_ISL_2876376, EPI_ISL_2876377, EPI_ISL_2876378, EPI_ISL_2876379, EPI_ISL_2876380, EPI_ISL_2876381, EPI_ISL_2876382, EPI_ISL_2876383, EPI_ISL_2876384, EPI_ISL_2876388, EPI_ISL_2876389, EPI_ISL_2876390, EPI_ISL_2876391, EPI_ISL_2876392, EPI_ISL_2876393, EPI_ISL_2876394, EPI_ISL_2876395 | | | |
| see above | Division of Medical Virology, National Health Laboratory Service (NHLS), Tygerberg Hospital / Stellenbosch University | Division of Medical Virology, National Health Laboratory Service (NHLS), Tygerberg Hospital / Stellenbosch University | Bronwyn Kleinhans; Gert van Zyl; Jean Maritz; Petra Raimond; San Emmanual James; Susan Engelbrecht; Tania Stander; Tongai Maponga; Tulio de Oliveira; Wolfgang Preiser |
| EPI_ISL_2621146, EPI_ISL_2621147 | Dysselsdorp Clinic wc DDC | NHLS/UCT | Arash Iranzadeh; Bruna Galvao; Carolyn Williamson; Deelan Doolabh; Diana Hardie; Gert Marais; Innocent Mudau; Lynn Tyers; Marvin Hsiao; Stephen Korsman |
| EPI_ISL_2086267 | Enthabeni Lancet Laboratories | KRISP, KZN Research Innovation and Sequencing Platform | Emmanuel SJ; Giandhari J; Khan S; Lessells R; Mdlalose K; Naidoo Y; Pillay S; Ramphal U; Tegally H; Wilkinson E; York D; de Oliveira T |
| EPI_ISL_2802122 | Florianville Clinic | NHLS/UCT | Arash Iranzadeh; Bruna Galvao; Carolyn Williamson; Deelan Doolabh; Diana Hardie; Gert Marais; Innocent Mudau; Lynn Tyers; Marvin Hsiao; Stephen Korsman |
| EPI_ISL_2547341 | Garies PHC Clinic | National Health Laboratory Service/University of Cape Town (NHLS/UCT) | Arash Iranzadeh; Bruna Galvao; Carolyn Williamson; Deelan Doolabh; Diana Hardie; Gert Marais; Innocent Mudau; Lynn Tyers; Marvin Hsiao; Stephen Korsman |
| EPI_ISL_2621137, EPI_ISL_2876951, EPI_ISL_2876959, EPI_ISL_2876984 | George Hospital wc GRH | NHLS/UCT | Arash Iranzadeh; Bruna Galvao; Carolyn Williamson; Deelan Doolabh; Diana Hardie; Gert Marais; Innocent Mudau; Lynn Tyers; Marvin Hsiao; Stephen Korsman |
| EPI_ISL_2621142, EPI_ISL_2693328, EPI_ISL_2802195, EPI_ISL_2876941, EPI_ISL_2876948, EPI_ISL_2876950, EPI_ISL_2876961, EPI_ISL_2876962, EPI_ISL_2876963, EPI_ISL_2876964, EPI_ISL_2876967, EPI_ISL_2876968, EPI_ISL_2876969 | | | |
| see above | Great Brak River Clinic wc GBC | NHLS/UCT | Arash Iranzadeh; Bruna Galvao; Carolyn Williamson; Deelan Doolabh; Diana Hardie; Gert Marais; Innocent Mudau; Lynn Tyers; Marvin Hsiao; Stephen Korsman |
| EPI_ISL_2547426 | Great Brak River Clinic wc GBC | National Health Laboratory Service/University of Cape Town (NHLS/UCT) | Arash Iranzadeh; Bruna Galvao; Carolyn Williamson; Deelan Doolabh; Diana Hardie; Gert Marais; Innocent Mudau; Lynn Tyers; Marvin Hsiao; Stephen Korsman |
| EPI_ISL_2621054, EPI_ISL_2621059, EPI_ISL_2621132, EPI_ISL_2621133, EPI_ISL_2693324, EPI_ISL_2693325, EPI_ISL_2693326, EPI_ISL_2693327, EPI_ISL_2802182, EPI_ISL_2802192, EPI_ISL_2802193, EPI_ISL_2802194, EPI_ISL_2802199, EPI_ISL_2876936, EPI_ISL_2876955, EPI_ISL_2876971, EPI_ISL_2876972, EPI_ISL_2876973, EPI_ISL_2876974, EPI_ISL_2876975, EPI_ISL_2876977, EPI_ISL_2876978, EPI_ISL_2876979, EPI_ISL_2876981, EPI_ISL_2876983, EPI_ISL_2876989, EPI_ISL_2876991, EPI_ISL_2876998 | | | |
| see above | Groote Schuur Hospital wc GSH | NHLS/UCT | Arash Iranzadeh; Bruna Galvao; Carolyn Williamson; Deelan Doolabh; Diana Hardie; Gert Marais; Innocent Mudau; Lynn Tyers; Marvin Hsiao; Stephen Korsman |
| EPI_ISL_2547418, EPI_ISL_2547419, EPI_ISL_2547424, EPI_ISL_2547425 | Groote Schuur Hospital wc GSH | National Health Laboratory Service/University of Cape Town (NHLS/UCT) | Arash Iranzadeh; Bruna Galvao; Carolyn Williamson; Deelan Doolabh; Diana Hardie; Gert Marais; Innocent Mudau; Lynn Tyers; Marvin Hsiao; Stephen Korsman |
| EPI_ISL_2695588, EPI_ISL_2695589, EPI_ISL_2695590, EPI_ISL_2695591, EPI_ISL_2695592, EPI_ISL_2695593, EPI_ISL_2695594, EPI_ISL_2695595, EPI_ISL_2695596, EPI_ISL_2695597, EPI_ISL_2695598, EPI_ISL_2695599, EPI_ISL_2695600, EPI_ISL_2695601, EPI_ISL_2695602, EPI_ISL_2695603, EPI_ISL_2695604, EPI_ISL_2695605, EPI_ISL_2695606, EPI_ISL_2695727, EPI_ISL_2695728, EPI_ISL_2695729, EPI_ISL_2695730, EPI_ISL_2695731, EPI_ISL_2695732, EPI_ISL_2695733, EPI_ISL_2695734, EPI_ISL_2695735, EPI_ISL_2695736, EPI_ISL_2695737, EPI_ISL_2695738, EPI_ISL_2695739, EPI_ISL_2695740, EPI_ISL_2695741, EPI_ISL_2695742, EPI_ISL_2695743, EPI_ISL_2695744, EPI_ISL_2695745, EPI_ISL_2695746, EPI_ISL_2695747, EPI_ISL_2695748, EPI_ISL_2695749, EPI_ISL_2695750, EPI_ISL_2695751, EPI_ISL_2709863, EPI_ISL_2709864, EPI_ISL_2709872, EPI_ISL_2709873, EPI_ISL_2709874, EPI_ISL_2709875, EPI_ISL_2709877, EPI_ISL_2709878, EPI_ISL_2709885, EPI_ISL_2709886 | | | |
| see above | HELEN JOSEPH | National Institute for Communicable Diseases of the National Health Laboratory Service | Amoako DG; Bhiman JN; Everatt J; Ismail A; Mahlangu B; Mnguni A; Mohale T; Ntuli N; Scheepers C |
| EPI_ISL_2827072, EPI_ISL_2827073, EPI_ISL_2827075, EPI_ISL_2827076, EPI_ISL_2827077, EPI_ISL_2827078, EPI_ISL_2827079, EPI_ISL_2827080, EPI_ISL_2827081, EPI_ISL_2827082, EPI_ISL_2827083, EPI_ISL_2827084, EPI_ISL_2827085, EPI_ISL_2827086, EPI_ISL_2827087, EPI_ISL_2827088, EPI_ISL_2827089, EPI_ISL_2827090, EPI_ISL_2827092, EPI_ISL_2827093, EPI_ISL_2827094, EPI_ISL_2827096, EPI_ISL_2827097, EPI_ISL_2827098, EPI_ISL_2827099, EPI_ISL_2827100, EPI_ISL_2827102, EPI_ISL_2827103, EPI_ISL_2827104, EPI_ISL_2827106, EPI_ISL_2827107, EPI_ISL_2827108, EPI_ISL_2827109, EPI_ISL_2827110, EPI_ISL_2827111, EPI_ISL_2827113, EPI_ISL_2827114, EPI_ISL_2827115, EPI_ISL_2827116, EPI_ISL_2827117, EPI_ISL_2827118, EPI_ISL_2827119, EPI_ISL_2827120, EPI_ISL_2827121, EPI_ISL_2827122, EPI_ISL_2827123, EPI_ISL_2827124, EPI_ISL_2827125, EPI_ISL_2827127, EPI_ISL_2827128, EPI_ISL_2827129, EPI_ISL_2827130, EPI_ISL_2827132, EPI_ISL_2895772, EPI_ISL_2895773, EPI_ISL_2895774, EPI_ISL_2895775, EPI_ISL_2895776, EPI_ISL_2895777, EPI_ISL_2895778, EPI_ISL_2895779, EPI_ISL_2895780, EPI_ISL_2895781, EPI_ISL_2895782, EPI_ISL_2895783, EPI_ISL_2895784, EPI_ISL_2895785, EPI_ISL_2895786, EPI_ISL_2895787, EPI_ISL_2895788, EPI_ISL_2895789, EPI_ISL_2895790, EPI_ISL_2895791, EPI_ISL_2895792, EPI_ISL_2895793, EPI_ISL_2895794, EPI_ISL_2895795, EPI_ISL_2895796, EPI_ISL_2895797, EPI_ISL_2942284, EPI_ISL_2942285, EPI_ISL_2942286, EPI_ISL_2942287 | | | |

| | | | |
|---|---|---|---|
| see above | HELEN JOSEPH LABORATORY | National Institute for Communicable Diseases of the National Health Laboratory Service | Amoako DG; Bhiman JN; Everatt J; Ismail A; Mahlangu B; Mnguni A; Mohale T; Ntuli N; Scheepers C |
| EPI_ISL_2876940 | Heidelberg Clinic wc HBC | NHLS/UCT | Arash Iranzadeh; Bruna Galvao; Carolyn Williamson; Deelan Doolabh; Diana Hardie; Gert Marais; Innocent Mudau; Lynn Tyers; Marvin Hsiao; Stephen Korsman |
| EPI_ISL_2877001 | Heideveld Emergency Centre | NHLS/UCT | Arash Iranzadeh; Bruna Galvao; Carolyn Williamson; Deelan Doolabh; Diana Hardie; Gert Marais; Innocent Mudau; Lynn Tyers; Marvin Hsiao; Stephen Korsman |
| EPI_ISL_2375927, EPI_ISL_2693320, EPI_ISL_2802188 | Helderberg Hospital wc HHH | NHLS/UCT | Arash Iranzadeh; Bruna Galvao; Carolyn Williamson; Deelan Doolabh; Diana Hardie; Gert Marais; Innocent Mudau; Kruger Marais; Lynn Tyers; Marvin Hsiao; Stephen Korsman |
| EPI_ISL_2547372 | Helderberg Hospital wc HHH | National Health Laboratory Service/University of Cape Town (NHLS/UCT) | Arash Iranzadeh; Bruna Galvao; Carolyn Williamson; Deelan Doolabh; Diana Hardie; Gert Marais; Innocent Mudau; Lynn Tyers; Marvin Hsiao; Stephen Korsman |
| EPI_ISL_2547352 | Hondeklipbaai Clinic | National Health Laboratory Service/University of Cape Town (NHLS/UCT) | Arash Iranzadeh; Bruna Galvao; Carolyn Williamson; Deelan Doolabh; Diana Hardie; Gert Marais; Innocent Mudau; Lynn Tyers; Marvin Hsiao; Stephen Korsman |
| EPI_ISL_2802117 | Hopetown RSC Clinic | NHLS/UCT | Arash Iranzadeh; Bruna Galvao; Carolyn Williamson; Deelan Doolabh; Diana Hardie; Gert Marais; Innocent Mudau; Lynn Tyers; Marvin Hsiao; Stephen Korsman |
| EPI_ISL_2877006 | Hornlee Clinic wc HLC | NHLS/UCT | Arash Iranzadeh; Bruna Galvao; Carolyn Williamson; Deelan Doolabh; Diana Hardie; Gert Marais; Innocent Mudau; Lynn Tyers; Marvin Hsiao; Stephen Korsman |
| EPI_ISL_2547427 | Hornlee Clinic wc HLC | National Health Laboratory Service/University of Cape Town (NHLS/UCT) | Arash Iranzadeh; Bruna Galvao; Carolyn Williamson; Deelan Doolabh; Diana Hardie; Gert Marais; Innocent Mudau; Lynn Tyers; Marvin Hsiao; Stephen Korsman |
| EPI_ISL_2802107 | Jan Kemp Hospital | NHLS/UCT | Arash Iranzadeh; Bruna Galvao; Carolyn Williamson; Deelan Doolabh; Diana Hardie; Gert Marais; Innocent Mudau; Lynn Tyers; Marvin Hsiao; Stephen Korsman |
| EPI_ISL_2547340 | Joe Slovo Hospital | National Health Laboratory Service/University of Cape Town (National Health Laboratory Service/University of Cape Town (NHLS/UCT)) | Arash Iranzadeh; Bruna Galvao; Carolyn Williamson; Deelan Doolabh; Diana Hardie; Gert Marais; Innocent Mudau; Lynn Tyers; Marvin Hsiao; Stephen Korsman |
| EPI_ISL_2382192, EPI_ISL_2382194, EPI_ISL_2382195, EPI_ISL_2382197, EPI_ISL_2382198, EPI_ISL_2382200, EPI_ISL_2382201, EPI_ISL_2382202, EPI_ISL_2382203, EPI_ISL_2382204, EPI_ISL_2382205, EPI_ISL_2382206, EPI_ISL_2382207, EPI_ISL_2382208, EPI_ISL_2382210, EPI_ISL_2382211, EPI_ISL_2382212, EPI_ISL_2382214, EPI_ISL_2382215, EPI_ISL_2382216, EPI_ISL_2382217, EPI_ISL_2382218, EPI_ISL_2382219, EPI_ISL_2382222, EPI_ISL_2382223, EPI_ISL_2382224, EPI_ISL_2382700, EPI_ISL_2382701, EPI_ISL_2382702, EPI_ISL_2382703, EPI_ISL_2382706, EPI_ISL_2382710, EPI_ISL_2582855, EPI_ISL_2582862, EPI_ISL_2582865, EPI_ISL_2582868, EPI_ISL_2582870, EPI_ISL_2582872, EPI_ISL_2582876, EPI_ISL_2582885, EPI_ISL_2582937, EPI_ISL_2582947, EPI_ISL_2582950, EPI_ISL_2662682, EPI_ISL_2662684, EPI_ISL_2662686, EPI_ISL_2662688, EPI_ISL_2662689, EPI_ISL_2662690, EPI_ISL_2662691, EPI_ISL_2662693, EPI_ISL_2662694, EPI_ISL_2662695, EPI_ISL_2662696, EPI_ISL_2662699, EPI_ISL_2662700, EPI_ISL_2662701, EPI_ISL_2662702, EPI_ISL_2662704, EPI_ISL_2662705, EPI_ISL_2662707 | | | |
| see above | KIMBERLEY LABORATORY | National Institute for Communicable Diseases of the National Health Laboratory Service | Amoako DG; Bhiman JN; Everatt J; Ismail A; Mahlangu B; Mnguni A; Mohale T; Ntuli N; Scheepers C |
| EPI_ISL_2694614, EPI_ISL_2694615, EPI_ISL_2695512, EPI_ISL_2695513, EPI_ISL_2695523, EPI_ISL_2695524 | KOPANONG | National Institute for Communicable Diseases of the National Health Laboratory Service | Amoako DG; Bhiman JN; Everatt J; Ismail A; Mahlangu B; Mnguni A; Mohale T; Ntuli N; Scheepers C |
| EPI_ISL_2382260, EPI_ISL_2382261, EPI_ISL_2382262, EPI_ISL_2382263, EPI_ISL_2382264, EPI_ISL_2382265, EPI_ISL_2382266, EPI_ISL_2582990, EPI_ISL_2582993, EPI_ISL_2582996, EPI_ISL_2582999, EPI_ISL_2583001, EPI_ISL_2583005, EPI_ISL_2662663, EPI_ISL_2662664, EPI_ISL_2695759, EPI_ISL_2695760, EPI_ISL_2695761, EPI_ISL_2895766, EPI_ISL_2895767, EPI_ISL_2895768, EPI_ISL_2895769, EPI_ISL_2895770, EPI_ISL_2895771, EPI_ISL_2895799, EPI_ISL_2895800, EPI_ISL_2942283 | | | |
| see above | KOPANONG LABORATORY | National Institute for Communicable Diseases of the National Health Laboratory Service | Amoako DG; Bhiman JN; Everatt J; Ismail A; Mahlangu B; Mnguni A; Mohale T; Ntuli N; Scheepers C |
| EPI_ISL_2688547, EPI_ISL_2688548 | KRISP, KZn Research Innovation and Sequencing Platform | KRISP, KZn Research Innovation and Sequencing Platform | Giandhari Jennifer; Naidoo Yeshnee; Pillay Sureshnee; San James; Tegally Houriiyah; Tshabuila Derek; Wilkinson Eduan; Yajna Ramphal; de Oliveira Tulio |
| EPI_ISL_2621145, EPI_ISL_2877003 | Knysna CDC wc WLC | NHLS/UCT | Arash Iranzadeh; Bruna Galvao; Carolyn Williamson; Deelan Doolabh; Diana Hardie; Gert Marais; Innocent Mudau; Lynn Tyers; Marvin Hsiao; Stephen Korsman |
| EPI_ISL_2877002 | Knysna Hospital wc KNY | NHLS/UCT | Arash Iranzadeh; Bruna Galvao; Carolyn Williamson; Deelan Doolabh; Diana Hardie; Gert Marais; Innocent Mudau; Lynn Tyers; Marvin Hsiao; Stephen Korsman |
| EPI_ISL_2547415 | Knysna Hospital wc KNY | National Health Laboratory Service/University of Cape Town (NHLS/UCT) | Arash Iranzadeh; Bruna Galvao; Carolyn Williamson; Deelan Doolabh; Diana Hardie; Gert Marais; Innocent Mudau; Lynn Tyers; Marvin Hsiao; Stephen Korsman |
| EPI_ISL_2802114 | Kraaifontein | NHLS/UCT | Arash Iranzadeh; Bruna Galvao; Carolyn Williamson; Deelan Doolabh; Diana Hardie; Gert Marais; Innocent Mudau; Lynn Tyers; Marvin Hsiao; Stephen Korsman |
| EPI_ISL_2802116 | Kuils river | NHLS/UCT | Arash Iranzadeh; Bruna Galvao; Carolyn Williamson; Deelan Doolabh; Diana Hardie; Gert Marais; Innocent Mudau; Lynn Tyers; Marvin Hsiao; Stephen Korsman |
| EPI_ISL_2547368, EPI_ISL_2547370 | Kuruman Hospital | National Health Laboratory Service/University of Cape Town (NHLS/UCT) | Arash Iranzadeh; Bruna Galvao; Carolyn Williamson; Deelan Doolabh; Diana Hardie; Gert Marais; Innocent Mudau; Lynn Tyers; Marvin Hsiao; Stephen Korsman |
| EPI_ISL_2877004, EPI_ISL_2877005 | Kwanokuthula CDC wc KWA | NHLS/UCT | Arash Iranzadeh; Bruna Galvao; Carolyn Williamson; Deelan Doolabh; Diana Hardie; Gert Marais; Innocent Mudau; Lynn Tyers; Marvin Hsiao; Stephen Korsman |
| EPI_ISL_2463301, EPI_ISL_2463302, EPI_ISL_2463303, EPI_ISL_2463304, EPI_ISL_2463305, EPI_ISL_2463306, EPI_ISL_2463307, EPI_ISL_2463308, EPI_ISL_2463309, EPI_ISL_2463310 | | | |
| see above | LANCET LABORATORY | National Institute for Communicable Diseases of the National Health Laboratory Service | Amoako DG; Bhiman JN; Everatt J; Glass A; Sisonke Team; Viana R |
| EPI_ISL_2828731, EPI_ISL_2828732 | Lancet | National Institute for Communicable Diseases of the National Health Laboratory Service | Amoako DG; Bhiman JN; Everatt J; Ismail A; Mahlangu B; Mnguni A; Mohale T; Ntuli N; Scheepers C |
| EPI_ISL_2013037, EPI_ISL_2013040, EPI_ISL_2013053 | Lancet Laboratories | National Institute for Communicable Diseases of the National Health Laboratory Service | Amoako DG; Bhiman JN; Glass A; Gottberg A; Mahlangu B; Mohale T; Ntuli N; Oliveira TD; Scheepers C; Tegally H; Viana R |
| EPI_ISL_2802123 | Lingelethu Clinic [Upington] | NHLS/UCT | Arash Iranzadeh; Bruna Galvao; Carolyn Williamson; Deelan Doolabh; Diana Hardie; Gert Marais; Innocent Mudau; Lynn Tyers; Marvin Hsiao; Stephen Korsman |
| EPI_ISL_2547366 | Lutzville Clinic wc LVP | National Health Laboratory Service/University of Cape Town (NHLS/UCT) | Arash Iranzadeh; Bruna Galvao; Carolyn Williamson; Deelan Doolabh; Diana Hardie; Gert Marais; Innocent Mudau; Lynn Tyers; Marvin Hsiao; Stephen Korsman |
| EPI_ISL_2709848, EPI_ISL_2827071, EPI_ISL_2827133, EPI_ISL_2827134, EPI_ISL_2827135, EPI_ISL_2827136, EPI_ISL_2827137, EPI_ISL_2827138, EPI_ISL_2827139, EPI_ISL_2827140, EPI_ISL_2827141, EPI_ISL_2827158, EPI_ISL_2827160, EPI_ISL_2827161, EPI_ISL_2827162, EPI_ISL_2827163, EPI_ISL_2827164, EPI_ISL_2827165, EPI_ISL_2827166, EPI_ISL_2827927, EPI_ISL_2827928, EPI_ISL_2827929, EPI_ISL_2827930, EPI_ISL_2827931, EPI_ISL_2827932, EPI_ISL_2827933, EPI_ISL_2827934, EPI_ISL_2827935, EPI_ISL_2827936, EPI_ISL_2827937, EPI_ISL_2827938, EPI_ISL_2827939, EPI_ISL_2827940, EPI_ISL_2827941, EPI_ISL_2827942, EPI_ISL_2827943, EPI_ISL_2827944, EPI_ISL_2827945, EPI_ISL_2827946, EPI_ISL_2827947, EPI_ISL_2827948, EPI_ISL_2827949, EPI_ISL_2827950, EPI_ISL_2827951 | | | |
| see above | MANKWENG LABORATORY | National Institute for Communicable Diseases of the National Health Laboratory Service | Amoako DG; Bhiman JN; Everatt J; Ismail A; Mahlangu B; Mnguni A; Mohale T; Ntuli N; Scheepers C |
| EPI_ISL_2895749, EPI_ISL_2895750, EPI_ISL_2895751, EPI_ISL_2895752, EPI_ISL_2895753, EPI_ISL_2895755, EPI_ISL_2895756, EPI_ISL_2895757, EPI_ISL_2895758, EPI_ISL_2895759, EPI_ISL_2895760, EPI_ISL_2895762, EPI_ISL_2895763, EPI_ISL_2895764, EPI_ISL_2895765, EPI_ISL_2942282 | | | |
| see above | MIDDELBURG LABORATORY | National Institute for Communicable Diseases of the National Health Laboratory Service | Amoako DG; Bhiman JN; Everatt J; Ismail A; Mahlangu B; Mnguni A; Mohale T; Ntuli N; Scheepers C |
| EPI_ISL_2547344 | Matjieskloof Clinic | National Health Laboratory Service/University of Cape Town (NHLS/UCT) | Arash Iranzadeh; Bruna Galvao; Carolyn Williamson; Deelan Doolabh; Diana Hardie; Gert Marais; Innocent Mudau; Lynn Tyers; Marvin Hsiao; Stephen Korsman |
| EPI_ISL_2547354 | Max Shapiro Clinic | National Health Laboratory Service/University of Cape Town (NHLS/UCT) | Arash Iranzadeh; Bruna Galvao; Carolyn Williamson; Deelan Doolabh; Diana Hardie; Gert Marais; Innocent Mudau; Lynn Tyers; Marvin Hsiao; Stephen Korsman |
| EPI_ISL_2876937, EPI_ISL_2877000 | Mitchells Plain Hospital wc MPH | NHLS/UCT | Arash Iranzadeh; Bruna Galvao; Carolyn Williamson; Deelan Doolabh; Diana Hardie; Gert Marais; Innocent Mudau; Lynn Tyers; Marvin Hsiao; Stephen Korsman |
| EPI_ISL_2547414 | Mitchells Plain Hospital wc MPH | National Health Laboratory Service/University of Cape Town (NHLS/UCT) | Arash Iranzadeh; Bruna Galvao; Carolyn Williamson; Deelan Doolabh; Diana Hardie; Gert Marais; Innocent Mudau; Lynn Tyers; Marvin Hsiao; Stephen Korsman |
| EPI_ISL_2710249, EPI_ISL_2710250, EPI_ISL_2710251, EPI_ISL_2710252, EPI_ISL_2710253, EPI_ISL_2710254, EPI_ISL_2710255, EPI_ISL_2710256, EPI_ISL_2710257, EPI_ISL_2710258, EPI_ISL_2710259, EPI_ISL_2710260, EPI_ISL_2710261, EPI_ISL_2710262, EPI_ISL_2710263, EPI_ISL_2710264, EPI_ISL_2710265, EPI_ISL_2710266, EPI_ISL_2710267, EPI_ISL_2710268, EPI_ISL_2710269, EPI_ISL_2710270, EPI_ISL_2710271, EPI_ISL_2710272, EPI_ISL_2710273, EPI_ISL_2710274, EPI_ISL_2710275, EPI_ISL_2710276, EPI_ISL_2710277, EPI_ISL_2710278, EPI_ISL_2710279, EPI_ISL_2710280, EPI_ISL_2710281, EPI_ISL_2710282, EPI_ISL_2710283, EPI_ISL_2710284, EPI_ISL_2710285, EPI_ISL_2710286, EPI_ISL_2710287, EPI_ISL_2710288, EPI_ISL_2710289, EPI_ISL_2710290, EPI_ISL_2710291, EPI_ISL_2710292, EPI_ISL_2710293, EPI_ISL_2710294, EPI_ISL_2710295, EPI_ISL_2710296, EPI_ISL_2710297, EPI_ISL_2710298, EPI_ISL_2710299, EPI_ISL_2710300, EPI_ISL_2710301, EPI_ISL_2710302, EPI_ISL_2710303, EPI_ISL_2710304, EPI_ISL_2710305, EPI_ISL_2726854, EPI_ISL_2726855, EPI_ISL_2726856, EPI_ISL_2770439, EPI_ISL_2770440, EPI_ISL_2770441, EPI_ISL_2770442, EPI_ISL_2770443, EPI_ISL_2770444, EPI_ISL_2770445, EPI_ISL_2770446, EPI_ISL_2770447, EPI_ISL_2770448, EPI_ISL_2770449, EPI_ISL_2770450, EPI_ISL_2770451, EPI_ISL_2770452, EPI_ISL_2770453, EPI_ISL_2770454, EPI_ISL_2770455, EPI_ISL_2770456, EPI_ISL_2770457, EPI_ISL_2770458, EPI_ISL_2770459, EPI_ISL_2770460, EPI_ISL_2770461, EPI_ISL_2770462, EPI_ISL_2770463, EPI_ISL_2770464, EPI_ISL_2770465, EPI_ISL_2770466, EPI_ISL_2770467, EPI_ISL_2770468, EPI_ISL_2770469, EPI_ISL_2770470, EPI_ISL_2770471, EPI_ISL_2770472, EPI_ISL_2770473, EPI_ISL_2770474, EPI_ISL_2770475, EPI_ISL_2770477, EPI_ISL_2770478, EPI_ISL_2770479, EPI_ISL_2770480, EPI_ISL_2770481, EPI_ISL_2770482, EPI_ISL_2770483, EPI_ISL_2770484, EPI_ISL_2770485, EPI_ISL_2770486, EPI_ISL_2770487, EPI_ISL_2770488 | | | |
| see above | NHLS Charlotte Maxeke Johannesburg Academic Hospital and the University of the Witwatersrand | KRISP, KZN Research Innovation and Sequencing Platform | Bulelani Manene; Florette Treurnicht; Giandhari Jennifer; Kathleen Subramoney; Naidoo Yeshnee; Pillay Sureshnee; San James; Sisonke; Tegally Houriiyah; Tshabuila Derek; Wilkinson Eduan; Yajna Ramphal; de Oliveira Tulio |
| EPI_ISL_2841643, EPI_ISL_2841644, EPI_ISL_2841645, EPI_ISL_2841646, EPI_ISL_2841647, EPI_ISL_2841648, EPI_ISL_2841649 | | | |
| see above | NHLS Charlotte Maxeke Johannesburg Academic Hospital and the University of the Witwatersrand | KRISP, KZn Research Innovation and Sequencing Platform | Bulelani Manene; Florette Treurnicht; Giandhari Jennifer; Kathleen Subramoney; Naidoo Yeshnee; Pillay Sureshnee; San James; Tegally Houriiyah; Tshabuila Derek; Wilkinson Eduan; Yajna Ramphal; de Oliveira Tulio |
| EPI_ISL_2162299, EPI_ISL_2162300, EPI_ISL_2162301, EPI_ISL_2162302, EPI_ISL_2162303, EPI_ISL_2162304, EPI_ISL_2162305, EPI_ISL_2162306, EPI_ISL_2162307, EPI_ISL_2162308, EPI_ISL_2162309, EPI_ISL_2162310, EPI_ISL_2162311, EPI_ISL_2162312, EPI_ISL_2162313, EPI_ISL_2162314, EPI_ISL_2162315, EPI_ISL_2162316, EPI_ISL_2162317, EPI_ISL_2162318, EPI_ISL_2162319, EPI_ISL_2162320, EPI_ISL_2162321, EPI_ISL_2162322, EPI_ISL_2162323, EPI_ISL_2162324, EPI_ISL_2162325, EPI_ISL_2162326, EPI_ISL_2162327, EPI_ISL_2162328, EPI_ISL_2162329, EPI_ISL_2162330, EPI_ISL_2162331, EPI_ISL_2162332, EPI_ISL_2162333, EPI_ISL_2162334, EPI_ISL_2162335, EPI_ISL_2162336, EPI_ISL_2162337, EPI_ISL_2162338 | | | |
| see above | NHLS Universitas Academic | UFS Virology | D Goedhals; Emmanuel Ogunbayo; MM Nyaga; MT Mogotsi; P Nthiga; PA Bester; T de Oliveira |
| EPI_ISL_2693316, EPI_ISL_2693317, EPI_ISL_2693329, EPI_ISL_2693330, EPI_ISL_2693331, EPI_ISL_2693332, EPI_ISL_2802183, EPI_ISL_2802184, EPI_ISL_2802185, EPI_ISL_2802196, EPI_ISL_2802197, EPI_ISL_2802198 | | | |
| see above | NHLS/UCT | NHLS/UCT | Arash Iranzadeh; Bruna Galvao; Carolyn Williamson; Deelan Doolabh; Diana Hardie; Gert Marais; Innocent Mudau; Lynn Tyers; Marvin Hsiao; Stephen Korsman |
| EPI_ISL_2693036, EPI_ISL_2693037, EPI_ISL_2693038, EPI_ISL_2693039, EPI_ISL_2693040, EPI_ISL_2693041, EPI_ISL_2693042, EPI_ISL_2693043, EPI_ISL_2693044, EPI_ISL_2693045, EPI_ISL_2693046, EPI_ISL_2693047, EPI_ISL_2693048, EPI_ISL_2693049, EPI_ISL_2693050, EPI_ISL_2693051, EPI_ISL_2693052, EPI_ISL_2693053, EPI_ISL_2693054, EPI_ISL_2693055, EPI_ISL_2693056, EPI_ISL_2693057, EPI_ISL_2693058, EPI_ISL_2693059, EPI_ISL_2693060, EPI_ISL_2693061, EPI_ISL_2693063, EPI_ISL_2693064, EPI_ISL_2693065, EPI_ISL_2693066, EPI_ISL_2693067, EPI_ISL_2693068, EPI_ISL_2693069, EPI_ISL_2693070, EPI_ISL_2693071, EPI_ISL_2693072, EPI_ISL_2693073, EPI_ISL_2693074, EPI_ISL_2693075, EPI_ISL_2693076, EPI_ISL_2693077, EPI_ISL_2693078, EPI_ISL_2693079, EPI_ISL_2693080, EPI_ISL_2693081, EPI_ISL_2693082, EPI_ISL_2693083, EPI_ISL_2693084, EPI_ISL_2693085, EPI_ISL_2693086, EPI_ISL_2693087, EPI_ISL_2693089, EPI_ISL_2693090, EPI_ISL_2693091, EPI_ISL_2693092, EPI_ISL_2693093, EPI_ISL_2693094, EPI_ISL_2693095 | | | |
| see above | NHLS_VIRO | KRISP, KZN Research Innovation and Sequencing Platform | Giandhari Jennifer; Naidoo Yeshnee; Pillay Sureshnee; San James; Tegally Houriiyah; Tshabuila Derek; Wilkinson Eduan; Yajna Ramphal; de Oliveira Tulio |
| EPI_ISL_2727380, EPI_ISL_2727381, EPI_ISL_2727382, EPI_ISL_2727383, EPI_ISL_2727384, EPI_ISL_2727385, EPI_ISL_2727386, EPI_ISL_2727387, EPI_ISL_2727388, EPI_ISL_2727389, EPI_ISL_2727390, EPI_ISL_2727391, EPI_ISL_2727392, EPI_ISL_2727393, EPI_ISL_2727394, EPI_ISL_2727395, EPI_ISL_2727396, EPI_ISL_2727397, EPI_ISL_2727398, EPI_ISL_2727399, EPI_ISL_2727400, EPI_ISL_2727401, EPI_ISL_2727402, EPI_ISL_2727403 | | | |
| see above | NHLS_VIRO | KRISP, KZn Research Innovation and Sequencing Platform | Giandhari Jennifer; Naidoo Yeshnee; Pillay Sureshnee; San James; Tegally Houriiyah; Tshabuila Derek; Wilkinson Eduan; Yajna Ramphal; de Oliveira Tulio |
| EPI_ISL_2693824, EPI_ISL_2693825, EPI_ISL_2693826, EPI_ISL_2693827, EPI_ISL_2693828, EPI_ISL_2693829, EPI_ISL_2693830, EPI_ISL_2693831, EPI_ISL_2693832, EPI_ISL_2693833, EPI_ISL_2693834, EPI_ISL_2693835, EPI_ISL_2693836, EPI_ISL_2693837, EPI_ISL_2693838, EPI_ISL_2693839, EPI_ISL_2693840, EPI_ISL_2693841, EPI_ISL_2693842, EPI_ISL_2693843, EPI_ISL_2693844, EPI_ISL_2693845, EPI_ISL_2693846, EPI_ISL_2693847, EPI_ISL_2693848, EPI_ISL_2693849, EPI_ISL_2693850, EPI_ISL_2693851, EPI_ISL_2693852, EPI_ISL_2693853, EPI_ISL_2693854, EPI_ISL_2693855, EPI_ISL_2693856, EPI_ISL_2693857, EPI_ISL_2693858, EPI_ISL_2693859, EPI_ISL_2693860, EPI_ISL_2693861, EPI_ISL_2693862, EPI_ISL_2693863, EPI_ISL_2693864, EPI_ISL_2693865, EPI_ISL_2693866, EPI_ISL_2693867, EPI_ISL_2693868, EPI_ISL_2693869, EPI_ISL_2693870, EPI_ISL_2693871, EPI_ISL_2693872, EPI_ISL_2693873, EPI_ISL_2693874, EPI_ISL_2693875, EPI_ISL_2693876, EPI_ISL_2693877, EPI_ISL_2693878, EPI_ISL_2693879, EPI_ISL_2693880, EPI_ISL_2693881, EPI_ISL_2693882, EPI_ISL_2693883, EPI_ISL_2693884, EPI_ISL_2693885, EPI_ISL_2693886, EPI_ISL_2693887, EPI_ISL_2693888, EPI_ISL_2693889, EPI_ISL_2693890, EPI_ISL_2693891, EPI_ISL_2693892, EPI_ISL_2693893, EPI_ISL_2693894, EPI_ISL_2693895, EPI_ISL_2693896, EPI_ISL_2693897, EPI_ISL_2693898, EPI_ISL_2693899, EPI_ISL_2693900, EPI_ISL_2693901, EPI_ISL_2893243, EPI_ISL_2893244, EPI_ISL_2893245, EPI_ISL_2893246, EPI_ISL_2893247, EPI_ISL_2893248, EPI_ISL_2893249, EPI_ISL_2893250, EPI_ISL_2893251, EPI_ISL_2893252, EPI_ISL_2893253, EPI_ISL_2893254, EPI_ISL_2893255, EPI_ISL_2893256, EPI_ISL_2893257, EPI_ISL_2893258, EPI_ISL_2893259, EPI_ISL_2893260, EPI_ISL_2893261, EPI_ISL_2893262, EPI_ISL_2893263, EPI_ISL_2893264, EPI_ISL_2893265, EPI_ISL_2893266, EPI_ISL_2893267, EPI_ISL_2893268, EPI_ISL_2893269, EPI_ISL_2893270, EPI_ISL_2893271, EPI_ISL_2893272, EPI_ISL_2893273, EPI_ISL_2893274, EPI_ISL_2893275, EPI_ISL_2893276, EPI_ISL_2893277, EPI_ISL_2893278, EPI_ISL_2893279, EPI_ISL_2893280, EPI_ISL_2893281, EPI_ISL_2893282, EPI_ISL_2893283, EPI_ISL_2893284, EPI_ISL_2893285, EPI_ISL_2893286, EPI_ISL_2893287, EPI_ISL_2893288, EPI_ISL_2893289, EPI_ISL_2893290, EPI_ISL_2893291, EPI_ISL_2893292, EPI_ISL_2893293, EPI_ISL_2893294, EPI_ISL_2893295, EPI_ISL_2893296, EPI_ISL_2893297, EPI_ISL_2893298, EPI_ISL_2893299 | | | |
| see above | NHLS_VIRO Eastern Cape | KRISP, KZN Research Innovation and Sequencing Platform | Emmanuel SJ; Giandhari J; Giandhari Jennifer; Khan S; Laguda-Akingba O; Lessells R; Mdlalose K; Naidoo Yeshnee; Pillay S; Pillay Sureshnee; San James; Tegally H; Tegally Houriiyah; Tshabuila Derek; Wilkinson E; Wilkinson Eduan; Yajna Ramphal; York D; de Oliveira T; de Oliveira Tulio |
| EPI_ISL_2727175, EPI_ISL_2727176, EPI_ISL_2727177, EPI_ISL_2727178, EPI_ISL_2727190, EPI_ISL_2727199, EPI_ISL_2727200, EPI_ISL_2727201, EPI_ISL_2727202, EPI_ISL_2727203, EPI_ISL_2727204, EPI_ISL_2727205, EPI_ISL_2727206, EPI_ISL_2727207, EPI_ISL_2727208, EPI_ISL_2727209, EPI_ISL_2727210, EPI_ISL_2727211, EPI_ISL_2727212, EPI_ISL_2727213, EPI_ISL_2727214, EPI_ISL_2727215, EPI_ISL_2727217, EPI_ISL_2727218, EPI_ISL_2727219, EPI_ISL_2727220, EPI_ISL_2727221, EPI_ISL_2727222, EPI_ISL_2727223, EPI_ISL_2727224, EPI_ISL_2727226, EPI_ISL_2727227, EPI_ISL_2727228, EPI_ISL_2727229, EPI_ISL_2727230, EPI_ISL_2727231, EPI_ISL_2727232, EPI_ISL_2955453, EPI_ISL_2955454, EPI_ISL_2955455, EPI_ISL_2955456, EPI_ISL_2955457, EPI_ISL_2955458, EPI_ISL_2955459, EPI_ISL_2955460, EPI_ISL_2955461, EPI_ISL_2955462, EPI_ISL_2955463, EPI_ISL_2955464, EPI_ISL_2955465, EPI_ISL_2955466, EPI_ISL_2955467, EPI_ISL_2955468, EPI_ISL_2955469, EPI_ISL_2955470, EPI_ISL_2955471, EPI_ISL_2955472, EPI_ISL_2955473, EPI_ISL_2955474, EPI_ISL_2955475, EPI_ISL_2955476, EPI_ISL_2955477, EPI_ISL_2955478, EPI_ISL_2955479, EPI_ISL_2955480, EPI_ISL_2955481, EPI_ISL_2955482, EPI_ISL_2955483, EPI_ISL_2955484, EPI_ISL_2955485, EPI_ISL_2955486, EPI_ISL_2955487, EPI_ISL_2955488, EPI_ISL_2955489, EPI_ISL_2955490, EPI_ISL_2955491, EPI_ISL_2955492, EPI_ISL_2955493, EPI_ISL_2955494, EPI_ISL_2955495, EPI_ISL_2955496, EPI_ISL_2955497, EPI_ISL_2955498, EPI_ISL_2955499 | | | |
| see above | NHLS_VIRO Eastern Cape | KRISP, KZn Research Innovation and Sequencing Platform | Emmanuel SJ; Giandhari J; Khan S; Laguda-Akingba O; Lessells R; Mdlalose K; Pillay S; Tegally H; Wilkinson E; York D; de Oliveira T |
| EPI_ISL_2547343 | Nababeep Hospital | National Health Laboratory Service/University of Cape Town (NHLS/UCT) | Arash Iranzadeh; Bruna Galvao; Carolyn Williamson; Deelan Doolabh; Diana Hardie; Gert Marais; Innocent Mudau; Lynn Tyers; Marvin Hsiao; Stephen Korsman |
| EPI_ISL_2617186, EPI_ISL_2617187, EPI_ISL_2617188, EPI_ISL_2617189, EPI_ISL_2617190, EPI_ISL_2617192, EPI_ISL_2617194, EPI_ISL_2617195, EPI_ISL_2617196, EPI_ISL_2617197, EPI_ISL_2617198, EPI_ISL_2617199, EPI_ISL_2617203, EPI_ISL_2617204, EPI_ISL_2617206, EPI_ISL_2617207, EPI_ISL_2617208, EPI_ISL_2617209, EPI_ISL_2617210, EPI_ISL_2617211, EPI_ISL_2617212, EPI_ISL_2617213, EPI_ISL_2617214, EPI_ISL_2617215, EPI_ISL_2617216, EPI_ISL_2688531, EPI_ISL_2688532, EPI_ISL_2688533, EPI_ISL_2688534, EPI_ISL_2688535, EPI_ISL_2688536, EPI_ISL_2688537, EPI_ISL_2688538, EPI_ISL_2688539, EPI_ISL_2688540, EPI_ISL_2688541, EPI_ISL_2688542, EPI_ISL_2688543, EPI_ISL_2688544, EPI_ISL_2688545, EPI_ISL_2688546, EPI_ISL_2688549, EPI_ISL_2688550, EPI_ISL_2688551, EPI_ISL_2688552, EPI_ISL_2688553, EPI_ISL_2688554, EPI_ISL_2688555, EPI_ISL_2688556, EPI_ISL_2688557, EPI_ISL_2688558, EPI_ISL_2688559, EPI_ISL_2688560, EPI_ISL_2688561, EPI_ISL_2688562, EPI_ISL_2688563, EPI_ISL_2688564, EPI_ISL_2688565, EPI_ISL_2688566, EPI_ISL_2688567, EPI_ISL_2688568, EPI_ISL_2688569, EPI_ISL_2688570, EPI_ISL_2688571, EPI_ISL_2688572, EPI_ISL_2688573, EPI_ISL_2688574, EPI_ISL_2688575, EPI_ISL_2899687, EPI_ISL_2899688, EPI_ISL_2899689, EPI_ISL_2899690, EPI_ISL_2899691, EPI_ISL_2899692, EPI_ISL_2899693, EPI_ISL_2899694, EPI_ISL_2899695, EPI_ISL_2899696, EPI_ISL_2899697, EPI_ISL_2899698, EPI_ISL_2899699, EPI_ISL_2899700, EPI_ISL_2899701, EPI_ISL_2899702, EPI_ISL_2899703, EPI_ISL_2899704, EPI_ISL_2899705, EPI_ISL_2899706, | | | |

| | | | |
|---|---|---|---|
| EPI_ISL_2899707, EPI_ISL_2899708, EPI_ISL_2899709, EPI_ISL_2899710, EPI_ISL_2899711, EPI_ISL_2899712, EPI_ISL_2899713, EPI_ISL_2899714, EPI_ISL_2899715, EPI_ISL_2899716, EPI_ISL_2899717, EPI_ISL_2899718, EPI_ISL_2899719, EPI_ISL_2899720, EPI_ISL_2899721, EPI_ISL_2899722, EPI_ISL_2899723, EPI_ISL_2899724, EPI_ISL_2899725, EPI_ISL_2899726, EPI_ISL_2899727, EPI_ISL_2899728, EPI_ISL_2899729, EPI_ISL_2899730, EPI_ISL_2899731, EPI_ISL_2899732, EPI_ISL_2899733, EPI_ISL_2899734, EPI_ISL_2899735, EPI_ISL_2899736, EPI_ISL_2899737, EPI_ISL_2899738, EPI_ISL_2899739, EPI_ISL_2899740 | | | |
| see above | National Health Laboratory Service, South Africa | KRISP, KZn Research Innovation and Sequencing Platform | Giandhari Jennifer; Naidoo Yeshnee; Pillay Sureshnee; San James; Tegally Houriiyah; Tshabuila Derek; Wilkinson Eduan; Yajna Ramphal; de Oliveira Tulio |
| EPI_ISL_2662708, EPI_ISL_2662709, EPI_ISL_2662710, EPI_ISL_2662711, EPI_ISL_2662712, EPI_ISL_2662713, EPI_ISL_2662714, EPI_ISL_2662715, EPI_ISL_2662716, EPI_ISL_2662717, EPI_ISL_2662718, EPI_ISL_2662719, EPI_ISL_2662720, EPI_ISL_2662721, EPI_ISL_2662722, EPI_ISL_2662723, EPI_ISL_2662724, EPI_ISL_2662725 | | | |
| see above | National Institute for Communicable Diseases of the National Health Laboratory Service | National Institute for Communicable Diseases of the National Health Laboratory Service | Amoako DG; Bhiman JN; Everatt J; Ismail A; Mahlangu B; Mnguni A; Mohale T; Ntuli N; Scheepers C |
| EPI_ISL_2547420 | Nelspoort Clinic wc NPP | National Health Laboratory Service/University of Cape Town (NHLS/UCT) | Arash Iranzadeh; Bruna Galvao; Carolyn Williamson; Deelan Doolabh; Diana Hardie; Gert Marais; Innocent Mudau; Lynn Tyers; Marvin Hsiao; Stephen Korsman |
| EPI_ISL_2770489 | Netcare Kingsway | KRISP, KZN Research Innovation and Sequencing Platform | Giandhari Jennifer; Naidoo Yeshnee; Pillay Sureshnee; San James; Tegally Houriiyah; Tshabuila Derek; Wilkinson Eduan; Yajna Ramphal; de Oliveira Tulio |
| EPI_ISL_2693062 | Netcare Kingsway Hospital | KRISP, KZN Research Innovation and Sequencing Platform | Giandhari Jennifer; Naidoo Yeshnee; Pillay Sureshnee; San James; Tegally Houriiyah; Tshabuila Derek; Wilkinson Eduan; Yajna Ramphal; de Oliveira Tulio |
| EPI_ISL_2841651 | Netcare Kingsway Hospital | KRISP, KZn Research Innovation and Sequencing Platform | Giandhari Jennifer; Naidoo Yeshnee; Pillay Sureshnee; San James; Tegally Houriiyah; Tshabuila Derek; Wilkinson Eduan; Yajna Ramphal; de Oliveira Tulio |
| EPI_ISL_2086211, EPI_ISL_2086212, EPI_ISL_2086213, EPI_ISL_2086214, EPI_ISL_2086215, EPI_ISL_2086216, EPI_ISL_2086217, EPI_ISL_2086218, EPI_ISL_2086219 | | | |
| see above | Netcare Linksfield Hospital | KRISP, KZN Research Innovation and Sequencing Platform | Emmanuel S; Emmanuel SJ; Giandhari J; Lessells R; Naidoo Y; Pillay S; Ramphal U; Tegally H; Wilkinson E; de Oliveira T |
| EPI_ISL_2494975, EPI_ISL_2501091, EPI_ISL_2501092, EPI_ISL_2501093, EPI_ISL_2501094, EPI_ISL_2501095 | Netcare St Augustines Hospital | KRISP, KZN Research Innovation and Sequencing Platform | Emmanuel SJ; Giandhari J; Khan S; Lessells R; Mdlalose K; Naidoo Y; Pillay S; Ramphal U; Sisonke Team; Tegally H; Wilkinson E; York D; de Oliveira T |
| EPI_ISL_2841650 | Netcare The Bay Hospital | KRISP, KZn Research Innovation and Sequencing Platform | Giandhari Jennifer; Naidoo Yeshnee; Pillay Sureshnee; San James; Tegally Houriiyah; Tshabuila Derek; Wilkinson Eduan; Yajna Ramphal; de Oliveira Tulio |
| EPI_ISL_2727404 | Netcare Umhlanga Hospital | KRISP, KZn Research Innovation and Sequencing Platform | Giandhari Jennifer; Naidoo Yeshnee; Pillay Sureshnee; San James; Tegally Houriiyah; Tshabuila Derek; Wilkinson Eduan; Yajna Ramphal; de Oliveira Tulio |
| EPI_ISL_2802124 | New Somerset Hospital wc NSH | NHLS/UCT | Arash Iranzadeh; Bruna Galvao; Carolyn Williamson; Deelan Doolabh; Diana Hardie; Gert Marais; Innocent Mudau; Lynn Tyers; Marvin Hsiao; Stephen Korsman |
| EPI_ISL_2547342, EPI_ISL_2547345, EPI_ISL_2547351, EPI_ISL_2547353 | Okiep CHC | National Health Laboratory Service/University of Cape Town (NHLS/UCT) | Arash Iranzadeh; Bruna Galvao; Carolyn Williamson; Deelan Doolabh; Diana Hardie; Gert Marais; Innocent Mudau; Lynn Tyers; Marvin Hsiao; Stephen Korsman |
| EPI_ISL_2695583, EPI_ISL_2695584, EPI_ISL_2695585, EPI_ISL_2695586, EPI_ISL_2695587 | PATHCARE LABORATORY | National Institute for Communicable Diseases of the National Health Laboratory Service | Amoako DG; Bhiman JN; Everatt J; Ismail A; Mahlangu B; Mnguni A; Mohale T; Ntuli N; Scheepers C |
| EPI_ISL_2709978 | PHOLOSONG LABORATORY | National Institute for Communicable Diseases of the National Health Laboratory Service | Amoako DG; Bhiman JN; Everatt J; Ismail A; Mahlangu B; Mnguni A; Mohale T; Ntuli N; Scheepers C |
| EPI_ISL_2827167, EPI_ISL_2827168, EPI_ISL_2827169, EPI_ISL_2827170 | POLOKWANE LABORATORY | National Institute for Communicable Diseases of the National Health Laboratory Service | Amoako DG; Bhiman JN; Everatt J; Ismail A; Mahlangu B; Mnguni A; Mohale T; Ntuli N; Scheepers C |
| EPI_ISL_2695530, EPI_ISL_2695531, EPI_ISL_2695532, EPI_ISL_2695533, EPI_ISL_2695535, EPI_ISL_2695536, EPI_ISL_2695538, EPI_ISL_2695539, EPI_ISL_2695540, EPI_ISL_2695541, EPI_ISL_2695542, EPI_ISL_2695543, EPI_ISL_2695544, EPI_ISL_2695545, EPI_ISL_2695546, EPI_ISL_2695547, EPI_ISL_2695548, EPI_ISL_2695550, EPI_ISL_2695551, EPI_ISL_2695552, EPI_ISL_2695553, EPI_ISL_2695554, EPI_ISL_2695555, EPI_ISL_2695556, EPI_ISL_2695558, EPI_ISL_2695559, EPI_ISL_2695560, EPI_ISL_2695561, EPI_ISL_2695562, EPI_ISL_2695563, EPI_ISL_2695565, EPI_ISL_2695566, EPI_ISL_2695567, EPI_ISL_2695568, EPI_ISL_2695569, EPI_ISL_2695570, EPI_ISL_2695571, EPI_ISL_2695572, EPI_ISL_2695573, EPI_ISL_2695574, EPI_ISL_2695575, EPI_ISL_2695576, EPI_ISL_2695577, EPI_ISL_2695578, EPI_ISL_2695579, EPI_ISL_2695580, EPI_ISL_2695581, EPI_ISL_2695582, EPI_ISL_2709849, EPI_ISL_2709850, EPI_ISL_2709851, EPI_ISL_2709852, EPI_ISL_2709853, EPI_ISL_2709854, EPI_ISL_2709855, EPI_ISL_2709856, EPI_ISL_2709857, EPI_ISL_2709858, EPI_ISL_2709859, EPI_ISL_2709860, EPI_ISL_2709861, EPI_ISL_2709862, EPI_ISL_2709865, EPI_ISL_2709867, EPI_ISL_2709869, EPI_ISL_2709871, EPI_ISL_2709916 | | | |
| see above | POLOKWANE MANKWENG HOSPITAL | National Institute for Communicable Diseases of the National Health Laboratory Service | Amoako DG; Bhiman JN; Everatt J; Ismail A; Mahlangu B; Mnguni A; Mohale T; Ntuli N; Scheepers C |
| EPI_ISL_2827899, EPI_ISL_2827900, EPI_ISL_2827901, EPI_ISL_2827902, EPI_ISL_2827903, EPI_ISL_2827904, EPI_ISL_2895717, EPI_ISL_2895718, EPI_ISL_2895719, EPI_ISL_2895720, EPI_ISL_2895721, EPI_ISL_2895722, EPI_ISL_2895723, EPI_ISL_2895724, EPI_ISL_2895725, EPI_ISL_2895726, EPI_ISL_2895727, EPI_ISL_2895728, EPI_ISL_2895729, EPI_ISL_2895730, EPI_ISL_2895731, EPI_ISL_2895732, EPI_ISL_2895733, EPI_ISL_2895734, EPI_ISL_2895735, EPI_ISL_2895736, EPI_ISL_2895737, EPI_ISL_2895738, EPI_ISL_2895739, EPI_ISL_2895740, EPI_ISL_2895741, EPI_ISL_2895742, EPI_ISL_2895743, EPI_ISL_2895744, EPI_ISL_2895745, EPI_ISL_2895746, EPI_ISL_2895747, EPI_ISL_2895748, EPI_ISL_2895798, EPI_ISL_2942288 | | | |
| see above | PORT ELIZABETH LABORATORY | National Institute for Communicable Diseases of the National Health Laboratory Service | Amoako DG; Bhiman JN; Everatt J; Ismail A; Mahlangu B; Mnguni A; Mohale T; Ntuli N; Scheepers C |
| EPI_ISL_2827909, EPI_ISL_2827917, EPI_ISL_2827918, EPI_ISL_2827920, EPI_ISL_2827921, EPI_ISL_2827922, EPI_ISL_2827926 | | | |
| see above | POTCHEFSTROOM LABORATORY | National Institute for Communicable Diseases of the National Health Laboratory Service | Amoako DG; Bhiman JN; Everatt J; Ismail A; Mahlangu B; Mnguni A; Mohale T; Ntuli N; Scheepers C |
| EPI_ISL_2876935, EPI_ISL_2876960, EPI_ISL_2876966, EPI_ISL_2876986 | Pacaltsdorp Clinic wc PAC | NHLS/UCT | Arash Iranzadeh; Bruna Galvao; Carolyn Williamson; Deelan Doolabh; Diana Hardie; Gert Marais; Innocent Mudau; Lynn Tyers; Marvin Hsiao; Stephen Korsman |
| EPI_ISL_2547416 | Pacaltsdorp Clinic wc PAC | National Health Laboratory Service/University of Cape Town (NHLS/UCT) | Arash Iranzadeh; Bruna Galvao; Carolyn Williamson; Deelan Doolabh; Diana Hardie; Gert Marais; Innocent Mudau; Lynn Tyers; Marvin Hsiao; Stephen Korsman |
| EPI_ISL_2876995, EPI_ISL_2876996 | Parkwood Clinic wc PRK | NHLS/UCT | Arash Iranzadeh; Bruna Galvao; Carolyn Williamson; Deelan Doolabh; Diana Hardie; Gert Marais; Innocent Mudau; Lynn Tyers; Marvin Hsiao; Stephen Korsman |
| EPI_ISL_2802115 | Parow | NHLS/UCT | Arash Iranzadeh; Bruna Galvao; Carolyn Williamson; Deelan Doolabh; Diana Hardie; Gert Marais; Innocent Mudau; Lynn Tyers; Marvin Hsiao; Stephen Korsman |
| EPI_ISL_2694593, EPI_ISL_2694594, EPI_ISL_2694595, EPI_ISL_2694596, EPI_ISL_2694597, EPI_ISL_2694598, EPI_ISL_2694599, EPI_ISL_2694600, EPI_ISL_2694601, EPI_ISL_2694602, EPI_ISL_2694603, EPI_ISL_2694604, EPI_ISL_2694605, EPI_ISL_2694606, EPI_ISL_2694607, EPI_ISL_2694608, EPI_ISL_2694609, EPI_ISL_2694610, EPI_ISL_2695811, EPI_ISL_2695812, EPI_ISL_2695813, EPI_ISL_2695814, EPI_ISL_2695815, EPI_ISL_2695816, EPI_ISL_2695817, EPI_ISL_2695818, EPI_ISL_2695819, EPI_ISL_2695820, EPI_ISL_2695821, EPI_ISL_2695822, EPI_ISL_2695823, EPI_ISL_2695824, EPI_ISL_2695825, EPI_ISL_2695826, EPI_ISL_2695827, EPI_ISL_2695828, EPI_ISL_2695829, EPI_ISL_2695830, EPI_ISL_2695831, EPI_ISL_2695832, EPI_ISL_2695833, EPI_ISL_2695834, EPI_ISL_2695835, EPI_ISL_2695836, EPI_ISL_2695837, EPI_ISL_2695838, EPI_ISL_2695839, EPI_ISL_2695840, EPI_ISL_2695841, EPI_ISL_2695842, EPI_ISL_2695843, EPI_ISL_2695844, EPI_ISL_2695845, EPI_ISL_2695846, EPI_ISL_2695847, EPI_ISL_2695848, EPI_ISL_2876315, EPI_ISL_2876316, EPI_ISL_2876327, EPI_ISL_2876328, EPI_ISL_2876338, EPI_ISL_2876339, EPI_ISL_2876352, EPI_ISL_2876353, EPI_ISL_2876363, EPI_ISL_2876364, EPI_ISL_2876374, EPI_ISL_2876375, EPI_ISL_2876385, EPI_ISL_2876386, EPI_ISL_2876387, EPI_ISL_2876396, EPI_ISL_2876397, EPI_ISL_2876398 | | | |
| see above | PathCare, Cape Town | Division of Medical Virology, National Health Laboratory Service (NHLS), Tygerberg Hospital / Stellenbosch University | Bronwyn Kleinhans; Gert van Zyl; Houriiyah Tegally; Jean Maritz; Petra Raimond; San Emmanual James; San Emmanuel James; Sisonke; Susan Engelbrecht; Tania Stander; Tongai Maponga; Tulio de Oliveira; Wolfgang Preiser |
| EPI_ISL_2828735, EPI_ISL_2828736, EPI_ISL_2828737, EPI_ISL_2828738, EPI_ISL_2828739, EPI_ISL_2828740, EPI_ISL_2828741, EPI_ISL_2828742, EPI_ISL_2828743, EPI_ISL_2828744, EPI_ISL_2828745, EPI_ISL_2828746, EPI_ISL_2828747, EPI_ISL_2828748, EPI_ISL_2828749, EPI_ISL_2828750, EPI_ISL_2828751, EPI_ISL_2828752, EPI_ISL_2828753, EPI_ISL_2828754, EPI_ISL_2828755, EPI_ISL_2828756, EPI_ISL_2828757, EPI_ISL_2828758 | | | |
| see above | Pathcare Vaal | National Institute for Communicable Diseases of the National Health Laboratory Service | Amoako DG; Bhiman JN; Everatt J; Ismail A; Mahlangu B; Mnguni A; Mohale T; Ntuli N; Scheepers C |
| EPI_ISL_2013038, EPI_ISL_2013042 | Pathcare-Vermaak Centurion | National Institute for Communicable Diseases of the National Health Laboratory Service | Amoako DG; Bhiman JN; Glass A; Gottberg A; Mahlangu B; Mohale T; Ntuli N; Oliveira TD; Scheepers C; Tegally H; Viana R |
| EPI_ISL_2876956, EPI_ISL_2876994 | Pelican Park CDC wc PAX | NHLS/UCT | Arash Iranzadeh; Bruna Galvao; Carolyn Williamson; Deelan Doolabh; Diana Hardie; Gert Marais; Innocent Mudau; Lynn Tyers; Marvin Hsiao; Stephen Korsman |
| EPI_ISL_2802119 | Phutanang Clinic | NHLS/UCT | Arash Iranzadeh; Bruna Galvao; Carolyn Williamson; Deelan Doolabh; Diana Hardie; Gert Marais; Innocent Mudau; Lynn Tyers; Marvin Hsiao; Stephen Korsman |
| EPI_ISL_2547371 | Postdene Clinic | National Health Laboratory Service/University of Cape Town (NHLS/UCT) | Arash Iranzadeh; Bruna Galvao; Carolyn Williamson; Deelan Doolabh; Diana Hardie; Gert Marais; Innocent Mudau; Lynn Tyers; Marvin Hsiao; Stephen Korsman |
| EPI_ISL_2802120 | Prieska Municipal Clinic | NHLS/UCT | Arash Iranzadeh; Bruna Galvao; Carolyn Williamson; Deelan Doolabh; Diana Hardie; Gert Marais; Innocent Mudau; Lynn Tyers; Marvin Hsiao; Stephen Korsman |
| EPI_ISL_2273063, EPI_ISL_2273064, EPI_ISL_2273068, EPI_ISL_2273069, EPI_ISL_2695607, EPI_ISL_2695608, EPI_ISL_2695609, EPI_ISL_2695610, EPI_ISL_2695611, EPI_ISL_2695612, EPI_ISL_2695613, EPI_ISL_2695614, EPI_ISL_2695615, EPI_ISL_2695616, EPI_ISL_2695617, EPI_ISL_2695618, EPI_ISL_2695619, EPI_ISL_2695620, EPI_ISL_2695621, EPI_ISL_2695623, EPI_ISL_2695624, EPI_ISL_2695625, EPI_ISL_2695626, EPI_ISL_2695627, EPI_ISL_2695628, EPI_ISL_2695629, EPI_ISL_2695630, EPI_ISL_2695631, EPI_ISL_2695632, EPI_ISL_2695633, EPI_ISL_2695725, EPI_ISL_2695726, EPI_ISL_2695752, EPI_ISL_2695753, EPI_ISL_2695754, EPI_ISL_2695755, EPI_ISL_2695756, EPI_ISL_2695757, EPI_ISL_2695758, EPI_ISL_2695763, EPI_ISL_2695764, EPI_ISL_2695765, EPI_ISL_2695766, EPI_ISL_2695767, EPI_ISL_2695768, EPI_ISL_2695769, EPI_ISL_2695770, EPI_ISL_2695771 | | | |
| see above | ROB FERREIRA LABORATORY | National Institute for Communicable Diseases of the National Health Laboratory Service | Amoako DG; Bhiman JN; Everatt J; Ismail A; Mahlangu B; Mnguni A; Mohale T; Ntuli N; Scheepers C |
| EPI_ISL_2827868, EPI_ISL_2827869, EPI_ISL_2827870, EPI_ISL_2827871, EPI_ISL_2827872, EPI_ISL_2827873, EPI_ISL_2827874, EPI_ISL_2827875, EPI_ISL_2827876, EPI_ISL_2827877, EPI_ISL_2827878, EPI_ISL_2827879, EPI_ISL_2827880, EPI_ISL_2827881, EPI_ISL_2827882, EPI_ISL_2827883, EPI_ISL_2827884, EPI_ISL_2827885, EPI_ISL_2827886, EPI_ISL_2827887, EPI_ISL_2827888, EPI_ISL_2827889, EPI_ISL_2827890, EPI_ISL_2827891, EPI_ISL_2827892, EPI_ISL_2827893, EPI_ISL_2827894, EPI_ISL_2827895, EPI_ISL_2827896, EPI_ISL_2827897, EPI_ISL_2827898, EPI_ISL_2895678, EPI_ISL_2895679, EPI_ISL_2895680, EPI_ISL_2895681, EPI_ISL_2895682, EPI_ISL_2895683, EPI_ISL_2895684, EPI_ISL_2895685, EPI_ISL_2895687, EPI_ISL_2895688, EPI_ISL_2895689, EPI_ISL_2895690, EPI_ISL_2895691, EPI_ISL_2895692, EPI_ISL_2895693, EPI_ISL_2895694, EPI_ISL_2895695, EPI_ISL_2895696, EPI_ISL_2895697, EPI_ISL_2895698, EPI_ISL_2895699, EPI_ISL_2895701, EPI_ISL_2895702, EPI_ISL_2895703, EPI_ISL_2895704, EPI_ISL_2895705, EPI_ISL_2895706, EPI_ISL_2895707, EPI_ISL_2895708, EPI_ISL_2895709, EPI_ISL_2895710, EPI_ISL_2895711, EPI_ISL_2895712, EPI_ISL_2895713, EPI_ISL_2895714, EPI_ISL_2895715, EPI_ISL_2895716, EPI_ISL_2942280, EPI_ISL_2942281 | | | |
| see above | RUSTENBURG LABORATORY | National Institute for Communicable Diseases of the National Health Laboratory Service | Amoako DG; Bhiman JN; Everatt J; Ismail A; Mahlangu B; Mnguni A; Mohale T; Ntuli N; Scheepers C |
| EPI_ISL_2876999 | Red Cross Children's Hospital wc RXH | NHLS/UCT | Arash Iranzadeh; Bruna Galvao; Carolyn Williamson; Deelan Doolabh; Diana Hardie; Gert Marais; Innocent Mudau; Lynn Tyers; Marvin Hsiao; Stephen Korsman |
| EPI_ISL_2547346, EPI_ISL_2547347, EPI_ISL_2547348, EPI_ISL_2547349, EPI_ISL_2547350 | Sanddrift nc Clinic | National Health Laboratory Service/University of Cape Town (NHLS/UCT) | Arash Iranzadeh; Bruna Galvao; Carolyn Williamson; Deelan Doolabh; Diana Hardie; Gert Marais; Innocent Mudau; Lynn Tyers; Marvin Hsiao; Stephen Korsman |
| EPI_ISL_2621135, EPI_ISL_2621138, EPI_ISL_2621139, EPI_ISL_2621140, EPI_ISL_2802180 | Sedgefield Clinic wc SGE | NHLS/UCT | Arash Iranzadeh; Bruna Galvao; Carolyn Williamson; Deelan Doolabh; Diana Hardie; Gert Marais; Innocent Mudau; Lynn Tyers; Marvin Hsiao; Stephen Korsman |
| EPI_ISL_2086220, EPI_ISL_2086221, EPI_ISL_2086222, EPI_ISL_2086223, EPI_ISL_2086224, EPI_ISL_2086225, EPI_ISL_2086226, EPI_ISL_2086227 | | | |
| see above | ShipMed Inc | KRISP, KZN Research Innovation and Sequencing Platform | Emmanuel SJ; Giandhari J; Lessells R; Naidoo Y; Pillay S; Ramphal U; Tegally H; Wilkinson E; de Oliveira T |
| EPI_ISL_2802118 | Stellenbosch Hospital wc STB | NHLS/UCT | Arash Iranzadeh; Bruna Galvao; Carolyn Williamson; Deelan Doolabh; Diana Hardie; Gert Marais; Innocent Mudau; Lynn Tyers; Marvin Hsiao; Stephen Korsman |
| EPI_ISL_2876938 | Strandfontein Clinic wc STR | NHLS/UCT | Arash Iranzadeh; Bruna Galvao; Carolyn Williamson; Deelan Doolabh; Diana Hardie; Gert Marais; Innocent Mudau; Lynn Tyers; Marvin Hsiao; Stephen Korsman |
| EPI_ISL_2693321, EPI_ISL_2693322, EPI_ISL_2802189, EPI_ISL_2802190 | Swellendam PHC Clinic wc SHC | NHLS/UCT | Arash Iranzadeh; Bruna Galvao; Carolyn Williamson; Deelan Doolabh; Diana Hardie; Gert Marais; Innocent Mudau; Lynn Tyers; Marvin Hsiao; Stephen Korsman |
| EPI_ISL_2694611, EPI_ISL_2694612, EPI_ISL_2694613, EPI_ISL_2694616, EPI_ISL_2694617, EPI_ISL_2694618, EPI_ISL_2694619, EPI_ISL_2694620, EPI_ISL_2694621, EPI_ISL_2694622, EPI_ISL_2694623, EPI_ISL_2694624, EPI_ISL_2694625, EPI_ISL_2694626, EPI_ISL_2694632, EPI_ISL_2694678, EPI_ISL_2694679, EPI_ISL_2694680, EPI_ISL_2694681, EPI_ISL_2694682, EPI_ISL_2694683, EPI_ISL_2694684, EPI_ISL_2694685, EPI_ISL_2694686, EPI_ISL_2694687, EPI_ISL_2694688, EPI_ISL_2694689, EPI_ISL_2694690, EPI_ISL_2694691, EPI_ISL_2694692, EPI_ISL_2694693, EPI_ISL_2694694, EPI_ISL_2694695, EPI_ISL_2694696, EPI_ISL_2694697, EPI_ISL_2694698, EPI_ISL_2694699, EPI_ISL_2694700, EPI_ISL_2694701, EPI_ISL_2694702, EPI_ISL_2694703, EPI_ISL_2694704, EPI_ISL_2694705, EPI_ISL_2694706, EPI_ISL_2694707, EPI_ISL_2694708, EPI_ISL_2694709, EPI_ISL_2694710, EPI_ISL_2694711, EPI_ISL_2694712, EPI_ISL_2694713, EPI_ISL_2694714, EPI_ISL_2694715, EPI_ISL_2694716, EPI_ISL_2694717, EPI_ISL_2694718, EPI_ISL_2694719, EPI_ISL_2694720, EPI_ISL_2694721, EPI_ISL_2694722, EPI_ISL_2694723, EPI_ISL_2694724, EPI_ISL_2694725, EPI_ISL_2694726, EPI_ISL_2695504, EPI_ISL_2695505, EPI_ISL_2695507, EPI_ISL_2695508, EPI_ISL_2695509, EPI_ISL_2695510, EPI_ISL_2695511, EPI_ISL_2695514, EPI_ISL_2695515, EPI_ISL_2695516, EPI_ISL_2695517, EPI_ISL_2695518, EPI_ISL_2695519, EPI_ISL_2695520, EPI_ISL_2695521, EPI_ISL_2695522, EPI_ISL_2695525, EPI_ISL_2695526, EPI_ISL_2695527, EPI_ISL_2695528 | | | |
| see above | TAMBO MEMORIAL | National Institute for Communicable Diseases of the National Health Laboratory Service | Amoako DG; Bhiman JN; Everatt J; Ismail A; Mahlangu B; Mnguni A; Mohale T; Ntuli N; Scheepers C |
| EPI_ISL_2662556, EPI_ISL_2662557, EPI_ISL_2662558, EPI_ISL_2662559, EPI_ISL_2662560, EPI_ISL_2662561, EPI_ISL_2662562, EPI_ISL_2662563, EPI_ISL_2662564, EPI_ISL_2662565, EPI_ISL_2662566, EPI_ISL_2662567, EPI_ISL_2662568, EPI_ISL_2662569, EPI_ISL_2662570, EPI_ISL_2662571, EPI_ISL_2662572, EPI_ISL_2662573, EPI_ISL_2662574, EPI_ISL_2662575, EPI_ISL_2662576, EPI_ISL_2662577, EPI_ISL_2662578, EPI_ISL_2662579, EPI_ISL_2662580, EPI_ISL_2662581, EPI_ISL_2662582, EPI_ISL_2662583, EPI_ISL_2662584, EPI_ISL_2662585, EPI_ISL_2662586, EPI_ISL_2662587, EPI_ISL_2662588, EPI_ISL_2662589, EPI_ISL_2662590, EPI_ISL_2662591, EPI_ISL_2662592, EPI_ISL_2662593, EPI_ISL_2662594, EPI_ISL_2662595, EPI_ISL_2662596, EPI_ISL_2662597, EPI_ISL_2662598, EPI_ISL_2662599, EPI_ISL_2662600, EPI_ISL_2662601, EPI_ISL_2662602, EPI_ISL_2662603, EPI_ISL_2662604, EPI_ISL_2662605, EPI_ISL_2662606, EPI_ISL_2662608, EPI_ISL_2662609, EPI_ISL_2662610, EPI_ISL_2662611, EPI_ISL_2662612, EPI_ISL_2662613, EPI_ISL_2662614, EPI_ISL_2662615, EPI_ISL_2662616, EPI_ISL_2662617, EPI_ISL_2695772, EPI_ISL_2695773, EPI_ISL_2695774, EPI_ISL_2695775, EPI_ISL_2695776, EPI_ISL_2695777, EPI_ISL_2695778, EPI_ISL_2695779, EPI_ISL_2695780, EPI_ISL_2695781, EPI_ISL_2695782, EPI_ISL_2695783, EPI_ISL_2695784, EPI_ISL_2709943, EPI_ISL_2709944, EPI_ISL_2709945, EPI_ISL_2709946, EPI_ISL_2709947, EPI_ISL_2709948, EPI_ISL_2709949, EPI_ISL_2709950, EPI_ISL_2709951, EPI_ISL_2709953, EPI_ISL_2709954, EPI_ISL_2709955, EPI_ISL_2709956, EPI_ISL_2709957, EPI_ISL_2709958, EPI_ISL_2709959, EPI_ISL_2709960, EPI_ISL_2709961, EPI_ISL_2709962, EPI_ISL_2709963, EPI_ISL_2709964, EPI_ISL_2709965, EPI_ISL_2709966, EPI_ISL_2709967, EPI_ISL_2709968, EPI_ISL_2709969, EPI_ISL_2709970, EPI_ISL_2709971, EPI_ISL_2709972, EPI_ISL_2709973, EPI_ISL_2709975, EPI_ISL_2709976, EPI_ISL_2709979, EPI_ISL_2709980, EPI_ISL_2709982, EPI_ISL_2709983, EPI_ISL_2709984, EPI_ISL_2709985, EPI_ISL_2709986, EPI_ISL_2709987, EPI_ISL_2709988, EPI_ISL_2709989, EPI_ISL_2709990, EPI_ISL_2709991, EPI_ISL_2709992, EPI_ISL_2709993, EPI_ISL_2709994, EPI_ISL_2827037, EPI_ISL_2827038, EPI_ISL_2827039, EPI_ISL_2827040, EPI_ISL_2827041, EPI_ISL_2827042, EPI_ISL_2827043, EPI_ISL_2827044, EPI_ISL_2827045, EPI_ISL_2827046, EPI_ISL_2827047, EPI_ISL_2827048, EPI_ISL_2827049, EPI_ISL_2827050, EPI_ISL_2827051, EPI_ISL_2827052, EPI_ISL_2827053, EPI_ISL_2827054, EPI_ISL_2827055, EPI_ISL_2827056, EPI_ISL_2827057, EPI_ISL_2827058, EPI_ISL_2827059, EPI_ISL_2827060, EPI_ISL_2827061, EPI_ISL_2827062, EPI_ISL_2827063, EPI_ISL_2827064, EPI_ISL_2827065, EPI_ISL_2827066, EPI_ISL_2827067, EPI_ISL_2827068, EPI_ISL_2827069, EPI_ISL_2827142, EPI_ISL_2827143, EPI_ISL_2827144, EPI_ISL_2827145, EPI_ISL_2827146, EPI_ISL_2827147, EPI_ISL_2827148, EPI_ISL_2827150, EPI_ISL_2827151, EPI_ISL_2827152, EPI_ISL_2827153, EPI_ISL_2827154, EPI_ISL_2827155 | | | |
| see above | TAMBO MEMORIAL LABORATORY | National Institute for Communicable Diseases of the National Health Laboratory Service | Amoako DG; Bhiman JN; Everatt J; Ismail A; Mahlangu B; Mnguni A; Mohale T; Ntuli N; Scheepers C |

| | | | |
|---|---|---|---|
| EPI_ISL_2693319, EPI_ISL_2693323, EPI_ISL_2802187, EPI_ISL_2802191 | TC Newman CDC wc TCN | NHLS/UCT | Arash Iranzadeh; Bruna Galvao; Carolyn Williamson; Deelan Doolabh; Diana Hardie; Gert Marais; Innocent Mudau; Lynn Tyers; Marvin Hsiao; Stephen Korsman |
| EPI_ISL_2827906, EPI_ISL_2827907, EPI_ISL_2827908, EPI_ISL_2827911, EPI_ISL_2827912, EPI_ISL_2827913, EPI_ISL_2827914, EPI_ISL_2827915, EPI_ISL_2827916, EPI_ISL_2827919, EPI_ISL_2827923, EPI_ISL_2827924, EPI_ISL_2827925 | | | |
| see above | TSHEPONG LABORATORY | National Institute for Communicable Diseases of the National Health Laboratory Service | Amoako DG; Bhiman JN; Everatt J; Ismail A; Mahlangu B; Mnguni A; Mohale T; Ntuli N; Scheepers C |
| EPI_ISL_2802181, EPI_ISL_2876985 | Thembalethu CDC wc THC | NHLS/UCT | Arash Iranzadeh; Bruna Galvao; Carolyn Williamson; Deelan Doolabh; Diana Hardie; Gert Marais; Innocent Mudau; Lynn Tyers; Marvin Hsiao; Stephen Korsman |
| EPI_ISL_2547369 | Tshwaragano Hospital | National Health Laboratory Service/University of Cape Town (NHLS/UCT) | Arash Iranzadeh; Bruna Galvao; Carolyn Williamson; Deelan Doolabh; Diana Hardie; Gert Marais; Innocent Mudau; Lynn Tyers; Marvin Hsiao; Stephen Korsman |
| EPI_ISL_2802121 | Tsineng Clinic | NHLS/UCT | Arash Iranzadeh; Bruna Galvao; Carolyn Williamson; Deelan Doolabh; Diana Hardie; Gert Marais; Innocent Mudau; Lynn Tyers; Marvin Hsiao; Stephen Korsman |
| EPI_ISL_2802112 | Tygerdal | NHLS/UCT | Arash Iranzadeh; Bruna Galvao; Carolyn Williamson; Deelan Doolabh; Diana Hardie; Gert Marais; Innocent Mudau; Lynn Tyers; Marvin Hsiao; Stephen Korsman |
| EPI_ISL_2812356, EPI_ISL_2812357, EPI_ISL_2812358, EPI_ISL_2812359, EPI_ISL_2812360, EPI_ISL_2812361, EPI_ISL_2812362, EPI_ISL_2812363, EPI_ISL_2812364, EPI_ISL_2812365, EPI_ISL_2812366, EPI_ISL_2812367, EPI_ISL_2812368, EPI_ISL_2812369, EPI_ISL_2812370, EPI_ISL_2812371, EPI_ISL_2812372, EPI_ISL_2812373, EPI_ISL_2812374, EPI_ISL_2812375, EPI_ISL_2812376, EPI_ISL_2812377, EPI_ISL_2812378, EPI_ISL_2812379, EPI_ISL_2812380, EPI_ISL_2812381, EPI_ISL_2812382, EPI_ISL_2812383, EPI_ISL_2812384, EPI_ISL_2812385, EPI_ISL_2812386, EPI_ISL_2812387, EPI_ISL_2812388, EPI_ISL_2812389, EPI_ISL_2812390, EPI_ISL_2812391, EPI_ISL_2812392, EPI_ISL_2812393, EPI_ISL_2812394, EPI_ISL_2812395, EPI_ISL_2812396, EPI_ISL_2812397, EPI_ISL_2812398, EPI_ISL_2812399, EPI_ISL_2812400, EPI_ISL_2812401, EPI_ISL_2812402, EPI_ISL_2812403, EPI_ISL_2812404, EPI_ISL_2812405, EPI_ISL_2812406, EPI_ISL_2812407, EPI_ISL_2812408, EPI_ISL_2812409, EPI_ISL_2812410, EPI_ISL_2812411, EPI_ISL_2812412, EPI_ISL_2812413, EPI_ISL_2812414, EPI_ISL_2812415, EPI_ISL_2812416, EPI_ISL_2812417, EPI_ISL_2812418, EPI_ISL_2812419, EPI_ISL_2812420, EPI_ISL_2812421, EPI_ISL_2812422, EPI_ISL_2812423, EPI_ISL_2812424, EPI_ISL_2812425, EPI_ISL_2812426, EPI_ISL_2812427, EPI_ISL_2812428, EPI_ISL_2812429, EPI_ISL_2812430, EPI_ISL_2812431, EPI_ISL_2812432, EPI_ISL_2812433, EPI_ISL_2812434, EPI_ISL_2812435, EPI_ISL_2812436, EPI_ISL_2812437, EPI_ISL_2812438, EPI_ISL_2841635, EPI_ISL_2841636, EPI_ISL_2841637, EPI_ISL_2841638, EPI_ISL_2841639, EPI_ISL_2841640, EPI_ISL_2841641, EPI_ISL_2841642 | | | |
| see above | UFS Virology | KRISP, KZn Research Innovation and Sequencing Platform | Giandhari Jennifer; Naidoo Yeshnee; Pillay Sureshnee; San James; Tegally Houriiyah; Tshabuila Derek; Wilkinson Eduan; Yajna Ramphal; de Oliveira Tulio |
| EPI_ISL_2375993, EPI_ISL_2375995, EPI_ISL_2621061, EPI_ISL_2802178, EPI_ISL_2802179, EPI_ISL_2876943, EPI_ISL_2876946, EPI_ISL_2876970, EPI_ISL_2876976 | | | |
| see above | Victoria Hospital wc VHW | NHLS/UCT | Arash Iranzadeh; Bruna Galvao; Carolyn Williamson; Deelan Doolabh; Diana Hardie; Gert Marais; Innocent Mudau; Kruger Marais; Lynn Tyers; Marvin Hsiao; Stephen Korsman |
| EPI_ISL_2547417 | Victoria Hospital wc VHW | National Health Laboratory Service/University of Cape Town (NHLS/UCT) | Arash Iranzadeh; Bruna Galvao; Carolyn Williamson; Deelan Doolabh; Diana Hardie; Gert Marais; Innocent Mudau; Lynn Tyers; Marvin Hsiao; Stephen Korsman |
| EPI_ISL_2802125, EPI_ISL_2802126, EPI_ISL_2802127 | Villiersdorp Clinic wc VDP | NHLS/UCT | Arash Iranzadeh; Bruna Galvao; Carolyn Williamson; Deelan Doolabh; Diana Hardie; Gert Marais; Innocent Mudau; Lynn Tyers; Marvin Hsiao; Stephen Korsman |
| EPI_ISL_2547363 | Vredendal Hospital wc VRE | National Health Laboratory Service/University of Cape Town (NHLS/UCT) | Arash Iranzadeh; Bruna Galvao; Carolyn Williamson; Deelan Doolabh; Diana Hardie; Gert Marais; Innocent Mudau; Lynn Tyers; Marvin Hsiao; Stephen Korsman |
| EPI_ISL_2375932 | Vredendal North Clinic wc VDN | NHLS/UCT | Arash Iranzadeh; Bruna Galvao; Carolyn Williamson; Deelan Doolabh; Diana Hardie; Innocent Mudau; Kruger Marais; Lynn Tyers; Marvin Hsiao; Stephen Korsman |
| EPI_ISL_2547367 | Vredendal North Clinic wc VDN | National Health Laboratory Service/University of Cape Town (NHLS/UCT) | Arash Iranzadeh; Bruna Galvao; Carolyn Williamson; Deelan Doolabh; Diana Hardie; Gert Marais; Innocent Mudau; Lynn Tyers; Marvin Hsiao; Stephen Korsman |
| EPI_ISL_2841660, EPI_ISL_2841661, EPI_ISL_2841662, EPI_ISL_2841663, EPI_ISL_2841664, EPI_ISL_2841665, EPI_ISL_2841666, EPI_ISL_2841667, EPI_ISL_2841668, EPI_ISL_2841669, EPI_ISL_2841670, EPI_ISL_2841671, EPI_ISL_2841672, EPI_ISL_2841673, EPI_ISL_2841674, EPI_ISL_2841675, EPI_ISL_2841676, EPI_ISL_2841677, EPI_ISL_2841678, EPI_ISL_2841679, EPI_ISL_2841680, EPI_ISL_2841681, EPI_ISL_2841682, EPI_ISL_2841683, EPI_ISL_2841684, EPI_ISL_2841685, EPI_ISL_2841686, EPI_ISL_2841687, EPI_ISL_2841688, EPI_ISL_2841689, EPI_ISL_2841690, EPI_ISL_2841691, EPI_ISL_2841692, EPI_ISL_2841693, EPI_ISL_2841694, EPI_ISL_2841695, EPI_ISL_2841696, EPI_ISL_2841697, EPI_ISL_2841698, EPI_ISL_2841699, EPI_ISL_2841700 | | | |
| see above | Wits Vaccines & Infectious Diseases Analytics (VIDA) Research Unit | KRISP, KZn Research Innovation and Sequencing Platform | Baillie Vicky; Giandhari Jennifer; Madhi Shabir; Naidoo Yeshnee; Pillay Sureshnee; San James; Tegally Houriiyah; Wilkinson Eduan; de Oliveira Tulio; du Plessis Jeanine |
| EPI_ISL_2375925, EPI_ISL_2375931 | Worcester Hospital wc WOC | NHLS/UCT | Arash Iranzadeh; Bruna Galvao; Carolyn Williamson; Deelan Doolabh; Diana Hardie; Innocent Mudau; Kruger Marais; Lynn Tyers; Marvin Hsiao; Stephen Korsman |
| EPI_ISL_2547364 | Worcester Hospital wc WOC | National Health Laboratory Service/University of Cape Town (NHLS/UCT) | Arash Iranzadeh; Bruna Galvao; Carolyn Williamson; Deelan Doolabh; Diana Hardie; Gert Marais; Innocent Mudau; Lynn Tyers; Marvin Hsiao; Stephen Korsman |
| EPI_ISL_2727328, EPI_ISL_2727329, EPI_ISL_2727330, EPI_ISL_2727331, EPI_ISL_2727332, EPI_ISL_2727333, EPI_ISL_2727334, EPI_ISL_2727335, EPI_ISL_2727336, EPI_ISL_2727337, EPI_ISL_2727338, EPI_ISL_2727339, EPI_ISL_2727340, EPI_ISL_2727341, EPI_ISL_2727342, EPI_ISL_2727343, EPI_ISL_2727344, EPI_ISL_2727345, EPI_ISL_2727346, EPI_ISL_2727347, EPI_ISL_2727348, EPI_ISL_2727351, EPI_ISL_2727352, EPI_ISL_2727353, EPI_ISL_2727354, EPI_ISL_2727355, EPI_ISL_2727356, EPI_ISL_2727357, EPI_ISL_2727358, EPI_ISL_2727359, EPI_ISL_2727360, EPI_ISL_2727361, EPI_ISL_2727362, EPI_ISL_2727363, EPI_ISL_2727364, EPI_ISL_2727365, EPI_ISL_2727366, EPI_ISL_2727367, EPI_ISL_2727368, EPI_ISL_2727369, EPI_ISL_2727370, EPI_ISL_2727371, EPI_ISL_2727372, EPI_ISL_2727373, EPI_ISL_2727374, EPI_ISL_2727375, EPI_ISL_2727376, EPI_ISL_2727377, EPI_ISL_2727378, EPI_ISL_2727379, EPI_ISL_2899741, EPI_ISL_2899742, EPI_ISL_2899743, EPI_ISL_2899744, EPI_ISL_2899745, EPI_ISL_2899746, EPI_ISL_2899747, EPI_ISL_2899748, EPI_ISL_2899749, EPI_ISL_2899750, EPI_ISL_2899751, EPI_ISL_2899752, EPI_ISL_2899753, EPI_ISL_2899754, EPI_ISL_2899755, EPI_ISL_2899756, EPI_ISL_2899757, EPI_ISL_2955421, EPI_ISL_2955422, EPI_ISL_2955423, EPI_ISL_2955424, EPI_ISL_2955425, EPI_ISL_2955426, EPI_ISL_2955427, EPI_ISL_2955428, EPI_ISL_2955429, EPI_ISL_2955430, EPI_ISL_2955431, EPI_ISL_2955432, EPI_ISL_2955433, EPI_ISL_2955434, EPI_ISL_2955435, EPI_ISL_2955436, EPI_ISL_2955437, EPI_ISL_2955438, EPI_ISL_2955439, EPI_ISL_2955440, EPI_ISL_2955441, EPI_ISL_2955442, EPI_ISL_2955443, EPI_ISL_2955444, EPI_ISL_2955445, EPI_ISL_2955446, EPI_ISL_2955447, EPI_ISL_2955448, EPI_ISL_2955449, EPI_ISL_2955450, EPI_ISL_2955451, EPI_ISL_2955452 | | | |
| see above | ZARV/NHLS, Department Medical Virology, University of Pretoria | KRISP, KZn Research Innovation and Sequencing Platform | Adriano Mendes; Amy Strydom; Emmanuel SJ; Giandhari J; Lessells R; Micheala Davids; Naidoo Y; Pillay S; Ramphal U; Sim Mayaphi and Marietjie Venter; Tegally H; Wilkinson E; de Oliveira T |

We gratefully acknowledge the following Authors from the Originating laboratories responsible for obtaining the specimens, as well as the Submitting laboratories where the genome data were generated and shared via GISAID, on which this research is based.

All Submitters of data may be contacted directly via www.gisaid.org

Authors are sorted alphabetically.

| Accession ID | Originating Laboratory | Submitting Laboratory | Authors |
|---|---|---|---|
| EPI_ISL_1178008, EPI_ISL_1474810, EPI_ISL_1474811, EPI_ISL_1474812, EPI_ISL_1474813, EPI_ISL_1474814, EPI_ISL_1474815, EPI_ISL_1474816, EPI_ISL_1474817, EPI_ISL_1474818, EPI_ISL_1474819, EPI_ISL_1474828, EPI_ISL_1730312 | | | |
| see above | Barts Health NHS Trust | COVID-19 Genomics UK (COG-UK) Consortium | BROAD; Beatrix; CUTINO-MOGUEL; Claire; David; Dola; HARRINGTON; KELE; KULASEGARAN-SHYLINI; Maria-Teresa; OWOYEMI; Raghavendran |
| EPI_ISL_1180134, EPI_ISL_1180135, EPI_ISL_1180136, EPI_ISL_1180137, EPI_ISL_1180138, EPI_ISL_1180139, EPI_ISL_1180140, EPI_ISL_1250312, EPI_ISL_1250314, EPI_ISL_1250315 | | | |
| see above | Bioinformatics and Biostatistics Lab, Advanced Sequencing Facility | COVID-19 Genomics UK (COG-UK) Consortium | Aengus Stewart; Chelsea Sawyer; Harshil Patel; Jerome Nicod; Laura Cubitt; Margaret Crawford |
| EPI_ISL_1249974, EPI_ISL_1249975, EPI_ISL_1249976, EPI_ISL_1249977, EPI_ISL_1249978, EPI_ISL_1250118, EPI_ISL_1250119, EPI_ISL_1310336, EPI_ISL_1310338, EPI_ISL_1310340, EPI_ISL_1310341, EPI_ISL_1310342, EPI_ISL_1310343, EPI_ISL_1310344, EPI_ISL_1310345, EPI_ISL_1310346, EPI_ISL_1310347, EPI_ISL_1310357, EPI_ISL_1310438, EPI_ISL_1310439, EPI_ISL_1310441, EPI_ISL_1310442, EPI_ISL_1310443, EPI_ISL_1310445, EPI_ISL_1310446, EPI_ISL_1310447, EPI_ISL_1310448, EPI_ISL_1310450, EPI_ISL_1310451, EPI_ISL_1310452, EPI_ISL_1388660, EPI_ISL_1388661, EPI_ISL_1388664, EPI_ISL_1388665, EPI_ISL_1388667, EPI_ISL_1388670, EPI_ISL_1388672, EPI_ISL_1388674, EPI_ISL_1388682, EPI_ISL_1388698, EPI_ISL_1388754, EPI_ISL_1388766, EPI_ISL_1388768, EPI_ISL_1388770, EPI_ISL_1476549, EPI_ISL_1476550, EPI_ISL_1476551, EPI_ISL_1476552, EPI_ISL_1476553, EPI_ISL_1476554, EPI_ISL_1476672 | | | |
| see above | Centre for Enzyme Innovation, University of Portsmouth / Translational Research Laboratory, Portsmouth Hospitals NHS Trust | COVID-19 Genomics UK (COG-UK) Consortium | Angela Beckett; Christopher Fearn; Kate Cook; Katie Loveson; Salman Goudarzi; Samuel Robson; Scott Elliott; Sharon Glaysher |
| EPI_ISL_1177683, EPI_ISL_1177684, EPI_ISL_1177685, EPI_ISL_1177686, EPI_ISL_1177687, EPI_ISL_1247289, EPI_ISL_1247290, EPI_ISL_1247291, EPI_ISL_1247292, EPI_ISL_1247293, EPI_ISL_1247294, EPI_ISL_1247295, EPI_ISL_1247296, EPI_ISL_1247297, EPI_ISL_1247298, EPI_ISL_1247299, EPI_ISL_1247300, EPI_ISL_1247301, EPI_ISL_1247302, EPI_ISL_1247303, EPI_ISL_1247304, EPI_ISL_1247305, EPI_ISL_1247306, EPI_ISL_1247307, EPI_ISL_1247308, EPI_ISL_1247309, EPI_ISL_1247310, EPI_ISL_1247311, EPI_ISL_1247312, EPI_ISL_1247313, EPI_ISL_1247314, EPI_ISL_1247315, EPI_ISL_1247316, EPI_ISL_1247317, EPI_ISL_1247318, EPI_ISL_1247319, EPI_ISL_1247320, EPI_ISL_1247321, EPI_ISL_1247322, EPI_ISL_1247323, EPI_ISL_1247324, EPI_ISL_1247325, EPI_ISL_1247326, EPI_ISL_1247327, EPI_ISL_1247328, EPI_ISL_1247329, EPI_ISL_1247330, EPI_ISL_1247331, EPI_ISL_1247332, EPI_ISL_1247333, EPI_ISL_1247334, EPI_ISL_1247335, EPI_ISL_1247336, EPI_ISL_1247337, EPI_ISL_1247338, EPI_ISL_1247339, EPI_ISL_1247340, EPI_ISL_1247341, EPI_ISL_1247342, EPI_ISL_1247343, EPI_ISL_1247344, EPI_ISL_1247345, EPI_ISL_1247346, EPI_ISL_1247347, EPI_ISL_1247348, EPI_ISL_1247349, EPI_ISL_1247350, EPI_ISL_1247352, EPI_ISL_1247353, EPI_ISL_1247354, EPI_ISL_1247355, EPI_ISL_1247356, EPI_ISL_1247357, EPI_ISL_1247358, EPI_ISL_1247360, EPI_ISL_1247361, EPI_ISL_1247362, EPI_ISL_1247363, EPI_ISL_1247364, EPI_ISL_1247365, EPI_ISL_1247366, EPI_ISL_1247367, EPI_ISL_1247368, EPI_ISL_1247612, EPI_ISL_1247613, EPI_ISL_1247614, EPI_ISL_1247615, EPI_ISL_1247616, EPI_ISL_1247618, EPI_ISL_1247631, EPI_ISL_1247632, EPI_ISL_1250124, EPI_ISL_1250125, EPI_ISL_1250126, EPI_ISL_1250127, EPI_ISL_1250128, EPI_ISL_1250129, EPI_ISL_1250130, EPI_ISL_1250131, EPI_ISL_1250132, EPI_ISL_1250133, EPI_ISL_1250134, EPI_ISL_1250135, EPI_ISL_1250136, EPI_ISL_1250137, EPI_ISL_1250138, EPI_ISL_1250139, EPI_ISL_1250140, EPI_ISL_1250141, EPI_ISL_1250142, EPI_ISL_1250143, EPI_ISL_1250144, EPI_ISL_1250145, EPI_ISL_1250146, EPI_ISL_1250147, EPI_ISL_1250148, EPI_ISL_1250149, EPI_ISL_1250150, EPI_ISL_1250151, EPI_ISL_1250152, EPI_ISL_1250153, EPI_ISL_1250154, EPI_ISL_1250155, EPI_ISL_1250156, EPI_ISL_1250157, EPI_ISL_1250158, EPI_ISL_1250159, EPI_ISL_1250160, EPI_ISL_1250161, EPI_ISL_1250162, EPI_ISL_1250163, EPI_ISL_1250164, EPI_ISL_1250165, EPI_ISL_1250166, EPI_ISL_1250167, EPI_ISL_1250168, EPI_ISL_1250169, EPI_ISL_1250170, EPI_ISL_1250171, EPI_ISL_1250172, EPI_ISL_1250173, EPI_ISL_1250174, EPI_ISL_1250175, EPI_ISL_1250176, EPI_ISL_1250177, EPI_ISL_1250178, EPI_ISL_1250179, EPI_ISL_1250180, EPI_ISL_1250181, EPI_ISL_1250182, EPI_ISL_1250183, EPI_ISL_1250184, EPI_ISL_1250185, EPI_ISL_1250186, EPI_ISL_1250187, EPI_ISL_1250188, EPI_ISL_1250189, EPI_ISL_1250190, EPI_ISL_1250191, EPI_ISL_1250192, EPI_ISL_1250193, EPI_ISL_1250194, EPI_ISL_1250195, EPI_ISL_1250196, EPI_ISL_1250197, EPI_ISL_1250198, EPI_ISL_1250199, EPI_ISL_1250200, EPI_ISL_1250201, EPI_ISL_1250202, EPI_ISL_1250203, EPI_ISL_1250204, EPI_ISL_1250205, EPI_ISL_1250206, EPI_ISL_1250207, EPI_ISL_1250208, EPI_ISL_1250209, EPI_ISL_1250210, EPI_ISL_1250211, EPI_ISL_1250212, EPI_ISL_1250213, EPI_ISL_1250214, EPI_ISL_1250215, EPI_ISL_1250216, EPI_ISL_1250218, EPI_ISL_1250219, EPI_ISL_1250220, EPI_ISL_1310455, EPI_ISL_1310456, EPI_ISL_1310457, EPI_ISL_1310458, EPI_ISL_1310459, EPI_ISL_1310460, EPI_ISL_1310461, EPI_ISL_1310463, EPI_ISL_1310464, EPI_ISL_1310465, EPI_ISL_1310466, EPI_ISL_1310467, EPI_ISL_1310468, EPI_ISL_1310469, EPI_ISL_1310470, EPI_ISL_1310471, EPI_ISL_1310472, EPI_ISL_1310473, EPI_ISL_1310474, EPI_ISL_1310475, EPI_ISL_1310476, EPI_ISL_1310477, EPI_ISL_1310478, EPI_ISL_1310479, EPI_ISL_1310480, EPI_ISL_1310481, EPI_ISL_1310482, EPI_ISL_1310483, EPI_ISL_1310484, EPI_ISL_1310485, EPI_ISL_1310486, EPI_ISL_1310487, EPI_ISL_1310488, EPI_ISL_1310489, EPI_ISL_1310490, EPI_ISL_1310491, EPI_ISL_1310492, EPI_ISL_1310494, EPI_ISL_1310495, EPI_ISL_1310496, EPI_ISL_1310497, EPI_ISL_1310498, EPI_ISL_1310499, EPI_ISL_1310500, EPI_ISL_1310501, EPI_ISL_1310502, EPI_ISL_1310503, EPI_ISL_1310504, EPI_ISL_1310505, EPI_ISL_1310506, EPI_ISL_1310507, EPI_ISL_1310508, EPI_ISL_1310509, EPI_ISL_1310510, EPI_ISL_1310511, EPI_ISL_1310512, EPI_ISL_1310513, EPI_ISL_1310514, EPI_ISL_1310515, EPI_ISL_1310516, EPI_ISL_1310517, EPI_ISL_1310518, EPI_ISL_1310519, EPI_ISL_1310521, EPI_ISL_1310522, EPI_ISL_1310523, EPI_ISL_1310524, EPI_ISL_1310525, EPI_ISL_1310526, EPI_ISL_1310527, EPI_ISL_1310528, EPI_ISL_1310529, EPI_ISL_1310530, EPI_ISL_1310531, EPI_ISL_1310532, EPI_ISL_1310533, EPI_ISL_1310534, EPI_ISL_1310535, EPI_ISL_1310536, EPI_ISL_1310537, EPI_ISL_1310538, EPI_ISL_1310539, EPI_ISL_1310540, EPI_ISL_1310541, EPI_ISL_1310542, EPI_ISL_1310543, EPI_ISL_1310544, EPI_ISL_1310545, EPI_ISL_1310546, EPI_ISL_1310547, EPI_ISL_1310548, EPI_ISL_1310549, EPI_ISL_1310550, EPI_ISL_1310551, EPI_ISL_1310552, EPI_ISL_1310553, EPI_ISL_1310554, EPI_ISL_1310555, EPI_ISL_1310556, EPI_ISL_1310557, EPI_ISL_1310558, EPI_ISL_1310559, EPI_ISL_1310560, EPI_ISL_1310561, EPI_ISL_1310562, EPI_ISL_1310563, EPI_ISL_1310564, EPI_ISL_1310565, EPI_ISL_1310566, EPI_ISL_1310567, EPI_ISL_1310568, EPI_ISL_1310569, EPI_ISL_1310570, EPI_ISL_1310571, EPI_ISL_1310572, EPI_ISL_1310573, EPI_ISL_1310574, EPI_ISL_1310575, EPI_ISL_1310576, EPI_ISL_1310577, EPI_ISL_1310578, EPI_ISL_1310579, EPI_ISL_1310580, EPI_ISL_1310581, EPI_ISL_1310582, EPI_ISL_1310583, EPI_ISL_1310584, EPI_ISL_1310585, EPI_ISL_1310586, EPI_ISL_1310587, EPI_ISL_1310588, EPI_ISL_1310589, EPI_ISL_1310590, EPI_ISL_1310591, EPI_ISL_1310592, EPI_ISL_1310593, EPI_ISL_1310594, EPI_ISL_1310595, EPI_ISL_1310596, EPI_ISL_1310597, EPI_ISL_1310598, EPI_ISL_1310599, EPI_ISL_1310600, EPI_ISL_1310601, EPI_ISL_1310602, EPI_ISL_1310603, EPI_ISL_1310604, EPI_ISL_1310605, EPI_ISL_1310606, EPI_ISL_1310607, EPI_ISL_1310608, EPI_ISL_1310609, EPI_ISL_1310610, EPI_ISL_1310611, EPI_ISL_1310612, EPI_ISL_1310613, EPI_ISL_1310614, EPI_ISL_1310615, EPI_ISL_1310616, EPI_ISL_1310617, EPI_ISL_1310618, EPI_ISL_1310619, EPI_ISL_1310620, EPI_ISL_1310621, EPI_ISL_1310625, EPI_ISL_1310626, EPI_ISL_1310628, EPI_ISL_1310629, EPI_ISL_1310632, EPI_ISL_1310633, EPI_ISL_1310634, EPI_ISL_1310636, EPI_ISL_1310637, EPI_ISL_1310638, EPI_ISL_1310639, EPI_ISL_1310643, EPI_ISL_1310644, EPI_ISL_1310645, EPI_ISL_1310647, EPI_ISL_1310648, EPI_ISL_1310649, EPI_ISL_1310650, EPI_ISL_1310653, EPI_ISL_1310654, EPI_ISL_1310655, EPI_ISL_1310656, EPI_ISL_1310657, EPI_ISL_1310658, EPI_ISL_1310659, EPI_ISL_1310660, EPI_ISL_1310664, EPI_ISL_1310665, EPI_ISL_1310666, EPI_ISL_1310668, EPI_ISL_1310669, EPI_ISL_1310670, EPI_ISL_1310674, EPI_ISL_1310675, EPI_ISL_1310678, EPI_ISL_1310679, EPI_ISL_1310680, EPI_ISL_1310683, EPI_ISL_1310684, EPI_ISL_1310687, EPI_ISL_1310688, EPI_ISL_1310689, EPI_ISL_1310690, EPI_ISL_1310692, EPI_ISL_1310693, EPI_ISL_1310694, EPI_ISL_1310695, EPI_ISL_1310696, EPI_ISL_1310697, EPI_ISL_1310698, EPI_ISL_1310699, EPI_ISL_1310700, EPI_ISL_1310701 | | | |
| see above | Department of Pathology, University of Cambridge | COVID-19 Genomics UK (COG-UK) Consortium | Aminu S. Jahun; Ian Goodfellow; Iliana Georgana; Martin D. Curran; Myra Hosmillo; Rhys Izuagbe; Surendra Parmar; William L. Hamilton; Yasmin Chaudhry |
| EPI_ISL_1186849, EPI_ISL_1187313, EPI_ISL_1205707, EPI_ISL_1205709, EPI_ISL_1205710, EPI_ISL_1205714, EPI_ISL_1205719, EPI_ISL_1205720, EPI_ISL_1205721, EPI_ISL_1205725, EPI_ISL_1205727, EPI_ISL_1205728, EPI_ISL_1205731, EPI_ISL_1205736, EPI_ISL_1205745, EPI_ISL_1205748, EPI_ISL_1205752, EPI_ISL_1205753, EPI_ISL_1205757, EPI_ISL_1205758, EPI_ISL_1205759, EPI_ISL_1205760, EPI_ISL_1205761, EPI_ISL_1205767, EPI_ISL_1205770, EPI_ISL_1205776, EPI_ISL_1205785, EPI_ISL_1205789, EPI_ISL_1205791, EPI_ISL_1205802, EPI_ISL_1205805, EPI_ISL_1205819, EPI_ISL_1205824, EPI_ISL_1205825, EPI_ISL_1205828, EPI_ISL_1205831, EPI_ISL_1205838, EPI_ISL_1205843, EPI_ISL_1205850, EPI_ISL_1205862, EPI_ISL_1205873, EPI_ISL_1205876, EPI_ISL_1205881, EPI_ISL_1205889, EPI_ISL_1205897, EPI_ISL_1205906, EPI_ISL_1205910, EPI_ISL_1205911, EPI_ISL_1205913, EPI_ISL_1205916, EPI_ISL_1205932, EPI_ISL_1205934, EPI_ISL_1205938, EPI_ISL_1205940, EPI_ISL_1205949, EPI_ISL_1205951, EPI_ISL_1205954, EPI_ISL_1205958, EPI_ISL_1205962, EPI_ISL_1205967, EPI_ISL_1205978, EPI_ISL_1205988, EPI_ISL_1205997, EPI_ISL_1206003, EPI_ISL_1206005, EPI_ISL_1206021, EPI_ISL_1206027, EPI_ISL_1206028, EPI_ISL_1206030, EPI_ISL_1206032, EPI_ISL_1223666, EPI_ISL_1223674, EPI_ISL_1223686, EPI_ISL_1223689, EPI_ISL_1223695, EPI_ISL_1223718, EPI_ISL_1223737, EPI_ISL_1223740, EPI_ISL_1223746, EPI_ISL_1223750, EPI_ISL_1223767, EPI_ISL_1223792, EPI_ISL_1223800, EPI_ISL_1223828, EPI_ISL_1223830, EPI_ISL_1223835, EPI_ISL_1223843, EPI_ISL_1223847, EPI_ISL_1223920, EPI_ISL_1223929, EPI_ISL_1223931, EPI_ISL_1242828, EPI_ISL_1242831, EPI_ISL_1242832, EPI_ISL_1242834, EPI_ISL_1242839, EPI_ISL_1242840, EPI_ISL_1242845, EPI_ISL_1242846, EPI_ISL_1242848, EPI_ISL_1242849, EPI_ISL_1242850, EPI_ISL_1242854, EPI_ISL_1242857, EPI_ISL_1242859, EPI_ISL_1242861, EPI_ISL_1242866, EPI_ISL_1242868, EPI_ISL_1242869, EPI_ISL_1242870, EPI_ISL_1242873, EPI_ISL_1242874, EPI_ISL_1242875, EPI_ISL_1242876, EPI_ISL_1242877, EPI_ISL_1242878, EPI_ISL_1242879, EPI_ISL_1242882, EPI_ISL_1242891, EPI_ISL_1242892, EPI_ISL_1242895, EPI_ISL_1242899, EPI_ISL_1242900, EPI_ISL_1242901, EPI_ISL_1242903, EPI_ISL_1242904, EPI_ISL_1242911, EPI_ISL_1242913, EPI_ISL_1242915, EPI_ISL_1242917, EPI_ISL_1242918, EPI_ISL_1242919, EPI_ISL_1242921, EPI_ISL_1242922, EPI_ISL_1242923, EPI_ISL_1242933, EPI_ISL_1242936, EPI_ISL_1242937, EPI_ISL_1242940, EPI_ISL_1242941, EPI_ISL_1242942, EPI_ISL_1242944, EPI_ISL_1242949, EPI_ISL_1242950, EPI_ISL_1242953, EPI_ISL_1242962, EPI_ISL_1242963, EPI_ISL_1242965, EPI_ISL_1242967, EPI_ISL_1242972, EPI_ISL_1242973, EPI_ISL_1242975, EPI_ISL_1242978, EPI_ISL_1242984, EPI_ISL_1242985, EPI_ISL_1242988, EPI_ISL_1242990, EPI_ISL_1242991, EPI_ISL_1242992, EPI_ISL_1242993, EPI_ISL_1242994, EPI_ISL_1242997, EPI_ISL_1243004, EPI_ISL_1243005, EPI_ISL_1243007, EPI_ISL_1243015, EPI_ISL_1243017, EPI_ISL_1243018, EPI_ISL_1243021, EPI_ISL_1243027, EPI_ISL_1243029, EPI_ISL_1243030, EPI_ISL_1243032, EPI_ISL_1243033, EPI_ISL_1243035, EPI_ISL_1243040, EPI_ISL_1243043, EPI_ISL_1243044, EPI_ISL_1243047, EPI_ISL_1243048, EPI_ISL_1243053, EPI_ISL_1243054, EPI_ISL_1243055, EPI_ISL_1243060, EPI_ISL_1243065, EPI_ISL_1243068, EPI_ISL_1243069, EPI_ISL_1243070, EPI_ISL_1243075, EPI_ISL_1243076, EPI_ISL_1243077, EPI_ISL_1243078, EPI_ISL_1243082, EPI_ISL_1243084, EPI_ISL_1243087, EPI_ISL_1243092, EPI_ISL_1243093, EPI_ISL_1243094, EPI_ISL_1243095, EPI_ISL_1243098, EPI_ISL_1243101, EPI_ISL_1243102, EPI_ISL_1243105, EPI_ISL_1243111, EPI_ISL_1243117, EPI_ISL_1243119, EPI_ISL_1243126, EPI_ISL_1243127, EPI_ISL_1243128, EPI_ISL_1243134, EPI_ISL_1243138, EPI_ISL_1243145, EPI_ISL_1243146, EPI_ISL_1243150, EPI_ISL_1243152, EPI_ISL_1243156, EPI_ISL_1243157, EPI_ISL_1243158, EPI_ISL_1243160, EPI_ISL_1243164, EPI_ISL_1243166, EPI_ISL_1243167, EPI_ISL_1243168, EPI_ISL_1243169, EPI_ISL_1243171, EPI_ISL_1243175, EPI_ISL_1243179, EPI_ISL_1243181, EPI_ISL_1243183, EPI_ISL_1243184, EPI_ISL_1243187, EPI_ISL_1243190, EPI_ISL_1243194, EPI_ISL_1243197, EPI_ISL_1243198, EPI_ISL_1243199, EPI_ISL_1243200, EPI_ISL_1243201, EPI_ISL_1243205, EPI_ISL_1243206, EPI_ISL_1243207, EPI_ISL_1243208, EPI_ISL_1243209, EPI_ISL_1243210, EPI_ISL_1243211, EPI_ISL_1243212, EPI_ISL_1243213, EPI_ISL_1243214, EPI_ISL_1243215, EPI_ISL_1243216, EPI_ISL_1243217, EPI_ISL_1243221, EPI_ISL_1243222, EPI_ISL_1243223, EPI_ISL_1243224, EPI_ISL_1243225, EPI_ISL_1243227, EPI_ISL_1243229, EPI_ISL_1243231, EPI_ISL_1243232, EPI_ISL_1243233, EPI_ISL_1243234, EPI_ISL_1243235, EPI_ISL_1243236, EPI_ISL_1243241, EPI_ISL_1243243, EPI_ISL_1243245, EPI_ISL_1243246, EPI_ISL_1243248, EPI_ISL_1243250, EPI_ISL_1243251, EPI_ISL_1243252, EPI_ISL_1243253, EPI_ISL_1243256, EPI_ISL_1243257, EPI_ISL_1243259, EPI_ISL_1243261, EPI_ISL_1243263, EPI_ISL_1243266, EPI_ISL_1243267, EPI_ISL_1243268, EPI_ISL_1243269, EPI_ISL_1243270, EPI_ISL_1243272, EPI_ISL_1243274, EPI_ISL_1243277, EPI_ISL_1243278, EPI_ISL_1243280, EPI_ISL_1243283, EPI_ISL_1243284, EPI_ISL_1243285, EPI_ISL_1243286, EPI_ISL_1243287, EPI_ISL_1243288, EPI_ISL_1243289, EPI_ISL_1243290, EPI_ISL_1243291, EPI_ISL_1243294, EPI_ISL_1243295, EPI_ISL_1243297, EPI_ISL_1243298, EPI_ISL_1243299, EPI_ISL_1243301, EPI_ISL_1243302, EPI_ISL_1243305, EPI_ISL_1243307, EPI_ISL_1243309, EPI_ISL_1243311, EPI_ISL_1243312, EPI_ISL_1243313, EPI_ISL_1243314, EPI_ISL_1243316, EPI_ISL_1243318, EPI_ISL_1243319, EPI_ISL_1243320, EPI_ISL_1243321, EPI_ISL_1243322, EPI_ISL_1243325, EPI_ISL_1243326, EPI_ISL_1243328, EPI_ISL_1243330, EPI_ISL_1243331, EPI_ISL_1243332, EPI_ISL_1243333, EPI_ISL_1243334, EPI_ISL_1243335, EPI_ISL_1243336, EPI_ISL_1243338, EPI_ISL_1243342, EPI_ISL_1243343, EPI_ISL_1243344, EPI_ISL_1243345, EPI_ISL_1243348, EPI_ISL_1243349, EPI_ISL_1243352, EPI_ISL_1243354, EPI_ISL_1243355, EPI_ISL_1243356, EPI_ISL_1243357, EPI_ISL_1243360, EPI_ISL_1243362, EPI_ISL_1243364, EPI_ISL_1243365, EPI_ISL_1243367, EPI_ISL_1243368, EPI_ISL_1243370, EPI_ISL_1243371, EPI_ISL_1243372, EPI_ISL_1243373, EPI_ISL_1243375, EPI_ISL_1243376, EPI_ISL_1243381, EPI_ISL_1243382, EPI_ISL_1243384, EPI_ISL_1243386, EPI_ISL_1243388, EPI_ISL_1243389, EPI_ISL_1243390, EPI_ISL_1243391, EPI_ISL_1243392, EPI_ISL_1243393, EPI_ISL_1243397, EPI_ISL_1243398, EPI_ISL_1243399, EPI_ISL_1243400, EPI_ISL_1243402, EPI_ISL_1243406, EPI_ISL_1243409, EPI_ISL_1243410, EPI_ISL_1243412, EPI_ISL_1243413, EPI_ISL_1243414, EPI_ISL_1243415, EPI_ISL_1243417, EPI_ISL_1243418, EPI_ISL_1243419, EPI_ISL_1243420, EPI_ISL_1243430, EPI_ISL_1243431, EPI_ISL_1243432, EPI_ISL_1243433, EPI_ISL_1243613, EPI_ISL_1243615, EPI_ISL_1243619, EPI_ISL_1243624, EPI_ISL_1243629, EPI_ISL_1243635, EPI_ISL_1243637, EPI_ISL_1243644, EPI_ISL_1243660, EPI_ISL_1243661, EPI_ISL_1243662, EPI_ISL_1243663, EPI_ISL_1243665, EPI_ISL_1243668, EPI_ISL_1243670, EPI_ISL_1243674, EPI_ISL_1243690, EPI_ISL_1243698, EPI_ISL_1243699, EPI_ISL_1243703, EPI_ISL_1243708, EPI_ISL_1243726, EPI_ISL_1243728, EPI_ISL_1243729, EPI_ISL_1243734, EPI_ISL_1243735, EPI_ISL_1243743, EPI_ISL_1243744, EPI_ISL_1243745, EPI_ISL_1243747, EPI_ISL_1243748, EPI_ISL_1243755, EPI_ISL_1243757, EPI_ISL_1243760, EPI_ISL_1243779, EPI_ISL_1243781, EPI_ISL_1243783, EPI_ISL_1243786, EPI_ISL_1243795, EPI_ISL_1243804, EPI_ISL_1243806, EPI_ISL_1243808, EPI_ISL_1243812, EPI_ISL_1243815, EPI_ISL_1243816, EPI_ISL_1243822, EPI_ISL_1243824, EPI_ISL_1243825, EPI_ISL_1243832, EPI_ISL_1243843, EPI_ISL_1243847, EPI_ISL_1243849, EPI_ISL_1243856, EPI_ISL_1243858, EPI_ISL_1243860, EPI_ISL_1243861, EPI_ISL_1243862, EPI_ISL_1243863, EPI_ISL_1243867, EPI_ISL_1243869, EPI_ISL_1243872, EPI_ISL_1243874, EPI_ISL_1243875, EPI_ISL_1243877, EPI_ISL_1243881, EPI_ISL_1243891, EPI_ISL_1243897, EPI_ISL_1243899, EPI_ISL_1243901, EPI_ISL_1243902, EPI_ISL_1243903, EPI_ISL_1243908, EPI_ISL_1243911, EPI_ISL_1243921, EPI_ISL_1243922, EPI_ISL_1243923, EPI_ISL_1243924, EPI_ISL_1243935, EPI_ISL_1243936, EPI_ISL_1243940, EPI_ISL_1243945, EPI_ISL_1243952, EPI_ISL_1243957, EPI_ISL_1243961, EPI_ISL_1243962, EPI_ISL_1243970, EPI_ISL_1243972, EPI_ISL_1243973, EPI_ISL_1243979, EPI_ISL_1243984, EPI_ISL_1243991, EPI_ISL_1243992, EPI_ISL_1243993, EPI_ISL_1243998, EPI_ISL_1244001, EPI_ISL_1244002, EPI_ISL_1244004, EPI_ISL_1244005, EPI_ISL_1244009, EPI_ISL_1244013, EPI_ISL_1244015, EPI_ISL_1244019, EPI_ISL_1244028, EPI_ISL_1244032, EPI_ISL_1244039, EPI_ISL_1244046, EPI_ISL_1244047, EPI_ISL_1244048, EPI_ISL_1244051, EPI_ISL_1244057, EPI_ISL_1244061, EPI_ISL_1244064, EPI_ISL_1244065, EPI_ISL_1244068, EPI_ISL_1244069, EPI_ISL_1244070, EPI_ISL_1244071, EPI_ISL_1244074, EPI_ISL_1244086, EPI_ISL_1244092, EPI_ISL_1244095, EPI_ISL_1244100, EPI_ISL_1244101, EPI_ISL_1244104, EPI_ISL_1244106, EPI_ISL_1244108, EPI_ISL_1244112, EPI_ISL_1244120, EPI_ISL_1244124, EPI_ISL_1244125, EPI_ISL_1244128, EPI_ISL_1244131, EPI_ISL_1244138, EPI_ISL_1244142, EPI_ISL_1244144, EPI_ISL_1244157, EPI_ISL_1244164, EPI_ISL_1244165, EPI_ISL_1244171, EPI_ISL_1244172, EPI_ISL_1244176, EPI_ISL_1244178, EPI_ISL_1244179, EPI_ISL_1244181, EPI_ISL_1244191, EPI_ISL_1244193, EPI_ISL_1244201, EPI_ISL_1244205, EPI_ISL_1244208, EPI_ISL_1244213, EPI_ISL_1244214, EPI_ISL_1244221, EPI_ISL_1244222, EPI_ISL_1244225, EPI_ISL_1244226, EPI_ISL_1244229, EPI_ISL_1244231, EPI_ISL_1244241, EPI_ISL_1244248, EPI_ISL_1244254, EPI_ISL_1244266, EPI_ISL_1244269, EPI_ISL_1244270, EPI_ISL_1244275, EPI_ISL_1244277, EPI_ISL_1244280, EPI_ISL_1244282, EPI_ISL_1244288, EPI_ISL_1244291, EPI_ISL_1244292, EPI_ISL_1244293, EPI_ISL_1244294, EPI_ISL_1244296, EPI_ISL_1244297, EPI_ISL_1244298, EPI_ISL_1244300, EPI_ISL_1244302, EPI_ISL_1244305, EPI_ISL_1244306, EPI_ISL_1244309, EPI_ISL_1244311, EPI_ISL_1244312, EPI_ISL_1244315, EPI_ISL_1244319, EPI_ISL_1244320, EPI_ISL_1244323, EPI_ISL_1244324, EPI_ISL_1244326, EPI_ISL_1244329, EPI_ISL_1244330, EPI_ISL_1244332, EPI_ISL_1244333, EPI_ISL_1244335, EPI_ISL_1244336, EPI_ISL_1244341, EPI_ISL_1244342, EPI_ISL_1244346, EPI_ISL_1244348, EPI_ISL_1244349, EPI_ISL_1244350, EPI_ISL_1244351, EPI_ISL_1244352, EPI_ISL_1244353, EPI_ISL_1244356, EPI_ISL_1244357, EPI_ISL_1244358, EPI_ISL_1244359, EPI_ISL_1244360, EPI_ISL_1244361, EPI_ISL_1244362, EPI_ISL_1244363, EPI_ISL_1244365, EPI_ISL_1244366, EPI_ISL_1244367, EPI_ISL_1244369, EPI_ISL_1244372, EPI_ISL_1244373, EPI_ISL_1244374, EPI_ISL_1244376, EPI_ISL_1244377, EPI_ISL_1244378, EPI_ISL_1244379, EPI_ISL_1244380, EPI_ISL_1244381, EPI_ISL_1244386, EPI_ISL_1244387, EPI_ISL_1244391, EPI_ISL_1244392, EPI_ISL_1244393, EPI_ISL_1244401, EPI_ISL_1244405, EPI_ISL_1244406, EPI_ISL_1244407, EPI_ISL_1244408, EPI_ISL_1244409, EPI_ISL_1244413, EPI_ISL_1244414, EPI_ISL_1244415, EPI_ISL_1244416, EPI_ISL_1244418, EPI_ISL_1244420, EPI_ISL_1244421, EPI_ISL_1244422, EPI_ISL_1244423, EPI_ISL_1244424, EPI_ISL_1244426, EPI_ISL_1244427, EPI_ISL_1244429, EPI_ISL_1244430, EPI_ISL_1244434, EPI_ISL_1244435, EPI_ISL_1244437, EPI_ISL_1244438, EPI_ISL_1244439, EPI_ISL_1244440, EPI_ISL_1244442, EPI_ISL_1244443, EPI_ISL_1244444, EPI_ISL_1244445, EPI_ISL_1244448, EPI_ISL_1244449, EPI_ISL_1244450, EPI_ISL_1244452, EPI_ISL_1244453, EPI_ISL_1244455, EPI_ISL_1244457, EPI_ISL_1244461, EPI_ISL_1244462, EPI_ISL_1244463, EPI_ISL_1244464, EPI_ISL_1244465, EPI_ISL_1244467, EPI_ISL_1244468, EPI_ISL_1244470, EPI_ISL_1244471, EPI_ISL_1244473, EPI_ISL_1244474, EPI_ISL_1244477, EPI_ISL_1244478, EPI_ISL_1244479, EPI_ISL_1244481, EPI_ISL_1244482, EPI_ISL_1244483, EPI_ISL_1244486, EPI_ISL_1244487, EPI_ISL_1244489, EPI_ISL_1244491, EPI_ISL_1244492, EPI_ISL_1244493, EPI_ISL_1244494, EPI_ISL_1244495, EPI_ISL_1244498, EPI_ISL_1244499, EPI_ISL_1244501, EPI_ISL_1244502, EPI_ISL_1244503, EPI_ISL_1244505, EPI_ISL_1244507, EPI_ISL_1244508, EPI_ISL_1244509, EPI_ISL_1244510, EPI_ISL_1244511, EPI_ISL_1244512, EPI_ISL_1244514, EPI_ISL_1244515, EPI_ISL_1244516, EPI_ISL_1244519, EPI_ISL_1244520, EPI_ISL_1244523, EPI_ISL_1244524, EPI_ISL_1244525, EPI_ISL_1244527, EPI_ISL_1244529, EPI_ISL_1244530, EPI_ISL_1244531, EPI_ISL_1244532, EPI_ISL_1244533, EPI_ISL_1244534, EPI_ISL_1244537, EPI_ISL_1244538, EPI_ISL_1244542, EPI_ISL_1244543, EPI_ISL_1244544, EPI_ISL_1244545, EPI_ISL_1244549, EPI_ISL_1244550, EPI_ISL_1244551, EPI_ISL_1244552, EPI_ISL_1244555, EPI_ISL_1244556, EPI_ISL_1244559, EPI_ISL_1244560, EPI_ISL_1244561, EPI_ISL_1244564, EPI_ISL_1244567, EPI_ISL_1244568, EPI_ISL_1244572, EPI_ISL_1244573, EPI_ISL_1244575, EPI_ISL_1244579, EPI_ISL_1244580, EPI_ISL_1244581, EPI_ISL_1244583, EPI_ISL_1244584, EPI_ISL_1244585, EPI_ISL_1244586, EPI_ISL_1244587, EPI_ISL_1244588, EPI_ISL_1244589, EPI_ISL_1244591, EPI_ISL_1244592, EPI_ISL_1244595, EPI_ISL_1244597, EPI_ISL_1244598, EPI_ISL_1244600, EPI_ISL_1244601, EPI_ISL_1244602, EPI_ISL_1244604, EPI_ISL_1244606, EPI_ISL_1244607, EPI_ISL_1244610, EPI_ISL_1244611, EPI_ISL_1245360, EPI_ISL_1245363, EPI_ISL_1245367, EPI_ISL_1245370, EPI_ISL_1245371, EPI_ISL_1245375, EPI_ISL_1245379, EPI_ISL_1245380, EPI_ISL_1245393, EPI_ISL_1245395, EPI_ISL_1245401, EPI_ISL_1245406, EPI_ISL_1245410, EPI_ISL_1245411, EPI_ISL_1245412, EPI_ISL_1245418, EPI_ISL_1245419, EPI_ISL_1245420, EPI_ISL_1245421, EPI_ISL_1245424, EPI_ISL_1245427, EPI_ISL_1245430, EPI_ISL_1245432, EPI_ISL_1245433, EPI_ISL_1245436, EPI_ISL_1245438, EPI_ISL_1245439, EPI_ISL_1245445, EPI_ISL_1245447, EPI_ISL_1245450, EPI_ISL_1245451, EPI_ISL_1245457, EPI_ISL_1245462, EPI_ISL_1245466, EPI_ISL_1245473, EPI_ISL_1245474, EPI_ISL_1245476, EPI_ISL_1245479, EPI_ISL_1245480, EPI_ISL_1245482, EPI_ISL_1245483, EPI_ISL_1245484, EPI_ISL_1245488, EPI_ISL_1245491, EPI_ISL_1245492, EPI_ISL_1245493, EPI_ISL_1245494, EPI_ISL_1245501, EPI_ISL_1245504, EPI_ISL_1245510, EPI_ISL_1245515, EPI_ISL_1245517, EPI_ISL_1245518, EPI_ISL_1245524, EPI_ISL_1245526, EPI_ISL_1245527, EPI_ISL_1245529, EPI_ISL_1245533, EPI_ISL_1245535, EPI_ISL_1245538, EPI_ISL_1245541, EPI_ISL_1245544, EPI_ISL_1245548, EPI_ISL_1245549, EPI_ISL_1245553, EPI_ISL_1245554, EPI_ISL_1245555, EPI_ISL_1245558, EPI_ISL_1245566, EPI_ISL_1245567, EPI_ISL_1245568, EPI_ISL_1245569, EPI_ISL_1245570, EPI_ISL_1245571, EPI_ISL_1245576, EPI_ISL_1245577, EPI_ISL_1245584, EPI_ISL_1245586, EPI_ISL_1245588, EPI_ISL_1245589, EPI_ISL_1245591, EPI_ISL_1245594, EPI_ISL_1245595, EPI_ISL_1245598, EPI_ISL_1245599, EPI_ISL_1245603, EPI_ISL_1245605, EPI_ISL_1245606, EPI_ISL_1245610, EPI_ISL_1245613, EPI_ISL_1245614, EPI_ISL_1245615, EPI_ISL_1245620, EPI_ISL_1245621, EPI_ISL_1245623, EPI_ISL_1245624, EPI_ISL_1245625, EPI_ISL_1245627, EPI_ISL_1245628, EPI_ISL_1245630, EPI_ISL_1245631, EPI_ISL_1245632, EPI_ISL_1245634, EPI_ISL_1245635, EPI_ISL_1245636, EPI_ISL_1245637, EPI_ISL_1245639, EPI_ISL_1245641, EPI_ISL_1245646, EPI_ISL_1245648, EPI_ISL_1245653, EPI_ISL_1245655, EPI_ISL_1245662, EPI_ISL_1245665, EPI_ISL_1245666, EPI_ISL_1245677, EPI_ISL_1245684, EPI_ISL_1245685, EPI_ISL_1245686, EPI_ISL_1245687, EPI_ISL_1245688, EPI_ISL_1245691, EPI_ISL_1245693, EPI_ISL_1245694, EPI_ISL_1245697, EPI_ISL_1245698, EPI_ISL_1245700, EPI_ISL_1245703, EPI_ISL_1245705, EPI_ISL_1245707, EPI_ISL_1245708, EPI_ISL_1245709, EPI_ISL_1245710, EPI_ISL_1245711, EPI_ISL_1245712, EPI_ISL_1245715, EPI_ISL_1245716, EPI_ISL_1245718, EPI_ISL_1245720, EPI_ISL_1245722, EPI_ISL_1245723, EPI_ISL_1245725, EPI_ISL_1245726, EPI_ISL_1245728, EPI_ISL_1245729, EPI_ISL_1245732, EPI_ISL_1245734, EPI_ISL_1245735, EPI_ISL_1245736, EPI_ISL_1245737, EPI_ISL_1245738, EPI_ISL_1245740, EPI_ISL_1245741, EPI_ISL_1245743, EPI_ISL_1245744, EPI_ISL_1245745, EPI_ISL_1245746, EPI_ISL_1245751, EPI_ISL_1245754, EPI_ISL_1245758, EPI_ISL_1245759, EPI_ISL_1245761, EPI_ISL_1245763, EPI_ISL_1245764, EPI_ISL_1245765, EPI_ISL_1245766, EPI_ISL_1245768, EPI_ISL_1245769, EPI_ISL_1245770, EPI_ISL_1245771, EPI_ISL_1245772, EPI_ISL_1245774, EPI_ISL_1245775, EPI_ISL_1245778, EPI_ISL_1245779, EPI_ISL_1245780, EPI_ISL_1245781, EPI_ISL_1245785, EPI_ISL_1245788, EPI_ISL_1245793, EPI_ISL_1245794, EPI_ISL_1245795, EPI_ISL_1245796, EPI_ISL_1245797, EPI_ISL_1245798, EPI_ISL_1245799, EPI_ISL_1245800, EPI_ISL_1245802, EPI_ISL_1245803, EPI_ISL_1245804, EPI_ISL_1245806, EPI_ISL_1245807, EPI_ISL_1245809, EPI_ISL_1245810, EPI_ISL_1245811, EPI_ISL_1245812, EPI_ISL_1245813, EPI_ISL_1245816, EPI_ISL_1245817, EPI_ISL_1245818, EPI_ISL_1245819, EPI_ISL_1245820, EPI_ISL_1245821, EPI_ISL_1245824, EPI_ISL_1245825, EPI_ISL_1245826, EPI_ISL_1245827, EPI_ISL_1245828, EPI_ISL_1245829, EPI_ISL_1245831, EPI_ISL_1245832, EPI_ISL_1245833, EPI_ISL_1245835, EPI_ISL_1245836, EPI_ISL_1245837, EPI_ISL_1245838, EPI_ISL_1245839, EPI_ISL_1245840, EPI_ISL_1245841, EPI_ISL_1245842, EPI_ISL_1245843, EPI_ISL_1245845, EPI_ISL_1245846, EPI_ISL_1245848, EPI_ISL_1245849, EPI_ISL_1245851, EPI_ISL_1245853, EPI_ISL_1245854, EPI_ISL_1245856, EPI_ISL_1245857, EPI_ISL_1245858, EPI_ISL_1245859, EPI_ISL_1245860, EPI_ISL_1245861, EPI_ISL_1245865, EPI_ISL_1245866, EPI_ISL_1245867, EPI_ISL_1245868, EPI_ISL_1245869, EPI_ISL_1245870, EPI_ISL_1245871, EPI_ISL_1245872, EPI_ISL_1245873, EPI_ISL_1245874, EPI_ISL_1245875, EPI_ISL_1245876, EPI_ISL_1245878, EPI_ISL_1245880, EPI_ISL_1245882, EPI_ISL_1245884, EPI_ISL_1245885, EPI_ISL_1245887, EPI_ISL_1245889, EPI_ISL_1245891, EPI_ISL_1245892, EPI_ISL_1245893, EPI_ISL_1245894, EPI_ISL_1245897, EPI_ISL_1245898, EPI_ISL_1245899, EPI_ISL_1245900, EPI_ISL_1245901, EPI_ISL_1245902, EPI_ISL_1245904, EPI_ISL_1245905, EPI_ISL_1245906, EPI_ISL_1245909, EPI_ISL_1245912, EPI_ISL_1245913, EPI_ISL_1245915, EPI_ISL_1245917, EPI_ISL_1245918, EPI_ISL_1245919, EPI_ISL_1245920, EPI_ISL_1245922, EPI_ISL_1245923, EPI_ISL_1245924, EPI_ISL_1245925, EPI_ISL_1245926, EPI_ISL_1245928, EPI_ISL_1245929, EPI_ISL_1245930, EPI_ISL_1245931, EPI_ISL_1245934, EPI_ISL_1245936, EPI_ISL_1245937, EPI_ISL_1245938, EPI_ISL_1245939, EPI_ISL_1245940, EPI_ISL_1245942, EPI_ISL_1245943, EPI_ISL_1245944, EPI_ISL_1245945, EPI_ISL_1245946, EPI_ISL_1245947, EPI_ISL_1245948, EPI_ISL_1245949, EPI_ISL_1245951, EPI_ISL_1245952, EPI_ISL_1245953, EPI_ISL_1245955, EPI_ISL_1245959, EPI_ISL_1245961, EPI_ISL_1245965, EPI_ISL_1245966, EPI_ISL_1245970, EPI_ISL_1245971, EPI_ISL_1245972, EPI_ISL_1245973, EPI_ISL_1245975, EPI_ISL_1245976, EPI_ISL_1245979, EPI_ISL_1245980, EPI_ISL_1245981, EPI_ISL_1245982, EPI_ISL_1245983, EPI_ISL_1245984, EPI_ISL_1245988, EPI_ISL_1245989, EPI_ISL_1245991, EPI_ISL_1245993, EPI_ISL_1245996, EPI_ISL_1246000, EPI_ISL_1246001, EPI_ISL_1246002, EPI_ISL_1246003, EPI_ISL_1246004, EPI_ISL_1246006, EPI_ISL_1246007, EPI_ISL_1246009, EPI_ISL_1246010, EPI_ISL_1246011, EPI_ISL_1246013, EPI_ISL_1246015, EPI_ISL_1246016, EPI_ISL_1246017, EPI_ISL_1246869, EPI_ISL_1255425, EPI_ISL_1255429, EPI_ISL_1255430, EPI_ISL_1255435, EPI_ISL_1255436, EPI_ISL_1255442, EPI_ISL_1255443, EPI_ISL_1255445, EPI_ISL_1255447, EPI_ISL_1255451, EPI_ISL_1255453, EPI_ISL_1255454, EPI_ISL_1255458, EPI_ISL_1255461, EPI_ISL_1255464, EPI_ISL_1255466, EPI_ISL_1255469, EPI_ISL_1255470, EPI_ISL_1255472, EPI_ISL_1255473, EPI_ISL_1255481, EPI_ISL_1255486, EPI_ISL_1255488, EPI_ISL_1255490, EPI_ISL_1255493, EPI_ISL_1255494, EPI_ISL_1255496, EPI_ISL_1255502, EPI_ISL_1255503, EPI_ISL_1255509, EPI_ISL_1255514, EPI_ISL_1255515, EPI_ISL_1255523, EPI_ISL_1255528, EPI_ISL_1255535, EPI_ISL_1255537, EPI_ISL_1255538, EPI_ISL_1255540, EPI_ISL_1255545, EPI_ISL_1255546, EPI_ISL_1255547, EPI_ISL_1255550, EPI_ISL_1255558, EPI_ISL_1255561, EPI_ISL_1255563, EPI_ISL_1255566, EPI_ISL_1255567, EPI_ISL_1255570, EPI_ISL_1255572, EPI_ISL_1255573, EPI_ISL_1255574, EPI_ISL_1255576, EPI_ISL_1255579, EPI_ISL_1255580, EPI_ISL_1255584, EPI_ISL_1255585, EPI_ISL_1255586, EPI_ISL_1255587, EPI_ISL_1255594, EPI_ISL_1255596, EPI_ISL_1255598, EPI_ISL_1255599, EPI_ISL_1255600, EPI_ISL_1255602, EPI_ISL_1255603, EPI_ISL_1255605, EPI_ISL_1255612, EPI_ISL_1255614, EPI_ISL_1255622, EPI_ISL_1255624, EPI_ISL_1255625, EPI_ISL_1255628, EPI_ISL_1255629, EPI_ISL_1255631, EPI_ISL_1255632, EPI_ISL_1255634, EPI_ISL_1255635, EPI_ISL_1255636, EPI_ISL_1255643, EPI_ISL_1255645, EPI_ISL_1255646, EPI_ISL_1255651, EPI_ISL_1255655, EPI_ISL_1255656, EPI_ISL_1255659, EPI_ISL_1255661, EPI_ISL_1255662, EPI_ISL_1255663, EPI_ISL_1255667, EPI_ISL_1255671, EPI_ISL_1255674, EPI_ISL_1255687, EPI_ISL_1255689, EPI_ISL_1255691, EPI_ISL_1255709, EPI_ISL_1255712, EPI_ISL_1255732, EPI_ISL_1255765, EPI_ISL_1255766, EPI_ISL_1255771, EPI_ISL_1255790, EPI_ISL_1255801, EPI_ISL_1255820, EPI_ISL_1255834, EPI_ISL_1255855, EPI_ISL_1255856, EPI_ISL_1255866, EPI_ISL_1255867, EPI_ISL_1255872, EPI_ISL_1255877, EPI_ISL_1255881, EPI_ISL_1255883, EPI_ISL_1255885, EPI_ISL_1255890, EPI_ISL_1255895, EPI_ISL_1255897, EPI_ISL_1255928, EPI_ISL_1255952, EPI_ISL_1255967, EPI_ISL_1255976, EPI_ISL_1256003, EPI_ISL_1256006, EPI_ISL_1256040, EPI_ISL_1256044, EPI_ISL_1256049, EPI_ISL_1256050, EPI_ISL_1256052, EPI_ISL_1256054, EPI_ISL_1256056, EPI_ISL_1256058, EPI_ISL_1256060, EPI_ISL_1256061, EPI_ISL_1256063, EPI_ISL_1256064, EPI_ISL_1256069, EPI_ISL_1256070, EPI_ISL_1256071, EPI_ISL_1256072, EPI_ISL_1256075, EPI_ISL_1256076, EPI_ISL_1256077, EPI_ISL_1256079, EPI_ISL_1256080, EPI_ISL_1256081, EPI_ISL_1256084, EPI_ISL_1256085, EPI_ISL_1256086, EPI_ISL_1256088, EPI_ISL_1256090, EPI_ISL_1256091, EPI_ISL_1256092, EPI_ISL_1256093, EPI_ISL_1256095, EPI_ISL_1256096, EPI_ISL_1256098, EPI_ISL_1256101, EPI_ISL_1256102, EPI_ISL_1256105, EPI_ISL_1256111, EPI_ISL_1256112, EPI_ISL_1256113, EPI_ISL_1256115, EPI_ISL_1256117, EPI_ISL_1256119, EPI_ISL_1256120, EPI_ISL_1256121, EPI_ISL_1256122, EPI_ISL_1256123, EPI_ISL_1256126, EPI_ISL_1256131, EPI_ISL_1256132, EPI_ISL_1256133, EPI_ISL_1256135, EPI_ISL_1256138, EPI_ISL_1256139, EPI_ISL_1256140, EPI_ISL_1256141, EPI_ISL_1256142, EPI_ISL_1256146, EPI_ISL_1256150, EPI_ISL_1256151, EPI_ISL_1256153, EPI_ISL_1256154, EPI_ISL_1256157, EPI_ISL_1256158, EPI_ISL_1256161, EPI_ISL_1256162, EPI_ISL_1256164, EPI_ISL_1256165, EPI_ISL_1256167, EPI_ISL_1256169, EPI_ISL_1256170, EPI_ISL_1256173, EPI_ISL_1256174, EPI_ISL_1256177, EPI_ISL_1256178, EPI_ISL_1256180, EPI_ISL_1256181, EPI_ISL_1256182, EPI_ISL_1256183, EPI_ISL_1256184, EPI_ISL_1256185, EPI_ISL_1256186, EPI_ISL_1256187, EPI_ISL_1256189, EPI_ISL_1256190, EPI_ISL_1256191, EPI_ISL_1256192, EPI_ISL_1256194, EPI_ISL_1256196, EPI_ISL_1256199, EPI_ISL_1256202, EPI_ISL_1256203, EPI_ISL_1256204, EPI_ISL_1256205, EPI_ISL_1256207, EPI_ISL_1256209, EPI_ISL_1256210, EPI_ISL_1256212, EPI_ISL_1256214, EPI_ISL_1256215, EPI_ISL_1256216, EPI_ISL_1256218, EPI_ISL_1256223, EPI_ISL_1256224, EPI_ISL_1256226, EPI_ISL_1256228, EPI_ISL_1256230, EPI_ISL_1256232, EPI_ISL_1256234, EPI_ISL_1256235, EPI_ISL_1256236, EPI_ISL_1256237, EPI_ISL_1256238, EPI_ISL_1256240, EPI_ISL_1256241, EPI_ISL_1256243, EPI_ISL_1256244, EPI_ISL_1256245, EPI_ISL_1256248, EPI_ISL_1256249, EPI_ISL_1256250, EPI_ISL_1256251, EPI_ISL_1256253, EPI_ISL_1256256, EPI_ISL_1256257, EPI_ISL_1256258, EPI_ISL_1256263, EPI_ISL_1256266, EPI_ISL_1256267, EPI_ISL_1256268, EPI_ISL_1256269, EPI_ISL_1256270, EPI_ISL_1256271, EPI_ISL_1256272, EPI_ISL_1256276, EPI_ISL_1256279, EPI_ISL_1256280, EPI_ISL_1256286, EPI_ISL_1256287, EPI_ISL_1256288, EPI_ISL_1256289, EPI_ISL_1256290, EPI_ISL_1256292, EPI_ISL_1256293, EPI_ISL_1256294, EPI_ISL_1256295, EPI_ISL_1256298, EPI_ISL_1256300, EPI_ISL_1256301, EPI_ISL_1256302, EPI_ISL_1256303, EPI_ISL_1256304, EPI_ISL_1256305, EPI_ISL_1256309, EPI_ISL_1256311, EPI_ISL_1256313, EPI_ISL_1256314, EPI_ISL_1256315, EPI_ISL_1256316, EPI_ISL_1256317, EPI_ISL_1256319, EPI_ISL_1256320, EPI_ISL_1256321, EPI_ISL_1256322, EPI_ISL_1256323, EPI_ISL_1256325, EPI_ISL_1256326, EPI_ISL_1256327, EPI_ISL_1256330, EPI_ISL_1256333, EPI_ISL_1256335, EPI_ISL_1256337, EPI_ISL_1256340, EPI_ISL_1256341, EPI_ISL_1256342, EPI_ISL_1256344, EPI_ISL_1256346, EPI_ISL_1256348, EPI_ISL_1256349, EPI_ISL_1256352, EPI_ISL_1256354, EPI_ISL_1256358, EPI_ISL_1256359, EPI_ISL_1256362, EPI_ISL_1256363, | | | |

EPI_ISL_1256683, EPI_ISL_1256710, EPI_ISL_1256720, EPI_ISL_1256730, EPI_ISL_1256740, EPI_ISL_1256743, EPI_ISL_1256773, EPI_ISL_1256780, EPI_ISL_1256794, EPI_ISL_1256799, EPI_ISL_1256803, EPI_ISL_1256814, EPI_ISL_1256831, EPI_ISL_1256836, EPI_ISL_1256845, EPI_ISL_1256848, EPI_ISL_1256860, EPI_ISL_1256890, EPI_ISL_1256901, EPI_ISL_1256918, EPI_ISL_1256932, EPI_ISL_1264099, EPI_ISL_1264143, EPI_ISL_1264166, EPI_ISL_1264233, EPI_ISL_1264415, EPI_ISL_1264439, EPI_ISL_1264616, EPI_ISL_1264629, EPI_ISL_1264638, EPI_ISL_1274059, EPI_ISL_1274061, EPI_ISL_1276351, EPI_ISL_1276732, EPI_ISL_1276761, EPI_ISL_1276796, EPI_ISL_1276798, EPI_ISL_1276805, EPI_ISL_1277212, EPI_ISL_1277235, EPI_ISL_1277242, EPI_ISL_1277253, EPI_ISL_1277270, EPI_ISL_1277303, EPI_ISL_1277311, EPI_ISL_1277316, EPI_ISL_1277319, EPI_ISL_1277320, EPI_ISL_1277325, EPI_ISL_1277328, EPI_ISL_1277329, EPI_ISL_1277330, EPI_ISL_1277331, EPI_ISL_1277333, EPI_ISL_1277334, EPI_ISL_1277336, EPI_ISL_1277337, EPI_ISL_1277339, EPI_ISL_1277344, EPI_ISL_1277347, EPI_ISL_1277613, EPI_ISL_1277966, EPI_ISL_1277968, EPI_ISL_1277969, EPI_ISL_1277970, EPI_ISL_1277971, EPI_ISL_1277972, EPI_ISL_1277973, EPI_ISL_1277974, EPI_ISL_1277975, EPI_ISL_1277976, EPI_ISL_1277977, EPI_ISL_1277978, EPI_ISL_1277979, EPI_ISL_1277981, EPI_ISL_1277982, EPI_ISL_1277983, EPI_ISL_1277984, EPI_ISL_1277985, EPI_ISL_1277986, EPI_ISL_1277987, EPI_ISL_1277989, EPI_ISL_1277990, EPI_ISL_1277992, EPI_ISL_1277994, EPI_ISL_1277995, EPI_ISL_1277996, EPI_ISL_1277997, EPI_ISL_1277998, EPI_ISL_1277999, EPI_ISL_1278000, EPI_ISL_1278001, EPI_ISL_1278002, EPI_ISL_1316427, EPI_ISL_1316431, EPI_ISL_1316455, EPI_ISL_1316466, EPI_ISL_1316495, EPI_ISL_1316511, EPI_ISL_1316535, EPI_ISL_1316574, EPI_ISL_1316577, EPI_ISL_1316594, EPI_ISL_1316598, EPI_ISL_1316639, EPI_ISL_1316659, EPI_ISL_1316678, EPI_ISL_1316683, EPI_ISL_1316687, EPI_ISL_1316689, EPI_ISL_1316739, EPI_ISL_1316757, EPI_ISL_1316764, EPI_ISL_1316777, EPI_ISL_1317211, EPI_ISL_1328899, EPI_ISL_1328900, EPI_ISL_1328901, EPI_ISL_1328905, EPI_ISL_1328906, EPI_ISL_1328908, EPI_ISL_1328912, EPI_ISL_1328915, EPI_ISL_1328920, EPI_ISL_1328923, EPI_ISL_1328928, EPI_ISL_1328934, EPI_ISL_1328937, EPI_ISL_1328952, EPI_ISL_1328958, EPI_ISL_1328960, EPI_ISL_1328962, EPI_ISL_1328963, EPI_ISL_1328967, EPI_ISL_1328971, EPI_ISL_1328972, EPI_ISL_1328982, EPI_ISL_1328983, EPI_ISL_1328984, EPI_ISL_1328986, EPI_ISL_1328991, EPI_ISL_1328993, EPI_ISL_1328994, EPI_ISL_1328995, EPI_ISL_1328999, EPI_ISL_1329001, EPI_ISL_1329004, EPI_ISL_1329005, EPI_ISL_1329007, EPI_ISL_1329018, EPI_ISL_1329030, EPI_ISL_1329031, EPI_ISL_1329032, EPI_ISL_1329033, EPI_ISL_1329035, EPI_ISL_1329036, EPI_ISL_1329037, EPI_ISL_1329038, EPI_ISL_1329045, EPI_ISL_1329048, EPI_ISL_1329052, EPI_ISL_1329055, EPI_ISL_1329058, EPI_ISL_1329060, EPI_ISL_1329062, EPI_ISL_1329064, EPI_ISL_1329066, EPI_ISL_1329068, EPI_ISL_1329069, EPI_ISL_1329070, EPI_ISL_1329072, EPI_ISL_1329074, EPI_ISL_1329086, EPI_ISL_1329093, EPI_ISL_1329099, EPI_ISL_1329101, EPI_ISL_1329102, EPI_ISL_1329104, EPI_ISL_1329108, EPI_ISL_1329109, EPI_ISL_1329111, EPI_ISL_1329112, EPI_ISL_1329113, EPI_ISL_1329115, EPI_ISL_1329116, EPI_ISL_1329121, EPI_ISL_1329124, EPI_ISL_1329125, EPI_ISL_1329130, EPI_ISL_1329131, EPI_ISL_1329136, EPI_ISL_1329138, EPI_ISL_1329139, EPI_ISL_1329140, EPI_ISL_1329142, EPI_ISL_1329143, EPI_ISL_1329144, EPI_ISL_1329145, EPI_ISL_1329147, EPI_ISL_1329148, EPI_ISL_1329149, EPI_ISL_1329156, EPI_ISL_1329159, EPI_ISL_1329161, EPI_ISL_1329163, EPI_ISL_1329172, EPI_ISL_1329175, EPI_ISL_1329181, EPI_ISL_1329183, EPI_ISL_1329185, EPI_ISL_1329187, EPI_ISL_1329189, EPI_ISL_1329194, EPI_ISL_1329199, EPI_ISL_1329200, EPI_ISL_1329980, EPI_ISL_1329981, EPI_ISL_1329982, EPI_ISL_1329985, EPI_ISL_1329986, EPI_ISL_1329987, EPI_ISL_1329988, EPI_ISL_1329989, EPI_ISL_1329990, EPI_ISL_1329991, EPI_ISL_1329993, EPI_ISL_1329996, EPI_ISL_1329998, EPI_ISL_1330001, EPI_ISL_1330002, EPI_ISL_1330003, EPI_ISL_1330004, EPI_ISL_1330005, EPI_ISL_1330006, EPI_ISL_1330007, EPI_ISL_1330009, EPI_ISL_1330010, EPI_ISL_1330014, EPI_ISL_1330015, EPI_ISL_1330016, EPI_ISL_1330017, EPI_ISL_1330018, EPI_ISL_1330019, EPI_ISL_1330020, EPI_ISL_1330022, EPI_ISL_1330024, EPI_ISL_1330025, EPI_ISL_1330026, EPI_ISL_1330028, EPI_ISL_1330031, EPI_ISL_1330032, EPI_ISL_1330033, EPI_ISL_1330036, EPI_ISL_1330037, EPI_ISL_1330038, EPI_ISL_1330039, EPI_ISL_1330040, EPI_ISL_1330042, EPI_ISL_1330043, EPI_ISL_1330044, EPI_ISL_1330045, EPI_ISL_1330048, EPI_ISL_1330049, EPI_ISL_1330052, EPI_ISL_1330053, EPI_ISL_1330055, EPI_ISL_1330056, EPI_ISL_1330058, EPI_ISL_1330060, EPI_ISL_1330061, EPI_ISL_1330062, EPI_ISL_1330063, EPI_ISL_1330064, EPI_ISL_1330065, EPI_ISL_1330066, EPI_ISL_1330068, EPI_ISL_1330070, EPI_ISL_1330072, EPI_ISL_1330073, EPI_ISL_1330074, EPI_ISL_1330076, EPI_ISL_1330077, EPI_ISL_1330079, EPI_ISL_1330082, EPI_ISL_1330083, EPI_ISL_1330084, EPI_ISL_1330086, EPI_ISL_1330087, EPI_ISL_1330088, EPI_ISL_1330089, EPI_ISL_1330090, EPI_ISL_1330091, EPI_ISL_1330093, EPI_ISL_1330098, EPI_ISL_1330099, EPI_ISL_1330101, EPI_ISL_1330103, EPI_ISL_1332830, EPI_ISL_1333335, EPI_ISL_1333476, EPI_ISL_1333479, EPI_ISL_1333511, EPI_ISL_1333516, EPI_ISL_1333611, EPI_ISL_1346365, EPI_ISL_1346400, EPI_ISL_1346401, EPI_ISL_1346402, EPI_ISL_1449651, EPI_ISL_1449654, EPI_ISL_1449655, EPI_ISL_1449656, EPI_ISL_1449657, EPI_ISL_1449658, EPI_ISL_1449660, EPI_ISL_1449661, EPI_ISL_1449663, EPI_ISL_1449664, EPI_ISL_1449665, EPI_ISL_1449666, EPI_ISL_1449675, EPI_ISL_1449677, EPI_ISL_1449678, EPI_ISL_1449679, EPI_ISL_1449680, EPI_ISL_1449683, EPI_ISL_1449685, EPI_ISL_1449686, EPI_ISL_1449687, EPI_ISL_1449688, EPI_ISL_1449689, EPI_ISL_1449691, EPI_ISL_1449694, EPI_ISL_1449696, EPI_ISL_1449699, EPI_ISL_1449700, EPI_ISL_1449701, EPI_ISL_1449703, EPI_ISL_1449704, EPI_ISL_1449707, EPI_ISL_1449711, EPI_ISL_1449712, EPI_ISL_1449714, EPI_ISL_1449717, EPI_ISL_1449720, EPI_ISL_1449723, EPI_ISL_1449726, EPI_ISL_1449727, EPI_ISL_1449729, EPI_ISL_1449732, EPI_ISL_1449735, EPI_ISL_1449742, EPI_ISL_1449743, EPI_ISL_1449745, EPI_ISL_1449747, EPI_ISL_1449748, EPI_ISL_1449752, EPI_ISL_1449753, EPI_ISL_1449754, EPI_ISL_1449755, EPI_ISL_1449758, EPI_ISL_1449759, EPI_ISL_1449760, EPI_ISL_1449761, EPI_ISL_1449762, EPI_ISL_1449763, EPI_ISL_1449764, EPI_ISL_1449768, EPI_ISL_1449771, EPI_ISL_1449772, EPI_ISL_1449773, EPI_ISL_1449774, EPI_ISL_1449775, EPI_ISL_1449776, EPI_ISL_1449777, EPI_ISL_1449780, EPI_ISL_1449785, EPI_ISL_1449787, EPI_ISL_1449789, EPI_ISL_1449791, EPI_ISL_1449792, EPI_ISL_1449794, EPI_ISL_1449796, EPI_ISL_1449798, EPI_ISL_1449799, EPI_ISL_1449804, EPI_ISL_1449806, EPI_ISL_1449807, EPI_ISL_1449810, EPI_ISL_1449812, EPI_ISL_1449813, EPI_ISL_1449816, EPI_ISL_1449818, EPI_ISL_1449819, EPI_ISL_1449820, EPI_ISL_1449821, EPI_ISL_1449822, EPI_ISL_1449823, EPI_ISL_1449825, EPI_ISL_1449827, EPI_ISL_1449828, EPI_ISL_1449829, EPI_ISL_1449830, EPI_ISL_1449831, EPI_ISL_1449833, EPI_ISL_1449834, EPI_ISL_1449835, EPI_ISL_1449836, EPI_ISL_1449837, EPI_ISL_1449838, EPI_ISL_1449840, EPI_ISL_1449841, EPI_ISL_1449842, EPI_ISL_1449843, EPI_ISL_1449845, EPI_ISL_1449848, EPI_ISL_1449851, EPI_ISL_1449852, EPI_ISL_1449853, EPI_ISL_1449854, EPI_ISL_1449855, EPI_ISL_1449856, EPI_ISL_1449858, EPI_ISL_1449859, EPI_ISL_1449860, EPI_ISL_1449861, EPI_ISL_1449864, EPI_ISL_1449865, EPI_ISL_1449867, EPI_ISL_1449868, EPI_ISL_1449869, EPI_ISL_1449873, EPI_ISL_1449874, EPI_ISL_1449877, EPI_ISL_1449878, EPI_ISL_1449879, EPI_ISL_1449881, EPI_ISL_1449884, EPI_ISL_1449890, EPI_ISL_1449891, EPI_ISL_1449892, EPI_ISL_1449893, EPI_ISL_1449895, EPI_ISL_1449896, EPI_ISL_1449897, EPI_ISL_1449898, EPI_ISL_1449899, EPI_ISL_1449900, EPI_ISL_1449902, EPI_ISL_1449903, EPI_ISL_1449904, EPI_ISL_1449907, EPI_ISL_1449910, EPI_ISL_1449911, EPI_ISL_1449912, EPI_ISL_1449914, EPI_ISL_1449915, EPI_ISL_1449917, EPI_ISL_1449920, EPI_ISL_1449921, EPI_ISL_1449922, EPI_ISL_1449925, EPI_ISL_1449926, EPI_ISL_1449927, EPI_ISL_1449928, EPI_ISL_1449932, EPI_ISL_1449933, EPI_ISL_1449935, EPI_ISL_1449936, EPI_ISL_1449938, EPI_ISL_1449939, EPI_ISL_1449943, EPI_ISL_1449945, EPI_ISL_1449947, EPI_ISL_1449948, EPI_ISL_1449949, EPI_ISL_1449950, EPI_ISL_1449952, EPI_ISL_1449953, EPI_ISL_1449955, EPI_ISL_1449958, EPI_ISL_1449959, EPI_ISL_1449960, EPI_ISL_1449961, EPI_ISL_1449963, EPI_ISL_1449965, EPI_ISL_1449967, EPI_ISL_1449969, EPI_ISL_1449971, EPI_ISL_1449977, EPI_ISL_1449978, EPI_ISL_1454545, EPI_ISL_1507796, EPI_ISL_1507805, EPI_ISL_1507807

| | | | |
|---|---|---|---|
| see above | Lighthouse Lab in Alderley Park | Wellcome Sanger Institute for the COVID-19 Genomics UK (COG-UK) Consortium | Cordelia Langford; David K. Jackson; Dominic Kwiatkowski; Ewan Harrison; Ian Johnston; Jacquelyn Wynn; Jeffrey Barrett; John Sillitoe on behalf of the Wellcome Sanger Institute COVID-19 Surveillance Team; Mairead Hyland; Roberto Amato; Sonia Goncalves; The Lighthouse Lab in Alderley Park and Alex Alderton |

EPI_ISL_1190101, EPI_ISL_1206407, EPI_ISL_1242048, EPI_ISL_1242055, EPI_ISL_1242058, EPI_ISL_1242061, EPI_ISL_1242074, EPI_ISL_1242077, EPI_ISL_1242079, EPI_ISL_1242086, EPI_ISL_1242098, EPI_ISL_1242104, EPI_ISL_1242108, EPI_ISL_1242111, EPI_ISL_1242113, EPI_ISL_1242119, EPI_ISL_1242120, EPI_ISL_1242123, EPI_ISL_1242129, EPI_ISL_1242133, EPI_ISL_1242140, EPI_ISL_1242148, EPI_ISL_1242155, EPI_ISL_1242164, EPI_ISL_1242172, EPI_ISL_1242173, EPI_ISL_1242184, EPI_ISL_1242185, EPI_ISL_1242190, EPI_ISL_1242191, EPI_ISL_1242193, EPI_ISL_1242195, EPI_ISL_1242199, EPI_ISL_1242203, EPI_ISL_1242204, EPI_ISL_1242205, EPI_ISL_1242213, EPI_ISL_1242215, EPI_ISL_1242219, EPI_ISL_1242222, EPI_ISL_1242230, EPI_ISL_1242231, EPI_ISL_1242232, EPI_ISL_1242233, EPI_ISL_1242235, EPI_ISL_1242240, EPI_ISL_1242246, EPI_ISL_1242251, EPI_ISL_1242255, EPI_ISL_1242256, EPI_ISL_1242257, EPI_ISL_1242258, EPI_ISL_1242269, EPI_ISL_1242270, EPI_ISL_1242272, EPI_ISL_1242278, EPI_ISL_1244852, EPI_ISL_1244853, EPI_ISL_1244854, EPI_ISL_1244855, EPI_ISL_1244856, EPI_ISL_1244857, EPI_ISL_1244859, EPI_ISL_1244860, EPI_ISL_1244861, EPI_ISL_1244865, EPI_ISL_1244868, EPI_ISL_1244873, EPI_ISL_1244874, EPI_ISL_1244876, EPI_ISL_1244878, EPI_ISL_1244879, EPI_ISL_1244881, EPI_ISL_1244883, EPI_ISL_1244884, EPI_ISL_1244885, EPI_ISL_1244887, EPI_ISL_1244889, EPI_ISL_1244890, EPI_ISL_1244891, EPI_ISL_1244893, EPI_ISL_1244895, EPI_ISL_1244896, EPI_ISL_1244897, EPI_ISL_1244899, EPI_ISL_1244900, EPI_ISL_1244901, EPI_ISL_1244902, EPI_ISL_1244903, EPI_ISL_1244907, EPI_ISL_1244908, EPI_ISL_1244909, EPI_ISL_1244910, EPI_ISL_1244911, EPI_ISL_1244914, EPI_ISL_1244916, EPI_ISL_1244917, EPI_ISL_1244918, EPI_ISL_1244923, EPI_ISL_1244925, EPI_ISL_1244926, EPI_ISL_1244927, EPI_ISL_1244928, EPI_ISL_1244930, EPI_ISL_1244934, EPI_ISL_1244935, EPI_ISL_1244937, EPI_ISL_1244938, EPI_ISL_1244942, EPI_ISL_1244943, EPI_ISL_1244944, EPI_ISL_1244945, EPI_ISL_1244947, EPI_ISL_1244948, EPI_ISL_1244951, EPI_ISL_1244952, EPI_ISL_1244953, EPI_ISL_1244955, EPI_ISL_1244956, EPI_ISL_1244957, EPI_ISL_1244958, EPI_ISL_1244959, EPI_ISL_1244960, EPI_ISL_1244962, EPI_ISL_1244965, EPI_ISL_1244966, EPI_ISL_1244967, EPI_ISL_1244968, EPI_ISL_1244970, EPI_ISL_1244972, EPI_ISL_1244973, EPI_ISL_1244974, EPI_ISL_1244975, EPI_ISL_1244976, EPI_ISL_1244978, EPI_ISL_1244980, EPI_ISL_1244981, EPI_ISL_1244985, EPI_ISL_1244986, EPI_ISL_1244987, EPI_ISL_1244989, EPI_ISL_1244990, EPI_ISL_1244996, EPI_ISL_1244998, EPI_ISL_1244999, EPI_ISL_1245000, EPI_ISL_1245001, EPI_ISL_1245005, EPI_ISL_1246271, EPI_ISL_1256366, EPI_ISL_1256367, EPI_ISL_1256372, EPI_ISL_1256373, EPI_ISL_1256374, EPI_ISL_1256377, EPI_ISL_1256379, EPI_ISL_1256382, EPI_ISL_1256383, EPI_ISL_1256385, EPI_ISL_1256386, EPI_ISL_1256388, EPI_ISL_1256398, EPI_ISL_1256406, EPI_ISL_1256409, EPI_ISL_1256411, EPI_ISL_1256412, EPI_ISL_1256420, EPI_ISL_1256422, EPI_ISL_1256429, EPI_ISL_1256430, EPI_ISL_1256432, EPI_ISL_1256436, EPI_ISL_1256438, EPI_ISL_1256444, EPI_ISL_1256446, EPI_ISL_1256449, EPI_ISL_1256453, EPI_ISL_1256458, EPI_ISL_1256462, EPI_ISL_1256464, EPI_ISL_1256944, EPI_ISL_1256945, EPI_ISL_1256949, EPI_ISL_1256951, EPI_ISL_1256952, EPI_ISL_1256953, EPI_ISL_1256955, EPI_ISL_1256956, EPI_ISL_1256957, EPI_ISL_1256959, EPI_ISL_1256961, EPI_ISL_1256962, EPI_ISL_1256963, EPI_ISL_1256964, EPI_ISL_1256965, EPI_ISL_1256966, EPI_ISL_1256969, EPI_ISL_1256970, EPI_ISL_1256972, EPI_ISL_1256973, EPI_ISL_1256974, EPI_ISL_1256975, EPI_ISL_1256976, EPI_ISL_1256978, EPI_ISL_1256981, EPI_ISL_1256985, EPI_ISL_1256987, EPI_ISL_1256988, EPI_ISL_1256989, EPI_ISL_1256990, EPI_ISL_1256991, EPI_ISL_1256992, EPI_ISL_1256993, EPI_ISL_1256994, EPI_ISL_1256995, EPI_ISL_1256996, EPI_ISL_1256998, EPI_ISL_1256999, EPI_ISL_1257000, EPI_ISL_1257001, EPI_ISL_1257002, EPI_ISL_1257004, EPI_ISL_1257005, EPI_ISL_1257006, EPI_ISL_1257007, EPI_ISL_1257009, EPI_ISL_1257011, EPI_ISL_1257012, EPI_ISL_1257013, EPI_ISL_1257014, EPI_ISL_1257015, EPI_ISL_1257016, EPI_ISL_1257017, EPI_ISL_1257018, EPI_ISL_1257019, EPI_ISL_1257021, EPI_ISL_1257022, EPI_ISL_1257023, EPI_ISL_1257024, EPI_ISL_1257026, EPI_ISL_1257028, EPI_ISL_1257029, EPI_ISL_1257030, EPI_ISL_1257031, EPI_ISL_1257033, EPI_ISL_1257037, EPI_ISL_1257038, EPI_ISL_1257040, EPI_ISL_1257041, EPI_ISL_1257042, EPI_ISL_1257043, EPI_ISL_1257044, EPI_ISL_1257045, EPI_ISL_1257046, EPI_ISL_1257047, EPI_ISL_1257049, EPI_ISL_1257050, EPI_ISL_1257053, EPI_ISL_1257055, EPI_ISL_1257056, EPI_ISL_1257057, EPI_ISL_1257058, EPI_ISL_1257059, EPI_ISL_1257060, EPI_ISL_1257061, EPI_ISL_1257062, EPI_ISL_1257643, EPI_ISL_1257646, EPI_ISL_1263856, EPI_ISL_1263862, EPI_ISL_1263872, EPI_ISL_1263888, EPI_ISL_1263889, EPI_ISL_1263896, EPI_ISL_1263897, EPI_ISL_1263898, EPI_ISL_1263899, EPI_ISL_1263902, EPI_ISL_1263905, EPI_ISL_1263911, EPI_ISL_1263915, EPI_ISL_1263917, EPI_ISL_1263918, EPI_ISL_1263920, EPI_ISL_1263921, EPI_ISL_1263928, EPI_ISL_1263933, EPI_ISL_1263936, EPI_ISL_1263938, EPI_ISL_1263943, EPI_ISL_1263946, EPI_ISL_1263950, EPI_ISL_1263951, EPI_ISL_1263952, EPI_ISL_1263956, EPI_ISL_1263961, EPI_ISL_1263965, EPI_ISL_1263966, EPI_ISL_1263969, EPI_ISL_1263973, EPI_ISL_1263975, EPI_ISL_1263977, EPI_ISL_1263978, EPI_ISL_1263980, EPI_ISL_1263986, EPI_ISL_1263990, EPI_ISL_1263993, EPI_ISL_1263996, EPI_ISL_1263997, EPI_ISL_1264001, EPI_ISL_1264012, EPI_ISL_1264016, EPI_ISL_1264025, EPI_ISL_1264027, EPI_ISL_1264028, EPI_ISL_1264032, EPI_ISL_1264033, EPI_ISL_1264035, EPI_ISL_1264037, EPI_ISL_1264045, EPI_ISL_1264046, EPI_ISL_1264050, EPI_ISL_1264051, EPI_ISL_1264054, EPI_ISL_1264056, EPI_ISL_1264060, EPI_ISL_1264064, EPI_ISL_1275565, EPI_ISL_1275566, EPI_ISL_1275568, EPI_ISL_1275573, EPI_ISL_1275574, EPI_ISL_1275575, EPI_ISL_1275579, EPI_ISL_1275582, EPI_ISL_1275586, EPI_ISL_1275588, EPI_ISL_1275591, EPI_ISL_1275592, EPI_ISL_1275600, EPI_ISL_1275604, EPI_ISL_1275607, EPI_ISL_1275609, EPI_ISL_1275614, EPI_ISL_1275616, EPI_ISL_1275619, EPI_ISL_1275620, EPI_ISL_1275621, EPI_ISL_1275625, EPI_ISL_1275628, EPI_ISL_1275630, EPI_ISL_1275632, EPI_ISL_1275634, EPI_ISL_1275635, EPI_ISL_1275638, EPI_ISL_1275640, EPI_ISL_1275642, EPI_ISL_1275643, EPI_ISL_1275650, EPI_ISL_1275651, EPI_ISL_1275653, EPI_ISL_1275655, EPI_ISL_1275657, EPI_ISL_1275659, EPI_ISL_1275662, EPI_ISL_1275663, EPI_ISL_1275667, EPI_ISL_1275669, EPI_ISL_1275677, EPI_ISL_1275678, EPI_ISL_1275679, EPI_ISL_1275680, EPI_ISL_1275681, EPI_ISL_1275690, EPI_ISL_1275694, EPI_ISL_1275695, EPI_ISL_1275697, EPI_ISL_1275702, EPI_ISL_1275705, EPI_ISL_1275716, EPI_ISL_1275719, EPI_ISL_1275724, EPI_ISL_1275725, EPI_ISL_1275730, EPI_ISL_1275733, EPI_ISL_1275740, EPI_ISL_1275745, EPI_ISL_1275754, EPI_ISL_1275756, EPI_ISL_1275778, EPI_ISL_1275780, EPI_ISL_1275801, EPI_ISL_1275810, EPI_ISL_1275818, EPI_ISL_1275826, EPI_ISL_1275828, EPI_ISL_1275832, EPI_ISL_1275835, EPI_ISL_1275846, EPI_ISL_1275847, EPI_ISL_1275849, EPI_ISL_1275850, EPI_ISL_1275863, EPI_ISL_1275869, EPI_ISL_1275877, EPI_ISL_1275878, EPI_ISL_1275888, EPI_ISL_1275891, EPI_ISL_1275901, EPI_ISL_1275908, EPI_ISL_1275911, EPI_ISL_1275917, EPI_ISL_1275919, EPI_ISL_1275920, EPI_ISL_1275923, EPI_ISL_1275931, EPI_ISL_1275934, EPI_ISL_1275940, EPI_ISL_1275943, EPI_ISL_1275951, EPI_ISL_1275953, EPI_ISL_1275967, EPI_ISL_1275972, EPI_ISL_1275973, EPI_ISL_1275975, EPI_ISL_1275976, EPI_ISL_1275986, EPI_ISL_1275990, EPI_ISL_1276003, EPI_ISL_1276009, EPI_ISL_1276011, EPI_ISL_1276014, EPI_ISL_1276017, EPI_ISL_1276022, EPI_ISL_1276040, EPI_ISL_1276047, EPI_ISL_1276049, EPI_ISL_1276069, EPI_ISL_1276072, EPI_ISL_1276079, EPI_ISL_1276081, EPI_ISL_1276086, EPI_ISL_1276088, EPI_ISL_1276097, EPI_ISL_1276099, EPI_ISL_1276106, EPI_ISL_1276109, EPI_ISL_1276893, EPI_ISL_1276897, EPI_ISL_1276903, EPI_ISL_1276904, EPI_ISL_1276906, EPI_ISL_1276907, EPI_ISL_1276908, EPI_ISL_1276912, EPI_ISL_1276919, EPI_ISL_1276929, EPI_ISL_1276933, EPI_ISL_1276935, EPI_ISL_1276936, EPI_ISL_1276941, EPI_ISL_1276945, EPI_ISL_1276947, EPI_ISL_1276953, EPI_ISL_1276970, EPI_ISL_1276973, EPI_ISL_1276975, EPI_ISL_1276976, EPI_ISL_1276977, EPI_ISL_1276983, EPI_ISL_1276997, EPI_ISL_1277003, EPI_ISL_1277008, EPI_ISL_1277009, EPI_ISL_1277017, EPI_ISL_1277022, EPI_ISL_1277024, EPI_ISL_1277034, EPI_ISL_1277036, EPI_ISL_1277037, EPI_ISL_1277044, EPI_ISL_1277045, EPI_ISL_1277051, EPI_ISL_1277052, EPI_ISL_1277058, EPI_ISL_1277077, EPI_ISL_1277085, EPI_ISL_1277089, EPI_ISL_1277099, EPI_ISL_1277101, EPI_ISL_1277104, EPI_ISL_1277109, EPI_ISL_1277125, EPI_ISL_1277126, EPI_ISL_1277143, EPI_ISL_1277145, EPI_ISL_1277155, EPI_ISL_1277161, EPI_ISL_1277162, EPI_ISL_1277164, EPI_ISL_1277182, EPI_ISL_1277185, EPI_ISL_1277190, EPI_ISL_1277201, EPI_ISL_1277865, EPI_ISL_1277866, EPI_ISL_1277993, EPI_ISL_1294618, EPI_ISL_1294623, EPI_ISL_1315996, EPI_ISL_1316004, EPI_ISL_1316005, EPI_ISL_1316008, EPI_ISL_1316010, EPI_ISL_1316013, EPI_ISL_1316022, EPI_ISL_1316113, EPI_ISL_1316137, EPI_ISL_1316161, EPI_ISL_1316174, EPI_ISL_1316188, EPI_ISL_1316192, EPI_ISL_1316195, EPI_ISL_1316207, EPI_ISL_1316223, EPI_ISL_1316230, EPI_ISL_1326985, EPI_ISL_1327037, EPI_ISL_1327242, EPI_ISL_1327275, EPI_ISL_1327392, EPI_ISL_1327393, EPI_ISL_1327451, EPI_ISL_1327498, EPI_ISL_1327503, EPI_ISL_1327541, EPI_ISL_1327551, EPI_ISL_1329337, EPI_ISL_1329373, EPI_ISL_1329999, EPI_ISL_1330047, EPI_ISL_1330057, EPI_ISL_1332656, EPI_ISL_1332657, EPI_ISL_1332659, EPI_ISL_1332662, EPI_ISL_1332663, EPI_ISL_1332667, EPI_ISL_1332668, EPI_ISL_1332670, EPI_ISL_1332673, EPI_ISL_1332675, EPI_ISL_1332682, EPI_ISL_1332686, EPI_ISL_1332689, EPI_ISL_1332690, EPI_ISL_1332692, EPI_ISL_1332693, EPI_ISL_1332694, EPI_ISL_1332697, EPI_ISL_1332700, EPI_ISL_1332701, EPI_ISL_1332702, EPI_ISL_1332833, EPI_ISL_1332835, EPI_ISL_1332836, EPI_ISL_1332837, EPI_ISL_1332850, EPI_ISL_1332853, EPI_ISL_1332875, EPI_ISL_1332876, EPI_ISL_1332896, EPI_ISL_1332902, EPI_ISL_1332907, EPI_ISL_1332910, EPI_ISL_1332919, EPI_ISL_1332922, EPI_ISL_1332925, EPI_ISL_1332931, EPI_ISL_1332934, EPI_ISL_1333176, EPI_ISL_1333227, EPI_ISL_1333269, EPI_ISL_1333280, EPI_ISL_1333289, EPI_ISL_1333304, EPI_ISL_1333307, EPI_ISL_1333313, EPI_ISL_1333339, EPI_ISL_1333340, EPI_ISL_1333349, EPI_ISL_1333355, EPI_ISL_1333376, EPI_ISL_1333383, EPI_ISL_1333392, EPI_ISL_1333409, EPI_ISL_1333415, EPI_ISL_1333417, EPI_ISL_1333482, EPI_ISL_1333483, EPI_ISL_1333484, EPI_ISL_1333485, EPI_ISL_1333493, EPI_ISL_1333494, EPI_ISL_1333495, EPI_ISL_1333499, EPI_ISL_1333501, EPI_ISL_1333503, EPI_ISL_1333506, EPI_ISL_1333509, EPI_ISL_1333522, EPI_ISL_1333523, EPI_ISL_1333537, EPI_ISL_1333539, EPI_ISL_1333543, EPI_ISL_1333547, EPI_ISL_1333548, EPI_ISL_1333549, EPI_ISL_1333564, EPI_ISL_1333565, EPI_ISL_1333566, EPI_ISL_1333567, EPI_ISL_1333568, EPI_ISL_1333569, EPI_ISL_1333575, EPI_ISL_1333577, EPI_ISL_1333579, EPI_ISL_1333581, EPI_ISL_1333582, EPI_ISL_1333587, EPI_ISL_1333588, EPI_ISL_1333591, EPI_ISL_1333597, EPI_ISL_1333599, EPI_ISL_1333600, EPI_ISL_1333604, EPI_ISL_1333609, EPI_ISL_1333874, EPI_ISL_1346305, EPI_ISL_1409510, EPI_ISL_1409649, EPI_ISL_1416183, EPI_ISL_1416185, EPI_ISL_1451152, EPI_ISL_1451153, EPI_ISL_1451163, EPI_ISL_1451164, EPI_ISL_1451165, EPI_ISL_1451167, EPI_ISL_1451169, EPI_ISL_1451173, EPI_ISL_1451175, EPI_ISL_1451176, EPI_ISL_1451178, EPI_ISL_1451180, EPI_ISL_1451181, EPI_ISL_1451192, EPI_ISL_1451193, EPI_ISL_1451194, EPI_ISL_1451195, EPI_ISL_1451198, EPI_ISL_1451201, EPI_ISL_1451203, EPI_ISL_1451206, EPI_ISL_1451210, EPI_ISL_1451216, EPI_ISL_1451221, EPI_ISL_1451225, EPI_ISL_1451227, EPI_ISL_1451228, EPI_ISL_1451231, EPI_ISL_1451233, EPI_ISL_1451237, EPI_ISL_1451240, EPI_ISL_1451243, EPI_ISL_1451244, EPI_ISL_1451254, EPI_ISL_1451255, EPI_ISL_1451260, EPI_ISL_1451263, EPI_ISL_1451264, EPI_ISL_1451266, EPI_ISL_1451267, EPI_ISL_1451268, EPI_ISL_1451269, EPI_ISL_1451270, EPI_ISL_1451274, EPI_ISL_1451276, EPI_ISL_1451280, EPI_ISL_1451284, EPI_ISL_1451286, EPI_ISL_1451290, EPI_ISL_1451291, EPI_ISL_1451295, EPI_ISL_1451296, EPI_ISL_1451300, EPI_ISL_1451304, EPI_ISL_1451309, EPI_ISL_1451310, EPI_ISL_1451314, EPI_ISL_1451316, EPI_ISL_1451317, EPI_ISL_1451320, EPI_ISL_1451322, EPI_ISL_1451326, EPI_ISL_1451327, EPI_ISL_1451332, EPI_ISL_1451334, EPI_ISL_1451339, EPI_ISL_1451342, EPI_ISL_1451343, EPI_ISL_1451344, EPI_ISL_1451345, EPI_ISL_1451347, EPI_ISL_1451349, EPI_ISL_1451350, EPI_ISL_1451351, EPI_ISL_1451354, EPI_ISL_1451356, EPI_ISL_1451357, EPI_ISL_1451358, EPI_ISL_1451360, EPI_ISL_1451362, EPI_ISL_1451367, EPI_ISL_1451369, EPI_ISL_1451370, EPI_ISL_1451371, EPI_ISL_1451376, EPI_ISL_1451380, EPI_ISL_1451382, EPI_ISL_1451384, EPI_ISL_1451387, EPI_ISL_1451388, EPI_ISL_1451390, EPI_ISL_1451393, EPI_ISL_1451394, EPI_ISL_1451399, EPI_ISL_1451402, EPI_ISL_1451406, EPI_ISL_1451408, EPI_ISL_1451411, EPI_ISL_1451412, EPI_ISL_1451413, EPI_ISL_1451416, EPI_ISL_1451417, EPI_ISL_1451420, EPI_ISL_1451424, EPI_ISL_1451428, EPI_ISL_1451433, EPI_ISL_1451435, EPI_ISL_1451436, EPI_ISL_1451437, EPI_ISL_1451440, EPI_ISL_1451443, EPI_ISL_1451444, EPI_ISL_1451446, EPI_ISL_1451459, EPI_ISL_1451460, EPI_ISL_1451461, EPI_ISL_1451463, EPI_ISL_1451465, EPI_ISL_1451466, EPI_ISL_1467588, EPI_ISL_1467591, EPI_ISL_1467595, EPI_ISL_1467597, EPI_ISL_1467601, EPI_ISL_1467602, EPI_ISL_1467605, EPI_ISL_1467607, EPI_ISL_1467610, EPI_ISL_1467624, EPI_ISL_1467627, EPI_ISL_1467628, EPI_ISL_1467630, EPI_ISL_1467632, EPI_ISL_1467634, EPI_ISL_1467635, EPI_ISL_1467640, EPI_ISL_1467642, EPI_ISL_1467645, EPI_ISL_1467647, EPI_ISL_1467650, EPI_ISL_1467652, EPI_ISL_1467656, EPI_ISL_1467662, EPI_ISL_1467664, EPI_ISL_1467670, EPI_ISL_1467671, EPI_ISL_1467673, EPI_ISL_1467676, EPI_ISL_1467678, EPI_ISL_1467679, EPI_ISL_1467681, EPI_ISL_1467682, EPI_ISL_1467688, EPI_ISL_1467690, EPI_ISL_1467696, EPI_ISL_1467699, EPI_ISL_1467701, EPI_ISL_1467707, EPI_ISL_1467713, EPI_ISL_1467715, EPI_ISL_1467718, EPI_ISL_1467720, EPI_ISL_1467721, EPI_ISL_1467730, EPI_ISL_1467732, EPI_ISL_1467736, EPI_ISL_1467744, EPI_ISL_1467745, EPI_ISL_1467746, EPI_ISL_1467747, EPI_ISL_1467748, EPI_ISL_1467751, EPI_ISL_1467755, EPI_ISL_1467763, EPI_ISL_1467764, EPI_ISL_1467765, EPI_ISL_1467782, EPI_ISL_1467788, EPI_ISL_1467789, EPI_ISL_1467791, EPI_ISL_1467796, EPI_ISL_1467797, EPI_ISL_1467799, EPI_ISL_1473448, EPI_ISL_1473450, EPI_ISL_1473456, EPI_ISL_1473458, EPI_ISL_1473462, EPI_ISL_1473463, EPI_ISL_1473467, EPI_ISL_1473470, EPI_ISL_1473472, EPI_ISL_1473473, EPI_ISL_1473474, EPI_ISL_1473476, EPI_ISL_1473477, EPI_ISL_1473478, EPI_ISL_1473480, EPI_ISL_1473482, EPI_ISL_1473484, EPI_ISL_1473487, EPI_ISL_1473488, EPI_ISL_1473490, EPI_ISL_1473491, EPI_ISL_1473493, EPI_ISL_1473496, EPI_ISL_1473501, EPI_ISL_1473502, EPI_ISL_1473503, EPI_ISL_1473505, EPI_ISL_1473507, EPI_ISL_1473509, EPI_ISL_1473510, EPI_ISL_1473511, EPI_ISL_1473513, EPI_ISL_1473515, EPI_ISL_1473517, EPI_ISL_1473518, EPI_ISL_1473520, EPI_ISL_1473521, EPI_ISL_1473522, EPI_ISL_1473523, EPI_ISL_1473524, EPI_ISL_1473526, EPI_ISL_1473528, EPI_ISL_1473530, EPI_ISL_1473534, EPI_ISL_1473535, EPI_ISL_1473537, EPI_ISL_1473539, EPI_ISL_1473540, EPI_ISL_1473541, EPI_ISL_1473542, EPI_ISL_1473543, EPI_ISL_1473544, EPI_ISL_1473547, EPI_ISL_1473550, EPI_ISL_1473552, EPI_ISL_1473554, EPI_ISL_1473556, EPI_ISL_1473557, EPI_ISL_1473558, EPI_ISL_1473561, EPI_ISL_1473566, EPI_ISL_1473567, EPI_ISL_1473568, EPI_ISL_1473571, EPI_ISL_1473572, EPI_ISL_1473573, EPI_ISL_1473575, EPI_ISL_1473578, EPI_ISL_1473579, EPI_ISL_1473580, EPI_ISL_1473581, EPI_ISL_1473583, EPI_ISL_1473584, EPI_ISL_1473588, EPI_ISL_1473589, EPI_ISL_1473590, EPI_ISL_1473591, EPI_ISL_1474165, EPI_ISL_1487775, EPI_ISL_1489499, EPI_ISL_1489503, EPI_ISL_1489505, EPI_ISL_1489507, EPI_ISL_1489510, EPI_ISL_1489511, EPI_ISL_1489515, EPI_ISL_1489518, EPI_ISL_1489520, EPI_ISL_1507597, EPI_ISL_1831430

| | | | |
|---|---|---|---|
| see above | Lighthouse Lab in Cambridge | Wellcome Sanger Institute for the COVID-19 Genomics UK (COG-UK) Consortium | Cordelia Langford; David K. Jackson; Dominic Kwiatkowski; Ewan Harrison; Ian Johnston; Jeffrey Barrett; John Sillitoe on behalf of the Wellcome Sanger Institute COVID-19 Surveillance Team; Rob Howes; Roberto Amato; Sonia Goncalves; The Lighthouse Lab in Cambridge and Alex Alderton |

EPI_ISL_1186308, EPI_ISL_1186349, EPI_ISL_1186354, EPI_ISL_1186391, EPI_ISL_1186409, EPI_ISL_1186415, EPI_ISL_1186453, EPI_ISL_1186468, EPI_ISL_1186508, EPI_ISL_1186509, EPI_ISL_1186522, EPI_ISL_1186525, EPI_ISL_1186567, EPI_ISL_1186574, EPI_ISL_1186576, EPI_ISL_1186577, EPI_ISL_1186579, EPI_ISL_1186580, EPI_ISL_1186581, EPI_ISL_1186583, EPI_ISL_1186590, EPI_ISL_1186593, EPI_ISL_1186601, EPI_ISL_1186602, EPI_ISL_1186603, EPI_ISL_1186604, EPI_ISL_1186605, EPI_ISL_1186606, EPI_ISL_1186609, EPI_ISL_1186610, EPI_ISL_1186611, EPI_ISL_1186614, EPI_ISL_1186620, EPI_ISL_1186622, EPI_ISL_1186623, EPI_ISL_1186627, EPI_ISL_1186628, EPI_ISL_1186629, EPI_ISL_1186633, EPI_ISL_1186635, EPI_ISL_1186643, EPI_ISL_1186647, EPI_ISL_1186652, EPI_ISL_1186653, EPI_ISL_1186654, EPI_ISL_1186656, EPI_ISL_1186661, EPI_ISL_1186672, EPI_ISL_1186673, EPI_ISL_1186675, EPI_ISL_1186679, EPI_ISL_1186682, EPI_ISL_1186684, EPI_ISL_1186686, EPI_ISL_1186695, EPI_ISL_1186697, EPI_ISL_1186703, EPI_ISL_1186704, EPI_ISL_1186709, EPI_ISL_1186713, EPI_ISL_1186714, EPI_ISL_1186719, EPI_ISL_1186726, EPI_ISL_1186729, EPI_ISL_1186730, EPI_ISL_1186732, EPI_ISL_1186737, EPI_ISL_1186738, EPI_ISL_1186743, EPI_ISL_1186750, EPI_ISL_1186753, EPI_ISL_1186754, EPI_ISL_1186755, EPI_ISL_1186756, EPI_ISL_1186759, EPI_ISL_1186763, EPI_ISL_1186764, EPI_ISL_1186766, EPI_ISL_1186768, EPI_ISL_1186772, EPI_ISL_1186773, EPI_ISL_1186775, EPI_ISL_1186776, EPI_ISL_1186778, EPI_ISL_1186779, EPI_ISL_1186780, EPI_ISL_1186781, EPI_ISL_1186785, EPI_ISL_1186787, EPI_ISL_1186790, EPI_ISL_1186793, EPI_ISL_1186794, EPI_ISL_1186799, EPI_ISL_1186801, EPI_ISL_1186806, EPI_ISL_1186809, EPI_ISL_1186810, EPI_ISL_1186813, EPI_ISL_1186815, EPI_ISL_1186817, EPI_ISL_1186824, EPI_ISL_1186825, EPI_ISL_1186827, EPI_ISL_1186835, EPI_ISL_1186839, EPI_ISL_1186840, EPI_ISL_1190342, EPI_ISL_1207155, EPI_ISL_1207157, EPI_ISL_1207167, EPI_ISL_1207169, EPI_ISL_1207172, EPI_ISL_1207173, EPI_ISL_1207185, EPI_ISL_1207191, EPI_ISL_1207193, EPI_ISL_1207195, EPI_ISL_1207197, EPI_ISL_1207198, EPI_ISL_1207203, EPI_ISL_1207208, EPI_ISL_1207216, EPI_ISL_1207218, EPI_ISL_1207235, EPI_ISL_1207238, EPI_ISL_1207243, EPI_ISL_1207248, EPI_ISL_1207249, EPI_ISL_1207258, EPI_ISL_1207261, EPI_ISL_1207265, EPI_ISL_1207266, EPI_ISL_1207270, EPI_ISL_1207273, EPI_ISL_1207276, EPI_ISL_1207283, EPI_ISL_1207288, EPI_ISL_1207289, EPI_ISL_1207296, EPI_ISL_1207304, EPI_ISL_1207309, EPI_ISL_1207315, EPI_ISL_1207320, EPI_ISL_1207324, EPI_ISL_1207325, EPI_ISL_1207328, EPI_ISL_1207329, EPI_ISL_1207338, EPI_ISL_1207343, EPI_ISL_1207345, EPI_ISL_1207348, EPI_ISL_1207356, EPI_ISL_1207366, EPI_ISL_1207375, EPI_ISL_1207376, EPI_ISL_1207381, EPI_ISL_1207384, EPI_ISL_1207387, EPI_ISL_1207388, EPI_ISL_1207390, EPI_ISL_1207391, EPI_ISL_1207397, EPI_ISL_1207400, EPI_ISL_1207404, EPI_ISL_1207406, EPI_ISL_1207407, EPI_ISL_1207410, EPI_ISL_1207411, EPI_ISL_1207416, EPI_ISL_1207420, EPI_ISL_1207422, EPI_ISL_1207424, EPI_ISL_1207425, EPI_ISL_1207431, EPI_ISL_1207443, EPI_ISL_1207448, EPI_ISL_1207451, EPI_ISL_1207815, EPI_ISL_1207822, EPI_ISL_1207846, EPI_ISL_1207848, EPI_ISL_1207849, EPI_ISL_1207852, EPI_ISL_1207853, EPI_ISL_1207854, EPI_ISL_1207856, EPI_ISL_1207857, EPI_ISL_1207858, EPI_ISL_1207861, EPI_ISL_1207862, EPI_ISL_1207866, EPI_ISL_1207867, EPI_ISL_1207869, EPI_ISL_1207871, EPI_ISL_1207873, EPI_ISL_1207874, EPI_ISL_1207875, EPI_ISL_1207876, EPI_ISL_1207877, EPI_ISL_1207878, EPI_ISL_1207879, EPI_ISL_1207880, EPI_ISL_1207883, EPI_ISL_1207884, EPI_ISL_1207886, EPI_ISL_1207887, EPI_ISL_1207889, EPI_ISL_1207893, EPI_ISL_1207895, EPI_ISL_1207897, EPI_ISL_1207899, EPI_ISL_1207903, EPI_ISL_1207907, EPI_ISL_1207908, EPI_ISL_1207909, EPI_ISL_1207910, EPI_ISL_1223438, EPI_ISL_1223439, EPI_ISL_1223442, EPI_ISL_1223445, EPI_ISL_1223446, EPI_ISL_1223447, EPI_ISL_1223448, EPI_ISL_1223451, EPI_ISL_1223452, EPI_ISL_1223453, EPI_ISL_1223454, EPI_ISL_1223456, EPI_ISL_1223457, EPI_ISL_1223458, EPI_ISL_1223459, EPI_ISL_1223460, EPI_ISL_1223461, EPI_ISL_1223462, EPI_ISL_1223463, EPI_ISL_1223466, EPI_ISL_1223467, EPI_ISL_1223468, EPI_ISL_1223469, EPI_ISL_1223470, EPI_ISL_1223471, EPI_ISL_1223473, EPI_ISL_1223474, EPI_ISL_1223475, EPI_ISL_1223476, EPI_ISL_1223477, EPI_ISL_1223478, EPI_ISL_1223480, EPI_ISL_1223481, EPI_ISL_1223482, EPI_ISL_1223483, EPI_ISL_1223484, EPI_ISL_1223486, EPI_ISL_1223487, EPI_ISL_1223488, EPI_ISL_1223489, EPI_ISL_1223491, EPI_ISL_1223492, EPI_ISL_1223493, EPI_ISL_1223494, EPI_ISL_1223496, EPI_ISL_1223503, EPI_ISL_1223504, EPI_ISL_1223505, EPI_ISL_1223507, EPI_ISL_1223508, EPI_ISL_1223509, EPI_ISL_1223510, EPI_ISL_1223511, EPI_ISL_1223512, EPI_ISL_1223513, EPI_ISL_1223514, EPI_ISL_1223516, EPI_ISL_1223517, EPI_ISL_1223519, EPI_ISL_1223520, EPI_ISL_1223521, EPI_ISL_1223523, EPI_ISL_1223524, EPI_ISL_1223525, EPI_ISL_1223526, EPI_ISL_1223528, EPI_ISL_1223529, EPI_ISL_1223530, EPI_ISL_1223531, EPI_ISL_1223532, EPI_ISL_1223533, EPI_ISL_1223535, EPI_ISL_1223536, EPI_ISL_1223538, EPI_ISL_1223539, EPI_ISL_1223541, EPI_ISL_1223542, EPI_ISL_1223544, EPI_ISL_1223546, EPI_ISL_1223547, EPI_ISL_1223548, EPI_ISL_1223549, EPI_ISL_1223551, EPI_ISL_1223553, EPI_ISL_1223554, EPI_ISL_1223555, EPI_ISL_1223557, EPI_ISL_1223558, EPI_ISL_1223559, EPI_ISL_1223560, EPI_ISL_1223562, EPI_ISL_1223563, EPI_ISL_1223565, EPI_ISL_1223567, EPI_ISL_1223568, EPI_ISL_1223570, EPI_ISL_1223572, EPI_ISL_1223573, EPI_ISL_1223574, EPI_ISL_1223575, EPI_ISL_1223577, EPI_ISL_1223578, EPI_ISL_1223579, EPI_ISL_1223581, EPI_ISL_1223582, EPI_ISL_1223583, EPI_ISL_1223585, EPI_ISL_1223586, EPI_ISL_1223589, EPI_ISL_1223590, EPI_ISL_1223591, EPI_ISL_1223592, EPI_ISL_1223593, EPI_ISL_1223594, EPI_ISL_1223595, EPI_ISL_1223596, EPI_ISL_1223597, EPI_ISL_1223598, EPI_ISL_1223599, EPI_ISL_1223600, EPI_ISL_1223602, EPI_ISL_1223603, EPI_ISL_1223604, EPI_ISL_1223606, EPI_ISL_1223608, EPI_ISL_1223609, EPI_ISL_1223610, EPI_ISL_1223613, EPI_ISL_1223615, EPI_ISL_1223616, EPI_ISL_1223617, EPI_ISL_1223618, EPI_ISL_1223619, EPI_ISL_1223621, EPI_ISL_1223622, EPI_ISL_1223623, EPI_ISL_1223625, EPI_ISL_1223626, EPI_ISL_1223627, EPI_ISL_1223628, EPI_ISL_1223629, EPI_ISL_1223630, EPI_ISL_1223632, EPI_ISL_1223633, EPI_ISL_1223634, EPI_ISL_1223635, EPI_ISL_1223636, EPI_ISL_1223637, EPI_ISL_1223638, EPI_ISL_1223640, EPI_ISL_1223642, EPI_ISL_1223643, EPI_ISL_1223644, EPI_ISL_1223645, EPI_ISL_1223646, EPI_ISL_1223647, EPI_ISL_1223649, EPI_ISL_1223650, EPI_ISL_1223651, EPI_ISL_1223652, EPI_ISL_1223653, EPI_ISL_1223662, EPI_ISL_1223663, EPI_ISL_1223676, EPI_ISL_1223678, EPI_ISL_1223698, EPI_ISL_1223702, EPI_ISL_1223722, EPI_ISL_1223727, EPI_ISL_1223728, EPI_ISL_1223729, EPI_ISL_1223754, EPI_ISL_1223757, EPI_ISL_1223758, EPI_ISL_1223759, EPI_ISL_1223763, EPI_ISL_1223764, EPI_ISL_1223773, EPI_ISL_1223778, EPI_ISL_1223779, EPI_ISL_1223781, EPI_ISL_1223798, EPI_ISL_1223805, EPI_ISL_1223806, EPI_ISL_1223808, EPI_ISL_1223826, EPI_ISL_1223832, EPI_ISL_1223846, EPI_ISL_1223849, EPI_ISL_1223853, EPI_ISL_1223854, EPI_ISL_1223861, EPI_ISL_1223871, EPI_ISL_1223887, EPI_ISL_1223902, EPI_ISL_1223904, EPI_ISL_1223919, EPI_ISL_1223925, EPI_ISL_1223928, EPI_ISL_1223940, EPI_ISL_1223946, EPI_ISL_1223947, EPI_ISL_1223948, EPI_ISL_1223949, EPI_ISL_1223950, EPI_ISL_1223951, EPI_ISL_1223952, EPI_ISL_1223953, EPI_ISL_1223954, EPI_ISL_1223955, EPI_ISL_1223956, EPI_ISL_1223957, EPI_ISL_1223958, EPI_ISL_1223959, EPI_ISL_1223960, EPI_ISL_1223961, EPI_ISL_1223962, EPI_ISL_1223963, EPI_ISL_1223964, EPI_ISL_1223965, EPI_ISL_1223966, EPI_ISL_1223967, EPI_ISL_1223969, EPI_ISL_1223970, EPI_ISL_1223971, EPI_ISL_1223972, EPI_ISL_1223973, EPI_ISL_1223974, EPI_ISL_1223975, EPI_ISL_1223976, EPI_ISL_1223977, EPI_ISL_1223978, EPI_ISL_1223979, EPI_ISL_1223980, EPI_ISL_1223981, EPI_ISL_1223982, EPI_ISL_1223983, EPI_ISL_1223984, EPI_ISL_1223985, EPI_ISL_1223986, EPI_ISL_1223987, EPI_ISL_1223988, EPI_ISL_1223989, EPI_ISL_1223990, EPI_ISL_1223991, EPI_ISL_1223992, EPI_ISL_1223993, EPI_ISL_1223994, EPI_ISL_1223995, EPI_ISL_1223996, EPI_ISL_1223997, EPI_ISL_1223998, EPI_ISL_1223999, EPI_ISL_1224000, EPI_ISL_1224001, EPI_ISL_1224002, EPI_ISL_1224003, EPI_ISL_1224004, EPI_ISL_1224005, EPI_ISL_1224006, EPI_ISL_1224007, EPI_ISL_1224008, EPI_ISL_1224009, EPI_ISL_1224010, EPI_ISL_1224011, EPI_ISL_1224012, EPI_ISL_1224013, EPI_ISL_1224014, EPI_ISL_1224015, EPI_ISL_1224016, EPI_ISL_1224017, EPI_ISL_1224019, EPI_ISL_1224020, EPI_ISL_1224021, EPI_ISL_1224022, EPI_ISL_1224023, EPI_ISL_1224024, EPI_ISL_1224026, EPI_ISL_1224027, EPI_ISL_1224028, EPI_ISL_1224029, EPI_ISL_1224030, EPI_ISL_1224031, EPI_ISL_1224032, EPI_ISL_1224033, EPI_ISL_1224034, EPI_ISL_1224035, EPI_ISL_1224036, EPI_ISL_1224037, EPI_ISL_1224038, EPI_ISL_1224039, EPI_ISL_1224040, EPI_ISL_1224041, EPI_ISL_1224042, EPI_ISL_1224043, EPI_ISL_1224044, EPI_ISL_1224045, EPI_ISL_1224046, EPI_ISL_1224047, EPI_ISL_1224048, EPI_ISL_1224049, EPI_ISL_1224050, EPI_ISL_1224051, EPI_ISL_1224052, EPI_ISL_1224053, EPI_ISL_1224054, EPI_ISL_1224055, EPI_ISL_1224056, EPI_ISL_1224057, EPI_ISL_1224058, EPI_ISL_1224059, EPI_ISL_1224060, EPI_ISL_1224061, EPI_ISL_1224062, EPI_ISL_1224063, EPI_ISL_1224064, EPI_ISL_1224065, EPI_ISL_1224066, EPI_ISL_1224067, EPI_ISL_1224068, EPI_ISL_1224069, EPI_ISL_1224070, EPI_ISL_1224071, EPI_ISL_1224072, EPI_ISL_1224073, EPI_ISL_1224074, EPI_ISL_1224075, EPI_ISL_1224076, EPI_ISL_1224077, EPI_ISL_1224078, EPI_ISL_1224079, EPI_ISL_1224080, EPI_ISL_1224081, EPI_ISL_1224082, EPI_ISL_1224083, EPI_ISL_1224084, EPI_ISL_1224085, EPI_ISL_1224086, EPI_ISL_1224087, EPI_ISL_1224088, EPI_ISL_1224089, EPI_ISL_1224090, EPI_ISL_1224091, EPI_ISL_1224092, EPI_ISL_1224093, EPI_ISL_1224094, EPI_ISL_1224095, EPI_ISL_1224096, EPI_ISL_1224097, EPI_ISL_1224098, EPI_ISL_1224099, EPI_ISL_1224100, EPI_ISL_1224101, EPI_ISL_1224102, EPI_ISL_1224103, EPI_ISL_1224104, EPI_ISL_1224105, EPI_ISL_1224106, EPI_ISL_1224107, EPI_ISL_1224108, EPI_ISL_1224109, EPI_ISL_1224110, EPI_ISL_1224111, EPI_ISL_1224112, EPI_ISL_1224113, EPI_ISL_1224114, EPI_ISL_1224115, EPI_ISL_1224116, EPI_ISL_1224117, EPI_ISL_1224118, EPI_ISL_1224119, EPI_ISL_1224120, EPI_ISL_1224121, EPI_ISL_1224122, EPI_ISL_1224123, EPI_ISL_1224124, EPI_ISL_1224125, EPI_ISL_1224126, EPI_ISL_1224127, EPI_ISL_1224128, EPI_ISL_1224129, EPI_ISL_1224130, EPI_ISL_1224131, EPI_ISL_1224132, EPI_ISL_1224133, EPI_ISL_1224134, EPI_ISL_1224135, EPI_ISL_1224136, EPI_ISL_1224137, EPI_ISL_1224138, EPI_ISL_1224139, EPI_ISL_1224140, EPI_ISL_1224141, EPI_ISL_1224142, EPI_ISL_1224143, EPI_ISL_1224144, EPI_ISL_1224145, EPI_ISL_1224146, EPI_ISL_1224147, EPI_ISL_1224148, EPI_ISL_1224149, EPI_ISL_1224150, EPI_ISL_1224151, EPI_ISL_1224152, EPI_ISL_1224153, EPI_ISL_1224154, EPI_ISL_1224156, EPI_ISL_1224157, EPI_ISL_1224158, EPI_ISL_1224159, EPI_ISL_1224160, EPI_ISL_1224161, EPI_ISL_1224162, EPI_ISL_1243147, EPI_ISL_1243148, EPI_ISL_1243149, EPI_ISL_1243151, EPI_ISL_1243153, EPI_ISL_1243154, EPI_ISL_1243155, EPI_ISL_1243159, EPI_ISL_1243161, EPI_ISL_1243162, EPI_ISL_1243163, EPI_ISL_1243165, EPI_ISL_1243170, EPI_ISL_1243172, EPI_ISL_1243173, EPI_ISL_1243174, EPI_ISL_1243176, EPI_ISL_1243177, EPI_ISL_1243178, EPI_ISL_1243180, EPI_ISL_1243182, EPI_ISL_1243185, EPI_ISL_1243186, EPI_ISL_1243188, EPI_ISL_1243189, EPI_ISL_1243191, EPI_ISL_1243192, EPI_ISL_1243193, EPI_ISL_1243195, EPI_ISL_1243196, EPI_ISL_1243202, EPI_ISL_1243203, EPI_ISL_1243204, EPI_ISL_1243218, EPI_ISL_1243219, EPI_ISL_1243220, EPI_ISL_1243226, EPI_ISL_1243228, EPI_ISL_1243230, EPI_ISL_1243237, EPI_ISL_1243238, EPI_ISL_1243239, EPI_ISL_1243240, EPI_ISL_1243242, EPI_ISL_1243244, EPI_ISL_1243247, EPI_ISL_1243249, EPI_ISL_1243254, EPI_ISL_1243255, EPI_ISL_1243258, EPI_ISL_1243260, EPI_ISL_1243262, EPI_ISL_1243264, EPI_ISL_1243265, EPI_ISL_1243271, EPI_ISL_1243273, EPI_ISL_1243275, EPI_ISL_1243276, EPI_ISL_1243279, EPI_ISL_1243281, EPI_ISL_1243282, EPI_ISL_1243292, EPI_ISL_1243293, EPI_ISL_1243296, EPI_ISL_1243300, EPI_ISL_1243303, EPI_ISL_1243304, EPI_ISL_1243306, EPI_ISL_1243308, EPI_ISL_1243310, EPI_ISL_1243315, EPI_ISL_1243317, EPI_ISL_1243323, EPI_ISL_1243324, EPI_ISL_1243327, EPI_ISL_1243329, EPI_ISL_1243337, EPI_ISL_1243339, EPI_ISL_1243340, EPI_ISL_1243341, EPI_ISL_1243346, EPI_ISL_1243347, EPI_ISL_1243350, EPI_ISL_1243351, EPI_ISL_1243353, EPI_ISL_1243358, EPI_ISL_1243359, EPI_ISL_1243361, EPI_ISL_1243363, EPI_ISL_1243366, EPI_ISL_1243369, EPI_ISL_1243374, EPI_ISL_1243377, EPI_ISL_1243378, EPI_ISL_1243379, EPI_ISL_1243380, EPI_ISL_1243383, EPI_ISL_1243385, EPI_ISL_1243387, EPI_ISL_1243394, EPI_ISL_1243395, EPI_ISL_1243396, EPI_ISL_1243401, EPI_ISL_1243403, EPI_ISL_1243404, EPI_ISL_1243405, EPI_ISL_1243407, EPI_ISL_1243408, EPI_ISL_1243411, EPI_ISL_1243416, EPI_ISL_1243421, EPI_ISL_1243422, EPI_ISL_1243423, EPI_ISL_1243424, EPI_ISL_1243425, EPI_ISL_1243426, EPI_ISL_1243427, EPI_ISL_1243428, EPI_ISL_1243429, EPI_ISL_1243434, EPI_ISL_1243614, EPI_ISL_1243616, EPI_ISL_1243617, EPI_ISL_1243620, EPI_ISL_1243621, EPI_ISL_1243622, EPI_ISL_1243623, EPI_ISL_1243625, EPI_ISL_1243626, EPI_ISL_1243627, EPI_ISL_1243628, EPI_ISL_1243631, EPI_ISL_1243632, EPI_ISL_1243633, EPI_ISL_1243634, EPI_ISL_1243638, EPI_ISL_1243639, EPI_ISL_1243640, EPI_ISL_1243641, EPI_ISL_1243643, EPI_ISL_1243646, EPI_ISL_1243647, EPI_ISL_1243648, EPI_ISL_1243649, EPI_ISL_1243650, EPI_ISL_1243651, EPI_ISL_1243653, EPI_ISL_1243655, EPI_ISL_1243658, EPI_ISL_1243659, EPI_ISL_1243666, EPI_ISL_1243667, EPI_ISL_1243669, EPI_ISL_1243671, EPI_ISL_1243676, EPI_ISL_1243678, EPI_ISL_1243679, EPI_ISL_1243680, EPI_ISL_1243681, EPI_ISL_1243682, EPI_ISL_1243683, EPI_ISL_1243685, EPI_ISL_1243686, EPI_ISL_1243689, EPI_ISL_1243691, EPI_ISL_1243692, EPI_ISL_1243693, EPI_ISL_1243694, EPI_ISL_1243695, EPI_ISL_1243697, EPI_ISL_1243701, EPI_ISL_1243702, EPI_ISL_1243704, EPI_ISL_1243705, EPI_ISL_1243706, EPI_ISL_1243709, EPI_ISL_1243710, EPI_ISL_1243711,

EPI_ISL_1243712, EPI_ISL_1243713, EPI_ISL_1243714, EPI_ISL_1243716, EPI_ISL_1243718, EPI_ISL_1243720, EPI_ISL_1243721, EPI_ISL_1243723, EPI_ISL_1243724, EPI_ISL_1243727, EPI_ISL_1243730, EPI_ISL_1243731, EPI_ISL_1243733, EPI_ISL_1243737, EPI_ISL_1243739, EPI_ISL_1243741, EPI_ISL_1243742, EPI_ISL_1243746, EPI_ISL_1243750, EPI_ISL_1243751, EPI_ISL_1243754, EPI_ISL_1243758,
EPI_ISL_1243761, EPI_ISL_1243762, EPI_ISL_1243763, EPI_ISL_1243764, EPI_ISL_1243765, EPI_ISL_1243767, EPI_ISL_1243768, EPI_ISL_1243769, EPI_ISL_1243770, EPI_ISL_1243771, EPI_ISL_1243772, EPI_ISL_1243773, EPI_ISL_1243774, EPI_ISL_1243775, EPI_ISL_1243776, EPI_ISL_1243778, EPI_ISL_1243780, EPI_ISL_1243784, EPI_ISL_1243785, EPI_ISL_1243789, EPI_ISL_1243790, EPI_ISL_1243793,
EPI_ISL_1243794, EPI_ISL_1243796, EPI_ISL_1243798, EPI_ISL_1243800, EPI_ISL_1243801, EPI_ISL_1243802, EPI_ISL_1243803, EPI_ISL_1243805, EPI_ISL_1243809, EPI_ISL_1243810, EPI_ISL_1243811, EPI_ISL_1243813, EPI_ISL_1243814, EPI_ISL_1243817, EPI_ISL_1243818, EPI_ISL_1243819, EPI_ISL_1243820, EPI_ISL_1243821, EPI_ISL_1243823, EPI_ISL_1243827, EPI_ISL_1243828, EPI_ISL_1243829,
EPI_ISL_1243831, EPI_ISL_1243833, EPI_ISL_1243834, EPI_ISL_1243835, EPI_ISL_1243836, EPI_ISL_1243838, EPI_ISL_1243839, EPI_ISL_1243840, EPI_ISL_1243841, EPI_ISL_1243842, EPI_ISL_1243846, EPI_ISL_1243850, EPI_ISL_1243851, EPI_ISL_1243852, EPI_ISL_1243853, EPI_ISL_1243855, EPI_ISL_1243857, EPI_ISL_1243866, EPI_ISL_1243868, EPI_ISL_1243870, EPI_ISL_1243871, EPI_ISL_1243873,
EPI_ISL_1243878, EPI_ISL_1243879, EPI_ISL_1243880, EPI_ISL_1243883, EPI_ISL_1243884, EPI_ISL_1243885, EPI_ISL_1243886, EPI_ISL_1243888, EPI_ISL_1243889, EPI_ISL_1243892, EPI_ISL_1243893, EPI_ISL_1243896, EPI_ISL_1243904, EPI_ISL_1243906, EPI_ISL_1243907, EPI_ISL_1243910, EPI_ISL_1243912, EPI_ISL_1243913, EPI_ISL_1243914, EPI_ISL_1243915, EPI_ISL_1243916, EPI_ISL_1243917,
EPI_ISL_1243919, EPI_ISL_1243920, EPI_ISL_1243925, EPI_ISL_1243927, EPI_ISL_1243928, EPI_ISL_1243930, EPI_ISL_1243931, EPI_ISL_1243932, EPI_ISL_1243933, EPI_ISL_1243934, EPI_ISL_1244276, EPI_ISL_1244278, EPI_ISL_1244322, EPI_ISL_1244331, EPI_ISL_1244340, EPI_ISL_1244472, EPI_ISL_1244475, EPI_ISL_1244518, EPI_ISL_1244521, EPI_ISL_1244603, EPI_ISL_1244612, EPI_ISL_1244613,
EPI_ISL_1244614, EPI_ISL_1244615, EPI_ISL_1244616, EPI_ISL_1244617, EPI_ISL_1244618, EPI_ISL_1244619, EPI_ISL_1244620, EPI_ISL_1244621, EPI_ISL_1244622, EPI_ISL_1244623, EPI_ISL_1244624, EPI_ISL_1244625, EPI_ISL_1244626, EPI_ISL_1244627, EPI_ISL_1244628, EPI_ISL_1244629, EPI_ISL_1244630, EPI_ISL_1244631, EPI_ISL_1244633, EPI_ISL_1244634, EPI_ISL_1244635,
EPI_ISL_1244636, EPI_ISL_1244637, EPI_ISL_1244639, EPI_ISL_1244640, EPI_ISL_1244641, EPI_ISL_1244642, EPI_ISL_1244643, EPI_ISL_1244644, EPI_ISL_1244645, EPI_ISL_1244646, EPI_ISL_1244647, EPI_ISL_1244648, EPI_ISL_1244649, EPI_ISL_1244650, EPI_ISL_1244651, EPI_ISL_1244652, EPI_ISL_1244653, EPI_ISL_1244654, EPI_ISL_1244655, EPI_ISL_1244656, EPI_ISL_1244657, EPI_ISL_1244658,
EPI_ISL_1244659, EPI_ISL_1244660, EPI_ISL_1244661, EPI_ISL_1244662, EPI_ISL_1244663, EPI_ISL_1244664, EPI_ISL_1244665, EPI_ISL_1244666, EPI_ISL_1244667, EPI_ISL_1244668, EPI_ISL_1244669, EPI_ISL_1244670, EPI_ISL_1244671, EPI_ISL_1244672, EPI_ISL_1244673, EPI_ISL_1244674, EPI_ISL_1244675, EPI_ISL_1244676, EPI_ISL_1244677, EPI_ISL_1244678, EPI_ISL_1244679, EPI_ISL_1244680,
EPI_ISL_1244681, EPI_ISL_1244682, EPI_ISL_1244683, EPI_ISL_1244684, EPI_ISL_1244685, EPI_ISL_1244686, EPI_ISL_1244687, EPI_ISL_1244688, EPI_ISL_1244689, EPI_ISL_1244690, EPI_ISL_1244691, EPI_ISL_1244692, EPI_ISL_1244693, EPI_ISL_1244694, EPI_ISL_1244695, EPI_ISL_1244696, EPI_ISL_1244697, EPI_ISL_1244698, EPI_ISL_1244699, EPI_ISL_1244700, EPI_ISL_1244701, EPI_ISL_1244702,
EPI_ISL_1244703, EPI_ISL_1244704, EPI_ISL_1244705, EPI_ISL_1244706, EPI_ISL_1244707, EPI_ISL_1244708, EPI_ISL_1244709, EPI_ISL_1244710, EPI_ISL_1244711, EPI_ISL_1244712, EPI_ISL_1244713, EPI_ISL_1244714, EPI_ISL_1244715, EPI_ISL_1244716, EPI_ISL_1244717, EPI_ISL_1244718, EPI_ISL_1244719, EPI_ISL_1244720, EPI_ISL_1244721, EPI_ISL_1244722, EPI_ISL_1244723, EPI_ISL_1244724,
EPI_ISL_1244725, EPI_ISL_1244726, EPI_ISL_1244727, EPI_ISL_1244728, EPI_ISL_1244729, EPI_ISL_1244730, EPI_ISL_1244731, EPI_ISL_1244732, EPI_ISL_1244733, EPI_ISL_1244734, EPI_ISL_1244735, EPI_ISL_1244736, EPI_ISL_1244737, EPI_ISL_1244738, EPI_ISL_1244739, EPI_ISL_1244740, EPI_ISL_1244741, EPI_ISL_1244742, EPI_ISL_1244743, EPI_ISL_1244744, EPI_ISL_1244745, EPI_ISL_1244746,
EPI_ISL_1244747, EPI_ISL_1244748, EPI_ISL_1244749, EPI_ISL_1244750, EPI_ISL_1244751, EPI_ISL_1244752, EPI_ISL_1244753, EPI_ISL_1244754, EPI_ISL_1244755, EPI_ISL_1244756, EPI_ISL_1244757, EPI_ISL_1244758, EPI_ISL_1244759, EPI_ISL_1244760, EPI_ISL_1244761, EPI_ISL_1244762, EPI_ISL_1244763, EPI_ISL_1244764, EPI_ISL_1244765, EPI_ISL_1244766, EPI_ISL_1244767, EPI_ISL_1244768,
EPI_ISL_1244769, EPI_ISL_1244770, EPI_ISL_1244771, EPI_ISL_1244772, EPI_ISL_1244773, EPI_ISL_1244774, EPI_ISL_1244775, EPI_ISL_1244776, EPI_ISL_1244777, EPI_ISL_1244778, EPI_ISL_1244779, EPI_ISL_1244780, EPI_ISL_1244781, EPI_ISL_1244782, EPI_ISL_1244783, EPI_ISL_1244784, EPI_ISL_1244785, EPI_ISL_1244786, EPI_ISL_1244787, EPI_ISL_1244788, EPI_ISL_1244789, EPI_ISL_1244790,
EPI_ISL_1244791, EPI_ISL_1244792, EPI_ISL_1244793, EPI_ISL_1244794, EPI_ISL_1244795, EPI_ISL_1244796, EPI_ISL_1244797, EPI_ISL_1244798, EPI_ISL_1244799, EPI_ISL_1244800, EPI_ISL_1244801, EPI_ISL_1244802, EPI_ISL_1244803, EPI_ISL_1244804, EPI_ISL_1244805, EPI_ISL_1244806, EPI_ISL_1244807, EPI_ISL_1244808, EPI_ISL_1244809, EPI_ISL_1244810, EPI_ISL_1244811, EPI_ISL_1244812,
EPI_ISL_1244813, EPI_ISL_1244814, EPI_ISL_1244815, EPI_ISL_1244816, EPI_ISL_1244817, EPI_ISL_1244818, EPI_ISL_1244819, EPI_ISL_1244820, EPI_ISL_1244821, EPI_ISL_1244822, EPI_ISL_1244823, EPI_ISL_1244824, EPI_ISL_1244825, EPI_ISL_1244826, EPI_ISL_1244827, EPI_ISL_1244828, EPI_ISL_1244829, EPI_ISL_1244830, EPI_ISL_1244831, EPI_ISL_1244832, EPI_ISL_1244833, EPI_ISL_1244834,
EPI_ISL_1244835, EPI_ISL_1244836, EPI_ISL_1244837, EPI_ISL_1244838, EPI_ISL_1244839, EPI_ISL_1244840, EPI_ISL_1244841, EPI_ISL_1244842, EPI_ISL_1244843, EPI_ISL_1244844, EPI_ISL_1244845, EPI_ISL_1244846, EPI_ISL_1244847, EPI_ISL_1244848, EPI_ISL_1244849, EPI_ISL_1244850, EPI_ISL_1244851, EPI_ISL_1245007, EPI_ISL_1245008, EPI_ISL_1245009, EPI_ISL_1245010, EPI_ISL_1245011,
EPI_ISL_1245012, EPI_ISL_1245013, EPI_ISL_1245014, EPI_ISL_1245015, EPI_ISL_1245016, EPI_ISL_1245017, EPI_ISL_1245018, EPI_ISL_1245019, EPI_ISL_1245020, EPI_ISL_1245021, EPI_ISL_1245022, EPI_ISL_1245023, EPI_ISL_1245024, EPI_ISL_1245025, EPI_ISL_1245026, EPI_ISL_1245027, EPI_ISL_1245028, EPI_ISL_1245029, EPI_ISL_1245030, EPI_ISL_1245031, EPI_ISL_1245032, EPI_ISL_1245033,
EPI_ISL_1245034, EPI_ISL_1245035, EPI_ISL_1245036, EPI_ISL_1245037, EPI_ISL_1245038, EPI_ISL_1245039, EPI_ISL_1245040, EPI_ISL_1245041, EPI_ISL_1245042, EPI_ISL_1245043, EPI_ISL_1245044, EPI_ISL_1245045, EPI_ISL_1245046, EPI_ISL_1245047, EPI_ISL_1245048, EPI_ISL_1245049, EPI_ISL_1245050, EPI_ISL_1245051, EPI_ISL_1245052, EPI_ISL_1245053, EPI_ISL_1245054, EPI_ISL_1245055,
EPI_ISL_1245056, EPI_ISL_1245057, EPI_ISL_1245058, EPI_ISL_1245059, EPI_ISL_1245060, EPI_ISL_1245061, EPI_ISL_1245062, EPI_ISL_1245063, EPI_ISL_1245064, EPI_ISL_1245065, EPI_ISL_1245066, EPI_ISL_1245067, EPI_ISL_1245068, EPI_ISL_1245069, EPI_ISL_1245070, EPI_ISL_1245071, EPI_ISL_1245072, EPI_ISL_1245073, EPI_ISL_1245074, EPI_ISL_1245075, EPI_ISL_1245076, EPI_ISL_1245077,
EPI_ISL_1245078, EPI_ISL_1245079, EPI_ISL_1245080, EPI_ISL_1245081, EPI_ISL_1245082, EPI_ISL_1245083, EPI_ISL_1245084, EPI_ISL_1245085, EPI_ISL_1245086, EPI_ISL_1245087, EPI_ISL_1245088, EPI_ISL_1245089, EPI_ISL_1245090, EPI_ISL_1245091, EPI_ISL_1245092, EPI_ISL_1245093, EPI_ISL_1245094, EPI_ISL_1245095, EPI_ISL_1245096, EPI_ISL_1245097, EPI_ISL_1245098, EPI_ISL_1245099,
EPI_ISL_1245100, EPI_ISL_1245101, EPI_ISL_1245102, EPI_ISL_1245103, EPI_ISL_1245104, EPI_ISL_1245105, EPI_ISL_1245106, EPI_ISL_1245107, EPI_ISL_1245108, EPI_ISL_1245109, EPI_ISL_1245110, EPI_ISL_1245111, EPI_ISL_1245112, EPI_ISL_1245113, EPI_ISL_1245114, EPI_ISL_1245115, EPI_ISL_1245116, EPI_ISL_1245117, EPI_ISL_1245118, EPI_ISL_1245119, EPI_ISL_1245120, EPI_ISL_1245121,
EPI_ISL_1245122, EPI_ISL_1245123, EPI_ISL_1245124, EPI_ISL_1245125, EPI_ISL_1245126, EPI_ISL_1245127, EPI_ISL_1245128, EPI_ISL_1245129, EPI_ISL_1245130, EPI_ISL_1245131, EPI_ISL_1245132, EPI_ISL_1245133, EPI_ISL_1245134, EPI_ISL_1245135, EPI_ISL_1245136, EPI_ISL_1245137, EPI_ISL_1245138, EPI_ISL_1245139, EPI_ISL_1245140, EPI_ISL_1245141, EPI_ISL_1245142, EPI_ISL_1245143,
EPI_ISL_1245144, EPI_ISL_1245145, EPI_ISL_1245146, EPI_ISL_1245147, EPI_ISL_1245148, EPI_ISL_1245149, EPI_ISL_1245150, EPI_ISL_1245151, EPI_ISL_1245152, EPI_ISL_1245153, EPI_ISL_1245154, EPI_ISL_1245155, EPI_ISL_1245156, EPI_ISL_1245157, EPI_ISL_1245158, EPI_ISL_1245159, EPI_ISL_1245160, EPI_ISL_1245161, EPI_ISL_1245162, EPI_ISL_1245163, EPI_ISL_1245164, EPI_ISL_1245165,
EPI_ISL_1245166, EPI_ISL_1245167, EPI_ISL_1245168, EPI_ISL_1245169, EPI_ISL_1245170, EPI_ISL_1245171, EPI_ISL_1245172, EPI_ISL_1245173, EPI_ISL_1245174, EPI_ISL_1245175, EPI_ISL_1245176, EPI_ISL_1245177, EPI_ISL_1245178, EPI_ISL_1245179, EPI_ISL_1245180, EPI_ISL_1245181, EPI_ISL_1245182, EPI_ISL_1245183, EPI_ISL_1245184, EPI_ISL_1245185, EPI_ISL_1245186, EPI_ISL_1245187,
EPI_ISL_1245188, EPI_ISL_1245189, EPI_ISL_1245190, EPI_ISL_1245191, EPI_ISL_1245192, EPI_ISL_1245193, EPI_ISL_1245194, EPI_ISL_1245195, EPI_ISL_1245196, EPI_ISL_1245197, EPI_ISL_1245198, EPI_ISL_1245199, EPI_ISL_1245200, EPI_ISL_1245201, EPI_ISL_1245202, EPI_ISL_1245203, EPI_ISL_1245204, EPI_ISL_1245205, EPI_ISL_1245206, EPI_ISL_1245207, EPI_ISL_1245208, EPI_ISL_1245209,
EPI_ISL_1245210, EPI_ISL_1245211, EPI_ISL_1245212, EPI_ISL_1245213, EPI_ISL_1245214, EPI_ISL_1245215, EPI_ISL_1245216, EPI_ISL_1245217, EPI_ISL_1245218, EPI_ISL_1245219, EPI_ISL_1245220, EPI_ISL_1245221, EPI_ISL_1245222, EPI_ISL_1245223, EPI_ISL_1245224, EPI_ISL_1245225, EPI_ISL_1245226, EPI_ISL_1245227, EPI_ISL_1245228, EPI_ISL_1245229, EPI_ISL_1245230, EPI_ISL_1245231,
EPI_ISL_1245232, EPI_ISL_1245233, EPI_ISL_1245235, EPI_ISL_1245236, EPI_ISL_1245237, EPI_ISL_1245238, EPI_ISL_1245239, EPI_ISL_1245240, EPI_ISL_1245241, EPI_ISL_1245242, EPI_ISL_1245243, EPI_ISL_1245244, EPI_ISL_1245245, EPI_ISL_1245246, EPI_ISL_1245247, EPI_ISL_1245248, EPI_ISL_1245249, EPI_ISL_1245250, EPI_ISL_1245251, EPI_ISL_1245252, EPI_ISL_1245253, EPI_ISL_1245254,
EPI_ISL_1245255, EPI_ISL_1245256, EPI_ISL_1245257, EPI_ISL_1245258, EPI_ISL_1245259, EPI_ISL_1245260, EPI_ISL_1245261, EPI_ISL_1245262, EPI_ISL_1245263, EPI_ISL_1245264, EPI_ISL_1245265, EPI_ISL_1245266, EPI_ISL_1245267, EPI_ISL_1245268, EPI_ISL_1245269, EPI_ISL_1245270, EPI_ISL_1245271, EPI_ISL_1245272, EPI_ISL_1245273, EPI_ISL_1245274, EPI_ISL_1245275, EPI_ISL_1245276,
EPI_ISL_1245277, EPI_ISL_1245278, EPI_ISL_1245279, EPI_ISL_1245280, EPI_ISL_1245281, EPI_ISL_1245282, EPI_ISL_1245283, EPI_ISL_1245284, EPI_ISL_1245285, EPI_ISL_1245286, EPI_ISL_1245287, EPI_ISL_1245288, EPI_ISL_1245289, EPI_ISL_1245290, EPI_ISL_1245291, EPI_ISL_1245292, EPI_ISL_1245293, EPI_ISL_1245294, EPI_ISL_1245295, EPI_ISL_1245296, EPI_ISL_1245297, EPI_ISL_1245298,
EPI_ISL_1245299, EPI_ISL_1245300, EPI_ISL_1245301, EPI_ISL_1245302, EPI_ISL_1245303, EPI_ISL_1245304, EPI_ISL_1245305, EPI_ISL_1245306, EPI_ISL_1245307, EPI_ISL_1245308, EPI_ISL_1245309, EPI_ISL_1245310, EPI_ISL_1245311, EPI_ISL_1245312, EPI_ISL_1245313, EPI_ISL_1245314, EPI_ISL_1245315, EPI_ISL_1245316, EPI_ISL_1245317, EPI_ISL_1245318, EPI_ISL_1245319, EPI_ISL_1245320,
EPI_ISL_1245321, EPI_ISL_1245322, EPI_ISL_1245323, EPI_ISL_1245324, EPI_ISL_1245325, EPI_ISL_1245326, EPI_ISL_1245327, EPI_ISL_1245328, EPI_ISL_1245329, EPI_ISL_1245330, EPI_ISL_1245331, EPI_ISL_1245332, EPI_ISL_1245333, EPI_ISL_1245334, EPI_ISL_1245335, EPI_ISL_1245336, EPI_ISL_1245338, EPI_ISL_1245339, EPI_ISL_1245340, EPI_ISL_1245341, EPI_ISL_1245342, EPI_ISL_1245343,
EPI_ISL_1245344, EPI_ISL_1245345, EPI_ISL_1245346, EPI_ISL_1245347, EPI_ISL_1245348, EPI_ISL_1245349, EPI_ISL_1245350, EPI_ISL_1245351, EPI_ISL_1245352, EPI_ISL_1245353, EPI_ISL_1245354, EPI_ISL_1245355, EPI_ISL_1245356, EPI_ISL_1245357, EPI_ISL_1246383, EPI_ISL_1246384, EPI_ISL_1246386, EPI_ISL_1246387, EPI_ISL_1246388, EPI_ISL_1246389, EPI_ISL_1246390, EPI_ISL_1246392,
EPI_ISL_1246393, EPI_ISL_1246394, EPI_ISL_1246395, EPI_ISL_1246396, EPI_ISL_1246397, EPI_ISL_1246399, EPI_ISL_1246400, EPI_ISL_1246401, EPI_ISL_1246402, EPI_ISL_1246403, EPI_ISL_1246404, EPI_ISL_1246405, EPI_ISL_1246406, EPI_ISL_1246408, EPI_ISL_1246409, EPI_ISL_1246410, EPI_ISL_1246412, EPI_ISL_1246413, EPI_ISL_1246415, EPI_ISL_1246416, EPI_ISL_1246419, EPI_ISL_1246421,
EPI_ISL_1246423, EPI_ISL_1246427, EPI_ISL_1246428, EPI_ISL_1246429, EPI_ISL_1246430, EPI_ISL_1246431, EPI_ISL_1246433, EPI_ISL_1246435, EPI_ISL_1246436, EPI_ISL_1246438, EPI_ISL_1246439, EPI_ISL_1246441, EPI_ISL_1246442, EPI_ISL_1246443, EPI_ISL_1246446, EPI_ISL_1246447, EPI_ISL_1246448, EPI_ISL_1246449, EPI_ISL_1246452, EPI_ISL_1246453, EPI_ISL_1246454, EPI_ISL_1246455,
EPI_ISL_1246456, EPI_ISL_1246457, EPI_ISL_1246458, EPI_ISL_1246461, EPI_ISL_1246462, EPI_ISL_1246464, EPI_ISL_1246466, EPI_ISL_1246467, EPI_ISL_1246468, EPI_ISL_1246469, EPI_ISL_1246470, EPI_ISL_1246471, EPI_ISL_1246472, EPI_ISL_1246475, EPI_ISL_1246478, EPI_ISL_1246479, EPI_ISL_1246480, EPI_ISL_1246481, EPI_ISL_1246482, EPI_ISL_1246484, EPI_ISL_1246487, EPI_ISL_1246488,
EPI_ISL_1246489, EPI_ISL_1246491, EPI_ISL_1246492, EPI_ISL_1246493, EPI_ISL_1246495, EPI_ISL_1246497, EPI_ISL_1246498, EPI_ISL_1246499, EPI_ISL_1246502, EPI_ISL_1246504, EPI_ISL_1246507, EPI_ISL_1246509, EPI_ISL_1246510, EPI_ISL_1246513, EPI_ISL_1246514, EPI_ISL_1246515, EPI_ISL_1246516, EPI_ISL_1246517, EPI_ISL_1246519, EPI_ISL_1246520, EPI_ISL_1246523, EPI_ISL_1246524,
EPI_ISL_1246525, EPI_ISL_1246527, EPI_ISL_1246529, EPI_ISL_1246530, EPI_ISL_1246531, EPI_ISL_1246533, EPI_ISL_1246535, EPI_ISL_1246536, EPI_ISL_1246537, EPI_ISL_1246538, EPI_ISL_1246539, EPI_ISL_1246540, EPI_ISL_1246541, EPI_ISL_1246542, EPI_ISL_1246543, EPI_ISL_1246544, EPI_ISL_1246545, EPI_ISL_1246546, EPI_ISL_1246547, EPI_ISL_1246548, EPI_ISL_1246549, EPI_ISL_1246550,
EPI_ISL_1246551, EPI_ISL_1246553, EPI_ISL_1246554, EPI_ISL_1246555, EPI_ISL_1246556, EPI_ISL_1246557, EPI_ISL_1246558, EPI_ISL_1246559, EPI_ISL_1246560, EPI_ISL_1246561, EPI_ISL_1246562, EPI_ISL_1246563, EPI_ISL_1246564, EPI_ISL_1246565, EPI_ISL_1246566, EPI_ISL_1246567, EPI_ISL_1246568, EPI_ISL_1246569, EPI_ISL_1246570, EPI_ISL_1246571, EPI_ISL_1246572, EPI_ISL_1246573,
EPI_ISL_1246574, EPI_ISL_1246575, EPI_ISL_1246576, EPI_ISL_1246577, EPI_ISL_1246578, EPI_ISL_1246579, EPI_ISL_1246581, EPI_ISL_1246582, EPI_ISL_1246583, EPI_ISL_1246584, EPI_ISL_1246587, EPI_ISL_1246588, EPI_ISL_1246589, EPI_ISL_1246590, EPI_ISL_1246593, EPI_ISL_1246594, EPI_ISL_1246595, EPI_ISL_1246596, EPI_ISL_1246597, EPI_ISL_1246598, EPI_ISL_1246599, EPI_ISL_1246600,
EPI_ISL_1246601, EPI_ISL_1246602, EPI_ISL_1246603, EPI_ISL_1246604, EPI_ISL_1246605, EPI_ISL_1246606, EPI_ISL_1246607, EPI_ISL_1246608, EPI_ISL_1246609, EPI_ISL_1246610, EPI_ISL_1246611, EPI_ISL_1246612, EPI_ISL_1246613, EPI_ISL_1246614, EPI_ISL_1246618, EPI_ISL_1246622, EPI_ISL_1246624, EPI_ISL_1246625, EPI_ISL_1246626, EPI_ISL_1246627, EPI_ISL_1246628, EPI_ISL_1246629,
EPI_ISL_1246630, EPI_ISL_1246631, EPI_ISL_1246632, EPI_ISL_1246635, EPI_ISL_1246636, EPI_ISL_1246637, EPI_ISL_1246638, EPI_ISL_1246639, EPI_ISL_1246641, EPI_ISL_1246642, EPI_ISL_1246643, EPI_ISL_1246644, EPI_ISL_1246645, EPI_ISL_1246646, EPI_ISL_1246647, EPI_ISL_1246648, EPI_ISL_1246649, EPI_ISL_1246650, EPI_ISL_1246651, EPI_ISL_1246654, EPI_ISL_1246655, EPI_ISL_1246656,
EPI_ISL_1246657, EPI_ISL_1246659, EPI_ISL_1246660, EPI_ISL_1246661, EPI_ISL_1246662, EPI_ISL_1246663, EPI_ISL_1246664, EPI_ISL_1246665, EPI_ISL_1246666, EPI_ISL_1246667, EPI_ISL_1246668, EPI_ISL_1246669, EPI_ISL_1246670, EPI_ISL_1246671, EPI_ISL_1246672, EPI_ISL_1246673, EPI_ISL_1246674, EPI_ISL_1246675, EPI_ISL_1246676, EPI_ISL_1246677, EPI_ISL_1246678, EPI_ISL_1246679,
EPI_ISL_1246680, EPI_ISL_1246681, EPI_ISL_1246683, EPI_ISL_1246684, EPI_ISL_1246685, EPI_ISL_1246686, EPI_ISL_1246687, EPI_ISL_1246688, EPI_ISL_1246689, EPI_ISL_1246690, EPI_ISL_1246691, EPI_ISL_1246693, EPI_ISL_1246694, EPI_ISL_1246695, EPI_ISL_1246696, EPI_ISL_1246697, EPI_ISL_1246698, EPI_ISL_1246699, EPI_ISL_1246701, EPI_ISL_1246702, EPI_ISL_1246703, EPI_ISL_1246704,
EPI_ISL_1246705, EPI_ISL_1246706, EPI_ISL_1246707, EPI_ISL_1246708, EPI_ISL_1246709, EPI_ISL_1246711, EPI_ISL_1246712, EPI_ISL_1246713, EPI_ISL_1246714, EPI_ISL_1246716, EPI_ISL_1246719, EPI_ISL_1246720, EPI_ISL_1246722, EPI_ISL_1246723, EPI_ISL_1246724, EPI_ISL_1246875, EPI_ISL_1246876, EPI_ISL_1246877, EPI_ISL_1246878, EPI_ISL_1246879, EPI_ISL_1246880, EPI_ISL_1246881,
EPI_ISL_1246882, EPI_ISL_1246883, EPI_ISL_1246884, EPI_ISL_1246885, EPI_ISL_1246886, EPI_ISL_1246887, EPI_ISL_1246888, EPI_ISL_1246889, EPI_ISL_1246890, EPI_ISL_1246891, EPI_ISL_1246892, EPI_ISL_1246893, EPI_ISL_1246894, EPI_ISL_1246895, EPI_ISL_1246896, EPI_ISL_1246897, EPI_ISL_1246898, EPI_ISL_1246899, EPI_ISL_1246901, EPI_ISL_1246902, EPI_ISL_1246903, EPI_ISL_1246904,
EPI_ISL_1246905, EPI_ISL_1246906, EPI_ISL_1246907, EPI_ISL_1246908, EPI_ISL_1246909, EPI_ISL_1246910, EPI_ISL_1246911, EPI_ISL_1246912, EPI_ISL_1246913, EPI_ISL_1246914, EPI_ISL_1246915, EPI_ISL_1246916, EPI_ISL_1246917, EPI_ISL_1246918, EPI_ISL_1246919, EPI_ISL_1246920, EPI_ISL_1246921, EPI_ISL_1246922, EPI_ISL_1246923, EPI_ISL_1246924, EPI_ISL_1246925, EPI_ISL_1246926,
EPI_ISL_1246927, EPI_ISL_1255465, EPI_ISL_1255491, EPI_ISL_1255505, EPI_ISL_1255507, EPI_ISL_1255557, EPI_ISL_1255597, EPI_ISL_1255611, EPI_ISL_1255616, EPI_ISL_1255653, EPI_ISL_1255665, EPI_ISL_1256097, EPI_ISL_1256124, EPI_ISL_1256136, EPI_ISL_1256188, EPI_ISL_1256278, EPI_ISL_1256365, EPI_ISL_1256817, EPI_ISL_1256830, EPI_ISL_1256899, EPI_ISL_1257364, EPI_ISL_1257365,
EPI_ISL_1257366, EPI_ISL_1257369, EPI_ISL_1257370, EPI_ISL_1257371, EPI_ISL_1257372, EPI_ISL_1257373, EPI_ISL_1257374, EPI_ISL_1257375, EPI_ISL_1257377, EPI_ISL_1257378, EPI_ISL_1257379, EPI_ISL_1257381, EPI_ISL_1257382, EPI_ISL_1257385, EPI_ISL_1257388, EPI_ISL_1257389, EPI_ISL_1257390, EPI_ISL_1257391, EPI_ISL_1257393, EPI_ISL_1257395, EPI_ISL_1257396,
EPI_ISL_1257398, EPI_ISL_1257399, EPI_ISL_1257400, EPI_ISL_1257402, EPI_ISL_1257403, EPI_ISL_1257404, EPI_ISL_1257405, EPI_ISL_1257408, EPI_ISL_1257410, EPI_ISL_1257413, EPI_ISL_1257414, EPI_ISL_1257416, EPI_ISL_1257417, EPI_ISL_1257418, EPI_ISL_1257419, EPI_ISL_1257420, EPI_ISL_1257421, EPI_ISL_1257422, EPI_ISL_1257423, EPI_ISL_1257486, EPI_ISL_1257487, EPI_ISL_1257488,
EPI_ISL_1257489, EPI_ISL_1257490, EPI_ISL_1257491, EPI_ISL_1257492, EPI_ISL_1257493, EPI_ISL_1257494, EPI_ISL_1257495, EPI_ISL_1257496, EPI_ISL_1257497, EPI_ISL_1257498, EPI_ISL_1257499, EPI_ISL_1257500, EPI_ISL_1257501, EPI_ISL_1257502, EPI_ISL_1257504, EPI_ISL_1257505, EPI_ISL_1257507, EPI_ISL_1257508, EPI_ISL_1257509, EPI_ISL_1257510, EPI_ISL_1257511, EPI_ISL_1257512,
EPI_ISL_1257513, EPI_ISL_1257514, EPI_ISL_1257515, EPI_ISL_1257516, EPI_ISL_1257517, EPI_ISL_1257518, EPI_ISL_1257519, EPI_ISL_1257520, EPI_ISL_1257521, EPI_ISL_1257522, EPI_ISL_1257523, EPI_ISL_1257524, EPI_ISL_1257525, EPI_ISL_1257526, EPI_ISL_1257527, EPI_ISL_1257528, EPI_ISL_1257529, EPI_ISL_1257530, EPI_ISL_1257531, EPI_ISL_1257532, EPI_ISL_1257533, EPI_ISL_1257534,
EPI_ISL_1257535, EPI_ISL_1257536, EPI_ISL_1257537, EPI_ISL_1257538, EPI_ISL_1257539, EPI_ISL_1257540, EPI_ISL_1257541, EPI_ISL_1257542, EPI_ISL_1257543, EPI_ISL_1257544, EPI_ISL_1257545, EPI_ISL_1257546, EPI_ISL_1257547, EPI_ISL_1257548, EPI_ISL_1257549, EPI_ISL_1257550, EPI_ISL_1257551, EPI_ISL_1257552, EPI_ISL_1257553, EPI_ISL_1257554, EPI_ISL_1257555, EPI_ISL_1257556,
EPI_ISL_1257557, EPI_ISL_1257558, EPI_ISL_1257559, EPI_ISL_1257560, EPI_ISL_1257561, EPI_ISL_1257562, EPI_ISL_1257563, EPI_ISL_1257564, EPI_ISL_1257565, EPI_ISL_1257566, EPI_ISL_1257567, EPI_ISL_1257568, EPI_ISL_1257569, EPI_ISL_1257570, EPI_ISL_1257571, EPI_ISL_1257572, EPI_ISL_1257573, EPI_ISL_1257574, EPI_ISL_1257575, EPI_ISL_1257576, EPI_ISL_1257577, EPI_ISL_1257578,
EPI_ISL_1257579, EPI_ISL_1257580, EPI_ISL_1257581, EPI_ISL_1257582, EPI_ISL_1257583, EPI_ISL_1257584, EPI_ISL_1257585, EPI_ISL_1257586, EPI_ISL_1257587, EPI_ISL_1257588, EPI_ISL_1257589, EPI_ISL_1257590, EPI_ISL_1257591, EPI_ISL_1257592, EPI_ISL_1257593, EPI_ISL_1257594, EPI_ISL_1257595, EPI_ISL_1257596, EPI_ISL_1257597, EPI_ISL_1257598, EPI_ISL_1257599, EPI_ISL_1257600,
EPI_ISL_1257601, EPI_ISL_1257602, EPI_ISL_1257603, EPI_ISL_1257604, EPI_ISL_1257605, EPI_ISL_1257606, EPI_ISL_1257607, EPI_ISL_1257608, EPI_ISL_1257609, EPI_ISL_1257610, EPI_ISL_1257611, EPI_ISL_1257612, EPI_ISL_1257613, EPI_ISL_1257614, EPI_ISL_1257615, EPI_ISL_1257616, EPI_ISL_1257617, EPI_ISL_1257618, EPI_ISL_1257619, EPI_ISL_1257620, EPI_ISL_1257621, EPI_ISL_1257622,
EPI_ISL_1257623, EPI_ISL_1257624, EPI_ISL_1257625, EPI_ISL_1257626, EPI_ISL_1257627, EPI_ISL_1257628, EPI_ISL_1257629, EPI_ISL_1257630, EPI_ISL_1257631, EPI_ISL_1257632, EPI_ISL_1257633, EPI_ISL_1257634, EPI_ISL_1257635, EPI_ISL_1257636, EPI_ISL_1257637, EPI_ISL_1257638, EPI_ISL_1257640, EPI_ISL_1257641, EPI_ISL_1257642, EPI_ISL_1257644, EPI_ISL_1257645, EPI_ISL_1257647,
EPI_ISL_1257648, EPI_ISL_1264087, EPI_ISL_1264142, EPI_ISL_1264258, EPI_ISL_1264325, EPI_ISL_1264332, EPI_ISL_1264643, EPI_ISL_1264644, EPI_ISL_1264645, EPI_ISL_1264646, EPI_ISL_1264647, EPI_ISL_1264648, EPI_ISL_1264649, EPI_ISL_1264650, EPI_ISL_1264651, EPI_ISL_1264652, EPI_ISL_1264653, EPI_ISL_1264654, EPI_ISL_1264655, EPI_ISL_1264656, EPI_ISL_1264657, EPI_ISL_1264658,
EPI_ISL_1264659, EPI_ISL_1264660, EPI_ISL_1264661, EPI_ISL_1264662, EPI_ISL_1264663, EPI_ISL_1264664, EPI_ISL_1264665, EPI_ISL_1264666, EPI_ISL_1264667, EPI_ISL_1264668, EPI_ISL_1264669, EPI_ISL_1264670, EPI_ISL_1264671, EPI_ISL_1264672, EPI_ISL_1264673, EPI_ISL_1264674, EPI_ISL_1264675, EPI_ISL_1264676, EPI_ISL_1264677, EPI_ISL_1264678, EPI_ISL_1264679, EPI_ISL_1264680,
EPI_ISL_1264681, EPI_ISL_1264682, EPI_ISL_1264683, EPI_ISL_1264684, EPI_ISL_1264685, EPI_ISL_1264686, EPI_ISL_1264687, EPI_ISL_1264688, EPI_ISL_1264689, EPI_ISL_1264690, EPI_ISL_1264691, EPI_ISL_1264692, EPI_ISL_1264693, EPI_ISL_1264694, EPI_ISL_1264695, EPI_ISL_1264696, EPI_ISL_1264697, EPI_ISL_1264698, EPI_ISL_1264699, EPI_ISL_1264700, EPI_ISL_1264701, EPI_ISL_1264702,
EPI_ISL_1264703, EPI_ISL_1264704, EPI_ISL_1264705, EPI_ISL_1264706, EPI_ISL_1264707, EPI_ISL_1264708, EPI_ISL_1264709, EPI_ISL_1264710, EPI_ISL_1264711, EPI_ISL_1264712, EPI_ISL_1264713, EPI_ISL_1264714, EPI_ISL_1264715, EPI_ISL_1264716, EPI_ISL_1264717, EPI_ISL_1264718, EPI_ISL_1264719, EPI_ISL_1264720, EPI_ISL_1264721, EPI_ISL_1264722, EPI_ISL_1264723, EPI_ISL_1264724,
EPI_ISL_1264725, EPI_ISL_1264726, EPI_ISL_1264727, EPI_ISL_1264728, EPI_ISL_1264729, EPI_ISL_1264730, EPI_ISL_1264731, EPI_ISL_1264732, EPI_ISL_1264733, EPI_ISL_1264734, EPI_ISL_1264735, EPI_ISL_1264736, EPI_ISL_1264737, EPI_ISL_1264738, EPI_ISL_1264739, EPI_ISL_1264740, EPI_ISL_1264741, EPI_ISL_1264742, EPI_ISL_1264743, EPI_ISL_1264744, EPI_ISL_1264745, EPI_ISL_1264746,
EPI_ISL_1264747, EPI_ISL_1264748, EPI_ISL_1264749, EPI_ISL_1264750, EPI_ISL_1264751, EPI_ISL_1264752, EPI_ISL_1264753, EPI_ISL_1264754, EPI_ISL_1264755, EPI_ISL_1264756, EPI_ISL_1264757, EPI_ISL_1264758, EPI_ISL_1264759, EPI_ISL_1264760, EPI_ISL_1264761, EPI_ISL_1264763, EPI_ISL_1264764, EPI_ISL_1264765, EPI_ISL_1264766, EPI_ISL_1264767, EPI_ISL_1264768, EPI_ISL_1264769,
EPI_ISL_1264770, EPI_ISL_1264771, EPI_ISL_1264772, EPI_ISL_1264773, EPI_ISL_1264774, EPI_ISL_1264775, EPI_ISL_1264776, EPI_ISL_1264777, EPI_ISL_1264778, EPI_ISL_1264779, EPI_ISL_1264780, EPI_ISL_1264781, EPI_ISL_1264782, EPI_ISL_1264783, EPI_ISL_1264784, EPI_ISL_1264785, EPI_ISL_1264786, EPI_ISL_1264787, EPI_ISL_1264788, EPI_ISL_1264789, EPI_ISL_1264790, EPI_ISL_1264791,
EPI_ISL_1264792, EPI_ISL_1264793, EPI_ISL_1264794, EPI_ISL_1264795, EPI_ISL_1264796, EPI_ISL_1264797, EPI_ISL_1264798, EPI_ISL_1264799, EPI_ISL_1264800, EPI_ISL_1264801, EPI_ISL_1264802, EPI_ISL_1264803, EPI_ISL_1264804, EPI_ISL_1264805, EPI_ISL_1264806, EPI_ISL_1264807, EPI_ISL_1264808, EPI_ISL_1264809, EPI_ISL_1264810, EPI_ISL_1264811, EPI_ISL_1264812, EPI_ISL_1264813,
EPI_ISL_1264814, EPI_ISL_1264815, EPI_ISL_1264816, EPI_ISL_1264817, EPI_ISL_1264818, EPI_ISL_1264819, EPI_ISL_1264820, EPI_ISL_1264821, EPI_ISL_1264822, EPI_ISL_1264823, EPI_ISL_1264824, EPI_ISL_1264825, EPI_ISL_1264826, EPI_ISL_1264827, EPI_ISL_1264828, EPI_ISL_1264829, EPI_ISL_1264830, EPI_ISL_1264831, EPI_ISL_1264832, EPI_ISL_1264833, EPI_ISL_1264834, EPI_ISL_1264835,
EPI_ISL_1264836, EPI_ISL_1264837, EPI_ISL_1264838, EPI_ISL_1264839, EPI_ISL_1264840, EPI_ISL_1264841, EPI_ISL_1264842, EPI_ISL_1264843, EPI_ISL_1264844, EPI_ISL_1264845, EPI_ISL_1264846, EPI_ISL_1264847, EPI_ISL_1264848, EPI_ISL_1264849, EPI_ISL_1264850, EPI_ISL_1264851, EPI_ISL_1264852, EPI_ISL_1264853, EPI_ISL_1264854, EPI_ISL_1264855, EPI_ISL_1264856, EPI_ISL_1264857,
EPI_ISL_1264858, EPI_ISL_1264859, EPI_ISL_1264860, EPI_ISL_1264861, EPI_ISL_1264862, EPI_ISL_1264863, EPI_ISL_1264864, EPI_ISL_1264865, EPI_ISL_1264866, EPI_ISL_1264867, EPI_ISL_1264868, EPI_ISL_1264869, EPI_ISL_1264870, EPI_ISL_1264871, EPI_ISL_1264872, EPI_ISL_1264873, EPI_ISL_1264874, EPI_ISL_1264875, EPI_ISL_1264876, EPI_ISL_1264877, EPI_ISL_1264878, EPI_ISL_1264879,
EPI_ISL_1264880, EPI_ISL_1264881, EPI_ISL_1264882, EPI_ISL_1264883, EPI_ISL_1264884, EPI_ISL_1264885, EPI_ISL_1264886, EPI_ISL_1264887, EPI_ISL_1264888, EPI_ISL_1264889, EPI_ISL_1264890, EPI_ISL_1264891, EPI_ISL_1264892, EPI_ISL_1264893, EPI_ISL_1264894, EPI_ISL_1264895, EPI_ISL_1264896, EPI_ISL_1264897, EPI_ISL_1264898, EPI_ISL_1264899, EPI_ISL_1264900, EPI_ISL_1264901,
EPI_ISL_1264902, EPI_ISL_1264903, EPI_ISL_1264904, EPI_ISL_1264905, EPI_ISL_1274586, EPI_ISL_1276823, EPI_ISL_1277221, EPI_ISL_1277226, EPI_ISL_1277227, EPI_ISL_1277248, EPI_ISL_1277268, EPI_ISL_1277605, EPI_ISL_1277748, EPI_ISL_1277755, EPI_ISL_1277763, EPI_ISL_1277781, EPI_ISL_1277811, EPI_ISL_1277829, EPI_ISL_1278006, EPI_ISL_1294617, EPI_ISL_1294619,
EPI_ISL_1294650, EPI_ISL_1294656, EPI_ISL_1294970, EPI_ISL_1295073, EPI_ISL_1295074, EPI_ISL_1295075, EPI_ISL_1295076, EPI_ISL_1295077, EPI_ISL_1295078, EPI_ISL_1295080, EPI_ISL_1295081, EPI_ISL_1295082, EPI_ISL_1295083, EPI_ISL_1295084, EPI_ISL_1295085, EPI_ISL_1295086, EPI_ISL_1295087, EPI_ISL_1295088, EPI_ISL_1295089, EPI_ISL_1295090, EPI_ISL_1295091, EPI_ISL_1295092,
EPI_ISL_1295093, EPI_ISL_1295094, EPI_ISL_1295095, EPI_ISL_1295096, EPI_ISL_1295097, EPI_ISL_1295098, EPI_ISL_1295099, EPI_ISL_1295100, EPI_ISL_1295101, EPI_ISL_1295102, EPI_ISL_1295103, EPI_ISL_1295104, EPI_ISL_1295105, EPI_ISL_1295106, EPI_ISL_1295107, EPI_ISL_1295108, EPI_ISL_1295109, EPI_ISL_1295110, EPI_ISL_1295111, EPI_ISL_1295112, EPI_ISL_1295113, EPI_ISL_1295114,
EPI_ISL_1295115, EPI_ISL_1295116, EPI_ISL_1295117, EPI_ISL_1295118, EPI_ISL_1295119, EPI_ISL_1295120, EPI_ISL_1295121, EPI_ISL_1295122, EPI_ISL_1295123, EPI_ISL_1295124, EPI_ISL_1295125, EPI_ISL_1295126, EPI_ISL_1295127, EPI_ISL_1295128, EPI_ISL_1295129, EPI_ISL_1295130, EPI_ISL_1295131, EPI_ISL_1295132, EPI_ISL_1295133, EPI_ISL_1295134, EPI_ISL_1295135, EPI_ISL_1295136,
EPI_ISL_1295137, EPI_ISL_1295138, EPI_ISL_1295139, EPI_ISL_1295140, EPI_ISL_1295141, EPI_ISL_1295142, EPI_ISL_1295143, EPI_ISL_1295144, EPI_ISL_1295145, EPI_ISL_1295146, EPI_ISL_1295147, EPI_ISL_1295148, EPI_ISL_1295149, EPI_ISL_1295150, EPI_ISL_1295151, EPI_ISL_1295152, EPI_ISL_1295153, EPI_ISL_1295154, EPI_ISL_1295155, EPI_ISL_1295156, EPI_ISL_1295157, EPI_ISL_1295158,
EPI_ISL_1295159, EPI_ISL_1295160, EPI_ISL_1295161, EPI_ISL_1295162, EPI_ISL_1295163, EPI_ISL_1295164, EPI_ISL_1295165, EPI_ISL_1295166, EPI_ISL_1316480, EPI_ISL_1316571, EPI_ISL_1316643, EPI_ISL_1316722, EPI_ISL_1328216, EPI_ISL_1332845, EPI_ISL_1332846, EPI_ISL_1332848, EPI_ISL_1332849, EPI_ISL_1332851, EPI_ISL_1332852, EPI_ISL_1332854, EPI_ISL_1332855, EPI_ISL_1332856,
EPI_ISL_1332857, EPI_ISL_1332858, EPI_ISL_1332859, EPI_ISL_1332860, EPI_ISL_1332861, EPI_ISL_1332862, EPI_ISL_1332863, EPI_ISL_1332864, EPI_ISL_1332865, EPI_ISL_1332867, EPI_ISL_1332868, EPI_ISL_1332869, EPI_ISL_1332870, EPI_ISL_1332871, EPI_ISL_1332872, EPI_ISL_1332873, EPI_ISL_1332874, EPI_ISL_1332877, EPI_ISL_1332878, EPI_ISL_1332879, EPI_ISL_1332880, EPI_ISL_1332882,
EPI_ISL_1332884, EPI_ISL_1332886, EPI_ISL_1332888, EPI_ISL_1332889, EPI_ISL_1332891, EPI_ISL_1332892, EPI_ISL_1332894, EPI_ISL_1332895, EPI_ISL_1332897, EPI_ISL_1332899, EPI_ISL_1332900, EPI_ISL_1332901, EPI_ISL_1332903, EPI_ISL_1332904, EPI_ISL_1332905, EPI_ISL_1332906, EPI_ISL_1332908, EPI_ISL_1332911, EPI_ISL_1332916, EPI_ISL_1332917, EPI_ISL_1332918, EPI_ISL_1332956,
EPI_ISL_1332958, EPI_ISL_1332960, EPI_ISL_1332963, EPI_ISL_1332964, EPI_ISL_1332966, EPI_ISL_1332971, EPI_ISL_1332972, EPI_ISL_1332973, EPI_ISL_1332975, EPI_ISL_1332977, EPI_ISL_1332980, EPI_ISL_1332982, EPI_ISL_1332984, EPI_ISL_1332985, EPI_ISL_1332987, EPI_ISL_1332990, EPI_ISL_1332991, EPI_ISL_1332992, EPI_ISL_1332994, EPI_ISL_1332995, EPI_ISL_1332997, EPI_ISL_1332999,
EPI_ISL_1333000, EPI_ISL_1333001, EPI_ISL_1333012, EPI_ISL_1333013, EPI_ISL_1333017, EPI_ISL_1333019, EPI_ISL_1333020, EPI_ISL_1333021, EPI_ISL_1333024, EPI_ISL_1333025, EPI_ISL_1333027, EPI_ISL_1333028, EPI_ISL_1333033, EPI_ISL_1333036, EPI_ISL_1333041, EPI_ISL_1333043, EPI_ISL_1333047, EPI_ISL_1333048, EPI_ISL_1333050, EPI_ISL_1333051, EPI_ISL_1333052, EPI_ISL_1333054,
EPI_ISL_1333055, EPI_ISL_1333056, EPI_ISL_1333057, EPI_ISL_1333059, EPI_ISL_1333062, EPI_ISL_1333066, EPI_ISL_1333068, EPI_ISL_1333071, EPI_ISL_1333072, EPI_ISL_1333076, EPI_ISL_1333080, EPI_ISL_1333081, EPI_ISL_1333082, EPI_ISL_1333085, EPI_ISL_1333086, EPI_ISL_1333087, EPI_ISL_1333088, EPI_ISL_1333090, EPI_ISL_1333094, EPI_ISL_1333097, EPI_ISL_1333098, EPI_ISL_1333100,
EPI_ISL_1333105, EPI_ISL_1333106, EPI_ISL_1333107, EPI_ISL_1333108, EPI_ISL_1333110, EPI_ISL_1333111, EPI_ISL_1333113, EPI_ISL_1333115, EPI_ISL_1333117, EPI_ISL_1333118, EPI_ISL_1333121, EPI_ISL_1333122, EPI_ISL_1333124, EPI_ISL_1333125, EPI_ISL_1333126, EPI_ISL_1333128, EPI_ISL_1333130, EPI_ISL_1333132, EPI_ISL_1333135, EPI_ISL_1333136, EPI_ISL_1333138, EPI_ISL_1333142,
EPI_ISL_1333145, EPI_ISL_1333146, EPI_ISL_1333147, EPI_ISL_1333149, EPI_ISL_1333153, EPI_ISL_1333160, EPI_ISL_1333162, EPI_ISL_1333163, EPI_ISL_1333164, EPI_ISL_1333165, EPI_ISL_1333166, EPI_ISL_1333169, EPI_ISL_1333170, EPI_ISL_1333173, EPI_ISL_1333175, EPI_ISL_1333336, EPI_ISL_1333341, EPI_ISL_1333343, EPI_ISL_1333350, EPI_ISL_1333357, EPI_ISL_1333359, EPI_ISL_1333361,
EPI_ISL_1333362, EPI_ISL_1333363, EPI_ISL_1333366, EPI_ISL_1333368, EPI_ISL_1333369, EPI_ISL_1333371, EPI_ISL_1333378, EPI_ISL_1333379, EPI_ISL_1333381, EPI_ISL_1333385, EPI_ISL_1333391, EPI_ISL_1333399, EPI_ISL_1333401, EPI_ISL_1333402, EPI_ISL_1333405, EPI_ISL_1333406, EPI_ISL_1333410, EPI_ISL_1333416, EPI_ISL_1333420, EPI_ISL_1333421, EPI_ISL_1333423, EPI_ISL_1333427,
EPI_ISL_1333435, EPI_ISL_1333436, EPI_ISL_1333438, EPI_ISL_1333439, EPI_ISL_1333443, EPI_ISL_1333447, EPI_ISL_1333448, EPI_ISL_1333449, EPI_ISL_1333450, EPI_ISL_1333451, EPI_ISL_1333453, EPI_ISL_1333457, EPI_ISL_1333458, EPI_ISL_1333466, EPI_ISL_1333474, EPI_ISL_1333514, EPI_ISL_1333827, EPI_ISL_1333990, EPI_ISL_1365313, EPI_ISL_1365314, EPI_ISL_1365315, EPI_ISL_1365316,
EPI_ISL_1365317, EPI_ISL_1365318, EPI_ISL_1365319, EPI_ISL_1365320, EPI_ISL_1365321, EPI_ISL_1365322, EPI_ISL_1365324, EPI_ISL_1365325, EPI_ISL_1365326, EPI_ISL_1365327, EPI_ISL_1365328, EPI_ISL_1365329, EPI_ISL_1365330, EPI_ISL_1365331, EPI_ISL_1365332, EPI_ISL_1365333, EPI_ISL_1365334, EPI_ISL_1365335, EPI_ISL_1365336, EPI_ISL_1365337, EPI_ISL_1365338, EPI_ISL_1365339,
EPI_ISL_1365340, EPI_ISL_1365341, EPI_ISL_1365342, EPI_ISL_1365343, EPI_ISL_1365344, EPI_ISL_1365345, EPI_ISL_1365346, EPI_ISL_1365347, EPI_ISL_1365348, EPI_ISL_1365349, EPI_ISL_1365350, EPI_ISL_1365351, EPI_ISL_1365352, EPI_ISL_1365353, EPI_ISL_1365354, EPI_ISL_1365355, EPI_ISL_1365356, EPI_ISL_1365358, EPI_ISL_1365359, EPI_ISL_1365360, EPI_ISL_1378428, EPI_ISL_1378429,
EPI_ISL_1378430, EPI_ISL_1378431, EPI_ISL_1378432, EPI_ISL_1378433, EPI_ISL_1378434, EPI_ISL_1378435, EPI_ISL_1378436, EPI_ISL_1378437, EPI_ISL_1378438, EPI_ISL_1378439, EPI_ISL_1378440, EPI_ISL_1378441, EPI_ISL_1378442, EPI_ISL_1378443, EPI_ISL_1378444, EPI_ISL_1378445, EPI_ISL_1378446, EPI_ISL_1378447, EPI_ISL_1449650, EPI_ISL_1449662, EPI_ISL_1449682, EPI_ISL_1449690,
EPI_ISL_1449692, EPI_ISL_1449693, EPI_ISL_1449695, EPI_ISL_1449697, EPI_ISL_1449698, EPI_ISL_1449702, EPI_ISL_1449705, EPI_ISL_1449709, EPI_ISL_1449716, EPI_ISL_1449721, EPI_ISL_1449722, EPI_ISL_1449725, EPI_ISL_1449730, EPI_ISL_1449733, EPI_ISL_1449737, EPI_ISL_1449741, EPI_ISL_1449749, EPI_ISL_1449750, EPI_ISL_1449751, EPI_ISL_1449769, EPI_ISL_1449782,
EPI_ISL_1449788, EPI_ISL_1449795, EPI_ISL_1449800, EPI_ISL_1449801, EPI_ISL_1449802, EPI_ISL_1449809, EPI_ISL_1449814, EPI_ISL_1449817, EPI_ISL_1449826, EPI_ISL_1449839, EPI_ISL_1449847, EPI_ISL_1449850, EPI_ISL_1449857, EPI_ISL_1449863, EPI_ISL_1449870, EPI_ISL_1449876, EPI_ISL_1449887, EPI_ISL_1449908, EPI_ISL_1449923, EPI_ISL_1449924, EPI_ISL_1449930, EPI_ISL_1449934,
EPI_ISL_1507795, EPI_ISL_1507797, EPI_ISL_1507798, EPI_ISL_1507801, EPI_ISL_1507803, EPI_ISL_1507804, EPI_ISL_1507810, EPI_ISL_1507811, EPI_ISL_1507812, EPI_ISL_1759944, EPI_ISL_1759945, EPI_ISL_1759946, EPI_ISL_1759947, EPI_ISL_1759948, EPI_ISL_1759949, EPI_ISL_1759950, EPI_ISL_1759951, EPI_ISL_1759952, EPI_ISL_1759953, EPI_ISL_1759954, EPI_ISL_1759955, EPI_ISL_1759956,
EPI_ISL_1759957, EPI_ISL_1759958, EPI_ISL_1759959, EPI_ISL_1759960, EPI_ISL_1759961, EPI_ISL_1759962, EPI_ISL_1759963, EPI_ISL_1759964, EPI_ISL_1759965, EPI_ISL_1759966, EPI_ISL_1759967, EPI_ISL_1759968, EPI_ISL_1759969, EPI_ISL_1759970, EPI_ISL_1759971, EPI_ISL_1759972, EPI_ISL_1759973, EPI_ISL_1759974, EPI_ISL_1759975, EPI_ISL_1759976, EPI_ISL_1759977, EPI_ISL_1759978,
EPI_ISL_1759979, EPI_ISL_1759980, EPI_ISL_1759981, EPI_ISL_1759982, EPI_ISL_1759983, EPI_ISL_1759984, EPI_ISL_1759985, EPI_ISL_1759986, EPI_ISL_1759987, EPI_ISL_1759988, EPI_ISL_1759989, EPI_ISL_1759990, EPI_ISL_1759991, EPI_ISL_1759992, EPI_ISL_1759993, EPI_ISL_1759994, EPI_ISL_1759995, EPI_ISL_1759996, EPI_ISL_1759997, EPI_ISL_1759998, EPI_ISL_1759999, EPI_ISL_1760000,
EPI_ISL_1760001, EPI_ISL_1760002

| | | | |
|---|---|---|---|
| see above | Lighthouse Lab in Glasgow | Wellcome Sanger Institute for the COVID-19 Genomics UK (COG-UK) Consortium | Anna Dominiczak and Alex Alderton; Carol Clugston; Cordelia Langford; David Gray; David K. Jackson; Dominic Kwiatkowski; Ewan Harrison; Harper VanSteenhouse; Ian Johnston; Jeffrey Barrett; John Sillitoe on behalf of the Wellcome Sanger Institute COVID-19 Surveillance Team; Roberto Amato; Sonia Goncalves; Yumi Kasai |

EPI_ISL_1205708, EPI_ISL_1205713, EPI_ISL_1205716, EPI_ISL_1205717, EPI_ISL_1205718, EPI_ISL_1205722, EPI_ISL_1205729, EPI_ISL_1205730, EPI_ISL_1205732, EPI_ISL_1205734, EPI_ISL_1205739, EPI_ISL_1205740, EPI_ISL_1205744, EPI_ISL_1205749, EPI_ISL_1205750, EPI_ISL_1205769, EPI_ISL_1205772, EPI_ISL_1205774, EPI_ISL_1205779, EPI_ISL_1205781, EPI_ISL_1205783, EPI_ISL_1205784,
EPI_ISL_1205786, EPI_ISL_1205787, EPI_ISL_1205790, EPI_ISL_1205793, EPI_ISL_1205795, EPI_ISL_1205807, EPI_ISL_1205810, EPI_ISL_1205813, EPI_ISL_1205818, EPI_ISL_1205820, EPI_ISL_1205821, EPI_ISL_1205823, EPI_ISL_1205827, EPI_ISL_1205829, EPI_ISL_1205835, EPI_ISL_1205836, EPI_ISL_1205837, EPI_ISL_1205841, EPI_ISL_1205849, EPI_ISL_1205851, EPI_ISL_1205863, EPI_ISL_1205866,
EPI_ISL_1205867, EPI_ISL_1205870, EPI_ISL_1205872, EPI_ISL_1205877, EPI_ISL_1205880, EPI_ISL_1205883, EPI_ISL_1205884, EPI_ISL_1205891, EPI_ISL_1205894, EPI_ISL_1205899, EPI_ISL_1205903, EPI_ISL_1205908, EPI_ISL_1205909, EPI_ISL_1205912, EPI_ISL_1205915, EPI_ISL_1205917, EPI_ISL_1205925, EPI_ISL_1205926, EPI_ISL_1205928, EPI_ISL_1205929, EPI_ISL_1205930, EPI_ISL_1205931,
EPI_ISL_1205933, EPI_ISL_1205936, EPI_ISL_1205937, EPI_ISL_1205942, EPI_ISL_1205943, EPI_ISL_1205945, EPI_ISL_1205947, EPI_ISL_1205950, EPI_ISL_1205952, EPI_ISL_1205959, EPI_ISL_1205966, EPI_ISL_1205968, EPI_ISL_1205971, EPI_ISL_1205972, EPI_ISL_1205973, EPI_ISL_1205974, EPI_ISL_1205975, EPI_ISL_1205976, EPI_ISL_1205977, EPI_ISL_1205980, EPI_ISL_1205981, EPI_ISL_1205983,
EPI_ISL_1205984, EPI_ISL_1205985, EPI_ISL_1205993, EPI_ISL_1205994, EPI_ISL_1205995, EPI_ISL_1205998, EPI_ISL_1206004, EPI_ISL_1206007, EPI_ISL_1206009, EPI_ISL_1206012, EPI_ISL_1206013, EPI_ISL_1206016, EPI_ISL_1206018, EPI_ISL_1206019, EPI_ISL_1206029, EPI_ISL_1206033, EPI_ISL_1206036, EPI_ISL_1207165, EPI_ISL_1207179, EPI_ISL_1207186, EPI_ISL_1207188, EPI_ISL_1207213,
EPI_ISL_1207220, EPI_ISL_1207224, EPI_ISL_1207226, EPI_ISL_1207254, EPI_ISL_1207262, EPI_ISL_1207264, EPI_ISL_1207268, EPI_ISL_1207272, EPI_ISL_1207274, EPI_ISL_1207282, EPI_ISL_1207294, EPI_ISL_1207302, EPI_ISL_1207303, EPI_ISL_1207310, EPI_ISL_1207313, EPI_ISL_1207333, EPI_ISL_1207335, EPI_ISL_1207340, EPI_ISL_1207344, EPI_ISL_1207361, EPI_ISL_1207402, EPI_ISL_1207429,
EPI_ISL_1207432, EPI_ISL_1207433, EPI_ISL_1207447, EPI_ISL_1207449, EPI_ISL_1207452, EPI_ISL_1223654, EPI_ISL_1223655, EPI_ISL_1223656, EPI_ISL_1223659, EPI_ISL_1223661, EPI_ISL_1223665, EPI_ISL_1223667, EPI_ISL_1223671, EPI_ISL_1223673, EPI_ISL_1223677, EPI_ISL_1223679, EPI_ISL_1223683, EPI_ISL_1223684, EPI_ISL_1223685, EPI_ISL_1223688, EPI_ISL_1223691, EPI_ISL_1223696,
EPI_ISL_1223697, EPI_ISL_1223703, EPI_ISL_1223705, EPI_ISL_1223710, EPI_ISL_1223713, EPI_ISL_1223714, EPI_ISL_1223715, EPI_ISL_1223716, EPI_ISL_1223720, EPI_ISL_1223723, EPI_ISL_1223724, EPI_ISL_1223731, EPI_ISL_1223736, EPI_ISL_1223738, EPI_ISL_1223739, EPI_ISL_1223741, EPI_ISL_1223744, EPI_ISL_1223745, EPI_ISL_1223747, EPI_ISL_1223749, EPI_ISL_1223751, EPI_ISL_1223752,
EPI_ISL_1223753, EPI_ISL_1223755, EPI_ISL_1223756, EPI_ISL_1223760, EPI_ISL_1223766, EPI_ISL_1223772, EPI_ISL_1223774, EPI_ISL_1223775, EPI_ISL_1223776, EPI_ISL_1223782, EPI_ISL_1223785, EPI_ISL_1223788, EPI_ISL_1223789, EPI_ISL_1223790, EPI_ISL_1223793, EPI_ISL_1223795, EPI_ISL_1223797, EPI_ISL_1223801, EPI_ISL_1223804, EPI_ISL_1223807, EPI_ISL_1223809, EPI_ISL_1223810,
EPI_ISL_1223812, EPI_ISL_1223821, EPI_ISL_1223824, EPI_ISL_1223825, EPI_ISL_1223829, EPI_ISL_1223837, EPI_ISL_1223848, EPI_ISL_1223851, EPI_ISL_1223852, EPI_ISL_1223855, EPI_ISL_1223857, EPI_ISL_1223859, EPI_ISL_1223860, EPI_ISL_1223864, EPI_ISL_1223868, EPI_ISL_1223872, EPI_ISL_1223873, EPI_ISL_1223877, EPI_ISL_1223882, EPI_ISL_1223888, EPI_ISL_1223895,
EPI_ISL_1223901, EPI_ISL_1223907, EPI_ISL_1223908, EPI_ISL_1223911, EPI_ISL_1223917, EPI_ISL_1223918, EPI_ISL_1223921, EPI_ISL_1223933, EPI_ISL_1223934, EPI_ISL_1223936, EPI_ISL_1223939, EPI_ISL_1223941, EPI_ISL_1223942, EPI_ISL_1223944, EPI_ISL_1223945, EPI_ISL_1224163, EPI_ISL_1224164, EPI_ISL_1224165, EPI_ISL_1224166, EPI_ISL_1224167, EPI_ISL_1224168, EPI_ISL_1224169,
EPI_ISL_1224170, EPI_ISL_1224171, EPI_ISL_1224172, EPI_ISL_1224173, EPI_ISL_1224174, EPI_ISL_1224175, EPI_ISL_1224176, EPI_ISL_1224177, EPI_ISL_1224178, EPI_ISL_1224179, EPI_ISL_1224180, EPI_ISL_1224181, EPI_ISL_1224182, EPI_ISL_1224183, EPI_ISL_1224184, EPI_ISL_1224185, EPI_ISL_1224186, EPI_ISL_1224187, EPI_ISL_1224188, EPI_ISL_1224189, EPI_ISL_1224190, EPI_ISL_1224191,
EPI_ISL_1224192, EPI_ISL_1224193, EPI_ISL_1224194, EPI_ISL_1224195, EPI_ISL_1224196, EPI_ISL_1224197, EPI_ISL_1224198, EPI_ISL_1224199, EPI_ISL_1224200, EPI_ISL_1224201, EPI_ISL_1224202, EPI_ISL_1224203, EPI_ISL_1224204, EPI_ISL_1224205, EPI_ISL_1224206, EPI_ISL_1224207, EPI_ISL_1224208, EPI_ISL_1224209, EPI_ISL_1224210, EPI_ISL_1224211, EPI_ISL_1224212, EPI_ISL_1224213,

EPI_ISL_1224214, EPI_ISL_1224215, EPI_ISL_1224216, EPI_ISL_1224217, EPI_ISL_1224218, EPI_ISL_1224219, EPI_ISL_1224220, EPI_ISL_1224221, EPI_ISL_1224222, EPI_ISL_1224223, EPI_ISL_1224224, EPI_ISL_1224225, EPI_ISL_1224226, EPI_ISL_1224227, EPI_ISL_1224228, EPI_ISL_1224229, EPI_ISL_1224230, EPI_ISL_1224231, EPI_ISL_1224232, EPI_ISL_1224233, EPI_ISL_1224234, EPI_ISL_1224235,
EPI_ISL_1224236, EPI_ISL_1224237, EPI_ISL_1224238, EPI_ISL_1224239, EPI_ISL_1224240, EPI_ISL_1224241, EPI_ISL_1224242, EPI_ISL_1224243, EPI_ISL_1224244, EPI_ISL_1224245, EPI_ISL_1224246, EPI_ISL_1224247, EPI_ISL_1224248, EPI_ISL_1224249, EPI_ISL_1224250, EPI_ISL_1224251, EPI_ISL_1224252, EPI_ISL_1224253, EPI_ISL_1224254, EPI_ISL_1224255, EPI_ISL_1224256, EPI_ISL_1224257,
EPI_ISL_1224258, EPI_ISL_1224259, EPI_ISL_1224260, EPI_ISL_1224261, EPI_ISL_1224262, EPI_ISL_1224263, EPI_ISL_1224264, EPI_ISL_1224265, EPI_ISL_1224266, EPI_ISL_1224267, EPI_ISL_1224268, EPI_ISL_1224269, EPI_ISL_1224270, EPI_ISL_1224271, EPI_ISL_1224272, EPI_ISL_1224273, EPI_ISL_1224274, EPI_ISL_1224275, EPI_ISL_1224276, EPI_ISL_1224277, EPI_ISL_1224278, EPI_ISL_1224279,
EPI_ISL_1224280, EPI_ISL_1224281, EPI_ISL_1224282, EPI_ISL_1224283, EPI_ISL_1224284, EPI_ISL_1224285, EPI_ISL_1224286, EPI_ISL_1224287, EPI_ISL_1224288, EPI_ISL_1224289, EPI_ISL_1224290, EPI_ISL_1224291, EPI_ISL_1224292, EPI_ISL_1224293, EPI_ISL_1224294, EPI_ISL_1224295, EPI_ISL_1224296, EPI_ISL_1224297, EPI_ISL_1224298, EPI_ISL_1224299, EPI_ISL_1224300, EPI_ISL_1224301,
EPI_ISL_1224302, EPI_ISL_1224303, EPI_ISL_1224304, EPI_ISL_1224305, EPI_ISL_1224306, EPI_ISL_1224307, EPI_ISL_1224308, EPI_ISL_1224309, EPI_ISL_1224310, EPI_ISL_1224311, EPI_ISL_1224312, EPI_ISL_1224313, EPI_ISL_1224314, EPI_ISL_1224315, EPI_ISL_1224316, EPI_ISL_1224317, EPI_ISL_1224318, EPI_ISL_1224319, EPI_ISL_1224320, EPI_ISL_1224321, EPI_ISL_1224322, EPI_ISL_1224323,
EPI_ISL_1224324, EPI_ISL_1224325, EPI_ISL_1224326, EPI_ISL_1224327, EPI_ISL_1224328, EPI_ISL_1224330, EPI_ISL_1224331, EPI_ISL_1224332, EPI_ISL_1224333, EPI_ISL_1224334, EPI_ISL_1224335, EPI_ISL_1224336, EPI_ISL_1224337, EPI_ISL_1224338, EPI_ISL_1224339, EPI_ISL_1224340, EPI_ISL_1224341, EPI_ISL_1224342, EPI_ISL_1224343, EPI_ISL_1224344, EPI_ISL_1224345, EPI_ISL_1224346,
EPI_ISL_1224347, EPI_ISL_1224348, EPI_ISL_1224349, EPI_ISL_1224350, EPI_ISL_1224351, EPI_ISL_1224352, EPI_ISL_1224353, EPI_ISL_1224354, EPI_ISL_1224355, EPI_ISL_1224356, EPI_ISL_1224357, EPI_ISL_1224358, EPI_ISL_1224359, EPI_ISL_1224360, EPI_ISL_1224361, EPI_ISL_1224362, EPI_ISL_1224363, EPI_ISL_1224364, EPI_ISL_1224365, EPI_ISL_1224366, EPI_ISL_1224367, EPI_ISL_1224368,
EPI_ISL_1224369, EPI_ISL_1224370, EPI_ISL_1224371, EPI_ISL_1224372, EPI_ISL_1224373, EPI_ISL_1224374, EPI_ISL_1224375, EPI_ISL_1224376, EPI_ISL_1224377, EPI_ISL_1224378, EPI_ISL_1224379, EPI_ISL_1224380, EPI_ISL_1224381, EPI_ISL_1224382, EPI_ISL_1224383, EPI_ISL_1224384, EPI_ISL_1224385, EPI_ISL_1224386, EPI_ISL_1224387, EPI_ISL_1224388, EPI_ISL_1224389, EPI_ISL_1224390,
EPI_ISL_1224391, EPI_ISL_1224392, EPI_ISL_1224393, EPI_ISL_1224394, EPI_ISL_1224396, EPI_ISL_1224397, EPI_ISL_1224398, EPI_ISL_1224399, EPI_ISL_1224400, EPI_ISL_1224401, EPI_ISL_1224402, EPI_ISL_1224403, EPI_ISL_1224404, EPI_ISL_1224405, EPI_ISL_1224406, EPI_ISL_1224407, EPI_ISL_1224408, EPI_ISL_1224409, EPI_ISL_1224410, EPI_ISL_1224411, EPI_ISL_1224412, EPI_ISL_1224413,
EPI_ISL_1224414, EPI_ISL_1224415, EPI_ISL_1224416, EPI_ISL_1224417, EPI_ISL_1224418, EPI_ISL_1224419, EPI_ISL_1224420, EPI_ISL_1224421, EPI_ISL_1224422, EPI_ISL_1224423, EPI_ISL_1224424, EPI_ISL_1224425, EPI_ISL_1224426, EPI_ISL_1224427, EPI_ISL_1224428, EPI_ISL_1224429, EPI_ISL_1224430, EPI_ISL_1224431, EPI_ISL_1224432, EPI_ISL_1224433, EPI_ISL_1224434, EPI_ISL_1224435,
EPI_ISL_1224436, EPI_ISL_1224437, EPI_ISL_1224438, EPI_ISL_1224439, EPI_ISL_1224440, EPI_ISL_1224441, EPI_ISL_1224442, EPI_ISL_1224443, EPI_ISL_1224444, EPI_ISL_1224445, EPI_ISL_1224446, EPI_ISL_1224447, EPI_ISL_1224448, EPI_ISL_1224449, EPI_ISL_1224450, EPI_ISL_1224451, EPI_ISL_1224452, EPI_ISL_1224453, EPI_ISL_1224454, EPI_ISL_1224455, EPI_ISL_1224457, EPI_ISL_1224458,
EPI_ISL_1224459, EPI_ISL_1224460, EPI_ISL_1224461, EPI_ISL_1224462, EPI_ISL_1224463, EPI_ISL_1224464, EPI_ISL_1224465, EPI_ISL_1224466, EPI_ISL_1224467, EPI_ISL_1224468, EPI_ISL_1224469, EPI_ISL_1224470, EPI_ISL_1224471, EPI_ISL_1224472, EPI_ISL_1224473, EPI_ISL_1224474, EPI_ISL_1224475, EPI_ISL_1224476, EPI_ISL_1224477, EPI_ISL_1224478, EPI_ISL_1224479, EPI_ISL_1224480,
EPI_ISL_1224481, EPI_ISL_1224482, EPI_ISL_1224483, EPI_ISL_1224484, EPI_ISL_1224485, EPI_ISL_1224486, EPI_ISL_1224487, EPI_ISL_1224488, EPI_ISL_1224489, EPI_ISL_1224490, EPI_ISL_1224491, EPI_ISL_1224492, EPI_ISL_1224493, EPI_ISL_1224494, EPI_ISL_1224495, EPI_ISL_1224496, EPI_ISL_1224497, EPI_ISL_1224498, EPI_ISL_1224499, EPI_ISL_1242820, EPI_ISL_1242821, EPI_ISL_1242822,
EPI_ISL_1242823, EPI_ISL_1242824, EPI_ISL_1242825, EPI_ISL_1242826, EPI_ISL_1242827, EPI_ISL_1242829, EPI_ISL_1242830, EPI_ISL_1242833, EPI_ISL_1242835, EPI_ISL_1242836, EPI_ISL_1242837, EPI_ISL_1242838, EPI_ISL_1242841, EPI_ISL_1242842, EPI_ISL_1242843, EPI_ISL_1242844, EPI_ISL_1242847, EPI_ISL_1242851, EPI_ISL_1242852, EPI_ISL_1242853, EPI_ISL_1242855, EPI_ISL_1242856,
EPI_ISL_1242858, EPI_ISL_1242860, EPI_ISL_1242862, EPI_ISL_1242863, EPI_ISL_1242864, EPI_ISL_1242865, EPI_ISL_1242867, EPI_ISL_1242871, EPI_ISL_1242872, EPI_ISL_1242880, EPI_ISL_1242881, EPI_ISL_1242883, EPI_ISL_1242884, EPI_ISL_1242885, EPI_ISL_1242886, EPI_ISL_1242887, EPI_ISL_1242888, EPI_ISL_1242889, EPI_ISL_1242890, EPI_ISL_1242893, EPI_ISL_1242894, EPI_ISL_1242896,
EPI_ISL_1242897, EPI_ISL_1242898, EPI_ISL_1242902, EPI_ISL_1242905, EPI_ISL_1242906, EPI_ISL_1242907, EPI_ISL_1242908, EPI_ISL_1242909, EPI_ISL_1242910, EPI_ISL_1242912, EPI_ISL_1242914, EPI_ISL_1242920, EPI_ISL_1242924, EPI_ISL_1242925, EPI_ISL_1242926, EPI_ISL_1242927, EPI_ISL_1242928, EPI_ISL_1242929, EPI_ISL_1242930, EPI_ISL_1242932, EPI_ISL_1242934, EPI_ISL_1242935,
EPI_ISL_1242938, EPI_ISL_1242939, EPI_ISL_1242943, EPI_ISL_1242945, EPI_ISL_1242946, EPI_ISL_1242947, EPI_ISL_1242948, EPI_ISL_1242951, EPI_ISL_1242952, EPI_ISL_1242954, EPI_ISL_1242955, EPI_ISL_1242956, EPI_ISL_1242957, EPI_ISL_1242958, EPI_ISL_1242959, EPI_ISL_1242960, EPI_ISL_1242961, EPI_ISL_1242964, EPI_ISL_1242966, EPI_ISL_1242968, EPI_ISL_1242969, EPI_ISL_1242970,
EPI_ISL_1242971, EPI_ISL_1242974, EPI_ISL_1242976, EPI_ISL_1242977, EPI_ISL_1242979, EPI_ISL_1242980, EPI_ISL_1242981, EPI_ISL_1242982, EPI_ISL_1242983, EPI_ISL_1242986, EPI_ISL_1242987, EPI_ISL_1242989, EPI_ISL_1242995, EPI_ISL_1242996, EPI_ISL_1242998, EPI_ISL_1242999, EPI_ISL_1243000, EPI_ISL_1243001, EPI_ISL_1243002, EPI_ISL_1243003, EPI_ISL_1243006, EPI_ISL_1243008,
EPI_ISL_1243009, EPI_ISL_1243010, EPI_ISL_1243011, EPI_ISL_1243012, EPI_ISL_1243013, EPI_ISL_1243014, EPI_ISL_1243016, EPI_ISL_1243019, EPI_ISL_1243020, EPI_ISL_1243022, EPI_ISL_1243023, EPI_ISL_1243024, EPI_ISL_1243025, EPI_ISL_1243026, EPI_ISL_1243028, EPI_ISL_1243031, EPI_ISL_1243034, EPI_ISL_1243036, EPI_ISL_1243037, EPI_ISL_1243038, EPI_ISL_1243039, EPI_ISL_1243041,
EPI_ISL_1243042, EPI_ISL_1243045, EPI_ISL_1243046, EPI_ISL_1243049, EPI_ISL_1243050, EPI_ISL_1243051, EPI_ISL_1243052, EPI_ISL_1243056, EPI_ISL_1243057, EPI_ISL_1243058, EPI_ISL_1243059, EPI_ISL_1243061, EPI_ISL_1243062, EPI_ISL_1243063, EPI_ISL_1243064, EPI_ISL_1243066, EPI_ISL_1243067, EPI_ISL_1243071, EPI_ISL_1243072, EPI_ISL_1243073, EPI_ISL_1243074, EPI_ISL_1243079,
EPI_ISL_1243080, EPI_ISL_1243081, EPI_ISL_1243083, EPI_ISL_1243085, EPI_ISL_1243086, EPI_ISL_1243088, EPI_ISL_1243089, EPI_ISL_1243090, EPI_ISL_1243091, EPI_ISL_1243096, EPI_ISL_1243097, EPI_ISL_1243099, EPI_ISL_1243100, EPI_ISL_1243103, EPI_ISL_1243104, EPI_ISL_1243106, EPI_ISL_1243107, EPI_ISL_1243108, EPI_ISL_1243109, EPI_ISL_1243110, EPI_ISL_1243112, EPI_ISL_1243113,
EPI_ISL_1243114, EPI_ISL_1243115, EPI_ISL_1243116, EPI_ISL_1243118, EPI_ISL_1243120, EPI_ISL_1243121, EPI_ISL_1243122, EPI_ISL_1243123, EPI_ISL_1243124, EPI_ISL_1243125, EPI_ISL_1243129, EPI_ISL_1243130, EPI_ISL_1243131, EPI_ISL_1243132, EPI_ISL_1243133, EPI_ISL_1243135, EPI_ISL_1243136, EPI_ISL_1243137, EPI_ISL_1243139, EPI_ISL_1243140, EPI_ISL_1243141, EPI_ISL_1243142,
EPI_ISL_1243143, EPI_ISL_1243937, EPI_ISL_1243938, EPI_ISL_1243939, EPI_ISL_1243941, EPI_ISL_1243942, EPI_ISL_1243943, EPI_ISL_1243946, EPI_ISL_1243947, EPI_ISL_1243949, EPI_ISL_1243950, EPI_ISL_1243951, EPI_ISL_1243953, EPI_ISL_1243954, EPI_ISL_1243955, EPI_ISL_1243958, EPI_ISL_1243960, EPI_ISL_1243963, EPI_ISL_1243964, EPI_ISL_1243965, EPI_ISL_1243967, EPI_ISL_1243969,
EPI_ISL_1243971, EPI_ISL_1243974, EPI_ISL_1243975, EPI_ISL_1243976, EPI_ISL_1243978, EPI_ISL_1243980, EPI_ISL_1243981, EPI_ISL_1243982, EPI_ISL_1243983, EPI_ISL_1243985, EPI_ISL_1243986, EPI_ISL_1243988, EPI_ISL_1243990, EPI_ISL_1243994, EPI_ISL_1243996, EPI_ISL_1243997, EPI_ISL_1243999, EPI_ISL_1244006, EPI_ISL_1244007, EPI_ISL_1244008, EPI_ISL_1244010, EPI_ISL_1244011,
EPI_ISL_1244012, EPI_ISL_1244016, EPI_ISL_1244017, EPI_ISL_1244018, EPI_ISL_1244020, EPI_ISL_1244021, EPI_ISL_1244022, EPI_ISL_1244024, EPI_ISL_1244026, EPI_ISL_1244027, EPI_ISL_1244029, EPI_ISL_1244030, EPI_ISL_1244033, EPI_ISL_1244034, EPI_ISL_1244035, EPI_ISL_1244036, EPI_ISL_1244040, EPI_ISL_1244041, EPI_ISL_1244042, EPI_ISL_1244044, EPI_ISL_1244045, EPI_ISL_1244050,
EPI_ISL_1244053, EPI_ISL_1244054, EPI_ISL_1244055, EPI_ISL_1244056, EPI_ISL_1244059, EPI_ISL_1244060, EPI_ISL_1244063, EPI_ISL_1244066, EPI_ISL_1244067, EPI_ISL_1244072, EPI_ISL_1244075, EPI_ISL_1244076, EPI_ISL_1244077, EPI_ISL_1244081, EPI_ISL_1244082, EPI_ISL_1244083, EPI_ISL_1244084, EPI_ISL_1244085, EPI_ISL_1244087, EPI_ISL_1244093, EPI_ISL_1244094, EPI_ISL_1244096,
EPI_ISL_1244102, EPI_ISL_1244103, EPI_ISL_1244105, EPI_ISL_1244107, EPI_ISL_1244109, EPI_ISL_1244110, EPI_ISL_1244111, EPI_ISL_1244113, EPI_ISL_1244115, EPI_ISL_1244117, EPI_ISL_1244118, EPI_ISL_1244119, EPI_ISL_1244121, EPI_ISL_1244122, EPI_ISL_1244126, EPI_ISL_1244127, EPI_ISL_1244129, EPI_ISL_1244130, EPI_ISL_1244132, EPI_ISL_1244133, EPI_ISL_1244135, EPI_ISL_1244136,
EPI_ISL_1244137, EPI_ISL_1244139, EPI_ISL_1244140, EPI_ISL_1244141, EPI_ISL_1244143, EPI_ISL_1244145, EPI_ISL_1244146, EPI_ISL_1244148, EPI_ISL_1244149, EPI_ISL_1244150, EPI_ISL_1244151, EPI_ISL_1244152, EPI_ISL_1244153, EPI_ISL_1244154, EPI_ISL_1244155, EPI_ISL_1244156, EPI_ISL_1244158, EPI_ISL_1244159, EPI_ISL_1244160, EPI_ISL_1244161, EPI_ISL_1244162, EPI_ISL_1244166,
EPI_ISL_1244167, EPI_ISL_1244168, EPI_ISL_1244169, EPI_ISL_1244170, EPI_ISL_1244173, EPI_ISL_1244174, EPI_ISL_1244175, EPI_ISL_1244177, EPI_ISL_1244180, EPI_ISL_1244182, EPI_ISL_1244183, EPI_ISL_1244184, EPI_ISL_1244185, EPI_ISL_1244186, EPI_ISL_1244189, EPI_ISL_1244190, EPI_ISL_1244192, EPI_ISL_1244194, EPI_ISL_1244195, EPI_ISL_1244196, EPI_ISL_1244198, EPI_ISL_1244199,
EPI_ISL_1244200, EPI_ISL_1244202, EPI_ISL_1244204, EPI_ISL_1244206, EPI_ISL_1244207, EPI_ISL_1244209, EPI_ISL_1244210, EPI_ISL_1244211, EPI_ISL_1244212, EPI_ISL_1244216, EPI_ISL_1244218, EPI_ISL_1244219, EPI_ISL_1244220, EPI_ISL_1244223, EPI_ISL_1244227, EPI_ISL_1244228, EPI_ISL_1244233, EPI_ISL_1244234, EPI_ISL_1244235, EPI_ISL_1244236, EPI_ISL_1244237, EPI_ISL_1244239,
EPI_ISL_1244240, EPI_ISL_1244243, EPI_ISL_1244244, EPI_ISL_1244245, EPI_ISL_1244247, EPI_ISL_1244250, EPI_ISL_1244251, EPI_ISL_1244253, EPI_ISL_1244255, EPI_ISL_1244256, EPI_ISL_1244257, EPI_ISL_1244259, EPI_ISL_1244260, EPI_ISL_1244261, EPI_ISL_1244263, EPI_ISL_1244264, EPI_ISL_1244267, EPI_ISL_1244268, EPI_ISL_1244271, EPI_ISL_1244272, EPI_ISL_1244273, EPI_ISL_1244274,
EPI_ISL_1244283, EPI_ISL_1244284, EPI_ISL_1244286, EPI_ISL_1244287, EPI_ISL_1244289, EPI_ISL_1244295, EPI_ISL_1244303, EPI_ISL_1244308, EPI_ISL_1244321, EPI_ISL_1244327, EPI_ISL_1244338, EPI_ISL_1244354, EPI_ISL_1244355, EPI_ISL_1244385, EPI_ISL_1244389, EPI_ISL_1244390, EPI_ISL_1244394, EPI_ISL_1244397, EPI_ISL_1244398, EPI_ISL_1244402, EPI_ISL_1244403, EPI_ISL_1244410,
EPI_ISL_1244411, EPI_ISL_1244425, EPI_ISL_1244432, EPI_ISL_1244446, EPI_ISL_1244447, EPI_ISL_1244459, EPI_ISL_1244460, EPI_ISL_1244466, EPI_ISL_1244476, EPI_ISL_1244485, EPI_ISL_1244496, EPI_ISL_1244497, EPI_ISL_1244506, EPI_ISL_1244513, EPI_ISL_1244528, EPI_ISL_1244541, EPI_ISL_1244557, EPI_ISL_1244562, EPI_ISL_1244574, EPI_ISL_1244576, EPI_ISL_1244582, EPI_ISL_1244596,
EPI_ISL_1244609, EPI_ISL_1245358, EPI_ISL_1245359, EPI_ISL_1245361, EPI_ISL_1245362, EPI_ISL_1245364, EPI_ISL_1245365, EPI_ISL_1245369, EPI_ISL_1245372, EPI_ISL_1245373, EPI_ISL_1245374, EPI_ISL_1245376, EPI_ISL_1245377, EPI_ISL_1245378, EPI_ISL_1245381, EPI_ISL_1245382, EPI_ISL_1245383, EPI_ISL_1245384, EPI_ISL_1245385, EPI_ISL_1245386, EPI_ISL_1245387, EPI_ISL_1245388,
EPI_ISL_1245389, EPI_ISL_1245390, EPI_ISL_1245391, EPI_ISL_1245394, EPI_ISL_1245396, EPI_ISL_1245397, EPI_ISL_1245399, EPI_ISL_1245402, EPI_ISL_1245403, EPI_ISL_1245404, EPI_ISL_1245405, EPI_ISL_1245407, EPI_ISL_1245408, EPI_ISL_1245413, EPI_ISL_1245414, EPI_ISL_1245415, EPI_ISL_1245416, EPI_ISL_1245417, EPI_ISL_1245422, EPI_ISL_1245423, EPI_ISL_1245425, EPI_ISL_1245428,
EPI_ISL_1245429, EPI_ISL_1245431, EPI_ISL_1245434, EPI_ISL_1245435, EPI_ISL_1245437, EPI_ISL_1245440, EPI_ISL_1245441, EPI_ISL_1245442, EPI_ISL_1245444, EPI_ISL_1245446, EPI_ISL_1245448, EPI_ISL_1245449, EPI_ISL_1245452, EPI_ISL_1245454, EPI_ISL_1245455, EPI_ISL_1245456, EPI_ISL_1245458, EPI_ISL_1245459, EPI_ISL_1245460, EPI_ISL_1245461, EPI_ISL_1245463, EPI_ISL_1245464,
EPI_ISL_1245465, EPI_ISL_1245467, EPI_ISL_1245468, EPI_ISL_1245469, EPI_ISL_1245470, EPI_ISL_1245471, EPI_ISL_1245472, EPI_ISL_1245475, EPI_ISL_1245477, EPI_ISL_1245478, EPI_ISL_1245481, EPI_ISL_1245485, EPI_ISL_1245486, EPI_ISL_1245487, EPI_ISL_1245489, EPI_ISL_1245490, EPI_ISL_1245495, EPI_ISL_1245496, EPI_ISL_1245497, EPI_ISL_1245498, EPI_ISL_1245499, EPI_ISL_1245500,
EPI_ISL_1245502, EPI_ISL_1245503, EPI_ISL_1245505, EPI_ISL_1245506, EPI_ISL_1245507, EPI_ISL_1245508, EPI_ISL_1245509, EPI_ISL_1245511, EPI_ISL_1245512, EPI_ISL_1245513, EPI_ISL_1245514, EPI_ISL_1245516, EPI_ISL_1245519, EPI_ISL_1245520, EPI_ISL_1245521, EPI_ISL_1245525, EPI_ISL_1245528, EPI_ISL_1245530, EPI_ISL_1245531, EPI_ISL_1245532, EPI_ISL_1245534, EPI_ISL_1245536,
EPI_ISL_1245537, EPI_ISL_1245539, EPI_ISL_1245540, EPI_ISL_1245542, EPI_ISL_1245543, EPI_ISL_1245545, EPI_ISL_1245546, EPI_ISL_1245550, EPI_ISL_1245551, EPI_ISL_1245552, EPI_ISL_1245556, EPI_ISL_1245557, EPI_ISL_1245561, EPI_ISL_1245562, EPI_ISL_1245563, EPI_ISL_1245564, EPI_ISL_1245565, EPI_ISL_1245572, EPI_ISL_1245574, EPI_ISL_1245575, EPI_ISL_1245578, EPI_ISL_1245579,
EPI_ISL_1245580, EPI_ISL_1245581, EPI_ISL_1245582, EPI_ISL_1245583, EPI_ISL_1245585, EPI_ISL_1245587, EPI_ISL_1245590, EPI_ISL_1245593, EPI_ISL_1245596, EPI_ISL_1245597, EPI_ISL_1245600, EPI_ISL_1245601, EPI_ISL_1245602, EPI_ISL_1245604, EPI_ISL_1245607, EPI_ISL_1245608, EPI_ISL_1245609, EPI_ISL_1245612, EPI_ISL_1245616, EPI_ISL_1245617, EPI_ISL_1245618, EPI_ISL_1245619,
EPI_ISL_1245622, EPI_ISL_1245626, EPI_ISL_1245629, EPI_ISL_1245633, EPI_ISL_1245638, EPI_ISL_1245640, EPI_ISL_1245643, EPI_ISL_1245644, EPI_ISL_1245647, EPI_ISL_1245649, EPI_ISL_1245650, EPI_ISL_1245651, EPI_ISL_1245652, EPI_ISL_1245654, EPI_ISL_1245656, EPI_ISL_1245657, EPI_ISL_1245658, EPI_ISL_1245659, EPI_ISL_1245660, EPI_ISL_1245661, EPI_ISL_1245663, EPI_ISL_1245664,
EPI_ISL_1245668, EPI_ISL_1245669, EPI_ISL_1245670, EPI_ISL_1245671, EPI_ISL_1245672, EPI_ISL_1245673, EPI_ISL_1245674, EPI_ISL_1245675, EPI_ISL_1245676, EPI_ISL_1245678, EPI_ISL_1245680, EPI_ISL_1245681, EPI_ISL_1245682, EPI_ISL_1245683, EPI_ISL_1245689, EPI_ISL_1245690, EPI_ISL_1245692, EPI_ISL_1245695, EPI_ISL_1245699, EPI_ISL_1245701, EPI_ISL_1245702, EPI_ISL_1245704,
EPI_ISL_1245706, EPI_ISL_1245713, EPI_ISL_1245714, EPI_ISL_1245717, EPI_ISL_1245719, EPI_ISL_1245721, EPI_ISL_1245724, EPI_ISL_1245727, EPI_ISL_1245730, EPI_ISL_1245731, EPI_ISL_1245733, EPI_ISL_1245739, EPI_ISL_1245742, EPI_ISL_1245747, EPI_ISL_1245748, EPI_ISL_1245749, EPI_ISL_1245750, EPI_ISL_1245752, EPI_ISL_1245753, EPI_ISL_1245755, EPI_ISL_1245756, EPI_ISL_1245757,
EPI_ISL_1245760, EPI_ISL_1245762, EPI_ISL_1245767, EPI_ISL_1245773, EPI_ISL_1245776, EPI_ISL_1245777, EPI_ISL_1245782, EPI_ISL_1245783, EPI_ISL_1245784, EPI_ISL_1245786, EPI_ISL_1245787, EPI_ISL_1245789, EPI_ISL_1245790, EPI_ISL_1245791, EPI_ISL_1245801, EPI_ISL_1245805, EPI_ISL_1245808, EPI_ISL_1245814, EPI_ISL_1245822, EPI_ISL_1245823, EPI_ISL_1245830, EPI_ISL_1245834,
EPI_ISL_1245847, EPI_ISL_1245850, EPI_ISL_1245852, EPI_ISL_1245862, EPI_ISL_1245863, EPI_ISL_1245864, EPI_ISL_1245877, EPI_ISL_1245879, EPI_ISL_1245881, EPI_ISL_1245883, EPI_ISL_1245886, EPI_ISL_1245888, EPI_ISL_1245890, EPI_ISL_1245895, EPI_ISL_1245896, EPI_ISL_1245903, EPI_ISL_1245908, EPI_ISL_1245910, EPI_ISL_1245911, EPI_ISL_1245914, EPI_ISL_1245916, EPI_ISL_1245921,
EPI_ISL_1245927, EPI_ISL_1245932, EPI_ISL_1245933, EPI_ISL_1245935, EPI_ISL_1245950, EPI_ISL_1245954, EPI_ISL_1245956, EPI_ISL_1245957, EPI_ISL_1245958, EPI_ISL_1245960, EPI_ISL_1245962, EPI_ISL_1245963, EPI_ISL_1245964, EPI_ISL_1245967, EPI_ISL_1245969, EPI_ISL_1245974, EPI_ISL_1245977, EPI_ISL_1245978, EPI_ISL_1245985, EPI_ISL_1245986, EPI_ISL_1245987, EPI_ISL_1245990,
EPI_ISL_1245992, EPI_ISL_1245994, EPI_ISL_1245995, EPI_ISL_1245997, EPI_ISL_1245998, EPI_ISL_1245999, EPI_ISL_1246005, EPI_ISL_1246012, EPI_ISL_1246014, EPI_ISL_1255431, EPI_ISL_1255434, EPI_ISL_1255439, EPI_ISL_1255446, EPI_ISL_1255456, EPI_ISL_1255468, EPI_ISL_1255475, EPI_ISL_1255477, EPI_ISL_1255492, EPI_ISL_1255500, EPI_ISL_1255518, EPI_ISL_1255531, EPI_ISL_1255544,
EPI_ISL_1255549, EPI_ISL_1255562, EPI_ISL_1255568, EPI_ISL_1255588, EPI_ISL_1255590, EPI_ISL_1255592, EPI_ISL_1255593, EPI_ISL_1255613, EPI_ISL_1255619, EPI_ISL_1255623, EPI_ISL_1255642, EPI_ISL_1255652, EPI_ISL_1255669, EPI_ISL_1255670, EPI_ISL_1255675, EPI_ISL_1255734, EPI_ISL_1255760, EPI_ISL_1255861, EPI_ISL_1256012, EPI_ISL_1256025, EPI_ISL_1256685, EPI_ISL_1256688,
EPI_ISL_1256698, EPI_ISL_1256700, EPI_ISL_1256702, EPI_ISL_1256714, EPI_ISL_1256715, EPI_ISL_1256718, EPI_ISL_1256719, EPI_ISL_1256724, EPI_ISL_1256741, EPI_ISL_1256744, EPI_ISL_1256756, EPI_ISL_1256758, EPI_ISL_1256765, EPI_ISL_1256770, EPI_ISL_1256771, EPI_ISL_1256779, EPI_ISL_1256782, EPI_ISL_1256789, EPI_ISL_1256796, EPI_ISL_1256824, EPI_ISL_1256837, EPI_ISL_1256870,
EPI_ISL_1256876, EPI_ISL_1256877, EPI_ISL_1256887, EPI_ISL_1256891, EPI_ISL_1256912, EPI_ISL_1256931, EPI_ISL_1256939, EPI_ISL_1264067, EPI_ISL_1264070, EPI_ISL_1264073, EPI_ISL_1264074, EPI_ISL_1264078, EPI_ISL_1264080, EPI_ISL_1264083, EPI_ISL_1264094, EPI_ISL_1264095, EPI_ISL_1264101, EPI_ISL_1264117, EPI_ISL_1264118, EPI_ISL_1264121, EPI_ISL_1264125, EPI_ISL_1264126,
EPI_ISL_1264128, EPI_ISL_1264133, EPI_ISL_1264136, EPI_ISL_1264144, EPI_ISL_1264147, EPI_ISL_1264153, EPI_ISL_1264154, EPI_ISL_1264156, EPI_ISL_1264158, EPI_ISL_1264160, EPI_ISL_1264176, EPI_ISL_1264180, EPI_ISL_1264184, EPI_ISL_1264189, EPI_ISL_1264200, EPI_ISL_1264207, EPI_ISL_1264208, EPI_ISL_1264220, EPI_ISL_1264227, EPI_ISL_1264228, EPI_ISL_1264229, EPI_ISL_1264236,
EPI_ISL_1264241, EPI_ISL_1264252, EPI_ISL_1264257, EPI_ISL_1264273, EPI_ISL_1264283, EPI_ISL_1264284, EPI_ISL_1264292, EPI_ISL_1264301, EPI_ISL_1264304, EPI_ISL_1264308, EPI_ISL_1264309, EPI_ISL_1264311, EPI_ISL_1264312, EPI_ISL_1264318, EPI_ISL_1264320, EPI_ISL_1264321, EPI_ISL_1264326, EPI_ISL_1264328, EPI_ISL_1264329, EPI_ISL_1264330, EPI_ISL_1264334, EPI_ISL_1264379,
EPI_ISL_1264490, EPI_ISL_1264517, EPI_ISL_1264571, EPI_ISL_1264574, EPI_ISL_1264591, EPI_ISL_1264602, EPI_ISL_1264634, EPI_ISL_1274311, EPI_ISL_1274354, EPI_ISL_1274363, EPI_ISL_1274430, EPI_ISL_1274438, EPI_ISL_1274522, EPI_ISL_1274761, EPI_ISL_1274836, EPI_ISL_1274892, EPI_ISL_1274930, EPI_ISL_1274983, EPI_ISL_1274985, EPI_ISL_1275020, EPI_ISL_1276176, EPI_ISL_1276207,
EPI_ISL_1276255, EPI_ISL_1276281, EPI_ISL_1276297, EPI_ISL_1276299, EPI_ISL_1276321, EPI_ISL_1276339, EPI_ISL_1276366, EPI_ISL_1276826, EPI_ISL_1324882, EPI_ISL_1325007, EPI_ISL_1325064, EPI_ISL_1327979, EPI_ISL_1330778, EPI_ISL_1333512, EPI_ISL_1333833, EPI_ISL_1449652, EPI_ISL_1449668, EPI_ISL_1449672, EPI_ISL_1449673, EPI_ISL_1449674, EPI_ISL_1449681, EPI_ISL_1449715,
EPI_ISL_1449718, EPI_ISL_1449719, EPI_ISL_1449738, EPI_ISL_1449740, EPI_ISL_1449746, EPI_ISL_1449757, EPI_ISL_1449766, EPI_ISL_1449781, EPI_ISL_1449805, EPI_ISL_1449844, EPI_ISL_1449846, EPI_ISL_1449849, EPI_ISL_1449862, EPI_ISL_1449866, EPI_ISL_1449880, EPI_ISL_1449883, EPI_ISL_1449886, EPI_ISL_1449906, EPI_ISL_1449909, EPI_ISL_1449937, EPI_ISL_1449941, EPI_ISL_1449942,
EPI_ISL_1449951, EPI_ISL_1449956, EPI_ISL_1449962, EPI_ISL_1449968, EPI_ISL_1449970, EPI_ISL_1449973, EPI_ISL_1449974, EPI_ISL_1449975, EPI_ISL_1473514, EPI_ISL_1473559

| | | | |
|---|---|---|---|
| see above | Lighthouse Lab in Milton Keynes | Wellcome Sanger Institute for the COVID-19 Genomics UK (COG-UK) Consortium | Cordelia Langford; David K. Jackson; Dominic Kwiatkowski; Ewan Harrison; Ian Johnston; Jeffrey Barrett; John Sillitoe on behalf of the Wellcome Sanger Institute COVID-19 Surveillance Team; Roberto Amato; Sonia Goncalves; The Lighthouse Lab in Milton Keynes and Alex Alderton |

EPI_ISL_1309711, EPI_ISL_1309712, EPI_ISL_1309713, EPI_ISL_1309714, EPI_ISL_1309715, EPI_ISL_1309716, EPI_ISL_1309717, EPI_ISL_1309718, EPI_ISL_1309719, EPI_ISL_1387206, EPI_ISL_1387208, EPI_ISL_1387209, EPI_ISL_1387210, EPI_ISL_1387211, EPI_ISL_1475496, EPI_ISL_1475497, EPI_ISL_1475498, EPI_ISL_1475499, EPI_ISL_1475500, EPI_ISL_1475501, EPI_ISL_1475502, EPI_ISL_1475503,
EPI_ISL_1978952

| | | | |
|---|---|---|---|
| see above | Lincolnshire Hospitals and DeepSeq Nottingham | COVID-19 Genomics UK (COG-UK) Consortium | Christopher Moore; Fei Sang; Joeseph Chappell; Johnny Debebe; Jonathan Ball; Matthew Carlisle; Matthew Loose; Nadine Holmes; Nichola Duckworth; Patrick McClure; Sarah Walsh; Tim Sloan; Victoria Wright |

EPI_ISL_1247828, EPI_ISL_1247829, EPI_ISL_1247830, EPI_ISL_1247831, EPI_ISL_1247838, EPI_ISL_1247839, EPI_ISL_1247840, EPI_ISL_1247841, EPI_ISL_1247842, EPI_ISL_1247843, EPI_ISL_1308891, EPI_ISL_1308919, EPI_ISL_1308920, EPI_ISL_1308921, EPI_ISL_1308922, EPI_ISL_1308923, EPI_ISL_1308924, EPI_ISL_1308925, EPI_ISL_1308926, EPI_ISL_1308927, EPI_ISL_1308928, EPI_ISL_1308929,
EPI_ISL_1308930, EPI_ISL_1308931, EPI_ISL_1308932, EPI_ISL_1308933, EPI_ISL_1308934, EPI_ISL_1308935, EPI_ISL_1308936, EPI_ISL_1308937, EPI_ISL_1308938, EPI_ISL_1308940, EPI_ISL_1308941, EPI_ISL_1309785, EPI_ISL_1309786, EPI_ISL_1309787, EPI_ISL_1309788, EPI_ISL_1309790, EPI_ISL_1386739, EPI_ISL_1386782, EPI_ISL_1386785, EPI_ISL_1386786, EPI_ISL_1386791, EPI_ISL_1386794,
EPI_ISL_1386795, EPI_ISL_1386798, EPI_ISL_1474662, EPI_ISL_1474663, EPI_ISL_1474687, EPI_ISL_1474697, EPI_ISL_1474718, EPI_ISL_1474721, EPI_ISL_1474722, EPI_ISL_1474724, EPI_ISL_1546150, EPI_ISL_1636825, EPI_ISL_1833111, EPI_ISL_1833112, EPI_ISL_1833115, EPI_ISL_1833116, EPI_ISL_1833117, EPI_ISL_1833118, EPI_ISL_1833120, EPI_ISL_1833123, EPI_ISL_1833124

| | | | |
|---|---|---|---|
| see above | Liverpool Clinical Laboratories | COVID-19 Genomics UK (COG-UK) Consortium | A Alrezaihi; Alessandro Gerada; Alistair Darby; Angela Cowell; Anita Lucaci; Anu Chawla; Cassie Olateju; Catherine Hartley; Charlotte Nelson; Ecaterina Vamos; Elaine O'Toole; Eleanor G Bentley; Ghada T Shawli; Hermione Webster; Isabel García-Dorival; James Johnson; James P Stewart; Jenifer Manson; Joanne Watts; Jones Benjamin; Jordan J Clark; Julian Hiscox; L Luu; Lucille Rainbow; M Almsaud; Margaret Hughes; Mark Whitehead; Matthew Gemmell; Miren Iturriza-Gomara; Muhannad Alruwaili; N.P Randle; Neil Swainston; PKF Gilmore; Parul Sharma; Rebekah Penrice-Randal; Richard Eccles; Richard Gregory; Sam Haldenby; Steve Paterson; Stuart D Armstrong; Trevor Ian Robinson; Ximeng Han |

EPI_ISL_1248818, EPI_ISL_1248819, EPI_ISL_1248820, EPI_ISL_1248821, EPI_ISL_1248896, EPI_ISL_1248897, EPI_ISL_1248898, EPI_ISL_1248899, EPI_ISL_1248900, EPI_ISL_1248915, EPI_ISL_1248916, EPI_ISL_1248917, EPI_ISL_1248918, EPI_ISL_1248919, EPI_ISL_1248920, EPI_ISL_1309506, EPI_ISL_1309532, EPI_ISL_1309536, EPI_ISL_1309537, EPI_ISL_1309538, EPI_ISL_1309539, EPI_ISL_1309540,
EPI_ISL_1309541, EPI_ISL_1309542, EPI_ISL_1309543, EPI_ISL_1309544, EPI_ISL_1309545, EPI_ISL_1309546, EPI_ISL_1309547, EPI_ISL_1309548, EPI_ISL_1309549, EPI_ISL_1475189, EPI_ISL_1475191, EPI_ISL_1475192, EPI_ISL_1475193, EPI_ISL_1475196, EPI_ISL_1475383, EPI_ISL_1475384, EPI_ISL_1475385, EPI_ISL_1475386, EPI_ISL_1475388, EPI_ISL_1475389, EPI_ISL_1475390, EPI_ISL_1475391,
EPI_ISL_1475392, EPI_ISL_1475446, EPI_ISL_1475447, EPI_ISL_1475448, EPI_ISL_1475450, EPI_ISL_1475452, EPI_ISL_1475453, EPI_ISL_1475454, EPI_ISL_1475455, EPI_ISL_1475480, EPI_ISL_1475481, EPI_ISL_1475482, EPI_ISL_1475484, EPI_ISL_1475485

| | | | |
|---|---|---|---|
| see above | Northumbria University / South Tees Hospitals NHS Foundation Trust / North Cumbria Integrated Care NHS Foundation Trust / North Tees and Hartlepool NHS Foundation Trust / Newcastle Hospitals NHS Foundation Trust | COVID-19 Genomics UK (COG-UK) Consortium | Andrew Nelson; Brendan Payne; Clive Graham; Darren L Smith; Debra Padgett; Edward Barton; Emma Swindells; Garren Scott; Gary Black; Gary Eltringham; Giles S Holt; Greg R Young; Jane Greenaway; Jennifer Collins; John Allan; Joshua Loh; Lynn Dover; Matthew Bashton; Mohammad A Tariq; Paul Baker; Sarah Essex; Steve Liggett; Wen C Yew; Yusri Taha |

EPI_ISL_1249297, EPI_ISL_1249298, EPI_ISL_1249306, EPI_ISL_1249389, EPI_ISL_1249391, EPI_ISL_1249395, EPI_ISL_1249397, EPI_ISL_1249398, EPI_ISL_1249407, EPI_ISL_1249408, EPI_ISL_1249411, EPI_ISL_1249413, EPI_ISL_1249414, EPI_ISL_1249415, EPI_ISL_1249416, EPI_ISL_1249419, EPI_ISL_1249420, EPI_ISL_1249421, EPI_ISL_1249426, EPI_ISL_1249427, EPI_ISL_1249430, EPI_ISL_1249431,
EPI_ISL_1249433, EPI_ISL_1249434, EPI_ISL_1249435, EPI_ISL_1249446, EPI_ISL_1249448, EPI_ISL_1249456, EPI_ISL_1249459, EPI_ISL_1249461, EPI_ISL_1249462, EPI_ISL_1249464, EPI_ISL_1249465, EPI_ISL_1249466, EPI_ISL_1249469, EPI_ISL_1249473, EPI_ISL_1249484, EPI_ISL_1249485, EPI_ISL_1249486, EPI_ISL_1249487, EPI_ISL_1249493, EPI_ISL_1249494, EPI_ISL_1249495, EPI_ISL_1249497,
EPI_ISL_1249500, EPI_ISL_1249503, EPI_ISL_1249504, EPI_ISL_1249505, EPI_ISL_1249506, EPI_ISL_1249507, EPI_ISL_1249509, EPI_ISL_1249510, EPI_ISL_1249511, EPI_ISL_1249513, EPI_ISL_1249514, EPI_ISL_1249516, EPI_ISL_1249518, EPI_ISL_1249519, EPI_ISL_1249520, EPI_ISL_1249521, EPI_ISL_1249522, EPI_ISL_1249523, EPI_ISL_1249525, EPI_ISL_1249530, EPI_ISL_1249531, EPI_ISL_1249532,
EPI_ISL_1249534, EPI_ISL_1249535, EPI_ISL_1249536, EPI_ISL_1249537, EPI_ISL_1249538, EPI_ISL_1249541, EPI_ISL_1249542, EPI_ISL_1249543, EPI_ISL_1249544, EPI_ISL_1249545, EPI_ISL_1249546, EPI_ISL_1249548, EPI_ISL_1249550, EPI_ISL_1249551, EPI_ISL_1249552, EPI_ISL_1249555, EPI_ISL_1249556, EPI_ISL_1249557, EPI_ISL_1249558, EPI_ISL_1249560, EPI_ISL_1249564, EPI_ISL_1249566,
EPI_ISL_1249570, EPI_ISL_1249577, EPI_ISL_1249580, EPI_ISL_1249581, EPI_ISL_1249582, EPI_ISL_1249583, EPI_ISL_1249584, EPI_ISL_1249586, EPI_ISL_1249587, EPI_ISL_1249588, EPI_ISL_1249589, EPI_ISL_1249590, EPI_ISL_1249592, EPI_ISL_1249604, EPI_ISL_1249605, EPI_ISL_1249606, EPI_ISL_1249608, EPI_ISL_1249612, EPI_ISL_1249621, EPI_ISL_1249622, EPI_ISL_1249623, EPI_ISL_1249625,
EPI_ISL_1249626, EPI_ISL_1249714, EPI_ISL_1249715, EPI_ISL_1249716, EPI_ISL_1249725, EPI_ISL_1249726, EPI_ISL_1249732, EPI_ISL_1249733, EPI_ISL_1249734, EPI_ISL_1249749, EPI_ISL_1249755, EPI_ISL_1249758, EPI_ISL_1249775, EPI_ISL_1249776, EPI_ISL_1249778, EPI_ISL_1249779, EPI_ISL_1249780, EPI_ISL_1249784, EPI_ISL_1249801, EPI_ISL_1249802, EPI_ISL_1249803, EPI_ISL_1249804,
EPI_ISL_1249805, EPI_ISL_1249806, EPI_ISL_1249838, EPI_ISL_1249839, EPI_ISL_1249841, EPI_ISL_1249844, EPI_ISL_1249845, EPI_ISL_1249846, EPI_ISL_1249847, EPI_ISL_1249852, EPI_ISL_1249853, EPI_ISL_1249854, EPI_ISL_1249857, EPI_ISL_1249882, EPI_ISL_1309792, EPI_ISL_1309794, EPI_ISL_1309959, EPI_ISL_1387669, EPI_ISL_1387671, EPI_ISL_1387674, EPI_ISL_1387676, EPI_ISL_1387678,
EPI_ISL_1387679, EPI_ISL_1387680, EPI_ISL_1387681, EPI_ISL_1387682, EPI_ISL_1387683, EPI_ISL_1387686, EPI_ISL_1387690, EPI_ISL_1387691, EPI_ISL_1387692, EPI_ISL_1387693, EPI_ISL_1387696, EPI_ISL_1387697, EPI_ISL_1387698, EPI_ISL_1387699, EPI_ISL_1387702, EPI_ISL_1387703, EPI_ISL_1387704, EPI_ISL_1387707, EPI_ISL_1387708, EPI_ISL_1387710, EPI_ISL_1387711, EPI_ISL_1387712,
EPI_ISL_1387713, EPI_ISL_1387714, EPI_ISL_1387715, EPI_ISL_1387721, EPI_ISL_1387723, EPI_ISL_1387725, EPI_ISL_1387730, EPI_ISL_1387731, EPI_ISL_1387732, EPI_ISL_1387735, EPI_ISL_1387736, EPI_ISL_1387739, EPI_ISL_1387743, EPI_ISL_1387745, EPI_ISL_1387746, EPI_ISL_1387749, EPI_ISL_1387750, EPI_ISL_1387762, EPI_ISL_1387765, EPI_ISL_1387791, EPI_ISL_1387792, EPI_ISL_1387795,
EPI_ISL_1387796, EPI_ISL_1546795, EPI_ISL_1547044, EPI_ISL_1547045, EPI_ISL_1547046, EPI_ISL_1547050, EPI_ISL_1547052, EPI_ISL_1547053, EPI_ISL_1547054

| | | | |
|---|---|---|---|
| see above | Originating lab: Wales Specialist Virology Centre Sequencing lab: Pathogen Genomics Unit | Public Health Wales Microbiology Cardiff Wales Specialist Virology Centre | Alec Birchley; Alexander Adams; Amy Gaskin; Angela Marchbank; Bree Gatica-Wilcox; Catherine Moore; Jason Coombes; Joanne Watkins; Joel Southgate; Johnathan Evans; Laura Gifford; Lauren Gilbert; Lee Graham; Malorie Perry; Matthew Bull; Nicole Pacchiarini; Sally Corden; Sara Kumziene-Summerhayes; Sara Rey; Sarah Taylor; Simon Cottrell; Sophie Jones; Tom Connor |

EPI_ISL_1475605, EPI_ISL_1475606, EPI_ISL_1475607, EPI_ISL_1475609, EPI_ISL_1475612, EPI_ISL_1475613, EPI_ISL_1475615, EPI_ISL_1475616, EPI_ISL_1475617, EPI_ISL_1475618, EPI_ISL_1475619, EPI_ISL_1475620, EPI_ISL_1475621, EPI_ISL_1475623, EPI_ISL_1475624, EPI_ISL_1475625, EPI_ISL_1475626, EPI_ISL_1475627, EPI_ISL_1475628, EPI_ISL_1475629, EPI_ISL_1475630, EPI_ISL_1475631,
EPI_ISL_1475632, EPI_ISL_1475633, EPI_ISL_1475635, EPI_ISL_1475638, EPI_ISL_1475639, EPI_ISL_1475640, EPI_ISL_1475641, EPI_ISL_1475642, EPI_ISL_1475644, EPI_ISL_1475648, EPI_ISL_1475651, EPI_ISL_1475652, EPI_ISL_1475676

| | | | |
|---|---|---|---|
| see above | Oxford Viromics, NDM, University of Oxford; Oxford University Hospitals; Basingstoke and North Hampshire Hospital | COVID-19 Genomics UK (COG-UK) Consortium | Alex Mobbs; Amy Trebes; Anita Justice; Catrin Moore; Christophe Fraser; David Bonsall; David Buck; Emma Wise; George Macintyre; Jessica Lynch; John Todd; Mariateresa de Cesare; Matilde Mori; Monique Andersson; Nathan Moore; Nick Cortes; Robert Shaw; Stephen Kidd; Tanya Golubchik; Timothy Peto |

EPI_ISL_1309661, EPI_ISL_1309665, EPI_ISL_1309687, EPI_ISL_1309693, EPI_ISL_1309694, EPI_ISL_1309695, EPI_ISL_1832973

| | | | |
|---|---|---|---|
| see above | Quadram Institute Bioscience | COVID-19 Genomics UK (COG-UK) Consortium | Alexander J Trotter; Alison E. Mather; Alp Aydin; Ana P. Tedim; Anastasia Kolyva; Andrew Bell; Andrew J. Page; Claire Stuart; Dave J. Baker; Gemma L. Kay; John Wain; Justin O'Grady; Leonardo de Oliveira Martins; Lizzie Meadows; Maria Diaz; Mark Webber; Muhammed Yasir; Nabil-Fareed Alikhan; Ngozi Elumogo; Nicholas M. Thomson; Rachael Stanley; Rachel Gilroy; Reenesh Prakash; Samir Dervisevic; Samuel Bloomfield; Steven Rudder; Thanh Le-Viet |

EPI_ISL_1178580, EPI_ISL_1178581, EPI_ISL_1178588, EPI_ISL_1178623, EPI_ISL_1178624, EPI_ISL_1249063, EPI_ISL_1249064, EPI_ISL_1249065, EPI_ISL_1249066, EPI_ISL_1249071, EPI_ISL_1387167, EPI_ISL_1387168, EPI_ISL_1387169, EPI_ISL_1387170, EPI_ISL_1387171, EPI_ISL_1387172, EPI_ISL_1387173, EPI_ISL_1387174, EPI_ISL_1387175, EPI_ISL_1387176, EPI_ISL_1387177, EPI_ISL_1387179,
EPI_ISL_1387180, EPI_ISL_1387181

| | | | |
|---|---|---|---|
| see above | Queens Medical Centre, Clinical Microbiology Department / DeepSeq Nottingham | COVID-19 Genomics UK (COG-UK) Consortium | Christopher Moore; Emily Park; Fei Sang; Gemma Clark; Hannah Howson-Wells; Jack Hill; Jayasree Dey; Johnny Debebe; Jonathan Ball; Joseph Chappell; Manjinder Khakh; Matthew Carlisle; Matthew Loose; Michelle M Lister; Nadine Holmes; Patrick McClure; Theocharis Tsoleridis; Timothy Byaruhanga; Vicki M Fleming; Victoria Wright; Wendy Smith |

EPI_ISL_1275873, EPI_ISL_1275879, EPI_ISL_1275880, EPI_ISL_1275902, EPI_ISL_1275969, EPI_ISL_1275981, EPI_ISL_1276008, EPI_ISL_1276048, EPI_ISL_1276051, EPI_ISL_1276056, EPI_ISL_1276064, EPI_ISL_1276084, EPI_ISL_1276103, EPI_ISL_1332939, EPI_ISL_1374072, EPI_ISL_1374074, EPI_ISL_1374075, EPI_ISL_1374076, EPI_ISL_1374077, EPI_ISL_1374078, EPI_ISL_1374079, EPI_ISL_1374080,
EPI_ISL_1374084, EPI_ISL_1374085, EPI_ISL_1374086, EPI_ISL_1374087, EPI_ISL_1374089, EPI_ISL_1374090, EPI_ISL_1374091, EPI_ISL_1374093, EPI_ISL_1374095, EPI_ISL_1374097, EPI_ISL_1374099, EPI_ISL_1374102, EPI_ISL_1374103, EPI_ISL_1374104, EPI_ISL_1374106, EPI_ISL_1374107, EPI_ISL_1374109, EPI_ISL_1374110, EPI_ISL_1374111, EPI_ISL_1374112, EPI_ISL_1374113, EPI_ISL_1374114,
EPI_ISL_1374116, EPI_ISL_1374117, EPI_ISL_1374118, EPI_ISL_1374120, EPI_ISL_1374121, EPI_ISL_1374124, EPI_ISL_1374125, EPI_ISL_1374126, EPI_ISL_1374127, EPI_ISL_1374128, EPI_ISL_1374129, EPI_ISL_1374131, EPI_ISL_1374132, EPI_ISL_1374133, EPI_ISL_1374135, EPI_ISL_1374136, EPI_ISL_1374138, EPI_ISL_1374145, EPI_ISL_1374146, EPI_ISL_1374147, EPI_ISL_1374149, EPI_ISL_1374151,

| | | | |
|---|---|---|---|
| EPI_ISL_1374153, EPI_ISL_1374154, EPI_ISL_1374155, EPI_ISL_1374156, EPI_ISL_1374157, EPI_ISL_1374158, EPI_ISL_1374160, EPI_ISL_1374161, EPI_ISL_1374166, EPI_ISL_1374167, EPI_ISL_1374168, EPI_ISL_1374170, EPI_ISL_1374171, EPI_ISL_1374173, EPI_ISL_1374176, EPI_ISL_1374177, EPI_ISL_1374178, EPI_ISL_1374179, EPI_ISL_1374181, EPI_ISL_1374184, EPI_ISL_1374185, EPI_ISL_1374190, EPI_ISL_1374191, EPI_ISL_1374192, EPI_ISL_1374196, EPI_ISL_1374198, EPI_ISL_1374199, EPI_ISL_1374201, EPI_ISL_1374202, EPI_ISL_1374203, EPI_ISL_1374205, EPI_ISL_1374207, EPI_ISL_1374208, EPI_ISL_1374210, EPI_ISL_1374212, EPI_ISL_1374213, EPI_ISL_1374215, EPI_ISL_1374216, EPI_ISL_1374218, EPI_ISL_1374220, EPI_ISL_1374222, EPI_ISL_1374225, EPI_ISL_1374226, EPI_ISL_1374227, EPI_ISL_1374229, EPI_ISL_1374231, EPI_ISL_1374232, EPI_ISL_1374235, EPI_ISL_1374237, EPI_ISL_1374238, EPI_ISL_1374239, EPI_ISL_1374241, EPI_ISL_1374245, EPI_ISL_1374246, EPI_ISL_1374247, EPI_ISL_1374251, EPI_ISL_1374252, EPI_ISL_1374253, EPI_ISL_1374254, EPI_ISL_1374255, EPI_ISL_1374257, EPI_ISL_1374258, EPI_ISL_1374262, EPI_ISL_1374263, EPI_ISL_1374266, EPI_ISL_1374267, EPI_ISL_1374268, EPI_ISL_1374269, EPI_ISL_1374270, EPI_ISL_1374271, EPI_ISL_1374272, EPI_ISL_1374273, EPI_ISL_1374275, EPI_ISL_1374276, EPI_ISL_1374277, EPI_ISL_1374278, EPI_ISL_1374280, EPI_ISL_1374282, EPI_ISL_1374283, EPI_ISL_1374284, EPI_ISL_1374289, EPI_ISL_1374292, EPI_ISL_1374293, EPI_ISL_1374294, EPI_ISL_1374296, EPI_ISL_1374297, EPI_ISL_1374300, EPI_ISL_1374301, EPI_ISL_1374302, EPI_ISL_1374304, EPI_ISL_1374308, EPI_ISL_1374309, EPI_ISL_1374311, EPI_ISL_1374312, EPI_ISL_1374313, EPI_ISL_1374316, EPI_ISL_1374317, EPI_ISL_1374318, EPI_ISL_1374320, EPI_ISL_1374321, EPI_ISL_1374322, EPI_ISL_1374323, EPI_ISL_1374324, EPI_ISL_1374325, EPI_ISL_1374327, EPI_ISL_1374328, EPI_ISL_1374329, EPI_ISL_1374330, EPI_ISL_1374331, EPI_ISL_1389400, EPI_ISL_1389401, EPI_ISL_1389403, EPI_ISL_1389404, EPI_ISL_1389405, EPI_ISL_1389406, EPI_ISL_1389407, EPI_ISL_1389408, EPI_ISL_1389409, EPI_ISL_1389410, EPI_ISL_1389412, EPI_ISL_1389413, EPI_ISL_1389414, EPI_ISL_1389415, EPI_ISL_1389416, EPI_ISL_1389417, EPI_ISL_1389418, EPI_ISL_1389419, EPI_ISL_1389421, EPI_ISL_1389422, EPI_ISL_1389424, EPI_ISL_1389425, EPI_ISL_1389426, EPI_ISL_1389427, EPI_ISL_1389428, EPI_ISL_1389429, EPI_ISL_1389430, EPI_ISL_1389431, EPI_ISL_1389432, EPI_ISL_1389433, EPI_ISL_1389434, EPI_ISL_1389435, EPI_ISL_1389436, EPI_ISL_1389437, EPI_ISL_1389438, EPI_ISL_1389439, EPI_ISL_1389440, EPI_ISL_1389441, EPI_ISL_1389442, EPI_ISL_1389443, EPI_ISL_1389445, EPI_ISL_1389446, EPI_ISL_1389447, EPI_ISL_1389448, EPI_ISL_1389450, EPI_ISL_1389451, EPI_ISL_1389452, EPI_ISL_1389453, EPI_ISL_1389454, EPI_ISL_1389455, EPI_ISL_1389456, EPI_ISL_1389457, EPI_ISL_1389458, EPI_ISL_1389459, EPI_ISL_1389460, EPI_ISL_1389461, EPI_ISL_1389463, EPI_ISL_1389465, EPI_ISL_1389466, EPI_ISL_1389467, EPI_ISL_1389468, EPI_ISL_1389470, EPI_ISL_1389471, EPI_ISL_1389472, EPI_ISL_1389474, EPI_ISL_1389475, EPI_ISL_1389476, EPI_ISL_1389477, EPI_ISL_1389478, EPI_ISL_1389482, EPI_ISL_1389483, EPI_ISL_1389484, EPI_ISL_1389485, EPI_ISL_1389486, EPI_ISL_1389488, EPI_ISL_1389490, EPI_ISL_1389491, EPI_ISL_1389492, EPI_ISL_1389493, EPI_ISL_1389494, EPI_ISL_1389495, EPI_ISL_1389496, EPI_ISL_1389497, EPI_ISL_1389498, EPI_ISL_1389499, EPI_ISL_1389500, EPI_ISL_1389501, EPI_ISL_1389502, EPI_ISL_1389503, EPI_ISL_1389504, EPI_ISL_1389506, EPI_ISL_1389507, EPI_ISL_1389508, EPI_ISL_1389509, EPI_ISL_1389511, EPI_ISL_1389512, EPI_ISL_1389513, EPI_ISL_1389514, EPI_ISL_1389515, EPI_ISL_1389516, EPI_ISL_1389517, EPI_ISL_1389519, EPI_ISL_1389520, EPI_ISL_1389522, EPI_ISL_1389523, EPI_ISL_1389524, EPI_ISL_1389525, EPI_ISL_1389526, EPI_ISL_1389527, EPI_ISL_1389528, EPI_ISL_1389530, EPI_ISL_1389531, EPI_ISL_1389532, EPI_ISL_1389533, EPI_ISL_1389535, EPI_ISL_1389538, EPI_ISL_1389539, EPI_ISL_1389540, EPI_ISL_1389541, EPI_ISL_1389542, EPI_ISL_1389543, EPI_ISL_1389544, EPI_ISL_1389545, EPI_ISL_1389546, EPI_ISL_1389547, EPI_ISL_1389548, EPI_ISL_1389550, EPI_ISL_1389551, EPI_ISL_1389553, EPI_ISL_1389554, EPI_ISL_1389555, EPI_ISL_1389556, EPI_ISL_1389557, EPI_ISL_1389560, EPI_ISL_1389561, EPI_ISL_1389562, EPI_ISL_1389563, EPI_ISL_1389564, EPI_ISL_1389565, EPI_ISL_1389566, EPI_ISL_1389567, EPI_ISL_1389568, EPI_ISL_1389569, EPI_ISL_1389571, EPI_ISL_1389572, EPI_ISL_1389573, EPI_ISL_1389574, EPI_ISL_1389575, EPI_ISL_1389577, EPI_ISL_1389578, EPI_ISL_1389579, EPI_ISL_1389580, EPI_ISL_1389582, EPI_ISL_1389583, EPI_ISL_1389584, EPI_ISL_1389585, EPI_ISL_1389586, EPI_ISL_1389587, EPI_ISL_1389588, EPI_ISL_1389592, EPI_ISL_1389593, EPI_ISL_1389595, EPI_ISL_1389596, EPI_ISL_1389597, EPI_ISL_1389601, EPI_ISL_1389603, EPI_ISL_1389604, EPI_ISL_1389605, EPI_ISL_1389606, EPI_ISL_1389607, EPI_ISL_1389608, EPI_ISL_1389609, EPI_ISL_1389611, EPI_ISL_1389612, EPI_ISL_1389613, EPI_ISL_1389614, EPI_ISL_1389615, EPI_ISL_1389616, EPI_ISL_1389617, EPI_ISL_1389618, EPI_ISL_1389619, EPI_ISL_1389620, EPI_ISL_1389621, EPI_ISL_1412934, EPI_ISL_1412938, EPI_ISL_1412940, EPI_ISL_1412943, EPI_ISL_1412946, EPI_ISL_1412947, EPI_ISL_1412952, EPI_ISL_1412954, EPI_ISL_1412959, EPI_ISL_1412963, EPI_ISL_1412968, EPI_ISL_1412973, EPI_ISL_1412975, EPI_ISL_1412976, EPI_ISL_1412996, EPI_ISL_1412999, EPI_ISL_1413000, EPI_ISL_1413010, EPI_ISL_1413012, EPI_ISL_1413020, EPI_ISL_1413021, EPI_ISL_1413023, EPI_ISL_1413024, EPI_ISL_1413036, EPI_ISL_1413037, EPI_ISL_1413042, EPI_ISL_1413048, EPI_ISL_1413049, EPI_ISL_1413052, EPI_ISL_1416129, EPI_ISL_1416132, EPI_ISL_1416137, EPI_ISL_1416139, EPI_ISL_1416149, EPI_ISL_1416154, EPI_ISL_1416155, EPI_ISL_1416156, EPI_ISL_1416167, EPI_ISL_1450803 | | | |
| see above | Randox Laboratories | Wellcome Sanger Institute for the COVID-19 Genomics UK (COG-UK) Consortium | Cordelia Langford; David K. Jackson; Dominic Kwiatkowski; Ewan Harrison; Ian Johnston; Jeffrey Barrett; John Sillitoe on behalf of the Wellcome Sanger Institute COVID-19 Surveillance Team; Randox Laboratories and Alex Alderton; Roberto Amato; Sonia Goncalves |
| EPI_ISL_1975185, EPI_ISL_1975186, EPI_ISL_1975188, EPI_ISL_1975190, EPI_ISL_1975191, EPI_ISL_1975193, EPI_ISL_1975194, EPI_ISL_1975196, EPI_ISL_1975200, EPI_ISL_1975202, EPI_ISL_1975203, EPI_ISL_1975205, EPI_ISL_1975207, EPI_ISL_1975208, EPI_ISL_1975210, EPI_ISL_1975215, EPI_ISL_1975217, EPI_ISL_1975218, EPI_ISL_1975220, EPI_ISL_1975222, EPI_ISL_1975224, EPI_ISL_1975225, EPI_ISL_1975227, EPI_ISL_1975228, EPI_ISL_1975230, EPI_ISL_1975232, EPI_ISL_1975234, EPI_ISL_1975237, EPI_ISL_1975239, EPI_ISL_1975240, EPI_ISL_1975246, EPI_ISL_1975248, EPI_ISL_1975249, EPI_ISL_1975251, EPI_ISL_1975254, EPI_ISL_1975256, EPI_ISL_1975257, EPI_ISL_1975259, EPI_ISL_1975261, EPI_ISL_1975262, EPI_ISL_1975264, EPI_ISL_1975265, EPI_ISL_1975267, EPI_ISL_1975269, EPI_ISL_1975270, EPI_ISL_1975272, EPI_ISL_1975274, EPI_ISL_1975275, EPI_ISL_1975277, EPI_ISL_1975279, EPI_ISL_1975285, EPI_ISL_1975287, EPI_ISL_1975288, EPI_ISL_1975290, EPI_ISL_1975291, EPI_ISL_1975293, EPI_ISL_1975295, EPI_ISL_1975296, EPI_ISL_1975298, EPI_ISL_1975302, EPI_ISL_1975714, EPI_ISL_1975716, EPI_ISL_1975718, EPI_ISL_1975719, EPI_ISL_1975721, EPI_ISL_1975723, EPI_ISL_1975725, EPI_ISL_1975726, EPI_ISL_1975728, EPI_ISL_1975729, EPI_ISL_1975731, EPI_ISL_1975733, EPI_ISL_1975736, EPI_ISL_1975740, EPI_ISL_1975742, EPI_ISL_1975745, EPI_ISL_1975748, EPI_ISL_1975750, EPI_ISL_1975751, EPI_ISL_1975753, EPI_ISL_1975755, EPI_ISL_1975757, EPI_ISL_1975758, EPI_ISL_1975760, EPI_ISL_1975765, EPI_ISL_1975767, EPI_ISL_1975768, EPI_ISL_1975769, EPI_ISL_1975770, EPI_ISL_1975771, EPI_ISL_1975772, EPI_ISL_1975773, EPI_ISL_1975774, EPI_ISL_1975776, EPI_ISL_1975777, EPI_ISL_1975778, EPI_ISL_1975779, EPI_ISL_1975780, EPI_ISL_1975781, EPI_ISL_1975782, EPI_ISL_1975784, EPI_ISL_1975785, EPI_ISL_1975789, EPI_ISL_1975790, EPI_ISL_1975795, EPI_ISL_1975797, EPI_ISL_1975799, EPI_ISL_1975802, EPI_ISL_1975804, EPI_ISL_1975805, EPI_ISL_1975807, EPI_ISL_1975808, EPI_ISL_1975809, EPI_ISL_1975810, EPI_ISL_1975811, EPI_ISL_1975812, EPI_ISL_1975813, EPI_ISL_1975814, EPI_ISL_1975816, EPI_ISL_1975817, EPI_ISL_1975818, EPI_ISL_1975819, EPI_ISL_1975820, EPI_ISL_1975821, EPI_ISL_1975822, EPI_ISL_1975823, EPI_ISL_1975825, EPI_ISL_1975830, EPI_ISL_1975832, EPI_ISL_1975833 | | | |
| see above | Regional Virus Laboratory, Belfast Health and Social Care Trust | COVID-19 Genomics UK (COG-UK) Consortium | Alison Watt; Ciara Cox; Conall McCaughey; David Simpson; Derek Fairley; James McKenna; Mairead Connor; Susan Feeney; Tanya Curran; Zoltan Molnar |
| EPI_ISL_1833807, EPI_ISL_1833821 | St George's University Hospitals NHS Foundation Trust | COVID-19 Genomics UK (COG-UK) Consortium | Adam Witney; Cassie Pope; Irene Monahan; Joshua Taylor; Ken Laing; NgeeKeong Tan |
| EPI_ISL_1248446, EPI_ISL_1248447, EPI_ISL_1248453, EPI_ISL_1248497, EPI_ISL_1637037, EPI_ISL_1637038 | University College London Hospital | COVID-19 Genomics UK (COG-UK) Consortium | Dr Catherine Houlihan; Dr Daniel Frampton; Dr Judith Heaney; Dr Moira Spyer; Dr Paul Grant and Dr Eleni Nastouli; Matthew Byott; Stuart Kirk |
| EPI_ISL_1178019, EPI_ISL_1178021, EPI_ISL_1178022, EPI_ISL_1178023, EPI_ISL_1178024, EPI_ISL_1178025, EPI_ISL_1178026, EPI_ISL_1178027, EPI_ISL_1178028, EPI_ISL_1178029, EPI_ISL_1178030, EPI_ISL_1178031, EPI_ISL_1178032, EPI_ISL_1178033, EPI_ISL_1178034, EPI_ISL_1178035, EPI_ISL_1178036, EPI_ISL_1178037, EPI_ISL_1247994, EPI_ISL_1248284, EPI_ISL_1248289, EPI_ISL_1248292, EPI_ISL_1248307, EPI_ISL_1248309, EPI_ISL_1248310, EPI_ISL_1248314, EPI_ISL_1248316, EPI_ISL_1248318, EPI_ISL_1248319, EPI_ISL_1248334, EPI_ISL_1248335, EPI_ISL_1248337, EPI_ISL_1248339, EPI_ISL_1248341, EPI_ISL_1248342, EPI_ISL_1248343, EPI_ISL_1248346, EPI_ISL_1248348, EPI_ISL_1248350, EPI_ISL_1248517, EPI_ISL_1248518, EPI_ISL_1248519, EPI_ISL_1248520, EPI_ISL_1248522, EPI_ISL_1248523, EPI_ISL_1248524, EPI_ISL_1248540, EPI_ISL_1248541, EPI_ISL_1248542, EPI_ISL_1309106, EPI_ISL_1309120, EPI_ISL_1386840, EPI_ISL_1386844, EPI_ISL_1386848, EPI_ISL_1386853, EPI_ISL_1386863, EPI_ISL_1386864, EPI_ISL_1386869, EPI_ISL_1637160, EPI_ISL_1637168 | | | |
| see above | University College London, Great Ormond Street Hospital for Children NHS Foundation Trust, Imperial College Healthcare NHS Trust | COVID-19 Genomics UK (COG-UK) Consortium | Alison Holmes; Charlotte Williams; Helena Tutill; Jacqueline Findlay; James Price; Judith Breuer; Julianne Brown; Kathryn Harris; Leysa Forrest; Mark Kristiansen; Paola Niola; Paola Resende Silva; Patricia Dyal; Paul Randell; Rachel Williams; Samuel Weeks; Sergi Castellano; Sunando Roy; Tony Brooks; Yasmin Panchbhaya |
| EPI_ISL_1247762, EPI_ISL_1247763, EPI_ISL_1247764, EPI_ISL_1247765, EPI_ISL_1308868, EPI_ISL_1308869, EPI_ISL_1308870, EPI_ISL_1308874, EPI_ISL_1308875, EPI_ISL_1308877, EPI_ISL_1308878, EPI_ISL_1308879, EPI_ISL_1308880, EPI_ISL_1308881, EPI_ISL_1308882, EPI_ISL_1308883, EPI_ISL_1730299 | | | |
| see above | University Hospitals Of Leicester NHS Trust and DeepSeq Nottingham | COVID-19 Genomics UK (COG-UK) Consortium | Christopher Holmes; Christopher Moore; Fei Sang; Joeseph Chappell; Johnny Debebe; Jonathan Ball; Julian Tang; Karlie Fallon; Matthew Carlisle; Matthew Loose; Nadine Holmes; Patrick McClure; Paul Bird; Thomas Helmer; Victoria Wright |
| EPI_ISL_1308488, EPI_ISL_1308490, EPI_ISL_1308491, EPI_ISL_1308495, EPI_ISL_1308499, EPI_ISL_1308501, EPI_ISL_1308502, EPI_ISL_1308503, EPI_ISL_1308506, EPI_ISL_1474399, EPI_ISL_1474400, EPI_ISL_1474466, EPI_ISL_1474484 | | | |
| see above | University of Birmingham | COVID-19 Genomics UK (COG-UK) Consortium | Alex Richter; Andrew Bosworth; Andrew Bosworth. Queen Elizabeth Hospital: Anna Casey; Andrew D Beggs PHE Heartlands Lab: Husam Osman; Charlotte Poxon; Institute of Microbiology; Joanne Stockton; Josh Quick; Kasun Wanigasooriya; Mike Kidd; Nicholas Loman. University of Birmingham Testing Laboratory: Celina M Whalley; Oliver Pickles; Radoslaw Poplawski; Samuel Nicholls; University of Birmingham: Claire McMurray; Will Rowe |
| EPI_ISL_1308646, EPI_ISL_1308647, EPI_ISL_1308648, EPI_ISL_1308654, EPI_ISL_1308666, EPI_ISL_1308672, EPI_ISL_1474533, EPI_ISL_1474539, EPI_ISL_1474548, EPI_ISL_1474549, EPI_ISL_1474550, EPI_ISL_1474551, EPI_ISL_1474552, EPI_ISL_1474554, EPI_ISL_1474555, EPI_ISL_1474556, EPI_ISL_1474557, EPI_ISL_1474559, EPI_ISL_1474561, EPI_ISL_1474562, EPI_ISL_1474563, EPI_ISL_1474614, EPI_ISL_1474615, EPI_ISL_1474616, EPI_ISL_1474617, EPI_ISL_1474618, EPI_ISL_1474619, EPI_ISL_1474620, EPI_ISL_1474621, EPI_ISL_1474622, EPI_ISL_1474623, EPI_ISL_1474624, EPI_ISL_1474625, EPI_ISL_1474626, EPI_ISL_1474627, EPI_ISL_1545766, EPI_ISL_1545772, EPI_ISL_1545775, EPI_ISL_1545777, EPI_ISL_1545778, EPI_ISL_1545780, EPI_ISL_1545781, EPI_ISL_1545783, EPI_ISL_1545786, EPI_ISL_1545787, EPI_ISL_1545788, EPI_ISL_1545789, EPI_ISL_1730171, EPI_ISL_1730172 | | | |
| see above | University of Exeter | COVID-19 Genomics UK (COG-UK) Consortium | Aaron Jeffries; Audrey Farbos; Ben Temperton; Jane Masoli; Joanna Warwick-Dugdale; Michelle Michelsen; Robyn Manley; Stephen Michell |
| EPI_ISL_1247725, EPI_ISL_1308719, EPI_ISL_1308774, EPI_ISL_1308775, EPI_ISL_1308776, EPI_ISL_1308777, EPI_ISL_1308807, EPI_ISL_1308808, EPI_ISL_1308809, EPI_ISL_1308810, EPI_ISL_1308811, EPI_ISL_1308816, EPI_ISL_1308817, EPI_ISL_1308864, EPI_ISL_1386653, EPI_ISL_1386654, EPI_ISL_1386655, EPI_ISL_1545911, EPI_ISL_1545913, EPI_ISL_1545916, EPI_ISL_1545919, EPI_ISL_1545929, EPI_ISL_1545934, EPI_ISL_1545939, EPI_ISL_1545940, EPI_ISL_1730080, EPI_ISL_1730235, EPI_ISL_1730236, EPI_ISL_1730250, EPI_ISL_1832236, EPI_ISL_1832237, EPI_ISL_1832238, EPI_ISL_1832239, EPI_ISL_1832250, EPI_ISL_1832251, EPI_ISL_1974811, EPI_ISL_1974812, EPI_ISL_2125629, EPI_ISL_2125631, EPI_ISL_2125632, EPI_ISL_2125634, EPI_ISL_2125736, EPI_ISL_2125737 | | | |
| see above | Virology Department, Royal Infirmary of Edinburgh, NHS Lothian / School of Biological Sciences, University of Edinburgh | COVID-19 Genomics UK (COG-UK) Consortium | Colquhoun R; Cotton S; Dewar R; Gallagher A; Hill V; Jackson B; Maloney D; McCrone JT; McHugh M; O'Toole Á; Rambaut A; Rooke S; Scher E; Templeton K; Yu X |
| EPI_ISL_1180057, EPI_ISL_1180067, EPI_ISL_1180077, EPI_ISL_1180085, EPI_ISL_1180092, EPI_ISL_1180094, EPI_ISL_1180096, EPI_ISL_1180100, EPI_ISL_1180101, EPI_ISL_1180103, EPI_ISL_1180104, EPI_ISL_1250258, EPI_ISL_1250264, EPI_ISL_1250266, EPI_ISL_1250270, EPI_ISL_1250271, EPI_ISL_1250272, EPI_ISL_1250275, EPI_ISL_1250277, EPI_ISL_1250284, EPI_ISL_1250287, EPI_ISL_1250288, EPI_ISL_1250289, EPI_ISL_1250290, EPI_ISL_1250293, EPI_ISL_1250295, EPI_ISL_1250301, EPI_ISL_1250302, EPI_ISL_1250305, EPI_ISL_1250307, EPI_ISL_1638021, EPI_ISL_1638024, EPI_ISL_1730951, EPI_ISL_1730985, EPI_ISL_1730995, EPI_ISL_1731002, EPI_ISL_1731012, EPI_ISL_1731026, EPI_ISL_1833841, EPI_ISL_1833844, EPI_ISL_1833865, EPI_ISL_1833881, EPI_ISL_1833894 | | | |
| see above | Virology Department, Sheffield Teaching Hospitals NHS Foundation Trust/Department of Infection, Immunity and Cardiovascular Disease, The Medical School, University of Sheffield | COVID-19 Genomics UK (COG-UK) Consortium | Adri Angyal; Alex Keeley; Benjamin Lindsey; Cariad Evans; Danielle Groves; Dave Partridge; Katie Johnson; Laura Carrilero; Luke Green; Matthew Parker; Matthew Wyles; Mehmet Yavuz; Mohammad Raza; Nikki Smith; Paul Parsons; Rachel Tucker; Rebecca Brown; Thushan de Silva |
| EPI_ISL_1249303, EPI_ISL_1249304, EPI_ISL_1249390, EPI_ISL_1249392, EPI_ISL_1249393, EPI_ISL_1249394, EPI_ISL_1249396, EPI_ISL_1249403, EPI_ISL_1249404, EPI_ISL_1249418, EPI_ISL_1249423, EPI_ISL_1249428, EPI_ISL_1249429, EPI_ISL_1249436, EPI_ISL_1249439, EPI_ISL_1249440, EPI_ISL_1249445, EPI_ISL_1249453, EPI_ISL_1249454, EPI_ISL_1249468, EPI_ISL_1249471, EPI_ISL_1249472, EPI_ISL_1249474, EPI_ISL_1249475, EPI_ISL_1249476, EPI_ISL_1249477, EPI_ISL_1249478, EPI_ISL_1249479, EPI_ISL_1249480, EPI_ISL_1249481, EPI_ISL_1249482, EPI_ISL_1249483, EPI_ISL_1249490, EPI_ISL_1249491, EPI_ISL_1249498, EPI_ISL_1249508, EPI_ISL_1249515, EPI_ISL_1249533, EPI_ISL_1249540, EPI_ISL_1249549, EPI_ISL_1249559, EPI_ISL_1249561, EPI_ISL_1249576, EPI_ISL_1249578, EPI_ISL_1249579, EPI_ISL_1249585, EPI_ISL_1249593, EPI_ISL_1249607, EPI_ISL_1249609, EPI_ISL_1249611, EPI_ISL_1249613, EPI_ISL_1249614, EPI_ISL_1249615, EPI_ISL_1249616, EPI_ISL_1249617, EPI_ISL_1249618, EPI_ISL_1249619, EPI_ISL_1249620, EPI_ISL_1249624, EPI_ISL_1249680, EPI_ISL_1249682, EPI_ISL_1249684, EPI_ISL_1249685, EPI_ISL_1249690, EPI_ISL_1249691, EPI_ISL_1249692, EPI_ISL_1249697, EPI_ISL_1249698, EPI_ISL_1249706, EPI_ISL_1249707, EPI_ISL_1249708, EPI_ISL_1249709, EPI_ISL_1249710, EPI_ISL_1249712, EPI_ISL_1249713, EPI_ISL_1249722, EPI_ISL_1249723, EPI_ISL_1249724, EPI_ISL_1249738, EPI_ISL_1249750, EPI_ISL_1249751, EPI_ISL_1249759, EPI_ISL_1249790, EPI_ISL_1249811, EPI_ISL_1249815, EPI_ISL_1249816, EPI_ISL_1249817, EPI_ISL_1249818, EPI_ISL_1249851, EPI_ISL_1249867, EPI_ISL_1387670, EPI_ISL_1387672, EPI_ISL_1387675, EPI_ISL_1387677, EPI_ISL_1387684, EPI_ISL_1387685, EPI_ISL_1387689, EPI_ISL_1387694, EPI_ISL_1387709, EPI_ISL_1387716, EPI_ISL_1387717, EPI_ISL_1387718, EPI_ISL_1387719, EPI_ISL_1387720, EPI_ISL_1387722, EPI_ISL_1387724, EPI_ISL_1387726, EPI_ISL_1387727, EPI_ISL_1387728, EPI_ISL_1387729, EPI_ISL_1387733, EPI_ISL_1387734, EPI_ISL_1387737, EPI_ISL_1387738, EPI_ISL_1387744, EPI_ISL_1387775, EPI_ISL_1387776, EPI_ISL_1387777, EPI_ISL_1387778, EPI_ISL_1387789, EPI_ISL_1387790, EPI_ISL_1387793, EPI_ISL_1387794, EPI_ISL_1833127 | | | |
| see above | Wales Specialist Virology Centre Sequencing lab: Pathogen Genomics Unit | Public Health Wales Microbiology Cardiff Wales Specialist Virology Centre | Alec Birchley; Alexander Adams; Amy Gaskin; Angela Marchbank; Bree Gatica-Wilcox; Catherine Moore; Jason Coombes; Joanne Watkins; Joel Southgate; Johnathan Evans; Laura Gifford; Lauren Gilbert; Lee Graham; Malorie Perry; Matthew Bull; Nicole Pacchiarini; Sally Corden; Sara Kumziene-Summerhayes; Sara Rey; Sarah Taylor; Simon Cottrell; Sophie Jones; Tom Connor |
| EPI_ISL_1247685, EPI_ISL_1386591, EPI_ISL_1386592, EPI_ISL_1386593, EPI_ISL_1386594, EPI_ISL_1386609, EPI_ISL_1545872, EPI_ISL_1545873, EPI_ISL_1545874, EPI_ISL_1545876, EPI_ISL_1545877, EPI_ISL_1545879, EPI_ISL_1545889, EPI_ISL_1636626 | | | |
| see above | West of Scotland Specialist Virology Centre, NHSGGC / MRC-University of Glasgow Centre for Virus Research | COVID-19 Genomics UK (COG-UK) Consortium | Alasdair MacLean; Alice Broos; Ana da Silva Filipe; Antonia Ho; Daniel Mair; David L Robertson; Emma Thomson; Guy Mollett; James Shepherd; Jenna Nichols; Joseph Hughes; Kathy Li; Kathy Smollett; Kyriaki Nomikou; Lily Tong; Matthew Holden; Natasha Johnson; Rachel Blacow; Richard Orton; Rory Gunson; Sarah McDonald; Sharif Shaaban; Sreenu Vattipally; Stephen Carmichael |

We gratefully acknowledge the following Authors from the Originating laboratories responsible for obtaining the specimens, as well as the Submitting laboratories where the genome data were generated and shared via GISAID, on which this research is based.

All Submitters of data may be contacted directly via www.gisaid.org

Authors are sorted alphabetically.

| Accession ID | Originating Laboratory | Submitting Laboratory | Authors |
| --- | --- | --- | --- |
| EPI_ISL_1308949, EPI_ISL_1308950, EPI_ISL_1474802, EPI_ISL_1474820, EPI_ISL_1474821, EPI_ISL_1474822, EPI_ISL_1474823, EPI_ISL_1730313, EPI_ISL_1730314, EPI_ISL_1730315 | | | |
| see above | Barts Health NHS Trust | COVID-19 Genomics UK (COG-UK) Consortium | BROAD; Beatrix; CUTINO-MOGUEL; Claire; David; Dola; HARRINGTON; KELE; KULASEGARAN-SHYLINI; Maria-Teresa; OWOYEMI; Raghavendran |
| EPI_ISL_1250310, EPI_ISL_1250311, EPI_ISL_1250313, EPI_ISL_1250316, EPI_ISL_1250317 | Bioinformatics and Biostatistics Lab, Advanced Sequencing Facility | COVID-19 Genomics UK (COG-UK) Consortium | Aengus Stewart; Chelsea Sawyer; Harshil Patel; Jerome Nicod; Laura Cubitt; Margaret Crawford |
| EPI_ISL_1310440, EPI_ISL_1310444, EPI_ISL_1388676, EPI_ISL_1388677, EPI_ISL_1388680, EPI_ISL_1388684, EPI_ISL_1388686, EPI_ISL_1388688, EPI_ISL_1388690, EPI_ISL_1388692, EPI_ISL_1388694, EPI_ISL_1388696, EPI_ISL_1388700, EPI_ISL_1388702, EPI_ISL_1388704, EPI_ISL_1388874, EPI_ISL_1388878, EPI_ISL_1388880, EPI_ISL_1388882, EPI_ISL_1388891, EPI_ISL_1388893, EPI_ISL_1388897, EPI_ISL_1388899, EPI_ISL_1388901, EPI_ISL_1388904, EPI_ISL_1388906, EPI_ISL_1388908, EPI_ISL_1388910, EPI_ISL_1388912, EPI_ISL_1388914, EPI_ISL_1476555, EPI_ISL_1476556, EPI_ISL_1476557, EPI_ISL_1476558, EPI_ISL_1476559, EPI_ISL_1476560, EPI_ISL_1476561, EPI_ISL_1476562, EPI_ISL_1476565, EPI_ISL_1637990, EPI_ISL_1637995 | | | |
| see above | Centre for Enzyme Innovation, University of Portsmouth / Translational Research Laboratory, Portsmouth Hospitals NHS Trust | COVID-19 Genomics UK (COG-UK) Consortium | Angela Beckett; Christopher Fearn; Kate Cook; Katie Loveson; Salman Goudarzi; Samuel Robson; Scott Elliott; Sharon Glaysher |
| EPI_ISL_1247351, EPI_ISL_1247617, EPI_ISL_1247619, EPI_ISL_1247620, EPI_ISL_1247621, EPI_ISL_1247622, EPI_ISL_1247624, EPI_ISL_1247625, EPI_ISL_1247626, EPI_ISL_1247627, EPI_ISL_1247628, EPI_ISL_1247629, EPI_ISL_1250217, EPI_ISL_1250221, EPI_ISL_1308683, EPI_ISL_1310462, EPI_ISL_1310493, EPI_ISL_1310520, EPI_ISL_1310622, EPI_ISL_1310623, EPI_ISL_1310624, EPI_ISL_1310627, EPI_ISL_1310630, EPI_ISL_1310631, EPI_ISL_1310635, EPI_ISL_1310640, EPI_ISL_1310641, EPI_ISL_1310642, EPI_ISL_1310646, EPI_ISL_1310651, EPI_ISL_1310652, EPI_ISL_1310661, EPI_ISL_1310662, EPI_ISL_1310663, EPI_ISL_1310667, EPI_ISL_1310671, EPI_ISL_1310672, EPI_ISL_1310673, EPI_ISL_1310676, EPI_ISL_1310677, EPI_ISL_1310681, EPI_ISL_1310682, EPI_ISL_1310685, EPI_ISL_1310686, EPI_ISL_1310691, EPI_ISL_1730020, EPI_ISL_1730034 | | | |
| see above | Department of Pathology, University of Cambridge | COVID-19 Genomics UK (COG-UK) Consortium | Aminu S. Jahun; Ian Goodfellow; Iliana Georgana; Martin D. Curran; Myra Hosmillo; Rhys Izuagbe; Surendra Parmar; William L. Hamilton; Yasmin Chaudhry |
| EPI_ISL_1246008, EPI_ISL_1255489, EPI_ISL_1255520, EPI_ISL_1255524, EPI_ISL_1255617, EPI_ISL_1255678, EPI_ISL_1255680, EPI_ISL_1255682, EPI_ISL_1255684, EPI_ISL_1255685, EPI_ISL_1255692, EPI_ISL_1255693, EPI_ISL_1255695, EPI_ISL_1255696, EPI_ISL_1255697, EPI_ISL_1255698, EPI_ISL_1255703, EPI_ISL_1255704, EPI_ISL_1255705, EPI_ISL_1255707, EPI_ISL_1255708, EPI_ISL_1255710, EPI_ISL_1255716, EPI_ISL_1255719, EPI_ISL_1255720, EPI_ISL_1255721, EPI_ISL_1255722, EPI_ISL_1255723, EPI_ISL_1255724, EPI_ISL_1255725, EPI_ISL_1255726, EPI_ISL_1255727, EPI_ISL_1255728, EPI_ISL_1255729, EPI_ISL_1255730, EPI_ISL_1255733, EPI_ISL_1255735, EPI_ISL_1255736, EPI_ISL_1255737, EPI_ISL_1255738, EPI_ISL_1255739, EPI_ISL_1255740, EPI_ISL_1255744, EPI_ISL_1255746, EPI_ISL_1255748, EPI_ISL_1255750, EPI_ISL_1255751, EPI_ISL_1255752, EPI_ISL_1255753, EPI_ISL_1255754, EPI_ISL_1255755, EPI_ISL_1255756, EPI_ISL_1255757, EPI_ISL_1255758, EPI_ISL_1255759, EPI_ISL_1255761, EPI_ISL_1255762, EPI_ISL_1255764, EPI_ISL_1255767, EPI_ISL_1255768, EPI_ISL_1255769, EPI_ISL_1255772, EPI_ISL_1255773, EPI_ISL_1255774, EPI_ISL_1255776, EPI_ISL_1255778, EPI_ISL_1255779, EPI_ISL_1255780, EPI_ISL_1255781, EPI_ISL_1255782, EPI_ISL_1255783, EPI_ISL_1255784, EPI_ISL_1255785, EPI_ISL_1255786, EPI_ISL_1255787, EPI_ISL_1255788, EPI_ISL_1255789, EPI_ISL_1255791, EPI_ISL_1255793, EPI_ISL_1255795, EPI_ISL_1255799, EPI_ISL_1255800, EPI_ISL_1255804, EPI_ISL_1255805, EPI_ISL_1255807, EPI_ISL_1255808, EPI_ISL_1255809, EPI_ISL_1255810, EPI_ISL_1255811, EPI_ISL_1255816, EPI_ISL_1255817, EPI_ISL_1255818, EPI_ISL_1255819, EPI_ISL_1255821, EPI_ISL_1255824, EPI_ISL_1255825, EPI_ISL_1255828, EPI_ISL_1255829, EPI_ISL_1255831, EPI_ISL_1255832, EPI_ISL_1255833, EPI_ISL_1255836, EPI_ISL_1255837, EPI_ISL_1255838, EPI_ISL_1255840, EPI_ISL_1255841, EPI_ISL_1255842, EPI_ISL_1255843, EPI_ISL_1255844, EPI_ISL_1255845, EPI_ISL_1255846, EPI_ISL_1255847, EPI_ISL_1255849, EPI_ISL_1255850, EPI_ISL_1255851, EPI_ISL_1255853, EPI_ISL_1255857, EPI_ISL_1255858, EPI_ISL_1255860, EPI_ISL_1255863, EPI_ISL_1255868, EPI_ISL_1255869, EPI_ISL_1255870, EPI_ISL_1255871, EPI_ISL_1255873, EPI_ISL_1255876, EPI_ISL_1255878, EPI_ISL_1255880, EPI_ISL_1255882, EPI_ISL_1255884, EPI_ISL_1255886, EPI_ISL_1255887, EPI_ISL_1255888, EPI_ISL_1255892, EPI_ISL_1255893, EPI_ISL_1255896, EPI_ISL_1255898, EPI_ISL_1255899, EPI_ISL_1255901, EPI_ISL_1255902, EPI_ISL_1255903, EPI_ISL_1255905, EPI_ISL_1255906, EPI_ISL_1255907, EPI_ISL_1255909, EPI_ISL_1255910, EPI_ISL_1255911, EPI_ISL_1255912, EPI_ISL_1255913, EPI_ISL_1255914, EPI_ISL_1255916, EPI_ISL_1255917, EPI_ISL_1255918, EPI_ISL_1255920, EPI_ISL_1255923, EPI_ISL_1255924, EPI_ISL_1255925, EPI_ISL_1255926, EPI_ISL_1255929, EPI_ISL_1255930, EPI_ISL_1255931, EPI_ISL_1255933, EPI_ISL_1255934, EPI_ISL_1255941, EPI_ISL_1255942, EPI_ISL_1255945, EPI_ISL_1255948, EPI_ISL_1255953, EPI_ISL_1255955, EPI_ISL_1255958, EPI_ISL_1255959, EPI_ISL_1255961, EPI_ISL_1255962, EPI_ISL_1255964, EPI_ISL_1255965, EPI_ISL_1255968, EPI_ISL_1255971, EPI_ISL_1255973, EPI_ISL_1255974, EPI_ISL_1255978, EPI_ISL_1255980, EPI_ISL_1255981, EPI_ISL_1255983, EPI_ISL_1255984, EPI_ISL_1255986, EPI_ISL_1255987, EPI_ISL_1255989, EPI_ISL_1255990, EPI_ISL_1255992, EPI_ISL_1255993, EPI_ISL_1255995, EPI_ISL_1255996, EPI_ISL_1255998, EPI_ISL_1256001, EPI_ISL_1256004, EPI_ISL_1256007, EPI_ISL_1256009, EPI_ISL_1256010, EPI_ISL_1256015, EPI_ISL_1256018, EPI_ISL_1256020, EPI_ISL_1256022, EPI_ISL_1256023, EPI_ISL_1256026, EPI_ISL_1256030, EPI_ISL_1256031, EPI_ISL_1256033, EPI_ISL_1256034, EPI_ISL_1256037, EPI_ISL_1256047, EPI_ISL_1256055, EPI_ISL_1256057, EPI_ISL_1256078, EPI_ISL_1256082, EPI_ISL_1256089, EPI_ISL_1256094, EPI_ISL_1256106, EPI_ISL_1256110, EPI_ISL_1256114, EPI_ISL_1256116, EPI_ISL_1256125, EPI_ISL_1256128, EPI_ISL_1256129, EPI_ISL_1256130, EPI_ISL_1256134, EPI_ISL_1256144, EPI_ISL_1256145, EPI_ISL_1256148, EPI_ISL_1256149, EPI_ISL_1256152, EPI_ISL_1256155, EPI_ISL_1256160, EPI_ISL_1256166, EPI_ISL_1256172, EPI_ISL_1256175, EPI_ISL_1256176, EPI_ISL_1256193, EPI_ISL_1256195, EPI_ISL_1256198, EPI_ISL_1256200, EPI_ISL_1256201, EPI_ISL_1256208, EPI_ISL_1256211, EPI_ISL_1256213, EPI_ISL_1256217, EPI_ISL_1256219, EPI_ISL_1256220, EPI_ISL_1256222, EPI_ISL_1256225, EPI_ISL_1256229, EPI_ISL_1256239, EPI_ISL_1256246, EPI_ISL_1256252, EPI_ISL_1256259, EPI_ISL_1256260, EPI_ISL_1256261, EPI_ISL_1256277, EPI_ISL_1256281, EPI_ISL_1256284, EPI_ISL_1256291, EPI_ISL_1256299, EPI_ISL_1256307, EPI_ISL_1256308, EPI_ISL_1256310, EPI_ISL_1256312, EPI_ISL_1256332, EPI_ISL_1256334, EPI_ISL_1256338, EPI_ISL_1256350, EPI_ISL_1256355, EPI_ISL_1256357, EPI_ISL_1256360, EPI_ISL_1256684, EPI_ISL_1256686, EPI_ISL_1256692, EPI_ISL_1256693, EPI_ISL_1256696, EPI_ISL_1256699, EPI_ISL_1256705, EPI_ISL_1256706, EPI_ISL_1256708, EPI_ISL_1256711, EPI_ISL_1256713, EPI_ISL_1256722, EPI_ISL_1256727, EPI_ISL_1256728, EPI_ISL_1256729, EPI_ISL_1256731, EPI_ISL_1256732, EPI_ISL_1256736, EPI_ISL_1256737, EPI_ISL_1256742, EPI_ISL_1256746, EPI_ISL_1256748, EPI_ISL_1256751, EPI_ISL_1256754, EPI_ISL_1256762, EPI_ISL_1256763, EPI_ISL_1256768, EPI_ISL_1256769, EPI_ISL_1256774, EPI_ISL_1256775, EPI_ISL_1256778, EPI_ISL_1256781, EPI_ISL_1256785, EPI_ISL_1256790, EPI_ISL_1256791, EPI_ISL_1256792, EPI_ISL_1256797, EPI_ISL_1256800, EPI_ISL_1256801, EPI_ISL_1256805, EPI_ISL_1256806, EPI_ISL_1256807, EPI_ISL_1256811, EPI_ISL_1256812, EPI_ISL_1256813, EPI_ISL_1256816, EPI_ISL_1256818, EPI_ISL_1256825, EPI_ISL_1256832, EPI_ISL_1256835, EPI_ISL_1256842, EPI_ISL_1256844, EPI_ISL_1256849, EPI_ISL_1256854, EPI_ISL_1256855, EPI_ISL_1256861, EPI_ISL_1256863, EPI_ISL_1256864, EPI_ISL_1256866, EPI_ISL_1256873, EPI_ISL_1256874, EPI_ISL_1256875, EPI_ISL_1256881, EPI_ISL_1256882, EPI_ISL_1256885, EPI_ISL_1256888, EPI_ISL_1256896, EPI_ISL_1256897, EPI_ISL_1256902, EPI_ISL_1256909, EPI_ISL_1256920, EPI_ISL_1256921, EPI_ISL_1256923, EPI_ISL_1256926, EPI_ISL_1256927, EPI_ISL_1256928, EPI_ISL_1256930, EPI_ISL_1256935, EPI_ISL_1256936, EPI_ISL_1256938, EPI_ISL_1256940, EPI_ISL_1264072, EPI_ISL_1264085, EPI_ISL_1264108, EPI_ISL_1264127, EPI_ISL_1264129, EPI_ISL_1264151, EPI_ISL_1264157, EPI_ISL_1264174, EPI_ISL_1264178, EPI_ISL_1264186, EPI_ISL_1264232, EPI_ISL_1264235, EPI_ISL_1264254, EPI_ISL_1264256, EPI_ISL_1264295, EPI_ISL_1264297, EPI_ISL_1264302, EPI_ISL_1264336, EPI_ISL_1264349, EPI_ISL_1264352, EPI_ISL_1264355, EPI_ISL_1264357, EPI_ISL_1264378, EPI_ISL_1264382, EPI_ISL_1264384, EPI_ISL_1264394, EPI_ISL_1264401, EPI_ISL_1264406, EPI_ISL_1264411, EPI_ISL_1264436, EPI_ISL_1264437, EPI_ISL_1264442, EPI_ISL_1264448, EPI_ISL_1264457, EPI_ISL_1264464, EPI_ISL_1264511, EPI_ISL_1264512, EPI_ISL_1264535, EPI_ISL_1264536, EPI_ISL_1264547, EPI_ISL_1264558, EPI_ISL_1264565, EPI_ISL_1264570, EPI_ISL_1264580, EPI_ISL_1264583, EPI_ISL_1264590, EPI_ISL_1264595, EPI_ISL_1264622, EPI_ISL_1273989, EPI_ISL_1273992, EPI_ISL_1273995, EPI_ISL_1273996, EPI_ISL_1273999, EPI_ISL_1274000, EPI_ISL_1274003, EPI_ISL_1274010, EPI_ISL_1274011, EPI_ISL_1274018, EPI_ISL_1274024, EPI_ISL_1274025, EPI_ISL_1274027, EPI_ISL_1274029, EPI_ISL_1274031, EPI_ISL_1274033, EPI_ISL_1274034, EPI_ISL_1274039, EPI_ISL_1274043, EPI_ISL_1274044, EPI_ISL_1274051, EPI_ISL_1274052, EPI_ISL_1274053, EPI_ISL_1274055, EPI_ISL_1274063, EPI_ISL_1274064, EPI_ISL_1274071, EPI_ISL_1274077, EPI_ISL_1274082, EPI_ISL_1274090, EPI_ISL_1274098, EPI_ISL_1274101, EPI_ISL_1274102, EPI_ISL_1274103, EPI_ISL_1274104, EPI_ISL_1274106, EPI_ISL_1274108, EPI_ISL_1274109, EPI_ISL_1274111, EPI_ISL_1274115, EPI_ISL_1274117, EPI_ISL_1274118, EPI_ISL_1274119, EPI_ISL_1274120, EPI_ISL_1274121, EPI_ISL_1274129, EPI_ISL_1274132, EPI_ISL_1274137, EPI_ISL_1274139, EPI_ISL_1274142, EPI_ISL_1274148, EPI_ISL_1274149, EPI_ISL_1274150, EPI_ISL_1274152, EPI_ISL_1274153, EPI_ISL_1274154, EPI_ISL_1274156, EPI_ISL_1274160, EPI_ISL_1274164, EPI_ISL_1274168, EPI_ISL_1274174, EPI_ISL_1274178, EPI_ISL_1274180, EPI_ISL_1274182, EPI_ISL_1274189, EPI_ISL_1274193, EPI_ISL_1274196, EPI_ISL_1274205, EPI_ISL_1274206, EPI_ISL_1274211, EPI_ISL_1274225, EPI_ISL_1274226, EPI_ISL_1274227, EPI_ISL_1274229, EPI_ISL_1274230, EPI_ISL_1274234, EPI_ISL_1274235, EPI_ISL_1274236, EPI_ISL_1274241, EPI_ISL_1274244, EPI_ISL_1274248, EPI_ISL_1274255, EPI_ISL_1274261, EPI_ISL_1274263, EPI_ISL_1274267, EPI_ISL_1274268, EPI_ISL_1274270, EPI_ISL_1274275, EPI_ISL_1274277, EPI_ISL_1274278, EPI_ISL_1274282, EPI_ISL_1274284, EPI_ISL_1274285, EPI_ISL_1274291, EPI_ISL_1274293, EPI_ISL_1274294, EPI_ISL_1274295, EPI_ISL_1274299, EPI_ISL_1276133, EPI_ISL_1276152, EPI_ISL_1276166, EPI_ISL_1276169, EPI_ISL_1276180, EPI_ISL_1276191, EPI_ISL_1276197, EPI_ISL_1276215, EPI_ISL_1276220, EPI_ISL_1276238, EPI_ISL_1276256, EPI_ISL_1276257, EPI_ISL_1276265, EPI_ISL_1276286, EPI_ISL_1276300, EPI_ISL_1276306, EPI_ISL_1276315, EPI_ISL_1276319, EPI_ISL_1276326, EPI_ISL_1276328, EPI_ISL_1276332, EPI_ISL_1276335, EPI_ISL_1276347, EPI_ISL_1276348, EPI_ISL_1276369, EPI_ISL_1276492, EPI_ISL_1276657, EPI_ISL_1276663, EPI_ISL_1276664, EPI_ISL_1276686, EPI_ISL_1276698, EPI_ISL_1276701, EPI_ISL_1276710, EPI_ISL_1276714, EPI_ISL_1276720, EPI_ISL_1276729, EPI_ISL_1276734, EPI_ISL_1276745, EPI_ISL_1276752, EPI_ISL_1276754, EPI_ISL_1276758, EPI_ISL_1276760, EPI_ISL_1276786, EPI_ISL_1276800, EPI_ISL_1276802, EPI_ISL_1276806, EPI_ISL_1276808, EPI_ISL_1276811, EPI_ISL_1276814, EPI_ISL_1276853, EPI_ISL_1276854, EPI_ISL_1276861, EPI_ISL_1276864, EPI_ISL_1276869, EPI_ISL_1276875, EPI_ISL_1276876, EPI_ISL_1276879, EPI_ISL_1276882, EPI_ISL_1276891, EPI_ISL_1277338, EPI_ISL_1277758, EPI_ISL_1277760, EPI_ISL_1315994, EPI_ISL_1316423, EPI_ISL_1316434, EPI_ISL_1316448, EPI_ISL_1316450, EPI_ISL_1316454, EPI_ISL_1316459, EPI_ISL_1316460, EPI_ISL_1316464, EPI_ISL_1316469, EPI_ISL_1316476, EPI_ISL_1316481, EPI_ISL_1316485, EPI_ISL_1316486, EPI_ISL_1316497, EPI_ISL_1316498, EPI_ISL_1316504, EPI_ISL_1316509, EPI_ISL_1316514, EPI_ISL_1316518, EPI_ISL_1316533, EPI_ISL_1316536, EPI_ISL_1316540, EPI_ISL_1316545, EPI_ISL_1316549, EPI_ISL_1316552, EPI_ISL_1316557, EPI_ISL_1316559, EPI_ISL_1316561, EPI_ISL_1316572, EPI_ISL_1316583, EPI_ISL_1316584, EPI_ISL_1316586, EPI_ISL_1316589, EPI_ISL_1316595, EPI_ISL_1316596, EPI_ISL_1316604, EPI_ISL_1316605, EPI_ISL_1316610, EPI_ISL_1316611, EPI_ISL_1316613, EPI_ISL_1316614, EPI_ISL_1316627, EPI_ISL_1316629, EPI_ISL_1316635, EPI_ISL_1316637, EPI_ISL_1316640, EPI_ISL_1316642, EPI_ISL_1316644, EPI_ISL_1316647, EPI_ISL_1316656, EPI_ISL_1316657, EPI_ISL_1316660, EPI_ISL_1316663, EPI_ISL_1316664, EPI_ISL_1316667, EPI_ISL_1316669, EPI_ISL_1316675, EPI_ISL_1316693, EPI_ISL_1316700, EPI_ISL_1316701, EPI_ISL_1316711, EPI_ISL_1316721, EPI_ISL_1316723, EPI_ISL_1316725, EPI_ISL_1316727, EPI_ISL_1316731, EPI_ISL_1316762, EPI_ISL_1316765, EPI_ISL_1316773, EPI_ISL_1326675, EPI_ISL_1326706, EPI_ISL_1326724, EPI_ISL_1326773, EPI_ISL_1326779, EPI_ISL_1326784, EPI_ISL_1326794, EPI_ISL_1326795, EPI_ISL_1326808, EPI_ISL_1326828, EPI_ISL_1326842, EPI_ISL_1326894, EPI_ISL_1326927, EPI_ISL_1326937, EPI_ISL_1326960, EPI_ISL_1326974, EPI_ISL_1326978, EPI_ISL_1326980, EPI_ISL_1326983, EPI_ISL_1326986, EPI_ISL_1326987, EPI_ISL_1326989, EPI_ISL_1327003, EPI_ISL_1327005, EPI_ISL_1327009, EPI_ISL_1327011, EPI_ISL_1327012, EPI_ISL_1327013, EPI_ISL_1327014, EPI_ISL_1327015, EPI_ISL_1327016, EPI_ISL_1327029, EPI_ISL_1327030, EPI_ISL_1327031, EPI_ISL_1327039, EPI_ISL_1327040, EPI_ISL_1327042, EPI_ISL_1327046, EPI_ISL_1327050, EPI_ISL_1327055, EPI_ISL_1327058, EPI_ISL_1327063, EPI_ISL_1327064, EPI_ISL_1327070, EPI_ISL_1327073, EPI_ISL_1327075, EPI_ISL_1327078, EPI_ISL_1327081, EPI_ISL_1327083, EPI_ISL_1327084, EPI_ISL_1327090, EPI_ISL_1327091, EPI_ISL_1327098, EPI_ISL_1327102, EPI_ISL_1327105, EPI_ISL_1327107, EPI_ISL_1327109, EPI_ISL_1327112, EPI_ISL_1327115, EPI_ISL_1327119, EPI_ISL_1327125, EPI_ISL_1327131, EPI_ISL_1327132, EPI_ISL_1327137, EPI_ISL_1327140, EPI_ISL_1327141, EPI_ISL_1327144, EPI_ISL_1327146, EPI_ISL_1327161, EPI_ISL_1327163, EPI_ISL_1327164, EPI_ISL_1327165, EPI_ISL_1327171, EPI_ISL_1327177, EPI_ISL_1327180, EPI_ISL_1327182, EPI_ISL_1327183, EPI_ISL_1327187, EPI_ISL_1327191, EPI_ISL_1327193, EPI_ISL_1327208, EPI_ISL_1327213, EPI_ISL_1327219, EPI_ISL_1327231, EPI_ISL_1327233, EPI_ISL_1327237, EPI_ISL_1328898, EPI_ISL_1328902, EPI_ISL_1328909, EPI_ISL_1328910, EPI_ISL_1328913, EPI_ISL_1328916, EPI_ISL_1328921, EPI_ISL_1328924, EPI_ISL_1328925, EPI_ISL_1328948, EPI_ISL_1328949, EPI_ISL_1328951, EPI_ISL_1328955, EPI_ISL_1328956, EPI_ISL_1328961, EPI_ISL_1328965, EPI_ISL_1328969, EPI_ISL_1328973, EPI_ISL_1328978, EPI_ISL_1329000, EPI_ISL_1329014, EPI_ISL_1329015, EPI_ISL_1329034, EPI_ISL_1329044, EPI_ISL_1329046, EPI_ISL_1329047, EPI_ISL_1329050, EPI_ISL_1329073, EPI_ISL_1329075, EPI_ISL_1329078, EPI_ISL_1329085, EPI_ISL_1329089, EPI_ISL_1329090, EPI_ISL_1329095, EPI_ISL_1329096, EPI_ISL_1329097, EPI_ISL_1329100, EPI_ISL_1329118, EPI_ISL_1329119, EPI_ISL_1329120, EPI_ISL_1329126, EPI_ISL_1329128, EPI_ISL_1329135, EPI_ISL_1329146, EPI_ISL_1329151, EPI_ISL_1329154, EPI_ISL_1329160, EPI_ISL_1329162, EPI_ISL_1329164, EPI_ISL_1329170, EPI_ISL_1329174, EPI_ISL_1329178, EPI_ISL_1329184, EPI_ISL_1329186, EPI_ISL_1329188, EPI_ISL_1329190, EPI_ISL_1329192, EPI_ISL_1329198, EPI_ISL_1329207, EPI_ISL_1329220, EPI_ISL_1329224, EPI_ISL_1329245, EPI_ISL_1329252, EPI_ISL_1329269, EPI_ISL_1329274, EPI_ISL_1329286, EPI_ISL_1329291, EPI_ISL_1329310, EPI_ISL_1329319, EPI_ISL_1329323, EPI_ISL_1329336, EPI_ISL_1329338, EPI_ISL_1329342, EPI_ISL_1329372, EPI_ISL_1330027, EPI_ISL_1330297, EPI_ISL_1330300, EPI_ISL_1330308, EPI_ISL_1330313, EPI_ISL_1330315, EPI_ISL_1330322, EPI_ISL_1330326, EPI_ISL_1330351, EPI_ISL_1330362, EPI_ISL_1330363, EPI_ISL_1330364, EPI_ISL_1330378, EPI_ISL_1330381, EPI_ISL_1330382, EPI_ISL_1330385, EPI_ISL_1330396, EPI_ISL_1330399, EPI_ISL_1330407, EPI_ISL_1330413, EPI_ISL_1330420, EPI_ISL_1330429, EPI_ISL_1330434, EPI_ISL_1330445, EPI_ISL_1330457, EPI_ISL_1330458, EPI_ISL_1330460, EPI_ISL_1330465, EPI_ISL_1330470, EPI_ISL_1330480, EPI_ISL_1330489, EPI_ISL_1330492, EPI_ISL_1330495, EPI_ISL_1330505, EPI_ISL_1330508, EPI_ISL_1330513, EPI_ISL_1330519, EPI_ISL_1330521, EPI_ISL_1330527, EPI_ISL_1330537, EPI_ISL_1330548, EPI_ISL_1330550, EPI_ISL_1330556, EPI_ISL_1330564, EPI_ISL_1330580, EPI_ISL_1330588, EPI_ISL_1330591, EPI_ISL_1330600, EPI_ISL_1330601, EPI_ISL_1330610, EPI_ISL_1331131, EPI_ISL_1331132, EPI_ISL_1331137, EPI_ISL_1331139, EPI_ISL_1331140, EPI_ISL_1331142, EPI_ISL_1331147, EPI_ISL_1331150, EPI_ISL_1331152, EPI_ISL_1331157, EPI_ISL_1331180, EPI_ISL_1331184, EPI_ISL_1331188, EPI_ISL_1331195, EPI_ISL_1331196, EPI_ISL_1331209, EPI_ISL_1331213, EPI_ISL_1331215, EPI_ISL_1331216, EPI_ISL_1331221, EPI_ISL_1331232, EPI_ISL_1331247, EPI_ISL_1331251, EPI_ISL_1331257, EPI_ISL_1331261, EPI_ISL_1331262, EPI_ISL_1331264, EPI_ISL_1331269, EPI_ISL_1331277, EPI_ISL_1331280, EPI_ISL_1331293, EPI_ISL_1331294, EPI_ISL_1331300, EPI_ISL_1331304, EPI_ISL_1331306, EPI_ISL_1331317, EPI_ISL_1331323, EPI_ISL_1331328, EPI_ISL_1331330, EPI_ISL_1331339, EPI_ISL_1331341, EPI_ISL_1331344, EPI_ISL_1331353, EPI_ISL_1331358, EPI_ISL_1331363, EPI_ISL_1331365, EPI_ISL_1331367, EPI_ISL_1331374, EPI_ISL_1331387, EPI_ISL_1331396, EPI_ISL_1331399, EPI_ISL_1331400, EPI_ISL_1331409, EPI_ISL_1331422, EPI_ISL_1331425, EPI_ISL_1331430, EPI_ISL_1331432, EPI_ISL_1331435, EPI_ISL_1331436, EPI_ISL_1331437, EPI_ISL_1331438, EPI_ISL_1331444, EPI_ISL_1331457, EPI_ISL_1331458, EPI_ISL_1331459, EPI_ISL_1331461, EPI_ISL_1331718, EPI_ISL_1331761, EPI_ISL_1331764, EPI_ISL_1331780, EPI_ISL_1331811, EPI_ISL_1331888, EPI_ISL_1331896, EPI_ISL_1331902, EPI_ISL_1331970, EPI_ISL_1331998, EPI_ISL_1332014, EPI_ISL_1332027, EPI_ISL_1332063, EPI_ISL_1332089, EPI_ISL_1332093, EPI_ISL_1332098, EPI_ISL_1332137, EPI_ISL_1332156, EPI_ISL_1332159, EPI_ISL_1332172, EPI_ISL_1332204, EPI_ISL_1332228, EPI_ISL_1332239, EPI_ISL_1332244, EPI_ISL_1332250, EPI_ISL_1332811, EPI_ISL_1332812, EPI_ISL_1332838, EPI_ISL_1332839, EPI_ISL_1332840, EPI_ISL_1332841, EPI_ISL_1332842, EPI_ISL_1333174, EPI_ISL_1333334, EPI_ISL_1333477, EPI_ISL_1346364, EPI_ISL_1346366, EPI_ISL_1346369, EPI_ISL_1346371, EPI_ISL_1346438, EPI_ISL_1346442, EPI_ISL_1346461, EPI_ISL_1346472, EPI_ISL_1374336, EPI_ISL_1374345, EPI_ISL_1374355, EPI_ISL_1374356, EPI_ISL_1374357, EPI_ISL_1374370, EPI_ISL_1374406, EPI_ISL_1374433, EPI_ISL_1374456, EPI_ISL_1374461, EPI_ISL_1374466, EPI_ISL_1374475, EPI_ISL_1374481, EPI_ISL_1374485, EPI_ISL_1374497, EPI_ISL_1374530, EPI_ISL_1374532, EPI_ISL_1374543, EPI_ISL_1374547, EPI_ISL_1374767, EPI_ISL_1378450, EPI_ISL_1410757, EPI_ISL_1410823, EPI_ISL_1410826, EPI_ISL_1410859, EPI_ISL_1410869, EPI_ISL_1411075, EPI_ISL_1411864, EPI_ISL_1411919, EPI_ISL_1412008, EPI_ISL_1412041, EPI_ISL_1412054, EPI_ISL_1412095, EPI_ISL_1412108, EPI_ISL_1413174, EPI_ISL_1413210, EPI_ISL_1413223, EPI_ISL_1413240, EPI_ISL_1413464, EPI_ISL_1413947, EPI_ISL_1413971, EPI_ISL_1414075, EPI_ISL_1414146, EPI_ISL_1414173, EPI_ISL_1414203, EPI_ISL_1414210, EPI_ISL_1414223, EPI_ISL_1414227, EPI_ISL_1414241, EPI_ISL_1414388, EPI_ISL_1414420, EPI_ISL_1414433, EPI_ISL_1414504, EPI_ISL_1414512, EPI_ISL_1414638, EPI_ISL_1414844, EPI_ISL_1415018, EPI_ISL_1454950, EPI_ISL_1455038, EPI_ISL_1455498, EPI_ISL_1455503, EPI_ISL_1455506, EPI_ISL_1455518, EPI_ISL_1455531, EPI_ISL_1455538 | | | |
| see above | Lighthouse Lab in Alderley Park | Wellcome Sanger Institute for the COVID-19 Genomics UK (COG-UK) Consortium | Cordelia Langford; David K. Jackson; Dominic Kwiatkowski; Ewan Harrison; Ian Johnston; Jacquelyn Wynn; Jeffrey Barrett; John Sillitoe on behalf of the Wellcome Sanger Institute COVID-19 Surveillance Team; Mairead Hyland; Roberto Amato; Sonia Goncalves; The Lighthouse Lab in Alderley Park and Alex Alderton |
| EPI_ISL_1263853, EPI_ISL_1263857, EPI_ISL_1263858, EPI_ISL_1263865, EPI_ISL_1263868, EPI_ISL_1263869, EPI_ISL_1263870, EPI_ISL_1263871, EPI_ISL_1263880, EPI_ISL_1263881, EPI_ISL_1263882, EPI_ISL_1263883, EPI_ISL_1263884, EPI_ISL_1263891, EPI_ISL_1263892, EPI_ISL_1263895, EPI_ISL_1263906, EPI_ISL_1263908, EPI_ISL_1263909, EPI_ISL_1263912, EPI_ISL_1263914, EPI_ISL_1263919, EPI_ISL_1263923, EPI_ISL_1263927, EPI_ISL_1263929, EPI_ISL_1263931, EPI_ISL_1263932, EPI_ISL_1263935, EPI_ISL_1263937, EPI_ISL_1263939, EPI_ISL_1263940, EPI_ISL_1263941, EPI_ISL_1263942, EPI_ISL_1263944, EPI_ISL_1263947, EPI_ISL_1263949, EPI_ISL_1263957, EPI_ISL_1263958, EPI_ISL_1263960, EPI_ISL_1263963, EPI_ISL_1263964, EPI_ISL_1263972, EPI_ISL_1263974, EPI_ISL_1263976, EPI_ISL_1263988, EPI_ISL_1263989, EPI_ISL_1263994, EPI_ISL_1263995, EPI_ISL_1263999, EPI_ISL_1264000, EPI_ISL_1264003, EPI_ISL_1264006, EPI_ISL_1264008, EPI_ISL_1264009, EPI_ISL_1264010, EPI_ISL_1264015, EPI_ISL_1264018, EPI_ISL_1264019, EPI_ISL_1264024, EPI_ISL_1264029, EPI_ISL_1264030, EPI_ISL_1264034, EPI_ISL_1264048, EPI_ISL_1264052, EPI_ISL_1264053, EPI_ISL_1264057, EPI_ISL_1264062, EPI_ISL_1275562, EPI_ISL_1275564, EPI_ISL_1275567, EPI_ISL_1275569, EPI_ISL_1275570, EPI_ISL_1275571, EPI_ISL_1275578, EPI_ISL_1275581, EPI_ISL_1275583, EPI_ISL_1275585, EPI_ISL_1275587, EPI_ISL_1275589, EPI_ISL_1275593, EPI_ISL_1275595, EPI_ISL_1275597, EPI_ISL_1275599, EPI_ISL_1275601, EPI_ISL_1275605, EPI_ISL_1275606, EPI_ISL_1275608, EPI_ISL_1275610, EPI_ISL_1275611, EPI_ISL_1275617, EPI_ISL_1275624, EPI_ISL_1275626, EPI_ISL_1275627, EPI_ISL_1275629, EPI_ISL_1275631, EPI_ISL_1275633, EPI_ISL_1275636, EPI_ISL_1275639, EPI_ISL_1275644, EPI_ISL_1275646, EPI_ISL_1275648, EPI_ISL_1275649, EPI_ISL_1275654, EPI_ISL_1275656, EPI_ISL_1275658, EPI_ISL_1275660, EPI_ISL_1275664, EPI_ISL_1275671, EPI_ISL_1275674, EPI_ISL_1275675, EPI_ISL_1275676, EPI_ISL_1275684, EPI_ISL_1275687, EPI_ISL_1275688, EPI_ISL_1275691, EPI_ISL_1275692, EPI_ISL_1275696, EPI_ISL_1275698, EPI_ISL_1275700, EPI_ISL_1275701, EPI_ISL_1275703, EPI_ISL_1275704, EPI_ISL_1275706, EPI_ISL_1275709, EPI_ISL_1275720, EPI_ISL_1275723, EPI_ISL_1275727, EPI_ISL_1275728, EPI_ISL_1275729, EPI_ISL_1275731, EPI_ISL_1275732, EPI_ISL_1275736, EPI_ISL_1275737, EPI_ISL_1275738, EPI_ISL_1275741, EPI_ISL_1275742, EPI_ISL_1275743, EPI_ISL_1275744, EPI_ISL_1275746, EPI_ISL_1275747, EPI_ISL_1275748, EPI_ISL_1275749, EPI_ISL_1275750, EPI_ISL_1275751, EPI_ISL_1275752, EPI_ISL_1275753, EPI_ISL_1275755, EPI_ISL_1275757, EPI_ISL_1275758, EPI_ISL_1275759, EPI_ISL_1275760, EPI_ISL_1275762, EPI_ISL_1275763, EPI_ISL_1275764, EPI_ISL_1275765, EPI_ISL_1275766, EPI_ISL_1275767, EPI_ISL_1275768, EPI_ISL_1275769, EPI_ISL_1275770, EPI_ISL_1275771, EPI_ISL_1275772, EPI_ISL_1275773, EPI_ISL_1275774, EPI_ISL_1275775, EPI_ISL_1275776, EPI_ISL_1275777, EPI_ISL_1275779, EPI_ISL_1275781, EPI_ISL_1275782, EPI_ISL_1275783, EPI_ISL_1275784, EPI_ISL_1275785, EPI_ISL_1275786, EPI_ISL_1275787, EPI_ISL_1275788, EPI_ISL_1275789, EPI_ISL_1275790, EPI_ISL_1275791, EPI_ISL_1275792, EPI_ISL_1275793, EPI_ISL_1275794, EPI_ISL_1275795, EPI_ISL_1275796, EPI_ISL_1275798, EPI_ISL_1275799, EPI_ISL_1275800, EPI_ISL_1275803, EPI_ISL_1275804, EPI_ISL_1275805, EPI_ISL_1275806, EPI_ISL_1275807, EPI_ISL_1275808, EPI_ISL_1275809, EPI_ISL_1275811, EPI_ISL_1275812, EPI_ISL_1275813, EPI_ISL_1275814, EPI_ISL_1275815, EPI_ISL_1275816, EPI_ISL_1275817, EPI_ISL_1275819, EPI_ISL_1275820, EPI_ISL_1275821, EPI_ISL_1275822, EPI_ISL_1275823, EPI_ISL_1275824, EPI_ISL_1275825, EPI_ISL_1275827, EPI_ISL_1275829, EPI_ISL_1275830, EPI_ISL_1275831, EPI_ISL_1275833, EPI_ISL_1275834, EPI_ISL_1275836, EPI_ISL_1275837, EPI_ISL_1275838, EPI_ISL_1275839, EPI_ISL_1275840, EPI_ISL_1275841, EPI_ISL_1275842, EPI_ISL_1275843, EPI_ISL_1275844, EPI_ISL_1275848, EPI_ISL_1275851, EPI_ISL_1275852, EPI_ISL_1275853, EPI_ISL_1275854, EPI_ISL_1275855, EPI_ISL_1275856, EPI_ISL_1275857, EPI_ISL_1275858, EPI_ISL_1275859, EPI_ISL_1275861, EPI_ISL_1275862, EPI_ISL_1275864, EPI_ISL_1275865, EPI_ISL_1275866, EPI_ISL_1275875, EPI_ISL_1275892, EPI_ISL_1275915, EPI_ISL_1275926, EPI_ISL_1275927, EPI_ISL_1275929, EPI_ISL_1275933, EPI_ISL_1275941, EPI_ISL_1275960, EPI_ISL_1275961, EPI_ISL_1275971, EPI_ISL_1275979, EPI_ISL_1275983, EPI_ISL_1275995, EPI_ISL_1275997, EPI_ISL_1276006, EPI_ISL_1276007, EPI_ISL_1276018, EPI_ISL_1276020, EPI_ISL_1276028, EPI_ISL_1276030, EPI_ISL_1276034, EPI_ISL_1276054, EPI_ISL_1276061, EPI_ISL_1276065, EPI_ISL_1276066, EPI_ISL_1276068, EPI_ISL_1276073, EPI_ISL_1276075, EPI_ISL_1276083, EPI_ISL_1276093, EPI_ISL_1276095, EPI_ISL_1276102, EPI_ISL_1276105, EPI_ISL_1276894, EPI_ISL_1276896, EPI_ISL_1276900, EPI_ISL_1276902, EPI_ISL_1276914, EPI_ISL_1276915, EPI_ISL_1276916, EPI_ISL_1276917, EPI_ISL_1276918, EPI_ISL_1276920, EPI_ISL_1276921, EPI_ISL_1276923, EPI_ISL_1276924, EPI_ISL_1276925, EPI_ISL_1276927, EPI_ISL_1276937, EPI_ISL_1276939, EPI_ISL_1276940, EPI_ISL_1276946, EPI_ISL_1276949, EPI_ISL_1276952, EPI_ISL_1276954, EPI_ISL_1276956, EPI_ISL_1276957, EPI_ISL_1276958, EPI_ISL_1276959, EPI_ISL_1276960, EPI_ISL_1276962, EPI_ISL_1276965, EPI_ISL_1276966, EPI_ISL_1276969, EPI_ISL_1276971, EPI_ISL_1276972, EPI_ISL_1276974, EPI_ISL_1276980, EPI_ISL_1276981, EPI_ISL_1276984, EPI_ISL_1276985, EPI_ISL_1276987, EPI_ISL_1276989, EPI_ISL_1276990, EPI_ISL_1276993, EPI_ISL_1276994, EPI_ISL_1276995, EPI_ISL_1276996, EPI_ISL_1276999, EPI_ISL_1277000, EPI_ISL_1277001, EPI_ISL_1277006, EPI_ISL_1277007, EPI_ISL_1277010, EPI_ISL_1277012, EPI_ISL_1277019, EPI_ISL_1277020, EPI_ISL_1277023, EPI_ISL_1277025, EPI_ISL_1277028, EPI_ISL_1277029, EPI_ISL_1277032, EPI_ISL_1277033, EPI_ISL_1277035, EPI_ISL_1277039, EPI_ISL_1277040, EPI_ISL_1277041, EPI_ISL_1277049, EPI_ISL_1277050, EPI_ISL_1277057, EPI_ISL_1277060, EPI_ISL_1277061, EPI_ISL_1277064, EPI_ISL_1277066, EPI_ISL_1277068, EPI_ISL_1277070, EPI_ISL_1277074, EPI_ISL_1277075, EPI_ISL_1277079, EPI_ISL_1277080, EPI_ISL_1277082, EPI_ISL_1277084, EPI_ISL_1277087, EPI_ISL_1277088, EPI_ISL_1277090, EPI_ISL_1277092, EPI_ISL_1277093, EPI_ISL_1277095, EPI_ISL_1277097, EPI_ISL_1277098, EPI_ISL_1277100, EPI_ISL_1277105, EPI_ISL_1277110, EPI_ISL_1277111, EPI_ISL_1277112, EPI_ISL_1277115, EPI_ISL_1277116, EPI_ISL_1277119, EPI_ISL_1277121, EPI_ISL_1277122, EPI_ISL_1277123, EPI_ISL_1277127, EPI_ISL_1277128, EPI_ISL_1277129, EPI_ISL_1277130, EPI_ISL_1277132, EPI_ISL_1277133, EPI_ISL_1277135, EPI_ISL_1277136, EPI_ISL_1277138, EPI_ISL_1277139, EPI_ISL_1277144, EPI_ISL_1277147, EPI_ISL_1277148, EPI_ISL_1277150, EPI_ISL_1277151, EPI_ISL_1277152, EPI_ISL_1277153, EPI_ISL_1277154, EPI_ISL_1277156, EPI_ISL_1277157, EPI_ISL_1277158, EPI_ISL_1277160, EPI_ISL_1277165, EPI_ISL_1277167, EPI_ISL_1277168, EPI_ISL_1277171, EPI_ISL_1277172, EPI_ISL_1277178, EPI_ISL_1277179, EPI_ISL_1277180, EPI_ISL_1277181, EPI_ISL_1277186, EPI_ISL_1277187, EPI_ISL_1277188, EPI_ISL_1277189, EPI_ISL_1277191, EPI_ISL_1277193, EPI_ISL_1277194, EPI_ISL_1277196, EPI_ISL_1277197, EPI_ISL_1277199, EPI_ISL_1277200, EPI_ISL_1294557, EPI_ISL_1294620, EPI_ISL_1294622, EPI_ISL_1315645, EPI_ISL_1315995, EPI_ISL_1315997, EPI_ISL_1315998, EPI_ISL_1315999, EPI_ISL_1316000, EPI_ISL_1316001, EPI_ISL_1316002, EPI_ISL_1316003, EPI_ISL_1316006, EPI_ISL_1316007, EPI_ISL_1316009, EPI_ISL_1316011, EPI_ISL_1316014, EPI_ISL_1316015, EPI_ISL_1316016, EPI_ISL_1316017, EPI_ISL_1316018, EPI_ISL_1316019, EPI_ISL_1316020, EPI_ISL_1316021, EPI_ISL_1316095, EPI_ISL_1316096, EPI_ISL_1316097, EPI_ISL_1316098, EPI_ISL_1316099, EPI_ISL_1316100, EPI_ISL_1316101, EPI_ISL_1316102, EPI_ISL_1316103, EPI_ISL_1316104, EPI_ISL_1316105, EPI_ISL_1316106, EPI_ISL_1316107, EPI_ISL_1316108, EPI_ISL_1316109, EPI_ISL_1316110, EPI_ISL_1316111, EPI_ISL_1316112, EPI_ISL_1316114, EPI_ISL_1316115, EPI_ISL_1316116, EPI_ISL_1316117, EPI_ISL_1316118, EPI_ISL_1316119, EPI_ISL_1316120, EPI_ISL_1316121, EPI_ISL_1316122, EPI_ISL_1316123, EPI_ISL_1316124, EPI_ISL_1316125, EPI_ISL_1316126, EPI_ISL_1316127, EPI_ISL_1316128, EPI_ISL_1316129, EPI_ISL_1316130, EPI_ISL_1316131, EPI_ISL_1316132, EPI_ISL_1316133, EPI_ISL_1316134, EPI_ISL_1316135, EPI_ISL_1316136, EPI_ISL_1316138, EPI_ISL_1316139, EPI_ISL_1316140, EPI_ISL_1316141, EPI_ISL_1316142, EPI_ISL_1316143, EPI_ISL_1316144, EPI_ISL_1316145, EPI_ISL_1316146, EPI_ISL_1316147, EPI_ISL_1316148, EPI_ISL_1316149, EPI_ISL_1316150, EPI_ISL_1316151, EPI_ISL_1316152, EPI_ISL_1316153, EPI_ISL_1316154, EPI_ISL_1316155, EPI_ISL_1316156, EPI_ISL_1316157, EPI_ISL_1316158, EPI_ISL_1316159, EPI_ISL_1316160, EPI_ISL_1316162, EPI_ISL_1316164, EPI_ISL_1316165, EPI_ISL_1316166, EPI_ISL_1316167, EPI_ISL_1316168, EPI_ISL_1316169, EPI_ISL_1316170, EPI_ISL_1316171, EPI_ISL_1316172, EPI_ISL_1316173, EPI_ISL_1316175, EPI_ISL_1316176, EPI_ISL_1316177, EPI_ISL_1316178, EPI_ISL_1316179, EPI_ISL_1316180, EPI_ISL_1316181, EPI_ISL_1316182, EPI_ISL_1316183, EPI_ISL_1316184, EPI_ISL_1316185, EPI_ISL_1316186, EPI_ISL_1316187, EPI_ISL_1316189, EPI_ISL_1316190, EPI_ISL_1316191, EPI_ISL_1316193, EPI_ISL_1316194, EPI_ISL_1316196, EPI_ISL_1316197, EPI_ISL_1316198, EPI_ISL_1316199, EPI_ISL_1316200, EPI_ISL_1316201, EPI_ISL_1316202, EPI_ISL_1316203, EPI_ISL_1316204, EPI_ISL_1316205, EPI_ISL_1316206, EPI_ISL_1316209, EPI_ISL_1316210, EPI_ISL_1316211, EPI_ISL_1316212, EPI_ISL_1316213, EPI_ISL_1316214, EPI_ISL_1316215, EPI_ISL_1316216, EPI_ISL_1316217, EPI_ISL_1316218, EPI_ISL_1316220, EPI_ISL_1316221, EPI_ISL_1316222, EPI_ISL_1316224, EPI_ISL_1316225, EPI_ISL_1316226, EPI_ISL_1316227, EPI_ISL_1316228, EPI_ISL_1316229, EPI_ISL_1316231, EPI_ISL_1326674, EPI_ISL_1326719, EPI_ISL_1326721, EPI_ISL_1326819, EPI_ISL_1326831, EPI_ISL_1326848, EPI_ISL_1326856, EPI_ISL_1326877, EPI_ISL_1326881, EPI_ISL_1326883, EPI_ISL_1326917, EPI_ISL_1326922, EPI_ISL_1326935, EPI_ISL_1326946, EPI_ISL_1326954, EPI_ISL_1326958, EPI_ISL_1326988, EPI_ISL_1326992, EPI_ISL_1327004, EPI_ISL_1327041, EPI_ISL_1327072, EPI_ISL_1327079, EPI_ISL_1327087, EPI_ISL_1327100, EPI_ISL_1327157, EPI_ISL_1327162, EPI_ISL_1327181, EPI_ISL_1327205, EPI_ISL_1327206, EPI_ISL_1327228, EPI_ISL_1327243, EPI_ISL_1327247, EPI_ISL_1327251, EPI_ISL_1327253, EPI_ISL_1327255, EPI_ISL_1327256, EPI_ISL_1327260, EPI_ISL_1327263, EPI_ISL_1327264, EPI_ISL_1327267, EPI_ISL_1327269, EPI_ISL_1327270, EPI_ISL_1327272, EPI_ISL_1327274, EPI_ISL_1327279, EPI_ISL_1327284, EPI_ISL_1327287, EPI_ISL_1327288, EPI_ISL_1327289, EPI_ISL_1327290, EPI_ISL_1327291, EPI_ISL_1327294, EPI_ISL_1327295, EPI_ISL_1327297, EPI_ISL_1327302, EPI_ISL_1327303, EPI_ISL_1327304, EPI_ISL_1327305, EPI_ISL_1327306, EPI_ISL_1327321, EPI_ISL_1327324, EPI_ISL_1327325, EPI_ISL_1327328, EPI_ISL_1327331, EPI_ISL_1327332, EPI_ISL_1327336, EPI_ISL_1327337, EPI_ISL_1327339, EPI_ISL_1327341, EPI_ISL_1327343, EPI_ISL_1327346, EPI_ISL_1327351, EPI_ISL_1327352, EPI_ISL_1327353, EPI_ISL_1327354, EPI_ISL_1327355, EPI_ISL_1327360, EPI_ISL_1327363, EPI_ISL_1327367, EPI_ISL_1327371, EPI_ISL_1327373, EPI_ISL_1327376, EPI_ISL_1327377, EPI_ISL_1327381, EPI_ISL_1327384, EPI_ISL_1327386, EPI_ISL_1327389, EPI_ISL_1327394, EPI_ISL_1327396, EPI_ISL_1327397, EPI_ISL_1327399, EPI_ISL_1327400, EPI_ISL_1327402, EPI_ISL_1327406, EPI_ISL_1327412, EPI_ISL_1327419, EPI_ISL_1327422, EPI_ISL_1327423, EPI_ISL_1327424, EPI_ISL_1327425, | | | |

EPI_ISL_1327429, EPI_ISL_1327435, EPI_ISL_1327436, EPI_ISL_1327437, EPI_ISL_1327449, EPI_ISL_1327450, EPI_ISL_1327454, EPI_ISL_1327464, EPI_ISL_1327482, EPI_ISL_1327485, EPI_ISL_1327502, EPI_ISL_1327506, EPI_ISL_1327509, EPI_ISL_1327537, EPI_ISL_1327599, EPI_ISL_1327600, EPI_ISL_1327604, EPI_ISL_1327610, EPI_ISL_1327659, EPI_ISL_1327669, EPI_ISL_1327683, EPI_ISL_1327697,
EPI_ISL_1328903, EPI_ISL_1328904, EPI_ISL_1328907, EPI_ISL_1328911, EPI_ISL_1328914, EPI_ISL_1328931, EPI_ISL_1328932, EPI_ISL_1328935, EPI_ISL_1328936, EPI_ISL_1328940, EPI_ISL_1328942, EPI_ISL_1328944, EPI_ISL_1328945, EPI_ISL_1328957, EPI_ISL_1328959, EPI_ISL_1328964, EPI_ISL_1328968, EPI_ISL_1328970, EPI_ISL_1328974, EPI_ISL_1328985, EPI_ISL_1328990, EPI_ISL_1328992,
EPI_ISL_1328996, EPI_ISL_1328997, EPI_ISL_1329002, EPI_ISL_1329003, EPI_ISL_1329009, EPI_ISL_1329010, EPI_ISL_1329026, EPI_ISL_1329027, EPI_ISL_1329029, EPI_ISL_1329039, EPI_ISL_1329040, EPI_ISL_1329042, EPI_ISL_1329043, EPI_ISL_1329051, EPI_ISL_1329053, EPI_ISL_1329056, EPI_ISL_1329057, EPI_ISL_1329059, EPI_ISL_1329061, EPI_ISL_1329067, EPI_ISL_1329079, EPI_ISL_1329083,
EPI_ISL_1329087, EPI_ISL_1329092, EPI_ISL_1329094, EPI_ISL_1329103, EPI_ISL_1329105, EPI_ISL_1329106, EPI_ISL_1329110, EPI_ISL_1329114, EPI_ISL_1329127, EPI_ISL_1329133, EPI_ISL_1329155, EPI_ISL_1329157, EPI_ISL_1329166, EPI_ISL_1329167, EPI_ISL_1329169, EPI_ISL_1329171, EPI_ISL_1329176, EPI_ISL_1329179, EPI_ISL_1329180, EPI_ISL_1329182, EPI_ISL_1329196, EPI_ISL_1329201,
EPI_ISL_1329202, EPI_ISL_1329203, EPI_ISL_1329204, EPI_ISL_1329206, EPI_ISL_1329208, EPI_ISL_1329211, EPI_ISL_1329213, EPI_ISL_1329216, EPI_ISL_1329219, EPI_ISL_1329221, EPI_ISL_1329222, EPI_ISL_1329225, EPI_ISL_1329227, EPI_ISL_1329228, EPI_ISL_1329229, EPI_ISL_1329231, EPI_ISL_1329233, EPI_ISL_1329235, EPI_ISL_1329237, EPI_ISL_1329239, EPI_ISL_1329240, EPI_ISL_1329244,
EPI_ISL_1329246, EPI_ISL_1329248, EPI_ISL_1329249, EPI_ISL_1329253, EPI_ISL_1329254, EPI_ISL_1329255, EPI_ISL_1329256, EPI_ISL_1329257, EPI_ISL_1329259, EPI_ISL_1329260, EPI_ISL_1329262, EPI_ISL_1329263, EPI_ISL_1329267, EPI_ISL_1329273, EPI_ISL_1329275, EPI_ISL_1329277, EPI_ISL_1329278, EPI_ISL_1329280, EPI_ISL_1329283, EPI_ISL_1329284, EPI_ISL_1329285, EPI_ISL_1329289,
EPI_ISL_1329292, EPI_ISL_1329293, EPI_ISL_1329294, EPI_ISL_1329296, EPI_ISL_1329301, EPI_ISL_1329304, EPI_ISL_1329306, EPI_ISL_1329307, EPI_ISL_1329308, EPI_ISL_1329309, EPI_ISL_1329313, EPI_ISL_1329315, EPI_ISL_1329317, EPI_ISL_1329322, EPI_ISL_1329326, EPI_ISL_1329330, EPI_ISL_1329331, EPI_ISL_1329332, EPI_ISL_1329333, EPI_ISL_1329335, EPI_ISL_1329340, EPI_ISL_1329341,
EPI_ISL_1329343, EPI_ISL_1329345, EPI_ISL_1329346, EPI_ISL_1329349, EPI_ISL_1329351, EPI_ISL_1329353, EPI_ISL_1329355, EPI_ISL_1329356, EPI_ISL_1329357, EPI_ISL_1329358, EPI_ISL_1329359, EPI_ISL_1329360, EPI_ISL_1329361, EPI_ISL_1329365, EPI_ISL_1329366, EPI_ISL_1329367, EPI_ISL_1329368, EPI_ISL_1329369, EPI_ISL_1329370, EPI_ISL_1329371, EPI_ISL_1329374, EPI_ISL_1329376,
EPI_ISL_1329377, EPI_ISL_1329983, EPI_ISL_1329984, EPI_ISL_1329992, EPI_ISL_1329994, EPI_ISL_1329995, EPI_ISL_1329997, EPI_ISL_1330000, EPI_ISL_1330008, EPI_ISL_1330011, EPI_ISL_1330013, EPI_ISL_1330021, EPI_ISL_1330023, EPI_ISL_1330029, EPI_ISL_1330030, EPI_ISL_1330034, EPI_ISL_1330035, EPI_ISL_1330041, EPI_ISL_1330046, EPI_ISL_1330050, EPI_ISL_1330051, EPI_ISL_1330054,
EPI_ISL_1330059, EPI_ISL_1330067, EPI_ISL_1330069, EPI_ISL_1330075, EPI_ISL_1330078, EPI_ISL_1330080, EPI_ISL_1330081, EPI_ISL_1330085, EPI_ISL_1330092, EPI_ISL_1330094, EPI_ISL_1330095, EPI_ISL_1330096, EPI_ISL_1330097, EPI_ISL_1330100, EPI_ISL_1330102, EPI_ISL_1332555, EPI_ISL_1332556, EPI_ISL_1332560, EPI_ISL_1332566, EPI_ISL_1332571, EPI_ISL_1332586, EPI_ISL_1332600,
EPI_ISL_1332602, EPI_ISL_1332605, EPI_ISL_1332612, EPI_ISL_1332614, EPI_ISL_1332616, EPI_ISL_1332622, EPI_ISL_1332625, EPI_ISL_1332640, EPI_ISL_1332655, EPI_ISL_1332658, EPI_ISL_1332660, EPI_ISL_1332661, EPI_ISL_1332664, EPI_ISL_1332665, EPI_ISL_1332666, EPI_ISL_1332669, EPI_ISL_1332671, EPI_ISL_1332672, EPI_ISL_1332674, EPI_ISL_1332676, EPI_ISL_1332677, EPI_ISL_1332678,
EPI_ISL_1332679, EPI_ISL_1332680, EPI_ISL_1332681, EPI_ISL_1332683, EPI_ISL_1332684, EPI_ISL_1332685, EPI_ISL_1332687, EPI_ISL_1332688, EPI_ISL_1332691, EPI_ISL_1332695, EPI_ISL_1332696, EPI_ISL_1332698, EPI_ISL_1332699, EPI_ISL_1332703, EPI_ISL_1332704, EPI_ISL_1332707, EPI_ISL_1332709, EPI_ISL_1332711, EPI_ISL_1332712, EPI_ISL_1332715, EPI_ISL_1332717, EPI_ISL_1332719,
EPI_ISL_1332720, EPI_ISL_1332721, EPI_ISL_1332722, EPI_ISL_1332725, EPI_ISL_1332727, EPI_ISL_1332734, EPI_ISL_1332739, EPI_ISL_1332740, EPI_ISL_1332746, EPI_ISL_1332758, EPI_ISL_1332764, EPI_ISL_1332765, EPI_ISL_1332767, EPI_ISL_1332770, EPI_ISL_1332772, EPI_ISL_1332781, EPI_ISL_1332790, EPI_ISL_1332793, EPI_ISL_1332807, EPI_ISL_1332808, EPI_ISL_1332809, EPI_ISL_1332831,
EPI_ISL_1332832, EPI_ISL_1332834, EPI_ISL_1332920, EPI_ISL_1332926, EPI_ISL_1332929, EPI_ISL_1332930, EPI_ISL_1332932, EPI_ISL_1332933, EPI_ISL_1332936, EPI_ISL_1332938, EPI_ISL_1332944, EPI_ISL_1332949, EPI_ISL_1332952, EPI_ISL_1344511, EPI_ISL_1344512, EPI_ISL_1344527, EPI_ISL_1346309, EPI_ISL_1346311, EPI_ISL_1346312, EPI_ISL_1346315, EPI_ISL_1346316, EPI_ISL_1346319,
EPI_ISL_1346327, EPI_ISL_1346361, EPI_ISL_1346362, EPI_ISL_1346363, EPI_ISL_1346367, EPI_ISL_1346368, EPI_ISL_1346373, EPI_ISL_1346375, EPI_ISL_1346377, EPI_ISL_1376978, EPI_ISL_1377060, EPI_ISL_1377121, EPI_ISL_1378491, EPI_ISL_1409586, EPI_ISL_1409731, EPI_ISL_1410151, EPI_ISL_1416182, EPI_ISL_1451215, EPI_ISL_1483817, EPI_ISL_1483834, EPI_ISL_1483843, EPI_ISL_1483900,
EPI_ISL_1483926, EPI_ISL_1483935, EPI_ISL_1483987, EPI_ISL_1484047, EPI_ISL_1484367, EPI_ISL_1484372, EPI_ISL_1484376, EPI_ISL_1484388, EPI_ISL_1484391, EPI_ISL_1484484, EPI_ISL_1484491, EPI_ISL_1484533, EPI_ISL_1484576, EPI_ISL_1484591, EPI_ISL_1484602, EPI_ISL_1506514, EPI_ISL_1584029

see above — Lighthouse Lab in Cambridge — Wellcome Sanger Institute for the COVID-19 Genomics UK (COG-UK) Consortium — Cordelia Langford; David K. Jackson; Dominic Kwiatkowski; Ewan Harrison; Ian Johnston; Jeffrey Barrett; John Sillitoe on behalf of the Wellcome Sanger Institute COVID-19 Surveillance Team; Rob Howes; Roberto Amato; Sonia Goncalves; The Lighthouse Lab in Cambridge and Alex Alderton

EPI_ISL_1243144, EPI_ISL_1255424, EPI_ISL_1255426, EPI_ISL_1255427, EPI_ISL_1255428, EPI_ISL_1255432, EPI_ISL_1255433, EPI_ISL_1255437, EPI_ISL_1255438, EPI_ISL_1255440, EPI_ISL_1255441, EPI_ISL_1255444, EPI_ISL_1255448, EPI_ISL_1255449, EPI_ISL_1255450, EPI_ISL_1255452, EPI_ISL_1255455, EPI_ISL_1255457, EPI_ISL_1255459, EPI_ISL_1255460, EPI_ISL_1255462, EPI_ISL_1255463,
EPI_ISL_1255467, EPI_ISL_1255471, EPI_ISL_1255474, EPI_ISL_1255476, EPI_ISL_1255478, EPI_ISL_1255479, EPI_ISL_1255480, EPI_ISL_1255482, EPI_ISL_1255483, EPI_ISL_1255484, EPI_ISL_1255485, EPI_ISL_1255487, EPI_ISL_1255495, EPI_ISL_1255497, EPI_ISL_1255498, EPI_ISL_1255499, EPI_ISL_1255501, EPI_ISL_1255504, EPI_ISL_1255506, EPI_ISL_1255508, EPI_ISL_1255510, EPI_ISL_1255511,
EPI_ISL_1255512, EPI_ISL_1255513, EPI_ISL_1255516, EPI_ISL_1255517, EPI_ISL_1255519, EPI_ISL_1255521, EPI_ISL_1255522, EPI_ISL_1255525, EPI_ISL_1255526, EPI_ISL_1255527, EPI_ISL_1255529, EPI_ISL_1255530, EPI_ISL_1255532, EPI_ISL_1255533, EPI_ISL_1255534, EPI_ISL_1255536, EPI_ISL_1255539, EPI_ISL_1255541, EPI_ISL_1255542, EPI_ISL_1255543, EPI_ISL_1255548, EPI_ISL_1255551,
EPI_ISL_1255552, EPI_ISL_1255553, EPI_ISL_1255554, EPI_ISL_1255555, EPI_ISL_1255556, EPI_ISL_1255559, EPI_ISL_1255560, EPI_ISL_1255564, EPI_ISL_1255565, EPI_ISL_1255569, EPI_ISL_1255571, EPI_ISL_1255575, EPI_ISL_1255577, EPI_ISL_1255578, EPI_ISL_1255581, EPI_ISL_1255582, EPI_ISL_1255583, EPI_ISL_1255589, EPI_ISL_1255591, EPI_ISL_1255595, EPI_ISL_1255601, EPI_ISL_1255604,
EPI_ISL_1255606, EPI_ISL_1255607, EPI_ISL_1255608, EPI_ISL_1255609, EPI_ISL_1255610, EPI_ISL_1255615, EPI_ISL_1255618, EPI_ISL_1255620, EPI_ISL_1255621, EPI_ISL_1255626, EPI_ISL_1255627, EPI_ISL_1255630, EPI_ISL_1255633, EPI_ISL_1255637, EPI_ISL_1255638, EPI_ISL_1255639, EPI_ISL_1255640, EPI_ISL_1255641, EPI_ISL_1255644, EPI_ISL_1255647, EPI_ISL_1255648, EPI_ISL_1255649,
EPI_ISL_1255650, EPI_ISL_1255654, EPI_ISL_1255657, EPI_ISL_1255658, EPI_ISL_1255660, EPI_ISL_1255664, EPI_ISL_1255666, EPI_ISL_1255668, EPI_ISL_1255672, EPI_ISL_1255673, EPI_ISL_1255676, EPI_ISL_1255677, EPI_ISL_1255681, EPI_ISL_1255683, EPI_ISL_1255694, EPI_ISL_1255700, EPI_ISL_1255702, EPI_ISL_1255713, EPI_ISL_1255714, EPI_ISL_1255717, EPI_ISL_1255731, EPI_ISL_1255742,
EPI_ISL_1255743, EPI_ISL_1255747, EPI_ISL_1255749, EPI_ISL_1255763, EPI_ISL_1255775, EPI_ISL_1255796, EPI_ISL_1255797, EPI_ISL_1255798, EPI_ISL_1255802, EPI_ISL_1255813, EPI_ISL_1255814, EPI_ISL_1255815, EPI_ISL_1255826, EPI_ISL_1255827, EPI_ISL_1255835, EPI_ISL_1255839, EPI_ISL_1255848, EPI_ISL_1255852, EPI_ISL_1255859, EPI_ISL_1255862, EPI_ISL_1255864, EPI_ISL_1255875,
EPI_ISL_1255889, EPI_ISL_1255891, EPI_ISL_1255894, EPI_ISL_1255919, EPI_ISL_1255927, EPI_ISL_1255936, EPI_ISL_1255938, EPI_ISL_1255939, EPI_ISL_1255944, EPI_ISL_1255947, EPI_ISL_1255950, EPI_ISL_1255956, EPI_ISL_1255970, EPI_ISL_1256000, EPI_ISL_1256028, EPI_ISL_1256036, EPI_ISL_1256041, EPI_ISL_1256043, EPI_ISL_1256046, EPI_ISL_1256059, EPI_ISL_1256062, EPI_ISL_1256065,
EPI_ISL_1256066, EPI_ISL_1256067, EPI_ISL_1256068, EPI_ISL_1256073, EPI_ISL_1256074, EPI_ISL_1256083, EPI_ISL_1256087, EPI_ISL_1256099, EPI_ISL_1256100, EPI_ISL_1256103, EPI_ISL_1256104, EPI_ISL_1256107, EPI_ISL_1256108, EPI_ISL_1256109, EPI_ISL_1256118, EPI_ISL_1256127, EPI_ISL_1256137, EPI_ISL_1256143, EPI_ISL_1256147, EPI_ISL_1256156, EPI_ISL_1256159, EPI_ISL_1256163,
EPI_ISL_1256168, EPI_ISL_1256171, EPI_ISL_1256179, EPI_ISL_1256197, EPI_ISL_1256206, EPI_ISL_1256221, EPI_ISL_1256227, EPI_ISL_1256231, EPI_ISL_1256233, EPI_ISL_1256242, EPI_ISL_1256247, EPI_ISL_1256254, EPI_ISL_1256255, EPI_ISL_1256262, EPI_ISL_1256264, EPI_ISL_1256265, EPI_ISL_1256273, EPI_ISL_1256274, EPI_ISL_1256275, EPI_ISL_1256282, EPI_ISL_1256283, EPI_ISL_1256285,
EPI_ISL_1256296, EPI_ISL_1256297, EPI_ISL_1256306, EPI_ISL_1256318, EPI_ISL_1256324, EPI_ISL_1256328, EPI_ISL_1256329, EPI_ISL_1256331, EPI_ISL_1256336, EPI_ISL_1256339, EPI_ISL_1256343, EPI_ISL_1256345, EPI_ISL_1256347, EPI_ISL_1256351, EPI_ISL_1256353, EPI_ISL_1256356, EPI_ISL_1256361, EPI_ISL_1256364, EPI_ISL_1256687, EPI_ISL_1256689, EPI_ISL_1256694, EPI_ISL_1256695,
EPI_ISL_1256697, EPI_ISL_1256701, EPI_ISL_1256704, EPI_ISL_1256717, EPI_ISL_1256725, EPI_ISL_1256735, EPI_ISL_1256739, EPI_ISL_1256747, EPI_ISL_1256752, EPI_ISL_1256755, EPI_ISL_1256757, EPI_ISL_1256761, EPI_ISL_1256764, EPI_ISL_1256766, EPI_ISL_1256772, EPI_ISL_1256783, EPI_ISL_1256787, EPI_ISL_1256793, EPI_ISL_1256795, EPI_ISL_1256798, EPI_ISL_1256802, EPI_ISL_1256804,
EPI_ISL_1256809, EPI_ISL_1256815, EPI_ISL_1256819, EPI_ISL_1256820, EPI_ISL_1256821, EPI_ISL_1256827, EPI_ISL_1256828, EPI_ISL_1256829, EPI_ISL_1256833, EPI_ISL_1256834, EPI_ISL_1256840, EPI_ISL_1256841, EPI_ISL_1256843, EPI_ISL_1256846, EPI_ISL_1256847, EPI_ISL_1256856, EPI_ISL_1256857, EPI_ISL_1256862, EPI_ISL_1256869, EPI_ISL_1256871, EPI_ISL_1256872, EPI_ISL_1256880,
EPI_ISL_1256884, EPI_ISL_1256892, EPI_ISL_1256893, EPI_ISL_1256894, EPI_ISL_1256895, EPI_ISL_1256900, EPI_ISL_1256903, EPI_ISL_1256904, EPI_ISL_1256905, EPI_ISL_1256906, EPI_ISL_1256907, EPI_ISL_1256913, EPI_ISL_1256922, EPI_ISL_1256925, EPI_ISL_1256933, EPI_ISL_1256934, EPI_ISL_1256941, EPI_ISL_1256942, EPI_ISL_1257367, EPI_ISL_1257368, EPI_ISL_1257376, EPI_ISL_1257380,
EPI_ISL_1257383, EPI_ISL_1257384, EPI_ISL_1257386, EPI_ISL_1257387, EPI_ISL_1257394, EPI_ISL_1257397, EPI_ISL_1257401, EPI_ISL_1257406, EPI_ISL_1257407, EPI_ISL_1257409, EPI_ISL_1257411, EPI_ISL_1257412, EPI_ISL_1257415, EPI_ISL_1263542, EPI_ISL_1263543, EPI_ISL_1263544, EPI_ISL_1263545, EPI_ISL_1263546, EPI_ISL_1263547, EPI_ISL_1263548, EPI_ISL_1263549, EPI_ISL_1263550,
EPI_ISL_1263551, EPI_ISL_1263552, EPI_ISL_1263553, EPI_ISL_1263554, EPI_ISL_1263555, EPI_ISL_1263556, EPI_ISL_1263557, EPI_ISL_1263558, EPI_ISL_1263559, EPI_ISL_1263560, EPI_ISL_1263561, EPI_ISL_1263562, EPI_ISL_1263563, EPI_ISL_1263564, EPI_ISL_1263565, EPI_ISL_1263566, EPI_ISL_1263567, EPI_ISL_1263568, EPI_ISL_1263569, EPI_ISL_1263570, EPI_ISL_1263571, EPI_ISL_1263572,
EPI_ISL_1263573, EPI_ISL_1263574, EPI_ISL_1263575, EPI_ISL_1263576, EPI_ISL_1263577, EPI_ISL_1263578, EPI_ISL_1263579, EPI_ISL_1263580, EPI_ISL_1263581, EPI_ISL_1263582, EPI_ISL_1263583, EPI_ISL_1263584, EPI_ISL_1263585, EPI_ISL_1263586, EPI_ISL_1263587, EPI_ISL_1263588, EPI_ISL_1263589, EPI_ISL_1263590, EPI_ISL_1263591, EPI_ISL_1263592, EPI_ISL_1263593, EPI_ISL_1263594,
EPI_ISL_1263595, EPI_ISL_1263596, EPI_ISL_1263597, EPI_ISL_1263598, EPI_ISL_1263599, EPI_ISL_1263600, EPI_ISL_1263601, EPI_ISL_1263602, EPI_ISL_1263603, EPI_ISL_1263604, EPI_ISL_1263605, EPI_ISL_1263606, EPI_ISL_1263607, EPI_ISL_1263608, EPI_ISL_1263609, EPI_ISL_1263610, EPI_ISL_1263611, EPI_ISL_1263612, EPI_ISL_1263613, EPI_ISL_1263614, EPI_ISL_1263615, EPI_ISL_1263616,
EPI_ISL_1263617, EPI_ISL_1263618, EPI_ISL_1263619, EPI_ISL_1263620, EPI_ISL_1263621, EPI_ISL_1263622, EPI_ISL_1263623, EPI_ISL_1263624, EPI_ISL_1263625, EPI_ISL_1263626, EPI_ISL_1263627, EPI_ISL_1263628, EPI_ISL_1263629, EPI_ISL_1263630, EPI_ISL_1263631, EPI_ISL_1263632, EPI_ISL_1263633, EPI_ISL_1263634, EPI_ISL_1263635, EPI_ISL_1263636, EPI_ISL_1263637, EPI_ISL_1263638,
EPI_ISL_1263639, EPI_ISL_1263640, EPI_ISL_1263641, EPI_ISL_1263642, EPI_ISL_1263643, EPI_ISL_1263644, EPI_ISL_1263645, EPI_ISL_1263646, EPI_ISL_1263647, EPI_ISL_1263648, EPI_ISL_1263649, EPI_ISL_1263650, EPI_ISL_1263651, EPI_ISL_1263652, EPI_ISL_1263653, EPI_ISL_1263654, EPI_ISL_1263655, EPI_ISL_1263656, EPI_ISL_1263657, EPI_ISL_1263658, EPI_ISL_1263659, EPI_ISL_1263660,
EPI_ISL_1263661, EPI_ISL_1263662, EPI_ISL_1263663, EPI_ISL_1263664, EPI_ISL_1263665, EPI_ISL_1263666, EPI_ISL_1263667, EPI_ISL_1263668, EPI_ISL_1263669, EPI_ISL_1263670, EPI_ISL_1263671, EPI_ISL_1263672, EPI_ISL_1263673, EPI_ISL_1263674, EPI_ISL_1263675, EPI_ISL_1263676, EPI_ISL_1263677, EPI_ISL_1263678, EPI_ISL_1263679, EPI_ISL_1263680, EPI_ISL_1263681, EPI_ISL_1263682,
EPI_ISL_1263683, EPI_ISL_1263684, EPI_ISL_1263685, EPI_ISL_1263686, EPI_ISL_1263687, EPI_ISL_1263688, EPI_ISL_1263689, EPI_ISL_1263690, EPI_ISL_1263691, EPI_ISL_1263692, EPI_ISL_1263693, EPI_ISL_1263694, EPI_ISL_1263695, EPI_ISL_1263696, EPI_ISL_1263697, EPI_ISL_1263698, EPI_ISL_1263699, EPI_ISL_1263700, EPI_ISL_1263701, EPI_ISL_1263702, EPI_ISL_1263703, EPI_ISL_1263704,
EPI_ISL_1263705, EPI_ISL_1263706, EPI_ISL_1263707, EPI_ISL_1263708, EPI_ISL_1263709, EPI_ISL_1263710, EPI_ISL_1263711, EPI_ISL_1263712, EPI_ISL_1263713, EPI_ISL_1263714, EPI_ISL_1263715, EPI_ISL_1263716, EPI_ISL_1263717, EPI_ISL_1263718, EPI_ISL_1263719, EPI_ISL_1263720, EPI_ISL_1263721, EPI_ISL_1263722, EPI_ISL_1263723, EPI_ISL_1263724, EPI_ISL_1263725, EPI_ISL_1263726,
EPI_ISL_1263727, EPI_ISL_1263728, EPI_ISL_1263729, EPI_ISL_1263730, EPI_ISL_1263731, EPI_ISL_1263732, EPI_ISL_1263733, EPI_ISL_1263734, EPI_ISL_1263735, EPI_ISL_1263736, EPI_ISL_1263737, EPI_ISL_1263738, EPI_ISL_1263739, EPI_ISL_1263740, EPI_ISL_1263741, EPI_ISL_1263742, EPI_ISL_1263743, EPI_ISL_1263744, EPI_ISL_1263745, EPI_ISL_1263746, EPI_ISL_1263747, EPI_ISL_1263748,
EPI_ISL_1263749, EPI_ISL_1263750, EPI_ISL_1263751, EPI_ISL_1263752, EPI_ISL_1263753, EPI_ISL_1263754, EPI_ISL_1263755, EPI_ISL_1263756, EPI_ISL_1263757, EPI_ISL_1263758, EPI_ISL_1263759, EPI_ISL_1263760, EPI_ISL_1263761, EPI_ISL_1263762, EPI_ISL_1263763, EPI_ISL_1263764, EPI_ISL_1263765, EPI_ISL_1263766, EPI_ISL_1263767, EPI_ISL_1263768, EPI_ISL_1263769, EPI_ISL_1263770,
EPI_ISL_1263771, EPI_ISL_1263772, EPI_ISL_1263773, EPI_ISL_1263774, EPI_ISL_1263775, EPI_ISL_1263776, EPI_ISL_1263777, EPI_ISL_1263778, EPI_ISL_1263779, EPI_ISL_1263780, EPI_ISL_1263781, EPI_ISL_1263782, EPI_ISL_1263783, EPI_ISL_1263784, EPI_ISL_1263785, EPI_ISL_1263786, EPI_ISL_1263787, EPI_ISL_1263788, EPI_ISL_1263789, EPI_ISL_1263790, EPI_ISL_1263791, EPI_ISL_1263792,
EPI_ISL_1263793, EPI_ISL_1263794, EPI_ISL_1263795, EPI_ISL_1263796, EPI_ISL_1263797, EPI_ISL_1263798, EPI_ISL_1263799, EPI_ISL_1263800, EPI_ISL_1263801, EPI_ISL_1263802, EPI_ISL_1263803, EPI_ISL_1263804, EPI_ISL_1263805, EPI_ISL_1263806, EPI_ISL_1263807, EPI_ISL_1263808, EPI_ISL_1263809, EPI_ISL_1263810, EPI_ISL_1263811, EPI_ISL_1263812, EPI_ISL_1263813, EPI_ISL_1263814,
EPI_ISL_1263815, EPI_ISL_1263816, EPI_ISL_1263817, EPI_ISL_1263818, EPI_ISL_1263819, EPI_ISL_1263820, EPI_ISL_1263821, EPI_ISL_1263822, EPI_ISL_1263823, EPI_ISL_1263824, EPI_ISL_1263825, EPI_ISL_1263826, EPI_ISL_1263827, EPI_ISL_1263828, EPI_ISL_1263829, EPI_ISL_1263830, EPI_ISL_1263831, EPI_ISL_1263832, EPI_ISL_1263833, EPI_ISL_1263834, EPI_ISL_1263835, EPI_ISL_1263836,
EPI_ISL_1263837, EPI_ISL_1263838, EPI_ISL_1263839, EPI_ISL_1263840, EPI_ISL_1263841, EPI_ISL_1263842, EPI_ISL_1263843, EPI_ISL_1263844, EPI_ISL_1263845, EPI_ISL_1263846, EPI_ISL_1263847, EPI_ISL_1263848, EPI_ISL_1263849, EPI_ISL_1263850, EPI_ISL_1263851, EPI_ISL_1263852, EPI_ISL_1264065, EPI_ISL_1264066, EPI_ISL_1264068, EPI_ISL_1264075, EPI_ISL_1264076, EPI_ISL_1264089,
EPI_ISL_1264090, EPI_ISL_1264096, EPI_ISL_1264097, EPI_ISL_1264103, EPI_ISL_1264105, EPI_ISL_1264106, EPI_ISL_1264109, EPI_ISL_1264111, EPI_ISL_1264113, EPI_ISL_1264115, EPI_ISL_1264116, EPI_ISL_1264122, EPI_ISL_1264123, EPI_ISL_1264130, EPI_ISL_1264132, EPI_ISL_1264134, EPI_ISL_1264138, EPI_ISL_1264140, EPI_ISL_1264145, EPI_ISL_1264146, EPI_ISL_1264150, EPI_ISL_1264152,
EPI_ISL_1264159, EPI_ISL_1264164, EPI_ISL_1264165, EPI_ISL_1264167, EPI_ISL_1264171, EPI_ISL_1264173, EPI_ISL_1264179, EPI_ISL_1264181, EPI_ISL_1264185, EPI_ISL_1264187, EPI_ISL_1264191, EPI_ISL_1264193, EPI_ISL_1264195, EPI_ISL_1264196, EPI_ISL_1264197, EPI_ISL_1264203, EPI_ISL_1264204, EPI_ISL_1264205, EPI_ISL_1264209, EPI_ISL_1264210, EPI_ISL_1264213,
EPI_ISL_1264217, EPI_ISL_1264218, EPI_ISL_1264221, EPI_ISL_1264222, EPI_ISL_1264223, EPI_ISL_1264224, EPI_ISL_1264226, EPI_ISL_1264231, EPI_ISL_1264237, EPI_ISL_1264240, EPI_ISL_1264242, EPI_ISL_1264244, EPI_ISL_1264245, EPI_ISL_1264246, EPI_ISL_1264248, EPI_ISL_1264249, EPI_ISL_1264250, EPI_ISL_1264253, EPI_ISL_1264255, EPI_ISL_1264259, EPI_ISL_1264260, EPI_ISL_1264263,
EPI_ISL_1264264, EPI_ISL_1264265, EPI_ISL_1264266, EPI_ISL_1264269, EPI_ISL_1264270, EPI_ISL_1264272, EPI_ISL_1264274, EPI_ISL_1264282, EPI_ISL_1264285, EPI_ISL_1264290, EPI_ISL_1264293, EPI_ISL_1264299, EPI_ISL_1264300, EPI_ISL_1264303, EPI_ISL_1264305, EPI_ISL_1264307, EPI_ISL_1264319, EPI_ISL_1264324, EPI_ISL_1264331, EPI_ISL_1264333, EPI_ISL_1264340, EPI_ISL_1264341,
EPI_ISL_1264342, EPI_ISL_1264343, EPI_ISL_1264345, EPI_ISL_1264348, EPI_ISL_1264350, EPI_ISL_1264353, EPI_ISL_1264362, EPI_ISL_1264363, EPI_ISL_1264365, EPI_ISL_1264366, EPI_ISL_1264367, EPI_ISL_1264368, EPI_ISL_1264369, EPI_ISL_1264372, EPI_ISL_1264376, EPI_ISL_1264377, EPI_ISL_1264383, EPI_ISL_1264386, EPI_ISL_1264389, EPI_ISL_1264396, EPI_ISL_1264398, EPI_ISL_1264399,
EPI_ISL_1264404, EPI_ISL_1264409, EPI_ISL_1264413, EPI_ISL_1264431, EPI_ISL_1264432, EPI_ISL_1264440, EPI_ISL_1264443, EPI_ISL_1264444, EPI_ISL_1264445, EPI_ISL_1264449, EPI_ISL_1264450, EPI_ISL_1264451, EPI_ISL_1264453, EPI_ISL_1264456, EPI_ISL_1264459, EPI_ISL_1264461, EPI_ISL_1264462, EPI_ISL_1264463, EPI_ISL_1264466, EPI_ISL_1264468, EPI_ISL_1264470, EPI_ISL_1264471,
EPI_ISL_1264472, EPI_ISL_1264474, EPI_ISL_1264475, EPI_ISL_1264476, EPI_ISL_1264479, EPI_ISL_1264482, EPI_ISL_1264484, EPI_ISL_1264485, EPI_ISL_1264486, EPI_ISL_1264487, EPI_ISL_1264488, EPI_ISL_1264489, EPI_ISL_1264491, EPI_ISL_1264492, EPI_ISL_1264496, EPI_ISL_1264498, EPI_ISL_1264499, EPI_ISL_1264502, EPI_ISL_1264504, EPI_ISL_1264505, EPI_ISL_1264507, EPI_ISL_1264508,
EPI_ISL_1264510, EPI_ISL_1264514, EPI_ISL_1264515, EPI_ISL_1264518, EPI_ISL_1264522, EPI_ISL_1264527, EPI_ISL_1264529, EPI_ISL_1264530, EPI_ISL_1264533, EPI_ISL_1264537, EPI_ISL_1264538, EPI_ISL_1264540, EPI_ISL_1264542, EPI_ISL_1264544, EPI_ISL_1264546, EPI_ISL_1264548, EPI_ISL_1264549, EPI_ISL_1264550, EPI_ISL_1264551, EPI_ISL_1264553, EPI_ISL_1264554, EPI_ISL_1264555,
EPI_ISL_1264556, EPI_ISL_1264563, EPI_ISL_1264564, EPI_ISL_1264566, EPI_ISL_1264567, EPI_ISL_1264569, EPI_ISL_1264575, EPI_ISL_1264576, EPI_ISL_1264577, EPI_ISL_1264578, EPI_ISL_1264582, EPI_ISL_1264585, EPI_ISL_1264587, EPI_ISL_1264588, EPI_ISL_1264592, EPI_ISL_1264594, EPI_ISL_1264596, EPI_ISL_1264597, EPI_ISL_1264598, EPI_ISL_1264599, EPI_ISL_1264600, EPI_ISL_1264601,
EPI_ISL_1264603, EPI_ISL_1264604, EPI_ISL_1264605, EPI_ISL_1264606, EPI_ISL_1264608, EPI_ISL_1264610, EPI_ISL_1264611, EPI_ISL_1264619, EPI_ISL_1264620, EPI_ISL_1264621, EPI_ISL_1264624, EPI_ISL_1264626, EPI_ISL_1264627, EPI_ISL_1264628, EPI_ISL_1264631, EPI_ISL_1264637, EPI_ISL_1273843, EPI_ISL_1273844, EPI_ISL_1273845, EPI_ISL_1273846, EPI_ISL_1273847, EPI_ISL_1273848,
EPI_ISL_1273849, EPI_ISL_1273850, EPI_ISL_1273851, EPI_ISL_1273852, EPI_ISL_1273853, EPI_ISL_1273854, EPI_ISL_1273855, EPI_ISL_1273856, EPI_ISL_1273857, EPI_ISL_1273858, EPI_ISL_1273859, EPI_ISL_1273860, EPI_ISL_1273861, EPI_ISL_1273862, EPI_ISL_1273863, EPI_ISL_1273864, EPI_ISL_1273865, EPI_ISL_1273866, EPI_ISL_1273867, EPI_ISL_1273868, EPI_ISL_1273869, EPI_ISL_1273870,
EPI_ISL_1273871, EPI_ISL_1273872, EPI_ISL_1273873, EPI_ISL_1273874, EPI_ISL_1273875, EPI_ISL_1273876, EPI_ISL_1273877, EPI_ISL_1273878, EPI_ISL_1273879, EPI_ISL_1273880, EPI_ISL_1273881, EPI_ISL_1273882, EPI_ISL_1273883, EPI_ISL_1273884, EPI_ISL_1273885, EPI_ISL_1273886, EPI_ISL_1273887, EPI_ISL_1273888, EPI_ISL_1273889, EPI_ISL_1273890, EPI_ISL_1273891, EPI_ISL_1273892,
EPI_ISL_1273893, EPI_ISL_1273894, EPI_ISL_1273895, EPI_ISL_1273896, EPI_ISL_1273897, EPI_ISL_1273898, EPI_ISL_1273899, EPI_ISL_1273900, EPI_ISL_1273901, EPI_ISL_1273902, EPI_ISL_1273903, EPI_ISL_1273904, EPI_ISL_1273905, EPI_ISL_1273906, EPI_ISL_1273907, EPI_ISL_1273908, EPI_ISL_1273909, EPI_ISL_1273910, EPI_ISL_1273911, EPI_ISL_1273912, EPI_ISL_1273913, EPI_ISL_1273914,
EPI_ISL_1273915, EPI_ISL_1273916, EPI_ISL_1273917, EPI_ISL_1273918, EPI_ISL_1273919, EPI_ISL_1273920, EPI_ISL_1273921, EPI_ISL_1273922, EPI_ISL_1273923, EPI_ISL_1273924, EPI_ISL_1273925, EPI_ISL_1273926, EPI_ISL_1273927, EPI_ISL_1273928, EPI_ISL_1273929, EPI_ISL_1273930, EPI_ISL_1273931, EPI_ISL_1273932, EPI_ISL_1273933, EPI_ISL_1273934, EPI_ISL_1273935, EPI_ISL_1273936,
EPI_ISL_1273937, EPI_ISL_1273938, EPI_ISL_1273939, EPI_ISL_1273940, EPI_ISL_1273941, EPI_ISL_1273942, EPI_ISL_1273943, EPI_ISL_1273944, EPI_ISL_1273945, EPI_ISL_1273946, EPI_ISL_1273947, EPI_ISL_1273948, EPI_ISL_1273949, EPI_ISL_1273950, EPI_ISL_1273951, EPI_ISL_1273952, EPI_ISL_1273953, EPI_ISL_1273954, EPI_ISL_1273955, EPI_ISL_1273956, EPI_ISL_1273957, EPI_ISL_1273958,
EPI_ISL_1273959, EPI_ISL_1273960, EPI_ISL_1273961, EPI_ISL_1273962, EPI_ISL_1273963, EPI_ISL_1273964, EPI_ISL_1273965, EPI_ISL_1273966, EPI_ISL_1273967, EPI_ISL_1273968, EPI_ISL_1273969, EPI_ISL_1273970, EPI_ISL_1273971, EPI_ISL_1273972, EPI_ISL_1273973, EPI_ISL_1273974, EPI_ISL_1273975, EPI_ISL_1273976, EPI_ISL_1273977, EPI_ISL_1273978, EPI_ISL_1273979, EPI_ISL_1273980,
EPI_ISL_1273981, EPI_ISL_1273982, EPI_ISL_1273983, EPI_ISL_1273984, EPI_ISL_1273985, EPI_ISL_1273986, EPI_ISL_1273991, EPI_ISL_1274001, EPI_ISL_1274009, EPI_ISL_1274012, EPI_ISL_1274015, EPI_ISL_1274019, EPI_ISL_1274022, EPI_ISL_1274023, EPI_ISL_1274040, EPI_ISL_1274046, EPI_ISL_1274047, EPI_ISL_1274048, EPI_ISL_1274056, EPI_ISL_1274068, EPI_ISL_1274072, EPI_ISL_1274074,
EPI_ISL_1274075, EPI_ISL_1274079, EPI_ISL_1274085, EPI_ISL_1274088, EPI_ISL_1274095, EPI_ISL_1274097, EPI_ISL_1274099, EPI_ISL_1274107, EPI_ISL_1274125, EPI_ISL_1274126, EPI_ISL_1274130, EPI_ISL_1274131, EPI_ISL_1274138, EPI_ISL_1274146, EPI_ISL_1274151, EPI_ISL_1274155, EPI_ISL_1274159, EPI_ISL_1274163, EPI_ISL_1274181, EPI_ISL_1274187, EPI_ISL_1274190, EPI_ISL_1274192,
EPI_ISL_1274209, EPI_ISL_1274210, EPI_ISL_1274213, EPI_ISL_1274217, EPI_ISL_1274221, EPI_ISL_1274232, EPI_ISL_1274238, EPI_ISL_1274239, EPI_ISL_1274242, EPI_ISL_1274246, EPI_ISL_1274254, EPI_ISL_1274260, EPI_ISL_1274271, EPI_ISL_1274272, EPI_ISL_1274276, EPI_ISL_1274286, EPI_ISL_1274290, EPI_ISL_1274298, EPI_ISL_1274303, EPI_ISL_1274304, EPI_ISL_1274305,
EPI_ISL_1274306, EPI_ISL_1274307, EPI_ISL_1274308, EPI_ISL_1274310, EPI_ISL_1274313, EPI_ISL_1274314, EPI_ISL_1274315, EPI_ISL_1274316, EPI_ISL_1274317, EPI_ISL_1274318, EPI_ISL_1274320, EPI_ISL_1274321, EPI_ISL_1274322, EPI_ISL_1274323, EPI_ISL_1274324, EPI_ISL_1274326, EPI_ISL_1274330, EPI_ISL_1274331, EPI_ISL_1274332, EPI_ISL_1274333, EPI_ISL_1274334, EPI_ISL_1274335,
EPI_ISL_1274336, EPI_ISL_1274337, EPI_ISL_1274339, EPI_ISL_1274341, EPI_ISL_1274342, EPI_ISL_1274345, EPI_ISL_1274347, EPI_ISL_1274350, EPI_ISL_1274352, EPI_ISL_1274355, EPI_ISL_1274361, EPI_ISL_1274364, EPI_ISL_1274365, EPI_ISL_1274367, EPI_ISL_1274368, EPI_ISL_1274375, EPI_ISL_1274377, EPI_ISL_1274378, EPI_ISL_1274380, EPI_ISL_1274381, EPI_ISL_1274382, EPI_ISL_1274383,
EPI_ISL_1274384, EPI_ISL_1274385, EPI_ISL_1274387, EPI_ISL_1274388, EPI_ISL_1274389, EPI_ISL_1274390, EPI_ISL_1274391, EPI_ISL_1274392, EPI_ISL_1274393, EPI_ISL_1274394, EPI_ISL_1274395, EPI_ISL_1274396, EPI_ISL_1274397, EPI_ISL_1274399, EPI_ISL_1274400, EPI_ISL_1274403, EPI_ISL_1274404, EPI_ISL_1274405, EPI_ISL_1274408, EPI_ISL_1274409, EPI_ISL_1274411, EPI_ISL_1274412,
EPI_ISL_1274413, EPI_ISL_1274414, EPI_ISL_1274418, EPI_ISL_1274419, EPI_ISL_1274420, EPI_ISL_1274421, EPI_ISL_1274422, EPI_ISL_1274423, EPI_ISL_1274426, EPI_ISL_1274427, EPI_ISL_1274428, EPI_ISL_1274432, EPI_ISL_1274434, EPI_ISL_1274435, EPI_ISL_1274437, EPI_ISL_1274440, EPI_ISL_1274441, EPI_ISL_1274443, EPI_ISL_1274444, EPI_ISL_1274445, EPI_ISL_1274447, EPI_ISL_1274448,
EPI_ISL_1274450, EPI_ISL_1274451, EPI_ISL_1274452, EPI_ISL_1274454, EPI_ISL_1274455, EPI_ISL_1274456, EPI_ISL_1274457, EPI_ISL_1274459, EPI_ISL_1274460, EPI_ISL_1274461, EPI_ISL_1274464, EPI_ISL_1274465, EPI_ISL_1274466, EPI_ISL_1274471, EPI_ISL_1274473, EPI_ISL_1274474, EPI_ISL_1274475, EPI_ISL_1274477, EPI_ISL_1274478, EPI_ISL_1274481, EPI_ISL_1274482, EPI_ISL_1274483,
EPI_ISL_1274487, EPI_ISL_1274488, EPI_ISL_1274489, EPI_ISL_1274491, EPI_ISL_1274493, EPI_ISL_1274494, EPI_ISL_1274495, EPI_ISL_1274497, EPI_ISL_1274498, EPI_ISL_1274500, EPI_ISL_1274501, EPI_ISL_1274502, EPI_ISL_1274503, EPI_ISL_1274504, EPI_ISL_1274506, EPI_ISL_1274508, EPI_ISL_1274509, EPI_ISL_1274513, EPI_ISL_1274516, EPI_ISL_1274518, EPI_ISL_1274519, EPI_ISL_1274521,
EPI_ISL_1274523, EPI_ISL_1274526, EPI_ISL_1274527, EPI_ISL_1274528, EPI_ISL_1274530, EPI_ISL_1274531, EPI_ISL_1274532, EPI_ISL_1274533, EPI_ISL_1274534, EPI_ISL_1274536, EPI_ISL_1274538, EPI_ISL_1274539, EPI_ISL_1274541, EPI_ISL_1274543, EPI_ISL_1274544, EPI_ISL_1274545, EPI_ISL_1274547, EPI_ISL_1274548, EPI_ISL_1274552, EPI_ISL_1274554, EPI_ISL_1274555, EPI_ISL_1274556,
EPI_ISL_1274557, EPI_ISL_1274558, EPI_ISL_1274560, EPI_ISL_1274564, EPI_ISL_1274565, EPI_ISL_1274567, EPI_ISL_1274570, EPI_ISL_1274571, EPI_ISL_1274573, EPI_ISL_1274577, EPI_ISL_1274578, EPI_ISL_1274581, EPI_ISL_1274585, EPI_ISL_1274587, EPI_ISL_1274589, EPI_ISL_1274591, EPI_ISL_1274594, EPI_ISL_1274596, EPI_ISL_1274597, EPI_ISL_1274598,
EPI_ISL_1274599, EPI_ISL_1274601, EPI_ISL_1274603, EPI_ISL_1274604, EPI_ISL_1274605, EPI_ISL_1274608, EPI_ISL_1274609, EPI_ISL_1274611, EPI_ISL_1274612, EPI_ISL_1274614, EPI_ISL_1274615, EPI_ISL_1274616, EPI_ISL_1274620, EPI_ISL_1274622, EPI_ISL_1274624, EPI_ISL_1274625, EPI_ISL_1274628, EPI_ISL_1274629, EPI_ISL_1274631, EPI_ISL_1274633, EPI_ISL_1274634, EPI_ISL_1274635,
EPI_ISL_1274636, EPI_ISL_1274638, EPI_ISL_1274639, EPI_ISL_1274641, EPI_ISL_1274642, EPI_ISL_1274644, EPI_ISL_1274645, EPI_ISL_1274646, EPI_ISL_1274647, EPI_ISL_1274649, EPI_ISL_1274650, EPI_ISL_1274651, EPI_ISL_1274652, EPI_ISL_1274654, EPI_ISL_1274656, EPI_ISL_1274657, EPI_ISL_1274660, EPI_ISL_1274662, EPI_ISL_1274663, EPI_ISL_1274664, EPI_ISL_1274670, EPI_ISL_1274672,
EPI_ISL_1274673, EPI_ISL_1274675, EPI_ISL_1274676, EPI_ISL_1274677, EPI_ISL_1274678, EPI_ISL_1274679, EPI_ISL_1274681, EPI_ISL_1274682, EPI_ISL_1274683, EPI_ISL_1274687, EPI_ISL_1274691, EPI_ISL_1274692, EPI_ISL_1274693, EPI_ISL_1274694, EPI_ISL_1274695, EPI_ISL_1274696, EPI_ISL_1274697, EPI_ISL_1274698, EPI_ISL_1274700, EPI_ISL_1274701, EPI_ISL_1274702,
EPI_ISL_1274704, EPI_ISL_1274705, EPI_ISL_1274706, EPI_ISL_1274707, EPI_ISL_1274710, EPI_ISL_1274711, EPI_ISL_1274713, EPI_ISL_1274715, EPI_ISL_1274716, EPI_ISL_1274717, EPI_ISL_1274721, EPI_ISL_1274727, EPI_ISL_1274729, EPI_ISL_1274730, EPI_ISL_1274732, EPI_ISL_1274733, EPI_ISL_1274736, EPI_ISL_1274738, EPI_ISL_1274739, EPI_ISL_1274741, EPI_ISL_1274742,
EPI_ISL_1274743, EPI_ISL_1274744, EPI_ISL_1274745, EPI_ISL_1274748, EPI_ISL_1274755, EPI_ISL_1274756, EPI_ISL_1274757, EPI_ISL_1274758, EPI_ISL_1274759, EPI_ISL_1274762, EPI_ISL_1274763, EPI_ISL_1274766, EPI_ISL_1274767, EPI_ISL_1274768, EPI_ISL_1274769, EPI_ISL_1274771, EPI_ISL_1274772, EPI_ISL_1274774, EPI_ISL_1274778, EPI_ISL_1274780, EPI_ISL_1274785, EPI_ISL_1274786,
EPI_ISL_1274787, EPI_ISL_1274790, EPI_ISL_1274792, EPI_ISL_1274794, EPI_ISL_1274795, EPI_ISL_1274796, EPI_ISL_1274797, EPI_ISL_1274798, EPI_ISL_1274799, EPI_ISL_1274805, EPI_ISL_1274806, EPI_ISL_1274808, EPI_ISL_1274810, EPI_ISL_1274811, EPI_ISL_1274813, EPI_ISL_1274814, EPI_ISL_1274815, EPI_ISL_1274816, EPI_ISL_1274817, EPI_ISL_1274818, EPI_ISL_1274820, EPI_ISL_1274822,
EPI_ISL_1274823, EPI_ISL_1274824, EPI_ISL_1274825, EPI_ISL_1274827, EPI_ISL_1274829, EPI_ISL_1274830, EPI_ISL_1274831, EPI_ISL_1274833, EPI_ISL_1274834, EPI_ISL_1274837, EPI_ISL_1274838, EPI_ISL_1274840, EPI_ISL_1274843, EPI_ISL_1274845, EPI_ISL_1274847, EPI_ISL_1274850, EPI_ISL_1274851, EPI_ISL_1274853, EPI_ISL_1274855, EPI_ISL_1274857, EPI_ISL_1274860, EPI_ISL_1274861,
EPI_ISL_1274862, EPI_ISL_1274863, EPI_ISL_1274864, EPI_ISL_1274865, EPI_ISL_1274867, EPI_ISL_1274869, EPI_ISL_1274870, EPI_ISL_1274871, EPI_ISL_1274873, EPI_ISL_1274874, EPI_ISL_1274876, EPI_ISL_1274879, EPI_ISL_1274880, EPI_ISL_1274882, EPI_ISL_1274884, EPI_ISL_1274887, EPI_ISL_1274889, EPI_ISL_1274890, EPI_ISL_1274891, EPI_ISL_1274893, EPI_ISL_1274894, EPI_ISL_1274898,
EPI_ISL_1274899, EPI_ISL_1274900, EPI_ISL_1274901, EPI_ISL_1274902, EPI_ISL_1274904, EPI_ISL_1274906, EPI_ISL_1274907, EPI_ISL_1274908, EPI_ISL_1274909, EPI_ISL_1274910, EPI_ISL_1274912, EPI_ISL_1274913, EPI_ISL_1274915, EPI_ISL_1274916, EPI_ISL_1274917, EPI_ISL_1274918, EPI_ISL_1274919, EPI_ISL_1274921, EPI_ISL_1274922, EPI_ISL_1274923, EPI_ISL_1274924, EPI_ISL_1274925,
EPI_ISL_1274928, EPI_ISL_1274929, EPI_ISL_1274931, EPI_ISL_1274932, EPI_ISL_1274933, EPI_ISL_1274938, EPI_ISL_1274939, EPI_ISL_1274940, EPI_ISL_1274941, EPI_ISL_1274942, EPI_ISL_1274944, EPI_ISL_1274945, EPI_ISL_1274946, EPI_ISL_1274947, EPI_ISL_1274948, EPI_ISL_1274949, EPI_ISL_1274950, EPI_ISL_1274951, EPI_ISL_1274952, EPI_ISL_1274953, EPI_ISL_1274954,
EPI_ISL_1274955, EPI_ISL_1274957, EPI_ISL_1274958, EPI_ISL_1274959, EPI_ISL_1274960, EPI_ISL_1274961, EPI_ISL_1274963, EPI_ISL_1274964, EPI_ISL_1274965, EPI_ISL_1274967, EPI_ISL_1274968, EPI_ISL_1274970, EPI_ISL_1274971, EPI_ISL_1274972, EPI_ISL_1274973, EPI_ISL_1274975, EPI_ISL_1274976, EPI_ISL_1274978, EPI_ISL_1274979, EPI_ISL_1274980, EPI_ISL_1274984, EPI_ISL_1274986,
EPI_ISL_1274987, EPI_ISL_1274988, EPI_ISL_1274989, EPI_ISL_1274990, EPI_ISL_1274991, EPI_ISL_1274992, EPI_ISL_1274995, EPI_ISL_1274998, EPI_ISL_1274999, EPI_ISL_1275000, EPI_ISL_1275002, EPI_ISL_1275005, EPI_ISL_1275008, EPI_ISL_1275013, EPI_ISL_1275018, EPI_ISL_1275024, EPI_ISL_1275027, EPI_ISL_1275030, EPI_ISL_1275048, EPI_ISL_1275064, EPI_ISL_1275071, EPI_ISL_1275072,
EPI_ISL_1275082, EPI_ISL_1275087, EPI_ISL_1275104, EPI_ISL_1275109, EPI_ISL_1275115, EPI_ISL_1275116, EPI_ISL_1275117, EPI_ISL_1275118, EPI_ISL_1275119, EPI_ISL_1275120, EPI_ISL_1275121, EPI_ISL_1275122, EPI_ISL_1275123, EPI_ISL_1275124, EPI_ISL_1275125, EPI_ISL_1275126, EPI_ISL_1275127, EPI_ISL_1275128, EPI_ISL_1275129, EPI_ISL_1275130, EPI_ISL_1275131, EPI_ISL_1275132,
EPI_ISL_1275133, EPI_ISL_1275134, EPI_ISL_1275135, EPI_ISL_1275136, EPI_ISL_1275137, EPI_ISL_1275138, EPI_ISL_1275139, EPI_ISL_1275140, EPI_ISL_1275141, EPI_ISL_1275142, EPI_ISL_1275143, EPI_ISL_1275144, EPI_ISL_1275145, EPI_ISL_1275146, EPI_ISL_1275147, EPI_ISL_1275148, EPI_ISL_1275149, EPI_ISL_1275150, EPI_ISL_1275151, EPI_ISL_1275152, EPI_ISL_1275153, EPI_ISL_1275154,
EPI_ISL_1275155, EPI_ISL_1275156, EPI_ISL_1275157, EPI_ISL_1275158, EPI_ISL_1275159, EPI_ISL_1275160, EPI_ISL_1275161, EPI_ISL_1275162, EPI_ISL_1275163, EPI_ISL_1275164, EPI_ISL_1275165, EPI_ISL_1275166, EPI_ISL_1275167, EPI_ISL_1275168, EPI_ISL_1275169, EPI_ISL_1275170, EPI_ISL_1275171, EPI_ISL_1275172, EPI_ISL_1275173, EPI_ISL_1275174, EPI_ISL_1275175, EPI_ISL_1275176,
EPI_ISL_1275177, EPI_ISL_1275178, EPI_ISL_1275179, EPI_ISL_1275180, EPI_ISL_1275181, EPI_ISL_1275182, EPI_ISL_1275183, EPI_ISL_1275184, EPI_ISL_1275185, EPI_ISL_1275186, EPI_ISL_1275187, EPI_ISL_1275188, EPI_ISL_1275189, EPI_ISL_1275190, EPI_ISL_1275191, EPI_ISL_1275192, EPI_ISL_1275193, EPI_ISL_1275194, EPI_ISL_1275195, EPI_ISL_1275196, EPI_ISL_1275197, EPI_ISL_1275198,
EPI_ISL_1275199, EPI_ISL_1275200, EPI_ISL_1275201, EPI_ISL_1275202, EPI_ISL_1275203, EPI_ISL_1275204, EPI_ISL_1275205, EPI_ISL_1275206, EPI_ISL_1275207, EPI_ISL_1275208, EPI_ISL_1275209, EPI_ISL_1275210, EPI_ISL_1275211, EPI_ISL_1275212, EPI_ISL_1275213, EPI_ISL_1275214, EPI_ISL_1275215, EPI_ISL_1275216, EPI_ISL_1275217, EPI_ISL_1275218, EPI_ISL_1275219, EPI_ISL_1275220,
EPI_ISL_1275221, EPI_ISL_1275222, EPI_ISL_1275223, EPI_ISL_1275224, EPI_ISL_1275225, EPI_ISL_1275226, EPI_ISL_1275227, EPI_ISL_1275228, EPI_ISL_1275229, EPI_ISL_1275230, EPI_ISL_1275231, EPI_ISL_1275232, EPI_ISL_1275233, EPI_ISL_1275234, EPI_ISL_1275235, EPI_ISL_1275236, EPI_ISL_1275237, EPI_ISL_1275238, EPI_ISL_1275239, EPI_ISL_1275240, EPI_ISL_1275241, EPI_ISL_1275242,
EPI_ISL_1275243, EPI_ISL_1275244, EPI_ISL_1275245, EPI_ISL_1275246, EPI_ISL_1275247, EPI_ISL_1275249, EPI_ISL_1275250, EPI_ISL_1275251, EPI_ISL_1275252, EPI_ISL_1275253, EPI_ISL_1275254, EPI_ISL_1275255, EPI_ISL_1275256, EPI_ISL_1275257, EPI_ISL_1275258, EPI_ISL_1275259, EPI_ISL_1275260, EPI_ISL_1275261, EPI_ISL_1275262, EPI_ISL_1275263, EPI_ISL_1275264,
EPI_ISL_1275265, EPI_ISL_1275266, EPI_ISL_1275267, EPI_ISL_1275268, EPI_ISL_1275269, EPI_ISL_1275270, EPI_ISL_1275271, EPI_ISL_1275272, EPI_ISL_1275273, EPI_ISL_1275274, EPI_ISL_1275275, EPI_ISL_1275276, EPI_ISL_1275277, EPI_ISL_1275278, EPI_ISL_1275279, EPI_ISL_1275280, EPI_ISL_1275281, EPI_ISL_1275282, EPI_ISL_1275283, EPI_ISL_1275284, EPI_ISL_1275285, EPI_ISL_1275286,
EPI_ISL_1275287, EPI_ISL_1275288, EPI_ISL_1275289, EPI_ISL_1275290, EPI_ISL_1275291, EPI_ISL_1275292, EPI_ISL_1275293, EPI_ISL_1275294, EPI_ISL_1275295, EPI_ISL_1275296, EPI_ISL_1275297, EPI_ISL_1275298, EPI_ISL_1275299, EPI_ISL_1275300, EPI_ISL_1275301, EPI_ISL_1275302, EPI_ISL_1275303, EPI_ISL_1275304, EPI_ISL_1275305, EPI_ISL_1275306, EPI_ISL_1275307, EPI_ISL_1275308,
EPI_ISL_1275309, EPI_ISL_1275310, EPI_ISL_1275311, EPI_ISL_1275312, EPI_ISL_1275313, EPI_ISL_1275314, EPI_ISL_1275315, EPI_ISL_1275316, EPI_ISL_1275317, EPI_ISL_1275318, EPI_ISL_1275319, EPI_ISL_1275320, EPI_ISL_1275321, EPI_ISL_1275322, EPI_ISL_1275323, EPI_ISL_1275324, EPI_ISL_1275325, EPI_ISL_1275326, EPI_ISL_1275327, EPI_ISL_1275328, EPI_ISL_1275329, EPI_ISL_1275330,
EPI_ISL_1275331, EPI_ISL_1275332, EPI_ISL_1275333, EPI_ISL_1275334, EPI_ISL_1275335, EPI_ISL_1275336, EPI_ISL_1275337, EPI_ISL_1275338, EPI_ISL_1275339, EPI_ISL_1275340, EPI_ISL_1275341, EPI_ISL_1275342, EPI_ISL_1275347, EPI_ISL_1275348, EPI_ISL_1275349, EPI_ISL_1275350, EPI_ISL_1275351, EPI_ISL_1275353, EPI_ISL_1275354, EPI_ISL_1275358, EPI_ISL_1275359, EPI_ISL_1275361,
EPI_ISL_1275362, EPI_ISL_1275363, EPI_ISL_1275365, EPI_ISL_1275366, EPI_ISL_1275367, EPI_ISL_1275369, EPI_ISL_1275373, EPI_ISL_1275376, EPI_ISL_1275377, EPI_ISL_1275378, EPI_ISL_1275379, EPI_ISL_1275380, EPI_ISL_1275382, EPI_ISL_1275383, EPI_ISL_1275384, EPI_ISL_1275385, EPI_ISL_1275388, EPI_ISL_1275390, EPI_ISL_1275392, EPI_ISL_1275394, EPI_ISL_1275395, EPI_ISL_1275396,
EPI_ISL_1275397, EPI_ISL_1275398, EPI_ISL_1275399, EPI_ISL_1275401, EPI_ISL_1275402, EPI_ISL_1275403, EPI_ISL_1275405, EPI_ISL_1275408, EPI_ISL_1275410, EPI_ISL_1275411, EPI_ISL_1275412, EPI_ISL_1275414, EPI_ISL_1275415, EPI_ISL_1275416, EPI_ISL_1275417, EPI_ISL_1275420, EPI_ISL_1275421, EPI_ISL_1275423, EPI_ISL_1275426, EPI_ISL_1275427, EPI_ISL_1275428, EPI_ISL_1275430,
EPI_ISL_1275431, EPI_ISL_1275435, EPI_ISL_1275436, EPI_ISL_1275437, EPI_ISL_1275439, EPI_ISL_1275440, EPI_ISL_1275442, EPI_ISL_1275444, EPI_ISL_1275446, EPI_ISL_1275447, EPI_ISL_1275448, EPI_ISL_1275449, EPI_ISL_1275450, EPI_ISL_1275451, EPI_ISL_1275452, EPI_ISL_1275453, EPI_ISL_1275454, EPI_ISL_1275456, EPI_ISL_1275461, EPI_ISL_1275462, EPI_ISL_1275463, EPI_ISL_1275464,
EPI_ISL_1275465, EPI_ISL_1275466, EPI_ISL_1275468, EPI_ISL_1275469, EPI_ISL_1275470, EPI_ISL_1275471, EPI_ISL_1275474, EPI_ISL_1275475, EPI_ISL_1275476, EPI_ISL_1275478, EPI_ISL_1275483, EPI_ISL_1275485, EPI_ISL_1275487, EPI_ISL_1275488, EPI_ISL_1275489, EPI_ISL_1275490, EPI_ISL_1275491, EPI_ISL_1275494, EPI_ISL_1275495, EPI_ISL_1275496, EPI_ISL_1275497, EPI_ISL_1275498,
EPI_ISL_1275499, EPI_ISL_1275502, EPI_ISL_1275503, EPI_ISL_1275505, EPI_ISL_1275506, EPI_ISL_1275507, EPI_ISL_1275508, EPI_ISL_1275509, EPI_ISL_1275511, EPI_ISL_1275512, EPI_ISL_1275513, EPI_ISL_1275514, EPI_ISL_1275515, EPI_ISL_1275516, EPI_ISL_1275518, EPI_ISL_1275520, EPI_ISL_1275521, EPI_ISL_1275522, EPI_ISL_1275525, EPI_ISL_1275527, EPI_ISL_1275528, EPI_ISL_1275529,
EPI_ISL_1275531, EPI_ISL_1275533, EPI_ISL_1275534, EPI_ISL_1275535, EPI_ISL_1275536, EPI_ISL_1275537, EPI_ISL_1275539, EPI_ISL_1275540, EPI_ISL_1275541, EPI_ISL_1275543, EPI_ISL_1275544, EPI_ISL_1275545, EPI_ISL_1275546, EPI_ISL_1275548, EPI_ISL_1275549, EPI_ISL_1275552, EPI_ISL_1275553, EPI_ISL_1275554, EPI_ISL_1275556, EPI_ISL_1275557, EPI_ISL_1275558, EPI_ISL_1275559,
EPI_ISL_1276115, EPI_ISL_1276124, EPI_ISL_1276125, EPI_ISL_1276126, EPI_ISL_1276128, EPI_ISL_1276130, EPI_ISL_1276131, EPI_ISL_1276137, EPI_ISL_1276139, EPI_ISL_1276141, EPI_ISL_1276143, EPI_ISL_1276146, EPI_ISL_1276148, EPI_ISL_1276151, EPI_ISL_1276155, EPI_ISL_1276156, EPI_ISL_1276160, EPI_ISL_1276163, EPI_ISL_1276165, EPI_ISL_1276167, EPI_ISL_1276168,
EPI_ISL_1276170, EPI_ISL_1276172, EPI_ISL_1276173, EPI_ISL_1276174, EPI_ISL_1276182, EPI_ISL_1276185, EPI_ISL_1276186, EPI_ISL_1276187, EPI_ISL_1276190, EPI_ISL_1276194, EPI_ISL_1276195, EPI_ISL_1276199, EPI_ISL_1276200, EPI_ISL_1276201, EPI_ISL_1276203, EPI_ISL_1276211, EPI_ISL_1276212, EPI_ISL_1276216, EPI_ISL_1276217, EPI_ISL_1276219, EPI_ISL_1276221, EPI_ISL_1276223,
EPI_ISL_1276231, EPI_ISL_1276234, EPI_ISL_1276237, EPI_ISL_1276240, EPI_ISL_1276243, EPI_ISL_1276245, EPI_ISL_1276246, EPI_ISL_1276247, EPI_ISL_1276250, EPI_ISL_1276252, EPI_ISL_1276254, EPI_ISL_1276260, EPI_ISL_1276262, EPI_ISL_1276264, EPI_ISL_1276267, EPI_ISL_1276268, EPI_ISL_1276269, EPI_ISL_1276274, EPI_ISL_1276278, EPI_ISL_1276279, EPI_ISL_1276280, EPI_ISL_1276283,
EPI_ISL_1276284, EPI_ISL_1276289, EPI_ISL_1276291, EPI_ISL_1276295, EPI_ISL_1276301, EPI_ISL_1276302, EPI_ISL_1276303, EPI_ISL_1276307, EPI_ISL_1276309, EPI_ISL_1276310, EPI_ISL_1276312, EPI_ISL_1276313, EPI_ISL_1276314, EPI_ISL_1276316, EPI_ISL_1276317, EPI_ISL_1276318, EPI_ISL_1276320, EPI_ISL_1276322, EPI_ISL_1276323, EPI_ISL_1276327, EPI_ISL_1276330, EPI_ISL_1276331,
EPI_ISL_1276333, EPI_ISL_1276336, EPI_ISL_1276338, EPI_ISL_1276340, EPI_ISL_1276341, EPI_ISL_1276342, EPI_ISL_1276345, EPI_ISL_1276346, EPI_ISL_1276349, EPI_ISL_1276350, EPI_ISL_1276352, EPI_ISL_1276353, EPI_ISL_1276354, EPI_ISL_1276355, EPI_ISL_1276356, EPI_ISL_1276357, EPI_ISL_1276359, EPI_ISL_1276361, EPI_ISL_1276362, EPI_ISL_1276364, EPI_ISL_1276371, EPI_ISL_1276375,

EPI_ISL_1276376, EPI_ISL_1276378, EPI_ISL_1276379, EPI_ISL_1276382, EPI_ISL_1276383, EPI_ISL_1276384, EPI_ISL_1276386, EPI_ISL_1276387, EPI_ISL_1276388, EPI_ISL_1276391, EPI_ISL_1276392, EPI_ISL_1276394, EPI_ISL_1276395, EPI_ISL_1276397, EPI_ISL_1276398, EPI_ISL_1276399, EPI_ISL_1276400, EPI_ISL_1276401, EPI_ISL_1276402, EPI_ISL_1276404, EPI_ISL_1276406, EPI_ISL_1276407,
EPI_ISL_1276408, EPI_ISL_1276409, EPI_ISL_1276411, EPI_ISL_1276413, EPI_ISL_1276414, EPI_ISL_1276415, EPI_ISL_1276416, EPI_ISL_1276417, EPI_ISL_1276418, EPI_ISL_1276419, EPI_ISL_1276420, EPI_ISL_1276421, EPI_ISL_1276422, EPI_ISL_1276424, EPI_ISL_1276425, EPI_ISL_1276426, EPI_ISL_1276427, EPI_ISL_1276429, EPI_ISL_1276430, EPI_ISL_1276431, EPI_ISL_1276432, EPI_ISL_1276433,
EPI_ISL_1276434, EPI_ISL_1276435, EPI_ISL_1276436, EPI_ISL_1276437, EPI_ISL_1276438, EPI_ISL_1276439, EPI_ISL_1276443, EPI_ISL_1276444, EPI_ISL_1276445, EPI_ISL_1276446, EPI_ISL_1276447, EPI_ISL_1276448, EPI_ISL_1276449, EPI_ISL_1276450, EPI_ISL_1276451, EPI_ISL_1276452, EPI_ISL_1276453, EPI_ISL_1276455, EPI_ISL_1276456, EPI_ISL_1276457, EPI_ISL_1276458, EPI_ISL_1276459,
EPI_ISL_1276460, EPI_ISL_1276462, EPI_ISL_1276463, EPI_ISL_1276464, EPI_ISL_1276466, EPI_ISL_1276467, EPI_ISL_1276470, EPI_ISL_1276472, EPI_ISL_1276473, EPI_ISL_1276474, EPI_ISL_1276475, EPI_ISL_1276476, EPI_ISL_1276477, EPI_ISL_1276479, EPI_ISL_1276480, EPI_ISL_1276482, EPI_ISL_1276483, EPI_ISL_1276484, EPI_ISL_1276485, EPI_ISL_1276486, EPI_ISL_1276487, EPI_ISL_1276488,
EPI_ISL_1276494, EPI_ISL_1276495, EPI_ISL_1276496, EPI_ISL_1276497, EPI_ISL_1276498, EPI_ISL_1276500, EPI_ISL_1276502, EPI_ISL_1276503, EPI_ISL_1276504, EPI_ISL_1276506, EPI_ISL_1276507, EPI_ISL_1276509, EPI_ISL_1276510, EPI_ISL_1276511, EPI_ISL_1276512, EPI_ISL_1276513, EPI_ISL_1276516, EPI_ISL_1276517, EPI_ISL_1276518, EPI_ISL_1276519, EPI_ISL_1276520, EPI_ISL_1276521,
EPI_ISL_1276522, EPI_ISL_1276524, EPI_ISL_1276525, EPI_ISL_1276526, EPI_ISL_1276528, EPI_ISL_1276529, EPI_ISL_1276530, EPI_ISL_1276531, EPI_ISL_1276533, EPI_ISL_1276534, EPI_ISL_1276535, EPI_ISL_1276536, EPI_ISL_1276538, EPI_ISL_1276539, EPI_ISL_1276540, EPI_ISL_1276541, EPI_ISL_1276542, EPI_ISL_1276543, EPI_ISL_1276545, EPI_ISL_1276546, EPI_ISL_1276547, EPI_ISL_1276548,
EPI_ISL_1276549, EPI_ISL_1276552, EPI_ISL_1276554, EPI_ISL_1276555, EPI_ISL_1276556, EPI_ISL_1276557, EPI_ISL_1276558, EPI_ISL_1276559, EPI_ISL_1276560, EPI_ISL_1276562, EPI_ISL_1276563, EPI_ISL_1276564, EPI_ISL_1276565, EPI_ISL_1276566, EPI_ISL_1276567, EPI_ISL_1276568, EPI_ISL_1276569, EPI_ISL_1276570, EPI_ISL_1276571, EPI_ISL_1276572, EPI_ISL_1276573, EPI_ISL_1276574,
EPI_ISL_1276575, EPI_ISL_1276578, EPI_ISL_1276579, EPI_ISL_1276580, EPI_ISL_1276581, EPI_ISL_1276582, EPI_ISL_1276583, EPI_ISL_1276585, EPI_ISL_1276586, EPI_ISL_1276587, EPI_ISL_1276588, EPI_ISL_1276590, EPI_ISL_1276591, EPI_ISL_1276592, EPI_ISL_1276593, EPI_ISL_1276595, EPI_ISL_1276596, EPI_ISL_1276598, EPI_ISL_1276599, EPI_ISL_1276600, EPI_ISL_1276601, EPI_ISL_1276603,
EPI_ISL_1276605, EPI_ISL_1276606, EPI_ISL_1276608, EPI_ISL_1276609, EPI_ISL_1276610, EPI_ISL_1276612, EPI_ISL_1276614, EPI_ISL_1276615, EPI_ISL_1276617, EPI_ISL_1276618, EPI_ISL_1276620, EPI_ISL_1276621, EPI_ISL_1276622, EPI_ISL_1276623, EPI_ISL_1276624, EPI_ISL_1276625, EPI_ISL_1276626, EPI_ISL_1276628, EPI_ISL_1276629, EPI_ISL_1276630, EPI_ISL_1276631, EPI_ISL_1276632,
EPI_ISL_1276633, EPI_ISL_1276634, EPI_ISL_1276635, EPI_ISL_1276637, EPI_ISL_1276638, EPI_ISL_1276639, EPI_ISL_1276640, EPI_ISL_1276641, EPI_ISL_1276642, EPI_ISL_1276644, EPI_ISL_1276645, EPI_ISL_1276646, EPI_ISL_1276647, EPI_ISL_1276650, EPI_ISL_1276651, EPI_ISL_1276652, EPI_ISL_1276653, EPI_ISL_1276655, EPI_ISL_1276656, EPI_ISL_1276658, EPI_ISL_1276659, EPI_ISL_1276660,
EPI_ISL_1276661, EPI_ISL_1276665, EPI_ISL_1276666, EPI_ISL_1276668, EPI_ISL_1276669, EPI_ISL_1276670, EPI_ISL_1276671, EPI_ISL_1276672, EPI_ISL_1276673, EPI_ISL_1276676, EPI_ISL_1276677, EPI_ISL_1276679, EPI_ISL_1276680, EPI_ISL_1276681, EPI_ISL_1276682, EPI_ISL_1276684, EPI_ISL_1276685, EPI_ISL_1276687, EPI_ISL_1276688, EPI_ISL_1276689, EPI_ISL_1276690, EPI_ISL_1276691,
EPI_ISL_1276692, EPI_ISL_1276693, EPI_ISL_1276694, EPI_ISL_1276695, EPI_ISL_1276697, EPI_ISL_1276699, EPI_ISL_1276700, EPI_ISL_1276703, EPI_ISL_1276704, EPI_ISL_1276706, EPI_ISL_1276707, EPI_ISL_1276709, EPI_ISL_1276711, EPI_ISL_1276713, EPI_ISL_1276716, EPI_ISL_1276717, EPI_ISL_1276719, EPI_ISL_1276722, EPI_ISL_1276724, EPI_ISL_1276726, EPI_ISL_1276728, EPI_ISL_1276730,
EPI_ISL_1276731, EPI_ISL_1276733, EPI_ISL_1276736, EPI_ISL_1276739, EPI_ISL_1276740, EPI_ISL_1276741, EPI_ISL_1276743, EPI_ISL_1276744, EPI_ISL_1276746, EPI_ISL_1276748, EPI_ISL_1276749, EPI_ISL_1276750, EPI_ISL_1276753, EPI_ISL_1276755, EPI_ISL_1276759, EPI_ISL_1276762, EPI_ISL_1276764, EPI_ISL_1276765, EPI_ISL_1276766, EPI_ISL_1276767, EPI_ISL_1276768, EPI_ISL_1276769,
EPI_ISL_1276771, EPI_ISL_1276772, EPI_ISL_1276773, EPI_ISL_1276774, EPI_ISL_1276776, EPI_ISL_1276777, EPI_ISL_1276779, EPI_ISL_1276781, EPI_ISL_1276782, EPI_ISL_1276783, EPI_ISL_1276784, EPI_ISL_1276785, EPI_ISL_1276787, EPI_ISL_1276788, EPI_ISL_1276790, EPI_ISL_1276791, EPI_ISL_1276792, EPI_ISL_1276793, EPI_ISL_1276794, EPI_ISL_1276799, EPI_ISL_1276801, EPI_ISL_1276803,
EPI_ISL_1276804, EPI_ISL_1276807, EPI_ISL_1276809, EPI_ISL_1276810, EPI_ISL_1276813, EPI_ISL_1276818, EPI_ISL_1276819, EPI_ISL_1276820, EPI_ISL_1276821, EPI_ISL_1276822, EPI_ISL_1276824, EPI_ISL_1276825, EPI_ISL_1276827, EPI_ISL_1276828, EPI_ISL_1276830, EPI_ISL_1276831, EPI_ISL_1276832, EPI_ISL_1276833, EPI_ISL_1276834, EPI_ISL_1276836, EPI_ISL_1276837, EPI_ISL_1276838,
EPI_ISL_1276839, EPI_ISL_1276840, EPI_ISL_1276841, EPI_ISL_1276842, EPI_ISL_1276843, EPI_ISL_1276844, EPI_ISL_1276845, EPI_ISL_1276846, EPI_ISL_1276847, EPI_ISL_1276848, EPI_ISL_1276849, EPI_ISL_1276850, EPI_ISL_1276852, EPI_ISL_1276855, EPI_ISL_1276856, EPI_ISL_1276857, EPI_ISL_1276858, EPI_ISL_1276859, EPI_ISL_1276860, EPI_ISL_1276863, EPI_ISL_1276865, EPI_ISL_1276866,
EPI_ISL_1276867, EPI_ISL_1276868, EPI_ISL_1276870, EPI_ISL_1276872, EPI_ISL_1276873, EPI_ISL_1276874, EPI_ISL_1276877, EPI_ISL_1276880, EPI_ISL_1276881, EPI_ISL_1276883, EPI_ISL_1276884, EPI_ISL_1276885, EPI_ISL_1276886, EPI_ISL_1276887, EPI_ISL_1276888, EPI_ISL_1276889, EPI_ISL_1276890, EPI_ISL_1277205, EPI_ISL_1277206, EPI_ISL_1277207, EPI_ISL_1277208, EPI_ISL_1277209,
EPI_ISL_1277210, EPI_ISL_1277211, EPI_ISL_1277213, EPI_ISL_1277214, EPI_ISL_1277215, EPI_ISL_1277216, EPI_ISL_1277217, EPI_ISL_1277218, EPI_ISL_1277219, EPI_ISL_1277220, EPI_ISL_1277222, EPI_ISL_1277223, EPI_ISL_1277224, EPI_ISL_1277225, EPI_ISL_1277228, EPI_ISL_1277229, EPI_ISL_1277230, EPI_ISL_1277231, EPI_ISL_1277232, EPI_ISL_1277233, EPI_ISL_1277234, EPI_ISL_1277236,
EPI_ISL_1277237, EPI_ISL_1277238, EPI_ISL_1277239, EPI_ISL_1277240, EPI_ISL_1277241, EPI_ISL_1277243, EPI_ISL_1277244, EPI_ISL_1277245, EPI_ISL_1277246, EPI_ISL_1277247, EPI_ISL_1277249, EPI_ISL_1277250, EPI_ISL_1277251, EPI_ISL_1277252, EPI_ISL_1277254, EPI_ISL_1277255, EPI_ISL_1277256, EPI_ISL_1277257, EPI_ISL_1277258, EPI_ISL_1277259, EPI_ISL_1277260, EPI_ISL_1277261,
EPI_ISL_1277262, EPI_ISL_1277263, EPI_ISL_1277264, EPI_ISL_1277265, EPI_ISL_1277266, EPI_ISL_1277267, EPI_ISL_1277269, EPI_ISL_1277271, EPI_ISL_1277272, EPI_ISL_1277273, EPI_ISL_1277274, EPI_ISL_1277275, EPI_ISL_1277276, EPI_ISL_1277277, EPI_ISL_1277279, EPI_ISL_1277280, EPI_ISL_1277281, EPI_ISL_1277282, EPI_ISL_1277283, EPI_ISL_1277284, EPI_ISL_1277285, EPI_ISL_1277286,
EPI_ISL_1277287, EPI_ISL_1277288, EPI_ISL_1277289, EPI_ISL_1277290, EPI_ISL_1277291, EPI_ISL_1277292, EPI_ISL_1277293, EPI_ISL_1277294, EPI_ISL_1277295, EPI_ISL_1277296, EPI_ISL_1277297, EPI_ISL_1277298, EPI_ISL_1277299, EPI_ISL_1277300, EPI_ISL_1277301, EPI_ISL_1277302, EPI_ISL_1277304, EPI_ISL_1277305, EPI_ISL_1277306, EPI_ISL_1277307, EPI_ISL_1277308, EPI_ISL_1277309,
EPI_ISL_1277310, EPI_ISL_1277312, EPI_ISL_1277313, EPI_ISL_1277314, EPI_ISL_1277315, EPI_ISL_1277317, EPI_ISL_1277318, EPI_ISL_1277321, EPI_ISL_1277322, EPI_ISL_1277323, EPI_ISL_1277324, EPI_ISL_1277326, EPI_ISL_1277327, EPI_ISL_1277332, EPI_ISL_1277335, EPI_ISL_1277340, EPI_ISL_1277341, EPI_ISL_1277342, EPI_ISL_1277343, EPI_ISL_1277345, EPI_ISL_1277346, EPI_ISL_1277348,
EPI_ISL_1277604, EPI_ISL_1277606, EPI_ISL_1277607, EPI_ISL_1277608, EPI_ISL_1277609, EPI_ISL_1277610, EPI_ISL_1277611, EPI_ISL_1277612, EPI_ISL_1277614, EPI_ISL_1277615, EPI_ISL_1277616, EPI_ISL_1277617, EPI_ISL_1277618, EPI_ISL_1277619, EPI_ISL_1277620, EPI_ISL_1277621, EPI_ISL_1277622, EPI_ISL_1277623, EPI_ISL_1277624, EPI_ISL_1277625, EPI_ISL_1277626, EPI_ISL_1277627,
EPI_ISL_1277628, EPI_ISL_1277736, EPI_ISL_1277737, EPI_ISL_1277738, EPI_ISL_1277739, EPI_ISL_1277740, EPI_ISL_1277741, EPI_ISL_1277742, EPI_ISL_1277743, EPI_ISL_1277744, EPI_ISL_1277745, EPI_ISL_1277746, EPI_ISL_1277747, EPI_ISL_1277749, EPI_ISL_1277750, EPI_ISL_1277751, EPI_ISL_1277752, EPI_ISL_1277753, EPI_ISL_1277754, EPI_ISL_1277756, EPI_ISL_1277757, EPI_ISL_1277759,
EPI_ISL_1277761, EPI_ISL_1277762, EPI_ISL_1277764, EPI_ISL_1277765, EPI_ISL_1277766, EPI_ISL_1277767, EPI_ISL_1277768, EPI_ISL_1277769, EPI_ISL_1277770, EPI_ISL_1277771, EPI_ISL_1277772, EPI_ISL_1277773, EPI_ISL_1277774, EPI_ISL_1277775, EPI_ISL_1277776, EPI_ISL_1277777, EPI_ISL_1277778, EPI_ISL_1277779, EPI_ISL_1277780, EPI_ISL_1277782, EPI_ISL_1277783, EPI_ISL_1277784,
EPI_ISL_1277785, EPI_ISL_1277786, EPI_ISL_1277787, EPI_ISL_1277788, EPI_ISL_1277789, EPI_ISL_1277790, EPI_ISL_1277791, EPI_ISL_1277792, EPI_ISL_1277793, EPI_ISL_1277794, EPI_ISL_1277795, EPI_ISL_1277796, EPI_ISL_1277797, EPI_ISL_1277798, EPI_ISL_1277799, EPI_ISL_1277800, EPI_ISL_1277801, EPI_ISL_1277802, EPI_ISL_1277803, EPI_ISL_1277804, EPI_ISL_1277805, EPI_ISL_1277806,
EPI_ISL_1277807, EPI_ISL_1277808, EPI_ISL_1277809, EPI_ISL_1277810, EPI_ISL_1277812, EPI_ISL_1277813, EPI_ISL_1277814, EPI_ISL_1277815, EPI_ISL_1277816, EPI_ISL_1277817, EPI_ISL_1277818, EPI_ISL_1277819, EPI_ISL_1277820, EPI_ISL_1277821, EPI_ISL_1277822, EPI_ISL_1277823, EPI_ISL_1277824, EPI_ISL_1277825, EPI_ISL_1277826, EPI_ISL_1277827, EPI_ISL_1277828, EPI_ISL_1277830,
EPI_ISL_1277831, EPI_ISL_1277832, EPI_ISL_1277833, EPI_ISL_1277834, EPI_ISL_1277835, EPI_ISL_1277836, EPI_ISL_1277837, EPI_ISL_1277838, EPI_ISL_1277839, EPI_ISL_1277841, EPI_ISL_1277842, EPI_ISL_1277843, EPI_ISL_1277844, EPI_ISL_1277845, EPI_ISL_1277846, EPI_ISL_1277847, EPI_ISL_1277848, EPI_ISL_1277849, EPI_ISL_1277850, EPI_ISL_1277851, EPI_ISL_1277852, EPI_ISL_1277853,
EPI_ISL_1277854, EPI_ISL_1277855, EPI_ISL_1277856, EPI_ISL_1277857, EPI_ISL_1277858, EPI_ISL_1277859, EPI_ISL_1277860, EPI_ISL_1277861, EPI_ISL_1277863, EPI_ISL_1277864, EPI_ISL_1293377, EPI_ISL_1293403, EPI_ISL_1293422, EPI_ISL_1293459, EPI_ISL_1293472, EPI_ISL_1293487, EPI_ISL_1293565, EPI_ISL_1293605, EPI_ISL_1293609, EPI_ISL_1293624, EPI_ISL_1293633, EPI_ISL_1293635,
EPI_ISL_1293636, EPI_ISL_1293639, EPI_ISL_1293640, EPI_ISL_1293641, EPI_ISL_1293644, EPI_ISL_1293645, EPI_ISL_1293646, EPI_ISL_1293647, EPI_ISL_1293649, EPI_ISL_1293650, EPI_ISL_1293651, EPI_ISL_1293652, EPI_ISL_1293654, EPI_ISL_1293655, EPI_ISL_1293658, EPI_ISL_1293660, EPI_ISL_1293662, EPI_ISL_1293663, EPI_ISL_1293664, EPI_ISL_1293665, EPI_ISL_1293666, EPI_ISL_1293667,
EPI_ISL_1293668, EPI_ISL_1293669, EPI_ISL_1293670, EPI_ISL_1293671, EPI_ISL_1293673, EPI_ISL_1293674, EPI_ISL_1293675, EPI_ISL_1293676, EPI_ISL_1293678, EPI_ISL_1293680, EPI_ISL_1293681, EPI_ISL_1293683, EPI_ISL_1293685, EPI_ISL_1293686, EPI_ISL_1293688, EPI_ISL_1293689, EPI_ISL_1293690, EPI_ISL_1293691, EPI_ISL_1293693, EPI_ISL_1293694, EPI_ISL_1293696, EPI_ISL_1293698,
EPI_ISL_1293699, EPI_ISL_1293700, EPI_ISL_1293701, EPI_ISL_1293702, EPI_ISL_1293703, EPI_ISL_1293704, EPI_ISL_1293706, EPI_ISL_1293708, EPI_ISL_1293709, EPI_ISL_1293710, EPI_ISL_1293712, EPI_ISL_1293713, EPI_ISL_1293715, EPI_ISL_1293718, EPI_ISL_1293719, EPI_ISL_1293720, EPI_ISL_1293722, EPI_ISL_1293723, EPI_ISL_1293724, EPI_ISL_1293725, EPI_ISL_1293726, EPI_ISL_1293733,
EPI_ISL_1293735, EPI_ISL_1293736, EPI_ISL_1293738, EPI_ISL_1293739, EPI_ISL_1293740, EPI_ISL_1293741, EPI_ISL_1293742, EPI_ISL_1293744, EPI_ISL_1293745, EPI_ISL_1293747, EPI_ISL_1293749, EPI_ISL_1293751, EPI_ISL_1293753, EPI_ISL_1293754, EPI_ISL_1293755, EPI_ISL_1293756, EPI_ISL_1293757, EPI_ISL_1293758, EPI_ISL_1293760, EPI_ISL_1293762, EPI_ISL_1293763, EPI_ISL_1293765,
EPI_ISL_1293768, EPI_ISL_1293771, EPI_ISL_1293773, EPI_ISL_1293775, EPI_ISL_1293779, EPI_ISL_1293781, EPI_ISL_1293783, EPI_ISL_1293784, EPI_ISL_1293785, EPI_ISL_1293786, EPI_ISL_1293787, EPI_ISL_1293788, EPI_ISL_1293789, EPI_ISL_1293790, EPI_ISL_1293791, EPI_ISL_1293792, EPI_ISL_1293793, EPI_ISL_1293794, EPI_ISL_1293795, EPI_ISL_1293796, EPI_ISL_1293797, EPI_ISL_1293798,
EPI_ISL_1293799, EPI_ISL_1293800, EPI_ISL_1293801, EPI_ISL_1293802, EPI_ISL_1293803, EPI_ISL_1293804, EPI_ISL_1293805, EPI_ISL_1293806, EPI_ISL_1293807, EPI_ISL_1293808, EPI_ISL_1293809, EPI_ISL_1293810, EPI_ISL_1293811, EPI_ISL_1293812, EPI_ISL_1293813, EPI_ISL_1293814, EPI_ISL_1293815, EPI_ISL_1293816, EPI_ISL_1293817, EPI_ISL_1293818, EPI_ISL_1293819, EPI_ISL_1293820,
EPI_ISL_1293821, EPI_ISL_1293822, EPI_ISL_1293823, EPI_ISL_1293824, EPI_ISL_1293825, EPI_ISL_1293826, EPI_ISL_1293827, EPI_ISL_1293828, EPI_ISL_1293829, EPI_ISL_1293830, EPI_ISL_1293831, EPI_ISL_1293832, EPI_ISL_1293833, EPI_ISL_1293834, EPI_ISL_1293835, EPI_ISL_1293836, EPI_ISL_1293837, EPI_ISL_1293838, EPI_ISL_1293839, EPI_ISL_1293840, EPI_ISL_1293841, EPI_ISL_1293842,
EPI_ISL_1293843, EPI_ISL_1293844, EPI_ISL_1293845, EPI_ISL_1293846, EPI_ISL_1293847, EPI_ISL_1293848, EPI_ISL_1293849, EPI_ISL_1293850, EPI_ISL_1293851, EPI_ISL_1293852, EPI_ISL_1293853, EPI_ISL_1293854, EPI_ISL_1293855, EPI_ISL_1293856, EPI_ISL_1293857, EPI_ISL_1293858, EPI_ISL_1293859, EPI_ISL_1293860, EPI_ISL_1293861, EPI_ISL_1293862, EPI_ISL_1293863, EPI_ISL_1293864,
EPI_ISL_1293865, EPI_ISL_1293866, EPI_ISL_1293867, EPI_ISL_1293868, EPI_ISL_1293869, EPI_ISL_1293870, EPI_ISL_1293871, EPI_ISL_1293872, EPI_ISL_1293873, EPI_ISL_1293874, EPI_ISL_1293875, EPI_ISL_1293876, EPI_ISL_1293877, EPI_ISL_1293878, EPI_ISL_1293879, EPI_ISL_1293880, EPI_ISL_1293881, EPI_ISL_1293882, EPI_ISL_1293883, EPI_ISL_1293884, EPI_ISL_1293885, EPI_ISL_1293886,
EPI_ISL_1293887, EPI_ISL_1293888, EPI_ISL_1293889, EPI_ISL_1293890, EPI_ISL_1293891, EPI_ISL_1293892, EPI_ISL_1293893, EPI_ISL_1293894, EPI_ISL_1293895, EPI_ISL_1293896, EPI_ISL_1293897, EPI_ISL_1293898, EPI_ISL_1293899, EPI_ISL_1293900, EPI_ISL_1293901, EPI_ISL_1293902, EPI_ISL_1293903, EPI_ISL_1293904, EPI_ISL_1293905, EPI_ISL_1293906, EPI_ISL_1293907, EPI_ISL_1293908,
EPI_ISL_1293909, EPI_ISL_1293910, EPI_ISL_1293911, EPI_ISL_1293912, EPI_ISL_1293913, EPI_ISL_1293914, EPI_ISL_1293915, EPI_ISL_1293916, EPI_ISL_1293917, EPI_ISL_1293918, EPI_ISL_1293919, EPI_ISL_1293920, EPI_ISL_1293921, EPI_ISL_1293922, EPI_ISL_1293923, EPI_ISL_1293924, EPI_ISL_1293925, EPI_ISL_1293926, EPI_ISL_1293927, EPI_ISL_1293928, EPI_ISL_1293929, EPI_ISL_1293930,
EPI_ISL_1293931, EPI_ISL_1293932, EPI_ISL_1293933, EPI_ISL_1293934, EPI_ISL_1293935, EPI_ISL_1293936, EPI_ISL_1293937, EPI_ISL_1293938, EPI_ISL_1293939, EPI_ISL_1293940, EPI_ISL_1293941, EPI_ISL_1293942, EPI_ISL_1293943, EPI_ISL_1293944, EPI_ISL_1293945, EPI_ISL_1293946, EPI_ISL_1293947, EPI_ISL_1293948, EPI_ISL_1293949, EPI_ISL_1293950, EPI_ISL_1293951, EPI_ISL_1293952,
EPI_ISL_1293953, EPI_ISL_1293954, EPI_ISL_1293955, EPI_ISL_1293956, EPI_ISL_1293957, EPI_ISL_1293958, EPI_ISL_1293959, EPI_ISL_1293960, EPI_ISL_1293961, EPI_ISL_1293962, EPI_ISL_1293963, EPI_ISL_1293964, EPI_ISL_1293965, EPI_ISL_1293966, EPI_ISL_1293967, EPI_ISL_1293968, EPI_ISL_1293969, EPI_ISL_1293970, EPI_ISL_1293971, EPI_ISL_1293972, EPI_ISL_1293973, EPI_ISL_1293974,
EPI_ISL_1293975, EPI_ISL_1293976, EPI_ISL_1293977, EPI_ISL_1293978, EPI_ISL_1293979, EPI_ISL_1293980, EPI_ISL_1293981, EPI_ISL_1293982, EPI_ISL_1293983, EPI_ISL_1293984, EPI_ISL_1293985, EPI_ISL_1293986, EPI_ISL_1293987, EPI_ISL_1293988, EPI_ISL_1293989, EPI_ISL_1293990, EPI_ISL_1293991, EPI_ISL_1293992, EPI_ISL_1293993, EPI_ISL_1293994, EPI_ISL_1293995, EPI_ISL_1293996,
EPI_ISL_1293997, EPI_ISL_1293998, EPI_ISL_1293999, EPI_ISL_1294000, EPI_ISL_1294001, EPI_ISL_1294002, EPI_ISL_1294003, EPI_ISL_1294004, EPI_ISL_1294005, EPI_ISL_1294006, EPI_ISL_1294007, EPI_ISL_1294008, EPI_ISL_1294009, EPI_ISL_1294010, EPI_ISL_1294011, EPI_ISL_1294012, EPI_ISL_1294013, EPI_ISL_1294014, EPI_ISL_1294015, EPI_ISL_1294016, EPI_ISL_1294017, EPI_ISL_1294018,
EPI_ISL_1294019, EPI_ISL_1294020, EPI_ISL_1294021, EPI_ISL_1294022, EPI_ISL_1294023, EPI_ISL_1294024, EPI_ISL_1294025, EPI_ISL_1294026, EPI_ISL_1294027, EPI_ISL_1294028, EPI_ISL_1294029, EPI_ISL_1294030, EPI_ISL_1294031, EPI_ISL_1294032, EPI_ISL_1294033, EPI_ISL_1294034, EPI_ISL_1294035, EPI_ISL_1294036, EPI_ISL_1294037, EPI_ISL_1294038, EPI_ISL_1294039, EPI_ISL_1294040,
EPI_ISL_1294041, EPI_ISL_1294042, EPI_ISL_1294043, EPI_ISL_1294044, EPI_ISL_1294045, EPI_ISL_1294046, EPI_ISL_1294047, EPI_ISL_1294048, EPI_ISL_1294049, EPI_ISL_1294050, EPI_ISL_1294051, EPI_ISL_1294052, EPI_ISL_1294053, EPI_ISL_1294054, EPI_ISL_1294055, EPI_ISL_1294056, EPI_ISL_1294057, EPI_ISL_1294058, EPI_ISL_1294059, EPI_ISL_1294060, EPI_ISL_1294061, EPI_ISL_1294062,
EPI_ISL_1294063, EPI_ISL_1294064, EPI_ISL_1294065, EPI_ISL_1294066, EPI_ISL_1294067, EPI_ISL_1294068, EPI_ISL_1294069, EPI_ISL_1294070, EPI_ISL_1294071, EPI_ISL_1294072, EPI_ISL_1294073, EPI_ISL_1294074, EPI_ISL_1294075, EPI_ISL_1294076, EPI_ISL_1294077, EPI_ISL_1294078, EPI_ISL_1294079, EPI_ISL_1294080, EPI_ISL_1294081, EPI_ISL_1294082, EPI_ISL_1294083, EPI_ISL_1294084,
EPI_ISL_1294085, EPI_ISL_1294086, EPI_ISL_1294087, EPI_ISL_1294088, EPI_ISL_1294089, EPI_ISL_1294090, EPI_ISL_1294091, EPI_ISL_1294092, EPI_ISL_1294093, EPI_ISL_1294094, EPI_ISL_1294095, EPI_ISL_1294096, EPI_ISL_1294097, EPI_ISL_1294098, EPI_ISL_1294099, EPI_ISL_1294100, EPI_ISL_1294101, EPI_ISL_1294102, EPI_ISL_1294103, EPI_ISL_1294104, EPI_ISL_1294105, EPI_ISL_1294106,
EPI_ISL_1294107, EPI_ISL_1294108, EPI_ISL_1294109, EPI_ISL_1294110, EPI_ISL_1294111, EPI_ISL_1294112, EPI_ISL_1294113, EPI_ISL_1294114, EPI_ISL_1294115, EPI_ISL_1294116, EPI_ISL_1294117, EPI_ISL_1294118, EPI_ISL_1294119, EPI_ISL_1294120, EPI_ISL_1294121, EPI_ISL_1294122, EPI_ISL_1294123, EPI_ISL_1294124, EPI_ISL_1294125, EPI_ISL_1294126, EPI_ISL_1294127, EPI_ISL_1294128,
EPI_ISL_1294129, EPI_ISL_1294130, EPI_ISL_1294131, EPI_ISL_1294132, EPI_ISL_1294133, EPI_ISL_1294134, EPI_ISL_1294135, EPI_ISL_1294136, EPI_ISL_1294137, EPI_ISL_1294138, EPI_ISL_1294139, EPI_ISL_1294140, EPI_ISL_1294141, EPI_ISL_1294142, EPI_ISL_1294143, EPI_ISL_1294145, EPI_ISL_1294147, EPI_ISL_1294152, EPI_ISL_1294154, EPI_ISL_1294155, EPI_ISL_1294157, EPI_ISL_1294158,
EPI_ISL_1294162, EPI_ISL_1294164, EPI_ISL_1294168, EPI_ISL_1294171, EPI_ISL_1294173, EPI_ISL_1294180, EPI_ISL_1294184, EPI_ISL_1294185, EPI_ISL_1294186, EPI_ISL_1294188, EPI_ISL_1294190, EPI_ISL_1294192, EPI_ISL_1294194, EPI_ISL_1294197, EPI_ISL_1294200, EPI_ISL_1294201, EPI_ISL_1294204, EPI_ISL_1294206, EPI_ISL_1294209, EPI_ISL_1294211, EPI_ISL_1294213, EPI_ISL_1294215,
EPI_ISL_1294216, EPI_ISL_1294218, EPI_ISL_1294223, EPI_ISL_1294225, EPI_ISL_1294232, EPI_ISL_1294233, EPI_ISL_1294234, EPI_ISL_1294236, EPI_ISL_1294239, EPI_ISL_1294243, EPI_ISL_1294245, EPI_ISL_1294248, EPI_ISL_1294260, EPI_ISL_1294263, EPI_ISL_1294264, EPI_ISL_1294265, EPI_ISL_1294266, EPI_ISL_1294271, EPI_ISL_1294275, EPI_ISL_1294278, EPI_ISL_1294279, EPI_ISL_1294282,
EPI_ISL_1294285, EPI_ISL_1294286, EPI_ISL_1294287, EPI_ISL_1294288, EPI_ISL_1294289, EPI_ISL_1294290, EPI_ISL_1294291, EPI_ISL_1294292, EPI_ISL_1294293, EPI_ISL_1294294, EPI_ISL_1294295, EPI_ISL_1294296, EPI_ISL_1294297, EPI_ISL_1294298, EPI_ISL_1294299, EPI_ISL_1294300, EPI_ISL_1294301, EPI_ISL_1294302, EPI_ISL_1294303, EPI_ISL_1294304, EPI_ISL_1294305, EPI_ISL_1294306,
EPI_ISL_1294307, EPI_ISL_1294308, EPI_ISL_1294309, EPI_ISL_1294310, EPI_ISL_1294311, EPI_ISL_1294312, EPI_ISL_1294313, EPI_ISL_1294314, EPI_ISL_1294315, EPI_ISL_1294316, EPI_ISL_1294317, EPI_ISL_1294318, EPI_ISL_1294319, EPI_ISL_1294320, EPI_ISL_1294321, EPI_ISL_1294322, EPI_ISL_1294323, EPI_ISL_1294324, EPI_ISL_1294325, EPI_ISL_1294326, EPI_ISL_1294327, EPI_ISL_1294328,
EPI_ISL_1294329, EPI_ISL_1294330, EPI_ISL_1294331, EPI_ISL_1294332, EPI_ISL_1294333, EPI_ISL_1294334, EPI_ISL_1294335, EPI_ISL_1294336, EPI_ISL_1294337, EPI_ISL_1294338, EPI_ISL_1294339, EPI_ISL_1294340, EPI_ISL_1294341, EPI_ISL_1294342, EPI_ISL_1294343, EPI_ISL_1294344, EPI_ISL_1294345, EPI_ISL_1294346, EPI_ISL_1294347, EPI_ISL_1294348, EPI_ISL_1294349, EPI_ISL_1294350,
EPI_ISL_1294351, EPI_ISL_1294352, EPI_ISL_1294353, EPI_ISL_1294354, EPI_ISL_1294355, EPI_ISL_1294356, EPI_ISL_1294357, EPI_ISL_1294359, EPI_ISL_1294360, EPI_ISL_1294361, EPI_ISL_1294362, EPI_ISL_1294363, EPI_ISL_1294364, EPI_ISL_1294365, EPI_ISL_1294366, EPI_ISL_1294367, EPI_ISL_1294368, EPI_ISL_1294369, EPI_ISL_1294370, EPI_ISL_1294371, EPI_ISL_1294372, EPI_ISL_1294373,
EPI_ISL_1294374, EPI_ISL_1294375, EPI_ISL_1294376, EPI_ISL_1294377, EPI_ISL_1294378, EPI_ISL_1294379, EPI_ISL_1294380, EPI_ISL_1294381, EPI_ISL_1294382, EPI_ISL_1294383, EPI_ISL_1294384, EPI_ISL_1294385, EPI_ISL_1294386, EPI_ISL_1294387, EPI_ISL_1294388, EPI_ISL_1294389, EPI_ISL_1294390, EPI_ISL_1294391, EPI_ISL_1294392, EPI_ISL_1294393, EPI_ISL_1294394, EPI_ISL_1294395,
EPI_ISL_1294396, EPI_ISL_1294397, EPI_ISL_1294398, EPI_ISL_1294399, EPI_ISL_1294400, EPI_ISL_1294401, EPI_ISL_1294402, EPI_ISL_1294403, EPI_ISL_1294404, EPI_ISL_1294405, EPI_ISL_1294406, EPI_ISL_1294407, EPI_ISL_1294408, EPI_ISL_1294409, EPI_ISL_1294410, EPI_ISL_1294411, EPI_ISL_1294412, EPI_ISL_1294413, EPI_ISL_1294414, EPI_ISL_1294415, EPI_ISL_1294416, EPI_ISL_1294417,
EPI_ISL_1294418, EPI_ISL_1294419, EPI_ISL_1294420, EPI_ISL_1294421, EPI_ISL_1294422, EPI_ISL_1294423, EPI_ISL_1294424, EPI_ISL_1294427, EPI_ISL_1294437, EPI_ISL_1294440, EPI_ISL_1294441, EPI_ISL_1294443, EPI_ISL_1294444, EPI_ISL_1294448, EPI_ISL_1294450, EPI_ISL_1294451, EPI_ISL_1294452, EPI_ISL_1294453, EPI_ISL_1294454, EPI_ISL_1294457, EPI_ISL_1294461, EPI_ISL_1294463,
EPI_ISL_1294464, EPI_ISL_1294466, EPI_ISL_1294467, EPI_ISL_1294468, EPI_ISL_1294469, EPI_ISL_1294474, EPI_ISL_1294475, EPI_ISL_1294478, EPI_ISL_1294479, EPI_ISL_1294480, EPI_ISL_1294482, EPI_ISL_1294483, EPI_ISL_1294485, EPI_ISL_1294486, EPI_ISL_1294487, EPI_ISL_1294488, EPI_ISL_1294489, EPI_ISL_1294490, EPI_ISL_1294491, EPI_ISL_1294492, EPI_ISL_1294493, EPI_ISL_1294494,
EPI_ISL_1294498, EPI_ISL_1294499, EPI_ISL_1294501, EPI_ISL_1294502, EPI_ISL_1294503, EPI_ISL_1294504, EPI_ISL_1294505, EPI_ISL_1294506, EPI_ISL_1294507, EPI_ISL_1294508, EPI_ISL_1294510, EPI_ISL_1294511, EPI_ISL_1294514, EPI_ISL_1294515, EPI_ISL_1294516, EPI_ISL_1294518, EPI_ISL_1294520, EPI_ISL_1294521, EPI_ISL_1294522, EPI_ISL_1294523, EPI_ISL_1294525, EPI_ISL_1294530,
EPI_ISL_1294533, EPI_ISL_1294535, EPI_ISL_1294537, EPI_ISL_1294538, EPI_ISL_1294539, EPI_ISL_1294540, EPI_ISL_1294541, EPI_ISL_1294543, EPI_ISL_1294544, EPI_ISL_1294545, EPI_ISL_1294546, EPI_ISL_1294548, EPI_ISL_1294550, EPI_ISL_1294551, EPI_ISL_1294554, EPI_ISL_1294556, EPI_ISL_1294558, EPI_ISL_1294559, EPI_ISL_1294560, EPI_ISL_1294561, EPI_ISL_1294562, EPI_ISL_1294563,
EPI_ISL_1294564, EPI_ISL_1294565, EPI_ISL_1294566, EPI_ISL_1294567, EPI_ISL_1294568, EPI_ISL_1294569, EPI_ISL_1294570, EPI_ISL_1294571, EPI_ISL_1294572, EPI_ISL_1294573, EPI_ISL_1294574, EPI_ISL_1294575, EPI_ISL_1294576, EPI_ISL_1294577, EPI_ISL_1294578, EPI_ISL_1294579, EPI_ISL_1294580, EPI_ISL_1294581, EPI_ISL_1294582, EPI_ISL_1294583, EPI_ISL_1294584, EPI_ISL_1294585,
EPI_ISL_1294586, EPI_ISL_1294587, EPI_ISL_1294588, EPI_ISL_1294589, EPI_ISL_1294590, EPI_ISL_1294591, EPI_ISL_1294592, EPI_ISL_1294593, EPI_ISL_1294594, EPI_ISL_1294595, EPI_ISL_1294596, EPI_ISL_1294597, EPI_ISL_1294598, EPI_ISL_1294599, EPI_ISL_1294600, EPI_ISL_1294601, EPI_ISL_1294602, EPI_ISL_1294603, EPI_ISL_1294604, EPI_ISL_1294605, EPI_ISL_1294606, EPI_ISL_1294607,
EPI_ISL_1294608, EPI_ISL_1294609, EPI_ISL_1294610, EPI_ISL_1294611, EPI_ISL_1294612, EPI_ISL_1294613, EPI_ISL_1294614, EPI_ISL_1294615, EPI_ISL_1294616, EPI_ISL_1294624, EPI_ISL_1294625, EPI_ISL_1294626, EPI_ISL_1294627, EPI_ISL_1294629, EPI_ISL_1294630, EPI_ISL_1294631, EPI_ISL_1294632, EPI_ISL_1294634, EPI_ISL_1294635, EPI_ISL_1294636, EPI_ISL_1294638, EPI_ISL_1294639,
EPI_ISL_1294640, EPI_ISL_1294641, EPI_ISL_1294643, EPI_ISL_1294644, EPI_ISL_1294645, EPI_ISL_1294646, EPI_ISL_1294648, EPI_ISL_1294649, EPI_ISL_1294651, EPI_ISL_1294652, EPI_ISL_1294653, EPI_ISL_1294654, EPI_ISL_1294657, EPI_ISL_1294658, EPI_ISL_1294659, EPI_ISL_1294660, EPI_ISL_1294661, EPI_ISL_1294662, EPI_ISL_1294663, EPI_ISL_1294664, EPI_ISL_1294665, EPI_ISL_1294666,
EPI_ISL_1294667, EPI_ISL_1294668, EPI_ISL_1294669, EPI_ISL_1294670, EPI_ISL_1294671, EPI_ISL_1294672, EPI_ISL_1294673, EPI_ISL_1294674, EPI_ISL_1294675, EPI_ISL_1294676, EPI_ISL_1294677, EPI_ISL_1294678, EPI_ISL_1294679, EPI_ISL_1294680, EPI_ISL_1294681, EPI_ISL_1294682, EPI_ISL_1294683, EPI_ISL_1294684, EPI_ISL_1294685, EPI_ISL_1294686, EPI_ISL_1294687, EPI_ISL_1294688,
EPI_ISL_1294689, EPI_ISL_1294690, EPI_ISL_1294691, EPI_ISL_1294692, EPI_ISL_1294693, EPI_ISL_1294694, EPI_ISL_1294695, EPI_ISL_1294696, EPI_ISL_1294697, EPI_ISL_1294698, EPI_ISL_1294699, EPI_ISL_1294700, EPI_ISL_1294701, EPI_ISL_1294702, EPI_ISL_1294703, EPI_ISL_1294704, EPI_ISL_1294705, EPI_ISL_1294706, EPI_ISL_1294707, EPI_ISL_1294708, EPI_ISL_1294709, EPI_ISL_1294710,
EPI_ISL_1294711, EPI_ISL_1294712, EPI_ISL_1294713, EPI_ISL_1294714, EPI_ISL_1294715, EPI_ISL_1294716, EPI_ISL_1294717, EPI_ISL_1294718, EPI_ISL_1294719, EPI_ISL_1294720, EPI_ISL_1294721, EPI_ISL_1294722, EPI_ISL_1294723, EPI_ISL_1294724, EPI_ISL_1294725, EPI_ISL_1294726, EPI_ISL_1294727, EPI_ISL_1294728, EPI_ISL_1294729, EPI_ISL_1294730, EPI_ISL_1294731, EPI_ISL_1294732,
EPI_ISL_1294733, EPI_ISL_1294734, EPI_ISL_1294735, EPI_ISL_1294736, EPI_ISL_1294737, EPI_ISL_1294738, EPI_ISL_1294739, EPI_ISL_1294740, EPI_ISL_1294741, EPI_ISL_1294742, EPI_ISL_1294743, EPI_ISL_1294744, EPI_ISL_1294745, EPI_ISL_1294746, EPI_ISL_1294747, EPI_ISL_1294748, EPI_ISL_1294749, EPI_ISL_1294750, EPI_ISL_1294751, EPI_ISL_1294752, EPI_ISL_1294753, EPI_ISL_1294754,
EPI_ISL_1294755, EPI_ISL_1294756, EPI_ISL_1294757, EPI_ISL_1294758, EPI_ISL_1294759, EPI_ISL_1294760, EPI_ISL_1294761, EPI_ISL_1294762, EPI_ISL_1294763, EPI_ISL_1294764, EPI_ISL_1294765, EPI_ISL_1294766, EPI_ISL_1294767, EPI_ISL_1294768, EPI_ISL_1294769, EPI_ISL_1294770, EPI_ISL_1294771, EPI_ISL_1294772, EPI_ISL_1294773, EPI_ISL_1294774, EPI_ISL_1294775, EPI_ISL_1294776,
EPI_ISL_1294777, EPI_ISL_1294778, EPI_ISL_1294779, EPI_ISL_1294780, EPI_ISL_1294781, EPI_ISL_1294782, EPI_ISL_1294783, EPI_ISL_1294784, EPI_ISL_1294785, EPI_ISL_1294786, EPI_ISL_1294787, EPI_ISL_1294788, EPI_ISL_1294789, EPI_ISL_1294790, EPI_ISL_1294791, EPI_ISL_1294792, EPI_ISL_1294793, EPI_ISL_1294794, EPI_ISL_1294795, EPI_ISL_1294796, EPI_ISL_1294797, EPI_ISL_1294798,
EPI_ISL_1294799, EPI_ISL_1294800, EPI_ISL_1294801, EPI_ISL_1294802, EPI_ISL_1294803, EPI_ISL_1294804, EPI_ISL_1294805, EPI_ISL_1294806, EPI_ISL_1294807, EPI_ISL_1294808, EPI_ISL_1294809, EPI_ISL_1294810, EPI_ISL_1294811, EPI_ISL_1294812, EPI_ISL_1294813, EPI_ISL_1294814, EPI_ISL_1294815, EPI_ISL_1294816, EPI_ISL_1294817, EPI_ISL_1294818, EPI_ISL_1294819, EPI_ISL_1294820,
EPI_ISL_1294821, EPI_ISL_1294822, EPI_ISL_1294823, EPI_ISL_1294824, EPI_ISL_1294825, EPI_ISL_1294826, EPI_ISL_1294827, EPI_ISL_1294828, EPI_ISL_1294829, EPI_ISL_1294830, EPI_ISL_1294831, EPI_ISL_1294832, EPI_ISL_1294833, EPI_ISL_1294834, EPI_ISL_1294835, EPI_ISL_1294836, EPI_ISL_1294837, EPI_ISL_1294838, EPI_ISL_1294839, EPI_ISL_1294840, EPI_ISL_1294841, EPI_ISL_1294842,
EPI_ISL_1294843, EPI_ISL_1294844, EPI_ISL_1294845, EPI_ISL_1294846, EPI_ISL_1294847, EPI_ISL_1294848, EPI_ISL_1294849, EPI_ISL_1294850, EPI_ISL_1294851, EPI_ISL_1294852, EPI_ISL_1294853, EPI_ISL_1294854, EPI_ISL_1294855, EPI_ISL_1294856, EPI_ISL_1294857, EPI_ISL_1294858, EPI_ISL_1294859, EPI_ISL_1294860, EPI_ISL_1294861, EPI_ISL_1294862, EPI_ISL_1294863, EPI_ISL_1294864,
EPI_ISL_1294865, EPI_ISL_1294866, EPI_ISL_1294867, EPI_ISL_1294868, EPI_ISL_1294869, EPI_ISL_1294870, EPI_ISL_1294871, EPI_ISL_1294872, EPI_ISL_1294873, EPI_ISL_1294874, EPI_ISL_1294875, EPI_ISL_1294876, EPI_ISL_1294877, EPI_ISL_1294878, EPI_ISL_1294879, EPI_ISL_1294880, EPI_ISL_1294881, EPI_ISL_1294882, EPI_ISL_1294883, EPI_ISL_1294884, EPI_ISL_1294885, EPI_ISL_1294886,
EPI_ISL_1294887, EPI_ISL_1294888, EPI_ISL_1294889, EPI_ISL_1294890, EPI_ISL_1294891, EPI_ISL_1294892, EPI_ISL_1294893, EPI_ISL_1294894, EPI_ISL_1294895, EPI_ISL_1294896, EPI_ISL_1294897, EPI_ISL_1294898, EPI_ISL_1294899, EPI_ISL_1294900, EPI_ISL_1294901, EPI_ISL_1294902, EPI_ISL_1294903, EPI_ISL_1294904, EPI_ISL_1294905, EPI_ISL_1294906, EPI_ISL_1294907, EPI_ISL_1294908,
EPI_ISL_1294909, EPI_ISL_1294910, EPI_ISL_1294911, EPI_ISL_1294912, EPI_ISL_1294913, EPI_ISL_1294914, EPI_ISL_1294915, EPI_ISL_1294916, EPI_ISL_1294917, EPI_ISL_1294918, EPI_ISL_1294919, EPI_ISL_1294920, EPI_ISL_1294921, EPI_ISL_1294922, EPI_ISL_1294923, EPI_ISL_1294924, EPI_ISL_1294925, EPI_ISL_1294926, EPI_ISL_1294927, EPI_ISL_1294928, EPI_ISL_1294929, EPI_ISL_1294930,
EPI_ISL_1294931, EPI_ISL_1294932, EPI_ISL_1294933, EPI_ISL_1294934, EPI_ISL_1294935, EPI_ISL_1294936, EPI_ISL_1294937, EPI_ISL_1294938, EPI_ISL_1294939, EPI_ISL_1294940, EPI_ISL_1294941, EPI_ISL_1294942, EPI_ISL_1294943, EPI_ISL_1294944, EPI_ISL_1294945, EPI_ISL_1294946, EPI_ISL_1294947, EPI_ISL_1294948, EPI_ISL_1294949, EPI_ISL_1294950, EPI_ISL_1294951, EPI_ISL_1294952,
EPI_ISL_1294953, EPI_ISL_1294954, EPI_ISL_1294955, EPI_ISL_1294956, EPI_ISL_1294957, EPI_ISL_1294958, EPI_ISL_1294959, EPI_ISL_1294960, EPI_ISL_1294961, EPI_ISL_1294962, EPI_ISL_1294963, EPI_ISL_1294964, EPI_ISL_1294965, EPI_ISL_1294966, EPI_ISL_1294967, EPI_ISL_1294968, EPI_ISL_1294969, EPI_ISL_1294971, EPI_ISL_1294972, EPI_ISL_1294973, EPI_ISL_1294974, EPI_ISL_1294975,
EPI_ISL_1294976, EPI_ISL_1294977, EPI_ISL_1294978, EPI_ISL_1294979, EPI_ISL_1294980, EPI_ISL_1294981, EPI_ISL_1294982, EPI_ISL_1294983, EPI_ISL_1294984, EPI_ISL_1294985, EPI_ISL_1294986, EPI_ISL_1294987, EPI_ISL_1294988, EPI_ISL_1294989, EPI_ISL_1294990, EPI_ISL_1294991, EPI_ISL_1294992, EPI_ISL_1294993, EPI_ISL_1294994, EPI_ISL_1294995, EPI_ISL_1294996, EPI_ISL_1294997,
EPI_ISL_1294998, EPI_ISL_1294999, EPI_ISL_1295000, EPI_ISL_1295001, EPI_ISL_1295002, EPI_ISL_1295003, EPI_ISL_1295004, EPI_ISL_1295005, EPI_ISL_1295006, EPI_ISL_1295007, EPI_ISL_1295008, EPI_ISL_1295009, EPI_ISL_1295010, EPI_ISL_1295011, EPI_ISL_1295012, EPI_ISL_1295014, EPI_ISL_1295015, EPI_ISL_1295016, EPI_ISL_1295017, EPI_ISL_1295018, EPI_ISL_1295019, EPI_ISL_1295020,
EPI_ISL_1295021, EPI_ISL_1295022, EPI_ISL_1295023, EPI_ISL_1295024, EPI_ISL_1295025, EPI_ISL_1295026, EPI_ISL_1295027, EPI_ISL_1295028, EPI_ISL_1295029, EPI_ISL_1295030, EPI_ISL_1295031, EPI_ISL_1295032, EPI_ISL_1295033, EPI_ISL_1295034, EPI_ISL_1295035, EPI_ISL_1295036, EPI_ISL_1295037, EPI_ISL_1295038, EPI_ISL_1295039, EPI_ISL_1295040, EPI_ISL_1295041, EPI_ISL_1295042,
EPI_ISL_1295043, EPI_ISL_1295044, EPI_ISL_1295045, EPI_ISL_1295046, EPI_ISL_1295047, EPI_ISL_1295048, EPI_ISL_1295049, EPI_ISL_1295050, EPI_ISL_1295051, EPI_ISL_1295052, EPI_ISL_1295053, EPI_ISL_1295054, EPI_ISL_1295055, EPI_ISL_1295056, EPI_ISL_1295057, EPI_ISL_1295058, EPI_ISL_1295059, EPI_ISL_1295060, EPI_ISL_1295061, EPI_ISL_1295062, EPI_ISL_1295063, EPI_ISL_1295064,
EPI_ISL_1295065, EPI_ISL_1295066, EPI_ISL_1295067, EPI_ISL_1295068, EPI_ISL_1295069, EPI_ISL_1295070, EPI_ISL_1295071, EPI_ISL_1295072, EPI_ISL_1315847, EPI_ISL_1315985, EPI_ISL_1316418, EPI_ISL_1316419, EPI_ISL_1316420, EPI_ISL_1316421, EPI_ISL_1316422, EPI_ISL_1316424, EPI_ISL_1316426, EPI_ISL_1316428, EPI_ISL_1316429, EPI_ISL_1316430, EPI_ISL_1316432, EPI_ISL_1316433,
EPI_ISL_1316435, EPI_ISL_1316436, EPI_ISL_1316437, EPI_ISL_1316438, EPI_ISL_1316439, EPI_ISL_1316440, EPI_ISL_1316441, EPI_ISL_1316442, EPI_ISL_1316444, EPI_ISL_1316445, EPI_ISL_1316446, EPI_ISL_1316447, EPI_ISL_1316449, EPI_ISL_1316451, EPI_ISL_1316452, EPI_ISL_1316453, EPI_ISL_1316456, EPI_ISL_1316457, EPI_ISL_1316458, EPI_ISL_1316463, EPI_ISL_1316465, EPI_ISL_1316467,
EPI_ISL_1316468, EPI_ISL_1316470, EPI_ISL_1316471, EPI_ISL_1316472, EPI_ISL_1316473, EPI_ISL_1316474, EPI_ISL_1316475, EPI_ISL_1316477, EPI_ISL_1316478, EPI_ISL_1316479, EPI_ISL_1316482, EPI_ISL_1316483, EPI_ISL_1316484, EPI_ISL_1316487, EPI_ISL_1316488, EPI_ISL_1316489, EPI_ISL_1316490, EPI_ISL_1316491, EPI_ISL_1316493, EPI_ISL_1316494, EPI_ISL_1316496, EPI_ISL_1316499,
EPI_ISL_1316500, EPI_ISL_1316501, EPI_ISL_1316502, EPI_ISL_1316503, EPI_ISL_1316505, EPI_ISL_1316506, EPI_ISL_1316507, EPI_ISL_1316508, EPI_ISL_1316510, EPI_ISL_1316513, EPI_ISL_1316517, EPI_ISL_1316519, EPI_ISL_1316520, EPI_ISL_1316521, EPI_ISL_1316523, EPI_ISL_1316524, EPI_ISL_1316525, EPI_ISL_1316526, EPI_ISL_1316527, EPI_ISL_1316528, EPI_ISL_1316529, EPI_ISL_1316530,
EPI_ISL_1316531, EPI_ISL_1316532, EPI_ISL_1316534, EPI_ISL_1316537, EPI_ISL_1316538, EPI_ISL_1316539, EPI_ISL_1316541, EPI_ISL_1316542, EPI_ISL_1316543, EPI_ISL_1316544, EPI_ISL_1316546, EPI_ISL_1316548, EPI_ISL_1316550, EPI_ISL_1316551, EPI_ISL_1316553, EPI_ISL_1316554, EPI_ISL_1316555, EPI_ISL_1316556, EPI_ISL_1316558, EPI_ISL_1316563, EPI_ISL_1316564, EPI_ISL_1316565,
EPI_ISL_1316567, EPI_ISL_1316568, EPI_ISL_1316569, EPI_ISL_1316570, EPI_ISL_1316573, EPI_ISL_1316576, EPI_ISL_1316578, EPI_ISL_1316580, EPI_ISL_1316581, EPI_ISL_1316582, EPI_ISL_1316585, EPI_ISL_1316587, EPI_ISL_1316588, EPI_ISL_1316591, EPI_ISL_1316592, EPI_ISL_1316593, EPI_ISL_1316597, EPI_ISL_1316599, EPI_ISL_1316600, EPI_ISL_1316601, EPI_ISL_1316603, EPI_ISL_1316606,
EPI_ISL_1316607, EPI_ISL_1316608, EPI_ISL_1316609, EPI_ISL_1316612, EPI_ISL_1316615, EPI_ISL_1316616, EPI_ISL_1316618, EPI_ISL_1316619, EPI_ISL_1316620, EPI_ISL_1316621, EPI_ISL_1316623, EPI_ISL_1316624, EPI_ISL_1316625, EPI_ISL_1316626, EPI_ISL_1316628, EPI_ISL_1316630, EPI_ISL_1316631, EPI_ISL_1316633, EPI_ISL_1316634, EPI_ISL_1316636, EPI_ISL_1316638, EPI_ISL_1316641,
EPI_ISL_1316645, EPI_ISL_1316646, EPI_ISL_1316648, EPI_ISL_1316649, EPI_ISL_1316650, EPI_ISL_1316651, EPI_ISL_1316652, EPI_ISL_1316653, EPI_ISL_1316654, EPI_ISL_1316655, EPI_ISL_1316658, EPI_ISL_1316661, EPI_ISL_1316662, EPI_ISL_1316665, EPI_ISL_1316666, EPI_ISL_1316670, EPI_ISL_1316671, EPI_ISL_1316672, EPI_ISL_1316673, EPI_ISL_1316674, EPI_ISL_1316676, EPI_ISL_1316677,
EPI_ISL_1316679, EPI_ISL_1316680, EPI_ISL_1316681, EPI_ISL_1316682, EPI_ISL_1316684, EPI_ISL_1316685, EPI_ISL_1316686, EPI_ISL_1316688, EPI_ISL_1316690, EPI_ISL_1316691, EPI_ISL_1316692, EPI_ISL_1316694, EPI_ISL_1316695, EPI_ISL_1316696, EPI_ISL_1316697, EPI_ISL_1316698, EPI_ISL_1316699, EPI_ISL_1316703, EPI_ISL_1316704, EPI_ISL_1316706, EPI_ISL_1316707, EPI_ISL_1316708,
EPI_ISL_1316710, EPI_ISL_1316712, EPI_ISL_1316713, EPI_ISL_1316714, EPI_ISL_1316715, EPI_ISL_1316716, EPI_ISL_1316717, EPI_ISL_1316718, EPI_ISL_1316720, EPI_ISL_1316724, EPI_ISL_1316726, EPI_ISL_1316728, EPI_ISL_1316729, EPI_ISL_1316730, EPI_ISL_1316732, EPI_ISL_1316733, EPI_ISL_1316734, EPI_ISL_1316735, EPI_ISL_1316736, EPI_ISL_1316737, EPI_ISL_1316738, EPI_ISL_1316740,
EPI_ISL_1316741, EPI_ISL_1316742, EPI_ISL_1316743, EPI_ISL_1316744, EPI_ISL_1316746, EPI_ISL_1316747, EPI_ISL_1316748, EPI_ISL_1316749, EPI_ISL_1316750, EPI_ISL_1316751, EPI_ISL_1316752, EPI_ISL_1316753, EPI_ISL_1316754, EPI_ISL_1316755, EPI_ISL_1316756, EPI_ISL_1316758, EPI_ISL_1316759, EPI_ISL_1316760, EPI_ISL_1316761, EPI_ISL_1316763, EPI_ISL_1316766, EPI_ISL_1316767,
EPI_ISL_1316768, EPI_ISL_1316769, EPI_ISL_1316770, EPI_ISL_1316771, EPI_ISL_1316772, EPI_ISL_1316774, EPI_ISL_1316775, EPI_ISL_1316776, EPI_ISL_1316778, EPI_ISL_1316779, EPI_ISL_1316780, EPI_ISL_1316781, EPI_ISL_1316782, EPI_ISL_1316978, EPI_ISL_1317001, EPI_ISL_1317003, EPI_ISL_1317006, EPI_ISL_1317020, EPI_ISL_1317062, EPI_ISL_1317080, EPI_ISL_1317210, EPI_ISL_1328801,
EPI_ISL_1328885, EPI_ISL_1328917, EPI_ISL_1328919, EPI_ISL_1328922, EPI_ISL_1328927, EPI_ISL_1328929, EPI_ISL_1328933, EPI_ISL_1328938, EPI_ISL_1328941, EPI_ISL_1328946, EPI_ISL_1328947, EPI_ISL_1328950, EPI_ISL_1328953, EPI_ISL_1328954, EPI_ISL_1328966, EPI_ISL_1328976, EPI_ISL_1328979, EPI_ISL_1328981, EPI_ISL_1328987, EPI_ISL_1328988, EPI_ISL_1328989, EPI_ISL_1329006,
EPI_ISL_1329011, EPI_ISL_1329016, EPI_ISL_1329017, EPI_ISL_1329020, EPI_ISL_1329021, EPI_ISL_1329023, EPI_ISL_1329024, EPI_ISL_1329025, EPI_ISL_1329028, EPI_ISL_1329041, EPI_ISL_1329049, EPI_ISL_1329063, EPI_ISL_1329065, EPI_ISL_1329071, EPI_ISL_1329077, EPI_ISL_1329080, EPI_ISL_1329081, EPI_ISL_1329084, EPI_ISL_1329088, EPI_ISL_1329091, EPI_ISL_1329107, EPI_ISL_1329122,
EPI_ISL_1329134, EPI_ISL_1329137, EPI_ISL_1329141, EPI_ISL_1329150, EPI_ISL_1329152, EPI_ISL_1329158, EPI_ISL_1329165, EPI_ISL_1329168, EPI_ISL_1329173, EPI_ISL_1329191, EPI_ISL_1329197, EPI_ISL_1331144, EPI_ISL_1331185, EPI_ISL_1331206, EPI_ISL_1331211, EPI_ISL_1331225, EPI_ISL_1331233, EPI_ISL_1331343, EPI_ISL_1331349, EPI_ISL_1331419, EPI_ISL_1331527, EPI_ISL_1331547,
EPI_ISL_1331559, EPI_ISL_1331587, EPI_ISL_1331685, EPI_ISL_1331702, EPI_ISL_1331708, EPI_ISL_1331725, EPI_ISL_1331772, EPI_ISL_1331825, EPI_ISL_1331833, EPI_ISL_1331835, EPI_ISL_1331841, EPI_ISL_1331856, EPI_ISL_1331870, EPI_ISL_1331886, EPI_ISL_1331891, EPI_ISL_1331893, EPI_ISL_1331937, EPI_ISL_1331946, EPI_ISL_1331950, EPI_ISL_1331969, EPI_ISL_1331981, EPI_ISL_1332043,
EPI_ISL_1332052, EPI_ISL_1332059, EPI_ISL_1332112, EPI_ISL_1332133, EPI_ISL_1332135, EPI_ISL_1332158, EPI_ISL_1332180, EPI_ISL_1332212, EPI_ISL_1332284, EPI_ISL_1332287, EPI_ISL_1332298, EPI_ISL_1332305, EPI_ISL_1332349, EPI_ISL_1332365, EPI_ISL_1332389, EPI_ISL_1332392, EPI_ISL_1332393, EPI_ISL_1332399, EPI_ISL_1332425, EPI_ISL_1332451, EPI_ISL_1332482, EPI_ISL_1332511,
EPI_ISL_1332512, EPI_ISL_1332545, EPI_ISL_1332603, EPI_ISL_1332615, EPI_ISL_1332620, EPI_ISL_1332647, EPI_ISL_1332705, EPI_ISL_1332706, EPI_ISL_1332708, EPI_ISL_1332710, EPI_ISL_1332713, EPI_ISL_1332714, EPI_ISL_1332716, EPI_ISL_1332718, EPI_ISL_1332723, EPI_ISL_1332724, EPI_ISL_1332726, EPI_ISL_1332728, EPI_ISL_1332729, EPI_ISL_1332730, EPI_ISL_1332731, EPI_ISL_1332732,
EPI_ISL_1332733, EPI_ISL_1332735, EPI_ISL_1332736, EPI_ISL_1332737, EPI_ISL_1332738, EPI_ISL_1332741, EPI_ISL_1332742, EPI_ISL_1332743, EPI_ISL_1332744, EPI_ISL_1332745, EPI_ISL_1332747, EPI_ISL_1332748, EPI_ISL_1332749, EPI_ISL_1332750, EPI_ISL_1332751, EPI_ISL_1332752, EPI_ISL_1332753, EPI_ISL_1332754, EPI_ISL_1332755, EPI_ISL_1332756, EPI_ISL_1332757, EPI_ISL_1332759,
EPI_ISL_1332760, EPI_ISL_1332761, EPI_ISL_1332762, EPI_ISL_1332763, EPI_ISL_1332766, EPI_ISL_1332768, EPI_ISL_1332769, EPI_ISL_1332771, EPI_ISL_1332773, EPI_ISL_1332774, EPI_ISL_1332775, EPI_ISL_1332776, EPI_ISL_1332777, EPI_ISL_1332778, EPI_ISL_1332779, EPI_ISL_1332780, EPI_ISL_1332782, EPI_ISL_1332783, EPI_ISL_1332784, EPI_ISL_1332785, EPI_ISL_1332786, EPI_ISL_1332787,
EPI_ISL_1332788, EPI_ISL_1332789, EPI_ISL_1332791, EPI_ISL_1332792, EPI_ISL_1332794, EPI_ISL_1332795, EPI_ISL_1332796, EPI_ISL_1332797, EPI_ISL_1332798, EPI_ISL_1332799, EPI_ISL_1332800, EPI_ISL_1332801, EPI_ISL_1332802, EPI_ISL_1332803, EPI_ISL_1332804, EPI_ISL_1332805, EPI_ISL_1332806, EPI_ISL_1332810, EPI_ISL_1332827, EPI_ISL_1332843, EPI_ISL_1332844, EPI_ISL_1332921,
EPI_ISL_1332923, EPI_ISL_1332924, EPI_ISL_1332935, EPI_ISL_1332937, EPI_ISL_1332940, EPI_ISL_1332941, EPI_ISL_1332942, EPI_ISL_1332945, EPI_ISL_1332946, EPI_ISL_1332947, EPI_ISL_1332948, EPI_ISL_1332951, EPI_ISL_1332953, EPI_ISL_1332954, EPI_ISL_1332955, EPI_ISL_1332957, EPI_ISL_1332959, EPI_ISL_1332962, EPI_ISL_1332968, EPI_ISL_1332969, EPI_ISL_1332970, EPI_ISL_1332974,
EPI_ISL_1332979, EPI_ISL_1332993, EPI_ISL_1333002, EPI_ISL_1333003, EPI_ISL_1333006, EPI_ISL_1333007, EPI_ISL_1333008, EPI_ISL_1333009, EPI_ISL_1333011, EPI_ISL_1333014, EPI_ISL_1333015, EPI_ISL_1333029, EPI_ISL_1333037, EPI_ISL_1333042, EPI_ISL_1333044, EPI_ISL_1333045, EPI_ISL_1333058, EPI_ISL_1333067, EPI_ISL_1333078, EPI_ISL_1333079, EPI_ISL_1333099, EPI_ISL_1333102,
EPI_ISL_1333104, EPI_ISL_1333109, EPI_ISL_1333112, EPI_ISL_1333114, EPI_ISL_1333116, EPI_ISL_1333119, EPI_ISL_1333123, EPI_ISL_1333134, EPI_ISL_1333137, EPI_ISL_1333143, EPI_ISL_1333144, EPI_ISL_1333151, EPI_ISL_1333155, EPI_ISL_1333157, EPI_ISL_1333161, EPI_ISL_1333167, EPI_ISL_1333171, EPI_ISL_1333337, EPI_ISL_1333338, EPI_ISL_1333342, EPI_ISL_1333344, EPI_ISL_1333345,
EPI_ISL_1333348, EPI_ISL_1333351, EPI_ISL_1333352, EPI_ISL_1333356, EPI_ISL_1333358, EPI_ISL_1333360, EPI_ISL_1333364, EPI_ISL_1333365, EPI_ISL_1333370, EPI_ISL_1333372, EPI_ISL_1333374, EPI_ISL_1333380, EPI_ISL_1333382, EPI_ISL_1333384, EPI_ISL_1333387, EPI_ISL_1333388, EPI_ISL_1333389, EPI_ISL_1333393, EPI_ISL_1333394, EPI_ISL_1333395, EPI_ISL_1333400, EPI_ISL_1333412,
EPI_ISL_1333414, EPI_ISL_1333422, EPI_ISL_1333426, EPI_ISL_1333429, EPI_ISL_1333433, EPI_ISL_1333434, EPI_ISL_1333440, EPI_ISL_1333441, EPI_ISL_1333442, EPI_ISL_1333444, EPI_ISL_1333445, EPI_ISL_1333446, EPI_ISL_1333454, EPI_ISL_1333455, EPI_ISL_1333456, EPI_ISL_1333459, EPI_ISL_1333460, EPI_ISL_1333461, EPI_ISL_1333464, EPI_ISL_1333468, EPI_ISL_1333469, EPI_ISL_1333471,

EPI_ISL_1333472, EPI_ISL_1333478, EPI_ISL_1346345, EPI_ISL_1346431, EPI_ISL_1346439, EPI_ISL_1346453, EPI_ISL_1346473, EPI_ISL_1365361, EPI_ISL_1378492, EPI_ISL_1378493, EPI_ISL_1378494, EPI_ISL_1378495, EPI_ISL_1378496, EPI_ISL_1378497, EPI_ISL_1378498

| | | | |
|---|---|---|---|
| see above | Lighthouse Lab in Glasgow | Wellcome Sanger Institute for the COVID-19 Genomics UK (COG-UK) Consortium | Anna Dominiczak and Alex Alderton; Carol Clugston; Cordelia Langford; David Gray; David K. Jackson; Dominic Kwiatkowski; Ewan Harrison; Harper VanSteenhouse; Ian Johnston; Jeffrey Barrett; John Sillitoe on behalf of the Wellcome Sanger Institute COVID-19 Surveillance Team; Roberto Amato; Sonia Goncalves; Yumi Kasai |

EPI_ISL_1246288, EPI_ISL_1255679, EPI_ISL_1255686, EPI_ISL_1255688, EPI_ISL_1255690, EPI_ISL_1255699, EPI_ISL_1255701, EPI_ISL_1255706, EPI_ISL_1255711, EPI_ISL_1255715, EPI_ISL_1255718, EPI_ISL_1255741, EPI_ISL_1255745, EPI_ISL_1255770, EPI_ISL_1255777, EPI_ISL_1255792, EPI_ISL_1255794, EPI_ISL_1255803, EPI_ISL_1255812, EPI_ISL_1255822, EPI_ISL_1255823, EPI_ISL_1255830, EPI_ISL_1255854, EPI_ISL_1255865, EPI_ISL_1255874, EPI_ISL_1255879, EPI_ISL_1255904, EPI_ISL_1255908, EPI_ISL_1255915, EPI_ISL_1255921, EPI_ISL_1255922, EPI_ISL_1256013, EPI_ISL_1256038, EPI_ISL_1256690, EPI_ISL_1256691, EPI_ISL_1256703, EPI_ISL_1256707, EPI_ISL_1256709, EPI_ISL_1256712, EPI_ISL_1256716, EPI_ISL_1256721, EPI_ISL_1256723, EPI_ISL_1256726, EPI_ISL_1256733, EPI_ISL_1256734, EPI_ISL_1256738, EPI_ISL_1256745, EPI_ISL_1256749, EPI_ISL_1256750, EPI_ISL_1256753, EPI_ISL_1256759, EPI_ISL_1256767, EPI_ISL_1256776, EPI_ISL_1256777, EPI_ISL_1256784, EPI_ISL_1256786, EPI_ISL_1256788, EPI_ISL_1256808, EPI_ISL_1256810, EPI_ISL_1256822, EPI_ISL_1256823, EPI_ISL_1256826, EPI_ISL_1256838, EPI_ISL_1256839, EPI_ISL_1256850, EPI_ISL_1256851, EPI_ISL_1256852, EPI_ISL_1256853, EPI_ISL_1256858, EPI_ISL_1256859, EPI_ISL_1256865, EPI_ISL_1256867, EPI_ISL_1256868, EPI_ISL_1256878, EPI_ISL_1256879, EPI_ISL_1256883, EPI_ISL_1256886, EPI_ISL_1256889, EPI_ISL_1256898, EPI_ISL_1256908, EPI_ISL_1256910, EPI_ISL_1256911, EPI_ISL_1256914, EPI_ISL_1256915, EPI_ISL_1256916, EPI_ISL_1256917, EPI_ISL_1256919, EPI_ISL_1256924, EPI_ISL_1256929, EPI_ISL_1256937, EPI_ISL_1264069, EPI_ISL_1264071, EPI_ISL_1264077, EPI_ISL_1264079, EPI_ISL_1264081, EPI_ISL_1264082, EPI_ISL_1264084, EPI_ISL_1264086, EPI_ISL_1264088, EPI_ISL_1264091, EPI_ISL_1264092, EPI_ISL_1264093, EPI_ISL_1264098, EPI_ISL_1264100, EPI_ISL_1264102, EPI_ISL_1264104, EPI_ISL_1264107, EPI_ISL_1264110, EPI_ISL_1264112, EPI_ISL_1264114, EPI_ISL_1264119, EPI_ISL_1264120, EPI_ISL_1264124, EPI_ISL_1264131, EPI_ISL_1264135, EPI_ISL_1264137, EPI_ISL_1264139, EPI_ISL_1264141, EPI_ISL_1264148, EPI_ISL_1264149, EPI_ISL_1264155, EPI_ISL_1264161, EPI_ISL_1264162, EPI_ISL_1264163, EPI_ISL_1264169, EPI_ISL_1264170, EPI_ISL_1264175, EPI_ISL_1264177, EPI_ISL_1264183, EPI_ISL_1264188, EPI_ISL_1264190, EPI_ISL_1264192, EPI_ISL_1264194, EPI_ISL_1264198, EPI_ISL_1264199, EPI_ISL_1264201, EPI_ISL_1264202, EPI_ISL_1264206, EPI_ISL_1264211, EPI_ISL_1264212, EPI_ISL_1264214, EPI_ISL_1264215, EPI_ISL_1264216, EPI_ISL_1264219, EPI_ISL_1264225, EPI_ISL_1264230, EPI_ISL_1264234, EPI_ISL_1264238, EPI_ISL_1264239, EPI_ISL_1264243, EPI_ISL_1264251, EPI_ISL_1264261, EPI_ISL_1264262, EPI_ISL_1264267, EPI_ISL_1264268, EPI_ISL_1264271, EPI_ISL_1264275, EPI_ISL_1264276, EPI_ISL_1264277, EPI_ISL_1264278, EPI_ISL_1264279, EPI_ISL_1264280, EPI_ISL_1264281, EPI_ISL_1264286, EPI_ISL_1264287, EPI_ISL_1264288, EPI_ISL_1264289, EPI_ISL_1264291, EPI_ISL_1264294, EPI_ISL_1264296, EPI_ISL_1264298, EPI_ISL_1264306, EPI_ISL_1264310, EPI_ISL_1264313, EPI_ISL_1264314, EPI_ISL_1264315, EPI_ISL_1264316, EPI_ISL_1264317, EPI_ISL_1264322, EPI_ISL_1264323, EPI_ISL_1264327, EPI_ISL_1264335, EPI_ISL_1264337, EPI_ISL_1264338, EPI_ISL_1264339, EPI_ISL_1264344, EPI_ISL_1264346, EPI_ISL_1264347, EPI_ISL_1264351, EPI_ISL_1264354, EPI_ISL_1264356, EPI_ISL_1264358, EPI_ISL_1264359, EPI_ISL_1264360, EPI_ISL_1264361, EPI_ISL_1264364, EPI_ISL_1264370, EPI_ISL_1264371, EPI_ISL_1264373, EPI_ISL_1264374, EPI_ISL_1264375, EPI_ISL_1264380, EPI_ISL_1264381, EPI_ISL_1264388, EPI_ISL_1264391, EPI_ISL_1264392, EPI_ISL_1264402, EPI_ISL_1264407, EPI_ISL_1264417, EPI_ISL_1264419, EPI_ISL_1264420, EPI_ISL_1264423, EPI_ISL_1264424, EPI_ISL_1264426, EPI_ISL_1264428, EPI_ISL_1264429, EPI_ISL_1264434, EPI_ISL_1264438, EPI_ISL_1264441, EPI_ISL_1264446, EPI_ISL_1264447, EPI_ISL_1264452, EPI_ISL_1264454, EPI_ISL_1264455, EPI_ISL_1264458, EPI_ISL_1264460, EPI_ISL_1264465, EPI_ISL_1264467, EPI_ISL_1264469, EPI_ISL_1264473, EPI_ISL_1264477, EPI_ISL_1264478, EPI_ISL_1264480, EPI_ISL_1264481, EPI_ISL_1264483, EPI_ISL_1264493, EPI_ISL_1264494, EPI_ISL_1264495, EPI_ISL_1264497, EPI_ISL_1264500, EPI_ISL_1264501, EPI_ISL_1264503, EPI_ISL_1264506, EPI_ISL_1264509, EPI_ISL_1264513, EPI_ISL_1264516, EPI_ISL_1264519, EPI_ISL_1264520, EPI_ISL_1264521, EPI_ISL_1264523, EPI_ISL_1264524, EPI_ISL_1264525, EPI_ISL_1264526, EPI_ISL_1264528, EPI_ISL_1264531, EPI_ISL_1264532, EPI_ISL_1264534, EPI_ISL_1264539, EPI_ISL_1264541, EPI_ISL_1264543, EPI_ISL_1264545, EPI_ISL_1264552, EPI_ISL_1264557, EPI_ISL_1264559, EPI_ISL_1264560, EPI_ISL_1264561, EPI_ISL_1264562, EPI_ISL_1264568, EPI_ISL_1264572, EPI_ISL_1264573, EPI_ISL_1264579, EPI_ISL_1264581, EPI_ISL_1264584, EPI_ISL_1264586, EPI_ISL_1264589, EPI_ISL_1264593, EPI_ISL_1264607, EPI_ISL_1264609, EPI_ISL_1264612, EPI_ISL_1264613, EPI_ISL_1264614, EPI_ISL_1264615, EPI_ISL_1264617, EPI_ISL_1264618, EPI_ISL_1264623, EPI_ISL_1264625, EPI_ISL_1264630, EPI_ISL_1264632, EPI_ISL_1264633, EPI_ISL_1264635, EPI_ISL_1264636, EPI_ISL_1264639, EPI_ISL_1264640, EPI_ISL_1264641, EPI_ISL_1264642, EPI_ISL_1273987, EPI_ISL_1273988, EPI_ISL_1273990, EPI_ISL_1273993, EPI_ISL_1273994, EPI_ISL_1273997, EPI_ISL_1273998, EPI_ISL_1274002, EPI_ISL_1274004, EPI_ISL_1274005, EPI_ISL_1274006, EPI_ISL_1274007, EPI_ISL_1274008, EPI_ISL_1274013, EPI_ISL_1274014, EPI_ISL_1274016, EPI_ISL_1274017, EPI_ISL_1274020, EPI_ISL_1274021, EPI_ISL_1274026, EPI_ISL_1274028, EPI_ISL_1274030, EPI_ISL_1274032, EPI_ISL_1274035, EPI_ISL_1274036, EPI_ISL_1274037, EPI_ISL_1274038, EPI_ISL_1274041, EPI_ISL_1274042, EPI_ISL_1274045, EPI_ISL_1274049, EPI_ISL_1274050, EPI_ISL_1274054, EPI_ISL_1274057, EPI_ISL_1274058, EPI_ISL_1274060, EPI_ISL_1274062, EPI_ISL_1274065, EPI_ISL_1274066, EPI_ISL_1274067, EPI_ISL_1274069, EPI_ISL_1274070, EPI_ISL_1274073, EPI_ISL_1274076, EPI_ISL_1274078, EPI_ISL_1274080, EPI_ISL_1274081, EPI_ISL_1274083, EPI_ISL_1274084, EPI_ISL_1274086, EPI_ISL_1274087, EPI_ISL_1274089, EPI_ISL_1274091, EPI_ISL_1274092, EPI_ISL_1274093, EPI_ISL_1274094, EPI_ISL_1274096, EPI_ISL_1274100, EPI_ISL_1274105, EPI_ISL_1274110, EPI_ISL_1274112, EPI_ISL_1274113, EPI_ISL_1274114, EPI_ISL_1274116, EPI_ISL_1274122, EPI_ISL_1274123, EPI_ISL_1274124, EPI_ISL_1274127, EPI_ISL_1274128, EPI_ISL_1274133, EPI_ISL_1274134, EPI_ISL_1274135, EPI_ISL_1274136, EPI_ISL_1274140, EPI_ISL_1274141, EPI_ISL_1274143, EPI_ISL_1274144, EPI_ISL_1274145, EPI_ISL_1274147, EPI_ISL_1274157, EPI_ISL_1274158, EPI_ISL_1274161, EPI_ISL_1274162, EPI_ISL_1274165, EPI_ISL_1274166, EPI_ISL_1274167, EPI_ISL_1274169, EPI_ISL_1274170, EPI_ISL_1274171, EPI_ISL_1274172, EPI_ISL_1274173, EPI_ISL_1274175, EPI_ISL_1274176, EPI_ISL_1274177, EPI_ISL_1274179, EPI_ISL_1274183, EPI_ISL_1274184, EPI_ISL_1274185, EPI_ISL_1274186, EPI_ISL_1274188, EPI_ISL_1274191, EPI_ISL_1274194, EPI_ISL_1274195, EPI_ISL_1274197, EPI_ISL_1274198, EPI_ISL_1274199, EPI_ISL_1274200, EPI_ISL_1274201, EPI_ISL_1274202, EPI_ISL_1274203, EPI_ISL_1274204, EPI_ISL_1274207, EPI_ISL_1274208, EPI_ISL_1274212, EPI_ISL_1274214, EPI_ISL_1274215, EPI_ISL_1274216, EPI_ISL_1274218, EPI_ISL_1274219, EPI_ISL_1274220, EPI_ISL_1274222, EPI_ISL_1274223, EPI_ISL_1274224, EPI_ISL_1274228, EPI_ISL_1274231, EPI_ISL_1274233, EPI_ISL_1274237, EPI_ISL_1274240, EPI_ISL_1274243, EPI_ISL_1274245, EPI_ISL_1274247, EPI_ISL_1274249, EPI_ISL_1274250, EPI_ISL_1274251, EPI_ISL_1274252, EPI_ISL_1274253, EPI_ISL_1274256, EPI_ISL_1274257, EPI_ISL_1274258, EPI_ISL_1274259, EPI_ISL_1274262, EPI_ISL_1274264, EPI_ISL_1274265, EPI_ISL_1274266, EPI_ISL_1274269, EPI_ISL_1274273, EPI_ISL_1274274, EPI_ISL_1274279, EPI_ISL_1274280, EPI_ISL_1274281, EPI_ISL_1274283, EPI_ISL_1274287, EPI_ISL_1274288, EPI_ISL_1274289, EPI_ISL_1274292, EPI_ISL_1274296, EPI_ISL_1274297, EPI_ISL_1274301, EPI_ISL_1274302, EPI_ISL_1274309, EPI_ISL_1274312, EPI_ISL_1274319, EPI_ISL_1274325, EPI_ISL_1274327, EPI_ISL_1274328, EPI_ISL_1274329, EPI_ISL_1274338, EPI_ISL_1274340, EPI_ISL_1274343, EPI_ISL_1274344, EPI_ISL_1274346, EPI_ISL_1274348, EPI_ISL_1274349, EPI_ISL_1274351, EPI_ISL_1274353, EPI_ISL_1274356, EPI_ISL_1274357, EPI_ISL_1274358, EPI_ISL_1274359, EPI_ISL_1274360, EPI_ISL_1274362, EPI_ISL_1274366, EPI_ISL_1274369, EPI_ISL_1274370, EPI_ISL_1274371, EPI_ISL_1274372, EPI_ISL_1274373, EPI_ISL_1274374, EPI_ISL_1274376, EPI_ISL_1274379, EPI_ISL_1274386, EPI_ISL_1274398, EPI_ISL_1274401, EPI_ISL_1274402, EPI_ISL_1274406, EPI_ISL_1274407, EPI_ISL_1274410, EPI_ISL_1274415, EPI_ISL_1274416, EPI_ISL_1274417, EPI_ISL_1274424, EPI_ISL_1274425, EPI_ISL_1274429, EPI_ISL_1274433, EPI_ISL_1274436, EPI_ISL_1274439, EPI_ISL_1274442, EPI_ISL_1274446, EPI_ISL_1274449, EPI_ISL_1274453, EPI_ISL_1274458, EPI_ISL_1274462, EPI_ISL_1274463, EPI_ISL_1274467, EPI_ISL_1274468, EPI_ISL_1274469, EPI_ISL_1274470, EPI_ISL_1274472, EPI_ISL_1274476, EPI_ISL_1274479, EPI_ISL_1274480, EPI_ISL_1274484, EPI_ISL_1274485, EPI_ISL_1274486, EPI_ISL_1274490, EPI_ISL_1274492, EPI_ISL_1274496, EPI_ISL_1274499, EPI_ISL_1274505, EPI_ISL_1274507, EPI_ISL_1274510, EPI_ISL_1274511, EPI_ISL_1274512, EPI_ISL_1274514, EPI_ISL_1274517, EPI_ISL_1274520, EPI_ISL_1274524, EPI_ISL_1274525, EPI_ISL_1274529, EPI_ISL_1274535, EPI_ISL_1274537, EPI_ISL_1274542, EPI_ISL_1274546, EPI_ISL_1274549, EPI_ISL_1274550, EPI_ISL_1274551, EPI_ISL_1274553, EPI_ISL_1274559, EPI_ISL_1274561, EPI_ISL_1274562, EPI_ISL_1274563, EPI_ISL_1274566, EPI_ISL_1274568, EPI_ISL_1274569, EPI_ISL_1274572, EPI_ISL_1274575, EPI_ISL_1274576, EPI_ISL_1274579, EPI_ISL_1274580, EPI_ISL_1274582, EPI_ISL_1274583, EPI_ISL_1274588, EPI_ISL_1274590, EPI_ISL_1274592, EPI_ISL_1274593, EPI_ISL_1274600, EPI_ISL_1274602, EPI_ISL_1274606, EPI_ISL_1274607, EPI_ISL_1274610, EPI_ISL_1274613, EPI_ISL_1274617, EPI_ISL_1274618, EPI_ISL_1274619, EPI_ISL_1274623, EPI_ISL_1274626, EPI_ISL_1274627, EPI_ISL_1274630, EPI_ISL_1274632, EPI_ISL_1274637, EPI_ISL_1274640, EPI_ISL_1274643, EPI_ISL_1274648, EPI_ISL_1274653, EPI_ISL_1274655, EPI_ISL_1274658, EPI_ISL_1274659, EPI_ISL_1274661, EPI_ISL_1274665, EPI_ISL_1274666, EPI_ISL_1274667, EPI_ISL_1274668, EPI_ISL_1274669, EPI_ISL_1274671, EPI_ISL_1274674, EPI_ISL_1274680, EPI_ISL_1274684, EPI_ISL_1274685, EPI_ISL_1274686, EPI_ISL_1274689, EPI_ISL_1274690, EPI_ISL_1274699, EPI_ISL_1274703, EPI_ISL_1274708, EPI_ISL_1274709, EPI_ISL_1274712, EPI_ISL_1274714, EPI_ISL_1274718, EPI_ISL_1274719, EPI_ISL_1274720, EPI_ISL_1274722, EPI_ISL_1274723, EPI_ISL_1274724, EPI_ISL_1274725, EPI_ISL_1274726, EPI_ISL_1274728, EPI_ISL_1274731, EPI_ISL_1274735, EPI_ISL_1274737, EPI_ISL_1274740, EPI_ISL_1274746, EPI_ISL_1274747, EPI_ISL_1274749, EPI_ISL_1274750, EPI_ISL_1274751, EPI_ISL_1274752, EPI_ISL_1274753, EPI_ISL_1274754, EPI_ISL_1274760, EPI_ISL_1274764, EPI_ISL_1274765, EPI_ISL_1274770, EPI_ISL_1274773, EPI_ISL_1274775, EPI_ISL_1274776, EPI_ISL_1274777, EPI_ISL_1274779, EPI_ISL_1274781, EPI_ISL_1274782, EPI_ISL_1274783, EPI_ISL_1274784, EPI_ISL_1274788, EPI_ISL_1274789, EPI_ISL_1274791, EPI_ISL_1274793, EPI_ISL_1274800, EPI_ISL_1274801, EPI_ISL_1274802, EPI_ISL_1274803, EPI_ISL_1274804, EPI_ISL_1274807, EPI_ISL_1274809, EPI_ISL_1274812, EPI_ISL_1274819, EPI_ISL_1274821, EPI_ISL_1274826, EPI_ISL_1274828, EPI_ISL_1274832, EPI_ISL_1274835, EPI_ISL_1274839, EPI_ISL_1274841, EPI_ISL_1274842, EPI_ISL_1274844, EPI_ISL_1274846, EPI_ISL_1274848, EPI_ISL_1274849, EPI_ISL_1274852, EPI_ISL_1274854, EPI_ISL_1274856, EPI_ISL_1274858, EPI_ISL_1274859, EPI_ISL_1274866, EPI_ISL_1274868, EPI_ISL_1274872, EPI_ISL_1274875, EPI_ISL_1274877, EPI_ISL_1274878, EPI_ISL_1274881, EPI_ISL_1274883, EPI_ISL_1274885, EPI_ISL_1274886, EPI_ISL_1274888, EPI_ISL_1274895, EPI_ISL_1274896, EPI_ISL_1274897, EPI_ISL_1274903, EPI_ISL_1274905, EPI_ISL_1274911, EPI_ISL_1274914, EPI_ISL_1274920, EPI_ISL_1274926, EPI_ISL_1274927, EPI_ISL_1274935, EPI_ISL_1274936, EPI_ISL_1274937, EPI_ISL_1274943, EPI_ISL_1274956, EPI_ISL_1274962, EPI_ISL_1274966, EPI_ISL_1274969, EPI_ISL_1274974, EPI_ISL_1274977, EPI_ISL_1274981, EPI_ISL_1274982, EPI_ISL_1274993, EPI_ISL_1274994, EPI_ISL_1274996, EPI_ISL_1274997, EPI_ISL_1275001, EPI_ISL_1275003, EPI_ISL_1275004, EPI_ISL_1275006, EPI_ISL_1275007, EPI_ISL_1275009, EPI_ISL_1275010, EPI_ISL_1275011, EPI_ISL_1275012, EPI_ISL_1275014, EPI_ISL_1275016, EPI_ISL_1275017, EPI_ISL_1275021, EPI_ISL_1275022, EPI_ISL_1275023, EPI_ISL_1275025, EPI_ISL_1275028, EPI_ISL_1275031, EPI_ISL_1275032, EPI_ISL_1275033, EPI_ISL_1275034, EPI_ISL_1275035, EPI_ISL_1275036, EPI_ISL_1275037, EPI_ISL_1275038, EPI_ISL_1275039, EPI_ISL_1275040, EPI_ISL_1275041, EPI_ISL_1275043, EPI_ISL_1275044, EPI_ISL_1275045, EPI_ISL_1275046, EPI_ISL_1275047, EPI_ISL_1275049, EPI_ISL_1275050, EPI_ISL_1275051, EPI_ISL_1275054, EPI_ISL_1275057, EPI_ISL_1275058, EPI_ISL_1275059, EPI_ISL_1275060, EPI_ISL_1275063, EPI_ISL_1275065, EPI_ISL_1275066, EPI_ISL_1275067, EPI_ISL_1275068, EPI_ISL_1275069, EPI_ISL_1275070, EPI_ISL_1275073, EPI_ISL_1275075, EPI_ISL_1275077, EPI_ISL_1275079, EPI_ISL_1275083, EPI_ISL_1275084, EPI_ISL_1275086, EPI_ISL_1275088, EPI_ISL_1275090, EPI_ISL_1275092, EPI_ISL_1275093, EPI_ISL_1275094, EPI_ISL_1275096, EPI_ISL_1275097, EPI_ISL_1275098, EPI_ISL_1275100, EPI_ISL_1275101, EPI_ISL_1275105, EPI_ISL_1275108, EPI_ISL_1275110, EPI_ISL_1275111, EPI_ISL_1275112, EPI_ISL_1275113, EPI_ISL_1275114, EPI_ISL_1276113, EPI_ISL_1276114, EPI_ISL_1276116, EPI_ISL_1276117, EPI_ISL_1276118, EPI_ISL_1276119, EPI_ISL_1276120, EPI_ISL_1276121, EPI_ISL_1276122, EPI_ISL_1276123, EPI_ISL_1276127, EPI_ISL_1276129, EPI_ISL_1276132, EPI_ISL_1276134, EPI_ISL_1276135, EPI_ISL_1276136, EPI_ISL_1276138, EPI_ISL_1276140, EPI_ISL_1276142, EPI_ISL_1276144, EPI_ISL_1276145, EPI_ISL_1276147, EPI_ISL_1276149, EPI_ISL_1276150, EPI_ISL_1276153, EPI_ISL_1276154, EPI_ISL_1276157, EPI_ISL_1276158, EPI_ISL_1276159, EPI_ISL_1276161, EPI_ISL_1276162, EPI_ISL_1276171, EPI_ISL_1276175, EPI_ISL_1276177, EPI_ISL_1276178, EPI_ISL_1276179, EPI_ISL_1276181, EPI_ISL_1276183, EPI_ISL_1276184, EPI_ISL_1276188, EPI_ISL_1276189, EPI_ISL_1276192, EPI_ISL_1276195, EPI_ISL_1276196, EPI_ISL_1276198, EPI_ISL_1276202, EPI_ISL_1276204, EPI_ISL_1276205, EPI_ISL_1276206, EPI_ISL_1276208, EPI_ISL_1276209, EPI_ISL_1276210, EPI_ISL_1276213, EPI_ISL_1276214, EPI_ISL_1276218, EPI_ISL_1276222, EPI_ISL_1276224, EPI_ISL_1276225, EPI_ISL_1276226, EPI_ISL_1276227, EPI_ISL_1276228, EPI_ISL_1276229, EPI_ISL_1276230, EPI_ISL_1276232, EPI_ISL_1276233, EPI_ISL_1276235, EPI_ISL_1276236, EPI_ISL_1276239, EPI_ISL_1276241, EPI_ISL_1276242, EPI_ISL_1276244, EPI_ISL_1276248, EPI_ISL_1276249, EPI_ISL_1276251, EPI_ISL_1276253, EPI_ISL_1276258, EPI_ISL_1276259, EPI_ISL_1276261, EPI_ISL_1276263, EPI_ISL_1276266, EPI_ISL_1276270, EPI_ISL_1276271, EPI_ISL_1276272, EPI_ISL_1276273, EPI_ISL_1276275, EPI_ISL_1276276, EPI_ISL_1276277, EPI_ISL_1276282, EPI_ISL_1276285, EPI_ISL_1276287, EPI_ISL_1276288, EPI_ISL_1276290, EPI_ISL_1276292, EPI_ISL_1276293, EPI_ISL_1276294, EPI_ISL_1276296, EPI_ISL_1276298, EPI_ISL_1276304, EPI_ISL_1276305, EPI_ISL_1276308, EPI_ISL_1276311, EPI_ISL_1276324, EPI_ISL_1276325, EPI_ISL_1276329, EPI_ISL_1276334, EPI_ISL_1276337, EPI_ISL_1276343, EPI_ISL_1276344, EPI_ISL_1276358, EPI_ISL_1276360, EPI_ISL_1276363, EPI_ISL_1276365, EPI_ISL_1276367, EPI_ISL_1276368, EPI_ISL_1276370, EPI_ISL_1276372, EPI_ISL_1276373, EPI_ISL_1276374, EPI_ISL_1276377, EPI_ISL_1276380, EPI_ISL_1276381, EPI_ISL_1276385, EPI_ISL_1276389, EPI_ISL_1276390, EPI_ISL_1276393, EPI_ISL_1276396, EPI_ISL_1276403, EPI_ISL_1276405, EPI_ISL_1276410, EPI_ISL_1276412, EPI_ISL_1276423, EPI_ISL_1276428, EPI_ISL_1276440, EPI_ISL_1276441, EPI_ISL_1276442, EPI_ISL_1276454, EPI_ISL_1276461, EPI_ISL_1276465, EPI_ISL_1276468, EPI_ISL_1276469, EPI_ISL_1276471, EPI_ISL_1276478, EPI_ISL_1276481, EPI_ISL_1276489, EPI_ISL_1276490, EPI_ISL_1276491, EPI_ISL_1276493, EPI_ISL_1276499, EPI_ISL_1276501, EPI_ISL_1276505, EPI_ISL_1276508, EPI_ISL_1276514, EPI_ISL_1276515, EPI_ISL_1276523, EPI_ISL_1276527, EPI_ISL_1276532, EPI_ISL_1276537, EPI_ISL_1276544, EPI_ISL_1276550, EPI_ISL_1276551, EPI_ISL_1276553, EPI_ISL_1276561, EPI_ISL_1276576, EPI_ISL_1276577, EPI_ISL_1276584, EPI_ISL_1276589, EPI_ISL_1276594, EPI_ISL_1276597, EPI_ISL_1276602, EPI_ISL_1276604, EPI_ISL_1276607, EPI_ISL_1276611, EPI_ISL_1276613, EPI_ISL_1276616, EPI_ISL_1276619, EPI_ISL_1276627, EPI_ISL_1276636, EPI_ISL_1276643, EPI_ISL_1276648, EPI_ISL_1276649, EPI_ISL_1276654, EPI_ISL_1276662, EPI_ISL_1276667, EPI_ISL_1276674, EPI_ISL_1276675, EPI_ISL_1276678, EPI_ISL_1276683, EPI_ISL_1276696, EPI_ISL_1276702, EPI_ISL_1276705, EPI_ISL_1276708, EPI_ISL_1276712, EPI_ISL_1276715, EPI_ISL_1276718, EPI_ISL_1276721, EPI_ISL_1276723, EPI_ISL_1276725, EPI_ISL_1276727, EPI_ISL_1276735, EPI_ISL_1276737, EPI_ISL_1276738, EPI_ISL_1276742, EPI_ISL_1276747, EPI_ISL_1276751, EPI_ISL_1276756, EPI_ISL_1276757, EPI_ISL_1276763, EPI_ISL_1276770, EPI_ISL_1276775, EPI_ISL_1276778, EPI_ISL_1276780, EPI_ISL_1276789, EPI_ISL_1276795, EPI_ISL_1276797, EPI_ISL_1276812, EPI_ISL_1276815, EPI_ISL_1276816, EPI_ISL_1276817, EPI_ISL_1276829, EPI_ISL_1276835, EPI_ISL_1276851, EPI_ISL_1276862, EPI_ISL_1276871, EPI_ISL_1276878, EPI_ISL_1316425, EPI_ISL_1316443, EPI_ISL_1316461, EPI_ISL_1316462, EPI_ISL_1316492, EPI_ISL_1316512, EPI_ISL_1316515, EPI_ISL_1316516, EPI_ISL_1316522, EPI_ISL_1316547, EPI_ISL_1316560, EPI_ISL_1316562, EPI_ISL_1316566, EPI_ISL_1316575, EPI_ISL_1316579, EPI_ISL_1316590, EPI_ISL_1316602, EPI_ISL_1316617, EPI_ISL_1316622, EPI_ISL_1316632, EPI_ISL_1316668, EPI_ISL_1316702, EPI_ISL_1316705, EPI_ISL_1316709, EPI_ISL_1316719, EPI_ISL_1316745, EPI_ISL_1317152, EPI_ISL_1324832, EPI_ISL_1324883, EPI_ISL_1324974, EPI_ISL_1325066, EPI_ISL_1325623, EPI_ISL_1326150, EPI_ISL_1326159, EPI_ISL_1326233, EPI_ISL_1326322, EPI_ISL_1326365, EPI_ISL_1326366, EPI_ISL_1326367, EPI_ISL_1326375, EPI_ISL_1326378, EPI_ISL_1326381, EPI_ISL_1326386, EPI_ISL_1326391, EPI_ISL_1326392, EPI_ISL_1326397, EPI_ISL_1326398, EPI_ISL_1326399, EPI_ISL_1326408, EPI_ISL_1326417, EPI_ISL_1326420, EPI_ISL_1326435, EPI_ISL_1326436, EPI_ISL_1326441, EPI_ISL_1326442, EPI_ISL_1326444, EPI_ISL_1326449, EPI_ISL_1326452, EPI_ISL_1326457, EPI_ISL_1326458, EPI_ISL_1326461, EPI_ISL_1326462, EPI_ISL_1326464, EPI_ISL_1326465, EPI_ISL_1326469, EPI_ISL_1326470, EPI_ISL_1326473, EPI_ISL_1326479, EPI_ISL_1326481, EPI_ISL_1326483, EPI_ISL_1326486, EPI_ISL_1326488, EPI_ISL_1326492, EPI_ISL_1326497, EPI_ISL_1326500, EPI_ISL_1326501, EPI_ISL_1326515, EPI_ISL_1326517, EPI_ISL_1326521, EPI_ISL_1326525, EPI_ISL_1326529, EPI_ISL_1326530, EPI_ISL_1326537, EPI_ISL_1326540, EPI_ISL_1326542, EPI_ISL_1326545, EPI_ISL_1326548, EPI_ISL_1326550, EPI_ISL_1326551, EPI_ISL_1326554, EPI_ISL_1326561, EPI_ISL_1326567, EPI_ISL_1326568, EPI_ISL_1326570, EPI_ISL_1326571, EPI_ISL_1326573, EPI_ISL_1326575, EPI_ISL_1326582, EPI_ISL_1326583, EPI_ISL_1326585, EPI_ISL_1326588, EPI_ISL_1326590, EPI_ISL_1326591, EPI_ISL_1326592, EPI_ISL_1326594, EPI_ISL_1326595, EPI_ISL_1326596, EPI_ISL_1326602, EPI_ISL_1326603, EPI_ISL_1326608, EPI_ISL_1326614, EPI_ISL_1326615, EPI_ISL_1326617, EPI_ISL_1326620, EPI_ISL_1326624, EPI_ISL_1326628, EPI_ISL_1326632, EPI_ISL_1326637, EPI_ISL_1326638, EPI_ISL_1326641, EPI_ISL_1326643, EPI_ISL_1326650, EPI_ISL_1326653, EPI_ISL_1326659, EPI_ISL_1328530, EPI_ISL_1328557, EPI_ISL_1328659, EPI_ISL_1328712, EPI_ISL_1328836, EPI_ISL_1329639, EPI_ISL_1329858, EPI_ISL_1330299, EPI_ISL_1330303, EPI_ISL_1330329, EPI_ISL_1330336, EPI_ISL_1330337, EPI_ISL_1330347, EPI_ISL_1330353, EPI_ISL_1330357, EPI_ISL_1330360, EPI_ISL_1330371, EPI_ISL_1330379, EPI_ISL_1330387, EPI_ISL_1330394, EPI_ISL_1330397, EPI_ISL_1330403, EPI_ISL_1330409, EPI_ISL_1330414, EPI_ISL_1330417, EPI_ISL_1330418, EPI_ISL_1330421, EPI_ISL_1330422, EPI_ISL_1330436, EPI_ISL_1330440, EPI_ISL_1330441, EPI_ISL_1330444, EPI_ISL_1330449, EPI_ISL_1330450, EPI_ISL_1330456, EPI_ISL_1330464, EPI_ISL_1330471, EPI_ISL_1330475, EPI_ISL_1330483, EPI_ISL_1330484, EPI_ISL_1330485, EPI_ISL_1330494, EPI_ISL_1330503, EPI_ISL_1330525, EPI_ISL_1330531, EPI_ISL_1330532, EPI_ISL_1330546, EPI_ISL_1330553, EPI_ISL_1330563, EPI_ISL_1330565, EPI_ISL_1330574, EPI_ISL_1330575, EPI_ISL_1330584, EPI_ISL_1330586, EPI_ISL_1330595, EPI_ISL_1330597, EPI_ISL_1330603, EPI_ISL_1330607, EPI_ISL_1330609, EPI_ISL_1330612, EPI_ISL_1330617, EPI_ISL_1330619, EPI_ISL_1330620, EPI_ISL_1330623, EPI_ISL_1330625, EPI_ISL_1330628, EPI_ISL_1330629, EPI_ISL_1330630, EPI_ISL_1330631, EPI_ISL_1330632, EPI_ISL_1330633, EPI_ISL_1330636, EPI_ISL_1330637, EPI_ISL_1330639, EPI_ISL_1330640, EPI_ISL_1330641, EPI_ISL_1330642, EPI_ISL_1330643, EPI_ISL_1330645, EPI_ISL_1330646, EPI_ISL_1330647, EPI_ISL_1330648, EPI_ISL_1330651, EPI_ISL_1330654, EPI_ISL_1330655, EPI_ISL_1330657, EPI_ISL_1330659, EPI_ISL_1330660, EPI_ISL_1330661, EPI_ISL_1330664, EPI_ISL_1330665, EPI_ISL_1330666, EPI_ISL_1330668, EPI_ISL_1330670, EPI_ISL_1330671, EPI_ISL_1330674, EPI_ISL_1330679, EPI_ISL_1330680, EPI_ISL_1330681, EPI_ISL_1330682, EPI_ISL_1330683, EPI_ISL_1330684, EPI_ISL_1330686, EPI_ISL_1330687, EPI_ISL_1330688, EPI_ISL_1330690, EPI_ISL_1330691, EPI_ISL_1330693, EPI_ISL_1330695, EPI_ISL_1330696, EPI_ISL_1330697, EPI_ISL_1330698, EPI_ISL_1330701, EPI_ISL_1330702, EPI_ISL_1330703, EPI_ISL_1330704, EPI_ISL_1330705, EPI_ISL_1330706, EPI_ISL_1330709, EPI_ISL_1330711, EPI_ISL_1330717, EPI_ISL_1330718, EPI_ISL_1330719, EPI_ISL_1330721, EPI_ISL_1330722, EPI_ISL_1330723, EPI_ISL_1330725, EPI_ISL_1330727, EPI_ISL_1330728, EPI_ISL_1330729, EPI_ISL_1330730, EPI_ISL_1330731, EPI_ISL_1330732, EPI_ISL_1330734, EPI_ISL_1330737, EPI_ISL_1330738, EPI_ISL_1330739, EPI_ISL_1330742, EPI_ISL_1330744, EPI_ISL_1330745, EPI_ISL_1330746, EPI_ISL_1330747, EPI_ISL_1330748, EPI_ISL_1330749, EPI_ISL_1330751, EPI_ISL_1330752, EPI_ISL_1330754, EPI_ISL_1330756, EPI_ISL_1330758, EPI_ISL_1330759, EPI_ISL_1330763, EPI_ISL_1330764, EPI_ISL_1330765, EPI_ISL_1330767, EPI_ISL_1330769, EPI_ISL_1330770, EPI_ISL_1330772, EPI_ISL_1330774, EPI_ISL_1330775, EPI_ISL_1330776, EPI_ISL_1330780, EPI_ISL_1330783, EPI_ISL_1330784, EPI_ISL_1330785, EPI_ISL_1330786, EPI_ISL_1330787, EPI_ISL_1330788, EPI_ISL_1330789, EPI_ISL_1330790, EPI_ISL_1330792, EPI_ISL_1330795, EPI_ISL_1330796, EPI_ISL_1330799, EPI_ISL_1330801, EPI_ISL_1330803, EPI_ISL_1330804, EPI_ISL_1330805, EPI_ISL_1330806, EPI_ISL_1330807, EPI_ISL_1330809, EPI_ISL_1330813, EPI_ISL_1330814, EPI_ISL_1330815, EPI_ISL_1330816, EPI_ISL_1330817, EPI_ISL_1330819, EPI_ISL_1330820, EPI_ISL_1330821, EPI_ISL_1330822, EPI_ISL_1330823, EPI_ISL_1330824, EPI_ISL_1330826, EPI_ISL_1330827, EPI_ISL_1330832, EPI_ISL_1330833, EPI_ISL_1330834, EPI_ISL_1330839, EPI_ISL_1330840, EPI_ISL_1330841, EPI_ISL_1330844, EPI_ISL_1330849, EPI_ISL_1330850, EPI_ISL_1330851, EPI_ISL_1330852, EPI_ISL_1330853, EPI_ISL_1330854, EPI_ISL_1330856, EPI_ISL_1330858, EPI_ISL_1330859, EPI_ISL_1330862, EPI_ISL_1330863, EPI_ISL_1330865, EPI_ISL_1330867, EPI_ISL_1330868, EPI_ISL_1330869, EPI_ISL_1330870, EPI_ISL_1330871, EPI_ISL_1330872, EPI_ISL_1330873, EPI_ISL_1330874, EPI_ISL_1330875, EPI_ISL_1330876, EPI_ISL_1330877, EPI_ISL_1330878, EPI_ISL_1330880, EPI_ISL_1330886, EPI_ISL_1330887, EPI_ISL_1330890, EPI_ISL_1330892, EPI_ISL_1330893, EPI_ISL_1330894, EPI_ISL_1330895, EPI_ISL_1330896, EPI_ISL_1330897, EPI_ISL_1330898, EPI_ISL_1330899, EPI_ISL_1330900, EPI_ISL_1330902, EPI_ISL_1330903, EPI_ISL_1330904, EPI_ISL_1330905, EPI_ISL_1330906, EPI_ISL_1330909, EPI_ISL_1330910, EPI_ISL_1330911, EPI_ISL_1330912, EPI_ISL_1330913, EPI_ISL_1330915, EPI_ISL_1330920, EPI_ISL_1330922, EPI_ISL_1330925, EPI_ISL_1330928, EPI_ISL_1330933, EPI_ISL_1330938, EPI_ISL_1330946, EPI_ISL_1330947, EPI_ISL_1330951, EPI_ISL_1330954, EPI_ISL_1330984, EPI_ISL_1330995, EPI_ISL_1330997, EPI_ISL_1331015, EPI_ISL_1331016, EPI_ISL_1331019, EPI_ISL_1331034, EPI_ISL_1331035, EPI_ISL_1331040, EPI_ISL_1331041, EPI_ISL_1331043, EPI_ISL_1331050, EPI_ISL_1331059, EPI_ISL_1331067, EPI_ISL_1331086, EPI_ISL_1331092, EPI_ISL_1331093, EPI_ISL_1331097, EPI_ISL_1331098, EPI_ISL_1331099, EPI_ISL_1331103, EPI_ISL_1331104, EPI_ISL_1331109, EPI_ISL_1331112, EPI_ISL_1331123, EPI_ISL_1331125, EPI_ISL_1331127, EPI_ISL_1332568, EPI_ISL_1332572, EPI_ISL_1332579, EPI_ISL_1332595, EPI_ISL_1332608, EPI_ISL_1332617, EPI_ISL_1332633, EPI_ISL_1332635, EPI_ISL_1332813, EPI_ISL_1332814, EPI_ISL_1332815, EPI_ISL_1332816, EPI_ISL_1332817, EPI_ISL_1332818, EPI_ISL_1332819, EPI_ISL_1332820, EPI_ISL_1332821, EPI_ISL_1332822, EPI_ISL_1332823, EPI_ISL_1332824, EPI_ISL_1332825, EPI_ISL_1332826, EPI_ISL_1332828, EPI_ISL_1332829, EPI_ISL_1343361, EPI_ISL_1343371, EPI_ISL_1343378, EPI_ISL_1343386, EPI_ISL_1343388, EPI_ISL_1343389, EPI_ISL_1343398, EPI_ISL_1343403, EPI_ISL_1343426, EPI_ISL_1343499, EPI_ISL_1343539, EPI_ISL_1343541, EPI_ISL_1343580, EPI_ISL_1346262, EPI_ISL_1346267, EPI_ISL_1346277, EPI_ISL_1346281, EPI_ISL_1346414, EPI_ISL_1346415, EPI_ISL_1346416, EPI_ISL_1346419, EPI_ISL_1346420, EPI_ISL_1346423, EPI_ISL_1346474, EPI_ISL_1375215, EPI_ISL_1467918, EPI_ISL_1468009, EPI_ISL_1468063, EPI_ISL_1986635, EPI_ISL_1986708

| | | | |
|---|---|---|---|
| see above | Lighthouse Lab in Milton Keynes | Wellcome Sanger Institute for the COVID-19 Genomics UK (COG-UK) Consortium | Cordelia Langford; David K. Jackson; Dominic Kwiatkowski; Ewan Harrison; Ian Johnston; Jeffrey Barrett; John Sillitoe on behalf of the Wellcome Sanger Institute COVID-19 Surveillance Team; Roberto Amato; Sonia Goncalves; The Lighthouse Lab in Milton Keynes and Alex Alderton |

EPI_ISL_1309720, EPI_ISL_1309721, EPI_ISL_1475504, EPI_ISL_1475505, EPI_ISL_1475506, EPI_ISL_1475507, EPI_ISL_1475508

| | | | |
|---|---|---|---|
| see above | Lincolnshire Hospitals and DeepSeq Nottingham | COVID-19 Genomics UK (COG-UK) Consortium | Christopher Moore; Fei Sang; Joeseph Chappell; Johnny Debebe; Jonathan Ball; Matthew Carlisle; Matthew Loose; Nadine Holmes; Nichola Duckworth; Patrick McClure; Sarah Walsh; Tim Sloan; Victoria Wright |

EPI_ISL_1308942, EPI_ISL_1308943, EPI_ISL_1308944, EPI_ISL_1308945, EPI_ISL_1308946, EPI_ISL_1308947, EPI_ISL_1308948, EPI_ISL_1386729, EPI_ISL_1386732, EPI_ISL_1386740, EPI_ISL_1386741, EPI_ISL_1386742, EPI_ISL_1386743, EPI_ISL_1386744, EPI_ISL_1386745, EPI_ISL_1386781, EPI_ISL_1386783, EPI_ISL_1386784, EPI_ISL_1386789, EPI_ISL_1386790, EPI_ISL_1386792, EPI_ISL_1386796, EPI_ISL_1386797, EPI_ISL_1386804, EPI_ISL_1474688, EPI_ISL_1474689, EPI_ISL_1474691, EPI_ISL_1474692, EPI_ISL_1474694, EPI_ISL_1474695, EPI_ISL_1474696, EPI_ISL_1474698, EPI_ISL_1474699, EPI_ISL_1474701, EPI_ISL_1474702, EPI_ISL_1474703, EPI_ISL_1474704, EPI_ISL_1474705, EPI_ISL_1474711, EPI_ISL_1474712, EPI_ISL_1474716, EPI_ISL_1474719, EPI_ISL_1474720, EPI_ISL_1474723, EPI_ISL_1833119, EPI_ISL_1833121, EPI_ISL_1833122, EPI_ISL_1833125, EPI_ISL_1833126

| | | | |
|---|---|---|---|
| see above | Liverpool Clinical Laboratories | COVID-19 Genomics UK (COG-UK) Consortium | A Alrezaihi; Alessandro Gerada; Alistair Darby; Angela Cowell; Anita Lucaci; Anu Chawla; Cassie Olateju; Catherine Hartley; Charlotte Nelson; Ecaterina Vamos; Elaine O'Toole; Eleanor G Bentley; Ghada T Shawli; Hermione Webster; Isabel García-Dorival; James Johnson; James P Stewart; Jenifer Manson; Joanne Watts; Jones Benjamin; Jordan J Clark; Julian Hiscox; L Luu; Lucille Rainbow; M Almsaud; Margaret Hughes; Mark Whitehead; Matthew Gemmell; Miren Iturriza-Gomara; Muhannad Alruwaili; N.P Randle; Neil Swainston; PKF Gilmore; Parul Sharma; Rebekah Penrice-Randal; Richard Eccles; Richard Gregory; Sam Haldenby; Steve Paterson; Stuart D Armstrong; Trevor Ian Robinson; Ximeng Han |

EPI_ISL_1309455, EPI_ISL_1309456, EPI_ISL_1309459, EPI_ISL_1309460, EPI_ISL_1309461, EPI_ISL_1309462, EPI_ISL_1309463, EPI_ISL_1309464, EPI_ISL_1309465, EPI_ISL_1309466, EPI_ISL_1309507, EPI_ISL_1309508, EPI_ISL_1309509, EPI_ISL_1309510, EPI_ISL_1309511, EPI_ISL_1309520, EPI_ISL_1309521, EPI_ISL_1309522, EPI_ISL_1309523, EPI_ISL_1309524, EPI_ISL_1309525, EPI_ISL_1309526, EPI_ISL_1309527, EPI_ISL_1309528, EPI_ISL_1309529, EPI_ISL_1309530, EPI_ISL_1309531, EPI_ISL_1309533, EPI_ISL_1309534, EPI_ISL_1309535, EPI_ISL_1475204, EPI_ISL_1475393, EPI_ISL_1475394, EPI_ISL_1475395, EPI_ISL_1475396, EPI_ISL_1475397, EPI_ISL_1475398, EPI_ISL_1475399, EPI_ISL_1475400, EPI_ISL_1475401, EPI_ISL_1475402, EPI_ISL_1475403, EPI_ISL_1475404, EPI_ISL_1475405, EPI_ISL_1475456, EPI_ISL_1475457, EPI_ISL_1475458, EPI_ISL_1475459, EPI_ISL_1475460, EPI_ISL_1475463, EPI_ISL_1475476, EPI_ISL_1475477, EPI_ISL_1475478, EPI_ISL_1475479, EPI_ISL_1475483

| | | | |
|---|---|---|---|
| see above | Northumbria University / South Tees Hospitals NHS Foundation Trust / North Cumbria Integrated Care NHS Foundation Trust / North Tees and Hartlepool NHS Foundation Trust / Newcastle Hospitals NHS Foundation Trust | COVID-19 Genomics UK (COG-UK) Consortium | Andrew Nelson; Brendan Payne; Clive Graham; Darren L Smith; Debra Padgett; Edward Barton; Emma Swindells; Garren Scott; Gary Black; Gary Eltringham; Giles S Holt; Greg R Young; Jane Greenaway; Jennifer Collins; John Allan; Joshua Loh; Lynn Dover; Matthew Bashton; Mohammad A Tariq; Paul Baker; Sarah Essex; Steve Liggett; Wen C Yew; Yusri Taha |

EPI_ISL_1249591, EPI_ISL_1249594, EPI_ISL_1249595, EPI_ISL_1249596, EPI_ISL_1249597, EPI_ISL_1249598, EPI_ISL_1249599, EPI_ISL_1249628, EPI_ISL_1249631, EPI_ISL_1249633, EPI_ISL_1249634, EPI_ISL_1249658, EPI_ISL_1249659, EPI_ISL_1249662, EPI_ISL_1249663, EPI_ISL_1249664, EPI_ISL_1249665, EPI_ISL_1249667, EPI_ISL_1249669, EPI_ISL_1249670, EPI_ISL_1249673, EPI_ISL_1249674, EPI_ISL_1249676, EPI_ISL_1249677, EPI_ISL_1249678, EPI_ISL_1249681, EPI_ISL_1249683, EPI_ISL_1249689, EPI_ISL_1249694, EPI_ISL_1249695, EPI_ISL_1249701, EPI_ISL_1249711, EPI_ISL_1249720, EPI_ISL_1249727, EPI_ISL_1249729, EPI_ISL_1249730, EPI_ISL_1249736, EPI_ISL_1249739, EPI_ISL_1249740, EPI_ISL_1249741, EPI_ISL_1249743, EPI_ISL_1249745, EPI_ISL_1249746, EPI_ISL_1249747, EPI_ISL_1249748, EPI_ISL_1249756, EPI_ISL_1249760, EPI_ISL_1249763, EPI_ISL_1249764, EPI_ISL_1249766, EPI_ISL_1249767, EPI_ISL_1249768, EPI_ISL_1249769, EPI_ISL_1249770, EPI_ISL_1249772, EPI_ISL_1249773, EPI_ISL_1249777, EPI_ISL_1249781, EPI_ISL_1249786, EPI_ISL_1249788, EPI_ISL_1249789, EPI_ISL_1249793, EPI_ISL_1249794, EPI_ISL_1249798, EPI_ISL_1249799, EPI_ISL_1249807, EPI_ISL_1249808, EPI_ISL_1249809, EPI_ISL_1249810, EPI_ISL_1249812, EPI_ISL_1249813, EPI_ISL_1249814, EPI_ISL_1249819, EPI_ISL_1249820, EPI_ISL_1249825, EPI_ISL_1249831, EPI_ISL_1249832, EPI_ISL_1249834, EPI_ISL_1249835, EPI_ISL_1249836, EPI_ISL_1249837, EPI_ISL_1249842, EPI_ISL_1249843, EPI_ISL_1249848, EPI_ISL_1249849, EPI_ISL_1249850, EPI_ISL_1249855, EPI_ISL_1249856, EPI_ISL_1249858, EPI_ISL_1249860, EPI_ISL_1249861, EPI_ISL_1249862, EPI_ISL_1249866, EPI_ISL_1249868, EPI_ISL_1249869, EPI_ISL_1249870, EPI_ISL_1249873, EPI_ISL_1249875, EPI_ISL_1249877, EPI_ISL_1249878, EPI_ISL_1249880, EPI_ISL_1249883, EPI_ISL_1249885, EPI_ISL_1249886, EPI_ISL_1249887, EPI_ISL_1309793, EPI_ISL_1309795, EPI_ISL_1309796, EPI_ISL_1309797, EPI_ISL_1309798, EPI_ISL_1309799, EPI_ISL_1309800, EPI_ISL_1309801, EPI_ISL_1309802, EPI_ISL_1309803, EPI_ISL_1309804, EPI_ISL_1309805, EPI_ISL_1309806, EPI_ISL_1309807, EPI_ISL_1309808, EPI_ISL_1309809, EPI_ISL_1309810, EPI_ISL_1309811, EPI_ISL_1309812, EPI_ISL_1309813, EPI_ISL_1309814, EPI_ISL_1309815, EPI_ISL_1309816, EPI_ISL_1309817, EPI_ISL_1309818, EPI_ISL_1309819, EPI_ISL_1309820, EPI_ISL_1309821, EPI_ISL_1309822, EPI_ISL_1309823, EPI_ISL_1309824, EPI_ISL_1309825, EPI_ISL_1309826, EPI_ISL_1309827, EPI_ISL_1309828, EPI_ISL_1309829, EPI_ISL_1309830, EPI_ISL_1309831, EPI_ISL_1309832, EPI_ISL_1309833, EPI_ISL_1309837, EPI_ISL_1309838, EPI_ISL_1309839, EPI_ISL_1309841, EPI_ISL_1309842, EPI_ISL_1309843, EPI_ISL_1309844, EPI_ISL_1309845, EPI_ISL_1309846, EPI_ISL_1309847, EPI_ISL_1309848, EPI_ISL_1309849, EPI_ISL_1309850, EPI_ISL_1309851, EPI_ISL_1309852, EPI_ISL_1309853, EPI_ISL_1309854, EPI_ISL_1309855, EPI_ISL_1309856, EPI_ISL_1309857, EPI_ISL_1309858, EPI_ISL_1309859, EPI_ISL_1309860, EPI_ISL_1309861, EPI_ISL_1309862, EPI_ISL_1309863, EPI_ISL_1309864, EPI_ISL_1309865, EPI_ISL_1309866, EPI_ISL_1309867, EPI_ISL_1309868, EPI_ISL_1309869, EPI_ISL_1309870, EPI_ISL_1309871, EPI_ISL_1309872, EPI_ISL_1309873, EPI_ISL_1309874, EPI_ISL_1309875, EPI_ISL_1309876, EPI_ISL_1309877, EPI_ISL_1309878, EPI_ISL_1309879, EPI_ISL_1309880, EPI_ISL_1309881, EPI_ISL_1309882, EPI_ISL_1309883, EPI_ISL_1309884, EPI_ISL_1309885, EPI_ISL_1309886, EPI_ISL_1309887, EPI_ISL_1309888, EPI_ISL_1309889, EPI_ISL_1309890, EPI_ISL_1309891, EPI_ISL_1309892, EPI_ISL_1309893, EPI_ISL_1309894, EPI_ISL_1309895, EPI_ISL_1309896, EPI_ISL_1309897, EPI_ISL_1309898, EPI_ISL_1309899, EPI_ISL_1309900, EPI_ISL_1309901, EPI_ISL_1309902, EPI_ISL_1309903, EPI_ISL_1309904, EPI_ISL_1309905, EPI_ISL_1309906, EPI_ISL_1309907, EPI_ISL_1309908, EPI_ISL_1309909, EPI_ISL_1309910, EPI_ISL_1309911, EPI_ISL_1309912, EPI_ISL_1309913, EPI_ISL_1309914, EPI_ISL_1309915, EPI_ISL_1309916, EPI_ISL_1309917, EPI_ISL_1309918, EPI_ISL_1309919, EPI_ISL_1309920, EPI_ISL_1309921, EPI_ISL_1309922, EPI_ISL_1309923, EPI_ISL_1309924, EPI_ISL_1309925, EPI_ISL_1309926, EPI_ISL_1309927, EPI_ISL_1309928, EPI_ISL_1309929, EPI_ISL_1309930, EPI_ISL_1309931, EPI_ISL_1309933, EPI_ISL_1309934, EPI_ISL_1309935, EPI_ISL_1309936, EPI_ISL_1309937, EPI_ISL_1309938, EPI_ISL_1309939, EPI_ISL_1309940, EPI_ISL_1309941, EPI_ISL_1309942, EPI_ISL_1309943, EPI_ISL_1309944, EPI_ISL_1309945, EPI_ISL_1309946, EPI_ISL_1309947, EPI_ISL_1309948, EPI_ISL_1309949, EPI_ISL_1309950, EPI_ISL_1309951, EPI_ISL_1309952, EPI_ISL_1309953, EPI_ISL_1309954, EPI_ISL_1309955, EPI_ISL_1309957, EPI_ISL_1309958, EPI_ISL_1309960, EPI_ISL_1309961, EPI_ISL_1309962, EPI_ISL_1309963, EPI_ISL_1309964, EPI_ISL_1309965, EPI_ISL_1309966, EPI_ISL_1309967, EPI_ISL_1309968, EPI_ISL_1309969, EPI_ISL_1309970, EPI_ISL_1309971, EPI_ISL_1309972, EPI_ISL_1309973, EPI_ISL_1309974, EPI_ISL_1309975, EPI_ISL_1309976, EPI_ISL_1309977, EPI_ISL_1309978, EPI_ISL_1309979, EPI_ISL_1309980, EPI_ISL_1309981, EPI_ISL_1309982, EPI_ISL_1309983, EPI_ISL_1309984, EPI_ISL_1309985, EPI_ISL_1309986, EPI_ISL_1309987, EPI_ISL_1309988, EPI_ISL_1309989, EPI_ISL_1309990, EPI_ISL_1309991, EPI_ISL_1309992, EPI_ISL_1309994, EPI_ISL_1309995, EPI_ISL_1309996, EPI_ISL_1309997, EPI_ISL_1309998, EPI_ISL_1309999, EPI_ISL_1310000, EPI_ISL_1310001, EPI_ISL_1310002, EPI_ISL_1310003, EPI_ISL_1310004, EPI_ISL_1310005, EPI_ISL_1310008, EPI_ISL_1310009, EPI_ISL_1310010, EPI_ISL_1310011, EPI_ISL_1310012, EPI_ISL_1310013, EPI_ISL_1310014, EPI_ISL_1310015, EPI_ISL_1310016, EPI_ISL_1310022, EPI_ISL_1310023, EPI_ISL_1310024, EPI_ISL_1310025, EPI_ISL_1310026, EPI_ISL_1310027, EPI_ISL_1310028, EPI_ISL_1310029, EPI_ISL_1310033, EPI_ISL_1310034, EPI_ISL_1310035, EPI_ISL_1310036, EPI_ISL_1310037, EPI_ISL_1310038, EPI_ISL_1310039, EPI_ISL_1310040, EPI_ISL_1310041, EPI_ISL_1310042, EPI_ISL_1310043, EPI_ISL_1310044, EPI_ISL_1310045, EPI_ISL_1310046, EPI_ISL_1310047, EPI_ISL_1310051, EPI_ISL_1310052, EPI_ISL_1310056, EPI_ISL_1310057, EPI_ISL_1310058, EPI_ISL_1310059, EPI_ISL_1310103, EPI_ISL_1310108, EPI_ISL_1310145, EPI_ISL_1310149, EPI_ISL_1310150, EPI_ISL_1310164, EPI_ISL_1310170, EPI_ISL_1310171, EPI_ISL_1310174, EPI_ISL_1310179, EPI_ISL_1310180, EPI_ISL_1310207, EPI_ISL_1310215, EPI_ISL_1310216, EPI_ISL_1310217, EPI_ISL_1310240, EPI_ISL_1310241, EPI_ISL_1310242, EPI_ISL_1310253, EPI_ISL_1310261, EPI_ISL_1310287, EPI_ISL_1310288, EPI_ISL_1387747, EPI_ISL_1387748,

| | | | |
|---|---|---|---|
| EPI_ISL_1387751, EPI_ISL_1387754, EPI_ISL_1387756, EPI_ISL_1387757, EPI_ISL_1387758, EPI_ISL_1387760, EPI_ISL_1387761, EPI_ISL_1387766, EPI_ISL_1387767, EPI_ISL_1387768, EPI_ISL_1387769, EPI_ISL_1387770, EPI_ISL_1387772, EPI_ISL_1387773, EPI_ISL_1387780, EPI_ISL_1387782, EPI_ISL_1387783, EPI_ISL_1387784, EPI_ISL_1387785, EPI_ISL_1387788, EPI_ISL_1387799, EPI_ISL_1387801, EPI_ISL_1387804, EPI_ISL_1387805, EPI_ISL_1387806, EPI_ISL_1387807, EPI_ISL_1387808, EPI_ISL_1387809, EPI_ISL_1387810, EPI_ISL_1387811, EPI_ISL_1387812, EPI_ISL_1387813, EPI_ISL_1387814, EPI_ISL_1387815, EPI_ISL_1387816, EPI_ISL_1387817, EPI_ISL_1387818, EPI_ISL_1387820, EPI_ISL_1387821, EPI_ISL_1387822, EPI_ISL_1387824, EPI_ISL_1387826, EPI_ISL_1387827, EPI_ISL_1387828, EPI_ISL_1387829, EPI_ISL_1387830, EPI_ISL_1387831, EPI_ISL_1387832, EPI_ISL_1388127, EPI_ISL_1388161, EPI_ISL_1388165, EPI_ISL_1388172, EPI_ISL_1388173, EPI_ISL_1546793, EPI_ISL_1547043, EPI_ISL_1547057 | | | |
| see above | Originating lab: Wales Specialist Virology Centre Sequencing lab: Pathogen Genomics Unit | Public Health Wales Microbiology Cardiff Wales Specialist Virology Centre | Alec Birchley; Alexander Adams; Amy Gaskin; Angela Marchbank; Bree Gatica-Wilcox; Catherine Moore; Jason Coombes; Joanne Watkins; Joel Southgate; Johnathan Evans; Laura Gifford; Lauren Gilbert; Lee Graham; Malorie Perry; Matthew Bull; Nicole Pacchiarini; Sally Corden; Sara Kumziene-Summerhayes; Sara Rey; Sarah Taylor; Simon Cottrell; Sophie Jones; Tom Connor |
| EPI_ISL_1475647, EPI_ISL_1475649, EPI_ISL_1475653, EPI_ISL_1475654, EPI_ISL_1475655, EPI_ISL_1475656, EPI_ISL_1475657, EPI_ISL_1475658, EPI_ISL_1475661, EPI_ISL_1475662, EPI_ISL_1475664, EPI_ISL_1475665, EPI_ISL_1475666, EPI_ISL_1475667, EPI_ISL_1475668, EPI_ISL_1475669, EPI_ISL_1475670, EPI_ISL_1475731, EPI_ISL_1730503, EPI_ISL_1730504, EPI_ISL_1730505 | | | |
| see above | Oxford Viromics, NDM, University of Oxford; Oxford University Hospitals; Basingstoke and North Hampshire Hospital | COVID-19 Genomics UK (COG-UK) Consortium | Alex Mobbs; Amy Trebes; Anita Justice; Catrin Moore; Christophe Fraser; David Bonsall; David Buck; Emma Wise; George Macintyre; Jessica Lynch; John Todd; Mariateresa de Cesare; Matilde Mori; Monique Andersson; Nathan Moore; Nick Cortes; Robert Shaw; Stephen Kidd; Tanya Golubchik; Timothy Peto |
| EPI_ISL_1309662, EPI_ISL_1309663, EPI_ISL_1309664, EPI_ISL_1309666, EPI_ISL_1309679, EPI_ISL_1309680, EPI_ISL_1309681, EPI_ISL_1309686, EPI_ISL_1309688, EPI_ISL_1309690, EPI_ISL_1309692, EPI_ISL_1387147, EPI_ISL_1387148, EPI_ISL_1387153, EPI_ISL_1832810, EPI_ISL_1978921 | | | |
| see above | Quadram Institute Bioscience | COVID-19 Genomics UK (COG-UK) Consortium | Alexander J Trotter; Alison E. Mather; Alp Aydin; Ana P. Tedim; Anastasia Kolyva; Andrew Bell; Andrew J. Page; Claire Stuart; Dave J. Baker; Gemma L. Kay; John Wain; Justin O'Grady; Leonardo de Oliveira Martins; Lizzie Meadows; Maria Diaz; Mark Webber; Muhammed Yasir; Nabil-Fareed Alikhan; Ngozi Elumogo; Nicholas M. Thomson; Rachael Stanley; Rachel Gilroy; Reenesh Prakash; Samir Dervisevic; Samuel Bloomfield; Steven Rudder; Thanh Le-Viet |
| EPI_ISL_1249067, EPI_ISL_1249068, EPI_ISL_1249069, EPI_ISL_1249070, EPI_ISL_1387182, EPI_ISL_1387183, EPI_ISL_1387184, EPI_ISL_1387185, EPI_ISL_1387186, EPI_ISL_1978955, EPI_ISL_1978956, EPI_ISL_1978957, EPI_ISL_1978964 | | | |
| see above | Queens Medical Centre, Clinical Microbiology Department / DeepSeq Nottingham | COVID-19 Genomics UK (COG-UK) Consortium | Christopher Moore; Emily Park; Fei Sang; Gemma Clark; Hannah Howson-Wells; Jack Hill; Jayasree Dey; Johnny Debebe; Jonathan Ball; Joseph Chappell; Manjinder Khakh; Matthew Carlisle; Matthew Loose; Michelle M Lister; Nadine Holmes; Patrick McClure; Theocharis Tsoleridis; Timothy Byaruhanga; Vicki M Fleming; Victoria Wright; Wendy Smith |
| EPI_ISL_1344469, EPI_ISL_1344470, EPI_ISL_1344531, EPI_ISL_1344536, EPI_ISL_1344542, EPI_ISL_1344550, EPI_ISL_1344555, EPI_ISL_1344556, EPI_ISL_1344576, EPI_ISL_1344586, EPI_ISL_1344599, EPI_ISL_1344603, EPI_ISL_1344614, EPI_ISL_1344618, EPI_ISL_1344620, EPI_ISL_1344639, EPI_ISL_1344641, EPI_ISL_1344649, EPI_ISL_1344656, EPI_ISL_1344677, EPI_ISL_1344684, EPI_ISL_1344690, EPI_ISL_1344696, EPI_ISL_1344701, EPI_ISL_1344711, EPI_ISL_1344717, EPI_ISL_1344719, EPI_ISL_1344720, EPI_ISL_1344724, EPI_ISL_1344729, EPI_ISL_1344731, EPI_ISL_1344732, EPI_ISL_1344734, EPI_ISL_1344735, EPI_ISL_1344744, EPI_ISL_1344746, EPI_ISL_1344748, EPI_ISL_1344750, EPI_ISL_1344753, EPI_ISL_1344757, EPI_ISL_1344758, EPI_ISL_1344759, EPI_ISL_1344760, EPI_ISL_1344761, EPI_ISL_1344762, EPI_ISL_1344766, EPI_ISL_1344768, EPI_ISL_1344773, EPI_ISL_1344780, EPI_ISL_1344785, EPI_ISL_1344786, EPI_ISL_1344788, EPI_ISL_1344789, EPI_ISL_1344791, EPI_ISL_1344797, EPI_ISL_1344800, EPI_ISL_1344801, EPI_ISL_1344802, EPI_ISL_1344804, EPI_ISL_1344805, EPI_ISL_1344806, EPI_ISL_1344815, EPI_ISL_1344820, EPI_ISL_1344821, EPI_ISL_1344822, EPI_ISL_1344824, EPI_ISL_1344826, EPI_ISL_1344830, EPI_ISL_1344831, EPI_ISL_1344835, EPI_ISL_1344837, EPI_ISL_1344839, EPI_ISL_1344841, EPI_ISL_1344842, EPI_ISL_1344845, EPI_ISL_1344849, EPI_ISL_1344852, EPI_ISL_1344854, EPI_ISL_1344855, EPI_ISL_1344857, EPI_ISL_1344859, EPI_ISL_1344861, EPI_ISL_1344862, EPI_ISL_1344865, EPI_ISL_1344870, EPI_ISL_1344874, EPI_ISL_1344875, EPI_ISL_1344878, EPI_ISL_1344881, EPI_ISL_1344884, EPI_ISL_1344887, EPI_ISL_1344892, EPI_ISL_1344894, EPI_ISL_1344897, EPI_ISL_1344898, EPI_ISL_1344899, EPI_ISL_1344906, EPI_ISL_1344907, EPI_ISL_1344908, EPI_ISL_1344910, EPI_ISL_1344911, EPI_ISL_1344913, EPI_ISL_1344918, EPI_ISL_1344920, EPI_ISL_1344921, EPI_ISL_1365037, EPI_ISL_1365038, EPI_ISL_1365039, EPI_ISL_1365040, EPI_ISL_1365041, EPI_ISL_1365042, EPI_ISL_1365043, EPI_ISL_1365044, EPI_ISL_1365045, EPI_ISL_1365046, EPI_ISL_1365047, EPI_ISL_1365048, EPI_ISL_1365049, EPI_ISL_1365050, EPI_ISL_1365051, EPI_ISL_1365052, EPI_ISL_1365053, EPI_ISL_1365054, EPI_ISL_1365055, EPI_ISL_1365056, EPI_ISL_1365057, EPI_ISL_1365058, EPI_ISL_1365059, EPI_ISL_1365060, EPI_ISL_1365061, EPI_ISL_1365062, EPI_ISL_1365063, EPI_ISL_1365064, EPI_ISL_1365065, EPI_ISL_1365066, EPI_ISL_1365067, EPI_ISL_1365068, EPI_ISL_1365069, EPI_ISL_1365070, EPI_ISL_1365071, EPI_ISL_1365072, EPI_ISL_1365073, EPI_ISL_1365074, EPI_ISL_1365075, EPI_ISL_1365076, EPI_ISL_1365077, EPI_ISL_1365078, EPI_ISL_1365079, EPI_ISL_1365080, EPI_ISL_1365081, EPI_ISL_1365082, EPI_ISL_1365083, EPI_ISL_1365084, EPI_ISL_1365085, EPI_ISL_1365086, EPI_ISL_1365087, EPI_ISL_1365088, EPI_ISL_1365089, EPI_ISL_1365090, EPI_ISL_1365091, EPI_ISL_1365092, EPI_ISL_1365093, EPI_ISL_1365094, EPI_ISL_1365095, EPI_ISL_1365096, EPI_ISL_1365097, EPI_ISL_1365098, EPI_ISL_1365099, EPI_ISL_1365100, EPI_ISL_1365101, EPI_ISL_1365102, EPI_ISL_1365103, EPI_ISL_1365104, EPI_ISL_1365105, EPI_ISL_1365106, EPI_ISL_1365107, EPI_ISL_1365108, EPI_ISL_1365109, EPI_ISL_1365110, EPI_ISL_1365111, EPI_ISL_1365112, EPI_ISL_1365113, EPI_ISL_1365114, EPI_ISL_1365115, EPI_ISL_1365116, EPI_ISL_1365117, EPI_ISL_1365118, EPI_ISL_1365119, EPI_ISL_1365120, EPI_ISL_1365121, EPI_ISL_1365122, EPI_ISL_1365123, EPI_ISL_1365124, EPI_ISL_1365125, EPI_ISL_1365126, EPI_ISL_1365127, EPI_ISL_1365128, EPI_ISL_1365129, EPI_ISL_1365130, EPI_ISL_1365131, EPI_ISL_1365132, EPI_ISL_1365133, EPI_ISL_1365134, EPI_ISL_1365135, EPI_ISL_1365136, EPI_ISL_1365137, EPI_ISL_1365138, EPI_ISL_1365139, EPI_ISL_1365140, EPI_ISL_1365141, EPI_ISL_1365142, EPI_ISL_1365143, EPI_ISL_1365144, EPI_ISL_1365145, EPI_ISL_1365146, EPI_ISL_1365147, EPI_ISL_1365148, EPI_ISL_1365183, EPI_ISL_1365185, EPI_ISL_1365211, EPI_ISL_1365227, EPI_ISL_1365228, EPI_ISL_1365229, EPI_ISL_1365233, EPI_ISL_1365241, EPI_ISL_1365242, EPI_ISL_1365243, EPI_ISL_1365244, EPI_ISL_1365246, EPI_ISL_1365256, EPI_ISL_1365262, EPI_ISL_1365267, EPI_ISL_1365269, EPI_ISL_1365272, EPI_ISL_1365275, EPI_ISL_1365276, EPI_ISL_1365279, EPI_ISL_1365281, EPI_ISL_1365285, EPI_ISL_1365286, EPI_ISL_1365289, EPI_ISL_1365301, EPI_ISL_1365303, EPI_ISL_1365305, EPI_ISL_1365307, EPI_ISL_1374082, EPI_ISL_1374083, EPI_ISL_1374092, EPI_ISL_1374096, EPI_ISL_1374098, EPI_ISL_1374100, EPI_ISL_1374101, EPI_ISL_1374122, EPI_ISL_1374134, EPI_ISL_1374139, EPI_ISL_1374140, EPI_ISL_1374141, EPI_ISL_1374143, EPI_ISL_1374144, EPI_ISL_1374148, EPI_ISL_1374162, EPI_ISL_1374163, EPI_ISL_1374164, EPI_ISL_1374165, EPI_ISL_1374172, EPI_ISL_1374180, EPI_ISL_1374182, EPI_ISL_1374186, EPI_ISL_1374187, EPI_ISL_1374189, EPI_ISL_1374193, EPI_ISL_1374194, EPI_ISL_1374195, EPI_ISL_1374206, EPI_ISL_1374209, EPI_ISL_1374211, EPI_ISL_1374219, EPI_ISL_1374221, EPI_ISL_1374228, EPI_ISL_1374233, EPI_ISL_1374234, EPI_ISL_1374236, EPI_ISL_1374248, EPI_ISL_1374259, EPI_ISL_1374260, EPI_ISL_1374264, EPI_ISL_1374265, EPI_ISL_1374274, EPI_ISL_1374281, EPI_ISL_1374286, EPI_ISL_1374290, EPI_ISL_1374291, EPI_ISL_1374295, EPI_ISL_1374298, EPI_ISL_1374303, EPI_ISL_1374305, EPI_ISL_1374306, EPI_ISL_1374310, EPI_ISL_1374315, EPI_ISL_1374319, EPI_ISL_1374326, EPI_ISL_1374332, EPI_ISL_1389402, EPI_ISL_1389411, EPI_ISL_1389420, EPI_ISL_1389423, EPI_ISL_1389444, EPI_ISL_1389449, EPI_ISL_1389462, EPI_ISL_1389464, EPI_ISL_1389469, EPI_ISL_1389473, EPI_ISL_1389479, EPI_ISL_1389480, EPI_ISL_1389481, EPI_ISL_1389487, EPI_ISL_1389489, EPI_ISL_1389505, EPI_ISL_1389510, EPI_ISL_1389518, EPI_ISL_1389521, EPI_ISL_1389529, EPI_ISL_1389534, EPI_ISL_1389536, EPI_ISL_1389537, EPI_ISL_1389549, EPI_ISL_1389552, EPI_ISL_1389558, EPI_ISL_1389559, EPI_ISL_1389570, EPI_ISL_1389576, EPI_ISL_1389581, EPI_ISL_1389589, EPI_ISL_1389590, EPI_ISL_1389591, EPI_ISL_1389594, EPI_ISL_1389598, EPI_ISL_1389599, EPI_ISL_1389600, EPI_ISL_1389602, EPI_ISL_1389610, EPI_ISL_1412928, EPI_ISL_1412930, EPI_ISL_1412936, EPI_ISL_1412941, EPI_ISL_1412950, EPI_ISL_1412951, EPI_ISL_1412957, EPI_ISL_1412961, EPI_ISL_1412972, EPI_ISL_1412974, EPI_ISL_1412977, EPI_ISL_1412982, EPI_ISL_1412983, EPI_ISL_1412984, EPI_ISL_1412985, EPI_ISL_1412990, EPI_ISL_1412991, EPI_ISL_1412992, EPI_ISL_1412993, EPI_ISL_1413001, EPI_ISL_1413003, EPI_ISL_1413004, EPI_ISL_1413011, EPI_ISL_1413022, EPI_ISL_1413030, EPI_ISL_1413031, EPI_ISL_1413040, EPI_ISL_1413041, EPI_ISL_1413045, EPI_ISL_1413050, EPI_ISL_1413058, EPI_ISL_1413060, EPI_ISL_1416126, EPI_ISL_1416127, EPI_ISL_1416128, EPI_ISL_1416130, EPI_ISL_1416134, EPI_ISL_1416135, EPI_ISL_1416136, EPI_ISL_1416138, EPI_ISL_1416140, EPI_ISL_1416141, EPI_ISL_1416142, EPI_ISL_1416143, EPI_ISL_1416144, EPI_ISL_1416145, EPI_ISL_1416146, EPI_ISL_1416147, EPI_ISL_1416148, EPI_ISL_1416150, EPI_ISL_1416151, EPI_ISL_1416152, EPI_ISL_1416153, EPI_ISL_1416157, EPI_ISL_1416158, EPI_ISL_1416159, EPI_ISL_1416160, EPI_ISL_1416161, EPI_ISL_1416162, EPI_ISL_1416163, EPI_ISL_1416164, EPI_ISL_1416166, EPI_ISL_1416168, EPI_ISL_1416169, EPI_ISL_1416170, EPI_ISL_1416171 | | | |
| see above | Randox Laboratories | Wellcome Sanger Institute for the COVID-19 Genomics UK (COG-UK) Consortium | Cordelia Langford; David K. Jackson; Dominic Kwiatkowski; Ewan Harrison; Ian Johnston; Jeffrey Barrett; John Sillitoe on behalf of the Wellcome Sanger Institute COVID-19 Surveillance Team; Randox Laboratories and Alex Alderton; Roberto Amato; Sonia Goncalves |
| EPI_ISL_1976143, EPI_ISL_1976144, EPI_ISL_1976146, EPI_ISL_1976147, EPI_ISL_1976149, EPI_ISL_1976153, EPI_ISL_1976154, EPI_ISL_1976156, EPI_ISL_1976163, EPI_ISL_1976164, EPI_ISL_1976166, EPI_ISL_1976168, EPI_ISL_1976170, EPI_ISL_1976172, EPI_ISL_1976173, EPI_ISL_1976175, EPI_ISL_1976177, EPI_ISL_1976179, EPI_ISL_1976182, EPI_ISL_1976184, EPI_ISL_1976185, EPI_ISL_1976187, EPI_ISL_1976188, EPI_ISL_1976190, EPI_ISL_1976192, EPI_ISL_1976195, EPI_ISL_1976197, EPI_ISL_1976199, EPI_ISL_1976200, EPI_ISL_1976202, EPI_ISL_1976203, EPI_ISL_1976207, EPI_ISL_1976209, EPI_ISL_1976211, EPI_ISL_1976212, EPI_ISL_1976214, EPI_ISL_1976215, EPI_ISL_1976217, EPI_ISL_1976219, EPI_ISL_1976220, EPI_ISL_1976222, EPI_ISL_1976223, EPI_ISL_1976225, EPI_ISL_1976227, EPI_ISL_1976232, EPI_ISL_1976233, EPI_ISL_1976235, EPI_ISL_1976237, EPI_ISL_1976238, EPI_ISL_1976240, EPI_ISL_1976241, EPI_ISL_1976243, EPI_ISL_1976244, EPI_ISL_1976245, EPI_ISL_1976247, EPI_ISL_1976249, EPI_ISL_1976251, EPI_ISL_1976252, EPI_ISL_1976254, EPI_ISL_1976256, EPI_ISL_1976257, EPI_ISL_1976259, EPI_ISL_1976261, EPI_ISL_1976263, EPI_ISL_1976264, EPI_ISL_1976268, EPI_ISL_1976270, EPI_ISL_1976271, EPI_ISL_1976273, EPI_ISL_1976275, EPI_ISL_1976276, EPI_ISL_1976278, EPI_ISL_1976280, EPI_ISL_1976282, EPI_ISL_1976283, EPI_ISL_1976286, EPI_ISL_1976288, EPI_ISL_1976290, EPI_ISL_1976291, EPI_ISL_1976293, EPI_ISL_1976295, EPI_ISL_1976296, EPI_ISL_1976298, EPI_ISL_1976300, EPI_ISL_1976301, EPI_ISL_1976303, EPI_ISL_1976304, EPI_ISL_1976306, EPI_ISL_1976307, EPI_ISL_1976309, EPI_ISL_1976310, EPI_ISL_1976312, EPI_ISL_1976313, EPI_ISL_1976315, EPI_ISL_1976317, EPI_ISL_1976319, EPI_ISL_1976321, EPI_ISL_1976322, EPI_ISL_1976324, EPI_ISL_1976332, EPI_ISL_1976334, EPI_ISL_1976335, EPI_ISL_1976337, EPI_ISL_1976339, EPI_ISL_1976347, EPI_ISL_1976350, EPI_ISL_1976352, EPI_ISL_1976354, EPI_ISL_1976355, EPI_ISL_1976357, EPI_ISL_1976358, EPI_ISL_1976360 | | | |
| see above | Regional Virus Laboratory, Belfast Health and Social Care Trust | COVID-19 Genomics UK (COG-UK) Consortium | Alison Watt; Ciara Cox; Conall McCaughey; David Simpson; Derek Fairley; James McKenna; Mairead Connor; Susan Feeney; Tanya Curran; Zoltan Molnar |
| EPI_ISL_1833810, EPI_ISL_1833820 | St George's University Hospitals NHS Foundation Trust | COVID-19 Genomics UK (COG-UK) Consortium | Adam Witney; Cassie Pope; Irene Monahan; Joshua Taylor; Ken Laing; NgeeKeong Tan |
| EPI_ISL_1637039, EPI_ISL_1637040, EPI_ISL_1637041, EPI_ISL_1637042, EPI_ISL_1637043, EPI_ISL_1637044 | University College London Hospital | COVID-19 Genomics UK (COG-UK) Consortium | Dr Catherine Houlihan; Dr Daniel Frampton; Dr Judith Heaney; Dr Moira Spyer; Dr Paul Grant and Dr Eleni Nastouli; Matthew Byott; Stuart Kirk |
| EPI_ISL_1247987, EPI_ISL_1247988, EPI_ISL_1248298, EPI_ISL_1248303, EPI_ISL_1248305, EPI_ISL_1248312, EPI_ISL_1248321, EPI_ISL_1248323, EPI_ISL_1248325, EPI_ISL_1248326, EPI_ISL_1248328, EPI_ISL_1248330, EPI_ISL_1248347, EPI_ISL_1248349, EPI_ISL_1248351, EPI_ISL_1248352, EPI_ISL_1248353, EPI_ISL_1248360, EPI_ISL_1248362, EPI_ISL_1248363, EPI_ISL_1248364, EPI_ISL_1248365, EPI_ISL_1248525, EPI_ISL_1248526, EPI_ISL_1248527, EPI_ISL_1248528, EPI_ISL_1248529, EPI_ISL_1248530, EPI_ISL_1248543, EPI_ISL_1248544, EPI_ISL_1309105, EPI_ISL_1309126, EPI_ISL_1386850, EPI_ISL_1386851, EPI_ISL_1386854, EPI_ISL_1386865, EPI_ISL_1386866 | | | |
| see above | University College London, Great Ormond Street Hospital for Children NHS Foundation Trust, Imperial College Healthcare NHS Trust | COVID-19 Genomics UK (COG-UK) Consortium | Alison Holmes; Charlotte Williams; Helena Tutill; Jacqueline Findlay; James Price; Judith Breuer; Julianne Brown; Kathryn Harris; Leysa Forrest; Mark Kristiansen; Paola Niola; Paola Resende Silva; Patricia Dyal; Paul Randell; Rachel Williams; Samuel Weeks; Sergi Castellano; Sunando Roy; Tony Brooks; Yasmin Panchbhaya |
| EPI_ISL_1308871, EPI_ISL_1308872, EPI_ISL_1308873, EPI_ISL_1308876, EPI_ISL_1308887, EPI_ISL_1308888, EPI_ISL_1308889, EPI_ISL_1730295, EPI_ISL_1730296, EPI_ISL_1730297, EPI_ISL_1730298, EPI_ISL_1730301 | | | |
| see above | University Hospitals Of Leicester NHS Trust and DeepSeq Nottingham | COVID-19 Genomics UK (COG-UK) Consortium | Christopher Holmes; Christopher Moore; Fei Sang; Joeseph Chappell; Johnny Debebe; Jonathan Ball; Julian Tang; Karlie Fallon; Matthew Carlisle; Matthew Loose; Nadine Holmes; Patrick McClure; Paul Bird; Thomas Helmer; Victoria Wright |
| EPI_ISL_1308489, EPI_ISL_1308500, EPI_ISL_1308505, EPI_ISL_1308509, EPI_ISL_1308527, EPI_ISL_1386543, EPI_ISL_1386544, EPI_ISL_1386545, EPI_ISL_1386546, EPI_ISL_1386547, EPI_ISL_1386548, EPI_ISL_1386551, EPI_ISL_1386552, EPI_ISL_1386554, EPI_ISL_1386555, EPI_ISL_1386556, EPI_ISL_1386557, EPI_ISL_1386558, EPI_ISL_1386559, EPI_ISL_1386560, EPI_ISL_1386562, EPI_ISL_1474401, EPI_ISL_1474402, EPI_ISL_1474404, EPI_ISL_1474406, EPI_ISL_1474412, EPI_ISL_1474439, EPI_ISL_1474490, EPI_ISL_1474491, EPI_ISL_1474493, EPI_ISL_1474496, EPI_ISL_1474498, EPI_ISL_1474514, EPI_ISL_1474515, EPI_ISL_1474516, EPI_ISL_1474517, EPI_ISL_1474518, EPI_ISL_1474520, EPI_ISL_1474521, EPI_ISL_1474522, EPI_ISL_1474523, EPI_ISL_1474524, EPI_ISL_1474525, EPI_ISL_1474526, EPI_ISL_1729970, EPI_ISL_1832091, EPI_ISL_1832095, EPI_ISL_1832097, EPI_ISL_1832099, EPI_ISL_1832102, EPI_ISL_1832104, EPI_ISL_1832106, EPI_ISL_1832108, EPI_ISL_1832110, EPI_ISL_1832114, EPI_ISL_1832117, EPI_ISL_1832119, EPI_ISL_1832121, EPI_ISL_1832125, EPI_ISL_1832127, EPI_ISL_1832129, EPI_ISL_1832131, EPI_ISL_1832133, EPI_ISL_1832135, EPI_ISL_1832161, EPI_ISL_1832163, EPI_ISL_1832165, EPI_ISL_1832167, EPI_ISL_1832169 | | | |
| see above | University of Birmingham | COVID-19 Genomics UK (COG-UK) Consortium | Alex Richter; Andrew Bosworth; Andrew Bosworth. Queen Elizabeth Hospital: Anna Casey; Andrew D Beggs PHE Heartlands Lab: Husam Osman; Charlotte Poxon; Institute of Microbiology: Joanne Stockton; Josh Quick; Kasun Wanigasooriya; Mike Kidd; Nicholas Loman. University of Birmingham Testing Laboratory: Celina M Whalley; Oliver Pickles; Radoslaw Poplawski; Samuel Nicholls; University of Birmingham: Claire McMurray; Will Rowe |
| EPI_ISL_1308640, EPI_ISL_1308644, EPI_ISL_1308645, EPI_ISL_1474530, EPI_ISL_1474531, EPI_ISL_1474532, EPI_ISL_1474534, EPI_ISL_1474535, EPI_ISL_1474536, EPI_ISL_1474540, EPI_ISL_1474541, EPI_ISL_1474543, EPI_ISL_1474544, EPI_ISL_1474545, EPI_ISL_1474546, EPI_ISL_1474547, EPI_ISL_1474560, EPI_ISL_1474603, EPI_ISL_1474604, EPI_ISL_1474605, EPI_ISL_1474606, EPI_ISL_1474607, EPI_ISL_1474608, EPI_ISL_1474628, EPI_ISL_1474629, EPI_ISL_1474630, EPI_ISL_1474631, EPI_ISL_1474632, EPI_ISL_1474633, EPI_ISL_1474634, EPI_ISL_1474635, EPI_ISL_1474636, EPI_ISL_1474637, EPI_ISL_1474638, EPI_ISL_1474639, EPI_ISL_1474642, EPI_ISL_1545944, EPI_ISL_1730161, EPI_ISL_1730166, EPI_ISL_1730167, EPI_ISL_1730168, EPI_ISL_1730169, EPI_ISL_1730170 | | | |
| see above | University of Exeter | COVID-19 Genomics UK (COG-UK) Consortium | Aaron Jeffries; Audrey Farbos; Ben Temperton; Jane Masoli; Joanna Warwick-Dugdale; Michelle Michelsen; Robyn Manley; Stephen Michell |
| EPI_ISL_1308778, EPI_ISL_1308779, EPI_ISL_1308812, EPI_ISL_1308813, EPI_ISL_1308814, EPI_ISL_1308815, EPI_ISL_1308856, EPI_ISL_1308857, EPI_ISL_1308858, EPI_ISL_1308865, EPI_ISL_1308866, EPI_ISL_1386656, EPI_ISL_1386657, EPI_ISL_1386658, EPI_ISL_1386659, EPI_ISL_1386660, EPI_ISL_1386662, EPI_ISL_1386663, EPI_ISL_1386664, EPI_ISL_1545917, EPI_ISL_1545918, EPI_ISL_1545926, EPI_ISL_1545933, EPI_ISL_1545938, EPI_ISL_1545941, EPI_ISL_1730081, EPI_ISL_1730082, EPI_ISL_1730083, EPI_ISL_1730084, EPI_ISL_1730085, EPI_ISL_1730238, EPI_ISL_1730239, EPI_ISL_1730240, EPI_ISL_1730241, EPI_ISL_1730248, EPI_ISL_1832240, EPI_ISL_1832241, EPI_ISL_1832242, EPI_ISL_1832244, EPI_ISL_1832245, EPI_ISL_1832246, EPI_ISL_2125734 | | | |
| see above | Virology Department, Royal Infirmary of Edinburgh, NHS Lothian / School of Biological Sciences, University of Edinburgh | COVID-19 Genomics UK (COG-UK) Consortium | Colquhoun R; Cotton S; Dewar R; Gallagher A; Hill V; Jackson B; Maloney D; McCrone JT; McHugh M; O'Toole Á; Rambaut A; Rooke S; Scher E; Templeton K; Yu X |
| EPI_ISL_1250260, EPI_ISL_1250262, EPI_ISL_1250263, EPI_ISL_1250265, EPI_ISL_1250267, EPI_ISL_1250268, EPI_ISL_1250269, EPI_ISL_1250273, EPI_ISL_1250274, EPI_ISL_1250279, EPI_ISL_1250280, EPI_ISL_1250281, EPI_ISL_1250283, EPI_ISL_1250285, EPI_ISL_1250286, EPI_ISL_1250291, EPI_ISL_1250292, EPI_ISL_1250294, EPI_ISL_1250297, EPI_ISL_1250298, EPI_ISL_1250299, EPI_ISL_1250300, EPI_ISL_1250304, EPI_ISL_1250306, EPI_ISL_1250308, EPI_ISL_1250309, EPI_ISL_1310714, EPI_ISL_1310721, EPI_ISL_1310722, EPI_ISL_1310724, EPI_ISL_1310726, EPI_ISL_1310727, EPI_ISL_1310737, EPI_ISL_1310739, EPI_ISL_1310740, EPI_ISL_1310741, EPI_ISL_1310743, EPI_ISL_1310745, EPI_ISL_1310747, EPI_ISL_1310753, EPI_ISL_1310759, EPI_ISL_1310761, EPI_ISL_1310763, EPI_ISL_1310764, EPI_ISL_1638035, EPI_ISL_1833830, EPI_ISL_1833846, EPI_ISL_1833847, EPI_ISL_1833849, EPI_ISL_1833866, EPI_ISL_1833872, EPI_ISL_1833880, EPI_ISL_1833888, EPI_ISL_1833892, EPI_ISL_1833904, EPI_ISL_1833913 | | | |
| see above | Virology Department, Sheffield Teaching Hospitals NHS Foundation Trust/Department of Infection, Immunity and Cardiovascular Disease, The Medical School, University of Sheffield | COVID-19 Genomics UK (COG-UK) Consortium | Adri Angyal; Alex Keeley; Benjamin Lindsey; Cariad Evans; Danielle Groves; Dave Partridge; Katie Johnson; Laura Carrilero; Luke Green; Matthew Parker; Matthew Wyles; Mehmet Yavuz; Mohammad Raza; Nikki Smith; Paul Parsons; Rachel Tucker; Rebecca Brown; Thushan de Silva |
| EPI_ISL_1249627, EPI_ISL_1249629, EPI_ISL_1249630, EPI_ISL_1249632, EPI_ISL_1249635, EPI_ISL_1249636, EPI_ISL_1249637, EPI_ISL_1249638, EPI_ISL_1249639, EPI_ISL_1249640, EPI_ISL_1249641, EPI_ISL_1249642, EPI_ISL_1249643, EPI_ISL_1249644, EPI_ISL_1249645, EPI_ISL_1249646, EPI_ISL_1249647, EPI_ISL_1249648, EPI_ISL_1249649, EPI_ISL_1249650, EPI_ISL_1249651, EPI_ISL_1249652, EPI_ISL_1249653, EPI_ISL_1249654, EPI_ISL_1249655, EPI_ISL_1249656, EPI_ISL_1249657, EPI_ISL_1249660, EPI_ISL_1249661, EPI_ISL_1249666, EPI_ISL_1249668, EPI_ISL_1249671, EPI_ISL_1249672, EPI_ISL_1249675, EPI_ISL_1249679, EPI_ISL_1249686, EPI_ISL_1249687, EPI_ISL_1249688, EPI_ISL_1249693, EPI_ISL_1249696, EPI_ISL_1249699, EPI_ISL_1249700, EPI_ISL_1249702, EPI_ISL_1249703, EPI_ISL_1249704, EPI_ISL_1249705, EPI_ISL_1249717, EPI_ISL_1249718, EPI_ISL_1249719, EPI_ISL_1249721, EPI_ISL_1249728, EPI_ISL_1249731, EPI_ISL_1249735, EPI_ISL_1249737, EPI_ISL_1249742, EPI_ISL_1249744, EPI_ISL_1249752, EPI_ISL_1249753, EPI_ISL_1249754, EPI_ISL_1249757, EPI_ISL_1249761, EPI_ISL_1249762, EPI_ISL_1249771, EPI_ISL_1249774, EPI_ISL_1249782, EPI_ISL_1249783, EPI_ISL_1249785, EPI_ISL_1249787, EPI_ISL_1249791, EPI_ISL_1249792, EPI_ISL_1249795, EPI_ISL_1249796, EPI_ISL_1249797, EPI_ISL_1249800, EPI_ISL_1249821, EPI_ISL_1249822, EPI_ISL_1249823, EPI_ISL_1249824, EPI_ISL_1249826, EPI_ISL_1249827, EPI_ISL_1249828, EPI_ISL_1249829, EPI_ISL_1249830, EPI_ISL_1249833, EPI_ISL_1249840, EPI_ISL_1249859, EPI_ISL_1249863, EPI_ISL_1249864, EPI_ISL_1249865, EPI_ISL_1249871, EPI_ISL_1249872, EPI_ISL_1249874, EPI_ISL_1249876, EPI_ISL_1249879, EPI_ISL_1249881, EPI_ISL_1249884, EPI_ISL_1309834, EPI_ISL_1309835, EPI_ISL_1309836, EPI_ISL_1309956, EPI_ISL_1387740, EPI_ISL_1387741, EPI_ISL_1387742, EPI_ISL_1387752, EPI_ISL_1387753, EPI_ISL_1387755, EPI_ISL_1387759, EPI_ISL_1387763, EPI_ISL_1387764, EPI_ISL_1387771, EPI_ISL_1387774, EPI_ISL_1387779, EPI_ISL_1387781, EPI_ISL_1387786, EPI_ISL_1387787, EPI_ISL_1387800, EPI_ISL_1387802, EPI_ISL_1387803, EPI_ISL_1387823, EPI_ISL_1387825, EPI_ISL_1476489, EPI_ISL_1637270, EPI_ISL_1637271, EPI_ISL_1637272, EPI_ISL_1637277, EPI_ISL_1637280, EPI_ISL_1637281, EPI_ISL_1637283, EPI_ISL_1637288 | | | |
| see above | Wales Specialist Virology Centre Sequencing lab: Pathogen Genomics Unit | Public Health Wales Microbiology Cardiff Wales Specialist Virology Centre | Alec Birchley; Alexander Adams; Amy Gaskin; Angela Marchbank; Bree Gatica-Wilcox; Catherine Moore; Jason Coombes; Joanne Watkins; Joel Southgate; Johnathan Evans; Laura Gifford; Lauren Gilbert; Lee Graham; Malorie Perry; Matthew Bull; Nicole Pacchiarini; Sally Corden; Sara Kumziene-Summerhayes; Sara Rey; Sarah Taylor; Simon Cottrell; Sophie Jones; Tom Connor |
| EPI_ISL_1386596, EPI_ISL_1386597, EPI_ISL_1386598, EPI_ISL_1386606, EPI_ISL_1386607, EPI_ISL_1386608, EPI_ISL_1386610, EPI_ISL_1386611, EPI_ISL_1386612, EPI_ISL_1386613, EPI_ISL_1386614, EPI_ISL_1386615, EPI_ISL_1386616, EPI_ISL_1386617, EPI_ISL_1386618, EPI_ISL_1386619, EPI_ISL_1386620, EPI_ISL_1386621, EPI_ISL_1386622, EPI_ISL_1386623, EPI_ISL_1386624, EPI_ISL_1386625, EPI_ISL_1386626, EPI_ISL_1386627, EPI_ISL_1386628, EPI_ISL_1386629, EPI_ISL_1386630, EPI_ISL_1386631, EPI_ISL_1386632, EPI_ISL_1386636, EPI_ISL_1386637, EPI_ISL_1386641, EPI_ISL_1386642, EPI_ISL_1386643, EPI_ISL_1386644, EPI_ISL_1386645, EPI_ISL_1386646, EPI_ISL_1545878, EPI_ISL_1545880, EPI_ISL_1545881, EPI_ISL_1545882, EPI_ISL_1545884, EPI_ISL_1545886, EPI_ISL_1545890, EPI_ISL_1545891, EPI_ISL_1545893, EPI_ISL_1545896, EPI_ISL_1636658, EPI_ISL_1636659, EPI_ISL_1636663, EPI_ISL_1636664, EPI_ISL_1636665, EPI_ISL_1636666 | | | |
| see above | West of Scotland Specialist Virology Centre, NHSGGC / MRC-University of Glasgow Centre for Virus Research | COVID-19 Genomics UK (COG-UK) Consortium | Alasdair MacLean; Alice Broos; Ana da Silva Filipe; Antonia Ho; Daniel Mair; David L Robertson; Emma Thomson; Guy Mollett; James Shepherd; Jenna Nichols; Joseph Hughes; Kathy Li; Kathy Smollett; Kyriaki Nomikou; Lily Tong; Matthew Holden; Natasha Johnson; Rachel Blacow; Richard Orton; Rory Gunson; Sarah McDonald; Sharif Shaaban; Sreenu Vattipally; Stephen Carmichael |

We gratefully acknowledge the following Authors from the Originating laboratories responsible for obtaining the specimens, as well as the Submitting laboratories where the genome data were generated and shared via GISAID, on which this research is based.

All Submitters of data may be contacted directly via www.gisaid.org

Authors are sorted alphabetically.

| Accession ID | Originating Laboratory | Submitting Laboratory | Authors |
|---|---|---|---|
| EPI_ISL_1474824 | Barts Health NHS Trust | COVID-19 Genomics UK (COG-UK) Consortium | BROAD; Beatrix; CUTINO-MOGUEL; Claire; David; Dola; HARRINGTON; KELE; KULASEGARAN-SHYLINI; Maria-Teresa; OWOYEMI; Raghavendran |
| EPI_ISL_1388782, EPI_ISL_1388784, EPI_ISL_1388870, EPI_ISL_1388872, EPI_ISL_1388876, EPI_ISL_1388888, EPI_ISL_1388890, EPI_ISL_1388896, EPI_ISL_1476564, EPI_ISL_1637984, EPI_ISL_1637991, EPI_ISL_1637992, EPI_ISL_1637993, EPI_ISL_1637994, EPI_ISL_1637998, EPI_ISL_1637999, EPI_ISL_1638001, EPI_ISL_2127989 | | | |
| see above | Centre for Enzyme Innovation, University of Portsmouth / Translational Research Laboratory, Portsmouth Hospitals NHS Trust | COVID-19 Genomics UK (COG-UK) Consortium | Angela Beckett; Christopher Fearn; Kate Cook; Katie Loveson; Salman Goudarzi; Samuel Robson; Scott Elliott; Sharon Glaysher |
| EPI_ISL_1308676, EPI_ISL_1308677, EPI_ISL_1308678, EPI_ISL_1308679, EPI_ISL_1308680, EPI_ISL_1308681, EPI_ISL_1308682, EPI_ISL_1308685, EPI_ISL_1308690, EPI_ISL_1308692, EPI_ISL_1308694, EPI_ISL_1308696, EPI_ISL_1308697, EPI_ISL_1308699, EPI_ISL_1730018 | | | |
| see above | Department of Pathology, University of Cambridge | COVID-19 Genomics UK (COG-UK) Consortium | Aminu S. Jahun; Ian Goodfellow; Iliana Georgana; Martin D. Curran; Myra Hosmillo; Rhys Izuagbe; Surendra Parmar; William L. Hamilton; Yasmin Chaudhry |
| EPI_ISL_1326670, EPI_ISL_1326673, EPI_ISL_1326693, EPI_ISL_1326695, EPI_ISL_1326699, EPI_ISL_1326701, EPI_ISL_1326704, EPI_ISL_1326707, EPI_ISL_1326713, EPI_ISL_1326723, EPI_ISL_1326736, EPI_ISL_1326740, EPI_ISL_1326741, EPI_ISL_1326747, EPI_ISL_1326752, EPI_ISL_1326754, EPI_ISL_1326757, EPI_ISL_1326759, EPI_ISL_1326760, EPI_ISL_1326761, EPI_ISL_1326763, EPI_ISL_1326771, EPI_ISL_1326774, EPI_ISL_1326777, EPI_ISL_1326782, EPI_ISL_1326783, EPI_ISL_1326785, EPI_ISL_1326788, EPI_ISL_1326790, EPI_ISL_1326797, EPI_ISL_1326798, EPI_ISL_1326805, EPI_ISL_1326807, EPI_ISL_1326810, EPI_ISL_1326813, EPI_ISL_1326817, EPI_ISL_1326830, EPI_ISL_1326840, EPI_ISL_1326846, EPI_ISL_1326847, EPI_ISL_1326850, EPI_ISL_1326855, EPI_ISL_1326859, EPI_ISL_1326861, EPI_ISL_1326868, EPI_ISL_1326875, EPI_ISL_1326876, EPI_ISL_1326879, EPI_ISL_1326895, EPI_ISL_1326901, EPI_ISL_1326903, EPI_ISL_1326904, EPI_ISL_1326905, EPI_ISL_1326914, EPI_ISL_1326929, EPI_ISL_1326932, EPI_ISL_1326942, EPI_ISL_1326950, EPI_ISL_1326952, EPI_ISL_1326956, EPI_ISL_1326957, EPI_ISL_1326963, EPI_ISL_1326966, EPI_ISL_1326968, EPI_ISL_1326970, EPI_ISL_1326982, EPI_ISL_1326984, EPI_ISL_1326998, EPI_ISL_1327001, EPI_ISL_1327008, EPI_ISL_1327017, EPI_ISL_1327025, EPI_ISL_1327028, EPI_ISL_1327034, EPI_ISL_1327048, EPI_ISL_1327076, EPI_ISL_1327082, EPI_ISL_1327085, EPI_ISL_1327089, EPI_ISL_1327096, EPI_ISL_1327097, EPI_ISL_1327106, EPI_ISL_1327114, EPI_ISL_1327116, EPI_ISL_1327120, EPI_ISL_1327127, EPI_ISL_1327129, EPI_ISL_1327134, EPI_ISL_1327142, EPI_ISL_1327148, EPI_ISL_1327147, EPI_ISL_1327148, EPI_ISL_1327149, EPI_ISL_1327151, EPI_ISL_1327156, EPI_ISL_1327170, EPI_ISL_1327176, EPI_ISL_1327184, EPI_ISL_1327188, EPI_ISL_1327190, EPI_ISL_1327192, EPI_ISL_1327198, EPI_ISL_1327201, EPI_ISL_1327203, EPI_ISL_1327209, EPI_ISL_1327230, EPI_ISL_1327232, EPI_ISL_1327234, EPI_ISL_1327236, EPI_ISL_1327238, EPI_ISL_1330071, EPI_ISL_1330302, EPI_ISL_1330316, EPI_ISL_1330324, EPI_ISL_1330333, EPI_ISL_1330334, EPI_ISL_1330340, EPI_ISL_1330341, EPI_ISL_1330345, EPI_ISL_1330350, EPI_ISL_1330354, EPI_ISL_1330355, EPI_ISL_1330359, EPI_ISL_1330366, EPI_ISL_1330369, EPI_ISL_1330376, EPI_ISL_1330386, EPI_ISL_1330388, EPI_ISL_1330389, EPI_ISL_1330393, EPI_ISL_1330395, EPI_ISL_1330402, EPI_ISL_1330416, EPI_ISL_1330424, EPI_ISL_1330431, EPI_ISL_1330442, EPI_ISL_1330443, EPI_ISL_1330446, EPI_ISL_1330448, EPI_ISL_1330451, EPI_ISL_1330463, EPI_ISL_1330466, EPI_ISL_1330467, EPI_ISL_1330468, EPI_ISL_1330473, EPI_ISL_1330474, EPI_ISL_1330477, EPI_ISL_1330478, EPI_ISL_1330479, EPI_ISL_1330481, EPI_ISL_1330502, EPI_ISL_1330506, EPI_ISL_1330507, EPI_ISL_1330511, EPI_ISL_1330512, EPI_ISL_1330517, EPI_ISL_1330520, EPI_ISL_1330523, EPI_ISL_1330526, EPI_ISL_1330528, EPI_ISL_1330535, EPI_ISL_1330538, EPI_ISL_1330541, EPI_ISL_1330542, EPI_ISL_1330543, EPI_ISL_1330547, EPI_ISL_1330555, EPI_ISL_1330560, EPI_ISL_1330567, EPI_ISL_1330570, EPI_ISL_1330577, EPI_ISL_1330578, EPI_ISL_1330581, EPI_ISL_1330585, EPI_ISL_1330590, EPI_ISL_1330594, EPI_ISL_1330596, EPI_ISL_1330602, EPI_ISL_1330605, EPI_ISL_1331133, EPI_ISL_1331134, EPI_ISL_1331135, EPI_ISL_1331136, EPI_ISL_1331138, EPI_ISL_1331141, EPI_ISL_1331143, EPI_ISL_1331145, EPI_ISL_1331146, EPI_ISL_1331148, EPI_ISL_1331149, EPI_ISL_1331151, EPI_ISL_1331153, EPI_ISL_1331155, EPI_ISL_1331156, EPI_ISL_1331159, EPI_ISL_1331160, EPI_ISL_1331161, EPI_ISL_1331163, EPI_ISL_1331164, EPI_ISL_1331165, EPI_ISL_1331166, EPI_ISL_1331167, EPI_ISL_1331168, EPI_ISL_1331169, EPI_ISL_1331170, EPI_ISL_1331171, EPI_ISL_1331172, EPI_ISL_1331173, EPI_ISL_1331174, EPI_ISL_1331175, EPI_ISL_1331176, EPI_ISL_1331177, EPI_ISL_1331178, EPI_ISL_1331179, EPI_ISL_1331181, EPI_ISL_1331182, EPI_ISL_1331183, EPI_ISL_1331186, EPI_ISL_1331187, EPI_ISL_1331189, EPI_ISL_1331191, EPI_ISL_1331192, EPI_ISL_1331193, EPI_ISL_1331194, EPI_ISL_1331197, EPI_ISL_1331198, EPI_ISL_1331201, EPI_ISL_1331202, EPI_ISL_1331204, EPI_ISL_1331205, EPI_ISL_1331207, EPI_ISL_1331208, EPI_ISL_1331210, EPI_ISL_1331212, EPI_ISL_1331214, EPI_ISL_1331217, EPI_ISL_1331218, EPI_ISL_1331219, EPI_ISL_1331220, EPI_ISL_1331222, EPI_ISL_1331223, EPI_ISL_1331227, EPI_ISL_1331228, EPI_ISL_1331229, EPI_ISL_1331230, EPI_ISL_1331231, EPI_ISL_1331234, EPI_ISL_1331235, EPI_ISL_1331236, EPI_ISL_1331237, EPI_ISL_1331238, EPI_ISL_1331239, EPI_ISL_1331240, EPI_ISL_1331241, EPI_ISL_1331242, EPI_ISL_1331243, EPI_ISL_1331244, EPI_ISL_1331245, EPI_ISL_1331246, EPI_ISL_1331248, EPI_ISL_1331249, EPI_ISL_1331250, EPI_ISL_1331252, EPI_ISL_1331253, EPI_ISL_1331254, EPI_ISL_1331255, EPI_ISL_1331258, EPI_ISL_1331259, EPI_ISL_1331260, EPI_ISL_1331263, EPI_ISL_1331266, EPI_ISL_1331267, EPI_ISL_1331268, EPI_ISL_1331271, EPI_ISL_1331272, EPI_ISL_1331273, EPI_ISL_1331274, EPI_ISL_1331275, EPI_ISL_1331276, EPI_ISL_1331278, EPI_ISL_1331279, EPI_ISL_1331281, EPI_ISL_1331282, EPI_ISL_1331283, EPI_ISL_1331284, EPI_ISL_1331285, EPI_ISL_1331286, EPI_ISL_1331287, EPI_ISL_1331288, EPI_ISL_1331290, EPI_ISL_1331291, EPI_ISL_1331292, EPI_ISL_1331295, EPI_ISL_1331296, EPI_ISL_1331297, EPI_ISL_1331299, EPI_ISL_1331302, EPI_ISL_1331303, EPI_ISL_1331305, EPI_ISL_1331307, EPI_ISL_1331308, EPI_ISL_1331309, EPI_ISL_1331310, EPI_ISL_1331312, EPI_ISL_1331314, EPI_ISL_1331315, EPI_ISL_1331316, EPI_ISL_1331319, EPI_ISL_1331320, EPI_ISL_1331321, EPI_ISL_1331325, EPI_ISL_1331326, EPI_ISL_1331327, EPI_ISL_1331329, EPI_ISL_1331331, EPI_ISL_1331332, EPI_ISL_1331333, EPI_ISL_1331334, EPI_ISL_1331335, EPI_ISL_1331336, EPI_ISL_1331337, EPI_ISL_1331340, EPI_ISL_1331342, EPI_ISL_1331345, EPI_ISL_1331346, EPI_ISL_1331347, EPI_ISL_1331348, EPI_ISL_1331350, EPI_ISL_1331351, EPI_ISL_1331352, EPI_ISL_1331354, EPI_ISL_1331356, EPI_ISL_1331357, EPI_ISL_1331360, EPI_ISL_1331361, EPI_ISL_1331362, EPI_ISL_1331364, EPI_ISL_1331366, EPI_ISL_1331368, EPI_ISL_1331369, EPI_ISL_1331370, EPI_ISL_1331371, EPI_ISL_1331373, EPI_ISL_1331375, EPI_ISL_1331376, EPI_ISL_1331377, EPI_ISL_1331378, EPI_ISL_1331379, EPI_ISL_1331380, EPI_ISL_1331381, EPI_ISL_1331385, EPI_ISL_1331386, EPI_ISL_1331388, EPI_ISL_1331390, EPI_ISL_1331392, EPI_ISL_1331395, EPI_ISL_1331397, EPI_ISL_1331398, EPI_ISL_1331402, EPI_ISL_1331403, EPI_ISL_1331404, EPI_ISL_1331405, EPI_ISL_1331406, EPI_ISL_1331407, EPI_ISL_1331408, EPI_ISL_1331410, EPI_ISL_1331411, EPI_ISL_1331413, EPI_ISL_1331414, EPI_ISL_1331415, EPI_ISL_1331416, EPI_ISL_1331417, EPI_ISL_1331418, EPI_ISL_1331420, EPI_ISL_1331421, EPI_ISL_1331423, EPI_ISL_1331424, EPI_ISL_1331426, EPI_ISL_1331427, EPI_ISL_1331428, EPI_ISL_1331429, EPI_ISL_1331431, EPI_ISL_1331433, EPI_ISL_1331434, EPI_ISL_1331439, EPI_ISL_1331440, EPI_ISL_1331442, EPI_ISL_1331443, EPI_ISL_1331445, EPI_ISL_1331446, EPI_ISL_1331448, EPI_ISL_1331449, EPI_ISL_1331450, EPI_ISL_1331451, EPI_ISL_1331452, EPI_ISL_1331453, EPI_ISL_1331454, EPI_ISL_1331455, EPI_ISL_1331456, EPI_ISL_1331460, EPI_ISL_1331462, EPI_ISL_1331697, EPI_ISL_1331701, EPI_ISL_1331706, EPI_ISL_1331712, EPI_ISL_1331714, EPI_ISL_1331717, EPI_ISL_1331719, EPI_ISL_1331720, EPI_ISL_1331721, EPI_ISL_1331724, EPI_ISL_1331726, EPI_ISL_1331727, EPI_ISL_1331733, EPI_ISL_1331735, EPI_ISL_1331736, EPI_ISL_1331740, EPI_ISL_1331744, EPI_ISL_1331759, EPI_ISL_1331760, EPI_ISL_1331766, EPI_ISL_1331769, EPI_ISL_1331774, EPI_ISL_1331776, EPI_ISL_1331779, EPI_ISL_1331782, EPI_ISL_1331785, EPI_ISL_1331788, EPI_ISL_1331789, EPI_ISL_1331792, EPI_ISL_1331794, EPI_ISL_1331802, EPI_ISL_1331806, EPI_ISL_1331813, EPI_ISL_1331818, EPI_ISL_1331819, EPI_ISL_1331822, EPI_ISL_1331824, EPI_ISL_1331842, EPI_ISL_1331843, EPI_ISL_1331847, EPI_ISL_1331848, EPI_ISL_1331852, EPI_ISL_1331859, EPI_ISL_1331865, EPI_ISL_1331867, EPI_ISL_1331871, EPI_ISL_1331873, EPI_ISL_1331877, EPI_ISL_1331887, EPI_ISL_1331899, EPI_ISL_1331900, EPI_ISL_1331901, EPI_ISL_1331903, EPI_ISL_1331905, EPI_ISL_1331908, EPI_ISL_1331909, EPI_ISL_1331910, EPI_ISL_1331915, EPI_ISL_1331919, EPI_ISL_1331920, EPI_ISL_1331924, EPI_ISL_1331926, EPI_ISL_1331927, EPI_ISL_1331929, EPI_ISL_1331934, EPI_ISL_1331935, EPI_ISL_1331936, EPI_ISL_1331944, EPI_ISL_1331945, EPI_ISL_1331947, EPI_ISL_1331948, EPI_ISL_1331952, EPI_ISL_1331953, EPI_ISL_1331955, EPI_ISL_1331961, EPI_ISL_1331965, EPI_ISL_1331966, EPI_ISL_1331968, EPI_ISL_1331985, EPI_ISL_1331994, EPI_ISL_1332000, EPI_ISL_1332003, EPI_ISL_1332013, EPI_ISL_1332016, EPI_ISL_1332021, EPI_ISL_1332029, EPI_ISL_1332034, EPI_ISL_1332037, EPI_ISL_1332038, EPI_ISL_1332041, EPI_ISL_1332047, EPI_ISL_1332050, EPI_ISL_1332051, EPI_ISL_1332055, EPI_ISL_1332061, EPI_ISL_1332064, EPI_ISL_1332065, EPI_ISL_1332068, EPI_ISL_1332071, EPI_ISL_1332075, EPI_ISL_1332078, EPI_ISL_1332085, EPI_ISL_1332099, EPI_ISL_1332104, EPI_ISL_1332119, EPI_ISL_1332123, EPI_ISL_1332127, EPI_ISL_1332128, EPI_ISL_1332136, EPI_ISL_1332141, EPI_ISL_1332144, EPI_ISL_1332145, EPI_ISL_1332146, EPI_ISL_1332148, EPI_ISL_1332160, EPI_ISL_1332163, EPI_ISL_1332165, EPI_ISL_1332166, EPI_ISL_1332170, EPI_ISL_1332181, EPI_ISL_1332187, EPI_ISL_1332188, EPI_ISL_1332192, EPI_ISL_1332193, EPI_ISL_1332196, EPI_ISL_1332201, EPI_ISL_1332205, EPI_ISL_1332206, EPI_ISL_1332213, EPI_ISL_1332214, EPI_ISL_1332215, EPI_ISL_1332217, EPI_ISL_1332222, EPI_ISL_1332237, EPI_ISL_1332238, EPI_ISL_1332240, EPI_ISL_1332247, EPI_ISL_1332260, EPI_ISL_1332261, EPI_ISL_1332266, EPI_ISL_1332267, EPI_ISL_1332268, EPI_ISL_1332269, EPI_ISL_1332271, EPI_ISL_1332274, EPI_ISL_1342419, EPI_ISL_1342480, EPI_ISL_1343042, EPI_ISL_1344466, EPI_ISL_1344494, EPI_ISL_1344508, EPI_ISL_1344528, EPI_ISL_1344552, EPI_ISL_1344569, EPI_ISL_1344575, EPI_ISL_1344592, EPI_ISL_1344627, EPI_ISL_1344660, EPI_ISL_1344679, EPI_ISL_1344688, EPI_ISL_1346283, EPI_ISL_1346284, EPI_ISL_1346287, EPI_ISL_1346294, EPI_ISL_1346295, EPI_ISL_1346300, EPI_ISL_1346302, EPI_ISL_1346408, EPI_ISL_1346426, EPI_ISL_1346427, EPI_ISL_1346429, EPI_ISL_1346430, EPI_ISL_1346432, EPI_ISL_1346433, EPI_ISL_1346434, EPI_ISL_1346435, EPI_ISL_1346436, EPI_ISL_1346437, EPI_ISL_1346440, EPI_ISL_1346441, EPI_ISL_1346443, EPI_ISL_1346462, EPI_ISL_1346464, EPI_ISL_1346469, EPI_ISL_1365216, EPI_ISL_1374338, EPI_ISL_1374339, EPI_ISL_1374340, EPI_ISL_1374341, EPI_ISL_1374342, EPI_ISL_1374344, EPI_ISL_1374346, EPI_ISL_1374347, EPI_ISL_1374348, EPI_ISL_1374349, EPI_ISL_1374350, EPI_ISL_1374352, EPI_ISL_1374353, EPI_ISL_1374359, EPI_ISL_1374360, EPI_ISL_1374362, EPI_ISL_1374363, EPI_ISL_1374364, EPI_ISL_1374367, EPI_ISL_1374368, EPI_ISL_1374369, EPI_ISL_1374371, EPI_ISL_1374372, EPI_ISL_1374373, EPI_ISL_1374376, EPI_ISL_1374377, EPI_ISL_1374378, EPI_ISL_1374379, EPI_ISL_1374383, EPI_ISL_1374384, EPI_ISL_1374385, EPI_ISL_1374386, EPI_ISL_1374387, EPI_ISL_1374388, EPI_ISL_1374390, EPI_ISL_1374392, EPI_ISL_1374394, EPI_ISL_1374395, EPI_ISL_1374396, EPI_ISL_1374397, EPI_ISL_1374398, EPI_ISL_1374399, EPI_ISL_1374401, EPI_ISL_1374402, EPI_ISL_1374403, EPI_ISL_1374404, EPI_ISL_1374405, EPI_ISL_1374409, EPI_ISL_1374410, EPI_ISL_1374412, EPI_ISL_1374413, EPI_ISL_1374414, EPI_ISL_1374418, EPI_ISL_1374420, EPI_ISL_1374421, EPI_ISL_1374422, EPI_ISL_1374423, EPI_ISL_1374424, EPI_ISL_1374425, EPI_ISL_1374426, EPI_ISL_1374427, EPI_ISL_1374428, EPI_ISL_1374429, EPI_ISL_1374432, EPI_ISL_1374435, EPI_ISL_1374436, EPI_ISL_1374437, EPI_ISL_1374438, EPI_ISL_1374439, EPI_ISL_1374440, EPI_ISL_1374441, EPI_ISL_1374442, EPI_ISL_1374443, EPI_ISL_1374444, EPI_ISL_1374446, EPI_ISL_1374447, EPI_ISL_1374450, EPI_ISL_1374451, EPI_ISL_1374454, EPI_ISL_1374455, EPI_ISL_1374457, EPI_ISL_1374459, EPI_ISL_1374463, EPI_ISL_1374464, EPI_ISL_1374465, EPI_ISL_1374467, EPI_ISL_1374468, EPI_ISL_1374469, EPI_ISL_1374470, EPI_ISL_1374471, EPI_ISL_1374472, EPI_ISL_1374473, EPI_ISL_1374476, EPI_ISL_1374478, EPI_ISL_1374479, EPI_ISL_1374480, EPI_ISL_1374482, EPI_ISL_1374483, EPI_ISL_1374484, EPI_ISL_1374486, EPI_ISL_1374487, EPI_ISL_1374488, EPI_ISL_1374489, EPI_ISL_1374493, EPI_ISL_1374494, EPI_ISL_1374495, EPI_ISL_1374496, EPI_ISL_1374499, EPI_ISL_1374500, EPI_ISL_1374503, EPI_ISL_1374504, EPI_ISL_1374505, EPI_ISL_1374506, EPI_ISL_1374507, EPI_ISL_1374508, EPI_ISL_1374509, EPI_ISL_1374510, EPI_ISL_1374511, EPI_ISL_1374512, EPI_ISL_1374513, EPI_ISL_1374514, EPI_ISL_1374515, EPI_ISL_1374516, EPI_ISL_1374517, EPI_ISL_1374518, EPI_ISL_1374519, EPI_ISL_1374520, EPI_ISL_1374521, EPI_ISL_1374523, EPI_ISL_1374525, EPI_ISL_1374527, EPI_ISL_1374528, EPI_ISL_1374529, EPI_ISL_1374533, EPI_ISL_1374535, EPI_ISL_1374538, EPI_ISL_1374540, EPI_ISL_1374541, EPI_ISL_1374542, EPI_ISL_1374544, EPI_ISL_1374548, EPI_ISL_1374549, EPI_ISL_1374552, EPI_ISL_1374555, EPI_ISL_1374556, EPI_ISL_1374557, EPI_ISL_1374559, EPI_ISL_1374563, EPI_ISL_1374564, EPI_ISL_1374566, EPI_ISL_1374567, EPI_ISL_1374569, EPI_ISL_1374570, EPI_ISL_1374571, EPI_ISL_1374572, EPI_ISL_1374574, EPI_ISL_1374575, EPI_ISL_1374576, EPI_ISL_1374577, EPI_ISL_1374578, EPI_ISL_1374579, EPI_ISL_1374580, EPI_ISL_1374581, EPI_ISL_1374582, EPI_ISL_1374584, EPI_ISL_1374585, EPI_ISL_1374586, EPI_ISL_1374588, EPI_ISL_1374590, EPI_ISL_1374591, EPI_ISL_1374596, EPI_ISL_1374598, EPI_ISL_1374599, EPI_ISL_1374600, EPI_ISL_1374604, EPI_ISL_1374606, EPI_ISL_1374607, EPI_ISL_1374609, EPI_ISL_1374610, EPI_ISL_1374611, EPI_ISL_1374612, EPI_ISL_1374613, EPI_ISL_1374617, EPI_ISL_1374618, EPI_ISL_1374620, EPI_ISL_1374621, EPI_ISL_1374624, EPI_ISL_1374625, EPI_ISL_1374628, EPI_ISL_1374631, EPI_ISL_1374632, EPI_ISL_1374633, EPI_ISL_1374635, EPI_ISL_1374637, EPI_ISL_1374638, EPI_ISL_1374640, EPI_ISL_1374641, EPI_ISL_1374645, EPI_ISL_1374646, EPI_ISL_1374648, EPI_ISL_1374649, EPI_ISL_1374650, EPI_ISL_1374651, EPI_ISL_1374653, EPI_ISL_1374657, EPI_ISL_1374659, EPI_ISL_1374660, EPI_ISL_1374661, EPI_ISL_1374662, EPI_ISL_1374663, EPI_ISL_1374664, EPI_ISL_1374666, EPI_ISL_1374668, EPI_ISL_1374670, EPI_ISL_1374672, EPI_ISL_1374675, EPI_ISL_1374676, EPI_ISL_1374677, EPI_ISL_1374678, EPI_ISL_1374679, EPI_ISL_1374680, EPI_ISL_1374681, EPI_ISL_1374682, EPI_ISL_1374684, EPI_ISL_1374685, EPI_ISL_1374686, EPI_ISL_1374688, EPI_ISL_1374689, EPI_ISL_1374691, EPI_ISL_1374692, EPI_ISL_1374694, EPI_ISL_1374695, EPI_ISL_1374696, EPI_ISL_1374700, EPI_ISL_1374703, EPI_ISL_1374704, EPI_ISL_1374706, EPI_ISL_1374707, EPI_ISL_1374708, EPI_ISL_1374711, EPI_ISL_1374713, EPI_ISL_1374715, EPI_ISL_1374717, EPI_ISL_1374720, EPI_ISL_1374723, EPI_ISL_1374725, EPI_ISL_1374727, EPI_ISL_1374728, EPI_ISL_1374729, EPI_ISL_1374730, EPI_ISL_1374732, EPI_ISL_1374733, EPI_ISL_1374734, EPI_ISL_1374735, EPI_ISL_1374736, EPI_ISL_1374737, EPI_ISL_1374738, EPI_ISL_1374740, EPI_ISL_1374741, EPI_ISL_1374744, EPI_ISL_1374745, EPI_ISL_1374746, EPI_ISL_1374750, EPI_ISL_1374751, EPI_ISL_1374752, EPI_ISL_1374753, EPI_ISL_1374755, EPI_ISL_1374756, EPI_ISL_1374759, EPI_ISL_1374760, EPI_ISL_1374761, EPI_ISL_1374762, EPI_ISL_1374763, EPI_ISL_1374764, EPI_ISL_1374765, EPI_ISL_1374768, EPI_ISL_1374769, EPI_ISL_1374771, EPI_ISL_1374772, EPI_ISL_1374773, EPI_ISL_1374774, EPI_ISL_1374775, EPI_ISL_1374777, EPI_ISL_1374778, EPI_ISL_1374779, EPI_ISL_1374780, EPI_ISL_1374781, EPI_ISL_1374783, EPI_ISL_1374785, EPI_ISL_1374786, EPI_ISL_1374788, EPI_ISL_1374793, EPI_ISL_1374798, EPI_ISL_1374800, EPI_ISL_1374801, EPI_ISL_1374802, EPI_ISL_1374805, EPI_ISL_1374806, EPI_ISL_1374807, EPI_ISL_1374808, EPI_ISL_1374810, EPI_ISL_1374812, EPI_ISL_1374814, EPI_ISL_1374815, EPI_ISL_1374817, EPI_ISL_1374818, EPI_ISL_1374819, EPI_ISL_1374820, EPI_ISL_1374821, EPI_ISL_1374823, EPI_ISL_1374824, EPI_ISL_1374825, EPI_ISL_1374826, EPI_ISL_1374827, EPI_ISL_1374828, EPI_ISL_1374829, EPI_ISL_1374830, EPI_ISL_1374833, EPI_ISL_1374835, EPI_ISL_1374836, EPI_ISL_1374837, EPI_ISL_1374838, EPI_ISL_1374839, EPI_ISL_1374840, EPI_ISL_1374841, EPI_ISL_1374844, EPI_ISL_1374846, EPI_ISL_1374847, EPI_ISL_1374848, EPI_ISL_1374849, EPI_ISL_1374850, EPI_ISL_1374851, EPI_ISL_1374852, EPI_ISL_1374853, EPI_ISL_1374854, EPI_ISL_1374855, EPI_ISL_1374856, EPI_ISL_1374857, EPI_ISL_1374861, EPI_ISL_1374862, EPI_ISL_1374868, EPI_ISL_1374874, EPI_ISL_1374883, EPI_ISL_1374887, EPI_ISL_1374894, EPI_ISL_1374896, EPI_ISL_1374901, EPI_ISL_1374902, EPI_ISL_1374904, EPI_ISL_1374906, EPI_ISL_1374907, EPI_ISL_1374910, EPI_ISL_1374912, EPI_ISL_1374914, EPI_ISL_1374922, EPI_ISL_1374943, EPI_ISL_1374951, EPI_ISL_1374956, EPI_ISL_1374959, EPI_ISL_1374960, EPI_ISL_1374966, EPI_ISL_1374974, EPI_ISL_1374981, EPI_ISL_1374982, EPI_ISL_1374985, EPI_ISL_1374988, EPI_ISL_1375005, EPI_ISL_1375011, EPI_ISL_1375013, EPI_ISL_1375019, EPI_ISL_1375023, EPI_ISL_1375025, EPI_ISL_1375028, EPI_ISL_1375031, EPI_ISL_1375033, EPI_ISL_1375036, EPI_ISL_1375045, EPI_ISL_1375055, EPI_ISL_1375056, EPI_ISL_1375058, EPI_ISL_1375060, EPI_ISL_1375085, EPI_ISL_1375089, EPI_ISL_1375094, EPI_ISL_1375100, EPI_ISL_1375101, EPI_ISL_1375103, EPI_ISL_1375104, EPI_ISL_1375107, EPI_ISL_1375112, EPI_ISL_1375113, EPI_ISL_1375117, EPI_ISL_1375119, EPI_ISL_1375122, EPI_ISL_1375125, EPI_ISL_1375127, EPI_ISL_1375130, EPI_ISL_1375137, EPI_ISL_1375141, EPI_ISL_1375142, EPI_ISL_1375150, EPI_ISL_1376762, EPI_ISL_1410718, EPI_ISL_1410740, EPI_ISL_1410777, EPI_ISL_1410796, EPI_ISL_1410809, EPI_ISL_1410832, EPI_ISL_1410848, EPI_ISL_1410855, EPI_ISL_1410938, EPI_ISL_1410947, EPI_ISL_1410983, EPI_ISL_1410988, EPI_ISL_1410994, EPI_ISL_1411002, EPI_ISL_1411029, EPI_ISL_1411039, EPI_ISL_1411060, EPI_ISL_1411062, EPI_ISL_1411070, EPI_ISL_1411854, EPI_ISL_1411857, EPI_ISL_1411863, EPI_ISL_1411869, EPI_ISL_1411875, EPI_ISL_1411899, EPI_ISL_1411902, EPI_ISL_1411904, EPI_ISL_1411908, EPI_ISL_1411913, EPI_ISL_1411924, EPI_ISL_1411929, EPI_ISL_1411938, EPI_ISL_1411941, EPI_ISL_1411949, EPI_ISL_1411950, EPI_ISL_1411954, EPI_ISL_1411959, EPI_ISL_1411973, EPI_ISL_1411975, EPI_ISL_1411977, EPI_ISL_1411982, EPI_ISL_1411990, EPI_ISL_1411992, EPI_ISL_1411996, EPI_ISL_1412004, EPI_ISL_1412006, EPI_ISL_1412013, EPI_ISL_1412020, EPI_ISL_1412021, EPI_ISL_1412028, EPI_ISL_1412032, EPI_ISL_1412033, EPI_ISL_1412034, EPI_ISL_1412036, EPI_ISL_1412058, EPI_ISL_1412064, EPI_ISL_1412068, EPI_ISL_1412073, EPI_ISL_1412081, EPI_ISL_1412088, EPI_ISL_1412101, EPI_ISL_1412103, EPI_ISL_1412104, EPI_ISL_1412106, EPI_ISL_1412107, EPI_ISL_1412118, EPI_ISL_1412121, EPI_ISL_1412122, EPI_ISL_1412133, EPI_ISL_1412137, EPI_ISL_1412143, EPI_ISL_1412148, EPI_ISL_1412149, EPI_ISL_1412150, EPI_ISL_1412164, EPI_ISL_1412167, EPI_ISL_1412168, EPI_ISL_1412172, EPI_ISL_1412174, EPI_ISL_1412187, EPI_ISL_1412197, EPI_ISL_1412231, EPI_ISL_1412262, EPI_ISL_1412287, EPI_ISL_1412310, EPI_ISL_1412324, EPI_ISL_1412352, EPI_ISL_1412364, EPI_ISL_1412401, EPI_ISL_1412427, EPI_ISL_1412446, EPI_ISL_1412534, EPI_ISL_1413064, EPI_ISL_1413067, EPI_ISL_1413073, EPI_ISL_1413074, EPI_ISL_1413079, EPI_ISL_1413082, EPI_ISL_1413084, EPI_ISL_1413086, EPI_ISL_1413090, EPI_ISL_1413091, EPI_ISL_1413107, EPI_ISL_1413114, EPI_ISL_1413119, EPI_ISL_1413120, EPI_ISL_1413123, EPI_ISL_1413125, EPI_ISL_1413134, EPI_ISL_1413138, EPI_ISL_1413145, EPI_ISL_1413149, EPI_ISL_1413167, EPI_ISL_1413169, EPI_ISL_1413178, EPI_ISL_1413183, EPI_ISL_1413185, EPI_ISL_1413189, EPI_ISL_1413208, EPI_ISL_1413222, EPI_ISL_1413227, EPI_ISL_1413236, EPI_ISL_1413245, EPI_ISL_1413248, EPI_ISL_1413253, EPI_ISL_1413255, EPI_ISL_1413258, EPI_ISL_1413265, EPI_ISL_1413273, EPI_ISL_1413292, EPI_ISL_1413305, EPI_ISL_1413315, EPI_ISL_1413323, EPI_ISL_1413329, EPI_ISL_1413332, EPI_ISL_1413343, EPI_ISL_1413348, EPI_ISL_1413351, EPI_ISL_1413353, EPI_ISL_1413359, EPI_ISL_1413360, EPI_ISL_1413365, EPI_ISL_1413370, EPI_ISL_1413373, EPI_ISL_1413376, EPI_ISL_1413384, EPI_ISL_1413387, EPI_ISL_1413389, EPI_ISL_1413398, EPI_ISL_1413412, EPI_ISL_1413443, EPI_ISL_1413457, EPI_ISL_1413459, EPI_ISL_1413480, EPI_ISL_1413500, EPI_ISL_1413505, EPI_ISL_1413510, EPI_ISL_1413518, EPI_ISL_1413539, EPI_ISL_1413550, EPI_ISL_1413552, EPI_ISL_1413555, EPI_ISL_1413562, EPI_ISL_1413570, EPI_ISL_1413577, EPI_ISL_1413586, EPI_ISL_1413595, EPI_ISL_1413604, EPI_ISL_1413606, EPI_ISL_1413609, EPI_ISL_1413617, EPI_ISL_1413629, EPI_ISL_1413640, EPI_ISL_1413673, EPI_ISL_1413679, EPI_ISL_1413691, EPI_ISL_1413692, EPI_ISL_1413694, EPI_ISL_1413716, EPI_ISL_1413738, EPI_ISL_1413740, EPI_ISL_1413741, EPI_ISL_1413747, EPI_ISL_1413752, EPI_ISL_1413770, EPI_ISL_1413800, EPI_ISL_1413801, EPI_ISL_1413806, EPI_ISL_1413811, EPI_ISL_1413825, EPI_ISL_1413834, EPI_ISL_1413841, EPI_ISL_1413843, EPI_ISL_1413849, EPI_ISL_1413867, EPI_ISL_1413869, EPI_ISL_1413873, EPI_ISL_1413874, EPI_ISL_1413883, EPI_ISL_1413891, EPI_ISL_1413894, EPI_ISL_1413896, EPI_ISL_1413903, EPI_ISL_1413905, EPI_ISL_1413913, EPI_ISL_1413921, EPI_ISL_1413924, EPI_ISL_1413931, EPI_ISL_1413934, EPI_ISL_1413944, EPI_ISL_1413959, EPI_ISL_1413967, EPI_ISL_1413969, EPI_ISL_1413978, EPI_ISL_1413979, EPI_ISL_1413980, EPI_ISL_1413983, EPI_ISL_1413997, EPI_ISL_1413999, EPI_ISL_1414000, EPI_ISL_1414004, EPI_ISL_1414006, EPI_ISL_1414007, EPI_ISL_1414014, EPI_ISL_1414032, EPI_ISL_1414035, EPI_ISL_1414043, EPI_ISL_1414048, EPI_ISL_1414054, EPI_ISL_1414062, EPI_ISL_1414063, EPI_ISL_1414064, EPI_ISL_1414066, EPI_ISL_1414067, EPI_ISL_1414068, EPI_ISL_1414069, EPI_ISL_1414070, EPI_ISL_1414071, EPI_ISL_1414072, EPI_ISL_1414073, EPI_ISL_1414074, EPI_ISL_1414076, EPI_ISL_1414079, EPI_ISL_1414080, EPI_ISL_1414081, EPI_ISL_1414082, EPI_ISL_1414083, EPI_ISL_1414085, EPI_ISL_1414086, EPI_ISL_1414088, EPI_ISL_1414090, EPI_ISL_1414091, EPI_ISL_1414093, EPI_ISL_1414094, EPI_ISL_1414095, EPI_ISL_1414096, EPI_ISL_1414097, EPI_ISL_1414098, EPI_ISL_1414100, EPI_ISL_1414102, EPI_ISL_1414103, EPI_ISL_1414104, EPI_ISL_1414105, EPI_ISL_1414109, EPI_ISL_1414110, EPI_ISL_1414111, EPI_ISL_1414112, EPI_ISL_1414113, EPI_ISL_1414116, EPI_ISL_1414117, EPI_ISL_1414118, EPI_ISL_1414119, EPI_ISL_1414121, EPI_ISL_1414123, EPI_ISL_1414124, EPI_ISL_1414125, EPI_ISL_1414126, EPI_ISL_1414127, EPI_ISL_1414130, EPI_ISL_1414131, EPI_ISL_1414132, EPI_ISL_1414133, EPI_ISL_1414134, EPI_ISL_1414135, EPI_ISL_1414137, EPI_ISL_1414138, EPI_ISL_1414139, EPI_ISL_1414141, EPI_ISL_1414142, EPI_ISL_1414143, EPI_ISL_1414145, EPI_ISL_1414147, EPI_ISL_1414148, EPI_ISL_1414149, EPI_ISL_1414152, EPI_ISL_1414153, EPI_ISL_1414154, EPI_ISL_1414155, EPI_ISL_1414156, EPI_ISL_1414158, EPI_ISL_1414159, EPI_ISL_1414160, EPI_ISL_1414161, EPI_ISL_1414162, EPI_ISL_1414164, EPI_ISL_1414165, EPI_ISL_1414166, EPI_ISL_1414168, EPI_ISL_1414169, EPI_ISL_1414170, EPI_ISL_1414171, EPI_ISL_1414172, EPI_ISL_1414175, EPI_ISL_1414176, EPI_ISL_1414177, EPI_ISL_1414178, EPI_ISL_1414180, EPI_ISL_1414181, EPI_ISL_1414182, EPI_ISL_1414184, EPI_ISL_1414185, EPI_ISL_1414186, EPI_ISL_1414189, EPI_ISL_1414190, EPI_ISL_1414191, EPI_ISL_1414193, EPI_ISL_1414194, EPI_ISL_1414195, EPI_ISL_1414197, EPI_ISL_1414200, EPI_ISL_1414201, EPI_ISL_1414202, EPI_ISL_1414204, EPI_ISL_1414205, EPI_ISL_1414206, EPI_ISL_1414207, EPI_ISL_1414208, EPI_ISL_1414209, EPI_ISL_1414212, EPI_ISL_1414213, EPI_ISL_1414215, EPI_ISL_1414216, EPI_ISL_1414217, EPI_ISL_1414218, EPI_ISL_1414219, EPI_ISL_1414224, EPI_ISL_1414225, EPI_ISL_1414226, EPI_ISL_1414229, EPI_ISL_1414231, EPI_ISL_1414233, EPI_ISL_1414234, EPI_ISL_1414236, EPI_ISL_1414237, EPI_ISL_1414238, EPI_ISL_1414239, EPI_ISL_1414240, EPI_ISL_1414242, EPI_ISL_1414243, EPI_ISL_1414245, EPI_ISL_1414246, EPI_ISL_1414247, EPI_ISL_1414249, EPI_ISL_1414250, EPI_ISL_1414251, EPI_ISL_1414252, EPI_ISL_1414253, EPI_ISL_1414254, EPI_ISL_1414257, EPI_ISL_1414260, EPI_ISL_1414261, EPI_ISL_1414264, EPI_ISL_1414265, EPI_ISL_1414267, EPI_ISL_1414271, EPI_ISL_1414272, EPI_ISL_1414274, EPI_ISL_1414276, EPI_ISL_1414278, EPI_ISL_1414280, EPI_ISL_1414281, EPI_ISL_1414282, EPI_ISL_1414284, EPI_ISL_1414286, EPI_ISL_1414287, EPI_ISL_1414290, EPI_ISL_1414291, EPI_ISL_1414296, EPI_ISL_1414297, EPI_ISL_1414298, EPI_ISL_1414299, EPI_ISL_1414300, EPI_ISL_1414301, EPI_ISL_1414302, EPI_ISL_1414303, EPI_ISL_1414304, EPI_ISL_1414305, EPI_ISL_1414306, EPI_ISL_1414307, EPI_ISL_1414309, EPI_ISL_1414310, EPI_ISL_1414311, EPI_ISL_1414313, EPI_ISL_1414315, EPI_ISL_1414316, EPI_ISL_1414317, EPI_ISL_1414318, EPI_ISL_1414319, EPI_ISL_1414320, EPI_ISL_1414321, EPI_ISL_1414322, EPI_ISL_1414324, EPI_ISL_1414326, EPI_ISL_1414327, EPI_ISL_1414329, EPI_ISL_1414330, EPI_ISL_1414331, EPI_ISL_1414334, EPI_ISL_1414335, EPI_ISL_1414337, EPI_ISL_1414338, EPI_ISL_1414339, EPI_ISL_1414340, EPI_ISL_1414341, EPI_ISL_1414342, EPI_ISL_1414343, EPI_ISL_1414344, EPI_ISL_1414345, EPI_ISL_1414347, EPI_ISL_1414349, EPI_ISL_1414350, EPI_ISL_1414351, EPI_ISL_1414352, EPI_ISL_1414353, EPI_ISL_1414354, EPI_ISL_1414355, EPI_ISL_1414356, EPI_ISL_1414357, EPI_ISL_1414359, EPI_ISL_1414360, EPI_ISL_1414361, EPI_ISL_1414362, EPI_ISL_1414363, EPI_ISL_1414365, EPI_ISL_1414367, EPI_ISL_1414368, EPI_ISL_1414370, EPI_ISL_1414371, EPI_ISL_1414372, EPI_ISL_1414373, EPI_ISL_1414375, EPI_ISL_1414377, EPI_ISL_1414378, EPI_ISL_1414379, EPI_ISL_1414380, EPI_ISL_1414381, EPI_ISL_1414382, EPI_ISL_1414383, EPI_ISL_1414384, EPI_ISL_1414385, EPI_ISL_1414386, EPI_ISL_1414387, EPI_ISL_1414389, EPI_ISL_1414390, EPI_ISL_1414391, EPI_ISL_1414392, EPI_ISL_1414393, EPI_ISL_1414394, EPI_ISL_1414397, EPI_ISL_1414398, EPI_ISL_1414399, EPI_ISL_1414400, EPI_ISL_1414401, EPI_ISL_1414402, EPI_ISL_1414403, EPI_ISL_1414405, EPI_ISL_1414406, EPI_ISL_1414407, EPI_ISL_1414408, EPI_ISL_1414409, EPI_ISL_1414411, EPI_ISL_1414413, EPI_ISL_1414414, EPI_ISL_1414415, EPI_ISL_1414417, EPI_ISL_1414418, EPI_ISL_1414419, EPI_ISL_1414422, EPI_ISL_1414423, EPI_ISL_1414424, EPI_ISL_1414425, EPI_ISL_1414426, EPI_ISL_1414429, EPI_ISL_1414430, EPI_ISL_1414432, EPI_ISL_1414436, EPI_ISL_1414440, EPI_ISL_1414441, EPI_ISL_1414443, EPI_ISL_1414446, EPI_ISL_1414447, EPI_ISL_1414448, EPI_ISL_1414449, EPI_ISL_1414451, EPI_ISL_1414452, EPI_ISL_1414453, EPI_ISL_1414454, EPI_ISL_1414457, EPI_ISL_1414458, EPI_ISL_1414459, EPI_ISL_1414462, EPI_ISL_1414465, EPI_ISL_1414466, EPI_ISL_1414471, EPI_ISL_1414472, EPI_ISL_1414473, EPI_ISL_1414476, EPI_ISL_1414478, EPI_ISL_1414480, EPI_ISL_1414481, EPI_ISL_1414483, EPI_ISL_1414485, EPI_ISL_1414487, EPI_ISL_1414488, EPI_ISL_1414490, EPI_ISL_1414492, EPI_ISL_1414494, EPI_ISL_1414495, EPI_ISL_1414496, EPI_ISL_1414497, EPI_ISL_1414499, EPI_ISL_1414501, EPI_ISL_1414503, EPI_ISL_1414505, EPI_ISL_1414508, EPI_ISL_1414509, EPI_ISL_1414511, EPI_ISL_1414513, EPI_ISL_1414515, EPI_ISL_1414517, EPI_ISL_1414520, EPI_ISL_1414522, EPI_ISL_1414523, EPI_ISL_1414524, EPI_ISL_1414533, EPI_ISL_1414535, EPI_ISL_1414537, EPI_ISL_1414538, EPI_ISL_1414539, EPI_ISL_1414540, EPI_ISL_1414541, EPI_ISL_1414543, EPI_ISL_1414544, EPI_ISL_1414545, EPI_ISL_1414546, EPI_ISL_1414547, EPI_ISL_1414548, EPI_ISL_1414549, EPI_ISL_1414551, EPI_ISL_1414553, EPI_ISL_1414554, EPI_ISL_1414555, EPI_ISL_1414557, EPI_ISL_1414560, EPI_ISL_1414562, EPI_ISL_1414563, EPI_ISL_1414564, EPI_ISL_1414566, EPI_ISL_1414568, EPI_ISL_1414569, EPI_ISL_1414570, EPI_ISL_1414573, EPI_ISL_1414575, EPI_ISL_1414576, EPI_ISL_1414577, EPI_ISL_1414578, EPI_ISL_1414581, EPI_ISL_1414582, EPI_ISL_1414584, EPI_ISL_1414585, EPI_ISL_1414588, EPI_ISL_1414589, EPI_ISL_1414590, EPI_ISL_1414591, EPI_ISL_1414592, EPI_ISL_1414596, EPI_ISL_1414598, EPI_ISL_1414599, EPI_ISL_1414600, EPI_ISL_1414601, EPI_ISL_1414602, EPI_ISL_1414604, EPI_ISL_1414605, EPI_ISL_1414607, EPI_ISL_1414608, EPI_ISL_1414610, EPI_ISL_1414611, EPI_ISL_1414612, EPI_ISL_1414613, EPI_ISL_1414614, EPI_ISL_1414616, EPI_ISL_1414617, EPI_ISL_1414618, EPI_ISL_1414619, EPI_ISL_1414620, EPI_ISL_1414623, EPI_ISL_1414624, EPI_ISL_1414626, EPI_ISL_1414629, EPI_ISL_1414630, EPI_ISL_1414631, EPI_ISL_1414633, EPI_ISL_1414642, EPI_ISL_1414643, EPI_ISL_1414645, EPI_ISL_1414646, EPI_ISL_1414648, EPI_ISL_1414651, EPI_ISL_1414652, EPI_ISL_1414654, EPI_ISL_1414655, EPI_ISL_1414658, EPI_ISL_1414659, EPI_ISL_1414660, EPI_ISL_1414661, EPI_ISL_1414662, EPI_ISL_1414665, EPI_ISL_1414667, EPI_ISL_1414668, EPI_ISL_1414669, EPI_ISL_1414670, EPI_ISL_1414672, EPI_ISL_1414673, EPI_ISL_1414674, EPI_ISL_1414677, EPI_ISL_1414679, EPI_ISL_1414680, EPI_ISL_1414682, EPI_ISL_1414683, EPI_ISL_1414686, EPI_ISL_1414687, EPI_ISL_1414688, EPI_ISL_1414689, EPI_ISL_1414691, EPI_ISL_1414693, EPI_ISL_1414696, EPI_ISL_1414697, EPI_ISL_1414698, EPI_ISL_1414699, EPI_ISL_1414700, EPI_ISL_1414701, EPI_ISL_1414702, EPI_ISL_1414703, EPI_ISL_1414704, EPI_ISL_1414705, EPI_ISL_1414709, EPI_ISL_1414710, EPI_ISL_1414711, EPI_ISL_1414712, EPI_ISL_1414713, EPI_ISL_1414714, EPI_ISL_1414715, EPI_ISL_1414716, EPI_ISL_1414717, EPI_ISL_1414719, EPI_ISL_1414720, EPI_ISL_1414724, EPI_ISL_1414725, EPI_ISL_1414727, EPI_ISL_1414728, EPI_ISL_1414731, EPI_ISL_1414733, EPI_ISL_1414734, EPI_ISL_1414736, EPI_ISL_1414740, EPI_ISL_1414743, EPI_ISL_1414746, EPI_ISL_1414747, EPI_ISL_1414748, EPI_ISL_1414751, EPI_ISL_1414753, EPI_ISL_1414754, EPI_ISL_1414756, EPI_ISL_1414757, EPI_ISL_1414759, EPI_ISL_1414761, EPI_ISL_1414762, EPI_ISL_1414763, EPI_ISL_1414764, EPI_ISL_1414766, EPI_ISL_1414767, EPI_ISL_1414768, EPI_ISL_1414769, EPI_ISL_1414770, EPI_ISL_1414772, EPI_ISL_1414773, EPI_ISL_1414774, EPI_ISL_1414776, EPI_ISL_1414778, EPI_ISL_1414779, EPI_ISL_1414780, EPI_ISL_1414781, EPI_ISL_1414782, EPI_ISL_1414783, EPI_ISL_1414785, EPI_ISL_1414786, EPI_ISL_1414787, EPI_ISL_1414788, EPI_ISL_1414789, EPI_ISL_1414798, EPI_ISL_1414799, EPI_ISL_1414801, EPI_ISL_1414802, EPI_ISL_1414805, EPI_ISL_1414808, EPI_ISL_1414809, EPI_ISL_1414810, EPI_ISL_1414812, EPI_ISL_1414814, EPI_ISL_1414815, EPI_ISL_1414816, EPI_ISL_1414818, EPI_ISL_1414819, EPI_ISL_1414821, EPI_ISL_1414822, EPI_ISL_1414826, EPI_ISL_1414827, EPI_ISL_1414828, EPI_ISL_1414831, EPI_ISL_1414832, EPI_ISL_1414834, EPI_ISL_1414835, EPI_ISL_1414837, EPI_ISL_1414838, EPI_ISL_1414840, EPI_ISL_1414842, EPI_ISL_1414843, EPI_ISL_1414845, EPI_ISL_1414849, EPI_ISL_1414850, EPI_ISL_1414851, EPI_ISL_1414852, EPI_ISL_1414853, EPI_ISL_1414854, EPI_ISL_1414856, EPI_ISL_1414858, EPI_ISL_1414860, EPI_ISL_1414861, EPI_ISL_1414862, EPI_ISL_1414863, EPI_ISL_1414864, EPI_ISL_1414865, EPI_ISL_1414868, EPI_ISL_1414869, EPI_ISL_1414872, EPI_ISL_1414873, EPI_ISL_1414877, EPI_ISL_1414878, EPI_ISL_1414879, EPI_ISL_1414880, EPI_ISL_1414881, EPI_ISL_1414882, EPI_ISL_1414883, EPI_ISL_1414884, EPI_ISL_1414886, EPI_ISL_1414887, EPI_ISL_1414888, EPI_ISL_1414889, EPI_ISL_1414891, EPI_ISL_1414892, EPI_ISL_1414893, EPI_ISL_1414897, EPI_ISL_1414901, EPI_ISL_1414902, EPI_ISL_1414903, EPI_ISL_1414904, EPI_ISL_1414906, EPI_ISL_1414908, EPI_ISL_1414914, EPI_ISL_1414915, EPI_ISL_1414916, EPI_ISL_1414917, EPI_ISL_1414918, EPI_ISL_1414921, EPI_ISL_1414923, EPI_ISL_1414925, EPI_ISL_1414926, EPI_ISL_1414927, EPI_ISL_1414928, EPI_ISL_1414931, EPI_ISL_1414933, EPI_ISL_1414936, EPI_ISL_1414937, EPI_ISL_1414938, EPI_ISL_1414939, EPI_ISL_1414940, EPI_ISL_1414941, EPI_ISL_1414942, EPI_ISL_1414944, EPI_ISL_1414945, EPI_ISL_1414947, EPI_ISL_1414948, EPI_ISL_1414949, EPI_ISL_1414950, EPI_ISL_1414951, EPI_ISL_1414952, EPI_ISL_1414955, EPI_ISL_1414957, EPI_ISL_1414959, EPI_ISL_1414961, EPI_ISL_1414962, EPI_ISL_1414963, EPI_ISL_1414964, EPI_ISL_1414965, EPI_ISL_1414969, EPI_ISL_1414971, EPI_ISL_1414973, EPI_ISL_1414974, EPI_ISL_1414975, EPI_ISL_1414976, EPI_ISL_1414977, EPI_ISL_1414978, EPI_ISL_1414979, EPI_ISL_1414981, EPI_ISL_1414983, EPI_ISL_1414984, EPI_ISL_1414986, EPI_ISL_1414988, EPI_ISL_1414989, EPI_ISL_1414992, EPI_ISL_1414993, EPI_ISL_1414994, EPI_ISL_1414995, EPI_ISL_1414996, EPI_ISL_1414997, EPI_ISL_1414999, EPI_ISL_1415003, EPI_ISL_1415004, EPI_ISL_1415005, EPI_ISL_1415007, EPI_ISL_1415008, EPI_ISL_1415010, EPI_ISL_1415012, EPI_ISL_1415013, EPI_ISL_1415014, EPI_ISL_1415016, EPI_ISL_1415017, EPI_ISL_1415019, EPI_ISL_1415021, EPI_ISL_1415022, EPI_ISL_1415023, EPI_ISL_1415024, EPI_ISL_1415025, EPI_ISL_1415027, EPI_ISL_1415029, EPI_ISL_1415031, EPI_ISL_1415032, EPI_ISL_1415033, EPI_ISL_1415034, EPI_ISL_1415035, EPI_ISL_1415037, EPI_ISL_1415038, EPI_ISL_1415039, EPI_ISL_1415040, EPI_ISL_1415042, EPI_ISL_1415043, EPI_ISL_1415044, EPI_ISL_1415045, EPI_ISL_1415047, EPI_ISL_1415048, EPI_ISL_1415049, EPI_ISL_1415050, EPI_ISL_1415051, EPI_ISL_1415052, EPI_ISL_1415053, EPI_ISL_1415056, EPI_ISL_1415060, EPI_ISL_1415429, EPI_ISL_1415430, EPI_ISL_1415431, EPI_ISL_1415433, EPI_ISL_1415434, EPI_ISL_1415438, EPI_ISL_1415440, EPI_ISL_1415441, EPI_ISL_1415445, EPI_ISL_1415447, EPI_ISL_1415448, EPI_ISL_1415451, EPI_ISL_1415453, EPI_ISL_1415454, EPI_ISL_1415456, EPI_ISL_1415457, EPI_ISL_1415458, EPI_ISL_1415459, EPI_ISL_1415460, EPI_ISL_1415465, EPI_ISL_1415515, EPI_ISL_1415566, EPI_ISL_1415685, EPI_ISL_1415733, EPI_ISL_1415758, EPI_ISL_1415777, EPI_ISL_1416172, EPI_ISL_1416173, EPI_ISL_1416175, EPI_ISL_1416176, EPI_ISL_1416177, EPI_ISL_1416178, EPI_ISL_1416179, EPI_ISL_1454326, EPI_ISL_1454327, EPI_ISL_1454329, EPI_ISL_1454330, EPI_ISL_1454331, EPI_ISL_1454332, EPI_ISL_1454334, EPI_ISL_1454337, EPI_ISL_1454338, EPI_ISL_1454339, EPI_ISL_1454340, EPI_ISL_1454341, EPI_ISL_1454342, EPI_ISL_1454343, EPI_ISL_1454345, EPI_ISL_1454346, EPI_ISL_1454350, EPI_ISL_1454351, EPI_ISL_1454353, EPI_ISL_1454356, EPI_ISL_1454358, EPI_ISL_1454359, EPI_ISL_1454361, EPI_ISL_1454362, EPI_ISL_1454363, EPI_ISL_1454365, EPI_ISL_1454366, EPI_ISL_1454367, EPI_ISL_1454369, EPI_ISL_1454370, EPI_ISL_1454371, EPI_ISL_1454372, EPI_ISL_1454373, EPI_ISL_1454374, EPI_ISL_1454376, EPI_ISL_1454377, EPI_ISL_1454378, EPI_ISL_1454380, EPI_ISL_1454381, EPI_ISL_1454384, EPI_ISL_1454385, EPI_ISL_1454386, EPI_ISL_1454387, EPI_ISL_1454388, EPI_ISL_1454389, EPI_ISL_1454392, | | | |

EPI_ISL_1454393, EPI_ISL_1454394, EPI_ISL_1454395, EPI_ISL_1454396, EPI_ISL_1454397, EPI_ISL_1454400, EPI_ISL_1454401, EPI_ISL_1454403, EPI_ISL_1454404, EPI_ISL_1454405, EPI_ISL_1454406, EPI_ISL_1454408, EPI_ISL_1454410, EPI_ISL_1454412, EPI_ISL_1454413, EPI_ISL_1454414, EPI_ISL_1454415, EPI_ISL_1454417, EPI_ISL_1454419, EPI_ISL_1454420, EPI_ISL_1454421, EPI_ISL_1454422,
EPI_ISL_1454423, EPI_ISL_1454424, EPI_ISL_1454425, EPI_ISL_1454426, EPI_ISL_1454427, EPI_ISL_1454428, EPI_ISL_1454430, EPI_ISL_1454431, EPI_ISL_1454432, EPI_ISL_1454433, EPI_ISL_1454434, EPI_ISL_1454437, EPI_ISL_1454438, EPI_ISL_1454439, EPI_ISL_1454441, EPI_ISL_1454443, EPI_ISL_1454444, EPI_ISL_1454447, EPI_ISL_1454449, EPI_ISL_1454450, EPI_ISL_1454451, EPI_ISL_1454452,
EPI_ISL_1454455, EPI_ISL_1454457, EPI_ISL_1454459, EPI_ISL_1454460, EPI_ISL_1454461, EPI_ISL_1454462, EPI_ISL_1454463, EPI_ISL_1454464, EPI_ISL_1454465, EPI_ISL_1454469, EPI_ISL_1454470, EPI_ISL_1454472, EPI_ISL_1454473, EPI_ISL_1454476, EPI_ISL_1454477, EPI_ISL_1454478, EPI_ISL_1454480, EPI_ISL_1454481, EPI_ISL_1454482, EPI_ISL_1454484, EPI_ISL_1454486, EPI_ISL_1454487,
EPI_ISL_1454488, EPI_ISL_1454489, EPI_ISL_1454490, EPI_ISL_1454492, EPI_ISL_1454493, EPI_ISL_1454498, EPI_ISL_1454499, EPI_ISL_1454500, EPI_ISL_1454501, EPI_ISL_1454503, EPI_ISL_1454505, EPI_ISL_1454506, EPI_ISL_1454508, EPI_ISL_1454509, EPI_ISL_1454510, EPI_ISL_1454511, EPI_ISL_1454512, EPI_ISL_1454514, EPI_ISL_1454515, EPI_ISL_1454516, EPI_ISL_1454517, EPI_ISL_1454518,
EPI_ISL_1454519, EPI_ISL_1454520, EPI_ISL_1454522, EPI_ISL_1454524, EPI_ISL_1454527, EPI_ISL_1454528, EPI_ISL_1454529, EPI_ISL_1454531, EPI_ISL_1454532, EPI_ISL_1454533, EPI_ISL_1454534, EPI_ISL_1454535, EPI_ISL_1454537, EPI_ISL_1454538, EPI_ISL_1454539, EPI_ISL_1454540, EPI_ISL_1454543, EPI_ISL_1454544, EPI_ISL_1454547, EPI_ISL_1454549, EPI_ISL_1454550, EPI_ISL_1454552,
EPI_ISL_1454554, EPI_ISL_1454555, EPI_ISL_1454556, EPI_ISL_1454557, EPI_ISL_1454560, EPI_ISL_1454561, EPI_ISL_1454562, EPI_ISL_1454564, EPI_ISL_1454565, EPI_ISL_1454566, EPI_ISL_1454567, EPI_ISL_1454569, EPI_ISL_1454570, EPI_ISL_1454571, EPI_ISL_1454573, EPI_ISL_1454574, EPI_ISL_1454576, EPI_ISL_1454577, EPI_ISL_1454579, EPI_ISL_1454580, EPI_ISL_1454581, EPI_ISL_1454583,
EPI_ISL_1454585, EPI_ISL_1454586, EPI_ISL_1454587, EPI_ISL_1454588, EPI_ISL_1454589, EPI_ISL_1454590, EPI_ISL_1454591, EPI_ISL_1454592, EPI_ISL_1454593, EPI_ISL_1454595, EPI_ISL_1454596, EPI_ISL_1454598, EPI_ISL_1454599, EPI_ISL_1454600, EPI_ISL_1454601, EPI_ISL_1454603, EPI_ISL_1454605, EPI_ISL_1454607, EPI_ISL_1454608, EPI_ISL_1454610, EPI_ISL_1454611, EPI_ISL_1454613,
EPI_ISL_1454614, EPI_ISL_1454615, EPI_ISL_1454616, EPI_ISL_1454617, EPI_ISL_1454635, EPI_ISL_1454653, EPI_ISL_1454679, EPI_ISL_1454687, EPI_ISL_1454735, EPI_ISL_1454738, EPI_ISL_1454749, EPI_ISL_1454770, EPI_ISL_1454786, EPI_ISL_1454808, EPI_ISL_1454810, EPI_ISL_1454812, EPI_ISL_1454821, EPI_ISL_1454824, EPI_ISL_1454832, EPI_ISL_1454837, EPI_ISL_1454859, EPI_ISL_1454860,
EPI_ISL_1454887, EPI_ISL_1454909, EPI_ISL_1454919, EPI_ISL_1454920, EPI_ISL_1454921, EPI_ISL_1454924, EPI_ISL_1454946, EPI_ISL_1454969, EPI_ISL_1454971, EPI_ISL_1455012, EPI_ISL_1455014, EPI_ISL_1455016, EPI_ISL_1455017, EPI_ISL_1455018, EPI_ISL_1455019, EPI_ISL_1455022, EPI_ISL_1455028, EPI_ISL_1455029, EPI_ISL_1455035, EPI_ISL_1455040, EPI_ISL_1455046, EPI_ISL_1455047,
EPI_ISL_1455051, EPI_ISL_1455053, EPI_ISL_1455055, EPI_ISL_1455056, EPI_ISL_1455060, EPI_ISL_1455067, EPI_ISL_1455070, EPI_ISL_1455072, EPI_ISL_1455073, EPI_ISL_1455075, EPI_ISL_1455079, EPI_ISL_1455080, EPI_ISL_1455081, EPI_ISL_1455084, EPI_ISL_1455085, EPI_ISL_1455086, EPI_ISL_1455090, EPI_ISL_1455092, EPI_ISL_1455094, EPI_ISL_1455095, EPI_ISL_1455099, EPI_ISL_1455100,
EPI_ISL_1455101, EPI_ISL_1455102, EPI_ISL_1455103, EPI_ISL_1455104, EPI_ISL_1455105, EPI_ISL_1455109, EPI_ISL_1455111, EPI_ISL_1455112, EPI_ISL_1455113, EPI_ISL_1455114, EPI_ISL_1455115, EPI_ISL_1455117, EPI_ISL_1455119, EPI_ISL_1455120, EPI_ISL_1455122, EPI_ISL_1455124, EPI_ISL_1455126, EPI_ISL_1455127, EPI_ISL_1455128, EPI_ISL_1455129, EPI_ISL_1455132, EPI_ISL_1455134,
EPI_ISL_1455135, EPI_ISL_1455136, EPI_ISL_1455137, EPI_ISL_1455138, EPI_ISL_1455139, EPI_ISL_1455142, EPI_ISL_1455144, EPI_ISL_1455147, EPI_ISL_1455148, EPI_ISL_1455150, EPI_ISL_1455151, EPI_ISL_1455152, EPI_ISL_1455153, EPI_ISL_1455158, EPI_ISL_1455161, EPI_ISL_1455162, EPI_ISL_1455163, EPI_ISL_1455165, EPI_ISL_1455167, EPI_ISL_1455168, EPI_ISL_1455170, EPI_ISL_1455173,
EPI_ISL_1455174, EPI_ISL_1455175, EPI_ISL_1455177, EPI_ISL_1455180, EPI_ISL_1455183, EPI_ISL_1455186, EPI_ISL_1455187, EPI_ISL_1455189, EPI_ISL_1455190, EPI_ISL_1455191, EPI_ISL_1455197, EPI_ISL_1455201, EPI_ISL_1455202, EPI_ISL_1455204, EPI_ISL_1455205, EPI_ISL_1455207, EPI_ISL_1455209, EPI_ISL_1455214, EPI_ISL_1455215, EPI_ISL_1455216, EPI_ISL_1455222, EPI_ISL_1455223,
EPI_ISL_1455224, EPI_ISL_1455225, EPI_ISL_1455226, EPI_ISL_1455228, EPI_ISL_1455231, EPI_ISL_1455236, EPI_ISL_1455265, EPI_ISL_1455279, EPI_ISL_1455323, EPI_ISL_1455414, EPI_ISL_1455492, EPI_ISL_1455496, EPI_ISL_1455499, EPI_ISL_1455500, EPI_ISL_1455501, EPI_ISL_1455502, EPI_ISL_1455505, EPI_ISL_1455507, EPI_ISL_1455508, EPI_ISL_1455509, EPI_ISL_1455510, EPI_ISL_1455511,
EPI_ISL_1455512, EPI_ISL_1455514, EPI_ISL_1455519, EPI_ISL_1455521, EPI_ISL_1455523, EPI_ISL_1455524, EPI_ISL_1455525, EPI_ISL_1455526, EPI_ISL_1455527, EPI_ISL_1455528, EPI_ISL_1455529, EPI_ISL_1455530, EPI_ISL_1455533

| | | | |
|---|---|---|---|
| see above | Lighthouse Lab in Alderley Park | Wellcome Sanger Institute for the COVID-19 Genomics UK (COG-UK) Consortium | Cordelia Langford; David K. Jackson; Dominic Kwiatkowski; Ewan Harrison; Ian Johnston; Jacquelyn Wynn; Jeffrey Barrett; John Sillitoe on behalf of the Wellcome Sanger Institute COVID-19 Surveillance Team; Mairead Hyland; Roberto Amato; Sonia Goncalves; The Lighthouse Lab in Alderley Park and Alex Alderton |

EPI_ISL_1275802, EPI_ISL_1315643, EPI_ISL_1315644, EPI_ISL_1315646, EPI_ISL_1315647, EPI_ISL_1315648, EPI_ISL_1315649, EPI_ISL_1315650, EPI_ISL_1315651, EPI_ISL_1315652, EPI_ISL_1316012, EPI_ISL_1326689, EPI_ISL_1326758, EPI_ISL_1326834, EPI_ISL_1326841, EPI_ISL_1326873, EPI_ISL_1326899, EPI_ISL_1326944, EPI_ISL_1326947, EPI_ISL_1327062, EPI_ISL_1327088, EPI_ISL_1327103,
EPI_ISL_1327110, EPI_ISL_1327118, EPI_ISL_1327150, EPI_ISL_1327167, EPI_ISL_1327202, EPI_ISL_1327241, EPI_ISL_1327244, EPI_ISL_1327245, EPI_ISL_1327246, EPI_ISL_1327248, EPI_ISL_1327249, EPI_ISL_1327250, EPI_ISL_1327252, EPI_ISL_1327254, EPI_ISL_1327257, EPI_ISL_1327258, EPI_ISL_1327259, EPI_ISL_1327261, EPI_ISL_1327265, EPI_ISL_1327266, EPI_ISL_1327268, EPI_ISL_1327271,
EPI_ISL_1327273, EPI_ISL_1327276, EPI_ISL_1327277, EPI_ISL_1327278, EPI_ISL_1327280, EPI_ISL_1327281, EPI_ISL_1327282, EPI_ISL_1327283, EPI_ISL_1327285, EPI_ISL_1327286, EPI_ISL_1327292, EPI_ISL_1327293, EPI_ISL_1327296, EPI_ISL_1327298, EPI_ISL_1327299, EPI_ISL_1327300, EPI_ISL_1327301, EPI_ISL_1327307, EPI_ISL_1327308, EPI_ISL_1327309, EPI_ISL_1327310, EPI_ISL_1327311,
EPI_ISL_1327312, EPI_ISL_1327313, EPI_ISL_1327314, EPI_ISL_1327315, EPI_ISL_1327316, EPI_ISL_1327317, EPI_ISL_1327318, EPI_ISL_1327319, EPI_ISL_1327320, EPI_ISL_1327322, EPI_ISL_1327323, EPI_ISL_1327326, EPI_ISL_1327327, EPI_ISL_1327329, EPI_ISL_1327330, EPI_ISL_1327333, EPI_ISL_1327334, EPI_ISL_1327335, EPI_ISL_1327338, EPI_ISL_1327340, EPI_ISL_1327342, EPI_ISL_1327344,
EPI_ISL_1327345, EPI_ISL_1327347, EPI_ISL_1327348, EPI_ISL_1327350, EPI_ISL_1327356, EPI_ISL_1327357, EPI_ISL_1327358, EPI_ISL_1327359, EPI_ISL_1327361, EPI_ISL_1327362, EPI_ISL_1327364, EPI_ISL_1327365, EPI_ISL_1327368, EPI_ISL_1327370, EPI_ISL_1327372, EPI_ISL_1327378, EPI_ISL_1327379, EPI_ISL_1327380, EPI_ISL_1327382, EPI_ISL_1327383, EPI_ISL_1327385, EPI_ISL_1327387,
EPI_ISL_1327388, EPI_ISL_1327390, EPI_ISL_1327391, EPI_ISL_1327395, EPI_ISL_1327398, EPI_ISL_1327401, EPI_ISL_1327403, EPI_ISL_1327404, EPI_ISL_1327407, EPI_ISL_1327408, EPI_ISL_1327409, EPI_ISL_1327410, EPI_ISL_1327411, EPI_ISL_1327413, EPI_ISL_1327414, EPI_ISL_1327415, EPI_ISL_1327416, EPI_ISL_1327417, EPI_ISL_1327418, EPI_ISL_1327421, EPI_ISL_1327426, EPI_ISL_1327427,
EPI_ISL_1327428, EPI_ISL_1327430, EPI_ISL_1327431, EPI_ISL_1327432, EPI_ISL_1327434, EPI_ISL_1327438, EPI_ISL_1327439, EPI_ISL_1327440, EPI_ISL_1327441, EPI_ISL_1327442, EPI_ISL_1327444, EPI_ISL_1327445, EPI_ISL_1327446, EPI_ISL_1327447, EPI_ISL_1327453, EPI_ISL_1327455, EPI_ISL_1327456, EPI_ISL_1327458, EPI_ISL_1327460, EPI_ISL_1327468, EPI_ISL_1327470, EPI_ISL_1327471,
EPI_ISL_1327472, EPI_ISL_1327475, EPI_ISL_1327476, EPI_ISL_1327477, EPI_ISL_1327478, EPI_ISL_1327481, EPI_ISL_1327484, EPI_ISL_1327486, EPI_ISL_1327489, EPI_ISL_1327490, EPI_ISL_1327492, EPI_ISL_1327493, EPI_ISL_1327494, EPI_ISL_1327495, EPI_ISL_1327497, EPI_ISL_1327499, EPI_ISL_1327500, EPI_ISL_1327501, EPI_ISL_1327505, EPI_ISL_1327507, EPI_ISL_1327508, EPI_ISL_1327510,
EPI_ISL_1327511, EPI_ISL_1327513, EPI_ISL_1327514, EPI_ISL_1327515, EPI_ISL_1327516, EPI_ISL_1327520, EPI_ISL_1327521, EPI_ISL_1327524, EPI_ISL_1327525, EPI_ISL_1327526, EPI_ISL_1327528, EPI_ISL_1327529, EPI_ISL_1327531, EPI_ISL_1327532, EPI_ISL_1327533, EPI_ISL_1327534, EPI_ISL_1327535, EPI_ISL_1327538, EPI_ISL_1327539, EPI_ISL_1327540, EPI_ISL_1327545, EPI_ISL_1327548,
EPI_ISL_1327553, EPI_ISL_1327555, EPI_ISL_1327556, EPI_ISL_1327558, EPI_ISL_1327561, EPI_ISL_1327562, EPI_ISL_1327563, EPI_ISL_1327564, EPI_ISL_1327565, EPI_ISL_1327566, EPI_ISL_1327567, EPI_ISL_1327568, EPI_ISL_1327569, EPI_ISL_1327570, EPI_ISL_1327572, EPI_ISL_1327573, EPI_ISL_1327575, EPI_ISL_1327576, EPI_ISL_1327577, EPI_ISL_1327578, EPI_ISL_1327579, EPI_ISL_1327581,
EPI_ISL_1327582, EPI_ISL_1327583, EPI_ISL_1327584, EPI_ISL_1327585, EPI_ISL_1327587, EPI_ISL_1327590, EPI_ISL_1327591, EPI_ISL_1327592, EPI_ISL_1327593, EPI_ISL_1327594, EPI_ISL_1327596, EPI_ISL_1327597, EPI_ISL_1327602, EPI_ISL_1327603, EPI_ISL_1327605, EPI_ISL_1327606, EPI_ISL_1327607, EPI_ISL_1327608, EPI_ISL_1327609, EPI_ISL_1327613, EPI_ISL_1327614, EPI_ISL_1327615,
EPI_ISL_1327616, EPI_ISL_1327617, EPI_ISL_1327618, EPI_ISL_1327619, EPI_ISL_1327620, EPI_ISL_1327621, EPI_ISL_1327622, EPI_ISL_1327624, EPI_ISL_1327628, EPI_ISL_1327630, EPI_ISL_1327631, EPI_ISL_1327634, EPI_ISL_1327635, EPI_ISL_1327636, EPI_ISL_1327638, EPI_ISL_1327639, EPI_ISL_1327640, EPI_ISL_1327646, EPI_ISL_1327647, EPI_ISL_1327648, EPI_ISL_1327650, EPI_ISL_1327651,
EPI_ISL_1327652, EPI_ISL_1327653, EPI_ISL_1327654, EPI_ISL_1327655, EPI_ISL_1327656, EPI_ISL_1327657, EPI_ISL_1327658, EPI_ISL_1327660, EPI_ISL_1327662, EPI_ISL_1327663, EPI_ISL_1327664, EPI_ISL_1327665, EPI_ISL_1327668, EPI_ISL_1327670, EPI_ISL_1327671, EPI_ISL_1327672, EPI_ISL_1327673, EPI_ISL_1327674, EPI_ISL_1327675, EPI_ISL_1327677, EPI_ISL_1327679, EPI_ISL_1327680,
EPI_ISL_1327682, EPI_ISL_1327684, EPI_ISL_1327685, EPI_ISL_1327686, EPI_ISL_1327687, EPI_ISL_1327689, EPI_ISL_1327691, EPI_ISL_1327693, EPI_ISL_1327694, EPI_ISL_1327695, EPI_ISL_1327696, EPI_ISL_1327698, EPI_ISL_1327699, EPI_ISL_1327700, EPI_ISL_1327701, EPI_ISL_1329205, EPI_ISL_1329209, EPI_ISL_1329210, EPI_ISL_1329212, EPI_ISL_1329214, EPI_ISL_1329215, EPI_ISL_1329217,
EPI_ISL_1329218, EPI_ISL_1329223, EPI_ISL_1329226, EPI_ISL_1329230, EPI_ISL_1329232, EPI_ISL_1329234, EPI_ISL_1329236, EPI_ISL_1329238, EPI_ISL_1329241, EPI_ISL_1329242, EPI_ISL_1329243, EPI_ISL_1329247, EPI_ISL_1329250, EPI_ISL_1329251, EPI_ISL_1329258, EPI_ISL_1329261, EPI_ISL_1329264, EPI_ISL_1329265, EPI_ISL_1329266, EPI_ISL_1329268, EPI_ISL_1329270, EPI_ISL_1329271,
EPI_ISL_1329272, EPI_ISL_1329276, EPI_ISL_1329279, EPI_ISL_1329281, EPI_ISL_1329282, EPI_ISL_1329287, EPI_ISL_1329288, EPI_ISL_1329290, EPI_ISL_1329295, EPI_ISL_1329297, EPI_ISL_1329298, EPI_ISL_1329299, EPI_ISL_1329300, EPI_ISL_1329302, EPI_ISL_1329303, EPI_ISL_1329305, EPI_ISL_1329311, EPI_ISL_1329312, EPI_ISL_1329314, EPI_ISL_1329316, EPI_ISL_1329318, EPI_ISL_1329320,
EPI_ISL_1329321, EPI_ISL_1329324, EPI_ISL_1329325, EPI_ISL_1329327, EPI_ISL_1329328, EPI_ISL_1329329, EPI_ISL_1329334, EPI_ISL_1329339, EPI_ISL_1329344, EPI_ISL_1329347, EPI_ISL_1329348, EPI_ISL_1329350, EPI_ISL_1329352, EPI_ISL_1329354, EPI_ISL_1329362, EPI_ISL_1329363, EPI_ISL_1329364, EPI_ISL_1329375, EPI_ISL_1329378, EPI_ISL_1329379, EPI_ISL_1329380, EPI_ISL_1329381,
EPI_ISL_1330301, EPI_ISL_1330309, EPI_ISL_1330311, EPI_ISL_1330312, EPI_ISL_1330319, EPI_ISL_1330320, EPI_ISL_1330321, EPI_ISL_1330330, EPI_ISL_1330331, EPI_ISL_1330342, EPI_ISL_1330349, EPI_ISL_1330358, EPI_ISL_1330361, EPI_ISL_1330367, EPI_ISL_1330373, EPI_ISL_1330375, EPI_ISL_1330380, EPI_ISL_1330404, EPI_ISL_1330410, EPI_ISL_1330411, EPI_ISL_1330412, EPI_ISL_1330415,
EPI_ISL_1330452, EPI_ISL_1330459, EPI_ISL_1330461, EPI_ISL_1330469, EPI_ISL_1330472, EPI_ISL_1330487, EPI_ISL_1330488, EPI_ISL_1330498, EPI_ISL_1330504, EPI_ISL_1330518, EPI_ISL_1330522, EPI_ISL_1330534, EPI_ISL_1330540, EPI_ISL_1330549, EPI_ISL_1330551, EPI_ISL_1330568, EPI_ISL_1330569, EPI_ISL_1330573, EPI_ISL_1330582, EPI_ISL_1330593, EPI_ISL_1330598, EPI_ISL_1330606,
EPI_ISL_1330608, EPI_ISL_1332559, EPI_ISL_1332561, EPI_ISL_1332564, EPI_ISL_1332567, EPI_ISL_1332573, EPI_ISL_1332576, EPI_ISL_1332588, EPI_ISL_1332589, EPI_ISL_1332592, EPI_ISL_1332594, EPI_ISL_1332596, EPI_ISL_1332598, EPI_ISL_1332604, EPI_ISL_1332607, EPI_ISL_1332610, EPI_ISL_1332611, EPI_ISL_1332613, EPI_ISL_1332618, EPI_ISL_1332621, EPI_ISL_1332623, EPI_ISL_1332626,
EPI_ISL_1332636, EPI_ISL_1332637, EPI_ISL_1332639, EPI_ISL_1332641, EPI_ISL_1332642, EPI_ISL_1332643, EPI_ISL_1332645, EPI_ISL_1332648, EPI_ISL_1332928, EPI_ISL_1333975, EPI_ISL_1344517, EPI_ISL_1344623, EPI_ISL_1344675, EPI_ISL_1344693, EPI_ISL_1344704, EPI_ISL_1346286, EPI_ISL_1346291, EPI_ISL_1346308, EPI_ISL_1346310, EPI_ISL_1346313, EPI_ISL_1346317, EPI_ISL_1346318,
EPI_ISL_1346320, EPI_ISL_1346321, EPI_ISL_1346322, EPI_ISL_1346323, EPI_ISL_1346324, EPI_ISL_1346325, EPI_ISL_1346326, EPI_ISL_1346328, EPI_ISL_1346370, EPI_ISL_1346372, EPI_ISL_1346374, EPI_ISL_1346478, EPI_ISL_1374159, EPI_ISL_1374183, EPI_ISL_1377048, EPI_ISL_1377093, EPI_ISL_1377162, EPI_ISL_1409500, EPI_ISL_1409501, EPI_ISL_1409506, EPI_ISL_1409508, EPI_ISL_1409517,
EPI_ISL_1409520, EPI_ISL_1409526, EPI_ISL_1409535, EPI_ISL_1409537, EPI_ISL_1409541, EPI_ISL_1409546, EPI_ISL_1409559, EPI_ISL_1409560, EPI_ISL_1409561, EPI_ISL_1409562, EPI_ISL_1409563, EPI_ISL_1409566, EPI_ISL_1409582, EPI_ISL_1409585, EPI_ISL_1409587, EPI_ISL_1409598, EPI_ISL_1409601, EPI_ISL_1409608, EPI_ISL_1409610, EPI_ISL_1409618, EPI_ISL_1409619, EPI_ISL_1409657,
EPI_ISL_1409661, EPI_ISL_1409673, EPI_ISL_1409674, EPI_ISL_1409677, EPI_ISL_1409682, EPI_ISL_1409683, EPI_ISL_1409690, EPI_ISL_1409693, EPI_ISL_1409697, EPI_ISL_1409702, EPI_ISL_1409705, EPI_ISL_1409713, EPI_ISL_1409715, EPI_ISL_1409721, EPI_ISL_1409722, EPI_ISL_1409732, EPI_ISL_1409742, EPI_ISL_1409746, EPI_ISL_1409760, EPI_ISL_1409763, EPI_ISL_1409766, EPI_ISL_1410164,
EPI_ISL_1410274, EPI_ISL_1412927, EPI_ISL_1412931, EPI_ISL_1412933, EPI_ISL_1412942, EPI_ISL_1412945, EPI_ISL_1412948, EPI_ISL_1412949, EPI_ISL_1412953, EPI_ISL_1412955, EPI_ISL_1412956, EPI_ISL_1412960, EPI_ISL_1412962, EPI_ISL_1412966, EPI_ISL_1412967, EPI_ISL_1412969, EPI_ISL_1412970, EPI_ISL_1412971, EPI_ISL_1412978, EPI_ISL_1412979, EPI_ISL_1412981, EPI_ISL_1412987,
EPI_ISL_1412989, EPI_ISL_1412995, EPI_ISL_1413005, EPI_ISL_1413007, EPI_ISL_1413008, EPI_ISL_1413013, EPI_ISL_1413015, EPI_ISL_1413016, EPI_ISL_1413018, EPI_ISL_1413027, EPI_ISL_1413028, EPI_ISL_1413029, EPI_ISL_1413032, EPI_ISL_1413033, EPI_ISL_1413038, EPI_ISL_1413039, EPI_ISL_1413043, EPI_ISL_1413044, EPI_ISL_1413046, EPI_ISL_1413051, EPI_ISL_1413053, EPI_ISL_1413054,
EPI_ISL_1413055, EPI_ISL_1413056, EPI_ISL_1413057, EPI_ISL_1413059, EPI_ISL_1416184, EPI_ISL_1473599, EPI_ISL_1473625, EPI_ISL_1473636, EPI_ISL_1473694, EPI_ISL_1473754, EPI_ISL_1473758, EPI_ISL_1473774, EPI_ISL_1473794, EPI_ISL_1484323, EPI_ISL_1484332, EPI_ISL_1484337, EPI_ISL_1484338, EPI_ISL_1484343, EPI_ISL_1484346, EPI_ISL_1484358, EPI_ISL_1484370, EPI_ISL_1484373,
EPI_ISL_1484386, EPI_ISL_1484397, EPI_ISL_1484404, EPI_ISL_1484411, EPI_ISL_1484413, EPI_ISL_1484422, EPI_ISL_1484434, EPI_ISL_1484436, EPI_ISL_1484440, EPI_ISL_1484443, EPI_ISL_1484445, EPI_ISL_1484448, EPI_ISL_1484457, EPI_ISL_1484467, EPI_ISL_1484474, EPI_ISL_1484477, EPI_ISL_1484478, EPI_ISL_1484494, EPI_ISL_1484499, EPI_ISL_1484502, EPI_ISL_1484503, EPI_ISL_1484504,
EPI_ISL_1484516, EPI_ISL_1484518, EPI_ISL_1484519, EPI_ISL_1484522, EPI_ISL_1484535, EPI_ISL_1484537, EPI_ISL_1484540, EPI_ISL_1484544, EPI_ISL_1484548, EPI_ISL_1484550, EPI_ISL_1484551, EPI_ISL_1484554, EPI_ISL_1484556, EPI_ISL_1484559, EPI_ISL_1484561, EPI_ISL_1484564, EPI_ISL_1484566, EPI_ISL_1484567, EPI_ISL_1484568, EPI_ISL_1484571, EPI_ISL_1484574, EPI_ISL_1484578,
EPI_ISL_1484586, EPI_ISL_1484588, EPI_ISL_1484603, EPI_ISL_1484607, EPI_ISL_1484609

| | | | |
|---|---|---|---|
| see above | Lighthouse Lab in Cambridge | Wellcome Sanger Institute for the COVID-19 Genomics UK (COG-UK) Consortium | Cordelia Langford; David K. Jackson; Dominic Kwiatkowski; Ewan Harrison; Ian Johnston; Jeffrey Barrett; John Sillitoe on behalf of the Wellcome Sanger Institute COVID-19 Surveillance Team; Rob Howes; Roberto Amato; Sonia Goncalves; The Lighthouse Lab in Cambridge and Alex Alderton |

EPI_ISL_1275015, EPI_ISL_1275019, EPI_ISL_1275026, EPI_ISL_1275029, EPI_ISL_1275042, EPI_ISL_1275052, EPI_ISL_1275053, EPI_ISL_1275055, EPI_ISL_1275056, EPI_ISL_1275061, EPI_ISL_1275062, EPI_ISL_1275074, EPI_ISL_1275076, EPI_ISL_1275078, EPI_ISL_1275080, EPI_ISL_1275081, EPI_ISL_1275085, EPI_ISL_1275089, EPI_ISL_1275091, EPI_ISL_1275095, EPI_ISL_1275099, EPI_ISL_1275102,
EPI_ISL_1275103, EPI_ISL_1275106, EPI_ISL_1275107, EPI_ISL_1275343, EPI_ISL_1275344, EPI_ISL_1275345, EPI_ISL_1275346, EPI_ISL_1275352, EPI_ISL_1275355, EPI_ISL_1275356, EPI_ISL_1275357, EPI_ISL_1275360, EPI_ISL_1275364, EPI_ISL_1275368, EPI_ISL_1275370, EPI_ISL_1275371, EPI_ISL_1275372, EPI_ISL_1275374, EPI_ISL_1275375, EPI_ISL_1275381, EPI_ISL_1275386, EPI_ISL_1275387,
EPI_ISL_1275389, EPI_ISL_1275391, EPI_ISL_1275393, EPI_ISL_1275400, EPI_ISL_1275404, EPI_ISL_1275406, EPI_ISL_1275407, EPI_ISL_1275409, EPI_ISL_1275413, EPI_ISL_1275418, EPI_ISL_1275419, EPI_ISL_1275422, EPI_ISL_1275424, EPI_ISL_1275425, EPI_ISL_1275429, EPI_ISL_1275432, EPI_ISL_1275433, EPI_ISL_1275434, EPI_ISL_1275438, EPI_ISL_1275441, EPI_ISL_1275443, EPI_ISL_1275445,
EPI_ISL_1275455, EPI_ISL_1275457, EPI_ISL_1275458, EPI_ISL_1275459, EPI_ISL_1275460, EPI_ISL_1275467, EPI_ISL_1275472, EPI_ISL_1275473, EPI_ISL_1275477, EPI_ISL_1275479, EPI_ISL_1275480, EPI_ISL_1275481, EPI_ISL_1275482, EPI_ISL_1275484, EPI_ISL_1275486, EPI_ISL_1275492, EPI_ISL_1275493, EPI_ISL_1275500, EPI_ISL_1275501, EPI_ISL_1275504, EPI_ISL_1275510, EPI_ISL_1275517,
EPI_ISL_1275519, EPI_ISL_1275523, EPI_ISL_1275524, EPI_ISL_1275526, EPI_ISL_1275530, EPI_ISL_1275532, EPI_ISL_1275538, EPI_ISL_1275542, EPI_ISL_1275547, EPI_ISL_1275550, EPI_ISL_1275551, EPI_ISL_1275555, EPI_ISL_1293357, EPI_ISL_1293358, EPI_ISL_1293359, EPI_ISL_1293360, EPI_ISL_1293361, EPI_ISL_1293362, EPI_ISL_1293363, EPI_ISL_1293364, EPI_ISL_1293365, EPI_ISL_1293366,
EPI_ISL_1293367, EPI_ISL_1293368, EPI_ISL_1293369, EPI_ISL_1293370, EPI_ISL_1293371, EPI_ISL_1293372, EPI_ISL_1293373, EPI_ISL_1293374, EPI_ISL_1293375, EPI_ISL_1293376, EPI_ISL_1293378, EPI_ISL_1293379, EPI_ISL_1293380, EPI_ISL_1293381, EPI_ISL_1293382, EPI_ISL_1293383, EPI_ISL_1293384, EPI_ISL_1293385, EPI_ISL_1293386, EPI_ISL_1293387, EPI_ISL_1293388, EPI_ISL_1293389,
EPI_ISL_1293390, EPI_ISL_1293391, EPI_ISL_1293392, EPI_ISL_1293393, EPI_ISL_1293394, EPI_ISL_1293395, EPI_ISL_1293396, EPI_ISL_1293397, EPI_ISL_1293398, EPI_ISL_1293399, EPI_ISL_1293400, EPI_ISL_1293401, EPI_ISL_1293402, EPI_ISL_1293404, EPI_ISL_1293405, EPI_ISL_1293406, EPI_ISL_1293407, EPI_ISL_1293408, EPI_ISL_1293409, EPI_ISL_1293410, EPI_ISL_1293411, EPI_ISL_1293412,
EPI_ISL_1293413, EPI_ISL_1293414, EPI_ISL_1293415, EPI_ISL_1293416, EPI_ISL_1293417, EPI_ISL_1293418, EPI_ISL_1293419, EPI_ISL_1293420, EPI_ISL_1293421, EPI_ISL_1293423, EPI_ISL_1293424, EPI_ISL_1293425, EPI_ISL_1293426, EPI_ISL_1293427, EPI_ISL_1293428, EPI_ISL_1293429, EPI_ISL_1293430, EPI_ISL_1293431, EPI_ISL_1293432, EPI_ISL_1293433, EPI_ISL_1293434, EPI_ISL_1293435,
EPI_ISL_1293436, EPI_ISL_1293437, EPI_ISL_1293438, EPI_ISL_1293439, EPI_ISL_1293440, EPI_ISL_1293441, EPI_ISL_1293442, EPI_ISL_1293443, EPI_ISL_1293444, EPI_ISL_1293445, EPI_ISL_1293446, EPI_ISL_1293447, EPI_ISL_1293448, EPI_ISL_1293449, EPI_ISL_1293450, EPI_ISL_1293451, EPI_ISL_1293452, EPI_ISL_1293453, EPI_ISL_1293454, EPI_ISL_1293455, EPI_ISL_1293456, EPI_ISL_1293457,
EPI_ISL_1293458, EPI_ISL_1293460, EPI_ISL_1293461, EPI_ISL_1293462, EPI_ISL_1293463, EPI_ISL_1293464, EPI_ISL_1293465, EPI_ISL_1293466, EPI_ISL_1293467, EPI_ISL_1293468, EPI_ISL_1293469, EPI_ISL_1293470, EPI_ISL_1293471, EPI_ISL_1293473, EPI_ISL_1293474, EPI_ISL_1293475, EPI_ISL_1293476, EPI_ISL_1293477, EPI_ISL_1293478, EPI_ISL_1293479, EPI_ISL_1293480, EPI_ISL_1293481,
EPI_ISL_1293482, EPI_ISL_1293483, EPI_ISL_1293484, EPI_ISL_1293485, EPI_ISL_1293486, EPI_ISL_1293488, EPI_ISL_1293489, EPI_ISL_1293490, EPI_ISL_1293491, EPI_ISL_1293492, EPI_ISL_1293493, EPI_ISL_1293494, EPI_ISL_1293495, EPI_ISL_1293496, EPI_ISL_1293497, EPI_ISL_1293498, EPI_ISL_1293499, EPI_ISL_1293500, EPI_ISL_1293501, EPI_ISL_1293502, EPI_ISL_1293503, EPI_ISL_1293504,
EPI_ISL_1293505, EPI_ISL_1293506, EPI_ISL_1293507, EPI_ISL_1293508, EPI_ISL_1293509, EPI_ISL_1293510, EPI_ISL_1293511, EPI_ISL_1293512, EPI_ISL_1293513, EPI_ISL_1293514, EPI_ISL_1293515, EPI_ISL_1293516, EPI_ISL_1293517, EPI_ISL_1293518, EPI_ISL_1293519, EPI_ISL_1293520, EPI_ISL_1293521, EPI_ISL_1293522, EPI_ISL_1293523, EPI_ISL_1293524, EPI_ISL_1293525, EPI_ISL_1293526,
EPI_ISL_1293527, EPI_ISL_1293528, EPI_ISL_1293529, EPI_ISL_1293530, EPI_ISL_1293531, EPI_ISL_1293532, EPI_ISL_1293533, EPI_ISL_1293534, EPI_ISL_1293535, EPI_ISL_1293536, EPI_ISL_1293537, EPI_ISL_1293538, EPI_ISL_1293539, EPI_ISL_1293540, EPI_ISL_1293541, EPI_ISL_1293542, EPI_ISL_1293543, EPI_ISL_1293544, EPI_ISL_1293545, EPI_ISL_1293546, EPI_ISL_1293547, EPI_ISL_1293548,
EPI_ISL_1293549, EPI_ISL_1293550, EPI_ISL_1293551, EPI_ISL_1293552, EPI_ISL_1293553, EPI_ISL_1293554, EPI_ISL_1293555, EPI_ISL_1293556, EPI_ISL_1293557, EPI_ISL_1293558, EPI_ISL_1293559, EPI_ISL_1293560, EPI_ISL_1293561, EPI_ISL_1293562, EPI_ISL_1293563, EPI_ISL_1293564, EPI_ISL_1293566, EPI_ISL_1293567, EPI_ISL_1293568, EPI_ISL_1293569, EPI_ISL_1293570, EPI_ISL_1293571,
EPI_ISL_1293572, EPI_ISL_1293573, EPI_ISL_1293574, EPI_ISL_1293575, EPI_ISL_1293576, EPI_ISL_1293577, EPI_ISL_1293578, EPI_ISL_1293579, EPI_ISL_1293580, EPI_ISL_1293581, EPI_ISL_1293582, EPI_ISL_1293583, EPI_ISL_1293584, EPI_ISL_1293585, EPI_ISL_1293586, EPI_ISL_1293587, EPI_ISL_1293588, EPI_ISL_1293589, EPI_ISL_1293590, EPI_ISL_1293591, EPI_ISL_1293592, EPI_ISL_1293593,
EPI_ISL_1293594, EPI_ISL_1293595, EPI_ISL_1293596, EPI_ISL_1293597, EPI_ISL_1293598, EPI_ISL_1293599, EPI_ISL_1293600, EPI_ISL_1293601, EPI_ISL_1293602, EPI_ISL_1293603, EPI_ISL_1293604, EPI_ISL_1293606, EPI_ISL_1293607, EPI_ISL_1293608, EPI_ISL_1293610, EPI_ISL_1293611, EPI_ISL_1293612, EPI_ISL_1293613, EPI_ISL_1293614, EPI_ISL_1293615, EPI_ISL_1293616, EPI_ISL_1293617,
EPI_ISL_1293618, EPI_ISL_1293619, EPI_ISL_1293620, EPI_ISL_1293621, EPI_ISL_1293622, EPI_ISL_1293623, EPI_ISL_1293625, EPI_ISL_1293626, EPI_ISL_1293627, EPI_ISL_1293628, EPI_ISL_1293629, EPI_ISL_1293630, EPI_ISL_1293631, EPI_ISL_1293632, EPI_ISL_1293634, EPI_ISL_1293637, EPI_ISL_1293638, EPI_ISL_1293643, EPI_ISL_1293648, EPI_ISL_1293653, EPI_ISL_1293656, EPI_ISL_1293657,
EPI_ISL_1293659, EPI_ISL_1293661, EPI_ISL_1293672, EPI_ISL_1293677, EPI_ISL_1293679, EPI_ISL_1293682, EPI_ISL_1293684, EPI_ISL_1293687, EPI_ISL_1293692, EPI_ISL_1293695, EPI_ISL_1293697, EPI_ISL_1293705, EPI_ISL_1293707, EPI_ISL_1293711, EPI_ISL_1293714, EPI_ISL_1293716, EPI_ISL_1293717, EPI_ISL_1293721, EPI_ISL_1293727, EPI_ISL_1293728, EPI_ISL_1293729, EPI_ISL_1293730,
EPI_ISL_1293731, EPI_ISL_1293732, EPI_ISL_1293734, EPI_ISL_1293737, EPI_ISL_1293743, EPI_ISL_1293746, EPI_ISL_1293748, EPI_ISL_1293750, EPI_ISL_1293752, EPI_ISL_1293759, EPI_ISL_1293761, EPI_ISL_1293764, EPI_ISL_1293766, EPI_ISL_1293767, EPI_ISL_1293769, EPI_ISL_1293770, EPI_ISL_1293772, EPI_ISL_1293774, EPI_ISL_1293776, EPI_ISL_1293777, EPI_ISL_1293778, EPI_ISL_1293780,
EPI_ISL_1293782, EPI_ISL_1294144, EPI_ISL_1294146, EPI_ISL_1294148, EPI_ISL_1294149, EPI_ISL_1294150, EPI_ISL_1294151, EPI_ISL_1294153, EPI_ISL_1294156, EPI_ISL_1294159, EPI_ISL_1294160, EPI_ISL_1294161, EPI_ISL_1294163, EPI_ISL_1294165, EPI_ISL_1294166, EPI_ISL_1294167, EPI_ISL_1294169, EPI_ISL_1294170, EPI_ISL_1294172, EPI_ISL_1294174, EPI_ISL_1294175, EPI_ISL_1294176,
EPI_ISL_1294177, EPI_ISL_1294178, EPI_ISL_1294179, EPI_ISL_1294181, EPI_ISL_1294182, EPI_ISL_1294183, EPI_ISL_1294187, EPI_ISL_1294189, EPI_ISL_1294191, EPI_ISL_1294193, EPI_ISL_1294195, EPI_ISL_1294196, EPI_ISL_1294198, EPI_ISL_1294199, EPI_ISL_1294202, EPI_ISL_1294203, EPI_ISL_1294205, EPI_ISL_1294207, EPI_ISL_1294208, EPI_ISL_1294210, EPI_ISL_1294212, EPI_ISL_1294214,
EPI_ISL_1294217, EPI_ISL_1294219, EPI_ISL_1294220, EPI_ISL_1294221, EPI_ISL_1294222, EPI_ISL_1294224, EPI_ISL_1294226, EPI_ISL_1294227, EPI_ISL_1294228, EPI_ISL_1294229, EPI_ISL_1294230, EPI_ISL_1294231, EPI_ISL_1294235, EPI_ISL_1294237, EPI_ISL_1294238, EPI_ISL_1294240, EPI_ISL_1294241, EPI_ISL_1294242, EPI_ISL_1294244, EPI_ISL_1294246, EPI_ISL_1294247, EPI_ISL_1294249,
EPI_ISL_1294250, EPI_ISL_1294251, EPI_ISL_1294252, EPI_ISL_1294253, EPI_ISL_1294254, EPI_ISL_1294255, EPI_ISL_1294256, EPI_ISL_1294257, EPI_ISL_1294258, EPI_ISL_1294259, EPI_ISL_1294261, EPI_ISL_1294262, EPI_ISL_1294267, EPI_ISL_1294268, EPI_ISL_1294269, EPI_ISL_1294270, EPI_ISL_1294272, EPI_ISL_1294273, EPI_ISL_1294274, EPI_ISL_1294276, EPI_ISL_1294277, EPI_ISL_1294280,
EPI_ISL_1294281, EPI_ISL_1294283, EPI_ISL_1294284, EPI_ISL_1294425, EPI_ISL_1294426, EPI_ISL_1294428, EPI_ISL_1294429, EPI_ISL_1294430, EPI_ISL_1294431, EPI_ISL_1294432, EPI_ISL_1294433, EPI_ISL_1294434, EPI_ISL_1294435, EPI_ISL_1294436, EPI_ISL_1294438, EPI_ISL_1294439, EPI_ISL_1294442, EPI_ISL_1294445, EPI_ISL_1294446, EPI_ISL_1294447, EPI_ISL_1294449, EPI_ISL_1294455,
EPI_ISL_1294456, EPI_ISL_1294458, EPI_ISL_1294459, EPI_ISL_1294460, EPI_ISL_1294462, EPI_ISL_1294465, EPI_ISL_1294470, EPI_ISL_1294471, EPI_ISL_1294472, EPI_ISL_1294473, EPI_ISL_1294476, EPI_ISL_1294477, EPI_ISL_1294481, EPI_ISL_1294484, EPI_ISL_1294495, EPI_ISL_1294496, EPI_ISL_1294497, EPI_ISL_1294500, EPI_ISL_1294509, EPI_ISL_1294512, EPI_ISL_1294513, EPI_ISL_1294517,
EPI_ISL_1294519, EPI_ISL_1294524, EPI_ISL_1294526, EPI_ISL_1294527, EPI_ISL_1294528, EPI_ISL_1294529, EPI_ISL_1294531, EPI_ISL_1294532, EPI_ISL_1294534, EPI_ISL_1294536, EPI_ISL_1294542, EPI_ISL_1294547, EPI_ISL_1294549, EPI_ISL_1294552, EPI_ISL_1294553, EPI_ISL_1294555, EPI_ISL_1315669, EPI_ISL_1315671, EPI_ISL_1315759, EPI_ISL_1315774, EPI_ISL_1315780, EPI_ISL_1315807,
EPI_ISL_1315808, EPI_ISL_1315832, EPI_ISL_1315834, EPI_ISL_1315835, EPI_ISL_1315836, EPI_ISL_1315837, EPI_ISL_1315838, EPI_ISL_1315839, EPI_ISL_1315840, EPI_ISL_1315841, EPI_ISL_1315842, EPI_ISL_1315843, EPI_ISL_1315844, EPI_ISL_1315845, EPI_ISL_1315846, EPI_ISL_1315848, EPI_ISL_1315849, EPI_ISL_1315850, EPI_ISL_1315851, EPI_ISL_1315852, EPI_ISL_1315853, EPI_ISL_1315854,
EPI_ISL_1315855, EPI_ISL_1315856, EPI_ISL_1315857, EPI_ISL_1315858, EPI_ISL_1315859, EPI_ISL_1315860, EPI_ISL_1315861, EPI_ISL_1315862, EPI_ISL_1315863, EPI_ISL_1315864, EPI_ISL_1315865, EPI_ISL_1315866, EPI_ISL_1315867, EPI_ISL_1315868, EPI_ISL_1315869, EPI_ISL_1315870, EPI_ISL_1315871, EPI_ISL_1315872, EPI_ISL_1315873, EPI_ISL_1315874, EPI_ISL_1315875, EPI_ISL_1315876,
EPI_ISL_1315877, EPI_ISL_1315878, EPI_ISL_1315879, EPI_ISL_1315880, EPI_ISL_1315881, EPI_ISL_1315882, EPI_ISL_1315883, EPI_ISL_1315884, EPI_ISL_1315885, EPI_ISL_1315886, EPI_ISL_1315887, EPI_ISL_1315888, EPI_ISL_1315889, EPI_ISL_1315890, EPI_ISL_1315891, EPI_ISL_1315892, EPI_ISL_1315893, EPI_ISL_1315894, EPI_ISL_1315895, EPI_ISL_1315896, EPI_ISL_1315897, EPI_ISL_1315898,
EPI_ISL_1315899, EPI_ISL_1315900, EPI_ISL_1315901, EPI_ISL_1315902, EPI_ISL_1315903, EPI_ISL_1315904, EPI_ISL_1315905, EPI_ISL_1315906, EPI_ISL_1315907, EPI_ISL_1315908, EPI_ISL_1315909, EPI_ISL_1315910, EPI_ISL_1315911, EPI_ISL_1315912, EPI_ISL_1315913, EPI_ISL_1315914, EPI_ISL_1315915, EPI_ISL_1315916, EPI_ISL_1315917, EPI_ISL_1315918, EPI_ISL_1315919, EPI_ISL_1315920,
EPI_ISL_1315921, EPI_ISL_1315922, EPI_ISL_1315923, EPI_ISL_1315924, EPI_ISL_1315925, EPI_ISL_1315926, EPI_ISL_1315927, EPI_ISL_1315928, EPI_ISL_1315929, EPI_ISL_1315930, EPI_ISL_1315931, EPI_ISL_1315932, EPI_ISL_1315933, EPI_ISL_1315934, EPI_ISL_1315935, EPI_ISL_1315936, EPI_ISL_1315937, EPI_ISL_1315938, EPI_ISL_1315939, EPI_ISL_1315940, EPI_ISL_1315941, EPI_ISL_1315942,
EPI_ISL_1315943, EPI_ISL_1315944, EPI_ISL_1315945, EPI_ISL_1315946, EPI_ISL_1315947, EPI_ISL_1315948, EPI_ISL_1315949, EPI_ISL_1315950, EPI_ISL_1315951, EPI_ISL_1315952, EPI_ISL_1315953, EPI_ISL_1315954, EPI_ISL_1315955, EPI_ISL_1315956, EPI_ISL_1315957, EPI_ISL_1315958, EPI_ISL_1315959, EPI_ISL_1315960, EPI_ISL_1315961, EPI_ISL_1315962, EPI_ISL_1315963, EPI_ISL_1315964,
EPI_ISL_1315965, EPI_ISL_1315966, EPI_ISL_1315967, EPI_ISL_1315968, EPI_ISL_1315969, EPI_ISL_1315970, EPI_ISL_1315971, EPI_ISL_1315972, EPI_ISL_1315973, EPI_ISL_1315974, EPI_ISL_1315975, EPI_ISL_1315976, EPI_ISL_1315977, EPI_ISL_1315978, EPI_ISL_1315979, EPI_ISL_1315980, EPI_ISL_1315981, EPI_ISL_1315982, EPI_ISL_1315983, EPI_ISL_1315984, EPI_ISL_1315986, EPI_ISL_1315987,
EPI_ISL_1315988, EPI_ISL_1315989, EPI_ISL_1315990, EPI_ISL_1315991, EPI_ISL_1315992, EPI_ISL_1315993, EPI_ISL_1316028, EPI_ISL_1316057, EPI_ISL_1316232, EPI_ISL_1316233, EPI_ISL_1316234, EPI_ISL_1316235, EPI_ISL_1316236, EPI_ISL_1316237, EPI_ISL_1316238, EPI_ISL_1316239, EPI_ISL_1316240, EPI_ISL_1316241, EPI_ISL_1316242, EPI_ISL_1316243, EPI_ISL_1316244, EPI_ISL_1316245,
EPI_ISL_1316246, EPI_ISL_1316247, EPI_ISL_1316248, EPI_ISL_1316249, EPI_ISL_1316250, EPI_ISL_1316251, EPI_ISL_1316252, EPI_ISL_1316253, EPI_ISL_1316254, EPI_ISL_1316255, EPI_ISL_1316256, EPI_ISL_1316257, EPI_ISL_1316258, EPI_ISL_1316259, EPI_ISL_1316260, EPI_ISL_1316261, EPI_ISL_1316262, EPI_ISL_1316263, EPI_ISL_1316264, EPI_ISL_1316265, EPI_ISL_1316266, EPI_ISL_1316267,
EPI_ISL_1316268, EPI_ISL_1316269, EPI_ISL_1316270, EPI_ISL_1316271, EPI_ISL_1316272, EPI_ISL_1316273, EPI_ISL_1316274, EPI_ISL_1316275, EPI_ISL_1316276, EPI_ISL_1316277, EPI_ISL_1316278, EPI_ISL_1316279, EPI_ISL_1316280, EPI_ISL_1316281, EPI_ISL_1316282, EPI_ISL_1316283, EPI_ISL_1316284, EPI_ISL_1316285, EPI_ISL_1316286, EPI_ISL_1316287, EPI_ISL_1316288, EPI_ISL_1316289,
EPI_ISL_1316290, EPI_ISL_1316291, EPI_ISL_1316292, EPI_ISL_1316293, EPI_ISL_1316294, EPI_ISL_1316295, EPI_ISL_1316296, EPI_ISL_1316297, EPI_ISL_1316298, EPI_ISL_1316299, EPI_ISL_1316300, EPI_ISL_1316301, EPI_ISL_1316302, EPI_ISL_1316303, EPI_ISL_1316304, EPI_ISL_1316305, EPI_ISL_1316306, EPI_ISL_1316307, EPI_ISL_1316308, EPI_ISL_1316309, EPI_ISL_1316310, EPI_ISL_1316311,
EPI_ISL_1316312, EPI_ISL_1316313, EPI_ISL_1316314, EPI_ISL_1316315, EPI_ISL_1316316, EPI_ISL_1316317, EPI_ISL_1316318, EPI_ISL_1316319, EPI_ISL_1316320, EPI_ISL_1316321, EPI_ISL_1316322, EPI_ISL_1316323, EPI_ISL_1316324, EPI_ISL_1316325, EPI_ISL_1316326, EPI_ISL_1316327, EPI_ISL_1316328, EPI_ISL_1316329, EPI_ISL_1316330, EPI_ISL_1316331, EPI_ISL_1316332, EPI_ISL_1316333,
EPI_ISL_1316334, EPI_ISL_1316335, EPI_ISL_1316336, EPI_ISL_1316337, EPI_ISL_1316338, EPI_ISL_1316339, EPI_ISL_1316340, EPI_ISL_1316341, EPI_ISL_1316342, EPI_ISL_1316343, EPI_ISL_1316344, EPI_ISL_1316345, EPI_ISL_1316346, EPI_ISL_1316347, EPI_ISL_1316348, EPI_ISL_1316349, EPI_ISL_1316350, EPI_ISL_1316351, EPI_ISL_1316352, EPI_ISL_1316353, EPI_ISL_1316354, EPI_ISL_1316355,
EPI_ISL_1316356, EPI_ISL_1316357, EPI_ISL_1316358, EPI_ISL_1316359, EPI_ISL_1316360, EPI_ISL_1316361, EPI_ISL_1316362, EPI_ISL_1316363, EPI_ISL_1316364, EPI_ISL_1316365, EPI_ISL_1316366, EPI_ISL_1316367, EPI_ISL_1316368, EPI_ISL_1316369, EPI_ISL_1316370, EPI_ISL_1316371, EPI_ISL_1316372, EPI_ISL_1316373, EPI_ISL_1316374, EPI_ISL_1316375, EPI_ISL_1316376, EPI_ISL_1316377,
EPI_ISL_1316378, EPI_ISL_1316379, EPI_ISL_1316380, EPI_ISL_1316381, EPI_ISL_1316382, EPI_ISL_1316383, EPI_ISL_1316384, EPI_ISL_1316385, EPI_ISL_1316386, EPI_ISL_1316387, EPI_ISL_1316388, EPI_ISL_1316389, EPI_ISL_1316390, EPI_ISL_1316391, EPI_ISL_1316392, EPI_ISL_1316393, EPI_ISL_1316394, EPI_ISL_1316395, EPI_ISL_1316396, EPI_ISL_1316397, EPI_ISL_1316398, EPI_ISL_1316399,
EPI_ISL_1316400, EPI_ISL_1316401, EPI_ISL_1316402, EPI_ISL_1316403, EPI_ISL_1316404, EPI_ISL_1316405, EPI_ISL_1316406, EPI_ISL_1316407, EPI_ISL_1316408, EPI_ISL_1316409, EPI_ISL_1316410, EPI_ISL_1316411, EPI_ISL_1316412, EPI_ISL_1316413, EPI_ISL_1316414, EPI_ISL_1316415, EPI_ISL_1316416, EPI_ISL_1316417, EPI_ISL_1316783, EPI_ISL_1316785, EPI_ISL_1316786, EPI_ISL_1316787,
EPI_ISL_1316788, EPI_ISL_1316790, EPI_ISL_1316791, EPI_ISL_1316793, EPI_ISL_1316794, EPI_ISL_1316796, EPI_ISL_1316797, EPI_ISL_1316798, EPI_ISL_1316799, EPI_ISL_1316800, EPI_ISL_1316802, EPI_ISL_1316804, EPI_ISL_1316805, EPI_ISL_1316806, EPI_ISL_1316808, EPI_ISL_1316809, EPI_ISL_1316813, EPI_ISL_1316814, EPI_ISL_1316815, EPI_ISL_1316816, EPI_ISL_1316818, EPI_ISL_1316819,
EPI_ISL_1316821, EPI_ISL_1316822, EPI_ISL_1316823, EPI_ISL_1316824, EPI_ISL_1316825, EPI_ISL_1316826, EPI_ISL_1316829, EPI_ISL_1316830, EPI_ISL_1316831, EPI_ISL_1316832, EPI_ISL_1316833, EPI_ISL_1316836, EPI_ISL_1316838, EPI_ISL_1316839, EPI_ISL_1316840, EPI_ISL_1316841, EPI_ISL_1316842, EPI_ISL_1316843, EPI_ISL_1316844, EPI_ISL_1316846, EPI_ISL_1316847, EPI_ISL_1316848,
EPI_ISL_1316852, EPI_ISL_1316854, EPI_ISL_1316855, EPI_ISL_1316858, EPI_ISL_1316859, EPI_ISL_1316860, EPI_ISL_1316861, EPI_ISL_1316862, EPI_ISL_1316863, EPI_ISL_1316864, EPI_ISL_1316865, EPI_ISL_1316867, EPI_ISL_1316869, EPI_ISL_1316871, EPI_ISL_1316872, EPI_ISL_1316874, EPI_ISL_1316875, EPI_ISL_1316878, EPI_ISL_1316880, EPI_ISL_1316881, EPI_ISL_1316883, EPI_ISL_1316885,
EPI_ISL_1316886, EPI_ISL_1316887, EPI_ISL_1316889, EPI_ISL_1316890, EPI_ISL_1316891, EPI_ISL_1316892, EPI_ISL_1316893, EPI_ISL_1316894, EPI_ISL_1316896, EPI_ISL_1316898, EPI_ISL_1316899, EPI_ISL_1316901, EPI_ISL_1316902, EPI_ISL_1316903, EPI_ISL_1316904, EPI_ISL_1316905, EPI_ISL_1316906, EPI_ISL_1316907, EPI_ISL_1316909, EPI_ISL_1316912, EPI_ISL_1316913, EPI_ISL_1316914,
EPI_ISL_1316915, EPI_ISL_1316916, EPI_ISL_1316918, EPI_ISL_1316919, EPI_ISL_1316920, EPI_ISL_1316921, EPI_ISL_1316922, EPI_ISL_1316924, EPI_ISL_1316925, EPI_ISL_1316926, EPI_ISL_1316927, EPI_ISL_1316930, EPI_ISL_1316931, EPI_ISL_1316934, EPI_ISL_1316935, EPI_ISL_1316936, EPI_ISL_1316937, EPI_ISL_1316939, EPI_ISL_1316940, EPI_ISL_1316944, EPI_ISL_1316945, EPI_ISL_1316946,
EPI_ISL_1316947, EPI_ISL_1316948, EPI_ISL_1316949, EPI_ISL_1316950, EPI_ISL_1316951, EPI_ISL_1316952, EPI_ISL_1316953, EPI_ISL_1316954, EPI_ISL_1316955, EPI_ISL_1316956, EPI_ISL_1316957, EPI_ISL_1316958, EPI_ISL_1316959, EPI_ISL_1316960, EPI_ISL_1316961, EPI_ISL_1316962, EPI_ISL_1316963, EPI_ISL_1316964, EPI_ISL_1316965, EPI_ISL_1316966, EPI_ISL_1316967, EPI_ISL_1316968,
EPI_ISL_1316969, EPI_ISL_1316970, EPI_ISL_1316971, EPI_ISL_1316972, EPI_ISL_1316973, EPI_ISL_1316974, EPI_ISL_1316975, EPI_ISL_1316976, EPI_ISL_1316977, EPI_ISL_1316979, EPI_ISL_1316980, EPI_ISL_1316981, EPI_ISL_1316982, EPI_ISL_1316983, EPI_ISL_1316984, EPI_ISL_1316985, EPI_ISL_1316986, EPI_ISL_1316987, EPI_ISL_1316988, EPI_ISL_1316989, EPI_ISL_1316990, EPI_ISL_1316991,
EPI_ISL_1316992, EPI_ISL_1316993, EPI_ISL_1316994, EPI_ISL_1316995, EPI_ISL_1316996, EPI_ISL_1316997, EPI_ISL_1316998, EPI_ISL_1316999, EPI_ISL_1317000, EPI_ISL_1317002, EPI_ISL_1317004, EPI_ISL_1317005, EPI_ISL_1317007, EPI_ISL_1317008, EPI_ISL_1317009, EPI_ISL_1317010, EPI_ISL_1317011, EPI_ISL_1317012, EPI_ISL_1317013, EPI_ISL_1317014, EPI_ISL_1317015, EPI_ISL_1317016,
EPI_ISL_1317017, EPI_ISL_1317018, EPI_ISL_1317019, EPI_ISL_1317021, EPI_ISL_1317022, EPI_ISL_1317023, EPI_ISL_1317024, EPI_ISL_1317025, EPI_ISL_1317026, EPI_ISL_1317027, EPI_ISL_1317028, EPI_ISL_1317029, EPI_ISL_1317030, EPI_ISL_1317031, EPI_ISL_1317032, EPI_ISL_1317033, EPI_ISL_1317034, EPI_ISL_1317035, EPI_ISL_1317036, EPI_ISL_1317037, EPI_ISL_1317038, EPI_ISL_1317039,
EPI_ISL_1317040, EPI_ISL_1317041, EPI_ISL_1317042, EPI_ISL_1317043, EPI_ISL_1317044, EPI_ISL_1317045, EPI_ISL_1317046, EPI_ISL_1317047, EPI_ISL_1317048, EPI_ISL_1317049, EPI_ISL_1317050, EPI_ISL_1317051, EPI_ISL_1317052, EPI_ISL_1317053, EPI_ISL_1317054, EPI_ISL_1317055, EPI_ISL_1317056, EPI_ISL_1317057, EPI_ISL_1317058, EPI_ISL_1317059, EPI_ISL_1317060, EPI_ISL_1317061,
EPI_ISL_1317063, EPI_ISL_1317064, EPI_ISL_1317065, EPI_ISL_1317066, EPI_ISL_1317067, EPI_ISL_1317068, EPI_ISL_1317069, EPI_ISL_1317070, EPI_ISL_1317071, EPI_ISL_1317072, EPI_ISL_1317073, EPI_ISL_1317074, EPI_ISL_1317075, EPI_ISL_1317076, EPI_ISL_1317077, EPI_ISL_1317078, EPI_ISL_1317079, EPI_ISL_1317081, EPI_ISL_1317082, EPI_ISL_1317083, EPI_ISL_1317084, EPI_ISL_1317085,
EPI_ISL_1317086, EPI_ISL_1317087, EPI_ISL_1317088, EPI_ISL_1317089, EPI_ISL_1317090, EPI_ISL_1317091, EPI_ISL_1317092, EPI_ISL_1317093, EPI_ISL_1317094, EPI_ISL_1317095, EPI_ISL_1317096, EPI_ISL_1317097, EPI_ISL_1317098, EPI_ISL_1317099, EPI_ISL_1317100, EPI_ISL_1317101, EPI_ISL_1317102, EPI_ISL_1317103, EPI_ISL_1317104, EPI_ISL_1317105, EPI_ISL_1317106, EPI_ISL_1317107,
EPI_ISL_1317108, EPI_ISL_1317109, EPI_ISL_1317110, EPI_ISL_1317111, EPI_ISL_1317112, EPI_ISL_1317113, EPI_ISL_1317114, EPI_ISL_1317115, EPI_ISL_1317116, EPI_ISL_1317117, EPI_ISL_1317118, EPI_ISL_1317119, EPI_ISL_1317120, EPI_ISL_1317121, EPI_ISL_1317122, EPI_ISL_1317123, EPI_ISL_1317124, EPI_ISL_1317125, EPI_ISL_1317126, EPI_ISL_1317127, EPI_ISL_1317128, EPI_ISL_1317129,
EPI_ISL_1317130, EPI_ISL_1317131, EPI_ISL_1317132, EPI_ISL_1317133, EPI_ISL_1317134, EPI_ISL_1317135, EPI_ISL_1317136, EPI_ISL_1317137, EPI_ISL_1317138, EPI_ISL_1317139, EPI_ISL_1317140, EPI_ISL_1317141, EPI_ISL_1317142, EPI_ISL_1317143, EPI_ISL_1317144, EPI_ISL_1317145, EPI_ISL_1317146, EPI_ISL_1317147, EPI_ISL_1317148, EPI_ISL_1317149, EPI_ISL_1317150, EPI_ISL_1317151,
EPI_ISL_1317153, EPI_ISL_1317154, EPI_ISL_1317155, EPI_ISL_1317156, EPI_ISL_1317157, EPI_ISL_1317158, EPI_ISL_1317159, EPI_ISL_1317160, EPI_ISL_1317161, EPI_ISL_1317162, EPI_ISL_1317163, EPI_ISL_1317164, EPI_ISL_1317165, EPI_ISL_1317166, EPI_ISL_1317167, EPI_ISL_1317168, EPI_ISL_1317169, EPI_ISL_1317170, EPI_ISL_1317171, EPI_ISL_1317172, EPI_ISL_1317173, EPI_ISL_1317174,
EPI_ISL_1317175, EPI_ISL_1317176, EPI_ISL_1317177, EPI_ISL_1317178, EPI_ISL_1317179, EPI_ISL_1317180, EPI_ISL_1317181, EPI_ISL_1317182, EPI_ISL_1317183, EPI_ISL_1317184, EPI_ISL_1317185, EPI_ISL_1317186, EPI_ISL_1317187, EPI_ISL_1317188, EPI_ISL_1317189, EPI_ISL_1317190, EPI_ISL_1317191, EPI_ISL_1317192, EPI_ISL_1317193, EPI_ISL_1317194, EPI_ISL_1317195, EPI_ISL_1317196,

EPI_ISL_1317197, EPI_ISL_1317198, EPI_ISL_1317199, EPI_ISL_1317200, EPI_ISL_1317201, EPI_ISL_1317202, EPI_ISL_1317203, EPI_ISL_1317204, EPI_ISL_1317205, EPI_ISL_1317206, EPI_ISL_1317207, EPI_ISL_1317208, EPI_ISL_1325782, EPI_ISL_1326126, EPI_ISL_1326127, EPI_ISL_1326128, EPI_ISL_1326129, EPI_ISL_1326130, EPI_ISL_1326131, EPI_ISL_1326132, EPI_ISL_1326133, EPI_ISL_1326134,
EPI_ISL_1326135, EPI_ISL_1326136, EPI_ISL_1326137, EPI_ISL_1326138, EPI_ISL_1326139, EPI_ISL_1326140, EPI_ISL_1326141, EPI_ISL_1326142, EPI_ISL_1326143, EPI_ISL_1326144, EPI_ISL_1326145, EPI_ISL_1326146, EPI_ISL_1326147, EPI_ISL_1326148, EPI_ISL_1326149, EPI_ISL_1326151, EPI_ISL_1326152, EPI_ISL_1326153, EPI_ISL_1326154, EPI_ISL_1326156, EPI_ISL_1326157, EPI_ISL_1326158,
EPI_ISL_1326160, EPI_ISL_1326161, EPI_ISL_1326162, EPI_ISL_1326163, EPI_ISL_1326164, EPI_ISL_1326165, EPI_ISL_1326166, EPI_ISL_1326167, EPI_ISL_1326168, EPI_ISL_1326169, EPI_ISL_1326170, EPI_ISL_1326171, EPI_ISL_1326173, EPI_ISL_1326174, EPI_ISL_1326175, EPI_ISL_1326176, EPI_ISL_1326177, EPI_ISL_1326178, EPI_ISL_1326179, EPI_ISL_1326180, EPI_ISL_1326181, EPI_ISL_1326182,
EPI_ISL_1326183, EPI_ISL_1326184, EPI_ISL_1326187, EPI_ISL_1326188, EPI_ISL_1326189, EPI_ISL_1326190, EPI_ISL_1326191, EPI_ISL_1326192, EPI_ISL_1326193, EPI_ISL_1326194, EPI_ISL_1326195, EPI_ISL_1326197, EPI_ISL_1326198, EPI_ISL_1326199, EPI_ISL_1326200, EPI_ISL_1326201, EPI_ISL_1326202, EPI_ISL_1326203, EPI_ISL_1326204, EPI_ISL_1326205, EPI_ISL_1326206, EPI_ISL_1326207,
EPI_ISL_1326208, EPI_ISL_1326210, EPI_ISL_1326211, EPI_ISL_1326212, EPI_ISL_1326213, EPI_ISL_1326214, EPI_ISL_1326215, EPI_ISL_1326216, EPI_ISL_1326217, EPI_ISL_1326218, EPI_ISL_1326219, EPI_ISL_1326220, EPI_ISL_1326222, EPI_ISL_1326223, EPI_ISL_1326224, EPI_ISL_1326225, EPI_ISL_1326226, EPI_ISL_1326227, EPI_ISL_1326228, EPI_ISL_1326230, EPI_ISL_1326231, EPI_ISL_1326232,
EPI_ISL_1326234, EPI_ISL_1326235, EPI_ISL_1326236, EPI_ISL_1326237, EPI_ISL_1326238, EPI_ISL_1326239, EPI_ISL_1326240, EPI_ISL_1326241, EPI_ISL_1326242, EPI_ISL_1326243, EPI_ISL_1326244, EPI_ISL_1326245, EPI_ISL_1326246, EPI_ISL_1326247, EPI_ISL_1326248, EPI_ISL_1326249, EPI_ISL_1326250, EPI_ISL_1326251, EPI_ISL_1326252, EPI_ISL_1326254, EPI_ISL_1326255, EPI_ISL_1326256,
EPI_ISL_1326257, EPI_ISL_1326258, EPI_ISL_1326259, EPI_ISL_1326260, EPI_ISL_1326261, EPI_ISL_1326264, EPI_ISL_1326266, EPI_ISL_1326267, EPI_ISL_1326268, EPI_ISL_1326269, EPI_ISL_1326270, EPI_ISL_1326271, EPI_ISL_1326272, EPI_ISL_1326273, EPI_ISL_1326274, EPI_ISL_1326276, EPI_ISL_1326277, EPI_ISL_1326278, EPI_ISL_1326279, EPI_ISL_1326280, EPI_ISL_1326281, EPI_ISL_1326282,
EPI_ISL_1326283, EPI_ISL_1326284, EPI_ISL_1326285, EPI_ISL_1326286, EPI_ISL_1326287, EPI_ISL_1326288, EPI_ISL_1326289, EPI_ISL_1326290, EPI_ISL_1326291, EPI_ISL_1326292, EPI_ISL_1326293, EPI_ISL_1326294, EPI_ISL_1326295, EPI_ISL_1326296, EPI_ISL_1326297, EPI_ISL_1326298, EPI_ISL_1326299, EPI_ISL_1326300, EPI_ISL_1326301, EPI_ISL_1326302, EPI_ISL_1326303, EPI_ISL_1326304,
EPI_ISL_1326305, EPI_ISL_1326306, EPI_ISL_1326307, EPI_ISL_1326308, EPI_ISL_1326309, EPI_ISL_1326310, EPI_ISL_1326312, EPI_ISL_1326313, EPI_ISL_1326314, EPI_ISL_1326315, EPI_ISL_1326316, EPI_ISL_1326317, EPI_ISL_1326318, EPI_ISL_1326319, EPI_ISL_1326320, EPI_ISL_1326321, EPI_ISL_1326323, EPI_ISL_1326324, EPI_ISL_1326325, EPI_ISL_1326326, EPI_ISL_1326327, EPI_ISL_1326328,
EPI_ISL_1326329, EPI_ISL_1326330, EPI_ISL_1326331, EPI_ISL_1326332, EPI_ISL_1326333, EPI_ISL_1326334, EPI_ISL_1326335, EPI_ISL_1326336, EPI_ISL_1326337, EPI_ISL_1326338, EPI_ISL_1326339, EPI_ISL_1326340, EPI_ISL_1326341, EPI_ISL_1326342, EPI_ISL_1326343, EPI_ISL_1326344, EPI_ISL_1326345, EPI_ISL_1326346, EPI_ISL_1326347, EPI_ISL_1326348, EPI_ISL_1326349, EPI_ISL_1326350,
EPI_ISL_1326351, EPI_ISL_1326352, EPI_ISL_1326353, EPI_ISL_1326354, EPI_ISL_1326355, EPI_ISL_1326356, EPI_ISL_1326357, EPI_ISL_1326358, EPI_ISL_1326359, EPI_ISL_1326360, EPI_ISL_1326361, EPI_ISL_1326362, EPI_ISL_1326363, EPI_ISL_1326364, EPI_ISL_1326374, EPI_ISL_1326379, EPI_ISL_1326380, EPI_ISL_1326387, EPI_ISL_1326389, EPI_ISL_1326400, EPI_ISL_1326409, EPI_ISL_1326413,
EPI_ISL_1326414, EPI_ISL_1326415, EPI_ISL_1326418, EPI_ISL_1326429, EPI_ISL_1326432, EPI_ISL_1326443, EPI_ISL_1326446, EPI_ISL_1326450, EPI_ISL_1326451, EPI_ISL_1326456, EPI_ISL_1326459, EPI_ISL_1326460, EPI_ISL_1326468, EPI_ISL_1326477, EPI_ISL_1326478, EPI_ISL_1326480, EPI_ISL_1326494, EPI_ISL_1326505, EPI_ISL_1326518, EPI_ISL_1326519, EPI_ISL_1326526, EPI_ISL_1326527,
EPI_ISL_1326536, EPI_ISL_1326538, EPI_ISL_1326541, EPI_ISL_1326543, EPI_ISL_1326553, EPI_ISL_1326564, EPI_ISL_1326566, EPI_ISL_1326569, EPI_ISL_1326574, EPI_ISL_1326576, EPI_ISL_1326580, EPI_ISL_1326581, EPI_ISL_1326589, EPI_ISL_1326610, EPI_ISL_1326611, EPI_ISL_1326613, EPI_ISL_1326630, EPI_ISL_1326631, EPI_ISL_1326636, EPI_ISL_1326646, EPI_ISL_1326652, EPI_ISL_1326656,
EPI_ISL_1326657, EPI_ISL_1326667, EPI_ISL_1326671, EPI_ISL_1326672, EPI_ISL_1326686, EPI_ISL_1326700, EPI_ISL_1326709, EPI_ISL_1326712, EPI_ISL_1326714, EPI_ISL_1326715, EPI_ISL_1326722, EPI_ISL_1326733, EPI_ISL_1326744, EPI_ISL_1326753, EPI_ISL_1326769, EPI_ISL_1326789, EPI_ISL_1326801, EPI_ISL_1326818, EPI_ISL_1326826, EPI_ISL_1326866, EPI_ISL_1326874, EPI_ISL_1326884,
EPI_ISL_1326888, EPI_ISL_1326891, EPI_ISL_1326896, EPI_ISL_1326911, EPI_ISL_1326912, EPI_ISL_1326913, EPI_ISL_1326919, EPI_ISL_1326920, EPI_ISL_1326925, EPI_ISL_1326941, EPI_ISL_1326962, EPI_ISL_1326973, EPI_ISL_1326975, EPI_ISL_1326981, EPI_ISL_1326991, EPI_ISL_1326995, EPI_ISL_1326999, EPI_ISL_1327000, EPI_ISL_1327010, EPI_ISL_1327018, EPI_ISL_1327019, EPI_ISL_1327020,
EPI_ISL_1327021, EPI_ISL_1327024, EPI_ISL_1327035, EPI_ISL_1327047, EPI_ISL_1327049, EPI_ISL_1327054, EPI_ISL_1327056, EPI_ISL_1327061, EPI_ISL_1327067, EPI_ISL_1327068, EPI_ISL_1327094, EPI_ISL_1327099, EPI_ISL_1327108, EPI_ISL_1327122, EPI_ISL_1327124, EPI_ISL_1327128, EPI_ISL_1327138, EPI_ISL_1327145, EPI_ISL_1327158, EPI_ISL_1327159, EPI_ISL_1327160, EPI_ISL_1327173,
EPI_ISL_1327175, EPI_ISL_1327195, EPI_ISL_1327196, EPI_ISL_1327197, EPI_ISL_1327200, EPI_ISL_1327204, EPI_ISL_1327212, EPI_ISL_1327214, EPI_ISL_1327221, EPI_ISL_1327222, EPI_ISL_1327223, EPI_ISL_1327224, EPI_ISL_1327240, EPI_ISL_1327452, EPI_ISL_1327457, EPI_ISL_1327461, EPI_ISL_1327462, EPI_ISL_1327463, EPI_ISL_1327466, EPI_ISL_1327467, EPI_ISL_1327469, EPI_ISL_1327473,
EPI_ISL_1327474, EPI_ISL_1327479, EPI_ISL_1327480, EPI_ISL_1327487, EPI_ISL_1327488, EPI_ISL_1327491, EPI_ISL_1327512, EPI_ISL_1327517, EPI_ISL_1327518, EPI_ISL_1327522, EPI_ISL_1327523, EPI_ISL_1327527, EPI_ISL_1327530, EPI_ISL_1327542, EPI_ISL_1327544, EPI_ISL_1327550, EPI_ISL_1327559, EPI_ISL_1327560, EPI_ISL_1327571, EPI_ISL_1327574, EPI_ISL_1327580, EPI_ISL_1327588,
EPI_ISL_1327595, EPI_ISL_1327598, EPI_ISL_1327601, EPI_ISL_1327611, EPI_ISL_1327612, EPI_ISL_1327623, EPI_ISL_1327625, EPI_ISL_1327626, EPI_ISL_1327627, EPI_ISL_1327629, EPI_ISL_1327632, EPI_ISL_1327633, EPI_ISL_1327637, EPI_ISL_1327641, EPI_ISL_1327642, EPI_ISL_1327643, EPI_ISL_1327644, EPI_ISL_1327645, EPI_ISL_1327661, EPI_ISL_1327666, EPI_ISL_1327667, EPI_ISL_1327676,
EPI_ISL_1327681, EPI_ISL_1327688, EPI_ISL_1327690, EPI_ISL_1327721, EPI_ISL_1327933, EPI_ISL_1327970, EPI_ISL_1328020, EPI_ISL_1328022, EPI_ISL_1328088, EPI_ISL_1328110, EPI_ISL_1328161, EPI_ISL_1328205, EPI_ISL_1328217, EPI_ISL_1328218, EPI_ISL_1328219, EPI_ISL_1328220, EPI_ISL_1328221, EPI_ISL_1328222, EPI_ISL_1328223, EPI_ISL_1328224, EPI_ISL_1328225, EPI_ISL_1328226,
EPI_ISL_1328227, EPI_ISL_1328228, EPI_ISL_1328229, EPI_ISL_1328230, EPI_ISL_1328231, EPI_ISL_1328232, EPI_ISL_1328233, EPI_ISL_1328234, EPI_ISL_1328235, EPI_ISL_1328236, EPI_ISL_1328237, EPI_ISL_1328238, EPI_ISL_1328239, EPI_ISL_1328240, EPI_ISL_1328241, EPI_ISL_1328242, EPI_ISL_1328243, EPI_ISL_1328244, EPI_ISL_1328245, EPI_ISL_1328247, EPI_ISL_1328248, EPI_ISL_1328249,
EPI_ISL_1328250, EPI_ISL_1328251, EPI_ISL_1328252, EPI_ISL_1328253, EPI_ISL_1328254, EPI_ISL_1328255, EPI_ISL_1328256, EPI_ISL_1328257, EPI_ISL_1328258, EPI_ISL_1328259, EPI_ISL_1328260, EPI_ISL_1328261, EPI_ISL_1328262, EPI_ISL_1328263, EPI_ISL_1328264, EPI_ISL_1328265, EPI_ISL_1328266, EPI_ISL_1328267, EPI_ISL_1328268, EPI_ISL_1328269, EPI_ISL_1328270, EPI_ISL_1328271,
EPI_ISL_1328272, EPI_ISL_1328273, EPI_ISL_1328274, EPI_ISL_1328275, EPI_ISL_1328276, EPI_ISL_1328277, EPI_ISL_1328278, EPI_ISL_1328279, EPI_ISL_1328280, EPI_ISL_1328281, EPI_ISL_1328282, EPI_ISL_1328283, EPI_ISL_1328284, EPI_ISL_1328285, EPI_ISL_1328286, EPI_ISL_1328287, EPI_ISL_1328288, EPI_ISL_1328289, EPI_ISL_1328290, EPI_ISL_1328291, EPI_ISL_1328292, EPI_ISL_1328293,
EPI_ISL_1328294, EPI_ISL_1328295, EPI_ISL_1328296, EPI_ISL_1328297, EPI_ISL_1328298, EPI_ISL_1328299, EPI_ISL_1328300, EPI_ISL_1328301, EPI_ISL_1328302, EPI_ISL_1328303, EPI_ISL_1328304, EPI_ISL_1328305, EPI_ISL_1328306, EPI_ISL_1328307, EPI_ISL_1328308, EPI_ISL_1328309, EPI_ISL_1328310, EPI_ISL_1328311, EPI_ISL_1328312, EPI_ISL_1328313, EPI_ISL_1328314, EPI_ISL_1328315,
EPI_ISL_1328316, EPI_ISL_1328317, EPI_ISL_1328318, EPI_ISL_1328319, EPI_ISL_1328320, EPI_ISL_1328321, EPI_ISL_1328322, EPI_ISL_1328323, EPI_ISL_1328324, EPI_ISL_1328325, EPI_ISL_1328326, EPI_ISL_1328327, EPI_ISL_1328328, EPI_ISL_1328329, EPI_ISL_1328330, EPI_ISL_1328331, EPI_ISL_1328332, EPI_ISL_1328333, EPI_ISL_1328334, EPI_ISL_1328335, EPI_ISL_1328336, EPI_ISL_1328338,
EPI_ISL_1328339, EPI_ISL_1328340, EPI_ISL_1328341, EPI_ISL_1328342, EPI_ISL_1328343, EPI_ISL_1328344, EPI_ISL_1328345, EPI_ISL_1328346, EPI_ISL_1328347, EPI_ISL_1328348, EPI_ISL_1328349, EPI_ISL_1328350, EPI_ISL_1328351, EPI_ISL_1328352, EPI_ISL_1328353, EPI_ISL_1328354, EPI_ISL_1328355, EPI_ISL_1328357, EPI_ISL_1328358, EPI_ISL_1328359, EPI_ISL_1328360, EPI_ISL_1328361,
EPI_ISL_1328362, EPI_ISL_1328363, EPI_ISL_1328364, EPI_ISL_1328365, EPI_ISL_1328366, EPI_ISL_1328367, EPI_ISL_1328368, EPI_ISL_1328369, EPI_ISL_1328370, EPI_ISL_1328371, EPI_ISL_1328372, EPI_ISL_1328373, EPI_ISL_1328374, EPI_ISL_1328375, EPI_ISL_1328376, EPI_ISL_1328377, EPI_ISL_1328378, EPI_ISL_1328379, EPI_ISL_1328380, EPI_ISL_1328381, EPI_ISL_1328382, EPI_ISL_1328383,
EPI_ISL_1328384, EPI_ISL_1328385, EPI_ISL_1328386, EPI_ISL_1328387, EPI_ISL_1328388, EPI_ISL_1328389, EPI_ISL_1328390, EPI_ISL_1328391, EPI_ISL_1328392, EPI_ISL_1328393, EPI_ISL_1328394, EPI_ISL_1328395, EPI_ISL_1328396, EPI_ISL_1328397, EPI_ISL_1328398, EPI_ISL_1328399, EPI_ISL_1328400, EPI_ISL_1328401, EPI_ISL_1328402, EPI_ISL_1328403, EPI_ISL_1328404, EPI_ISL_1328405,
EPI_ISL_1328406, EPI_ISL_1328407, EPI_ISL_1328408, EPI_ISL_1328420, EPI_ISL_1328433, EPI_ISL_1328490, EPI_ISL_1328508, EPI_ISL_1328525, EPI_ISL_1328527, EPI_ISL_1328558, EPI_ISL_1328586, EPI_ISL_1328594, EPI_ISL_1328601, EPI_ISL_1328614, EPI_ISL_1328643, EPI_ISL_1328680, EPI_ISL_1328687, EPI_ISL_1328696, EPI_ISL_1328708, EPI_ISL_1328713, EPI_ISL_1328715, EPI_ISL_1328716,
EPI_ISL_1328717, EPI_ISL_1328721, EPI_ISL_1328722, EPI_ISL_1328724, EPI_ISL_1328726, EPI_ISL_1328727, EPI_ISL_1328730, EPI_ISL_1328732, EPI_ISL_1328733, EPI_ISL_1328734, EPI_ISL_1328735, EPI_ISL_1328737, EPI_ISL_1328738, EPI_ISL_1328740, EPI_ISL_1328741, EPI_ISL_1328742, EPI_ISL_1328743, EPI_ISL_1328744, EPI_ISL_1328745, EPI_ISL_1328749, EPI_ISL_1328750, EPI_ISL_1328752,
EPI_ISL_1328754, EPI_ISL_1328757, EPI_ISL_1328758, EPI_ISL_1328760, EPI_ISL_1328761, EPI_ISL_1328762, EPI_ISL_1328763, EPI_ISL_1328765, EPI_ISL_1328766, EPI_ISL_1328767, EPI_ISL_1328770, EPI_ISL_1328771, EPI_ISL_1328772, EPI_ISL_1328773, EPI_ISL_1328774, EPI_ISL_1328775, EPI_ISL_1328776, EPI_ISL_1328777, EPI_ISL_1328778, EPI_ISL_1328780, EPI_ISL_1328781, EPI_ISL_1328783,
EPI_ISL_1328784, EPI_ISL_1328785, EPI_ISL_1328786, EPI_ISL_1328787, EPI_ISL_1328788, EPI_ISL_1328789, EPI_ISL_1328790, EPI_ISL_1328791, EPI_ISL_1328792, EPI_ISL_1328793, EPI_ISL_1328794, EPI_ISL_1328795, EPI_ISL_1328796, EPI_ISL_1328798, EPI_ISL_1328804, EPI_ISL_1328815, EPI_ISL_1328816, EPI_ISL_1328822, EPI_ISL_1328823, EPI_ISL_1328825, EPI_ISL_1328827, EPI_ISL_1328828,
EPI_ISL_1328829, EPI_ISL_1328831, EPI_ISL_1328832, EPI_ISL_1328834, EPI_ISL_1328835, EPI_ISL_1328837, EPI_ISL_1328838, EPI_ISL_1328839, EPI_ISL_1328842, EPI_ISL_1328843, EPI_ISL_1328844, EPI_ISL_1328845, EPI_ISL_1328847, EPI_ISL_1328848, EPI_ISL_1328849, EPI_ISL_1328851, EPI_ISL_1328853, EPI_ISL_1328857, EPI_ISL_1328858, EPI_ISL_1328862, EPI_ISL_1328864, EPI_ISL_1328865,
EPI_ISL_1328866, EPI_ISL_1328867, EPI_ISL_1328868, EPI_ISL_1328870, EPI_ISL_1328871, EPI_ISL_1328873, EPI_ISL_1328874, EPI_ISL_1328880, EPI_ISL_1328881, EPI_ISL_1328883, EPI_ISL_1328884, EPI_ISL_1328886, EPI_ISL_1328888, EPI_ISL_1328890, EPI_ISL_1328891, EPI_ISL_1328894, EPI_ISL_1328895, EPI_ISL_1328897, EPI_ISL_1328918, EPI_ISL_1328926, EPI_ISL_1328930, EPI_ISL_1328939,
EPI_ISL_1328943, EPI_ISL_1328975, EPI_ISL_1328977, EPI_ISL_1328980, EPI_ISL_1328998, EPI_ISL_1329008, EPI_ISL_1329012, EPI_ISL_1329019, EPI_ISL_1329022, EPI_ISL_1329054, EPI_ISL_1329076, EPI_ISL_1329082, EPI_ISL_1329098, EPI_ISL_1329117, EPI_ISL_1329123, EPI_ISL_1329129, EPI_ISL_1329132, EPI_ISL_1329153, EPI_ISL_1329177, EPI_ISL_1329193, EPI_ISL_1329195, EPI_ISL_1329536,
EPI_ISL_1329669, EPI_ISL_1330104, EPI_ISL_1330105, EPI_ISL_1330106, EPI_ISL_1330107, EPI_ISL_1330108, EPI_ISL_1330109, EPI_ISL_1330110, EPI_ISL_1330111, EPI_ISL_1330112, EPI_ISL_1330113, EPI_ISL_1330114, EPI_ISL_1330115, EPI_ISL_1330116, EPI_ISL_1330117, EPI_ISL_1330118, EPI_ISL_1330119, EPI_ISL_1330120, EPI_ISL_1330121, EPI_ISL_1330122, EPI_ISL_1330123, EPI_ISL_1330124,
EPI_ISL_1330125, EPI_ISL_1330126, EPI_ISL_1330127, EPI_ISL_1330128, EPI_ISL_1330129, EPI_ISL_1330130, EPI_ISL_1330131, EPI_ISL_1330132, EPI_ISL_1330133, EPI_ISL_1330134, EPI_ISL_1330135, EPI_ISL_1330136, EPI_ISL_1330137, EPI_ISL_1330138, EPI_ISL_1330139, EPI_ISL_1330140, EPI_ISL_1330141, EPI_ISL_1330142, EPI_ISL_1330143, EPI_ISL_1330144, EPI_ISL_1330145, EPI_ISL_1330146,
EPI_ISL_1330147, EPI_ISL_1330148, EPI_ISL_1330149, EPI_ISL_1330150, EPI_ISL_1330151, EPI_ISL_1330153, EPI_ISL_1330155, EPI_ISL_1330156, EPI_ISL_1330157, EPI_ISL_1330159, EPI_ISL_1330160, EPI_ISL_1330161, EPI_ISL_1330162, EPI_ISL_1330163, EPI_ISL_1330164, EPI_ISL_1330165, EPI_ISL_1330167, EPI_ISL_1330168, EPI_ISL_1330169, EPI_ISL_1330170, EPI_ISL_1330171, EPI_ISL_1330172,
EPI_ISL_1330173, EPI_ISL_1330174, EPI_ISL_1330175, EPI_ISL_1330177, EPI_ISL_1330178, EPI_ISL_1330179, EPI_ISL_1330180, EPI_ISL_1330181, EPI_ISL_1330182, EPI_ISL_1330184, EPI_ISL_1330185, EPI_ISL_1330186, EPI_ISL_1330187, EPI_ISL_1330188, EPI_ISL_1330189, EPI_ISL_1330190, EPI_ISL_1330192, EPI_ISL_1330193, EPI_ISL_1330194, EPI_ISL_1330195, EPI_ISL_1330196, EPI_ISL_1330197,
EPI_ISL_1330198, EPI_ISL_1330200, EPI_ISL_1330202, EPI_ISL_1330203, EPI_ISL_1330205, EPI_ISL_1330207, EPI_ISL_1330208, EPI_ISL_1330209, EPI_ISL_1330210, EPI_ISL_1330211, EPI_ISL_1330212, EPI_ISL_1330213, EPI_ISL_1330215, EPI_ISL_1330216, EPI_ISL_1330217, EPI_ISL_1330218, EPI_ISL_1330219, EPI_ISL_1330220, EPI_ISL_1330221, EPI_ISL_1330222, EPI_ISL_1330223, EPI_ISL_1330225,
EPI_ISL_1330226, EPI_ISL_1330227, EPI_ISL_1330228, EPI_ISL_1330229, EPI_ISL_1330230, EPI_ISL_1330231, EPI_ISL_1330233, EPI_ISL_1330234, EPI_ISL_1330235, EPI_ISL_1330236, EPI_ISL_1330237, EPI_ISL_1330238, EPI_ISL_1330239, EPI_ISL_1330241, EPI_ISL_1330242, EPI_ISL_1330243, EPI_ISL_1330244, EPI_ISL_1330245, EPI_ISL_1330246, EPI_ISL_1330247, EPI_ISL_1330249, EPI_ISL_1330250,
EPI_ISL_1330252, EPI_ISL_1330253, EPI_ISL_1330255, EPI_ISL_1330256, EPI_ISL_1330258, EPI_ISL_1330259, EPI_ISL_1330261, EPI_ISL_1330262, EPI_ISL_1330263, EPI_ISL_1330264, EPI_ISL_1330265, EPI_ISL_1330266, EPI_ISL_1330267, EPI_ISL_1330268, EPI_ISL_1330269, EPI_ISL_1330270, EPI_ISL_1330271, EPI_ISL_1330272, EPI_ISL_1330273, EPI_ISL_1330274, EPI_ISL_1330275, EPI_ISL_1330276,
EPI_ISL_1330277, EPI_ISL_1330278, EPI_ISL_1330279, EPI_ISL_1330280, EPI_ISL_1330281, EPI_ISL_1330282, EPI_ISL_1330283, EPI_ISL_1330284, EPI_ISL_1330285, EPI_ISL_1330286, EPI_ISL_1330287, EPI_ISL_1330288, EPI_ISL_1330289, EPI_ISL_1330290, EPI_ISL_1330291, EPI_ISL_1330292, EPI_ISL_1330293, EPI_ISL_1330294, EPI_ISL_1330295, EPI_ISL_1330296, EPI_ISL_1330924, EPI_ISL_1330926,
EPI_ISL_1330929, EPI_ISL_1330930, EPI_ISL_1330931, EPI_ISL_1330932, EPI_ISL_1330934, EPI_ISL_1330936, EPI_ISL_1330940, EPI_ISL_1330941, EPI_ISL_1330943, EPI_ISL_1330944, EPI_ISL_1330945, EPI_ISL_1330948, EPI_ISL_1330949, EPI_ISL_1330950, EPI_ISL_1330952, EPI_ISL_1330953, EPI_ISL_1330955, EPI_ISL_1330956, EPI_ISL_1330957, EPI_ISL_1330960, EPI_ISL_1330962, EPI_ISL_1330964,
EPI_ISL_1330965, EPI_ISL_1330966, EPI_ISL_1330967, EPI_ISL_1330969, EPI_ISL_1330970, EPI_ISL_1330971, EPI_ISL_1330972, EPI_ISL_1330973, EPI_ISL_1330974, EPI_ISL_1330975, EPI_ISL_1330976, EPI_ISL_1330977, EPI_ISL_1330978, EPI_ISL_1330979, EPI_ISL_1330981, EPI_ISL_1330982, EPI_ISL_1330985, EPI_ISL_1330986, EPI_ISL_1330987, EPI_ISL_1330988, EPI_ISL_1330990, EPI_ISL_1330992,
EPI_ISL_1330993, EPI_ISL_1330994, EPI_ISL_1330996, EPI_ISL_1330998, EPI_ISL_1330999, EPI_ISL_1331000, EPI_ISL_1331001, EPI_ISL_1331002, EPI_ISL_1331003, EPI_ISL_1331004, EPI_ISL_1331005, EPI_ISL_1331006, EPI_ISL_1331007, EPI_ISL_1331008, EPI_ISL_1331009, EPI_ISL_1331010, EPI_ISL_1331011, EPI_ISL_1331012, EPI_ISL_1331013, EPI_ISL_1331017, EPI_ISL_1331018, EPI_ISL_1331020,
EPI_ISL_1331021, EPI_ISL_1331023, EPI_ISL_1331024, EPI_ISL_1331025, EPI_ISL_1331026, EPI_ISL_1331027, EPI_ISL_1331028, EPI_ISL_1331029, EPI_ISL_1331030, EPI_ISL_1331032, EPI_ISL_1331033, EPI_ISL_1331036, EPI_ISL_1331038, EPI_ISL_1331039, EPI_ISL_1331044, EPI_ISL_1331046, EPI_ISL_1331047, EPI_ISL_1331048, EPI_ISL_1331052, EPI_ISL_1331053, EPI_ISL_1331054, EPI_ISL_1331055,
EPI_ISL_1331056, EPI_ISL_1331057, EPI_ISL_1331058, EPI_ISL_1331060, EPI_ISL_1331061, EPI_ISL_1331062, EPI_ISL_1331063, EPI_ISL_1331064, EPI_ISL_1331065, EPI_ISL_1331066, EPI_ISL_1331068, EPI_ISL_1331069, EPI_ISL_1331070, EPI_ISL_1331071, EPI_ISL_1331072, EPI_ISL_1331073, EPI_ISL_1331074, EPI_ISL_1331075, EPI_ISL_1331076, EPI_ISL_1331077, EPI_ISL_1331078, EPI_ISL_1331079,
EPI_ISL_1331080, EPI_ISL_1331081, EPI_ISL_1331082, EPI_ISL_1331083, EPI_ISL_1331084, EPI_ISL_1331085, EPI_ISL_1331087, EPI_ISL_1331088, EPI_ISL_1331089, EPI_ISL_1331091, EPI_ISL_1331095, EPI_ISL_1331096, EPI_ISL_1331100, EPI_ISL_1331101, EPI_ISL_1331102, EPI_ISL_1331105, EPI_ISL_1331106, EPI_ISL_1331107, EPI_ISL_1331108, EPI_ISL_1331111, EPI_ISL_1331113, EPI_ISL_1331115,
EPI_ISL_1331117, EPI_ISL_1331118, EPI_ISL_1331120, EPI_ISL_1331121, EPI_ISL_1331122, EPI_ISL_1331124, EPI_ISL_1331126, EPI_ISL_1331128, EPI_ISL_1331129, EPI_ISL_1331130, EPI_ISL_1331154, EPI_ISL_1331158, EPI_ISL_1331162, EPI_ISL_1331190, EPI_ISL_1331199, EPI_ISL_1331200, EPI_ISL_1331203, EPI_ISL_1331224, EPI_ISL_1331226, EPI_ISL_1331256, EPI_ISL_1331265, EPI_ISL_1331270,
EPI_ISL_1331289, EPI_ISL_1331298, EPI_ISL_1331301, EPI_ISL_1331311, EPI_ISL_1331313, EPI_ISL_1331318, EPI_ISL_1331322, EPI_ISL_1331324, EPI_ISL_1331338, EPI_ISL_1331355, EPI_ISL_1331359, EPI_ISL_1331372, EPI_ISL_1331382, EPI_ISL_1331383, EPI_ISL_1331384, EPI_ISL_1331389, EPI_ISL_1331391, EPI_ISL_1331393, EPI_ISL_1331394, EPI_ISL_1331401, EPI_ISL_1331412, EPI_ISL_1331441,
EPI_ISL_1331447, EPI_ISL_1331463, EPI_ISL_1331464, EPI_ISL_1331465, EPI_ISL_1331466, EPI_ISL_1331467, EPI_ISL_1331468, EPI_ISL_1331469, EPI_ISL_1331470, EPI_ISL_1331471, EPI_ISL_1331472, EPI_ISL_1331473, EPI_ISL_1331474, EPI_ISL_1331475, EPI_ISL_1331476, EPI_ISL_1331477, EPI_ISL_1331478, EPI_ISL_1331479, EPI_ISL_1331480, EPI_ISL_1331481, EPI_ISL_1331482, EPI_ISL_1331483,
EPI_ISL_1331484, EPI_ISL_1331485, EPI_ISL_1331486, EPI_ISL_1331487, EPI_ISL_1331488, EPI_ISL_1331489, EPI_ISL_1331490, EPI_ISL_1331491, EPI_ISL_1331492, EPI_ISL_1331493, EPI_ISL_1331494, EPI_ISL_1331495, EPI_ISL_1331496, EPI_ISL_1331497, EPI_ISL_1331498, EPI_ISL_1331499, EPI_ISL_1331500, EPI_ISL_1331501, EPI_ISL_1331502, EPI_ISL_1331503, EPI_ISL_1331504, EPI_ISL_1331505,
EPI_ISL_1331506, EPI_ISL_1331507, EPI_ISL_1331508, EPI_ISL_1331509, EPI_ISL_1331510, EPI_ISL_1331511, EPI_ISL_1331512, EPI_ISL_1331513, EPI_ISL_1331514, EPI_ISL_1331515, EPI_ISL_1331516, EPI_ISL_1331517, EPI_ISL_1331518, EPI_ISL_1331519, EPI_ISL_1331520, EPI_ISL_1331521, EPI_ISL_1331522, EPI_ISL_1331523, EPI_ISL_1331524, EPI_ISL_1331525, EPI_ISL_1331526, EPI_ISL_1331528,
EPI_ISL_1331529, EPI_ISL_1331530, EPI_ISL_1331531, EPI_ISL_1331532, EPI_ISL_1331533, EPI_ISL_1331534, EPI_ISL_1331535, EPI_ISL_1331536, EPI_ISL_1331537, EPI_ISL_1331538, EPI_ISL_1331539, EPI_ISL_1331540, EPI_ISL_1331541, EPI_ISL_1331542, EPI_ISL_1331543, EPI_ISL_1331544, EPI_ISL_1331545, EPI_ISL_1331546, EPI_ISL_1331548, EPI_ISL_1331549, EPI_ISL_1331550, EPI_ISL_1331551,
EPI_ISL_1331552, EPI_ISL_1331553, EPI_ISL_1331554, EPI_ISL_1331555, EPI_ISL_1331556, EPI_ISL_1331557, EPI_ISL_1331558, EPI_ISL_1331560, EPI_ISL_1331561, EPI_ISL_1331562, EPI_ISL_1331563, EPI_ISL_1331564, EPI_ISL_1331565, EPI_ISL_1331566, EPI_ISL_1331567, EPI_ISL_1331568, EPI_ISL_1331569, EPI_ISL_1331570, EPI_ISL_1331571, EPI_ISL_1331572, EPI_ISL_1331573, EPI_ISL_1331574,
EPI_ISL_1331575, EPI_ISL_1331576, EPI_ISL_1331577, EPI_ISL_1331578, EPI_ISL_1331579, EPI_ISL_1331580, EPI_ISL_1331581, EPI_ISL_1331582, EPI_ISL_1331583, EPI_ISL_1331584, EPI_ISL_1331585, EPI_ISL_1331586, EPI_ISL_1331588, EPI_ISL_1331589, EPI_ISL_1331590, EPI_ISL_1331591, EPI_ISL_1331592, EPI_ISL_1331593, EPI_ISL_1331594, EPI_ISL_1331595, EPI_ISL_1331596, EPI_ISL_1331597,
EPI_ISL_1331598, EPI_ISL_1331599, EPI_ISL_1331600, EPI_ISL_1331601, EPI_ISL_1331602, EPI_ISL_1331603, EPI_ISL_1331604, EPI_ISL_1331605, EPI_ISL_1331606, EPI_ISL_1331607, EPI_ISL_1331608, EPI_ISL_1331609, EPI_ISL_1331610, EPI_ISL_1331611, EPI_ISL_1331612, EPI_ISL_1331613, EPI_ISL_1331614, EPI_ISL_1331615, EPI_ISL_1331616, EPI_ISL_1331617, EPI_ISL_1331618, EPI_ISL_1331619,
EPI_ISL_1331620, EPI_ISL_1331621, EPI_ISL_1331622, EPI_ISL_1331623, EPI_ISL_1331624, EPI_ISL_1331625, EPI_ISL_1331626, EPI_ISL_1331627, EPI_ISL_1331628, EPI_ISL_1331629, EPI_ISL_1331630, EPI_ISL_1331631, EPI_ISL_1331632, EPI_ISL_1331633, EPI_ISL_1331634, EPI_ISL_1331635, EPI_ISL_1331636, EPI_ISL_1331637, EPI_ISL_1331638, EPI_ISL_1331639, EPI_ISL_1331640, EPI_ISL_1331641,
EPI_ISL_1331642, EPI_ISL_1331643, EPI_ISL_1331644, EPI_ISL_1331645, EPI_ISL_1331646, EPI_ISL_1331647, EPI_ISL_1331648, EPI_ISL_1331649, EPI_ISL_1331650, EPI_ISL_1331651, EPI_ISL_1331652, EPI_ISL_1331653, EPI_ISL_1331654, EPI_ISL_1331655, EPI_ISL_1331656, EPI_ISL_1331657, EPI_ISL_1331658, EPI_ISL_1331659, EPI_ISL_1331660, EPI_ISL_1331661, EPI_ISL_1331662, EPI_ISL_1331663,
EPI_ISL_1331664, EPI_ISL_1331665, EPI_ISL_1331666, EPI_ISL_1331667, EPI_ISL_1331668, EPI_ISL_1331669, EPI_ISL_1331670, EPI_ISL_1331671, EPI_ISL_1331672, EPI_ISL_1331673, EPI_ISL_1331674, EPI_ISL_1331675, EPI_ISL_1331676, EPI_ISL_1331677, EPI_ISL_1331678, EPI_ISL_1331679, EPI_ISL_1331680, EPI_ISL_1331681, EPI_ISL_1331682, EPI_ISL_1331683, EPI_ISL_1331684, EPI_ISL_1331686,
EPI_ISL_1331687, EPI_ISL_1331688, EPI_ISL_1331689, EPI_ISL_1331690, EPI_ISL_1331691, EPI_ISL_1331692, EPI_ISL_1331693, EPI_ISL_1331694, EPI_ISL_1331695, EPI_ISL_1331696, EPI_ISL_1331698, EPI_ISL_1331699, EPI_ISL_1331700, EPI_ISL_1331703, EPI_ISL_1331704, EPI_ISL_1331705, EPI_ISL_1331707, EPI_ISL_1331709, EPI_ISL_1331710, EPI_ISL_1331711, EPI_ISL_1331713, EPI_ISL_1331715,
EPI_ISL_1331716, EPI_ISL_1331722, EPI_ISL_1331723, EPI_ISL_1331728, EPI_ISL_1331729, EPI_ISL_1331730, EPI_ISL_1331731, EPI_ISL_1331732, EPI_ISL_1331734, EPI_ISL_1331737, EPI_ISL_1331738, EPI_ISL_1331739, EPI_ISL_1331741, EPI_ISL_1331742, EPI_ISL_1331743, EPI_ISL_1331745, EPI_ISL_1331746, EPI_ISL_1331747, EPI_ISL_1331748, EPI_ISL_1331749, EPI_ISL_1331750, EPI_ISL_1331751,
EPI_ISL_1331752, EPI_ISL_1331753, EPI_ISL_1331754, EPI_ISL_1331755, EPI_ISL_1331756, EPI_ISL_1331757, EPI_ISL_1331758, EPI_ISL_1331762, EPI_ISL_1331763, EPI_ISL_1331765, EPI_ISL_1331767, EPI_ISL_1331768, EPI_ISL_1331770, EPI_ISL_1331771, EPI_ISL_1331773, EPI_ISL_1331775, EPI_ISL_1331777, EPI_ISL_1331778, EPI_ISL_1331781, EPI_ISL_1331783, EPI_ISL_1331784, EPI_ISL_1331786,
EPI_ISL_1331787, EPI_ISL_1331790, EPI_ISL_1331791, EPI_ISL_1331793, EPI_ISL_1331795, EPI_ISL_1331796, EPI_ISL_1331797, EPI_ISL_1331798, EPI_ISL_1331799, EPI_ISL_1331800, EPI_ISL_1331801, EPI_ISL_1331803, EPI_ISL_1331804, EPI_ISL_1331805, EPI_ISL_1331807, EPI_ISL_1331808, EPI_ISL_1331809, EPI_ISL_1331810, EPI_ISL_1331812, EPI_ISL_1331814, EPI_ISL_1331815, EPI_ISL_1331816,
EPI_ISL_1331817, EPI_ISL_1331820, EPI_ISL_1331821, EPI_ISL_1331823, EPI_ISL_1331826, EPI_ISL_1331827, EPI_ISL_1331828, EPI_ISL_1331829, EPI_ISL_1331830, EPI_ISL_1331831, EPI_ISL_1331832, EPI_ISL_1331834, EPI_ISL_1331836, EPI_ISL_1331837, EPI_ISL_1331838, EPI_ISL_1331839, EPI_ISL_1331840, EPI_ISL_1331844, EPI_ISL_1331845, EPI_ISL_1331846, EPI_ISL_1331849, EPI_ISL_1331850,
EPI_ISL_1331851, EPI_ISL_1331853, EPI_ISL_1331854, EPI_ISL_1331855, EPI_ISL_1331857, EPI_ISL_1331858, EPI_ISL_1331860, EPI_ISL_1331861, EPI_ISL_1331862, EPI_ISL_1331863, EPI_ISL_1331864, EPI_ISL_1331866, EPI_ISL_1331868, EPI_ISL_1331869, EPI_ISL_1331872, EPI_ISL_1331874, EPI_ISL_1331875, EPI_ISL_1331876, EPI_ISL_1331878, EPI_ISL_1331879, EPI_ISL_1331880, EPI_ISL_1331881,
EPI_ISL_1331882, EPI_ISL_1331883, EPI_ISL_1331884, EPI_ISL_1331885, EPI_ISL_1331889, EPI_ISL_1331890, EPI_ISL_1331892, EPI_ISL_1331894, EPI_ISL_1331895, EPI_ISL_1331897, EPI_ISL_1331898, EPI_ISL_1331904, EPI_ISL_1331906, EPI_ISL_1331907, EPI_ISL_1331911, EPI_ISL_1331912, EPI_ISL_1331913, EPI_ISL_1331914, EPI_ISL_1331916, EPI_ISL_1331917, EPI_ISL_1331918, EPI_ISL_1331921,
EPI_ISL_1331922, EPI_ISL_1331923, EPI_ISL_1331925, EPI_ISL_1331928, EPI_ISL_1331930, EPI_ISL_1331931, EPI_ISL_1331932, EPI_ISL_1331933, EPI_ISL_1331938, EPI_ISL_1331939, EPI_ISL_1331940, EPI_ISL_1331941, EPI_ISL_1331942, EPI_ISL_1331943, EPI_ISL_1331949, EPI_ISL_1331951, EPI_ISL_1331954, EPI_ISL_1331956, EPI_ISL_1331957, EPI_ISL_1331958, EPI_ISL_1331959, EPI_ISL_1331960,
EPI_ISL_1331962, EPI_ISL_1331963, EPI_ISL_1331964, EPI_ISL_1331967, EPI_ISL_1331971, EPI_ISL_1331972, EPI_ISL_1331973, EPI_ISL_1331974, EPI_ISL_1331975, EPI_ISL_1331976, EPI_ISL_1331977, EPI_ISL_1331978, EPI_ISL_1331979, EPI_ISL_1331980, EPI_ISL_1331982, EPI_ISL_1331983, EPI_ISL_1331984, EPI_ISL_1331986, EPI_ISL_1331987, EPI_ISL_1331988, EPI_ISL_1331989, EPI_ISL_1331990,
EPI_ISL_1331991, EPI_ISL_1331992, EPI_ISL_1331993, EPI_ISL_1331995, EPI_ISL_1331996, EPI_ISL_1331997, EPI_ISL_1331999, EPI_ISL_1332001, EPI_ISL_1332002, EPI_ISL_1332004, EPI_ISL_1332005, EPI_ISL_1332006, EPI_ISL_1332007, EPI_ISL_1332008, EPI_ISL_1332009, EPI_ISL_1332010, EPI_ISL_1332011, EPI_ISL_1332012, EPI_ISL_1332015, EPI_ISL_1332017, EPI_ISL_1332018, EPI_ISL_1332019,
EPI_ISL_1332020, EPI_ISL_1332022, EPI_ISL_1332023, EPI_ISL_1332024, EPI_ISL_1332025, EPI_ISL_1332026, EPI_ISL_1332028, EPI_ISL_1332030, EPI_ISL_1332031, EPI_ISL_1332032, EPI_ISL_1332033, EPI_ISL_1332035, EPI_ISL_1332036, EPI_ISL_1332039, EPI_ISL_1332040, EPI_ISL_1332042, EPI_ISL_1332044, EPI_ISL_1332045, EPI_ISL_1332046, EPI_ISL_1332048, EPI_ISL_1332049, EPI_ISL_1332053,
EPI_ISL_1332054, EPI_ISL_1332056, EPI_ISL_1332057, EPI_ISL_1332058, EPI_ISL_1332060, EPI_ISL_1332062, EPI_ISL_1332066, EPI_ISL_1332067, EPI_ISL_1332069, EPI_ISL_1332070, EPI_ISL_1332072, EPI_ISL_1332073, EPI_ISL_1332074, EPI_ISL_1332076, EPI_ISL_1332077, EPI_ISL_1332079, EPI_ISL_1332080, EPI_ISL_1332081, EPI_ISL_1332082, EPI_ISL_1332083, EPI_ISL_1332084, EPI_ISL_1332086,
EPI_ISL_1332087, EPI_ISL_1332088, EPI_ISL_1332090, EPI_ISL_1332091, EPI_ISL_1332092, EPI_ISL_1332094, EPI_ISL_1332095, EPI_ISL_1332096, EPI_ISL_1332097, EPI_ISL_1332100, EPI_ISL_1332101, EPI_ISL_1332102, EPI_ISL_1332103, EPI_ISL_1332105, EPI_ISL_1332106, EPI_ISL_1332107, EPI_ISL_1332108, EPI_ISL_1332109, EPI_ISL_1332110, EPI_ISL_1332111, EPI_ISL_1332113, EPI_ISL_1332114,
EPI_ISL_1332115, EPI_ISL_1332116, EPI_ISL_1332117, EPI_ISL_1332118, EPI_ISL_1332120, EPI_ISL_1332121, EPI_ISL_1332122, EPI_ISL_1332124, EPI_ISL_1332125, EPI_ISL_1332126, EPI_ISL_1332129, EPI_ISL_1332130, EPI_ISL_1332131, EPI_ISL_1332132, EPI_ISL_1332134, EPI_ISL_1332138, EPI_ISL_1332139, EPI_ISL_1332140, EPI_ISL_1332142, EPI_ISL_1332143, EPI_ISL_1332147, EPI_ISL_1332149,
EPI_ISL_1332150, EPI_ISL_1332151, EPI_ISL_1332152, EPI_ISL_1332153, EPI_ISL_1332154, EPI_ISL_1332155, EPI_ISL_1332157, EPI_ISL_1332161, EPI_ISL_1332162, EPI_ISL_1332164, EPI_ISL_1332167, EPI_ISL_1332168, EPI_ISL_1332169, EPI_ISL_1332171, EPI_ISL_1332173, EPI_ISL_1332174, EPI_ISL_1332175, EPI_ISL_1332176, EPI_ISL_1332177, EPI_ISL_1332178, EPI_ISL_1332179, EPI_ISL_1332182,
EPI_ISL_1332183, EPI_ISL_1332184, EPI_ISL_1332185, EPI_ISL_1332186, EPI_ISL_1332189, EPI_ISL_1332190, EPI_ISL_1332191, EPI_ISL_1332194, EPI_ISL_1332195, EPI_ISL_1332197, EPI_ISL_1332198, EPI_ISL_1332199, EPI_ISL_1332200, EPI_ISL_1332202, EPI_ISL_1332203, EPI_ISL_1332207, EPI_ISL_1332208, EPI_ISL_1332209, EPI_ISL_1332210, EPI_ISL_1332211, EPI_ISL_1332216, EPI_ISL_1332218,
EPI_ISL_1332219, EPI_ISL_1332220, EPI_ISL_1332221, EPI_ISL_1332223, EPI_ISL_1332224, EPI_ISL_1332225, EPI_ISL_1332226, EPI_ISL_1332227, EPI_ISL_1332229, EPI_ISL_1332230, EPI_ISL_1332231, EPI_ISL_1332232, EPI_ISL_1332233, EPI_ISL_1332234, EPI_ISL_1332235, EPI_ISL_1332236, EPI_ISL_1332241, EPI_ISL_1332242, EPI_ISL_1332243, EPI_ISL_1332245, EPI_ISL_1332246, EPI_ISL_1332248,
EPI_ISL_1332249, EPI_ISL_1332251, EPI_ISL_1332252, EPI_ISL_1332253, EPI_ISL_1332254, EPI_ISL_1332255, EPI_ISL_1332256, EPI_ISL_1332257, EPI_ISL_1332258, EPI_ISL_1332259, EPI_ISL_1332262, EPI_ISL_1332263, EPI_ISL_1332264, EPI_ISL_1332265, EPI_ISL_1332270, EPI_ISL_1332272, EPI_ISL_1332273, EPI_ISL_1332275, EPI_ISL_1332276, EPI_ISL_1332277, EPI_ISL_1332278, EPI_ISL_1332279,
EPI_ISL_1332280, EPI_ISL_1332281, EPI_ISL_1332282, EPI_ISL_1332283, EPI_ISL_1332285, EPI_ISL_1332286, EPI_ISL_1332288, EPI_ISL_1332289, EPI_ISL_1332290, EPI_ISL_1332291, EPI_ISL_1332292, EPI_ISL_1332293, EPI_ISL_1332294, EPI_ISL_1332295, EPI_ISL_1332296, EPI_ISL_1332297, EPI_ISL_1332299, EPI_ISL_1332300, EPI_ISL_1332301, EPI_ISL_1332302, EPI_ISL_1332303, EPI_ISL_1332304,
EPI_ISL_1332306, EPI_ISL_1332307, EPI_ISL_1332308, EPI_ISL_1332309, EPI_ISL_1332310, EPI_ISL_1332311, EPI_ISL_1332312, EPI_ISL_1332313, EPI_ISL_1332314, EPI_ISL_1332315, EPI_ISL_1332316, EPI_ISL_1332317, EPI_ISL_1332318, EPI_ISL_1332319, EPI_ISL_1332320, EPI_ISL_1332321, EPI_ISL_1332322, EPI_ISL_1332323, EPI_ISL_1332324, EPI_ISL_1332325, EPI_ISL_1332326, EPI_ISL_1332327,
EPI_ISL_1332328, EPI_ISL_1332329, EPI_ISL_1332330, EPI_ISL_1332331, EPI_ISL_1332332, EPI_ISL_1332333, EPI_ISL_1332334, EPI_ISL_1332335, EPI_ISL_1332336, EPI_ISL_1332337, EPI_ISL_1332338, EPI_ISL_1332339, EPI_ISL_1332340, EPI_ISL_1332341, EPI_ISL_1332342, EPI_ISL_1332343, EPI_ISL_1332344, EPI_ISL_1332345, EPI_ISL_1332346, EPI_ISL_1332347, EPI_ISL_1332348, EPI_ISL_1332350,
EPI_ISL_1332351, EPI_ISL_1332352, EPI_ISL_1332353, EPI_ISL_1332354, EPI_ISL_1332355, EPI_ISL_1332356, EPI_ISL_1332357, EPI_ISL_1332358, EPI_ISL_1332359, EPI_ISL_1332360, EPI_ISL_1332361, EPI_ISL_1332362, EPI_ISL_1332363, EPI_ISL_1332364, EPI_ISL_1332366, EPI_ISL_1332367, EPI_ISL_1332368, EPI_ISL_1332369, EPI_ISL_1332370, EPI_ISL_1332371, EPI_ISL_1332372, EPI_ISL_1332373,
EPI_ISL_1332374, EPI_ISL_1332375, EPI_ISL_1332376, EPI_ISL_1332377, EPI_ISL_1332378, EPI_ISL_1332379, EPI_ISL_1332380, EPI_ISL_1332381, EPI_ISL_1332382, EPI_ISL_1332383, EPI_ISL_1332384, EPI_ISL_1332385, EPI_ISL_1332386, EPI_ISL_1332387, EPI_ISL_1332388, EPI_ISL_1332390, EPI_ISL_1332391, EPI_ISL_1332394, EPI_ISL_1332395, EPI_ISL_1332396, EPI_ISL_1332397, EPI_ISL_1332398,
EPI_ISL_1332400, EPI_ISL_1332401, EPI_ISL_1332402, EPI_ISL_1332403, EPI_ISL_1332404, EPI_ISL_1332405, EPI_ISL_1332406, EPI_ISL_1332407, EPI_ISL_1332408, EPI_ISL_1332409, EPI_ISL_1332410, EPI_ISL_1332411, EPI_ISL_1332412, EPI_ISL_1332413, EPI_ISL_1332414, EPI_ISL_1332415, EPI_ISL_1332416, EPI_ISL_1332417, EPI_ISL_1332418, EPI_ISL_1332419, EPI_ISL_1332420, EPI_ISL_1332421,
EPI_ISL_1332422, EPI_ISL_1332423, EPI_ISL_1332424, EPI_ISL_1332426, EPI_ISL_1332427, EPI_ISL_1332428, EPI_ISL_1332429, EPI_ISL_1332430, EPI_ISL_1332431, EPI_ISL_1332432, EPI_ISL_1332433, EPI_ISL_1332434, EPI_ISL_1332435, EPI_ISL_1332436, EPI_ISL_1332437, EPI_ISL_1332438, EPI_ISL_1332439, EPI_ISL_1332440, EPI_ISL_1332441, EPI_ISL_1332442, EPI_ISL_1332443, EPI_ISL_1332444,
EPI_ISL_1332445, EPI_ISL_1332446, EPI_ISL_1332447, EPI_ISL_1332448, EPI_ISL_1332449, EPI_ISL_1332450, EPI_ISL_1332452, EPI_ISL_1332453, EPI_ISL_1332454, EPI_ISL_1332455, EPI_ISL_1332456, EPI_ISL_1332457, EPI_ISL_1332458, EPI_ISL_1332459, EPI_ISL_1332460, EPI_ISL_1332461, EPI_ISL_1332462, EPI_ISL_1332463, EPI_ISL_1332464, EPI_ISL_1332465, EPI_ISL_1332466, EPI_ISL_1332467,
EPI_ISL_1332468, EPI_ISL_1332469, EPI_ISL_1332470, EPI_ISL_1332471, EPI_ISL_1332472, EPI_ISL_1332473, EPI_ISL_1332474, EPI_ISL_1332475, EPI_ISL_1332476, EPI_ISL_1332477, EPI_ISL_1332478, EPI_ISL_1332479, EPI_ISL_1332480, EPI_ISL_1332481, EPI_ISL_1332483, EPI_ISL_1332484, EPI_ISL_1332485, EPI_ISL_1332486, EPI_ISL_1332487, EPI_ISL_1332488, EPI_ISL_1332489, EPI_ISL_1332490,
EPI_ISL_1332491, EPI_ISL_1332492, EPI_ISL_1332493, EPI_ISL_1332494, EPI_ISL_1332495, EPI_ISL_1332496, EPI_ISL_1332497, EPI_ISL_1332498, EPI_ISL_1332499, EPI_ISL_1332500, EPI_ISL_1332501, EPI_ISL_1332502, EPI_ISL_1332503, EPI_ISL_1332504, EPI_ISL_1332505, EPI_ISL_1332506, EPI_ISL_1332507, EPI_ISL_1332508, EPI_ISL_1332509, EPI_ISL_1332510, EPI_ISL_1332513, EPI_ISL_1332514,
EPI_ISL_1332515, EPI_ISL_1332516, EPI_ISL_1332517, EPI_ISL_1332518, EPI_ISL_1332519, EPI_ISL_1332520, EPI_ISL_1332521, EPI_ISL_1332522, EPI_ISL_1332523, EPI_ISL_1332524, EPI_ISL_1332525, EPI_ISL_1332526, EPI_ISL_1332527, EPI_ISL_1332528, EPI_ISL_1332529, EPI_ISL_1332530, EPI_ISL_1332531, EPI_ISL_1332532, EPI_ISL_1332533, EPI_ISL_1332534, EPI_ISL_1332535, EPI_ISL_1332536,
EPI_ISL_1332537, EPI_ISL_1332538, EPI_ISL_1332539, EPI_ISL_1332540, EPI_ISL_1332541, EPI_ISL_1332542, EPI_ISL_1332543, EPI_ISL_1332544, EPI_ISL_1332546, EPI_ISL_1332547, EPI_ISL_1332548, EPI_ISL_1332549, EPI_ISL_1332550, EPI_ISL_1332551, EPI_ISL_1332552, EPI_ISL_1332553, EPI_ISL_1332554, EPI_ISL_1332557, EPI_ISL_1332558, EPI_ISL_1332562, EPI_ISL_1332563, EPI_ISL_1332565,
EPI_ISL_1332569, EPI_ISL_1332570, EPI_ISL_1332574, EPI_ISL_1332575, EPI_ISL_1332577, EPI_ISL_1332578, EPI_ISL_1332580, EPI_ISL_1332581, EPI_ISL_1332582, EPI_ISL_1332583, EPI_ISL_1332584, EPI_ISL_1332585, EPI_ISL_1332587, EPI_ISL_1332590, EPI_ISL_1332591, EPI_ISL_1332593, EPI_ISL_1332597, EPI_ISL_1332599, EPI_ISL_1332601, EPI_ISL_1332609, EPI_ISL_1332619, EPI_ISL_1332624,
EPI_ISL_1332627, EPI_ISL_1332628, EPI_ISL_1332629, EPI_ISL_1332630, EPI_ISL_1332631, EPI_ISL_1332632, EPI_ISL_1332634, EPI_ISL_1332638, EPI_ISL_1332644, EPI_ISL_1332646, EPI_ISL_1332649, EPI_ISL_1332650, EPI_ISL_1332651, EPI_ISL_1332652, EPI_ISL_1332653, EPI_ISL_1332654, EPI_ISL_1333186, EPI_ISL_1344965, EPI_ISL_1345004, EPI_ISL_1345055, EPI_ISL_1345057, EPI_ISL_1345058,
EPI_ISL_1345062, EPI_ISL_1345073, EPI_ISL_1345119, EPI_ISL_1345146, EPI_ISL_1345174, EPI_ISL_1345177, EPI_ISL_1345208, EPI_ISL_1345284, EPI_ISL_1345988, EPI_ISL_1346103, EPI_ISL_1346105, EPI_ISL_1346106, EPI_ISL_1346107, EPI_ISL_1346109, EPI_ISL_1346110, EPI_ISL_1346112, EPI_ISL_1346113, EPI_ISL_1346114, EPI_ISL_1346115, EPI_ISL_1346116, EPI_ISL_1346117, EPI_ISL_1346118,
EPI_ISL_1346119, EPI_ISL_1346120, EPI_ISL_1346121, EPI_ISL_1346122, EPI_ISL_1346123, EPI_ISL_1346124, EPI_ISL_1346125, EPI_ISL_1346126, EPI_ISL_1346127, EPI_ISL_1346128, EPI_ISL_1346129, EPI_ISL_1346130, EPI_ISL_1346131, EPI_ISL_1346132, EPI_ISL_1346135, EPI_ISL_1346136, EPI_ISL_1346137, EPI_ISL_1346139, EPI_ISL_1346140, EPI_ISL_1346141, EPI_ISL_1346142, EPI_ISL_1346144,
EPI_ISL_1346145, EPI_ISL_1346146, EPI_ISL_1346148, EPI_ISL_1346149, EPI_ISL_1346150, EPI_ISL_1346151, EPI_ISL_1346152, EPI_ISL_1346153, EPI_ISL_1346154, EPI_ISL_1346155, EPI_ISL_1346157, EPI_ISL_1346159, EPI_ISL_1346160, EPI_ISL_1346161, EPI_ISL_1346163, EPI_ISL_1346166, EPI_ISL_1346167, EPI_ISL_1346168, EPI_ISL_1346169, EPI_ISL_1346171, EPI_ISL_1346172, EPI_ISL_1346173,
EPI_ISL_1346174, EPI_ISL_1346175, EPI_ISL_1346176, EPI_ISL_1346177, EPI_ISL_1346178, EPI_ISL_1346180, EPI_ISL_1346181, EPI_ISL_1346182, EPI_ISL_1346183, EPI_ISL_1346184, EPI_ISL_1346186, EPI_ISL_1346187, EPI_ISL_1346190, EPI_ISL_1346192, EPI_ISL_1346195, EPI_ISL_1346196, EPI_ISL_1346198, EPI_ISL_1346200, EPI_ISL_1346201, EPI_ISL_1346202, EPI_ISL_1346203, EPI_ISL_1346204,
EPI_ISL_1346205, EPI_ISL_1346206, EPI_ISL_1346207, EPI_ISL_1346208, EPI_ISL_1346209, EPI_ISL_1346210, EPI_ISL_1346211, EPI_ISL_1346212, EPI_ISL_1346213, EPI_ISL_1346214, EPI_ISL_1346215, EPI_ISL_1346216, EPI_ISL_1346217, EPI_ISL_1346218, EPI_ISL_1346219, EPI_ISL_1346220, EPI_ISL_1346222, EPI_ISL_1346226, EPI_ISL_1346227, EPI_ISL_1346228, EPI_ISL_1346229, EPI_ISL_1346230,
EPI_ISL_1346232, EPI_ISL_1346234, EPI_ISL_1346235, EPI_ISL_1346237, EPI_ISL_1346239, EPI_ISL_1346240, EPI_ISL_1346241, EPI_ISL_1346242, EPI_ISL_1346243, EPI_ISL_1346244, EPI_ISL_1346245, EPI_ISL_1346246, EPI_ISL_1346247, EPI_ISL_1346248, EPI_ISL_1346249, EPI_ISL_1346250, EPI_ISL_1346251, EPI_ISL_1346252, EPI_ISL_1346254, EPI_ISL_1346255, EPI_ISL_1346256, EPI_ISL_1346257,
EPI_ISL_1346258, EPI_ISL_1346259, EPI_ISL_1346263, EPI_ISL_1346264, EPI_ISL_1346266, EPI_ISL_1346268, EPI_ISL_1346269, EPI_ISL_1346270, EPI_ISL_1346271, EPI_ISL_1346272, EPI_ISL_1346273, EPI_ISL_1346274, EPI_ISL_1346275, EPI_ISL_1346276, EPI_ISL_1346278, EPI_ISL_1346279, EPI_ISL_1346280, EPI_ISL_1346282, EPI_ISL_1346292, EPI_ISL_1346296, EPI_ISL_1346297, EPI_ISL_1346299,
EPI_ISL_1346303, EPI_ISL_1346304, EPI_ISL_1346306, EPI_ISL_1346307, EPI_ISL_1346333, EPI_ISL_1346336, EPI_ISL_1346343, EPI_ISL_1346346, EPI_ISL_1346347, EPI_ISL_1346348, EPI_ISL_1346359, EPI_ISL_1346403, EPI_ISL_1346404, EPI_ISL_1346405, EPI_ISL_1346406, EPI_ISL_1346407, EPI_ISL_1346421, EPI_ISL_1346422, EPI_ISL_1346424, EPI_ISL_1346425, EPI_ISL_1346428, EPI_ISL_1346444,
EPI_ISL_1346445, EPI_ISL_1346446, EPI_ISL_1346447, EPI_ISL_1346448, EPI_ISL_1346449, EPI_ISL_1346450, EPI_ISL_1346451, EPI_ISL_1346452, EPI_ISL_1346454, EPI_ISL_1346455, EPI_ISL_1346456, EPI_ISL_1346457, EPI_ISL_1346458, EPI_ISL_1346459, EPI_ISL_1346460, EPI_ISL_1346463, EPI_ISL_1346465, EPI_ISL_1346466, EPI_ISL_1346467, EPI_ISL_1346468, EPI_ISL_1346470, EPI_ISL_1346471,
EPI_ISL_1346475, EPI_ISL_1346476, EPI_ISL_1346477, EPI_ISL_1346479, EPI_ISL_1346480, EPI_ISL_1376085, EPI_ISL_1377399, EPI_ISL_1378299, EPI_ISL_1697743, EPI_ISL_1697760, EPI_ISL_1697764, EPI_ISL_1697915, EPI_ISL_1698558, EPI_ISL_1698566, EPI_ISL_1698590, EPI_ISL_1698610, EPI_ISL_1698624, EPI_ISL_1698626, EPI_ISL_1698636, EPI_ISL_1698653, EPI_ISL_1698759, EPI_ISL_1699404,
EPI_ISL_1699476, EPI_ISL_1699479, EPI_ISL_1699524, EPI_ISL_1699541, EPI_ISL_1699560, EPI_ISL_1742205

| | | | |
|---|---|---|---|
| see above | Lighthouse Lab in Glasgow | Wellcome Sanger Institute for the COVID-19 Genomics UK (COG-UK) Consortium | Anna Dominiczak and Alex Alderton; Carol Clugston; Cordelia Langford; David Gray; David K. Jackson; Dominic Kwiatkowski; Ewan Harrison; Harper VanSteenhouse; Ian Johnston; Jeffrey Barrett; John Sillitoe on behalf of the Wellcome Sanger Institute COVID-19 Surveillance Team; Roberto Amato; Sonia Goncalves; Yumi Kasai |

EPI_ISL_1315831, EPI_ISL_1315833, EPI_ISL_1316077, EPI_ISL_1324807, EPI_ISL_1324823, EPI_ISL_1324830, EPI_ISL_1324831, EPI_ISL_1324835, EPI_ISL_1324838, EPI_ISL_1324858, EPI_ISL_1324860, EPI_ISL_1324865, EPI_ISL_1324866, EPI_ISL_1324881, EPI_ISL_1324884, EPI_ISL_1324890, EPI_ISL_1324891, EPI_ISL_1324899, EPI_ISL_1324901, EPI_ISL_1324905, EPI_ISL_1324910, EPI_ISL_1324920,

EPI_ISL_1324934, EPI_ISL_1324940, EPI_ISL_1324945, EPI_ISL_1324970, EPI_ISL_1324975, EPI_ISL_1324995, EPI_ISL_1324997, EPI_ISL_1325000, EPI_ISL_1325002, EPI_ISL_1325004, EPI_ISL_1325005, EPI_ISL_1325012, EPI_ISL_1325019, EPI_ISL_1325020, EPI_ISL_1325022, EPI_ISL_1325025, EPI_ISL_1325031, EPI_ISL_1325039, EPI_ISL_1325042, EPI_ISL_1325046, EPI_ISL_1325048, EPI_ISL_1325050, EPI_ISL_1325052, EPI_ISL_1325075, EPI_ISL_1325077, EPI_ISL_1325083, EPI_ISL_1325086, EPI_ISL_1325095, EPI_ISL_1325096, EPI_ISL_1325097, EPI_ISL_1325101, EPI_ISL_1325102, EPI_ISL_1325108, EPI_ISL_1325109, EPI_ISL_1325110, EPI_ISL_1325121, EPI_ISL_1325558, EPI_ISL_1325615, EPI_ISL_1325628, EPI_ISL_1325634, EPI_ISL_1325873, EPI_ISL_1325880, EPI_ISL_1325913, EPI_ISL_1325916, EPI_ISL_1325931, EPI_ISL_1325954, EPI_ISL_1325967, EPI_ISL_1325968, EPI_ISL_1325974, EPI_ISL_1325976, EPI_ISL_1325977, EPI_ISL_1326001, EPI_ISL_1326005, EPI_ISL_1326034, EPI_ISL_1326066, EPI_ISL_1326070, EPI_ISL_1326073, EPI_ISL_1326082, EPI_ISL_1326114, EPI_ISL_1326119, EPI_ISL_1326155, EPI_ISL_1326172, EPI_ISL_1326185, EPI_ISL_1326186, EPI_ISL_1326196, EPI_ISL_1326209, EPI_ISL_1326221, EPI_ISL_1326229, EPI_ISL_1326253, EPI_ISL_1326262, EPI_ISL_1326263, EPI_ISL_1326265, EPI_ISL_1326275, EPI_ISL_1326311, EPI_ISL_1326368, EPI_ISL_1326369, EPI_ISL_1326370, EPI_ISL_1326371, EPI_ISL_1326372, EPI_ISL_1326376, EPI_ISL_1326377, EPI_ISL_1326382, EPI_ISL_1326383, EPI_ISL_1326384, EPI_ISL_1326385, EPI_ISL_1326388, EPI_ISL_1326390, EPI_ISL_1326393, EPI_ISL_1326394, EPI_ISL_1326395, EPI_ISL_1326396, EPI_ISL_1326401, EPI_ISL_1326402, EPI_ISL_1326403, EPI_ISL_1326404, EPI_ISL_1326405, EPI_ISL_1326406, EPI_ISL_1326407, EPI_ISL_1326410, EPI_ISL_1326411, EPI_ISL_1326412, EPI_ISL_1326416, EPI_ISL_1326419, EPI_ISL_1326421, EPI_ISL_1326422, EPI_ISL_1326423, EPI_ISL_1326424, EPI_ISL_1326425, EPI_ISL_1326426, EPI_ISL_1326427, EPI_ISL_1326428, EPI_ISL_1326430, EPI_ISL_1326431, EPI_ISL_1326433, EPI_ISL_1326434, EPI_ISL_1326437, EPI_ISL_1326438, EPI_ISL_1326439, EPI_ISL_1326440, EPI_ISL_1326445, EPI_ISL_1326447, EPI_ISL_1326448, EPI_ISL_1326453, EPI_ISL_1326454, EPI_ISL_1326455, EPI_ISL_1326463, EPI_ISL_1326466, EPI_ISL_1326467, EPI_ISL_1326471, EPI_ISL_1326472, EPI_ISL_1326474, EPI_ISL_1326475, EPI_ISL_1326476, EPI_ISL_1326482, EPI_ISL_1326484, EPI_ISL_1326485, EPI_ISL_1326487, EPI_ISL_1326489, EPI_ISL_1326490, EPI_ISL_1326491, EPI_ISL_1326493, EPI_ISL_1326495, EPI_ISL_1326496, EPI_ISL_1326498, EPI_ISL_1326499, EPI_ISL_1326502, EPI_ISL_1326503, EPI_ISL_1326504, EPI_ISL_1326506, EPI_ISL_1326507, EPI_ISL_1326508, EPI_ISL_1326509, EPI_ISL_1326510, EPI_ISL_1326511, EPI_ISL_1326512, EPI_ISL_1326513, EPI_ISL_1326514, EPI_ISL_1326516, EPI_ISL_1326520, EPI_ISL_1326522, EPI_ISL_1326523, EPI_ISL_1326524, EPI_ISL_1326528, EPI_ISL_1326531, EPI_ISL_1326532, EPI_ISL_1326533, EPI_ISL_1326534, EPI_ISL_1326535, EPI_ISL_1326539, EPI_ISL_1326544, EPI_ISL_1326546, EPI_ISL_1326547, EPI_ISL_1326549, EPI_ISL_1326552, EPI_ISL_1326555, EPI_ISL_1326556, EPI_ISL_1326557, EPI_ISL_1326558, EPI_ISL_1326559, EPI_ISL_1326560, EPI_ISL_1326562, EPI_ISL_1326563, EPI_ISL_1326565, EPI_ISL_1326572, EPI_ISL_1326577, EPI_ISL_1326578, EPI_ISL_1326579, EPI_ISL_1326584, EPI_ISL_1326586, EPI_ISL_1326587, EPI_ISL_1326593, EPI_ISL_1326597, EPI_ISL_1326598, EPI_ISL_1326599, EPI_ISL_1326600, EPI_ISL_1326601, EPI_ISL_1326604, EPI_ISL_1326605, EPI_ISL_1326606, EPI_ISL_1326607, EPI_ISL_1326609, EPI_ISL_1326612, EPI_ISL_1326616, EPI_ISL_1326618, EPI_ISL_1326619, EPI_ISL_1326621, EPI_ISL_1326622, EPI_ISL_1326623, EPI_ISL_1326625, EPI_ISL_1326626, EPI_ISL_1326627, EPI_ISL_1326629, EPI_ISL_1326633, EPI_ISL_1326634, EPI_ISL_1326635, EPI_ISL_1326639, EPI_ISL_1326640, EPI_ISL_1326642, EPI_ISL_1326644, EPI_ISL_1326645, EPI_ISL_1326647, EPI_ISL_1326648, EPI_ISL_1326649, EPI_ISL_1326651, EPI_ISL_1326655, EPI_ISL_1326658, EPI_ISL_1326660, EPI_ISL_1326661, EPI_ISL_1327719, EPI_ISL_1327720, EPI_ISL_1327724, EPI_ISL_1327728, EPI_ISL_1327729, EPI_ISL_1327731, EPI_ISL_1327734, EPI_ISL_1327737, EPI_ISL_1327738, EPI_ISL_1327739, EPI_ISL_1327740, EPI_ISL_1327741, EPI_ISL_1327742, EPI_ISL_1327743, EPI_ISL_1327751, EPI_ISL_1327753, EPI_ISL_1327754, EPI_ISL_1327755, EPI_ISL_1327756, EPI_ISL_1327758, EPI_ISL_1327761, EPI_ISL_1327762, EPI_ISL_1327770, EPI_ISL_1327772, EPI_ISL_1327775, EPI_ISL_1327789, EPI_ISL_1327791, EPI_ISL_1327811, EPI_ISL_1327815, EPI_ISL_1327816, EPI_ISL_1327821, EPI_ISL_1327823, EPI_ISL_1327827, EPI_ISL_1327835, EPI_ISL_1327836, EPI_ISL_1327838, EPI_ISL_1327839, EPI_ISL_1327840, EPI_ISL_1327841, EPI_ISL_1327849, EPI_ISL_1327852, EPI_ISL_1327863, EPI_ISL_1327864, EPI_ISL_1327866, EPI_ISL_1327868, EPI_ISL_1327870, EPI_ISL_1327881, EPI_ISL_1327883, EPI_ISL_1327892, EPI_ISL_1327896, EPI_ISL_1327898, EPI_ISL_1327911, EPI_ISL_1327913, EPI_ISL_1327923, EPI_ISL_1327924, EPI_ISL_1327928, EPI_ISL_1327934, EPI_ISL_1327936, EPI_ISL_1327939, EPI_ISL_1327943, EPI_ISL_1327948, EPI_ISL_1327956, EPI_ISL_1327973, EPI_ISL_1327995, EPI_ISL_1328000, EPI_ISL_1328001, EPI_ISL_1328021, EPI_ISL_1328027, EPI_ISL_1328035, EPI_ISL_1328036, EPI_ISL_1328040, EPI_ISL_1328043, EPI_ISL_1328054, EPI_ISL_1328056, EPI_ISL_1328061, EPI_ISL_1328079, EPI_ISL_1328083, EPI_ISL_1328101, EPI_ISL_1328105, EPI_ISL_1328109, EPI_ISL_1328126, EPI_ISL_1328128, EPI_ISL_1328138, EPI_ISL_1328148, EPI_ISL_1328154, EPI_ISL_1328160, EPI_ISL_1328162, EPI_ISL_1328194, EPI_ISL_1328204, EPI_ISL_1328209, EPI_ISL_1328409, EPI_ISL_1328410, EPI_ISL_1328411, EPI_ISL_1328412, EPI_ISL_1328413, EPI_ISL_1328414, EPI_ISL_1328415, EPI_ISL_1328416, EPI_ISL_1328417, EPI_ISL_1328418, EPI_ISL_1328419, EPI_ISL_1328421, EPI_ISL_1328422, EPI_ISL_1328423, EPI_ISL_1328424, EPI_ISL_1328425, EPI_ISL_1328426, EPI_ISL_1328427, EPI_ISL_1328428, EPI_ISL_1328429, EPI_ISL_1328430, EPI_ISL_1328431, EPI_ISL_1328432, EPI_ISL_1328434, EPI_ISL_1328435, EPI_ISL_1328436, EPI_ISL_1328437, EPI_ISL_1328438, EPI_ISL_1328439, EPI_ISL_1328440, EPI_ISL_1328441, EPI_ISL_1328443, EPI_ISL_1328444, EPI_ISL_1328445, EPI_ISL_1328446, EPI_ISL_1328447, EPI_ISL_1328448, EPI_ISL_1328449, EPI_ISL_1328450, EPI_ISL_1328451, EPI_ISL_1328452, EPI_ISL_1328453, EPI_ISL_1328454, EPI_ISL_1328455, EPI_ISL_1328456, EPI_ISL_1328457, EPI_ISL_1328458, EPI_ISL_1328459, EPI_ISL_1328461, EPI_ISL_1328462, EPI_ISL_1328463, EPI_ISL_1328464, EPI_ISL_1328465, EPI_ISL_1328466, EPI_ISL_1328467, EPI_ISL_1328468, EPI_ISL_1328469, EPI_ISL_1328470, EPI_ISL_1328471, EPI_ISL_1328472, EPI_ISL_1328473, EPI_ISL_1328474, EPI_ISL_1328475, EPI_ISL_1328476, EPI_ISL_1328477, EPI_ISL_1328478, EPI_ISL_1328479, EPI_ISL_1328480, EPI_ISL_1328481, EPI_ISL_1328482, EPI_ISL_1328483, EPI_ISL_1328484, EPI_ISL_1328485, EPI_ISL_1328486, EPI_ISL_1328487, EPI_ISL_1328488, EPI_ISL_1328489, EPI_ISL_1328491, EPI_ISL_1328492, EPI_ISL_1328493, EPI_ISL_1328494, EPI_ISL_1328495, EPI_ISL_1328496, EPI_ISL_1328497, EPI_ISL_1328498, EPI_ISL_1328499, EPI_ISL_1328500, EPI_ISL_1328501, EPI_ISL_1328502, EPI_ISL_1328503, EPI_ISL_1328504, EPI_ISL_1328505, EPI_ISL_1328506, EPI_ISL_1328507, EPI_ISL_1328509, EPI_ISL_1328510, EPI_ISL_1328511, EPI_ISL_1328512, EPI_ISL_1328513, EPI_ISL_1328514, EPI_ISL_1328515, EPI_ISL_1328516, EPI_ISL_1328517, EPI_ISL_1328518, EPI_ISL_1328519, EPI_ISL_1328520, EPI_ISL_1328521, EPI_ISL_1328522, EPI_ISL_1328523, EPI_ISL_1328524, EPI_ISL_1328526, EPI_ISL_1328528, EPI_ISL_1328529, EPI_ISL_1328531, EPI_ISL_1328532, EPI_ISL_1328533, EPI_ISL_1328534, EPI_ISL_1328535, EPI_ISL_1328536, EPI_ISL_1328537, EPI_ISL_1328538, EPI_ISL_1328539, EPI_ISL_1328540, EPI_ISL_1328541, EPI_ISL_1328542, EPI_ISL_1328543, EPI_ISL_1328544, EPI_ISL_1328545, EPI_ISL_1328546, EPI_ISL_1328547, EPI_ISL_1328548, EPI_ISL_1328549, EPI_ISL_1328550, EPI_ISL_1328551, EPI_ISL_1328552, EPI_ISL_1328553, EPI_ISL_1328554, EPI_ISL_1328555, EPI_ISL_1328556, EPI_ISL_1328559, EPI_ISL_1328560, EPI_ISL_1328561, EPI_ISL_1328562, EPI_ISL_1328563, EPI_ISL_1328564, EPI_ISL_1328565, EPI_ISL_1328566, EPI_ISL_1328567, EPI_ISL_1328568, EPI_ISL_1328569, EPI_ISL_1328570, EPI_ISL_1328571, EPI_ISL_1328572, EPI_ISL_1328573, EPI_ISL_1328574, EPI_ISL_1328575, EPI_ISL_1328576, EPI_ISL_1328577, EPI_ISL_1328578, EPI_ISL_1328579, EPI_ISL_1328580, EPI_ISL_1328581, EPI_ISL_1328582, EPI_ISL_1328583, EPI_ISL_1328584, EPI_ISL_1328585, EPI_ISL_1328587, EPI_ISL_1328588, EPI_ISL_1328589, EPI_ISL_1328590, EPI_ISL_1328592, EPI_ISL_1328593, EPI_ISL_1328595, EPI_ISL_1328596, EPI_ISL_1328597, EPI_ISL_1328598, EPI_ISL_1328600, EPI_ISL_1328602, EPI_ISL_1328603, EPI_ISL_1328604, EPI_ISL_1328605, EPI_ISL_1328606, EPI_ISL_1328607, EPI_ISL_1328608, EPI_ISL_1328609, EPI_ISL_1328610, EPI_ISL_1328612, EPI_ISL_1328613, EPI_ISL_1328615, EPI_ISL_1328616, EPI_ISL_1328617, EPI_ISL_1328618, EPI_ISL_1328619, EPI_ISL_1328620, EPI_ISL_1328621, EPI_ISL_1328622, EPI_ISL_1328624, EPI_ISL_1328625, EPI_ISL_1328626, EPI_ISL_1328627, EPI_ISL_1328628, EPI_ISL_1328629, EPI_ISL_1328630, EPI_ISL_1328631, EPI_ISL_1328632, EPI_ISL_1328633, EPI_ISL_1328634, EPI_ISL_1328635, EPI_ISL_1328636, EPI_ISL_1328637, EPI_ISL_1328638, EPI_ISL_1328639, EPI_ISL_1328640, EPI_ISL_1328641, EPI_ISL_1328642, EPI_ISL_1328644, EPI_ISL_1328645, EPI_ISL_1328646, EPI_ISL_1328647, EPI_ISL_1328648, EPI_ISL_1328649, EPI_ISL_1328650, EPI_ISL_1328651, EPI_ISL_1328652, EPI_ISL_1328653, EPI_ISL_1328654, EPI_ISL_1328655, EPI_ISL_1328656, EPI_ISL_1328657, EPI_ISL_1328658, EPI_ISL_1328660, EPI_ISL_1328661, EPI_ISL_1328662, EPI_ISL_1328663, EPI_ISL_1328664, EPI_ISL_1328665, EPI_ISL_1328666, EPI_ISL_1328667, EPI_ISL_1328668, EPI_ISL_1328669, EPI_ISL_1328670, EPI_ISL_1328671, EPI_ISL_1328672, EPI_ISL_1328673, EPI_ISL_1328675, EPI_ISL_1328676, EPI_ISL_1328677, EPI_ISL_1328678, EPI_ISL_1328679, EPI_ISL_1328681, EPI_ISL_1328682, EPI_ISL_1328683, EPI_ISL_1328684, EPI_ISL_1328685, EPI_ISL_1328686, EPI_ISL_1328688, EPI_ISL_1328689, EPI_ISL_1328690, EPI_ISL_1328691, EPI_ISL_1328692, EPI_ISL_1328693, EPI_ISL_1328694, EPI_ISL_1328695, EPI_ISL_1328697, EPI_ISL_1328698, EPI_ISL_1328699, EPI_ISL_1328700, EPI_ISL_1328701, EPI_ISL_1328702, EPI_ISL_1328703, EPI_ISL_1328704, EPI_ISL_1328705, EPI_ISL_1328706, EPI_ISL_1328707, EPI_ISL_1328709, EPI_ISL_1328710, EPI_ISL_1328711, EPI_ISL_1328714, EPI_ISL_1328718, EPI_ISL_1328723, EPI_ISL_1328739, EPI_ISL_1328746, EPI_ISL_1328747, EPI_ISL_1328748, EPI_ISL_1328751, EPI_ISL_1328753, EPI_ISL_1328755, EPI_ISL_1328756, EPI_ISL_1328759, EPI_ISL_1328768, EPI_ISL_1328769, EPI_ISL_1328779, EPI_ISL_1328782, EPI_ISL_1328803, EPI_ISL_1328805, EPI_ISL_1328806, EPI_ISL_1328807, EPI_ISL_1328809, EPI_ISL_1328810, EPI_ISL_1328812, EPI_ISL_1328814, EPI_ISL_1328817, EPI_ISL_1328818, EPI_ISL_1328820, EPI_ISL_1328830, EPI_ISL_1328840, EPI_ISL_1328852, EPI_ISL_1328854, EPI_ISL_1328855, EPI_ISL_1328856, EPI_ISL_1328859, EPI_ISL_1328860, EPI_ISL_1328861, EPI_ISL_1328872, EPI_ISL_1328875, EPI_ISL_1328876, EPI_ISL_1328877, EPI_ISL_1328878, EPI_ISL_1328879, EPI_ISL_1328882, EPI_ISL_1328887, EPI_ISL_1328889, EPI_ISL_1328892, EPI_ISL_1328896, EPI_ISL_1329385, EPI_ISL_1329386, EPI_ISL_1329387, EPI_ISL_1329388, EPI_ISL_1329389, EPI_ISL_1329390, EPI_ISL_1329392, EPI_ISL_1329394, EPI_ISL_1329396, EPI_ISL_1329398, EPI_ISL_1329400, EPI_ISL_1329403, EPI_ISL_1329408, EPI_ISL_1329409, EPI_ISL_1329411, EPI_ISL_1329420, EPI_ISL_1329422, EPI_ISL_1329425, EPI_ISL_1329426, EPI_ISL_1329427, EPI_ISL_1329430, EPI_ISL_1329432, EPI_ISL_1329434, EPI_ISL_1329435, EPI_ISL_1329436, EPI_ISL_1329438, EPI_ISL_1329439, EPI_ISL_1329441, EPI_ISL_1329445, EPI_ISL_1329448, EPI_ISL_1329449, EPI_ISL_1329450, EPI_ISL_1329454, EPI_ISL_1329458, EPI_ISL_1329459, EPI_ISL_1329460, EPI_ISL_1329461, EPI_ISL_1329462, EPI_ISL_1329463, EPI_ISL_1329467, EPI_ISL_1329469, EPI_ISL_1329470, EPI_ISL_1329471, EPI_ISL_1329472, EPI_ISL_1329474, EPI_ISL_1329475, EPI_ISL_1329476, EPI_ISL_1329477, EPI_ISL_1329478, EPI_ISL_1329480, EPI_ISL_1329481, EPI_ISL_1329483, EPI_ISL_1329484, EPI_ISL_1329486, EPI_ISL_1329487, EPI_ISL_1329489, EPI_ISL_1329492, EPI_ISL_1329494, EPI_ISL_1329496, EPI_ISL_1329498, EPI_ISL_1329502, EPI_ISL_1329503, EPI_ISL_1329504, EPI_ISL_1329505, EPI_ISL_1329507, EPI_ISL_1329509, EPI_ISL_1329510, EPI_ISL_1329517, EPI_ISL_1329520, EPI_ISL_1329521, EPI_ISL_1329523, EPI_ISL_1329527, EPI_ISL_1329529, EPI_ISL_1329530, EPI_ISL_1329531, EPI_ISL_1329532, EPI_ISL_1329534, EPI_ISL_1329535, EPI_ISL_1329538, EPI_ISL_1329539, EPI_ISL_1329540, EPI_ISL_1329542, EPI_ISL_1329543, EPI_ISL_1329545, EPI_ISL_1329549, EPI_ISL_1329550, EPI_ISL_1329554, EPI_ISL_1329555, EPI_ISL_1329558, EPI_ISL_1329559, EPI_ISL_1329560, EPI_ISL_1329561, EPI_ISL_1329562, EPI_ISL_1329563, EPI_ISL_1329566, EPI_ISL_1329568, EPI_ISL_1329570, EPI_ISL_1329572, EPI_ISL_1329573, EPI_ISL_1329574, EPI_ISL_1329575, EPI_ISL_1329578, EPI_ISL_1329579, EPI_ISL_1329580, EPI_ISL_1329581, EPI_ISL_1329583, EPI_ISL_1329584, EPI_ISL_1329588, EPI_ISL_1329589, EPI_ISL_1329591, EPI_ISL_1329592, EPI_ISL_1329593, EPI_ISL_1329594, EPI_ISL_1329595, EPI_ISL_1329597, EPI_ISL_1329599, EPI_ISL_1329600, EPI_ISL_1329601, EPI_ISL_1329603, EPI_ISL_1329604, EPI_ISL_1329606, EPI_ISL_1329607, EPI_ISL_1329608, EPI_ISL_1329609, EPI_ISL_1329612, EPI_ISL_1329613, EPI_ISL_1329614, EPI_ISL_1329615, EPI_ISL_1329618, EPI_ISL_1329619, EPI_ISL_1329620, EPI_ISL_1329621, EPI_ISL_1329622, EPI_ISL_1329623, EPI_ISL_1329626, EPI_ISL_1329628, EPI_ISL_1329631, EPI_ISL_1329633, EPI_ISL_1329635, EPI_ISL_1329636, EPI_ISL_1329638, EPI_ISL_1329651, EPI_ISL_1329652, EPI_ISL_1329653, EPI_ISL_1329656, EPI_ISL_1329658, EPI_ISL_1329659, EPI_ISL_1329662, EPI_ISL_1329663, EPI_ISL_1329664, EPI_ISL_1329666, EPI_ISL_1329667, EPI_ISL_1329668, EPI_ISL_1329673, EPI_ISL_1329674, EPI_ISL_1329677, EPI_ISL_1329678, EPI_ISL_1329679, EPI_ISL_1329681, EPI_ISL_1329682, EPI_ISL_1329683, EPI_ISL_1329685, EPI_ISL_1329695, EPI_ISL_1329701, EPI_ISL_1329702, EPI_ISL_1329705, EPI_ISL_1329706, EPI_ISL_1329707, EPI_ISL_1329709, EPI_ISL_1329710, EPI_ISL_1329713, EPI_ISL_1329715, EPI_ISL_1329718, EPI_ISL_1329722, EPI_ISL_1329728, EPI_ISL_1329729, EPI_ISL_1329736, EPI_ISL_1329737, EPI_ISL_1329741, EPI_ISL_1329744, EPI_ISL_1329748, EPI_ISL_1329754, EPI_ISL_1329755, EPI_ISL_1329758, EPI_ISL_1329759, EPI_ISL_1329760, EPI_ISL_1329767, EPI_ISL_1329768, EPI_ISL_1329769, EPI_ISL_1329775, EPI_ISL_1329777, EPI_ISL_1329781, EPI_ISL_1329782, EPI_ISL_1329784, EPI_ISL_1329787, EPI_ISL_1329788, EPI_ISL_1329789, EPI_ISL_1329790, EPI_ISL_1329793, EPI_ISL_1329807, EPI_ISL_1329808, EPI_ISL_1329813, EPI_ISL_1329823, EPI_ISL_1329835, EPI_ISL_1329836, EPI_ISL_1329837, EPI_ISL_1329838, EPI_ISL_1329840, EPI_ISL_1329841, EPI_ISL_1329843, EPI_ISL_1329845, EPI_ISL_1329846, EPI_ISL_1329847, EPI_ISL_1329848, EPI_ISL_1329851, EPI_ISL_1329853, EPI_ISL_1329854, EPI_ISL_1329856, EPI_ISL_1329864, EPI_ISL_1329867, EPI_ISL_1329869, EPI_ISL_1329870, EPI_ISL_1329873, EPI_ISL_1329877, EPI_ISL_1329878, EPI_ISL_1329879, EPI_ISL_1329881, EPI_ISL_1329882, EPI_ISL_1329883, EPI_ISL_1329887, EPI_ISL_1329889, EPI_ISL_1329890, EPI_ISL_1329894, EPI_ISL_1329898, EPI_ISL_1329905, EPI_ISL_1329907, EPI_ISL_1329909, EPI_ISL_1329911, EPI_ISL_1329918, EPI_ISL_1329919, EPI_ISL_1329921, EPI_ISL_1329924, EPI_ISL_1329925, EPI_ISL_1329926, EPI_ISL_1329930, EPI_ISL_1329932, EPI_ISL_1329934, EPI_ISL_1329935, EPI_ISL_1329945, EPI_ISL_1329950, EPI_ISL_1329952, EPI_ISL_1329959, EPI_ISL_1329960, EPI_ISL_1329962, EPI_ISL_1329966, EPI_ISL_1329967, EPI_ISL_1330154, EPI_ISL_1330304, EPI_ISL_1330306, EPI_ISL_1330307, EPI_ISL_1330310, EPI_ISL_1330314, EPI_ISL_1330317, EPI_ISL_1330318, EPI_ISL_1330323, EPI_ISL_1330325, EPI_ISL_1330327, EPI_ISL_1330328, EPI_ISL_1330332, EPI_ISL_1330338, EPI_ISL_1330339, EPI_ISL_1330344, EPI_ISL_1330346, EPI_ISL_1330348, EPI_ISL_1330356, EPI_ISL_1330365, EPI_ISL_1330368, EPI_ISL_1330374, EPI_ISL_1330377, EPI_ISL_1330384, EPI_ISL_1330391, EPI_ISL_1330392, EPI_ISL_1330398, EPI_ISL_1330400, EPI_ISL_1330401, EPI_ISL_1330405, EPI_ISL_1330406, EPI_ISL_1330408, EPI_ISL_1330419, EPI_ISL_1330423, EPI_ISL_1330427, EPI_ISL_1330430, EPI_ISL_1330433, EPI_ISL_1330437, EPI_ISL_1330438, EPI_ISL_1330439, EPI_ISL_1330453, EPI_ISL_1330455, EPI_ISL_1330462, EPI_ISL_1330476, EPI_ISL_1330482, EPI_ISL_1330486, EPI_ISL_1330490, EPI_ISL_1330491, EPI_ISL_1330493, EPI_ISL_1330497, EPI_ISL_1330499, EPI_ISL_1330500, EPI_ISL_1330501, EPI_ISL_1330509, EPI_ISL_1330510, EPI_ISL_1330515, EPI_ISL_1330516, EPI_ISL_1330524, EPI_ISL_1330529, EPI_ISL_1330530, EPI_ISL_1330533, EPI_ISL_1330536, EPI_ISL_1330539, EPI_ISL_1330544, EPI_ISL_1330545, EPI_ISL_1330552, EPI_ISL_1330559, EPI_ISL_1330561, EPI_ISL_1330562, EPI_ISL_1330566, EPI_ISL_1330571, EPI_ISL_1330572, EPI_ISL_1330576, EPI_ISL_1330579, EPI_ISL_1330583, EPI_ISL_1330587, EPI_ISL_1330589, EPI_ISL_1330599, EPI_ISL_1330604, EPI_ISL_1330611, EPI_ISL_1330613, EPI_ISL_1330614, EPI_ISL_1330615, EPI_ISL_1330616, EPI_ISL_1330618, EPI_ISL_1330621, EPI_ISL_1330622, EPI_ISL_1330624, EPI_ISL_1330626, EPI_ISL_1330634, EPI_ISL_1330635, EPI_ISL_1330638, EPI_ISL_1330644, EPI_ISL_1330649, EPI_ISL_1330650, EPI_ISL_1330652, EPI_ISL_1330653, EPI_ISL_1330656, EPI_ISL_1330658, EPI_ISL_1330662, EPI_ISL_1330663, EPI_ISL_1330667, EPI_ISL_1330669, EPI_ISL_1330672, EPI_ISL_1330673, EPI_ISL_1330675, EPI_ISL_1330676, EPI_ISL_1330677, EPI_ISL_1330678, EPI_ISL_1330685, EPI_ISL_1330689, EPI_ISL_1330692, EPI_ISL_1330694, EPI_ISL_1330699, EPI_ISL_1330700, EPI_ISL_1330707, EPI_ISL_1330708, EPI_ISL_1330710, EPI_ISL_1330712, EPI_ISL_1330713, EPI_ISL_1330714, EPI_ISL_1330715, EPI_ISL_1330716, EPI_ISL_1330720, EPI_ISL_1330724, EPI_ISL_1330726, EPI_ISL_1330733, EPI_ISL_1330735, EPI_ISL_1330736, EPI_ISL_1330740, EPI_ISL_1330741, EPI_ISL_1330743, EPI_ISL_1330750, EPI_ISL_1330753, EPI_ISL_1330755, EPI_ISL_1330757, EPI_ISL_1330760, EPI_ISL_1330761, EPI_ISL_1330762, EPI_ISL_1330766, EPI_ISL_1330768, EPI_ISL_1330771, EPI_ISL_1330773, EPI_ISL_1330777, EPI_ISL_1330779, EPI_ISL_1330781, EPI_ISL_1330782, EPI_ISL_1330791, EPI_ISL_1330793, EPI_ISL_1330794, EPI_ISL_1330797, EPI_ISL_1330798, EPI_ISL_1330800, EPI_ISL_1330802, EPI_ISL_1330808, EPI_ISL_1330810, EPI_ISL_1330811, EPI_ISL_1330812, EPI_ISL_1330818, EPI_ISL_1330825, EPI_ISL_1330828, EPI_ISL_1330829, EPI_ISL_1330830, EPI_ISL_1330831, EPI_ISL_1330835, EPI_ISL_1330836, EPI_ISL_1330837, EPI_ISL_1330838, EPI_ISL_1330842, EPI_ISL_1330843, EPI_ISL_1330845, EPI_ISL_1330846, EPI_ISL_1330847, EPI_ISL_1330848, EPI_ISL_1330855, EPI_ISL_1330857, EPI_ISL_1330860, EPI_ISL_1330861, EPI_ISL_1330864, EPI_ISL_1330866, EPI_ISL_1330879, EPI_ISL_1330881, EPI_ISL_1330882, EPI_ISL_1330883, EPI_ISL_1330884, EPI_ISL_1330885, EPI_ISL_1330888, EPI_ISL_1330889, EPI_ISL_1330891, EPI_ISL_1330901, EPI_ISL_1330907, EPI_ISL_1330908, EPI_ISL_1330914, EPI_ISL_1330916, EPI_ISL_1330917, EPI_ISL_1330918, EPI_ISL_1330919, EPI_ISL_1330923, EPI_ISL_1330927, EPI_ISL_1330935, EPI_ISL_1330937, EPI_ISL_1330939, EPI_ISL_1330942, EPI_ISL_1330958, EPI_ISL_1330959, EPI_ISL_1330961, EPI_ISL_1330963, EPI_ISL_1330968, EPI_ISL_1330980, EPI_ISL_1330983, EPI_ISL_1330989, EPI_ISL_1330991, EPI_ISL_1331014, EPI_ISL_1331022, EPI_ISL_1331031, EPI_ISL_1331037, EPI_ISL_1331042, EPI_ISL_1331045, EPI_ISL_1331049, EPI_ISL_1331051, EPI_ISL_1331090, EPI_ISL_1331094, EPI_ISL_1331110, EPI_ISL_1331114, EPI_ISL_1331116, EPI_ISL_1331119, EPI_ISL_1343368, EPI_ISL_1343372, EPI_ISL_1343375, EPI_ISL_1343429, EPI_ISL_1343437, EPI_ISL_1343445, EPI_ISL_1343446, EPI_ISL_1343454, EPI_ISL_1343467, EPI_ISL_1343481, EPI_ISL_1343498, EPI_ISL_1343511, EPI_ISL_1343516, EPI_ISL_1343540, EPI_ISL_1343553, EPI_ISL_1343595, EPI_ISL_1343687, EPI_ISL_1344108, EPI_ISL_1344144, EPI_ISL_1345280, EPI_ISL_1345654, EPI_ISL_1345680, EPI_ISL_1345871, EPI_ISL_1346238, EPI_ISL_1346253, EPI_ISL_1346260, EPI_ISL_1346261, EPI_ISL_1346265, EPI_ISL_1346340, EPI_ISL_1346349, EPI_ISL_1346351, EPI_ISL_1346352, EPI_ISL_1346353, EPI_ISL_1346354, EPI_ISL_1346355, EPI_ISL_1346356, EPI_ISL_1346357, EPI_ISL_1346360, EPI_ISL_1346380, EPI_ISL_1346382, EPI_ISL_1346383, EPI_ISL_1346385, EPI_ISL_1346387, EPI_ISL_1346393, EPI_ISL_1346396, EPI_ISL_1346410, EPI_ISL_1346411, EPI_ISL_1346412, EPI_ISL_1346413, EPI_ISL_1346417, EPI_ISL_1346418, EPI_ISL_1377926, EPI_ISL_1415867, EPI_ISL_1415889, EPI_ISL_1415923, EPI_ISL_1415975, EPI_ISL_1415980, EPI_ISL_1416017, EPI_ISL_1416028, EPI_ISL_1416070

| | | | |
|---|---|---|---|
| see above | Lighthouse Lab in Milton Keynes | Wellcome Sanger Institute for the COVID-19 Genomics UK (COG-UK) Consortium | Cordelia Langford; David K. Jackson; Dominic Kwiatkowski; Ewan Harrison; Ian Johnston; Jeffrey Barrett; John Sillitoe on behalf of the Wellcome Sanger Institute COVID-19 Surveillance Team; Roberto Amato; Sonia Goncalves; The Lighthouse Lab in Milton Keynes and Alex Alderton |

EPI_ISL_1475509, EPI_ISL_1475510, EPI_ISL_1475511, EPI_ISL_1475512, EPI_ISL_1475516, EPI_ISL_1475517, EPI_ISL_1475518, EPI_ISL_1475519, EPI_ISL_1475520, EPI_ISL_1475522

| | | | |
|---|---|---|---|
| see above | Lincolnshire Hospitals and DeepSeq Nottingham | COVID-19 Genomics UK (COG-UK) Consortium | Christopher Moore; Fei Sang; Joeseph Chappell; Johnny Debebe; Jonathan Ball; Matthew Carlisle; Matthew Loose; Nadine Holmes; Nichola Duckworth; Patrick McClure; Sarah Walsh; Tim Sloan; Victoria Wright |

EPI_ISL_1386736, EPI_ISL_1386737, EPI_ISL_1386738, EPI_ISL_1386760, EPI_ISL_1386762, EPI_ISL_1386763, EPI_ISL_1386765, EPI_ISL_1386768, EPI_ISL_1386769, EPI_ISL_1386770, EPI_ISL_1386772, EPI_ISL_1386774, EPI_ISL_1386776, EPI_ISL_1386777, EPI_ISL_1386778, EPI_ISL_1386779, EPI_ISL_1386780, EPI_ISL_1386793, EPI_ISL_1386803, EPI_ISL_1386805, EPI_ISL_1474708, EPI_ISL_1474709, EPI_ISL_1474710, EPI_ISL_1474713, EPI_ISL_1474717, EPI_ISL_1474725, EPI_ISL_1474729, EPI_ISL_1474732, EPI_ISL_1474733, EPI_ISL_1474748, EPI_ISL_1474752, EPI_ISL_1475753, EPI_ISL_1475754, EPI_ISL_1475755, EPI_ISL_1475756, EPI_ISL_1546151, EPI_ISL_1546152

| | | | |
|---|---|---|---|
| see above | Liverpool Clinical Laboratories | COVID-19 Genomics UK (COG-UK) Consortium | A Alrezaihi; Alessandro Gerada; Alistair Darby; Angela Cowell; Anita Lucaci; Anu Chawla; Cassie Olateju; Catherine Hartley; Charlotte Nelson; Ecaterina Vamos; Elaine O'Toole; Eleanor G Bentley; Ghada T Shawli; Hermione Webster; Isabel García-Dorival; James Johnson; James P Stewart; Jenifer Manson; Joanne Watts; Jones Benjamin; Jordan J Clark; Julian Hiscox; L Luu; Lucille Rainbow; M Almsaud; Margaret Hughes; Mark Whitehead; Matthew Gemmell; Miren Iturriza-Gomara; Muhannad Alruwaili; N.P Randle; Neil Swainston; PKF Gilmore; Parul Sharma; Rebekah Penrice-Randal; Richard Eccles; Richard Gregory; Sam Haldenby; Steve Paterson; Stuart D Armstrong; Trevor Ian Robinson; Ximeng Han |

EPI_ISL_1309512, EPI_ISL_1309513, EPI_ISL_1309514, EPI_ISL_1309635, EPI_ISL_1309636, EPI_ISL_1387088, EPI_ISL_1387090, EPI_ISL_1475220, EPI_ISL_1475221, EPI_ISL_1475222, EPI_ISL_1475306, EPI_ISL_1475307, EPI_ISL_1475308, EPI_ISL_1475309, EPI_ISL_1475310, EPI_ISL_1475314, EPI_ISL_1475315, EPI_ISL_1475316, EPI_ISL_1475317, EPI_ISL_1475327, EPI_ISL_1475328, EPI_ISL_1475329, EPI_ISL_1475330, EPI_ISL_1475331, EPI_ISL_1475332, EPI_ISL_1475406, EPI_ISL_1475407, EPI_ISL_1475461, EPI_ISL_1475462, EPI_ISL_1475464, EPI_ISL_1475465, EPI_ISL_1475466, EPI_ISL_1475467, EPI_ISL_1475472, EPI_ISL_1475473, EPI_ISL_1475474, EPI_ISL_1475475

| | | | |
|---|---|---|---|
| see above | Northumbria University / South Tees Hospitals NHS Foundation Trust / North Cumbria Integrated Care NHS Foundation Trust / North Tees and Hartlepool NHS Foundation Trust / Newcastle Hospitals NHS Foundation Trust | COVID-19 Genomics UK (COG-UK) Consortium | Andrew Nelson; Brendan Payne; Clive Graham; Darren L Smith; Debra Padgett; Edward Barton; Emma Swindells; Garren Scott; Gary Black; Gary Eltringham; Giles S Holt; Greg R Young; Jane Greenaway; Jennifer Collins; John Allan; Joshua Loh; Lynn Dover; Matthew Bashton; Mohammad A Tariq; Paul Baker; Sarah Essex; Steve Liggett; Wen C Yew; Yusri Taha |

EPI_ISL_1309840, EPI_ISL_1309932, EPI_ISL_1309993, EPI_ISL_1310006, EPI_ISL_1310007, EPI_ISL_1310017, EPI_ISL_1310018, EPI_ISL_1310019, EPI_ISL_1310020, EPI_ISL_1310021, EPI_ISL_1310030, EPI_ISL_1310031, EPI_ISL_1310049, EPI_ISL_1310050, EPI_ISL_1310053, EPI_ISL_1310054, EPI_ISL_1310055, EPI_ISL_1310060, EPI_ISL_1310061, EPI_ISL_1310062, EPI_ISL_1310063, EPI_ISL_1310064, EPI_ISL_1310065, EPI_ISL_1310066, EPI_ISL_1310067, EPI_ISL_1310068, EPI_ISL_1310069, EPI_ISL_1310070, EPI_ISL_1310071, EPI_ISL_1310072, EPI_ISL_1310073, EPI_ISL_1310074, EPI_ISL_1310075, EPI_ISL_1310076, EPI_ISL_1310077, EPI_ISL_1310078, EPI_ISL_1310079, EPI_ISL_1310080, EPI_ISL_1310081, EPI_ISL_1310082, EPI_ISL_1310083, EPI_ISL_1310084, EPI_ISL_1310085, EPI_ISL_1310086, EPI_ISL_1310087, EPI_ISL_1310088, EPI_ISL_1310089, EPI_ISL_1310090, EPI_ISL_1310094, EPI_ISL_1310095, EPI_ISL_1310096, EPI_ISL_1310097, EPI_ISL_1310101, EPI_ISL_1310102, EPI_ISL_1310104, EPI_ISL_1310105, EPI_ISL_1310106, EPI_ISL_1310107, EPI_ISL_1310113, EPI_ISL_1310114, EPI_ISL_1310115, EPI_ISL_1310116, EPI_ISL_1310117, EPI_ISL_1310118, EPI_ISL_1310119, EPI_ISL_1310120, EPI_ISL_1310121, EPI_ISL_1310122, EPI_ISL_1310125, EPI_ISL_1310128, EPI_ISL_1310129, EPI_ISL_1310135, EPI_ISL_1310136, EPI_ISL_1310137, EPI_ISL_1310138, EPI_ISL_1310139, EPI_ISL_1310140, EPI_ISL_1310143, EPI_ISL_1310144, EPI_ISL_1310146, EPI_ISL_1310151, EPI_ISL_1310152, EPI_ISL_1310153, EPI_ISL_1310154, EPI_ISL_1310155, EPI_ISL_1310156, EPI_ISL_1310158, EPI_ISL_1310159, EPI_ISL_1310160, EPI_ISL_1310161, EPI_ISL_1310166, EPI_ISL_1310167, EPI_ISL_1310168, EPI_ISL_1310169, EPI_ISL_1310173, EPI_ISL_1310176, EPI_ISL_1310177, EPI_ISL_1310178, EPI_ISL_1310181, EPI_ISL_1310182, EPI_ISL_1310183, EPI_ISL_1310184, EPI_ISL_1310185, EPI_ISL_1310186, EPI_ISL_1310187, EPI_ISL_1310188, EPI_ISL_1310189, EPI_ISL_1310190, EPI_ISL_1310195, EPI_ISL_1310200, EPI_ISL_1310201, EPI_ISL_1310202, EPI_ISL_1310203, EPI_ISL_1310204, EPI_ISL_1310205, EPI_ISL_1310206, EPI_ISL_1310208, EPI_ISL_1310209, EPI_ISL_1310210, EPI_ISL_1310211, EPI_ISL_1310212, EPI_ISL_1310213, EPI_ISL_1310214, EPI_ISL_1310218, EPI_ISL_1310219, EPI_ISL_1310220, EPI_ISL_1310221, EPI_ISL_1310222, EPI_ISL_1310223, EPI_ISL_1310224, EPI_ISL_1310225, EPI_ISL_1310226, EPI_ISL_1310227, EPI_ISL_1310228, EPI_ISL_1310229, EPI_ISL_1310230, EPI_ISL_1310231, EPI_ISL_1310232, EPI_ISL_1310233, EPI_ISL_1310234, EPI_ISL_1310235, EPI_ISL_1310236, EPI_ISL_1310237, EPI_ISL_1310239, EPI_ISL_1310243, EPI_ISL_1310244, EPI_ISL_1310246, EPI_ISL_1310247, EPI_ISL_1310248, EPI_ISL_1310254, EPI_ISL_1310255, EPI_ISL_1310256, EPI_ISL_1310257, EPI_ISL_1310258, EPI_ISL_1310259, EPI_ISL_1310260, EPI_ISL_1310262, EPI_ISL_1310263, EPI_ISL_1310264, EPI_ISL_1310265, EPI_ISL_1310266, EPI_ISL_1310267, EPI_ISL_1310268, EPI_ISL_1310277, EPI_ISL_1310278, EPI_ISL_1310279, EPI_ISL_1310280, EPI_ISL_1310283, EPI_ISL_1310284, EPI_ISL_1310285, EPI_ISL_1310286, EPI_ISL_1387797, EPI_ISL_1387798, EPI_ISL_1387819, EPI_ISL_1387833, EPI_ISL_1387834, EPI_ISL_1387835, EPI_ISL_1387836, EPI_ISL_1387838, EPI_ISL_1387839, EPI_ISL_1387841, EPI_ISL_1387855, EPI_ISL_1387857, EPI_ISL_1387858, EPI_ISL_1387859, EPI_ISL_1387860, EPI_ISL_1387862, EPI_ISL_1387872, EPI_ISL_1387873, EPI_ISL_1387874, EPI_ISL_1387875, EPI_ISL_1387876, EPI_ISL_1387903, EPI_ISL_1387914, EPI_ISL_1387918, EPI_ISL_1387923, EPI_ISL_1387926, EPI_ISL_1387927, EPI_ISL_1387931, EPI_ISL_1387934, EPI_ISL_1387942, EPI_ISL_1387947, EPI_ISL_1387954, EPI_ISL_1387955, EPI_ISL_1387962, EPI_ISL_1387963, EPI_ISL_1387965, EPI_ISL_1387972, EPI_ISL_1387973, EPI_ISL_1387974, EPI_ISL_1387983, EPI_ISL_1387994, EPI_ISL_1388018, EPI_ISL_1388023, EPI_ISL_1388024, EPI_ISL_1388029, EPI_ISL_1388031, EPI_ISL_1388035, EPI_ISL_1388036, EPI_ISL_1388037, EPI_ISL_1388040, EPI_ISL_1388041, EPI_ISL_1388045, EPI_ISL_1388046, EPI_ISL_1388047, EPI_ISL_1388048, EPI_ISL_1388049, EPI_ISL_1388053, EPI_ISL_1388059, EPI_ISL_1388060, EPI_ISL_1388061, EPI_ISL_1388062, EPI_ISL_1388092, EPI_ISL_1388093, EPI_ISL_1388095, EPI_ISL_1388096, EPI_ISL_1388098, EPI_ISL_1388100, EPI_ISL_1388111, EPI_ISL_1388115, EPI_ISL_1388120, EPI_ISL_1388125, EPI_ISL_1388129, EPI_ISL_1388135, EPI_ISL_1388140, EPI_ISL_1388142, EPI_ISL_1388148, EPI_ISL_1388151, EPI_ISL_1388155, EPI_ISL_1388159, EPI_ISL_1388176, EPI_ISL_1388190, EPI_ISL_1388192, EPI_ISL_1388205, EPI_ISL_1388207, EPI_ISL_1388217, EPI_ISL_1388219, EPI_ISL_1388223, EPI_ISL_1388231, EPI_ISL_1388236, EPI_ISL_1388237, EPI_ISL_1388241, EPI_ISL_1388246, EPI_ISL_1388250, EPI_ISL_1388411, EPI_ISL_1388424, EPI_ISL_1388426, EPI_ISL_1388549, EPI_ISL_1388551, EPI_ISL_1388553, EPI_ISL_1388557, EPI_ISL_1388563, EPI_ISL_1388567, EPI_ISL_1388569, EPI_ISL_1388577, EPI_ISL_1388581, EPI_ISL_1388582, EPI_ISL_1388584, EPI_ISL_1388589, EPI_ISL_1388593, EPI_ISL_1476492, EPI_ISL_1476493, EPI_ISL_1546788, EPI_ISL_1546797, EPI_ISL_1546798, EPI_ISL_1546799, EPI_ISL_1546800, EPI_ISL_1547031, EPI_ISL_1547032, EPI_ISL_1547033, EPI_ISL_1547037

| | | | |
|---|---|---|---|
| see above | Originating lab: Wales Specialist Virology Centre Sequencing lab: Pathogen Genomics Unit | Public Health Wales Microbiology Cardiff Wales Specialist Virology Centre | Alec Birchley; Alexander Adams; Amy Gaskin; Angela Marchbank; Bree Gatica-Wilcox; Catherine Moore; Jason Coombes; Joanne Watkins; Joel Southgate; Johnathan Evans; Laura Gifford; Lauren Gilbert; Lee Graham; Malorie Perry; Matthew Bull; Nicole Pacchiarini; Sally Corden; Sara Kumziene-Summerhayes; Sara Rey; Sarah Taylor; Simon Cottrell; Sophie Jones; Tom Connor |

EPI_ISL_1475672, EPI_ISL_1475679, EPI_ISL_1475683, EPI_ISL_1475684, EPI_ISL_1475686, EPI_ISL_1475688, EPI_ISL_1475689, EPI_ISL_1475692, EPI_ISL_1475693, EPI_ISL_1475721, EPI_ISL_1475723, EPI_ISL_1475724, EPI_ISL_1475725, EPI_ISL_1475726, EPI_ISL_1475727, EPI_ISL_1475728, EPI_ISL_1475729, EPI_ISL_1475730, EPI_ISL_1475733, EPI_ISL_1475734, EPI_ISL_1475737, EPI_ISL_1833067, EPI_ISL_1833097

| | | | |
|---|---|---|---|
| see above | Oxford Viromics, NDM, University of Oxford; Oxford University Hospitals; Basingstoke and North Hampshire Hospital | COVID-19 Genomics UK (COG-UK) Consortium | Alex Mobbs; Amy Trebes; Anita Justice; Catrin Moore; Christophe Fraser; David Bonsall; David Buck; Emma Wise; George Macintyre; Jessica Lynch; John Todd; Mariateresa de Cesare; Matilde Mori; Monique Andersson; Nathan Moore; Nick Cortes; Robert Shaw; Stephen Kidd; Tanya Golubchik; Timothy Peto |
| EPI_ISL_1387143 | Quadram Institute Bioscience | COVID-19 Genomics UK (COG-UK) Consortium | Alexander J Trotter; Alison E. Mather; Alp Aydin; Ana P. Tedim; Anastasia Kolyva; Andrew Bell; Andrew J. Page; Claire Stuart; Dave J. Baker; Gemma L. Kay; John Wain; Justin O'Grady; Leonardo de Oliveira Martins; Lizzie Meadows; Maria Diaz; Mark Webber; Muhammed Yasir; Nabil-Fareed Alikhan; Ngozi Elumogo; Nicholas M. Thomson; Rachael Stanley; Rachel Gilroy; Reenesh Prakash; Samir Dervisevic; Samuel Bloomfield; Steven Rudder; Thanh Le-Viet |
| EPI_ISL_1309696, EPI_ISL_1309722, EPI_ISL_1309723, EPI_ISL_1309728, EPI_ISL_1309729, EPI_ISL_1387187 | Queens Medical Centre, Clinical Microbiology Department / DeepSeq Nottingham | COVID-19 Genomics UK (COG-UK) Consortium | Christopher Moore; Emily Park; Fei Sang; Gemma Clark; Hannah Howson-Wells; Jack Hill; Jayasree Dey; Johnny Debebe; Jonathan Ball; Joseph Chappell; Manjinder Khakh; Matthew Carlisle; Matthew Loose; Michelle M Lister; Nadine Holmes; Patrick McClure; Theocharis Tsoleridis; Timothy Byaruhanga; Vicki M Fleming; Victoria Wright; Wendy Smith |

EPI_ISL_1326663, EPI_ISL_1326664, EPI_ISL_1326665, EPI_ISL_1326668, EPI_ISL_1326676, EPI_ISL_1326677, EPI_ISL_1326680, EPI_ISL_1326681, EPI_ISL_1326684, EPI_ISL_1326685, EPI_ISL_1326690, EPI_ISL_1326691, EPI_ISL_1326692, EPI_ISL_1326694, EPI_ISL_1326696, EPI_ISL_1326697, EPI_ISL_1326705, EPI_ISL_1326708, EPI_ISL_1326716, EPI_ISL_1326717, EPI_ISL_1326718, EPI_ISL_1326730, EPI_ISL_1326742, EPI_ISL_1326748, EPI_ISL_1326749, EPI_ISL_1326755, EPI_ISL_1326756, EPI_ISL_1326764, EPI_ISL_1326765, EPI_ISL_1326766, EPI_ISL_1326768, EPI_ISL_1326772, EPI_ISL_1326776, EPI_ISL_1326778, EPI_ISL_1326780, EPI_ISL_1326786, EPI_ISL_1326792, EPI_ISL_1326793, EPI_ISL_1326796, EPI_ISL_1326799, EPI_ISL_1326800, EPI_ISL_1326802, EPI_ISL_1326806, EPI_ISL_1326809, EPI_ISL_1326812, EPI_ISL_1326814, EPI_ISL_1326821, EPI_ISL_1326827, EPI_ISL_1326833, EPI_ISL_1326835, EPI_ISL_1326836, EPI_ISL_1326839, EPI_ISL_1326843, EPI_ISL_1326845, EPI_ISL_1326849, EPI_ISL_1326851, EPI_ISL_1326857, EPI_ISL_1326863, EPI_ISL_1326865, EPI_ISL_1326869, EPI_ISL_1326878, EPI_ISL_1326880, EPI_ISL_1326889, EPI_ISL_1326890, EPI_ISL_1326897, EPI_ISL_1326906, EPI_ISL_1326907, EPI_ISL_1326918, EPI_ISL_1326921, EPI_ISL_1326923, EPI_ISL_1326926, EPI_ISL_1326928, EPI_ISL_1326930, EPI_ISL_1326933, EPI_ISL_1326934, EPI_ISL_1326936, EPI_ISL_1326939, EPI_ISL_1326943, EPI_ISL_1326945, EPI_ISL_1326948, EPI_ISL_1326949, EPI_ISL_1326955, EPI_ISL_1326961, EPI_ISL_1326965, EPI_ISL_1326967, EPI_ISL_1326969, EPI_ISL_1326971, EPI_ISL_1326972, EPI_ISL_1326976, EPI_ISL_1326979, EPI_ISL_1326997, EPI_ISL_1327006, EPI_ISL_1327045, EPI_ISL_1327051, EPI_ISL_1327057, EPI_ISL_1327059, EPI_ISL_1327066, EPI_ISL_1327071, EPI_ISL_1327077, EPI_ISL_1327093, EPI_ISL_1327101, EPI_ISL_1327104, EPI_ISL_1327113, EPI_ISL_1327117, EPI_ISL_1327123, EPI_ISL_1327130, EPI_ISL_1327139, EPI_ISL_1327152, EPI_ISL_1327154, EPI_ISL_1327166, EPI_ISL_1327169, EPI_ISL_1327186, EPI_ISL_1327210, EPI_ISL_1327211, EPI_ISL_1327215, EPI_ISL_1327217, EPI_ISL_1327220, EPI_ISL_1327226, EPI_ISL_1327239, EPI_ISL_1344464, EPI_ISL_1344465, EPI_ISL_1344467, EPI_ISL_1344473, EPI_ISL_1344474, EPI_ISL_1344475, EPI_ISL_1344476, EPI_ISL_1344477, EPI_ISL_1344478, EPI_ISL_1344481, EPI_ISL_1344484, EPI_ISL_1344486, EPI_ISL_1344487, EPI_ISL_1344488, EPI_ISL_1344490, EPI_ISL_1344492, EPI_ISL_1344493, EPI_ISL_1344497, EPI_ISL_1344498, EPI_ISL_1344500, EPI_ISL_1344503, EPI_ISL_1344504, EPI_ISL_1344505, EPI_ISL_1344506, EPI_ISL_1344507, EPI_ISL_1344509, EPI_ISL_1344510, EPI_ISL_1344513, EPI_ISL_1344516, EPI_ISL_1344521, EPI_ISL_1344529, EPI_ISL_1344530, EPI_ISL_1344532, EPI_ISL_1344533, EPI_ISL_1344534, EPI_ISL_1344535, EPI_ISL_1344538, EPI_ISL_1344539, EPI_ISL_1344545, EPI_ISL_1344546, EPI_ISL_1344547, EPI_ISL_1344549, EPI_ISL_1344551, EPI_ISL_1344554, EPI_ISL_1344559, EPI_ISL_1344561, EPI_ISL_1344562, EPI_ISL_1344564, EPI_ISL_1344565, EPI_ISL_1344567, EPI_ISL_1344568, EPI_ISL_1344570, EPI_ISL_1344574, EPI_ISL_1344578, EPI_ISL_1344580, EPI_ISL_1344581, EPI_ISL_1344583,

| | | | |
|---|---|---|---|
| EPI_ISL_1344584, EPI_ISL_1344585, EPI_ISL_1344588, EPI_ISL_1344591, EPI_ISL_1344593, EPI_ISL_1344594, EPI_ISL_1344595, EPI_ISL_1344596, EPI_ISL_1344597, EPI_ISL_1344600, EPI_ISL_1344601, EPI_ISL_1344602, EPI_ISL_1344605, EPI_ISL_1344606, EPI_ISL_1344608, EPI_ISL_1344609, EPI_ISL_1344610, EPI_ISL_1344613, EPI_ISL_1344615, EPI_ISL_1344617, EPI_ISL_1344626, EPI_ISL_1344628, EPI_ISL_1344634, EPI_ISL_1344635, EPI_ISL_1344636, EPI_ISL_1344638, EPI_ISL_1344640, EPI_ISL_1344642, EPI_ISL_1344643, EPI_ISL_1344644, EPI_ISL_1344647, EPI_ISL_1344650, EPI_ISL_1344651, EPI_ISL_1344654, EPI_ISL_1344655, EPI_ISL_1344657, EPI_ISL_1344661, EPI_ISL_1344662, EPI_ISL_1344664, EPI_ISL_1344666, EPI_ISL_1344670, EPI_ISL_1344671, EPI_ISL_1344672, EPI_ISL_1344673, EPI_ISL_1344676, EPI_ISL_1344678, EPI_ISL_1344680, EPI_ISL_1344681, EPI_ISL_1344683, EPI_ISL_1344689, EPI_ISL_1344692, EPI_ISL_1344697, EPI_ISL_1344698, EPI_ISL_1344700, EPI_ISL_1344706, EPI_ISL_1344707, EPI_ISL_1344708, EPI_ISL_1344709, EPI_ISL_1344710, EPI_ISL_1344713, EPI_ISL_1344716, EPI_ISL_1344718, EPI_ISL_1344721, EPI_ISL_1344725, EPI_ISL_1344727, EPI_ISL_1344728, EPI_ISL_1344730, EPI_ISL_1344736, EPI_ISL_1344738, EPI_ISL_1344740, EPI_ISL_1344742, EPI_ISL_1344743, EPI_ISL_1344745, EPI_ISL_1344751, EPI_ISL_1344752, EPI_ISL_1344755, EPI_ISL_1344763, EPI_ISL_1344769, EPI_ISL_1344775, EPI_ISL_1344778, EPI_ISL_1344781, EPI_ISL_1344783, EPI_ISL_1344784, EPI_ISL_1344790, EPI_ISL_1344793, EPI_ISL_1344794, EPI_ISL_1344798, EPI_ISL_1344799, EPI_ISL_1344803, EPI_ISL_1344807, EPI_ISL_1344809, EPI_ISL_1344811, EPI_ISL_1344812, EPI_ISL_1344813, EPI_ISL_1344816, EPI_ISL_1344827, EPI_ISL_1344829, EPI_ISL_1344833, EPI_ISL_1344834, EPI_ISL_1344836, EPI_ISL_1344838, EPI_ISL_1344840, EPI_ISL_1344848, EPI_ISL_1344850, EPI_ISL_1344851, EPI_ISL_1344853, EPI_ISL_1344856, EPI_ISL_1344858, EPI_ISL_1344860, EPI_ISL_1344869, EPI_ISL_1344871, EPI_ISL_1344872, EPI_ISL_1344879, EPI_ISL_1344880, EPI_ISL_1344882, EPI_ISL_1344888, EPI_ISL_1344895, EPI_ISL_1344896, EPI_ISL_1344902, EPI_ISL_1344904, EPI_ISL_1344909, EPI_ISL_1344912, EPI_ISL_1344914, EPI_ISL_1344915, EPI_ISL_1344916, EPI_ISL_1344917, EPI_ISL_1344919, EPI_ISL_1346288, EPI_ISL_1346298, EPI_ISL_1365178, EPI_ISL_1365179, EPI_ISL_1365181, EPI_ISL_1365191, EPI_ISL_1365192, EPI_ISL_1365193, EPI_ISL_1365194, EPI_ISL_1365195, EPI_ISL_1365196, EPI_ISL_1365197, EPI_ISL_1365200, EPI_ISL_1365202, EPI_ISL_1365205, EPI_ISL_1365207, EPI_ISL_1365208, EPI_ISL_1365210, EPI_ISL_1365212, EPI_ISL_1365213, EPI_ISL_1365215, EPI_ISL_1365219, EPI_ISL_1365220, EPI_ISL_1365221, EPI_ISL_1365222, EPI_ISL_1365226, EPI_ISL_1365231, EPI_ISL_1365232, EPI_ISL_1365236, EPI_ISL_1365249, EPI_ISL_1365251, EPI_ISL_1365252, EPI_ISL_1365253, EPI_ISL_1365254, EPI_ISL_1365255, EPI_ISL_1365263, EPI_ISL_1365264, EPI_ISL_1365265, EPI_ISL_1365266, EPI_ISL_1365271, EPI_ISL_1365278, EPI_ISL_1365288, EPI_ISL_1365291, EPI_ISL_1365294, EPI_ISL_1365295, EPI_ISL_1365296, EPI_ISL_1365297, EPI_ISL_1365298, EPI_ISL_1365299, EPI_ISL_1365306, EPI_ISL_1376789, EPI_ISL_1376894 | | | |
| see above | Randox Laboratories | Wellcome Sanger Institute for the COVID-19 Genomics UK (COG-UK) Consortium | Cordelia Langford; David K. Jackson; Dominic Kwiatkowski; Ewan Harrison; Ian Johnston; Jeffrey Barrett; John Sillitoe on behalf of the Wellcome Sanger Institute COVID-19 Surveillance Team; Randox Laboratories and Alex Alderton; Roberto Amato; Sonia Goncalves |
| EPI_ISL_1976285, EPI_ISL_1976340, EPI_ISL_1976342, EPI_ISL_1976343, EPI_ISL_1976345, EPI_ISL_1976403, EPI_ISL_1976407, EPI_ISL_1976415, EPI_ISL_1976417, EPI_ISL_1976418, EPI_ISL_1976420, EPI_ISL_1976422, EPI_ISL_1976423, EPI_ISL_1976434, EPI_ISL_1976435, EPI_ISL_1976437, EPI_ISL_1977452, EPI_ISL_1977454, EPI_ISL_1977456, EPI_ISL_1977458, EPI_ISL_1977459, EPI_ISL_1977461, EPI_ISL_1977463, EPI_ISL_1977466, EPI_ISL_1977468, EPI_ISL_1977471, EPI_ISL_1977473, EPI_ISL_1977475, EPI_ISL_1977477, EPI_ISL_1977478, EPI_ISL_1977480, EPI_ISL_1977481, EPI_ISL_1977483, EPI_ISL_1977485, EPI_ISL_1977486, EPI_ISL_1977503, EPI_ISL_1977505, EPI_ISL_1977507, EPI_ISL_1977508, EPI_ISL_1977510, EPI_ISL_1977511, EPI_ISL_1977513, EPI_ISL_1977514, EPI_ISL_1977516, EPI_ISL_1977530, EPI_ISL_1977531, EPI_ISL_1977535, EPI_ISL_1977536, EPI_ISL_1977538, EPI_ISL_1977550, EPI_ISL_1977552, EPI_ISL_1977554, EPI_ISL_1977556, EPI_ISL_1977557, EPI_ISL_1977560, EPI_ISL_1977564, EPI_ISL_1977566, EPI_ISL_1977569, EPI_ISL_1977583, EPI_ISL_1977589, EPI_ISL_1977591, EPI_ISL_1977592, EPI_ISL_1977594, EPI_ISL_1977596, EPI_ISL_1977598, EPI_ISL_1977599, EPI_ISL_1977601, EPI_ISL_1977603, EPI_ISL_1977606, EPI_ISL_1977609, EPI_ISL_1977611, EPI_ISL_1977612, EPI_ISL_1977616, EPI_ISL_1977618, EPI_ISL_1977620, EPI_ISL_1977622, EPI_ISL_1977683, EPI_ISL_1977685, EPI_ISL_1977687, EPI_ISL_1977688, EPI_ISL_1977690, EPI_ISL_1977692, EPI_ISL_1977725, EPI_ISL_1977727, EPI_ISL_1977729, EPI_ISL_1977731, EPI_ISL_1977736, EPI_ISL_1977738, EPI_ISL_1977739, EPI_ISL_1977741, EPI_ISL_1977743, EPI_ISL_1977745, EPI_ISL_1977747, EPI_ISL_1977748, EPI_ISL_1977750, EPI_ISL_1977751, EPI_ISL_1977753, EPI_ISL_1977813, EPI_ISL_1977814, EPI_ISL_1977816, EPI_ISL_1977818, EPI_ISL_1977819, EPI_ISL_1977821, EPI_ISL_1977823, EPI_ISL_1977824, EPI_ISL_1977826, EPI_ISL_1977828, EPI_ISL_1977830, EPI_ISL_1977832, EPI_ISL_1977845, EPI_ISL_1977846, EPI_ISL_1977848, EPI_ISL_1977850, EPI_ISL_1977865, EPI_ISL_1977866 | | | |
| see above | Regional Virus Laboratory, Belfast Health and Social Care Trust | COVID-19 Genomics UK (COG-UK) Consortium | Alison Watt; Ciara Cox; Conall McCaughey; David Simpson; Derek Fairley; James McKenna; Mairead Connor; Susan Feeney; Tanya Curran; Zoltan Molnar |
| EPI_ISL_1637045 | University College London Hospital | COVID-19 Genomics UK (COG-UK) Consortium | Dr Catherine Houlihan; Dr Daniel Frampton; Dr Judith Heaney; Dr Moira Spyer; Dr Paul Grant and Dr Eleni Nastouli; Matthew Byott; Stuart Kirk |
| EPI_ISL_1248361, EPI_ISL_1308952, EPI_ISL_1308957, EPI_ISL_1308968, EPI_ISL_1308969, EPI_ISL_1308970, EPI_ISL_1309128, EPI_ISL_1309129, EPI_ISL_1309130, EPI_ISL_1309136, EPI_ISL_1309137, EPI_ISL_1309139, EPI_ISL_1309140, EPI_ISL_1386845, EPI_ISL_1386867 | | | |
| see above | University College London, Great Ormond Street Hospital for Children NHS Foundation Trust, Imperial College Healthcare NHS Trust | COVID-19 Genomics UK (COG-UK) Consortium | Alison Holmes; Charlotte Williams; Helena Tutill; Jacqueline Findlay; James Price; Judith Breuer; Julianne Brown; Kathryn Harris; Leysa Forrest; Mark Kristiansen; Paola Niola; Paola Resende Silva; Patricia Dyal; Paul Randell; Rachel Williams; Samuel Weeks; Sergi Castellano; Sunando Roy; Tony Brooks; Yasmin Panchbhaya |
| EPI_ISL_1308884, EPI_ISL_1308885, EPI_ISL_1308886, EPI_ISL_1308890, EPI_ISL_1474649, EPI_ISL_1730300, EPI_ISL_1730302, EPI_ISL_1730303 | | | |
| see above | University Hospitals Of Leicester NHS Trust and DeepSeq Nottingham | COVID-19 Genomics UK (COG-UK) Consortium | Christopher Holmes; Christopher Moore; Fei Sang; Joeseph Chappell; Johnny Debebe; Jonathan Ball; Julian Tang; Karlie Fallon; Matthew Carlisle; Matthew Loose; Nadine Holmes; Patrick McClure; Paul Bird; Thomas Helmer; Victoria Wright |
| EPI_ISL_1308472, EPI_ISL_1308473, EPI_ISL_1308475, EPI_ISL_1308476, EPI_ISL_1308478, EPI_ISL_1308479, EPI_ISL_1308482, EPI_ISL_1308483, EPI_ISL_1308486, EPI_ISL_1308487, EPI_ISL_1308510, EPI_ISL_1308514, EPI_ISL_1308515, EPI_ISL_1308519, EPI_ISL_1308520, EPI_ISL_1308521, EPI_ISL_1308522, EPI_ISL_1308526, EPI_ISL_1308529, EPI_ISL_1308530, EPI_ISL_1308532, EPI_ISL_1386476, EPI_ISL_1386511, EPI_ISL_1386512, EPI_ISL_1386513, EPI_ISL_1386514, EPI_ISL_1386515, EPI_ISL_1386549, EPI_ISL_1386550, EPI_ISL_1386553, EPI_ISL_1386561, EPI_ISL_1474417, EPI_ISL_1474418 | | | |
| see above | University of Birmingham | COVID-19 Genomics UK (COG-UK) Consortium | Alex Richter; Andrew Bosworth; Andrew Bosworth. Queen Elizabeth Hospital: Anna Casey; Andrew D Beggs PHE Heartlands Lab: Husam Osman; Charlotte Poxon; Institute of Microbiology; Joanne Stockton; Josh Quick; Kasun Wanigasooriya; Mike Kidd; Nicholas Loman. University of Birmingham Testing Laboratory: Celina M Whalley; Oliver Pickles; Radoslaw Poplawski; Samuel Nicholls; University of Birmingham: Claire McMurray; Will Rowe |
| EPI_ISL_1474609, EPI_ISL_1474610, EPI_ISL_1474640, EPI_ISL_1474641, EPI_ISL_1474643, EPI_ISL_1545948, EPI_ISL_1545949, EPI_ISL_1545950 | | | |
| see above | University of Exeter | COVID-19 Genomics UK (COG-UK) Consortium | Aaron Jeffries; Audrey Farbos; Ben Temperton; Jane Masoli; Joanna Warwick-Dugdale; Michelle Michelsen; Robyn Manley; Stephen Michell |
| EPI_ISL_1386665, EPI_ISL_1386666, EPI_ISL_1386667, EPI_ISL_1386668, EPI_ISL_1386669, EPI_ISL_1386670, EPI_ISL_1386671, EPI_ISL_1386674, EPI_ISL_1474577, EPI_ISL_1545904, EPI_ISL_1545914, EPI_ISL_1545915, EPI_ISL_1545928, EPI_ISL_1545931, EPI_ISL_1545936, EPI_ISL_1636698, EPI_ISL_1636699, EPI_ISL_1730086, EPI_ISL_1730087, EPI_ISL_1730088, EPI_ISL_1730089, EPI_ISL_1730097, EPI_ISL_2125738 | | | |
| see above | Virology Department, Royal Infirmary of Edinburgh, NHS Lothian / School of Biological Sciences, University of Edinburgh | COVID-19 Genomics UK (COG-UK) Consortium | Colquhoun R; Cotton S; Dewar R; Gallagher A; Hill V; Jackson B; Maloney D; McCrone JT; McHugh M; O'Toole Á; Rambaut A; Rooke S; Scher E; Templeton K; Yu X |
| EPI_ISL_1310704, EPI_ISL_1310705, EPI_ISL_1310706, EPI_ISL_1310707, EPI_ISL_1310708, EPI_ISL_1310713, EPI_ISL_1310715, EPI_ISL_1310717, EPI_ISL_1310719, EPI_ISL_1310720, EPI_ISL_1310729, EPI_ISL_1310731, EPI_ISL_1310733, EPI_ISL_1310734, EPI_ISL_1310735, EPI_ISL_1310736, EPI_ISL_1310738, EPI_ISL_1310742, EPI_ISL_1310746, EPI_ISL_1310750, EPI_ISL_1310752, EPI_ISL_1310755, EPI_ISL_1310756, EPI_ISL_1310757, EPI_ISL_1310760, EPI_ISL_1310765, EPI_ISL_1310768, EPI_ISL_1310770, EPI_ISL_1310771, EPI_ISL_1310772, EPI_ISL_1310774, EPI_ISL_1310776, EPI_ISL_1389090, EPI_ISL_1389100, EPI_ISL_1389112, EPI_ISL_1389117, EPI_ISL_1389126, EPI_ISL_1638017, EPI_ISL_1638018, EPI_ISL_1638027, EPI_ISL_1833840 | | | |
| see above | Virology Department, Sheffield Teaching Hospitals NHS Foundation Trust/Department of Infection, Immunity and Cardiovascular Disease, The Medical School, University of Sheffield | COVID-19 Genomics UK (COG-UK) Consortium | Adri Angyal; Alex Keeley; Benjamin Lindsey; Cariad Evans; Danielle Groves; Dave Partridge; Katie Johnson; Laura Carrilero; Luke Green; Matthew Parker; Matthew Wyles; Mehmet Yavuz; Mohammad Raza; Nikki Smith; Paul Parsons; Rachel Tucker; Rebecca Brown; Thushan de Silva |
| EPI_ISL_1310157, EPI_ISL_1387837, EPI_ISL_1387840, EPI_ISL_1387856, EPI_ISL_1387864, EPI_ISL_1387879, EPI_ISL_1387884, EPI_ISL_1387885, EPI_ISL_1387886, EPI_ISL_1387887, EPI_ISL_1387891, EPI_ISL_1387892, EPI_ISL_1387895, EPI_ISL_1387896, EPI_ISL_1387904, EPI_ISL_1387913, EPI_ISL_1387922, EPI_ISL_1387925, EPI_ISL_1387932, EPI_ISL_1387933, EPI_ISL_1387940, EPI_ISL_1387941, EPI_ISL_1387943, EPI_ISL_1387944, EPI_ISL_1387946, EPI_ISL_1387948, EPI_ISL_1387953, EPI_ISL_1387968, EPI_ISL_1387969, EPI_ISL_1387976, EPI_ISL_1387978, EPI_ISL_1388021, EPI_ISL_1388022, EPI_ISL_1388025, EPI_ISL_1388026, EPI_ISL_1388027, EPI_ISL_1388028, EPI_ISL_1388032, EPI_ISL_1388039, EPI_ISL_1388050, EPI_ISL_1388051, EPI_ISL_1388054, EPI_ISL_1388055, EPI_ISL_1388057, EPI_ISL_1388058, EPI_ISL_1388063, EPI_ISL_1388094, EPI_ISL_1388105, EPI_ISL_1388106, EPI_ISL_1388107, EPI_ISL_1388108, EPI_ISL_1388109, EPI_ISL_1388121, EPI_ISL_1388131, EPI_ISL_1388133, EPI_ISL_1388146, EPI_ISL_1388167, EPI_ISL_1388189, EPI_ISL_1388194, EPI_ISL_1388203, EPI_ISL_1388229, EPI_ISL_1388407, EPI_ISL_1388409, EPI_ISL_1388413, EPI_ISL_1388420, EPI_ISL_1388561, EPI_ISL_1388575, EPI_ISL_1388578, EPI_ISL_1637273, EPI_ISL_1637274, EPI_ISL_1637275, EPI_ISL_1637276, EPI_ISL_1637278, EPI_ISL_1637279, EPI_ISL_1637282, EPI_ISL_1637284, EPI_ISL_1637286, EPI_ISL_1637289, EPI_ISL_1637290, EPI_ISL_1637291, EPI_ISL_1637292, EPI_ISL_1637293, EPI_ISL_1637295, EPI_ISL_1637296, EPI_ISL_1637297, EPI_ISL_1637299, EPI_ISL_1637300, EPI_ISL_1637301, EPI_ISL_1637303, EPI_ISL_1637304, EPI_ISL_1637305, EPI_ISL_1637887, EPI_ISL_1637901, EPI_ISL_1637903, EPI_ISL_1637905, EPI_ISL_1637907, EPI_ISL_1637910, EPI_ISL_1637911, EPI_ISL_1637913, EPI_ISL_1637914, EPI_ISL_1637915, EPI_ISL_1637916, EPI_ISL_1637919, EPI_ISL_1637933, EPI_ISL_1637934, EPI_ISL_1637937, EPI_ISL_1637939, EPI_ISL_1637941, EPI_ISL_1637943, EPI_ISL_1637945 | | | |
| see above | Wales Specialist Virology Centre Sequencing lab: Pathogen Genomics Unit | Public Health Wales Microbiology Cardiff Wales Specialist Virology Centre | Alec Birchley; Alexander Adams; Amy Gaskin; Angela Marchbank; Bree Gatica-Wilcox; Catherine Moore; Jason Coombes; Joanne Watkins; Joel Southgate; Johnathan Evans; Laura Gifford; Lauren Gilbert; Lee Graham; Malorie Perry; Matthew Bull; Nicole Pacchiarini; Sally Corden; Sara Kumziene-Summerhayes; Sara Rey; Sarah Taylor; Simon Cottrell; Sophie Jones; Tom Connor |
| EPI_ISL_1386599, EPI_ISL_1386600, EPI_ISL_1386601, EPI_ISL_1386602, EPI_ISL_1386603, EPI_ISL_1386633, EPI_ISL_1386634, EPI_ISL_1386635, EPI_ISL_1386638, EPI_ISL_1386639, EPI_ISL_1386640, EPI_ISL_1545883, EPI_ISL_1545885, EPI_ISL_1545887, EPI_ISL_1545888, EPI_ISL_1545892, EPI_ISL_1636627, EPI_ISL_1636628, EPI_ISL_1636629, EPI_ISL_1636630, EPI_ISL_1636631, EPI_ISL_1636632, EPI_ISL_1636633, EPI_ISL_1636634, EPI_ISL_1636635, EPI_ISL_1636636, EPI_ISL_1636637, EPI_ISL_1636638, EPI_ISL_1636640, EPI_ISL_1636644, EPI_ISL_1636657, EPI_ISL_1636660, EPI_ISL_1636661, EPI_ISL_1636662, EPI_ISL_1636667, EPI_ISL_1636668, EPI_ISL_1636669, EPI_ISL_1636670 | | | |
| see above | West of Scotland Specialist Virology Centre, NHSGGC / MRC-University of Glasgow Centre for Virus Research | COVID-19 Genomics UK (COG-UK) Consortium | Alasdair MacLean; Alice Broos; Ana da Silva Filipe; Antonia Ho; Daniel Mair; David L Robertson; Emma Thomson; Guy Mollett; James Shepherd; Jenna Nichols; Joseph Hughes; Kathy Li; Kathy Smollett; Kyriaki Nomikou; Lily Tong; Matthew Holden; Natasha Johnson; Rachel Blacow; Richard Orton; Rory Gunson; Sarah McDonald; Sharif Shaaban; Sreenu Vattipally; Stephen Carmichael |

We gratefully acknowledge the following Authors from the Originating laboratories responsible for obtaining the specimens, as well as the Submitting laboratories where the genome data were generated and shared via GISAID, on which this research is based.

All Submitters of data may be contacted directly via www.gisaid.org

Authors are sorted alphabetically.

| Accession ID | Originating Laboratory | Submitting Laboratory | Authors |
|---|---|---|---|
| EPI_ISL_1474825 | Barts Health NHS Trust | COVID-19 Genomics UK (COG-UK) Consortium | BROAD; Beatrix; CUTINO-MOGUEL; Claire; David; Dola; HARRINGTON; KELE; KULASEGARAN-SHYLINI; Maria-Teresa; OWOYEMI; Raghavendran |
| EPI_ISL_1388772, EPI_ISL_1388775, EPI_ISL_1388777, EPI_ISL_1388779, EPI_ISL_1388786, EPI_ISL_1388788, EPI_ISL_1388790, EPI_ISL_1388863, EPI_ISL_1388884, EPI_ISL_1388886, EPI_ISL_1388923, EPI_ISL_1388925, EPI_ISL_1388927, EPI_ISL_1638003, EPI_ISL_1730861, EPI_ISL_2128017 | | | |
| see above | Centre for Enzyme Innovation, University of Portsmouth / Translational Research Laboratory, Portsmouth Hospitals NHS Trust | COVID-19 Genomics UK (COG-UK) Consortium | Angela Beckett; Christopher Fearn; Kate Cook; Katie Loveson; Salman Goudarzi; Samuel Robson; Scott Elliott; Sharon Glaysher |
| EPI_ISL_1308684, EPI_ISL_1308686, EPI_ISL_1308687, EPI_ISL_1308688, EPI_ISL_1308689, EPI_ISL_1308691, EPI_ISL_1308693, EPI_ISL_1308695, EPI_ISL_1386589 | | | |
| see above | Department of Pathology, University of Cambridge | COVID-19 Genomics UK (COG-UK) Consortium | Aminu S. Jahun; Ian Goodfellow; Iliana Georgana; Martin D. Curran; Myra Hosmillo; Rhys Izuagbe; Surendra Parmar; William L. Hamilton; Yasmin Chaudhry |

EPI_ISL_1326996, EPI_ISL_1342235, EPI_ISL_1342239, EPI_ISL_1342245, EPI_ISL_1342246, EPI_ISL_1342247, EPI_ISL_1342250, EPI_ISL_1342252, EPI_ISL_1342254, EPI_ISL_1342256, EPI_ISL_1342257, EPI_ISL_1342261, EPI_ISL_1342262, EPI_ISL_1342263, EPI_ISL_1342265, EPI_ISL_1342268, EPI_ISL_1342269, EPI_ISL_1342270, EPI_ISL_1342271, EPI_ISL_1342274, EPI_ISL_1342275, EPI_ISL_1342276, EPI_ISL_1342277, EPI_ISL_1342281, EPI_ISL_1342282, EPI_ISL_1342283, EPI_ISL_1342285, EPI_ISL_1342286, EPI_ISL_1342290, EPI_ISL_1342292, EPI_ISL_1342295, EPI_ISL_1342297, EPI_ISL_1342298, EPI_ISL_1342300, EPI_ISL_1342304, EPI_ISL_1342307, EPI_ISL_1342308, EPI_ISL_1342310, EPI_ISL_1342311, EPI_ISL_1342312, EPI_ISL_1342314, EPI_ISL_1342315, EPI_ISL_1342321, EPI_ISL_1342322, EPI_ISL_1342324, EPI_ISL_1342329, EPI_ISL_1342330, EPI_ISL_1342331, EPI_ISL_1342334, EPI_ISL_1342335, EPI_ISL_1342336, EPI_ISL_1342340, EPI_ISL_1342343, EPI_ISL_1342351, EPI_ISL_1342352, EPI_ISL_1342359, EPI_ISL_1342363, EPI_ISL_1342365, EPI_ISL_1342366, EPI_ISL_1342367, EPI_ISL_1342374, EPI_ISL_1342379, EPI_ISL_1342382, EPI_ISL_1342383, EPI_ISL_1342384, EPI_ISL_1342387, EPI_ISL_1342393, EPI_ISL_1342394, EPI_ISL_1342396, EPI_ISL_1342399, EPI_ISL_1342402, EPI_ISL_1342404, EPI_ISL_1342406, EPI_ISL_1342410, EPI_ISL_1342412, EPI_ISL_1342413, EPI_ISL_1342415, EPI_ISL_1342417, EPI_ISL_1342418, EPI_ISL_1342420, EPI_ISL_1342421, EPI_ISL_1342423, EPI_ISL_1342424, EPI_ISL_1342429, EPI_ISL_1342430, EPI_ISL_1342431, EPI_ISL_1342432, EPI_ISL_1342435, EPI_ISL_1342436, EPI_ISL_1342439, EPI_ISL_1342441, EPI_ISL_1342445, EPI_ISL_1342447, EPI_ISL_1342450, EPI_ISL_1342452, EPI_ISL_1342457, EPI_ISL_1342459, EPI_ISL_1342463, EPI_ISL_1342464, EPI_ISL_1342465, EPI_ISL_1342473, EPI_ISL_1342475, EPI_ISL_1342476, EPI_ISL_1342478, EPI_ISL_1342479, EPI_ISL_1342482, EPI_ISL_1342483, EPI_ISL_1342484, EPI_ISL_1342486, EPI_ISL_1342487, EPI_ISL_1342490, EPI_ISL_1342493, EPI_ISL_1342494, EPI_ISL_1342498, EPI_ISL_1342499, EPI_ISL_1342502, EPI_ISL_1342508, EPI_ISL_1342509, EPI_ISL_1342510, EPI_ISL_1342511, EPI_ISL_1342514, EPI_ISL_1342516, EPI_ISL_1342517, EPI_ISL_1342518, EPI_ISL_1342519, EPI_ISL_1342520, EPI_ISL_1342521, EPI_ISL_1342523, EPI_ISL_1342525, EPI_ISL_1342528, EPI_ISL_1342529, EPI_ISL_1342532, EPI_ISL_1342534, EPI_ISL_1342535, EPI_ISL_1342536, EPI_ISL_1342537, EPI_ISL_1342539, EPI_ISL_1342540, EPI_ISL_1342545, EPI_ISL_1342546, EPI_ISL_1342549, EPI_ISL_1342550, EPI_ISL_1342551, EPI_ISL_1342556, EPI_ISL_1342558, EPI_ISL_1342559, EPI_ISL_1342564, EPI_ISL_1342565, EPI_ISL_1342566, EPI_ISL_1342567, EPI_ISL_1342568, EPI_ISL_1342569, EPI_ISL_1342570, EPI_ISL_1342573, EPI_ISL_1342574, EPI_ISL_1342576, EPI_ISL_1342577, EPI_ISL_1342578, EPI_ISL_1342579, EPI_ISL_1342580, EPI_ISL_1342581, EPI_ISL_1342582, EPI_ISL_1342583, EPI_ISL_1342584, EPI_ISL_1342587, EPI_ISL_1342588, EPI_ISL_1342589, EPI_ISL_1342590, EPI_ISL_1342592, EPI_ISL_1342593, EPI_ISL_1342595, EPI_ISL_1342598, EPI_ISL_1342600, EPI_ISL_1342603, EPI_ISL_1342605, EPI_ISL_1342606, EPI_ISL_1342607, EPI_ISL_1342608, EPI_ISL_1342610, EPI_ISL_1342611, EPI_ISL_1342612, EPI_ISL_1342615, EPI_ISL_1342617, EPI_ISL_1342618, EPI_ISL_1342619, EPI_ISL_1342622, EPI_ISL_1342623, EPI_ISL_1342624, EPI_ISL_1342625, EPI_ISL_1342627, EPI_ISL_1342628, EPI_ISL_1342630, EPI_ISL_1342631, EPI_ISL_1342632, EPI_ISL_1342633, EPI_ISL_1342635, EPI_ISL_1342637, EPI_ISL_1342638, EPI_ISL_1342639, EPI_ISL_1342642, EPI_ISL_1342646, EPI_ISL_1342647, EPI_ISL_1342648, EPI_ISL_1342649, EPI_ISL_1342650, EPI_ISL_1342651, EPI_ISL_1342652, EPI_ISL_1342653, EPI_ISL_1342654, EPI_ISL_1342656, EPI_ISL_1342657, EPI_ISL_1342660, EPI_ISL_1342661, EPI_ISL_1342662, EPI_ISL_1342663, EPI_ISL_1342664, EPI_ISL_1342665, EPI_ISL_1342667, EPI_ISL_1342668, EPI_ISL_1342672, EPI_ISL_1342674, EPI_ISL_1342675, EPI_ISL_1342679, EPI_ISL_1342681, EPI_ISL_1342682, EPI_ISL_1342683, EPI_ISL_1342684, EPI_ISL_1342687, EPI_ISL_1342688, EPI_ISL_1342690, EPI_ISL_1342691, EPI_ISL_1342692, EPI_ISL_1342694, EPI_ISL_1342696, EPI_ISL_1342697, EPI_ISL_1342698, EPI_ISL_1342699, EPI_ISL_1342700, EPI_ISL_1342703, EPI_ISL_1342704, EPI_ISL_1342706, EPI_ISL_1342707, EPI_ISL_1342708, EPI_ISL_1342709, EPI_ISL_1342710, EPI_ISL_1342711, EPI_ISL_1342715, EPI_ISL_1342716, EPI_ISL_1342717, EPI_ISL_1342719, EPI_ISL_1342721, EPI_ISL_1342722, EPI_ISL_1342723, EPI_ISL_1342725, EPI_ISL_1342727, EPI_ISL_1342729, EPI_ISL_1342730, EPI_ISL_1342731, EPI_ISL_1342732, EPI_ISL_1342734, EPI_ISL_1342735, EPI_ISL_1342737, EPI_ISL_1342738, EPI_ISL_1342739, EPI_ISL_1342740, EPI_ISL_1342741, EPI_ISL_1342742, EPI_ISL_1342744, EPI_ISL_1342745, EPI_ISL_1342746, EPI_ISL_1342747, EPI_ISL_1342748, EPI_ISL_1342749, EPI_ISL_1342750, EPI_ISL_1342751, EPI_ISL_1342753, EPI_ISL_1342755, EPI_ISL_1342758, EPI_ISL_1342760, EPI_ISL_1342761, EPI_ISL_1342762, EPI_ISL_1342763, EPI_ISL_1342764, EPI_ISL_1342765, EPI_ISL_1342767, EPI_ISL_1342769, EPI_ISL_1342770, EPI_ISL_1342774, EPI_ISL_1342775, EPI_ISL_1342776, EPI_ISL_1342779, EPI_ISL_1342780, EPI_ISL_1342781, EPI_ISL_1342782, EPI_ISL_1342783, EPI_ISL_1342784, EPI_ISL_1342785, EPI_ISL_1342787, EPI_ISL_1342790, EPI_ISL_1342791, EPI_ISL_1342792, EPI_ISL_1342793, EPI_ISL_1342795, EPI_ISL_1342796, EPI_ISL_1342798, EPI_ISL_1342799, EPI_ISL_1342800, EPI_ISL_1342801, EPI_ISL_1342802, EPI_ISL_1342803, EPI_ISL_1342804, EPI_ISL_1342805, EPI_ISL_1342808, EPI_ISL_1342809, EPI_ISL_1342811, EPI_ISL_1342812, EPI_ISL_1342818, EPI_ISL_1342819, EPI_ISL_1342820, EPI_ISL_1342830, EPI_ISL_1342831, EPI_ISL_1342841, EPI_ISL_1342844, EPI_ISL_1342850, EPI_ISL_1342851, EPI_ISL_1342855, EPI_ISL_1342857, EPI_ISL_1342864, EPI_ISL_1342869, EPI_ISL_1342888, EPI_ISL_1342893, EPI_ISL_1342898, EPI_ISL_1342900, EPI_ISL_1342902, EPI_ISL_1342903, EPI_ISL_1342907, EPI_ISL_1342917, EPI_ISL_1342919, EPI_ISL_1342922, EPI_ISL_1342935, EPI_ISL_1342946, EPI_ISL_1342950, EPI_ISL_1342955, EPI_ISL_1342960, EPI_ISL_1342963, EPI_ISL_1342969, EPI_ISL_1342971, EPI_ISL_1342972, EPI_ISL_1342978, EPI_ISL_1342982, EPI_ISL_1343010, EPI_ISL_1343023, EPI_ISL_1343034, EPI_ISL_1343036, EPI_ISL_1343044, EPI_ISL_1343050, EPI_ISL_1343053, EPI_ISL_1343054, EPI_ISL_1343055, EPI_ISL_1343061, EPI_ISL_1343065, EPI_ISL_1343069, EPI_ISL_1343073, EPI_ISL_1343079, EPI_ISL_1343081, EPI_ISL_1343085, EPI_ISL_1343087, EPI_ISL_1343089, EPI_ISL_1343090, EPI_ISL_1343091, EPI_ISL_1343093, EPI_ISL_1343097, EPI_ISL_1343098, EPI_ISL_1343099, EPI_ISL_1343101, EPI_ISL_1343104, EPI_ISL_1343106, EPI_ISL_1343107, EPI_ISL_1343108, EPI_ISL_1343109, EPI_ISL_1343110, EPI_ISL_1343111, EPI_ISL_1343112, EPI_ISL_1343114, EPI_ISL_1343115, EPI_ISL_1343116, EPI_ISL_1343117, EPI_ISL_1343119, EPI_ISL_1343120, EPI_ISL_1343121, EPI_ISL_1343122, EPI_ISL_1343123, EPI_ISL_1343124, EPI_ISL_1343126, EPI_ISL_1343132, EPI_ISL_1343134, EPI_ISL_1343135, EPI_ISL_1343137, EPI_ISL_1343138, EPI_ISL_1343139, EPI_ISL_1343141, EPI_ISL_1343142, EPI_ISL_1343147, EPI_ISL_1343152, EPI_ISL_1343154, EPI_ISL_1343155, EPI_ISL_1343156, EPI_ISL_1343159, EPI_ISL_1343161, EPI_ISL_1343162, EPI_ISL_1343164, EPI_ISL_1343165, EPI_ISL_1343166, EPI_ISL_1343167, EPI_ISL_1343168, EPI_ISL_1343169, EPI_ISL_1343170, EPI_ISL_1343171, EPI_ISL_1343172, EPI_ISL_1343173, EPI_ISL_1343174, EPI_ISL_1343176, EPI_ISL_1343177, EPI_ISL_1343178, EPI_ISL_1343180, EPI_ISL_1343182, EPI_ISL_1343183, EPI_ISL_1343184, EPI_ISL_1343186, EPI_ISL_1343187, EPI_ISL_1343189, EPI_ISL_1343192, EPI_ISL_1343194, EPI_ISL_1343196, EPI_ISL_1343199, EPI_ISL_1343202, EPI_ISL_1343203, EPI_ISL_1343205, EPI_ISL_1343206, EPI_ISL_1343208, EPI_ISL_1343212, EPI_ISL_1343213, EPI_ISL_1343214, EPI_ISL_1343215, EPI_ISL_1343216, EPI_ISL_1343217, EPI_ISL_1343219, EPI_ISL_1343221, EPI_ISL_1343223, EPI_ISL_1343224, EPI_ISL_1343225, EPI_ISL_1343230, EPI_ISL_1343234, EPI_ISL_1343236, EPI_ISL_1343237, EPI_ISL_1343238, EPI_ISL_1343239, EPI_ISL_1343242, EPI_ISL_1343243, EPI_ISL_1343244, EPI_ISL_1343251, EPI_ISL_1343252, EPI_ISL_1343254, EPI_ISL_1343256, EPI_ISL_1343257, EPI_ISL_1343260, EPI_ISL_1343261, EPI_ISL_1343262, EPI_ISL_1343264, EPI_ISL_1343266, EPI_ISL_1343267, EPI_ISL_1343270, EPI_ISL_1343271, EPI_ISL_1343273, EPI_ISL_1343274, EPI_ISL_1343275, EPI_ISL_1343277, EPI_ISL_1343278, EPI_ISL_1343281, EPI_ISL_1343282, EPI_ISL_1343284, EPI_ISL_1343285, EPI_ISL_1343286, EPI_ISL_1343287, EPI_ISL_1343290, EPI_ISL_1343292, EPI_ISL_1343293, EPI_ISL_1343295, EPI_ISL_1343296, EPI_ISL_1343297, EPI_ISL_1343298, EPI_ISL_1343299, EPI_ISL_1343300, EPI_ISL_1343302, EPI_ISL_1343303, EPI_ISL_1343307, EPI_ISL_1343311, EPI_ISL_1343316, EPI_ISL_1343317, EPI_ISL_1343318, EPI_ISL_1343319, EPI_ISL_1343320, EPI_ISL_1343323, EPI_ISL_1343324, EPI_ISL_1343328, EPI_ISL_1343329, EPI_ISL_1343331, EPI_ISL_1343333, EPI_ISL_1343337, EPI_ISL_1343338, EPI_ISL_1343339, EPI_ISL_1343340, EPI_ISL_1343342, EPI_ISL_1343343, EPI_ISL_1343344, EPI_ISL_1343346, EPI_ISL_1343348, EPI_ISL_1343349, EPI_ISL_1343350, EPI_ISL_1343351, EPI_ISL_1343354, EPI_ISL_1344468, EPI_ISL_1344472, EPI_ISL_1344479, EPI_ISL_1344480, EPI_ISL_1344483, EPI_ISL_1344485, EPI_ISL_1344489, EPI_ISL_1344495, EPI_ISL_1344501, EPI_ISL_1344502, EPI_ISL_1344515, EPI_ISL_1344518, EPI_ISL_1344522, EPI_ISL_1344523, EPI_ISL_1344540, EPI_ISL_1344541, EPI_ISL_1344543, EPI_ISL_1344548, EPI_ISL_1344553, EPI_ISL_1344558, EPI_ISL_1344563, EPI_ISL_1344566, EPI_ISL_1344611, EPI_ISL_1344616, EPI_ISL_1344621, EPI_ISL_1344629, EPI_ISL_1344632, EPI_ISL_1344633, EPI_ISL_1344637, EPI_ISL_1344652, EPI_ISL_1344658, EPI_ISL_1344667, EPI_ISL_1344685, EPI_ISL_1344691, EPI_ISL_1344694, EPI_ISL_1344695, EPI_ISL_1344703, EPI_ISL_1344705, EPI_ISL_1345296, EPI_ISL_1345302, EPI_ISL_1345304, EPI_ISL_1345306, EPI_ISL_1345310, EPI_ISL_1345313, EPI_ISL_1345324, EPI_ISL_1345326, EPI_ISL_1345328, EPI_ISL_1345335, EPI_ISL_1345338, EPI_ISL_1345343, EPI_ISL_1345350, EPI_ISL_1345351, EPI_ISL_1345352, EPI_ISL_1345353, EPI_ISL_1345356, EPI_ISL_1345360, EPI_ISL_1345364, EPI_ISL_1345366, EPI_ISL_1345369, EPI_ISL_1345373, EPI_ISL_1345376, EPI_ISL_1345380, EPI_ISL_1345384, EPI_ISL_1345385, EPI_ISL_1345390, EPI_ISL_1345391, EPI_ISL_1345395, EPI_ISL_1345396, EPI_ISL_1345398, EPI_ISL_1345402, EPI_ISL_1345405, EPI_ISL_1345407, EPI_ISL_1345410, EPI_ISL_1345411, EPI_ISL_1345412, EPI_ISL_1345415, EPI_ISL_1345418, EPI_ISL_1345421, EPI_ISL_1345422, EPI_ISL_1345423, EPI_ISL_1345424, EPI_ISL_1345427, EPI_ISL_1345428, EPI_ISL_1345429, EPI_ISL_1345431, EPI_ISL_1345434, EPI_ISL_1345441, EPI_ISL_1345443, EPI_ISL_1345444, EPI_ISL_1345446, EPI_ISL_1345447, EPI_ISL_1345455, EPI_ISL_1345459, EPI_ISL_1345461, EPI_ISL_1345462, EPI_ISL_1345464, EPI_ISL_1345466, EPI_ISL_1345468, EPI_ISL_1345475, EPI_ISL_1345479, EPI_ISL_1345481, EPI_ISL_1345486, EPI_ISL_1345487, EPI_ISL_1345488, EPI_ISL_1345497, EPI_ISL_1345500, EPI_ISL_1345501, EPI_ISL_1345504, EPI_ISL_1345506, EPI_ISL_1345507, EPI_ISL_1345511, EPI_ISL_1345515, EPI_ISL_1345519, EPI_ISL_1345522, EPI_ISL_1345523, EPI_ISL_1345524, EPI_ISL_1345525, EPI_ISL_1345526, EPI_ISL_1345527, EPI_ISL_1345531, EPI_ISL_1345532, EPI_ISL_1345533, EPI_ISL_1345534, EPI_ISL_1345537, EPI_ISL_1345538, EPI_ISL_1345542, EPI_ISL_1345543, EPI_ISL_1345547, EPI_ISL_1345548, EPI_ISL_1345553, EPI_ISL_1345556, EPI_ISL_1345557, EPI_ISL_1345563, EPI_ISL_1345569, EPI_ISL_1345570, EPI_ISL_1345571, EPI_ISL_1345575, EPI_ISL_1345576, EPI_ISL_1345582, EPI_ISL_1345583, EPI_ISL_1345587, EPI_ISL_1345589, EPI_ISL_1345590, EPI_ISL_1345595, EPI_ISL_1345596, EPI_ISL_1345597, EPI_ISL_1345599, EPI_ISL_1345602, EPI_ISL_1345603, EPI_ISL_1345604, EPI_ISL_1345605, EPI_ISL_1345607, EPI_ISL_1345611, EPI_ISL_1345614, EPI_ISL_1345617, EPI_ISL_1345620, EPI_ISL_1345621, EPI_ISL_1345624, EPI_ISL_1345629, EPI_ISL_1345632, EPI_ISL_1345635, EPI_ISL_1345639, EPI_ISL_1345641, EPI_ISL_1345644, EPI_ISL_1345646, EPI_ISL_1345647, EPI_ISL_1345649, EPI_ISL_1365149, EPI_ISL_1365150, EPI_ISL_1365151, EPI_ISL_1365152, EPI_ISL_1365153, EPI_ISL_1365154, EPI_ISL_1365155, EPI_ISL_1365156, EPI_ISL_1365157, EPI_ISL_1365158, EPI_ISL_1365159, EPI_ISL_1365160, EPI_ISL_1365161, EPI_ISL_1365162, EPI_ISL_1365163, EPI_ISL_1365164, EPI_ISL_1365165, EPI_ISL_1365166, EPI_ISL_1365167, EPI_ISL_1365168, EPI_ISL_1365169, EPI_ISL_1365170, EPI_ISL_1365171, EPI_ISL_1365172, EPI_ISL_1365173, EPI_ISL_1365174, EPI_ISL_1365175, EPI_ISL_1365176, EPI_ISL_1365177, EPI_ISL_1365218, EPI_ISL_1374337, EPI_ISL_1374343, EPI_ISL_1374361, EPI_ISL_1374365, EPI_ISL_1374389, EPI_ISL_1374458, EPI_ISL_1374462, EPI_ISL_1374501, EPI_ISL_1374587, EPI_ISL_1374859, EPI_ISL_1374863, EPI_ISL_1374865, EPI_ISL_1374866, EPI_ISL_1374871, EPI_ISL_1374872, EPI_ISL_1374876, EPI_ISL_1374877, EPI_ISL_1374878, EPI_ISL_1374886, EPI_ISL_1374891, EPI_ISL_1374893, EPI_ISL_1374898, EPI_ISL_1374900, EPI_ISL_1374909, EPI_ISL_1374911, EPI_ISL_1374913, EPI_ISL_1374915, EPI_ISL_1374919, EPI_ISL_1374921, EPI_ISL_1374923, EPI_ISL_1374924, EPI_ISL_1374925, EPI_ISL_1374929, EPI_ISL_1374930, EPI_ISL_1374931, EPI_ISL_1374933, EPI_ISL_1374936, EPI_ISL_1374937, EPI_ISL_1374938, EPI_ISL_1374939, EPI_ISL_1374942, EPI_ISL_1374944, EPI_ISL_1374947, EPI_ISL_1374948, EPI_ISL_1374950, EPI_ISL_1374957, EPI_ISL_1374961, EPI_ISL_1374962, EPI_ISL_1374963, EPI_ISL_1374965, EPI_ISL_1374967, EPI_ISL_1374969, EPI_ISL_1374971, EPI_ISL_1374975, EPI_ISL_1374976, EPI_ISL_1374977, EPI_ISL_1374979, EPI_ISL_1374980, EPI_ISL_1374984, EPI_ISL_1374991, EPI_ISL_1374992, EPI_ISL_1374993, EPI_ISL_1374994, EPI_ISL_1374997, EPI_ISL_1374999, EPI_ISL_1375001, EPI_ISL_1375002, EPI_ISL_1375004, EPI_ISL_1375006, EPI_ISL_1375007, EPI_ISL_1375008, EPI_ISL_1375010, EPI_ISL_1375018, EPI_ISL_1375024, EPI_ISL_1375026, EPI_ISL_1375027, EPI_ISL_1375029, EPI_ISL_1375032, EPI_ISL_1375034, EPI_ISL_1375035, EPI_ISL_1375039, EPI_ISL_1375040, EPI_ISL_1375046, EPI_ISL_1375048, EPI_ISL_1375049, EPI_ISL_1375054, EPI_ISL_1375062, EPI_ISL_1375063, EPI_ISL_1375064, EPI_ISL_1375073, EPI_ISL_1375075, EPI_ISL_1375079, EPI_ISL_1375083, EPI_ISL_1375084, EPI_ISL_1375086, EPI_ISL_1375088, EPI_ISL_1375091, EPI_ISL_1375092, EPI_ISL_1375093, EPI_ISL_1375097, EPI_ISL_1375098, EPI_ISL_1375099, EPI_ISL_1375102, EPI_ISL_1375109, EPI_ISL_1375110, EPI_ISL_1375115, EPI_ISL_1375121, EPI_ISL_1375124, EPI_ISL_1375129, EPI_ISL_1375132, EPI_ISL_1375133, EPI_ISL_1375136, EPI_ISL_1375140, EPI_ISL_1375143, EPI_ISL_1375144, EPI_ISL_1375148, EPI_ISL_1375149, EPI_ISL_1375151, EPI_ISL_1375154, EPI_ISL_1375155, EPI_ISL_1375157, EPI_ISL_1375160, EPI_ISL_1375168, EPI_ISL_1375171, EPI_ISL_1375180, EPI_ISL_1375183, EPI_ISL_1375200, EPI_ISL_1375202, EPI_ISL_1375207, EPI_ISL_1375210, EPI_ISL_1375213, EPI_ISL_1375217, EPI_ISL_1375220, EPI_ISL_1375225, EPI_ISL_1375230, EPI_ISL_1375239, EPI_ISL_1375260, EPI_ISL_1375261, EPI_ISL_1375268, EPI_ISL_1375272, EPI_ISL_1375275, EPI_ISL_1375282, EPI_ISL_1375284, EPI_ISL_1375288, EPI_ISL_1375291, EPI_ISL_1375296, EPI_ISL_1375299, EPI_ISL_1375300, EPI_ISL_1375302, EPI_ISL_1375304, EPI_ISL_1375305, EPI_ISL_1375312, EPI_ISL_1375313, EPI_ISL_1375314, EPI_ISL_1375318, EPI_ISL_1375323, EPI_ISL_1375338, EPI_ISL_1375340, EPI_ISL_1375346, EPI_ISL_1375355, EPI_ISL_1375362, EPI_ISL_1375368, EPI_ISL_1375374, EPI_ISL_1375375, EPI_ISL_1375381, EPI_ISL_1375382, EPI_ISL_1375387, EPI_ISL_1375409, EPI_ISL_1375410, EPI_ISL_1375411, EPI_ISL_1375434, EPI_ISL_1375435, EPI_ISL_1375436, EPI_ISL_1375444, EPI_ISL_1375447, EPI_ISL_1375448, EPI_ISL_1375450, EPI_ISL_1375451, EPI_ISL_1375469, EPI_ISL_1375474, EPI_ISL_1375489, EPI_ISL_1375527, EPI_ISL_1375533, EPI_ISL_1375562, EPI_ISL_1375641, EPI_ISL_1375691, EPI_ISL_1375706, EPI_ISL_1375719, EPI_ISL_1375755, EPI_ISL_1375764, EPI_ISL_1376262, EPI_ISL_1376289, EPI_ISL_1376316, EPI_ISL_1376341, EPI_ISL_1376392, EPI_ISL_1376403, EPI_ISL_1376405, EPI_ISL_1376425, EPI_ISL_1376487, EPI_ISL_1376523, EPI_ISL_1376579, EPI_ISL_1376611, EPI_ISL_1376622, EPI_ISL_1376729, EPI_ISL_1377191, EPI_ISL_1377208, EPI_ISL_1377229, EPI_ISL_1377257, EPI_ISL_1377348, EPI_ISL_1377357, EPI_ISL_1377368, EPI_ISL_1377386, EPI_ISL_1377409, EPI_ISL_1377414, EPI_ISL_1377415, EPI_ISL_1377418, EPI_ISL_1377419, EPI_ISL_1377422, EPI_ISL_1377423, EPI_ISL_1377427, EPI_ISL_1377431, EPI_ISL_1377440, EPI_ISL_1377441, EPI_ISL_1377450, EPI_ISL_1377455, EPI_ISL_1377460, EPI_ISL_1377461, EPI_ISL_1377462, EPI_ISL_1377463, EPI_ISL_1377464, EPI_ISL_1377468, EPI_ISL_1377469, EPI_ISL_1377473, EPI_ISL_1377480, EPI_ISL_1377481, EPI_ISL_1377489, EPI_ISL_1377491, EPI_ISL_1377494, EPI_ISL_1377497, EPI_ISL_1377506, EPI_ISL_1377508, EPI_ISL_1377511, EPI_ISL_1377512, EPI_ISL_1377515, EPI_ISL_1377516, EPI_ISL_1377523, EPI_ISL_1377527, EPI_ISL_1377530, EPI_ISL_1377536, EPI_ISL_1377544, EPI_ISL_1377545, EPI_ISL_1377551, EPI_ISL_1377552, EPI_ISL_1377557, EPI_ISL_1377559, EPI_ISL_1377562, EPI_ISL_1377564, EPI_ISL_1377567, EPI_ISL_1377573, EPI_ISL_1377578, EPI_ISL_1377583, EPI_ISL_1377586, EPI_ISL_1377594, EPI_ISL_1377595, EPI_ISL_1377597, EPI_ISL_1377599, EPI_ISL_1377600, EPI_ISL_1377604, EPI_ISL_1377613, EPI_ISL_1377623, EPI_ISL_1377628, EPI_ISL_1377633, EPI_ISL_1377634, EPI_ISL_1377636, EPI_ISL_1377638, EPI_ISL_1377640, EPI_ISL_1377643, EPI_ISL_1377646, EPI_ISL_1377648, EPI_ISL_1377652, EPI_ISL_1377654, EPI_ISL_1377665, EPI_ISL_1377668, EPI_ISL_1377674, EPI_ISL_1377675, EPI_ISL_1377677, EPI_ISL_1377678, EPI_ISL_1377679, EPI_ISL_1377681, EPI_ISL_1377683, EPI_ISL_1377684, EPI_ISL_1377685, EPI_ISL_1377686, EPI_ISL_1377690, EPI_ISL_1377694, EPI_ISL_1377697, EPI_ISL_1377700, EPI_ISL_1377701, EPI_ISL_1377702, EPI_ISL_1377703, EPI_ISL_1377704, EPI_ISL_1377706, EPI_ISL_1377707, EPI_ISL_1377708, EPI_ISL_1377709, EPI_ISL_1377712, EPI_ISL_1377720, EPI_ISL_1377724, EPI_ISL_1377725, EPI_ISL_1377727, EPI_ISL_1377730, EPI_ISL_1377735, EPI_ISL_1377736, EPI_ISL_1377738, EPI_ISL_1377741, EPI_ISL_1377744, EPI_ISL_1377745, EPI_ISL_1377748, EPI_ISL_1377749, EPI_ISL_1377750, EPI_ISL_1377751, EPI_ISL_1377752, EPI_ISL_1377753, EPI_ISL_1377754, EPI_ISL_1377755, EPI_ISL_1377757, EPI_ISL_1377758, EPI_ISL_1377760, EPI_ISL_1377762, EPI_ISL_1377763, EPI_ISL_1377764, EPI_ISL_1377766, EPI_ISL_1377770, EPI_ISL_1377772, EPI_ISL_1377775, EPI_ISL_1377776, EPI_ISL_1377778, EPI_ISL_1377779, EPI_ISL_1377780, EPI_ISL_1377787, EPI_ISL_1377788, EPI_ISL_1377791, EPI_ISL_1377792, EPI_ISL_1377793, EPI_ISL_1377794, EPI_ISL_1377796, EPI_ISL_1377798, EPI_ISL_1377799, EPI_ISL_1377804, EPI_ISL_1377807, EPI_ISL_1377808, EPI_ISL_1377809, EPI_ISL_1377814, EPI_ISL_1377816, EPI_ISL_1377818, EPI_ISL_1377822, EPI_ISL_1377824, EPI_ISL_1377827, EPI_ISL_1377828, EPI_ISL_1377831, EPI_ISL_1377832, EPI_ISL_1377834, EPI_ISL_1377835, EPI_ISL_1377840, EPI_ISL_1377841, EPI_ISL_1377843, EPI_ISL_1377844, EPI_ISL_1377845, EPI_ISL_1377846, EPI_ISL_1377847, EPI_ISL_1377848, EPI_ISL_1377852, EPI_ISL_1377856, EPI_ISL_1377857, EPI_ISL_1377858, EPI_ISL_1377863, EPI_ISL_1377864, EPI_ISL_1377865, EPI_ISL_1377868, EPI_ISL_1377871, EPI_ISL_1377873, EPI_ISL_1377876, EPI_ISL_1377878, EPI_ISL_1377883, EPI_ISL_1377884, EPI_ISL_1377885, EPI_ISL_1377886, EPI_ISL_1377887, EPI_ISL_1377891, EPI_ISL_1377892, EPI_ISL_1377897, EPI_ISL_1377898, EPI_ISL_1377901, EPI_ISL_1377903, EPI_ISL_1377905, EPI_ISL_1377908, EPI_ISL_1377909, EPI_ISL_1377911, EPI_ISL_1377914, EPI_ISL_1377917, EPI_ISL_1377920, EPI_ISL_1377922, EPI_ISL_1377924, EPI_ISL_1377925, EPI_ISL_1377927, EPI_ISL_1377930, EPI_ISL_1377931, EPI_ISL_1377933, EPI_ISL_1377934, EPI_ISL_1377935, EPI_ISL_1377936, EPI_ISL_1377937, EPI_ISL_1377938, EPI_ISL_1377940, EPI_ISL_1377943, EPI_ISL_1377945, EPI_ISL_1377947, EPI_ISL_1377948, EPI_ISL_1377951, EPI_ISL_1377953, EPI_ISL_1377954, EPI_ISL_1377960, EPI_ISL_1377962, EPI_ISL_1377963, EPI_ISL_1377971, EPI_ISL_1377975, EPI_ISL_1377977, EPI_ISL_1377980, EPI_ISL_1377982, EPI_ISL_1377983, EPI_ISL_1377987, EPI_ISL_1377992, EPI_ISL_1377993, EPI_ISL_1377994, EPI_ISL_1377995, EPI_ISL_1377996, EPI_ISL_1377998, EPI_ISL_1378490, EPI_ISL_1411868, EPI_ISL_1411895, EPI_ISL_1411943, EPI_ISL_1411962, EPI_ISL_1411966, EPI_ISL_1411970, EPI_ISL_1411978, EPI_ISL_1411984, EPI_ISL_1411993, EPI_ISL_1412000, EPI_ISL_1412009, EPI_ISL_1412089, EPI_ISL_1412100, EPI_ISL_1412109, EPI_ISL_1412125, EPI_ISL_1412154, EPI_ISL_1412175, EPI_ISL_1412213, EPI_ISL_1412218, EPI_ISL_1412225, EPI_ISL_1412246, EPI_ISL_1412267, EPI_ISL_1412284, EPI_ISL_1412286, EPI_ISL_1412299, EPI_ISL_1412312, EPI_ISL_1412329, EPI_ISL_1412355, EPI_ISL_1412362, EPI_ISL_1412377, EPI_ISL_1412429, EPI_ISL_1412447, EPI_ISL_1412470, EPI_ISL_1412482, EPI_ISL_1412517, EPI_ISL_1412543, EPI_ISL_1412549, EPI_ISL_1412562, EPI_ISL_1413115, EPI_ISL_1413128, EPI_ISL_1413140, EPI_ISL_1413182, EPI_ISL_1413203, EPI_ISL_1413214, EPI_ISL_1413250, EPI_ISL_1413284, EPI_ISL_1413297, EPI_ISL_1413375, EPI_ISL_1413383, EPI_ISL_1413402, EPI_ISL_1413415, EPI_ISL_1413484, EPI_ISL_1413495, EPI_ISL_1413507, EPI_ISL_1413560, EPI_ISL_1413958, EPI_ISL_1414065, EPI_ISL_1414077, EPI_ISL_1414078, EPI_ISL_1414084, EPI_ISL_1414087, EPI_ISL_1414089, EPI_ISL_1414092, EPI_ISL_1414099, EPI_ISL_1414101, EPI_ISL_1414106, EPI_ISL_1414107, EPI_ISL_1414108, EPI_ISL_1414114, EPI_ISL_1414115, EPI_ISL_1414120, EPI_ISL_1414122, EPI_ISL_1414128, EPI_ISL_1414129, EPI_ISL_1414136, EPI_ISL_1414140, EPI_ISL_1414144, EPI_ISL_1414150, EPI_ISL_1414151, EPI_ISL_1414157, EPI_ISL_1414167, EPI_ISL_1414174, EPI_ISL_1414179, EPI_ISL_1414183, EPI_ISL_1414187, EPI_ISL_1414188, EPI_ISL_1414192, EPI_ISL_1414196, EPI_ISL_1414198, EPI_ISL_1414199, EPI_ISL_1414211, EPI_ISL_1414214, EPI_ISL_1414220, EPI_ISL_1414221, EPI_ISL_1414222, EPI_ISL_1414228, EPI_ISL_1414230, EPI_ISL_1414232, EPI_ISL_1414235, EPI_ISL_1414244, EPI_ISL_1414248, EPI_ISL_1414255, EPI_ISL_1414256, EPI_ISL_1414258, EPI_ISL_1414259, EPI_ISL_1414262, EPI_ISL_1414263, EPI_ISL_1414266, EPI_ISL_1414268, EPI_ISL_1414269, EPI_ISL_1414270, EPI_ISL_1414273, EPI_ISL_1414275, EPI_ISL_1414277, EPI_ISL_1414279, EPI_ISL_1414283, EPI_ISL_1414285, EPI_ISL_1414289, EPI_ISL_1414292, EPI_ISL_1414294, EPI_ISL_1414295, EPI_ISL_1414308, EPI_ISL_1414312, EPI_ISL_1414314, EPI_ISL_1414323, EPI_ISL_1414325, EPI_ISL_1414328, EPI_ISL_1414332, EPI_ISL_1414333, EPI_ISL_1414336, EPI_ISL_1414346, EPI_ISL_1414358, EPI_ISL_1414364, EPI_ISL_1414366, EPI_ISL_1414369, EPI_ISL_1414374, EPI_ISL_1414376, EPI_ISL_1414395, EPI_ISL_1414396, EPI_ISL_1414404, EPI_ISL_1414410, EPI_ISL_1414412, EPI_ISL_1414416, EPI_ISL_1414421, EPI_ISL_1414427, EPI_ISL_1414428, EPI_ISL_1414431, EPI_ISL_1414435, EPI_ISL_1414438, EPI_ISL_1414439, EPI_ISL_1414442, EPI_ISL_1414450, EPI_ISL_1414455, EPI_ISL_1414456, EPI_ISL_1414460, EPI_ISL_1414461, EPI_ISL_1414463, EPI_ISL_1414464, EPI_ISL_1414467, EPI_ISL_1414468, EPI_ISL_1414469, EPI_ISL_1414470, EPI_ISL_1414474, EPI_ISL_1414475, EPI_ISL_1414477, EPI_ISL_1414479, EPI_ISL_1414484, EPI_ISL_1414486, EPI_ISL_1414489, EPI_ISL_1414493, EPI_ISL_1414498, EPI_ISL_1414502, EPI_ISL_1414506, EPI_ISL_1414507, EPI_ISL_1414510, EPI_ISL_1414514, EPI_ISL_1414516, EPI_ISL_1414518, EPI_ISL_1414519, EPI_ISL_1414525, EPI_ISL_1414526, EPI_ISL_1414527, EPI_ISL_1414528, EPI_ISL_1414529, EPI_ISL_1414530, EPI_ISL_1414531, EPI_ISL_1414532, EPI_ISL_1414534, EPI_ISL_1414536, EPI_ISL_1414550, EPI_ISL_1414552, EPI_ISL_1414556, EPI_ISL_1414558, EPI_ISL_1414559, EPI_ISL_1414561, EPI_ISL_1414565, EPI_ISL_1414567, EPI_ISL_1414571, EPI_ISL_1414572, EPI_ISL_1414579, EPI_ISL_1414580, EPI_ISL_1414583, EPI_ISL_1414586, EPI_ISL_1414587, EPI_ISL_1414593, EPI_ISL_1414594, EPI_ISL_1414595, EPI_ISL_1414603, EPI_ISL_1414606, EPI_ISL_1414609, EPI_ISL_1414615, EPI_ISL_1414622, EPI_ISL_1414625, EPI_ISL_1414627, EPI_ISL_1414632, EPI_ISL_1414634, EPI_ISL_1414635, EPI_ISL_1414636, EPI_ISL_1414637, EPI_ISL_1414639, EPI_ISL_1414640, EPI_ISL_1414641, EPI_ISL_1414644, EPI_ISL_1414649, EPI_ISL_1414650, EPI_ISL_1414653, EPI_ISL_1414656, EPI_ISL_1414657, EPI_ISL_1414663, EPI_ISL_1414664, EPI_ISL_1414666, EPI_ISL_1414671, EPI_ISL_1414675, EPI_ISL_1414676, EPI_ISL_1414678, EPI_ISL_1414681, EPI_ISL_1414684, EPI_ISL_1414685, EPI_ISL_1414690, EPI_ISL_1414692, EPI_ISL_1414694, EPI_ISL_1414695, EPI_ISL_1414706, EPI_ISL_1414707, EPI_ISL_1414708, EPI_ISL_1414718, EPI_ISL_1414721, EPI_ISL_1414722, EPI_ISL_1414723, EPI_ISL_1414726, EPI_ISL_1414729, EPI_ISL_1414730, EPI_ISL_1414732, EPI_ISL_1414737, EPI_ISL_1414738, EPI_ISL_1414739, EPI_ISL_1414741, EPI_ISL_1414742, EPI_ISL_1414744, EPI_ISL_1414745, EPI_ISL_1414749, EPI_ISL_1414750, EPI_ISL_1414752, EPI_ISL_1414755, EPI_ISL_1414758, EPI_ISL_1414760, EPI_ISL_1414765, EPI_ISL_1414771, EPI_ISL_1414775, EPI_ISL_1414784, EPI_ISL_1414790, EPI_ISL_1414791, EPI_ISL_1414792, EPI_ISL_1414794, EPI_ISL_1414795, EPI_ISL_1414796, EPI_ISL_1414797, EPI_ISL_1414803, EPI_ISL_1414804, EPI_ISL_1414806, EPI_ISL_1414811, EPI_ISL_1414813, EPI_ISL_1414817, EPI_ISL_1414820, EPI_ISL_1414823, EPI_ISL_1414824, EPI_ISL_1414825, EPI_ISL_1414833, EPI_ISL_1414836, EPI_ISL_1414839, EPI_ISL_1414848, EPI_ISL_1414855, EPI_ISL_1414857, EPI_ISL_1414859, EPI_ISL_1414870, EPI_ISL_1414871, EPI_ISL_1414874, EPI_ISL_1414875, EPI_ISL_1414876, EPI_ISL_1414885, EPI_ISL_1414890, EPI_ISL_1414894, EPI_ISL_1414896, EPI_ISL_1414898, EPI_ISL_1414900, EPI_ISL_1414905, EPI_ISL_1414909, EPI_ISL_1414910, EPI_ISL_1414911, EPI_ISL_1414912, EPI_ISL_1414913, EPI_ISL_1414922, EPI_ISL_1414924, EPI_ISL_1414929, EPI_ISL_1414932, EPI_ISL_1414943, EPI_ISL_1414946, EPI_ISL_1414953, EPI_ISL_1414958, EPI_ISL_1414967, EPI_ISL_1414968, EPI_ISL_1414970, EPI_ISL_1414985, EPI_ISL_1414987, EPI_ISL_1414990, EPI_ISL_1414991, EPI_ISL_1414998, EPI_ISL_1415000, EPI_ISL_1415001, EPI_ISL_1415002, EPI_ISL_1415006, EPI_ISL_1415009, EPI_ISL_1415010, EPI_ISL_1415020, EPI_ISL_1415036, EPI_ISL_1415041, EPI_ISL_1415046, EPI_ISL_1415054, EPI_ISL_1415055, EPI_ISL_1415057, EPI_ISL_1415058, EPI_ISL_1415059, EPI_ISL_1415432, EPI_ISL_1415435, EPI_ISL_1415436, EPI_ISL_1415439, EPI_ISL_1415442, EPI_ISL_1415443, EPI_ISL_1415444, EPI_ISL_1415446, EPI_ISL_1415450, EPI_ISL_1415452, EPI_ISL_1415455, EPI_ISL_1415461, EPI_ISL_1415462, EPI_ISL_1415463, EPI_ISL_1415464, EPI_ISL_1415466, EPI_ISL_1415467, EPI_ISL_1415469, EPI_ISL_1415470, EPI_ISL_1415471, EPI_ISL_1415472, EPI_ISL_1415475, EPI_ISL_1415476, EPI_ISL_1415477, EPI_ISL_1415478, EPI_ISL_1415479, EPI_ISL_1415480, EPI_ISL_1415481, EPI_ISL_1415482, EPI_ISL_1415483, EPI_ISL_1415484, EPI_ISL_1415485, EPI_ISL_1415487, EPI_ISL_1415488, EPI_ISL_1415489, EPI_ISL_1415490, EPI_ISL_1415491, EPI_ISL_1415492, EPI_ISL_1415493, EPI_ISL_1415494, EPI_ISL_1415496, EPI_ISL_1415497, EPI_ISL_1415498, EPI_ISL_1415500, EPI_ISL_1415501, EPI_ISL_1415502, EPI_ISL_1415503, EPI_ISL_1415505, EPI_ISL_1415506, EPI_ISL_1415507, EPI_ISL_1415508, EPI_ISL_1415509, EPI_ISL_1415510, EPI_ISL_1415511, EPI_ISL_1415512, EPI_ISL_1415513, EPI_ISL_1415516, EPI_ISL_1415517, EPI_ISL_1415518, EPI_ISL_1415519, EPI_ISL_1415520, EPI_ISL_1415521, EPI_ISL_1415522, EPI_ISL_1415523, EPI_ISL_1415524, EPI_ISL_1415525, EPI_ISL_1415526, EPI_ISL_1415527, EPI_ISL_1415528, EPI_ISL_1415529, EPI_ISL_1415530, EPI_ISL_1415531, EPI_ISL_1415532, EPI_ISL_1415533, EPI_ISL_1415534, EPI_ISL_1415535, EPI_ISL_1415536, EPI_ISL_1415537, EPI_ISL_1415538, EPI_ISL_1415539, EPI_ISL_1415540, EPI_ISL_1415541, EPI_ISL_1415542, EPI_ISL_1415543, EPI_ISL_1415544, EPI_ISL_1415545, EPI_ISL_1415546, EPI_ISL_1415547, EPI_ISL_1415548, EPI_ISL_1415550, EPI_ISL_1415551, EPI_ISL_1415552, EPI_ISL_1415553, EPI_ISL_1415554, EPI_ISL_1415555, EPI_ISL_1415556, EPI_ISL_1415557, EPI_ISL_1415558, EPI_ISL_1415559, EPI_ISL_1415560, EPI_ISL_1415561, EPI_ISL_1415562, EPI_ISL_1415563, EPI_ISL_1415564, EPI_ISL_1415565, EPI_ISL_1415567, EPI_ISL_1415569, EPI_ISL_1415570, EPI_ISL_1415571, EPI_ISL_1415572, EPI_ISL_1415573, EPI_ISL_1415574, EPI_ISL_1415575, EPI_ISL_1415576, EPI_ISL_1415577, EPI_ISL_1415578, EPI_ISL_1415580, EPI_ISL_1415581, EPI_ISL_1415582, EPI_ISL_1415583, EPI_ISL_1415584, EPI_ISL_1415586, EPI_ISL_1415587, EPI_ISL_1415588, EPI_ISL_1415589, EPI_ISL_1415590, EPI_ISL_1415591, EPI_ISL_1415592, EPI_ISL_1415593, EPI_ISL_1415594, EPI_ISL_1415595, EPI_ISL_1415596, EPI_ISL_1415597, EPI_ISL_1415598, EPI_ISL_1415599, EPI_ISL_1415600, EPI_ISL_1415601, EPI_ISL_1415602, EPI_ISL_1415603, EPI_ISL_1415604, EPI_ISL_1415605, EPI_ISL_1415606, EPI_ISL_1415607, EPI_ISL_1415608, EPI_ISL_1415609, EPI_ISL_1415610, EPI_ISL_1415611, EPI_ISL_1415612, EPI_ISL_1415613, EPI_ISL_1415614, EPI_ISL_1415615, EPI_ISL_1415616, EPI_ISL_1415617, EPI_ISL_1415618, EPI_ISL_1415619, EPI_ISL_1415620, EPI_ISL_1415621, EPI_ISL_1415622, EPI_ISL_1415623, EPI_ISL_1415624, EPI_ISL_1415625, EPI_ISL_1415626, EPI_ISL_1415627, EPI_ISL_1415628, EPI_ISL_1415629, EPI_ISL_1415630, EPI_ISL_1415632, EPI_ISL_1415633, EPI_ISL_1415634, EPI_ISL_1415636, EPI_ISL_1415637, EPI_ISL_1415638, EPI_ISL_1415639, EPI_ISL_1415640, EPI_ISL_1415641, EPI_ISL_1415642, EPI_ISL_1415643, EPI_ISL_1415644, EPI_ISL_1415645, EPI_ISL_1415648, EPI_ISL_1415649, EPI_ISL_1415650, EPI_ISL_1415651, EPI_ISL_1415652, EPI_ISL_1415653, EPI_ISL_1415654, EPI_ISL_1415655, EPI_ISL_1415656, EPI_ISL_1415658, EPI_ISL_1415659, EPI_ISL_1415660, EPI_ISL_1415661, EPI_ISL_1415662, EPI_ISL_1415663, EPI_ISL_1415664, EPI_ISL_1415665, EPI_ISL_1415666, EPI_ISL_1415667, EPI_ISL_1415668, EPI_ISL_1415669, EPI_ISL_1415670, EPI_ISL_1415671, EPI_ISL_1415672, EPI_ISL_1415673, EPI_ISL_1415674, EPI_ISL_1415676, EPI_ISL_1415677, EPI_ISL_1415678, EPI_ISL_1415679, EPI_ISL_1415680, EPI_ISL_1415681, EPI_ISL_1415682, EPI_ISL_1415683, EPI_ISL_1415684, EPI_ISL_1415686, EPI_ISL_1415687, EPI_ISL_1415688, EPI_ISL_1415689, EPI_ISL_1415690, EPI_ISL_1415691, EPI_ISL_1415692, EPI_ISL_1415694, EPI_ISL_1415695, EPI_ISL_1415696, EPI_ISL_1415697, EPI_ISL_1415698, EPI_ISL_1415699, EPI_ISL_1415700, EPI_ISL_1415701, EPI_ISL_1415702, EPI_ISL_1415703, EPI_ISL_1415704, EPI_ISL_1415705, EPI_ISL_1415706, EPI_ISL_1415707, EPI_ISL_1415708, EPI_ISL_1415709, EPI_ISL_1415710, EPI_ISL_1415711, EPI_ISL_1415712, EPI_ISL_1415713, EPI_ISL_1415714, EPI_ISL_1415715, EPI_ISL_1415716, EPI_ISL_1415717, EPI_ISL_1415718, EPI_ISL_1415719, EPI_ISL_1415721, EPI_ISL_1415722, EPI_ISL_1415723, EPI_ISL_1415724, EPI_ISL_1415725, EPI_ISL_1415726, EPI_ISL_1415727, EPI_ISL_1415728, EPI_ISL_1415729, EPI_ISL_1415730, EPI_ISL_1415731, EPI_ISL_1415732, EPI_ISL_1415734, EPI_ISL_1415735, EPI_ISL_1415736, EPI_ISL_1415737, EPI_ISL_1415738, EPI_ISL_1415739, EPI_ISL_1415740, EPI_ISL_1415741, EPI_ISL_1415742, EPI_ISL_1415743, EPI_ISL_1415745, EPI_ISL_1415746, EPI_ISL_1415747, EPI_ISL_1415748, EPI_ISL_1415749, EPI_ISL_1415750, EPI_ISL_1415751, EPI_ISL_1415752, EPI_ISL_1415753, EPI_ISL_1415754, EPI_ISL_1415755, EPI_ISL_1415756, EPI_ISL_1415757, EPI_ISL_1415759, EPI_ISL_1415760, EPI_ISL_1415761, EPI_ISL_1415762, EPI_ISL_1415763, EPI_ISL_1415764, EPI_ISL_1415765, EPI_ISL_1415766, EPI_ISL_1415767, EPI_ISL_1415768, EPI_ISL_1415769, EPI_ISL_1415770, EPI_ISL_1415771, EPI_ISL_1415773, EPI_ISL_1415775, EPI_ISL_1415776, EPI_ISL_1415778, EPI_ISL_1415779, EPI_ISL_1415780, EPI_ISL_1415781, EPI_ISL_1415782, EPI_ISL_1415783, EPI_ISL_1415784, EPI_ISL_1415785, EPI_ISL_1415786, EPI_ISL_1415787, EPI_ISL_1415788, EPI_ISL_1415789, EPI_ISL_1415790, EPI_ISL_1415791, EPI_ISL_1415792, EPI_ISL_1415793, EPI_ISL_1415794, EPI_ISL_1415795, EPI_ISL_1415796, EPI_ISL_1415797, EPI_ISL_1415798, EPI_ISL_1415799, EPI_ISL_1415800, EPI_ISL_1415801, EPI_ISL_1415803, EPI_ISL_1415804, EPI_ISL_1415805, EPI_ISL_1415806, EPI_ISL_1415807, EPI_ISL_1415808, EPI_ISL_1415809, EPI_ISL_1415810, EPI_ISL_1415811, EPI_ISL_1415812, EPI_ISL_1415813, EPI_ISL_1415814, EPI_ISL_1415815, EPI_ISL_1415820, EPI_ISL_1415829, EPI_ISL_1415831, EPI_ISL_1415837, EPI_ISL_1415851, EPI_ISL_1415855, EPI_ISL_1415862, EPI_ISL_1415876, EPI_ISL_1415877, EPI_ISL_1415879, EPI_ISL_1415882, EPI_ISL_1415883, EPI_ISL_1415886, EPI_ISL_1415891, EPI_ISL_1415893, EPI_ISL_1415894, EPI_ISL_1415901, EPI_ISL_1415904, EPI_ISL_1415914, EPI_ISL_1415926, EPI_ISL_1415928, EPI_ISL_1415929, EPI_ISL_1415931, EPI_ISL_1415932, EPI_ISL_1415938, EPI_ISL_1415941, EPI_ISL_1415942, EPI_ISL_1415954, EPI_ISL_1415960, EPI_ISL_1415966, EPI_ISL_1415969, EPI_ISL_1415991, EPI_ISL_1415997, EPI_ISL_1416004, EPI_ISL_1416007, EPI_ISL_1416029, EPI_ISL_1416032, EPI_ISL_1416033, EPI_ISL_1416034, EPI_ISL_1416036, EPI_ISL_1416044, EPI_ISL_1416045, EPI_ISL_1416055, EPI_ISL_1416057, EPI_ISL_1416063, EPI_ISL_1416072, EPI_ISL_1416073, EPI_ISL_1416076, EPI_ISL_1416081, EPI_ISL_1416082, EPI_ISL_1416083, EPI_ISL_1416090, EPI_ISL_1416091, EPI_ISL_1416095, EPI_ISL_1416096, EPI_ISL_1416108, EPI_ISL_1416118, EPI_ISL_1416122, EPI_ISL_1454645, EPI_ISL_1454648, EPI_ISL_1454661, EPI_ISL_1454663, EPI_ISL_1454670, EPI_ISL_1454672, EPI_ISL_1454678, EPI_ISL_1454682, EPI_ISL_1454698, EPI_ISL_1454705, EPI_ISL_1454709, EPI_ISL_1454713, EPI_ISL_1454717, EPI_ISL_1454725, EPI_ISL_1454726, EPI_ISL_1454728, EPI_ISL_1454734, EPI_ISL_1454750, EPI_ISL_1454756, EPI_ISL_1454765, EPI_ISL_1454766, EPI_ISL_1454768, EPI_ISL_1454772, EPI_ISL_1454777, EPI_ISL_1454779, EPI_ISL_1454780, EPI_ISL_1454781, EPI_ISL_1454783, EPI_ISL_1454784, EPI_ISL_1454789, EPI_ISL_1454791, EPI_ISL_1454794, EPI_ISL_1454796, EPI_ISL_1454798, EPI_ISL_1454803, EPI_ISL_1454809, EPI_ISL_1454815, EPI_ISL_1454816, EPI_ISL_1454818, EPI_ISL_1454820, EPI_ISL_1454826, EPI_ISL_1454829, EPI_ISL_1454830, EPI_ISL_1454835, EPI_ISL_1454838, EPI_ISL_1454839, EPI_ISL_1454840, EPI_ISL_1454847, EPI_ISL_1454857, EPI_ISL_1454862, EPI_ISL_1454868, EPI_ISL_1454870, EPI_ISL_1454883, EPI_ISL_1454884, EPI_ISL_1454886, EPI_ISL_1454898, EPI_ISL_1454899, EPI_ISL_1454901, EPI_ISL_1454913,

EPI_ISL_1454916, EPI_ISL_1454922, EPI_ISL_1454930, EPI_ISL_1454934, EPI_ISL_1454937, EPI_ISL_1454940, EPI_ISL_1454941, EPI_ISL_1454944, EPI_ISL_1455013, EPI_ISL_1455015, EPI_ISL_1455020, EPI_ISL_1455050, EPI_ISL_1455219, EPI_ISL_1455239, EPI_ISL_1455245, EPI_ISL_1455252, EPI_ISL_1455253, EPI_ISL_1455254, EPI_ISL_1455259, EPI_ISL_1455260, EPI_ISL_1455261, EPI_ISL_1455264,
EPI_ISL_1455269, EPI_ISL_1455271, EPI_ISL_1455285, EPI_ISL_1455290, EPI_ISL_1455292, EPI_ISL_1455296, EPI_ISL_1455297, EPI_ISL_1455303, EPI_ISL_1455304, EPI_ISL_1455307, EPI_ISL_1455308, EPI_ISL_1455309, EPI_ISL_1455310, EPI_ISL_1455328, EPI_ISL_1455330, EPI_ISL_1455336, EPI_ISL_1455338, EPI_ISL_1455347, EPI_ISL_1455348, EPI_ISL_1455353, EPI_ISL_1455360, EPI_ISL_1455363,
EPI_ISL_1455366, EPI_ISL_1455377, EPI_ISL_1455392, EPI_ISL_1455393, EPI_ISL_1455406, EPI_ISL_1455408, EPI_ISL_1455412, EPI_ISL_1455415, EPI_ISL_1455420, EPI_ISL_1455426, EPI_ISL_1455430, EPI_ISL_1455436, EPI_ISL_1455443, EPI_ISL_1455448, EPI_ISL_1455463, EPI_ISL_1455465, EPI_ISL_1455466, EPI_ISL_1455468, EPI_ISL_1455473, EPI_ISL_1455475, EPI_ISL_1455477, EPI_ISL_1455478,
EPI_ISL_1455479, EPI_ISL_1455485, EPI_ISL_1455487, EPI_ISL_1455488, EPI_ISL_1455493, EPI_ISL_1455494, EPI_ISL_1455495, EPI_ISL_1455497, EPI_ISL_1455504, EPI_ISL_1455522, EPI_ISL_1455535, EPI_ISL_1455539, EPI_ISL_1455540, EPI_ISL_1455542, EPI_ISL_1455543, EPI_ISL_1455544, EPI_ISL_1455545, EPI_ISL_1455547, EPI_ISL_1455548, EPI_ISL_1455549, EPI_ISL_1455550, EPI_ISL_1455551,
EPI_ISL_1455553, EPI_ISL_1455557, EPI_ISL_1455561, EPI_ISL_1455563, EPI_ISL_1455564, EPI_ISL_1455576, EPI_ISL_1455579, EPI_ISL_1455615, EPI_ISL_1455617, EPI_ISL_1455625, EPI_ISL_1455631, EPI_ISL_1455632

| | | | |
|---|---|---|---|
| see above | Lighthouse Lab in Alderley Park | Wellcome Sanger Institute for the COVID-19 Genomics UK (COG-UK) Consortium | Cordelia Langford; David K. Jackson; Dominic Kwiatkowski; Ewan Harrison; Ian Johnston; Jacquelyn Wynn; Jeffrey Barrett; John Sillitoe on behalf of the Wellcome Sanger Institute COVID-19 Surveillance Team; Mairead Hyland; Roberto Amato; Sonia Goncalves; The Lighthouse Lab in Alderley Park and Alex Alderton |

EPI_ISL_1326662, EPI_ISL_1326666, EPI_ISL_1326678, EPI_ISL_1326682, EPI_ISL_1326688, EPI_ISL_1326698, EPI_ISL_1326703, EPI_ISL_1326710, EPI_ISL_1326711, EPI_ISL_1326720, EPI_ISL_1326725, EPI_ISL_1326726, EPI_ISL_1326727, EPI_ISL_1326728, EPI_ISL_1326731, EPI_ISL_1326734, EPI_ISL_1326746, EPI_ISL_1326770, EPI_ISL_1326804, EPI_ISL_1326811, EPI_ISL_1326822, EPI_ISL_1326824,
EPI_ISL_1326832, EPI_ISL_1326838, EPI_ISL_1326844, EPI_ISL_1326852, EPI_ISL_1326860, EPI_ISL_1326862, EPI_ISL_1326864, EPI_ISL_1326882, EPI_ISL_1326892, EPI_ISL_1326893, EPI_ISL_1326900, EPI_ISL_1326908, EPI_ISL_1326910, EPI_ISL_1326915, EPI_ISL_1326931, EPI_ISL_1326959, EPI_ISL_1326977, EPI_ISL_1326990, EPI_ISL_1326993, EPI_ISL_1326994, EPI_ISL_1327002, EPI_ISL_1327007,
EPI_ISL_1327022, EPI_ISL_1327023, EPI_ISL_1327026, EPI_ISL_1327027, EPI_ISL_1327032, EPI_ISL_1327033, EPI_ISL_1327036, EPI_ISL_1327038, EPI_ISL_1327043, EPI_ISL_1327044, EPI_ISL_1327052, EPI_ISL_1327053, EPI_ISL_1327060, EPI_ISL_1327065, EPI_ISL_1327069, EPI_ISL_1327074, EPI_ISL_1327080, EPI_ISL_1327086, EPI_ISL_1327092, EPI_ISL_1327095, EPI_ISL_1327111, EPI_ISL_1327121,
EPI_ISL_1327126, EPI_ISL_1327133, EPI_ISL_1327135, EPI_ISL_1327136, EPI_ISL_1327143, EPI_ISL_1327153, EPI_ISL_1327155, EPI_ISL_1327168, EPI_ISL_1327172, EPI_ISL_1327174, EPI_ISL_1327178, EPI_ISL_1327179, EPI_ISL_1327185, EPI_ISL_1327189, EPI_ISL_1327194, EPI_ISL_1327199, EPI_ISL_1327207, EPI_ISL_1327216, EPI_ISL_1327218, EPI_ISL_1327225, EPI_ISL_1327227, EPI_ISL_1327229,
EPI_ISL_1327235, EPI_ISL_1327374, EPI_ISL_1327433, EPI_ISL_1327448, EPI_ISL_1327459, EPI_ISL_1327465, EPI_ISL_1327483, EPI_ISL_1327496, EPI_ISL_1327504, EPI_ISL_1327519, EPI_ISL_1327547, EPI_ISL_1327586, EPI_ISL_1327589, EPI_ISL_1327649, EPI_ISL_1327678, EPI_ISL_1327692, EPI_ISL_1328442, EPI_ISL_1328460, EPI_ISL_1328591, EPI_ISL_1328599, EPI_ISL_1328611, EPI_ISL_1328623,
EPI_ISL_1328674, EPI_ISL_1328719, EPI_ISL_1328720, EPI_ISL_1328725, EPI_ISL_1328728, EPI_ISL_1328729, EPI_ISL_1328731, EPI_ISL_1328736, EPI_ISL_1328764, EPI_ISL_1328797, EPI_ISL_1328799, EPI_ISL_1328800, EPI_ISL_1328802, EPI_ISL_1328808, EPI_ISL_1328811, EPI_ISL_1328813, EPI_ISL_1328833, EPI_ISL_1328841, EPI_ISL_1328846, EPI_ISL_1328850, EPI_ISL_1328863, EPI_ISL_1328869,
EPI_ISL_1328893, EPI_ISL_1329691, EPI_ISL_1329698, EPI_ISL_1329700, EPI_ISL_1329727, EPI_ISL_1329747, EPI_ISL_1329750, EPI_ISL_1329762, EPI_ISL_1329765, EPI_ISL_1329772, EPI_ISL_1329792, EPI_ISL_1329794, EPI_ISL_1329799, EPI_ISL_1329812, EPI_ISL_1329815, EPI_ISL_1329816, EPI_ISL_1329828, EPI_ISL_1329833, EPI_ISL_1329861, EPI_ISL_1329871, EPI_ISL_1329897, EPI_ISL_1329903,
EPI_ISL_1329914, EPI_ISL_1329922, EPI_ISL_1329928, EPI_ISL_1329931, EPI_ISL_1329937, EPI_ISL_1329940, EPI_ISL_1329947, EPI_ISL_1329948, EPI_ISL_1329965, EPI_ISL_1329971, EPI_ISL_1329973, EPI_ISL_1329976, EPI_ISL_1330298, EPI_ISL_1330305, EPI_ISL_1330335, EPI_ISL_1330343, EPI_ISL_1330352, EPI_ISL_1330370, EPI_ISL_1330372, EPI_ISL_1330383, EPI_ISL_1330390, EPI_ISL_1330425,
EPI_ISL_1330426, EPI_ISL_1330428, EPI_ISL_1330432, EPI_ISL_1330435, EPI_ISL_1330447, EPI_ISL_1330454, EPI_ISL_1330496, EPI_ISL_1330514, EPI_ISL_1330554, EPI_ISL_1330557, EPI_ISL_1330558, EPI_ISL_1330592, EPI_ISL_1344471, EPI_ISL_1344482, EPI_ISL_1344491, EPI_ISL_1344496, EPI_ISL_1344499, EPI_ISL_1344514, EPI_ISL_1344519, EPI_ISL_1344520, EPI_ISL_1344524, EPI_ISL_1344526,
EPI_ISL_1344537, EPI_ISL_1344544, EPI_ISL_1344557, EPI_ISL_1344560, EPI_ISL_1344573, EPI_ISL_1344577, EPI_ISL_1344579, EPI_ISL_1344582, EPI_ISL_1344589, EPI_ISL_1344590, EPI_ISL_1344598, EPI_ISL_1344604, EPI_ISL_1344607, EPI_ISL_1344619, EPI_ISL_1344622, EPI_ISL_1344624, EPI_ISL_1344630, EPI_ISL_1344631, EPI_ISL_1344646, EPI_ISL_1344648, EPI_ISL_1344659, EPI_ISL_1344663,
EPI_ISL_1344665, EPI_ISL_1344668, EPI_ISL_1344674, EPI_ISL_1344682, EPI_ISL_1344686, EPI_ISL_1344687, EPI_ISL_1344699, EPI_ISL_1344702, EPI_ISL_1344739, EPI_ISL_1344774, EPI_ISL_1346285, EPI_ISL_1346289, EPI_ISL_1346290, EPI_ISL_1346293, EPI_ISL_1346301, EPI_ISL_1346350, EPI_ISL_1346358, EPI_ISL_1346394, EPI_ISL_1346397, EPI_ISL_1346409, EPI_ISL_1365180, EPI_ISL_1365182,
EPI_ISL_1365184, EPI_ISL_1365186, EPI_ISL_1365187, EPI_ISL_1365188, EPI_ISL_1365189, EPI_ISL_1365190, EPI_ISL_1365198, EPI_ISL_1365199, EPI_ISL_1365201, EPI_ISL_1365203, EPI_ISL_1365204, EPI_ISL_1365206, EPI_ISL_1365209, EPI_ISL_1365214, EPI_ISL_1365217, EPI_ISL_1376970, EPI_ISL_1376971, EPI_ISL_1376972, EPI_ISL_1376973, EPI_ISL_1376974, EPI_ISL_1376975, EPI_ISL_1376976,
EPI_ISL_1376979, EPI_ISL_1376980, EPI_ISL_1376981, EPI_ISL_1376983, EPI_ISL_1376984, EPI_ISL_1376987, EPI_ISL_1376988, EPI_ISL_1376990, EPI_ISL_1376991, EPI_ISL_1376992, EPI_ISL_1376993, EPI_ISL_1376994, EPI_ISL_1376996, EPI_ISL_1376997, EPI_ISL_1376998, EPI_ISL_1376999, EPI_ISL_1377000, EPI_ISL_1377001, EPI_ISL_1377002, EPI_ISL_1377003, EPI_ISL_1377004, EPI_ISL_1377005,
EPI_ISL_1377006, EPI_ISL_1377007, EPI_ISL_1377008, EPI_ISL_1377009, EPI_ISL_1377010, EPI_ISL_1377012, EPI_ISL_1377013, EPI_ISL_1377014, EPI_ISL_1377015, EPI_ISL_1377017, EPI_ISL_1377018, EPI_ISL_1377019, EPI_ISL_1377020, EPI_ISL_1377021, EPI_ISL_1377022, EPI_ISL_1377023, EPI_ISL_1377024, EPI_ISL_1377025, EPI_ISL_1377026, EPI_ISL_1377028, EPI_ISL_1377029, EPI_ISL_1377030,
EPI_ISL_1377033, EPI_ISL_1377034, EPI_ISL_1377035, EPI_ISL_1377036, EPI_ISL_1377037, EPI_ISL_1377038, EPI_ISL_1377039, EPI_ISL_1377041, EPI_ISL_1377042, EPI_ISL_1377043, EPI_ISL_1377044, EPI_ISL_1377045, EPI_ISL_1377046, EPI_ISL_1377047, EPI_ISL_1377049, EPI_ISL_1377051, EPI_ISL_1377052, EPI_ISL_1377054, EPI_ISL_1377055, EPI_ISL_1377056, EPI_ISL_1377059, EPI_ISL_1377061,
EPI_ISL_1377062, EPI_ISL_1377063, EPI_ISL_1377064, EPI_ISL_1377065, EPI_ISL_1377066, EPI_ISL_1377067, EPI_ISL_1377068, EPI_ISL_1377070, EPI_ISL_1377072, EPI_ISL_1377073, EPI_ISL_1377074, EPI_ISL_1377075, EPI_ISL_1377076, EPI_ISL_1377079, EPI_ISL_1377082, EPI_ISL_1377083, EPI_ISL_1377084, EPI_ISL_1377086, EPI_ISL_1377087, EPI_ISL_1377088, EPI_ISL_1377089, EPI_ISL_1377090,
EPI_ISL_1377092, EPI_ISL_1377094, EPI_ISL_1377095, EPI_ISL_1377096, EPI_ISL_1377098, EPI_ISL_1377099, EPI_ISL_1377100, EPI_ISL_1377101, EPI_ISL_1377103, EPI_ISL_1377104, EPI_ISL_1377105, EPI_ISL_1377106, EPI_ISL_1377107, EPI_ISL_1377108, EPI_ISL_1377109, EPI_ISL_1377110, EPI_ISL_1377111, EPI_ISL_1377112, EPI_ISL_1377113, EPI_ISL_1377114, EPI_ISL_1377116, EPI_ISL_1377118,
EPI_ISL_1377119, EPI_ISL_1377120, EPI_ISL_1377122, EPI_ISL_1377125, EPI_ISL_1377126, EPI_ISL_1377128, EPI_ISL_1377129, EPI_ISL_1377130, EPI_ISL_1377131, EPI_ISL_1377132, EPI_ISL_1377134, EPI_ISL_1377136, EPI_ISL_1377137, EPI_ISL_1377138, EPI_ISL_1377140, EPI_ISL_1377141, EPI_ISL_1377143, EPI_ISL_1377145, EPI_ISL_1377147, EPI_ISL_1377148, EPI_ISL_1377149, EPI_ISL_1377150,
EPI_ISL_1377151, EPI_ISL_1377152, EPI_ISL_1377153, EPI_ISL_1377154, EPI_ISL_1377156, EPI_ISL_1377157, EPI_ISL_1377158, EPI_ISL_1377159, EPI_ISL_1377160, EPI_ISL_1377161, EPI_ISL_1377163, EPI_ISL_1377164, EPI_ISL_1377165, EPI_ISL_1377166, EPI_ISL_1377167, EPI_ISL_1377168, EPI_ISL_1377170, EPI_ISL_1377171, EPI_ISL_1377172, EPI_ISL_1377173, EPI_ISL_1377174, EPI_ISL_1377175,
EPI_ISL_1377176, EPI_ISL_1377177, EPI_ISL_1377178, EPI_ISL_1377179, EPI_ISL_1378448, EPI_ISL_1378449, EPI_ISL_1390314, EPI_ISL_1390315, EPI_ISL_1390316, EPI_ISL_1390317, EPI_ISL_1390319, EPI_ISL_1409552, EPI_ISL_1410086, EPI_ISL_1410096, EPI_ISL_1410101, EPI_ISL_1410106, EPI_ISL_1410116, EPI_ISL_1410117, EPI_ISL_1410119, EPI_ISL_1410128, EPI_ISL_1410142, EPI_ISL_1410143,
EPI_ISL_1410161, EPI_ISL_1410182, EPI_ISL_1410191, EPI_ISL_1410211, EPI_ISL_1410216, EPI_ISL_1410218, EPI_ISL_1410238, EPI_ISL_1410246, EPI_ISL_1410256, EPI_ISL_1410264, EPI_ISL_1410269, EPI_ISL_1410280, EPI_ISL_1410281, EPI_ISL_1410283, EPI_ISL_1410286, EPI_ISL_1412994, EPI_ISL_1416181, EPI_ISL_1454185, EPI_ISL_1454218, EPI_ISL_1454997, EPI_ISL_1455638, EPI_ISL_1455640,
EPI_ISL_1455642, EPI_ISL_1455643, EPI_ISL_1455644, EPI_ISL_1455645, EPI_ISL_1473790, EPI_ISL_1759943

| | | | |
|---|---|---|---|
| see above | Lighthouse Lab in Cambridge | Wellcome Sanger Institute for the COVID-19 Genomics UK (COG-UK) Consortium | Cordelia Langford; David K. Jackson; Dominic Kwiatkowski; Ewan Harrison; Ian Johnston; Jeffrey Barrett; John Sillitoe on behalf of the Wellcome Sanger Institute COVID-19 Surveillance Team; Rob Howes; Roberto Amato; Sonia Goncalves; The Lighthouse Lab in Cambridge and Alex Alderton |

EPI_ISL_1295167, EPI_ISL_1315653, EPI_ISL_1315654, EPI_ISL_1315655, EPI_ISL_1315656, EPI_ISL_1315657, EPI_ISL_1315658, EPI_ISL_1315659, EPI_ISL_1315660, EPI_ISL_1315661, EPI_ISL_1315662, EPI_ISL_1315663, EPI_ISL_1315664, EPI_ISL_1315665, EPI_ISL_1315666, EPI_ISL_1315667, EPI_ISL_1315668, EPI_ISL_1315670, EPI_ISL_1315672, EPI_ISL_1315673, EPI_ISL_1315674, EPI_ISL_1315675,
EPI_ISL_1315676, EPI_ISL_1315677, EPI_ISL_1315678, EPI_ISL_1315679, EPI_ISL_1315680, EPI_ISL_1315681, EPI_ISL_1315682, EPI_ISL_1315683, EPI_ISL_1315684, EPI_ISL_1315685, EPI_ISL_1315686, EPI_ISL_1315687, EPI_ISL_1315688, EPI_ISL_1315689, EPI_ISL_1315690, EPI_ISL_1315691, EPI_ISL_1315692, EPI_ISL_1315693, EPI_ISL_1315694, EPI_ISL_1315695, EPI_ISL_1315696, EPI_ISL_1315697,
EPI_ISL_1315698, EPI_ISL_1315699, EPI_ISL_1315700, EPI_ISL_1315701, EPI_ISL_1315702, EPI_ISL_1315703, EPI_ISL_1315704, EPI_ISL_1315705, EPI_ISL_1315706, EPI_ISL_1315707, EPI_ISL_1315708, EPI_ISL_1315709, EPI_ISL_1315710, EPI_ISL_1315711, EPI_ISL_1315712, EPI_ISL_1315713, EPI_ISL_1315714, EPI_ISL_1315715, EPI_ISL_1315716, EPI_ISL_1315717, EPI_ISL_1315718, EPI_ISL_1315719,
EPI_ISL_1315720, EPI_ISL_1315721, EPI_ISL_1315722, EPI_ISL_1315723, EPI_ISL_1315724, EPI_ISL_1315725, EPI_ISL_1315726, EPI_ISL_1315727, EPI_ISL_1315728, EPI_ISL_1315729, EPI_ISL_1315730, EPI_ISL_1315731, EPI_ISL_1315732, EPI_ISL_1315733, EPI_ISL_1315734, EPI_ISL_1315735, EPI_ISL_1315736, EPI_ISL_1315737, EPI_ISL_1315738, EPI_ISL_1315739, EPI_ISL_1315740, EPI_ISL_1315741,
EPI_ISL_1315742, EPI_ISL_1315743, EPI_ISL_1315744, EPI_ISL_1315745, EPI_ISL_1315746, EPI_ISL_1315747, EPI_ISL_1315748, EPI_ISL_1315749, EPI_ISL_1315750, EPI_ISL_1315751, EPI_ISL_1315752, EPI_ISL_1315753, EPI_ISL_1315754, EPI_ISL_1315755, EPI_ISL_1315756, EPI_ISL_1315757, EPI_ISL_1315758, EPI_ISL_1315760, EPI_ISL_1315761, EPI_ISL_1315762, EPI_ISL_1315763, EPI_ISL_1315764,
EPI_ISL_1315765, EPI_ISL_1315766, EPI_ISL_1315767, EPI_ISL_1315768, EPI_ISL_1315769, EPI_ISL_1315770, EPI_ISL_1315771, EPI_ISL_1315772, EPI_ISL_1315773, EPI_ISL_1315775, EPI_ISL_1315776, EPI_ISL_1315777, EPI_ISL_1315778, EPI_ISL_1315779, EPI_ISL_1315781, EPI_ISL_1315782, EPI_ISL_1315783, EPI_ISL_1315784, EPI_ISL_1315785, EPI_ISL_1315786, EPI_ISL_1315787, EPI_ISL_1315788,
EPI_ISL_1315789, EPI_ISL_1315790, EPI_ISL_1315791, EPI_ISL_1315792, EPI_ISL_1315793, EPI_ISL_1315794, EPI_ISL_1315795, EPI_ISL_1315796, EPI_ISL_1315797, EPI_ISL_1315798, EPI_ISL_1315799, EPI_ISL_1315800, EPI_ISL_1315801, EPI_ISL_1315802, EPI_ISL_1315803, EPI_ISL_1315804, EPI_ISL_1315805, EPI_ISL_1315806, EPI_ISL_1315809, EPI_ISL_1315810, EPI_ISL_1315811, EPI_ISL_1315812,
EPI_ISL_1315813, EPI_ISL_1315814, EPI_ISL_1315815, EPI_ISL_1315816, EPI_ISL_1315817, EPI_ISL_1315818, EPI_ISL_1315819, EPI_ISL_1315820, EPI_ISL_1315821, EPI_ISL_1315822, EPI_ISL_1315823, EPI_ISL_1315824, EPI_ISL_1315825, EPI_ISL_1315826, EPI_ISL_1315827, EPI_ISL_1315828, EPI_ISL_1315829, EPI_ISL_1315830, EPI_ISL_1316023, EPI_ISL_1316024, EPI_ISL_1316025, EPI_ISL_1316026,
EPI_ISL_1316027, EPI_ISL_1316029, EPI_ISL_1316030, EPI_ISL_1316031, EPI_ISL_1316032, EPI_ISL_1316033, EPI_ISL_1316034, EPI_ISL_1316035, EPI_ISL_1316036, EPI_ISL_1316037, EPI_ISL_1316038, EPI_ISL_1316039, EPI_ISL_1316040, EPI_ISL_1316041, EPI_ISL_1316042, EPI_ISL_1316043, EPI_ISL_1316044, EPI_ISL_1316045, EPI_ISL_1316046, EPI_ISL_1316047, EPI_ISL_1316048, EPI_ISL_1316049,
EPI_ISL_1316050, EPI_ISL_1316051, EPI_ISL_1316052, EPI_ISL_1316053, EPI_ISL_1316054, EPI_ISL_1316055, EPI_ISL_1316056, EPI_ISL_1316058, EPI_ISL_1316059, EPI_ISL_1316060, EPI_ISL_1316061, EPI_ISL_1316062, EPI_ISL_1316063, EPI_ISL_1316064, EPI_ISL_1316065, EPI_ISL_1316066, EPI_ISL_1316067, EPI_ISL_1316068, EPI_ISL_1316069, EPI_ISL_1316070, EPI_ISL_1316071, EPI_ISL_1316072,
EPI_ISL_1316073, EPI_ISL_1316074, EPI_ISL_1316075, EPI_ISL_1316076, EPI_ISL_1316078, EPI_ISL_1316079, EPI_ISL_1316080, EPI_ISL_1316081, EPI_ISL_1316082, EPI_ISL_1316083, EPI_ISL_1316084, EPI_ISL_1316085, EPI_ISL_1316086, EPI_ISL_1316087, EPI_ISL_1316088, EPI_ISL_1316089, EPI_ISL_1316090, EPI_ISL_1316091, EPI_ISL_1316092, EPI_ISL_1316093, EPI_ISL_1316094, EPI_ISL_1316784,
EPI_ISL_1316789, EPI_ISL_1316792, EPI_ISL_1316795, EPI_ISL_1316801, EPI_ISL_1316803, EPI_ISL_1316807, EPI_ISL_1316810, EPI_ISL_1316811, EPI_ISL_1316812, EPI_ISL_1316817, EPI_ISL_1316820, EPI_ISL_1316827, EPI_ISL_1316828, EPI_ISL_1316834, EPI_ISL_1316835, EPI_ISL_1316837, EPI_ISL_1316845, EPI_ISL_1316849, EPI_ISL_1316850, EPI_ISL_1316851, EPI_ISL_1316853, EPI_ISL_1316856,
EPI_ISL_1316857, EPI_ISL_1316866, EPI_ISL_1316868, EPI_ISL_1316870, EPI_ISL_1316873, EPI_ISL_1316876, EPI_ISL_1316877, EPI_ISL_1316879, EPI_ISL_1316882, EPI_ISL_1316884, EPI_ISL_1316888, EPI_ISL_1316895, EPI_ISL_1316897, EPI_ISL_1316900, EPI_ISL_1316908, EPI_ISL_1316910, EPI_ISL_1316911, EPI_ISL_1316917, EPI_ISL_1316923, EPI_ISL_1316928, EPI_ISL_1316929, EPI_ISL_1316932,
EPI_ISL_1316933, EPI_ISL_1316938, EPI_ISL_1316941, EPI_ISL_1316942, EPI_ISL_1316943, EPI_ISL_1324804, EPI_ISL_1324806, EPI_ISL_1324809, EPI_ISL_1324810, EPI_ISL_1324811, EPI_ISL_1324812, EPI_ISL_1324813, EPI_ISL_1324814, EPI_ISL_1324816, EPI_ISL_1324817, EPI_ISL_1324820, EPI_ISL_1324821, EPI_ISL_1324822, EPI_ISL_1324826, EPI_ISL_1324828, EPI_ISL_1324829, EPI_ISL_1324833,
EPI_ISL_1324834, EPI_ISL_1324839, EPI_ISL_1324841, EPI_ISL_1324842, EPI_ISL_1324843, EPI_ISL_1324844, EPI_ISL_1324846, EPI_ISL_1324847, EPI_ISL_1324848, EPI_ISL_1324851, EPI_ISL_1324852, EPI_ISL_1324853, EPI_ISL_1324854, EPI_ISL_1324855, EPI_ISL_1324856, EPI_ISL_1324862, EPI_ISL_1324864, EPI_ISL_1324867, EPI_ISL_1324869, EPI_ISL_1324870, EPI_ISL_1324871, EPI_ISL_1324872,
EPI_ISL_1324873, EPI_ISL_1324875, EPI_ISL_1324876, EPI_ISL_1324878, EPI_ISL_1324879, EPI_ISL_1324880, EPI_ISL_1324885, EPI_ISL_1324886, EPI_ISL_1324893, EPI_ISL_1324894, EPI_ISL_1324895, EPI_ISL_1324904, EPI_ISL_1324906, EPI_ISL_1324907, EPI_ISL_1324908, EPI_ISL_1324909, EPI_ISL_1324913, EPI_ISL_1324916, EPI_ISL_1324917, EPI_ISL_1324918, EPI_ISL_1324919, EPI_ISL_1324921,
EPI_ISL_1324925, EPI_ISL_1324931, EPI_ISL_1324932, EPI_ISL_1324935, EPI_ISL_1324937, EPI_ISL_1324938, EPI_ISL_1324939, EPI_ISL_1324942, EPI_ISL_1324947, EPI_ISL_1324948, EPI_ISL_1324951, EPI_ISL_1324952, EPI_ISL_1324953, EPI_ISL_1324954, EPI_ISL_1324956, EPI_ISL_1324957, EPI_ISL_1324958, EPI_ISL_1324962, EPI_ISL_1324966, EPI_ISL_1324967, EPI_ISL_1324968, EPI_ISL_1324972,
EPI_ISL_1324973, EPI_ISL_1324976, EPI_ISL_1324977, EPI_ISL_1324980, EPI_ISL_1324981, EPI_ISL_1324983, EPI_ISL_1324984, EPI_ISL_1324985, EPI_ISL_1324986, EPI_ISL_1324987, EPI_ISL_1324988, EPI_ISL_1324989, EPI_ISL_1324990, EPI_ISL_1324991, EPI_ISL_1324992, EPI_ISL_1324993, EPI_ISL_1325006, EPI_ISL_1325008, EPI_ISL_1325010, EPI_ISL_1325017, EPI_ISL_1325018, EPI_ISL_1325021,
EPI_ISL_1325023, EPI_ISL_1325024, EPI_ISL_1325028, EPI_ISL_1325029, EPI_ISL_1325034, EPI_ISL_1325038, EPI_ISL_1325041, EPI_ISL_1325043, EPI_ISL_1325045, EPI_ISL_1325047, EPI_ISL_1325051, EPI_ISL_1325053, EPI_ISL_1325054, EPI_ISL_1325055, EPI_ISL_1325058, EPI_ISL_1325060, EPI_ISL_1325062, EPI_ISL_1325063, EPI_ISL_1325065, EPI_ISL_1325067, EPI_ISL_1325073, EPI_ISL_1325074,
EPI_ISL_1325076, EPI_ISL_1325079, EPI_ISL_1325082, EPI_ISL_1325084, EPI_ISL_1325085, EPI_ISL_1325087, EPI_ISL_1325090, EPI_ISL_1325091, EPI_ISL_1325092, EPI_ISL_1325093, EPI_ISL_1325094, EPI_ISL_1325103, EPI_ISL_1325105, EPI_ISL_1325106, EPI_ISL_1325107, EPI_ISL_1325111, EPI_ISL_1325113, EPI_ISL_1325114, EPI_ISL_1325116, EPI_ISL_1325118, EPI_ISL_1325119, EPI_ISL_1325120,
EPI_ISL_1325122, EPI_ISL_1325125, EPI_ISL_1325126, EPI_ISL_1325127, EPI_ISL_1325128, EPI_ISL_1325129, EPI_ISL_1325130, EPI_ISL_1325131, EPI_ISL_1325132, EPI_ISL_1325133, EPI_ISL_1325134, EPI_ISL_1325135, EPI_ISL_1325136, EPI_ISL_1325137, EPI_ISL_1325138, EPI_ISL_1325139, EPI_ISL_1325140, EPI_ISL_1325141, EPI_ISL_1325142, EPI_ISL_1325143, EPI_ISL_1325144, EPI_ISL_1325145,
EPI_ISL_1325146, EPI_ISL_1325147, EPI_ISL_1325148, EPI_ISL_1325149, EPI_ISL_1325150, EPI_ISL_1325151, EPI_ISL_1325152, EPI_ISL_1325153, EPI_ISL_1325154, EPI_ISL_1325155, EPI_ISL_1325156, EPI_ISL_1325157, EPI_ISL_1325158, EPI_ISL_1325159, EPI_ISL_1325160, EPI_ISL_1325161, EPI_ISL_1325162, EPI_ISL_1325163, EPI_ISL_1325164, EPI_ISL_1325165, EPI_ISL_1325166, EPI_ISL_1325167,
EPI_ISL_1325168, EPI_ISL_1325169, EPI_ISL_1325170, EPI_ISL_1325171, EPI_ISL_1325172, EPI_ISL_1325173, EPI_ISL_1325174, EPI_ISL_1325175, EPI_ISL_1325176, EPI_ISL_1325177, EPI_ISL_1325178, EPI_ISL_1325179, EPI_ISL_1325180, EPI_ISL_1325181, EPI_ISL_1325182, EPI_ISL_1325183, EPI_ISL_1325184, EPI_ISL_1325185, EPI_ISL_1325186, EPI_ISL_1325187, EPI_ISL_1325188, EPI_ISL_1325189,
EPI_ISL_1325190, EPI_ISL_1325191, EPI_ISL_1325192, EPI_ISL_1325193, EPI_ISL_1325194, EPI_ISL_1325195, EPI_ISL_1325196, EPI_ISL_1325197, EPI_ISL_1325198, EPI_ISL_1325199, EPI_ISL_1325200, EPI_ISL_1325201, EPI_ISL_1325202, EPI_ISL_1325203, EPI_ISL_1325204, EPI_ISL_1325205, EPI_ISL_1325206, EPI_ISL_1325207, EPI_ISL_1325208, EPI_ISL_1325209, EPI_ISL_1325210, EPI_ISL_1325211,
EPI_ISL_1325212, EPI_ISL_1325213, EPI_ISL_1325214, EPI_ISL_1325215, EPI_ISL_1325216, EPI_ISL_1325217, EPI_ISL_1325218, EPI_ISL_1325219, EPI_ISL_1325220, EPI_ISL_1325221, EPI_ISL_1325222, EPI_ISL_1325223, EPI_ISL_1325224, EPI_ISL_1325225, EPI_ISL_1325226, EPI_ISL_1325227, EPI_ISL_1325228, EPI_ISL_1325229, EPI_ISL_1325230, EPI_ISL_1325231, EPI_ISL_1325232, EPI_ISL_1325233,
EPI_ISL_1325234, EPI_ISL_1325235, EPI_ISL_1325236, EPI_ISL_1325237, EPI_ISL_1325238, EPI_ISL_1325239, EPI_ISL_1325240, EPI_ISL_1325241, EPI_ISL_1325242, EPI_ISL_1325243, EPI_ISL_1325244, EPI_ISL_1325245, EPI_ISL_1325246, EPI_ISL_1325247, EPI_ISL_1325248, EPI_ISL_1325249, EPI_ISL_1325250, EPI_ISL_1325251, EPI_ISL_1325252, EPI_ISL_1325253, EPI_ISL_1325254, EPI_ISL_1325255,
EPI_ISL_1325256, EPI_ISL_1325257, EPI_ISL_1325258, EPI_ISL_1325259, EPI_ISL_1325260, EPI_ISL_1325261, EPI_ISL_1325262, EPI_ISL_1325263, EPI_ISL_1325264, EPI_ISL_1325265, EPI_ISL_1325266, EPI_ISL_1325267, EPI_ISL_1325268, EPI_ISL_1325269, EPI_ISL_1325270, EPI_ISL_1325271, EPI_ISL_1325272, EPI_ISL_1325273, EPI_ISL_1325274, EPI_ISL_1325275, EPI_ISL_1325276, EPI_ISL_1325277,
EPI_ISL_1325278, EPI_ISL_1325279, EPI_ISL_1325280, EPI_ISL_1325281, EPI_ISL_1325282, EPI_ISL_1325283, EPI_ISL_1325284, EPI_ISL_1325285, EPI_ISL_1325286, EPI_ISL_1325287, EPI_ISL_1325288, EPI_ISL_1325289, EPI_ISL_1325290, EPI_ISL_1325291, EPI_ISL_1325292, EPI_ISL_1325293, EPI_ISL_1325294, EPI_ISL_1325295, EPI_ISL_1325296, EPI_ISL_1325297, EPI_ISL_1325298, EPI_ISL_1325299,
EPI_ISL_1325300, EPI_ISL_1325301, EPI_ISL_1325302, EPI_ISL_1325303, EPI_ISL_1325304, EPI_ISL_1325305, EPI_ISL_1325306, EPI_ISL_1325307, EPI_ISL_1325308, EPI_ISL_1325309, EPI_ISL_1325310, EPI_ISL_1325311, EPI_ISL_1325312, EPI_ISL_1325313, EPI_ISL_1325314, EPI_ISL_1325315, EPI_ISL_1325316, EPI_ISL_1325317, EPI_ISL_1325318, EPI_ISL_1325319, EPI_ISL_1325320, EPI_ISL_1325321,
EPI_ISL_1325322, EPI_ISL_1325323, EPI_ISL_1325324, EPI_ISL_1325325, EPI_ISL_1325326, EPI_ISL_1325327, EPI_ISL_1325328, EPI_ISL_1325329, EPI_ISL_1325330, EPI_ISL_1325331, EPI_ISL_1325332, EPI_ISL_1325333, EPI_ISL_1325334, EPI_ISL_1325335, EPI_ISL_1325336, EPI_ISL_1325337, EPI_ISL_1325338, EPI_ISL_1325339, EPI_ISL_1325340, EPI_ISL_1325341, EPI_ISL_1325342, EPI_ISL_1325343,
EPI_ISL_1325344, EPI_ISL_1325345, EPI_ISL_1325346, EPI_ISL_1325347, EPI_ISL_1325348, EPI_ISL_1325349, EPI_ISL_1325350, EPI_ISL_1325351, EPI_ISL_1325352, EPI_ISL_1325353, EPI_ISL_1325354, EPI_ISL_1325355, EPI_ISL_1325356, EPI_ISL_1325357, EPI_ISL_1325358, EPI_ISL_1325359, EPI_ISL_1325360, EPI_ISL_1325361, EPI_ISL_1325362, EPI_ISL_1325363, EPI_ISL_1325364, EPI_ISL_1325365,
EPI_ISL_1325366, EPI_ISL_1325367, EPI_ISL_1325368, EPI_ISL_1325369, EPI_ISL_1325370, EPI_ISL_1325372, EPI_ISL_1325373, EPI_ISL_1325374, EPI_ISL_1325376, EPI_ISL_1325377, EPI_ISL_1325378, EPI_ISL_1325380, EPI_ISL_1325381, EPI_ISL_1325385, EPI_ISL_1325386, EPI_ISL_1325387, EPI_ISL_1325388, EPI_ISL_1325389, EPI_ISL_1325390, EPI_ISL_1325392, EPI_ISL_1325393, EPI_ISL_1325394,
EPI_ISL_1325396, EPI_ISL_1325398, EPI_ISL_1325399, EPI_ISL_1325400, EPI_ISL_1325401, EPI_ISL_1325402, EPI_ISL_1325403, EPI_ISL_1325405, EPI_ISL_1325406, EPI_ISL_1325412, EPI_ISL_1325414, EPI_ISL_1325415, EPI_ISL_1325416, EPI_ISL_1325418, EPI_ISL_1325420, EPI_ISL_1325422, EPI_ISL_1325425, EPI_ISL_1325427, EPI_ISL_1325428, EPI_ISL_1325430, EPI_ISL_1325432, EPI_ISL_1325435,
EPI_ISL_1325437, EPI_ISL_1325439, EPI_ISL_1325440, EPI_ISL_1325445, EPI_ISL_1325446, EPI_ISL_1325449, EPI_ISL_1325452, EPI_ISL_1325454, EPI_ISL_1325455, EPI_ISL_1325456, EPI_ISL_1325457, EPI_ISL_1325458, EPI_ISL_1325459, EPI_ISL_1325460, EPI_ISL_1325461, EPI_ISL_1325462, EPI_ISL_1325464, EPI_ISL_1325466, EPI_ISL_1325467, EPI_ISL_1325468, EPI_ISL_1325469, EPI_ISL_1325470,
EPI_ISL_1325472, EPI_ISL_1325473, EPI_ISL_1325475, EPI_ISL_1325476, EPI_ISL_1325478, EPI_ISL_1325480, EPI_ISL_1325481, EPI_ISL_1325482, EPI_ISL_1325485, EPI_ISL_1325486, EPI_ISL_1325488, EPI_ISL_1325489, EPI_ISL_1325490, EPI_ISL_1325491, EPI_ISL_1325492, EPI_ISL_1325493, EPI_ISL_1325496, EPI_ISL_1325498, EPI_ISL_1325499, EPI_ISL_1325500, EPI_ISL_1325505, EPI_ISL_1325506,
EPI_ISL_1325507, EPI_ISL_1325508, EPI_ISL_1325509, EPI_ISL_1325511, EPI_ISL_1325512, EPI_ISL_1325515, EPI_ISL_1325516, EPI_ISL_1325517, EPI_ISL_1325520, EPI_ISL_1325521, EPI_ISL_1325525, EPI_ISL_1325526, EPI_ISL_1325527, EPI_ISL_1325528, EPI_ISL_1325530, EPI_ISL_1325531, EPI_ISL_1325533, EPI_ISL_1325534, EPI_ISL_1325536, EPI_ISL_1325537, EPI_ISL_1325538, EPI_ISL_1325539,
EPI_ISL_1325540, EPI_ISL_1325542, EPI_ISL_1325543, EPI_ISL_1325544, EPI_ISL_1325547, EPI_ISL_1325548, EPI_ISL_1325549, EPI_ISL_1325551, EPI_ISL_1325552, EPI_ISL_1325554, EPI_ISL_1325555, EPI_ISL_1325556, EPI_ISL_1325557, EPI_ISL_1325559, EPI_ISL_1325560, EPI_ISL_1325561, EPI_ISL_1325562, EPI_ISL_1325565, EPI_ISL_1325566, EPI_ISL_1325568, EPI_ISL_1325570, EPI_ISL_1325571,
EPI_ISL_1325574, EPI_ISL_1325575, EPI_ISL_1325576, EPI_ISL_1325577, EPI_ISL_1325578, EPI_ISL_1325579, EPI_ISL_1325580, EPI_ISL_1325581, EPI_ISL_1325582, EPI_ISL_1325583, EPI_ISL_1325585, EPI_ISL_1325586, EPI_ISL_1325588, EPI_ISL_1325589, EPI_ISL_1325591, EPI_ISL_1325592, EPI_ISL_1325593, EPI_ISL_1325594, EPI_ISL_1325595, EPI_ISL_1325596, EPI_ISL_1325597, EPI_ISL_1325598,
EPI_ISL_1325599, EPI_ISL_1325603, EPI_ISL_1325604, EPI_ISL_1325606, EPI_ISL_1325607, EPI_ISL_1325610, EPI_ISL_1325611, EPI_ISL_1325612, EPI_ISL_1325614, EPI_ISL_1325618, EPI_ISL_1325620, EPI_ISL_1325621, EPI_ISL_1325622, EPI_ISL_1325624, EPI_ISL_1325625, EPI_ISL_1325627, EPI_ISL_1325629, EPI_ISL_1325630, EPI_ISL_1325631, EPI_ISL_1325632, EPI_ISL_1325633, EPI_ISL_1325635,
EPI_ISL_1325637, EPI_ISL_1325639, EPI_ISL_1325640, EPI_ISL_1325641, EPI_ISL_1325642, EPI_ISL_1325643, EPI_ISL_1325644, EPI_ISL_1325645, EPI_ISL_1325646, EPI_ISL_1325647, EPI_ISL_1325648, EPI_ISL_1325649, EPI_ISL_1325650, EPI_ISL_1325651, EPI_ISL_1325652, EPI_ISL_1325653, EPI_ISL_1325654, EPI_ISL_1325655, EPI_ISL_1325656, EPI_ISL_1325657, EPI_ISL_1325658, EPI_ISL_1325659,
EPI_ISL_1325660, EPI_ISL_1325661, EPI_ISL_1325662, EPI_ISL_1325663, EPI_ISL_1325664, EPI_ISL_1325665, EPI_ISL_1325666, EPI_ISL_1325667, EPI_ISL_1325668, EPI_ISL_1325669, EPI_ISL_1325670, EPI_ISL_1325671, EPI_ISL_1325672, EPI_ISL_1325673, EPI_ISL_1325674, EPI_ISL_1325675, EPI_ISL_1325676, EPI_ISL_1325677, EPI_ISL_1325678, EPI_ISL_1325679, EPI_ISL_1325680, EPI_ISL_1325681,
EPI_ISL_1325682, EPI_ISL_1325683, EPI_ISL_1325684, EPI_ISL_1325685, EPI_ISL_1325686, EPI_ISL_1325687, EPI_ISL_1325688, EPI_ISL_1325689, EPI_ISL_1325690, EPI_ISL_1325691, EPI_ISL_1325692, EPI_ISL_1325693, EPI_ISL_1325694, EPI_ISL_1325695, EPI_ISL_1325696, EPI_ISL_1325697, EPI_ISL_1325698, EPI_ISL_1325699, EPI_ISL_1325700, EPI_ISL_1325701, EPI_ISL_1325702, EPI_ISL_1325703,
EPI_ISL_1325704, EPI_ISL_1325705, EPI_ISL_1325706, EPI_ISL_1325707, EPI_ISL_1325708, EPI_ISL_1325709, EPI_ISL_1325710, EPI_ISL_1325711, EPI_ISL_1325712, EPI_ISL_1325713, EPI_ISL_1325714, EPI_ISL_1325715, EPI_ISL_1325716, EPI_ISL_1325717, EPI_ISL_1325718, EPI_ISL_1325719, EPI_ISL_1325720, EPI_ISL_1325721, EPI_ISL_1325722, EPI_ISL_1325723, EPI_ISL_1325724, EPI_ISL_1325725,
EPI_ISL_1325726, EPI_ISL_1325727, EPI_ISL_1325728, EPI_ISL_1325729, EPI_ISL_1325730, EPI_ISL_1325731, EPI_ISL_1325732, EPI_ISL_1325733, EPI_ISL_1325734, EPI_ISL_1325735, EPI_ISL_1325736, EPI_ISL_1325737, EPI_ISL_1325738, EPI_ISL_1325739, EPI_ISL_1325740, EPI_ISL_1325741, EPI_ISL_1325742, EPI_ISL_1325743, EPI_ISL_1325744, EPI_ISL_1325745, EPI_ISL_1325746, EPI_ISL_1325747,
EPI_ISL_1325748, EPI_ISL_1325749, EPI_ISL_1325750, EPI_ISL_1325751, EPI_ISL_1325752, EPI_ISL_1325753, EPI_ISL_1325754, EPI_ISL_1325755, EPI_ISL_1325756, EPI_ISL_1325757, EPI_ISL_1325758, EPI_ISL_1325759, EPI_ISL_1325760, EPI_ISL_1325761, EPI_ISL_1325762, EPI_ISL_1325763, EPI_ISL_1325764, EPI_ISL_1325765, EPI_ISL_1325766, EPI_ISL_1325767, EPI_ISL_1325768, EPI_ISL_1325769,
EPI_ISL_1325770, EPI_ISL_1325771, EPI_ISL_1325772, EPI_ISL_1325773, EPI_ISL_1325774, EPI_ISL_1325775, EPI_ISL_1325776, EPI_ISL_1325777, EPI_ISL_1325778, EPI_ISL_1325779, EPI_ISL_1325780, EPI_ISL_1325781, EPI_ISL_1325783, EPI_ISL_1325784, EPI_ISL_1325785, EPI_ISL_1325786, EPI_ISL_1325787, EPI_ISL_1325788, EPI_ISL_1325789, EPI_ISL_1325790, EPI_ISL_1325791, EPI_ISL_1325792,
EPI_ISL_1325793, EPI_ISL_1325794, EPI_ISL_1325795, EPI_ISL_1325796, EPI_ISL_1325797, EPI_ISL_1325798, EPI_ISL_1325799, EPI_ISL_1325800, EPI_ISL_1325801, EPI_ISL_1325802, EPI_ISL_1325803, EPI_ISL_1325804, EPI_ISL_1325805, EPI_ISL_1325806, EPI_ISL_1325807, EPI_ISL_1325808, EPI_ISL_1325809, EPI_ISL_1325810, EPI_ISL_1325811, EPI_ISL_1325812, EPI_ISL_1325813, EPI_ISL_1325814,
EPI_ISL_1325815, EPI_ISL_1325816, EPI_ISL_1325817, EPI_ISL_1325818, EPI_ISL_1325819, EPI_ISL_1325820, EPI_ISL_1325821, EPI_ISL_1325822, EPI_ISL_1325823, EPI_ISL_1325824, EPI_ISL_1325825, EPI_ISL_1325826, EPI_ISL_1325827, EPI_ISL_1325828, EPI_ISL_1325829, EPI_ISL_1325830, EPI_ISL_1325831, EPI_ISL_1325832, EPI_ISL_1325833, EPI_ISL_1325834, EPI_ISL_1325835, EPI_ISL_1325836,
EPI_ISL_1325837, EPI_ISL_1325838, EPI_ISL_1325839, EPI_ISL_1325840, EPI_ISL_1325841, EPI_ISL_1325842, EPI_ISL_1325843, EPI_ISL_1325844, EPI_ISL_1325845, EPI_ISL_1325846, EPI_ISL_1325847, EPI_ISL_1325848, EPI_ISL_1325849, EPI_ISL_1325850, EPI_ISL_1325851, EPI_ISL_1325852, EPI_ISL_1325853, EPI_ISL_1325854, EPI_ISL_1325855, EPI_ISL_1325856, EPI_ISL_1325857, EPI_ISL_1325858,
EPI_ISL_1325859, EPI_ISL_1325861, EPI_ISL_1325862, EPI_ISL_1325865, EPI_ISL_1325868, EPI_ISL_1325872, EPI_ISL_1325883, EPI_ISL_1325896, EPI_ISL_1325897, EPI_ISL_1325907, EPI_ISL_1325919, EPI_ISL_1325925, EPI_ISL_1325934, EPI_ISL_1325939, EPI_ISL_1325946, EPI_ISL_1325949, EPI_ISL_1325951, EPI_ISL_1325957, EPI_ISL_1325962, EPI_ISL_1325966, EPI_ISL_1325971, EPI_ISL_1325975,
EPI_ISL_1325978, EPI_ISL_1325992, EPI_ISL_1326000, EPI_ISL_1326010, EPI_ISL_1326019, EPI_ISL_1326029, EPI_ISL_1326031, EPI_ISL_1326032, EPI_ISL_1326037, EPI_ISL_1326038, EPI_ISL_1326040, EPI_ISL_1326041, EPI_ISL_1326043, EPI_ISL_1326081, EPI_ISL_1326102, EPI_ISL_1326107, EPI_ISL_1326124, EPI_ISL_1326669, EPI_ISL_1326679, EPI_ISL_1326683, EPI_ISL_1326687, EPI_ISL_1326702,
EPI_ISL_1326729, EPI_ISL_1326732, EPI_ISL_1326735, EPI_ISL_1326737, EPI_ISL_1326738, EPI_ISL_1326739, EPI_ISL_1326743, EPI_ISL_1326745, EPI_ISL_1326750, EPI_ISL_1326751, EPI_ISL_1326762, EPI_ISL_1326767, EPI_ISL_1326775, EPI_ISL_1326781, EPI_ISL_1326787, EPI_ISL_1326791, EPI_ISL_1326803, EPI_ISL_1326815, EPI_ISL_1326816, EPI_ISL_1326820, EPI_ISL_1326823, EPI_ISL_1326825,
EPI_ISL_1326829, EPI_ISL_1326837, EPI_ISL_1326853, EPI_ISL_1326854, EPI_ISL_1326858, EPI_ISL_1326867, EPI_ISL_1326870, EPI_ISL_1326871, EPI_ISL_1326872, EPI_ISL_1326885, EPI_ISL_1326886, EPI_ISL_1326887, EPI_ISL_1326898, EPI_ISL_1326902, EPI_ISL_1326909, EPI_ISL_1326916, EPI_ISL_1326924, EPI_ISL_1326938, EPI_ISL_1326940, EPI_ISL_1326951, EPI_ISL_1326953, EPI_ISL_1326964,
EPI_ISL_1327702, EPI_ISL_1327703, EPI_ISL_1327704, EPI_ISL_1327705, EPI_ISL_1327706, EPI_ISL_1327707, EPI_ISL_1327708, EPI_ISL_1327709, EPI_ISL_1327710, EPI_ISL_1327711, EPI_ISL_1327712, EPI_ISL_1327713, EPI_ISL_1327714, EPI_ISL_1327715, EPI_ISL_1327716, EPI_ISL_1327717, EPI_ISL_1327718, EPI_ISL_1327722, EPI_ISL_1327723, EPI_ISL_1327725, EPI_ISL_1327726, EPI_ISL_1327727,
EPI_ISL_1327730, EPI_ISL_1327732, EPI_ISL_1327733, EPI_ISL_1327735, EPI_ISL_1327736, EPI_ISL_1327744, EPI_ISL_1327745, EPI_ISL_1327746, EPI_ISL_1327747, EPI_ISL_1327748, EPI_ISL_1327749, EPI_ISL_1327750, EPI_ISL_1327752, EPI_ISL_1327757, EPI_ISL_1327759, EPI_ISL_1327760, EPI_ISL_1327763, EPI_ISL_1327764, EPI_ISL_1327765, EPI_ISL_1327766, EPI_ISL_1327767, EPI_ISL_1327768,
EPI_ISL_1327769, EPI_ISL_1327771, EPI_ISL_1327773, EPI_ISL_1327774, EPI_ISL_1327776, EPI_ISL_1327777, EPI_ISL_1327778, EPI_ISL_1327779, EPI_ISL_1327780, EPI_ISL_1327781, EPI_ISL_1327782, EPI_ISL_1327783, EPI_ISL_1327784, EPI_ISL_1327785, EPI_ISL_1327786, EPI_ISL_1327787, EPI_ISL_1327788, EPI_ISL_1327790, EPI_ISL_1327792, EPI_ISL_1327793, EPI_ISL_1327794, EPI_ISL_1327795,
EPI_ISL_1327796, EPI_ISL_1327797, EPI_ISL_1327798, EPI_ISL_1327799, EPI_ISL_1327800, EPI_ISL_1327801, EPI_ISL_1327802, EPI_ISL_1327803, EPI_ISL_1327804, EPI_ISL_1327805, EPI_ISL_1327806, EPI_ISL_1327807, EPI_ISL_1327808, EPI_ISL_1327809, EPI_ISL_1327810, EPI_ISL_1327812, EPI_ISL_1327813, EPI_ISL_1327814, EPI_ISL_1327817, EPI_ISL_1327818, EPI_ISL_1327819, EPI_ISL_1327820,
EPI_ISL_1327822, EPI_ISL_1327824, EPI_ISL_1327825, EPI_ISL_1327826, EPI_ISL_1327828, EPI_ISL_1327829, EPI_ISL_1327830, EPI_ISL_1327831, EPI_ISL_1327832, EPI_ISL_1327833, EPI_ISL_1327834, EPI_ISL_1327837, EPI_ISL_1327842, EPI_ISL_1327843, EPI_ISL_1327844, EPI_ISL_1327845, EPI_ISL_1327846, EPI_ISL_1327847, EPI_ISL_1327848, EPI_ISL_1327850, EPI_ISL_1327851, EPI_ISL_1327853,
EPI_ISL_1327854, EPI_ISL_1327856, EPI_ISL_1327857, EPI_ISL_1327858, EPI_ISL_1327859, EPI_ISL_1327860, EPI_ISL_1327861, EPI_ISL_1327862, EPI_ISL_1327865, EPI_ISL_1327867, EPI_ISL_1327869, EPI_ISL_1327871, EPI_ISL_1327872, EPI_ISL_1327873, EPI_ISL_1327874, EPI_ISL_1327875, EPI_ISL_1327876, EPI_ISL_1327877, EPI_ISL_1327878, EPI_ISL_1327879, EPI_ISL_1327880, EPI_ISL_1327882,
EPI_ISL_1327884, EPI_ISL_1327885, EPI_ISL_1327886, EPI_ISL_1327887, EPI_ISL_1327888, EPI_ISL_1327889, EPI_ISL_1327890, EPI_ISL_1327891, EPI_ISL_1327893, EPI_ISL_1327894, EPI_ISL_1327895, EPI_ISL_1327897, EPI_ISL_1327899, EPI_ISL_1327900, EPI_ISL_1327901, EPI_ISL_1327902, EPI_ISL_1327903, EPI_ISL_1327904, EPI_ISL_1327905, EPI_ISL_1327906, EPI_ISL_1327907, EPI_ISL_1327908,
EPI_ISL_1327909, EPI_ISL_1327910, EPI_ISL_1327912, EPI_ISL_1327914, EPI_ISL_1327915, EPI_ISL_1327916, EPI_ISL_1327917, EPI_ISL_1327918, EPI_ISL_1327919, EPI_ISL_1327920, EPI_ISL_1327921, EPI_ISL_1327922, EPI_ISL_1327925, EPI_ISL_1327926, EPI_ISL_1327927, EPI_ISL_1327929, EPI_ISL_1327930, EPI_ISL_1327931, EPI_ISL_1327932, EPI_ISL_1327935, EPI_ISL_1327937, EPI_ISL_1327938,
EPI_ISL_1327940, EPI_ISL_1327941, EPI_ISL_1327942, EPI_ISL_1327944, EPI_ISL_1327945, EPI_ISL_1327946, EPI_ISL_1327947, EPI_ISL_1327949, EPI_ISL_1327950, EPI_ISL_1327951, EPI_ISL_1327952, EPI_ISL_1327953, EPI_ISL_1327954, EPI_ISL_1327955, EPI_ISL_1327957, EPI_ISL_1327958, EPI_ISL_1327959, EPI_ISL_1327960, EPI_ISL_1327961, EPI_ISL_1327962, EPI_ISL_1327963, EPI_ISL_1327964,
EPI_ISL_1327965, EPI_ISL_1327966, EPI_ISL_1327967, EPI_ISL_1327968, EPI_ISL_1327969, EPI_ISL_1327971, EPI_ISL_1327972, EPI_ISL_1327974, EPI_ISL_1327975, EPI_ISL_1327976, EPI_ISL_1327977, EPI_ISL_1327978, EPI_ISL_1327980, EPI_ISL_1327981, EPI_ISL_1327982, EPI_ISL_1327983, EPI_ISL_1327984, EPI_ISL_1327985, EPI_ISL_1327986, EPI_ISL_1327987, EPI_ISL_1327988, EPI_ISL_1327989,
EPI_ISL_1327990, EPI_ISL_1327991, EPI_ISL_1327992, EPI_ISL_1327993, EPI_ISL_1327994, EPI_ISL_1327996, EPI_ISL_1327997, EPI_ISL_1327998, EPI_ISL_1327999, EPI_ISL_1328002, EPI_ISL_1328003, EPI_ISL_1328004, EPI_ISL_1328005, EPI_ISL_1328006, EPI_ISL_1328007, EPI_ISL_1328008, EPI_ISL_1328009, EPI_ISL_1328010, EPI_ISL_1328011, EPI_ISL_1328012, EPI_ISL_1328013, EPI_ISL_1328014,
EPI_ISL_1328015, EPI_ISL_1328016, EPI_ISL_1328017, EPI_ISL_1328018, EPI_ISL_1328019, EPI_ISL_1328023, EPI_ISL_1328024, EPI_ISL_1328025, EPI_ISL_1328026, EPI_ISL_1328028, EPI_ISL_1328029, EPI_ISL_1328030, EPI_ISL_1328031, EPI_ISL_1328032, EPI_ISL_1328033, EPI_ISL_1328034, EPI_ISL_1328037, EPI_ISL_1328038, EPI_ISL_1328039, EPI_ISL_1328041, EPI_ISL_1328042, EPI_ISL_1328044,
EPI_ISL_1328045, EPI_ISL_1328046, EPI_ISL_1328047, EPI_ISL_1328048, EPI_ISL_1328049, EPI_ISL_1328050, EPI_ISL_1328051, EPI_ISL_1328052, EPI_ISL_1328053, EPI_ISL_1328055, EPI_ISL_1328057, EPI_ISL_1328058, EPI_ISL_1328059, EPI_ISL_1328060, EPI_ISL_1328062, EPI_ISL_1328063, EPI_ISL_1328064, EPI_ISL_1328065, EPI_ISL_1328066, EPI_ISL_1328067, EPI_ISL_1328068, EPI_ISL_1328069,
EPI_ISL_1328070, EPI_ISL_1328071, EPI_ISL_1328072, EPI_ISL_1328073, EPI_ISL_1328074, EPI_ISL_1328075, EPI_ISL_1328076, EPI_ISL_1328077, EPI_ISL_1328078, EPI_ISL_1328080, EPI_ISL_1328081, EPI_ISL_1328082, EPI_ISL_1328084, EPI_ISL_1328085, EPI_ISL_1328086, EPI_ISL_1328087, EPI_ISL_1328089, EPI_ISL_1328090, EPI_ISL_1328091, EPI_ISL_1328092, EPI_ISL_1328093, EPI_ISL_1328094,
EPI_ISL_1328095, EPI_ISL_1328096, EPI_ISL_1328097, EPI_ISL_1328098, EPI_ISL_1328099, EPI_ISL_1328100, EPI_ISL_1328102, EPI_ISL_1328103, EPI_ISL_1328104, EPI_ISL_1328106, EPI_ISL_1328107, EPI_ISL_1328108, EPI_ISL_1328111, EPI_ISL_1328112, EPI_ISL_1328113, EPI_ISL_1328114, EPI_ISL_1328115, EPI_ISL_1328116, EPI_ISL_1328117, EPI_ISL_1328118, EPI_ISL_1328119, EPI_ISL_1328120,
EPI_ISL_1328121, EPI_ISL_1328122, EPI_ISL_1328123, EPI_ISL_1328124, EPI_ISL_1328125, EPI_ISL_1328127, EPI_ISL_1328129, EPI_ISL_1328130, EPI_ISL_1328131, EPI_ISL_1328132, EPI_ISL_1328133, EPI_ISL_1328134, EPI_ISL_1328135, EPI_ISL_1328136, EPI_ISL_1328137, EPI_ISL_1328139, EPI_ISL_1328140, EPI_ISL_1328141, EPI_ISL_1328142, EPI_ISL_1328143, EPI_ISL_1328144, EPI_ISL_1328145,
EPI_ISL_1328146, EPI_ISL_1328147, EPI_ISL_1328149, EPI_ISL_1328150, EPI_ISL_1328151, EPI_ISL_1328152, EPI_ISL_1328153, EPI_ISL_1328155, EPI_ISL_1328156, EPI_ISL_1328157, EPI_ISL_1328158, EPI_ISL_1328159, EPI_ISL_1328163, EPI_ISL_1328164, EPI_ISL_1328165, EPI_ISL_1328166, EPI_ISL_1328167, EPI_ISL_1328168, EPI_ISL_1328169, EPI_ISL_1328170, EPI_ISL_1328171, EPI_ISL_1328172,
EPI_ISL_1328173, EPI_ISL_1328174, EPI_ISL_1328175, EPI_ISL_1328176, EPI_ISL_1328177, EPI_ISL_1328178, EPI_ISL_1328179, EPI_ISL_1328180, EPI_ISL_1328181, EPI_ISL_1328182, EPI_ISL_1328183, EPI_ISL_1328184, EPI_ISL_1328185, EPI_ISL_1328186, EPI_ISL_1328187, EPI_ISL_1328188, EPI_ISL_1328189, EPI_ISL_1328190, EPI_ISL_1328191, EPI_ISL_1328192, EPI_ISL_1328193, EPI_ISL_1328195,
EPI_ISL_1328196, EPI_ISL_1328197, EPI_ISL_1328198, EPI_ISL_1328199, EPI_ISL_1328200, EPI_ISL_1328201, EPI_ISL_1328202, EPI_ISL_1328203, EPI_ISL_1328206, EPI_ISL_1328207, EPI_ISL_1328208, EPI_ISL_1328210, EPI_ISL_1328211, EPI_ISL_1328212, EPI_ISL_1328213, EPI_ISL_1328214, EPI_ISL_1328215, EPI_ISL_1328246, EPI_ISL_1328337, EPI_ISL_1328356, EPI_ISL_1329382, EPI_ISL_1329383,
EPI_ISL_1329384, EPI_ISL_1329391, EPI_ISL_1329393, EPI_ISL_1329395, EPI_ISL_1329397, EPI_ISL_1329399, EPI_ISL_1329401, EPI_ISL_1329402, EPI_ISL_1329404, EPI_ISL_1329405, EPI_ISL_1329406, EPI_ISL_1329407, EPI_ISL_1329410, EPI_ISL_1329412, EPI_ISL_1329413, EPI_ISL_1329414, EPI_ISL_1329415, EPI_ISL_1329416, EPI_ISL_1329417, EPI_ISL_1329418, EPI_ISL_1329419, EPI_ISL_1329421,
EPI_ISL_1329423, EPI_ISL_1329424, EPI_ISL_1329428, EPI_ISL_1329429, EPI_ISL_1329431, EPI_ISL_1329433, EPI_ISL_1329437, EPI_ISL_1329440, EPI_ISL_1329442, EPI_ISL_1329443, EPI_ISL_1329444, EPI_ISL_1329446, EPI_ISL_1329447, EPI_ISL_1329451, EPI_ISL_1329452, EPI_ISL_1329453, EPI_ISL_1329455, EPI_ISL_1329456, EPI_ISL_1329457, EPI_ISL_1329464, EPI_ISL_1329465, EPI_ISL_1329466,
EPI_ISL_1329468, EPI_ISL_1329473, EPI_ISL_1329479, EPI_ISL_1329482, EPI_ISL_1329485, EPI_ISL_1329488, EPI_ISL_1329490, EPI_ISL_1329491, EPI_ISL_1329493, EPI_ISL_1329495, EPI_ISL_1329497, EPI_ISL_1329499, EPI_ISL_1329500, EPI_ISL_1329501, EPI_ISL_1329506, EPI_ISL_1329508, EPI_ISL_1329511, EPI_ISL_1329512, EPI_ISL_1329513, EPI_ISL_1329514, EPI_ISL_1329515, EPI_ISL_1329516,

EPI_ISL_1329518, EPI_ISL_1329519, EPI_ISL_1329522, EPI_ISL_1329524, EPI_ISL_1329525, EPI_ISL_1329526, EPI_ISL_1329528, EPI_ISL_1329533, EPI_ISL_1329537, EPI_ISL_1329541, EPI_ISL_1329544, EPI_ISL_1329546, EPI_ISL_1329547, EPI_ISL_1329548, EPI_ISL_1329551, EPI_ISL_1329552, EPI_ISL_1329553, EPI_ISL_1329556, EPI_ISL_1329557, EPI_ISL_1329564, EPI_ISL_1329565, EPI_ISL_1329567, EPI_ISL_1329569, EPI_ISL_1329571, EPI_ISL_1329576, EPI_ISL_1329577, EPI_ISL_1329582, EPI_ISL_1329585, EPI_ISL_1329586, EPI_ISL_1329587, EPI_ISL_1329590, EPI_ISL_1329596, EPI_ISL_1329598, EPI_ISL_1329602, EPI_ISL_1329605, EPI_ISL_1329610, EPI_ISL_1329611, EPI_ISL_1329616, EPI_ISL_1329617, EPI_ISL_1329624, EPI_ISL_1329625, EPI_ISL_1329627, EPI_ISL_1329629, EPI_ISL_1329630, EPI_ISL_1329632, EPI_ISL_1329634, EPI_ISL_1329637, EPI_ISL_1329640, EPI_ISL_1329641, EPI_ISL_1329642, EPI_ISL_1329643, EPI_ISL_1329644, EPI_ISL_1329645, EPI_ISL_1329646, EPI_ISL_1329647, EPI_ISL_1329648, EPI_ISL_1329649, EPI_ISL_1329650, EPI_ISL_1329654, EPI_ISL_1329655, EPI_ISL_1329657, EPI_ISL_1329660, EPI_ISL_1329661, EPI_ISL_1329665, EPI_ISL_1329670, EPI_ISL_1329671, EPI_ISL_1329672, EPI_ISL_1329675, EPI_ISL_1329676, EPI_ISL_1329680, EPI_ISL_1329684, EPI_ISL_1329686, EPI_ISL_1329692, EPI_ISL_1329694, EPI_ISL_1329696, EPI_ISL_1329699, EPI_ISL_1329711, EPI_ISL_1329716, EPI_ISL_1329719, EPI_ISL_1329721, EPI_ISL_1329723, EPI_ISL_1329724, EPI_ISL_1329734, EPI_ISL_1329745, EPI_ISL_1329749, EPI_ISL_1329751, EPI_ISL_1329753, EPI_ISL_1329757, EPI_ISL_1329764, EPI_ISL_1329770, EPI_ISL_1329773, EPI_ISL_1329776, EPI_ISL_1329780, EPI_ISL_1329786, EPI_ISL_1329795, EPI_ISL_1329798, EPI_ISL_1329800, EPI_ISL_1329811, EPI_ISL_1329818, EPI_ISL_1329822, EPI_ISL_1329824, EPI_ISL_1329829, EPI_ISL_1329831, EPI_ISL_1329832, EPI_ISL_1329849, EPI_ISL_1329857, EPI_ISL_1329868, EPI_ISL_1329876, EPI_ISL_1329880, EPI_ISL_1329884, EPI_ISL_1329885, EPI_ISL_1329886, EPI_ISL_1329888, EPI_ISL_1329892, EPI_ISL_1329904, EPI_ISL_1329908, EPI_ISL_1329910, EPI_ISL_1329913, EPI_ISL_1329917, EPI_ISL_1329920, EPI_ISL_1329923, EPI_ISL_1329927, EPI_ISL_1329939, EPI_ISL_1329942, EPI_ISL_1329949, EPI_ISL_1329953, EPI_ISL_1329958, EPI_ISL_1329968, EPI_ISL_1329979, EPI_ISL_1341940, EPI_ISL_1342004, EPI_ISL_1342395, EPI_ISL_1342437, EPI_ISL_1342848, EPI_ISL_1343247, EPI_ISL_1343335, EPI_ISL_1343357, EPI_ISL_1343364, EPI_ISL_1343365, EPI_ISL_1343367, EPI_ISL_1343380, EPI_ISL_1343385, EPI_ISL_1343391, EPI_ISL_1343400, EPI_ISL_1343414, EPI_ISL_1343421, EPI_ISL_1343449, EPI_ISL_1343457, EPI_ISL_1343468, EPI_ISL_1343489, EPI_ISL_1343507, EPI_ISL_1343508, EPI_ISL_1343524, EPI_ISL_1343554, EPI_ISL_1343564, EPI_ISL_1343571, EPI_ISL_1343582, EPI_ISL_1343585, EPI_ISL_1343586, EPI_ISL_1343587, EPI_ISL_1343589, EPI_ISL_1343590, EPI_ISL_1343592, EPI_ISL_1343593, EPI_ISL_1343594, EPI_ISL_1343597, EPI_ISL_1343598, EPI_ISL_1343599, EPI_ISL_1343600, EPI_ISL_1343601, EPI_ISL_1343602, EPI_ISL_1343605, EPI_ISL_1343607, EPI_ISL_1343608, EPI_ISL_1343609, EPI_ISL_1343610, EPI_ISL_1343611, EPI_ISL_1343614, EPI_ISL_1343615, EPI_ISL_1343616, EPI_ISL_1343617, EPI_ISL_1343620, EPI_ISL_1343621, EPI_ISL_1343622, EPI_ISL_1343623, EPI_ISL_1343627, EPI_ISL_1343629, EPI_ISL_1343631, EPI_ISL_1343633, EPI_ISL_1343634, EPI_ISL_1343636, EPI_ISL_1343639, EPI_ISL_1343640, EPI_ISL_1343643, EPI_ISL_1343644, EPI_ISL_1343650, EPI_ISL_1343651, EPI_ISL_1343652, EPI_ISL_1343653, EPI_ISL_1343654, EPI_ISL_1343655, EPI_ISL_1343658, EPI_ISL_1343659, EPI_ISL_1343662, EPI_ISL_1343667, EPI_ISL_1343669, EPI_ISL_1343672, EPI_ISL_1343674, EPI_ISL_1343676, EPI_ISL_1343677, EPI_ISL_1343678, EPI_ISL_1343679, EPI_ISL_1343680, EPI_ISL_1343684, EPI_ISL_1343686, EPI_ISL_1343688, EPI_ISL_1343689, EPI_ISL_1343690, EPI_ISL_1343691, EPI_ISL_1343693, EPI_ISL_1343694, EPI_ISL_1343695, EPI_ISL_1343698, EPI_ISL_1343699, EPI_ISL_1343701, EPI_ISL_1343702, EPI_ISL_1343703, EPI_ISL_1343706, EPI_ISL_1343708, EPI_ISL_1343710, EPI_ISL_1343711, EPI_ISL_1343712, EPI_ISL_1343714, EPI_ISL_1343715, EPI_ISL_1343716, EPI_ISL_1343717, EPI_ISL_1343719, EPI_ISL_1343720, EPI_ISL_1343723, EPI_ISL_1343724, EPI_ISL_1343725, EPI_ISL_1343726, EPI_ISL_1343728, EPI_ISL_1343729, EPI_ISL_1343730, EPI_ISL_1343732, EPI_ISL_1343733, EPI_ISL_1343734, EPI_ISL_1343737, EPI_ISL_1343738, EPI_ISL_1343741, EPI_ISL_1343743, EPI_ISL_1343745, EPI_ISL_1343746, EPI_ISL_1343749, EPI_ISL_1343750, EPI_ISL_1343754, EPI_ISL_1343758, EPI_ISL_1343760, EPI_ISL_1343762, EPI_ISL_1343764, EPI_ISL_1343765, EPI_ISL_1343766, EPI_ISL_1343767, EPI_ISL_1343768, EPI_ISL_1343770, EPI_ISL_1343771, EPI_ISL_1343772, EPI_ISL_1343773, EPI_ISL_1343774, EPI_ISL_1343775, EPI_ISL_1343778, EPI_ISL_1343779, EPI_ISL_1343780, EPI_ISL_1343781, EPI_ISL_1343783, EPI_ISL_1343785, EPI_ISL_1343790, EPI_ISL_1343791, EPI_ISL_1343792, EPI_ISL_1343793, EPI_ISL_1343795, EPI_ISL_1343800, EPI_ISL_1343801, EPI_ISL_1343802, EPI_ISL_1343803, EPI_ISL_1343806, EPI_ISL_1343807, EPI_ISL_1343815, EPI_ISL_1343819, EPI_ISL_1343820, EPI_ISL_1343824, EPI_ISL_1343830, EPI_ISL_1343841, EPI_ISL_1343848, EPI_ISL_1343850, EPI_ISL_1343854, EPI_ISL_1343855, EPI_ISL_1343867, EPI_ISL_1343868, EPI_ISL_1343869, EPI_ISL_1343878, EPI_ISL_1343879, EPI_ISL_1343880, EPI_ISL_1343885, EPI_ISL_1343887, EPI_ISL_1343891, EPI_ISL_1343899, EPI_ISL_1343911, EPI_ISL_1343915, EPI_ISL_1343917, EPI_ISL_1343924, EPI_ISL_1343927, EPI_ISL_1343932, EPI_ISL_1343937, EPI_ISL_1343953, EPI_ISL_1343957, EPI_ISL_1343961, EPI_ISL_1343964, EPI_ISL_1343969, EPI_ISL_1343973, EPI_ISL_1343978, EPI_ISL_1343983, EPI_ISL_1344001, EPI_ISL_1344003, EPI_ISL_1344004, EPI_ISL_1344006, EPI_ISL_1344009, EPI_ISL_1344010, EPI_ISL_1344014, EPI_ISL_1344030, EPI_ISL_1344034, EPI_ISL_1344042, EPI_ISL_1344043, EPI_ISL_1344052, EPI_ISL_1344053, EPI_ISL_1344059, EPI_ISL_1344071, EPI_ISL_1344074, EPI_ISL_1344076, EPI_ISL_1344084, EPI_ISL_1344085, EPI_ISL_1344112, EPI_ISL_1344113, EPI_ISL_1344120, EPI_ISL_1344129, EPI_ISL_1344134, EPI_ISL_1344146, EPI_ISL_1344150, EPI_ISL_1344156, EPI_ISL_1344162, EPI_ISL_1344163, EPI_ISL_1344173, EPI_ISL_1344174, EPI_ISL_1344176, EPI_ISL_1344177, EPI_ISL_1344178, EPI_ISL_1344179, EPI_ISL_1344180, EPI_ISL_1344181, EPI_ISL_1344183, EPI_ISL_1344189, EPI_ISL_1344190, EPI_ISL_1344192, EPI_ISL_1344193, EPI_ISL_1344195, EPI_ISL_1344200, EPI_ISL_1344202, EPI_ISL_1344204, EPI_ISL_1344206, EPI_ISL_1344211, EPI_ISL_1344212, EPI_ISL_1344215, EPI_ISL_1344216, EPI_ISL_1344221, EPI_ISL_1344227, EPI_ISL_1344231, EPI_ISL_1344233, EPI_ISL_1344234, EPI_ISL_1344235, EPI_ISL_1344236, EPI_ISL_1344239, EPI_ISL_1344241, EPI_ISL_1344242, EPI_ISL_1344243, EPI_ISL_1344246, EPI_ISL_1344247, EPI_ISL_1344249, EPI_ISL_1344251, EPI_ISL_1344256, EPI_ISL_1344259, EPI_ISL_1344262, EPI_ISL_1344269, EPI_ISL_1344273, EPI_ISL_1344278, EPI_ISL_1344281, EPI_ISL_1344283, EPI_ISL_1344285, EPI_ISL_1344286, EPI_ISL_1344288, EPI_ISL_1344295, EPI_ISL_1344296, EPI_ISL_1344297, EPI_ISL_1344301, EPI_ISL_1344305, EPI_ISL_1344306, EPI_ISL_1344307, EPI_ISL_1344308, EPI_ISL_1344309, EPI_ISL_1344310, EPI_ISL_1344313, EPI_ISL_1344314, EPI_ISL_1344315, EPI_ISL_1344317, EPI_ISL_1344321, EPI_ISL_1344322, EPI_ISL_1344326, EPI_ISL_1344327, EPI_ISL_1344330, EPI_ISL_1344334, EPI_ISL_1344335, EPI_ISL_1344337, EPI_ISL_1344339, EPI_ISL_1344344, EPI_ISL_1344345, EPI_ISL_1344346, EPI_ISL_1344347, EPI_ISL_1344352, EPI_ISL_1344353, EPI_ISL_1344355, EPI_ISL_1344356, EPI_ISL_1344359, EPI_ISL_1344361, EPI_ISL_1344362, EPI_ISL_1344365, EPI_ISL_1344366, EPI_ISL_1344367, EPI_ISL_1344370, EPI_ISL_1344372, EPI_ISL_1344373, EPI_ISL_1344375, EPI_ISL_1344376, EPI_ISL_1344377, EPI_ISL_1344378, EPI_ISL_1344379, EPI_ISL_1344380, EPI_ISL_1344381, EPI_ISL_1344382, EPI_ISL_1344383, EPI_ISL_1344384, EPI_ISL_1344385, EPI_ISL_1344386, EPI_ISL_1344387, EPI_ISL_1344390, EPI_ISL_1344391, EPI_ISL_1344394, EPI_ISL_1344395, EPI_ISL_1344396, EPI_ISL_1344398, EPI_ISL_1344399, EPI_ISL_1344400, EPI_ISL_1344401, EPI_ISL_1344403, EPI_ISL_1344405, EPI_ISL_1344406, EPI_ISL_1344408, EPI_ISL_1344412, EPI_ISL_1344413, EPI_ISL_1344415, EPI_ISL_1344416, EPI_ISL_1344421, EPI_ISL_1344425, EPI_ISL_1344427, EPI_ISL_1344430, EPI_ISL_1344432, EPI_ISL_1344434, EPI_ISL_1344439, EPI_ISL_1344440, EPI_ISL_1344441, EPI_ISL_1344443, EPI_ISL_1344446, EPI_ISL_1344449, EPI_ISL_1344452, EPI_ISL_1344453, EPI_ISL_1344454, EPI_ISL_1344455, EPI_ISL_1344456, EPI_ISL_1344457, EPI_ISL_1344459, EPI_ISL_1344461, EPI_ISL_1344462, EPI_ISL_1344924, EPI_ISL_1344925, EPI_ISL_1344926, EPI_ISL_1344927, EPI_ISL_1344928, EPI_ISL_1344929, EPI_ISL_1344930, EPI_ISL_1344931, EPI_ISL_1344932, EPI_ISL_1344933, EPI_ISL_1344934, EPI_ISL_1344935, EPI_ISL_1344936, EPI_ISL_1344937, EPI_ISL_1344938, EPI_ISL_1344939, EPI_ISL_1344940, EPI_ISL_1344941, EPI_ISL_1344942, EPI_ISL_1344943, EPI_ISL_1344944, EPI_ISL_1344945, EPI_ISL_1344946, EPI_ISL_1344947, EPI_ISL_1344948, EPI_ISL_1344949, EPI_ISL_1344950, EPI_ISL_1344951, EPI_ISL_1344952, EPI_ISL_1344953, EPI_ISL_1344954, EPI_ISL_1344955, EPI_ISL_1344956, EPI_ISL_1344957, EPI_ISL_1344958, EPI_ISL_1344959, EPI_ISL_1344960, EPI_ISL_1344961, EPI_ISL_1344962, EPI_ISL_1344963, EPI_ISL_1344964, EPI_ISL_1344966, EPI_ISL_1344967, EPI_ISL_1344968, EPI_ISL_1344969, EPI_ISL_1344970, EPI_ISL_1344971, EPI_ISL_1344972, EPI_ISL_1344973, EPI_ISL_1344974, EPI_ISL_1344975, EPI_ISL_1344976, EPI_ISL_1344977, EPI_ISL_1344978, EPI_ISL_1344979, EPI_ISL_1344980, EPI_ISL_1344981, EPI_ISL_1344982, EPI_ISL_1344983, EPI_ISL_1344984, EPI_ISL_1344985, EPI_ISL_1344986, EPI_ISL_1344987, EPI_ISL_1344988, EPI_ISL_1344989, EPI_ISL_1344990, EPI_ISL_1344991, EPI_ISL_1344992, EPI_ISL_1344993, EPI_ISL_1344994, EPI_ISL_1344995, EPI_ISL_1344996, EPI_ISL_1344997, EPI_ISL_1344998, EPI_ISL_1344999, EPI_ISL_1345000, EPI_ISL_1345001, EPI_ISL_1345002, EPI_ISL_1345003, EPI_ISL_1345005, EPI_ISL_1345006, EPI_ISL_1345007, EPI_ISL_1345008, EPI_ISL_1345009, EPI_ISL_1345010, EPI_ISL_1345011, EPI_ISL_1345012, EPI_ISL_1345013, EPI_ISL_1345014, EPI_ISL_1345015, EPI_ISL_1345016, EPI_ISL_1345017, EPI_ISL_1345018, EPI_ISL_1345019, EPI_ISL_1345020, EPI_ISL_1345021, EPI_ISL_1345022, EPI_ISL_1345023, EPI_ISL_1345024, EPI_ISL_1345025, EPI_ISL_1345026, EPI_ISL_1345027, EPI_ISL_1345028, EPI_ISL_1345029, EPI_ISL_1345030, EPI_ISL_1345031, EPI_ISL_1345032, EPI_ISL_1345033, EPI_ISL_1345034, EPI_ISL_1345035, EPI_ISL_1345036, EPI_ISL_1345037, EPI_ISL_1345038, EPI_ISL_1345039, EPI_ISL_1345040, EPI_ISL_1345041, EPI_ISL_1345042, EPI_ISL_1345043, EPI_ISL_1345044, EPI_ISL_1345045, EPI_ISL_1345046, EPI_ISL_1345047, EPI_ISL_1345048, EPI_ISL_1345049, EPI_ISL_1345050, EPI_ISL_1345051, EPI_ISL_1345052, EPI_ISL_1345053, EPI_ISL_1345054, EPI_ISL_1345055, EPI_ISL_1345056, EPI_ISL_1345059, EPI_ISL_1345060, EPI_ISL_1345061, EPI_ISL_1345063, EPI_ISL_1345064, EPI_ISL_1345065, EPI_ISL_1345066, EPI_ISL_1345067, EPI_ISL_1345068, EPI_ISL_1345069, EPI_ISL_1345070, EPI_ISL_1345071, EPI_ISL_1345072, EPI_ISL_1345074, EPI_ISL_1345075, EPI_ISL_1345076, EPI_ISL_1345077, EPI_ISL_1345078, EPI_ISL_1345079, EPI_ISL_1345080, EPI_ISL_1345081, EPI_ISL_1345082, EPI_ISL_1345083, EPI_ISL_1345084, EPI_ISL_1345085, EPI_ISL_1345086, EPI_ISL_1345087, EPI_ISL_1345088, EPI_ISL_1345089, EPI_ISL_1345090, EPI_ISL_1345091, EPI_ISL_1345092, EPI_ISL_1345093, EPI_ISL_1345094, EPI_ISL_1345095, EPI_ISL_1345096, EPI_ISL_1345097, EPI_ISL_1345098, EPI_ISL_1345099, EPI_ISL_1345100, EPI_ISL_1345101, EPI_ISL_1345102, EPI_ISL_1345103, EPI_ISL_1345104, EPI_ISL_1345105, EPI_ISL_1345106, EPI_ISL_1345107, EPI_ISL_1345108, EPI_ISL_1345109, EPI_ISL_1345110, EPI_ISL_1345111, EPI_ISL_1345112, EPI_ISL_1345113, EPI_ISL_1345114, EPI_ISL_1345115, EPI_ISL_1345116, EPI_ISL_1345117, EPI_ISL_1345118, EPI_ISL_1345120, EPI_ISL_1345121, EPI_ISL_1345122, EPI_ISL_1345123, EPI_ISL_1345124, EPI_ISL_1345125, EPI_ISL_1345126, EPI_ISL_1345127, EPI_ISL_1345128, EPI_ISL_1345129, EPI_ISL_1345130, EPI_ISL_1345131, EPI_ISL_1345132, EPI_ISL_1345133, EPI_ISL_1345134, EPI_ISL_1345135, EPI_ISL_1345136, EPI_ISL_1345137, EPI_ISL_1345138, EPI_ISL_1345139, EPI_ISL_1345140, EPI_ISL_1345141, EPI_ISL_1345142, EPI_ISL_1345143, EPI_ISL_1345144, EPI_ISL_1345145, EPI_ISL_1345147, EPI_ISL_1345148, EPI_ISL_1345149, EPI_ISL_1345150, EPI_ISL_1345151, EPI_ISL_1345152, EPI_ISL_1345153, EPI_ISL_1345154, EPI_ISL_1345155, EPI_ISL_1345156, EPI_ISL_1345157, EPI_ISL_1345158, EPI_ISL_1345159, EPI_ISL_1345160, EPI_ISL_1345161, EPI_ISL_1345162, EPI_ISL_1345163, EPI_ISL_1345164, EPI_ISL_1345165, EPI_ISL_1345166, EPI_ISL_1345167, EPI_ISL_1345168, EPI_ISL_1345169, EPI_ISL_1345170, EPI_ISL_1345171, EPI_ISL_1345172, EPI_ISL_1345173, EPI_ISL_1345175, EPI_ISL_1345176, EPI_ISL_1345178, EPI_ISL_1345179, EPI_ISL_1345180, EPI_ISL_1345181, EPI_ISL_1345182, EPI_ISL_1345183, EPI_ISL_1345184, EPI_ISL_1345185, EPI_ISL_1345186, EPI_ISL_1345187, EPI_ISL_1345188, EPI_ISL_1345189, EPI_ISL_1345190, EPI_ISL_1345191, EPI_ISL_1345192, EPI_ISL_1345193, EPI_ISL_1345194, EPI_ISL_1345195, EPI_ISL_1345196, EPI_ISL_1345197, EPI_ISL_1345198, EPI_ISL_1345199, EPI_ISL_1345200, EPI_ISL_1345201, EPI_ISL_1345202, EPI_ISL_1345203, EPI_ISL_1345204, EPI_ISL_1345205, EPI_ISL_1345206, EPI_ISL_1345207, EPI_ISL_1345209, EPI_ISL_1345210, EPI_ISL_1345211, EPI_ISL_1345212, EPI_ISL_1345213, EPI_ISL_1345214, EPI_ISL_1345215, EPI_ISL_1345216, EPI_ISL_1345217, EPI_ISL_1345218, EPI_ISL_1345219, EPI_ISL_1345220, EPI_ISL_1345221, EPI_ISL_1345222, EPI_ISL_1345223, EPI_ISL_1345224, EPI_ISL_1345225, EPI_ISL_1345226, EPI_ISL_1345227, EPI_ISL_1345228, EPI_ISL_1345229, EPI_ISL_1345230, EPI_ISL_1345231, EPI_ISL_1345233, EPI_ISL_1345234, EPI_ISL_1345235, EPI_ISL_1345236, EPI_ISL_1345237, EPI_ISL_1345238, EPI_ISL_1345239, EPI_ISL_1345240, EPI_ISL_1345241, EPI_ISL_1345242, EPI_ISL_1345243, EPI_ISL_1345244, EPI_ISL_1345245, EPI_ISL_1345246, EPI_ISL_1345247, EPI_ISL_1345248, EPI_ISL_1345249, EPI_ISL_1345250, EPI_ISL_1345251, EPI_ISL_1345252, EPI_ISL_1345253, EPI_ISL_1345254, EPI_ISL_1345255, EPI_ISL_1345256, EPI_ISL_1345257, EPI_ISL_1345258, EPI_ISL_1345259, EPI_ISL_1345260, EPI_ISL_1345261, EPI_ISL_1345262, EPI_ISL_1345263, EPI_ISL_1345264, EPI_ISL_1345265, EPI_ISL_1345266, EPI_ISL_1345267, EPI_ISL_1345268, EPI_ISL_1345269, EPI_ISL_1345270, EPI_ISL_1345271, EPI_ISL_1345272, EPI_ISL_1345273, EPI_ISL_1345274, EPI_ISL_1345275, EPI_ISL_1345276, EPI_ISL_1345277, EPI_ISL_1345278, EPI_ISL_1345279, EPI_ISL_1345281, EPI_ISL_1345282, EPI_ISL_1345283, EPI_ISL_1345285, EPI_ISL_1345286, EPI_ISL_1345287, EPI_ISL_1345288, EPI_ISL_1345289, EPI_ISL_1345290, EPI_ISL_1345291, EPI_ISL_1345292, EPI_ISL_1345293, EPI_ISL_1345474, EPI_ISL_1345650, EPI_ISL_1345651, EPI_ISL_1345652, EPI_ISL_1345653, EPI_ISL_1345655, EPI_ISL_1345656, EPI_ISL_1345657, EPI_ISL_1345658, EPI_ISL_1345659, EPI_ISL_1345660, EPI_ISL_1345661, EPI_ISL_1345662, EPI_ISL_1345663, EPI_ISL_1345664, EPI_ISL_1345665, EPI_ISL_1345666, EPI_ISL_1345667, EPI_ISL_1345668, EPI_ISL_1345669, EPI_ISL_1345670, EPI_ISL_1345671, EPI_ISL_1345672, EPI_ISL_1345673, EPI_ISL_1345674, EPI_ISL_1345676, EPI_ISL_1345677, EPI_ISL_1345678, EPI_ISL_1345679, EPI_ISL_1345681, EPI_ISL_1345682, EPI_ISL_1345683, EPI_ISL_1345684, EPI_ISL_1345685, EPI_ISL_1345686, EPI_ISL_1345687, EPI_ISL_1345688, EPI_ISL_1345689, EPI_ISL_1345691, EPI_ISL_1345692, EPI_ISL_1345693, EPI_ISL_1345694, EPI_ISL_1345695, EPI_ISL_1345696, EPI_ISL_1345697, EPI_ISL_1345698, EPI_ISL_1345699, EPI_ISL_1345700, EPI_ISL_1345701, EPI_ISL_1345702, EPI_ISL_1345703, EPI_ISL_1345704, EPI_ISL_1345705, EPI_ISL_1345706, EPI_ISL_1345707, EPI_ISL_1345708, EPI_ISL_1345709, EPI_ISL_1345710, EPI_ISL_1345711, EPI_ISL_1345712, EPI_ISL_1345713, EPI_ISL_1345714, EPI_ISL_1345715, EPI_ISL_1345716, EPI_ISL_1345717, EPI_ISL_1345718, EPI_ISL_1345719, EPI_ISL_1345720, EPI_ISL_1345721, EPI_ISL_1345722, EPI_ISL_1345723, EPI_ISL_1345724, EPI_ISL_1345725, EPI_ISL_1345726, EPI_ISL_1345727, EPI_ISL_1345728, EPI_ISL_1345729, EPI_ISL_1345730, EPI_ISL_1345731, EPI_ISL_1345732, EPI_ISL_1345733, EPI_ISL_1345734, EPI_ISL_1345735, EPI_ISL_1345736, EPI_ISL_1345737, EPI_ISL_1345738, EPI_ISL_1345739, EPI_ISL_1345740, EPI_ISL_1345741, EPI_ISL_1345742, EPI_ISL_1345743, EPI_ISL_1345744, EPI_ISL_1345745, EPI_ISL_1345746, EPI_ISL_1345747, EPI_ISL_1345748, EPI_ISL_1345749, EPI_ISL_1345750, EPI_ISL_1345751, EPI_ISL_1345752, EPI_ISL_1345753, EPI_ISL_1345754, EPI_ISL_1345755, EPI_ISL_1345756, EPI_ISL_1345757, EPI_ISL_1345758, EPI_ISL_1345759, EPI_ISL_1345760, EPI_ISL_1345761, EPI_ISL_1345762, EPI_ISL_1345763, EPI_ISL_1345764, EPI_ISL_1345765, EPI_ISL_1345766, EPI_ISL_1345767, EPI_ISL_1345768, EPI_ISL_1345769, EPI_ISL_1345770, EPI_ISL_1345771, EPI_ISL_1345772, EPI_ISL_1345773, EPI_ISL_1345774, EPI_ISL_1345775, EPI_ISL_1345776, EPI_ISL_1345777, EPI_ISL_1345778, EPI_ISL_1345779, EPI_ISL_1345780, EPI_ISL_1345781, EPI_ISL_1345782, EPI_ISL_1345783, EPI_ISL_1345784, EPI_ISL_1345785, EPI_ISL_1345786, EPI_ISL_1345787, EPI_ISL_1345788, EPI_ISL_1345789, EPI_ISL_1345790, EPI_ISL_1345791, EPI_ISL_1345792, EPI_ISL_1345793, EPI_ISL_1345794, EPI_ISL_1345795, EPI_ISL_1345796, EPI_ISL_1345797, EPI_ISL_1345798, EPI_ISL_1345799, EPI_ISL_1345800, EPI_ISL_1345801, EPI_ISL_1345802, EPI_ISL_1345803, EPI_ISL_1345804, EPI_ISL_1345805, EPI_ISL_1345806, EPI_ISL_1345807, EPI_ISL_1345808, EPI_ISL_1345809, EPI_ISL_1345810, EPI_ISL_1345811, EPI_ISL_1345812, EPI_ISL_1345813, EPI_ISL_1345814, EPI_ISL_1345815, EPI_ISL_1345816, EPI_ISL_1345817, EPI_ISL_1345818, EPI_ISL_1345819, EPI_ISL_1345820, EPI_ISL_1345821, EPI_ISL_1345822, EPI_ISL_1345823, EPI_ISL_1345824, EPI_ISL_1345825, EPI_ISL_1345826, EPI_ISL_1345827, EPI_ISL_1345828, EPI_ISL_1345829, EPI_ISL_1345830, EPI_ISL_1345831, EPI_ISL_1345832, EPI_ISL_1345833, EPI_ISL_1345834, EPI_ISL_1345835, EPI_ISL_1345836, EPI_ISL_1345837, EPI_ISL_1345838, EPI_ISL_1345839, EPI_ISL_1345840, EPI_ISL_1345841, EPI_ISL_1345842, EPI_ISL_1345843, EPI_ISL_1345844, EPI_ISL_1345845, EPI_ISL_1345846, EPI_ISL_1345847, EPI_ISL_1345848, EPI_ISL_1345849, EPI_ISL_1345850, EPI_ISL_1345851, EPI_ISL_1345852, EPI_ISL_1345853, EPI_ISL_1345854, EPI_ISL_1345855, EPI_ISL_1345857, EPI_ISL_1345858, EPI_ISL_1345859, EPI_ISL_1345860, EPI_ISL_1345861, EPI_ISL_1345862, EPI_ISL_1345863, EPI_ISL_1345864, EPI_ISL_1345865, EPI_ISL_1345866, EPI_ISL_1345867, EPI_ISL_1345868, EPI_ISL_1345870, EPI_ISL_1345872, EPI_ISL_1345873, EPI_ISL_1345874, EPI_ISL_1345875, EPI_ISL_1345876, EPI_ISL_1345877, EPI_ISL_1345878, EPI_ISL_1345879, EPI_ISL_1345880, EPI_ISL_1345881, EPI_ISL_1345882, EPI_ISL_1345883, EPI_ISL_1345885, EPI_ISL_1345886, EPI_ISL_1345887, EPI_ISL_1345888, EPI_ISL_1345889, EPI_ISL_1345890, EPI_ISL_1345891, EPI_ISL_1345892, EPI_ISL_1345893, EPI_ISL_1345894, EPI_ISL_1345895, EPI_ISL_1345896, EPI_ISL_1345897, EPI_ISL_1345898, EPI_ISL_1345899, EPI_ISL_1345900, EPI_ISL_1345901, EPI_ISL_1345902, EPI_ISL_1345903, EPI_ISL_1345904, EPI_ISL_1345905, EPI_ISL_1345906, EPI_ISL_1345907, EPI_ISL_1345908, EPI_ISL_1345909, EPI_ISL_1345910, EPI_ISL_1345911, EPI_ISL_1345912, EPI_ISL_1345913, EPI_ISL_1345914, EPI_ISL_1345915, EPI_ISL_1345916, EPI_ISL_1345917, EPI_ISL_1345918, EPI_ISL_1345919, EPI_ISL_1345920, EPI_ISL_1345921, EPI_ISL_1345922, EPI_ISL_1345923, EPI_ISL_1345924, EPI_ISL_1345925, EPI_ISL_1345926, EPI_ISL_1345927, EPI_ISL_1345928, EPI_ISL_1345929, EPI_ISL_1345930, EPI_ISL_1345931, EPI_ISL_1345932, EPI_ISL_1345933, EPI_ISL_1345934, EPI_ISL_1345935, EPI_ISL_1345936, EPI_ISL_1345937, EPI_ISL_1345938, EPI_ISL_1345939, EPI_ISL_1345940, EPI_ISL_1345941, EPI_ISL_1345942, EPI_ISL_1345943, EPI_ISL_1345944, EPI_ISL_1345945, EPI_ISL_1345946, EPI_ISL_1345947, EPI_ISL_1345948, EPI_ISL_1345949, EPI_ISL_1345950, EPI_ISL_1345951, EPI_ISL_1345952, EPI_ISL_1345953, EPI_ISL_1345954, EPI_ISL_1345955, EPI_ISL_1345956, EPI_ISL_1345957, EPI_ISL_1345958, EPI_ISL_1345959, EPI_ISL_1345960, EPI_ISL_1345961, EPI_ISL_1345962, EPI_ISL_1345963, EPI_ISL_1345964, EPI_ISL_1345965, EPI_ISL_1345966, EPI_ISL_1345967, EPI_ISL_1345968, EPI_ISL_1345969, EPI_ISL_1345970, EPI_ISL_1345971, EPI_ISL_1345972, EPI_ISL_1345973, EPI_ISL_1345974, EPI_ISL_1345975, EPI_ISL_1345976, EPI_ISL_1345977, EPI_ISL_1345978, EPI_ISL_1345979, EPI_ISL_1345980, EPI_ISL_1345981, EPI_ISL_1345983, EPI_ISL_1345984, EPI_ISL_1345985, EPI_ISL_1345986, EPI_ISL_1345987, EPI_ISL_1345989, EPI_ISL_1345990, EPI_ISL_1345991, EPI_ISL_1345992, EPI_ISL_1345993, EPI_ISL_1345994, EPI_ISL_1345995, EPI_ISL_1345996, EPI_ISL_1345997, EPI_ISL_1345998, EPI_ISL_1345999, EPI_ISL_1346000, EPI_ISL_1346001, EPI_ISL_1346002, EPI_ISL_1346003, EPI_ISL_1346004, EPI_ISL_1346005, EPI_ISL_1346006, EPI_ISL_1346007, EPI_ISL_1346008, EPI_ISL_1346009, EPI_ISL_1346010, EPI_ISL_1346011, EPI_ISL_1346012, EPI_ISL_1346013, EPI_ISL_1346014, EPI_ISL_1346015, EPI_ISL_1346016, EPI_ISL_1346017, EPI_ISL_1346018, EPI_ISL_1346019, EPI_ISL_1346020, EPI_ISL_1346021, EPI_ISL_1346022, EPI_ISL_1346023, EPI_ISL_1346024, EPI_ISL_1346025, EPI_ISL_1346026, EPI_ISL_1346027, EPI_ISL_1346028, EPI_ISL_1346029, EPI_ISL_1346030, EPI_ISL_1346031, EPI_ISL_1346032, EPI_ISL_1346033, EPI_ISL_1346034, EPI_ISL_1346035, EPI_ISL_1346036, EPI_ISL_1346037, EPI_ISL_1346038, EPI_ISL_1346039, EPI_ISL_1346040, EPI_ISL_1346041, EPI_ISL_1346042, EPI_ISL_1346043, EPI_ISL_1346044, EPI_ISL_1346045, EPI_ISL_1346046, EPI_ISL_1346048, EPI_ISL_1346049, EPI_ISL_1346050, EPI_ISL_1346051, EPI_ISL_1346052, EPI_ISL_1346053, EPI_ISL_1346054, EPI_ISL_1346055, EPI_ISL_1346056, EPI_ISL_1346057, EPI_ISL_1346058, EPI_ISL_1346059, EPI_ISL_1346060, EPI_ISL_1346061, EPI_ISL_1346062, EPI_ISL_1346063, EPI_ISL_1346064, EPI_ISL_1346065, EPI_ISL_1346066, EPI_ISL_1346068, EPI_ISL_1346069, EPI_ISL_1346071, EPI_ISL_1346072, EPI_ISL_1346073, EPI_ISL_1346074, EPI_ISL_1346077, EPI_ISL_1346078, EPI_ISL_1346079, EPI_ISL_1346080, EPI_ISL_1346081, EPI_ISL_1346082, EPI_ISL_1346083, EPI_ISL_1346084, EPI_ISL_1346086, EPI_ISL_1346087, EPI_ISL_1346088, EPI_ISL_1346089, EPI_ISL_1346090, EPI_ISL_1346092, EPI_ISL_1346093, EPI_ISL_1346095, EPI_ISL_1346096, EPI_ISL_1346097, EPI_ISL_1346098, EPI_ISL_1346101, EPI_ISL_1346102, EPI_ISL_1346104, EPI_ISL_1346108, EPI_ISL_1346111, EPI_ISL_1346133, EPI_ISL_1346134, EPI_ISL_1346138, EPI_ISL_1346143, EPI_ISL_1346147, EPI_ISL_1346156, EPI_ISL_1346158, EPI_ISL_1346162, EPI_ISL_1346164, EPI_ISL_1346165, EPI_ISL_1346170, EPI_ISL_1346179, EPI_ISL_1346185, EPI_ISL_1346188, EPI_ISL_1346189, EPI_ISL_1346191, EPI_ISL_1346193, EPI_ISL_1346194, EPI_ISL_1346197, EPI_ISL_1346199, EPI_ISL_1346221, EPI_ISL_1346223, EPI_ISL_1346224, EPI_ISL_1346225, EPI_ISL_1346231, EPI_ISL_1346233, EPI_ISL_1346236, EPI_ISL_1346329, EPI_ISL_1346330, EPI_ISL_1346331, EPI_ISL_1346332, EPI_ISL_1346334, EPI_ISL_1346335, EPI_ISL_1346337, EPI_ISL_1346338, EPI_ISL_1346339, EPI_ISL_1346341, EPI_ISL_1346342, EPI_ISL_1346344, EPI_ISL_1346378, EPI_ISL_1346379, EPI_ISL_1346381, EPI_ISL_1346384, EPI_ISL_1346386, EPI_ISL_1346388, EPI_ISL_1346389, EPI_ISL_1346392, EPI_ISL_1365223, EPI_ISL_1365224, EPI_ISL_1365225, EPI_ISL_1365230, EPI_ISL_1365234, EPI_ISL_1365235, EPI_ISL_1365237, EPI_ISL_1365238, EPI_ISL_1365239, EPI_ISL_1365240, EPI_ISL_1365245, EPI_ISL_1365247, EPI_ISL_1365248, EPI_ISL_1365250, EPI_ISL_1365257, EPI_ISL_1365258, EPI_ISL_1365259, EPI_ISL_1365260, EPI_ISL_1365261, EPI_ISL_1365268, EPI_ISL_1365270, EPI_ISL_1365273, EPI_ISL_1365274, EPI_ISL_1365277, EPI_ISL_1365280, EPI_ISL_1365282, EPI_ISL_1365283, EPI_ISL_1365284, EPI_ISL_1365287, EPI_ISL_1365290, EPI_ISL_1365292, EPI_ISL_1365293, EPI_ISL_1365300, EPI_ISL_1365302, EPI_ISL_1365304, EPI_ISL_1365308, EPI_ISL_1365309, EPI_ISL_1365310, EPI_ISL_1365311, EPI_ISL_1365312, EPI_ISL_1375786, EPI_ISL_1375845, EPI_ISL_1375871, EPI_ISL_1375907, EPI_ISL_1375990, EPI_ISL_1375998, EPI_ISL_1376051, EPI_ISL_1376076, EPI_ISL_1376117, EPI_ISL_1376779, EPI_ISL_1376785, EPI_ISL_1376790, EPI_ISL_1376794, EPI_ISL_1376799, EPI_ISL_1376800, EPI_ISL_1376818, EPI_ISL_1376820, EPI_ISL_1376835, EPI_ISL_1376843, EPI_ISL_1376846, EPI_ISL_1376852, EPI_ISL_1376854, EPI_ISL_1376866, EPI_ISL_1376873, EPI_ISL_1376875, EPI_ISL_1376876, EPI_ISL_1376881, EPI_ISL_1376882, EPI_ISL_1376885, EPI_ISL_1376891, EPI_ISL_1376893, EPI_ISL_1376902, EPI_ISL_1376909, EPI_ISL_1376911, EPI_ISL_1376920, EPI_ISL_1376922, EPI_ISL_1376926, EPI_ISL_1376928, EPI_ISL_1376929, EPI_ISL_1376931, EPI_ISL_1376935, EPI_ISL_1376941, EPI_ISL_1376950, EPI_ISL_1376955, EPI_ISL_1376962, EPI_ISL_1376965, EPI_ISL_1377195, EPI_ISL_1377231, EPI_ISL_1377265, EPI_ISL_1377347, EPI_ISL_1377352, EPI_ISL_1377448, EPI_ISL_1378297, EPI_ISL_1378300, EPI_ISL_1378305, EPI_ISL_1378307, EPI_ISL_1378308, EPI_ISL_1378319, EPI_ISL_1378320, EPI_ISL_1378324, EPI_ISL_1378328, EPI_ISL_1378330, EPI_ISL_1378333, EPI_ISL_1378334, EPI_ISL_1378339, EPI_ISL_1378340, EPI_ISL_1378341, EPI_ISL_1378343, EPI_ISL_1378349, EPI_ISL_1378350, EPI_ISL_1378354, EPI_ISL_1378355, EPI_ISL_1378356, EPI_ISL_1378357, EPI_ISL_1378360, EPI_ISL_1378361, EPI_ISL_1378362, EPI_ISL_1378367, EPI_ISL_1378368, EPI_ISL_1378369, EPI_ISL_1378370, EPI_ISL_1378371, EPI_ISL_1378372, EPI_ISL_1378373, EPI_ISL_1378375, EPI_ISL_1378376, EPI_ISL_1378377, EPI_ISL_1378381, EPI_ISL_1378382, EPI_ISL_1378386, EPI_ISL_1378387, EPI_ISL_1378388, EPI_ISL_1378390, EPI_ISL_1378391, EPI_ISL_1378392, EPI_ISL_1378393, EPI_ISL_1378395, EPI_ISL_1378396, EPI_ISL_1378397, EPI_ISL_1378398, EPI_ISL_1378401, EPI_ISL_1378402, EPI_ISL_1378403, EPI_ISL_1378404, EPI_ISL_1378405, EPI_ISL_1378406, EPI_ISL_1378407, EPI_ISL_1378408, EPI_ISL_1378409, EPI_ISL_1378410, EPI_ISL_1378412, EPI_ISL_1378413, EPI_ISL_1378415, EPI_ISL_1378416, EPI_ISL_1378418, EPI_ISL_1378419, EPI_ISL_1378420, EPI_ISL_1378421, EPI_ISL_1378422, EPI_ISL_1378423, EPI_ISL_1378424, EPI_ISL_1378425, EPI_ISL_1378427, EPI_ISL_1378451, EPI_ISL_1378452, EPI_ISL_1378453, EPI_ISL_1378454, EPI_ISL_1378455, EPI_ISL_1378456, EPI_ISL_1378457, EPI_ISL_1378458, EPI_ISL_1378459, EPI_ISL_1378460, EPI_ISL_1378461, EPI_ISL_1378462, EPI_ISL_1378463, EPI_ISL_1378464, EPI_ISL_1378465, EPI_ISL_1378466, EPI_ISL_1378467, EPI_ISL_1378468, EPI_ISL_1378469, EPI_ISL_1378470, EPI_ISL_1378471, EPI_ISL_1378472, EPI_ISL_1378473, EPI_ISL_1378474, EPI_ISL_1378475, EPI_ISL_1378476, EPI_ISL_1378477, EPI_ISL_1378478, EPI_ISL_1378479, EPI_ISL_1378480, EPI_ISL_1378481, EPI_ISL_1378482, EPI_ISL_1378483, EPI_ISL_1378484, EPI_ISL_1378485, EPI_ISL_1378486, EPI_ISL_1378487, EPI_ISL_1378488, EPI_ISL_1378489

| | | | |
|---|---|---|---|
| see above | Lighthouse Lab in Glasgow | Wellcome Sanger Institute for the COVID-19 Genomics UK (COG-UK) Consortium | Anna Dominiczak and Alex Alderton; Carol Clugston; Cordelia Langford; David Gray; David K. Jackson; Dominic Kwiatkowski; Ewan Harrison; Harper VanSteenhouse; Ian Johnston; Jeffrey Barrett; John Sillitoe on behalf of the Wellcome Sanger Institute COVID-19 Surveillance Team; Roberto Amato; Sonia Goncalves; Yumi Kasai |

EPI_ISL_1324805, EPI_ISL_1324808, EPI_ISL_1324815, EPI_ISL_1324818, EPI_ISL_1324819, EPI_ISL_1324824, EPI_ISL_1324825, EPI_ISL_1324827, EPI_ISL_1324836, EPI_ISL_1324837, EPI_ISL_1324840, EPI_ISL_1324845, EPI_ISL_1324849, EPI_ISL_1324850, EPI_ISL_1324857, EPI_ISL_1324859, EPI_ISL_1324861, EPI_ISL_1324863, EPI_ISL_1324868, EPI_ISL_1324874, EPI_ISL_1324877, EPI_ISL_1324887, EPI_ISL_1324888, EPI_ISL_1324889, EPI_ISL_1324892, EPI_ISL_1324896, EPI_ISL_1324897, EPI_ISL_1324898, EPI_ISL_1324900, EPI_ISL_1324902, EPI_ISL_1324903, EPI_ISL_1324911, EPI_ISL_1324912, EPI_ISL_1324914, EPI_ISL_1324915, EPI_ISL_1324922, EPI_ISL_1324923, EPI_ISL_1324924, EPI_ISL_1324926, EPI_ISL_1324927, EPI_ISL_1324928, EPI_ISL_1324929, EPI_ISL_1324930, EPI_ISL_1324933, EPI_ISL_1324936, EPI_ISL_1324941, EPI_ISL_1324943, EPI_ISL_1324944, EPI_ISL_1324946, EPI_ISL_1324949, EPI_ISL_1324950, EPI_ISL_1324955, EPI_ISL_1324959, EPI_ISL_1324960, EPI_ISL_1324961, EPI_ISL_1324963, EPI_ISL_1324964, EPI_ISL_1324965, EPI_ISL_1324969, EPI_ISL_1324971, EPI_ISL_1324978, EPI_ISL_1324979, EPI_ISL_1324982, EPI_ISL_1324994, EPI_ISL_1324996, EPI_ISL_1324998, EPI_ISL_1324999, EPI_ISL_1325001, EPI_ISL_1325009, EPI_ISL_1325011, EPI_ISL_1325013, EPI_ISL_1325014, EPI_ISL_1325015, EPI_ISL_1325016, EPI_ISL_1325026, EPI_ISL_1325027, EPI_ISL_1325030, EPI_ISL_1325032, EPI_ISL_1325033, EPI_ISL_1325035, EPI_ISL_1325036, EPI_ISL_1325037, EPI_ISL_1325040, EPI_ISL_1325044, EPI_ISL_1325049, EPI_ISL_1325056, EPI_ISL_1325057, EPI_ISL_1325059, EPI_ISL_1325061, EPI_ISL_1325068, EPI_ISL_1325069, EPI_ISL_1325070, EPI_ISL_1325071, EPI_ISL_1325072, EPI_ISL_1325078, EPI_ISL_1325080, EPI_ISL_1325081, EPI_ISL_1325088, EPI_ISL_1325089, EPI_ISL_1325098, EPI_ISL_1325099, EPI_ISL_1325100, EPI_ISL_1325104, EPI_ISL_1325112, EPI_ISL_1325115, EPI_ISL_1325117, EPI_ISL_1325123, EPI_ISL_1325124, EPI_ISL_1325371, EPI_ISL_1325375, EPI_ISL_1325379, EPI_ISL_1325382, EPI_ISL_1325383, EPI_ISL_1325384, EPI_ISL_1325391, EPI_ISL_1325395, EPI_ISL_1325397, EPI_ISL_1325404, EPI_ISL_1325407, EPI_ISL_1325408, EPI_ISL_1325409, EPI_ISL_1325410, EPI_ISL_1325411, EPI_ISL_1325413, EPI_ISL_1325417, EPI_ISL_1325419, EPI_ISL_1325421, EPI_ISL_1325423, EPI_ISL_1325424, EPI_ISL_1325426, EPI_ISL_1325429, EPI_ISL_1325431, EPI_ISL_1325433, EPI_ISL_1325434, EPI_ISL_1325436, EPI_ISL_1325438, EPI_ISL_1325441, EPI_ISL_1325442, EPI_ISL_1325443, EPI_ISL_1325444, EPI_ISL_1325447, EPI_ISL_1325448, EPI_ISL_1325450, EPI_ISL_1325451, EPI_ISL_1325453, EPI_ISL_1325463, EPI_ISL_1325465, EPI_ISL_1325471, EPI_ISL_1325474, EPI_ISL_1325477, EPI_ISL_1325479, EPI_ISL_1325483, EPI_ISL_1325484, EPI_ISL_1325487, EPI_ISL_1325494, EPI_ISL_1325495, EPI_ISL_1325497, EPI_ISL_1325501, EPI_ISL_1325502, EPI_ISL_1325503, EPI_ISL_1325504, EPI_ISL_1325510, EPI_ISL_1325513, EPI_ISL_1325514, EPI_ISL_1325518, EPI_ISL_1325519, EPI_ISL_1325522, EPI_ISL_1325523, EPI_ISL_1325524, EPI_ISL_1325529, EPI_ISL_1325532, EPI_ISL_1325535, EPI_ISL_1325541, EPI_ISL_1325545, EPI_ISL_1325546, EPI_ISL_1325550, EPI_ISL_1325553, EPI_ISL_1325563, EPI_ISL_1325564, EPI_ISL_1325567, EPI_ISL_1325569, EPI_ISL_1325572, EPI_ISL_1325573, EPI_ISL_1325584, EPI_ISL_1325587, EPI_ISL_1325590, EPI_ISL_1325600, EPI_ISL_1325601, EPI_ISL_1325602, EPI_ISL_1325605, EPI_ISL_1325608, EPI_ISL_1325609, EPI_ISL_1325613, EPI_ISL_1325616, EPI_ISL_1325617, EPI_ISL_1325619, EPI_ISL_1325626, EPI_ISL_1325636, EPI_ISL_1325638, EPI_ISL_1325860, EPI_ISL_1325863, EPI_ISL_1325864, EPI_ISL_1325866, EPI_ISL_1325867, EPI_ISL_1325869, EPI_ISL_1325870, EPI_ISL_1325871, EPI_ISL_1325874, EPI_ISL_1325875, EPI_ISL_1325876, EPI_ISL_1325877, EPI_ISL_1325878, EPI_ISL_1325879, EPI_ISL_1325881, EPI_ISL_1325882, EPI_ISL_1325884, EPI_ISL_1325885, EPI_ISL_1325886, EPI_ISL_1325887, EPI_ISL_1325888, EPI_ISL_1325889, EPI_ISL_1325890, EPI_ISL_1325891, EPI_ISL_1325892, EPI_ISL_1325893, EPI_ISL_1325894, EPI_ISL_1325895, EPI_ISL_1325898, EPI_ISL_1325899, EPI_ISL_1325900, EPI_ISL_1325901, EPI_ISL_1325902, EPI_ISL_1325903, EPI_ISL_1325904, EPI_ISL_1325905, EPI_ISL_1325906, EPI_ISL_1325908, EPI_ISL_1325909, EPI_ISL_1325910, EPI_ISL_1325911, EPI_ISL_1325912, EPI_ISL_1325914, EPI_ISL_1325915, EPI_ISL_1325917, EPI_ISL_1325918, EPI_ISL_1325920, EPI_ISL_1325921, EPI_ISL_1325922, EPI_ISL_1325923, EPI_ISL_1325924, EPI_ISL_1325926, EPI_ISL_1325927, EPI_ISL_1325928, EPI_ISL_1325929, EPI_ISL_1325930, EPI_ISL_1325932, EPI_ISL_1325933, EPI_ISL_1325935, EPI_ISL_1325936, EPI_ISL_1325937, EPI_ISL_1325938, EPI_ISL_1325940, EPI_ISL_1325941, EPI_ISL_1325942, EPI_ISL_1325943, EPI_ISL_1325944, EPI_ISL_1325945, EPI_ISL_1325947, EPI_ISL_1325948, EPI_ISL_1325950, EPI_ISL_1325952, EPI_ISL_1325953, EPI_ISL_1325955, EPI_ISL_1325956, EPI_ISL_1325958, EPI_ISL_1325959, EPI_ISL_1325960, EPI_ISL_1325961, EPI_ISL_1325963, EPI_ISL_1325964, EPI_ISL_1325965, EPI_ISL_1325969, EPI_ISL_1325970, EPI_ISL_1325972, EPI_ISL_1325973, EPI_ISL_1325979, EPI_ISL_1325980, EPI_ISL_1325982, EPI_ISL_1325983, EPI_ISL_1325984, EPI_ISL_1325985, EPI_ISL_1325986, EPI_ISL_1325987, EPI_ISL_1325989, EPI_ISL_1325990, EPI_ISL_1325991, EPI_ISL_1325993, EPI_ISL_1325994, EPI_ISL_1325995, EPI_ISL_1325996, EPI_ISL_1325997, EPI_ISL_1325999, EPI_ISL_1326002, EPI_ISL_1326003, EPI_ISL_1326004, EPI_ISL_1326006, EPI_ISL_1326007, EPI_ISL_1326008, EPI_ISL_1326009, EPI_ISL_1326011, EPI_ISL_1326012, EPI_ISL_1326013, EPI_ISL_1326014, EPI_ISL_1326015, EPI_ISL_1326016, EPI_ISL_1326017, EPI_ISL_1326018, EPI_ISL_1326020, EPI_ISL_1326021, EPI_ISL_1326022, EPI_ISL_1326023, EPI_ISL_1326024, EPI_ISL_1326025, EPI_ISL_1326026, EPI_ISL_1326027, EPI_ISL_1326028, EPI_ISL_1326030, EPI_ISL_1326033, EPI_ISL_1326035, EPI_ISL_1326036, EPI_ISL_1326039, EPI_ISL_1326042, EPI_ISL_1326044, EPI_ISL_1326045, EPI_ISL_1326046, EPI_ISL_1326047, EPI_ISL_1326048, EPI_ISL_1326049, EPI_ISL_1326050, EPI_ISL_1326051, EPI_ISL_1326052, EPI_ISL_1326053, EPI_ISL_1326055, EPI_ISL_1326056, EPI_ISL_1326057, EPI_ISL_1326058, EPI_ISL_1326059, EPI_ISL_1326060, EPI_ISL_1326061, EPI_ISL_1326062, EPI_ISL_1326063, EPI_ISL_1326064, EPI_ISL_1326065, EPI_ISL_1326067, EPI_ISL_1326068, EPI_ISL_1326069, EPI_ISL_1326071, EPI_ISL_1326072, EPI_ISL_1326074, EPI_ISL_1326075, EPI_ISL_1326076, EPI_ISL_1326077, EPI_ISL_1326078, EPI_ISL_1326079, EPI_ISL_1326080, EPI_ISL_1326083, EPI_ISL_1326084, EPI_ISL_1326085, EPI_ISL_1326086, EPI_ISL_1326087, EPI_ISL_1326088, EPI_ISL_1326089, EPI_ISL_1326090, EPI_ISL_1326091, EPI_ISL_1326092, EPI_ISL_1326093, EPI_ISL_1326094, EPI_ISL_1326095, EPI_ISL_1326096, EPI_ISL_1326097, EPI_ISL_1326098, EPI_ISL_1326099, EPI_ISL_1326100, EPI_ISL_1326101, EPI_ISL_1326103, EPI_ISL_1326104, EPI_ISL_1326105, EPI_ISL_1326106, EPI_ISL_1326108, EPI_ISL_1326109, EPI_ISL_1326110, EPI_ISL_1326111, EPI_ISL_1326112, EPI_ISL_1326113, EPI_ISL_1326115, EPI_ISL_1326116, EPI_ISL_1326117, EPI_ISL_1326118, EPI_ISL_1326120, EPI_ISL_1326121, EPI_ISL_1326122, EPI_ISL_1326123, EPI_ISL_1326125, EPI_ISL_1327855, EPI_ISL_1329687, EPI_ISL_1329688, EPI_ISL_1329689, EPI_ISL_1329690, EPI_ISL_1329693, EPI_ISL_1329697, EPI_ISL_1329703, EPI_ISL_1329704, EPI_ISL_1329708, EPI_ISL_1329712, EPI_ISL_1329714, EPI_ISL_1329717, EPI_ISL_1329720, EPI_ISL_1329725, EPI_ISL_1329726, EPI_ISL_1329730, EPI_ISL_1329731, EPI_ISL_1329732, EPI_ISL_1329733, EPI_ISL_1329735, EPI_ISL_1329738, EPI_ISL_1329739, EPI_ISL_1329740, EPI_ISL_1329742, EPI_ISL_1329743, EPI_ISL_1329746, EPI_ISL_1329752, EPI_ISL_1329756, EPI_ISL_1329761, EPI_ISL_1329763, EPI_ISL_1329766, EPI_ISL_1329771, EPI_ISL_1329774, EPI_ISL_1329778, EPI_ISL_1329779, EPI_ISL_1329783, EPI_ISL_1329785, EPI_ISL_1329791, EPI_ISL_1329796, EPI_ISL_1329797, EPI_ISL_1329801, EPI_ISL_1329802, EPI_ISL_1329803, EPI_ISL_1329804, EPI_ISL_1329805, EPI_ISL_1329806, EPI_ISL_1329809, EPI_ISL_1329810, EPI_ISL_1329814, EPI_ISL_1329817, EPI_ISL_1329819, EPI_ISL_1329820, EPI_ISL_1329821, EPI_ISL_1329825, EPI_ISL_1329826, EPI_ISL_1329827, EPI_ISL_1329830, EPI_ISL_1329834, EPI_ISL_1329839, EPI_ISL_1329842, EPI_ISL_1329844, EPI_ISL_1329850, EPI_ISL_1329852, EPI_ISL_1329855, EPI_ISL_1329859, EPI_ISL_1329860, EPI_ISL_1329862, EPI_ISL_1329863, EPI_ISL_1329865, EPI_ISL_1329866, EPI_ISL_1329872, EPI_ISL_1329874, EPI_ISL_1329875, EPI_ISL_1329891, EPI_ISL_1329893, EPI_ISL_1329895, EPI_ISL_1329896, EPI_ISL_1329899, EPI_ISL_1329900, EPI_ISL_1329901, EPI_ISL_1329902, EPI_ISL_1329906, EPI_ISL_1329912, EPI_ISL_1329915, EPI_ISL_1329916, EPI_ISL_1329929, EPI_ISL_1329933, EPI_ISL_1329936, EPI_ISL_1329938, EPI_ISL_1329941, EPI_ISL_1329943, EPI_ISL_1329944, EPI_ISL_1329946, EPI_ISL_1329951, EPI_ISL_1329954, EPI_ISL_1329955, EPI_ISL_1329956, EPI_ISL_1329957, EPI_ISL_1329961, EPI_ISL_1329963, EPI_ISL_1329964, EPI_ISL_1329969, EPI_ISL_1329970, EPI_ISL_1329972, EPI_ISL_1329974, EPI_ISL_1329975, EPI_ISL_1329977, EPI_ISL_1329978, EPI_ISL_1342238, EPI_ISL_1342244, EPI_ISL_1342259, EPI_ISL_1342299, EPI_ISL_1342302, EPI_ISL_1342326, EPI_ISL_1342328, EPI_ISL_1342362, EPI_ISL_1342385, EPI_ISL_1342389, EPI_ISL_1342401, EPI_ISL_1342408, EPI_ISL_1342466, EPI_ISL_1342471, EPI_ISL_1342495, EPI_ISL_1342815, EPI_ISL_1342829, EPI_ISL_1342834, EPI_ISL_1342835, EPI_ISL_1342861, EPI_ISL_1342873, EPI_ISL_1342874, EPI_ISL_1342879, EPI_ISL_1342910, EPI_ISL_1342924, EPI_ISL_1342930, EPI_ISL_1342941, EPI_ISL_1342942, EPI_ISL_1342945, EPI_ISL_1342957, EPI_ISL_1342959, EPI_ISL_1342979, EPI_ISL_1342980, EPI_ISL_1342987, EPI_ISL_1342993, EPI_ISL_1342994, EPI_ISL_1342995, EPI_ISL_1343002, EPI_ISL_1343007, EPI_ISL_1343009, EPI_ISL_1343020, EPI_ISL_1343025, EPI_ISL_1343035, EPI_ISL_1343040, EPI_ISL_1343048, EPI_ISL_1343059, EPI_ISL_1343064, EPI_ISL_1343084, EPI_ISL_1343092, EPI_ISL_1343359, EPI_ISL_1343370, EPI_ISL_1343376, EPI_ISL_1343387, EPI_ISL_1343392, EPI_ISL_1343404, EPI_ISL_1343409, EPI_ISL_1343411, EPI_ISL_1343435, EPI_ISL_1343438, EPI_ISL_1343441, EPI_ISL_1343450, EPI_ISL_1343451, EPI_ISL_1343452, EPI_ISL_1343456, EPI_ISL_1343460, EPI_ISL_1343475, EPI_ISL_1343482, EPI_ISL_1343488, EPI_ISL_1343492, EPI_ISL_1343493, EPI_ISL_1343494, EPI_ISL_1343497, EPI_ISL_1343501, EPI_ISL_1343506, EPI_ISL_1343520, EPI_ISL_1343523, EPI_ISL_1343526, EPI_ISL_1343529, EPI_ISL_1343530, EPI_ISL_1343531, EPI_ISL_1343536, EPI_ISL_1343537, EPI_ISL_1343543, EPI_ISL_1343544, EPI_ISL_1343547, EPI_ISL_1343548, EPI_ISL_1343549, EPI_ISL_1343550, EPI_ISL_1343551, EPI_ISL_1343557, EPI_ISL_1343566, EPI_ISL_1343573, EPI_ISL_1343575, EPI_ISL_1343576, EPI_ISL_1343583, EPI_ISL_1343588, EPI_ISL_1343591, EPI_ISL_1343596, EPI_ISL_1343603, EPI_ISL_1343604, EPI_ISL_1343606, EPI_ISL_1343612, EPI_ISL_1343613, EPI_ISL_1343618, EPI_ISL_1343619, EPI_ISL_1343625, EPI_ISL_1343626, EPI_ISL_1343628, EPI_ISL_1343630, EPI_ISL_1343632, EPI_ISL_1343635, EPI_ISL_1343637, EPI_ISL_1343638, EPI_ISL_1343641, EPI_ISL_1343642, EPI_ISL_1343646, EPI_ISL_1343647, EPI_ISL_1343648, EPI_ISL_1343649, EPI_ISL_1343656, EPI_ISL_1343657, EPI_ISL_1343660, EPI_ISL_1343663, EPI_ISL_1343664, EPI_ISL_1343665, EPI_ISL_1343666, EPI_ISL_1343668, EPI_ISL_1343670, EPI_ISL_1343673, EPI_ISL_1343675, EPI_ISL_1343681, EPI_ISL_1343682, EPI_ISL_1343685, EPI_ISL_1343692, EPI_ISL_1343696, EPI_ISL_1343697, EPI_ISL_1343700, EPI_ISL_1343705, EPI_ISL_1343707, EPI_ISL_1343709, EPI_ISL_1343713, EPI_ISL_1343718, EPI_ISL_1343721, EPI_ISL_1343727, EPI_ISL_1343731, EPI_ISL_1343735, EPI_ISL_1343736, EPI_ISL_1343739, EPI_ISL_1343744, EPI_ISL_1343747, EPI_ISL_1343748, EPI_ISL_1343751, EPI_ISL_1343752, EPI_ISL_1343755, EPI_ISL_1343757, EPI_ISL_1343759, EPI_ISL_1343761, EPI_ISL_1343763, EPI_ISL_1343769, EPI_ISL_1343776, EPI_ISL_1343777, EPI_ISL_1343782, EPI_ISL_1343784, EPI_ISL_1343786, EPI_ISL_1343787, EPI_ISL_1343788, EPI_ISL_1343794, EPI_ISL_1343796, EPI_ISL_1343797, EPI_ISL_1343799, EPI_ISL_1343804, EPI_ISL_1343810, EPI_ISL_1343811, EPI_ISL_1343812, EPI_ISL_1343816, EPI_ISL_1343817, EPI_ISL_1343822, EPI_ISL_1343825, EPI_ISL_1343826, EPI_ISL_1343833, EPI_ISL_1343834, EPI_ISL_1343835, EPI_ISL_1343836, EPI_ISL_1343839, EPI_ISL_1343842, EPI_ISL_1343846, EPI_ISL_1343847, EPI_ISL_1343851, EPI_ISL_1343852, EPI_ISL_1343853, EPI_ISL_1343857, EPI_ISL_1343858, EPI_ISL_1343860, EPI_ISL_1343861, EPI_ISL_1343863, EPI_ISL_1343865, EPI_ISL_1343866, EPI_ISL_1343870, EPI_ISL_1343871, EPI_ISL_1343872, EPI_ISL_1343873, EPI_ISL_1343876, EPI_ISL_1343877, EPI_ISL_1343881, EPI_ISL_1343882, EPI_ISL_1343883, EPI_ISL_1343886, EPI_ISL_1343892, EPI_ISL_1343894, EPI_ISL_1343896, EPI_ISL_1343897, EPI_ISL_1343898, EPI_ISL_1343900, EPI_ISL_1343903, EPI_ISL_1343904, EPI_ISL_1343905, EPI_ISL_1343906, EPI_ISL_1343907, EPI_ISL_1343909, EPI_ISL_1343910, EPI_ISL_1343912, EPI_ISL_1343914, EPI_ISL_1343918, EPI_ISL_1343919, EPI_ISL_1343923, EPI_ISL_1343925, EPI_ISL_1343928, EPI_ISL_1343931, EPI_ISL_1343933, EPI_ISL_1343938, EPI_ISL_1343940, EPI_ISL_1343943, EPI_ISL_1343944, EPI_ISL_1343945, EPI_ISL_1343947, EPI_ISL_1343948, EPI_ISL_1343950, EPI_ISL_1343951, EPI_ISL_1343952, EPI_ISL_1343954, EPI_ISL_1343955, EPI_ISL_1343956, EPI_ISL_1343958, EPI_ISL_1343959, EPI_ISL_1343960, EPI_ISL_1343962, EPI_ISL_1343963, EPI_ISL_1343965, EPI_ISL_1343966, EPI_ISL_1343967, EPI_ISL_1343970, EPI_ISL_1343971, EPI_ISL_1343974, EPI_ISL_1343975, EPI_ISL_1343976, EPI_ISL_1343979, EPI_ISL_1343984, EPI_ISL_1343986, EPI_ISL_1343987, EPI_ISL_1343994, EPI_ISL_1343995, EPI_ISL_1343996, EPI_ISL_1343998, EPI_ISL_1344002, EPI_ISL_1344016, EPI_ISL_1344017, EPI_ISL_1344018, EPI_ISL_1344019, EPI_ISL_1344020, EPI_ISL_1344021, EPI_ISL_1344025, EPI_ISL_1344026, EPI_ISL_1344027,

| | | | |
|---|---|---|---|
| EPI_ISL_1344028, EPI_ISL_1344033, EPI_ISL_1344035, EPI_ISL_1344037, EPI_ISL_1344038, EPI_ISL_1344040, EPI_ISL_1344041, EPI_ISL_1344047, EPI_ISL_1344048, EPI_ISL_1344049, EPI_ISL_1344050, EPI_ISL_1344054, EPI_ISL_1344056, EPI_ISL_1344057, EPI_ISL_1344060, EPI_ISL_1344062, EPI_ISL_1344064, EPI_ISL_1344065, EPI_ISL_1344067, EPI_ISL_1344068, EPI_ISL_1344075, EPI_ISL_1344080, EPI_ISL_1344081, EPI_ISL_1344083, EPI_ISL_1344087, EPI_ISL_1344088, EPI_ISL_1344089, EPI_ISL_1344093, EPI_ISL_1344094, EPI_ISL_1344095, EPI_ISL_1344097, EPI_ISL_1344098, EPI_ISL_1344099, EPI_ISL_1344100, EPI_ISL_1344104, EPI_ISL_1344105, EPI_ISL_1344107, EPI_ISL_1344109, EPI_ISL_1344110, EPI_ISL_1344116, EPI_ISL_1344117, EPI_ISL_1344121, EPI_ISL_1344122, EPI_ISL_1344123, EPI_ISL_1344127, EPI_ISL_1344128, EPI_ISL_1344130, EPI_ISL_1344131, EPI_ISL_1344133, EPI_ISL_1344135, EPI_ISL_1344140, EPI_ISL_1344141, EPI_ISL_1344143, EPI_ISL_1344147, EPI_ISL_1344148, EPI_ISL_1344153, EPI_ISL_1344158, EPI_ISL_1344165, EPI_ISL_1344167, EPI_ISL_1344170, EPI_ISL_1344172, EPI_ISL_1344184, EPI_ISL_1344185, EPI_ISL_1344187, EPI_ISL_1344188, EPI_ISL_1344191, EPI_ISL_1344196, EPI_ISL_1344197, EPI_ISL_1344198, EPI_ISL_1344203, EPI_ISL_1344205, EPI_ISL_1344213, EPI_ISL_1344217, EPI_ISL_1344218, EPI_ISL_1344220, EPI_ISL_1344222, EPI_ISL_1344225, EPI_ISL_1344228, EPI_ISL_1344232, EPI_ISL_1344237, EPI_ISL_1344240, EPI_ISL_1344244, EPI_ISL_1344248, EPI_ISL_1344250, EPI_ISL_1344252, EPI_ISL_1344255, EPI_ISL_1344263, EPI_ISL_1344264, EPI_ISL_1344265, EPI_ISL_1344266, EPI_ISL_1344267, EPI_ISL_1344274, EPI_ISL_1344275, EPI_ISL_1344276, EPI_ISL_1344279, EPI_ISL_1344282, EPI_ISL_1344287, EPI_ISL_1344289, EPI_ISL_1344298, EPI_ISL_1344303, EPI_ISL_1344304, EPI_ISL_1344311, EPI_ISL_1344312, EPI_ISL_1344320, EPI_ISL_1344324, EPI_ISL_1344329, EPI_ISL_1344331, EPI_ISL_1344332, EPI_ISL_1344333, EPI_ISL_1344336, EPI_ISL_1344338, EPI_ISL_1344342, EPI_ISL_1344343, EPI_ISL_1344351, EPI_ISL_1344358, EPI_ISL_1344360, EPI_ISL_1344363, EPI_ISL_1344389, EPI_ISL_1344392, EPI_ISL_1344393, EPI_ISL_1344402, EPI_ISL_1344404, EPI_ISL_1344407, EPI_ISL_1344409, EPI_ISL_1344414, EPI_ISL_1344417, EPI_ISL_1344418, EPI_ISL_1344424, EPI_ISL_1344431, EPI_ISL_1344433, EPI_ISL_1344436, EPI_ISL_1344437, EPI_ISL_1344438, EPI_ISL_1344444, EPI_ISL_1344447, EPI_ISL_1344448, EPI_ISL_1344460, EPI_ISL_1344463, EPI_ISL_1345675, EPI_ISL_1345690, EPI_ISL_1345856, EPI_ISL_1345869, EPI_ISL_1345884, EPI_ISL_1346067, EPI_ISL_1346070, EPI_ISL_1346075, EPI_ISL_1346076, EPI_ISL_1346085, EPI_ISL_1346091, EPI_ISL_1346094, EPI_ISL_1346099, EPI_ISL_1346100, EPI_ISL_1346390, EPI_ISL_1346391, EPI_ISL_1346395, EPI_ISL_1346398, EPI_ISL_1346399, EPI_ISL_1375174, EPI_ISL_1375199, EPI_ISL_1375252, EPI_ISL_1375271, EPI_ISL_1375292, EPI_ISL_1375342, EPI_ISL_1375352, EPI_ISL_1375357, EPI_ISL_1375386, EPI_ISL_1375391, EPI_ISL_1375402, EPI_ISL_1375425, EPI_ISL_1375426, EPI_ISL_1375467, EPI_ISL_1376476, EPI_ISL_1376749, EPI_ISL_1377416, EPI_ISL_1377421, EPI_ISL_1377428, EPI_ISL_1377430, EPI_ISL_1377432, EPI_ISL_1377436, EPI_ISL_1377437, EPI_ISL_1377438, EPI_ISL_1377447, EPI_ISL_1377449, EPI_ISL_1377451, EPI_ISL_1377458, EPI_ISL_1377466, EPI_ISL_1377467, EPI_ISL_1377470, EPI_ISL_1377471, EPI_ISL_1377476, EPI_ISL_1377487, EPI_ISL_1377488, EPI_ISL_1377490, EPI_ISL_1377493, EPI_ISL_1377496, EPI_ISL_1377502, EPI_ISL_1377503, EPI_ISL_1377505, EPI_ISL_1377509, EPI_ISL_1377510, EPI_ISL_1377518, EPI_ISL_1377529, EPI_ISL_1377533, EPI_ISL_1377540, EPI_ISL_1377560, EPI_ISL_1377561, EPI_ISL_1377565, EPI_ISL_1377569, EPI_ISL_1377572, EPI_ISL_1377576, EPI_ISL_1377585, EPI_ISL_1377587, EPI_ISL_1377588, EPI_ISL_1377591, EPI_ISL_1377602, EPI_ISL_1377605, EPI_ISL_1377610, EPI_ISL_1377614, EPI_ISL_1377616, EPI_ISL_1377619, EPI_ISL_1377624, EPI_ISL_1377626, EPI_ISL_1377641, EPI_ISL_1377642, EPI_ISL_1377644, EPI_ISL_1377656, EPI_ISL_1377657, EPI_ISL_1377663, EPI_ISL_1377667, EPI_ISL_1377688, EPI_ISL_1377696, EPI_ISL_1377698, EPI_ISL_1377711, EPI_ISL_1377715, EPI_ISL_1377719, EPI_ISL_1377722, EPI_ISL_1377726, EPI_ISL_1377729, EPI_ISL_1377756, EPI_ISL_1377759, EPI_ISL_1377767, EPI_ISL_1377768, EPI_ISL_1377771, EPI_ISL_1377777, EPI_ISL_1377783, EPI_ISL_1377789, EPI_ISL_1377795, EPI_ISL_1377800, EPI_ISL_1377802, EPI_ISL_1377811, EPI_ISL_1377812, EPI_ISL_1377815, EPI_ISL_1377820, EPI_ISL_1377821, EPI_ISL_1377823, EPI_ISL_1377825, EPI_ISL_1377826, EPI_ISL_1377830, EPI_ISL_1377838, EPI_ISL_1377842, EPI_ISL_1377850, EPI_ISL_1377851, EPI_ISL_1377853, EPI_ISL_1377854, EPI_ISL_1377855, EPI_ISL_1377859, EPI_ISL_1377866, EPI_ISL_1377870, EPI_ISL_1377872, EPI_ISL_1377880, EPI_ISL_1377882, EPI_ISL_1377889, EPI_ISL_1377890, EPI_ISL_1377895, EPI_ISL_1377896, EPI_ISL_1377902, EPI_ISL_1377910, EPI_ISL_1377915, EPI_ISL_1377916, EPI_ISL_1377918, EPI_ISL_1377923, EPI_ISL_1377942, EPI_ISL_1377944, EPI_ISL_1377946, EPI_ISL_1377950, EPI_ISL_1377961, EPI_ISL_1377964, EPI_ISL_1377965, EPI_ISL_1377966, EPI_ISL_1377973, EPI_ISL_1377978, EPI_ISL_1377979, EPI_ISL_1377981, EPI_ISL_1377984, EPI_ISL_1377986, EPI_ISL_1377997, EPI_ISL_1415976, EPI_ISL_1455568, EPI_ISL_1455570, EPI_ISL_1455612, EPI_ISL_1455619, EPI_ISL_1455624, EPI_ISL_1455628, EPI_ISL_1455629 | | | |
| see above | Lighthouse Lab in Milton Keynes | Wellcome Sanger Institute for the COVID-19 Genomics UK (COG-UK) Consortium | Cordelia Langford; David K. Jackson; Dominic Kwiatkowski; Ewan Harrison; Ian Johnston; Jeffrey Barrett; John Sillitoe on behalf of the Wellcome Sanger Institute COVID-19 Surveillance Team; Roberto Amato; Sonia Goncalves; The Lighthouse Lab in Milton Keynes and Alex Alderton |
| EPI_ISL_1475513, EPI_ISL_1475514, EPI_ISL_1475521, EPI_ISL_1475523, EPI_ISL_1475524, EPI_ISL_1475525, EPI_ISL_1475526, EPI_ISL_1475527, EPI_ISL_1475528 | | | |
| see above | Lincolnshire Hospitals and DeepSeq Nottingham | COVID-19 Genomics UK (COG-UK) Consortium | Christopher Moore; Fei Sang; Joeseph Chappell; Johnny Debebe; Jonathan Ball; Matthew Carlisle; Matthew Loose; Nadine Holmes; Nichola Duckworth; Patrick McClure; Sarah Walsh; Tim Sloan; Victoria Wright |
| EPI_ISL_1386759, EPI_ISL_1386761, EPI_ISL_1386764, EPI_ISL_1386766, EPI_ISL_1386767, EPI_ISL_1386771, EPI_ISL_1386773, EPI_ISL_1386775, EPI_ISL_1386807, EPI_ISL_1386808, EPI_ISL_1386809, EPI_ISL_1386810, EPI_ISL_1386811, EPI_ISL_1386812, EPI_ISL_1474726, EPI_ISL_1474727, EPI_ISL_1474728, EPI_ISL_1474730, EPI_ISL_1474731, EPI_ISL_1474747, EPI_ISL_1474750, EPI_ISL_1474751, EPI_ISL_1474769, EPI_ISL_1474770, EPI_ISL_1474772, EPI_ISL_1474779, EPI_ISL_1474781, EPI_ISL_1475751, EPI_ISL_1475752, EPI_ISL_1475757, EPI_ISL_1546153, EPI_ISL_1546222, EPI_ISL_1546224, EPI_ISL_1636827, EPI_ISL_1637251 | | | |
| see above | Liverpool Clinical Laboratories | COVID-19 Genomics UK (COG-UK) Consortium | A Alrezaihi; Alessandro Gerada; Alistair Darby; Angela Cowell; Anita Lucaci; Anu Chawla; Cassie Olateju; Catherine Hartley; Charlotte Nelson; Ecaterina Vamos; Elaine O'Toole; Eleanor G Bentley; Ghada T Shawli; Hermione Webster; Isabel García-Dorival; James Johnson; James P Stewart; Jenifer Manson; Joanne Watts; Jones Benjamin; Jordan J Clark; Julian Hiscox; L Luu; Lucille Rainbow; M Almsaud; Margaret Hughes; Mark Whitehead; Matthew Gemmell; Miren Iturriza-Gomara; Muhannad Alruwaili; N.P Randle; Neil Swainston; PKF Gilmore; Parul Sharma; Rebekah Penrice-Randal; Richard Eccles; Richard Gregory; Sam Haldenby; Steve Paterson; Stuart D Armstrong; Trevor Ian Robinson; Ximeng Han |
| EPI_ISL_1309515, EPI_ISL_1309516, EPI_ISL_1309517, EPI_ISL_1309518, EPI_ISL_1309519, EPI_ISL_1387094, EPI_ISL_1387096, EPI_ISL_1387097, EPI_ISL_1387098, EPI_ISL_1387130, EPI_ISL_1475311, EPI_ISL_1475312, EPI_ISL_1475313, EPI_ISL_1475318, EPI_ISL_1475319, EPI_ISL_1475320, EPI_ISL_1475321, EPI_ISL_1475322, EPI_ISL_1475323, EPI_ISL_1475324, EPI_ISL_1475325, EPI_ISL_1475326, EPI_ISL_1475333, EPI_ISL_1475334, EPI_ISL_1475335, EPI_ISL_1475468, EPI_ISL_1475469, EPI_ISL_1475470, EPI_ISL_1475471 | | | |
| see above | Northumbria University / South Tees Hospitals NHS Foundation Trust / North Cumbria Integrated Care NHS Foundation Trust / North Tees and Hartlepool NHS Foundation Trust / Newcastle Hospitals NHS Foundation Trust | COVID-19 Genomics UK (COG-UK) Consortium | Andrew Nelson; Brendan Payne; Clive Graham; Darren L Smith; Debra Padgett; Edward Barton; Emma Swindells; Garren Scott; Gary Black; Gary Eltringham; Giles S Holt; Greg R Young; Jane Greenaway; Jennifer Collins; John Allan; Joshua Loh; Lynn Dover; Matthew Bashton; Mohammad A Tariq; Paul Baker; Sarah Essex; Steve Liggett; Wen C Yew; Yusri Taha |
| EPI_ISL_1310032, EPI_ISL_1310091, EPI_ISL_1310092, EPI_ISL_1310093, EPI_ISL_1310098, EPI_ISL_1310099, EPI_ISL_1310100, EPI_ISL_1310109, EPI_ISL_1310110, EPI_ISL_1310111, EPI_ISL_1310112, EPI_ISL_1310123, EPI_ISL_1310124, EPI_ISL_1310126, EPI_ISL_1310127, EPI_ISL_1310130, EPI_ISL_1310131, EPI_ISL_1310132, EPI_ISL_1310133, EPI_ISL_1310134, EPI_ISL_1310141, EPI_ISL_1310142, EPI_ISL_1310147, EPI_ISL_1310148, EPI_ISL_1310162, EPI_ISL_1310163, EPI_ISL_1310165, EPI_ISL_1310175, EPI_ISL_1310191, EPI_ISL_1310192, EPI_ISL_1310193, EPI_ISL_1310194, EPI_ISL_1310196, EPI_ISL_1310197, EPI_ISL_1310198, EPI_ISL_1310199, EPI_ISL_1310238, EPI_ISL_1310245, EPI_ISL_1310249, EPI_ISL_1310250, EPI_ISL_1310251, EPI_ISL_1310252, EPI_ISL_1310269, EPI_ISL_1310270, EPI_ISL_1310271, EPI_ISL_1310273, EPI_ISL_1310274, EPI_ISL_1310275, EPI_ISL_1310276, EPI_ISL_1310281, EPI_ISL_1310282, EPI_ISL_1387843, EPI_ISL_1387845, EPI_ISL_1387846, EPI_ISL_1387847, EPI_ISL_1387849, EPI_ISL_1387850, EPI_ISL_1387851, EPI_ISL_1387852, EPI_ISL_1387853, EPI_ISL_1387861, EPI_ISL_1387863, EPI_ISL_1387866, EPI_ISL_1387877, EPI_ISL_1387881, EPI_ISL_1387882, EPI_ISL_1387883, EPI_ISL_1387889, EPI_ISL_1387893, EPI_ISL_1387898, EPI_ISL_1387899, EPI_ISL_1387900, EPI_ISL_1387901, EPI_ISL_1387902, EPI_ISL_1387905, EPI_ISL_1387907, EPI_ISL_1387908, EPI_ISL_1387916, EPI_ISL_1387917, EPI_ISL_1387919, EPI_ISL_1387921, EPI_ISL_1387928, EPI_ISL_1387929, EPI_ISL_1387930, EPI_ISL_1387938, EPI_ISL_1387945, EPI_ISL_1387950, EPI_ISL_1387956, EPI_ISL_1387957, EPI_ISL_1387958, EPI_ISL_1387959, EPI_ISL_1387960, EPI_ISL_1387961, EPI_ISL_1387964, EPI_ISL_1387967, EPI_ISL_1387970, EPI_ISL_1387971, EPI_ISL_1387975, EPI_ISL_1387979, EPI_ISL_1387981, EPI_ISL_1387982, EPI_ISL_1387984, EPI_ISL_1387989, EPI_ISL_1387991, EPI_ISL_1387992, EPI_ISL_1387993, EPI_ISL_1387996, EPI_ISL_1387997, EPI_ISL_1388002, EPI_ISL_1388003, EPI_ISL_1388006, EPI_ISL_1388011, EPI_ISL_1388015, EPI_ISL_1388019, EPI_ISL_1388020, EPI_ISL_1388030, EPI_ISL_1388033, EPI_ISL_1388034, EPI_ISL_1388042, EPI_ISL_1388044, EPI_ISL_1388052, EPI_ISL_1388056, EPI_ISL_1388065, EPI_ISL_1388068, EPI_ISL_1388071, EPI_ISL_1388072, EPI_ISL_1388074, EPI_ISL_1388077, EPI_ISL_1388078, EPI_ISL_1388079, EPI_ISL_1388081, EPI_ISL_1388090, EPI_ISL_1388097, EPI_ISL_1388099, EPI_ISL_1388101, EPI_ISL_1388102, EPI_ISL_1388103, EPI_ISL_1388104, EPI_ISL_1388113, EPI_ISL_1388123, EPI_ISL_1388138, EPI_ISL_1388153, EPI_ISL_1388157, EPI_ISL_1388169, EPI_ISL_1388178, EPI_ISL_1388180, EPI_ISL_1388186, EPI_ISL_1388196, EPI_ISL_1388209, EPI_ISL_1388211, EPI_ISL_1388213, EPI_ISL_1388215, EPI_ISL_1388221, EPI_ISL_1388225, EPI_ISL_1388243, EPI_ISL_1388248, EPI_ISL_1388254, EPI_ISL_1388256, EPI_ISL_1388268, EPI_ISL_1388274, EPI_ISL_1388282, EPI_ISL_1388286, EPI_ISL_1388288, EPI_ISL_1388294, EPI_ISL_1388296, EPI_ISL_1388302, EPI_ISL_1388303, EPI_ISL_1388307, EPI_ISL_1388309, EPI_ISL_1388311, EPI_ISL_1388313, EPI_ISL_1388331, EPI_ISL_1388335, EPI_ISL_1388337, EPI_ISL_1388343, EPI_ISL_1388361, EPI_ISL_1388374, EPI_ISL_1388381, EPI_ISL_1388387, EPI_ISL_1388391, EPI_ISL_1388415, EPI_ISL_1388418, EPI_ISL_1388433, EPI_ISL_1388435, EPI_ISL_1388441, EPI_ISL_1388443, EPI_ISL_1388446, EPI_ISL_1388450, EPI_ISL_1388452, EPI_ISL_1388454, EPI_ISL_1388459, EPI_ISL_1388463, EPI_ISL_1388465, EPI_ISL_1388468, EPI_ISL_1388470, EPI_ISL_1388479, EPI_ISL_1388485, EPI_ISL_1388489, EPI_ISL_1388495, EPI_ISL_1388496, EPI_ISL_1388498, EPI_ISL_1388508, EPI_ISL_1388510, EPI_ISL_1388513, EPI_ISL_1388515, EPI_ISL_1388519, EPI_ISL_1388527, EPI_ISL_1388533, EPI_ISL_1388537, EPI_ISL_1388539, EPI_ISL_1388541, EPI_ISL_1388545, EPI_ISL_1388555, EPI_ISL_1388559, EPI_ISL_1388565, EPI_ISL_1388573, EPI_ISL_1388591, EPI_ISL_1388594, EPI_ISL_1388601, EPI_ISL_1388602, EPI_ISL_1388611, EPI_ISL_1388613, EPI_ISL_1388617, EPI_ISL_1388619, EPI_ISL_1388621, EPI_ISL_1388623, EPI_ISL_1388625, EPI_ISL_1388627, EPI_ISL_1388632, EPI_ISL_1388634, EPI_ISL_1388636, EPI_ISL_1388638, EPI_ISL_1476190, EPI_ISL_1476253, EPI_ISL_1476267, EPI_ISL_1476485, EPI_ISL_1476494, EPI_ISL_1476498, EPI_ISL_1476503, EPI_ISL_1546758, EPI_ISL_1546760, EPI_ISL_1546770, EPI_ISL_1546775, EPI_ISL_1546777, EPI_ISL_1546778, EPI_ISL_1546779, EPI_ISL_1546780, EPI_ISL_1546787, EPI_ISL_1546801 | | | |
| see above | Originating lab: Wales Specialist Virology Centre Sequencing lab: Pathogen Genomics Unit | Public Health Wales Microbiology Cardiff Wales Specialist Virology Centre | Alec Birchley; Alexander Adams; Amy Gaskin; Angela Marchbank; Bree Gatica-Wilcox; Catherine Moore; Jason Coombes; Joanne Watkins; Joel Southgate; Johnathan Evans; Laura Gifford; Lauren Gilbert; Lee Graham; Malorie Perry; Matthew Bull; Nicole Pacchiarini; Sally Corden; Sara Kumziene-Summerhayes; Sara Rey; Sarah Taylor; Simon Cottrell; Sophie Jones; Tom Connor |
| EPI_ISL_1475671, EPI_ISL_1475678, EPI_ISL_1475697, EPI_ISL_1475698, EPI_ISL_1475699, EPI_ISL_1475700, EPI_ISL_1475703, EPI_ISL_1475704, EPI_ISL_1475706, EPI_ISL_1475708, EPI_ISL_1475709, EPI_ISL_1475710, EPI_ISL_1475711, EPI_ISL_1475712, EPI_ISL_1475718, EPI_ISL_1475719, EPI_ISL_1475720, EPI_ISL_1475738, EPI_ISL_1475740, EPI_ISL_1475741, EPI_ISL_1475742, EPI_ISL_1475743, EPI_ISL_1475744, EPI_ISL_1730579 | | | |
| see above | Oxford Viromics, NDM, University of Oxford; Oxford University Hospitals; Basingstoke and North Hampshire Hospital | COVID-19 Genomics UK (COG-UK) Consortium | Alex Mobbs; Amy Trebes; Anita Justice; Catrin Moore; Christophe Fraser; David Bonsall; David Buck; Emma Wise; George Macintyre; Jessica Lynch; John Todd; Mariateresa de Cesare; Matilde Mori; Monique Andersson; Nathan Moore; Nick Cortes; Robert Shaw; Stephen Kidd; Tanya Golubchik; Timothy Peto |
| EPI_ISL_1387137, EPI_ISL_1387138, EPI_ISL_1387144 | Quadram Institute Bioscience | COVID-19 Genomics UK (COG-UK) Consortium | Alexander J Trotter; Alison E. Mather; Alp Aydin; Ana P. Tedim; Anastasia Kolyva; Andrew Bell; Andrew J. Page; Claire Stuart; Dave J. Baker; Gemma L. Kay; John Wain; Justin O'Grady; Leonardo de Oliveira Martins; Lizzie Meadows; Maria Diaz; Mark Webber; Muhammed Yasir; Nabil-Fareed Alikhan; Ngozi Elumogo; Nicholas M. Thomson; Rachael Stanley; Rachel Gilroy; Reenesh Prakash; Samir Dervisevic; Samuel Bloomfield; Steven Rudder; Thanh Le-Viet |
| EPI_ISL_1309724, EPI_ISL_1309725, EPI_ISL_1309726, EPI_ISL_1309727, EPI_ISL_1309730 | Queens Medical Centre, Clinical Microbiology Department / DeepSeq Nottingham | COVID-19 Genomics UK (COG-UK) Consortium | Christopher Moore; Emily Park; Fei Sang; Gemma Clark; Hannah Howson-Wells; Jack Hill; Jayasree Dey; Johnny Debebe; Jonathan Ball; Joseph Chappell; Manjinder Khakh; Matthew Carlisle; Matthew Loose; Michelle M Lister; Nadine Holmes; Patrick McClure; Theocharis Tsoleridis; Timothy Byaruhanga; Vicki M Fleming; Victoria Wright; Wendy Smith |
| EPI_ISL_1344712, EPI_ISL_1344714, EPI_ISL_1344715, EPI_ISL_1344722, EPI_ISL_1344723, EPI_ISL_1344726, EPI_ISL_1344733, EPI_ISL_1344737, EPI_ISL_1344741, EPI_ISL_1344747, EPI_ISL_1344749, EPI_ISL_1344754, EPI_ISL_1344756, EPI_ISL_1344764, EPI_ISL_1344765, EPI_ISL_1344767, EPI_ISL_1344770, EPI_ISL_1344771, EPI_ISL_1344772, EPI_ISL_1344776, EPI_ISL_1344777, EPI_ISL_1344779, EPI_ISL_1344782, EPI_ISL_1344787, EPI_ISL_1344792, EPI_ISL_1344795, EPI_ISL_1344796, EPI_ISL_1344808, EPI_ISL_1344810, EPI_ISL_1344814, EPI_ISL_1344817, EPI_ISL_1344818, EPI_ISL_1344819, EPI_ISL_1344823, EPI_ISL_1344825, EPI_ISL_1344828, EPI_ISL_1344832, EPI_ISL_1344843, EPI_ISL_1344844, EPI_ISL_1344846, EPI_ISL_1344847, EPI_ISL_1344863, EPI_ISL_1344864, EPI_ISL_1344866, EPI_ISL_1344867, EPI_ISL_1344868, EPI_ISL_1344873, EPI_ISL_1344876, EPI_ISL_1344877, EPI_ISL_1344883, EPI_ISL_1344885, EPI_ISL_1344886, EPI_ISL_1344889, EPI_ISL_1344890, EPI_ISL_1344891, EPI_ISL_1344893, EPI_ISL_1344900, EPI_ISL_1344901, EPI_ISL_1344903, EPI_ISL_1344905, EPI_ISL_1344922, EPI_ISL_1344923, EPI_ISL_1376777, EPI_ISL_1376778, EPI_ISL_1376780, EPI_ISL_1376781, EPI_ISL_1376782, EPI_ISL_1376784, EPI_ISL_1376786, EPI_ISL_1376787, EPI_ISL_1376788, EPI_ISL_1376791, EPI_ISL_1376792, EPI_ISL_1376793, EPI_ISL_1376795, EPI_ISL_1376796, EPI_ISL_1376797, EPI_ISL_1376798, EPI_ISL_1376801, EPI_ISL_1376802, EPI_ISL_1376803, EPI_ISL_1376804, EPI_ISL_1376805, EPI_ISL_1376806, EPI_ISL_1376807, EPI_ISL_1376808, EPI_ISL_1376809, EPI_ISL_1376810, EPI_ISL_1376811, EPI_ISL_1376812, EPI_ISL_1376813, EPI_ISL_1376814, EPI_ISL_1376815, EPI_ISL_1376816, EPI_ISL_1376817, EPI_ISL_1376819, EPI_ISL_1376821, EPI_ISL_1376822, EPI_ISL_1376824, EPI_ISL_1376826, EPI_ISL_1376827, EPI_ISL_1376833, EPI_ISL_1376834, EPI_ISL_1376836, EPI_ISL_1376837, EPI_ISL_1376838, EPI_ISL_1376839, EPI_ISL_1376841, EPI_ISL_1376842, EPI_ISL_1376844, EPI_ISL_1376845, EPI_ISL_1376847, EPI_ISL_1376848, EPI_ISL_1376849, EPI_ISL_1376851, EPI_ISL_1376855, EPI_ISL_1376856, EPI_ISL_1376857, EPI_ISL_1376858, EPI_ISL_1376859, EPI_ISL_1376860, EPI_ISL_1376861, EPI_ISL_1376862, EPI_ISL_1376863, EPI_ISL_1376865, EPI_ISL_1376867, EPI_ISL_1376869, EPI_ISL_1376870, EPI_ISL_1376871, EPI_ISL_1376874, EPI_ISL_1376877, EPI_ISL_1376878, EPI_ISL_1376879, EPI_ISL_1376880, EPI_ISL_1376883, EPI_ISL_1376884, EPI_ISL_1376886, EPI_ISL_1376888, EPI_ISL_1376889, EPI_ISL_1376890, EPI_ISL_1376892, EPI_ISL_1376895, EPI_ISL_1376896, EPI_ISL_1376897, EPI_ISL_1376898, EPI_ISL_1376899, EPI_ISL_1376900, EPI_ISL_1376901, EPI_ISL_1376903, EPI_ISL_1376905, EPI_ISL_1376907, EPI_ISL_1376908, EPI_ISL_1376910, EPI_ISL_1376912, EPI_ISL_1376914, EPI_ISL_1376916, EPI_ISL_1376917, EPI_ISL_1376918, EPI_ISL_1376919, EPI_ISL_1376921, EPI_ISL_1376923, EPI_ISL_1376925, EPI_ISL_1376930, EPI_ISL_1376932, EPI_ISL_1376934, EPI_ISL_1376938, EPI_ISL_1376939, EPI_ISL_1376940, EPI_ISL_1376942, EPI_ISL_1376943, EPI_ISL_1376944, EPI_ISL_1376945, EPI_ISL_1376946, EPI_ISL_1376947, EPI_ISL_1376948, EPI_ISL_1376951, EPI_ISL_1376954, EPI_ISL_1376956, EPI_ISL_1376957, EPI_ISL_1376958, EPI_ISL_1376959, EPI_ISL_1376960, EPI_ISL_1376961, EPI_ISL_1376964, EPI_ISL_1376967, EPI_ISL_1411116, EPI_ISL_1411171, EPI_ISL_1411184, EPI_ISL_1411209, EPI_ISL_1411213, EPI_ISL_1411239, EPI_ISL_1411245, EPI_ISL_1411246, EPI_ISL_1411309, EPI_ISL_1411320, EPI_ISL_1411322, EPI_ISL_1411334, EPI_ISL_1411391, EPI_ISL_1411400, EPI_ISL_1411402, EPI_ISL_1411411, EPI_ISL_1411429, EPI_ISL_1412212, EPI_ISL_1412239, EPI_ISL_1412241, EPI_ISL_1412265, EPI_ISL_1412317, EPI_ISL_1412339, EPI_ISL_1412340, EPI_ISL_1412346, EPI_ISL_1412374, EPI_ISL_1412392, EPI_ISL_1412464, EPI_ISL_1412527, EPI_ISL_1412547, EPI_ISL_1455633, EPI_ISL_1455634, EPI_ISL_1455635, EPI_ISL_1455636 | | | |
| see above | Randox Laboratories | Wellcome Sanger Institute for the COVID-19 Genomics UK (COG-UK) Consortium | Cordelia Langford; David K. Jackson; Dominic Kwiatkowski; Ewan Harrison; Ian Johnston; Jeffrey Barrett; John Sillitoe on behalf of the Wellcome Sanger Institute COVID-19 Surveillance Team; Randox Laboratories and Alex Alderton; Roberto Amato; Sonia Goncalves |
| EPI_ISL_1976362, EPI_ISL_1976363, EPI_ISL_1976365, EPI_ISL_1976366, EPI_ISL_1976367, EPI_ISL_1976369, EPI_ISL_1976371, EPI_ISL_1976374, EPI_ISL_1976376, EPI_ISL_1976380, EPI_ISL_1976385, EPI_ISL_1976387, EPI_ISL_1976388, EPI_ISL_1976390, EPI_ISL_1976391, EPI_ISL_1976395, EPI_ISL_1976396, EPI_ISL_1976398, EPI_ISL_1976400, EPI_ISL_1976401, EPI_ISL_1976411, EPI_ISL_1976412, EPI_ISL_1976413, EPI_ISL_1976414, EPI_ISL_1976427, EPI_ISL_1976430, EPI_ISL_1976432, EPI_ISL_1976440, EPI_ISL_1976442, EPI_ISL_1976443, EPI_ISL_1977487, EPI_ISL_1977489, EPI_ISL_1977492, EPI_ISL_1977494, EPI_ISL_1977495, EPI_ISL_1977497, EPI_ISL_1977499, EPI_ISL_1977500, EPI_ISL_1977502, EPI_ISL_1977518, EPI_ISL_1977519, EPI_ISL_1977523, EPI_ISL_1977524, EPI_ISL_1977526, EPI_ISL_1977528, EPI_ISL_1977540, EPI_ISL_1977541, EPI_ISL_1977543, EPI_ISL_1977545, EPI_ISL_1977549, EPI_ISL_1977571, EPI_ISL_1977573, EPI_ISL_1977574, EPI_ISL_1977579, EPI_ISL_1977581, EPI_ISL_1977586, EPI_ISL_1977587, EPI_ISL_1977625, EPI_ISL_1977627, EPI_ISL_1977629, EPI_ISL_1977630, EPI_ISL_1977632, EPI_ISL_1977634, EPI_ISL_1977636, EPI_ISL_1977639, EPI_ISL_1977642, EPI_ISL_1977644, EPI_ISL_1977646, EPI_ISL_1977648, EPI_ISL_1977649, EPI_ISL_1977651, EPI_ISL_1977652, EPI_ISL_1977654, EPI_ISL_1977656, EPI_ISL_1977658, EPI_ISL_1977662, EPI_ISL_1977663, EPI_ISL_1977665, EPI_ISL_1977667, EPI_ISL_1977668, EPI_ISL_1977670, EPI_ISL_1977671, EPI_ISL_1977673, EPI_ISL_1977675, EPI_ISL_1977676, EPI_ISL_1977678, EPI_ISL_1977680, EPI_ISL_1977682, EPI_ISL_1977694, EPI_ISL_1977696, EPI_ISL_1977698, EPI_ISL_1977700, EPI_ISL_1977701, EPI_ISL_1977703, EPI_ISL_1977704, EPI_ISL_1977706, EPI_ISL_1977708, EPI_ISL_1977710, EPI_ISL_1977712, EPI_ISL_1977713, EPI_ISL_1977715, EPI_ISL_1977717, EPI_ISL_1977718, EPI_ISL_1977720, EPI_ISL_1977722, EPI_ISL_1977723, EPI_ISL_1977732, EPI_ISL_1977734, EPI_ISL_1977755, EPI_ISL_1977756, EPI_ISL_1977758, EPI_ISL_1977760, EPI_ISL_1977761, EPI_ISL_1977763, EPI_ISL_1977765, EPI_ISL_1977767, EPI_ISL_1977768, EPI_ISL_1977770, EPI_ISL_1977772, EPI_ISL_1977773, EPI_ISL_1977775, EPI_ISL_1977777, EPI_ISL_1977779, EPI_ISL_1977780, EPI_ISL_1977782, EPI_ISL_1977784, EPI_ISL_1977786, EPI_ISL_1977787, EPI_ISL_1977789, EPI_ISL_1977791, EPI_ISL_1977793, EPI_ISL_1977794, EPI_ISL_1977796, EPI_ISL_1977798, EPI_ISL_1977800, EPI_ISL_1977801, EPI_ISL_1977803, EPI_ISL_1977805, EPI_ISL_1977806, EPI_ISL_1977808, EPI_ISL_1977810, EPI_ISL_1977811, EPI_ISL_1977834, EPI_ISL_1977836, EPI_ISL_1977838, EPI_ISL_1977839, EPI_ISL_1977842, EPI_ISL_1977843, EPI_ISL_1977851, EPI_ISL_1977853, EPI_ISL_1977854, EPI_ISL_1977856, EPI_ISL_1977857, EPI_ISL_1977859, EPI_ISL_1977860, EPI_ISL_1977862, EPI_ISL_1977863, EPI_ISL_1977868, EPI_ISL_1977870, EPI_ISL_1977955, EPI_ISL_1977957, EPI_ISL_1977959, EPI_ISL_1977960, EPI_ISL_1977962, EPI_ISL_1977964, EPI_ISL_1978048, EPI_ISL_1978050, EPI_ISL_1978052, EPI_ISL_1978054, EPI_ISL_1978055, EPI_ISL_1978057, EPI_ISL_1978066, EPI_ISL_1978067, EPI_ISL_1978069, EPI_ISL_1978089 | | | |
| see above | Regional Virus Laboratory, Belfast Health and Social Care Trust | COVID-19 Genomics UK (COG-UK) Consortium | Alison Watt; Ciara Cox; Conall McCaughey; David Simpson; Derek Fairley; James McKenna; Mairead Connor; Susan Feeney; Tanya Curran; Zoltan Molnar |
| EPI_ISL_1637046, EPI_ISL_1637047, EPI_ISL_1637048 | University College London Hospital | COVID-19 Genomics UK (COG-UK) Consortium | Dr Catherine Houlihan; Dr Daniel Frampton; Dr Judith Heaney; Dr Moira Spyer; Dr Paul Grant and Dr Eleni Nastouli; Matthew Byott; Stuart Kirk |
| EPI_ISL_1308953, EPI_ISL_1308954, EPI_ISL_1308955, EPI_ISL_1308956, EPI_ISL_1308958, EPI_ISL_1308959, EPI_ISL_1308967, EPI_ISL_1309071, EPI_ISL_1309131, EPI_ISL_1309132, EPI_ISL_1309133, EPI_ISL_1309135, EPI_ISL_1309138, EPI_ISL_1309141, EPI_ISL_1309142, EPI_ISL_1310702, EPI_ISL_1310703, EPI_ISL_1386846, EPI_ISL_1386847, EPI_ISL_1386852, EPI_ISL_1386868, EPI_ISL_1386870, EPI_ISL_1386871, EPI_ISL_1386882, EPI_ISL_1832647 | | | |
| see above | University College London, Great Ormond Street Hospital for Children NHS Foundation Trust, Imperial College Healthcare NHS Trust | COVID-19 Genomics UK (COG-UK) Consortium | Alison Holmes; Charlotte Williams; Helena Tutill; Jacqueline Findlay; James Price; Judith Breuer; Julianne Brown; Kathryn Harris; Leysa Forrest; Mark Kristiansen; Paola Niola; Paola Resende Silva; Patricia Dyal; Paul Randell; Rachel Williams; Samuel Weeks; Sergi Castellano; Sunando Roy; Tony Brooks; Yasmin Panchbhaya |
| EPI_ISL_1474648 | University Hospitals Of Leicester NHS Trust and DeepSeq Nottingham | COVID-19 Genomics UK (COG-UK) Consortium | Christopher Holmes; Christopher Moore; Fei Sang; Joseph Chappell; Johnny Debebe; Jonathan Ball; Julian Tang; Karlie Fallon; Matthew Carlisle; Matthew Loose; Nadine Holmes; Patrick McClure; Paul Bird; Thomas Helmer; Victoria Wright |
| EPI_ISL_1308471, EPI_ISL_1308474, EPI_ISL_1308477, EPI_ISL_1308481, EPI_ISL_1308484, EPI_ISL_1308485, EPI_ISL_1308511, EPI_ISL_1308512, EPI_ISL_1308513, EPI_ISL_1308516, EPI_ISL_1308517, EPI_ISL_1308518, EPI_ISL_1308523, EPI_ISL_1308524, EPI_ISL_1308525, EPI_ISL_1308533, EPI_ISL_1386480, EPI_ISL_1386481, EPI_ISL_1386482, EPI_ISL_1386483, EPI_ISL_1386484, EPI_ISL_1386485, EPI_ISL_1386486, EPI_ISL_1386510, EPI_ISL_1386516, EPI_ISL_1386517, EPI_ISL_1386518, EPI_ISL_1386540, EPI_ISL_1386541, EPI_ISL_1474411, EPI_ISL_1474414, EPI_ISL_1474419, EPI_ISL_1474456, EPI_ISL_1474459, EPI_ISL_1474487 | | | |
| see above | University of Birmingham | COVID-19 Genomics UK (COG-UK) Consortium | Alex Richter; Andrew Bosworth; Andrew Bosworth. Queen Elizabeth Hospital: Anna Casey; Andrew D Beggs PHE Heartlands Lab: Husam Osman; Charlotte Poxon; Institute of Microbiology; Joanne Stockton; Josh Quick; Kasun Wanigasooriya; Mike Kidd; Nicholas Loman. University of Birmingham Testing Laboratory: Celina M Whalley; Oliver Pickles; Radoslaw Poplawski; Samuel Nicholls; University of Birmingham: Claire McMurray; Will Rowe |
| EPI_ISL_1474611, EPI_ISL_1474612, EPI_ISL_1545823, EPI_ISL_1545825, EPI_ISL_1545829, EPI_ISL_1545830, EPI_ISL_1545837, EPI_ISL_1545844, EPI_ISL_1545946, EPI_ISL_1545947, EPI_ISL_1730163, EPI_ISL_1730164, EPI_ISL_1730165 | | | |
| see above | University of Exeter | COVID-19 Genomics UK (COG-UK) Consortium | Aaron Jeffries; Audrey Farbos; Ben Temperton; Jane Masoli; Joanna Warwick-Dugdale; Michelle Michelsen; Robyn Manley; Stephen Michell |
| EPI_ISL_1308861, EPI_ISL_1308862, EPI_ISL_1308863, EPI_ISL_1386672, EPI_ISL_1386673, EPI_ISL_1386692, EPI_ISL_1386693, EPI_ISL_1386697, EPI_ISL_1386703, EPI_ISL_1545900, EPI_ISL_1545905, EPI_ISL_1545906, EPI_ISL_1545924, EPI_ISL_1545925, EPI_ISL_1636695, EPI_ISL_1636709, EPI_ISL_1636710, EPI_ISL_1730090, EPI_ISL_1730099, EPI_ISL_1730101, EPI_ISL_1730103, EPI_ISL_1730105, EPI_ISL_1832265, EPI_ISL_1832266, EPI_ISL_1832267, EPI_ISL_1832268, EPI_ISL_1832269, EPI_ISL_1832271, EPI_ISL_2125739 | | | |

see above | Virology Department, Royal Infirmary of Edinburgh, NHS Lothian / School of Biological Sciences, University of Edinburgh | COVID-19 Genomics UK (COG-UK) Consortium | Colquhoun R; Cotton S; Dewar R; Gallagher A; Hill V; Jackson B; Maloney D; McCrone JT; McHugh M; O'Toole Á; Rambaut A; Rooke S; Scher E; Templeton K; Yu X

EPI_ISL_1310709, EPI_ISL_1310710, EPI_ISL_1310712, EPI_ISL_1310716, EPI_ISL_1310718, EPI_ISL_1310723, EPI_ISL_1310728, EPI_ISL_1310730, EPI_ISL_1310732, EPI_ISL_1310748, EPI_ISL_1310749, EPI_ISL_1310751, EPI_ISL_1310754, EPI_ISL_1310758, EPI_ISL_1310762, EPI_ISL_1310766, EPI_ISL_1310767, EPI_ISL_1310769, EPI_ISL_1310773, EPI_ISL_1310775, EPI_ISL_1310777, EPI_ISL_1389084, EPI_ISL_1389086, EPI_ISL_1389098, EPI_ISL_1389102, EPI_ISL_1389110, EPI_ISL_1389132, EPI_ISL_1833871

see above | Virology Department, Sheffield Teaching Hospitals NHS Foundation Trust/Department of Infection, Immunity and Cardiovascular Disease, The Medical School, University of Sheffield | COVID-19 Genomics UK (COG-UK) Consortium | Adri Angyal; Alex Keeley; Benjamin Lindsey; Cariad Evans; Danielle Groves; Dave Partridge; Katie Johnson; Laura Carrilero; Luke Green; Matthew Parker; Matthew Wyles; Mehmet Yavuz; Mohammad Raza; Nikki Smith; Paul Parsons; Rachel Tucker; Rebecca Brown; Thushan de Silva

EPI_ISL_1310272, EPI_ISL_1387842, EPI_ISL_1387844, EPI_ISL_1387848, EPI_ISL_1387854, EPI_ISL_1387865, EPI_ISL_1387867, EPI_ISL_1387868, EPI_ISL_1387869, EPI_ISL_1387870, EPI_ISL_1387871, EPI_ISL_1387878, EPI_ISL_1387880, EPI_ISL_1387888, EPI_ISL_1387890, EPI_ISL_1387894, EPI_ISL_1387897, EPI_ISL_1387906, EPI_ISL_1387909, EPI_ISL_1387910, EPI_ISL_1387911, EPI_ISL_1387912, EPI_ISL_1387915, EPI_ISL_1387920, EPI_ISL_1387924, EPI_ISL_1387935, EPI_ISL_1387936, EPI_ISL_1387937, EPI_ISL_1387939, EPI_ISL_1387949, EPI_ISL_1387951, EPI_ISL_1387952, EPI_ISL_1387966, EPI_ISL_1387977, EPI_ISL_1387980, EPI_ISL_1387985, EPI_ISL_1387986, EPI_ISL_1387987, EPI_ISL_1387990, EPI_ISL_1388012, EPI_ISL_1388038, EPI_ISL_1388043, EPI_ISL_1388069, EPI_ISL_1388073, EPI_ISL_1388083, EPI_ISL_1388091, EPI_ISL_1388117, EPI_ISL_1388144, EPI_ISL_1388182, EPI_ISL_1388184, EPI_ISL_1388198, EPI_ISL_1388200, EPI_ISL_1388227, EPI_ISL_1388233, EPI_ISL_1388239, EPI_ISL_1388245, EPI_ISL_1388252, EPI_ISL_1388272, EPI_ISL_1388276, EPI_ISL_1388278, EPI_ISL_1388280, EPI_ISL_1388290, EPI_ISL_1388292, EPI_ISL_1388298, EPI_ISL_1388299, EPI_ISL_1388305, EPI_ISL_1388319, EPI_ISL_1388323, EPI_ISL_1388333, EPI_ISL_1388339, EPI_ISL_1388341, EPI_ISL_1388349, EPI_ISL_1388365, EPI_ISL_1388389, EPI_ISL_1388398, EPI_ISL_1388401, EPI_ISL_1388403, EPI_ISL_1388405, EPI_ISL_1388416, EPI_ISL_1388422, EPI_ISL_1388437, EPI_ISL_1388481, EPI_ISL_1388483, EPI_ISL_1388517, EPI_ISL_1388571, EPI_ISL_1388586, EPI_ISL_1476497, EPI_ISL_1546767, EPI_ISL_1546781, EPI_ISL_1637285, EPI_ISL_1637287, EPI_ISL_1637294, EPI_ISL_1637298, EPI_ISL_1637302, EPI_ISL_1637306, EPI_ISL_1637866, EPI_ISL_1637867, EPI_ISL_1637868, EPI_ISL_1637886, EPI_ISL_1637902, EPI_ISL_1637904, EPI_ISL_1637906, EPI_ISL_1637908, EPI_ISL_1637909, EPI_ISL_1637912, EPI_ISL_1637917, EPI_ISL_1637918, EPI_ISL_1637920, EPI_ISL_1637921, EPI_ISL_1637922, EPI_ISL_1637927, EPI_ISL_1637928, EPI_ISL_1637930, EPI_ISL_1637931, EPI_ISL_1637932, EPI_ISL_1637935, EPI_ISL_1637936, EPI_ISL_1637938, EPI_ISL_1637940, EPI_ISL_1637942, EPI_ISL_1637944, EPI_ISL_1637946, EPI_ISL_1637947, EPI_ISL_1637948, EPI_ISL_1637949, EPI_ISL_1637950, EPI_ISL_1637951, EPI_ISL_1637954, EPI_ISL_1637956, EPI_ISL_1637958, EPI_ISL_1637959, EPI_ISL_1637960, EPI_ISL_1637961, EPI_ISL_1637962, EPI_ISL_1637963, EPI_ISL_1637964, EPI_ISL_1637965, EPI_ISL_1637966, EPI_ISL_1637967, EPI_ISL_1637968, EPI_ISL_1637971

see above | Wales Specialist Virology Centre Sequencing lab: Pathogen Genomics Unit | Public Health Wales Microbiology Cardiff Wales Specialist Virology Centre | Alec Birchley; Alexander Adams; Amy Gaskin; Angela Marchbank; Bree Gatica-Wilcox; Catherine Moore; Jason Coombes; Joanne Watkins; Joel Southgate; Johnathan Evans; Laura Gifford; Lauren Gilbert; Lee Graham; Malorie Perry; Matthew Bull; Nicole Pacchiarini; Sally Corden; Sara Kumziene-Summerhayes; Sara Rey; Sarah Taylor; Simon Cottrell; Sophie Jones; Tom Connor

EPI_ISL_1386604, EPI_ISL_1386605, EPI_ISL_1545857, EPI_ISL_1545894, EPI_ISL_1545895, EPI_ISL_1636639, EPI_ISL_1636641, EPI_ISL_1636642, EPI_ISL_1636643, EPI_ISL_1636645, EPI_ISL_1636646, EPI_ISL_1636648, EPI_ISL_1636649, EPI_ISL_1636651, EPI_ISL_1636654

see above | West of Scotland Specialist Virology Centre, NHSGGC / MRC-University of Glasgow Centre for Virus Research | COVID-19 Genomics UK (COG-UK) Consortium | Alasdair MacLean; Alice Broos; Ana da Silva Filipe; Antonia Ho; Daniel Mair; David L Robertson; Emma Thomson; Guy Mollett; James Shepherd; Jenna Nichols; Joseph Hughes; Kathy Li; Kathy Smollett; Kyriaki Nomikou; Lily Tong; Matthew Holden; Natasha Johnson; Rachel Blacow; Richard Orton; Rory Gunson; Sarah McDonald; Sharif Shaaban; Sreenu Vattipally; Stephen Carmichael

We gratefully acknowledge the following Authors from the Originating laboratories responsible for obtaining the specimens, as well as the Submitting laboratories where the genome data were generated and shared via GISAID, on which this research is based.

All Submitters of data may be contacted directly via www.gisaid.org

Authors are sorted alphabetically.

| Accession ID | Originating Laboratory | Submitting Laboratory | Authors |
|---|---|---|---|
| EPI_ISL_1474827, EPI_ISL_1730316 | Barts Health NHS Trust | COVID-19 Genomics UK (COG-UK) Consortium | BROAD; Beatrix; CUTINO-MOGUEL; Claire; David; Dola; HARRINGTON; KELE; KULASEGARAN-SHYLINI; Maria-Teresa; OWOYEMI; Raghavendran |
| EPI_ISL_1389140, EPI_ISL_1389146 | Bioinformatics and Biostatistics Lab, Advanced Sequencing Facility | COVID-19 Genomics UK (COG-UK) Consortium | Aengus Stewart; Chelsea Sawyer; Harshil Patel; Jerome Nicod; Laura Cubitt; Margaret Crawford |
| EPI_ISL_1388792, EPI_ISL_1388794, EPI_ISL_1388796, EPI_ISL_1388800, EPI_ISL_1388917, EPI_ISL_1388919, EPI_ISL_1388921, EPI_ISL_1388929, EPI_ISL_1476678, EPI_ISL_1638004, EPI_ISL_1638005, EPI_ISL_1638006, EPI_ISL_1638007, EPI_ISL_1638008, EPI_ISL_1638010 | | | |
| see above | Centre for Enzyme Innovation, University of Portsmouth / Translational Research Laboratory, Portsmouth Hospitals NHS Trust | COVID-19 Genomics UK (COG-UK) Consortium | Angela Beckett; Christopher Fearn; Kate Cook; Katie Loveson; Salman Goudarzi; Samuel Robson; Scott Elliott; Sharon Glaysher |
| EPI_ISL_1308700, EPI_ISL_1308701, EPI_ISL_1308702, EPI_ISL_1386574, EPI_ISL_1386577, EPI_ISL_1386578, EPI_ISL_1386579, EPI_ISL_1386583, EPI_ISL_1386586, EPI_ISL_1386588, EPI_ISL_1474564, EPI_ISL_1474565, EPI_ISL_1474566, EPI_ISL_1474567, EPI_ISL_1474568, EPI_ISL_1474569, EPI_ISL_1636606, EPI_ISL_1730026 | | | |
| see above | Department of Pathology, University of Cambridge | COVID-19 Genomics UK (COG-UK) Consortium | Aminu S. Jahun; Ian Goodfellow; Iliana Georgana; Martin D. Curran; Myra Hosmillo; Rhys Izuagbe; Surendra Parmar; William L. Hamilton; Yasmin Chaudhry |
| EPI_ISL_1333832, EPI_ISL_1342240, EPI_ISL_1342253, EPI_ISL_1342266, EPI_ISL_1342272, EPI_ISL_1342273, EPI_ISL_1342284, EPI_ISL_1342291, EPI_ISL_1342294, EPI_ISL_1342296, EPI_ISL_1342313, EPI_ISL_1342316, EPI_ISL_1342318, EPI_ISL_1342332, EPI_ISL_1342337, EPI_ISL_1342339, EPI_ISL_1342345, EPI_ISL_1342347, EPI_ISL_1342353, EPI_ISL_1342361, EPI_ISL_1342369, EPI_ISL_1342372, EPI_ISL_1342376, EPI_ISL_1342378, EPI_ISL_1342380, EPI_ISL_1342386, EPI_ISL_1342388, EPI_ISL_1342390, EPI_ISL_1342398, EPI_ISL_1342403, EPI_ISL_1342409, EPI_ISL_1342411, EPI_ISL_1342422, EPI_ISL_1342443, EPI_ISL_1342449, EPI_ISL_1342451, EPI_ISL_1342460, EPI_ISL_1342468, EPI_ISL_1342481, EPI_ISL_1342492, EPI_ISL_1342501, EPI_ISL_1342507, EPI_ISL_1342513, EPI_ISL_1342526, EPI_ISL_1342548, EPI_ISL_1342561, EPI_ISL_1342562, EPI_ISL_1342604, EPI_ISL_1342613, EPI_ISL_1342659, EPI_ISL_1342666, EPI_ISL_1342678, EPI_ISL_1342680, EPI_ISL_1342689, EPI_ISL_1342701, EPI_ISL_1342702, EPI_ISL_1342712, EPI_ISL_1342733, EPI_ISL_1342759, EPI_ISL_1342771, EPI_ISL_1342773, EPI_ISL_1342786, EPI_ISL_1342788, EPI_ISL_1342814, EPI_ISL_1342816, EPI_ISL_1342821, EPI_ISL_1342823, EPI_ISL_1342824, EPI_ISL_1342825, EPI_ISL_1342826, EPI_ISL_1342828, EPI_ISL_1342837, EPI_ISL_1342838, EPI_ISL_1342840, EPI_ISL_1342842, EPI_ISL_1342843, EPI_ISL_1342846, EPI_ISL_1342849, EPI_ISL_1342852, EPI_ISL_1342853, EPI_ISL_1342854, EPI_ISL_1342856, EPI_ISL_1342858, EPI_ISL_1342859, EPI_ISL_1342860, EPI_ISL_1342862, EPI_ISL_1342863, EPI_ISL_1342867, EPI_ISL_1342872, EPI_ISL_1342875, EPI_ISL_1342877, EPI_ISL_1342878, EPI_ISL_1342880, EPI_ISL_1342882, EPI_ISL_1342883, EPI_ISL_1342884, EPI_ISL_1342886, EPI_ISL_1342887, EPI_ISL_1342889, EPI_ISL_1342890, EPI_ISL_1342892, EPI_ISL_1342894, EPI_ISL_1342895, EPI_ISL_1342897, EPI_ISL_1342899, EPI_ISL_1342901, EPI_ISL_1342904, EPI_ISL_1342905, EPI_ISL_1342908, EPI_ISL_1342909, EPI_ISL_1342911, EPI_ISL_1342912, EPI_ISL_1342913, EPI_ISL_1342915, EPI_ISL_1342916, EPI_ISL_1342920, EPI_ISL_1342925, EPI_ISL_1342927, EPI_ISL_1342928, EPI_ISL_1342929, EPI_ISL_1342931, EPI_ISL_1342934, EPI_ISL_1342938, EPI_ISL_1342939, EPI_ISL_1342940, EPI_ISL_1342943, EPI_ISL_1342944, EPI_ISL_1342947, EPI_ISL_1342949, EPI_ISL_1342951, EPI_ISL_1342953, EPI_ISL_1342956, EPI_ISL_1342958, EPI_ISL_1342961, EPI_ISL_1342962, EPI_ISL_1342964, EPI_ISL_1342966, EPI_ISL_1342967, EPI_ISL_1342968, EPI_ISL_1342973, EPI_ISL_1342974, EPI_ISL_1342975, EPI_ISL_1342976, EPI_ISL_1342977, EPI_ISL_1342983, EPI_ISL_1342986, EPI_ISL_1342988, EPI_ISL_1342989, EPI_ISL_1342996, EPI_ISL_1343000, EPI_ISL_1343001, EPI_ISL_1343003, EPI_ISL_1343005, EPI_ISL_1343011, EPI_ISL_1343012, EPI_ISL_1343013, EPI_ISL_1343015, EPI_ISL_1343017, EPI_ISL_1343018, EPI_ISL_1343019, EPI_ISL_1343021, EPI_ISL_1343022, EPI_ISL_1343024, EPI_ISL_1343027, EPI_ISL_1343037, EPI_ISL_1343038, EPI_ISL_1343039, EPI_ISL_1343041, EPI_ISL_1343045, EPI_ISL_1343047, EPI_ISL_1343049, EPI_ISL_1343051, EPI_ISL_1343052, EPI_ISL_1343056, EPI_ISL_1343057, EPI_ISL_1343058, EPI_ISL_1343063, EPI_ISL_1343066, EPI_ISL_1343067, EPI_ISL_1343068, EPI_ISL_1343070, EPI_ISL_1343071, EPI_ISL_1343075, EPI_ISL_1343077, EPI_ISL_1343078, EPI_ISL_1343080, EPI_ISL_1343082, EPI_ISL_1343086, EPI_ISL_1343088, EPI_ISL_1343094, EPI_ISL_1343096, EPI_ISL_1343100, EPI_ISL_1343103, EPI_ISL_1343105, EPI_ISL_1343113, EPI_ISL_1343125, EPI_ISL_1343127, EPI_ISL_1343128, EPI_ISL_1343129, EPI_ISL_1343131, EPI_ISL_1343133, EPI_ISL_1343136, EPI_ISL_1343140, EPI_ISL_1343143, EPI_ISL_1343144, EPI_ISL_1343145, EPI_ISL_1343146, EPI_ISL_1343148, EPI_ISL_1343149, EPI_ISL_1343151, EPI_ISL_1343153, EPI_ISL_1343157, EPI_ISL_1343158, EPI_ISL_1343160, EPI_ISL_1343163, EPI_ISL_1343175, EPI_ISL_1343179, EPI_ISL_1343181, EPI_ISL_1343188, EPI_ISL_1343190, EPI_ISL_1343191, EPI_ISL_1343193, EPI_ISL_1343195, EPI_ISL_1343197, EPI_ISL_1343198, EPI_ISL_1343200, EPI_ISL_1343207, EPI_ISL_1343209, EPI_ISL_1343211, EPI_ISL_1343220, EPI_ISL_1343227, EPI_ISL_1343228, EPI_ISL_1343229, EPI_ISL_1343232, EPI_ISL_1343233, EPI_ISL_1343235, EPI_ISL_1343240, EPI_ISL_1343241, EPI_ISL_1343245, EPI_ISL_1343246, EPI_ISL_1343250, EPI_ISL_1343253, EPI_ISL_1343258, EPI_ISL_1343259, EPI_ISL_1343263, EPI_ISL_1343265, EPI_ISL_1343268, EPI_ISL_1343269, EPI_ISL_1343272, EPI_ISL_1343283, EPI_ISL_1343288, EPI_ISL_1343289, EPI_ISL_1343294, EPI_ISL_1343301, EPI_ISL_1343304, EPI_ISL_1343308, EPI_ISL_1343309, EPI_ISL_1343310, EPI_ISL_1343312, EPI_ISL_1343313, EPI_ISL_1343314, EPI_ISL_1343315, EPI_ISL_1343321, EPI_ISL_1343325, EPI_ISL_1343326, EPI_ISL_1343327, EPI_ISL_1343334, EPI_ISL_1343336, EPI_ISL_1343341, EPI_ISL_1343345, EPI_ISL_1343347, EPI_ISL_1343353, EPI_ISL_1345297, EPI_ISL_1345298, EPI_ISL_1345301, EPI_ISL_1345305, EPI_ISL_1345316, EPI_ISL_1345318, EPI_ISL_1345377, EPI_ISL_1345382, EPI_ISL_1345388, EPI_ISL_1345389, EPI_ISL_1345404, EPI_ISL_1345413, EPI_ISL_1345416, EPI_ISL_1345417, EPI_ISL_1345433, EPI_ISL_1345435, EPI_ISL_1345436, EPI_ISL_1345450, EPI_ISL_1345452, EPI_ISL_1345454, EPI_ISL_1345456, EPI_ISL_1345460, EPI_ISL_1345463, EPI_ISL_1345465, EPI_ISL_1345478, EPI_ISL_1345483, EPI_ISL_1345484, EPI_ISL_1345491, EPI_ISL_1345493, EPI_ISL_1345495, EPI_ISL_1345512, EPI_ISL_1345516, EPI_ISL_1345521, EPI_ISL_1345530, EPI_ISL_1345545, EPI_ISL_1345546, EPI_ISL_1345551, EPI_ISL_1345566, EPI_ISL_1345567, EPI_ISL_1345568, EPI_ISL_1345640, EPI_ISL_1375166, EPI_ISL_1375172, EPI_ISL_1375173, EPI_ISL_1375175, EPI_ISL_1375176, EPI_ISL_1375181, EPI_ISL_1375184, EPI_ISL_1375186, EPI_ISL_1375188, EPI_ISL_1375190, EPI_ISL_1375191, EPI_ISL_1375192, EPI_ISL_1375194, EPI_ISL_1375196, EPI_ISL_1375197, EPI_ISL_1375203, EPI_ISL_1375209, EPI_ISL_1375211, EPI_ISL_1375212, EPI_ISL_1375214, EPI_ISL_1375222, EPI_ISL_1375223, EPI_ISL_1375229, EPI_ISL_1375237, EPI_ISL_1375244, EPI_ISL_1375247, EPI_ISL_1375248, EPI_ISL_1375250, EPI_ISL_1375253, EPI_ISL_1375256, EPI_ISL_1375265, EPI_ISL_1375274, EPI_ISL_1375276, EPI_ISL_1375277, EPI_ISL_1375295, EPI_ISL_1375298, EPI_ISL_1375301, EPI_ISL_1375306, EPI_ISL_1375307, EPI_ISL_1375310, EPI_ISL_1375316, EPI_ISL_1375317, EPI_ISL_1375320, EPI_ISL_1375322, EPI_ISL_1375326, EPI_ISL_1375328, EPI_ISL_1375331, EPI_ISL_1375332, EPI_ISL_1375333, EPI_ISL_1375335, EPI_ISL_1375337, EPI_ISL_1375339, EPI_ISL_1375343, EPI_ISL_1375344, EPI_ISL_1375347, EPI_ISL_1375349, EPI_ISL_1375350, EPI_ISL_1375351, EPI_ISL_1375353, EPI_ISL_1375354, EPI_ISL_1375365, EPI_ISL_1375366, EPI_ISL_1375372, EPI_ISL_1375376, EPI_ISL_1375383, EPI_ISL_1375385, EPI_ISL_1375388, EPI_ISL_1375395, EPI_ISL_1375396, EPI_ISL_1375399, EPI_ISL_1375401, EPI_ISL_1375408, EPI_ISL_1375414, EPI_ISL_1375417, EPI_ISL_1375422, EPI_ISL_1375423, EPI_ISL_1375429, EPI_ISL_1375433, EPI_ISL_1375445, EPI_ISL_1375456, EPI_ISL_1375458, EPI_ISL_1375459, EPI_ISL_1375461, EPI_ISL_1375462, EPI_ISL_1375464, EPI_ISL_1375465, EPI_ISL_1375466, EPI_ISL_1375470, EPI_ISL_1375471, EPI_ISL_1375472, EPI_ISL_1375475, EPI_ISL_1375476, EPI_ISL_1375477, EPI_ISL_1375478, EPI_ISL_1375479, EPI_ISL_1375480, EPI_ISL_1375482, EPI_ISL_1375483, EPI_ISL_1375485, EPI_ISL_1375486, EPI_ISL_1375488, EPI_ISL_1375490, EPI_ISL_1375491, EPI_ISL_1375493, EPI_ISL_1375494, EPI_ISL_1375495, EPI_ISL_1375496, EPI_ISL_1375497, EPI_ISL_1375498, EPI_ISL_1375499, EPI_ISL_1375501, EPI_ISL_1375502, EPI_ISL_1375503, EPI_ISL_1375504, EPI_ISL_1375505, EPI_ISL_1375506, EPI_ISL_1375509, EPI_ISL_1375511, EPI_ISL_1375512, EPI_ISL_1375514, EPI_ISL_1375515, EPI_ISL_1375516, EPI_ISL_1375517, EPI_ISL_1375518, EPI_ISL_1375519, EPI_ISL_1375521, EPI_ISL_1375522, EPI_ISL_1375523, EPI_ISL_1375525, EPI_ISL_1375526, EPI_ISL_1375528, EPI_ISL_1375529, EPI_ISL_1375530, EPI_ISL_1375531, EPI_ISL_1375535, EPI_ISL_1375536, EPI_ISL_1375537, EPI_ISL_1375538, EPI_ISL_1375539, EPI_ISL_1375542, EPI_ISL_1375543, EPI_ISL_1375544, EPI_ISL_1375548, EPI_ISL_1375549, EPI_ISL_1375550, EPI_ISL_1375552, EPI_ISL_1375554, EPI_ISL_1375555, EPI_ISL_1375556, EPI_ISL_1375559, EPI_ISL_1375560, EPI_ISL_1375561, EPI_ISL_1375563, EPI_ISL_1375571, EPI_ISL_1375573, EPI_ISL_1375575, EPI_ISL_1375577, EPI_ISL_1375579, EPI_ISL_1375580, EPI_ISL_1375581, EPI_ISL_1375584, EPI_ISL_1375585, EPI_ISL_1375587, EPI_ISL_1375589, EPI_ISL_1375593, EPI_ISL_1375595, EPI_ISL_1375596, EPI_ISL_1375598, EPI_ISL_1375599, EPI_ISL_1375600, EPI_ISL_1375602, EPI_ISL_1375604, EPI_ISL_1375606, EPI_ISL_1375607, EPI_ISL_1375608, EPI_ISL_1375609, EPI_ISL_1375610, EPI_ISL_1375611, EPI_ISL_1375613, EPI_ISL_1375615, EPI_ISL_1375616, EPI_ISL_1375617, EPI_ISL_1375618, EPI_ISL_1375619, EPI_ISL_1375622, EPI_ISL_1375623, EPI_ISL_1375624, EPI_ISL_1375627, EPI_ISL_1375628, EPI_ISL_1375629, EPI_ISL_1375630, EPI_ISL_1375631, EPI_ISL_1375633, EPI_ISL_1375634, EPI_ISL_1375635, EPI_ISL_1375637, EPI_ISL_1375638, EPI_ISL_1375639, EPI_ISL_1375640, EPI_ISL_1375642, EPI_ISL_1375644, EPI_ISL_1375645, EPI_ISL_1375647, EPI_ISL_1375648, EPI_ISL_1375649, EPI_ISL_1375650, EPI_ISL_1375651, EPI_ISL_1375652, EPI_ISL_1375653, EPI_ISL_1375654, EPI_ISL_1375656, EPI_ISL_1375657, EPI_ISL_1375658, EPI_ISL_1375659, EPI_ISL_1375660, EPI_ISL_1375662, EPI_ISL_1375663, EPI_ISL_1375664, EPI_ISL_1375668, EPI_ISL_1375669, EPI_ISL_1375670, EPI_ISL_1375671, EPI_ISL_1375672, EPI_ISL_1375674, EPI_ISL_1375675, EPI_ISL_1375676, EPI_ISL_1375677, EPI_ISL_1375679, EPI_ISL_1375680, EPI_ISL_1375682, EPI_ISL_1375684, EPI_ISL_1375685, EPI_ISL_1375687, EPI_ISL_1375688, EPI_ISL_1375689, EPI_ISL_1375692, EPI_ISL_1375693, EPI_ISL_1375694, EPI_ISL_1375695, EPI_ISL_1375696, EPI_ISL_1375697, EPI_ISL_1375699, EPI_ISL_1375702, EPI_ISL_1375703, EPI_ISL_1375708, EPI_ISL_1375709, EPI_ISL_1375710, EPI_ISL_1375711, EPI_ISL_1375713, EPI_ISL_1375714, EPI_ISL_1375715, EPI_ISL_1375717, EPI_ISL_1375718, EPI_ISL_1375722, EPI_ISL_1375723, EPI_ISL_1375724, EPI_ISL_1375725, EPI_ISL_1375727, EPI_ISL_1375728, EPI_ISL_1375731, EPI_ISL_1375732, EPI_ISL_1375733, EPI_ISL_1375734, EPI_ISL_1375735, EPI_ISL_1375736, EPI_ISL_1375737, EPI_ISL_1375738, EPI_ISL_1375739, EPI_ISL_1375742, EPI_ISL_1375744, EPI_ISL_1375746, EPI_ISL_1375747, EPI_ISL_1375748, EPI_ISL_1375749, EPI_ISL_1375750, EPI_ISL_1375751, EPI_ISL_1375752, EPI_ISL_1375753, EPI_ISL_1375754, EPI_ISL_1375756, EPI_ISL_1375757, EPI_ISL_1375758, EPI_ISL_1375759, EPI_ISL_1375761, EPI_ISL_1375763, EPI_ISL_1376224, EPI_ISL_1376226, EPI_ISL_1376228, EPI_ISL_1376232, EPI_ISL_1376237, EPI_ISL_1376238, EPI_ISL_1376240, EPI_ISL_1376241, EPI_ISL_1376242, EPI_ISL_1376243, EPI_ISL_1376246, EPI_ISL_1376250, EPI_ISL_1376253, EPI_ISL_1376254, EPI_ISL_1376256, EPI_ISL_1376260, EPI_ISL_1376261, EPI_ISL_1376263, EPI_ISL_1376264, EPI_ISL_1376265, EPI_ISL_1376273, EPI_ISL_1376276, EPI_ISL_1376278, EPI_ISL_1376279, EPI_ISL_1376283, EPI_ISL_1376290, EPI_ISL_1376291, EPI_ISL_1376292, EPI_ISL_1376296, EPI_ISL_1376300, EPI_ISL_1376310, EPI_ISL_1376312, EPI_ISL_1376313, EPI_ISL_1376325, EPI_ISL_1376332, EPI_ISL_1376335, EPI_ISL_1376336, EPI_ISL_1376337, EPI_ISL_1376345, EPI_ISL_1376346, EPI_ISL_1376348, EPI_ISL_1376351, EPI_ISL_1376352, EPI_ISL_1376355, EPI_ISL_1376357, EPI_ISL_1376371, EPI_ISL_1376375, EPI_ISL_1376378, EPI_ISL_1376382, EPI_ISL_1376384, EPI_ISL_1376388, EPI_ISL_1376389, EPI_ISL_1376391, EPI_ISL_1376393, EPI_ISL_1376394, EPI_ISL_1376395, EPI_ISL_1376396, EPI_ISL_1376400, EPI_ISL_1376401, EPI_ISL_1376406, EPI_ISL_1376408, EPI_ISL_1376409, EPI_ISL_1376412, EPI_ISL_1376414, EPI_ISL_1376415, EPI_ISL_1376416, EPI_ISL_1376417, EPI_ISL_1376429, EPI_ISL_1376432, EPI_ISL_1376434, EPI_ISL_1376435, EPI_ISL_1376440, EPI_ISL_1376442, EPI_ISL_1376446, EPI_ISL_1376447, EPI_ISL_1376454, EPI_ISL_1376455, EPI_ISL_1376456, EPI_ISL_1376460, EPI_ISL_1376461, EPI_ISL_1376463, EPI_ISL_1376468, EPI_ISL_1376472, EPI_ISL_1376484, EPI_ISL_1376486, EPI_ISL_1376489, EPI_ISL_1376490, EPI_ISL_1376493, EPI_ISL_1376498, EPI_ISL_1376500, EPI_ISL_1376502, EPI_ISL_1376503, EPI_ISL_1376506, EPI_ISL_1376511, EPI_ISL_1376513, EPI_ISL_1376514, EPI_ISL_1376515, EPI_ISL_1376516, EPI_ISL_1376517, EPI_ISL_1376520, EPI_ISL_1376524, EPI_ISL_1376529, EPI_ISL_1376531, EPI_ISL_1376532, EPI_ISL_1376534, EPI_ISL_1376535, EPI_ISL_1376538, EPI_ISL_1376541, EPI_ISL_1376542, EPI_ISL_1376547, EPI_ISL_1376548, EPI_ISL_1376554, EPI_ISL_1376558, EPI_ISL_1376559, EPI_ISL_1376560, EPI_ISL_1376564, EPI_ISL_1376573, EPI_ISL_1376574, EPI_ISL_1376585, EPI_ISL_1376589, EPI_ISL_1376590, EPI_ISL_1376598, EPI_ISL_1376600, EPI_ISL_1376603, EPI_ISL_1376621, EPI_ISL_1376623, EPI_ISL_1376627, EPI_ISL_1376631, EPI_ISL_1376632, EPI_ISL_1376639, EPI_ISL_1376640, EPI_ISL_1376641, EPI_ISL_1376650, EPI_ISL_1376654, EPI_ISL_1376655, EPI_ISL_1376657, EPI_ISL_1376660, EPI_ISL_1376664, EPI_ISL_1376665, EPI_ISL_1376666, EPI_ISL_1376670, EPI_ISL_1376676, EPI_ISL_1376688, EPI_ISL_1376689, EPI_ISL_1376695, EPI_ISL_1376698, EPI_ISL_1376700, EPI_ISL_1376703, EPI_ISL_1376708, EPI_ISL_1376709, EPI_ISL_1376710, EPI_ISL_1376711, EPI_ISL_1376713, EPI_ISL_1376714, EPI_ISL_1376715, EPI_ISL_1376722, EPI_ISL_1376723, EPI_ISL_1376727, EPI_ISL_1376732, EPI_ISL_1376733, EPI_ISL_1376734, EPI_ISL_1376737, EPI_ISL_1376739, EPI_ISL_1376740, EPI_ISL_1376741, EPI_ISL_1376742, EPI_ISL_1376743, EPI_ISL_1376745, EPI_ISL_1376752, EPI_ISL_1376755, EPI_ISL_1376760, EPI_ISL_1376761, EPI_ISL_1376763, EPI_ISL_1376769, EPI_ISL_1377269, EPI_ISL_1377286, EPI_ISL_1377417, EPI_ISL_1377492, EPI_ISL_1377554, EPI_ISL_1377606, EPI_ISL_1377673, EPI_ISL_1377680, EPI_ISL_1377691, EPI_ISL_1377716, EPI_ISL_1377761, EPI_ISL_1377765, EPI_ISL_1377769, EPI_ISL_1377773, EPI_ISL_1377781, EPI_ISL_1377801, EPI_ISL_1377817, EPI_ISL_1377837, EPI_ISL_1377875, EPI_ISL_1377894, EPI_ISL_1377907, EPI_ISL_1377941, EPI_ISL_1377949, EPI_ISL_1377952, EPI_ISL_1377959, EPI_ISL_1377974, EPI_ISL_1377985, EPI_ISL_1377988, EPI_ISL_1408911, EPI_ISL_1408953, EPI_ISL_1408959, EPI_ISL_1408988, EPI_ISL_1409082, EPI_ISL_1409114, EPI_ISL_1409503, EPI_ISL_1409504, EPI_ISL_1409505, EPI_ISL_1409507, EPI_ISL_1409511, EPI_ISL_1409512, EPI_ISL_1409515, EPI_ISL_1409521, EPI_ISL_1409528, EPI_ISL_1409534, EPI_ISL_1409538, EPI_ISL_1409542, EPI_ISL_1409548, EPI_ISL_1409554, EPI_ISL_1409555, EPI_ISL_1409556, EPI_ISL_1409565, EPI_ISL_1409570, EPI_ISL_1409573, EPI_ISL_1409575, EPI_ISL_1409578, EPI_ISL_1409581, EPI_ISL_1409595, EPI_ISL_1409603, EPI_ISL_1409613, EPI_ISL_1409614, EPI_ISL_1409621, EPI_ISL_1409626, EPI_ISL_1409627, EPI_ISL_1409628, EPI_ISL_1409631, EPI_ISL_1409635, EPI_ISL_1409636, EPI_ISL_1409644, EPI_ISL_1409645, EPI_ISL_1409648, EPI_ISL_1409650, EPI_ISL_1409653, EPI_ISL_1409667, EPI_ISL_1409676, EPI_ISL_1409678, EPI_ISL_1409688, EPI_ISL_1409695, EPI_ISL_1409703, EPI_ISL_1409704, EPI_ISL_1409706, EPI_ISL_1409714, EPI_ISL_1409720, EPI_ISL_1409725, EPI_ISL_1409726, EPI_ISL_1409729, EPI_ISL_1409740, EPI_ISL_1409750, EPI_ISL_1409755, EPI_ISL_1409756, EPI_ISL_1409759, EPI_ISL_1409762, EPI_ISL_1409764, EPI_ISL_1409765, EPI_ISL_1409768, EPI_ISL_1409769, EPI_ISL_1409770, EPI_ISL_1409788, EPI_ISL_1409789, EPI_ISL_1409808, EPI_ISL_1409811, EPI_ISL_1409815, EPI_ISL_1409820, EPI_ISL_1409824, EPI_ISL_1409849, EPI_ISL_1409871, EPI_ISL_1409885, EPI_ISL_1409895, EPI_ISL_1409903, EPI_ISL_1409905, EPI_ISL_1409917, EPI_ISL_1409925, EPI_ISL_1409928, EPI_ISL_1409930, EPI_ISL_1409946, EPI_ISL_1409962, EPI_ISL_1409963, EPI_ISL_1409964, EPI_ISL_1409965, EPI_ISL_1409974, EPI_ISL_1409992, EPI_ISL_1410007, EPI_ISL_1410024, EPI_ISL_1410043, EPI_ISL_1410049, EPI_ISL_1410054, EPI_ISL_1410058, EPI_ISL_1410305, EPI_ISL_1410363, EPI_ISL_1410367, EPI_ISL_1410382, EPI_ISL_1410392, EPI_ISL_1410430, EPI_ISL_1410438, EPI_ISL_1410439, EPI_ISL_1410455, EPI_ISL_1410512, EPI_ISL_1410524, EPI_ISL_1410542, EPI_ISL_1410554, EPI_ISL_1410563, EPI_ISL_1410573, EPI_ISL_1410596, EPI_ISL_1410615, EPI_ISL_1410625, EPI_ISL_1410632, EPI_ISL_1410659, EPI_ISL_1410678, EPI_ISL_1410680, EPI_ISL_1410683, EPI_ISL_1410708, EPI_ISL_1410710, EPI_ISL_1410713, EPI_ISL_1410714, EPI_ISL_1410724, EPI_ISL_1410727, EPI_ISL_1410729, EPI_ISL_1410732, EPI_ISL_1410739, EPI_ISL_1410742, EPI_ISL_1410748, EPI_ISL_1410752, EPI_ISL_1410754, EPI_ISL_1410762, EPI_ISL_1410763, EPI_ISL_1410765, EPI_ISL_1410766, EPI_ISL_1410768, EPI_ISL_1410770, EPI_ISL_1410771, EPI_ISL_1410775, EPI_ISL_1410780, EPI_ISL_1410781, EPI_ISL_1410783, EPI_ISL_1410786, EPI_ISL_1410789, EPI_ISL_1410793, EPI_ISL_1410798, EPI_ISL_1410802, EPI_ISL_1410803, EPI_ISL_1410804, EPI_ISL_1410805, EPI_ISL_1410806, EPI_ISL_1410808, EPI_ISL_1410815, EPI_ISL_1410817, EPI_ISL_1410818, EPI_ISL_1410821, EPI_ISL_1410824, EPI_ISL_1410825, EPI_ISL_1410828, EPI_ISL_1410830, EPI_ISL_1410831, EPI_ISL_1410834, EPI_ISL_1410836, EPI_ISL_1410837, EPI_ISL_1410838, EPI_ISL_1410846, EPI_ISL_1410851, EPI_ISL_1410852, EPI_ISL_1410857, EPI_ISL_1410860, EPI_ISL_1410862, EPI_ISL_1410863, EPI_ISL_1410866, EPI_ISL_1410868, EPI_ISL_1410870, EPI_ISL_1410871, EPI_ISL_1410872, EPI_ISL_1410874, EPI_ISL_1410875, EPI_ISL_1410876, EPI_ISL_1410877, EPI_ISL_1410883, EPI_ISL_1410885, EPI_ISL_1410886, EPI_ISL_1410887, EPI_ISL_1410888, EPI_ISL_1410889, EPI_ISL_1410891, EPI_ISL_1410893, EPI_ISL_1410894, EPI_ISL_1410896, EPI_ISL_1410897, EPI_ISL_1410899, EPI_ISL_1410901, EPI_ISL_1410902, EPI_ISL_1410904, EPI_ISL_1410907, EPI_ISL_1410910, EPI_ISL_1410911, EPI_ISL_1410913, EPI_ISL_1410915, EPI_ISL_1410918, EPI_ISL_1410919, EPI_ISL_1410924, EPI_ISL_1410928, EPI_ISL_1410930, EPI_ISL_1410931, EPI_ISL_1410935, EPI_ISL_1410940, EPI_ISL_1410942, EPI_ISL_1410944, EPI_ISL_1410946, EPI_ISL_1410949, EPI_ISL_1410951, EPI_ISL_1410953, EPI_ISL_1410954, EPI_ISL_1410957, EPI_ISL_1410958, EPI_ISL_1410961, EPI_ISL_1410962, EPI_ISL_1410963, EPI_ISL_1410971, EPI_ISL_1410972, EPI_ISL_1410973, EPI_ISL_1410974, EPI_ISL_1410977, EPI_ISL_1410981, EPI_ISL_1410985, EPI_ISL_1410986, EPI_ISL_1410987, EPI_ISL_1410990, EPI_ISL_1410995, EPI_ISL_1410997, EPI_ISL_1410999, EPI_ISL_1411000, EPI_ISL_1411001, EPI_ISL_1411010, EPI_ISL_1411015, EPI_ISL_1411019, EPI_ISL_1411022, EPI_ISL_1411024, EPI_ISL_1411028, EPI_ISL_1411030, EPI_ISL_1411032, EPI_ISL_1411035, EPI_ISL_1411036, EPI_ISL_1411037, EPI_ISL_1411038, EPI_ISL_1411041, EPI_ISL_1411044, EPI_ISL_1411045, EPI_ISL_1411051, EPI_ISL_1411052, EPI_ISL_1411053, EPI_ISL_1411054, EPI_ISL_1411056, EPI_ISL_1411058, EPI_ISL_1411061, EPI_ISL_1411067, EPI_ISL_1411069, EPI_ISL_1411071, EPI_ISL_1411072, EPI_ISL_1411073, EPI_ISL_1411080, EPI_ISL_1411085, EPI_ISL_1411086, EPI_ISL_1411089, EPI_ISL_1411091, EPI_ISL_1411094, EPI_ISL_1411100, EPI_ISL_1411458, EPI_ISL_1411477, EPI_ISL_1411497, EPI_ISL_1411569, EPI_ISL_1411582, EPI_ISL_1411592, EPI_ISL_1411657, EPI_ISL_1411671, EPI_ISL_1411697, EPI_ISL_1411708, EPI_ISL_1411729, EPI_ISL_1411740, EPI_ISL_1411774, EPI_ISL_1411794, EPI_ISL_1411797, EPI_ISL_1411806, EPI_ISL_1411816, EPI_ISL_1411827, EPI_ISL_1411852, EPI_ISL_1411853, EPI_ISL_1411855, EPI_ISL_1411856, EPI_ISL_1411858, EPI_ISL_1411859, EPI_ISL_1411860, EPI_ISL_1411862, EPI_ISL_1411865, EPI_ISL_1411866, EPI_ISL_1411867, EPI_ISL_1411870, EPI_ISL_1411874, EPI_ISL_1411877, EPI_ISL_1411878, EPI_ISL_1411880, EPI_ISL_1411882, EPI_ISL_1411884, EPI_ISL_1411886, EPI_ISL_1411888, EPI_ISL_1411889, EPI_ISL_1411893, EPI_ISL_1411896, EPI_ISL_1411897, EPI_ISL_1411898, EPI_ISL_1411900, EPI_ISL_1411901, EPI_ISL_1411903, EPI_ISL_1411905, EPI_ISL_1411906, EPI_ISL_1411907, EPI_ISL_1411910, EPI_ISL_1411911, EPI_ISL_1411916, EPI_ISL_1411920, EPI_ISL_1411921, EPI_ISL_1411922, EPI_ISL_1411923, EPI_ISL_1411925, EPI_ISL_1411926, EPI_ISL_1411928, EPI_ISL_1411931, EPI_ISL_1411933, EPI_ISL_1411934, EPI_ISL_1411935, EPI_ISL_1411936, EPI_ISL_1411937, EPI_ISL_1411939, EPI_ISL_1411944, EPI_ISL_1411945, EPI_ISL_1411947, EPI_ISL_1411948, EPI_ISL_1411951, EPI_ISL_1411953, EPI_ISL_1411955, EPI_ISL_1411956, EPI_ISL_1411960, EPI_ISL_1411961, EPI_ISL_1411963, EPI_ISL_1411964, EPI_ISL_1411965, EPI_ISL_1411967, EPI_ISL_1411968, EPI_ISL_1411969, EPI_ISL_1411974, EPI_ISL_1411976, EPI_ISL_1411979, EPI_ISL_1411980, EPI_ISL_1411981, EPI_ISL_1411983, EPI_ISL_1411985, EPI_ISL_1411986, EPI_ISL_1411987, EPI_ISL_1411988, EPI_ISL_1411991, EPI_ISL_1411994, EPI_ISL_1411995, EPI_ISL_1411998, EPI_ISL_1412001, EPI_ISL_1412002, EPI_ISL_1412003, EPI_ISL_1412010, EPI_ISL_1412012, EPI_ISL_1412014, EPI_ISL_1412015, EPI_ISL_1412017, EPI_ISL_1412018, EPI_ISL_1412019, EPI_ISL_1412024, EPI_ISL_1412025, EPI_ISL_1412027, EPI_ISL_1412029, EPI_ISL_1412030, EPI_ISL_1412038, EPI_ISL_1412042, EPI_ISL_1412046, EPI_ISL_1412048, EPI_ISL_1412050, EPI_ISL_1412053, EPI_ISL_1412056, EPI_ISL_1412057, EPI_ISL_1412060, EPI_ISL_1412061, EPI_ISL_1412062, EPI_ISL_1412063, EPI_ISL_1412065, EPI_ISL_1412066, EPI_ISL_1412067, EPI_ISL_1412069, EPI_ISL_1412072, EPI_ISL_1412074, EPI_ISL_1412079, EPI_ISL_1412080, EPI_ISL_1412082, EPI_ISL_1412083, EPI_ISL_1412084, EPI_ISL_1412085, EPI_ISL_1412087, EPI_ISL_1412090, EPI_ISL_1412094, EPI_ISL_1412096, EPI_ISL_1412097, EPI_ISL_1412105, EPI_ISL_1412110, EPI_ISL_1412111, EPI_ISL_1412113, EPI_ISL_1412115, EPI_ISL_1412116, EPI_ISL_1412119, EPI_ISL_1412120, EPI_ISL_1412126, EPI_ISL_1412127, EPI_ISL_1412128, EPI_ISL_1412131, EPI_ISL_1412132, EPI_ISL_1412134, EPI_ISL_1412138, EPI_ISL_1412141, EPI_ISL_1412145, EPI_ISL_1412146, EPI_ISL_1412151, EPI_ISL_1412153, EPI_ISL_1412155, EPI_ISL_1412156, EPI_ISL_1412158, EPI_ISL_1412159, EPI_ISL_1412160, EPI_ISL_1412162, EPI_ISL_1412165, EPI_ISL_1412166, EPI_ISL_1412171, EPI_ISL_1412177, EPI_ISL_1412178, EPI_ISL_1412179, EPI_ISL_1412180, EPI_ISL_1412182, EPI_ISL_1412184, EPI_ISL_1412185, EPI_ISL_1412188, EPI_ISL_1412189, EPI_ISL_1412190, EPI_ISL_1412191, EPI_ISL_1412192, EPI_ISL_1412193, EPI_ISL_1412194, EPI_ISL_1412196, EPI_ISL_1412198, EPI_ISL_1412199, EPI_ISL_1412200, EPI_ISL_1412201, EPI_ISL_1412202, EPI_ISL_1412203, EPI_ISL_1412204, EPI_ISL_1412205, EPI_ISL_1412207, EPI_ISL_1412208, EPI_ISL_1412209, EPI_ISL_1412210, EPI_ISL_1412211, EPI_ISL_1412214, EPI_ISL_1412215, EPI_ISL_1412216, EPI_ISL_1412217, EPI_ISL_1412219, EPI_ISL_1412220, EPI_ISL_1412221, EPI_ISL_1412222, EPI_ISL_1412223, EPI_ISL_1412226, EPI_ISL_1412227, EPI_ISL_1412228, EPI_ISL_1412230, EPI_ISL_1412232, EPI_ISL_1412233, EPI_ISL_1412234, EPI_ISL_1412236, EPI_ISL_1412237, EPI_ISL_1412238, EPI_ISL_1412242, EPI_ISL_1412243, EPI_ISL_1412244, EPI_ISL_1412245, EPI_ISL_1412247, EPI_ISL_1412248, EPI_ISL_1412249, EPI_ISL_1412250, EPI_ISL_1412251, EPI_ISL_1412252, EPI_ISL_1412253, EPI_ISL_1412254, EPI_ISL_1412255, EPI_ISL_1412256, EPI_ISL_1412257, EPI_ISL_1412258, EPI_ISL_1412259, EPI_ISL_1412263, EPI_ISL_1412266, EPI_ISL_1412269, EPI_ISL_1412271, EPI_ISL_1412272, EPI_ISL_1412273, EPI_ISL_1412274, EPI_ISL_1412275, EPI_ISL_1412276, EPI_ISL_1412277, EPI_ISL_1412278, EPI_ISL_1412279, EPI_ISL_1412280, EPI_ISL_1412282, EPI_ISL_1412283, EPI_ISL_1412285, EPI_ISL_1412289, EPI_ISL_1412290, EPI_ISL_1412291, EPI_ISL_1412292, EPI_ISL_1412293, EPI_ISL_1412294, EPI_ISL_1412295, EPI_ISL_1412296, EPI_ISL_1412297, EPI_ISL_1412300, EPI_ISL_1412302, EPI_ISL_1412303, EPI_ISL_1412304, EPI_ISL_1412305, EPI_ISL_1412307, EPI_ISL_1412309, EPI_ISL_1412313, EPI_ISL_1412314, EPI_ISL_1412315, EPI_ISL_1412316, EPI_ISL_1412318, EPI_ISL_1412319, EPI_ISL_1412322, EPI_ISL_1412323, EPI_ISL_1412326, EPI_ISL_1412327, EPI_ISL_1412334, EPI_ISL_1412336, EPI_ISL_1412341, EPI_ISL_1412342, EPI_ISL_1412343, EPI_ISL_1412344, EPI_ISL_1412347, EPI_ISL_1412348, EPI_ISL_1412349, EPI_ISL_1412350, EPI_ISL_1412351, EPI_ISL_1412353, EPI_ISL_1412357, EPI_ISL_1412359, EPI_ISL_1412360, EPI_ISL_1412363, EPI_ISL_1412366, EPI_ISL_1412367, EPI_ISL_1412370, EPI_ISL_1412371, EPI_ISL_1412373, EPI_ISL_1412375, EPI_ISL_1412378, EPI_ISL_1412379, EPI_ISL_1412380, EPI_ISL_1412381, EPI_ISL_1412382, EPI_ISL_1412384, EPI_ISL_1412385, EPI_ISL_1412387, EPI_ISL_1412389, EPI_ISL_1412390, EPI_ISL_1412391, EPI_ISL_1412393, EPI_ISL_1412395, EPI_ISL_1412396, EPI_ISL_1412397, EPI_ISL_1412398, EPI_ISL_1412400, EPI_ISL_1412402, EPI_ISL_1412403, EPI_ISL_1412406, EPI_ISL_1412407, EPI_ISL_1412408, EPI_ISL_1412409, EPI_ISL_1412412, EPI_ISL_1412413, EPI_ISL_1412415, EPI_ISL_1412418, EPI_ISL_1412419, EPI_ISL_1412421, EPI_ISL_1412422, EPI_ISL_1412423, EPI_ISL_1412424, EPI_ISL_1412425, EPI_ISL_1412426, EPI_ISL_1412428, EPI_ISL_1412430, EPI_ISL_1412432, EPI_ISL_1412433, EPI_ISL_1412434, EPI_ISL_1412436, EPI_ISL_1412438, EPI_ISL_1412441, EPI_ISL_1412442, EPI_ISL_1412443, EPI_ISL_1412444, EPI_ISL_1412445, EPI_ISL_1412448, EPI_ISL_1412451, EPI_ISL_1412452, EPI_ISL_1412453, EPI_ISL_1412455, EPI_ISL_1412456, EPI_ISL_1412457, EPI_ISL_1412458, EPI_ISL_1412459, EPI_ISL_1412460, EPI_ISL_1412461, EPI_ISL_1412465, EPI_ISL_1412467, EPI_ISL_1412469, EPI_ISL_1412471, EPI_ISL_1412472, EPI_ISL_1412473, EPI_ISL_1412474, EPI_ISL_1412477, EPI_ISL_1412479, EPI_ISL_1412480, EPI_ISL_1412483, EPI_ISL_1412485, EPI_ISL_1412488, EPI_ISL_1412489, EPI_ISL_1412491, EPI_ISL_1412492, EPI_ISL_1412494, EPI_ISL_1412497, EPI_ISL_1412498, EPI_ISL_1412500, EPI_ISL_1412501, EPI_ISL_1412503, EPI_ISL_1412504, EPI_ISL_1412506, EPI_ISL_1412507, EPI_ISL_1412511, EPI_ISL_1412512, EPI_ISL_1412514, EPI_ISL_1412515, EPI_ISL_1412518, EPI_ISL_1412520, EPI_ISL_1412521, EPI_ISL_1412523, EPI_ISL_1412524, EPI_ISL_1412526, EPI_ISL_1412529, EPI_ISL_1412531, EPI_ISL_1412532, EPI_ISL_1412533, EPI_ISL_1412535, EPI_ISL_1412536, EPI_ISL_1412537, EPI_ISL_1412538, EPI_ISL_1412540, EPI_ISL_1412541, EPI_ISL_1412542, EPI_ISL_1412544, EPI_ISL_1412546, EPI_ISL_1412548, EPI_ISL_1412551, EPI_ISL_1412552, EPI_ISL_1412553, EPI_ISL_1412554, EPI_ISL_1412556, EPI_ISL_1412557, EPI_ISL_1412558, EPI_ISL_1412559, EPI_ISL_1412560, EPI_ISL_1412561, EPI_ISL_1412563, EPI_ISL_1412565, EPI_ISL_1412566, EPI_ISL_1412567, EPI_ISL_1412577, EPI_ISL_1412596, EPI_ISL_1412610, EPI_ISL_1412681, EPI_ISL_1412689, EPI_ISL_1412730, EPI_ISL_1412749, EPI_ISL_1412760, EPI_ISL_1412774, EPI_ISL_1412803, EPI_ISL_1412836, EPI_ISL_1412843, EPI_ISL_1412845, EPI_ISL_1412864, EPI_ISL_1412870, EPI_ISL_1412871, EPI_ISL_1412887, EPI_ISL_1412913, EPI_ISL_1412925, EPI_ISL_1413061, EPI_ISL_1413066, EPI_ISL_1413078, EPI_ISL_1413093, EPI_ISL_1413096, EPI_ISL_1413099, EPI_ISL_1413104, EPI_ISL_1413111, EPI_ISL_1413117, EPI_ISL_1413118, EPI_ISL_1413127, EPI_ISL_1413129, EPI_ISL_1413131, EPI_ISL_1413132, EPI_ISL_1413146, EPI_ISL_1413155, EPI_ISL_1413156, EPI_ISL_1413164, EPI_ISL_1413186, EPI_ISL_1413195, EPI_ISL_1413197, EPI_ISL_1413206, EPI_ISL_1413213, EPI_ISL_1413216, EPI_ISL_1413225, EPI_ISL_1413231, EPI_ISL_1413234, EPI_ISL_1413239, EPI_ISL_1413261, EPI_ISL_1413268, EPI_ISL_1413270, EPI_ISL_1413278, EPI_ISL_1413281, EPI_ISL_1413286, EPI_ISL_1413289, EPI_ISL_1413300, EPI_ISL_1413301, EPI_ISL_1413306, EPI_ISL_1413308, EPI_ISL_1413313, EPI_ISL_1413317, EPI_ISL_1413321, EPI_ISL_1413327, EPI_ISL_1413337, EPI_ISL_1413345, EPI_ISL_1413349, EPI_ISL_1413356, EPI_ISL_1413357, EPI_ISL_1413371, EPI_ISL_1413381, EPI_ISL_1413395, EPI_ISL_1413409, EPI_ISL_1413417, EPI_ISL_1413419, EPI_ISL_1413420, EPI_ISL_1413428, EPI_ISL_1413438, EPI_ISL_1413444, EPI_ISL_1413446, EPI_ISL_1413461, EPI_ISL_1413462, EPI_ISL_1413466, EPI_ISL_1413472, EPI_ISL_1413474, EPI_ISL_1413481, EPI_ISL_1413487, EPI_ISL_1413492, EPI_ISL_1413502, EPI_ISL_1413503, EPI_ISL_1413517, EPI_ISL_1413521, EPI_ISL_1413526, EPI_ISL_1413543, EPI_ISL_1413545, EPI_ISL_1413548, EPI_ISL_1413558, EPI_ISL_1413565, EPI_ISL_1413567, EPI_ISL_1413573, EPI_ISL_1413578, EPI_ISL_1413581, EPI_ISL_1413584, EPI_ISL_1413668, EPI_ISL_1413717, EPI_ISL_1413756, EPI_ISL_1413817, EPI_ISL_1413858, EPI_ISL_1413871, EPI_ISL_1413889, EPI_ISL_1413920, EPI_ISL_1413933, EPI_ISL_1413985, EPI_ISL_1413990, EPI_ISL_1414001, EPI_ISL_1414018, EPI_ISL_1414025, EPI_ISL_1414050, EPI_ISL_1414057, EPI_ISL_1414482, EPI_ISL_1414491, EPI_ISL_1414500, EPI_ISL_1414542, EPI_ISL_1414597, EPI_ISL_1414621, EPI_ISL_1414628, EPI_ISL_1414735, EPI_ISL_1414777, EPI_ISL_1414793, EPI_ISL_1414800, EPI_ISL_1414807, EPI_ISL_1414829, EPI_ISL_1414830, EPI_ISL_1414841, EPI_ISL_1414846, EPI_ISL_1414847, EPI_ISL_1414867, EPI_ISL_1414899, EPI_ISL_1414907, EPI_ISL_1414919, EPI_ISL_1414920, EPI_ISL_1414930, EPI_ISL_1414934, EPI_ISL_1414954, EPI_ISL_1414956, EPI_ISL_1414960, EPI_ISL_1414966, EPI_ISL_1414972, EPI_ISL_1414980, EPI_ISL_1414982, EPI_ISL_1415026, EPI_ISL_1415028, EPI_ISL_1415030, EPI_ISL_1415437, EPI_ISL_1415468, EPI_ISL_1415473, EPI_ISL_1415474, EPI_ISL_1415486, EPI_ISL_1415495, EPI_ISL_1415499, EPI_ISL_1415504, EPI_ISL_1415514, EPI_ISL_1415549, EPI_ISL_1415568, EPI_ISL_1415631, EPI_ISL_1415635, EPI_ISL_1415646, EPI_ISL_1415647, EPI_ISL_1415657, EPI_ISL_1415675, EPI_ISL_1415693, EPI_ISL_1415720, EPI_ISL_1415744, EPI_ISL_1415772, EPI_ISL_1415802, EPI_ISL_1415817, EPI_ISL_1415819, EPI_ISL_1415821, EPI_ISL_1415825, EPI_ISL_1415826, EPI_ISL_1415830, EPI_ISL_1415832, EPI_ISL_1415833, EPI_ISL_1415836, EPI_ISL_1415841, EPI_ISL_1415851, EPI_ISL_1415853, EPI_ISL_1415856, EPI_ISL_1415860, EPI_ISL_1415875, EPI_ISL_1415880, EPI_ISL_1415881, EPI_ISL_1415910, EPI_ISL_1415911, EPI_ISL_1415915, EPI_ISL_1415916, EPI_ISL_1415917, EPI_ISL_1415918, EPI_ISL_1415924, EPI_ISL_1415934, EPI_ISL_1415937, EPI_ISL_1415945, EPI_ISL_1415947, EPI_ISL_1415948, EPI_ISL_1415949, EPI_ISL_1415958, EPI_ISL_1415963, EPI_ISL_1415964, EPI_ISL_1415968, EPI_ISL_1415972, EPI_ISL_1415978, EPI_ISL_1415979, EPI_ISL_1415985, EPI_ISL_1415988, EPI_ISL_1416002, EPI_ISL_1416003, EPI_ISL_1416011, EPI_ISL_1416012, EPI_ISL_1416015, EPI_ISL_1416016, EPI_ISL_1416019, EPI_ISL_1416021, EPI_ISL_1416030, EPI_ISL_1416035, EPI_ISL_1416039, EPI_ISL_1416041, EPI_ISL_1416047, EPI_ISL_1416052, EPI_ISL_1416058, EPI_ISL_1416066, EPI_ISL_1416077, EPI_ISL_1416078, EPI_ISL_1416080, EPI_ISL_1416084, EPI_ISL_1416087, EPI_ISL_1416088, EPI_ISL_1416089, EPI_ISL_1416103, EPI_ISL_1416106, EPI_ISL_1416107, EPI_ISL_1416111, EPI_ISL_1416115, EPI_ISL_1416119, EPI_ISL_1416124, EPI_ISL_1416125, EPI_ISL_1449067, EPI_ISL_1453243, EPI_ISL_1453372, EPI_ISL_1453473, EPI_ISL_1453478, EPI_ISL_1453494, EPI_ISL_1453496, EPI_ISL_1453497, EPI_ISL_1453499, EPI_ISL_1453501, EPI_ISL_1453503, EPI_ISL_1453508, EPI_ISL_1453523, EPI_ISL_1453525, EPI_ISL_1453531, EPI_ISL_1453542, EPI_ISL_1453544, EPI_ISL_1453546, EPI_ISL_1453552, EPI_ISL_1453554, EPI_ISL_1453556, EPI_ISL_1453561, EPI_ISL_1453574, EPI_ISL_1453575, EPI_ISL_1453578, EPI_ISL_1453581, EPI_ISL_1453588, EPI_ISL_1453594, EPI_ISL_1453602, EPI_ISL_1453605, EPI_ISL_1453630, EPI_ISL_1453631, EPI_ISL_1453635, EPI_ISL_1453636, EPI_ISL_1453637, EPI_ISL_1453638, EPI_ISL_1453647, EPI_ISL_1453651, EPI_ISL_1453654, EPI_ISL_1453656, EPI_ISL_1453658, EPI_ISL_1453663, EPI_ISL_1453666, EPI_ISL_1453667, EPI_ISL_1453679, EPI_ISL_1453681, EPI_ISL_1453683, EPI_ISL_1453689, EPI_ISL_1453693, EPI_ISL_1453709, EPI_ISL_1453723, EPI_ISL_1453727, EPI_ISL_1453738, EPI_ISL_1453741, EPI_ISL_1453744, EPI_ISL_1453745, EPI_ISL_1453755, EPI_ISL_1453760, EPI_ISL_1453762, EPI_ISL_1453765, EPI_ISL_1453768, EPI_ISL_1453770, EPI_ISL_1453774, EPI_ISL_1453784, EPI_ISL_1453785, EPI_ISL_1453792, EPI_ISL_1453801, EPI_ISL_1453811, EPI_ISL_1453817, EPI_ISL_1453820, EPI_ISL_1453829, EPI_ISL_1453836, EPI_ISL_1453840, EPI_ISL_1453844, EPI_ISL_1453845, EPI_ISL_1453848, EPI_ISL_1453861, EPI_ISL_1453864, EPI_ISL_1453877, EPI_ISL_1453878, EPI_ISL_1453901, EPI_ISL_1453915, EPI_ISL_1453932, EPI_ISL_1453955, EPI_ISL_1453956, EPI_ISL_1453967, EPI_ISL_1453979, EPI_ISL_1453985, EPI_ISL_1454009, EPI_ISL_1454010, EPI_ISL_1454026, EPI_ISL_1454041, EPI_ISL_1454050, EPI_ISL_1454077, EPI_ISL_1454103, EPI_ISL_1454112, EPI_ISL_1454131, EPI_ISL_1454143, EPI_ISL_1454416, EPI_ISL_1454541, EPI_ISL_1454621, EPI_ISL_1454622, EPI_ISL_1454623, EPI_ISL_1454624, EPI_ISL_1454627, EPI_ISL_1454629, EPI_ISL_1454637, EPI_ISL_1454643, EPI_ISL_1454647, EPI_ISL_1454650, EPI_ISL_1454652, EPI_ISL_1454655, EPI_ISL_1454656, EPI_ISL_1454657, EPI_ISL_1454659, EPI_ISL_1454660, EPI_ISL_1454662, EPI_ISL_1454664, EPI_ISL_1454666, EPI_ISL_1454667, EPI_ISL_1454668, EPI_ISL_1454669, EPI_ISL_1454673, EPI_ISL_1454674, EPI_ISL_1454676, EPI_ISL_1454677, EPI_ISL_1454691, EPI_ISL_1454696, EPI_ISL_1454699, EPI_ISL_1454700, EPI_ISL_1454703, EPI_ISL_1454704, EPI_ISL_1454706, EPI_ISL_1454708, EPI_ISL_1454712, EPI_ISL_1454714, EPI_ISL_1454716, EPI_ISL_1454727, EPI_ISL_1454729, EPI_ISL_1454737, EPI_ISL_1454739, EPI_ISL_1454741, | | | |

EPI_ISL_1454743, EPI_ISL_1454744, EPI_ISL_1454746, EPI_ISL_1454747, EPI_ISL_1454754, EPI_ISL_1454755, EPI_ISL_1454762, EPI_ISL_1454767, EPI_ISL_1454774, EPI_ISL_1454775, EPI_ISL_1454782, EPI_ISL_1454787, EPI_ISL_1454788, EPI_ISL_1454790, EPI_ISL_1454792, EPI_ISL_1454793, EPI_ISL_1454795, EPI_ISL_1454800, EPI_ISL_1454801, EPI_ISL_1454806, EPI_ISL_1454807, EPI_ISL_1454815, EPI_ISL_1454822, EPI_ISL_1454823, EPI_ISL_1454825, EPI_ISL_1454828, EPI_ISL_1454831, EPI_ISL_1454834, EPI_ISL_1454836, EPI_ISL_1454841, EPI_ISL_1454845, EPI_ISL_1454848, EPI_ISL_1454851, EPI_ISL_1454853, EPI_ISL_1454858, EPI_ISL_1454869, EPI_ISL_1454871, EPI_ISL_1454873, EPI_ISL_1454878, EPI_ISL_1454879, EPI_ISL_1454881, EPI_ISL_1454885, EPI_ISL_1454892, EPI_ISL_1454894, EPI_ISL_1454895, EPI_ISL_1454902, EPI_ISL_1454905, EPI_ISL_1454906, EPI_ISL_1454908, EPI_ISL_1454912, EPI_ISL_1454914, EPI_ISL_1454918, EPI_ISL_1454923, EPI_ISL_1454931, EPI_ISL_1454936, EPI_ISL_1454951, EPI_ISL_1454952, EPI_ISL_1454954, EPI_ISL_1454970, EPI_ISL_1454972, EPI_ISL_1454973, EPI_ISL_1454975, EPI_ISL_1454976, EPI_ISL_1454977, EPI_ISL_1454978, EPI_ISL_1454979, EPI_ISL_1454980, EPI_ISL_1454981, EPI_ISL_1454982, EPI_ISL_1454983, EPI_ISL_1454984, EPI_ISL_1455023, EPI_ISL_1455031, EPI_ISL_1455032, EPI_ISL_1455513, EPI_ISL_1455546, EPI_ISL_1455552, EPI_ISL_1455556, EPI_ISL_1455558, EPI_ISL_1455562, EPI_ISL_1455565, EPI_ISL_1455567, EPI_ISL_1455573, EPI_ISL_1455575, EPI_ISL_1455578, EPI_ISL_1455613, EPI_ISL_1455622, EPI_ISL_1455627, EPI_ISL_1467216, EPI_ISL_1467988, EPI_ISL_1468160, EPI_ISL_1472501, EPI_ISL_1635428

| | | | |
|---|---|---|---|
| see above | Lighthouse Lab in Alderley Park | Wellcome Sanger Institute for the COVID-19 Genomics UK (COG-UK) Consortium | Cordelia Langford; David K. Jackson; Dominic Kwiatkowski; Ewan Harrison; Ian Johnston; Jacquelyn Wynn; Jeffrey Barrett; John Sillitoe on behalf of the Wellcome Sanger Institute COVID-19 Surveillance Team; Mairead Hyland; Roberto Amato; Sonia Goncalves; The Lighthouse Lab in Alderley Park and Alex Alderton |

EPI_ISL_1327536, EPI_ISL_1342515, EPI_ISL_1342530, EPI_ISL_1342538, EPI_ISL_1342542, EPI_ISL_1342544, EPI_ISL_1342547, EPI_ISL_1342563, EPI_ISL_1342571, EPI_ISL_1342586, EPI_ISL_1342596, EPI_ISL_1342597, EPI_ISL_1342599, EPI_ISL_1342601, EPI_ISL_1342609, EPI_ISL_1342614, EPI_ISL_1342626, EPI_ISL_1342634, EPI_ISL_1342640, EPI_ISL_1342645, EPI_ISL_1342655, EPI_ISL_1342669, EPI_ISL_1342673, EPI_ISL_1342676, EPI_ISL_1342685, EPI_ISL_1342726, EPI_ISL_1342752, EPI_ISL_1342756, EPI_ISL_1342757, EPI_ISL_1342772, EPI_ISL_1342794, EPI_ISL_1342810, EPI_ISL_1346314, EPI_ISL_1374071, EPI_ISL_1374073, EPI_ISL_1374081, EPI_ISL_1374088, EPI_ISL_1374094, EPI_ISL_1374105, EPI_ISL_1374108, EPI_ISL_1374115, EPI_ISL_1374119, EPI_ISL_1374123, EPI_ISL_1374130, EPI_ISL_1374137, EPI_ISL_1374142, EPI_ISL_1374150, EPI_ISL_1374152, EPI_ISL_1374169, EPI_ISL_1374174, EPI_ISL_1374175, EPI_ISL_1374188, EPI_ISL_1374197, EPI_ISL_1374200, EPI_ISL_1374204, EPI_ISL_1374214, EPI_ISL_1374217, EPI_ISL_1374223, EPI_ISL_1374224, EPI_ISL_1374230, EPI_ISL_1374240, EPI_ISL_1374242, EPI_ISL_1374243, EPI_ISL_1374244, EPI_ISL_1374249, EPI_ISL_1374250, EPI_ISL_1374256, EPI_ISL_1374261, EPI_ISL_1374279, EPI_ISL_1374285, EPI_ISL_1374287, EPI_ISL_1374288, EPI_ISL_1374299, EPI_ISL_1374307, EPI_ISL_1374314, EPI_ISL_1376783, EPI_ISL_1376823, EPI_ISL_1376828, EPI_ISL_1376829, EPI_ISL_1376830, EPI_ISL_1376831, EPI_ISL_1376832, EPI_ISL_1376840, EPI_ISL_1376850, EPI_ISL_1376864, EPI_ISL_1376868, EPI_ISL_1376872, EPI_ISL_1376887, EPI_ISL_1376904, EPI_ISL_1376906, EPI_ISL_1376913, EPI_ISL_1376915, EPI_ISL_1376924, EPI_ISL_1376927, EPI_ISL_1376933, EPI_ISL_1376936, EPI_ISL_1376937, EPI_ISL_1376949, EPI_ISL_1376952, EPI_ISL_1376953, EPI_ISL_1376963, EPI_ISL_1376966, EPI_ISL_1376968, EPI_ISL_1376969, EPI_ISL_1376977, EPI_ISL_1376982, EPI_ISL_1376985, EPI_ISL_1376986, EPI_ISL_1376989, EPI_ISL_1376995, EPI_ISL_1377011, EPI_ISL_1377016, EPI_ISL_1377031, EPI_ISL_1377032, EPI_ISL_1377040, EPI_ISL_1377050, EPI_ISL_1377053, EPI_ISL_1377057, EPI_ISL_1377058, EPI_ISL_1377069, EPI_ISL_1377071, EPI_ISL_1377077, EPI_ISL_1377078, EPI_ISL_1377080, EPI_ISL_1377081, EPI_ISL_1377085, EPI_ISL_1377091, EPI_ISL_1377097, EPI_ISL_1377102, EPI_ISL_1377115, EPI_ISL_1377117, EPI_ISL_1377123, EPI_ISL_1377124, EPI_ISL_1377127, EPI_ISL_1377133, EPI_ISL_1377135, EPI_ISL_1377139, EPI_ISL_1377142, EPI_ISL_1377144, EPI_ISL_1377146, EPI_ISL_1377155, EPI_ISL_1377169, EPI_ISL_1390313, EPI_ISL_1390318, EPI_ISL_1409497, EPI_ISL_1409502, EPI_ISL_1409509, EPI_ISL_1409514, EPI_ISL_1409519, EPI_ISL_1409522, EPI_ISL_1409545, EPI_ISL_1409549, EPI_ISL_1409564, EPI_ISL_1409569, EPI_ISL_1409584, EPI_ISL_1409590, EPI_ISL_1409591, EPI_ISL_1409592, EPI_ISL_1409605, EPI_ISL_1409606, EPI_ISL_1409617, EPI_ISL_1409622, EPI_ISL_1409623, EPI_ISL_1409643, EPI_ISL_1409646, EPI_ISL_1409647, EPI_ISL_1409656, EPI_ISL_1409659, EPI_ISL_1409666, EPI_ISL_1409668, EPI_ISL_1409672, EPI_ISL_1409679, EPI_ISL_1409694, EPI_ISL_1409699, EPI_ISL_1409701, EPI_ISL_1409710, EPI_ISL_1409712, EPI_ISL_1409716, EPI_ISL_1409718, EPI_ISL_1409728, EPI_ISL_1409730, EPI_ISL_1409734, EPI_ISL_1409735, EPI_ISL_1409739, EPI_ISL_1409748, EPI_ISL_1409749, EPI_ISL_1409752, EPI_ISL_1409761, EPI_ISL_1409767, EPI_ISL_1409773, EPI_ISL_1410079, EPI_ISL_1410080, EPI_ISL_1410081, EPI_ISL_1410082, EPI_ISL_1410083, EPI_ISL_1410084, EPI_ISL_1410085, EPI_ISL_1410087, EPI_ISL_1410088, EPI_ISL_1410089, EPI_ISL_1410090, EPI_ISL_1410091, EPI_ISL_1410092, EPI_ISL_1410093, EPI_ISL_1410094, EPI_ISL_1410095, EPI_ISL_1410097, EPI_ISL_1410098, EPI_ISL_1410099, EPI_ISL_1410100, EPI_ISL_1410102, EPI_ISL_1410103, EPI_ISL_1410104, EPI_ISL_1410105, EPI_ISL_1410107, EPI_ISL_1410108, EPI_ISL_1410109, EPI_ISL_1410110, EPI_ISL_1410111, EPI_ISL_1410112, EPI_ISL_1410113, EPI_ISL_1410114, EPI_ISL_1410115, EPI_ISL_1410118, EPI_ISL_1410120, EPI_ISL_1410121, EPI_ISL_1410122, EPI_ISL_1410123, EPI_ISL_1410124, EPI_ISL_1410125, EPI_ISL_1410126, EPI_ISL_1410127, EPI_ISL_1410129, EPI_ISL_1410130, EPI_ISL_1410131, EPI_ISL_1410132, EPI_ISL_1410133, EPI_ISL_1410134, EPI_ISL_1410135, EPI_ISL_1410136, EPI_ISL_1410137, EPI_ISL_1410138, EPI_ISL_1410139, EPI_ISL_1410140, EPI_ISL_1410141, EPI_ISL_1410144, EPI_ISL_1410145, EPI_ISL_1410146, EPI_ISL_1410147, EPI_ISL_1410148, EPI_ISL_1410149, EPI_ISL_1410150, EPI_ISL_1410152, EPI_ISL_1410153, EPI_ISL_1410154, EPI_ISL_1410155, EPI_ISL_1410156, EPI_ISL_1410157, EPI_ISL_1410158, EPI_ISL_1410159, EPI_ISL_1410160, EPI_ISL_1410162, EPI_ISL_1410163, EPI_ISL_1410165, EPI_ISL_1410166, EPI_ISL_1410167, EPI_ISL_1410168, EPI_ISL_1410169, EPI_ISL_1410170, EPI_ISL_1410171, EPI_ISL_1410172, EPI_ISL_1410173, EPI_ISL_1410174, EPI_ISL_1410175, EPI_ISL_1410176, EPI_ISL_1410177, EPI_ISL_1410178, EPI_ISL_1410179, EPI_ISL_1410180, EPI_ISL_1410181, EPI_ISL_1410183, EPI_ISL_1410184, EPI_ISL_1410185, EPI_ISL_1410186, EPI_ISL_1410187, EPI_ISL_1410188, EPI_ISL_1410189, EPI_ISL_1410190, EPI_ISL_1410192, EPI_ISL_1410193, EPI_ISL_1410194, EPI_ISL_1410195, EPI_ISL_1410196, EPI_ISL_1410197, EPI_ISL_1410198, EPI_ISL_1410199, EPI_ISL_1410200, EPI_ISL_1410201, EPI_ISL_1410202, EPI_ISL_1410203, EPI_ISL_1410204, EPI_ISL_1410205, EPI_ISL_1410206, EPI_ISL_1410207, EPI_ISL_1410208, EPI_ISL_1410209, EPI_ISL_1410210, EPI_ISL_1410212, EPI_ISL_1410213, EPI_ISL_1410214, EPI_ISL_1410215, EPI_ISL_1410217, EPI_ISL_1410219, EPI_ISL_1410220, EPI_ISL_1410221, EPI_ISL_1410222, EPI_ISL_1410223, EPI_ISL_1410224, EPI_ISL_1410225, EPI_ISL_1410226, EPI_ISL_1410227, EPI_ISL_1410228, EPI_ISL_1410229, EPI_ISL_1410230, EPI_ISL_1410231, EPI_ISL_1410232, EPI_ISL_1410233, EPI_ISL_1410234, EPI_ISL_1410235, EPI_ISL_1410236, EPI_ISL_1410237, EPI_ISL_1410239, EPI_ISL_1410240, EPI_ISL_1410241, EPI_ISL_1410242, EPI_ISL_1410243, EPI_ISL_1410244, EPI_ISL_1410245, EPI_ISL_1410247, EPI_ISL_1410248, EPI_ISL_1410249, EPI_ISL_1410250, EPI_ISL_1410251, EPI_ISL_1410252, EPI_ISL_1410253, EPI_ISL_1410254, EPI_ISL_1410255, EPI_ISL_1410257, EPI_ISL_1410258, EPI_ISL_1410259, EPI_ISL_1410260, EPI_ISL_1410261, EPI_ISL_1410262, EPI_ISL_1410263, EPI_ISL_1410265, EPI_ISL_1410266, EPI_ISL_1410267, EPI_ISL_1410268, EPI_ISL_1410270, EPI_ISL_1410271, EPI_ISL_1410272, EPI_ISL_1410273, EPI_ISL_1410275, EPI_ISL_1410276, EPI_ISL_1410277, EPI_ISL_1410278, EPI_ISL_1410279, EPI_ISL_1410282, EPI_ISL_1410284, EPI_ISL_1410285, EPI_ISL_1410287, EPI_ISL_1412937, EPI_ISL_1412939, EPI_ISL_1412944, EPI_ISL_1412958, EPI_ISL_1412964, EPI_ISL_1412965, EPI_ISL_1412980, EPI_ISL_1412986, EPI_ISL_1412988, EPI_ISL_1412997, EPI_ISL_1413002, EPI_ISL_1413006, EPI_ISL_1413009, EPI_ISL_1413014, EPI_ISL_1413017, EPI_ISL_1413019, EPI_ISL_1413025, EPI_ISL_1413026, EPI_ISL_1413034, EPI_ISL_1413035, EPI_ISL_1413047, EPI_ISL_1415847, EPI_ISL_1415890, EPI_ISL_1415962, EPI_ISL_1416008, EPI_ISL_1416046, EPI_ISL_1416131, EPI_ISL_1416133, EPI_ISL_1454161, EPI_ISL_1454181, EPI_ISL_1454194, EPI_ISL_1454205, EPI_ISL_1454210, EPI_ISL_1454221, EPI_ISL_1454223, EPI_ISL_1454236, EPI_ISL_1454253, EPI_ISL_1454258, EPI_ISL_1454272, EPI_ISL_1454281, EPI_ISL_1454282, EPI_ISL_1454292, EPI_ISL_1454309, EPI_ISL_1454318, EPI_ISL_1454324, EPI_ISL_1454357, EPI_ISL_1454379, EPI_ISL_1454440, EPI_ISL_1454446, EPI_ISL_1454448, EPI_ISL_1454454, EPI_ISL_1454467, EPI_ISL_1454471, EPI_ISL_1454497, EPI_ISL_1454523, EPI_ISL_1454526, EPI_ISL_1454536, EPI_ISL_1454551, EPI_ISL_1454572, EPI_ISL_1454582, EPI_ISL_1454602, EPI_ISL_1454606, EPI_ISL_1454609, EPI_ISL_1454618, EPI_ISL_1454619, EPI_ISL_1454620, EPI_ISL_1454625, EPI_ISL_1454628, EPI_ISL_1454993, EPI_ISL_1454994, EPI_ISL_1454995, EPI_ISL_1454996, EPI_ISL_1454998, EPI_ISL_1454999, EPI_ISL_1455000, EPI_ISL_1455001, EPI_ISL_1455002, EPI_ISL_1455241, EPI_ISL_1455305, EPI_ISL_1455306, EPI_ISL_1455318, EPI_ISL_1455320, EPI_ISL_1455356, EPI_ISL_1455358, EPI_ISL_1455384, EPI_ISL_1455399, EPI_ISL_1455422, EPI_ISL_1455428, EPI_ISL_1455433, EPI_ISL_1455434, EPI_ISL_1455438, EPI_ISL_1455515, EPI_ISL_1455516, EPI_ISL_1455517, EPI_ISL_1455639, EPI_ISL_1455641, EPI_ISL_1473619, EPI_ISL_1473623, EPI_ISL_1473660, EPI_ISL_1473675, EPI_ISL_1473741, EPI_ISL_1473750, EPI_ISL_1473752, EPI_ISL_1473756, EPI_ISL_1473792, EPI_ISL_1484083, EPI_ISL_1484328, EPI_ISL_1484335, EPI_ISL_1484349, EPI_ISL_1484352, EPI_ISL_1484361, EPI_ISL_1484366, EPI_ISL_1484375, EPI_ISL_1484377, EPI_ISL_1484393, EPI_ISL_1484410, EPI_ISL_1484417, EPI_ISL_1484449, EPI_ISL_1484453, EPI_ISL_1484458, EPI_ISL_1484465, EPI_ISL_1484490, EPI_ISL_1484493, EPI_ISL_1484497, EPI_ISL_1484498, EPI_ISL_1484500, EPI_ISL_1484509, EPI_ISL_1484520, EPI_ISL_1484525, EPI_ISL_1484527, EPI_ISL_1484534, EPI_ISL_1484545, EPI_ISL_1484546, EPI_ISL_1484549, EPI_ISL_1484553, EPI_ISL_1484555, EPI_ISL_1484558, EPI_ISL_1484570, EPI_ISL_1484590, EPI_ISL_1484595, EPI_ISL_1484600, EPI_ISL_1508561, EPI_ISL_1508648, EPI_ISL_1517787, EPI_ISL_1517788, EPI_ISL_1517794, EPI_ISL_1517902, EPI_ISL_1518032, EPI_ISL_1518057, EPI_ISL_1537316

| | | | |
|---|---|---|---|
| see above | Lighthouse Lab in Cambridge | Wellcome Sanger Institute for the COVID-19 Genomics UK (COG-UK) Consortium | Cordelia Langford; David K. Jackson; Dominic Kwiatkowski; Ewan Harrison; Ian Johnston; Jeffrey Barrett; John Sillitoe on behalf of the Wellcome Sanger Institute COVID-19 Surveillance Team; Rob Howes; Roberto Amato; Sonia Goncalves; The Lighthouse Lab in Cambridge and Alex Alderton |

EPI_ISL_1341803, EPI_ISL_1341804, EPI_ISL_1341805, EPI_ISL_1341806, EPI_ISL_1341807, EPI_ISL_1341808, EPI_ISL_1341809, EPI_ISL_1341810, EPI_ISL_1341811, EPI_ISL_1341812, EPI_ISL_1341813, EPI_ISL_1341814, EPI_ISL_1341815, EPI_ISL_1341816, EPI_ISL_1341817, EPI_ISL_1341818, EPI_ISL_1341819, EPI_ISL_1341820, EPI_ISL_1341821, EPI_ISL_1341822, EPI_ISL_1341823, EPI_ISL_1341824, EPI_ISL_1341825, EPI_ISL_1341826, EPI_ISL_1341827, EPI_ISL_1341828, EPI_ISL_1341829, EPI_ISL_1341830, EPI_ISL_1341831, EPI_ISL_1341832, EPI_ISL_1341833, EPI_ISL_1341834, EPI_ISL_1341835, EPI_ISL_1341836, EPI_ISL_1341837, EPI_ISL_1341838, EPI_ISL_1341839, EPI_ISL_1341840, EPI_ISL_1341841, EPI_ISL_1341842, EPI_ISL_1341843, EPI_ISL_1341844, EPI_ISL_1341845, EPI_ISL_1341846, EPI_ISL_1341847, EPI_ISL_1341848, EPI_ISL_1341849, EPI_ISL_1341850, EPI_ISL_1341851, EPI_ISL_1341852, EPI_ISL_1341853, EPI_ISL_1341854, EPI_ISL_1341855, EPI_ISL_1341856, EPI_ISL_1341857, EPI_ISL_1341858, EPI_ISL_1341859, EPI_ISL_1341860, EPI_ISL_1341861, EPI_ISL_1341862, EPI_ISL_1341863, EPI_ISL_1341864, EPI_ISL_1341865, EPI_ISL_1341866, EPI_ISL_1341867, EPI_ISL_1341868, EPI_ISL_1341869, EPI_ISL_1341870, EPI_ISL_1341871, EPI_ISL_1341872, EPI_ISL_1341873, EPI_ISL_1341874, EPI_ISL_1341875, EPI_ISL_1341876, EPI_ISL_1341877, EPI_ISL_1341878, EPI_ISL_1341879, EPI_ISL_1341880, EPI_ISL_1341881, EPI_ISL_1341882, EPI_ISL_1341883, EPI_ISL_1341884, EPI_ISL_1341885, EPI_ISL_1341886, EPI_ISL_1341887, EPI_ISL_1341888, EPI_ISL_1341889, EPI_ISL_1341890, EPI_ISL_1341891, EPI_ISL_1341892, EPI_ISL_1341893, EPI_ISL_1341894, EPI_ISL_1341895, EPI_ISL_1341896, EPI_ISL_1341897, EPI_ISL_1341898, EPI_ISL_1341899, EPI_ISL_1341900, EPI_ISL_1341901, EPI_ISL_1341902, EPI_ISL_1341903, EPI_ISL_1341904, EPI_ISL_1341905, EPI_ISL_1341906, EPI_ISL_1341908, EPI_ISL_1341909, EPI_ISL_1341910, EPI_ISL_1341911, EPI_ISL_1341912, EPI_ISL_1341913, EPI_ISL_1341914, EPI_ISL_1341915, EPI_ISL_1341916, EPI_ISL_1341917, EPI_ISL_1341918, EPI_ISL_1341919, EPI_ISL_1341920, EPI_ISL_1341921, EPI_ISL_1341922, EPI_ISL_1341923, EPI_ISL_1341924, EPI_ISL_1341925, EPI_ISL_1341926, EPI_ISL_1341927, EPI_ISL_1341928, EPI_ISL_1341929, EPI_ISL_1341930, EPI_ISL_1341931, EPI_ISL_1341932, EPI_ISL_1341933, EPI_ISL_1341934, EPI_ISL_1341935, EPI_ISL_1341936, EPI_ISL_1341937, EPI_ISL_1341938, EPI_ISL_1341939, EPI_ISL_1341941, EPI_ISL_1341942, EPI_ISL_1341943, EPI_ISL_1341944, EPI_ISL_1341945, EPI_ISL_1341946, EPI_ISL_1341947, EPI_ISL_1341948, EPI_ISL_1341949, EPI_ISL_1341950, EPI_ISL_1341951, EPI_ISL_1341952, EPI_ISL_1341953, EPI_ISL_1341954, EPI_ISL_1341955, EPI_ISL_1341956, EPI_ISL_1341957, EPI_ISL_1341958, EPI_ISL_1341959, EPI_ISL_1341960, EPI_ISL_1341961, EPI_ISL_1341962, EPI_ISL_1341963, EPI_ISL_1341964, EPI_ISL_1341965, EPI_ISL_1341966, EPI_ISL_1341967, EPI_ISL_1341968, EPI_ISL_1341969, EPI_ISL_1341970, EPI_ISL_1341971, EPI_ISL_1341972, EPI_ISL_1341973, EPI_ISL_1341974, EPI_ISL_1341975, EPI_ISL_1341976, EPI_ISL_1341977, EPI_ISL_1341978, EPI_ISL_1341979, EPI_ISL_1341980, EPI_ISL_1341981, EPI_ISL_1341982, EPI_ISL_1341983, EPI_ISL_1341984, EPI_ISL_1341985, EPI_ISL_1341986, EPI_ISL_1341987, EPI_ISL_1341988, EPI_ISL_1341989, EPI_ISL_1341990, EPI_ISL_1341991, EPI_ISL_1341992, EPI_ISL_1341993, EPI_ISL_1341994, EPI_ISL_1341995, EPI_ISL_1341996, EPI_ISL_1341997, EPI_ISL_1341998, EPI_ISL_1341999, EPI_ISL_1342000, EPI_ISL_1342001, EPI_ISL_1342002, EPI_ISL_1342003, EPI_ISL_1342005, EPI_ISL_1342006, EPI_ISL_1342007, EPI_ISL_1342008, EPI_ISL_1342009, EPI_ISL_1342010, EPI_ISL_1342011, EPI_ISL_1342012, EPI_ISL_1342013, EPI_ISL_1342014, EPI_ISL_1342015, EPI_ISL_1342016, EPI_ISL_1342017, EPI_ISL_1342018, EPI_ISL_1342019, EPI_ISL_1342020, EPI_ISL_1342021, EPI_ISL_1342022, EPI_ISL_1342023, EPI_ISL_1342024, EPI_ISL_1342025, EPI_ISL_1342026, EPI_ISL_1342027, EPI_ISL_1342028, EPI_ISL_1342029, EPI_ISL_1342030, EPI_ISL_1342031, EPI_ISL_1342032, EPI_ISL_1342033, EPI_ISL_1342034, EPI_ISL_1342035, EPI_ISL_1342036, EPI_ISL_1342037, EPI_ISL_1342038, EPI_ISL_1342039, EPI_ISL_1342040, EPI_ISL_1342041, EPI_ISL_1342042, EPI_ISL_1342043, EPI_ISL_1342044, EPI_ISL_1342045, EPI_ISL_1342046, EPI_ISL_1342047, EPI_ISL_1342048, EPI_ISL_1342049, EPI_ISL_1342050, EPI_ISL_1342051, EPI_ISL_1342052, EPI_ISL_1342053, EPI_ISL_1342054, EPI_ISL_1342055, EPI_ISL_1342056, EPI_ISL_1342057, EPI_ISL_1342058, EPI_ISL_1342059, EPI_ISL_1342060, EPI_ISL_1342061, EPI_ISL_1342062, EPI_ISL_1342063, EPI_ISL_1342064, EPI_ISL_1342065, EPI_ISL_1342066, EPI_ISL_1342067, EPI_ISL_1342068, EPI_ISL_1342069, EPI_ISL_1342070, EPI_ISL_1342071, EPI_ISL_1342072, EPI_ISL_1342073, EPI_ISL_1342074, EPI_ISL_1342075, EPI_ISL_1342076, EPI_ISL_1342077, EPI_ISL_1342078, EPI_ISL_1342079, EPI_ISL_1342080, EPI_ISL_1342081, EPI_ISL_1342082, EPI_ISL_1342083, EPI_ISL_1342084, EPI_ISL_1342085, EPI_ISL_1342086, EPI_ISL_1342087, EPI_ISL_1342088, EPI_ISL_1342089, EPI_ISL_1342090, EPI_ISL_1342091, EPI_ISL_1342092, EPI_ISL_1342093, EPI_ISL_1342094, EPI_ISL_1342095, EPI_ISL_1342096, EPI_ISL_1342097, EPI_ISL_1342098, EPI_ISL_1342099, EPI_ISL_1342100, EPI_ISL_1342101, EPI_ISL_1342102, EPI_ISL_1342103, EPI_ISL_1342104, EPI_ISL_1342105, EPI_ISL_1342106, EPI_ISL_1342107, EPI_ISL_1342108, EPI_ISL_1342109, EPI_ISL_1342110, EPI_ISL_1342111, EPI_ISL_1342112, EPI_ISL_1342113, EPI_ISL_1342114, EPI_ISL_1342115, EPI_ISL_1342116, EPI_ISL_1342117, EPI_ISL_1342118, EPI_ISL_1342119, EPI_ISL_1342120, EPI_ISL_1342121, EPI_ISL_1342122, EPI_ISL_1342123, EPI_ISL_1342124, EPI_ISL_1342125, EPI_ISL_1342126, EPI_ISL_1342127, EPI_ISL_1342128, EPI_ISL_1342129, EPI_ISL_1342130, EPI_ISL_1342131, EPI_ISL_1342132, EPI_ISL_1342133, EPI_ISL_1342134, EPI_ISL_1342135, EPI_ISL_1342136, EPI_ISL_1342137, EPI_ISL_1342138, EPI_ISL_1342139, EPI_ISL_1342140, EPI_ISL_1342141, EPI_ISL_1342142, EPI_ISL_1342143, EPI_ISL_1342144, EPI_ISL_1342145, EPI_ISL_1342146, EPI_ISL_1342147, EPI_ISL_1342148, EPI_ISL_1342149, EPI_ISL_1342150, EPI_ISL_1342151, EPI_ISL_1342152, EPI_ISL_1342153, EPI_ISL_1342154, EPI_ISL_1342155, EPI_ISL_1342156, EPI_ISL_1342157, EPI_ISL_1342158, EPI_ISL_1342159, EPI_ISL_1342160, EPI_ISL_1342161, EPI_ISL_1342162, EPI_ISL_1342163, EPI_ISL_1342164, EPI_ISL_1342165, EPI_ISL_1342166, EPI_ISL_1342167, EPI_ISL_1342168, EPI_ISL_1342169, EPI_ISL_1342170, EPI_ISL_1342171, EPI_ISL_1342172, EPI_ISL_1342173, EPI_ISL_1342174, EPI_ISL_1342175, EPI_ISL_1342176, EPI_ISL_1342177, EPI_ISL_1342178, EPI_ISL_1342179, EPI_ISL_1342180, EPI_ISL_1342181, EPI_ISL_1342182, EPI_ISL_1342183, EPI_ISL_1342184, EPI_ISL_1342185, EPI_ISL_1342186, EPI_ISL_1342187, EPI_ISL_1342188, EPI_ISL_1342189, EPI_ISL_1342190, EPI_ISL_1342191, EPI_ISL_1342192, EPI_ISL_1342193, EPI_ISL_1342194, EPI_ISL_1342195, EPI_ISL_1342196, EPI_ISL_1342197, EPI_ISL_1342198, EPI_ISL_1342199, EPI_ISL_1342200, EPI_ISL_1342201, EPI_ISL_1342202, EPI_ISL_1342203, EPI_ISL_1342204, EPI_ISL_1342205, EPI_ISL_1342206, EPI_ISL_1342207, EPI_ISL_1342208, EPI_ISL_1342209, EPI_ISL_1342210, EPI_ISL_1342211, EPI_ISL_1342212, EPI_ISL_1342213, EPI_ISL_1342214, EPI_ISL_1342215, EPI_ISL_1342216, EPI_ISL_1342217, EPI_ISL_1342218, EPI_ISL_1342219, EPI_ISL_1342220, EPI_ISL_1342221, EPI_ISL_1342222, EPI_ISL_1342223, EPI_ISL_1342224, EPI_ISL_1342225, EPI_ISL_1342226, EPI_ISL_1342227, EPI_ISL_1342228, EPI_ISL_1342229, EPI_ISL_1342230, EPI_ISL_1342231, EPI_ISL_1342232, EPI_ISL_1342233, EPI_ISL_1342234, EPI_ISL_1342237, EPI_ISL_1342241, EPI_ISL_1342248, EPI_ISL_1342260, EPI_ISL_1342279, EPI_ISL_1342289, EPI_ISL_1342293, EPI_ISL_1342301, EPI_ISL_1342303, EPI_ISL_1342306, EPI_ISL_1342320, EPI_ISL_1342327, EPI_ISL_1342341, EPI_ISL_1342342, EPI_ISL_1342344, EPI_ISL_1342350, EPI_ISL_1342357, EPI_ISL_1342358, EPI_ISL_1342368, EPI_ISL_1342381, EPI_ISL_1342414, EPI_ISL_1342416, EPI_ISL_1342425, EPI_ISL_1342426, EPI_ISL_1342427, EPI_ISL_1342444, EPI_ISL_1342448, EPI_ISL_1342453, EPI_ISL_1342455, EPI_ISL_1342456, EPI_ISL_1342458, EPI_ISL_1342461, EPI_ISL_1342469, EPI_ISL_1342496, EPI_ISL_1342500, EPI_ISL_1342503, EPI_ISL_1342504, EPI_ISL_1342522, EPI_ISL_1342524, EPI_ISL_1342527, EPI_ISL_1342531, EPI_ISL_1342533, EPI_ISL_1342541, EPI_ISL_1342543, EPI_ISL_1342552, EPI_ISL_1342553, EPI_ISL_1342554, EPI_ISL_1342555, EPI_ISL_1342557, EPI_ISL_1342560, EPI_ISL_1342572, EPI_ISL_1342575, EPI_ISL_1342585, EPI_ISL_1342591, EPI_ISL_1342594, EPI_ISL_1342602, EPI_ISL_1342616, EPI_ISL_1342620, EPI_ISL_1342621, EPI_ISL_1342629, EPI_ISL_1342636, EPI_ISL_1342641, EPI_ISL_1342643, EPI_ISL_1342644, EPI_ISL_1342658, EPI_ISL_1342670, EPI_ISL_1342671, EPI_ISL_1342677, EPI_ISL_1342686, EPI_ISL_1342693, EPI_ISL_1342695, EPI_ISL_1342705, EPI_ISL_1342713, EPI_ISL_1342714, EPI_ISL_1342718, EPI_ISL_1342720, EPI_ISL_1342724, EPI_ISL_1342728, EPI_ISL_1342736, EPI_ISL_1342743, EPI_ISL_1342754, EPI_ISL_1342766, EPI_ISL_1342768, EPI_ISL_1342777, EPI_ISL_1342778, EPI_ISL_1342789, EPI_ISL_1342797, EPI_ISL_1342806, EPI_ISL_1342807, EPI_ISL_1342813, EPI_ISL_1342833, EPI_ISL_1342839, EPI_ISL_1342845, EPI_ISL_1342870, EPI_ISL_1342871, EPI_ISL_1342881, EPI_ISL_1342885, EPI_ISL_1342926, EPI_ISL_1342932, EPI_ISL_1342936, EPI_ISL_1342948, EPI_ISL_1342965, EPI_ISL_1342981, EPI_ISL_1342985, EPI_ISL_1342991, EPI_ISL_1342998, EPI_ISL_1343014, EPI_ISL_1343031, EPI_ISL_1343046, EPI_ISL_1343060, EPI_ISL_1343076, EPI_ISL_1343102, EPI_ISL_1343118, EPI_ISL_1343130, EPI_ISL_1343150, EPI_ISL_1343185, EPI_ISL_1343201, EPI_ISL_1343204, EPI_ISL_1343210, EPI_ISL_1343222, EPI_ISL_1343226, EPI_ISL_1343231, EPI_ISL_1343248, EPI_ISL_1343249, EPI_ISL_1343255, EPI_ISL_1343276, EPI_ISL_1343279, EPI_ISL_1343280, EPI_ISL_1343291, EPI_ISL_1343305, EPI_ISL_1343322, EPI_ISL_1343330, EPI_ISL_1343352, EPI_ISL_1343356, EPI_ISL_1343358, EPI_ISL_1343360, EPI_ISL_1343362, EPI_ISL_1343363, EPI_ISL_1343366, EPI_ISL_1343369, EPI_ISL_1343373, EPI_ISL_1343374, EPI_ISL_1343377, EPI_ISL_1343379, EPI_ISL_1343381, EPI_ISL_1343382, EPI_ISL_1343383, EPI_ISL_1343384, EPI_ISL_1343390, EPI_ISL_1343393, EPI_ISL_1343394, EPI_ISL_1343395, EPI_ISL_1343396, EPI_ISL_1343397, EPI_ISL_1343399, EPI_ISL_1343401, EPI_ISL_1343402, EPI_ISL_1343405, EPI_ISL_1343406, EPI_ISL_1343407, EPI_ISL_1343408, EPI_ISL_1343410, EPI_ISL_1343412, EPI_ISL_1343413, EPI_ISL_1343415, EPI_ISL_1343416, EPI_ISL_1343417, EPI_ISL_1343418, EPI_ISL_1343419, EPI_ISL_1343420, EPI_ISL_1343422, EPI_ISL_1343423, EPI_ISL_1343424, EPI_ISL_1343425, EPI_ISL_1343427, EPI_ISL_1343428, EPI_ISL_1343430, EPI_ISL_1343431, EPI_ISL_1343432, EPI_ISL_1343433, EPI_ISL_1343434, EPI_ISL_1343436, EPI_ISL_1343439, EPI_ISL_1343440, EPI_ISL_1343442, EPI_ISL_1343443, EPI_ISL_1343444, EPI_ISL_1343447, EPI_ISL_1343448, EPI_ISL_1343453, EPI_ISL_1343455, EPI_ISL_1343458, EPI_ISL_1343459, EPI_ISL_1343461, EPI_ISL_1343462, EPI_ISL_1343463, EPI_ISL_1343464, EPI_ISL_1343465, EPI_ISL_1343466, EPI_ISL_1343469, EPI_ISL_1343470, EPI_ISL_1343471, EPI_ISL_1343472, EPI_ISL_1343473, EPI_ISL_1343474, EPI_ISL_1343476, EPI_ISL_1343477, EPI_ISL_1343478, EPI_ISL_1343479, EPI_ISL_1343480, EPI_ISL_1343483, EPI_ISL_1343484, EPI_ISL_1343485, EPI_ISL_1343486, EPI_ISL_1343487, EPI_ISL_1343490, EPI_ISL_1343491, EPI_ISL_1343495, EPI_ISL_1343496, EPI_ISL_1343500, EPI_ISL_1343502, EPI_ISL_1343503, EPI_ISL_1343504, EPI_ISL_1343505, EPI_ISL_1343509, EPI_ISL_1343510, EPI_ISL_1343512, EPI_ISL_1343513, EPI_ISL_1343514, EPI_ISL_1343515, EPI_ISL_1343517, EPI_ISL_1343518, EPI_ISL_1343519, EPI_ISL_1343521, EPI_ISL_1343522, EPI_ISL_1343525, EPI_ISL_1343527, EPI_ISL_1343528, EPI_ISL_1343532, EPI_ISL_1343533, EPI_ISL_1343534, EPI_ISL_1343535, EPI_ISL_1343538, EPI_ISL_1343542, EPI_ISL_1343545, EPI_ISL_1343546, EPI_ISL_1343552, EPI_ISL_1343555, EPI_ISL_1343556, EPI_ISL_1343558, EPI_ISL_1343559, EPI_ISL_1343560, EPI_ISL_1343561, EPI_ISL_1343562, EPI_ISL_1343563, EPI_ISL_1343565, EPI_ISL_1343567, EPI_ISL_1343568, EPI_ISL_1343569, EPI_ISL_1343570, EPI_ISL_1343572, EPI_ISL_1343574, EPI_ISL_1343577, EPI_ISL_1343578, EPI_ISL_1343579, EPI_ISL_1343581, EPI_ISL_1343624, EPI_ISL_1343645, EPI_ISL_1343661, EPI_ISL_1343671, EPI_ISL_1343683, EPI_ISL_1343704, EPI_ISL_1343722, EPI_ISL_1343740, EPI_ISL_1343742, EPI_ISL_1343753, EPI_ISL_1343756, EPI_ISL_1343798, EPI_ISL_1343805, EPI_ISL_1343808, EPI_ISL_1343809, EPI_ISL_1343813, EPI_ISL_1343814, EPI_ISL_1343821, EPI_ISL_1343823, EPI_ISL_1343828, EPI_ISL_1343829, EPI_ISL_1343831, EPI_ISL_1343832, EPI_ISL_1343837, EPI_ISL_1343843, EPI_ISL_1343845, EPI_ISL_1343849, EPI_ISL_1343856, EPI_ISL_1343859, EPI_ISL_1343862, EPI_ISL_1343864, EPI_ISL_1343874, EPI_ISL_1343875, EPI_ISL_1343884, EPI_ISL_1343888, EPI_ISL_1343889, EPI_ISL_1343890, EPI_ISL_1343893, EPI_ISL_1343895, EPI_ISL_1343901, EPI_ISL_1343902, EPI_ISL_1343908, EPI_ISL_1343913, EPI_ISL_1343916, EPI_ISL_1343920, EPI_ISL_1343921, EPI_ISL_1343922, EPI_ISL_1343926, EPI_ISL_1343929, EPI_ISL_1343930, EPI_ISL_1343934, EPI_ISL_1343935, EPI_ISL_1343936, EPI_ISL_1343939, EPI_ISL_1343941, EPI_ISL_1343942, EPI_ISL_1343946, EPI_ISL_1343949, EPI_ISL_1343968, EPI_ISL_1343972, EPI_ISL_1343977, EPI_ISL_1343980, EPI_ISL_1343981, EPI_ISL_1343982, EPI_ISL_1343985, EPI_ISL_1343988, EPI_ISL_1343989, EPI_ISL_1343990, EPI_ISL_1343991, EPI_ISL_1343992, EPI_ISL_1343993, EPI_ISL_1343997, EPI_ISL_1343999, EPI_ISL_1344000, EPI_ISL_1344005, EPI_ISL_1344007, EPI_ISL_1344008, EPI_ISL_1344011, EPI_ISL_1344012, EPI_ISL_1344013, EPI_ISL_1344022, EPI_ISL_1344023, EPI_ISL_1344024, EPI_ISL_1344029, EPI_ISL_1344031, EPI_ISL_1344032, EPI_ISL_1344039, EPI_ISL_1344044, EPI_ISL_1344045, EPI_ISL_1344046, EPI_ISL_1344051, EPI_ISL_1344055, EPI_ISL_1344058, EPI_ISL_1344061, EPI_ISL_1344063, EPI_ISL_1344066, EPI_ISL_1344069, EPI_ISL_1344072, EPI_ISL_1344073, EPI_ISL_1344077, EPI_ISL_1344078, EPI_ISL_1344079, EPI_ISL_1344082, EPI_ISL_1344086, EPI_ISL_1344090, EPI_ISL_1344092, EPI_ISL_1344096, EPI_ISL_1344101, EPI_ISL_1344102, EPI_ISL_1344103, EPI_ISL_1344106, EPI_ISL_1344111, EPI_ISL_1344114, EPI_ISL_1344115, EPI_ISL_1344118, EPI_ISL_1344119, EPI_ISL_1344124, EPI_ISL_1344125, EPI_ISL_1344126, EPI_ISL_1344132, EPI_ISL_1344136, EPI_ISL_1344137, EPI_ISL_1344138, EPI_ISL_1344139, EPI_ISL_1344142, EPI_ISL_1344145, EPI_ISL_1344149, EPI_ISL_1344151, EPI_ISL_1344152, EPI_ISL_1344154, EPI_ISL_1344155, EPI_ISL_1344157, EPI_ISL_1344159, EPI_ISL_1344160, EPI_ISL_1344161, EPI_ISL_1344164, EPI_ISL_1344166, EPI_ISL_1344168, EPI_ISL_1344169, EPI_ISL_1344171, EPI_ISL_1344175, EPI_ISL_1344182, EPI_ISL_1344186, EPI_ISL_1344194, EPI_ISL_1344199, EPI_ISL_1344201, EPI_ISL_1344207, EPI_ISL_1344208, EPI_ISL_1344209, EPI_ISL_1344210, EPI_ISL_1344214, EPI_ISL_1344219, EPI_ISL_1344223, EPI_ISL_1344224, EPI_ISL_1344226, EPI_ISL_1344229, EPI_ISL_1344230, EPI_ISL_1344238, EPI_ISL_1344245, EPI_ISL_1344253, EPI_ISL_1344254, EPI_ISL_1344257, EPI_ISL_1344258, EPI_ISL_1344260, EPI_ISL_1344261, EPI_ISL_1344268, EPI_ISL_1344270, EPI_ISL_1344271, EPI_ISL_1344272, EPI_ISL_1344277, EPI_ISL_1344280, EPI_ISL_1344284, EPI_ISL_1344290, EPI_ISL_1344291, EPI_ISL_1344292, EPI_ISL_1344293, EPI_ISL_1344294, EPI_ISL_1344299, EPI_ISL_1344300, EPI_ISL_1344302, EPI_ISL_1344316, EPI_ISL_1344318, EPI_ISL_1344319, EPI_ISL_1344323, EPI_ISL_1344325, EPI_ISL_1344328, EPI_ISL_1344340, EPI_ISL_1344341, EPI_ISL_1344348, EPI_ISL_1344349, EPI_ISL_1344350, EPI_ISL_1344354, EPI_ISL_1344357, EPI_ISL_1344364, EPI_ISL_1344368, EPI_ISL_1344369, EPI_ISL_1344371, EPI_ISL_1344374, EPI_ISL_1344388, EPI_ISL_1344397, EPI_ISL_1344410, EPI_ISL_1344411, EPI_ISL_1344419, EPI_ISL_1344420, EPI_ISL_1344422, EPI_ISL_1344423, EPI_ISL_1344426, EPI_ISL_1344428, EPI_ISL_1344429, EPI_ISL_1344435, EPI_ISL_1344442, EPI_ISL_1344445, EPI_ISL_1344450, EPI_ISL_1344451, EPI_ISL_1344458, EPI_ISL_1345294, EPI_ISL_1345295, EPI_ISL_1345299, EPI_ISL_1345300, EPI_ISL_1345303, EPI_ISL_1345307, EPI_ISL_1345308, EPI_ISL_1345309, EPI_ISL_1345311, EPI_ISL_1345312, EPI_ISL_1345314, EPI_ISL_1345315, EPI_ISL_1345317, EPI_ISL_1345319, EPI_ISL_1345320, EPI_ISL_1345321, EPI_ISL_1345322, EPI_ISL_1345323, EPI_ISL_1345325, EPI_ISL_1345327, EPI_ISL_1345329, EPI_ISL_1345330, EPI_ISL_1345331, EPI_ISL_1345332, EPI_ISL_1345333, EPI_ISL_1345334, EPI_ISL_1345336, EPI_ISL_1345337, EPI_ISL_1345339, EPI_ISL_1345340, EPI_ISL_1345341, EPI_ISL_1345342, EPI_ISL_1345344, EPI_ISL_1345345, EPI_ISL_1345346, EPI_ISL_1345347, EPI_ISL_1345348, EPI_ISL_1345349, EPI_ISL_1345354, EPI_ISL_1345355, EPI_ISL_1345357, EPI_ISL_1345358, EPI_ISL_1345359, EPI_ISL_1345361, EPI_ISL_1345362, EPI_ISL_1345363, EPI_ISL_1345365, EPI_ISL_1345367, EPI_ISL_1345368, EPI_ISL_1345370, EPI_ISL_1345371, EPI_ISL_1345372, EPI_ISL_1345374, EPI_ISL_1345375, EPI_ISL_1345378, EPI_ISL_1345379, EPI_ISL_1345381, EPI_ISL_1345383, EPI_ISL_1345386, EPI_ISL_1345387, EPI_ISL_1345392, EPI_ISL_1345393, EPI_ISL_1345394, EPI_ISL_1345397, EPI_ISL_1345399, EPI_ISL_1345400, EPI_ISL_1345401, EPI_ISL_1345403, EPI_ISL_1345406, EPI_ISL_1345408, EPI_ISL_1345409, EPI_ISL_1345414, EPI_ISL_1345419, EPI_ISL_1345420, EPI_ISL_1345425, EPI_ISL_1345426, EPI_ISL_1345430, EPI_ISL_1345432, EPI_ISL_1345437, EPI_ISL_1345438, EPI_ISL_1345439, EPI_ISL_1345440, EPI_ISL_1345442, EPI_ISL_1345445, EPI_ISL_1345448, EPI_ISL_1345449, EPI_ISL_1345451, EPI_ISL_1345453, EPI_ISL_1345457, EPI_ISL_1345458, EPI_ISL_1345467, EPI_ISL_1345469, EPI_ISL_1345470, EPI_ISL_1345471, EPI_ISL_1345472, EPI_ISL_1345473, EPI_ISL_1345476, EPI_ISL_1345477, EPI_ISL_1345480, EPI_ISL_1345482, EPI_ISL_1345485, EPI_ISL_1345489, EPI_ISL_1345490, EPI_ISL_1345492, EPI_ISL_1345494, EPI_ISL_1345496, EPI_ISL_1345498, EPI_ISL_1345499, EPI_ISL_1345502, EPI_ISL_1345503, EPI_ISL_1345505, EPI_ISL_1345508, EPI_ISL_1345509, EPI_ISL_1345510, EPI_ISL_1345513, EPI_ISL_1345514, EPI_ISL_1345517, EPI_ISL_1345518, EPI_ISL_1345520, EPI_ISL_1345528, EPI_ISL_1345529, EPI_ISL_1345535, EPI_ISL_1345536, EPI_ISL_1345539, EPI_ISL_1345540, EPI_ISL_1345541, EPI_ISL_1345544, EPI_ISL_1345549, EPI_ISL_1345550, EPI_ISL_1345552, EPI_ISL_1345554, EPI_ISL_1345555, EPI_ISL_1345558, EPI_ISL_1345559, EPI_ISL_1345560, EPI_ISL_1345561, EPI_ISL_1345562, EPI_ISL_1345564, EPI_ISL_1345565, EPI_ISL_1345572, EPI_ISL_1345573, EPI_ISL_1345574, EPI_ISL_1345577, EPI_ISL_1345578, EPI_ISL_1345579, EPI_ISL_1345580, EPI_ISL_1345581, EPI_ISL_1345584, EPI_ISL_1345585, EPI_ISL_1345586, EPI_ISL_1345588, EPI_ISL_1345591, EPI_ISL_1345592, EPI_ISL_1345593, EPI_ISL_1345594, EPI_ISL_1345598, EPI_ISL_1345600, EPI_ISL_1345601, EPI_ISL_1345606, EPI_ISL_1345608, EPI_ISL_1345609, EPI_ISL_1345610, EPI_ISL_1345612, EPI_ISL_1345613, EPI_ISL_1345615, EPI_ISL_1345616, EPI_ISL_1345618, EPI_ISL_1345619, EPI_ISL_1345622, EPI_ISL_1345623, EPI_ISL_1345625, EPI_ISL_1345626, EPI_ISL_1345627, EPI_ISL_1345628, EPI_ISL_1345630, EPI_ISL_1345631, EPI_ISL_1345633, EPI_ISL_1345634, EPI_ISL_1345636, EPI_ISL_1345637, EPI_ISL_1345638, EPI_ISL_1345642, EPI_ISL_1345643, EPI_ISL_1345645, EPI_ISL_1345648, EPI_ISL_1365357, EPI_ISL_1374333, EPI_ISL_1374334, EPI_ISL_1374335, EPI_ISL_1374358, EPI_ISL_1374375, EPI_ISL_1374380, EPI_ISL_1374381, EPI_ISL_1374382, EPI_ISL_1374400, EPI_ISL_1374407, EPI_ISL_1374408, EPI_ISL_1374411, EPI_ISL_1374415, EPI_ISL_1374416, EPI_ISL_1374417, EPI_ISL_1374419, EPI_ISL_1374430, EPI_ISL_1374434, EPI_ISL_1374448, EPI_ISL_1374449, EPI_ISL_1374452, EPI_ISL_1374460, EPI_ISL_1374477, EPI_ISL_1374490, EPI_ISL_1374491, EPI_ISL_1374498, EPI_ISL_1374502, EPI_ISL_1374536, EPI_ISL_1374550, EPI_ISL_1374551, EPI_ISL_1374553, EPI_ISL_1374554, EPI_ISL_1374558, EPI_ISL_1374560, EPI_ISL_1374561, EPI_ISL_1374562, EPI_ISL_1374565, EPI_ISL_1374568, EPI_ISL_1374573, EPI_ISL_1374583, EPI_ISL_1374589, EPI_ISL_1374592, EPI_ISL_1374593, EPI_ISL_1374594, EPI_ISL_1374595, EPI_ISL_1374597, EPI_ISL_1374601, EPI_ISL_1374603, EPI_ISL_1374605, EPI_ISL_1374608, EPI_ISL_1374614, EPI_ISL_1374615, EPI_ISL_1374616, EPI_ISL_1374619, EPI_ISL_1374622, EPI_ISL_1374623, EPI_ISL_1374626, EPI_ISL_1374627, EPI_ISL_1374629, EPI_ISL_1374630, EPI_ISL_1374634, EPI_ISL_1374636, EPI_ISL_1374639, EPI_ISL_1374642, EPI_ISL_1374643, EPI_ISL_1374644, EPI_ISL_1374647, EPI_ISL_1374652, EPI_ISL_1374654, EPI_ISL_1374655, EPI_ISL_1374656, EPI_ISL_1374658, EPI_ISL_1374665, EPI_ISL_1374667, EPI_ISL_1374669, EPI_ISL_1374671, EPI_ISL_1374673, EPI_ISL_1374674, EPI_ISL_1374683, EPI_ISL_1374687, EPI_ISL_1374690, EPI_ISL_1374693, EPI_ISL_1374697, EPI_ISL_1374698, EPI_ISL_1374699, EPI_ISL_1374701, EPI_ISL_1374702, EPI_ISL_1374705, EPI_ISL_1374709, EPI_ISL_1374710, EPI_ISL_1374712, EPI_ISL_1374714, EPI_ISL_1374716, EPI_ISL_1374718, EPI_ISL_1374719, EPI_ISL_1374721, EPI_ISL_1374722, EPI_ISL_1374724, EPI_ISL_1374726, EPI_ISL_1374731, EPI_ISL_1374739, EPI_ISL_1374742, EPI_ISL_1374743, EPI_ISL_1374747, EPI_ISL_1374748, EPI_ISL_1374749, EPI_ISL_1374754, EPI_ISL_1374757, EPI_ISL_1374758, EPI_ISL_1374766, EPI_ISL_1374770, EPI_ISL_1374776, EPI_ISL_1374782, EPI_ISL_1374784, EPI_ISL_1374787, EPI_ISL_1374789, EPI_ISL_1374790, EPI_ISL_1374791, EPI_ISL_1374792, EPI_ISL_1374794, EPI_ISL_1374795, EPI_ISL_1374796, EPI_ISL_1374797, EPI_ISL_1374799, EPI_ISL_1374803, EPI_ISL_1374804, EPI_ISL_1374809, EPI_ISL_1374811, EPI_ISL_1374813, EPI_ISL_1374816, EPI_ISL_1374822, EPI_ISL_1374831, EPI_ISL_1374832, EPI_ISL_1374834, EPI_ISL_1374842, EPI_ISL_1374843, EPI_ISL_1374845, EPI_ISL_1374858, EPI_ISL_1374860, EPI_ISL_1374864, EPI_ISL_1374867, EPI_ISL_1374869, EPI_ISL_1374870, EPI_ISL_1374873, EPI_ISL_1374875, EPI_ISL_1374879, EPI_ISL_1374880, EPI_ISL_1374881, EPI_ISL_1374884, EPI_ISL_1374885, EPI_ISL_1374888, EPI_ISL_1374889, EPI_ISL_1374890, EPI_ISL_1374892, EPI_ISL_1374895, EPI_ISL_1374897, EPI_ISL_1374899, EPI_ISL_1374903, EPI_ISL_1374905, EPI_ISL_1374908, EPI_ISL_1374916, EPI_ISL_1374917, EPI_ISL_1374918, EPI_ISL_1374920, EPI_ISL_1374926, EPI_ISL_1374927, EPI_ISL_1374928, EPI_ISL_1374932, EPI_ISL_1374934, EPI_ISL_1374935, EPI_ISL_1374940, EPI_ISL_1374946, EPI_ISL_1374949, EPI_ISL_1374953, EPI_ISL_1374954, EPI_ISL_1374955, EPI_ISL_1374964, EPI_ISL_1374968, EPI_ISL_1374970, EPI_ISL_1374972, EPI_ISL_1374973, EPI_ISL_1374978, EPI_ISL_1374983, EPI_ISL_1374986, EPI_ISL_1374987, EPI_ISL_1374990, EPI_ISL_1374995, EPI_ISL_1374996, EPI_ISL_1374998, EPI_ISL_1375000, EPI_ISL_1375003, EPI_ISL_1375009, EPI_ISL_1375012, EPI_ISL_1375014, EPI_ISL_1375015, EPI_ISL_1375016, EPI_ISL_1375017, EPI_ISL_1375020, EPI_ISL_1375021, EPI_ISL_1375022, EPI_ISL_1375030, EPI_ISL_1375037, EPI_ISL_1375038, EPI_ISL_1375041, EPI_ISL_1375042, EPI_ISL_1375043, EPI_ISL_1375044, EPI_ISL_1375047, EPI_ISL_1375050, EPI_ISL_1375052, EPI_ISL_1375053, EPI_ISL_1375057, EPI_ISL_1375059, EPI_ISL_1375061, EPI_ISL_1375065, EPI_ISL_1375066, EPI_ISL_1375067, EPI_ISL_1375068, EPI_ISL_1375069, EPI_ISL_1375070, EPI_ISL_1375071, EPI_ISL_1375072, EPI_ISL_1375074, EPI_ISL_1375076, EPI_ISL_1375077, EPI_ISL_1375078, EPI_ISL_1375080, EPI_ISL_1375081, EPI_ISL_1375082, EPI_ISL_1375087, EPI_ISL_1375090, EPI_ISL_1375095, EPI_ISL_1375096, EPI_ISL_1375105, EPI_ISL_1375106, EPI_ISL_1375108, EPI_ISL_1375111, EPI_ISL_1375114, EPI_ISL_1375116, EPI_ISL_1375120, EPI_ISL_1375123, EPI_ISL_1375126, EPI_ISL_1375128, EPI_ISL_1375131, EPI_ISL_1375134, EPI_ISL_1375135, EPI_ISL_1375138, EPI_ISL_1375145, EPI_ISL_1375146, EPI_ISL_1375147, EPI_ISL_1375152, EPI_ISL_1375153, EPI_ISL_1375156, EPI_ISL_1375158, EPI_ISL_1375159, EPI_ISL_1375254, EPI_ISL_1375473, EPI_ISL_1375481, EPI_ISL_1375484, EPI_ISL_1375487, EPI_ISL_1375492, EPI_ISL_1375500, EPI_ISL_1375507, EPI_ISL_1375508, EPI_ISL_1375510, EPI_ISL_1375513, EPI_ISL_1375520, EPI_ISL_1375524, EPI_ISL_1375532, EPI_ISL_1375534, EPI_ISL_1375540, EPI_ISL_1375541, EPI_ISL_1375545, EPI_ISL_1375546, EPI_ISL_1375547, EPI_ISL_1375551, EPI_ISL_1375553, EPI_ISL_1375557, EPI_ISL_1375558, EPI_ISL_1375564, EPI_ISL_1375565, EPI_ISL_1375566, EPI_ISL_1375567, EPI_ISL_1375568, EPI_ISL_1375569, EPI_ISL_1375570, EPI_ISL_1375572, EPI_ISL_1375574, EPI_ISL_1375576, EPI_ISL_1375578, EPI_ISL_1375582, EPI_ISL_1375583, EPI_ISL_1375586, EPI_ISL_1375590, EPI_ISL_1375591, EPI_ISL_1375592, EPI_ISL_1375594, EPI_ISL_1375597, EPI_ISL_1375601, EPI_ISL_1375603, EPI_ISL_1375605, EPI_ISL_1375612, EPI_ISL_1375614, EPI_ISL_1375620, EPI_ISL_1375621, EPI_ISL_1375625, EPI_ISL_1375626, EPI_ISL_1375632, EPI_ISL_1375636, EPI_ISL_1375643, EPI_ISL_1375646, EPI_ISL_1375655, EPI_ISL_1375661, EPI_ISL_1375665, EPI_ISL_1375666, EPI_ISL_1375667, EPI_ISL_1375673, EPI_ISL_1375678, EPI_ISL_1375681, EPI_ISL_1375683, EPI_ISL_1375686, EPI_ISL_1375690, EPI_ISL_1375698, EPI_ISL_1375700, EPI_ISL_1375701, EPI_ISL_1375704, EPI_ISL_1375705, EPI_ISL_1375707, EPI_ISL_1375712, EPI_ISL_1375716, EPI_ISL_1375720, EPI_ISL_1375721, EPI_ISL_1375726, EPI_ISL_1375729, EPI_ISL_1375730, EPI_ISL_1375740, EPI_ISL_1375741, EPI_ISL_1375743, EPI_ISL_1375745, EPI_ISL_1375760, EPI_ISL_1375762, EPI_ISL_1375765, EPI_ISL_1375766, EPI_ISL_1375767, EPI_ISL_1375768, EPI_ISL_1375769, EPI_ISL_1375770, EPI_ISL_1375771, EPI_ISL_1375772, EPI_ISL_1375773, EPI_ISL_1375774, EPI_ISL_1375775, EPI_ISL_1375776, EPI_ISL_1375777, EPI_ISL_1375778, EPI_ISL_1375779, EPI_ISL_1375780, EPI_ISL_1375781, EPI_ISL_1375782, EPI_ISL_1375783, EPI_ISL_1375784, EPI_ISL_1375785, EPI_ISL_1375787, EPI_ISL_1375788, EPI_ISL_1375789, EPI_ISL_1375790, EPI_ISL_1375791, EPI_ISL_1375792, EPI_ISL_1375793, EPI_ISL_1375794, EPI_ISL_1375795, EPI_ISL_1375796, EPI_ISL_1375797, EPI_ISL_1375798, EPI_ISL_1375799, EPI_ISL_1375800, EPI_ISL_1375801, EPI_ISL_1375802, EPI_ISL_1375803, EPI_ISL_1375804, EPI_ISL_1375805, EPI_ISL_1375806, EPI_ISL_1375807, EPI_ISL_1375808, EPI_ISL_1375809, EPI_ISL_1375810, EPI_ISL_1375811, EPI_ISL_1375812, EPI_ISL_1375813, EPI_ISL_1375814, EPI_ISL_1375815, EPI_ISL_1375816, EPI_ISL_1375817, EPI_ISL_1375818, EPI_ISL_1375819, EPI_ISL_1375820, EPI_ISL_1375821, EPI_ISL_1375822, EPI_ISL_1375823, EPI_ISL_1375824, EPI_ISL_1375825, EPI_ISL_1375826, EPI_ISL_1375827, EPI_ISL_1375828, EPI_ISL_1375829, EPI_ISL_1375830, EPI_ISL_1375831, EPI_ISL_1375832, EPI_ISL_1375833, EPI_ISL_1375834, EPI_ISL_1375835, EPI_ISL_1375836, EPI_ISL_1375837, EPI_ISL_1375838, EPI_ISL_1375839, EPI_ISL_1375840, EPI_ISL_1375841, EPI_ISL_1375842, EPI_ISL_1375843, EPI_ISL_1375844, EPI_ISL_1375846, EPI_ISL_1375847, EPI_ISL_1375848, EPI_ISL_1375849, EPI_ISL_1375850, EPI_ISL_1375851, EPI_ISL_1375852, EPI_ISL_1375853, EPI_ISL_1375854, EPI_ISL_1375855, EPI_ISL_1375856, EPI_ISL_1375857, EPI_ISL_1375858, EPI_ISL_1375859, EPI_ISL_1375860, EPI_ISL_1375861, EPI_ISL_1375862, EPI_ISL_1375863, EPI_ISL_1375864, EPI_ISL_1375865, EPI_ISL_1375866, EPI_ISL_1375867, EPI_ISL_1375868, EPI_ISL_1375869, EPI_ISL_1375870, EPI_ISL_1375872, EPI_ISL_1375873, EPI_ISL_1375874, EPI_ISL_1375875, EPI_ISL_1375876, EPI_ISL_1375877, EPI_ISL_1375878, EPI_ISL_1375879, EPI_ISL_1375880, EPI_ISL_1375881, EPI_ISL_1375882, EPI_ISL_1375883, EPI_ISL_1375884, EPI_ISL_1375885, EPI_ISL_1375886, EPI_ISL_1375887, EPI_ISL_1375888, EPI_ISL_1375889, EPI_ISL_1375890, EPI_ISL_1375891, EPI_ISL_1375892, EPI_ISL_1375893, EPI_ISL_1375894, EPI_ISL_1375895, EPI_ISL_1375896, EPI_ISL_1375897, EPI_ISL_1375898, EPI_ISL_1375899, EPI_ISL_1375900, EPI_ISL_1375901, EPI_ISL_1375902, EPI_ISL_1375903, EPI_ISL_1375904, EPI_ISL_1375905, EPI_ISL_1375906, EPI_ISL_1375908, EPI_ISL_1375909, EPI_ISL_1375910, EPI_ISL_1375911, EPI_ISL_1375912, EPI_ISL_1375913, EPI_ISL_1375914, EPI_ISL_1375915, EPI_ISL_1375916, EPI_ISL_1375917, EPI_ISL_1375918, EPI_ISL_1375919, EPI_ISL_1375920, EPI_ISL_1375921, EPI_ISL_1375922, EPI_ISL_1375923, EPI_ISL_1375924, EPI_ISL_1375925, EPI_ISL_1375926, EPI_ISL_1375927, EPI_ISL_1375928, EPI_ISL_1375929, EPI_ISL_1375930, EPI_ISL_1375931, EPI_ISL_1375932, EPI_ISL_1375933, EPI_ISL_1375934, EPI_ISL_1375935, EPI_ISL_1375936, EPI_ISL_1375937,

EPI_ISL_1375938, EPI_ISL_1375939, EPI_ISL_1375940, EPI_ISL_1375941, EPI_ISL_1375942, EPI_ISL_1375943, EPI_ISL_1375944, EPI_ISL_1375945, EPI_ISL_1375946, EPI_ISL_1375947, EPI_ISL_1375948, EPI_ISL_1375949, EPI_ISL_1375950, EPI_ISL_1375951, EPI_ISL_1375952, EPI_ISL_1375953, EPI_ISL_1375954, EPI_ISL_1375955, EPI_ISL_1375956, EPI_ISL_1375957, EPI_ISL_1375958, EPI_ISL_1375959,
EPI_ISL_1375960, EPI_ISL_1375961, EPI_ISL_1375962, EPI_ISL_1375963, EPI_ISL_1375964, EPI_ISL_1375965, EPI_ISL_1375966, EPI_ISL_1375967, EPI_ISL_1375968, EPI_ISL_1375969, EPI_ISL_1375970, EPI_ISL_1375971, EPI_ISL_1375972, EPI_ISL_1375973, EPI_ISL_1375974, EPI_ISL_1375975, EPI_ISL_1375976, EPI_ISL_1375977, EPI_ISL_1375978, EPI_ISL_1375979, EPI_ISL_1375980, EPI_ISL_1375981,
EPI_ISL_1375982, EPI_ISL_1375983, EPI_ISL_1375984, EPI_ISL_1375985, EPI_ISL_1375986, EPI_ISL_1375987, EPI_ISL_1375988, EPI_ISL_1375989, EPI_ISL_1375991, EPI_ISL_1375992, EPI_ISL_1375993, EPI_ISL_1375994, EPI_ISL_1375995, EPI_ISL_1375996, EPI_ISL_1375997, EPI_ISL_1375999, EPI_ISL_1376000, EPI_ISL_1376001, EPI_ISL_1376002, EPI_ISL_1376003, EPI_ISL_1376004, EPI_ISL_1376005,
EPI_ISL_1376006, EPI_ISL_1376007, EPI_ISL_1376008, EPI_ISL_1376009, EPI_ISL_1376010, EPI_ISL_1376011, EPI_ISL_1376012, EPI_ISL_1376013, EPI_ISL_1376014, EPI_ISL_1376015, EPI_ISL_1376016, EPI_ISL_1376017, EPI_ISL_1376018, EPI_ISL_1376019, EPI_ISL_1376020, EPI_ISL_1376021, EPI_ISL_1376022, EPI_ISL_1376023, EPI_ISL_1376024, EPI_ISL_1376025, EPI_ISL_1376026, EPI_ISL_1376027,
EPI_ISL_1376028, EPI_ISL_1376029, EPI_ISL_1376030, EPI_ISL_1376031, EPI_ISL_1376032, EPI_ISL_1376033, EPI_ISL_1376034, EPI_ISL_1376035, EPI_ISL_1376036, EPI_ISL_1376037, EPI_ISL_1376038, EPI_ISL_1376039, EPI_ISL_1376040, EPI_ISL_1376041, EPI_ISL_1376042, EPI_ISL_1376043, EPI_ISL_1376044, EPI_ISL_1376045, EPI_ISL_1376046, EPI_ISL_1376047, EPI_ISL_1376048, EPI_ISL_1376049,
EPI_ISL_1376050, EPI_ISL_1376052, EPI_ISL_1376053, EPI_ISL_1376054, EPI_ISL_1376055, EPI_ISL_1376056, EPI_ISL_1376057, EPI_ISL_1376058, EPI_ISL_1376059, EPI_ISL_1376060, EPI_ISL_1376061, EPI_ISL_1376062, EPI_ISL_1376063, EPI_ISL_1376064, EPI_ISL_1376065, EPI_ISL_1376066, EPI_ISL_1376067, EPI_ISL_1376068, EPI_ISL_1376069, EPI_ISL_1376070, EPI_ISL_1376071, EPI_ISL_1376072,
EPI_ISL_1376073, EPI_ISL_1376074, EPI_ISL_1376075, EPI_ISL_1376077, EPI_ISL_1376078, EPI_ISL_1376079, EPI_ISL_1376080, EPI_ISL_1376081, EPI_ISL_1376082, EPI_ISL_1376083, EPI_ISL_1376084, EPI_ISL_1376086, EPI_ISL_1376087, EPI_ISL_1376088, EPI_ISL_1376089, EPI_ISL_1376090, EPI_ISL_1376091, EPI_ISL_1376092, EPI_ISL_1376093, EPI_ISL_1376094, EPI_ISL_1376095, EPI_ISL_1376096,
EPI_ISL_1376097, EPI_ISL_1376098, EPI_ISL_1376099, EPI_ISL_1376100, EPI_ISL_1376101, EPI_ISL_1376102, EPI_ISL_1376103, EPI_ISL_1376104, EPI_ISL_1376105, EPI_ISL_1376106, EPI_ISL_1376107, EPI_ISL_1376108, EPI_ISL_1376109, EPI_ISL_1376110, EPI_ISL_1376111, EPI_ISL_1376112, EPI_ISL_1376113, EPI_ISL_1376114, EPI_ISL_1376115, EPI_ISL_1376116, EPI_ISL_1376118, EPI_ISL_1376119,
EPI_ISL_1376120, EPI_ISL_1376121, EPI_ISL_1376122, EPI_ISL_1376123, EPI_ISL_1376124, EPI_ISL_1376125, EPI_ISL_1376126, EPI_ISL_1376127, EPI_ISL_1376128, EPI_ISL_1376129, EPI_ISL_1376130, EPI_ISL_1376131, EPI_ISL_1376132, EPI_ISL_1376133, EPI_ISL_1376134, EPI_ISL_1376135, EPI_ISL_1376136, EPI_ISL_1376137, EPI_ISL_1376138, EPI_ISL_1376139, EPI_ISL_1376140, EPI_ISL_1376141,
EPI_ISL_1376142, EPI_ISL_1376143, EPI_ISL_1376144, EPI_ISL_1376145, EPI_ISL_1376146, EPI_ISL_1376147, EPI_ISL_1376148, EPI_ISL_1376149, EPI_ISL_1376150, EPI_ISL_1376151, EPI_ISL_1376152, EPI_ISL_1376153, EPI_ISL_1376154, EPI_ISL_1376155, EPI_ISL_1376156, EPI_ISL_1376157, EPI_ISL_1376158, EPI_ISL_1376159, EPI_ISL_1376160, EPI_ISL_1376161, EPI_ISL_1376162, EPI_ISL_1376163,
EPI_ISL_1376164, EPI_ISL_1376165, EPI_ISL_1376166, EPI_ISL_1376167, EPI_ISL_1376168, EPI_ISL_1376169, EPI_ISL_1376170, EPI_ISL_1376171, EPI_ISL_1376172, EPI_ISL_1376173, EPI_ISL_1376174, EPI_ISL_1376175, EPI_ISL_1376176, EPI_ISL_1376177, EPI_ISL_1376178, EPI_ISL_1376179, EPI_ISL_1376180, EPI_ISL_1376181, EPI_ISL_1376182, EPI_ISL_1376183, EPI_ISL_1376184, EPI_ISL_1376185,
EPI_ISL_1376186, EPI_ISL_1376187, EPI_ISL_1376188, EPI_ISL_1376189, EPI_ISL_1376190, EPI_ISL_1376191, EPI_ISL_1376192, EPI_ISL_1376193, EPI_ISL_1376194, EPI_ISL_1376195, EPI_ISL_1376196, EPI_ISL_1376197, EPI_ISL_1376198, EPI_ISL_1376199, EPI_ISL_1376200, EPI_ISL_1376201, EPI_ISL_1376202, EPI_ISL_1376203, EPI_ISL_1376204, EPI_ISL_1376205, EPI_ISL_1376206, EPI_ISL_1376207,
EPI_ISL_1376208, EPI_ISL_1376209, EPI_ISL_1376210, EPI_ISL_1376211, EPI_ISL_1376212, EPI_ISL_1376213, EPI_ISL_1376214, EPI_ISL_1376215, EPI_ISL_1376216, EPI_ISL_1376217, EPI_ISL_1376218, EPI_ISL_1376219, EPI_ISL_1376220, EPI_ISL_1376221, EPI_ISL_1376222, EPI_ISL_1376227, EPI_ISL_1376230, EPI_ISL_1376233, EPI_ISL_1376235, EPI_ISL_1376239, EPI_ISL_1376245, EPI_ISL_1376247,
EPI_ISL_1376249, EPI_ISL_1376267, EPI_ISL_1376268, EPI_ISL_1376270, EPI_ISL_1376272, EPI_ISL_1376277, EPI_ISL_1376281, EPI_ISL_1376284, EPI_ISL_1376286, EPI_ISL_1376287, EPI_ISL_1376288, EPI_ISL_1376294, EPI_ISL_1376302, EPI_ISL_1376304, EPI_ISL_1376311, EPI_ISL_1376315, EPI_ISL_1376318, EPI_ISL_1376321, EPI_ISL_1376324, EPI_ISL_1376326, EPI_ISL_1376329, EPI_ISL_1376330,
EPI_ISL_1376333, EPI_ISL_1376340, EPI_ISL_1376342, EPI_ISL_1376347, EPI_ISL_1376349, EPI_ISL_1376354, EPI_ISL_1376360, EPI_ISL_1376361, EPI_ISL_1376362, EPI_ISL_1376366, EPI_ISL_1376367, EPI_ISL_1376373, EPI_ISL_1376377, EPI_ISL_1376379, EPI_ISL_1376381, EPI_ISL_1376385, EPI_ISL_1376386, EPI_ISL_1376399, EPI_ISL_1376410, EPI_ISL_1376411, EPI_ISL_1376413, EPI_ISL_1376418,
EPI_ISL_1376419, EPI_ISL_1376420, EPI_ISL_1376421, EPI_ISL_1376422, EPI_ISL_1376423, EPI_ISL_1376426, EPI_ISL_1376428, EPI_ISL_1376430, EPI_ISL_1376438, EPI_ISL_1376441, EPI_ISL_1376443, EPI_ISL_1376444, EPI_ISL_1376449, EPI_ISL_1376457, EPI_ISL_1376458, EPI_ISL_1376459, EPI_ISL_1376465, EPI_ISL_1376466, EPI_ISL_1376470, EPI_ISL_1376473, EPI_ISL_1376475, EPI_ISL_1376477,
EPI_ISL_1376478, EPI_ISL_1376481, EPI_ISL_1376482, EPI_ISL_1376483, EPI_ISL_1376485, EPI_ISL_1376488, EPI_ISL_1376491, EPI_ISL_1376495, EPI_ISL_1376496, EPI_ISL_1376499, EPI_ISL_1376504, EPI_ISL_1376505, EPI_ISL_1376522, EPI_ISL_1376525, EPI_ISL_1376527, EPI_ISL_1376533, EPI_ISL_1376536, EPI_ISL_1376539, EPI_ISL_1376540, EPI_ISL_1376543, EPI_ISL_1376544, EPI_ISL_1376550,
EPI_ISL_1376552, EPI_ISL_1376557, EPI_ISL_1376561, EPI_ISL_1376571, EPI_ISL_1376577, EPI_ISL_1376581, EPI_ISL_1376583, EPI_ISL_1376588, EPI_ISL_1376592, EPI_ISL_1376594, EPI_ISL_1376597, EPI_ISL_1376601, EPI_ISL_1376605, EPI_ISL_1376608, EPI_ISL_1376609, EPI_ISL_1376612, EPI_ISL_1376617, EPI_ISL_1376625, EPI_ISL_1376629, EPI_ISL_1376634, EPI_ISL_1376637, EPI_ISL_1376638,
EPI_ISL_1376642, EPI_ISL_1376647, EPI_ISL_1376656, EPI_ISL_1376658, EPI_ISL_1376661, EPI_ISL_1376662, EPI_ISL_1376667, EPI_ISL_1376671, EPI_ISL_1376672, EPI_ISL_1376678, EPI_ISL_1376679, EPI_ISL_1376684, EPI_ISL_1376690, EPI_ISL_1376691, EPI_ISL_1376694, EPI_ISL_1376712, EPI_ISL_1376718, EPI_ISL_1376721, EPI_ISL_1376724, EPI_ISL_1376725, EPI_ISL_1376726, EPI_ISL_1376731,
EPI_ISL_1376736, EPI_ISL_1376744, EPI_ISL_1376746, EPI_ISL_1376748, EPI_ISL_1376751, EPI_ISL_1376756, EPI_ISL_1376758, EPI_ISL_1376759, EPI_ISL_1376765, EPI_ISL_1376768, EPI_ISL_1376771, EPI_ISL_1376773, EPI_ISL_1376776, EPI_ISL_1377180, EPI_ISL_1377181, EPI_ISL_1377182, EPI_ISL_1377183, EPI_ISL_1377184, EPI_ISL_1377185, EPI_ISL_1377186, EPI_ISL_1377187, EPI_ISL_1377188,
EPI_ISL_1377189, EPI_ISL_1377190, EPI_ISL_1377192, EPI_ISL_1377193, EPI_ISL_1377194, EPI_ISL_1377196, EPI_ISL_1377197, EPI_ISL_1377198, EPI_ISL_1377199, EPI_ISL_1377200, EPI_ISL_1377201, EPI_ISL_1377202, EPI_ISL_1377203, EPI_ISL_1377204, EPI_ISL_1377205, EPI_ISL_1377206, EPI_ISL_1377207, EPI_ISL_1377209, EPI_ISL_1377210, EPI_ISL_1377211, EPI_ISL_1377212, EPI_ISL_1377213,
EPI_ISL_1377214, EPI_ISL_1377215, EPI_ISL_1377216, EPI_ISL_1377217, EPI_ISL_1377218, EPI_ISL_1377219, EPI_ISL_1377220, EPI_ISL_1377221, EPI_ISL_1377222, EPI_ISL_1377223, EPI_ISL_1377224, EPI_ISL_1377225, EPI_ISL_1377226, EPI_ISL_1377227, EPI_ISL_1377228, EPI_ISL_1377230, EPI_ISL_1377232, EPI_ISL_1377233, EPI_ISL_1377234, EPI_ISL_1377235, EPI_ISL_1377236, EPI_ISL_1377237,
EPI_ISL_1377238, EPI_ISL_1377239, EPI_ISL_1377240, EPI_ISL_1377241, EPI_ISL_1377242, EPI_ISL_1377243, EPI_ISL_1377244, EPI_ISL_1377245, EPI_ISL_1377246, EPI_ISL_1377247, EPI_ISL_1377248, EPI_ISL_1377249, EPI_ISL_1377250, EPI_ISL_1377251, EPI_ISL_1377252, EPI_ISL_1377253, EPI_ISL_1377254, EPI_ISL_1377255, EPI_ISL_1377256, EPI_ISL_1377258, EPI_ISL_1377259, EPI_ISL_1377260,
EPI_ISL_1377261, EPI_ISL_1377262, EPI_ISL_1377263, EPI_ISL_1377264, EPI_ISL_1377266, EPI_ISL_1377267, EPI_ISL_1377268, EPI_ISL_1377270, EPI_ISL_1377271, EPI_ISL_1377272, EPI_ISL_1377273, EPI_ISL_1377274, EPI_ISL_1377275, EPI_ISL_1377276, EPI_ISL_1377277, EPI_ISL_1377278, EPI_ISL_1377279, EPI_ISL_1377280, EPI_ISL_1377281, EPI_ISL_1377282, EPI_ISL_1377283, EPI_ISL_1377284,
EPI_ISL_1377285, EPI_ISL_1377287, EPI_ISL_1377288, EPI_ISL_1377289, EPI_ISL_1377290, EPI_ISL_1377291, EPI_ISL_1377292, EPI_ISL_1377293, EPI_ISL_1377294, EPI_ISL_1377295, EPI_ISL_1377296, EPI_ISL_1377297, EPI_ISL_1377298, EPI_ISL_1377299, EPI_ISL_1377300, EPI_ISL_1377301, EPI_ISL_1377302, EPI_ISL_1377303, EPI_ISL_1377304, EPI_ISL_1377305, EPI_ISL_1377306, EPI_ISL_1377307,
EPI_ISL_1377308, EPI_ISL_1377309, EPI_ISL_1377310, EPI_ISL_1377311, EPI_ISL_1377312, EPI_ISL_1377313, EPI_ISL_1377314, EPI_ISL_1377315, EPI_ISL_1377316, EPI_ISL_1377317, EPI_ISL_1377318, EPI_ISL_1377319, EPI_ISL_1377320, EPI_ISL_1377321, EPI_ISL_1377322, EPI_ISL_1377323, EPI_ISL_1377324, EPI_ISL_1377325, EPI_ISL_1377326, EPI_ISL_1377327, EPI_ISL_1377328, EPI_ISL_1377329,
EPI_ISL_1377330, EPI_ISL_1377331, EPI_ISL_1377332, EPI_ISL_1377333, EPI_ISL_1377334, EPI_ISL_1377335, EPI_ISL_1377336, EPI_ISL_1377337, EPI_ISL_1377338, EPI_ISL_1377339, EPI_ISL_1377340, EPI_ISL_1377341, EPI_ISL_1377342, EPI_ISL_1377343, EPI_ISL_1377344, EPI_ISL_1377345, EPI_ISL_1377346, EPI_ISL_1377349, EPI_ISL_1377350, EPI_ISL_1377351, EPI_ISL_1377353, EPI_ISL_1377354,
EPI_ISL_1377355, EPI_ISL_1377356, EPI_ISL_1377358, EPI_ISL_1377359, EPI_ISL_1377360, EPI_ISL_1377361, EPI_ISL_1377362, EPI_ISL_1377363, EPI_ISL_1377364, EPI_ISL_1377365, EPI_ISL_1377366, EPI_ISL_1377367, EPI_ISL_1377369, EPI_ISL_1377370, EPI_ISL_1377371, EPI_ISL_1377372, EPI_ISL_1377373, EPI_ISL_1377374, EPI_ISL_1377375, EPI_ISL_1377376, EPI_ISL_1377377, EPI_ISL_1377378,
EPI_ISL_1377379, EPI_ISL_1377380, EPI_ISL_1377381, EPI_ISL_1377382, EPI_ISL_1377383, EPI_ISL_1377384, EPI_ISL_1377385, EPI_ISL_1377387, EPI_ISL_1377388, EPI_ISL_1377389, EPI_ISL_1377390, EPI_ISL_1377391, EPI_ISL_1377392, EPI_ISL_1377393, EPI_ISL_1377394, EPI_ISL_1377395, EPI_ISL_1377396, EPI_ISL_1377397, EPI_ISL_1377398, EPI_ISL_1377400, EPI_ISL_1377401, EPI_ISL_1377402,
EPI_ISL_1377403, EPI_ISL_1377404, EPI_ISL_1377405, EPI_ISL_1377406, EPI_ISL_1377407, EPI_ISL_1377408, EPI_ISL_1377410, EPI_ISL_1377411, EPI_ISL_1377412, EPI_ISL_1377413, EPI_ISL_1377420, EPI_ISL_1377424, EPI_ISL_1377425, EPI_ISL_1377426, EPI_ISL_1377429, EPI_ISL_1377433, EPI_ISL_1377434, EPI_ISL_1377435, EPI_ISL_1377439, EPI_ISL_1377442, EPI_ISL_1377443, EPI_ISL_1377444,
EPI_ISL_1377445, EPI_ISL_1377446, EPI_ISL_1377452, EPI_ISL_1377453, EPI_ISL_1377454, EPI_ISL_1377456, EPI_ISL_1377457, EPI_ISL_1377465, EPI_ISL_1377474, EPI_ISL_1377475, EPI_ISL_1377477, EPI_ISL_1377478, EPI_ISL_1377479, EPI_ISL_1377482, EPI_ISL_1377483, EPI_ISL_1377484, EPI_ISL_1377485, EPI_ISL_1377486, EPI_ISL_1377495, EPI_ISL_1377498, EPI_ISL_1377499, EPI_ISL_1377500,
EPI_ISL_1377501, EPI_ISL_1377504, EPI_ISL_1377507, EPI_ISL_1377513, EPI_ISL_1377514, EPI_ISL_1377517, EPI_ISL_1377519, EPI_ISL_1377520, EPI_ISL_1377521, EPI_ISL_1377522, EPI_ISL_1377524, EPI_ISL_1377525, EPI_ISL_1377526, EPI_ISL_1377528, EPI_ISL_1377531, EPI_ISL_1377532, EPI_ISL_1377534, EPI_ISL_1377535, EPI_ISL_1377537, EPI_ISL_1377538, EPI_ISL_1377539, EPI_ISL_1377541,
EPI_ISL_1377542, EPI_ISL_1377543, EPI_ISL_1377546, EPI_ISL_1377547, EPI_ISL_1377548, EPI_ISL_1377549, EPI_ISL_1377550, EPI_ISL_1377553, EPI_ISL_1377555, EPI_ISL_1377556, EPI_ISL_1377558, EPI_ISL_1377563, EPI_ISL_1377566, EPI_ISL_1377570, EPI_ISL_1377574, EPI_ISL_1377575, EPI_ISL_1377577, EPI_ISL_1377579, EPI_ISL_1377580, EPI_ISL_1377581, EPI_ISL_1377582, EPI_ISL_1377584,
EPI_ISL_1377589, EPI_ISL_1377590, EPI_ISL_1377592, EPI_ISL_1377593, EPI_ISL_1377596, EPI_ISL_1377598, EPI_ISL_1377601, EPI_ISL_1377603, EPI_ISL_1377607, EPI_ISL_1377608, EPI_ISL_1377609, EPI_ISL_1377611, EPI_ISL_1377612, EPI_ISL_1377615, EPI_ISL_1377617, EPI_ISL_1377618, EPI_ISL_1377620, EPI_ISL_1377621, EPI_ISL_1377625, EPI_ISL_1377627, EPI_ISL_1377629, EPI_ISL_1377630,
EPI_ISL_1377631, EPI_ISL_1377635, EPI_ISL_1377639, EPI_ISL_1377645, EPI_ISL_1377647, EPI_ISL_1377649, EPI_ISL_1377651, EPI_ISL_1377653, EPI_ISL_1377655, EPI_ISL_1377659, EPI_ISL_1377660, EPI_ISL_1377661, EPI_ISL_1377662, EPI_ISL_1377664, EPI_ISL_1377666, EPI_ISL_1377669, EPI_ISL_1377670, EPI_ISL_1377999, EPI_ISL_1378000, EPI_ISL_1378001, EPI_ISL_1378002, EPI_ISL_1378003,
EPI_ISL_1378004, EPI_ISL_1378005, EPI_ISL_1378006, EPI_ISL_1378007, EPI_ISL_1378008, EPI_ISL_1378009, EPI_ISL_1378010, EPI_ISL_1378011, EPI_ISL_1378012, EPI_ISL_1378013, EPI_ISL_1378014, EPI_ISL_1378015, EPI_ISL_1378016, EPI_ISL_1378017, EPI_ISL_1378018, EPI_ISL_1378019, EPI_ISL_1378020, EPI_ISL_1378021, EPI_ISL_1378022, EPI_ISL_1378023, EPI_ISL_1378024, EPI_ISL_1378025,
EPI_ISL_1378026, EPI_ISL_1378027, EPI_ISL_1378028, EPI_ISL_1378029, EPI_ISL_1378030, EPI_ISL_1378031, EPI_ISL_1378032, EPI_ISL_1378033, EPI_ISL_1378034, EPI_ISL_1378035, EPI_ISL_1378036, EPI_ISL_1378037, EPI_ISL_1378038, EPI_ISL_1378039, EPI_ISL_1378040, EPI_ISL_1378041, EPI_ISL_1378042, EPI_ISL_1378043, EPI_ISL_1378044, EPI_ISL_1378045, EPI_ISL_1378046, EPI_ISL_1378047,
EPI_ISL_1378049, EPI_ISL_1378050, EPI_ISL_1378051, EPI_ISL_1378052, EPI_ISL_1378053, EPI_ISL_1378054, EPI_ISL_1378055, EPI_ISL_1378056, EPI_ISL_1378057, EPI_ISL_1378058, EPI_ISL_1378059, EPI_ISL_1378060, EPI_ISL_1378061, EPI_ISL_1378062, EPI_ISL_1378063, EPI_ISL_1378064, EPI_ISL_1378065, EPI_ISL_1378066, EPI_ISL_1378067, EPI_ISL_1378068, EPI_ISL_1378069, EPI_ISL_1378070,
EPI_ISL_1378071, EPI_ISL_1378072, EPI_ISL_1378073, EPI_ISL_1378074, EPI_ISL_1378075, EPI_ISL_1378076, EPI_ISL_1378077, EPI_ISL_1378078, EPI_ISL_1378079, EPI_ISL_1378080, EPI_ISL_1378081, EPI_ISL_1378082, EPI_ISL_1378083, EPI_ISL_1378084, EPI_ISL_1378085, EPI_ISL_1378086, EPI_ISL_1378087, EPI_ISL_1378088, EPI_ISL_1378089, EPI_ISL_1378090, EPI_ISL_1378091, EPI_ISL_1378092,
EPI_ISL_1378093, EPI_ISL_1378094, EPI_ISL_1378095, EPI_ISL_1378096, EPI_ISL_1378097, EPI_ISL_1378098, EPI_ISL_1378099, EPI_ISL_1378100, EPI_ISL_1378101, EPI_ISL_1378102, EPI_ISL_1378103, EPI_ISL_1378104, EPI_ISL_1378105, EPI_ISL_1378106, EPI_ISL_1378107, EPI_ISL_1378108, EPI_ISL_1378109, EPI_ISL_1378110, EPI_ISL_1378111, EPI_ISL_1378112, EPI_ISL_1378113, EPI_ISL_1378114,
EPI_ISL_1378115, EPI_ISL_1378116, EPI_ISL_1378117, EPI_ISL_1378118, EPI_ISL_1378119, EPI_ISL_1378120, EPI_ISL_1378121, EPI_ISL_1378122, EPI_ISL_1378123, EPI_ISL_1378124, EPI_ISL_1378125, EPI_ISL_1378126, EPI_ISL_1378127, EPI_ISL_1378128, EPI_ISL_1378129, EPI_ISL_1378130, EPI_ISL_1378131, EPI_ISL_1378132, EPI_ISL_1378133, EPI_ISL_1378134, EPI_ISL_1378135, EPI_ISL_1378136,
EPI_ISL_1378137, EPI_ISL_1378138, EPI_ISL_1378139, EPI_ISL_1378140, EPI_ISL_1378141, EPI_ISL_1378142, EPI_ISL_1378143, EPI_ISL_1378144, EPI_ISL_1378145, EPI_ISL_1378146, EPI_ISL_1378147, EPI_ISL_1378148, EPI_ISL_1378149, EPI_ISL_1378150, EPI_ISL_1378151, EPI_ISL_1378152, EPI_ISL_1378153, EPI_ISL_1378154, EPI_ISL_1378155, EPI_ISL_1378156, EPI_ISL_1378157, EPI_ISL_1378158,
EPI_ISL_1378159, EPI_ISL_1378160, EPI_ISL_1378161, EPI_ISL_1378162, EPI_ISL_1378163, EPI_ISL_1378164, EPI_ISL_1378165, EPI_ISL_1378166, EPI_ISL_1378167, EPI_ISL_1378168, EPI_ISL_1378169, EPI_ISL_1378170, EPI_ISL_1378171, EPI_ISL_1378172, EPI_ISL_1378173, EPI_ISL_1378174, EPI_ISL_1378175, EPI_ISL_1378176, EPI_ISL_1378177, EPI_ISL_1378178, EPI_ISL_1378179, EPI_ISL_1378180,
EPI_ISL_1378181, EPI_ISL_1378182, EPI_ISL_1378183, EPI_ISL_1378184, EPI_ISL_1378185, EPI_ISL_1378186, EPI_ISL_1378187, EPI_ISL_1378188, EPI_ISL_1378189, EPI_ISL_1378190, EPI_ISL_1378191, EPI_ISL_1378192, EPI_ISL_1378193, EPI_ISL_1378194, EPI_ISL_1378195, EPI_ISL_1378196, EPI_ISL_1378197, EPI_ISL_1378198, EPI_ISL_1378199, EPI_ISL_1378200, EPI_ISL_1378201, EPI_ISL_1378202,
EPI_ISL_1378203, EPI_ISL_1378204, EPI_ISL_1378205, EPI_ISL_1378206, EPI_ISL_1378207, EPI_ISL_1378208, EPI_ISL_1378209, EPI_ISL_1378210, EPI_ISL_1378211, EPI_ISL_1378212, EPI_ISL_1378213, EPI_ISL_1378214, EPI_ISL_1378215, EPI_ISL_1378216, EPI_ISL_1378217, EPI_ISL_1378218, EPI_ISL_1378219, EPI_ISL_1378220, EPI_ISL_1378221, EPI_ISL_1378222, EPI_ISL_1378223, EPI_ISL_1378224,
EPI_ISL_1378225, EPI_ISL_1378226, EPI_ISL_1378227, EPI_ISL_1378228, EPI_ISL_1378229, EPI_ISL_1378230, EPI_ISL_1378231, EPI_ISL_1378232, EPI_ISL_1378233, EPI_ISL_1378234, EPI_ISL_1378235, EPI_ISL_1378236, EPI_ISL_1378237, EPI_ISL_1378238, EPI_ISL_1378239, EPI_ISL_1378240, EPI_ISL_1378241, EPI_ISL_1378242, EPI_ISL_1378243, EPI_ISL_1378244, EPI_ISL_1378245, EPI_ISL_1378246,
EPI_ISL_1378247, EPI_ISL_1378248, EPI_ISL_1378249, EPI_ISL_1378250, EPI_ISL_1378251, EPI_ISL_1378252, EPI_ISL_1378253, EPI_ISL_1378254, EPI_ISL_1378255, EPI_ISL_1378256, EPI_ISL_1378257, EPI_ISL_1378258, EPI_ISL_1378259, EPI_ISL_1378260, EPI_ISL_1378261, EPI_ISL_1378262, EPI_ISL_1378263, EPI_ISL_1378264, EPI_ISL_1378265, EPI_ISL_1378266, EPI_ISL_1378267, EPI_ISL_1378268,
EPI_ISL_1378269, EPI_ISL_1378270, EPI_ISL_1378271, EPI_ISL_1378272, EPI_ISL_1378273, EPI_ISL_1378274, EPI_ISL_1378275, EPI_ISL_1378276, EPI_ISL_1378277, EPI_ISL_1378278, EPI_ISL_1378279, EPI_ISL_1378280, EPI_ISL_1378281, EPI_ISL_1378282, EPI_ISL_1378283, EPI_ISL_1378284, EPI_ISL_1378285, EPI_ISL_1378286, EPI_ISL_1378287, EPI_ISL_1378288, EPI_ISL_1378289, EPI_ISL_1378290,
EPI_ISL_1378291, EPI_ISL_1378292, EPI_ISL_1378293, EPI_ISL_1378294, EPI_ISL_1378295, EPI_ISL_1378296, EPI_ISL_1378298, EPI_ISL_1378301, EPI_ISL_1378302, EPI_ISL_1378303, EPI_ISL_1378304, EPI_ISL_1378306, EPI_ISL_1378309, EPI_ISL_1378310, EPI_ISL_1378311, EPI_ISL_1378312, EPI_ISL_1378313, EPI_ISL_1378314, EPI_ISL_1378315, EPI_ISL_1378316, EPI_ISL_1378317, EPI_ISL_1378318,
EPI_ISL_1378321, EPI_ISL_1378322, EPI_ISL_1378323, EPI_ISL_1378325, EPI_ISL_1378326, EPI_ISL_1378327, EPI_ISL_1378329, EPI_ISL_1378331, EPI_ISL_1378332, EPI_ISL_1378335, EPI_ISL_1378336, EPI_ISL_1378337, EPI_ISL_1378338, EPI_ISL_1378342, EPI_ISL_1378344, EPI_ISL_1378345, EPI_ISL_1378346, EPI_ISL_1378347, EPI_ISL_1378348, EPI_ISL_1378351, EPI_ISL_1378352, EPI_ISL_1378353,
EPI_ISL_1378358, EPI_ISL_1378359, EPI_ISL_1378363, EPI_ISL_1378364, EPI_ISL_1378365, EPI_ISL_1378366, EPI_ISL_1378374, EPI_ISL_1378378, EPI_ISL_1378379, EPI_ISL_1378380, EPI_ISL_1378383, EPI_ISL_1378384, EPI_ISL_1378385, EPI_ISL_1378389, EPI_ISL_1378394, EPI_ISL_1378399, EPI_ISL_1378400, EPI_ISL_1378411, EPI_ISL_1378414, EPI_ISL_1378417, EPI_ISL_1378426, EPI_ISL_1389622,
EPI_ISL_1389623, EPI_ISL_1389624, EPI_ISL_1389625, EPI_ISL_1389626, EPI_ISL_1389627, EPI_ISL_1389628, EPI_ISL_1389629, EPI_ISL_1389630, EPI_ISL_1389631, EPI_ISL_1389632, EPI_ISL_1389633, EPI_ISL_1389634, EPI_ISL_1389635, EPI_ISL_1389636, EPI_ISL_1389637, EPI_ISL_1389638, EPI_ISL_1389639, EPI_ISL_1389640, EPI_ISL_1389641, EPI_ISL_1389642, EPI_ISL_1389643, EPI_ISL_1389644,
EPI_ISL_1389645, EPI_ISL_1389646, EPI_ISL_1389647, EPI_ISL_1389650, EPI_ISL_1389651, EPI_ISL_1389658, EPI_ISL_1389661, EPI_ISL_1389664, EPI_ISL_1389668, EPI_ISL_1389672, EPI_ISL_1389674, EPI_ISL_1389676, EPI_ISL_1389678, EPI_ISL_1389680, EPI_ISL_1389681, EPI_ISL_1389689, EPI_ISL_1389691, EPI_ISL_1389692, EPI_ISL_1389693, EPI_ISL_1389695, EPI_ISL_1389696, EPI_ISL_1389700,
EPI_ISL_1389703, EPI_ISL_1389705, EPI_ISL_1389707, EPI_ISL_1389711, EPI_ISL_1389727, EPI_ISL_1389728, EPI_ISL_1389731, EPI_ISL_1389732, EPI_ISL_1389734, EPI_ISL_1389735, EPI_ISL_1389736, EPI_ISL_1389737, EPI_ISL_1389740, EPI_ISL_1389742, EPI_ISL_1389744, EPI_ISL_1389746, EPI_ISL_1389749, EPI_ISL_1389751, EPI_ISL_1389756, EPI_ISL_1389765, EPI_ISL_1389766, EPI_ISL_1389770,
EPI_ISL_1389771, EPI_ISL_1389772, EPI_ISL_1389773, EPI_ISL_1389774, EPI_ISL_1389775, EPI_ISL_1389776, EPI_ISL_1389777, EPI_ISL_1389778, EPI_ISL_1389779, EPI_ISL_1389780, EPI_ISL_1389781, EPI_ISL_1389782, EPI_ISL_1389783, EPI_ISL_1389784, EPI_ISL_1389785, EPI_ISL_1389786, EPI_ISL_1389787, EPI_ISL_1389788, EPI_ISL_1389789, EPI_ISL_1389790, EPI_ISL_1389791, EPI_ISL_1389792,
EPI_ISL_1389793, EPI_ISL_1389794, EPI_ISL_1389795, EPI_ISL_1389796, EPI_ISL_1389797, EPI_ISL_1389798, EPI_ISL_1389799, EPI_ISL_1389800, EPI_ISL_1389801, EPI_ISL_1389802, EPI_ISL_1389803, EPI_ISL_1389804, EPI_ISL_1389805, EPI_ISL_1389806, EPI_ISL_1389807, EPI_ISL_1389808, EPI_ISL_1389809, EPI_ISL_1389810, EPI_ISL_1389811, EPI_ISL_1389812, EPI_ISL_1389813, EPI_ISL_1389814,
EPI_ISL_1389815, EPI_ISL_1389816, EPI_ISL_1389817, EPI_ISL_1389818, EPI_ISL_1389820, EPI_ISL_1389824, EPI_ISL_1389826, EPI_ISL_1389827, EPI_ISL_1389828, EPI_ISL_1389830, EPI_ISL_1389831, EPI_ISL_1389832, EPI_ISL_1389833, EPI_ISL_1389834, EPI_ISL_1389835, EPI_ISL_1389836, EPI_ISL_1389837, EPI_ISL_1389838, EPI_ISL_1389839, EPI_ISL_1389841, EPI_ISL_1389842, EPI_ISL_1389845,
EPI_ISL_1389846, EPI_ISL_1389847, EPI_ISL_1389848, EPI_ISL_1389849, EPI_ISL_1389850, EPI_ISL_1389851, EPI_ISL_1389852, EPI_ISL_1389853, EPI_ISL_1389854, EPI_ISL_1389856, EPI_ISL_1389857, EPI_ISL_1389858, EPI_ISL_1389859, EPI_ISL_1389860, EPI_ISL_1389861, EPI_ISL_1389863, EPI_ISL_1389864, EPI_ISL_1389865, EPI_ISL_1389866, EPI_ISL_1389867, EPI_ISL_1389868, EPI_ISL_1389869,
EPI_ISL_1389870, EPI_ISL_1389871, EPI_ISL_1389872, EPI_ISL_1389873, EPI_ISL_1389874, EPI_ISL_1389875, EPI_ISL_1389876, EPI_ISL_1389877, EPI_ISL_1389878, EPI_ISL_1389879, EPI_ISL_1389880, EPI_ISL_1389881, EPI_ISL_1389882, EPI_ISL_1389883, EPI_ISL_1389884, EPI_ISL_1389885, EPI_ISL_1389886, EPI_ISL_1389887, EPI_ISL_1389888, EPI_ISL_1389889, EPI_ISL_1389890, EPI_ISL_1389891,
EPI_ISL_1389892, EPI_ISL_1389893, EPI_ISL_1389894, EPI_ISL_1389895, EPI_ISL_1389896, EPI_ISL_1389897, EPI_ISL_1389898, EPI_ISL_1389899, EPI_ISL_1389900, EPI_ISL_1389901, EPI_ISL_1389902, EPI_ISL_1389903, EPI_ISL_1389904, EPI_ISL_1389905, EPI_ISL_1389906, EPI_ISL_1389907, EPI_ISL_1389908, EPI_ISL_1389909, EPI_ISL_1389910, EPI_ISL_1389911, EPI_ISL_1389912, EPI_ISL_1389913,
EPI_ISL_1389914, EPI_ISL_1389915, EPI_ISL_1389916, EPI_ISL_1389917, EPI_ISL_1389918, EPI_ISL_1389919, EPI_ISL_1389920, EPI_ISL_1389921, EPI_ISL_1389922, EPI_ISL_1389923, EPI_ISL_1389924, EPI_ISL_1389925, EPI_ISL_1389926, EPI_ISL_1389927, EPI_ISL_1389928, EPI_ISL_1389929, EPI_ISL_1389930, EPI_ISL_1389931, EPI_ISL_1389932, EPI_ISL_1389933, EPI_ISL_1389934, EPI_ISL_1389935,
EPI_ISL_1389936, EPI_ISL_1389937, EPI_ISL_1389938, EPI_ISL_1389939, EPI_ISL_1389940, EPI_ISL_1389941, EPI_ISL_1389942, EPI_ISL_1389943, EPI_ISL_1389944, EPI_ISL_1389945, EPI_ISL_1389946, EPI_ISL_1389947, EPI_ISL_1389948, EPI_ISL_1389949, EPI_ISL_1389950, EPI_ISL_1389951, EPI_ISL_1389952, EPI_ISL_1389953, EPI_ISL_1389954, EPI_ISL_1389955, EPI_ISL_1389956, EPI_ISL_1389957,
EPI_ISL_1389958, EPI_ISL_1389959, EPI_ISL_1389960, EPI_ISL_1389961, EPI_ISL_1389962, EPI_ISL_1389963, EPI_ISL_1389964, EPI_ISL_1389965, EPI_ISL_1389966, EPI_ISL_1389967, EPI_ISL_1389968, EPI_ISL_1389969, EPI_ISL_1389970, EPI_ISL_1389971, EPI_ISL_1389972, EPI_ISL_1389973, EPI_ISL_1389974, EPI_ISL_1389975, EPI_ISL_1389976, EPI_ISL_1389977, EPI_ISL_1389978, EPI_ISL_1389979,
EPI_ISL_1389980, EPI_ISL_1389981, EPI_ISL_1389982, EPI_ISL_1389983, EPI_ISL_1389984, EPI_ISL_1389985, EPI_ISL_1389986, EPI_ISL_1389987, EPI_ISL_1389988, EPI_ISL_1389989, EPI_ISL_1389990, EPI_ISL_1389991, EPI_ISL_1389992, EPI_ISL_1389993, EPI_ISL_1389994, EPI_ISL_1389995, EPI_ISL_1389996, EPI_ISL_1389997, EPI_ISL_1389998, EPI_ISL_1389999, EPI_ISL_1390000, EPI_ISL_1390001,
EPI_ISL_1390002, EPI_ISL_1390003, EPI_ISL_1390004, EPI_ISL_1390005, EPI_ISL_1390006, EPI_ISL_1390007, EPI_ISL_1390008, EPI_ISL_1390009, EPI_ISL_1390010, EPI_ISL_1390011, EPI_ISL_1390012, EPI_ISL_1390013, EPI_ISL_1390014, EPI_ISL_1390015, EPI_ISL_1390016, EPI_ISL_1390017, EPI_ISL_1390018, EPI_ISL_1390019, EPI_ISL_1390020, EPI_ISL_1390021, EPI_ISL_1390022, EPI_ISL_1390023,
EPI_ISL_1390024, EPI_ISL_1390025, EPI_ISL_1390026, EPI_ISL_1390027, EPI_ISL_1390028, EPI_ISL_1390029, EPI_ISL_1390030, EPI_ISL_1390031, EPI_ISL_1390032, EPI_ISL_1390033, EPI_ISL_1390034, EPI_ISL_1390035, EPI_ISL_1390036, EPI_ISL_1390037, EPI_ISL_1390038, EPI_ISL_1390039, EPI_ISL_1390040, EPI_ISL_1390041, EPI_ISL_1390042, EPI_ISL_1390043, EPI_ISL_1390044, EPI_ISL_1390045,
EPI_ISL_1390046, EPI_ISL_1390047, EPI_ISL_1390048, EPI_ISL_1390049, EPI_ISL_1390050, EPI_ISL_1390051, EPI_ISL_1390052, EPI_ISL_1390053, EPI_ISL_1390054, EPI_ISL_1390055, EPI_ISL_1390056, EPI_ISL_1390057, EPI_ISL_1390058, EPI_ISL_1390059, EPI_ISL_1390060, EPI_ISL_1390061, EPI_ISL_1390062, EPI_ISL_1390063, EPI_ISL_1390064, EPI_ISL_1390065, EPI_ISL_1390066, EPI_ISL_1390067,
EPI_ISL_1390068, EPI_ISL_1390069, EPI_ISL_1390070, EPI_ISL_1390071, EPI_ISL_1390072, EPI_ISL_1390073, EPI_ISL_1390074, EPI_ISL_1390075, EPI_ISL_1390076, EPI_ISL_1390077, EPI_ISL_1390078, EPI_ISL_1390079, EPI_ISL_1390080, EPI_ISL_1390081, EPI_ISL_1390082, EPI_ISL_1390083, EPI_ISL_1390084, EPI_ISL_1390085, EPI_ISL_1390086, EPI_ISL_1390087, EPI_ISL_1390088, EPI_ISL_1390089,
EPI_ISL_1390090, EPI_ISL_1390091, EPI_ISL_1390092, EPI_ISL_1390093, EPI_ISL_1390094, EPI_ISL_1390095, EPI_ISL_1390096, EPI_ISL_1390097, EPI_ISL_1390098, EPI_ISL_1390099, EPI_ISL_1390100, EPI_ISL_1390101, EPI_ISL_1390102, EPI_ISL_1390103, EPI_ISL_1390104, EPI_ISL_1390105, EPI_ISL_1390106, EPI_ISL_1390107, EPI_ISL_1390108, EPI_ISL_1390109, EPI_ISL_1390110, EPI_ISL_1390111,
EPI_ISL_1390112, EPI_ISL_1390113, EPI_ISL_1390114, EPI_ISL_1390115, EPI_ISL_1390116, EPI_ISL_1390117, EPI_ISL_1390118, EPI_ISL_1390119, EPI_ISL_1390120, EPI_ISL_1390121, EPI_ISL_1390122, EPI_ISL_1390123, EPI_ISL_1390124, EPI_ISL_1390125, EPI_ISL_1390126, EPI_ISL_1390127, EPI_ISL_1390128, EPI_ISL_1390129, EPI_ISL_1390130, EPI_ISL_1390131, EPI_ISL_1390132, EPI_ISL_1390133,
EPI_ISL_1390134, EPI_ISL_1390135, EPI_ISL_1390136, EPI_ISL_1390137, EPI_ISL_1390138, EPI_ISL_1390139, EPI_ISL_1390140, EPI_ISL_1390141, EPI_ISL_1390142, EPI_ISL_1390143, EPI_ISL_1390144, EPI_ISL_1390145, EPI_ISL_1390146, EPI_ISL_1390147, EPI_ISL_1390148, EPI_ISL_1390149, EPI_ISL_1390150, EPI_ISL_1390151, EPI_ISL_1390152, EPI_ISL_1390153, EPI_ISL_1390154, EPI_ISL_1390155,
EPI_ISL_1390156, EPI_ISL_1390157, EPI_ISL_1390158, EPI_ISL_1390159, EPI_ISL_1390160, EPI_ISL_1390161, EPI_ISL_1390162, EPI_ISL_1390163, EPI_ISL_1390164, EPI_ISL_1390165, EPI_ISL_1390166, EPI_ISL_1390167, EPI_ISL_1390168, EPI_ISL_1390169, EPI_ISL_1390170, EPI_ISL_1390171, EPI_ISL_1390172, EPI_ISL_1390173, EPI_ISL_1390174, EPI_ISL_1390175, EPI_ISL_1390176, EPI_ISL_1390177,
EPI_ISL_1390178, EPI_ISL_1390179, EPI_ISL_1390180, EPI_ISL_1390181, EPI_ISL_1390182, EPI_ISL_1390183, EPI_ISL_1390184, EPI_ISL_1390185, EPI_ISL_1390186, EPI_ISL_1390187, EPI_ISL_1390188, EPI_ISL_1390189, EPI_ISL_1390190, EPI_ISL_1390191, EPI_ISL_1390192, EPI_ISL_1390193, EPI_ISL_1390194, EPI_ISL_1390195, EPI_ISL_1390196, EPI_ISL_1390197, EPI_ISL_1390198, EPI_ISL_1390199,
EPI_ISL_1390200, EPI_ISL_1390201, EPI_ISL_1390202, EPI_ISL_1390203, EPI_ISL_1390204, EPI_ISL_1390205, EPI_ISL_1390206, EPI_ISL_1390207, EPI_ISL_1390208, EPI_ISL_1390209, EPI_ISL_1390210, EPI_ISL_1390211, EPI_ISL_1390212, EPI_ISL_1390213, EPI_ISL_1390214, EPI_ISL_1390215, EPI_ISL_1390216, EPI_ISL_1390217, EPI_ISL_1390218, EPI_ISL_1390219, EPI_ISL_1390220, EPI_ISL_1390221,
EPI_ISL_1390222, EPI_ISL_1390223, EPI_ISL_1390224, EPI_ISL_1390225, EPI_ISL_1390226, EPI_ISL_1390227, EPI_ISL_1390228, EPI_ISL_1390229, EPI_ISL_1390230, EPI_ISL_1390231, EPI_ISL_1390232, EPI_ISL_1390233, EPI_ISL_1390234, EPI_ISL_1390235, EPI_ISL_1390236, EPI_ISL_1390237, EPI_ISL_1390238, EPI_ISL_1390239, EPI_ISL_1390240, EPI_ISL_1390241, EPI_ISL_1390242, EPI_ISL_1390243,
EPI_ISL_1390244, EPI_ISL_1390245, EPI_ISL_1390246, EPI_ISL_1390247, EPI_ISL_1390248, EPI_ISL_1390249, EPI_ISL_1390250, EPI_ISL_1390251, EPI_ISL_1390252, EPI_ISL_1390253, EPI_ISL_1390254, EPI_ISL_1390255, EPI_ISL_1390256, EPI_ISL_1390257, EPI_ISL_1390258, EPI_ISL_1390259, EPI_ISL_1390260, EPI_ISL_1390261, EPI_ISL_1390262, EPI_ISL_1390263, EPI_ISL_1390264, EPI_ISL_1390265,
EPI_ISL_1390266, EPI_ISL_1390267, EPI_ISL_1390268, EPI_ISL_1390269, EPI_ISL_1390270, EPI_ISL_1390271, EPI_ISL_1390272, EPI_ISL_1390273, EPI_ISL_1390274, EPI_ISL_1390275, EPI_ISL_1390276, EPI_ISL_1390277, EPI_ISL_1390278, EPI_ISL_1390279, EPI_ISL_1390280, EPI_ISL_1390281, EPI_ISL_1390282, EPI_ISL_1390283, EPI_ISL_1390284, EPI_ISL_1390285, EPI_ISL_1390286, EPI_ISL_1390287,
EPI_ISL_1390288, EPI_ISL_1390289, EPI_ISL_1390290, EPI_ISL_1390291, EPI_ISL_1390292, EPI_ISL_1390293, EPI_ISL_1390294, EPI_ISL_1390295, EPI_ISL_1390296, EPI_ISL_1390297, EPI_ISL_1390298, EPI_ISL_1390299, EPI_ISL_1390300, EPI_ISL_1390301, EPI_ISL_1390302, EPI_ISL_1390303, EPI_ISL_1390304, EPI_ISL_1390305, EPI_ISL_1390306, EPI_ISL_1390307, EPI_ISL_1390308, EPI_ISL_1390309,
EPI_ISL_1390310, EPI_ISL_1390311, EPI_ISL_1390312, EPI_ISL_1390320, EPI_ISL_1390321, EPI_ISL_1390322, EPI_ISL_1390323, EPI_ISL_1390324, EPI_ISL_1390325, EPI_ISL_1390326, EPI_ISL_1390327, EPI_ISL_1390328, EPI_ISL_1390329, EPI_ISL_1390330, EPI_ISL_1390331, EPI_ISL_1390332, EPI_ISL_1390333, EPI_ISL_1390334, EPI_ISL_1390335, EPI_ISL_1390336, EPI_ISL_1390337, EPI_ISL_1390338,
EPI_ISL_1390339, EPI_ISL_1390340, EPI_ISL_1390341, EPI_ISL_1390342, EPI_ISL_1390343, EPI_ISL_1390344, EPI_ISL_1390345, EPI_ISL_1390346, EPI_ISL_1390347, EPI_ISL_1390348, EPI_ISL_1390349, EPI_ISL_1390350, EPI_ISL_1390351, EPI_ISL_1390352, EPI_ISL_1390353, EPI_ISL_1390354, EPI_ISL_1390355, EPI_ISL_1390356, EPI_ISL_1390357, EPI_ISL_1390358, EPI_ISL_1390359, EPI_ISL_1390360,
EPI_ISL_1390361, EPI_ISL_1390362, EPI_ISL_1390363, EPI_ISL_1390364, EPI_ISL_1390365, EPI_ISL_1390366, EPI_ISL_1390367, EPI_ISL_1390368, EPI_ISL_1390369, EPI_ISL_1390370, EPI_ISL_1390371, EPI_ISL_1390372, EPI_ISL_1390373, EPI_ISL_1390374, EPI_ISL_1390375, EPI_ISL_1390376, EPI_ISL_1409518, EPI_ISL_1409524, EPI_ISL_1409529, EPI_ISL_1409531, EPI_ISL_1409547, EPI_ISL_1409579,
EPI_ISL_1409583, EPI_ISL_1409597, EPI_ISL_1409609, EPI_ISL_1409615, EPI_ISL_1409638, EPI_ISL_1409652, EPI_ISL_1409670, EPI_ISL_1409717, EPI_ISL_1409723, EPI_ISL_1409738, EPI_ISL_1410730, EPI_ISL_1410799, EPI_ISL_1410819, EPI_ISL_1410932, EPI_ISL_1410989, EPI_ISL_1411013, EPI_ISL_1411055, EPI_ISL_1411145, EPI_ISL_1411176, EPI_ISL_1411271, EPI_ISL_1411338, EPI_ISL_1411363,
EPI_ISL_1411909, EPI_ISL_1411917, EPI_ISL_1411952, EPI_ISL_1411971, EPI_ISL_1411989, EPI_ISL_1412035, EPI_ISL_1412075, EPI_ISL_1412092, EPI_ISL_1412139, EPI_ISL_1412147, EPI_ISL_1413211, EPI_ISL_1413276, EPI_ISL_1413291, EPI_ISL_1413354, EPI_ISL_1413537, EPI_ISL_1413541, EPI_ISL_1413569, EPI_ISL_1413689, EPI_ISL_1413721, EPI_ISL_1413847, EPI_ISL_1413926, EPI_ISL_1414024,
EPI_ISL_1414040, EPI_ISL_1414045, EPI_ISL_1414061, EPI_ISL_1415816, EPI_ISL_1415818, EPI_ISL_1415827, EPI_ISL_1415834, EPI_ISL_1415845, EPI_ISL_1415852, EPI_ISL_1415854, EPI_ISL_1415863, EPI_ISL_1415864, EPI_ISL_1415865, EPI_ISL_1415870, EPI_ISL_1415874, EPI_ISL_1415878, EPI_ISL_1415884, EPI_ISL_1415885, EPI_ISL_1415888, EPI_ISL_1415892, EPI_ISL_1415905, EPI_ISL_1415907,
EPI_ISL_1415908, EPI_ISL_1415921, EPI_ISL_1415925, EPI_ISL_1415930, EPI_ISL_1415935, EPI_ISL_1415940, EPI_ISL_1415943, EPI_ISL_1415946, EPI_ISL_1415957, EPI_ISL_1415970, EPI_ISL_1415984, EPI_ISL_1415986, EPI_ISL_1415987, EPI_ISL_1415995, EPI_ISL_1415999, EPI_ISL_1416001, EPI_ISL_1416013, EPI_ISL_1416014, EPI_ISL_1416018, EPI_ISL_1416024, EPI_ISL_1416025, EPI_ISL_1416042,
EPI_ISL_1416043, EPI_ISL_1416050, EPI_ISL_1416071, EPI_ISL_1416074, EPI_ISL_1416075, EPI_ISL_1416086, EPI_ISL_1416094, EPI_ISL_1416098, EPI_ISL_1416102, EPI_ISL_1416109, EPI_ISL_1416110, EPI_ISL_1416117, EPI_ISL_1416123, EPI_ISL_1416180, EPI_ISL_1453897, EPI_ISL_1453948, EPI_ISL_1454158, EPI_ISL_1454165, EPI_ISL_1454167, EPI_ISL_1454169, EPI_ISL_1454172, EPI_ISL_1454174,
EPI_ISL_1454179, EPI_ISL_1454180, EPI_ISL_1454184, EPI_ISL_1454187, EPI_ISL_1454193, EPI_ISL_1454199, EPI_ISL_1454203, EPI_ISL_1454211, EPI_ISL_1454213, EPI_ISL_1454214, EPI_ISL_1454215, EPI_ISL_1454216, EPI_ISL_1454217, EPI_ISL_1454219, EPI_ISL_1454224, EPI_ISL_1454226, EPI_ISL_1454228, EPI_ISL_1454229, EPI_ISL_1454231, EPI_ISL_1454232, EPI_ISL_1454233, EPI_ISL_1454235,
EPI_ISL_1454237, EPI_ISL_1454239, EPI_ISL_1454240, EPI_ISL_1454244, EPI_ISL_1454245, EPI_ISL_1454246, EPI_ISL_1454247, EPI_ISL_1454260, EPI_ISL_1454261, EPI_ISL_1454262, EPI_ISL_1454264, EPI_ISL_1454265, EPI_ISL_1454266, EPI_ISL_1454268, EPI_ISL_1454276, EPI_ISL_1454278, EPI_ISL_1454279, EPI_ISL_1454284, EPI_ISL_1454285, EPI_ISL_1454286, EPI_ISL_1454287, EPI_ISL_1454290,
EPI_ISL_1454291, EPI_ISL_1454296, EPI_ISL_1454297, EPI_ISL_1454298, EPI_ISL_1454299, EPI_ISL_1454301, EPI_ISL_1454302, EPI_ISL_1454304, EPI_ISL_1454305, EPI_ISL_1454308, EPI_ISL_1454310, EPI_ISL_1454312, EPI_ISL_1454313, EPI_ISL_1454314, EPI_ISL_1454322, EPI_ISL_1454865, EPI_ISL_1455043, EPI_ISL_1455044, EPI_ISL_1455045, EPI_ISL_1455048, EPI_ISL_1455049, EPI_ISL_1455052,
EPI_ISL_1455054, EPI_ISL_1455057, EPI_ISL_1455058, EPI_ISL_1455059, EPI_ISL_1455061, EPI_ISL_1455062, EPI_ISL_1455063, EPI_ISL_1455064, EPI_ISL_1455065, EPI_ISL_1455066, EPI_ISL_1455068, EPI_ISL_1455069, EPI_ISL_1455071, EPI_ISL_1455074, EPI_ISL_1455076, EPI_ISL_1455077, EPI_ISL_1455078, EPI_ISL_1455082, EPI_ISL_1455083, EPI_ISL_1455087, EPI_ISL_1455088, EPI_ISL_1455089,
EPI_ISL_1455091, EPI_ISL_1455093, EPI_ISL_1455096, EPI_ISL_1455097, EPI_ISL_1455098, EPI_ISL_1455106, EPI_ISL_1455107, EPI_ISL_1455108, EPI_ISL_1455110, EPI_ISL_1455116, EPI_ISL_1455118, EPI_ISL_1455121, EPI_ISL_1455123, EPI_ISL_1455125, EPI_ISL_1455130, EPI_ISL_1455131, EPI_ISL_1455133, EPI_ISL_1455140, EPI_ISL_1455141, EPI_ISL_1455143, EPI_ISL_1455145, EPI_ISL_1455146,
EPI_ISL_1455149, EPI_ISL_1455154, EPI_ISL_1455155, EPI_ISL_1455156, EPI_ISL_1455157, EPI_ISL_1455159, EPI_ISL_1455160, EPI_ISL_1455164, EPI_ISL_1455166, EPI_ISL_1455169, EPI_ISL_1455171, EPI_ISL_1455172, EPI_ISL_1455176, EPI_ISL_1455178, EPI_ISL_1455179, EPI_ISL_1455181, EPI_ISL_1455182, EPI_ISL_1455184, EPI_ISL_1455185, EPI_ISL_1455188, EPI_ISL_1455192, EPI_ISL_1455193,
EPI_ISL_1455194, EPI_ISL_1455195, EPI_ISL_1455196, EPI_ISL_1455198, EPI_ISL_1455199, EPI_ISL_1455200, EPI_ISL_1455203, EPI_ISL_1455206, EPI_ISL_1455208, EPI_ISL_1455210, EPI_ISL_1455211, EPI_ISL_1455212, EPI_ISL_1455213, EPI_ISL_1455217, EPI_ISL_1455218, EPI_ISL_1455220, EPI_ISL_1455221, EPI_ISL_1455227, EPI_ISL_1455229, EPI_ISL_1455230, EPI_ISL_1455232, EPI_ISL_1455233,
EPI_ISL_1455234, EPI_ISL_1455235, EPI_ISL_1455237, EPI_ISL_1455243, EPI_ISL_1455249, EPI_ISL_1455251, EPI_ISL_1455255, EPI_ISL_1455262, EPI_ISL_1455263, EPI_ISL_1455273, EPI_ISL_1455282, EPI_ISL_1455283, EPI_ISL_1455286, EPI_ISL_1455287, EPI_ISL_1455288, EPI_ISL_1455293, EPI_ISL_1455299, EPI_ISL_1455312, EPI_ISL_1455314, EPI_ISL_1455319, EPI_ISL_1455321, EPI_ISL_1455325,
EPI_ISL_1455332, EPI_ISL_1455334, EPI_ISL_1455335, EPI_ISL_1455343, EPI_ISL_1455346, EPI_ISL_1455351, EPI_ISL_1455352, EPI_ISL_1455354, EPI_ISL_1455357, EPI_ISL_1455359, EPI_ISL_1455367, EPI_ISL_1455368, EPI_ISL_1455370, EPI_ISL_1455372, EPI_ISL_1455381, EPI_ISL_1455385, EPI_ISL_1455389, EPI_ISL_1455398, EPI_ISL_1455402, EPI_ISL_1455403, EPI_ISL_1455404, EPI_ISL_1455407,
EPI_ISL_1455409, EPI_ISL_1455413, EPI_ISL_1455447, EPI_ISL_1455449, EPI_ISL_1455451, EPI_ISL_1455452, EPI_ISL_1455454, EPI_ISL_1455458, EPI_ISL_1455464, EPI_ISL_1455472, EPI_ISL_1455474, EPI_ISL_1455480, EPI_ISL_1455481, EPI_ISL_1455520, EPI_ISL_1455532, EPI_ISL_1455534, EPI_ISL_1455536, EPI_ISL_1455537, EPI_ISL_1455541, EPI_ISL_1455566, EPI_ISL_1455572, EPI_ISL_1455574,
EPI_ISL_1455577, EPI_ISL_1455580, EPI_ISL_1455581, EPI_ISL_1455582, EPI_ISL_1455583, EPI_ISL_1455584, EPI_ISL_1455585, EPI_ISL_1455586, EPI_ISL_1455587, EPI_ISL_1455588, EPI_ISL_1455589, EPI_ISL_1455590, EPI_ISL_1455591, EPI_ISL_1455592, EPI_ISL_1455594, EPI_ISL_1455595, EPI_ISL_1455596, EPI_ISL_1455599, EPI_ISL_1455603, EPI_ISL_1455604, EPI_ISL_1455605, EPI_ISL_1455608,
EPI_ISL_1455609, EPI_ISL_1455610, EPI_ISL_1455611, EPI_ISL_1455616, EPI_ISL_1455618, EPI_ISL_1455620, EPI_ISL_1455623, EPI_ISL_1508158, EPI_ISL_1508291, EPI_ISL_1508312, EPI_ISL_1508447, EPI_ISL_1508728, EPI_ISL_1508730, EPI_ISL_1508731, EPI_ISL_1508732, EPI_ISL_1508733, EPI_ISL_1508734, EPI_ISL_1508735, EPI_ISL_1508738, EPI_ISL_1508739, EPI_ISL_1508740, EPI_ISL_1508741,
EPI_ISL_1508742, EPI_ISL_1508743, EPI_ISL_1508744, EPI_ISL_1508745, EPI_ISL_1508746, EPI_ISL_1508748, EPI_ISL_1508749, EPI_ISL_1508750, EPI_ISL_1508751, EPI_ISL_1508752, EPI_ISL_1508753, EPI_ISL_1508754, EPI_ISL_1508756, EPI_ISL_1508757, EPI_ISL_1508758, EPI_ISL_1508759, EPI_ISL_1508760, EPI_ISL_1508763, EPI_ISL_1508764, EPI_ISL_1508765, EPI_ISL_1508766, EPI_ISL_1508767,
EPI_ISL_1508768, EPI_ISL_1508769, EPI_ISL_1508770, EPI_ISL_1508771, EPI_ISL_1508773, EPI_ISL_1508774, EPI_ISL_1508775, EPI_ISL_1508776, EPI_ISL_1508778, EPI_ISL_1508779, EPI_ISL_1508780, EPI_ISL_1508781, EPI_ISL_1508782, EPI_ISL_1508783, EPI_ISL_1508784, EPI_ISL_1508785, EPI_ISL_1508786, EPI_ISL_1508787, EPI_ISL_1508788, EPI_ISL_1508789, EPI_ISL_1508790, EPI_ISL_1508791,
EPI_ISL_1508792, EPI_ISL_1508793, EPI_ISL_1508794, EPI_ISL_1508795, EPI_ISL_1508796, EPI_ISL_1508797, EPI_ISL_1508798, EPI_ISL_1508799, EPI_ISL_1508800, EPI_ISL_1508801, EPI_ISL_1508802, EPI_ISL_1508803, EPI_ISL_1508804, EPI_ISL_1508805, EPI_ISL_1508806, EPI_ISL_1508807, EPI_ISL_1508808, EPI_ISL_1508809, EPI_ISL_1508810, EPI_ISL_1508811, EPI_ISL_1508812, EPI_ISL_1508813,
EPI_ISL_1508814, EPI_ISL_1508815, EPI_ISL_1508816, EPI_ISL_1508817, EPI_ISL_1508818, EPI_ISL_1508819, EPI_ISL_1635429

| | | | |
|---|---|---|---|
| see above | Lighthouse Lab in Glasgow | Wellcome Sanger Institute for the COVID-19 Genomics UK (COG-UK) Consortium | Anna Dominiczak and Alex Alderton; Carol Clugston; Cordelia Langford; David Gray; David K. Jackson; Dominic Kwiatkowski; Ewan Harrison; Harper VanSteenhouse; Ian Johnston; Jeffrey Barrett; John Sillitoe on behalf of the Wellcome Sanger Institute COVID-19 Surveillance Team; Roberto Amato; Sonia Goncalves; Yumi Kasai |

EPI_ISL_1342236, EPI_ISL_1342242, EPI_ISL_1342243, EPI_ISL_1342249, EPI_ISL_1342251, EPI_ISL_1342255, EPI_ISL_1342258, EPI_ISL_1342264, EPI_ISL_1342267, EPI_ISL_1342278, EPI_ISL_1342280, EPI_ISL_1342287, EPI_ISL_1342288, EPI_ISL_1342305, EPI_ISL_1342309, EPI_ISL_1342317, EPI_ISL_1342319, EPI_ISL_1342323, EPI_ISL_1342325, EPI_ISL_1342333, EPI_ISL_1342338, EPI_ISL_1342346,
EPI_ISL_1342348, EPI_ISL_1342349, EPI_ISL_1342354, EPI_ISL_1342355, EPI_ISL_1342356, EPI_ISL_1342360, EPI_ISL_1342364, EPI_ISL_1342370, EPI_ISL_1342371, EPI_ISL_1342373, EPI_ISL_1342375, EPI_ISL_1342377, EPI_ISL_1342391, EPI_ISL_1342392, EPI_ISL_1342397, EPI_ISL_1342400, EPI_ISL_1342405, EPI_ISL_1342407, EPI_ISL_1342428, EPI_ISL_1342433, EPI_ISL_1342434, EPI_ISL_1342438,
EPI_ISL_1342440, EPI_ISL_1342442, EPI_ISL_1342446, EPI_ISL_1342454, EPI_ISL_1342462, EPI_ISL_1342467, EPI_ISL_1342470, EPI_ISL_1342472, EPI_ISL_1342474, EPI_ISL_1342477, EPI_ISL_1342485, EPI_ISL_1342488, EPI_ISL_1342489, EPI_ISL_1342491, EPI_ISL_1342497, EPI_ISL_1342505, EPI_ISL_1342506, EPI_ISL_1342512, EPI_ISL_1342817, EPI_ISL_1342822, EPI_ISL_1342827, EPI_ISL_1342832,
EPI_ISL_1342836, EPI_ISL_1342847, EPI_ISL_1342865, EPI_ISL_1342866, EPI_ISL_1342868, EPI_ISL_1342876, EPI_ISL_1342891, EPI_ISL_1342896, EPI_ISL_1342906, EPI_ISL_1342918, EPI_ISL_1342921, EPI_ISL_1342923, EPI_ISL_1342933, EPI_ISL_1342937, EPI_ISL_1342952, EPI_ISL_1342954, EPI_ISL_1342970, EPI_ISL_1342984, EPI_ISL_1342990, EPI_ISL_1342992, EPI_ISL_1342997, EPI_ISL_1342999,

EPI_ISL_1343004, EPI_ISL_1343006, EPI_ISL_1343008, EPI_ISL_1343016, EPI_ISL_1343026, EPI_ISL_1343028, EPI_ISL_1343029, EPI_ISL_1343030, EPI_ISL_1343032, EPI_ISL_1343033, EPI_ISL_1343043, EPI_ISL_1343062, EPI_ISL_1343072, EPI_ISL_1343074, EPI_ISL_1343083, EPI_ISL_1343095, EPI_ISL_1343827, EPI_ISL_1343838, EPI_ISL_1343840, EPI_ISL_1343844, EPI_ISL_1344015, EPI_ISL_1344036, EPI_ISL_1344070, EPI_ISL_1344091, EPI_ISL_1375161, EPI_ISL_1375162, EPI_ISL_1375163, EPI_ISL_1375164, EPI_ISL_1375165, EPI_ISL_1375167, EPI_ISL_1375169, EPI_ISL_1375170, EPI_ISL_1375177, EPI_ISL_1375178, EPI_ISL_1375179, EPI_ISL_1375182, EPI_ISL_1375185, EPI_ISL_1375187, EPI_ISL_1375189, EPI_ISL_1375193, EPI_ISL_1375195, EPI_ISL_1375198, EPI_ISL_1375201, EPI_ISL_1375204, EPI_ISL_1375205, EPI_ISL_1375206, EPI_ISL_1375208, EPI_ISL_1375216, EPI_ISL_1375218, EPI_ISL_1375219, EPI_ISL_1375221, EPI_ISL_1375224, EPI_ISL_1375226, EPI_ISL_1375227, EPI_ISL_1375228, EPI_ISL_1375232, EPI_ISL_1375233, EPI_ISL_1375234, EPI_ISL_1375235, EPI_ISL_1375236, EPI_ISL_1375238, EPI_ISL_1375240, EPI_ISL_1375241, EPI_ISL_1375242, EPI_ISL_1375243, EPI_ISL_1375245, EPI_ISL_1375246, EPI_ISL_1375249, EPI_ISL_1375251, EPI_ISL_1375255, EPI_ISL_1375257, EPI_ISL_1375258, EPI_ISL_1375259, EPI_ISL_1375262, EPI_ISL_1375263, EPI_ISL_1375264, EPI_ISL_1375266, EPI_ISL_1375267, EPI_ISL_1375269, EPI_ISL_1375270, EPI_ISL_1375273, EPI_ISL_1375278, EPI_ISL_1375279, EPI_ISL_1375280, EPI_ISL_1375281, EPI_ISL_1375283, EPI_ISL_1375285, EPI_ISL_1375286, EPI_ISL_1375287, EPI_ISL_1375289, EPI_ISL_1375290, EPI_ISL_1375293, EPI_ISL_1375294, EPI_ISL_1375297, EPI_ISL_1375303, EPI_ISL_1375308, EPI_ISL_1375309, EPI_ISL_1375311, EPI_ISL_1375315, EPI_ISL_1375319, EPI_ISL_1375321, EPI_ISL_1375324, EPI_ISL_1375325, EPI_ISL_1375327, EPI_ISL_1375329, EPI_ISL_1375330, EPI_ISL_1375334, EPI_ISL_1375336, EPI_ISL_1375341, EPI_ISL_1375345, EPI_ISL_1375348, EPI_ISL_1375356, EPI_ISL_1375358, EPI_ISL_1375359, EPI_ISL_1375360, EPI_ISL_1375361, EPI_ISL_1375363, EPI_ISL_1375364, EPI_ISL_1375367, EPI_ISL_1375369, EPI_ISL_1375370, EPI_ISL_1375371, EPI_ISL_1375373, EPI_ISL_1375377, EPI_ISL_1375378, EPI_ISL_1375379, EPI_ISL_1375380, EPI_ISL_1375384, EPI_ISL_1375389, EPI_ISL_1375390, EPI_ISL_1375392, EPI_ISL_1375393, EPI_ISL_1375394, EPI_ISL_1375397, EPI_ISL_1375398, EPI_ISL_1375400, EPI_ISL_1375403, EPI_ISL_1375404, EPI_ISL_1375405, EPI_ISL_1375406, EPI_ISL_1375407, EPI_ISL_1375412, EPI_ISL_1375413, EPI_ISL_1375415, EPI_ISL_1375416, EPI_ISL_1375418, EPI_ISL_1375419, EPI_ISL_1375420, EPI_ISL_1375421, EPI_ISL_1375424, EPI_ISL_1375427, EPI_ISL_1375428, EPI_ISL_1375430, EPI_ISL_1375431, EPI_ISL_1375432, EPI_ISL_1375437, EPI_ISL_1375438, EPI_ISL_1375439, EPI_ISL_1375440, EPI_ISL_1375441, EPI_ISL_1375442, EPI_ISL_1375443, EPI_ISL_1375446, EPI_ISL_1375449, EPI_ISL_1375452, EPI_ISL_1375453, EPI_ISL_1375454, EPI_ISL_1375455, EPI_ISL_1375457, EPI_ISL_1375460, EPI_ISL_1375463, EPI_ISL_1375468, EPI_ISL_1376223, EPI_ISL_1376225, EPI_ISL_1376229, EPI_ISL_1376231, EPI_ISL_1376234, EPI_ISL_1376236, EPI_ISL_1376244, EPI_ISL_1376248, EPI_ISL_1376251, EPI_ISL_1376252, EPI_ISL_1376255, EPI_ISL_1376257, EPI_ISL_1376258, EPI_ISL_1376259, EPI_ISL_1376266, EPI_ISL_1376269, EPI_ISL_1376271, EPI_ISL_1376274, EPI_ISL_1376275, EPI_ISL_1376280, EPI_ISL_1376282, EPI_ISL_1376285, EPI_ISL_1376293, EPI_ISL_1376295, EPI_ISL_1376297, EPI_ISL_1376298, EPI_ISL_1376299, EPI_ISL_1376301, EPI_ISL_1376303, EPI_ISL_1376305, EPI_ISL_1376306, EPI_ISL_1376307, EPI_ISL_1376308, EPI_ISL_1376309, EPI_ISL_1376314, EPI_ISL_1376317, EPI_ISL_1376319, EPI_ISL_1376320, EPI_ISL_1376322, EPI_ISL_1376323, EPI_ISL_1376327, EPI_ISL_1376328, EPI_ISL_1376331, EPI_ISL_1376334, EPI_ISL_1376338, EPI_ISL_1376339, EPI_ISL_1376343, EPI_ISL_1376344, EPI_ISL_1376350, EPI_ISL_1376353, EPI_ISL_1376356, EPI_ISL_1376358, EPI_ISL_1376359, EPI_ISL_1376363, EPI_ISL_1376364, EPI_ISL_1376365, EPI_ISL_1376368, EPI_ISL_1376369, EPI_ISL_1376370, EPI_ISL_1376372, EPI_ISL_1376374, EPI_ISL_1376376, EPI_ISL_1376380, EPI_ISL_1376383, EPI_ISL_1376387, EPI_ISL_1376390, EPI_ISL_1376397, EPI_ISL_1376398, EPI_ISL_1376402, EPI_ISL_1376404, EPI_ISL_1376407, EPI_ISL_1376424, EPI_ISL_1376427, EPI_ISL_1376431, EPI_ISL_1376433, EPI_ISL_1376436, EPI_ISL_1376437, EPI_ISL_1376439, EPI_ISL_1376445, EPI_ISL_1376448, EPI_ISL_1376450, EPI_ISL_1376451, EPI_ISL_1376452, EPI_ISL_1376453, EPI_ISL_1376462, EPI_ISL_1376464, EPI_ISL_1376467, EPI_ISL_1376471, EPI_ISL_1376474, EPI_ISL_1376479, EPI_ISL_1376480, EPI_ISL_1376492, EPI_ISL_1376494, EPI_ISL_1376497, EPI_ISL_1376501, EPI_ISL_1376507, EPI_ISL_1376508, EPI_ISL_1376509, EPI_ISL_1376510, EPI_ISL_1376512, EPI_ISL_1376519, EPI_ISL_1376521, EPI_ISL_1376526, EPI_ISL_1376528, EPI_ISL_1376530, EPI_ISL_1376537, EPI_ISL_1376545, EPI_ISL_1376546, EPI_ISL_1376549, EPI_ISL_1376551, EPI_ISL_1376553, EPI_ISL_1376555, EPI_ISL_1376556, EPI_ISL_1376562, EPI_ISL_1376563, EPI_ISL_1376565, EPI_ISL_1376566, EPI_ISL_1376567, EPI_ISL_1376568, EPI_ISL_1376569, EPI_ISL_1376570, EPI_ISL_1376572, EPI_ISL_1376575, EPI_ISL_1376576, EPI_ISL_1376578, EPI_ISL_1376580, EPI_ISL_1376582, EPI_ISL_1376584, EPI_ISL_1376586, EPI_ISL_1376587, EPI_ISL_1376591, EPI_ISL_1376593, EPI_ISL_1376595, EPI_ISL_1376596, EPI_ISL_1376599, EPI_ISL_1376602, EPI_ISL_1376604, EPI_ISL_1376606, EPI_ISL_1376607, EPI_ISL_1376610, EPI_ISL_1376613, EPI_ISL_1376614, EPI_ISL_1376615, EPI_ISL_1376616, EPI_ISL_1376618, EPI_ISL_1376619, EPI_ISL_1376620, EPI_ISL_1376624, EPI_ISL_1376628, EPI_ISL_1376630, EPI_ISL_1376633, EPI_ISL_1376635, EPI_ISL_1376636, EPI_ISL_1376643, EPI_ISL_1376644, EPI_ISL_1376645, EPI_ISL_1376646, EPI_ISL_1376648, EPI_ISL_1376649, EPI_ISL_1376651, EPI_ISL_1376652, EPI_ISL_1376653, EPI_ISL_1376659, EPI_ISL_1376663, EPI_ISL_1376668, EPI_ISL_1376669, EPI_ISL_1376674, EPI_ISL_1376675, EPI_ISL_1376677, EPI_ISL_1376680, EPI_ISL_1376681, EPI_ISL_1376682, EPI_ISL_1376683, EPI_ISL_1376685, EPI_ISL_1376686, EPI_ISL_1376687, EPI_ISL_1376692, EPI_ISL_1376693, EPI_ISL_1376696, EPI_ISL_1376697, EPI_ISL_1376699, EPI_ISL_1376701, EPI_ISL_1376702, EPI_ISL_1376704, EPI_ISL_1376705, EPI_ISL_1376706, EPI_ISL_1376707, EPI_ISL_1376716, EPI_ISL_1376717, EPI_ISL_1376719, EPI_ISL_1376720, EPI_ISL_1376728, EPI_ISL_1376730, EPI_ISL_1376735, EPI_ISL_1376738, EPI_ISL_1376747, EPI_ISL_1376750, EPI_ISL_1376753, EPI_ISL_1376754, EPI_ISL_1376757, EPI_ISL_1376764, EPI_ISL_1376766, EPI_ISL_1376767, EPI_ISL_1376770, EPI_ISL_1376772, EPI_ISL_1376774, EPI_ISL_1376775, EPI_ISL_1377459, EPI_ISL_1377472, EPI_ISL_1377568, EPI_ISL_1377622, EPI_ISL_1377632, EPI_ISL_1377637, EPI_ISL_1377650, EPI_ISL_1377658, EPI_ISL_1377671, EPI_ISL_1377672, EPI_ISL_1377676, EPI_ISL_1377682, EPI_ISL_1377687, EPI_ISL_1377689, EPI_ISL_1377692, EPI_ISL_1377693, EPI_ISL_1377695, EPI_ISL_1377699, EPI_ISL_1377705, EPI_ISL_1377710, EPI_ISL_1377713, EPI_ISL_1377714, EPI_ISL_1377717, EPI_ISL_1377718, EPI_ISL_1377721, EPI_ISL_1377723, EPI_ISL_1377728, EPI_ISL_1377731, EPI_ISL_1377732, EPI_ISL_1377733, EPI_ISL_1377734, EPI_ISL_1377737, EPI_ISL_1377739, EPI_ISL_1377740, EPI_ISL_1377742, EPI_ISL_1377743, EPI_ISL_1377746, EPI_ISL_1377747, EPI_ISL_1377774, EPI_ISL_1377782, EPI_ISL_1377784, EPI_ISL_1377785, EPI_ISL_1377786, EPI_ISL_1377790, EPI_ISL_1377797, EPI_ISL_1377803, EPI_ISL_1377805, EPI_ISL_1377806, EPI_ISL_1377810, EPI_ISL_1377813, EPI_ISL_1377819, EPI_ISL_1377829, EPI_ISL_1377833, EPI_ISL_1377836, EPI_ISL_1377839, EPI_ISL_1377849, EPI_ISL_1377860, EPI_ISL_1377861, EPI_ISL_1377862, EPI_ISL_1377867, EPI_ISL_1377869, EPI_ISL_1377874, EPI_ISL_1377877, EPI_ISL_1377879, EPI_ISL_1377881, EPI_ISL_1377888, EPI_ISL_1377893, EPI_ISL_1377899, EPI_ISL_1377900, EPI_ISL_1377904, EPI_ISL_1377906, EPI_ISL_1377912, EPI_ISL_1377913, EPI_ISL_1377919, EPI_ISL_1377921, EPI_ISL_1377928, EPI_ISL_1377929, EPI_ISL_1377932, EPI_ISL_1377939, EPI_ISL_1377955, EPI_ISL_1377956, EPI_ISL_1377957, EPI_ISL_1377958, EPI_ISL_1377967, EPI_ISL_1377968, EPI_ISL_1377969, EPI_ISL_1377970, EPI_ISL_1377972, EPI_ISL_1377976, EPI_ISL_1377989, EPI_ISL_1377990, EPI_ISL_1377991, EPI_ISL_1408956, EPI_ISL_1409022, EPI_ISL_1409023, EPI_ISL_1409042, EPI_ISL_1409045, EPI_ISL_1409054, EPI_ISL_1409065, EPI_ISL_1409077, EPI_ISL_1409089, EPI_ISL_1409098, EPI_ISL_1409106, EPI_ISL_1409227, EPI_ISL_1409303, EPI_ISL_1411793, EPI_ISL_1415822, EPI_ISL_1415823, EPI_ISL_1415824, EPI_ISL_1415828, EPI_ISL_1415835, EPI_ISL_1415838, EPI_ISL_1415839, EPI_ISL_1415840, EPI_ISL_1415843, EPI_ISL_1415844, EPI_ISL_1415846, EPI_ISL_1415848, EPI_ISL_1415849, EPI_ISL_1415850, EPI_ISL_1415857, EPI_ISL_1415858, EPI_ISL_1415859, EPI_ISL_1415861, EPI_ISL_1415866, EPI_ISL_1415868, EPI_ISL_1415869, EPI_ISL_1415871, EPI_ISL_1415872, EPI_ISL_1415873, EPI_ISL_1415887, EPI_ISL_1415896, EPI_ISL_1415897, EPI_ISL_1415899, EPI_ISL_1415900, EPI_ISL_1415902, EPI_ISL_1415903, EPI_ISL_1415906, EPI_ISL_1415909, EPI_ISL_1415912, EPI_ISL_1415913, EPI_ISL_1415919, EPI_ISL_1415920, EPI_ISL_1415922, EPI_ISL_1415927, EPI_ISL_1415933, EPI_ISL_1415936, EPI_ISL_1415939, EPI_ISL_1415944, EPI_ISL_1415950, EPI_ISL_1415951, EPI_ISL_1415952, EPI_ISL_1415953, EPI_ISL_1415955, EPI_ISL_1415956, EPI_ISL_1415959, EPI_ISL_1415961, EPI_ISL_1415965, EPI_ISL_1415967, EPI_ISL_1415971, EPI_ISL_1415973, EPI_ISL_1415974, EPI_ISL_1415977, EPI_ISL_1415981, EPI_ISL_1415982, EPI_ISL_1415983, EPI_ISL_1415989, EPI_ISL_1415990, EPI_ISL_1415992, EPI_ISL_1415994, EPI_ISL_1415996, EPI_ISL_1415998, EPI_ISL_1416000, EPI_ISL_1416005, EPI_ISL_1416006, EPI_ISL_1416009, EPI_ISL_1416010, EPI_ISL_1416020, EPI_ISL_1416022, EPI_ISL_1416023, EPI_ISL_1416026, EPI_ISL_1416027, EPI_ISL_1416031, EPI_ISL_1416037, EPI_ISL_1416038, EPI_ISL_1416040, EPI_ISL_1416048, EPI_ISL_1416049, EPI_ISL_1416051, EPI_ISL_1416054, EPI_ISL_1416059, EPI_ISL_1416060, EPI_ISL_1416061, EPI_ISL_1416062, EPI_ISL_1416064, EPI_ISL_1416065, EPI_ISL_1416067, EPI_ISL_1416068, EPI_ISL_1416069, EPI_ISL_1416079, EPI_ISL_1416085, EPI_ISL_1416092, EPI_ISL_1416093, EPI_ISL_1416097, EPI_ISL_1416099, EPI_ISL_1416100, EPI_ISL_1416101, EPI_ISL_1416104, EPI_ISL_1416105, EPI_ISL_1416113, EPI_ISL_1416114, EPI_ISL_1416116, EPI_ISL_1416120, EPI_ISL_1416121, EPI_ISL_1448798, EPI_ISL_1450018, EPI_ISL_1450054, EPI_ISL_1450115, EPI_ISL_1450246, EPI_ISL_1450254, EPI_ISL_1450269, EPI_ISL_1450903, EPI_ISL_1450999, EPI_ISL_1451128, EPI_ISL_1451133, EPI_ISL_1452103, EPI_ISL_1452111, EPI_ISL_1452213, EPI_ISL_1452277, EPI_ISL_1452281, EPI_ISL_1452313, EPI_ISL_1453480, EPI_ISL_1454665, EPI_ISL_1454690, EPI_ISL_1454711, EPI_ISL_1454723, EPI_ISL_1454736, EPI_ISL_1454771, EPI_ISL_1454819, EPI_ISL_1454866, EPI_ISL_1455238, EPI_ISL_1455240, EPI_ISL_1455242, EPI_ISL_1455244, EPI_ISL_1455246, EPI_ISL_1455248, EPI_ISL_1455250, EPI_ISL_1455257, EPI_ISL_1455258, EPI_ISL_1455266, EPI_ISL_1455267, EPI_ISL_1455268, EPI_ISL_1455270, EPI_ISL_1455274, EPI_ISL_1455275, EPI_ISL_1455276, EPI_ISL_1455277, EPI_ISL_1455278, EPI_ISL_1455280, EPI_ISL_1455281, EPI_ISL_1455284, EPI_ISL_1455291, EPI_ISL_1455294, EPI_ISL_1455295, EPI_ISL_1455298, EPI_ISL_1455301, EPI_ISL_1455302, EPI_ISL_1455311, EPI_ISL_1455313, EPI_ISL_1455315, EPI_ISL_1455316, EPI_ISL_1455317, EPI_ISL_1455322, EPI_ISL_1455324, EPI_ISL_1455326, EPI_ISL_1455329, EPI_ISL_1455331, EPI_ISL_1455333, EPI_ISL_1455337, EPI_ISL_1455339, EPI_ISL_1455340, EPI_ISL_1455342, EPI_ISL_1455344, EPI_ISL_1455345, EPI_ISL_1455349, EPI_ISL_1455350, EPI_ISL_1455355, EPI_ISL_1455361, EPI_ISL_1455362, EPI_ISL_1455364, EPI_ISL_1455365, EPI_ISL_1455369, EPI_ISL_1455371, EPI_ISL_1455373, EPI_ISL_1455374, EPI_ISL_1455375, EPI_ISL_1455376, EPI_ISL_1455378, EPI_ISL_1455379, EPI_ISL_1455380, EPI_ISL_1455382, EPI_ISL_1455383, EPI_ISL_1455386, EPI_ISL_1455387, EPI_ISL_1455388, EPI_ISL_1455390, EPI_ISL_1455391, EPI_ISL_1455394, EPI_ISL_1455395, EPI_ISL_1455396, EPI_ISL_1455397, EPI_ISL_1455400, EPI_ISL_1455401, EPI_ISL_1455405, EPI_ISL_1455410, EPI_ISL_1455411, EPI_ISL_1455416, EPI_ISL_1455417, EPI_ISL_1455418, EPI_ISL_1455419, EPI_ISL_1455421, EPI_ISL_1455423, EPI_ISL_1455424, EPI_ISL_1455425, EPI_ISL_1455427, EPI_ISL_1455429, EPI_ISL_1455432, EPI_ISL_1455435, EPI_ISL_1455437, EPI_ISL_1455439, EPI_ISL_1455440, EPI_ISL_1455441, EPI_ISL_1455442, EPI_ISL_1455444, EPI_ISL_1455445, EPI_ISL_1455446, EPI_ISL_1455450, EPI_ISL_1455453, EPI_ISL_1455455, EPI_ISL_1455456, EPI_ISL_1455457, EPI_ISL_1455459, EPI_ISL_1455460, EPI_ISL_1455461, EPI_ISL_1455462, EPI_ISL_1455467, EPI_ISL_1455469, EPI_ISL_1455470, EPI_ISL_1455471, EPI_ISL_1455476, EPI_ISL_1455482, EPI_ISL_1455483, EPI_ISL_1455484, EPI_ISL_1455486, EPI_ISL_1455489, EPI_ISL_1455490, EPI_ISL_1455491, EPI_ISL_1455554, EPI_ISL_1455555, EPI_ISL_1455559, EPI_ISL_1455560, EPI_ISL_1455569, EPI_ISL_1455571, EPI_ISL_1455593, EPI_ISL_1455597, EPI_ISL_1455598, EPI_ISL_1455600, EPI_ISL_1455601, EPI_ISL_1455602, EPI_ISL_1455606, EPI_ISL_1455607, EPI_ISL_1455614, EPI_ISL_1455621, EPI_ISL_1455626, EPI_ISL_1455630, EPI_ISL_1467858, EPI_ISL_1518071, EPI_ISL_1518083, EPI_ISL_1518088, EPI_ISL_1518145, EPI_ISL_1518167, EPI_ISL_1518221, EPI_ISL_1518222, EPI_ISL_1518260, EPI_ISL_1518312, EPI_ISL_1518316, EPI_ISL_1518322, EPI_ISL_1518369, EPI_ISL_1518830, EPI_ISL_1537331, EPI_ISL_1537332, EPI_ISL_1537363, EPI_ISL_1537373, EPI_ISL_1537377, EPI_ISL_1537382, EPI_ISL_1537386, EPI_ISL_1537405, EPI_ISL_1537431, EPI_ISL_1537466, EPI_ISL_1537539, EPI_ISL_1537583, EPI_ISL_1537584, EPI_ISL_1537645, EPI_ISL_1537753, EPI_ISL_1537785, EPI_ISL_1537788, EPI_ISL_1537834, EPI_ISL_1564060, EPI_ISL_1564070, EPI_ISL_1564071, EPI_ISL_1564081, EPI_ISL_1564092, EPI_ISL_1564093, EPI_ISL_1564094, EPI_ISL_1564098, EPI_ISL_1564099, EPI_ISL_1564103, EPI_ISL_1564108, EPI_ISL_1564116, EPI_ISL_1564129, EPI_ISL_1564131, EPI_ISL_1564150, EPI_ISL_1564153, EPI_ISL_1564160, EPI_ISL_1564173, EPI_ISL_1564205, EPI_ISL_1564210, EPI_ISL_1564251, EPI_ISL_1564291, EPI_ISL_1564324, EPI_ISL_1584172, EPI_ISL_1584175, EPI_ISL_1584178, EPI_ISL_1584193, EPI_ISL_1584197, EPI_ISL_1584201, EPI_ISL_1584223, EPI_ISL_1584264, EPI_ISL_1584275, EPI_ISL_1584282, EPI_ISL_1584297, EPI_ISL_1584311, EPI_ISL_1584316, EPI_ISL_1584328, EPI_ISL_1584338, EPI_ISL_1584389, EPI_ISL_1584417, EPI_ISL_1594997, EPI_ISL_1595000, EPI_ISL_1595001, EPI_ISL_1595003, EPI_ISL_1595006, EPI_ISL_1595007, EPI_ISL_1595013, EPI_ISL_1595016, EPI_ISL_1595019, EPI_ISL_1595024, EPI_ISL_1595033, EPI_ISL_1595042, EPI_ISL_1595043, EPI_ISL_1595044, EPI_ISL_1595048, EPI_ISL_1595059, EPI_ISL_1595070, EPI_ISL_1595083, EPI_ISL_1595092, EPI_ISL_1595101, EPI_ISL_1595106, EPI_ISL_1595107, EPI_ISL_1595111, EPI_ISL_1595120, EPI_ISL_1595123, EPI_ISL_1595128, EPI_ISL_1595140, EPI_ISL_1595144, EPI_ISL_1595146, EPI_ISL_1595160, EPI_ISL_1595167, EPI_ISL_1595168, EPI_ISL_1595172, EPI_ISL_1595173, EPI_ISL_1595175, EPI_ISL_1595185, EPI_ISL_1595195, EPI_ISL_1595196, EPI_ISL_1595197, EPI_ISL_1595199

| | | | |
|---|---|---|---|
| see above | Lighthouse Lab in Milton Keynes | Wellcome Sanger Institute for the COVID-19 Genomics UK (COG-UK) Consortium | Cordelia Langford; David K. Jackson; Dominic Kwiatkowski; Ewan Harrison; Ian Johnston; Jeffrey Barrett; John Sillitoe on behalf of the Wellcome Sanger Institute COVID-19 Surveillance Team; Roberto Amato; Sonia Goncalves; The Lighthouse Lab in Milton Keynes and Alex Alderton |

EPI_ISL_1475515, EPI_ISL_1475529, EPI_ISL_1475530, EPI_ISL_1475531, EPI_ISL_1475532, EPI_ISL_1475533, EPI_ISL_1475534, EPI_ISL_1475535, EPI_ISL_1475536, EPI_ISL_1475537

| | | | |
|---|---|---|---|
| see above | Lincolnshire Hospitals and DeepSeq Nottingham | COVID-19 Genomics UK (COG-UK) Consortium | Christopher Moore; Fei Sang; Joeseph Chappell; Johnny Debebe; Jonathan Ball; Matthew Carlisle; Matthew Loose; Nadine Holmes; Nichola Duckworth; Patrick McClure; Sarah Walsh; Tim Sloan; Victoria Wright |

EPI_ISL_1386806, EPI_ISL_1474735, EPI_ISL_1474736, EPI_ISL_1474737, EPI_ISL_1474744, EPI_ISL_1474753, EPI_ISL_1474756, EPI_ISL_1474758, EPI_ISL_1474759, EPI_ISL_1474760, EPI_ISL_1474761, EPI_ISL_1474763, EPI_ISL_1474765, EPI_ISL_1474766, EPI_ISL_1474767, EPI_ISL_1474768, EPI_ISL_1474771, EPI_ISL_1474775, EPI_ISL_1474777, EPI_ISL_1474778, EPI_ISL_1474780, EPI_ISL_1474782, EPI_ISL_1474790, EPI_ISL_1474791, EPI_ISL_1474792, EPI_ISL_1474793, EPI_ISL_1474794, EPI_ISL_1474795, EPI_ISL_1546154, EPI_ISL_1546221, EPI_ISL_1546223, EPI_ISL_1546225, EPI_ISL_1546227, EPI_ISL_1637242, EPI_ISL_1637243, EPI_ISL_1637244, EPI_ISL_1637245, EPI_ISL_1637247, EPI_ISL_1637248, EPI_ISL_1637268

| | | | |
|---|---|---|---|
| see above | Liverpool Clinical Laboratories | COVID-19 Genomics UK (COG-UK) Consortium | A Alrezaihi; Alessandro Gerada; Alistair Darby; Angela Cowell; Anita Lucaci; Anu Chawla; Cassie Olateju; Catherine Hartley; Charlotte Nelson; Ecaterina Vamos; Elaine O'Toole; Eleanor G Bentley; Ghada T Shawli; Hermione Webster; Isabel García-Dorival; James Johnson; James P Stewart; Jenifer Manson; Joanne Watts; Jones Benjamin; Jordan J Clark; Julian Hiscox; L Luu; Lucille Rainbow; M Almsaud; Margaret Hughes; Mark Whitehead; Matthew Gemmell; Miren Iturriza-Gomara; Muhannad Alruwaili; N.P Randle; Neil Swainston; PKF Gilmore; Parul Sharma; Rebekah Penrice-Randal; Richard Eccles; Richard Gregory; Sam Haldenby; Steve Paterson; Stuart D Armstrong; Trevor Ian Robinson; Ximeng Han |

EPI_ISL_1387079, EPI_ISL_1387080, EPI_ISL_1387091, EPI_ISL_1387099, EPI_ISL_1387100, EPI_ISL_1475206, EPI_ISL_1475207, EPI_ISL_1475336, EPI_ISL_1475337, EPI_ISL_1475338

| | | | |
|---|---|---|---|
| see above | Northumbria University / South Tees Hospitals NHS Foundation Trust / North Cumbria Integrated Care NHS Foundation Trust / North Tees and Hartlepool NHS Foundation Trust / Newcastle Hospitals NHS Foundation Trust | COVID-19 Genomics UK (COG-UK) Consortium | Andrew Nelson; Brendan Payne; Clive Graham; Darren L Smith; Debra Padgett; Edward Barton; Emma Swindells; Garren Scott; Gary Black; Gary Eltringham; Giles S Holt; Greg R Young; Jane Greenaway; Jennifer Collins; John Allan; Joshua Loh; Lynn Dover; Matthew Bashton; Mohammad A Tariq; Paul Baker; Sarah Essex; Steve Liggett; Wen C Yew; Yusri Taha |

EPI_ISL_1387995, EPI_ISL_1387998, EPI_ISL_1387999, EPI_ISL_1388000, EPI_ISL_1388001, EPI_ISL_1388005, EPI_ISL_1388007, EPI_ISL_1388008, EPI_ISL_1388009, EPI_ISL_1388010, EPI_ISL_1388013, EPI_ISL_1388014, EPI_ISL_1388017, EPI_ISL_1388064, EPI_ISL_1388076, EPI_ISL_1388080, EPI_ISL_1388082, EPI_ISL_1388084, EPI_ISL_1388085, EPI_ISL_1388086, EPI_ISL_1388089, EPI_ISL_1388163, EPI_ISL_1388260, EPI_ISL_1388266, EPI_ISL_1388284, EPI_ISL_1388315, EPI_ISL_1388327, EPI_ISL_1388345, EPI_ISL_1388347, EPI_ISL_1388353, EPI_ISL_1388357, EPI_ISL_1388359, EPI_ISL_1388363, EPI_ISL_1388369, EPI_ISL_1388371, EPI_ISL_1388378, EPI_ISL_1388383, EPI_ISL_1388395, EPI_ISL_1388399, EPI_ISL_1388428, EPI_ISL_1388430, EPI_ISL_1388431, EPI_ISL_1388439, EPI_ISL_1388445, EPI_ISL_1388448, EPI_ISL_1388456, EPI_ISL_1388458, EPI_ISL_1388461, EPI_ISL_1388467, EPI_ISL_1388475, EPI_ISL_1388477, EPI_ISL_1388487, EPI_ISL_1388491, EPI_ISL_1388493, EPI_ISL_1388500, EPI_ISL_1388502, EPI_ISL_1388505, EPI_ISL_1388506, EPI_ISL_1388521, EPI_ISL_1388523, EPI_ISL_1388525, EPI_ISL_1388531, EPI_ISL_1388535, EPI_ISL_1388596, EPI_ISL_1388599, EPI_ISL_1388605, EPI_ISL_1388607, EPI_ISL_1388609, EPI_ISL_1388615, EPI_ISL_1388628, EPI_ISL_1388630, EPI_ISL_1388640, EPI_ISL_1388641, EPI_ISL_1475787, EPI_ISL_1475847, EPI_ISL_1475857, EPI_ISL_1475859, EPI_ISL_1475866, EPI_ISL_1475874, EPI_ISL_1475877, EPI_ISL_1475878, EPI_ISL_1475880, EPI_ISL_1475890, EPI_ISL_1475901, EPI_ISL_1475904, EPI_ISL_1475907, EPI_ISL_1475908, EPI_ISL_1475913, EPI_ISL_1475918, EPI_ISL_1475921, EPI_ISL_1475922, EPI_ISL_1475923, EPI_ISL_1475925, EPI_ISL_1475933, EPI_ISL_1475948, EPI_ISL_1475949, EPI_ISL_1475950, EPI_ISL_1475951, EPI_ISL_1475953, EPI_ISL_1475954, EPI_ISL_1475960, EPI_ISL_1475961, EPI_ISL_1475967, EPI_ISL_1475968, EPI_ISL_1475970, EPI_ISL_1475996, EPI_ISL_1476000, EPI_ISL_1476002, EPI_ISL_1476013, EPI_ISL_1476014, EPI_ISL_1476019, EPI_ISL_1476020, EPI_ISL_1476026, EPI_ISL_1476029, EPI_ISL_1476030, EPI_ISL_1476034, EPI_ISL_1476035, EPI_ISL_1476036, EPI_ISL_1476038, EPI_ISL_1476042, EPI_ISL_1476044, EPI_ISL_1476049, EPI_ISL_1476052, EPI_ISL_1476053, EPI_ISL_1476055, EPI_ISL_1476057, EPI_ISL_1476058, EPI_ISL_1476059, EPI_ISL_1476060, EPI_ISL_1476061, EPI_ISL_1476066, EPI_ISL_1476070, EPI_ISL_1476073, EPI_ISL_1476106, EPI_ISL_1476108, EPI_ISL_1476111, EPI_ISL_1476112, EPI_ISL_1476113, EPI_ISL_1476116, EPI_ISL_1476117, EPI_ISL_1476122, EPI_ISL_1476125, EPI_ISL_1476127, EPI_ISL_1476128, EPI_ISL_1476130, EPI_ISL_1476133, EPI_ISL_1476135, EPI_ISL_1476152, EPI_ISL_1476153, EPI_ISL_1476154, EPI_ISL_1476155, EPI_ISL_1476235, EPI_ISL_1476244, EPI_ISL_1476251, EPI_ISL_1476257, EPI_ISL_1476258, EPI_ISL_1476259, EPI_ISL_1476260, EPI_ISL_1476261, EPI_ISL_1476262, EPI_ISL_1476263, EPI_ISL_1476264, EPI_ISL_1476265, EPI_ISL_1476266, EPI_ISL_1476269, EPI_ISL_1476270, EPI_ISL_1476271, EPI_ISL_1476272, EPI_ISL_1476273, EPI_ISL_1476274, EPI_ISL_1476276, EPI_ISL_1476277, EPI_ISL_1476294, EPI_ISL_1476295, EPI_ISL_1476299, EPI_ISL_1476300, EPI_ISL_1476301, EPI_ISL_1476302, EPI_ISL_1476304, EPI_ISL_1476305, EPI_ISL_1476306, EPI_ISL_1476307, EPI_ISL_1476308, EPI_ISL_1476311, EPI_ISL_1476312, EPI_ISL_1476313, EPI_ISL_1476314, EPI_ISL_1476316, EPI_ISL_1476319, EPI_ISL_1476323, EPI_ISL_1476324, EPI_ISL_1476325, EPI_ISL_1476327, EPI_ISL_1476328, EPI_ISL_1476329, EPI_ISL_1476330, EPI_ISL_1476332, EPI_ISL_1476333, EPI_ISL_1476334, EPI_ISL_1476335, EPI_ISL_1476336, EPI_ISL_1476337, EPI_ISL_1476340, EPI_ISL_1476341, EPI_ISL_1476342, EPI_ISL_1476344, EPI_ISL_1476345, EPI_ISL_1476346, EPI_ISL_1476347, EPI_ISL_1476348, EPI_ISL_1476349, EPI_ISL_1476350, EPI_ISL_1476351, EPI_ISL_1476352, EPI_ISL_1476353, EPI_ISL_1476354, EPI_ISL_1476355, EPI_ISL_1476356, EPI_ISL_1476357, EPI_ISL_1476358, EPI_ISL_1476359, EPI_ISL_1476360, EPI_ISL_1476361, EPI_ISL_1476362, EPI_ISL_1476363, EPI_ISL_1476365, EPI_ISL_1476366, EPI_ISL_1476370, EPI_ISL_1476372, EPI_ISL_1476373, EPI_ISL_1476375, EPI_ISL_1476376, EPI_ISL_1476378, EPI_ISL_1476379, EPI_ISL_1476381, EPI_ISL_1476382, EPI_ISL_1476383, EPI_ISL_1476384, EPI_ISL_1476385, EPI_ISL_1476386, EPI_ISL_1476387, EPI_ISL_1476389, EPI_ISL_1476391, EPI_ISL_1476392, EPI_ISL_1476395, EPI_ISL_1476400, EPI_ISL_1476401, EPI_ISL_1476402, EPI_ISL_1476403, EPI_ISL_1476404, EPI_ISL_1476405, EPI_ISL_1476406, EPI_ISL_1476407, EPI_ISL_1476411, EPI_ISL_1476414, EPI_ISL_1476415, EPI_ISL_1476416, EPI_ISL_1476418, EPI_ISL_1476419, EPI_ISL_1476421, EPI_ISL_1476422, EPI_ISL_1476424, EPI_ISL_1476426, EPI_ISL_1476430, EPI_ISL_1476431, EPI_ISL_1476432, EPI_ISL_1476435, EPI_ISL_1476436, EPI_ISL_1476438, EPI_ISL_1476441, EPI_ISL_1476443, EPI_ISL_1476446, EPI_ISL_1476480, EPI_ISL_1476481, EPI_ISL_1476482, EPI_ISL_1476483, EPI_ISL_1476487, EPI_ISL_1476490, EPI_ISL_1476491, EPI_ISL_1476496, EPI_ISL_1476500, EPI_ISL_1476501, EPI_ISL_1476502, EPI_ISL_1476505, EPI_ISL_1476515, EPI_ISL_1476516, EPI_ISL_1547068

| | | | |
|---|---|---|---|
| see above | Originating lab: Wales Specialist Virology Centre Sequencing lab: Pathogen Genomics Unit | Public Health Wales Microbiology Cardiff Wales Specialist Virology Centre | Alec Birchley; Alexander Adams; Amy Gaskin; Angela Marchbank; Bree Gatica-Wilcox; Catherine Moore; Jason Coombes; Joanne Watkins; Joel Southgate; Johnathan Evans; Laura Gifford; Lauren Gilbert; Lee Graham; Malorie Perry; Matthew Bull; Nicole Pacchiarini; Sally Corden; Sara Kumziene-Summerhayes; Sara Rey; Sarah Taylor; Simon Cottrell; Sophie Jones; Tom Connor |

EPI_ISL_1475701, EPI_ISL_1475716, EPI_ISL_1475746, EPI_ISL_1475747, EPI_ISL_1475749, EPI_ISL_1475750, EPI_ISL_1730432, EPI_ISL_1730433, EPI_ISL_1730435, EPI_ISL_1730436, EPI_ISL_1730437, EPI_ISL_1730438, EPI_ISL_1730439, EPI_ISL_1730441, EPI_ISL_1730444, EPI_ISL_1730446, EPI_ISL_1730469, EPI_ISL_1730578

| | | | |
|---|---|---|---|
| see above | Oxford Viromics, NDM, University of Oxford; Oxford University Hospitals; Basingstoke and North Hampshire Hospital | COVID-19 Genomics UK (COG-UK) Consortium | Alex Mobbs; Amy Trebes; Anita Justice; Catrin Moore; Christophe Fraser; David Bonsall; David Buck; Emma Wise; George Macintyre; Jessica Lynch; John Todd; Mariateresa de Cesare; Matilde Mori; Monique Andersson; Nathan Moore; Nick Cortes; Robert Shaw; Stephen Kidd; Tanya Golubchik; Timothy Peto |

EPI_ISL_1387133, EPI_ISL_1387134, EPI_ISL_1387135, EPI_ISL_1387139, EPI_ISL_1387140, EPI_ISL_1832744, EPI_ISL_1832756, EPI_ISL_1832760

| | | | |
|---|---|---|---|
| see above | Quadram Institute Bioscience | COVID-19 Genomics UK (COG-UK) Consortium | Alexander J Trotter; Alison E. Mather; Alp Aydin; Ana P. Tedim; Anastasia Kolyva; Andrew Bell; Andrew J. Page; Claire Stuart; Dave J. Baker; Gemma L. Kay; John Wain; Justin O'Grady; Leonardo de Oliveira Martins; Lizzie Meadows; Maria Diaz; Mark Webber; Muhammed Yasir; Nabil-Fareed Alikhan; Ngozi Elumogo; Nicholas M. Thomson; Rachael Stanley; Rachel Gilroy; Reenesh Prakash; Samir Dervisevic; Samuel Bloomfield; Steven Rudder; Thanh Le-Viet |

EPI_ISL_1309731, EPI_ISL_1309732, EPI_ISL_1387157, EPI_ISL_1387158, EPI_ISL_1387159, EPI_ISL_1387160, EPI_ISL_1387161, EPI_ISL_1387162, EPI_ISL_1387188, EPI_ISL_1387189, EPI_ISL_1387193

| | | | |
|---|---|---|---|
| see above | Queens Medical Centre, Clinical Microbiology Department / DeepSeq Nottingham | COVID-19 Genomics UK (COG-UK) Consortium | Christopher Moore; Emily Park; Fei Sang; Gemma Clark; Hannah Howson-Wells; Jack Hill; Jayasree Dey; Johnny Debebe; Jonathan Ball; Joseph Chappell; Manjinder Khakh; Matthew Carlisle; Matthew Loose; Michelle M Lister; Nadine Holmes; Patrick McClure; Theocharis Tsoleridis; Timothy Byaruhanga; Vicki M Fleming; Victoria Wright; Wendy Smith |

EPI_ISL_1376853, EPI_ISL_1409224, EPI_ISL_1409278, EPI_ISL_1409294, EPI_ISL_1409420, EPI_ISL_1409443, EPI_ISL_1409483, EPI_ISL_1411101, EPI_ISL_1411102, EPI_ISL_1411105, EPI_ISL_1411112, EPI_ISL_1411113, EPI_ISL_1411114, EPI_ISL_1411115, EPI_ISL_1411118, EPI_ISL_1411122, EPI_ISL_1411123, EPI_ISL_1411125, EPI_ISL_1411127, EPI_ISL_1411128, EPI_ISL_1411131, EPI_ISL_1411133, EPI_ISL_1411136, EPI_ISL_1411138, EPI_ISL_1411142, EPI_ISL_1411147, EPI_ISL_1411150, EPI_ISL_1411153, EPI_ISL_1411156, EPI_ISL_1411158, EPI_ISL_1411161, EPI_ISL_1411163, EPI_ISL_1411168, EPI_ISL_1411170, EPI_ISL_1411172, EPI_ISL_1411179, EPI_ISL_1411180, EPI_ISL_1411183, EPI_ISL_1411185, EPI_ISL_1411189, EPI_ISL_1411190, EPI_ISL_1411194, EPI_ISL_1411195, EPI_ISL_1411196, EPI_ISL_1411197, EPI_ISL_1411204, EPI_ISL_1411205, EPI_ISL_1411218, EPI_ISL_1411219, EPI_ISL_1411223, EPI_ISL_1411225, EPI_ISL_1411229, EPI_ISL_1411230, EPI_ISL_1411231, EPI_ISL_1411232, EPI_ISL_1411233, EPI_ISL_1411234, EPI_ISL_1411236, EPI_ISL_1411237, EPI_ISL_1411238, EPI_ISL_1411240, EPI_ISL_1411243, EPI_ISL_1411244, EPI_ISL_1411250, EPI_ISL_1411256, EPI_ISL_1411258, EPI_ISL_1411260, EPI_ISL_1411261, EPI_ISL_1411268, EPI_ISL_1411272, EPI_ISL_1411274, EPI_ISL_1411278, EPI_ISL_1411279, EPI_ISL_1411280, EPI_ISL_1411282, EPI_ISL_1411283, EPI_ISL_1411285, EPI_ISL_1411286, EPI_ISL_1411287, EPI_ISL_1411295, EPI_ISL_1411296, EPI_ISL_1411297, EPI_ISL_1411299, EPI_ISL_1411302, EPI_ISL_1411303, EPI_ISL_1411304, EPI_ISL_1411306, EPI_ISL_1411308, EPI_ISL_1411310, EPI_ISL_1411311, EPI_ISL_1411313, EPI_ISL_1411315, EPI_ISL_1411316, EPI_ISL_1411317, EPI_ISL_1411319, EPI_ISL_1411325, EPI_ISL_1411328, EPI_ISL_1411329, EPI_ISL_1411335, EPI_ISL_1411337, EPI_ISL_1411339, EPI_ISL_1411345, EPI_ISL_1411346, EPI_ISL_1411348, EPI_ISL_1411352, EPI_ISL_1411356, EPI_ISL_1411357, EPI_ISL_1411358, EPI_ISL_1411360, EPI_ISL_1411362, EPI_ISL_1411365, EPI_ISL_1411369, EPI_ISL_1411370, EPI_ISL_1411371, EPI_ISL_1411372, EPI_ISL_1411374, EPI_ISL_1411375, EPI_ISL_1411377, EPI_ISL_1411378, EPI_ISL_1411380, EPI_ISL_1411382, EPI_ISL_1411384, EPI_ISL_1411385, EPI_ISL_1411386, EPI_ISL_1411387, EPI_ISL_1411389, EPI_ISL_1411393, EPI_ISL_1411394, EPI_ISL_1411395, EPI_ISL_1411401, EPI_ISL_1411404, EPI_ISL_1411405, EPI_ISL_1411406, EPI_ISL_1411409, EPI_ISL_1411412, EPI_ISL_1411413, EPI_ISL_1411415, EPI_ISL_1411419, EPI_ISL_1411422, EPI_ISL_1411423, EPI_ISL_1411424, EPI_ISL_1411425, EPI_ISL_1411426, EPI_ISL_1411428, EPI_ISL_1411430, EPI_ISL_1411432, EPI_ISL_1411436, EPI_ISL_1411437, EPI_ISL_1411438, EPI_ISL_1411440, EPI_ISL_1411441, EPI_ISL_1411445, EPI_ISL_1411446, EPI_ISL_1411448, EPI_ISL_1411452, EPI_ISL_1411455, EPI_ISL_1411456, EPI_ISL_1411464, EPI_ISL_1411467, EPI_ISL_1411480, EPI_ISL_1411483, EPI_ISL_1411487, EPI_ISL_1411489, EPI_ISL_1411490, EPI_ISL_1411493, EPI_ISL_1411495, EPI_ISL_1411496, EPI_ISL_1411555, EPI_ISL_1411575, EPI_ISL_1411586, EPI_ISL_1411605, EPI_ISL_1411613, EPI_ISL_1411615, EPI_ISL_1411620, EPI_ISL_1411628, EPI_ISL_1411634, EPI_ISL_1411637, EPI_ISL_1411642, EPI_ISL_1411655, EPI_ISL_1411663, EPI_ISL_1411664, EPI_ISL_1411673, EPI_ISL_1411676, EPI_ISL_1411678, EPI_ISL_1411688, EPI_ISL_1411699, EPI_ISL_1411702, EPI_ISL_1411707, EPI_ISL_1411712, EPI_ISL_1411716, EPI_ISL_1411724, EPI_ISL_1411726, EPI_ISL_1411728, EPI_ISL_1411730, EPI_ISL_1411742, EPI_ISL_1411745, EPI_ISL_1411747, EPI_ISL_1411748, EPI_ISL_1411749, EPI_ISL_1411756, EPI_ISL_1411760, EPI_ISL_1411763, EPI_ISL_1411770, EPI_ISL_1411782, EPI_ISL_1411786, EPI_ISL_1411787, EPI_ISL_1411789, EPI_ISL_1411802, EPI_ISL_1411813, EPI_ISL_1411822, EPI_ISL_1411823, EPI_ISL_1411828, EPI_ISL_1411830, EPI_ISL_1411839, EPI_ISL_1411843, EPI_ISL_1411846, EPI_ISL_1411847, EPI_ISL_1411851, EPI_ISL_1412206, EPI_ISL_1412229, EPI_ISL_1412240, EPI_ISL_1412270, EPI_ISL_1412281, EPI_ISL_1412301, EPI_ISL_1412338, EPI_ISL_1412354, EPI_ISL_1412365, EPI_ISL_1412388, EPI_ISL_1412410, EPI_ISL_1412437, EPI_ISL_1412450, EPI_ISL_1412454, EPI_ISL_1412462, EPI_ISL_1412486, EPI_ISL_1451554, EPI_ISL_1454955, EPI_ISL_1454959, EPI_ISL_1454962

| | | | |
|---|---|---|---|
| see above | Randox Laboratories | Wellcome Sanger Institute for the COVID-19 Genomics UK (COG-UK) Consortium | Cordelia Langford; David K. Jackson; Dominic Kwiatkowski; Ewan Harrison; Ian Johnston; Jeffrey Barrett; John Sillitoe on behalf of the Wellcome Sanger Institute COVID-19 Surveillance Team; Randox Laboratories and Alex Alderton; Roberto Amato; Sonia Goncalves |

EPI_ISL_1977235, EPI_ISL_1977237, EPI_ISL_1977239, EPI_ISL_1977241, EPI_ISL_1977242, EPI_ISL_1977244, EPI_ISL_1977245, EPI_ISL_1977247, EPI_ISL_1977249, EPI_ISL_1977251, EPI_ISL_1977252, EPI_ISL_1977254, EPI_ISL_1977258, EPI_ISL_1977259, EPI_ISL_1977264, EPI_ISL_1977265, EPI_ISL_1977267, EPI_ISL_1977280, EPI_ISL_1977282, EPI_ISL_1977285, EPI_ISL_1977286, EPI_ISL_1977288, EPI_ISL_1977290, EPI_ISL_1977292, EPI_ISL_1977293, EPI_ISL_1977295, EPI_ISL_1977296, EPI_ISL_1977314, EPI_ISL_1977316, EPI_ISL_1977325, EPI_ISL_1977326, EPI_ISL_1977329, EPI_ISL_1977330, EPI_ISL_1977332, EPI_ISL_1977335, EPI_ISL_1977337, EPI_ISL_1977338, EPI_ISL_1977340, EPI_ISL_1977342, EPI_ISL_1977344, EPI_ISL_1977346, EPI_ISL_1977347, EPI_ISL_1977349, EPI_ISL_1977351, EPI_ISL_1977352, EPI_ISL_1977354, EPI_ISL_1977355, EPI_ISL_1977357, EPI_ISL_1977370, EPI_ISL_1977371, EPI_ISL_1977372, EPI_ISL_1977374, EPI_ISL_1977377, EPI_ISL_1977389, EPI_ISL_1977391, EPI_ISL_1977392, EPI_ISL_1977394, EPI_ISL_1977401, EPI_ISL_1977403, EPI_ISL_1977404, EPI_ISL_1977406, EPI_ISL_1977412, EPI_ISL_1977414, EPI_ISL_1977416, EPI_ISL_1977417, EPI_ISL_1977419, EPI_ISL_1977420, EPI_ISL_1977424, EPI_ISL_1977426, EPI_ISL_1977427, EPI_ISL_1977429, EPI_ISL_1977431, EPI_ISL_1977433, EPI_ISL_1977434, EPI_ISL_1977435, EPI_ISL_1977437, EPI_ISL_1977440, EPI_ISL_1977442, EPI_ISL_1977444, EPI_ISL_1977447, EPI_ISL_1977871, EPI_ISL_1977873, EPI_ISL_1977874, EPI_ISL_1977876, EPI_ISL_1977878, EPI_ISL_1977880, EPI_ISL_1977881, EPI_ISL_1977883, EPI_ISL_1977884, EPI_ISL_1977886, EPI_ISL_1977888, EPI_ISL_1977890, EPI_ISL_1977892, EPI_ISL_1977893, EPI_ISL_1977895, EPI_ISL_1977897, EPI_ISL_1977899, EPI_ISL_1977901, EPI_ISL_1977903, EPI_ISL_1977905, EPI_ISL_1977906, EPI_ISL_1977908, EPI_ISL_1977909, EPI_ISL_1977911, EPI_ISL_1977913, EPI_ISL_1977915, EPI_ISL_1977917, EPI_ISL_1977919, EPI_ISL_1977920, EPI_ISL_1977922, EPI_ISL_1977924, EPI_ISL_1977925, EPI_ISL_1977927, EPI_ISL_1977929, EPI_ISL_1977931, EPI_ISL_1977933, EPI_ISL_1977934, EPI_ISL_1977936, EPI_ISL_1977938, EPI_ISL_1977940, EPI_ISL_1977941, EPI_ISL_1977943, EPI_ISL_1977945, EPI_ISL_1977947, EPI_ISL_1977948, EPI_ISL_1977950, EPI_ISL_1977951, EPI_ISL_1977953, EPI_ISL_1977965, EPI_ISL_1977967, EPI_ISL_1977969, EPI_ISL_1977971, EPI_ISL_1977972, EPI_ISL_1977974, EPI_ISL_1977976, EPI_ISL_1977977, EPI_ISL_1977979, EPI_ISL_1977981, EPI_ISL_1977982, EPI_ISL_1977984, EPI_ISL_1977986, EPI_ISL_1977987, EPI_ISL_1977989, EPI_ISL_1977991, EPI_ISL_1977992, EPI_ISL_1977994, EPI_ISL_1977996, EPI_ISL_1977997, EPI_ISL_1977999, EPI_ISL_1978000, EPI_ISL_1978002, EPI_ISL_1978004, EPI_ISL_1978006, EPI_ISL_1978007,

| | | | |
|---|---|---|---|
| EPI_ISL_1978009, EPI_ISL_1978011, EPI_ISL_1978013, EPI_ISL_1978014, EPI_ISL_1978016, EPI_ISL_1978017, EPI_ISL_1978019, EPI_ISL_1978021, EPI_ISL_1978023, EPI_ISL_1978025, EPI_ISL_1978026, EPI_ISL_1978028, EPI_ISL_1978030, EPI_ISL_1978032, EPI_ISL_1978033, EPI_ISL_1978035, EPI_ISL_1978037, EPI_ISL_1978038, EPI_ISL_1978040, EPI_ISL_1978042, EPI_ISL_1978043, EPI_ISL_1978045, EPI_ISL_1978046, EPI_ISL_1978059, EPI_ISL_1978061, EPI_ISL_1978063, EPI_ISL_1978064, EPI_ISL_1978071, EPI_ISL_1978072, EPI_ISL_1978074, EPI_ISL_1978075, EPI_ISL_1978077, EPI_ISL_1978079, EPI_ISL_1978081, EPI_ISL_1978083, EPI_ISL_1978084, EPI_ISL_1978086, EPI_ISL_1978087, EPI_ISL_1978091 | | | |
| see above | Regional Virus Laboratory, Belfast Health and Social Care Trust | COVID-19 Genomics UK (COG-UK) Consortium | Alison Watt; Ciara Cox; Conall McCaughey; David Simpson; Derek Fairley; James McKenna; Mairead Connor; Susan Feeney; Tanya Curran; Zoltan Molnar |
| EPI_ISL_1637049, EPI_ISL_1637050, EPI_ISL_1637051, EPI_ISL_1637085, EPI_ISL_1637086 | University College London Hospital | COVID-19 Genomics UK (COG-UK) Consortium | Dr Catherine Houlihan; Dr Daniel Frampton; Dr Judith Heaney; Dr Moira Spyer; Dr Paul Grant and Dr Eleni Nastouli; Matthew Byott; Stuart Kirk |
| EPI_ISL_1308951, EPI_ISL_1309134, EPI_ISL_1309143, EPI_ISL_1309144, EPI_ISL_1309145, EPI_ISL_1386814, EPI_ISL_1386815, EPI_ISL_1386816, EPI_ISL_1386817, EPI_ISL_1386818, EPI_ISL_1386819, EPI_ISL_1386820, EPI_ISL_1386821, EPI_ISL_1386837, EPI_ISL_1386838, EPI_ISL_1386839, EPI_ISL_1386842, EPI_ISL_1386849, EPI_ISL_1386914, EPI_ISL_1386915, EPI_ISL_1386916, EPI_ISL_1386917, EPI_ISL_1474839, EPI_ISL_1474842, EPI_ISL_1474843, EPI_ISL_1474856, EPI_ISL_1474868, EPI_ISL_1474882, EPI_ISL_1474883, EPI_ISL_1474884, EPI_ISL_1474885, EPI_ISL_1474886, EPI_ISL_1474887, EPI_ISL_1637176 | | | |
| see above | University College London, Great Ormond Street Hospital for Children NHS Foundation Trust, Imperial College Healthcare NHS Trust | COVID-19 Genomics UK (COG-UK) Consortium | Alison Holmes; Charlotte Williams; Helena Tutill; Jacqueline Findlay; James Price; Judith Breuer; Julianne Brown; Kathryn Harris; Leysa Forrest; Mark Kristiansen; Paola Niola; Paola Resende Silva; Patricia Dyal; Paul Randell; Rachel Williams; Samuel Weeks; Sergi Castellano; Sunando Roy; Tony Brooks; Yasmin Panchbhaya |
| EPI_ISL_1474645, EPI_ISL_1474646, EPI_ISL_1474647, EPI_ISL_1474650, EPI_ISL_1474651, EPI_ISL_1474652, EPI_ISL_1474653 | | | |
| see above | University Hospitals Of Leicester NHS Trust and DeepSeq Nottingham | COVID-19 Genomics UK (COG-UK) Consortium | Christopher Holmes; Christopher Moore; Fei Sang; Joeseph Chappell; Johnny Debebe; Jonathan Ball; Julian Tang; Karlie Fallon; Matthew Carlisle; Matthew Loose; Nadine Holmes; Patrick McClure; Paul Bird; Thomas Helmer; Victoria Wright |
| EPI_ISL_1308534, EPI_ISL_1386477, EPI_ISL_1386478, EPI_ISL_1386479, EPI_ISL_1386487, EPI_ISL_1386488, EPI_ISL_1386489, EPI_ISL_1386490, EPI_ISL_1386491, EPI_ISL_1386492, EPI_ISL_1386493, EPI_ISL_1386494, EPI_ISL_1386495, EPI_ISL_1386496, EPI_ISL_1386497, EPI_ISL_1386498, EPI_ISL_1386499, EPI_ISL_1386500, EPI_ISL_1386501, EPI_ISL_1386502, EPI_ISL_1386503, EPI_ISL_1386504, EPI_ISL_1386505, EPI_ISL_1386506, EPI_ISL_1386507, EPI_ISL_1386508, EPI_ISL_1386521, EPI_ISL_1386542, EPI_ISL_1386563, EPI_ISL_1386564, EPI_ISL_1474413, EPI_ISL_1474415, EPI_ISL_1474416, EPI_ISL_1474433, EPI_ISL_1474440, EPI_ISL_1474444, EPI_ISL_1474446, EPI_ISL_1474447, EPI_ISL_1474448, EPI_ISL_1474449, EPI_ISL_1474450, EPI_ISL_1474451, EPI_ISL_1474457, EPI_ISL_1474458, EPI_ISL_1474460, EPI_ISL_1474461, EPI_ISL_1729971, EPI_ISL_1729972, EPI_ISL_1729982, EPI_ISL_1729984 | | | |
| see above | University of Birmingham | COVID-19 Genomics UK (COG-UK) Consortium | Alex Richter; Andrew Bosworth; Andrew Bosworth. Queen Elizabeth Hospital: Anna Casey; Andrew D Beggs PHE Heartlands Lab: Husam Osman; Charlotte Poxon; Institute of Microbiology; Joanne Stockton; Josh Quick; Kasun Wanigasooriya; Mike Kidd; Nicholas Loman. University of Birmingham Testing Laboratory: Celina M Whalley; Oliver Pickles; Radoslaw Poplawski; Samuel Nicholls; University of Birmingham: Claire McMurray; Will Rowe |
| EPI_ISL_1545824, EPI_ISL_1545826, EPI_ISL_1545827, EPI_ISL_1545828, EPI_ISL_1545831, EPI_ISL_1545832, EPI_ISL_1545833, EPI_ISL_1545834, EPI_ISL_1545835, EPI_ISL_1545836, EPI_ISL_1545838, EPI_ISL_1545839, EPI_ISL_1545840, EPI_ISL_1545841, EPI_ISL_1545845, EPI_ISL_1545952, EPI_ISL_1545953, EPI_ISL_1545954, EPI_ISL_1545955, EPI_ISL_1545956, EPI_ISL_1545957, EPI_ISL_1545958, EPI_ISL_1545959 | | | |
| see above | University of Exeter | COVID-19 Genomics UK (COG-UK) Consortium | Aaron Jeffries; Audrey Farbos; Ben Temperton; Jane Masoli; Joanna Warwick-Dugdale; Michelle Michelsen; Robyn Manley; Stephen Michell |
| EPI_ISL_1386694, EPI_ISL_1386695, EPI_ISL_1386696, EPI_ISL_1386698, EPI_ISL_1386699, EPI_ISL_1386700, EPI_ISL_1386702, EPI_ISL_1386704, EPI_ISL_1386705, EPI_ISL_1474582, EPI_ISL_1474583, EPI_ISL_1474586, EPI_ISL_1474587, EPI_ISL_1545912, EPI_ISL_1545921, EPI_ISL_1545922, EPI_ISL_1545923, EPI_ISL_1545927, EPI_ISL_1545930, EPI_ISL_1545935, EPI_ISL_1545942, EPI_ISL_1636678, EPI_ISL_1636679, EPI_ISL_1636680, EPI_ISL_1636681, EPI_ISL_1636682, EPI_ISL_1636683, EPI_ISL_1636693, EPI_ISL_1636711, EPI_ISL_1730091, EPI_ISL_1730098, EPI_ISL_1730100, EPI_ISL_1730242, EPI_ISL_1730243, EPI_ISL_1730244, EPI_ISL_1730245, EPI_ISL_1730246, EPI_ISL_1730247, EPI_ISL_1730251, EPI_ISL_1730252, EPI_ISL_1730254 | | | |
| see above | Virology Department, Royal Infirmary of Edinburgh, NHS Lothian / School of Biological Sciences, University of Edinburgh | COVID-19 Genomics UK (COG-UK) Consortium | Colquhoun R; Cotton S; Dewar R; Gallagher A; Hill V; Jackson B; Maloney D; McCrone JT; McHugh M; O'Toole Á; Rambaut A; Rooke S; Scher E; Templeton K; Yu X |
| EPI_ISL_1310711, EPI_ISL_1310725, EPI_ISL_1310744, EPI_ISL_1389038, EPI_ISL_1389040, EPI_ISL_1389042, EPI_ISL_1389044, EPI_ISL_1389046, EPI_ISL_1389048, EPI_ISL_1389050, EPI_ISL_1389052, EPI_ISL_1389054, EPI_ISL_1389056, EPI_ISL_1389058, EPI_ISL_1389059, EPI_ISL_1389062, EPI_ISL_1389064, EPI_ISL_1389066, EPI_ISL_1389068, EPI_ISL_1389069, EPI_ISL_1389071, EPI_ISL_1389075, EPI_ISL_1389077, EPI_ISL_1389080, EPI_ISL_1389094, EPI_ISL_1389104, EPI_ISL_1389115, EPI_ISL_1389118, EPI_ISL_1389125, EPI_ISL_1389130, EPI_ISL_1389135, EPI_ISL_1389138, EPI_ISL_1476723, EPI_ISL_1476725, EPI_ISL_1476726, EPI_ISL_1476735, EPI_ISL_1476739, EPI_ISL_1731020 | | | |
| see above | Virology Department, Sheffield Teaching Hospitals NHS Foundation Trust/Department of Infection, Immunity and Cardiovascular Disease, The Medical School, University of Sheffield | COVID-19 Genomics UK (COG-UK) Consortium | Adri Angyal; Alex Keeley; Benjamin Lindsey; Cariad Evans; Danielle Groves; Dave Partridge; Katie Johnson; Laura Carrilero; Luke Green; Matthew Parker; Matthew Wyles; Mehmet Yavuz; Mohammad Raza; Nikki Smith; Paul Parsons; Rachel Tucker; Rebecca Brown; Thushan de Silva |
| EPI_ISL_1388004, EPI_ISL_1388016, EPI_ISL_1388066, EPI_ISL_1388067, EPI_ISL_1388070, EPI_ISL_1388075, EPI_ISL_1388087, EPI_ISL_1388088, EPI_ISL_1388258, EPI_ISL_1388262, EPI_ISL_1388264, EPI_ISL_1388270, EPI_ISL_1388317, EPI_ISL_1388321, EPI_ISL_1388325, EPI_ISL_1388329, EPI_ISL_1388350, EPI_ISL_1388367, EPI_ISL_1388376, EPI_ISL_1388379, EPI_ISL_1388385, EPI_ISL_1388393, EPI_ISL_1388472, EPI_ISL_1388512, EPI_ISL_1388529, EPI_ISL_1388543, EPI_ISL_1475763, EPI_ISL_1475765, EPI_ISL_1475771, EPI_ISL_1475846, EPI_ISL_1475848, EPI_ISL_1475849, EPI_ISL_1475851, EPI_ISL_1475856, EPI_ISL_1475862, EPI_ISL_1475867, EPI_ISL_1475873, EPI_ISL_1475891, EPI_ISL_1475892, EPI_ISL_1475920, EPI_ISL_1475927, EPI_ISL_1475936, EPI_ISL_1475941, EPI_ISL_1475943, EPI_ISL_1475945, EPI_ISL_1475969, EPI_ISL_1475998, EPI_ISL_1475999, EPI_ISL_1476009, EPI_ISL_1476021, EPI_ISL_1476022, EPI_ISL_1476031, EPI_ISL_1476032, EPI_ISL_1476040, EPI_ISL_1476047, EPI_ISL_1476051, EPI_ISL_1476056, EPI_ISL_1476069, EPI_ISL_1476076, EPI_ISL_1476107, EPI_ISL_1476110, EPI_ISL_1476115, EPI_ISL_1476132, EPI_ISL_1476136, EPI_ISL_1476145, EPI_ISL_1476148, EPI_ISL_1476180, EPI_ISL_1476181, EPI_ISL_1476183, EPI_ISL_1476296, EPI_ISL_1476297, EPI_ISL_1476298, EPI_ISL_1476303, EPI_ISL_1476309, EPI_ISL_1476310, EPI_ISL_1476315, EPI_ISL_1476317, EPI_ISL_1476318, EPI_ISL_1476320, EPI_ISL_1476321, EPI_ISL_1476322, EPI_ISL_1476326, EPI_ISL_1476331, EPI_ISL_1476338, EPI_ISL_1476339, EPI_ISL_1476343, EPI_ISL_1476364, EPI_ISL_1476367, EPI_ISL_1476368, EPI_ISL_1476369, EPI_ISL_1476371, EPI_ISL_1476374, EPI_ISL_1476377, EPI_ISL_1476380, EPI_ISL_1476390, EPI_ISL_1476393, EPI_ISL_1476394, EPI_ISL_1476396, EPI_ISL_1476399, EPI_ISL_1476410, EPI_ISL_1476412, EPI_ISL_1476420, EPI_ISL_1476423, EPI_ISL_1476425, EPI_ISL_1476427, EPI_ISL_1476428, EPI_ISL_1476429, EPI_ISL_1476433, EPI_ISL_1476434, EPI_ISL_1476437, EPI_ISL_1476442, EPI_ISL_1476444, EPI_ISL_1476445, EPI_ISL_1476447, EPI_ISL_1476448, EPI_ISL_1476478, EPI_ISL_1476479, EPI_ISL_1637328, EPI_ISL_1637330, EPI_ISL_1637331, EPI_ISL_1637332, EPI_ISL_1637333, EPI_ISL_1637334, EPI_ISL_1637337, EPI_ISL_1637339, EPI_ISL_1637342, EPI_ISL_1637344, EPI_ISL_1637347, EPI_ISL_1637348, EPI_ISL_1637349, EPI_ISL_1637356, EPI_ISL_1637361, EPI_ISL_1637362, EPI_ISL_1637363, EPI_ISL_1637364, EPI_ISL_1637369, EPI_ISL_1637370, EPI_ISL_1637379, EPI_ISL_1637380, EPI_ISL_1637381, EPI_ISL_1637382, EPI_ISL_1637387, EPI_ISL_1637388, EPI_ISL_1637389, EPI_ISL_1637390, EPI_ISL_1637391, EPI_ISL_1637392, EPI_ISL_1637393, EPI_ISL_1637394, EPI_ISL_1637396, EPI_ISL_1637397, EPI_ISL_1637424, EPI_ISL_1637435, EPI_ISL_1637438, EPI_ISL_1637439, EPI_ISL_1637440, EPI_ISL_1637441, EPI_ISL_1637442, EPI_ISL_1637443, EPI_ISL_1637444, EPI_ISL_1637869, EPI_ISL_1637870, EPI_ISL_1637871, EPI_ISL_1637872, EPI_ISL_1637873, EPI_ISL_1637874, EPI_ISL_1637875, EPI_ISL_1637876, EPI_ISL_1637877, EPI_ISL_1637878, EPI_ISL_1637879, EPI_ISL_1637880, EPI_ISL_1637881, EPI_ISL_1637882, EPI_ISL_1637883, EPI_ISL_1637884, EPI_ISL_1637885, EPI_ISL_1637888, EPI_ISL_1637889, EPI_ISL_1637890, EPI_ISL_1637891, EPI_ISL_1637892, EPI_ISL_1637893, EPI_ISL_1637894, EPI_ISL_1637895, EPI_ISL_1637896, EPI_ISL_1637897, EPI_ISL_1637898, EPI_ISL_1637899, EPI_ISL_1637900, EPI_ISL_1637923, EPI_ISL_1637924, EPI_ISL_1637925, EPI_ISL_1637926, EPI_ISL_1637929, EPI_ISL_1637952, EPI_ISL_1637953, EPI_ISL_1637955, EPI_ISL_1637957, EPI_ISL_1637969, EPI_ISL_1637970, EPI_ISL_1637972, EPI_ISL_1637973, EPI_ISL_1637974, EPI_ISL_1637975, EPI_ISL_1637976, EPI_ISL_1833153, EPI_ISL_1833154, EPI_ISL_1833159, EPI_ISL_1833164, EPI_ISL_1833169, EPI_ISL_1833172, EPI_ISL_1833173, EPI_ISL_1833174, EPI_ISL_1833175, EPI_ISL_1833176, EPI_ISL_1833178 | | | |
| see above | Wales Specialist Virology Centre Sequencing lab: Pathogen Genomics Unit | Public Health Wales Microbiology Cardiff Wales Specialist Virology Centre | Alec Birchley; Alexander Adams; Amy Gaskin; Angela Marchbank; Bree Gatica-Wilcox; Catherine Moore; Jason Coombes; Joanne Watkins; Joel Southgate; Johnathan Evans; Laura Gifford; Lauren Gilbert; Lee Graham; Malorie Perry; Matthew Bull; Nicole Pacchiarini; Sally Corden; Sara Kumziene-Summerhayes; Sara Rey; Sarah Taylor; Simon Cottrell; Sophie Jones; Tom Connor |
| EPI_ISL_1545852, EPI_ISL_1545853, EPI_ISL_1545854, EPI_ISL_1545855, EPI_ISL_1545856, EPI_ISL_1545858, EPI_ISL_1545859, EPI_ISL_1545860, EPI_ISL_1545861, EPI_ISL_1545862, EPI_ISL_1545863, EPI_ISL_1545864, EPI_ISL_1545865, EPI_ISL_1545866, EPI_ISL_1545897, EPI_ISL_1636647, EPI_ISL_1636650, EPI_ISL_1636652, EPI_ISL_1636653, EPI_ISL_1636655, EPI_ISL_1636656 | | | |
| see above | West of Scotland Specialist Virology Centre, NHSGGC / MRC-University of Glasgow Centre for Virus Research | COVID-19 Genomics UK (COG-UK) Consortium | Alasdair MacLean; Alice Broos; Ana da Silva Filipe; Antonia Ho; Daniel Mair; David L Robertson; Emma Thomson; Guy Mollett; James Shepherd; Jenna Nichols; Joseph Hughes; Kathy Li; Kathy Smollett; Kyriaki Nomikou; Lily Tong; Matthew Holden; Natasha Johnson; Rachel Blacow; Richard Orton; Rory Gunson; Sarah McDonald; Sharif Shaaban; Sreenu Vattipally; Stephen Carmichael |

We gratefully acknowledge the following Authors from the Originating laboratories responsible for obtaining the specimens, as well as the Submitting laboratories where the genome data were generated and shared via GISAID, on which this research is based.

All Submitters of data may be contacted directly via www.gisaid.org

Authors are sorted alphabetically.

| Accession ID | Originating Laboratory | Submitting Laboratory | Authors |
| --- | --- | --- | --- |
| EPI_ISL_1474826, EPI_ISL_1636963 | Barts Health NHS Trust | COVID-19 Genomics UK (COG-UK) Consortium | BROAD; Beatrix; CUTINO-MOGUEL; Claire; David; Dola; HARRINGTON; KELE; KULASEGARAN-SHYLINI; Maria-Teresa; OWOYEMI; Raghavendran |
| EPI_ISL_1389142 | Bioinformatics and Biostatistics Lab, Advanced Sequencing Facility | COVID-19 Genomics UK (COG-UK) Consortium | Aengus Stewart; Chelsea Sawyer; Harshil Patel; Jerome Nicod; Laura Cubitt; Margaret Crawford |
| EPI_ISL_1388798, EPI_ISL_1388802, EPI_ISL_1388804, EPI_ISL_1388806, EPI_ISL_1388865, EPI_ISL_1388867, EPI_ISL_1388931, EPI_ISL_1476675, EPI_ISL_1476683, EPI_ISL_1476684, EPI_ISL_1476685, EPI_ISL_1638009, EPI_ISL_1638011, EPI_ISL_1638012, EPI_ISL_1638013 | | | |
| see above | Centre for Enzyme Innovation, University of Portsmouth / Translational Research Laboratory, Portsmouth Hospitals NHS Trust | COVID-19 Genomics UK (COG-UK) Consortium | Angela Beckett; Christopher Fearn; Kate Cook; Katie Loveson; Salman Goudarzi; Samuel Robson; Scott Elliott; Sharon Glaysher |
| EPI_ISL_1386571, EPI_ISL_1386572, EPI_ISL_1386573, EPI_ISL_1386575, EPI_ISL_1386580 | Department of Pathology, University of Cambridge | COVID-19 Genomics UK (COG-UK) Consortium | Aminu S. Jahun; Ian Goodfellow; Iliana Georgana; Martin D. Curran; Myra Hosmillo; Rhys Izuagbe; Surendra Parmar; William L. Hamilton; Yasmin Chaudhry |

EPI_ISL_1408908, EPI_ISL_1408913, EPI_ISL_1408932, EPI_ISL_1408941, EPI_ISL_1408948, EPI_ISL_1408958, EPI_ISL_1408961, EPI_ISL_1408966, EPI_ISL_1408973, EPI_ISL_1408990, EPI_ISL_1409004, EPI_ISL_1409006, EPI_ISL_1409011, EPI_ISL_1409016, EPI_ISL_1409031, EPI_ISL_1409075, EPI_ISL_1409092, EPI_ISL_1409104, EPI_ISL_1409124, EPI_ISL_1409146, EPI_ISL_1409151, EPI_ISL_1409164, EPI_ISL_1409180, EPI_ISL_1409187, EPI_ISL_1409189, EPI_ISL_1409200, EPI_ISL_1409208, EPI_ISL_1409211, EPI_ISL_1409216, EPI_ISL_1409218, EPI_ISL_1409223, EPI_ISL_1409228, EPI_ISL_1409229, EPI_ISL_1409232, EPI_ISL_1409236, EPI_ISL_1409238, EPI_ISL_1409239, EPI_ISL_1409241, EPI_ISL_1409243, EPI_ISL_1409244, EPI_ISL_1409257, EPI_ISL_1409273, EPI_ISL_1409276, EPI_ISL_1409281, EPI_ISL_1409284, EPI_ISL_1409285, EPI_ISL_1409286, EPI_ISL_1409288, EPI_ISL_1409293, EPI_ISL_1409299, EPI_ISL_1409300, EPI_ISL_1409306, EPI_ISL_1409308, EPI_ISL_1409312, EPI_ISL_1409319, EPI_ISL_1409320, EPI_ISL_1409323, EPI_ISL_1409328, EPI_ISL_1409329, EPI_ISL_1409334, EPI_ISL_1409346, EPI_ISL_1409348, EPI_ISL_1409351, EPI_ISL_1409353, EPI_ISL_1409355, EPI_ISL_1409363, EPI_ISL_1409364, EPI_ISL_1409365, EPI_ISL_1409374, EPI_ISL_1409376, EPI_ISL_1409381, EPI_ISL_1409382, EPI_ISL_1409385, EPI_ISL_1409387, EPI_ISL_1409397, EPI_ISL_1409398, EPI_ISL_1409400, EPI_ISL_1409402, EPI_ISL_1409405, EPI_ISL_1409408, EPI_ISL_1409412, EPI_ISL_1409415, EPI_ISL_1409418, EPI_ISL_1409419, EPI_ISL_1409424, EPI_ISL_1409426, EPI_ISL_1409431, EPI_ISL_1409434, EPI_ISL_1409435, EPI_ISL_1409436, EPI_ISL_1409438, EPI_ISL_1409440, EPI_ISL_1409445, EPI_ISL_1409451, EPI_ISL_1409455, EPI_ISL_1409459, EPI_ISL_1409463, EPI_ISL_1409464, EPI_ISL_1409466, EPI_ISL_1409467, EPI_ISL_1409473, EPI_ISL_1409476, EPI_ISL_1409480, EPI_ISL_1409486, EPI_ISL_1409487, EPI_ISL_1409488, EPI_ISL_1409489, EPI_ISL_1409490, EPI_ISL_1409513, EPI_ISL_1409588, EPI_ISL_1409658, EPI_ISL_1409681, EPI_ISL_1409719, EPI_ISL_1409775, EPI_ISL_1409776, EPI_ISL_1409777, EPI_ISL_1409779, EPI_ISL_1409780, EPI_ISL_1409781, EPI_ISL_1409782, EPI_ISL_1409784, EPI_ISL_1409785, EPI_ISL_1409786, EPI_ISL_1409787, EPI_ISL_1409790, EPI_ISL_1409791, EPI_ISL_1409792, EPI_ISL_1409793, EPI_ISL_1409795, EPI_ISL_1409796, EPI_ISL_1409797, EPI_ISL_1409798, EPI_ISL_1409799, EPI_ISL_1409800, EPI_ISL_1409801, EPI_ISL_1409802, EPI_ISL_1409803, EPI_ISL_1409804, EPI_ISL_1409805, EPI_ISL_1409806, EPI_ISL_1409807, EPI_ISL_1409810, EPI_ISL_1409812, EPI_ISL_1409814, EPI_ISL_1409816, EPI_ISL_1409818, EPI_ISL_1409819, EPI_ISL_1409821, EPI_ISL_1409822, EPI_ISL_1409823, EPI_ISL_1409825, EPI_ISL_1409826, EPI_ISL_1409827, EPI_ISL_1409828, EPI_ISL_1409829, EPI_ISL_1409830, EPI_ISL_1409831, EPI_ISL_1409832, EPI_ISL_1409833, EPI_ISL_1409834, EPI_ISL_1409835, EPI_ISL_1409837, EPI_ISL_1409838, EPI_ISL_1409839, EPI_ISL_1409840, EPI_ISL_1409841, EPI_ISL_1409842, EPI_ISL_1409844, EPI_ISL_1409845, EPI_ISL_1409846, EPI_ISL_1409847, EPI_ISL_1409848, EPI_ISL_1409850, EPI_ISL_1409851, EPI_ISL_1409852, EPI_ISL_1409853, EPI_ISL_1409854, EPI_ISL_1409855, EPI_ISL_1409856, EPI_ISL_1409857, EPI_ISL_1409860, EPI_ISL_1409862, EPI_ISL_1409864, EPI_ISL_1409865, EPI_ISL_1409866, EPI_ISL_1409867, EPI_ISL_1409869, EPI_ISL_1409872, EPI_ISL_1409874, EPI_ISL_1409876, EPI_ISL_1409877, EPI_ISL_1409878, EPI_ISL_1409879, EPI_ISL_1409880, EPI_ISL_1409881, EPI_ISL_1409882, EPI_ISL_1409884, EPI_ISL_1409887, EPI_ISL_1409888, EPI_ISL_1409890, EPI_ISL_1409892, EPI_ISL_1409893, EPI_ISL_1409896, EPI_ISL_1409897, EPI_ISL_1409898, EPI_ISL_1409899, EPI_ISL_1409900, EPI_ISL_1409901, EPI_ISL_1409902, EPI_ISL_1409904, EPI_ISL_1409908, EPI_ISL_1409909, EPI_ISL_1409910, EPI_ISL_1409911, EPI_ISL_1409914, EPI_ISL_1409915, EPI_ISL_1409916, EPI_ISL_1409919, EPI_ISL_1409920, EPI_ISL_1409921, EPI_ISL_1409922, EPI_ISL_1409923, EPI_ISL_1409924, EPI_ISL_1409927, EPI_ISL_1409929, EPI_ISL_1409931, EPI_ISL_1409933, EPI_ISL_1409934, EPI_ISL_1409936, EPI_ISL_1409937, EPI_ISL_1409939, EPI_ISL_1409940, EPI_ISL_1409941, EPI_ISL_1409942, EPI_ISL_1409943, EPI_ISL_1409944, EPI_ISL_1409945, EPI_ISL_1409947, EPI_ISL_1409948, EPI_ISL_1409949, EPI_ISL_1409950, EPI_ISL_1409952, EPI_ISL_1409953, EPI_ISL_1409954, EPI_ISL_1409955, EPI_ISL_1409956, EPI_ISL_1409957, EPI_ISL_1409958, EPI_ISL_1409959, EPI_ISL_1409966, EPI_ISL_1409968, EPI_ISL_1409969, EPI_ISL_1409971, EPI_ISL_1409972, EPI_ISL_1409973, EPI_ISL_1409975, EPI_ISL_1409977, EPI_ISL_1409978, EPI_ISL_1409979, EPI_ISL_1409980, EPI_ISL_1409982, EPI_ISL_1409983, EPI_ISL_1409984, EPI_ISL_1409985, EPI_ISL_1409986, EPI_ISL_1409988, EPI_ISL_1409989, EPI_ISL_1409990, EPI_ISL_1409991, EPI_ISL_1409994, EPI_ISL_1409995, EPI_ISL_1409997, EPI_ISL_1409998, EPI_ISL_1409999, EPI_ISL_1410000, EPI_ISL_1410003, EPI_ISL_1410004, EPI_ISL_1410005, EPI_ISL_1410008, EPI_ISL_1410010, EPI_ISL_1410011, EPI_ISL_1410012, EPI_ISL_1410014, EPI_ISL_1410015, EPI_ISL_1410016, EPI_ISL_1410017, EPI_ISL_1410018, EPI_ISL_1410019, EPI_ISL_1410020, EPI_ISL_1410021, EPI_ISL_1410022, EPI_ISL_1410023, EPI_ISL_1410025, EPI_ISL_1410026, EPI_ISL_1410027, EPI_ISL_1410028, EPI_ISL_1410029, EPI_ISL_1410030, EPI_ISL_1410031, EPI_ISL_1410033, EPI_ISL_1410034, EPI_ISL_1410035, EPI_ISL_1410036, EPI_ISL_1410038, EPI_ISL_1410039, EPI_ISL_1410040, EPI_ISL_1410041, EPI_ISL_1410042, EPI_ISL_1410044, EPI_ISL_1410045, EPI_ISL_1410050, EPI_ISL_1410052, EPI_ISL_1410055, EPI_ISL_1410057, EPI_ISL_1410059, EPI_ISL_1410060, EPI_ISL_1410061, EPI_ISL_1410062, EPI_ISL_1410063, EPI_ISL_1410064, EPI_ISL_1410065, EPI_ISL_1410066, EPI_ISL_1410068, EPI_ISL_1410069, EPI_ISL_1410070, EPI_ISL_1410072, EPI_ISL_1410074, EPI_ISL_1410075, EPI_ISL_1410077, EPI_ISL_1410078, EPI_ISL_1410288, EPI_ISL_1410294, EPI_ISL_1410296, EPI_ISL_1410301, EPI_ISL_1410302, EPI_ISL_1410303, EPI_ISL_1410307, EPI_ISL_1410308, EPI_ISL_1410309, EPI_ISL_1410310, EPI_ISL_1410311, EPI_ISL_1410312, EPI_ISL_1410313, EPI_ISL_1410317, EPI_ISL_1410319, EPI_ISL_1410321, EPI_ISL_1410329, EPI_ISL_1410331, EPI_ISL_1410332, EPI_ISL_1410335, EPI_ISL_1410336, EPI_ISL_1410337, EPI_ISL_1410339, EPI_ISL_1410340, EPI_ISL_1410343, EPI_ISL_1410344, EPI_ISL_1410346, EPI_ISL_1410348, EPI_ISL_1410350, EPI_ISL_1410351, EPI_ISL_1410352, EPI_ISL_1410353, EPI_ISL_1410354, EPI_ISL_1410359, EPI_ISL_1410362, EPI_ISL_1410365, EPI_ISL_1410369, EPI_ISL_1410370, EPI_ISL_1410371, EPI_ISL_1410372, EPI_ISL_1410376, EPI_ISL_1410377, EPI_ISL_1410378, EPI_ISL_1410379, EPI_ISL_1410380, EPI_ISL_1410381, EPI_ISL_1410386, EPI_ISL_1410388, EPI_ISL_1410389, EPI_ISL_1410393, EPI_ISL_1410398, EPI_ISL_1410399, EPI_ISL_1410401, EPI_ISL_1410402, EPI_ISL_1410404, EPI_ISL_1410405, EPI_ISL_1410407, EPI_ISL_1410408, EPI_ISL_1410411, EPI_ISL_1410414, EPI_ISL_1410416, EPI_ISL_1410418, EPI_ISL_1410419, EPI_ISL_1410420, EPI_ISL_1410423, EPI_ISL_1410429, EPI_ISL_1410431, EPI_ISL_1410436, EPI_ISL_1410440, EPI_ISL_1410442, EPI_ISL_1410443, EPI_ISL_1410444, EPI_ISL_1410445, EPI_ISL_1410446, EPI_ISL_1410447, EPI_ISL_1410449, EPI_ISL_1410452, EPI_ISL_1410454, EPI_ISL_1410456, EPI_ISL_1410457, EPI_ISL_1410458, EPI_ISL_1410463, EPI_ISL_1410466, EPI_ISL_1410468, EPI_ISL_1410470, EPI_ISL_1410472, EPI_ISL_1410474, EPI_ISL_1410478, EPI_ISL_1410479, EPI_ISL_1410481, EPI_ISL_1410497, EPI_ISL_1410498, EPI_ISL_1410500, EPI_ISL_1410503, EPI_ISL_1410504, EPI_ISL_1410505, EPI_ISL_1410506, EPI_ISL_1410510, EPI_ISL_1410513, EPI_ISL_1410520, EPI_ISL_1410523, EPI_ISL_1410527, EPI_ISL_1410530, EPI_ISL_1410531, EPI_ISL_1410532, EPI_ISL_1410533, EPI_ISL_1410541, EPI_ISL_1410549, EPI_ISL_1410550, EPI_ISL_1410557, EPI_ISL_1410559, EPI_ISL_1410560, EPI_ISL_1410565, EPI_ISL_1410570, EPI_ISL_1410575, EPI_ISL_1410580, EPI_ISL_1410588, EPI_ISL_1410591, EPI_ISL_1410593, EPI_ISL_1410594, EPI_ISL_1410597, EPI_ISL_1410602, EPI_ISL_1410607, EPI_ISL_1410612, EPI_ISL_1410620, EPI_ISL_1410622, EPI_ISL_1410627, EPI_ISL_1410628, EPI_ISL_1410630, EPI_ISL_1410639, EPI_ISL_1410641, EPI_ISL_1410642, EPI_ISL_1410644, EPI_ISL_1410646, EPI_ISL_1410649, EPI_ISL_1410651, EPI_ISL_1410652, EPI_ISL_1410662, EPI_ISL_1410664, EPI_ISL_1410665, EPI_ISL_1410667, EPI_ISL_1410668, EPI_ISL_1410670, EPI_ISL_1410672, EPI_ISL_1410681, EPI_ISL_1410688, EPI_ISL_1410689, EPI_ISL_1410691, EPI_ISL_1410698, EPI_ISL_1410704, EPI_ISL_1410706, EPI_ISL_1411457, EPI_ISL_1411462, EPI_ISL_1411465, EPI_ISL_1411468, EPI_ISL_1411470, EPI_ISL_1411473, EPI_ISL_1411476, EPI_ISL_1411482, EPI_ISL_1411492, EPI_ISL_1411498, EPI_ISL_1411501, EPI_ISL_1411503, EPI_ISL_1411508, EPI_ISL_1411556, EPI_ISL_1411558, EPI_ISL_1411567, EPI_ISL_1411579, EPI_ISL_1411581, EPI_ISL_1411587, EPI_ISL_1411589, EPI_ISL_1411600, EPI_ISL_1411618, EPI_ISL_1411625, EPI_ISL_1411651, EPI_ISL_1411652, EPI_ISL_1411656, EPI_ISL_1411658, EPI_ISL_1411665, EPI_ISL_1411675, EPI_ISL_1411680, EPI_ISL_1411682, EPI_ISL_1411683, EPI_ISL_1411684, EPI_ISL_1411687, EPI_ISL_1411689, EPI_ISL_1411691, EPI_ISL_1411694, EPI_ISL_1411698, EPI_ISL_1411701, EPI_ISL_1411705, EPI_ISL_1411710, EPI_ISL_1411711, EPI_ISL_1411727, EPI_ISL_1411731, EPI_ISL_1411735, EPI_ISL_1411741, EPI_ISL_1411753, EPI_ISL_1411758, EPI_ISL_1411765, EPI_ISL_1411766, EPI_ISL_1411775, EPI_ISL_1411779, EPI_ISL_1411781, EPI_ISL_1411788, EPI_ISL_1411791, EPI_ISL_1411804, EPI_ISL_1411810, EPI_ISL_1411811, EPI_ISL_1411819, EPI_ISL_1411820, EPI_ISL_1411826, EPI_ISL_1411840, EPI_ISL_1411848, EPI_ISL_1411849, EPI_ISL_1412574, EPI_ISL_1412579, EPI_ISL_1412588, EPI_ISL_1412590, EPI_ISL_1412591, EPI_ISL_1412592, EPI_ISL_1412593, EPI_ISL_1412599, EPI_ISL_1412601, EPI_ISL_1412603, EPI_ISL_1412604, EPI_ISL_1412606, EPI_ISL_1412607, EPI_ISL_1412611, EPI_ISL_1412613, EPI_ISL_1412618, EPI_ISL_1412619, EPI_ISL_1412620, EPI_ISL_1412623, EPI_ISL_1412627, EPI_ISL_1412631, EPI_ISL_1412633, EPI_ISL_1412634, EPI_ISL_1412635, EPI_ISL_1412636, EPI_ISL_1412638, EPI_ISL_1412639, EPI_ISL_1412642, EPI_ISL_1412645, EPI_ISL_1412649, EPI_ISL_1412651, EPI_ISL_1412652, EPI_ISL_1412653, EPI_ISL_1412655, EPI_ISL_1412656, EPI_ISL_1412660, EPI_ISL_1412661, EPI_ISL_1412665, EPI_ISL_1412667, EPI_ISL_1412668, EPI_ISL_1412669, EPI_ISL_1412670, EPI_ISL_1412671, EPI_ISL_1412677, EPI_ISL_1412679, EPI_ISL_1412680, EPI_ISL_1412684, EPI_ISL_1412685, EPI_ISL_1412686, EPI_ISL_1412687, EPI_ISL_1412695, EPI_ISL_1412696, EPI_ISL_1412697, EPI_ISL_1412699, EPI_ISL_1412700, EPI_ISL_1412707, EPI_ISL_1412709, EPI_ISL_1412710, EPI_ISL_1412714, EPI_ISL_1412716, EPI_ISL_1412718, EPI_ISL_1412721, EPI_ISL_1412726, EPI_ISL_1412728, EPI_ISL_1412731, EPI_ISL_1412732, EPI_ISL_1412733, EPI_ISL_1412737, EPI_ISL_1412738, EPI_ISL_1412742, EPI_ISL_1412746, EPI_ISL_1412750, EPI_ISL_1412752, EPI_ISL_1412753, EPI_ISL_1412756, EPI_ISL_1412758, EPI_ISL_1412759, EPI_ISL_1412761, EPI_ISL_1412766, EPI_ISL_1412769, EPI_ISL_1412770, EPI_ISL_1412772, EPI_ISL_1412773, EPI_ISL_1412778, EPI_ISL_1412780, EPI_ISL_1412784, EPI_ISL_1412785, EPI_ISL_1412788, EPI_ISL_1412789, EPI_ISL_1412791, EPI_ISL_1412795, EPI_ISL_1412796, EPI_ISL_1412799, EPI_ISL_1412800, EPI_ISL_1412801, EPI_ISL_1412808, EPI_ISL_1412810, EPI_ISL_1412814, EPI_ISL_1412815, EPI_ISL_1412818, EPI_ISL_1412819, EPI_ISL_1412821, EPI_ISL_1412822, EPI_ISL_1412824, EPI_ISL_1412825, EPI_ISL_1412826, EPI_ISL_1412829, EPI_ISL_1412832, EPI_ISL_1412833, EPI_ISL_1412835, EPI_ISL_1412838, EPI_ISL_1412839, EPI_ISL_1412840, EPI_ISL_1412841, EPI_ISL_1412842, EPI_ISL_1412844, EPI_ISL_1412849, EPI_ISL_1412850, EPI_ISL_1412851, EPI_ISL_1412853, EPI_ISL_1412855, EPI_ISL_1412856, EPI_ISL_1412860, EPI_ISL_1412862, EPI_ISL_1412869, EPI_ISL_1412872, EPI_ISL_1412873, EPI_ISL_1412874, EPI_ISL_1412878, EPI_ISL_1412880, EPI_ISL_1412882, EPI_ISL_1412884, EPI_ISL_1412886, EPI_ISL_1412888, EPI_ISL_1412890, EPI_ISL_1412892, EPI_ISL_1412895, EPI_ISL_1412896, EPI_ISL_1412897, EPI_ISL_1412900, EPI_ISL_1412904, EPI_ISL_1412905, EPI_ISL_1412906, EPI_ISL_1412907, EPI_ISL_1412908, EPI_ISL_1412910, EPI_ISL_1412911, EPI_ISL_1412912, EPI_ISL_1412915, EPI_ISL_1412917, EPI_ISL_1412919, EPI_ISL_1412923, EPI_ISL_1412926, EPI_ISL_1414288, EPI_ISL_1449033, EPI_ISL_1449038, EPI_ISL_1449044, EPI_ISL_1449054, EPI_ISL_1449062, EPI_ISL_1449068, EPI_ISL_1449081, EPI_ISL_1449090, EPI_ISL_1449111, EPI_ISL_1449112, EPI_ISL_1449114, EPI_ISL_1449130, EPI_ISL_1449131, EPI_ISL_1449149, EPI_ISL_1449155, EPI_ISL_1449188, EPI_ISL_1449190, EPI_ISL_1449200, EPI_ISL_1449205, EPI_ISL_1449212, EPI_ISL_1449215, EPI_ISL_1449229, EPI_ISL_1449235, EPI_ISL_1449243, EPI_ISL_1449271, EPI_ISL_1449284, EPI_ISL_1449295, EPI_ISL_1449299, EPI_ISL_1449311, EPI_ISL_1449332, EPI_ISL_1449348, EPI_ISL_1449349, EPI_ISL_1449352, EPI_ISL_1449357, EPI_ISL_1449358, EPI_ISL_1449360, EPI_ISL_1449370, EPI_ISL_1449371, EPI_ISL_1449373, EPI_ISL_1449375, EPI_ISL_1449380, EPI_ISL_1449382, EPI_ISL_1449384, EPI_ISL_1449386, EPI_ISL_1449392, EPI_ISL_1449393, EPI_ISL_1449394, EPI_ISL_1449399, EPI_ISL_1449401, EPI_ISL_1449402, EPI_ISL_1449408, EPI_ISL_1449411, EPI_ISL_1449413, EPI_ISL_1449414, EPI_ISL_1449415, EPI_ISL_1449417, EPI_ISL_1449419, EPI_ISL_1449421, EPI_ISL_1449422, EPI_ISL_1449423, EPI_ISL_1449424, EPI_ISL_1449426, EPI_ISL_1449432, EPI_ISL_1449438, EPI_ISL_1449442, EPI_ISL_1449447, EPI_ISL_1449451, EPI_ISL_1449453, EPI_ISL_1449455, EPI_ISL_1449456, EPI_ISL_1449458, EPI_ISL_1449460, EPI_ISL_1449463, EPI_ISL_1449464, EPI_ISL_1449465, EPI_ISL_1449466, EPI_ISL_1449467, EPI_ISL_1449469, EPI_ISL_1449473, EPI_ISL_1449475, EPI_ISL_1449479, EPI_ISL_1449480, EPI_ISL_1449487, EPI_ISL_1449488, EPI_ISL_1449490, EPI_ISL_1449495, EPI_ISL_1449496, EPI_ISL_1449497, EPI_ISL_1449499, EPI_ISL_1449500, EPI_ISL_1449502, EPI_ISL_1449504, EPI_ISL_1449505, EPI_ISL_1449507, EPI_ISL_1449510, EPI_ISL_1449512, EPI_ISL_1449513, EPI_ISL_1449514, EPI_ISL_1449519, EPI_ISL_1449525, EPI_ISL_1449531, EPI_ISL_1449534, EPI_ISL_1449539, EPI_ISL_1449549, EPI_ISL_1449552, EPI_ISL_1449555, EPI_ISL_1449565, EPI_ISL_1449568, EPI_ISL_1449569, EPI_ISL_1449574, EPI_ISL_1449577, EPI_ISL_1449578, EPI_ISL_1449579, EPI_ISL_1449580, EPI_ISL_1449584, EPI_ISL_1449587, EPI_ISL_1449589, EPI_ISL_1449596, EPI_ISL_1449599, EPI_ISL_1449610, EPI_ISL_1449612, EPI_ISL_1449613, EPI_ISL_1449615, EPI_ISL_1449622, EPI_ISL_1449625, EPI_ISL_1449629, EPI_ISL_1449633, EPI_ISL_1449639, EPI_ISL_1449642, EPI_ISL_1449979, EPI_ISL_1449980, EPI_ISL_1449983, EPI_ISL_1449984, EPI_ISL_1449985, EPI_ISL_1449987, EPI_ISL_1449988, EPI_ISL_1449990, EPI_ISL_1449992, EPI_ISL_1449994, EPI_ISL_1449995, EPI_ISL_1449996, EPI_ISL_1449998, EPI_ISL_1450000, EPI_ISL_1450002, EPI_ISL_1450003, EPI_ISL_1450005, EPI_ISL_1450007, EPI_ISL_1450009, EPI_ISL_1450010, EPI_ISL_1450011, EPI_ISL_1450012, EPI_ISL_1450013, EPI_ISL_1450014, EPI_ISL_1450017, EPI_ISL_1450021, EPI_ISL_1450023, EPI_ISL_1450027, EPI_ISL_1450030, EPI_ISL_1450032, EPI_ISL_1450035, EPI_ISL_1450036, EPI_ISL_1450037, EPI_ISL_1450038, EPI_ISL_1450039, EPI_ISL_1450040, EPI_ISL_1450041, EPI_ISL_1450043, EPI_ISL_1450046, EPI_ISL_1450048, EPI_ISL_1450049, EPI_ISL_1450050, EPI_ISL_1450052, EPI_ISL_1450057, EPI_ISL_1450062, EPI_ISL_1450064, EPI_ISL_1450067, EPI_ISL_1450068, EPI_ISL_1450069, EPI_ISL_1450070, EPI_ISL_1450072, EPI_ISL_1450074, EPI_ISL_1450076, EPI_ISL_1450077, EPI_ISL_1450078, EPI_ISL_1450079, EPI_ISL_1450081, EPI_ISL_1450082, EPI_ISL_1450084, EPI_ISL_1450089, EPI_ISL_1450090, EPI_ISL_1450091, EPI_ISL_1450092, EPI_ISL_1450093, EPI_ISL_1450094, EPI_ISL_1450096, EPI_ISL_1450097, EPI_ISL_1450102, EPI_ISL_1450103, EPI_ISL_1450104, EPI_ISL_1450107, EPI_ISL_1450108, EPI_ISL_1450111, EPI_ISL_1450112, EPI_ISL_1450114, EPI_ISL_1450116, EPI_ISL_1450117, EPI_ISL_1450118, EPI_ISL_1450119, EPI_ISL_1450120, EPI_ISL_1450121, EPI_ISL_1450123, EPI_ISL_1450126, EPI_ISL_1450127, EPI_ISL_1450129, EPI_ISL_1450130, EPI_ISL_1450131, EPI_ISL_1450132, EPI_ISL_1450133, EPI_ISL_1450134, EPI_ISL_1450135, EPI_ISL_1450137, EPI_ISL_1450141, EPI_ISL_1450142, EPI_ISL_1450143, EPI_ISL_1450146, EPI_ISL_1450148, EPI_ISL_1450149, EPI_ISL_1450150, EPI_ISL_1450152, EPI_ISL_1450155, EPI_ISL_1450156, EPI_ISL_1450164, EPI_ISL_1450165, EPI_ISL_1450169, EPI_ISL_1450170, EPI_ISL_1450172, EPI_ISL_1450177, EPI_ISL_1450178, EPI_ISL_1450181, EPI_ISL_1450182, EPI_ISL_1450183, EPI_ISL_1450187, EPI_ISL_1450188, EPI_ISL_1450189, EPI_ISL_1450193, EPI_ISL_1450197, EPI_ISL_1450198, EPI_ISL_1450200, EPI_ISL_1450201, EPI_ISL_1450203, EPI_ISL_1450204, EPI_ISL_1450205, EPI_ISL_1450206, EPI_ISL_1450208, EPI_ISL_1450209, EPI_ISL_1450212, EPI_ISL_1450213, EPI_ISL_1450216, EPI_ISL_1450217, EPI_ISL_1450218, EPI_ISL_1450219, EPI_ISL_1450221, EPI_ISL_1450222, EPI_ISL_1450224, EPI_ISL_1450226, EPI_ISL_1450228, EPI_ISL_1450229, EPI_ISL_1450230, EPI_ISL_1450231, EPI_ISL_1450237, EPI_ISL_1450238, EPI_ISL_1450239, EPI_ISL_1450240, EPI_ISL_1450241, EPI_ISL_1450243, EPI_ISL_1450244, EPI_ISL_1450247, EPI_ISL_1450248, EPI_ISL_1450249, EPI_ISL_1450250, EPI_ISL_1450253, EPI_ISL_1450256, EPI_ISL_1450257, EPI_ISL_1450258, EPI_ISL_1450268, EPI_ISL_1450270, EPI_ISL_1450271, EPI_ISL_1450272, EPI_ISL_1450276, EPI_ISL_1450277, EPI_ISL_1450278, EPI_ISL_1450279, EPI_ISL_1450280, EPI_ISL_1450281, EPI_ISL_1450282, EPI_ISL_1450283, EPI_ISL_1450286, EPI_ISL_1450287, EPI_ISL_1450293, EPI_ISL_1450294, EPI_ISL_1450300, EPI_ISL_1450301, EPI_ISL_1450305, EPI_ISL_1450307, EPI_ISL_1450308, EPI_ISL_1450309, EPI_ISL_1450311, EPI_ISL_1450318, EPI_ISL_1450319, EPI_ISL_1450320, EPI_ISL_1450322, EPI_ISL_1450323, EPI_ISL_1450324, EPI_ISL_1450326, EPI_ISL_1450330, EPI_ISL_1450332, EPI_ISL_1450336, EPI_ISL_1450338, EPI_ISL_1450345, EPI_ISL_1450346, EPI_ISL_1450348, EPI_ISL_1450350, EPI_ISL_1450352, EPI_ISL_1450354, EPI_ISL_1450356, EPI_ISL_1450358, EPI_ISL_1450361, EPI_ISL_1450363, EPI_ISL_1450367, EPI_ISL_1450368, EPI_ISL_1450371, EPI_ISL_1450374, EPI_ISL_1450376, EPI_ISL_1450377, EPI_ISL_1450379, EPI_ISL_1450380, EPI_ISL_1450381, EPI_ISL_1450382, EPI_ISL_1450384, EPI_ISL_1450388, EPI_ISL_1450389, EPI_ISL_1450390, EPI_ISL_1450391, EPI_ISL_1450392, EPI_ISL_1450393, EPI_ISL_1450395, EPI_ISL_1450398, EPI_ISL_1450399, EPI_ISL_1450401, EPI_ISL_1450402, EPI_ISL_1450404, EPI_ISL_1450405, EPI_ISL_1450406, EPI_ISL_1450409, EPI_ISL_1450410, EPI_ISL_1450412, EPI_ISL_1450415, EPI_ISL_1450416, EPI_ISL_1450417, EPI_ISL_1450419, EPI_ISL_1450420, EPI_ISL_1450421, EPI_ISL_1450422, EPI_ISL_1450424, EPI_ISL_1450428, EPI_ISL_1450429, EPI_ISL_1450430, EPI_ISL_1450431, EPI_ISL_1450432, EPI_ISL_1450434, EPI_ISL_1450435, EPI_ISL_1450436, EPI_ISL_1450440, EPI_ISL_1450442, EPI_ISL_1450444, EPI_ISL_1450445, EPI_ISL_1450446, EPI_ISL_1450449, EPI_ISL_1450454, EPI_ISL_1450455, EPI_ISL_1450456, EPI_ISL_1450457, EPI_ISL_1450459, EPI_ISL_1450461, EPI_ISL_1450462, EPI_ISL_1450465, EPI_ISL_1450467, EPI_ISL_1450470, EPI_ISL_1450471, EPI_ISL_1450473, EPI_ISL_1450474, EPI_ISL_1450475, EPI_ISL_1450479, EPI_ISL_1450480, EPI_ISL_1450481, EPI_ISL_1450483, EPI_ISL_1450484, EPI_ISL_1450486, EPI_ISL_1450489, EPI_ISL_1450490, EPI_ISL_1450491, EPI_ISL_1450492, EPI_ISL_1450495, EPI_ISL_1450499, EPI_ISL_1450501, EPI_ISL_1450505, EPI_ISL_1450506, EPI_ISL_1450507, EPI_ISL_1450509, EPI_ISL_1450510, EPI_ISL_1450513, EPI_ISL_1450514, EPI_ISL_1450518, EPI_ISL_1450521, EPI_ISL_1450523, EPI_ISL_1450525, EPI_ISL_1450527, EPI_ISL_1450528, EPI_ISL_1450531, EPI_ISL_1450533, EPI_ISL_1450534, EPI_ISL_1450535, EPI_ISL_1450536, EPI_ISL_1450537, EPI_ISL_1450542, EPI_ISL_1450544, EPI_ISL_1450545, EPI_ISL_1450546, EPI_ISL_1450547, EPI_ISL_1450548, EPI_ISL_1450549, EPI_ISL_1450551, EPI_ISL_1450554, EPI_ISL_1450555, EPI_ISL_1450556, EPI_ISL_1450557, EPI_ISL_1450560, EPI_ISL_1450562, EPI_ISL_1450566, EPI_ISL_1450568, EPI_ISL_1450569, EPI_ISL_1450574, EPI_ISL_1450575, EPI_ISL_1450576, EPI_ISL_1450577, EPI_ISL_1450578, EPI_ISL_1450579, EPI_ISL_1450580, EPI_ISL_1450581, EPI_ISL_1450583, EPI_ISL_1450585, EPI_ISL_1450586, EPI_ISL_1450588, EPI_ISL_1450589, EPI_ISL_1450591, EPI_ISL_1450592, EPI_ISL_1450593, EPI_ISL_1450595, EPI_ISL_1450597, EPI_ISL_1450600, EPI_ISL_1450601, EPI_ISL_1450602, EPI_ISL_1450603, EPI_ISL_1451483, EPI_ISL_1451484, EPI_ISL_1451486, EPI_ISL_1451492, EPI_ISL_1451501, EPI_ISL_1451504, EPI_ISL_1451507, EPI_ISL_1451510, EPI_ISL_1451514, EPI_ISL_1451516, EPI_ISL_1451517, EPI_ISL_1451522, EPI_ISL_1451524, EPI_ISL_1451526, EPI_ISL_1451527, EPI_ISL_1451531, EPI_ISL_1451535, EPI_ISL_1451544, EPI_ISL_1451545, EPI_ISL_1451546, EPI_ISL_1451548, EPI_ISL_1451549, EPI_ISL_1451552, EPI_ISL_1451553, EPI_ISL_1451560, EPI_ISL_1451562, EPI_ISL_1451564, EPI_ISL_1451566, EPI_ISL_1451569, EPI_ISL_1451570, EPI_ISL_1451575, EPI_ISL_1451588, EPI_ISL_1451591, EPI_ISL_1451595, EPI_ISL_1451596, EPI_ISL_1451597, EPI_ISL_1451606, EPI_ISL_1451611, EPI_ISL_1451618, EPI_ISL_1451621, EPI_ISL_1451623, EPI_ISL_1451625, EPI_ISL_1451626, EPI_ISL_1451636, EPI_ISL_1451638, EPI_ISL_1451639, EPI_ISL_1451645, EPI_ISL_1451646, EPI_ISL_1451647, EPI_ISL_1451655, EPI_ISL_1451656, EPI_ISL_1451662, EPI_ISL_1451669, EPI_ISL_1451672, EPI_ISL_1451673, EPI_ISL_1451674, EPI_ISL_1451680, EPI_ISL_1451686, EPI_ISL_1451688, EPI_ISL_1451691, EPI_ISL_1451692, EPI_ISL_1451695, EPI_ISL_1451698, EPI_ISL_1451702, EPI_ISL_1451709, EPI_ISL_1451710, EPI_ISL_1451711, EPI_ISL_1451715, EPI_ISL_1451720, EPI_ISL_1451724, EPI_ISL_1451728, EPI_ISL_1451730, EPI_ISL_1451731, EPI_ISL_1451734, EPI_ISL_1451735, EPI_ISL_1451737, EPI_ISL_1451739, EPI_ISL_1451740, EPI_ISL_1451741, EPI_ISL_1451742, EPI_ISL_1451744, EPI_ISL_1451745, EPI_ISL_1451746, EPI_ISL_1451751, EPI_ISL_1451753, EPI_ISL_1451762, EPI_ISL_1451766, EPI_ISL_1451773, EPI_ISL_1451775, EPI_ISL_1451776, EPI_ISL_1451784, EPI_ISL_1451788, EPI_ISL_1451792, EPI_ISL_1451794, EPI_ISL_1451795, EPI_ISL_1451797, EPI_ISL_1451798, EPI_ISL_1451800, EPI_ISL_1451801, EPI_ISL_1451803, EPI_ISL_1451804, EPI_ISL_1451807, EPI_ISL_1451808, EPI_ISL_1451809, EPI_ISL_1451810, EPI_ISL_1451813, EPI_ISL_1451814, EPI_ISL_1451816, EPI_ISL_1451817, EPI_ISL_1451819, EPI_ISL_1451820, EPI_ISL_1451821, EPI_ISL_1451822, EPI_ISL_1451823, EPI_ISL_1451824, EPI_ISL_1451825, EPI_ISL_1451826, EPI_ISL_1451827, EPI_ISL_1451828, EPI_ISL_1451830, EPI_ISL_1451831, EPI_ISL_1451833, EPI_ISL_1451835, EPI_ISL_1451836, EPI_ISL_1451839, EPI_ISL_1451843, EPI_ISL_1451844, EPI_ISL_1451846, EPI_ISL_1451847, EPI_ISL_1451848, EPI_ISL_1451849, EPI_ISL_1451851, EPI_ISL_1451853, EPI_ISL_1451854, EPI_ISL_1451856, EPI_ISL_1451857, EPI_ISL_1451860, EPI_ISL_1451861, EPI_ISL_1451863, EPI_ISL_1451864, EPI_ISL_1451865, EPI_ISL_1451866, EPI_ISL_1451867, EPI_ISL_1451868, EPI_ISL_1451871, EPI_ISL_1451873, EPI_ISL_1451874, EPI_ISL_1451877, EPI_ISL_1451878, EPI_ISL_1451880, EPI_ISL_1451884, EPI_ISL_1451885, EPI_ISL_1451889, EPI_ISL_1451890, EPI_ISL_1451891, EPI_ISL_1451893, EPI_ISL_1451894, EPI_ISL_1451895, EPI_ISL_1451897, EPI_ISL_1451898, EPI_ISL_1451899, EPI_ISL_1451901, EPI_ISL_1451902, EPI_ISL_1451905, EPI_ISL_1451906, EPI_ISL_1451908, EPI_ISL_1451910, EPI_ISL_1451911, EPI_ISL_1451912, EPI_ISL_1451913, EPI_ISL_1451915, EPI_ISL_1451916, EPI_ISL_1451920, EPI_ISL_1451921, EPI_ISL_1451923, EPI_ISL_1451924, EPI_ISL_1451925, EPI_ISL_1451926, EPI_ISL_1451927, EPI_ISL_1451928, EPI_ISL_1451929, EPI_ISL_1451933, EPI_ISL_1451934, EPI_ISL_1451938, EPI_ISL_1451939, EPI_ISL_1451941, EPI_ISL_1451943, EPI_ISL_1451944, EPI_ISL_1451945, EPI_ISL_1451946, EPI_ISL_1451947, EPI_ISL_1451948, EPI_ISL_1451949, EPI_ISL_1451950, EPI_ISL_1451951, EPI_ISL_1451952, EPI_ISL_1451953, EPI_ISL_1451954, EPI_ISL_1451955, EPI_ISL_1451959, EPI_ISL_1451960, EPI_ISL_1451961, EPI_ISL_1451962, EPI_ISL_1451963, EPI_ISL_1451964, EPI_ISL_1451965, EPI_ISL_1451968, EPI_ISL_1451971, EPI_ISL_1451972, EPI_ISL_1451973, EPI_ISL_1451974, EPI_ISL_1451976, EPI_ISL_1451978, EPI_ISL_1451981, EPI_ISL_1451982, EPI_ISL_1451983, EPI_ISL_1451984, EPI_ISL_1451985, EPI_ISL_1451986, EPI_ISL_1451989, EPI_ISL_1451990, EPI_ISL_1451991, EPI_ISL_1451993, EPI_ISL_1451994, EPI_ISL_1451995, EPI_ISL_1451996, EPI_ISL_1451997, EPI_ISL_1452000, EPI_ISL_1452001, EPI_ISL_1452004, EPI_ISL_1452006, EPI_ISL_1452007, EPI_ISL_1452009, EPI_ISL_1452012, EPI_ISL_1452013, EPI_ISL_1452015, EPI_ISL_1452018, EPI_ISL_1452020, EPI_ISL_1452022, EPI_ISL_1452023, EPI_ISL_1452024, EPI_ISL_1452025, EPI_ISL_1452026, EPI_ISL_1452027, EPI_ISL_1452028, EPI_ISL_1452029, EPI_ISL_1452031, EPI_ISL_1452033, EPI_ISL_1452034, EPI_ISL_1452037, EPI_ISL_1452038, EPI_ISL_1452042, EPI_ISL_1452044, EPI_ISL_1452045, EPI_ISL_1452046, EPI_ISL_1452047, EPI_ISL_1452048, EPI_ISL_1452049, EPI_ISL_1452055, EPI_ISL_1452057, EPI_ISL_1452058, EPI_ISL_1452059, EPI_ISL_1452064, EPI_ISL_1452065, EPI_ISL_1452066, EPI_ISL_1452067, EPI_ISL_1452068, EPI_ISL_1452069, EPI_ISL_1452070, EPI_ISL_1452071, EPI_ISL_1452072, EPI_ISL_1452074, EPI_ISL_1452077, EPI_ISL_1452086, EPI_ISL_1452090, EPI_ISL_1452091, EPI_ISL_1452092, EPI_ISL_1452093, EPI_ISL_1452095, EPI_ISL_1452104, EPI_ISL_1452105, EPI_ISL_1452107, EPI_ISL_1452109, EPI_ISL_1452113, EPI_ISL_1452114, EPI_ISL_1452115, EPI_ISL_1452119, EPI_ISL_1452122, EPI_ISL_1452123, EPI_ISL_1452126, EPI_ISL_1452128, EPI_ISL_1452131, EPI_ISL_1452135, EPI_ISL_1452138, EPI_ISL_1452141, EPI_ISL_1452142, EPI_ISL_1452143, EPI_ISL_1452144, EPI_ISL_1452146, EPI_ISL_1452147, EPI_ISL_1452158, EPI_ISL_1452160, EPI_ISL_1452162, EPI_ISL_1452163, EPI_ISL_1452169, EPI_ISL_1452172, EPI_ISL_1452174, EPI_ISL_1452177, EPI_ISL_1452178, EPI_ISL_1452187, EPI_ISL_1452189, EPI_ISL_1452191, EPI_ISL_1452194, EPI_ISL_1452198, EPI_ISL_1452204, EPI_ISL_1452205, EPI_ISL_1452208, EPI_ISL_1452210, EPI_ISL_1452212, EPI_ISL_1452215, EPI_ISL_1452216, EPI_ISL_1452220, EPI_ISL_1452228, EPI_ISL_1452229, EPI_ISL_1452238, EPI_ISL_1452239, EPI_ISL_1452240, EPI_ISL_1452242, EPI_ISL_1452245, EPI_ISL_1452246, EPI_ISL_1452252, EPI_ISL_1452253, EPI_ISL_1452256, EPI_ISL_1452257, EPI_ISL_1452258, EPI_ISL_1452260, EPI_ISL_1452278, EPI_ISL_1452280, EPI_ISL_1449622, EPI_ISL_1452284, EPI_ISL_1452289, EPI_ISL_1452295, EPI_ISL_1452299, EPI_ISL_1452302, EPI_ISL_1452306, EPI_ISL_1452308, EPI_ISL_1452309, EPI_ISL_1452315, EPI_ISL_1452317, EPI_ISL_1452318, EPI_ISL_1452320, EPI_ISL_1452322, EPI_ISL_1452325, EPI_ISL_1452326, EPI_ISL_1452344, EPI_ISL_1452626, EPI_ISL_1452631, EPI_ISL_1452632, EPI_ISL_1452638, EPI_ISL_1452644, EPI_ISL_1452647, EPI_ISL_1452649, EPI_ISL_1452650, EPI_ISL_1452665, EPI_ISL_1452674, EPI_ISL_1452680, EPI_ISL_1452682, EPI_ISL_1452684, EPI_ISL_1452685, EPI_ISL_1452692, EPI_ISL_1452693, EPI_ISL_1452694, EPI_ISL_1452697, EPI_ISL_1452699, EPI_ISL_1452702, EPI_ISL_1452703, EPI_ISL_1452704, EPI_ISL_1452707, EPI_ISL_1452708, EPI_ISL_1452710, EPI_ISL_1452711, EPI_ISL_1452712, EPI_ISL_1452715, EPI_ISL_1452716, EPI_ISL_1452718, EPI_ISL_1452720, EPI_ISL_1452722, EPI_ISL_1452728, EPI_ISL_1452729, EPI_ISL_1452730, EPI_ISL_1452738, EPI_ISL_1452740, EPI_ISL_1452743, EPI_ISL_1452745, EPI_ISL_1452750, EPI_ISL_1452751, EPI_ISL_1452753, EPI_ISL_1452755, EPI_ISL_1452758, EPI_ISL_1452761, EPI_ISL_1452764, EPI_ISL_1452767, EPI_ISL_1452768, EPI_ISL_1452772, EPI_ISL_1452777, EPI_ISL_1452778, EPI_ISL_1452780, EPI_ISL_1452781, EPI_ISL_1452783, EPI_ISL_1452786, EPI_ISL_1452787, EPI_ISL_1452790, EPI_ISL_1452792, EPI_ISL_1452794, EPI_ISL_1452795, EPI_ISL_1452798, EPI_ISL_1452800, EPI_ISL_1452808, EPI_ISL_1452812, EPI_ISL_1452815, EPI_ISL_1452817, EPI_ISL_1452819, EPI_ISL_1452826, EPI_ISL_1452831, EPI_ISL_1452833, EPI_ISL_1452835, EPI_ISL_1452836, EPI_ISL_1452839, EPI_ISL_1452842, EPI_ISL_1452851, EPI_ISL_1452854, EPI_ISL_1452856, EPI_ISL_1452857, EPI_ISL_1452860, EPI_ISL_1452862, EPI_ISL_1452866, EPI_ISL_1452870, EPI_ISL_1452871, EPI_ISL_1452872, EPI_ISL_1452873, EPI_ISL_1452874, EPI_ISL_1452877, EPI_ISL_1452882, EPI_ISL_1452887, EPI_ISL_1452890, EPI_ISL_1452891, EPI_ISL_1452892, EPI_ISL_1452893, EPI_ISL_1452899, EPI_ISL_1452901, EPI_ISL_1452904, EPI_ISL_1452905, EPI_ISL_1452910, EPI_ISL_1452911, EPI_ISL_1452914, EPI_ISL_1452917, EPI_ISL_1452920, EPI_ISL_1452922, EPI_ISL_1452923, EPI_ISL_1452925, EPI_ISL_1452929, EPI_ISL_1452932, EPI_ISL_1452935, EPI_ISL_1452945, EPI_ISL_1452949, EPI_ISL_1452950, EPI_ISL_1452951, EPI_ISL_1452954, EPI_ISL_1452957, EPI_ISL_1452959, EPI_ISL_1452961, EPI_ISL_1452963, EPI_ISL_1452967, EPI_ISL_1452976, EPI_ISL_1452982, EPI_ISL_1452986, EPI_ISL_1452987, EPI_ISL_1452992, EPI_ISL_1452994, EPI_ISL_1453001, EPI_ISL_1453004, EPI_ISL_1453007, EPI_ISL_1453009, EPI_ISL_1453010, EPI_ISL_1453017, EPI_ISL_1453018, EPI_ISL_1453025, EPI_ISL_1453026, EPI_ISL_1453029, EPI_ISL_1453040, EPI_ISL_1453046, EPI_ISL_1453048, EPI_ISL_1453050, EPI_ISL_1453051, EPI_ISL_1453052, EPI_ISL_1453053, EPI_ISL_1453055, EPI_ISL_1453061, EPI_ISL_1453062, EPI_ISL_1453063, EPI_ISL_1453069, EPI_ISL_1453070, EPI_ISL_1453071, EPI_ISL_1453073, EPI_ISL_1453079, EPI_ISL_1453104, EPI_ISL_1453107, EPI_ISL_1453108, EPI_ISL_1453111, EPI_ISL_1453113, EPI_ISL_1453115, EPI_ISL_1453116, EPI_ISL_1453124, EPI_ISL_1453128, EPI_ISL_1453137, EPI_ISL_1453142, EPI_ISL_1453147, EPI_ISL_1453148, EPI_ISL_1453152, EPI_ISL_1453158, EPI_ISL_1453160, EPI_ISL_1453162, EPI_ISL_1453166, EPI_ISL_1453178, EPI_ISL_1453182, EPI_ISL_1453183, EPI_ISL_1453184, EPI_ISL_1453185, EPI_ISL_1453189, EPI_ISL_1453190, EPI_ISL_1453191, EPI_ISL_1453193, EPI_ISL_1453194, EPI_ISL_1453195, EPI_ISL_1453198, EPI_ISL_1453200, EPI_ISL_1453201, EPI_ISL_1453202, EPI_ISL_1453203, EPI_ISL_1453205, EPI_ISL_1453207, EPI_ISL_1453208, EPI_ISL_1453209, EPI_ISL_1453211, EPI_ISL_1453216, EPI_ISL_1453217, EPI_ISL_1453218, EPI_ISL_1453223, EPI_ISL_1453224, EPI_ISL_1453225, EPI_ISL_1453226, EPI_ISL_1453227, EPI_ISL_1453229, EPI_ISL_1453230, EPI_ISL_1453231, EPI_ISL_1453232, EPI_ISL_1453233, EPI_ISL_1453235, EPI_ISL_1453236, EPI_ISL_1453237, EPI_ISL_1453238, EPI_ISL_1453239, EPI_ISL_1453240, EPI_ISL_1453242, EPI_ISL_1453244, EPI_ISL_1453245, EPI_ISL_1453247, EPI_ISL_1453248, EPI_ISL_1453251, EPI_ISL_1453252, EPI_ISL_1453253, EPI_ISL_1453256, EPI_ISL_1453257, EPI_ISL_1453259, EPI_ISL_1453261, EPI_ISL_1453262, EPI_ISL_1453263, EPI_ISL_1453264, EPI_ISL_1453269, EPI_ISL_1453270, EPI_ISL_1453272, EPI_ISL_1453274, EPI_ISL_1453275, EPI_ISL_1453277, EPI_ISL_1453282, EPI_ISL_1453284, EPI_ISL_1453285, EPI_ISL_1453286, EPI_ISL_1453287, EPI_ISL_1453289, EPI_ISL_1453290, EPI_ISL_1453293, EPI_ISL_1453295, EPI_ISL_1453296, EPI_ISL_1453299, EPI_ISL_1453301, EPI_ISL_1453302, EPI_ISL_1453304, EPI_ISL_1453305, EPI_ISL_1453306, EPI_ISL_1453307, EPI_ISL_1453308, EPI_ISL_1453309, EPI_ISL_1453310, EPI_ISL_1453312, EPI_ISL_1453318, EPI_ISL_1453321, EPI_ISL_1453322, EPI_ISL_1453324, EPI_ISL_1453326, EPI_ISL_1453328, EPI_ISL_1453330, EPI_ISL_1453332, EPI_ISL_1453333, EPI_ISL_1453334, EPI_ISL_1453336, EPI_ISL_1453339, EPI_ISL_1453340, EPI_ISL_1453342, EPI_ISL_1453343, EPI_ISL_1453344, EPI_ISL_1453345, EPI_ISL_1453348, EPI_ISL_1453349, EPI_ISL_1453350, EPI_ISL_1453352, EPI_ISL_1453353, EPI_ISL_1453354, EPI_ISL_1453356, EPI_ISL_1453357, EPI_ISL_1453358, EPI_ISL_1453363, EPI_ISL_1453365, EPI_ISL_1453366, EPI_ISL_1453367, EPI_ISL_1453368, EPI_ISL_1453374, EPI_ISL_1453376, EPI_ISL_1453377, EPI_ISL_1453379, EPI_ISL_1453380, EPI_ISL_1453381, EPI_ISL_1453382, EPI_ISL_1453383, EPI_ISL_1453384, EPI_ISL_1453385, EPI_ISL_1453386, EPI_ISL_1453387, EPI_ISL_1453388, EPI_ISL_1453389, EPI_ISL_1453390, EPI_ISL_1453391, EPI_ISL_1453397, EPI_ISL_1453399, EPI_ISL_1453401, EPI_ISL_1453402, EPI_ISL_1453403, EPI_ISL_1453406, EPI_ISL_1453408, EPI_ISL_1453410, EPI_ISL_1453412, EPI_ISL_1453414, EPI_ISL_1453416, EPI_ISL_1453417, EPI_ISL_1453419, EPI_ISL_1453421, EPI_ISL_1453422, EPI_ISL_1453423, EPI_ISL_1453425, EPI_ISL_1453428, EPI_ISL_1453429, EPI_ISL_1453430, EPI_ISL_1453431, EPI_ISL_1453432, EPI_ISL_1453438, EPI_ISL_1453439, EPI_ISL_1453441, EPI_ISL_1453443, EPI_ISL_1453446, EPI_ISL_1453450, EPI_ISL_1453451, EPI_ISL_1453452, EPI_ISL_1453454, EPI_ISL_1453455, EPI_ISL_1453456, EPI_ISL_1453457, EPI_ISL_1453458, EPI_ISL_1453460, EPI_ISL_1453461, EPI_ISL_1453462, EPI_ISL_1453464, EPI_ISL_1453465, EPI_ISL_1453466, EPI_ISL_1453467, EPI_ISL_1453468, EPI_ISL_1453469, EPI_ISL_1453487, EPI_ISL_1453490,

EPI_ISL_1453491, EPI_ISL_1453492, EPI_ISL_1453493, EPI_ISL_1453495, EPI_ISL_1453502, EPI_ISL_1453505, EPI_ISL_1453509, EPI_ISL_1453510, EPI_ISL_1453512, EPI_ISL_1453514, EPI_ISL_1453515, EPI_ISL_1453516, EPI_ISL_1453517, EPI_ISL_1453520, EPI_ISL_1453522, EPI_ISL_1453524, EPI_ISL_1453527, EPI_ISL_1453528, EPI_ISL_1453529, EPI_ISL_1453530, EPI_ISL_1453532, EPI_ISL_1453533,
EPI_ISL_1453535, EPI_ISL_1453538, EPI_ISL_1453539, EPI_ISL_1453541, EPI_ISL_1453543, EPI_ISL_1453545, EPI_ISL_1453547, EPI_ISL_1453549, EPI_ISL_1453551, EPI_ISL_1453553, EPI_ISL_1453557, EPI_ISL_1453558, EPI_ISL_1453560, EPI_ISL_1453562, EPI_ISL_1453563, EPI_ISL_1453565, EPI_ISL_1453566, EPI_ISL_1453568, EPI_ISL_1453570, EPI_ISL_1453576, EPI_ISL_1453580, EPI_ISL_1453585,
EPI_ISL_1453587, EPI_ISL_1453589, EPI_ISL_1453590, EPI_ISL_1453591, EPI_ISL_1453592, EPI_ISL_1453593, EPI_ISL_1453596, EPI_ISL_1453597, EPI_ISL_1453598, EPI_ISL_1453599, EPI_ISL_1453600, EPI_ISL_1453601, EPI_ISL_1453606, EPI_ISL_1453607, EPI_ISL_1453608, EPI_ISL_1453609, EPI_ISL_1453610, EPI_ISL_1453611, EPI_ISL_1453612, EPI_ISL_1453613, EPI_ISL_1453614, EPI_ISL_1453616,
EPI_ISL_1453617, EPI_ISL_1453618, EPI_ISL_1453619, EPI_ISL_1453620, EPI_ISL_1453621, EPI_ISL_1453622, EPI_ISL_1453623, EPI_ISL_1453624, EPI_ISL_1453625, EPI_ISL_1453627, EPI_ISL_1453628, EPI_ISL_1453629, EPI_ISL_1453632, EPI_ISL_1453633, EPI_ISL_1453634, EPI_ISL_1453639, EPI_ISL_1453640, EPI_ISL_1453641, EPI_ISL_1453642, EPI_ISL_1453643, EPI_ISL_1453645, EPI_ISL_1453649,
EPI_ISL_1453650, EPI_ISL_1453652, EPI_ISL_1453653, EPI_ISL_1453657, EPI_ISL_1453661, EPI_ISL_1453662, EPI_ISL_1453664, EPI_ISL_1453668, EPI_ISL_1453669, EPI_ISL_1453670, EPI_ISL_1453671, EPI_ISL_1453672, EPI_ISL_1453673, EPI_ISL_1453674, EPI_ISL_1453675, EPI_ISL_1453676, EPI_ISL_1453680, EPI_ISL_1453682, EPI_ISL_1453684, EPI_ISL_1453685, EPI_ISL_1453686, EPI_ISL_1453687,
EPI_ISL_1453688, EPI_ISL_1453690, EPI_ISL_1453691, EPI_ISL_1453694, EPI_ISL_1453695, EPI_ISL_1453696, EPI_ISL_1453697, EPI_ISL_1453698, EPI_ISL_1453701, EPI_ISL_1453703, EPI_ISL_1453704, EPI_ISL_1453707, EPI_ISL_1453710, EPI_ISL_1453713, EPI_ISL_1453715, EPI_ISL_1453716, EPI_ISL_1453717, EPI_ISL_1453719, EPI_ISL_1453720, EPI_ISL_1453721, EPI_ISL_1453722, EPI_ISL_1453724,
EPI_ISL_1453725, EPI_ISL_1453726, EPI_ISL_1453728, EPI_ISL_1453729, EPI_ISL_1453730, EPI_ISL_1453731, EPI_ISL_1453732, EPI_ISL_1453733, EPI_ISL_1453734, EPI_ISL_1453735, EPI_ISL_1453737, EPI_ISL_1453739, EPI_ISL_1453740, EPI_ISL_1453742, EPI_ISL_1453743, EPI_ISL_1453746, EPI_ISL_1453747, EPI_ISL_1453748, EPI_ISL_1453750, EPI_ISL_1453751, EPI_ISL_1453752, EPI_ISL_1453754,
EPI_ISL_1453756, EPI_ISL_1453757, EPI_ISL_1453758, EPI_ISL_1453759, EPI_ISL_1453761, EPI_ISL_1453763, EPI_ISL_1453766, EPI_ISL_1453767, EPI_ISL_1453769, EPI_ISL_1453771, EPI_ISL_1453773, EPI_ISL_1453775, EPI_ISL_1453776, EPI_ISL_1453778, EPI_ISL_1453780, EPI_ISL_1453786, EPI_ISL_1453788, EPI_ISL_1453789, EPI_ISL_1453790, EPI_ISL_1453795, EPI_ISL_1453797, EPI_ISL_1453798,
EPI_ISL_1453800, EPI_ISL_1453805, EPI_ISL_1453806, EPI_ISL_1453807, EPI_ISL_1453809, EPI_ISL_1453810, EPI_ISL_1453812, EPI_ISL_1453813, EPI_ISL_1453815, EPI_ISL_1453816, EPI_ISL_1453822, EPI_ISL_1453823, EPI_ISL_1453824, EPI_ISL_1453825, EPI_ISL_1453826, EPI_ISL_1453828, EPI_ISL_1453831, EPI_ISL_1453835, EPI_ISL_1453837, EPI_ISL_1453839, EPI_ISL_1453841, EPI_ISL_1453842,
EPI_ISL_1453843, EPI_ISL_1453849, EPI_ISL_1453851, EPI_ISL_1453852, EPI_ISL_1453857, EPI_ISL_1453859, EPI_ISL_1453860, EPI_ISL_1453862, EPI_ISL_1453867, EPI_ISL_1453873, EPI_ISL_1453874, EPI_ISL_1453876, EPI_ISL_1453881, EPI_ISL_1453882, EPI_ISL_1453883, EPI_ISL_1453886, EPI_ISL_1453889, EPI_ISL_1453891, EPI_ISL_1453892, EPI_ISL_1453895, EPI_ISL_1453896, EPI_ISL_1453899,
EPI_ISL_1453900, EPI_ISL_1453902, EPI_ISL_1453903, EPI_ISL_1453904, EPI_ISL_1453905, EPI_ISL_1453907, EPI_ISL_1453911, EPI_ISL_1453912, EPI_ISL_1453917, EPI_ISL_1453918, EPI_ISL_1453919, EPI_ISL_1453920, EPI_ISL_1453922, EPI_ISL_1453923, EPI_ISL_1453925, EPI_ISL_1453930, EPI_ISL_1453933, EPI_ISL_1453934, EPI_ISL_1453935, EPI_ISL_1453936, EPI_ISL_1453937, EPI_ISL_1453938,
EPI_ISL_1453939, EPI_ISL_1453943, EPI_ISL_1453944, EPI_ISL_1453945, EPI_ISL_1453946, EPI_ISL_1453949, EPI_ISL_1453950, EPI_ISL_1453953, EPI_ISL_1453954, EPI_ISL_1453959, EPI_ISL_1453960, EPI_ISL_1453961, EPI_ISL_1453962, EPI_ISL_1453963, EPI_ISL_1453964, EPI_ISL_1453969, EPI_ISL_1453971, EPI_ISL_1453972, EPI_ISL_1453973, EPI_ISL_1453974, EPI_ISL_1453976, EPI_ISL_1453977,
EPI_ISL_1453980, EPI_ISL_1453981, EPI_ISL_1453982, EPI_ISL_1453983, EPI_ISL_1453984, EPI_ISL_1453986, EPI_ISL_1453987, EPI_ISL_1453989, EPI_ISL_1453990, EPI_ISL_1453991, EPI_ISL_1453992, EPI_ISL_1453996, EPI_ISL_1453998, EPI_ISL_1453999, EPI_ISL_1454001, EPI_ISL_1454003, EPI_ISL_1454006, EPI_ISL_1454012, EPI_ISL_1454014, EPI_ISL_1454015, EPI_ISL_1454016, EPI_ISL_1454018,
EPI_ISL_1454019, EPI_ISL_1454024, EPI_ISL_1454027, EPI_ISL_1454029, EPI_ISL_1454030, EPI_ISL_1454031, EPI_ISL_1454032, EPI_ISL_1454036, EPI_ISL_1454037, EPI_ISL_1454039, EPI_ISL_1454040, EPI_ISL_1454043, EPI_ISL_1454046, EPI_ISL_1454053, EPI_ISL_1454054, EPI_ISL_1454055, EPI_ISL_1454059, EPI_ISL_1454062, EPI_ISL_1454063, EPI_ISL_1454064, EPI_ISL_1454066, EPI_ISL_1454068,
EPI_ISL_1454070, EPI_ISL_1454071, EPI_ISL_1454072, EPI_ISL_1454073, EPI_ISL_1454078, EPI_ISL_1454081, EPI_ISL_1454082, EPI_ISL_1454084, EPI_ISL_1454088, EPI_ISL_1454090, EPI_ISL_1454091, EPI_ISL_1454092, EPI_ISL_1454093, EPI_ISL_1454094, EPI_ISL_1454098, EPI_ISL_1454099, EPI_ISL_1454106, EPI_ISL_1454108, EPI_ISL_1454110, EPI_ISL_1454111, EPI_ISL_1454113, EPI_ISL_1454114,
EPI_ISL_1454116, EPI_ISL_1454118, EPI_ISL_1454119, EPI_ISL_1454120, EPI_ISL_1454121, EPI_ISL_1454122, EPI_ISL_1454124, EPI_ISL_1454126, EPI_ISL_1454127, EPI_ISL_1454128, EPI_ISL_1454129, EPI_ISL_1454130, EPI_ISL_1454138, EPI_ISL_1454139, EPI_ISL_1454142, EPI_ISL_1454146, EPI_ISL_1454148, EPI_ISL_1454150, EPI_ISL_1454151, EPI_ISL_1454153, EPI_ISL_1454154, EPI_ISL_1454155,
EPI_ISL_1454156, EPI_ISL_1454333, EPI_ISL_1454335, EPI_ISL_1454336, EPI_ISL_1454352, EPI_ISL_1454354, EPI_ISL_1454360, EPI_ISL_1454382, EPI_ISL_1454390, EPI_ISL_1454402, EPI_ISL_1454407, EPI_ISL_1454436, EPI_ISL_1454445, EPI_ISL_1454453, EPI_ISL_1454456, EPI_ISL_1454458, EPI_ISL_1454466, EPI_ISL_1454474, EPI_ISL_1454483, EPI_ISL_1454491, EPI_ISL_1454495, EPI_ISL_1454496,
EPI_ISL_1454513, EPI_ISL_1454542, EPI_ISL_1454548, EPI_ISL_1454559, EPI_ISL_1454563, EPI_ISL_1454568, EPI_ISL_1454597, EPI_ISL_1454604, EPI_ISL_1454630, EPI_ISL_1454631, EPI_ISL_1454632, EPI_ISL_1454633, EPI_ISL_1454634, EPI_ISL_1454636, EPI_ISL_1454642, EPI_ISL_1454960, EPI_ISL_1454963, EPI_ISL_1454985, EPI_ISL_1454986, EPI_ISL_1454987, EPI_ISL_1454988, EPI_ISL_1454990,
EPI_ISL_1454991, EPI_ISL_1454992, EPI_ISL_1455005, EPI_ISL_1455008, EPI_ISL_1455009, EPI_ISL_1455010, EPI_ISL_1455011, EPI_ISL_1466251, EPI_ISL_1466323, EPI_ISL_1466364, EPI_ISL_1466402, EPI_ISL_1466453, EPI_ISL_1466462, EPI_ISL_1466476, EPI_ISL_1466486, EPI_ISL_1466493, EPI_ISL_1466531, EPI_ISL_1466557, EPI_ISL_1466590, EPI_ISL_1466602, EPI_ISL_1466628, EPI_ISL_1466639,
EPI_ISL_1466642, EPI_ISL_1466666, EPI_ISL_1466668, EPI_ISL_1466688, EPI_ISL_1466719, EPI_ISL_1466721, EPI_ISL_1466724, EPI_ISL_1466730, EPI_ISL_1466939, EPI_ISL_1466959, EPI_ISL_1466960, EPI_ISL_1466969, EPI_ISL_1466995, EPI_ISL_1467002, EPI_ISL_1467010, EPI_ISL_1467015, EPI_ISL_1467052, EPI_ISL_1467054, EPI_ISL_1467058, EPI_ISL_1467061, EPI_ISL_1467064, EPI_ISL_1467072,
EPI_ISL_1467082, EPI_ISL_1467083, EPI_ISL_1467100, EPI_ISL_1467112, EPI_ISL_1467165, EPI_ISL_1467178, EPI_ISL_1467184, EPI_ISL_1467192, EPI_ISL_1467196, EPI_ISL_1467213, EPI_ISL_1467219, EPI_ISL_1467234, EPI_ISL_1467246, EPI_ISL_1467273, EPI_ISL_1467302, EPI_ISL_1467386, EPI_ISL_1467397, EPI_ISL_1467408, EPI_ISL_1467417, EPI_ISL_1467468, EPI_ISL_1467475, EPI_ISL_1467481,
EPI_ISL_1467487, EPI_ISL_1467534, EPI_ISL_1467545, EPI_ISL_1467565, EPI_ISL_1467829, EPI_ISL_1467848, EPI_ISL_1467851, EPI_ISL_1467883, EPI_ISL_1467890, EPI_ISL_1467902, EPI_ISL_1467912, EPI_ISL_1467922, EPI_ISL_1467934, EPI_ISL_1467976, EPI_ISL_1467977, EPI_ISL_1467982, EPI_ISL_1467989, EPI_ISL_1468018, EPI_ISL_1468020, EPI_ISL_1468021, EPI_ISL_1468024, EPI_ISL_1468047,
EPI_ISL_1468050, EPI_ISL_1468086, EPI_ISL_1468107, EPI_ISL_1468117, EPI_ISL_1468119, EPI_ISL_1468132, EPI_ISL_1468133, EPI_ISL_1468135, EPI_ISL_1468138, EPI_ISL_1468140, EPI_ISL_1468142, EPI_ISL_1468143, EPI_ISL_1468145, EPI_ISL_1468148, EPI_ISL_1468149, EPI_ISL_1468151, EPI_ISL_1468152, EPI_ISL_1468153, EPI_ISL_1468154, EPI_ISL_1468155, EPI_ISL_1468158, EPI_ISL_1468161,
EPI_ISL_1468162, EPI_ISL_1468163, EPI_ISL_1468167, EPI_ISL_1468168, EPI_ISL_1468171, EPI_ISL_1468175, EPI_ISL_1468177, EPI_ISL_1468179, EPI_ISL_1468180, EPI_ISL_1468182, EPI_ISL_1468184, EPI_ISL_1468186, EPI_ISL_1468187, EPI_ISL_1468189, EPI_ISL_1468190, EPI_ISL_1468191, EPI_ISL_1468194, EPI_ISL_1468195, EPI_ISL_1468197, EPI_ISL_1468198, EPI_ISL_1468200, EPI_ISL_1468203,
EPI_ISL_1468204, EPI_ISL_1468205, EPI_ISL_1468206, EPI_ISL_1468209, EPI_ISL_1468211, EPI_ISL_1468216, EPI_ISL_1468217, EPI_ISL_1468219, EPI_ISL_1468222, EPI_ISL_1468223, EPI_ISL_1468224, EPI_ISL_1468225, EPI_ISL_1468227, EPI_ISL_1468231, EPI_ISL_1468233, EPI_ISL_1468238, EPI_ISL_1468239, EPI_ISL_1468240, EPI_ISL_1468243, EPI_ISL_1468245, EPI_ISL_1468247, EPI_ISL_1468250,
EPI_ISL_1468251, EPI_ISL_1468252, EPI_ISL_1468253, EPI_ISL_1468254, EPI_ISL_1468257, EPI_ISL_1468258, EPI_ISL_1468259, EPI_ISL_1468260, EPI_ISL_1468262, EPI_ISL_1468263, EPI_ISL_1468264, EPI_ISL_1468265, EPI_ISL_1468272, EPI_ISL_1468273, EPI_ISL_1468274, EPI_ISL_1468276, EPI_ISL_1468279, EPI_ISL_1468280, EPI_ISL_1468281, EPI_ISL_1468283, EPI_ISL_1468289, EPI_ISL_1468290,
EPI_ISL_1468291, EPI_ISL_1468293, EPI_ISL_1468294, EPI_ISL_1468295, EPI_ISL_1468298, EPI_ISL_1468299, EPI_ISL_1468300, EPI_ISL_1468301, EPI_ISL_1468307, EPI_ISL_1468308, EPI_ISL_1468310, EPI_ISL_1468311, EPI_ISL_1468312, EPI_ISL_1468316, EPI_ISL_1468317, EPI_ISL_1468320, EPI_ISL_1468324, EPI_ISL_1468327, EPI_ISL_1468328, EPI_ISL_1468332, EPI_ISL_1468333, EPI_ISL_1468337,
EPI_ISL_1468338, EPI_ISL_1468340, EPI_ISL_1468346, EPI_ISL_1468347, EPI_ISL_1468348, EPI_ISL_1468350, EPI_ISL_1468351, EPI_ISL_1468356, EPI_ISL_1468357, EPI_ISL_1468359, EPI_ISL_1468360, EPI_ISL_1468361, EPI_ISL_1468362, EPI_ISL_1468363, EPI_ISL_1468365, EPI_ISL_1468368, EPI_ISL_1468369, EPI_ISL_1468372, EPI_ISL_1468373, EPI_ISL_1468375, EPI_ISL_1468377, EPI_ISL_1468379,
EPI_ISL_1468380, EPI_ISL_1468381, EPI_ISL_1468383, EPI_ISL_1468385, EPI_ISL_1468387, EPI_ISL_1468389, EPI_ISL_1468391, EPI_ISL_1468392, EPI_ISL_1468395, EPI_ISL_1468396, EPI_ISL_1468397, EPI_ISL_1468403, EPI_ISL_1472425, EPI_ISL_1472438, EPI_ISL_1472450, EPI_ISL_1472488, EPI_ISL_1472509, EPI_ISL_1472534, EPI_ISL_1472537, EPI_ISL_1472585, EPI_ISL_1472595, EPI_ISL_1472599,
EPI_ISL_1472636, EPI_ISL_1472648, EPI_ISL_1472659, EPI_ISL_1472682, EPI_ISL_1472689, EPI_ISL_1473803, EPI_ISL_1473936, EPI_ISL_1474041, EPI_ISL_1474080, EPI_ISL_1474125, EPI_ISL_1474159, EPI_ISL_1474160, EPI_ISL_1484315, EPI_ISL_1484321, EPI_ISL_1484326, EPI_ISL_1484329, EPI_ISL_1484334, EPI_ISL_1484348, EPI_ISL_1484351, EPI_ISL_1484355, EPI_ISL_1484359, EPI_ISL_1484363,
EPI_ISL_1484364, EPI_ISL_1484383, EPI_ISL_1484390, EPI_ISL_1484396, EPI_ISL_1484399, EPI_ISL_1484402, EPI_ISL_1484405, EPI_ISL_1484407, EPI_ISL_1484408, EPI_ISL_1484419, EPI_ISL_1484420, EPI_ISL_1484423, EPI_ISL_1484425, EPI_ISL_1484429, EPI_ISL_1484432, EPI_ISL_1484437, EPI_ISL_1484439, EPI_ISL_1484442, EPI_ISL_1484456, EPI_ISL_1484461, EPI_ISL_1484463, EPI_ISL_1484464,
EPI_ISL_1484468, EPI_ISL_1484469, EPI_ISL_1484470, EPI_ISL_1484475, EPI_ISL_1484489, EPI_ISL_1484492, EPI_ISL_1484495, EPI_ISL_1484506, EPI_ISL_1484507, EPI_ISL_1484508, EPI_ISL_1484512, EPI_ISL_1484514, EPI_ISL_1484521, EPI_ISL_1484524, EPI_ISL_1484526, EPI_ISL_1484529, EPI_ISL_1484530, EPI_ISL_1484536, EPI_ISL_1484541, EPI_ISL_1484543, EPI_ISL_1484563, EPI_ISL_1484565,
EPI_ISL_1484569, EPI_ISL_1484577, EPI_ISL_1484580, EPI_ISL_1484581, EPI_ISL_1484584, EPI_ISL_1484585, EPI_ISL_1484589, EPI_ISL_1484605, EPI_ISL_1484608, EPI_ISL_1484610, EPI_ISL_1486742, EPI_ISL_1487512, EPI_ISL_1487883, EPI_ISL_1487908, EPI_ISL_1487914, EPI_ISL_1487944, EPI_ISL_1487951, EPI_ISL_1488054, EPI_ISL_1488071, EPI_ISL_1488097, EPI_ISL_1488101, EPI_ISL_1488945,
EPI_ISL_1489042, EPI_ISL_1489117, EPI_ISL_1489331, EPI_ISL_1489404, EPI_ISL_1489466, EPI_ISL_1489484, EPI_ISL_1489491, EPI_ISL_1489528, EPI_ISL_1507575, EPI_ISL_1507766, EPI_ISL_1507815, EPI_ISL_1507816, EPI_ISL_1507817, EPI_ISL_1507818, EPI_ISL_1507852, EPI_ISL_1507950, EPI_ISL_1507988, EPI_ISL_1508008, EPI_ISL_1508438, EPI_ISL_1508491

| | | | |
|---|---|---|---|
| see above | Lighthouse Lab in Alderley Park | Wellcome Sanger Institute for the COVID-19 Genomics UK (COG-UK) Consortium | Cordelia Langford; David K. Jackson; Dominic Kwiatkowski; Ewan Harrison; Ian Johnston; Jacquelyn Wynn; Jeffrey Barrett; John Sillitoe on behalf of the Wellcome Sanger Institute COVID-19 Surveillance Team; Mairead Hyland; Roberto Amato; Sonia Goncalves; The Lighthouse Lab in Alderley Park and Alex Alderton |

EPI_ISL_1377027, EPI_ISL_1409495, EPI_ISL_1409496, EPI_ISL_1409499, EPI_ISL_1409523, EPI_ISL_1409533, EPI_ISL_1409567, EPI_ISL_1409568, EPI_ISL_1409571, EPI_ISL_1409580, EPI_ISL_1409602, EPI_ISL_1409630, EPI_ISL_1409641, EPI_ISL_1409655, EPI_ISL_1409660, EPI_ISL_1409671, EPI_ISL_1409684, EPI_ISL_1409686, EPI_ISL_1409696, EPI_ISL_1409698, EPI_ISL_1409709, EPI_ISL_1409741,
EPI_ISL_1409747, EPI_ISL_1409772, EPI_ISL_1411119, EPI_ISL_1411135, EPI_ISL_1411154, EPI_ISL_1411174, EPI_ISL_1411207, EPI_ISL_1411220, EPI_ISL_1411267, EPI_ISL_1411273, EPI_ISL_1411289, EPI_ISL_1412224, EPI_ISL_1412261, EPI_ISL_1412268, EPI_ISL_1412298, EPI_ISL_1412306, EPI_ISL_1412308, EPI_ISL_1412311, EPI_ISL_1412320, EPI_ISL_1412321, EPI_ISL_1412328, EPI_ISL_1412331,
EPI_ISL_1412332, EPI_ISL_1412333, EPI_ISL_1412337, EPI_ISL_1412345, EPI_ISL_1412361, EPI_ISL_1412368, EPI_ISL_1412369, EPI_ISL_1412386, EPI_ISL_1412394, EPI_ISL_1412404, EPI_ISL_1412405, EPI_ISL_1412411, EPI_ISL_1412417, EPI_ISL_1412420, EPI_ISL_1412435, EPI_ISL_1412439, EPI_ISL_1412440, EPI_ISL_1412468, EPI_ISL_1412539, EPI_ISL_1412564, EPI_ISL_1415895, EPI_ISL_1415993,
EPI_ISL_1416053, EPI_ISL_1416056, EPI_ISL_1416112, EPI_ISL_1451158, EPI_ISL_1451182, EPI_ISL_1451191, EPI_ISL_1451208, EPI_ISL_1451214, EPI_ISL_1451224, EPI_ISL_1451232, EPI_ISL_1451239, EPI_ISL_1451279, EPI_ISL_1451287, EPI_ISL_1451288, EPI_ISL_1451311, EPI_ISL_1451315, EPI_ISL_1451329, EPI_ISL_1451346, EPI_ISL_1451442, EPI_ISL_1451457, EPI_ISL_1454157, EPI_ISL_1454159,
EPI_ISL_1454160, EPI_ISL_1454162, EPI_ISL_1454163, EPI_ISL_1454164, EPI_ISL_1454166, EPI_ISL_1454168, EPI_ISL_1454170, EPI_ISL_1454171, EPI_ISL_1454173, EPI_ISL_1454175, EPI_ISL_1454176, EPI_ISL_1454177, EPI_ISL_1454178, EPI_ISL_1454182, EPI_ISL_1454183, EPI_ISL_1454186, EPI_ISL_1454188, EPI_ISL_1454189, EPI_ISL_1454190, EPI_ISL_1454191, EPI_ISL_1454192, EPI_ISL_1454195,
EPI_ISL_1454196, EPI_ISL_1454197, EPI_ISL_1454198, EPI_ISL_1454200, EPI_ISL_1454201, EPI_ISL_1454202, EPI_ISL_1454204, EPI_ISL_1454206, EPI_ISL_1454207, EPI_ISL_1454208, EPI_ISL_1454209, EPI_ISL_1454212, EPI_ISL_1454220, EPI_ISL_1454222, EPI_ISL_1454225, EPI_ISL_1454227, EPI_ISL_1454230, EPI_ISL_1454234, EPI_ISL_1454238, EPI_ISL_1454241, EPI_ISL_1454242, EPI_ISL_1454243,
EPI_ISL_1454248, EPI_ISL_1454249, EPI_ISL_1454250, EPI_ISL_1454251, EPI_ISL_1454252, EPI_ISL_1454254, EPI_ISL_1454255, EPI_ISL_1454256, EPI_ISL_1454257, EPI_ISL_1454259, EPI_ISL_1454263, EPI_ISL_1454267, EPI_ISL_1454269, EPI_ISL_1454270, EPI_ISL_1454271, EPI_ISL_1454273, EPI_ISL_1454274, EPI_ISL_1454275, EPI_ISL_1454277, EPI_ISL_1454280, EPI_ISL_1454283, EPI_ISL_1454288,
EPI_ISL_1454289, EPI_ISL_1454293, EPI_ISL_1454294, EPI_ISL_1454295, EPI_ISL_1454300, EPI_ISL_1454306, EPI_ISL_1454307, EPI_ISL_1454311, EPI_ISL_1454315, EPI_ISL_1454316, EPI_ISL_1454319, EPI_ISL_1454320, EPI_ISL_1454321, EPI_ISL_1454323, EPI_ISL_1454325, EPI_ISL_1454328, EPI_ISL_1454347, EPI_ISL_1454348, EPI_ISL_1454349, EPI_ISL_1454355, EPI_ISL_1454368, EPI_ISL_1454375,
EPI_ISL_1454383, EPI_ISL_1454391, EPI_ISL_1454398, EPI_ISL_1454399, EPI_ISL_1454409, EPI_ISL_1454411, EPI_ISL_1454418, EPI_ISL_1454429, EPI_ISL_1454435, EPI_ISL_1454442, EPI_ISL_1454468, EPI_ISL_1454475, EPI_ISL_1454479, EPI_ISL_1454485, EPI_ISL_1454494, EPI_ISL_1454502, EPI_ISL_1454504, EPI_ISL_1454507, EPI_ISL_1454521, EPI_ISL_1454525, EPI_ISL_1454530, EPI_ISL_1454546,
EPI_ISL_1454553, EPI_ISL_1454558, EPI_ISL_1454575, EPI_ISL_1454594, EPI_ISL_1454612, EPI_ISL_1454626, EPI_ISL_1455247, EPI_ISL_1455256, EPI_ISL_1455272, EPI_ISL_1455289, EPI_ISL_1455300, EPI_ISL_1455327, EPI_ISL_1455341, EPI_ISL_1455431, EPI_ISL_1468404, EPI_ISL_1473447, EPI_ISL_1473451, EPI_ISL_1473481, EPI_ISL_1473533, EPI_ISL_1473576, EPI_ISL_1473585, EPI_ISL_1473594,
EPI_ISL_1473595, EPI_ISL_1473602, EPI_ISL_1473603, EPI_ISL_1473604, EPI_ISL_1473606, EPI_ISL_1473607, EPI_ISL_1473609, EPI_ISL_1473611, EPI_ISL_1473616, EPI_ISL_1473617, EPI_ISL_1473621, EPI_ISL_1473626, EPI_ISL_1473628, EPI_ISL_1473630, EPI_ISL_1473631, EPI_ISL_1473637, EPI_ISL_1473640, EPI_ISL_1473641, EPI_ISL_1473645, EPI_ISL_1473653, EPI_ISL_1473658, EPI_ISL_1473661,
EPI_ISL_1473662, EPI_ISL_1473663, EPI_ISL_1473666, EPI_ISL_1473667, EPI_ISL_1473671, EPI_ISL_1473673, EPI_ISL_1473677, EPI_ISL_1473679, EPI_ISL_1473680, EPI_ISL_1473681, EPI_ISL_1473689, EPI_ISL_1473698, EPI_ISL_1473700, EPI_ISL_1473708, EPI_ISL_1473713, EPI_ISL_1473717, EPI_ISL_1473718, EPI_ISL_1473724, EPI_ISL_1473726, EPI_ISL_1473734, EPI_ISL_1473739, EPI_ISL_1473745,
EPI_ISL_1473746, EPI_ISL_1473747, EPI_ISL_1473748, EPI_ISL_1473753, EPI_ISL_1473759, EPI_ISL_1473760, EPI_ISL_1473761, EPI_ISL_1473764, EPI_ISL_1473765, EPI_ISL_1473766, EPI_ISL_1473768, EPI_ISL_1473775, EPI_ISL_1473776, EPI_ISL_1473781, EPI_ISL_1473783, EPI_ISL_1473785, EPI_ISL_1473786, EPI_ISL_1473791, EPI_ISL_1484317, EPI_ISL_1484318, EPI_ISL_1484320, EPI_ISL_1484325,
EPI_ISL_1484331, EPI_ISL_1484340, EPI_ISL_1484341, EPI_ISL_1484344, EPI_ISL_1484354, EPI_ISL_1484356, EPI_ISL_1484369, EPI_ISL_1484379, EPI_ISL_1484380, EPI_ISL_1484382, EPI_ISL_1484385, EPI_ISL_1484394, EPI_ISL_1484400, EPI_ISL_1484414, EPI_ISL_1484416, EPI_ISL_1484428, EPI_ISL_1484431, EPI_ISL_1484446, EPI_ISL_1484451, EPI_ISL_1484454, EPI_ISL_1484455, EPI_ISL_1484459,
EPI_ISL_1484460, EPI_ISL_1484462, EPI_ISL_1484466, EPI_ISL_1484471, EPI_ISL_1484472, EPI_ISL_1484473, EPI_ISL_1484476, EPI_ISL_1484479, EPI_ISL_1484480, EPI_ISL_1484481, EPI_ISL_1484482, EPI_ISL_1484483, EPI_ISL_1484485, EPI_ISL_1484486, EPI_ISL_1484487, EPI_ISL_1484488, EPI_ISL_1484496, EPI_ISL_1484505, EPI_ISL_1484510, EPI_ISL_1484511, EPI_ISL_1484513, EPI_ISL_1484515,
EPI_ISL_1484523, EPI_ISL_1484528, EPI_ISL_1484531, EPI_ISL_1484538, EPI_ISL_1484539, EPI_ISL_1484542, EPI_ISL_1484557, EPI_ISL_1484560, EPI_ISL_1484562, EPI_ISL_1484572, EPI_ISL_1484573, EPI_ISL_1484575, EPI_ISL_1484579, EPI_ISL_1484582, EPI_ISL_1484583, EPI_ISL_1484592, EPI_ISL_1484593, EPI_ISL_1484594, EPI_ISL_1484596, EPI_ISL_1484597, EPI_ISL_1484599, EPI_ISL_1484604,
EPI_ISL_1484606, EPI_ISL_1487858, EPI_ISL_1506108, EPI_ISL_1506109, EPI_ISL_1506110, EPI_ISL_1506128, EPI_ISL_1506130, EPI_ISL_1506131, EPI_ISL_1506132, EPI_ISL_1508560, EPI_ISL_1519929, EPI_ISL_1519930, EPI_ISL_1519931, EPI_ISL_1519968

| | | | |
|---|---|---|---|
| see above | Lighthouse Lab in Cambridge | Wellcome Sanger Institute for the COVID-19 Genomics UK (COG-UK) Consortium | Cordelia Langford; David K. Jackson; Dominic Kwiatkowski; Ewan Harrison; Ian Johnston; Jeffrey Barrett; John Sillitoe on behalf of the Wellcome Sanger Institute COVID-19 Surveillance Team; Rob Howes; Roberto Amato; Sonia Goncalves; The Lighthouse Lab in Cambridge and Alex Alderton |

EPI_ISL_1374351, EPI_ISL_1374354, EPI_ISL_1374366, EPI_ISL_1374374, EPI_ISL_1374391, EPI_ISL_1374393, EPI_ISL_1374431, EPI_ISL_1374445, EPI_ISL_1374453, EPI_ISL_1374474, EPI_ISL_1374492, EPI_ISL_1374522, EPI_ISL_1374524, EPI_ISL_1374526, EPI_ISL_1374531, EPI_ISL_1374534, EPI_ISL_1374537, EPI_ISL_1374539, EPI_ISL_1374545, EPI_ISL_1374546, EPI_ISL_1374882, EPI_ISL_1374941,
EPI_ISL_1374952, EPI_ISL_1374958, EPI_ISL_1374989, EPI_ISL_1375051, EPI_ISL_1375118, EPI_ISL_1375139, EPI_ISL_1389648, EPI_ISL_1389649, EPI_ISL_1389652, EPI_ISL_1389653, EPI_ISL_1389654, EPI_ISL_1389655, EPI_ISL_1389656, EPI_ISL_1389657, EPI_ISL_1389659, EPI_ISL_1389660, EPI_ISL_1389662, EPI_ISL_1389663, EPI_ISL_1389665, EPI_ISL_1389666, EPI_ISL_1389667, EPI_ISL_1389669,
EPI_ISL_1389670, EPI_ISL_1389671, EPI_ISL_1389673, EPI_ISL_1389675, EPI_ISL_1389677, EPI_ISL_1389679, EPI_ISL_1389682, EPI_ISL_1389683, EPI_ISL_1389684, EPI_ISL_1389685, EPI_ISL_1389686, EPI_ISL_1389687, EPI_ISL_1389688, EPI_ISL_1389690, EPI_ISL_1389694, EPI_ISL_1389697, EPI_ISL_1389698, EPI_ISL_1389699, EPI_ISL_1389701, EPI_ISL_1389702, EPI_ISL_1389704, EPI_ISL_1389706,
EPI_ISL_1389708, EPI_ISL_1389709, EPI_ISL_1389710, EPI_ISL_1389712, EPI_ISL_1389713, EPI_ISL_1389714, EPI_ISL_1389715, EPI_ISL_1389716, EPI_ISL_1389717, EPI_ISL_1389718, EPI_ISL_1389719, EPI_ISL_1389720, EPI_ISL_1389721, EPI_ISL_1389722, EPI_ISL_1389723, EPI_ISL_1389724, EPI_ISL_1389725, EPI_ISL_1389726, EPI_ISL_1389729, EPI_ISL_1389730, EPI_ISL_1389733, EPI_ISL_1389738,
EPI_ISL_1389739, EPI_ISL_1389741, EPI_ISL_1389743, EPI_ISL_1389745, EPI_ISL_1389747, EPI_ISL_1389748, EPI_ISL_1389750, EPI_ISL_1389752, EPI_ISL_1389753, EPI_ISL_1389754, EPI_ISL_1389755, EPI_ISL_1389757, EPI_ISL_1389758, EPI_ISL_1389759, EPI_ISL_1389760, EPI_ISL_1389761, EPI_ISL_1389762, EPI_ISL_1389763, EPI_ISL_1389764, EPI_ISL_1389767, EPI_ISL_1389768, EPI_ISL_1389769,
EPI_ISL_1389819, EPI_ISL_1389821, EPI_ISL_1389822, EPI_ISL_1389823, EPI_ISL_1389825, EPI_ISL_1389829, EPI_ISL_1389840, EPI_ISL_1389843, EPI_ISL_1389844, EPI_ISL_1389855, EPI_ISL_1389862, EPI_ISL_1408909, EPI_ISL_1408915, EPI_ISL_1408918, EPI_ISL_1408921, EPI_ISL_1408922, EPI_ISL_1408928, EPI_ISL_1408929, EPI_ISL_1408934, EPI_ISL_1408937, EPI_ISL_1408939, EPI_ISL_1408940,
EPI_ISL_1408944, EPI_ISL_1408945, EPI_ISL_1408946, EPI_ISL_1408947, EPI_ISL_1408950, EPI_ISL_1408954, EPI_ISL_1408962, EPI_ISL_1408963, EPI_ISL_1408965, EPI_ISL_1408968, EPI_ISL_1408969, EPI_ISL_1408972, EPI_ISL_1408974, EPI_ISL_1408978, EPI_ISL_1408980, EPI_ISL_1408981, EPI_ISL_1408982, EPI_ISL_1408983, EPI_ISL_1408986, EPI_ISL_1408992, EPI_ISL_1408996, EPI_ISL_1408997,
EPI_ISL_1409000, EPI_ISL_1409002, EPI_ISL_1409005, EPI_ISL_1409008, EPI_ISL_1409009, EPI_ISL_1409010, EPI_ISL_1409012, EPI_ISL_1409013, EPI_ISL_1409014, EPI_ISL_1409015, EPI_ISL_1409017, EPI_ISL_1409018, EPI_ISL_1409020, EPI_ISL_1409021, EPI_ISL_1409024, EPI_ISL_1409027, EPI_ISL_1409029, EPI_ISL_1409032, EPI_ISL_1409034, EPI_ISL_1409037, EPI_ISL_1409038, EPI_ISL_1409039,
EPI_ISL_1409040, EPI_ISL_1409043, EPI_ISL_1409044, EPI_ISL_1409047, EPI_ISL_1409048, EPI_ISL_1409049, EPI_ISL_1409050, EPI_ISL_1409051, EPI_ISL_1409055, EPI_ISL_1409063, EPI_ISL_1409066, EPI_ISL_1409068, EPI_ISL_1409069, EPI_ISL_1409071, EPI_ISL_1409073, EPI_ISL_1409074, EPI_ISL_1409076, EPI_ISL_1409078, EPI_ISL_1409080, EPI_ISL_1409081, EPI_ISL_1409085, EPI_ISL_1409087,
EPI_ISL_1409088, EPI_ISL_1409090, EPI_ISL_1409091, EPI_ISL_1409095, EPI_ISL_1409096, EPI_ISL_1409097, EPI_ISL_1409099, EPI_ISL_1409100, EPI_ISL_1409103, EPI_ISL_1409109, EPI_ISL_1409110, EPI_ISL_1409113, EPI_ISL_1409115, EPI_ISL_1409116, EPI_ISL_1409117, EPI_ISL_1409118, EPI_ISL_1409123, EPI_ISL_1409129, EPI_ISL_1409130, EPI_ISL_1409131, EPI_ISL_1409136, EPI_ISL_1409137,
EPI_ISL_1409138, EPI_ISL_1409142, EPI_ISL_1409143, EPI_ISL_1409144, EPI_ISL_1409145, EPI_ISL_1409148, EPI_ISL_1409155, EPI_ISL_1409156, EPI_ISL_1409157, EPI_ISL_1409159, EPI_ISL_1409160, EPI_ISL_1409161, EPI_ISL_1409163, EPI_ISL_1409165, EPI_ISL_1409170, EPI_ISL_1409172, EPI_ISL_1409173, EPI_ISL_1409174, EPI_ISL_1409178, EPI_ISL_1409181, EPI_ISL_1409182, EPI_ISL_1409184,
EPI_ISL_1409198, EPI_ISL_1409199, EPI_ISL_1409204, EPI_ISL_1409205, EPI_ISL_1409210, EPI_ISL_1409214, EPI_ISL_1409220, EPI_ISL_1409221, EPI_ISL_1409225, EPI_ISL_1409231, EPI_ISL_1409233, EPI_ISL_1409234, EPI_ISL_1409237, EPI_ISL_1409247, EPI_ISL_1409256, EPI_ISL_1409261, EPI_ISL_1409263, EPI_ISL_1409264, EPI_ISL_1409270, EPI_ISL_1409277, EPI_ISL_1409280, EPI_ISL_1409309,
EPI_ISL_1409314, EPI_ISL_1409318, EPI_ISL_1409324, EPI_ISL_1409327, EPI_ISL_1409331, EPI_ISL_1409332, EPI_ISL_1409336, EPI_ISL_1409342, EPI_ISL_1409344, EPI_ISL_1409345, EPI_ISL_1409347, EPI_ISL_1409352, EPI_ISL_1409354, EPI_ISL_1409357, EPI_ISL_1409358, EPI_ISL_1409361, EPI_ISL_1409366, EPI_ISL_1409370, EPI_ISL_1409371, EPI_ISL_1409377, EPI_ISL_1409379, EPI_ISL_1409393,
EPI_ISL_1409396, EPI_ISL_1409399, EPI_ISL_1409403, EPI_ISL_1409406, EPI_ISL_1409407, EPI_ISL_1409414, EPI_ISL_1409421, EPI_ISL_1409429, EPI_ISL_1409441, EPI_ISL_1409444, EPI_ISL_1409457, EPI_ISL_1409458, EPI_ISL_1409461, EPI_ISL_1409468, EPI_ISL_1409469, EPI_ISL_1409471, EPI_ISL_1409477, EPI_ISL_1409484, EPI_ISL_1409494, EPI_ISL_1409516, EPI_ISL_1409532, EPI_ISL_1409539,
EPI_ISL_1409550, EPI_ISL_1409551, EPI_ISL_1409557, EPI_ISL_1409572, EPI_ISL_1409577, EPI_ISL_1409593, EPI_ISL_1409594, EPI_ISL_1409596, EPI_ISL_1409604, EPI_ISL_1409607, EPI_ISL_1409611, EPI_ISL_1409612, EPI_ISL_1409624, EPI_ISL_1409632, EPI_ISL_1409634, EPI_ISL_1409642, EPI_ISL_1409654, EPI_ISL_1409663, EPI_ISL_1409664, EPI_ISL_1409665, EPI_ISL_1409669, EPI_ISL_1409685,
EPI_ISL_1409689, EPI_ISL_1409691, EPI_ISL_1409692, EPI_ISL_1409727, EPI_ISL_1409733, EPI_ISL_1409736, EPI_ISL_1409754, EPI_ISL_1409758, EPI_ISL_1409771, EPI_ISL_1409774, EPI_ISL_1409778, EPI_ISL_1409783, EPI_ISL_1409794, EPI_ISL_1409809, EPI_ISL_1409813, EPI_ISL_1409817, EPI_ISL_1409836, EPI_ISL_1409843, EPI_ISL_1409858, EPI_ISL_1409859, EPI_ISL_1409861, EPI_ISL_1409863,
EPI_ISL_1409868, EPI_ISL_1409870, EPI_ISL_1409873, EPI_ISL_1409875, EPI_ISL_1409883, EPI_ISL_1409886, EPI_ISL_1409889, EPI_ISL_1409891, EPI_ISL_1409894, EPI_ISL_1409906, EPI_ISL_1409907, EPI_ISL_1409912, EPI_ISL_1409913, EPI_ISL_1409918, EPI_ISL_1409926, EPI_ISL_1409932, EPI_ISL_1409935, EPI_ISL_1409938, EPI_ISL_1409951, EPI_ISL_1409960, EPI_ISL_1409961, EPI_ISL_1409967,
EPI_ISL_1409970, EPI_ISL_1409976, EPI_ISL_1409981, EPI_ISL_1409987, EPI_ISL_1409993, EPI_ISL_1409996, EPI_ISL_1410001, EPI_ISL_1410002, EPI_ISL_1410006, EPI_ISL_1410009, EPI_ISL_1410013, EPI_ISL_1410032, EPI_ISL_1410037, EPI_ISL_1410046, EPI_ISL_1410047, EPI_ISL_1410048, EPI_ISL_1410051, EPI_ISL_1410053, EPI_ISL_1410056, EPI_ISL_1410067, EPI_ISL_1410071, EPI_ISL_1410073,
EPI_ISL_1410076, EPI_ISL_1410289, EPI_ISL_1410290, EPI_ISL_1410291, EPI_ISL_1410292, EPI_ISL_1410293, EPI_ISL_1410295, EPI_ISL_1410297, EPI_ISL_1410298, EPI_ISL_1410299, EPI_ISL_1410300, EPI_ISL_1410304, EPI_ISL_1410306, EPI_ISL_1410314, EPI_ISL_1410315, EPI_ISL_1410316, EPI_ISL_1410318, EPI_ISL_1410320, EPI_ISL_1410322, EPI_ISL_1410323, EPI_ISL_1410324, EPI_ISL_1410325,
EPI_ISL_1410326, EPI_ISL_1410327, EPI_ISL_1410328, EPI_ISL_1410330, EPI_ISL_1410333, EPI_ISL_1410334, EPI_ISL_1410338, EPI_ISL_1410341, EPI_ISL_1410342, EPI_ISL_1410345, EPI_ISL_1410347, EPI_ISL_1410349, EPI_ISL_1410355, EPI_ISL_1410356, EPI_ISL_1410357, EPI_ISL_1410358, EPI_ISL_1410360, EPI_ISL_1410361, EPI_ISL_1410364, EPI_ISL_1410366, EPI_ISL_1410368, EPI_ISL_1410373,
EPI_ISL_1410374, EPI_ISL_1410375, EPI_ISL_1410383, EPI_ISL_1410384, EPI_ISL_1410385, EPI_ISL_1410387, EPI_ISL_1410390, EPI_ISL_1410391, EPI_ISL_1410394, EPI_ISL_1410395, EPI_ISL_1410396, EPI_ISL_1410397, EPI_ISL_1410400, EPI_ISL_1410403, EPI_ISL_1410406, EPI_ISL_1410409, EPI_ISL_1410410, EPI_ISL_1410412, EPI_ISL_1410413, EPI_ISL_1410415, EPI_ISL_1410417, EPI_ISL_1410421,
EPI_ISL_1410422, EPI_ISL_1410424, EPI_ISL_1410425, EPI_ISL_1410426, EPI_ISL_1410427, EPI_ISL_1410428, EPI_ISL_1410432, EPI_ISL_1410433, EPI_ISL_1410434, EPI_ISL_1410435, EPI_ISL_1410437, EPI_ISL_1410441, EPI_ISL_1410448, EPI_ISL_1410450, EPI_ISL_1410451, EPI_ISL_1410453, EPI_ISL_1410459, EPI_ISL_1410460, EPI_ISL_1410461, EPI_ISL_1410462, EPI_ISL_1410464, EPI_ISL_1410465,
EPI_ISL_1410467, EPI_ISL_1410469, EPI_ISL_1410471, EPI_ISL_1410473, EPI_ISL_1410475, EPI_ISL_1410476, EPI_ISL_1410477, EPI_ISL_1410480, EPI_ISL_1410482, EPI_ISL_1410483, EPI_ISL_1410484, EPI_ISL_1410485, EPI_ISL_1410486, EPI_ISL_1410487, EPI_ISL_1410488, EPI_ISL_1410489, EPI_ISL_1410490, EPI_ISL_1410491, EPI_ISL_1410492, EPI_ISL_1410493, EPI_ISL_1410494, EPI_ISL_1410495,
EPI_ISL_1410496, EPI_ISL_1410499, EPI_ISL_1410501, EPI_ISL_1410502, EPI_ISL_1410507, EPI_ISL_1410508, EPI_ISL_1410509, EPI_ISL_1410511, EPI_ISL_1410514, EPI_ISL_1410515, EPI_ISL_1410516, EPI_ISL_1410517, EPI_ISL_1410518, EPI_ISL_1410519, EPI_ISL_1410521, EPI_ISL_1410522, EPI_ISL_1410525, EPI_ISL_1410526, EPI_ISL_1410528, EPI_ISL_1410529, EPI_ISL_1410534, EPI_ISL_1410535,
EPI_ISL_1410536, EPI_ISL_1410537, EPI_ISL_1410539, EPI_ISL_1410544, EPI_ISL_1410546, EPI_ISL_1410547, EPI_ISL_1410552, EPI_ISL_1410555, EPI_ISL_1410562, EPI_ISL_1410567, EPI_ISL_1410568, EPI_ISL_1410572, EPI_ISL_1410576, EPI_ISL_1410578, EPI_ISL_1410581, EPI_ISL_1410583, EPI_ISL_1410584, EPI_ISL_1410586, EPI_ISL_1410589, EPI_ISL_1410599, EPI_ISL_1410601, EPI_ISL_1410604,
EPI_ISL_1410606, EPI_ISL_1410609, EPI_ISL_1410610, EPI_ISL_1410614, EPI_ISL_1410617, EPI_ISL_1410619, EPI_ISL_1410623, EPI_ISL_1410633, EPI_ISL_1410635, EPI_ISL_1410636, EPI_ISL_1410638, EPI_ISL_1410647, EPI_ISL_1410654, EPI_ISL_1410656, EPI_ISL_1410657, EPI_ISL_1410661, EPI_ISL_1410673, EPI_ISL_1410675, EPI_ISL_1410676, EPI_ISL_1410684, EPI_ISL_1410686, EPI_ISL_1410692,
EPI_ISL_1410694, EPI_ISL_1410695, EPI_ISL_1410697, EPI_ISL_1410700, EPI_ISL_1410702, EPI_ISL_1410703, EPI_ISL_1410711, EPI_ISL_1410716, EPI_ISL_1410719, EPI_ISL_1410721, EPI_ISL_1410722, EPI_ISL_1410726, EPI_ISL_1410734, EPI_ISL_1410735, EPI_ISL_1410737, EPI_ISL_1410744, EPI_ISL_1410745, EPI_ISL_1410747, EPI_ISL_1410750, EPI_ISL_1410755, EPI_ISL_1410759, EPI_ISL_1410760,
EPI_ISL_1410773, EPI_ISL_1410778, EPI_ISL_1410784, EPI_ISL_1410787, EPI_ISL_1410788, EPI_ISL_1410790, EPI_ISL_1410791, EPI_ISL_1410792, EPI_ISL_1410794, EPI_ISL_1410795, EPI_ISL_1410797, EPI_ISL_1410800, EPI_ISL_1410801, EPI_ISL_1410807, EPI_ISL_1410810, EPI_ISL_1410811, EPI_ISL_1410812, EPI_ISL_1410813, EPI_ISL_1410814, EPI_ISL_1410820, EPI_ISL_1410822, EPI_ISL_1410827,
EPI_ISL_1410829, EPI_ISL_1410833, EPI_ISL_1410835, EPI_ISL_1410839, EPI_ISL_1410840, EPI_ISL_1410841, EPI_ISL_1410842, EPI_ISL_1410843, EPI_ISL_1410844, EPI_ISL_1410845, EPI_ISL_1410847, EPI_ISL_1410849, EPI_ISL_1410850, EPI_ISL_1410853, EPI_ISL_1410854, EPI_ISL_1410856, EPI_ISL_1410858, EPI_ISL_1410861, EPI_ISL_1410864, EPI_ISL_1410865, EPI_ISL_1410867, EPI_ISL_1410873,
EPI_ISL_1410878, EPI_ISL_1410879, EPI_ISL_1410880, EPI_ISL_1410881, EPI_ISL_1410882, EPI_ISL_1410884, EPI_ISL_1410890, EPI_ISL_1410892, EPI_ISL_1410895, EPI_ISL_1410898, EPI_ISL_1410900, EPI_ISL_1410903, EPI_ISL_1410905, EPI_ISL_1410906, EPI_ISL_1410908, EPI_ISL_1410909, EPI_ISL_1410912, EPI_ISL_1410914, EPI_ISL_1410916, EPI_ISL_1410917, EPI_ISL_1410920, EPI_ISL_1410921,
EPI_ISL_1410922, EPI_ISL_1410923, EPI_ISL_1410925, EPI_ISL_1410926, EPI_ISL_1410927, EPI_ISL_1410929, EPI_ISL_1410933, EPI_ISL_1410934, EPI_ISL_1410936, EPI_ISL_1410937, EPI_ISL_1410939, EPI_ISL_1410941, EPI_ISL_1410943, EPI_ISL_1410945, EPI_ISL_1410948, EPI_ISL_1410950, EPI_ISL_1410952, EPI_ISL_1410955, EPI_ISL_1410956, EPI_ISL_1410959, EPI_ISL_1410960, EPI_ISL_1410964,
EPI_ISL_1410965, EPI_ISL_1410966, EPI_ISL_1410967, EPI_ISL_1410968, EPI_ISL_1410969, EPI_ISL_1410970, EPI_ISL_1410975, EPI_ISL_1410976, EPI_ISL_1410978, EPI_ISL_1410979, EPI_ISL_1410980, EPI_ISL_1410982, EPI_ISL_1410984, EPI_ISL_1410991, EPI_ISL_1410992, EPI_ISL_1410993, EPI_ISL_1410996, EPI_ISL_1410998, EPI_ISL_1411003, EPI_ISL_1411008, EPI_ISL_1411009, EPI_ISL_1411011,
EPI_ISL_1411012, EPI_ISL_1411014, EPI_ISL_1411016, EPI_ISL_1411017, EPI_ISL_1411018, EPI_ISL_1411020, EPI_ISL_1411021, EPI_ISL_1411023, EPI_ISL_1411025, EPI_ISL_1411026, EPI_ISL_1411027, EPI_ISL_1411031, EPI_ISL_1411033, EPI_ISL_1411034, EPI_ISL_1411040, EPI_ISL_1411042, EPI_ISL_1411043, EPI_ISL_1411046, EPI_ISL_1411047, EPI_ISL_1411048, EPI_ISL_1411049, EPI_ISL_1411050,
EPI_ISL_1411057, EPI_ISL_1411059, EPI_ISL_1411063, EPI_ISL_1411064, EPI_ISL_1411065, EPI_ISL_1411066, EPI_ISL_1411068, EPI_ISL_1411074, EPI_ISL_1411076, EPI_ISL_1411077, EPI_ISL_1411078, EPI_ISL_1411079, EPI_ISL_1411081, EPI_ISL_1411082, EPI_ISL_1411083, EPI_ISL_1411084, EPI_ISL_1411087, EPI_ISL_1411088, EPI_ISL_1411090, EPI_ISL_1411092, EPI_ISL_1411093, EPI_ISL_1411095,
EPI_ISL_1411096, EPI_ISL_1411097, EPI_ISL_1411098, EPI_ISL_1411099, EPI_ISL_1411103, EPI_ISL_1411104, EPI_ISL_1411106, EPI_ISL_1411107, EPI_ISL_1411108, EPI_ISL_1411109, EPI_ISL_1411110, EPI_ISL_1411111, EPI_ISL_1411117, EPI_ISL_1411120, EPI_ISL_1411121, EPI_ISL_1411126, EPI_ISL_1411129, EPI_ISL_1411130, EPI_ISL_1411132, EPI_ISL_1411134, EPI_ISL_1411137, EPI_ISL_1411139,
EPI_ISL_1411140, EPI_ISL_1411141, EPI_ISL_1411143, EPI_ISL_1411144, EPI_ISL_1411146, EPI_ISL_1411148, EPI_ISL_1411149, EPI_ISL_1411151, EPI_ISL_1411152, EPI_ISL_1411155, EPI_ISL_1411157, EPI_ISL_1411160, EPI_ISL_1411162, EPI_ISL_1411164, EPI_ISL_1411165, EPI_ISL_1411166, EPI_ISL_1411167, EPI_ISL_1411169, EPI_ISL_1411173, EPI_ISL_1411175, EPI_ISL_1411177, EPI_ISL_1411178,
EPI_ISL_1411181, EPI_ISL_1411182, EPI_ISL_1411186, EPI_ISL_1411187, EPI_ISL_1411188, EPI_ISL_1411191, EPI_ISL_1411192, EPI_ISL_1411193, EPI_ISL_1411198, EPI_ISL_1411199, EPI_ISL_1411200, EPI_ISL_1411201, EPI_ISL_1411202, EPI_ISL_1411203, EPI_ISL_1411206, EPI_ISL_1411208, EPI_ISL_1411210, EPI_ISL_1411212, EPI_ISL_1411214, EPI_ISL_1411215, EPI_ISL_1411216, EPI_ISL_1411217,
EPI_ISL_1411221, EPI_ISL_1411222, EPI_ISL_1411224, EPI_ISL_1411226, EPI_ISL_1411227, EPI_ISL_1411228, EPI_ISL_1411235, EPI_ISL_1411241, EPI_ISL_1411242, EPI_ISL_1411247, EPI_ISL_1411248, EPI_ISL_1411249, EPI_ISL_1411251, EPI_ISL_1411252, EPI_ISL_1411253, EPI_ISL_1411254, EPI_ISL_1411255, EPI_ISL_1411257, EPI_ISL_1411259, EPI_ISL_1411262, EPI_ISL_1411263, EPI_ISL_1411264,
EPI_ISL_1411265, EPI_ISL_1411269, EPI_ISL_1411270, EPI_ISL_1411275, EPI_ISL_1411276, EPI_ISL_1411277, EPI_ISL_1411281, EPI_ISL_1411284, EPI_ISL_1411288, EPI_ISL_1411290, EPI_ISL_1411291, EPI_ISL_1411292, EPI_ISL_1411293, EPI_ISL_1411294, EPI_ISL_1411298, EPI_ISL_1411300, EPI_ISL_1411301, EPI_ISL_1411305, EPI_ISL_1411307, EPI_ISL_1411312, EPI_ISL_1411314, EPI_ISL_1411318,
EPI_ISL_1411321, EPI_ISL_1411323, EPI_ISL_1411324, EPI_ISL_1411326, EPI_ISL_1411327, EPI_ISL_1411330, EPI_ISL_1411331, EPI_ISL_1411332, EPI_ISL_1411333, EPI_ISL_1411336, EPI_ISL_1411340, EPI_ISL_1411341, EPI_ISL_1411342, EPI_ISL_1411343, EPI_ISL_1411344, EPI_ISL_1411347, EPI_ISL_1411349, EPI_ISL_1411350, EPI_ISL_1411351, EPI_ISL_1411353, EPI_ISL_1411354, EPI_ISL_1411355,
EPI_ISL_1411359, EPI_ISL_1411361, EPI_ISL_1411364, EPI_ISL_1411366, EPI_ISL_1411367, EPI_ISL_1411368, EPI_ISL_1411373, EPI_ISL_1411376, EPI_ISL_1411381, EPI_ISL_1411383, EPI_ISL_1411388, EPI_ISL_1411390, EPI_ISL_1411392, EPI_ISL_1411396, EPI_ISL_1411397, EPI_ISL_1411398, EPI_ISL_1411399, EPI_ISL_1411403, EPI_ISL_1411407, EPI_ISL_1411408, EPI_ISL_1411410, EPI_ISL_1411414,
EPI_ISL_1411417, EPI_ISL_1411418, EPI_ISL_1411420, EPI_ISL_1411421, EPI_ISL_1411427, EPI_ISL_1411431, EPI_ISL_1411433, EPI_ISL_1411434, EPI_ISL_1411435, EPI_ISL_1411439, EPI_ISL_1411442, EPI_ISL_1411443, EPI_ISL_1411444, EPI_ISL_1411447, EPI_ISL_1411449, EPI_ISL_1411450, EPI_ISL_1411451, EPI_ISL_1411453, EPI_ISL_1411460, EPI_ISL_1411461, EPI_ISL_1411463, EPI_ISL_1411466,
EPI_ISL_1411469, EPI_ISL_1411474, EPI_ISL_1411475, EPI_ISL_1411478, EPI_ISL_1411479, EPI_ISL_1411484, EPI_ISL_1411486, EPI_ISL_1411494, EPI_ISL_1411499, EPI_ISL_1411502, EPI_ISL_1411507, EPI_ISL_1411563, EPI_ISL_1411572, EPI_ISL_1411577, EPI_ISL_1411596, EPI_ISL_1411603, EPI_ISL_1411607, EPI_ISL_1411608, EPI_ISL_1411621, EPI_ISL_1411630, EPI_ISL_1411635, EPI_ISL_1411638,
EPI_ISL_1411639, EPI_ISL_1411640, EPI_ISL_1411641, EPI_ISL_1411643, EPI_ISL_1411646, EPI_ISL_1411647, EPI_ISL_1411649, EPI_ISL_1411661, EPI_ISL_1411666, EPI_ISL_1411668, EPI_ISL_1411672, EPI_ISL_1411677, EPI_ISL_1411679, EPI_ISL_1411681, EPI_ISL_1411685, EPI_ISL_1411686, EPI_ISL_1411690, EPI_ISL_1411693, EPI_ISL_1411696, EPI_ISL_1411706, EPI_ISL_1411713, EPI_ISL_1411714,
EPI_ISL_1411715, EPI_ISL_1411718, EPI_ISL_1411719, EPI_ISL_1411721, EPI_ISL_1411722, EPI_ISL_1411723, EPI_ISL_1411736, EPI_ISL_1411738, EPI_ISL_1411739, EPI_ISL_1411743, EPI_ISL_1411744, EPI_ISL_1411750, EPI_ISL_1411751, EPI_ISL_1411752, EPI_ISL_1411761, EPI_ISL_1411768, EPI_ISL_1411772, EPI_ISL_1411776, EPI_ISL_1411778, EPI_ISL_1411784, EPI_ISL_1411785, EPI_ISL_1411795,
EPI_ISL_1411796, EPI_ISL_1411798, EPI_ISL_1411799, EPI_ISL_1411803, EPI_ISL_1411807, EPI_ISL_1411812, EPI_ISL_1411814, EPI_ISL_1411815, EPI_ISL_1411817, EPI_ISL_1411829, EPI_ISL_1411833, EPI_ISL_1411834, EPI_ISL_1411836, EPI_ISL_1411837, EPI_ISL_1411838, EPI_ISL_1411841, EPI_ISL_1411842, EPI_ISL_1411861, EPI_ISL_1411871, EPI_ISL_1411872, EPI_ISL_1411873, EPI_ISL_1411876,
EPI_ISL_1411879, EPI_ISL_1411881, EPI_ISL_1411883, EPI_ISL_1411885, EPI_ISL_1411887, EPI_ISL_1411890, EPI_ISL_1411891, EPI_ISL_1411892, EPI_ISL_1411894, EPI_ISL_1411912, EPI_ISL_1411914, EPI_ISL_1411915, EPI_ISL_1411918, EPI_ISL_1411927, EPI_ISL_1411932, EPI_ISL_1411940, EPI_ISL_1411946, EPI_ISL_1411957, EPI_ISL_1411958, EPI_ISL_1411972, EPI_ISL_1411997, EPI_ISL_1411999,
EPI_ISL_1412005, EPI_ISL_1412007, EPI_ISL_1412011, EPI_ISL_1412016, EPI_ISL_1412022, EPI_ISL_1412023, EPI_ISL_1412026, EPI_ISL_1412031, EPI_ISL_1412037, EPI_ISL_1412039, EPI_ISL_1412040, EPI_ISL_1412043, EPI_ISL_1412044, EPI_ISL_1412045, EPI_ISL_1412047, EPI_ISL_1412049, EPI_ISL_1412051, EPI_ISL_1412052, EPI_ISL_1412055, EPI_ISL_1412059, EPI_ISL_1412070, EPI_ISL_1412071,
EPI_ISL_1412076, EPI_ISL_1412077, EPI_ISL_1412078, EPI_ISL_1412086, EPI_ISL_1412091, EPI_ISL_1412093, EPI_ISL_1412098, EPI_ISL_1412099, EPI_ISL_1412102, EPI_ISL_1412112, EPI_ISL_1412114, EPI_ISL_1412117, EPI_ISL_1412123, EPI_ISL_1412124, EPI_ISL_1412129, EPI_ISL_1412130, EPI_ISL_1412135, EPI_ISL_1412136, EPI_ISL_1412140, EPI_ISL_1412142, EPI_ISL_1412144, EPI_ISL_1412152,
EPI_ISL_1412157, EPI_ISL_1412161, EPI_ISL_1412163, EPI_ISL_1412169, EPI_ISL_1412170, EPI_ISL_1412173, EPI_ISL_1412176, EPI_ISL_1412181, EPI_ISL_1412183, EPI_ISL_1412186, EPI_ISL_1412235, EPI_ISL_1412260, EPI_ISL_1412264, EPI_ISL_1412288, EPI_ISL_1412325, EPI_ISL_1412330, EPI_ISL_1412356, EPI_ISL_1412358, EPI_ISL_1412372, EPI_ISL_1412376, EPI_ISL_1412383, EPI_ISL_1412399,
EPI_ISL_1412414, EPI_ISL_1412416, EPI_ISL_1412431, EPI_ISL_1412449, EPI_ISL_1412463, EPI_ISL_1412466, EPI_ISL_1412476, EPI_ISL_1412495, EPI_ISL_1412509, EPI_ISL_1412545, EPI_ISL_1412550, EPI_ISL_1412568, EPI_ISL_1412569, EPI_ISL_1412570, EPI_ISL_1412571, EPI_ISL_1412572, EPI_ISL_1412573, EPI_ISL_1412575, EPI_ISL_1412576, EPI_ISL_1412578, EPI_ISL_1412580, EPI_ISL_1412581,
EPI_ISL_1412582, EPI_ISL_1412583, EPI_ISL_1412584, EPI_ISL_1412585, EPI_ISL_1412586, EPI_ISL_1412587, EPI_ISL_1412589, EPI_ISL_1412594, EPI_ISL_1412595, EPI_ISL_1412597, EPI_ISL_1412598, EPI_ISL_1412600, EPI_ISL_1412602, EPI_ISL_1412605, EPI_ISL_1412608, EPI_ISL_1412609, EPI_ISL_1412612, EPI_ISL_1412614, EPI_ISL_1412615, EPI_ISL_1412616, EPI_ISL_1412617, EPI_ISL_1412621,

EPI_ISL_1412622, EPI_ISL_1412624, EPI_ISL_1412625, EPI_ISL_1412626, EPI_ISL_1412628, EPI_ISL_1412629, EPI_ISL_1412630, EPI_ISL_1412632, EPI_ISL_1412637, EPI_ISL_1412640, EPI_ISL_1412641, EPI_ISL_1412643, EPI_ISL_1412644, EPI_ISL_1412646, EPI_ISL_1412647, EPI_ISL_1412648, EPI_ISL_1412650, EPI_ISL_1412654, EPI_ISL_1412657, EPI_ISL_1412658, EPI_ISL_1412659, EPI_ISL_1412662,
EPI_ISL_1412663, EPI_ISL_1412664, EPI_ISL_1412666, EPI_ISL_1412672, EPI_ISL_1412673, EPI_ISL_1412674, EPI_ISL_1412675, EPI_ISL_1412676, EPI_ISL_1412678, EPI_ISL_1412682, EPI_ISL_1412683, EPI_ISL_1412688, EPI_ISL_1412690, EPI_ISL_1412691, EPI_ISL_1412692, EPI_ISL_1412693, EPI_ISL_1412694, EPI_ISL_1412698, EPI_ISL_1412701, EPI_ISL_1412702, EPI_ISL_1412703, EPI_ISL_1412704,
EPI_ISL_1412705, EPI_ISL_1412706, EPI_ISL_1412708, EPI_ISL_1412711, EPI_ISL_1412712, EPI_ISL_1412713, EPI_ISL_1412715, EPI_ISL_1412717, EPI_ISL_1412719, EPI_ISL_1412720, EPI_ISL_1412722, EPI_ISL_1412723, EPI_ISL_1412724, EPI_ISL_1412725, EPI_ISL_1412727, EPI_ISL_1412729, EPI_ISL_1412734, EPI_ISL_1412735, EPI_ISL_1412739, EPI_ISL_1412740, EPI_ISL_1412741,
EPI_ISL_1412743, EPI_ISL_1412744, EPI_ISL_1412745, EPI_ISL_1412747, EPI_ISL_1412748, EPI_ISL_1412751, EPI_ISL_1412754, EPI_ISL_1412755, EPI_ISL_1412757, EPI_ISL_1412762, EPI_ISL_1412763, EPI_ISL_1412764, EPI_ISL_1412765, EPI_ISL_1412767, EPI_ISL_1412768, EPI_ISL_1412771, EPI_ISL_1412775, EPI_ISL_1412776, EPI_ISL_1412777, EPI_ISL_1412779, EPI_ISL_1412781, EPI_ISL_1412782,
EPI_ISL_1412783, EPI_ISL_1412786, EPI_ISL_1412787, EPI_ISL_1412790, EPI_ISL_1412792, EPI_ISL_1412793, EPI_ISL_1412794, EPI_ISL_1412797, EPI_ISL_1412798, EPI_ISL_1412802, EPI_ISL_1412804, EPI_ISL_1412805, EPI_ISL_1412806, EPI_ISL_1412807, EPI_ISL_1412809, EPI_ISL_1412811, EPI_ISL_1412812, EPI_ISL_1412813, EPI_ISL_1412816, EPI_ISL_1412817, EPI_ISL_1412820, EPI_ISL_1412823,
EPI_ISL_1412827, EPI_ISL_1412828, EPI_ISL_1412830, EPI_ISL_1412831, EPI_ISL_1412834, EPI_ISL_1412837, EPI_ISL_1412846, EPI_ISL_1412847, EPI_ISL_1412848, EPI_ISL_1412852, EPI_ISL_1412857, EPI_ISL_1412858, EPI_ISL_1412859, EPI_ISL_1412861, EPI_ISL_1412863, EPI_ISL_1412865, EPI_ISL_1412866, EPI_ISL_1412867, EPI_ISL_1412868, EPI_ISL_1412875, EPI_ISL_1412876, EPI_ISL_1412877,
EPI_ISL_1412879, EPI_ISL_1412881, EPI_ISL_1412883, EPI_ISL_1412885, EPI_ISL_1412889, EPI_ISL_1412891, EPI_ISL_1412893, EPI_ISL_1412894, EPI_ISL_1412898, EPI_ISL_1412899, EPI_ISL_1412901, EPI_ISL_1412902, EPI_ISL_1412903, EPI_ISL_1412909, EPI_ISL_1412914, EPI_ISL_1412916, EPI_ISL_1412918, EPI_ISL_1412920, EPI_ISL_1412921, EPI_ISL_1412922, EPI_ISL_1412924, EPI_ISL_1413062,
EPI_ISL_1413063, EPI_ISL_1413065, EPI_ISL_1413068, EPI_ISL_1413069, EPI_ISL_1413070, EPI_ISL_1413071, EPI_ISL_1413072, EPI_ISL_1413075, EPI_ISL_1413076, EPI_ISL_1413077, EPI_ISL_1413080, EPI_ISL_1413081, EPI_ISL_1413083, EPI_ISL_1413085, EPI_ISL_1413087, EPI_ISL_1413088, EPI_ISL_1413089, EPI_ISL_1413092, EPI_ISL_1413094, EPI_ISL_1413095, EPI_ISL_1413097, EPI_ISL_1413098,
EPI_ISL_1413100, EPI_ISL_1413101, EPI_ISL_1413102, EPI_ISL_1413103, EPI_ISL_1413105, EPI_ISL_1413106, EPI_ISL_1413108, EPI_ISL_1413109, EPI_ISL_1413110, EPI_ISL_1413112, EPI_ISL_1413113, EPI_ISL_1413121, EPI_ISL_1413122, EPI_ISL_1413124, EPI_ISL_1413126, EPI_ISL_1413130, EPI_ISL_1413135, EPI_ISL_1413137, EPI_ISL_1413143, EPI_ISL_1413148, EPI_ISL_1413151, EPI_ISL_1413153,
EPI_ISL_1413158, EPI_ISL_1413159, EPI_ISL_1413161, EPI_ISL_1413162, EPI_ISL_1413166, EPI_ISL_1413170, EPI_ISL_1413172, EPI_ISL_1413175, EPI_ISL_1413177, EPI_ISL_1413180, EPI_ISL_1413188, EPI_ISL_1413190, EPI_ISL_1413192, EPI_ISL_1413194, EPI_ISL_1413198, EPI_ISL_1413200, EPI_ISL_1413201, EPI_ISL_1413205, EPI_ISL_1413217, EPI_ISL_1413219, EPI_ISL_1413220, EPI_ISL_1413228,
EPI_ISL_1413230, EPI_ISL_1413233, EPI_ISL_1413237, EPI_ISL_1413242, EPI_ISL_1413244, EPI_ISL_1413247, EPI_ISL_1413252, EPI_ISL_1413256, EPI_ISL_1413259, EPI_ISL_1413262, EPI_ISL_1413264, EPI_ISL_1413267, EPI_ISL_1413271, EPI_ISL_1413275, EPI_ISL_1413279, EPI_ISL_1413283, EPI_ISL_1413288, EPI_ISL_1413294, EPI_ISL_1413295, EPI_ISL_1413298, EPI_ISL_1413303, EPI_ISL_1413310,
EPI_ISL_1413312, EPI_ISL_1413318, EPI_ISL_1413320, EPI_ISL_1413324, EPI_ISL_1413326, EPI_ISL_1413331, EPI_ISL_1413334, EPI_ISL_1413335, EPI_ISL_1413338, EPI_ISL_1413340, EPI_ISL_1413342, EPI_ISL_1413346, EPI_ISL_1413362, EPI_ISL_1413364, EPI_ISL_1413367, EPI_ISL_1413368, EPI_ISL_1413378, EPI_ISL_1413379, EPI_ISL_1413386, EPI_ISL_1413391, EPI_ISL_1413392, EPI_ISL_1413394,
EPI_ISL_1413397, EPI_ISL_1413400, EPI_ISL_1413403, EPI_ISL_1413405, EPI_ISL_1413406, EPI_ISL_1413408, EPI_ISL_1413411, EPI_ISL_1413414, EPI_ISL_1413422, EPI_ISL_1413423, EPI_ISL_1413425, EPI_ISL_1413430, EPI_ISL_1413432, EPI_ISL_1413433, EPI_ISL_1413435, EPI_ISL_1413436, EPI_ISL_1413440, EPI_ISL_1413441, EPI_ISL_1413448, EPI_ISL_1413449, EPI_ISL_1413451, EPI_ISL_1413453,
EPI_ISL_1413454, EPI_ISL_1413456, EPI_ISL_1413467, EPI_ISL_1413469, EPI_ISL_1413470, EPI_ISL_1413475, EPI_ISL_1413477, EPI_ISL_1413478, EPI_ISL_1413483, EPI_ISL_1413486, EPI_ISL_1413489, EPI_ISL_1413491, EPI_ISL_1413494, EPI_ISL_1413497, EPI_ISL_1413499, EPI_ISL_1413508, EPI_ISL_1413512, EPI_ISL_1413513, EPI_ISL_1413515, EPI_ISL_1413520, EPI_ISL_1413523, EPI_ISL_1413524,
EPI_ISL_1413527, EPI_ISL_1413529, EPI_ISL_1413531, EPI_ISL_1413532, EPI_ISL_1413534, EPI_ISL_1413535, EPI_ISL_1413540, EPI_ISL_1413546, EPI_ISL_1413553, EPI_ISL_1413557, EPI_ISL_1413564, EPI_ISL_1413572, EPI_ISL_1413575, EPI_ISL_1413580, EPI_ISL_1413583, EPI_ISL_1413588, EPI_ISL_1413590, EPI_ISL_1413591, EPI_ISL_1413593, EPI_ISL_1413596, EPI_ISL_1413598, EPI_ISL_1413599,
EPI_ISL_1413601, EPI_ISL_1413603, EPI_ISL_1413608, EPI_ISL_1413611, EPI_ISL_1413612, EPI_ISL_1413614, EPI_ISL_1413616, EPI_ISL_1413619, EPI_ISL_1413620, EPI_ISL_1413622, EPI_ISL_1413624, EPI_ISL_1413625, EPI_ISL_1413627, EPI_ISL_1413630, EPI_ISL_1413632, EPI_ISL_1413633, EPI_ISL_1413635, EPI_ISL_1413636, EPI_ISL_1413638, EPI_ISL_1413641, EPI_ISL_1413643, EPI_ISL_1413644,
EPI_ISL_1413646, EPI_ISL_1413648, EPI_ISL_1413649, EPI_ISL_1413650, EPI_ISL_1413652, EPI_ISL_1413654, EPI_ISL_1413655, EPI_ISL_1413657, EPI_ISL_1413659, EPI_ISL_1413660, EPI_ISL_1413662, EPI_ISL_1413663, EPI_ISL_1413665, EPI_ISL_1413666, EPI_ISL_1413670, EPI_ISL_1413671, EPI_ISL_1413674, EPI_ISL_1413676, EPI_ISL_1413681, EPI_ISL_1413683, EPI_ISL_1413684, EPI_ISL_1413686,
EPI_ISL_1413688, EPI_ISL_1413695, EPI_ISL_1413697, EPI_ISL_1413698, EPI_ISL_1413700, EPI_ISL_1413701, EPI_ISL_1413703, EPI_ISL_1413705, EPI_ISL_1413706, EPI_ISL_1413708, EPI_ISL_1413709, EPI_ISL_1413711, EPI_ISL_1413713, EPI_ISL_1413714, EPI_ISL_1413719, EPI_ISL_1413722, EPI_ISL_1413724, EPI_ISL_1413725, EPI_ISL_1413727, EPI_ISL_1413729, EPI_ISL_1413730, EPI_ISL_1413732,
EPI_ISL_1413734, EPI_ISL_1413735, EPI_ISL_1413737, EPI_ISL_1413743, EPI_ISL_1413744, EPI_ISL_1413746, EPI_ISL_1413749, EPI_ISL_1413751, EPI_ISL_1413754, EPI_ISL_1413757, EPI_ISL_1413759, EPI_ISL_1413760, EPI_ISL_1413762, EPI_ISL_1413764, EPI_ISL_1413765, EPI_ISL_1413767, EPI_ISL_1413768, EPI_ISL_1413772, EPI_ISL_1413773, EPI_ISL_1413775, EPI_ISL_1413777, EPI_ISL_1413778,
EPI_ISL_1413780, EPI_ISL_1413782, EPI_ISL_1413783, EPI_ISL_1413785, EPI_ISL_1413786, EPI_ISL_1413788, EPI_ISL_1413790, EPI_ISL_1413791, EPI_ISL_1413793, EPI_ISL_1413795, EPI_ISL_1413796, EPI_ISL_1413798, EPI_ISL_1413802, EPI_ISL_1413804, EPI_ISL_1413808, EPI_ISL_1413809, EPI_ISL_1413812, EPI_ISL_1413814, EPI_ISL_1413816, EPI_ISL_1413819, EPI_ISL_1413820, EPI_ISL_1413822,
EPI_ISL_1413824, EPI_ISL_1413827, EPI_ISL_1413828, EPI_ISL_1413830, EPI_ISL_1413832, EPI_ISL_1413833, EPI_ISL_1413836, EPI_ISL_1413838, EPI_ISL_1413839, EPI_ISL_1413844, EPI_ISL_1413846, EPI_ISL_1413850, EPI_ISL_1413852, EPI_ISL_1413854, EPI_ISL_1413855, EPI_ISL_1413857, EPI_ISL_1413860, EPI_ISL_1413862, EPI_ISL_1413863, EPI_ISL_1413865, EPI_ISL_1413866, EPI_ISL_1413868,
EPI_ISL_1413870, EPI_ISL_1413872, EPI_ISL_1413875, EPI_ISL_1413876, EPI_ISL_1413877, EPI_ISL_1413878, EPI_ISL_1413879, EPI_ISL_1413880, EPI_ISL_1413881, EPI_ISL_1413882, EPI_ISL_1413884, EPI_ISL_1413885, EPI_ISL_1413886, EPI_ISL_1413887, EPI_ISL_1413892, EPI_ISL_1413898, EPI_ISL_1413900, EPI_ISL_1413901, EPI_ISL_1413906, EPI_ISL_1413908, EPI_ISL_1413910, EPI_ISL_1413911,
EPI_ISL_1413915, EPI_ISL_1413916, EPI_ISL_1413918, EPI_ISL_1413923, EPI_ISL_1413928, EPI_ISL_1413930, EPI_ISL_1413936, EPI_ISL_1413938, EPI_ISL_1413939, EPI_ISL_1413941, EPI_ISL_1413942, EPI_ISL_1413945, EPI_ISL_1413946, EPI_ISL_1413948, EPI_ISL_1413949, EPI_ISL_1413950, EPI_ISL_1413951, EPI_ISL_1413952, EPI_ISL_1413953, EPI_ISL_1413954, EPI_ISL_1413955, EPI_ISL_1413956,
EPI_ISL_1413957, EPI_ISL_1413960, EPI_ISL_1413961, EPI_ISL_1413962, EPI_ISL_1413963, EPI_ISL_1413964, EPI_ISL_1413965, EPI_ISL_1413966, EPI_ISL_1413968, EPI_ISL_1413970, EPI_ISL_1413972, EPI_ISL_1413973, EPI_ISL_1413974, EPI_ISL_1413975, EPI_ISL_1413976, EPI_ISL_1413977, EPI_ISL_1413981, EPI_ISL_1413982, EPI_ISL_1413984, EPI_ISL_1413986, EPI_ISL_1413987, EPI_ISL_1413988,
EPI_ISL_1413989, EPI_ISL_1413991, EPI_ISL_1413992, EPI_ISL_1413994, EPI_ISL_1413995, EPI_ISL_1413996, EPI_ISL_1413998, EPI_ISL_1414002, EPI_ISL_1414003, EPI_ISL_1414005, EPI_ISL_1414008, EPI_ISL_1414009, EPI_ISL_1414010, EPI_ISL_1414011, EPI_ISL_1414012, EPI_ISL_1414013, EPI_ISL_1414015, EPI_ISL_1414016, EPI_ISL_1414017, EPI_ISL_1414019, EPI_ISL_1414020, EPI_ISL_1414021,
EPI_ISL_1414022, EPI_ISL_1414023, EPI_ISL_1414026, EPI_ISL_1414027, EPI_ISL_1414028, EPI_ISL_1414029, EPI_ISL_1414030, EPI_ISL_1414031, EPI_ISL_1414033, EPI_ISL_1414034, EPI_ISL_1414036, EPI_ISL_1414037, EPI_ISL_1414038, EPI_ISL_1414039, EPI_ISL_1414041, EPI_ISL_1414042, EPI_ISL_1414044, EPI_ISL_1414046, EPI_ISL_1414047, EPI_ISL_1414049, EPI_ISL_1414051, EPI_ISL_1414052,
EPI_ISL_1414053, EPI_ISL_1414055, EPI_ISL_1414056, EPI_ISL_1414058, EPI_ISL_1414059, EPI_ISL_1414060, EPI_ISL_1449561, EPI_ISL_1450303, EPI_ISL_1450304, EPI_ISL_1450310, EPI_ISL_1450314, EPI_ISL_1450315, EPI_ISL_1450321, EPI_ISL_1450327, EPI_ISL_1450328, EPI_ISL_1450331, EPI_ISL_1450333, EPI_ISL_1450334, EPI_ISL_1450337, EPI_ISL_1450340, EPI_ISL_1450342, EPI_ISL_1450343,
EPI_ISL_1450344, EPI_ISL_1450349, EPI_ISL_1450353, EPI_ISL_1450362, EPI_ISL_1450366, EPI_ISL_1450370, EPI_ISL_1450372, EPI_ISL_1450373, EPI_ISL_1450378, EPI_ISL_1450385, EPI_ISL_1450386, EPI_ISL_1450387, EPI_ISL_1450394, EPI_ISL_1450397, EPI_ISL_1450403, EPI_ISL_1450408, EPI_ISL_1450411, EPI_ISL_1450414, EPI_ISL_1450418, EPI_ISL_1450433, EPI_ISL_1450438, EPI_ISL_1450441,
EPI_ISL_1450450, EPI_ISL_1450451, EPI_ISL_1450453, EPI_ISL_1450463, EPI_ISL_1450464, EPI_ISL_1450466, EPI_ISL_1450469, EPI_ISL_1450472, EPI_ISL_1450476, EPI_ISL_1450477, EPI_ISL_1450482, EPI_ISL_1450485, EPI_ISL_1450487, EPI_ISL_1450488, EPI_ISL_1450500, EPI_ISL_1450503, EPI_ISL_1450508, EPI_ISL_1450519, EPI_ISL_1450522, EPI_ISL_1450524, EPI_ISL_1450526, EPI_ISL_1450529,
EPI_ISL_1450538, EPI_ISL_1450552, EPI_ISL_1450553, EPI_ISL_1450558, EPI_ISL_1450559, EPI_ISL_1450565, EPI_ISL_1450567, EPI_ISL_1450570, EPI_ISL_1450582, EPI_ISL_1450584, EPI_ISL_1450590, EPI_ISL_1450596, EPI_ISL_1450598, EPI_ISL_1450599, EPI_ISL_1450604, EPI_ISL_1450605, EPI_ISL_1450606, EPI_ISL_1451154, EPI_ISL_1451157, EPI_ISL_1451159, EPI_ISL_1451162, EPI_ISL_1451170,
EPI_ISL_1451174, EPI_ISL_1451179, EPI_ISL_1451183, EPI_ISL_1451185, EPI_ISL_1451187, EPI_ISL_1451188, EPI_ISL_1451189, EPI_ISL_1451196, EPI_ISL_1451197, EPI_ISL_1451211, EPI_ISL_1451212, EPI_ISL_1451220, EPI_ISL_1451235, EPI_ISL_1451241, EPI_ISL_1451246, EPI_ISL_1451251, EPI_ISL_1451271, EPI_ISL_1451272, EPI_ISL_1451275, EPI_ISL_1451277, EPI_ISL_1451285, EPI_ISL_1451293,
EPI_ISL_1451301, EPI_ISL_1451302, EPI_ISL_1451303, EPI_ISL_1451313, EPI_ISL_1451321, EPI_ISL_1451323, EPI_ISL_1451324, EPI_ISL_1451325, EPI_ISL_1451333, EPI_ISL_1451335, EPI_ISL_1451353, EPI_ISL_1451355, EPI_ISL_1451359, EPI_ISL_1451363, EPI_ISL_1451366, EPI_ISL_1451372, EPI_ISL_1451375, EPI_ISL_1451378, EPI_ISL_1451385, EPI_ISL_1451389, EPI_ISL_1451392, EPI_ISL_1451397,
EPI_ISL_1451398, EPI_ISL_1451400, EPI_ISL_1451404, EPI_ISL_1451407, EPI_ISL_1451410, EPI_ISL_1451418, EPI_ISL_1451421, EPI_ISL_1451423, EPI_ISL_1451429, EPI_ISL_1451434, EPI_ISL_1451439, EPI_ISL_1451441, EPI_ISL_1451445, EPI_ISL_1451447, EPI_ISL_1451448, EPI_ISL_1451449, EPI_ISL_1451453, EPI_ISL_1451602, EPI_ISL_1451940, EPI_ISL_1452076, EPI_ISL_1452078, EPI_ISL_1452080,
EPI_ISL_1452082, EPI_ISL_1452083, EPI_ISL_1452088, EPI_ISL_1452089, EPI_ISL_1452094, EPI_ISL_1452098, EPI_ISL_1452101, EPI_ISL_1452108, EPI_ISL_1452110, EPI_ISL_1452112, EPI_ISL_1452117, EPI_ISL_1452121, EPI_ISL_1452124, EPI_ISL_1452127, EPI_ISL_1452129, EPI_ISL_1452132, EPI_ISL_1452136, EPI_ISL_1452139, EPI_ISL_1452148, EPI_ISL_1452151, EPI_ISL_1452152, EPI_ISL_1452153,
EPI_ISL_1452156, EPI_ISL_1452165, EPI_ISL_1452167, EPI_ISL_1452170, EPI_ISL_1452171, EPI_ISL_1452179, EPI_ISL_1452181, EPI_ISL_1452183, EPI_ISL_1452190, EPI_ISL_1452201, EPI_ISL_1452203, EPI_ISL_1452209, EPI_ISL_1452211, EPI_ISL_1452214, EPI_ISL_1452217, EPI_ISL_1452219, EPI_ISL_1452221, EPI_ISL_1452227, EPI_ISL_1452230, EPI_ISL_1452234, EPI_ISL_1452235, EPI_ISL_1452237,
EPI_ISL_1452241, EPI_ISL_1452243, EPI_ISL_1452244, EPI_ISL_1452247, EPI_ISL_1452248, EPI_ISL_1452249, EPI_ISL_1452251, EPI_ISL_1452255, EPI_ISL_1452261, EPI_ISL_1452263, EPI_ISL_1452264, EPI_ISL_1452265, EPI_ISL_1452267, EPI_ISL_1452268, EPI_ISL_1452270, EPI_ISL_1452271, EPI_ISL_1452274, EPI_ISL_1452275, EPI_ISL_1452276, EPI_ISL_1452283, EPI_ISL_1452286, EPI_ISL_1452287,
EPI_ISL_1452290, EPI_ISL_1452291, EPI_ISL_1452292, EPI_ISL_1452294, EPI_ISL_1452296, EPI_ISL_1452300, EPI_ISL_1452301, EPI_ISL_1452303, EPI_ISL_1452305, EPI_ISL_1452307, EPI_ISL_1452310, EPI_ISL_1452312, EPI_ISL_1452314, EPI_ISL_1452316, EPI_ISL_1452319, EPI_ISL_1452323, EPI_ISL_1452324, EPI_ISL_1452327, EPI_ISL_1452328, EPI_ISL_1452329, EPI_ISL_1452330, EPI_ISL_1452331,
EPI_ISL_1452332, EPI_ISL_1452333, EPI_ISL_1452335, EPI_ISL_1452336, EPI_ISL_1452337, EPI_ISL_1452339, EPI_ISL_1452341, EPI_ISL_1452343, EPI_ISL_1452345, EPI_ISL_1452346, EPI_ISL_1452351, EPI_ISL_1452352, EPI_ISL_1452353, EPI_ISL_1452354, EPI_ISL_1452355, EPI_ISL_1452359, EPI_ISL_1452360, EPI_ISL_1452362, EPI_ISL_1452363, EPI_ISL_1452364, EPI_ISL_1452365, EPI_ISL_1452366,
EPI_ISL_1452367, EPI_ISL_1452368, EPI_ISL_1452370, EPI_ISL_1452371, EPI_ISL_1452372, EPI_ISL_1452374, EPI_ISL_1452376, EPI_ISL_1452377, EPI_ISL_1452378, EPI_ISL_1452379, EPI_ISL_1452380, EPI_ISL_1452381, EPI_ISL_1452382, EPI_ISL_1452385, EPI_ISL_1452386, EPI_ISL_1452387, EPI_ISL_1452388, EPI_ISL_1452389, EPI_ISL_1452390, EPI_ISL_1452391, EPI_ISL_1452394,
EPI_ISL_1452395, EPI_ISL_1452398, EPI_ISL_1452400, EPI_ISL_1452402, EPI_ISL_1452403, EPI_ISL_1452404, EPI_ISL_1452407, EPI_ISL_1452409, EPI_ISL_1452412, EPI_ISL_1452413, EPI_ISL_1452414, EPI_ISL_1452417, EPI_ISL_1452418, EPI_ISL_1452419, EPI_ISL_1452420, EPI_ISL_1452421, EPI_ISL_1452422, EPI_ISL_1452423, EPI_ISL_1452424, EPI_ISL_1452425, EPI_ISL_1452426, EPI_ISL_1452427,
EPI_ISL_1452428, EPI_ISL_1452429, EPI_ISL_1452430, EPI_ISL_1452432, EPI_ISL_1452434, EPI_ISL_1452435, EPI_ISL_1452437, EPI_ISL_1452438, EPI_ISL_1452439, EPI_ISL_1452440, EPI_ISL_1452442, EPI_ISL_1452444, EPI_ISL_1452446, EPI_ISL_1452448, EPI_ISL_1452450, EPI_ISL_1452451, EPI_ISL_1452452, EPI_ISL_1452453, EPI_ISL_1452455, EPI_ISL_1452456, EPI_ISL_1452459, EPI_ISL_1452460,
EPI_ISL_1452461, EPI_ISL_1452462, EPI_ISL_1452463, EPI_ISL_1452464, EPI_ISL_1452470, EPI_ISL_1452473, EPI_ISL_1452474, EPI_ISL_1452476, EPI_ISL_1452477, EPI_ISL_1452478, EPI_ISL_1452480, EPI_ISL_1452481, EPI_ISL_1452483, EPI_ISL_1452485, EPI_ISL_1452487, EPI_ISL_1452490, EPI_ISL_1452492, EPI_ISL_1452494, EPI_ISL_1452495, EPI_ISL_1452496, EPI_ISL_1452497, EPI_ISL_1452498,
EPI_ISL_1452502, EPI_ISL_1452503, EPI_ISL_1452505, EPI_ISL_1452506, EPI_ISL_1452507, EPI_ISL_1452508, EPI_ISL_1452510, EPI_ISL_1452511, EPI_ISL_1452512, EPI_ISL_1452514, EPI_ISL_1452515, EPI_ISL_1452518, EPI_ISL_1452519, EPI_ISL_1452520, EPI_ISL_1452522, EPI_ISL_1452523, EPI_ISL_1452526, EPI_ISL_1452527, EPI_ISL_1452528, EPI_ISL_1452529, EPI_ISL_1452530, EPI_ISL_1452531,
EPI_ISL_1452532, EPI_ISL_1452533, EPI_ISL_1452534, EPI_ISL_1452535, EPI_ISL_1452536, EPI_ISL_1452538, EPI_ISL_1452539, EPI_ISL_1452541, EPI_ISL_1452543, EPI_ISL_1452546, EPI_ISL_1452548, EPI_ISL_1452549, EPI_ISL_1452550, EPI_ISL_1452552, EPI_ISL_1452554, EPI_ISL_1452556, EPI_ISL_1452557, EPI_ISL_1452558, EPI_ISL_1452559, EPI_ISL_1452560, EPI_ISL_1452561, EPI_ISL_1452562,
EPI_ISL_1452563, EPI_ISL_1452564, EPI_ISL_1452565, EPI_ISL_1452567, EPI_ISL_1452568, EPI_ISL_1452569, EPI_ISL_1452571, EPI_ISL_1452573, EPI_ISL_1452574, EPI_ISL_1452575, EPI_ISL_1452577, EPI_ISL_1452582, EPI_ISL_1452583, EPI_ISL_1452584, EPI_ISL_1452585, EPI_ISL_1452586, EPI_ISL_1452587, EPI_ISL_1452588, EPI_ISL_1452589, EPI_ISL_1452590, EPI_ISL_1452592, EPI_ISL_1452593,
EPI_ISL_1452595, EPI_ISL_1452597, EPI_ISL_1452598, EPI_ISL_1452599, EPI_ISL_1452600, EPI_ISL_1452602, EPI_ISL_1452603, EPI_ISL_1452606, EPI_ISL_1452608, EPI_ISL_1452609, EPI_ISL_1452612, EPI_ISL_1452613, EPI_ISL_1452614, EPI_ISL_1452615, EPI_ISL_1452616, EPI_ISL_1452618, EPI_ISL_1452619, EPI_ISL_1452620, EPI_ISL_1452621, EPI_ISL_1452622, EPI_ISL_1452623, EPI_ISL_1452624,
EPI_ISL_1452627, EPI_ISL_1452628, EPI_ISL_1452629, EPI_ISL_1452630, EPI_ISL_1452633, EPI_ISL_1452634, EPI_ISL_1452635, EPI_ISL_1452636, EPI_ISL_1452637, EPI_ISL_1452639, EPI_ISL_1452640, EPI_ISL_1452641, EPI_ISL_1452642, EPI_ISL_1452643, EPI_ISL_1452645, EPI_ISL_1452646, EPI_ISL_1452648, EPI_ISL_1452651, EPI_ISL_1452652, EPI_ISL_1452653, EPI_ISL_1452654, EPI_ISL_1452655,
EPI_ISL_1452656, EPI_ISL_1452657, EPI_ISL_1452658, EPI_ISL_1452659, EPI_ISL_1452660, EPI_ISL_1452661, EPI_ISL_1452662, EPI_ISL_1452663, EPI_ISL_1452664, EPI_ISL_1452666, EPI_ISL_1452667, EPI_ISL_1452669, EPI_ISL_1452670, EPI_ISL_1452671, EPI_ISL_1452672, EPI_ISL_1452673, EPI_ISL_1452676, EPI_ISL_1452677, EPI_ISL_1452678, EPI_ISL_1452679, EPI_ISL_1452681, EPI_ISL_1452683,
EPI_ISL_1452686, EPI_ISL_1452687, EPI_ISL_1452688, EPI_ISL_1452689, EPI_ISL_1452690, EPI_ISL_1452691, EPI_ISL_1452695, EPI_ISL_1452696, EPI_ISL_1452698, EPI_ISL_1452700, EPI_ISL_1452701, EPI_ISL_1452705, EPI_ISL_1452706, EPI_ISL_1452709, EPI_ISL_1452713, EPI_ISL_1452714, EPI_ISL_1452717, EPI_ISL_1452719, EPI_ISL_1452721, EPI_ISL_1452723, EPI_ISL_1452724, EPI_ISL_1452725,
EPI_ISL_1452727, EPI_ISL_1452731, EPI_ISL_1452732, EPI_ISL_1452733, EPI_ISL_1452734, EPI_ISL_1452735, EPI_ISL_1452736, EPI_ISL_1452737, EPI_ISL_1452739, EPI_ISL_1452741, EPI_ISL_1452742, EPI_ISL_1452744, EPI_ISL_1452746, EPI_ISL_1452747, EPI_ISL_1452748, EPI_ISL_1452749, EPI_ISL_1452752, EPI_ISL_1452754, EPI_ISL_1452756, EPI_ISL_1452757, EPI_ISL_1452759, EPI_ISL_1452760,
EPI_ISL_1452762, EPI_ISL_1452763, EPI_ISL_1452765, EPI_ISL_1452766, EPI_ISL_1452769, EPI_ISL_1452770, EPI_ISL_1452771, EPI_ISL_1452773, EPI_ISL_1452774, EPI_ISL_1452775, EPI_ISL_1452776, EPI_ISL_1452779, EPI_ISL_1452782, EPI_ISL_1452784, EPI_ISL_1452785, EPI_ISL_1452788, EPI_ISL_1452789, EPI_ISL_1452791, EPI_ISL_1452793, EPI_ISL_1452796, EPI_ISL_1452797, EPI_ISL_1452799,
EPI_ISL_1452801, EPI_ISL_1452802, EPI_ISL_1452803, EPI_ISL_1452804, EPI_ISL_1452805, EPI_ISL_1452806, EPI_ISL_1452807, EPI_ISL_1452809, EPI_ISL_1452810, EPI_ISL_1452811, EPI_ISL_1452813, EPI_ISL_1452814, EPI_ISL_1452816, EPI_ISL_1452818, EPI_ISL_1452820, EPI_ISL_1452821, EPI_ISL_1452822, EPI_ISL_1452823, EPI_ISL_1452824, EPI_ISL_1452825, EPI_ISL_1452827, EPI_ISL_1452828,
EPI_ISL_1452829, EPI_ISL_1452830, EPI_ISL_1452832, EPI_ISL_1452834, EPI_ISL_1452837, EPI_ISL_1452838, EPI_ISL_1452840, EPI_ISL_1452841, EPI_ISL_1452843, EPI_ISL_1452844, EPI_ISL_1452845, EPI_ISL_1452846, EPI_ISL_1452847, EPI_ISL_1452848, EPI_ISL_1452849, EPI_ISL_1452850, EPI_ISL_1452852, EPI_ISL_1452853, EPI_ISL_1452855, EPI_ISL_1452858, EPI_ISL_1452859, EPI_ISL_1452861,
EPI_ISL_1452863, EPI_ISL_1452864, EPI_ISL_1452865, EPI_ISL_1452867, EPI_ISL_1452868, EPI_ISL_1452869, EPI_ISL_1452875, EPI_ISL_1452876, EPI_ISL_1452878, EPI_ISL_1452879, EPI_ISL_1452880, EPI_ISL_1452881, EPI_ISL_1452883, EPI_ISL_1452884, EPI_ISL_1452885, EPI_ISL_1452886, EPI_ISL_1452889, EPI_ISL_1452896, EPI_ISL_1452900, EPI_ISL_1452906, EPI_ISL_1452908, EPI_ISL_1452909,
EPI_ISL_1452912, EPI_ISL_1452913, EPI_ISL_1452915, EPI_ISL_1452916, EPI_ISL_1452919, EPI_ISL_1452921, EPI_ISL_1452924, EPI_ISL_1452926, EPI_ISL_1452928, EPI_ISL_1452930, EPI_ISL_1452936, EPI_ISL_1452938, EPI_ISL_1452939, EPI_ISL_1452943, EPI_ISL_1452946, EPI_ISL_1452955, EPI_ISL_1452960, EPI_ISL_1452962, EPI_ISL_1452965, EPI_ISL_1452968, EPI_ISL_1452969, EPI_ISL_1452970,
EPI_ISL_1452971, EPI_ISL_1452973, EPI_ISL_1452974, EPI_ISL_1452975, EPI_ISL_1452977, EPI_ISL_1452978, EPI_ISL_1452981, EPI_ISL_1452983, EPI_ISL_1452985, EPI_ISL_1452991, EPI_ISL_1452993, EPI_ISL_1452996, EPI_ISL_1452999, EPI_ISL_1453000, EPI_ISL_1453002, EPI_ISL_1453003, EPI_ISL_1453005, EPI_ISL_1453008, EPI_ISL_1453011, EPI_ISL_1453012, EPI_ISL_1453013, EPI_ISL_1453015,
EPI_ISL_1453016, EPI_ISL_1453021, EPI_ISL_1453022, EPI_ISL_1453023, EPI_ISL_1453027, EPI_ISL_1453030, EPI_ISL_1453031, EPI_ISL_1453032, EPI_ISL_1453034, EPI_ISL_1453035, EPI_ISL_1453036, EPI_ISL_1453037, EPI_ISL_1453038, EPI_ISL_1453039, EPI_ISL_1453042, EPI_ISL_1453043, EPI_ISL_1453047, EPI_ISL_1453056, EPI_ISL_1453057, EPI_ISL_1453058, EPI_ISL_1453060, EPI_ISL_1453065,
EPI_ISL_1453066, EPI_ISL_1453068, EPI_ISL_1453072, EPI_ISL_1453075, EPI_ISL_1453076, EPI_ISL_1453078, EPI_ISL_1453080, EPI_ISL_1453081, EPI_ISL_1453082, EPI_ISL_1453083, EPI_ISL_1453084, EPI_ISL_1453085, EPI_ISL_1453086, EPI_ISL_1453087, EPI_ISL_1453088, EPI_ISL_1453089, EPI_ISL_1453092, EPI_ISL_1453093, EPI_ISL_1453094, EPI_ISL_1453095, EPI_ISL_1453097, EPI_ISL_1453098,
EPI_ISL_1453100, EPI_ISL_1453101, EPI_ISL_1453103, EPI_ISL_1453105, EPI_ISL_1453109, EPI_ISL_1453110, EPI_ISL_1453114, EPI_ISL_1453117, EPI_ISL_1453118, EPI_ISL_1453120, EPI_ISL_1453122, EPI_ISL_1453123, EPI_ISL_1453125, EPI_ISL_1453126, EPI_ISL_1453127, EPI_ISL_1453130, EPI_ISL_1453132, EPI_ISL_1453135, EPI_ISL_1453136, EPI_ISL_1453138, EPI_ISL_1453139, EPI_ISL_1453143,
EPI_ISL_1453144, EPI_ISL_1453146, EPI_ISL_1453153, EPI_ISL_1453154, EPI_ISL_1453155, EPI_ISL_1453156, EPI_ISL_1453157, EPI_ISL_1453159, EPI_ISL_1453163, EPI_ISL_1453165, EPI_ISL_1453167, EPI_ISL_1453169, EPI_ISL_1453174, EPI_ISL_1453175, EPI_ISL_1453179, EPI_ISL_1453181, EPI_ISL_1453186, EPI_ISL_1453187, EPI_ISL_1453188, EPI_ISL_1453192, EPI_ISL_1453196, EPI_ISL_1453197,
EPI_ISL_1453199, EPI_ISL_1453204, EPI_ISL_1453206, EPI_ISL_1453210, EPI_ISL_1453212, EPI_ISL_1453213, EPI_ISL_1453214, EPI_ISL_1453215, EPI_ISL_1453219, EPI_ISL_1453220, EPI_ISL_1453221, EPI_ISL_1453222, EPI_ISL_1453228, EPI_ISL_1453234, EPI_ISL_1453241, EPI_ISL_1453246, EPI_ISL_1453249, EPI_ISL_1453250, EPI_ISL_1453254, EPI_ISL_1453255, EPI_ISL_1453258, EPI_ISL_1453260,
EPI_ISL_1453265, EPI_ISL_1453266, EPI_ISL_1453268, EPI_ISL_1453271, EPI_ISL_1453273, EPI_ISL_1453276, EPI_ISL_1453278, EPI_ISL_1453279, EPI_ISL_1453280, EPI_ISL_1453281, EPI_ISL_1453283, EPI_ISL_1453291, EPI_ISL_1453292, EPI_ISL_1453294, EPI_ISL_1453297, EPI_ISL_1453298, EPI_ISL_1453300, EPI_ISL_1453303, EPI_ISL_1453311, EPI_ISL_1453313, EPI_ISL_1453314,
EPI_ISL_1453315, EPI_ISL_1453316, EPI_ISL_1453317, EPI_ISL_1453319, EPI_ISL_1453320, EPI_ISL_1453323, EPI_ISL_1453325, EPI_ISL_1453327, EPI_ISL_1453329, EPI_ISL_1453331, EPI_ISL_1453335, EPI_ISL_1453337, EPI_ISL_1453338, EPI_ISL_1453341, EPI_ISL_1453346, EPI_ISL_1453347, EPI_ISL_1453351, EPI_ISL_1453355, EPI_ISL_1453359, EPI_ISL_1453360, EPI_ISL_1453361, EPI_ISL_1453362,
EPI_ISL_1453364, EPI_ISL_1453369, EPI_ISL_1453370, EPI_ISL_1453371, EPI_ISL_1453373, EPI_ISL_1453375, EPI_ISL_1453378, EPI_ISL_1453392, EPI_ISL_1453393, EPI_ISL_1453394, EPI_ISL_1453395, EPI_ISL_1453396, EPI_ISL_1453398, EPI_ISL_1453400, EPI_ISL_1453404, EPI_ISL_1453405, EPI_ISL_1453407, EPI_ISL_1453409, EPI_ISL_1453411, EPI_ISL_1453413, EPI_ISL_1453415, EPI_ISL_1453420,
EPI_ISL_1453424, EPI_ISL_1453426, EPI_ISL_1453427, EPI_ISL_1453433, EPI_ISL_1453434, EPI_ISL_1453437, EPI_ISL_1453440, EPI_ISL_1453442, EPI_ISL_1453444, EPI_ISL_1453447, EPI_ISL_1453448, EPI_ISL_1453449, EPI_ISL_1453453, EPI_ISL_1453459, EPI_ISL_1453463, EPI_ISL_1453470, EPI_ISL_1453471, EPI_ISL_1453475, EPI_ISL_1453476, EPI_ISL_1453479, EPI_ISL_1453481, EPI_ISL_1453482,
EPI_ISL_1453484, EPI_ISL_1453485, EPI_ISL_1453850, EPI_ISL_1453853, EPI_ISL_1453854, EPI_ISL_1453855, EPI_ISL_1453856, EPI_ISL_1453858, EPI_ISL_1453863, EPI_ISL_1453865, EPI_ISL_1453866, EPI_ISL_1453868, EPI_ISL_1453869, EPI_ISL_1453870, EPI_ISL_1453871, EPI_ISL_1453872, EPI_ISL_1453875, EPI_ISL_1453879, EPI_ISL_1453880, EPI_ISL_1453884, EPI_ISL_1453885, EPI_ISL_1453887,
EPI_ISL_1453888, EPI_ISL_1453890, EPI_ISL_1453893, EPI_ISL_1453898, EPI_ISL_1453906, EPI_ISL_1453908, EPI_ISL_1453909, EPI_ISL_1453910, EPI_ISL_1453913, EPI_ISL_1453914, EPI_ISL_1453916, EPI_ISL_1453921, EPI_ISL_1453924, EPI_ISL_1453926, EPI_ISL_1453927, EPI_ISL_1453928, EPI_ISL_1453929, EPI_ISL_1453931, EPI_ISL_1453940, EPI_ISL_1453941, EPI_ISL_1453942, EPI_ISL_1453947,
EPI_ISL_1453951, EPI_ISL_1453952, EPI_ISL_1453957, EPI_ISL_1453958, EPI_ISL_1453965, EPI_ISL_1453966, EPI_ISL_1453968, EPI_ISL_1453970, EPI_ISL_1453975, EPI_ISL_1453978, EPI_ISL_1453988, EPI_ISL_1453993, EPI_ISL_1453994, EPI_ISL_1453995, EPI_ISL_1453997, EPI_ISL_1454000, EPI_ISL_1454002, EPI_ISL_1454004, EPI_ISL_1454005, EPI_ISL_1454007, EPI_ISL_1454008, EPI_ISL_1454011,
EPI_ISL_1454013, EPI_ISL_1454017, EPI_ISL_1454020, EPI_ISL_1454021, EPI_ISL_1454022, EPI_ISL_1454023, EPI_ISL_1454025, EPI_ISL_1454028, EPI_ISL_1454033, EPI_ISL_1454034, EPI_ISL_1454035, EPI_ISL_1454038, EPI_ISL_1454042, EPI_ISL_1454044, EPI_ISL_1454045, EPI_ISL_1454047, EPI_ISL_1454048, EPI_ISL_1454049, EPI_ISL_1454051, EPI_ISL_1454052, EPI_ISL_1454056, EPI_ISL_1454057,
EPI_ISL_1454058, EPI_ISL_1454060, EPI_ISL_1454061, EPI_ISL_1454065, EPI_ISL_1454067, EPI_ISL_1454069, EPI_ISL_1454074, EPI_ISL_1454075, EPI_ISL_1454076, EPI_ISL_1454079, EPI_ISL_1454080, EPI_ISL_1454083, EPI_ISL_1454085, EPI_ISL_1454086, EPI_ISL_1454087, EPI_ISL_1454089, EPI_ISL_1454095, EPI_ISL_1454096, EPI_ISL_1454097, EPI_ISL_1454100, EPI_ISL_1454101, EPI_ISL_1454102,
EPI_ISL_1454104, EPI_ISL_1454105, EPI_ISL_1454107, EPI_ISL_1454115, EPI_ISL_1454117, EPI_ISL_1454123, EPI_ISL_1454125, EPI_ISL_1454132, EPI_ISL_1454133, EPI_ISL_1454134, EPI_ISL_1454135, EPI_ISL_1454136, EPI_ISL_1454137, EPI_ISL_1454140, EPI_ISL_1454141, EPI_ISL_1454144, EPI_ISL_1454145, EPI_ISL_1454147, EPI_ISL_1454149, EPI_ISL_1454152, EPI_ISL_1454638, EPI_ISL_1454639,
EPI_ISL_1454640, EPI_ISL_1454641, EPI_ISL_1454644, EPI_ISL_1454646, EPI_ISL_1454649, EPI_ISL_1454675, EPI_ISL_1454683, EPI_ISL_1454684, EPI_ISL_1454685, EPI_ISL_1454686, EPI_ISL_1454695, EPI_ISL_1454701, EPI_ISL_1454702, EPI_ISL_1454719, EPI_ISL_1454722, EPI_ISL_1454730, EPI_ISL_1454732, EPI_ISL_1454745, EPI_ISL_1454752, EPI_ISL_1454757, EPI_ISL_1454758, EPI_ISL_1454761,
EPI_ISL_1454763, EPI_ISL_1454773, EPI_ISL_1454778, EPI_ISL_1454797, EPI_ISL_1454799, EPI_ISL_1454827, EPI_ISL_1454844, EPI_ISL_1454846, EPI_ISL_1454852, EPI_ISL_1454854, EPI_ISL_1454856, EPI_ISL_1454867, EPI_ISL_1454875, EPI_ISL_1454877, EPI_ISL_1454880, EPI_ISL_1454890, EPI_ISL_1454891, EPI_ISL_1454896, EPI_ISL_1454900, EPI_ISL_1454903, EPI_ISL_1454915, EPI_ISL_1454925,
EPI_ISL_1454926, EPI_ISL_1454927, EPI_ISL_1454928, EPI_ISL_1454939, EPI_ISL_1454943, EPI_ISL_1454948, EPI_ISL_1454949, EPI_ISL_1454953, EPI_ISL_1454956, EPI_ISL_1454957, EPI_ISL_1454958, EPI_ISL_1454965, EPI_ISL_1454967, EPI_ISL_1454968, EPI_ISL_1454974, EPI_ISL_1454989, EPI_ISL_1455003, EPI_ISL_1455004, EPI_ISL_1455006, EPI_ISL_1455007, EPI_ISL_1455021, EPI_ISL_1455024,
EPI_ISL_1455025, EPI_ISL_1455026, EPI_ISL_1455027, EPI_ISL_1455030, EPI_ISL_1455033, EPI_ISL_1455034, EPI_ISL_1455036, EPI_ISL_1455037, EPI_ISL_1455039, EPI_ISL_1455041, EPI_ISL_1455042, EPI_ISL_1468237, EPI_ISL_1487825, EPI_ISL_1507867, EPI_ISL_1507883, EPI_ISL_1507929, EPI_ISL_1507941, EPI_ISL_1507943, EPI_ISL_1508012, EPI_ISL_1508017, EPI_ISL_1508041, EPI_ISL_1508045,
EPI_ISL_1508141, EPI_ISL_1508142, EPI_ISL_1508143, EPI_ISL_1508144, EPI_ISL_1508145, EPI_ISL_1508146, EPI_ISL_1508147, EPI_ISL_1508148, EPI_ISL_1508149, EPI_ISL_1508150, EPI_ISL_1508151, EPI_ISL_1508153, EPI_ISL_1508154, EPI_ISL_1508155, EPI_ISL_1508156, EPI_ISL_1508157, EPI_ISL_1508159, EPI_ISL_1508160, EPI_ISL_1508161, EPI_ISL_1508162, EPI_ISL_1508163, EPI_ISL_1508165,
EPI_ISL_1508166, EPI_ISL_1508167, EPI_ISL_1508169, EPI_ISL_1508170, EPI_ISL_1508172, EPI_ISL_1508173, EPI_ISL_1508175, EPI_ISL_1508176, EPI_ISL_1508177, EPI_ISL_1508178, EPI_ISL_1508179, EPI_ISL_1508180, EPI_ISL_1508181, EPI_ISL_1508182, EPI_ISL_1508183, EPI_ISL_1508184, EPI_ISL_1508185, EPI_ISL_1508186, EPI_ISL_1508187, EPI_ISL_1508188, EPI_ISL_1508190, EPI_ISL_1508191,
EPI_ISL_1508192, EPI_ISL_1508194, EPI_ISL_1508195, EPI_ISL_1508197, EPI_ISL_1508198, EPI_ISL_1508199, EPI_ISL_1508200, EPI_ISL_1508201, EPI_ISL_1508202, EPI_ISL_1508203, EPI_ISL_1508204, EPI_ISL_1508205, EPI_ISL_1508206, EPI_ISL_1508207, EPI_ISL_1508208, EPI_ISL_1508209, EPI_ISL_1508210, EPI_ISL_1508211, EPI_ISL_1508212, EPI_ISL_1508213, EPI_ISL_1508214, EPI_ISL_1508215,
EPI_ISL_1508216, EPI_ISL_1508217, EPI_ISL_1508218, EPI_ISL_1508219, EPI_ISL_1508220, EPI_ISL_1508221, EPI_ISL_1508222, EPI_ISL_1508223, EPI_ISL_1508225, EPI_ISL_1508226, EPI_ISL_1508227, EPI_ISL_1508228, EPI_ISL_1508229, EPI_ISL_1508230, EPI_ISL_1508232, EPI_ISL_1508233, EPI_ISL_1508234, EPI_ISL_1508235, EPI_ISL_1508236, EPI_ISL_1508237, EPI_ISL_1508238, EPI_ISL_1508239,
EPI_ISL_1508241, EPI_ISL_1508242, EPI_ISL_1508243, EPI_ISL_1508245, EPI_ISL_1508246, EPI_ISL_1508247, EPI_ISL_1508248, EPI_ISL_1508249, EPI_ISL_1508250, EPI_ISL_1508252, EPI_ISL_1508253, EPI_ISL_1508254, EPI_ISL_1508255, EPI_ISL_1508256, EPI_ISL_1508257, EPI_ISL_1508258, EPI_ISL_1508259, EPI_ISL_1508260, EPI_ISL_1508261, EPI_ISL_1508262, EPI_ISL_1508263, EPI_ISL_1508265,
EPI_ISL_1508266, EPI_ISL_1508267, EPI_ISL_1508268, EPI_ISL_1508269, EPI_ISL_1508272, EPI_ISL_1508273, EPI_ISL_1508274, EPI_ISL_1508275, EPI_ISL_1508276, EPI_ISL_1508277, EPI_ISL_1508278, EPI_ISL_1508279, EPI_ISL_1508280, EPI_ISL_1508281, EPI_ISL_1508282, EPI_ISL_1508283, EPI_ISL_1508284, EPI_ISL_1508285, EPI_ISL_1508286, EPI_ISL_1508287, EPI_ISL_1508289, EPI_ISL_1508290,
EPI_ISL_1508292, EPI_ISL_1508294, EPI_ISL_1508295, EPI_ISL_1508296, EPI_ISL_1508297, EPI_ISL_1508298, EPI_ISL_1508299, EPI_ISL_1508300, EPI_ISL_1508301, EPI_ISL_1508302, EPI_ISL_1508303, EPI_ISL_1508304, EPI_ISL_1508305, EPI_ISL_1508306, EPI_ISL_1508307, EPI_ISL_1508308, EPI_ISL_1508309, EPI_ISL_1508310, EPI_ISL_1508311, EPI_ISL_1508314, EPI_ISL_1508315, EPI_ISL_1508316,
EPI_ISL_1508318, EPI_ISL_1508319, EPI_ISL_1508320, EPI_ISL_1508321, EPI_ISL_1508322, EPI_ISL_1508323, EPI_ISL_1508324, EPI_ISL_1508325, EPI_ISL_1508326, EPI_ISL_1508329, EPI_ISL_1508330, EPI_ISL_1508331, EPI_ISL_1508332, EPI_ISL_1508333, EPI_ISL_1508334, EPI_ISL_1508335, EPI_ISL_1508336, EPI_ISL_1508338, EPI_ISL_1508339, EPI_ISL_1508340, EPI_ISL_1508342, EPI_ISL_1508343,
EPI_ISL_1508344, EPI_ISL_1508345, EPI_ISL_1508346, EPI_ISL_1508347, EPI_ISL_1508348, EPI_ISL_1508349, EPI_ISL_1508350, EPI_ISL_1508351, EPI_ISL_1508354, EPI_ISL_1508355, EPI_ISL_1508356, EPI_ISL_1508357, EPI_ISL_1508358, EPI_ISL_1508360, EPI_ISL_1508361, EPI_ISL_1508362, EPI_ISL_1508363, EPI_ISL_1508367, EPI_ISL_1508368, EPI_ISL_1508369, EPI_ISL_1508370, EPI_ISL_1508372,
EPI_ISL_1508373, EPI_ISL_1508374, EPI_ISL_1508376, EPI_ISL_1508377, EPI_ISL_1508378, EPI_ISL_1508379, EPI_ISL_1508380, EPI_ISL_1508382, EPI_ISL_1508383, EPI_ISL_1508384, EPI_ISL_1508385, EPI_ISL_1508386, EPI_ISL_1508387, EPI_ISL_1508388, EPI_ISL_1508389, EPI_ISL_1508390, EPI_ISL_1508391, EPI_ISL_1508392, EPI_ISL_1508393, EPI_ISL_1508394, EPI_ISL_1508395, EPI_ISL_1508396,
EPI_ISL_1508397, EPI_ISL_1508398, EPI_ISL_1508399, EPI_ISL_1508400, EPI_ISL_1508401, EPI_ISL_1508402, EPI_ISL_1508403, EPI_ISL_1508404, EPI_ISL_1508405, EPI_ISL_1508406, EPI_ISL_1508407, EPI_ISL_1508408, EPI_ISL_1508409, EPI_ISL_1508410, EPI_ISL_1508411, EPI_ISL_1508412, EPI_ISL_1508413, EPI_ISL_1508414, EPI_ISL_1508415, EPI_ISL_1508416, EPI_ISL_1508417, EPI_ISL_1508418,
EPI_ISL_1508419, EPI_ISL_1508420, EPI_ISL_1508421, EPI_ISL_1508422, EPI_ISL_1508423, EPI_ISL_1508424, EPI_ISL_1508425, EPI_ISL_1508426, EPI_ISL_1508427, EPI_ISL_1508428, EPI_ISL_1508429, EPI_ISL_1508430, EPI_ISL_1508431, EPI_ISL_1508432, EPI_ISL_1508433, EPI_ISL_1508434, EPI_ISL_1508435, EPI_ISL_1508436, EPI_ISL_1508437, EPI_ISL_1508439, EPI_ISL_1508440, EPI_ISL_1508441,
EPI_ISL_1508442, EPI_ISL_1508443, EPI_ISL_1508444, EPI_ISL_1508445, EPI_ISL_1508446, EPI_ISL_1508448, EPI_ISL_1508449, EPI_ISL_1508450, EPI_ISL_1508451, EPI_ISL_1508452, EPI_ISL_1508453, EPI_ISL_1508454, EPI_ISL_1508455, EPI_ISL_1508456, EPI_ISL_1508457, EPI_ISL_1508458, EPI_ISL_1508459, EPI_ISL_1508460, EPI_ISL_1508461, EPI_ISL_1508462, EPI_ISL_1508463, EPI_ISL_1508464,
EPI_ISL_1508466, EPI_ISL_1508467, EPI_ISL_1508468, EPI_ISL_1508469, EPI_ISL_1508470, EPI_ISL_1508471, EPI_ISL_1508472, EPI_ISL_1508473, EPI_ISL_1508474, EPI_ISL_1508475, EPI_ISL_1508476, EPI_ISL_1508477, EPI_ISL_1508478, EPI_ISL_1508479, EPI_ISL_1508480, EPI_ISL_1508481, EPI_ISL_1508482, EPI_ISL_1508483, EPI_ISL_1508484, EPI_ISL_1508485, EPI_ISL_1508486, EPI_ISL_1508487,
EPI_ISL_1508488, EPI_ISL_1508489, EPI_ISL_1508490, EPI_ISL_1508492, EPI_ISL_1508493, EPI_ISL_1508494, EPI_ISL_1508495, EPI_ISL_1508496, EPI_ISL_1508497, EPI_ISL_1508498, EPI_ISL_1508499, EPI_ISL_1508500, EPI_ISL_1508501, EPI_ISL_1508502, EPI_ISL_1508503, EPI_ISL_1508504, EPI_ISL_1508505, EPI_ISL_1508506, EPI_ISL_1508507, EPI_ISL_1508508, EPI_ISL_1508509, EPI_ISL_1508510,
EPI_ISL_1508511, EPI_ISL_1508512, EPI_ISL_1508513, EPI_ISL_1508514, EPI_ISL_1508516, EPI_ISL_1508517, EPI_ISL_1508518, EPI_ISL_1508519, EPI_ISL_1508520, EPI_ISL_1508521, EPI_ISL_1508522, EPI_ISL_1508523, EPI_ISL_1508524, EPI_ISL_1508525, EPI_ISL_1508526, EPI_ISL_1508527, EPI_ISL_1508528, EPI_ISL_1508529, EPI_ISL_1508530, EPI_ISL_1508531, EPI_ISL_1508532, EPI_ISL_1508533,
EPI_ISL_1508534, EPI_ISL_1508535, EPI_ISL_1508536, EPI_ISL_1508537, EPI_ISL_1508538, EPI_ISL_1508539, EPI_ISL_1508540, EPI_ISL_1508541, EPI_ISL_1508542, EPI_ISL_1508543, EPI_ISL_1508544, EPI_ISL_1508545, EPI_ISL_1508546, EPI_ISL_1508547, EPI_ISL_1508548, EPI_ISL_1508549, EPI_ISL_1508550, EPI_ISL_1508551, EPI_ISL_1508552, EPI_ISL_1508553, EPI_ISL_1508554, EPI_ISL_1508555,
EPI_ISL_1508556, EPI_ISL_1508557, EPI_ISL_1508558, EPI_ISL_1508559, EPI_ISL_1508562, EPI_ISL_1508563, EPI_ISL_1508564, EPI_ISL_1508565, EPI_ISL_1508566, EPI_ISL_1508567, EPI_ISL_1508568, EPI_ISL_1508569, EPI_ISL_1508570, EPI_ISL_1508571, EPI_ISL_1508572, EPI_ISL_1508573, EPI_ISL_1508574, EPI_ISL_1508575, EPI_ISL_1508576, EPI_ISL_1508577, EPI_ISL_1508578, EPI_ISL_1508579,
EPI_ISL_1508580, EPI_ISL_1508581, EPI_ISL_1508582, EPI_ISL_1508583, EPI_ISL_1508584, EPI_ISL_1508585, EPI_ISL_1508586, EPI_ISL_1508587, EPI_ISL_1508588, EPI_ISL_1508589, EPI_ISL_1508590, EPI_ISL_1508591, EPI_ISL_1508592, EPI_ISL_1508593, EPI_ISL_1508594, EPI_ISL_1508595, EPI_ISL_1508596, EPI_ISL_1508597, EPI_ISL_1508598, EPI_ISL_1508599, EPI_ISL_1508600, EPI_ISL_1508601,
EPI_ISL_1508602, EPI_ISL_1508603, EPI_ISL_1508604, EPI_ISL_1508605, EPI_ISL_1508606, EPI_ISL_1508607, EPI_ISL_1508608, EPI_ISL_1508609, EPI_ISL_1508610, EPI_ISL_1508611, EPI_ISL_1508612, EPI_ISL_1508613, EPI_ISL_1508614, EPI_ISL_1508615, EPI_ISL_1508616, EPI_ISL_1508617, EPI_ISL_1508618, EPI_ISL_1508619, EPI_ISL_1508620, EPI_ISL_1508621, EPI_ISL_1508622, EPI_ISL_1508623,
EPI_ISL_1508624, EPI_ISL_1508625, EPI_ISL_1508626, EPI_ISL_1508627, EPI_ISL_1508628, EPI_ISL_1508629, EPI_ISL_1508630, EPI_ISL_1508631, EPI_ISL_1508632, EPI_ISL_1508633, EPI_ISL_1508634, EPI_ISL_1508635, EPI_ISL_1508636, EPI_ISL_1508637, EPI_ISL_1508638, EPI_ISL_1508639, EPI_ISL_1508640, EPI_ISL_1508641, EPI_ISL_1508642, EPI_ISL_1508643, EPI_ISL_1508644, EPI_ISL_1508645,
EPI_ISL_1508646, EPI_ISL_1508647, EPI_ISL_1508649, EPI_ISL_1508650, EPI_ISL_1508651, EPI_ISL_1508652, EPI_ISL_1508653, EPI_ISL_1508654, EPI_ISL_1508655, EPI_ISL_1508656, EPI_ISL_1508657, EPI_ISL_1508658, EPI_ISL_1508659, EPI_ISL_1508660, EPI_ISL_1508661, EPI_ISL_1508662, EPI_ISL_1508663, EPI_ISL_1508664, EPI_ISL_1508665, EPI_ISL_1508666, EPI_ISL_1508667, EPI_ISL_1508668,
EPI_ISL_1508669, EPI_ISL_1508670, EPI_ISL_1508671, EPI_ISL_1508672, EPI_ISL_1508673, EPI_ISL_1508674, EPI_ISL_1508675, EPI_ISL_1508676, EPI_ISL_1508677, EPI_ISL_1508679, EPI_ISL_1508680, EPI_ISL_1508681, EPI_ISL_1508682, EPI_ISL_1508683, EPI_ISL_1508684, EPI_ISL_1508685, EPI_ISL_1508686, EPI_ISL_1508687, EPI_ISL_1508688, EPI_ISL_1508689, EPI_ISL_1508690,
EPI_ISL_1508691, EPI_ISL_1508692, EPI_ISL_1508693, EPI_ISL_1508694, EPI_ISL_1508695, EPI_ISL_1508696, EPI_ISL_1508697, EPI_ISL_1508698, EPI_ISL_1508699, EPI_ISL_1508700, EPI_ISL_1508701, EPI_ISL_1508702, EPI_ISL_1508703, EPI_ISL_1508704, EPI_ISL_1508705, EPI_ISL_1508706, EPI_ISL_1508707, EPI_ISL_1508708, EPI_ISL_1508709, EPI_ISL_1508710, EPI_ISL_1508712, EPI_ISL_1508713,
EPI_ISL_1508714, EPI_ISL_1508715, EPI_ISL_1508716, EPI_ISL_1508717, EPI_ISL_1508718, EPI_ISL_1508719, EPI_ISL_1508720, EPI_ISL_1508721, EPI_ISL_1508722, EPI_ISL_1508723, EPI_ISL_1508724, EPI_ISL_1508725, EPI_ISL_1508726, EPI_ISL_1508727, EPI_ISL_1508729, EPI_ISL_1508736, EPI_ISL_1508737, EPI_ISL_1508747, EPI_ISL_1508755, EPI_ISL_1508761, EPI_ISL_1508762, EPI_ISL_1508772,
EPI_ISL_1508777, EPI_ISL_1519845, EPI_ISL_1519854, EPI_ISL_1635425, EPI_ISL_1635426, EPI_ISL_1635427

| | | | |
|---|---|---|---|
| see above | Lighthouse Lab in Glasgow | Wellcome Sanger Institute for the COVID-19 Genomics UK (COG-UK) Consortium | Anna Dominiczak and Alex Alderton; Carol Clugston; Cordelia Langford; David Gray; David K. Jackson; Dominic Kwiatkowski; Ewan Harrison; Harper VanSteenhouse; Ian Johnston; Jeffrey Barrett; John Sillitoe on behalf of the Wellcome Sanger Institute COVID-19 Surveillance Team; Roberto Amato; Sonia Goncalves; Yumi Kasai |

EPI_ISL_1408910, EPI_ISL_1408912, EPI_ISL_1408914, EPI_ISL_1408916, EPI_ISL_1408917, EPI_ISL_1408919, EPI_ISL_1408920, EPI_ISL_1408923, EPI_ISL_1408924, EPI_ISL_1408925, EPI_ISL_1408926, EPI_ISL_1408927, EPI_ISL_1408930, EPI_ISL_1408931, EPI_ISL_1408933, EPI_ISL_1408935, EPI_ISL_1408936, EPI_ISL_1408938, EPI_ISL_1408942, EPI_ISL_1408943, EPI_ISL_1408949, EPI_ISL_1408951,
EPI_ISL_1408952, EPI_ISL_1408955, EPI_ISL_1408957, EPI_ISL_1408960, EPI_ISL_1408964, EPI_ISL_1408967, EPI_ISL_1408970, EPI_ISL_1408971, EPI_ISL_1408975, EPI_ISL_1408976, EPI_ISL_1408977, EPI_ISL_1408979, EPI_ISL_1408984, EPI_ISL_1408985, EPI_ISL_1408987, EPI_ISL_1408989, EPI_ISL_1408991, EPI_ISL_1408993, EPI_ISL_1408995, EPI_ISL_1408998, EPI_ISL_1408999,
EPI_ISL_1409001, EPI_ISL_1409003, EPI_ISL_1409007, EPI_ISL_1409019, EPI_ISL_1409025, EPI_ISL_1409026, EPI_ISL_1409028, EPI_ISL_1409030, EPI_ISL_1409033, EPI_ISL_1409035, EPI_ISL_1409036, EPI_ISL_1409041, EPI_ISL_1409046, EPI_ISL_1409052, EPI_ISL_1409056, EPI_ISL_1409057, EPI_ISL_1409058, EPI_ISL_1409059, EPI_ISL_1409060, EPI_ISL_1409061, EPI_ISL_1409062, EPI_ISL_1409064,
EPI_ISL_1409067, EPI_ISL_1409070, EPI_ISL_1409072, EPI_ISL_1409079, EPI_ISL_1409083, EPI_ISL_1409084, EPI_ISL_1409086, EPI_ISL_1409093, EPI_ISL_1409094, EPI_ISL_1409101, EPI_ISL_1409102, EPI_ISL_1409105, EPI_ISL_1409107, EPI_ISL_1409108, EPI_ISL_1409111, EPI_ISL_1409112, EPI_ISL_1409119, EPI_ISL_1409120, EPI_ISL_1409121, EPI_ISL_1409122, EPI_ISL_1409125, EPI_ISL_1409126,

| | | | |
|---|---|---|---|
| EPI_ISL_1409127, EPI_ISL_1409128, EPI_ISL_1409132, EPI_ISL_1409133, EPI_ISL_1409134, EPI_ISL_1409135, EPI_ISL_1409139, EPI_ISL_1409140, EPI_ISL_1409141, EPI_ISL_1409147, EPI_ISL_1409149, EPI_ISL_1409150, EPI_ISL_1409152, EPI_ISL_1409153, EPI_ISL_1409154, EPI_ISL_1409158, EPI_ISL_1409162, EPI_ISL_1409166, EPI_ISL_1409167, EPI_ISL_1409168, EPI_ISL_1409169, EPI_ISL_1409171, EPI_ISL_1409175, EPI_ISL_1409176, EPI_ISL_1409177, EPI_ISL_1409179, EPI_ISL_1409183, EPI_ISL_1409186, EPI_ISL_1409190, EPI_ISL_1409191, EPI_ISL_1409192, EPI_ISL_1409193, EPI_ISL_1409194, EPI_ISL_1409195, EPI_ISL_1409202, EPI_ISL_1409203, EPI_ISL_1409206, EPI_ISL_1409207, EPI_ISL_1409215, EPI_ISL_1409219, EPI_ISL_1409222, EPI_ISL_1409245, EPI_ISL_1409246, EPI_ISL_1409248, EPI_ISL_1409251, EPI_ISL_1409253, EPI_ISL_1409254, EPI_ISL_1409260, EPI_ISL_1409262, EPI_ISL_1409265, EPI_ISL_1409267, EPI_ISL_1409268, EPI_ISL_1409269, EPI_ISL_1409272, EPI_ISL_1409275, EPI_ISL_1409279, EPI_ISL_1409282, EPI_ISL_1409287, EPI_ISL_1409289, EPI_ISL_1409290, EPI_ISL_1409291, EPI_ISL_1409295, EPI_ISL_1409296, EPI_ISL_1409298, EPI_ISL_1409301, EPI_ISL_1409302, EPI_ISL_1409305, EPI_ISL_1409310, EPI_ISL_1409313, EPI_ISL_1409315, EPI_ISL_1409322, EPI_ISL_1409325, EPI_ISL_1409326, EPI_ISL_1409335, EPI_ISL_1409350, EPI_ISL_1409360, EPI_ISL_1409368, EPI_ISL_1409369, EPI_ISL_1409372, EPI_ISL_1409375, EPI_ISL_1409383, EPI_ISL_1409391, EPI_ISL_1409395, EPI_ISL_1409409, EPI_ISL_1409410, EPI_ISL_1409413, EPI_ISL_1409422, EPI_ISL_1409423, EPI_ISL_1409425, EPI_ISL_1409427, EPI_ISL_1409439, EPI_ISL_1409442, EPI_ISL_1409446, EPI_ISL_1409447, EPI_ISL_1409453, EPI_ISL_1409454, EPI_ISL_1409460, EPI_ISL_1409478, EPI_ISL_1409479, EPI_ISL_1409481, EPI_ISL_1409482, EPI_ISL_1409491, EPI_ISL_1409492, EPI_ISL_1409493, EPI_ISL_1411454, EPI_ISL_1411471, EPI_ISL_1411472, EPI_ISL_1411481, EPI_ISL_1411488, EPI_ISL_1411491, EPI_ISL_1411504, EPI_ISL_1411506, EPI_ISL_1411560, EPI_ISL_1411561, EPI_ISL_1411565, EPI_ISL_1411570, EPI_ISL_1411584, EPI_ISL_1411597, EPI_ISL_1411599, EPI_ISL_1411602, EPI_ISL_1411610, EPI_ISL_1411612, EPI_ISL_1411623, EPI_ISL_1411627, EPI_ISL_1411632, EPI_ISL_1411645, EPI_ISL_1411648, EPI_ISL_1411650, EPI_ISL_1411653, EPI_ISL_1411654, EPI_ISL_1411659, EPI_ISL_1411660, EPI_ISL_1411662, EPI_ISL_1411667, EPI_ISL_1411669, EPI_ISL_1411670, EPI_ISL_1411695, EPI_ISL_1411700, EPI_ISL_1411703, EPI_ISL_1411717, EPI_ISL_1411725, EPI_ISL_1411732, EPI_ISL_1411737, EPI_ISL_1411746, EPI_ISL_1411754, EPI_ISL_1411755, EPI_ISL_1411757, EPI_ISL_1411759, EPI_ISL_1411762, EPI_ISL_1411764, EPI_ISL_1411769, EPI_ISL_1411771, EPI_ISL_1411777, EPI_ISL_1411790, EPI_ISL_1411800, EPI_ISL_1411801, EPI_ISL_1411805, EPI_ISL_1411818, EPI_ISL_1411821, EPI_ISL_1411825, EPI_ISL_1411832, EPI_ISL_1411835, EPI_ISL_1411844, EPI_ISL_1448788, EPI_ISL_1448791, EPI_ISL_1448797, EPI_ISL_1448799, EPI_ISL_1448801, EPI_ISL_1448804, EPI_ISL_1448807, EPI_ISL_1448809, EPI_ISL_1448818, EPI_ISL_1448829, EPI_ISL_1448840, EPI_ISL_1448843, EPI_ISL_1448847, EPI_ISL_1448854, EPI_ISL_1448859, EPI_ISL_1448860, EPI_ISL_1448863, EPI_ISL_1448868, EPI_ISL_1448874, EPI_ISL_1448899, EPI_ISL_1448919, EPI_ISL_1448920, EPI_ISL_1448947, EPI_ISL_1448949, EPI_ISL_1448952, EPI_ISL_1448965, EPI_ISL_1448972, EPI_ISL_1448976, EPI_ISL_1448977, EPI_ISL_1448979, EPI_ISL_1448989, EPI_ISL_1448990, EPI_ISL_1448994, EPI_ISL_1449005, EPI_ISL_1449017, EPI_ISL_1449018, EPI_ISL_1449019, EPI_ISL_1449024, EPI_ISL_1449026, EPI_ISL_1449459, EPI_ISL_1449538, EPI_ISL_1449570, EPI_ISL_1449989, EPI_ISL_1449991, EPI_ISL_1449997, EPI_ISL_1450001, EPI_ISL_1450004, EPI_ISL_1450016, EPI_ISL_1450019, EPI_ISL_1450022, EPI_ISL_1450029, EPI_ISL_1450031, EPI_ISL_1450034, EPI_ISL_1450042, EPI_ISL_1450044, EPI_ISL_1450045, EPI_ISL_1450053, EPI_ISL_1450056, EPI_ISL_1450058, EPI_ISL_1450060, EPI_ISL_1450073, EPI_ISL_1450075, EPI_ISL_1450080, EPI_ISL_1450083, EPI_ISL_1450085, EPI_ISL_1450086, EPI_ISL_1450088, EPI_ISL_1450099, EPI_ISL_1450101, EPI_ISL_1450105, EPI_ISL_1450109, EPI_ISL_1450113, EPI_ISL_1450125, EPI_ISL_1450144, EPI_ISL_1450145, EPI_ISL_1450151, EPI_ISL_1450154, EPI_ISL_1450157, EPI_ISL_1450161, EPI_ISL_1450162, EPI_ISL_1450166, EPI_ISL_1450180, EPI_ISL_1450190, EPI_ISL_1450191, EPI_ISL_1450192, EPI_ISL_1450194, EPI_ISL_1450196, EPI_ISL_1450202, EPI_ISL_1450207, EPI_ISL_1450210, EPI_ISL_1450211, EPI_ISL_1450214, EPI_ISL_1450220, EPI_ISL_1450223, EPI_ISL_1450225, EPI_ISL_1450232, EPI_ISL_1450233, EPI_ISL_1450234, EPI_ISL_1450261, EPI_ISL_1450263, EPI_ISL_1450266, EPI_ISL_1450267, EPI_ISL_1450275, EPI_ISL_1450288, EPI_ISL_1450289, EPI_ISL_1450290, EPI_ISL_1450297, EPI_ISL_1450302, EPI_ISL_1450306, EPI_ISL_1450312, EPI_ISL_1450313, EPI_ISL_1450316, EPI_ISL_1450317, EPI_ISL_1450325, EPI_ISL_1450329, EPI_ISL_1450335, EPI_ISL_1450339, EPI_ISL_1450341, EPI_ISL_1450347, EPI_ISL_1450351, EPI_ISL_1450355, EPI_ISL_1450357, EPI_ISL_1450359, EPI_ISL_1450360, EPI_ISL_1450364, EPI_ISL_1450365, EPI_ISL_1450369, EPI_ISL_1450375, EPI_ISL_1450383, EPI_ISL_1450396, EPI_ISL_1450400, EPI_ISL_1450407, EPI_ISL_1450413, EPI_ISL_1450423, EPI_ISL_1450425, EPI_ISL_1450426, EPI_ISL_1450427, EPI_ISL_1450437, EPI_ISL_1450439, EPI_ISL_1450443, EPI_ISL_1450447, EPI_ISL_1450448, EPI_ISL_1450452, EPI_ISL_1450458, EPI_ISL_1450460, EPI_ISL_1450468, EPI_ISL_1450478, EPI_ISL_1450493, EPI_ISL_1450494, EPI_ISL_1450496, EPI_ISL_1450497, EPI_ISL_1450498, EPI_ISL_1450502, EPI_ISL_1450504, EPI_ISL_1450511, EPI_ISL_1450512, EPI_ISL_1450515, EPI_ISL_1450516, EPI_ISL_1450517, EPI_ISL_1450520, EPI_ISL_1450530, EPI_ISL_1450532, EPI_ISL_1450539, EPI_ISL_1450540, EPI_ISL_1450541, EPI_ISL_1450543, EPI_ISL_1450550, EPI_ISL_1450561, EPI_ISL_1450563, EPI_ISL_1450564, EPI_ISL_1450571, EPI_ISL_1450572, EPI_ISL_1450573, EPI_ISL_1450587, EPI_ISL_1450594, EPI_ISL_1450890, EPI_ISL_1450891, EPI_ISL_1450892, EPI_ISL_1450893, EPI_ISL_1450894, EPI_ISL_1450900, EPI_ISL_1450902, EPI_ISL_1450905, EPI_ISL_1450907, EPI_ISL_1450908, EPI_ISL_1450909, EPI_ISL_1450911, EPI_ISL_1450912, EPI_ISL_1450913, EPI_ISL_1450914, EPI_ISL_1450916, EPI_ISL_1450917, EPI_ISL_1450921, EPI_ISL_1450923, EPI_ISL_1450924, EPI_ISL_1450925, EPI_ISL_1450926, EPI_ISL_1450928, EPI_ISL_1450929, EPI_ISL_1450931, EPI_ISL_1450933, EPI_ISL_1450938, EPI_ISL_1450939, EPI_ISL_1450940, EPI_ISL_1450942, EPI_ISL_1450943, EPI_ISL_1450944, EPI_ISL_1450946, EPI_ISL_1450948, EPI_ISL_1450951, EPI_ISL_1450952, EPI_ISL_1450953, EPI_ISL_1450955, EPI_ISL_1450957, EPI_ISL_1450959, EPI_ISL_1450960, EPI_ISL_1450961, EPI_ISL_1450962, EPI_ISL_1450963, EPI_ISL_1450964, EPI_ISL_1450966, EPI_ISL_1450968, EPI_ISL_1450971, EPI_ISL_1450972, EPI_ISL_1450975, EPI_ISL_1450976, EPI_ISL_1450977, EPI_ISL_1450979, EPI_ISL_1450980, EPI_ISL_1450983, EPI_ISL_1450984, EPI_ISL_1450985, EPI_ISL_1450986, EPI_ISL_1450987, EPI_ISL_1450988, EPI_ISL_1450990, EPI_ISL_1450992, EPI_ISL_1450994, EPI_ISL_1450995, EPI_ISL_1450996, EPI_ISL_1450998, EPI_ISL_1451000, EPI_ISL_1451002, EPI_ISL_1451003, EPI_ISL_1451004, EPI_ISL_1451005, EPI_ISL_1451006, EPI_ISL_1451007, EPI_ISL_1451008, EPI_ISL_1451013, EPI_ISL_1451017, EPI_ISL_1451018, EPI_ISL_1451019, EPI_ISL_1451020, EPI_ISL_1451021, EPI_ISL_1451022, EPI_ISL_1451023, EPI_ISL_1451026, EPI_ISL_1451027, EPI_ISL_1451031, EPI_ISL_1451032, EPI_ISL_1451033, EPI_ISL_1451036, EPI_ISL_1451037, EPI_ISL_1451038, EPI_ISL_1451040, EPI_ISL_1451041, EPI_ISL_1451046, EPI_ISL_1451047, EPI_ISL_1451049, EPI_ISL_1451050, EPI_ISL_1451051, EPI_ISL_1451053, EPI_ISL_1451054, EPI_ISL_1451055, EPI_ISL_1451057, EPI_ISL_1451060, EPI_ISL_1451061, EPI_ISL_1451062, EPI_ISL_1451063, EPI_ISL_1451065, EPI_ISL_1451066, EPI_ISL_1451069, EPI_ISL_1451070, EPI_ISL_1451073, EPI_ISL_1451074, EPI_ISL_1451075, EPI_ISL_1451077, EPI_ISL_1451078, EPI_ISL_1451079, EPI_ISL_1451080, EPI_ISL_1451082, EPI_ISL_1451083, EPI_ISL_1451084, EPI_ISL_1451085, EPI_ISL_1451086, EPI_ISL_1451087, EPI_ISL_1451088, EPI_ISL_1451091, EPI_ISL_1451092, EPI_ISL_1451093, EPI_ISL_1451094, EPI_ISL_1451096, EPI_ISL_1451098, EPI_ISL_1451099, EPI_ISL_1451100, EPI_ISL_1451101, EPI_ISL_1451104, EPI_ISL_1451108, EPI_ISL_1451109, EPI_ISL_1451110, EPI_ISL_1451111, EPI_ISL_1451112, EPI_ISL_1451113, EPI_ISL_1451115, EPI_ISL_1451121, EPI_ISL_1451123, EPI_ISL_1451124, EPI_ISL_1451125, EPI_ISL_1451127, EPI_ISL_1451132, EPI_ISL_1451134, EPI_ISL_1451135, EPI_ISL_1451136, EPI_ISL_1451137, EPI_ISL_1451138, EPI_ISL_1451139, EPI_ISL_1451140, EPI_ISL_1451141, EPI_ISL_1451142, EPI_ISL_1451143, EPI_ISL_1451144, EPI_ISL_1451145, EPI_ISL_1451146, EPI_ISL_1451148, EPI_ISL_1451150, EPI_ISL_1451480, EPI_ISL_1451495, EPI_ISL_1451525, EPI_ISL_1451558, EPI_ISL_1451559, EPI_ISL_1451586, EPI_ISL_1451637, EPI_ISL_1451649, EPI_ISL_1451675, EPI_ISL_1451681, EPI_ISL_1451718, EPI_ISL_1451726, EPI_ISL_1451765, EPI_ISL_1451770, EPI_ISL_1451772, EPI_ISL_1451777, EPI_ISL_1452075, EPI_ISL_1452079, EPI_ISL_1452081, EPI_ISL_1452084, EPI_ISL_1452085, EPI_ISL_1452087, EPI_ISL_1452096, EPI_ISL_1452097, EPI_ISL_1452099, EPI_ISL_1452100, EPI_ISL_1452102, EPI_ISL_1452106, EPI_ISL_1452116, EPI_ISL_1452118, EPI_ISL_1452120, EPI_ISL_1452125, EPI_ISL_1452130, EPI_ISL_1452133, EPI_ISL_1452134, EPI_ISL_1452137, EPI_ISL_1452140, EPI_ISL_1452145, EPI_ISL_1452149, EPI_ISL_1452150, EPI_ISL_1452154, EPI_ISL_1452155, EPI_ISL_1452157, EPI_ISL_1452159, EPI_ISL_1452161, EPI_ISL_1452164, EPI_ISL_1452166, EPI_ISL_1452168, EPI_ISL_1452173, EPI_ISL_1452175, EPI_ISL_1452176, EPI_ISL_1452180, EPI_ISL_1452182, EPI_ISL_1452184, EPI_ISL_1452185, EPI_ISL_1452186, EPI_ISL_1452188, EPI_ISL_1452192, EPI_ISL_1452193, EPI_ISL_1452195, EPI_ISL_1452196, EPI_ISL_1452197, EPI_ISL_1452199, EPI_ISL_1452200, EPI_ISL_1452202, EPI_ISL_1452206, EPI_ISL_1452207, EPI_ISL_1452218, EPI_ISL_1452222, EPI_ISL_1452223, EPI_ISL_1452224, EPI_ISL_1452225, EPI_ISL_1452226, EPI_ISL_1452231, EPI_ISL_1452232, EPI_ISL_1452236, EPI_ISL_1452250, EPI_ISL_1452254, EPI_ISL_1452259, EPI_ISL_1452262, EPI_ISL_1452266, EPI_ISL_1452269, EPI_ISL_1452272, EPI_ISL_1452273, EPI_ISL_1452279, EPI_ISL_1452282, EPI_ISL_1452285, EPI_ISL_1452288, EPI_ISL_1452293, EPI_ISL_1452297, EPI_ISL_1452298, EPI_ISL_1452304, EPI_ISL_1452311, EPI_ISL_1452321, EPI_ISL_1452334, EPI_ISL_1452338, EPI_ISL_1452340, EPI_ISL_1452342, EPI_ISL_1452347, EPI_ISL_1452348, EPI_ISL_1452349, EPI_ISL_1452350, EPI_ISL_1452356, EPI_ISL_1452357, EPI_ISL_1452358, EPI_ISL_1452361, EPI_ISL_1452369, EPI_ISL_1452375, EPI_ISL_1452383, EPI_ISL_1452384, EPI_ISL_1452392, EPI_ISL_1452393, EPI_ISL_1452396, EPI_ISL_1452397, EPI_ISL_1452399, EPI_ISL_1452401, EPI_ISL_1452405, EPI_ISL_1452406, EPI_ISL_1452408, EPI_ISL_1452410, EPI_ISL_1452411, EPI_ISL_1452415, EPI_ISL_1452416, EPI_ISL_1452431, EPI_ISL_1452433, EPI_ISL_1452436, EPI_ISL_1452441, EPI_ISL_1452443, EPI_ISL_1452445, EPI_ISL_1452447, EPI_ISL_1452449, EPI_ISL_1452454, EPI_ISL_1452457, EPI_ISL_1452458, EPI_ISL_1452465, EPI_ISL_1452466, EPI_ISL_1452467, EPI_ISL_1452468, EPI_ISL_1452469, EPI_ISL_1452471, EPI_ISL_1452472, EPI_ISL_1452475, EPI_ISL_1452479, EPI_ISL_1452482, EPI_ISL_1452484, EPI_ISL_1452486, EPI_ISL_1452488, EPI_ISL_1452489, EPI_ISL_1452491, EPI_ISL_1452493, EPI_ISL_1452499, EPI_ISL_1452501, EPI_ISL_1452504, EPI_ISL_1452509, EPI_ISL_1452513, EPI_ISL_1452516, EPI_ISL_1452517, EPI_ISL_1452521, EPI_ISL_1452524, EPI_ISL_1452525, EPI_ISL_1452537, EPI_ISL_1452540, EPI_ISL_1452542, EPI_ISL_1452544, EPI_ISL_1452545, EPI_ISL_1452547, EPI_ISL_1452551, EPI_ISL_1452553, EPI_ISL_1452555, EPI_ISL_1452566, EPI_ISL_1452570, EPI_ISL_1452572, EPI_ISL_1452576, EPI_ISL_1452578, EPI_ISL_1452579, EPI_ISL_1452580, EPI_ISL_1452581, EPI_ISL_1452591, EPI_ISL_1452594, EPI_ISL_1452596, EPI_ISL_1452601, EPI_ISL_1452604, EPI_ISL_1452605, EPI_ISL_1452607, EPI_ISL_1452610, EPI_ISL_1452611, EPI_ISL_1452617, EPI_ISL_1452625, EPI_ISL_1452888, EPI_ISL_1452894, EPI_ISL_1452895, EPI_ISL_1452897, EPI_ISL_1452898, EPI_ISL_1452902, EPI_ISL_1452903, EPI_ISL_1452907, EPI_ISL_1452918, EPI_ISL_1452927, EPI_ISL_1452931, EPI_ISL_1452933, EPI_ISL_1452934, EPI_ISL_1452937, EPI_ISL_1452940, EPI_ISL_1452941, EPI_ISL_1452942, EPI_ISL_1452944, EPI_ISL_1452947, EPI_ISL_1452948, EPI_ISL_1452952, EPI_ISL_1452953, EPI_ISL_1452956, EPI_ISL_1452958, EPI_ISL_1452964, EPI_ISL_1452966, EPI_ISL_1452972, EPI_ISL_1452979, EPI_ISL_1452980, EPI_ISL_1452984, EPI_ISL_1452988, EPI_ISL_1452989, EPI_ISL_1452990, EPI_ISL_1452995, EPI_ISL_1452997, EPI_ISL_1452998, EPI_ISL_1453006, EPI_ISL_1453014, EPI_ISL_1453019, EPI_ISL_1453020, EPI_ISL_1453024, EPI_ISL_1453028, EPI_ISL_1453033, EPI_ISL_1453041, EPI_ISL_1453044, EPI_ISL_1453045, EPI_ISL_1453049, EPI_ISL_1453054, EPI_ISL_1453059, EPI_ISL_1453064, EPI_ISL_1453067, EPI_ISL_1453074, EPI_ISL_1453077, EPI_ISL_1453090, EPI_ISL_1453091, EPI_ISL_1453096, EPI_ISL_1453099, EPI_ISL_1453102, EPI_ISL_1453106, EPI_ISL_1453112, EPI_ISL_1453119, EPI_ISL_1453121, EPI_ISL_1453129, EPI_ISL_1453131, EPI_ISL_1453133, EPI_ISL_1453134, EPI_ISL_1453140, EPI_ISL_1453141, EPI_ISL_1453145, EPI_ISL_1453149, EPI_ISL_1453150, EPI_ISL_1453151, EPI_ISL_1453161, EPI_ISL_1453164, EPI_ISL_1453168, EPI_ISL_1453170, EPI_ISL_1453171, EPI_ISL_1453172, EPI_ISL_1453173, EPI_ISL_1453176, EPI_ISL_1453177, EPI_ISL_1453180, EPI_ISL_1453472, EPI_ISL_1453474, EPI_ISL_1453477, EPI_ISL_1453483, EPI_ISL_1453486, EPI_ISL_1453488, EPI_ISL_1453498, EPI_ISL_1453500, EPI_ISL_1453504, EPI_ISL_1453506, EPI_ISL_1453507, EPI_ISL_1453511, EPI_ISL_1453513, EPI_ISL_1453518, EPI_ISL_1453519, EPI_ISL_1453521, EPI_ISL_1453526, EPI_ISL_1453534, EPI_ISL_1453536, EPI_ISL_1453537, EPI_ISL_1453540, EPI_ISL_1453548, EPI_ISL_1453550, EPI_ISL_1453555, EPI_ISL_1453559, EPI_ISL_1453564, EPI_ISL_1453567, EPI_ISL_1453569, EPI_ISL_1453571, EPI_ISL_1453572, EPI_ISL_1453573, EPI_ISL_1453577, EPI_ISL_1453579, EPI_ISL_1453582, EPI_ISL_1453583, EPI_ISL_1453584, EPI_ISL_1453586, EPI_ISL_1453595, EPI_ISL_1453603, EPI_ISL_1453604, EPI_ISL_1453615, EPI_ISL_1453626, EPI_ISL_1453644, EPI_ISL_1453646, EPI_ISL_1453648, EPI_ISL_1453655, EPI_ISL_1453659, EPI_ISL_1453660, EPI_ISL_1453665, EPI_ISL_1453677, EPI_ISL_1453678, EPI_ISL_1453692, EPI_ISL_1453699, EPI_ISL_1453700, EPI_ISL_1453702, EPI_ISL_1453705, EPI_ISL_1453706, EPI_ISL_1453708, EPI_ISL_1453711, EPI_ISL_1453712, EPI_ISL_1453714, EPI_ISL_1453718, EPI_ISL_1453736, EPI_ISL_1453749, EPI_ISL_1453753, EPI_ISL_1453764, EPI_ISL_1453772, EPI_ISL_1453777, EPI_ISL_1453779, EPI_ISL_1453781, EPI_ISL_1453782, EPI_ISL_1453783, EPI_ISL_1453787, EPI_ISL_1453791, EPI_ISL_1453793, EPI_ISL_1453794, EPI_ISL_1453796, EPI_ISL_1453799, EPI_ISL_1453802, EPI_ISL_1453803, EPI_ISL_1453804, EPI_ISL_1453808, EPI_ISL_1453814, EPI_ISL_1453818, EPI_ISL_1453819, EPI_ISL_1453821, EPI_ISL_1453827, EPI_ISL_1453830, EPI_ISL_1453832, EPI_ISL_1453833, EPI_ISL_1453834, EPI_ISL_1453838, EPI_ISL_1453846, EPI_ISL_1453847, EPI_ISL_1454651, EPI_ISL_1454654, EPI_ISL_1454658, EPI_ISL_1454671, EPI_ISL_1454680, EPI_ISL_1454681, EPI_ISL_1454689, EPI_ISL_1454692, EPI_ISL_1454693, EPI_ISL_1454694, EPI_ISL_1454697, EPI_ISL_1454707, EPI_ISL_1454710, EPI_ISL_1454715, EPI_ISL_1454718, EPI_ISL_1454720, EPI_ISL_1454721, EPI_ISL_1454724, EPI_ISL_1454731, EPI_ISL_1454733, EPI_ISL_1454740, EPI_ISL_1454742, EPI_ISL_1454748, EPI_ISL_1454751, EPI_ISL_1454753, EPI_ISL_1454759, EPI_ISL_1454760, EPI_ISL_1454764, EPI_ISL_1454769, EPI_ISL_1454776, EPI_ISL_1454802, EPI_ISL_1454804, EPI_ISL_1454805, EPI_ISL_1454811, EPI_ISL_1454813, EPI_ISL_1454817, EPI_ISL_1454833, EPI_ISL_1454842, EPI_ISL_1454843, EPI_ISL_1454849, EPI_ISL_1454850, EPI_ISL_1454855, EPI_ISL_1454861, EPI_ISL_1454863, EPI_ISL_1454864, EPI_ISL_1454872, EPI_ISL_1454874, EPI_ISL_1454876, EPI_ISL_1454882, EPI_ISL_1454888, EPI_ISL_1454889, EPI_ISL_1454893, EPI_ISL_1454897, EPI_ISL_1454904, EPI_ISL_1454907, EPI_ISL_1454910, EPI_ISL_1454911, EPI_ISL_1454917, EPI_ISL_1454929, EPI_ISL_1454932, EPI_ISL_1454933, EPI_ISL_1454935, EPI_ISL_1454938, EPI_ISL_1454942, EPI_ISL_1454945, EPI_ISL_1454947, EPI_ISL_1454966, EPI_ISL_1466444, EPI_ISL_1466469, EPI_ISL_1466621, EPI_ISL_1466641, EPI_ISL_1466653, EPI_ISL_1466786, EPI_ISL_1466788, EPI_ISL_1466795, EPI_ISL_1466829, EPI_ISL_1466832, EPI_ISL_1466896, EPI_ISL_1466916, EPI_ISL_1466917, EPI_ISL_1467297, EPI_ISL_1467928, EPI_ISL_1467965, EPI_ISL_1468022, EPI_ISL_1468123, EPI_ISL_1472627, EPI_ISL_1472660, EPI_ISL_1519182 | | | |
| see above | Lighthouse Lab in Milton Keynes | Wellcome Sanger Institute for the COVID-19 Genomics UK (COG-UK) Consortium | Cordelia Langford; David K. Jackson; Dominic Kwiatkowski; Ewan Harrison; Ian Johnston; Jeffrey Barrett; John Sillitoe on behalf of the Wellcome Sanger Institute COVID-19 Surveillance Team; Roberto Amato; Sonia Goncalves; The Lighthouse Lab in Milton Keynes and Alex Alderton |
| EPI_ISL_1475538, EPI_ISL_1475539, EPI_ISL_1475540, EPI_ISL_1475541, EPI_ISL_1475542 | Lincolnshire Hospitals and DeepSeq Nottingham | COVID-19 Genomics UK (COG-UK) Consortium | Christopher Moore; Fei Sang; Joeseph Chappell; Johnny Debebe; Jonathan Ball; Matthew Carlisle; Matthew Loose; Nadine Holmes; Nichola Duckworth; Patrick McClure; Sarah Walsh; Tim Sloan; Victoria Wright |
| EPI_ISL_1474738, EPI_ISL_1474740, EPI_ISL_1474741, EPI_ISL_1474742, EPI_ISL_1474743, EPI_ISL_1474754, EPI_ISL_1474755, EPI_ISL_1474757, EPI_ISL_1474762, EPI_ISL_1474764, EPI_ISL_1474773, EPI_ISL_1474774, EPI_ISL_1474776, EPI_ISL_1474783, EPI_ISL_1474784, EPI_ISL_1474786, EPI_ISL_1474787, EPI_ISL_1474788, EPI_ISL_1546175, EPI_ISL_1546176, EPI_ISL_1546177, EPI_ISL_1546178, EPI_ISL_1546179, EPI_ISL_1546180, EPI_ISL_1546181, EPI_ISL_1546185, EPI_ISL_1546186, EPI_ISL_1546187, EPI_ISL_1546188, EPI_ISL_1546189, EPI_ISL_1636826, EPI_ISL_1636838, EPI_ISL_1637246, EPI_ISL_1637249, EPI_ISL_1637250, EPI_ISL_1637252, EPI_ISL_1637253 | | | |
| see above | Liverpool Clinical Laboratories | COVID-19 Genomics UK (COG-UK) Consortium | A Alrezaihi; Alessandro Gerada; Alistair Darby; Angela Cowell; Anita Lucaci; Anu Chawla; Cassie Olateju; Catherine Hartley; Charlotte Nelson; Ecaterina Vamos; Elaine O'Toole; Eleanor G Bentley; Ghada T Shawli; Hermione Webster; Isabel García-Dorival; James Johnson; James P Stewart; Jenifer Manson; Joanne Watts; Jones Benjamin; Jordan J Clark; Julian Hiscox; L Luu; Lucille Rainbow; M Almsaud; Margaret Hughes; Mark Whitehead; Matthew Gemmell; Miren Iturriza-Gomara; Muhannad Alruwaili; N.P Randle; Neil Swainston; PKF Gilmore; Parul Sharma; Rebekah Penrice-Randal; Richard Eccles; Richard Gregory; Sam Haldenby; Steve Paterson; Stuart D Armstrong; Trevor Ian Robinson; Ximeng Han |
| EPI_ISL_1387081, EPI_ISL_1387082, EPI_ISL_1387083, EPI_ISL_1387108, EPI_ISL_1387128, EPI_ISL_1387129, EPI_ISL_1387131, EPI_ISL_1387132, EPI_ISL_1475208, EPI_ISL_1475209 | | | |
| see above | Northumbria University / South Tees Hospitals NHS Foundation Trust / North Cumbria Integrated Care NHS Foundation Trust / North Tees and Hartlepool NHS Foundation Trust / Newcastle Hospitals NHS Foundation Trust | COVID-19 Genomics UK (COG-UK) Consortium | Andrew Nelson; Brendan Payne; Clive Graham; Darren L Smith; Debra Padgett; Edward Barton; Emma Swindells; Garren Scott; Gary Black; Gary Eltringham; Giles S Holt; Greg R Young; Jane Greenaway; Jennifer Collins; John Allan; Joshua Loh; Lynn Dover; Matthew Bashton; Mohammad A Tariq; Paul Baker; Sarah Essex; Steve Liggett; Wen C Yew; Yusri Taha |
| EPI_ISL_1475759, EPI_ISL_1475760, EPI_ISL_1475767, EPI_ISL_1475768, EPI_ISL_1475772, EPI_ISL_1475773, EPI_ISL_1475776, EPI_ISL_1475779, EPI_ISL_1475781, EPI_ISL_1475782, EPI_ISL_1475783, EPI_ISL_1475784, EPI_ISL_1475786, EPI_ISL_1475788, EPI_ISL_1475789, EPI_ISL_1475791, EPI_ISL_1475792, EPI_ISL_1475794, EPI_ISL_1475795, EPI_ISL_1475798, EPI_ISL_1475799, EPI_ISL_1475801, EPI_ISL_1475810, EPI_ISL_1475818, EPI_ISL_1475823, EPI_ISL_1475824, EPI_ISL_1475825, EPI_ISL_1475827, EPI_ISL_1475830, EPI_ISL_1475833, EPI_ISL_1475836, EPI_ISL_1475837, EPI_ISL_1475838, EPI_ISL_1475840, EPI_ISL_1475841, EPI_ISL_1475844, EPI_ISL_1475850, EPI_ISL_1475852, EPI_ISL_1475853, EPI_ISL_1475854, EPI_ISL_1475855, EPI_ISL_1475858, EPI_ISL_1475861, EPI_ISL_1475863, EPI_ISL_1475865, EPI_ISL_1475868, EPI_ISL_1475869, EPI_ISL_1475870, EPI_ISL_1475871, EPI_ISL_1475872, EPI_ISL_1475876, EPI_ISL_1475879, EPI_ISL_1475881, EPI_ISL_1475882, EPI_ISL_1475883, EPI_ISL_1475884, EPI_ISL_1475885, EPI_ISL_1475886, EPI_ISL_1475887, EPI_ISL_1475888, EPI_ISL_1475889, EPI_ISL_1475894, EPI_ISL_1475895, EPI_ISL_1475896, EPI_ISL_1475897, EPI_ISL_1475902, EPI_ISL_1475903, EPI_ISL_1475909, EPI_ISL_1475910, EPI_ISL_1475911, EPI_ISL_1475912, EPI_ISL_1475914, EPI_ISL_1475915, EPI_ISL_1475916, EPI_ISL_1475917, EPI_ISL_1475919, EPI_ISL_1475924, EPI_ISL_1475926, EPI_ISL_1475928, EPI_ISL_1475930, EPI_ISL_1475931, EPI_ISL_1475932, EPI_ISL_1475934, EPI_ISL_1475935, EPI_ISL_1475937, EPI_ISL_1475938, EPI_ISL_1475942, EPI_ISL_1475947, EPI_ISL_1475956, EPI_ISL_1475963, EPI_ISL_1475964, EPI_ISL_1475965, EPI_ISL_1475971, EPI_ISL_1475972, EPI_ISL_1475977, EPI_ISL_1475978, EPI_ISL_1475983, EPI_ISL_1475987, EPI_ISL_1475988, EPI_ISL_1475989, EPI_ISL_1475990, EPI_ISL_1475991, EPI_ISL_1475993, EPI_ISL_1475994, EPI_ISL_1475995, EPI_ISL_1475997, EPI_ISL_1476001, EPI_ISL_1476004, EPI_ISL_1476005, EPI_ISL_1476006, EPI_ISL_1476007, EPI_ISL_1476008, EPI_ISL_1476010, EPI_ISL_1476012, EPI_ISL_1476015, EPI_ISL_1476016, EPI_ISL_1476018, EPI_ISL_1476023, EPI_ISL_1476024, EPI_ISL_1476025, EPI_ISL_1476027, EPI_ISL_1476028, EPI_ISL_1476033, EPI_ISL_1476037, EPI_ISL_1476039, EPI_ISL_1476041, EPI_ISL_1476043, EPI_ISL_1476046, EPI_ISL_1476050, EPI_ISL_1476054, EPI_ISL_1476062, EPI_ISL_1476065, EPI_ISL_1476072, EPI_ISL_1476074, EPI_ISL_1476103, EPI_ISL_1476104, EPI_ISL_1476105, EPI_ISL_1476109, EPI_ISL_1476114, EPI_ISL_1476118, EPI_ISL_1476119, EPI_ISL_1476120, EPI_ISL_1476121, EPI_ISL_1476123, EPI_ISL_1476124, EPI_ISL_1476126, EPI_ISL_1476129, EPI_ISL_1476131, EPI_ISL_1476134, EPI_ISL_1476137, EPI_ISL_1476138, EPI_ISL_1476140, EPI_ISL_1476141, EPI_ISL_1476142, EPI_ISL_1476143, EPI_ISL_1476149, EPI_ISL_1476151, EPI_ISL_1476156, EPI_ISL_1476157, EPI_ISL_1476168, EPI_ISL_1476173, EPI_ISL_1476174, EPI_ISL_1476207, EPI_ISL_1476223, EPI_ISL_1476227, EPI_ISL_1476228, EPI_ISL_1476243, EPI_ISL_1476245, EPI_ISL_1476246, EPI_ISL_1476249, EPI_ISL_1476250, EPI_ISL_1476252, EPI_ISL_1476254, EPI_ISL_1476268, EPI_ISL_1476275, EPI_ISL_1476278, EPI_ISL_1476279, EPI_ISL_1476284, EPI_ISL_1476285, EPI_ISL_1476287, EPI_ISL_1476289, EPI_ISL_1476290, EPI_ISL_1476292, EPI_ISL_1476388, EPI_ISL_1476397, EPI_ISL_1476398, EPI_ISL_1476439, EPI_ISL_1476440, EPI_ISL_1546762, EPI_ISL_1546764, EPI_ISL_1546786, EPI_ISL_1547035, EPI_ISL_1547073 | | | |
| see above | Originating lab: Wales Specialist Virology Centre Sequencing lab: Pathogen Genomics Unit | Public Health Wales Microbiology Cardiff Wales Specialist Virology Centre | Alec Birchley; Alexander Adams; Amy Gaskin; Angela Marchbank; Bree Gatica-Wilcox; Catherine Moore; Jason Coombes; Joanne Watkins; Joel Southgate; Johnathan Evans; Laura Gifford; Lauren Gilbert; Lee Graham; Malorie Perry; Matthew Bull; Nicole Pacchiarini; Sally Corden; Sara Kumziene-Summerhayes; Sara Rey; Sarah Taylor; Simon Cottrell; Sophie Jones; Tom Connor |
| EPI_ISL_1730426, EPI_ISL_1730431, EPI_ISL_1730451, EPI_ISL_1730452, EPI_ISL_1730454, EPI_ISL_1730456, EPI_ISL_1730457, EPI_ISL_1730458, EPI_ISL_1730459, EPI_ISL_1730460, EPI_ISL_1730461, EPI_ISL_1730462, EPI_ISL_1730463, EPI_ISL_1730464, EPI_ISL_1730465, EPI_ISL_1730466, EPI_ISL_1730468 | | | |
| see above | Oxford Viromics, NDM, University of Oxford; Oxford University Hospitals; Basingstoke and North Hampshire Hospital | COVID-19 Genomics UK (COG-UK) Consortium | Alex Mobbs; Amy Trebes; Anita Justice; Catrin Moore; Christophe Fraser; David Bonsall; David Buck; Emma Wise; George Macintyre; Jessica Lynch; John Todd; Mariateresa de Cesare; Matilde Mori; Monique Andersson; Nathan Moore; Nick Cortes; Robert Shaw; Stephen Kidd; Tanya Golubchik; Timothy Peto |
| EPI_ISL_1387136, EPI_ISL_1387141, EPI_ISL_1387142, EPI_ISL_1832738, EPI_ISL_1832740, EPI_ISL_1832750, EPI_ISL_1832753, EPI_ISL_1832758, EPI_ISL_1832767 | | | |
| see above | Quadram Institute Bioscience | COVID-19 Genomics UK (COG-UK) Consortium | Alexander J Trotter; Alison E. Mather; Alp Aydin; Ana P. Tedim; Anastasia Kolyva; Andrew Bell; Andrew J. Page; Claire Stuart; Dave J. Baker; Gemma L. Kay; John Wain; Justin O'Grady; Leonardo de Oliveira Martins; Lizzie Meadows; Maria Diaz; Mark Webber; Muhammed Yasir; Nabil-Fareed Alikhan; Ngozi Elumogo; Nicholas M. Thomson; Rachael Stanley; Rachel Gilroy; Reenesh Prakash; Samir Dervisevic; Samuel Bloomfield; Steven Rudder; Thanh Le-Viet |
| EPI_ISL_1387163, EPI_ISL_1387164, EPI_ISL_1387165, EPI_ISL_1387190, EPI_ISL_1387191, EPI_ISL_1387192, EPI_ISL_1387195, EPI_ISL_1546242 | | | |
| see above | Queens Medical Centre, Clinical Microbiology Department / DeepSeq Nottingham | COVID-19 Genomics UK (COG-UK) Consortium | Christopher Moore; Emily Park; Fei Sang; Gemma Clark; Hannah Howson-Wells; Jack Hill; Jayasree Dey; Johnny Debebe; Jonathan Ball; Joseph Chappell; Manjinder Khakh; Matthew Carlisle; Matthew Loose; Michelle M Lister; Nadine Holmes; Patrick McClure; Theocharis Tsoleridis; Timothy Byaruhanga; Vicki M Fleming; Victoria Wright; Wendy Smith |
| EPI_ISL_1409185, EPI_ISL_1409188, EPI_ISL_1409201, EPI_ISL_1409209, EPI_ISL_1409212, EPI_ISL_1409217, EPI_ISL_1409230, EPI_ISL_1409235, EPI_ISL_1409240, EPI_ISL_1409242, EPI_ISL_1409252, EPI_ISL_1409255, EPI_ISL_1409258, EPI_ISL_1409259, EPI_ISL_1409271, EPI_ISL_1409283, EPI_ISL_1409297, EPI_ISL_1409304, EPI_ISL_1409311, EPI_ISL_1409316, EPI_ISL_1409321, EPI_ISL_1409330, EPI_ISL_1409333, EPI_ISL_1409338, EPI_ISL_1409340, EPI_ISL_1409343, EPI_ISL_1409359, EPI_ISL_1409362, EPI_ISL_1409367, EPI_ISL_1409373, EPI_ISL_1409386, EPI_ISL_1409388, EPI_ISL_1409392, EPI_ISL_1409394, EPI_ISL_1409401, EPI_ISL_1409411, EPI_ISL_1409416, EPI_ISL_1409428, EPI_ISL_1409430, EPI_ISL_1409432, EPI_ISL_1409433, EPI_ISL_1409437, EPI_ISL_1409448, EPI_ISL_1409449, EPI_ISL_1409456, EPI_ISL_1409462, EPI_ISL_1409475, EPI_ISL_1409485, EPI_ISL_1411124, EPI_ISL_1411211, EPI_ISL_1411266, EPI_ISL_1411379, EPI_ISL_1411416, EPI_ISL_1411459, EPI_ISL_1411485, EPI_ISL_1411500, EPI_ISL_1411505, EPI_ISL_1411591, EPI_ISL_1411594, EPI_ISL_1411617, EPI_ISL_1411644, EPI_ISL_1411674, EPI_ISL_1411692, EPI_ISL_1411704, EPI_ISL_1411720, EPI_ISL_1411733, EPI_ISL_1411734, EPI_ISL_1411767, EPI_ISL_1411773, EPI_ISL_1411780, EPI_ISL_1411783, EPI_ISL_1411792, EPI_ISL_1411808, EPI_ISL_1411824, EPI_ISL_1411831, EPI_ISL_1411845, EPI_ISL_1451469, EPI_ISL_1451474, EPI_ISL_1451477, EPI_ISL_1451478, EPI_ISL_1451489, EPI_ISL_1451490, EPI_ISL_1451493, EPI_ISL_1451498, EPI_ISL_1451499, EPI_ISL_1451500, EPI_ISL_1451506, EPI_ISL_1451511, EPI_ISL_1451513, EPI_ISL_1451515, EPI_ISL_1451518, EPI_ISL_1451521, EPI_ISL_1451523, EPI_ISL_1451528, EPI_ISL_1451529, EPI_ISL_1451530, EPI_ISL_1451532, EPI_ISL_1451536, EPI_ISL_1451542, EPI_ISL_1451543, EPI_ISL_1451550, EPI_ISL_1451551, EPI_ISL_1451557, EPI_ISL_1451563, EPI_ISL_1451565, EPI_ISL_1451568, EPI_ISL_1451574, EPI_ISL_1451578, EPI_ISL_1451582, EPI_ISL_1451583, EPI_ISL_1451584, EPI_ISL_1451587, EPI_ISL_1451589, EPI_ISL_1451592, EPI_ISL_1451593, EPI_ISL_1451594, EPI_ISL_1451598, EPI_ISL_1451603, EPI_ISL_1451604, EPI_ISL_1451605, EPI_ISL_1451608, EPI_ISL_1451610, EPI_ISL_1451612, EPI_ISL_1451614, EPI_ISL_1451616, EPI_ISL_1451617, EPI_ISL_1451619, EPI_ISL_1451622, EPI_ISL_1451624, EPI_ISL_1451629, EPI_ISL_1451630, EPI_ISL_1451631, EPI_ISL_1451632, EPI_ISL_1451635, EPI_ISL_1451642, EPI_ISL_1451643, EPI_ISL_1451644, EPI_ISL_1451650, EPI_ISL_1451652, EPI_ISL_1451658, EPI_ISL_1451659, EPI_ISL_1451660, EPI_ISL_1451661, EPI_ISL_1451664, EPI_ISL_1451665, EPI_ISL_1451679, EPI_ISL_1451696, EPI_ISL_1451699, EPI_ISL_1451700, EPI_ISL_1451701, EPI_ISL_1451703, EPI_ISL_1451704, EPI_ISL_1451705, EPI_ISL_1451712, EPI_ISL_1451714, EPI_ISL_1451716, EPI_ISL_1451721, EPI_ISL_1451722, EPI_ISL_1451723, EPI_ISL_1451725, EPI_ISL_1451727, EPI_ISL_1451733, EPI_ISL_1451738, EPI_ISL_1451743, EPI_ISL_1451748, EPI_ISL_1451749, EPI_ISL_1451755, EPI_ISL_1451758, EPI_ISL_1451759, EPI_ISL_1451763, EPI_ISL_1451764, EPI_ISL_1451768, EPI_ISL_1451771, EPI_ISL_1451774, EPI_ISL_1451780, EPI_ISL_1451781, EPI_ISL_1451782, EPI_ISL_1451783, EPI_ISL_1454961, EPI_ISL_1454964, EPI_ISL_1466987, EPI_ISL_1467038, EPI_ISL_1467206, EPI_ISL_1467252, EPI_ISL_1467293, EPI_ISL_1467418, EPI_ISL_1467460, EPI_ISL_1489493, EPI_ISL_1520019 | | | |
| see above | Randox Laboratories | Wellcome Sanger Institute for the COVID-19 Genomics UK (COG-UK) Consortium | Cordelia Langford; David K. Jackson; Dominic Kwiatkowski; Ewan Harrison; Ian Johnston; Jeffrey Barrett; John Sillitoe on behalf of the Wellcome Sanger Institute COVID-19 Surveillance Team; Randox Laboratories and Alex Alderton; Roberto Amato; Sonia Goncalves |
| EPI_ISL_1976753, EPI_ISL_1976756, EPI_ISL_1976770, EPI_ISL_1976783, EPI_ISL_1976788, EPI_ISL_1976791, EPI_ISL_1976793, EPI_ISL_1976797, EPI_ISL_1976798, EPI_ISL_1976800, EPI_ISL_1976801, EPI_ISL_1976835, EPI_ISL_1976843, EPI_ISL_1976844, EPI_ISL_1976868, EPI_ISL_1976869, EPI_ISL_1976870, EPI_ISL_1976871, EPI_ISL_1976872, EPI_ISL_1976887, EPI_ISL_1976889, EPI_ISL_1976891, EPI_ISL_1976904, EPI_ISL_1977211, EPI_ISL_1977213, EPI_ISL_1977215, EPI_ISL_1977216, EPI_ISL_1977218, EPI_ISL_1977219, EPI_ISL_1977222, EPI_ISL_1977224, EPI_ISL_1977226, EPI_ISL_1977228, EPI_ISL_1977229, EPI_ISL_1977231, EPI_ISL_1977232, EPI_ISL_1977234, EPI_ISL_1977268, EPI_ISL_1977270, EPI_ISL_1977271, EPI_ISL_1977272, EPI_ISL_1977275, EPI_ISL_1977298, EPI_ISL_1977300, EPI_ISL_1977301, EPI_ISL_1977303, EPI_ISL_1977304, EPI_ISL_1977306, EPI_ISL_1977308, EPI_ISL_1977309, EPI_ISL_1977312, EPI_ISL_1977317, EPI_ISL_1977322, EPI_ISL_1977324, EPI_ISL_1977358, EPI_ISL_1977360, EPI_ISL_1977362, EPI_ISL_1977363, EPI_ISL_1977365, EPI_ISL_1977366, EPI_ISL_1977368, EPI_ISL_1977369, EPI_ISL_1977379, EPI_ISL_1977381, EPI_ISL_1977386, EPI_ISL_1977387, EPI_ISL_1977449, EPI_ISL_1977451, EPI_ISL_1978093, EPI_ISL_1978094, EPI_ISL_1978096, EPI_ISL_1978098, EPI_ISL_1978100, EPI_ISL_1978101, EPI_ISL_1978103, EPI_ISL_1978105, EPI_ISL_1978106, EPI_ISL_1978108, EPI_ISL_1978110, EPI_ISL_1978112, EPI_ISL_1978113, EPI_ISL_1978115, EPI_ISL_1978117, EPI_ISL_1978118, EPI_ISL_1978120, EPI_ISL_1978122, EPI_ISL_1978123, EPI_ISL_1978125, EPI_ISL_1978127, EPI_ISL_1978129, EPI_ISL_1978130, EPI_ISL_1978132, EPI_ISL_1978134, EPI_ISL_1978135, EPI_ISL_1978137, EPI_ISL_1978139, EPI_ISL_1978141, EPI_ISL_1978142, EPI_ISL_1978144, EPI_ISL_1978145, EPI_ISL_1978147, EPI_ISL_1978234, EPI_ISL_1978235, EPI_ISL_1978253 | | | |
| see above | Regional Virus Laboratory, Belfast Health and Social Care Trust | COVID-19 Genomics UK (COG-UK) Consortium | Alison Watt; Ciara Cox; Conall McCaughey; David Simpson; Derek Fairley; James McKenna; Mairead Connor; Susan Feeney; Tanya Curran; Zoltan Molnar |
| EPI_ISL_1637052 | University College London Hospital | COVID-19 Genomics UK | Dr Catherine Houlihan; Dr Daniel Frampton; Dr Judith Heaney; Dr Moira Spyer; Dr Paul Grant and Dr Eleni Nastouli; Matthew Byott; Stuart Kirk |

| | | | |
|---|---|---|---|
| | | (COG-UK) Consortium | |
| EPI_ISL_1386813, EPI_ISL_1386824, EPI_ISL_1386825, EPI_ISL_1386827, EPI_ISL_1386828, EPI_ISL_1386829, EPI_ISL_1386841, EPI_ISL_1386843, EPI_ISL_1386928, EPI_ISL_1386929, EPI_ISL_1386930, EPI_ISL_1386931, EPI_ISL_1474833, EPI_ISL_1474835, EPI_ISL_1474840, EPI_ISL_1474841, EPI_ISL_1474869, EPI_ISL_1474881, EPI_ISL_1637166 | | | |
| see above | University College London, Great Ormond Street Hospital for Children NHS Foundation Trust, Imperial College Healthcare NHS Trust | COVID-19 Genomics UK (COG-UK) Consortium | Alison Holmes; Charlotte Williams; Helena Tutill; Jacqueline Findlay; James Price; Judith Breuer; Julianne Brown; Kathryn Harris; Leysa Forrest; Mark Kristiansen; Paola Niola; Paola Resende Silva; Patricia Dyal; Paul Randell; Rachel Williams; Samuel Weeks; Sergi Castellano; Sunando Roy; Tony Brooks; Yasmin Panchbhaya |
| EPI_ISL_1545974, EPI_ISL_1545982 | University Hospitals Of Leicester NHS Trust and DeepSeq Nottingham | COVID-19 Genomics UK (COG-UK) Consortium | Christopher Holmes; Christopher Moore; Fei Sang; Joeseph Chappell; Johnny Debebe; Jonathan Ball; Julian Tang; Karlie Fallon; Matthew Carlisle; Matthew Loose; Nadine Holmes; Patrick McClure; Paul Bird; Thomas Helmer; Victoria Wright |
| EPI_ISL_1386519, EPI_ISL_1386520, EPI_ISL_1386522, EPI_ISL_1386524, EPI_ISL_1386525, EPI_ISL_1386526, EPI_ISL_1386530, EPI_ISL_1386531, EPI_ISL_1386532, EPI_ISL_1386538, EPI_ISL_1386565, EPI_ISL_1386566, EPI_ISL_1386567, EPI_ISL_1386568, EPI_ISL_1386569, EPI_ISL_1386570, EPI_ISL_1474434, EPI_ISL_1474435, EPI_ISL_1474442, EPI_ISL_1729967, EPI_ISL_1729968, EPI_ISL_1729974, EPI_ISL_1729975, EPI_ISL_1729979, EPI_ISL_1729980, EPI_ISL_1729983 | | | |
| see above | University of Birmingham | COVID-19 Genomics UK (COG-UK) Consortium | Alex Richter; Andrew Bosworth; Andrew Bosworth. Queen Elizabeth Hospital: Anna Casey; Andrew D Beggs PHE Heartlands Lab: Husam Osman; Charlotte Poxon; Institute of Microbiology; Joanne Stockton; Josh Quick; Kasun Wanigasooriya; Mike Kidd; Nicholas Loman. University of Birmingham Testing Laboratory: Celina M Whalley; Oliver Pickles; Radoslaw Poplawski; Samuel Nicholls; University of Birmingham: Claire McMurray; Will Rowe |
| EPI_ISL_1545951 | University of Exeter | COVID-19 Genomics UK (COG-UK) Consortium | Aaron Jeffries; Audrey Farbos; Ben Temperton; Jane Masoli; Joanna Warwick-Dugdale; Michelle Michelsen; Robyn Manley; Stephen Michell |
| EPI_ISL_1386706, EPI_ISL_1474584, EPI_ISL_1474585, EPI_ISL_1545907, EPI_ISL_1545908, EPI_ISL_1545909, EPI_ISL_1545910, EPI_ISL_1545932, EPI_ISL_1545937, EPI_ISL_1636671, EPI_ISL_1636672, EPI_ISL_1636684, EPI_ISL_1636685, EPI_ISL_1636696, EPI_ISL_1730076, EPI_ISL_1974892, EPI_ISL_1974893, EPI_ISL_1974894, EPI_ISL_1974907, EPI_ISL_1974908, EPI_ISL_1974914, EPI_ISL_2125863, EPI_ISL_2125885 | | | |
| see above | Virology Department, Royal Infirmary of Edinburgh, NHS Lothian / School of Biological Sciences, University of Edinburgh | COVID-19 Genomics UK (COG-UK) Consortium | Colquhoun R; Cotton S; Dewar R; Gallagher A; Hill V; Jackson B; Maloney D; McCrone JT; McHugh M; O'Toole Á; Rambaut A; Rooke S; Scher E; Templeton K; Yu X |
| EPI_ISL_1389073, EPI_ISL_1389079, EPI_ISL_1389082, EPI_ISL_1389088, EPI_ISL_1389096, EPI_ISL_1389122, EPI_ISL_1389134, EPI_ISL_1476695, EPI_ISL_1476708, EPI_ISL_1476713, EPI_ISL_1476721, EPI_ISL_1476724, EPI_ISL_1476727, EPI_ISL_1476729, EPI_ISL_1476732, EPI_ISL_1476734, EPI_ISL_1476736, EPI_ISL_1476737, EPI_ISL_1476738, EPI_ISL_1476740, EPI_ISL_1547346, EPI_ISL_1547356, EPI_ISL_1731029, EPI_ISL_1731034, EPI_ISL_1833885 | | | |
| see above | Virology Department, Sheffield Teaching Hospitals NHS Foundation Trust/Department of Infection, Immunity and Cardiovascular Disease, The Medical School, University of Sheffield | COVID-19 Genomics UK (COG-UK) Consortium | Adri Angyal; Alex Keeley; Benjamin Lindsey; Cariad Evans; Danielle Groves; Dave Partridge; Katie Johnson; Laura Carrilero; Luke Green; Matthew Parker; Matthew Wyles; Mehmet Yavuz; Mohammad Raza; Nikki Smith; Paul Parsons; Rachel Tucker; Rebecca Brown; Thushan de Silva |
| EPI_ISL_1475761, EPI_ISL_1475764, EPI_ISL_1475766, EPI_ISL_1475769, EPI_ISL_1475770, EPI_ISL_1475774, EPI_ISL_1475785, EPI_ISL_1475790, EPI_ISL_1475860, EPI_ISL_1475864, EPI_ISL_1475875, EPI_ISL_1475893, EPI_ISL_1475898, EPI_ISL_1475899, EPI_ISL_1475900, EPI_ISL_1475905, EPI_ISL_1475906, EPI_ISL_1475929, EPI_ISL_1475939, EPI_ISL_1475940, EPI_ISL_1475944, EPI_ISL_1475946, EPI_ISL_1475952, EPI_ISL_1475955, EPI_ISL_1475957, EPI_ISL_1475958, EPI_ISL_1475959, EPI_ISL_1475962, EPI_ISL_1475966, EPI_ISL_1476003, EPI_ISL_1476011, EPI_ISL_1476017, EPI_ISL_1476045, EPI_ISL_1476048, EPI_ISL_1476063, EPI_ISL_1476064, EPI_ISL_1476067, EPI_ISL_1476068, EPI_ISL_1476071, EPI_ISL_1476075, EPI_ISL_1476139, EPI_ISL_1476144, EPI_ISL_1476146, EPI_ISL_1476147, EPI_ISL_1476150, EPI_ISL_1476158, EPI_ISL_1476159, EPI_ISL_1476161, EPI_ISL_1476162, EPI_ISL_1476163, EPI_ISL_1476167, EPI_ISL_1476209, EPI_ISL_1476210, EPI_ISL_1476213, EPI_ISL_1476214, EPI_ISL_1476216, EPI_ISL_1476217, EPI_ISL_1476220, EPI_ISL_1476221, EPI_ISL_1476229, EPI_ISL_1476230, EPI_ISL_1476234, EPI_ISL_1476283, EPI_ISL_1476408, EPI_ISL_1476409, EPI_ISL_1476413, EPI_ISL_1476417, EPI_ISL_1546761, EPI_ISL_1546765, EPI_ISL_1546766, EPI_ISL_1637307, EPI_ISL_1637322, EPI_ISL_1637324, EPI_ISL_1637325, EPI_ISL_1637326, EPI_ISL_1637327, EPI_ISL_1637329, EPI_ISL_1637335, EPI_ISL_1637336, EPI_ISL_1637338, EPI_ISL_1637340, EPI_ISL_1637341, EPI_ISL_1637343, EPI_ISL_1637345, EPI_ISL_1637346, EPI_ISL_1637350, EPI_ISL_1637351, EPI_ISL_1637352, EPI_ISL_1637353, EPI_ISL_1637354, EPI_ISL_1637355, EPI_ISL_1637357, EPI_ISL_1637358, EPI_ISL_1637359, EPI_ISL_1637360, EPI_ISL_1637368, EPI_ISL_1637371, EPI_ISL_1637372, EPI_ISL_1637373, EPI_ISL_1637374, EPI_ISL_1637375, EPI_ISL_1637376, EPI_ISL_1637377, EPI_ISL_1637378, EPI_ISL_1637383, EPI_ISL_1637384, EPI_ISL_1637385, EPI_ISL_1637386, EPI_ISL_1637395, EPI_ISL_1637398, EPI_ISL_1637401, EPI_ISL_1637404, EPI_ISL_1637406, EPI_ISL_1637407, EPI_ISL_1637408, EPI_ISL_1637409, EPI_ISL_1637415, EPI_ISL_1637416, EPI_ISL_1637418, EPI_ISL_1637423, EPI_ISL_1637425, EPI_ISL_1637427, EPI_ISL_1637429, EPI_ISL_1637432, EPI_ISL_1637433, EPI_ISL_1637434, EPI_ISL_1637436, EPI_ISL_1637437, EPI_ISL_1637446, EPI_ISL_1637447, EPI_ISL_1637450, EPI_ISL_1637451, EPI_ISL_1637454, EPI_ISL_1637455, EPI_ISL_1833155, EPI_ISL_1833156, EPI_ISL_1833163 | | | |
| see above | Wales Specialist Virology Centre Sequencing lab: Pathogen Genomics Unit | Public Health Wales Microbiology Cardiff Wales Specialist Virology Centre | Alec Birchley; Alexander Adams; Amy Gaskin; Angela Marchbank; Bree Gatica-Wilcox; Catherine Moore; Jason Coombes; Joanne Watkins; Joel Southgate; Johnathan Evans; Laura Gifford; Lauren Gilbert; Lee Graham; Malorie Perry; Matthew Bull; Nicole Pacchiarini; Sally Corden; Sara Kumziene-Summerhayes; Sara Rey; Sarah Taylor; Simon Cottrell; Sophie Jones; Tom Connor |
| EPI_ISL_1545867, EPI_ISL_1545868 | West of Scotland Specialist Virology Centre, NHSGGC / MRC-University of Glasgow Centre for Virus Research | COVID-19 Genomics UK (COG-UK) Consortium | Alasdair MacLean; Alice Broos; Ana da Silva Filipe; Antonia Ho; Daniel Mair; David L Robertson; Emma Thomson; Guy Mollett; James Shepherd; Jenna Nichols; Joseph Hughes; Kathy Li; Kathy Smollett; Kyriaki Nomikou; Lily Tong; Matthew Holden; Natasha Johnson; Rachel Blacow; Richard Orton; Rory Gunson; Sarah McDonald; Sharif Shaaban; Sreenu Vattipally; Stephen Carmichael |

We gratefully acknowledge the following Authors from the Originating laboratories responsible for obtaining the specimens, as well as the Submitting laboratories where the genome data were generated and shared via GISAID, on which this research is based.

All Submitters of data may be contacted directly via www.gisaid.org

Authors are sorted alphabetically.

| Accession ID | Originating Laboratory | Submitting Laboratory | Authors |
|---|---|---|---|
| EPI_ISL_1636961, EPI_ISL_1636964, EPI_ISL_2126021 | Barts Health NHS Trust | COVID-19 Genomics UK (COG-UK) Consortium | BROAD; Beatrix; CUTINO-MOGUEL; Claire; David; Dola; HARRINGTON; KELE; KULASEGARAN-SHYLINI; Maria-Teresa; OWOYEMI; Raghavendran |
| EPI_ISL_1389139 | Bioinformatics and Biostatistics Lab, Advanced Sequencing Facility | COVID-19 Genomics UK (COG-UK) Consortium | Aengus Stewart; Chelsea Sawyer; Harshil Patel; Jerome Nicod; Laura Cubitt; Margaret Crawford |
| EPI_ISL_1388808, EPI_ISL_1476674, EPI_ISL_1476676, EPI_ISL_1476677, EPI_ISL_1476679, EPI_ISL_1476680, EPI_ISL_1476681, EPI_ISL_1476682, EPI_ISL_1547232, EPI_ISL_1547233, EPI_ISL_1547234, EPI_ISL_1547235, EPI_ISL_1547236, EPI_ISL_1547237, EPI_ISL_1547242, EPI_ISL_1637985, EPI_ISL_1637988, EPI_ISL_1637989, EPI_ISL_1637997, EPI_ISL_1638014, EPI_ISL_1638015, EPI_ISL_1638016 | | | |
| see above | Centre for Enzyme Innovation, University of Portsmouth / Translational Research Laboratory, Portsmouth Hospitals NHS Trust | COVID-19 Genomics UK (COG-UK) Consortium | Angela Beckett; Christopher Fearn; Kate Cook; Katie Loveson; Salman Goudarzi; Samuel Robson; Scott Elliott; Sharon Glaysher |
| EPI_ISL_1678813 | Department of Medical Microbiology & Infection prevention, Amsterdam University Medical Centers location AMC | Department of Medical Microbiology & Infection prevention, Amsterdam University Medical Centers location AMC | Fokla Zorgdrager; Janke Schinkel; Marcel Jonges; Matthijs Welkers; Menno de Jong; Robin van Houdt; Sebastien Matamoros; Sjoerd Rebers |
| EPI_ISL_1386581, EPI_ISL_1474572, EPI_ISL_1636603, EPI_ISL_1636604, EPI_ISL_1636610, EPI_ISL_1636611, EPI_ISL_1636616, EPI_ISL_1636619, EPI_ISL_1636620, EPI_ISL_1730024, EPI_ISL_1730032 | | | |
| see above | Department of Pathology, University of Cambridge | COVID-19 Genomics UK (COG-UK) Consortium | Aminu S. Jahun; Ian Goodfellow; Iliana Georgana; Martin D. Curran; Myra Hosmillo; Rhys Izuagbe; Surendra Parmar; William L. Hamilton; Yasmin Chaudhry |
| EPI_ISL_1449029, EPI_ISL_1449030, EPI_ISL_1449031, EPI_ISL_1449032, EPI_ISL_1449034, EPI_ISL_1449035, EPI_ISL_1449036, EPI_ISL_1449037, EPI_ISL_1449039, EPI_ISL_1449040, EPI_ISL_1449041, EPI_ISL_1449042, EPI_ISL_1449043, EPI_ISL_1449045, EPI_ISL_1449046, EPI_ISL_1449047, EPI_ISL_1449048, EPI_ISL_1449049, EPI_ISL_1449050, EPI_ISL_1449051, EPI_ISL_1449052, EPI_ISL_1449053, EPI_ISL_1449055, EPI_ISL_1449056, EPI_ISL_1449057, EPI_ISL_1449058, EPI_ISL_1449059, EPI_ISL_1449060, EPI_ISL_1449061, EPI_ISL_1449063, EPI_ISL_1449064, EPI_ISL_1449065, EPI_ISL_1449066, EPI_ISL_1449069, EPI_ISL_1449070, EPI_ISL_1449071, EPI_ISL_1449072, EPI_ISL_1449073, EPI_ISL_1449074, EPI_ISL_1449075, EPI_ISL_1449076, EPI_ISL_1449077, EPI_ISL_1449078, EPI_ISL_1449079, EPI_ISL_1449080, EPI_ISL_1449082, EPI_ISL_1449083, EPI_ISL_1449084, EPI_ISL_1449085, EPI_ISL_1449086, EPI_ISL_1449087, EPI_ISL_1449088, EPI_ISL_1449089, EPI_ISL_1449091, EPI_ISL_1449092, EPI_ISL_1449093, EPI_ISL_1449094, EPI_ISL_1449095, EPI_ISL_1449096, EPI_ISL_1449097, EPI_ISL_1449098, EPI_ISL_1449099, EPI_ISL_1449100, EPI_ISL_1449101, EPI_ISL_1449102, EPI_ISL_1449103, EPI_ISL_1449104, EPI_ISL_1449105, EPI_ISL_1449106, EPI_ISL_1449107, EPI_ISL_1449108, EPI_ISL_1449109, EPI_ISL_1449110, EPI_ISL_1449113, EPI_ISL_1449115, EPI_ISL_1449116, EPI_ISL_1449117, EPI_ISL_1449118, EPI_ISL_1449119, EPI_ISL_1449120, EPI_ISL_1449121, EPI_ISL_1449122, EPI_ISL_1449123, EPI_ISL_1449124, EPI_ISL_1449125, EPI_ISL_1449126, EPI_ISL_1449127, EPI_ISL_1449128, EPI_ISL_1449129, EPI_ISL_1449132, EPI_ISL_1449133, EPI_ISL_1449134, EPI_ISL_1449135, EPI_ISL_1449136, EPI_ISL_1449137, EPI_ISL_1449138, EPI_ISL_1449139, EPI_ISL_1449140, EPI_ISL_1449141, EPI_ISL_1449142, EPI_ISL_1449143, EPI_ISL_1449144, EPI_ISL_1449145, EPI_ISL_1449146, EPI_ISL_1449147, EPI_ISL_1449148, EPI_ISL_1449150, EPI_ISL_1449151, EPI_ISL_1449152, EPI_ISL_1449153, EPI_ISL_1449154, EPI_ISL_1449156, EPI_ISL_1449157, EPI_ISL_1449158, EPI_ISL_1449159, EPI_ISL_1449160, EPI_ISL_1449161, EPI_ISL_1449162, EPI_ISL_1449163, EPI_ISL_1449164, EPI_ISL_1449165, EPI_ISL_1449166, EPI_ISL_1449167, EPI_ISL_1449168, EPI_ISL_1449169, EPI_ISL_1449170, EPI_ISL_1449171, EPI_ISL_1449172, EPI_ISL_1449173, EPI_ISL_1449174, EPI_ISL_1449175, EPI_ISL_1449176, EPI_ISL_1449177, EPI_ISL_1449178, EPI_ISL_1449179, EPI_ISL_1449180, EPI_ISL_1449181, EPI_ISL_1449182, EPI_ISL_1449183, EPI_ISL_1449184, EPI_ISL_1449185, EPI_ISL_1449186, EPI_ISL_1449187, EPI_ISL_1449189, EPI_ISL_1449191, EPI_ISL_1449192, EPI_ISL_1449193, EPI_ISL_1449194, EPI_ISL_1449195, EPI_ISL_1449196, EPI_ISL_1449197, EPI_ISL_1449198, EPI_ISL_1449199, EPI_ISL_1449201, EPI_ISL_1449202, EPI_ISL_1449203, EPI_ISL_1449204, EPI_ISL_1449206, EPI_ISL_1449207, EPI_ISL_1449208, EPI_ISL_1449209, EPI_ISL_1449210, EPI_ISL_1449211, EPI_ISL_1449213, EPI_ISL_1449214, EPI_ISL_1449216, EPI_ISL_1449217, EPI_ISL_1449218, EPI_ISL_1449219, EPI_ISL_1449220, EPI_ISL_1449221, EPI_ISL_1449222, EPI_ISL_1449223, EPI_ISL_1449224, EPI_ISL_1449225, EPI_ISL_1449226, EPI_ISL_1449227, EPI_ISL_1449228, EPI_ISL_1449230, EPI_ISL_1449231, EPI_ISL_1449232, EPI_ISL_1449233, EPI_ISL_1449234, EPI_ISL_1449236, EPI_ISL_1449237, EPI_ISL_1449238, EPI_ISL_1449239, EPI_ISL_1449240, EPI_ISL_1449241, EPI_ISL_1449242, EPI_ISL_1449244, EPI_ISL_1449245, EPI_ISL_1449246, EPI_ISL_1449247, EPI_ISL_1449248, EPI_ISL_1449249, EPI_ISL_1449250, EPI_ISL_1449251, EPI_ISL_1449252, EPI_ISL_1449253, EPI_ISL_1449254, EPI_ISL_1449255, EPI_ISL_1449256, EPI_ISL_1449257, EPI_ISL_1449258, EPI_ISL_1449259, EPI_ISL_1449260, EPI_ISL_1449261, EPI_ISL_1449262, EPI_ISL_1449263, EPI_ISL_1449264, EPI_ISL_1449265, EPI_ISL_1449266, EPI_ISL_1449267, EPI_ISL_1449268, EPI_ISL_1449269, EPI_ISL_1449270, EPI_ISL_1449272, EPI_ISL_1449273, EPI_ISL_1449274, EPI_ISL_1449275, EPI_ISL_1449276, EPI_ISL_1449277, EPI_ISL_1449278, EPI_ISL_1449279, EPI_ISL_1449280, EPI_ISL_1449281, EPI_ISL_1449282, EPI_ISL_1449283, EPI_ISL_1449285, EPI_ISL_1449286, EPI_ISL_1449287, EPI_ISL_1449288, EPI_ISL_1449289, EPI_ISL_1449290, EPI_ISL_1449291, EPI_ISL_1449292, EPI_ISL_1449293, EPI_ISL_1449294, EPI_ISL_1449296, EPI_ISL_1449297, EPI_ISL_1449298, EPI_ISL_1449300, EPI_ISL_1449301, EPI_ISL_1449302, EPI_ISL_1449303, EPI_ISL_1449304, EPI_ISL_1449305, EPI_ISL_1449306, EPI_ISL_1449307, EPI_ISL_1449308, EPI_ISL_1449309, EPI_ISL_1449310, EPI_ISL_1449312, EPI_ISL_1449313, EPI_ISL_1449314, EPI_ISL_1449315, EPI_ISL_1449316, EPI_ISL_1449317, EPI_ISL_1449318, EPI_ISL_1449319, EPI_ISL_1449320, EPI_ISL_1449321, EPI_ISL_1449322, EPI_ISL_1449323, EPI_ISL_1449324, EPI_ISL_1449325, EPI_ISL_1449326, EPI_ISL_1449327, EPI_ISL_1449328, EPI_ISL_1449329, EPI_ISL_1449330, EPI_ISL_1449331, EPI_ISL_1449333, EPI_ISL_1449334, EPI_ISL_1449335, EPI_ISL_1449336, EPI_ISL_1449337, EPI_ISL_1449338, EPI_ISL_1449339, EPI_ISL_1449340, EPI_ISL_1449341, EPI_ISL_1449342, EPI_ISL_1449343, EPI_ISL_1449344, EPI_ISL_1449345, EPI_ISL_1449346, EPI_ISL_1449347, EPI_ISL_1449470, EPI_ISL_1450167, EPI_ISL_1451855, EPI_ISL_1453894, EPI_ISL_1466161, EPI_ISL_1466163, EPI_ISL_1466164, EPI_ISL_1466165, EPI_ISL_1466166, EPI_ISL_1466167, EPI_ISL_1466168, EPI_ISL_1466169, EPI_ISL_1466170, EPI_ISL_1466171, EPI_ISL_1466172, EPI_ISL_1466174, EPI_ISL_1466175, EPI_ISL_1466176, EPI_ISL_1466177, EPI_ISL_1466178, EPI_ISL_1466179, EPI_ISL_1466180, EPI_ISL_1466181, EPI_ISL_1466182, EPI_ISL_1466183, EPI_ISL_1466185, EPI_ISL_1466186, EPI_ISL_1466187, EPI_ISL_1466188, EPI_ISL_1466189, EPI_ISL_1466190, EPI_ISL_1466191, EPI_ISL_1466192, EPI_ISL_1466193, EPI_ISL_1466197, EPI_ISL_1466198, EPI_ISL_1466199, EPI_ISL_1466200, EPI_ISL_1466201, EPI_ISL_1466202, EPI_ISL_1466203, EPI_ISL_1466204, EPI_ISL_1466205, EPI_ISL_1466206, EPI_ISL_1466207, EPI_ISL_1466208, EPI_ISL_1466209, EPI_ISL_1466210, EPI_ISL_1466211, EPI_ISL_1466212, EPI_ISL_1466213, EPI_ISL_1466214, EPI_ISL_1466215, EPI_ISL_1466216, EPI_ISL_1466217, EPI_ISL_1466218, EPI_ISL_1466219, EPI_ISL_1466220, EPI_ISL_1466221, EPI_ISL_1466222, EPI_ISL_1466223, EPI_ISL_1466224, EPI_ISL_1466225, EPI_ISL_1466226, EPI_ISL_1466227, EPI_ISL_1466228, EPI_ISL_1466230, EPI_ISL_1466231, EPI_ISL_1466232, EPI_ISL_1466233, EPI_ISL_1466234, EPI_ISL_1466236, EPI_ISL_1466237, EPI_ISL_1466238, EPI_ISL_1466240, EPI_ISL_1466241, EPI_ISL_1466243, EPI_ISL_1466244, EPI_ISL_1466245, EPI_ISL_1466248, EPI_ISL_1466249, EPI_ISL_1466250, EPI_ISL_1466253, EPI_ISL_1466255, EPI_ISL_1466256, EPI_ISL_1466257, EPI_ISL_1466258, EPI_ISL_1466259, EPI_ISL_1466261, EPI_ISL_1466262, EPI_ISL_1466263, EPI_ISL_1466264, EPI_ISL_1466265, EPI_ISL_1466266, EPI_ISL_1466267, EPI_ISL_1466268, EPI_ISL_1466269, EPI_ISL_1466272, EPI_ISL_1466273, EPI_ISL_1466274, EPI_ISL_1466275, EPI_ISL_1466276, EPI_ISL_1466277, EPI_ISL_1466278, EPI_ISL_1466279, EPI_ISL_1466280, EPI_ISL_1466282, EPI_ISL_1466285, EPI_ISL_1466286, EPI_ISL_1466288, EPI_ISL_1466289, EPI_ISL_1466291, EPI_ISL_1466292, EPI_ISL_1466293, EPI_ISL_1466294, EPI_ISL_1466295, EPI_ISL_1466297, EPI_ISL_1466298, EPI_ISL_1466299, EPI_ISL_1466300, EPI_ISL_1466301, EPI_ISL_1466302, EPI_ISL_1466303, EPI_ISL_1466304, EPI_ISL_1466305, EPI_ISL_1466307, EPI_ISL_1466308, EPI_ISL_1466310, EPI_ISL_1466311, EPI_ISL_1466312, EPI_ISL_1466313, EPI_ISL_1466315, EPI_ISL_1466317, EPI_ISL_1466318, EPI_ISL_1466319, EPI_ISL_1466320, EPI_ISL_1466321, EPI_ISL_1466324, EPI_ISL_1466327, EPI_ISL_1466328, EPI_ISL_1466329, EPI_ISL_1466330, EPI_ISL_1466331, EPI_ISL_1466332, EPI_ISL_1466333, EPI_ISL_1466336, EPI_ISL_1466337, EPI_ISL_1466339, EPI_ISL_1466340, EPI_ISL_1466341, EPI_ISL_1466342, EPI_ISL_1466343, EPI_ISL_1466345, EPI_ISL_1466346, EPI_ISL_1466347, EPI_ISL_1466348, EPI_ISL_1466349, EPI_ISL_1466350, EPI_ISL_1466351, EPI_ISL_1466352, EPI_ISL_1466353, EPI_ISL_1466354, EPI_ISL_1466355, EPI_ISL_1466356, EPI_ISL_1466357, EPI_ISL_1466358, EPI_ISL_1466359, EPI_ISL_1466360, EPI_ISL_1466361, EPI_ISL_1466363, EPI_ISL_1466365, EPI_ISL_1466366, EPI_ISL_1466367, EPI_ISL_1466368, EPI_ISL_1466369, EPI_ISL_1466370, EPI_ISL_1466371, EPI_ISL_1466372, EPI_ISL_1466373, EPI_ISL_1466376, EPI_ISL_1466377, EPI_ISL_1466378, EPI_ISL_1466379, EPI_ISL_1466380, EPI_ISL_1466382, EPI_ISL_1466383, EPI_ISL_1466384, EPI_ISL_1466385, EPI_ISL_1466386, EPI_ISL_1466387, EPI_ISL_1466388, EPI_ISL_1466389, EPI_ISL_1466390, EPI_ISL_1466391, EPI_ISL_1466392, EPI_ISL_1466393, EPI_ISL_1466394, EPI_ISL_1466395, EPI_ISL_1466396, EPI_ISL_1466397, EPI_ISL_1466399, EPI_ISL_1466400, EPI_ISL_1466401, EPI_ISL_1466403, EPI_ISL_1466404, EPI_ISL_1466405, EPI_ISL_1466406, EPI_ISL_1466407, EPI_ISL_1466408, EPI_ISL_1466410, EPI_ISL_1466411, EPI_ISL_1466412, EPI_ISL_1466413, EPI_ISL_1466414, EPI_ISL_1466415, EPI_ISL_1466416, EPI_ISL_1466418, EPI_ISL_1466419, EPI_ISL_1466421, EPI_ISL_1466422, EPI_ISL_1466423, EPI_ISL_1466424, EPI_ISL_1466425, EPI_ISL_1466427, EPI_ISL_1466429, EPI_ISL_1466430, EPI_ISL_1466431, EPI_ISL_1466432, EPI_ISL_1466433, EPI_ISL_1466434, EPI_ISL_1466435, EPI_ISL_1466436, EPI_ISL_1466438, EPI_ISL_1466439, EPI_ISL_1466441, EPI_ISL_1466442, EPI_ISL_1466443, EPI_ISL_1466445, EPI_ISL_1466447, EPI_ISL_1466450, EPI_ISL_1466451, EPI_ISL_1466452, EPI_ISL_1466454, EPI_ISL_1466455, EPI_ISL_1466456, EPI_ISL_1466458, EPI_ISL_1466459, EPI_ISL_1466460, EPI_ISL_1466461, EPI_ISL_1466464, EPI_ISL_1466465, EPI_ISL_1466466, EPI_ISL_1466467, EPI_ISL_1466468, EPI_ISL_1466470, EPI_ISL_1466472, EPI_ISL_1466473, EPI_ISL_1466475, EPI_ISL_1466477, EPI_ISL_1466479, EPI_ISL_1466481, EPI_ISL_1466483, EPI_ISL_1466485, EPI_ISL_1466488, EPI_ISL_1466489, EPI_ISL_1466490, EPI_ISL_1466494, EPI_ISL_1466497, EPI_ISL_1466500, EPI_ISL_1466503, EPI_ISL_1466504, EPI_ISL_1466507, EPI_ISL_1466510, EPI_ISL_1466512, EPI_ISL_1466514, EPI_ISL_1466515, EPI_ISL_1466518, EPI_ISL_1466519, EPI_ISL_1466520, EPI_ISL_1466522, EPI_ISL_1466523, EPI_ISL_1466525, EPI_ISL_1466526, EPI_ISL_1466529, EPI_ISL_1466530, EPI_ISL_1466534, EPI_ISL_1466535, EPI_ISL_1466537, EPI_ISL_1466539, EPI_ISL_1466540, EPI_ISL_1466541, EPI_ISL_1466544, EPI_ISL_1466545, EPI_ISL_1466546, EPI_ISL_1466547, EPI_ISL_1466548, EPI_ISL_1466549, EPI_ISL_1466552, EPI_ISL_1466554, EPI_ISL_1466555, EPI_ISL_1466558, EPI_ISL_1466562, EPI_ISL_1466563, EPI_ISL_1466566, EPI_ISL_1466567, EPI_ISL_1466568, EPI_ISL_1466570, EPI_ISL_1466571, EPI_ISL_1466572, EPI_ISL_1466573, EPI_ISL_1466574, EPI_ISL_1466575, EPI_ISL_1466576, EPI_ISL_1466578, EPI_ISL_1466579, EPI_ISL_1466582, EPI_ISL_1466583, EPI_ISL_1466585, EPI_ISL_1466586, EPI_ISL_1466588, EPI_ISL_1466591, EPI_ISL_1466592, EPI_ISL_1466603, EPI_ISL_1466605, EPI_ISL_1466607, EPI_ISL_1466608, EPI_ISL_1466609, EPI_ISL_1466611, EPI_ISL_1466613, EPI_ISL_1466614, EPI_ISL_1466615, EPI_ISL_1466618, EPI_ISL_1466619, EPI_ISL_1466623, EPI_ISL_1466624, EPI_ISL_1466625, EPI_ISL_1466629, EPI_ISL_1466631, EPI_ISL_1466632, EPI_ISL_1466633, EPI_ISL_1466635, EPI_ISL_1466636, EPI_ISL_1466637, EPI_ISL_1466638, EPI_ISL_1466640, EPI_ISL_1466643, EPI_ISL_1466646, EPI_ISL_1466647, EPI_ISL_1466651, EPI_ISL_1466657, EPI_ISL_1466662, EPI_ISL_1466664, EPI_ISL_1466669, EPI_ISL_1466670, EPI_ISL_1466671, EPI_ISL_1466672, EPI_ISL_1466676, EPI_ISL_1466677, EPI_ISL_1466681, EPI_ISL_1466684, EPI_ISL_1466689, EPI_ISL_1466690, EPI_ISL_1466692, EPI_ISL_1466699, EPI_ISL_1466700, EPI_ISL_1466703, EPI_ISL_1466704, EPI_ISL_1466705, EPI_ISL_1466706, EPI_ISL_1466712, EPI_ISL_1466713, EPI_ISL_1466714, EPI_ISL_1466715, EPI_ISL_1466716, EPI_ISL_1466717, EPI_ISL_1466718, EPI_ISL_1466720, EPI_ISL_1466723, EPI_ISL_1466726, EPI_ISL_1466727, EPI_ISL_1466729, EPI_ISL_1466731, EPI_ISL_1466732, EPI_ISL_1466734, EPI_ISL_1466735, EPI_ISL_1466740, EPI_ISL_1466743, EPI_ISL_1466923, EPI_ISL_1466924, EPI_ISL_1466925, EPI_ISL_1466926, EPI_ISL_1466927, EPI_ISL_1466928, EPI_ISL_1466929, EPI_ISL_1466930, EPI_ISL_1466931, EPI_ISL_1466932, EPI_ISL_1466933, EPI_ISL_1466935, EPI_ISL_1466936, EPI_ISL_1466937, EPI_ISL_1466938, EPI_ISL_1466940, EPI_ISL_1466941, EPI_ISL_1466942, EPI_ISL_1466943, EPI_ISL_1466944, EPI_ISL_1466945, EPI_ISL_1466947, EPI_ISL_1466948, EPI_ISL_1466950, EPI_ISL_1466952, EPI_ISL_1466953, EPI_ISL_1466954, EPI_ISL_1466961, EPI_ISL_1466963, EPI_ISL_1466964, EPI_ISL_1466965, EPI_ISL_1466966, EPI_ISL_1466967, EPI_ISL_1466968, EPI_ISL_1466970, EPI_ISL_1466971, EPI_ISL_1466972, EPI_ISL_1466973, EPI_ISL_1466974, EPI_ISL_1466976, EPI_ISL_1466977, EPI_ISL_1466978, EPI_ISL_1466979, EPI_ISL_1466980, EPI_ISL_1466981, EPI_ISL_1466982, EPI_ISL_1466984, EPI_ISL_1466985, EPI_ISL_1466986, EPI_ISL_1466988, EPI_ISL_1466989, EPI_ISL_1466994, EPI_ISL_1466996, EPI_ISL_1466998, EPI_ISL_1466999, EPI_ISL_1467001, EPI_ISL_1467003, EPI_ISL_1467004, EPI_ISL_1467005, EPI_ISL_1467006, EPI_ISL_1467007, EPI_ISL_1467008, EPI_ISL_1467011, EPI_ISL_1467012, EPI_ISL_1467013, EPI_ISL_1467014, EPI_ISL_1467016, EPI_ISL_1467020, EPI_ISL_1467021, EPI_ISL_1467022, EPI_ISL_1467023, EPI_ISL_1467024, EPI_ISL_1467025, EPI_ISL_1467027, EPI_ISL_1467028, EPI_ISL_1467030, EPI_ISL_1467031, EPI_ISL_1467032, EPI_ISL_1467033, EPI_ISL_1467035, EPI_ISL_1467036, EPI_ISL_1467037, EPI_ISL_1467039, EPI_ISL_1467040, EPI_ISL_1467042, EPI_ISL_1467043, EPI_ISL_1467045, EPI_ISL_1467047, EPI_ISL_1467048, EPI_ISL_1467050, EPI_ISL_1467053, EPI_ISL_1467056, EPI_ISL_1467057, EPI_ISL_1467060, EPI_ISL_1467062, EPI_ISL_1467063, EPI_ISL_1467065, EPI_ISL_1467066, EPI_ISL_1467069, EPI_ISL_1467070, EPI_ISL_1467071, EPI_ISL_1467073, EPI_ISL_1467074, EPI_ISL_1467076, EPI_ISL_1467078, EPI_ISL_1467079, EPI_ISL_1467081, EPI_ISL_1467084, EPI_ISL_1467085, EPI_ISL_1467086, EPI_ISL_1467087, EPI_ISL_1467089, EPI_ISL_1467090, EPI_ISL_1467091, EPI_ISL_1467092, EPI_ISL_1467093, EPI_ISL_1467094, EPI_ISL_1467095, EPI_ISL_1467096, EPI_ISL_1467097, EPI_ISL_1467099, EPI_ISL_1467101, EPI_ISL_1467102, EPI_ISL_1467103, EPI_ISL_1467104, EPI_ISL_1467105, EPI_ISL_1467106, EPI_ISL_1467107, EPI_ISL_1467108, EPI_ISL_1467109, EPI_ISL_1467110, EPI_ISL_1467111, EPI_ISL_1467114, EPI_ISL_1467116, EPI_ISL_1467117, EPI_ISL_1467118, EPI_ISL_1467119, EPI_ISL_1467120, EPI_ISL_1467121, EPI_ISL_1467123, EPI_ISL_1467124, EPI_ISL_1467125, EPI_ISL_1467126, EPI_ISL_1467127, EPI_ISL_1467129, EPI_ISL_1467130, EPI_ISL_1467132, EPI_ISL_1467133, EPI_ISL_1467135, EPI_ISL_1467136, EPI_ISL_1467137, EPI_ISL_1467141, EPI_ISL_1467142, EPI_ISL_1467143, EPI_ISL_1467144, EPI_ISL_1467145, EPI_ISL_1467146, EPI_ISL_1467148, EPI_ISL_1467149, EPI_ISL_1467150, EPI_ISL_1467151, EPI_ISL_1467152, EPI_ISL_1467153, EPI_ISL_1467154, EPI_ISL_1467155, EPI_ISL_1467156, EPI_ISL_1467157, EPI_ISL_1467158, EPI_ISL_1467159, EPI_ISL_1467160, EPI_ISL_1467161, EPI_ISL_1467162, EPI_ISL_1467163, EPI_ISL_1467164, EPI_ISL_1467166, EPI_ISL_1467168, EPI_ISL_1467169, EPI_ISL_1467172, EPI_ISL_1467173, EPI_ISL_1467174, EPI_ISL_1467175, EPI_ISL_1467176, EPI_ISL_1467177, EPI_ISL_1467179, EPI_ISL_1467181, EPI_ISL_1467183, EPI_ISL_1467185, EPI_ISL_1467186, EPI_ISL_1467189, EPI_ISL_1467191, EPI_ISL_1467193, EPI_ISL_1467194, EPI_ISL_1467195, EPI_ISL_1467197, EPI_ISL_1467198, EPI_ISL_1467199, EPI_ISL_1467200, EPI_ISL_1467201, EPI_ISL_1467202, EPI_ISL_1467203, EPI_ISL_1467204, EPI_ISL_1467205, EPI_ISL_1467207, EPI_ISL_1467209, EPI_ISL_1467210, EPI_ISL_1467211, EPI_ISL_1467212, EPI_ISL_1467214, EPI_ISL_1467215, EPI_ISL_1467217, EPI_ISL_1467218, EPI_ISL_1467220, EPI_ISL_1467221, EPI_ISL_1467222, EPI_ISL_1467223, EPI_ISL_1467224, EPI_ISL_1467225, EPI_ISL_1467226, EPI_ISL_1467227, EPI_ISL_1467228, EPI_ISL_1467232, EPI_ISL_1467235, EPI_ISL_1467237, EPI_ISL_1467238, EPI_ISL_1467240, EPI_ISL_1467241, EPI_ISL_1467242, EPI_ISL_1467243, EPI_ISL_1467244, EPI_ISL_1467245, EPI_ISL_1467247, EPI_ISL_1467248, EPI_ISL_1467249, EPI_ISL_1467250, EPI_ISL_1467251, EPI_ISL_1467254, EPI_ISL_1467255, EPI_ISL_1467256, EPI_ISL_1467257, EPI_ISL_1467262, EPI_ISL_1467265, EPI_ISL_1467268, EPI_ISL_1467270, EPI_ISL_1467271, EPI_ISL_1467272, EPI_ISL_1467278, EPI_ISL_1467281, EPI_ISL_1467285, EPI_ISL_1467286, EPI_ISL_1467289, EPI_ISL_1467294, EPI_ISL_1467299, EPI_ISL_1467308, EPI_ISL_1467309, EPI_ISL_1467312, EPI_ISL_1467315, EPI_ISL_1467318, EPI_ISL_1467322, EPI_ISL_1467323, EPI_ISL_1467329, EPI_ISL_1467331, EPI_ISL_1467332, EPI_ISL_1467334, EPI_ISL_1467350, EPI_ISL_1467355, EPI_ISL_1467366, EPI_ISL_1467369, EPI_ISL_1467373, EPI_ISL_1467379, EPI_ISL_1467380, EPI_ISL_1467383, EPI_ISL_1467388, EPI_ISL_1467401, EPI_ISL_1467406, EPI_ISL_1467409, EPI_ISL_1467412, EPI_ISL_1467428, EPI_ISL_1467429, EPI_ISL_1467430, EPI_ISL_1467431, EPI_ISL_1467434, EPI_ISL_1467437, EPI_ISL_1467444, EPI_ISL_1467451, EPI_ISL_1467463, EPI_ISL_1467467, EPI_ISL_1467470, EPI_ISL_1467471, EPI_ISL_1467479, EPI_ISL_1467486, EPI_ISL_1467491, EPI_ISL_1467493, EPI_ISL_1467499, EPI_ISL_1467510, EPI_ISL_1467517, EPI_ISL_1467525, EPI_ISL_1467531, EPI_ISL_1467535, EPI_ISL_1467538, EPI_ISL_1467546, EPI_ISL_1467549, EPI_ISL_1467553, EPI_ISL_1467554, EPI_ISL_1467563, EPI_ISL_1467567, EPI_ISL_1467568, EPI_ISL_1467576, EPI_ISL_1467804, EPI_ISL_1467805, EPI_ISL_1467806, EPI_ISL_1467807, EPI_ISL_1467808, EPI_ISL_1467809, EPI_ISL_1467810, EPI_ISL_1467811, EPI_ISL_1467812, EPI_ISL_1467813, EPI_ISL_1467814, EPI_ISL_1467815, EPI_ISL_1467816, EPI_ISL_1467817, EPI_ISL_1467818, EPI_ISL_1467819, EPI_ISL_1467820, EPI_ISL_1467821, EPI_ISL_1467822, EPI_ISL_1467823, EPI_ISL_1467824, EPI_ISL_1467825, EPI_ISL_1467826, EPI_ISL_1467827, EPI_ISL_1467828, EPI_ISL_1467830, EPI_ISL_1467831, EPI_ISL_1467832, EPI_ISL_1467833, EPI_ISL_1467834, EPI_ISL_1467839, EPI_ISL_1467842, EPI_ISL_1467846, EPI_ISL_1467849, EPI_ISL_1467850, EPI_ISL_1467855, EPI_ISL_1467856, EPI_ISL_1467861, EPI_ISL_1467863, EPI_ISL_1467864, EPI_ISL_1467865, EPI_ISL_1467866, EPI_ISL_1467867, EPI_ISL_1467869, EPI_ISL_1467870, EPI_ISL_1467871, EPI_ISL_1467872, EPI_ISL_1467873, EPI_ISL_1467878, EPI_ISL_1467882, EPI_ISL_1467884, EPI_ISL_1467891, EPI_ISL_1467893, EPI_ISL_1467894, EPI_ISL_1467895, EPI_ISL_1467896, EPI_ISL_1467901, EPI_ISL_1467903, EPI_ISL_1467904, EPI_ISL_1467906, EPI_ISL_1467907, EPI_ISL_1467909, EPI_ISL_1467910, EPI_ISL_1467911, EPI_ISL_1467915, EPI_ISL_1467916, EPI_ISL_1467920, EPI_ISL_1467923, EPI_ISL_1467931, EPI_ISL_1467935, EPI_ISL_1467936, EPI_ISL_1467938, EPI_ISL_1467940, EPI_ISL_1467942, EPI_ISL_1467944, EPI_ISL_1467945, EPI_ISL_1467950, EPI_ISL_1467951, EPI_ISL_1467954, EPI_ISL_1467958, EPI_ISL_1467959, EPI_ISL_1467960, EPI_ISL_1467961, EPI_ISL_1467962, EPI_ISL_1467964, EPI_ISL_1467971, EPI_ISL_1467974, EPI_ISL_1467983, EPI_ISL_1467984, EPI_ISL_1467985, EPI_ISL_1467990, EPI_ISL_1467992, EPI_ISL_1467997, EPI_ISL_1467998, EPI_ISL_1468003, EPI_ISL_1468004, EPI_ISL_1468005, EPI_ISL_1468011, EPI_ISL_1468013, EPI_ISL_1468015, EPI_ISL_1468023, EPI_ISL_1468027, EPI_ISL_1468030, EPI_ISL_1468033, EPI_ISL_1468034, EPI_ISL_1468035, EPI_ISL_1468036, EPI_ISL_1468038, EPI_ISL_1468039, EPI_ISL_1468040, EPI_ISL_1468042, EPI_ISL_1468044, EPI_ISL_1468046, EPI_ISL_1468054, EPI_ISL_1468057, EPI_ISL_1468058, EPI_ISL_1468061, EPI_ISL_1468064, EPI_ISL_1468067, EPI_ISL_1468068, EPI_ISL_1468070, EPI_ISL_1468072, EPI_ISL_1468078, EPI_ISL_1468079, EPI_ISL_1468082, EPI_ISL_1468083, EPI_ISL_1468090, EPI_ISL_1468092, EPI_ISL_1468093, EPI_ISL_1468101, EPI_ISL_1468108, EPI_ISL_1468111, EPI_ISL_1468124, EPI_ISL_1468127, EPI_ISL_1468128, EPI_ISL_1468129, EPI_ISL_1468141, EPI_ISL_1468164, EPI_ISL_1468169, EPI_ISL_1468183, EPI_ISL_1468188, EPI_ISL_1468192, EPI_ISL_1468199, EPI_ISL_1468202, EPI_ISL_1468212, EPI_ISL_1468214, EPI_ISL_1468226, EPI_ISL_1468228, EPI_ISL_1468229, EPI_ISL_1468230, EPI_ISL_1468242, EPI_ISL_1468244, EPI_ISL_1468255, EPI_ISL_1468284, EPI_ISL_1468285, EPI_ISL_1468286, EPI_ISL_1468288, EPI_ISL_1468302, EPI_ISL_1468315, EPI_ISL_1468322, EPI_ISL_1468323, EPI_ISL_1468326, EPI_ISL_1468330, EPI_ISL_1468335, EPI_ISL_1468343, EPI_ISL_1468349, EPI_ISL_1468378, EPI_ISL_1468390, EPI_ISL_1468393, EPI_ISL_1468398, EPI_ISL_1468399, EPI_ISL_1468400, EPI_ISL_1472409, EPI_ISL_1472412, EPI_ISL_1472413, EPI_ISL_1472416, EPI_ISL_1472417, EPI_ISL_1472419, EPI_ISL_1472420, EPI_ISL_1472423, EPI_ISL_1472424, EPI_ISL_1472429, EPI_ISL_1472430, EPI_ISL_1472432, EPI_ISL_1472433, EPI_ISL_1472437, EPI_ISL_1472439, EPI_ISL_1472440, EPI_ISL_1472444, EPI_ISL_1472445, EPI_ISL_1472447, EPI_ISL_1472449, EPI_ISL_1472457, EPI_ISL_1472459, EPI_ISL_1472461, EPI_ISL_1472466, EPI_ISL_1472467, EPI_ISL_1472469, EPI_ISL_1472470, EPI_ISL_1472471, EPI_ISL_1472472, EPI_ISL_1472473, EPI_ISL_1472475, EPI_ISL_1472476, EPI_ISL_1472477, EPI_ISL_1472479, EPI_ISL_1472480, EPI_ISL_1472482, EPI_ISL_1472484, EPI_ISL_1472486, EPI_ISL_1472487, EPI_ISL_1472489, EPI_ISL_1472490, EPI_ISL_1472491, EPI_ISL_1472493, EPI_ISL_1472495, EPI_ISL_1472497, EPI_ISL_1472498, EPI_ISL_1472499, EPI_ISL_1472500, EPI_ISL_1472503, EPI_ISL_1472504, EPI_ISL_1472505, EPI_ISL_1472506, EPI_ISL_1472507, EPI_ISL_1472510, EPI_ISL_1472514, EPI_ISL_1472515, EPI_ISL_1472516, EPI_ISL_1472517, EPI_ISL_1472518, EPI_ISL_1472519, EPI_ISL_1472520, EPI_ISL_1472521, EPI_ISL_1472522, EPI_ISL_1472523, EPI_ISL_1472524, EPI_ISL_1472525, EPI_ISL_1472529, EPI_ISL_1472531, EPI_ISL_1472538, EPI_ISL_1472539, EPI_ISL_1472540, EPI_ISL_1472541, EPI_ISL_1472542, EPI_ISL_1472543, EPI_ISL_1472544, EPI_ISL_1472545, EPI_ISL_1472546, EPI_ISL_1472548, EPI_ISL_1472550, EPI_ISL_1472553, EPI_ISL_1472555, EPI_ISL_1472559, EPI_ISL_1472561, EPI_ISL_1472563, EPI_ISL_1472564, EPI_ISL_1472565, EPI_ISL_1472566, EPI_ISL_1472567, EPI_ISL_1472570, EPI_ISL_1472571, EPI_ISL_1472573, EPI_ISL_1472574, EPI_ISL_1472575, EPI_ISL_1472576, EPI_ISL_1472577, EPI_ISL_1472581, EPI_ISL_1472586, EPI_ISL_1472587, EPI_ISL_1472588, EPI_ISL_1472589, EPI_ISL_1472590, EPI_ISL_1472591, EPI_ISL_1472592, EPI_ISL_1472598, EPI_ISL_1472601, EPI_ISL_1472602, EPI_ISL_1472603, EPI_ISL_1472607, EPI_ISL_1472609, EPI_ISL_1472610, EPI_ISL_1472614, EPI_ISL_1472615, EPI_ISL_1472616, EPI_ISL_1472618, EPI_ISL_1472620, EPI_ISL_1472623, EPI_ISL_1472624, EPI_ISL_1472630, EPI_ISL_1472632, EPI_ISL_1472635, EPI_ISL_1472637, EPI_ISL_1472639, EPI_ISL_1472640, EPI_ISL_1472641, EPI_ISL_1472642, EPI_ISL_1472645, EPI_ISL_1472649, EPI_ISL_1472651, EPI_ISL_1472654, EPI_ISL_1472666, EPI_ISL_1472671, EPI_ISL_1472673, EPI_ISL_1472674, EPI_ISL_1472675, EPI_ISL_1472678, EPI_ISL_1472679, EPI_ISL_1472683, EPI_ISL_1472685, EPI_ISL_1472687, EPI_ISL_1472688, EPI_ISL_1472690, EPI_ISL_1472691, EPI_ISL_1472693, EPI_ISL_1472694, EPI_ISL_1472695, EPI_ISL_1472699, EPI_ISL_1472700, EPI_ISL_1472706, EPI_ISL_1472707, EPI_ISL_1472715, EPI_ISL_1472727, EPI_ISL_1472742, EPI_ISL_1472756, EPI_ISL_1472766, EPI_ISL_1472769, EPI_ISL_1472777, EPI_ISL_1472778, EPI_ISL_1472780, EPI_ISL_1472786, EPI_ISL_1472797, EPI_ISL_1472809, EPI_ISL_1472819, EPI_ISL_1472830, EPI_ISL_1472840, EPI_ISL_1472867, EPI_ISL_1472869, EPI_ISL_1472870, EPI_ISL_1472876, EPI_ISL_1472880, EPI_ISL_1472898, EPI_ISL_1472911, EPI_ISL_1472925, EPI_ISL_1472929, EPI_ISL_1472931, EPI_ISL_1472932, EPI_ISL_1472943, EPI_ISL_1472944, EPI_ISL_1473804, EPI_ISL_1473807, EPI_ISL_1473808, EPI_ISL_1473809, EPI_ISL_1473812, EPI_ISL_1473813, EPI_ISL_1473814, EPI_ISL_1473815, EPI_ISL_1473816, EPI_ISL_1473817, EPI_ISL_1473818, EPI_ISL_1473819, EPI_ISL_1473820, EPI_ISL_1473821, EPI_ISL_1473822, EPI_ISL_1473823, EPI_ISL_1473824, EPI_ISL_1473825, EPI_ISL_1473826, EPI_ISL_1473827, EPI_ISL_1473828, EPI_ISL_1473829, EPI_ISL_1473830, EPI_ISL_1473831, EPI_ISL_1473832, EPI_ISL_1473833, EPI_ISL_1473834, EPI_ISL_1473835, EPI_ISL_1473836, EPI_ISL_1473837, EPI_ISL_1473838, EPI_ISL_1473839, EPI_ISL_1473840, EPI_ISL_1473841, EPI_ISL_1473842, EPI_ISL_1473843, EPI_ISL_1473844, EPI_ISL_1473845, EPI_ISL_1473846, EPI_ISL_1473847, EPI_ISL_1473848, EPI_ISL_1473849, EPI_ISL_1473850, EPI_ISL_1473851, EPI_ISL_1473852, EPI_ISL_1473853, EPI_ISL_1473854, EPI_ISL_1473855, EPI_ISL_1473856, EPI_ISL_1473857, EPI_ISL_1473858, EPI_ISL_1473859, EPI_ISL_1473860, EPI_ISL_1473861, EPI_ISL_1473862, EPI_ISL_1473863, EPI_ISL_1473864, EPI_ISL_1473865, EPI_ISL_1473866, EPI_ISL_1473867, EPI_ISL_1473868, EPI_ISL_1473869, EPI_ISL_1473870, EPI_ISL_1473871, EPI_ISL_1473872, EPI_ISL_1473873, EPI_ISL_1473874, EPI_ISL_1473875, EPI_ISL_1473876, EPI_ISL_1473877, EPI_ISL_1473878, EPI_ISL_1473879, EPI_ISL_1473880, EPI_ISL_1473881, EPI_ISL_1473882, EPI_ISL_1473883, EPI_ISL_1473884, EPI_ISL_1473885, EPI_ISL_1473886, EPI_ISL_1473887, EPI_ISL_1473888, EPI_ISL_1473889, EPI_ISL_1473890, EPI_ISL_1473891, EPI_ISL_1473892, EPI_ISL_1473893, EPI_ISL_1473894, EPI_ISL_1473895, EPI_ISL_1473896, EPI_ISL_1473897, EPI_ISL_1473898, EPI_ISL_1473899, EPI_ISL_1473900, EPI_ISL_1473901, EPI_ISL_1473902, EPI_ISL_1473903, EPI_ISL_1473904, EPI_ISL_1473905, EPI_ISL_1473906, EPI_ISL_1473907, EPI_ISL_1473908, EPI_ISL_1473909, EPI_ISL_1473910, EPI_ISL_1473911, EPI_ISL_1473912, EPI_ISL_1473913, EPI_ISL_1473914, EPI_ISL_1473915, EPI_ISL_1473916, EPI_ISL_1473917, EPI_ISL_1473918, EPI_ISL_1473919, EPI_ISL_1473920, EPI_ISL_1473921, EPI_ISL_1473922, EPI_ISL_1473923, EPI_ISL_1473924, EPI_ISL_1473925, EPI_ISL_1473926, EPI_ISL_1473927, EPI_ISL_1473928, EPI_ISL_1473929, EPI_ISL_1473930, EPI_ISL_1473931, EPI_ISL_1473932, EPI_ISL_1473933, EPI_ISL_1473934, EPI_ISL_1473935, EPI_ISL_1473937, EPI_ISL_1473938, EPI_ISL_1473939, EPI_ISL_1473940, EPI_ISL_1473941, EPI_ISL_1473942, EPI_ISL_1473943, EPI_ISL_1473944, EPI_ISL_1473945, EPI_ISL_1473946, EPI_ISL_1473947, EPI_ISL_1473948, EPI_ISL_1473949, EPI_ISL_1473950, EPI_ISL_1473951, EPI_ISL_1473952, EPI_ISL_1473953, EPI_ISL_1473954, EPI_ISL_1473955, EPI_ISL_1473956, EPI_ISL_1473957, EPI_ISL_1473958, EPI_ISL_1473959, EPI_ISL_1473960, EPI_ISL_1473961, EPI_ISL_1473962, EPI_ISL_1473963, EPI_ISL_1473964, EPI_ISL_1473965, EPI_ISL_1473966, EPI_ISL_1473967, EPI_ISL_1473968, EPI_ISL_1473969, EPI_ISL_1473970, EPI_ISL_1473971, EPI_ISL_1473972, EPI_ISL_1473973, EPI_ISL_1473974, EPI_ISL_1473975, EPI_ISL_1473976, EPI_ISL_1473977, EPI_ISL_1473978, EPI_ISL_1473979, EPI_ISL_1473980, EPI_ISL_1473981, EPI_ISL_1473982, EPI_ISL_1473983, EPI_ISL_1473984, EPI_ISL_1473985, EPI_ISL_1473986, EPI_ISL_1473987, EPI_ISL_1473988, EPI_ISL_1473989, EPI_ISL_1473990, EPI_ISL_1473991, EPI_ISL_1473992, EPI_ISL_1473993, EPI_ISL_1473994, EPI_ISL_1473995, EPI_ISL_1473996, EPI_ISL_1473997, EPI_ISL_1473998, EPI_ISL_1473999, EPI_ISL_1474000, EPI_ISL_1474001, EPI_ISL_1474002, EPI_ISL_1474003, EPI_ISL_1474004, EPI_ISL_1474005, EPI_ISL_1474006, EPI_ISL_1474007, EPI_ISL_1474008, EPI_ISL_1474009, EPI_ISL_1474010, EPI_ISL_1474011, EPI_ISL_1474012, EPI_ISL_1474013, EPI_ISL_1474014, EPI_ISL_1474015, EPI_ISL_1474016, EPI_ISL_1474017, EPI_ISL_1474018, EPI_ISL_1474019, EPI_ISL_1474020, EPI_ISL_1474021, EPI_ISL_1474022, EPI_ISL_1474023, EPI_ISL_1474024, EPI_ISL_1474025, EPI_ISL_1474026, EPI_ISL_1474027, EPI_ISL_1474028, EPI_ISL_1474029, EPI_ISL_1474030, EPI_ISL_1474031, EPI_ISL_1474032, EPI_ISL_1474033, EPI_ISL_1474034, EPI_ISL_1474035, EPI_ISL_1474036, EPI_ISL_1474037, EPI_ISL_1474038, EPI_ISL_1474039, EPI_ISL_1474040, EPI_ISL_1474042, EPI_ISL_1474043, EPI_ISL_1474044, EPI_ISL_1474045, EPI_ISL_1474046, EPI_ISL_1474047, EPI_ISL_1474048, EPI_ISL_1474049, EPI_ISL_1474050, EPI_ISL_1474051, EPI_ISL_1474052, EPI_ISL_1474053, EPI_ISL_1474054, EPI_ISL_1474055, EPI_ISL_1474056, EPI_ISL_1474057, EPI_ISL_1474058, EPI_ISL_1474059, EPI_ISL_1474060, EPI_ISL_1474061, EPI_ISL_1474062, EPI_ISL_1474063, EPI_ISL_1474064, EPI_ISL_1474065, EPI_ISL_1474066, EPI_ISL_1474067, EPI_ISL_1474068, EPI_ISL_1474069, EPI_ISL_1474070, EPI_ISL_1474071, EPI_ISL_1474072, EPI_ISL_1474073, EPI_ISL_1474074, EPI_ISL_1474075, EPI_ISL_1474076, EPI_ISL_1474077, EPI_ISL_1474078, EPI_ISL_1474079, EPI_ISL_1474081, EPI_ISL_1474082, EPI_ISL_1474083, EPI_ISL_1474084, EPI_ISL_1474085, EPI_ISL_1474086, EPI_ISL_1474087, EPI_ISL_1474088, EPI_ISL_1474089, EPI_ISL_1474090, EPI_ISL_1474091, EPI_ISL_1474092, EPI_ISL_1474093, EPI_ISL_1474094, EPI_ISL_1474095, EPI_ISL_1474096, EPI_ISL_1474097, EPI_ISL_1474098, EPI_ISL_1474099, EPI_ISL_1474100, EPI_ISL_1474101, EPI_ISL_1474102, EPI_ISL_1474103, EPI_ISL_1474104, EPI_ISL_1474105, EPI_ISL_1474106, EPI_ISL_1474107, EPI_ISL_1474108, EPI_ISL_1474109, EPI_ISL_1474110, EPI_ISL_1474111, EPI_ISL_1474112, EPI_ISL_1474113, EPI_ISL_1474114, EPI_ISL_1474115, EPI_ISL_1474116, EPI_ISL_1474117, EPI_ISL_1474118, EPI_ISL_1474119, EPI_ISL_1474120, EPI_ISL_1474121, EPI_ISL_1474122, EPI_ISL_1474123, EPI_ISL_1474124, EPI_ISL_1474126, EPI_ISL_1474127, EPI_ISL_1474128, EPI_ISL_1474129, EPI_ISL_1474130, EPI_ISL_1474131, EPI_ISL_1474132, EPI_ISL_1474133, EPI_ISL_1474134, EPI_ISL_1474135, EPI_ISL_1474136, EPI_ISL_1474137, EPI_ISL_1474138, EPI_ISL_1474139, EPI_ISL_1474140, EPI_ISL_1474141, EPI_ISL_1474142, EPI_ISL_1474143, EPI_ISL_1474144, EPI_ISL_1474145, EPI_ISL_1474146, EPI_ISL_1474147, EPI_ISL_1474148, EPI_ISL_1474149, EPI_ISL_1474150, EPI_ISL_1474151, EPI_ISL_1474152, EPI_ISL_1474153, EPI_ISL_1474154, EPI_ISL_1474155, EPI_ISL_1474158, EPI_ISL_1483863, EPI_ISL_1483865, EPI_ISL_1483975, EPI_ISL_1483996, EPI_ISL_1484037, EPI_ISL_1484094, EPI_ISL_1484106, EPI_ISL_1484193, EPI_ISL_1484194, EPI_ISL_1484840, EPI_ISL_1484844, EPI_ISL_1484850, EPI_ISL_1484862, EPI_ISL_1484878, EPI_ISL_1484887, EPI_ISL_1484892, EPI_ISL_1484898, EPI_ISL_1484901, EPI_ISL_1484907, EPI_ISL_1484915, EPI_ISL_1484916, EPI_ISL_1484936, EPI_ISL_1484939, EPI_ISL_1484963, EPI_ISL_1484972, EPI_ISL_1484980, EPI_ISL_1484990, EPI_ISL_1484997, EPI_ISL_1485002, EPI_ISL_1485008, EPI_ISL_1485015, EPI_ISL_1485020, EPI_ISL_1485021, EPI_ISL_1485033, EPI_ISL_1485046, EPI_ISL_1485054, EPI_ISL_1485061, EPI_ISL_1485064, EPI_ISL_1485069, EPI_ISL_1485074, EPI_ISL_1485083, EPI_ISL_1485095, EPI_ISL_1485110, EPI_ISL_1485116, EPI_ISL_1485123, EPI_ISL_1485135, EPI_ISL_1485136, EPI_ISL_1485142, EPI_ISL_1485144, EPI_ISL_1485149, EPI_ISL_1485155, EPI_ISL_1485156, EPI_ISL_1485158, EPI_ISL_1485160, EPI_ISL_1485174, EPI_ISL_1485180, EPI_ISL_1485193, EPI_ISL_1485197, EPI_ISL_1485209, EPI_ISL_1485217, EPI_ISL_1485225, EPI_ISL_1485231, EPI_ISL_1485235, EPI_ISL_1485236, EPI_ISL_1485245, EPI_ISL_1485251, EPI_ISL_1485257, EPI_ISL_1485267, EPI_ISL_1485268, EPI_ISL_1485269, EPI_ISL_1485275, EPI_ISL_1485285, EPI_ISL_1485286, EPI_ISL_1485293, EPI_ISL_1485294, EPI_ISL_1485305, EPI_ISL_1485308, EPI_ISL_1485312, EPI_ISL_1485317, EPI_ISL_1485319, EPI_ISL_1485320, EPI_ISL_1485338, EPI_ISL_1485340, EPI_ISL_1485349, EPI_ISL_1485354, EPI_ISL_1485364, EPI_ISL_1485366, EPI_ISL_1485368, EPI_ISL_1485384, EPI_ISL_1485397, EPI_ISL_1485400, EPI_ISL_1485410, EPI_ISL_1485421, EPI_ISL_1485434, EPI_ISL_1485436, EPI_ISL_1485443, EPI_ISL_1485444, EPI_ISL_1485446, EPI_ISL_1485449, EPI_ISL_1485453, EPI_ISL_1485456, EPI_ISL_1485463, EPI_ISL_1485465, EPI_ISL_1485468, EPI_ISL_1485471, EPI_ISL_1485478, EPI_ISL_1485479, EPI_ISL_1485483, EPI_ISL_1485484, EPI_ISL_1485485, EPI_ISL_1485488, EPI_ISL_1485489, EPI_ISL_1485490, EPI_ISL_1485494, EPI_ISL_1485495, EPI_ISL_1485497, EPI_ISL_1485499, EPI_ISL_1485502, EPI_ISL_1485503, EPI_ISL_1485505, EPI_ISL_1485506, EPI_ISL_1485507, EPI_ISL_1485510, | | | |

EPI_ISL_1485511, EPI_ISL_1485512, EPI_ISL_1485513, EPI_ISL_1485515, EPI_ISL_1485516, EPI_ISL_1485517, EPI_ISL_1485519, EPI_ISL_1485520, EPI_ISL_1485522, EPI_ISL_1485523, EPI_ISL_1485526, EPI_ISL_1485527, EPI_ISL_1485528, EPI_ISL_1485529, EPI_ISL_1485533, EPI_ISL_1485534, EPI_ISL_1485535, EPI_ISL_1485539, EPI_ISL_1485540, EPI_ISL_1485542, EPI_ISL_1485547, EPI_ISL_1485548,
EPI_ISL_1485549, EPI_ISL_1485552, EPI_ISL_1485555, EPI_ISL_1485557, EPI_ISL_1485558, EPI_ISL_1485561, EPI_ISL_1485565, EPI_ISL_1485566, EPI_ISL_1485567, EPI_ISL_1485569, EPI_ISL_1485570, EPI_ISL_1485571, EPI_ISL_1485572, EPI_ISL_1485573, EPI_ISL_1485576, EPI_ISL_1485580, EPI_ISL_1485581, EPI_ISL_1485585, EPI_ISL_1485586, EPI_ISL_1485587, EPI_ISL_1485588, EPI_ISL_1485589,
EPI_ISL_1485590, EPI_ISL_1485595, EPI_ISL_1485597, EPI_ISL_1485598, EPI_ISL_1485604, EPI_ISL_1485607, EPI_ISL_1485608, EPI_ISL_1485610, EPI_ISL_1485611, EPI_ISL_1485615, EPI_ISL_1485619, EPI_ISL_1485620, EPI_ISL_1485622, EPI_ISL_1485624, EPI_ISL_1485626, EPI_ISL_1485629, EPI_ISL_1485632, EPI_ISL_1485633, EPI_ISL_1485634, EPI_ISL_1485636, EPI_ISL_1485637, EPI_ISL_1485638,
EPI_ISL_1485639, EPI_ISL_1485640, EPI_ISL_1485643, EPI_ISL_1485645, EPI_ISL_1485647, EPI_ISL_1485650, EPI_ISL_1485654, EPI_ISL_1485656, EPI_ISL_1485657, EPI_ISL_1485660, EPI_ISL_1485661, EPI_ISL_1485663, EPI_ISL_1485664, EPI_ISL_1485665, EPI_ISL_1485666, EPI_ISL_1485668, EPI_ISL_1485669, EPI_ISL_1485671, EPI_ISL_1485673, EPI_ISL_1485674, EPI_ISL_1485675, EPI_ISL_1485678,
EPI_ISL_1485682, EPI_ISL_1485685, EPI_ISL_1485687, EPI_ISL_1485688, EPI_ISL_1485689, EPI_ISL_1485690, EPI_ISL_1485691, EPI_ISL_1485692, EPI_ISL_1485693, EPI_ISL_1485696, EPI_ISL_1485698, EPI_ISL_1485702, EPI_ISL_1485704, EPI_ISL_1485705, EPI_ISL_1485706, EPI_ISL_1485708, EPI_ISL_1485709, EPI_ISL_1485710, EPI_ISL_1485715, EPI_ISL_1485716, EPI_ISL_1485717, EPI_ISL_1485718,
EPI_ISL_1485720, EPI_ISL_1485721, EPI_ISL_1485724, EPI_ISL_1485725, EPI_ISL_1485729, EPI_ISL_1485730, EPI_ISL_1485732, EPI_ISL_1485735, EPI_ISL_1485736, EPI_ISL_1485737, EPI_ISL_1485738, EPI_ISL_1485741, EPI_ISL_1485742, EPI_ISL_1485743, EPI_ISL_1485744, EPI_ISL_1485745, EPI_ISL_1485750, EPI_ISL_1485760, EPI_ISL_1485762, EPI_ISL_1485766, EPI_ISL_1485767, EPI_ISL_1485769,
EPI_ISL_1485775, EPI_ISL_1485776, EPI_ISL_1485777, EPI_ISL_1485780, EPI_ISL_1485781, EPI_ISL_1485782, EPI_ISL_1485784, EPI_ISL_1485785, EPI_ISL_1485786, EPI_ISL_1485787, EPI_ISL_1485791, EPI_ISL_1485795, EPI_ISL_1485797, EPI_ISL_1485798, EPI_ISL_1485800, EPI_ISL_1485801, EPI_ISL_1485803, EPI_ISL_1485804, EPI_ISL_1485806, EPI_ISL_1485814, EPI_ISL_1485816, EPI_ISL_1485817,
EPI_ISL_1485819, EPI_ISL_1485821, EPI_ISL_1485825, EPI_ISL_1485829, EPI_ISL_1485830, EPI_ISL_1485831, EPI_ISL_1485833, EPI_ISL_1485834, EPI_ISL_1485837, EPI_ISL_1485838, EPI_ISL_1485839, EPI_ISL_1485840, EPI_ISL_1485841, EPI_ISL_1485842, EPI_ISL_1485843, EPI_ISL_1485845, EPI_ISL_1485850, EPI_ISL_1485851, EPI_ISL_1485858, EPI_ISL_1485861, EPI_ISL_1485862, EPI_ISL_1485867,
EPI_ISL_1485868, EPI_ISL_1485869, EPI_ISL_1485870, EPI_ISL_1485874, EPI_ISL_1485878, EPI_ISL_1485880, EPI_ISL_1485881, EPI_ISL_1485883, EPI_ISL_1485884, EPI_ISL_1485887, EPI_ISL_1485890, EPI_ISL_1485891, EPI_ISL_1485901, EPI_ISL_1485902, EPI_ISL_1485903, EPI_ISL_1485907, EPI_ISL_1485908, EPI_ISL_1485911, EPI_ISL_1485912, EPI_ISL_1485913, EPI_ISL_1485914, EPI_ISL_1485915,
EPI_ISL_1485917, EPI_ISL_1485919, EPI_ISL_1485920, EPI_ISL_1485921, EPI_ISL_1485922, EPI_ISL_1485929, EPI_ISL_1485931, EPI_ISL_1485932, EPI_ISL_1485944, EPI_ISL_1485948, EPI_ISL_1485949, EPI_ISL_1485951, EPI_ISL_1485953, EPI_ISL_1485957, EPI_ISL_1485958, EPI_ISL_1485959, EPI_ISL_1485961, EPI_ISL_1485963, EPI_ISL_1485967, EPI_ISL_1485968, EPI_ISL_1485970, EPI_ISL_1485971,
EPI_ISL_1485972, EPI_ISL_1485974, EPI_ISL_1485975, EPI_ISL_1485981, EPI_ISL_1485982, EPI_ISL_1485984, EPI_ISL_1485987, EPI_ISL_1485988, EPI_ISL_1485989, EPI_ISL_1485991, EPI_ISL_1485995, EPI_ISL_1485997, EPI_ISL_1486000, EPI_ISL_1486001, EPI_ISL_1486002, EPI_ISL_1486004, EPI_ISL_1486006, EPI_ISL_1486010, EPI_ISL_1486011, EPI_ISL_1486013, EPI_ISL_1486016, EPI_ISL_1486017,
EPI_ISL_1486018, EPI_ISL_1486019, EPI_ISL_1486020, EPI_ISL_1486027, EPI_ISL_1486029, EPI_ISL_1486031, EPI_ISL_1486032, EPI_ISL_1486035, EPI_ISL_1486037, EPI_ISL_1486039, EPI_ISL_1486042, EPI_ISL_1486044, EPI_ISL_1486047, EPI_ISL_1486048, EPI_ISL_1486049, EPI_ISL_1486051, EPI_ISL_1486052, EPI_ISL_1486053, EPI_ISL_1486055, EPI_ISL_1486057, EPI_ISL_1486061, EPI_ISL_1486062,
EPI_ISL_1486064, EPI_ISL_1486066, EPI_ISL_1486070, EPI_ISL_1486072, EPI_ISL_1486073, EPI_ISL_1486076, EPI_ISL_1486078, EPI_ISL_1486080, EPI_ISL_1486081, EPI_ISL_1486083, EPI_ISL_1486085, EPI_ISL_1486086, EPI_ISL_1486093, EPI_ISL_1486095, EPI_ISL_1486098, EPI_ISL_1486099, EPI_ISL_1486102, EPI_ISL_1486104, EPI_ISL_1486394, EPI_ISL_1486395, EPI_ISL_1486396, EPI_ISL_1486397,
EPI_ISL_1486398, EPI_ISL_1486401, EPI_ISL_1486402, EPI_ISL_1486405, EPI_ISL_1486409, EPI_ISL_1486412, EPI_ISL_1486415, EPI_ISL_1486418, EPI_ISL_1486421, EPI_ISL_1486424, EPI_ISL_1486425, EPI_ISL_1486426, EPI_ISL_1486427, EPI_ISL_1486428, EPI_ISL_1486430, EPI_ISL_1486431, EPI_ISL_1486434, EPI_ISL_1486436, EPI_ISL_1486439, EPI_ISL_1486440, EPI_ISL_1486441, EPI_ISL_1486442,
EPI_ISL_1486448, EPI_ISL_1486449, EPI_ISL_1486451, EPI_ISL_1486452, EPI_ISL_1486455, EPI_ISL_1486458, EPI_ISL_1486459, EPI_ISL_1486465, EPI_ISL_1486470, EPI_ISL_1486474, EPI_ISL_1486476, EPI_ISL_1486477, EPI_ISL_1486481, EPI_ISL_1486483, EPI_ISL_1486490, EPI_ISL_1486493, EPI_ISL_1486494, EPI_ISL_1486497, EPI_ISL_1486505, EPI_ISL_1486507, EPI_ISL_1486513, EPI_ISL_1486515,
EPI_ISL_1486520, EPI_ISL_1486521, EPI_ISL_1486529, EPI_ISL_1486533, EPI_ISL_1486538, EPI_ISL_1486542, EPI_ISL_1486554, EPI_ISL_1486556, EPI_ISL_1486557, EPI_ISL_1486559, EPI_ISL_1486560, EPI_ISL_1486566, EPI_ISL_1486570, EPI_ISL_1486579, EPI_ISL_1486581, EPI_ISL_1486582, EPI_ISL_1486588, EPI_ISL_1486591, EPI_ISL_1486592, EPI_ISL_1486594, EPI_ISL_1486597, EPI_ISL_1486598,
EPI_ISL_1486611, EPI_ISL_1486617, EPI_ISL_1486618, EPI_ISL_1486621, EPI_ISL_1486623, EPI_ISL_1486629, EPI_ISL_1486630, EPI_ISL_1486633, EPI_ISL_1486635, EPI_ISL_1486636, EPI_ISL_1486638, EPI_ISL_1486639, EPI_ISL_1486650, EPI_ISL_1486654, EPI_ISL_1486663, EPI_ISL_1486664, EPI_ISL_1486666, EPI_ISL_1486670, EPI_ISL_1486672, EPI_ISL_1486679, EPI_ISL_1486683, EPI_ISL_1486688,
EPI_ISL_1486690, EPI_ISL_1486692, EPI_ISL_1486697, EPI_ISL_1486700, EPI_ISL_1486701, EPI_ISL_1486708, EPI_ISL_1486711, EPI_ISL_1486715, EPI_ISL_1486717, EPI_ISL_1486720, EPI_ISL_1486721, EPI_ISL_1486723, EPI_ISL_1486725, EPI_ISL_1486728, EPI_ISL_1486729, EPI_ISL_1486734, EPI_ISL_1486735, EPI_ISL_1486738, EPI_ISL_1486750, EPI_ISL_1486754, EPI_ISL_1486759, EPI_ISL_1486761,
EPI_ISL_1486762, EPI_ISL_1486765, EPI_ISL_1486780, EPI_ISL_1486786, EPI_ISL_1486790, EPI_ISL_1486794, EPI_ISL_1486797, EPI_ISL_1486804, EPI_ISL_1486807, EPI_ISL_1486809, EPI_ISL_1486810, EPI_ISL_1486811, EPI_ISL_1486814, EPI_ISL_1486816, EPI_ISL_1486817, EPI_ISL_1486819, EPI_ISL_1486820, EPI_ISL_1486827, EPI_ISL_1486829, EPI_ISL_1486830, EPI_ISL_1486831, EPI_ISL_1486834,
EPI_ISL_1486838, EPI_ISL_1486843, EPI_ISL_1486845, EPI_ISL_1486846, EPI_ISL_1486848, EPI_ISL_1486850, EPI_ISL_1486851, EPI_ISL_1486852, EPI_ISL_1486853, EPI_ISL_1486854, EPI_ISL_1486855, EPI_ISL_1486858, EPI_ISL_1486861, EPI_ISL_1486862, EPI_ISL_1486863, EPI_ISL_1486864, EPI_ISL_1486865, EPI_ISL_1486867, EPI_ISL_1486869, EPI_ISL_1486870, EPI_ISL_1486874, EPI_ISL_1486879,
EPI_ISL_1486883, EPI_ISL_1486886, EPI_ISL_1486887, EPI_ISL_1486888, EPI_ISL_1486894, EPI_ISL_1486897, EPI_ISL_1486898, EPI_ISL_1486900, EPI_ISL_1486901, EPI_ISL_1486902, EPI_ISL_1486903, EPI_ISL_1486904, EPI_ISL_1486906, EPI_ISL_1486907, EPI_ISL_1486910, EPI_ISL_1486911, EPI_ISL_1486915, EPI_ISL_1486917, EPI_ISL_1486918, EPI_ISL_1486920, EPI_ISL_1486921, EPI_ISL_1486922,
EPI_ISL_1486923, EPI_ISL_1486927, EPI_ISL_1486928, EPI_ISL_1486929, EPI_ISL_1486930, EPI_ISL_1486931, EPI_ISL_1486932, EPI_ISL_1486936, EPI_ISL_1486938, EPI_ISL_1486940, EPI_ISL_1486941, EPI_ISL_1486942, EPI_ISL_1486945, EPI_ISL_1486946, EPI_ISL_1486947, EPI_ISL_1486948, EPI_ISL_1486949, EPI_ISL_1486950, EPI_ISL_1486954, EPI_ISL_1486957, EPI_ISL_1486961, EPI_ISL_1486962,
EPI_ISL_1486966, EPI_ISL_1486967, EPI_ISL_1486970, EPI_ISL_1486972, EPI_ISL_1486974, EPI_ISL_1486975, EPI_ISL_1486978, EPI_ISL_1486979, EPI_ISL_1486980, EPI_ISL_1486981, EPI_ISL_1486984, EPI_ISL_1486986, EPI_ISL_1486988, EPI_ISL_1486992, EPI_ISL_1486998, EPI_ISL_1487002, EPI_ISL_1487004, EPI_ISL_1487007, EPI_ISL_1487010, EPI_ISL_1487012, EPI_ISL_1487014, EPI_ISL_1487015,
EPI_ISL_1487018, EPI_ISL_1487019, EPI_ISL_1487020, EPI_ISL_1487021, EPI_ISL_1487023, EPI_ISL_1487025, EPI_ISL_1487026, EPI_ISL_1487027, EPI_ISL_1487028, EPI_ISL_1487031, EPI_ISL_1487033, EPI_ISL_1487035, EPI_ISL_1487038, EPI_ISL_1487040, EPI_ISL_1487041, EPI_ISL_1487043, EPI_ISL_1487046, EPI_ISL_1487047, EPI_ISL_1487048, EPI_ISL_1487049, EPI_ISL_1487053, EPI_ISL_1487054,
EPI_ISL_1487055, EPI_ISL_1487058, EPI_ISL_1487064, EPI_ISL_1487065, EPI_ISL_1487066, EPI_ISL_1487067, EPI_ISL_1487069, EPI_ISL_1487071, EPI_ISL_1487075, EPI_ISL_1487076, EPI_ISL_1487079, EPI_ISL_1487081, EPI_ISL_1487085, EPI_ISL_1487087, EPI_ISL_1487088, EPI_ISL_1487092, EPI_ISL_1487093, EPI_ISL_1487099, EPI_ISL_1487101, EPI_ISL_1487102, EPI_ISL_1487104, EPI_ISL_1487105,
EPI_ISL_1487111, EPI_ISL_1487112, EPI_ISL_1487115, EPI_ISL_1487116, EPI_ISL_1487119, EPI_ISL_1487121, EPI_ISL_1487128, EPI_ISL_1487130, EPI_ISL_1487134, EPI_ISL_1487138, EPI_ISL_1487139, EPI_ISL_1487140, EPI_ISL_1487141, EPI_ISL_1487144, EPI_ISL_1487145, EPI_ISL_1487147, EPI_ISL_1487150, EPI_ISL_1487154, EPI_ISL_1487155, EPI_ISL_1487156, EPI_ISL_1487157, EPI_ISL_1487158,
EPI_ISL_1487378, EPI_ISL_1487379, EPI_ISL_1487380, EPI_ISL_1487381, EPI_ISL_1487382, EPI_ISL_1487383, EPI_ISL_1487384, EPI_ISL_1487385, EPI_ISL_1487389, EPI_ISL_1487390, EPI_ISL_1487391, EPI_ISL_1487392, EPI_ISL_1487395, EPI_ISL_1487397, EPI_ISL_1487398, EPI_ISL_1487401, EPI_ISL_1487403, EPI_ISL_1487406, EPI_ISL_1487407, EPI_ISL_1487408, EPI_ISL_1487409, EPI_ISL_1487410,
EPI_ISL_1487411, EPI_ISL_1487412, EPI_ISL_1487413, EPI_ISL_1487414, EPI_ISL_1487417, EPI_ISL_1487418, EPI_ISL_1487420, EPI_ISL_1487421, EPI_ISL_1487422, EPI_ISL_1487425, EPI_ISL_1487426, EPI_ISL_1487427, EPI_ISL_1487428, EPI_ISL_1487429, EPI_ISL_1487430, EPI_ISL_1487431, EPI_ISL_1487432, EPI_ISL_1487436, EPI_ISL_1487438, EPI_ISL_1487439, EPI_ISL_1487442, EPI_ISL_1487443,
EPI_ISL_1487444, EPI_ISL_1487445, EPI_ISL_1487447, EPI_ISL_1487449, EPI_ISL_1487450, EPI_ISL_1487451, EPI_ISL_1487454, EPI_ISL_1487455, EPI_ISL_1487456, EPI_ISL_1487459, EPI_ISL_1487461, EPI_ISL_1487463, EPI_ISL_1487465, EPI_ISL_1487466, EPI_ISL_1487467, EPI_ISL_1487469, EPI_ISL_1487470, EPI_ISL_1487471, EPI_ISL_1487472, EPI_ISL_1487473, EPI_ISL_1487476, EPI_ISL_1487478,
EPI_ISL_1487481, EPI_ISL_1487482, EPI_ISL_1487483, EPI_ISL_1487484, EPI_ISL_1487486, EPI_ISL_1487487, EPI_ISL_1487488, EPI_ISL_1487489, EPI_ISL_1487492, EPI_ISL_1487493, EPI_ISL_1487494, EPI_ISL_1487495, EPI_ISL_1487498, EPI_ISL_1487500, EPI_ISL_1487503, EPI_ISL_1487505, EPI_ISL_1487506, EPI_ISL_1487508, EPI_ISL_1487509, EPI_ISL_1487510, EPI_ISL_1487513, EPI_ISL_1487514,
EPI_ISL_1487515, EPI_ISL_1487516, EPI_ISL_1487517, EPI_ISL_1487519, EPI_ISL_1487523, EPI_ISL_1487524, EPI_ISL_1487527, EPI_ISL_1487528, EPI_ISL_1487532, EPI_ISL_1487533, EPI_ISL_1487534, EPI_ISL_1487535, EPI_ISL_1487536, EPI_ISL_1487538, EPI_ISL_1487540, EPI_ISL_1487542, EPI_ISL_1487543, EPI_ISL_1487544, EPI_ISL_1487545, EPI_ISL_1487548, EPI_ISL_1487553, EPI_ISL_1487554,
EPI_ISL_1487555, EPI_ISL_1487556, EPI_ISL_1487557, EPI_ISL_1487558, EPI_ISL_1487559, EPI_ISL_1487560, EPI_ISL_1487561, EPI_ISL_1487563, EPI_ISL_1487564, EPI_ISL_1487565, EPI_ISL_1487567, EPI_ISL_1487568, EPI_ISL_1487569, EPI_ISL_1487570, EPI_ISL_1487571, EPI_ISL_1487574, EPI_ISL_1487575, EPI_ISL_1487576, EPI_ISL_1487578, EPI_ISL_1487579, EPI_ISL_1487580, EPI_ISL_1487582,
EPI_ISL_1487584, EPI_ISL_1487586, EPI_ISL_1487587, EPI_ISL_1487588, EPI_ISL_1487590, EPI_ISL_1487591, EPI_ISL_1487593, EPI_ISL_1487594, EPI_ISL_1487598, EPI_ISL_1487599, EPI_ISL_1487600, EPI_ISL_1487601, EPI_ISL_1487602, EPI_ISL_1487604, EPI_ISL_1487605, EPI_ISL_1487606, EPI_ISL_1487608, EPI_ISL_1487610, EPI_ISL_1487611, EPI_ISL_1487612, EPI_ISL_1487615, EPI_ISL_1487618,
EPI_ISL_1487619, EPI_ISL_1487620, EPI_ISL_1487622, EPI_ISL_1487623, EPI_ISL_1487626, EPI_ISL_1487627, EPI_ISL_1487628, EPI_ISL_1487630, EPI_ISL_1487631, EPI_ISL_1487632, EPI_ISL_1487633, EPI_ISL_1487636, EPI_ISL_1487638, EPI_ISL_1487639, EPI_ISL_1487642, EPI_ISL_1487644, EPI_ISL_1487645, EPI_ISL_1487646, EPI_ISL_1487647, EPI_ISL_1487649, EPI_ISL_1487650, EPI_ISL_1487651,
EPI_ISL_1487652, EPI_ISL_1487654, EPI_ISL_1487656, EPI_ISL_1487659, EPI_ISL_1487661, EPI_ISL_1487662, EPI_ISL_1487663, EPI_ISL_1487664, EPI_ISL_1487665, EPI_ISL_1487666, EPI_ISL_1487667, EPI_ISL_1487668, EPI_ISL_1487669, EPI_ISL_1487671, EPI_ISL_1487672, EPI_ISL_1487673, EPI_ISL_1487674, EPI_ISL_1487675, EPI_ISL_1487676, EPI_ISL_1487678, EPI_ISL_1487679, EPI_ISL_1487680,
EPI_ISL_1487681, EPI_ISL_1487682, EPI_ISL_1487683, EPI_ISL_1487684, EPI_ISL_1487685, EPI_ISL_1487686, EPI_ISL_1487687, EPI_ISL_1487688, EPI_ISL_1487689, EPI_ISL_1487690, EPI_ISL_1487691, EPI_ISL_1487692, EPI_ISL_1487693, EPI_ISL_1487694, EPI_ISL_1487695, EPI_ISL_1487697, EPI_ISL_1487699, EPI_ISL_1487702, EPI_ISL_1487703, EPI_ISL_1487705, EPI_ISL_1487706, EPI_ISL_1487707,
EPI_ISL_1487709, EPI_ISL_1487710, EPI_ISL_1487711, EPI_ISL_1487713, EPI_ISL_1487714, EPI_ISL_1487715, EPI_ISL_1487717, EPI_ISL_1487718, EPI_ISL_1487720, EPI_ISL_1487721, EPI_ISL_1487722, EPI_ISL_1487723, EPI_ISL_1487724, EPI_ISL_1487873, EPI_ISL_1487874, EPI_ISL_1487875, EPI_ISL_1487876, EPI_ISL_1487877, EPI_ISL_1487878, EPI_ISL_1487879, EPI_ISL_1487880, EPI_ISL_1487882,
EPI_ISL_1487884, EPI_ISL_1487885, EPI_ISL_1487886, EPI_ISL_1487887, EPI_ISL_1487888, EPI_ISL_1487889, EPI_ISL_1487890, EPI_ISL_1487891, EPI_ISL_1487892, EPI_ISL_1487893, EPI_ISL_1487894, EPI_ISL_1487895, EPI_ISL_1487896, EPI_ISL_1487897, EPI_ISL_1487898, EPI_ISL_1487900, EPI_ISL_1487902, EPI_ISL_1487904, EPI_ISL_1487906, EPI_ISL_1487907, EPI_ISL_1487909, EPI_ISL_1487910,
EPI_ISL_1487911, EPI_ISL_1487912, EPI_ISL_1487913, EPI_ISL_1487915, EPI_ISL_1487916, EPI_ISL_1487917, EPI_ISL_1487918, EPI_ISL_1487919, EPI_ISL_1487920, EPI_ISL_1487922, EPI_ISL_1487923, EPI_ISL_1487924, EPI_ISL_1487925, EPI_ISL_1487926, EPI_ISL_1487927, EPI_ISL_1487928, EPI_ISL_1487929, EPI_ISL_1487930, EPI_ISL_1487931, EPI_ISL_1487932, EPI_ISL_1487933, EPI_ISL_1487934,
EPI_ISL_1487935, EPI_ISL_1487936, EPI_ISL_1487937, EPI_ISL_1487938, EPI_ISL_1487939, EPI_ISL_1487940, EPI_ISL_1487941, EPI_ISL_1487942, EPI_ISL_1487943, EPI_ISL_1487945, EPI_ISL_1487946, EPI_ISL_1487947, EPI_ISL_1487948, EPI_ISL_1487949, EPI_ISL_1487950, EPI_ISL_1487952, EPI_ISL_1487953, EPI_ISL_1487954, EPI_ISL_1487955, EPI_ISL_1487956, EPI_ISL_1487957, EPI_ISL_1487958,
EPI_ISL_1487959, EPI_ISL_1487960, EPI_ISL_1487962, EPI_ISL_1487963, EPI_ISL_1487964, EPI_ISL_1487965, EPI_ISL_1487966, EPI_ISL_1487968, EPI_ISL_1487969, EPI_ISL_1487971, EPI_ISL_1487972, EPI_ISL_1487973, EPI_ISL_1487974, EPI_ISL_1487975, EPI_ISL_1487976, EPI_ISL_1487977, EPI_ISL_1487978, EPI_ISL_1487979, EPI_ISL_1487980, EPI_ISL_1487981, EPI_ISL_1487982, EPI_ISL_1487983,
EPI_ISL_1487984, EPI_ISL_1487985, EPI_ISL_1487986, EPI_ISL_1487987, EPI_ISL_1487988, EPI_ISL_1487989, EPI_ISL_1487990, EPI_ISL_1487991, EPI_ISL_1487992, EPI_ISL_1487993, EPI_ISL_1487994, EPI_ISL_1487995, EPI_ISL_1487996, EPI_ISL_1487998, EPI_ISL_1488000, EPI_ISL_1488001, EPI_ISL_1488002, EPI_ISL_1488003, EPI_ISL_1488004, EPI_ISL_1488005, EPI_ISL_1488006, EPI_ISL_1488007,
EPI_ISL_1488008, EPI_ISL_1488009, EPI_ISL_1488010, EPI_ISL_1488011, EPI_ISL_1488012, EPI_ISL_1488013, EPI_ISL_1488014, EPI_ISL_1488015, EPI_ISL_1488016, EPI_ISL_1488017, EPI_ISL_1488018, EPI_ISL_1488019, EPI_ISL_1488020, EPI_ISL_1488021, EPI_ISL_1488022, EPI_ISL_1488023, EPI_ISL_1488024, EPI_ISL_1488025, EPI_ISL_1488026, EPI_ISL_1488027, EPI_ISL_1488028, EPI_ISL_1488029,
EPI_ISL_1488030, EPI_ISL_1488031, EPI_ISL_1488032, EPI_ISL_1488033, EPI_ISL_1488034, EPI_ISL_1488035, EPI_ISL_1488036, EPI_ISL_1488037, EPI_ISL_1488038, EPI_ISL_1488039, EPI_ISL_1488040, EPI_ISL_1488041, EPI_ISL_1488042, EPI_ISL_1488043, EPI_ISL_1488044, EPI_ISL_1488045, EPI_ISL_1488046, EPI_ISL_1488047, EPI_ISL_1488048, EPI_ISL_1488049, EPI_ISL_1488050, EPI_ISL_1488051,
EPI_ISL_1488052, EPI_ISL_1488053, EPI_ISL_1488055, EPI_ISL_1488057, EPI_ISL_1488058, EPI_ISL_1488059, EPI_ISL_1488060, EPI_ISL_1488061, EPI_ISL_1488062, EPI_ISL_1488063, EPI_ISL_1488064, EPI_ISL_1488065, EPI_ISL_1488066, EPI_ISL_1488067, EPI_ISL_1488068, EPI_ISL_1488069, EPI_ISL_1488070, EPI_ISL_1488072, EPI_ISL_1488073, EPI_ISL_1488074, EPI_ISL_1488075, EPI_ISL_1488076,
EPI_ISL_1488077, EPI_ISL_1488078, EPI_ISL_1488079, EPI_ISL_1488080, EPI_ISL_1488081, EPI_ISL_1488082, EPI_ISL_1488083, EPI_ISL_1488084, EPI_ISL_1488085, EPI_ISL_1488086, EPI_ISL_1488087, EPI_ISL_1488088, EPI_ISL_1488089, EPI_ISL_1488090, EPI_ISL_1488091, EPI_ISL_1488092, EPI_ISL_1488093, EPI_ISL_1488094, EPI_ISL_1488095, EPI_ISL_1488096, EPI_ISL_1488098, EPI_ISL_1488099,
EPI_ISL_1488100, EPI_ISL_1488102, EPI_ISL_1488103, EPI_ISL_1488104, EPI_ISL_1488105, EPI_ISL_1488106, EPI_ISL_1488107, EPI_ISL_1488108, EPI_ISL_1488109, EPI_ISL_1488110, EPI_ISL_1488111, EPI_ISL_1488112, EPI_ISL_1488113, EPI_ISL_1488114, EPI_ISL_1488115, EPI_ISL_1488116, EPI_ISL_1488117, EPI_ISL_1488118, EPI_ISL_1488119, EPI_ISL_1488120, EPI_ISL_1488121, EPI_ISL_1488122,
EPI_ISL_1488123, EPI_ISL_1488124, EPI_ISL_1488125, EPI_ISL_1488126, EPI_ISL_1488127, EPI_ISL_1488129, EPI_ISL_1488130, EPI_ISL_1488131, EPI_ISL_1488133, EPI_ISL_1488134, EPI_ISL_1488135, EPI_ISL_1488136, EPI_ISL_1488137, EPI_ISL_1488138, EPI_ISL_1488139, EPI_ISL_1488140, EPI_ISL_1488141, EPI_ISL_1488142, EPI_ISL_1488143, EPI_ISL_1488144, EPI_ISL_1488145, EPI_ISL_1488147,
EPI_ISL_1488148, EPI_ISL_1488149, EPI_ISL_1488150, EPI_ISL_1488151, EPI_ISL_1488152, EPI_ISL_1488154, EPI_ISL_1488155, EPI_ISL_1488156, EPI_ISL_1488157, EPI_ISL_1488158, EPI_ISL_1488159, EPI_ISL_1488160, EPI_ISL_1488161, EPI_ISL_1488162, EPI_ISL_1488163, EPI_ISL_1488164, EPI_ISL_1488166, EPI_ISL_1488167, EPI_ISL_1488169, EPI_ISL_1488170, EPI_ISL_1488171, EPI_ISL_1488172,
EPI_ISL_1488173, EPI_ISL_1488174, EPI_ISL_1488175, EPI_ISL_1488176, EPI_ISL_1488177, EPI_ISL_1488178, EPI_ISL_1488179, EPI_ISL_1488180, EPI_ISL_1488181, EPI_ISL_1488182, EPI_ISL_1488183, EPI_ISL_1488184, EPI_ISL_1488185, EPI_ISL_1488186, EPI_ISL_1488187, EPI_ISL_1488188, EPI_ISL_1488189, EPI_ISL_1488190, EPI_ISL_1488191, EPI_ISL_1488192, EPI_ISL_1488193, EPI_ISL_1488194,
EPI_ISL_1488195, EPI_ISL_1488196, EPI_ISL_1488197, EPI_ISL_1488198, EPI_ISL_1488199, EPI_ISL_1488200, EPI_ISL_1488201, EPI_ISL_1488203, EPI_ISL_1488204, EPI_ISL_1488205, EPI_ISL_1488206, EPI_ISL_1488207, EPI_ISL_1488208, EPI_ISL_1488209, EPI_ISL_1488210, EPI_ISL_1488211, EPI_ISL_1488212, EPI_ISL_1488213, EPI_ISL_1488214, EPI_ISL_1488215, EPI_ISL_1488216, EPI_ISL_1488217,
EPI_ISL_1488218, EPI_ISL_1488219, EPI_ISL_1488220, EPI_ISL_1488221, EPI_ISL_1488222, EPI_ISL_1488224, EPI_ISL_1488225, EPI_ISL_1488228, EPI_ISL_1488235, EPI_ISL_1488237, EPI_ISL_1488238, EPI_ISL_1488247, EPI_ISL_1488256, EPI_ISL_1488258, EPI_ISL_1488262, EPI_ISL_1488274, EPI_ISL_1488275, EPI_ISL_1488276, EPI_ISL_1488278, EPI_ISL_1488279, EPI_ISL_1488281, EPI_ISL_1488282,
EPI_ISL_1488287, EPI_ISL_1488299, EPI_ISL_1488302, EPI_ISL_1488305, EPI_ISL_1488310, EPI_ISL_1488318, EPI_ISL_1488319, EPI_ISL_1488327, EPI_ISL_1488330, EPI_ISL_1488334, EPI_ISL_1488340, EPI_ISL_1488350, EPI_ISL_1488352, EPI_ISL_1488357, EPI_ISL_1488358, EPI_ISL_1488363, EPI_ISL_1488370, EPI_ISL_1488371, EPI_ISL_1488372, EPI_ISL_1488374, EPI_ISL_1488378,
EPI_ISL_1488381, EPI_ISL_1488386, EPI_ISL_1488396, EPI_ISL_1488402, EPI_ISL_1488407, EPI_ISL_1488408, EPI_ISL_1488409, EPI_ISL_1488411, EPI_ISL_1488412, EPI_ISL_1488413, EPI_ISL_1488414, EPI_ISL_1488421, EPI_ISL_1488429, EPI_ISL_1488431, EPI_ISL_1488432, EPI_ISL_1488436, EPI_ISL_1488439, EPI_ISL_1488442, EPI_ISL_1488452, EPI_ISL_1488453, EPI_ISL_1488454, EPI_ISL_1488456,
EPI_ISL_1488457, EPI_ISL_1488460, EPI_ISL_1488462, EPI_ISL_1488465, EPI_ISL_1488470, EPI_ISL_1488471, EPI_ISL_1488475, EPI_ISL_1488480, EPI_ISL_1488484, EPI_ISL_1488487, EPI_ISL_1488491, EPI_ISL_1488492, EPI_ISL_1488493, EPI_ISL_1488497, EPI_ISL_1488499, EPI_ISL_1488505, EPI_ISL_1488506, EPI_ISL_1488507, EPI_ISL_1488508, EPI_ISL_1488511, EPI_ISL_1488512, EPI_ISL_1488515,
EPI_ISL_1488518, EPI_ISL_1488520, EPI_ISL_1488524, EPI_ISL_1488526, EPI_ISL_1488527, EPI_ISL_1488528, EPI_ISL_1488530, EPI_ISL_1488531, EPI_ISL_1488532, EPI_ISL_1488533, EPI_ISL_1488534, EPI_ISL_1488536, EPI_ISL_1488537, EPI_ISL_1488538, EPI_ISL_1488539, EPI_ISL_1488540, EPI_ISL_1488542, EPI_ISL_1488543, EPI_ISL_1488546, EPI_ISL_1488547, EPI_ISL_1488548, EPI_ISL_1488549,
EPI_ISL_1488551, EPI_ISL_1488553, EPI_ISL_1488554, EPI_ISL_1488556, EPI_ISL_1488559, EPI_ISL_1488562, EPI_ISL_1488564, EPI_ISL_1488567, EPI_ISL_1488569, EPI_ISL_1488570, EPI_ISL_1488572, EPI_ISL_1488573, EPI_ISL_1488577, EPI_ISL_1488578, EPI_ISL_1488583, EPI_ISL_1488584, EPI_ISL_1488586, EPI_ISL_1488587, EPI_ISL_1488588, EPI_ISL_1488589, EPI_ISL_1488590, EPI_ISL_1488591,
EPI_ISL_1488592, EPI_ISL_1488593, EPI_ISL_1488595, EPI_ISL_1488596, EPI_ISL_1488597, EPI_ISL_1488599, EPI_ISL_1488602, EPI_ISL_1488603, EPI_ISL_1488604, EPI_ISL_1488606, EPI_ISL_1488607, EPI_ISL_1488610, EPI_ISL_1488611, EPI_ISL_1488612, EPI_ISL_1488616, EPI_ISL_1488619, EPI_ISL_1488620, EPI_ISL_1488621, EPI_ISL_1488622, EPI_ISL_1488623, EPI_ISL_1488624, EPI_ISL_1488626,
EPI_ISL_1488628, EPI_ISL_1488629, EPI_ISL_1488631, EPI_ISL_1488634, EPI_ISL_1488635, EPI_ISL_1488636, EPI_ISL_1488638, EPI_ISL_1488639, EPI_ISL_1488642, EPI_ISL_1488643, EPI_ISL_1488645, EPI_ISL_1488648, EPI_ISL_1488650, EPI_ISL_1488652, EPI_ISL_1488653, EPI_ISL_1488655, EPI_ISL_1488656, EPI_ISL_1488657, EPI_ISL_1488658, EPI_ISL_1488660, EPI_ISL_1488661, EPI_ISL_1488662,
EPI_ISL_1488663, EPI_ISL_1488664, EPI_ISL_1488665, EPI_ISL_1488666, EPI_ISL_1488668, EPI_ISL_1488669, EPI_ISL_1488670, EPI_ISL_1488671, EPI_ISL_1488672, EPI_ISL_1488673, EPI_ISL_1488676, EPI_ISL_1488677, EPI_ISL_1488679, EPI_ISL_1488681, EPI_ISL_1488682, EPI_ISL_1488683, EPI_ISL_1488685, EPI_ISL_1488686, EPI_ISL_1488687, EPI_ISL_1488688, EPI_ISL_1488689, EPI_ISL_1488690,
EPI_ISL_1488691, EPI_ISL_1488692, EPI_ISL_1488693, EPI_ISL_1488694, EPI_ISL_1488696, EPI_ISL_1488697, EPI_ISL_1488698, EPI_ISL_1488699, EPI_ISL_1488700, EPI_ISL_1488701, EPI_ISL_1488702, EPI_ISL_1488703, EPI_ISL_1488705, EPI_ISL_1488707, EPI_ISL_1488708, EPI_ISL_1488710, EPI_ISL_1488711, EPI_ISL_1488712, EPI_ISL_1488713, EPI_ISL_1488715, EPI_ISL_1488718, EPI_ISL_1488720,
EPI_ISL_1488721, EPI_ISL_1488724, EPI_ISL_1488725, EPI_ISL_1488727, EPI_ISL_1488729, EPI_ISL_1488730, EPI_ISL_1488732, EPI_ISL_1488734, EPI_ISL_1488735, EPI_ISL_1488736, EPI_ISL_1488739, EPI_ISL_1488742, EPI_ISL_1488743, EPI_ISL_1488744, EPI_ISL_1488745, EPI_ISL_1488746, EPI_ISL_1488747, EPI_ISL_1488748, EPI_ISL_1488749, EPI_ISL_1488750, EPI_ISL_1488754, EPI_ISL_1488755,
EPI_ISL_1488758, EPI_ISL_1488759, EPI_ISL_1488760, EPI_ISL_1488766, EPI_ISL_1488767, EPI_ISL_1488768, EPI_ISL_1488769, EPI_ISL_1488771, EPI_ISL_1488772, EPI_ISL_1488773, EPI_ISL_1488775, EPI_ISL_1488776, EPI_ISL_1488778, EPI_ISL_1488779, EPI_ISL_1488780, EPI_ISL_1488781, EPI_ISL_1488783, EPI_ISL_1488786, EPI_ISL_1488787, EPI_ISL_1488788, EPI_ISL_1488790, EPI_ISL_1488791,
EPI_ISL_1488792, EPI_ISL_1488793, EPI_ISL_1488794, EPI_ISL_1488796, EPI_ISL_1488799, EPI_ISL_1488800, EPI_ISL_1488801, EPI_ISL_1488803, EPI_ISL_1488804, EPI_ISL_1488805, EPI_ISL_1488807, EPI_ISL_1488811, EPI_ISL_1488812, EPI_ISL_1488816, EPI_ISL_1488817, EPI_ISL_1488818, EPI_ISL_1488819, EPI_ISL_1488820, EPI_ISL_1488822, EPI_ISL_1488825, EPI_ISL_1488826, EPI_ISL_1488828,
EPI_ISL_1488829, EPI_ISL_1488831, EPI_ISL_1488833, EPI_ISL_1488837, EPI_ISL_1488840, EPI_ISL_1488842, EPI_ISL_1488843, EPI_ISL_1488844, EPI_ISL_1488845, EPI_ISL_1488846, EPI_ISL_1488847, EPI_ISL_1488848, EPI_ISL_1488849, EPI_ISL_1488850, EPI_ISL_1488851, EPI_ISL_1488852, EPI_ISL_1488853, EPI_ISL_1488854, EPI_ISL_1488855, EPI_ISL_1488856, EPI_ISL_1488860, EPI_ISL_1488863,
EPI_ISL_1488864, EPI_ISL_1488865, EPI_ISL_1488869, EPI_ISL_1488870, EPI_ISL_1488873, EPI_ISL_1488875, EPI_ISL_1488879, EPI_ISL_1488884, EPI_ISL_1488888, EPI_ISL_1488891, EPI_ISL_1488896, EPI_ISL_1488899, EPI_ISL_1488904, EPI_ISL_1488906, EPI_ISL_1488910, EPI_ISL_1488911, EPI_ISL_1488912, EPI_ISL_1488917, EPI_ISL_1488920, EPI_ISL_1488922, EPI_ISL_1488924, EPI_ISL_1488925,
EPI_ISL_1488931, EPI_ISL_1488937, EPI_ISL_1488939, EPI_ISL_1488942, EPI_ISL_1488943, EPI_ISL_1488946, EPI_ISL_1488947, EPI_ISL_1488949, EPI_ISL_1488951, EPI_ISL_1488952, EPI_ISL_1488962, EPI_ISL_1488966, EPI_ISL_1488968, EPI_ISL_1488970, EPI_ISL_1488972, EPI_ISL_1488973, EPI_ISL_1488975, EPI_ISL_1488978, EPI_ISL_1488983, EPI_ISL_1488993, EPI_ISL_1489001, EPI_ISL_1489003,
EPI_ISL_1489004, EPI_ISL_1489009, EPI_ISL_1489010, EPI_ISL_1489012, EPI_ISL_1489016, EPI_ISL_1489017, EPI_ISL_1489020, EPI_ISL_1489021, EPI_ISL_1489025, EPI_ISL_1489028, EPI_ISL_1489033, EPI_ISL_1489035, EPI_ISL_1489036, EPI_ISL_1489041, EPI_ISL_1489046, EPI_ISL_1489048, EPI_ISL_1489050, EPI_ISL_1489054, EPI_ISL_1489055, EPI_ISL_1489058, EPI_ISL_1489063,
EPI_ISL_1489072, EPI_ISL_1489075, EPI_ISL_1489078, EPI_ISL_1489087, EPI_ISL_1489089, EPI_ISL_1489093, EPI_ISL_1489094, EPI_ISL_1489097, EPI_ISL_1489100, EPI_ISL_1489101, EPI_ISL_1489104, EPI_ISL_1489108, EPI_ISL_1489109, EPI_ISL_1489111, EPI_ISL_1489113, EPI_ISL_1489115, EPI_ISL_1489116, EPI_ISL_1489122, EPI_ISL_1489126, EPI_ISL_1489138, EPI_ISL_1489140, EPI_ISL_1489141,
EPI_ISL_1489142, EPI_ISL_1489151, EPI_ISL_1489155, EPI_ISL_1489180, EPI_ISL_1489190, EPI_ISL_1489195, EPI_ISL_1489215, EPI_ISL_1489227, EPI_ISL_1489240, EPI_ISL_1489246, EPI_ISL_1489249, EPI_ISL_1489252, EPI_ISL_1489271, EPI_ISL_1489273, EPI_ISL_1489299, EPI_ISL_1489300, EPI_ISL_1489301, EPI_ISL_1489308, EPI_ISL_1489312, EPI_ISL_1489329, EPI_ISL_1489340,
EPI_ISL_1489355, EPI_ISL_1489365, EPI_ISL_1489381, EPI_ISL_1489383, EPI_ISL_1489386, EPI_ISL_1489398, EPI_ISL_1489408, EPI_ISL_1489425, EPI_ISL_1489428, EPI_ISL_1489435, EPI_ISL_1489457, EPI_ISL_1489458, EPI_ISL_1489459, EPI_ISL_1489460, EPI_ISL_1489461, EPI_ISL_1489463, EPI_ISL_1489464, EPI_ISL_1489465, EPI_ISL_1489467, EPI_ISL_1489468, EPI_ISL_1489469, EPI_ISL_1489470,
EPI_ISL_1489471, EPI_ISL_1489473, EPI_ISL_1489474, EPI_ISL_1489475, EPI_ISL_1489476, EPI_ISL_1489480, EPI_ISL_1489481, EPI_ISL_1489482, EPI_ISL_1489483, EPI_ISL_1489485, EPI_ISL_1489486, EPI_ISL_1489487, EPI_ISL_1489488, EPI_ISL_1489489, EPI_ISL_1489490, EPI_ISL_1489492, EPI_ISL_1489526, EPI_ISL_1505863, EPI_ISL_1506426, EPI_ISL_1506427, EPI_ISL_1506440, EPI_ISL_1506461,
EPI_ISL_1506480, EPI_ISL_1506532, EPI_ISL_1506533, EPI_ISL_1506534, EPI_ISL_1506535, EPI_ISL_1506536, EPI_ISL_1506549, EPI_ISL_1506564, EPI_ISL_1506571, EPI_ISL_1506573, EPI_ISL_1506574, EPI_ISL_1506668, EPI_ISL_1507042, EPI_ISL_1507043, EPI_ISL_1507044, EPI_ISL_1507196, EPI_ISL_1507197, EPI_ISL_1507211, EPI_ISL_1507213, EPI_ISL_1507232, EPI_ISL_1507519, EPI_ISL_1507573,
EPI_ISL_1507574, EPI_ISL_1507576, EPI_ISL_1507577, EPI_ISL_1507578, EPI_ISL_1507579, EPI_ISL_1507580, EPI_ISL_1507707, EPI_ISL_1507708, EPI_ISL_1507709, EPI_ISL_1518624, EPI_ISL_1518776, EPI_ISL_1518829, EPI_ISL_1518850, EPI_ISL_1536153, EPI_ISL_1536169, EPI_ISL_1565342, EPI_ISL_1630518, EPI_ISL_1630519, EPI_ISL_1630527, EPI_ISL_1630529, EPI_ISL_1630534,
EPI_ISL_1630537, EPI_ISL_1630538, EPI_ISL_1630539, EPI_ISL_1630540, EPI_ISL_1630541, EPI_ISL_1630545, EPI_ISL_1630547, EPI_ISL_1630549, EPI_ISL_1630551, EPI_ISL_1630554, EPI_ISL_1630559, EPI_ISL_1630560, EPI_ISL_1630562, EPI_ISL_1630565, EPI_ISL_1630570, EPI_ISL_1630573, EPI_ISL_1630574, EPI_ISL_1630575, EPI_ISL_1630578, EPI_ISL_1630581, EPI_ISL_1630585, EPI_ISL_1630588,
EPI_ISL_1630593, EPI_ISL_1630595, EPI_ISL_1630596, EPI_ISL_1630602, EPI_ISL_1630603, EPI_ISL_1630605, EPI_ISL_1630609, EPI_ISL_1630611, EPI_ISL_1630613, EPI_ISL_1630615, EPI_ISL_1630619, EPI_ISL_1630625, EPI_ISL_1630628, EPI_ISL_1630632, EPI_ISL_1630633, EPI_ISL_1630642, EPI_ISL_1630644, EPI_ISL_1630645, EPI_ISL_1630646, EPI_ISL_1630647, EPI_ISL_1630649,
EPI_ISL_1630654, EPI_ISL_1630655, EPI_ISL_1630656, EPI_ISL_1630657, EPI_ISL_1630658, EPI_ISL_1630660, EPI_ISL_1630662, EPI_ISL_1630666, EPI_ISL_1630669, EPI_ISL_1630673, EPI_ISL_1630680, EPI_ISL_1630681, EPI_ISL_1630685, EPI_ISL_1630686, EPI_ISL_1630689, EPI_ISL_1630693, EPI_ISL_1630695, EPI_ISL_1630701, EPI_ISL_1630703, EPI_ISL_1630705, EPI_ISL_1630707, EPI_ISL_1630709,
EPI_ISL_1630712, EPI_ISL_1630716, EPI_ISL_1630718, EPI_ISL_1630720, EPI_ISL_1630722, EPI_ISL_1630729, EPI_ISL_1630731, EPI_ISL_1630736, EPI_ISL_1630737, EPI_ISL_1630738, EPI_ISL_1630740, EPI_ISL_1630743, EPI_ISL_1630746, EPI_ISL_1630751, EPI_ISL_1630755, EPI_ISL_1630757, EPI_ISL_1630759, EPI_ISL_1630764, EPI_ISL_1630766, EPI_ISL_1630768,
EPI_ISL_1630769, EPI_ISL_1630777, EPI_ISL_1630780, EPI_ISL_1630781, EPI_ISL_1630782, EPI_ISL_1630783, EPI_ISL_1630785, EPI_ISL_1630787, EPI_ISL_1630791, EPI_ISL_1630794, EPI_ISL_1630795, EPI_ISL_1630796, EPI_ISL_1630797, EPI_ISL_1630798, EPI_ISL_1630799, EPI_ISL_1630801, EPI_ISL_1630810, EPI_ISL_1630811, EPI_ISL_1630812, EPI_ISL_1630813, EPI_ISL_1630821, EPI_ISL_1653738

| | | | |
|---|---|---|---|
| see above | Lighthouse Lab in Alderley Park | Wellcome Sanger Institute for the COVID-19 Genomics UK (COG-UK) Consortium | Cordelia Langford; David K. Jackson; Dominic Kwiatkowski; Ewan Harrison; Ian Johnston; Jacquelyn Wynn; Jeffrey Barrett; John Sillitoe on behalf of the Wellcome Sanger Institute COVID-19 Surveillance Team; Mairead Hyland; Roberto Amato; Sonia Goncalves; The Lighthouse Lab in Alderley Park and Alex Alderton |

EPI_ISL_1409196, EPI_ISL_1409197, EPI_ISL_1409213, EPI_ISL_1409226, EPI_ISL_1409249, EPI_ISL_1409250, EPI_ISL_1409266, EPI_ISL_1409274, EPI_ISL_1409292, EPI_ISL_1409307, EPI_ISL_1409317, EPI_ISL_1409337, EPI_ISL_1409341, EPI_ISL_1409349, EPI_ISL_1409356, EPI_ISL_1409378, EPI_ISL_1409380, EPI_ISL_1409384, EPI_ISL_1409389, EPI_ISL_1409390, EPI_ISL_1409404, EPI_ISL_1409417,
EPI_ISL_1409450, EPI_ISL_1409452, EPI_ISL_1409465, EPI_ISL_1409470, EPI_ISL_1409472, EPI_ISL_1409474, EPI_ISL_1411573, EPI_ISL_1411850, EPI_ISL_1412998, EPI_ISL_1451160, EPI_ISL_1451166, EPI_ISL_1451199, EPI_ISL_1451200, EPI_ISL_1451222, EPI_ISL_1451226, EPI_ISL_1451238, EPI_ISL_1451249, EPI_ISL_1451252, EPI_ISL_1451261, EPI_ISL_1451281, EPI_ISL_1451294, EPI_ISL_1451308,
EPI_ISL_1451364, EPI_ISL_1451368, EPI_ISL_1451373, EPI_ISL_1451383, EPI_ISL_1451386, EPI_ISL_1451395, EPI_ISL_1451409, EPI_ISL_1451415, EPI_ISL_1451425, EPI_ISL_1451450, EPI_ISL_1451451, EPI_ISL_1451452, EPI_ISL_1451454, EPI_ISL_1451471, EPI_ISL_1451508, EPI_ISL_1451537, EPI_ISL_1451541, EPI_ISL_1451555, EPI_ISL_1451556, EPI_ISL_1451576, EPI_ISL_1451579, EPI_ISL_1451581,
EPI_ISL_1451600, EPI_ISL_1451613, EPI_ISL_1451620, EPI_ISL_1451628, EPI_ISL_1451641, EPI_ISL_1451653, EPI_ISL_1451657, EPI_ISL_1451663, EPI_ISL_1451670, EPI_ISL_1451682, EPI_ISL_1451685, EPI_ISL_1451707, EPI_ISL_1451708, EPI_ISL_1451719, EPI_ISL_1451752, EPI_ISL_1451778, EPI_ISL_1451779, EPI_ISL_1453489, EPI_ISL_1454303, EPI_ISL_1454317, EPI_ISL_1455637, EPI_ISL_1455646,
EPI_ISL_1455647, EPI_ISL_1465925, EPI_ISL_1465927, EPI_ISL_1465936, EPI_ISL_1465941, EPI_ISL_1465952, EPI_ISL_1465971, EPI_ISL_1465974, EPI_ISL_1466009, EPI_ISL_1466010, EPI_ISL_1466011, EPI_ISL_1466016, EPI_ISL_1466020, EPI_ISL_1466024, EPI_ISL_1466044, EPI_ISL_1466055, EPI_ISL_1466101, EPI_ISL_1466113, EPI_ISL_1466121, EPI_ISL_1466125, EPI_ISL_1466148, EPI_ISL_1473449,
EPI_ISL_1473452, EPI_ISL_1473453, EPI_ISL_1473457, EPI_ISL_1473459, EPI_ISL_1473460, EPI_ISL_1473461, EPI_ISL_1473464, EPI_ISL_1473465, EPI_ISL_1473466, EPI_ISL_1473468, EPI_ISL_1473469, EPI_ISL_1473471, EPI_ISL_1473475, EPI_ISL_1473479, EPI_ISL_1473483, EPI_ISL_1473485, EPI_ISL_1473486, EPI_ISL_1473489, EPI_ISL_1473492, EPI_ISL_1473494, EPI_ISL_1473495, EPI_ISL_1473497,
EPI_ISL_1473498, EPI_ISL_1473499, EPI_ISL_1473500, EPI_ISL_1473504, EPI_ISL_1473505, EPI_ISL_1473508, EPI_ISL_1473512, EPI_ISL_1473516, EPI_ISL_1473519, EPI_ISL_1473525, EPI_ISL_1473527, EPI_ISL_1473529, EPI_ISL_1473531, EPI_ISL_1473532, EPI_ISL_1473536, EPI_ISL_1473538, EPI_ISL_1473545, EPI_ISL_1473546, EPI_ISL_1473548, EPI_ISL_1473551, EPI_ISL_1473553, EPI_ISL_1473555,
EPI_ISL_1473562, EPI_ISL_1473563, EPI_ISL_1473564, EPI_ISL_1473565, EPI_ISL_1473569, EPI_ISL_1473570, EPI_ISL_1473574, EPI_ISL_1473577, EPI_ISL_1473587, EPI_ISL_1473592, EPI_ISL_1473593, EPI_ISL_1473596, EPI_ISL_1473598, EPI_ISL_1473600, EPI_ISL_1473608, EPI_ISL_1473618, EPI_ISL_1473620, EPI_ISL_1473632, EPI_ISL_1473634, EPI_ISL_1473639, EPI_ISL_1473642, EPI_ISL_1473643,
EPI_ISL_1473644, EPI_ISL_1473646, EPI_ISL_1473648, EPI_ISL_1473651, EPI_ISL_1473655, EPI_ISL_1473656, EPI_ISL_1473657, EPI_ISL_1473664, EPI_ISL_1473668, EPI_ISL_1473678, EPI_ISL_1473683, EPI_ISL_1473685, EPI_ISL_1473686, EPI_ISL_1473688, EPI_ISL_1473693, EPI_ISL_1473697, EPI_ISL_1473703, EPI_ISL_1473704, EPI_ISL_1473706, EPI_ISL_1473711, EPI_ISL_1473714, EPI_ISL_1473715,
EPI_ISL_1473716, EPI_ISL_1473719, EPI_ISL_1473721, EPI_ISL_1473723, EPI_ISL_1473725, EPI_ISL_1473728, EPI_ISL_1473733, EPI_ISL_1473737, EPI_ISL_1473740, EPI_ISL_1473742, EPI_ISL_1473743, EPI_ISL_1473757, EPI_ISL_1473763, EPI_ISL_1473767, EPI_ISL_1473769, EPI_ISL_1473770, EPI_ISL_1473771, EPI_ISL_1473773, EPI_ISL_1473778, EPI_ISL_1473779, EPI_ISL_1473782, EPI_ISL_1473787,
EPI_ISL_1473788, EPI_ISL_1473795, EPI_ISL_1473797, EPI_ISL_1473799, EPI_ISL_1483829, EPI_ISL_1484065, EPI_ISL_1484426, EPI_ISL_1484501, EPI_ISL_1484517, EPI_ISL_1484532, EPI_ISL_1484547, EPI_ISL_1484552, EPI_ISL_1484587, EPI_ISL_1484598, EPI_ISL_1484601, EPI_ISL_1487726, EPI_ISL_1487744, EPI_ISL_1487791, EPI_ISL_1487813, EPI_ISL_1487831, EPI_ISL_1487832, EPI_ISL_1487852,
EPI_ISL_1487853, EPI_ISL_1487854, EPI_ISL_1487855, EPI_ISL_1487856, EPI_ISL_1487857, EPI_ISL_1487859, EPI_ISL_1487860, EPI_ISL_1487861, EPI_ISL_1487862, EPI_ISL_1487863, EPI_ISL_1487864, EPI_ISL_1487865, EPI_ISL_1487866, EPI_ISL_1487867, EPI_ISL_1487868, EPI_ISL_1487869, EPI_ISL_1487870, EPI_ISL_1487871, EPI_ISL_1487872, EPI_ISL_1504713, EPI_ISL_1504717, EPI_ISL_1504832,
EPI_ISL_1504848, EPI_ISL_1506105, EPI_ISL_1506106, EPI_ISL_1506107, EPI_ISL_1506111, EPI_ISL_1506112, EPI_ISL_1506113, EPI_ISL_1506129, EPI_ISL_1506133, EPI_ISL_1506520, EPI_ISL_1506528, EPI_ISL_1506529, EPI_ISL_1506530, EPI_ISL_1506531, EPI_ISL_1507942, EPI_ISL_1519277, EPI_ISL_1519291, EPI_ISL_1519301, EPI_ISL_1519306, EPI_ISL_1584081

| | | | |
|---|---|---|---|
| see above | Lighthouse Lab in Cambridge | Wellcome Sanger Institute for the COVID-19 Genomics UK (COG-UK) Consortium | Cordelia Langford; David K. Jackson; Dominic Kwiatkowski; Ewan Harrison; Ian Johnston; Jeffrey Barrett; John Sillitoe on behalf of the Wellcome Sanger Institute COVID-19 Surveillance Team; Rob Howes; Roberto Amato; Sonia Goncalves; The Lighthouse Lab in Cambridge and Alex Alderton |

EPI_ISL_1378048, EPI_ISL_1416174, EPI_ISL_1448789, EPI_ISL_1448790, EPI_ISL_1448792, EPI_ISL_1448793, EPI_ISL_1448794, EPI_ISL_1448795, EPI_ISL_1448796, EPI_ISL_1448800, EPI_ISL_1448802, EPI_ISL_1448803, EPI_ISL_1448805, EPI_ISL_1448806, EPI_ISL_1448808, EPI_ISL_1448810, EPI_ISL_1448811, EPI_ISL_1448812, EPI_ISL_1448813, EPI_ISL_1448814, EPI_ISL_1448815, EPI_ISL_1448816,
EPI_ISL_1448817, EPI_ISL_1448819, EPI_ISL_1448820, EPI_ISL_1448821, EPI_ISL_1448822, EPI_ISL_1448823, EPI_ISL_1448824, EPI_ISL_1448825, EPI_ISL_1448826, EPI_ISL_1448827, EPI_ISL_1448828, EPI_ISL_1448830, EPI_ISL_1448831, EPI_ISL_1448832, EPI_ISL_1448833, EPI_ISL_1448834, EPI_ISL_1448835, EPI_ISL_1448836, EPI_ISL_1448837, EPI_ISL_1448839, EPI_ISL_1448841, EPI_ISL_1448842,
EPI_ISL_1448844, EPI_ISL_1448845, EPI_ISL_1448846, EPI_ISL_1448848, EPI_ISL_1448849, EPI_ISL_1448850, EPI_ISL_1448851, EPI_ISL_1448852, EPI_ISL_1448853, EPI_ISL_1448855, EPI_ISL_1448856, EPI_ISL_1448858, EPI_ISL_1448861, EPI_ISL_1448862, EPI_ISL_1448864, EPI_ISL_1448865, EPI_ISL_1448866, EPI_ISL_1448867, EPI_ISL_1448869, EPI_ISL_1448870, EPI_ISL_1448871, EPI_ISL_1448872,
EPI_ISL_1448873, EPI_ISL_1448875, EPI_ISL_1448877, EPI_ISL_1448878, EPI_ISL_1448879, EPI_ISL_1448880, EPI_ISL_1448881, EPI_ISL_1448882, EPI_ISL_1448883, EPI_ISL_1448884, EPI_ISL_1448885, EPI_ISL_1448886, EPI_ISL_1448887, EPI_ISL_1448888, EPI_ISL_1448889, EPI_ISL_1448890, EPI_ISL_1448891, EPI_ISL_1448892, EPI_ISL_1448893, EPI_ISL_1448894, EPI_ISL_1448895, EPI_ISL_1448897,
EPI_ISL_1448898, EPI_ISL_1448900, EPI_ISL_1448901, EPI_ISL_1448902, EPI_ISL_1448903, EPI_ISL_1448904, EPI_ISL_1448906, EPI_ISL_1448907, EPI_ISL_1448910, EPI_ISL_1448911, EPI_ISL_1448913, EPI_ISL_1448914, EPI_ISL_1448915, EPI_ISL_1448916, EPI_ISL_1448918, EPI_ISL_1448921, EPI_ISL_1448922, EPI_ISL_1448923, EPI_ISL_1448924, EPI_ISL_1448925, EPI_ISL_1448926,
EPI_ISL_1448928, EPI_ISL_1448929, EPI_ISL_1448930, EPI_ISL_1448931, EPI_ISL_1448932, EPI_ISL_1448934, EPI_ISL_1448935, EPI_ISL_1448936, EPI_ISL_1448937, EPI_ISL_1448938, EPI_ISL_1448940, EPI_ISL_1448941, EPI_ISL_1448942, EPI_ISL_1448943, EPI_ISL_1448944, EPI_ISL_1448945, EPI_ISL_1448946, EPI_ISL_1448950, EPI_ISL_1448953, EPI_ISL_1448955, EPI_ISL_1448956, EPI_ISL_1448957,
EPI_ISL_1448958, EPI_ISL_1448959, EPI_ISL_1448960, EPI_ISL_1448961, EPI_ISL_1448962, EPI_ISL_1448963, EPI_ISL_1448964, EPI_ISL_1448966, EPI_ISL_1448967, EPI_ISL_1448969, EPI_ISL_1448973, EPI_ISL_1448974, EPI_ISL_1448978, EPI_ISL_1448980, EPI_ISL_1448981, EPI_ISL_1448983, EPI_ISL_1448985, EPI_ISL_1448987, EPI_ISL_1448988,
EPI_ISL_1448991, EPI_ISL_1448992, EPI_ISL_1448995, EPI_ISL_1448996, EPI_ISL_1448997, EPI_ISL_1448998, EPI_ISL_1448999, EPI_ISL_1449000, EPI_ISL_1449001, EPI_ISL_1449002, EPI_ISL_1449003, EPI_ISL_1449004, EPI_ISL_1449006, EPI_ISL_1449007, EPI_ISL_1449008, EPI_ISL_1449009, EPI_ISL_1449010, EPI_ISL_1449011, EPI_ISL_1449012, EPI_ISL_1449013, EPI_ISL_1449014, EPI_ISL_1449016,
EPI_ISL_1449020, EPI_ISL_1449022, EPI_ISL_1449027, EPI_ISL_1449028, EPI_ISL_1449353, EPI_ISL_1449354, EPI_ISL_1449355, EPI_ISL_1449356, EPI_ISL_1449359, EPI_ISL_1449361, EPI_ISL_1449363, EPI_ISL_1449366, EPI_ISL_1449367, EPI_ISL_1449368, EPI_ISL_1449369, EPI_ISL_1449372, EPI_ISL_1449377, EPI_ISL_1449378, EPI_ISL_1449379, EPI_ISL_1449388, EPI_ISL_1449389,
EPI_ISL_1449390, EPI_ISL_1449395, EPI_ISL_1449396, EPI_ISL_1449397, EPI_ISL_1449398, EPI_ISL_1449400, EPI_ISL_1449404, EPI_ISL_1449405, EPI_ISL_1449406, EPI_ISL_1449409, EPI_ISL_1449410, EPI_ISL_1449416, EPI_ISL_1449418, EPI_ISL_1449420, EPI_ISL_1449425, EPI_ISL_1449429, EPI_ISL_1449430, EPI_ISL_1449433, EPI_ISL_1449434, EPI_ISL_1449435, EPI_ISL_1449437, EPI_ISL_1449441,

EPI_ISL_1449443, EPI_ISL_1449444, EPI_ISL_1449445, EPI_ISL_1449446, EPI_ISL_1449448, EPI_ISL_1449461, EPI_ISL_1449462, EPI_ISL_1449468, EPI_ISL_1449471, EPI_ISL_1449472, EPI_ISL_1449474, EPI_ISL_1449478, EPI_ISL_1449482, EPI_ISL_1449483, EPI_ISL_1449484, EPI_ISL_1449486, EPI_ISL_1449492, EPI_ISL_1449493, EPI_ISL_1449498, EPI_ISL_1449501, EPI_ISL_1449503, EPI_ISL_1449509,
EPI_ISL_1449516, EPI_ISL_1449517, EPI_ISL_1449518, EPI_ISL_1449520, EPI_ISL_1449524, EPI_ISL_1449527, EPI_ISL_1449528, EPI_ISL_1449529, EPI_ISL_1449533, EPI_ISL_1449535, EPI_ISL_1449537, EPI_ISL_1449541, EPI_ISL_1449542, EPI_ISL_1449546, EPI_ISL_1449548, EPI_ISL_1449550, EPI_ISL_1449551, EPI_ISL_1449553, EPI_ISL_1449554, EPI_ISL_1449556, EPI_ISL_1449558, EPI_ISL_1449560,
EPI_ISL_1449562, EPI_ISL_1449563, EPI_ISL_1449564, EPI_ISL_1449571, EPI_ISL_1449572, EPI_ISL_1449575, EPI_ISL_1449576, EPI_ISL_1449581, EPI_ISL_1449582, EPI_ISL_1449583, EPI_ISL_1449585, EPI_ISL_1449586, EPI_ISL_1449588, EPI_ISL_1449591, EPI_ISL_1449592, EPI_ISL_1449593, EPI_ISL_1449594, EPI_ISL_1449595, EPI_ISL_1449597, EPI_ISL_1449598, EPI_ISL_1449600, EPI_ISL_1449601,
EPI_ISL_1449602, EPI_ISL_1449603, EPI_ISL_1449604, EPI_ISL_1449606, EPI_ISL_1449607, EPI_ISL_1449608, EPI_ISL_1449609, EPI_ISL_1449611, EPI_ISL_1449614, EPI_ISL_1449616, EPI_ISL_1449617, EPI_ISL_1449619, EPI_ISL_1449623, EPI_ISL_1449624, EPI_ISL_1449626, EPI_ISL_1449628, EPI_ISL_1449631, EPI_ISL_1449632, EPI_ISL_1449635, EPI_ISL_1449636, EPI_ISL_1449638, EPI_ISL_1449644,
EPI_ISL_1449645, EPI_ISL_1449646, EPI_ISL_1449648, EPI_ISL_1449649, EPI_ISL_1450895, EPI_ISL_1450896, EPI_ISL_1450897, EPI_ISL_1450898, EPI_ISL_1450899, EPI_ISL_1450901, EPI_ISL_1450904, EPI_ISL_1450906, EPI_ISL_1450910, EPI_ISL_1450915, EPI_ISL_1450918, EPI_ISL_1450919, EPI_ISL_1450920, EPI_ISL_1450922, EPI_ISL_1450927, EPI_ISL_1450930, EPI_ISL_1450932, EPI_ISL_1450934,
EPI_ISL_1450935, EPI_ISL_1450936, EPI_ISL_1450937, EPI_ISL_1450941, EPI_ISL_1450945, EPI_ISL_1450947, EPI_ISL_1450949, EPI_ISL_1450950, EPI_ISL_1450954, EPI_ISL_1450956, EPI_ISL_1450958, EPI_ISL_1450965, EPI_ISL_1450967, EPI_ISL_1450969, EPI_ISL_1450970, EPI_ISL_1450973, EPI_ISL_1450974, EPI_ISL_1450978, EPI_ISL_1450981, EPI_ISL_1450982, EPI_ISL_1450989, EPI_ISL_1450991,
EPI_ISL_1450993, EPI_ISL_1450997, EPI_ISL_1451001, EPI_ISL_1451009, EPI_ISL_1451010, EPI_ISL_1451011, EPI_ISL_1451012, EPI_ISL_1451014, EPI_ISL_1451015, EPI_ISL_1451016, EPI_ISL_1451024, EPI_ISL_1451025, EPI_ISL_1451028, EPI_ISL_1451029, EPI_ISL_1451030, EPI_ISL_1451034, EPI_ISL_1451035, EPI_ISL_1451039, EPI_ISL_1451042, EPI_ISL_1451043, EPI_ISL_1451044, EPI_ISL_1451045,
EPI_ISL_1451048, EPI_ISL_1451052, EPI_ISL_1451056, EPI_ISL_1451058, EPI_ISL_1451059, EPI_ISL_1451064, EPI_ISL_1451067, EPI_ISL_1451068, EPI_ISL_1451071, EPI_ISL_1451072, EPI_ISL_1451076, EPI_ISL_1451081, EPI_ISL_1451089, EPI_ISL_1451090, EPI_ISL_1451095, EPI_ISL_1451097, EPI_ISL_1451102, EPI_ISL_1451103, EPI_ISL_1451105, EPI_ISL_1451106, EPI_ISL_1451107, EPI_ISL_1451114,
EPI_ISL_1451116, EPI_ISL_1451117, EPI_ISL_1451118, EPI_ISL_1451119, EPI_ISL_1451120, EPI_ISL_1451122, EPI_ISL_1451126, EPI_ISL_1451129, EPI_ISL_1451130, EPI_ISL_1451131, EPI_ISL_1451147, EPI_ISL_1451149, EPI_ISL_1451472, EPI_ISL_1451475, EPI_ISL_1451481, EPI_ISL_1451487, EPI_ISL_1451496, EPI_ISL_1451497, EPI_ISL_1451502, EPI_ISL_1451503, EPI_ISL_1451505, EPI_ISL_1451509,
EPI_ISL_1451519, EPI_ISL_1451520, EPI_ISL_1451533, EPI_ISL_1451534, EPI_ISL_1451538, EPI_ISL_1451539, EPI_ISL_1451540, EPI_ISL_1451547, EPI_ISL_1451561, EPI_ISL_1451567, EPI_ISL_1451571, EPI_ISL_1451572, EPI_ISL_1451573, EPI_ISL_1451577, EPI_ISL_1451580, EPI_ISL_1451585, EPI_ISL_1451590, EPI_ISL_1451599, EPI_ISL_1451601, EPI_ISL_1451607, EPI_ISL_1451609, EPI_ISL_1451615,
EPI_ISL_1451633, EPI_ISL_1451634, EPI_ISL_1451640, EPI_ISL_1451648, EPI_ISL_1451651, EPI_ISL_1451654, EPI_ISL_1451666, EPI_ISL_1451667, EPI_ISL_1451668, EPI_ISL_1451671, EPI_ISL_1451676, EPI_ISL_1451677, EPI_ISL_1451678, EPI_ISL_1451683, EPI_ISL_1451684, EPI_ISL_1451687, EPI_ISL_1451690, EPI_ISL_1451693, EPI_ISL_1451694, EPI_ISL_1451697, EPI_ISL_1451706, EPI_ISL_1451713,
EPI_ISL_1451717, EPI_ISL_1451729, EPI_ISL_1451732, EPI_ISL_1451736, EPI_ISL_1451747, EPI_ISL_1451750, EPI_ISL_1451754, EPI_ISL_1451756, EPI_ISL_1451757, EPI_ISL_1451760, EPI_ISL_1451761, EPI_ISL_1451766, EPI_ISL_1451769, EPI_ISL_1451785, EPI_ISL_1451786, EPI_ISL_1451787, EPI_ISL_1451789, EPI_ISL_1451790, EPI_ISL_1451791, EPI_ISL_1451793, EPI_ISL_1451796, EPI_ISL_1451799,
EPI_ISL_1451802, EPI_ISL_1451805, EPI_ISL_1451806, EPI_ISL_1451811, EPI_ISL_1451812, EPI_ISL_1451815, EPI_ISL_1451818, EPI_ISL_1451829, EPI_ISL_1451832, EPI_ISL_1451834, EPI_ISL_1451837, EPI_ISL_1451838, EPI_ISL_1451840, EPI_ISL_1451841, EPI_ISL_1451842, EPI_ISL_1451845, EPI_ISL_1451850, EPI_ISL_1451852, EPI_ISL_1451858, EPI_ISL_1451859, EPI_ISL_1451862, EPI_ISL_1451869,
EPI_ISL_1451870, EPI_ISL_1451872, EPI_ISL_1451875, EPI_ISL_1451876, EPI_ISL_1451879, EPI_ISL_1451881, EPI_ISL_1451882, EPI_ISL_1451883, EPI_ISL_1451886, EPI_ISL_1451887, EPI_ISL_1451888, EPI_ISL_1451892, EPI_ISL_1451896, EPI_ISL_1451900, EPI_ISL_1451903, EPI_ISL_1451904, EPI_ISL_1451907, EPI_ISL_1451909, EPI_ISL_1451914, EPI_ISL_1451917, EPI_ISL_1451918, EPI_ISL_1451919,
EPI_ISL_1451922, EPI_ISL_1451930, EPI_ISL_1451931, EPI_ISL_1451932, EPI_ISL_1451935, EPI_ISL_1451936, EPI_ISL_1451937, EPI_ISL_1451942, EPI_ISL_1451956, EPI_ISL_1451957, EPI_ISL_1451958, EPI_ISL_1451966, EPI_ISL_1451967, EPI_ISL_1451969, EPI_ISL_1451970, EPI_ISL_1451975, EPI_ISL_1451977, EPI_ISL_1451979, EPI_ISL_1451980, EPI_ISL_1451987, EPI_ISL_1451988, EPI_ISL_1451992,
EPI_ISL_1451998, EPI_ISL_1451999, EPI_ISL_1452002, EPI_ISL_1452003, EPI_ISL_1452005, EPI_ISL_1452008, EPI_ISL_1452010, EPI_ISL_1452011, EPI_ISL_1452014, EPI_ISL_1452016, EPI_ISL_1452017, EPI_ISL_1452019, EPI_ISL_1452030, EPI_ISL_1452032, EPI_ISL_1452035, EPI_ISL_1452036, EPI_ISL_1452039, EPI_ISL_1452040, EPI_ISL_1452041, EPI_ISL_1452043, EPI_ISL_1452050, EPI_ISL_1452051,
EPI_ISL_1452052, EPI_ISL_1452053, EPI_ISL_1452054, EPI_ISL_1452056, EPI_ISL_1452060, EPI_ISL_1452061, EPI_ISL_1452062, EPI_ISL_1452063, EPI_ISL_1452073, EPI_ISL_1452500, EPI_ISL_1452668, EPI_ISL_1452675, EPI_ISL_1452726, EPI_ISL_1453445, EPI_ISL_1465897, EPI_ISL_1465898, EPI_ISL_1465899, EPI_ISL_1465900, EPI_ISL_1465901, EPI_ISL_1465902, EPI_ISL_1465903, EPI_ISL_1465908,
EPI_ISL_1465909, EPI_ISL_1465910, EPI_ISL_1465911, EPI_ISL_1465912, EPI_ISL_1465914, EPI_ISL_1465916, EPI_ISL_1465917, EPI_ISL_1465918, EPI_ISL_1465919, EPI_ISL_1465920, EPI_ISL_1465921, EPI_ISL_1465922, EPI_ISL_1465923, EPI_ISL_1465924, EPI_ISL_1465928, EPI_ISL_1465931, EPI_ISL_1465932, EPI_ISL_1465933, EPI_ISL_1465934, EPI_ISL_1465935, EPI_ISL_1465937, EPI_ISL_1465938,
EPI_ISL_1465939, EPI_ISL_1465940, EPI_ISL_1465942, EPI_ISL_1465943, EPI_ISL_1465945, EPI_ISL_1465946, EPI_ISL_1465947, EPI_ISL_1465948, EPI_ISL_1465950, EPI_ISL_1465951, EPI_ISL_1465953, EPI_ISL_1465954, EPI_ISL_1465955, EPI_ISL_1465956, EPI_ISL_1465958, EPI_ISL_1465959, EPI_ISL_1465960, EPI_ISL_1465961, EPI_ISL_1465962, EPI_ISL_1465963, EPI_ISL_1465964, EPI_ISL_1465965,
EPI_ISL_1465966, EPI_ISL_1465968, EPI_ISL_1465969, EPI_ISL_1465972, EPI_ISL_1465976, EPI_ISL_1465977, EPI_ISL_1465978, EPI_ISL_1465979, EPI_ISL_1465980, EPI_ISL_1465981, EPI_ISL_1465983, EPI_ISL_1465984, EPI_ISL_1465985, EPI_ISL_1465986, EPI_ISL_1465987, EPI_ISL_1465988, EPI_ISL_1465989, EPI_ISL_1465990, EPI_ISL_1465991, EPI_ISL_1465993, EPI_ISL_1465994, EPI_ISL_1465995,
EPI_ISL_1465996, EPI_ISL_1465997, EPI_ISL_1465998, EPI_ISL_1465999, EPI_ISL_1466000, EPI_ISL_1466001, EPI_ISL_1466002, EPI_ISL_1466003, EPI_ISL_1466004, EPI_ISL_1466005, EPI_ISL_1466007, EPI_ISL_1466008, EPI_ISL_1466012, EPI_ISL_1466014, EPI_ISL_1466015, EPI_ISL_1466017, EPI_ISL_1466018, EPI_ISL_1466021, EPI_ISL_1466022, EPI_ISL_1466025, EPI_ISL_1466026, EPI_ISL_1466027,
EPI_ISL_1466028, EPI_ISL_1466030, EPI_ISL_1466031, EPI_ISL_1466032, EPI_ISL_1466033, EPI_ISL_1466035, EPI_ISL_1466036, EPI_ISL_1466037, EPI_ISL_1466038, EPI_ISL_1466040, EPI_ISL_1466041, EPI_ISL_1466042, EPI_ISL_1466043, EPI_ISL_1466045, EPI_ISL_1466047, EPI_ISL_1466048, EPI_ISL_1466049, EPI_ISL_1466050, EPI_ISL_1466051, EPI_ISL_1466052, EPI_ISL_1466053, EPI_ISL_1466056,
EPI_ISL_1466057, EPI_ISL_1466058, EPI_ISL_1466059, EPI_ISL_1466060, EPI_ISL_1466061, EPI_ISL_1466062, EPI_ISL_1466063, EPI_ISL_1466064, EPI_ISL_1466065, EPI_ISL_1466066, EPI_ISL_1466068, EPI_ISL_1466069, EPI_ISL_1466070, EPI_ISL_1466071, EPI_ISL_1466072, EPI_ISL_1466073, EPI_ISL_1466074, EPI_ISL_1466075, EPI_ISL_1466077, EPI_ISL_1466078, EPI_ISL_1466080, EPI_ISL_1466081,
EPI_ISL_1466082, EPI_ISL_1466085, EPI_ISL_1466086, EPI_ISL_1466087, EPI_ISL_1466089, EPI_ISL_1466090, EPI_ISL_1466091, EPI_ISL_1466092, EPI_ISL_1466093, EPI_ISL_1466094, EPI_ISL_1466095, EPI_ISL_1466096, EPI_ISL_1466097, EPI_ISL_1466098, EPI_ISL_1466099, EPI_ISL_1466103, EPI_ISL_1466104, EPI_ISL_1466105, EPI_ISL_1466106, EPI_ISL_1466107, EPI_ISL_1466108, EPI_ISL_1466110,
EPI_ISL_1466114, EPI_ISL_1466116, EPI_ISL_1466118, EPI_ISL_1466119, EPI_ISL_1466120, EPI_ISL_1466122, EPI_ISL_1466123, EPI_ISL_1466124, EPI_ISL_1466126, EPI_ISL_1466127, EPI_ISL_1466128, EPI_ISL_1466130, EPI_ISL_1466131, EPI_ISL_1466132, EPI_ISL_1466134, EPI_ISL_1466135, EPI_ISL_1466136, EPI_ISL_1466137, EPI_ISL_1466138, EPI_ISL_1466139, EPI_ISL_1466140, EPI_ISL_1466142,
EPI_ISL_1466144, EPI_ISL_1466145, EPI_ISL_1466146, EPI_ISL_1466147, EPI_ISL_1466149, EPI_ISL_1466150, EPI_ISL_1466151, EPI_ISL_1466152, EPI_ISL_1466153, EPI_ISL_1466154, EPI_ISL_1466155, EPI_ISL_1466156, EPI_ISL_1466157, EPI_ISL_1466159, EPI_ISL_1466160, EPI_ISL_1466162, EPI_ISL_1466173, EPI_ISL_1466184, EPI_ISL_1466194, EPI_ISL_1466195, EPI_ISL_1466196, EPI_ISL_1466229,
EPI_ISL_1466235, EPI_ISL_1466239, EPI_ISL_1466242, EPI_ISL_1466246, EPI_ISL_1466247, EPI_ISL_1466252, EPI_ISL_1466254, EPI_ISL_1466260, EPI_ISL_1466270, EPI_ISL_1466271, EPI_ISL_1466281, EPI_ISL_1466283, EPI_ISL_1466284, EPI_ISL_1466287, EPI_ISL_1466296, EPI_ISL_1466306, EPI_ISL_1466309, EPI_ISL_1466314, EPI_ISL_1466316, EPI_ISL_1466322, EPI_ISL_1466325, EPI_ISL_1466326,
EPI_ISL_1466334, EPI_ISL_1466335, EPI_ISL_1466338, EPI_ISL_1466344, EPI_ISL_1466362, EPI_ISL_1466374, EPI_ISL_1466375, EPI_ISL_1466381, EPI_ISL_1466398, EPI_ISL_1466409, EPI_ISL_1466417, EPI_ISL_1466420, EPI_ISL_1466426, EPI_ISL_1466428, EPI_ISL_1466437, EPI_ISL_1466440, EPI_ISL_1466449, EPI_ISL_1466471, EPI_ISL_1466480, EPI_ISL_1466498, EPI_ISL_1466502, EPI_ISL_1466506,
EPI_ISL_1466524, EPI_ISL_1466527, EPI_ISL_1466533, EPI_ISL_1466538, EPI_ISL_1466542, EPI_ISL_1466543, EPI_ISL_1466553, EPI_ISL_1466556, EPI_ISL_1466559, EPI_ISL_1466561, EPI_ISL_1466564, EPI_ISL_1466565, EPI_ISL_1466580, EPI_ISL_1466587, EPI_ISL_1466589, EPI_ISL_1466596, EPI_ISL_1466597, EPI_ISL_1466600, EPI_ISL_1466601, EPI_ISL_1466604, EPI_ISL_1466612,
EPI_ISL_1466620, EPI_ISL_1466622, EPI_ISL_1466627, EPI_ISL_1466644, EPI_ISL_1466645, EPI_ISL_1466648, EPI_ISL_1466650, EPI_ISL_1466654, EPI_ISL_1466658, EPI_ISL_1466659, EPI_ISL_1466661, EPI_ISL_1466665, EPI_ISL_1466667, EPI_ISL_1466673, EPI_ISL_1466674, EPI_ISL_1466679, EPI_ISL_1466687, EPI_ISL_1466694, EPI_ISL_1466701, EPI_ISL_1466702, EPI_ISL_1466707, EPI_ISL_1466710,
EPI_ISL_1466728, EPI_ISL_1466736, EPI_ISL_1466744, EPI_ISL_1466745, EPI_ISL_1466747, EPI_ISL_1466748, EPI_ISL_1466749, EPI_ISL_1466750, EPI_ISL_1466751, EPI_ISL_1466753, EPI_ISL_1466754, EPI_ISL_1466755, EPI_ISL_1466756, EPI_ISL_1466757, EPI_ISL_1466758, EPI_ISL_1466759, EPI_ISL_1466760, EPI_ISL_1466761, EPI_ISL_1466762, EPI_ISL_1466763, EPI_ISL_1466765, EPI_ISL_1466766,
EPI_ISL_1466767, EPI_ISL_1466769, EPI_ISL_1466770, EPI_ISL_1466772, EPI_ISL_1466773, EPI_ISL_1466774, EPI_ISL_1466776, EPI_ISL_1466777, EPI_ISL_1466778, EPI_ISL_1466780, EPI_ISL_1466781, EPI_ISL_1466782, EPI_ISL_1466784, EPI_ISL_1466790, EPI_ISL_1466791, EPI_ISL_1466792, EPI_ISL_1466796, EPI_ISL_1466797, EPI_ISL_1466799, EPI_ISL_1466800, EPI_ISL_1466801, EPI_ISL_1466802,
EPI_ISL_1466803, EPI_ISL_1466804, EPI_ISL_1466806, EPI_ISL_1466807, EPI_ISL_1466808, EPI_ISL_1466810, EPI_ISL_1466811, EPI_ISL_1466814, EPI_ISL_1466815, EPI_ISL_1466816, EPI_ISL_1466818, EPI_ISL_1466819, EPI_ISL_1466821, EPI_ISL_1466823, EPI_ISL_1466825, EPI_ISL_1466826, EPI_ISL_1466827, EPI_ISL_1466828, EPI_ISL_1466830, EPI_ISL_1466833, EPI_ISL_1466834, EPI_ISL_1466835,
EPI_ISL_1466836, EPI_ISL_1466837, EPI_ISL_1466839, EPI_ISL_1466841, EPI_ISL_1466842, EPI_ISL_1466843, EPI_ISL_1466845, EPI_ISL_1466848, EPI_ISL_1466849, EPI_ISL_1466851, EPI_ISL_1466853, EPI_ISL_1466855, EPI_ISL_1466857, EPI_ISL_1466858, EPI_ISL_1466859, EPI_ISL_1466860, EPI_ISL_1466861, EPI_ISL_1466862, EPI_ISL_1466863, EPI_ISL_1466864, EPI_ISL_1466865, EPI_ISL_1466869,
EPI_ISL_1466871, EPI_ISL_1466873, EPI_ISL_1466874, EPI_ISL_1466875, EPI_ISL_1466876, EPI_ISL_1466877, EPI_ISL_1466879, EPI_ISL_1466880, EPI_ISL_1466882, EPI_ISL_1466885, EPI_ISL_1466887, EPI_ISL_1466891, EPI_ISL_1466893, EPI_ISL_1466895, EPI_ISL_1466898, EPI_ISL_1466899, EPI_ISL_1466900, EPI_ISL_1466901, EPI_ISL_1466902, EPI_ISL_1466903, EPI_ISL_1466904, EPI_ISL_1466905,
EPI_ISL_1466907, EPI_ISL_1466908, EPI_ISL_1466909, EPI_ISL_1466911, EPI_ISL_1466912, EPI_ISL_1466914, EPI_ISL_1466915, EPI_ISL_1466918, EPI_ISL_1466919, EPI_ISL_1466921, EPI_ISL_1467279, EPI_ISL_1467291, EPI_ISL_1467360, EPI_ISL_1467374, EPI_ISL_1467404, EPI_ISL_1467421, EPI_ISL_1467423, EPI_ISL_1467425, EPI_ISL_1467441, EPI_ISL_1467474, EPI_ISL_1467483, EPI_ISL_1467516,
EPI_ISL_1467536, EPI_ISL_1467544, EPI_ISL_1467552, EPI_ISL_1467579, EPI_ISL_1468134, EPI_ISL_1468136, EPI_ISL_1468137, EPI_ISL_1468139, EPI_ISL_1468144, EPI_ISL_1468146, EPI_ISL_1468147, EPI_ISL_1468150, EPI_ISL_1468156, EPI_ISL_1468157, EPI_ISL_1468159, EPI_ISL_1468165, EPI_ISL_1468166, EPI_ISL_1468170, EPI_ISL_1468172, EPI_ISL_1468173, EPI_ISL_1468174, EPI_ISL_1468176,
EPI_ISL_1468178, EPI_ISL_1468181, EPI_ISL_1468185, EPI_ISL_1468193, EPI_ISL_1468196, EPI_ISL_1468201, EPI_ISL_1468207, EPI_ISL_1468208, EPI_ISL_1468210, EPI_ISL_1468213, EPI_ISL_1468215, EPI_ISL_1468218, EPI_ISL_1468220, EPI_ISL_1468221, EPI_ISL_1468232, EPI_ISL_1468234, EPI_ISL_1468235, EPI_ISL_1468236, EPI_ISL_1468241, EPI_ISL_1468246, EPI_ISL_1468248, EPI_ISL_1468249,
EPI_ISL_1468256, EPI_ISL_1468261, EPI_ISL_1468266, EPI_ISL_1468267, EPI_ISL_1468268, EPI_ISL_1468269, EPI_ISL_1468270, EPI_ISL_1468271, EPI_ISL_1468275, EPI_ISL_1468277, EPI_ISL_1468278, EPI_ISL_1468282, EPI_ISL_1468287, EPI_ISL_1468292, EPI_ISL_1468296, EPI_ISL_1468297, EPI_ISL_1468303, EPI_ISL_1468304, EPI_ISL_1468305, EPI_ISL_1468306, EPI_ISL_1468309, EPI_ISL_1468313,
EPI_ISL_1468314, EPI_ISL_1468318, EPI_ISL_1468319, EPI_ISL_1468321, EPI_ISL_1468325, EPI_ISL_1468329, EPI_ISL_1468331, EPI_ISL_1468334, EPI_ISL_1468336, EPI_ISL_1468339, EPI_ISL_1468341, EPI_ISL_1468342, EPI_ISL_1468344, EPI_ISL_1468345, EPI_ISL_1468352, EPI_ISL_1468353, EPI_ISL_1468354, EPI_ISL_1468355, EPI_ISL_1468358, EPI_ISL_1468364, EPI_ISL_1468366, EPI_ISL_1468367,
EPI_ISL_1468370, EPI_ISL_1468371, EPI_ISL_1468374, EPI_ISL_1468376, EPI_ISL_1468382, EPI_ISL_1468384, EPI_ISL_1468386, EPI_ISL_1468388, EPI_ISL_1468394, EPI_ISL_1468401, EPI_ISL_1468402, EPI_ISL_1472414, EPI_ISL_1472421, EPI_ISL_1472434, EPI_ISL_1472441, EPI_ISL_1472443, EPI_ISL_1472454, EPI_ISL_1472460, EPI_ISL_1472463, EPI_ISL_1472478, EPI_ISL_1472481,
EPI_ISL_1472485, EPI_ISL_1472526, EPI_ISL_1472527, EPI_ISL_1472530, EPI_ISL_1472533, EPI_ISL_1472535, EPI_ISL_1472549, EPI_ISL_1472556, EPI_ISL_1472557, EPI_ISL_1472558, EPI_ISL_1472568, EPI_ISL_1472579, EPI_ISL_1472583, EPI_ISL_1472597, EPI_ISL_1472604, EPI_ISL_1472605, EPI_ISL_1472606, EPI_ISL_1472612, EPI_ISL_1472617, EPI_ISL_1472619, EPI_ISL_1472625, EPI_ISL_1472629,
EPI_ISL_1472631, EPI_ISL_1472633, EPI_ISL_1472643, EPI_ISL_1472644, EPI_ISL_1472647, EPI_ISL_1472652, EPI_ISL_1472653, EPI_ISL_1472656, EPI_ISL_1472661, EPI_ISL_1472662, EPI_ISL_1472664, EPI_ISL_1472665, EPI_ISL_1472668, EPI_ISL_1472676, EPI_ISL_1472677, EPI_ISL_1472697, EPI_ISL_1472698, EPI_ISL_1472703, EPI_ISL_1472704, EPI_ISL_1472708, EPI_ISL_1472709, EPI_ISL_1472710,
EPI_ISL_1472712, EPI_ISL_1472713, EPI_ISL_1472714, EPI_ISL_1472718, EPI_ISL_1472720, EPI_ISL_1472721, EPI_ISL_1472723, EPI_ISL_1472724, EPI_ISL_1472725, EPI_ISL_1472726, EPI_ISL_1472728, EPI_ISL_1472730, EPI_ISL_1472732, EPI_ISL_1472733, EPI_ISL_1472735, EPI_ISL_1472736, EPI_ISL_1472737, EPI_ISL_1472738, EPI_ISL_1472739, EPI_ISL_1472740, EPI_ISL_1472743, EPI_ISL_1472744,
EPI_ISL_1472745, EPI_ISL_1472746, EPI_ISL_1472748, EPI_ISL_1472749, EPI_ISL_1472750, EPI_ISL_1472751, EPI_ISL_1472752, EPI_ISL_1472755, EPI_ISL_1472758, EPI_ISL_1472759, EPI_ISL_1472760, EPI_ISL_1472761, EPI_ISL_1472762, EPI_ISL_1472763, EPI_ISL_1472768, EPI_ISL_1472770, EPI_ISL_1472771, EPI_ISL_1472773, EPI_ISL_1472774, EPI_ISL_1472775, EPI_ISL_1472776, EPI_ISL_1472779,
EPI_ISL_1472781, EPI_ISL_1472783, EPI_ISL_1472784, EPI_ISL_1472787, EPI_ISL_1472788, EPI_ISL_1472790, EPI_ISL_1472791, EPI_ISL_1472792, EPI_ISL_1472793, EPI_ISL_1472795, EPI_ISL_1472796, EPI_ISL_1472798, EPI_ISL_1472799, EPI_ISL_1472803, EPI_ISL_1472804, EPI_ISL_1472805, EPI_ISL_1472806, EPI_ISL_1472808, EPI_ISL_1472810, EPI_ISL_1472811, EPI_ISL_1472812, EPI_ISL_1472813,
EPI_ISL_1472816, EPI_ISL_1472817, EPI_ISL_1472818, EPI_ISL_1472820, EPI_ISL_1472821, EPI_ISL_1472822, EPI_ISL_1472823, EPI_ISL_1472826, EPI_ISL_1472828, EPI_ISL_1472829, EPI_ISL_1472831, EPI_ISL_1472833, EPI_ISL_1472834, EPI_ISL_1472835, EPI_ISL_1472836, EPI_ISL_1472838, EPI_ISL_1472842, EPI_ISL_1472844, EPI_ISL_1472845, EPI_ISL_1472846, EPI_ISL_1472847, EPI_ISL_1472848,
EPI_ISL_1472849, EPI_ISL_1472850, EPI_ISL_1472852, EPI_ISL_1472853, EPI_ISL_1472855, EPI_ISL_1472856, EPI_ISL_1472857, EPI_ISL_1472858, EPI_ISL_1472859, EPI_ISL_1472861, EPI_ISL_1472862, EPI_ISL_1472864, EPI_ISL_1472865, EPI_ISL_1472868, EPI_ISL_1472871, EPI_ISL_1472872, EPI_ISL_1472873, EPI_ISL_1472874, EPI_ISL_1472875, EPI_ISL_1472877, EPI_ISL_1472878, EPI_ISL_1472879,
EPI_ISL_1472882, EPI_ISL_1472883, EPI_ISL_1472884, EPI_ISL_1472886, EPI_ISL_1472887, EPI_ISL_1472888, EPI_ISL_1472889, EPI_ISL_1472890, EPI_ISL_1472891, EPI_ISL_1472892, EPI_ISL_1472894, EPI_ISL_1472895, EPI_ISL_1472896, EPI_ISL_1472897, EPI_ISL_1472899, EPI_ISL_1472902, EPI_ISL_1472903, EPI_ISL_1472904, EPI_ISL_1472905, EPI_ISL_1472906, EPI_ISL_1472908, EPI_ISL_1472909,
EPI_ISL_1472912, EPI_ISL_1472913, EPI_ISL_1472914, EPI_ISL_1472915, EPI_ISL_1472916, EPI_ISL_1472917, EPI_ISL_1472918, EPI_ISL_1472919, EPI_ISL_1472921, EPI_ISL_1472922, EPI_ISL_1472923, EPI_ISL_1472924, EPI_ISL_1472926, EPI_ISL_1472930, EPI_ISL_1472933, EPI_ISL_1472936, EPI_ISL_1472938, EPI_ISL_1472939, EPI_ISL_1472942, EPI_ISL_1472945, EPI_ISL_1472946, EPI_ISL_1472947,
EPI_ISL_1472949, EPI_ISL_1472950, EPI_ISL_1472951, EPI_ISL_1472952, EPI_ISL_1472953, EPI_ISL_1472955, EPI_ISL_1472956, EPI_ISL_1472959, EPI_ISL_1472960, EPI_ISL_1472961, EPI_ISL_1472962, EPI_ISL_1472963, EPI_ISL_1472964, EPI_ISL_1472965, EPI_ISL_1472966, EPI_ISL_1472967, EPI_ISL_1472968, EPI_ISL_1472969, EPI_ISL_1472970, EPI_ISL_1472972, EPI_ISL_1472973, EPI_ISL_1472975,
EPI_ISL_1472976, EPI_ISL_1472977, EPI_ISL_1472978, EPI_ISL_1472980, EPI_ISL_1472981, EPI_ISL_1472982, EPI_ISL_1472983, EPI_ISL_1472984, EPI_ISL_1472985, EPI_ISL_1472986, EPI_ISL_1472987, EPI_ISL_1472988, EPI_ISL_1472989, EPI_ISL_1472990, EPI_ISL_1472991, EPI_ISL_1472992, EPI_ISL_1472993, EPI_ISL_1472994, EPI_ISL_1472995, EPI_ISL_1472996, EPI_ISL_1472997, EPI_ISL_1472998,
EPI_ISL_1472999, EPI_ISL_1473000, EPI_ISL_1473001, EPI_ISL_1473003, EPI_ISL_1473004, EPI_ISL_1473005, EPI_ISL_1473006, EPI_ISL_1473007, EPI_ISL_1473008, EPI_ISL_1473009, EPI_ISL_1473010, EPI_ISL_1473011, EPI_ISL_1473012, EPI_ISL_1473014, EPI_ISL_1473015, EPI_ISL_1473016, EPI_ISL_1473017, EPI_ISL_1473018, EPI_ISL_1473020, EPI_ISL_1473021, EPI_ISL_1473022, EPI_ISL_1473023,
EPI_ISL_1473024, EPI_ISL_1473026, EPI_ISL_1473027, EPI_ISL_1473028, EPI_ISL_1473029, EPI_ISL_1473031, EPI_ISL_1473032, EPI_ISL_1473033, EPI_ISL_1473034, EPI_ISL_1473036, EPI_ISL_1473037, EPI_ISL_1473038, EPI_ISL_1473039, EPI_ISL_1473040, EPI_ISL_1473041, EPI_ISL_1473042, EPI_ISL_1473043, EPI_ISL_1473044, EPI_ISL_1473045, EPI_ISL_1473046, EPI_ISL_1473047, EPI_ISL_1473048,
EPI_ISL_1473049, EPI_ISL_1473050, EPI_ISL_1473052, EPI_ISL_1473053, EPI_ISL_1473054, EPI_ISL_1473055, EPI_ISL_1473056, EPI_ISL_1473057, EPI_ISL_1473058, EPI_ISL_1473059, EPI_ISL_1473060, EPI_ISL_1473061, EPI_ISL_1473062, EPI_ISL_1473063, EPI_ISL_1473065, EPI_ISL_1473066, EPI_ISL_1473067, EPI_ISL_1473068, EPI_ISL_1473069, EPI_ISL_1473070, EPI_ISL_1473071, EPI_ISL_1473072,
EPI_ISL_1473073, EPI_ISL_1473074, EPI_ISL_1473075, EPI_ISL_1473076, EPI_ISL_1473077, EPI_ISL_1473079, EPI_ISL_1473081, EPI_ISL_1473083, EPI_ISL_1473084, EPI_ISL_1473085, EPI_ISL_1473086, EPI_ISL_1473088, EPI_ISL_1473089, EPI_ISL_1473090, EPI_ISL_1473091, EPI_ISL_1473092, EPI_ISL_1473093, EPI_ISL_1473094, EPI_ISL_1473095, EPI_ISL_1473097, EPI_ISL_1473098, EPI_ISL_1473099,
EPI_ISL_1473100, EPI_ISL_1473101, EPI_ISL_1473103, EPI_ISL_1473104, EPI_ISL_1473105, EPI_ISL_1473106, EPI_ISL_1473107, EPI_ISL_1473108, EPI_ISL_1473109, EPI_ISL_1473110, EPI_ISL_1473111, EPI_ISL_1473112, EPI_ISL_1473113, EPI_ISL_1473114, EPI_ISL_1473115, EPI_ISL_1473116, EPI_ISL_1473117, EPI_ISL_1473118, EPI_ISL_1473119, EPI_ISL_1473120, EPI_ISL_1473121, EPI_ISL_1473122,
EPI_ISL_1473123, EPI_ISL_1473124, EPI_ISL_1473126, EPI_ISL_1473127, EPI_ISL_1473128, EPI_ISL_1473130, EPI_ISL_1473131, EPI_ISL_1473132, EPI_ISL_1473134, EPI_ISL_1473135, EPI_ISL_1473136, EPI_ISL_1473137, EPI_ISL_1473138, EPI_ISL_1473139, EPI_ISL_1473140, EPI_ISL_1473141, EPI_ISL_1473142, EPI_ISL_1473143, EPI_ISL_1473144, EPI_ISL_1473145, EPI_ISL_1473146, EPI_ISL_1473147,
EPI_ISL_1473148, EPI_ISL_1473149, EPI_ISL_1473152, EPI_ISL_1473153, EPI_ISL_1473154, EPI_ISL_1473156, EPI_ISL_1473157, EPI_ISL_1473158, EPI_ISL_1473159, EPI_ISL_1473160, EPI_ISL_1473161, EPI_ISL_1473162, EPI_ISL_1473163, EPI_ISL_1473165, EPI_ISL_1473166, EPI_ISL_1473168, EPI_ISL_1473169, EPI_ISL_1473170, EPI_ISL_1473171, EPI_ISL_1473172, EPI_ISL_1473173, EPI_ISL_1473174,
EPI_ISL_1473175, EPI_ISL_1473176, EPI_ISL_1473177, EPI_ISL_1473178, EPI_ISL_1473179, EPI_ISL_1473180, EPI_ISL_1473181, EPI_ISL_1473182, EPI_ISL_1473183, EPI_ISL_1473184, EPI_ISL_1473185, EPI_ISL_1473186, EPI_ISL_1473187, EPI_ISL_1473188, EPI_ISL_1473189, EPI_ISL_1473190, EPI_ISL_1473191, EPI_ISL_1473192, EPI_ISL_1473193, EPI_ISL_1473194, EPI_ISL_1473196, EPI_ISL_1473198,
EPI_ISL_1473199, EPI_ISL_1473200, EPI_ISL_1473201, EPI_ISL_1473202, EPI_ISL_1473203, EPI_ISL_1473205, EPI_ISL_1473206, EPI_ISL_1473209, EPI_ISL_1473212, EPI_ISL_1473213, EPI_ISL_1473214, EPI_ISL_1473215, EPI_ISL_1473216, EPI_ISL_1473219, EPI_ISL_1473221, EPI_ISL_1473222, EPI_ISL_1473223, EPI_ISL_1473227, EPI_ISL_1473229, EPI_ISL_1473230, EPI_ISL_1473231, EPI_ISL_1473233,
EPI_ISL_1473237, EPI_ISL_1473238, EPI_ISL_1473239, EPI_ISL_1473240, EPI_ISL_1473241, EPI_ISL_1473242, EPI_ISL_1473243, EPI_ISL_1473245, EPI_ISL_1473246, EPI_ISL_1473247, EPI_ISL_1473248, EPI_ISL_1473249, EPI_ISL_1473251, EPI_ISL_1473252, EPI_ISL_1473254, EPI_ISL_1473256, EPI_ISL_1473258, EPI_ISL_1473259, EPI_ISL_1473263, EPI_ISL_1473266, EPI_ISL_1473267, EPI_ISL_1473268,
EPI_ISL_1473269, EPI_ISL_1473271, EPI_ISL_1473272, EPI_ISL_1473273, EPI_ISL_1473274, EPI_ISL_1473275, EPI_ISL_1473276, EPI_ISL_1473277, EPI_ISL_1473278, EPI_ISL_1473279, EPI_ISL_1473280, EPI_ISL_1473283, EPI_ISL_1473284, EPI_ISL_1473285, EPI_ISL_1473286, EPI_ISL_1473288, EPI_ISL_1473291, EPI_ISL_1473298, EPI_ISL_1473299, EPI_ISL_1473302, EPI_ISL_1473303, EPI_ISL_1473305,
EPI_ISL_1473307, EPI_ISL_1473308, EPI_ISL_1473309, EPI_ISL_1473311, EPI_ISL_1473313, EPI_ISL_1473316, EPI_ISL_1473317, EPI_ISL_1473318, EPI_ISL_1473321, EPI_ISL_1473323, EPI_ISL_1473324, EPI_ISL_1473328, EPI_ISL_1473330, EPI_ISL_1473331, EPI_ISL_1473332, EPI_ISL_1473333, EPI_ISL_1473334, EPI_ISL_1473335, EPI_ISL_1473337, EPI_ISL_1473339, EPI_ISL_1473340, EPI_ISL_1473341,
EPI_ISL_1473342, EPI_ISL_1473343, EPI_ISL_1473345, EPI_ISL_1473346, EPI_ISL_1473349, EPI_ISL_1473354, EPI_ISL_1473355, EPI_ISL_1473357, EPI_ISL_1473360, EPI_ISL_1473361, EPI_ISL_1473363, EPI_ISL_1473366, EPI_ISL_1473367, EPI_ISL_1473368, EPI_ISL_1473369, EPI_ISL_1473370, EPI_ISL_1473371, EPI_ISL_1473372, EPI_ISL_1473373, EPI_ISL_1473376, EPI_ISL_1473377, EPI_ISL_1473378,
EPI_ISL_1473379, EPI_ISL_1473381, EPI_ISL_1473383, EPI_ISL_1473384, EPI_ISL_1473387, EPI_ISL_1473388, EPI_ISL_1473389, EPI_ISL_1473390, EPI_ISL_1473392, EPI_ISL_1473395, EPI_ISL_1473396, EPI_ISL_1473398, EPI_ISL_1473399, EPI_ISL_1473400, EPI_ISL_1473402, EPI_ISL_1473406, EPI_ISL_1473407, EPI_ISL_1473408, EPI_ISL_1473410, EPI_ISL_1473411, EPI_ISL_1473412, EPI_ISL_1473415,
EPI_ISL_1473417, EPI_ISL_1473418, EPI_ISL_1473422, EPI_ISL_1473423, EPI_ISL_1473424, EPI_ISL_1473425, EPI_ISL_1473426, EPI_ISL_1473428, EPI_ISL_1473430, EPI_ISL_1473431, EPI_ISL_1473434, EPI_ISL_1473435, EPI_ISL_1473436, EPI_ISL_1473437, EPI_ISL_1473438, EPI_ISL_1473439, EPI_ISL_1473441, EPI_ISL_1473442, EPI_ISL_1473443, EPI_ISL_1473444, EPI_ISL_1473800, EPI_ISL_1473801,
EPI_ISL_1473802, EPI_ISL_1473805, EPI_ISL_1473806, EPI_ISL_1473811, EPI_ISL_1474157, EPI_ISL_1485300, EPI_ISL_1487168, EPI_ISL_1487192, EPI_ISL_1487201, EPI_ISL_1487211, EPI_ISL_1487213, EPI_ISL_1487214, EPI_ISL_1487248, EPI_ISL_1487250, EPI_ISL_1487263, EPI_ISL_1487279, EPI_ISL_1487285, EPI_ISL_1487290, EPI_ISL_1487313, EPI_ISL_1487352, EPI_ISL_1487360, EPI_ISL_1487725,
EPI_ISL_1487727, EPI_ISL_1487728, EPI_ISL_1487729, EPI_ISL_1487730, EPI_ISL_1487731, EPI_ISL_1487732, EPI_ISL_1487733, EPI_ISL_1487734, EPI_ISL_1487735, EPI_ISL_1487736, EPI_ISL_1487737, EPI_ISL_1487738, EPI_ISL_1487739, EPI_ISL_1487740, EPI_ISL_1487741, EPI_ISL_1487742, EPI_ISL_1487743, EPI_ISL_1487745, EPI_ISL_1487746, EPI_ISL_1487747, EPI_ISL_1487748, EPI_ISL_1487749,
EPI_ISL_1487750, EPI_ISL_1487751, EPI_ISL_1487752, EPI_ISL_1487753, EPI_ISL_1487754, EPI_ISL_1487755, EPI_ISL_1487756, EPI_ISL_1487757, EPI_ISL_1487758, EPI_ISL_1487759, EPI_ISL_1487760, EPI_ISL_1487761, EPI_ISL_1487762, EPI_ISL_1487763, EPI_ISL_1487764, EPI_ISL_1487765, EPI_ISL_1487766, EPI_ISL_1487767, EPI_ISL_1487768, EPI_ISL_1487769, EPI_ISL_1487770, EPI_ISL_1487771,
EPI_ISL_1487772, EPI_ISL_1487773, EPI_ISL_1487774, EPI_ISL_1487776, EPI_ISL_1487777, EPI_ISL_1487778, EPI_ISL_1487779, EPI_ISL_1487780, EPI_ISL_1487781, EPI_ISL_1487782, EPI_ISL_1487783, EPI_ISL_1487784, EPI_ISL_1487785, EPI_ISL_1487786, EPI_ISL_1487787, EPI_ISL_1487788, EPI_ISL_1487789, EPI_ISL_1487790, EPI_ISL_1487792, EPI_ISL_1487793, EPI_ISL_1487794, EPI_ISL_1487795,
EPI_ISL_1487796, EPI_ISL_1487797, EPI_ISL_1487798, EPI_ISL_1487799, EPI_ISL_1487800, EPI_ISL_1487801, EPI_ISL_1487802, EPI_ISL_1487803, EPI_ISL_1487804, EPI_ISL_1487805, EPI_ISL_1487806, EPI_ISL_1487807, EPI_ISL_1487808, EPI_ISL_1487809, EPI_ISL_1487810, EPI_ISL_1487811, EPI_ISL_1487812, EPI_ISL_1487814, EPI_ISL_1487815, EPI_ISL_1487816, EPI_ISL_1487817, EPI_ISL_1487818,
EPI_ISL_1487819, EPI_ISL_1487820, EPI_ISL_1487821, EPI_ISL_1487822, EPI_ISL_1487823, EPI_ISL_1487824, EPI_ISL_1487826, EPI_ISL_1487827, EPI_ISL_1487828, EPI_ISL_1487829, EPI_ISL_1487830, EPI_ISL_1487833, EPI_ISL_1487834, EPI_ISL_1487835, EPI_ISL_1487836, EPI_ISL_1487837, EPI_ISL_1487838, EPI_ISL_1487839, EPI_ISL_1487840, EPI_ISL_1487841, EPI_ISL_1487842, EPI_ISL_1487843,
EPI_ISL_1487844, EPI_ISL_1487845, EPI_ISL_1487846, EPI_ISL_1487847, EPI_ISL_1487848, EPI_ISL_1487849, EPI_ISL_1487850, EPI_ISL_1487851, EPI_ISL_1488866, EPI_ISL_1488867, EPI_ISL_1488868, EPI_ISL_1488872, EPI_ISL_1488874, EPI_ISL_1488876, EPI_ISL_1488877, EPI_ISL_1488878, EPI_ISL_1488881, EPI_ISL_1488882, EPI_ISL_1488885, EPI_ISL_1488886, EPI_ISL_1488889, EPI_ISL_1488890,
EPI_ISL_1488892, EPI_ISL_1488894, EPI_ISL_1488895, EPI_ISL_1488897, EPI_ISL_1488898, EPI_ISL_1488900, EPI_ISL_1488901, EPI_ISL_1488902, EPI_ISL_1488903, EPI_ISL_1488908, EPI_ISL_1488909, EPI_ISL_1488913, EPI_ISL_1488915, EPI_ISL_1488918, EPI_ISL_1488921, EPI_ISL_1488923, EPI_ISL_1488926, EPI_ISL_1488929, EPI_ISL_1488933, EPI_ISL_1488934, EPI_ISL_1488936, EPI_ISL_1488940,
EPI_ISL_1488941, EPI_ISL_1488950, EPI_ISL_1488955, EPI_ISL_1488956, EPI_ISL_1488957, EPI_ISL_1488958, EPI_ISL_1488963, EPI_ISL_1488964, EPI_ISL_1488967, EPI_ISL_1488969, EPI_ISL_1488979, EPI_ISL_1488981, EPI_ISL_1488982, EPI_ISL_1488984, EPI_ISL_1488985, EPI_ISL_1488986, EPI_ISL_1488988, EPI_ISL_1488992, EPI_ISL_1488994, EPI_ISL_1488995, EPI_ISL_1488996, EPI_ISL_1488997,
EPI_ISL_1488998, EPI_ISL_1488999, EPI_ISL_1489000, EPI_ISL_1489005, EPI_ISL_1489011, EPI_ISL_1489013, EPI_ISL_1489018, EPI_ISL_1489022, EPI_ISL_1489023, EPI_ISL_1489024, EPI_ISL_1489026, EPI_ISL_1489031, EPI_ISL_1489032, EPI_ISL_1489034, EPI_ISL_1489037, EPI_ISL_1489040, EPI_ISL_1489043, EPI_ISL_1489044, EPI_ISL_1489045, EPI_ISL_1489047, EPI_ISL_1489049, EPI_ISL_1489053,
EPI_ISL_1489056, EPI_ISL_1489060, EPI_ISL_1489061, EPI_ISL_1489062, EPI_ISL_1489064, EPI_ISL_1489065, EPI_ISL_1489068, EPI_ISL_1489071, EPI_ISL_1489074, EPI_ISL_1489076, EPI_ISL_1489082, EPI_ISL_1489083, EPI_ISL_1489084, EPI_ISL_1489085, EPI_ISL_1489088, EPI_ISL_1489091, EPI_ISL_1489096, EPI_ISL_1489098, EPI_ISL_1489099, EPI_ISL_1489102, EPI_ISL_1489105, EPI_ISL_1489106,
EPI_ISL_1489107, EPI_ISL_1489118, EPI_ISL_1489119, EPI_ISL_1489123, EPI_ISL_1489125, EPI_ISL_1489128, EPI_ISL_1489129, EPI_ISL_1489133, EPI_ISL_1489136, EPI_ISL_1489137, EPI_ISL_1489144, EPI_ISL_1489145, EPI_ISL_1489146, EPI_ISL_1489148, EPI_ISL_1489150, EPI_ISL_1489152, EPI_ISL_1489156, EPI_ISL_1489158, EPI_ISL_1489188, EPI_ISL_1489211, EPI_ISL_1489260, EPI_ISL_1489276,
EPI_ISL_1489281, EPI_ISL_1489290, EPI_ISL_1489314, EPI_ISL_1489345, EPI_ISL_1489347, EPI_ISL_1489397, EPI_ISL_1489402, EPI_ISL_1489418, EPI_ISL_1489444, EPI_ISL_1489477, EPI_ISL_1489478, EPI_ISL_1489479, EPI_ISL_1506512, EPI_ISL_1506513, EPI_ISL_1506515, EPI_ISL_1506517, EPI_ISL_1506518, EPI_ISL_1506519, EPI_ISL_1506521, EPI_ISL_1506522, EPI_ISL_1506523, EPI_ISL_1506524,
EPI_ISL_1506525, EPI_ISL_1506526, EPI_ISL_1506527, EPI_ISL_1506575, EPI_ISL_1506576, EPI_ISL_1506578, EPI_ISL_1506579, EPI_ISL_1506580, EPI_ISL_1506581, EPI_ISL_1506582, EPI_ISL_1506584, EPI_ISL_1506585, EPI_ISL_1506587, EPI_ISL_1506589, EPI_ISL_1506591, EPI_ISL_1506593, EPI_ISL_1506594, EPI_ISL_1506595, EPI_ISL_1506596, EPI_ISL_1506598, EPI_ISL_1506599, EPI_ISL_1506600,
EPI_ISL_1506602, EPI_ISL_1506603, EPI_ISL_1506606, EPI_ISL_1506607, EPI_ISL_1506610, EPI_ISL_1506611, EPI_ISL_1506613, EPI_ISL_1506614, EPI_ISL_1506615, EPI_ISL_1506617, EPI_ISL_1506618, EPI_ISL_1506620, EPI_ISL_1506623, EPI_ISL_1506624, EPI_ISL_1506627, EPI_ISL_1506628, EPI_ISL_1506629, EPI_ISL_1506630, EPI_ISL_1506631, EPI_ISL_1506632, EPI_ISL_1506633, EPI_ISL_1506634,
EPI_ISL_1506638, EPI_ISL_1506641, EPI_ISL_1506642, EPI_ISL_1506643, EPI_ISL_1506645, EPI_ISL_1506653, EPI_ISL_1506686, EPI_ISL_1506698, EPI_ISL_1506699, EPI_ISL_1506700, EPI_ISL_1506701, EPI_ISL_1506702, EPI_ISL_1506703, EPI_ISL_1506705, EPI_ISL_1506706, EPI_ISL_1506707, EPI_ISL_1506708, EPI_ISL_1506709, EPI_ISL_1506710, EPI_ISL_1506711, EPI_ISL_1506712, EPI_ISL_1506714,
EPI_ISL_1506715, EPI_ISL_1506716, EPI_ISL_1506717, EPI_ISL_1506718, EPI_ISL_1506720, EPI_ISL_1506721, EPI_ISL_1506722, EPI_ISL_1506723, EPI_ISL_1506724, EPI_ISL_1506725, EPI_ISL_1506726, EPI_ISL_1506727, EPI_ISL_1506728, EPI_ISL_1506729, EPI_ISL_1506730, EPI_ISL_1506731, EPI_ISL_1506732, EPI_ISL_1506733, EPI_ISL_1506734, EPI_ISL_1506735, EPI_ISL_1506736,
EPI_ISL_1506737, EPI_ISL_1506738, EPI_ISL_1506739, EPI_ISL_1506741, EPI_ISL_1506742, EPI_ISL_1506743, EPI_ISL_1506744, EPI_ISL_1506745, EPI_ISL_1506746, EPI_ISL_1506747, EPI_ISL_1506748, EPI_ISL_1506749, EPI_ISL_1506750, EPI_ISL_1506752, EPI_ISL_1506753, EPI_ISL_1506754, EPI_ISL_1506755, EPI_ISL_1506756, EPI_ISL_1506757, EPI_ISL_1506758, EPI_ISL_1506759, EPI_ISL_1506760,
EPI_ISL_1506761, EPI_ISL_1506762, EPI_ISL_1506763, EPI_ISL_1506764, EPI_ISL_1506765, EPI_ISL_1506766, EPI_ISL_1506767, EPI_ISL_1506768, EPI_ISL_1506769, EPI_ISL_1506770, EPI_ISL_1506771, EPI_ISL_1506772, EPI_ISL_1506773, EPI_ISL_1506774, EPI_ISL_1506775, EPI_ISL_1506776, EPI_ISL_1506777, EPI_ISL_1506778, EPI_ISL_1506779, EPI_ISL_1506780, EPI_ISL_1506781, EPI_ISL_1506782,
EPI_ISL_1506783, EPI_ISL_1506784, EPI_ISL_1506785, EPI_ISL_1506786, EPI_ISL_1506787, EPI_ISL_1506788, EPI_ISL_1506789, EPI_ISL_1506790, EPI_ISL_1506791, EPI_ISL_1506792, EPI_ISL_1506793, EPI_ISL_1506794, EPI_ISL_1506795, EPI_ISL_1506796, EPI_ISL_1506797, EPI_ISL_1506798, EPI_ISL_1506799, EPI_ISL_1506800, EPI_ISL_1506801, EPI_ISL_1506802, EPI_ISL_1506803, EPI_ISL_1506804,
EPI_ISL_1506805, EPI_ISL_1506806, EPI_ISL_1506807, EPI_ISL_1506808, EPI_ISL_1506809, EPI_ISL_1506810, EPI_ISL_1506811, EPI_ISL_1506812, EPI_ISL_1506813, EPI_ISL_1506814, EPI_ISL_1506815, EPI_ISL_1506816, EPI_ISL_1506817, EPI_ISL_1506818, EPI_ISL_1506819, EPI_ISL_1506820, EPI_ISL_1506821, EPI_ISL_1506822, EPI_ISL_1506823, EPI_ISL_1506824, EPI_ISL_1506826, EPI_ISL_1506839,
EPI_ISL_1506847, EPI_ISL_1506853, EPI_ISL_1506859, EPI_ISL_1506871, EPI_ISL_1506888, EPI_ISL_1506889, EPI_ISL_1506890, EPI_ISL_1506898, EPI_ISL_1506909, EPI_ISL_1506910, EPI_ISL_1506915, EPI_ISL_1506924, EPI_ISL_1506928, EPI_ISL_1506936, EPI_ISL_1506943, EPI_ISL_1506944, EPI_ISL_1506945, EPI_ISL_1506956, EPI_ISL_1506966, EPI_ISL_1506972, EPI_ISL_1506974, EPI_ISL_1506976,
EPI_ISL_1506994, EPI_ISL_1506996, EPI_ISL_1507021, EPI_ISL_1507025, EPI_ISL_1507036, EPI_ISL_1507198, EPI_ISL_1507199, EPI_ISL_1507200, EPI_ISL_1507201, EPI_ISL_1507202, EPI_ISL_1507203, EPI_ISL_1507204, EPI_ISL_1507205, EPI_ISL_1507206, EPI_ISL_1507207, EPI_ISL_1507208, EPI_ISL_1507209, EPI_ISL_1507210, EPI_ISL_1507214, EPI_ISL_1507215, EPI_ISL_1507216,
EPI_ISL_1507217, EPI_ISL_1507218, EPI_ISL_1507219, EPI_ISL_1507220, EPI_ISL_1507221, EPI_ISL_1507222, EPI_ISL_1507223, EPI_ISL_1507224, EPI_ISL_1507225, EPI_ISL_1507226, EPI_ISL_1507227, EPI_ISL_1507228, EPI_ISL_1507229, EPI_ISL_1507230, EPI_ISL_1507231, EPI_ISL_1507233, EPI_ISL_1507235, EPI_ISL_1507236, EPI_ISL_1507237, EPI_ISL_1507238, EPI_ISL_1507239, EPI_ISL_1507240,
EPI_ISL_1507241, EPI_ISL_1507242, EPI_ISL_1507243, EPI_ISL_1507244, EPI_ISL_1507245, EPI_ISL_1507246, EPI_ISL_1507247, EPI_ISL_1507248, EPI_ISL_1507249, EPI_ISL_1507250, EPI_ISL_1507251, EPI_ISL_1507253, EPI_ISL_1507254, EPI_ISL_1507256, EPI_ISL_1507257, EPI_ISL_1507258, EPI_ISL_1507259, EPI_ISL_1507260, EPI_ISL_1507261, EPI_ISL_1507262, EPI_ISL_1507263,
EPI_ISL_1507264, EPI_ISL_1507265, EPI_ISL_1507266, EPI_ISL_1507267, EPI_ISL_1507268, EPI_ISL_1507269, EPI_ISL_1507270, EPI_ISL_1507271, EPI_ISL_1507272, EPI_ISL_1507273, EPI_ISL_1507274, EPI_ISL_1507275, EPI_ISL_1507276, EPI_ISL_1507277, EPI_ISL_1507278, EPI_ISL_1507279, EPI_ISL_1507280, EPI_ISL_1507281, EPI_ISL_1507282, EPI_ISL_1507283, EPI_ISL_1507284, EPI_ISL_1507285,
EPI_ISL_1507286, EPI_ISL_1507287, EPI_ISL_1507288, EPI_ISL_1507289, EPI_ISL_1507290, EPI_ISL_1507291, EPI_ISL_1507292, EPI_ISL_1507293, EPI_ISL_1507294, EPI_ISL_1507295, EPI_ISL_1507296, EPI_ISL_1507297, EPI_ISL_1507298, EPI_ISL_1507299, EPI_ISL_1507300, EPI_ISL_1507301, EPI_ISL_1507302, EPI_ISL_1507303, EPI_ISL_1507304, EPI_ISL_1507305, EPI_ISL_1507306, EPI_ISL_1507307,
EPI_ISL_1507308, EPI_ISL_1507309, EPI_ISL_1507310, EPI_ISL_1507311, EPI_ISL_1507312, EPI_ISL_1507313, EPI_ISL_1507314, EPI_ISL_1507315, EPI_ISL_1507316, EPI_ISL_1507317, EPI_ISL_1507318, EPI_ISL_1507319, EPI_ISL_1507320, EPI_ISL_1507321, EPI_ISL_1507322, EPI_ISL_1507323, EPI_ISL_1507324, EPI_ISL_1507325, EPI_ISL_1507326, EPI_ISL_1507327, EPI_ISL_1507328, EPI_ISL_1507329, EPI_ISL_1507330,
EPI_ISL_1507331, EPI_ISL_1507332, EPI_ISL_1507333, EPI_ISL_1507334, EPI_ISL_1507335, EPI_ISL_1507336, EPI_ISL_1507337, EPI_ISL_1507338, EPI_ISL_1507339, EPI_ISL_1507340, EPI_ISL_1507341, EPI_ISL_1507342, EPI_ISL_1507343, EPI_ISL_1507344, EPI_ISL_1507345, EPI_ISL_1507346, EPI_ISL_1507347, EPI_ISL_1507348, EPI_ISL_1507349, EPI_ISL_1507350, EPI_ISL_1507351, EPI_ISL_1507352,
EPI_ISL_1507353, EPI_ISL_1507354, EPI_ISL_1507355, EPI_ISL_1507356, EPI_ISL_1507357, EPI_ISL_1507358, EPI_ISL_1507359, EPI_ISL_1507360, EPI_ISL_1507361, EPI_ISL_1507362, EPI_ISL_1507363, EPI_ISL_1507364, EPI_ISL_1507365, EPI_ISL_1507366, EPI_ISL_1507367, EPI_ISL_1507368, EPI_ISL_1507369, EPI_ISL_1507370, EPI_ISL_1507371, EPI_ISL_1507372, EPI_ISL_1507373, EPI_ISL_1507374,
EPI_ISL_1507375, EPI_ISL_1507376, EPI_ISL_1507377, EPI_ISL_1507378, EPI_ISL_1507379, EPI_ISL_1507380, EPI_ISL_1507381, EPI_ISL_1507382, EPI_ISL_1507383, EPI_ISL_1507384, EPI_ISL_1507385, EPI_ISL_1507386, EPI_ISL_1507387, EPI_ISL_1507388, EPI_ISL_1507389, EPI_ISL_1507390, EPI_ISL_1507391, EPI_ISL_1507392, EPI_ISL_1507393, EPI_ISL_1507394, EPI_ISL_1507395, EPI_ISL_1507396,
EPI_ISL_1507397, EPI_ISL_1507398, EPI_ISL_1507399, EPI_ISL_1507400, EPI_ISL_1507401, EPI_ISL_1507402, EPI_ISL_1507403, EPI_ISL_1507404, EPI_ISL_1507405, EPI_ISL_1507406, EPI_ISL_1507407, EPI_ISL_1507408, EPI_ISL_1507409, EPI_ISL_1507410, EPI_ISL_1507411, EPI_ISL_1507412, EPI_ISL_1507413, EPI_ISL_1507414, EPI_ISL_1507415, EPI_ISL_1507416, EPI_ISL_1507417, EPI_ISL_1507418,
EPI_ISL_1507419, EPI_ISL_1507420, EPI_ISL_1507421, EPI_ISL_1507423, EPI_ISL_1507424, EPI_ISL_1507425, EPI_ISL_1507426, EPI_ISL_1507427, EPI_ISL_1507428, EPI_ISL_1507429, EPI_ISL_1507430, EPI_ISL_1507432, EPI_ISL_1507433, EPI_ISL_1507434, EPI_ISL_1507435, EPI_ISL_1507436, EPI_ISL_1507437, EPI_ISL_1507438, EPI_ISL_1507439, EPI_ISL_1507440, EPI_ISL_1507441,
EPI_ISL_1507442, EPI_ISL_1507443, EPI_ISL_1507444, EPI_ISL_1507445, EPI_ISL_1507446, EPI_ISL_1507447, EPI_ISL_1507448, EPI_ISL_1507449, EPI_ISL_1507450, EPI_ISL_1507451, EPI_ISL_1507452, EPI_ISL_1507453, EPI_ISL_1507454, EPI_ISL_1507455, EPI_ISL_1507456, EPI_ISL_1507457, EPI_ISL_1507458, EPI_ISL_1507459, EPI_ISL_1507460, EPI_ISL_1507461, EPI_ISL_1507462, EPI_ISL_1507463,
EPI_ISL_1507464, EPI_ISL_1507465, EPI_ISL_1507466, EPI_ISL_1507467, EPI_ISL_1507468, EPI_ISL_1507469, EPI_ISL_1507470, EPI_ISL_1507471, EPI_ISL_1507472, EPI_ISL_1507473, EPI_ISL_1507474, EPI_ISL_1507475, EPI_ISL_1507476, EPI_ISL_1507477, EPI_ISL_1507478, EPI_ISL_1507479, EPI_ISL_1507480, EPI_ISL_1507481, EPI_ISL_1507482, EPI_ISL_1507484, EPI_ISL_1507485, EPI_ISL_1507486,
EPI_ISL_1507487, EPI_ISL_1507488, EPI_ISL_1507489, EPI_ISL_1507490, EPI_ISL_1507491, EPI_ISL_1507492, EPI_ISL_1507493, EPI_ISL_1507494, EPI_ISL_1507495, EPI_ISL_1507496, EPI_ISL_1507497, EPI_ISL_1507498, EPI_ISL_1507499, EPI_ISL_1507500, EPI_ISL_1507501, EPI_ISL_1507502, EPI_ISL_1507503, EPI_ISL_1507504, EPI_ISL_1507505, EPI_ISL_1507506, EPI_ISL_1507507, EPI_ISL_1507508,
EPI_ISL_1507509, EPI_ISL_1507510, EPI_ISL_1507511, EPI_ISL_1507512, EPI_ISL_1507513, EPI_ISL_1507516, EPI_ISL_1507518, EPI_ISL_1507520, EPI_ISL_1507521, EPI_ISL_1507522, EPI_ISL_1507523, EPI_ISL_1507524, EPI_ISL_1507525, EPI_ISL_1507527, EPI_ISL_1507529, EPI_ISL_1507530, EPI_ISL_1507531, EPI_ISL_1507532, EPI_ISL_1507534, EPI_ISL_1507535,
EPI_ISL_1507536, EPI_ISL_1507537, EPI_ISL_1507538, EPI_ISL_1507539, EPI_ISL_1507540, EPI_ISL_1507542, EPI_ISL_1507543, EPI_ISL_1507544, EPI_ISL_1507545, EPI_ISL_1507547, EPI_ISL_1507548, EPI_ISL_1507549, EPI_ISL_1507550, EPI_ISL_1507551, EPI_ISL_1507552, EPI_ISL_1507553, EPI_ISL_1507554, EPI_ISL_1507555, EPI_ISL_1507557, EPI_ISL_1507558, EPI_ISL_1507559, EPI_ISL_1507560,
EPI_ISL_1507561, EPI_ISL_1507562, EPI_ISL_1507563, EPI_ISL_1507565, EPI_ISL_1507566, EPI_ISL_1507567, EPI_ISL_1507568, EPI_ISL_1507569, EPI_ISL_1507570, EPI_ISL_1507571, EPI_ISL_1507581, EPI_ISL_1507582, EPI_ISL_1507583, EPI_ISL_1507584, EPI_ISL_1507585, EPI_ISL_1507586, EPI_ISL_1507587, EPI_ISL_1507588, EPI_ISL_1507589, EPI_ISL_1507590, EPI_ISL_1507591, EPI_ISL_1507592,
EPI_ISL_1507593, EPI_ISL_1507594, EPI_ISL_1507595, EPI_ISL_1507596, EPI_ISL_1507598, EPI_ISL_1507599, EPI_ISL_1507600, EPI_ISL_1507601, EPI_ISL_1507602, EPI_ISL_1507603, EPI_ISL_1507604, EPI_ISL_1507605, EPI_ISL_1507606, EPI_ISL_1507607, EPI_ISL_1507608, EPI_ISL_1507609, EPI_ISL_1507610, EPI_ISL_1507611, EPI_ISL_1507612, EPI_ISL_1507613, EPI_ISL_1507614, EPI_ISL_1507615,
EPI_ISL_1507616, EPI_ISL_1507617, EPI_ISL_1507618, EPI_ISL_1507619, EPI_ISL_1507620, EPI_ISL_1507621, EPI_ISL_1507622, EPI_ISL_1507623, EPI_ISL_1507624, EPI_ISL_1507626, EPI_ISL_1507627, EPI_ISL_1507628, EPI_ISL_1507629, EPI_ISL_1507630, EPI_ISL_1507631, EPI_ISL_1507632, EPI_ISL_1507633, EPI_ISL_1507634, EPI_ISL_1507635, EPI_ISL_1507636, EPI_ISL_1507637,
EPI_ISL_1507638, EPI_ISL_1507639, EPI_ISL_1507640, EPI_ISL_1507641, EPI_ISL_1507642, EPI_ISL_1507643, EPI_ISL_1507644, EPI_ISL_1507645, EPI_ISL_1507646, EPI_ISL_1507648, EPI_ISL_1507649, EPI_ISL_1507650, EPI_ISL_1507651, EPI_ISL_1507652, EPI_ISL_1507653, EPI_ISL_1507654, EPI_ISL_1507655, EPI_ISL_1507656, EPI_ISL_1507657, EPI_ISL_1507658, EPI_ISL_1507659, EPI_ISL_1507660,
EPI_ISL_1507661, EPI_ISL_1507662, EPI_ISL_1507663, EPI_ISL_1507664, EPI_ISL_1507665, EPI_ISL_1507666, EPI_ISL_1507667, EPI_ISL_1507668, EPI_ISL_1507669, EPI_ISL_1507670, EPI_ISL_1507671, EPI_ISL_1507672, EPI_ISL_1507673, EPI_ISL_1507674, EPI_ISL_1507675, EPI_ISL_1507676, EPI_ISL_1507677, EPI_ISL_1507678, EPI_ISL_1507679, EPI_ISL_1507680, EPI_ISL_1507681, EPI_ISL_1507682,

EPI_ISL_1507683, EPI_ISL_1507684, EPI_ISL_1507685, EPI_ISL_1507686, EPI_ISL_1507687, EPI_ISL_1507688, EPI_ISL_1507689, EPI_ISL_1507690, EPI_ISL_1507691, EPI_ISL_1507692, EPI_ISL_1507693, EPI_ISL_1507694, EPI_ISL_1507695, EPI_ISL_1507696, EPI_ISL_1507697, EPI_ISL_1507698, EPI_ISL_1507699, EPI_ISL_1507700, EPI_ISL_1507701, EPI_ISL_1507702, EPI_ISL_1507703, EPI_ISL_1507704,
EPI_ISL_1507705, EPI_ISL_1507706, EPI_ISL_1507710, EPI_ISL_1507711, EPI_ISL_1507712, EPI_ISL_1507713, EPI_ISL_1507714, EPI_ISL_1507715, EPI_ISL_1507716, EPI_ISL_1507717, EPI_ISL_1507718, EPI_ISL_1507719, EPI_ISL_1507720, EPI_ISL_1507721, EPI_ISL_1507722, EPI_ISL_1507723, EPI_ISL_1507724, EPI_ISL_1507725, EPI_ISL_1507726, EPI_ISL_1507727, EPI_ISL_1507728, EPI_ISL_1507729,
EPI_ISL_1507730, EPI_ISL_1507731, EPI_ISL_1507732, EPI_ISL_1507733, EPI_ISL_1507734, EPI_ISL_1507735, EPI_ISL_1507736, EPI_ISL_1507737, EPI_ISL_1507738, EPI_ISL_1507739, EPI_ISL_1507740, EPI_ISL_1507741, EPI_ISL_1507742, EPI_ISL_1507743, EPI_ISL_1507744, EPI_ISL_1507745, EPI_ISL_1507746, EPI_ISL_1507747, EPI_ISL_1507748, EPI_ISL_1507749, EPI_ISL_1507750, EPI_ISL_1507751,
EPI_ISL_1507752, EPI_ISL_1507753, EPI_ISL_1507754, EPI_ISL_1507755, EPI_ISL_1507756, EPI_ISL_1507757, EPI_ISL_1507758, EPI_ISL_1507759, EPI_ISL_1507760, EPI_ISL_1507761, EPI_ISL_1507762, EPI_ISL_1507763, EPI_ISL_1507764, EPI_ISL_1507765, EPI_ISL_1507767, EPI_ISL_1507768, EPI_ISL_1507769, EPI_ISL_1507770, EPI_ISL_1507771, EPI_ISL_1507772, EPI_ISL_1507773, EPI_ISL_1507774,
EPI_ISL_1507775, EPI_ISL_1507776, EPI_ISL_1507777, EPI_ISL_1507778, EPI_ISL_1507779, EPI_ISL_1507780, EPI_ISL_1507781, EPI_ISL_1507782, EPI_ISL_1507783, EPI_ISL_1507784, EPI_ISL_1507785, EPI_ISL_1507786, EPI_ISL_1507787, EPI_ISL_1507788, EPI_ISL_1507789, EPI_ISL_1507790, EPI_ISL_1507791, EPI_ISL_1507792, EPI_ISL_1507793, EPI_ISL_1507819, EPI_ISL_1507820, EPI_ISL_1507821,
EPI_ISL_1507822, EPI_ISL_1507823, EPI_ISL_1507824, EPI_ISL_1507825, EPI_ISL_1507826, EPI_ISL_1507827, EPI_ISL_1507828, EPI_ISL_1507829, EPI_ISL_1507830, EPI_ISL_1507831, EPI_ISL_1507832, EPI_ISL_1507833, EPI_ISL_1507834, EPI_ISL_1507835, EPI_ISL_1507836, EPI_ISL_1507837, EPI_ISL_1507838, EPI_ISL_1507839, EPI_ISL_1507840, EPI_ISL_1507841, EPI_ISL_1507842, EPI_ISL_1507843,
EPI_ISL_1507844, EPI_ISL_1507845, EPI_ISL_1507846, EPI_ISL_1507847, EPI_ISL_1507848, EPI_ISL_1507849, EPI_ISL_1507850, EPI_ISL_1507851, EPI_ISL_1507853, EPI_ISL_1507854, EPI_ISL_1507855, EPI_ISL_1507856, EPI_ISL_1507857, EPI_ISL_1507858, EPI_ISL_1507859, EPI_ISL_1507860, EPI_ISL_1507861, EPI_ISL_1507862, EPI_ISL_1507863, EPI_ISL_1507864, EPI_ISL_1507865, EPI_ISL_1507866,
EPI_ISL_1507868, EPI_ISL_1507869, EPI_ISL_1507870, EPI_ISL_1507871, EPI_ISL_1507872, EPI_ISL_1507873, EPI_ISL_1507874, EPI_ISL_1507875, EPI_ISL_1507876, EPI_ISL_1507878, EPI_ISL_1507879, EPI_ISL_1507880, EPI_ISL_1507881, EPI_ISL_1507882, EPI_ISL_1507884, EPI_ISL_1507885, EPI_ISL_1507886, EPI_ISL_1507887, EPI_ISL_1507888, EPI_ISL_1507889, EPI_ISL_1507890, EPI_ISL_1507891,
EPI_ISL_1507892, EPI_ISL_1507893, EPI_ISL_1507894, EPI_ISL_1507895, EPI_ISL_1507896, EPI_ISL_1507897, EPI_ISL_1507898, EPI_ISL_1507899, EPI_ISL_1507900, EPI_ISL_1507901, EPI_ISL_1507902, EPI_ISL_1507903, EPI_ISL_1507904, EPI_ISL_1507905, EPI_ISL_1507906, EPI_ISL_1507907, EPI_ISL_1507908, EPI_ISL_1507909, EPI_ISL_1507910, EPI_ISL_1507911, EPI_ISL_1507912, EPI_ISL_1507913,
EPI_ISL_1507914, EPI_ISL_1507915, EPI_ISL_1507916, EPI_ISL_1507917, EPI_ISL_1507918, EPI_ISL_1507919, EPI_ISL_1507920, EPI_ISL_1507921, EPI_ISL_1507922, EPI_ISL_1507923, EPI_ISL_1507924, EPI_ISL_1507925, EPI_ISL_1507926, EPI_ISL_1507927, EPI_ISL_1507928, EPI_ISL_1507930, EPI_ISL_1507931, EPI_ISL_1507932, EPI_ISL_1507933, EPI_ISL_1507934, EPI_ISL_1507935, EPI_ISL_1507936,
EPI_ISL_1507937, EPI_ISL_1507938, EPI_ISL_1507939, EPI_ISL_1507940, EPI_ISL_1507944, EPI_ISL_1507945, EPI_ISL_1507946, EPI_ISL_1507947, EPI_ISL_1507948, EPI_ISL_1507949, EPI_ISL_1507951, EPI_ISL_1507952, EPI_ISL_1507953, EPI_ISL_1507954, EPI_ISL_1507955, EPI_ISL_1507956, EPI_ISL_1507957, EPI_ISL_1507958, EPI_ISL_1507959, EPI_ISL_1507960, EPI_ISL_1507961, EPI_ISL_1507962,
EPI_ISL_1507963, EPI_ISL_1507964, EPI_ISL_1507965, EPI_ISL_1507966, EPI_ISL_1507967, EPI_ISL_1507968, EPI_ISL_1507969, EPI_ISL_1507970, EPI_ISL_1507971, EPI_ISL_1507972, EPI_ISL_1507973, EPI_ISL_1507974, EPI_ISL_1507975, EPI_ISL_1507976, EPI_ISL_1507977, EPI_ISL_1507978, EPI_ISL_1507979, EPI_ISL_1507980, EPI_ISL_1507981, EPI_ISL_1507982, EPI_ISL_1507983, EPI_ISL_1507984,
EPI_ISL_1507985, EPI_ISL_1507986, EPI_ISL_1507987, EPI_ISL_1507989, EPI_ISL_1507990, EPI_ISL_1507991, EPI_ISL_1507992, EPI_ISL_1507993, EPI_ISL_1507994, EPI_ISL_1507995, EPI_ISL_1507996, EPI_ISL_1507997, EPI_ISL_1507998, EPI_ISL_1507999, EPI_ISL_1508000, EPI_ISL_1508001, EPI_ISL_1508002, EPI_ISL_1508003, EPI_ISL_1508004, EPI_ISL_1508005, EPI_ISL_1508006, EPI_ISL_1508007,
EPI_ISL_1508009, EPI_ISL_1508010, EPI_ISL_1508011, EPI_ISL_1508013, EPI_ISL_1508014, EPI_ISL_1508015, EPI_ISL_1508016, EPI_ISL_1508018, EPI_ISL_1508019, EPI_ISL_1508020, EPI_ISL_1508021, EPI_ISL_1508022, EPI_ISL_1508023, EPI_ISL_1508024, EPI_ISL_1508025, EPI_ISL_1508026, EPI_ISL_1508027, EPI_ISL_1508028, EPI_ISL_1508029, EPI_ISL_1508030, EPI_ISL_1508031, EPI_ISL_1508032,
EPI_ISL_1508033, EPI_ISL_1508034, EPI_ISL_1508035, EPI_ISL_1508036, EPI_ISL_1508037, EPI_ISL_1508038, EPI_ISL_1508039, EPI_ISL_1508040, EPI_ISL_1508042, EPI_ISL_1508043, EPI_ISL_1508044, EPI_ISL_1508046, EPI_ISL_1508047, EPI_ISL_1508048, EPI_ISL_1508049, EPI_ISL_1508050, EPI_ISL_1508051, EPI_ISL_1508052, EPI_ISL_1508053, EPI_ISL_1508054, EPI_ISL_1508055, EPI_ISL_1508056,
EPI_ISL_1508057, EPI_ISL_1508058, EPI_ISL_1508059, EPI_ISL_1508060, EPI_ISL_1508061, EPI_ISL_1508062, EPI_ISL_1508063, EPI_ISL_1508064, EPI_ISL_1508065, EPI_ISL_1508066, EPI_ISL_1508067, EPI_ISL_1508068, EPI_ISL_1508069, EPI_ISL_1508070, EPI_ISL_1508071, EPI_ISL_1508072, EPI_ISL_1508073, EPI_ISL_1508074, EPI_ISL_1508075, EPI_ISL_1508076, EPI_ISL_1508077, EPI_ISL_1508078,
EPI_ISL_1508079, EPI_ISL_1508080, EPI_ISL_1508081, EPI_ISL_1508082, EPI_ISL_1508083, EPI_ISL_1508084, EPI_ISL_1508085, EPI_ISL_1508086, EPI_ISL_1508087, EPI_ISL_1508088, EPI_ISL_1508089, EPI_ISL_1508090, EPI_ISL_1508091, EPI_ISL_1508092, EPI_ISL_1508093, EPI_ISL_1508094, EPI_ISL_1508095, EPI_ISL_1508096, EPI_ISL_1508097, EPI_ISL_1508098, EPI_ISL_1508099, EPI_ISL_1508100,
EPI_ISL_1508101, EPI_ISL_1508102, EPI_ISL_1508103, EPI_ISL_1508104, EPI_ISL_1508105, EPI_ISL_1508106, EPI_ISL_1508107, EPI_ISL_1508108, EPI_ISL_1508109, EPI_ISL_1508110, EPI_ISL_1508111, EPI_ISL_1508112, EPI_ISL_1508113, EPI_ISL_1508114, EPI_ISL_1508115, EPI_ISL_1508116, EPI_ISL_1508117, EPI_ISL_1508118, EPI_ISL_1508119, EPI_ISL_1508120, EPI_ISL_1508121, EPI_ISL_1508122,
EPI_ISL_1508123, EPI_ISL_1508124, EPI_ISL_1508125, EPI_ISL_1508126, EPI_ISL_1508127, EPI_ISL_1508128, EPI_ISL_1508129, EPI_ISL_1508130, EPI_ISL_1508131, EPI_ISL_1508132, EPI_ISL_1508133, EPI_ISL_1508134, EPI_ISL_1508135, EPI_ISL_1508136, EPI_ISL_1508137, EPI_ISL_1508138, EPI_ISL_1508139, EPI_ISL_1508140, EPI_ISL_1508152, EPI_ISL_1508164, EPI_ISL_1508168, EPI_ISL_1508171,
EPI_ISL_1508174, EPI_ISL_1508189, EPI_ISL_1508193, EPI_ISL_1508196, EPI_ISL_1508224, EPI_ISL_1508231, EPI_ISL_1508240, EPI_ISL_1508244, EPI_ISL_1508251, EPI_ISL_1508264, EPI_ISL_1508270, EPI_ISL_1508271, EPI_ISL_1508288, EPI_ISL_1508293, EPI_ISL_1508313, EPI_ISL_1508317, EPI_ISL_1508327, EPI_ISL_1508328, EPI_ISL_1508337, EPI_ISL_1508341, EPI_ISL_1508352, EPI_ISL_1508353,
EPI_ISL_1508359, EPI_ISL_1508364, EPI_ISL_1508365, EPI_ISL_1508366, EPI_ISL_1508371, EPI_ISL_1508375, EPI_ISL_1508381, EPI_ISL_1508465, EPI_ISL_1508515, EPI_ISL_1565339, EPI_ISL_1565340, EPI_ISL_1630544, EPI_ISL_1630579, EPI_ISL_1630678, EPI_ISL_1630690, EPI_ISL_1635082, EPI_ISL_1635093, EPI_ISL_1635097, EPI_ISL_1635109, EPI_ISL_1635117, EPI_ISL_1741784, EPI_ISL_1741796,
EPI_ISL_1741814, EPI_ISL_1741824, EPI_ISL_1741888

| | | | |
|---|---|---|---|
| see above | Lighthouse Lab in Glasgow | Wellcome Sanger Institute for the COVID-19 Genomics UK (COG-UK) Consortium | Anna Dominiczak and Alex Alderton; Carol Clugston; Cordelia Langford; David Gray; David K. Jackson; Dominic Kwiatkowski; Ewan Harrison; Harper VanSteenhouse; Ian Johnston; Jeffrey Barrett; John Sillitoe on behalf of the Wellcome Sanger Institute COVID-19 Surveillance Team; Roberto Amato; Sonia Goncalves; Yumi Kasai |

EPI_ISL_1409053, EPI_ISL_1448838, EPI_ISL_1448857, EPI_ISL_1448876, EPI_ISL_1448896, EPI_ISL_1448905, EPI_ISL_1448908, EPI_ISL_1448909, EPI_ISL_1448917, EPI_ISL_1448927, EPI_ISL_1448933, EPI_ISL_1448939, EPI_ISL_1448948, EPI_ISL_1448951, EPI_ISL_1448954, EPI_ISL_1448968, EPI_ISL_1448970, EPI_ISL_1448971, EPI_ISL_1448984, EPI_ISL_1448993, EPI_ISL_1449015, EPI_ISL_1449021,
EPI_ISL_1449023, EPI_ISL_1449025, EPI_ISL_1449351, EPI_ISL_1449362, EPI_ISL_1449364, EPI_ISL_1449365, EPI_ISL_1449374, EPI_ISL_1449376, EPI_ISL_1449381, EPI_ISL_1449383, EPI_ISL_1449385, EPI_ISL_1449387, EPI_ISL_1449391, EPI_ISL_1449403, EPI_ISL_1449407, EPI_ISL_1449412, EPI_ISL_1449427, EPI_ISL_1449428, EPI_ISL_1449431, EPI_ISL_1449436, EPI_ISL_1449439, EPI_ISL_1449440,
EPI_ISL_1449449, EPI_ISL_1449450, EPI_ISL_1449452, EPI_ISL_1449454, EPI_ISL_1449457, EPI_ISL_1449476, EPI_ISL_1449477, EPI_ISL_1449481, EPI_ISL_1449485, EPI_ISL_1449489, EPI_ISL_1449491, EPI_ISL_1449494, EPI_ISL_1449506, EPI_ISL_1449508, EPI_ISL_1449511, EPI_ISL_1449515, EPI_ISL_1449521, EPI_ISL_1449522, EPI_ISL_1449523, EPI_ISL_1449530, EPI_ISL_1449532, EPI_ISL_1449536,
EPI_ISL_1449540, EPI_ISL_1449543, EPI_ISL_1449544, EPI_ISL_1449545, EPI_ISL_1449547, EPI_ISL_1449557, EPI_ISL_1449559, EPI_ISL_1449566, EPI_ISL_1449567, EPI_ISL_1449573, EPI_ISL_1449590, EPI_ISL_1449605, EPI_ISL_1449618, EPI_ISL_1449620, EPI_ISL_1449621, EPI_ISL_1449627, EPI_ISL_1449630, EPI_ISL_1449634, EPI_ISL_1449637, EPI_ISL_1449640, EPI_ISL_1449641, EPI_ISL_1449643,
EPI_ISL_1449647, EPI_ISL_1449981, EPI_ISL_1449982, EPI_ISL_1449986, EPI_ISL_1449993, EPI_ISL_1449999, EPI_ISL_1450006, EPI_ISL_1450008, EPI_ISL_1450015, EPI_ISL_1450020, EPI_ISL_1450024, EPI_ISL_1450025, EPI_ISL_1450026, EPI_ISL_1450028, EPI_ISL_1450033, EPI_ISL_1450047, EPI_ISL_1450051, EPI_ISL_1450055, EPI_ISL_1450059, EPI_ISL_1450061, EPI_ISL_1450063, EPI_ISL_1450065,
EPI_ISL_1450066, EPI_ISL_1450071, EPI_ISL_1450087, EPI_ISL_1450095, EPI_ISL_1450100, EPI_ISL_1450106, EPI_ISL_1450110, EPI_ISL_1450122, EPI_ISL_1450124, EPI_ISL_1450128, EPI_ISL_1450136, EPI_ISL_1450138, EPI_ISL_1450139, EPI_ISL_1450140, EPI_ISL_1450153, EPI_ISL_1450158, EPI_ISL_1450159, EPI_ISL_1450160, EPI_ISL_1450163, EPI_ISL_1450168, EPI_ISL_1450171, EPI_ISL_1450173,
EPI_ISL_1450174, EPI_ISL_1450175, EPI_ISL_1450176, EPI_ISL_1450179, EPI_ISL_1450184, EPI_ISL_1450185, EPI_ISL_1450186, EPI_ISL_1450195, EPI_ISL_1450199, EPI_ISL_1450215, EPI_ISL_1450227, EPI_ISL_1450235, EPI_ISL_1450236, EPI_ISL_1450242, EPI_ISL_1450245, EPI_ISL_1450251, EPI_ISL_1450252, EPI_ISL_1450255, EPI_ISL_1450259, EPI_ISL_1450260, EPI_ISL_1450262, EPI_ISL_1450264,
EPI_ISL_1450265, EPI_ISL_1450273, EPI_ISL_1450274, EPI_ISL_1450284, EPI_ISL_1450285, EPI_ISL_1450291, EPI_ISL_1450292, EPI_ISL_1450295, EPI_ISL_1450296, EPI_ISL_1450298, EPI_ISL_1450299, EPI_ISL_1466446, EPI_ISL_1466448, EPI_ISL_1466457, EPI_ISL_1466463, EPI_ISL_1466474, EPI_ISL_1466478, EPI_ISL_1466482, EPI_ISL_1466484, EPI_ISL_1466487, EPI_ISL_1466491, EPI_ISL_1466492,
EPI_ISL_1466495, EPI_ISL_1466496, EPI_ISL_1466499, EPI_ISL_1466501, EPI_ISL_1466505, EPI_ISL_1466508, EPI_ISL_1466509, EPI_ISL_1466511, EPI_ISL_1466513, EPI_ISL_1466516, EPI_ISL_1466517, EPI_ISL_1466521, EPI_ISL_1466528, EPI_ISL_1466536, EPI_ISL_1466550, EPI_ISL_1466551, EPI_ISL_1466560, EPI_ISL_1466569, EPI_ISL_1466577, EPI_ISL_1466581, EPI_ISL_1466584, EPI_ISL_1466593,
EPI_ISL_1466594, EPI_ISL_1466595, EPI_ISL_1466598, EPI_ISL_1466599, EPI_ISL_1466606, EPI_ISL_1466610, EPI_ISL_1466616, EPI_ISL_1466617, EPI_ISL_1466626, EPI_ISL_1466630, EPI_ISL_1466634, EPI_ISL_1466649, EPI_ISL_1466652, EPI_ISL_1466655, EPI_ISL_1466656, EPI_ISL_1466660, EPI_ISL_1466663, EPI_ISL_1466675, EPI_ISL_1466678, EPI_ISL_1466680, EPI_ISL_1466683, EPI_ISL_1466685,
EPI_ISL_1466686, EPI_ISL_1466691, EPI_ISL_1466693, EPI_ISL_1466695, EPI_ISL_1466696, EPI_ISL_1466697, EPI_ISL_1466698, EPI_ISL_1466708, EPI_ISL_1466709, EPI_ISL_1466711, EPI_ISL_1466722, EPI_ISL_1466725, EPI_ISL_1466733, EPI_ISL_1466737, EPI_ISL_1466738, EPI_ISL_1466739, EPI_ISL_1466741, EPI_ISL_1466742, EPI_ISL_1466746, EPI_ISL_1466752, EPI_ISL_1466764, EPI_ISL_1466768,
EPI_ISL_1466771, EPI_ISL_1466775, EPI_ISL_1466779, EPI_ISL_1466783, EPI_ISL_1466785, EPI_ISL_1466787, EPI_ISL_1466789, EPI_ISL_1466793, EPI_ISL_1466794, EPI_ISL_1466798, EPI_ISL_1466805, EPI_ISL_1466809, EPI_ISL_1466812, EPI_ISL_1466813, EPI_ISL_1466817, EPI_ISL_1466820, EPI_ISL_1466822, EPI_ISL_1466824, EPI_ISL_1466831, EPI_ISL_1466838, EPI_ISL_1466840, EPI_ISL_1466844,
EPI_ISL_1466846, EPI_ISL_1466847, EPI_ISL_1466850, EPI_ISL_1466852, EPI_ISL_1466854, EPI_ISL_1466856, EPI_ISL_1466866, EPI_ISL_1466868, EPI_ISL_1466870, EPI_ISL_1466872, EPI_ISL_1466878, EPI_ISL_1466881, EPI_ISL_1466883, EPI_ISL_1466884, EPI_ISL_1466886, EPI_ISL_1466888, EPI_ISL_1466889, EPI_ISL_1466890, EPI_ISL_1466892, EPI_ISL_1466894, EPI_ISL_1466897, EPI_ISL_1466906,
EPI_ISL_1466910, EPI_ISL_1466913, EPI_ISL_1466920, EPI_ISL_1466922, EPI_ISL_1467259, EPI_ISL_1467266, EPI_ISL_1467274, EPI_ISL_1467277, EPI_ISL_1467280, EPI_ISL_1467284, EPI_ISL_1467298, EPI_ISL_1467301, EPI_ISL_1467306, EPI_ISL_1467307, EPI_ISL_1467310, EPI_ISL_1467314, EPI_ISL_1467316, EPI_ISL_1467320, EPI_ISL_1467324, EPI_ISL_1467328, EPI_ISL_1467333, EPI_ISL_1467338,
EPI_ISL_1467339, EPI_ISL_1467344, EPI_ISL_1467345, EPI_ISL_1467346, EPI_ISL_1467347, EPI_ISL_1467353, EPI_ISL_1467357, EPI_ISL_1467359, EPI_ISL_1467361, EPI_ISL_1467368, EPI_ISL_1467375, EPI_ISL_1467376, EPI_ISL_1467378, EPI_ISL_1467381, EPI_ISL_1467382, EPI_ISL_1467384, EPI_ISL_1467391, EPI_ISL_1467393, EPI_ISL_1467394, EPI_ISL_1467396, EPI_ISL_1467398, EPI_ISL_1467400,
EPI_ISL_1467415, EPI_ISL_1467416, EPI_ISL_1467419, EPI_ISL_1467420, EPI_ISL_1467422, EPI_ISL_1467432, EPI_ISL_1467436, EPI_ISL_1467438, EPI_ISL_1467439, EPI_ISL_1467445, EPI_ISL_1467446, EPI_ISL_1467448, EPI_ISL_1467452, EPI_ISL_1467453, EPI_ISL_1467457, EPI_ISL_1467464, EPI_ISL_1467465, EPI_ISL_1467469, EPI_ISL_1467472, EPI_ISL_1467477, EPI_ISL_1467484, EPI_ISL_1467492,
EPI_ISL_1467495, EPI_ISL_1467496, EPI_ISL_1467497, EPI_ISL_1467503, EPI_ISL_1467504, EPI_ISL_1467506, EPI_ISL_1467509, EPI_ISL_1467511, EPI_ISL_1467513, EPI_ISL_1467515, EPI_ISL_1467518, EPI_ISL_1467520, EPI_ISL_1467522, EPI_ISL_1467523, EPI_ISL_1467524, EPI_ISL_1467528, EPI_ISL_1467529, EPI_ISL_1467533, EPI_ISL_1467537, EPI_ISL_1467539, EPI_ISL_1467540, EPI_ISL_1467541,
EPI_ISL_1467550, EPI_ISL_1467559, EPI_ISL_1467564, EPI_ISL_1467573, EPI_ISL_1467574, EPI_ISL_1467577, EPI_ISL_1467578, EPI_ISL_1467803, EPI_ISL_1467835, EPI_ISL_1467836, EPI_ISL_1467837, EPI_ISL_1467838, EPI_ISL_1467840, EPI_ISL_1467841, EPI_ISL_1467843, EPI_ISL_1467844, EPI_ISL_1467845, EPI_ISL_1467847, EPI_ISL_1467852, EPI_ISL_1467853, EPI_ISL_1467854, EPI_ISL_1467857,
EPI_ISL_1467859, EPI_ISL_1467860, EPI_ISL_1467862, EPI_ISL_1467868, EPI_ISL_1467874, EPI_ISL_1467875, EPI_ISL_1467876, EPI_ISL_1467877, EPI_ISL_1467879, EPI_ISL_1467880, EPI_ISL_1467881, EPI_ISL_1467885, EPI_ISL_1467886, EPI_ISL_1467887, EPI_ISL_1467888, EPI_ISL_1467889, EPI_ISL_1467892, EPI_ISL_1467897, EPI_ISL_1467898, EPI_ISL_1467899, EPI_ISL_1467900, EPI_ISL_1467905,
EPI_ISL_1467908, EPI_ISL_1467913, EPI_ISL_1467914, EPI_ISL_1467917, EPI_ISL_1467919, EPI_ISL_1467921, EPI_ISL_1467924, EPI_ISL_1467925, EPI_ISL_1467926, EPI_ISL_1467927, EPI_ISL_1467929, EPI_ISL_1467930, EPI_ISL_1467932, EPI_ISL_1467933, EPI_ISL_1467937, EPI_ISL_1467939, EPI_ISL_1467941, EPI_ISL_1467943, EPI_ISL_1467946, EPI_ISL_1467947, EPI_ISL_1467948, EPI_ISL_1467949,
EPI_ISL_1467952, EPI_ISL_1467953, EPI_ISL_1467955, EPI_ISL_1467956, EPI_ISL_1467957, EPI_ISL_1467963, EPI_ISL_1467966, EPI_ISL_1467967, EPI_ISL_1467968, EPI_ISL_1467969, EPI_ISL_1467970, EPI_ISL_1467972, EPI_ISL_1467973, EPI_ISL_1467975, EPI_ISL_1467978, EPI_ISL_1467979, EPI_ISL_1467980, EPI_ISL_1467981, EPI_ISL_1467986, EPI_ISL_1467987, EPI_ISL_1467991, EPI_ISL_1467993,
EPI_ISL_1467994, EPI_ISL_1467995, EPI_ISL_1467996, EPI_ISL_1467999, EPI_ISL_1468000, EPI_ISL_1468001, EPI_ISL_1468002, EPI_ISL_1468006, EPI_ISL_1468007, EPI_ISL_1468008, EPI_ISL_1468010, EPI_ISL_1468012, EPI_ISL_1468014, EPI_ISL_1468016, EPI_ISL_1468017, EPI_ISL_1468019, EPI_ISL_1468025, EPI_ISL_1468026, EPI_ISL_1468028, EPI_ISL_1468029, EPI_ISL_1468031, EPI_ISL_1468032,
EPI_ISL_1468037, EPI_ISL_1468041, EPI_ISL_1468043, EPI_ISL_1468045, EPI_ISL_1468048, EPI_ISL_1468049, EPI_ISL_1468051, EPI_ISL_1468052, EPI_ISL_1468053, EPI_ISL_1468055, EPI_ISL_1468056, EPI_ISL_1468059, EPI_ISL_1468060, EPI_ISL_1468062, EPI_ISL_1468065, EPI_ISL_1468066, EPI_ISL_1468069, EPI_ISL_1468071, EPI_ISL_1468073, EPI_ISL_1468074, EPI_ISL_1468075, EPI_ISL_1468076,
EPI_ISL_1468077, EPI_ISL_1468080, EPI_ISL_1468081, EPI_ISL_1468084, EPI_ISL_1468085, EPI_ISL_1468087, EPI_ISL_1468088, EPI_ISL_1468089, EPI_ISL_1468091, EPI_ISL_1468094, EPI_ISL_1468095, EPI_ISL_1468096, EPI_ISL_1468097, EPI_ISL_1468098, EPI_ISL_1468099, EPI_ISL_1468100, EPI_ISL_1468102, EPI_ISL_1468103, EPI_ISL_1468104, EPI_ISL_1468105, EPI_ISL_1468106, EPI_ISL_1468109,
EPI_ISL_1468110, EPI_ISL_1468112, EPI_ISL_1468113, EPI_ISL_1468114, EPI_ISL_1468115, EPI_ISL_1468116, EPI_ISL_1468118, EPI_ISL_1468120, EPI_ISL_1468121, EPI_ISL_1468122, EPI_ISL_1468125, EPI_ISL_1468126, EPI_ISL_1468130, EPI_ISL_1468131, EPI_ISL_1472408, EPI_ISL_1472410, EPI_ISL_1472411, EPI_ISL_1472415, EPI_ISL_1472418, EPI_ISL_1472422, EPI_ISL_1472426, EPI_ISL_1472427,
EPI_ISL_1472428, EPI_ISL_1472431, EPI_ISL_1472435, EPI_ISL_1472436, EPI_ISL_1472442, EPI_ISL_1472446, EPI_ISL_1472448, EPI_ISL_1472451, EPI_ISL_1472452, EPI_ISL_1472453, EPI_ISL_1472455, EPI_ISL_1472456, EPI_ISL_1472458, EPI_ISL_1472462, EPI_ISL_1472464, EPI_ISL_1472465, EPI_ISL_1472474, EPI_ISL_1472483, EPI_ISL_1472492, EPI_ISL_1472494, EPI_ISL_1472496, EPI_ISL_1472502,
EPI_ISL_1472508, EPI_ISL_1472511, EPI_ISL_1472512, EPI_ISL_1472513, EPI_ISL_1472528, EPI_ISL_1472532, EPI_ISL_1472536, EPI_ISL_1472547, EPI_ISL_1472551, EPI_ISL_1472552, EPI_ISL_1472554, EPI_ISL_1472560, EPI_ISL_1472562, EPI_ISL_1472569, EPI_ISL_1472572, EPI_ISL_1472578, EPI_ISL_1472580, EPI_ISL_1472582, EPI_ISL_1472584, EPI_ISL_1472593, EPI_ISL_1472594, EPI_ISL_1472596,
EPI_ISL_1472600, EPI_ISL_1472608, EPI_ISL_1472611, EPI_ISL_1472613, EPI_ISL_1472621, EPI_ISL_1472622, EPI_ISL_1472626, EPI_ISL_1472628, EPI_ISL_1472634, EPI_ISL_1472638, EPI_ISL_1472646, EPI_ISL_1472650, EPI_ISL_1472655, EPI_ISL_1472657, EPI_ISL_1472658, EPI_ISL_1472663, EPI_ISL_1472667, EPI_ISL_1472669, EPI_ISL_1472670, EPI_ISL_1472672, EPI_ISL_1472680, EPI_ISL_1472681,
EPI_ISL_1472684, EPI_ISL_1472686, EPI_ISL_1472692, EPI_ISL_1472696, EPI_ISL_1473810, EPI_ISL_1474156, EPI_ISL_1474161, EPI_ISL_1485501, EPI_ISL_1485563, EPI_ISL_1485574, EPI_ISL_1485584, EPI_ISL_1486404, EPI_ISL_1486420, EPI_ISL_1486433, EPI_ISL_1486444, EPI_ISL_1486446, EPI_ISL_1486450, EPI_ISL_1486457, EPI_ISL_1486461, EPI_ISL_1486466, EPI_ISL_1486473, EPI_ISL_1486480,
EPI_ISL_1486489, EPI_ISL_1486498, EPI_ISL_1486510, EPI_ISL_1486524, EPI_ISL_1486527, EPI_ISL_1486536, EPI_ISL_1486541, EPI_ISL_1486544, EPI_ISL_1486563, EPI_ISL_1486567, EPI_ISL_1486569, EPI_ISL_1486595, EPI_ISL_1486604, EPI_ISL_1486612, EPI_ISL_1486632, EPI_ISL_1486641, EPI_ISL_1486644, EPI_ISL_1486659, EPI_ISL_1486661, EPI_ISL_1486673, EPI_ISL_1486676, EPI_ISL_1486694,
EPI_ISL_1486756, EPI_ISL_1486757, EPI_ISL_1486763, EPI_ISL_1486768, EPI_ISL_1486777, EPI_ISL_1486798, EPI_ISL_1486802, EPI_ISL_1486866, EPI_ISL_1486876, EPI_ISL_1486989, EPI_ISL_1487016, EPI_ISL_1487127, EPI_ISL_1487388, EPI_ISL_1487393, EPI_ISL_1487396, EPI_ISL_1487405, EPI_ISL_1487433, EPI_ISL_1487434, EPI_ISL_1487437, EPI_ISL_1487440, EPI_ISL_1487452, EPI_ISL_1487458,
EPI_ISL_1487462, EPI_ISL_1487474, EPI_ISL_1487477, EPI_ISL_1487491, EPI_ISL_1487497, EPI_ISL_1487499, EPI_ISL_1487501, EPI_ISL_1487502, EPI_ISL_1487511, EPI_ISL_1487518, EPI_ISL_1487521, EPI_ISL_1487522, EPI_ISL_1487530, EPI_ISL_1487537, EPI_ISL_1487541, EPI_ISL_1487546, EPI_ISL_1487549, EPI_ISL_1487551, EPI_ISL_1487566, EPI_ISL_1487573, EPI_ISL_1487577, EPI_ISL_1487589,
EPI_ISL_1487595, EPI_ISL_1487603, EPI_ISL_1487609, EPI_ISL_1487613, EPI_ISL_1487614, EPI_ISL_1487617, EPI_ISL_1487624, EPI_ISL_1487648, EPI_ISL_1487653, EPI_ISL_1487655, EPI_ISL_1487657, EPI_ISL_1487658, EPI_ISL_1487670, EPI_ISL_1487704, EPI_ISL_1487716, EPI_ISL_1487719, EPI_ISL_1488259, EPI_ISL_1488292, EPI_ISL_1488387, EPI_ISL_1488438, EPI_ISL_1489462, EPI_ISL_1489472,
EPI_ISL_1504911, EPI_ISL_1504915, EPI_ISL_1504924, EPI_ISL_1504925, EPI_ISL_1504926, EPI_ISL_1504927, EPI_ISL_1504928, EPI_ISL_1504929, EPI_ISL_1504932, EPI_ISL_1504936, EPI_ISL_1504937, EPI_ISL_1504938, EPI_ISL_1504942, EPI_ISL_1504945, EPI_ISL_1504949, EPI_ISL_1504956, EPI_ISL_1504962, EPI_ISL_1504967, EPI_ISL_1504973, EPI_ISL_1504984, EPI_ISL_1504989, EPI_ISL_1504993,
EPI_ISL_1504994, EPI_ISL_1504995, EPI_ISL_1504997, EPI_ISL_1504998, EPI_ISL_1505000, EPI_ISL_1505003, EPI_ISL_1505007, EPI_ISL_1505009, EPI_ISL_1505019, EPI_ISL_1505020, EPI_ISL_1505026, EPI_ISL_1505030, EPI_ISL_1505031, EPI_ISL_1505032, EPI_ISL_1505035, EPI_ISL_1505041, EPI_ISL_1505042, EPI_ISL_1505047, EPI_ISL_1505052, EPI_ISL_1505053, EPI_ISL_1505063, EPI_ISL_1505067,
EPI_ISL_1505071, EPI_ISL_1505079, EPI_ISL_1505081, EPI_ISL_1505088, EPI_ISL_1505099, EPI_ISL_1505100, EPI_ISL_1505102, EPI_ISL_1505109, EPI_ISL_1505112, EPI_ISL_1505114, EPI_ISL_1505118, EPI_ISL_1505121, EPI_ISL_1505123, EPI_ISL_1505127, EPI_ISL_1505131, EPI_ISL_1505133, EPI_ISL_1505134, EPI_ISL_1505137, EPI_ISL_1505141, EPI_ISL_1505149, EPI_ISL_1505151, EPI_ISL_1505155,
EPI_ISL_1505156, EPI_ISL_1505161, EPI_ISL_1505162, EPI_ISL_1505163, EPI_ISL_1505166, EPI_ISL_1505169, EPI_ISL_1505171, EPI_ISL_1518066, EPI_ISL_1518067, EPI_ISL_1518084, EPI_ISL_1518085, EPI_ISL_1518087, EPI_ISL_1518093, EPI_ISL_1518094, EPI_ISL_1518101, EPI_ISL_1518103, EPI_ISL_1518111, EPI_ISL_1518118, EPI_ISL_1518121, EPI_ISL_1518133, EPI_ISL_1518135, EPI_ISL_1518136,
EPI_ISL_1518138, EPI_ISL_1518140, EPI_ISL_1518149, EPI_ISL_1518157, EPI_ISL_1518160, EPI_ISL_1518161, EPI_ISL_1518162, EPI_ISL_1518164, EPI_ISL_1518172, EPI_ISL_1518173, EPI_ISL_1518174, EPI_ISL_1518175, EPI_ISL_1518176, EPI_ISL_1518178, EPI_ISL_1518179, EPI_ISL_1518182, EPI_ISL_1518185, EPI_ISL_1518186, EPI_ISL_1518189, EPI_ISL_1518194, EPI_ISL_1518198, EPI_ISL_1518203,
EPI_ISL_1518205, EPI_ISL_1518207, EPI_ISL_1518214, EPI_ISL_1518223, EPI_ISL_1518228, EPI_ISL_1518230, EPI_ISL_1518231, EPI_ISL_1518249, EPI_ISL_1518251, EPI_ISL_1518254, EPI_ISL_1518255, EPI_ISL_1518257, EPI_ISL_1518263, EPI_ISL_1518266, EPI_ISL_1518273, EPI_ISL_1518277, EPI_ISL_1518278, EPI_ISL_1518281, EPI_ISL_1518289, EPI_ISL_1518291, EPI_ISL_1518294, EPI_ISL_1518297,
EPI_ISL_1518303, EPI_ISL_1518308, EPI_ISL_1518311, EPI_ISL_1518314, EPI_ISL_1518318, EPI_ISL_1518328, EPI_ISL_1518330, EPI_ISL_1518332, EPI_ISL_1518334, EPI_ISL_1518336, EPI_ISL_1518343, EPI_ISL_1518344, EPI_ISL_1518349, EPI_ISL_1518355, EPI_ISL_1518358, EPI_ISL_1518363, EPI_ISL_1518374, EPI_ISL_1518382, EPI_ISL_1518385, EPI_ISL_1518388, EPI_ISL_1518392, EPI_ISL_1518687,
EPI_ISL_1518690, EPI_ISL_1518691, EPI_ISL_1518695, EPI_ISL_1518696, EPI_ISL_1518701, EPI_ISL_1518713, EPI_ISL_1518715, EPI_ISL_1518718, EPI_ISL_1518725, EPI_ISL_1518727, EPI_ISL_1518730, EPI_ISL_1518736, EPI_ISL_1518746, EPI_ISL_1518753, EPI_ISL_1518757, EPI_ISL_1518759, EPI_ISL_1518762, EPI_ISL_1518763, EPI_ISL_1518771, EPI_ISL_1518777, EPI_ISL_1518786, EPI_ISL_1518791,
EPI_ISL_1518792, EPI_ISL_1518793, EPI_ISL_1518803, EPI_ISL_1518810, EPI_ISL_1518811, EPI_ISL_1518817, EPI_ISL_1518818, EPI_ISL_1518819, EPI_ISL_1518821, EPI_ISL_1518822, EPI_ISL_1518836, EPI_ISL_1518838, EPI_ISL_1518848, EPI_ISL_1518853, EPI_ISL_1518861, EPI_ISL_1518862, EPI_ISL_1518865, EPI_ISL_1518873, EPI_ISL_1518877, EPI_ISL_1518881, EPI_ISL_1518888, EPI_ISL_1518889,
EPI_ISL_1518901, EPI_ISL_1518903, EPI_ISL_1518907, EPI_ISL_1518913, EPI_ISL_1518916, EPI_ISL_1518920, EPI_ISL_1518921, EPI_ISL_1518927, EPI_ISL_1518935, EPI_ISL_1518943, EPI_ISL_1518946, EPI_ISL_1518957, EPI_ISL_1518959, EPI_ISL_1518969, EPI_ISL_1518978, EPI_ISL_1518984, EPI_ISL_1518988, EPI_ISL_1518989, EPI_ISL_1518996, EPI_ISL_1518997, EPI_ISL_1518999, EPI_ISL_1519000,
EPI_ISL_1519004, EPI_ISL_1519005, EPI_ISL_1519009, EPI_ISL_1519010, EPI_ISL_1519011, EPI_ISL_1519012, EPI_ISL_1519013, EPI_ISL_1519014, EPI_ISL_1519015, EPI_ISL_1519017, EPI_ISL_1519018, EPI_ISL_1519020, EPI_ISL_1519022, EPI_ISL_1519023, EPI_ISL_1519024, EPI_ISL_1519025, EPI_ISL_1519026, EPI_ISL_1519027, EPI_ISL_1519028, EPI_ISL_1519033, EPI_ISL_1519034, EPI_ISL_1519035,
EPI_ISL_1519037, EPI_ISL_1519038, EPI_ISL_1519040, EPI_ISL_1519041, EPI_ISL_1519042, EPI_ISL_1519043, EPI_ISL_1519044, EPI_ISL_1519045, EPI_ISL_1519047, EPI_ISL_1519049, EPI_ISL_1519051, EPI_ISL_1519052, EPI_ISL_1519053, EPI_ISL_1519054, EPI_ISL_1519055, EPI_ISL_1519057, EPI_ISL_1519061, EPI_ISL_1519063, EPI_ISL_1519066, EPI_ISL_1519068, EPI_ISL_1519070, EPI_ISL_1519071,
EPI_ISL_1519076, EPI_ISL_1519079, EPI_ISL_1519080, EPI_ISL_1519081, EPI_ISL_1519082, EPI_ISL_1519083, EPI_ISL_1519084, EPI_ISL_1519085, EPI_ISL_1519088, EPI_ISL_1519090, EPI_ISL_1519091, EPI_ISL_1519093, EPI_ISL_1519094, EPI_ISL_1519097, EPI_ISL_1519098, EPI_ISL_1519099, EPI_ISL_1519102, EPI_ISL_1519103, EPI_ISL_1519105, EPI_ISL_1519106, EPI_ISL_1519108, EPI_ISL_1519110,
EPI_ISL_1519111, EPI_ISL_1519115, EPI_ISL_1519117, EPI_ISL_1519118, EPI_ISL_1519119, EPI_ISL_1519121, EPI_ISL_1519122, EPI_ISL_1519123, EPI_ISL_1519124, EPI_ISL_1519126, EPI_ISL_1519128, EPI_ISL_1519129, EPI_ISL_1519130, EPI_ISL_1519131, EPI_ISL_1519132, EPI_ISL_1519134, EPI_ISL_1519135, EPI_ISL_1519136, EPI_ISL_1519137, EPI_ISL_1519140, EPI_ISL_1519141, EPI_ISL_1519142,
EPI_ISL_1519143, EPI_ISL_1519144, EPI_ISL_1519148, EPI_ISL_1519149, EPI_ISL_1519151, EPI_ISL_1519152, EPI_ISL_1519153, EPI_ISL_1519158, EPI_ISL_1519159, EPI_ISL_1519160, EPI_ISL_1519161, EPI_ISL_1519163, EPI_ISL_1519164, EPI_ISL_1519165, EPI_ISL_1519166, EPI_ISL_1519167, EPI_ISL_1519168, EPI_ISL_1519170, EPI_ISL_1519171, EPI_ISL_1519172, EPI_ISL_1519173, EPI_ISL_1519174,
EPI_ISL_1519176, EPI_ISL_1519177, EPI_ISL_1519179, EPI_ISL_1519180, EPI_ISL_1519181, EPI_ISL_1519183, EPI_ISL_1519184, EPI_ISL_1519186, EPI_ISL_1519187, EPI_ISL_1519188, EPI_ISL_1519191, EPI_ISL_1519194, EPI_ISL_1519195, EPI_ISL_1519198, EPI_ISL_1519199, EPI_ISL_1519201, EPI_ISL_1519203, EPI_ISL_1519205, EPI_ISL_1519206, EPI_ISL_1519207, EPI_ISL_1519209, EPI_ISL_1519210,
EPI_ISL_1519211, EPI_ISL_1519213, EPI_ISL_1519214, EPI_ISL_1519217, EPI_ISL_1519219, EPI_ISL_1519220, EPI_ISL_1519221, EPI_ISL_1519223, EPI_ISL_1519229, EPI_ISL_1519231, EPI_ISL_1519232, EPI_ISL_1519234, EPI_ISL_1519235, EPI_ISL_1519236, EPI_ISL_1519238, EPI_ISL_1519239, EPI_ISL_1519241, EPI_ISL_1519242, EPI_ISL_1519245, EPI_ISL_1519248, EPI_ISL_1519251, EPI_ISL_1519256,
EPI_ISL_1519257, EPI_ISL_1519258, EPI_ISL_1519262, EPI_ISL_1519265, EPI_ISL_1519295, EPI_ISL_1535603, EPI_ISL_1535650, EPI_ISL_1535694, EPI_ISL_1535700, EPI_ISL_1535817, EPI_ISL_1537360, EPI_ISL_1537374, EPI_ISL_1537380, EPI_ISL_1537394, EPI_ISL_1537404, EPI_ISL_1537409, EPI_ISL_1537410, EPI_ISL_1537418, EPI_ISL_1537422, EPI_ISL_1537435, EPI_ISL_1537472, EPI_ISL_1537480,
EPI_ISL_1537486, EPI_ISL_1537508, EPI_ISL_1537558, EPI_ISL_1537573, EPI_ISL_1537588, EPI_ISL_1537590, EPI_ISL_1537595, EPI_ISL_1537597, EPI_ISL_1537600, EPI_ISL_1537601, EPI_ISL_1537602, EPI_ISL_1537603, EPI_ISL_1537610, EPI_ISL_1537611, EPI_ISL_1537612, EPI_ISL_1537613, EPI_ISL_1537624, EPI_ISL_1537627, EPI_ISL_1537628, EPI_ISL_1537629, EPI_ISL_1537630, EPI_ISL_1537633,
EPI_ISL_1537634, EPI_ISL_1537637, EPI_ISL_1537659, EPI_ISL_1537666, EPI_ISL_1537671, EPI_ISL_1537674, EPI_ISL_1537692, EPI_ISL_1537700, EPI_ISL_1537704, EPI_ISL_1537705, EPI_ISL_1537729, EPI_ISL_1537736, EPI_ISL_1537738, EPI_ISL_1537747, EPI_ISL_1537748, EPI_ISL_1537749, EPI_ISL_1537763, EPI_ISL_1537767, EPI_ISL_1537768, EPI_ISL_1537770, EPI_ISL_1537774, EPI_ISL_1537775,
EPI_ISL_1537776, EPI_ISL_1537778, EPI_ISL_1537786, EPI_ISL_1537789, EPI_ISL_1537794, EPI_ISL_1537801, EPI_ISL_1537802, EPI_ISL_1537807, EPI_ISL_1537814, EPI_ISL_1537816, EPI_ISL_1537819, EPI_ISL_1537823, EPI_ISL_1537827, EPI_ISL_1537828, EPI_ISL_1537830, EPI_ISL_1537837, EPI_ISL_1537838, EPI_ISL_1537843, EPI_ISL_1538373, EPI_ISL_1564077, EPI_ISL_1564147, EPI_ISL_1564188,
EPI_ISL_1564194, EPI_ISL_1564202, EPI_ISL_1564234, EPI_ISL_1584171, EPI_ISL_1584173, EPI_ISL_1584257, EPI_ISL_1584299, EPI_ISL_1584314, EPI_ISL_1584333, EPI_ISL_1584363, EPI_ISL_1584367, EPI_ISL_1584382, EPI_ISL_1584394, EPI_ISL_1584403, EPI_ISL_1584431, EPI_ISL_1594994, EPI_ISL_1594998, EPI_ISL_1595029, EPI_ISL_1595073, EPI_ISL_1595075, EPI_ISL_1595084, EPI_ISL_1595098,
EPI_ISL_1595122, EPI_ISL_1595132, EPI_ISL_1595135, EPI_ISL_1595141, EPI_ISL_1595142, EPI_ISL_1595147, EPI_ISL_1595151, EPI_ISL_1595161, EPI_ISL_1595165, EPI_ISL_1595180, EPI_ISL_1595198

| | | | |
|---|---|---|---|
| see above | Lighthouse Lab in Milton Keynes | Wellcome Sanger Institute for the COVID-19 Genomics UK (COG-UK) Consortium | Cordelia Langford; David K. Jackson; Dominic Kwiatkowski; Ewan Harrison; Ian Johnston; Jeffrey Barrett; John Sillitoe on behalf of the Wellcome Sanger Institute COVID-19 Surveillance Team; Roberto Amato; Sonia Goncalves; The Lighthouse Lab in Milton Keynes and Alex Alderton |
| EPI_ISL_1833004, EPI_ISL_1833005, EPI_ISL_1833006, EPI_ISL_1833007, EPI_ISL_1833008, EPI_ISL_1833014 | Lincolnshire Hospitals and DeepSeq Nottingham | COVID-19 Genomics UK (COG-UK) Consortium | Christopher Moore; Fei Sang; Joeseph Chappell; Johnny Debebe; Jonathan Ball; Matthew Carlisle; Matthew Loose; Nadine Holmes; Nichola Duckworth; Patrick McClure; Sarah Walsh; Tim Sloan; Victoria Wright |

EPI_ISL_1474785, EPI_ISL_1474789, EPI_ISL_1474796, EPI_ISL_1474797, EPI_ISL_1474798, EPI_ISL_1474799, EPI_ISL_1546155, EPI_ISL_1546156, EPI_ISL_1546157, EPI_ISL_1546158, EPI_ISL_1546159, EPI_ISL_1546160, EPI_ISL_1546161, EPI_ISL_1546162, EPI_ISL_1546163, EPI_ISL_1546164, EPI_ISL_1546165, EPI_ISL_1546166, EPI_ISL_1546167, EPI_ISL_1546168, EPI_ISL_1546169, EPI_ISL_1546170,
EPI_ISL_1546171, EPI_ISL_1546172, EPI_ISL_1546173, EPI_ISL_1546182, EPI_ISL_1546183, EPI_ISL_1546184, EPI_ISL_1546192, EPI_ISL_1546193, EPI_ISL_1546194, EPI_ISL_1546195, EPI_ISL_1546196, EPI_ISL_1546197, EPI_ISL_1546198, EPI_ISL_1546199, EPI_ISL_1546200, EPI_ISL_1546201, EPI_ISL_1546202, EPI_ISL_1546204, EPI_ISL_1546205, EPI_ISL_1546206, EPI_ISL_1546212, EPI_ISL_1546213,
EPI_ISL_1546214, EPI_ISL_1546215, EPI_ISL_1546216, EPI_ISL_1546217, EPI_ISL_1546219, EPI_ISL_1546220, EPI_ISL_1546226, EPI_ISL_1546228, EPI_ISL_1636828, EPI_ISL_1636835, EPI_ISL_1636836, EPI_ISL_1636837, EPI_ISL_1637254, EPI_ISL_1637255, EPI_ISL_1637256, EPI_ISL_1637257, EPI_ISL_1637258, EPI_ISL_1637259, EPI_ISL_1637260, EPI_ISL_1637261, EPI_ISL_1637262, EPI_ISL_1637264,
EPI_ISL_1637265, EPI_ISL_1637266, EPI_ISL_1832385, EPI_ISL_1832387, EPI_ISL_1832391

| | | | |
|---|---|---|---|
| see above | Liverpool Clinical Laboratories | COVID-19 Genomics UK (COG-UK) Consortium | A Alrezaihi; Alessandro Gerada; Alistair Darby; Angela Cowell; Anita Lucaci; Anu Chawla; Cassie Olateju; Catherine Hartley; Charlotte Nelson; Ecaterina Vamos; Elaine O'Toole; Eleanor G Bentley; Ghada T Shawli; Hermione Webster; Isabel García-Dorival; James Johnson; James P Stewart; Jenifer Manson; Joanne Watts; Jones Benjamin; Jordan J Clark; Julian Hiscox; L Luu; Lucille Rainbow; M Almsaud; Margaret Hughes; Mark Whitehead; Matthew Gemmell; Miren Iturriza-Gomara; Muhannad Alruwaili; N.P Randle; Neil Swainston; PKF Gilmore; Parul Sharma; Rebekah Penrice-Randal; Richard Eccles; Richard Gregory; Sam Haldenby; Steve Paterson; Stuart D Armstrong; Trevor Ian Robinson; Ximeng Han |

EPI_ISL_1387084, EPI_ISL_1387085, EPI_ISL_1475210, EPI_ISL_1475211, EPI_ISL_1475212, EPI_ISL_1475213, EPI_ISL_1475214, EPI_ISL_1475215

| | | | |
|---|---|---|---|
| see above | Northumbria University / South Tees Hospitals NHS Foundation Trust / North Cumbria Integrated Care NHS Foundation Trust / North Tees and Hartlepool NHS Foundation Trust / Newcastle Hospitals NHS Foundation Trust | COVID-19 Genomics UK (COG-UK) Consortium | Andrew Nelson; Brendan Payne; Clive Graham; Darren L Smith; Debra Padgett; Edward Barton; Emma Swindells; Garren Scott; Gary Black; Gary Eltringham; Giles S Holt; Greg R Young; Jane Greenaway; Jennifer Collins; John Allan; Joshua Loh; Lynn Dover; Matthew Bashton; Mohammad A Tariq; Paul Baker; Sarah Essex; Steve Liggett; Wen C Yew; Yusri Taha |

EPI_ISL_1475762, EPI_ISL_1475780, EPI_ISL_1475793, EPI_ISL_1475796, EPI_ISL_1475800, EPI_ISL_1475802, EPI_ISL_1475804, EPI_ISL_1475805, EPI_ISL_1475808, EPI_ISL_1475809, EPI_ISL_1475811, EPI_ISL_1475812, EPI_ISL_1475813, EPI_ISL_1475814, EPI_ISL_1475815, EPI_ISL_1475816, EPI_ISL_1475817, EPI_ISL_1475819, EPI_ISL_1475821, EPI_ISL_1475822, EPI_ISL_1475828, EPI_ISL_1475832,
EPI_ISL_1475843, EPI_ISL_1475973, EPI_ISL_1475979, EPI_ISL_1475980, EPI_ISL_1475982, EPI_ISL_1475985, EPI_ISL_1475986, EPI_ISL_1476160, EPI_ISL_1476165, EPI_ISL_1476166, EPI_ISL_1476171, EPI_ISL_1476206, EPI_ISL_1476208, EPI_ISL_1476215, EPI_ISL_1476219, EPI_ISL_1476222, EPI_ISL_1476224, EPI_ISL_1476225, EPI_ISL_1476231, EPI_ISL_1476232, EPI_ISL_1476233, EPI_ISL_1476280,
EPI_ISL_1476281, EPI_ISL_1476282, EPI_ISL_1476286, EPI_ISL_1476288, EPI_ISL_1476293, EPI_ISL_1546282, EPI_ISL_1546290, EPI_ISL_1546293, EPI_ISL_1546299, EPI_ISL_1546326, EPI_ISL_1546327, EPI_ISL_1546331, EPI_ISL_1546332, EPI_ISL_1546333, EPI_ISL_1546341, EPI_ISL_1546345, EPI_ISL_1546348, EPI_ISL_1546350, EPI_ISL_1546351, EPI_ISL_1546356, EPI_ISL_1546373, EPI_ISL_1546377,
EPI_ISL_1546408, EPI_ISL_1546409, EPI_ISL_1546496, EPI_ISL_1546502, EPI_ISL_1546511, EPI_ISL_1546512, EPI_ISL_1546515, EPI_ISL_1546577, EPI_ISL_1546583, EPI_ISL_1546641, EPI_ISL_1546648, EPI_ISL_1546649, EPI_ISL_1546663, EPI_ISL_1546664, EPI_ISL_1546665, EPI_ISL_1546666, EPI_ISL_1546668, EPI_ISL_1546670, EPI_ISL_1546671, EPI_ISL_1546672, EPI_ISL_1546675, EPI_ISL_1546676,
EPI_ISL_1546677, EPI_ISL_1546681, EPI_ISL_1546686, EPI_ISL_1546687, EPI_ISL_1546689, EPI_ISL_1546690, EPI_ISL_1546692, EPI_ISL_1546693, EPI_ISL_1546695, EPI_ISL_1546696, EPI_ISL_1546697, EPI_ISL_1546698, EPI_ISL_1546700, EPI_ISL_1546702, EPI_ISL_1546703, EPI_ISL_1546704, EPI_ISL_1546706, EPI_ISL_1546709, EPI_ISL_1546712, EPI_ISL_1546713, EPI_ISL_1546715, EPI_ISL_1546716,
EPI_ISL_1546717, EPI_ISL_1546718, EPI_ISL_1546719, EPI_ISL_1546721, EPI_ISL_1546723, EPI_ISL_1546724, EPI_ISL_1546725, EPI_ISL_1546727, EPI_ISL_1546730, EPI_ISL_1546731, EPI_ISL_1546732, EPI_ISL_1546733, EPI_ISL_1546734, EPI_ISL_1546735, EPI_ISL_1546737, EPI_ISL_1546738, EPI_ISL_1546739, EPI_ISL_1546742, EPI_ISL_1546744, EPI_ISL_1546748, EPI_ISL_1546751, EPI_ISL_1546753,
EPI_ISL_1546754, EPI_ISL_1546755, EPI_ISL_1546763, EPI_ISL_1546768, EPI_ISL_1546769, EPI_ISL_1546771, EPI_ISL_1546772, EPI_ISL_1546773, EPI_ISL_1546776, EPI_ISL_1546782, EPI_ISL_1546785, EPI_ISL_1546789, EPI_ISL_1546790, EPI_ISL_1546846, EPI_ISL_1546847, EPI_ISL_1546852, EPI_ISL_1546853, EPI_ISL_1546854, EPI_ISL_1546855, EPI_ISL_1546857, EPI_ISL_1546858, EPI_ISL_1546859,
EPI_ISL_1546860, EPI_ISL_1546861, EPI_ISL_1546864, EPI_ISL_1546865, EPI_ISL_1546866, EPI_ISL_1546867, EPI_ISL_1546868, EPI_ISL_1546869, EPI_ISL_1546870, EPI_ISL_1546871, EPI_ISL_1546872, EPI_ISL_1546873, EPI_ISL_1546874, EPI_ISL_1546875, EPI_ISL_1546876, EPI_ISL_1546877, EPI_ISL_1546878, EPI_ISL_1546879, EPI_ISL_1546880, EPI_ISL_1546882, EPI_ISL_1546883, EPI_ISL_1546884,

| | | | |
|---|---|---|---|
| EPI_ISL_1546885, EPI_ISL_1546922, EPI_ISL_1546947, EPI_ISL_1546948, EPI_ISL_1546949, EPI_ISL_1546950, EPI_ISL_1546954, EPI_ISL_1546956, EPI_ISL_1546957, EPI_ISL_1546958, EPI_ISL_1546959, EPI_ISL_1546962, EPI_ISL_1546964, EPI_ISL_1546967, EPI_ISL_1546969, EPI_ISL_1546970, EPI_ISL_1546972, EPI_ISL_1546973, EPI_ISL_1546974, EPI_ISL_1546975, EPI_ISL_1546976, EPI_ISL_1546977, EPI_ISL_1546978, EPI_ISL_1546979, EPI_ISL_1546980, EPI_ISL_1546981, EPI_ISL_1546986, EPI_ISL_1546991, EPI_ISL_1546992, EPI_ISL_1546993, EPI_ISL_1546995, EPI_ISL_1546996, EPI_ISL_1546998, EPI_ISL_1547034, EPI_ISL_1547058, EPI_ISL_1547063, EPI_ISL_1547064, EPI_ISL_1547066, EPI_ISL_1547067, EPI_ISL_1547069, EPI_ISL_1547070, EPI_ISL_1547071, EPI_ISL_1547072, EPI_ISL_1547074, EPI_ISL_1547075, EPI_ISL_1547076, EPI_ISL_1547077, EPI_ISL_1547078, EPI_ISL_1547079, EPI_ISL_1547080, EPI_ISL_1547081, EPI_ISL_1547082, EPI_ISL_1547083, EPI_ISL_1547084, EPI_ISL_1547085, EPI_ISL_1547086, EPI_ISL_1547087, EPI_ISL_1547088, EPI_ISL_1547089, EPI_ISL_1547090, EPI_ISL_1547091, EPI_ISL_1547092, EPI_ISL_1547093, EPI_ISL_1547094, EPI_ISL_1547095, EPI_ISL_1547096, EPI_ISL_1547097, EPI_ISL_1547098, EPI_ISL_1547099, EPI_ISL_1547100, EPI_ISL_1547101, EPI_ISL_1547102, EPI_ISL_1547103, EPI_ISL_1547104, EPI_ISL_1547105, EPI_ISL_1547107, EPI_ISL_1547108, EPI_ISL_1547111, EPI_ISL_1547112, EPI_ISL_1547113, EPI_ISL_1547115, EPI_ISL_1547118, EPI_ISL_1547119, EPI_ISL_1547121, EPI_ISL_1547122, EPI_ISL_1547123, EPI_ISL_1547124, EPI_ISL_1547126, EPI_ISL_1547127, EPI_ISL_1547128, EPI_ISL_1547129, EPI_ISL_1547130, EPI_ISL_1547134, EPI_ISL_1547135, EPI_ISL_1547136, EPI_ISL_1547137, EPI_ISL_1547138, EPI_ISL_1547141, EPI_ISL_1547143, EPI_ISL_1547144, EPI_ISL_1547145, EPI_ISL_1547146, EPI_ISL_1547147, EPI_ISL_1547148, EPI_ISL_1547149, EPI_ISL_1547151, EPI_ISL_1547152, EPI_ISL_1547153, EPI_ISL_1547157, EPI_ISL_1547158, EPI_ISL_1547159, EPI_ISL_1547160, EPI_ISL_1547162, EPI_ISL_1547163, EPI_ISL_1547164, EPI_ISL_1547166, EPI_ISL_1547167, EPI_ISL_1547171, EPI_ISL_1547172, EPI_ISL_1547173, EPI_ISL_1547175, EPI_ISL_1547179, EPI_ISL_1547180, EPI_ISL_1547182, EPI_ISL_1547185, EPI_ISL_1547186, EPI_ISL_1547187, EPI_ISL_1547188, EPI_ISL_1547189, EPI_ISL_1547190, EPI_ISL_1547191, EPI_ISL_1547192, EPI_ISL_1547193, EPI_ISL_1547194, EPI_ISL_1547195, EPI_ISL_1547196, EPI_ISL_1547197, EPI_ISL_1547198, EPI_ISL_1547199, EPI_ISL_1547200, EPI_ISL_1547201, EPI_ISL_1547202, EPI_ISL_1547203, EPI_ISL_1547204, EPI_ISL_1547205, EPI_ISL_1547206, EPI_ISL_1547207, EPI_ISL_1547208, EPI_ISL_1547209, EPI_ISL_1547210, EPI_ISL_1547211, EPI_ISL_1547212, EPI_ISL_1547213, EPI_ISL_1547214, EPI_ISL_1547215, EPI_ISL_1547216, EPI_ISL_1547217, EPI_ISL_1547218, EPI_ISL_1547219, EPI_ISL_1547220, EPI_ISL_1547222, EPI_ISL_1547223, EPI_ISL_1547224, EPI_ISL_1547225 | | | |
| see above | Originating lab: Wales Specialist Virology Centre Sequencing lab: Pathogen Genomics Unit | Public Health Wales Microbiology Cardiff Wales Specialist Virology Centre | Alec Birchley; Alexander Adams; Amy Gaskin; Angela Marchbank; Bree Gatica-Wilcox; Catherine Moore; Jason Coombes; Joanne Watkins; Joel Southgate; Johnathan Evans; Laura Gifford; Lauren Gilbert; Lee Graham; Malorie Perry; Matthew Bull; Nicole Pacchiarini; Sally Corden; Sara Kumziene-Summerhayes; Sara Rey; Sarah Taylor; Simon Cottrell; Sophie Jones; Tom Connor |
| EPI_ISL_1730450, EPI_ISL_1730472, EPI_ISL_1730473, EPI_ISL_1730474, EPI_ISL_1730475, EPI_ISL_1730476, EPI_ISL_1730477, EPI_ISL_1730478, EPI_ISL_1730479, EPI_ISL_1730480, EPI_ISL_1730481, EPI_ISL_1730482, EPI_ISL_1730483, EPI_ISL_1730484, EPI_ISL_1730485, EPI_ISL_1730486, EPI_ISL_1730487, EPI_ISL_1730488, EPI_ISL_1730489, EPI_ISL_1730490, EPI_ISL_1730491, EPI_ISL_1730492, EPI_ISL_1730494, EPI_ISL_1730495, EPI_ISL_1730496, EPI_ISL_1730497, EPI_ISL_1730498, EPI_ISL_1730513, EPI_ISL_1730520, EPI_ISL_1833070 | | | |
| see above | Oxford Viromics, NDM, University of Oxford; Oxford University Hospitals; Basingstoke and North Hampshire Hospital | COVID-19 Genomics UK (COG-UK) Consortium | Alex Mobbs; Amy Trebes; Anita Justice; Catrin Moore; Christophe Fraser; David Bonsall; David Buck; Emma Wise; George Macintyre; Jessica Lynch; John Todd; Mariateresa de Cesare; Matilde Mori; Monique Andersson; Nathan Moore; Nick Cortes; Robert Shaw; Stephen Kidd; Tanya Golubchik; Timothy Peto |
| EPI_ISL_1832728, EPI_ISL_1832730, EPI_ISL_1832732, EPI_ISL_1832734, EPI_ISL_1832736, EPI_ISL_1832748 | Quadram Institute Bioscience | COVID-19 Genomics UK (COG-UK) Consortium | Alexander J Trotter; Alison E. Mather; Alp Aydin; Ana P. Tedim; Anastasia Kolyva; Andrew Bell; Andrew J. Page; Claire Stuart; Dave J. Baker; Gemma L. Kay; John Wain; Justin O'Grady; Leonardo de Oliveira Martins; Lizzie Meadows; Maria Diaz; Mark Webber; Muhammed Yasir; Nabil-Fareed Alikhan; Ngozi Elumogo; Nicholas M. Thomson; Rachael Stanley; Rachel Gilroy; Reenesh Prakash; Samir Dervisevic; Samuel Bloomfield; Steven Rudder; Thanh Le-Viet |
| EPI_ISL_1387194, EPI_ISL_1475486, EPI_ISL_1475487, EPI_ISL_1475488, EPI_ISL_1475493, EPI_ISL_1475494, EPI_ISL_1546243, EPI_ISL_1546244, EPI_ISL_1546245, EPI_ISL_1546246, EPI_ISL_1546247, EPI_ISL_1546248, EPI_ISL_1978954 | | | |
| see above | Queens Medical Centre, Clinical Microbiology Department / DeepSeq Nottingham | COVID-19 Genomics UK (COG-UK) Consortium | Christopher Moore; Emily Park; Fei Sang; Gemma Clark; Hannah Howson-Wells; Jack Hill; Jayasree Dey; Johnny Debebe; Jonathan Ball; Joseph Chappell; Manjinder Khakh; Matthew Carlisle; Matthew Loose; Michelle M Lister; Nadine Holmes; Patrick McClure; Theocharis Tsoleridis; Timothy Byaruhanga; Vicki M Fleming; Victoria Wright; Wendy Smith |
| EPI_ISL_1451512, EPI_ISL_1451627, EPI_ISL_1466934, EPI_ISL_1466946, EPI_ISL_1466949, EPI_ISL_1466951, EPI_ISL_1466955, EPI_ISL_1466956, EPI_ISL_1466957, EPI_ISL_1466958, EPI_ISL_1466962, EPI_ISL_1466975, EPI_ISL_1466983, EPI_ISL_1466990, EPI_ISL_1466991, EPI_ISL_1466992, EPI_ISL_1466993, EPI_ISL_1466997, EPI_ISL_1467000, EPI_ISL_1467009, EPI_ISL_1467017, EPI_ISL_1467018, EPI_ISL_1467019, EPI_ISL_1467026, EPI_ISL_1467029, EPI_ISL_1467034, EPI_ISL_1467041, EPI_ISL_1467044, EPI_ISL_1467046, EPI_ISL_1467049, EPI_ISL_1467051, EPI_ISL_1467055, EPI_ISL_1467059, EPI_ISL_1467067, EPI_ISL_1467068, EPI_ISL_1467075, EPI_ISL_1467077, EPI_ISL_1467080, EPI_ISL_1467088, EPI_ISL_1467098, EPI_ISL_1467113, EPI_ISL_1467115, EPI_ISL_1467122, EPI_ISL_1467128, EPI_ISL_1467131, EPI_ISL_1467134, EPI_ISL_1467138, EPI_ISL_1467139, EPI_ISL_1467140, EPI_ISL_1467147, EPI_ISL_1467167, EPI_ISL_1467170, EPI_ISL_1467171, EPI_ISL_1467180, EPI_ISL_1467182, EPI_ISL_1467187, EPI_ISL_1467188, EPI_ISL_1467190, EPI_ISL_1467208, EPI_ISL_1467229, EPI_ISL_1467230, EPI_ISL_1467231, EPI_ISL_1467233, EPI_ISL_1467236, EPI_ISL_1467239, EPI_ISL_1467253, EPI_ISL_1467258, EPI_ISL_1467260, EPI_ISL_1467261, EPI_ISL_1467263, EPI_ISL_1467264, EPI_ISL_1467267, EPI_ISL_1467269, EPI_ISL_1467275, EPI_ISL_1467276, EPI_ISL_1467282, EPI_ISL_1467283, EPI_ISL_1467287, EPI_ISL_1467288, EPI_ISL_1467290, EPI_ISL_1467292, EPI_ISL_1467295, EPI_ISL_1467296, EPI_ISL_1467300, EPI_ISL_1467303, EPI_ISL_1467304, EPI_ISL_1467305, EPI_ISL_1467311, EPI_ISL_1467313, EPI_ISL_1467317, EPI_ISL_1467319, EPI_ISL_1467321, EPI_ISL_1467325, EPI_ISL_1467326, EPI_ISL_1467327, EPI_ISL_1467330, EPI_ISL_1467335, EPI_ISL_1467336, EPI_ISL_1467337, EPI_ISL_1467340, EPI_ISL_1467341, EPI_ISL_1467342, EPI_ISL_1467343, EPI_ISL_1467348, EPI_ISL_1467349, EPI_ISL_1467351, EPI_ISL_1467352, EPI_ISL_1467354, EPI_ISL_1467356, EPI_ISL_1467358, EPI_ISL_1467362, EPI_ISL_1467363, EPI_ISL_1467364, EPI_ISL_1467365, EPI_ISL_1467367, EPI_ISL_1467370, EPI_ISL_1467371, EPI_ISL_1467372, EPI_ISL_1467377, EPI_ISL_1467385, EPI_ISL_1467387, EPI_ISL_1467389, EPI_ISL_1467390, EPI_ISL_1467392, EPI_ISL_1467395, EPI_ISL_1467399, EPI_ISL_1467402, EPI_ISL_1467403, EPI_ISL_1467405, EPI_ISL_1467407, EPI_ISL_1467410, EPI_ISL_1467411, EPI_ISL_1467413, EPI_ISL_1467414, EPI_ISL_1467424, EPI_ISL_1467426, EPI_ISL_1467427, EPI_ISL_1467433, EPI_ISL_1467435, EPI_ISL_1467440, EPI_ISL_1467442, EPI_ISL_1467443, EPI_ISL_1467447, EPI_ISL_1467449, EPI_ISL_1467450, EPI_ISL_1467454, EPI_ISL_1467455, EPI_ISL_1467456, EPI_ISL_1467458, EPI_ISL_1467459, EPI_ISL_1467461, EPI_ISL_1467462, EPI_ISL_1467466, EPI_ISL_1467473, EPI_ISL_1467476, EPI_ISL_1467478, EPI_ISL_1467480, EPI_ISL_1467482, EPI_ISL_1467485, EPI_ISL_1467488, EPI_ISL_1467489, EPI_ISL_1467490, EPI_ISL_1467494, EPI_ISL_1467498, EPI_ISL_1467500, EPI_ISL_1467501, EPI_ISL_1467502, EPI_ISL_1467505, EPI_ISL_1467507, EPI_ISL_1467508, EPI_ISL_1467512, EPI_ISL_1467514, EPI_ISL_1467519, EPI_ISL_1467521, EPI_ISL_1467526, EPI_ISL_1467527, EPI_ISL_1467530, EPI_ISL_1467532, EPI_ISL_1467542, EPI_ISL_1467543, EPI_ISL_1467547, EPI_ISL_1467548, EPI_ISL_1467551, EPI_ISL_1467555, EPI_ISL_1467556, EPI_ISL_1467557, EPI_ISL_1467558, EPI_ISL_1467560, EPI_ISL_1467561, EPI_ISL_1467566, EPI_ISL_1467569, EPI_ISL_1467570, EPI_ISL_1467571, EPI_ISL_1467572, EPI_ISL_1467575, EPI_ISL_1467580, EPI_ISL_1472701, EPI_ISL_1472702, EPI_ISL_1472705, EPI_ISL_1472711, EPI_ISL_1472716, EPI_ISL_1472717, EPI_ISL_1472719, EPI_ISL_1472722, EPI_ISL_1472729, EPI_ISL_1472731, EPI_ISL_1472734, EPI_ISL_1472741, EPI_ISL_1472747, EPI_ISL_1472753, EPI_ISL_1472754, EPI_ISL_1472757, EPI_ISL_1472764, EPI_ISL_1472765, EPI_ISL_1472767, EPI_ISL_1472772, EPI_ISL_1472782, EPI_ISL_1472785, EPI_ISL_1472789, EPI_ISL_1472794, EPI_ISL_1472800, EPI_ISL_1472801, EPI_ISL_1472802, EPI_ISL_1472807, EPI_ISL_1472814, EPI_ISL_1472815, EPI_ISL_1472824, EPI_ISL_1472825, EPI_ISL_1472827, EPI_ISL_1472832, EPI_ISL_1472837, EPI_ISL_1472839, EPI_ISL_1472841, EPI_ISL_1472843, EPI_ISL_1472851, EPI_ISL_1472854, EPI_ISL_1472860, EPI_ISL_1472863, EPI_ISL_1472866, EPI_ISL_1472881, EPI_ISL_1472885, EPI_ISL_1472893, EPI_ISL_1472900, EPI_ISL_1472901, EPI_ISL_1472907, EPI_ISL_1472910, EPI_ISL_1472920, EPI_ISL_1472927, EPI_ISL_1472928, EPI_ISL_1472934, EPI_ISL_1472935, EPI_ISL_1472937, EPI_ISL_1472940, EPI_ISL_1472941, EPI_ISL_1483883, EPI_ISL_1485482, EPI_ISL_1485536, EPI_ISL_1485562, EPI_ISL_1485609, EPI_ISL_1485614, EPI_ISL_1485623, EPI_ISL_1485628, EPI_ISL_1485646, EPI_ISL_1485679, EPI_ISL_1485711, EPI_ISL_1485731, EPI_ISL_1486399, EPI_ISL_1486429, EPI_ISL_1486437, EPI_ISL_1486438, EPI_ISL_1486445, EPI_ISL_1486499, EPI_ISL_1486545, EPI_ISL_1486548, EPI_ISL_1486573, EPI_ISL_1486642, EPI_ISL_1486660, EPI_ISL_1486680, EPI_ISL_1486707, EPI_ISL_1486732, EPI_ISL_1486744, EPI_ISL_1486781, EPI_ISL_1486788, EPI_ISL_1486796, EPI_ISL_1486805, EPI_ISL_1487881, EPI_ISL_1487899, EPI_ISL_1487901, EPI_ISL_1487903, EPI_ISL_1487905, EPI_ISL_1487921, EPI_ISL_1487961, EPI_ISL_1487967, EPI_ISL_1487970, EPI_ISL_1487997, EPI_ISL_1487999, EPI_ISL_1488056, EPI_ISL_1488128, EPI_ISL_1488132, EPI_ISL_1488146, EPI_ISL_1488153, EPI_ISL_1488165, EPI_ISL_1488168, EPI_ISL_1488202, EPI_ISL_1488223, EPI_ISL_1488226, EPI_ISL_1488579, EPI_ISL_1488600, EPI_ISL_1488651, EPI_ISL_1488680, EPI_ISL_1488733, EPI_ISL_1488763, EPI_ISL_1488798, EPI_ISL_1488813, EPI_ISL_1488814, EPI_ISL_1488830, EPI_ISL_1489161, EPI_ISL_1489165, EPI_ISL_1489171, EPI_ISL_1489175, EPI_ISL_1489183, EPI_ISL_1489185, EPI_ISL_1489187, EPI_ISL_1489189, EPI_ISL_1489192, EPI_ISL_1489200, EPI_ISL_1489205, EPI_ISL_1489206, EPI_ISL_1489207, EPI_ISL_1489208, EPI_ISL_1489212, EPI_ISL_1489214, EPI_ISL_1489217, EPI_ISL_1489219, EPI_ISL_1489221, EPI_ISL_1489222, EPI_ISL_1489223, EPI_ISL_1489225, EPI_ISL_1489226, EPI_ISL_1489228, EPI_ISL_1489236, EPI_ISL_1489239, EPI_ISL_1489242, EPI_ISL_1489243, EPI_ISL_1489244, EPI_ISL_1489245, EPI_ISL_1489253, EPI_ISL_1489261, EPI_ISL_1489262, EPI_ISL_1489263, EPI_ISL_1489267, EPI_ISL_1489274, EPI_ISL_1489278, EPI_ISL_1489280, EPI_ISL_1489284, EPI_ISL_1489285, EPI_ISL_1489293, EPI_ISL_1489296, EPI_ISL_1489304, EPI_ISL_1489305, EPI_ISL_1489317, EPI_ISL_1489318, EPI_ISL_1489319, EPI_ISL_1489321, EPI_ISL_1489332, EPI_ISL_1489336, EPI_ISL_1489350, EPI_ISL_1489352, EPI_ISL_1489353, EPI_ISL_1489358, EPI_ISL_1489362, EPI_ISL_1489368, EPI_ISL_1489375, EPI_ISL_1489376, EPI_ISL_1489389, EPI_ISL_1489393, EPI_ISL_1489395, EPI_ISL_1489396, EPI_ISL_1489400, EPI_ISL_1489416, EPI_ISL_1489420, EPI_ISL_1489423, EPI_ISL_1489424, EPI_ISL_1489426, EPI_ISL_1489437, EPI_ISL_1489438, EPI_ISL_1489441, EPI_ISL_1489445, EPI_ISL_1489446, EPI_ISL_1489448, EPI_ISL_1489450, EPI_ISL_1489452, EPI_ISL_1489454, EPI_ISL_1489494, EPI_ISL_1506070, EPI_ISL_1519932, EPI_ISL_1519933, EPI_ISL_1519934, EPI_ISL_1519996, EPI_ISL_1519997, EPI_ISL_1519998, EPI_ISL_1519999, EPI_ISL_1520000, EPI_ISL_1520001, EPI_ISL_1520002, EPI_ISL_1520003, EPI_ISL_1520004, EPI_ISL_1520005, EPI_ISL_1520006, EPI_ISL_1520007, EPI_ISL_1520008, EPI_ISL_1520009, EPI_ISL_1520010, EPI_ISL_1520011, EPI_ISL_1520012, EPI_ISL_1520013, EPI_ISL_1520014, EPI_ISL_1520015, EPI_ISL_1520016, EPI_ISL_1520017, EPI_ISL_1520018, EPI_ISL_1520020 | | | |
| see above | Randox Laboratories | Wellcome Sanger Institute for the COVID-19 Genomics UK (COG-UK) Consortium | Cordelia Langford; David K. Jackson; Dominic Kwiatkowski; Ewan Harrison; Ian Johnston; Jeffrey Barrett; John Sillitoe on behalf of the Wellcome Sanger Institute COVID-19 Surveillance Team; Randox Laboratories and Alex Alderton; Roberto Amato; Sonia Goncalves |
| EPI_ISL_1976658, EPI_ISL_1976660, EPI_ISL_1976667, EPI_ISL_1976671, EPI_ISL_1976673, EPI_ISL_1976674, EPI_ISL_1976675, EPI_ISL_1976677, EPI_ISL_1976681, EPI_ISL_1976683, EPI_ISL_1976684, EPI_ISL_1976687, EPI_ISL_1976691, EPI_ISL_1976692, EPI_ISL_1976694, EPI_ISL_1976697, EPI_ISL_1976699, EPI_ISL_1976701, EPI_ISL_1976704, EPI_ISL_1976706, EPI_ISL_1976707, EPI_ISL_1976708, EPI_ISL_1976711, EPI_ISL_1976712, EPI_ISL_1976714, EPI_ISL_1976715, EPI_ISL_1976717, EPI_ISL_1976718, EPI_ISL_1976720, EPI_ISL_1976721, EPI_ISL_1976725, EPI_ISL_1976730, EPI_ISL_1976732, EPI_ISL_1976734, EPI_ISL_1976735, EPI_ISL_1976737, EPI_ISL_1976739, EPI_ISL_1976741, EPI_ISL_1976743, EPI_ISL_1976744, EPI_ISL_1976746, EPI_ISL_1976748, EPI_ISL_1976749, EPI_ISL_1976751, EPI_ISL_1976758, EPI_ISL_1976760, EPI_ISL_1976762, EPI_ISL_1976763, EPI_ISL_1976767, EPI_ISL_1976774, EPI_ISL_1976776, EPI_ISL_1976779, EPI_ISL_1976781, EPI_ISL_1976784, EPI_ISL_1976786, EPI_ISL_1976803, EPI_ISL_1976805, EPI_ISL_1976807, EPI_ISL_1976809, EPI_ISL_1976814, EPI_ISL_1976816, EPI_ISL_1976818, EPI_ISL_1976819, EPI_ISL_1976821, EPI_ISL_1976823, EPI_ISL_1976826, EPI_ISL_1976828, EPI_ISL_1976830, EPI_ISL_1976832, EPI_ISL_1976833, EPI_ISL_1976848, EPI_ISL_1976850, EPI_ISL_1976851, EPI_ISL_1976853, EPI_ISL_1976855, EPI_ISL_1976858, EPI_ISL_1976860, EPI_ISL_1976863, EPI_ISL_1976864, EPI_ISL_1976865, EPI_ISL_1976866, EPI_ISL_1976867, EPI_ISL_1976873, EPI_ISL_1976874, EPI_ISL_1976876, EPI_ISL_1976877, EPI_ISL_1976878, EPI_ISL_1976880, EPI_ISL_1976883, EPI_ISL_1976884, EPI_ISL_1976886, EPI_ISL_1976894, EPI_ISL_1976896, EPI_ISL_1976897, EPI_ISL_1976899, EPI_ISL_1976901, EPI_ISL_1976905, EPI_ISL_1976908, EPI_ISL_1976909, EPI_ISL_1976910, EPI_ISL_1976912, EPI_ISL_1976913, EPI_ISL_1976915, EPI_ISL_1976916, EPI_ISL_1976918, EPI_ISL_1976920, EPI_ISL_1976921, EPI_ISL_1976923, EPI_ISL_1976925, EPI_ISL_1976926, EPI_ISL_1976928, EPI_ISL_1976931, EPI_ISL_1976933, EPI_ISL_1976935, EPI_ISL_1976936, EPI_ISL_1976938, EPI_ISL_1976940, EPI_ISL_1976944, EPI_ISL_1976966, EPI_ISL_1976968, EPI_ISL_1976969, EPI_ISL_1976971, EPI_ISL_1976972, EPI_ISL_1976976, EPI_ISL_1976979, EPI_ISL_1976981, EPI_ISL_1976983, EPI_ISL_1976985, EPI_ISL_1976986, EPI_ISL_1976988, EPI_ISL_1976990, EPI_ISL_1976993, EPI_ISL_1976995, EPI_ISL_1976997, EPI_ISL_1976998, EPI_ISL_1977000, EPI_ISL_1977002, EPI_ISL_1977003, EPI_ISL_1977007, EPI_ISL_1977009, EPI_ISL_1977010, EPI_ISL_1977014, EPI_ISL_1977016, EPI_ISL_1977019, EPI_ISL_1977021, EPI_ISL_1977023, EPI_ISL_1977024, EPI_ISL_1977026, EPI_ISL_1977030, EPI_ISL_1977032, EPI_ISL_1977033, EPI_ISL_1977035, EPI_ISL_1977037, EPI_ISL_1977052, EPI_ISL_1977054, EPI_ISL_1977055, EPI_ISL_1977066, EPI_ISL_1977084, EPI_ISL_1977086, EPI_ISL_1977088, EPI_ISL_1977089, EPI_ISL_1977091, EPI_ISL_1977093, EPI_ISL_1977096, EPI_ISL_1977098, EPI_ISL_1977099, EPI_ISL_1977118, EPI_ISL_1977120, EPI_ISL_1977123, EPI_ISL_1977125, EPI_ISL_1977127, EPI_ISL_1977128, EPI_ISL_1977130, EPI_ISL_1977132, EPI_ISL_1977133, EPI_ISL_1977135, EPI_ISL_1977137, EPI_ISL_1977138, EPI_ISL_1977150, EPI_ISL_1977151, EPI_ISL_1977153, EPI_ISL_1977154, EPI_ISL_1977156, EPI_ISL_1977165, EPI_ISL_1977167, EPI_ISL_1977169, EPI_ISL_1977170, EPI_ISL_1977185, EPI_ISL_1977187, EPI_ISL_1977189, EPI_ISL_1977201, EPI_ISL_1977203, EPI_ISL_1977205, EPI_ISL_1977206, EPI_ISL_1977208, EPI_ISL_1977209, EPI_ISL_1978149, EPI_ISL_1978151, EPI_ISL_1978152, EPI_ISL_1978154, EPI_ISL_1978155, EPI_ISL_1978157, EPI_ISL_1978159, EPI_ISL_1978160, EPI_ISL_1978162, EPI_ISL_1978163, EPI_ISL_1978165, EPI_ISL_1978167, EPI_ISL_1978168, EPI_ISL_1978170, EPI_ISL_1978172, EPI_ISL_1978173, EPI_ISL_1978175, EPI_ISL_1978177, EPI_ISL_1978179, EPI_ISL_1978181, EPI_ISL_1978183, EPI_ISL_1978184, EPI_ISL_1978186, EPI_ISL_1978188, EPI_ISL_1978190, EPI_ISL_1978192, EPI_ISL_1978193, EPI_ISL_1978195, EPI_ISL_1978197, EPI_ISL_1978198, EPI_ISL_1978200, EPI_ISL_1978201, EPI_ISL_1978203, EPI_ISL_1978205, EPI_ISL_1978206, EPI_ISL_1978207, EPI_ISL_1978208, EPI_ISL_1978209, EPI_ISL_1978211, EPI_ISL_1978212, EPI_ISL_1978214, EPI_ISL_1978215, EPI_ISL_1978217, EPI_ISL_1978218, EPI_ISL_1978220, EPI_ISL_1978222, EPI_ISL_1978224, EPI_ISL_1978225, EPI_ISL_1978227, EPI_ISL_1978229, EPI_ISL_1978230, EPI_ISL_1978232, EPI_ISL_1978237, EPI_ISL_1978239, EPI_ISL_1978240, EPI_ISL_1978241, EPI_ISL_1978242, EPI_ISL_1978243, EPI_ISL_1978245, EPI_ISL_1978247, EPI_ISL_1978249, EPI_ISL_1978251, EPI_ISL_1978254, EPI_ISL_1978256 | | | |
| see above | Regional Virus Laboratory, Belfast Health and Social Care Trust | COVID-19 Genomics UK (COG-UK) Consortium | Alison Watt; Ciara Cox; Conall McCaughey; David Simpson; Derek Fairley; James McKenna; Mairead Connor; Susan Feeney; Tanya Curran; Zoltan Molnar |
| EPI_ISL_1833818 | St George's University Hospitals NHS Foundation Trust | COVID-19 Genomics UK (COG-UK) Consortium | Adam Witney; Cassie Pope; Irene Monahan; Joshua Taylor; Ken Laing; NgeeKeong Tan |
| EPI_ISL_1386822, EPI_ISL_1386823, EPI_ISL_1386826, EPI_ISL_1386830, EPI_ISL_1474834, EPI_ISL_1474844, EPI_ISL_1474852, EPI_ISL_1474854, EPI_ISL_1474860, EPI_ISL_1474861, EPI_ISL_1474870, EPI_ISL_1474871, EPI_ISL_1474872, EPI_ISL_1474874, EPI_ISL_1474876, EPI_ISL_1474877, EPI_ISL_1474878, EPI_ISL_1474880, EPI_ISL_1474888, EPI_ISL_1637090, EPI_ISL_1637165, EPI_ISL_1637173, EPI_ISL_1637174, EPI_ISL_1637175, EPI_ISL_1730362 | | | |
| see above | University College London, Great Ormond Street Hospital for Children NHS Foundation Trust, Imperial College Healthcare NHS Trust | COVID-19 Genomics UK (COG-UK) Consortium | Alison Holmes; Charlotte Williams; Helena Tutill; Jacqueline Findlay; James Price; Judith Breuer; Julianne Brown; Kathryn Harris; Leysa Forrest; Mark Kristiansen; Paola Niola; Paola Resende Silva; Patricia Dyal; Paul Randell; Rachel Williams; Samuel Weeks; Sergi Castellano; Sunando Roy; Tony Brooks; Yasmin Panchbhaya |
| EPI_ISL_1545973, EPI_ISL_1545975, EPI_ISL_1545976, EPI_ISL_1545978, EPI_ISL_1545980, EPI_ISL_1545983 | University Hospitals Of Leicester NHS Trust and DeepSeq Nottingham | COVID-19 Genomics UK (COG-UK) Consortium | Christopher Holmes; Christopher Moore; Fei Sang; Joeseph Chappell; Johnny Debebe; Jonathan Ball; Julian Tang; Karlie Fallon; Matthew Carlisle; Matthew Loose; Nadine Holmes; Patrick McClure; Paul Bird; Thomas Helmer; Victoria Wright |
| EPI_ISL_1386527, EPI_ISL_1386528, EPI_ISL_1386533, EPI_ISL_1474420, EPI_ISL_1474421, EPI_ISL_1474422, EPI_ISL_1474423, EPI_ISL_1474424, EPI_ISL_1474425, EPI_ISL_1474426, EPI_ISL_1474427, EPI_ISL_1474428, EPI_ISL_1474429, EPI_ISL_1474430, EPI_ISL_1474432, EPI_ISL_1474436, EPI_ISL_1474437, EPI_ISL_1474438, EPI_ISL_1474464, EPI_ISL_1474494, EPI_ISL_1474527, EPI_ISL_1474528, EPI_ISL_1474529, EPI_ISL_1636554, EPI_ISL_1636555, EPI_ISL_1636556, EPI_ISL_1636557, EPI_ISL_1636587, EPI_ISL_1636588, EPI_ISL_1636590, EPI_ISL_1729978 | | | |
| see above | University of Birmingham | COVID-19 Genomics UK (COG-UK) Consortium | Alex Richter; Andrew Bosworth; Andrew Bosworth. Queen Elizabeth Hospital: Anna Casey; Andrew D Beggs PHE Heartlands Lab: Husam Osman; Charlotte Poxon; Institute of Microbiology; Joanne Stockton; Josh Quick; Kasun Wanigasooriya; Mike Kidd; Nicholas Loman. University of Birmingham Testing Laboratory: Celina M Whalley; Oliver Pickles; Radoslaw Poplawski; Samuel Nicholls; University of Birmingham: Claire McMurray; Will Rowe |
| EPI_ISL_1545969 | University of Exeter | COVID-19 Genomics UK (COG-UK) Consortium | Aaron Jeffries; Audrey Farbos; Ben Temperton; Jane Masoli; Joanna Warwick-Dugdale; Michelle Michelsen; Robyn Manley; Stephen Michell |
| EPI_ISL_1474588, EPI_ISL_1474595, EPI_ISL_1474596, EPI_ISL_1474597, EPI_ISL_1474598, EPI_ISL_1474599, EPI_ISL_1474600, EPI_ISL_1474601, EPI_ISL_1545901, EPI_ISL_1545920, EPI_ISL_1636673, EPI_ISL_1636686, EPI_ISL_1636702, EPI_ISL_1730077, EPI_ISL_1730078, EPI_ISL_1730079, EPI_ISL_1730092, EPI_ISL_1730093, EPI_ISL_1730094, EPI_ISL_1730285, EPI_ISL_1974910, EPI_ISL_2125733, EPI_ISL_2125851, EPI_ISL_2125853, EPI_ISL_2125857, EPI_ISL_2125859, EPI_ISL_2125861 | | | |
| see above | Virology Department, Royal Infirmary of Edinburgh, NHS Lothian / School of Biological Sciences, University of Edinburgh | COVID-19 Genomics UK (COG-UK) Consortium | Colquhoun R; Cotton S; Dewar R; Gallagher A; Hill V; Jackson B; Maloney D; McCrone JT; McHugh M; O'Toole Á; Rambaut A; Rooke S; Scher E; Templeton K; Yu X |
| EPI_ISL_1476691, EPI_ISL_1476692, EPI_ISL_1476693, EPI_ISL_1476694, EPI_ISL_1476696, EPI_ISL_1476697, EPI_ISL_1476698, EPI_ISL_1476699, EPI_ISL_1476700, EPI_ISL_1476701, EPI_ISL_1476702, EPI_ISL_1476703, EPI_ISL_1476704, EPI_ISL_1476705, EPI_ISL_1476706, EPI_ISL_1476711, EPI_ISL_1476714, EPI_ISL_1476715, EPI_ISL_1476717, EPI_ISL_1476718, EPI_ISL_1476719, EPI_ISL_1476728, EPI_ISL_1476730, EPI_ISL_1476731, EPI_ISL_1476733, EPI_ISL_1547312, EPI_ISL_1547317, EPI_ISL_1547318, EPI_ISL_1547322, EPI_ISL_1547325, EPI_ISL_1547326, EPI_ISL_1547327, EPI_ISL_1547330, EPI_ISL_1547332, EPI_ISL_1547337, EPI_ISL_1547340, EPI_ISL_1547341, EPI_ISL_1547342, EPI_ISL_1547347, EPI_ISL_1547349, EPI_ISL_1547351, EPI_ISL_1547352, EPI_ISL_1547354, EPI_ISL_1547355, EPI_ISL_1547357, EPI_ISL_1547358, EPI_ISL_1638075, EPI_ISL_1730931, EPI_ISL_1730935, EPI_ISL_1730969, EPI_ISL_1730978, EPI_ISL_1730987, EPI_ISL_1731042, EPI_ISL_1833854, EPI_ISL_1833896, EPI_ISL_1833903 | | | |
| see above | Virology Department, Sheffield Teaching Hospitals NHS Foundation Trust/Department of Infection, Immunity and Cardiovascular Disease, The Medical School, University of Sheffield | COVID-19 Genomics UK (COG-UK) Consortium | Adri Angyal; Alex Keeley; Benjamin Lindsey; Cariad Evans; Danielle Groves; Dave Partridge; Katie Johnson; Laura Carrilero; Luke Green; Matthew Parker; Matthew Wyles; Mehmet Yavuz; Mohammad Raza; Nikki Smith; Paul Parsons; Rachel Tucker; Rebecca Brown; Thushan de Silva |
| EPI_ISL_1475775, EPI_ISL_1475777, EPI_ISL_1475778, EPI_ISL_1475797, EPI_ISL_1475803, EPI_ISL_1475806, EPI_ISL_1475807, EPI_ISL_1475820, EPI_ISL_1475826, EPI_ISL_1475829, EPI_ISL_1475831, EPI_ISL_1475834, EPI_ISL_1475835, EPI_ISL_1475839, EPI_ISL_1475842, EPI_ISL_1475845, EPI_ISL_1475974, EPI_ISL_1475975, EPI_ISL_1475976, EPI_ISL_1475981, EPI_ISL_1475984, EPI_ISL_1475992, EPI_ISL_1476164, EPI_ISL_1476169, EPI_ISL_1476170, EPI_ISL_1476172, EPI_ISL_1476175, EPI_ISL_1476205, EPI_ISL_1476211, EPI_ISL_1476212, EPI_ISL_1476218, EPI_ISL_1476226, EPI_ISL_1476291, EPI_ISL_1546519, EPI_ISL_1546542, EPI_ISL_1546667, EPI_ISL_1546673, EPI_ISL_1546694, EPI_ISL_1546729, EPI_ISL_1546756, EPI_ISL_1546757, EPI_ISL_1546759, EPI_ISL_1546774, EPI_ISL_1546792, EPI_ISL_1546845, EPI_ISL_1546862, EPI_ISL_1546863, EPI_ISL_1637308, EPI_ISL_1637309, EPI_ISL_1637310, EPI_ISL_1637311, EPI_ISL_1637312, EPI_ISL_1637313, EPI_ISL_1637314, EPI_ISL_1637315, EPI_ISL_1637316, EPI_ISL_1637317, EPI_ISL_1637318, EPI_ISL_1637319, EPI_ISL_1637320, EPI_ISL_1637321, EPI_ISL_1637323, EPI_ISL_1637365, EPI_ISL_1637366, EPI_ISL_1637367, EPI_ISL_1637400, EPI_ISL_1637402, EPI_ISL_1637403, EPI_ISL_1637405, EPI_ISL_1637410, EPI_ISL_1637411, EPI_ISL_1637412, EPI_ISL_1637413, EPI_ISL_1637414, EPI_ISL_1637417, EPI_ISL_1637419, EPI_ISL_1637420, EPI_ISL_1637421, EPI_ISL_1637422, EPI_ISL_1637426, EPI_ISL_1637428, EPI_ISL_1637430, EPI_ISL_1637431, EPI_ISL_1637445, EPI_ISL_1637448, EPI_ISL_1637449, EPI_ISL_1637452, EPI_ISL_1637453, EPI_ISL_1637550, EPI_ISL_1637584, EPI_ISL_1637587, EPI_ISL_1637593, EPI_ISL_1637594, EPI_ISL_1637598, EPI_ISL_1637660, EPI_ISL_1637667, EPI_ISL_1637770, EPI_ISL_1637771, EPI_ISL_1637772, EPI_ISL_1637774, EPI_ISL_1637775, EPI_ISL_1637777, EPI_ISL_1637778, EPI_ISL_1637780, EPI_ISL_1637781, EPI_ISL_1637782, EPI_ISL_1637786, EPI_ISL_1637787, EPI_ISL_1637791, EPI_ISL_1637793, EPI_ISL_1637794, EPI_ISL_1637797, EPI_ISL_1637801, EPI_ISL_1637803, EPI_ISL_1637804, EPI_ISL_1637805, EPI_ISL_1637806, EPI_ISL_1637807, EPI_ISL_1637808, EPI_ISL_1637809, EPI_ISL_1637810, EPI_ISL_1637811, EPI_ISL_1637812, EPI_ISL_1637813, EPI_ISL_1637814, EPI_ISL_1637815, EPI_ISL_1637816, EPI_ISL_1637817, EPI_ISL_1637818, EPI_ISL_1637819, EPI_ISL_1637820, EPI_ISL_1637821, EPI_ISL_1637822, EPI_ISL_1637823, EPI_ISL_1637824, EPI_ISL_1637826, EPI_ISL_1637827, EPI_ISL_1637828, EPI_ISL_1637829, EPI_ISL_1637830, EPI_ISL_1637831, EPI_ISL_1637832, EPI_ISL_1637833, EPI_ISL_1637834, EPI_ISL_1637836, EPI_ISL_1637837, EPI_ISL_1637838, EPI_ISL_1637840, EPI_ISL_1637841, EPI_ISL_1637842, EPI_ISL_1637843, EPI_ISL_1637844, EPI_ISL_1637845, EPI_ISL_1637846, EPI_ISL_1637848, EPI_ISL_1637849, EPI_ISL_1637850, EPI_ISL_1637851, EPI_ISL_1637853, EPI_ISL_1637854, EPI_ISL_1637855, EPI_ISL_1637856, EPI_ISL_1637858, EPI_ISL_1637859, EPI_ISL_1637860, EPI_ISL_1637861, EPI_ISL_1637862, EPI_ISL_1637864, EPI_ISL_1833158 | | | |
| see above | Wales Specialist Virology Centre Sequencing lab: Pathogen Genomics Unit | Public Health Wales Microbiology Cardiff Wales Specialist Virology Centre | Alec Birchley; Alexander Adams; Amy Gaskin; Angela Marchbank; Bree Gatica-Wilcox; Catherine Moore; Jason Coombes; Joanne Watkins; Joel Southgate; Johnathan Evans; Laura Gifford; Lauren Gilbert; Lee Graham; Malorie Perry; Matthew Bull; Nicole Pacchiarini; Sally Corden; Sara Kumziene-Summerhayes; Sara Rey; Sarah Taylor; Simon Cottrell; Sophie Jones; Tom Connor |
| EPI_ISL_1545869, EPI_ISL_1545870, EPI_ISL_1545898, EPI_ISL_1545899 | West of Scotland Specialist Virology Centre, NHSGGC / MRC-University of Glasgow Centre for Virus Research | COVID-19 Genomics UK (COG-UK) Consortium | Alasdair MacLean; Alice Broos; Ana da Silva Filipe; Antonia Ho; Daniel Mair; David L Robertson; Emma Thomson; Guy Mollett; James Shepherd; Jenna Nichols; Joseph Hughes; Kathy Li; Kathy Smollett; Kyriaki Nomikou; Lily Tong; Matthew Holden; Natasha Johnson; Rachel Blacow; Richard Orton; Rory Gunson; Sarah McDonald; Sharif Shaaban; Sreenu Vattipally; Stephen Carmichael |

We gratefully acknowledge the following Authors from the Originating laboratories responsible for obtaining the specimens, as well as the Submitting laboratories where the genome data were generated and shared via GISAID, on which this research is based.

All Submitters of data may be contacted directly via www.gisaid.org

Authors are sorted alphabetically.

| Accession ID | Originating Laboratory | Submitting Laboratory | Authors |
|---|---|---|---|
| EPI_ISL_1636962, EPI_ISL_1636965 | Barts Health NHS Trust | COVID-19 Genomics UK (COG-UK) Consortium | BROAD; Beatrix; CUTINO-MOGUEL; Claire; David; Dola; HARRINGTON; KELE; KULASEGARAN-SHYLINI; Maria-Teresa; OWOYEMI; Raghavendran |
| EPI_ISL_1476673, EPI_ISL_1547238, EPI_ISL_1547239, EPI_ISL_1547240, EPI_ISL_1547241, EPI_ISL_1547243, EPI_ISL_1547244, EPI_ISL_1547245, EPI_ISL_1547246, EPI_ISL_1547248, EPI_ISL_1547249, EPI_ISL_1547300, EPI_ISL_1547304, EPI_ISL_1547307, EPI_ISL_1547309, EPI_ISL_1637978, EPI_ISL_1637983, EPI_ISL_1637987, EPI_ISL_1637996 | | | |
| see above | Centre for Enzyme Innovation, University of Portsmouth / Translational Research Laboratory, Portsmouth Hospitals NHS Trust | COVID-19 Genomics UK (COG-UK) Consortium | Angela Beckett; Christopher Fearn; Kate Cook; Katie Loveson; Salman Goudarzi; Samuel Robson; Scott Elliott; Sharon Glaysher |
| EPI_ISL_1474570, EPI_ISL_1474571, EPI_ISL_1474573, EPI_ISL_1545847, EPI_ISL_1545848, EPI_ISL_1636605, EPI_ISL_1636607, EPI_ISL_1636608, EPI_ISL_1636609, EPI_ISL_1636617, EPI_ISL_1636618, EPI_ISL_1636621, EPI_ISL_1730030, EPI_ISL_1730037 | | | |
| see above | Department of Pathology, University of Cambridge | COVID-19 Genomics UK (COG-UK) Consortium | Aminu S. Jahun; Ian Goodfellow; Iliana Georgana; Martin D. Curran; Myra Hosmillo; Rhys Izuagbe; Surendra Parmar; William L. Hamilton; Yasmin Chaudhry |
| EPI_ISL_1451689, EPI_ISL_1466290, EPI_ISL_1483803, EPI_ISL_1483809, EPI_ISL_1483812, EPI_ISL_1483815, EPI_ISL_1483816, EPI_ISL_1483840, EPI_ISL_1483846, EPI_ISL_1483848, EPI_ISL_1483850, EPI_ISL_1483864, EPI_ISL_1483869, EPI_ISL_1483879, EPI_ISL_1483893, EPI_ISL_1483908, EPI_ISL_1483909, EPI_ISL_1483913, EPI_ISL_1483914, EPI_ISL_1483921, EPI_ISL_1483924, EPI_ISL_1483940, EPI_ISL_1483942, EPI_ISL_1483943, EPI_ISL_1483944, EPI_ISL_1483947, EPI_ISL_1483948, EPI_ISL_1483950, EPI_ISL_1483952, EPI_ISL_1483956, EPI_ISL_1483958, EPI_ISL_1483961, EPI_ISL_1483966, EPI_ISL_1483967, EPI_ISL_1483969, EPI_ISL_1483973, EPI_ISL_1483976, EPI_ISL_1483978, EPI_ISL_1483980, EPI_ISL_1483982, EPI_ISL_1483985, EPI_ISL_1483990, EPI_ISL_1483991, EPI_ISL_1483992, EPI_ISL_1483993, EPI_ISL_1483994, EPI_ISL_1483998, EPI_ISL_1484002, EPI_ISL_1484007, EPI_ISL_1484012, EPI_ISL_1484017, EPI_ISL_1484022, EPI_ISL_1484026, EPI_ISL_1484030, EPI_ISL_1484033, EPI_ISL_1484039, EPI_ISL_1484052, EPI_ISL_1484055, EPI_ISL_1484061, EPI_ISL_1484066, EPI_ISL_1484067, EPI_ISL_1484072, EPI_ISL_1484077, EPI_ISL_1484090, EPI_ISL_1484091, EPI_ISL_1484095, EPI_ISL_1484097, EPI_ISL_1484098, EPI_ISL_1484100, EPI_ISL_1484112, EPI_ISL_1484113, EPI_ISL_1484114, EPI_ISL_1484115, EPI_ISL_1484119, EPI_ISL_1484122, EPI_ISL_1484124, EPI_ISL_1484125, EPI_ISL_1484127, EPI_ISL_1484129, EPI_ISL_1484130, EPI_ISL_1484134, EPI_ISL_1484135, EPI_ISL_1484149, EPI_ISL_1484155, EPI_ISL_1484160, EPI_ISL_1484165, EPI_ISL_1484169, EPI_ISL_1484172, EPI_ISL_1484180, EPI_ISL_1484186, EPI_ISL_1484187, EPI_ISL_1484188, EPI_ISL_1484190, EPI_ISL_1484191, EPI_ISL_1484195, EPI_ISL_1484198, EPI_ISL_1484199, EPI_ISL_1484204, EPI_ISL_1484207, EPI_ISL_1484213, EPI_ISL_1484214, EPI_ISL_1484215, EPI_ISL_1484216, EPI_ISL_1484217, EPI_ISL_1484218, EPI_ISL_1484219, EPI_ISL_1484221, EPI_ISL_1484222, EPI_ISL_1484227, EPI_ISL_1484228, EPI_ISL_1484229, EPI_ISL_1484246, EPI_ISL_1484252, EPI_ISL_1484254, EPI_ISL_1484256, EPI_ISL_1484257, EPI_ISL_1484264, EPI_ISL_1484265, EPI_ISL_1484269, EPI_ISL_1484270, EPI_ISL_1484271, EPI_ISL_1484275, EPI_ISL_1484283, EPI_ISL_1484289, EPI_ISL_1484297, EPI_ISL_1484299, EPI_ISL_1484302, EPI_ISL_1484308, EPI_ISL_1484311, EPI_ISL_1484612, EPI_ISL_1484613, EPI_ISL_1484616, EPI_ISL_1484618, EPI_ISL_1484620, EPI_ISL_1484622, EPI_ISL_1484623, EPI_ISL_1484624, EPI_ISL_1484625, EPI_ISL_1484626, EPI_ISL_1484628, EPI_ISL_1484630, EPI_ISL_1484632, EPI_ISL_1484635, EPI_ISL_1484636, EPI_ISL_1484638, EPI_ISL_1484639, EPI_ISL_1484640, EPI_ISL_1484642, EPI_ISL_1484643, EPI_ISL_1484644, EPI_ISL_1484646, EPI_ISL_1484647, EPI_ISL_1484652, EPI_ISL_1484653, EPI_ISL_1484655, EPI_ISL_1484657, EPI_ISL_1484660, EPI_ISL_1484661, EPI_ISL_1484662, EPI_ISL_1484663, EPI_ISL_1484664, EPI_ISL_1484666, EPI_ISL_1484668, EPI_ISL_1484669, EPI_ISL_1484670, EPI_ISL_1484671, EPI_ISL_1484672, EPI_ISL_1484673, EPI_ISL_1484674, EPI_ISL_1484676, EPI_ISL_1484677, EPI_ISL_1484678, EPI_ISL_1484679, EPI_ISL_1484681, EPI_ISL_1484682, EPI_ISL_1484683, EPI_ISL_1484684, EPI_ISL_1484686, EPI_ISL_1484688, EPI_ISL_1484689, EPI_ISL_1484690, EPI_ISL_1484692, EPI_ISL_1484693, EPI_ISL_1484694, EPI_ISL_1484695, EPI_ISL_1484696, EPI_ISL_1484697, EPI_ISL_1484699, EPI_ISL_1484700, EPI_ISL_1484701, EPI_ISL_1484702, EPI_ISL_1484703, EPI_ISL_1484704, EPI_ISL_1484705, EPI_ISL_1484707, EPI_ISL_1484709, EPI_ISL_1484711, EPI_ISL_1484713, EPI_ISL_1484714, EPI_ISL_1484715, EPI_ISL_1484716, EPI_ISL_1484721, EPI_ISL_1484722, EPI_ISL_1484723, EPI_ISL_1484724, EPI_ISL_1484725, EPI_ISL_1484726, EPI_ISL_1484727, EPI_ISL_1484729, EPI_ISL_1484730, EPI_ISL_1484731, EPI_ISL_1484732, EPI_ISL_1484734, EPI_ISL_1484735, EPI_ISL_1484736, EPI_ISL_1484737, EPI_ISL_1484738, EPI_ISL_1484740, EPI_ISL_1484741, EPI_ISL_1484742, EPI_ISL_1484743, EPI_ISL_1484744, EPI_ISL_1484745, EPI_ISL_1484746, EPI_ISL_1484748, EPI_ISL_1484749, EPI_ISL_1484750, EPI_ISL_1484751, EPI_ISL_1484752, EPI_ISL_1484753, EPI_ISL_1484755, EPI_ISL_1484756, EPI_ISL_1484757, EPI_ISL_1484758, EPI_ISL_1484760, EPI_ISL_1484761, EPI_ISL_1484763, EPI_ISL_1484766, EPI_ISL_1484767, EPI_ISL_1484768, EPI_ISL_1484769, EPI_ISL_1484770, EPI_ISL_1484774, EPI_ISL_1484775, EPI_ISL_1484776, EPI_ISL_1484777, EPI_ISL_1484778, EPI_ISL_1484779, EPI_ISL_1484780, EPI_ISL_1484782, EPI_ISL_1484783, EPI_ISL_1484784, EPI_ISL_1484785, EPI_ISL_1484786, EPI_ISL_1484787, EPI_ISL_1484790, EPI_ISL_1484791, EPI_ISL_1484792, EPI_ISL_1484793, EPI_ISL_1484796, EPI_ISL_1484798, EPI_ISL_1484799, EPI_ISL_1484801, EPI_ISL_1484802, EPI_ISL_1484804, EPI_ISL_1484805, EPI_ISL_1484806, EPI_ISL_1484808, EPI_ISL_1484809, EPI_ISL_1484810, EPI_ISL_1484811, EPI_ISL_1484812, EPI_ISL_1484814, EPI_ISL_1484816, EPI_ISL_1484817, EPI_ISL_1484819, EPI_ISL_1484820, EPI_ISL_1484821, EPI_ISL_1484822, EPI_ISL_1484823, EPI_ISL_1484824, EPI_ISL_1484825, EPI_ISL_1484826, EPI_ISL_1484828, EPI_ISL_1484829, EPI_ISL_1484831, EPI_ISL_1484832, EPI_ISL_1484833, EPI_ISL_1484835, EPI_ISL_1484837, EPI_ISL_1484838, EPI_ISL_1484839, EPI_ISL_1484841, EPI_ISL_1484843, EPI_ISL_1484845, EPI_ISL_1484846, EPI_ISL_1484847, EPI_ISL_1484848, EPI_ISL_1484849, EPI_ISL_1484851, EPI_ISL_1484852, EPI_ISL_1484853, EPI_ISL_1484855, EPI_ISL_1484856, EPI_ISL_1484858, EPI_ISL_1484859, EPI_ISL_1484860, EPI_ISL_1484861, EPI_ISL_1484864, EPI_ISL_1484865, EPI_ISL_1484866, EPI_ISL_1484868, EPI_ISL_1484869, EPI_ISL_1484870, EPI_ISL_1484871, EPI_ISL_1484873, EPI_ISL_1484874, EPI_ISL_1484875, EPI_ISL_1484876, EPI_ISL_1484877, EPI_ISL_1484879, EPI_ISL_1484880, EPI_ISL_1484881, EPI_ISL_1484882, EPI_ISL_1484884, EPI_ISL_1484885, EPI_ISL_1484886, EPI_ISL_1484888, EPI_ISL_1484889, EPI_ISL_1484890, EPI_ISL_1484893, EPI_ISL_1484897, EPI_ISL_1484899, EPI_ISL_1484900, EPI_ISL_1484903, EPI_ISL_1484904, EPI_ISL_1484905, EPI_ISL_1484906, EPI_ISL_1484908, EPI_ISL_1484909, EPI_ISL_1484910, EPI_ISL_1484912, EPI_ISL_1484914, EPI_ISL_1484917, EPI_ISL_1484918, EPI_ISL_1484920, EPI_ISL_1484921, EPI_ISL_1484924, EPI_ISL_1484926, EPI_ISL_1484928, EPI_ISL_1484929, EPI_ISL_1484930, EPI_ISL_1484931, EPI_ISL_1484932, EPI_ISL_1484933, EPI_ISL_1484934, EPI_ISL_1484937, EPI_ISL_1484940, EPI_ISL_1484941, EPI_ISL_1484943, EPI_ISL_1484944, EPI_ISL_1484945, EPI_ISL_1484946, EPI_ISL_1484947, EPI_ISL_1484949, EPI_ISL_1484950, EPI_ISL_1484951, EPI_ISL_1484954, EPI_ISL_1484956, EPI_ISL_1484959, EPI_ISL_1484962, EPI_ISL_1484964, EPI_ISL_1484965, EPI_ISL_1484966, EPI_ISL_1484967, EPI_ISL_1484968, EPI_ISL_1484969, EPI_ISL_1484970, EPI_ISL_1484971, EPI_ISL_1484974, EPI_ISL_1484975, EPI_ISL_1484976, EPI_ISL_1484977, EPI_ISL_1484978, EPI_ISL_1484979, EPI_ISL_1484981, EPI_ISL_1484982, EPI_ISL_1484983, EPI_ISL_1484987, EPI_ISL_1484988, EPI_ISL_1484989, EPI_ISL_1484992, EPI_ISL_1484993, EPI_ISL_1484994, EPI_ISL_1484995, EPI_ISL_1484996, EPI_ISL_1484999, EPI_ISL_1485000, EPI_ISL_1485001, EPI_ISL_1485003, EPI_ISL_1485004, EPI_ISL_1485005, EPI_ISL_1485006, EPI_ISL_1485010, EPI_ISL_1485011, EPI_ISL_1485012, EPI_ISL_1485013, EPI_ISL_1485014, EPI_ISL_1485016, EPI_ISL_1485017, EPI_ISL_1485022, EPI_ISL_1485023, EPI_ISL_1485024, EPI_ISL_1485025, EPI_ISL_1485027, EPI_ISL_1485028, EPI_ISL_1485029, EPI_ISL_1485030, EPI_ISL_1485031, EPI_ISL_1485032, EPI_ISL_1485034, EPI_ISL_1485035, EPI_ISL_1485036, EPI_ISL_1485037, EPI_ISL_1485038, EPI_ISL_1485039, EPI_ISL_1485040, EPI_ISL_1485041, EPI_ISL_1485043, EPI_ISL_1485045, EPI_ISL_1485047, EPI_ISL_1485049, EPI_ISL_1485051, EPI_ISL_1485052, EPI_ISL_1485053, EPI_ISL_1485055, EPI_ISL_1485057, EPI_ISL_1485059, EPI_ISL_1485060, EPI_ISL_1485062, EPI_ISL_1485063, EPI_ISL_1485065, EPI_ISL_1485066, EPI_ISL_1485068, EPI_ISL_1485070, EPI_ISL_1485071, EPI_ISL_1485072, EPI_ISL_1485073, EPI_ISL_1485075, EPI_ISL_1485077, EPI_ISL_1485078, EPI_ISL_1485081, EPI_ISL_1485085, EPI_ISL_1485086, EPI_ISL_1485087, EPI_ISL_1485088, EPI_ISL_1485089, EPI_ISL_1485090, EPI_ISL_1485091, EPI_ISL_1485092, EPI_ISL_1485094, EPI_ISL_1485097, EPI_ISL_1485099, EPI_ISL_1485100, EPI_ISL_1485102, EPI_ISL_1485103, EPI_ISL_1485104, EPI_ISL_1485105, EPI_ISL_1485108, EPI_ISL_1485109, EPI_ISL_1485111, EPI_ISL_1485113, EPI_ISL_1485114, EPI_ISL_1485115, EPI_ISL_1485117, EPI_ISL_1485118, EPI_ISL_1485119, EPI_ISL_1485120, EPI_ISL_1485122, EPI_ISL_1485124, EPI_ISL_1485125, EPI_ISL_1485127, EPI_ISL_1485128, EPI_ISL_1485129, EPI_ISL_1485131, EPI_ISL_1485137, EPI_ISL_1485139, EPI_ISL_1485140, EPI_ISL_1485143, EPI_ISL_1485146, EPI_ISL_1485148, EPI_ISL_1485150, EPI_ISL_1485151, EPI_ISL_1485152, EPI_ISL_1485153, EPI_ISL_1485154, EPI_ISL_1485157, EPI_ISL_1485159, EPI_ISL_1485161, EPI_ISL_1485162, EPI_ISL_1485163, EPI_ISL_1485164, EPI_ISL_1485165, EPI_ISL_1485167, EPI_ISL_1485168, EPI_ISL_1485169, EPI_ISL_1485170, EPI_ISL_1485171, EPI_ISL_1485172, EPI_ISL_1485173, EPI_ISL_1485176, EPI_ISL_1485177, EPI_ISL_1485179, EPI_ISL_1485181, EPI_ISL_1485186, EPI_ISL_1485187, EPI_ISL_1485188, EPI_ISL_1485189, EPI_ISL_1485190, EPI_ISL_1485192, EPI_ISL_1485194, EPI_ISL_1485198, EPI_ISL_1485200, EPI_ISL_1485201, EPI_ISL_1485202, EPI_ISL_1485203, EPI_ISL_1485205, EPI_ISL_1485206, EPI_ISL_1485207, EPI_ISL_1485208, EPI_ISL_1485210, EPI_ISL_1485212, EPI_ISL_1485215, EPI_ISL_1485219, EPI_ISL_1485220, EPI_ISL_1485221, EPI_ISL_1485222, EPI_ISL_1485223, EPI_ISL_1485229, EPI_ISL_1485232, EPI_ISL_1485233, EPI_ISL_1485239, EPI_ISL_1485242, EPI_ISL_1485243, EPI_ISL_1485244, EPI_ISL_1485246, EPI_ISL_1485247, EPI_ISL_1485248, EPI_ISL_1485250, EPI_ISL_1485253, EPI_ISL_1485254, EPI_ISL_1485255, EPI_ISL_1485256, EPI_ISL_1485258, EPI_ISL_1485259, EPI_ISL_1485261, EPI_ISL_1485265, EPI_ISL_1485271, EPI_ISL_1485272, EPI_ISL_1485274, EPI_ISL_1485277, EPI_ISL_1485278, EPI_ISL_1485279, EPI_ISL_1485280, EPI_ISL_1485281, EPI_ISL_1485282, EPI_ISL_1485283, EPI_ISL_1485284, EPI_ISL_1485287, EPI_ISL_1485288, EPI_ISL_1485289, EPI_ISL_1485291, EPI_ISL_1485295, EPI_ISL_1485296, EPI_ISL_1485297, EPI_ISL_1485299, EPI_ISL_1485302, EPI_ISL_1485304, EPI_ISL_1485306, EPI_ISL_1485309, EPI_ISL_1485310, EPI_ISL_1485311, EPI_ISL_1485314, EPI_ISL_1485316, EPI_ISL_1485318, EPI_ISL_1485322, EPI_ISL_1485323, EPI_ISL_1485324, EPI_ISL_1485330, EPI_ISL_1485331, EPI_ISL_1485337, EPI_ISL_1485339, EPI_ISL_1485341, EPI_ISL_1485342, EPI_ISL_1485345, EPI_ISL_1485346, EPI_ISL_1485350, EPI_ISL_1485352, EPI_ISL_1485353, EPI_ISL_1485356, EPI_ISL_1485357, EPI_ISL_1485358, EPI_ISL_1485359, EPI_ISL_1485360, EPI_ISL_1485362, EPI_ISL_1485365, EPI_ISL_1485370, EPI_ISL_1485373, EPI_ISL_1485374, EPI_ISL_1485376, EPI_ISL_1485377, EPI_ISL_1485380, EPI_ISL_1485383, EPI_ISL_1485386, EPI_ISL_1485389, EPI_ISL_1485390, EPI_ISL_1485391, EPI_ISL_1485392, EPI_ISL_1485393, EPI_ISL_1485394, EPI_ISL_1485396, EPI_ISL_1485401, EPI_ISL_1485402, EPI_ISL_1485403, EPI_ISL_1485404, EPI_ISL_1485405, EPI_ISL_1485407, EPI_ISL_1485408, EPI_ISL_1485409, EPI_ISL_1485411, EPI_ISL_1485413, EPI_ISL_1485415, EPI_ISL_1485416, EPI_ISL_1485417, EPI_ISL_1485420, EPI_ISL_1485427, EPI_ISL_1485430, EPI_ISL_1485431, EPI_ISL_1485432, EPI_ISL_1485435, EPI_ISL_1485437, EPI_ISL_1485439, EPI_ISL_1485440, EPI_ISL_1485441, EPI_ISL_1485442, EPI_ISL_1485445, EPI_ISL_1485448, EPI_ISL_1485450, EPI_ISL_1485451, EPI_ISL_1485452, EPI_ISL_1485454, EPI_ISL_1485459, EPI_ISL_1485464, EPI_ISL_1485466, EPI_ISL_1485469, EPI_ISL_1485473, EPI_ISL_1485474, EPI_ISL_1485476, EPI_ISL_1485477, EPI_ISL_1485480, EPI_ISL_1485486, EPI_ISL_1485491, EPI_ISL_1485492, EPI_ISL_1485500, EPI_ISL_1485509, EPI_ISL_1485514, EPI_ISL_1485518, EPI_ISL_1485521, EPI_ISL_1485525, EPI_ISL_1485532, EPI_ISL_1485537, EPI_ISL_1485538, EPI_ISL_1485541, EPI_ISL_1485543, EPI_ISL_1485545, EPI_ISL_1485546, EPI_ISL_1485550, EPI_ISL_1485551, EPI_ISL_1485553, EPI_ISL_1485554, EPI_ISL_1485559, EPI_ISL_1485560, EPI_ISL_1485564, EPI_ISL_1485568, EPI_ISL_1485575, EPI_ISL_1485577, EPI_ISL_1485579, EPI_ISL_1485582, EPI_ISL_1485583, EPI_ISL_1485592, EPI_ISL_1485596, EPI_ISL_1485605, EPI_ISL_1485612, EPI_ISL_1485616, EPI_ISL_1485617, EPI_ISL_1485618, EPI_ISL_1485621, EPI_ISL_1485627, EPI_ISL_1485631, EPI_ISL_1485644, EPI_ISL_1485648, EPI_ISL_1485652, EPI_ISL_1485655, EPI_ISL_1485662, EPI_ISL_1485667, EPI_ISL_1485672, EPI_ISL_1485676, EPI_ISL_1485677, EPI_ISL_1485680, EPI_ISL_1485683, EPI_ISL_1485686, EPI_ISL_1485697, EPI_ISL_1485700, EPI_ISL_1485707, EPI_ISL_1485719, EPI_ISL_1485722, EPI_ISL_1485726, EPI_ISL_1485734, EPI_ISL_1485739, EPI_ISL_1485746, EPI_ISL_1485747, EPI_ISL_1485748, EPI_ISL_1485752, EPI_ISL_1485754, EPI_ISL_1485757, EPI_ISL_1485765, EPI_ISL_1485768, EPI_ISL_1485770, EPI_ISL_1485772, EPI_ISL_1485773, EPI_ISL_1485778, EPI_ISL_1485779, EPI_ISL_1485783, EPI_ISL_1485788, EPI_ISL_1485796, EPI_ISL_1485799, EPI_ISL_1485805, EPI_ISL_1485807, EPI_ISL_1485808, EPI_ISL_1485809, EPI_ISL_1485811, EPI_ISL_1485812, EPI_ISL_1485818, EPI_ISL_1485820, EPI_ISL_1485822, EPI_ISL_1485823, EPI_ISL_1485824, EPI_ISL_1485826, EPI_ISL_1485827, EPI_ISL_1485828, EPI_ISL_1485836, EPI_ISL_1485844, EPI_ISL_1485846, EPI_ISL_1485847, EPI_ISL_1485848, EPI_ISL_1485849, EPI_ISL_1485852, EPI_ISL_1485853, EPI_ISL_1485854, EPI_ISL_1485855, EPI_ISL_1485857, EPI_ISL_1485859, EPI_ISL_1485860, EPI_ISL_1485864, EPI_ISL_1485865, EPI_ISL_1485873, EPI_ISL_1485875, EPI_ISL_1485877, EPI_ISL_1485882, EPI_ISL_1485885, EPI_ISL_1485886, EPI_ISL_1485888, EPI_ISL_1485889, EPI_ISL_1485893, EPI_ISL_1485894, EPI_ISL_1485895, EPI_ISL_1485896, EPI_ISL_1485897, EPI_ISL_1485899, EPI_ISL_1485900, EPI_ISL_1485904, EPI_ISL_1485905, EPI_ISL_1485906, EPI_ISL_1485909, EPI_ISL_1485916, EPI_ISL_1485918, EPI_ISL_1485923, EPI_ISL_1485924, EPI_ISL_1485925, EPI_ISL_1485926, EPI_ISL_1485927, EPI_ISL_1485928, EPI_ISL_1485930, EPI_ISL_1485933, EPI_ISL_1485934, EPI_ISL_1485935, EPI_ISL_1485937, EPI_ISL_1485938, EPI_ISL_1485939, EPI_ISL_1485940, EPI_ISL_1485941, EPI_ISL_1485942, EPI_ISL_1485943, EPI_ISL_1485945, EPI_ISL_1485946, EPI_ISL_1485950, EPI_ISL_1485952, EPI_ISL_1485954, EPI_ISL_1485955, EPI_ISL_1485956, EPI_ISL_1485960, EPI_ISL_1485964, EPI_ISL_1485965, EPI_ISL_1485969, EPI_ISL_1485976, EPI_ISL_1485977, EPI_ISL_1485978, EPI_ISL_1485979, EPI_ISL_1485980, EPI_ISL_1485983, EPI_ISL_1485986, EPI_ISL_1485990, EPI_ISL_1485993, EPI_ISL_1485994, EPI_ISL_1485996, EPI_ISL_1485998, EPI_ISL_1486003, EPI_ISL_1486005, EPI_ISL_1486007, EPI_ISL_1486008, EPI_ISL_1486009, EPI_ISL_1486014, EPI_ISL_1486015, EPI_ISL_1486021, EPI_ISL_1486022, EPI_ISL_1486023, EPI_ISL_1486025, EPI_ISL_1486026, EPI_ISL_1486033, EPI_ISL_1486034, EPI_ISL_1486036, EPI_ISL_1486038, EPI_ISL_1486040, EPI_ISL_1486041, EPI_ISL_1486043, EPI_ISL_1486046, EPI_ISL_1486050, EPI_ISL_1486056, EPI_ISL_1486063, EPI_ISL_1486068, EPI_ISL_1486069, EPI_ISL_1486074, EPI_ISL_1486077, EPI_ISL_1486082, EPI_ISL_1486084, EPI_ISL_1486088, EPI_ISL_1486089, EPI_ISL_1486090, EPI_ISL_1486094, EPI_ISL_1486096, EPI_ISL_1486100, EPI_ISL_1486101, EPI_ISL_1486103, EPI_ISL_1486105, EPI_ISL_1486106, EPI_ISL_1486108, EPI_ISL_1486109, EPI_ISL_1486110, EPI_ISL_1486111, EPI_ISL_1486112, EPI_ISL_1486114, EPI_ISL_1486115, EPI_ISL_1486116, EPI_ISL_1486119, EPI_ISL_1486121, EPI_ISL_1486122, EPI_ISL_1486123, EPI_ISL_1486126, EPI_ISL_1486128, EPI_ISL_1486129, EPI_ISL_1486130, EPI_ISL_1486131, EPI_ISL_1486133, EPI_ISL_1486134, EPI_ISL_1486136, EPI_ISL_1486137, EPI_ISL_1486138, EPI_ISL_1486143, EPI_ISL_1486145, EPI_ISL_1486146, EPI_ISL_1486147, EPI_ISL_1486148, EPI_ISL_1486150, EPI_ISL_1486151, EPI_ISL_1486154, EPI_ISL_1486155, EPI_ISL_1486157, EPI_ISL_1486158, EPI_ISL_1486161, EPI_ISL_1486162, EPI_ISL_1486163, EPI_ISL_1486164, EPI_ISL_1486165, EPI_ISL_1486167, EPI_ISL_1486171, EPI_ISL_1486174, EPI_ISL_1486177, EPI_ISL_1486178, EPI_ISL_1486180, EPI_ISL_1486183, EPI_ISL_1486184, EPI_ISL_1486185, EPI_ISL_1486186, EPI_ISL_1486187, EPI_ISL_1486188, EPI_ISL_1486193, EPI_ISL_1486194, EPI_ISL_1486195, EPI_ISL_1486196, EPI_ISL_1486197, EPI_ISL_1486200, EPI_ISL_1486201, EPI_ISL_1486202, EPI_ISL_1486203, EPI_ISL_1486207, EPI_ISL_1486208, EPI_ISL_1486209, EPI_ISL_1486210, EPI_ISL_1486212, EPI_ISL_1486214, EPI_ISL_1486215, EPI_ISL_1486216, EPI_ISL_1486217, EPI_ISL_1486219, EPI_ISL_1486220, EPI_ISL_1486221, EPI_ISL_1486223, EPI_ISL_1486224, EPI_ISL_1486225, EPI_ISL_1486227, EPI_ISL_1486228, EPI_ISL_1486230, EPI_ISL_1486231, EPI_ISL_1486232, EPI_ISL_1486235, EPI_ISL_1486236, EPI_ISL_1486239, EPI_ISL_1486240, EPI_ISL_1486241, EPI_ISL_1486243, EPI_ISL_1486244, EPI_ISL_1486247, EPI_ISL_1486249, EPI_ISL_1486251, EPI_ISL_1486252, EPI_ISL_1486255, EPI_ISL_1486256, EPI_ISL_1486257, EPI_ISL_1486258, EPI_ISL_1486260, EPI_ISL_1486261, EPI_ISL_1486264, EPI_ISL_1486265, EPI_ISL_1486266, EPI_ISL_1486267, EPI_ISL_1486268, EPI_ISL_1486269, EPI_ISL_1486273, EPI_ISL_1486274, EPI_ISL_1486278, EPI_ISL_1486280, EPI_ISL_1486281, EPI_ISL_1486282, EPI_ISL_1486283, EPI_ISL_1486284, EPI_ISL_1486286, EPI_ISL_1486287, EPI_ISL_1486289, EPI_ISL_1486290, EPI_ISL_1486294, EPI_ISL_1486295, EPI_ISL_1486296, EPI_ISL_1486302, EPI_ISL_1486303, EPI_ISL_1486304, EPI_ISL_1486308, EPI_ISL_1486309, EPI_ISL_1486310, EPI_ISL_1486312, EPI_ISL_1486314, EPI_ISL_1486315, EPI_ISL_1486316, EPI_ISL_1486318, EPI_ISL_1486320, EPI_ISL_1486321, EPI_ISL_1486322, EPI_ISL_1486323, EPI_ISL_1486324, EPI_ISL_1486325, EPI_ISL_1486328, EPI_ISL_1486331, EPI_ISL_1486332, EPI_ISL_1486333, EPI_ISL_1486335, EPI_ISL_1486336, EPI_ISL_1486337, EPI_ISL_1486338, EPI_ISL_1486339, EPI_ISL_1486340, EPI_ISL_1486342, EPI_ISL_1486343, EPI_ISL_1486345, EPI_ISL_1486346, EPI_ISL_1486347, EPI_ISL_1486349, EPI_ISL_1486352, EPI_ISL_1486353, EPI_ISL_1486354, EPI_ISL_1486355, EPI_ISL_1486356, EPI_ISL_1486357, EPI_ISL_1486358, EPI_ISL_1486360, EPI_ISL_1486362, EPI_ISL_1486363, EPI_ISL_1486364, EPI_ISL_1486365, EPI_ISL_1486366, EPI_ISL_1486368, EPI_ISL_1486370, EPI_ISL_1486371, EPI_ISL_1486373, EPI_ISL_1486374, EPI_ISL_1486377, EPI_ISL_1486378, EPI_ISL_1486380, EPI_ISL_1486382, EPI_ISL_1486384, EPI_ISL_1486386, EPI_ISL_1486387, EPI_ISL_1486389, EPI_ISL_1486390, EPI_ISL_1486393, EPI_ISL_1486411, EPI_ISL_1486443, EPI_ISL_1486472, EPI_ISL_1486475, EPI_ISL_1486484, EPI_ISL_1486526, EPI_ISL_1486530, EPI_ISL_1486600, EPI_ISL_1486601, EPI_ISL_1486603, EPI_ISL_1486620, EPI_ISL_1486646, EPI_ISL_1486667, EPI_ISL_1486689, EPI_ISL_1486752, EPI_ISL_1486792, EPI_ISL_1486795, EPI_ISL_1486808, EPI_ISL_1486813, EPI_ISL_1486821, EPI_ISL_1486822, EPI_ISL_1486823, EPI_ISL_1486824, EPI_ISL_1486826, EPI_ISL_1486828, EPI_ISL_1486833, EPI_ISL_1486835, EPI_ISL_1486837, EPI_ISL_1486841, EPI_ISL_1486842, EPI_ISL_1486844, EPI_ISL_1486847, EPI_ISL_1486849, EPI_ISL_1486856, EPI_ISL_1486859, EPI_ISL_1486860, EPI_ISL_1486871, EPI_ISL_1486875, EPI_ISL_1486877, EPI_ISL_1486878, EPI_ISL_1486881, EPI_ISL_1486882, EPI_ISL_1486889, EPI_ISL_1486892, EPI_ISL_1486893, EPI_ISL_1486895, EPI_ISL_1486905, EPI_ISL_1486908, EPI_ISL_1486909, EPI_ISL_1486912, EPI_ISL_1486914, EPI_ISL_1486924, EPI_ISL_1486934, EPI_ISL_1486935, EPI_ISL_1486943, EPI_ISL_1486955, EPI_ISL_1486958, EPI_ISL_1486963, EPI_ISL_1486969, EPI_ISL_1486971, EPI_ISL_1486973, EPI_ISL_1486976, EPI_ISL_1486983, EPI_ISL_1486985, EPI_ISL_1486990, EPI_ISL_1486991, EPI_ISL_1486995, EPI_ISL_1487006, EPI_ISL_1487008, EPI_ISL_1487009, EPI_ISL_1487011, EPI_ISL_1487029, EPI_ISL_1487030, EPI_ISL_1487039, EPI_ISL_1487050, EPI_ISL_1487051, EPI_ISL_1487057, EPI_ISL_1487059, EPI_ISL_1487061, EPI_ISL_1487073, EPI_ISL_1487078, EPI_ISL_1487080, EPI_ISL_1487082, EPI_ISL_1487084, EPI_ISL_1487091, EPI_ISL_1487097, EPI_ISL_1487108, EPI_ISL_1487109, EPI_ISL_1487110, EPI_ISL_1487113, EPI_ISL_1487120, EPI_ISL_1487122, EPI_ISL_1487133, EPI_ISL_1487137, EPI_ISL_1487142, EPI_ISL_1487148, EPI_ISL_1487152, EPI_ISL_1487153, EPI_ISL_1487386, EPI_ISL_1487387, EPI_ISL_1487400, EPI_ISL_1487404, EPI_ISL_1487419, EPI_ISL_1487448, EPI_ISL_1487460, EPI_ISL_1487504, EPI_ISL_1487520, EPI_ISL_1487529, EPI_ISL_1487583, EPI_ISL_1487596, EPI_ISL_1487634, EPI_ISL_1487643, EPI_ISL_1487677, EPI_ISL_1488231, EPI_ISL_1488232, EPI_ISL_1488233, EPI_ISL_1488234, EPI_ISL_1488236, EPI_ISL_1488239, EPI_ISL_1488240, EPI_ISL_1488241, EPI_ISL_1488242, EPI_ISL_1488243, EPI_ISL_1488244, EPI_ISL_1488248, EPI_ISL_1488250, EPI_ISL_1488257, EPI_ISL_1488260, EPI_ISL_1488263, EPI_ISL_1488265, EPI_ISL_1488272, EPI_ISL_1488277, EPI_ISL_1488288, EPI_ISL_1488290, EPI_ISL_1488293, EPI_ISL_1488295, EPI_ISL_1488301, EPI_ISL_1488303, EPI_ISL_1488306, EPI_ISL_1488309, EPI_ISL_1488311, EPI_ISL_1488312, EPI_ISL_1488315, EPI_ISL_1488321, EPI_ISL_1488324, EPI_ISL_1488331, EPI_ISL_1488332, EPI_ISL_1488336, EPI_ISL_1488339, EPI_ISL_1488342, EPI_ISL_1488345, EPI_ISL_1488349, EPI_ISL_1488356, EPI_ISL_1488360, EPI_ISL_1488365, EPI_ISL_1488368, EPI_ISL_1488373, EPI_ISL_1488375, EPI_ISL_1488377, EPI_ISL_1488379, EPI_ISL_1488380, EPI_ISL_1488383, EPI_ISL_1488385, EPI_ISL_1488401, EPI_ISL_1488403, EPI_ISL_1488404, EPI_ISL_1488415, EPI_ISL_1488417, EPI_ISL_1488419, EPI_ISL_1488425, EPI_ISL_1488430, EPI_ISL_1488434, EPI_ISL_1488437, EPI_ISL_1488440, EPI_ISL_1488441, EPI_ISL_1488443, EPI_ISL_1488445, EPI_ISL_1488447, EPI_ISL_1488448, EPI_ISL_1488449, EPI_ISL_1488461, EPI_ISL_1488464, EPI_ISL_1488466, EPI_ISL_1488467, EPI_ISL_1488468, EPI_ISL_1488469, EPI_ISL_1488473, EPI_ISL_1488474, EPI_ISL_1488478, EPI_ISL_1488482, EPI_ISL_1488485, EPI_ISL_1488486, EPI_ISL_1488488, EPI_ISL_1488490, EPI_ISL_1488501, EPI_ISL_1488503, EPI_ISL_1488504, EPI_ISL_1488509, EPI_ISL_1488514, EPI_ISL_1488517, EPI_ISL_1488519, EPI_ISL_1488522, EPI_ISL_1488545, EPI_ISL_1488557, EPI_ISL_1488560, EPI_ISL_1488561, EPI_ISL_1488563, EPI_ISL_1488566, EPI_ISL_1488568, EPI_ISL_1488571, EPI_ISL_1488574, EPI_ISL_1488580, EPI_ISL_1488582, EPI_ISL_1488601, EPI_ISL_1488609, EPI_ISL_1488618, EPI_ISL_1488625, EPI_ISL_1488637, EPI_ISL_1488644, EPI_ISL_1488646, EPI_ISL_1488649, EPI_ISL_1488667, EPI_ISL_1488706, EPI_ISL_1488709, EPI_ISL_1488716, EPI_ISL_1488717, EPI_ISL_1488726, EPI_ISL_1488731, EPI_ISL_1488738, EPI_ISL_1488741, EPI_ISL_1488753, EPI_ISL_1488757, EPI_ISL_1488761, EPI_ISL_1488765, EPI_ISL_1488782, EPI_ISL_1488784, EPI_ISL_1488785, EPI_ISL_1488797, EPI_ISL_1488808, EPI_ISL_1488824, EPI_ISL_1488832, EPI_ISL_1488838, EPI_ISL_1488857, EPI_ISL_1504117, EPI_ISL_1504145, EPI_ISL_1504155, EPI_ISL_1504158, EPI_ISL_1504168, EPI_ISL_1504185, EPI_ISL_1504186, EPI_ISL_1504205, EPI_ISL_1504207, EPI_ISL_1504213, EPI_ISL_1504218, EPI_ISL_1504219, EPI_ISL_1504223, EPI_ISL_1504227, EPI_ISL_1504229, EPI_ISL_1504233, EPI_ISL_1504247, EPI_ISL_1504256, EPI_ISL_1504260, EPI_ISL_1504263, EPI_ISL_1504272, EPI_ISL_1504317, EPI_ISL_1504318, EPI_ISL_1504324, EPI_ISL_1504328, EPI_ISL_1504332, EPI_ISL_1504336, EPI_ISL_1504339, EPI_ISL_1504341, EPI_ISL_1504345, EPI_ISL_1504351, EPI_ISL_1504357, EPI_ISL_1504363, EPI_ISL_1504377, EPI_ISL_1504385, EPI_ISL_1504395, EPI_ISL_1504405, EPI_ISL_1504433, EPI_ISL_1504450, EPI_ISL_1504455, EPI_ISL_1504462, EPI_ISL_1504494, EPI_ISL_1504500, EPI_ISL_1504508, EPI_ISL_1504553, EPI_ISL_1504578, EPI_ISL_1504579, EPI_ISL_1504603, EPI_ISL_1504605, EPI_ISL_1504612, EPI_ISL_1504617, EPI_ISL_1504634, EPI_ISL_1504652, EPI_ISL_1504721, EPI_ISL_1504761, EPI_ISL_1504766, EPI_ISL_1504775, EPI_ISL_1504788, EPI_ISL_1504789, EPI_ISL_1504792, EPI_ISL_1504813, EPI_ISL_1504822, EPI_ISL_1504838, EPI_ISL_1504840, EPI_ISL_1504844, EPI_ISL_1504847, EPI_ISL_1504849, EPI_ISL_1504858, EPI_ISL_1504859, EPI_ISL_1504883, EPI_ISL_1504902, EPI_ISL_1504903, EPI_ISL_1504918, EPI_ISL_1504921, EPI_ISL_1504922, EPI_ISL_1504951, EPI_ISL_1504965, EPI_ISL_1504977, EPI_ISL_1504990, EPI_ISL_1504992, EPI_ISL_1505066, EPI_ISL_1505074, EPI_ISL_1505091, EPI_ISL_1505125, EPI_ISL_1505153, EPI_ISL_1505168, EPI_ISL_1505175, EPI_ISL_1505176, EPI_ISL_1505177, EPI_ISL_1505178, EPI_ISL_1505179, EPI_ISL_1505180, EPI_ISL_1505183, EPI_ISL_1505185, EPI_ISL_1505186, EPI_ISL_1505188, EPI_ISL_1505189, EPI_ISL_1505193, EPI_ISL_1505195, EPI_ISL_1505196, EPI_ISL_1505197, EPI_ISL_1505198, EPI_ISL_1505200, EPI_ISL_1505203, EPI_ISL_1505204, EPI_ISL_1505205, EPI_ISL_1505209, EPI_ISL_1505212, EPI_ISL_1505213, EPI_ISL_1505214, EPI_ISL_1505216, EPI_ISL_1505217, EPI_ISL_1505222, EPI_ISL_1505224, EPI_ISL_1505228, EPI_ISL_1505231, EPI_ISL_1505232, EPI_ISL_1505234, EPI_ISL_1505236, EPI_ISL_1505237, EPI_ISL_1505239, EPI_ISL_1505242, EPI_ISL_1505244, EPI_ISL_1505245, EPI_ISL_1505246, EPI_ISL_1505250, EPI_ISL_1505253, EPI_ISL_1505254, EPI_ISL_1505255, EPI_ISL_1505256, EPI_ISL_1505257, EPI_ISL_1505259, EPI_ISL_1505263, EPI_ISL_1505264, EPI_ISL_1505266, EPI_ISL_1505268, EPI_ISL_1505269, EPI_ISL_1505271, EPI_ISL_1505272, EPI_ISL_1505278, EPI_ISL_1505279, EPI_ISL_1505280, EPI_ISL_1505281, EPI_ISL_1505283, EPI_ISL_1505284, EPI_ISL_1505286, EPI_ISL_1505290, EPI_ISL_1505292, EPI_ISL_1505296, EPI_ISL_1505297, EPI_ISL_1505299, EPI_ISL_1505300, EPI_ISL_1505303, EPI_ISL_1505305, EPI_ISL_1505308, EPI_ISL_1505309, EPI_ISL_1505310, EPI_ISL_1505312, EPI_ISL_1505313, EPI_ISL_1505316, EPI_ISL_1505317, EPI_ISL_1505318, EPI_ISL_1505319, EPI_ISL_1505321, EPI_ISL_1505322, EPI_ISL_1505324, EPI_ISL_1505326, EPI_ISL_1505328, EPI_ISL_1505329, EPI_ISL_1505330, EPI_ISL_1505331, EPI_ISL_1505332, EPI_ISL_1505333, EPI_ISL_1505334, EPI_ISL_1505335, EPI_ISL_1505338, EPI_ISL_1505339, EPI_ISL_1505343, EPI_ISL_1505344, EPI_ISL_1505345, EPI_ISL_1505346, EPI_ISL_1505347, EPI_ISL_1505348, EPI_ISL_1505349, EPI_ISL_1505354, EPI_ISL_1505355, EPI_ISL_1505357, EPI_ISL_1505358, EPI_ISL_1505359, EPI_ISL_1505364, EPI_ISL_1505366, EPI_ISL_1505368, EPI_ISL_1505369, EPI_ISL_1505373, EPI_ISL_1505374, EPI_ISL_1505375, EPI_ISL_1505376, EPI_ISL_1505377, EPI_ISL_1505378, EPI_ISL_1505379, EPI_ISL_1505381, EPI_ISL_1505382, EPI_ISL_1505388, EPI_ISL_1505389, EPI_ISL_1505390, EPI_ISL_1505394, EPI_ISL_1505395, EPI_ISL_1505397, EPI_ISL_1505398, EPI_ISL_1505400, EPI_ISL_1505401, EPI_ISL_1505403, EPI_ISL_1505404, EPI_ISL_1505407, EPI_ISL_1505408, EPI_ISL_1505409, EPI_ISL_1505410, EPI_ISL_1505411, EPI_ISL_1505412, EPI_ISL_1505413, EPI_ISL_1505414, EPI_ISL_1505415, EPI_ISL_1505417, EPI_ISL_1505419, EPI_ISL_1505422, EPI_ISL_1505423, EPI_ISL_1505424, EPI_ISL_1505425, EPI_ISL_1505429, EPI_ISL_1505430, EPI_ISL_1505431, EPI_ISL_1505432, EPI_ISL_1505433, EPI_ISL_1505435, EPI_ISL_1505437, EPI_ISL_1505438, EPI_ISL_1505441, EPI_ISL_1505443, EPI_ISL_1505444, EPI_ISL_1505445, EPI_ISL_1505447, EPI_ISL_1505449, EPI_ISL_1505450, EPI_ISL_1505451, EPI_ISL_1505455, EPI_ISL_1505462, EPI_ISL_1505463, EPI_ISL_1505464, EPI_ISL_1505466, EPI_ISL_1505468, EPI_ISL_1505469, EPI_ISL_1505473, EPI_ISL_1505474, EPI_ISL_1505475, EPI_ISL_1505477, EPI_ISL_1505478, EPI_ISL_1505480, EPI_ISL_1505481, EPI_ISL_1505482, EPI_ISL_1505483, EPI_ISL_1505484, EPI_ISL_1505485, EPI_ISL_1505486, EPI_ISL_1505490, EPI_ISL_1505494, EPI_ISL_1505495, EPI_ISL_1505498, EPI_ISL_1505500, EPI_ISL_1505501, EPI_ISL_1505502, EPI_ISL_1505503, EPI_ISL_1505504, EPI_ISL_1505505, EPI_ISL_1505506, EPI_ISL_1505507, EPI_ISL_1505508, EPI_ISL_1505510, EPI_ISL_1505511, EPI_ISL_1505512, EPI_ISL_1505513, EPI_ISL_1505514, EPI_ISL_1505515, EPI_ISL_1505516, EPI_ISL_1505517, EPI_ISL_1505519, EPI_ISL_1505521, EPI_ISL_1505522, EPI_ISL_1505523, EPI_ISL_1505524, EPI_ISL_1505526, EPI_ISL_1505527, EPI_ISL_1505528, EPI_ISL_1505529, EPI_ISL_1505530, EPI_ISL_1505532, EPI_ISL_1505536, EPI_ISL_1505537, EPI_ISL_1505538, EPI_ISL_1505540, EPI_ISL_1505542, EPI_ISL_1505544, EPI_ISL_1505546, EPI_ISL_1505549, EPI_ISL_1505553, EPI_ISL_1505555, EPI_ISL_1505557, EPI_ISL_1505558, EPI_ISL_1505559, EPI_ISL_1505560, EPI_ISL_1505562, EPI_ISL_1505564, EPI_ISL_1505566, EPI_ISL_1505567, EPI_ISL_1505568, EPI_ISL_1505570, EPI_ISL_1505571, EPI_ISL_1505572, EPI_ISL_1505573, EPI_ISL_1505574, EPI_ISL_1505575, EPI_ISL_1505576, EPI_ISL_1505577, EPI_ISL_1505579, EPI_ISL_1505581, EPI_ISL_1505582, EPI_ISL_1505585, EPI_ISL_1505586, EPI_ISL_1505587, EPI_ISL_1505588, EPI_ISL_1505590, EPI_ISL_1505592, EPI_ISL_1505596, EPI_ISL_1505597, EPI_ISL_1505598, EPI_ISL_1505602, EPI_ISL_1505603, EPI_ISL_1505604, EPI_ISL_1505605, EPI_ISL_1505606, EPI_ISL_1505607, EPI_ISL_1505609, EPI_ISL_1505611, EPI_ISL_1505615, EPI_ISL_1505616, EPI_ISL_1505617, EPI_ISL_1505618, EPI_ISL_1505619, EPI_ISL_1505620, EPI_ISL_1505623, EPI_ISL_1505624, EPI_ISL_1505626, EPI_ISL_1505628, EPI_ISL_1505629, EPI_ISL_1505630, EPI_ISL_1505633, EPI_ISL_1505634, EPI_ISL_1505636, EPI_ISL_1505639, EPI_ISL_1505640, EPI_ISL_1505641, EPI_ISL_1505642, EPI_ISL_1505644, EPI_ISL_1505645, EPI_ISL_1505646, EPI_ISL_1505647, EPI_ISL_1505648, EPI_ISL_1505649, EPI_ISL_1505652, EPI_ISL_1505654, EPI_ISL_1505660, EPI_ISL_1505661, EPI_ISL_1505662, EPI_ISL_1505663, EPI_ISL_1505664, EPI_ISL_1505666, EPI_ISL_1505667, EPI_ISL_1505669, EPI_ISL_1505670, EPI_ISL_1505671, EPI_ISL_1505672, EPI_ISL_1505673, EPI_ISL_1505674, EPI_ISL_1505675, EPI_ISL_1505676, EPI_ISL_1505678, EPI_ISL_1505679, EPI_ISL_1505680, EPI_ISL_1505682, EPI_ISL_1505683, EPI_ISL_1505684, EPI_ISL_1505685, EPI_ISL_1505686, EPI_ISL_1505687, EPI_ISL_1505689, EPI_ISL_1505690, EPI_ISL_1505691, EPI_ISL_1505692, EPI_ISL_1505696, EPI_ISL_1505697, EPI_ISL_1505699, EPI_ISL_1505700, EPI_ISL_1505701, EPI_ISL_1505702, EPI_ISL_1505703, EPI_ISL_1505704, EPI_ISL_1505705, EPI_ISL_1505707, EPI_ISL_1505708, EPI_ISL_1505709, EPI_ISL_1505710, EPI_ISL_1505713, EPI_ISL_1505714, EPI_ISL_1505715, EPI_ISL_1505717, EPI_ISL_1505719, EPI_ISL_1505720, EPI_ISL_1505721, EPI_ISL_1505722, EPI_ISL_1505724, EPI_ISL_1505725, EPI_ISL_1505726, EPI_ISL_1505729, EPI_ISL_1505730, EPI_ISL_1505731, EPI_ISL_1505732, EPI_ISL_1505733, EPI_ISL_1505734, EPI_ISL_1505737, EPI_ISL_1505740, EPI_ISL_1505741, EPI_ISL_1505744, EPI_ISL_1505745, EPI_ISL_1505746, EPI_ISL_1505747, EPI_ISL_1505748, EPI_ISL_1505749, EPI_ISL_1505751, EPI_ISL_1505752, EPI_ISL_1505753, EPI_ISL_1505755, EPI_ISL_1505758, EPI_ISL_1505759, EPI_ISL_1505760, EPI_ISL_1505761, EPI_ISL_1505762, EPI_ISL_1505763, EPI_ISL_1505764, EPI_ISL_1505766, EPI_ISL_1505769, EPI_ISL_1505773, EPI_ISL_1505774, EPI_ISL_1505775, EPI_ISL_1505776, EPI_ISL_1505777, EPI_ISL_1505778, EPI_ISL_1505779, EPI_ISL_1505780, EPI_ISL_1505781, EPI_ISL_1505782, EPI_ISL_1505783, EPI_ISL_1505785, EPI_ISL_1505787, EPI_ISL_1505789, EPI_ISL_1505790, EPI_ISL_1505791, EPI_ISL_1505792, EPI_ISL_1505795, EPI_ISL_1505796, EPI_ISL_1505798, EPI_ISL_1505799, EPI_ISL_1505801, EPI_ISL_1505802, EPI_ISL_1505803, EPI_ISL_1505806, EPI_ISL_1505809, EPI_ISL_1505810, EPI_ISL_1505813, EPI_ISL_1505815, EPI_ISL_1505816, EPI_ISL_1505817, EPI_ISL_1505818, EPI_ISL_1505820, EPI_ISL_1505823, EPI_ISL_1505824, EPI_ISL_1505825, EPI_ISL_1505828, EPI_ISL_1505829, EPI_ISL_1505830, EPI_ISL_1505832, EPI_ISL_1505833, EPI_ISL_1505835, EPI_ISL_1505836, EPI_ISL_1505837, EPI_ISL_1505838, EPI_ISL_1505840, EPI_ISL_1505844, EPI_ISL_1505846, EPI_ISL_1505847, EPI_ISL_1505848, EPI_ISL_1505849, EPI_ISL_1505852, EPI_ISL_1505853, EPI_ISL_1505854, EPI_ISL_1505855, EPI_ISL_1505860, EPI_ISL_1505867, EPI_ISL_1505868, EPI_ISL_1505869, EPI_ISL_1505870, EPI_ISL_1505871, EPI_ISL_1505874, EPI_ISL_1505875, EPI_ISL_1505876, EPI_ISL_1505878, EPI_ISL_1505879, EPI_ISL_1505880, EPI_ISL_1505881, EPI_ISL_1505883, EPI_ISL_1505887, EPI_ISL_1505888, EPI_ISL_1505890, EPI_ISL_1505892, EPI_ISL_1505893, EPI_ISL_1505895, EPI_ISL_1505896, EPI_ISL_1505897, EPI_ISL_1505900, EPI_ISL_1505901, EPI_ISL_1505908, EPI_ISL_1505910, EPI_ISL_1505912, EPI_ISL_1505913, EPI_ISL_1505915, EPI_ISL_1505917, EPI_ISL_1505918, EPI_ISL_1505919, EPI_ISL_1505921, EPI_ISL_1505922, EPI_ISL_1505924, EPI_ISL_1505925, EPI_ISL_1505926, EPI_ISL_1505927, EPI_ISL_1505928, EPI_ISL_1505933, EPI_ISL_1505934, EPI_ISL_1505935, EPI_ISL_1505937, EPI_ISL_1505938, EPI_ISL_1505939, EPI_ISL_1505941, EPI_ISL_1505942, EPI_ISL_1505943, EPI_ISL_1505945, EPI_ISL_1505949, EPI_ISL_1505951, EPI_ISL_1505952, EPI_ISL_1505953, EPI_ISL_1505954, EPI_ISL_1505956, EPI_ISL_1505958, EPI_ISL_1505959, EPI_ISL_1505964, EPI_ISL_1505965, EPI_ISL_1505969, EPI_ISL_1505970, EPI_ISL_1505971, EPI_ISL_1505974, EPI_ISL_1505975, EPI_ISL_1505979, EPI_ISL_1505980, EPI_ISL_1505986, EPI_ISL_1505987, EPI_ISL_1505989, EPI_ISL_1505990, EPI_ISL_1505991, EPI_ISL_1505993, EPI_ISL_1505994, EPI_ISL_1505996, EPI_ISL_1506000, EPI_ISL_1506001, EPI_ISL_1506004, EPI_ISL_1506005, EPI_ISL_1506007, EPI_ISL_1506008, EPI_ISL_1506009, EPI_ISL_1506010, EPI_ISL_1506014, EPI_ISL_1506016, EPI_ISL_1506017, EPI_ISL_1506018, EPI_ISL_1506019, EPI_ISL_1506020, EPI_ISL_1506023, EPI_ISL_1506025, EPI_ISL_1506027, EPI_ISL_1506029, EPI_ISL_1506030, EPI_ISL_1506033, EPI_ISL_1506034, EPI_ISL_1506035, EPI_ISL_1506036, EPI_ISL_1506040, EPI_ISL_1506042, EPI_ISL_1506043, EPI_ISL_1506044, EPI_ISL_1506047, EPI_ISL_1506049, EPI_ISL_1506051, EPI_ISL_1506052, EPI_ISL_1506053, EPI_ISL_1506054, EPI_ISL_1506055, EPI_ISL_1506057, EPI_ISL_1506058, EPI_ISL_1506059, EPI_ISL_1506062, EPI_ISL_1506063, EPI_ISL_1506067, EPI_ISL_1506069, EPI_ISL_1506071, EPI_ISL_1506074, EPI_ISL_1506075, EPI_ISL_1506076, EPI_ISL_1506079, EPI_ISL_1506080, EPI_ISL_1506086, EPI_ISL_1506088, EPI_ISL_1506090, EPI_ISL_1506091, EPI_ISL_1506093, EPI_ISL_1506097, EPI_ISL_1506099, EPI_ISL_1506100, EPI_ISL_1506102, EPI_ISL_1506103, EPI_ISL_1506144, EPI_ISL_1506163, EPI_ISL_1506192, EPI_ISL_1506266, EPI_ISL_1506277, EPI_ISL_1506278, EPI_ISL_1506292, EPI_ISL_1506305, EPI_ISL_1506353, EPI_ISL_1506407, EPI_ISL_1506424, EPI_ISL_1506434, EPI_ISL_1506562, EPI_ISL_1506572, EPI_ISL_1517496, EPI_ISL_1517498, EPI_ISL_1517500, EPI_ISL_1517501, EPI_ISL_1517502, EPI_ISL_1517503, EPI_ISL_1517505, EPI_ISL_1517508, EPI_ISL_1517509, EPI_ISL_1517512, EPI_ISL_1517516, EPI_ISL_1517517, EPI_ISL_1517518, EPI_ISL_1517521, EPI_ISL_1517523, EPI_ISL_1517527, EPI_ISL_1517528, EPI_ISL_1517529, EPI_ISL_1517530, EPI_ISL_1517532, EPI_ISL_1517537, EPI_ISL_1517538, EPI_ISL_1517539, | | | |

EPI_ISL_1517540, EPI_ISL_1517541, EPI_ISL_1517544, EPI_ISL_1517546, EPI_ISL_1517548, EPI_ISL_1517550, EPI_ISL_1517553, EPI_ISL_1517555, EPI_ISL_1517556, EPI_ISL_1517557, EPI_ISL_1517558, EPI_ISL_1517560, EPI_ISL_1517563, EPI_ISL_1517570, EPI_ISL_1517573, EPI_ISL_1517576, EPI_ISL_1517577, EPI_ISL_1517579, EPI_ISL_1517580, EPI_ISL_1517584, EPI_ISL_1517592, EPI_ISL_1517603, EPI_ISL_1517604, EPI_ISL_1517607, EPI_ISL_1517615, EPI_ISL_1517623, EPI_ISL_1517630, EPI_ISL_1517635, EPI_ISL_1517640, EPI_ISL_1517645, EPI_ISL_1517647, EPI_ISL_1517649, EPI_ISL_1517654, EPI_ISL_1517666, EPI_ISL_1517668, EPI_ISL_1517671, EPI_ISL_1517675, EPI_ISL_1517676, EPI_ISL_1517677, EPI_ISL_1517678, EPI_ISL_1517679, EPI_ISL_1517681, EPI_ISL_1517685, EPI_ISL_1517688, EPI_ISL_1517697, EPI_ISL_1517698, EPI_ISL_1517700, EPI_ISL_1517702, EPI_ISL_1517706, EPI_ISL_1517708, EPI_ISL_1517711, EPI_ISL_1517717, EPI_ISL_1517718, EPI_ISL_1517719, EPI_ISL_1517723, EPI_ISL_1517725, EPI_ISL_1517729, EPI_ISL_1517731, EPI_ISL_1517734, EPI_ISL_1517735, EPI_ISL_1517736, EPI_ISL_1517740, EPI_ISL_1517743, EPI_ISL_1517746, EPI_ISL_1517748, EPI_ISL_1517751, EPI_ISL_1517776, EPI_ISL_1517779, EPI_ISL_1517796, EPI_ISL_1517804, EPI_ISL_1517813, EPI_ISL_1517826, EPI_ISL_1517836, EPI_ISL_1517841, EPI_ISL_1517852, EPI_ISL_1517853, EPI_ISL_1517859, EPI_ISL_1517870, EPI_ISL_1517873, EPI_ISL_1517881, EPI_ISL_1517896, EPI_ISL_1517916, EPI_ISL_1517937, EPI_ISL_1517957, EPI_ISL_1517963, EPI_ISL_1517977, EPI_ISL_1517978, EPI_ISL_1517988, EPI_ISL_1517994, EPI_ISL_1518004, EPI_ISL_1518011, EPI_ISL_1518016, EPI_ISL_1518022, EPI_ISL_1518028, EPI_ISL_1518056, EPI_ISL_1518070, EPI_ISL_1518077, EPI_ISL_1518126, EPI_ISL_1518134, EPI_ISL_1518142, EPI_ISL_1518151, EPI_ISL_1518153, EPI_ISL_1518181, EPI_ISL_1518187, EPI_ISL_1518188, EPI_ISL_1518220, EPI_ISL_1518233, EPI_ISL_1518275, EPI_ISL_1518279, EPI_ISL_1518299, EPI_ISL_1518304, EPI_ISL_1518305, EPI_ISL_1518310, EPI_ISL_1518321, EPI_ISL_1518327, EPI_ISL_1518338, EPI_ISL_1518341, EPI_ISL_1518356, EPI_ISL_1518370, EPI_ISL_1518377, EPI_ISL_1518383, EPI_ISL_1518395, EPI_ISL_1518397, EPI_ISL_1518401, EPI_ISL_1518403, EPI_ISL_1518404, EPI_ISL_1518408, EPI_ISL_1518412, EPI_ISL_1518415, EPI_ISL_1518428, EPI_ISL_1518430, EPI_ISL_1518433, EPI_ISL_1518435, EPI_ISL_1518437, EPI_ISL_1518438, EPI_ISL_1518441, EPI_ISL_1518442, EPI_ISL_1518445, EPI_ISL_1518446, EPI_ISL_1518447, EPI_ISL_1518450, EPI_ISL_1518453, EPI_ISL_1518454, EPI_ISL_1518457, EPI_ISL_1518460, EPI_ISL_1518464, EPI_ISL_1518466, EPI_ISL_1518467, EPI_ISL_1518468, EPI_ISL_1518469, EPI_ISL_1518470, EPI_ISL_1518472, EPI_ISL_1518475, EPI_ISL_1518478, EPI_ISL_1518482, EPI_ISL_1518485, EPI_ISL_1518490, EPI_ISL_1518494, EPI_ISL_1518497, EPI_ISL_1518498, EPI_ISL_1518504, EPI_ISL_1518505, EPI_ISL_1518509, EPI_ISL_1518516, EPI_ISL_1518520, EPI_ISL_1518522, EPI_ISL_1518540, EPI_ISL_1518541, EPI_ISL_1518543, EPI_ISL_1518545, EPI_ISL_1518546, EPI_ISL_1518551, EPI_ISL_1518552, EPI_ISL_1518555, EPI_ISL_1518560, EPI_ISL_1518566, EPI_ISL_1518567, EPI_ISL_1518568, EPI_ISL_1518569, EPI_ISL_1518571, EPI_ISL_1518574, EPI_ISL_1518575, EPI_ISL_1518577, EPI_ISL_1518578, EPI_ISL_1518583, EPI_ISL_1518587, EPI_ISL_1518588, EPI_ISL_1518591, EPI_ISL_1518597, EPI_ISL_1518599, EPI_ISL_1518600, EPI_ISL_1518605, EPI_ISL_1518606, EPI_ISL_1518609, EPI_ISL_1518611, EPI_ISL_1518615, EPI_ISL_1518617, EPI_ISL_1518629, EPI_ISL_1518630, EPI_ISL_1518633, EPI_ISL_1518640, EPI_ISL_1518642, EPI_ISL_1518643, EPI_ISL_1518655, EPI_ISL_1518661, EPI_ISL_1518662, EPI_ISL_1518668, EPI_ISL_1518675, EPI_ISL_1518676, EPI_ISL_1518677, EPI_ISL_1518678, EPI_ISL_1518680, EPI_ISL_1518683, EPI_ISL_1518684, EPI_ISL_1518685, EPI_ISL_1518686, EPI_ISL_1518688, EPI_ISL_1518689, EPI_ISL_1518692, EPI_ISL_1518693, EPI_ISL_1518694, EPI_ISL_1518697, EPI_ISL_1518698, EPI_ISL_1518699, EPI_ISL_1518700, EPI_ISL_1518702, EPI_ISL_1518703, EPI_ISL_1518704, EPI_ISL_1518705, EPI_ISL_1518706, EPI_ISL_1518707, EPI_ISL_1518708, EPI_ISL_1518709, EPI_ISL_1518710, EPI_ISL_1518711, EPI_ISL_1518712, EPI_ISL_1518714, EPI_ISL_1518716, EPI_ISL_1518717, EPI_ISL_1518719, EPI_ISL_1518720, EPI_ISL_1518721, EPI_ISL_1518722, EPI_ISL_1518723, EPI_ISL_1518724, EPI_ISL_1518726, EPI_ISL_1518728, EPI_ISL_1518729, EPI_ISL_1518731, EPI_ISL_1518732, EPI_ISL_1518733, EPI_ISL_1518734, EPI_ISL_1518735, EPI_ISL_1518737, EPI_ISL_1518738, EPI_ISL_1518739, EPI_ISL_1518740, EPI_ISL_1518741, EPI_ISL_1518742, EPI_ISL_1518743, EPI_ISL_1518744, EPI_ISL_1518745, EPI_ISL_1518747, EPI_ISL_1518748, EPI_ISL_1518749, EPI_ISL_1518750, EPI_ISL_1518751, EPI_ISL_1518752, EPI_ISL_1518754, EPI_ISL_1518755, EPI_ISL_1518756, EPI_ISL_1518758, EPI_ISL_1518760, EPI_ISL_1518761, EPI_ISL_1518764, EPI_ISL_1518765, EPI_ISL_1518766, EPI_ISL_1518767, EPI_ISL_1518768, EPI_ISL_1518769, EPI_ISL_1518770, EPI_ISL_1518772, EPI_ISL_1518773, EPI_ISL_1518774, EPI_ISL_1518775, EPI_ISL_1518778, EPI_ISL_1518779, EPI_ISL_1518780, EPI_ISL_1518781, EPI_ISL_1518782, EPI_ISL_1518783, EPI_ISL_1518784, EPI_ISL_1518785, EPI_ISL_1518787, EPI_ISL_1518788, EPI_ISL_1518789, EPI_ISL_1518790, EPI_ISL_1518794, EPI_ISL_1518795, EPI_ISL_1518796, EPI_ISL_1518797, EPI_ISL_1518798, EPI_ISL_1518799, EPI_ISL_1518800, EPI_ISL_1518801, EPI_ISL_1518802, EPI_ISL_1518804, EPI_ISL_1518805, EPI_ISL_1518806, EPI_ISL_1518807, EPI_ISL_1518808, EPI_ISL_1518809, EPI_ISL_1518812, EPI_ISL_1518813, EPI_ISL_1518814, EPI_ISL_1518815, EPI_ISL_1518816, EPI_ISL_1518820, EPI_ISL_1518823, EPI_ISL_1518824, EPI_ISL_1518825, EPI_ISL_1518826, EPI_ISL_1518827, EPI_ISL_1518828, EPI_ISL_1518831, EPI_ISL_1518832, EPI_ISL_1518833, EPI_ISL_1518834, EPI_ISL_1518835, EPI_ISL_1518837, EPI_ISL_1518839, EPI_ISL_1518840, EPI_ISL_1518841, EPI_ISL_1518842, EPI_ISL_1518843, EPI_ISL_1518844, EPI_ISL_1518845, EPI_ISL_1518846, EPI_ISL_1518847, EPI_ISL_1518849, EPI_ISL_1518851, EPI_ISL_1518852, EPI_ISL_1518854, EPI_ISL_1518855, EPI_ISL_1518856, EPI_ISL_1518857, EPI_ISL_1518858, EPI_ISL_1518859, EPI_ISL_1518860, EPI_ISL_1518863, EPI_ISL_1518864, EPI_ISL_1518866, EPI_ISL_1518867, EPI_ISL_1518868, EPI_ISL_1518869, EPI_ISL_1518870, EPI_ISL_1518871, EPI_ISL_1518872, EPI_ISL_1518874, EPI_ISL_1518875, EPI_ISL_1518876, EPI_ISL_1518878, EPI_ISL_1518879, EPI_ISL_1518880, EPI_ISL_1518882, EPI_ISL_1518883, EPI_ISL_1518884, EPI_ISL_1518885, EPI_ISL_1518886, EPI_ISL_1518887, EPI_ISL_1518890, EPI_ISL_1518891, EPI_ISL_1518892, EPI_ISL_1518893, EPI_ISL_1518894, EPI_ISL_1518895, EPI_ISL_1518896, EPI_ISL_1518897, EPI_ISL_1518898, EPI_ISL_1518899, EPI_ISL_1518900, EPI_ISL_1518902, EPI_ISL_1518904, EPI_ISL_1518905, EPI_ISL_1518906, EPI_ISL_1518908, EPI_ISL_1518909, EPI_ISL_1518910, EPI_ISL_1518911, EPI_ISL_1518912, EPI_ISL_1518914, EPI_ISL_1518915, EPI_ISL_1518917, EPI_ISL_1518918, EPI_ISL_1518919, EPI_ISL_1518922, EPI_ISL_1518923, EPI_ISL_1518924, EPI_ISL_1518925, EPI_ISL_1518926, EPI_ISL_1518929, EPI_ISL_1518930, EPI_ISL_1518931, EPI_ISL_1518932, EPI_ISL_1518933, EPI_ISL_1518934, EPI_ISL_1518936, EPI_ISL_1518937, EPI_ISL_1518938, EPI_ISL_1518939, EPI_ISL_1518940, EPI_ISL_1518941, EPI_ISL_1518942, EPI_ISL_1518944, EPI_ISL_1518945, EPI_ISL_1518947, EPI_ISL_1518948, EPI_ISL_1518949, EPI_ISL_1518950, EPI_ISL_1518951, EPI_ISL_1518952, EPI_ISL_1518953, EPI_ISL_1518954, EPI_ISL_1518955, EPI_ISL_1518956, EPI_ISL_1518958, EPI_ISL_1518960, EPI_ISL_1518961, EPI_ISL_1518962, EPI_ISL_1518963, EPI_ISL_1518964, EPI_ISL_1518965, EPI_ISL_1518966, EPI_ISL_1518967, EPI_ISL_1518968, EPI_ISL_1518970, EPI_ISL_1518971, EPI_ISL_1518972, EPI_ISL_1518973, EPI_ISL_1518974, EPI_ISL_1518975, EPI_ISL_1518976, EPI_ISL_1518977, EPI_ISL_1518979, EPI_ISL_1518980, EPI_ISL_1518981, EPI_ISL_1518982, EPI_ISL_1518983, EPI_ISL_1518985, EPI_ISL_1518986, EPI_ISL_1518987, EPI_ISL_1518990, EPI_ISL_1518991, EPI_ISL_1518992, EPI_ISL_1518993, EPI_ISL_1518994, EPI_ISL_1519307, EPI_ISL_1519308, EPI_ISL_1519311, EPI_ISL_1519314, EPI_ISL_1519315, EPI_ISL_1519317, EPI_ISL_1519319, EPI_ISL_1519320, EPI_ISL_1519321, EPI_ISL_1519324, EPI_ISL_1519325, EPI_ISL_1519327, EPI_ISL_1519329, EPI_ISL_1519331, EPI_ISL_1519332, EPI_ISL_1519333, EPI_ISL_1519334, EPI_ISL_1519335, EPI_ISL_1519337, EPI_ISL_1519339, EPI_ISL_1519340, EPI_ISL_1519341, EPI_ISL_1519342, EPI_ISL_1519343, EPI_ISL_1519344, EPI_ISL_1519346, EPI_ISL_1519347, EPI_ISL_1519349, EPI_ISL_1519350, EPI_ISL_1519351, EPI_ISL_1519389, EPI_ISL_1519390, EPI_ISL_1519392, EPI_ISL_1519394, EPI_ISL_1519395, EPI_ISL_1519397, EPI_ISL_1519399, EPI_ISL_1519400, EPI_ISL_1519403, EPI_ISL_1519404, EPI_ISL_1519406, EPI_ISL_1519408, EPI_ISL_1519409, EPI_ISL_1519411, EPI_ISL_1519412, EPI_ISL_1519414, EPI_ISL_1519417, EPI_ISL_1519418, EPI_ISL_1519423, EPI_ISL_1519425, EPI_ISL_1519426, EPI_ISL_1519427, EPI_ISL_1519428, EPI_ISL_1519431, EPI_ISL_1519432, EPI_ISL_1519433, EPI_ISL_1519435, EPI_ISL_1519436, EPI_ISL_1519437, EPI_ISL_1519438, EPI_ISL_1519439, EPI_ISL_1519441, EPI_ISL_1519442, EPI_ISL_1519443, EPI_ISL_1519444, EPI_ISL_1519445, EPI_ISL_1519446, EPI_ISL_1519447, EPI_ISL_1519449, EPI_ISL_1519450, EPI_ISL_1519452, EPI_ISL_1519453, EPI_ISL_1519454, EPI_ISL_1519456, EPI_ISL_1519457, EPI_ISL_1519458, EPI_ISL_1519460, EPI_ISL_1519461, EPI_ISL_1519462, EPI_ISL_1519463, EPI_ISL_1519464, EPI_ISL_1519465, EPI_ISL_1519466, EPI_ISL_1519467, EPI_ISL_1519468, EPI_ISL_1519470, EPI_ISL_1519471, EPI_ISL_1519472, EPI_ISL_1519474, EPI_ISL_1519476, EPI_ISL_1519477, EPI_ISL_1519478, EPI_ISL_1519480, EPI_ISL_1519482, EPI_ISL_1519483, EPI_ISL_1519485, EPI_ISL_1519490, EPI_ISL_1519491, EPI_ISL_1519492, EPI_ISL_1519494, EPI_ISL_1519496, EPI_ISL_1519497, EPI_ISL_1519498, EPI_ISL_1519501, EPI_ISL_1519502, EPI_ISL_1519503, EPI_ISL_1519504, EPI_ISL_1519506, EPI_ISL_1519507, EPI_ISL_1519508, EPI_ISL_1519509, EPI_ISL_1519512, EPI_ISL_1519513, EPI_ISL_1519515, EPI_ISL_1519517, EPI_ISL_1519518, EPI_ISL_1519519, EPI_ISL_1519520, EPI_ISL_1519521, EPI_ISL_1519522, EPI_ISL_1519524, EPI_ISL_1519525, EPI_ISL_1519526, EPI_ISL_1519527, EPI_ISL_1519528, EPI_ISL_1519530, EPI_ISL_1519533, EPI_ISL_1519534, EPI_ISL_1519535, EPI_ISL_1519537, EPI_ISL_1519539, EPI_ISL_1519540, EPI_ISL_1519541, EPI_ISL_1519544, EPI_ISL_1519545, EPI_ISL_1519546, EPI_ISL_1519548, EPI_ISL_1519549, EPI_ISL_1519550, EPI_ISL_1519551, EPI_ISL_1519555, EPI_ISL_1519556, EPI_ISL_1519558, EPI_ISL_1519559, EPI_ISL_1519560, EPI_ISL_1519561, EPI_ISL_1519562, EPI_ISL_1519563, EPI_ISL_1519564, EPI_ISL_1519565, EPI_ISL_1519566, EPI_ISL_1519567, EPI_ISL_1519569, EPI_ISL_1519570, EPI_ISL_1519571, EPI_ISL_1519573, EPI_ISL_1519574, EPI_ISL_1519576, EPI_ISL_1519578, EPI_ISL_1519579, EPI_ISL_1519580, EPI_ISL_1519581, EPI_ISL_1519582, EPI_ISL_1519583, EPI_ISL_1519584, EPI_ISL_1519585, EPI_ISL_1519587, EPI_ISL_1519590, EPI_ISL_1519591, EPI_ISL_1519592, EPI_ISL_1519595, EPI_ISL_1519596, EPI_ISL_1519599, EPI_ISL_1519600, EPI_ISL_1519603, EPI_ISL_1519604, EPI_ISL_1519605, EPI_ISL_1519606, EPI_ISL_1519608, EPI_ISL_1519610, EPI_ISL_1519611, EPI_ISL_1519612, EPI_ISL_1519613, EPI_ISL_1519614, EPI_ISL_1519615, EPI_ISL_1519617, EPI_ISL_1519618, EPI_ISL_1519619, EPI_ISL_1519620, EPI_ISL_1519621, EPI_ISL_1519622, EPI_ISL_1519623, EPI_ISL_1519624, EPI_ISL_1519625, EPI_ISL_1519626, EPI_ISL_1519628, EPI_ISL_1519629, EPI_ISL_1519630, EPI_ISL_1519631, EPI_ISL_1519632, EPI_ISL_1519633, EPI_ISL_1519634, EPI_ISL_1519635, EPI_ISL_1519636, EPI_ISL_1519637, EPI_ISL_1519638, EPI_ISL_1519640, EPI_ISL_1519641, EPI_ISL_1519642, EPI_ISL_1519643, EPI_ISL_1519644, EPI_ISL_1519646, EPI_ISL_1519647, EPI_ISL_1519648, EPI_ISL_1519649, EPI_ISL_1519918, EPI_ISL_1519919, EPI_ISL_1519921, EPI_ISL_1519922, EPI_ISL_1519923, EPI_ISL_1519924, EPI_ISL_1519925, EPI_ISL_1535712, EPI_ISL_1536078, EPI_ISL_1536356, EPI_ISL_1536387, EPI_ISL_1536480, EPI_ISL_1536541, EPI_ISL_1536545, EPI_ISL_1536546, EPI_ISL_1536547, EPI_ISL_1536548, EPI_ISL_1536549, EPI_ISL_1536550, EPI_ISL_1536551, EPI_ISL_1536553, EPI_ISL_1536555, EPI_ISL_1536556, EPI_ISL_1536567, EPI_ISL_1536571, EPI_ISL_1536572, EPI_ISL_1536575, EPI_ISL_1536576, EPI_ISL_1536577, EPI_ISL_1536583, EPI_ISL_1536585, EPI_ISL_1536586, EPI_ISL_1536596, EPI_ISL_1536602, EPI_ISL_1536606, EPI_ISL_1536618, EPI_ISL_1536619, EPI_ISL_1536625, EPI_ISL_1536629, EPI_ISL_1536630, EPI_ISL_1536635, EPI_ISL_1536637, EPI_ISL_1536639, EPI_ISL_1536640, EPI_ISL_1536641, EPI_ISL_1536643, EPI_ISL_1536644, EPI_ISL_1536647, EPI_ISL_1536651, EPI_ISL_1536652, EPI_ISL_1536653, EPI_ISL_1536656, EPI_ISL_1536657, EPI_ISL_1536667, EPI_ISL_1536673, EPI_ISL_1536675, EPI_ISL_1536678, EPI_ISL_1536686, EPI_ISL_1536687, EPI_ISL_1536695, EPI_ISL_1536696, EPI_ISL_1536699, EPI_ISL_1536706, EPI_ISL_1536709, EPI_ISL_1536718, EPI_ISL_1536720, EPI_ISL_1536721, EPI_ISL_1536724, EPI_ISL_1536727, EPI_ISL_1536728, EPI_ISL_1536731, EPI_ISL_1536735, EPI_ISL_1536738, EPI_ISL_1536741, EPI_ISL_1536751, EPI_ISL_1536757, EPI_ISL_1536758, EPI_ISL_1536759, EPI_ISL_1536762, EPI_ISL_1536764, EPI_ISL_1536765, EPI_ISL_1536769, EPI_ISL_1536771, EPI_ISL_1536774, EPI_ISL_1536777, EPI_ISL_1536781, EPI_ISL_1536785, EPI_ISL_1536786, EPI_ISL_1536787, EPI_ISL_1536791, EPI_ISL_1536797, EPI_ISL_1536798, EPI_ISL_1536803, EPI_ISL_1536812, EPI_ISL_1536815, EPI_ISL_1536820, EPI_ISL_1536830, EPI_ISL_1536834, EPI_ISL_1536836, EPI_ISL_1536840, EPI_ISL_1537059, EPI_ISL_1537132, EPI_ISL_1537152, EPI_ISL_1537193, EPI_ISL_1537226, EPI_ISL_1537236, EPI_ISL_1537240, EPI_ISL_1537328, EPI_ISL_1538102, EPI_ISL_1538153, EPI_ISL_1538273, EPI_ISL_1538277, EPI_ISL_1538278, EPI_ISL_1538283, EPI_ISL_1538288, EPI_ISL_1538294, EPI_ISL_1538297, EPI_ISL_1538300, EPI_ISL_1538304, EPI_ISL_1538397, EPI_ISL_1544934, EPI_ISL_1544939, EPI_ISL_1630517, EPI_ISL_1630523, EPI_ISL_1630530, EPI_ISL_1630533, EPI_ISL_1630535, EPI_ISL_1630548, EPI_ISL_1630557, EPI_ISL_1630564, EPI_ISL_1630569, EPI_ISL_1630571, EPI_ISL_1630572, EPI_ISL_1630580, EPI_ISL_1630582, EPI_ISL_1630583, EPI_ISL_1630584, EPI_ISL_1630587, EPI_ISL_1630589, EPI_ISL_1630591, EPI_ISL_1630594, EPI_ISL_1630597, EPI_ISL_1630600, EPI_ISL_1630606, EPI_ISL_1630607, EPI_ISL_1630610, EPI_ISL_1630616, EPI_ISL_1630621, EPI_ISL_1630634, EPI_ISL_1630636, EPI_ISL_1630638, EPI_ISL_1630640, EPI_ISL_1630641, EPI_ISL_1630643, EPI_ISL_1630650, EPI_ISL_1630659, EPI_ISL_1630663, EPI_ISL_1630664, EPI_ISL_1630668, EPI_ISL_1630670, EPI_ISL_1630677, EPI_ISL_1630682, EPI_ISL_1630687, EPI_ISL_1630691, EPI_ISL_1630692, EPI_ISL_1630697, EPI_ISL_1630698, EPI_ISL_1630699, EPI_ISL_1630702, EPI_ISL_1630706, EPI_ISL_1630711, EPI_ISL_1630713, EPI_ISL_1630714, EPI_ISL_1630717, EPI_ISL_1630721, EPI_ISL_1630723, EPI_ISL_1630733, EPI_ISL_1630734, EPI_ISL_1630741, EPI_ISL_1630744, EPI_ISL_1630745, EPI_ISL_1630747, EPI_ISL_1630748, EPI_ISL_1630750, EPI_ISL_1630752, EPI_ISL_1630753, EPI_ISL_1630756, EPI_ISL_1630758, EPI_ISL_1630760, EPI_ISL_1630761, EPI_ISL_1630767, EPI_ISL_1630770, EPI_ISL_1630772, EPI_ISL_1630773, EPI_ISL_1630778, EPI_ISL_1630784, EPI_ISL_1630788, EPI_ISL_1630790, EPI_ISL_1630792, EPI_ISL_1630793, EPI_ISL_1630800, EPI_ISL_1630802, EPI_ISL_1630803, EPI_ISL_1630804, EPI_ISL_1630819, EPI_ISL_1635088

| | | | |
|---|---|---|---|
| see above | Lighthouse Lab in Alderley Park | Wellcome Sanger Institute for the COVID-19 Genomics UK (COG-UK) Consortium | Cordelia Langford; David K. Jackson; Dominic Kwiatkowski; Ewan Harrison; Ian Johnston; Jacquelyn Wynn; Jeffrey Barrett; John Sillitoe on behalf of the Wellcome Sanger Institute COVID-19 Surveillance Team; Mairead Hyland; Roberto Amato; Sonia Goncalves; The Lighthouse Lab in Alderley Park and Alex Alderton |

EPI_ISL_1465904, EPI_ISL_1465905, EPI_ISL_1465906, EPI_ISL_1465907, EPI_ISL_1465913, EPI_ISL_1465915, EPI_ISL_1465926, EPI_ISL_1465929, EPI_ISL_1465930, EPI_ISL_1465944, EPI_ISL_1465949, EPI_ISL_1465957, EPI_ISL_1465967, EPI_ISL_1465970, EPI_ISL_1465973, EPI_ISL_1465975, EPI_ISL_1465982, EPI_ISL_1465992, EPI_ISL_1466006, EPI_ISL_1466013, EPI_ISL_1466019, EPI_ISL_1466023, EPI_ISL_1466029, EPI_ISL_1466034, EPI_ISL_1466039, EPI_ISL_1466046, EPI_ISL_1466054, EPI_ISL_1466067, EPI_ISL_1466076, EPI_ISL_1466079, EPI_ISL_1466083, EPI_ISL_1466084, EPI_ISL_1466088, EPI_ISL_1466100, EPI_ISL_1466102, EPI_ISL_1466111, EPI_ISL_1466112, EPI_ISL_1466115, EPI_ISL_1466117, EPI_ISL_1466129, EPI_ISL_1466133, EPI_ISL_1466141, EPI_ISL_1466143, EPI_ISL_1466158, EPI_ISL_1473549, EPI_ISL_1483806, EPI_ISL_1483807, EPI_ISL_1483810, EPI_ISL_1483811, EPI_ISL_1483813, EPI_ISL_1483814, EPI_ISL_1483818, EPI_ISL_1483819, EPI_ISL_1483821, EPI_ISL_1483822, EPI_ISL_1483823, EPI_ISL_1483824, EPI_ISL_1483826, EPI_ISL_1483832, EPI_ISL_1483836, EPI_ISL_1483842, EPI_ISL_1483845, EPI_ISL_1483847, EPI_ISL_1483849, EPI_ISL_1483851, EPI_ISL_1483852, EPI_ISL_1483854, EPI_ISL_1483856, EPI_ISL_1483857, EPI_ISL_1483859, EPI_ISL_1483867, EPI_ISL_1483873, EPI_ISL_1483874, EPI_ISL_1483876, EPI_ISL_1483878, EPI_ISL_1483881, EPI_ISL_1483885, EPI_ISL_1483888, EPI_ISL_1483890, EPI_ISL_1483891, EPI_ISL_1483894, EPI_ISL_1483898, EPI_ISL_1483899, EPI_ISL_1483901, EPI_ISL_1483904, EPI_ISL_1483906, EPI_ISL_1483911, EPI_ISL_1483917, EPI_ISL_1483918, EPI_ISL_1483920, EPI_ISL_1483929, EPI_ISL_1483938, EPI_ISL_1483954, EPI_ISL_1483957, EPI_ISL_1483959, EPI_ISL_1483963, EPI_ISL_1483964, EPI_ISL_1483965, EPI_ISL_1483968, EPI_ISL_1483970, EPI_ISL_1483972, EPI_ISL_1483974, EPI_ISL_1483983, EPI_ISL_1483984, EPI_ISL_1483986, EPI_ISL_1483999, EPI_ISL_1484003, EPI_ISL_1484011, EPI_ISL_1484015, EPI_ISL_1484024, EPI_ISL_1484029, EPI_ISL_1484031, EPI_ISL_1484034, EPI_ISL_1484045, EPI_ISL_1484050, EPI_ISL_1484053, EPI_ISL_1484054, EPI_ISL_1484058, EPI_ISL_1484059, EPI_ISL_1484063, EPI_ISL_1484068, EPI_ISL_1484069, EPI_ISL_1484070, EPI_ISL_1484074, EPI_ISL_1484075, EPI_ISL_1484076, EPI_ISL_1484080, EPI_ISL_1484081, EPI_ISL_1484082, EPI_ISL_1484087, EPI_ISL_1484089, EPI_ISL_1484452, EPI_ISL_1487163, EPI_ISL_1487176, EPI_ISL_1487181, EPI_ISL_1487206, EPI_ISL_1487208, EPI_ISL_1487210, EPI_ISL_1487255, EPI_ISL_1487270, EPI_ISL_1487288, EPI_ISL_1487291, EPI_ISL_1487306, EPI_ISL_1487309, EPI_ISL_1487310, EPI_ISL_1487316, EPI_ISL_1487318, EPI_ISL_1487320, EPI_ISL_1487355, EPI_ISL_1487376, EPI_ISL_1487394, EPI_ISL_1487399, EPI_ISL_1487402, EPI_ISL_1487416, EPI_ISL_1487423, EPI_ISL_1487424, EPI_ISL_1487435, EPI_ISL_1487441, EPI_ISL_1487446, EPI_ISL_1487453, EPI_ISL_1487457, EPI_ISL_1487464, EPI_ISL_1487468, EPI_ISL_1487475, EPI_ISL_1487479, EPI_ISL_1487480, EPI_ISL_1487485, EPI_ISL_1487490, EPI_ISL_1487496, EPI_ISL_1487507, EPI_ISL_1487525, EPI_ISL_1487526, EPI_ISL_1487531, EPI_ISL_1487539, EPI_ISL_1487547, EPI_ISL_1487550, EPI_ISL_1487562, EPI_ISL_1487572, EPI_ISL_1487581, EPI_ISL_1487585, EPI_ISL_1487592, EPI_ISL_1487597, EPI_ISL_1487607, EPI_ISL_1487616, EPI_ISL_1487621, EPI_ISL_1487625, EPI_ISL_1487629, EPI_ISL_1487635, EPI_ISL_1487637, EPI_ISL_1487640, EPI_ISL_1487641, EPI_ISL_1487660, EPI_ISL_1487696, EPI_ISL_1487698, EPI_ISL_1487700, EPI_ISL_1487701, EPI_ISL_1487708, EPI_ISL_1487712, EPI_ISL_1488521, EPI_ISL_1488529, EPI_ISL_1488535, EPI_ISL_1488541, EPI_ISL_1488550, EPI_ISL_1488555, EPI_ISL_1488565, EPI_ISL_1488581, EPI_ISL_1488585, EPI_ISL_1488594, EPI_ISL_1488598, EPI_ISL_1488613, EPI_ISL_1488614, EPI_ISL_1488632, EPI_ISL_1488640, EPI_ISL_1488647, EPI_ISL_1488684, EPI_ISL_1488695, EPI_ISL_1488714, EPI_ISL_1488719, EPI_ISL_1488722, EPI_ISL_1488728, EPI_ISL_1488737, EPI_ISL_1488751, EPI_ISL_1488752, EPI_ISL_1488756, EPI_ISL_1488764, EPI_ISL_1488770, EPI_ISL_1488810, EPI_ISL_1488823, EPI_ISL_1488827, EPI_ISL_1488834, EPI_ISL_1488835, EPI_ISL_1488858, EPI_ISL_1489163, EPI_ISL_1489167, EPI_ISL_1489173, EPI_ISL_1489174, EPI_ISL_1489181, EPI_ISL_1489184, EPI_ISL_1489191, EPI_ISL_1489194, EPI_ISL_1489197, EPI_ISL_1489199, EPI_ISL_1489202, EPI_ISL_1489204, EPI_ISL_1489220, EPI_ISL_1489231, EPI_ISL_1489232, EPI_ISL_1489233, EPI_ISL_1489237, EPI_ISL_1489248, EPI_ISL_1489251, EPI_ISL_1489256, EPI_ISL_1489257, EPI_ISL_1489264, EPI_ISL_1489272, EPI_ISL_1489275, EPI_ISL_1489288, EPI_ISL_1489295, EPI_ISL_1489307, EPI_ISL_1489311, EPI_ISL_1489313, EPI_ISL_1489323, EPI_ISL_1489325, EPI_ISL_1489330, EPI_ISL_1489334, EPI_ISL_1489338, EPI_ISL_1489344, EPI_ISL_1489346, EPI_ISL_1489356, EPI_ISL_1489359, EPI_ISL_1489360, EPI_ISL_1489369, EPI_ISL_1489378, EPI_ISL_1489379, EPI_ISL_1489382, EPI_ISL_1489388, EPI_ISL_1489391, EPI_ISL_1489392, EPI_ISL_1489394, EPI_ISL_1489401, EPI_ISL_1489403, EPI_ISL_1489407, EPI_ISL_1489413, EPI_ISL_1489421, EPI_ISL_1489422, EPI_ISL_1489430, EPI_ISL_1489436, EPI_ISL_1489439, EPI_ISL_1489440, EPI_ISL_1489443, EPI_ISL_1489451, EPI_ISL_1489455, EPI_ISL_1504668, EPI_ISL_1504669, EPI_ISL_1504670, EPI_ISL_1504671, EPI_ISL_1504673, EPI_ISL_1504674, EPI_ISL_1504675, EPI_ISL_1504678, EPI_ISL_1504679, EPI_ISL_1504688, EPI_ISL_1504689, EPI_ISL_1504691, EPI_ISL_1504695, EPI_ISL_1504699, EPI_ISL_1504701, EPI_ISL_1504702, EPI_ISL_1504704, EPI_ISL_1504705, EPI_ISL_1504708, EPI_ISL_1504715, EPI_ISL_1504718, EPI_ISL_1504719, EPI_ISL_1504720, EPI_ISL_1504722, EPI_ISL_1504725, EPI_ISL_1504726, EPI_ISL_1504733, EPI_ISL_1504735, EPI_ISL_1504737, EPI_ISL_1504738, EPI_ISL_1504739, EPI_ISL_1504746, EPI_ISL_1504759, EPI_ISL_1504760, EPI_ISL_1504774, EPI_ISL_1504777, EPI_ISL_1504780, EPI_ISL_1504781, EPI_ISL_1504782, EPI_ISL_1504785, EPI_ISL_1504787, EPI_ISL_1504793, EPI_ISL_1504794, EPI_ISL_1504796, EPI_ISL_1504797, EPI_ISL_1504798, EPI_ISL_1504799, EPI_ISL_1504800, EPI_ISL_1504802, EPI_ISL_1504803, EPI_ISL_1504809, EPI_ISL_1504810, EPI_ISL_1504811, EPI_ISL_1504812, EPI_ISL_1504818, EPI_ISL_1504821, EPI_ISL_1504823, EPI_ISL_1504825, EPI_ISL_1504828, EPI_ISL_1504830, EPI_ISL_1504837, EPI_ISL_1504839, EPI_ISL_1504841, EPI_ISL_1504842, EPI_ISL_1504843, EPI_ISL_1504845, EPI_ISL_1504850, EPI_ISL_1504856, EPI_ISL_1504857, EPI_ISL_1504860, EPI_ISL_1504862, EPI_ISL_1504865, EPI_ISL_1504866, EPI_ISL_1504867, EPI_ISL_1504869, EPI_ISL_1504870, EPI_ISL_1504872, EPI_ISL_1504873, EPI_ISL_1504874, EPI_ISL_1504876, EPI_ISL_1504881, EPI_ISL_1504884, EPI_ISL_1504885, EPI_ISL_1504891, EPI_ISL_1504896, EPI_ISL_1504897, EPI_ISL_1504898, EPI_ISL_1504899, EPI_ISL_1504901, EPI_ISL_1504904, EPI_ISL_1504907, EPI_ISL_1504908, EPI_ISL_1504909, EPI_ISL_1504917, EPI_ISL_1504923, EPI_ISL_1504933, EPI_ISL_1504939, EPI_ISL_1504941, EPI_ISL_1504943, EPI_ISL_1504948, EPI_ISL_1504957, EPI_ISL_1504959, EPI_ISL_1504961, EPI_ISL_1504964, EPI_ISL_1504966, EPI_ISL_1504979, EPI_ISL_1504980, EPI_ISL_1504981, EPI_ISL_1504987, EPI_ISL_1504988, EPI_ISL_1504991, EPI_ISL_1504996, EPI_ISL_1505004, EPI_ISL_1505014, EPI_ISL_1505017, EPI_ISL_1505018, EPI_ISL_1505028, EPI_ISL_1505034, EPI_ISL_1505036, EPI_ISL_1505037, EPI_ISL_1505043, EPI_ISL_1505045, EPI_ISL_1505049, EPI_ISL_1505050, EPI_ISL_1505061, EPI_ISL_1505070, EPI_ISL_1505077, EPI_ISL_1505080, EPI_ISL_1505082, EPI_ISL_1505084, EPI_ISL_1505090, EPI_ISL_1505095, EPI_ISL_1505096, EPI_ISL_1505108, EPI_ISL_1505111, EPI_ISL_1505129, EPI_ISL_1505138, EPI_ISL_1505139, EPI_ISL_1505140, EPI_ISL_1505144, EPI_ISL_1505147, EPI_ISL_1505159, EPI_ISL_1505165, EPI_ISL_1505170, EPI_ISL_1506139, EPI_ISL_1506145, EPI_ISL_1506157, EPI_ISL_1506158, EPI_ISL_1506159, EPI_ISL_1506165, EPI_ISL_1506175, EPI_ISL_1517894, EPI_ISL_1517921, EPI_ISL_1517932, EPI_ISL_1517949, EPI_ISL_1517966, EPI_ISL_1518006, EPI_ISL_1518012, EPI_ISL_1518014, EPI_ISL_1518044, EPI_ISL_1519271, EPI_ISL_1519274, EPI_ISL_1519278, EPI_ISL_1519285, EPI_ISL_1519288, EPI_ISL_1519289, EPI_ISL_1519290, EPI_ISL_1519292, EPI_ISL_1519297, EPI_ISL_1519298, EPI_ISL_1519299, EPI_ISL_1519303, EPI_ISL_1519304, EPI_ISL_1519305, EPI_ISL_1534917, EPI_ISL_1537321, EPI_ISL_1537324, EPI_ISL_1538020, EPI_ISL_1538029, EPI_ISL_1538036, EPI_ISL_1538039, EPI_ISL_1538043, EPI_ISL_1538053, EPI_ISL_1538057, EPI_ISL_1538067, EPI_ISL_1538069, EPI_ISL_1538073, EPI_ISL_1538078, EPI_ISL_1538080, EPI_ISL_1538088, EPI_ISL_1538090, EPI_ISL_1538091, EPI_ISL_1538093, EPI_ISL_1544783, EPI_ISL_1583993, EPI_ISL_1742308, EPI_ISL_1742309

| | | | |
|---|---|---|---|
| see above | Lighthouse Lab in Cambridge | Wellcome Sanger Institute for the COVID-19 Genomics UK (COG-UK) Consortium | Cordelia Langford; David K. Jackson; Dominic Kwiatkowski; Ewan Harrison; Ian Johnston; Jeffrey Barrett; John Sillitoe on behalf of the Wellcome Sanger Institute COVID-19 Surveillance Team; Rob Howes; Roberto Amato; Sonia Goncalves; The Lighthouse Lab in Cambridge and Alex Alderton |

EPI_ISL_1466867, EPI_ISL_1472948, EPI_ISL_1472954, EPI_ISL_1472957, EPI_ISL_1472958, EPI_ISL_1472971, EPI_ISL_1472974, EPI_ISL_1472979, EPI_ISL_1473002, EPI_ISL_1473013, EPI_ISL_1473019, EPI_ISL_1473025, EPI_ISL_1473030, EPI_ISL_1473035, EPI_ISL_1473051, EPI_ISL_1473064, EPI_ISL_1473078, EPI_ISL_1473080, EPI_ISL_1473082, EPI_ISL_1473087, EPI_ISL_1473096, EPI_ISL_1473102, EPI_ISL_1473125, EPI_ISL_1473129, EPI_ISL_1473133, EPI_ISL_1473150, EPI_ISL_1473151, EPI_ISL_1473155, EPI_ISL_1473164, EPI_ISL_1473167, EPI_ISL_1473195, EPI_ISL_1473197, EPI_ISL_1473204, EPI_ISL_1473207, EPI_ISL_1473208, EPI_ISL_1473210, EPI_ISL_1473211, EPI_ISL_1473217, EPI_ISL_1473218, EPI_ISL_1473220, EPI_ISL_1473224, EPI_ISL_1473225, EPI_ISL_1473226, EPI_ISL_1473228, EPI_ISL_1473232, EPI_ISL_1473234, EPI_ISL_1473235, EPI_ISL_1473236, EPI_ISL_1473244, EPI_ISL_1473250, EPI_ISL_1473253, EPI_ISL_1473255, EPI_ISL_1473257, EPI_ISL_1473260, EPI_ISL_1473261, EPI_ISL_1473262, EPI_ISL_1473264, EPI_ISL_1473265, EPI_ISL_1473270, EPI_ISL_1473281, EPI_ISL_1473282, EPI_ISL_1473287, EPI_ISL_1473289, EPI_ISL_1473290, EPI_ISL_1473292, EPI_ISL_1473293, EPI_ISL_1473294, EPI_ISL_1473295, EPI_ISL_1473296, EPI_ISL_1473297, EPI_ISL_1473300, EPI_ISL_1473301, EPI_ISL_1473304, EPI_ISL_1473306, EPI_ISL_1473310, EPI_ISL_1473312, EPI_ISL_1473314, EPI_ISL_1473315, EPI_ISL_1473319, EPI_ISL_1473320, EPI_ISL_1473322, EPI_ISL_1473325, EPI_ISL_1473326, EPI_ISL_1473327, EPI_ISL_1473329, EPI_ISL_1473336, EPI_ISL_1473338, EPI_ISL_1473344, EPI_ISL_1473347, EPI_ISL_1473348, EPI_ISL_1473350, EPI_ISL_1473351, EPI_ISL_1473352, EPI_ISL_1473353, EPI_ISL_1473356, EPI_ISL_1473358, EPI_ISL_1473359, EPI_ISL_1473362, EPI_ISL_1473364, EPI_ISL_1473365, EPI_ISL_1473374, EPI_ISL_1473375, EPI_ISL_1473380, EPI_ISL_1473382, EPI_ISL_1473385, EPI_ISL_1473386, EPI_ISL_1473391, EPI_ISL_1473393, EPI_ISL_1473394, EPI_ISL_1473397, EPI_ISL_1473401, EPI_ISL_1473403, EPI_ISL_1473405, EPI_ISL_1473409, EPI_ISL_1473413, EPI_ISL_1473414, EPI_ISL_1473416, EPI_ISL_1473419, EPI_ISL_1473420, EPI_ISL_1473421, EPI_ISL_1473427, EPI_ISL_1473429, EPI_ISL_1473432, EPI_ISL_1473433, EPI_ISL_1473440, EPI_ISL_1473445, EPI_ISL_1473446, EPI_ISL_1483804, EPI_ISL_1483805, EPI_ISL_1483808, EPI_ISL_1483820, EPI_ISL_1483825, EPI_ISL_1483835, EPI_ISL_1483838, EPI_ISL_1483841, EPI_ISL_1483844, EPI_ISL_1483853, EPI_ISL_1483855, EPI_ISL_1483861, EPI_ISL_1483862, EPI_ISL_1483870, EPI_ISL_1483871, EPI_ISL_1483872, EPI_ISL_1483875, EPI_ISL_1483877, EPI_ISL_1483887, EPI_ISL_1483889, EPI_ISL_1483892, EPI_ISL_1483896, EPI_ISL_1483902, EPI_ISL_1483910, EPI_ISL_1483916, EPI_ISL_1483919, EPI_ISL_1483922, EPI_ISL_1483925, EPI_ISL_1483928, EPI_ISL_1483930, EPI_ISL_1483933, EPI_ISL_1483937, EPI_ISL_1483939, EPI_ISL_1483941, EPI_ISL_1483946, EPI_ISL_1483951, EPI_ISL_1483955, EPI_ISL_1483977, EPI_ISL_1483988, EPI_ISL_1483989, EPI_ISL_1484000, EPI_ISL_1484005, EPI_ISL_1484008, EPI_ISL_1484009, EPI_ISL_1484018, EPI_ISL_1484021, EPI_ISL_1484027, EPI_ISL_1484028, EPI_ISL_1484035, EPI_ISL_1484040, EPI_ISL_1484046, EPI_ISL_1484049, EPI_ISL_1484051, EPI_ISL_1484056, EPI_ISL_1484060, EPI_ISL_1484062, EPI_ISL_1484064, EPI_ISL_1484078, EPI_ISL_1484085, EPI_ISL_1484088, EPI_ISL_1484092, EPI_ISL_1484093, EPI_ISL_1484096, EPI_ISL_1484099, EPI_ISL_1484101, EPI_ISL_1484102, EPI_ISL_1484104, EPI_ISL_1484105, EPI_ISL_1484107, EPI_ISL_1484108, EPI_ISL_1484109, EPI_ISL_1484110, EPI_ISL_1484111, EPI_ISL_1484116, EPI_ISL_1484117, EPI_ISL_1484120, EPI_ISL_1484121, EPI_ISL_1484123, EPI_ISL_1484128, EPI_ISL_1484132, EPI_ISL_1484136, EPI_ISL_1484138, EPI_ISL_1484139, EPI_ISL_1484140, EPI_ISL_1484141, EPI_ISL_1484143, EPI_ISL_1484144, EPI_ISL_1484145, EPI_ISL_1484146, EPI_ISL_1484147, EPI_ISL_1484148, EPI_ISL_1484150, EPI_ISL_1484152, EPI_ISL_1484153, EPI_ISL_1484154, EPI_ISL_1484156, EPI_ISL_1484157, EPI_ISL_1484158, EPI_ISL_1484159, EPI_ISL_1484161, EPI_ISL_1484162, EPI_ISL_1484163, EPI_ISL_1484164, EPI_ISL_1484166, EPI_ISL_1484167, EPI_ISL_1484168, EPI_ISL_1484171, EPI_ISL_1484174, EPI_ISL_1484176, EPI_ISL_1484177, EPI_ISL_1484178, EPI_ISL_1484179, EPI_ISL_1484181, EPI_ISL_1484182, EPI_ISL_1484183, EPI_ISL_1484184, EPI_ISL_1484185, EPI_ISL_1484189, EPI_ISL_1484192, EPI_ISL_1484196, EPI_ISL_1484197, EPI_ISL_1484200, EPI_ISL_1484201, EPI_ISL_1484205, EPI_ISL_1484206, EPI_ISL_1484208, EPI_ISL_1484209, EPI_ISL_1484211, EPI_ISL_1484212, EPI_ISL_1484220, EPI_ISL_1484223, EPI_ISL_1484224, EPI_ISL_1484225, EPI_ISL_1484226, EPI_ISL_1484230, EPI_ISL_1484231, EPI_ISL_1484232, EPI_ISL_1484234, EPI_ISL_1484235, EPI_ISL_1484236, EPI_ISL_1484237, EPI_ISL_1484238, EPI_ISL_1484239, EPI_ISL_1484240, EPI_ISL_1484241, EPI_ISL_1484242, EPI_ISL_1484243, EPI_ISL_1484244, EPI_ISL_1484245, EPI_ISL_1484247, EPI_ISL_1484248, EPI_ISL_1484249, EPI_ISL_1484250, EPI_ISL_1484253, EPI_ISL_1484255, EPI_ISL_1484259, EPI_ISL_1484262, EPI_ISL_1484263, EPI_ISL_1484266, EPI_ISL_1484267, EPI_ISL_1484268, EPI_ISL_1484272, EPI_ISL_1484273, EPI_ISL_1484274, EPI_ISL_1484276, EPI_ISL_1484277, EPI_ISL_1484278, EPI_ISL_1484279, EPI_ISL_1484280, EPI_ISL_1484281, EPI_ISL_1484284, EPI_ISL_1484285, EPI_ISL_1484286, EPI_ISL_1484287, EPI_ISL_1484288, EPI_ISL_1484290, EPI_ISL_1484291, EPI_ISL_1484292, EPI_ISL_1484294, EPI_ISL_1484295, EPI_ISL_1484296, EPI_ISL_1484298, EPI_ISL_1484300, EPI_ISL_1484301, EPI_ISL_1484303, EPI_ISL_1484304, EPI_ISL_1484305, EPI_ISL_1484306, EPI_ISL_1484307, EPI_ISL_1484309, EPI_ISL_1484310, EPI_ISL_1484312, EPI_ISL_1484313, EPI_ISL_1484611, EPI_ISL_1484614, EPI_ISL_1484615, EPI_ISL_1484617, EPI_ISL_1484619, EPI_ISL_1484621, EPI_ISL_1484627, EPI_ISL_1484629, EPI_ISL_1484631, EPI_ISL_1484633, EPI_ISL_1484634, EPI_ISL_1484637, EPI_ISL_1484641, EPI_ISL_1484645, EPI_ISL_1484648, EPI_ISL_1484649, EPI_ISL_1484650, EPI_ISL_1484651, EPI_ISL_1484654, EPI_ISL_1484656, EPI_ISL_1484658, EPI_ISL_1484659, EPI_ISL_1484665, EPI_ISL_1484667, EPI_ISL_1484675, EPI_ISL_1484680, EPI_ISL_1484685, EPI_ISL_1484687, EPI_ISL_1484691, EPI_ISL_1484698, EPI_ISL_1484706, EPI_ISL_1484708, EPI_ISL_1484710, EPI_ISL_1484712, EPI_ISL_1484717, EPI_ISL_1484718, EPI_ISL_1484719, EPI_ISL_1484720, EPI_ISL_1484728, EPI_ISL_1484733, EPI_ISL_1484739, EPI_ISL_1484747, EPI_ISL_1484754, EPI_ISL_1484759, EPI_ISL_1484762, EPI_ISL_1484764, EPI_ISL_1484765, EPI_ISL_1484771, EPI_ISL_1484772, EPI_ISL_1484773, EPI_ISL_1484781, EPI_ISL_1484788, EPI_ISL_1484789, EPI_ISL_1484794, EPI_ISL_1484795, EPI_ISL_1484797, EPI_ISL_1484800, EPI_ISL_1484803, EPI_ISL_1484807, EPI_ISL_1484813, EPI_ISL_1484815, EPI_ISL_1484818, EPI_ISL_1484827, EPI_ISL_1484830, EPI_ISL_1484834, EPI_ISL_1484836, EPI_ISL_1485182, EPI_ISL_1485183, EPI_ISL_1485184, EPI_ISL_1485191, EPI_ISL_1485195, EPI_ISL_1485196, EPI_ISL_1485199, EPI_ISL_1485204, EPI_ISL_1485211, EPI_ISL_1485213, EPI_ISL_1485214, EPI_ISL_1485216, EPI_ISL_1485218, EPI_ISL_1485224, EPI_ISL_1485226, EPI_ISL_1485227, EPI_ISL_1485228, EPI_ISL_1485230, EPI_ISL_1485234, EPI_ISL_1485237, EPI_ISL_1485238, EPI_ISL_1485240, EPI_ISL_1485241, EPI_ISL_1485249, EPI_ISL_1485252, EPI_ISL_1485260, EPI_ISL_1485262, EPI_ISL_1485263, EPI_ISL_1485264, EPI_ISL_1485266, EPI_ISL_1485270, EPI_ISL_1485273, EPI_ISL_1485276, EPI_ISL_1485290, EPI_ISL_1485292, EPI_ISL_1485298, EPI_ISL_1485301, EPI_ISL_1485303, EPI_ISL_1485307, EPI_ISL_1485313, EPI_ISL_1485315, EPI_ISL_1485321, EPI_ISL_1485325, EPI_ISL_1485326, EPI_ISL_1485327, EPI_ISL_1485328, EPI_ISL_1485329, EPI_ISL_1485332, EPI_ISL_1485333, EPI_ISL_1485334, EPI_ISL_1485335, EPI_ISL_1485336, EPI_ISL_1485343, EPI_ISL_1485344, EPI_ISL_1485347, EPI_ISL_1485348, EPI_ISL_1485351, EPI_ISL_1485355, EPI_ISL_1485361, EPI_ISL_1485363, EPI_ISL_1485367, EPI_ISL_1485371, EPI_ISL_1485372, EPI_ISL_1485375, EPI_ISL_1485378, EPI_ISL_1485379, EPI_ISL_1485381, EPI_ISL_1485382, EPI_ISL_1485385, EPI_ISL_1485388, EPI_ISL_1485395, EPI_ISL_1485398, EPI_ISL_1485399, EPI_ISL_1485406, EPI_ISL_1485412, EPI_ISL_1485414, EPI_ISL_1485418, EPI_ISL_1485419, EPI_ISL_1485422, EPI_ISL_1485423, EPI_ISL_1485424, EPI_ISL_1485425, EPI_ISL_1485426, EPI_ISL_1485428, EPI_ISL_1485429, EPI_ISL_1485433, EPI_ISL_1485438, EPI_ISL_1485447, EPI_ISL_1485455, EPI_ISL_1485456, EPI_ISL_1485457, EPI_ISL_1485458, EPI_ISL_1485460, EPI_ISL_1485461, EPI_ISL_1485467, EPI_ISL_1485470, EPI_ISL_1485472, EPI_ISL_1485475, EPI_ISL_1485810, EPI_ISL_1485813, EPI_ISL_1485815, EPI_ISL_1485832, EPI_ISL_1485835, EPI_ISL_1485856, EPI_ISL_1485863, EPI_ISL_1485866, EPI_ISL_1485871, EPI_ISL_1485872, EPI_ISL_1485876, EPI_ISL_1485879, EPI_ISL_1485892, EPI_ISL_1485898, EPI_ISL_1485910, EPI_ISL_1485936, EPI_ISL_1485947, EPI_ISL_1485962, EPI_ISL_1485966, EPI_ISL_1485973, EPI_ISL_1485985, EPI_ISL_1485992, EPI_ISL_1485999, EPI_ISL_1486012, EPI_ISL_1486024, EPI_ISL_1486028, EPI_ISL_1486030, EPI_ISL_1486045, EPI_ISL_1486054, EPI_ISL_1486058, EPI_ISL_1486059, EPI_ISL_1486060, EPI_ISL_1486065, EPI_ISL_1486067, EPI_ISL_1486071, EPI_ISL_1486075, EPI_ISL_1486079, EPI_ISL_1486087, EPI_ISL_1486091, EPI_ISL_1486092, EPI_ISL_1486097, EPI_ISL_1486107, EPI_ISL_1486118, EPI_ISL_1486125, EPI_ISL_1486127, EPI_ISL_1486140, EPI_ISL_1486142, EPI_ISL_1486149, EPI_ISL_1486152, EPI_ISL_1486153, EPI_ISL_1486160, EPI_ISL_1486168, EPI_ISL_1486172, EPI_ISL_1486175, EPI_ISL_1486176, EPI_ISL_1486181, EPI_ISL_1486182, EPI_ISL_1486190, EPI_ISL_1486198, EPI_ISL_1486199, EPI_ISL_1486204, EPI_ISL_1486211, EPI_ISL_1486213, EPI_ISL_1486218, EPI_ISL_1486229, EPI_ISL_1486234, EPI_ISL_1486237, EPI_ISL_1486238, EPI_ISL_1486245, EPI_ISL_1486246, EPI_ISL_1486248, EPI_ISL_1486250, EPI_ISL_1486253, EPI_ISL_1486263, EPI_ISL_1486272, EPI_ISL_1486275, EPI_ISL_1486277, EPI_ISL_1486279, EPI_ISL_1486285, EPI_ISL_1486291, EPI_ISL_1486292, EPI_ISL_1486297, EPI_ISL_1486299, EPI_ISL_1486300, EPI_ISL_1486305, EPI_ISL_1486317, EPI_ISL_1486319, EPI_ISL_1486326, EPI_ISL_1486329, EPI_ISL_1486330, EPI_ISL_1486334, EPI_ISL_1486350, EPI_ISL_1486361, EPI_ISL_1486372, EPI_ISL_1486375, EPI_ISL_1486379, EPI_ISL_1486388, EPI_ISL_1486391, EPI_ISL_1486392, EPI_ISL_1487159, EPI_ISL_1487160, EPI_ISL_1487161, EPI_ISL_1487162, EPI_ISL_1487164, EPI_ISL_1487165, EPI_ISL_1487166, EPI_ISL_1487167, EPI_ISL_1487169, EPI_ISL_1487170, EPI_ISL_1487171, EPI_ISL_1487172, EPI_ISL_1487173, EPI_ISL_1487174, EPI_ISL_1487175, EPI_ISL_1487177, EPI_ISL_1487178, EPI_ISL_1487179, EPI_ISL_1487180, EPI_ISL_1487182, EPI_ISL_1487183, EPI_ISL_1487184, EPI_ISL_1487185, EPI_ISL_1487186, EPI_ISL_1487187, EPI_ISL_1487188, EPI_ISL_1487189, EPI_ISL_1487190, EPI_ISL_1487191, EPI_ISL_1487193, EPI_ISL_1487194, EPI_ISL_1487195, EPI_ISL_1487196, EPI_ISL_1487197, EPI_ISL_1487198, EPI_ISL_1487199, EPI_ISL_1487200, EPI_ISL_1487202, EPI_ISL_1487203, EPI_ISL_1487204, EPI_ISL_1487205, EPI_ISL_1487207, EPI_ISL_1487209, EPI_ISL_1487212, EPI_ISL_1487215, EPI_ISL_1487216, EPI_ISL_1487217, EPI_ISL_1487218, EPI_ISL_1487219, EPI_ISL_1487220, EPI_ISL_1487221, EPI_ISL_1487222, EPI_ISL_1487223, EPI_ISL_1487224, EPI_ISL_1487226, EPI_ISL_1487227, EPI_ISL_1487228, EPI_ISL_1487229, EPI_ISL_1487230, EPI_ISL_1487231, EPI_ISL_1487232, EPI_ISL_1487233, EPI_ISL_1487234, EPI_ISL_1487235, EPI_ISL_1487236, EPI_ISL_1487237, EPI_ISL_1487238, EPI_ISL_1487239, EPI_ISL_1487240, EPI_ISL_1487241, EPI_ISL_1487242, EPI_ISL_1487243, EPI_ISL_1487244, EPI_ISL_1487245, EPI_ISL_1487246, EPI_ISL_1487247, EPI_ISL_1487249, EPI_ISL_1487251, EPI_ISL_1487252, EPI_ISL_1487253, EPI_ISL_1487254, EPI_ISL_1487256, EPI_ISL_1487257, EPI_ISL_1487258, EPI_ISL_1487259, EPI_ISL_1487260, EPI_ISL_1487261, EPI_ISL_1487262, EPI_ISL_1487265, EPI_ISL_1487266, EPI_ISL_1487267, EPI_ISL_1487268, EPI_ISL_1487269, EPI_ISL_1487271, EPI_ISL_1487272, EPI_ISL_1487273, EPI_ISL_1487274, EPI_ISL_1487275, EPI_ISL_1487276, EPI_ISL_1487277, EPI_ISL_1487278, EPI_ISL_1487280, EPI_ISL_1487281, EPI_ISL_1487282, EPI_ISL_1487283, EPI_ISL_1487284, EPI_ISL_1487286, EPI_ISL_1487287, EPI_ISL_1487289, EPI_ISL_1487292, EPI_ISL_1487293, EPI_ISL_1487294, EPI_ISL_1487295, EPI_ISL_1487296, EPI_ISL_1487297, EPI_ISL_1487298, EPI_ISL_1487299, EPI_ISL_1487300, EPI_ISL_1487301, EPI_ISL_1487302, EPI_ISL_1487303, EPI_ISL_1487304, EPI_ISL_1487305, EPI_ISL_1487307, EPI_ISL_1487308, EPI_ISL_1487311, EPI_ISL_1487312, EPI_ISL_1487314, EPI_ISL_1487315, EPI_ISL_1487317, EPI_ISL_1487319, EPI_ISL_1487321, EPI_ISL_1487322, EPI_ISL_1487323, EPI_ISL_1487324, EPI_ISL_1487325, EPI_ISL_1487326, EPI_ISL_1487327, EPI_ISL_1487328, EPI_ISL_1487329, EPI_ISL_1487330, EPI_ISL_1487331, EPI_ISL_1487332, EPI_ISL_1487333, EPI_ISL_1487334, EPI_ISL_1487335, EPI_ISL_1487336, EPI_ISL_1487337, EPI_ISL_1487338, EPI_ISL_1487339, EPI_ISL_1487340, EPI_ISL_1487341, EPI_ISL_1487342, EPI_ISL_1487343, EPI_ISL_1487344, EPI_ISL_1487345, EPI_ISL_1487346, EPI_ISL_1487347, EPI_ISL_1487348, EPI_ISL_1487349, EPI_ISL_1487350, EPI_ISL_1487351, EPI_ISL_1487353, EPI_ISL_1487354, EPI_ISL_1487356, EPI_ISL_1487357, EPI_ISL_1487358, EPI_ISL_1487359, EPI_ISL_1487361, EPI_ISL_1487362, EPI_ISL_1487363, EPI_ISL_1487364, EPI_ISL_1487365, EPI_ISL_1487366, EPI_ISL_1487367, EPI_ISL_1487368, EPI_ISL_1487369, EPI_ISL_1487370, EPI_ISL_1487371, EPI_ISL_1487372, EPI_ISL_1487373, EPI_ISL_1487374, EPI_ISL_1487375, EPI_ISL_1487377, EPI_ISL_1488229, EPI_ISL_1488245, EPI_ISL_1488246, EPI_ISL_1488249, EPI_ISL_1488251, EPI_ISL_1488252, EPI_ISL_1488253, EPI_ISL_1488254, EPI_ISL_1488255, EPI_ISL_1488264, EPI_ISL_1488266, EPI_ISL_1488267, EPI_ISL_1488268, EPI_ISL_1488269, EPI_ISL_1488271, EPI_ISL_1488273, EPI_ISL_1488280, EPI_ISL_1488283, EPI_ISL_1488284, EPI_ISL_1488285, EPI_ISL_1488289, EPI_ISL_1488291, EPI_ISL_1488296, EPI_ISL_1488297, EPI_ISL_1488300, EPI_ISL_1488304, EPI_ISL_1488307, EPI_ISL_1488308, EPI_ISL_1488313, EPI_ISL_1488314, EPI_ISL_1488316, EPI_ISL_1488317, EPI_ISL_1488320, EPI_ISL_1488322, EPI_ISL_1488323, EPI_ISL_1488325, EPI_ISL_1488326, EPI_ISL_1488328, EPI_ISL_1488329, EPI_ISL_1488335, EPI_ISL_1488341, EPI_ISL_1488344, EPI_ISL_1488347, EPI_ISL_1488348, EPI_ISL_1488353, EPI_ISL_1488355, EPI_ISL_1488359, EPI_ISL_1488362, EPI_ISL_1488367, EPI_ISL_1488369, EPI_ISL_1488382, EPI_ISL_1488384, EPI_ISL_1488389, EPI_ISL_1488390, EPI_ISL_1488391, EPI_ISL_1488392, EPI_ISL_1488393, EPI_ISL_1488394, EPI_ISL_1488395, EPI_ISL_1488398, EPI_ISL_1488399, EPI_ISL_1488400, EPI_ISL_1488405, EPI_ISL_1488410, EPI_ISL_1488416, EPI_ISL_1488418, EPI_ISL_1488420, EPI_ISL_1488424, EPI_ISL_1488426, EPI_ISL_1488444, EPI_ISL_1488446, EPI_ISL_1488455, EPI_ISL_1488459, EPI_ISL_1488463, EPI_ISL_1488472, EPI_ISL_1488479, EPI_ISL_1488481, EPI_ISL_1488489, EPI_ISL_1488495, EPI_ISL_1488496, EPI_ISL_1488498, EPI_ISL_1488500, EPI_ISL_1488502, EPI_ISL_1488510, EPI_ISL_1488513, EPI_ISL_1488516, EPI_ISL_1488871, EPI_ISL_1488880, EPI_ISL_1488885, EPI_ISL_1488887, EPI_ISL_1488893, EPI_ISL_1488905, EPI_ISL_1488907, EPI_ISL_1488914, EPI_ISL_1488916, EPI_ISL_1488919, EPI_ISL_1488927, EPI_ISL_1488928, EPI_ISL_1488930, EPI_ISL_1488932, EPI_ISL_1488935, EPI_ISL_1488938,

EPI_ISL_1488944, EPI_ISL_1488948, EPI_ISL_1488953, EPI_ISL_1488954, EPI_ISL_1488959, EPI_ISL_1488960, EPI_ISL_1488961, EPI_ISL_1488965, EPI_ISL_1488971, EPI_ISL_1488974, EPI_ISL_1488976, EPI_ISL_1488977, EPI_ISL_1488980, EPI_ISL_1488987, EPI_ISL_1488989, EPI_ISL_1488990, EPI_ISL_1488991, EPI_ISL_1489002, EPI_ISL_1489006, EPI_ISL_1489007, EPI_ISL_1489008, EPI_ISL_1489015,
EPI_ISL_1489019, EPI_ISL_1489027, EPI_ISL_1489029, EPI_ISL_1489030, EPI_ISL_1489038, EPI_ISL_1489039, EPI_ISL_1489051, EPI_ISL_1489052, EPI_ISL_1489057, EPI_ISL_1489059, EPI_ISL_1489066, EPI_ISL_1489067, EPI_ISL_1489069, EPI_ISL_1489070, EPI_ISL_1489073, EPI_ISL_1489077, EPI_ISL_1489079, EPI_ISL_1489080, EPI_ISL_1489081, EPI_ISL_1489086, EPI_ISL_1489090, EPI_ISL_1489092,
EPI_ISL_1489095, EPI_ISL_1489103, EPI_ISL_1489110, EPI_ISL_1489112, EPI_ISL_1489114, EPI_ISL_1489120, EPI_ISL_1489121, EPI_ISL_1489124, EPI_ISL_1489127, EPI_ISL_1489130, EPI_ISL_1489131, EPI_ISL_1489132, EPI_ISL_1489134, EPI_ISL_1489135, EPI_ISL_1489139, EPI_ISL_1489143, EPI_ISL_1489147, EPI_ISL_1489149, EPI_ISL_1489154, EPI_ISL_1489157, EPI_ISL_1489159, EPI_ISL_1489160,
EPI_ISL_1489162, EPI_ISL_1489164, EPI_ISL_1489166, EPI_ISL_1489168, EPI_ISL_1489169, EPI_ISL_1489170, EPI_ISL_1489172, EPI_ISL_1489176, EPI_ISL_1489177, EPI_ISL_1489178, EPI_ISL_1489179, EPI_ISL_1489182, EPI_ISL_1489186, EPI_ISL_1489193, EPI_ISL_1489196, EPI_ISL_1489198, EPI_ISL_1489201, EPI_ISL_1489203, EPI_ISL_1489209, EPI_ISL_1489210, EPI_ISL_1489213, EPI_ISL_1489216,
EPI_ISL_1489218, EPI_ISL_1489224, EPI_ISL_1489229, EPI_ISL_1489230, EPI_ISL_1489234, EPI_ISL_1489235, EPI_ISL_1489238, EPI_ISL_1489241, EPI_ISL_1489247, EPI_ISL_1489250, EPI_ISL_1489254, EPI_ISL_1489255, EPI_ISL_1489258, EPI_ISL_1489259, EPI_ISL_1489265, EPI_ISL_1489266, EPI_ISL_1489268, EPI_ISL_1489269, EPI_ISL_1489270, EPI_ISL_1489277, EPI_ISL_1489279, EPI_ISL_1489282,
EPI_ISL_1489283, EPI_ISL_1489286, EPI_ISL_1489287, EPI_ISL_1489289, EPI_ISL_1489291, EPI_ISL_1489292, EPI_ISL_1489294, EPI_ISL_1489297, EPI_ISL_1489298, EPI_ISL_1489302, EPI_ISL_1489303, EPI_ISL_1489306, EPI_ISL_1489309, EPI_ISL_1489310, EPI_ISL_1489315, EPI_ISL_1489316, EPI_ISL_1489320, EPI_ISL_1489322, EPI_ISL_1489324, EPI_ISL_1489326, EPI_ISL_1489327, EPI_ISL_1489328,
EPI_ISL_1489333, EPI_ISL_1489335, EPI_ISL_1489337, EPI_ISL_1489339, EPI_ISL_1489341, EPI_ISL_1489342, EPI_ISL_1489348, EPI_ISL_1489349, EPI_ISL_1489351, EPI_ISL_1489354, EPI_ISL_1489357, EPI_ISL_1489361, EPI_ISL_1489363, EPI_ISL_1489364, EPI_ISL_1489366, EPI_ISL_1489367, EPI_ISL_1489370, EPI_ISL_1489371, EPI_ISL_1489372, EPI_ISL_1489373, EPI_ISL_1489374, EPI_ISL_1489377,
EPI_ISL_1489380, EPI_ISL_1489384, EPI_ISL_1489385, EPI_ISL_1489387, EPI_ISL_1489390, EPI_ISL_1489399, EPI_ISL_1489405, EPI_ISL_1489406, EPI_ISL_1489409, EPI_ISL_1489410, EPI_ISL_1489411, EPI_ISL_1489412, EPI_ISL_1489414, EPI_ISL_1489415, EPI_ISL_1489417, EPI_ISL_1489419, EPI_ISL_1489427, EPI_ISL_1489429, EPI_ISL_1489431, EPI_ISL_1489432, EPI_ISL_1489433, EPI_ISL_1489434,
EPI_ISL_1489442, EPI_ISL_1489447, EPI_ISL_1489449, EPI_ISL_1489453, EPI_ISL_1489456, EPI_ISL_1504120, EPI_ISL_1504130, EPI_ISL_1504160, EPI_ISL_1504193, EPI_ISL_1504236, EPI_ISL_1504241, EPI_ISL_1504246, EPI_ISL_1504258, EPI_ISL_1504270, EPI_ISL_1504278, EPI_ISL_1504390, EPI_ISL_1504676, EPI_ISL_1504677, EPI_ISL_1504680, EPI_ISL_1504681, EPI_ISL_1504686, EPI_ISL_1504687,
EPI_ISL_1504697, EPI_ISL_1504700, EPI_ISL_1504709, EPI_ISL_1504711, EPI_ISL_1504714, EPI_ISL_1504727, EPI_ISL_1504729, EPI_ISL_1504731, EPI_ISL_1504736, EPI_ISL_1504740, EPI_ISL_1504741, EPI_ISL_1504747, EPI_ISL_1504752, EPI_ISL_1504754, EPI_ISL_1504762, EPI_ISL_1504763, EPI_ISL_1504764, EPI_ISL_1504765, EPI_ISL_1504770, EPI_ISL_1504778, EPI_ISL_1504783, EPI_ISL_1504786,
EPI_ISL_1504801, EPI_ISL_1504807, EPI_ISL_1504815, EPI_ISL_1504819, EPI_ISL_1504824, EPI_ISL_1504836, EPI_ISL_1504852, EPI_ISL_1504853, EPI_ISL_1504854, EPI_ISL_1504855, EPI_ISL_1504863, EPI_ISL_1504864, EPI_ISL_1504877, EPI_ISL_1504880, EPI_ISL_1504886, EPI_ISL_1504889, EPI_ISL_1504890, EPI_ISL_1504900, EPI_ISL_1504906, EPI_ISL_1505261, EPI_ISL_1505306, EPI_ISL_1505399,
EPI_ISL_1505807, EPI_ISL_1505821, EPI_ISL_1505822, EPI_ISL_1505834, EPI_ISL_1505839, EPI_ISL_1505843, EPI_ISL_1505877, EPI_ISL_1505885, EPI_ISL_1505886, EPI_ISL_1505903, EPI_ISL_1505920, EPI_ISL_1505929, EPI_ISL_1505931, EPI_ISL_1505936, EPI_ISL_1505972, EPI_ISL_1505976, EPI_ISL_1505999, EPI_ISL_1506012, EPI_ISL_1506022, EPI_ISL_1506037, EPI_ISL_1506038, EPI_ISL_1506048,
EPI_ISL_1506056, EPI_ISL_1506072, EPI_ISL_1506083, EPI_ISL_1506134, EPI_ISL_1506135, EPI_ISL_1506136, EPI_ISL_1506137, EPI_ISL_1506138, EPI_ISL_1506140, EPI_ISL_1506141, EPI_ISL_1506142, EPI_ISL_1506143, EPI_ISL_1506146, EPI_ISL_1506147, EPI_ISL_1506148, EPI_ISL_1506149, EPI_ISL_1506150, EPI_ISL_1506151, EPI_ISL_1506152, EPI_ISL_1506153, EPI_ISL_1506154, EPI_ISL_1506155,
EPI_ISL_1506156, EPI_ISL_1506160, EPI_ISL_1506161, EPI_ISL_1506162, EPI_ISL_1506164, EPI_ISL_1506166, EPI_ISL_1506167, EPI_ISL_1506168, EPI_ISL_1506169, EPI_ISL_1506170, EPI_ISL_1506171, EPI_ISL_1506173, EPI_ISL_1506174, EPI_ISL_1506176, EPI_ISL_1506177, EPI_ISL_1506178, EPI_ISL_1506179, EPI_ISL_1506180, EPI_ISL_1506181, EPI_ISL_1506182, EPI_ISL_1506183, EPI_ISL_1506184,
EPI_ISL_1506185, EPI_ISL_1506186, EPI_ISL_1506187, EPI_ISL_1506188, EPI_ISL_1506189, EPI_ISL_1506190, EPI_ISL_1506191, EPI_ISL_1506193, EPI_ISL_1506194, EPI_ISL_1506195, EPI_ISL_1506196, EPI_ISL_1506197, EPI_ISL_1506198, EPI_ISL_1506199, EPI_ISL_1506200, EPI_ISL_1506201, EPI_ISL_1506202, EPI_ISL_1506203, EPI_ISL_1506204, EPI_ISL_1506205, EPI_ISL_1506206, EPI_ISL_1506207,
EPI_ISL_1506208, EPI_ISL_1506209, EPI_ISL_1506210, EPI_ISL_1506211, EPI_ISL_1506212, EPI_ISL_1506213, EPI_ISL_1506214, EPI_ISL_1506215, EPI_ISL_1506216, EPI_ISL_1506217, EPI_ISL_1506218, EPI_ISL_1506219, EPI_ISL_1506220, EPI_ISL_1506221, EPI_ISL_1506222, EPI_ISL_1506223, EPI_ISL_1506224, EPI_ISL_1506225, EPI_ISL_1506226, EPI_ISL_1506227, EPI_ISL_1506228, EPI_ISL_1506229,
EPI_ISL_1506230, EPI_ISL_1506231, EPI_ISL_1506232, EPI_ISL_1506233, EPI_ISL_1506234, EPI_ISL_1506236, EPI_ISL_1506237, EPI_ISL_1506238, EPI_ISL_1506239, EPI_ISL_1506240, EPI_ISL_1506241, EPI_ISL_1506242, EPI_ISL_1506243, EPI_ISL_1506244, EPI_ISL_1506245, EPI_ISL_1506246, EPI_ISL_1506247, EPI_ISL_1506248, EPI_ISL_1506249, EPI_ISL_1506250, EPI_ISL_1506251, EPI_ISL_1506252,
EPI_ISL_1506253, EPI_ISL_1506254, EPI_ISL_1506255, EPI_ISL_1506256, EPI_ISL_1506257, EPI_ISL_1506258, EPI_ISL_1506259, EPI_ISL_1506260, EPI_ISL_1506261, EPI_ISL_1506262, EPI_ISL_1506263, EPI_ISL_1506264, EPI_ISL_1506265, EPI_ISL_1506267, EPI_ISL_1506268, EPI_ISL_1506269, EPI_ISL_1506270, EPI_ISL_1506271, EPI_ISL_1506272, EPI_ISL_1506273, EPI_ISL_1506274, EPI_ISL_1506275,
EPI_ISL_1506276, EPI_ISL_1506279, EPI_ISL_1506280, EPI_ISL_1506281, EPI_ISL_1506282, EPI_ISL_1506283, EPI_ISL_1506284, EPI_ISL_1506285, EPI_ISL_1506286, EPI_ISL_1506287, EPI_ISL_1506288, EPI_ISL_1506289, EPI_ISL_1506290, EPI_ISL_1506291, EPI_ISL_1506293, EPI_ISL_1506294, EPI_ISL_1506295, EPI_ISL_1506296, EPI_ISL_1506297, EPI_ISL_1506298, EPI_ISL_1506299, EPI_ISL_1506300,
EPI_ISL_1506301, EPI_ISL_1506303, EPI_ISL_1506304, EPI_ISL_1506306, EPI_ISL_1506307, EPI_ISL_1506308, EPI_ISL_1506309, EPI_ISL_1506310, EPI_ISL_1506311, EPI_ISL_1506312, EPI_ISL_1506313, EPI_ISL_1506314, EPI_ISL_1506315, EPI_ISL_1506316, EPI_ISL_1506317, EPI_ISL_1506318, EPI_ISL_1506319, EPI_ISL_1506320, EPI_ISL_1506321, EPI_ISL_1506322, EPI_ISL_1506323, EPI_ISL_1506324,
EPI_ISL_1506325, EPI_ISL_1506326, EPI_ISL_1506327, EPI_ISL_1506328, EPI_ISL_1506329, EPI_ISL_1506330, EPI_ISL_1506331, EPI_ISL_1506332, EPI_ISL_1506333, EPI_ISL_1506334, EPI_ISL_1506335, EPI_ISL_1506336, EPI_ISL_1506337, EPI_ISL_1506338, EPI_ISL_1506339, EPI_ISL_1506340, EPI_ISL_1506341, EPI_ISL_1506342, EPI_ISL_1506343, EPI_ISL_1506344, EPI_ISL_1506345, EPI_ISL_1506346,
EPI_ISL_1506347, EPI_ISL_1506348, EPI_ISL_1506349, EPI_ISL_1506350, EPI_ISL_1506351, EPI_ISL_1506352, EPI_ISL_1506354, EPI_ISL_1506355, EPI_ISL_1506356, EPI_ISL_1506357, EPI_ISL_1506358, EPI_ISL_1506359, EPI_ISL_1506360, EPI_ISL_1506361, EPI_ISL_1506362, EPI_ISL_1506363, EPI_ISL_1506364, EPI_ISL_1506365, EPI_ISL_1506366, EPI_ISL_1506368, EPI_ISL_1506369, EPI_ISL_1506370,
EPI_ISL_1506371, EPI_ISL_1506372, EPI_ISL_1506373, EPI_ISL_1506374, EPI_ISL_1506375, EPI_ISL_1506376, EPI_ISL_1506377, EPI_ISL_1506378, EPI_ISL_1506379, EPI_ISL_1506380, EPI_ISL_1506381, EPI_ISL_1506382, EPI_ISL_1506383, EPI_ISL_1506384, EPI_ISL_1506385, EPI_ISL_1506386, EPI_ISL_1506387, EPI_ISL_1506388, EPI_ISL_1506389, EPI_ISL_1506390, EPI_ISL_1506391, EPI_ISL_1506392,
EPI_ISL_1506393, EPI_ISL_1506394, EPI_ISL_1506395, EPI_ISL_1506396, EPI_ISL_1506397, EPI_ISL_1506398, EPI_ISL_1506399, EPI_ISL_1506400, EPI_ISL_1506401, EPI_ISL_1506402, EPI_ISL_1506403, EPI_ISL_1506404, EPI_ISL_1506405, EPI_ISL_1506406, EPI_ISL_1506408, EPI_ISL_1506409, EPI_ISL_1506410, EPI_ISL_1506411, EPI_ISL_1506412, EPI_ISL_1506413, EPI_ISL_1506414, EPI_ISL_1506415,
EPI_ISL_1506416, EPI_ISL_1506417, EPI_ISL_1506418, EPI_ISL_1506419, EPI_ISL_1506420, EPI_ISL_1506421, EPI_ISL_1506422, EPI_ISL_1506423, EPI_ISL_1506428, EPI_ISL_1506429, EPI_ISL_1506430, EPI_ISL_1506431, EPI_ISL_1506432, EPI_ISL_1506433, EPI_ISL_1506435, EPI_ISL_1506436, EPI_ISL_1506437, EPI_ISL_1506438, EPI_ISL_1506439, EPI_ISL_1506441, EPI_ISL_1506442, EPI_ISL_1506443,
EPI_ISL_1506444, EPI_ISL_1506445, EPI_ISL_1506446, EPI_ISL_1506447, EPI_ISL_1506448, EPI_ISL_1506450, EPI_ISL_1506451, EPI_ISL_1506452, EPI_ISL_1506453, EPI_ISL_1506454, EPI_ISL_1506455, EPI_ISL_1506456, EPI_ISL_1506457, EPI_ISL_1506458, EPI_ISL_1506459, EPI_ISL_1506460, EPI_ISL_1506462, EPI_ISL_1506463, EPI_ISL_1506464, EPI_ISL_1506465, EPI_ISL_1506466, EPI_ISL_1506467,
EPI_ISL_1506468, EPI_ISL_1506469, EPI_ISL_1506470, EPI_ISL_1506471, EPI_ISL_1506472, EPI_ISL_1506473, EPI_ISL_1506474, EPI_ISL_1506475, EPI_ISL_1506476, EPI_ISL_1506477, EPI_ISL_1506478, EPI_ISL_1506479, EPI_ISL_1506481, EPI_ISL_1506482, EPI_ISL_1506483, EPI_ISL_1506484, EPI_ISL_1506485, EPI_ISL_1506486, EPI_ISL_1506487, EPI_ISL_1506488, EPI_ISL_1506489, EPI_ISL_1506490,
EPI_ISL_1506491, EPI_ISL_1506492, EPI_ISL_1506493, EPI_ISL_1506494, EPI_ISL_1506495, EPI_ISL_1506496, EPI_ISL_1506497, EPI_ISL_1506498, EPI_ISL_1506499, EPI_ISL_1506500, EPI_ISL_1506501, EPI_ISL_1506503, EPI_ISL_1506504, EPI_ISL_1506505, EPI_ISL_1506506, EPI_ISL_1506507, EPI_ISL_1506508, EPI_ISL_1506509, EPI_ISL_1506510, EPI_ISL_1506511, EPI_ISL_1506537, EPI_ISL_1506538,
EPI_ISL_1506539, EPI_ISL_1506540, EPI_ISL_1506541, EPI_ISL_1506542, EPI_ISL_1506543, EPI_ISL_1506544, EPI_ISL_1506545, EPI_ISL_1506546, EPI_ISL_1506547, EPI_ISL_1506550, EPI_ISL_1506551, EPI_ISL_1506552, EPI_ISL_1506553, EPI_ISL_1506554, EPI_ISL_1506555, EPI_ISL_1506556, EPI_ISL_1506557, EPI_ISL_1506558, EPI_ISL_1506559, EPI_ISL_1506560, EPI_ISL_1506561, EPI_ISL_1506563,
EPI_ISL_1506565, EPI_ISL_1506566, EPI_ISL_1506567, EPI_ISL_1506568, EPI_ISL_1506569, EPI_ISL_1506570, EPI_ISL_1506577, EPI_ISL_1506583, EPI_ISL_1506586, EPI_ISL_1506588, EPI_ISL_1506590, EPI_ISL_1506592, EPI_ISL_1506597, EPI_ISL_1506601, EPI_ISL_1506604, EPI_ISL_1506605, EPI_ISL_1506608, EPI_ISL_1506609, EPI_ISL_1506612, EPI_ISL_1506616, EPI_ISL_1506619, EPI_ISL_1506621,
EPI_ISL_1506622, EPI_ISL_1506625, EPI_ISL_1506626, EPI_ISL_1506635, EPI_ISL_1506636, EPI_ISL_1506637, EPI_ISL_1506639, EPI_ISL_1506640, EPI_ISL_1506644, EPI_ISL_1506646, EPI_ISL_1506647, EPI_ISL_1506648, EPI_ISL_1506649, EPI_ISL_1506650, EPI_ISL_1506651, EPI_ISL_1506652, EPI_ISL_1506654, EPI_ISL_1506655, EPI_ISL_1506657, EPI_ISL_1506658, EPI_ISL_1506659, EPI_ISL_1506660,
EPI_ISL_1506661, EPI_ISL_1506662, EPI_ISL_1506663, EPI_ISL_1506664, EPI_ISL_1506665, EPI_ISL_1506666, EPI_ISL_1506667, EPI_ISL_1506669, EPI_ISL_1506670, EPI_ISL_1506671, EPI_ISL_1506672, EPI_ISL_1506673, EPI_ISL_1506674, EPI_ISL_1506675, EPI_ISL_1506676, EPI_ISL_1506677, EPI_ISL_1506678, EPI_ISL_1506679, EPI_ISL_1506680, EPI_ISL_1506681, EPI_ISL_1506682, EPI_ISL_1506683,
EPI_ISL_1506684, EPI_ISL_1506685, EPI_ISL_1506687, EPI_ISL_1506688, EPI_ISL_1506689, EPI_ISL_1506690, EPI_ISL_1506691, EPI_ISL_1506692, EPI_ISL_1506693, EPI_ISL_1506694, EPI_ISL_1506695, EPI_ISL_1506696, EPI_ISL_1506697, EPI_ISL_1506704, EPI_ISL_1506713, EPI_ISL_1506740, EPI_ISL_1506751, EPI_ISL_1506825, EPI_ISL_1506827, EPI_ISL_1506828, EPI_ISL_1506829, EPI_ISL_1506830,
EPI_ISL_1506831, EPI_ISL_1506832, EPI_ISL_1506833, EPI_ISL_1506834, EPI_ISL_1506835, EPI_ISL_1506836, EPI_ISL_1506837, EPI_ISL_1506838, EPI_ISL_1506840, EPI_ISL_1506841, EPI_ISL_1506842, EPI_ISL_1506843, EPI_ISL_1506844, EPI_ISL_1506845, EPI_ISL_1506846, EPI_ISL_1506848, EPI_ISL_1506849, EPI_ISL_1506850, EPI_ISL_1506851, EPI_ISL_1506852, EPI_ISL_1506854, EPI_ISL_1506855,
EPI_ISL_1506856, EPI_ISL_1506857, EPI_ISL_1506858, EPI_ISL_1506860, EPI_ISL_1506861, EPI_ISL_1506862, EPI_ISL_1506863, EPI_ISL_1506864, EPI_ISL_1506865, EPI_ISL_1506866, EPI_ISL_1506867, EPI_ISL_1506868, EPI_ISL_1506869, EPI_ISL_1506870, EPI_ISL_1506872, EPI_ISL_1506873, EPI_ISL_1506874, EPI_ISL_1506875, EPI_ISL_1506876, EPI_ISL_1506877, EPI_ISL_1506878, EPI_ISL_1506879,
EPI_ISL_1506880, EPI_ISL_1506881, EPI_ISL_1506882, EPI_ISL_1506883, EPI_ISL_1506884, EPI_ISL_1506885, EPI_ISL_1506886, EPI_ISL_1506887, EPI_ISL_1506891, EPI_ISL_1506892, EPI_ISL_1506893, EPI_ISL_1506894, EPI_ISL_1506895, EPI_ISL_1506896, EPI_ISL_1506897, EPI_ISL_1506899, EPI_ISL_1506900, EPI_ISL_1506901, EPI_ISL_1506902, EPI_ISL_1506903, EPI_ISL_1506904, EPI_ISL_1506905,
EPI_ISL_1506906, EPI_ISL_1506907, EPI_ISL_1506908, EPI_ISL_1506911, EPI_ISL_1506912, EPI_ISL_1506913, EPI_ISL_1506914, EPI_ISL_1506916, EPI_ISL_1506917, EPI_ISL_1506918, EPI_ISL_1506919, EPI_ISL_1506920, EPI_ISL_1506921, EPI_ISL_1506922, EPI_ISL_1506923, EPI_ISL_1506925, EPI_ISL_1506926, EPI_ISL_1506927, EPI_ISL_1506929, EPI_ISL_1506930, EPI_ISL_1506931, EPI_ISL_1506932,
EPI_ISL_1506933, EPI_ISL_1506934, EPI_ISL_1506935, EPI_ISL_1506937, EPI_ISL_1506938, EPI_ISL_1506939, EPI_ISL_1506940, EPI_ISL_1506941, EPI_ISL_1506942, EPI_ISL_1506946, EPI_ISL_1506947, EPI_ISL_1506948, EPI_ISL_1506949, EPI_ISL_1506950, EPI_ISL_1506951, EPI_ISL_1506952, EPI_ISL_1506953, EPI_ISL_1506954, EPI_ISL_1506955, EPI_ISL_1506957, EPI_ISL_1506958, EPI_ISL_1506959,
EPI_ISL_1506960, EPI_ISL_1506961, EPI_ISL_1506962, EPI_ISL_1506963, EPI_ISL_1506964, EPI_ISL_1506965, EPI_ISL_1506967, EPI_ISL_1506968, EPI_ISL_1506969, EPI_ISL_1506970, EPI_ISL_1506971, EPI_ISL_1506973, EPI_ISL_1506975, EPI_ISL_1506977, EPI_ISL_1506978, EPI_ISL_1506979, EPI_ISL_1506980, EPI_ISL_1506981, EPI_ISL_1506982, EPI_ISL_1506983, EPI_ISL_1506984, EPI_ISL_1506985,
EPI_ISL_1506986, EPI_ISL_1506987, EPI_ISL_1506988, EPI_ISL_1506989, EPI_ISL_1506990, EPI_ISL_1506991, EPI_ISL_1506992, EPI_ISL_1506993, EPI_ISL_1506995, EPI_ISL_1506997, EPI_ISL_1506998, EPI_ISL_1506999, EPI_ISL_1507000, EPI_ISL_1507001, EPI_ISL_1507002, EPI_ISL_1507003, EPI_ISL_1507004, EPI_ISL_1507005, EPI_ISL_1507006, EPI_ISL_1507007, EPI_ISL_1507008, EPI_ISL_1507009,
EPI_ISL_1507010, EPI_ISL_1507011, EPI_ISL_1507012, EPI_ISL_1507013, EPI_ISL_1507014, EPI_ISL_1507015, EPI_ISL_1507016, EPI_ISL_1507017, EPI_ISL_1507018, EPI_ISL_1507019, EPI_ISL_1507020, EPI_ISL_1507022, EPI_ISL_1507023, EPI_ISL_1507024, EPI_ISL_1507026, EPI_ISL_1507027, EPI_ISL_1507028, EPI_ISL_1507029, EPI_ISL_1507030, EPI_ISL_1507031, EPI_ISL_1507032, EPI_ISL_1507033,
EPI_ISL_1507034, EPI_ISL_1507035, EPI_ISL_1507037, EPI_ISL_1507038, EPI_ISL_1507039, EPI_ISL_1507040, EPI_ISL_1507045, EPI_ISL_1507046, EPI_ISL_1507047, EPI_ISL_1507048, EPI_ISL_1507049, EPI_ISL_1507050, EPI_ISL_1507051, EPI_ISL_1507052, EPI_ISL_1507053, EPI_ISL_1507054, EPI_ISL_1507055, EPI_ISL_1507056, EPI_ISL_1507057, EPI_ISL_1507058, EPI_ISL_1507059, EPI_ISL_1507060,
EPI_ISL_1507061, EPI_ISL_1507062, EPI_ISL_1507063, EPI_ISL_1507064, EPI_ISL_1507065, EPI_ISL_1507066, EPI_ISL_1507067, EPI_ISL_1507068, EPI_ISL_1507069, EPI_ISL_1507070, EPI_ISL_1507071, EPI_ISL_1507072, EPI_ISL_1507073, EPI_ISL_1507074, EPI_ISL_1507075, EPI_ISL_1507076, EPI_ISL_1507077, EPI_ISL_1507078, EPI_ISL_1507079, EPI_ISL_1507080, EPI_ISL_1507081, EPI_ISL_1507082,
EPI_ISL_1507083, EPI_ISL_1507084, EPI_ISL_1507085, EPI_ISL_1507086, EPI_ISL_1507087, EPI_ISL_1507088, EPI_ISL_1507089, EPI_ISL_1507090, EPI_ISL_1507091, EPI_ISL_1507092, EPI_ISL_1507093, EPI_ISL_1507094, EPI_ISL_1507095, EPI_ISL_1507096, EPI_ISL_1507097, EPI_ISL_1507098, EPI_ISL_1507099, EPI_ISL_1507100, EPI_ISL_1507101, EPI_ISL_1507102, EPI_ISL_1507103, EPI_ISL_1507104,
EPI_ISL_1507105, EPI_ISL_1507106, EPI_ISL_1507107, EPI_ISL_1507108, EPI_ISL_1507109, EPI_ISL_1507110, EPI_ISL_1507111, EPI_ISL_1507112, EPI_ISL_1507113, EPI_ISL_1507114, EPI_ISL_1507115, EPI_ISL_1507116, EPI_ISL_1507117, EPI_ISL_1507118, EPI_ISL_1507119, EPI_ISL_1507120, EPI_ISL_1507121, EPI_ISL_1507122, EPI_ISL_1507123, EPI_ISL_1507124, EPI_ISL_1507125, EPI_ISL_1507126,
EPI_ISL_1507127, EPI_ISL_1507128, EPI_ISL_1507129, EPI_ISL_1507130, EPI_ISL_1507131, EPI_ISL_1507132, EPI_ISL_1507133, EPI_ISL_1507134, EPI_ISL_1507135, EPI_ISL_1507136, EPI_ISL_1507137, EPI_ISL_1507138, EPI_ISL_1507139, EPI_ISL_1507140, EPI_ISL_1507141, EPI_ISL_1507142, EPI_ISL_1507143, EPI_ISL_1507144, EPI_ISL_1507145, EPI_ISL_1507146, EPI_ISL_1507147, EPI_ISL_1507148,
EPI_ISL_1507149, EPI_ISL_1507150, EPI_ISL_1507151, EPI_ISL_1507152, EPI_ISL_1507153, EPI_ISL_1507154, EPI_ISL_1507155, EPI_ISL_1507156, EPI_ISL_1507157, EPI_ISL_1507158, EPI_ISL_1507159, EPI_ISL_1507160, EPI_ISL_1507161, EPI_ISL_1507162, EPI_ISL_1507163, EPI_ISL_1507164, EPI_ISL_1507165, EPI_ISL_1507166, EPI_ISL_1507167, EPI_ISL_1507168, EPI_ISL_1507169, EPI_ISL_1507170,
EPI_ISL_1507171, EPI_ISL_1507172, EPI_ISL_1507173, EPI_ISL_1507174, EPI_ISL_1507175, EPI_ISL_1507176, EPI_ISL_1507177, EPI_ISL_1507178, EPI_ISL_1507179, EPI_ISL_1507180, EPI_ISL_1507181, EPI_ISL_1507182, EPI_ISL_1507183, EPI_ISL_1507184, EPI_ISL_1507185, EPI_ISL_1507186, EPI_ISL_1507187, EPI_ISL_1507188, EPI_ISL_1507189, EPI_ISL_1507190, EPI_ISL_1507191, EPI_ISL_1507192,
EPI_ISL_1507193, EPI_ISL_1507194, EPI_ISL_1507195, EPI_ISL_1507255, EPI_ISL_1507310, EPI_ISL_1507431, EPI_ISL_1507515, EPI_ISL_1507517, EPI_ISL_1507526, EPI_ISL_1507528, EPI_ISL_1507541, EPI_ISL_1507546, EPI_ISL_1507556, EPI_ISL_1507564, EPI_ISL_1507572, EPI_ISL_1507647, EPI_ISL_1517499, EPI_ISL_1517504, EPI_ISL_1517506, EPI_ISL_1517507, EPI_ISL_1517510, EPI_ISL_1517511,
EPI_ISL_1517513, EPI_ISL_1517514, EPI_ISL_1517515, EPI_ISL_1517522, EPI_ISL_1517524, EPI_ISL_1517525, EPI_ISL_1517526, EPI_ISL_1517531, EPI_ISL_1517534, EPI_ISL_1517542, EPI_ISL_1517545, EPI_ISL_1517547, EPI_ISL_1517549, EPI_ISL_1517552, EPI_ISL_1517559, EPI_ISL_1517561, EPI_ISL_1517564, EPI_ISL_1517565, EPI_ISL_1517566, EPI_ISL_1517567, EPI_ISL_1517569, EPI_ISL_1517571,
EPI_ISL_1517572, EPI_ISL_1517574, EPI_ISL_1517575, EPI_ISL_1517578, EPI_ISL_1517581, EPI_ISL_1517582, EPI_ISL_1517583, EPI_ISL_1517585, EPI_ISL_1517586, EPI_ISL_1517587, EPI_ISL_1517589, EPI_ISL_1517590, EPI_ISL_1517591, EPI_ISL_1517593, EPI_ISL_1517594, EPI_ISL_1517595, EPI_ISL_1517596, EPI_ISL_1517597, EPI_ISL_1517601, EPI_ISL_1517602, EPI_ISL_1517605, EPI_ISL_1517608,
EPI_ISL_1517609, EPI_ISL_1517611, EPI_ISL_1517612, EPI_ISL_1517613, EPI_ISL_1517616, EPI_ISL_1517617, EPI_ISL_1517618, EPI_ISL_1517619, EPI_ISL_1517620, EPI_ISL_1517624, EPI_ISL_1517626, EPI_ISL_1517627, EPI_ISL_1517633, EPI_ISL_1517634, EPI_ISL_1517636, EPI_ISL_1517637, EPI_ISL_1517638, EPI_ISL_1517641, EPI_ISL_1517642, EPI_ISL_1517643, EPI_ISL_1517644, EPI_ISL_1517646,
EPI_ISL_1517650, EPI_ISL_1517651, EPI_ISL_1517652, EPI_ISL_1517653, EPI_ISL_1517655, EPI_ISL_1517657, EPI_ISL_1517659, EPI_ISL_1517660, EPI_ISL_1517661, EPI_ISL_1517662, EPI_ISL_1517663, EPI_ISL_1517664, EPI_ISL_1517665, EPI_ISL_1517667, EPI_ISL_1517669, EPI_ISL_1517670, EPI_ISL_1517674, EPI_ISL_1517680, EPI_ISL_1517682, EPI_ISL_1517683, EPI_ISL_1517684, EPI_ISL_1517689,
EPI_ISL_1517690, EPI_ISL_1517693, EPI_ISL_1517699, EPI_ISL_1517703, EPI_ISL_1517704, EPI_ISL_1517705, EPI_ISL_1517707, EPI_ISL_1517710, EPI_ISL_1517713, EPI_ISL_1517714, EPI_ISL_1517716, EPI_ISL_1517720, EPI_ISL_1517721, EPI_ISL_1517722, EPI_ISL_1517724, EPI_ISL_1517726, EPI_ISL_1517727, EPI_ISL_1517728, EPI_ISL_1517730, EPI_ISL_1517737, EPI_ISL_1517738, EPI_ISL_1517739,
EPI_ISL_1517744, EPI_ISL_1517745, EPI_ISL_1517747, EPI_ISL_1517749, EPI_ISL_1517752, EPI_ISL_1518394, EPI_ISL_1518399, EPI_ISL_1518405, EPI_ISL_1518406, EPI_ISL_1518413, EPI_ISL_1518414, EPI_ISL_1518421, EPI_ISL_1518426, EPI_ISL_1518431, EPI_ISL_1518432, EPI_ISL_1518439, EPI_ISL_1518444, EPI_ISL_1518448, EPI_ISL_1518451, EPI_ISL_1518455, EPI_ISL_1518456, EPI_ISL_1518458,
EPI_ISL_1518465, EPI_ISL_1518476, EPI_ISL_1518478, EPI_ISL_1518484, EPI_ISL_1518488, EPI_ISL_1518499, EPI_ISL_1518501, EPI_ISL_1518503, EPI_ISL_1518507, EPI_ISL_1518511, EPI_ISL_1518515, EPI_ISL_1518519, EPI_ISL_1518521, EPI_ISL_1518528, EPI_ISL_1518530, EPI_ISL_1518531, EPI_ISL_1518532, EPI_ISL_1518534, EPI_ISL_1518535, EPI_ISL_1518536, EPI_ISL_1518548, EPI_ISL_1518549,
EPI_ISL_1518557, EPI_ISL_1518562, EPI_ISL_1518563, EPI_ISL_1518565, EPI_ISL_1518579, EPI_ISL_1518582, EPI_ISL_1518590, EPI_ISL_1518608, EPI_ISL_1518610, EPI_ISL_1518620, EPI_ISL_1518622, EPI_ISL_1518626, EPI_ISL_1518636, EPI_ISL_1518638, EPI_ISL_1518639, EPI_ISL_1518641, EPI_ISL_1518645, EPI_ISL_1518646, EPI_ISL_1518649, EPI_ISL_1518653, EPI_ISL_1518654, EPI_ISL_1518657,
EPI_ISL_1518658, EPI_ISL_1518665, EPI_ISL_1518666, EPI_ISL_1518672, EPI_ISL_1518674, EPI_ISL_1518995, EPI_ISL_1518998, EPI_ISL_1519001, EPI_ISL_1519002, EPI_ISL_1519003, EPI_ISL_1519006, EPI_ISL_1519007, EPI_ISL_1519008, EPI_ISL_1519016, EPI_ISL_1519019, EPI_ISL_1519021, EPI_ISL_1519029, EPI_ISL_1519030, EPI_ISL_1519031, EPI_ISL_1519032, EPI_ISL_1519036, EPI_ISL_1519039,
EPI_ISL_1519046, EPI_ISL_1519048, EPI_ISL_1519050, EPI_ISL_1519056, EPI_ISL_1519058, EPI_ISL_1519059, EPI_ISL_1519060, EPI_ISL_1519062, EPI_ISL_1519064, EPI_ISL_1519065, EPI_ISL_1519067, EPI_ISL_1519069, EPI_ISL_1519072, EPI_ISL_1519073, EPI_ISL_1519074, EPI_ISL_1519075, EPI_ISL_1519077, EPI_ISL_1519078, EPI_ISL_1519086, EPI_ISL_1519087, EPI_ISL_1519089, EPI_ISL_1519092,
EPI_ISL_1519095, EPI_ISL_1519096, EPI_ISL_1519100, EPI_ISL_1519101, EPI_ISL_1519104, EPI_ISL_1519107, EPI_ISL_1519109, EPI_ISL_1519112, EPI_ISL_1519113, EPI_ISL_1519114, EPI_ISL_1519116, EPI_ISL_1519120, EPI_ISL_1519125, EPI_ISL_1519127, EPI_ISL_1519133, EPI_ISL_1519138, EPI_ISL_1519139, EPI_ISL_1519145, EPI_ISL_1519146, EPI_ISL_1519147, EPI_ISL_1519150, EPI_ISL_1519154,
EPI_ISL_1519155, EPI_ISL_1519156, EPI_ISL_1519157, EPI_ISL_1519162, EPI_ISL_1519169, EPI_ISL_1519175, EPI_ISL_1519178, EPI_ISL_1519185, EPI_ISL_1519189, EPI_ISL_1519190, EPI_ISL_1519192, EPI_ISL_1519193, EPI_ISL_1519196, EPI_ISL_1519197, EPI_ISL_1519200, EPI_ISL_1519202, EPI_ISL_1519204, EPI_ISL_1519208, EPI_ISL_1519212, EPI_ISL_1519215, EPI_ISL_1519216, EPI_ISL_1519218,
EPI_ISL_1519222, EPI_ISL_1519224, EPI_ISL_1519225, EPI_ISL_1519226, EPI_ISL_1519228, EPI_ISL_1519230, EPI_ISL_1519233, EPI_ISL_1519237, EPI_ISL_1519240, EPI_ISL_1519243, EPI_ISL_1519244, EPI_ISL_1519247, EPI_ISL_1519249, EPI_ISL_1519250, EPI_ISL_1519252, EPI_ISL_1519253, EPI_ISL_1519254, EPI_ISL_1519255, EPI_ISL_1519259, EPI_ISL_1519260, EPI_ISL_1519261, EPI_ISL_1519263,
EPI_ISL_1519266, EPI_ISL_1519267, EPI_ISL_1519309, EPI_ISL_1519310, EPI_ISL_1519312, EPI_ISL_1519313, EPI_ISL_1519318, EPI_ISL_1519322, EPI_ISL_1519323, EPI_ISL_1519326, EPI_ISL_1519330, EPI_ISL_1519336, EPI_ISL_1519338, EPI_ISL_1519348, EPI_ISL_1519387, EPI_ISL_1519420, EPI_ISL_1519421, EPI_ISL_1519422, EPI_ISL_1519429, EPI_ISL_1519430, EPI_ISL_1519448, EPI_ISL_1519451,
EPI_ISL_1519455, EPI_ISL_1519473, EPI_ISL_1519479, EPI_ISL_1519484, EPI_ISL_1519486, EPI_ISL_1519488, EPI_ISL_1519489, EPI_ISL_1519495, EPI_ISL_1519500, EPI_ISL_1519511, EPI_ISL_1519514, EPI_ISL_1519531, EPI_ISL_1519532, EPI_ISL_1519538, EPI_ISL_1519552, EPI_ISL_1519553, EPI_ISL_1519554, EPI_ISL_1519557, EPI_ISL_1519568, EPI_ISL_1519572, EPI_ISL_1519575, EPI_ISL_1519577,
EPI_ISL_1519586, EPI_ISL_1519588, EPI_ISL_1519589, EPI_ISL_1519593, EPI_ISL_1519594, EPI_ISL_1519597, EPI_ISL_1519598, EPI_ISL_1519601, EPI_ISL_1519602, EPI_ISL_1519607, EPI_ISL_1519609, EPI_ISL_1519616, EPI_ISL_1519627, EPI_ISL_1519639, EPI_ISL_1519645, EPI_ISL_1519650, EPI_ISL_1519651, EPI_ISL_1519652, EPI_ISL_1519653, EPI_ISL_1519654, EPI_ISL_1519655, EPI_ISL_1519656,
EPI_ISL_1519657, EPI_ISL_1519658, EPI_ISL_1519659, EPI_ISL_1519660, EPI_ISL_1519661, EPI_ISL_1519662, EPI_ISL_1519663, EPI_ISL_1519664, EPI_ISL_1519665, EPI_ISL_1519666, EPI_ISL_1519667, EPI_ISL_1519668, EPI_ISL_1519669, EPI_ISL_1519670, EPI_ISL_1519671, EPI_ISL_1519672, EPI_ISL_1519673, EPI_ISL_1519674, EPI_ISL_1519675, EPI_ISL_1519676, EPI_ISL_1519677, EPI_ISL_1519678,
EPI_ISL_1519679, EPI_ISL_1519680, EPI_ISL_1519681, EPI_ISL_1519682, EPI_ISL_1519683, EPI_ISL_1519684, EPI_ISL_1519685, EPI_ISL_1519686, EPI_ISL_1519687, EPI_ISL_1519688, EPI_ISL_1519689, EPI_ISL_1519690, EPI_ISL_1519691, EPI_ISL_1519692, EPI_ISL_1519693, EPI_ISL_1519694, EPI_ISL_1519695, EPI_ISL_1519696, EPI_ISL_1519697, EPI_ISL_1519698, EPI_ISL_1519699, EPI_ISL_1519700,
EPI_ISL_1519701, EPI_ISL_1519702, EPI_ISL_1519703, EPI_ISL_1519704, EPI_ISL_1519705, EPI_ISL_1519706, EPI_ISL_1519707, EPI_ISL_1519708, EPI_ISL_1519709, EPI_ISL_1519710, EPI_ISL_1519711, EPI_ISL_1519712, EPI_ISL_1519713, EPI_ISL_1519714, EPI_ISL_1519715, EPI_ISL_1519716, EPI_ISL_1519717, EPI_ISL_1519718, EPI_ISL_1519719, EPI_ISL_1519720, EPI_ISL_1519721, EPI_ISL_1519722,
EPI_ISL_1519723, EPI_ISL_1519724, EPI_ISL_1519725, EPI_ISL_1519726, EPI_ISL_1519727, EPI_ISL_1519728, EPI_ISL_1519729, EPI_ISL_1519730, EPI_ISL_1519731, EPI_ISL_1519732, EPI_ISL_1519733, EPI_ISL_1519734, EPI_ISL_1519735, EPI_ISL_1519736, EPI_ISL_1519737, EPI_ISL_1519738, EPI_ISL_1519739, EPI_ISL_1519740, EPI_ISL_1519741, EPI_ISL_1519742, EPI_ISL_1519743, EPI_ISL_1519744,
EPI_ISL_1519745, EPI_ISL_1519746, EPI_ISL_1519747, EPI_ISL_1519748, EPI_ISL_1519749, EPI_ISL_1519750, EPI_ISL_1519751, EPI_ISL_1519752, EPI_ISL_1519753, EPI_ISL_1519754, EPI_ISL_1519755, EPI_ISL_1519756, EPI_ISL_1519757, EPI_ISL_1519758, EPI_ISL_1519759, EPI_ISL_1519760, EPI_ISL_1519761, EPI_ISL_1519762, EPI_ISL_1519763, EPI_ISL_1519764, EPI_ISL_1519765, EPI_ISL_1519766,
EPI_ISL_1519767, EPI_ISL_1519768, EPI_ISL_1519769, EPI_ISL_1519770, EPI_ISL_1519771, EPI_ISL_1519772, EPI_ISL_1519773, EPI_ISL_1519774, EPI_ISL_1519775, EPI_ISL_1519776, EPI_ISL_1519777, EPI_ISL_1519778, EPI_ISL_1519779, EPI_ISL_1519780, EPI_ISL_1519781, EPI_ISL_1519782, EPI_ISL_1519783, EPI_ISL_1519784, EPI_ISL_1519785, EPI_ISL_1519786, EPI_ISL_1519787, EPI_ISL_1519788,
EPI_ISL_1519789, EPI_ISL_1519790, EPI_ISL_1519791, EPI_ISL_1519792, EPI_ISL_1519793, EPI_ISL_1519794, EPI_ISL_1519795, EPI_ISL_1519796, EPI_ISL_1519797, EPI_ISL_1519798, EPI_ISL_1519799, EPI_ISL_1519800, EPI_ISL_1519801, EPI_ISL_1519802, EPI_ISL_1519803, EPI_ISL_1519804, EPI_ISL_1519805, EPI_ISL_1519806, EPI_ISL_1519807, EPI_ISL_1519808, EPI_ISL_1519809, EPI_ISL_1519810,
EPI_ISL_1519811, EPI_ISL_1519812, EPI_ISL_1519813, EPI_ISL_1519814, EPI_ISL_1519815, EPI_ISL_1519816, EPI_ISL_1519817, EPI_ISL_1519818, EPI_ISL_1519819, EPI_ISL_1519820, EPI_ISL_1519821, EPI_ISL_1519822, EPI_ISL_1519823, EPI_ISL_1519824, EPI_ISL_1519825, EPI_ISL_1519826, EPI_ISL_1519827, EPI_ISL_1519828, EPI_ISL_1519829, EPI_ISL_1519830, EPI_ISL_1519831, EPI_ISL_1519832,
EPI_ISL_1519833, EPI_ISL_1519834, EPI_ISL_1519835, EPI_ISL_1519836, EPI_ISL_1519837, EPI_ISL_1519838, EPI_ISL_1519839, EPI_ISL_1519840, EPI_ISL_1519841, EPI_ISL_1519842, EPI_ISL_1519843, EPI_ISL_1519844, EPI_ISL_1519846, EPI_ISL_1519847, EPI_ISL_1519848, EPI_ISL_1519850, EPI_ISL_1519851, EPI_ISL_1519852, EPI_ISL_1519853, EPI_ISL_1519855, EPI_ISL_1519856, EPI_ISL_1519857,
EPI_ISL_1519858, EPI_ISL_1519859, EPI_ISL_1519860, EPI_ISL_1519861, EPI_ISL_1519862, EPI_ISL_1519863, EPI_ISL_1519864, EPI_ISL_1519865, EPI_ISL_1519866, EPI_ISL_1519867, EPI_ISL_1519868, EPI_ISL_1519869, EPI_ISL_1519870, EPI_ISL_1519871, EPI_ISL_1519872, EPI_ISL_1519873, EPI_ISL_1519874, EPI_ISL_1519875, EPI_ISL_1519876, EPI_ISL_1519877, EPI_ISL_1519878, EPI_ISL_1519879,
EPI_ISL_1519880, EPI_ISL_1519881, EPI_ISL_1519882, EPI_ISL_1519883, EPI_ISL_1519885, EPI_ISL_1519886, EPI_ISL_1519887, EPI_ISL_1519888, EPI_ISL_1519889, EPI_ISL_1519890, EPI_ISL_1519891, EPI_ISL_1519892, EPI_ISL_1519893, EPI_ISL_1519894, EPI_ISL_1519895, EPI_ISL_1519896, EPI_ISL_1519897, EPI_ISL_1519898, EPI_ISL_1519899, EPI_ISL_1519900, EPI_ISL_1519901, EPI_ISL_1519902,
EPI_ISL_1519903, EPI_ISL_1519904, EPI_ISL_1519905, EPI_ISL_1519906, EPI_ISL_1519907, EPI_ISL_1519908, EPI_ISL_1519909, EPI_ISL_1535530, EPI_ISL_1537146, EPI_ISL_1537154, EPI_ISL_1537158, EPI_ISL_1537160, EPI_ISL_1537167, EPI_ISL_1537168, EPI_ISL_1537175, EPI_ISL_1537179, EPI_ISL_1537188, EPI_ISL_1537201, EPI_ISL_1537210, EPI_ISL_1537211, EPI_ISL_1537212, EPI_ISL_1537213,
EPI_ISL_1537217, EPI_ISL_1537218, EPI_ISL_1537219, EPI_ISL_1537221, EPI_ISL_1537222, EPI_ISL_1537223, EPI_ISL_1537224, EPI_ISL_1537225, EPI_ISL_1537227, EPI_ISL_1537228, EPI_ISL_1537229, EPI_ISL_1537230, EPI_ISL_1537231, EPI_ISL_1537232, EPI_ISL_1537233, EPI_ISL_1537234, EPI_ISL_1537235, EPI_ISL_1537237, EPI_ISL_1537238, EPI_ISL_1537239, EPI_ISL_1537241, EPI_ISL_1537243,
EPI_ISL_1537244, EPI_ISL_1537245, EPI_ISL_1537246, EPI_ISL_1537247, EPI_ISL_1537248, EPI_ISL_1537251, EPI_ISL_1537252, EPI_ISL_1537253, EPI_ISL_1537254, EPI_ISL_1537255, EPI_ISL_1537256, EPI_ISL_1537257, EPI_ISL_1537264, EPI_ISL_1537265, EPI_ISL_1537266, EPI_ISL_1537267, EPI_ISL_1537269, EPI_ISL_1537270, EPI_ISL_1537271, EPI_ISL_1537272, EPI_ISL_1537275, EPI_ISL_1537276,
EPI_ISL_1537277, EPI_ISL_1537278, EPI_ISL_1537279, EPI_ISL_1537280, EPI_ISL_1537281, EPI_ISL_1537282, EPI_ISL_1537284, EPI_ISL_1537285, EPI_ISL_1537287, EPI_ISL_1537288, EPI_ISL_1537291, EPI_ISL_1537292, EPI_ISL_1537293, EPI_ISL_1537294, EPI_ISL_1537295, EPI_ISL_1537296, EPI_ISL_1537297, EPI_ISL_1537298, EPI_ISL_1537299, EPI_ISL_1537300, EPI_ISL_1537303, EPI_ISL_1537305,
EPI_ISL_1537306, EPI_ISL_1537307, EPI_ISL_1537308, EPI_ISL_1537309, EPI_ISL_1537312, EPI_ISL_1537353, EPI_ISL_1537358, EPI_ISL_1537413, EPI_ISL_1537443, EPI_ISL_1537462, EPI_ISL_1537531, EPI_ISL_1537567, EPI_ISL_1537906, EPI_ISL_1537961, EPI_ISL_1538015, EPI_ISL_1538016, EPI_ISL_1538017, EPI_ISL_1538018, EPI_ISL_1538019, EPI_ISL_1538021, EPI_ISL_1538022, EPI_ISL_1538023,
EPI_ISL_1538024, EPI_ISL_1538025, EPI_ISL_1538026, EPI_ISL_1538027, EPI_ISL_1538028, EPI_ISL_1538030, EPI_ISL_1538032, EPI_ISL_1538033, EPI_ISL_1538034, EPI_ISL_1538035, EPI_ISL_1538037, EPI_ISL_1538038, EPI_ISL_1538040, EPI_ISL_1538041, EPI_ISL_1538042, EPI_ISL_1538044, EPI_ISL_1538045, EPI_ISL_1538046, EPI_ISL_1538047, EPI_ISL_1538048, EPI_ISL_1538050, EPI_ISL_1538051,
EPI_ISL_1538052, EPI_ISL_1538054, EPI_ISL_1538055, EPI_ISL_1538056, EPI_ISL_1538058, EPI_ISL_1538059, EPI_ISL_1538060, EPI_ISL_1538061, EPI_ISL_1538062, EPI_ISL_1538064, EPI_ISL_1538065, EPI_ISL_1538066, EPI_ISL_1538094, EPI_ISL_1538095, EPI_ISL_1538097, EPI_ISL_1538099, EPI_ISL_1538100, EPI_ISL_1538103, EPI_ISL_1538105, EPI_ISL_1538109, EPI_ISL_1538110, EPI_ISL_1538111,
EPI_ISL_1538113, EPI_ISL_1538114, EPI_ISL_1538115, EPI_ISL_1538117, EPI_ISL_1538118, EPI_ISL_1538119, EPI_ISL_1538121, EPI_ISL_1538123, EPI_ISL_1538125, EPI_ISL_1538126, EPI_ISL_1538127, EPI_ISL_1538129, EPI_ISL_1538130, EPI_ISL_1538132, EPI_ISL_1538133, EPI_ISL_1538135, EPI_ISL_1538138, EPI_ISL_1538139, EPI_ISL_1538140, EPI_ISL_1538141, EPI_ISL_1538142, EPI_ISL_1538143,
EPI_ISL_1538146, EPI_ISL_1538147, EPI_ISL_1538148, EPI_ISL_1538150, EPI_ISL_1538152, EPI_ISL_1538155, EPI_ISL_1538156, EPI_ISL_1538157, EPI_ISL_1538158, EPI_ISL_1538159, EPI_ISL_1538160, EPI_ISL_1538161, EPI_ISL_1538162, EPI_ISL_1538163, EPI_ISL_1538164, EPI_ISL_1538165, EPI_ISL_1538167, EPI_ISL_1538168, EPI_ISL_1538169, EPI_ISL_1538170, EPI_ISL_1538172, EPI_ISL_1538173,
EPI_ISL_1538174, EPI_ISL_1538176, EPI_ISL_1538177, EPI_ISL_1538178, EPI_ISL_1538179, EPI_ISL_1538180, EPI_ISL_1538181, EPI_ISL_1538182, EPI_ISL_1538183, EPI_ISL_1538184, EPI_ISL_1538186, EPI_ISL_1538187, EPI_ISL_1538188, EPI_ISL_1538189, EPI_ISL_1538190, EPI_ISL_1538191, EPI_ISL_1538192, EPI_ISL_1538193, EPI_ISL_1538194, EPI_ISL_1538195, EPI_ISL_1538197, EPI_ISL_1538198,
EPI_ISL_1538199, EPI_ISL_1538200, EPI_ISL_1538201, EPI_ISL_1538202, EPI_ISL_1538203, EPI_ISL_1538204, EPI_ISL_1538205, EPI_ISL_1538207, EPI_ISL_1538210, EPI_ISL_1538211, EPI_ISL_1538213, EPI_ISL_1538215, EPI_ISL_1538217, EPI_ISL_1538220, EPI_ISL_1538221, EPI_ISL_1538223, EPI_ISL_1538224, EPI_ISL_1538225, EPI_ISL_1538226, EPI_ISL_1538227, EPI_ISL_1538228, EPI_ISL_1538229,
EPI_ISL_1538230, EPI_ISL_1538231, EPI_ISL_1538232, EPI_ISL_1538233, EPI_ISL_1538234, EPI_ISL_1538236, EPI_ISL_1538237, EPI_ISL_1538238, EPI_ISL_1538239, EPI_ISL_1538240, EPI_ISL_1538242, EPI_ISL_1538243, EPI_ISL_1538244, EPI_ISL_1538245, EPI_ISL_1538246, EPI_ISL_1538248, EPI_ISL_1538250, EPI_ISL_1538251, EPI_ISL_1538255, EPI_ISL_1538256, EPI_ISL_1538259, EPI_ISL_1538260,
EPI_ISL_1538261, EPI_ISL_1538262, EPI_ISL_1538264, EPI_ISL_1538298, EPI_ISL_1538299, EPI_ISL_1538302, EPI_ISL_1538303, EPI_ISL_1538305, EPI_ISL_1538306, EPI_ISL_1538307, EPI_ISL_1538308, EPI_ISL_1538309, EPI_ISL_1538310, EPI_ISL_1538311, EPI_ISL_1538312, EPI_ISL_1538313, EPI_ISL_1538314, EPI_ISL_1538315, EPI_ISL_1538316, EPI_ISL_1538317, EPI_ISL_1538318, EPI_ISL_1538319,
EPI_ISL_1538320, EPI_ISL_1538321, EPI_ISL_1538322, EPI_ISL_1538323, EPI_ISL_1538324, EPI_ISL_1538325, EPI_ISL_1538326, EPI_ISL_1538327, EPI_ISL_1538328, EPI_ISL_1538329, EPI_ISL_1538330, EPI_ISL_1538331, EPI_ISL_1538332, EPI_ISL_1538333, EPI_ISL_1538334, EPI_ISL_1538335, EPI_ISL_1538336, EPI_ISL_1538337, EPI_ISL_1538338, EPI_ISL_1538339, EPI_ISL_1538340, EPI_ISL_1538341,
EPI_ISL_1538342, EPI_ISL_1538343, EPI_ISL_1538344, EPI_ISL_1538345, EPI_ISL_1538346, EPI_ISL_1538347, EPI_ISL_1538348, EPI_ISL_1538349, EPI_ISL_1538350, EPI_ISL_1538351, EPI_ISL_1538352, EPI_ISL_1538353, EPI_ISL_1538354, EPI_ISL_1538355, EPI_ISL_1538356, EPI_ISL_1538357, EPI_ISL_1538358, EPI_ISL_1538359, EPI_ISL_1538360, EPI_ISL_1538361, EPI_ISL_1538362, EPI_ISL_1538363,
EPI_ISL_1538364, EPI_ISL_1538365, EPI_ISL_1538366, EPI_ISL_1538367, EPI_ISL_1538368, EPI_ISL_1538369, EPI_ISL_1538370, EPI_ISL_1538371, EPI_ISL_1538372, EPI_ISL_1538374, EPI_ISL_1538375, EPI_ISL_1538376, EPI_ISL_1538377, EPI_ISL_1538378, EPI_ISL_1538379, EPI_ISL_1538380, EPI_ISL_1538381, EPI_ISL_1538382, EPI_ISL_1538383, EPI_ISL_1538384, EPI_ISL_1538385, EPI_ISL_1538386,
EPI_ISL_1538387, EPI_ISL_1538388, EPI_ISL_1538389, EPI_ISL_1538390, EPI_ISL_1538391, EPI_ISL_1538392, EPI_ISL_1538393, EPI_ISL_1538394, EPI_ISL_1538395, EPI_ISL_1538396, EPI_ISL_1538398, EPI_ISL_1538399, EPI_ISL_1538400, EPI_ISL_1538401, EPI_ISL_1564884, EPI_ISL_1565035, EPI_ISL_1565132, EPI_ISL_1565247, EPI_ISL_1584853, EPI_ISL_1595208, EPI_ISL_1595209, EPI_ISL_1595213,
EPI_ISL_1595214, EPI_ISL_1595216, EPI_ISL_1595217, EPI_ISL_1595222, EPI_ISL_1595224, EPI_ISL_1595225, EPI_ISL_1595227, EPI_ISL_1595231, EPI_ISL_1595234, EPI_ISL_1595237, EPI_ISL_1595238, EPI_ISL_1595242, EPI_ISL_1595243, EPI_ISL_1595244, EPI_ISL_1595245, EPI_ISL_1595248, EPI_ISL_1595252, EPI_ISL_1595253, EPI_ISL_1595256, EPI_ISL_1595257, EPI_ISL_1595267, EPI_ISL_1595268,
EPI_ISL_1595269, EPI_ISL_1595270, EPI_ISL_1595273, EPI_ISL_1595275, EPI_ISL_1595276, EPI_ISL_1595278, EPI_ISL_1595281, EPI_ISL_1595282, EPI_ISL_1595285, EPI_ISL_1595286, EPI_ISL_1595289, EPI_ISL_1595290, EPI_ISL_1595291, EPI_ISL_1595297, EPI_ISL_1595298, EPI_ISL_1595300, EPI_ISL_1595302, EPI_ISL_1595303, EPI_ISL_1595305, EPI_ISL_1595306, EPI_ISL_1595307, EPI_ISL_1595308,
EPI_ISL_1595314, EPI_ISL_1595317, EPI_ISL_1595318, EPI_ISL_1595319, EPI_ISL_1595320, EPI_ISL_1595321, EPI_ISL_1595325, EPI_ISL_1595326, EPI_ISL_1595327, EPI_ISL_1595330, EPI_ISL_1595331, EPI_ISL_1595332, EPI_ISL_1595333, EPI_ISL_1595336, EPI_ISL_1595337, EPI_ISL_1595338, EPI_ISL_1595344, EPI_ISL_1595345, EPI_ISL_1595347, EPI_ISL_1595348, EPI_ISL_1595349, EPI_ISL_1595350,
EPI_ISL_1595354, EPI_ISL_1595356, EPI_ISL_1595357, EPI_ISL_1595360, EPI_ISL_1595361, EPI_ISL_1595366, EPI_ISL_1595368, EPI_ISL_1595370, EPI_ISL_1595374, EPI_ISL_1595375, EPI_ISL_1595377, EPI_ISL_1595379, EPI_ISL_1595380, EPI_ISL_1595385, EPI_ISL_1595388, EPI_ISL_1595391, EPI_ISL_1595394, EPI_ISL_1595397, EPI_ISL_1595398, EPI_ISL_1595399, EPI_ISL_1595405, EPI_ISL_1595407,
EPI_ISL_1595409, EPI_ISL_1595414, EPI_ISL_1595420, EPI_ISL_1595423, EPI_ISL_1595424, EPI_ISL_1595427, EPI_ISL_1595428, EPI_ISL_1595434, EPI_ISL_1595436, EPI_ISL_1595439, EPI_ISL_1595444, EPI_ISL_1595448, EPI_ISL_1595449, EPI_ISL_1595451, EPI_ISL_1595452, EPI_ISL_1595455, EPI_ISL_1595456, EPI_ISL_1595459, EPI_ISL_1595460, EPI_ISL_1595462, EPI_ISL_1595467, EPI_ISL_1616571,
EPI_ISL_1630516, EPI_ISL_1630520, EPI_ISL_1630521, EPI_ISL_1630522, EPI_ISL_1630524, EPI_ISL_1630525, EPI_ISL_1630528, EPI_ISL_1630531, EPI_ISL_1630532, EPI_ISL_1630536, EPI_ISL_1630542, EPI_ISL_1630543, EPI_ISL_1630546, EPI_ISL_1630550, EPI_ISL_1630552, EPI_ISL_1630553, EPI_ISL_1630555, EPI_ISL_1630556, EPI_ISL_1630558, EPI_ISL_1630561, EPI_ISL_1630563, EPI_ISL_1630566,
EPI_ISL_1630567, EPI_ISL_1630568, EPI_ISL_1630576, EPI_ISL_1630577, EPI_ISL_1630586, EPI_ISL_1630590, EPI_ISL_1630592, EPI_ISL_1630598, EPI_ISL_1630599, EPI_ISL_1630601, EPI_ISL_1630604, EPI_ISL_1630608, EPI_ISL_1630612, EPI_ISL_1630614, EPI_ISL_1630617, EPI_ISL_1630618, EPI_ISL_1630620, EPI_ISL_1630622, EPI_ISL_1630623, EPI_ISL_1630624, EPI_ISL_1630627, EPI_ISL_1630629,
EPI_ISL_1630630, EPI_ISL_1630631, EPI_ISL_1630635, EPI_ISL_1630637, EPI_ISL_1630639, EPI_ISL_1630648, EPI_ISL_1630651, EPI_ISL_1630652, EPI_ISL_1630653, EPI_ISL_1630661, EPI_ISL_1630665, EPI_ISL_1630667, EPI_ISL_1630671, EPI_ISL_1630672, EPI_ISL_1630674, EPI_ISL_1630675, EPI_ISL_1630676, EPI_ISL_1630679, EPI_ISL_1630683, EPI_ISL_1630684, EPI_ISL_1630688, EPI_ISL_1630694,
EPI_ISL_1630696, EPI_ISL_1630700, EPI_ISL_1630704, EPI_ISL_1630708, EPI_ISL_1630710, EPI_ISL_1630715, EPI_ISL_1630719, EPI_ISL_1630724, EPI_ISL_1630725, EPI_ISL_1630726, EPI_ISL_1630728, EPI_ISL_1630730, EPI_ISL_1630732, EPI_ISL_1630735, EPI_ISL_1630739, EPI_ISL_1630742, EPI_ISL_1630749, EPI_ISL_1630754, EPI_ISL_1630763, EPI_ISL_1630765, EPI_ISL_1630771, EPI_ISL_1630774,
EPI_ISL_1630775, EPI_ISL_1630776, EPI_ISL_1630779, EPI_ISL_1630786, EPI_ISL_1630789, EPI_ISL_1630805, EPI_ISL_1630806, EPI_ISL_1630807, EPI_ISL_1630808, EPI_ISL_1630809, EPI_ISL_1630814, EPI_ISL_1630815, EPI_ISL_1630816, EPI_ISL_1630817, EPI_ISL_1630818, EPI_ISL_1630820, EPI_ISL_1635070, EPI_ISL_1635071, EPI_ISL_1635072, EPI_ISL_1635073, EPI_ISL_1635074, EPI_ISL_1635075,
EPI_ISL_1635076, EPI_ISL_1635077, EPI_ISL_1635078, EPI_ISL_1635079, EPI_ISL_1635080, EPI_ISL_1635081, EPI_ISL_1635083, EPI_ISL_1635084, EPI_ISL_1635085, EPI_ISL_1635086, EPI_ISL_1635087, EPI_ISL_1635089, EPI_ISL_1635090, EPI_ISL_1635091, EPI_ISL_1635092, EPI_ISL_1635094, EPI_ISL_1635095, EPI_ISL_1635096, EPI_ISL_1635098, EPI_ISL_1635099, EPI_ISL_1635100, EPI_ISL_1635101,
EPI_ISL_1635102, EPI_ISL_1635103, EPI_ISL_1635104, EPI_ISL_1635105, EPI_ISL_1635106, EPI_ISL_1635107, EPI_ISL_1635108, EPI_ISL_1635110, EPI_ISL_1635111, EPI_ISL_1635112, EPI_ISL_1635113, EPI_ISL_1635114, EPI_ISL_1635115, EPI_ISL_1635116, EPI_ISL_1635118, EPI_ISL_1635423, EPI_ISL_1635424

| | | | |
|---|---|---|---|
| see above | Lighthouse Lab in Glasgow | Wellcome Sanger Institute for the COVID-19 Genomics UK (COG-UK) Consortium | Anna Dominiczak and Alex Alderton; Carol Clugston; Cordelia Langford; David Gray; David K. Jackson; Dominic Kwiatkowski; Ewan Harrison; Harper VanSteenhouse; Ian Johnston; Jeffrey Barrett; John Sillitoe on behalf of the Wellcome Sanger Institute COVID-19 Surveillance Team; Roberto Amato; Sonia Goncalves; Yumi Kasai |

EPI_ISL_1484103, EPI_ISL_1484118, EPI_ISL_1484126, EPI_ISL_1484131, EPI_ISL_1484133, EPI_ISL_1484137, EPI_ISL_1484142, EPI_ISL_1484151, EPI_ISL_1484170, EPI_ISL_1484173, EPI_ISL_1484175, EPI_ISL_1484202, EPI_ISL_1484203, EPI_ISL_1484210, EPI_ISL_1484251, EPI_ISL_1484258, EPI_ISL_1484260, EPI_ISL_1484261, EPI_ISL_1484282, EPI_ISL_1484293, EPI_ISL_1484842, EPI_ISL_1484854,

| | | | |
|---|---|---|---|
| EPI_ISL_1484857, EPI_ISL_1484863, EPI_ISL_1484867, EPI_ISL_1484872, EPI_ISL_1484883, EPI_ISL_1484891, EPI_ISL_1484894, EPI_ISL_1484895, EPI_ISL_1484896, EPI_ISL_1484902, EPI_ISL_1484911, EPI_ISL_1484913, EPI_ISL_1484919, EPI_ISL_1484922, EPI_ISL_1484923, EPI_ISL_1484925, EPI_ISL_1484927, EPI_ISL_1484935, EPI_ISL_1484938, EPI_ISL_1484942, EPI_ISL_1484948, EPI_ISL_1484952, EPI_ISL_1484953, EPI_ISL_1484955, EPI_ISL_1484957, EPI_ISL_1484958, EPI_ISL_1484960, EPI_ISL_1484961, EPI_ISL_1484973, EPI_ISL_1484984, EPI_ISL_1484985, EPI_ISL_1484986, EPI_ISL_1484991, EPI_ISL_1484998, EPI_ISL_1485007, EPI_ISL_1485009, EPI_ISL_1485018, EPI_ISL_1485019, EPI_ISL_1485026, EPI_ISL_1485042, EPI_ISL_1485044, EPI_ISL_1485048, EPI_ISL_1485050, EPI_ISL_1485056, EPI_ISL_1485058, EPI_ISL_1485067, EPI_ISL_1485076, EPI_ISL_1485079, EPI_ISL_1485080, EPI_ISL_1485082, EPI_ISL_1485084, EPI_ISL_1485093, EPI_ISL_1485096, EPI_ISL_1485098, EPI_ISL_1485101, EPI_ISL_1485106, EPI_ISL_1485107, EPI_ISL_1485112, EPI_ISL_1485121, EPI_ISL_1485126, EPI_ISL_1485130, EPI_ISL_1485132, EPI_ISL_1485133, EPI_ISL_1485134, EPI_ISL_1485138, EPI_ISL_1485141, EPI_ISL_1485145, EPI_ISL_1485147, EPI_ISL_1485166, EPI_ISL_1485175, EPI_ISL_1485524, EPI_ISL_1485544, EPI_ISL_1485599, EPI_ISL_1485603, EPI_ISL_1485606, EPI_ISL_1485701, EPI_ISL_1485728, EPI_ISL_1485733, EPI_ISL_1485740, EPI_ISL_1485756, EPI_ISL_1485761, EPI_ISL_1485763, EPI_ISL_1486113, EPI_ISL_1486117, EPI_ISL_1486120, EPI_ISL_1486132, EPI_ISL_1486135, EPI_ISL_1486139, EPI_ISL_1486141, EPI_ISL_1486144, EPI_ISL_1486156, EPI_ISL_1486159, EPI_ISL_1486166, EPI_ISL_1486169, EPI_ISL_1486170, EPI_ISL_1486173, EPI_ISL_1486179, EPI_ISL_1486189, EPI_ISL_1486192, EPI_ISL_1486205, EPI_ISL_1486206, EPI_ISL_1486222, EPI_ISL_1486226, EPI_ISL_1486233, EPI_ISL_1486242, EPI_ISL_1486254, EPI_ISL_1486259, EPI_ISL_1486262, EPI_ISL_1486270, EPI_ISL_1486271, EPI_ISL_1486276, EPI_ISL_1486288, EPI_ISL_1486293, EPI_ISL_1486298, EPI_ISL_1486301, EPI_ISL_1486306, EPI_ISL_1486307, EPI_ISL_1486313, EPI_ISL_1486327, EPI_ISL_1486341, EPI_ISL_1486344, EPI_ISL_1486348, EPI_ISL_1486351, EPI_ISL_1486359, EPI_ISL_1486367, EPI_ISL_1486369, EPI_ISL_1486376, EPI_ISL_1486381, EPI_ISL_1486383, EPI_ISL_1486385, EPI_ISL_1486400, EPI_ISL_1486403, EPI_ISL_1486406, EPI_ISL_1486407, EPI_ISL_1486408, EPI_ISL_1486410, EPI_ISL_1486413, EPI_ISL_1486414, EPI_ISL_1486416, EPI_ISL_1486417, EPI_ISL_1486419, EPI_ISL_1486423, EPI_ISL_1486432, EPI_ISL_1486435, EPI_ISL_1486447, EPI_ISL_1486453, EPI_ISL_1486454, EPI_ISL_1486456, EPI_ISL_1486460, EPI_ISL_1486462, EPI_ISL_1486463, EPI_ISL_1486464, EPI_ISL_1486467, EPI_ISL_1486468, EPI_ISL_1486469, EPI_ISL_1486471, EPI_ISL_1486479, EPI_ISL_1486482, EPI_ISL_1486485, EPI_ISL_1486486, EPI_ISL_1486487, EPI_ISL_1486488, EPI_ISL_1486491, EPI_ISL_1486492, EPI_ISL_1486495, EPI_ISL_1486496, EPI_ISL_1486500, EPI_ISL_1486502, EPI_ISL_1486503, EPI_ISL_1486504, EPI_ISL_1486506, EPI_ISL_1486509, EPI_ISL_1486511, EPI_ISL_1486512, EPI_ISL_1486516, EPI_ISL_1486518, EPI_ISL_1486523, EPI_ISL_1486532, EPI_ISL_1486535, EPI_ISL_1486539, EPI_ISL_1486547, EPI_ISL_1486551, EPI_ISL_1486553, EPI_ISL_1486561, EPI_ISL_1486564, EPI_ISL_1486572, EPI_ISL_1486576, EPI_ISL_1486578, EPI_ISL_1486584, EPI_ISL_1486585, EPI_ISL_1486586, EPI_ISL_1486589, EPI_ISL_1486606, EPI_ISL_1486607, EPI_ISL_1486609, EPI_ISL_1486614, EPI_ISL_1486615, EPI_ISL_1486624, EPI_ISL_1486626, EPI_ISL_1486645, EPI_ISL_1486648, EPI_ISL_1486651, EPI_ISL_1486653, EPI_ISL_1486656, EPI_ISL_1486657, EPI_ISL_1486669, EPI_ISL_1486675, EPI_ISL_1486677, EPI_ISL_1486682, EPI_ISL_1486685, EPI_ISL_1486686, EPI_ISL_1486695, EPI_ISL_1486698, EPI_ISL_1486702, EPI_ISL_1486704, EPI_ISL_1486705, EPI_ISL_1486712, EPI_ISL_1486714, EPI_ISL_1486718, EPI_ISL_1486726, EPI_ISL_1486731, EPI_ISL_1486737, EPI_ISL_1486740, EPI_ISL_1486741, EPI_ISL_1486745, EPI_ISL_1486747, EPI_ISL_1486749, EPI_ISL_1486753, EPI_ISL_1486771, EPI_ISL_1486772, EPI_ISL_1486774, EPI_ISL_1486775, EPI_ISL_1486778, EPI_ISL_1486783, EPI_ISL_1486784, EPI_ISL_1486789, EPI_ISL_1486799, EPI_ISL_1486800, EPI_ISL_1486801, EPI_ISL_1486803, EPI_ISL_1486806, EPI_ISL_1486812, EPI_ISL_1486815, EPI_ISL_1486818, EPI_ISL_1486825, EPI_ISL_1486832, EPI_ISL_1486836, EPI_ISL_1486839, EPI_ISL_1486840, EPI_ISL_1486857, EPI_ISL_1486868, EPI_ISL_1486872, EPI_ISL_1486873, EPI_ISL_1486880, EPI_ISL_1486884, EPI_ISL_1486885, EPI_ISL_1486890, EPI_ISL_1486891, EPI_ISL_1486896, EPI_ISL_1486899, EPI_ISL_1486913, EPI_ISL_1486916, EPI_ISL_1486919, EPI_ISL_1486925, EPI_ISL_1486926, EPI_ISL_1486933, EPI_ISL_1486937, EPI_ISL_1486939, EPI_ISL_1486944, EPI_ISL_1486951, EPI_ISL_1486952, EPI_ISL_1486953, EPI_ISL_1486956, EPI_ISL_1486959, EPI_ISL_1486960, EPI_ISL_1486964, EPI_ISL_1486965, EPI_ISL_1486968, EPI_ISL_1486977, EPI_ISL_1486982, EPI_ISL_1486987, EPI_ISL_1486993, EPI_ISL_1486994, EPI_ISL_1486996, EPI_ISL_1486997, EPI_ISL_1486999, EPI_ISL_1487000, EPI_ISL_1487001, EPI_ISL_1487003, EPI_ISL_1487005, EPI_ISL_1487013, EPI_ISL_1487017, EPI_ISL_1487022, EPI_ISL_1487024, EPI_ISL_1487032, EPI_ISL_1487034, EPI_ISL_1487036, EPI_ISL_1487037, EPI_ISL_1487042, EPI_ISL_1487044, EPI_ISL_1487045, EPI_ISL_1487052, EPI_ISL_1487056, EPI_ISL_1487060, EPI_ISL_1487062, EPI_ISL_1487063, EPI_ISL_1487068, EPI_ISL_1487070, EPI_ISL_1487072, EPI_ISL_1487074, EPI_ISL_1487077, EPI_ISL_1487083, EPI_ISL_1487086, EPI_ISL_1487089, EPI_ISL_1487090, EPI_ISL_1487094, EPI_ISL_1487095, EPI_ISL_1487096, EPI_ISL_1487098, EPI_ISL_1487100, EPI_ISL_1487103, EPI_ISL_1487106, EPI_ISL_1487107, EPI_ISL_1487114, EPI_ISL_1487117, EPI_ISL_1487118, EPI_ISL_1487123, EPI_ISL_1487124, EPI_ISL_1487125, EPI_ISL_1487126, EPI_ISL_1487129, EPI_ISL_1487131, EPI_ISL_1487132, EPI_ISL_1487135, EPI_ISL_1487136, EPI_ISL_1487143, EPI_ISL_1487146, EPI_ISL_1487149, EPI_ISL_1487151, EPI_ISL_1487415, EPI_ISL_1488227, EPI_ISL_1488230, EPI_ISL_1488261, EPI_ISL_1488270, EPI_ISL_1488286, EPI_ISL_1488294, EPI_ISL_1488298, EPI_ISL_1488333, EPI_ISL_1488337, EPI_ISL_1488338, EPI_ISL_1488343, EPI_ISL_1488346, EPI_ISL_1488354, EPI_ISL_1488361, EPI_ISL_1488364, EPI_ISL_1488366, EPI_ISL_1488376, EPI_ISL_1488388, EPI_ISL_1488397, EPI_ISL_1488406, EPI_ISL_1488422, EPI_ISL_1488423, EPI_ISL_1488427, EPI_ISL_1488428, EPI_ISL_1488433, EPI_ISL_1488435, EPI_ISL_1488450, EPI_ISL_1488451, EPI_ISL_1488476, EPI_ISL_1488477, EPI_ISL_1504115, EPI_ISL_1504116, EPI_ISL_1504119, EPI_ISL_1504125, EPI_ISL_1504128, EPI_ISL_1504132, EPI_ISL_1504133, EPI_ISL_1504138, EPI_ISL_1504141, EPI_ISL_1504147, EPI_ISL_1504149, EPI_ISL_1504162, EPI_ISL_1504163, EPI_ISL_1504166, EPI_ISL_1504167, EPI_ISL_1504174, EPI_ISL_1504177, EPI_ISL_1504180, EPI_ISL_1504181, EPI_ISL_1504183, EPI_ISL_1504184, EPI_ISL_1504195, EPI_ISL_1504196, EPI_ISL_1504200, EPI_ISL_1504208, EPI_ISL_1504209, EPI_ISL_1504222, EPI_ISL_1504225, EPI_ISL_1504226, EPI_ISL_1504235, EPI_ISL_1504239, EPI_ISL_1504240, EPI_ISL_1504243, EPI_ISL_1504244, EPI_ISL_1504248, EPI_ISL_1504249, EPI_ISL_1504265, EPI_ISL_1504276, EPI_ISL_1504280, EPI_ISL_1504284, EPI_ISL_1504288, EPI_ISL_1504289, EPI_ISL_1504291, EPI_ISL_1504294, EPI_ISL_1504297, EPI_ISL_1504298, EPI_ISL_1504303, EPI_ISL_1504306, EPI_ISL_1504313, EPI_ISL_1504316, EPI_ISL_1504320, EPI_ISL_1504321, EPI_ISL_1504322, EPI_ISL_1504323, EPI_ISL_1504325, EPI_ISL_1504327, EPI_ISL_1504329, EPI_ISL_1504333, EPI_ISL_1504335, EPI_ISL_1504340, EPI_ISL_1504347, EPI_ISL_1504348, EPI_ISL_1504352, EPI_ISL_1504355, EPI_ISL_1504361, EPI_ISL_1504365, EPI_ISL_1504368, EPI_ISL_1504369, EPI_ISL_1504373, EPI_ISL_1504374, EPI_ISL_1504375, EPI_ISL_1504380, EPI_ISL_1504381, EPI_ISL_1504386, EPI_ISL_1504388, EPI_ISL_1504392, EPI_ISL_1504394, EPI_ISL_1504969, EPI_ISL_1504970, EPI_ISL_1505002, EPI_ISL_1505010, EPI_ISL_1505016, EPI_ISL_1505055, EPI_ISL_1505143, EPI_ISL_1505154, EPI_ISL_1505157, EPI_ISL_1505182, EPI_ISL_1505192, EPI_ISL_1505194, EPI_ISL_1505199, EPI_ISL_1505202, EPI_ISL_1505210, EPI_ISL_1505219, EPI_ISL_1505223, EPI_ISL_1505226, EPI_ISL_1505235, EPI_ISL_1505238, EPI_ISL_1505241, EPI_ISL_1505249, EPI_ISL_1505251, EPI_ISL_1505252, EPI_ISL_1505262, EPI_ISL_1505274, EPI_ISL_1505275, EPI_ISL_1505276, EPI_ISL_1505282, EPI_ISL_1505285, EPI_ISL_1505287, EPI_ISL_1505288, EPI_ISL_1505298, EPI_ISL_1505302, EPI_ISL_1505315, EPI_ISL_1505327, EPI_ISL_1505336, EPI_ISL_1505337, EPI_ISL_1505340, EPI_ISL_1505341, EPI_ISL_1505342, EPI_ISL_1505351, EPI_ISL_1505352, EPI_ISL_1505353, EPI_ISL_1505360, EPI_ISL_1505365, EPI_ISL_1505372, EPI_ISL_1505380, EPI_ISL_1505384, EPI_ISL_1505386, EPI_ISL_1505387, EPI_ISL_1505391, EPI_ISL_1505396, EPI_ISL_1505402, EPI_ISL_1505406, EPI_ISL_1505416, EPI_ISL_1505418, EPI_ISL_1505426, EPI_ISL_1505427, EPI_ISL_1505428, EPI_ISL_1505439, EPI_ISL_1505440, EPI_ISL_1505456, EPI_ISL_1505797, EPI_ISL_1505800, EPI_ISL_1505804, EPI_ISL_1505805, EPI_ISL_1505819, EPI_ISL_1505826, EPI_ISL_1505831, EPI_ISL_1505841, EPI_ISL_1505842, EPI_ISL_1505850, EPI_ISL_1505851, EPI_ISL_1505857, EPI_ISL_1505861, EPI_ISL_1505864, EPI_ISL_1505872, EPI_ISL_1505882, EPI_ISL_1505884, EPI_ISL_1505891, EPI_ISL_1505894, EPI_ISL_1505898, EPI_ISL_1505902, EPI_ISL_1505904, EPI_ISL_1505905, EPI_ISL_1505906, EPI_ISL_1505909, EPI_ISL_1505911, EPI_ISL_1505914, EPI_ISL_1505916, EPI_ISL_1505923, EPI_ISL_1505930, EPI_ISL_1505932, EPI_ISL_1505946, EPI_ISL_1505947, EPI_ISL_1505948, EPI_ISL_1505955, EPI_ISL_1505957, EPI_ISL_1505962, EPI_ISL_1505966, EPI_ISL_1505967, EPI_ISL_1505968, EPI_ISL_1505978, EPI_ISL_1505981, EPI_ISL_1505982, EPI_ISL_1505983, EPI_ISL_1505984, EPI_ISL_1505985, EPI_ISL_1505988, EPI_ISL_1505992, EPI_ISL_1505997, EPI_ISL_1505998, EPI_ISL_1506011, EPI_ISL_1506013, EPI_ISL_1506021, EPI_ISL_1506024, EPI_ISL_1506028, EPI_ISL_1506031, EPI_ISL_1506032, EPI_ISL_1506061, EPI_ISL_1506064, EPI_ISL_1506068, EPI_ISL_1506073, EPI_ISL_1506077, EPI_ISL_1506081, EPI_ISL_1506082, EPI_ISL_1506084, EPI_ISL_1506087, EPI_ISL_1506095, EPI_ISL_1506096, EPI_ISL_1506101, EPI_ISL_1535348, EPI_ISL_1537342, EPI_ISL_1537348, EPI_ISL_1537412, EPI_ISL_1537478, EPI_ISL_1537483, EPI_ISL_1537488, EPI_ISL_1537540, EPI_ISL_1537582, EPI_ISL_1537586, EPI_ISL_1537647, EPI_ISL_1537701, EPI_ISL_1631116, EPI_ISL_1631123, EPI_ISL_1631183, EPI_ISL_1631200, EPI_ISL_1631250, EPI_ISL_1631274, EPI_ISL_1631427, EPI_ISL_1631432, EPI_ISL_1631515, EPI_ISL_1631545, EPI_ISL_1631569, EPI_ISL_1631588, EPI_ISL_1631606, EPI_ISL_1631617, EPI_ISL_1631618, EPI_ISL_1631626, EPI_ISL_1631636, EPI_ISL_1631711, EPI_ISL_1631720, EPI_ISL_1631737 | | | |
| see above | Lighthouse Lab in Milton Keynes | Wellcome Sanger Institute for the COVID-19 Genomics UK (COG-UK) Consortium | Cordelia Langford; David K. Jackson; Dominic Kwiatkowski; Ewan Harrison; Ian Johnston; Jeffrey Barrett; John Sillitoe on behalf of the Wellcome Sanger Institute COVID-19 Surveillance Team; Roberto Amato; Sonia Goncalves; The Lighthouse Lab in Milton Keynes and Alex Alderton |
| EPI_ISL_1833009, EPI_ISL_1833010, EPI_ISL_1833015, EPI_ISL_1833016 | Lincolnshire Hospitals and DeepSeq Nottingham | COVID-19 Genomics UK (COG-UK) Consortium | Christopher Moore; Fei Sang; Joeseph Chappell; Johnny Debebe; Jonathan Ball; Matthew Carlisle; Matthew Loose; Nadine Holmes; Nichola Duckworth; Patrick McClure; Sarah Walsh; Tim Sloan; Victoria Wright |
| EPI_ISL_1546174, EPI_ISL_1546190, EPI_ISL_1546191, EPI_ISL_1546203, EPI_ISL_1546207, EPI_ISL_1546208, EPI_ISL_1546209, EPI_ISL_1546210, EPI_ISL_1546211, EPI_ISL_1546218, EPI_ISL_1636830, EPI_ISL_1636831, EPI_ISL_1636832, EPI_ISL_1636833, EPI_ISL_1636850, EPI_ISL_1636854, EPI_ISL_1636855, EPI_ISL_1636856, EPI_ISL_1636857, EPI_ISL_1636858, EPI_ISL_1636875, EPI_ISL_1636882, EPI_ISL_1637263, EPI_ISL_1637267, EPI_ISL_1637269, EPI_ISL_1832359, EPI_ISL_1832389, EPI_ISL_1832401, EPI_ISL_1832403, EPI_ISL_1832405, EPI_ISL_1832408, EPI_ISL_1979098, EPI_ISL_1979104 | | | |
| see above | Liverpool Clinical Laboratories | COVID-19 Genomics UK (COG-UK) Consortium | A Alrezaihi; Alessandro Gerada; Alistair Darby; Angela Cowell; Anita Lucaci; Anu Chawla; Cassie Olateju; Catherine Hartley; Charlotte Nelson; Ecaterina Vamos; Elaine O'Toole; Eleanor G Bentley; Ghada T Shawli; Hermione Webster; Isabel García-Dorival; James Johnson; James P Stewart; Jenifer Manson; Joanne Watts; Jones Benjamin; Jordan J Clark; Julian Hiscox; L Luu; Lucille Rainbow; M Almsaud; Margaret Hughes; Mark Whitehead; Matthew Gemmell; Miren Iturriza-Gomara; Muhannad Alruwaili; N.P Randle; Neil Swainston; PKF Gilmore; Parul Sharma; Rebekah Penrice-Randal; Richard Eccles; Richard Gregory; Sam Haldenby; Steve Paterson; Stuart D Armstrong; Trevor Ian Robinson; Ximeng Han |
| EPI_ISL_1475216, EPI_ISL_1475217, EPI_ISL_1475218, EPI_ISL_1475219, EPI_ISL_1637194, EPI_ISL_1637195, EPI_ISL_1637226, EPI_ISL_1637227, EPI_ISL_1637228, EPI_ISL_1637229 | | | |
| see above | Northumbria University / South Tees Hospitals NHS Foundation Trust / North Cumbria Integrated Care NHS Foundation Trust / North Tees and Hartlepool NHS Foundation Trust / Newcastle Hospitals NHS Foundation Trust | COVID-19 Genomics UK (COG-UK) Consortium | Andrew Nelson; Brendan Payne; Clive Graham; Darren L Smith; Debra Padgett; Edward Barton; Emma Swindells; Garren Scott; Gary Black; Gary Eltringham; Giles S Holt; Greg R Young; Jane Greenaway; Jennifer Collins; John Allan; Joshua Loh; Lynn Dover; Matthew Bashton; Mohammad A Tariq; Paul Baker; Sarah Essex; Steve Liggett; Wen C Yew; Yusri Taha |
| EPI_ISL_1546273, EPI_ISL_1546274, EPI_ISL_1546275, EPI_ISL_1546276, EPI_ISL_1546279, EPI_ISL_1546280, EPI_ISL_1546281, EPI_ISL_1546285, EPI_ISL_1546289, EPI_ISL_1546291, EPI_ISL_1546296, EPI_ISL_1546298, EPI_ISL_1546300, EPI_ISL_1546334, EPI_ISL_1546335, EPI_ISL_1546336, EPI_ISL_1546340, EPI_ISL_1546342, EPI_ISL_1546344, EPI_ISL_1546352, EPI_ISL_1546354, EPI_ISL_1546355, EPI_ISL_1546358, EPI_ISL_1546360, EPI_ISL_1546362, EPI_ISL_1546363, EPI_ISL_1546364, EPI_ISL_1546365, EPI_ISL_1546370, EPI_ISL_1546374, EPI_ISL_1546375, EPI_ISL_1546379, EPI_ISL_1546381, EPI_ISL_1546382, EPI_ISL_1546383, EPI_ISL_1546384, EPI_ISL_1546388, EPI_ISL_1546390, EPI_ISL_1546394, EPI_ISL_1546395, EPI_ISL_1546399, EPI_ISL_1546400, EPI_ISL_1546401, EPI_ISL_1546402, EPI_ISL_1546405, EPI_ISL_1546411, EPI_ISL_1546412, EPI_ISL_1546413, EPI_ISL_1546414, EPI_ISL_1546415, EPI_ISL_1546417, EPI_ISL_1546431, EPI_ISL_1546432, EPI_ISL_1546436, EPI_ISL_1546437, EPI_ISL_1546497, EPI_ISL_1546499, EPI_ISL_1546503, EPI_ISL_1546508, EPI_ISL_1546518, EPI_ISL_1546579, EPI_ISL_1546581, EPI_ISL_1546588, EPI_ISL_1546640, EPI_ISL_1546642, EPI_ISL_1546644, EPI_ISL_1546652, EPI_ISL_1546654, EPI_ISL_1546657, EPI_ISL_1546660, EPI_ISL_1546661, EPI_ISL_1546669, EPI_ISL_1546674, EPI_ISL_1546678, EPI_ISL_1546679, EPI_ISL_1546680, EPI_ISL_1546683, EPI_ISL_1546684, EPI_ISL_1546688, EPI_ISL_1546691, EPI_ISL_1546699, EPI_ISL_1546701, EPI_ISL_1546705, EPI_ISL_1546707, EPI_ISL_1546708, EPI_ISL_1546710, EPI_ISL_1546711, EPI_ISL_1546720, EPI_ISL_1546722, EPI_ISL_1546726, EPI_ISL_1546728, EPI_ISL_1546736, EPI_ISL_1546740, EPI_ISL_1546741, EPI_ISL_1546745, EPI_ISL_1546749, EPI_ISL_1546752, EPI_ISL_1546784, EPI_ISL_1546796, EPI_ISL_1546824, EPI_ISL_1546828, EPI_ISL_1546829, EPI_ISL_1546840, EPI_ISL_1546842, EPI_ISL_1546844, EPI_ISL_1546848, EPI_ISL_1546849, EPI_ISL_1546850, EPI_ISL_1546851, EPI_ISL_1546856, EPI_ISL_1546881, EPI_ISL_1546951, EPI_ISL_1546952, EPI_ISL_1546953, EPI_ISL_1546955, EPI_ISL_1546960, EPI_ISL_1546961, EPI_ISL_1546963, EPI_ISL_1546965, EPI_ISL_1546966, EPI_ISL_1546968, EPI_ISL_1546971, EPI_ISL_1546982, EPI_ISL_1546983, EPI_ISL_1546984, EPI_ISL_1546985, EPI_ISL_1546987, EPI_ISL_1546988, EPI_ISL_1546989, EPI_ISL_1546990, EPI_ISL_1546994, EPI_ISL_1546997, EPI_ISL_1547059, EPI_ISL_1547060, EPI_ISL_1547061, EPI_ISL_1547062, EPI_ISL_1547065, EPI_ISL_1547106, EPI_ISL_1547109, EPI_ISL_1547110, EPI_ISL_1547114, EPI_ISL_1547116, EPI_ISL_1547117, EPI_ISL_1547120, EPI_ISL_1547125, EPI_ISL_1547131, EPI_ISL_1547132, EPI_ISL_1547133, EPI_ISL_1547139, EPI_ISL_1547140, EPI_ISL_1547142, EPI_ISL_1547150, EPI_ISL_1547154, EPI_ISL_1547155, EPI_ISL_1547156, EPI_ISL_1547161, EPI_ISL_1547165, EPI_ISL_1547168, EPI_ISL_1547169, EPI_ISL_1547170, EPI_ISL_1547174, EPI_ISL_1547176, EPI_ISL_1547177, EPI_ISL_1547178, EPI_ISL_1547181, EPI_ISL_1547183, EPI_ISL_1547184, EPI_ISL_1547221, EPI_ISL_1547226, EPI_ISL_1547227 | | | |
| see above | Originating lab: Wales Specialist Virology Centre Sequencing lab: Pathogen Genomics Unit | Public Health Wales Microbiology Cardiff Wales Specialist Virology Centre | Alec Birchley; Alexander Adams; Amy Gaskin; Angela Marchbank; Bree Gatica-Wilcox; Catherine Moore; Jason Coombes; Joanne Watkins; Joel Southgate; Johnathan Evans; Laura Gifford; Lauren Gilbert; Lee Graham; Malorie Perry; Matthew Bull; Nicole Pacchiarini; Sally Corden; Sara Kumziene-Summerhayes; Sara Rey; Sarah Taylor; Simon Cottrell; Sophie Jones; Tom Connor |
| EPI_ISL_1730470, EPI_ISL_1730507, EPI_ISL_1730508, EPI_ISL_1730509, EPI_ISL_1730510, EPI_ISL_1730512, EPI_ISL_1730514, EPI_ISL_1730516, EPI_ISL_1730517, EPI_ISL_1730518, EPI_ISL_1730519, EPI_ISL_1730521, EPI_ISL_1730539, EPI_ISL_1730540, EPI_ISL_1730541, EPI_ISL_1730542, EPI_ISL_1730544, EPI_ISL_1730549, EPI_ISL_1730554, EPI_ISL_1730556, EPI_ISL_1730557, EPI_ISL_1730566, EPI_ISL_1730571, EPI_ISL_1730576 | | | |
| see above | Oxford Viromics, NDM, University of Oxford; Oxford University Hospitals; Basingstoke and North Hampshire Hospital | COVID-19 Genomics UK (COG-UK) Consortium | Alex Mobbs; Amy Trebes; Anita Justice; Catrin Moore; Christophe Fraser; David Bonsall; David Buck; Emma Wise; George Macintyre; Jessica Lynch; John Todd; Mariateresa de Cesare; Matilde Mori; Monique Andersson; Nathan Moore; Nick Cortes; Robert Shaw; Stephen Kidd; Tanya Golubchik; Timothy Peto |
| EPI_ISL_1475489, EPI_ISL_1475490, EPI_ISL_1475491, EPI_ISL_1475492, EPI_ISL_1546239, EPI_ISL_1546240, EPI_ISL_1546241, EPI_ISL_1546249, EPI_ISL_1546250, EPI_ISL_1730419 | | | |
| see above | Queens Medical Centre, Clinical Microbiology Department / DeepSeq Nottingham | COVID-19 Genomics UK (COG-UK) Consortium | Christopher Moore; Emily Park; Fei Sang; Gemma Clark; Hannah Howson-Wells; Jack Hill; Jayasree Dey; Johnny Debebe; Jonathan Ball; Joseph Chappell; Manjinder Khakh; Matthew Carlisle; Matthew Loose; Michelle M Lister; Nadine Holmes; Patrick McClure; Theocharis Tsoleridis; Timothy Byaruhanga; Vicki M Fleming; Victoria Wright; Wendy Smith |
| EPI_ISL_1467562, EPI_ISL_1483827, EPI_ISL_1483830, EPI_ISL_1483831, EPI_ISL_1483833, EPI_ISL_1483839, EPI_ISL_1483858, EPI_ISL_1483860, EPI_ISL_1483868, EPI_ISL_1483880, EPI_ISL_1483882, EPI_ISL_1483884, EPI_ISL_1483886, EPI_ISL_1483895, EPI_ISL_1483903, EPI_ISL_1483905, EPI_ISL_1483907, EPI_ISL_1483912, EPI_ISL_1483915, EPI_ISL_1483923, EPI_ISL_1483927, EPI_ISL_1483931, EPI_ISL_1483932, EPI_ISL_1483934, EPI_ISL_1483945, EPI_ISL_1483949, EPI_ISL_1483953, EPI_ISL_1483960, EPI_ISL_1483962, EPI_ISL_1483971, EPI_ISL_1483979, EPI_ISL_1483981, EPI_ISL_1483997, EPI_ISL_1484001, EPI_ISL_1484004, EPI_ISL_1484006, EPI_ISL_1484010, EPI_ISL_1484014, EPI_ISL_1484016, EPI_ISL_1484019, EPI_ISL_1484023, EPI_ISL_1484032, EPI_ISL_1484038, EPI_ISL_1484041, EPI_ISL_1484043, EPI_ISL_1484044, EPI_ISL_1484048, EPI_ISL_1484071, EPI_ISL_1484073, EPI_ISL_1484079, EPI_ISL_1484084, EPI_ISL_1484086, EPI_ISL_1485481, EPI_ISL_1485487, EPI_ISL_1485493, EPI_ISL_1485496, EPI_ISL_1485498, EPI_ISL_1485504, EPI_ISL_1485508, EPI_ISL_1485530, EPI_ISL_1485531, EPI_ISL_1485556, EPI_ISL_1485578, EPI_ISL_1485591, EPI_ISL_1485593, EPI_ISL_1485594, EPI_ISL_1485600, EPI_ISL_1485601, EPI_ISL_1485602, EPI_ISL_1485613, EPI_ISL_1485625, EPI_ISL_1485630, EPI_ISL_1485635, EPI_ISL_1485641, EPI_ISL_1485642, EPI_ISL_1485649, EPI_ISL_1485651, EPI_ISL_1485653, EPI_ISL_1485658, EPI_ISL_1485659, EPI_ISL_1485670, EPI_ISL_1485681, EPI_ISL_1485684, EPI_ISL_1485694, EPI_ISL_1485695, EPI_ISL_1485699, EPI_ISL_1485703, EPI_ISL_1485712, EPI_ISL_1485713, EPI_ISL_1485714, EPI_ISL_1485723, EPI_ISL_1485727, EPI_ISL_1485749, EPI_ISL_1485751, EPI_ISL_1485753, EPI_ISL_1485755, EPI_ISL_1485758, EPI_ISL_1485759, EPI_ISL_1485764, EPI_ISL_1485771, EPI_ISL_1485774, EPI_ISL_1485789, EPI_ISL_1485790, EPI_ISL_1485792, EPI_ISL_1485793, EPI_ISL_1485794, EPI_ISL_1485802, EPI_ISL_1486422, EPI_ISL_1486478, EPI_ISL_1486501, EPI_ISL_1486508, EPI_ISL_1486550, EPI_ISL_1486575, EPI_ISL_1486627, EPI_ISL_1486710, EPI_ISL_1486766, EPI_ISL_1486769, EPI_ISL_1488523, EPI_ISL_1488525, EPI_ISL_1488544, EPI_ISL_1488552, EPI_ISL_1488558, EPI_ISL_1488575, EPI_ISL_1488576, EPI_ISL_1488605, EPI_ISL_1488608, EPI_ISL_1488615, EPI_ISL_1488617, EPI_ISL_1488627, EPI_ISL_1488630, EPI_ISL_1488633, EPI_ISL_1488641, EPI_ISL_1488654, EPI_ISL_1488659, EPI_ISL_1488674, EPI_ISL_1488675, EPI_ISL_1488678, EPI_ISL_1488704, EPI_ISL_1488723, EPI_ISL_1488740, EPI_ISL_1488762, EPI_ISL_1488774, EPI_ISL_1488777, EPI_ISL_1488789, EPI_ISL_1488795, EPI_ISL_1488802, EPI_ISL_1488806, EPI_ISL_1488809, EPI_ISL_1488815, EPI_ISL_1488821, EPI_ISL_1488836, EPI_ISL_1488839, EPI_ISL_1488841, EPI_ISL_1488859, EPI_ISL_1488861, EPI_ISL_1488862, EPI_ISL_1504667, EPI_ISL_1504682, EPI_ISL_1504683, EPI_ISL_1504684, EPI_ISL_1504685, EPI_ISL_1504690, EPI_ISL_1504694, EPI_ISL_1504696, EPI_ISL_1504698, EPI_ISL_1504703, EPI_ISL_1504706, EPI_ISL_1504707, EPI_ISL_1504710, EPI_ISL_1504712, EPI_ISL_1504716, EPI_ISL_1504723, EPI_ISL_1504724, EPI_ISL_1504728, EPI_ISL_1504730, EPI_ISL_1504732, EPI_ISL_1504734, EPI_ISL_1504742, EPI_ISL_1504743, EPI_ISL_1504744, EPI_ISL_1504745, EPI_ISL_1504749, EPI_ISL_1504750, EPI_ISL_1504751, EPI_ISL_1504753, EPI_ISL_1504756, EPI_ISL_1504758, EPI_ISL_1504767, EPI_ISL_1504768, EPI_ISL_1504769, EPI_ISL_1504771, EPI_ISL_1504772, EPI_ISL_1504773, EPI_ISL_1504776, EPI_ISL_1504784, EPI_ISL_1504791, EPI_ISL_1504795, EPI_ISL_1504804, EPI_ISL_1504805, EPI_ISL_1504806, EPI_ISL_1504808, EPI_ISL_1504814, EPI_ISL_1504817, EPI_ISL_1504820, EPI_ISL_1504826, EPI_ISL_1504827, EPI_ISL_1504829, EPI_ISL_1504831, EPI_ISL_1504834, EPI_ISL_1504835, EPI_ISL_1504846, EPI_ISL_1504851, EPI_ISL_1504861, EPI_ISL_1504868, EPI_ISL_1504871, EPI_ISL_1504875, EPI_ISL_1504878, EPI_ISL_1504879, EPI_ISL_1504882, EPI_ISL_1504888, EPI_ISL_1504893, EPI_ISL_1504894, EPI_ISL_1504895, EPI_ISL_1504905, EPI_ISL_1504913, EPI_ISL_1504940, EPI_ISL_1504974, EPI_ISL_1504978, EPI_ISL_1504985, EPI_ISL_1505085, EPI_ISL_1505092, EPI_ISL_1505097, EPI_ISL_1505101, EPI_ISL_1505107, EPI_ISL_1505113, EPI_ISL_1505117, EPI_ISL_1505145, EPI_ISL_1505152, EPI_ISL_1505470, EPI_ISL_1505491, EPI_ISL_1505492, EPI_ISL_1505496, EPI_ISL_1505497, EPI_ISL_1505520, EPI_ISL_1505525, EPI_ISL_1505531, EPI_ISL_1505534, EPI_ISL_1505535, EPI_ISL_1505539, EPI_ISL_1505545, EPI_ISL_1505550, EPI_ISL_1505569, EPI_ISL_1505580, EPI_ISL_1505583, EPI_ISL_1505589, EPI_ISL_1505591, EPI_ISL_1505599, EPI_ISL_1505600, EPI_ISL_1505608, EPI_ISL_1505612, EPI_ISL_1505621, EPI_ISL_1505625, EPI_ISL_1505637, EPI_ISL_1505643, EPI_ISL_1505650, EPI_ISL_1505653, EPI_ISL_1505655, EPI_ISL_1505681, EPI_ISL_1505694, EPI_ISL_1505695, EPI_ISL_1505698, EPI_ISL_1505706, EPI_ISL_1505718, EPI_ISL_1505728, EPI_ISL_1505735, EPI_ISL_1505739, EPI_ISL_1505750, EPI_ISL_1505767, EPI_ISL_1505768, EPI_ISL_1505770, EPI_ISL_1505771, EPI_ISL_1505786, EPI_ISL_1505793, EPI_ISL_1505808, EPI_ISL_1505811, EPI_ISL_1505812, EPI_ISL_1505814, EPI_ISL_1505827, EPI_ISL_1505845, EPI_ISL_1505856, EPI_ISL_1505858, EPI_ISL_1505859, EPI_ISL_1505862, EPI_ISL_1505866, EPI_ISL_1505873, EPI_ISL_1505889, EPI_ISL_1505899, EPI_ISL_1505907, EPI_ISL_1505940, EPI_ISL_1505944, EPI_ISL_1505950, EPI_ISL_1505960, EPI_ISL_1505961, EPI_ISL_1505963, EPI_ISL_1505973, EPI_ISL_1505977, EPI_ISL_1505995, EPI_ISL_1506002, EPI_ISL_1506003, EPI_ISL_1506006, EPI_ISL_1506015, EPI_ISL_1506026, EPI_ISL_1506039, EPI_ISL_1506041, EPI_ISL_1506045, EPI_ISL_1506046, EPI_ISL_1506050, EPI_ISL_1506060, EPI_ISL_1506065, EPI_ISL_1506066, EPI_ISL_1506078, EPI_ISL_1506085, EPI_ISL_1506089, EPI_ISL_1506092, EPI_ISL_1506094, EPI_ISL_1506104, EPI_ISL_1518092, EPI_ISL_1518239, EPI_ISL_1518331, EPI_ISL_1519270, EPI_ISL_1519272, EPI_ISL_1519273, EPI_ISL_1519279, EPI_ISL_1519281, EPI_ISL_1519282, EPI_ISL_1519286, EPI_ISL_1519302, EPI_ISL_1519910, EPI_ISL_1519911, EPI_ISL_1519912, EPI_ISL_1519913, EPI_ISL_1519914, EPI_ISL_1519915, EPI_ISL_1519916, EPI_ISL_1519917, EPI_ISL_1519920, EPI_ISL_1519926, EPI_ISL_1519927, EPI_ISL_1519928, EPI_ISL_1519935, EPI_ISL_1519993, EPI_ISL_1519994, EPI_ISL_1519995, EPI_ISL_1535868, EPI_ISL_1535878, EPI_ISL_1537138, EPI_ISL_1564811 | | | |
| see above | Randox Laboratories | Wellcome Sanger Institute for the COVID-19 Genomics UK (COG-UK) Consortium | Cordelia Langford; David K. Jackson; Dominic Kwiatkowski; Ewan Harrison; Ian Johnston; Jeffrey Barrett; John Sillitoe on behalf of the Wellcome Sanger Institute COVID-19 Surveillance Team; Randox Laboratories and Alex Alderton; Roberto Amato; Sonia Goncalves |
| EPI_ISL_1976463, EPI_ISL_1976465, EPI_ISL_1976466, EPI_ISL_1976468, EPI_ISL_1976469, EPI_ISL_1976471, EPI_ISL_1976473, EPI_ISL_1976475, EPI_ISL_1976476, EPI_ISL_1976478, EPI_ISL_1976483, EPI_ISL_1976486, EPI_ISL_1976488, EPI_ISL_1976490, EPI_ISL_1976492, EPI_ISL_1976493, EPI_ISL_1976495, EPI_ISL_1976498, EPI_ISL_1976500, EPI_ISL_1976501, EPI_ISL_1976512, EPI_ISL_1976513, EPI_ISL_1976515, EPI_ISL_1976517, EPI_ISL_1976521, EPI_ISL_1976522, EPI_ISL_1976524, EPI_ISL_1976526, EPI_ISL_1976528, EPI_ISL_1976530, EPI_ISL_1976531, EPI_ISL_1976533, EPI_ISL_1976534, EPI_ISL_1976537, EPI_ISL_1976543, EPI_ISL_1976545, EPI_ISL_1976547, EPI_ISL_1976549, EPI_ISL_1976551, EPI_ISL_1976945, EPI_ISL_1976946, EPI_ISL_1976948, EPI_ISL_1976950, EPI_ISL_1976951, EPI_ISL_1976953, EPI_ISL_1976957, EPI_ISL_1976959, EPI_ISL_1976960, EPI_ISL_1976962, EPI_ISL_1977038, EPI_ISL_1977040, EPI_ISL_1977042, EPI_ISL_1977044, EPI_ISL_1977046, EPI_ISL_1977047, EPI_ISL_1977048, EPI_ISL_1977050, EPI_ISL_1977057, EPI_ISL_1977058, EPI_ISL_1977059, EPI_ISL_1977060, EPI_ISL_1977061, EPI_ISL_1977062, EPI_ISL_1977063, EPI_ISL_1977064, EPI_ISL_1977065, EPI_ISL_1977067, EPI_ISL_1977068, EPI_ISL_1977069, EPI_ISL_1977070, EPI_ISL_1977071, EPI_ISL_1977073, EPI_ISL_1977075, EPI_ISL_1977076, EPI_ISL_1977077, EPI_ISL_1977078, EPI_ISL_1977080, EPI_ISL_1977081, EPI_ISL_1977083, EPI_ISL_1977103, EPI_ISL_1977104, EPI_ISL_1977108, EPI_ISL_1977109, EPI_ISL_1977111, EPI_ISL_1977112, EPI_ISL_1977114, EPI_ISL_1977116, EPI_ISL_1977140, EPI_ISL_1977141, EPI_ISL_1977143, EPI_ISL_1977145, EPI_ISL_1977146, EPI_ISL_1977148, EPI_ISL_1977157, EPI_ISL_1977161, EPI_ISL_1977162, EPI_ISL_1977172, EPI_ISL_1977175, EPI_ISL_1977179, EPI_ISL_1977180, EPI_ISL_1977182, EPI_ISL_1977184, EPI_ISL_1977190, EPI_ISL_1977192, EPI_ISL_1977194, EPI_ISL_1977196, EPI_ISL_1977197, EPI_ISL_1977199 | | | |
| see above | Regional Virus Laboratory, Belfast Health and Social Care Trust | COVID-19 Genomics UK (COG-UK) Consortium | Alison Watt; Ciara Cox; Conall McCaughey; David Simpson; Derek Fairley; James McKenna; Mairead Connor; Susan Feeney; Tanya Curran; Zoltan Molnar |
| EPI_ISL_1474829, EPI_ISL_1474830, EPI_ISL_1474831, EPI_ISL_1474832, EPI_ISL_1474845, EPI_ISL_1474846, EPI_ISL_1474847, EPI_ISL_1474873, EPI_ISL_1474875, EPI_ISL_1474879, EPI_ISL_1474889, EPI_ISL_1474890, EPI_ISL_1474891, EPI_ISL_1637000, EPI_ISL_1637170, EPI_ISL_1637172, EPI_ISL_1730367, EPI_ISL_1730368, EPI_ISL_1730381, EPI_ISL_1730383 | | | |
| see above | University College London, Great Ormond Street Hospital for Children NHS Foundation Trust, Imperial College Healthcare NHS Trust | COVID-19 Genomics UK (COG-UK) Consortium | Alison Holmes; Charlotte Williams; Helena Tutill; Jacqueline Findlay; James Price; Judith Breuer; Julianne Brown; Kathryn Harris; Leysa Forrest; Mark Kristiansen; Paola Niola; Paola Resende Silva; Patricia Dyal; Paul Randell; Rachel Williams; Samuel Weeks; Sergi Castellano; Sunando Roy; Tony Brooks; Yasmin Panchbhaya |
| EPI_ISL_1545977, EPI_ISL_1545979, EPI_ISL_1545981, EPI_ISL_1636714, EPI_ISL_1636716 | University Hospitals Of Leicester NHS Trust and DeepSeq Nottingham | COVID-19 Genomics UK (COG-UK) Consortium | Christopher Holmes; Christopher Moore; Fei Sang; Joeseph Chappell; Johnny Debebe; Jonathan Ball; Julian Tang; Karlie Fallon; Matthew Carlisle; Matthew Loose; Nadine Holmes; Patrick McClure; Paul Bird; Thomas Helmer; Victoria Wright |
| EPI_ISL_1474452, EPI_ISL_1474453, EPI_ISL_1636551, EPI_ISL_1636552, EPI_ISL_1636553, EPI_ISL_1636561, EPI_ISL_1636562, EPI_ISL_1636563, EPI_ISL_1636565, EPI_ISL_1636566, EPI_ISL_1636567, EPI_ISL_1636569, EPI_ISL_1636575, EPI_ISL_1636579, EPI_ISL_1636581, EPI_ISL_1636583, EPI_ISL_1636585, EPI_ISL_1636586, EPI_ISL_1636589, EPI_ISL_1636591, EPI_ISL_1729977, EPI_ISL_2125450 | | | |
| see above | University of Birmingham | COVID-19 Genomics UK (COG-UK) Consortium | Alex Richter; Andrew Bosworth; Andrew Bosworth. Queen Elizabeth Hospital: Anna Casey; Andrew D Beggs PHE Heartlands Lab: Husam Osman; Charlotte Poxon; Institute of Microbiology; Joanne Stockton; Josh Quick; Kasun Wanigasooriya; Mike Kidd; Nicholas Loman. University of Birmingham Testing Laboratory: Celina M Whalley; Oliver Pickles; Radoslaw Poplawski; Samuel Nicholls; University of Birmingham: Claire McMurray; Will Rowe |
| EPI_ISL_1730142, | University of Exeter | COVID-19 Genomics UK | Aaron Jeffries; Audrey Farbos; Ben Temperton; Jane Masoli; Joanna Warwick-Dugdale; Michelle Michelsen; Robyn Manley; Stephen Michell |

| | | | |
|---|---|---|---|
| EPI_ISL_1730143 | | (COG-UK) Consortium | |
| EPI_ISL_1474602, EPI_ISL_1545902, EPI_ISL_1545903, EPI_ISL_1636687, EPI_ISL_1636688, EPI_ISL_1636689, EPI_ISL_1636690, EPI_ISL_1636704, EPI_ISL_1730095, EPI_ISL_1730096, EPI_ISL_1730257, EPI_ISL_1730258, EPI_ISL_1730259, EPI_ISL_1730260, EPI_ISL_1730261, EPI_ISL_1730262, EPI_ISL_1730263, EPI_ISL_1730264, EPI_ISL_1730265, EPI_ISL_1730266, EPI_ISL_1730267, EPI_ISL_1730268, EPI_ISL_1730269, EPI_ISL_1730286, EPI_ISL_1730287, EPI_ISL_2125855, EPI_ISL_2125872 | | | |
| see above | Virology Department, Royal Infirmary of Edinburgh, NHS Lothian / School of Biological Sciences, University of Edinburgh | COVID-19 Genomics UK (COG-UK) Consortium | Colquhoun R; Cotton S; Dewar R; Gallagher A; Hill V; Jackson B; Maloney D; McCrone JT; McHugh M; O'Toole Á; Rambaut A; Rooke S; Scher E; Templeton K; Yu X |
| EPI_ISL_1476707, EPI_ISL_1476710, EPI_ISL_1476712, EPI_ISL_1476716, EPI_ISL_1476720, EPI_ISL_1476722, EPI_ISL_1547311, EPI_ISL_1547315, EPI_ISL_1547343, EPI_ISL_1547353, EPI_ISL_1638068, EPI_ISL_1638076, EPI_ISL_1730926, EPI_ISL_1730927, EPI_ISL_1730928, EPI_ISL_1730929, EPI_ISL_1730930, EPI_ISL_1730965, EPI_ISL_1730976, EPI_ISL_1731024, EPI_ISL_1731036, EPI_ISL_1833858, EPI_ISL_1833869 | | | |
| see above | Virology Department, Sheffield Teaching Hospitals NHS Foundation Trust/Department of Infection, Immunity and Cardiovascular Disease, The Medical School, University of Sheffield | COVID-19 Genomics UK (COG-UK) Consortium | Adri Angyal; Alex Keeley; Benjamin Lindsey; Cariad Evans; Danielle Groves; Dave Partridge; Katie Johnson; Laura Carrilero; Luke Green; Matthew Parker; Matthew Wyles; Mehmet Yavuz; Mohammad Raza; Nikki Smith; Paul Parsons; Rachel Tucker; Rebecca Brown; Thushan de Silva |
| EPI_ISL_1546271, EPI_ISL_1546278, EPI_ISL_1546287, EPI_ISL_1546288, EPI_ISL_1546292, EPI_ISL_1546294, EPI_ISL_1546330, EPI_ISL_1546337, EPI_ISL_1546338, EPI_ISL_1546366, EPI_ISL_1546367, EPI_ISL_1546368, EPI_ISL_1546369, EPI_ISL_1546372, EPI_ISL_1546391, EPI_ISL_1546393, EPI_ISL_1546397, EPI_ISL_1546404, EPI_ISL_1546406, EPI_ISL_1546416, EPI_ISL_1546418, EPI_ISL_1546429, EPI_ISL_1546504, EPI_ISL_1546506, EPI_ISL_1546509, EPI_ISL_1546510, EPI_ISL_1546513, EPI_ISL_1546532, EPI_ISL_1546543, EPI_ISL_1546643, EPI_ISL_1546659, EPI_ISL_1546662, EPI_ISL_1546682, EPI_ISL_1546685, EPI_ISL_1546714, EPI_ISL_1546743, EPI_ISL_1546746, EPI_ISL_1546747, EPI_ISL_1546750, EPI_ISL_1546826, EPI_ISL_1546827, EPI_ISL_1546830, EPI_ISL_1546831, EPI_ISL_1546832, EPI_ISL_1546833, EPI_ISL_1546835, EPI_ISL_1546836, EPI_ISL_1546837, EPI_ISL_1546838, EPI_ISL_1546839, EPI_ISL_1637551, EPI_ISL_1637582, EPI_ISL_1637583, EPI_ISL_1637589, EPI_ISL_1637590, EPI_ISL_1637591, EPI_ISL_1637595, EPI_ISL_1637597, EPI_ISL_1637599, EPI_ISL_1637600, EPI_ISL_1637655, EPI_ISL_1637656, EPI_ISL_1637657, EPI_ISL_1637659, EPI_ISL_1637662, EPI_ISL_1637663, EPI_ISL_1637664, EPI_ISL_1637742, EPI_ISL_1637769, EPI_ISL_1637773, EPI_ISL_1637776, EPI_ISL_1637779, EPI_ISL_1637783, EPI_ISL_1637784, EPI_ISL_1637785, EPI_ISL_1637788, EPI_ISL_1637792, EPI_ISL_1637795, EPI_ISL_1637796, EPI_ISL_1637798, EPI_ISL_1637799, EPI_ISL_1637800, EPI_ISL_1637802, EPI_ISL_1637825, EPI_ISL_1637835, EPI_ISL_1637839, EPI_ISL_1637847, EPI_ISL_1637852, EPI_ISL_1637857, EPI_ISL_1637863, EPI_ISL_1637865, EPI_ISL_1833189, EPI_ISL_1833205 | | | |
| see above | Wales Specialist Virology Centre Sequencing lab: Pathogen Genomics Unit | Public Health Wales Microbiology Cardiff Wales Specialist Virology Centre | Alec Birchley; Alexander Adams; Amy Gaskin; Angela Marchbank; Bree Gatica-Wilcox; Catherine Moore; Jason Coombes; Joanne Watkins; Joel Southgate; Johnathan Evans; Laura Gifford; Lauren Gilbert; Lee Graham; Malorie Perry; Matthew Bull; Nicole Pacchiarini; Sally Corden; Sara Kumziene-Summerhayes; Sara Rey; Sarah Taylor; Simon Cottrell; Sophie Jones; Tom Connor |
| EPI_ISL_2125495 | West of Scotland Specialist Virology Centre, NHSGGC / MRC-University of Glasgow Centre for Virus Research | COVID-19 Genomics UK (COG-UK) Consortium | Alasdair MacLean; Alice Broos; Ana da Silva Filipe; Antonia Ho; Daniel Mair; David L Robertson; Emma Thomson; Guy Mollett; Ioulia Tsatsani; James Shepherd; Jenna Nichols; Joseph Hughes; Kathy Li; Kathy Smollett; Kyriaki Nomikou; Lily Tong; Matthew Holden; Natasha Johnson; Rachel Blacow; Richard Orton; Rory Gunson; Sarah McDonald; Sharif Shaaban; Sreenu Vattipally; Stephen Carmichael |

We gratefully acknowledge the following Authors from the Originating laboratories responsible for obtaining the specimens, as well as the Submitting laboratories where the genome data were generated and shared via GISAID, on which this research is based.

All Submitters of data may be contacted directly via www.gisaid.org

Authors are sorted alphabetically.

| Accession ID | Originating Laboratory | Submitting Laboratory | Authors |
| --- | --- | --- | --- |
| EPI_ISL_1636998, EPI_ISL_1636999, EPI_ISL_1730318, EPI_ISL_1730319 | Barts Health NHS Trust | COVID-19 Genomics UK (COG-UK) Consortium | BROAD; Beatrix; CUTINO-MOGUEL; Claire; David; Dola; HARRINGTON; KELE; KULASEGARAN-SHYLINI; Maria-Teresa; OWOYEMI; Raghavendran |
| EPI_ISL_1638261, EPI_ISL_1638264, EPI_ISL_1638265 | Bioinformatics and Biostatistics Lab, Advanced Sequencing Facility | COVID-19 Genomics UK (COG-UK) Consortium | Aengus Stewart; Chelsea Sawyer; Harshil Patel; Jerome Nicod; Laura Cubitt; Margaret Crawford |

EPI_ISL_1547247, EPI_ISL_1547250, EPI_ISL_1547251, EPI_ISL_1547252, EPI_ISL_1547253, EPI_ISL_1547254, EPI_ISL_1547255, EPI_ISL_1547256, EPI_ISL_1547257, EPI_ISL_1547258, EPI_ISL_1547306, EPI_ISL_1547308, EPI_ISL_1637979, EPI_ISL_1637980, EPI_ISL_1730917, EPI_ISL_1730918, EPI_ISL_1730919, EPI_ISL_1730920, EPI_ISL_1730921

| Accession ID | Originating Laboratory | Submitting Laboratory | Authors |
| --- | --- | --- | --- |
| see above | Centre for Enzyme Innovation, University of Portsmouth / Translational Research Laboratory, Portsmouth Hospitals NHS Trust | COVID-19 Genomics UK (COG-UK) Consortium | Angela Beckett; Christopher Fearn; Kate Cook; Katie Loveson; Salman Goudarzi; Samuel Robson; Scott Elliott; Sharon Glaysher |

EPI_ISL_1545849, EPI_ISL_1545850, EPI_ISL_1545851, EPI_ISL_1636612, EPI_ISL_1636613, EPI_ISL_1636614, EPI_ISL_1636615, EPI_ISL_1730041, EPI_ISL_1730055, EPI_ISL_1730057, EPI_ISL_1730059

| Accession ID | Originating Laboratory | Submitting Laboratory | Authors |
| --- | --- | --- | --- |
| see above | Department of Pathology, University of Cambridge | COVID-19 Genomics UK (COG-UK) Consortium | Aminu S. Jahun; Ian Goodfellow; Iliana Georgana; Martin D. Curran; Myra Hosmillo; Rhys Izuagbe; Surendra Parmar; William L. Hamilton; Yasmin Chaudhry |

EPI_ISL_1504112, EPI_ISL_1504122, EPI_ISL_1504126, EPI_ISL_1504127, EPI_ISL_1504129, EPI_ISL_1504137, EPI_ISL_1504140, EPI_ISL_1504143, EPI_ISL_1504146, EPI_ISL_1504151, EPI_ISL_1504153, EPI_ISL_1504154, EPI_ISL_1504165, EPI_ISL_1504169, EPI_ISL_1504175, EPI_ISL_1504188, EPI_ISL_1504203, EPI_ISL_1504204, EPI_ISL_1504206, EPI_ISL_1504216, EPI_ISL_1504228, EPI_ISL_1504231, EPI_ISL_1504234, EPI_ISL_1504237, EPI_ISL_1504245, EPI_ISL_1504250, EPI_ISL_1504254, EPI_ISL_1504261, EPI_ISL_1504262, EPI_ISL_1504271, EPI_ISL_1504273, EPI_ISL_1504274, EPI_ISL_1504275, EPI_ISL_1504285, EPI_ISL_1504290, EPI_ISL_1504292, EPI_ISL_1504295, EPI_ISL_1504296, EPI_ISL_1504301, EPI_ISL_1504311, EPI_ISL_1504342, EPI_ISL_1504344, EPI_ISL_1504349, EPI_ISL_1504350, EPI_ISL_1504353, EPI_ISL_1504359, EPI_ISL_1504360, EPI_ISL_1504367, EPI_ISL_1504370, EPI_ISL_1504371, EPI_ISL_1504372, EPI_ISL_1504376, EPI_ISL_1504384, EPI_ISL_1504387, EPI_ISL_1504396, EPI_ISL_1504406, EPI_ISL_1504408, EPI_ISL_1504411, EPI_ISL_1504412, EPI_ISL_1504413, EPI_ISL_1504414, EPI_ISL_1504415, EPI_ISL_1504423, EPI_ISL_1504431, EPI_ISL_1504434, EPI_ISL_1504435, EPI_ISL_1504436, EPI_ISL_1504441, EPI_ISL_1504444, EPI_ISL_1504445, EPI_ISL_1504456, EPI_ISL_1504457, EPI_ISL_1504458, EPI_ISL_1504459, EPI_ISL_1504465, EPI_ISL_1504468, EPI_ISL_1504470, EPI_ISL_1504480, EPI_ISL_1504483, EPI_ISL_1504488, EPI_ISL_1504497, EPI_ISL_1504498, EPI_ISL_1504499, EPI_ISL_1504501, EPI_ISL_1504503, EPI_ISL_1504518, EPI_ISL_1504520, EPI_ISL_1504522, EPI_ISL_1504524, EPI_ISL_1504527, EPI_ISL_1504529, EPI_ISL_1504531, EPI_ISL_1504536, EPI_ISL_1504538, EPI_ISL_1504539, EPI_ISL_1504543, EPI_ISL_1504547, EPI_ISL_1504549, EPI_ISL_1504552, EPI_ISL_1504560, EPI_ISL_1504562, EPI_ISL_1504565, EPI_ISL_1504566, EPI_ISL_1504568, EPI_ISL_1504571, EPI_ISL_1504572, EPI_ISL_1504574, EPI_ISL_1504576, EPI_ISL_1504577, EPI_ISL_1504580, EPI_ISL_1504582, EPI_ISL_1504595, EPI_ISL_1504596, EPI_ISL_1504609, EPI_ISL_1504616, EPI_ISL_1504621, EPI_ISL_1504622, EPI_ISL_1504626, EPI_ISL_1504630, EPI_ISL_1504631, EPI_ISL_1504639, EPI_ISL_1504642, EPI_ISL_1504645, EPI_ISL_1504650, EPI_ISL_1504651, EPI_ISL_1504653, EPI_ISL_1504654, EPI_ISL_1504656, EPI_ISL_1504660, EPI_ISL_1504661, EPI_ISL_1504665, EPI_ISL_1504999, EPI_ISL_1505008, EPI_ISL_1505057, EPI_ISL_1505087, EPI_ISL_1505094, EPI_ISL_1505122, EPI_ISL_1505142, EPI_ISL_1505461, EPI_ISL_1505465, EPI_ISL_1505476, EPI_ISL_1505479, EPI_ISL_1505487, EPI_ISL_1505488, EPI_ISL_1505499, EPI_ISL_1505518, EPI_ISL_1505533, EPI_ISL_1505547, EPI_ISL_1505552, EPI_ISL_1505554, EPI_ISL_1505556, EPI_ISL_1505561, EPI_ISL_1505565, EPI_ISL_1505584, EPI_ISL_1505593, EPI_ISL_1505601, EPI_ISL_1505613, EPI_ISL_1505622, EPI_ISL_1505627, EPI_ISL_1505632, EPI_ISL_1505635, EPI_ISL_1505638, EPI_ISL_1505651, EPI_ISL_1505659, EPI_ISL_1505665, EPI_ISL_1505668, EPI_ISL_1505688, EPI_ISL_1505711, EPI_ISL_1505736, EPI_ISL_1505738, EPI_ISL_1505754, EPI_ISL_1505757, EPI_ISL_1505772, EPI_ISL_1505784, EPI_ISL_1507041, EPI_ISL_1517753, EPI_ISL_1517754, EPI_ISL_1517755, EPI_ISL_1517756, EPI_ISL_1517757, EPI_ISL_1517758, EPI_ISL_1517759, EPI_ISL_1517760, EPI_ISL_1517761, EPI_ISL_1517762, EPI_ISL_1517763, EPI_ISL_1517764, EPI_ISL_1517766, EPI_ISL_1517768, EPI_ISL_1517769, EPI_ISL_1517770, EPI_ISL_1517771, EPI_ISL_1517773, EPI_ISL_1517774, EPI_ISL_1517775, EPI_ISL_1517780, EPI_ISL_1517781, EPI_ISL_1517782, EPI_ISL_1517783, EPI_ISL_1517784, EPI_ISL_1517785, EPI_ISL_1517786, EPI_ISL_1517790, EPI_ISL_1517792, EPI_ISL_1517793, EPI_ISL_1517795, EPI_ISL_1517797, EPI_ISL_1517798, EPI_ISL_1517799, EPI_ISL_1517800, EPI_ISL_1517801, EPI_ISL_1517802, EPI_ISL_1517803, EPI_ISL_1517805, EPI_ISL_1517806, EPI_ISL_1517808, EPI_ISL_1517809, EPI_ISL_1517810, EPI_ISL_1517812, EPI_ISL_1517815, EPI_ISL_1517816, EPI_ISL_1517817, EPI_ISL_1517818, EPI_ISL_1517819, EPI_ISL_1517820, EPI_ISL_1517821, EPI_ISL_1517822, EPI_ISL_1517823, EPI_ISL_1517824, EPI_ISL_1517825, EPI_ISL_1517827, EPI_ISL_1517828, EPI_ISL_1517829, EPI_ISL_1517830, EPI_ISL_1517831, EPI_ISL_1517832, EPI_ISL_1517833, EPI_ISL_1517834, EPI_ISL_1517835, EPI_ISL_1517837, EPI_ISL_1517838, EPI_ISL_1517839, EPI_ISL_1517840, EPI_ISL_1517842, EPI_ISL_1517843, EPI_ISL_1517845, EPI_ISL_1517846, EPI_ISL_1517847, EPI_ISL_1517848, EPI_ISL_1517849, EPI_ISL_1517850, EPI_ISL_1517851, EPI_ISL_1517854, EPI_ISL_1517855, EPI_ISL_1517856, EPI_ISL_1517857, EPI_ISL_1517858, EPI_ISL_1517860, EPI_ISL_1517861, EPI_ISL_1517862, EPI_ISL_1517863, EPI_ISL_1517865, EPI_ISL_1517866, EPI_ISL_1517867, EPI_ISL_1517868, EPI_ISL_1517869, EPI_ISL_1517872, EPI_ISL_1517874, EPI_ISL_1517876, EPI_ISL_1517877, EPI_ISL_1517878, EPI_ISL_1517879, EPI_ISL_1517880, EPI_ISL_1517882, EPI_ISL_1517883, EPI_ISL_1517884, EPI_ISL_1517885, EPI_ISL_1517886, EPI_ISL_1517887, EPI_ISL_1517888, EPI_ISL_1517890, EPI_ISL_1517892, EPI_ISL_1517893, EPI_ISL_1517895, EPI_ISL_1517897, EPI_ISL_1517899, EPI_ISL_1517903, EPI_ISL_1517904, EPI_ISL_1517905, EPI_ISL_1517906, EPI_ISL_1517907, EPI_ISL_1517908, EPI_ISL_1517909, EPI_ISL_1517910, EPI_ISL_1517911, EPI_ISL_1517912, EPI_ISL_1517914, EPI_ISL_1517915, EPI_ISL_1517917, EPI_ISL_1517919, EPI_ISL_1517922, EPI_ISL_1517923, EPI_ISL_1517924, EPI_ISL_1517926, EPI_ISL_1517927, EPI_ISL_1517928, EPI_ISL_1517929, EPI_ISL_1517931, EPI_ISL_1517933, EPI_ISL_1517934, EPI_ISL_1517935, EPI_ISL_1517939, EPI_ISL_1517940, EPI_ISL_1517941, EPI_ISL_1517942, EPI_ISL_1517943, EPI_ISL_1517944, EPI_ISL_1517945, EPI_ISL_1517946, EPI_ISL_1517947, EPI_ISL_1517948, EPI_ISL_1517950, EPI_ISL_1517954, EPI_ISL_1517955, EPI_ISL_1517959, EPI_ISL_1517960, EPI_ISL_1517961, EPI_ISL_1517962, EPI_ISL_1517964, EPI_ISL_1517965, EPI_ISL_1517967, EPI_ISL_1517968, EPI_ISL_1517969, EPI_ISL_1517970, EPI_ISL_1517971, EPI_ISL_1517972, EPI_ISL_1517973, EPI_ISL_1517974, EPI_ISL_1517979, EPI_ISL_1517981, EPI_ISL_1517982, EPI_ISL_1517983, EPI_ISL_1517984, EPI_ISL_1517985, EPI_ISL_1517987, EPI_ISL_1517990, EPI_ISL_1517991, EPI_ISL_1517992, EPI_ISL_1517993, EPI_ISL_1517995, EPI_ISL_1517996, EPI_ISL_1517997, EPI_ISL_1517998, EPI_ISL_1518000, EPI_ISL_1518001, EPI_ISL_1518002, EPI_ISL_1518003, EPI_ISL_1518005, EPI_ISL_1518009, EPI_ISL_1518010, EPI_ISL_1518013, EPI_ISL_1518015, EPI_ISL_1518017, EPI_ISL_1518018, EPI_ISL_1518019, EPI_ISL_1518020, EPI_ISL_1518021, EPI_ISL_1518023, EPI_ISL_1518025, EPI_ISL_1518026, EPI_ISL_1518029, EPI_ISL_1518030, EPI_ISL_1518033, EPI_ISL_1518034, EPI_ISL_1518035, EPI_ISL_1518036, EPI_ISL_1518037, EPI_ISL_1518038, EPI_ISL_1518039, EPI_ISL_1518040, EPI_ISL_1518041, EPI_ISL_1518042, EPI_ISL_1518045, EPI_ISL_1518047, EPI_ISL_1518048, EPI_ISL_1518049, EPI_ISL_1518050, EPI_ISL_1518051, EPI_ISL_1518052, EPI_ISL_1518053, EPI_ISL_1518055, EPI_ISL_1518058, EPI_ISL_1518059, EPI_ISL_1518060, EPI_ISL_1518061, EPI_ISL_1518062, EPI_ISL_1518063, EPI_ISL_1518064, EPI_ISL_1518065, EPI_ISL_1518068, EPI_ISL_1518069, EPI_ISL_1518072, EPI_ISL_1518074, EPI_ISL_1518075, EPI_ISL_1518076, EPI_ISL_1518079, EPI_ISL_1518080, EPI_ISL_1518081, EPI_ISL_1518086, EPI_ISL_1518089, EPI_ISL_1518090, EPI_ISL_1518091, EPI_ISL_1518095, EPI_ISL_1518096, EPI_ISL_1518098, EPI_ISL_1518099, EPI_ISL_1518100, EPI_ISL_1518102, EPI_ISL_1518104, EPI_ISL_1518105, EPI_ISL_1518106, EPI_ISL_1518107, EPI_ISL_1518108, EPI_ISL_1518109, EPI_ISL_1518110, EPI_ISL_1518113, EPI_ISL_1518115, EPI_ISL_1518116, EPI_ISL_1518117, EPI_ISL_1518119, EPI_ISL_1518120, EPI_ISL_1518122, EPI_ISL_1518123, EPI_ISL_1518124, EPI_ISL_1518125, EPI_ISL_1518127, EPI_ISL_1518128, EPI_ISL_1518129, EPI_ISL_1518131, EPI_ISL_1518132, EPI_ISL_1518137, EPI_ISL_1518139, EPI_ISL_1518141, EPI_ISL_1518143, EPI_ISL_1518144, EPI_ISL_1518146, EPI_ISL_1518147, EPI_ISL_1518148, EPI_ISL_1518150, EPI_ISL_1518154, EPI_ISL_1518155, EPI_ISL_1518156, EPI_ISL_1518158, EPI_ISL_1518159, EPI_ISL_1518163, EPI_ISL_1518165, EPI_ISL_1518166, EPI_ISL_1518168, EPI_ISL_1518169, EPI_ISL_1518170, EPI_ISL_1518171, EPI_ISL_1518177, EPI_ISL_1518180, EPI_ISL_1518184, EPI_ISL_1518190, EPI_ISL_1518191, EPI_ISL_1518192, EPI_ISL_1518193, EPI_ISL_1518197, EPI_ISL_1518199, EPI_ISL_1518200, EPI_ISL_1518201, EPI_ISL_1518202, EPI_ISL_1518204, EPI_ISL_1518206, EPI_ISL_1518208, EPI_ISL_1518210, EPI_ISL_1518211, EPI_ISL_1518212, EPI_ISL_1518213, EPI_ISL_1518218, EPI_ISL_1518219, EPI_ISL_1518224, EPI_ISL_1518225, EPI_ISL_1518226, EPI_ISL_1518227, EPI_ISL_1518229, EPI_ISL_1518234, EPI_ISL_1518235, EPI_ISL_1518237, EPI_ISL_1518238, EPI_ISL_1518240, EPI_ISL_1518242, EPI_ISL_1518243, EPI_ISL_1518245, EPI_ISL_1518247, EPI_ISL_1518248, EPI_ISL_1518250, EPI_ISL_1518252, EPI_ISL_1518253, EPI_ISL_1518258, EPI_ISL_1518259, EPI_ISL_1518261, EPI_ISL_1518262, EPI_ISL_1518264, EPI_ISL_1518265, EPI_ISL_1518267, EPI_ISL_1518268, EPI_ISL_1518269, EPI_ISL_1518270, EPI_ISL_1518271, EPI_ISL_1518272, EPI_ISL_1518274, EPI_ISL_1518276, EPI_ISL_1518282, EPI_ISL_1518283, EPI_ISL_1518284, EPI_ISL_1518285, EPI_ISL_1518286, EPI_ISL_1518287, EPI_ISL_1518290, EPI_ISL_1518292, EPI_ISL_1518293, EPI_ISL_1518295, EPI_ISL_1518296, EPI_ISL_1518298, EPI_ISL_1518300, EPI_ISL_1518301, EPI_ISL_1518302, EPI_ISL_1518306, EPI_ISL_1518307, EPI_ISL_1518309, EPI_ISL_1518313, EPI_ISL_1518315, EPI_ISL_1518317, EPI_ISL_1518320, EPI_ISL_1518323, EPI_ISL_1518324, EPI_ISL_1518325, EPI_ISL_1518326, EPI_ISL_1518329, EPI_ISL_1518333, EPI_ISL_1518335, EPI_ISL_1518339, EPI_ISL_1518340, EPI_ISL_1518345, EPI_ISL_1518346, EPI_ISL_1518347, EPI_ISL_1518351, EPI_ISL_1518352, EPI_ISL_1518353, EPI_ISL_1518354, EPI_ISL_1518357, EPI_ISL_1518359, EPI_ISL_1518360, EPI_ISL_1518361, EPI_ISL_1518362, EPI_ISL_1518364, EPI_ISL_1518365, EPI_ISL_1518366, EPI_ISL_1518367, EPI_ISL_1518368, EPI_ISL_1518371, EPI_ISL_1518372, EPI_ISL_1518375, EPI_ISL_1518376, EPI_ISL_1518378, EPI_ISL_1518379, EPI_ISL_1518380, EPI_ISL_1518381, EPI_ISL_1518384, EPI_ISL_1518386, EPI_ISL_1518387, EPI_ISL_1518389, EPI_ISL_1518390, EPI_ISL_1518391, EPI_ISL_1518398, EPI_ISL_1518400, EPI_ISL_1518402, EPI_ISL_1518407, EPI_ISL_1518409, EPI_ISL_1518416, EPI_ISL_1518417, EPI_ISL_1518422, EPI_ISL_1518424, EPI_ISL_1518427, EPI_ISL_1518429, EPI_ISL_1518436, EPI_ISL_1518440, EPI_ISL_1518443, EPI_ISL_1518452, EPI_ISL_1518463, EPI_ISL_1518471, EPI_ISL_1518473, EPI_ISL_1518474, EPI_ISL_1518477, EPI_ISL_1518480, EPI_ISL_1518487, EPI_ISL_1518491, EPI_ISL_1518492, EPI_ISL_1518495, EPI_ISL_1518496, EPI_ISL_1518500, EPI_ISL_1518502, EPI_ISL_1518506, EPI_ISL_1518508, EPI_ISL_1518510, EPI_ISL_1518512, EPI_ISL_1518514, EPI_ISL_1518517, EPI_ISL_1518518, EPI_ISL_1518523, EPI_ISL_1518524, EPI_ISL_1518526, EPI_ISL_1518527, EPI_ISL_1518529, EPI_ISL_1518533, EPI_ISL_1518544, EPI_ISL_1518547, EPI_ISL_1518550, EPI_ISL_1518561, EPI_ISL_1518570, EPI_ISL_1518572, EPI_ISL_1518573, EPI_ISL_1518580, EPI_ISL_1518581, EPI_ISL_1518585, EPI_ISL_1518586, EPI_ISL_1518592, EPI_ISL_1518595, EPI_ISL_1518596, EPI_ISL_1518601, EPI_ISL_1518602, EPI_ISL_1518603, EPI_ISL_1518614, EPI_ISL_1518616, EPI_ISL_1518618, EPI_ISL_1518619, EPI_ISL_1518625, EPI_ISL_1518627, EPI_ISL_1518631, EPI_ISL_1518632, EPI_ISL_1518634, EPI_ISL_1518635, EPI_ISL_1518637, EPI_ISL_1518644, EPI_ISL_1518648, EPI_ISL_1518651, EPI_ISL_1518652, EPI_ISL_1518656, EPI_ISL_1518659, EPI_ISL_1518660, EPI_ISL_1518663, EPI_ISL_1518664, EPI_ISL_1518669, EPI_ISL_1518670, EPI_ISL_1518671, EPI_ISL_1518673, EPI_ISL_1518928, EPI_ISL_1519269, EPI_ISL_1534749, EPI_ISL_1534750, EPI_ISL_1534752, EPI_ISL_1534753, EPI_ISL_1534754, EPI_ISL_1534755, EPI_ISL_1534756, EPI_ISL_1534759, EPI_ISL_1534760, EPI_ISL_1534763, EPI_ISL_1534764, EPI_ISL_1534765, EPI_ISL_1534766, EPI_ISL_1534767, EPI_ISL_1534769, EPI_ISL_1534770, EPI_ISL_1534771, EPI_ISL_1534772, EPI_ISL_1534773, EPI_ISL_1534780, EPI_ISL_1534782, EPI_ISL_1534784, EPI_ISL_1534785, EPI_ISL_1534787, EPI_ISL_1534788, EPI_ISL_1534790, EPI_ISL_1534792, EPI_ISL_1534793, EPI_ISL_1534794, EPI_ISL_1534796, EPI_ISL_1534797, EPI_ISL_1534798, EPI_ISL_1534799, EPI_ISL_1534800, EPI_ISL_1534802, EPI_ISL_1534805, EPI_ISL_1534807, EPI_ISL_1534808, EPI_ISL_1534809, EPI_ISL_1534810, EPI_ISL_1534811, EPI_ISL_1534812, EPI_ISL_1534813, EPI_ISL_1534814, EPI_ISL_1534815, EPI_ISL_1534816, EPI_ISL_1534817, EPI_ISL_1534819, EPI_ISL_1534822, EPI_ISL_1534824, EPI_ISL_1534825, EPI_ISL_1534826, EPI_ISL_1534827, EPI_ISL_1534828, EPI_ISL_1534829, EPI_ISL_1534830, EPI_ISL_1534831, EPI_ISL_1534832, EPI_ISL_1534833, EPI_ISL_1534835, EPI_ISL_1534838, EPI_ISL_1534840, EPI_ISL_1534841, EPI_ISL_1534844, EPI_ISL_1534846, EPI_ISL_1534847, EPI_ISL_1534849, EPI_ISL_1534851, EPI_ISL_1534853, EPI_ISL_1534855, EPI_ISL_1534856, EPI_ISL_1534857, EPI_ISL_1534859, EPI_ISL_1534860, EPI_ISL_1534861, EPI_ISL_1534862, EPI_ISL_1534864, EPI_ISL_1534865, EPI_ISL_1534866, EPI_ISL_1534867, EPI_ISL_1534869, EPI_ISL_1534870, EPI_ISL_1534871, EPI_ISL_1534873, EPI_ISL_1534875, EPI_ISL_1534876, EPI_ISL_1534877, EPI_ISL_1534878, EPI_ISL_1534881, EPI_ISL_1534882, EPI_ISL_1534883, EPI_ISL_1534884, EPI_ISL_1534885, EPI_ISL_1534887, EPI_ISL_1534888, EPI_ISL_1534889, EPI_ISL_1534890, EPI_ISL_1534892, EPI_ISL_1534893, EPI_ISL_1534896, EPI_ISL_1534897, EPI_ISL_1534898, EPI_ISL_1534899, EPI_ISL_1534900, EPI_ISL_1534901, EPI_ISL_1534902, EPI_ISL_1534903, EPI_ISL_1534904, EPI_ISL_1534905, EPI_ISL_1534906, EPI_ISL_1534907, EPI_ISL_1534908, EPI_ISL_1534909, EPI_ISL_1534910, EPI_ISL_1534912, EPI_ISL_1534913, EPI_ISL_1534915, EPI_ISL_1534919, EPI_ISL_1534920, EPI_ISL_1534922, EPI_ISL_1534923, EPI_ISL_1534924, EPI_ISL_1534926, EPI_ISL_1534929, EPI_ISL_1534931, EPI_ISL_1534932, EPI_ISL_1534933, EPI_ISL_1534934, EPI_ISL_1534935, EPI_ISL_1534936, EPI_ISL_1534937, EPI_ISL_1534939, EPI_ISL_1534940, EPI_ISL_1534941, EPI_ISL_1534946, EPI_ISL_1534947, EPI_ISL_1534948, EPI_ISL_1534951, EPI_ISL_1534952, EPI_ISL_1534954, EPI_ISL_1534955, EPI_ISL_1534956, EPI_ISL_1534959, EPI_ISL_1534962, EPI_ISL_1534964, EPI_ISL_1534966, EPI_ISL_1534968, EPI_ISL_1534969, EPI_ISL_1534970, EPI_ISL_1534971, EPI_ISL_1534972, EPI_ISL_1534973, EPI_ISL_1534979, EPI_ISL_1534980, EPI_ISL_1534982, EPI_ISL_1534985, EPI_ISL_1534986, EPI_ISL_1534988, EPI_ISL_1534989, EPI_ISL_1534991, EPI_ISL_1534994, EPI_ISL_1534995, EPI_ISL_1534997, EPI_ISL_1534998, EPI_ISL_1534999, EPI_ISL_1535000, EPI_ISL_1535002, EPI_ISL_1535004, EPI_ISL_1535005, EPI_ISL_1535007, EPI_ISL_1535009, EPI_ISL_1535010, EPI_ISL_1535011, EPI_ISL_1535012, EPI_ISL_1535014, EPI_ISL_1535019, EPI_ISL_1535021, EPI_ISL_1535023, EPI_ISL_1535024, EPI_ISL_1535028, EPI_ISL_1535029, EPI_ISL_1535030, EPI_ISL_1535031, EPI_ISL_1535032, EPI_ISL_1535034, EPI_ISL_1535036, EPI_ISL_1535037, EPI_ISL_1535038, EPI_ISL_1535043, EPI_ISL_1535044, EPI_ISL_1535046, EPI_ISL_1535048, EPI_ISL_1535050, EPI_ISL_1535053, EPI_ISL_1535054, EPI_ISL_1535055, EPI_ISL_1535059, EPI_ISL_1535061, EPI_ISL_1535062, EPI_ISL_1535065, EPI_ISL_1535067, EPI_ISL_1535071, EPI_ISL_1535074, EPI_ISL_1535077, EPI_ISL_1535078, EPI_ISL_1535080, EPI_ISL_1535082, EPI_ISL_1535084, EPI_ISL_1535085, EPI_ISL_1535090, EPI_ISL_1535091, EPI_ISL_1535092, EPI_ISL_1535095, EPI_ISL_1535096, EPI_ISL_1535098, EPI_ISL_1535099, EPI_ISL_1535105, EPI_ISL_1535107, EPI_ISL_1535110, EPI_ISL_1535113, EPI_ISL_1535116, EPI_ISL_1535117, EPI_ISL_1535118, EPI_ISL_1535119, EPI_ISL_1535120, EPI_ISL_1535125, EPI_ISL_1535126, EPI_ISL_1535128, EPI_ISL_1535129, EPI_ISL_1535130, EPI_ISL_1535132, EPI_ISL_1535133, EPI_ISL_1535139, EPI_ISL_1535140, EPI_ISL_1535143, EPI_ISL_1535144, EPI_ISL_1535146, EPI_ISL_1535151, EPI_ISL_1535153, EPI_ISL_1535154, EPI_ISL_1535156, EPI_ISL_1535159, EPI_ISL_1535161, EPI_ISL_1535162, EPI_ISL_1535164, EPI_ISL_1535167, EPI_ISL_1535169, EPI_ISL_1535174, EPI_ISL_1535177, EPI_ISL_1535180, EPI_ISL_1535181, EPI_ISL_1535182, EPI_ISL_1535186, EPI_ISL_1535190, EPI_ISL_1535191, EPI_ISL_1535194, EPI_ISL_1535195, EPI_ISL_1535198, EPI_ISL_1535203, EPI_ISL_1535204, EPI_ISL_1535206, EPI_ISL_1535207, EPI_ISL_1535210, EPI_ISL_1535215, EPI_ISL_1535216, EPI_ISL_1535218, EPI_ISL_1535219, EPI_ISL_1535222, EPI_ISL_1535223, EPI_ISL_1535227, EPI_ISL_1535229, EPI_ISL_1535231, EPI_ISL_1535234, EPI_ISL_1535238, EPI_ISL_1535241, EPI_ISL_1535242, EPI_ISL_1535243, EPI_ISL_1535244, EPI_ISL_1535246, EPI_ISL_1535248, EPI_ISL_1535250, EPI_ISL_1535251, EPI_ISL_1535252, EPI_ISL_1535253, EPI_ISL_1535254, EPI_ISL_1535255, EPI_ISL_1535262, EPI_ISL_1535263, EPI_ISL_1535264, EPI_ISL_1535265, EPI_ISL_1535266, EPI_ISL_1535271, EPI_ISL_1535274, EPI_ISL_1535275, EPI_ISL_1535276, EPI_ISL_1535279, EPI_ISL_1535280, EPI_ISL_1535281, EPI_ISL_1535282, EPI_ISL_1535283, EPI_ISL_1535285, EPI_ISL_1535287, EPI_ISL_1535288, EPI_ISL_1535291, EPI_ISL_1535292, EPI_ISL_1535295, EPI_ISL_1535296, EPI_ISL_1535297, EPI_ISL_1535302, EPI_ISL_1535303, EPI_ISL_1535306, EPI_ISL_1535310, EPI_ISL_1535312, EPI_ISL_1535313, EPI_ISL_1535314, EPI_ISL_1535315, EPI_ISL_1535318, EPI_ISL_1535319, EPI_ISL_1535322, EPI_ISL_1535325, EPI_ISL_1535326, EPI_ISL_1535327, EPI_ISL_1535331, EPI_ISL_1535333, EPI_ISL_1535602, EPI_ISL_1535604, EPI_ISL_1535606, EPI_ISL_1535608, EPI_ISL_1535613, EPI_ISL_1535615, EPI_ISL_1535617, EPI_ISL_1535618, EPI_ISL_1535621, EPI_ISL_1535624, EPI_ISL_1535625, EPI_ISL_1535626, EPI_ISL_1535630, EPI_ISL_1535631, EPI_ISL_1535636, EPI_ISL_1535637, EPI_ISL_1535638, EPI_ISL_1535639, EPI_ISL_1535641, EPI_ISL_1535643, EPI_ISL_1535645, EPI_ISL_1535646, EPI_ISL_1535647, EPI_ISL_1535648, EPI_ISL_1535651, EPI_ISL_1535652, EPI_ISL_1535653, EPI_ISL_1535654, EPI_ISL_1535657, EPI_ISL_1535658, EPI_ISL_1535659, EPI_ISL_1535660, EPI_ISL_1535662, EPI_ISL_1535668, EPI_ISL_1535669, EPI_ISL_1535671, EPI_ISL_1535674, EPI_ISL_1535675, EPI_ISL_1535678, EPI_ISL_1535680, EPI_ISL_1535683, EPI_ISL_1535686, EPI_ISL_1535688, EPI_ISL_1535689, EPI_ISL_1535690, EPI_ISL_1535691, EPI_ISL_1535693, EPI_ISL_1535695, EPI_ISL_1535696, EPI_ISL_1535697, EPI_ISL_1535702, EPI_ISL_1535704, EPI_ISL_1535705, EPI_ISL_1535706, EPI_ISL_1535713, EPI_ISL_1535717, EPI_ISL_1535718, EPI_ISL_1535719, EPI_ISL_1535723, EPI_ISL_1535727, EPI_ISL_1535728, EPI_ISL_1535729, EPI_ISL_1535732, EPI_ISL_1535734, EPI_ISL_1535735, EPI_ISL_1535737, EPI_ISL_1535740, EPI_ISL_1535741, EPI_ISL_1535746, EPI_ISL_1535747, EPI_ISL_1535748, EPI_ISL_1535749, EPI_ISL_1535752, EPI_ISL_1535755, EPI_ISL_1535760, EPI_ISL_1535762, EPI_ISL_1535763, EPI_ISL_1535765, EPI_ISL_1535770, EPI_ISL_1535771, EPI_ISL_1535775, EPI_ISL_1535776, EPI_ISL_1535777, EPI_ISL_1535778, EPI_ISL_1535781, EPI_ISL_1535784, EPI_ISL_1535785, EPI_ISL_1535787, EPI_ISL_1535790, EPI_ISL_1535791, EPI_ISL_1535793, EPI_ISL_1535794, EPI_ISL_1535796, EPI_ISL_1535797, EPI_ISL_1535798, EPI_ISL_1535799, EPI_ISL_1535800, EPI_ISL_1535805, EPI_ISL_1535808, EPI_ISL_1535809, EPI_ISL_1535810, EPI_ISL_1535812, EPI_ISL_1535813, EPI_ISL_1535819, EPI_ISL_1535823, EPI_ISL_1535831, EPI_ISL_1535834, EPI_ISL_1535835, EPI_ISL_1535836, EPI_ISL_1535837, EPI_ISL_1535839, EPI_ISL_1535840, EPI_ISL_1535842, EPI_ISL_1535843, EPI_ISL_1535845, EPI_ISL_1535847, EPI_ISL_1535848, EPI_ISL_1535853, EPI_ISL_1535854, EPI_ISL_1535855, EPI_ISL_1535858, EPI_ISL_1535863, EPI_ISL_1535864, EPI_ISL_1535865, EPI_ISL_1535873, EPI_ISL_1535876, EPI_ISL_1535877, EPI_ISL_1535882, EPI_ISL_1535883, EPI_ISL_1535884, EPI_ISL_1535885, EPI_ISL_1535889, EPI_ISL_1535890, EPI_ISL_1535891, EPI_ISL_1535893, EPI_ISL_1535894, EPI_ISL_1535895, EPI_ISL_1535896, EPI_ISL_1535899, EPI_ISL_1535900, EPI_ISL_1535901, EPI_ISL_1535902, EPI_ISL_1535904, EPI_ISL_1535905, EPI_ISL_1535906, EPI_ISL_1535908, EPI_ISL_1535913, EPI_ISL_1535914, EPI_ISL_1535915, EPI_ISL_1535917, EPI_ISL_1535918, EPI_ISL_1535922, EPI_ISL_1535923, EPI_ISL_1535925, EPI_ISL_1535926, EPI_ISL_1535927, EPI_ISL_1535928, EPI_ISL_1535929, EPI_ISL_1535930, EPI_ISL_1535931, EPI_ISL_1535932, EPI_ISL_1535933, EPI_ISL_1535936, EPI_ISL_1535940, EPI_ISL_1535942, EPI_ISL_1535943, EPI_ISL_1535945, EPI_ISL_1535946, EPI_ISL_1535947, EPI_ISL_1535948, EPI_ISL_1535949, EPI_ISL_1535952, EPI_ISL_1535953, EPI_ISL_1535954, EPI_ISL_1535955, EPI_ISL_1535956, EPI_ISL_1535958, EPI_ISL_1535959, EPI_ISL_1535960, EPI_ISL_1535961, EPI_ISL_1535962, EPI_ISL_1535964, EPI_ISL_1535967, EPI_ISL_1535968, EPI_ISL_1535970, EPI_ISL_1535971, EPI_ISL_1535972, EPI_ISL_1535973, EPI_ISL_1535974, EPI_ISL_1535977, EPI_ISL_1535978, EPI_ISL_1535979, EPI_ISL_1535981, EPI_ISL_1535986, EPI_ISL_1535988, EPI_ISL_1535990, EPI_ISL_1535993, EPI_ISL_1535995, EPI_ISL_1535996, EPI_ISL_1535997, EPI_ISL_1535998, EPI_ISL_1536000, EPI_ISL_1536002, EPI_ISL_1536003, EPI_ISL_1536004, EPI_ISL_1536007, EPI_ISL_1536009, EPI_ISL_1536010, EPI_ISL_1536011, EPI_ISL_1536012, EPI_ISL_1536014, EPI_ISL_1536015, EPI_ISL_1536016, EPI_ISL_1536019, EPI_ISL_1536020, EPI_ISL_1536022, EPI_ISL_1536023, EPI_ISL_1536026, EPI_ISL_1536027, EPI_ISL_1536031, EPI_ISL_1536032, EPI_ISL_1536033, EPI_ISL_1536035, EPI_ISL_1536038, EPI_ISL_1536039, EPI_ISL_1536040, EPI_ISL_1536044, EPI_ISL_1536045, EPI_ISL_1536046, EPI_ISL_1536047, EPI_ISL_1536050, EPI_ISL_1536051, EPI_ISL_1536052, EPI_ISL_1536053, EPI_ISL_1536054, EPI_ISL_1536055, EPI_ISL_1536056, EPI_ISL_1536057, EPI_ISL_1536058, EPI_ISL_1536059, EPI_ISL_1536060, EPI_ISL_1536061, EPI_ISL_1536063, EPI_ISL_1536064, EPI_ISL_1536066, EPI_ISL_1536067, EPI_ISL_1536069, EPI_ISL_1536070, EPI_ISL_1536072, EPI_ISL_1536073, EPI_ISL_1536075, EPI_ISL_1536076, EPI_ISL_1536077, EPI_ISL_1536079, EPI_ISL_1536080, EPI_ISL_1536081, EPI_ISL_1536082, EPI_ISL_1536083, EPI_ISL_1536085, EPI_ISL_1536086, EPI_ISL_1536087, EPI_ISL_1536089, EPI_ISL_1536090, EPI_ISL_1536091, EPI_ISL_1536094, EPI_ISL_1536097, EPI_ISL_1536098, EPI_ISL_1536099, EPI_ISL_1536100, EPI_ISL_1536101, EPI_ISL_1536102, EPI_ISL_1536103, EPI_ISL_1536104, EPI_ISL_1536105, EPI_ISL_1536106, EPI_ISL_1536107, EPI_ISL_1536109, EPI_ISL_1536110, EPI_ISL_1536112, EPI_ISL_1536113, EPI_ISL_1536114, EPI_ISL_1536117, EPI_ISL_1536118, EPI_ISL_1536119, EPI_ISL_1536121, EPI_ISL_1536122, EPI_ISL_1536123, EPI_ISL_1536124, EPI_ISL_1536127, EPI_ISL_1536129, EPI_ISL_1536130, EPI_ISL_1536133, EPI_ISL_1536134, EPI_ISL_1536135, EPI_ISL_1536136, EPI_ISL_1536139, EPI_ISL_1536140, EPI_ISL_1536141, EPI_ISL_1536142, EPI_ISL_1536143, EPI_ISL_1536146, EPI_ISL_1536147, EPI_ISL_1536148, EPI_ISL_1536150, EPI_ISL_1536154, EPI_ISL_1536155, EPI_ISL_1536157, EPI_ISL_1536158, EPI_ISL_1536162, EPI_ISL_1536163, EPI_ISL_1536164, EPI_ISL_1536165, EPI_ISL_1536166, EPI_ISL_1536167, EPI_ISL_1536168, EPI_ISL_1536170, EPI_ISL_1536173, EPI_ISL_1536174, EPI_ISL_1536176, EPI_ISL_1536177, EPI_ISL_1536178, EPI_ISL_1536180, EPI_ISL_1536181, EPI_ISL_1536182, EPI_ISL_1536183, EPI_ISL_1536184, EPI_ISL_1536185, EPI_ISL_1536186, EPI_ISL_1536187, EPI_ISL_1536188, EPI_ISL_1536189, EPI_ISL_1536190, EPI_ISL_1536191, EPI_ISL_1536192, EPI_ISL_1536193, EPI_ISL_1536195, EPI_ISL_1536196, EPI_ISL_1536197, EPI_ISL_1536198, EPI_ISL_1536199, EPI_ISL_1536200, EPI_ISL_1536201, EPI_ISL_1536203, EPI_ISL_1536204, EPI_ISL_1536205, EPI_ISL_1536206, EPI_ISL_1536207, EPI_ISL_1536208, EPI_ISL_1536209, EPI_ISL_1536210, EPI_ISL_1536211, EPI_ISL_1536212, EPI_ISL_1536213, EPI_ISL_1536215, EPI_ISL_1536216, EPI_ISL_1536218, EPI_ISL_1536219, EPI_ISL_1536220, EPI_ISL_1536221, EPI_ISL_1536222, EPI_ISL_1536223, EPI_ISL_1536224, EPI_ISL_1536225, EPI_ISL_1536226, EPI_ISL_1536227, EPI_ISL_1536229, EPI_ISL_1536231, EPI_ISL_1536232, EPI_ISL_1536234, EPI_ISL_1536235, EPI_ISL_1536236, EPI_ISL_1536238, EPI_ISL_1536239, EPI_ISL_1536240, EPI_ISL_1536241, EPI_ISL_1536242, EPI_ISL_1536243, EPI_ISL_1536244, EPI_ISL_1536245, EPI_ISL_1536246, EPI_ISL_1536247, EPI_ISL_1536248, EPI_ISL_1536249, EPI_ISL_1536250, EPI_ISL_1536251, EPI_ISL_1536252, EPI_ISL_1536254, EPI_ISL_1536255, EPI_ISL_1536256, EPI_ISL_1536257, EPI_ISL_1536258, EPI_ISL_1536259, EPI_ISL_1536260, EPI_ISL_1536261, EPI_ISL_1536262, EPI_ISL_1536263, EPI_ISL_1536265, EPI_ISL_1536266, EPI_ISL_1536268, EPI_ISL_1536269, EPI_ISL_1536270, EPI_ISL_1536271, EPI_ISL_1536272, EPI_ISL_1536273, EPI_ISL_1536274, EPI_ISL_1536275, EPI_ISL_1536277, EPI_ISL_1536278, EPI_ISL_1536279, EPI_ISL_1536280, EPI_ISL_1536281, EPI_ISL_1536282, EPI_ISL_1536283, EPI_ISL_1536284, EPI_ISL_1536285, EPI_ISL_1536286, EPI_ISL_1536288, EPI_ISL_1536289, EPI_ISL_1536290, EPI_ISL_1536291, EPI_ISL_1536292, EPI_ISL_1536293, EPI_ISL_1536294, EPI_ISL_1536295, EPI_ISL_1536297, EPI_ISL_1536298, EPI_ISL_1536299, EPI_ISL_1536300, EPI_ISL_1536301, EPI_ISL_1536303, EPI_ISL_1536304, EPI_ISL_1536305, EPI_ISL_1536306, EPI_ISL_1536307, EPI_ISL_1536308, EPI_ISL_1536309, EPI_ISL_1536311, EPI_ISL_1536313, EPI_ISL_1536314, EPI_ISL_1536315, EPI_ISL_1536316, EPI_ISL_1536317, EPI_ISL_1536318, EPI_ISL_1536319, EPI_ISL_1536321, EPI_ISL_1536323, EPI_ISL_1536324, EPI_ISL_1536325, EPI_ISL_1536326, EPI_ISL_1536327, EPI_ISL_1536328, EPI_ISL_1536330, EPI_ISL_1536331, EPI_ISL_1536332, EPI_ISL_1536333, EPI_ISL_1536334, EPI_ISL_1536335, EPI_ISL_1536336, EPI_ISL_1536337, EPI_ISL_1536338, EPI_ISL_1536339, EPI_ISL_1536340, EPI_ISL_1536341, EPI_ISL_1536342, EPI_ISL_1536344, EPI_ISL_1536345, EPI_ISL_1536346, EPI_ISL_1536347, EPI_ISL_1536349, EPI_ISL_1536352, EPI_ISL_1536353, EPI_ISL_1536354, EPI_ISL_1536355, EPI_ISL_1536358, EPI_ISL_1536359, EPI_ISL_1536360, EPI_ISL_1536361, EPI_ISL_1536363, EPI_ISL_1536364, EPI_ISL_1536365, EPI_ISL_1536366, EPI_ISL_1536367, EPI_ISL_1536368, EPI_ISL_1536369, EPI_ISL_1536371, EPI_ISL_1536372, EPI_ISL_1536374, EPI_ISL_1536375, EPI_ISL_1536376, EPI_ISL_1536377, EPI_ISL_1536378, EPI_ISL_1536379, EPI_ISL_1536381, EPI_ISL_1536382, EPI_ISL_1536383, EPI_ISL_1536384, EPI_ISL_1536385, EPI_ISL_1536386, EPI_ISL_1536388, EPI_ISL_1536389, EPI_ISL_1536390, EPI_ISL_1536391, EPI_ISL_1536392, EPI_ISL_1536393, EPI_ISL_1536394, EPI_ISL_1536397, EPI_ISL_1536398, EPI_ISL_1536399, EPI_ISL_1536400, EPI_ISL_1536401, EPI_ISL_1536403, EPI_ISL_1536405, EPI_ISL_1536406, EPI_ISL_1536407, EPI_ISL_1536408, EPI_ISL_1536410, EPI_ISL_1536412, EPI_ISL_1536413, EPI_ISL_1536414, EPI_ISL_1536415, EPI_ISL_1536416, EPI_ISL_1536417, EPI_ISL_1536418, EPI_ISL_1536419, EPI_ISL_1536420, EPI_ISL_1536424, EPI_ISL_1536426, EPI_ISL_1536427, EPI_ISL_1536428, EPI_ISL_1536430, EPI_ISL_1536431, EPI_ISL_1536432, EPI_ISL_1536434, EPI_ISL_1536435, EPI_ISL_1536436, EPI_ISL_1536437, EPI_ISL_1536438, EPI_ISL_1536439, EPI_ISL_1536440, EPI_ISL_1536441, EPI_ISL_1536443, EPI_ISL_1536444, EPI_ISL_1536445, EPI_ISL_1536447, EPI_ISL_1536448, EPI_ISL_1536449, EPI_ISL_1536450, EPI_ISL_1536451, EPI_ISL_1536452, EPI_ISL_1536454, EPI_ISL_1536455, EPI_ISL_1536456, EPI_ISL_1536457, EPI_ISL_1536459, EPI_ISL_1536460, EPI_ISL_1536461, EPI_ISL_1536462, EPI_ISL_1536463, EPI_ISL_1536464, EPI_ISL_1536465, EPI_ISL_1536466, EPI_ISL_1536468, EPI_ISL_1536469, EPI_ISL_1536470, EPI_ISL_1536471, EPI_ISL_1536472, EPI_ISL_1536473, EPI_ISL_1536474, EPI_ISL_1536475, EPI_ISL_1536476, EPI_ISL_1536477, EPI_ISL_1536478, EPI_ISL_1536479, EPI_ISL_1536481, EPI_ISL_1536482, EPI_ISL_1536483, EPI_ISL_1536484, EPI_ISL_1536485, EPI_ISL_1536487, EPI_ISL_1536488, EPI_ISL_1536489, EPI_ISL_1536490, EPI_ISL_1536491, EPI_ISL_1536492, EPI_ISL_1536493, EPI_ISL_1536494, EPI_ISL_1536495, EPI_ISL_1536496, EPI_ISL_1536497, EPI_ISL_1536498, EPI_ISL_1536499, EPI_ISL_1536500, EPI_ISL_1536501, EPI_ISL_1536502, EPI_ISL_1536504, EPI_ISL_1536505, EPI_ISL_1536506, EPI_ISL_1536507, EPI_ISL_1536509, EPI_ISL_1536510, EPI_ISL_1536511, EPI_ISL_1536512, EPI_ISL_1536513, EPI_ISL_1536514, EPI_ISL_1536515, EPI_ISL_1536516, EPI_ISL_1536517, EPI_ISL_1536518, EPI_ISL_1536519, EPI_ISL_1536522, EPI_ISL_1536523, EPI_ISL_1536524, EPI_ISL_1536526, EPI_ISL_1536528, EPI_ISL_1536529, EPI_ISL_1536530, EPI_ISL_1536531, EPI_ISL_1536532, EPI_ISL_1536534, EPI_ISL_1536538, EPI_ISL_1536843, EPI_ISL_1536846, EPI_ISL_1536848, EPI_ISL_1536850, EPI_ISL_1536853, EPI_ISL_1536856, EPI_ISL_1536864, EPI_ISL_1536865, EPI_ISL_1536866, EPI_ISL_1536867, EPI_ISL_1536868, EPI_ISL_1536871, EPI_ISL_1536875, EPI_ISL_1536877, EPI_ISL_1536883, EPI_ISL_1536889, EPI_ISL_1536890, EPI_ISL_1536895, EPI_ISL_1536896, EPI_ISL_1536899, EPI_ISL_1536900, EPI_ISL_1536901, EPI_ISL_1536907, EPI_ISL_1536908, EPI_ISL_1536910, EPI_ISL_1536913, EPI_ISL_1536915, EPI_ISL_1536916, EPI_ISL_1536922, EPI_ISL_1536924, EPI_ISL_1536927, EPI_ISL_1536928, EPI_ISL_1536932, EPI_ISL_1536935, EPI_ISL_1536936, EPI_ISL_1536940, EPI_ISL_1536941, EPI_ISL_1536943, EPI_ISL_1536946, EPI_ISL_1536949, EPI_ISL_1536950, EPI_ISL_1536952, EPI_ISL_1536955, EPI_ISL_1536958, EPI_ISL_1536960, EPI_ISL_1536961, EPI_ISL_1536965, EPI_ISL_1536970, EPI_ISL_1536980, EPI_ISL_1536982, EPI_ISL_1536983, EPI_ISL_1536988, EPI_ISL_1536989, EPI_ISL_1536992, EPI_ISL_1536999, EPI_ISL_1537006, EPI_ISL_1537010, EPI_ISL_1537016, EPI_ISL_1537018, EPI_ISL_1537021, EPI_ISL_1537023, EPI_ISL_1537027, EPI_ISL_1537033, EPI_ISL_1537035, EPI_ISL_1537038, EPI_ISL_1537039, EPI_ISL_1537040, EPI_ISL_1537042, EPI_ISL_1537044, EPI_ISL_1537047, EPI_ISL_1537051, EPI_ISL_1537052, EPI_ISL_1537061, EPI_ISL_1537062, EPI_ISL_1537065, EPI_ISL_1537066, EPI_ISL_1537067, EPI_ISL_1537070, EPI_ISL_1537072, EPI_ISL_1537075, EPI_ISL_1537078, EPI_ISL_1537079, EPI_ISL_1537088, EPI_ISL_1537094, EPI_ISL_1537099, EPI_ISL_1537104, EPI_ISL_1537113, EPI_ISL_1537115, EPI_ISL_1537121, EPI_ISL_1537127, EPI_ISL_1537128, EPI_ISL_1537133, EPI_ISL_1537137, EPI_ISL_1537315, EPI_ISL_1537318, EPI_ISL_1537319, EPI_ISL_1537320, EPI_ISL_1537322, EPI_ISL_1537323, EPI_ISL_1537327, EPI_ISL_1537620, EPI_ISL_1537621, EPI_ISL_1537642, EPI_ISL_1537651, EPI_ISL_1537655, EPI_ISL_1537661, EPI_ISL_1537663, EPI_ISL_1537665, EPI_ISL_1537667, EPI_ISL_1537669, EPI_ISL_1537670, EPI_ISL_1537673, EPI_ISL_1537687, EPI_ISL_1537689, EPI_ISL_1537690, EPI_ISL_1537693, EPI_ISL_1537695, EPI_ISL_1537702, EPI_ISL_1537711, EPI_ISL_1537715, EPI_ISL_1537719, EPI_ISL_1537726, EPI_ISL_1537728, EPI_ISL_1537758, EPI_ISL_1537759, EPI_ISL_1537760, EPI_ISL_1537790, EPI_ISL_1537793, EPI_ISL_1537804, EPI_ISL_1537808, EPI_ISL_1537813, EPI_ISL_1537820, EPI_ISL_1537822, EPI_ISL_1537826, EPI_ISL_1537840, EPI_ISL_1538301, EPI_ISL_1544864, EPI_ISL_1564334, EPI_ISL_1564335, EPI_ISL_1564336, EPI_ISL_1564337, EPI_ISL_1564338, EPI_ISL_1564339, EPI_ISL_1564343, EPI_ISL_1564344, EPI_ISL_1564345, EPI_ISL_1564346, EPI_ISL_1564348, EPI_ISL_1564350, EPI_ISL_1564351, EPI_ISL_1564354, EPI_ISL_1564360, EPI_ISL_1564364, EPI_ISL_1564365, EPI_ISL_1564366, EPI_ISL_1564370, EPI_ISL_1564371, EPI_ISL_1564372, EPI_ISL_1564374, EPI_ISL_1564375, EPI_ISL_1564376, EPI_ISL_1564379, EPI_ISL_1564380, EPI_ISL_1564383, EPI_ISL_1564384, EPI_ISL_1564386, EPI_ISL_1564387, EPI_ISL_1564391, EPI_ISL_1564392, EPI_ISL_1564393, EPI_ISL_1564396, EPI_ISL_1564398, EPI_ISL_1564399, EPI_ISL_1564400, EPI_ISL_1564403, EPI_ISL_1564404, EPI_ISL_1564405, EPI_ISL_1564409, EPI_ISL_1564412, EPI_ISL_1564413, EPI_ISL_1564420, EPI_ISL_1564421, EPI_ISL_1564422, EPI_ISL_1564425, EPI_ISL_1564426, EPI_ISL_1564428, EPI_ISL_1564431, EPI_ISL_1564437, EPI_ISL_1564443, EPI_ISL_1564447, EPI_ISL_1564450, EPI_ISL_1564452, EPI_ISL_1564456, EPI_ISL_1564462, EPI_ISL_1564464, EPI_ISL_1564467, EPI_ISL_1564468, EPI_ISL_1564471, EPI_ISL_1564472, EPI_ISL_1564474, EPI_ISL_1564477, EPI_ISL_1564480, EPI_ISL_1564481, EPI_ISL_1564482, EPI_ISL_1564484, EPI_ISL_1564488, EPI_ISL_1564491, EPI_ISL_1564492,

EPI_ISL_1564494, EPI_ISL_1564495, EPI_ISL_1564496, EPI_ISL_1564497, EPI_ISL_1564499, EPI_ISL_1564500, EPI_ISL_1564502, EPI_ISL_1564505, EPI_ISL_1564506, EPI_ISL_1564508, EPI_ISL_1564511, EPI_ISL_1564512, EPI_ISL_1564513, EPI_ISL_1564514, EPI_ISL_1564518, EPI_ISL_1564520, EPI_ISL_1564523, EPI_ISL_1564524, EPI_ISL_1564525, EPI_ISL_1564527, EPI_ISL_1564529, EPI_ISL_1564531, EPI_ISL_1564533, EPI_ISL_1564535, EPI_ISL_1564538, EPI_ISL_1564540, EPI_ISL_1564541, EPI_ISL_1564544, EPI_ISL_1564545, EPI_ISL_1564546, EPI_ISL_1564548, EPI_ISL_1564553, EPI_ISL_1564555, EPI_ISL_1564556, EPI_ISL_1564560, EPI_ISL_1564562, EPI_ISL_1564563, EPI_ISL_1564564, EPI_ISL_1564565, EPI_ISL_1564566, EPI_ISL_1564570, EPI_ISL_1564576, EPI_ISL_1564577, EPI_ISL_1564579, EPI_ISL_1564581, EPI_ISL_1564583, EPI_ISL_1564585, EPI_ISL_1564586, EPI_ISL_1564587, EPI_ISL_1564588, EPI_ISL_1564591, EPI_ISL_1564593, EPI_ISL_1564596, EPI_ISL_1564597, EPI_ISL_1564598, EPI_ISL_1564601, EPI_ISL_1564602, EPI_ISL_1564603, EPI_ISL_1564604, EPI_ISL_1564605, EPI_ISL_1564607, EPI_ISL_1564608, EPI_ISL_1564609, EPI_ISL_1564629, EPI_ISL_1564630, EPI_ISL_1564631, EPI_ISL_1564632, EPI_ISL_1564635, EPI_ISL_1564636, EPI_ISL_1564639, EPI_ISL_1564644, EPI_ISL_1564645, EPI_ISL_1564646, EPI_ISL_1564650, EPI_ISL_1564651, EPI_ISL_1564653, EPI_ISL_1564654, EPI_ISL_1564655, EPI_ISL_1564657, EPI_ISL_1564659, EPI_ISL_1564660, EPI_ISL_1564661, EPI_ISL_1564662, EPI_ISL_1564663, EPI_ISL_1564668, EPI_ISL_1564669, EPI_ISL_1564670, EPI_ISL_1564672, EPI_ISL_1564673, EPI_ISL_1564675, EPI_ISL_1564676, EPI_ISL_1564677, EPI_ISL_1564679, EPI_ISL_1564680, EPI_ISL_1564681, EPI_ISL_1564682, EPI_ISL_1564685, EPI_ISL_1564687, EPI_ISL_1564689, EPI_ISL_1564690, EPI_ISL_1564694, EPI_ISL_1564695, EPI_ISL_1564696, EPI_ISL_1564697, EPI_ISL_1564698, EPI_ISL_1564700, EPI_ISL_1564701, EPI_ISL_1564702, EPI_ISL_1564704, EPI_ISL_1564705, EPI_ISL_1564709, EPI_ISL_1564710, EPI_ISL_1564711, EPI_ISL_1564712, EPI_ISL_1564716, EPI_ISL_1564717, EPI_ISL_1564722, EPI_ISL_1564723, EPI_ISL_1564727, EPI_ISL_1564728, EPI_ISL_1564732, EPI_ISL_1564734, EPI_ISL_1564740, EPI_ISL_1564741, EPI_ISL_1564744, EPI_ISL_1564749, EPI_ISL_1564755, EPI_ISL_1564756, EPI_ISL_1564757, EPI_ISL_1564758, EPI_ISL_1564761, EPI_ISL_1564773, EPI_ISL_1564783, EPI_ISL_1564787, EPI_ISL_1564796, EPI_ISL_1564797, EPI_ISL_1564799, EPI_ISL_1583685, EPI_ISL_1583693, EPI_ISL_1583696, EPI_ISL_1583698, EPI_ISL_1583699, EPI_ISL_1583709, EPI_ISL_1583712, EPI_ISL_1583720, EPI_ISL_1583723, EPI_ISL_1583728, EPI_ISL_1583742, EPI_ISL_1583743, EPI_ISL_1583744, EPI_ISL_1583748, EPI_ISL_1583750, EPI_ISL_1583752, EPI_ISL_1583753, EPI_ISL_1583755, EPI_ISL_1583756, EPI_ISL_1583757, EPI_ISL_1583758, EPI_ISL_1583759, EPI_ISL_1583763, EPI_ISL_1583764, EPI_ISL_1583769, EPI_ISL_1583770, EPI_ISL_1583771, EPI_ISL_1583773, EPI_ISL_1583774, EPI_ISL_1583776, EPI_ISL_1583777, EPI_ISL_1583780, EPI_ISL_1583781, EPI_ISL_1583782, EPI_ISL_1583783, EPI_ISL_1583785, EPI_ISL_1583787, EPI_ISL_1583789, EPI_ISL_1583790, EPI_ISL_1583791, EPI_ISL_1583792, EPI_ISL_1583797, EPI_ISL_1583799, EPI_ISL_1583807, EPI_ISL_1583808, EPI_ISL_1583811, EPI_ISL_1583813, EPI_ISL_1583816, EPI_ISL_1583817, EPI_ISL_1583819, EPI_ISL_1583824, EPI_ISL_1583826, EPI_ISL_1583828, EPI_ISL_1583829, EPI_ISL_1583832, EPI_ISL_1583844, EPI_ISL_1583845, EPI_ISL_1583852, EPI_ISL_1583853, EPI_ISL_1583857, EPI_ISL_1583858, EPI_ISL_1583859, EPI_ISL_1583861, EPI_ISL_1583864, EPI_ISL_1583865, EPI_ISL_1583869, EPI_ISL_1583870, EPI_ISL_1583872, EPI_ISL_1583875, EPI_ISL_1583878, EPI_ISL_1583879, EPI_ISL_1583880, EPI_ISL_1583884, EPI_ISL_1583886, EPI_ISL_1583889, EPI_ISL_1583891, EPI_ISL_1583892, EPI_ISL_1583896, EPI_ISL_1583897, EPI_ISL_1583898, EPI_ISL_1583900, EPI_ISL_1583902, EPI_ISL_1583905, EPI_ISL_1583906, EPI_ISL_1583910, EPI_ISL_1583912, EPI_ISL_1583914, EPI_ISL_1583916, EPI_ISL_1583917, EPI_ISL_1583919, EPI_ISL_1583935, EPI_ISL_1583939, EPI_ISL_1583943, EPI_ISL_1583975, EPI_ISL_1583982, EPI_ISL_1583989, EPI_ISL_1584019, EPI_ISL_1584021, EPI_ISL_1584030, EPI_ISL_1584044, EPI_ISL_1584054, EPI_ISL_1584064, EPI_ISL_1584079, EPI_ISL_1584095, EPI_ISL_1584097, EPI_ISL_1584124, EPI_ISL_1584134, EPI_ISL_1584137, EPI_ISL_1584167, EPI_ISL_1584169, EPI_ISL_1584170, EPI_ISL_1584174, EPI_ISL_1584176, EPI_ISL_1584184, EPI_ISL_1584194, EPI_ISL_1584196, EPI_ISL_1584198, EPI_ISL_1584202, EPI_ISL_1584214, EPI_ISL_1584220, EPI_ISL_1584225, EPI_ISL_1584230, EPI_ISL_1584231, EPI_ISL_1584236, EPI_ISL_1584237, EPI_ISL_1584242, EPI_ISL_1584243, EPI_ISL_1584244, EPI_ISL_1584247, EPI_ISL_1584249, EPI_ISL_1584251, EPI_ISL_1584254, EPI_ISL_1584261, EPI_ISL_1584262, EPI_ISL_1584265, EPI_ISL_1584267, EPI_ISL_1584268, EPI_ISL_1584270, EPI_ISL_1584273, EPI_ISL_1584276, EPI_ISL_1584278, EPI_ISL_1584280, EPI_ISL_1584281, EPI_ISL_1584285, EPI_ISL_1584291, EPI_ISL_1584296, EPI_ISL_1584303, EPI_ISL_1584304, EPI_ISL_1584312, EPI_ISL_1584319, EPI_ISL_1584320, EPI_ISL_1584321, EPI_ISL_1584323, EPI_ISL_1584324, EPI_ISL_1584332, EPI_ISL_1584339, EPI_ISL_1584343, EPI_ISL_1584344, EPI_ISL_1584347, EPI_ISL_1584349, EPI_ISL_1584351, EPI_ISL_1584352, EPI_ISL_1584355, EPI_ISL_1584359, EPI_ISL_1584361, EPI_ISL_1584362, EPI_ISL_1584365, EPI_ISL_1584366, EPI_ISL_1584372, EPI_ISL_1584373, EPI_ISL_1584376, EPI_ISL_1584377, EPI_ISL_1584379, EPI_ISL_1584381, EPI_ISL_1584384, EPI_ISL_1584393, EPI_ISL_1584397, EPI_ISL_1584400, EPI_ISL_1584401, EPI_ISL_1584405, EPI_ISL_1584410, EPI_ISL_1584411, EPI_ISL_1584412, EPI_ISL_1584413, EPI_ISL_1584423, EPI_ISL_1584427, EPI_ISL_1584433, EPI_ISL_1584435, EPI_ISL_1584438, EPI_ISL_1584439, EPI_ISL_1584529, EPI_ISL_1594722, EPI_ISL_1594745, EPI_ISL_1594751, EPI_ISL_1594785, EPI_ISL_1594789, EPI_ISL_1594793, EPI_ISL_1594797, EPI_ISL_1594814, EPI_ISL_1594815, EPI_ISL_1594821, EPI_ISL_1594824, EPI_ISL_1594836, EPI_ISL_1594857, EPI_ISL_1594861, EPI_ISL_1594865, EPI_ISL_1594873, EPI_ISL_1594877, EPI_ISL_1594899, EPI_ISL_1594915, EPI_ISL_1594948, EPI_ISL_1594957, EPI_ISL_1594967, EPI_ISL_1594969, EPI_ISL_1594970, EPI_ISL_1594980, EPI_ISL_1594981, EPI_ISL_1595470, EPI_ISL_1615697, EPI_ISL_1615713, EPI_ISL_1615735, EPI_ISL_1615815, EPI_ISL_1615843, EPI_ISL_1615901, EPI_ISL_1615919, EPI_ISL_1616593, EPI_ISL_1631171, EPI_ISL_1631287, EPI_ISL_1631772, EPI_ISL_1631783, EPI_ISL_1631804, EPI_ISL_1631819, EPI_ISL_1631852, EPI_ISL_1631864, EPI_ISL_1631870, EPI_ISL_1631896, EPI_ISL_1631963, EPI_ISL_1631966, EPI_ISL_1631978, EPI_ISL_1631985, EPI_ISL_1631995, EPI_ISL_1632093, EPI_ISL_1632214, EPI_ISL_1632273, EPI_ISL_1635124, EPI_ISL_1635149, EPI_ISL_1653486, EPI_ISL_1653504, EPI_ISL_1653509, EPI_ISL_1653520, EPI_ISL_1653521, EPI_ISL_1653533, EPI_ISL_1653555, EPI_ISL_1653558, EPI_ISL_1653563, EPI_ISL_1653572, EPI_ISL_1653594, EPI_ISL_1653597, EPI_ISL_1653615, EPI_ISL_1653628, EPI_ISL_1653629, EPI_ISL_1653634, EPI_ISL_1653638, EPI_ISL_1653642, EPI_ISL_1653654, EPI_ISL_1653673, EPI_ISL_1653682, EPI_ISL_1653703, EPI_ISL_1653708

| | | | |
|---|---|---|---|
| see above | Lighthouse Lab in Alderley Park | Wellcome Sanger Institute for the COVID-19 Genomics UK (COG-UK) Consortium | Cordelia Langford; David K. Jackson; Dominic Kwiatkowski; Ewan Harrison; Ian Johnston; Jacquelyn Wynn; Jeffrey Barrett; John Sillitoe on behalf of the Wellcome Sanger Institute COVID-19 Surveillance Team; Mairead Hyland; Roberto Amato; Sonia Goncalves; The Lighthouse Lab in Alderley Park and Alex Alderton |

EPI_ISL_1483828, EPI_ISL_1483837, EPI_ISL_1483866, EPI_ISL_1483936, EPI_ISL_1484013, EPI_ISL_1484020, EPI_ISL_1484025, EPI_ISL_1484036, EPI_ISL_1484042, EPI_ISL_1484057, EPI_ISL_1504403, EPI_ISL_1504409, EPI_ISL_1504419, EPI_ISL_1504421, EPI_ISL_1504425, EPI_ISL_1504427, EPI_ISL_1504428, EPI_ISL_1504437, EPI_ISL_1504440, EPI_ISL_1504447, EPI_ISL_1504451, EPI_ISL_1504453, EPI_ISL_1504472, EPI_ISL_1504473, EPI_ISL_1504476, EPI_ISL_1504477, EPI_ISL_1504484, EPI_ISL_1504486, EPI_ISL_1504492, EPI_ISL_1504496, EPI_ISL_1504506, EPI_ISL_1504513, EPI_ISL_1504514, EPI_ISL_1504515, EPI_ISL_1504517, EPI_ISL_1504525, EPI_ISL_1504526, EPI_ISL_1504530, EPI_ISL_1504534, EPI_ISL_1504548, EPI_ISL_1504551, EPI_ISL_1504555, EPI_ISL_1504556, EPI_ISL_1504561, EPI_ISL_1504569, EPI_ISL_1504583, EPI_ISL_1504584, EPI_ISL_1504587, EPI_ISL_1504588, EPI_ISL_1504590, EPI_ISL_1504591, EPI_ISL_1504598, EPI_ISL_1504607, EPI_ISL_1504610, EPI_ISL_1504614, EPI_ISL_1504615, EPI_ISL_1504620, EPI_ISL_1504625, EPI_ISL_1504629, EPI_ISL_1504636, EPI_ISL_1504637, EPI_ISL_1504643, EPI_ISL_1504644, EPI_ISL_1504662, EPI_ISL_1504663, EPI_ISL_1504672, EPI_ISL_1504692, EPI_ISL_1504693, EPI_ISL_1504748, EPI_ISL_1504755, EPI_ISL_1504757, EPI_ISL_1504779, EPI_ISL_1504790, EPI_ISL_1504816, EPI_ISL_1504833, EPI_ISL_1504887, EPI_ISL_1504892, EPI_ISL_1504920, EPI_ISL_1504955, EPI_ISL_1504968, EPI_ISL_1505012, EPI_ISL_1505013, EPI_ISL_1505025, EPI_ISL_1505038, EPI_ISL_1505046, EPI_ISL_1505064, EPI_ISL_1505098, EPI_ISL_1505103, EPI_ISL_1505110, EPI_ISL_1505116, EPI_ISL_1505124, EPI_ISL_1505126, EPI_ISL_1505136, EPI_ISL_1505150, EPI_ISL_1505158, EPI_ISL_1505164, EPI_ISL_1505173, EPI_ISL_1506172, EPI_ISL_1517765, EPI_ISL_1517767, EPI_ISL_1517772, EPI_ISL_1517777, EPI_ISL_1517778, EPI_ISL_1517789, EPI_ISL_1517791, EPI_ISL_1517807, EPI_ISL_1517811, EPI_ISL_1517814, EPI_ISL_1517844, EPI_ISL_1517864, EPI_ISL_1517871, EPI_ISL_1517875, EPI_ISL_1517889, EPI_ISL_1517898, EPI_ISL_1517900, EPI_ISL_1517901, EPI_ISL_1517913, EPI_ISL_1517918, EPI_ISL_1517920, EPI_ISL_1517925, EPI_ISL_1517930, EPI_ISL_1517936, EPI_ISL_1517938, EPI_ISL_1517951, EPI_ISL_1517952, EPI_ISL_1517953, EPI_ISL_1517956, EPI_ISL_1517958, EPI_ISL_1517975, EPI_ISL_1517976, EPI_ISL_1517980, EPI_ISL_1517986, EPI_ISL_1517999, EPI_ISL_1518007, EPI_ISL_1518008, EPI_ISL_1518024, EPI_ISL_1518027, EPI_ISL_1518031, EPI_ISL_1518043, EPI_ISL_1518046, EPI_ISL_1518054, EPI_ISL_1519276, EPI_ISL_1519280, EPI_ISL_1519293, EPI_ISL_1519294, EPI_ISL_1519300, EPI_ISL_1534757, EPI_ISL_1534758, EPI_ISL_1534762, EPI_ISL_1534775, EPI_ISL_1534777, EPI_ISL_1534778, EPI_ISL_1534779, EPI_ISL_1534781, EPI_ISL_1534783, EPI_ISL_1534786, EPI_ISL_1534789, EPI_ISL_1534791, EPI_ISL_1534795, EPI_ISL_1534803, EPI_ISL_1534804, EPI_ISL_1534818, EPI_ISL_1534820, EPI_ISL_1534821, EPI_ISL_1534823, EPI_ISL_1534834, EPI_ISL_1534836, EPI_ISL_1534837, EPI_ISL_1534839, EPI_ISL_1534850, EPI_ISL_1534854, EPI_ISL_1534858, EPI_ISL_1534863, EPI_ISL_1534874, EPI_ISL_1534879, EPI_ISL_1534886, EPI_ISL_1534895, EPI_ISL_1534914, EPI_ISL_1534918, EPI_ISL_1534921, EPI_ISL_1534927, EPI_ISL_1534928, EPI_ISL_1534930, EPI_ISL_1534938, EPI_ISL_1534943, EPI_ISL_1534945, EPI_ISL_1534949, EPI_ISL_1534957, EPI_ISL_1534958, EPI_ISL_1534961, EPI_ISL_1534963, EPI_ISL_1534965, EPI_ISL_1534967, EPI_ISL_1534975, EPI_ISL_1534976, EPI_ISL_1534978, EPI_ISL_1534981, EPI_ISL_1534983, EPI_ISL_1534984, EPI_ISL_1534992, EPI_ISL_1534993, EPI_ISL_1534996, EPI_ISL_1535001, EPI_ISL_1535003, EPI_ISL_1535006, EPI_ISL_1535008, EPI_ISL_1535016, EPI_ISL_1535027, EPI_ISL_1535033, EPI_ISL_1535035, EPI_ISL_1535042, EPI_ISL_1535070, EPI_ISL_1535100, EPI_ISL_1535157, EPI_ISL_1535178, EPI_ISL_1535258, EPI_ISL_1535267, EPI_ISL_1535272, EPI_ISL_1535293, EPI_ISL_1535301, EPI_ISL_1535329, EPI_ISL_1535330, EPI_ISL_1535332, EPI_ISL_1537317, EPI_ISL_1537325, EPI_ISL_1537326, EPI_ISL_1538031, EPI_ISL_1538063, EPI_ISL_1538071, EPI_ISL_1538072, EPI_ISL_1538084, EPI_ISL_1544778, EPI_ISL_1544792, EPI_ISL_1564038, EPI_ISL_1564053, EPI_ISL_1564062, EPI_ISL_1564078, EPI_ISL_1564086, EPI_ISL_1564115, EPI_ISL_1564125, EPI_ISL_1564132, EPI_ISL_1564172, EPI_ISL_1564179, EPI_ISL_1564181, EPI_ISL_1564187, EPI_ISL_1564200, EPI_ISL_1564206, EPI_ISL_1564241, EPI_ISL_1564256, EPI_ISL_1564270, EPI_ISL_1564276, EPI_ISL_1564284, EPI_ISL_1564286, EPI_ISL_1564287, EPI_ISL_1564296, EPI_ISL_1564300, EPI_ISL_1564314, EPI_ISL_1564326, EPI_ISL_1564772, EPI_ISL_1564775, EPI_ISL_1564776, EPI_ISL_1564777, EPI_ISL_1564778, EPI_ISL_1564779, EPI_ISL_1564780, EPI_ISL_1564782, EPI_ISL_1564784, EPI_ISL_1564785, EPI_ISL_1583760, EPI_ISL_1583786, EPI_ISL_1583922, EPI_ISL_1583923, EPI_ISL_1583927, EPI_ISL_1583928, EPI_ISL_1583929, EPI_ISL_1583930, EPI_ISL_1583932, EPI_ISL_1583933, EPI_ISL_1583934, EPI_ISL_1583936, EPI_ISL_1583941, EPI_ISL_1583948, EPI_ISL_1583949, EPI_ISL_1583952, EPI_ISL_1583953, EPI_ISL_1583956, EPI_ISL_1583957, EPI_ISL_1583958, EPI_ISL_1583959, EPI_ISL_1583961, EPI_ISL_1583964, EPI_ISL_1583965, EPI_ISL_1583968, EPI_ISL_1583971, EPI_ISL_1583973, EPI_ISL_1583976, EPI_ISL_1583977, EPI_ISL_1583978, EPI_ISL_1583979, EPI_ISL_1583980, EPI_ISL_1583988, EPI_ISL_1583990, EPI_ISL_1583992, EPI_ISL_1583995, EPI_ISL_1583996, EPI_ISL_1583997, EPI_ISL_1584000, EPI_ISL_1584001, EPI_ISL_1584003, EPI_ISL_1584004, EPI_ISL_1584005, EPI_ISL_1584009, EPI_ISL_1584013, EPI_ISL_1584014, EPI_ISL_1584018, EPI_ISL_1584020, EPI_ISL_1584023, EPI_ISL_1584024, EPI_ISL_1584025, EPI_ISL_1584027, EPI_ISL_1584028, EPI_ISL_1584031, EPI_ISL_1584035, EPI_ISL_1584037, EPI_ISL_1584038, EPI_ISL_1584041, EPI_ISL_1584042, EPI_ISL_1584045, EPI_ISL_1584048, EPI_ISL_1584049, EPI_ISL_1584051, EPI_ISL_1584057, EPI_ISL_1584059, EPI_ISL_1584060, EPI_ISL_1584061, EPI_ISL_1584063, EPI_ISL_1584065, EPI_ISL_1584068, EPI_ISL_1584069, EPI_ISL_1584074, EPI_ISL_1584075, EPI_ISL_1584078, EPI_ISL_1584080, EPI_ISL_1584085, EPI_ISL_1584086, EPI_ISL_1584088, EPI_ISL_1584092, EPI_ISL_1584093, EPI_ISL_1584098, EPI_ISL_1584099, EPI_ISL_1584105, EPI_ISL_1584107, EPI_ISL_1584109, EPI_ISL_1584111, EPI_ISL_1584113, EPI_ISL_1584117, EPI_ISL_1584120, EPI_ISL_1584126, EPI_ISL_1584129, EPI_ISL_1584133, EPI_ISL_1584136, EPI_ISL_1584138, EPI_ISL_1584139, EPI_ISL_1584140, EPI_ISL_1584141, EPI_ISL_1584142, EPI_ISL_1584143, EPI_ISL_1584144, EPI_ISL_1584145, EPI_ISL_1584146, EPI_ISL_1584147, EPI_ISL_1584148, EPI_ISL_1584149, EPI_ISL_1584150, EPI_ISL_1584151, EPI_ISL_1584152, EPI_ISL_1584153, EPI_ISL_1584154, EPI_ISL_1584155, EPI_ISL_1584156, EPI_ISL_1584157, EPI_ISL_1584158, EPI_ISL_1584159, EPI_ISL_1584160, EPI_ISL_1584161, EPI_ISL_1584162, EPI_ISL_1584163, EPI_ISL_1584164, EPI_ISL_1584444, EPI_ISL_1584460, EPI_ISL_1584470, EPI_ISL_1584476, EPI_ISL_1584497, EPI_ISL_1584511, EPI_ISL_1584513, EPI_ISL_1584515, EPI_ISL_1594464, EPI_ISL_1594482, EPI_ISL_1594484, EPI_ISL_1594512, EPI_ISL_1594590, EPI_ISL_1594613, EPI_ISL_1594730, EPI_ISL_1594736, EPI_ISL_1594748, EPI_ISL_1594749, EPI_ISL_1594770, EPI_ISL_1594807, EPI_ISL_1594810, EPI_ISL_1594848, EPI_ISL_1594866, EPI_ISL_1594901, EPI_ISL_1594929, EPI_ISL_1594934, EPI_ISL_1594956, EPI_ISL_1594962, EPI_ISL_1595201, EPI_ISL_1595202, EPI_ISL_1595203, EPI_ISL_1595204, EPI_ISL_1595205, EPI_ISL_1595206, EPI_ISL_1595259, EPI_ISL_1595313, EPI_ISL_1595334, EPI_ISL_1595378, EPI_ISL_1595404, EPI_ISL_1595419, EPI_ISL_1595450, EPI_ISL_1595457, EPI_ISL_1595472, EPI_ISL_1615752, EPI_ISL_1615766, EPI_ISL_1615808, EPI_ISL_1615869, EPI_ISL_1615879, EPI_ISL_1616211, EPI_ISL_1616262, EPI_ISL_1616265, EPI_ISL_1616279, EPI_ISL_1616287, EPI_ISL_1616523, EPI_ISL_1616534, EPI_ISL_1616540, EPI_ISL_1616552, EPI_ISL_1616564, EPI_ISL_1616567, EPI_ISL_1616569, EPI_ISL_1616570, EPI_ISL_1616572, EPI_ISL_1616573, EPI_ISL_1616574, EPI_ISL_1630372, EPI_ISL_1631094, EPI_ISL_1631102, EPI_ISL_1631106, EPI_ISL_1631117, EPI_ISL_1631134, EPI_ISL_1631149, EPI_ISL_1631155, EPI_ISL_1631177, EPI_ISL_1631185, EPI_ISL_1631189, EPI_ISL_1631196, EPI_ISL_1631286, EPI_ISL_1631303, EPI_ISL_1631330, EPI_ISL_1632310, EPI_ISL_1632315, EPI_ISL_1632354, EPI_ISL_1632355, EPI_ISL_1632360, EPI_ISL_1632386, EPI_ISL_1632389, EPI_ISL_1632443, EPI_ISL_1632451, EPI_ISL_1632452, EPI_ISL_1634718, EPI_ISL_1652954, EPI_ISL_1652964, EPI_ISL_1652967, EPI_ISL_1652982, EPI_ISL_1653006, EPI_ISL_1653018, EPI_ISL_1653027, EPI_ISL_1653059, EPI_ISL_1653074, EPI_ISL_1653078, EPI_ISL_1653100, EPI_ISL_1653102, EPI_ISL_1653104, EPI_ISL_1653109, EPI_ISL_1653126, EPI_ISL_1653136, EPI_ISL_1653601, EPI_ISL_1653680, EPI_ISL_1653681, EPI_ISL_1653717, EPI_ISL_1653721, EPI_ISL_1653754, EPI_ISL_1673100, EPI_ISL_1673114, EPI_ISL_1719747

| | | | |
|---|---|---|---|
| see above | Lighthouse Lab in Cambridge | Wellcome Sanger Institute for the COVID-19 Genomics UK (COG-UK) Consortium | Cordelia Langford; David K. Jackson; Dominic Kwiatkowski; Ewan Harrison; Ian Johnston; Jeffrey Barrett; John Sillitoe on behalf of the Wellcome Sanger Institute COVID-19 Surveillance Team; Rob Howes; Roberto Amato; Sonia Goncalves; The Lighthouse Lab in Cambridge and Alex Alderton |

EPI_ISL_1504113, EPI_ISL_1504114, EPI_ISL_1504118, EPI_ISL_1504121, EPI_ISL_1504123, EPI_ISL_1504124, EPI_ISL_1504131, EPI_ISL_1504134, EPI_ISL_1504135, EPI_ISL_1504136, EPI_ISL_1504139, EPI_ISL_1504142, EPI_ISL_1504144, EPI_ISL_1504148, EPI_ISL_1504150, EPI_ISL_1504152, EPI_ISL_1504156, EPI_ISL_1504157, EPI_ISL_1504159, EPI_ISL_1504161, EPI_ISL_1504164, EPI_ISL_1504170, EPI_ISL_1504171, EPI_ISL_1504172, EPI_ISL_1504173, EPI_ISL_1504176, EPI_ISL_1504178, EPI_ISL_1504179, EPI_ISL_1504182, EPI_ISL_1504187, EPI_ISL_1504189, EPI_ISL_1504190, EPI_ISL_1504191, EPI_ISL_1504192, EPI_ISL_1504194, EPI_ISL_1504197, EPI_ISL_1504198, EPI_ISL_1504199, EPI_ISL_1504201, EPI_ISL_1504202, EPI_ISL_1504211, EPI_ISL_1504212, EPI_ISL_1504214, EPI_ISL_1504215, EPI_ISL_1504217, EPI_ISL_1504220, EPI_ISL_1504221, EPI_ISL_1504224, EPI_ISL_1504230, EPI_ISL_1504238, EPI_ISL_1504242, EPI_ISL_1504251, EPI_ISL_1504252, EPI_ISL_1504253, EPI_ISL_1504255, EPI_ISL_1504257, EPI_ISL_1504259, EPI_ISL_1504264, EPI_ISL_1504266, EPI_ISL_1504267, EPI_ISL_1504268, EPI_ISL_1504269, EPI_ISL_1504277, EPI_ISL_1504279, EPI_ISL_1504281, EPI_ISL_1504282, EPI_ISL_1504283, EPI_ISL_1504286, EPI_ISL_1504287, EPI_ISL_1504293, EPI_ISL_1504299, EPI_ISL_1504300, EPI_ISL_1504302, EPI_ISL_1504304, EPI_ISL_1504305, EPI_ISL_1504307, EPI_ISL_1504308, EPI_ISL_1504309, EPI_ISL_1504310, EPI_ISL_1504312, EPI_ISL_1504319, EPI_ISL_1504326, EPI_ISL_1504330, EPI_ISL_1504331, EPI_ISL_1504334, EPI_ISL_1504337, EPI_ISL_1504338, EPI_ISL_1504343, EPI_ISL_1504346, EPI_ISL_1504354, EPI_ISL_1504356, EPI_ISL_1504358, EPI_ISL_1504362, EPI_ISL_1504364, EPI_ISL_1504366, EPI_ISL_1504378, EPI_ISL_1504379, EPI_ISL_1504382, EPI_ISL_1504383, EPI_ISL_1504389, EPI_ISL_1504391, EPI_ISL_1504393, EPI_ISL_1504397, EPI_ISL_1504398, EPI_ISL_1504399, EPI_ISL_1504400, EPI_ISL_1504401, EPI_ISL_1504402, EPI_ISL_1504404, EPI_ISL_1504407, EPI_ISL_1504410, EPI_ISL_1504416, EPI_ISL_1504417, EPI_ISL_1504418, EPI_ISL_1504420, EPI_ISL_1504422, EPI_ISL_1504424, EPI_ISL_1504426, EPI_ISL_1504429, EPI_ISL_1504430, EPI_ISL_1504432, EPI_ISL_1504438, EPI_ISL_1504439, EPI_ISL_1504442, EPI_ISL_1504443, EPI_ISL_1504446, EPI_ISL_1504448, EPI_ISL_1504449, EPI_ISL_1504452, EPI_ISL_1504454, EPI_ISL_1504460, EPI_ISL_1504461, EPI_ISL_1504463, EPI_ISL_1504464, EPI_ISL_1504466, EPI_ISL_1504467, EPI_ISL_1504469, EPI_ISL_1504471, EPI_ISL_1504474, EPI_ISL_1504475, EPI_ISL_1504478, EPI_ISL_1504479, EPI_ISL_1504482, EPI_ISL_1504485, EPI_ISL_1504487, EPI_ISL_1504489, EPI_ISL_1504490, EPI_ISL_1504493, EPI_ISL_1504495, EPI_ISL_1504502, EPI_ISL_1504504, EPI_ISL_1504505, EPI_ISL_1504507, EPI_ISL_1504509, EPI_ISL_1504510, EPI_ISL_1504511, EPI_ISL_1504512, EPI_ISL_1504516, EPI_ISL_1504519, EPI_ISL_1504521, EPI_ISL_1504523, EPI_ISL_1504528, EPI_ISL_1504532, EPI_ISL_1504533, EPI_ISL_1504535, EPI_ISL_1504537, EPI_ISL_1504541, EPI_ISL_1504542, EPI_ISL_1504544, EPI_ISL_1504545, EPI_ISL_1504546, EPI_ISL_1504550, EPI_ISL_1504554, EPI_ISL_1504557, EPI_ISL_1504558, EPI_ISL_1504559, EPI_ISL_1504563, EPI_ISL_1504564, EPI_ISL_1504567, EPI_ISL_1504570, EPI_ISL_1504573, EPI_ISL_1504575, EPI_ISL_1504581, EPI_ISL_1504585, EPI_ISL_1504586, EPI_ISL_1504589, EPI_ISL_1504592, EPI_ISL_1504593, EPI_ISL_1504594, EPI_ISL_1504597, EPI_ISL_1504599, EPI_ISL_1504600, EPI_ISL_1504601, EPI_ISL_1504602, EPI_ISL_1504604, EPI_ISL_1504606, EPI_ISL_1504608, EPI_ISL_1504611, EPI_ISL_1504613, EPI_ISL_1504618, EPI_ISL_1504619, EPI_ISL_1504623, EPI_ISL_1504624, EPI_ISL_1504627, EPI_ISL_1504628, EPI_ISL_1504632, EPI_ISL_1504633, EPI_ISL_1504635, EPI_ISL_1504638, EPI_ISL_1504640, EPI_ISL_1504641, EPI_ISL_1504646, EPI_ISL_1504647, EPI_ISL_1504648, EPI_ISL_1504649, EPI_ISL_1504655, EPI_ISL_1504657, EPI_ISL_1504658, EPI_ISL_1504659, EPI_ISL_1504664, EPI_ISL_1504666, EPI_ISL_1504910, EPI_ISL_1504912, EPI_ISL_1504914, EPI_ISL_1504916, EPI_ISL_1504919, EPI_ISL_1504931, EPI_ISL_1504934, EPI_ISL_1504935, EPI_ISL_1504946, EPI_ISL_1504947, EPI_ISL_1504950, EPI_ISL_1504952, EPI_ISL_1504958, EPI_ISL_1504963, EPI_ISL_1504971, EPI_ISL_1504972, EPI_ISL_1504975, EPI_ISL_1504976, EPI_ISL_1504982, EPI_ISL_1504983, EPI_ISL_1504986, EPI_ISL_1505001, EPI_ISL_1505005, EPI_ISL_1505006, EPI_ISL_1505011, EPI_ISL_1505015, EPI_ISL_1505021, EPI_ISL_1505022, EPI_ISL_1505024, EPI_ISL_1505027, EPI_ISL_1505029, EPI_ISL_1505033, EPI_ISL_1505039, EPI_ISL_1505040, EPI_ISL_1505044, EPI_ISL_1505048, EPI_ISL_1505054, EPI_ISL_1505056, EPI_ISL_1505058, EPI_ISL_1505060, EPI_ISL_1505062, EPI_ISL_1505065, EPI_ISL_1505068, EPI_ISL_1505069, EPI_ISL_1505072, EPI_ISL_1505073, EPI_ISL_1505076, EPI_ISL_1505083, EPI_ISL_1505086, EPI_ISL_1505089, EPI_ISL_1505093, EPI_ISL_1505104, EPI_ISL_1505105, EPI_ISL_1505106, EPI_ISL_1505115, EPI_ISL_1505119, EPI_ISL_1505120, EPI_ISL_1505128, EPI_ISL_1505130, EPI_ISL_1505132, EPI_ISL_1505148, EPI_ISL_1505160, EPI_ISL_1505167, EPI_ISL_1505172, EPI_ISL_1505174, EPI_ISL_1505181, EPI_ISL_1505184, EPI_ISL_1505187, EPI_ISL_1505190, EPI_ISL_1505191, EPI_ISL_1505201, EPI_ISL_1505206, EPI_ISL_1505207, EPI_ISL_1505208, EPI_ISL_1505211, EPI_ISL_1505215, EPI_ISL_1505218, EPI_ISL_1505220, EPI_ISL_1505221, EPI_ISL_1505225, EPI_ISL_1505227, EPI_ISL_1505229, EPI_ISL_1505230, EPI_ISL_1505233, EPI_ISL_1505240, EPI_ISL_1505243, EPI_ISL_1505247, EPI_ISL_1505248, EPI_ISL_1505258, EPI_ISL_1505260, EPI_ISL_1505265, EPI_ISL_1505267, EPI_ISL_1505270, EPI_ISL_1505273, EPI_ISL_1505277, EPI_ISL_1505289, EPI_ISL_1505291, EPI_ISL_1505293, EPI_ISL_1505294, EPI_ISL_1505295, EPI_ISL_1505301, EPI_ISL_1505304, EPI_ISL_1505307, EPI_ISL_1505311, EPI_ISL_1505314, EPI_ISL_1505320, EPI_ISL_1505323, EPI_ISL_1505325, EPI_ISL_1505350, EPI_ISL_1505356, EPI_ISL_1505361, EPI_ISL_1505362, EPI_ISL_1505363, EPI_ISL_1505367, EPI_ISL_1505370, EPI_ISL_1505371, EPI_ISL_1505383, EPI_ISL_1505385, EPI_ISL_1505392, EPI_ISL_1505393, EPI_ISL_1505405, EPI_ISL_1505420, EPI_ISL_1505421, EPI_ISL_1505434, EPI_ISL_1505436, EPI_ISL_1505442, EPI_ISL_1505446, EPI_ISL_1505448, EPI_ISL_1505452, EPI_ISL_1505453, EPI_ISL_1505454, EPI_ISL_1505457, EPI_ISL_1505458, EPI_ISL_1506235, EPI_ISL_1506302, EPI_ISL_1506367, EPI_ISL_1506449, EPI_ISL_1506502, EPI_ISL_1506548, EPI_ISL_1506656, EPI_ISL_1507234, EPI_ISL_1517497, EPI_ISL_1517519, EPI_ISL_1517520, EPI_ISL_1517533, EPI_ISL_1517535, EPI_ISL_1517536, EPI_ISL_1517543, EPI_ISL_1517551, EPI_ISL_1517554, EPI_ISL_1517562, EPI_ISL_1517568, EPI_ISL_1517588, EPI_ISL_1517598, EPI_ISL_1517599, EPI_ISL_1517600, EPI_ISL_1517606, EPI_ISL_1517610, EPI_ISL_1517614, EPI_ISL_1517621, EPI_ISL_1517622, EPI_ISL_1517625, EPI_ISL_1517628, EPI_ISL_1517629, EPI_ISL_1517631, EPI_ISL_1517632, EPI_ISL_1517639, EPI_ISL_1517648, EPI_ISL_1517656, EPI_ISL_1517658, EPI_ISL_1517672, EPI_ISL_1517673, EPI_ISL_1517686, EPI_ISL_1517687, EPI_ISL_1517691, EPI_ISL_1517692, EPI_ISL_1517694, EPI_ISL_1517695, EPI_ISL_1517696, EPI_ISL_1517701, EPI_ISL_1517709, EPI_ISL_1517712, EPI_ISL_1517715, EPI_ISL_1517732, EPI_ISL_1517733, EPI_ISL_1517741, EPI_ISL_1517742, EPI_ISL_1517750, EPI_ISL_1518393, EPI_ISL_1518396, EPI_ISL_1518410, EPI_ISL_1518411, EPI_ISL_1518418, EPI_ISL_1518419, EPI_ISL_1518420, EPI_ISL_1518423, EPI_ISL_1518425, EPI_ISL_1518434, EPI_ISL_1518449, EPI_ISL_1518459, EPI_ISL_1518461, EPI_ISL_1518462, EPI_ISL_1518481, EPI_ISL_1518483, EPI_ISL_1518486, EPI_ISL_1518489, EPI_ISL_1518493, EPI_ISL_1518513, EPI_ISL_1518525, EPI_ISL_1518537, EPI_ISL_1518538, EPI_ISL_1518539, EPI_ISL_1518542, EPI_ISL_1518553, EPI_ISL_1518554, EPI_ISL_1518556, EPI_ISL_1518558, EPI_ISL_1518559, EPI_ISL_1518564, EPI_ISL_1518576, EPI_ISL_1518584, EPI_ISL_1518589, EPI_ISL_1518593, EPI_ISL_1518594, EPI_ISL_1518598, EPI_ISL_1518604, EPI_ISL_1518607, EPI_ISL_1518612, EPI_ISL_1518613, EPI_ISL_1518621, EPI_ISL_1518623, EPI_ISL_1518628, EPI_ISL_1518647, EPI_ISL_1518650, EPI_ISL_1518667, EPI_ISL_1519268, EPI_ISL_1519434, EPI_ISL_1519849, EPI_ISL_1519884, EPI_ISL_1534751, EPI_ISL_1534761, EPI_ISL_1534774, EPI_ISL_1534776, EPI_ISL_1534801, EPI_ISL_1534806, EPI_ISL_1534842, EPI_ISL_1534845, EPI_ISL_1534848, EPI_ISL_1534852, EPI_ISL_1534868, EPI_ISL_1534872, EPI_ISL_1534880, EPI_ISL_1534891, EPI_ISL_1534894, EPI_ISL_1534911, EPI_ISL_1534925, EPI_ISL_1534942, EPI_ISL_1534950, EPI_ISL_1534953, EPI_ISL_1534960, EPI_ISL_1534974, EPI_ISL_1534977, EPI_ISL_1534987, EPI_ISL_1534990, EPI_ISL_1535337, EPI_ISL_1535357, EPI_ISL_1535360, EPI_ISL_1535364, EPI_ISL_1535366, EPI_ISL_1535377, EPI_ISL_1535378, EPI_ISL_1535380, EPI_ISL_1535381, EPI_ISL_1535385, EPI_ISL_1535387, EPI_ISL_1535388, EPI_ISL_1535392, EPI_ISL_1535399, EPI_ISL_1535403, EPI_ISL_1535404, EPI_ISL_1535406, EPI_ISL_1535409, EPI_ISL_1535410, EPI_ISL_1535413, EPI_ISL_1535416, EPI_ISL_1535418, EPI_ISL_1535419, EPI_ISL_1535422, EPI_ISL_1535423, EPI_ISL_1535429, EPI_ISL_1535430, EPI_ISL_1535432, EPI_ISL_1535435, EPI_ISL_1535437, EPI_ISL_1535446, EPI_ISL_1535448, EPI_ISL_1535457, EPI_ISL_1535460, EPI_ISL_1535462, EPI_ISL_1535465, EPI_ISL_1535466, EPI_ISL_1535467, EPI_ISL_1535470, EPI_ISL_1535471, EPI_ISL_1535473, EPI_ISL_1535474, EPI_ISL_1535475, EPI_ISL_1535481, EPI_ISL_1535483, EPI_ISL_1535485, EPI_ISL_1535490, EPI_ISL_1535492, EPI_ISL_1535494, EPI_ISL_1535498, EPI_ISL_1535501, EPI_ISL_1535502, EPI_ISL_1535504, EPI_ISL_1535517, EPI_ISL_1535518, EPI_ISL_1535519, EPI_ISL_1535522, EPI_ISL_1535523, EPI_ISL_1535528, EPI_ISL_1535529, EPI_ISL_1535537, EPI_ISL_1535540, EPI_ISL_1535541, EPI_ISL_1535542, EPI_ISL_1535544, EPI_ISL_1535546, EPI_ISL_1535549, EPI_ISL_1535553, EPI_ISL_1535555, EPI_ISL_1535556, EPI_ISL_1535559, EPI_ISL_1535560, EPI_ISL_1535564, EPI_ISL_1535568, EPI_ISL_1535573, EPI_ISL_1535578, EPI_ISL_1535580, EPI_ISL_1535583, EPI_ISL_1535585, EPI_ISL_1535590, EPI_ISL_1535594, EPI_ISL_1535596, EPI_ISL_1535599, EPI_ISL_1535610, EPI_ISL_1535611, EPI_ISL_1535614, EPI_ISL_1535616, EPI_ISL_1535622, EPI_ISL_1535634, EPI_ISL_1535649, EPI_ISL_1535655, EPI_ISL_1535656, EPI_ISL_1535661, EPI_ISL_1535667, EPI_ISL_1535672, EPI_ISL_1535673, EPI_ISL_1535677, EPI_ISL_1535679, EPI_ISL_1535682, EPI_ISL_1535685, EPI_ISL_1535687, EPI_ISL_1535692, EPI_ISL_1535698, EPI_ISL_1535708, EPI_ISL_1535714, EPI_ISL_1535716, EPI_ISL_1535722, EPI_ISL_1535725, EPI_ISL_1535730, EPI_ISL_1535736, EPI_ISL_1535742, EPI_ISL_1535744, EPI_ISL_1535751, EPI_ISL_1535754, EPI_ISL_1535764, EPI_ISL_1535766, EPI_ISL_1535773, EPI_ISL_1535774, EPI_ISL_1535782, EPI_ISL_1535783, EPI_ISL_1535802, EPI_ISL_1535803, EPI_ISL_1535807, EPI_ISL_1535814, EPI_ISL_1535816, EPI_ISL_1535824, EPI_ISL_1535828, EPI_ISL_1535829, EPI_ISL_1535838, EPI_ISL_1535841, EPI_ISL_1535846, EPI_ISL_1535851, EPI_ISL_1535852, EPI_ISL_1535857, EPI_ISL_1535861, EPI_ISL_1535867, EPI_ISL_1535870, EPI_ISL_1535874, EPI_ISL_1535875, EPI_ISL_1535879, EPI_ISL_1535886, EPI_ISL_1535892, EPI_ISL_1535897, EPI_ISL_1535903, EPI_ISL_1535907, EPI_ISL_1535910, EPI_ISL_1535912, EPI_ISL_1535916, EPI_ISL_1535919, EPI_ISL_1535920, EPI_ISL_1535921, EPI_ISL_1535924, EPI_ISL_1535934, EPI_ISL_1535937, EPI_ISL_1535939, EPI_ISL_1535944, EPI_ISL_1535950, EPI_ISL_1535951, EPI_ISL_1535963, EPI_ISL_1535966, EPI_ISL_1535969, EPI_ISL_1535975, EPI_ISL_1535976, EPI_ISL_1535982, EPI_ISL_1535983, EPI_ISL_1535984, EPI_ISL_1535985, EPI_ISL_1535991, EPI_ISL_1535992, EPI_ISL_1535994, EPI_ISL_1535999, EPI_ISL_1536001, EPI_ISL_1536005, EPI_ISL_1536006, EPI_ISL_1536008, EPI_ISL_1536024, EPI_ISL_1536025, EPI_ISL_1536028, EPI_ISL_1536029, EPI_ISL_1536030, EPI_ISL_1536036, EPI_ISL_1536041, EPI_ISL_1536042, EPI_ISL_1536043, EPI_ISL_1536049, EPI_ISL_1536062, EPI_ISL_1536068, EPI_ISL_1536071, EPI_ISL_1536108, EPI_ISL_1536111, EPI_ISL_1536116, EPI_ISL_1536120, EPI_ISL_1536125, EPI_ISL_1536126, EPI_ISL_1536131, EPI_ISL_1536132, EPI_ISL_1536137, EPI_ISL_1536138, EPI_ISL_1536144, EPI_ISL_1536149, EPI_ISL_1536151, EPI_ISL_1536152, EPI_ISL_1536156, EPI_ISL_1536160, EPI_ISL_1536161, EPI_ISL_1536171, EPI_ISL_1536172, EPI_ISL_1536175, EPI_ISL_1536179, EPI_ISL_1536539, EPI_ISL_1536542, EPI_ISL_1536543, EPI_ISL_1536544, EPI_ISL_1536558, EPI_ISL_1536559, EPI_ISL_1536560, EPI_ISL_1536568, EPI_ISL_1536570, EPI_ISL_1536574, EPI_ISL_1536580, EPI_ISL_1536581, EPI_ISL_1536584, EPI_ISL_1536588, EPI_ISL_1536590, EPI_ISL_1536597, EPI_ISL_1536599, EPI_ISL_1536600, EPI_ISL_1536603, EPI_ISL_1536604, EPI_ISL_1536605, EPI_ISL_1536609, EPI_ISL_1536613, EPI_ISL_1536616, EPI_ISL_1536617, EPI_ISL_1536626, EPI_ISL_1536628, EPI_ISL_1536631, EPI_ISL_1536632, EPI_ISL_1536634, EPI_ISL_1536638, EPI_ISL_1536646, EPI_ISL_1536654, EPI_ISL_1536655, EPI_ISL_1536660, EPI_ISL_1536665, EPI_ISL_1536671, EPI_ISL_1536680, EPI_ISL_1536683, EPI_ISL_1536685, EPI_ISL_1536689, EPI_ISL_1536690, EPI_ISL_1536693, EPI_ISL_1536694, EPI_ISL_1536698, EPI_ISL_1536701, EPI_ISL_1536702, EPI_ISL_1536703, EPI_ISL_1536708, EPI_ISL_1536710, EPI_ISL_1536715, EPI_ISL_1536716, EPI_ISL_1536719, EPI_ISL_1536725, EPI_ISL_1536729, EPI_ISL_1536733, EPI_ISL_1536742, EPI_ISL_1536743, EPI_ISL_1536745, EPI_ISL_1536747, EPI_ISL_1536753, EPI_ISL_1536754, EPI_ISL_1536760, EPI_ISL_1536766, EPI_ISL_1536768, EPI_ISL_1536770, EPI_ISL_1536773, EPI_ISL_1536775, EPI_ISL_1536778, EPI_ISL_1536780, EPI_ISL_1536782, EPI_ISL_1536783, EPI_ISL_1536788, EPI_ISL_1536789, EPI_ISL_1536790, EPI_ISL_1536792, EPI_ISL_1536793, EPI_ISL_1536794, EPI_ISL_1536795, EPI_ISL_1536796, EPI_ISL_1536799, EPI_ISL_1536800, EPI_ISL_1536802, EPI_ISL_1536804, EPI_ISL_1536806, EPI_ISL_1536809, EPI_ISL_1536810, EPI_ISL_1536811, EPI_ISL_1536813, EPI_ISL_1536816, EPI_ISL_1536817, EPI_ISL_1536821, EPI_ISL_1536823, EPI_ISL_1536824, EPI_ISL_1536827, EPI_ISL_1536829, EPI_ISL_1536831, EPI_ISL_1536835, EPI_ISL_1536837, EPI_ISL_1536842, EPI_ISL_1536844, EPI_ISL_1536845, EPI_ISL_1536847, EPI_ISL_1536849, EPI_ISL_1536851, EPI_ISL_1536854, EPI_ISL_1536855, EPI_ISL_1536857, EPI_ISL_1536858, EPI_ISL_1536859, EPI_ISL_1536861, EPI_ISL_1536869, EPI_ISL_1536870, EPI_ISL_1536872, EPI_ISL_1536873, EPI_ISL_1536874, EPI_ISL_1536878, EPI_ISL_1536879, EPI_ISL_1536880, EPI_ISL_1536881, EPI_ISL_1536882, EPI_ISL_1536886, EPI_ISL_1536888, EPI_ISL_1536891, EPI_ISL_1536893, EPI_ISL_1536897, EPI_ISL_1536903, EPI_ISL_1536905, EPI_ISL_1536906, EPI_ISL_1536911, EPI_ISL_1536912, EPI_ISL_1536914, EPI_ISL_1536917, EPI_ISL_1536918, EPI_ISL_1536919, EPI_ISL_1536923, EPI_ISL_1536925, EPI_ISL_1536926, EPI_ISL_1536930, EPI_ISL_1536933, EPI_ISL_1536937, EPI_ISL_1536938, EPI_ISL_1536942, EPI_ISL_1536944, EPI_ISL_1536945, EPI_ISL_1536947, EPI_ISL_1536948, EPI_ISL_1536951, EPI_ISL_1536954, EPI_ISL_1536956, EPI_ISL_1536959, EPI_ISL_1536962, EPI_ISL_1536963, EPI_ISL_1536964, EPI_ISL_1536966, EPI_ISL_1536967, EPI_ISL_1536968, EPI_ISL_1536969, EPI_ISL_1536972, EPI_ISL_1536973, EPI_ISL_1536974, EPI_ISL_1536975, EPI_ISL_1536976, EPI_ISL_1536978, EPI_ISL_1536979, EPI_ISL_1536981, EPI_ISL_1536984, EPI_ISL_1536985, EPI_ISL_1536987, EPI_ISL_1536990, EPI_ISL_1536991, EPI_ISL_1536993, EPI_ISL_1536994, EPI_ISL_1536995, EPI_ISL_1536998, EPI_ISL_1537002, EPI_ISL_1537003, EPI_ISL_1537004, EPI_ISL_1537007, EPI_ISL_1537008, EPI_ISL_1537009, EPI_ISL_1537013, EPI_ISL_1537015, EPI_ISL_1537017, EPI_ISL_1537020, EPI_ISL_1537022, EPI_ISL_1537024, EPI_ISL_1537026, EPI_ISL_1537029, EPI_ISL_1537030, EPI_ISL_1537032, EPI_ISL_1537034, EPI_ISL_1537036, EPI_ISL_1537043, EPI_ISL_1537045, EPI_ISL_1537046, EPI_ISL_1537050, EPI_ISL_1537053, EPI_ISL_1537054, EPI_ISL_1537055, EPI_ISL_1537057, EPI_ISL_1537058, EPI_ISL_1537060, EPI_ISL_1537063, EPI_ISL_1537064, EPI_ISL_1537068, EPI_ISL_1537069, EPI_ISL_1537073, EPI_ISL_1537074, EPI_ISL_1537076, EPI_ISL_1537080, EPI_ISL_1537081, EPI_ISL_1537082, EPI_ISL_1537083, EPI_ISL_1537085, EPI_ISL_1537086, EPI_ISL_1537087, EPI_ISL_1537089, EPI_ISL_1537095, EPI_ISL_1537096, EPI_ISL_1537097, EPI_ISL_1537098, EPI_ISL_1537100, EPI_ISL_1537102, EPI_ISL_1537103, EPI_ISL_1537107, EPI_ISL_1537110, EPI_ISL_1537111, EPI_ISL_1537112, EPI_ISL_1537116, EPI_ISL_1537117, EPI_ISL_1537118, EPI_ISL_1537119, EPI_ISL_1537120, EPI_ISL_1537123, EPI_ISL_1537125, EPI_ISL_1537129, EPI_ISL_1537130, EPI_ISL_1537141, EPI_ISL_1537142, EPI_ISL_1537143, EPI_ISL_1537144, EPI_ISL_1537145, EPI_ISL_1537147, EPI_ISL_1537148, EPI_ISL_1537149, EPI_ISL_1537150, EPI_ISL_1537151, EPI_ISL_1537153, EPI_ISL_1537155, EPI_ISL_1537156, EPI_ISL_1537157, EPI_ISL_1537159, EPI_ISL_1537161, EPI_ISL_1537162, EPI_ISL_1537163, EPI_ISL_1537164, EPI_ISL_1537165, EPI_ISL_1537166, EPI_ISL_1537169, EPI_ISL_1537170, EPI_ISL_1537171, EPI_ISL_1537172, EPI_ISL_1537173, EPI_ISL_1537176, EPI_ISL_1537177, EPI_ISL_1537178, EPI_ISL_1537180, EPI_ISL_1537181, EPI_ISL_1537182, EPI_ISL_1537183, EPI_ISL_1537184, EPI_ISL_1537185, EPI_ISL_1537186, EPI_ISL_1537187, EPI_ISL_1537189, EPI_ISL_1537190, EPI_ISL_1537192, EPI_ISL_1537194, EPI_ISL_1537195, EPI_ISL_1537196, EPI_ISL_1537197, EPI_ISL_1537198, EPI_ISL_1537199, EPI_ISL_1537200, EPI_ISL_1537202, EPI_ISL_1537203, EPI_ISL_1537204, EPI_ISL_1537205, EPI_ISL_1537206, EPI_ISL_1537207, EPI_ISL_1537208, EPI_ISL_1537209, EPI_ISL_1537214, EPI_ISL_1537215, EPI_ISL_1537216, EPI_ISL_1537220, EPI_ISL_1537242, EPI_ISL_1537249, EPI_ISL_1537250, EPI_ISL_1537258, EPI_ISL_1537259, EPI_ISL_1537260, EPI_ISL_1537261, EPI_ISL_1537262, EPI_ISL_1537263, EPI_ISL_1537268, EPI_ISL_1537273, EPI_ISL_1537274, EPI_ISL_1537283, EPI_ISL_1537286, EPI_ISL_1537289, EPI_ISL_1537290, EPI_ISL_1537301, EPI_ISL_1537302, EPI_ISL_1537304, EPI_ISL_1537310, EPI_ISL_1537311, EPI_ISL_1537313, EPI_ISL_1537314, EPI_ISL_1537329, EPI_ISL_1537333, EPI_ISL_1537336, EPI_ISL_1537337, EPI_ISL_1537341, EPI_ISL_1537343, EPI_ISL_1537344, EPI_ISL_1537346, EPI_ISL_1537349, EPI_ISL_1537350, EPI_ISL_1537352, EPI_ISL_1537354, EPI_ISL_1537356, EPI_ISL_1537357, EPI_ISL_1537362, EPI_ISL_1537366, EPI_ISL_1537368, EPI_ISL_1537371, EPI_ISL_1537379, EPI_ISL_1537381, EPI_ISL_1537383, EPI_ISL_1537384, EPI_ISL_1537389, EPI_ISL_1537391, EPI_ISL_1537392, EPI_ISL_1537393, EPI_ISL_1537396, EPI_ISL_1537397, EPI_ISL_1537398, EPI_ISL_1537400, EPI_ISL_1537401, EPI_ISL_1537402, EPI_ISL_1537408, EPI_ISL_1537417, EPI_ISL_1537423, EPI_ISL_1537424, EPI_ISL_1537427, EPI_ISL_1537433, EPI_ISL_1537436, EPI_ISL_1537438, EPI_ISL_1537441, EPI_ISL_1537442, EPI_ISL_1537445, EPI_ISL_1537447, EPI_ISL_1537449, EPI_ISL_1537450, EPI_ISL_1537451, EPI_ISL_1537452, EPI_ISL_1537455, EPI_ISL_1537457, EPI_ISL_1537458, EPI_ISL_1537459, EPI_ISL_1537460, EPI_ISL_1537461, EPI_ISL_1537464, EPI_ISL_1537465, EPI_ISL_1537468, EPI_ISL_1537470, EPI_ISL_1537471, EPI_ISL_1537473, EPI_ISL_1537474, EPI_ISL_1537477, EPI_ISL_1537479, EPI_ISL_1537482, EPI_ISL_1537487, EPI_ISL_1537489, EPI_ISL_1537490, EPI_ISL_1537491, EPI_ISL_1537492, EPI_ISL_1537493, EPI_ISL_1537494, EPI_ISL_1537497, EPI_ISL_1537498, EPI_ISL_1537500, EPI_ISL_1537501, EPI_ISL_1537504, EPI_ISL_1537505, EPI_ISL_1537506, EPI_ISL_1537507, EPI_ISL_1537509, EPI_ISL_1537510, EPI_ISL_1537514, EPI_ISL_1537515, EPI_ISL_1537516, EPI_ISL_1537518, EPI_ISL_1537522, EPI_ISL_1537524, EPI_ISL_1537525, EPI_ISL_1537527, EPI_ISL_1537529, EPI_ISL_1537530, EPI_ISL_1537533, EPI_ISL_1537535, EPI_ISL_1537537, EPI_ISL_1537538, EPI_ISL_1537542, EPI_ISL_1537546, EPI_ISL_1537548, EPI_ISL_1537550, EPI_ISL_1537552, EPI_ISL_1537554, EPI_ISL_1537555, EPI_ISL_1537556, EPI_ISL_1537559, EPI_ISL_1537561, EPI_ISL_1537565, EPI_ISL_1537566, EPI_ISL_1537568, EPI_ISL_1537569, EPI_ISL_1537570, EPI_ISL_1537575, EPI_ISL_1537578, EPI_ISL_1537579, EPI_ISL_1537580, EPI_ISL_1537581, EPI_ISL_1537585, EPI_ISL_1537587, EPI_ISL_1537592, EPI_ISL_1537593, EPI_ISL_1537594, EPI_ISL_1537596, EPI_ISL_1537599, EPI_ISL_1537605, EPI_ISL_1537606, EPI_ISL_1537607, EPI_ISL_1537614, EPI_ISL_1537617, EPI_ISL_1537622, EPI_ISL_1537625, EPI_ISL_1537636, EPI_ISL_1537639, EPI_ISL_1537640, EPI_ISL_1537641, EPI_ISL_1537643, EPI_ISL_1537644, EPI_ISL_1537646, EPI_ISL_1537648, EPI_ISL_1537649, EPI_ISL_1537650, EPI_ISL_1537652, EPI_ISL_1537656, EPI_ISL_1537657, EPI_ISL_1537660, EPI_ISL_1537662, EPI_ISL_1537668, EPI_ISL_1537672, EPI_ISL_1537675, EPI_ISL_1537676, EPI_ISL_1537677, EPI_ISL_1537680, EPI_ISL_1537681, EPI_ISL_1537683, EPI_ISL_1537684, EPI_ISL_1537685, EPI_ISL_1537688, EPI_ISL_1537691, EPI_ISL_1537696, EPI_ISL_1537697, EPI_ISL_1537698, EPI_ISL_1537699, EPI_ISL_1537703, EPI_ISL_1537706, EPI_ISL_1537712, EPI_ISL_1537713, EPI_ISL_1537716, EPI_ISL_1537717, EPI_ISL_1537721, EPI_ISL_1537725, EPI_ISL_1537727, EPI_ISL_1537730, EPI_ISL_1537733, EPI_ISL_1537734, EPI_ISL_1537735, EPI_ISL_1537737, EPI_ISL_1537739, EPI_ISL_1537740, EPI_ISL_1537741, EPI_ISL_1537743, EPI_ISL_1537744, EPI_ISL_1537745, EPI_ISL_1537746, EPI_ISL_1537750, EPI_ISL_1537751, EPI_ISL_1537752, EPI_ISL_1537755, EPI_ISL_1537756, EPI_ISL_1537757, EPI_ISL_1537761, EPI_ISL_1537762, EPI_ISL_1537764, EPI_ISL_1537765, EPI_ISL_1537769, EPI_ISL_1537771, EPI_ISL_1537773, EPI_ISL_1537777, EPI_ISL_1537779, EPI_ISL_1537780, EPI_ISL_1537782, EPI_ISL_1537783, EPI_ISL_1537784, EPI_ISL_1537787, EPI_ISL_1537792, EPI_ISL_1537795, EPI_ISL_1537796, EPI_ISL_1537797, EPI_ISL_1537799, EPI_ISL_1537803, EPI_ISL_1537805, EPI_ISL_1537809, EPI_ISL_1537811, EPI_ISL_1537812, EPI_ISL_1537815, EPI_ISL_1537817, EPI_ISL_1537821, EPI_ISL_1537824, EPI_ISL_1537825, EPI_ISL_1537831, EPI_ISL_1537832, EPI_ISL_1537833, EPI_ISL_1537835, EPI_ISL_1537836, EPI_ISL_1537839, EPI_ISL_1537841, EPI_ISL_1537844, EPI_ISL_1537845, EPI_ISL_1537846, EPI_ISL_1537847, EPI_ISL_1537848, EPI_ISL_1537849, EPI_ISL_1537850, EPI_ISL_1537851, EPI_ISL_1537852, EPI_ISL_1537853, EPI_ISL_1537854, EPI_ISL_1537855, EPI_ISL_1537856, EPI_ISL_1537857, EPI_ISL_1537858, EPI_ISL_1537859, EPI_ISL_1537860, EPI_ISL_1537861, EPI_ISL_1537862, EPI_ISL_1537863, EPI_ISL_1537864, EPI_ISL_1537866, EPI_ISL_1537867, EPI_ISL_1537868, EPI_ISL_1537869, EPI_ISL_1537871, EPI_ISL_1537872, EPI_ISL_1537873, EPI_ISL_1537874, EPI_ISL_1537875, EPI_ISL_1537876, EPI_ISL_1537878, EPI_ISL_1537879, EPI_ISL_1537880, EPI_ISL_1537881, EPI_ISL_1537882, EPI_ISL_1537883, EPI_ISL_1537884, EPI_ISL_1537885, EPI_ISL_1537886, EPI_ISL_1537887, EPI_ISL_1537888, EPI_ISL_1537889, EPI_ISL_1537890, EPI_ISL_1537891, EPI_ISL_1537892, EPI_ISL_1537893, EPI_ISL_1537894, EPI_ISL_1537895, EPI_ISL_1537896, EPI_ISL_1537897, EPI_ISL_1537898, EPI_ISL_1537899, EPI_ISL_1537900, EPI_ISL_1537901,

EPI_ISL_1537902, EPI_ISL_1537903, EPI_ISL_1537904, EPI_ISL_1537905, EPI_ISL_1537907, EPI_ISL_1537908, EPI_ISL_1537909, EPI_ISL_1537910, EPI_ISL_1537911, EPI_ISL_1537912, EPI_ISL_1537913, EPI_ISL_1537914, EPI_ISL_1537915, EPI_ISL_1537916, EPI_ISL_1537917, EPI_ISL_1537918, EPI_ISL_1537919, EPI_ISL_1537920, EPI_ISL_1537921, EPI_ISL_1537922, EPI_ISL_1537923, EPI_ISL_1537924, EPI_ISL_1537925, EPI_ISL_1537926, EPI_ISL_1537927, EPI_ISL_1537928, EPI_ISL_1537929, EPI_ISL_1537930, EPI_ISL_1537931, EPI_ISL_1537932, EPI_ISL_1537933, EPI_ISL_1537934, EPI_ISL_1537935, EPI_ISL_1537936, EPI_ISL_1537937, EPI_ISL_1537938, EPI_ISL_1537939, EPI_ISL_1537940, EPI_ISL_1537941, EPI_ISL_1537942, EPI_ISL_1537943, EPI_ISL_1537944, EPI_ISL_1537945, EPI_ISL_1537946, EPI_ISL_1537947, EPI_ISL_1537948, EPI_ISL_1537949, EPI_ISL_1537950, EPI_ISL_1537951, EPI_ISL_1537952, EPI_ISL_1537953, EPI_ISL_1537954, EPI_ISL_1537955, EPI_ISL_1537956, EPI_ISL_1537957, EPI_ISL_1537958, EPI_ISL_1537959, EPI_ISL_1537960, EPI_ISL_1537962, EPI_ISL_1537963, EPI_ISL_1537964, EPI_ISL_1537965, EPI_ISL_1537966, EPI_ISL_1537967, EPI_ISL_1537968, EPI_ISL_1537969, EPI_ISL_1537970, EPI_ISL_1537971, EPI_ISL_1537972, EPI_ISL_1537973, EPI_ISL_1537974, EPI_ISL_1537975, EPI_ISL_1537976, EPI_ISL_1537977, EPI_ISL_1537978, EPI_ISL_1537979, EPI_ISL_1537980, EPI_ISL_1537981, EPI_ISL_1537982, EPI_ISL_1537983, EPI_ISL_1537984, EPI_ISL_1537985, EPI_ISL_1537986, EPI_ISL_1537987, EPI_ISL_1537988, EPI_ISL_1537989, EPI_ISL_1537990, EPI_ISL_1537991, EPI_ISL_1537992, EPI_ISL_1537993, EPI_ISL_1537994, EPI_ISL_1537995, EPI_ISL_1537996, EPI_ISL_1537997, EPI_ISL_1537998, EPI_ISL_1537999, EPI_ISL_1538000, EPI_ISL_1538001, EPI_ISL_1538002, EPI_ISL_1538003, EPI_ISL_1538004, EPI_ISL_1538005, EPI_ISL_1538006, EPI_ISL_1538007, EPI_ISL_1538008, EPI_ISL_1538009, EPI_ISL_1538011, EPI_ISL_1538012, EPI_ISL_1538013, EPI_ISL_1538014, EPI_ISL_1538068, EPI_ISL_1538070, EPI_ISL_1538074, EPI_ISL_1538075, EPI_ISL_1538076, EPI_ISL_1538077, EPI_ISL_1538079, EPI_ISL_1538081, EPI_ISL_1538082, EPI_ISL_1538083, EPI_ISL_1538085, EPI_ISL_1538086, EPI_ISL_1538087, EPI_ISL_1538089, EPI_ISL_1538092, EPI_ISL_1538096, EPI_ISL_1538098, EPI_ISL_1538101, EPI_ISL_1538104, EPI_ISL_1538106, EPI_ISL_1538107, EPI_ISL_1538108, EPI_ISL_1538112, EPI_ISL_1538116, EPI_ISL_1538120, EPI_ISL_1538122, EPI_ISL_1538124, EPI_ISL_1538128, EPI_ISL_1538131, EPI_ISL_1538134, EPI_ISL_1538136, EPI_ISL_1538137, EPI_ISL_1538144, EPI_ISL_1538145, EPI_ISL_1538149, EPI_ISL_1538151, EPI_ISL_1538154, EPI_ISL_1538166, EPI_ISL_1538171, EPI_ISL_1538175, EPI_ISL_1538185, EPI_ISL_1538196, EPI_ISL_1538206, EPI_ISL_1538208, EPI_ISL_1538209, EPI_ISL_1538212, EPI_ISL_1538214, EPI_ISL_1538216, EPI_ISL_1538218, EPI_ISL_1538219, EPI_ISL_1538222, EPI_ISL_1538235, EPI_ISL_1538241, EPI_ISL_1538247, EPI_ISL_1538249, EPI_ISL_1538252, EPI_ISL_1538253, EPI_ISL_1538254, EPI_ISL_1538257, EPI_ISL_1538258, EPI_ISL_1538263, EPI_ISL_1538265, EPI_ISL_1538266, EPI_ISL_1538267, EPI_ISL_1538268, EPI_ISL_1538269, EPI_ISL_1538270, EPI_ISL_1538271, EPI_ISL_1538272, EPI_ISL_1538274, EPI_ISL_1538275, EPI_ISL_1538276, EPI_ISL_1538279, EPI_ISL_1538280, EPI_ISL_1538281, EPI_ISL_1538282, EPI_ISL_1538284, EPI_ISL_1538285, EPI_ISL_1538286, EPI_ISL_1538287, EPI_ISL_1538289, EPI_ISL_1538290, EPI_ISL_1538291, EPI_ISL_1538292, EPI_ISL_1538293, EPI_ISL_1538295, EPI_ISL_1538296, EPI_ISL_1544197, EPI_ISL_1544201, EPI_ISL_1544204, EPI_ISL_1544205, EPI_ISL_1544207, EPI_ISL_1544208, EPI_ISL_1544209, EPI_ISL_1544211, EPI_ISL_1544214, EPI_ISL_1544215, EPI_ISL_1544216, EPI_ISL_1544217, EPI_ISL_1544219, EPI_ISL_1544221, EPI_ISL_1544225, EPI_ISL_1544226, EPI_ISL_1544228, EPI_ISL_1544231, EPI_ISL_1544232, EPI_ISL_1544234, EPI_ISL_1544235, EPI_ISL_1544236, EPI_ISL_1544242, EPI_ISL_1544243, EPI_ISL_1544246, EPI_ISL_1544249, EPI_ISL_1544250, EPI_ISL_1544253, EPI_ISL_1544254, EPI_ISL_1544262, EPI_ISL_1544263, EPI_ISL_1544270, EPI_ISL_1544273, EPI_ISL_1544276, EPI_ISL_1544277, EPI_ISL_1544280, EPI_ISL_1544282, EPI_ISL_1544285, EPI_ISL_1544286, EPI_ISL_1544290, EPI_ISL_1544297, EPI_ISL_1544300, EPI_ISL_1544302, EPI_ISL_1544304, EPI_ISL_1544305, EPI_ISL_1544306, EPI_ISL_1544310, EPI_ISL_1544311, EPI_ISL_1544313, EPI_ISL_1544315, EPI_ISL_1544320, EPI_ISL_1544321, EPI_ISL_1544322, EPI_ISL_1544323, EPI_ISL_1544324, EPI_ISL_1544325, EPI_ISL_1544326, EPI_ISL_1544328, EPI_ISL_1544332, EPI_ISL_1544333, EPI_ISL_1544336, EPI_ISL_1544337, EPI_ISL_1544338, EPI_ISL_1544340, EPI_ISL_1544341, EPI_ISL_1544343, EPI_ISL_1544346, EPI_ISL_1544347, EPI_ISL_1544351, EPI_ISL_1544352, EPI_ISL_1544357, EPI_ISL_1544359, EPI_ISL_1544360, EPI_ISL_1544361, EPI_ISL_1544362, EPI_ISL_1544363, EPI_ISL_1544364, EPI_ISL_1544365, EPI_ISL_1544371, EPI_ISL_1544372, EPI_ISL_1544375, EPI_ISL_1544376, EPI_ISL_1544377, EPI_ISL_1544378, EPI_ISL_1544380, EPI_ISL_1544381, EPI_ISL_1544387, EPI_ISL_1544389, EPI_ISL_1544394, EPI_ISL_1544395, EPI_ISL_1544398, EPI_ISL_1544399, EPI_ISL_1544400, EPI_ISL_1544401, EPI_ISL_1544403, EPI_ISL_1544404, EPI_ISL_1544406, EPI_ISL_1544410, EPI_ISL_1544411, EPI_ISL_1544412, EPI_ISL_1544413, EPI_ISL_1544417, EPI_ISL_1544418, EPI_ISL_1544420, EPI_ISL_1544423, EPI_ISL_1544431, EPI_ISL_1544434, EPI_ISL_1544435, EPI_ISL_1544436, EPI_ISL_1544438, EPI_ISL_1544440, EPI_ISL_1544441, EPI_ISL_1544443, EPI_ISL_1544444, EPI_ISL_1544446, EPI_ISL_1544447, EPI_ISL_1544448, EPI_ISL_1544449, EPI_ISL_1544454, EPI_ISL_1544456, EPI_ISL_1544458, EPI_ISL_1544459, EPI_ISL_1544461, EPI_ISL_1544463, EPI_ISL_1544467, EPI_ISL_1544468, EPI_ISL_1544469, EPI_ISL_1544470, EPI_ISL_1544475, EPI_ISL_1544477, EPI_ISL_1544478, EPI_ISL_1544484, EPI_ISL_1544486, EPI_ISL_1544489, EPI_ISL_1544491, EPI_ISL_1544492, EPI_ISL_1544494, EPI_ISL_1544495, EPI_ISL_1544496, EPI_ISL_1544497, EPI_ISL_1544498, EPI_ISL_1544499, EPI_ISL_1544500, EPI_ISL_1544501, EPI_ISL_1544502, EPI_ISL_1544503, EPI_ISL_1544504, EPI_ISL_1544505, EPI_ISL_1544506, EPI_ISL_1544507, EPI_ISL_1544508, EPI_ISL_1544509, EPI_ISL_1544510, EPI_ISL_1544511, EPI_ISL_1544512, EPI_ISL_1544513, EPI_ISL_1544514, EPI_ISL_1544515, EPI_ISL_1544516, EPI_ISL_1544517, EPI_ISL_1544518, EPI_ISL_1544519, EPI_ISL_1544520, EPI_ISL_1544521, EPI_ISL_1544522, EPI_ISL_1544523, EPI_ISL_1544524, EPI_ISL_1544525, EPI_ISL_1544526, EPI_ISL_1544527, EPI_ISL_1544528, EPI_ISL_1544529, EPI_ISL_1544530, EPI_ISL_1544531, EPI_ISL_1544532, EPI_ISL_1544533, EPI_ISL_1544534, EPI_ISL_1544535, EPI_ISL_1544536, EPI_ISL_1544537, EPI_ISL_1544538, EPI_ISL_1544539, EPI_ISL_1544540, EPI_ISL_1544541, EPI_ISL_1544542, EPI_ISL_1544543, EPI_ISL_1544544, EPI_ISL_1544545, EPI_ISL_1544546, EPI_ISL_1544547, EPI_ISL_1544548, EPI_ISL_1544549, EPI_ISL_1544550, EPI_ISL_1544551, EPI_ISL_1544552, EPI_ISL_1544553, EPI_ISL_1544554, EPI_ISL_1544555, EPI_ISL_1544556, EPI_ISL_1544557, EPI_ISL_1544558, EPI_ISL_1544559, EPI_ISL_1544560, EPI_ISL_1544561, EPI_ISL_1544562, EPI_ISL_1544563, EPI_ISL_1544564, EPI_ISL_1544565, EPI_ISL_1544566, EPI_ISL_1544567, EPI_ISL_1544568, EPI_ISL_1544569, EPI_ISL_1544570, EPI_ISL_1544571, EPI_ISL_1544572, EPI_ISL_1544573, EPI_ISL_1544574, EPI_ISL_1544575, EPI_ISL_1544576, EPI_ISL_1544577, EPI_ISL_1544578, EPI_ISL_1544579, EPI_ISL_1544580, EPI_ISL_1544581, EPI_ISL_1544582, EPI_ISL_1544583, EPI_ISL_1544584, EPI_ISL_1544585, EPI_ISL_1544586, EPI_ISL_1544587, EPI_ISL_1544588, EPI_ISL_1544589, EPI_ISL_1544590, EPI_ISL_1544591, EPI_ISL_1544592, EPI_ISL_1544593, EPI_ISL_1544594, EPI_ISL_1544595, EPI_ISL_1544596, EPI_ISL_1544597, EPI_ISL_1544598, EPI_ISL_1544599, EPI_ISL_1544600, EPI_ISL_1544601, EPI_ISL_1544602, EPI_ISL_1544603, EPI_ISL_1544604, EPI_ISL_1544605, EPI_ISL_1544606, EPI_ISL_1544607, EPI_ISL_1544608, EPI_ISL_1544609, EPI_ISL_1544610, EPI_ISL_1544611, EPI_ISL_1544612, EPI_ISL_1544613, EPI_ISL_1544614, EPI_ISL_1544615, EPI_ISL_1544616, EPI_ISL_1544617, EPI_ISL_1544618, EPI_ISL_1544619, EPI_ISL_1544620, EPI_ISL_1544621, EPI_ISL_1544622, EPI_ISL_1544623, EPI_ISL_1544624, EPI_ISL_1544625, EPI_ISL_1544626, EPI_ISL_1544627, EPI_ISL_1544628, EPI_ISL_1544629, EPI_ISL_1544630, EPI_ISL_1544631, EPI_ISL_1544632, EPI_ISL_1544633, EPI_ISL_1544634, EPI_ISL_1544635, EPI_ISL_1544636, EPI_ISL_1544637, EPI_ISL_1544638, EPI_ISL_1544639, EPI_ISL_1544640, EPI_ISL_1544641, EPI_ISL_1544642, EPI_ISL_1544643, EPI_ISL_1544644, EPI_ISL_1544645, EPI_ISL_1544646, EPI_ISL_1544647, EPI_ISL_1544648, EPI_ISL_1544649, EPI_ISL_1544650, EPI_ISL_1544651, EPI_ISL_1544652, EPI_ISL_1544653, EPI_ISL_1544654, EPI_ISL_1544655, EPI_ISL_1544656, EPI_ISL_1544657, EPI_ISL_1544658, EPI_ISL_1544659, EPI_ISL_1544660, EPI_ISL_1544661, EPI_ISL_1544662, EPI_ISL_1544663, EPI_ISL_1544664, EPI_ISL_1544665, EPI_ISL_1544666, EPI_ISL_1544667, EPI_ISL_1544668, EPI_ISL_1544669, EPI_ISL_1544670, EPI_ISL_1544671, EPI_ISL_1544672, EPI_ISL_1544673, EPI_ISL_1544674, EPI_ISL_1544675, EPI_ISL_1544676, EPI_ISL_1544677, EPI_ISL_1544678, EPI_ISL_1544679, EPI_ISL_1544680, EPI_ISL_1544681, EPI_ISL_1544682, EPI_ISL_1544683, EPI_ISL_1544684, EPI_ISL_1544685, EPI_ISL_1544686, EPI_ISL_1544687, EPI_ISL_1544688, EPI_ISL_1544689, EPI_ISL_1544690, EPI_ISL_1544691, EPI_ISL_1544692, EPI_ISL_1544693, EPI_ISL_1544694, EPI_ISL_1544695, EPI_ISL_1544696, EPI_ISL_1544697, EPI_ISL_1544698, EPI_ISL_1544699, EPI_ISL_1544700, EPI_ISL_1544701, EPI_ISL_1544702, EPI_ISL_1544703, EPI_ISL_1544704, EPI_ISL_1544705, EPI_ISL_1544706, EPI_ISL_1544707, EPI_ISL_1544708, EPI_ISL_1544709, EPI_ISL_1544710, EPI_ISL_1544711, EPI_ISL_1544712, EPI_ISL_1544713, EPI_ISL_1544714, EPI_ISL_1544715, EPI_ISL_1544716, EPI_ISL_1544717, EPI_ISL_1544718, EPI_ISL_1544719, EPI_ISL_1544720, EPI_ISL_1544721, EPI_ISL_1544722, EPI_ISL_1544723, EPI_ISL_1544724, EPI_ISL_1544725, EPI_ISL_1544726, EPI_ISL_1544727, EPI_ISL_1544728, EPI_ISL_1544729, EPI_ISL_1544730, EPI_ISL_1544731, EPI_ISL_1544732, EPI_ISL_1544733, EPI_ISL_1544734, EPI_ISL_1544735, EPI_ISL_1544736, EPI_ISL_1544737, EPI_ISL_1544738, EPI_ISL_1544739, EPI_ISL_1544740, EPI_ISL_1544741, EPI_ISL_1544742, EPI_ISL_1544743, EPI_ISL_1544744, EPI_ISL_1544745, EPI_ISL_1544746, EPI_ISL_1544747, EPI_ISL_1544748, EPI_ISL_1544749, EPI_ISL_1544750, EPI_ISL_1544751, EPI_ISL_1544752, EPI_ISL_1544753, EPI_ISL_1544754, EPI_ISL_1544755, EPI_ISL_1544756, EPI_ISL_1544757, EPI_ISL_1544758, EPI_ISL_1544759, EPI_ISL_1544760, EPI_ISL_1544761, EPI_ISL_1544762, EPI_ISL_1544763, EPI_ISL_1544764, EPI_ISL_1544765, EPI_ISL_1544766, EPI_ISL_1544767, EPI_ISL_1544768, EPI_ISL_1544769, EPI_ISL_1544770, EPI_ISL_1544771, EPI_ISL_1544772, EPI_ISL_1544773, EPI_ISL_1544774, EPI_ISL_1544775, EPI_ISL_1544776, EPI_ISL_1544777, EPI_ISL_1544779, EPI_ISL_1544780, EPI_ISL_1544781, EPI_ISL_1544782, EPI_ISL_1544784, EPI_ISL_1544785, EPI_ISL_1544786, EPI_ISL_1544787, EPI_ISL_1544788, EPI_ISL_1544789, EPI_ISL_1544790, EPI_ISL_1544791, EPI_ISL_1544793, EPI_ISL_1544794, EPI_ISL_1544795, EPI_ISL_1544796, EPI_ISL_1544797, EPI_ISL_1544798, EPI_ISL_1544799, EPI_ISL_1544800, EPI_ISL_1544801, EPI_ISL_1544802, EPI_ISL_1544803, EPI_ISL_1544804, EPI_ISL_1544805, EPI_ISL_1544806, EPI_ISL_1544807, EPI_ISL_1544808, EPI_ISL_1544809, EPI_ISL_1544810, EPI_ISL_1544811, EPI_ISL_1544812, EPI_ISL_1544813, EPI_ISL_1544814, EPI_ISL_1544815, EPI_ISL_1544816, EPI_ISL_1544817, EPI_ISL_1544818, EPI_ISL_1544819, EPI_ISL_1544820, EPI_ISL_1544821, EPI_ISL_1544822, EPI_ISL_1544823, EPI_ISL_1544824, EPI_ISL_1544825, EPI_ISL_1544826, EPI_ISL_1544827, EPI_ISL_1544828, EPI_ISL_1544829, EPI_ISL_1544830, EPI_ISL_1544831, EPI_ISL_1544832, EPI_ISL_1544833, EPI_ISL_1544834, EPI_ISL_1544835, EPI_ISL_1544836, EPI_ISL_1544837, EPI_ISL_1544838, EPI_ISL_1544839, EPI_ISL_1544840, EPI_ISL_1544841, EPI_ISL_1544842, EPI_ISL_1544843, EPI_ISL_1544844, EPI_ISL_1544845, EPI_ISL_1544846, EPI_ISL_1544847, EPI_ISL_1544848, EPI_ISL_1544849, EPI_ISL_1544850, EPI_ISL_1544851, EPI_ISL_1544852, EPI_ISL_1544853, EPI_ISL_1544854, EPI_ISL_1544855, EPI_ISL_1544856, EPI_ISL_1544857, EPI_ISL_1544858, EPI_ISL_1544859, EPI_ISL_1544860, EPI_ISL_1544861, EPI_ISL_1544862, EPI_ISL_1544863, EPI_ISL_1544865, EPI_ISL_1544866, EPI_ISL_1544867, EPI_ISL_1544868, EPI_ISL_1544869, EPI_ISL_1544870, EPI_ISL_1544871, EPI_ISL_1544872, EPI_ISL_1544873, EPI_ISL_1544874, EPI_ISL_1544875, EPI_ISL_1544876, EPI_ISL_1544877, EPI_ISL_1544878, EPI_ISL_1544879, EPI_ISL_1544880, EPI_ISL_1544881, EPI_ISL_1544882, EPI_ISL_1544883, EPI_ISL_1544884, EPI_ISL_1544885, EPI_ISL_1544886, EPI_ISL_1544887, EPI_ISL_1544888, EPI_ISL_1544889, EPI_ISL_1544890, EPI_ISL_1544891, EPI_ISL_1544892, EPI_ISL_1544893, EPI_ISL_1544894, EPI_ISL_1544895, EPI_ISL_1544896, EPI_ISL_1544897, EPI_ISL_1544898, EPI_ISL_1544899, EPI_ISL_1544900, EPI_ISL_1544901, EPI_ISL_1544902, EPI_ISL_1544903, EPI_ISL_1544904, EPI_ISL_1544905, EPI_ISL_1544906, EPI_ISL_1544907, EPI_ISL_1544908, EPI_ISL_1544909, EPI_ISL_1544910, EPI_ISL_1544911, EPI_ISL_1544912, EPI_ISL_1544913, EPI_ISL_1544914, EPI_ISL_1544915, EPI_ISL_1544916, EPI_ISL_1544917, EPI_ISL_1544918, EPI_ISL_1544919, EPI_ISL_1544920, EPI_ISL_1544921, EPI_ISL_1544922, EPI_ISL_1544923, EPI_ISL_1544924, EPI_ISL_1544925, EPI_ISL_1544926, EPI_ISL_1544927, EPI_ISL_1544928, EPI_ISL_1544929, EPI_ISL_1544930, EPI_ISL_1544931, EPI_ISL_1544932, EPI_ISL_1544933, EPI_ISL_1544935, EPI_ISL_1544936, EPI_ISL_1544937, EPI_ISL_1544938, EPI_ISL_1544940, EPI_ISL_1544941, EPI_ISL_1544942, EPI_ISL_1544943, EPI_ISL_1544944, EPI_ISL_1544945, EPI_ISL_1544946, EPI_ISL_1544947, EPI_ISL_1544948, EPI_ISL_1544949, EPI_ISL_1544950, EPI_ISL_1544951, EPI_ISL_1544952, EPI_ISL_1544953, EPI_ISL_1544954, EPI_ISL_1544955, EPI_ISL_1544956, EPI_ISL_1544957, EPI_ISL_1544958, EPI_ISL_1544959, EPI_ISL_1544960, EPI_ISL_1544961, EPI_ISL_1544962, EPI_ISL_1544963, EPI_ISL_1544964, EPI_ISL_1544965, EPI_ISL_1544966, EPI_ISL_1544967, EPI_ISL_1544968, EPI_ISL_1544969, EPI_ISL_1544970, EPI_ISL_1544971, EPI_ISL_1544972, EPI_ISL_1544973, EPI_ISL_1544974, EPI_ISL_1544975, EPI_ISL_1544976, EPI_ISL_1544977, EPI_ISL_1544978, EPI_ISL_1544979, EPI_ISL_1544980, EPI_ISL_1544981, EPI_ISL_1544982, EPI_ISL_1544983, EPI_ISL_1544984, EPI_ISL_1544985, EPI_ISL_1544986, EPI_ISL_1544987, EPI_ISL_1544988, EPI_ISL_1544989, EPI_ISL_1544990, EPI_ISL_1544991, EPI_ISL_1544992, EPI_ISL_1544993, EPI_ISL_1544994, EPI_ISL_1544995, EPI_ISL_1544996, EPI_ISL_1544997, EPI_ISL_1544998, EPI_ISL_1544999, EPI_ISL_1545000, EPI_ISL_1545001, EPI_ISL_1545002, EPI_ISL_1545003, EPI_ISL_1545004, EPI_ISL_1545005, EPI_ISL_1545006, EPI_ISL_1545007, EPI_ISL_1545008, EPI_ISL_1545009, EPI_ISL_1545010, EPI_ISL_1545011, EPI_ISL_1545012, EPI_ISL_1545013, EPI_ISL_1545015, EPI_ISL_1545016, EPI_ISL_1545017, EPI_ISL_1545018, EPI_ISL_1545019, EPI_ISL_1545020, EPI_ISL_1545021, EPI_ISL_1545022, EPI_ISL_1545023, EPI_ISL_1545024, EPI_ISL_1545025, EPI_ISL_1545026, EPI_ISL_1545027, EPI_ISL_1545028, EPI_ISL_1545029, EPI_ISL_1545030, EPI_ISL_1545031, EPI_ISL_1545032, EPI_ISL_1545033, EPI_ISL_1545034, EPI_ISL_1545035, EPI_ISL_1545036, EPI_ISL_1545037, EPI_ISL_1545038, EPI_ISL_1545039, EPI_ISL_1545040, EPI_ISL_1545041, EPI_ISL_1545042, EPI_ISL_1545043, EPI_ISL_1545044, EPI_ISL_1545045, EPI_ISL_1545046, EPI_ISL_1545047, EPI_ISL_1545048, EPI_ISL_1545049, EPI_ISL_1545050, EPI_ISL_1545051, EPI_ISL_1545052, EPI_ISL_1545053, EPI_ISL_1545054, EPI_ISL_1545055, EPI_ISL_1545056, EPI_ISL_1545057, EPI_ISL_1545058, EPI_ISL_1545059, EPI_ISL_1545060, EPI_ISL_1545061, EPI_ISL_1545062, EPI_ISL_1545063, EPI_ISL_1545064, EPI_ISL_1545065, EPI_ISL_1545066, EPI_ISL_1545067, EPI_ISL_1545068, EPI_ISL_1545069, EPI_ISL_1545070, EPI_ISL_1545071, EPI_ISL_1545072, EPI_ISL_1545073, EPI_ISL_1545074, EPI_ISL_1545075, EPI_ISL_1545076, EPI_ISL_1545077, EPI_ISL_1545078, EPI_ISL_1545079, EPI_ISL_1545080, EPI_ISL_1545081, EPI_ISL_1545082, EPI_ISL_1545083, EPI_ISL_1545084, EPI_ISL_1545085, EPI_ISL_1545086, EPI_ISL_1545087, EPI_ISL_1545088, EPI_ISL_1545089, EPI_ISL_1545090, EPI_ISL_1545091, EPI_ISL_1545092, EPI_ISL_1545093, EPI_ISL_1545094, EPI_ISL_1545095, EPI_ISL_1545096, EPI_ISL_1545097, EPI_ISL_1545098, EPI_ISL_1545099, EPI_ISL_1545100, EPI_ISL_1545101, EPI_ISL_1564026, EPI_ISL_1564027, EPI_ISL_1564028, EPI_ISL_1564029, EPI_ISL_1564031, EPI_ISL_1564032, EPI_ISL_1564033, EPI_ISL_1564034, EPI_ISL_1564036, EPI_ISL_1564037, EPI_ISL_1564041, EPI_ISL_1564042, EPI_ISL_1564044, EPI_ISL_1564048, EPI_ISL_1564056, EPI_ISL_1564059, EPI_ISL_1564069, EPI_ISL_1564075, EPI_ISL_1564079, EPI_ISL_1564083, EPI_ISL_1564105, EPI_ISL_1564117, EPI_ISL_1564122, EPI_ISL_1564128, EPI_ISL_1564134, EPI_ISL_1564137, EPI_ISL_1564138, EPI_ISL_1564141, EPI_ISL_1564145, EPI_ISL_1564146, EPI_ISL_1564149, EPI_ISL_1564154, EPI_ISL_1564158, EPI_ISL_1564161, EPI_ISL_1564165, EPI_ISL_1564167, EPI_ISL_1564169, EPI_ISL_1564170, EPI_ISL_1564174, EPI_ISL_1564180, EPI_ISL_1564184, EPI_ISL_1564189, EPI_ISL_1564195, EPI_ISL_1564196, EPI_ISL_1564198, EPI_ISL_1564207, EPI_ISL_1564208, EPI_ISL_1564209, EPI_ISL_1564212, EPI_ISL_1564217, EPI_ISL_1564225, EPI_ISL_1564226, EPI_ISL_1564227, EPI_ISL_1564230, EPI_ISL_1564233, EPI_ISL_1564242, EPI_ISL_1564243, EPI_ISL_1564245, EPI_ISL_1564249, EPI_ISL_1564252, EPI_ISL_1564255, EPI_ISL_1564263, EPI_ISL_1564264, EPI_ISL_1564267, EPI_ISL_1564268, EPI_ISL_1564269, EPI_ISL_1564273, EPI_ISL_1564274, EPI_ISL_1564275, EPI_ISL_1564278, EPI_ISL_1564279, EPI_ISL_1564283, EPI_ISL_1564305, EPI_ISL_1564308, EPI_ISL_1564311, EPI_ISL_1564312, EPI_ISL_1564313, EPI_ISL_1564317, EPI_ISL_1564318, EPI_ISL_1564321, EPI_ISL_1564325, EPI_ISL_1564327, EPI_ISL_1564329, EPI_ISL_1564804, EPI_ISL_1564805, EPI_ISL_1564806, EPI_ISL_1564807, EPI_ISL_1564808, EPI_ISL_1564812, EPI_ISL_1564813, EPI_ISL_1564814, EPI_ISL_1564815, EPI_ISL_1564816, EPI_ISL_1564818, EPI_ISL_1564819, EPI_ISL_1564820, EPI_ISL_1564821, EPI_ISL_1564822, EPI_ISL_1564823, EPI_ISL_1564824, EPI_ISL_1564826, EPI_ISL_1564827, EPI_ISL_1564828, EPI_ISL_1564829, EPI_ISL_1564831, EPI_ISL_1564833, EPI_ISL_1564834, EPI_ISL_1564835, EPI_ISL_1564836, EPI_ISL_1564838, EPI_ISL_1564839, EPI_ISL_1564840, EPI_ISL_1564841, EPI_ISL_1564843, EPI_ISL_1564844, EPI_ISL_1564845, EPI_ISL_1564846, EPI_ISL_1564847, EPI_ISL_1564848, EPI_ISL_1564850, EPI_ISL_1564851, EPI_ISL_1564852, EPI_ISL_1564853, EPI_ISL_1564855, EPI_ISL_1564857, EPI_ISL_1564858, EPI_ISL_1564861, EPI_ISL_1564862, EPI_ISL_1564863, EPI_ISL_1564864, EPI_ISL_1564865, EPI_ISL_1564866, EPI_ISL_1564868, EPI_ISL_1564869, EPI_ISL_1564870, EPI_ISL_1564871, EPI_ISL_1564872, EPI_ISL_1564873, EPI_ISL_1564875, EPI_ISL_1564876, EPI_ISL_1564877, EPI_ISL_1564878, EPI_ISL_1564879, EPI_ISL_1564880, EPI_ISL_1564882, EPI_ISL_1564883, EPI_ISL_1564885, EPI_ISL_1564886, EPI_ISL_1564888, EPI_ISL_1564889, EPI_ISL_1564890, EPI_ISL_1564891, EPI_ISL_1564892, EPI_ISL_1564893, EPI_ISL_1564894, EPI_ISL_1564895, EPI_ISL_1564896, EPI_ISL_1564898, EPI_ISL_1564900, EPI_ISL_1564901, EPI_ISL_1564903, EPI_ISL_1564905, EPI_ISL_1564908, EPI_ISL_1564909, EPI_ISL_1564910, EPI_ISL_1564911, EPI_ISL_1564912, EPI_ISL_1564914, EPI_ISL_1564915, EPI_ISL_1564916, EPI_ISL_1564917, EPI_ISL_1564918, EPI_ISL_1564919, EPI_ISL_1564920, EPI_ISL_1564922, EPI_ISL_1564923, EPI_ISL_1564924, EPI_ISL_1564925, EPI_ISL_1564927, EPI_ISL_1564929, EPI_ISL_1564930, EPI_ISL_1564931, EPI_ISL_1564933, EPI_ISL_1564935, EPI_ISL_1564936, EPI_ISL_1564937, EPI_ISL_1564939, EPI_ISL_1564941, EPI_ISL_1564942, EPI_ISL_1564943, EPI_ISL_1564944, EPI_ISL_1564945, EPI_ISL_1564947, EPI_ISL_1564948, EPI_ISL_1564950, EPI_ISL_1564951, EPI_ISL_1564952, EPI_ISL_1564954, EPI_ISL_1564955, EPI_ISL_1564956, EPI_ISL_1564957, EPI_ISL_1564958, EPI_ISL_1564960, EPI_ISL_1564961, EPI_ISL_1564962, EPI_ISL_1564963, EPI_ISL_1564964, EPI_ISL_1564966, EPI_ISL_1564967, EPI_ISL_1564968, EPI_ISL_1564969, EPI_ISL_1564971, EPI_ISL_1564972, EPI_ISL_1564973, EPI_ISL_1564974, EPI_ISL_1564975, EPI_ISL_1564976, EPI_ISL_1564979, EPI_ISL_1564980, EPI_ISL_1564981, EPI_ISL_1564982, EPI_ISL_1564983, EPI_ISL_1564984, EPI_ISL_1564986, EPI_ISL_1564987, EPI_ISL_1564989, EPI_ISL_1564990, EPI_ISL_1564991, EPI_ISL_1564993, EPI_ISL_1564994, EPI_ISL_1564995, EPI_ISL_1564996, EPI_ISL_1564997, EPI_ISL_1564998, EPI_ISL_1564999, EPI_ISL_1565000, EPI_ISL_1565002, EPI_ISL_1565003, EPI_ISL_1565004, EPI_ISL_1565007, EPI_ISL_1565008, EPI_ISL_1565009, EPI_ISL_1565010, EPI_ISL_1565011, EPI_ISL_1565012, EPI_ISL_1565013, EPI_ISL_1565015, EPI_ISL_1565016, EPI_ISL_1565017, EPI_ISL_1565018, EPI_ISL_1565019, EPI_ISL_1565020, EPI_ISL_1565021, EPI_ISL_1565022, EPI_ISL_1565023, EPI_ISL_1565024, EPI_ISL_1565025, EPI_ISL_1565026, EPI_ISL_1565027, EPI_ISL_1565028, EPI_ISL_1565030, EPI_ISL_1565031, EPI_ISL_1565032, EPI_ISL_1565033, EPI_ISL_1565034, EPI_ISL_1565036, EPI_ISL_1565037, EPI_ISL_1565039, EPI_ISL_1565040, EPI_ISL_1565041, EPI_ISL_1565042, EPI_ISL_1565043, EPI_ISL_1565044, EPI_ISL_1565046, EPI_ISL_1565048, EPI_ISL_1565049, EPI_ISL_1565050, EPI_ISL_1565052, EPI_ISL_1565053, EPI_ISL_1565054, EPI_ISL_1565055, EPI_ISL_1565056, EPI_ISL_1565057, EPI_ISL_1565058, EPI_ISL_1565059, EPI_ISL_1565060, EPI_ISL_1565061, EPI_ISL_1565063, EPI_ISL_1565065, EPI_ISL_1565066, EPI_ISL_1565067, EPI_ISL_1565068, EPI_ISL_1565069, EPI_ISL_1565070, EPI_ISL_1565071, EPI_ISL_1565073, EPI_ISL_1565074, EPI_ISL_1565075, EPI_ISL_1565076, EPI_ISL_1565077, EPI_ISL_1565078, EPI_ISL_1565079, EPI_ISL_1565080, EPI_ISL_1565081, EPI_ISL_1565082, EPI_ISL_1565083, EPI_ISL_1565084, EPI_ISL_1565086, EPI_ISL_1565087, EPI_ISL_1565088, EPI_ISL_1565089, EPI_ISL_1565090, EPI_ISL_1565091, EPI_ISL_1565093, EPI_ISL_1565094, EPI_ISL_1565095, EPI_ISL_1565096, EPI_ISL_1565097, EPI_ISL_1565098, EPI_ISL_1565099, EPI_ISL_1565101, EPI_ISL_1565102, EPI_ISL_1565103, EPI_ISL_1565104, EPI_ISL_1565105, EPI_ISL_1565107, EPI_ISL_1565108, EPI_ISL_1565109, EPI_ISL_1565110, EPI_ISL_1565111, EPI_ISL_1565113, EPI_ISL_1565114, EPI_ISL_1565115, EPI_ISL_1565116, EPI_ISL_1565117, EPI_ISL_1565118, EPI_ISL_1565119, EPI_ISL_1565120, EPI_ISL_1565122, EPI_ISL_1565123, EPI_ISL_1565124, EPI_ISL_1565125, EPI_ISL_1565126, EPI_ISL_1565127, EPI_ISL_1565128, EPI_ISL_1565130, EPI_ISL_1565131, EPI_ISL_1565133, EPI_ISL_1565134, EPI_ISL_1565135, EPI_ISL_1565137, EPI_ISL_1565138, EPI_ISL_1565139, EPI_ISL_1565140, EPI_ISL_1565141, EPI_ISL_1565143, EPI_ISL_1565144, EPI_ISL_1565145, EPI_ISL_1565146, EPI_ISL_1565147, EPI_ISL_1565148, EPI_ISL_1565149, EPI_ISL_1565150, EPI_ISL_1565151, EPI_ISL_1565152, EPI_ISL_1565153, EPI_ISL_1565155, EPI_ISL_1565156, EPI_ISL_1565157, EPI_ISL_1565158, EPI_ISL_1565159, EPI_ISL_1565160, EPI_ISL_1565161, EPI_ISL_1565162, EPI_ISL_1565163, EPI_ISL_1565165, EPI_ISL_1565166, EPI_ISL_1565167, EPI_ISL_1565168, EPI_ISL_1565169, EPI_ISL_1565170, EPI_ISL_1565171, EPI_ISL_1565173, EPI_ISL_1565174, EPI_ISL_1565175, EPI_ISL_1565176, EPI_ISL_1565177, EPI_ISL_1565179, EPI_ISL_1565180, EPI_ISL_1565181, EPI_ISL_1565182, EPI_ISL_1565183, EPI_ISL_1565184, EPI_ISL_1565185, EPI_ISL_1565186, EPI_ISL_1565188, EPI_ISL_1565189, EPI_ISL_1565190, EPI_ISL_1565191, EPI_ISL_1565192, EPI_ISL_1565193, EPI_ISL_1565195, EPI_ISL_1565196, EPI_ISL_1565197, EPI_ISL_1565198, EPI_ISL_1565199, EPI_ISL_1565200, EPI_ISL_1565201, EPI_ISL_1565216, EPI_ISL_1565217, EPI_ISL_1565219, EPI_ISL_1565220, EPI_ISL_1565221, EPI_ISL_1565222, EPI_ISL_1565223, EPI_ISL_1565224, EPI_ISL_1565226, EPI_ISL_1565227, EPI_ISL_1565228, EPI_ISL_1565229, EPI_ISL_1565231, EPI_ISL_1565232, EPI_ISL_1565233, EPI_ISL_1565234, EPI_ISL_1565235, EPI_ISL_1565236, EPI_ISL_1565238, EPI_ISL_1565239, EPI_ISL_1565240, EPI_ISL_1565241, EPI_ISL_1565243, EPI_ISL_1565244, EPI_ISL_1565245, EPI_ISL_1565246, EPI_ISL_1565249, EPI_ISL_1565250, EPI_ISL_1565251, EPI_ISL_1565252, EPI_ISL_1565254, EPI_ISL_1565255, EPI_ISL_1565256, EPI_ISL_1565257, EPI_ISL_1565258, EPI_ISL_1565260, EPI_ISL_1565261, EPI_ISL_1565262, EPI_ISL_1565264, EPI_ISL_1565266, EPI_ISL_1565267, EPI_ISL_1565269, EPI_ISL_1565270, EPI_ISL_1565271, EPI_ISL_1565272, EPI_ISL_1565274, EPI_ISL_1565275, EPI_ISL_1565276, EPI_ISL_1565277, EPI_ISL_1565278, EPI_ISL_1565279, EPI_ISL_1565281, EPI_ISL_1565282, EPI_ISL_1565283, EPI_ISL_1565284, EPI_ISL_1565285, EPI_ISL_1565286, EPI_ISL_1565288, EPI_ISL_1565289, EPI_ISL_1565290, EPI_ISL_1565291, EPI_ISL_1565293, EPI_ISL_1565294, EPI_ISL_1565295, EPI_ISL_1565296, EPI_ISL_1565298, EPI_ISL_1565299, EPI_ISL_1565300, EPI_ISL_1565301, EPI_ISL_1565302, EPI_ISL_1565304, EPI_ISL_1565305, EPI_ISL_1565306, EPI_ISL_1565308, EPI_ISL_1565309, EPI_ISL_1565310, EPI_ISL_1565311, EPI_ISL_1565312, EPI_ISL_1565314, EPI_ISL_1565315, EPI_ISL_1565316, EPI_ISL_1565317, EPI_ISL_1565318, EPI_ISL_1565320, EPI_ISL_1565321, EPI_ISL_1565323, EPI_ISL_1565324, EPI_ISL_1565326, EPI_ISL_1565327, EPI_ISL_1565328, EPI_ISL_1565329, EPI_ISL_1565330, EPI_ISL_1565331, EPI_ISL_1565332, EPI_ISL_1565333, EPI_ISL_1565335, EPI_ISL_1565336, EPI_ISL_1565337, EPI_ISL_1583684, EPI_ISL_1583687, EPI_ISL_1583688, EPI_ISL_1583690, EPI_ISL_1583692, EPI_ISL_1583695, EPI_ISL_1583701, EPI_ISL_1583702, EPI_ISL_1583704, EPI_ISL_1583706, EPI_ISL_1583707, EPI_ISL_1583711, EPI_ISL_1583713, EPI_ISL_1583715, EPI_ISL_1583717, EPI_ISL_1583718, EPI_ISL_1583721, EPI_ISL_1583724, EPI_ISL_1583726, EPI_ISL_1583729, EPI_ISL_1583731, EPI_ISL_1583732, EPI_ISL_1583734, EPI_ISL_1583735, EPI_ISL_1583736, EPI_ISL_1583737, EPI_ISL_1583738, EPI_ISL_1583739, EPI_ISL_1583740, EPI_ISL_1583741, EPI_ISL_1583745, EPI_ISL_1583746, EPI_ISL_1583747, EPI_ISL_1583749, EPI_ISL_1583751, EPI_ISL_1583754, EPI_ISL_1583761, EPI_ISL_1583762, EPI_ISL_1583765, EPI_ISL_1583766, EPI_ISL_1583767, EPI_ISL_1583768, EPI_ISL_1583772, EPI_ISL_1583775, EPI_ISL_1583778, EPI_ISL_1583779, EPI_ISL_1583784, EPI_ISL_1583793, EPI_ISL_1583794, EPI_ISL_1583795, EPI_ISL_1583796, EPI_ISL_1583798, EPI_ISL_1583800, EPI_ISL_1583801, EPI_ISL_1583802, EPI_ISL_1583803, EPI_ISL_1583804, EPI_ISL_1583805, EPI_ISL_1583806, EPI_ISL_1583809, EPI_ISL_1583810, EPI_ISL_1583812, EPI_ISL_1583814, EPI_ISL_1583815, EPI_ISL_1583818, EPI_ISL_1583820, EPI_ISL_1583821, EPI_ISL_1583822, EPI_ISL_1583823, EPI_ISL_1583825, EPI_ISL_1583827, EPI_ISL_1583830, EPI_ISL_1583831, EPI_ISL_1583834, EPI_ISL_1583835, EPI_ISL_1583836, EPI_ISL_1583837, EPI_ISL_1583838, EPI_ISL_1583839, EPI_ISL_1583840, EPI_ISL_1583841, EPI_ISL_1583842, EPI_ISL_1583843, EPI_ISL_1583846, EPI_ISL_1583847, EPI_ISL_1583848, EPI_ISL_1583849, EPI_ISL_1583850, EPI_ISL_1583851, EPI_ISL_1583854, EPI_ISL_1583855, EPI_ISL_1583856, EPI_ISL_1583860, EPI_ISL_1583862, EPI_ISL_1583863, EPI_ISL_1583866, EPI_ISL_1583867, EPI_ISL_1583868, EPI_ISL_1583871, EPI_ISL_1583873, EPI_ISL_1583874, EPI_ISL_1583876, EPI_ISL_1583877, EPI_ISL_1583881, EPI_ISL_1583882, EPI_ISL_1583883, EPI_ISL_1583885, EPI_ISL_1583887, EPI_ISL_1583888, EPI_ISL_1583890, EPI_ISL_1583893, EPI_ISL_1583894, EPI_ISL_1583895, EPI_ISL_1583899, EPI_ISL_1583901, EPI_ISL_1583903, EPI_ISL_1583904, EPI_ISL_1583907, EPI_ISL_1583908, EPI_ISL_1583909, EPI_ISL_1583911, EPI_ISL_1583913, EPI_ISL_1583915, EPI_ISL_1583918, EPI_ISL_1583920, EPI_ISL_1583921, EPI_ISL_1583926, EPI_ISL_1583937, EPI_ISL_1583945, EPI_ISL_1583946, EPI_ISL_1583951, EPI_ISL_1583954, EPI_ISL_1583960, EPI_ISL_1583963, EPI_ISL_1583967, EPI_ISL_1583969, EPI_ISL_1583983, EPI_ISL_1583994, EPI_ISL_1583999, EPI_ISL_1584002, EPI_ISL_1584006, EPI_ISL_1584007, EPI_ISL_1584012, EPI_ISL_1584015, EPI_ISL_1584016, EPI_ISL_1584022, EPI_ISL_1584026, EPI_ISL_1584033, EPI_ISL_1584034, EPI_ISL_1584039, EPI_ISL_1584040, EPI_ISL_1584046, EPI_ISL_1584050, EPI_ISL_1584053, EPI_ISL_1584056, EPI_ISL_1584058, EPI_ISL_1584062, EPI_ISL_1584066, EPI_ISL_1584073, EPI_ISL_1584083, EPI_ISL_1584084, EPI_ISL_1584087, EPI_ISL_1584094, EPI_ISL_1584102, EPI_ISL_1584103, EPI_ISL_1584108, EPI_ISL_1584110, EPI_ISL_1584112, EPI_ISL_1584116, EPI_ISL_1584119, EPI_ISL_1584122, EPI_ISL_1584127, EPI_ISL_1584128, EPI_ISL_1584130, EPI_ISL_1584132, EPI_ISL_1584177, EPI_ISL_1584185, EPI_ISL_1584195, EPI_ISL_1584200, EPI_ISL_1584204, EPI_ISL_1584216, EPI_ISL_1584219, EPI_ISL_1584233, EPI_ISL_1584253, EPI_ISL_1584255, EPI_ISL_1584277, EPI_ISL_1584288, EPI_ISL_1584293, EPI_ISL_1584308, EPI_ISL_1584329, EPI_ISL_1584346, EPI_ISL_1584369, EPI_ISL_1584391, EPI_ISL_1584408, EPI_ISL_1584414, EPI_ISL_1584415, EPI_ISL_1584418, EPI_ISL_1584419, EPI_ISL_1584421, EPI_ISL_1584457, EPI_ISL_1584467, EPI_ISL_1584473, EPI_ISL_1584483, EPI_ISL_1584489, EPI_ISL_1584498, EPI_ISL_1584503, EPI_ISL_1584508, EPI_ISL_1584517, EPI_ISL_1584522, EPI_ISL_1584524, EPI_ISL_1584527, EPI_ISL_1584530, EPI_ISL_1584531, EPI_ISL_1584532, EPI_ISL_1584533, EPI_ISL_1584534, EPI_ISL_1584535, EPI_ISL_1584536, EPI_ISL_1584537, EPI_ISL_1584538, EPI_ISL_1584539, EPI_ISL_1584540, EPI_ISL_1584541, EPI_ISL_1584542, EPI_ISL_1584543, EPI_ISL_1584544, EPI_ISL_1584545, EPI_ISL_1584547, EPI_ISL_1584548, EPI_ISL_1584549, EPI_ISL_1584550, EPI_ISL_1584551, EPI_ISL_1584552, EPI_ISL_1584553, EPI_ISL_1584554, EPI_ISL_1584555, EPI_ISL_1584556, EPI_ISL_1584557, EPI_ISL_1584558, EPI_ISL_1584560, EPI_ISL_1584561, EPI_ISL_1584562, EPI_ISL_1584563, EPI_ISL_1584564, EPI_ISL_1584565, EPI_ISL_1584566, EPI_ISL_1584567, EPI_ISL_1584568, EPI_ISL_1584569, EPI_ISL_1584570, EPI_ISL_1584571, EPI_ISL_1584572, EPI_ISL_1584573, EPI_ISL_1584574, EPI_ISL_1584575, EPI_ISL_1584576, EPI_ISL_1584577, EPI_ISL_1584578, EPI_ISL_1584579, EPI_ISL_1584580, EPI_ISL_1584581, EPI_ISL_1584582, EPI_ISL_1584583, EPI_ISL_1584584, EPI_ISL_1584585, EPI_ISL_1584586, EPI_ISL_1584587, EPI_ISL_1584588, EPI_ISL_1584589, EPI_ISL_1584590, EPI_ISL_1584591, EPI_ISL_1584592, EPI_ISL_1584593, EPI_ISL_1584594, EPI_ISL_1584595, EPI_ISL_1584596, EPI_ISL_1584597, EPI_ISL_1584598, EPI_ISL_1584599, EPI_ISL_1584600, EPI_ISL_1584601, EPI_ISL_1584602, EPI_ISL_1584603, EPI_ISL_1584604, EPI_ISL_1584605, EPI_ISL_1584606, EPI_ISL_1584607, EPI_ISL_1584608, EPI_ISL_1584609, EPI_ISL_1584610, EPI_ISL_1584611, EPI_ISL_1584612, EPI_ISL_1584614, EPI_ISL_1584615, EPI_ISL_1584616, EPI_ISL_1584617, EPI_ISL_1584618, EPI_ISL_1584619, EPI_ISL_1584620, EPI_ISL_1584621, EPI_ISL_1584622, EPI_ISL_1584623, EPI_ISL_1584624, EPI_ISL_1584625, EPI_ISL_1584626, EPI_ISL_1584627, EPI_ISL_1584628, EPI_ISL_1584629, EPI_ISL_1584630, EPI_ISL_1584631, EPI_ISL_1584632, EPI_ISL_1584633, EPI_ISL_1584634, EPI_ISL_1584635, EPI_ISL_1584636, EPI_ISL_1584637, EPI_ISL_1584638, EPI_ISL_1584639, EPI_ISL_1584640, EPI_ISL_1584641, EPI_ISL_1584642, EPI_ISL_1584643, EPI_ISL_1584644, EPI_ISL_1584645, EPI_ISL_1584646, EPI_ISL_1584647, EPI_ISL_1584648, EPI_ISL_1584649, EPI_ISL_1584650, EPI_ISL_1584651, EPI_ISL_1584652, EPI_ISL_1584653, EPI_ISL_1584654, EPI_ISL_1584655, EPI_ISL_1584656, EPI_ISL_1584657, EPI_ISL_1584658, EPI_ISL_1584659, EPI_ISL_1584660, EPI_ISL_1584661, EPI_ISL_1584662, EPI_ISL_1584663, EPI_ISL_1584664, EPI_ISL_1584665, EPI_ISL_1584666, EPI_ISL_1584667, EPI_ISL_1584668, EPI_ISL_1584669, EPI_ISL_1584670, EPI_ISL_1584671, EPI_ISL_1584672, EPI_ISL_1584673, EPI_ISL_1584674, EPI_ISL_1584675, EPI_ISL_1584676, EPI_ISL_1584677, EPI_ISL_1584678, EPI_ISL_1584679, EPI_ISL_1584680, EPI_ISL_1584681, EPI_ISL_1584682, EPI_ISL_1584683, EPI_ISL_1584684, EPI_ISL_1584685, EPI_ISL_1584686, EPI_ISL_1584687, EPI_ISL_1584688, EPI_ISL_1584689, EPI_ISL_1584690, EPI_ISL_1584691, EPI_ISL_1584692, EPI_ISL_1584693, EPI_ISL_1584694, EPI_ISL_1584695, EPI_ISL_1584696, EPI_ISL_1584697, EPI_ISL_1584698, EPI_ISL_1584699, EPI_ISL_1584700, EPI_ISL_1584701, EPI_ISL_1584702, EPI_ISL_1584703, EPI_ISL_1584704, EPI_ISL_1584705, EPI_ISL_1584706, EPI_ISL_1584707, EPI_ISL_1584708, EPI_ISL_1584709, EPI_ISL_1584710, EPI_ISL_1584711, EPI_ISL_1584712, EPI_ISL_1584714, EPI_ISL_1584715, EPI_ISL_1584716, EPI_ISL_1584717, EPI_ISL_1584718, EPI_ISL_1584719, EPI_ISL_1584720, EPI_ISL_1584721, EPI_ISL_1584722, EPI_ISL_1584723, EPI_ISL_1584724, EPI_ISL_1584725, EPI_ISL_1584726, EPI_ISL_1584727, EPI_ISL_1584728, EPI_ISL_1584729, EPI_ISL_1584730, EPI_ISL_1584731, EPI_ISL_1584732, EPI_ISL_1584733, EPI_ISL_1584734, EPI_ISL_1584735, EPI_ISL_1584736, EPI_ISL_1584737, EPI_ISL_1584738, EPI_ISL_1584739, EPI_ISL_1584740, EPI_ISL_1584741, EPI_ISL_1584742, EPI_ISL_1584743, EPI_ISL_1584744, EPI_ISL_1584745, EPI_ISL_1584746, EPI_ISL_1584747, EPI_ISL_1584748, EPI_ISL_1584749, EPI_ISL_1584750, EPI_ISL_1584751, EPI_ISL_1584752, EPI_ISL_1584753, EPI_ISL_1584754, EPI_ISL_1584755, EPI_ISL_1584756, EPI_ISL_1584757, EPI_ISL_1584758, EPI_ISL_1584759, EPI_ISL_1584760, EPI_ISL_1584761, EPI_ISL_1584762, EPI_ISL_1584763, EPI_ISL_1584764, EPI_ISL_1584765, EPI_ISL_1584766, EPI_ISL_1584767, EPI_ISL_1584768, EPI_ISL_1584769, EPI_ISL_1584770, EPI_ISL_1584771, EPI_ISL_1584772, EPI_ISL_1584773, EPI_ISL_1584774, EPI_ISL_1584775, EPI_ISL_1584776, EPI_ISL_1584777, EPI_ISL_1584778, EPI_ISL_1584779, EPI_ISL_1584780, EPI_ISL_1584781, EPI_ISL_1584782, EPI_ISL_1584783, EPI_ISL_1584784, EPI_ISL_1584785, EPI_ISL_1584786, EPI_ISL_1584787, EPI_ISL_1584788, EPI_ISL_1584789, EPI_ISL_1584790, EPI_ISL_1584791, EPI_ISL_1584792, EPI_ISL_1584793, EPI_ISL_1584794, EPI_ISL_1584795, EPI_ISL_1584796, EPI_ISL_1584797, EPI_ISL_1584798, EPI_ISL_1584799, EPI_ISL_1584800, EPI_ISL_1584801, EPI_ISL_1584802, EPI_ISL_1584803, EPI_ISL_1584804, EPI_ISL_1584805, EPI_ISL_1584806, EPI_ISL_1584807, EPI_ISL_1584808, EPI_ISL_1584809, EPI_ISL_1584810, EPI_ISL_1584811, EPI_ISL_1584812, EPI_ISL_1584813, EPI_ISL_1584814, EPI_ISL_1584815, EPI_ISL_1584816, EPI_ISL_1584817, EPI_ISL_1584818, EPI_ISL_1584819, EPI_ISL_1584820, EPI_ISL_1584821, EPI_ISL_1584822, EPI_ISL_1584823, EPI_ISL_1584824, EPI_ISL_1584825, EPI_ISL_1584826, EPI_ISL_1584827, EPI_ISL_1584828, EPI_ISL_1584829, EPI_ISL_1584830, EPI_ISL_1584831, EPI_ISL_1584832, EPI_ISL_1584833, EPI_ISL_1584834, EPI_ISL_1584835, EPI_ISL_1584836, EPI_ISL_1584837, EPI_ISL_1584838, EPI_ISL_1584839, EPI_ISL_1584840, EPI_ISL_1584841, EPI_ISL_1584842, EPI_ISL_1584843, EPI_ISL_1584844, EPI_ISL_1584845, EPI_ISL_1584846, EPI_ISL_1584847, EPI_ISL_1584848, EPI_ISL_1584849, EPI_ISL_1584850, EPI_ISL_1584851, EPI_ISL_1584852, EPI_ISL_1584854, EPI_ISL_1584855, EPI_ISL_1584856, EPI_ISL_1584857, EPI_ISL_1584858, EPI_ISL_1584859, EPI_ISL_1584860, EPI_ISL_1584861, EPI_ISL_1584862, EPI_ISL_1584863, EPI_ISL_1584864, EPI_ISL_1584865, EPI_ISL_1584866, EPI_ISL_1584867, EPI_ISL_1584868, EPI_ISL_1584869, EPI_ISL_1584870, EPI_ISL_1584871, EPI_ISL_1584872, EPI_ISL_1584873, EPI_ISL_1584874, EPI_ISL_1584875, EPI_ISL_1584876, EPI_ISL_1584877, EPI_ISL_1584878, EPI_ISL_1584879, EPI_ISL_1584880, EPI_ISL_1584881, EPI_ISL_1584882, EPI_ISL_1584883, EPI_ISL_1584884, EPI_ISL_1584885, EPI_ISL_1584886, EPI_ISL_1584887, EPI_ISL_1584888, EPI_ISL_1584889, EPI_ISL_1584890, EPI_ISL_1584891, EPI_ISL_1584893, EPI_ISL_1584894, EPI_ISL_1584895, EPI_ISL_1584896, EPI_ISL_1584897, EPI_ISL_1584898, EPI_ISL_1584899, EPI_ISL_1584900, EPI_ISL_1584901, EPI_ISL_1584902, EPI_ISL_1584903, EPI_ISL_1584904, EPI_ISL_1584905, EPI_ISL_1584906, EPI_ISL_1584907, EPI_ISL_1584908, EPI_ISL_1584909, EPI_ISL_1584910, EPI_ISL_1584911, EPI_ISL_1584912, EPI_ISL_1584913, EPI_ISL_1584914, EPI_ISL_1584915, EPI_ISL_1584916, EPI_ISL_1584917, EPI_ISL_1584918, EPI_ISL_1584919, EPI_ISL_1584920, EPI_ISL_1584921, EPI_ISL_1584922, EPI_ISL_1584923, EPI_ISL_1584924, EPI_ISL_1584925, EPI_ISL_1584926, EPI_ISL_1584927, EPI_ISL_1584928, EPI_ISL_1584929, EPI_ISL_1584930, EPI_ISL_1584931, EPI_ISL_1584932, EPI_ISL_1584933, EPI_ISL_1584934, EPI_ISL_1584935, EPI_ISL_1584936, EPI_ISL_1584937, EPI_ISL_1584938, EPI_ISL_1584939, EPI_ISL_1584940, EPI_ISL_1584941, EPI_ISL_1584942, EPI_ISL_1584943, EPI_ISL_1584944, EPI_ISL_1584945, EPI_ISL_1584946, EPI_ISL_1584947, EPI_ISL_1584948, EPI_ISL_1584949, EPI_ISL_1584950, EPI_ISL_1584951, EPI_ISL_1584952, EPI_ISL_1584953, EPI_ISL_1584954, EPI_ISL_1584955, EPI_ISL_1584956, EPI_ISL_1584957, EPI_ISL_1584958, EPI_ISL_1584959, EPI_ISL_1584960, EPI_ISL_1584961, EPI_ISL_1584962, EPI_ISL_1584963, EPI_ISL_1584964, EPI_ISL_1584965, EPI_ISL_1584966, EPI_ISL_1584967, EPI_ISL_1584968, EPI_ISL_1584969, EPI_ISL_1584970, EPI_ISL_1584971, EPI_ISL_1584972, EPI_ISL_1584973, EPI_ISL_1584974, EPI_ISL_1584975, EPI_ISL_1584976, EPI_ISL_1584977, EPI_ISL_1584978, EPI_ISL_1584979, EPI_ISL_1584980, EPI_ISL_1584981, EPI_ISL_1584982, EPI_ISL_1584983, EPI_ISL_1584984, EPI_ISL_1584985, EPI_ISL_1584986, EPI_ISL_1584987, EPI_ISL_1584988, EPI_ISL_1584989, EPI_ISL_1584990, EPI_ISL_1584991, EPI_ISL_1584992, EPI_ISL_1584993, EPI_ISL_1584994, EPI_ISL_1584995, EPI_ISL_1584996, EPI_ISL_1595014, EPI_ISL_1595015, EPI_ISL_1595021, EPI_ISL_1595025, EPI_ISL_1595028, EPI_ISL_1595030, EPI_ISL_1595032, EPI_ISL_1595034, EPI_ISL_1595035, EPI_ISL_1595036, EPI_ISL_1595038, EPI_ISL_1595039, EPI_ISL_1595047, EPI_ISL_1595052, EPI_ISL_1595063, EPI_ISL_1595079, EPI_ISL_1595082, EPI_ISL_1595086, EPI_ISL_1595099, EPI_ISL_1595104, EPI_ISL_1595108, EPI_ISL_1595119, EPI_ISL_1595124, EPI_ISL_1595130, EPI_ISL_1595131, EPI_ISL_1595134, EPI_ISL_1595139, EPI_ISL_1595148, EPI_ISL_1595152, EPI_ISL_1595153, EPI_ISL_1595154, EPI_ISL_1595155, EPI_ISL_1595159, EPI_ISL_1595163, EPI_ISL_1595184, EPI_ISL_1595187, EPI_ISL_1595190, EPI_ISL_1595191, EPI_ISL_1595193, EPI_ISL_1595200, EPI_ISL_1595207, EPI_ISL_1595210, EPI_ISL_1595211, EPI_ISL_1595212, EPI_ISL_1595215, EPI_ISL_1595218, EPI_ISL_1595219, EPI_ISL_1595220, EPI_ISL_1595221, EPI_ISL_1595223, EPI_ISL_1595226, EPI_ISL_1595228, EPI_ISL_1595229, EPI_ISL_1595230, EPI_ISL_1595232, EPI_ISL_1595235, EPI_ISL_1595236, EPI_ISL_1595239, EPI_ISL_1595240, EPI_ISL_1595241, EPI_ISL_1595246, EPI_ISL_1595247, EPI_ISL_1595249, EPI_ISL_1595250, EPI_ISL_1595251, EPI_ISL_1595254, EPI_ISL_1595255, EPI_ISL_1595258, EPI_ISL_1595260, EPI_ISL_1595261, EPI_ISL_1595262, EPI_ISL_1595263, EPI_ISL_1595264, EPI_ISL_1595265, EPI_ISL_1595266, EPI_ISL_1595271, EPI_ISL_1595272, EPI_ISL_1595274, EPI_ISL_1595277, EPI_ISL_1595279, EPI_ISL_1595280, EPI_ISL_1595283, EPI_ISL_1595284, EPI_ISL_1595287, EPI_ISL_1595288, EPI_ISL_1595292, EPI_ISL_1595293, EPI_ISL_1595294, EPI_ISL_1595295, EPI_ISL_1595296, EPI_ISL_1595299, EPI_ISL_1595301, EPI_ISL_1595304, EPI_ISL_1595309, EPI_ISL_1595310, EPI_ISL_1595311, EPI_ISL_1595312, EPI_ISL_1595315, EPI_ISL_1595316, EPI_ISL_1595322, EPI_ISL_1595323, EPI_ISL_1595324, EPI_ISL_1595328, EPI_ISL_1595329, EPI_ISL_1595335, EPI_ISL_1595339, EPI_ISL_1595340, EPI_ISL_1595341, EPI_ISL_1595342, EPI_ISL_1595343, EPI_ISL_1595346, EPI_ISL_1595351, EPI_ISL_1595352, EPI_ISL_1595353, EPI_ISL_1595355, EPI_ISL_1595358, EPI_ISL_1595359, EPI_ISL_1595362, EPI_ISL_1595363, EPI_ISL_1595364, EPI_ISL_1595365, EPI_ISL_1595367, EPI_ISL_1595369, EPI_ISL_1595371, EPI_ISL_1595372, EPI_ISL_1595373, EPI_ISL_1595376, EPI_ISL_1595381, EPI_ISL_1595382, EPI_ISL_1595383, EPI_ISL_1595384, EPI_ISL_1595386, EPI_ISL_1595387, EPI_ISL_1595389, EPI_ISL_1595390, EPI_ISL_1595392, EPI_ISL_1595393, EPI_ISL_1595395, EPI_ISL_1595396, EPI_ISL_1595400, EPI_ISL_1595401, EPI_ISL_1595402, EPI_ISL_1595403, EPI_ISL_1595406, EPI_ISL_1595408, EPI_ISL_1595410, EPI_ISL_1595411, EPI_ISL_1595412, EPI_ISL_1595413, EPI_ISL_1595415, EPI_ISL_1595416, EPI_ISL_1595417, EPI_ISL_1595418, EPI_ISL_1595421, EPI_ISL_1595425, EPI_ISL_1595426, EPI_ISL_1595429, EPI_ISL_1595430, EPI_ISL_1595431, EPI_ISL_1595432, EPI_ISL_1595433, EPI_ISL_1595435, EPI_ISL_1595437, EPI_ISL_1595438, EPI_ISL_1595440, EPI_ISL_1595441, EPI_ISL_1595442, EPI_ISL_1595443, EPI_ISL_1595445, EPI_ISL_1595446, EPI_ISL_1595447, EPI_ISL_1595453, EPI_ISL_1595454, EPI_ISL_1595458, EPI_ISL_1595461, EPI_ISL_1595463, EPI_ISL_1595464, EPI_ISL_1595465, EPI_ISL_1595466, EPI_ISL_1595468, EPI_ISL_1595469, EPI_ISL_1595473, EPI_ISL_1615933, EPI_ISL_1615936, EPI_ISL_1615941, EPI_ISL_1615965, EPI_ISL_1615978, EPI_ISL_1615987, EPI_ISL_1615990, EPI_ISL_1615993, EPI_ISL_1616017, EPI_ISL_1616023, EPI_ISL_1616024, EPI_ISL_1616025, EPI_ISL_1616029, EPI_ISL_1616031, EPI_ISL_1616044, EPI_ISL_1616073, EPI_ISL_1616097, EPI_ISL_1616100, EPI_ISL_1616102, EPI_ISL_1616103, EPI_ISL_1616107, EPI_ISL_1616109, EPI_ISL_1616112, EPI_ISL_1616113, EPI_ISL_1616114, EPI_ISL_1616128, EPI_ISL_1616139, EPI_ISL_1616146, EPI_ISL_1616152, EPI_ISL_1616155, EPI_ISL_1616156, EPI_ISL_1616157, EPI_ISL_1616160, EPI_ISL_1616167, EPI_ISL_1616170, EPI_ISL_1616171, EPI_ISL_1616292, EPI_ISL_1616295, EPI_ISL_1616297, EPI_ISL_1616300, EPI_ISL_1616304, EPI_ISL_1616308, EPI_ISL_1616319, EPI_ISL_1616326, EPI_ISL_1616327, EPI_ISL_1616337, EPI_ISL_1616342, EPI_ISL_1616344, EPI_ISL_1616345, EPI_ISL_1616347, EPI_ISL_1616360, EPI_ISL_1616361, EPI_ISL_1616362, EPI_ISL_1616366, EPI_ISL_1616372, EPI_ISL_1616373, EPI_ISL_1616375, EPI_ISL_1616379, EPI_ISL_1616380, EPI_ISL_1616382, EPI_ISL_1616390, EPI_ISL_1616391, EPI_ISL_1616403, EPI_ISL_1616405, EPI_ISL_1616415, EPI_ISL_1616419, EPI_ISL_1616423, EPI_ISL_1616425, EPI_ISL_1616426, EPI_ISL_1616427, EPI_ISL_1616428, EPI_ISL_1616429, EPI_ISL_1616430, EPI_ISL_1616431, EPI_ISL_1616432, EPI_ISL_1616433, EPI_ISL_1616434, EPI_ISL_1616435, EPI_ISL_1616436, EPI_ISL_1616437, EPI_ISL_1616438, EPI_ISL_1616439, EPI_ISL_1616440, EPI_ISL_1616441, EPI_ISL_1616442, EPI_ISL_1616443, EPI_ISL_1616444, EPI_ISL_1616445, EPI_ISL_1616446, EPI_ISL_1616447, EPI_ISL_1616448, EPI_ISL_1616449, EPI_ISL_1616450, EPI_ISL_1616451, EPI_ISL_1616452, EPI_ISL_1616453, EPI_ISL_1616454, EPI_ISL_1616455, EPI_ISL_1616456, EPI_ISL_1616457, EPI_ISL_1616458, EPI_ISL_1616459, EPI_ISL_1616460, EPI_ISL_1616461, EPI_ISL_1616462, EPI_ISL_1616463, EPI_ISL_1616464, EPI_ISL_1616465, EPI_ISL_1616466, EPI_ISL_1616467, EPI_ISL_1616468,

| | | | |
|---|---|---|---|
| EPI_ISL_1616469, EPI_ISL_1616470, EPI_ISL_1616471, EPI_ISL_1616472, EPI_ISL_1616473, EPI_ISL_1616474, EPI_ISL_1616475, EPI_ISL_1616476, EPI_ISL_1616477, EPI_ISL_1616478, EPI_ISL_1616479, EPI_ISL_1616480, EPI_ISL_1616481, EPI_ISL_1616482, EPI_ISL_1616483, EPI_ISL_1616484, EPI_ISL_1616485, EPI_ISL_1616486, EPI_ISL_1616487, EPI_ISL_1616488, EPI_ISL_1616489, EPI_ISL_1616490, EPI_ISL_1616491, EPI_ISL_1616492, EPI_ISL_1616493, EPI_ISL_1616494, EPI_ISL_1616495, EPI_ISL_1616496, EPI_ISL_1616497, EPI_ISL_1616498, EPI_ISL_1616499, EPI_ISL_1616500, EPI_ISL_1616501, EPI_ISL_1616502, EPI_ISL_1616503, EPI_ISL_1616504, EPI_ISL_1616505, EPI_ISL_1616506, EPI_ISL_1616507, EPI_ISL_1616508, EPI_ISL_1616509, EPI_ISL_1616510, EPI_ISL_1616511, EPI_ISL_1616512, EPI_ISL_1616513, EPI_ISL_1616514, EPI_ISL_1616515, EPI_ISL_1616516, EPI_ISL_1616517, EPI_ISL_1616518, EPI_ISL_1616519, EPI_ISL_1616520, EPI_ISL_1616521, EPI_ISL_1616522, EPI_ISL_1616524, EPI_ISL_1616525, EPI_ISL_1616526, EPI_ISL_1616527, EPI_ISL_1616528, EPI_ISL_1616529, EPI_ISL_1616530, EPI_ISL_1616531, EPI_ISL_1616532, EPI_ISL_1616533, EPI_ISL_1616535, EPI_ISL_1616536, EPI_ISL_1616537, EPI_ISL_1616538, EPI_ISL_1616539, EPI_ISL_1616541, EPI_ISL_1616542, EPI_ISL_1616543, EPI_ISL_1616544, EPI_ISL_1616545, EPI_ISL_1616546, EPI_ISL_1616547, EPI_ISL_1616548, EPI_ISL_1616549, EPI_ISL_1616550, EPI_ISL_1616551, EPI_ISL_1616553, EPI_ISL_1616554, EPI_ISL_1616555, EPI_ISL_1616556, EPI_ISL_1616557, EPI_ISL_1616558, EPI_ISL_1616559, EPI_ISL_1616560, EPI_ISL_1616561, EPI_ISL_1616562, EPI_ISL_1616563, EPI_ISL_1616565, EPI_ISL_1616566, EPI_ISL_1616568, EPI_ISL_1616575, EPI_ISL_1616576, EPI_ISL_1616577, EPI_ISL_1616579, EPI_ISL_1616582, EPI_ISL_1616583, EPI_ISL_1616584, EPI_ISL_1616585, EPI_ISL_1616588, EPI_ISL_1616591, EPI_ISL_1616596, EPI_ISL_1616600, EPI_ISL_1616601, EPI_ISL_1616603, EPI_ISL_1631426, EPI_ISL_1631904, EPI_ISL_1631934, EPI_ISL_1631959, EPI_ISL_1631968, EPI_ISL_1632398, EPI_ISL_1634509, EPI_ISL_1634778, EPI_ISL_1634783, EPI_ISL_1634784, EPI_ISL_1634786, EPI_ISL_1634791, EPI_ISL_1634794, EPI_ISL_1634796, EPI_ISL_1634798, EPI_ISL_1634801, EPI_ISL_1634802, EPI_ISL_1634806, EPI_ISL_1634807, EPI_ISL_1634808, EPI_ISL_1634813, EPI_ISL_1634814, EPI_ISL_1634816, EPI_ISL_1634818, EPI_ISL_1634821, EPI_ISL_1634825, EPI_ISL_1634826, EPI_ISL_1634830, EPI_ISL_1634831, EPI_ISL_1634832, EPI_ISL_1634836, EPI_ISL_1634839, EPI_ISL_1634844, EPI_ISL_1634846, EPI_ISL_1634848, EPI_ISL_1634850, EPI_ISL_1634852, EPI_ISL_1634855, EPI_ISL_1634858, EPI_ISL_1634861, EPI_ISL_1634864, EPI_ISL_1634868, EPI_ISL_1634872, EPI_ISL_1634879, EPI_ISL_1634882, EPI_ISL_1634889, EPI_ISL_1634892, EPI_ISL_1634894, EPI_ISL_1634896, EPI_ISL_1634897, EPI_ISL_1634898, EPI_ISL_1634900, EPI_ISL_1634902, EPI_ISL_1634903, EPI_ISL_1634913, EPI_ISL_1634914, EPI_ISL_1634921, EPI_ISL_1634926, EPI_ISL_1634929, EPI_ISL_1634932, EPI_ISL_1634935, EPI_ISL_1634936, EPI_ISL_1634938, EPI_ISL_1634940, EPI_ISL_1634947, EPI_ISL_1634950, EPI_ISL_1634954, EPI_ISL_1634956, EPI_ISL_1634957, EPI_ISL_1634962, EPI_ISL_1634967, EPI_ISL_1634969, EPI_ISL_1634970, EPI_ISL_1634975, EPI_ISL_1634977, EPI_ISL_1634986, EPI_ISL_1634988, EPI_ISL_1635121, EPI_ISL_1635128, EPI_ISL_1635131, EPI_ISL_1635135, EPI_ISL_1635150, EPI_ISL_1635151, EPI_ISL_1635225, EPI_ISL_1635234, EPI_ISL_1635237, EPI_ISL_1635240, EPI_ISL_1635245, EPI_ISL_1635246, EPI_ISL_1635248, EPI_ISL_1635249, EPI_ISL_1635256, EPI_ISL_1635263, EPI_ISL_1635264, EPI_ISL_1635270, EPI_ISL_1635278, EPI_ISL_1635284, EPI_ISL_1635288, EPI_ISL_1635293, EPI_ISL_1635295, EPI_ISL_1635299, EPI_ISL_1635300, EPI_ISL_1635301, EPI_ISL_1635302, EPI_ISL_1697806, EPI_ISL_1697897, EPI_ISL_1697926, EPI_ISL_1697927, EPI_ISL_1697945, EPI_ISL_1699581, EPI_ISL_1699585, EPI_ISL_1699597, EPI_ISL_1699612, EPI_ISL_1699631, EPI_ISL_1699637, EPI_ISL_1699655, EPI_ISL_1699671, EPI_ISL_1699679, EPI_ISL_1699685, EPI_ISL_1699687, EPI_ISL_1699694, EPI_ISL_1699701, EPI_ISL_1699734, EPI_ISL_1699745, EPI_ISL_1699755, EPI_ISL_1699789, EPI_ISL_1699792, EPI_ISL_1699793, EPI_ISL_1699795, EPI_ISL_1699799, EPI_ISL_1742089, EPI_ISL_1742121, EPI_ISL_1742136, EPI_ISL_1742222 | | | |
| see above | Lighthouse Lab in Glasgow | Wellcome Sanger Institute for the COVID-19 Genomics UK (COG-UK) Consortium | Anna Dominiczak and Alex Alderton; Carol Clugston; Cordelia Langford; David Gray; David K. Jackson; Dominic Kwiatkowski; Ewan Harrison; Harper VanSteenhouse; Ian Johnston; Jeffrey Barrett; John Sillitoe on behalf of the Wellcome Sanger Institute COVID-19 Surveillance Team; Roberto Amato; Sonia Goncalves; Yumi Kasai |
| EPI_ISL_1506098, EPI_ISL_1535013, EPI_ISL_1535015, EPI_ISL_1535017, EPI_ISL_1535018, EPI_ISL_1535020, EPI_ISL_1535022, EPI_ISL_1535025, EPI_ISL_1535026, EPI_ISL_1535039, EPI_ISL_1535040, EPI_ISL_1535041, EPI_ISL_1535046, EPI_ISL_1535047, EPI_ISL_1535049, EPI_ISL_1535051, EPI_ISL_1535052, EPI_ISL_1535056, EPI_ISL_1535057, EPI_ISL_1535058, EPI_ISL_1535060, EPI_ISL_1535063, EPI_ISL_1535064, EPI_ISL_1535066, EPI_ISL_1535068, EPI_ISL_1535069, EPI_ISL_1535072, EPI_ISL_1535073, EPI_ISL_1535075, EPI_ISL_1535076, EPI_ISL_1535079, EPI_ISL_1535081, EPI_ISL_1535083, EPI_ISL_1535086, EPI_ISL_1535087, EPI_ISL_1535088, EPI_ISL_1535089, EPI_ISL_1535093, EPI_ISL_1535094, EPI_ISL_1535097, EPI_ISL_1535101, EPI_ISL_1535102, EPI_ISL_1535103, EPI_ISL_1535104, EPI_ISL_1535106, EPI_ISL_1535108, EPI_ISL_1535109, EPI_ISL_1535111, EPI_ISL_1535112, EPI_ISL_1535114, EPI_ISL_1535115, EPI_ISL_1535121, EPI_ISL_1535122, EPI_ISL_1535124, EPI_ISL_1535127, EPI_ISL_1535131, EPI_ISL_1535134, EPI_ISL_1535135, EPI_ISL_1535136, EPI_ISL_1535137, EPI_ISL_1535138, EPI_ISL_1535141, EPI_ISL_1535142, EPI_ISL_1535145, EPI_ISL_1535147, EPI_ISL_1535148, EPI_ISL_1535149, EPI_ISL_1535150, EPI_ISL_1535152, EPI_ISL_1535155, EPI_ISL_1535158, EPI_ISL_1535160, EPI_ISL_1535163, EPI_ISL_1535165, EPI_ISL_1535166, EPI_ISL_1535168, EPI_ISL_1535170, EPI_ISL_1535171, EPI_ISL_1535172, EPI_ISL_1535173, EPI_ISL_1535175, EPI_ISL_1535176, EPI_ISL_1535179, EPI_ISL_1535183, EPI_ISL_1535184, EPI_ISL_1535185, EPI_ISL_1535187, EPI_ISL_1535188, EPI_ISL_1535189, EPI_ISL_1535192, EPI_ISL_1535193, EPI_ISL_1535196, EPI_ISL_1535197, EPI_ISL_1535199, EPI_ISL_1535200, EPI_ISL_1535201, EPI_ISL_1535202, EPI_ISL_1535205, EPI_ISL_1535208, EPI_ISL_1535209, EPI_ISL_1535211, EPI_ISL_1535212, EPI_ISL_1535213, EPI_ISL_1535214, EPI_ISL_1535217, EPI_ISL_1535220, EPI_ISL_1535221, EPI_ISL_1535224, EPI_ISL_1535225, EPI_ISL_1535226, EPI_ISL_1535228, EPI_ISL_1535230, EPI_ISL_1535232, EPI_ISL_1535233, EPI_ISL_1535235, EPI_ISL_1535236, EPI_ISL_1535237, EPI_ISL_1535239, EPI_ISL_1535240, EPI_ISL_1535245, EPI_ISL_1535247, EPI_ISL_1535249, EPI_ISL_1535256, EPI_ISL_1535257, EPI_ISL_1535259, EPI_ISL_1535260, EPI_ISL_1535261, EPI_ISL_1535268, EPI_ISL_1535269, EPI_ISL_1535270, EPI_ISL_1535273, EPI_ISL_1535277, EPI_ISL_1535278, EPI_ISL_1535284, EPI_ISL_1535286, EPI_ISL_1535289, EPI_ISL_1535290, EPI_ISL_1535294, EPI_ISL_1535298, EPI_ISL_1535299, EPI_ISL_1535300, EPI_ISL_1535304, EPI_ISL_1535305, EPI_ISL_1535307, EPI_ISL_1535308, EPI_ISL_1535309, EPI_ISL_1535311, EPI_ISL_1535316, EPI_ISL_1535317, EPI_ISL_1535320, EPI_ISL_1535321, EPI_ISL_1535323, EPI_ISL_1535324, EPI_ISL_1535328, EPI_ISL_1535334, EPI_ISL_1535335, EPI_ISL_1535336, EPI_ISL_1535338, EPI_ISL_1535339, EPI_ISL_1535340, EPI_ISL_1535342, EPI_ISL_1535343, EPI_ISL_1535344, EPI_ISL_1535345, EPI_ISL_1535346, EPI_ISL_1535347, EPI_ISL_1535349, EPI_ISL_1535350, EPI_ISL_1535351, EPI_ISL_1535352, EPI_ISL_1535353, EPI_ISL_1535354, EPI_ISL_1535355, EPI_ISL_1535356, EPI_ISL_1535358, EPI_ISL_1535359, EPI_ISL_1535361, EPI_ISL_1535362, EPI_ISL_1535363, EPI_ISL_1535365, EPI_ISL_1535367, EPI_ISL_1535368, EPI_ISL_1535369, EPI_ISL_1535370, EPI_ISL_1535371, EPI_ISL_1535372, EPI_ISL_1535373, EPI_ISL_1535374, EPI_ISL_1535375, EPI_ISL_1535376, EPI_ISL_1535379, EPI_ISL_1535382, EPI_ISL_1535383, EPI_ISL_1535384, EPI_ISL_1535386, EPI_ISL_1535389, EPI_ISL_1535390, EPI_ISL_1535391, EPI_ISL_1535393, EPI_ISL_1535394, EPI_ISL_1535395, EPI_ISL_1535396, EPI_ISL_1535397, EPI_ISL_1535398, EPI_ISL_1535400, EPI_ISL_1535401, EPI_ISL_1535402, EPI_ISL_1535405, EPI_ISL_1535407, EPI_ISL_1535408, EPI_ISL_1535411, EPI_ISL_1535412, EPI_ISL_1535414, EPI_ISL_1535415, EPI_ISL_1535417, EPI_ISL_1535420, EPI_ISL_1535421, EPI_ISL_1535424, EPI_ISL_1535425, EPI_ISL_1535426, EPI_ISL_1535427, EPI_ISL_1535431, EPI_ISL_1535433, EPI_ISL_1535434, EPI_ISL_1535436, EPI_ISL_1535438, EPI_ISL_1535439, EPI_ISL_1535440, EPI_ISL_1535441, EPI_ISL_1535442, EPI_ISL_1535443, EPI_ISL_1535444, EPI_ISL_1535445, EPI_ISL_1535447, EPI_ISL_1535449, EPI_ISL_1535450, EPI_ISL_1535451, EPI_ISL_1535452, EPI_ISL_1535453, EPI_ISL_1535454, EPI_ISL_1535455, EPI_ISL_1535456, EPI_ISL_1535458, EPI_ISL_1535459, EPI_ISL_1535461, EPI_ISL_1535463, EPI_ISL_1535464, EPI_ISL_1535468, EPI_ISL_1535469, EPI_ISL_1535472, EPI_ISL_1535476, EPI_ISL_1535477, EPI_ISL_1535478, EPI_ISL_1535479, EPI_ISL_1535480, EPI_ISL_1535482, EPI_ISL_1535484, EPI_ISL_1535486, EPI_ISL_1535487, EPI_ISL_1535488, EPI_ISL_1535489, EPI_ISL_1535491, EPI_ISL_1535493, EPI_ISL_1535495, EPI_ISL_1535496, EPI_ISL_1535497, EPI_ISL_1535499, EPI_ISL_1535500, EPI_ISL_1535503, EPI_ISL_1535505, EPI_ISL_1535506, EPI_ISL_1535507, EPI_ISL_1535508, EPI_ISL_1535509, EPI_ISL_1535510, EPI_ISL_1535511, EPI_ISL_1535512, EPI_ISL_1535513, EPI_ISL_1535514, EPI_ISL_1535515, EPI_ISL_1535516, EPI_ISL_1535520, EPI_ISL_1535521, EPI_ISL_1535524, EPI_ISL_1535525, EPI_ISL_1535526, EPI_ISL_1535527, EPI_ISL_1535531, EPI_ISL_1535532, EPI_ISL_1535533, EPI_ISL_1535534, EPI_ISL_1535535, EPI_ISL_1535536, EPI_ISL_1535538, EPI_ISL_1535539, EPI_ISL_1535543, EPI_ISL_1535545, EPI_ISL_1535547, EPI_ISL_1535548, EPI_ISL_1535550, EPI_ISL_1535551, EPI_ISL_1535552, EPI_ISL_1535554, EPI_ISL_1535557, EPI_ISL_1535558, EPI_ISL_1535561, EPI_ISL_1535562, EPI_ISL_1535563, EPI_ISL_1535565, EPI_ISL_1535566, EPI_ISL_1535567, EPI_ISL_1535569, EPI_ISL_1535570, EPI_ISL_1535571, EPI_ISL_1535572, EPI_ISL_1535574, EPI_ISL_1535575, EPI_ISL_1535576, EPI_ISL_1535577, EPI_ISL_1535579, EPI_ISL_1535581, EPI_ISL_1535582, EPI_ISL_1535584, EPI_ISL_1535586, EPI_ISL_1535587, EPI_ISL_1535588, EPI_ISL_1535589, EPI_ISL_1535591, EPI_ISL_1535592, EPI_ISL_1535593, EPI_ISL_1535595, EPI_ISL_1535597, EPI_ISL_1535598, EPI_ISL_1535600, EPI_ISL_1535601, EPI_ISL_1535605, EPI_ISL_1535607, EPI_ISL_1535609, EPI_ISL_1535619, EPI_ISL_1535620, EPI_ISL_1535623, EPI_ISL_1535629, EPI_ISL_1535635, EPI_ISL_1535640, EPI_ISL_1535644, EPI_ISL_1535663, EPI_ISL_1535665, EPI_ISL_1535670, EPI_ISL_1535676, EPI_ISL_1535699, EPI_ISL_1535703, EPI_ISL_1535709, EPI_ISL_1535711, EPI_ISL_1535715, EPI_ISL_1535721, EPI_ISL_1535726, EPI_ISL_1535731, EPI_ISL_1535733, EPI_ISL_1535738, EPI_ISL_1535743, EPI_ISL_1535745, EPI_ISL_1535750, EPI_ISL_1535756, EPI_ISL_1535757, EPI_ISL_1535768, EPI_ISL_1535779, EPI_ISL_1535780, EPI_ISL_1535786, EPI_ISL_1535795, EPI_ISL_1535801, EPI_ISL_1535811, EPI_ISL_1535815, EPI_ISL_1535818, EPI_ISL_1535822, EPI_ISL_1535827, EPI_ISL_1535830, EPI_ISL_1535832, EPI_ISL_1535833, EPI_ISL_1535844, EPI_ISL_1535849, EPI_ISL_1535850, EPI_ISL_1535860, EPI_ISL_1535866, EPI_ISL_1535869, EPI_ISL_1535880, EPI_ISL_1535887, EPI_ISL_1535888, EPI_ISL_1535909, EPI_ISL_1535911, EPI_ISL_1535938, EPI_ISL_1535941, EPI_ISL_1535957, EPI_ISL_1535965, EPI_ISL_1535980, EPI_ISL_1535987, EPI_ISL_1535989, EPI_ISL_1536013, EPI_ISL_1536017, EPI_ISL_1536018, EPI_ISL_1536021, EPI_ISL_1536034, EPI_ISL_1536037, EPI_ISL_1536048, EPI_ISL_1536065, EPI_ISL_1536074, EPI_ISL_1536084, EPI_ISL_1536088, EPI_ISL_1536092, EPI_ISL_1536093, EPI_ISL_1536095, EPI_ISL_1536096, EPI_ISL_1536115, EPI_ISL_1536128, EPI_ISL_1536159, EPI_ISL_1536202, EPI_ISL_1536214, EPI_ISL_1536228, EPI_ISL_1536230, EPI_ISL_1536233, EPI_ISL_1536253, EPI_ISL_1536264, EPI_ISL_1536276, EPI_ISL_1536296, EPI_ISL_1536302, EPI_ISL_1536310, EPI_ISL_1536312, EPI_ISL_1536320, EPI_ISL_1536329, EPI_ISL_1536343, EPI_ISL_1536348, EPI_ISL_1536350, EPI_ISL_1536357, EPI_ISL_1536380, EPI_ISL_1536396, EPI_ISL_1536402, EPI_ISL_1536404, EPI_ISL_1536409, EPI_ISL_1536411, EPI_ISL_1536423, EPI_ISL_1536425, EPI_ISL_1536429, EPI_ISL_1536442, EPI_ISL_1536446, EPI_ISL_1536453, EPI_ISL_1536467, EPI_ISL_1536503, EPI_ISL_1536508, EPI_ISL_1536521, EPI_ISL_1536527, EPI_ISL_1536535, EPI_ISL_1536536, EPI_ISL_1536540, EPI_ISL_1536552, EPI_ISL_1536554, EPI_ISL_1536557, EPI_ISL_1536561, EPI_ISL_1536562, EPI_ISL_1536563, EPI_ISL_1536564, EPI_ISL_1536565, EPI_ISL_1536566, EPI_ISL_1536569, EPI_ISL_1536573, EPI_ISL_1536578, EPI_ISL_1536579, EPI_ISL_1536582, EPI_ISL_1536587, EPI_ISL_1536589, EPI_ISL_1536591, EPI_ISL_1536592, EPI_ISL_1536593, EPI_ISL_1536594, EPI_ISL_1536595, EPI_ISL_1536598, EPI_ISL_1536601, EPI_ISL_1536608, EPI_ISL_1536610, EPI_ISL_1536611, EPI_ISL_1536612, EPI_ISL_1536615, EPI_ISL_1536616, EPI_ISL_1536620, EPI_ISL_1536621, EPI_ISL_1536622, EPI_ISL_1536623, EPI_ISL_1536624, EPI_ISL_1536627, EPI_ISL_1536633, EPI_ISL_1536636, EPI_ISL_1536642, EPI_ISL_1536645, EPI_ISL_1536648, EPI_ISL_1536649, EPI_ISL_1536650, EPI_ISL_1536658, EPI_ISL_1536659, EPI_ISL_1536661, EPI_ISL_1536662, EPI_ISL_1536663, EPI_ISL_1536664, EPI_ISL_1536666, EPI_ISL_1536668, EPI_ISL_1536669, EPI_ISL_1536670, EPI_ISL_1536672, EPI_ISL_1536674, EPI_ISL_1536676, EPI_ISL_1536677, EPI_ISL_1536679, EPI_ISL_1536681, EPI_ISL_1536682, EPI_ISL_1536684, EPI_ISL_1536688, EPI_ISL_1536691, EPI_ISL_1536692, EPI_ISL_1536697, EPI_ISL_1536700, EPI_ISL_1536704, EPI_ISL_1536705, EPI_ISL_1536707, EPI_ISL_1536711, EPI_ISL_1536712, EPI_ISL_1536713, EPI_ISL_1536717, EPI_ISL_1536722, EPI_ISL_1536723, EPI_ISL_1536726, EPI_ISL_1536730, EPI_ISL_1536732, EPI_ISL_1536734, EPI_ISL_1536736, EPI_ISL_1536737, EPI_ISL_1536740, EPI_ISL_1536744, EPI_ISL_1536746, EPI_ISL_1536748, EPI_ISL_1536749, EPI_ISL_1536750, EPI_ISL_1536752, EPI_ISL_1536755, EPI_ISL_1536761, EPI_ISL_1536763, EPI_ISL_1536767, EPI_ISL_1536772, EPI_ISL_1536776, EPI_ISL_1536779, EPI_ISL_1536784, EPI_ISL_1536801, EPI_ISL_1536805, EPI_ISL_1536807, EPI_ISL_1536808, EPI_ISL_1536814, EPI_ISL_1536818, EPI_ISL_1536819, EPI_ISL_1536822, EPI_ISL_1536825, EPI_ISL_1536826, EPI_ISL_1536828, EPI_ISL_1536832, EPI_ISL_1536833, EPI_ISL_1536838, EPI_ISL_1536839, EPI_ISL_1536852, EPI_ISL_1536860, EPI_ISL_1536876, EPI_ISL_1536884, EPI_ISL_1536885, EPI_ISL_1536887, EPI_ISL_1536892, EPI_ISL_1536894, EPI_ISL_1536902, EPI_ISL_1536909, EPI_ISL_1536921, EPI_ISL_1536929, EPI_ISL_1536931, EPI_ISL_1536939, EPI_ISL_1536957, EPI_ISL_1536971, EPI_ISL_1536977, EPI_ISL_1536986, EPI_ISL_1537000, EPI_ISL_1537011, EPI_ISL_1537012, EPI_ISL_1537014, EPI_ISL_1537019, EPI_ISL_1537028, EPI_ISL_1537037, EPI_ISL_1537041, EPI_ISL_1537048, EPI_ISL_1537056, EPI_ISL_1537077, EPI_ISL_1537084, EPI_ISL_1537090, EPI_ISL_1537093, EPI_ISL_1537105, EPI_ISL_1537108, EPI_ISL_1537109, EPI_ISL_1537114, EPI_ISL_1537330, EPI_ISL_1537334, EPI_ISL_1537335, EPI_ISL_1537338, EPI_ISL_1537339, EPI_ISL_1537340, EPI_ISL_1537345, EPI_ISL_1537347, EPI_ISL_1537351, EPI_ISL_1537355, EPI_ISL_1537359, EPI_ISL_1537361, EPI_ISL_1537364, EPI_ISL_1537365, EPI_ISL_1537367, EPI_ISL_1537369, EPI_ISL_1537370, EPI_ISL_1537372, EPI_ISL_1537375, EPI_ISL_1537376, EPI_ISL_1537378, EPI_ISL_1537385, EPI_ISL_1537387, EPI_ISL_1537388, EPI_ISL_1537390, EPI_ISL_1537395, EPI_ISL_1537399, EPI_ISL_1537403, EPI_ISL_1537406, EPI_ISL_1537407, EPI_ISL_1537411, EPI_ISL_1537414, EPI_ISL_1537415, EPI_ISL_1537419, EPI_ISL_1537420, EPI_ISL_1537421, EPI_ISL_1537426, EPI_ISL_1537428, EPI_ISL_1537429, EPI_ISL_1537430, EPI_ISL_1537432, EPI_ISL_1537434, EPI_ISL_1537437, EPI_ISL_1537439, EPI_ISL_1537440, EPI_ISL_1537446, EPI_ISL_1537448, EPI_ISL_1537453, EPI_ISL_1537454, EPI_ISL_1537456, EPI_ISL_1537463, EPI_ISL_1537467, EPI_ISL_1537469, EPI_ISL_1537475, EPI_ISL_1537476, EPI_ISL_1537481, EPI_ISL_1537484, EPI_ISL_1537485, EPI_ISL_1537495, EPI_ISL_1537496, EPI_ISL_1537502, EPI_ISL_1537503, EPI_ISL_1537511, EPI_ISL_1537512, EPI_ISL_1537513, EPI_ISL_1537517, EPI_ISL_1537519, EPI_ISL_1537520, EPI_ISL_1537521, EPI_ISL_1537523, EPI_ISL_1537526, EPI_ISL_1537528, EPI_ISL_1537532, EPI_ISL_1537534, EPI_ISL_1537536, EPI_ISL_1537541, EPI_ISL_1537543, EPI_ISL_1537544, EPI_ISL_1537545, EPI_ISL_1537547, EPI_ISL_1537549, EPI_ISL_1537551, EPI_ISL_1537553, EPI_ISL_1537557, EPI_ISL_1537560, EPI_ISL_1537562, EPI_ISL_1537563, EPI_ISL_1537564, EPI_ISL_1537571, EPI_ISL_1537572, EPI_ISL_1537574, EPI_ISL_1537576, EPI_ISL_1537577, EPI_ISL_1537589, EPI_ISL_1537591, EPI_ISL_1537598, EPI_ISL_1537604, EPI_ISL_1537608, EPI_ISL_1537609, EPI_ISL_1537615, EPI_ISL_1537616, EPI_ISL_1537618, EPI_ISL_1537619, EPI_ISL_1537623, EPI_ISL_1537626, EPI_ISL_1537635, EPI_ISL_1537638, EPI_ISL_1537653, EPI_ISL_1537654, EPI_ISL_1537658, EPI_ISL_1537664, EPI_ISL_1537679, EPI_ISL_1537682, EPI_ISL_1537686, EPI_ISL_1537694, EPI_ISL_1537708, EPI_ISL_1537709, EPI_ISL_1537710, EPI_ISL_1537714, EPI_ISL_1537718, EPI_ISL_1537720, EPI_ISL_1537722, EPI_ISL_1537723, EPI_ISL_1537724, EPI_ISL_1537732, EPI_ISL_1537742, EPI_ISL_1537754, EPI_ISL_1537766, EPI_ISL_1537772, EPI_ISL_1537791, EPI_ISL_1537798, EPI_ISL_1537800, EPI_ISL_1537806, EPI_ISL_1537810, EPI_ISL_1537829, EPI_ISL_1537842, EPI_ISL_1544198, EPI_ISL_1544199, EPI_ISL_1544200, EPI_ISL_1544202, EPI_ISL_1544206, EPI_ISL_1544212, EPI_ISL_1544213, EPI_ISL_1544220, EPI_ISL_1544230, EPI_ISL_1544233, EPI_ISL_1544237, EPI_ISL_1544238, EPI_ISL_1544241, EPI_ISL_1544244, EPI_ISL_1544245, EPI_ISL_1544247, EPI_ISL_1544251, EPI_ISL_1544252, EPI_ISL_1544255, EPI_ISL_1544256, EPI_ISL_1544257, EPI_ISL_1544258, EPI_ISL_1544265, EPI_ISL_1544266, EPI_ISL_1544268, EPI_ISL_1544269, EPI_ISL_1544271, EPI_ISL_1544272, EPI_ISL_1544275, EPI_ISL_1544278, EPI_ISL_1544279, EPI_ISL_1544283, EPI_ISL_1544284, EPI_ISL_1544287, EPI_ISL_1544289, EPI_ISL_1544291, EPI_ISL_1544292, EPI_ISL_1544298, EPI_ISL_1544301, EPI_ISL_1544303, EPI_ISL_1544307, EPI_ISL_1544312, EPI_ISL_1544316, EPI_ISL_1544318, EPI_ISL_1544327, EPI_ISL_1544329, EPI_ISL_1544330, EPI_ISL_1544334, EPI_ISL_1544335, EPI_ISL_1544342, EPI_ISL_1544344, EPI_ISL_1544345, EPI_ISL_1544348, EPI_ISL_1544350, EPI_ISL_1544353, EPI_ISL_1544355, EPI_ISL_1544356, EPI_ISL_1544358, EPI_ISL_1544366, EPI_ISL_1544367, EPI_ISL_1544368, EPI_ISL_1544369, EPI_ISL_1544373, EPI_ISL_1544374, EPI_ISL_1544379, EPI_ISL_1544383, EPI_ISL_1544384, EPI_ISL_1544386, EPI_ISL_1544391, EPI_ISL_1544392, EPI_ISL_1544393, EPI_ISL_1544402, EPI_ISL_1544405, EPI_ISL_1544408, EPI_ISL_1544421, EPI_ISL_1544422, EPI_ISL_1544427, EPI_ISL_1544428, EPI_ISL_1544429, EPI_ISL_1544430, EPI_ISL_1544439, EPI_ISL_1544442, EPI_ISL_1544445, EPI_ISL_1544450, EPI_ISL_1544451, EPI_ISL_1544452, EPI_ISL_1544455, EPI_ISL_1544457, EPI_ISL_1544460, EPI_ISL_1544471, EPI_ISL_1544472, EPI_ISL_1544479, EPI_ISL_1544481, EPI_ISL_1544482, EPI_ISL_1544483, EPI_ISL_1544488, EPI_ISL_1564040, EPI_ISL_1564045, EPI_ISL_1564046, EPI_ISL_1564050, EPI_ISL_1564052, EPI_ISL_1564061, EPI_ISL_1564063, EPI_ISL_1564067, EPI_ISL_1564087, EPI_ISL_1564100, EPI_ISL_1564106, EPI_ISL_1564110, EPI_ISL_1564118, EPI_ISL_1564121, EPI_ISL_1564123, EPI_ISL_1564126, EPI_ISL_1564140, EPI_ISL_1564159, EPI_ISL_1564175, EPI_ISL_1564183, EPI_ISL_1564190, EPI_ISL_1564193, EPI_ISL_1564222, EPI_ISL_1564228, EPI_ISL_1564229, EPI_ISL_1564236, EPI_ISL_1564237, EPI_ISL_1564240, EPI_ISL_1564244, EPI_ISL_1564246, EPI_ISL_1564254, EPI_ISL_1564262, EPI_ISL_1564285, EPI_ISL_1564294, EPI_ISL_1564297, EPI_ISL_1564298, EPI_ISL_1564299, EPI_ISL_1564302, EPI_ISL_1564303, EPI_ISL_1564307, EPI_ISL_1564310, EPI_ISL_1564316, EPI_ISL_1564770, EPI_ISL_1564771, EPI_ISL_1584165, EPI_ISL_1584166, EPI_ISL_1584179, EPI_ISL_1584180, EPI_ISL_1584181, EPI_ISL_1584186, EPI_ISL_1584190, EPI_ISL_1584199, EPI_ISL_1584203, EPI_ISL_1584205, EPI_ISL_1584207, EPI_ISL_1584209, EPI_ISL_1584210, EPI_ISL_1584211, EPI_ISL_1584212, EPI_ISL_1584218, EPI_ISL_1584221, EPI_ISL_1584222, EPI_ISL_1584224, EPI_ISL_1584226, EPI_ISL_1584227, EPI_ISL_1584228, EPI_ISL_1584229, EPI_ISL_1584234, EPI_ISL_1584238, EPI_ISL_1584246, EPI_ISL_1584252, EPI_ISL_1584263, EPI_ISL_1584266, EPI_ISL_1584269, EPI_ISL_1584271, EPI_ISL_1584272, EPI_ISL_1584283, EPI_ISL_1584284, EPI_ISL_1584286, EPI_ISL_1584287, EPI_ISL_1584289, EPI_ISL_1584292, EPI_ISL_1584294, EPI_ISL_1584295, EPI_ISL_1584300, EPI_ISL_1584301, EPI_ISL_1584302, EPI_ISL_1584305, EPI_ISL_1584306, EPI_ISL_1584307, EPI_ISL_1584313, EPI_ISL_1584315, EPI_ISL_1584317, EPI_ISL_1584318, EPI_ISL_1584325, EPI_ISL_1584331, EPI_ISL_1584335, EPI_ISL_1584340, EPI_ISL_1584341, EPI_ISL_1584342, EPI_ISL_1584350, EPI_ISL_1584354, EPI_ISL_1584358, EPI_ISL_1584360, EPI_ISL_1584364, EPI_ISL_1584370, EPI_ISL_1584374, EPI_ISL_1584378, EPI_ISL_1584385, EPI_ISL_1584387, EPI_ISL_1584388, EPI_ISL_1584390, EPI_ISL_1584392, EPI_ISL_1584395, EPI_ISL_1584396, EPI_ISL_1584399, EPI_ISL_1584404, EPI_ISL_1584406, EPI_ISL_1584407, EPI_ISL_1584409, EPI_ISL_1584416, EPI_ISL_1584420, EPI_ISL_1584424, EPI_ISL_1584425, EPI_ISL_1584426, EPI_ISL_1584428, EPI_ISL_1584429, EPI_ISL_1584430, EPI_ISL_1584432, EPI_ISL_1584434, EPI_ISL_1594625, EPI_ISL_1594679, EPI_ISL_1595009, EPI_ISL_1595054, EPI_ISL_1595058, EPI_ISL_1595090, EPI_ISL_1595100, EPI_ISL_1595154, EPI_ISL_1595164, EPI_ISL_1595169, EPI_ISL_1595189, EPI_ISL_1595471, EPI_ISL_1615934, EPI_ISL_1615937, EPI_ISL_1615939, EPI_ISL_1615942, EPI_ISL_1615943, EPI_ISL_1615945, EPI_ISL_1615946, EPI_ISL_1615951, EPI_ISL_1615952, EPI_ISL_1615955, EPI_ISL_1615958, EPI_ISL_1615962, EPI_ISL_1615964, EPI_ISL_1615966, EPI_ISL_1615971, EPI_ISL_1615972, EPI_ISL_1615976, EPI_ISL_1615980, EPI_ISL_1615983, EPI_ISL_1615984, EPI_ISL_1615991, EPI_ISL_1615992, EPI_ISL_1616003, EPI_ISL_1616005, EPI_ISL_1616006, EPI_ISL_1616008, EPI_ISL_1616010, EPI_ISL_1616011, EPI_ISL_1616012, EPI_ISL_1616015, EPI_ISL_1616016, EPI_ISL_1616018, EPI_ISL_1616019, EPI_ISL_1616021, EPI_ISL_1616028, EPI_ISL_1616030, EPI_ISL_1616033, EPI_ISL_1616034, EPI_ISL_1616035, EPI_ISL_1616037, EPI_ISL_1616040, EPI_ISL_1616041, EPI_ISL_1616043, EPI_ISL_1616045, EPI_ISL_1616046, EPI_ISL_1616052, EPI_ISL_1616054, EPI_ISL_1616056, EPI_ISL_1616058, EPI_ISL_1616066, EPI_ISL_1616067, EPI_ISL_1616070, EPI_ISL_1616071, EPI_ISL_1616074, EPI_ISL_1616078, EPI_ISL_1616081, EPI_ISL_1616086, EPI_ISL_1616088, EPI_ISL_1616090, EPI_ISL_1616093, EPI_ISL_1616095, EPI_ISL_1616098, EPI_ISL_1616105, EPI_ISL_1616108, EPI_ISL_1616116, EPI_ISL_1616118, EPI_ISL_1616124, EPI_ISL_1616126, EPI_ISL_1616129, EPI_ISL_1616131, EPI_ISL_1616136, EPI_ISL_1616145, EPI_ISL_1616147, EPI_ISL_1616151, EPI_ISL_1616154, EPI_ISL_1616166, EPI_ISL_1616168, EPI_ISL_1616169, EPI_ISL_1616172, EPI_ISL_1616173, EPI_ISL_1616174, EPI_ISL_1616180, EPI_ISL_1616182, EPI_ISL_1616184, EPI_ISL_1631336, EPI_ISL_1631425, EPI_ISL_1631428, EPI_ISL_1631429, EPI_ISL_1631430, EPI_ISL_1631431, EPI_ISL_1631433, EPI_ISL_1631435, EPI_ISL_1631436, EPI_ISL_1631437, EPI_ISL_1631438, EPI_ISL_1631439, EPI_ISL_1631440, EPI_ISL_1631441, EPI_ISL_1631442, EPI_ISL_1631443, EPI_ISL_1631444, EPI_ISL_1631445, EPI_ISL_1631446, EPI_ISL_1631447, EPI_ISL_1631448, EPI_ISL_1631449, EPI_ISL_1631450, EPI_ISL_1631451, EPI_ISL_1631452, EPI_ISL_1631453, EPI_ISL_1631454, EPI_ISL_1631455, EPI_ISL_1631456, EPI_ISL_1631457, EPI_ISL_1631458, EPI_ISL_1631459, EPI_ISL_1631460, EPI_ISL_1631461, EPI_ISL_1631462, EPI_ISL_1631463, EPI_ISL_1631464, EPI_ISL_1631465, EPI_ISL_1631466, EPI_ISL_1631467, EPI_ISL_1631468, EPI_ISL_1631469, EPI_ISL_1631470, EPI_ISL_1631471, EPI_ISL_1631472, EPI_ISL_1631473, EPI_ISL_1631474, EPI_ISL_1631475, EPI_ISL_1631476, EPI_ISL_1631477, EPI_ISL_1631478, EPI_ISL_1631479, EPI_ISL_1631480, EPI_ISL_1631481, EPI_ISL_1631482, EPI_ISL_1631483, EPI_ISL_1631484, EPI_ISL_1631485, EPI_ISL_1631486, EPI_ISL_1631487, EPI_ISL_1631488, EPI_ISL_1631489, EPI_ISL_1631490, EPI_ISL_1631491, EPI_ISL_1631492, EPI_ISL_1631493, EPI_ISL_1631494, EPI_ISL_1631495, EPI_ISL_1631496, EPI_ISL_1631497, EPI_ISL_1631498, EPI_ISL_1631499, EPI_ISL_1631500, EPI_ISL_1631501, EPI_ISL_1631502, EPI_ISL_1631503, EPI_ISL_1631504, EPI_ISL_1631505, EPI_ISL_1631506, EPI_ISL_1631507, EPI_ISL_1631508, EPI_ISL_1631509, EPI_ISL_1631510, EPI_ISL_1631511, EPI_ISL_1631512, EPI_ISL_1631513, EPI_ISL_1631514, EPI_ISL_1631516, EPI_ISL_1631517, EPI_ISL_1631518, EPI_ISL_1631519, EPI_ISL_1631520, EPI_ISL_1631521, EPI_ISL_1631522, EPI_ISL_1631523, EPI_ISL_1631524, EPI_ISL_1631525, EPI_ISL_1631526, EPI_ISL_1631527, EPI_ISL_1631528, EPI_ISL_1631529, EPI_ISL_1631530, EPI_ISL_1631531, EPI_ISL_1631532, EPI_ISL_1631533, EPI_ISL_1631534, EPI_ISL_1631535, EPI_ISL_1631536, EPI_ISL_1631537, EPI_ISL_1631538, EPI_ISL_1631539, EPI_ISL_1631540, EPI_ISL_1631541, EPI_ISL_1631542, EPI_ISL_1631543, EPI_ISL_1631544, EPI_ISL_1631546, EPI_ISL_1631547, EPI_ISL_1631548, EPI_ISL_1631549, EPI_ISL_1631550, EPI_ISL_1631551, EPI_ISL_1631552, EPI_ISL_1631553, EPI_ISL_1631554, EPI_ISL_1631555, EPI_ISL_1631556, EPI_ISL_1631557, EPI_ISL_1631558, EPI_ISL_1631559, EPI_ISL_1631560, EPI_ISL_1631561, EPI_ISL_1631562, EPI_ISL_1631563, EPI_ISL_1631564, EPI_ISL_1631565, EPI_ISL_1631566, EPI_ISL_1631567, EPI_ISL_1631568, EPI_ISL_1631570, EPI_ISL_1631571, EPI_ISL_1631572, EPI_ISL_1631573, EPI_ISL_1631574, EPI_ISL_1631575, EPI_ISL_1631576, EPI_ISL_1631577, EPI_ISL_1631578, EPI_ISL_1631579, EPI_ISL_1631580, EPI_ISL_1631581, EPI_ISL_1631582, EPI_ISL_1631583, EPI_ISL_1631584, EPI_ISL_1631585, EPI_ISL_1631586, EPI_ISL_1631587, EPI_ISL_1631589, EPI_ISL_1631590, EPI_ISL_1631591, EPI_ISL_1631592, EPI_ISL_1631593, EPI_ISL_1631594, EPI_ISL_1631595, EPI_ISL_1631596, EPI_ISL_1631597, EPI_ISL_1631598, EPI_ISL_1631599, EPI_ISL_1631600, EPI_ISL_1631601, EPI_ISL_1631602, EPI_ISL_1631603, EPI_ISL_1631604, EPI_ISL_1631605, EPI_ISL_1631607, EPI_ISL_1631608, EPI_ISL_1631609, EPI_ISL_1631610, EPI_ISL_1631611, EPI_ISL_1631612, EPI_ISL_1631613, EPI_ISL_1631614, EPI_ISL_1631615, EPI_ISL_1631616, EPI_ISL_1631619, EPI_ISL_1631620, EPI_ISL_1631621, EPI_ISL_1631622, EPI_ISL_1631623, EPI_ISL_1631624, EPI_ISL_1631625, EPI_ISL_1631627, EPI_ISL_1631628, EPI_ISL_1631629, EPI_ISL_1631630, EPI_ISL_1631631, EPI_ISL_1631632, EPI_ISL_1631633, EPI_ISL_1631634, EPI_ISL_1631635, EPI_ISL_1631637, EPI_ISL_1631638, EPI_ISL_1631639, EPI_ISL_1631640, EPI_ISL_1631641, EPI_ISL_1631642, EPI_ISL_1631643, EPI_ISL_1631644, EPI_ISL_1631645, EPI_ISL_1631646, EPI_ISL_1631647, EPI_ISL_1631648, EPI_ISL_1631649, EPI_ISL_1631650, EPI_ISL_1631651, EPI_ISL_1631652, EPI_ISL_1631653, EPI_ISL_1631654, EPI_ISL_1631655, EPI_ISL_1631656, EPI_ISL_1631657, EPI_ISL_1631658, EPI_ISL_1631659, EPI_ISL_1631660, EPI_ISL_1631661, EPI_ISL_1631662, EPI_ISL_1631663, EPI_ISL_1631664, EPI_ISL_1631665, EPI_ISL_1631666, EPI_ISL_1631667, EPI_ISL_1631668, EPI_ISL_1631669, EPI_ISL_1631670, EPI_ISL_1631671, EPI_ISL_1631672, EPI_ISL_1631673, EPI_ISL_1631674, EPI_ISL_1631675, EPI_ISL_1631676, EPI_ISL_1631677, EPI_ISL_1631678, EPI_ISL_1631679, EPI_ISL_1631680, EPI_ISL_1631681, EPI_ISL_1631682, EPI_ISL_1631683, EPI_ISL_1631684, EPI_ISL_1631685, EPI_ISL_1631686, EPI_ISL_1631687, EPI_ISL_1631688, EPI_ISL_1631689, EPI_ISL_1631690, EPI_ISL_1631691, EPI_ISL_1631692, EPI_ISL_1631693, EPI_ISL_1631694, EPI_ISL_1631695, EPI_ISL_1631696, EPI_ISL_1631697, EPI_ISL_1631698, EPI_ISL_1631699, EPI_ISL_1631700, EPI_ISL_1631701, EPI_ISL_1631702, EPI_ISL_1631703, EPI_ISL_1631704, EPI_ISL_1631705, EPI_ISL_1631706, EPI_ISL_1631707, EPI_ISL_1631708, EPI_ISL_1631709, EPI_ISL_1631710, EPI_ISL_1631712, EPI_ISL_1631713, EPI_ISL_1631714, EPI_ISL_1631715, EPI_ISL_1631716, EPI_ISL_1631717, EPI_ISL_1631718, EPI_ISL_1631719, EPI_ISL_1631721, EPI_ISL_1631722, EPI_ISL_1631723, EPI_ISL_1631724, EPI_ISL_1631725, EPI_ISL_1631726, EPI_ISL_1631727, EPI_ISL_1631728, EPI_ISL_1631729, EPI_ISL_1631730, EPI_ISL_1631731, EPI_ISL_1631732, EPI_ISL_1631733, EPI_ISL_1631734, EPI_ISL_1631735, EPI_ISL_1631736, EPI_ISL_1631738, EPI_ISL_1631739, EPI_ISL_1631740, EPI_ISL_1631741, EPI_ISL_1631742, EPI_ISL_1631743, EPI_ISL_1631744, EPI_ISL_1631745, EPI_ISL_1631746, EPI_ISL_1631747, EPI_ISL_1631748, EPI_ISL_1631749, EPI_ISL_1631750, EPI_ISL_1631751, EPI_ISL_1631752, EPI_ISL_1631781, EPI_ISL_1631797, EPI_ISL_1631808, EPI_ISL_1631820, EPI_ISL_1631837, EPI_ISL_1631880, EPI_ISL_1632033, EPI_ISL_1632035, EPI_ISL_1632090, EPI_ISL_1632142, EPI_ISL_1632146, EPI_ISL_1632168, EPI_ISL_1632170, EPI_ISL_1632181, EPI_ISL_1632228, EPI_ISL_1632229, EPI_ISL_1632232, EPI_ISL_1632252, EPI_ISL_1632254, EPI_ISL_1632289, EPI_ISL_1632391, EPI_ISL_1632466, EPI_ISL_1632480, EPI_ISL_1635119, EPI_ISL_1635313, EPI_ISL_1653739, EPI_ISL_1758191, EPI_ISL_1758210, EPI_ISL_1758213, EPI_ISL_1758215, EPI_ISL_1758221, EPI_ISL_1758222, EPI_ISL_1758231, EPI_ISL_1758293, EPI_ISL_1758300, EPI_ISL_1758309, EPI_ISL_1758310, EPI_ISL_1758320, EPI_ISL_1758327, EPI_ISL_1758329, EPI_ISL_1758341, EPI_ISL_1758344, EPI_ISL_1758347, EPI_ISL_1758383, EPI_ISL_1758390, EPI_ISL_1758411, EPI_ISL_1758416, EPI_ISL_1758429, EPI_ISL_1758461, EPI_ISL_1758473, EPI_ISL_1758501, EPI_ISL_1758515, EPI_ISL_1758520, EPI_ISL_1758521, EPI_ISL_1758545, EPI_ISL_1758548, EPI_ISL_1758556, EPI_ISL_1758562, EPI_ISL_1758581, EPI_ISL_1758583, EPI_ISL_1758584, EPI_ISL_1758590, EPI_ISL_1758596, EPI_ISL_1758599, EPI_ISL_1758603, EPI_ISL_1758604, EPI_ISL_1758605, EPI_ISL_1758627, EPI_ISL_1758642, EPI_ISL_1758649, EPI_ISL_1758657, EPI_ISL_1758661, EPI_ISL_1759246, EPI_ISL_1759327, EPI_ISL_1791019, EPI_ISL_1806239, EPI_ISL_1806355, EPI_ISL_1806389, EPI_ISL_1830560, EPI_ISL_1831192, EPI_ISL_2005574, EPI_ISL_2005627, EPI_ISL_2005719, EPI_ISL_2005751, EPI_ISL_2005758 | | | |
| see above | Lighthouse Lab in Milton Keynes | Wellcome Sanger Institute for the COVID-19 Genomics UK (COG-UK) Consortium | Cordelia Langford; David K. Jackson; Dominic Kwiatkowski; Ewan Harrison; Ian Johnston; Jeffrey Barrett; John Sillitoe on behalf of the Wellcome Sanger Institute COVID-19 Surveillance Team; Roberto Amato; Sonia Goncalves; The Lighthouse Lab in Milton Keynes and Alex Alderton |
| EPI_ISL_1833017, EPI_ISL_1978953 | Lincolnshire Hospitals and DeepSeq Nottingham | COVID-19 Genomics UK (COG-UK) Consortium | Christopher Moore; Fei Sang; Joeseph Chappell; Johnny Debebe; Jonathan Ball; Matthew Carlisle; Matthew Loose; Nadine Holmes; Nichola Duckworth; Patrick McClure; Sarah Walsh; Tim Sloan; Victoria Wright |
| EPI_ISL_1636834, EPI_ISL_1636839, EPI_ISL_1636840, EPI_ISL_1636841, EPI_ISL_1636842, EPI_ISL_1636843, EPI_ISL_1636845, EPI_ISL_1636846, EPI_ISL_1636847, EPI_ISL_1636848, EPI_ISL_1636849, EPI_ISL_1636851, EPI_ISL_1636852, EPI_ISL_1636853, EPI_ISL_1636859, EPI_ISL_1636860, EPI_ISL_1636861, EPI_ISL_1636862, EPI_ISL_1636863, EPI_ISL_1636864, EPI_ISL_1636865, EPI_ISL_1636866, EPI_ISL_1636867, EPI_ISL_1636868, EPI_ISL_1636873, EPI_ISL_1636874, EPI_ISL_1636876, EPI_ISL_1636878, EPI_ISL_1636879, EPI_ISL_1636880, EPI_ISL_1636881, EPI_ISL_1832357, EPI_ISL_1832361, EPI_ISL_1832368, EPI_ISL_1832376, EPI_ISL_1832378, EPI_ISL_1832484, EPI_ISL_1832486, EPI_ISL_1832488, EPI_ISL_1832492, EPI_ISL_1832494, EPI_ISL_1832502, EPI_ISL_1974934, EPI_ISL_1974936, EPI_ISL_1974939, EPI_ISL_1974941, EPI_ISL_1974942, EPI_ISL_1979105, EPI_ISL_1979107, EPI_ISL_1979109, EPI_ISL_1979110, EPI_ISL_1979112, EPI_ISL_1979113, EPI_ISL_1979115, EPI_ISL_1979119, EPI_ISL_1979121, EPI_ISL_1979126, EPI_ISL_1979127, EPI_ISL_1979129 | | | |
| see above | Liverpool Clinical Laboratories | COVID-19 Genomics UK (COG-UK) Consortium | A Alrezaihi; Alessandro Gerada; Alistair Darby; Angela Cowell; Anita Lucaci; Anu Chawla; Cassie Olateju; Catherine Hartley; Charlotte Nelson; Ecaterina Vamos; Elaine O'Toole; Eleanor G Bentley; Ghada T Shawli; Hermione Webster; Isabel García-Dorival; James Johnson; James P Stewart; Jenifer Manson; Joanne Watts; Jones Benjamin; Jordan J Clark; Julian Hiscox; L Luu; Lucille Rainbow; M Almsaud; Margaret Hughes; Mark Whitehead; Matthew Gemmell; Miren Iturriza-Gomara; Muhannad Alruwaili; N.P Randle; Neil Swainston; PKF Gilmore; Parul Sharma; Rebekah Penrice-Randal; Richard Eccles; Richard Gregory; Sam Haldenby; Steve Paterson; Stuart D Armstrong; Trevor Ian Robinson; Ximeng Han |
| EPI_ISL_1637182, EPI_ISL_1637183, EPI_ISL_1637184, EPI_ISL_1637185, EPI_ISL_1637188, EPI_ISL_1637196, EPI_ISL_1637230, EPI_ISL_1637231, EPI_ISL_1637234, EPI_ISL_1978535 | | | |
| see above | Northumbria University / South Tees Hospitals NHS Foundation Trust / North Cumbria Integrated Care NHS Foundation Trust / North Tees and Hartlepool NHS Foundation Trust / Newcastle Hospitals NHS Foundation Trust | COVID-19 Genomics UK (COG-UK) Consortium | Andrew Nelson; Brendan Payne; Clive Graham; Darren L Smith; Debra Padgett; Edward Barton; Emma Swindells; Garren Scott; Gary Black; Gary Eltringham; Giles S Holt; Greg R Young; Jane Greenaway; Jennifer Collins; John Allan; Joshua Loh; Lynn Dover; Matthew Bashton; Mohammad A Tariq; Paul Baker; Sarah Essex; Steve Liggett; Wen C Yew; Yusri Taha |
| EPI_ISL_1546254, EPI_ISL_1546255, EPI_ISL_1546257, EPI_ISL_1546259, EPI_ISL_1546261, EPI_ISL_1546262, EPI_ISL_1546263, EPI_ISL_1546265, EPI_ISL_1546267, EPI_ISL_1546268, EPI_ISL_1546270, EPI_ISL_1546277, EPI_ISL_1546283, EPI_ISL_1546284, EPI_ISL_1546286, EPI_ISL_1546295, EPI_ISL_1546297, EPI_ISL_1546301, EPI_ISL_1546302, EPI_ISL_1546303, EPI_ISL_1546304, EPI_ISL_1546305, EPI_ISL_1546306, EPI_ISL_1546307, EPI_ISL_1546308, EPI_ISL_1546309, EPI_ISL_1546310, EPI_ISL_1546311, EPI_ISL_1546312, EPI_ISL_1546313, EPI_ISL_1546314, EPI_ISL_1546315, EPI_ISL_1546316, EPI_ISL_1546317, EPI_ISL_1546318, EPI_ISL_1546319, EPI_ISL_1546320, EPI_ISL_1546321, EPI_ISL_1546322, EPI_ISL_1546323, EPI_ISL_1546324, EPI_ISL_1546325, EPI_ISL_1546328, EPI_ISL_1546329, EPI_ISL_1546339, EPI_ISL_1546347, EPI_ISL_1546349, EPI_ISL_1546353, EPI_ISL_1546359, EPI_ISL_1546361, EPI_ISL_1546371, EPI_ISL_1546376, EPI_ISL_1546378, EPI_ISL_1546380, EPI_ISL_1546385, EPI_ISL_1546387, EPI_ISL_1546392, EPI_ISL_1546396, EPI_ISL_1546398, EPI_ISL_1546403, EPI_ISL_1546407, EPI_ISL_1546410, EPI_ISL_1546419, EPI_ISL_1546421, EPI_ISL_1546422, EPI_ISL_1546423, EPI_ISL_1546426, EPI_ISL_1546428, EPI_ISL_1546430, EPI_ISL_1546433, EPI_ISL_1546434, EPI_ISL_1546435, EPI_ISL_1546438, EPI_ISL_1546439, EPI_ISL_1546440, EPI_ISL_1546441, EPI_ISL_1546442, EPI_ISL_1546443, EPI_ISL_1546444, EPI_ISL_1546445, EPI_ISL_1546446, EPI_ISL_1546447, EPI_ISL_1546448, EPI_ISL_1546449, EPI_ISL_1546450, EPI_ISL_1546451, EPI_ISL_1546452, EPI_ISL_1546453, EPI_ISL_1546454, EPI_ISL_1546455, EPI_ISL_1546456, EPI_ISL_1546457, EPI_ISL_1546458, EPI_ISL_1546459, EPI_ISL_1546460, EPI_ISL_1546461, EPI_ISL_1546463, EPI_ISL_1546464, EPI_ISL_1546465, EPI_ISL_1546466, EPI_ISL_1546467, EPI_ISL_1546468, EPI_ISL_1546469, EPI_ISL_1546470, EPI_ISL_1546472, EPI_ISL_1546473, EPI_ISL_1546474, EPI_ISL_1546475, EPI_ISL_1546476, EPI_ISL_1546477, EPI_ISL_1546478, EPI_ISL_1546480, EPI_ISL_1546482, EPI_ISL_1546483, EPI_ISL_1546484, EPI_ISL_1546486, EPI_ISL_1546487, EPI_ISL_1546488, EPI_ISL_1546489, EPI_ISL_1546491, EPI_ISL_1546492, EPI_ISL_1546493, EPI_ISL_1546494, EPI_ISL_1546495, EPI_ISL_1546498, EPI_ISL_1546501, EPI_ISL_1546505, EPI_ISL_1546507, EPI_ISL_1546514, EPI_ISL_1546516, EPI_ISL_1546517, EPI_ISL_1546520, EPI_ISL_1546523, EPI_ISL_1546524, EPI_ISL_1546525, EPI_ISL_1546526, EPI_ISL_1546527, EPI_ISL_1546530, EPI_ISL_1546533, EPI_ISL_1546534, EPI_ISL_1546535, EPI_ISL_1546536, EPI_ISL_1546537, EPI_ISL_1546538, EPI_ISL_1546539, EPI_ISL_1546544, EPI_ISL_1546545, EPI_ISL_1546546, EPI_ISL_1546558, EPI_ISL_1546564, EPI_ISL_1546567, EPI_ISL_1546571, EPI_ISL_1546572, EPI_ISL_1546573, EPI_ISL_1546574, EPI_ISL_1546576, EPI_ISL_1546578, EPI_ISL_1546580, EPI_ISL_1546582, EPI_ISL_1546585, EPI_ISL_1546587, EPI_ISL_1546589, EPI_ISL_1546595, EPI_ISL_1546598, EPI_ISL_1546599, EPI_ISL_1546600, EPI_ISL_1546601, EPI_ISL_1546602, EPI_ISL_1546606, EPI_ISL_1546608, EPI_ISL_1546610, EPI_ISL_1546618, EPI_ISL_1546619, EPI_ISL_1546620, EPI_ISL_1546622, EPI_ISL_1546625, EPI_ISL_1546629, EPI_ISL_1546631, EPI_ISL_1546633, EPI_ISL_1546635, EPI_ISL_1546638, EPI_ISL_1546639, EPI_ISL_1546645, EPI_ISL_1546651, EPI_ISL_1546653, EPI_ISL_1546655, EPI_ISL_1546656, EPI_ISL_1546825, EPI_ISL_1546834, EPI_ISL_1546843, EPI_ISL_1730595, EPI_ISL_1730668, EPI_ISL_1730673, EPI_ISL_1730674, EPI_ISL_1730675, EPI_ISL_1730676, EPI_ISL_1730677, EPI_ISL_1730678, EPI_ISL_1730683, EPI_ISL_1730684, EPI_ISL_1730686, EPI_ISL_1730687, EPI_ISL_1730688, EPI_ISL_1730689 | | | |

| | | | |
|---|---|---|---|
| see above | Originating lab: Wales Specialist Virology Centre Sequencing lab: Pathogen Genomics Unit | Public Health Wales Microbiology Cardiff Wales Specialist Virology Centre | Alec Birchley; Alexander Adams; Amy Gaskin; Angela Marchbank; Bree Gatica-Wilcox; Catherine Moore; Jason Coombes; Joanne Watkins; Joel Southgate; Johnathan Evans; Laura Gifford; Lauren Gilbert; Lee Graham; Malorie Perry; Matthew Bull; Nicole Pacchiarini; Sally Corden; Sara Kumziene-Summerhayes; Sara Rey; Sarah Taylor; Simon Cottrell; Sophie Jones; Tom Connor |
| EPI_ISL_1730532, EPI_ISL_1730533, EPI_ISL_1730535, EPI_ISL_1730536, EPI_ISL_1730543, EPI_ISL_1730545, EPI_ISL_1730546, EPI_ISL_1730547, EPI_ISL_1730551, EPI_ISL_1730552, EPI_ISL_1730558, EPI_ISL_1730562, EPI_ISL_1730569, EPI_ISL_1730570, EPI_ISL_1730582, EPI_ISL_1730583, EPI_ISL_1730584, EPI_ISL_1730586, EPI_ISL_2127688 | | | |
| see above | Oxford Viromics, NDM, University of Oxford; Oxford University Hospitals; Basingstoke and North Hampshire Hospital | COVID-19 Genomics UK (COG-UK) Consortium | Alex Mobbs; Amy Trebes; Anita Justice; Catrin Moore; Christophe Fraser; David Bonsall; David Buck; Emma Wise; George Macintyre; Jessica Lynch; John Todd; Mariateresa de Cesare; Matilde Mori; Monique Andersson; Nathan Moore; Nick Cortes; Robert Shaw; Stephen Kidd; Tanya Golubchik; Timothy Peto |
| EPI_ISL_2080492 | POOLE MICROBIOLOGY LABORATORY | COVID-19 Genomics UK (COG-UK) Consortium | PHE Covid Sequencing Team |
| EPI_ISL_1978945 | Quadram Institute Bioscience | COVID-19 Genomics UK (COG-UK) Consortium | Alexander J Trotter; Alison E. Mather; Alp Aydin; Ana P. Tedim; Anastasia Kolyva; Andrew Bell; Andrew J. Page; Claire Stuart; Dave J. Baker; Gemma L. Kay; John Wain; Justin O'Grady; Leonardo de Oliveira Martins; Lizzie Meadows; Maria Diaz; Mark Webber; Muhammed Yasir; Nabil-Fareed Alikhan; Ngozi Elumogo; Nicholas M. Thomson; Rachael Stanley; Rachel Gilroy; Reenesh Prakash; Samir Dervisevic; Samuel Bloomfield; Steven Rudder; Thanh Le-Viet |
| EPI_ISL_1637235, EPI_ISL_1637236, EPI_ISL_1730420, EPI_ISL_1730421, EPI_ISL_1730422, EPI_ISL_1833025, EPI_ISL_1833026, EPI_ISL_1833027 | | | |
| see above | Queens Medical Centre, Clinical Microbiology Department / DeepSeq Nottingham | COVID-19 Genomics UK (COG-UK) Consortium | Christopher Moore; Emily Park; Fei Sang; Gemma Clark; Hannah Howson-Wells; Jack Hill; Jayasree Dey; Johnny Debebe; Jonathan Ball; Joseph Chappell; Manjinder Khakh; Matthew Carlisle; Matthew Loose; Michelle M Lister; Nadine Holmes; Patrick McClure; Theocharis Tsoleridis; Timothy Byaruhanga; Vicki M Fleming; Victoria Wright; Wendy Smith |
| EPI_ISL_1504930, EPI_ISL_1504944, EPI_ISL_1504953, EPI_ISL_1504954, EPI_ISL_1504960, EPI_ISL_1505023, EPI_ISL_1505051, EPI_ISL_1505059, EPI_ISL_1505075, EPI_ISL_1505078, EPI_ISL_1505135, EPI_ISL_1505146, EPI_ISL_1505459, EPI_ISL_1505460, EPI_ISL_1505467, EPI_ISL_1505471, EPI_ISL_1505472, EPI_ISL_1505489, EPI_ISL_1505493, EPI_ISL_1505509, EPI_ISL_1505541, EPI_ISL_1505543, EPI_ISL_1505548, EPI_ISL_1505551, EPI_ISL_1505563, EPI_ISL_1505578, EPI_ISL_1505594, EPI_ISL_1505595, EPI_ISL_1505610, EPI_ISL_1505614, EPI_ISL_1505631, EPI_ISL_1505656, EPI_ISL_1505657, EPI_ISL_1505658, EPI_ISL_1505677, EPI_ISL_1505693, EPI_ISL_1505712, EPI_ISL_1505716, EPI_ISL_1505723, EPI_ISL_1505727, EPI_ISL_1505742, EPI_ISL_1505743, EPI_ISL_1505756, EPI_ISL_1505788, EPI_ISL_1505794, EPI_ISL_1506425, EPI_ISL_1518073, EPI_ISL_1518078, EPI_ISL_1518082, EPI_ISL_1518097, EPI_ISL_1518112, EPI_ISL_1518114, EPI_ISL_1518130, EPI_ISL_1518152, EPI_ISL_1518183, EPI_ISL_1518195, EPI_ISL_1518196, EPI_ISL_1518209, EPI_ISL_1518215, EPI_ISL_1518216, EPI_ISL_1518217, EPI_ISL_1518232, EPI_ISL_1518241, EPI_ISL_1518246, EPI_ISL_1518256, EPI_ISL_1518280, EPI_ISL_1518288, EPI_ISL_1518319, EPI_ISL_1518342, EPI_ISL_1518348, EPI_ISL_1518350, EPI_ISL_1518373, EPI_ISL_1518679, EPI_ISL_1518681, EPI_ISL_1518682, EPI_ISL_1519275, EPI_ISL_1519284, EPI_ISL_1519287, EPI_ISL_1535612, EPI_ISL_1535627, EPI_ISL_1535628, EPI_ISL_1535632, EPI_ISL_1535633, EPI_ISL_1535642, EPI_ISL_1535664, EPI_ISL_1535666, EPI_ISL_1535684, EPI_ISL_1535701, EPI_ISL_1535707, EPI_ISL_1535710, EPI_ISL_1535720, EPI_ISL_1535724, EPI_ISL_1535739, EPI_ISL_1535753, EPI_ISL_1535758, EPI_ISL_1535759, EPI_ISL_1535761, EPI_ISL_1535769, EPI_ISL_1535772, EPI_ISL_1535788, EPI_ISL_1535789, EPI_ISL_1535792, EPI_ISL_1535804, EPI_ISL_1535806, EPI_ISL_1535820, EPI_ISL_1535821, EPI_ISL_1535825, EPI_ISL_1535826, EPI_ISL_1535856, EPI_ISL_1535862, EPI_ISL_1535872, EPI_ISL_1535881, EPI_ISL_1535898, EPI_ISL_1536194, EPI_ISL_1536217, EPI_ISL_1536237, EPI_ISL_1536267, EPI_ISL_1536287, EPI_ISL_1536322, EPI_ISL_1536351, EPI_ISL_1536362, EPI_ISL_1536370, EPI_ISL_1536373, EPI_ISL_1536395, EPI_ISL_1536421, EPI_ISL_1536422, EPI_ISL_1536433, EPI_ISL_1536458, EPI_ISL_1536486, EPI_ISL_1536520, EPI_ISL_1536525, EPI_ISL_1536533, EPI_ISL_1536537, EPI_ISL_1536862, EPI_ISL_1536863, EPI_ISL_1536898, EPI_ISL_1536904, EPI_ISL_1536920, EPI_ISL_1536934, EPI_ISL_1536996, EPI_ISL_1536997, EPI_ISL_1537005, EPI_ISL_1537025, EPI_ISL_1537031, EPI_ISL_1537049, EPI_ISL_1537071, EPI_ISL_1537091, EPI_ISL_1537092, EPI_ISL_1537101, EPI_ISL_1537106, EPI_ISL_1537122, EPI_ISL_1537124, EPI_ISL_1537126, EPI_ISL_1537131, EPI_ISL_1537134, EPI_ISL_1537135, EPI_ISL_1537136, EPI_ISL_1537139, EPI_ISL_1537140, EPI_ISL_1544203, EPI_ISL_1544210, EPI_ISL_1544218, EPI_ISL_1544222, EPI_ISL_1544223, EPI_ISL_1544224, EPI_ISL_1544227, EPI_ISL_1544229, EPI_ISL_1544239, EPI_ISL_1544240, EPI_ISL_1544248, EPI_ISL_1544259, EPI_ISL_1544260, EPI_ISL_1544261, EPI_ISL_1544264, EPI_ISL_1544267, EPI_ISL_1544274, EPI_ISL_1544281, EPI_ISL_1544288, EPI_ISL_1544293, EPI_ISL_1544294, EPI_ISL_1544295, EPI_ISL_1544296, EPI_ISL_1544299, EPI_ISL_1544308, EPI_ISL_1544309, EPI_ISL_1544314, EPI_ISL_1544317, EPI_ISL_1544319, EPI_ISL_1544331, EPI_ISL_1544339, EPI_ISL_1544349, EPI_ISL_1544354, EPI_ISL_1544370, EPI_ISL_1544382, EPI_ISL_1544385, EPI_ISL_1544388, EPI_ISL_1544390, EPI_ISL_1544396, EPI_ISL_1544397, EPI_ISL_1544407, EPI_ISL_1544409, EPI_ISL_1544414, EPI_ISL_1544415, EPI_ISL_1544416, EPI_ISL_1544419, EPI_ISL_1544424, EPI_ISL_1544425, EPI_ISL_1544426, EPI_ISL_1544432, EPI_ISL_1544433, EPI_ISL_1544437, EPI_ISL_1544453, EPI_ISL_1544462, EPI_ISL_1544464, EPI_ISL_1544465, EPI_ISL_1544466, EPI_ISL_1544473, EPI_ISL_1544474, EPI_ISL_1544476, EPI_ISL_1544480, EPI_ISL_1544485, EPI_ISL_1544487, EPI_ISL_1544490, EPI_ISL_1544493, EPI_ISL_1564356, EPI_ISL_1564357, EPI_ISL_1564385, EPI_ISL_1564401, EPI_ISL_1564419, EPI_ISL_1564438, EPI_ISL_1564489, EPI_ISL_1564530, EPI_ISL_1564536, EPI_ISL_1564558, EPI_ISL_1564559, EPI_ISL_1564584, EPI_ISL_1564599, EPI_ISL_1564684, EPI_ISL_1564706, EPI_ISL_1564764, EPI_ISL_1564765, EPI_ISL_1564767, EPI_ISL_1564768, EPI_ISL_1564769, EPI_ISL_1564786, EPI_ISL_1564788, EPI_ISL_1564789, EPI_ISL_1564791, EPI_ISL_1564792, EPI_ISL_1564793, EPI_ISL_1564794, EPI_ISL_1564795, EPI_ISL_1564798, EPI_ISL_1564800, EPI_ISL_1564802, EPI_ISL_1564803, EPI_ISL_1564810, EPI_ISL_1564830, EPI_ISL_1564856, EPI_ISL_1564860, EPI_ISL_1564899, EPI_ISL_1564904, EPI_ISL_1564906, EPI_ISL_1564926, EPI_ISL_1564934, EPI_ISL_1564940, EPI_ISL_1564949, EPI_ISL_1564978, EPI_ISL_1565001, EPI_ISL_1565006, EPI_ISL_1565047, EPI_ISL_1631763, EPI_ISL_1631766, EPI_ISL_1631769, EPI_ISL_1631789, EPI_ISL_1631809, EPI_ISL_1631810, EPI_ISL_1631829, EPI_ISL_1631834, EPI_ISL_1631839, EPI_ISL_1631844, EPI_ISL_1631847, EPI_ISL_1631850, EPI_ISL_1631871, EPI_ISL_1631878, EPI_ISL_1631910, EPI_ISL_1631921, EPI_ISL_1631928, EPI_ISL_1631973, EPI_ISL_1631986 | | | |
| see above | Randox Laboratories | Wellcome Sanger Institute for the COVID-19 Genomics UK (COG-UK) Consortium | Cordelia Langford; David K. Jackson; Dominic Kwiatkowski; Ewan Harrison; Ian Johnston; Jeffrey Barrett; John Sillitoe on behalf of the Wellcome Sanger Institute COVID-19 Surveillance Team; Randox Laboratories and Alex Alderton; Roberto Amato; Sonia Goncalves |
| EPI_ISL_1976445, EPI_ISL_1976447, EPI_ISL_1976448, EPI_ISL_1976450, EPI_ISL_1976452, EPI_ISL_1976453, EPI_ISL_1976455, EPI_ISL_1976457, EPI_ISL_1976459, EPI_ISL_1976460, EPI_ISL_1976462, EPI_ISL_1976503, EPI_ISL_1976506, EPI_ISL_1976508, EPI_ISL_1976510, EPI_ISL_1976538, EPI_ISL_1976540, EPI_ISL_1976542, EPI_ISL_1976552, EPI_ISL_1976554, EPI_ISL_1976619, EPI_ISL_1976621, EPI_ISL_1976623, EPI_ISL_1976626, EPI_ISL_1976628, EPI_ISL_1976650 | | | |
| see above | Regional Virus Laboratory, Belfast Health and Social Care Trust | COVID-19 Genomics UK (COG-UK) Consortium | Alison Watt; Ciara Cox; Conall McCaughey; David Simpson; Derek Fairley; James McKenna; Mairead Connor; Susan Feeney; Tanya Curran; Zoltan Molnar |
| EPI_ISL_1833814, EPI_ISL_1833815, EPI_ISL_1833822, EPI_ISL_1833823 | St George's University Hospitals NHS Foundation Trust | COVID-19 Genomics UK (COG-UK) Consortium | Adam Witney; Cassie Pope; Irene Monahan; Joshua Taylor; Ken Laing; NgeeKeong Tan |
| EPI_ISL_1474892, EPI_ISL_1546229, EPI_ISL_1546230, EPI_ISL_1546231, EPI_ISL_1546232, EPI_ISL_1546233, EPI_ISL_1546234, EPI_ISL_1546235, EPI_ISL_1546236, EPI_ISL_1546237, EPI_ISL_1546238, EPI_ISL_1637001, EPI_ISL_1637014, EPI_ISL_1637017, EPI_ISL_1637019, EPI_ISL_1637092, EPI_ISL_1637178, EPI_ISL_1637179, EPI_ISL_1637180, EPI_ISL_1730338, EPI_ISL_1730375, EPI_ISL_1730380 | | | |
| see above | University College London, Great Ormond Street Hospital for Children NHS Foundation Trust, Imperial College Healthcare NHS Trust | COVID-19 Genomics UK (COG-UK) Consortium | Alison Holmes; Charlotte Williams; Helena Tutill; Jacqueline Findlay; James Price; Judith Breuer; Julianne Brown; Kathryn Harris; Leysa Forrest; Mark Kristiansen; Paola Niola; Paola Resende Silva; Patricia Dyal; Paul Randell; Rachel Williams; Samuel Weeks; Sergi Castellano; Sunando Roy; Tony Brooks; Yasmin Panchbhaya |
| EPI_ISL_1636713, EPI_ISL_1636715, EPI_ISL_1636717, EPI_ISL_1636718, EPI_ISL_1636719, EPI_ISL_1636720, EPI_ISL_1636721, EPI_ISL_1636722, EPI_ISL_1636723, EPI_ISL_1636724 | | | |
| see above | University Hospitals Of Leicester NHS Trust and DeepSeq Nottingham | COVID-19 Genomics UK (COG-UK) Consortium | Christopher Holmes; Christopher Moore; Fei Sang; Joeseph Chappell; Johnny Debebe; Jonathan Ball; Julian Tang; Karlie Fallon; Matthew Carlisle; Matthew Loose; Nadine Holmes; Patrick McClure; Paul Bird; Thomas Helmer; Victoria Wright |
| EPI_ISL_1636531, EPI_ISL_1636533, EPI_ISL_1636534, EPI_ISL_1636535, EPI_ISL_1636536, EPI_ISL_1636537, EPI_ISL_1636539, EPI_ISL_1636540, EPI_ISL_1636541, EPI_ISL_1636542, EPI_ISL_1636543, EPI_ISL_1636544, EPI_ISL_1636545, EPI_ISL_1636546, EPI_ISL_1636547, EPI_ISL_1636548, EPI_ISL_1636550, EPI_ISL_1636558, EPI_ISL_1636559, EPI_ISL_1636560, EPI_ISL_1636568, EPI_ISL_1636570, EPI_ISL_1636571, EPI_ISL_1636572, EPI_ISL_1636573, EPI_ISL_1636576, EPI_ISL_1636578, EPI_ISL_1636580, EPI_ISL_1636599 | | | |
| see above | University of Birmingham | COVID-19 Genomics UK (COG-UK) Consortium | Alex Richter; Andrew Bosworth; Andrew Bosworth. Queen Elizabeth Hospital: Anna Casey; Andrew D Beggs PHE Heartlands Lab: Husam Osman; Charlotte Poxon; Institute of Microbiology; Joanne Stockton; Josh Quick; Kasun Wanigasooriya; Mike Kidd; Nicholas Loman. University of Birmingham Testing Laboratory: Celina M Whalley; Oliver Pickles; Radoslaw Poplawski; Samuel Nicholls; University of Birmingham: Claire McMurray; Will Rowe |
| EPI_ISL_1730144, EPI_ISL_1730153, EPI_ISL_1730154, EPI_ISL_1730155 | University of Exeter | COVID-19 Genomics UK (COG-UK) Consortium | Aaron Jeffries; Audrey Farbos; Ben Temperton; Jane Masoli; Joanna Warwick-Dugdale; Michelle Michelsen; Robyn Manley; Stephen Michell |
| EPI_ISL_1636674, EPI_ISL_1636675, EPI_ISL_1636676, EPI_ISL_1636677, EPI_ISL_1636691, EPI_ISL_1636694, EPI_ISL_1636703, EPI_ISL_1730270, EPI_ISL_1730271, EPI_ISL_1730272, EPI_ISL_1730273, EPI_ISL_1730274, EPI_ISL_1730275, EPI_ISL_1730276, EPI_ISL_1730277, EPI_ISL_1730278, EPI_ISL_1730279, EPI_ISL_1730280, EPI_ISL_1730281, EPI_ISL_1730282, EPI_ISL_1730283, EPI_ISL_1730284, EPI_ISL_1730288, EPI_ISL_1730289, EPI_ISL_1730290, EPI_ISL_1730291, EPI_ISL_1730292, EPI_ISL_1730293, EPI_ISL_1730294, EPI_ISL_1974896, EPI_ISL_1974897, EPI_ISL_1974911, EPI_ISL_2125691, EPI_ISL_2125693, EPI_ISL_2125846, EPI_ISL_2125848, EPI_ISL_2125850, EPI_ISL_2125868 | | | |
| see above | Virology Department, Royal Infirmary of Edinburgh, NHS Lothian / School of Biological Sciences, University of Edinburgh | COVID-19 Genomics UK (COG-UK) Consortium | Colquhoun R; Cotton S; Dewar R; Gallagher A; Hill V; Jackson B; Maloney D; McCrone JT; McHugh M; O'Toole Á; Rambaut A; Rooke S; Scher E; Templeton K; Yu X |
| EPI_ISL_1547313, EPI_ISL_1547319, EPI_ISL_1547320, EPI_ISL_1547323, EPI_ISL_1547324, EPI_ISL_1547328, EPI_ISL_1547329, EPI_ISL_1547331, EPI_ISL_1547333, EPI_ISL_1547334, EPI_ISL_1547335, EPI_ISL_1547338, EPI_ISL_1547339, EPI_ISL_1547344, EPI_ISL_1547345, EPI_ISL_1547348, EPI_ISL_1547350, EPI_ISL_1638041, EPI_ISL_1638051, EPI_ISL_1638058, EPI_ISL_1638062, EPI_ISL_1638063, EPI_ISL_1638064, EPI_ISL_1638069, EPI_ISL_1638070, EPI_ISL_1638079, EPI_ISL_1730948, EPI_ISL_1730955, EPI_ISL_1730974, EPI_ISL_1730979, EPI_ISL_1730990, EPI_ISL_1731018 | | | |
| see above | Virology Department, Sheffield Teaching Hospitals NHS Foundation Trust/Department of Infection, Immunity and Cardiovascular Disease, The Medical School, University of Sheffield | COVID-19 Genomics UK (COG-UK) Consortium | Adri Angyal; Alex Keeley; Benjamin Lindsey; Cariad Evans; Danielle Groves; Dave Partridge; Katie Johnson; Laura Carrilero; Luke Green; Matthew Parker; Matthew Wyles; Mehmet Yavuz; Mohammad Raza; Nikki Smith; Paul Parsons; Rachel Tucker; Rebecca Brown; Thushan de Silva |
| EPI_ISL_1546252, EPI_ISL_1546256, EPI_ISL_1546269, EPI_ISL_1546272, EPI_ISL_1546343, EPI_ISL_1546346, EPI_ISL_1546357, EPI_ISL_1546386, EPI_ISL_1546420, EPI_ISL_1546424, EPI_ISL_1546425, EPI_ISL_1546427, EPI_ISL_1546462, EPI_ISL_1546471, EPI_ISL_1546479, EPI_ISL_1546481, EPI_ISL_1546485, EPI_ISL_1546490, EPI_ISL_1546500, EPI_ISL_1546521, EPI_ISL_1546522, EPI_ISL_1546528, EPI_ISL_1546529, EPI_ISL_1546531, EPI_ISL_1546540, EPI_ISL_1546550, EPI_ISL_1546551, EPI_ISL_1546554, EPI_ISL_1546556, EPI_ISL_1546563, EPI_ISL_1546575, EPI_ISL_1546584, EPI_ISL_1546586, EPI_ISL_1546630, EPI_ISL_1546646, EPI_ISL_1546647, EPI_ISL_1546650, EPI_ISL_1546658, EPI_ISL_1546841, EPI_ISL_1637456, EPI_ISL_1637457, EPI_ISL_1637480, EPI_ISL_1637585, EPI_ISL_1637586, EPI_ISL_1637588, EPI_ISL_1637592, EPI_ISL_1637596, EPI_ISL_1637654, EPI_ISL_1637658, EPI_ISL_1637661, EPI_ISL_1637665, EPI_ISL_1637666, EPI_ISL_1637691, EPI_ISL_1637789, EPI_ISL_1637790, EPI_ISL_1833196 | | | |
| see above | Wales Specialist Virology Centre Sequencing lab: Pathogen Genomics Unit | Public Health Wales Microbiology Cardiff Wales Specialist Virology Centre | Alec Birchley; Alexander Adams; Amy Gaskin; Angela Marchbank; Bree Gatica-Wilcox; Catherine Moore; Jason Coombes; Joanne Watkins; Joel Southgate; Johnathan Evans; Laura Gifford; Lauren Gilbert; Lee Graham; Malorie Perry; Matthew Bull; Nicole Pacchiarini; Sally Corden; Sara Kumziene-Summerhayes; Sara Rey; Sarah Taylor; Simon Cottrell; Sophie Jones; Tom Connor |
| EPI_ISL_1832277, EPI_ISL_1832278, EPI_ISL_1832279, EPI_ISL_1832280, EPI_ISL_1832281, EPI_ISL_1832282, EPI_ISL_1832283, EPI_ISL_1832285, EPI_ISL_1832286, EPI_ISL_1832287 | | | |
| see above | West of Scotland Specialist Virology Centre, NHSGGC / MRC-University of Glasgow Centre for Virus Research | COVID-19 Genomics UK (COG-UK) Consortium | Alasdair MacLean; Alice Broos; Ana da Silva Filipe; Antonia Ho; Daniel Mair; David L Robertson; Emma Thomson; Guy Mollett; James Shepherd; Jenna Nichols; Joseph Hughes; Kathy Li; Kathy Smollett; Kyriaki Nomikou; Lily Tong; Matthew Holden; Natasha Johnson; Rachel Blacow; Richard Orton; Rory Gunson; Sarah McDonald; Sharif Shaaban; Sreenu Vattipally; Stephen Carmichael |

We gratefully acknowledge the following Authors from the Originating laboratories responsible for obtaining the specimens, as well as the Submitting laboratories where the genome data were generated and shared via GISAID, on which this research is based.

All Submitters of data may be contacted directly via www.gisaid.org

Authors are sorted alphabetically.

| Accession ID | Originating Laboratory | Submitting Laboratory | Authors |
|---|---|---|---|
| EPI_ISL_1730317, EPI_ISL_1730320, EPI_ISL_1730321, EPI_ISL_1730322, EPI_ISL_1730323, EPI_ISL_1832547 | Barts Health NHS Trust | COVID-19 Genomics UK (COG-UK) Consortium | BROAD; Beatrix; CUTINO-MOGUEL; Claire; David; Dola; HARRINGTON; KELE; KULASEGARAN-SHYLINI; Maria-Teresa; OWOYEMI; Raghavendran |
| EPI_ISL_1638255, EPI_ISL_1638257, EPI_ISL_1638258, EPI_ISL_1833927, EPI_ISL_1833947 | Bioinformatics and Biostatistics Lab, Advanced Sequencing Facility | COVID-19 Genomics UK (COG-UK) Consortium | Aengus Stewart; Chelsea Sawyer; Harshil Patel; Jerome Nicod; Laura Cubitt; Margaret Crawford |
| EPI_ISL_1730822, EPI_ISL_1730823, EPI_ISL_1730824, EPI_ISL_1730825, EPI_ISL_1730826, EPI_ISL_1730827, EPI_ISL_1730828, EPI_ISL_1730829, EPI_ISL_1730830, EPI_ISL_1730889, EPI_ISL_1730890, EPI_ISL_1730891, EPI_ISL_1730892, EPI_ISL_1730893, EPI_ISL_1730894, EPI_ISL_1730895, EPI_ISL_1730897, EPI_ISL_1730900, EPI_ISL_1730901, EPI_ISL_1730902, EPI_ISL_1730903, EPI_ISL_1730905, EPI_ISL_1730906, EPI_ISL_1730907, EPI_ISL_1730908, EPI_ISL_1730914, EPI_ISL_1730915, EPI_ISL_1730916, EPI_ISL_1730925 | | | |
| see above | Centre for Enzyme Innovation, University of Portsmouth / Translational Research Laboratory, Portsmouth Hospitals NHS Trust | COVID-19 Genomics UK (COG-UK) Consortium | Angela Beckett; Christopher Fearn; Kate Cook; Katie Loveson; Salman Goudarzi; Samuel Robson; Scott Elliott; Sharon Glaysher |
| EPI_ISL_1636622, EPI_ISL_1636623, EPI_ISL_1636624, EPI_ISL_1730043, EPI_ISL_1730045, EPI_ISL_1730046, EPI_ISL_1730047, EPI_ISL_1730048, EPI_ISL_1730049, EPI_ISL_1730050, EPI_ISL_1730051, EPI_ISL_1730052, EPI_ISL_1730053, EPI_ISL_1730054, EPI_ISL_1730056, EPI_ISL_1730058, EPI_ISL_1730060, EPI_ISL_1730061, EPI_ISL_1730062, EPI_ISL_1730063, EPI_ISL_1730065, EPI_ISL_1730066, EPI_ISL_1730068, EPI_ISL_1832184, EPI_ISL_1832187, EPI_ISL_1832189 | | | |
| see above | Department of Pathology, University of Cambridge | COVID-19 Genomics UK (COG-UK) Consortium | Aminu S. Jahun; Ian Goodfellow; Iliana Georgana; Martin D. Curran; Myra Hosmillo; Rhys Izuagbe; Surendra Parmar; William L. Hamilton; Yasmin Chaudhry |
| EPI_ISL_1831313, EPI_ISL_1831315, EPI_ISL_1831317, EPI_ISL_1831322, EPI_ISL_1831323, EPI_ISL_1831326, EPI_ISL_1831329, EPI_ISL_1831330, EPI_ISL_1831334, EPI_ISL_1831341, EPI_ISL_1831342, EPI_ISL_1831343, EPI_ISL_1831344, EPI_ISL_1831345, EPI_ISL_1831348, EPI_ISL_1831349, EPI_ISL_1831350, EPI_ISL_1831354, EPI_ISL_1831357, EPI_ISL_1831360, EPI_ISL_1831362, EPI_ISL_1831363, EPI_ISL_1831364, EPI_ISL_1831365, EPI_ISL_1831366, EPI_ISL_1831376, EPI_ISL_1831379, EPI_ISL_1831381, EPI_ISL_1831382, EPI_ISL_1831383, EPI_ISL_1831385 | | | |
| see above | Health Services Laboratories | Wellcome Sanger Institute for the COVID-19 Genomics UK (COG-UK) Consortium | Cordelia Langford; David K. Jackson; Dominic Kwiatkowski; Ewan Harrison; Health Services Laboratories and Alex Alderton; Ian Johnston; Jeffrey Barrett; John Sillitoe on behalf of the Wellcome Sanger Institute COVID-19 Surveillance Team; Roberto Amato; Sonia Goncalves |
| EPI_ISL_1564333, EPI_ISL_1564340, EPI_ISL_1564342, EPI_ISL_1564347, EPI_ISL_1564352, EPI_ISL_1564353, EPI_ISL_1564358, EPI_ISL_1564359, EPI_ISL_1564361, EPI_ISL_1564363, EPI_ISL_1564367, EPI_ISL_1564368, EPI_ISL_1564369, EPI_ISL_1564381, EPI_ISL_1564382, EPI_ISL_1564388, EPI_ISL_1564390, EPI_ISL_1564394, EPI_ISL_1564395, EPI_ISL_1564402, EPI_ISL_1564406, EPI_ISL_1564408, EPI_ISL_1564410, EPI_ISL_1564411, EPI_ISL_1564414, EPI_ISL_1564415, EPI_ISL_1564416, EPI_ISL_1564417, EPI_ISL_1564418, EPI_ISL_1564424, EPI_ISL_1564427, EPI_ISL_1564429, EPI_ISL_1564432, EPI_ISL_1564433, EPI_ISL_1564434, EPI_ISL_1564435, EPI_ISL_1564439, EPI_ISL_1564440, EPI_ISL_1564442, EPI_ISL_1564444, EPI_ISL_1564445, EPI_ISL_1564446, EPI_ISL_1564448, EPI_ISL_1564451, EPI_ISL_1564453, EPI_ISL_1564455, EPI_ISL_1564457, EPI_ISL_1564458, EPI_ISL_1564459, EPI_ISL_1564461, EPI_ISL_1564463, EPI_ISL_1564466, EPI_ISL_1564469, EPI_ISL_1564473, EPI_ISL_1564476, EPI_ISL_1564478, EPI_ISL_1564483, EPI_ISL_1564485, EPI_ISL_1564486, EPI_ISL_1564490, EPI_ISL_1564501, EPI_ISL_1564504, EPI_ISL_1564507, EPI_ISL_1564509, EPI_ISL_1564515, EPI_ISL_1564517, EPI_ISL_1564519, EPI_ISL_1564521, EPI_ISL_1564526, EPI_ISL_1564528, EPI_ISL_1564534, EPI_ISL_1564539, EPI_ISL_1564542, EPI_ISL_1564547, EPI_ISL_1564549, EPI_ISL_1564551, EPI_ISL_1564552, EPI_ISL_1564554, EPI_ISL_1564568, EPI_ISL_1564569, EPI_ISL_1564571, EPI_ISL_1564572, EPI_ISL_1564573, EPI_ISL_1564574, EPI_ISL_1564578, EPI_ISL_1564580, EPI_ISL_1564589, EPI_ISL_1564592, EPI_ISL_1564594, EPI_ISL_1564627, EPI_ISL_1564628, EPI_ISL_1564634, EPI_ISL_1564637, EPI_ISL_1564638, EPI_ISL_1564640, EPI_ISL_1564642, EPI_ISL_1564643, EPI_ISL_1564647, EPI_ISL_1564648, EPI_ISL_1564652, EPI_ISL_1564656, EPI_ISL_1564664, EPI_ISL_1564665, EPI_ISL_1564666, EPI_ISL_1564671, EPI_ISL_1564678, EPI_ISL_1564686, EPI_ISL_1564688, EPI_ISL_1564691, EPI_ISL_1564692, EPI_ISL_1564703, EPI_ISL_1564708, EPI_ISL_1564713, EPI_ISL_1564714, EPI_ISL_1564718, EPI_ISL_1564719, EPI_ISL_1564720, EPI_ISL_1564724, EPI_ISL_1564726, EPI_ISL_1564729, EPI_ISL_1564730, EPI_ISL_1564733, EPI_ISL_1564735, EPI_ISL_1564736, EPI_ISL_1564737, EPI_ISL_1564738, EPI_ISL_1564742, EPI_ISL_1564743, EPI_ISL_1564745, EPI_ISL_1564747, EPI_ISL_1564748, EPI_ISL_1564750, EPI_ISL_1564751, EPI_ISL_1564752, EPI_ISL_1564753, EPI_ISL_1564760, EPI_ISL_1564762, EPI_ISL_1564763, EPI_ISL_1565338, EPI_ISL_1584386, EPI_ISL_1584528, EPI_ISL_1594714, EPI_ISL_1594715, EPI_ISL_1594716, EPI_ISL_1594717, EPI_ISL_1594724, EPI_ISL_1594725, EPI_ISL_1594732, EPI_ISL_1594733, EPI_ISL_1594734, EPI_ISL_1594737, EPI_ISL_1594738, EPI_ISL_1594739, EPI_ISL_1594740, EPI_ISL_1594741, EPI_ISL_1594742, EPI_ISL_1594747, EPI_ISL_1594750, EPI_ISL_1594762, EPI_ISL_1594763, EPI_ISL_1594764, EPI_ISL_1594766, EPI_ISL_1594767, EPI_ISL_1594768, EPI_ISL_1594773, EPI_ISL_1594774, EPI_ISL_1594775, EPI_ISL_1594776, EPI_ISL_1594778, EPI_ISL_1594780, EPI_ISL_1594781, EPI_ISL_1594782, EPI_ISL_1594784, EPI_ISL_1594786, EPI_ISL_1594791, EPI_ISL_1594792, EPI_ISL_1594800, EPI_ISL_1594802, EPI_ISL_1594804, EPI_ISL_1594811, EPI_ISL_1594812, EPI_ISL_1594813, EPI_ISL_1594817, EPI_ISL_1594818, EPI_ISL_1594822, EPI_ISL_1594825, EPI_ISL_1594827, EPI_ISL_1594828, EPI_ISL_1594830, EPI_ISL_1594834, EPI_ISL_1594835, EPI_ISL_1594838, EPI_ISL_1594841, EPI_ISL_1594846, EPI_ISL_1594849, EPI_ISL_1594850, EPI_ISL_1594853, EPI_ISL_1594855, EPI_ISL_1594860, EPI_ISL_1594863, EPI_ISL_1594864, EPI_ISL_1594867, EPI_ISL_1594868, EPI_ISL_1594869, EPI_ISL_1594870, EPI_ISL_1594872, EPI_ISL_1594875, EPI_ISL_1594879, EPI_ISL_1594881, EPI_ISL_1594882, EPI_ISL_1594883, EPI_ISL_1594884, EPI_ISL_1594885, EPI_ISL_1594886, EPI_ISL_1594887, EPI_ISL_1594889, EPI_ISL_1594890, EPI_ISL_1594891, EPI_ISL_1594893, EPI_ISL_1594894, EPI_ISL_1594897, EPI_ISL_1594902, EPI_ISL_1594905, EPI_ISL_1594906, EPI_ISL_1594907, EPI_ISL_1594908, EPI_ISL_1594914, EPI_ISL_1594917, EPI_ISL_1594919, EPI_ISL_1594921, EPI_ISL_1594926, EPI_ISL_1594931, EPI_ISL_1594935, EPI_ISL_1594937, EPI_ISL_1594941, EPI_ISL_1594944, EPI_ISL_1594946, EPI_ISL_1594949, EPI_ISL_1594950, EPI_ISL_1594951, EPI_ISL_1594954, EPI_ISL_1594955, EPI_ISL_1594958, EPI_ISL_1594961, EPI_ISL_1594963, EPI_ISL_1594964, EPI_ISL_1594965, EPI_ISL_1594973, EPI_ISL_1594974, EPI_ISL_1594976, EPI_ISL_1594977, EPI_ISL_1594982, EPI_ISL_1594985, EPI_ISL_1594987, EPI_ISL_1594989, EPI_ISL_1594990, EPI_ISL_1594992, EPI_ISL_1594993, EPI_ISL_1615688, EPI_ISL_1615698, EPI_ISL_1615706, EPI_ISL_1615707, EPI_ISL_1615716, EPI_ISL_1615729, EPI_ISL_1615733, EPI_ISL_1615736, EPI_ISL_1615747, EPI_ISL_1615749, EPI_ISL_1615757, EPI_ISL_1615759, EPI_ISL_1615763, EPI_ISL_1615778, EPI_ISL_1615783, EPI_ISL_1615790, EPI_ISL_1615792, EPI_ISL_1615794, EPI_ISL_1615796, EPI_ISL_1615805, EPI_ISL_1615817, EPI_ISL_1615837, EPI_ISL_1615847, EPI_ISL_1615855, EPI_ISL_1615858, EPI_ISL_1615864, EPI_ISL_1615872, EPI_ISL_1615881, EPI_ISL_1615882, EPI_ISL_1615885, EPI_ISL_1615899, EPI_ISL_1615905, EPI_ISL_1615918, EPI_ISL_1616267, EPI_ISL_1616283, EPI_ISL_1630822, EPI_ISL_1630823, EPI_ISL_1630824, EPI_ISL_1630825, EPI_ISL_1630826, EPI_ISL_1630827, EPI_ISL_1630828, EPI_ISL_1630829, EPI_ISL_1630831, EPI_ISL_1630832, EPI_ISL_1630833, EPI_ISL_1630835, EPI_ISL_1630836, EPI_ISL_1630837, EPI_ISL_1630838, EPI_ISL_1630839, EPI_ISL_1630840, EPI_ISL_1630841, EPI_ISL_1630842, EPI_ISL_1630843, EPI_ISL_1630844, EPI_ISL_1630845, EPI_ISL_1630849, EPI_ISL_1630851, EPI_ISL_1630852, EPI_ISL_1630854, EPI_ISL_1630856, EPI_ISL_1630857, EPI_ISL_1630858, EPI_ISL_1630859, EPI_ISL_1630860, EPI_ISL_1630861, EPI_ISL_1630862, EPI_ISL_1630863, EPI_ISL_1630864, EPI_ISL_1630865, EPI_ISL_1630866, EPI_ISL_1630867, EPI_ISL_1630868, EPI_ISL_1630869, EPI_ISL_1630870, EPI_ISL_1630873, EPI_ISL_1630874, EPI_ISL_1630875, EPI_ISL_1630876, EPI_ISL_1630877, EPI_ISL_1630878, EPI_ISL_1630880, EPI_ISL_1630881, EPI_ISL_1630882, EPI_ISL_1630883, EPI_ISL_1630884, EPI_ISL_1630885, EPI_ISL_1630886, EPI_ISL_1630887, EPI_ISL_1630888, EPI_ISL_1630890, EPI_ISL_1630891, EPI_ISL_1630892, EPI_ISL_1630893, EPI_ISL_1630894, EPI_ISL_1630895, EPI_ISL_1630897, EPI_ISL_1630898, EPI_ISL_1630899, EPI_ISL_1630900, EPI_ISL_1630901, EPI_ISL_1630902, EPI_ISL_1630903, EPI_ISL_1630904, EPI_ISL_1630905, EPI_ISL_1630906, EPI_ISL_1630907, EPI_ISL_1630909, EPI_ISL_1630911, EPI_ISL_1630912, EPI_ISL_1630913, EPI_ISL_1630914, EPI_ISL_1630915, EPI_ISL_1630916, EPI_ISL_1630917, EPI_ISL_1630918, EPI_ISL_1630919, EPI_ISL_1630920, EPI_ISL_1630921, EPI_ISL_1630922, EPI_ISL_1630923, EPI_ISL_1630924, EPI_ISL_1630925, EPI_ISL_1630926, EPI_ISL_1630927, EPI_ISL_1630928, EPI_ISL_1630929, EPI_ISL_1630930, EPI_ISL_1630931, EPI_ISL_1630932, EPI_ISL_1630934, EPI_ISL_1630935, EPI_ISL_1630936, EPI_ISL_1630937, EPI_ISL_1630938, EPI_ISL_1630939, EPI_ISL_1630940, EPI_ISL_1630941, EPI_ISL_1630942, EPI_ISL_1630943, EPI_ISL_1630945, EPI_ISL_1630946, EPI_ISL_1630947, EPI_ISL_1630950, EPI_ISL_1630951, EPI_ISL_1630952, EPI_ISL_1630953, EPI_ISL_1630954, EPI_ISL_1630955, EPI_ISL_1630956, EPI_ISL_1630957, EPI_ISL_1630959, EPI_ISL_1630960, EPI_ISL_1630961, EPI_ISL_1630962, EPI_ISL_1630963, EPI_ISL_1630964, EPI_ISL_1630965, EPI_ISL_1630966, EPI_ISL_1630967, EPI_ISL_1630969, EPI_ISL_1630970, EPI_ISL_1630971, EPI_ISL_1630972, EPI_ISL_1630973, EPI_ISL_1630974, EPI_ISL_1630975, EPI_ISL_1630976, EPI_ISL_1630977, EPI_ISL_1630979, EPI_ISL_1630980, EPI_ISL_1630981, EPI_ISL_1630982, EPI_ISL_1630983, EPI_ISL_1630985, EPI_ISL_1630986, EPI_ISL_1630987, EPI_ISL_1630990, EPI_ISL_1630991, EPI_ISL_1630992, EPI_ISL_1630993, EPI_ISL_1630994, EPI_ISL_1630995, EPI_ISL_1630996, EPI_ISL_1630997, EPI_ISL_1630998, EPI_ISL_1630999, EPI_ISL_1631001, EPI_ISL_1631002, EPI_ISL_1631003, EPI_ISL_1631004, EPI_ISL_1631005, EPI_ISL_1631006, EPI_ISL_1631007, EPI_ISL_1631010, EPI_ISL_1631011, EPI_ISL_1631012, EPI_ISL_1631013, EPI_ISL_1631014, EPI_ISL_1631015, EPI_ISL_1631016, EPI_ISL_1631019, EPI_ISL_1631020, EPI_ISL_1631021, EPI_ISL_1631022, EPI_ISL_1631023, EPI_ISL_1631025, EPI_ISL_1631026, EPI_ISL_1631027, EPI_ISL_1631029, EPI_ISL_1631031, EPI_ISL_1631032, EPI_ISL_1631033, EPI_ISL_1631034, EPI_ISL_1631035, EPI_ISL_1631036, EPI_ISL_1631037, EPI_ISL_1631038, EPI_ISL_1631040, EPI_ISL_1631041, EPI_ISL_1631042, EPI_ISL_1631043, EPI_ISL_1631044, EPI_ISL_1631045, EPI_ISL_1631046, EPI_ISL_1631048, EPI_ISL_1631049, EPI_ISL_1631050, EPI_ISL_1631052, EPI_ISL_1631053, EPI_ISL_1631054, EPI_ISL_1631055, EPI_ISL_1631057, EPI_ISL_1631058, EPI_ISL_1631059, EPI_ISL_1631060, EPI_ISL_1631061, EPI_ISL_1631062, EPI_ISL_1631063, EPI_ISL_1631064, EPI_ISL_1631065, EPI_ISL_1631066, EPI_ISL_1631068, EPI_ISL_1631069, EPI_ISL_1631070, EPI_ISL_1631071, EPI_ISL_1631072, EPI_ISL_1631074, EPI_ISL_1631075, EPI_ISL_1631076, EPI_ISL_1631077, EPI_ISL_1631078, EPI_ISL_1631079, EPI_ISL_1631080, EPI_ISL_1631081, EPI_ISL_1631082, EPI_ISL_1631084, EPI_ISL_1631085, EPI_ISL_1631086, EPI_ISL_1631087, EPI_ISL_1631088, EPI_ISL_1631096, EPI_ISL_1631098, EPI_ISL_1631099, EPI_ISL_1631103, EPI_ISL_1631115, EPI_ISL_1631120, EPI_ISL_1631121, EPI_ISL_1631127, EPI_ISL_1631130, EPI_ISL_1631132, EPI_ISL_1631135, EPI_ISL_1631137, EPI_ISL_1631139, EPI_ISL_1631143, EPI_ISL_1631148, EPI_ISL_1631152, EPI_ISL_1631154, EPI_ISL_1631163, EPI_ISL_1631166, EPI_ISL_1631182, EPI_ISL_1631184, EPI_ISL_1631186, EPI_ISL_1631191, EPI_ISL_1631198, EPI_ISL_1631207, EPI_ISL_1631208, EPI_ISL_1631221, EPI_ISL_1631223, EPI_ISL_1631228, EPI_ISL_1631231, EPI_ISL_1631236, EPI_ISL_1631239, EPI_ISL_1631240, EPI_ISL_1631241, EPI_ISL_1631251, EPI_ISL_1631255, EPI_ISL_1631260, EPI_ISL_1631267, EPI_ISL_1631273, EPI_ISL_1631280, EPI_ISL_1631281, EPI_ISL_1631283, EPI_ISL_1631289, EPI_ISL_1631293, EPI_ISL_1631299, EPI_ISL_1631300, EPI_ISL_1631313, EPI_ISL_1631316, EPI_ISL_1631321, EPI_ISL_1631326, EPI_ISL_1631333, EPI_ISL_1631758, EPI_ISL_1631778, EPI_ISL_1631780, EPI_ISL_1631784, EPI_ISL_1631813, EPI_ISL_1631854, EPI_ISL_1631873, EPI_ISL_1631895, EPI_ISL_1631903, EPI_ISL_1631930, EPI_ISL_1631942, EPI_ISL_1631956, EPI_ISL_1631965, EPI_ISL_1631972, EPI_ISL_1631990, EPI_ISL_1631999, EPI_ISL_1632000, EPI_ISL_1632002, EPI_ISL_1632003, EPI_ISL_1632010, EPI_ISL_1632017, EPI_ISL_1632019, EPI_ISL_1632020, EPI_ISL_1632029, EPI_ISL_1632038, EPI_ISL_1632040, EPI_ISL_1632044, EPI_ISL_1632047, EPI_ISL_1632054, EPI_ISL_1632055, EPI_ISL_1632060, EPI_ISL_1632066, EPI_ISL_1632073, EPI_ISL_1632075, EPI_ISL_1632077, EPI_ISL_1632080, EPI_ISL_1632082, EPI_ISL_1632086, EPI_ISL_1632087, EPI_ISL_1632088, EPI_ISL_1632098, EPI_ISL_1632100, EPI_ISL_1632101, EPI_ISL_1632102, EPI_ISL_1632103, EPI_ISL_1632105, EPI_ISL_1632107, EPI_ISL_1632108, EPI_ISL_1632109, EPI_ISL_1632110, EPI_ISL_1632113, EPI_ISL_1632116, EPI_ISL_1632120, EPI_ISL_1632121, EPI_ISL_1632124, EPI_ISL_1632125, EPI_ISL_1632129, EPI_ISL_1632132, EPI_ISL_1632133, EPI_ISL_1632134, EPI_ISL_1632137, EPI_ISL_1632139, EPI_ISL_1632147, EPI_ISL_1632148, EPI_ISL_1632149, EPI_ISL_1632153, EPI_ISL_1632165, EPI_ISL_1632169, EPI_ISL_1632176, EPI_ISL_1632180, EPI_ISL_1632184, EPI_ISL_1632186, EPI_ISL_1632187, EPI_ISL_1632188, EPI_ISL_1632189, EPI_ISL_1632190, EPI_ISL_1632192, EPI_ISL_1632194, EPI_ISL_1632197, EPI_ISL_1632199, EPI_ISL_1632202, EPI_ISL_1632204, EPI_ISL_1632208, EPI_ISL_1632209, EPI_ISL_1632211, EPI_ISL_1632221, EPI_ISL_1632227, EPI_ISL_1632236, EPI_ISL_1632241, EPI_ISL_1632259, EPI_ISL_1632267, EPI_ISL_1632280, EPI_ISL_1632281, EPI_ISL_1632284, EPI_ISL_1632285, EPI_ISL_1632292, EPI_ISL_1632296, EPI_ISL_1632297, EPI_ISL_1634714, EPI_ISL_1634716, EPI_ISL_1635122, EPI_ISL_1635155, EPI_ISL_1635321, EPI_ISL_1652694, EPI_ISL_1652697, EPI_ISL_1652701, EPI_ISL_1652703, EPI_ISL_1652708, EPI_ISL_1652713, EPI_ISL_1652715, EPI_ISL_1652719, EPI_ISL_1652720, EPI_ISL_1652722, EPI_ISL_1652724, EPI_ISL_1652725, EPI_ISL_1652727, EPI_ISL_1652728, EPI_ISL_1652729, EPI_ISL_1652733, EPI_ISL_1652734, EPI_ISL_1652735, EPI_ISL_1652736, EPI_ISL_1652738, EPI_ISL_1652739, EPI_ISL_1652743, EPI_ISL_1652745, EPI_ISL_1652746, EPI_ISL_1652747, EPI_ISL_1652748, EPI_ISL_1652750, EPI_ISL_1652753, EPI_ISL_1652756, EPI_ISL_1652757, EPI_ISL_1652760, EPI_ISL_1652761, EPI_ISL_1652763, EPI_ISL_1652764, EPI_ISL_1652766, EPI_ISL_1652769, EPI_ISL_1652770, EPI_ISL_1652771, EPI_ISL_1652773, EPI_ISL_1652774, EPI_ISL_1652775, EPI_ISL_1652777, EPI_ISL_1652779, EPI_ISL_1652781, EPI_ISL_1652788, EPI_ISL_1652789, EPI_ISL_1652791, EPI_ISL_1652793, EPI_ISL_1652794, EPI_ISL_1652796, EPI_ISL_1652797, EPI_ISL_1652802, EPI_ISL_1652808, EPI_ISL_1652809, EPI_ISL_1652811, EPI_ISL_1652813, EPI_ISL_1652814, EPI_ISL_1652818, EPI_ISL_1652819, EPI_ISL_1652821, EPI_ISL_1652825, EPI_ISL_1652826, EPI_ISL_1652831, EPI_ISL_1652834, EPI_ISL_1652836, EPI_ISL_1652837, EPI_ISL_1652839, EPI_ISL_1652840, EPI_ISL_1652841, EPI_ISL_1652843, EPI_ISL_1652845, EPI_ISL_1652846, EPI_ISL_1652848, EPI_ISL_1652849, EPI_ISL_1652850, EPI_ISL_1652853, EPI_ISL_1652854, EPI_ISL_1652856, EPI_ISL_1652857, EPI_ISL_1652861, EPI_ISL_1652862, EPI_ISL_1652864, EPI_ISL_1652865, EPI_ISL_1652867, EPI_ISL_1652869, EPI_ISL_1652871, EPI_ISL_1652873, EPI_ISL_1652874, EPI_ISL_1652875, EPI_ISL_1652877, EPI_ISL_1652878, EPI_ISL_1652879, EPI_ISL_1652886, EPI_ISL_1652888, EPI_ISL_1652890, EPI_ISL_1652898, EPI_ISL_1652900, EPI_ISL_1652901, EPI_ISL_1652903, EPI_ISL_1652904, EPI_ISL_1652907, EPI_ISL_1652908, EPI_ISL_1652910, EPI_ISL_1652912, EPI_ISL_1652913, EPI_ISL_1652915, EPI_ISL_1652916, EPI_ISL_1652917, EPI_ISL_1652920, EPI_ISL_1652924, EPI_ISL_1652929, EPI_ISL_1652930, EPI_ISL_1652933, EPI_ISL_1652937, EPI_ISL_1652938, EPI_ISL_1652939, EPI_ISL_1652952, EPI_ISL_1652979, EPI_ISL_1652986, EPI_ISL_1653009, EPI_ISL_1653011, EPI_ISL_1653024, EPI_ISL_1653025, EPI_ISL_1653040, EPI_ISL_1653058, EPI_ISL_1653108, EPI_ISL_1653125, EPI_ISL_1653175, EPI_ISL_1653179, EPI_ISL_1653180, EPI_ISL_1653187, EPI_ISL_1653195, EPI_ISL_1653225, EPI_ISL_1653227, EPI_ISL_1653229, EPI_ISL_1653230, EPI_ISL_1653232, EPI_ISL_1653234, EPI_ISL_1653235, EPI_ISL_1653236, EPI_ISL_1653242, EPI_ISL_1653243, EPI_ISL_1653245, EPI_ISL_1653247, EPI_ISL_1653250, EPI_ISL_1653259, EPI_ISL_1653260, EPI_ISL_1653269, EPI_ISL_1653270, EPI_ISL_1653271, EPI_ISL_1653277, EPI_ISL_1653279, EPI_ISL_1653280, EPI_ISL_1653284, EPI_ISL_1653288, EPI_ISL_1653290, EPI_ISL_1653297, EPI_ISL_1653298, EPI_ISL_1653301, EPI_ISL_1653317, EPI_ISL_1653327, EPI_ISL_1653330, EPI_ISL_1653332, EPI_ISL_1653334, EPI_ISL_1653338, EPI_ISL_1653342, EPI_ISL_1653346, EPI_ISL_1653347, EPI_ISL_1653348, EPI_ISL_1653349, EPI_ISL_1653351, EPI_ISL_1653352, EPI_ISL_1653358, EPI_ISL_1653361, EPI_ISL_1653363, EPI_ISL_1653364, EPI_ISL_1653370, EPI_ISL_1653377, EPI_ISL_1653379, EPI_ISL_1653380, EPI_ISL_1653387, EPI_ISL_1653391, EPI_ISL_1653392, EPI_ISL_1653394, EPI_ISL_1653396, EPI_ISL_1653400, EPI_ISL_1653401, EPI_ISL_1653405, EPI_ISL_1653413, EPI_ISL_1653415, EPI_ISL_1653421, EPI_ISL_1653433, EPI_ISL_1653435, EPI_ISL_1653438, EPI_ISL_1653439, EPI_ISL_1653440, EPI_ISL_1653441, EPI_ISL_1653446, EPI_ISL_1653447, EPI_ISL_1653453, EPI_ISL_1653458, EPI_ISL_1653460, EPI_ISL_1653461, EPI_ISL_1653468, EPI_ISL_1653474, EPI_ISL_1653479, EPI_ISL_1653481, EPI_ISL_1653483, EPI_ISL_1653484, EPI_ISL_1653487, EPI_ISL_1653489, EPI_ISL_1653490, EPI_ISL_1653492, EPI_ISL_1653494, EPI_ISL_1653495, EPI_ISL_1653496, EPI_ISL_1653497, EPI_ISL_1653499, EPI_ISL_1653500, EPI_ISL_1653501, EPI_ISL_1653503, EPI_ISL_1653508, EPI_ISL_1653510, EPI_ISL_1653511, EPI_ISL_1653512, EPI_ISL_1653513, EPI_ISL_1653514, EPI_ISL_1653515, EPI_ISL_1653517, EPI_ISL_1653518, EPI_ISL_1653519, EPI_ISL_1653522, EPI_ISL_1653525, EPI_ISL_1653531, EPI_ISL_1653532, EPI_ISL_1653534, EPI_ISL_1653535, EPI_ISL_1653537, EPI_ISL_1653538, EPI_ISL_1653541, EPI_ISL_1653542, EPI_ISL_1653543, EPI_ISL_1653544, EPI_ISL_1653545, EPI_ISL_1653546, EPI_ISL_1653547, EPI_ISL_1653548, EPI_ISL_1653549, EPI_ISL_1653551, EPI_ISL_1653554, EPI_ISL_1653556, EPI_ISL_1653557, EPI_ISL_1653559, EPI_ISL_1653560, EPI_ISL_1653561, EPI_ISL_1653564, EPI_ISL_1653565, EPI_ISL_1653567, EPI_ISL_1653569, EPI_ISL_1653570, EPI_ISL_1653571, EPI_ISL_1653573, EPI_ISL_1653574, EPI_ISL_1653575, EPI_ISL_1653577, EPI_ISL_1653578, EPI_ISL_1653579, EPI_ISL_1653580, EPI_ISL_1653581, EPI_ISL_1653582, EPI_ISL_1653583, EPI_ISL_1653584, EPI_ISL_1653585, EPI_ISL_1653589, EPI_ISL_1653590, EPI_ISL_1653591, EPI_ISL_1653592, EPI_ISL_1653593, EPI_ISL_1653595, EPI_ISL_1653596, EPI_ISL_1653599, EPI_ISL_1653600, EPI_ISL_1653603, EPI_ISL_1653604, EPI_ISL_1653605, EPI_ISL_1653606, EPI_ISL_1653607, EPI_ISL_1653611, EPI_ISL_1653613, EPI_ISL_1653614, EPI_ISL_1653616, EPI_ISL_1653618, EPI_ISL_1653619, EPI_ISL_1653620, EPI_ISL_1653623, EPI_ISL_1653624, EPI_ISL_1653625, EPI_ISL_1653626, EPI_ISL_1653627, EPI_ISL_1653630, EPI_ISL_1653631, EPI_ISL_1653632, EPI_ISL_1653633, EPI_ISL_1653635, EPI_ISL_1653636, EPI_ISL_1653637, EPI_ISL_1653640, EPI_ISL_1653641, EPI_ISL_1653643, EPI_ISL_1653644, EPI_ISL_1653645, EPI_ISL_1653646, EPI_ISL_1653647, EPI_ISL_1653649, EPI_ISL_1653652, EPI_ISL_1653655, EPI_ISL_1653656, EPI_ISL_1653657, EPI_ISL_1653658, EPI_ISL_1653659, EPI_ISL_1653660, EPI_ISL_1653661, EPI_ISL_1653662, EPI_ISL_1653663, EPI_ISL_1653668, EPI_ISL_1653669, EPI_ISL_1653670, EPI_ISL_1653672, EPI_ISL_1653676, EPI_ISL_1653677, EPI_ISL_1653679, EPI_ISL_1653683, EPI_ISL_1653686, EPI_ISL_1653687, EPI_ISL_1653688, EPI_ISL_1653690, EPI_ISL_1653697, EPI_ISL_1653698, EPI_ISL_1653700, EPI_ISL_1653702, EPI_ISL_1653704, EPI_ISL_1653705, EPI_ISL_1653706, EPI_ISL_1653707, EPI_ISL_1653709, EPI_ISL_1653710, EPI_ISL_1653711, EPI_ISL_1653712, EPI_ISL_1653713, EPI_ISL_1653714, EPI_ISL_1653715, EPI_ISL_1653716, EPI_ISL_1653728, EPI_ISL_1653758, EPI_ISL_1672977, EPI_ISL_1673041, EPI_ISL_1697444, EPI_ISL_1697445, EPI_ISL_1697446, EPI_ISL_1697447, EPI_ISL_1697452, EPI_ISL_1697454, EPI_ISL_1697457, EPI_ISL_1697459, EPI_ISL_1697460, EPI_ISL_1697461, EPI_ISL_1697462, EPI_ISL_1697463, EPI_ISL_1697464, EPI_ISL_1697467, EPI_ISL_1697470, EPI_ISL_1697474, EPI_ISL_1697475, EPI_ISL_1697476, EPI_ISL_1697478, EPI_ISL_1697479, EPI_ISL_1697481, EPI_ISL_1697483, EPI_ISL_1697487, EPI_ISL_1697488, EPI_ISL_1697489, EPI_ISL_1697490, EPI_ISL_1697492, EPI_ISL_1697498, EPI_ISL_1697500, EPI_ISL_1697502, EPI_ISL_1697504, EPI_ISL_1697505, EPI_ISL_1697507, EPI_ISL_1697509, EPI_ISL_1697510, EPI_ISL_1697511, EPI_ISL_1697512, EPI_ISL_1697513, EPI_ISL_1697515, EPI_ISL_1697520, EPI_ISL_1697521, EPI_ISL_1697524, EPI_ISL_1697528, EPI_ISL_1697533, EPI_ISL_1697535, EPI_ISL_1697537, EPI_ISL_1697541, EPI_ISL_1697542, EPI_ISL_1697545, EPI_ISL_1697550, EPI_ISL_1697551, EPI_ISL_1697553, EPI_ISL_1697556, EPI_ISL_1697557, EPI_ISL_1697558, EPI_ISL_1697559, EPI_ISL_1697560, EPI_ISL_1697561, EPI_ISL_1697563, EPI_ISL_1697567, EPI_ISL_1697569, EPI_ISL_1697570, EPI_ISL_1697572, EPI_ISL_1697575, EPI_ISL_1697581, EPI_ISL_1697582, EPI_ISL_1697583, EPI_ISL_1697584, EPI_ISL_1697585, EPI_ISL_1697586, EPI_ISL_1697590, EPI_ISL_1697597, EPI_ISL_1697600, EPI_ISL_1697602, EPI_ISL_1697604, EPI_ISL_1697608, EPI_ISL_1697609, EPI_ISL_1697610, EPI_ISL_1697611, EPI_ISL_1697614, EPI_ISL_1697615, EPI_ISL_1697616, EPI_ISL_1697618, EPI_ISL_1697619, EPI_ISL_1697621, EPI_ISL_1697626, EPI_ISL_1697629, EPI_ISL_1697631, EPI_ISL_1697633, EPI_ISL_1697634, EPI_ISL_1697637, EPI_ISL_1697638, EPI_ISL_1697641, EPI_ISL_1697642, EPI_ISL_1697643, EPI_ISL_1697646, EPI_ISL_1697647, EPI_ISL_1697649, EPI_ISL_1697651, EPI_ISL_1697653, EPI_ISL_1697658, EPI_ISL_1697662, EPI_ISL_1697663, EPI_ISL_1697665, EPI_ISL_1697668, EPI_ISL_1697670, EPI_ISL_1697672, EPI_ISL_1697675, EPI_ISL_1697676, EPI_ISL_1697679, EPI_ISL_1697680, EPI_ISL_1697681, EPI_ISL_1697682, EPI_ISL_1697683, EPI_ISL_1697686, EPI_ISL_1697688, EPI_ISL_1697692, EPI_ISL_1697694, EPI_ISL_1697697, EPI_ISL_1697700, EPI_ISL_1697701, EPI_ISL_1697703, EPI_ISL_1697705, EPI_ISL_1697706, EPI_ISL_1697707, EPI_ISL_1697713, EPI_ISL_1697714, EPI_ISL_1697716, EPI_ISL_1697718, EPI_ISL_1697720, EPI_ISL_1697721, EPI_ISL_1697724, EPI_ISL_1697727, EPI_ISL_1697732, EPI_ISL_1697733, EPI_ISL_1697737, EPI_ISL_1697738, EPI_ISL_1697739, EPI_ISL_1697741, EPI_ISL_1697742, EPI_ISL_1698075, EPI_ISL_1698209, EPI_ISL_1698256, EPI_ISL_1698277, EPI_ISL_1698294, EPI_ISL_1698302, EPI_ISL_1698318, EPI_ISL_1698321, EPI_ISL_1698338, EPI_ISL_1698341, EPI_ISL_1698371, EPI_ISL_1698372, EPI_ISL_1698388, EPI_ISL_1698411, EPI_ISL_1698423, EPI_ISL_1698446, EPI_ISL_1698451, EPI_ISL_1698472, EPI_ISL_1698483, EPI_ISL_1698512, EPI_ISL_1698541, EPI_ISL_1698764, EPI_ISL_1698771, EPI_ISL_1698775, EPI_ISL_1698778, EPI_ISL_1698783, EPI_ISL_1698790, EPI_ISL_1698792, EPI_ISL_1698795, EPI_ISL_1698798, EPI_ISL_1698802, EPI_ISL_1698816, EPI_ISL_1698817, EPI_ISL_1698818, EPI_ISL_1698825, EPI_ISL_1698826, EPI_ISL_1698828, EPI_ISL_1698830, EPI_ISL_1698831, EPI_ISL_1698834, EPI_ISL_1698835, EPI_ISL_1698843, EPI_ISL_1698844, EPI_ISL_1698849, EPI_ISL_1698850, EPI_ISL_1698852, EPI_ISL_1698858, EPI_ISL_1698859, EPI_ISL_1698867, EPI_ISL_1698868, EPI_ISL_1698869, EPI_ISL_1698871, EPI_ISL_1698875, EPI_ISL_1698878, EPI_ISL_1698884, EPI_ISL_1698894, EPI_ISL_1698896, EPI_ISL_1698898, EPI_ISL_1698905, EPI_ISL_1698909, EPI_ISL_1698910, EPI_ISL_1698913, EPI_ISL_1698923, EPI_ISL_1698924, EPI_ISL_1698931, EPI_ISL_1698942, EPI_ISL_1698946, EPI_ISL_1698949, EPI_ISL_1698955, EPI_ISL_1698963, EPI_ISL_1698968, EPI_ISL_1698978, EPI_ISL_1698979, EPI_ISL_1698985, EPI_ISL_1698986, EPI_ISL_1698987, EPI_ISL_1698988, EPI_ISL_1698993, EPI_ISL_1698995, EPI_ISL_1698996, EPI_ISL_1698998, EPI_ISL_1698999, EPI_ISL_1699001, EPI_ISL_1699013, EPI_ISL_1699017, EPI_ISL_1699022, EPI_ISL_1699027, EPI_ISL_1699033, EPI_ISL_1699035, EPI_ISL_1699036, EPI_ISL_1699037, EPI_ISL_1699039, EPI_ISL_1699040, EPI_ISL_1699045, EPI_ISL_1699047, EPI_ISL_1699048, EPI_ISL_1699050, EPI_ISL_1699052, EPI_ISL_1699053, EPI_ISL_1699056, EPI_ISL_1699060, EPI_ISL_1699061, EPI_ISL_1699063, EPI_ISL_1699067, EPI_ISL_1699068, EPI_ISL_1699069, EPI_ISL_1699070, EPI_ISL_1699071, EPI_ISL_1699072, EPI_ISL_1699079, EPI_ISL_1699081, EPI_ISL_1699084, EPI_ISL_1699088, EPI_ISL_1699089, EPI_ISL_1699094, EPI_ISL_1699099, EPI_ISL_1699101, EPI_ISL_1699104, EPI_ISL_1699105, EPI_ISL_1699106, EPI_ISL_1699107, EPI_ISL_1699108, EPI_ISL_1699110, EPI_ISL_1699112, EPI_ISL_1699118, EPI_ISL_1699119, EPI_ISL_1699120, EPI_ISL_1699122, EPI_ISL_1699126, EPI_ISL_1699127, EPI_ISL_1699133, EPI_ISL_1699134, EPI_ISL_1699141, EPI_ISL_1699142, EPI_ISL_1699143, EPI_ISL_1699148, EPI_ISL_1699150, EPI_ISL_1699155, EPI_ISL_1699156, EPI_ISL_1699157, EPI_ISL_1699172, EPI_ISL_1699180, EPI_ISL_1699182, EPI_ISL_1699192, EPI_ISL_1699195, EPI_ISL_1699196, EPI_ISL_1699197, EPI_ISL_1699199, EPI_ISL_1699203, EPI_ISL_1699207, EPI_ISL_1699209, EPI_ISL_1699210, EPI_ISL_1699211, EPI_ISL_1699214, EPI_ISL_1699215, EPI_ISL_1699218, EPI_ISL_1699220, EPI_ISL_1699228, EPI_ISL_1699229, EPI_ISL_1699231, EPI_ISL_1699232, EPI_ISL_1699238, EPI_ISL_1699239, EPI_ISL_1699240, EPI_ISL_1699242, EPI_ISL_1699246, EPI_ISL_1699250, EPI_ISL_1699252, EPI_ISL_1699254, EPI_ISL_1699266, EPI_ISL_1699269, EPI_ISL_1699270, EPI_ISL_1699282, EPI_ISL_1699290, EPI_ISL_1699292, EPI_ISL_1699294, EPI_ISL_1699296, EPI_ISL_1699298, EPI_ISL_1699300, EPI_ISL_1699303, EPI_ISL_1699305, EPI_ISL_1699308, EPI_ISL_1699310, EPI_ISL_1699314, EPI_ISL_1699315, EPI_ISL_1699320, EPI_ISL_1699323, EPI_ISL_1699324, EPI_ISL_1699331, EPI_ISL_1699332, EPI_ISL_1699334, EPI_ISL_1699337, EPI_ISL_1699341, EPI_ISL_1699344, EPI_ISL_1699351, EPI_ISL_1699357, EPI_ISL_1699359, EPI_ISL_1699362, EPI_ISL_1699364, EPI_ISL_1699366, EPI_ISL_1699369, EPI_ISL_1699371, EPI_ISL_1699372, EPI_ISL_1699378, EPI_ISL_1699380, EPI_ISL_1699381, EPI_ISL_1699387, EPI_ISL_1699391, EPI_ISL_1699392, EPI_ISL_1699394, EPI_ISL_1699395, EPI_ISL_1699399, EPI_ISL_1699401, EPI_ISL_1699402, EPI_ISL_1699406, EPI_ISL_1699410, EPI_ISL_1699411, EPI_ISL_1699412, EPI_ISL_1699416, EPI_ISL_1699418, EPI_ISL_1699421, EPI_ISL_1699422, EPI_ISL_1699424, EPI_ISL_1699425, EPI_ISL_1699426, EPI_ISL_1699432, EPI_ISL_1699436, EPI_ISL_1699437, EPI_ISL_1699438, EPI_ISL_1699440, EPI_ISL_1699441, EPI_ISL_1699453, EPI_ISL_1699461, EPI_ISL_1699465, EPI_ISL_1699466, EPI_ISL_1699467, EPI_ISL_1699474, EPI_ISL_1699485, EPI_ISL_1699494, EPI_ISL_1699500, EPI_ISL_1699501, EPI_ISL_1699503, EPI_ISL_1699508, EPI_ISL_1699509, EPI_ISL_1699512, EPI_ISL_1699513, EPI_ISL_1699516, EPI_ISL_1699521, EPI_ISL_1699526, EPI_ISL_1699528, EPI_ISL_1699529, EPI_ISL_1699532, EPI_ISL_1699534, EPI_ISL_1699535, EPI_ISL_1699536, EPI_ISL_1699540, EPI_ISL_1699542, EPI_ISL_1699546, EPI_ISL_1699550, EPI_ISL_1699551, EPI_ISL_1699552, EPI_ISL_1699553, EPI_ISL_1699554, EPI_ISL_1699556, EPI_ISL_1699557, EPI_ISL_1699569, EPI_ISL_1699586, EPI_ISL_1699589, EPI_ISL_1699603, EPI_ISL_1699616, EPI_ISL_1699623, EPI_ISL_1699629, EPI_ISL_1699646, EPI_ISL_1699650, EPI_ISL_1699660, EPI_ISL_1699709, EPI_ISL_1699713, EPI_ISL_1699717, EPI_ISL_1699721, EPI_ISL_1699723, EPI_ISL_1699738, EPI_ISL_1699751, EPI_ISL_1699752, EPI_ISL_1699774, EPI_ISL_1699775, EPI_ISL_1699817, EPI_ISL_1699820, EPI_ISL_1699824, EPI_ISL_1699836, EPI_ISL_1699838, EPI_ISL_1699842, EPI_ISL_1699844, EPI_ISL_1699845, EPI_ISL_1699849, EPI_ISL_1699851, EPI_ISL_1699853, EPI_ISL_1699854, EPI_ISL_1699856, EPI_ISL_1699857, EPI_ISL_1699859, EPI_ISL_1699863, EPI_ISL_1699864, EPI_ISL_1699865, EPI_ISL_1699866, EPI_ISL_1699870, EPI_ISL_1699871, EPI_ISL_1699873, EPI_ISL_1699875, EPI_ISL_1699877, EPI_ISL_1699879, EPI_ISL_1699881, EPI_ISL_1699882, EPI_ISL_1699884, EPI_ISL_1699886, EPI_ISL_1699887, EPI_ISL_1699888, EPI_ISL_1699891, EPI_ISL_1699892, EPI_ISL_1699893, EPI_ISL_1699896, EPI_ISL_1699900, EPI_ISL_1699901, EPI_ISL_1699903, EPI_ISL_1699904, EPI_ISL_1699905, EPI_ISL_1699906, EPI_ISL_1699908, EPI_ISL_1699909, EPI_ISL_1699911, EPI_ISL_1699912, EPI_ISL_1699918, EPI_ISL_1699919, EPI_ISL_1699920, EPI_ISL_1699923, EPI_ISL_1699924, EPI_ISL_1699925, EPI_ISL_1699926, EPI_ISL_1699935, EPI_ISL_1699936, EPI_ISL_1699938, EPI_ISL_1699939, EPI_ISL_1699940, EPI_ISL_1699944, EPI_ISL_1699945, EPI_ISL_1699946, EPI_ISL_1699947, EPI_ISL_1699948, EPI_ISL_1699949, EPI_ISL_1699951, EPI_ISL_1699952, EPI_ISL_1699953, EPI_ISL_1699954, EPI_ISL_1699956, EPI_ISL_1699957, EPI_ISL_1699958, EPI_ISL_1699959, EPI_ISL_1699961, EPI_ISL_1699962, EPI_ISL_1699963, EPI_ISL_1699965, EPI_ISL_1699966, EPI_ISL_1699968, EPI_ISL_1699969, EPI_ISL_1699970, EPI_ISL_1699971, EPI_ISL_1699972, EPI_ISL_1699973, EPI_ISL_1699975, EPI_ISL_1699976, EPI_ISL_1699978, EPI_ISL_1699981, EPI_ISL_1699982, EPI_ISL_1699983, EPI_ISL_1699985, EPI_ISL_1699987, EPI_ISL_1699988, EPI_ISL_1699989, EPI_ISL_1699992, EPI_ISL_1699993, EPI_ISL_1699996, EPI_ISL_1699999, EPI_ISL_1700000, EPI_ISL_1700001, EPI_ISL_1700002, EPI_ISL_1700003, EPI_ISL_1700004, EPI_ISL_1700005, EPI_ISL_1700008, EPI_ISL_1700009, EPI_ISL_1700011, EPI_ISL_1700012, EPI_ISL_1700014, EPI_ISL_1700016, EPI_ISL_1700017, EPI_ISL_1700018, EPI_ISL_1700019, EPI_ISL_1700023, EPI_ISL_1700024, EPI_ISL_1700026, EPI_ISL_1700032, EPI_ISL_1700033, EPI_ISL_1700035, EPI_ISL_1700038, EPI_ISL_1700039, EPI_ISL_1700040, EPI_ISL_1700043, EPI_ISL_1700045, EPI_ISL_1700046, EPI_ISL_1700049, EPI_ISL_1700051, EPI_ISL_1700054, EPI_ISL_1700055, EPI_ISL_1700056, EPI_ISL_1700058, EPI_ISL_1700061, EPI_ISL_1700062, EPI_ISL_1700064, EPI_ISL_1700065, EPI_ISL_1700066, EPI_ISL_1700072, EPI_ISL_1700073, EPI_ISL_1700075, EPI_ISL_1700077, EPI_ISL_1700082, EPI_ISL_1700083, EPI_ISL_1700084, EPI_ISL_1700086, EPI_ISL_1700087, EPI_ISL_1700088, EPI_ISL_1700089, EPI_ISL_1700091, EPI_ISL_1700092, EPI_ISL_1700093, EPI_ISL_1700094, EPI_ISL_1700095, EPI_ISL_1700097, EPI_ISL_1700099, EPI_ISL_1700101, EPI_ISL_1700104, EPI_ISL_1700105, EPI_ISL_1700106, EPI_ISL_1700107, EPI_ISL_1700110, EPI_ISL_1700111, EPI_ISL_1700114, EPI_ISL_1700115, EPI_ISL_1700117, EPI_ISL_1700119, EPI_ISL_1700120, EPI_ISL_1700121, EPI_ISL_1700122, EPI_ISL_1700123, EPI_ISL_1700124, EPI_ISL_1700125, EPI_ISL_1700126, EPI_ISL_1700128, EPI_ISL_1700129, EPI_ISL_1700131, EPI_ISL_1700132, EPI_ISL_1700133, EPI_ISL_1700134, EPI_ISL_1700137, EPI_ISL_1700142, EPI_ISL_1700146, EPI_ISL_1700147, EPI_ISL_1700150, EPI_ISL_1700155, EPI_ISL_1700156, EPI_ISL_1700158, EPI_ISL_1700160, EPI_ISL_1700161, EPI_ISL_1700162, EPI_ISL_1700163, EPI_ISL_1700168, EPI_ISL_1700171, EPI_ISL_1700172, EPI_ISL_1700175, EPI_ISL_1700176, EPI_ISL_1700178, EPI_ISL_1700180, EPI_ISL_1700181, EPI_ISL_1700186, EPI_ISL_1700194, EPI_ISL_1700195, EPI_ISL_1700197, EPI_ISL_1700199, EPI_ISL_1700206, EPI_ISL_1700210, EPI_ISL_1700211, EPI_ISL_1700215, EPI_ISL_1700218, EPI_ISL_1700219, EPI_ISL_1700225, EPI_ISL_1700230, EPI_ISL_1700231, EPI_ISL_1700232, EPI_ISL_1700234, EPI_ISL_1700235, EPI_ISL_1700236, EPI_ISL_1700239, EPI_ISL_1700242, EPI_ISL_1700247, | | | |

EPI_ISL_1700249, EPI_ISL_1700252, EPI_ISL_1700259, EPI_ISL_1700260, EPI_ISL_1700261, EPI_ISL_1700264, EPI_ISL_1700266, EPI_ISL_1700267, EPI_ISL_1700271, EPI_ISL_1700273, EPI_ISL_1700276, EPI_ISL_1700279, EPI_ISL_1700281, EPI_ISL_1700282, EPI_ISL_1700284, EPI_ISL_1700287, EPI_ISL_1700289, EPI_ISL_1700290, EPI_ISL_1700291, EPI_ISL_1700298, EPI_ISL_1700304, EPI_ISL_1700305, EPI_ISL_1700306, EPI_ISL_1700309, EPI_ISL_1700311, EPI_ISL_1700314, EPI_ISL_1700315, EPI_ISL_1700316, EPI_ISL_1700321, EPI_ISL_1700322, EPI_ISL_1700323, EPI_ISL_1700324, EPI_ISL_1700327, EPI_ISL_1700328, EPI_ISL_1700330, EPI_ISL_1700334, EPI_ISL_1700335, EPI_ISL_1700337, EPI_ISL_1700339, EPI_ISL_1700341, EPI_ISL_1700344, EPI_ISL_1700345, EPI_ISL_1700346, EPI_ISL_1700349, EPI_ISL_1700350, EPI_ISL_1700351, EPI_ISL_1700352, EPI_ISL_1700354, EPI_ISL_1700358, EPI_ISL_1700362, EPI_ISL_1700368, EPI_ISL_1700372, EPI_ISL_1700375, EPI_ISL_1700376, EPI_ISL_1700377, EPI_ISL_1700379, EPI_ISL_1700381, EPI_ISL_1700383, EPI_ISL_1700384, EPI_ISL_1700388, EPI_ISL_1700390, EPI_ISL_1700391, EPI_ISL_1700395, EPI_ISL_1700398, EPI_ISL_1700400, EPI_ISL_1700402, EPI_ISL_1700403, EPI_ISL_1700404, EPI_ISL_1700405, EPI_ISL_1700407, EPI_ISL_1700412, EPI_ISL_1700416, EPI_ISL_1700418, EPI_ISL_1700419, EPI_ISL_1700420, EPI_ISL_1700422, EPI_ISL_1700424, EPI_ISL_1700427, EPI_ISL_1700428, EPI_ISL_1700429, EPI_ISL_1700432, EPI_ISL_1700434, EPI_ISL_1700437, EPI_ISL_1700438, EPI_ISL_1700439, EPI_ISL_1700440, EPI_ISL_1700445, EPI_ISL_1700446, EPI_ISL_1700447, EPI_ISL_1700450, EPI_ISL_1700451, EPI_ISL_1700452, EPI_ISL_1700453, EPI_ISL_1700455, EPI_ISL_1700462, EPI_ISL_1700463, EPI_ISL_1700464, EPI_ISL_1700465, EPI_ISL_1700467, EPI_ISL_1700472, EPI_ISL_1700474, EPI_ISL_1700477, EPI_ISL_1700478, EPI_ISL_1700481, EPI_ISL_1700482, EPI_ISL_1700483, EPI_ISL_1700489, EPI_ISL_1700491, EPI_ISL_1700495, EPI_ISL_1700496, EPI_ISL_1700497, EPI_ISL_1700498, EPI_ISL_1700499, EPI_ISL_1700500, EPI_ISL_1700501, EPI_ISL_1700502, EPI_ISL_1700503, EPI_ISL_1700504, EPI_ISL_1700505, EPI_ISL_1700506, EPI_ISL_1700507, EPI_ISL_1700508, EPI_ISL_1700509, EPI_ISL_1700512, EPI_ISL_1700513, EPI_ISL_1718330, EPI_ISL_1718732, EPI_ISL_1719451, EPI_ISL_1719500, EPI_ISL_1719513, EPI_ISL_1719571, EPI_ISL_1719743, EPI_ISL_1742171, EPI_ISL_1759033

| | | | |
|---|---|---|---|
| see above | Lighthouse Lab in Alderley Park | Wellcome Sanger Institute for the COVID-19 Genomics UK (COG-UK) Consortium | Cordelia Langford; David K. Jackson; Dominic Kwiatkowski; Ewan Harrison; Ian Johnston; Jacquelyn Wynn; Jeffrey Barrett; John Sillitoe on behalf of the Wellcome Sanger Institute COVID-19 Surveillance Team; Mairead Hyland; Roberto Amato; Sonia Goncalves; The Lighthouse Lab in Alderley Park and Alex Alderton |

EPI_ISL_1564047, EPI_ISL_1564065, EPI_ISL_1564066, EPI_ISL_1564073, EPI_ISL_1564088, EPI_ISL_1564097, EPI_ISL_1564102, EPI_ISL_1564112, EPI_ISL_1564124, EPI_ISL_1564130, EPI_ISL_1564139, EPI_ISL_1564178, EPI_ISL_1564185, EPI_ISL_1564203, EPI_ISL_1564215, EPI_ISL_1564221, EPI_ISL_1564223, EPI_ISL_1564224, EPI_ISL_1564258, EPI_ISL_1564271, EPI_ISL_1564281, EPI_ISL_1564304, EPI_ISL_1564319, EPI_ISL_1564328, EPI_ISL_1583924, EPI_ISL_1583925, EPI_ISL_1583931, EPI_ISL_1583938, EPI_ISL_1583940, EPI_ISL_1583942, EPI_ISL_1583944, EPI_ISL_1583947, EPI_ISL_1583950, EPI_ISL_1583955, EPI_ISL_1583962, EPI_ISL_1583966, EPI_ISL_1583970, EPI_ISL_1583972, EPI_ISL_1583974, EPI_ISL_1583981, EPI_ISL_1583984, EPI_ISL_1583985, EPI_ISL_1583986, EPI_ISL_1583987, EPI_ISL_1583998, EPI_ISL_1584008, EPI_ISL_1584010, EPI_ISL_1584011, EPI_ISL_1584017, EPI_ISL_1584032, EPI_ISL_1584036, EPI_ISL_1584043, EPI_ISL_1584047, EPI_ISL_1584055, EPI_ISL_1584067, EPI_ISL_1584070, EPI_ISL_1584071, EPI_ISL_1584072, EPI_ISL_1584076, EPI_ISL_1584077, EPI_ISL_1584082, EPI_ISL_1584089, EPI_ISL_1584090, EPI_ISL_1584091, EPI_ISL_1584096, EPI_ISL_1584100, EPI_ISL_1584101, EPI_ISL_1584106, EPI_ISL_1584114, EPI_ISL_1584115, EPI_ISL_1584118, EPI_ISL_1584121, EPI_ISL_1584123, EPI_ISL_1584125, EPI_ISL_1584131, EPI_ISL_1584135, EPI_ISL_1584440, EPI_ISL_1594466, EPI_ISL_1594471, EPI_ISL_1594500, EPI_ISL_1594520, EPI_ISL_1594522, EPI_ISL_1594528, EPI_ISL_1594540, EPI_ISL_1594569, EPI_ISL_1594596, EPI_ISL_1594635, EPI_ISL_1594641, EPI_ISL_1594650, EPI_ISL_1594659, EPI_ISL_1594660, EPI_ISL_1594703, EPI_ISL_1594719, EPI_ISL_1594726, EPI_ISL_1594728, EPI_ISL_1594731, EPI_ISL_1594756, EPI_ISL_1594758, EPI_ISL_1594759, EPI_ISL_1594760, EPI_ISL_1594765, EPI_ISL_1594769, EPI_ISL_1594787, EPI_ISL_1594788, EPI_ISL_1594794, EPI_ISL_1594798, EPI_ISL_1594801, EPI_ISL_1594803, EPI_ISL_1594819, EPI_ISL_1594820, EPI_ISL_1594823, EPI_ISL_1594826, EPI_ISL_1594832, EPI_ISL_1594837, EPI_ISL_1594842, EPI_ISL_1594843, EPI_ISL_1594847, EPI_ISL_1594854, EPI_ISL_1594862, EPI_ISL_1594871, EPI_ISL_1594874, EPI_ISL_1594876, EPI_ISL_1594892, EPI_ISL_1594895, EPI_ISL_1594898, EPI_ISL_1594900, EPI_ISL_1594904, EPI_ISL_1594911, EPI_ISL_1594918, EPI_ISL_1594924, EPI_ISL_1594927, EPI_ISL_1594933, EPI_ISL_1594936, EPI_ISL_1594966, EPI_ISL_1594983, EPI_ISL_1594988, EPI_ISL_1615689, EPI_ISL_1615692, EPI_ISL_1615693, EPI_ISL_1615694, EPI_ISL_1615695, EPI_ISL_1615699, EPI_ISL_1615700, EPI_ISL_1615702, EPI_ISL_1615703, EPI_ISL_1615705, EPI_ISL_1615709, EPI_ISL_1615714, EPI_ISL_1615717, EPI_ISL_1615719, EPI_ISL_1615723, EPI_ISL_1615726, EPI_ISL_1615728, EPI_ISL_1615731, EPI_ISL_1615732, EPI_ISL_1615734, EPI_ISL_1615738, EPI_ISL_1615740, EPI_ISL_1615742, EPI_ISL_1615744, EPI_ISL_1615745, EPI_ISL_1615751, EPI_ISL_1615753, EPI_ISL_1615754, EPI_ISL_1615755, EPI_ISL_1615756, EPI_ISL_1615761, EPI_ISL_1615762, EPI_ISL_1615764, EPI_ISL_1615765, EPI_ISL_1615768, EPI_ISL_1615769, EPI_ISL_1615771, EPI_ISL_1615775, EPI_ISL_1615776, EPI_ISL_1615780, EPI_ISL_1615781, EPI_ISL_1615784, EPI_ISL_1615787, EPI_ISL_1615789, EPI_ISL_1615793, EPI_ISL_1615797, EPI_ISL_1615798, EPI_ISL_1615800, EPI_ISL_1615807, EPI_ISL_1615812, EPI_ISL_1615816, EPI_ISL_1615818, EPI_ISL_1615820, EPI_ISL_1615822, EPI_ISL_1615823, EPI_ISL_1615827, EPI_ISL_1615828, EPI_ISL_1615829, EPI_ISL_1615830, EPI_ISL_1615831, EPI_ISL_1615832, EPI_ISL_1615836, EPI_ISL_1615838, EPI_ISL_1615839, EPI_ISL_1615842, EPI_ISL_1615848, EPI_ISL_1615854, EPI_ISL_1615856, EPI_ISL_1615857, EPI_ISL_1615861, EPI_ISL_1615862, EPI_ISL_1615863, EPI_ISL_1615865, EPI_ISL_1615866, EPI_ISL_1615870, EPI_ISL_1615875, EPI_ISL_1615877, EPI_ISL_1615878, EPI_ISL_1615880, EPI_ISL_1615883, EPI_ISL_1615884, EPI_ISL_1615886, EPI_ISL_1615888, EPI_ISL_1615889, EPI_ISL_1615890, EPI_ISL_1615891, EPI_ISL_1615893, EPI_ISL_1615894, EPI_ISL_1615896, EPI_ISL_1615897, EPI_ISL_1615900, EPI_ISL_1615904, EPI_ISL_1615906, EPI_ISL_1615907, EPI_ISL_1615909, EPI_ISL_1615910, EPI_ISL_1615911, EPI_ISL_1615912, EPI_ISL_1615914, EPI_ISL_1615915, EPI_ISL_1615916, EPI_ISL_1615921, EPI_ISL_1615922, EPI_ISL_1615923, EPI_ISL_1615926, EPI_ISL_1615928, EPI_ISL_1615929, EPI_ISL_1615932, EPI_ISL_1616188, EPI_ISL_1616261, EPI_ISL_1616272, EPI_ISL_1616273, EPI_ISL_1616281, EPI_ISL_1616285, EPI_ISL_1616286, EPI_ISL_1630292, EPI_ISL_1630314, EPI_ISL_1630333, EPI_ISL_1630366, EPI_ISL_1630387, EPI_ISL_1630416, EPI_ISL_1630432, EPI_ISL_1630435, EPI_ISL_1631097, EPI_ISL_1631100, EPI_ISL_1631124, EPI_ISL_1631158, EPI_ISL_1631162, EPI_ISL_1631179, EPI_ISL_1631190, EPI_ISL_1631201, EPI_ISL_1631215, EPI_ISL_1631219, EPI_ISL_1631227, EPI_ISL_1631246, EPI_ISL_1631263, EPI_ISL_1631279, EPI_ISL_1631284, EPI_ISL_1631291, EPI_ISL_1631305, EPI_ISL_1631332, EPI_ISL_1631424, EPI_ISL_1631997, EPI_ISL_1631998, EPI_ISL_1632001, EPI_ISL_1632004, EPI_ISL_1632008, EPI_ISL_1632023, EPI_ISL_1632069, EPI_ISL_1632152, EPI_ISL_1632155, EPI_ISL_1632185, EPI_ISL_1632237, EPI_ISL_1632299, EPI_ISL_1632305, EPI_ISL_1632306, EPI_ISL_1632311, EPI_ISL_1632314, EPI_ISL_1632316, EPI_ISL_1632318, EPI_ISL_1632320, EPI_ISL_1632325, EPI_ISL_1632326, EPI_ISL_1632332, EPI_ISL_1632333, EPI_ISL_1632335, EPI_ISL_1632343, EPI_ISL_1632359, EPI_ISL_1632361, EPI_ISL_1632365, EPI_ISL_1632368, EPI_ISL_1632371, EPI_ISL_1632372, EPI_ISL_1632379, EPI_ISL_1632381, EPI_ISL_1632383, EPI_ISL_1632384, EPI_ISL_1632390, EPI_ISL_1632394, EPI_ISL_1632395, EPI_ISL_1632401, EPI_ISL_1632405, EPI_ISL_1632407, EPI_ISL_1632410, EPI_ISL_1632412, EPI_ISL_1632419, EPI_ISL_1632426, EPI_ISL_1632433, EPI_ISL_1632434, EPI_ISL_1632437, EPI_ISL_1632448, EPI_ISL_1632449, EPI_ISL_1632450, EPI_ISL_1632461, EPI_ISL_1632463, EPI_ISL_1632467, EPI_ISL_1632469, EPI_ISL_1632471, EPI_ISL_1632475, EPI_ISL_1632476, EPI_ISL_1632481, EPI_ISL_1634743, EPI_ISL_1635163, EPI_ISL_1635165, EPI_ISL_1635195, EPI_ISL_1635207, EPI_ISL_1635223, EPI_ISL_1635344, EPI_ISL_1635351, EPI_ISL_1635356, EPI_ISL_1635359, EPI_ISL_1635361, EPI_ISL_1635371, EPI_ISL_1635379, EPI_ISL_1635392, EPI_ISL_1635400, EPI_ISL_1635403, EPI_ISL_1635404, EPI_ISL_1652958, EPI_ISL_1652968, EPI_ISL_1652970, EPI_ISL_1652971, EPI_ISL_1652976, EPI_ISL_1652978, EPI_ISL_1652980, EPI_ISL_1652981, EPI_ISL_1652983, EPI_ISL_1652984, EPI_ISL_1652989, EPI_ISL_1652993, EPI_ISL_1652994, EPI_ISL_1652995, EPI_ISL_1652996, EPI_ISL_1653001, EPI_ISL_1653005, EPI_ISL_1653010, EPI_ISL_1653015, EPI_ISL_1653020, EPI_ISL_1653021, EPI_ISL_1653028, EPI_ISL_1653033, EPI_ISL_1653042, EPI_ISL_1653043, EPI_ISL_1653045, EPI_ISL_1653050, EPI_ISL_1653051, EPI_ISL_1653061, EPI_ISL_1653069, EPI_ISL_1653073, EPI_ISL_1653081, EPI_ISL_1653083, EPI_ISL_1653090, EPI_ISL_1653093, EPI_ISL_1653096, EPI_ISL_1653097, EPI_ISL_1653099, EPI_ISL_1653110, EPI_ISL_1653111, EPI_ISL_1653114, EPI_ISL_1653116, EPI_ISL_1653119, EPI_ISL_1653120, EPI_ISL_1653121, EPI_ISL_1653130, EPI_ISL_1653132, EPI_ISL_1653133, EPI_ISL_1653141, EPI_ISL_1653146, EPI_ISL_1653147, EPI_ISL_1653156, EPI_ISL_1653157, EPI_ISL_1653159, EPI_ISL_1653167, EPI_ISL_1653174, EPI_ISL_1653178, EPI_ISL_1653181, EPI_ISL_1653191, EPI_ISL_1653197, EPI_ISL_1653200, EPI_ISL_1653201, EPI_ISL_1653233, EPI_ISL_1653244, EPI_ISL_1653251, EPI_ISL_1653257, EPI_ISL_1653268, EPI_ISL_1653272, EPI_ISL_1653276, EPI_ISL_1653278, EPI_ISL_1653281, EPI_ISL_1653282, EPI_ISL_1653286, EPI_ISL_1653287, EPI_ISL_1653289, EPI_ISL_1653300, EPI_ISL_1653305, EPI_ISL_1653310, EPI_ISL_1653315, EPI_ISL_1653316, EPI_ISL_1653320, EPI_ISL_1653326, EPI_ISL_1653345, EPI_ISL_1653350, EPI_ISL_1653359, EPI_ISL_1653362, EPI_ISL_1653368, EPI_ISL_1653371, EPI_ISL_1653372, EPI_ISL_1653374, EPI_ISL_1653375, EPI_ISL_1653378, EPI_ISL_1653382, EPI_ISL_1653386, EPI_ISL_1653389, EPI_ISL_1653390, EPI_ISL_1653403, EPI_ISL_1653406, EPI_ISL_1653417, EPI_ISL_1653419, EPI_ISL_1653422, EPI_ISL_1653423, EPI_ISL_1653424, EPI_ISL_1653426, EPI_ISL_1653428, EPI_ISL_1653429, EPI_ISL_1653431, EPI_ISL_1653449, EPI_ISL_1653455, EPI_ISL_1653457, EPI_ISL_1653463, EPI_ISL_1653469, EPI_ISL_1653472, EPI_ISL_1653473, EPI_ISL_1653480, EPI_ISL_1653498, EPI_ISL_1653553, EPI_ISL_1653598, EPI_ISL_1653639, EPI_ISL_1653650, EPI_ISL_1653666, EPI_ISL_1653675, EPI_ISL_1653719, EPI_ISL_1653755, EPI_ISL_1653757, EPI_ISL_1653759, EPI_ISL_1653769, EPI_ISL_1653770, EPI_ISL_1673053, EPI_ISL_1673055, EPI_ISL_1673056, EPI_ISL_1673058, EPI_ISL_1673061, EPI_ISL_1673087, EPI_ISL_1673094, EPI_ISL_1673103, EPI_ISL_1673115, EPI_ISL_1673132, EPI_ISL_1673140, EPI_ISL_1673141, EPI_ISL_1673142, EPI_ISL_1673147, EPI_ISL_1697443, EPI_ISL_1697453, EPI_ISL_1697468, EPI_ISL_1697469, EPI_ISL_1697472, EPI_ISL_1697482, EPI_ISL_1697484, EPI_ISL_1697486, EPI_ISL_1697491, EPI_ISL_1697495, EPI_ISL_1697496, EPI_ISL_1697503, EPI_ISL_1697508, EPI_ISL_1697514, EPI_ISL_1697517, EPI_ISL_1697519, EPI_ISL_1697523, EPI_ISL_1697525, EPI_ISL_1697529, EPI_ISL_1697531, EPI_ISL_1697532, EPI_ISL_1697534, EPI_ISL_1697536, EPI_ISL_1697546, EPI_ISL_1697564, EPI_ISL_1697573, EPI_ISL_1697576, EPI_ISL_1697588, EPI_ISL_1697595, EPI_ISL_1697599, EPI_ISL_1697606, EPI_ISL_1697607, EPI_ISL_1697613, EPI_ISL_1697620, EPI_ISL_1697623, EPI_ISL_1697625, EPI_ISL_1697630, EPI_ISL_1697635, EPI_ISL_1697636, EPI_ISL_1697639, EPI_ISL_1697640, EPI_ISL_1697648, EPI_ISL_1697652, EPI_ISL_1697655, EPI_ISL_1697657, EPI_ISL_1697660, EPI_ISL_1697664, EPI_ISL_1697666, EPI_ISL_1697673, EPI_ISL_1697677, EPI_ISL_1697678, EPI_ISL_1697693, EPI_ISL_1697695, EPI_ISL_1697698, EPI_ISL_1697704, EPI_ISL_1697709, EPI_ISL_1697710, EPI_ISL_1697717, EPI_ISL_1697722, EPI_ISL_1697723, EPI_ISL_1697725, EPI_ISL_1697735, EPI_ISL_1697736, EPI_ISL_1697749, EPI_ISL_1697761, EPI_ISL_1697772, EPI_ISL_1697779, EPI_ISL_1697787, EPI_ISL_1697802, EPI_ISL_1697803, EPI_ISL_1697807, EPI_ISL_1697813, EPI_ISL_1697823, EPI_ISL_1697829, EPI_ISL_1697837, EPI_ISL_1697851, EPI_ISL_1697852, EPI_ISL_1697864, EPI_ISL_1697867, EPI_ISL_1697886, EPI_ISL_1697887, EPI_ISL_1697898, EPI_ISL_1697907, EPI_ISL_1697908, EPI_ISL_1697912, EPI_ISL_1697914, EPI_ISL_1697918, EPI_ISL_1697919, EPI_ISL_1697923, EPI_ISL_1697925, EPI_ISL_1697932, EPI_ISL_1697942, EPI_ISL_1697946, EPI_ISL_1697956, EPI_ISL_1697960, EPI_ISL_1697962, EPI_ISL_1697963, EPI_ISL_1697964, EPI_ISL_1697965, EPI_ISL_1698255, EPI_ISL_1698261, EPI_ISL_1698262, EPI_ISL_1698264, EPI_ISL_1698270, EPI_ISL_1698272, EPI_ISL_1698283, EPI_ISL_1698314, EPI_ISL_1698315, EPI_ISL_1698324, EPI_ISL_1698347, EPI_ISL_1698361, EPI_ISL_1698364, EPI_ISL_1698370, EPI_ISL_1698378, EPI_ISL_1698392, EPI_ISL_1698394, EPI_ISL_1698405, EPI_ISL_1698409, EPI_ISL_1698410, EPI_ISL_1698412, EPI_ISL_1698431, EPI_ISL_1698439, EPI_ISL_1698447, EPI_ISL_1698458, EPI_ISL_1698461, EPI_ISL_1698463, EPI_ISL_1698486, EPI_ISL_1698493, EPI_ISL_1698498, EPI_ISL_1698501, EPI_ISL_1698502, EPI_ISL_1698508, EPI_ISL_1698510, EPI_ISL_1698520, EPI_ISL_1698534, EPI_ISL_1700177, EPI_ISL_1700182, EPI_ISL_1700183, EPI_ISL_1700187, EPI_ISL_1700203, EPI_ISL_1700217, EPI_ISL_1700220, EPI_ISL_1700237, EPI_ISL_1700238, EPI_ISL_1700245, EPI_ISL_1700246, EPI_ISL_1700248, EPI_ISL_1700256, EPI_ISL_1700262, EPI_ISL_1700263, EPI_ISL_1700268, EPI_ISL_1700283, EPI_ISL_1700288, EPI_ISL_1700292, EPI_ISL_1700293, EPI_ISL_1700297, EPI_ISL_1700301, EPI_ISL_1700318, EPI_ISL_1700325, EPI_ISL_1700326, EPI_ISL_1700336, EPI_ISL_1700342, EPI_ISL_1700360, EPI_ISL_1700365, EPI_ISL_1700366, EPI_ISL_1700367, EPI_ISL_1700371, EPI_ISL_1700393, EPI_ISL_1700399, EPI_ISL_1700408, EPI_ISL_1700411, EPI_ISL_1700414, EPI_ISL_1700421, EPI_ISL_1700423, EPI_ISL_1700426, EPI_ISL_1700442, EPI_ISL_1700443, EPI_ISL_1700444, EPI_ISL_1700449, EPI_ISL_1700458, EPI_ISL_1700460, EPI_ISL_1700466, EPI_ISL_1700475, EPI_ISL_1700485, EPI_ISL_1700510, EPI_ISL_1700511, EPI_ISL_1700514, EPI_ISL_1700515, EPI_ISL_1719144, EPI_ISL_1719439, EPI_ISL_1719487, EPI_ISL_1719569, EPI_ISL_1719574, EPI_ISL_1719592, EPI_ISL_1719635, EPI_ISL_1719645, EPI_ISL_1719697, EPI_ISL_1719727, EPI_ISL_1719732, EPI_ISL_1719734, EPI_ISL_1719735, EPI_ISL_1719736, EPI_ISL_1719737, EPI_ISL_1719738, EPI_ISL_1719739, EPI_ISL_1719744, EPI_ISL_1742168, EPI_ISL_1742176, EPI_ISL_1742178, EPI_ISL_1742179, EPI_ISL_1742183, EPI_ISL_1742203, EPI_ISL_1742229, EPI_ISL_1742242, EPI_ISL_1742288, EPI_ISL_1742292, EPI_ISL_1987383

| | | | |
|---|---|---|---|
| see above | Lighthouse Lab in Cambridge | Wellcome Sanger Institute for the COVID-19 Genomics UK (COG-UK) Consortium | Cordelia Langford; David K. Jackson; Dominic Kwiatkowski; Ewan Harrison; Ian Johnston; Jeffrey Barrett; John Sillitoe on behalf of the Wellcome Sanger Institute COVID-19 Surveillance Team; Rob Howes; Roberto Amato; Sonia Goncalves; The Lighthouse Lab in Cambridge and Alex Alderton |

EPI_ISL_1564039, EPI_ISL_1564049, EPI_ISL_1564054, EPI_ISL_1564055, EPI_ISL_1564058, EPI_ISL_1564074, EPI_ISL_1564082, EPI_ISL_1564085, EPI_ISL_1564090, EPI_ISL_1564091, EPI_ISL_1564096, EPI_ISL_1564104, EPI_ISL_1564109, EPI_ISL_1564111, EPI_ISL_1564114, EPI_ISL_1564120, EPI_ISL_1564133, EPI_ISL_1564135, EPI_ISL_1564142, EPI_ISL_1564143, EPI_ISL_1564148, EPI_ISL_1564151, EPI_ISL_1564155, EPI_ISL_1564156, EPI_ISL_1564157, EPI_ISL_1564162, EPI_ISL_1564164, EPI_ISL_1564168, EPI_ISL_1564177, EPI_ISL_1564186, EPI_ISL_1564191, EPI_ISL_1564192, EPI_ISL_1564199, EPI_ISL_1564201, EPI_ISL_1564213, EPI_ISL_1564214, EPI_ISL_1564216, EPI_ISL_1564218, EPI_ISL_1564220, EPI_ISL_1564232, EPI_ISL_1564235, EPI_ISL_1564238, EPI_ISL_1564248, EPI_ISL_1564250, EPI_ISL_1564257, EPI_ISL_1564260, EPI_ISL_1564261, EPI_ISL_1564266, EPI_ISL_1564282, EPI_ISL_1564288, EPI_ISL_1564290, EPI_ISL_1564292, EPI_ISL_1564293, EPI_ISL_1564306, EPI_ISL_1564309, EPI_ISL_1564320, EPI_ISL_1564322, EPI_ISL_1564330, EPI_ISL_1564331, EPI_ISL_1584168, EPI_ISL_1584182, EPI_ISL_1584183, EPI_ISL_1584187, EPI_ISL_1584188, EPI_ISL_1584189, EPI_ISL_1584191, EPI_ISL_1584192, EPI_ISL_1584206, EPI_ISL_1584208, EPI_ISL_1584213, EPI_ISL_1584215, EPI_ISL_1584217, EPI_ISL_1584232, EPI_ISL_1584235, EPI_ISL_1584239, EPI_ISL_1584240, EPI_ISL_1584241, EPI_ISL_1584245, EPI_ISL_1584248, EPI_ISL_1584250, EPI_ISL_1584256, EPI_ISL_1584258, EPI_ISL_1584259, EPI_ISL_1584260, EPI_ISL_1584279, EPI_ISL_1584290, EPI_ISL_1584298, EPI_ISL_1584309, EPI_ISL_1584310, EPI_ISL_1584327, EPI_ISL_1584330, EPI_ISL_1584334, EPI_ISL_1584337, EPI_ISL_1584345, EPI_ISL_1584348, EPI_ISL_1584353, EPI_ISL_1584356, EPI_ISL_1584357, EPI_ISL_1584368, EPI_ISL_1584371, EPI_ISL_1584375, EPI_ISL_1584380, EPI_ISL_1584383, EPI_ISL_1584398, EPI_ISL_1584402, EPI_ISL_1584422, EPI_ISL_1584436, EPI_ISL_1584437, EPI_ISL_1584441, EPI_ISL_1584442, EPI_ISL_1584443, EPI_ISL_1584445, EPI_ISL_1584446, EPI_ISL_1584447, EPI_ISL_1584448, EPI_ISL_1584449, EPI_ISL_1584450, EPI_ISL_1584451, EPI_ISL_1584452, EPI_ISL_1584453, EPI_ISL_1584454, EPI_ISL_1584455, EPI_ISL_1584456, EPI_ISL_1584458, EPI_ISL_1584459, EPI_ISL_1584461, EPI_ISL_1584462, EPI_ISL_1584463, EPI_ISL_1584464, EPI_ISL_1584465, EPI_ISL_1584468, EPI_ISL_1584471, EPI_ISL_1584474, EPI_ISL_1584477, EPI_ISL_1584479, EPI_ISL_1584480, EPI_ISL_1584482, EPI_ISL_1584485, EPI_ISL_1584486, EPI_ISL_1584491, EPI_ISL_1584492, EPI_ISL_1584494, EPI_ISL_1584495, EPI_ISL_1584496, EPI_ISL_1584499, EPI_ISL_1584500, EPI_ISL_1584501, EPI_ISL_1584502, EPI_ISL_1584504, EPI_ISL_1584505, EPI_ISL_1584506, EPI_ISL_1584507, EPI_ISL_1584509, EPI_ISL_1584510, EPI_ISL_1584512, EPI_ISL_1584514, EPI_ISL_1584516, EPI_ISL_1584518, EPI_ISL_1584519, EPI_ISL_1584520, EPI_ISL_1584521, EPI_ISL_1584523, EPI_ISL_1584525, EPI_ISL_1584526, EPI_ISL_1594453, EPI_ISL_1594454, EPI_ISL_1594455, EPI_ISL_1594456, EPI_ISL_1594458, EPI_ISL_1594459, EPI_ISL_1594460, EPI_ISL_1594461, EPI_ISL_1594462, EPI_ISL_1594467, EPI_ISL_1594468, EPI_ISL_1594469, EPI_ISL_1594470, EPI_ISL_1594472, EPI_ISL_1594473, EPI_ISL_1594474, EPI_ISL_1594475, EPI_ISL_1594476, EPI_ISL_1594477, EPI_ISL_1594479, EPI_ISL_1594480, EPI_ISL_1594481, EPI_ISL_1594483, EPI_ISL_1594485, EPI_ISL_1594486, EPI_ISL_1594487, EPI_ISL_1594488, EPI_ISL_1594489, EPI_ISL_1594490, EPI_ISL_1594491, EPI_ISL_1594493, EPI_ISL_1594495, EPI_ISL_1594496, EPI_ISL_1594497, EPI_ISL_1594498, EPI_ISL_1594499, EPI_ISL_1594501, EPI_ISL_1594502, EPI_ISL_1594503, EPI_ISL_1594504, EPI_ISL_1594505, EPI_ISL_1594506, EPI_ISL_1594508, EPI_ISL_1594509, EPI_ISL_1594510, EPI_ISL_1594511, EPI_ISL_1594513, EPI_ISL_1594514, EPI_ISL_1594515, EPI_ISL_1594516, EPI_ISL_1594517, EPI_ISL_1594518, EPI_ISL_1594519, EPI_ISL_1594521, EPI_ISL_1594523, EPI_ISL_1594524, EPI_ISL_1594525, EPI_ISL_1594526, EPI_ISL_1594527, EPI_ISL_1594529, EPI_ISL_1594531, EPI_ISL_1594532, EPI_ISL_1594533, EPI_ISL_1594534, EPI_ISL_1594535, EPI_ISL_1594537, EPI_ISL_1594538, EPI_ISL_1594539, EPI_ISL_1594542, EPI_ISL_1594543, EPI_ISL_1594544, EPI_ISL_1594545, EPI_ISL_1594546, EPI_ISL_1594548, EPI_ISL_1594550, EPI_ISL_1594552, EPI_ISL_1594553, EPI_ISL_1594554, EPI_ISL_1594555, EPI_ISL_1594556, EPI_ISL_1594558, EPI_ISL_1594559, EPI_ISL_1594560, EPI_ISL_1594561, EPI_ISL_1594562, EPI_ISL_1594564, EPI_ISL_1594565, EPI_ISL_1594566, EPI_ISL_1594568, EPI_ISL_1594570, EPI_ISL_1594572, EPI_ISL_1594573, EPI_ISL_1594575, EPI_ISL_1594576, EPI_ISL_1594577, EPI_ISL_1594578, EPI_ISL_1594579, EPI_ISL_1594580, EPI_ISL_1594581, EPI_ISL_1594583, EPI_ISL_1594584, EPI_ISL_1594585, EPI_ISL_1594586, EPI_ISL_1594587, EPI_ISL_1594589, EPI_ISL_1594591, EPI_ISL_1594592, EPI_ISL_1594593, EPI_ISL_1594594, EPI_ISL_1594595, EPI_ISL_1594597, EPI_ISL_1594598, EPI_ISL_1594599, EPI_ISL_1594600, EPI_ISL_1594601, EPI_ISL_1594602, EPI_ISL_1594603, EPI_ISL_1594604, EPI_ISL_1594605, EPI_ISL_1594606, EPI_ISL_1594607, EPI_ISL_1594608, EPI_ISL_1594609, EPI_ISL_1594610, EPI_ISL_1594611, EPI_ISL_1594614, EPI_ISL_1594616, EPI_ISL_1594617, EPI_ISL_1594618, EPI_ISL_1594619, EPI_ISL_1594620, EPI_ISL_1594621, EPI_ISL_1594623, EPI_ISL_1594626, EPI_ISL_1594627, EPI_ISL_1594628, EPI_ISL_1594629, EPI_ISL_1594630, EPI_ISL_1594631, EPI_ISL_1594632, EPI_ISL_1594633, EPI_ISL_1594634, EPI_ISL_1594636, EPI_ISL_1594637, EPI_ISL_1594638, EPI_ISL_1594640, EPI_ISL_1594642, EPI_ISL_1594644, EPI_ISL_1594645, EPI_ISL_1594646, EPI_ISL_1594647, EPI_ISL_1594648, EPI_ISL_1594651, EPI_ISL_1594652, EPI_ISL_1594653, EPI_ISL_1594654, EPI_ISL_1594656, EPI_ISL_1594657, EPI_ISL_1594658, EPI_ISL_1594663, EPI_ISL_1594664, EPI_ISL_1594665, EPI_ISL_1594666, EPI_ISL_1594670, EPI_ISL_1594671, EPI_ISL_1594672, EPI_ISL_1594673, EPI_ISL_1594674, EPI_ISL_1594676, EPI_ISL_1594677, EPI_ISL_1594683, EPI_ISL_1594684, EPI_ISL_1594685, EPI_ISL_1594686, EPI_ISL_1594688, EPI_ISL_1594690, EPI_ISL_1594691, EPI_ISL_1594692, EPI_ISL_1594693, EPI_ISL_1594695, EPI_ISL_1594696, EPI_ISL_1594697, EPI_ISL_1594698, EPI_ISL_1594699, EPI_ISL_1594701, EPI_ISL_1594702, EPI_ISL_1594704, EPI_ISL_1594705, EPI_ISL_1594706, EPI_ISL_1594707, EPI_ISL_1594708, EPI_ISL_1594709, EPI_ISL_1594710, EPI_ISL_1594711, EPI_ISL_1594713, EPI_ISL_1594718, EPI_ISL_1594720, EPI_ISL_1594721, EPI_ISL_1594723, EPI_ISL_1594727, EPI_ISL_1594729, EPI_ISL_1594735, EPI_ISL_1594743, EPI_ISL_1594744, EPI_ISL_1594746, EPI_ISL_1594752, EPI_ISL_1594754, EPI_ISL_1594755, EPI_ISL_1594757, EPI_ISL_1594761, EPI_ISL_1594771, EPI_ISL_1594772, EPI_ISL_1594777, EPI_ISL_1594779, EPI_ISL_1594783, EPI_ISL_1594790, EPI_ISL_1594795, EPI_ISL_1594796, EPI_ISL_1594805, EPI_ISL_1594806, EPI_ISL_1594808, EPI_ISL_1594809, EPI_ISL_1594816, EPI_ISL_1594829, EPI_ISL_1594839, EPI_ISL_1594840, EPI_ISL_1594844, EPI_ISL_1594845, EPI_ISL_1594851, EPI_ISL_1594852, EPI_ISL_1594856, EPI_ISL_1594858, EPI_ISL_1594859, EPI_ISL_1594878, EPI_ISL_1594880, EPI_ISL_1594896, EPI_ISL_1594903, EPI_ISL_1594909, EPI_ISL_1594912, EPI_ISL_1594913, EPI_ISL_1594916, EPI_ISL_1594920, EPI_ISL_1594922, EPI_ISL_1594923, EPI_ISL_1594925, EPI_ISL_1594928, EPI_ISL_1594930, EPI_ISL_1594932, EPI_ISL_1594939, EPI_ISL_1594940, EPI_ISL_1594942, EPI_ISL_1594943, EPI_ISL_1594952, EPI_ISL_1594959, EPI_ISL_1594960, EPI_ISL_1594968, EPI_ISL_1594971, EPI_ISL_1594972, EPI_ISL_1594978, EPI_ISL_1594979, EPI_ISL_1594984, EPI_ISL_1594986, EPI_ISL_1594991, EPI_ISL_1594995, EPI_ISL_1594996, EPI_ISL_1594999, EPI_ISL_1595002, EPI_ISL_1595004, EPI_ISL_1595005, EPI_ISL_1595008, EPI_ISL_1595010, EPI_ISL_1595011, EPI_ISL_1595012, EPI_ISL_1595017, EPI_ISL_1595018, EPI_ISL_1595020, EPI_ISL_1595022, EPI_ISL_1595023, EPI_ISL_1595026, EPI_ISL_1595027, EPI_ISL_1595031, EPI_ISL_1595037, EPI_ISL_1595040, EPI_ISL_1595041, EPI_ISL_1595045, EPI_ISL_1595046, EPI_ISL_1595049, EPI_ISL_1595050, EPI_ISL_1595051, EPI_ISL_1595053, EPI_ISL_1595055, EPI_ISL_1595056, EPI_ISL_1595057, EPI_ISL_1595060, EPI_ISL_1595061, EPI_ISL_1595062, EPI_ISL_1595064, EPI_ISL_1595065, EPI_ISL_1595066, EPI_ISL_1595067, EPI_ISL_1595068, EPI_ISL_1595069, EPI_ISL_1595071, EPI_ISL_1595072, EPI_ISL_1595074, EPI_ISL_1595076, EPI_ISL_1595077, EPI_ISL_1595078, EPI_ISL_1595080, EPI_ISL_1595081, EPI_ISL_1595085, EPI_ISL_1595087, EPI_ISL_1595088, EPI_ISL_1595089, EPI_ISL_1595091, EPI_ISL_1595093, EPI_ISL_1595094, EPI_ISL_1595095, EPI_ISL_1595096, EPI_ISL_1595097, EPI_ISL_1595102, EPI_ISL_1595103, EPI_ISL_1595105, EPI_ISL_1595109, EPI_ISL_1595110, EPI_ISL_1595113, EPI_ISL_1595114, EPI_ISL_1595115, EPI_ISL_1595116, EPI_ISL_1595117, EPI_ISL_1595118, EPI_ISL_1595121, EPI_ISL_1595125, EPI_ISL_1595126, EPI_ISL_1595127, EPI_ISL_1595129, EPI_ISL_1595133, EPI_ISL_1595136, EPI_ISL_1595137, EPI_ISL_1595138, EPI_ISL_1595143, EPI_ISL_1595145, EPI_ISL_1595149, EPI_ISL_1595150, EPI_ISL_1595156, EPI_ISL_1595157, EPI_ISL_1595158, EPI_ISL_1595162, EPI_ISL_1595166, EPI_ISL_1595170, EPI_ISL_1595171, EPI_ISL_1595174, EPI_ISL_1595176, EPI_ISL_1595177, EPI_ISL_1595178, EPI_ISL_1595179, EPI_ISL_1595181, EPI_ISL_1595182, EPI_ISL_1595183, EPI_ISL_1595186, EPI_ISL_1595188, EPI_ISL_1595192, EPI_ISL_1595194, EPI_ISL_1615691, EPI_ISL_1615721, EPI_ISL_1615727, EPI_ISL_1615739, EPI_ISL_1615741, EPI_ISL_1615746, EPI_ISL_1615750, EPI_ISL_1615801, EPI_ISL_1615803, EPI_ISL_1615814, EPI_ISL_1615833, EPI_ISL_1615844, EPI_ISL_1615852, EPI_ISL_1615868, EPI_ISL_1615873, EPI_ISL_1615887, EPI_ISL_1615895, EPI_ISL_1615898, EPI_ISL_1615902, EPI_ISL_1615908, EPI_ISL_1615917, EPI_ISL_1615924, EPI_ISL_1615931, EPI_ISL_1615935, EPI_ISL_1615938, EPI_ISL_1615940, EPI_ISL_1615947, EPI_ISL_1615948, EPI_ISL_1615953, EPI_ISL_1615956, EPI_ISL_1615977, EPI_ISL_1615982, EPI_ISL_1615986, EPI_ISL_1615988, EPI_ISL_1615989, EPI_ISL_1615994, EPI_ISL_1615996, EPI_ISL_1615998, EPI_ISL_1615999, EPI_ISL_1616001, EPI_ISL_1616004, EPI_ISL_1616007, EPI_ISL_1616020, EPI_ISL_1616022, EPI_ISL_1616027, EPI_ISL_1616032, EPI_ISL_1616039, EPI_ISL_1616048, EPI_ISL_1616049, EPI_ISL_1616051, EPI_ISL_1616055, EPI_ISL_1616057, EPI_ISL_1616059, EPI_ISL_1616060, EPI_ISL_1616061, EPI_ISL_1616063, EPI_ISL_1616064, EPI_ISL_1616069, EPI_ISL_1616075, EPI_ISL_1616076, EPI_ISL_1616077, EPI_ISL_1616079, EPI_ISL_1616082, EPI_ISL_1616087, EPI_ISL_1616091, EPI_ISL_1616092, EPI_ISL_1616099, EPI_ISL_1616101, EPI_ISL_1616104, EPI_ISL_1616106, EPI_ISL_1616110, EPI_ISL_1616111, EPI_ISL_1616117, EPI_ISL_1616120, EPI_ISL_1616121, EPI_ISL_1616123, EPI_ISL_1616127, EPI_ISL_1616130, EPI_ISL_1616134, EPI_ISL_1616137, EPI_ISL_1616140, EPI_ISL_1616143, EPI_ISL_1616150, EPI_ISL_1616153, EPI_ISL_1616159, EPI_ISL_1616162, EPI_ISL_1616163, EPI_ISL_1616165, EPI_ISL_1616177, EPI_ISL_1616179, EPI_ISL_1616181, EPI_ISL_1616186, EPI_ISL_1616187, EPI_ISL_1616189, EPI_ISL_1616190, EPI_ISL_1616191, EPI_ISL_1616192, EPI_ISL_1616193, EPI_ISL_1616194, EPI_ISL_1616195, EPI_ISL_1616196, EPI_ISL_1616197, EPI_ISL_1616198, EPI_ISL_1616199, EPI_ISL_1616200, EPI_ISL_1616201, EPI_ISL_1616202, EPI_ISL_1616203, EPI_ISL_1616204, EPI_ISL_1616205, EPI_ISL_1616206, EPI_ISL_1616207, EPI_ISL_1616208, EPI_ISL_1616209, EPI_ISL_1616210, EPI_ISL_1616212, EPI_ISL_1616213, EPI_ISL_1616214, EPI_ISL_1616215, EPI_ISL_1616216, EPI_ISL_1616217, EPI_ISL_1616218, EPI_ISL_1616219, EPI_ISL_1616220, EPI_ISL_1616221, EPI_ISL_1616222, EPI_ISL_1616223, EPI_ISL_1616224, EPI_ISL_1616225, EPI_ISL_1616226, EPI_ISL_1616227, EPI_ISL_1616228, EPI_ISL_1616229, EPI_ISL_1616230, EPI_ISL_1616231, EPI_ISL_1616232, EPI_ISL_1616233, EPI_ISL_1616234, EPI_ISL_1616235, EPI_ISL_1616236, EPI_ISL_1616237, EPI_ISL_1616238, EPI_ISL_1616239, EPI_ISL_1616240, EPI_ISL_1616241, EPI_ISL_1616242, EPI_ISL_1616243, EPI_ISL_1616244, EPI_ISL_1616245, EPI_ISL_1616246, EPI_ISL_1616247, EPI_ISL_1616248, EPI_ISL_1616249, EPI_ISL_1616250, EPI_ISL_1616251, EPI_ISL_1616252, EPI_ISL_1616253, EPI_ISL_1616254, EPI_ISL_1616255, EPI_ISL_1616256, EPI_ISL_1616257, EPI_ISL_1616258, EPI_ISL_1616259, EPI_ISL_1616260, EPI_ISL_1616263, EPI_ISL_1616264, EPI_ISL_1616266, EPI_ISL_1616268, EPI_ISL_1616269, EPI_ISL_1616270, EPI_ISL_1616271, EPI_ISL_1616274, EPI_ISL_1616275, EPI_ISL_1616276, EPI_ISL_1616277, EPI_ISL_1616278, EPI_ISL_1616280, EPI_ISL_1616282, EPI_ISL_1616284, EPI_ISL_1616288, EPI_ISL_1616289, EPI_ISL_1616290, EPI_ISL_1616291, EPI_ISL_1616293, EPI_ISL_1616294, EPI_ISL_1616296, EPI_ISL_1616298, EPI_ISL_1616299, EPI_ISL_1616301, EPI_ISL_1616302, EPI_ISL_1616305, EPI_ISL_1616306, EPI_ISL_1616307, EPI_ISL_1616309, EPI_ISL_1616310, EPI_ISL_1616311, EPI_ISL_1616312, EPI_ISL_1616313, EPI_ISL_1616314, EPI_ISL_1616315, EPI_ISL_1616316, EPI_ISL_1616317, EPI_ISL_1616318, EPI_ISL_1616320, EPI_ISL_1616321, EPI_ISL_1616322, EPI_ISL_1616323, EPI_ISL_1616324, EPI_ISL_1616325, EPI_ISL_1616328, EPI_ISL_1616329, EPI_ISL_1616330, EPI_ISL_1616331, EPI_ISL_1616332, EPI_ISL_1616333, EPI_ISL_1616334, EPI_ISL_1616335, EPI_ISL_1616336, EPI_ISL_1616338, EPI_ISL_1616339, EPI_ISL_1616340, EPI_ISL_1616341, EPI_ISL_1616343, EPI_ISL_1616346, EPI_ISL_1616348, EPI_ISL_1616349, EPI_ISL_1616350, EPI_ISL_1616351, EPI_ISL_1616352, EPI_ISL_1616353, EPI_ISL_1616354, EPI_ISL_1616355, EPI_ISL_1616356, EPI_ISL_1616357, EPI_ISL_1616358, EPI_ISL_1616359, EPI_ISL_1616363, EPI_ISL_1616364, EPI_ISL_1616365, EPI_ISL_1616367, EPI_ISL_1616368, EPI_ISL_1616369, EPI_ISL_1616370, EPI_ISL_1616371, EPI_ISL_1616374, EPI_ISL_1616376, EPI_ISL_1616377, EPI_ISL_1616378, EPI_ISL_1616381, EPI_ISL_1616383, EPI_ISL_1616384, EPI_ISL_1616385, EPI_ISL_1616386, EPI_ISL_1616387, EPI_ISL_1616388, EPI_ISL_1616389, EPI_ISL_1616392, EPI_ISL_1616393, EPI_ISL_1616394, EPI_ISL_1616395, EPI_ISL_1616396, EPI_ISL_1616397, EPI_ISL_1616398, EPI_ISL_1616399, EPI_ISL_1616400, EPI_ISL_1616401, EPI_ISL_1616402, EPI_ISL_1616404, EPI_ISL_1616406, EPI_ISL_1616407, EPI_ISL_1616408, EPI_ISL_1616409, EPI_ISL_1616410, EPI_ISL_1616411, EPI_ISL_1616412, EPI_ISL_1616413, EPI_ISL_1616414, EPI_ISL_1616416, EPI_ISL_1616417, EPI_ISL_1616418, EPI_ISL_1616420, EPI_ISL_1616421, EPI_ISL_1616422, EPI_ISL_1616424, EPI_ISL_1616578, EPI_ISL_1616580, EPI_ISL_1616581, EPI_ISL_1616586, EPI_ISL_1616587, EPI_ISL_1616589, EPI_ISL_1616590, EPI_ISL_1616592, EPI_ISL_1616594, EPI_ISL_1616595, EPI_ISL_1616597, EPI_ISL_1616598, EPI_ISL_1616599, EPI_ISL_1616602, EPI_ISL_1630282, EPI_ISL_1630283, EPI_ISL_1630285, EPI_ISL_1630286, EPI_ISL_1630287, EPI_ISL_1630288, EPI_ISL_1630290, EPI_ISL_1630296, EPI_ISL_1630297, EPI_ISL_1630300, EPI_ISL_1630301, EPI_ISL_1630302, EPI_ISL_1630303, EPI_ISL_1630306, EPI_ISL_1630307, EPI_ISL_1630308, EPI_ISL_1630309, EPI_ISL_1630310, EPI_ISL_1630315, EPI_ISL_1630316, EPI_ISL_1630317, EPI_ISL_1630318, EPI_ISL_1630321, EPI_ISL_1630322, EPI_ISL_1630323, EPI_ISL_1630325, EPI_ISL_1630327, EPI_ISL_1630330, EPI_ISL_1630331, EPI_ISL_1630332, EPI_ISL_1630334, EPI_ISL_1630335, EPI_ISL_1630337, EPI_ISL_1630340, EPI_ISL_1630341, EPI_ISL_1630343, EPI_ISL_1630344, EPI_ISL_1630345, EPI_ISL_1630346, EPI_ISL_1630347, EPI_ISL_1630350, EPI_ISL_1630351, EPI_ISL_1630354, EPI_ISL_1630355, EPI_ISL_1630356, EPI_ISL_1630359, EPI_ISL_1630361, EPI_ISL_1630362, EPI_ISL_1630364, EPI_ISL_1630365, EPI_ISL_1630369, EPI_ISL_1630370, EPI_ISL_1630371, EPI_ISL_1630373, EPI_ISL_1630376, EPI_ISL_1630379, EPI_ISL_1630381, EPI_ISL_1630382, EPI_ISL_1630385, EPI_ISL_1630389, EPI_ISL_1630392, EPI_ISL_1630393, EPI_ISL_1630394, EPI_ISL_1630396, EPI_ISL_1630398, EPI_ISL_1630399, EPI_ISL_1630400, EPI_ISL_1630405, EPI_ISL_1630407, EPI_ISL_1630408, EPI_ISL_1630409, EPI_ISL_1630411, EPI_ISL_1630413, EPI_ISL_1630415, EPI_ISL_1630417, EPI_ISL_1630418, EPI_ISL_1630419, EPI_ISL_1630421, EPI_ISL_1630422, EPI_ISL_1630423, EPI_ISL_1630424, EPI_ISL_1630425, EPI_ISL_1630426, EPI_ISL_1630428, EPI_ISL_1630429, EPI_ISL_1630430, EPI_ISL_1630433, EPI_ISL_1630436, EPI_ISL_1630437, EPI_ISL_1630440, EPI_ISL_1630441, EPI_ISL_1630442, EPI_ISL_1630443, EPI_ISL_1630445, EPI_ISL_1630446, EPI_ISL_1630447, EPI_ISL_1630448, EPI_ISL_1630449, EPI_ISL_1630452, EPI_ISL_1630453, EPI_ISL_1630454, EPI_ISL_1630456, EPI_ISL_1630457, EPI_ISL_1630458, EPI_ISL_1630459, EPI_ISL_1630460, EPI_ISL_1630462, EPI_ISL_1630466, EPI_ISL_1630467, EPI_ISL_1630468, EPI_ISL_1630470, EPI_ISL_1630471, EPI_ISL_1630472, EPI_ISL_1630473, EPI_ISL_1630477, EPI_ISL_1630481, EPI_ISL_1630482, EPI_ISL_1630483, EPI_ISL_1630484, EPI_ISL_1630485, EPI_ISL_1630487, EPI_ISL_1630495, EPI_ISL_1630496, EPI_ISL_1630498, EPI_ISL_1630499, EPI_ISL_1630501, EPI_ISL_1630502, EPI_ISL_1630504, EPI_ISL_1630505, EPI_ISL_1630507, EPI_ISL_1630509, EPI_ISL_1630511, EPI_ISL_1630512, EPI_ISL_1630514, EPI_ISL_1630515, EPI_ISL_1630830, EPI_ISL_1630834, EPI_ISL_1630846, EPI_ISL_1630847, EPI_ISL_1630848, EPI_ISL_1630850, EPI_ISL_1630853, EPI_ISL_1630855, EPI_ISL_1630871, EPI_ISL_1630872, EPI_ISL_1630879, EPI_ISL_1630889, EPI_ISL_1630896, EPI_ISL_1630908, EPI_ISL_1630910, EPI_ISL_1630933, EPI_ISL_1630944, EPI_ISL_1630948, EPI_ISL_1630949, EPI_ISL_1630958, EPI_ISL_1630968, EPI_ISL_1630978, EPI_ISL_1630984, EPI_ISL_1630988, EPI_ISL_1630989, EPI_ISL_1631000, EPI_ISL_1631008, EPI_ISL_1631009, EPI_ISL_1631017, EPI_ISL_1631018, EPI_ISL_1631024, EPI_ISL_1631028, EPI_ISL_1631030, EPI_ISL_1631039, EPI_ISL_1631047, EPI_ISL_1631051, EPI_ISL_1631056, EPI_ISL_1631067, EPI_ISL_1631073, EPI_ISL_1631083, EPI_ISL_1631089, EPI_ISL_1631090, EPI_ISL_1631091, EPI_ISL_1631095, EPI_ISL_1631104, EPI_ISL_1631107, EPI_ISL_1631108, EPI_ISL_1631109, EPI_ISL_1631110, EPI_ISL_1631111, EPI_ISL_1631112, EPI_ISL_1631113, EPI_ISL_1631114, EPI_ISL_1631119, EPI_ISL_1631125, EPI_ISL_1631128, EPI_ISL_1631131, EPI_ISL_1631133, EPI_ISL_1631136, EPI_ISL_1631138, EPI_ISL_1631140, EPI_ISL_1631145, EPI_ISL_1631147, EPI_ISL_1631150, EPI_ISL_1631151, EPI_ISL_1631153, EPI_ISL_1631156, EPI_ISL_1631157, EPI_ISL_1631165, EPI_ISL_1631167, EPI_ISL_1631168, EPI_ISL_1631169, EPI_ISL_1631172, EPI_ISL_1631173, EPI_ISL_1631174, EPI_ISL_1631175, EPI_ISL_1631176, EPI_ISL_1631178, EPI_ISL_1631180, EPI_ISL_1631187, EPI_ISL_1631192, EPI_ISL_1631194, EPI_ISL_1631195, EPI_ISL_1631204, EPI_ISL_1631205, EPI_ISL_1631206, EPI_ISL_1631209, EPI_ISL_1631211, EPI_ISL_1631214, EPI_ISL_1631216, EPI_ISL_1631220, EPI_ISL_1631222, EPI_ISL_1631224, EPI_ISL_1631226, EPI_ISL_1631229, EPI_ISL_1631232, EPI_ISL_1631235, EPI_ISL_1631237, EPI_ISL_1631242, EPI_ISL_1631243, EPI_ISL_1631244, EPI_ISL_1631245, EPI_ISL_1631247, EPI_ISL_1631248, EPI_ISL_1631253, EPI_ISL_1631254, EPI_ISL_1631256, EPI_ISL_1631258, EPI_ISL_1631259, EPI_ISL_1631261, EPI_ISL_1631262, EPI_ISL_1631264, EPI_ISL_1631265, EPI_ISL_1631266, EPI_ISL_1631268, EPI_ISL_1631269, EPI_ISL_1631270, EPI_ISL_1631272, EPI_ISL_1631275, EPI_ISL_1631278, EPI_ISL_1631282, EPI_ISL_1631285, EPI_ISL_1631288, EPI_ISL_1631294, EPI_ISL_1631295, EPI_ISL_1631296, EPI_ISL_1631297, EPI_ISL_1631298, EPI_ISL_1631301, EPI_ISL_1631302, EPI_ISL_1631304, EPI_ISL_1631306, EPI_ISL_1631307, EPI_ISL_1631308, EPI_ISL_1631309, EPI_ISL_1631310, EPI_ISL_1631311, EPI_ISL_1631312, EPI_ISL_1631317, EPI_ISL_1631318, EPI_ISL_1631320, EPI_ISL_1631322, EPI_ISL_1631323, EPI_ISL_1631324, EPI_ISL_1631325, EPI_ISL_1631327, EPI_ISL_1631328, EPI_ISL_1631329, EPI_ISL_1631335, EPI_ISL_1631337, EPI_ISL_1631338, EPI_ISL_1631339, EPI_ISL_1631340, EPI_ISL_1631341, EPI_ISL_1631342, EPI_ISL_1631343, EPI_ISL_1631344, EPI_ISL_1631345, EPI_ISL_1631346, EPI_ISL_1631347, EPI_ISL_1631348, EPI_ISL_1631349, EPI_ISL_1631350, EPI_ISL_1631351, EPI_ISL_1631352, EPI_ISL_1631353, EPI_ISL_1631354, EPI_ISL_1631355, EPI_ISL_1631356, EPI_ISL_1631357, EPI_ISL_1631358, EPI_ISL_1631359, EPI_ISL_1631360, EPI_ISL_1631361, EPI_ISL_1631362, EPI_ISL_1631363, EPI_ISL_1631364, EPI_ISL_1631365, EPI_ISL_1631366, EPI_ISL_1631367, EPI_ISL_1631368, EPI_ISL_1631369, EPI_ISL_1631370, EPI_ISL_1631371, EPI_ISL_1631372, EPI_ISL_1631373, EPI_ISL_1631374, EPI_ISL_1631375, EPI_ISL_1631376, EPI_ISL_1631377, EPI_ISL_1631378, EPI_ISL_1631379, EPI_ISL_1631380, EPI_ISL_1631381, EPI_ISL_1631382, EPI_ISL_1631383, EPI_ISL_1631384, EPI_ISL_1631385, EPI_ISL_1631386, EPI_ISL_1631387, EPI_ISL_1631388, EPI_ISL_1631389, EPI_ISL_1631390, EPI_ISL_1631391, EPI_ISL_1631392, EPI_ISL_1631393, EPI_ISL_1631394, EPI_ISL_1631395, EPI_ISL_1631396, EPI_ISL_1631397, EPI_ISL_1631398, EPI_ISL_1631399, EPI_ISL_1631400, EPI_ISL_1631401, EPI_ISL_1631402, EPI_ISL_1631403, EPI_ISL_1631404, EPI_ISL_1631405, EPI_ISL_1631406, EPI_ISL_1631407, EPI_ISL_1631408, EPI_ISL_1631409, EPI_ISL_1631410, EPI_ISL_1631411, EPI_ISL_1631412, EPI_ISL_1631413, EPI_ISL_1631414, EPI_ISL_1631415, EPI_ISL_1631416, EPI_ISL_1631417, EPI_ISL_1631418, EPI_ISL_1631419, EPI_ISL_1631420, EPI_ISL_1631421, EPI_ISL_1631422, EPI_ISL_1631423, EPI_ISL_1631764, EPI_ISL_1631771, EPI_ISL_1631774, EPI_ISL_1631777, EPI_ISL_1631787, EPI_ISL_1631794, EPI_ISL_1631816, EPI_ISL_1631825, EPI_ISL_1631828, EPI_ISL_1631831, EPI_ISL_1631848, EPI_ISL_1631849, EPI_ISL_1631853, EPI_ISL_1631855, EPI_ISL_1631857, EPI_ISL_1631861, EPI_ISL_1631862, EPI_ISL_1631874, EPI_ISL_1631881, EPI_ISL_1631884, EPI_ISL_1631889, EPI_ISL_1631890, EPI_ISL_1631893, EPI_ISL_1631898, EPI_ISL_1631899, EPI_ISL_1631905, EPI_ISL_1631907, EPI_ISL_1631908, EPI_ISL_1631915, EPI_ISL_1631916, EPI_ISL_1631919, EPI_ISL_1631925, EPI_ISL_1631926, EPI_ISL_1631927, EPI_ISL_1631931, EPI_ISL_1631935, EPI_ISL_1631937, EPI_ISL_1631938, EPI_ISL_1631939, EPI_ISL_1631940, EPI_ISL_1631947, EPI_ISL_1631948, EPI_ISL_1631952, EPI_ISL_1631954, EPI_ISL_1631955, EPI_ISL_1631957, EPI_ISL_1631960, EPI_ISL_1631974, EPI_ISL_1631975, EPI_ISL_1631979, EPI_ISL_1631980, EPI_ISL_1631984, EPI_ISL_1631987, EPI_ISL_1631988, EPI_ISL_1631989, EPI_ISL_1631993, EPI_ISL_1632007, EPI_ISL_1632013, EPI_ISL_1632014, EPI_ISL_1632015, EPI_ISL_1632022, EPI_ISL_1632024, EPI_ISL_1632025, EPI_ISL_1632026, EPI_ISL_1632027, EPI_ISL_1632028, EPI_ISL_1632036, EPI_ISL_1632037, EPI_ISL_1632039, EPI_ISL_1632041, EPI_ISL_1632042, EPI_ISL_1632043, EPI_ISL_1632046, EPI_ISL_1632048, EPI_ISL_1632049, EPI_ISL_1632051, EPI_ISL_1632052, EPI_ISL_1632058, EPI_ISL_1632059, EPI_ISL_1632065, EPI_ISL_1632068, EPI_ISL_1632070, EPI_ISL_1632071, EPI_ISL_1632072, EPI_ISL_1632074, EPI_ISL_1632085, EPI_ISL_1632089, EPI_ISL_1632092, EPI_ISL_1632094, EPI_ISL_1632095, EPI_ISL_1632097, EPI_ISL_1632112, EPI_ISL_1632114, EPI_ISL_1632115, EPI_ISL_1632135, EPI_ISL_1632138, EPI_ISL_1632140, EPI_ISL_1632143, EPI_ISL_1632150, EPI_ISL_1632151, EPI_ISL_1632154, EPI_ISL_1632157, EPI_ISL_1632163, EPI_ISL_1632166, EPI_ISL_1632167, EPI_ISL_1632171, EPI_ISL_1632172, EPI_ISL_1632173, EPI_ISL_1632179, EPI_ISL_1632182, EPI_ISL_1632195, EPI_ISL_1632196, EPI_ISL_1632201, EPI_ISL_1632205, EPI_ISL_1632207, EPI_ISL_1632212, EPI_ISL_1632213, EPI_ISL_1632216, EPI_ISL_1632218, EPI_ISL_1632220, EPI_ISL_1632222, EPI_ISL_1632224, EPI_ISL_1632226, EPI_ISL_1632233, EPI_ISL_1632234, EPI_ISL_1632238, EPI_ISL_1632240, EPI_ISL_1632242, EPI_ISL_1632244, EPI_ISL_1632246, EPI_ISL_1632248, EPI_ISL_1632249, EPI_ISL_1632250, EPI_ISL_1632255, EPI_ISL_1632258, EPI_ISL_1632260, EPI_ISL_1632265, EPI_ISL_1632266, EPI_ISL_1632269, EPI_ISL_1632274, EPI_ISL_1632276, EPI_ISL_1632283, EPI_ISL_1632290, EPI_ISL_1632291, EPI_ISL_1632294, EPI_ISL_1632298, EPI_ISL_1632300, EPI_ISL_1632301, EPI_ISL_1632302, EPI_ISL_1632303, EPI_ISL_1632304, EPI_ISL_1632307, EPI_ISL_1632308, EPI_ISL_1632309, EPI_ISL_1632312, EPI_ISL_1632313, EPI_ISL_1632317, EPI_ISL_1632319, EPI_ISL_1632321, EPI_ISL_1632322, EPI_ISL_1632323, EPI_ISL_1632324, EPI_ISL_1632327, EPI_ISL_1632329, EPI_ISL_1632330, EPI_ISL_1632334, EPI_ISL_1632336, EPI_ISL_1632337, EPI_ISL_1632338, EPI_ISL_1632339, EPI_ISL_1632340, EPI_ISL_1632341, EPI_ISL_1632344, EPI_ISL_1632345, EPI_ISL_1632346, EPI_ISL_1632347, EPI_ISL_1632349, EPI_ISL_1632350, EPI_ISL_1632351, EPI_ISL_1632352, EPI_ISL_1632356, EPI_ISL_1632357, EPI_ISL_1632358, EPI_ISL_1632362, EPI_ISL_1632363, EPI_ISL_1632364, EPI_ISL_1632366, EPI_ISL_1632367, EPI_ISL_1632369, EPI_ISL_1632370,

EPI_ISL_1632373, EPI_ISL_1632374, EPI_ISL_1632375, EPI_ISL_1632376, EPI_ISL_1632378, EPI_ISL_1632380, EPI_ISL_1632382, EPI_ISL_1632385, EPI_ISL_1632387, EPI_ISL_1632388, EPI_ISL_1632392, EPI_ISL_1632393, EPI_ISL_1632396, EPI_ISL_1632397, EPI_ISL_1632399, EPI_ISL_1632400, EPI_ISL_1632402, EPI_ISL_1632403, EPI_ISL_1632404, EPI_ISL_1632406, EPI_ISL_1632408, EPI_ISL_1632409,
EPI_ISL_1632411, EPI_ISL_1632413, EPI_ISL_1632414, EPI_ISL_1632415, EPI_ISL_1632416, EPI_ISL_1632417, EPI_ISL_1632418, EPI_ISL_1632420, EPI_ISL_1632421, EPI_ISL_1632422, EPI_ISL_1632423, EPI_ISL_1632424, EPI_ISL_1632427, EPI_ISL_1632428, EPI_ISL_1632429, EPI_ISL_1632430, EPI_ISL_1632431, EPI_ISL_1632432, EPI_ISL_1632435, EPI_ISL_1632438, EPI_ISL_1632440, EPI_ISL_1632441,
EPI_ISL_1632442, EPI_ISL_1632444, EPI_ISL_1632445, EPI_ISL_1632446, EPI_ISL_1632447, EPI_ISL_1632453, EPI_ISL_1632455, EPI_ISL_1632456, EPI_ISL_1632457, EPI_ISL_1632458, EPI_ISL_1632460, EPI_ISL_1632464, EPI_ISL_1632465, EPI_ISL_1632468, EPI_ISL_1632470, EPI_ISL_1632473, EPI_ISL_1632474, EPI_ISL_1632477, EPI_ISL_1632478, EPI_ISL_1632479, EPI_ISL_1634489,
EPI_ISL_1634490, EPI_ISL_1634492, EPI_ISL_1634493, EPI_ISL_1634495, EPI_ISL_1634496, EPI_ISL_1634497, EPI_ISL_1634498, EPI_ISL_1634501, EPI_ISL_1634502, EPI_ISL_1634503, EPI_ISL_1634504, EPI_ISL_1634508, EPI_ISL_1634512, EPI_ISL_1634513, EPI_ISL_1634518, EPI_ISL_1634522, EPI_ISL_1634527, EPI_ISL_1634528, EPI_ISL_1634532, EPI_ISL_1634537, EPI_ISL_1634539, EPI_ISL_1634540,
EPI_ISL_1634543, EPI_ISL_1634544, EPI_ISL_1634545, EPI_ISL_1634547, EPI_ISL_1634548, EPI_ISL_1634550, EPI_ISL_1634554, EPI_ISL_1634555, EPI_ISL_1634558, EPI_ISL_1634559, EPI_ISL_1634566, EPI_ISL_1634568, EPI_ISL_1634571, EPI_ISL_1634572, EPI_ISL_1634574, EPI_ISL_1634575, EPI_ISL_1634577, EPI_ISL_1634578, EPI_ISL_1634582, EPI_ISL_1634583, EPI_ISL_1634584, EPI_ISL_1634586,
EPI_ISL_1634587, EPI_ISL_1634589, EPI_ISL_1634595, EPI_ISL_1634600, EPI_ISL_1634601, EPI_ISL_1634602, EPI_ISL_1634603, EPI_ISL_1634604, EPI_ISL_1634605, EPI_ISL_1634607, EPI_ISL_1634610, EPI_ISL_1634613, EPI_ISL_1634615, EPI_ISL_1634617, EPI_ISL_1634620, EPI_ISL_1634622, EPI_ISL_1634623, EPI_ISL_1634624, EPI_ISL_1634626, EPI_ISL_1634628, EPI_ISL_1634631, EPI_ISL_1634632,
EPI_ISL_1634633, EPI_ISL_1634634, EPI_ISL_1634635, EPI_ISL_1634643, EPI_ISL_1634649, EPI_ISL_1634650, EPI_ISL_1634651, EPI_ISL_1634653, EPI_ISL_1634656, EPI_ISL_1634661, EPI_ISL_1634664, EPI_ISL_1634665, EPI_ISL_1634667, EPI_ISL_1634669, EPI_ISL_1634674, EPI_ISL_1634676, EPI_ISL_1634677, EPI_ISL_1634680, EPI_ISL_1634681, EPI_ISL_1634683, EPI_ISL_1634685, EPI_ISL_1634690,
EPI_ISL_1634691, EPI_ISL_1634695, EPI_ISL_1634696, EPI_ISL_1634697, EPI_ISL_1634700, EPI_ISL_1634701, EPI_ISL_1634702, EPI_ISL_1634703, EPI_ISL_1634704, EPI_ISL_1634705, EPI_ISL_1634706, EPI_ISL_1634707, EPI_ISL_1634708, EPI_ISL_1634709, EPI_ISL_1634710, EPI_ISL_1634711, EPI_ISL_1634712, EPI_ISL_1634713, EPI_ISL_1634715, EPI_ISL_1634717, EPI_ISL_1634719, EPI_ISL_1634720,
EPI_ISL_1634721, EPI_ISL_1634722, EPI_ISL_1634723, EPI_ISL_1634724, EPI_ISL_1634725, EPI_ISL_1634726, EPI_ISL_1634727, EPI_ISL_1634728, EPI_ISL_1634729, EPI_ISL_1634730, EPI_ISL_1634731, EPI_ISL_1634732, EPI_ISL_1634733, EPI_ISL_1634734, EPI_ISL_1634735, EPI_ISL_1634736, EPI_ISL_1634738, EPI_ISL_1634739, EPI_ISL_1634740, EPI_ISL_1634741, EPI_ISL_1634742, EPI_ISL_1634744,
EPI_ISL_1634745, EPI_ISL_1634746, EPI_ISL_1634747, EPI_ISL_1634748, EPI_ISL_1634749, EPI_ISL_1634750, EPI_ISL_1634751, EPI_ISL_1634752, EPI_ISL_1634753, EPI_ISL_1634754, EPI_ISL_1634755, EPI_ISL_1634756, EPI_ISL_1634757, EPI_ISL_1634758, EPI_ISL_1634759, EPI_ISL_1634760, EPI_ISL_1634761, EPI_ISL_1634762, EPI_ISL_1634763, EPI_ISL_1634764, EPI_ISL_1634765, EPI_ISL_1634766,
EPI_ISL_1634767, EPI_ISL_1634768, EPI_ISL_1634769, EPI_ISL_1634770, EPI_ISL_1634771, EPI_ISL_1634772, EPI_ISL_1634773, EPI_ISL_1634774, EPI_ISL_1634775, EPI_ISL_1634776, EPI_ISL_1634777, EPI_ISL_1634779, EPI_ISL_1634780, EPI_ISL_1634781, EPI_ISL_1634782, EPI_ISL_1634785, EPI_ISL_1634787, EPI_ISL_1634788, EPI_ISL_1634789, EPI_ISL_1634790, EPI_ISL_1634792, EPI_ISL_1634793,
EPI_ISL_1634795, EPI_ISL_1634797, EPI_ISL_1634799, EPI_ISL_1634800, EPI_ISL_1634803, EPI_ISL_1634804, EPI_ISL_1634805, EPI_ISL_1634809, EPI_ISL_1634810, EPI_ISL_1634811, EPI_ISL_1634812, EPI_ISL_1634815, EPI_ISL_1634817, EPI_ISL_1634819, EPI_ISL_1634820, EPI_ISL_1634822, EPI_ISL_1634823, EPI_ISL_1634824, EPI_ISL_1634827, EPI_ISL_1634828, EPI_ISL_1634829, EPI_ISL_1634833,
EPI_ISL_1634834, EPI_ISL_1634835, EPI_ISL_1634837, EPI_ISL_1634838, EPI_ISL_1634840, EPI_ISL_1634841, EPI_ISL_1634842, EPI_ISL_1634843, EPI_ISL_1634845, EPI_ISL_1634847, EPI_ISL_1634849, EPI_ISL_1634851, EPI_ISL_1634853, EPI_ISL_1634854, EPI_ISL_1634856, EPI_ISL_1634857, EPI_ISL_1634859, EPI_ISL_1634860, EPI_ISL_1634862, EPI_ISL_1634863, EPI_ISL_1634865, EPI_ISL_1634866,
EPI_ISL_1634867, EPI_ISL_1634869, EPI_ISL_1634870, EPI_ISL_1634871, EPI_ISL_1634873, EPI_ISL_1634874, EPI_ISL_1634875, EPI_ISL_1634876, EPI_ISL_1634877, EPI_ISL_1634878, EPI_ISL_1634880, EPI_ISL_1634881, EPI_ISL_1634883, EPI_ISL_1634884, EPI_ISL_1634885, EPI_ISL_1634886, EPI_ISL_1634887, EPI_ISL_1634888, EPI_ISL_1634890, EPI_ISL_1634891, EPI_ISL_1634893, EPI_ISL_1634895,
EPI_ISL_1634899, EPI_ISL_1634901, EPI_ISL_1634904, EPI_ISL_1634905, EPI_ISL_1634906, EPI_ISL_1634907, EPI_ISL_1634908, EPI_ISL_1634909, EPI_ISL_1634910, EPI_ISL_1634911, EPI_ISL_1634912, EPI_ISL_1634915, EPI_ISL_1634916, EPI_ISL_1634917, EPI_ISL_1634918, EPI_ISL_1634919, EPI_ISL_1634920, EPI_ISL_1634922, EPI_ISL_1634923, EPI_ISL_1634924, EPI_ISL_1634925, EPI_ISL_1634927,
EPI_ISL_1634928, EPI_ISL_1634930, EPI_ISL_1634931, EPI_ISL_1634933, EPI_ISL_1634934, EPI_ISL_1634937, EPI_ISL_1634939, EPI_ISL_1634941, EPI_ISL_1634942, EPI_ISL_1634943, EPI_ISL_1634944, EPI_ISL_1634945, EPI_ISL_1634946, EPI_ISL_1634948, EPI_ISL_1634949, EPI_ISL_1634951, EPI_ISL_1634952, EPI_ISL_1634953, EPI_ISL_1634955, EPI_ISL_1634958, EPI_ISL_1634959, EPI_ISL_1634960,
EPI_ISL_1634961, EPI_ISL_1634963, EPI_ISL_1634964, EPI_ISL_1634965, EPI_ISL_1634966, EPI_ISL_1634968, EPI_ISL_1634971, EPI_ISL_1634972, EPI_ISL_1634973, EPI_ISL_1634974, EPI_ISL_1634976, EPI_ISL_1634978, EPI_ISL_1634979, EPI_ISL_1634980, EPI_ISL_1634981, EPI_ISL_1634982, EPI_ISL_1634983, EPI_ISL_1634984, EPI_ISL_1634985, EPI_ISL_1634989, EPI_ISL_1634990, EPI_ISL_1634991,
EPI_ISL_1634992, EPI_ISL_1634993, EPI_ISL_1634994, EPI_ISL_1634995, EPI_ISL_1634996, EPI_ISL_1634997, EPI_ISL_1634998, EPI_ISL_1634999, EPI_ISL_1635000, EPI_ISL_1635001, EPI_ISL_1635002, EPI_ISL_1635003, EPI_ISL_1635004, EPI_ISL_1635005, EPI_ISL_1635006, EPI_ISL_1635007, EPI_ISL_1635008, EPI_ISL_1635009, EPI_ISL_1635010, EPI_ISL_1635011, EPI_ISL_1635012, EPI_ISL_1635013,
EPI_ISL_1635014, EPI_ISL_1635015, EPI_ISL_1635016, EPI_ISL_1635017, EPI_ISL_1635018, EPI_ISL_1635019, EPI_ISL_1635020, EPI_ISL_1635021, EPI_ISL_1635022, EPI_ISL_1635023, EPI_ISL_1635024, EPI_ISL_1635025, EPI_ISL_1635026, EPI_ISL_1635027, EPI_ISL_1635028, EPI_ISL_1635029, EPI_ISL_1635030, EPI_ISL_1635031, EPI_ISL_1635032, EPI_ISL_1635033, EPI_ISL_1635034, EPI_ISL_1635035,
EPI_ISL_1635036, EPI_ISL_1635037, EPI_ISL_1635038, EPI_ISL_1635039, EPI_ISL_1635040, EPI_ISL_1635041, EPI_ISL_1635042, EPI_ISL_1635043, EPI_ISL_1635044, EPI_ISL_1635045, EPI_ISL_1635046, EPI_ISL_1635047, EPI_ISL_1635048, EPI_ISL_1635049, EPI_ISL_1635050, EPI_ISL_1635051, EPI_ISL_1635052, EPI_ISL_1635053, EPI_ISL_1635054, EPI_ISL_1635055, EPI_ISL_1635056, EPI_ISL_1635057,
EPI_ISL_1635058, EPI_ISL_1635059, EPI_ISL_1635060, EPI_ISL_1635061, EPI_ISL_1635062, EPI_ISL_1635063, EPI_ISL_1635064, EPI_ISL_1635065, EPI_ISL_1635066, EPI_ISL_1635067, EPI_ISL_1635068, EPI_ISL_1635069, EPI_ISL_1635120, EPI_ISL_1635123, EPI_ISL_1635125, EPI_ISL_1635126, EPI_ISL_1635127, EPI_ISL_1635129, EPI_ISL_1635130, EPI_ISL_1635132, EPI_ISL_1635133, EPI_ISL_1635134,
EPI_ISL_1635136, EPI_ISL_1635137, EPI_ISL_1635138, EPI_ISL_1635139, EPI_ISL_1635140, EPI_ISL_1635141, EPI_ISL_1635142, EPI_ISL_1635143, EPI_ISL_1635144, EPI_ISL_1635145, EPI_ISL_1635146, EPI_ISL_1635147, EPI_ISL_1635148, EPI_ISL_1635152, EPI_ISL_1635153, EPI_ISL_1635154, EPI_ISL_1635156, EPI_ISL_1635157, EPI_ISL_1635158, EPI_ISL_1635159, EPI_ISL_1635160, EPI_ISL_1635161,
EPI_ISL_1635162, EPI_ISL_1635164, EPI_ISL_1635166, EPI_ISL_1635167, EPI_ISL_1635168, EPI_ISL_1635169, EPI_ISL_1635170, EPI_ISL_1635171, EPI_ISL_1635172, EPI_ISL_1635173, EPI_ISL_1635174, EPI_ISL_1635175, EPI_ISL_1635176, EPI_ISL_1635177, EPI_ISL_1635178, EPI_ISL_1635179, EPI_ISL_1635180, EPI_ISL_1635181, EPI_ISL_1635182, EPI_ISL_1635183, EPI_ISL_1635184, EPI_ISL_1635185,
EPI_ISL_1635186, EPI_ISL_1635187, EPI_ISL_1635188, EPI_ISL_1635189, EPI_ISL_1635190, EPI_ISL_1635191, EPI_ISL_1635192, EPI_ISL_1635193, EPI_ISL_1635194, EPI_ISL_1635196, EPI_ISL_1635197, EPI_ISL_1635198, EPI_ISL_1635199, EPI_ISL_1635200, EPI_ISL_1635201, EPI_ISL_1635202, EPI_ISL_1635203, EPI_ISL_1635204, EPI_ISL_1635205, EPI_ISL_1635206, EPI_ISL_1635208, EPI_ISL_1635209,
EPI_ISL_1635210, EPI_ISL_1635211, EPI_ISL_1635212, EPI_ISL_1635213, EPI_ISL_1635214, EPI_ISL_1635215, EPI_ISL_1635216, EPI_ISL_1635217, EPI_ISL_1635218, EPI_ISL_1635219, EPI_ISL_1635220, EPI_ISL_1635221, EPI_ISL_1635222, EPI_ISL_1635224, EPI_ISL_1635226, EPI_ISL_1635227, EPI_ISL_1635228, EPI_ISL_1635229, EPI_ISL_1635230, EPI_ISL_1635231, EPI_ISL_1635232, EPI_ISL_1635233,
EPI_ISL_1635235, EPI_ISL_1635236, EPI_ISL_1635238, EPI_ISL_1635239, EPI_ISL_1635241, EPI_ISL_1635242, EPI_ISL_1635243, EPI_ISL_1635244, EPI_ISL_1635247, EPI_ISL_1635250, EPI_ISL_1635251, EPI_ISL_1635252, EPI_ISL_1635253, EPI_ISL_1635254, EPI_ISL_1635255, EPI_ISL_1635257, EPI_ISL_1635258, EPI_ISL_1635259, EPI_ISL_1635260, EPI_ISL_1635261, EPI_ISL_1635262, EPI_ISL_1635265,
EPI_ISL_1635266, EPI_ISL_1635267, EPI_ISL_1635268, EPI_ISL_1635269, EPI_ISL_1635271, EPI_ISL_1635272, EPI_ISL_1635273, EPI_ISL_1635274, EPI_ISL_1635275, EPI_ISL_1635276, EPI_ISL_1635277, EPI_ISL_1635279, EPI_ISL_1635280, EPI_ISL_1635281, EPI_ISL_1635282, EPI_ISL_1635283, EPI_ISL_1635285, EPI_ISL_1635286, EPI_ISL_1635287, EPI_ISL_1635289, EPI_ISL_1635290, EPI_ISL_1635291,
EPI_ISL_1635292, EPI_ISL_1635294, EPI_ISL_1635296, EPI_ISL_1635297, EPI_ISL_1635298, EPI_ISL_1635303, EPI_ISL_1635304, EPI_ISL_1635305, EPI_ISL_1635306, EPI_ISL_1635307, EPI_ISL_1635308, EPI_ISL_1635309, EPI_ISL_1635310, EPI_ISL_1635311, EPI_ISL_1635312, EPI_ISL_1635314, EPI_ISL_1635315, EPI_ISL_1635316, EPI_ISL_1635317, EPI_ISL_1635318, EPI_ISL_1635319, EPI_ISL_1635320,
EPI_ISL_1635322, EPI_ISL_1635323, EPI_ISL_1635324, EPI_ISL_1635326, EPI_ISL_1635327, EPI_ISL_1635328, EPI_ISL_1635329, EPI_ISL_1635330, EPI_ISL_1635331, EPI_ISL_1635332, EPI_ISL_1635333, EPI_ISL_1635334, EPI_ISL_1635335, EPI_ISL_1635336, EPI_ISL_1635337, EPI_ISL_1635338, EPI_ISL_1635339, EPI_ISL_1635340, EPI_ISL_1635341, EPI_ISL_1635342, EPI_ISL_1635343, EPI_ISL_1635345,
EPI_ISL_1635346, EPI_ISL_1635347, EPI_ISL_1635348, EPI_ISL_1635349, EPI_ISL_1635350, EPI_ISL_1635352, EPI_ISL_1635353, EPI_ISL_1635354, EPI_ISL_1635355, EPI_ISL_1635357, EPI_ISL_1635358, EPI_ISL_1635360, EPI_ISL_1635362, EPI_ISL_1635363, EPI_ISL_1635364, EPI_ISL_1635365, EPI_ISL_1635366, EPI_ISL_1635367, EPI_ISL_1635368, EPI_ISL_1635369, EPI_ISL_1635370, EPI_ISL_1635372,
EPI_ISL_1635373, EPI_ISL_1635374, EPI_ISL_1635375, EPI_ISL_1635376, EPI_ISL_1635377, EPI_ISL_1635378, EPI_ISL_1635380, EPI_ISL_1635381, EPI_ISL_1635382, EPI_ISL_1635383, EPI_ISL_1635384, EPI_ISL_1635385, EPI_ISL_1635386, EPI_ISL_1635387, EPI_ISL_1635388, EPI_ISL_1635389, EPI_ISL_1635390, EPI_ISL_1635391, EPI_ISL_1635393, EPI_ISL_1635394, EPI_ISL_1635395, EPI_ISL_1635396,
EPI_ISL_1635397, EPI_ISL_1635398, EPI_ISL_1635399, EPI_ISL_1635401, EPI_ISL_1635402, EPI_ISL_1635405, EPI_ISL_1635406, EPI_ISL_1635407, EPI_ISL_1635408, EPI_ISL_1635409, EPI_ISL_1635410, EPI_ISL_1635411, EPI_ISL_1635412, EPI_ISL_1635413, EPI_ISL_1635414, EPI_ISL_1635415, EPI_ISL_1635416, EPI_ISL_1635417, EPI_ISL_1635418, EPI_ISL_1635419, EPI_ISL_1635420, EPI_ISL_1635421,
EPI_ISL_1635422, EPI_ISL_1652691, EPI_ISL_1652702, EPI_ISL_1652704, EPI_ISL_1652721, EPI_ISL_1652723, EPI_ISL_1652726, EPI_ISL_1652731, EPI_ISL_1652737, EPI_ISL_1652740, EPI_ISL_1652741, EPI_ISL_1652752, EPI_ISL_1652776, EPI_ISL_1652778, EPI_ISL_1652787, EPI_ISL_1652792, EPI_ISL_1652798, EPI_ISL_1652801, EPI_ISL_1652815, EPI_ISL_1652820, EPI_ISL_1652832, EPI_ISL_1652844,
EPI_ISL_1652851, EPI_ISL_1652858, EPI_ISL_1652866, EPI_ISL_1652872, EPI_ISL_1652876, EPI_ISL_1652894, EPI_ISL_1652895, EPI_ISL_1652896, EPI_ISL_1652899, EPI_ISL_1652918, EPI_ISL_1652919, EPI_ISL_1652922, EPI_ISL_1652926, EPI_ISL_1652928, EPI_ISL_1652935, EPI_ISL_1652936, EPI_ISL_1652943, EPI_ISL_1652944, EPI_ISL_1652945, EPI_ISL_1652946, EPI_ISL_1652947, EPI_ISL_1652948,
EPI_ISL_1652949, EPI_ISL_1652950, EPI_ISL_1652953, EPI_ISL_1652955, EPI_ISL_1652956, EPI_ISL_1652957, EPI_ISL_1652959, EPI_ISL_1652960, EPI_ISL_1652961, EPI_ISL_1652962, EPI_ISL_1652963, EPI_ISL_1652965, EPI_ISL_1652966, EPI_ISL_1652969, EPI_ISL_1652972, EPI_ISL_1652974, EPI_ISL_1652975, EPI_ISL_1652977, EPI_ISL_1652987, EPI_ISL_1652988, EPI_ISL_1652990,
EPI_ISL_1652992, EPI_ISL_1652997, EPI_ISL_1652999, EPI_ISL_1653000, EPI_ISL_1653002, EPI_ISL_1653003, EPI_ISL_1653004, EPI_ISL_1653007, EPI_ISL_1653008, EPI_ISL_1653013, EPI_ISL_1653014, EPI_ISL_1653016, EPI_ISL_1653017, EPI_ISL_1653019, EPI_ISL_1653023, EPI_ISL_1653026, EPI_ISL_1653029, EPI_ISL_1653030, EPI_ISL_1653031, EPI_ISL_1653032, EPI_ISL_1653034, EPI_ISL_1653035,
EPI_ISL_1653036, EPI_ISL_1653037, EPI_ISL_1653038, EPI_ISL_1653039, EPI_ISL_1653041, EPI_ISL_1653046, EPI_ISL_1653047, EPI_ISL_1653048, EPI_ISL_1653049, EPI_ISL_1653054, EPI_ISL_1653055, EPI_ISL_1653056, EPI_ISL_1653057, EPI_ISL_1653060, EPI_ISL_1653063, EPI_ISL_1653064, EPI_ISL_1653065, EPI_ISL_1653066, EPI_ISL_1653067, EPI_ISL_1653068, EPI_ISL_1653070,
EPI_ISL_1653071, EPI_ISL_1653072, EPI_ISL_1653075, EPI_ISL_1653076, EPI_ISL_1653077, EPI_ISL_1653079, EPI_ISL_1653082, EPI_ISL_1653084, EPI_ISL_1653085, EPI_ISL_1653086, EPI_ISL_1653087, EPI_ISL_1653088, EPI_ISL_1653089, EPI_ISL_1653091, EPI_ISL_1653092, EPI_ISL_1653094, EPI_ISL_1653095, EPI_ISL_1653098, EPI_ISL_1653101, EPI_ISL_1653103, EPI_ISL_1653105, EPI_ISL_1653106,
EPI_ISL_1653107, EPI_ISL_1653112, EPI_ISL_1653113, EPI_ISL_1653115, EPI_ISL_1653117, EPI_ISL_1653118, EPI_ISL_1653122, EPI_ISL_1653123, EPI_ISL_1653124, EPI_ISL_1653127, EPI_ISL_1653128, EPI_ISL_1653129, EPI_ISL_1653131, EPI_ISL_1653134, EPI_ISL_1653135, EPI_ISL_1653137, EPI_ISL_1653139, EPI_ISL_1653140, EPI_ISL_1653142, EPI_ISL_1653143, EPI_ISL_1653144, EPI_ISL_1653145,
EPI_ISL_1653148, EPI_ISL_1653150, EPI_ISL_1653151, EPI_ISL_1653152, EPI_ISL_1653153, EPI_ISL_1653154, EPI_ISL_1653155, EPI_ISL_1653158, EPI_ISL_1653160, EPI_ISL_1653161, EPI_ISL_1653162, EPI_ISL_1653163, EPI_ISL_1653164, EPI_ISL_1653165, EPI_ISL_1653166, EPI_ISL_1653168, EPI_ISL_1653169, EPI_ISL_1653170, EPI_ISL_1653171, EPI_ISL_1653172, EPI_ISL_1653173, EPI_ISL_1653177,
EPI_ISL_1653182, EPI_ISL_1653184, EPI_ISL_1653186, EPI_ISL_1653188, EPI_ISL_1653189, EPI_ISL_1653190, EPI_ISL_1653192, EPI_ISL_1653193, EPI_ISL_1653194, EPI_ISL_1653196, EPI_ISL_1653198, EPI_ISL_1653199, EPI_ISL_1653202, EPI_ISL_1653203, EPI_ISL_1653204, EPI_ISL_1653205, EPI_ISL_1653206, EPI_ISL_1653207, EPI_ISL_1653208, EPI_ISL_1653211, EPI_ISL_1653212, EPI_ISL_1653215,
EPI_ISL_1653217, EPI_ISL_1653220, EPI_ISL_1653221, EPI_ISL_1653224, EPI_ISL_1653226, EPI_ISL_1653231, EPI_ISL_1653237, EPI_ISL_1653239, EPI_ISL_1653241, EPI_ISL_1653249, EPI_ISL_1653252, EPI_ISL_1653253, EPI_ISL_1653255, EPI_ISL_1653256, EPI_ISL_1653262, EPI_ISL_1653264, EPI_ISL_1653273, EPI_ISL_1653293, EPI_ISL_1653294, EPI_ISL_1653296, EPI_ISL_1653299, EPI_ISL_1653302,
EPI_ISL_1653303, EPI_ISL_1653304, EPI_ISL_1653307, EPI_ISL_1653308, EPI_ISL_1653309, EPI_ISL_1653312, EPI_ISL_1653313, EPI_ISL_1653314, EPI_ISL_1653319, EPI_ISL_1653321, EPI_ISL_1653325, EPI_ISL_1653329, EPI_ISL_1653331, EPI_ISL_1653333, EPI_ISL_1653335, EPI_ISL_1653337, EPI_ISL_1653339, EPI_ISL_1653340, EPI_ISL_1653341, EPI_ISL_1653343, EPI_ISL_1653353, EPI_ISL_1653354,
EPI_ISL_1653356, EPI_ISL_1653357, EPI_ISL_1653360, EPI_ISL_1653365, EPI_ISL_1653369, EPI_ISL_1653381, EPI_ISL_1653385, EPI_ISL_1653388, EPI_ISL_1653395, EPI_ISL_1653402, EPI_ISL_1653404, EPI_ISL_1653408, EPI_ISL_1653409, EPI_ISL_1653412, EPI_ISL_1653414, EPI_ISL_1653418, EPI_ISL_1653427, EPI_ISL_1653432, EPI_ISL_1653436, EPI_ISL_1653445, EPI_ISL_1653456, EPI_ISL_1653464,
EPI_ISL_1653465, EPI_ISL_1653471, EPI_ISL_1653476, EPI_ISL_1653477, EPI_ISL_1653478, EPI_ISL_1653482, EPI_ISL_1653488, EPI_ISL_1653491, EPI_ISL_1653493, EPI_ISL_1653502, EPI_ISL_1653505, EPI_ISL_1653506, EPI_ISL_1653507, EPI_ISL_1653516, EPI_ISL_1653523, EPI_ISL_1653524, EPI_ISL_1653526, EPI_ISL_1653527, EPI_ISL_1653528, EPI_ISL_1653529, EPI_ISL_1653530,
EPI_ISL_1653536, EPI_ISL_1653539, EPI_ISL_1653540, EPI_ISL_1653550, EPI_ISL_1653552, EPI_ISL_1653562, EPI_ISL_1653566, EPI_ISL_1653568, EPI_ISL_1653576, EPI_ISL_1653586, EPI_ISL_1653587, EPI_ISL_1653588, EPI_ISL_1653602, EPI_ISL_1653608, EPI_ISL_1653609, EPI_ISL_1653610, EPI_ISL_1653612, EPI_ISL_1653617, EPI_ISL_1653621, EPI_ISL_1653622, EPI_ISL_1653648, EPI_ISL_1653651,
EPI_ISL_1653653, EPI_ISL_1653664, EPI_ISL_1653665, EPI_ISL_1653671, EPI_ISL_1653674, EPI_ISL_1653678, EPI_ISL_1653684, EPI_ISL_1653685, EPI_ISL_1653689, EPI_ISL_1653699, EPI_ISL_1653701, EPI_ISL_1653729, EPI_ISL_1672972, EPI_ISL_1672984, EPI_ISL_1672986, EPI_ISL_1672988, EPI_ISL_1672990, EPI_ISL_1672997, EPI_ISL_1673002, EPI_ISL_1673005, EPI_ISL_1673006, EPI_ISL_1673008,
EPI_ISL_1673010, EPI_ISL_1673017, EPI_ISL_1673018, EPI_ISL_1673019, EPI_ISL_1673021, EPI_ISL_1673023, EPI_ISL_1673024, EPI_ISL_1673031, EPI_ISL_1673035, EPI_ISL_1673042, EPI_ISL_1673044, EPI_ISL_1673045, EPI_ISL_1673046, EPI_ISL_1673047, EPI_ISL_1673048, EPI_ISL_1673049, EPI_ISL_1673050, EPI_ISL_1673051, EPI_ISL_1673052, EPI_ISL_1673054, EPI_ISL_1673057, EPI_ISL_1673059,
EPI_ISL_1673060, EPI_ISL_1673062, EPI_ISL_1673063, EPI_ISL_1673064, EPI_ISL_1673065, EPI_ISL_1673066, EPI_ISL_1673067, EPI_ISL_1673068, EPI_ISL_1673069, EPI_ISL_1673070, EPI_ISL_1673071, EPI_ISL_1673072, EPI_ISL_1673073, EPI_ISL_1673074, EPI_ISL_1673075, EPI_ISL_1673076, EPI_ISL_1673077, EPI_ISL_1673078, EPI_ISL_1673079, EPI_ISL_1673080, EPI_ISL_1673081, EPI_ISL_1673082,
EPI_ISL_1673083, EPI_ISL_1673084, EPI_ISL_1673085, EPI_ISL_1673086, EPI_ISL_1673088, EPI_ISL_1673089, EPI_ISL_1673090, EPI_ISL_1673091, EPI_ISL_1673092, EPI_ISL_1673093, EPI_ISL_1673095, EPI_ISL_1673096, EPI_ISL_1673097, EPI_ISL_1673098, EPI_ISL_1673099, EPI_ISL_1673101, EPI_ISL_1673102, EPI_ISL_1673104, EPI_ISL_1673105, EPI_ISL_1673106, EPI_ISL_1673107, EPI_ISL_1673108,
EPI_ISL_1673109, EPI_ISL_1673110, EPI_ISL_1673111, EPI_ISL_1673112, EPI_ISL_1673113, EPI_ISL_1673116, EPI_ISL_1673117, EPI_ISL_1673118, EPI_ISL_1673119, EPI_ISL_1673120, EPI_ISL_1673122, EPI_ISL_1673123, EPI_ISL_1673124, EPI_ISL_1673125, EPI_ISL_1673126, EPI_ISL_1673127, EPI_ISL_1673128, EPI_ISL_1673129, EPI_ISL_1673130, EPI_ISL_1673131, EPI_ISL_1673133, EPI_ISL_1673134,
EPI_ISL_1673135, EPI_ISL_1673136, EPI_ISL_1673137, EPI_ISL_1673138, EPI_ISL_1673139, EPI_ISL_1673143, EPI_ISL_1673144, EPI_ISL_1673145, EPI_ISL_1673146, EPI_ISL_1673148, EPI_ISL_1673149, EPI_ISL_1673150, EPI_ISL_1673151, EPI_ISL_1673152, EPI_ISL_1673153, EPI_ISL_1673154, EPI_ISL_1673155, EPI_ISL_1673156, EPI_ISL_1673157, EPI_ISL_1673158, EPI_ISL_1673159, EPI_ISL_1673160,
EPI_ISL_1673161, EPI_ISL_1673162, EPI_ISL_1673163, EPI_ISL_1673164, EPI_ISL_1673165, EPI_ISL_1673166, EPI_ISL_1673167, EPI_ISL_1673168, EPI_ISL_1673169, EPI_ISL_1673170, EPI_ISL_1673171, EPI_ISL_1673173, EPI_ISL_1673174, EPI_ISL_1673175, EPI_ISL_1673176, EPI_ISL_1673177, EPI_ISL_1673178, EPI_ISL_1673180, EPI_ISL_1673181, EPI_ISL_1673182,
EPI_ISL_1673183, EPI_ISL_1673184, EPI_ISL_1673185, EPI_ISL_1673186, EPI_ISL_1673187, EPI_ISL_1673188, EPI_ISL_1673189, EPI_ISL_1673190, EPI_ISL_1673191, EPI_ISL_1673192, EPI_ISL_1673193, EPI_ISL_1673194, EPI_ISL_1673195, EPI_ISL_1673196, EPI_ISL_1673197, EPI_ISL_1673198, EPI_ISL_1673199, EPI_ISL_1673200, EPI_ISL_1673201, EPI_ISL_1673202, EPI_ISL_1673203, EPI_ISL_1673204,
EPI_ISL_1673205, EPI_ISL_1673206, EPI_ISL_1673207, EPI_ISL_1673208, EPI_ISL_1673209, EPI_ISL_1673210, EPI_ISL_1673211, EPI_ISL_1673212, EPI_ISL_1673213, EPI_ISL_1673214, EPI_ISL_1673215, EPI_ISL_1673216, EPI_ISL_1673217, EPI_ISL_1673218, EPI_ISL_1673219, EPI_ISL_1673220, EPI_ISL_1673221, EPI_ISL_1673222, EPI_ISL_1673223, EPI_ISL_1673224, EPI_ISL_1673225, EPI_ISL_1673226,
EPI_ISL_1673227, EPI_ISL_1673228, EPI_ISL_1673229, EPI_ISL_1673230, EPI_ISL_1673231, EPI_ISL_1673232, EPI_ISL_1673233, EPI_ISL_1673234, EPI_ISL_1673235, EPI_ISL_1673236, EPI_ISL_1673237, EPI_ISL_1673238, EPI_ISL_1673239, EPI_ISL_1673240, EPI_ISL_1673241, EPI_ISL_1673242, EPI_ISL_1673243, EPI_ISL_1673244, EPI_ISL_1673245, EPI_ISL_1673246, EPI_ISL_1673247, EPI_ISL_1673248,
EPI_ISL_1673249, EPI_ISL_1673250, EPI_ISL_1673251, EPI_ISL_1673252, EPI_ISL_1673253, EPI_ISL_1673254, EPI_ISL_1697744, EPI_ISL_1697746, EPI_ISL_1697751, EPI_ISL_1697753, EPI_ISL_1697766, EPI_ISL_1697767, EPI_ISL_1697771, EPI_ISL_1697781, EPI_ISL_1697784, EPI_ISL_1697785, EPI_ISL_1697786, EPI_ISL_1697790, EPI_ISL_1697794, EPI_ISL_1697796, EPI_ISL_1697797, EPI_ISL_1697798,
EPI_ISL_1697808, EPI_ISL_1697809, EPI_ISL_1697810, EPI_ISL_1697811, EPI_ISL_1697815, EPI_ISL_1697822, EPI_ISL_1697825, EPI_ISL_1697827, EPI_ISL_1697830, EPI_ISL_1697833, EPI_ISL_1697834, EPI_ISL_1697836, EPI_ISL_1697838, EPI_ISL_1697840, EPI_ISL_1697842, EPI_ISL_1697845, EPI_ISL_1697860, EPI_ISL_1697865, EPI_ISL_1697869, EPI_ISL_1697872, EPI_ISL_1697873, EPI_ISL_1697879,
EPI_ISL_1697880, EPI_ISL_1697888, EPI_ISL_1697889, EPI_ISL_1697890, EPI_ISL_1697891, EPI_ISL_1697892, EPI_ISL_1697894, EPI_ISL_1697901, EPI_ISL_1697905, EPI_ISL_1697906, EPI_ISL_1697913, EPI_ISL_1697916, EPI_ISL_1697929, EPI_ISL_1697930, EPI_ISL_1697931, EPI_ISL_1697933, EPI_ISL_1697935, EPI_ISL_1697937, EPI_ISL_1697938, EPI_ISL_1697939, EPI_ISL_1697940, EPI_ISL_1697952,
EPI_ISL_1697955, EPI_ISL_1697957, EPI_ISL_1697958, EPI_ISL_1697966, EPI_ISL_1697968, EPI_ISL_1698113, EPI_ISL_1698548, EPI_ISL_1698549, EPI_ISL_1698551, EPI_ISL_1698552, EPI_ISL_1698553, EPI_ISL_1698554, EPI_ISL_1698557, EPI_ISL_1698561, EPI_ISL_1698562, EPI_ISL_1698563, EPI_ISL_1698564, EPI_ISL_1698565, EPI_ISL_1698567, EPI_ISL_1698568, EPI_ISL_1698570, EPI_ISL_1698571,
EPI_ISL_1698574, EPI_ISL_1698575, EPI_ISL_1698576, EPI_ISL_1698577, EPI_ISL_1698579, EPI_ISL_1698580, EPI_ISL_1698581, EPI_ISL_1698582, EPI_ISL_1698583, EPI_ISL_1698584, EPI_ISL_1698585, EPI_ISL_1698586, EPI_ISL_1698587, EPI_ISL_1698591, EPI_ISL_1698592, EPI_ISL_1698594, EPI_ISL_1698595, EPI_ISL_1698596, EPI_ISL_1698597, EPI_ISL_1698598, EPI_ISL_1698599, EPI_ISL_1698600,
EPI_ISL_1698604, EPI_ISL_1698605, EPI_ISL_1698606, EPI_ISL_1698607, EPI_ISL_1698608, EPI_ISL_1698611, EPI_ISL_1698613, EPI_ISL_1698615, EPI_ISL_1698616, EPI_ISL_1698618, EPI_ISL_1698620, EPI_ISL_1698621, EPI_ISL_1698622, EPI_ISL_1698627, EPI_ISL_1698628, EPI_ISL_1698629, EPI_ISL_1698630, EPI_ISL_1698631, EPI_ISL_1698632, EPI_ISL_1698633, EPI_ISL_1698637, EPI_ISL_1698638,
EPI_ISL_1698639, EPI_ISL_1698640, EPI_ISL_1698641, EPI_ISL_1698643, EPI_ISL_1698647, EPI_ISL_1698648, EPI_ISL_1698649, EPI_ISL_1698650, EPI_ISL_1698651, EPI_ISL_1698652, EPI_ISL_1698655, EPI_ISL_1698656, EPI_ISL_1698657, EPI_ISL_1698659, EPI_ISL_1698660, EPI_ISL_1698661, EPI_ISL_1698662, EPI_ISL_1698663, EPI_ISL_1698664, EPI_ISL_1698665, EPI_ISL_1698667, EPI_ISL_1698668,
EPI_ISL_1698669, EPI_ISL_1698671, EPI_ISL_1698672, EPI_ISL_1698673, EPI_ISL_1698674, EPI_ISL_1698675, EPI_ISL_1698677, EPI_ISL_1698679, EPI_ISL_1698680, EPI_ISL_1698681, EPI_ISL_1698682, EPI_ISL_1698683, EPI_ISL_1698684, EPI_ISL_1698686, EPI_ISL_1698687, EPI_ISL_1698688, EPI_ISL_1698689, EPI_ISL_1698690, EPI_ISL_1698691, EPI_ISL_1698692, EPI_ISL_1698693, EPI_ISL_1698694,
EPI_ISL_1698695, EPI_ISL_1698696, EPI_ISL_1698697, EPI_ISL_1698700, EPI_ISL_1698701, EPI_ISL_1698702, EPI_ISL_1698704, EPI_ISL_1698705, EPI_ISL_1698706, EPI_ISL_1698707, EPI_ISL_1698708, EPI_ISL_1698709, EPI_ISL_1698710, EPI_ISL_1698712, EPI_ISL_1698713, EPI_ISL_1698714, EPI_ISL_1698715, EPI_ISL_1698716, EPI_ISL_1698717, EPI_ISL_1698718, EPI_ISL_1698719, EPI_ISL_1698720,
EPI_ISL_1698721, EPI_ISL_1698723, EPI_ISL_1698724, EPI_ISL_1698727, EPI_ISL_1698728, EPI_ISL_1698729, EPI_ISL_1698730, EPI_ISL_1698731, EPI_ISL_1698732, EPI_ISL_1698734, EPI_ISL_1698735, EPI_ISL_1698736, EPI_ISL_1698738, EPI_ISL_1698739, EPI_ISL_1698740, EPI_ISL_1698742, EPI_ISL_1698743, EPI_ISL_1698744, EPI_ISL_1698745, EPI_ISL_1698747, EPI_ISL_1698748, EPI_ISL_1698749,
EPI_ISL_1698750, EPI_ISL_1698752, EPI_ISL_1698754, EPI_ISL_1698756, EPI_ISL_1698757, EPI_ISL_1698758, EPI_ISL_1698761, EPI_ISL_1698762, EPI_ISL_1698768, EPI_ISL_1698773, EPI_ISL_1698774, EPI_ISL_1698779, EPI_ISL_1698782, EPI_ISL_1698803, EPI_ISL_1698810, EPI_ISL_1698819, EPI_ISL_1698820, EPI_ISL_1698822, EPI_ISL_1698827, EPI_ISL_1698841, EPI_ISL_1698842, EPI_ISL_1698845,
EPI_ISL_1698847, EPI_ISL_1698851, EPI_ISL_1698853, EPI_ISL_1698854, EPI_ISL_1698856, EPI_ISL_1698863, EPI_ISL_1698866, EPI_ISL_1698881, EPI_ISL_1698887, EPI_ISL_1698892, EPI_ISL_1698920, EPI_ISL_1698922, EPI_ISL_1698925, EPI_ISL_1698926, EPI_ISL_1698928, EPI_ISL_1698932, EPI_ISL_1698937, EPI_ISL_1698938, EPI_ISL_1698940, EPI_ISL_1698941, EPI_ISL_1698947, EPI_ISL_1698948,
EPI_ISL_1698951, EPI_ISL_1698953, EPI_ISL_1698961, EPI_ISL_1698973, EPI_ISL_1698980, EPI_ISL_1698984, EPI_ISL_1698991, EPI_ISL_1699003, EPI_ISL_1699006, EPI_ISL_1699010, EPI_ISL_1699014, EPI_ISL_1699019, EPI_ISL_1699023, EPI_ISL_1699024, EPI_ISL_1699025, EPI_ISL_1699026, EPI_ISL_1699029, EPI_ISL_1699031, EPI_ISL_1699032, EPI_ISL_1699034, EPI_ISL_1699038, EPI_ISL_1699041,
EPI_ISL_1699043, EPI_ISL_1699046, EPI_ISL_1699049, EPI_ISL_1699051, EPI_ISL_1699055, EPI_ISL_1699057, EPI_ISL_1699064, EPI_ISL_1699066, EPI_ISL_1699073, EPI_ISL_1699074, EPI_ISL_1699076, EPI_ISL_1699077, EPI_ISL_1699078, EPI_ISL_1699085, EPI_ISL_1699096, EPI_ISL_1699100, EPI_ISL_1699102, EPI_ISL_1699109, EPI_ISL_1699111, EPI_ISL_1699113, EPI_ISL_1699114, EPI_ISL_1699115,
EPI_ISL_1699117, EPI_ISL_1699123, EPI_ISL_1699124, EPI_ISL_1699125, EPI_ISL_1699129, EPI_ISL_1699132, EPI_ISL_1699136, EPI_ISL_1699140, EPI_ISL_1699145, EPI_ISL_1699146, EPI_ISL_1699147, EPI_ISL_1699149, EPI_ISL_1699151, EPI_ISL_1699152, EPI_ISL_1699153, EPI_ISL_1699158, EPI_ISL_1699159, EPI_ISL_1699161, EPI_ISL_1699162, EPI_ISL_1699164, EPI_ISL_1699166, EPI_ISL_1699167,
EPI_ISL_1699169, EPI_ISL_1699171, EPI_ISL_1699173, EPI_ISL_1699175, EPI_ISL_1699177, EPI_ISL_1699184, EPI_ISL_1699188, EPI_ISL_1699189, EPI_ISL_1699194, EPI_ISL_1699198, EPI_ISL_1699200, EPI_ISL_1699201, EPI_ISL_1699202, EPI_ISL_1699212, EPI_ISL_1699213, EPI_ISL_1699217, EPI_ISL_1699219, EPI_ISL_1699222, EPI_ISL_1699224, EPI_ISL_1699225, EPI_ISL_1699226, EPI_ISL_1699233,
EPI_ISL_1699235, EPI_ISL_1699236, EPI_ISL_1699243, EPI_ISL_1699244, EPI_ISL_1699245, EPI_ISL_1699247, EPI_ISL_1699249, EPI_ISL_1699251, EPI_ISL_1699253, EPI_ISL_1699256, EPI_ISL_1699257, EPI_ISL_1699258, EPI_ISL_1699259, EPI_ISL_1699260, EPI_ISL_1699261, EPI_ISL_1699262, EPI_ISL_1699265, EPI_ISL_1699268, EPI_ISL_1699274, EPI_ISL_1699275, EPI_ISL_1699277, EPI_ISL_1699280,
EPI_ISL_1699283, EPI_ISL_1699285, EPI_ISL_1699286, EPI_ISL_1699307, EPI_ISL_1699326, EPI_ISL_1699365, EPI_ISL_1699451, EPI_ISL_1699452, EPI_ISL_1699456, EPI_ISL_1699478, EPI_ISL_1699481, EPI_ISL_1699496, EPI_ISL_1699504, EPI_ISL_1699519, EPI_ISL_1699530, EPI_ISL_1699531, EPI_ISL_1699575, EPI_ISL_1699595, EPI_ISL_1699611, EPI_ISL_1699618, EPI_ISL_1699668, EPI_ISL_1699683,
EPI_ISL_1699700, EPI_ISL_1699708, EPI_ISL_1699763, EPI_ISL_1699816, EPI_ISL_1699846, EPI_ISL_1699848, EPI_ISL_1699850, EPI_ISL_1699852, EPI_ISL_1699860, EPI_ISL_1699861, EPI_ISL_1699862, EPI_ISL_1699868, EPI_ISL_1699874, EPI_ISL_1699876, EPI_ISL_1699878, EPI_ISL_1699889, EPI_ISL_1699890, EPI_ISL_1699894, EPI_ISL_1699897, EPI_ISL_1699899, EPI_ISL_1699907, EPI_ISL_1699910,
EPI_ISL_1699915, EPI_ISL_1699916, EPI_ISL_1699917, EPI_ISL_1699921, EPI_ISL_1699922, EPI_ISL_1699928, EPI_ISL_1699929, EPI_ISL_1699930, EPI_ISL_1699932, EPI_ISL_1699933, EPI_ISL_1699934, EPI_ISL_1699941, EPI_ISL_1699942, EPI_ISL_1699943, EPI_ISL_1699950, EPI_ISL_1699955, EPI_ISL_1699964, EPI_ISL_1699974, EPI_ISL_1699986, EPI_ISL_1699991, EPI_ISL_1699994, EPI_ISL_1699995,
EPI_ISL_1699998, EPI_ISL_1700006, EPI_ISL_1700007, EPI_ISL_1700010, EPI_ISL_1700015, EPI_ISL_1700027, EPI_ISL_1700028, EPI_ISL_1700030, EPI_ISL_1700031, EPI_ISL_1700034, EPI_ISL_1700036, EPI_ISL_1700037, EPI_ISL_1700041, EPI_ISL_1700042, EPI_ISL_1700047, EPI_ISL_1700048, EPI_ISL_1700052, EPI_ISL_1700053, EPI_ISL_1700057, EPI_ISL_1700059, EPI_ISL_1700060, EPI_ISL_1700063,
EPI_ISL_1700067, EPI_ISL_1700068, EPI_ISL_1700069, EPI_ISL_1700070, EPI_ISL_1700079, EPI_ISL_1700081, EPI_ISL_1700085, EPI_ISL_1700090, EPI_ISL_1700096, EPI_ISL_1700098, EPI_ISL_1700100, EPI_ISL_1700103, EPI_ISL_1700109, EPI_ISL_1700112, EPI_ISL_1700113, EPI_ISL_1700118, EPI_ISL_1700127, EPI_ISL_1700135, EPI_ISL_1700136, EPI_ISL_1700139, EPI_ISL_1700141, EPI_ISL_1700143,
EPI_ISL_1700144, EPI_ISL_1700145, EPI_ISL_1700151, EPI_ISL_1700154, EPI_ISL_1700159, EPI_ISL_1700164, EPI_ISL_1700166, EPI_ISL_1700170, EPI_ISL_1700173, EPI_ISL_1700174, EPI_ISL_1700179, EPI_ISL_1700184, EPI_ISL_1700190, EPI_ISL_1700191, EPI_ISL_1700192, EPI_ISL_1700193, EPI_ISL_1700196, EPI_ISL_1700200, EPI_ISL_1700201, EPI_ISL_1700204, EPI_ISL_1700207, EPI_ISL_1700208,
EPI_ISL_1700209, EPI_ISL_1700212, EPI_ISL_1700213, EPI_ISL_1700214, EPI_ISL_1700216, EPI_ISL_1700221, EPI_ISL_1700223, EPI_ISL_1700227, EPI_ISL_1700228, EPI_ISL_1700240, EPI_ISL_1700241, EPI_ISL_1700243, EPI_ISL_1700244, EPI_ISL_1700250, EPI_ISL_1700251, EPI_ISL_1700253, EPI_ISL_1700254, EPI_ISL_1700255, EPI_ISL_1700257, EPI_ISL_1700258, EPI_ISL_1700265, EPI_ISL_1700269,
EPI_ISL_1700270, EPI_ISL_1700274, EPI_ISL_1700275, EPI_ISL_1700277, EPI_ISL_1700278, EPI_ISL_1700286, EPI_ISL_1700294, EPI_ISL_1700300, EPI_ISL_1700302, EPI_ISL_1700307, EPI_ISL_1700308, EPI_ISL_1700312, EPI_ISL_1700319, EPI_ISL_1700320, EPI_ISL_1700331, EPI_ISL_1700332, EPI_ISL_1700333, EPI_ISL_1700338, EPI_ISL_1700343, EPI_ISL_1700347, EPI_ISL_1700355, EPI_ISL_1700356,
EPI_ISL_1700357, EPI_ISL_1700361, EPI_ISL_1700363, EPI_ISL_1700364, EPI_ISL_1700369, EPI_ISL_1700370, EPI_ISL_1700373, EPI_ISL_1700378, EPI_ISL_1700380, EPI_ISL_1700385, EPI_ISL_1700386, EPI_ISL_1700389, EPI_ISL_1700392, EPI_ISL_1700396, EPI_ISL_1700410, EPI_ISL_1700413, EPI_ISL_1700415, EPI_ISL_1700433, EPI_ISL_1700436, EPI_ISL_1700441, EPI_ISL_1700456, EPI_ISL_1700457,
EPI_ISL_1700459, EPI_ISL_1700461, EPI_ISL_1700469, EPI_ISL_1700470, EPI_ISL_1700473, EPI_ISL_1700476, EPI_ISL_1700479, EPI_ISL_1700484, EPI_ISL_1700487, EPI_ISL_1700490, EPI_ISL_1700492, EPI_ISL_1700493, EPI_ISL_1741139, EPI_ISL_1741231, EPI_ISL_1741774, EPI_ISL_1741775, EPI_ISL_1741777, EPI_ISL_1741778, EPI_ISL_1741779, EPI_ISL_1741780, EPI_ISL_1741781, EPI_ISL_1741782,
EPI_ISL_1741785, EPI_ISL_1741786, EPI_ISL_1741788, EPI_ISL_1741789, EPI_ISL_1741790, EPI_ISL_1741793, EPI_ISL_1741794, EPI_ISL_1741795, EPI_ISL_1741799, EPI_ISL_1741800, EPI_ISL_1741801, EPI_ISL_1741803, EPI_ISL_1741804, EPI_ISL_1741806, EPI_ISL_1741807, EPI_ISL_1741808, EPI_ISL_1741809, EPI_ISL_1741812, EPI_ISL_1741813, EPI_ISL_1741815,
EPI_ISL_1741816, EPI_ISL_1741817, EPI_ISL_1741818, EPI_ISL_1741819, EPI_ISL_1741820, EPI_ISL_1741822, EPI_ISL_1741823, EPI_ISL_1741825, EPI_ISL_1741826, EPI_ISL_1741827, EPI_ISL_1741828, EPI_ISL_1741829, EPI_ISL_1741830, EPI_ISL_1741831, EPI_ISL_1741832, EPI_ISL_1741834, EPI_ISL_1741835, EPI_ISL_1741836, EPI_ISL_1741837, EPI_ISL_1741838, EPI_ISL_1741839, EPI_ISL_1741840,
EPI_ISL_1741842, EPI_ISL_1741843, EPI_ISL_1741845, EPI_ISL_1741846, EPI_ISL_1741847, EPI_ISL_1741848, EPI_ISL_1741850, EPI_ISL_1741853, EPI_ISL_1741854, EPI_ISL_1741855, EPI_ISL_1741857, EPI_ISL_1741859, EPI_ISL_1741860, EPI_ISL_1741861, EPI_ISL_1741862, EPI_ISL_1741863, EPI_ISL_1741865, EPI_ISL_1741866, EPI_ISL_1741869, EPI_ISL_1741870, EPI_ISL_1741871,
EPI_ISL_1741873, EPI_ISL_1741874, EPI_ISL_1741875, EPI_ISL_1741878, EPI_ISL_1741879, EPI_ISL_1741881, EPI_ISL_1741882, EPI_ISL_1741883, EPI_ISL_1741884, EPI_ISL_1741885, EPI_ISL_1741886, EPI_ISL_1741887, EPI_ISL_1741889, EPI_ISL_1741890, EPI_ISL_1741891, EPI_ISL_1741892, EPI_ISL_1741893, EPI_ISL_1741894, EPI_ISL_1741897, EPI_ISL_1741899, EPI_ISL_1741901, EPI_ISL_1741902,
EPI_ISL_1741903, EPI_ISL_1741904, EPI_ISL_1741905, EPI_ISL_1741906, EPI_ISL_1741907, EPI_ISL_1741909, EPI_ISL_1741910, EPI_ISL_1741911, EPI_ISL_1741913, EPI_ISL_1741914, EPI_ISL_1741915, EPI_ISL_1741916, EPI_ISL_1741917, EPI_ISL_1741918, EPI_ISL_1741919, EPI_ISL_1741920, EPI_ISL_1741921, EPI_ISL_1741922, EPI_ISL_1741923, EPI_ISL_1741924, EPI_ISL_1741926, EPI_ISL_1741927,
EPI_ISL_1741929, EPI_ISL_1741930, EPI_ISL_1741931, EPI_ISL_1741934, EPI_ISL_1741936, EPI_ISL_1741937, EPI_ISL_1741939, EPI_ISL_1741940, EPI_ISL_1741941, EPI_ISL_1741943, EPI_ISL_1741945, EPI_ISL_1741947, EPI_ISL_1741950, EPI_ISL_1741951, EPI_ISL_1741952, EPI_ISL_1741954, EPI_ISL_1741956, EPI_ISL_1741959, EPI_ISL_1741960, EPI_ISL_1741962, EPI_ISL_1741963, EPI_ISL_1741964,
EPI_ISL_1741965, EPI_ISL_1741966, EPI_ISL_1741967, EPI_ISL_1741968, EPI_ISL_1741969, EPI_ISL_1741971, EPI_ISL_1741973, EPI_ISL_1741974, EPI_ISL_1741976, EPI_ISL_1741978, EPI_ISL_1741979, EPI_ISL_1741982, EPI_ISL_1741983, EPI_ISL_1741984, EPI_ISL_1741986, EPI_ISL_1741988, EPI_ISL_1741990, EPI_ISL_1741993, EPI_ISL_1741994, EPI_ISL_1741995, EPI_ISL_1741996, EPI_ISL_1742002,
EPI_ISL_1742003, EPI_ISL_1742004, EPI_ISL_1742005, EPI_ISL_1742006, EPI_ISL_1742007, EPI_ISL_1742011, EPI_ISL_1742013, EPI_ISL_1742014, EPI_ISL_1742015, EPI_ISL_1742016, EPI_ISL_1742017, EPI_ISL_1742018, EPI_ISL_1742020, EPI_ISL_1742021, EPI_ISL_1742022, EPI_ISL_1742023, EPI_ISL_1742024, EPI_ISL_1742025, EPI_ISL_1742026, EPI_ISL_1742027, EPI_ISL_1742028, EPI_ISL_1742029,
EPI_ISL_1742030, EPI_ISL_1742031, EPI_ISL_1742032, EPI_ISL_1742033, EPI_ISL_1742035, EPI_ISL_1742036, EPI_ISL_1742037, EPI_ISL_1742038, EPI_ISL_1742039, EPI_ISL_1742040, EPI_ISL_1742041, EPI_ISL_1742042, EPI_ISL_1742043, EPI_ISL_1742044, EPI_ISL_1742045, EPI_ISL_1742046, EPI_ISL_1742047, EPI_ISL_1742048, EPI_ISL_1742049, EPI_ISL_1742050, EPI_ISL_1742051, EPI_ISL_1742052,
EPI_ISL_1742053, EPI_ISL_1742054, EPI_ISL_1742056, EPI_ISL_1742057, EPI_ISL_1742058, EPI_ISL_1742059, EPI_ISL_1742061, EPI_ISL_1742063, EPI_ISL_1742064, EPI_ISL_1742065, EPI_ISL_1742066, EPI_ISL_1742067, EPI_ISL_1742068, EPI_ISL_1742069, EPI_ISL_1742070, EPI_ISL_1742071, EPI_ISL_1742072, EPI_ISL_1742073, EPI_ISL_1742074, EPI_ISL_1742075, EPI_ISL_1742076, EPI_ISL_1742077,
EPI_ISL_1742078, EPI_ISL_1742079, EPI_ISL_1742080, EPI_ISL_1742081, EPI_ISL_1742082, EPI_ISL_1742083, EPI_ISL_1742084, EPI_ISL_1742085, EPI_ISL_1742086, EPI_ISL_1742091, EPI_ISL_1742093, EPI_ISL_1742101, EPI_ISL_1742102, EPI_ISL_1742104, EPI_ISL_1742122, EPI_ISL_1742125, EPI_ISL_1742127, EPI_ISL_1742132, EPI_ISL_1742133, EPI_ISL_1742146, EPI_ISL_1742147, EPI_ISL_1742152,
EPI_ISL_1742153, EPI_ISL_1742156, EPI_ISL_1742157, EPI_ISL_1742163, EPI_ISL_1742164, EPI_ISL_1742170, EPI_ISL_1742181, EPI_ISL_1742182, EPI_ISL_1742184, EPI_ISL_1742185, EPI_ISL_1742186, EPI_ISL_1742187, EPI_ISL_1742188, EPI_ISL_1742189, EPI_ISL_1742190, EPI_ISL_1742191, EPI_ISL_1742192, EPI_ISL_1742193, EPI_ISL_1742194, EPI_ISL_1742195, EPI_ISL_1742196, EPI_ISL_1742197,
EPI_ISL_1742198, EPI_ISL_1742199, EPI_ISL_1742200, EPI_ISL_1742201, EPI_ISL_1742202, EPI_ISL_1742204, EPI_ISL_1742206, EPI_ISL_1742207, EPI_ISL_1742208, EPI_ISL_1742209, EPI_ISL_1742210, EPI_ISL_1742211, EPI_ISL_1742212, EPI_ISL_1742213, EPI_ISL_1742214, EPI_ISL_1742216, EPI_ISL_1742217, EPI_ISL_1742218, EPI_ISL_1742219, EPI_ISL_1742220, EPI_ISL_1742221, EPI_ISL_1742223,
EPI_ISL_1742224, EPI_ISL_1742225, EPI_ISL_1742226, EPI_ISL_1742227, EPI_ISL_1742228, EPI_ISL_1742230, EPI_ISL_1742231, EPI_ISL_1742232, EPI_ISL_1742233, EPI_ISL_1742234, EPI_ISL_1742235, EPI_ISL_1742236, EPI_ISL_1742237, EPI_ISL_1742238, EPI_ISL_1742239, EPI_ISL_1742240, EPI_ISL_1742241, EPI_ISL_1742243, EPI_ISL_1742244, EPI_ISL_1742245, EPI_ISL_1742246, EPI_ISL_1742247,
EPI_ISL_1742248, EPI_ISL_1742249, EPI_ISL_1742250, EPI_ISL_1742251, EPI_ISL_1742253, EPI_ISL_1742254, EPI_ISL_1742255, EPI_ISL_1742256, EPI_ISL_1742257, EPI_ISL_1742258, EPI_ISL_1742259, EPI_ISL_1742260, EPI_ISL_1742261, EPI_ISL_1758948

| | | | |
|---|---|---|---|
| see above | Lighthouse Lab in Glasgow | Wellcome Sanger Institute for the COVID-19 Genomics UK (COG-UK) Consortium | Anna Dominiczak and Alex Alderton; Carol Clugston; Cordelia Langford; David Gray; David K. Jackson; Dominic Kwiatkowski; Ewan Harrison; Harper VanSteenhouse; Ian Johnston; Jeffrey Barrett; John Sillitoe on behalf of the Wellcome Sanger Institute COVID-19 Surveillance Team; Roberto Amato; Sonia Goncalves; Yumi Kasai |

EPI_ISL_1538402, EPI_ISL_1584274, EPI_ISL_1584322, EPI_ISL_1594457, EPI_ISL_1594463, EPI_ISL_1594465, EPI_ISL_1594478, EPI_ISL_1594492, EPI_ISL_1594494, EPI_ISL_1594507, EPI_ISL_1594530, EPI_ISL_1594536, EPI_ISL_1594541, EPI_ISL_1594547, EPI_ISL_1594549, EPI_ISL_1594551, EPI_ISL_1594557, EPI_ISL_1594563, EPI_ISL_1594567, EPI_ISL_1594571, EPI_ISL_1594574, EPI_ISL_1594582,
EPI_ISL_1594588, EPI_ISL_1594612, EPI_ISL_1594615, EPI_ISL_1594622, EPI_ISL_1594624, EPI_ISL_1594639, EPI_ISL_1594643, EPI_ISL_1594649, EPI_ISL_1594655, EPI_ISL_1594661, EPI_ISL_1594662, EPI_ISL_1594667, EPI_ISL_1594668, EPI_ISL_1594669, EPI_ISL_1594675, EPI_ISL_1594678, EPI_ISL_1594680, EPI_ISL_1594681, EPI_ISL_1594682, EPI_ISL_1594687, EPI_ISL_1594689, EPI_ISL_1594694,
EPI_ISL_1594700, EPI_ISL_1594712, EPI_ISL_1594753, EPI_ISL_1594799, EPI_ISL_1594831, EPI_ISL_1594833, EPI_ISL_1594888, EPI_ISL_1594910, EPI_ISL_1594938, EPI_ISL_1594945, EPI_ISL_1594947, EPI_ISL_1594953, EPI_ISL_1594975, EPI_ISL_1615690, EPI_ISL_1615696, EPI_ISL_1615701, EPI_ISL_1615704, EPI_ISL_1615708, EPI_ISL_1615710, EPI_ISL_1615711, EPI_ISL_1615712, EPI_ISL_1615715,
EPI_ISL_1615718, EPI_ISL_1615720, EPI_ISL_1615724, EPI_ISL_1615725, EPI_ISL_1615730, EPI_ISL_1615737, EPI_ISL_1615743, EPI_ISL_1615748, EPI_ISL_1615758, EPI_ISL_1615760, EPI_ISL_1615767, EPI_ISL_1615770, EPI_ISL_1615772, EPI_ISL_1615773, EPI_ISL_1615774, EPI_ISL_1615777, EPI_ISL_1615779, EPI_ISL_1615782, EPI_ISL_1615785, EPI_ISL_1615786, EPI_ISL_1615788,
EPI_ISL_1615791, EPI_ISL_1615795, EPI_ISL_1615799, EPI_ISL_1615802, EPI_ISL_1615804, EPI_ISL_1615806, EPI_ISL_1615809, EPI_ISL_1615810, EPI_ISL_1615811, EPI_ISL_1615813, EPI_ISL_1615819, EPI_ISL_1615821, EPI_ISL_1615824, EPI_ISL_1615825, EPI_ISL_1615826, EPI_ISL_1615834, EPI_ISL_1615835, EPI_ISL_1615840, EPI_ISL_1615841, EPI_ISL_1615845, EPI_ISL_1615846, EPI_ISL_1615849,
EPI_ISL_1615850, EPI_ISL_1615851, EPI_ISL_1615853, EPI_ISL_1615859, EPI_ISL_1615860, EPI_ISL_1615867, EPI_ISL_1615871, EPI_ISL_1615874, EPI_ISL_1615876, EPI_ISL_1615892, EPI_ISL_1615903, EPI_ISL_1615913, EPI_ISL_1615920, EPI_ISL_1615925, EPI_ISL_1615927, EPI_ISL_1615930, EPI_ISL_1615944, EPI_ISL_1615949, EPI_ISL_1615950, EPI_ISL_1615954, EPI_ISL_1615957, EPI_ISL_1615959,
EPI_ISL_1615960, EPI_ISL_1615961, EPI_ISL_1615963, EPI_ISL_1615967, EPI_ISL_1615968, EPI_ISL_1615969, EPI_ISL_1615970, EPI_ISL_1615973, EPI_ISL_1615974, EPI_ISL_1615975, EPI_ISL_1615979, EPI_ISL_1615981, EPI_ISL_1615985, EPI_ISL_1615995, EPI_ISL_1615997, EPI_ISL_1616002, EPI_ISL_1616009, EPI_ISL_1616013, EPI_ISL_1616014, EPI_ISL_1616026, EPI_ISL_1616036, EPI_ISL_1616038,
EPI_ISL_1616042, EPI_ISL_1616047, EPI_ISL_1616050, EPI_ISL_1616053, EPI_ISL_1616062, EPI_ISL_1616065, EPI_ISL_1616068, EPI_ISL_1616072, EPI_ISL_1616080, EPI_ISL_1616083, EPI_ISL_1616084, EPI_ISL_1616085, EPI_ISL_1616089, EPI_ISL_1616094, EPI_ISL_1616096, EPI_ISL_1616115, EPI_ISL_1616119, EPI_ISL_1616122, EPI_ISL_1616125, EPI_ISL_1616132, EPI_ISL_1616133, EPI_ISL_1616135,

| | | | |
|---|---|---|---|
| EPI_ISL_1616138, EPI_ISL_1616141, EPI_ISL_1616142, EPI_ISL_1616144, EPI_ISL_1616148, EPI_ISL_1616149, EPI_ISL_1616158, EPI_ISL_1616161, EPI_ISL_1616164, EPI_ISL_1616175, EPI_ISL_1616176, EPI_ISL_1616178, EPI_ISL_1616183, EPI_ISL_1616185, EPI_ISL_1630284, EPI_ISL_1630289, EPI_ISL_1630291, EPI_ISL_1630298, EPI_ISL_1630305, EPI_ISL_1630320, EPI_ISL_1630326, EPI_ISL_1630336, EPI_ISL_1630339, EPI_ISL_1630349, EPI_ISL_1630357, EPI_ISL_1630358, EPI_ISL_1630360, EPI_ISL_1630363, EPI_ISL_1630374, EPI_ISL_1630375, EPI_ISL_1630377, EPI_ISL_1630378, EPI_ISL_1630380, EPI_ISL_1630383, EPI_ISL_1630388, EPI_ISL_1630390, EPI_ISL_1630395, EPI_ISL_1630402, EPI_ISL_1630406, EPI_ISL_1630412, EPI_ISL_1630444, EPI_ISL_1630455, EPI_ISL_1630461, EPI_ISL_1630463, EPI_ISL_1630464, EPI_ISL_1630469, EPI_ISL_1630474, EPI_ISL_1630475, EPI_ISL_1630478, EPI_ISL_1630479, EPI_ISL_1630488, EPI_ISL_1630493, EPI_ISL_1630494, EPI_ISL_1630508, EPI_ISL_1631101, EPI_ISL_1631118, EPI_ISL_1631122, EPI_ISL_1631126, EPI_ISL_1631129, EPI_ISL_1631141, EPI_ISL_1631142, EPI_ISL_1631159, EPI_ISL_1631161, EPI_ISL_1631181, EPI_ISL_1631188, EPI_ISL_1631193, EPI_ISL_1631197, EPI_ISL_1631199, EPI_ISL_1631218, EPI_ISL_1631233, EPI_ISL_1631234, EPI_ISL_1631252, EPI_ISL_1631277, EPI_ISL_1631290, EPI_ISL_1631292, EPI_ISL_1631315, EPI_ISL_1631754, EPI_ISL_1631755, EPI_ISL_1631756, EPI_ISL_1631757, EPI_ISL_1631759, EPI_ISL_1631760, EPI_ISL_1631767, EPI_ISL_1631775, EPI_ISL_1631776, EPI_ISL_1631792, EPI_ISL_1631793, EPI_ISL_1631798, EPI_ISL_1631799, EPI_ISL_1631801, EPI_ISL_1631802, EPI_ISL_1631803, EPI_ISL_1631806, EPI_ISL_1631811, EPI_ISL_1631812, EPI_ISL_1631814, EPI_ISL_1631815, EPI_ISL_1631818, EPI_ISL_1631821, EPI_ISL_1631824, EPI_ISL_1631826, EPI_ISL_1631827, EPI_ISL_1631832, EPI_ISL_1631838, EPI_ISL_1631856, EPI_ISL_1631858, EPI_ISL_1631859, EPI_ISL_1631860, EPI_ISL_1631865, EPI_ISL_1631866, EPI_ISL_1631869, EPI_ISL_1631876, EPI_ISL_1631877, EPI_ISL_1631882, EPI_ISL_1631887, EPI_ISL_1631891, EPI_ISL_1631892, EPI_ISL_1631900, EPI_ISL_1631902, EPI_ISL_1631912, EPI_ISL_1631913, EPI_ISL_1631918, EPI_ISL_1631922, EPI_ISL_1631923, EPI_ISL_1631924, EPI_ISL_1631929, EPI_ISL_1631941, EPI_ISL_1631943, EPI_ISL_1631945, EPI_ISL_1631949, EPI_ISL_1631953, EPI_ISL_1631961, EPI_ISL_1631970, EPI_ISL_1631981, EPI_ISL_1631991, EPI_ISL_1631994, EPI_ISL_1631996, EPI_ISL_1632011, EPI_ISL_1632021, EPI_ISL_1632031, EPI_ISL_1632032, EPI_ISL_1632034, EPI_ISL_1632081, EPI_ISL_1632106, EPI_ISL_1632111, EPI_ISL_1632117, EPI_ISL_1632118, EPI_ISL_1632126, EPI_ISL_1632127, EPI_ISL_1632136, EPI_ISL_1632161, EPI_ISL_1632174, EPI_ISL_1632175, EPI_ISL_1632191, EPI_ISL_1632198, EPI_ISL_1632200, EPI_ISL_1632215, EPI_ISL_1632217, EPI_ISL_1632223, EPI_ISL_1632231, EPI_ISL_1632235, EPI_ISL_1632245, EPI_ISL_1632257, EPI_ISL_1632264, EPI_ISL_1632268, EPI_ISL_1632271, EPI_ISL_1632279, EPI_ISL_1632286, EPI_ISL_1632288, EPI_ISL_1632293, EPI_ISL_1632295, EPI_ISL_1632328, EPI_ISL_1632348, EPI_ISL_1632377, EPI_ISL_1632436, EPI_ISL_1632439, EPI_ISL_1632459, EPI_ISL_1632472, EPI_ISL_1634487, EPI_ISL_1634488, EPI_ISL_1634491, EPI_ISL_1634494, EPI_ISL_1634499, EPI_ISL_1634500, EPI_ISL_1634505, EPI_ISL_1634506, EPI_ISL_1634507, EPI_ISL_1634510, EPI_ISL_1634511, EPI_ISL_1634514, EPI_ISL_1634515, EPI_ISL_1634516, EPI_ISL_1634517, EPI_ISL_1634519, EPI_ISL_1634520, EPI_ISL_1634521, EPI_ISL_1634523, EPI_ISL_1634524, EPI_ISL_1634525, EPI_ISL_1634526, EPI_ISL_1634529, EPI_ISL_1634530, EPI_ISL_1634531, EPI_ISL_1634533, EPI_ISL_1634534, EPI_ISL_1634535, EPI_ISL_1634536, EPI_ISL_1634538, EPI_ISL_1634541, EPI_ISL_1634542, EPI_ISL_1634546, EPI_ISL_1634549, EPI_ISL_1634551, EPI_ISL_1634552, EPI_ISL_1634553, EPI_ISL_1634556, EPI_ISL_1634557, EPI_ISL_1634560, EPI_ISL_1634561, EPI_ISL_1634562, EPI_ISL_1634563, EPI_ISL_1634564, EPI_ISL_1634565, EPI_ISL_1634567, EPI_ISL_1634569, EPI_ISL_1634570, EPI_ISL_1634573, EPI_ISL_1634576, EPI_ISL_1634579, EPI_ISL_1634580, EPI_ISL_1634581, EPI_ISL_1634585, EPI_ISL_1634588, EPI_ISL_1634590, EPI_ISL_1634591, EPI_ISL_1634592, EPI_ISL_1634593, EPI_ISL_1634594, EPI_ISL_1634596, EPI_ISL_1634597, EPI_ISL_1634598, EPI_ISL_1634599, EPI_ISL_1634606, EPI_ISL_1634608, EPI_ISL_1634609, EPI_ISL_1634611, EPI_ISL_1634612, EPI_ISL_1634614, EPI_ISL_1634616, EPI_ISL_1634618, EPI_ISL_1634619, EPI_ISL_1634621, EPI_ISL_1634625, EPI_ISL_1634627, EPI_ISL_1634629, EPI_ISL_1634630, EPI_ISL_1634636, EPI_ISL_1634637, EPI_ISL_1634638, EPI_ISL_1634639, EPI_ISL_1634640, EPI_ISL_1634641, EPI_ISL_1634642, EPI_ISL_1634644, EPI_ISL_1634645, EPI_ISL_1634646, EPI_ISL_1634647, EPI_ISL_1634648, EPI_ISL_1634652, EPI_ISL_1634654, EPI_ISL_1634655, EPI_ISL_1634657, EPI_ISL_1634658, EPI_ISL_1634659, EPI_ISL_1634660, EPI_ISL_1634662, EPI_ISL_1634663, EPI_ISL_1634666, EPI_ISL_1634668, EPI_ISL_1634670, EPI_ISL_1634671, EPI_ISL_1634672, EPI_ISL_1634673, EPI_ISL_1634675, EPI_ISL_1634678, EPI_ISL_1634679, EPI_ISL_1634682, EPI_ISL_1634684, EPI_ISL_1634686, EPI_ISL_1634687, EPI_ISL_1634688, EPI_ISL_1634689, EPI_ISL_1634692, EPI_ISL_1634693, EPI_ISL_1634694, EPI_ISL_1634698, EPI_ISL_1634699, EPI_ISL_1652998, EPI_ISL_1653176, EPI_ISL_1653183, EPI_ISL_1653185, EPI_ISL_1653209, EPI_ISL_1653210, EPI_ISL_1653213, EPI_ISL_1653214, EPI_ISL_1653216, EPI_ISL_1653218, EPI_ISL_1653219, EPI_ISL_1653222, EPI_ISL_1653223, EPI_ISL_1653228, EPI_ISL_1653238, EPI_ISL_1653240, EPI_ISL_1653246, EPI_ISL_1653248, EPI_ISL_1653254, EPI_ISL_1653258, EPI_ISL_1653261, EPI_ISL_1653263, EPI_ISL_1653265, EPI_ISL_1653266, EPI_ISL_1653267, EPI_ISL_1653274, EPI_ISL_1653275, EPI_ISL_1653283, EPI_ISL_1653285, EPI_ISL_1653291, EPI_ISL_1653292, EPI_ISL_1653295, EPI_ISL_1653306, EPI_ISL_1653311, EPI_ISL_1653318, EPI_ISL_1653322, EPI_ISL_1653323, EPI_ISL_1653324, EPI_ISL_1653328, EPI_ISL_1653336, EPI_ISL_1653344, EPI_ISL_1653355, EPI_ISL_1653366, EPI_ISL_1653367, EPI_ISL_1653373, EPI_ISL_1653376, EPI_ISL_1653383, EPI_ISL_1653384, EPI_ISL_1653393, EPI_ISL_1653397, EPI_ISL_1653398, EPI_ISL_1653399, EPI_ISL_1653407, EPI_ISL_1653410, EPI_ISL_1653411, EPI_ISL_1653416, EPI_ISL_1653420, EPI_ISL_1653425, EPI_ISL_1653430, EPI_ISL_1653434, EPI_ISL_1653437, EPI_ISL_1653442, EPI_ISL_1653443, EPI_ISL_1653444, EPI_ISL_1653448, EPI_ISL_1653450, EPI_ISL_1653451, EPI_ISL_1653452, EPI_ISL_1653454, EPI_ISL_1653459, EPI_ISL_1653462, EPI_ISL_1653466, EPI_ISL_1653467, EPI_ISL_1653470, EPI_ISL_1653485, EPI_ISL_1673121, EPI_ISL_1697456, EPI_ISL_1697497, EPI_ISL_1697516, EPI_ISL_1697518, EPI_ISL_1697522, EPI_ISL_1697526, EPI_ISL_1697530, EPI_ISL_1697539, EPI_ISL_1697547, EPI_ISL_1697552, EPI_ISL_1697565, EPI_ISL_1697571, EPI_ISL_1697577, EPI_ISL_1697579, EPI_ISL_1697592, EPI_ISL_1697594, EPI_ISL_1697596, EPI_ISL_1697601, EPI_ISL_1697603, EPI_ISL_1697617, EPI_ISL_1697622, EPI_ISL_1697645, EPI_ISL_1697650, EPI_ISL_1697654, EPI_ISL_1697656, EPI_ISL_1697684, EPI_ISL_1697690, EPI_ISL_1697748, EPI_ISL_1697754, EPI_ISL_1697755, EPI_ISL_1697756, EPI_ISL_1697757, EPI_ISL_1697758, EPI_ISL_1697759, EPI_ISL_1697762, EPI_ISL_1697763, EPI_ISL_1697768, EPI_ISL_1697769, EPI_ISL_1697775, EPI_ISL_1697776, EPI_ISL_1697777, EPI_ISL_1697783, EPI_ISL_1697788, EPI_ISL_1697791, EPI_ISL_1697792, EPI_ISL_1697795, EPI_ISL_1697799, EPI_ISL_1697800, EPI_ISL_1697805, EPI_ISL_1697817, EPI_ISL_1697819, EPI_ISL_1697821, EPI_ISL_1697828, EPI_ISL_1697835, EPI_ISL_1697839, EPI_ISL_1697843, EPI_ISL_1697844, EPI_ISL_1697846, EPI_ISL_1697847, EPI_ISL_1697849, EPI_ISL_1697853, EPI_ISL_1697854, EPI_ISL_1697856, EPI_ISL_1697857, EPI_ISL_1697859, EPI_ISL_1697863, EPI_ISL_1697866, EPI_ISL_1697871, EPI_ISL_1697874, EPI_ISL_1697875, EPI_ISL_1697882, EPI_ISL_1697883, EPI_ISL_1697884, EPI_ISL_1697885, EPI_ISL_1697896, EPI_ISL_1697899, EPI_ISL_1697900, EPI_ISL_1697902, EPI_ISL_1697903, EPI_ISL_1697904, EPI_ISL_1697917, EPI_ISL_1697921, EPI_ISL_1697924, EPI_ISL_1697928, EPI_ISL_1697934, EPI_ISL_1697941, EPI_ISL_1697947, EPI_ISL_1697948, EPI_ISL_1697949, EPI_ISL_1697959, EPI_ISL_1697961, EPI_ISL_1697969, EPI_ISL_1697979, EPI_ISL_1698000, EPI_ISL_1698025, EPI_ISL_1698029, EPI_ISL_1698031, EPI_ISL_1698054, EPI_ISL_1698073, EPI_ISL_1698102, EPI_ISL_1698172, EPI_ISL_1698193, EPI_ISL_1698215, EPI_ISL_1698767, EPI_ISL_1698770, EPI_ISL_1698777, EPI_ISL_1698780, EPI_ISL_1698784, EPI_ISL_1698786, EPI_ISL_1698789, EPI_ISL_1698791, EPI_ISL_1698793, EPI_ISL_1698794, EPI_ISL_1698796, EPI_ISL_1698797, EPI_ISL_1698799, EPI_ISL_1698804, EPI_ISL_1698805, EPI_ISL_1698806, EPI_ISL_1698808, EPI_ISL_1698809, EPI_ISL_1698814, EPI_ISL_1698815, EPI_ISL_1698821, EPI_ISL_1698824, EPI_ISL_1698829, EPI_ISL_1698832, EPI_ISL_1698833, EPI_ISL_1698838, EPI_ISL_1698840, EPI_ISL_1698846, EPI_ISL_1698855, EPI_ISL_1698857, EPI_ISL_1698860, EPI_ISL_1698861, EPI_ISL_1698862, EPI_ISL_1698864, EPI_ISL_1698865, EPI_ISL_1698866, EPI_ISL_1698870, EPI_ISL_1698873, EPI_ISL_1698876, EPI_ISL_1698877, EPI_ISL_1698879, EPI_ISL_1698880, EPI_ISL_1698883, EPI_ISL_1698886, EPI_ISL_1698888, EPI_ISL_1698889, EPI_ISL_1698890, EPI_ISL_1698891, EPI_ISL_1698895, EPI_ISL_1698899, EPI_ISL_1698901, EPI_ISL_1698902, EPI_ISL_1698906, EPI_ISL_1698907, EPI_ISL_1698908, EPI_ISL_1698911, EPI_ISL_1698912, EPI_ISL_1698914, EPI_ISL_1698916, EPI_ISL_1698917, EPI_ISL_1698919, EPI_ISL_1698921, EPI_ISL_1698927, EPI_ISL_1698929, EPI_ISL_1698934, EPI_ISL_1698943, EPI_ISL_1698944, EPI_ISL_1698945, EPI_ISL_1698950, EPI_ISL_1698956, EPI_ISL_1698959, EPI_ISL_1698965, EPI_ISL_1698966, EPI_ISL_1698967, EPI_ISL_1698971, EPI_ISL_1698972, EPI_ISL_1698974, EPI_ISL_1698975, EPI_ISL_1698977, EPI_ISL_1698983, EPI_ISL_1698989, EPI_ISL_1698990, EPI_ISL_1698994, EPI_ISL_1699000, EPI_ISL_1699007, EPI_ISL_1699008, EPI_ISL_1699009, EPI_ISL_1699012, EPI_ISL_1699015, EPI_ISL_1699016, EPI_ISL_1699018, EPI_ISL_1699020, EPI_ISL_1699021, EPI_ISL_1699028, EPI_ISL_1699030, EPI_ISL_1699042, EPI_ISL_1699054, EPI_ISL_1699059, EPI_ISL_1699062, EPI_ISL_1699065, EPI_ISL_1699075, EPI_ISL_1699083, EPI_ISL_1699086, EPI_ISL_1699087, EPI_ISL_1699090, EPI_ISL_1699092, EPI_ISL_1699093, EPI_ISL_1699095, EPI_ISL_1699097, EPI_ISL_1699098, EPI_ISL_1699103, EPI_ISL_1699116, EPI_ISL_1699121, EPI_ISL_1699130, EPI_ISL_1699135, EPI_ISL_1699137, EPI_ISL_1699139, EPI_ISL_1699144, EPI_ISL_1699154, EPI_ISL_1699160, EPI_ISL_1699163, EPI_ISL_1699165, EPI_ISL_1699168, EPI_ISL_1699170, EPI_ISL_1699174, EPI_ISL_1699176, EPI_ISL_1699178, EPI_ISL_1699179, EPI_ISL_1699181, EPI_ISL_1699183, EPI_ISL_1699185, EPI_ISL_1699186, EPI_ISL_1699187, EPI_ISL_1699190, EPI_ISL_1699191, EPI_ISL_1699193, EPI_ISL_1699205, EPI_ISL_1699206, EPI_ISL_1699208, EPI_ISL_1699216, EPI_ISL_1699221, EPI_ISL_1699223, EPI_ISL_1699227, EPI_ISL_1699230, EPI_ISL_1699234, EPI_ISL_1699241, EPI_ISL_1699255, EPI_ISL_1699264, EPI_ISL_1699267, EPI_ISL_1699272, EPI_ISL_1699273, EPI_ISL_1699276, EPI_ISL_1699278, EPI_ISL_1699279, EPI_ISL_1699289, EPI_ISL_1699319, EPI_ISL_1699322, EPI_ISL_1699328, EPI_ISL_1699339, EPI_ISL_1699348, EPI_ISL_1699385, EPI_ISL_1699413, EPI_ISL_1699414, EPI_ISL_1699417, EPI_ISL_1699450, EPI_ISL_1699475, EPI_ISL_1699571, EPI_ISL_1699572, EPI_ISL_1699573, EPI_ISL_1699574, EPI_ISL_1699576, EPI_ISL_1699577, EPI_ISL_1699579, EPI_ISL_1699583, EPI_ISL_1699584, EPI_ISL_1699588, EPI_ISL_1699590, EPI_ISL_1699591, EPI_ISL_1699592, EPI_ISL_1699593, EPI_ISL_1699594, EPI_ISL_1699596, EPI_ISL_1699599, EPI_ISL_1699600, EPI_ISL_1699601, EPI_ISL_1699602, EPI_ISL_1699604, EPI_ISL_1699607, EPI_ISL_1699608, EPI_ISL_1699610, EPI_ISL_1699613, EPI_ISL_1699614, EPI_ISL_1699615, EPI_ISL_1699617, EPI_ISL_1699619, EPI_ISL_1699624, EPI_ISL_1699625, EPI_ISL_1699626, EPI_ISL_1699627, EPI_ISL_1699632, EPI_ISL_1699634, EPI_ISL_1699639, EPI_ISL_1699640, EPI_ISL_1699641, EPI_ISL_1699642, EPI_ISL_1699643, EPI_ISL_1699644, EPI_ISL_1699645, EPI_ISL_1699647, EPI_ISL_1699648, EPI_ISL_1699651, EPI_ISL_1699652, EPI_ISL_1699653, EPI_ISL_1699654, EPI_ISL_1699656, EPI_ISL_1699657, EPI_ISL_1699658, EPI_ISL_1699662, EPI_ISL_1699663, EPI_ISL_1699664, EPI_ISL_1699665, EPI_ISL_1699669, EPI_ISL_1699672, EPI_ISL_1699673, EPI_ISL_1699674, EPI_ISL_1699678, EPI_ISL_1699681, EPI_ISL_1699682, EPI_ISL_1699684, EPI_ISL_1699686, EPI_ISL_1699689, EPI_ISL_1699690, EPI_ISL_1699693, EPI_ISL_1699695, EPI_ISL_1699699, EPI_ISL_1699702, EPI_ISL_1699703, EPI_ISL_1699705, EPI_ISL_1699706, EPI_ISL_1699707, EPI_ISL_1699716, EPI_ISL_1699720, EPI_ISL_1699725, EPI_ISL_1699726, EPI_ISL_1699728, EPI_ISL_1699731, EPI_ISL_1699733, EPI_ISL_1699737, EPI_ISL_1699739, EPI_ISL_1699742, EPI_ISL_1699743, EPI_ISL_1699744, EPI_ISL_1699746, EPI_ISL_1699747, EPI_ISL_1699750, EPI_ISL_1699753, EPI_ISL_1699754, EPI_ISL_1699756, EPI_ISL_1699757, EPI_ISL_1699758, EPI_ISL_1699760, EPI_ISL_1699761, EPI_ISL_1699765, EPI_ISL_1699766, EPI_ISL_1699768, EPI_ISL_1699771, EPI_ISL_1699773, EPI_ISL_1699776, EPI_ISL_1699777, EPI_ISL_1699779, EPI_ISL_1699780, EPI_ISL_1699781, EPI_ISL_1699782, EPI_ISL_1699784, EPI_ISL_1699785, EPI_ISL_1699786, EPI_ISL_1699787, EPI_ISL_1699791, EPI_ISL_1699797, EPI_ISL_1699801, EPI_ISL_1699802, EPI_ISL_1699804, EPI_ISL_1699806, EPI_ISL_1699807, EPI_ISL_1699809, EPI_ISL_1699810, EPI_ISL_1699812, EPI_ISL_1699814, EPI_ISL_1699818, EPI_ISL_1699819, EPI_ISL_1699821, EPI_ISL_1699823, EPI_ISL_1699825, EPI_ISL_1699826, EPI_ISL_1699827, EPI_ISL_1699828, EPI_ISL_1699829, EPI_ISL_1699830, EPI_ISL_1699831, EPI_ISL_1699833, EPI_ISL_1699834, EPI_ISL_1699835, EPI_ISL_1699837, EPI_ISL_1699839, EPI_ISL_1699841, EPI_ISL_1699843, EPI_ISL_1699847, EPI_ISL_1699855, EPI_ISL_1699858, EPI_ISL_1699867, EPI_ISL_1699869, EPI_ISL_1699872, EPI_ISL_1699880, EPI_ISL_1699883, EPI_ISL_1699885, EPI_ISL_1699895, EPI_ISL_1699898, EPI_ISL_1699902, EPI_ISL_1699913, EPI_ISL_1699914, EPI_ISL_1699927, EPI_ISL_1699931, EPI_ISL_1699937, EPI_ISL_1699960, EPI_ISL_1699967, EPI_ISL_1699977, EPI_ISL_1699979, EPI_ISL_1699980, EPI_ISL_1699984, EPI_ISL_1699990, EPI_ISL_1699997, EPI_ISL_1700013, EPI_ISL_1700020, EPI_ISL_1700021, EPI_ISL_1700022, EPI_ISL_1700025, EPI_ISL_1700029, EPI_ISL_1700044, EPI_ISL_1700050, EPI_ISL_1700071, EPI_ISL_1700074, EPI_ISL_1700076, EPI_ISL_1700078, EPI_ISL_1700080, EPI_ISL_1700102, EPI_ISL_1700108, EPI_ISL_1700116, EPI_ISL_1700130, EPI_ISL_1700138, EPI_ISL_1700140, EPI_ISL_1700148, EPI_ISL_1700149, EPI_ISL_1700152, EPI_ISL_1700153, EPI_ISL_1700157, EPI_ISL_1700165, EPI_ISL_1700167, EPI_ISL_1700169, EPI_ISL_1718365, EPI_ISL_1718644, EPI_ISL_1719009, EPI_ISL_1719197, EPI_ISL_1719730, EPI_ISL_1719731, EPI_ISL_1741991, EPI_ISL_1758205, EPI_ISL_1758208, EPI_ISL_1758209, EPI_ISL_1758224, EPI_ISL_1758226, EPI_ISL_1758228, EPI_ISL_1758232, EPI_ISL_1758253, EPI_ISL_1758258, EPI_ISL_1758260, EPI_ISL_1758262, EPI_ISL_1758263, EPI_ISL_1758265, EPI_ISL_1758269, EPI_ISL_1758274, EPI_ISL_1758276, EPI_ISL_1758277, EPI_ISL_1758278, EPI_ISL_1758281, EPI_ISL_1758282, EPI_ISL_1758283, EPI_ISL_1758288, EPI_ISL_1758302, EPI_ISL_1758323, EPI_ISL_1758331, EPI_ISL_1758335, EPI_ISL_1758350, EPI_ISL_1758351, EPI_ISL_1758354, EPI_ISL_1758362, EPI_ISL_1758363, EPI_ISL_1758367, EPI_ISL_1758375, EPI_ISL_1758388, EPI_ISL_1758389, EPI_ISL_1758398, EPI_ISL_1758402, EPI_ISL_1758405, EPI_ISL_1758417, EPI_ISL_1758419, EPI_ISL_1758422, EPI_ISL_1758425, EPI_ISL_1758426, EPI_ISL_1758427, EPI_ISL_1758428, EPI_ISL_1758430, EPI_ISL_1758453, EPI_ISL_1758454, EPI_ISL_1758457, EPI_ISL_1758458, EPI_ISL_1758459, EPI_ISL_1758464, EPI_ISL_1758467, EPI_ISL_1758468, EPI_ISL_1758471, EPI_ISL_1758474, EPI_ISL_1758475, EPI_ISL_1758477, EPI_ISL_1758478, EPI_ISL_1758480, EPI_ISL_1758481, EPI_ISL_1758487, EPI_ISL_1758488, EPI_ISL_1758489, EPI_ISL_1758490, EPI_ISL_1758491, EPI_ISL_1758494, EPI_ISL_1758498, EPI_ISL_1758499, EPI_ISL_1758500, EPI_ISL_1758502, EPI_ISL_1758504, EPI_ISL_1758506, EPI_ISL_1758508, EPI_ISL_1758512, EPI_ISL_1758513, EPI_ISL_1758514, EPI_ISL_1758516, EPI_ISL_1758517, EPI_ISL_1758519, EPI_ISL_1758524, EPI_ISL_1758528, EPI_ISL_1758534, EPI_ISL_1758535, EPI_ISL_1758537, EPI_ISL_1758539, EPI_ISL_1758542, EPI_ISL_1758543, EPI_ISL_1758544, EPI_ISL_1758546, EPI_ISL_1758551, EPI_ISL_1758552, EPI_ISL_1758553, EPI_ISL_1758554, EPI_ISL_1758557, EPI_ISL_1758560, EPI_ISL_1758561, EPI_ISL_1758564, EPI_ISL_1758565, EPI_ISL_1758566, EPI_ISL_1758571, EPI_ISL_1758574, EPI_ISL_1758575, EPI_ISL_1758576, EPI_ISL_1758577, EPI_ISL_1758579, EPI_ISL_1758580, EPI_ISL_1758585, EPI_ISL_1758586, EPI_ISL_1758588, EPI_ISL_1758589, EPI_ISL_1758591, EPI_ISL_1758592, EPI_ISL_1758593, EPI_ISL_1758594, EPI_ISL_1758595, EPI_ISL_1758600, EPI_ISL_1758601, EPI_ISL_1758602, EPI_ISL_1758607, EPI_ISL_1758610, EPI_ISL_1758611, EPI_ISL_1758613, EPI_ISL_1758615, EPI_ISL_1758617, EPI_ISL_1758618, EPI_ISL_1758620, EPI_ISL_1758621, EPI_ISL_1758622, EPI_ISL_1758623, EPI_ISL_1758626, EPI_ISL_1758629, EPI_ISL_1758631, EPI_ISL_1758633, EPI_ISL_1758634, EPI_ISL_1758637, EPI_ISL_1758638, EPI_ISL_1758639, EPI_ISL_1758640, EPI_ISL_1758641, EPI_ISL_1758643, EPI_ISL_1758644, EPI_ISL_1758646, EPI_ISL_1758647, EPI_ISL_1758648, EPI_ISL_1758650, EPI_ISL_1758651, EPI_ISL_1758652, EPI_ISL_1758653, EPI_ISL_1758654, EPI_ISL_1758655, EPI_ISL_1758659, EPI_ISL_1758663, EPI_ISL_1758665, EPI_ISL_1759194, EPI_ISL_1759199, EPI_ISL_1759204, EPI_ISL_1759216, EPI_ISL_1759218, EPI_ISL_1759229, EPI_ISL_1759231, EPI_ISL_1759234, EPI_ISL_1759243, EPI_ISL_1759245, EPI_ISL_1759261, EPI_ISL_1759275, EPI_ISL_1759277, EPI_ISL_1759280, EPI_ISL_1759302, EPI_ISL_1759315, EPI_ISL_1759322, EPI_ISL_1759331, EPI_ISL_1759333, EPI_ISL_1759337, EPI_ISL_1759341, EPI_ISL_1759813, EPI_ISL_1759831, EPI_ISL_1759848, EPI_ISL_1759854, EPI_ISL_1759865, EPI_ISL_1759866, EPI_ISL_1759869, EPI_ISL_1759878, EPI_ISL_1759883, EPI_ISL_1759891, EPI_ISL_1759892, EPI_ISL_1790214, EPI_ISL_1790218, EPI_ISL_1790219, EPI_ISL_1790220, EPI_ISL_1790221, EPI_ISL_1790223, EPI_ISL_1790224, EPI_ISL_1790225, EPI_ISL_1790226, EPI_ISL_1790227, EPI_ISL_1790233, EPI_ISL_1790234, EPI_ISL_1790236, EPI_ISL_1790237, EPI_ISL_1790239, EPI_ISL_1790240, EPI_ISL_1790241, EPI_ISL_1790242, EPI_ISL_1790243, EPI_ISL_1790244, EPI_ISL_1790245, EPI_ISL_1790246, EPI_ISL_1790247, EPI_ISL_1790250, EPI_ISL_1790251, EPI_ISL_1790254, EPI_ISL_1790257, EPI_ISL_1790259, EPI_ISL_1790261, EPI_ISL_1790262, EPI_ISL_1790264, EPI_ISL_1790265, EPI_ISL_1790267, EPI_ISL_1790268, EPI_ISL_1790270, EPI_ISL_1790271, EPI_ISL_1790273, EPI_ISL_1790276, EPI_ISL_1790277, EPI_ISL_1790278, EPI_ISL_1790279, EPI_ISL_1790280, EPI_ISL_1790281, EPI_ISL_1790282, EPI_ISL_1790283, EPI_ISL_1790284, EPI_ISL_1790288, EPI_ISL_1790291, EPI_ISL_1790293, EPI_ISL_1790294, EPI_ISL_1790296, EPI_ISL_1790297, EPI_ISL_1790298, EPI_ISL_1790299, EPI_ISL_1790301, EPI_ISL_1790302, EPI_ISL_1790304, EPI_ISL_1790305, EPI_ISL_1790309, EPI_ISL_1790310, EPI_ISL_1790311, EPI_ISL_1790312, EPI_ISL_1790313, EPI_ISL_1790317, EPI_ISL_1790318, EPI_ISL_1790319, EPI_ISL_1790320, EPI_ISL_1790326, EPI_ISL_1790329, EPI_ISL_1790330, EPI_ISL_1790334, EPI_ISL_1790335, EPI_ISL_1790338, EPI_ISL_1790339, EPI_ISL_1790341, EPI_ISL_1790343, EPI_ISL_1790345, EPI_ISL_1790346, EPI_ISL_1790348, EPI_ISL_1790350, EPI_ISL_1790352, EPI_ISL_1790353, EPI_ISL_1790354, EPI_ISL_1790355, EPI_ISL_1790356, EPI_ISL_1790358, EPI_ISL_1790360, EPI_ISL_1790363, EPI_ISL_1790365, EPI_ISL_1790366, EPI_ISL_1790368, EPI_ISL_1790374, EPI_ISL_1790376, EPI_ISL_1790377, EPI_ISL_1790379, EPI_ISL_1790383, EPI_ISL_1790384, EPI_ISL_1790385, EPI_ISL_1790386, EPI_ISL_1790389, EPI_ISL_1790390, EPI_ISL_1790391, EPI_ISL_1790392, EPI_ISL_1790396, EPI_ISL_1790400, EPI_ISL_1790401, EPI_ISL_1790403, EPI_ISL_1790404, EPI_ISL_1790405, EPI_ISL_1790406, EPI_ISL_1790407, EPI_ISL_1790410, EPI_ISL_1790412, EPI_ISL_1790416, EPI_ISL_1790417, EPI_ISL_1790418, EPI_ISL_1790420, EPI_ISL_1790421, EPI_ISL_1790422, EPI_ISL_1790424, EPI_ISL_1790425, EPI_ISL_1790429, EPI_ISL_1790430, EPI_ISL_1806217, EPI_ISL_1806224, EPI_ISL_1806225, EPI_ISL_1806228, EPI_ISL_1806233, EPI_ISL_1806242, EPI_ISL_1806245, EPI_ISL_1806248, EPI_ISL_1806249, EPI_ISL_1806252, EPI_ISL_1806254, EPI_ISL_1806273, EPI_ISL_1806282, EPI_ISL_1806284, EPI_ISL_1806285, EPI_ISL_1806297, EPI_ISL_1806299, EPI_ISL_1806304, EPI_ISL_1806309, EPI_ISL_1806312, EPI_ISL_1806316, EPI_ISL_1806319, EPI_ISL_1806321, EPI_ISL_1806328, EPI_ISL_1806330, EPI_ISL_1806336, EPI_ISL_1806337, EPI_ISL_1806339, EPI_ISL_1806346, EPI_ISL_1806348, EPI_ISL_1806354, EPI_ISL_1806357, EPI_ISL_1806358, EPI_ISL_1806359, EPI_ISL_1806365, EPI_ISL_1806369, EPI_ISL_1806370, EPI_ISL_1806383, EPI_ISL_1806386, EPI_ISL_1806387, EPI_ISL_1806396, EPI_ISL_1806401, EPI_ISL_1806415, EPI_ISL_1806416, EPI_ISL_1806418, EPI_ISL_1806424, EPI_ISL_1806425, EPI_ISL_1806431, EPI_ISL_1806434, EPI_ISL_1806448, EPI_ISL_1806449, EPI_ISL_1806451, EPI_ISL_1806456, EPI_ISL_1806458, EPI_ISL_1806459, EPI_ISL_1806462, EPI_ISL_1806463, EPI_ISL_1806464, EPI_ISL_1806466, EPI_ISL_1806467, EPI_ISL_1806472, EPI_ISL_1806478, EPI_ISL_1806488, EPI_ISL_1806496, EPI_ISL_1806500, EPI_ISL_1806501, EPI_ISL_1829215, EPI_ISL_1829269, EPI_ISL_1829327, EPI_ISL_1829363, EPI_ISL_1829432, EPI_ISL_1829442, EPI_ISL_1829461, EPI_ISL_1829500, EPI_ISL_1829525, EPI_ISL_1829532, EPI_ISL_1829540, EPI_ISL_1829561, EPI_ISL_1829570, EPI_ISL_1829593, EPI_ISL_1829601, EPI_ISL_1829643, EPI_ISL_1829666, EPI_ISL_1829673, EPI_ISL_1829700, EPI_ISL_1829703, EPI_ISL_1829723, EPI_ISL_1829801, EPI_ISL_1830117, EPI_ISL_1830142, EPI_ISL_1830152, EPI_ISL_1830210, EPI_ISL_1830226, EPI_ISL_1830230, EPI_ISL_1830231, EPI_ISL_1830235, EPI_ISL_1830249, EPI_ISL_1830260, EPI_ISL_1830262, EPI_ISL_1830284, EPI_ISL_1830344, EPI_ISL_1830370, EPI_ISL_1830405, EPI_ISL_1830409, EPI_ISL_1830414, EPI_ISL_1830416, EPI_ISL_1830429, EPI_ISL_1830440, EPI_ISL_1830441, EPI_ISL_1830448, EPI_ISL_1830461, EPI_ISL_1830470, EPI_ISL_1830476, EPI_ISL_1830488, EPI_ISL_1830517, EPI_ISL_1830556, EPI_ISL_1830557, EPI_ISL_1830580, EPI_ISL_1830581, EPI_ISL_1830583, EPI_ISL_1830584, EPI_ISL_1830596, EPI_ISL_1830605, EPI_ISL_1830621, EPI_ISL_1830632, EPI_ISL_1830634, EPI_ISL_1830644, EPI_ISL_1830664, EPI_ISL_1830669, EPI_ISL_1830687, EPI_ISL_1830694, EPI_ISL_1830703, EPI_ISL_1830709, EPI_ISL_1830763, EPI_ISL_1830998, EPI_ISL_1831194, EPI_ISL_1831261, EPI_ISL_1831262, EPI_ISL_1912023, EPI_ISL_1912056, EPI_ISL_1912057, EPI_ISL_1912068, EPI_ISL_1912099, EPI_ISL_1912102, EPI_ISL_1912107, EPI_ISL_1912142, EPI_ISL_1912167, EPI_ISL_1912200, EPI_ISL_1912220, EPI_ISL_1912273, EPI_ISL_1912292, EPI_ISL_1912303, EPI_ISL_1912317, EPI_ISL_1986586, EPI_ISL_1986588, EPI_ISL_1986596, EPI_ISL_1986600, EPI_ISL_1986602, EPI_ISL_1986608, EPI_ISL_1986615, EPI_ISL_1986621, EPI_ISL_1986622, EPI_ISL_1986624, EPI_ISL_1986636, EPI_ISL_1986640, EPI_ISL_1986641, EPI_ISL_1986642, EPI_ISL_1986643, EPI_ISL_1986645, EPI_ISL_1986646, EPI_ISL_1986647, EPI_ISL_1986650, EPI_ISL_1986657, EPI_ISL_1986670, EPI_ISL_1986672, EPI_ISL_1986683, EPI_ISL_1986685, EPI_ISL_1986694, EPI_ISL_1986696, EPI_ISL_1986700, EPI_ISL_1986709, EPI_ISL_1986713, EPI_ISL_1986719, EPI_ISL_1986723, EPI_ISL_1986728, EPI_ISL_1986734, EPI_ISL_1986738, EPI_ISL_1986745, EPI_ISL_1986765, EPI_ISL_1986766, EPI_ISL_1986767, EPI_ISL_1986776, EPI_ISL_1986777, EPI_ISL_1986784, EPI_ISL_1986788, EPI_ISL_1986789, EPI_ISL_1986796, EPI_ISL_1986799, EPI_ISL_1986800, EPI_ISL_1986808, EPI_ISL_1986815, EPI_ISL_1986818, EPI_ISL_1986826, EPI_ISL_1986828, EPI_ISL_1986835, EPI_ISL_1986836, EPI_ISL_1986838, EPI_ISL_1986839, EPI_ISL_1986843, EPI_ISL_1986844, EPI_ISL_1986849, EPI_ISL_1986852, EPI_ISL_1986872, EPI_ISL_1986883, EPI_ISL_1986884, EPI_ISL_1986892, EPI_ISL_1986898, EPI_ISL_1986905, EPI_ISL_1986910, EPI_ISL_1986911, EPI_ISL_1986913, EPI_ISL_1986915, EPI_ISL_1986917, EPI_ISL_1986918, EPI_ISL_1986919, EPI_ISL_1986926, EPI_ISL_1986934, EPI_ISL_1986938, EPI_ISL_1986940, EPI_ISL_1986942, EPI_ISL_2005346, EPI_ISL_2005348, EPI_ISL_2005351, EPI_ISL_2005352, EPI_ISL_2005371, EPI_ISL_2005374, EPI_ISL_2005390, EPI_ISL_2005397, EPI_ISL_2005398, EPI_ISL_2005401, EPI_ISL_2005405, EPI_ISL_2005408, EPI_ISL_2005410, EPI_ISL_2005411, EPI_ISL_2005427, EPI_ISL_2005431, EPI_ISL_2005433, EPI_ISL_2005436, EPI_ISL_2005454, EPI_ISL_2005456, EPI_ISL_2005474, EPI_ISL_2005482, EPI_ISL_2005485, EPI_ISL_2005488, EPI_ISL_2005491, EPI_ISL_2005492, EPI_ISL_2005500, EPI_ISL_2005505, EPI_ISL_2005508, EPI_ISL_2005526, EPI_ISL_2005528, EPI_ISL_2005530, EPI_ISL_2005541, EPI_ISL_2005543, EPI_ISL_2005547, EPI_ISL_2005549, EPI_ISL_2005565, EPI_ISL_2005570, EPI_ISL_2005576, EPI_ISL_2005580, EPI_ISL_2005586, EPI_ISL_2005595, EPI_ISL_2005596, EPI_ISL_2005602, EPI_ISL_2005615, EPI_ISL_2005621, EPI_ISL_2005623, EPI_ISL_2005626, EPI_ISL_2005628, EPI_ISL_2005634, EPI_ISL_2005636, EPI_ISL_2005644, EPI_ISL_2005645, EPI_ISL_2005648, EPI_ISL_2005653, EPI_ISL_2005654, EPI_ISL_2005658, EPI_ISL_2005662, EPI_ISL_2005665, EPI_ISL_2005667, EPI_ISL_2005669, EPI_ISL_2005670, EPI_ISL_2005673, EPI_ISL_2005674, EPI_ISL_2005677, EPI_ISL_2005678, EPI_ISL_2005680, EPI_ISL_2005681, EPI_ISL_2005686, EPI_ISL_2005687, EPI_ISL_2005688, EPI_ISL_2005689, EPI_ISL_2005693, EPI_ISL_2005696, EPI_ISL_2005699, EPI_ISL_2005701, EPI_ISL_2005704, EPI_ISL_2005708, EPI_ISL_2005713, EPI_ISL_2005717, EPI_ISL_2005718, EPI_ISL_2005720, EPI_ISL_2005723, EPI_ISL_2005724, EPI_ISL_2005726, EPI_ISL_2005727, EPI_ISL_2005731, EPI_ISL_2005735, EPI_ISL_2005737, EPI_ISL_2005738, EPI_ISL_2005742, EPI_ISL_2005744, EPI_ISL_2005745, EPI_ISL_2005750, EPI_ISL_2005752, EPI_ISL_2005753, EPI_ISL_2005755, EPI_ISL_2005759, EPI_ISL_2005762, EPI_ISL_2005763, EPI_ISL_2005765, EPI_ISL_2005766, EPI_ISL_2005767, EPI_ISL_2005768, EPI_ISL_2005770, EPI_ISL_2005772, EPI_ISL_2005773, EPI_ISL_2005774, EPI_ISL_2005775, EPI_ISL_2005776, EPI_ISL_2005777, EPI_ISL_2005778, EPI_ISL_2005779, EPI_ISL_2005780, EPI_ISL_2005781, EPI_ISL_2005782, EPI_ISL_2005783, EPI_ISL_2005787, EPI_ISL_2005788, EPI_ISL_2005789, EPI_ISL_2005790, EPI_ISL_2005791, EPI_ISL_2005793, EPI_ISL_2005794, EPI_ISL_2005798, EPI_ISL_2005800, EPI_ISL_2005801, EPI_ISL_2005805, EPI_ISL_2005806, EPI_ISL_2005807, EPI_ISL_2005809, EPI_ISL_2006302, EPI_ISL_2006312, EPI_ISL_2006328, EPI_ISL_2006333, EPI_ISL_2006334, EPI_ISL_2006345, EPI_ISL_2006358, EPI_ISL_2006369, EPI_ISL_2006392, EPI_ISL_2006405, EPI_ISL_2006438, EPI_ISL_2006442, EPI_ISL_2006452, EPI_ISL_2006462, EPI_ISL_2006471, EPI_ISL_2006485, EPI_ISL_2006487, EPI_ISL_2006493, EPI_ISL_2006508, EPI_ISL_2022179, EPI_ISL_2022278, EPI_ISL_2022346, EPI_ISL_2022432, EPI_ISL_2172124, EPI_ISL_2172172, EPI_ISL_2172205, EPI_ISL_2172216, EPI_ISL_2172234, EPI_ISL_2172272, EPI_ISL_2172344 | | | |
| see above | Lighthouse Lab in Milton Keynes | Wellcome Sanger Institute for the COVID-19 Genomics UK (COG-UK) Consortium | Cordelia Langford; David K. Jackson; Dominic Kwiatkowski; Ewan Harrison; Ian Johnston; Jeffrey Barrett; John Sillitoe on behalf of the Wellcome Sanger Institute COVID-19 Surveillance Team; Roberto Amato; Sonia Goncalves; The Lighthouse Lab in Milton Keynes and Alex Alderton |
| EPI_ISL_1833011, EPI_ISL_1833012, EPI_ISL_1833018, EPI_ISL_1833019, EPI_ISL_1833020 | Lincolnshire Hospitals and DeepSeq Nottingham | COVID-19 Genomics UK (COG-UK) Consortium | Christopher Moore; Fei Sang; Joeseph Chappell; Johnny Debebe; Jonathan Ball; Matthew Carlisle; Matthew Loose; Nadine Holmes; Nichola Duckworth; Patrick McClure; Sarah Walsh; Tim Sloan; Victoria Wright |
| EPI_ISL_1636869, EPI_ISL_1636870, EPI_ISL_1636871, EPI_ISL_1636872, EPI_ISL_1636877, EPI_ISL_1636883, EPI_ISL_1832353, EPI_ISL_1832355, EPI_ISL_1832363, EPI_ISL_1832366, EPI_ISL_1832370, EPI_ISL_1832372, EPI_ISL_1832374, EPI_ISL_1832380, EPI_ISL_1832383, EPI_ISL_1832393, EPI_ISL_1832395, EPI_ISL_1832397, EPI_ISL_1832410, EPI_ISL_1832412, EPI_ISL_1832414, EPI_ISL_1832416, EPI_ISL_1832418, EPI_ISL_1832420, EPI_ISL_1832422, EPI_ISL_1832424, EPI_ISL_1832426, EPI_ISL_1832429, EPI_ISL_1832431, EPI_ISL_1832433, EPI_ISL_1832435, EPI_ISL_1832466, EPI_ISL_1832468, EPI_ISL_1832470, EPI_ISL_1832478, EPI_ISL_1832480, EPI_ISL_1832482, EPI_ISL_1832490, EPI_ISL_1832496, EPI_ISL_1832498, EPI_ISL_1832500, EPI_ISL_1832507, EPI_ISL_1832510, EPI_ISL_1832512, EPI_ISL_1832520, EPI_ISL_1974929, EPI_ISL_1974931, EPI_ISL_1974938, EPI_ISL_1974947, EPI_ISL_1974949, EPI_ISL_1974960, EPI_ISL_1974962, EPI_ISL_1974963, EPI_ISL_1974965, EPI_ISL_1974971, EPI_ISL_1974976 | | | |
| see above | Liverpool Clinical Laboratories | COVID-19 Genomics UK (COG-UK) Consortium | A Alrezaihi; Alessandro Gerada; Alistair Darby; Angela Cowell; Anita Lucaci; Anu Chawla; Cassie Olateju; Catherine Hartley; Charlotte Nelson; Ecaterina Vamos; Elaine O'Toole; Eleanor G Bentley; Ghada T Shawli; Hermione Webster; Isabel García-Dorival; James Johnson; James P Stewart; Jenifer Manson; Joanne Watts; Jones Benjamin; Jordan J Clark; Julian Hiscox; L Luu; Lucille Rainbow; M Almsaud; Margaret Hughes; Mark Whitehead; Matthew Gemmell; Miren Iturriza-Gomara; Muhannad Alruwaili; N.P Randle; Neil Swainston; PKF Gilmore; Parul Sharma; Rebekah Penrice-Randal; Richard Eccles; Richard Gregory; Sam Haldenby; Steve Paterson; Stuart D Armstrong; Trevor Ian Robinson; Ximeng Han |
| EPI_ISL_1637186, EPI_ISL_1637187, EPI_ISL_1637189, EPI_ISL_1637190, EPI_ISL_1637191, EPI_ISL_1637192, EPI_ISL_1637193, EPI_ISL_1637225, EPI_ISL_1637232, EPI_ISL_1637233, EPI_ISL_1730399, EPI_ISL_1730400, EPI_ISL_1730405, EPI_ISL_1730406, EPI_ISL_1730407, EPI_ISL_1832707, EPI_ISL_1978536, EPI_ISL_1978538, EPI_ISL_1978540, EPI_ISL_1978542, EPI_ISL_1978543, EPI_ISL_1978545, EPI_ISL_1978546, EPI_ISL_1978547, EPI_ISL_1978548, EPI_ISL_1978551, EPI_ISL_1978552, EPI_ISL_1978553, EPI_ISL_1978554, EPI_ISL_1978555, EPI_ISL_1978557, EPI_ISL_1978559, EPI_ISL_1978560, EPI_ISL_1978562, EPI_ISL_1978624, EPI_ISL_1978630, EPI_ISL_1978633, EPI_ISL_1978635, EPI_ISL_1978637, EPI_ISL_1978639, EPI_ISL_1978640, EPI_ISL_1978643, EPI_ISL_1978645, EPI_ISL_1978647, EPI_ISL_1978648, EPI_ISL_1978650, EPI_ISL_1978652, EPI_ISL_1978655, EPI_ISL_1978657, EPI_ISL_1978737, EPI_ISL_1978739, EPI_ISL_1978744, EPI_ISL_1978746, EPI_ISL_1978893 | | | |
| see above | Northumbria University / South Tees Hospitals NHS Foundation Trust / North Cumbria Integrated Care NHS Foundation Trust / North Tees and Hartlepool NHS Foundation Trust / Newcastle Hospitals NHS Foundation Trust | COVID-19 Genomics UK (COG-UK) Consortium | Andrew Nelson; Brendan Payne; Clive Graham; Darren L Smith; Debra Padgett; Edward Barton; Emma Swindells; Garren Scott; Gary Black; Gary Eltringham; Giles S Holt; Greg R Young; Jane Greenaway; Jennifer Collins; John Allan; Joshua Loh; Lynn Dover; Matthew Bashton; Mohammad A Tariq; Paul Baker; Sarah Essex; Steve Liggett; Wen C Yew; Yusri Taha |
| EPI_ISL_1546251, EPI_ISL_1546253, EPI_ISL_1546258, EPI_ISL_1546260, EPI_ISL_1546264, EPI_ISL_1546266, EPI_ISL_1546541, EPI_ISL_1546547, EPI_ISL_1546549, EPI_ISL_1546553, EPI_ISL_1546557, EPI_ISL_1546559, EPI_ISL_1546560, EPI_ISL_1546561, EPI_ISL_1546562, EPI_ISL_1546565, EPI_ISL_1546566, EPI_ISL_1546568, EPI_ISL_1546569, EPI_ISL_1546570, EPI_ISL_1546590, EPI_ISL_1546591, EPI_ISL_1546593, EPI_ISL_1546594, EPI_ISL_1546596, EPI_ISL_1546597, EPI_ISL_1546603, EPI_ISL_1546604, EPI_ISL_1546605, EPI_ISL_1546607, EPI_ISL_1546609, EPI_ISL_1546611, EPI_ISL_1546612, EPI_ISL_1546613, EPI_ISL_1546614, EPI_ISL_1546615, EPI_ISL_1546616, EPI_ISL_1546617, EPI_ISL_1546621, EPI_ISL_1546623, EPI_ISL_1546626, EPI_ISL_1546627, EPI_ISL_1546628, EPI_ISL_1546632, EPI_ISL_1546637, EPI_ISL_1730596, EPI_ISL_1730597, EPI_ISL_1730598, EPI_ISL_1730599, EPI_ISL_1730600, EPI_ISL_1730601, EPI_ISL_1730602, EPI_ISL_1730603, EPI_ISL_1730604, EPI_ISL_1730605, EPI_ISL_1730606, EPI_ISL_1730624, EPI_ISL_1730631, EPI_ISL_1730632, EPI_ISL_1730636, EPI_ISL_1730638, EPI_ISL_1730639, EPI_ISL_1730667, EPI_ISL_1730669, EPI_ISL_1730670, EPI_ISL_1730671, EPI_ISL_1730672, EPI_ISL_1730679, EPI_ISL_1730680, EPI_ISL_1730681, EPI_ISL_1730682, EPI_ISL_1730685, EPI_ISL_1730765, EPI_ISL_1730766, EPI_ISL_1730773, EPI_ISL_1730784, EPI_ISL_1730786, EPI_ISL_1730789, EPI_ISL_1730797 | | | |
| see above | Originating lab: Wales Specialist Virology Centre Sequencing lab: Pathogen Genomics Unit | Public Health Wales Microbiology Cardiff Wales Specialist Virology Centre | Alec Birchley; Alexander Adams; Amy Gaskin; Angela Marchbank; Bree Gatica-Wilcox; Catherine Moore; Jason Coombes; Joanne Watkins; Joel Southgate; Johnathan Evans; Laura Gifford; Lauren Gilbert; Lee Graham; Malorie Perry; Matthew Bull; Nicole Pacchiarini; Sally Corden; Sara Kumziene-Summerhayes; Sara Rey; Sarah Taylor; Simon Cottrell; Sophie Jones; Tom Connor |
| EPI_ISL_1730561, EPI_ISL_1730567, EPI_ISL_1730568, EPI_ISL_1730573, EPI_ISL_1730574, EPI_ISL_1730575, EPI_ISL_1730580, EPI_ISL_1730581, EPI_ISL_1730588, EPI_ISL_1730589, EPI_ISL_1730591, EPI_ISL_1730592, EPI_ISL_1730593, EPI_ISL_1979029, EPI_ISL_1979032, EPI_ISL_1979033, EPI_ISL_1979040, EPI_ISL_1979041, EPI_ISL_1979043, EPI_ISL_1979048, EPI_ISL_1979053, EPI_ISL_1979054, EPI_ISL_1979056, EPI_ISL_1979063, EPI_ISL_1979064, EPI_ISL_1979072, EPI_ISL_1979074, EPI_ISL_1979079, EPI_ISL_1979080, EPI_ISL_1979087, EPI_ISL_1979088, EPI_ISL_1979094, EPI_ISL_2127687 | | | |
| see above | Oxford Viromics, NDM, University of Oxford; Oxford University Hospitals; Basingstoke and North Hampshire Hospital | COVID-19 Genomics UK (COG-UK) Consortium | Alex Mobbs; Amy Trebes; Anita Justice; Catrin Moore; Christophe Fraser; David Bonsall; David Buck; Emma Wise; George Macintyre; Jessica Lynch; John Todd; Mariateresa de Cesare; Matilde Mori; Monique Andersson; Nathan Moore; Nick Cortes; Robert Shaw; Stephen Kidd; Tanya Golubchik; Timothy Peto |
| EPI_ISL_1832746, EPI_ISL_1832755, EPI_ISL_1832763, EPI_ISL_1832764, EPI_ISL_1832842, EPI_ISL_1832846, EPI_ISL_1832848, EPI_ISL_1832852, EPI_ISL_1832854, EPI_ISL_1832856, EPI_ISL_1832858, EPI_ISL_1832860, EPI_ISL_1832862, EPI_ISL_1832863, EPI_ISL_1832864, EPI_ISL_1832865, EPI_ISL_1832866, EPI_ISL_1832868, EPI_ISL_1832869, EPI_ISL_1832870, EPI_ISL_1832875, EPI_ISL_1832891, EPI_ISL_1832898, EPI_ISL_1832899, EPI_ISL_1832900, EPI_ISL_1832901, EPI_ISL_1832902, EPI_ISL_1832903, EPI_ISL_1832906, EPI_ISL_1832907, EPI_ISL_1832909, EPI_ISL_1832910, EPI_ISL_1832912, EPI_ISL_1832915 | | | |
| see above | Quadram Institute Bioscience | COVID-19 Genomics UK (COG-UK) Consortium | Alexander J Trotter; Alison E. Mather; Alp Aydin; Ana P. Tedim; Anastasia Kolyva; Andrew Bell; Andrew J. Page; Claire Stuart; Dave J. Baker; Gemma L. Kay; John Wain; Justin O'Grady; Leonardo de Oliveira Martins; Lizzie Meadows; Maria Diaz; Mark Webber; Muhammed Yasir; Nabil-Fareed Alikhan; Ngozi Elumogo; Nicholas M. Thomson; Rachael Stanley; Rachel Gilroy; Reenesh Prakash; Samir Dervisevic; Samuel Bloomfield; Steven Rudder; Thanh Le-Viet |
| EPI_ISL_1637237, EPI_ISL_1637238, EPI_ISL_1637239, EPI_ISL_1637240, EPI_ISL_1730411, EPI_ISL_1730412, EPI_ISL_1730413, EPI_ISL_1730414, EPI_ISL_1730415, EPI_ISL_1730416, EPI_ISL_1730417, EPI_ISL_1730418, EPI_ISL_1833028 | | | |

| | | | |
|---|---|---|---|
| see above | Queens Medical Centre, Clinical Microbiology Department / DeepSeq Nottingham | COVID-19 Genomics UK (COG-UK) Consortium | Christopher Moore; Emily Park; Fei Sang; Gemma Clark; Hannah Howson-Wells; Jack Hill; Jayasree Dey; Johnny Debebe; Jonathan Ball; Joseph Chappell; Manjinder Khakh; Matthew Carlisle; Matthew Loose; Michelle M Lister; Nadine Holmes; Patrick McClure; Theocharis Tsoleridis; Timothy Byaruhanga; Vicki M Fleming; Victoria Wright; Wendy Smith |
| EPI_ISL_1538049, EPI_ISL_1584559, EPI_ISL_1630293, EPI_ISL_1630294, EPI_ISL_1630295, EPI_ISL_1630299, EPI_ISL_1630304, EPI_ISL_1630311, EPI_ISL_1630312, EPI_ISL_1630313, EPI_ISL_1630319, EPI_ISL_1630324, EPI_ISL_1630328, EPI_ISL_1630329, EPI_ISL_1630338, EPI_ISL_1630342, EPI_ISL_1630348, EPI_ISL_1630352, EPI_ISL_1630353, EPI_ISL_1630367, EPI_ISL_1630368, EPI_ISL_1630384, EPI_ISL_1630386, EPI_ISL_1630391, EPI_ISL_1630397, EPI_ISL_1630401, EPI_ISL_1630403, EPI_ISL_1630404, EPI_ISL_1630410, EPI_ISL_1630414, EPI_ISL_1630420, EPI_ISL_1630427, EPI_ISL_1630431, EPI_ISL_1630434, EPI_ISL_1630438, EPI_ISL_1630439, EPI_ISL_1630450, EPI_ISL_1630451, EPI_ISL_1630465, EPI_ISL_1630476, EPI_ISL_1630480, EPI_ISL_1630486, EPI_ISL_1630489, EPI_ISL_1630490, EPI_ISL_1630491, EPI_ISL_1630492, EPI_ISL_1630497, EPI_ISL_1630500, EPI_ISL_1630503, EPI_ISL_1630506, EPI_ISL_1630510, EPI_ISL_1630513, EPI_ISL_1631092, EPI_ISL_1631093, EPI_ISL_1631105, EPI_ISL_1631144, EPI_ISL_1631146, EPI_ISL_1631160, EPI_ISL_1631164, EPI_ISL_1631170, EPI_ISL_1631202, EPI_ISL_1631203, EPI_ISL_1631210, EPI_ISL_1631212, EPI_ISL_1631213, EPI_ISL_1631217, EPI_ISL_1631225, EPI_ISL_1631230, EPI_ISL_1631238, EPI_ISL_1631249, EPI_ISL_1631257, EPI_ISL_1631271, EPI_ISL_1631276, EPI_ISL_1631314, EPI_ISL_1631319, EPI_ISL_1631331, EPI_ISL_1631334, EPI_ISL_1631753, EPI_ISL_1631761, EPI_ISL_1631762, EPI_ISL_1631765, EPI_ISL_1631768, EPI_ISL_1631770, EPI_ISL_1631773, EPI_ISL_1631779, EPI_ISL_1631782, EPI_ISL_1631785, EPI_ISL_1631786, EPI_ISL_1631788, EPI_ISL_1631790, EPI_ISL_1631791, EPI_ISL_1631795, EPI_ISL_1631796, EPI_ISL_1631800, EPI_ISL_1631805, EPI_ISL_1631807, EPI_ISL_1631817, EPI_ISL_1631822, EPI_ISL_1631823, EPI_ISL_1631830, EPI_ISL_1631833, EPI_ISL_1631835, EPI_ISL_1631836, EPI_ISL_1631840, EPI_ISL_1631841, EPI_ISL_1631842, EPI_ISL_1631843, EPI_ISL_1631845, EPI_ISL_1631846, EPI_ISL_1631851, EPI_ISL_1631863, EPI_ISL_1631867, EPI_ISL_1631868, EPI_ISL_1631872, EPI_ISL_1631875, EPI_ISL_1631879, EPI_ISL_1631883, EPI_ISL_1631885, EPI_ISL_1631886, EPI_ISL_1631888, EPI_ISL_1631894, EPI_ISL_1631897, EPI_ISL_1631901, EPI_ISL_1631906, EPI_ISL_1631909, EPI_ISL_1631911, EPI_ISL_1631914, EPI_ISL_1631917, EPI_ISL_1631920, EPI_ISL_1631932, EPI_ISL_1631933, EPI_ISL_1631936, EPI_ISL_1631944, EPI_ISL_1631946, EPI_ISL_1631950, EPI_ISL_1631951, EPI_ISL_1631958, EPI_ISL_1631962, EPI_ISL_1631964, EPI_ISL_1631967, EPI_ISL_1631969, EPI_ISL_1631971, EPI_ISL_1631976, EPI_ISL_1631977, EPI_ISL_1631982, EPI_ISL_1631983, EPI_ISL_1631992, EPI_ISL_1632005, EPI_ISL_1632006, EPI_ISL_1632009, EPI_ISL_1632012, EPI_ISL_1632016, EPI_ISL_1632018, EPI_ISL_1632030, EPI_ISL_1632045, EPI_ISL_1632050, EPI_ISL_1632053, EPI_ISL_1632056, EPI_ISL_1632057, EPI_ISL_1632061, EPI_ISL_1632062, EPI_ISL_1632063, EPI_ISL_1632064, EPI_ISL_1632067, EPI_ISL_1632076, EPI_ISL_1632078, EPI_ISL_1632079, EPI_ISL_1632083, EPI_ISL_1632084, EPI_ISL_1632091, EPI_ISL_1632096, EPI_ISL_1632099, EPI_ISL_1632104, EPI_ISL_1632119, EPI_ISL_1632122, EPI_ISL_1632123, EPI_ISL_1632128, EPI_ISL_1632130, EPI_ISL_1632131, EPI_ISL_1632141, EPI_ISL_1632144, EPI_ISL_1632145, EPI_ISL_1632156, EPI_ISL_1632158, EPI_ISL_1632159, EPI_ISL_1632160, EPI_ISL_1632162, EPI_ISL_1632164, EPI_ISL_1632177, EPI_ISL_1632178, EPI_ISL_1632183, EPI_ISL_1632193, EPI_ISL_1632203, EPI_ISL_1632206, EPI_ISL_1632210, EPI_ISL_1632219, EPI_ISL_1632225, EPI_ISL_1632230, EPI_ISL_1632239, EPI_ISL_1632243, EPI_ISL_1632247, EPI_ISL_1632251, EPI_ISL_1632253, EPI_ISL_1632256, EPI_ISL_1632261, EPI_ISL_1632262, EPI_ISL_1632263, EPI_ISL_1632270, EPI_ISL_1632272, EPI_ISL_1632275, EPI_ISL_1632277, EPI_ISL_1632278, EPI_ISL_1632282, EPI_ISL_1632287, EPI_ISL_1632331, EPI_ISL_1632342, EPI_ISL_1632353, EPI_ISL_1632425, EPI_ISL_1632462, EPI_ISL_1634737, EPI_ISL_1635325, EPI_ISL_1652692, EPI_ISL_1652693, EPI_ISL_1652698, EPI_ISL_1652700, EPI_ISL_1652705, EPI_ISL_1652711, EPI_ISL_1652712, EPI_ISL_1652714, EPI_ISL_1652716, EPI_ISL_1652717, EPI_ISL_1652718, EPI_ISL_1652732, EPI_ISL_1652742, EPI_ISL_1652744, EPI_ISL_1652754, EPI_ISL_1652755, EPI_ISL_1652759, EPI_ISL_1652762, EPI_ISL_1652765, EPI_ISL_1652767, EPI_ISL_1652768, EPI_ISL_1652772, EPI_ISL_1652782, EPI_ISL_1652790, EPI_ISL_1652799, EPI_ISL_1652804, EPI_ISL_1652805, EPI_ISL_1652807, EPI_ISL_1652812, EPI_ISL_1652822, EPI_ISL_1652824, EPI_ISL_1652827, EPI_ISL_1652828, EPI_ISL_1652830, EPI_ISL_1652833, EPI_ISL_1652835, EPI_ISL_1652838, EPI_ISL_1652847, EPI_ISL_1652852, EPI_ISL_1652855, EPI_ISL_1652859, EPI_ISL_1652860, EPI_ISL_1652863, EPI_ISL_1652870, EPI_ISL_1652880, EPI_ISL_1652881, EPI_ISL_1652882, EPI_ISL_1652883, EPI_ISL_1652884, EPI_ISL_1652887, EPI_ISL_1652891, EPI_ISL_1652892, EPI_ISL_1652893, EPI_ISL_1652897, EPI_ISL_1652902, EPI_ISL_1652905, EPI_ISL_1652906, EPI_ISL_1652921, EPI_ISL_1652923, EPI_ISL_1652927, EPI_ISL_1652931, EPI_ISL_1652941, EPI_ISL_1652942, EPI_ISL_1653718, EPI_ISL_1653720, EPI_ISL_1653722, EPI_ISL_1653723, EPI_ISL_1653724, EPI_ISL_1653725, EPI_ISL_1653726, EPI_ISL_1653727, EPI_ISL_1653730, EPI_ISL_1653731, EPI_ISL_1653732, EPI_ISL_1653733, EPI_ISL_1653734, EPI_ISL_1653735, EPI_ISL_1653736, EPI_ISL_1653737, EPI_ISL_1653740, EPI_ISL_1653741, EPI_ISL_1653742, EPI_ISL_1653743, EPI_ISL_1653744, EPI_ISL_1653745, EPI_ISL_1653746, EPI_ISL_1653747, EPI_ISL_1653748, EPI_ISL_1653749, EPI_ISL_1653750, EPI_ISL_1653751, EPI_ISL_1653752, EPI_ISL_1653753, EPI_ISL_1653756, EPI_ISL_1653760, EPI_ISL_1653761, EPI_ISL_1653762, EPI_ISL_1653763, EPI_ISL_1653764, EPI_ISL_1653765, EPI_ISL_1653766, EPI_ISL_1653767, EPI_ISL_1653768, EPI_ISL_1673255, EPI_ISL_1697975, EPI_ISL_1697986, EPI_ISL_1698012, EPI_ISL_1698016, EPI_ISL_1698036, EPI_ISL_1698037, EPI_ISL_1698062, EPI_ISL_1698066, EPI_ISL_1698090, EPI_ISL_1698103, EPI_ISL_1698176, EPI_ISL_1698197, EPI_ISL_1698226, EPI_ISL_1698228, EPI_ISL_1699044, EPI_ISL_1699058, EPI_ISL_1699080, EPI_ISL_1699082, EPI_ISL_1699091, EPI_ISL_1699128, EPI_ISL_1699131, EPI_ISL_1699138, EPI_ISL_1699204, EPI_ISL_1699237, EPI_ISL_1699248, EPI_ISL_1699263, EPI_ISL_1699271, EPI_ISL_1699281, EPI_ISL_1699284, EPI_ISL_1700185, EPI_ISL_1700188, EPI_ISL_1700189, EPI_ISL_1700198, EPI_ISL_1700202, EPI_ISL_1700205, EPI_ISL_1700222, EPI_ISL_1700224, EPI_ISL_1700226, EPI_ISL_1700229, EPI_ISL_1700233, EPI_ISL_1700272, EPI_ISL_1700280, EPI_ISL_1700285, EPI_ISL_1700295, EPI_ISL_1700296, EPI_ISL_1700299, EPI_ISL_1700303, EPI_ISL_1700310, EPI_ISL_1700313, EPI_ISL_1700317, EPI_ISL_1700329, EPI_ISL_1700340, EPI_ISL_1700348, EPI_ISL_1700353, EPI_ISL_1700359, EPI_ISL_1700374, EPI_ISL_1700382, EPI_ISL_1700387, EPI_ISL_1700394, EPI_ISL_1700397, EPI_ISL_1700401, EPI_ISL_1700406, EPI_ISL_1700409, EPI_ISL_1700417, EPI_ISL_1700425, EPI_ISL_1700430, EPI_ISL_1700431, EPI_ISL_1700435, EPI_ISL_1700448, EPI_ISL_1700454, EPI_ISL_1700468, EPI_ISL_1700471, EPI_ISL_1700480, EPI_ISL_1700486, EPI_ISL_1700488, EPI_ISL_1700494, EPI_ISL_1742012, EPI_ISL_1742019, EPI_ISL_1742034, EPI_ISL_1742055, EPI_ISL_1742060, EPI_ISL_1742062, EPI_ISL_1742294, EPI_ISL_1742295, EPI_ISL_1742296, EPI_ISL_1742297, EPI_ISL_1742298, EPI_ISL_1742299, EPI_ISL_1742300, EPI_ISL_1742301, EPI_ISL_1742302, EPI_ISL_1742303, EPI_ISL_1742304, EPI_ISL_1742305, EPI_ISL_1742306, EPI_ISL_1742307, EPI_ISL_1830131, EPI_ISL_1830141, EPI_ISL_1830165, EPI_ISL_1830206, EPI_ISL_1830224, EPI_ISL_1830245, EPI_ISL_1830248, EPI_ISL_1830275, EPI_ISL_1830286, EPI_ISL_1830289, EPI_ISL_1830291, EPI_ISL_1830296, EPI_ISL_1830304, EPI_ISL_1830382, EPI_ISL_1830386, EPI_ISL_1912774 | | | |
| see above | Randox Laboratories | Wellcome Sanger Institute for the COVID-19 Genomics UK (COG-UK) Consortium | Cordelia Langford; David K. Jackson; Dominic Kwiatkowski; Ewan Harrison; Ian Johnston; Jeffrey Barrett; John Sillitoe on behalf of the Wellcome Sanger Institute COVID-19 Surveillance Team; Randox Laboratories and Alex Alderton; Roberto Amato; Sonia Goncalves |
| EPI_ISL_1976555, EPI_ISL_1976557, EPI_ISL_1976559, EPI_ISL_1976563, EPI_ISL_1976568, EPI_ISL_1976570, EPI_ISL_1976572, EPI_ISL_1976573, EPI_ISL_1976575, EPI_ISL_1976577, EPI_ISL_1976579, EPI_ISL_1976580, EPI_ISL_1976582, EPI_ISL_1976584, EPI_ISL_1976586, EPI_ISL_1976589, EPI_ISL_1976590, EPI_ISL_1976594, EPI_ISL_1976596, EPI_ISL_1976599, EPI_ISL_1976601, EPI_ISL_1976602, EPI_ISL_1976604, EPI_ISL_1976606, EPI_ISL_1976609, EPI_ISL_1976611, EPI_ISL_1976612, EPI_ISL_1976616, EPI_ISL_1976631, EPI_ISL_1976633, EPI_ISL_1976635, EPI_ISL_1976636, EPI_ISL_1976638, EPI_ISL_1976640, EPI_ISL_1976642, EPI_ISL_1976644, EPI_ISL_1976645, EPI_ISL_1976647, EPI_ISL_1976652, EPI_ISL_1976655, EPI_ISL_1976657 | | | |
| see above | Regional Virus Laboratory, Belfast Health and Social Care Trust | COVID-19 Genomics UK (COG-UK) Consortium | Alison Watt; Ciara Cox; Conall McCaughey; David Simpson; Derek Fairley; James McKenna; Mairead Connor; Susan Feeney; Tanya Curran; Zoltan Molnar |
| EPI_ISL_1833824, EPI_ISL_1833825, EPI_ISL_1833826 | St George's University Hospitals NHS Foundation Trust | COVID-19 Genomics UK (COG-UK) Consortium | Adam Witney; Cassie Pope; Irene Monahan; Joshua Taylor; Ken Laing; NgeeKeong Tan |
| EPI_ISL_1637002, EPI_ISL_1637003, EPI_ISL_1637004, EPI_ISL_1637005, EPI_ISL_1637006, EPI_ISL_1637007, EPI_ISL_1637008, EPI_ISL_1637009, EPI_ISL_1637010, EPI_ISL_1637011, EPI_ISL_1637012, EPI_ISL_1637013, EPI_ISL_1637015, EPI_ISL_1637016, EPI_ISL_1637018, EPI_ISL_1637091, EPI_ISL_1637093, EPI_ISL_1637094, EPI_ISL_1637095, EPI_ISL_1637096, EPI_ISL_1637097, EPI_ISL_1637098, EPI_ISL_1637099, EPI_ISL_1637100, EPI_ISL_1637101, EPI_ISL_1637103, EPI_ISL_1637104, EPI_ISL_1637177, EPI_ISL_1637181, EPI_ISL_1730324, EPI_ISL_1730325, EPI_ISL_1730326, EPI_ISL_1730328, EPI_ISL_1730329, EPI_ISL_1730341, EPI_ISL_1730342, EPI_ISL_1730343, EPI_ISL_1730344, EPI_ISL_1730345, EPI_ISL_1730369, EPI_ISL_1730370, EPI_ISL_1730371, EPI_ISL_1730372, EPI_ISL_1730373, EPI_ISL_1730377, EPI_ISL_1730379, EPI_ISL_1730382, EPI_ISL_1730384, EPI_ISL_1730386, EPI_ISL_1730387, EPI_ISL_1730388, EPI_ISL_1730389, EPI_ISL_1730390, EPI_ISL_1832659 | | | |
| see above | University College London, Great Ormond Street Hospital for Children NHS Foundation Trust, Imperial College Healthcare NHS Trust | COVID-19 Genomics UK (COG-UK) Consortium | Alison Holmes; Charlotte Williams; Helena Tutill; Jacqueline Findlay; James Price; Judith Breuer; Julianne Brown; Kathryn Harris; Leysa Forrest; Mark Kristiansen; Paola Niola; Paola Resende Silva; Patricia Dyal; Paul Randell; Rachel Williams; Samuel Weeks; Sergi Castellano; Sunando Roy; Tony Brooks; Yasmin Panchbhaya |
| EPI_ISL_1730304 | University Hospitals Of Leicester NHS Trust and DeepSeq Nottingham | COVID-19 Genomics UK (COG-UK) Consortium | Christopher Holmes; Christopher Moore; Fei Sang; Joeseph Chappell; Johnny Debebe; Jonathan Ball; Julian Tang; Karlie Fallon; Matthew Carlisle; Matthew Loose; Nadine Holmes; Patrick McClure; Paul Bird; Thomas Helmer; Victoria Wright |
| EPI_ISL_1636529, EPI_ISL_1636530, EPI_ISL_1636538, EPI_ISL_1636549, EPI_ISL_1636574, EPI_ISL_1636577, EPI_ISL_1636592, EPI_ISL_1636597, EPI_ISL_1636598, EPI_ISL_1636600, EPI_ISL_1729990, EPI_ISL_1729991, EPI_ISL_1729992, EPI_ISL_1729994, EPI_ISL_1729995, EPI_ISL_1729996, EPI_ISL_1729997, EPI_ISL_1729998, EPI_ISL_1729999, EPI_ISL_1730000, EPI_ISL_1730008, EPI_ISL_1730009, EPI_ISL_1730016 | | | |
| see above | University of Birmingham | COVID-19 Genomics UK (COG-UK) Consortium | Alex Richter; Andrew Bosworth; Andrew Bosworth. Queen Elizabeth Hospital: Anna Casey; Andrew D Beggs PHE Heartlands Lab: Husam Osman; Charlotte Poxon; Institute of Microbiology; Joanne Stockton; Josh Quick; Kasun Wanigasooriya; Mike Kidd; Nicholas Loman. University of Birmingham Testing Laboratory: Celina M Whalley; Oliver Pickles; Radoslaw Poplawski; Samuel Nicholls; University of Birmingham: Claire McMurray; Will Rowe |
| EPI_ISL_1730149, EPI_ISL_1730150, EPI_ISL_1730151, EPI_ISL_1730152 | University of Exeter | COVID-19 Genomics UK (COG-UK) Consortium | Aaron Jeffries; Audrey Farbos; Ben Temperton; Jane Masoli; Joanna Warwick-Dugdale; Michelle Michelsen; Robyn Manley; Stephen Michell |
| EPI_ISL_1636697, EPI_ISL_1636700, EPI_ISL_1636705, EPI_ISL_1636706, EPI_ISL_1636707, EPI_ISL_1636708, EPI_ISL_1730106, EPI_ISL_1730107, EPI_ISL_1730108, EPI_ISL_1730109, EPI_ISL_1730110, EPI_ISL_1730111, EPI_ISL_1730112, EPI_ISL_1730113, EPI_ISL_1730114, EPI_ISL_1730115, EPI_ISL_1730116, EPI_ISL_1730117, EPI_ISL_1730118, EPI_ISL_1730119, EPI_ISL_1730124, EPI_ISL_1730125, EPI_ISL_1730126, EPI_ISL_1730130, EPI_ISL_1730131, EPI_ISL_1730132, EPI_ISL_1832247, EPI_ISL_1832248, EPI_ISL_1832272, EPI_ISL_1832273, EPI_ISL_1974814, EPI_ISL_1974890, EPI_ISL_1974898, EPI_ISL_2125694, EPI_ISL_2125696, EPI_ISL_2125700, EPI_ISL_2125701, EPI_ISL_2125870, EPI_ISL_2125878, EPI_ISL_2125879, EPI_ISL_2125881 | | | |
| see above | Virology Department, Royal Infirmary of Edinburgh, NHS Lothian / School of Biological Sciences, University of Edinburgh | COVID-19 Genomics UK (COG-UK) Consortium | Colquhoun R; Cotton S; Dewar R; Gallagher A; Hill V; Jackson B; Maloney D; McCrone JT; McHugh M; O'Toole Á; Rambaut A; Rooke S; Scher E; Templeton K; Yu X |
| EPI_ISL_1547321, EPI_ISL_1547336, EPI_ISL_1638036, EPI_ISL_1638037, EPI_ISL_1638039, EPI_ISL_1638040, EPI_ISL_1638042, EPI_ISL_1638043, EPI_ISL_1638044, EPI_ISL_1638045, EPI_ISL_1638046, EPI_ISL_1638047, EPI_ISL_1638048, EPI_ISL_1638049, EPI_ISL_1638050, EPI_ISL_1638053, EPI_ISL_1638054, EPI_ISL_1638055, EPI_ISL_1638056, EPI_ISL_1638057, EPI_ISL_1638059, EPI_ISL_1638061, EPI_ISL_1638065, EPI_ISL_1638066, EPI_ISL_1638067, EPI_ISL_1638071, EPI_ISL_1638072, EPI_ISL_1638073, EPI_ISL_1638077, EPI_ISL_1638078, EPI_ISL_1638080, EPI_ISL_1730937, EPI_ISL_1730939, EPI_ISL_1730940, EPI_ISL_1730946, EPI_ISL_1730950, EPI_ISL_1730952, EPI_ISL_1730953, EPI_ISL_1730954, EPI_ISL_1730957, EPI_ISL_1730958, EPI_ISL_1730959, EPI_ISL_1730961, EPI_ISL_1730962, EPI_ISL_1730963, EPI_ISL_1730964, EPI_ISL_1730972, EPI_ISL_1730973, EPI_ISL_1730975, EPI_ISL_1730981, EPI_ISL_1730983, EPI_ISL_1730984, EPI_ISL_1730986, EPI_ISL_1730988, EPI_ISL_1730989, EPI_ISL_1730992, EPI_ISL_1730993, EPI_ISL_1730994, EPI_ISL_1730996, EPI_ISL_1730997, EPI_ISL_1730998, EPI_ISL_1730999, EPI_ISL_1731000, EPI_ISL_1731001, EPI_ISL_1731004, EPI_ISL_1731005, EPI_ISL_1731007, EPI_ISL_1731009, EPI_ISL_1731010, EPI_ISL_1731011, EPI_ISL_1731016, EPI_ISL_1731019, EPI_ISL_1731021, EPI_ISL_1731022, EPI_ISL_1731025, EPI_ISL_1731028, EPI_ISL_1731030, EPI_ISL_1731031, EPI_ISL_1731032, EPI_ISL_1731033, EPI_ISL_1731039, EPI_ISL_1833835, EPI_ISL_1833856, EPI_ISL_1833859, EPI_ISL_1833875, EPI_ISL_1833878, EPI_ISL_1833879, EPI_ISL_1833898, EPI_ISL_1833909 | | | |
| see above | Virology Department, Sheffield Teaching Hospitals NHS Foundation Trust/Department of Infection, Immunity and Cardiovascular Disease, The Medical School, University of Sheffield | COVID-19 Genomics UK (COG-UK) Consortium | Adri Angyal; Alex Keeley; Benjamin Lindsey; Cariad Evans; Danielle Groves; Dave Partridge; Katie Johnson; Laura Carrilero; Luke Green; Matthew Parker; Matthew Wyles; Mehmet Yavuz; Mohammad Raza; Nikki Smith; Paul Parsons; Rachel Tucker; Rebecca Brown; Thushan de Silva |
| EPI_ISL_1546548, EPI_ISL_1546552, EPI_ISL_1546555, EPI_ISL_1546592, EPI_ISL_1546624, EPI_ISL_1546634, EPI_ISL_1546636, EPI_ISL_1637458, EPI_ISL_1637459, EPI_ISL_1637460, EPI_ISL_1637461, EPI_ISL_1637462, EPI_ISL_1637463, EPI_ISL_1637464, EPI_ISL_1637465, EPI_ISL_1637466, EPI_ISL_1637467, EPI_ISL_1637468, EPI_ISL_1637469, EPI_ISL_1637470, EPI_ISL_1637471, EPI_ISL_1637472, EPI_ISL_1637473, EPI_ISL_1637474, EPI_ISL_1637475, EPI_ISL_1637476, EPI_ISL_1637477, EPI_ISL_1637478, EPI_ISL_1637479, EPI_ISL_1637481, EPI_ISL_1637482, EPI_ISL_1637483, EPI_ISL_1637484, EPI_ISL_1637485, EPI_ISL_1637486, EPI_ISL_1637488, EPI_ISL_1637489, EPI_ISL_1637490, EPI_ISL_1637491, EPI_ISL_1637492, EPI_ISL_1637493, EPI_ISL_1637494, EPI_ISL_1637495, EPI_ISL_1637496, EPI_ISL_1637497, EPI_ISL_1637498, EPI_ISL_1637499, EPI_ISL_1637501, EPI_ISL_1637504, EPI_ISL_1637505, EPI_ISL_1637506, EPI_ISL_1637507, EPI_ISL_1637508, EPI_ISL_1637509, EPI_ISL_1637510, EPI_ISL_1637511, EPI_ISL_1637513, EPI_ISL_1637514, EPI_ISL_1637515, EPI_ISL_1637516, EPI_ISL_1637517, EPI_ISL_1637518, EPI_ISL_1637519, EPI_ISL_1637520, EPI_ISL_1637521, EPI_ISL_1637522, EPI_ISL_1637523, EPI_ISL_1637524, EPI_ISL_1637525, EPI_ISL_1637526, EPI_ISL_1637527, EPI_ISL_1637528, EPI_ISL_1637529, EPI_ISL_1637530, EPI_ISL_1637531, EPI_ISL_1637532, EPI_ISL_1637533, EPI_ISL_1637534, EPI_ISL_1637535, EPI_ISL_1637536, EPI_ISL_1637537, EPI_ISL_1637538, EPI_ISL_1637539, EPI_ISL_1637540, EPI_ISL_1637541, EPI_ISL_1637542, EPI_ISL_1637543, EPI_ISL_1637544, EPI_ISL_1637545, EPI_ISL_1637546, EPI_ISL_1637547, EPI_ISL_1637548, EPI_ISL_1637549, EPI_ISL_1637552, EPI_ISL_1637553, EPI_ISL_1637554, EPI_ISL_1637555, EPI_ISL_1637556, EPI_ISL_1637557, EPI_ISL_1637558, EPI_ISL_1637559, EPI_ISL_1637560, EPI_ISL_1637561, EPI_ISL_1637562, EPI_ISL_1637563, EPI_ISL_1637564, EPI_ISL_1637565, EPI_ISL_1637566, EPI_ISL_1637567, EPI_ISL_1637568, EPI_ISL_1637569, EPI_ISL_1637570, EPI_ISL_1637571, EPI_ISL_1637572, EPI_ISL_1637573, EPI_ISL_1637574, EPI_ISL_1637575, EPI_ISL_1637576, EPI_ISL_1637577, EPI_ISL_1637578, EPI_ISL_1637579, EPI_ISL_1637580, EPI_ISL_1637581, EPI_ISL_1637601, EPI_ISL_1637602, EPI_ISL_1637603, EPI_ISL_1637604, EPI_ISL_1637605, EPI_ISL_1637606, EPI_ISL_1637607, EPI_ISL_1637608, EPI_ISL_1637609, EPI_ISL_1637610, EPI_ISL_1637611, EPI_ISL_1637612, EPI_ISL_1637613, EPI_ISL_1637614, EPI_ISL_1637615, EPI_ISL_1637616, EPI_ISL_1637617, EPI_ISL_1637618, EPI_ISL_1637619, EPI_ISL_1637620, EPI_ISL_1637621, EPI_ISL_1637622, EPI_ISL_1637623, EPI_ISL_1637624, EPI_ISL_1637625, EPI_ISL_1637626, EPI_ISL_1637627, EPI_ISL_1637628, EPI_ISL_1637629, EPI_ISL_1637630, EPI_ISL_1637631, EPI_ISL_1637632, EPI_ISL_1637633, EPI_ISL_1637634, EPI_ISL_1637635, EPI_ISL_1637636, EPI_ISL_1637637, EPI_ISL_1637638, EPI_ISL_1637639, EPI_ISL_1637640, EPI_ISL_1637641, EPI_ISL_1637642, EPI_ISL_1637643, EPI_ISL_1637644, EPI_ISL_1637645, EPI_ISL_1637646, EPI_ISL_1637647, EPI_ISL_1637648, EPI_ISL_1637649, EPI_ISL_1637650, EPI_ISL_1637651, EPI_ISL_1637652, EPI_ISL_1637653, EPI_ISL_1637668, EPI_ISL_1637669, EPI_ISL_1637670, EPI_ISL_1637671, EPI_ISL_1637672, EPI_ISL_1637673, EPI_ISL_1637674, EPI_ISL_1637675, EPI_ISL_1637676, EPI_ISL_1637677, EPI_ISL_1637678, EPI_ISL_1637679, EPI_ISL_1637680, EPI_ISL_1637681, EPI_ISL_1637682, EPI_ISL_1637683, EPI_ISL_1637684, EPI_ISL_1637685, EPI_ISL_1637686, EPI_ISL_1637687, EPI_ISL_1637688, EPI_ISL_1637689, EPI_ISL_1637690, EPI_ISL_1637692, EPI_ISL_1637693, EPI_ISL_1637694, EPI_ISL_1637695, EPI_ISL_1637696, EPI_ISL_1637698, EPI_ISL_1637699, EPI_ISL_1637700, EPI_ISL_1637701, EPI_ISL_1637702, EPI_ISL_1637703, EPI_ISL_1637704, EPI_ISL_1637705, EPI_ISL_1637706, EPI_ISL_1637707, EPI_ISL_1637708, EPI_ISL_1637709, EPI_ISL_1637710, EPI_ISL_1637711, EPI_ISL_1637712, EPI_ISL_1637713, EPI_ISL_1637714, EPI_ISL_1637715, EPI_ISL_1637716, EPI_ISL_1637717, EPI_ISL_1637718, EPI_ISL_1637719, EPI_ISL_1637720, EPI_ISL_1637721, EPI_ISL_1637722, EPI_ISL_1637723, EPI_ISL_1637724, EPI_ISL_1637725, EPI_ISL_1637726, EPI_ISL_1637727, EPI_ISL_1637728, EPI_ISL_1637729, EPI_ISL_1637730, EPI_ISL_1637731, EPI_ISL_1637732, EPI_ISL_1637733, EPI_ISL_1637734, EPI_ISL_1637735, EPI_ISL_1637736, EPI_ISL_1637737, EPI_ISL_1637738, EPI_ISL_1637739, EPI_ISL_1637740, EPI_ISL_1637741, EPI_ISL_1637743, EPI_ISL_1637744, EPI_ISL_1637745, EPI_ISL_1637746, EPI_ISL_1637747, EPI_ISL_1637748, EPI_ISL_1637749, EPI_ISL_1637750, EPI_ISL_1637751, EPI_ISL_1637752, EPI_ISL_1637753, EPI_ISL_1637754, EPI_ISL_1637755, EPI_ISL_1637756, EPI_ISL_1637757, EPI_ISL_1637758, EPI_ISL_1637759, EPI_ISL_1637760, EPI_ISL_1637761, EPI_ISL_1637762, EPI_ISL_1637763, EPI_ISL_1637764, EPI_ISL_1637765, EPI_ISL_1637766, EPI_ISL_1637767, EPI_ISL_1637768, EPI_ISL_1833186, EPI_ISL_1833190, EPI_ISL_1833200 | | | |
| see above | Wales Specialist Virology Centre Sequencing lab: Pathogen Genomics Unit | Public Health Wales Microbiology Cardiff Wales Specialist Virology Centre | Alec Birchley; Alexander Adams; Amy Gaskin; Angela Marchbank; Bree Gatica-Wilcox; Catherine Moore; Jason Coombes; Joanne Watkins; Joel Southgate; Johnathan Evans; Laura Gifford; Lauren Gilbert; Lee Graham; Malorie Perry; Matthew Bull; Nicole Pacchiarini; Sally Corden; Sara Kumziene-Summerhayes; Sara Rey; Sarah Taylor; Simon Cottrell; Sophie Jones; Tom Connor |
| EPI_ISL_1832284, EPI_ISL_1832288, EPI_ISL_1832289, EPI_ISL_1832290, EPI_ISL_1832291, EPI_ISL_1832292, EPI_ISL_1832293, EPI_ISL_1832294, EPI_ISL_1832295, EPI_ISL_1832296, EPI_ISL_1832298, EPI_ISL_1832299, EPI_ISL_1832308 | | | |
| see above | West of Scotland Specialist Virology Centre, NHSGGC / MRC-University of Glasgow Centre for Virus Research | COVID-19 Genomics UK (COG-UK) Consortium | Alasdair MacLean; Alice Broos; Ana da Silva Filipe; Antonia Ho; Daniel Mair; David L Robertson; Emma Thomson; Guy Mollett; James Shepherd; Jenna Nichols; Joseph Hughes; Kathy Li; Kathy Smollett; Kyriaki Nomikou; Lily Tong; Matthew Holden; Natasha Johnson; Rachel Blacow; Richard Orton; Rory Gunson; Sarah McDonald; Sharif Shaaban; Sreenu Vattipally; Stephen Carmichael |

We gratefully acknowledge the following Authors from the Originating laboratories responsible for obtaining the specimens, as well as the Submitting laboratories where the genome data were generated and shared via GISAID, on which this research is based.

All Submitters of data may be contacted directly via www.gisaid.org

Authors are sorted alphabetically.

| Accession ID | Originating Laboratory | Submitting Laboratory | Authors |
|---|---|---|---|
| EPI_ISL_1832539, EPI_ISL_1832543, EPI_ISL_1832545, EPI_ISL_1832549, EPI_ISL_1832551, EPI_ISL_1832553, EPI_ISL_1832555, EPI_ISL_1832557, EPI_ISL_1832559, EPI_ISL_1832562, EPI_ISL_1832564, EPI_ISL_1832566, EPI_ISL_1832568, EPI_ISL_1832570, EPI_ISL_1975069, EPI_ISL_1975070, EPI_ISL_1975072, EPI_ISL_1975074, EPI_ISL_1975075, EPI_ISL_1975101, EPI_ISL_1975102 | | | |
| see above | Barts Health NHS Trust | COVID-19 Genomics UK (COG-UK) Consortium | BROAD; Beatrix; CUTINO-MOGUEL; Claire; David; Dola; HARRINGTON; KELE; KULASEGARAN-SHYLINI; Maria-Teresa; OWOYEMI; Raghavendran |
| EPI_ISL_1833923, EPI_ISL_1833931, EPI_ISL_1833933, EPI_ISL_1833946 | Bioinformatics and Biostatistics Lab, Advanced Sequencing Facility | COVID-19 Genomics UK (COG-UK) Consortium | Aengus Stewart; Chelsea Sawyer; Harshil Patel; Jerome Nicod; Laura Cubitt; Margaret Crawford |
| EPI_ISL_1730896, EPI_ISL_1730898, EPI_ISL_1730899, EPI_ISL_1730904, EPI_ISL_1730909, EPI_ISL_1730910, EPI_ISL_1730912, EPI_ISL_1730922, EPI_ISL_1730923, EPI_ISL_1730924, EPI_ISL_1833279, EPI_ISL_1833280, EPI_ISL_1833281, EPI_ISL_1833284, EPI_ISL_1833285, EPI_ISL_1833287, EPI_ISL_1833340, EPI_ISL_1833347, EPI_ISL_1833348, EPI_ISL_1833349, EPI_ISL_1833350, EPI_ISL_1979798, EPI_ISL_2127895 | | | |
| see above | Centre for Enzyme Innovation, University of Portsmouth / Translational Research Laboratory, Portsmouth Hospitals NHS Trust | COVID-19 Genomics UK (COG-UK) Consortium | Angela Beckett; Christopher Fearn; Kate Cook; Katie Loveson; Salman Goudarzi; Samuel Robson; Scott Elliott; Sharon Glaysher |
| EPI_ISL_1636625, EPI_ISL_1730064, EPI_ISL_1730067, EPI_ISL_1730070, EPI_ISL_1730071, EPI_ISL_1730072, EPI_ISL_1832176, EPI_ISL_1832178, EPI_ISL_1832180, EPI_ISL_1832182, EPI_ISL_1832191, EPI_ISL_1832193, EPI_ISL_1832194, EPI_ISL_1832195, EPI_ISL_1832196, EPI_ISL_1832197, EPI_ISL_1832198, EPI_ISL_1832199, EPI_ISL_1832200, EPI_ISL_1832201, EPI_ISL_1832202, EPI_ISL_1832216, EPI_ISL_1832226, EPI_ISL_1974789 | | | |
| see above | Department of Pathology, University of Cambridge | COVID-19 Genomics UK (COG-UK) Consortium | Aminu S. Jahun; Ian Goodfellow; Iliana Georgana; Martin D. Curran; Myra Hosmillo; Rhys Izuagbe; Surendra Parmar; William L. Hamilton; Yasmin Chaudhry |
| EPI_ISL_1831284, EPI_ISL_1831285, EPI_ISL_1831286, EPI_ISL_1831287, EPI_ISL_1831288, EPI_ISL_1831289, EPI_ISL_1831290, EPI_ISL_1831291, EPI_ISL_1831292, EPI_ISL_1831293, EPI_ISL_1831294, EPI_ISL_1831295, EPI_ISL_1831296, EPI_ISL_1831297, EPI_ISL_1831298, EPI_ISL_1831299, EPI_ISL_1831300, EPI_ISL_1831301, EPI_ISL_1831302, EPI_ISL_1831303, EPI_ISL_1831304, EPI_ISL_1831305, EPI_ISL_1831306, EPI_ISL_1831307, EPI_ISL_1831308, EPI_ISL_1831309, EPI_ISL_1831310, EPI_ISL_1831311, EPI_ISL_1831312, EPI_ISL_1831314, EPI_ISL_1831316, EPI_ISL_1831318, EPI_ISL_1831319, EPI_ISL_1831320, EPI_ISL_1831321, EPI_ISL_1831324, EPI_ISL_1831325, EPI_ISL_1831327, EPI_ISL_1831328, EPI_ISL_1831331, EPI_ISL_1831332, EPI_ISL_1831333, EPI_ISL_1831335, EPI_ISL_1831336, EPI_ISL_1831337, EPI_ISL_1831338, EPI_ISL_1831339, EPI_ISL_1831340, EPI_ISL_1831346, EPI_ISL_1831347, EPI_ISL_1831351, EPI_ISL_1831352, EPI_ISL_1831353, EPI_ISL_1831355, EPI_ISL_1831356, EPI_ISL_1831358, EPI_ISL_1831359, EPI_ISL_1831361, EPI_ISL_1831367, EPI_ISL_1831368, EPI_ISL_1831369, EPI_ISL_1831370, EPI_ISL_1831371, EPI_ISL_1831372, EPI_ISL_1831373, EPI_ISL_1831374, EPI_ISL_1831375, EPI_ISL_1831377, EPI_ISL_1831378, EPI_ISL_1831380, EPI_ISL_1831384, EPI_ISL_1831386, EPI_ISL_2006541, EPI_ISL_2006542, EPI_ISL_2006544, EPI_ISL_2006545, EPI_ISL_2006546, EPI_ISL_2006547, EPI_ISL_2006548, EPI_ISL_2006549, EPI_ISL_2006551, EPI_ISL_2006553, EPI_ISL_2006554, EPI_ISL_2006555, EPI_ISL_2006556, EPI_ISL_2006557, EPI_ISL_2006558, EPI_ISL_2006559, EPI_ISL_2006560, EPI_ISL_2006561, EPI_ISL_2006562, EPI_ISL_2006563, EPI_ISL_2006564, EPI_ISL_2006565, EPI_ISL_2006566, EPI_ISL_2006567, EPI_ISL_2006568, EPI_ISL_2006569, EPI_ISL_2006570, EPI_ISL_2006571, EPI_ISL_2006572, EPI_ISL_2006573, EPI_ISL_2006574, EPI_ISL_2006575, EPI_ISL_2006576, EPI_ISL_2006579, EPI_ISL_2006580, EPI_ISL_2006581, EPI_ISL_2006583, EPI_ISL_2006584, EPI_ISL_2006585, EPI_ISL_2006586, EPI_ISL_2006587, EPI_ISL_2006588, EPI_ISL_2006589, EPI_ISL_2006590, EPI_ISL_2006591, EPI_ISL_2006593, EPI_ISL_2006594, EPI_ISL_2006596, EPI_ISL_2006598, EPI_ISL_2006599, EPI_ISL_2006600, EPI_ISL_2006601, EPI_ISL_2006604, EPI_ISL_2006605, EPI_ISL_2006606, EPI_ISL_2006607, EPI_ISL_2006609, EPI_ISL_2006610, EPI_ISL_2006611, EPI_ISL_2006612, EPI_ISL_2006614, EPI_ISL_2006615, EPI_ISL_2006617, EPI_ISL_2006619, EPI_ISL_2006620, EPI_ISL_2006622, EPI_ISL_2006624, EPI_ISL_2006625, EPI_ISL_2006627, EPI_ISL_2006628, EPI_ISL_2006629, EPI_ISL_2006630, EPI_ISL_2006631, EPI_ISL_2006633, EPI_ISL_2006634, EPI_ISL_2006635, EPI_ISL_2006636, EPI_ISL_2006637, EPI_ISL_2006638, EPI_ISL_2006639, EPI_ISL_2006640, EPI_ISL_2006641, EPI_ISL_2006642, EPI_ISL_2006643, EPI_ISL_2006644, EPI_ISL_2006645, EPI_ISL_2006646, EPI_ISL_2006647, EPI_ISL_2006648, EPI_ISL_2006649, EPI_ISL_2006650, EPI_ISL_2006651, EPI_ISL_2006652, EPI_ISL_2006654, EPI_ISL_2006655, EPI_ISL_2006657, EPI_ISL_2006658, EPI_ISL_2006659, EPI_ISL_2006660, EPI_ISL_2006661, EPI_ISL_2006662, EPI_ISL_2006663, EPI_ISL_2006665, EPI_ISL_2006666 | | | |
| see above | Health Services Laboratories | Wellcome Sanger Institute for the COVID-19 Genomics UK (COG-UK) Consortium | Cordelia Langford; David K. Jackson; Dominic Kwiatkowski; Ewan Harrison; Health Services Laboratories and Alex Alderton; Ian Johnston; Jeffrey Barrett; John Sillitoe on behalf of the Wellcome Sanger Institute COVID-19 Surveillance Team; Roberto Amato; Sonia Goncalves |
| EPI_ISL_1697449, EPI_ISL_1697450, EPI_ISL_1697451, EPI_ISL_1697458, EPI_ISL_1697471, EPI_ISL_1697473, EPI_ISL_1697477, EPI_ISL_1697499, EPI_ISL_1697501, EPI_ISL_1697540, EPI_ISL_1697548, EPI_ISL_1697555, EPI_ISL_1697562, EPI_ISL_1697574, EPI_ISL_1697589, EPI_ISL_1697593, EPI_ISL_1697598, EPI_ISL_1697605, EPI_ISL_1697612, EPI_ISL_1697624, EPI_ISL_1697628, EPI_ISL_1697659, EPI_ISL_1697661, EPI_ISL_1697669, EPI_ISL_1697671, EPI_ISL_1697687, EPI_ISL_1697699, EPI_ISL_1697708, EPI_ISL_1697712, EPI_ISL_1697715, EPI_ISL_1697730, EPI_ISL_1697731, EPI_ISL_1697740, EPI_ISL_1697971, EPI_ISL_1697980, EPI_ISL_1697998, EPI_ISL_1698019, EPI_ISL_1698026, EPI_ISL_1698030, EPI_ISL_1698045, EPI_ISL_1698049, EPI_ISL_1698056, EPI_ISL_1698071, EPI_ISL_1698080, EPI_ISL_1698091, EPI_ISL_1698100, EPI_ISL_1698117, EPI_ISL_1698130, EPI_ISL_1698146, EPI_ISL_1698157, EPI_ISL_1698165, EPI_ISL_1698169, EPI_ISL_1698173, EPI_ISL_1698186, EPI_ISL_1698188, EPI_ISL_1698190, EPI_ISL_1698203, EPI_ISL_1698204, EPI_ISL_1698205, EPI_ISL_1698213, EPI_ISL_1698219, EPI_ISL_1698227, EPI_ISL_1698229, EPI_ISL_1698231, EPI_ISL_1698247, EPI_ISL_1698252, EPI_ISL_1698254, EPI_ISL_1698257, EPI_ISL_1698259, EPI_ISL_1698263, EPI_ISL_1698265, EPI_ISL_1698266, EPI_ISL_1698269, EPI_ISL_1698271, EPI_ISL_1698275, EPI_ISL_1698276, EPI_ISL_1698278, EPI_ISL_1698281, EPI_ISL_1698282, EPI_ISL_1698286, EPI_ISL_1698287, EPI_ISL_1698288, EPI_ISL_1698289, EPI_ISL_1698290, EPI_ISL_1698291, EPI_ISL_1698292, EPI_ISL_1698293, EPI_ISL_1698296, EPI_ISL_1698297, EPI_ISL_1698298, EPI_ISL_1698299, EPI_ISL_1698300, EPI_ISL_1698301, EPI_ISL_1698303, EPI_ISL_1698304, EPI_ISL_1698306, EPI_ISL_1698308, EPI_ISL_1698309, EPI_ISL_1698310, EPI_ISL_1698311, EPI_ISL_1698312, EPI_ISL_1698313, EPI_ISL_1698316, EPI_ISL_1698319, EPI_ISL_1698320, EPI_ISL_1698323, EPI_ISL_1698325, EPI_ISL_1698326, EPI_ISL_1698329, EPI_ISL_1698330, EPI_ISL_1698331, EPI_ISL_1698333, EPI_ISL_1698334, EPI_ISL_1698335, EPI_ISL_1698336, EPI_ISL_1698337, EPI_ISL_1698340, EPI_ISL_1698343, EPI_ISL_1698344, EPI_ISL_1698345, EPI_ISL_1698348, EPI_ISL_1698350, EPI_ISL_1698351, EPI_ISL_1698352, EPI_ISL_1698353, EPI_ISL_1698354, EPI_ISL_1698355, EPI_ISL_1698356, EPI_ISL_1698358, EPI_ISL_1698360, EPI_ISL_1698362, EPI_ISL_1698363, EPI_ISL_1698365, EPI_ISL_1698367, EPI_ISL_1698368, EPI_ISL_1698369, EPI_ISL_1698374, EPI_ISL_1698375, EPI_ISL_1698376, EPI_ISL_1698377, EPI_ISL_1698380, EPI_ISL_1698381, EPI_ISL_1698382, EPI_ISL_1698385, EPI_ISL_1698386, EPI_ISL_1698389, EPI_ISL_1698390, EPI_ISL_1698395, EPI_ISL_1698396, EPI_ISL_1698397, EPI_ISL_1698399, EPI_ISL_1698400, EPI_ISL_1698401, EPI_ISL_1698402, EPI_ISL_1698404, EPI_ISL_1698408, EPI_ISL_1698413, EPI_ISL_1698414, EPI_ISL_1698415, EPI_ISL_1698416, EPI_ISL_1698419, EPI_ISL_1698420, EPI_ISL_1698421, EPI_ISL_1698422, EPI_ISL_1698424, EPI_ISL_1698425, EPI_ISL_1698427, EPI_ISL_1698433, EPI_ISL_1698434, EPI_ISL_1698437, EPI_ISL_1698438, EPI_ISL_1698440, EPI_ISL_1698441, EPI_ISL_1698443, EPI_ISL_1698445, EPI_ISL_1698448, EPI_ISL_1698449, EPI_ISL_1698453, EPI_ISL_1698454, EPI_ISL_1698455, EPI_ISL_1698456, EPI_ISL_1698457, EPI_ISL_1698460, EPI_ISL_1698462, EPI_ISL_1698464, EPI_ISL_1698466, EPI_ISL_1698467, EPI_ISL_1698468, EPI_ISL_1698470, EPI_ISL_1698471, EPI_ISL_1698473, EPI_ISL_1698474, EPI_ISL_1698477, EPI_ISL_1698478, EPI_ISL_1698480, EPI_ISL_1698481, EPI_ISL_1698484, EPI_ISL_1698485, EPI_ISL_1698488, EPI_ISL_1698489, EPI_ISL_1698491, EPI_ISL_1698494, EPI_ISL_1698495, EPI_ISL_1698496, EPI_ISL_1698497, EPI_ISL_1698499, EPI_ISL_1698500, EPI_ISL_1698503, EPI_ISL_1698507, EPI_ISL_1698509, EPI_ISL_1698513, EPI_ISL_1698514, EPI_ISL_1698515, EPI_ISL_1698516, EPI_ISL_1698517, EPI_ISL_1698518, EPI_ISL_1698519, EPI_ISL_1698522, EPI_ISL_1698523, EPI_ISL_1698525, EPI_ISL_1698528, EPI_ISL_1698529, EPI_ISL_1698530, EPI_ISL_1698531, EPI_ISL_1698535, EPI_ISL_1698536, EPI_ISL_1698538, EPI_ISL_1698540, EPI_ISL_1698542, EPI_ISL_1698543, EPI_ISL_1698544, EPI_ISL_1698545, EPI_ISL_1698546, EPI_ISL_1698918, EPI_ISL_1699291, EPI_ISL_1699295, EPI_ISL_1699299, EPI_ISL_1699301, EPI_ISL_1699304, EPI_ISL_1699306, EPI_ISL_1699311, EPI_ISL_1699312, EPI_ISL_1699313, EPI_ISL_1699316, EPI_ISL_1699317, EPI_ISL_1699325, EPI_ISL_1699329, EPI_ISL_1699330, EPI_ISL_1699338, EPI_ISL_1699340, EPI_ISL_1699345, EPI_ISL_1699346, EPI_ISL_1699347, EPI_ISL_1699350, EPI_ISL_1699352, EPI_ISL_1699361, EPI_ISL_1699368, EPI_ISL_1699370, EPI_ISL_1699375, EPI_ISL_1699376, EPI_ISL_1699377, EPI_ISL_1699384, EPI_ISL_1699386, EPI_ISL_1699388, EPI_ISL_1699389, EPI_ISL_1699393, EPI_ISL_1699396, EPI_ISL_1699398, EPI_ISL_1699400, EPI_ISL_1699403, EPI_ISL_1699405, EPI_ISL_1699407, EPI_ISL_1699409, EPI_ISL_1699415, EPI_ISL_1699419, EPI_ISL_1699428, EPI_ISL_1699429, EPI_ISL_1699431, EPI_ISL_1699433, EPI_ISL_1699435, EPI_ISL_1699439, EPI_ISL_1699442, EPI_ISL_1699455, EPI_ISL_1699457, EPI_ISL_1699458, EPI_ISL_1699462, EPI_ISL_1699470, EPI_ISL_1699477, EPI_ISL_1699482, EPI_ISL_1699483, EPI_ISL_1699488, EPI_ISL_1699489, EPI_ISL_1699491, EPI_ISL_1699493, EPI_ISL_1699495, EPI_ISL_1699497, EPI_ISL_1699502, EPI_ISL_1699506, EPI_ISL_1699510, EPI_ISL_1699511, EPI_ISL_1699514, EPI_ISL_1699515, EPI_ISL_1699518, EPI_ISL_1699523, EPI_ISL_1699525, EPI_ISL_1699537, EPI_ISL_1699539, EPI_ISL_1699543, EPI_ISL_1699544, EPI_ISL_1699545, EPI_ISL_1699547, EPI_ISL_1699558, EPI_ISL_1699561, EPI_ISL_1699562, EPI_ISL_1699563, EPI_ISL_1699564, EPI_ISL_1699565, EPI_ISL_1699568, EPI_ISL_1699570, EPI_ISL_1718322, EPI_ISL_1718324, EPI_ISL_1718325, EPI_ISL_1718326, EPI_ISL_1718327, EPI_ISL_1718329, EPI_ISL_1718331, EPI_ISL_1718335, EPI_ISL_1718339, EPI_ISL_1718340, EPI_ISL_1718341, EPI_ISL_1718342, EPI_ISL_1718345, EPI_ISL_1718346, EPI_ISL_1718348, EPI_ISL_1718352, EPI_ISL_1718354, EPI_ISL_1718356, EPI_ISL_1718357, EPI_ISL_1718363, EPI_ISL_1718367, EPI_ISL_1718368, EPI_ISL_1718369, EPI_ISL_1718370, EPI_ISL_1718372, EPI_ISL_1718373, EPI_ISL_1718375, EPI_ISL_1718376, EPI_ISL_1718380, EPI_ISL_1718381, EPI_ISL_1718383, EPI_ISL_1718389, EPI_ISL_1718390, EPI_ISL_1718392, EPI_ISL_1718393, EPI_ISL_1718394, EPI_ISL_1718396, EPI_ISL_1718398, EPI_ISL_1718399, EPI_ISL_1718400, EPI_ISL_1718406, EPI_ISL_1718409, EPI_ISL_1718411, EPI_ISL_1718413, EPI_ISL_1718414, EPI_ISL_1718419, EPI_ISL_1718421, EPI_ISL_1718422, EPI_ISL_1718425, EPI_ISL_1718426, EPI_ISL_1718428, EPI_ISL_1718429, EPI_ISL_1718430, EPI_ISL_1718431, EPI_ISL_1718433, EPI_ISL_1718434, EPI_ISL_1718436, EPI_ISL_1718437, EPI_ISL_1718438, EPI_ISL_1718442, EPI_ISL_1718444, EPI_ISL_1718446, EPI_ISL_1718450, EPI_ISL_1718452, EPI_ISL_1718454, EPI_ISL_1718455, EPI_ISL_1718458, EPI_ISL_1718459, EPI_ISL_1718460, EPI_ISL_1718465, EPI_ISL_1718469, EPI_ISL_1718470, EPI_ISL_1718476, EPI_ISL_1718479, EPI_ISL_1718480, EPI_ISL_1718482, EPI_ISL_1718483, EPI_ISL_1718484, EPI_ISL_1718486, EPI_ISL_1718490, EPI_ISL_1718496, EPI_ISL_1718497, EPI_ISL_1718500, EPI_ISL_1718501, EPI_ISL_1718504, EPI_ISL_1718507, EPI_ISL_1718510, EPI_ISL_1718511, EPI_ISL_1718517, EPI_ISL_1718518, EPI_ISL_1718519, EPI_ISL_1718527, EPI_ISL_1718528, EPI_ISL_1718531, EPI_ISL_1718532, EPI_ISL_1718540, EPI_ISL_1718541, EPI_ISL_1718542, EPI_ISL_1718543, EPI_ISL_1718544, EPI_ISL_1718545, EPI_ISL_1718548, EPI_ISL_1718549, EPI_ISL_1718555, EPI_ISL_1718560, EPI_ISL_1718561, EPI_ISL_1718565, EPI_ISL_1718569, EPI_ISL_1718576, EPI_ISL_1718578, EPI_ISL_1718580, EPI_ISL_1718582, EPI_ISL_1718583, EPI_ISL_1718584, EPI_ISL_1718585, EPI_ISL_1718588, EPI_ISL_1718589, EPI_ISL_1718592, EPI_ISL_1718593, EPI_ISL_1718595, EPI_ISL_1718597, EPI_ISL_1718598, EPI_ISL_1718599, EPI_ISL_1718601, EPI_ISL_1718603, EPI_ISL_1718604, EPI_ISL_1718605, EPI_ISL_1718607, EPI_ISL_1718609, EPI_ISL_1718614, EPI_ISL_1718615, EPI_ISL_1718616, EPI_ISL_1718619, EPI_ISL_1718620, EPI_ISL_1718621, EPI_ISL_1718623, EPI_ISL_1718624, EPI_ISL_1718625, EPI_ISL_1718626, EPI_ISL_1718627, EPI_ISL_1718628, EPI_ISL_1718629, EPI_ISL_1718630, EPI_ISL_1718631, EPI_ISL_1718632, EPI_ISL_1718633, EPI_ISL_1718634, EPI_ISL_1718635, EPI_ISL_1718636, EPI_ISL_1718637, EPI_ISL_1718638, EPI_ISL_1718639, EPI_ISL_1718641, EPI_ISL_1718643, EPI_ISL_1718646, EPI_ISL_1718647, EPI_ISL_1718648, EPI_ISL_1718650, EPI_ISL_1718651, EPI_ISL_1718652, EPI_ISL_1718653, EPI_ISL_1718654, EPI_ISL_1718655, EPI_ISL_1718656, EPI_ISL_1718657, EPI_ISL_1718658, EPI_ISL_1718659, EPI_ISL_1718660, EPI_ISL_1718661, EPI_ISL_1718662, EPI_ISL_1718664, EPI_ISL_1718665, EPI_ISL_1718666, EPI_ISL_1718667, EPI_ISL_1718669, EPI_ISL_1718670, EPI_ISL_1718671, EPI_ISL_1718673, EPI_ISL_1718674, EPI_ISL_1718675, EPI_ISL_1718676, EPI_ISL_1718677, EPI_ISL_1718678, EPI_ISL_1718680, EPI_ISL_1718681, EPI_ISL_1718682, EPI_ISL_1718683, EPI_ISL_1718684, EPI_ISL_1718687, EPI_ISL_1718689, EPI_ISL_1718690, EPI_ISL_1718693, EPI_ISL_1718694, EPI_ISL_1718696, EPI_ISL_1718698, EPI_ISL_1718699, EPI_ISL_1718702, EPI_ISL_1718705, EPI_ISL_1718706, EPI_ISL_1718708, EPI_ISL_1718709, EPI_ISL_1718710, EPI_ISL_1718711, EPI_ISL_1718712, EPI_ISL_1718713, EPI_ISL_1718714, EPI_ISL_1718715, EPI_ISL_1718716, EPI_ISL_1718717, EPI_ISL_1718718, EPI_ISL_1718719, EPI_ISL_1718720, EPI_ISL_1718721, EPI_ISL_1718722, EPI_ISL_1718723, EPI_ISL_1718724, EPI_ISL_1718725, EPI_ISL_1718726, EPI_ISL_1718728, EPI_ISL_1718729, EPI_ISL_1718730, EPI_ISL_1718731, EPI_ISL_1718733, EPI_ISL_1718734, EPI_ISL_1718735, EPI_ISL_1718736, EPI_ISL_1718737, EPI_ISL_1718738, EPI_ISL_1718739, EPI_ISL_1718740, EPI_ISL_1718741, EPI_ISL_1718743, EPI_ISL_1718745, EPI_ISL_1718747, EPI_ISL_1718748, EPI_ISL_1718749, EPI_ISL_1718752, EPI_ISL_1718753, EPI_ISL_1718754, EPI_ISL_1718755, EPI_ISL_1718756, EPI_ISL_1718757, EPI_ISL_1718758, EPI_ISL_1718759, EPI_ISL_1718761, EPI_ISL_1718762, EPI_ISL_1718763, EPI_ISL_1718764, EPI_ISL_1718765, EPI_ISL_1718766, EPI_ISL_1718767, EPI_ISL_1718768, EPI_ISL_1718769, EPI_ISL_1718770, EPI_ISL_1718771, EPI_ISL_1718772, EPI_ISL_1718773, EPI_ISL_1718774, EPI_ISL_1718775, EPI_ISL_1718776, EPI_ISL_1718777, EPI_ISL_1718778, EPI_ISL_1718779, EPI_ISL_1718780, EPI_ISL_1718781, EPI_ISL_1718782, EPI_ISL_1718785, EPI_ISL_1718786, EPI_ISL_1718787, EPI_ISL_1718788, EPI_ISL_1718790, EPI_ISL_1718792, EPI_ISL_1718793, EPI_ISL_1718794, EPI_ISL_1718795, EPI_ISL_1718796, EPI_ISL_1718797, EPI_ISL_1718798, EPI_ISL_1718799, EPI_ISL_1718800, EPI_ISL_1718802, EPI_ISL_1718803, EPI_ISL_1718804, EPI_ISL_1718805, EPI_ISL_1718807, EPI_ISL_1718808, EPI_ISL_1718809, EPI_ISL_1718811, EPI_ISL_1718812, EPI_ISL_1718813, EPI_ISL_1718814, EPI_ISL_1718815, EPI_ISL_1718816, EPI_ISL_1718817, EPI_ISL_1718819, EPI_ISL_1718820, EPI_ISL_1718821, EPI_ISL_1718823, EPI_ISL_1718824, EPI_ISL_1718825, EPI_ISL_1718826, EPI_ISL_1718827, EPI_ISL_1718828, EPI_ISL_1718829, EPI_ISL_1718830, EPI_ISL_1718831, EPI_ISL_1718832, EPI_ISL_1718833, EPI_ISL_1718834, EPI_ISL_1718835, EPI_ISL_1718836, EPI_ISL_1718837, EPI_ISL_1718839, EPI_ISL_1718840, EPI_ISL_1718841, EPI_ISL_1718842, EPI_ISL_1718843, EPI_ISL_1718844, EPI_ISL_1718845, EPI_ISL_1718847, EPI_ISL_1718848, EPI_ISL_1718849, EPI_ISL_1718850, EPI_ISL_1718851, EPI_ISL_1718852, EPI_ISL_1718853, EPI_ISL_1718854, EPI_ISL_1718855, EPI_ISL_1718856, EPI_ISL_1718859, EPI_ISL_1718861, EPI_ISL_1718862, EPI_ISL_1718863, EPI_ISL_1718864, EPI_ISL_1718865, EPI_ISL_1718866, EPI_ISL_1718867, EPI_ISL_1718868, EPI_ISL_1718870, EPI_ISL_1718871, EPI_ISL_1718873, EPI_ISL_1718874, EPI_ISL_1718877, EPI_ISL_1718878, EPI_ISL_1718879, EPI_ISL_1718880, EPI_ISL_1718881, EPI_ISL_1718882, EPI_ISL_1718883, EPI_ISL_1718884, EPI_ISL_1719433, EPI_ISL_1719443, EPI_ISL_1719446, EPI_ISL_1719449, EPI_ISL_1719450, EPI_ISL_1719454, EPI_ISL_1719455, EPI_ISL_1719459, EPI_ISL_1719461, EPI_ISL_1719465, EPI_ISL_1719469, EPI_ISL_1719470, EPI_ISL_1719480, EPI_ISL_1719481, EPI_ISL_1719488, EPI_ISL_1719489, EPI_ISL_1719496, EPI_ISL_1719498, EPI_ISL_1719501, EPI_ISL_1719505, EPI_ISL_1719506, EPI_ISL_1719507, EPI_ISL_1719512, EPI_ISL_1719514, EPI_ISL_1719519, EPI_ISL_1719526, EPI_ISL_1719533, EPI_ISL_1719542, EPI_ISL_1719544, EPI_ISL_1719547, EPI_ISL_1719548, EPI_ISL_1719549, EPI_ISL_1719558, EPI_ISL_1719560, EPI_ISL_1719576, EPI_ISL_1719582, EPI_ISL_1719588, EPI_ISL_1719591, EPI_ISL_1719593, EPI_ISL_1719595, EPI_ISL_1719598, EPI_ISL_1719599, EPI_ISL_1719600, EPI_ISL_1719603, EPI_ISL_1719604, EPI_ISL_1719605, EPI_ISL_1719606, EPI_ISL_1719609, EPI_ISL_1719613, EPI_ISL_1719616, EPI_ISL_1719624, EPI_ISL_1719625, EPI_ISL_1719639, EPI_ISL_1719643, EPI_ISL_1719647, EPI_ISL_1719650, EPI_ISL_1719656, EPI_ISL_1719659, EPI_ISL_1719665, EPI_ISL_1719666, EPI_ISL_1719671, EPI_ISL_1719679, EPI_ISL_1719685, EPI_ISL_1719688, EPI_ISL_1719691, EPI_ISL_1719699, EPI_ISL_1719710, EPI_ISL_1719711, EPI_ISL_1719712, EPI_ISL_1719713, EPI_ISL_1719717, EPI_ISL_1719721, EPI_ISL_1719726, EPI_ISL_1719733, EPI_ISL_1719741, EPI_ISL_1719742, EPI_ISL_1740532, EPI_ISL_1740538, EPI_ISL_1740552, EPI_ISL_1740553, EPI_ISL_1740555, EPI_ISL_1740556, EPI_ISL_1740571, EPI_ISL_1740572, EPI_ISL_1740574, EPI_ISL_1740576, EPI_ISL_1740585, EPI_ISL_1740590, EPI_ISL_1740592, EPI_ISL_1740595, EPI_ISL_1740599, EPI_ISL_1740600, EPI_ISL_1740604, EPI_ISL_1740605, EPI_ISL_1740607, EPI_ISL_1740609, EPI_ISL_1740616, EPI_ISL_1740629, EPI_ISL_1740630, EPI_ISL_1740632, EPI_ISL_1740637, EPI_ISL_1740641, EPI_ISL_1740642, EPI_ISL_1740651, EPI_ISL_1740652, EPI_ISL_1740654, EPI_ISL_1740655, EPI_ISL_1740657, EPI_ISL_1740660, EPI_ISL_1740662, EPI_ISL_1740664, EPI_ISL_1740666, EPI_ISL_1740669, EPI_ISL_1740671, EPI_ISL_1740674, EPI_ISL_1740675, EPI_ISL_1740681, EPI_ISL_1740682, EPI_ISL_1740688, EPI_ISL_1740697, EPI_ISL_1740703, EPI_ISL_1740704, EPI_ISL_1740706, EPI_ISL_1740707, EPI_ISL_1740714, EPI_ISL_1740715, EPI_ISL_1740722, EPI_ISL_1740725, EPI_ISL_1740726, EPI_ISL_1740731, EPI_ISL_1740732, EPI_ISL_1740736, EPI_ISL_1740746, EPI_ISL_1740748, EPI_ISL_1740750, EPI_ISL_1740751, EPI_ISL_1740753, EPI_ISL_1740760, EPI_ISL_1740761, EPI_ISL_1740763, EPI_ISL_1740764, EPI_ISL_1740769, EPI_ISL_1740778, EPI_ISL_1740783, EPI_ISL_1740786, EPI_ISL_1740787, EPI_ISL_1740788, EPI_ISL_1740790, EPI_ISL_1740792, EPI_ISL_1740794, EPI_ISL_1740796, EPI_ISL_1740797, EPI_ISL_1740798, EPI_ISL_1740800, EPI_ISL_1740801, EPI_ISL_1740803, EPI_ISL_1740804, EPI_ISL_1740805, EPI_ISL_1740808, EPI_ISL_1740810, EPI_ISL_1740811, EPI_ISL_1740812, EPI_ISL_1740815, EPI_ISL_1740816, EPI_ISL_1740818, EPI_ISL_1740819, EPI_ISL_1740820, EPI_ISL_1740823, EPI_ISL_1740826, EPI_ISL_1740827, EPI_ISL_1740829, EPI_ISL_1740830, EPI_ISL_1740831, EPI_ISL_1740832, EPI_ISL_1740833, EPI_ISL_1740834, EPI_ISL_1740835, EPI_ISL_1740836, EPI_ISL_1740837, EPI_ISL_1740838, EPI_ISL_1740839, EPI_ISL_1740840, EPI_ISL_1740841, EPI_ISL_1740845, EPI_ISL_1740846, EPI_ISL_1740847, EPI_ISL_1740848, EPI_ISL_1740850, EPI_ISL_1740852, EPI_ISL_1740853, EPI_ISL_1740857, EPI_ISL_1740859, EPI_ISL_1740863, EPI_ISL_1740866, EPI_ISL_1740867, EPI_ISL_1740868, EPI_ISL_1740869, EPI_ISL_1740870, EPI_ISL_1740871, EPI_ISL_1740872, EPI_ISL_1740873, EPI_ISL_1740876, EPI_ISL_1740877, EPI_ISL_1740878, EPI_ISL_1740879, EPI_ISL_1740880, EPI_ISL_1740881, EPI_ISL_1740883, EPI_ISL_1740884, EPI_ISL_1740885, EPI_ISL_1740886, EPI_ISL_1740889, EPI_ISL_1740890, EPI_ISL_1740894, EPI_ISL_1740895, EPI_ISL_1740896, EPI_ISL_1740899, EPI_ISL_1740902, EPI_ISL_1740903, EPI_ISL_1740904, EPI_ISL_1740906, EPI_ISL_1740908, EPI_ISL_1740910, EPI_ISL_1740911, EPI_ISL_1740912, EPI_ISL_1740913, EPI_ISL_1740914, EPI_ISL_1740916, EPI_ISL_1740917, EPI_ISL_1740918, EPI_ISL_1740919, EPI_ISL_1740921, EPI_ISL_1740923, EPI_ISL_1740925, EPI_ISL_1740926, EPI_ISL_1740927, EPI_ISL_1740928, EPI_ISL_1740930, EPI_ISL_1740932, EPI_ISL_1740933, EPI_ISL_1740938, EPI_ISL_1740940, EPI_ISL_1740942, EPI_ISL_1740946, EPI_ISL_1740949, EPI_ISL_1740950, EPI_ISL_1740952, EPI_ISL_1740955, EPI_ISL_1740959, EPI_ISL_1740960, EPI_ISL_1740961, EPI_ISL_1740966, EPI_ISL_1740967, EPI_ISL_1740968, EPI_ISL_1740973, EPI_ISL_1740977, EPI_ISL_1740978, EPI_ISL_1740980, EPI_ISL_1740983, EPI_ISL_1740984, EPI_ISL_1740985, EPI_ISL_1740986, EPI_ISL_1740987, EPI_ISL_1740988, EPI_ISL_1740990, EPI_ISL_1740991, EPI_ISL_1740992, EPI_ISL_1740995, EPI_ISL_1740996, EPI_ISL_1740997, EPI_ISL_1741000, EPI_ISL_1741001, EPI_ISL_1741002, EPI_ISL_1741003, EPI_ISL_1741005, EPI_ISL_1741006, EPI_ISL_1741010, EPI_ISL_1741011, EPI_ISL_1741012, EPI_ISL_1741013, EPI_ISL_1741015, EPI_ISL_1741017, EPI_ISL_1741018, EPI_ISL_1741019, EPI_ISL_1741022, EPI_ISL_1741023, EPI_ISL_1741025, EPI_ISL_1741026, EPI_ISL_1741027, EPI_ISL_1741028, EPI_ISL_1741029, EPI_ISL_1741030, EPI_ISL_1741033, EPI_ISL_1741034, EPI_ISL_1741037, EPI_ISL_1741038, EPI_ISL_1741039, EPI_ISL_1741041, EPI_ISL_1741043, EPI_ISL_1741046, EPI_ISL_1741047, EPI_ISL_1741049, EPI_ISL_1741050, EPI_ISL_1741051, EPI_ISL_1741052, EPI_ISL_1741053, EPI_ISL_1741056, EPI_ISL_1741059, EPI_ISL_1741062, EPI_ISL_1741063, EPI_ISL_1741066, EPI_ISL_1741074, EPI_ISL_1741076, EPI_ISL_1741083, EPI_ISL_1741085, EPI_ISL_1741087, EPI_ISL_1741088, EPI_ISL_1741090, EPI_ISL_1741093, EPI_ISL_1741094, EPI_ISL_1741101, EPI_ISL_1741102, EPI_ISL_1741104, EPI_ISL_1741110, EPI_ISL_1741111, EPI_ISL_1741114, EPI_ISL_1741115, EPI_ISL_1741116, EPI_ISL_1741117, EPI_ISL_1741120, EPI_ISL_1741122, EPI_ISL_1741125, EPI_ISL_1741127, EPI_ISL_1741130, EPI_ISL_1741131, EPI_ISL_1741133, EPI_ISL_1741134, EPI_ISL_1741135, EPI_ISL_1741140, EPI_ISL_1741148, EPI_ISL_1741150, EPI_ISL_1741151, EPI_ISL_1741154, EPI_ISL_1741156, EPI_ISL_1741159, EPI_ISL_1741160, EPI_ISL_1741161, EPI_ISL_1741163, EPI_ISL_1741164, EPI_ISL_1741165, EPI_ISL_1741166, EPI_ISL_1741167, EPI_ISL_1741168, EPI_ISL_1741169, EPI_ISL_1741175, EPI_ISL_1741177, EPI_ISL_1741180, EPI_ISL_1741186, EPI_ISL_1741187, EPI_ISL_1741189, EPI_ISL_1741195, EPI_ISL_1741197, EPI_ISL_1741198, EPI_ISL_1741202, EPI_ISL_1741207, EPI_ISL_1741208, EPI_ISL_1741210, EPI_ISL_1741218, EPI_ISL_1741226, EPI_ISL_1741227, EPI_ISL_1741229, EPI_ISL_1741233, EPI_ISL_1741234, EPI_ISL_1741236, EPI_ISL_1741238, EPI_ISL_1741239, EPI_ISL_1741241, EPI_ISL_1741245, EPI_ISL_1741246, EPI_ISL_1741247, EPI_ISL_1741248, EPI_ISL_1741249, EPI_ISL_1741251, EPI_ISL_1741252, EPI_ISL_1741254, EPI_ISL_1741255, EPI_ISL_1741256, EPI_ISL_1741260, EPI_ISL_1741261, EPI_ISL_1741263, EPI_ISL_1741268, EPI_ISL_1741272, EPI_ISL_1741514, EPI_ISL_1741524, EPI_ISL_1741542, EPI_ISL_1741545, EPI_ISL_1741547, EPI_ISL_1741548, EPI_ISL_1741725, EPI_ISL_1741737, EPI_ISL_1741765, EPI_ISL_1742285, EPI_ISL_1742290, EPI_ISL_1758188, EPI_ISL_1758190, EPI_ISL_1758195, EPI_ISL_1758196, EPI_ISL_1758198, EPI_ISL_1758202, EPI_ISL_1758203, EPI_ISL_1758206, EPI_ISL_1758212, EPI_ISL_1758214, EPI_ISL_1758217, EPI_ISL_1758218, EPI_ISL_1758233, EPI_ISL_1758234, EPI_ISL_1758237, EPI_ISL_1758238, EPI_ISL_1758241, EPI_ISL_1758243, EPI_ISL_1758245, EPI_ISL_1758247, EPI_ISL_1758252, EPI_ISL_1758254, EPI_ISL_1758256, EPI_ISL_1758259, EPI_ISL_1758261, EPI_ISL_1758266, EPI_ISL_1758268, EPI_ISL_1758270, EPI_ISL_1758273, EPI_ISL_1758284, EPI_ISL_1758289, EPI_ISL_1758290, EPI_ISL_1758292, EPI_ISL_1758294, EPI_ISL_1758296, EPI_ISL_1758297, EPI_ISL_1758301, EPI_ISL_1758303, EPI_ISL_1758306, EPI_ISL_1758308, EPI_ISL_1758313, EPI_ISL_1758314, EPI_ISL_1758316, EPI_ISL_1758317, EPI_ISL_1758319, EPI_ISL_1758322, EPI_ISL_1758324, EPI_ISL_1758325, EPI_ISL_1758328, EPI_ISL_1758332, EPI_ISL_1758342, EPI_ISL_1758343, EPI_ISL_1758348, EPI_ISL_1758349, EPI_ISL_1758352, EPI_ISL_1758353, EPI_ISL_1758355, EPI_ISL_1758356, EPI_ISL_1758360, EPI_ISL_1758361, EPI_ISL_1758364, EPI_ISL_1758368, EPI_ISL_1758372, EPI_ISL_1758373, EPI_ISL_1758376, EPI_ISL_1758378, EPI_ISL_1758379, EPI_ISL_1758387, EPI_ISL_1758397, EPI_ISL_1758400, EPI_ISL_1758401, EPI_ISL_1758404, EPI_ISL_1758406, EPI_ISL_1758412, EPI_ISL_1758414, EPI_ISL_1758415, EPI_ISL_1758418, EPI_ISL_1758420, EPI_ISL_1758423, EPI_ISL_1758424, EPI_ISL_1758433, EPI_ISL_1758437, EPI_ISL_1758440, EPI_ISL_1758442, EPI_ISL_1758443, EPI_ISL_1758444, EPI_ISL_1758446, EPI_ISL_1758448, EPI_ISL_1758449, EPI_ISL_1758450, EPI_ISL_1758451, EPI_ISL_1758879, EPI_ISL_1758880, EPI_ISL_1758881, EPI_ISL_1758884, EPI_ISL_1758885, EPI_ISL_1758888, EPI_ISL_1758890, EPI_ISL_1758891, EPI_ISL_1758892, EPI_ISL_1758895, EPI_ISL_1758896, EPI_ISL_1758898, EPI_ISL_1758899, EPI_ISL_1758900, EPI_ISL_1758903, EPI_ISL_1758905, EPI_ISL_1758906, EPI_ISL_1758907, EPI_ISL_1758908, EPI_ISL_1758909, EPI_ISL_1758912, EPI_ISL_1758914, EPI_ISL_1758915, EPI_ISL_1758917, EPI_ISL_1758918, EPI_ISL_1758922, EPI_ISL_1758923, EPI_ISL_1758925, EPI_ISL_1758926, EPI_ISL_1758927, EPI_ISL_1758929, EPI_ISL_1758930, EPI_ISL_1758931, EPI_ISL_1758934, EPI_ISL_1758935, EPI_ISL_1758936, EPI_ISL_1758937, EPI_ISL_1758941, EPI_ISL_1758944, EPI_ISL_1758950, EPI_ISL_1758955, EPI_ISL_1758956, EPI_ISL_1758958, EPI_ISL_1758959, EPI_ISL_1758960, EPI_ISL_1758962, EPI_ISL_1758963, EPI_ISL_1758965, EPI_ISL_1758967, EPI_ISL_1758968, EPI_ISL_1758970, EPI_ISL_1758971, EPI_ISL_1758972, EPI_ISL_1758973, EPI_ISL_1758974, EPI_ISL_1758975, EPI_ISL_1758977, EPI_ISL_1758978, EPI_ISL_1758983, EPI_ISL_1758984, EPI_ISL_1758985, EPI_ISL_1758986, EPI_ISL_1758988, EPI_ISL_1758989, EPI_ISL_1758991, EPI_ISL_1758992, EPI_ISL_1758995, EPI_ISL_1758997, EPI_ISL_1758998, EPI_ISL_1759001, EPI_ISL_1759002, EPI_ISL_1759004, EPI_ISL_1759006, EPI_ISL_1759007, EPI_ISL_1759008, EPI_ISL_1759009, EPI_ISL_1759010, EPI_ISL_1759013, EPI_ISL_1759014, EPI_ISL_1759015, EPI_ISL_1759016, EPI_ISL_1759017, EPI_ISL_1759018, EPI_ISL_1759019, EPI_ISL_1759020, EPI_ISL_1759023, EPI_ISL_1759024, EPI_ISL_1759025, EPI_ISL_1759027, EPI_ISL_1759028, EPI_ISL_1759029, EPI_ISL_1759031, EPI_ISL_1759032, EPI_ISL_1759034, EPI_ISL_1759035, EPI_ISL_1759038, EPI_ISL_1759039, EPI_ISL_1759042, EPI_ISL_1759043, EPI_ISL_1759045, EPI_ISL_1759048, EPI_ISL_1759049, EPI_ISL_1759050, EPI_ISL_1759053, EPI_ISL_1759054, EPI_ISL_1759056, EPI_ISL_1759057, EPI_ISL_1759058, EPI_ISL_1759060, EPI_ISL_1759062, EPI_ISL_1759063, EPI_ISL_1759064, EPI_ISL_1759069, EPI_ISL_1759070, EPI_ISL_1759071, EPI_ISL_1759073, EPI_ISL_1759075, EPI_ISL_1759077, EPI_ISL_1759079, EPI_ISL_1759080, EPI_ISL_1759081, EPI_ISL_1759083, EPI_ISL_1759086, EPI_ISL_1759087, EPI_ISL_1759088, EPI_ISL_1759089, EPI_ISL_1759090, EPI_ISL_1759091, EPI_ISL_1759092, EPI_ISL_1759093, EPI_ISL_1759094, EPI_ISL_1759097, EPI_ISL_1759098, EPI_ISL_1759101, EPI_ISL_1759102, EPI_ISL_1759104, EPI_ISL_1759105, EPI_ISL_1759107, EPI_ISL_1759109, EPI_ISL_1759110, EPI_ISL_1759111, EPI_ISL_1759112, EPI_ISL_1759113, EPI_ISL_1759115, EPI_ISL_1759118, EPI_ISL_1759122, EPI_ISL_1759124, EPI_ISL_1759127, EPI_ISL_1759129, EPI_ISL_1759130, EPI_ISL_1759131, EPI_ISL_1759133, EPI_ISL_1759134, EPI_ISL_1759135, EPI_ISL_1759136, EPI_ISL_1759137, EPI_ISL_1759139, EPI_ISL_1759140, EPI_ISL_1759142, EPI_ISL_1759143, EPI_ISL_1759146, EPI_ISL_1759148, EPI_ISL_1759149, EPI_ISL_1759150, EPI_ISL_1759153, EPI_ISL_1759155, EPI_ISL_1759156, EPI_ISL_1759157, EPI_ISL_1759161, EPI_ISL_1759165, EPI_ISL_1759167, EPI_ISL_1759168, EPI_ISL_1759171, EPI_ISL_1759173, EPI_ISL_1759174, EPI_ISL_1759187, EPI_ISL_1759190, EPI_ISL_1759266, EPI_ISL_1759274, EPI_ISL_1759286, EPI_ISL_1759293, EPI_ISL_1759301, EPI_ISL_1759320, EPI_ISL_1759325, EPI_ISL_1759812, EPI_ISL_1759815, EPI_ISL_1759828, EPI_ISL_1759842, EPI_ISL_1759879, EPI_ISL_1790432, EPI_ISL_1790438, EPI_ISL_1790439, EPI_ISL_1790442, EPI_ISL_1790445, EPI_ISL_1790451, EPI_ISL_1790457, EPI_ISL_1790461, EPI_ISL_1790462, EPI_ISL_1790463, EPI_ISL_1790464, EPI_ISL_1790465, EPI_ISL_1790472, EPI_ISL_1790474, EPI_ISL_1790482, EPI_ISL_1790483, EPI_ISL_1790484, EPI_ISL_1790497, EPI_ISL_1790500, EPI_ISL_1790504, EPI_ISL_1790506, EPI_ISL_1790525, EPI_ISL_1790534, EPI_ISL_1790538, EPI_ISL_1790542, EPI_ISL_1790543, EPI_ISL_1790546, EPI_ISL_1790547, EPI_ISL_1790548, EPI_ISL_1790549, EPI_ISL_1790554, EPI_ISL_1790563, EPI_ISL_1790571, EPI_ISL_1790576, EPI_ISL_1790577, EPI_ISL_1790579, EPI_ISL_1790584, EPI_ISL_1790585, EPI_ISL_1790589, EPI_ISL_1790591, EPI_ISL_1790596, EPI_ISL_1790603, EPI_ISL_1790616, EPI_ISL_1790617, EPI_ISL_1790618, EPI_ISL_1790621, EPI_ISL_1790626, EPI_ISL_1790628, EPI_ISL_1790630, EPI_ISL_1790632, EPI_ISL_1790639, EPI_ISL_1790644, EPI_ISL_1790651, EPI_ISL_1790653, EPI_ISL_1790660, EPI_ISL_1790662, EPI_ISL_1790663, EPI_ISL_1790665, EPI_ISL_1790666, EPI_ISL_1790670, EPI_ISL_1790682, EPI_ISL_1790685, EPI_ISL_1790692, EPI_ISL_1790693, EPI_ISL_1790704, EPI_ISL_1790718, EPI_ISL_1790723, EPI_ISL_1790726, EPI_ISL_1790731, EPI_ISL_1790736, EPI_ISL_1790745, EPI_ISL_1790747, EPI_ISL_1790748, EPI_ISL_1790750, EPI_ISL_1790759, EPI_ISL_1790760, EPI_ISL_1790762, EPI_ISL_1790765, EPI_ISL_1790767, EPI_ISL_1790771, EPI_ISL_1790775, EPI_ISL_1790776, EPI_ISL_1790777, EPI_ISL_1790787, EPI_ISL_1790788, EPI_ISL_1790790, EPI_ISL_1790791, EPI_ISL_1790793, EPI_ISL_1790794, EPI_ISL_1790797, EPI_ISL_1790805, EPI_ISL_1790806, EPI_ISL_1790807, EPI_ISL_1790808, EPI_ISL_1790809, EPI_ISL_1790813, EPI_ISL_1790823, EPI_ISL_1790828, EPI_ISL_1790830, EPI_ISL_1790831, EPI_ISL_1790835, EPI_ISL_1790842, EPI_ISL_1790843, EPI_ISL_1790844, EPI_ISL_1790846, EPI_ISL_1790851, EPI_ISL_1790854, EPI_ISL_1790855, EPI_ISL_1790858, EPI_ISL_1790860, EPI_ISL_1790863, EPI_ISL_1790872, EPI_ISL_1790874, EPI_ISL_1790875, EPI_ISL_1790876, EPI_ISL_1790880, EPI_ISL_1790882, EPI_ISL_1790884, EPI_ISL_1790887, EPI_ISL_1790889, EPI_ISL_1790890, EPI_ISL_1790891, EPI_ISL_1790892, EPI_ISL_1790897, EPI_ISL_1790906, EPI_ISL_1790912, EPI_ISL_1790913, EPI_ISL_1790916, EPI_ISL_1790918, EPI_ISL_1790919, EPI_ISL_1790921, EPI_ISL_1790922, EPI_ISL_1790926, EPI_ISL_1790929, EPI_ISL_1790930, EPI_ISL_1790934, EPI_ISL_1790936, EPI_ISL_1790937, EPI_ISL_1790938, EPI_ISL_1790939, EPI_ISL_1790941, EPI_ISL_1790943, EPI_ISL_1790945, EPI_ISL_1790946, EPI_ISL_1790947, EPI_ISL_1790951, EPI_ISL_1790953, EPI_ISL_1790955, EPI_ISL_1790965, EPI_ISL_1790966, EPI_ISL_1790968, EPI_ISL_1790969, EPI_ISL_1790970, EPI_ISL_1790971, EPI_ISL_1790973, EPI_ISL_1790975, EPI_ISL_1790978, EPI_ISL_1790982, EPI_ISL_1790986, EPI_ISL_1790987, EPI_ISL_1790988, EPI_ISL_1790990, EPI_ISL_1790992, EPI_ISL_1790995, EPI_ISL_1791001, EPI_ISL_1791008, | | | |

EPI_ISL_1791010, EPI_ISL_1791011, EPI_ISL_1791014, EPI_ISL_1791016, EPI_ISL_1791017, EPI_ISL_1791021, EPI_ISL_1791022, EPI_ISL_1805964, EPI_ISL_1805965, EPI_ISL_1805969, EPI_ISL_1805970, EPI_ISL_1805974, EPI_ISL_1805979, EPI_ISL_1805982, EPI_ISL_1805986, EPI_ISL_1805988, EPI_ISL_1805993, EPI_ISL_1805994, EPI_ISL_1805995, EPI_ISL_1805997, EPI_ISL_1805999, EPI_ISL_1806001, EPI_ISL_1806004, EPI_ISL_1806007, EPI_ISL_1806008, EPI_ISL_1806010, EPI_ISL_1806013, EPI_ISL_1806017, EPI_ISL_1806019, EPI_ISL_1806021, EPI_ISL_1806027, EPI_ISL_1806033, EPI_ISL_1806034, EPI_ISL_1806038, EPI_ISL_1806043, EPI_ISL_1806045, EPI_ISL_1806046, EPI_ISL_1806048, EPI_ISL_1806051, EPI_ISL_1806052, EPI_ISL_1806053, EPI_ISL_1806063, EPI_ISL_1806064, EPI_ISL_1806069, EPI_ISL_1806073, EPI_ISL_1806075, EPI_ISL_1806076, EPI_ISL_1806080, EPI_ISL_1806082, EPI_ISL_1806083, EPI_ISL_1806087, EPI_ISL_1806089, EPI_ISL_1806093, EPI_ISL_1806095, EPI_ISL_1806096, EPI_ISL_1806100, EPI_ISL_1806102, EPI_ISL_1806104, EPI_ISL_1806106, EPI_ISL_1806108, EPI_ISL_1806111, EPI_ISL_1806112, EPI_ISL_1806119, EPI_ISL_1806121, EPI_ISL_1806122, EPI_ISL_1806123, EPI_ISL_1806124, EPI_ISL_1806125, EPI_ISL_1806131, EPI_ISL_1806142, EPI_ISL_1806143, EPI_ISL_1806144, EPI_ISL_1806149, EPI_ISL_1806152, EPI_ISL_1806153, EPI_ISL_1806156, EPI_ISL_1806157, EPI_ISL_1806158, EPI_ISL_1806161, EPI_ISL_1806167, EPI_ISL_1806169, EPI_ISL_1806170, EPI_ISL_1806171, EPI_ISL_1806172, EPI_ISL_1806174, EPI_ISL_1806175, EPI_ISL_1806176, EPI_ISL_1806189, EPI_ISL_1806195, EPI_ISL_1806202, EPI_ISL_1806205, EPI_ISL_1806504, EPI_ISL_1806505, EPI_ISL_1806507, EPI_ISL_1806509, EPI_ISL_1806510, EPI_ISL_1806511, EPI_ISL_1806512, EPI_ISL_1806513, EPI_ISL_1806514, EPI_ISL_1806515, EPI_ISL_1806516, EPI_ISL_1806517, EPI_ISL_1806518, EPI_ISL_1806519, EPI_ISL_1806520, EPI_ISL_1806521, EPI_ISL_1806522, EPI_ISL_1806523, EPI_ISL_1806525, EPI_ISL_1806527, EPI_ISL_1806528, EPI_ISL_1806529, EPI_ISL_1806530, EPI_ISL_1806531, EPI_ISL_1806532, EPI_ISL_1806533, EPI_ISL_1806534, EPI_ISL_1806535, EPI_ISL_1806537, EPI_ISL_1806538, EPI_ISL_1806539, EPI_ISL_1806540, EPI_ISL_1806541, EPI_ISL_1806542, EPI_ISL_1806543, EPI_ISL_1806544, EPI_ISL_1806545, EPI_ISL_1806546, EPI_ISL_1806547, EPI_ISL_1806548, EPI_ISL_1806549, EPI_ISL_1806550, EPI_ISL_1806551, EPI_ISL_1806552, EPI_ISL_1806553, EPI_ISL_1806554, EPI_ISL_1806555, EPI_ISL_1806556, EPI_ISL_1806557, EPI_ISL_1806559, EPI_ISL_1806560, EPI_ISL_1806561, EPI_ISL_1806562, EPI_ISL_1806563, EPI_ISL_1806564, EPI_ISL_1806565, EPI_ISL_1806567, EPI_ISL_1806568, EPI_ISL_1806570, EPI_ISL_1806571, EPI_ISL_1806572, EPI_ISL_1806573, EPI_ISL_1806574, EPI_ISL_1806575, EPI_ISL_1806576, EPI_ISL_1806577, EPI_ISL_1806578, EPI_ISL_1806579, EPI_ISL_1806580, EPI_ISL_1806581, EPI_ISL_1806582, EPI_ISL_1806583, EPI_ISL_1806584, EPI_ISL_1806585, EPI_ISL_1806586, EPI_ISL_1806588, EPI_ISL_1806589, EPI_ISL_1806590, EPI_ISL_1806591, EPI_ISL_1806593, EPI_ISL_1806594, EPI_ISL_1806595, EPI_ISL_1806597, EPI_ISL_1806598, EPI_ISL_1806599, EPI_ISL_1806600, EPI_ISL_1806602, EPI_ISL_1806603, EPI_ISL_1806604, EPI_ISL_1806605, EPI_ISL_1806607, EPI_ISL_1806608, EPI_ISL_1806609, EPI_ISL_1806610, EPI_ISL_1806612, EPI_ISL_1806613, EPI_ISL_1806614, EPI_ISL_1806615, EPI_ISL_1806616, EPI_ISL_1806617, EPI_ISL_1806618, EPI_ISL_1806619, EPI_ISL_1806620, EPI_ISL_1806622, EPI_ISL_1806623, EPI_ISL_1806624, EPI_ISL_1806625, EPI_ISL_1806626, EPI_ISL_1806627, EPI_ISL_1806628, EPI_ISL_1806629, EPI_ISL_1806630, EPI_ISL_1806633, EPI_ISL_1806634, EPI_ISL_1806635, EPI_ISL_1806637, EPI_ISL_1806638, EPI_ISL_1806640, EPI_ISL_1806641, EPI_ISL_1806644, EPI_ISL_1806645, EPI_ISL_1806647, EPI_ISL_1806648, EPI_ISL_1806649, EPI_ISL_1806650, EPI_ISL_1806651, EPI_ISL_1806654, EPI_ISL_1806655, EPI_ISL_1806656, EPI_ISL_1806657, EPI_ISL_1806658, EPI_ISL_1806659, EPI_ISL_1806660, EPI_ISL_1806661, EPI_ISL_1806662, EPI_ISL_1806663, EPI_ISL_1806664, EPI_ISL_1806665, EPI_ISL_1806666, EPI_ISL_1806668, EPI_ISL_1806669, EPI_ISL_1806670, EPI_ISL_1806671, EPI_ISL_1806672, EPI_ISL_1806676, EPI_ISL_1806677, EPI_ISL_1806678, EPI_ISL_1806679, EPI_ISL_1806680, EPI_ISL_1806682, EPI_ISL_1806683, EPI_ISL_1806684, EPI_ISL_1806685, EPI_ISL_1806686, EPI_ISL_1806688, EPI_ISL_1806689, EPI_ISL_1806690, EPI_ISL_1806691, EPI_ISL_1806692, EPI_ISL_1806693, EPI_ISL_1806694, EPI_ISL_1806695, EPI_ISL_1806696, EPI_ISL_1806697, EPI_ISL_1806698, EPI_ISL_1806699, EPI_ISL_1806700, EPI_ISL_1806701, EPI_ISL_1806702, EPI_ISL_1806703, EPI_ISL_1806704, EPI_ISL_1806705, EPI_ISL_1806706, EPI_ISL_1806707, EPI_ISL_1806708, EPI_ISL_1806710, EPI_ISL_1806711, EPI_ISL_1806712, EPI_ISL_1806713, EPI_ISL_1806714, EPI_ISL_1806715, EPI_ISL_1806716, EPI_ISL_1806717, EPI_ISL_1806718, EPI_ISL_1806719, EPI_ISL_1806721, EPI_ISL_1806722, EPI_ISL_1806724, EPI_ISL_1806726, EPI_ISL_1806727, EPI_ISL_1806728, EPI_ISL_1806729, EPI_ISL_1806730, EPI_ISL_1806731, EPI_ISL_1806732, EPI_ISL_1806733, EPI_ISL_1806734, EPI_ISL_1806736, EPI_ISL_1806737, EPI_ISL_1806738, EPI_ISL_1806739, EPI_ISL_1806740, EPI_ISL_1806742, EPI_ISL_1806743, EPI_ISL_1806744, EPI_ISL_1806745, EPI_ISL_1806746, EPI_ISL_1806747, EPI_ISL_1806749, EPI_ISL_1806750, EPI_ISL_1806751, EPI_ISL_1806752, EPI_ISL_1806754, EPI_ISL_1806755, EPI_ISL_1806756, EPI_ISL_1806757, EPI_ISL_1806758, EPI_ISL_1806759, EPI_ISL_1806760, EPI_ISL_1806762, EPI_ISL_1806763, EPI_ISL_1806764, EPI_ISL_1806765, EPI_ISL_1806766, EPI_ISL_1806768, EPI_ISL_1806769, EPI_ISL_1806770, EPI_ISL_1806771, EPI_ISL_1806773, EPI_ISL_1806774, EPI_ISL_1806775, EPI_ISL_1806776, EPI_ISL_1806778, EPI_ISL_1806779, EPI_ISL_1806780, EPI_ISL_1806781, EPI_ISL_1806782, EPI_ISL_1806783, EPI_ISL_1806784, EPI_ISL_1806785, EPI_ISL_1806786, EPI_ISL_1806787, EPI_ISL_1806788, EPI_ISL_1806789, EPI_ISL_1806790, EPI_ISL_1806791, EPI_ISL_1806793, EPI_ISL_1806794, EPI_ISL_1806795, EPI_ISL_1806796, EPI_ISL_1806797, EPI_ISL_1806799, EPI_ISL_1806800, EPI_ISL_1806801, EPI_ISL_1806803, EPI_ISL_1806804, EPI_ISL_1806805, EPI_ISL_1806807, EPI_ISL_1806809, EPI_ISL_1806810, EPI_ISL_1806811, EPI_ISL_1806812, EPI_ISL_1806813, EPI_ISL_1806814, EPI_ISL_1806815, EPI_ISL_1806816, EPI_ISL_1806817, EPI_ISL_1806818, EPI_ISL_1806819, EPI_ISL_1806820, EPI_ISL_1806822, EPI_ISL_1806823, EPI_ISL_1806824, EPI_ISL_1806825, EPI_ISL_1806826, EPI_ISL_1806827, EPI_ISL_1806829, EPI_ISL_1806831, EPI_ISL_1806833, EPI_ISL_1806834, EPI_ISL_1806836, EPI_ISL_1806837, EPI_ISL_1806838, EPI_ISL_1806839, EPI_ISL_1806841, EPI_ISL_1806842, EPI_ISL_1806843, EPI_ISL_1806844, EPI_ISL_1806845, EPI_ISL_1806846, EPI_ISL_1806849, EPI_ISL_1806850, EPI_ISL_1806851, EPI_ISL_1806852, EPI_ISL_1806853, EPI_ISL_1806898, EPI_ISL_1807081, EPI_ISL_1829828, EPI_ISL_1829835, EPI_ISL_1829865, EPI_ISL_1829872, EPI_ISL_1829899, EPI_ISL_1829916, EPI_ISL_1829934, EPI_ISL_1829946, EPI_ISL_1830026, EPI_ISL_1830048, EPI_ISL_1830092, EPI_ISL_1830101, EPI_ISL_1830109, EPI_ISL_1830110, EPI_ISL_1830114, EPI_ISL_1830121, EPI_ISL_1830122, EPI_ISL_1830125, EPI_ISL_1830126, EPI_ISL_1830128, EPI_ISL_1830133, EPI_ISL_1830137, EPI_ISL_1830140, EPI_ISL_1830150, EPI_ISL_1830154, EPI_ISL_1830158, EPI_ISL_1830159, EPI_ISL_1830166, EPI_ISL_1830167, EPI_ISL_1830170, EPI_ISL_1830171, EPI_ISL_1830173, EPI_ISL_1830174, EPI_ISL_1830175, EPI_ISL_1830176, EPI_ISL_1830181, EPI_ISL_1830182, EPI_ISL_1830183, EPI_ISL_1830184, EPI_ISL_1830185, EPI_ISL_1830188, EPI_ISL_1830192, EPI_ISL_1830195, EPI_ISL_1830203, EPI_ISL_1830208, EPI_ISL_1830214, EPI_ISL_1830217, EPI_ISL_1830225, EPI_ISL_1830234, EPI_ISL_1830237, EPI_ISL_1830241, EPI_ISL_1830251, EPI_ISL_1830252, EPI_ISL_1830255, EPI_ISL_1830259, EPI_ISL_1830271, EPI_ISL_1830298, EPI_ISL_1830300, EPI_ISL_1830303, EPI_ISL_1830306, EPI_ISL_1830308, EPI_ISL_1830312, EPI_ISL_1830315, EPI_ISL_1830316, EPI_ISL_1830319, EPI_ISL_1830325, EPI_ISL_1830326, EPI_ISL_1830330, EPI_ISL_1830338, EPI_ISL_1830339, EPI_ISL_1830340, EPI_ISL_1830347, EPI_ISL_1830350, EPI_ISL_1830353, EPI_ISL_1830356, EPI_ISL_1830357, EPI_ISL_1830360, EPI_ISL_1830361, EPI_ISL_1830364, EPI_ISL_1830365, EPI_ISL_1830368, EPI_ISL_1830383, EPI_ISL_1830388, EPI_ISL_1830391, EPI_ISL_1830397, EPI_ISL_1830398, EPI_ISL_1830402, EPI_ISL_1830422, EPI_ISL_1830464, EPI_ISL_1830490, EPI_ISL_1830505, EPI_ISL_1830539, EPI_ISL_1830552, EPI_ISL_1830590, EPI_ISL_1830598, EPI_ISL_1830613, EPI_ISL_1830658, EPI_ISL_1830690, EPI_ISL_1830731, EPI_ISL_1830735, EPI_ISL_1830748, EPI_ISL_1830758, EPI_ISL_1830769, EPI_ISL_1830772, EPI_ISL_1830791, EPI_ISL_1830805, EPI_ISL_1830806, EPI_ISL_1830807, EPI_ISL_1830811, EPI_ISL_1830817, EPI_ISL_1830819, EPI_ISL_1830823, EPI_ISL_1830828, EPI_ISL_1830836, EPI_ISL_1830840, EPI_ISL_1830844, EPI_ISL_1830846, EPI_ISL_1830848, EPI_ISL_1830850, EPI_ISL_1830856, EPI_ISL_1830868, EPI_ISL_1830869, EPI_ISL_1830870, EPI_ISL_1830878, EPI_ISL_1830883, EPI_ISL_1830896, EPI_ISL_1830911, EPI_ISL_1830914, EPI_ISL_1830944, EPI_ISL_1830947, EPI_ISL_1830949, EPI_ISL_1830956, EPI_ISL_1830959, EPI_ISL_1830966, EPI_ISL_1830968, EPI_ISL_1830970, EPI_ISL_1830979, EPI_ISL_1830983, EPI_ISL_1831001, EPI_ISL_1831005, EPI_ISL_1831006, EPI_ISL_1831008, EPI_ISL_1831011, EPI_ISL_1831015, EPI_ISL_1831023, EPI_ISL_1831027, EPI_ISL_1831029, EPI_ISL_1831039, EPI_ISL_1831045, EPI_ISL_1831048, EPI_ISL_1831049, EPI_ISL_1831052, EPI_ISL_1831054, EPI_ISL_1831055, EPI_ISL_1831057, EPI_ISL_1831060, EPI_ISL_1831061, EPI_ISL_1831070, EPI_ISL_1831074, EPI_ISL_1831079, EPI_ISL_1831080, EPI_ISL_1831082, EPI_ISL_1831084, EPI_ISL_1831263, EPI_ISL_1831264, EPI_ISL_1831265, EPI_ISL_1831266, EPI_ISL_1831267, EPI_ISL_1831268, EPI_ISL_1858060, EPI_ISL_1912481, EPI_ISL_1912491, EPI_ISL_1912506, EPI_ISL_1912545, EPI_ISL_1912582, EPI_ISL_1912785, EPI_ISL_1912786, EPI_ISL_1912787, EPI_ISL_1912788, EPI_ISL_1912789, EPI_ISL_1985195, EPI_ISL_1985199, EPI_ISL_1985208, EPI_ISL_1985209, EPI_ISL_1985217, EPI_ISL_1985219, EPI_ISL_1985221, EPI_ISL_1985222, EPI_ISL_1985229, EPI_ISL_1985236, EPI_ISL_1985237, EPI_ISL_1985240, EPI_ISL_1985246, EPI_ISL_1985249, EPI_ISL_1985250, EPI_ISL_1985255, EPI_ISL_1985257, EPI_ISL_1985258, EPI_ISL_1985259, EPI_ISL_1985263, EPI_ISL_1985269, EPI_ISL_1985272, EPI_ISL_1985280, EPI_ISL_1985291, EPI_ISL_1985293, EPI_ISL_1985301, EPI_ISL_1985306, EPI_ISL_1985307, EPI_ISL_1985309, EPI_ISL_1985311, EPI_ISL_1985314, EPI_ISL_1985318, EPI_ISL_1985320, EPI_ISL_1985323, EPI_ISL_1985324, EPI_ISL_1985336, EPI_ISL_1985337, EPI_ISL_1985338, EPI_ISL_1985340, EPI_ISL_1985342, EPI_ISL_1985347, EPI_ISL_1985348, EPI_ISL_1985349, EPI_ISL_1985354, EPI_ISL_1985357, EPI_ISL_1985359, EPI_ISL_1985360, EPI_ISL_1985362, EPI_ISL_1985364, EPI_ISL_1985370, EPI_ISL_1985373, EPI_ISL_1985383, EPI_ISL_1985384, EPI_ISL_1985387, EPI_ISL_1985389, EPI_ISL_1985390, EPI_ISL_1985394, EPI_ISL_1985396, EPI_ISL_1985398, EPI_ISL_1985401, EPI_ISL_1985402, EPI_ISL_1985411, EPI_ISL_1985413, EPI_ISL_1985414, EPI_ISL_1985419, EPI_ISL_1985420, EPI_ISL_1985421, EPI_ISL_1985427, EPI_ISL_1985429, EPI_ISL_1985431, EPI_ISL_1985436, EPI_ISL_1985437, EPI_ISL_1985454, EPI_ISL_1985461, EPI_ISL_1985462, EPI_ISL_1985470, EPI_ISL_1985473, EPI_ISL_1985474, EPI_ISL_1985477, EPI_ISL_1985479, EPI_ISL_1985488, EPI_ISL_1985491, EPI_ISL_1985494, EPI_ISL_1985495, EPI_ISL_1985499, EPI_ISL_1985503, EPI_ISL_1985504, EPI_ISL_2005614, EPI_ISL_2005620, EPI_ISL_2005622, EPI_ISL_2005629, EPI_ISL_2005630, EPI_ISL_2005632, EPI_ISL_2005633, EPI_ISL_2005635, EPI_ISL_2005637, EPI_ISL_2005639, EPI_ISL_2005640, EPI_ISL_2005641, EPI_ISL_2005643, EPI_ISL_2005655, EPI_ISL_2005657, EPI_ISL_2005661, EPI_ISL_2005668, EPI_ISL_2005682, EPI_ISL_2005684, EPI_ISL_2005691, EPI_ISL_2005692, EPI_ISL_2005694, EPI_ISL_2005697, EPI_ISL_2005698, EPI_ISL_2005702, EPI_ISL_2005707, EPI_ISL_2005709, EPI_ISL_2005710, EPI_ISL_2005711, EPI_ISL_2005722, EPI_ISL_2005728, EPI_ISL_2005730, EPI_ISL_2005741, EPI_ISL_2005743, EPI_ISL_2005746, EPI_ISL_2005747, EPI_ISL_2005748, EPI_ISL_2005749, EPI_ISL_2005756, EPI_ISL_2005760, EPI_ISL_2005771, EPI_ISL_2005785, EPI_ISL_2005786, EPI_ISL_2005797, EPI_ISL_2006072, EPI_ISL_2006074, EPI_ISL_2006075, EPI_ISL_2006078, EPI_ISL_2006080, EPI_ISL_2006090, EPI_ISL_2006096, EPI_ISL_2006102, EPI_ISL_2006105, EPI_ISL_2006112, EPI_ISL_2006113, EPI_ISL_2006114, EPI_ISL_2006120, EPI_ISL_2006121, EPI_ISL_2006124, EPI_ISL_2006126, EPI_ISL_2006128, EPI_ISL_2006132, EPI_ISL_2006134, EPI_ISL_2006135, EPI_ISL_2006138, EPI_ISL_2006139, EPI_ISL_2006149, EPI_ISL_2006150, EPI_ISL_2006153, EPI_ISL_2006159, EPI_ISL_2006163, EPI_ISL_2006166, EPI_ISL_2006167, EPI_ISL_2006169, EPI_ISL_2006176, EPI_ISL_2006185, EPI_ISL_2006187, EPI_ISL_2006188, EPI_ISL_2006191, EPI_ISL_2006192, EPI_ISL_2006197, EPI_ISL_2006200, EPI_ISL_2006203, EPI_ISL_2006206, EPI_ISL_2006208, EPI_ISL_2006209, EPI_ISL_2006210, EPI_ISL_2006214, EPI_ISL_2006216, EPI_ISL_2006220, EPI_ISL_2006221, EPI_ISL_2006228, EPI_ISL_2006234, EPI_ISL_2006236, EPI_ISL_2006242, EPI_ISL_2006247, EPI_ISL_2006248, EPI_ISL_2006251, EPI_ISL_2006252, EPI_ISL_2006255, EPI_ISL_2006257, EPI_ISL_2006259, EPI_ISL_2006260, EPI_ISL_2006261, EPI_ISL_2006263, EPI_ISL_2006268, EPI_ISL_2006269, EPI_ISL_2006270, EPI_ISL_2006272, EPI_ISL_2006274, EPI_ISL_2006278, EPI_ISL_2006282, EPI_ISL_2006289, EPI_ISL_2006291, EPI_ISL_2006294, EPI_ISL_2006536, EPI_ISL_2022645, EPI_ISL_2022667, EPI_ISL_2080485, EPI_ISL_2172092, EPI_ISL_2172112, EPI_ISL_2172148, EPI_ISL_2172170, EPI_ISL_2172242, EPI_ISL_2172286, EPI_ISL_2172293, EPI_ISL_2172301, EPI_ISL_2172335, EPI_ISL_2172389

| | | | |
|---|---|---|---|
| see above | Lighthouse Lab in Alderley Park | Wellcome Sanger Institute for the COVID-19 Genomics UK (COG-UK) Consortium | Cordelia Langford; David K. Jackson; Dominic Kwiatkowski; Ewan Harrison; Ian Johnston; Jacquelyn Wynn; Jeffrey Barrett; John Sillitoe on behalf of the Wellcome Sanger Institute COVID-19 Surveillance Team; Mairead Hyland; Roberto Amato; Sonia Goncalves; The Lighthouse Lab in Alderley Park and Alex Alderton |

EPI_ISL_1652951, EPI_ISL_1652973, EPI_ISL_1652991, EPI_ISL_1653012, EPI_ISL_1653022, EPI_ISL_1653052, EPI_ISL_1653053, EPI_ISL_1653062, EPI_ISL_1653080, EPI_ISL_1653138, EPI_ISL_1653149, EPI_ISL_1697554, EPI_ISL_1697719, EPI_ISL_1697773, EPI_ISL_1697818, EPI_ISL_1697824, EPI_ISL_1697826, EPI_ISL_1697848, EPI_ISL_1697855, EPI_ISL_1697858, EPI_ISL_1697868, EPI_ISL_1697870, EPI_ISL_1697911, EPI_ISL_1697922, EPI_ISL_1697967, EPI_ISL_1698258, EPI_ISL_1698260, EPI_ISL_1698267, EPI_ISL_1698268, EPI_ISL_1698273, EPI_ISL_1698274, EPI_ISL_1698279, EPI_ISL_1698280, EPI_ISL_1698284, EPI_ISL_1698285, EPI_ISL_1698295, EPI_ISL_1698305, EPI_ISL_1698307, EPI_ISL_1698317, EPI_ISL_1698327, EPI_ISL_1698328, EPI_ISL_1698332, EPI_ISL_1698339, EPI_ISL_1698342, EPI_ISL_1698346, EPI_ISL_1698349, EPI_ISL_1698357, EPI_ISL_1698359, EPI_ISL_1698366, EPI_ISL_1698373, EPI_ISL_1698379, EPI_ISL_1698383, EPI_ISL_1698384, EPI_ISL_1698387, EPI_ISL_1698391, EPI_ISL_1698393, EPI_ISL_1698398, EPI_ISL_1698403, EPI_ISL_1698406, EPI_ISL_1698407, EPI_ISL_1698417, EPI_ISL_1698418, EPI_ISL_1698426, EPI_ISL_1698428, EPI_ISL_1698429, EPI_ISL_1698430, EPI_ISL_1698432, EPI_ISL_1698435, EPI_ISL_1698436, EPI_ISL_1698442, EPI_ISL_1698444, EPI_ISL_1698450, EPI_ISL_1698452, EPI_ISL_1698459, EPI_ISL_1698465, EPI_ISL_1698469, EPI_ISL_1698475, EPI_ISL_1698476, EPI_ISL_1698479, EPI_ISL_1698482, EPI_ISL_1698487, EPI_ISL_1698490, EPI_ISL_1698492, EPI_ISL_1698504, EPI_ISL_1698505, EPI_ISL_1698506, EPI_ISL_1698511, EPI_ISL_1698521, EPI_ISL_1698524, EPI_ISL_1698526, EPI_ISL_1698527, EPI_ISL_1698532, EPI_ISL_1698533, EPI_ISL_1698537, EPI_ISL_1698539, EPI_ISL_1718897, EPI_ISL_1718904, EPI_ISL_1718912, EPI_ISL_1718913, EPI_ISL_1718921, EPI_ISL_1718927, EPI_ISL_1718928, EPI_ISL_1718929, EPI_ISL_1718930, EPI_ISL_1718931, EPI_ISL_1718932, EPI_ISL_1718943, EPI_ISL_1718947, EPI_ISL_1718959, EPI_ISL_1718968, EPI_ISL_1718971, EPI_ISL_1718973, EPI_ISL_1718979, EPI_ISL_1718980, EPI_ISL_1718981, EPI_ISL_1718989, EPI_ISL_1718991, EPI_ISL_1718992, EPI_ISL_1718997, EPI_ISL_1719004, EPI_ISL_1719007, EPI_ISL_1719010, EPI_ISL_1719011, EPI_ISL_1719013, EPI_ISL_1719017, EPI_ISL_1719022, EPI_ISL_1719026, EPI_ISL_1719027, EPI_ISL_1719033, EPI_ISL_1719036, EPI_ISL_1719042, EPI_ISL_1719045, EPI_ISL_1719048, EPI_ISL_1719051, EPI_ISL_1719062, EPI_ISL_1719065, EPI_ISL_1719066, EPI_ISL_1719071, EPI_ISL_1719084, EPI_ISL_1719088, EPI_ISL_1719089, EPI_ISL_1719099, EPI_ISL_1719107, EPI_ISL_1719115, EPI_ISL_1719118, EPI_ISL_1719120, EPI_ISL_1719127, EPI_ISL_1719128, EPI_ISL_1719134, EPI_ISL_1719142, EPI_ISL_1719145, EPI_ISL_1719150, EPI_ISL_1719157, EPI_ISL_1719159, EPI_ISL_1719167, EPI_ISL_1719168, EPI_ISL_1719175, EPI_ISL_1719177, EPI_ISL_1719191, EPI_ISL_1719195, EPI_ISL_1719196, EPI_ISL_1719214, EPI_ISL_1719216, EPI_ISL_1719434, EPI_ISL_1719435, EPI_ISL_1719438, EPI_ISL_1719441, EPI_ISL_1719444, EPI_ISL_1719445, EPI_ISL_1719447, EPI_ISL_1719452, EPI_ISL_1719458, EPI_ISL_1719460, EPI_ISL_1719466, EPI_ISL_1719467, EPI_ISL_1719468, EPI_ISL_1719479, EPI_ISL_1719482, EPI_ISL_1719484, EPI_ISL_1719485, EPI_ISL_1719486, EPI_ISL_1719493, EPI_ISL_1719499, EPI_ISL_1719503, EPI_ISL_1719504, EPI_ISL_1719508, EPI_ISL_1719515, EPI_ISL_1719517, EPI_ISL_1719518, EPI_ISL_1719521, EPI_ISL_1719522, EPI_ISL_1719524, EPI_ISL_1719527, EPI_ISL_1719528, EPI_ISL_1719529, EPI_ISL_1719534, EPI_ISL_1719536, EPI_ISL_1719539, EPI_ISL_1719543, EPI_ISL_1719546, EPI_ISL_1719551, EPI_ISL_1719561, EPI_ISL_1719570, EPI_ISL_1719575, EPI_ISL_1719583, EPI_ISL_1719584, EPI_ISL_1719585, EPI_ISL_1719587, EPI_ISL_1719594, EPI_ISL_1719596, EPI_ISL_1719607, EPI_ISL_1719618, EPI_ISL_1719623, EPI_ISL_1719629, EPI_ISL_1719630, EPI_ISL_1719636, EPI_ISL_1719640, EPI_ISL_1719642, EPI_ISL_1719648, EPI_ISL_1719651, EPI_ISL_1719655, EPI_ISL_1719658, EPI_ISL_1719660, EPI_ISL_1719668, EPI_ISL_1719676, EPI_ISL_1719680, EPI_ISL_1719684, EPI_ISL_1719687, EPI_ISL_1719690, EPI_ISL_1719692, EPI_ISL_1719696, EPI_ISL_1719703, EPI_ISL_1719709, EPI_ISL_1719719, EPI_ISL_1719725, EPI_ISL_1719729, EPI_ISL_1719740, EPI_ISL_1719745, EPI_ISL_1719746, EPI_ISL_1740534, EPI_ISL_1740541, EPI_ISL_1740550, EPI_ISL_1740565, EPI_ISL_1740580, EPI_ISL_1740618, EPI_ISL_1740643, EPI_ISL_1740658, EPI_ISL_1740672, EPI_ISL_1740687, EPI_ISL_1740689, EPI_ISL_1740698, EPI_ISL_1740699, EPI_ISL_1740713, EPI_ISL_1740729, EPI_ISL_1740734, EPI_ISL_1740737, EPI_ISL_1740741, EPI_ISL_1740758, EPI_ISL_1740768, EPI_ISL_1740779, EPI_ISL_1740780, EPI_ISL_1740781, EPI_ISL_1741274, EPI_ISL_1741283, EPI_ISL_1741336, EPI_ISL_1741358, EPI_ISL_1741363, EPI_ISL_1741390, EPI_ISL_1741405, EPI_ISL_1741412, EPI_ISL_1741427, EPI_ISL_1741433, EPI_ISL_1741440, EPI_ISL_1741446, EPI_ISL_1741550, EPI_ISL_1741551, EPI_ISL_1741553, EPI_ISL_1741755, EPI_ISL_1742215, EPI_ISL_1742284, EPI_ISL_1742286, EPI_ISL_1742287, EPI_ISL_1742289, EPI_ISL_1742291, EPI_ISL_1742293, EPI_ISL_1758455, EPI_ISL_1758463, EPI_ISL_1758509, EPI_ISL_1758530, EPI_ISL_1758532, EPI_ISL_1758536, EPI_ISL_1758547, EPI_ISL_1758558, EPI_ISL_1758625, EPI_ISL_1758645, EPI_ISL_1759180, EPI_ISL_1759202, EPI_ISL_1759215, EPI_ISL_1759219, EPI_ISL_1759242, EPI_ISL_1759257, EPI_ISL_1759265, EPI_ISL_1759267, EPI_ISL_1759268, EPI_ISL_1759290, EPI_ISL_1759295, EPI_ISL_1759306, EPI_ISL_1759307, EPI_ISL_1759308, EPI_ISL_1759340, EPI_ISL_1759346, EPI_ISL_1759350, EPI_ISL_1759823, EPI_ISL_1759824, EPI_ISL_1759827, EPI_ISL_1759832, EPI_ISL_1759844, EPI_ISL_1759850, EPI_ISL_1759863, EPI_ISL_1759864, EPI_ISL_1759884, EPI_ISL_1759888, EPI_ISL_1759894, EPI_ISL_1791020, EPI_ISL_1807055, EPI_ISL_1807068, EPI_ISL_1807155, EPI_ISL_1807184, EPI_ISL_1912016, EPI_ISL_1912019, EPI_ISL_1912026, EPI_ISL_1912027, EPI_ISL_1912028, EPI_ISL_1912029, EPI_ISL_1912036, EPI_ISL_1912038, EPI_ISL_1912041, EPI_ISL_1912049, EPI_ISL_1912064, EPI_ISL_1912066, EPI_ISL_1912074, EPI_ISL_1912076, EPI_ISL_1912078, EPI_ISL_1912085, EPI_ISL_1912086, EPI_ISL_1912088, EPI_ISL_1912091, EPI_ISL_1912093, EPI_ISL_1912111, EPI_ISL_1912114, EPI_ISL_1912119, EPI_ISL_1912124, EPI_ISL_1912157, EPI_ISL_1912173, EPI_ISL_1912174, EPI_ISL_1912175, EPI_ISL_1912203, EPI_ISL_1912205, EPI_ISL_1912218, EPI_ISL_1912237, EPI_ISL_1912251, EPI_ISL_1912253, EPI_ISL_1912267, EPI_ISL_1912300, EPI_ISL_1912306, EPI_ISL_1912322, EPI_ISL_1912337, EPI_ISL_1912338, EPI_ISL_1939720

| | | | |
|---|---|---|---|
| see above | Lighthouse Lab in Cambridge | Wellcome Sanger Institute for the COVID-19 Genomics UK (COG-UK) Consortium | Cordelia Langford; David K. Jackson; Dominic Kwiatkowski; Ewan Harrison; Ian Johnston; Jeffrey Barrett; John Sillitoe on behalf of the Wellcome Sanger Institute COVID-19 Surveillance Team; Rob Howes; Roberto Amato; Sonia Goncalves; The Lighthouse Lab in Cambridge and Alex Alderton |

EPI_ISL_1652695, EPI_ISL_1652696, EPI_ISL_1652699, EPI_ISL_1652706, EPI_ISL_1652707, EPI_ISL_1652709, EPI_ISL_1652710, EPI_ISL_1652730, EPI_ISL_1652749, EPI_ISL_1652751, EPI_ISL_1652758, EPI_ISL_1652780, EPI_ISL_1652783, EPI_ISL_1652784, EPI_ISL_1652785, EPI_ISL_1652786, EPI_ISL_1652795, EPI_ISL_1652800, EPI_ISL_1652803, EPI_ISL_1652806, EPI_ISL_1652810, EPI_ISL_1652816, EPI_ISL_1652817, EPI_ISL_1652823, EPI_ISL_1652829, EPI_ISL_1652842, EPI_ISL_1652868, EPI_ISL_1652885, EPI_ISL_1652889, EPI_ISL_1652909, EPI_ISL_1652911, EPI_ISL_1652914, EPI_ISL_1652925, EPI_ISL_1652932, EPI_ISL_1652934, EPI_ISL_1652940, EPI_ISL_1672971, EPI_ISL_1672973, EPI_ISL_1672974, EPI_ISL_1672975, EPI_ISL_1672976, EPI_ISL_1672978, EPI_ISL_1672979, EPI_ISL_1672980, EPI_ISL_1672981, EPI_ISL_1672982, EPI_ISL_1672983, EPI_ISL_1672985, EPI_ISL_1672987, EPI_ISL_1672989, EPI_ISL_1672991, EPI_ISL_1672992, EPI_ISL_1672993, EPI_ISL_1672994, EPI_ISL_1672995, EPI_ISL_1672996, EPI_ISL_1672998, EPI_ISL_1672999, EPI_ISL_1673000, EPI_ISL_1673001, EPI_ISL_1673003, EPI_ISL_1673004, EPI_ISL_1673007, EPI_ISL_1673009, EPI_ISL_1673011, EPI_ISL_1673012, EPI_ISL_1673013, EPI_ISL_1673014, EPI_ISL_1673015, EPI_ISL_1673020, EPI_ISL_1673022, EPI_ISL_1673025, EPI_ISL_1673026, EPI_ISL_1673027, EPI_ISL_1673028, EPI_ISL_1673029, EPI_ISL_1673030, EPI_ISL_1673032, EPI_ISL_1673033, EPI_ISL_1673034, EPI_ISL_1673036, EPI_ISL_1673037, EPI_ISL_1673038, EPI_ISL_1673039, EPI_ISL_1673040, EPI_ISL_1673043, EPI_ISL_1697448, EPI_ISL_1697455, EPI_ISL_1697465, EPI_ISL_1697466, EPI_ISL_1697485, EPI_ISL_1697493, EPI_ISL_1697494, EPI_ISL_1697527, EPI_ISL_1697538, EPI_ISL_1697543, EPI_ISL_1697544, EPI_ISL_1697566, EPI_ISL_1697578, EPI_ISL_1697580, EPI_ISL_1697587, EPI_ISL_1697632, EPI_ISL_1697644, EPI_ISL_1697667, EPI_ISL_1697674, EPI_ISL_1697689, EPI_ISL_1697691, EPI_ISL_1697726, EPI_ISL_1697728, EPI_ISL_1697729, EPI_ISL_1697734, EPI_ISL_1697745, EPI_ISL_1697750, EPI_ISL_1697765, EPI_ISL_1697774, EPI_ISL_1697778, EPI_ISL_1697780, EPI_ISL_1697793, EPI_ISL_1697816, EPI_ISL_1697832, EPI_ISL_1697841, EPI_ISL_1697850, EPI_ISL_1697862, EPI_ISL_1697893, EPI_ISL_1697910, EPI_ISL_1697936, EPI_ISL_1697944, EPI_ISL_1697950, EPI_ISL_1697951, EPI_ISL_1698011, EPI_ISL_1698046, EPI_ISL_1698061, EPI_ISL_1698078, EPI_ISL_1698115, EPI_ISL_1698116, EPI_ISL_1698132, EPI_ISL_1698182, EPI_ISL_1698185, EPI_ISL_1698194, EPI_ISL_1698195, EPI_ISL_1698199, EPI_ISL_1698207, EPI_ISL_1698225, EPI_ISL_1698242, EPI_ISL_1698547, EPI_ISL_1698550, EPI_ISL_1698555, EPI_ISL_1698556, EPI_ISL_1698559, EPI_ISL_1698560, EPI_ISL_1698569, EPI_ISL_1698572, EPI_ISL_1698573, EPI_ISL_1698578, EPI_ISL_1698588, EPI_ISL_1698589, EPI_ISL_1698593, EPI_ISL_1698601, EPI_ISL_1698602, EPI_ISL_1698603, EPI_ISL_1698609, EPI_ISL_1698612, EPI_ISL_1698614, EPI_ISL_1698617, EPI_ISL_1698619, EPI_ISL_1698623, EPI_ISL_1698625, EPI_ISL_1698634, EPI_ISL_1698635, EPI_ISL_1698642, EPI_ISL_1698644, EPI_ISL_1698645, EPI_ISL_1698646, EPI_ISL_1698654, EPI_ISL_1698658, EPI_ISL_1698666, EPI_ISL_1698670, EPI_ISL_1698676, EPI_ISL_1698678, EPI_ISL_1698685, EPI_ISL_1698698, EPI_ISL_1698699, EPI_ISL_1698703, EPI_ISL_1698711, EPI_ISL_1698722, EPI_ISL_1698725, EPI_ISL_1698726, EPI_ISL_1698733, EPI_ISL_1698737, EPI_ISL_1698741, EPI_ISL_1698746, EPI_ISL_1698753, EPI_ISL_1698755, EPI_ISL_1698760, EPI_ISL_1698763, EPI_ISL_1698765, EPI_ISL_1698766, EPI_ISL_1698769, EPI_ISL_1698772, EPI_ISL_1698776, EPI_ISL_1698781, EPI_ISL_1698785, EPI_ISL_1698787, EPI_ISL_1698788, EPI_ISL_1698800, EPI_ISL_1698801, EPI_ISL_1698807, EPI_ISL_1698811, EPI_ISL_1698812, EPI_ISL_1698823, EPI_ISL_1698836, EPI_ISL_1698837, EPI_ISL_1698839, EPI_ISL_1698848, EPI_ISL_1698872, EPI_ISL_1698885, EPI_ISL_1698893, EPI_ISL_1698897, EPI_ISL_1698900, EPI_ISL_1698903, EPI_ISL_1698904, EPI_ISL_1698915, EPI_ISL_1698930, EPI_ISL_1698933, EPI_ISL_1698935, EPI_ISL_1698936, EPI_ISL_1698939, EPI_ISL_1698952, EPI_ISL_1698954, EPI_ISL_1698957, EPI_ISL_1698958, EPI_ISL_1698960, EPI_ISL_1698962, EPI_ISL_1698964, EPI_ISL_1698969, EPI_ISL_1698970, EPI_ISL_1698976, EPI_ISL_1698981, EPI_ISL_1698982, EPI_ISL_1698992, EPI_ISL_1698997, EPI_ISL_1699002, EPI_ISL_1699004, EPI_ISL_1699005, EPI_ISL_1699011, EPI_ISL_1699287, EPI_ISL_1699288, EPI_ISL_1699293, EPI_ISL_1699309, EPI_ISL_1699318, EPI_ISL_1699327, EPI_ISL_1699333, EPI_ISL_1699342, EPI_ISL_1699343, EPI_ISL_1699349, EPI_ISL_1699353, EPI_ISL_1699354, EPI_ISL_1699356, EPI_ISL_1699358, EPI_ISL_1699360, EPI_ISL_1699367, EPI_ISL_1699373, EPI_ISL_1699374, EPI_ISL_1699379, EPI_ISL_1699382, EPI_ISL_1699383, EPI_ISL_1699390, EPI_ISL_1699397, EPI_ISL_1699408, EPI_ISL_1699423, EPI_ISL_1699427, EPI_ISL_1699430, EPI_ISL_1699434, EPI_ISL_1699454, EPI_ISL_1699459, EPI_ISL_1699460, EPI_ISL_1699463, EPI_ISL_1699464, EPI_ISL_1699468, EPI_ISL_1699469, EPI_ISL_1699471, EPI_ISL_1699472, EPI_ISL_1699473, EPI_ISL_1699480, EPI_ISL_1699484, EPI_ISL_1699487, EPI_ISL_1699490, EPI_ISL_1699492, EPI_ISL_1699498, EPI_ISL_1699499, EPI_ISL_1699505, EPI_ISL_1699507, EPI_ISL_1699517, EPI_ISL_1699520, EPI_ISL_1699522, EPI_ISL_1699527, EPI_ISL_1699533, EPI_ISL_1699538, EPI_ISL_1699548, EPI_ISL_1699549, EPI_ISL_1699567, EPI_ISL_1699621, EPI_ISL_1699633, EPI_ISL_1699649, EPI_ISL_1699670, EPI_ISL_1699677, EPI_ISL_1699688, EPI_ISL_1699697, EPI_ISL_1699711, EPI_ISL_1699714, EPI_ISL_1699718, EPI_ISL_1699719, EPI_ISL_1699732, EPI_ISL_1699741, EPI_ISL_1699748, EPI_ISL_1699749, EPI_ISL_1699762, EPI_ISL_1699764, EPI_ISL_1699778, EPI_ISL_1699783, EPI_ISL_1699790, EPI_ISL_1699808, EPI_ISL_1699813, EPI_ISL_1718332, EPI_ISL_1718336, EPI_ISL_1718338, EPI_ISL_1718343, EPI_ISL_1718351, EPI_ISL_1718360, EPI_ISL_1718361, EPI_ISL_1718364, EPI_ISL_1718366, EPI_ISL_1718379, EPI_ISL_1718384, EPI_ISL_1718385, EPI_ISL_1718387, EPI_ISL_1718388, EPI_ISL_1718401, EPI_ISL_1718416, EPI_ISL_1718417, EPI_ISL_1718423, EPI_ISL_1718439, EPI_ISL_1718453, EPI_ISL_1718464, EPI_ISL_1718466, EPI_ISL_1718471, EPI_ISL_1718474, EPI_ISL_1718492, EPI_ISL_1718493, EPI_ISL_1718498, EPI_ISL_1718499, EPI_ISL_1718505, EPI_ISL_1718512, EPI_ISL_1718521, EPI_ISL_1718522, EPI_ISL_1718535, EPI_ISL_1718536, EPI_ISL_1718538, EPI_ISL_1718557, EPI_ISL_1718563, EPI_ISL_1718567, EPI_ISL_1718570, EPI_ISL_1718579, EPI_ISL_1718591, EPI_ISL_1718608, EPI_ISL_1718679, EPI_ISL_1718727, EPI_ISL_1718784, EPI_ISL_1718806, EPI_ISL_1719218, EPI_ISL_1719219, EPI_ISL_1719220, EPI_ISL_1719221, EPI_ISL_1719222, EPI_ISL_1719223, EPI_ISL_1719224, EPI_ISL_1719225, EPI_ISL_1719226, EPI_ISL_1719227, EPI_ISL_1719228, EPI_ISL_1719229, EPI_ISL_1719230, EPI_ISL_1719231, EPI_ISL_1719232, EPI_ISL_1719233, EPI_ISL_1719234, EPI_ISL_1719235, EPI_ISL_1719236, EPI_ISL_1719237, EPI_ISL_1719238, EPI_ISL_1719239, EPI_ISL_1719240, EPI_ISL_1719241, EPI_ISL_1719243, EPI_ISL_1719244, EPI_ISL_1719245, EPI_ISL_1719246, EPI_ISL_1719247, EPI_ISL_1719248, EPI_ISL_1719249, EPI_ISL_1719250, EPI_ISL_1719251, EPI_ISL_1719252, EPI_ISL_1719253, EPI_ISL_1719254, EPI_ISL_1719255, EPI_ISL_1719256, EPI_ISL_1719257, EPI_ISL_1719258, EPI_ISL_1719259, EPI_ISL_1719260, EPI_ISL_1719261, EPI_ISL_1719262, EPI_ISL_1719263, EPI_ISL_1719264, EPI_ISL_1719265, EPI_ISL_1719266, EPI_ISL_1719267, EPI_ISL_1719268, EPI_ISL_1719269, EPI_ISL_1719270, EPI_ISL_1719271, EPI_ISL_1719272, EPI_ISL_1719273, EPI_ISL_1719274, EPI_ISL_1719275, EPI_ISL_1719276, EPI_ISL_1719277, EPI_ISL_1719278, EPI_ISL_1719279, EPI_ISL_1719280, EPI_ISL_1719281, EPI_ISL_1719282, EPI_ISL_1719283, EPI_ISL_1719284, EPI_ISL_1719285, EPI_ISL_1719286, EPI_ISL_1719287, EPI_ISL_1719288, EPI_ISL_1719289, EPI_ISL_1719290, EPI_ISL_1719291, EPI_ISL_1719292, EPI_ISL_1719293, EPI_ISL_1719294, EPI_ISL_1719295, EPI_ISL_1719296, EPI_ISL_1719297, EPI_ISL_1719298, EPI_ISL_1719299, EPI_ISL_1719300, EPI_ISL_1719301, EPI_ISL_1719302, EPI_ISL_1719303, EPI_ISL_1719304, EPI_ISL_1719305, EPI_ISL_1719306, EPI_ISL_1719307, EPI_ISL_1719308, EPI_ISL_1719309, EPI_ISL_1719310, EPI_ISL_1719311, EPI_ISL_1719312, EPI_ISL_1719313, EPI_ISL_1719314, EPI_ISL_1719315, EPI_ISL_1719316, EPI_ISL_1719317, EPI_ISL_1719318, EPI_ISL_1719319, EPI_ISL_1719320, EPI_ISL_1719321, EPI_ISL_1719322, EPI_ISL_1719323, EPI_ISL_1719324, EPI_ISL_1719325, EPI_ISL_1719326, EPI_ISL_1719327, EPI_ISL_1719328, EPI_ISL_1719329, EPI_ISL_1719330, EPI_ISL_1719331, EPI_ISL_1719332, EPI_ISL_1719333, EPI_ISL_1719334, EPI_ISL_1719335, EPI_ISL_1719336, EPI_ISL_1719337, EPI_ISL_1719338, EPI_ISL_1719339, EPI_ISL_1719340, EPI_ISL_1719341, EPI_ISL_1719342, EPI_ISL_1719343, EPI_ISL_1719344, EPI_ISL_1719345, EPI_ISL_1719346, EPI_ISL_1719347, EPI_ISL_1719348, EPI_ISL_1719349, EPI_ISL_1719350, EPI_ISL_1719351, EPI_ISL_1719352, EPI_ISL_1719353, EPI_ISL_1719354, EPI_ISL_1719355, EPI_ISL_1719356, EPI_ISL_1719357, EPI_ISL_1719358, EPI_ISL_1719359, EPI_ISL_1719360, EPI_ISL_1719361, EPI_ISL_1719362, EPI_ISL_1719363, EPI_ISL_1719364, EPI_ISL_1719365, EPI_ISL_1719366, EPI_ISL_1719367, EPI_ISL_1719368, EPI_ISL_1719369, EPI_ISL_1719370, EPI_ISL_1719371, EPI_ISL_1719372, EPI_ISL_1719373, EPI_ISL_1719374, EPI_ISL_1719375, EPI_ISL_1719376, EPI_ISL_1719377, EPI_ISL_1719378, EPI_ISL_1719379, EPI_ISL_1719380, EPI_ISL_1719381, EPI_ISL_1719382, EPI_ISL_1719383, EPI_ISL_1719384, EPI_ISL_1719385, EPI_ISL_1719386, EPI_ISL_1719387, EPI_ISL_1719388, EPI_ISL_1719389, EPI_ISL_1719390, EPI_ISL_1719391, EPI_ISL_1719392, EPI_ISL_1719393, EPI_ISL_1719394, EPI_ISL_1719395, EPI_ISL_1719396, EPI_ISL_1719397, EPI_ISL_1719398, EPI_ISL_1719399, EPI_ISL_1719400, EPI_ISL_1719401, EPI_ISL_1719402, EPI_ISL_1719403, EPI_ISL_1719404, EPI_ISL_1719405, EPI_ISL_1719406, EPI_ISL_1719407, EPI_ISL_1719408, EPI_ISL_1719409, EPI_ISL_1719410, EPI_ISL_1719411, EPI_ISL_1719412, EPI_ISL_1719413, EPI_ISL_1719414, EPI_ISL_1719415, EPI_ISL_1719416, EPI_ISL_1719417, EPI_ISL_1719418, EPI_ISL_1719419, EPI_ISL_1719420, EPI_ISL_1719421, EPI_ISL_1719422, EPI_ISL_1719423, EPI_ISL_1719424, EPI_ISL_1719425, EPI_ISL_1719426, EPI_ISL_1719427, EPI_ISL_1719428, EPI_ISL_1719429, EPI_ISL_1719430, EPI_ISL_1719431, EPI_ISL_1719432, EPI_ISL_1719436, EPI_ISL_1719440, EPI_ISL_1719448, EPI_ISL_1719453, EPI_ISL_1719456, EPI_ISL_1719457, EPI_ISL_1719462, EPI_ISL_1719463, EPI_ISL_1719464, EPI_ISL_1719471, EPI_ISL_1719472, EPI_ISL_1719473, EPI_ISL_1719474, EPI_ISL_1719475, EPI_ISL_1719476, EPI_ISL_1719477, EPI_ISL_1719478, EPI_ISL_1719483, EPI_ISL_1719490, EPI_ISL_1719491, EPI_ISL_1719492, EPI_ISL_1719494, EPI_ISL_1719495, EPI_ISL_1719497, EPI_ISL_1719502, EPI_ISL_1719509, EPI_ISL_1719510, EPI_ISL_1719511, EPI_ISL_1719516, EPI_ISL_1719520, EPI_ISL_1719523, EPI_ISL_1719525, EPI_ISL_1719530, EPI_ISL_1719531, EPI_ISL_1719532, EPI_ISL_1719535, EPI_ISL_1719537, EPI_ISL_1719538, EPI_ISL_1719540, EPI_ISL_1719541, EPI_ISL_1719545, EPI_ISL_1719550, EPI_ISL_1719552, EPI_ISL_1719553, EPI_ISL_1719555, EPI_ISL_1719556, EPI_ISL_1719557, EPI_ISL_1719559, EPI_ISL_1719562, EPI_ISL_1719563, EPI_ISL_1719564, EPI_ISL_1719565, EPI_ISL_1719566, EPI_ISL_1719567, EPI_ISL_1719568, EPI_ISL_1719572, EPI_ISL_1719573, EPI_ISL_1719577, EPI_ISL_1719579, EPI_ISL_1719580, EPI_ISL_1719581, EPI_ISL_1719586, EPI_ISL_1719589, EPI_ISL_1719590, EPI_ISL_1719597, EPI_ISL_1719601, EPI_ISL_1719602, EPI_ISL_1719608, EPI_ISL_1719610, EPI_ISL_1719611, EPI_ISL_1719612, EPI_ISL_1719614, EPI_ISL_1719615, EPI_ISL_1719617, EPI_ISL_1719619, EPI_ISL_1719620, EPI_ISL_1719621, EPI_ISL_1719622, EPI_ISL_1719626, EPI_ISL_1719627, EPI_ISL_1719628, EPI_ISL_1719631, EPI_ISL_1719632, EPI_ISL_1719633, EPI_ISL_1719634, EPI_ISL_1719637, EPI_ISL_1719638, EPI_ISL_1719641, EPI_ISL_1719644, EPI_ISL_1719646, EPI_ISL_1719649, EPI_ISL_1719652, EPI_ISL_1719653, EPI_ISL_1719654, EPI_ISL_1719657, EPI_ISL_1719661, EPI_ISL_1719662, EPI_ISL_1719663, EPI_ISL_1719664, EPI_ISL_1719667, EPI_ISL_1719669, EPI_ISL_1719672, EPI_ISL_1719673, EPI_ISL_1719674, EPI_ISL_1719675, EPI_ISL_1719677, EPI_ISL_1719678, EPI_ISL_1719681, EPI_ISL_1719682, EPI_ISL_1719683, EPI_ISL_1719689, EPI_ISL_1719693, EPI_ISL_1719694, EPI_ISL_1719695, EPI_ISL_1719698, EPI_ISL_1719700, EPI_ISL_1719701, EPI_ISL_1719702, EPI_ISL_1719704, EPI_ISL_1719705, EPI_ISL_1719706, EPI_ISL_1719707, EPI_ISL_1719708, EPI_ISL_1719714, EPI_ISL_1719715, EPI_ISL_1719716, EPI_ISL_1719718, EPI_ISL_1719720, EPI_ISL_1719722, EPI_ISL_1719723, EPI_ISL_1719724, EPI_ISL_1719728, EPI_ISL_1740536, EPI_ISL_1740537, EPI_ISL_1740539, EPI_ISL_1740540, EPI_ISL_1740542, EPI_ISL_1740543, EPI_ISL_1740544, EPI_ISL_1740545, EPI_ISL_1740546, EPI_ISL_1740547, EPI_ISL_1740548, EPI_ISL_1740549, EPI_ISL_1740554, EPI_ISL_1740557, EPI_ISL_1740559, EPI_ISL_1740560, EPI_ISL_1740561, EPI_ISL_1740562, EPI_ISL_1740563, EPI_ISL_1740566, EPI_ISL_1740568, EPI_ISL_1740569, EPI_ISL_1740570, EPI_ISL_1740573, EPI_ISL_1740575, EPI_ISL_1740577, EPI_ISL_1740581, EPI_ISL_1740582, EPI_ISL_1740583, EPI_ISL_1740584, EPI_ISL_1740586, EPI_ISL_1740587, EPI_ISL_1740591, EPI_ISL_1740593, EPI_ISL_1740594, EPI_ISL_1740596, EPI_ISL_1740602, EPI_ISL_1740603, EPI_ISL_1740606, EPI_ISL_1740608, EPI_ISL_1740612, EPI_ISL_1740613, EPI_ISL_1740615, EPI_ISL_1740617, EPI_ISL_1740620, EPI_ISL_1740622, EPI_ISL_1740623, EPI_ISL_1740625, EPI_ISL_1740626, EPI_ISL_1740627, EPI_ISL_1740628, EPI_ISL_1740631, EPI_ISL_1740634, EPI_ISL_1740635, EPI_ISL_1740638, EPI_ISL_1740639, EPI_ISL_1740640, EPI_ISL_1740644, EPI_ISL_1740645, EPI_ISL_1740647, EPI_ISL_1740648, EPI_ISL_1740649, EPI_ISL_1740650, EPI_ISL_1740653, EPI_ISL_1740659, EPI_ISL_1740665, EPI_ISL_1740667, EPI_ISL_1740670, EPI_ISL_1740676, EPI_ISL_1740679, EPI_ISL_1740680, EPI_ISL_1740683, EPI_ISL_1740684, EPI_ISL_1740685, EPI_ISL_1740686, EPI_ISL_1740692, EPI_ISL_1740693, EPI_ISL_1740694, EPI_ISL_1740695, EPI_ISL_1740696, EPI_ISL_1740700, EPI_ISL_1740702, EPI_ISL_1740705, EPI_ISL_1740709, EPI_ISL_1740711, EPI_ISL_1740717, EPI_ISL_1740718, EPI_ISL_1740720, EPI_ISL_1740721, EPI_ISL_1740723, EPI_ISL_1740724, EPI_ISL_1740727, EPI_ISL_1740728, EPI_ISL_1740733, EPI_ISL_1740735, EPI_ISL_1740738, EPI_ISL_1740739, EPI_ISL_1740742, EPI_ISL_1740743, EPI_ISL_1740747, EPI_ISL_1740749, EPI_ISL_1740752, EPI_ISL_1740755, EPI_ISL_1740756, EPI_ISL_1740757, EPI_ISL_1740765, EPI_ISL_1740767, EPI_ISL_1740770, EPI_ISL_1740771, EPI_ISL_1740775, EPI_ISL_1740776, EPI_ISL_1740777, EPI_ISL_1740789, EPI_ISL_1740791, EPI_ISL_1740795, EPI_ISL_1740806, EPI_ISL_1740809, EPI_ISL_1740817, EPI_ISL_1740825, EPI_ISL_1740828, EPI_ISL_1740849, EPI_ISL_1740854, EPI_ISL_1740855, EPI_ISL_1740861, EPI_ISL_1740865, EPI_ISL_1740892, EPI_ISL_1740897, EPI_ISL_1740901, EPI_ISL_1740905, EPI_ISL_1740907, EPI_ISL_1740920, EPI_ISL_1740924, EPI_ISL_1740929, EPI_ISL_1740934, EPI_ISL_1740937, EPI_ISL_1740939, EPI_ISL_1740941, EPI_ISL_1740943, EPI_ISL_1740944, EPI_ISL_1740948, EPI_ISL_1740951, EPI_ISL_1740956, EPI_ISL_1740957, EPI_ISL_1740958, EPI_ISL_1740962, EPI_ISL_1740969, EPI_ISL_1740974, EPI_ISL_1740975, EPI_ISL_1740979, EPI_ISL_1740981, EPI_ISL_1740982, EPI_ISL_1740993, EPI_ISL_1740994, EPI_ISL_1740998, EPI_ISL_1741004, EPI_ISL_1741007, EPI_ISL_1741009, EPI_ISL_1741014, EPI_ISL_1741031, EPI_ISL_1741032, EPI_ISL_1741035, EPI_ISL_1741045, EPI_ISL_1741055, EPI_ISL_1741058, EPI_ISL_1741060, EPI_ISL_1741061, EPI_ISL_1741064, EPI_ISL_1741065, EPI_ISL_1741067, EPI_ISL_1741068, EPI_ISL_1741069, EPI_ISL_1741070, EPI_ISL_1741071, EPI_ISL_1741072, EPI_ISL_1741073, EPI_ISL_1741075, EPI_ISL_1741077, EPI_ISL_1741078, EPI_ISL_1741079, EPI_ISL_1741080, EPI_ISL_1741081, EPI_ISL_1741082, EPI_ISL_1741084, EPI_ISL_1741086, EPI_ISL_1741089, EPI_ISL_1741091, EPI_ISL_1741092, EPI_ISL_1741095, EPI_ISL_1741096, EPI_ISL_1741097, EPI_ISL_1741098, EPI_ISL_1741099, EPI_ISL_1741100, EPI_ISL_1741103, EPI_ISL_1741105, EPI_ISL_1741106, EPI_ISL_1741107, EPI_ISL_1741108, EPI_ISL_1741109, EPI_ISL_1741112, EPI_ISL_1741118, EPI_ISL_1741119, EPI_ISL_1741121, EPI_ISL_1741123, EPI_ISL_1741124, EPI_ISL_1741126, EPI_ISL_1741128, EPI_ISL_1741129, EPI_ISL_1741132, EPI_ISL_1741136, EPI_ISL_1741137, EPI_ISL_1741138, EPI_ISL_1741141, EPI_ISL_1741142, EPI_ISL_1741143, EPI_ISL_1741144, EPI_ISL_1741146, EPI_ISL_1741147, EPI_ISL_1741149, EPI_ISL_1741152, EPI_ISL_1741153, EPI_ISL_1741155, EPI_ISL_1741157, EPI_ISL_1741158, EPI_ISL_1741162, EPI_ISL_1741170, EPI_ISL_1741171, EPI_ISL_1741172, EPI_ISL_1741173, EPI_ISL_1741174, EPI_ISL_1741176, EPI_ISL_1741178, EPI_ISL_1741179, EPI_ISL_1741181, EPI_ISL_1741182, EPI_ISL_1741183, EPI_ISL_1741184, EPI_ISL_1741185, EPI_ISL_1741188, EPI_ISL_1741190, EPI_ISL_1741191, EPI_ISL_1741192, EPI_ISL_1741193, EPI_ISL_1741194, EPI_ISL_1741196, EPI_ISL_1741199, EPI_ISL_1741200, EPI_ISL_1741201, EPI_ISL_1741203, EPI_ISL_1741204, EPI_ISL_1741205, EPI_ISL_1741206, EPI_ISL_1741209, EPI_ISL_1741211, EPI_ISL_1741213, EPI_ISL_1741214, EPI_ISL_1741215, EPI_ISL_1741217, EPI_ISL_1741219, EPI_ISL_1741220, EPI_ISL_1741221, EPI_ISL_1741222, EPI_ISL_1741223, EPI_ISL_1741224, EPI_ISL_1741225, EPI_ISL_1741228, EPI_ISL_1741230, EPI_ISL_1741232, EPI_ISL_1741235, EPI_ISL_1741237, EPI_ISL_1741240, EPI_ISL_1741242, EPI_ISL_1741243, EPI_ISL_1741244, EPI_ISL_1741250, EPI_ISL_1741253, EPI_ISL_1741257, EPI_ISL_1741258, EPI_ISL_1741259, EPI_ISL_1741262, EPI_ISL_1741264, EPI_ISL_1741265, EPI_ISL_1741266, EPI_ISL_1741267, EPI_ISL_1741269, EPI_ISL_1741270, EPI_ISL_1741271, EPI_ISL_1741287, EPI_ISL_1741299, EPI_ISL_1741303, EPI_ISL_1741304, EPI_ISL_1741305, EPI_ISL_1741308, EPI_ISL_1741309, EPI_ISL_1741311, EPI_ISL_1741312, EPI_ISL_1741314, EPI_ISL_1741323, EPI_ISL_1741326, EPI_ISL_1741331, EPI_ISL_1741338, EPI_ISL_1741368, EPI_ISL_1741370, EPI_ISL_1741371, EPI_ISL_1741372, EPI_ISL_1741373, EPI_ISL_1741374, EPI_ISL_1741375, EPI_ISL_1741382, EPI_ISL_1741392, EPI_ISL_1741393, EPI_ISL_1741394, EPI_ISL_1741404, EPI_ISL_1741406, EPI_ISL_1741407, EPI_ISL_1741410, EPI_ISL_1741417, EPI_ISL_1741420, EPI_ISL_1741421, EPI_ISL_1741438, EPI_ISL_1741439, EPI_ISL_1741451, EPI_ISL_1741452, EPI_ISL_1741453, EPI_ISL_1741463, EPI_ISL_1741469, EPI_ISL_1741482, EPI_ISL_1741486, EPI_ISL_1741487, EPI_ISL_1741495, EPI_ISL_1741496, EPI_ISL_1741497, EPI_ISL_1741498, EPI_ISL_1741499, EPI_ISL_1741500, EPI_ISL_1741501, EPI_ISL_1741502, EPI_ISL_1741504, EPI_ISL_1741505, EPI_ISL_1741506, EPI_ISL_1741507, EPI_ISL_1741508, EPI_ISL_1741510, EPI_ISL_1741511, EPI_ISL_1741512, EPI_ISL_1741513, EPI_ISL_1741515, EPI_ISL_1741516, EPI_ISL_1741517, EPI_ISL_1741518, EPI_ISL_1741519, EPI_ISL_1741520, EPI_ISL_1741521, EPI_ISL_1741522, EPI_ISL_1741523, EPI_ISL_1741525, EPI_ISL_1741526, EPI_ISL_1741527, EPI_ISL_1741528, EPI_ISL_1741529, EPI_ISL_1741530, EPI_ISL_1741531, EPI_ISL_1741532, EPI_ISL_1741534, EPI_ISL_1741535, EPI_ISL_1741536, EPI_ISL_1741537,

EPI_ISL_1741538, EPI_ISL_1741539, EPI_ISL_1741540, EPI_ISL_1741541, EPI_ISL_1741554, EPI_ISL_1741556, EPI_ISL_1741557, EPI_ISL_1741559, EPI_ISL_1741561, EPI_ISL_1741562, EPI_ISL_1741564, EPI_ISL_1741565, EPI_ISL_1741567, EPI_ISL_1741568, EPI_ISL_1741570, EPI_ISL_1741571, EPI_ISL_1741573, EPI_ISL_1741574, EPI_ISL_1741576, EPI_ISL_1741577, EPI_ISL_1741579, EPI_ISL_1741580, EPI_ISL_1741582, EPI_ISL_1741584, EPI_ISL_1741585, EPI_ISL_1741587, EPI_ISL_1741588, EPI_ISL_1741590, EPI_ISL_1741591, EPI_ISL_1741593, EPI_ISL_1741594, EPI_ISL_1741596, EPI_ISL_1741597, EPI_ISL_1741599, EPI_ISL_1741600, EPI_ISL_1741602, EPI_ISL_1741603, EPI_ISL_1741605, EPI_ISL_1741606, EPI_ISL_1741608, EPI_ISL_1741609, EPI_ISL_1741611, EPI_ISL_1741612, EPI_ISL_1741613, EPI_ISL_1741614, EPI_ISL_1741615, EPI_ISL_1741616, EPI_ISL_1741617, EPI_ISL_1741618, EPI_ISL_1741619, EPI_ISL_1741620, EPI_ISL_1741621, EPI_ISL_1741622, EPI_ISL_1741623, EPI_ISL_1741624, EPI_ISL_1741625, EPI_ISL_1741626, EPI_ISL_1741627, EPI_ISL_1741628, EPI_ISL_1741629, EPI_ISL_1741630, EPI_ISL_1741631, EPI_ISL_1741632, EPI_ISL_1741633, EPI_ISL_1741634, EPI_ISL_1741635, EPI_ISL_1741636, EPI_ISL_1741637, EPI_ISL_1741638, EPI_ISL_1741639, EPI_ISL_1741640, EPI_ISL_1741641, EPI_ISL_1741642, EPI_ISL_1741643, EPI_ISL_1741644, EPI_ISL_1741645, EPI_ISL_1741646, EPI_ISL_1741647, EPI_ISL_1741648, EPI_ISL_1741649, EPI_ISL_1741650, EPI_ISL_1741651, EPI_ISL_1741652, EPI_ISL_1741653, EPI_ISL_1741654, EPI_ISL_1741655, EPI_ISL_1741656, EPI_ISL_1741657, EPI_ISL_1741658, EPI_ISL_1741659, EPI_ISL_1741660, EPI_ISL_1741661, EPI_ISL_1741662, EPI_ISL_1741663, EPI_ISL_1741664, EPI_ISL_1741665, EPI_ISL_1741666, EPI_ISL_1741667, EPI_ISL_1741668, EPI_ISL_1741669, EPI_ISL_1741670, EPI_ISL_1741671, EPI_ISL_1741672, EPI_ISL_1741673, EPI_ISL_1741674, EPI_ISL_1741675, EPI_ISL_1741676, EPI_ISL_1741677, EPI_ISL_1741678, EPI_ISL_1741679, EPI_ISL_1741680, EPI_ISL_1741681, EPI_ISL_1741682, EPI_ISL_1741683, EPI_ISL_1741684, EPI_ISL_1741685, EPI_ISL_1741686, EPI_ISL_1741687, EPI_ISL_1741688, EPI_ISL_1741689, EPI_ISL_1741690, EPI_ISL_1741691, EPI_ISL_1741692, EPI_ISL_1741693, EPI_ISL_1741694, EPI_ISL_1741695, EPI_ISL_1741696, EPI_ISL_1741697, EPI_ISL_1741698, EPI_ISL_1741699, EPI_ISL_1741700, EPI_ISL_1741701, EPI_ISL_1741702, EPI_ISL_1741703, EPI_ISL_1741704, EPI_ISL_1741705, EPI_ISL_1741706, EPI_ISL_1741707, EPI_ISL_1741708, EPI_ISL_1741709, EPI_ISL_1741710, EPI_ISL_1741711, EPI_ISL_1741712, EPI_ISL_1741713, EPI_ISL_1741714, EPI_ISL_1741715, EPI_ISL_1741716, EPI_ISL_1741717, EPI_ISL_1741718, EPI_ISL_1741719, EPI_ISL_1741720, EPI_ISL_1741721, EPI_ISL_1741722, EPI_ISL_1741723, EPI_ISL_1741724, EPI_ISL_1741726, EPI_ISL_1741727, EPI_ISL_1741728, EPI_ISL_1741729, EPI_ISL_1741730, EPI_ISL_1741731, EPI_ISL_1741732, EPI_ISL_1741733, EPI_ISL_1741734, EPI_ISL_1741735, EPI_ISL_1741736, EPI_ISL_1741738, EPI_ISL_1741739, EPI_ISL_1741740, EPI_ISL_1741741, EPI_ISL_1741742, EPI_ISL_1741743, EPI_ISL_1741744, EPI_ISL_1741745, EPI_ISL_1741746, EPI_ISL_1741747, EPI_ISL_1741748, EPI_ISL_1741749, EPI_ISL_1741750, EPI_ISL_1741751, EPI_ISL_1741752, EPI_ISL_1741753, EPI_ISL_1741754, EPI_ISL_1741756, EPI_ISL_1741757, EPI_ISL_1741758, EPI_ISL_1741759, EPI_ISL_1741760, EPI_ISL_1741761, EPI_ISL_1741762, EPI_ISL_1741763, EPI_ISL_1741764, EPI_ISL_1741766, EPI_ISL_1741767, EPI_ISL_1741768, EPI_ISL_1741769, EPI_ISL_1741770, EPI_ISL_1741771, EPI_ISL_1741772, EPI_ISL_1741773, EPI_ISL_1741776, EPI_ISL_1741783, EPI_ISL_1741791, EPI_ISL_1741792, EPI_ISL_1741797, EPI_ISL_1741798, EPI_ISL_1741802, EPI_ISL_1741805, EPI_ISL_1741810, EPI_ISL_1741821, EPI_ISL_1741833, EPI_ISL_1741841, EPI_ISL_1741844, EPI_ISL_1741849, EPI_ISL_1741851, EPI_ISL_1741852, EPI_ISL_1741856, EPI_ISL_1741858, EPI_ISL_1741867, EPI_ISL_1741868, EPI_ISL_1741872, EPI_ISL_1741876, EPI_ISL_1741877, EPI_ISL_1741880, EPI_ISL_1741895, EPI_ISL_1741896, EPI_ISL_1741898, EPI_ISL_1741900, EPI_ISL_1741908, EPI_ISL_1741912, EPI_ISL_1741925, EPI_ISL_1741928, EPI_ISL_1741932, EPI_ISL_1741933, EPI_ISL_1741935, EPI_ISL_1741938, EPI_ISL_1741942, EPI_ISL_1741944, EPI_ISL_1741946, EPI_ISL_1741948, EPI_ISL_1741949, EPI_ISL_1741953, EPI_ISL_1741955, EPI_ISL_1741957, EPI_ISL_1741958, EPI_ISL_1741961, EPI_ISL_1741970, EPI_ISL_1741972, EPI_ISL_1741975, EPI_ISL_1741977, EPI_ISL_1741980, EPI_ISL_1741981, EPI_ISL_1741985, EPI_ISL_1741987, EPI_ISL_1741989, EPI_ISL_1741992, EPI_ISL_1741997, EPI_ISL_1741998, EPI_ISL_1741999, EPI_ISL_1742000, EPI_ISL_1742001, EPI_ISL_1742008, EPI_ISL_1742009, EPI_ISL_1742010, EPI_ISL_1742087, EPI_ISL_1742088, EPI_ISL_1742090, EPI_ISL_1742092, EPI_ISL_1742094, EPI_ISL_1742095, EPI_ISL_1742096, EPI_ISL_1742097, EPI_ISL_1742098, EPI_ISL_1742099, EPI_ISL_1742100, EPI_ISL_1742103, EPI_ISL_1742105, EPI_ISL_1742106, EPI_ISL_1742107, EPI_ISL_1742108, EPI_ISL_1742109, EPI_ISL_1742110, EPI_ISL_1742111, EPI_ISL_1742112, EPI_ISL_1742113, EPI_ISL_1742114, EPI_ISL_1742115, EPI_ISL_1742116, EPI_ISL_1742117, EPI_ISL_1742118, EPI_ISL_1742119, EPI_ISL_1742120, EPI_ISL_1742123, EPI_ISL_1742124, EPI_ISL_1742126, EPI_ISL_1742128, EPI_ISL_1742129, EPI_ISL_1742130, EPI_ISL_1742131, EPI_ISL_1742134, EPI_ISL_1742135, EPI_ISL_1742137, EPI_ISL_1742138, EPI_ISL_1742139, EPI_ISL_1742140, EPI_ISL_1742141, EPI_ISL_1742142, EPI_ISL_1742143, EPI_ISL_1742144, EPI_ISL_1742145, EPI_ISL_1742148, EPI_ISL_1742149, EPI_ISL_1742150, EPI_ISL_1742151, EPI_ISL_1742154, EPI_ISL_1742158, EPI_ISL_1742159, EPI_ISL_1742160, EPI_ISL_1742161, EPI_ISL_1742165, EPI_ISL_1742166, EPI_ISL_1742167, EPI_ISL_1742169, EPI_ISL_1742172, EPI_ISL_1742173, EPI_ISL_1742174, EPI_ISL_1742175, EPI_ISL_1742177, EPI_ISL_1742180, EPI_ISL_1742262, EPI_ISL_1742263, EPI_ISL_1742264, EPI_ISL_1742265, EPI_ISL_1742266, EPI_ISL_1742267, EPI_ISL_1742268, EPI_ISL_1742269, EPI_ISL_1742270, EPI_ISL_1742271, EPI_ISL_1742272, EPI_ISL_1742273, EPI_ISL_1742275, EPI_ISL_1742276, EPI_ISL_1742277, EPI_ISL_1742278, EPI_ISL_1742279, EPI_ISL_1742280, EPI_ISL_1742281, EPI_ISL_1742282, EPI_ISL_1742283, EPI_ISL_1758187, EPI_ISL_1758189, EPI_ISL_1758192, EPI_ISL_1758193, EPI_ISL_1758194, EPI_ISL_1758197, EPI_ISL_1758199, EPI_ISL_1758200, EPI_ISL_1758201, EPI_ISL_1758204, EPI_ISL_1758207, EPI_ISL_1758211, EPI_ISL_1758216, EPI_ISL_1758219, EPI_ISL_1758220, EPI_ISL_1758223, EPI_ISL_1758225, EPI_ISL_1758227, EPI_ISL_1758229, EPI_ISL_1758230, EPI_ISL_1758235, EPI_ISL_1758236, EPI_ISL_1758239, EPI_ISL_1758240, EPI_ISL_1758242, EPI_ISL_1758244, EPI_ISL_1758246, EPI_ISL_1758248, EPI_ISL_1758249, EPI_ISL_1758250, EPI_ISL_1758251, EPI_ISL_1758255, EPI_ISL_1758257, EPI_ISL_1758264, EPI_ISL_1758267, EPI_ISL_1758271, EPI_ISL_1758272, EPI_ISL_1758275, EPI_ISL_1758279, EPI_ISL_1758280, EPI_ISL_1758285, EPI_ISL_1758286, EPI_ISL_1758287, EPI_ISL_1758291, EPI_ISL_1758295, EPI_ISL_1758298, EPI_ISL_1758299, EPI_ISL_1758304, EPI_ISL_1758305, EPI_ISL_1758307, EPI_ISL_1758311, EPI_ISL_1758312, EPI_ISL_1758315, EPI_ISL_1758318, EPI_ISL_1758321, EPI_ISL_1758326, EPI_ISL_1758330, EPI_ISL_1758333, EPI_ISL_1758336, EPI_ISL_1758337, EPI_ISL_1758338, EPI_ISL_1758339, EPI_ISL_1758340, EPI_ISL_1758345, EPI_ISL_1758346, EPI_ISL_1758357, EPI_ISL_1758358, EPI_ISL_1758359, EPI_ISL_1758365, EPI_ISL_1758366, EPI_ISL_1758369, EPI_ISL_1758370, EPI_ISL_1758371, EPI_ISL_1758374, EPI_ISL_1758377, EPI_ISL_1758380, EPI_ISL_1758381, EPI_ISL_1758382, EPI_ISL_1758384, EPI_ISL_1758385, EPI_ISL_1758386, EPI_ISL_1758391, EPI_ISL_1758392, EPI_ISL_1758393, EPI_ISL_1758394, EPI_ISL_1758395, EPI_ISL_1758396, EPI_ISL_1758399, EPI_ISL_1758403, EPI_ISL_1758407, EPI_ISL_1758408, EPI_ISL_1758409, EPI_ISL_1758410, EPI_ISL_1758413, EPI_ISL_1758421, EPI_ISL_1758431, EPI_ISL_1758432, EPI_ISL_1758434, EPI_ISL_1758435, EPI_ISL_1758436, EPI_ISL_1758438, EPI_ISL_1758439, EPI_ISL_1758441, EPI_ISL_1758445, EPI_ISL_1758447, EPI_ISL_1758452, EPI_ISL_1758460, EPI_ISL_1758462, EPI_ISL_1758465, EPI_ISL_1758469, EPI_ISL_1758470, EPI_ISL_1758479, EPI_ISL_1758482, EPI_ISL_1758483, EPI_ISL_1758486, EPI_ISL_1758493, EPI_ISL_1758496, EPI_ISL_1758505, EPI_ISL_1758507, EPI_ISL_1758510, EPI_ISL_1758511, EPI_ISL_1758518, EPI_ISL_1758522, EPI_ISL_1758525, EPI_ISL_1758527, EPI_ISL_1758529, EPI_ISL_1758531, EPI_ISL_1758538, EPI_ISL_1758541, EPI_ISL_1758555, EPI_ISL_1758563, EPI_ISL_1758567, EPI_ISL_1758568, EPI_ISL_1758572, EPI_ISL_1758573, EPI_ISL_1758578, EPI_ISL_1758582, EPI_ISL_1758597, EPI_ISL_1758606, EPI_ISL_1758609, EPI_ISL_1758612, EPI_ISL_1758614, EPI_ISL_1758624, EPI_ISL_1758630, EPI_ISL_1758635, EPI_ISL_1758656, EPI_ISL_1758658, EPI_ISL_1758660, EPI_ISL_1758666, EPI_ISL_1758667, EPI_ISL_1758668, EPI_ISL_1758669, EPI_ISL_1758670, EPI_ISL_1758671, EPI_ISL_1758672, EPI_ISL_1758673, EPI_ISL_1758674, EPI_ISL_1758675, EPI_ISL_1758676, EPI_ISL_1758677, EPI_ISL_1758678, EPI_ISL_1758679, EPI_ISL_1758680, EPI_ISL_1758681, EPI_ISL_1758682, EPI_ISL_1758683, EPI_ISL_1758684, EPI_ISL_1758685, EPI_ISL_1758686, EPI_ISL_1758687, EPI_ISL_1758688, EPI_ISL_1758689, EPI_ISL_1758690, EPI_ISL_1758691, EPI_ISL_1758692, EPI_ISL_1758693, EPI_ISL_1758694, EPI_ISL_1758695, EPI_ISL_1758696, EPI_ISL_1758697, EPI_ISL_1758698, EPI_ISL_1758699, EPI_ISL_1758700, EPI_ISL_1758701, EPI_ISL_1758702, EPI_ISL_1758703, EPI_ISL_1758704, EPI_ISL_1758705, EPI_ISL_1758706, EPI_ISL_1758707, EPI_ISL_1758708, EPI_ISL_1758709, EPI_ISL_1758710, EPI_ISL_1758711, EPI_ISL_1758712, EPI_ISL_1758713, EPI_ISL_1758714, EPI_ISL_1758715, EPI_ISL_1758716, EPI_ISL_1758717, EPI_ISL_1758718, EPI_ISL_1758719, EPI_ISL_1758720, EPI_ISL_1758721, EPI_ISL_1758722, EPI_ISL_1758723, EPI_ISL_1758724, EPI_ISL_1758725, EPI_ISL_1758726, EPI_ISL_1758727, EPI_ISL_1758728, EPI_ISL_1758729, EPI_ISL_1758730, EPI_ISL_1758731, EPI_ISL_1758732, EPI_ISL_1758733, EPI_ISL_1758734, EPI_ISL_1758735, EPI_ISL_1758736, EPI_ISL_1758737, EPI_ISL_1758738, EPI_ISL_1758739, EPI_ISL_1758740, EPI_ISL_1758742, EPI_ISL_1758743, EPI_ISL_1758744, EPI_ISL_1758745, EPI_ISL_1758746, EPI_ISL_1758747, EPI_ISL_1758748, EPI_ISL_1758749, EPI_ISL_1758750, EPI_ISL_1758751, EPI_ISL_1758752, EPI_ISL_1758753, EPI_ISL_1758754, EPI_ISL_1758755, EPI_ISL_1758756, EPI_ISL_1758757, EPI_ISL_1758758, EPI_ISL_1758759, EPI_ISL_1758760, EPI_ISL_1758761, EPI_ISL_1758762, EPI_ISL_1758763, EPI_ISL_1758764, EPI_ISL_1758765, EPI_ISL_1758766, EPI_ISL_1758767, EPI_ISL_1758768, EPI_ISL_1758769, EPI_ISL_1758770, EPI_ISL_1758771, EPI_ISL_1758772, EPI_ISL_1758773, EPI_ISL_1758774, EPI_ISL_1758775, EPI_ISL_1758776, EPI_ISL_1758777, EPI_ISL_1758779, EPI_ISL_1758780, EPI_ISL_1758782, EPI_ISL_1758783, EPI_ISL_1758785, EPI_ISL_1758786, EPI_ISL_1758787, EPI_ISL_1758789, EPI_ISL_1758791, EPI_ISL_1758793, EPI_ISL_1758794, EPI_ISL_1758796, EPI_ISL_1758797, EPI_ISL_1758798, EPI_ISL_1758800, EPI_ISL_1758801, EPI_ISL_1758802, EPI_ISL_1758803, EPI_ISL_1758804, EPI_ISL_1758805, EPI_ISL_1758806, EPI_ISL_1758807, EPI_ISL_1758808, EPI_ISL_1758809, EPI_ISL_1758810, EPI_ISL_1758811, EPI_ISL_1758812, EPI_ISL_1758813, EPI_ISL_1758814, EPI_ISL_1758815, EPI_ISL_1758816, EPI_ISL_1758817, EPI_ISL_1758818, EPI_ISL_1758819, EPI_ISL_1758820, EPI_ISL_1758821, EPI_ISL_1758822, EPI_ISL_1758823, EPI_ISL_1758825, EPI_ISL_1758826, EPI_ISL_1758827, EPI_ISL_1758828, EPI_ISL_1758829, EPI_ISL_1758830, EPI_ISL_1758832, EPI_ISL_1758833, EPI_ISL_1758834, EPI_ISL_1758835, EPI_ISL_1758836, EPI_ISL_1758837, EPI_ISL_1758838, EPI_ISL_1758839, EPI_ISL_1758840, EPI_ISL_1758841, EPI_ISL_1758842, EPI_ISL_1758844, EPI_ISL_1758845, EPI_ISL_1758846, EPI_ISL_1758847, EPI_ISL_1758849, EPI_ISL_1758850, EPI_ISL_1758851, EPI_ISL_1758852, EPI_ISL_1758853, EPI_ISL_1758854, EPI_ISL_1758855, EPI_ISL_1758856, EPI_ISL_1758857, EPI_ISL_1758858, EPI_ISL_1758859, EPI_ISL_1758860, EPI_ISL_1758861, EPI_ISL_1758862, EPI_ISL_1758863, EPI_ISL_1758864, EPI_ISL_1758865, EPI_ISL_1758866, EPI_ISL_1758867, EPI_ISL_1758868, EPI_ISL_1758870, EPI_ISL_1758871, EPI_ISL_1758872, EPI_ISL_1758873, EPI_ISL_1758874, EPI_ISL_1758875, EPI_ISL_1758876, EPI_ISL_1758877, EPI_ISL_1758883, EPI_ISL_1758887, EPI_ISL_1758889, EPI_ISL_1758893, EPI_ISL_1758894, EPI_ISL_1758897, EPI_ISL_1758901, EPI_ISL_1758913, EPI_ISL_1758916, EPI_ISL_1758920, EPI_ISL_1758924, EPI_ISL_1758928, EPI_ISL_1758933, EPI_ISL_1758938, EPI_ISL_1758939, EPI_ISL_1758940, EPI_ISL_1758942, EPI_ISL_1758943, EPI_ISL_1758945, EPI_ISL_1758947, EPI_ISL_1758949, EPI_ISL_1758951, EPI_ISL_1758952, EPI_ISL_1758953, EPI_ISL_1758954, EPI_ISL_1758957, EPI_ISL_1758961, EPI_ISL_1758966, EPI_ISL_1758969, EPI_ISL_1758976, EPI_ISL_1758979, EPI_ISL_1758981, EPI_ISL_1758987, EPI_ISL_1758990, EPI_ISL_1758993, EPI_ISL_1758994, EPI_ISL_1758996, EPI_ISL_1758999, EPI_ISL_1759003, EPI_ISL_1759011, EPI_ISL_1759012, EPI_ISL_1759021, EPI_ISL_1759022, EPI_ISL_1759030, EPI_ISL_1759036, EPI_ISL_1759037, EPI_ISL_1759040, EPI_ISL_1759044, EPI_ISL_1759047, EPI_ISL_1759051, EPI_ISL_1759052, EPI_ISL_1759061, EPI_ISL_1759066, EPI_ISL_1759067, EPI_ISL_1759068, EPI_ISL_1759072, EPI_ISL_1759074, EPI_ISL_1759076, EPI_ISL_1759078, EPI_ISL_1759082, EPI_ISL_1759085, EPI_ISL_1759096, EPI_ISL_1759099, EPI_ISL_1759116, EPI_ISL_1759119, EPI_ISL_1759120, EPI_ISL_1759125, EPI_ISL_1759126, EPI_ISL_1759128, EPI_ISL_1759138, EPI_ISL_1759141, EPI_ISL_1759144, EPI_ISL_1759145, EPI_ISL_1759147, EPI_ISL_1759152, EPI_ISL_1759154, EPI_ISL_1759159, EPI_ISL_1759160, EPI_ISL_1759162, EPI_ISL_1759163, EPI_ISL_1759166, EPI_ISL_1759169, EPI_ISL_1759172, EPI_ISL_1759184, EPI_ISL_1759186, EPI_ISL_1759208, EPI_ISL_1759209, EPI_ISL_1759221, EPI_ISL_1759222, EPI_ISL_1759227, EPI_ISL_1759237, EPI_ISL_1759248, EPI_ISL_1759251, EPI_ISL_1759269, EPI_ISL_1759270, EPI_ISL_1759282, EPI_ISL_1759287, EPI_ISL_1759288, EPI_ISL_1759289, EPI_ISL_1759294, EPI_ISL_1759297, EPI_ISL_1759300, EPI_ISL_1759305, EPI_ISL_1759309, EPI_ISL_1759314, EPI_ISL_1759317, EPI_ISL_1759329, EPI_ISL_1759339, EPI_ISL_1759347, EPI_ISL_1759814, EPI_ISL_1759817, EPI_ISL_1759829, EPI_ISL_1759833, EPI_ISL_1759840, EPI_ISL_1759859, EPI_ISL_1759861, EPI_ISL_1759867, EPI_ISL_1759873, EPI_ISL_1759877, EPI_ISL_1759881, EPI_ISL_1759890, EPI_ISL_1759896, EPI_ISL_1759897, EPI_ISL_1759898, EPI_ISL_1759899, EPI_ISL_1759900, EPI_ISL_1759901, EPI_ISL_1759902, EPI_ISL_1759903, EPI_ISL_1759904, EPI_ISL_1759905, EPI_ISL_1759906, EPI_ISL_1759907, EPI_ISL_1759908, EPI_ISL_1759909, EPI_ISL_1759910, EPI_ISL_1759911, EPI_ISL_1759912, EPI_ISL_1759913, EPI_ISL_1759914, EPI_ISL_1759915, EPI_ISL_1759916, EPI_ISL_1759917, EPI_ISL_1759918, EPI_ISL_1759919, EPI_ISL_1759920, EPI_ISL_1759921, EPI_ISL_1759922, EPI_ISL_1759923, EPI_ISL_1759924, EPI_ISL_1759925, EPI_ISL_1759926, EPI_ISL_1759927, EPI_ISL_1759928, EPI_ISL_1759929, EPI_ISL_1759930, EPI_ISL_1759931, EPI_ISL_1759932, EPI_ISL_1759933, EPI_ISL_1759934, EPI_ISL_1759935, EPI_ISL_1759936, EPI_ISL_1759937, EPI_ISL_1759938, EPI_ISL_1759939, EPI_ISL_1759940, EPI_ISL_1759941, EPI_ISL_1759942, EPI_ISL_1790216, EPI_ISL_1790222, EPI_ISL_1790228, EPI_ISL_1790229, EPI_ISL_1790230, EPI_ISL_1790231, EPI_ISL_1790235, EPI_ISL_1790238, EPI_ISL_1790249, EPI_ISL_1790252, EPI_ISL_1790253, EPI_ISL_1790255, EPI_ISL_1790256, EPI_ISL_1790263, EPI_ISL_1790266, EPI_ISL_1790274, EPI_ISL_1790275, EPI_ISL_1790285, EPI_ISL_1790290, EPI_ISL_1790292, EPI_ISL_1790295, EPI_ISL_1790306, EPI_ISL_1790314, EPI_ISL_1790315, EPI_ISL_1790316, EPI_ISL_1790321, EPI_ISL_1790327, EPI_ISL_1790331, EPI_ISL_1790332, EPI_ISL_1790336, EPI_ISL_1790337, EPI_ISL_1790342, EPI_ISL_1790344, EPI_ISL_1790351, EPI_ISL_1790357, EPI_ISL_1790359, EPI_ISL_1790362, EPI_ISL_1790371, EPI_ISL_1790373, EPI_ISL_1790375, EPI_ISL_1790380, EPI_ISL_1790387, EPI_ISL_1790394, EPI_ISL_1790395, EPI_ISL_1790397, EPI_ISL_1790398, EPI_ISL_1790399, EPI_ISL_1790402, EPI_ISL_1790408, EPI_ISL_1790409, EPI_ISL_1790411, EPI_ISL_1790413, EPI_ISL_1790419, EPI_ISL_1790426, EPI_ISL_1790447, EPI_ISL_1790448, EPI_ISL_1790456, EPI_ISL_1790459, EPI_ISL_1790467, EPI_ISL_1790480, EPI_ISL_1790491, EPI_ISL_1790493, EPI_ISL_1790502, EPI_ISL_1790505, EPI_ISL_1790515, EPI_ISL_1790524, EPI_ISL_1790531, EPI_ISL_1790533, EPI_ISL_1790536, EPI_ISL_1790544, EPI_ISL_1790552, EPI_ISL_1790553, EPI_ISL_1790555, EPI_ISL_1790557, EPI_ISL_1790558, EPI_ISL_1790567, EPI_ISL_1790569, EPI_ISL_1790574, EPI_ISL_1790581, EPI_ISL_1790601, EPI_ISL_1790606, EPI_ISL_1790615, EPI_ISL_1790627, EPI_ISL_1790634, EPI_ISL_1790635, EPI_ISL_1790636, EPI_ISL_1790647, EPI_ISL_1790650, EPI_ISL_1790652, EPI_ISL_1790659, EPI_ISL_1790667, EPI_ISL_1790676, EPI_ISL_1790684, EPI_ISL_1790691, EPI_ISL_1790694, EPI_ISL_1790705, EPI_ISL_1790710, EPI_ISL_1790717, EPI_ISL_1790721, EPI_ISL_1790722, EPI_ISL_1790739, EPI_ISL_1790755, EPI_ISL_1790756, EPI_ISL_1790772, EPI_ISL_1790773, EPI_ISL_1790795, EPI_ISL_1790811, EPI_ISL_1790838, EPI_ISL_1790849, EPI_ISL_1790859, EPI_ISL_1790871, EPI_ISL_1790873, EPI_ISL_1790895, EPI_ISL_1790920, EPI_ISL_1790932, EPI_ISL_1790933, EPI_ISL_1790950, EPI_ISL_1790952, EPI_ISL_1790956, EPI_ISL_1790958, EPI_ISL_1790963, EPI_ISL_1790981, EPI_ISL_1790983, EPI_ISL_1790994, EPI_ISL_1791004, EPI_ISL_1791015, EPI_ISL_1805968, EPI_ISL_1805973, EPI_ISL_1805978, EPI_ISL_1805981, EPI_ISL_1805983, EPI_ISL_1805985, EPI_ISL_1805987, EPI_ISL_1805991, EPI_ISL_1805992, EPI_ISL_1806002, EPI_ISL_1806005, EPI_ISL_1806015, EPI_ISL_1806016, EPI_ISL_1806020, EPI_ISL_1806023, EPI_ISL_1806025, EPI_ISL_1806026, EPI_ISL_1806029, EPI_ISL_1806030, EPI_ISL_1806035, EPI_ISL_1806039, EPI_ISL_1806044, EPI_ISL_1806047, EPI_ISL_1806049, EPI_ISL_1806050, EPI_ISL_1806055, EPI_ISL_1806056, EPI_ISL_1806058, EPI_ISL_1806061, EPI_ISL_1806062, EPI_ISL_1806065, EPI_ISL_1806067, EPI_ISL_1806071, EPI_ISL_1806074, EPI_ISL_1806077, EPI_ISL_1806078, EPI_ISL_1806085, EPI_ISL_1806086, EPI_ISL_1806088, EPI_ISL_1806090, EPI_ISL_1806091, EPI_ISL_1806092, EPI_ISL_1806094, EPI_ISL_1806099, EPI_ISL_1806109, EPI_ISL_1806126, EPI_ISL_1806128, EPI_ISL_1806134, EPI_ISL_1806135, EPI_ISL_1806136, EPI_ISL_1806139, EPI_ISL_1806140, EPI_ISL_1806141, EPI_ISL_1806145, EPI_ISL_1806146, EPI_ISL_1806150, EPI_ISL_1806154, EPI_ISL_1806155, EPI_ISL_1806159, EPI_ISL_1806162, EPI_ISL_1806163, EPI_ISL_1806166, EPI_ISL_1806168, EPI_ISL_1806178, EPI_ISL_1806180, EPI_ISL_1806181, EPI_ISL_1806182, EPI_ISL_1806185, EPI_ISL_1806199, EPI_ISL_1806201, EPI_ISL_1806204, EPI_ISL_1806207, EPI_ISL_1806208, EPI_ISL_1806210, EPI_ISL_1806211, EPI_ISL_1806213, EPI_ISL_1806214, EPI_ISL_1806219, EPI_ISL_1806227, EPI_ISL_1806231, EPI_ISL_1806234, EPI_ISL_1806236, EPI_ISL_1806237, EPI_ISL_1806246, EPI_ISL_1806257, EPI_ISL_1806258, EPI_ISL_1806260, EPI_ISL_1806261, EPI_ISL_1806263, EPI_ISL_1806267, EPI_ISL_1806269, EPI_ISL_1806275, EPI_ISL_1806278, EPI_ISL_1806279, EPI_ISL_1806281, EPI_ISL_1806287, EPI_ISL_1806288, EPI_ISL_1806293, EPI_ISL_1806295, EPI_ISL_1806300, EPI_ISL_1806302, EPI_ISL_1806303, EPI_ISL_1806308, EPI_ISL_1806311, EPI_ISL_1806318, EPI_ISL_1806322, EPI_ISL_1806324, EPI_ISL_1806325, EPI_ISL_1806331, EPI_ISL_1806333, EPI_ISL_1806334, EPI_ISL_1806340, EPI_ISL_1806349, EPI_ISL_1806350, EPI_ISL_1806351, EPI_ISL_1806353, EPI_ISL_1806356, EPI_ISL_1806360, EPI_ISL_1806361, EPI_ISL_1806362, EPI_ISL_1806364, EPI_ISL_1806368, EPI_ISL_1806371, EPI_ISL_1806372, EPI_ISL_1806373, EPI_ISL_1806374, EPI_ISL_1806375, EPI_ISL_1806376, EPI_ISL_1806378, EPI_ISL_1806381, EPI_ISL_1806382, EPI_ISL_1806384, EPI_ISL_1806385, EPI_ISL_1806388, EPI_ISL_1806391, EPI_ISL_1806395, EPI_ISL_1806398, EPI_ISL_1806399, EPI_ISL_1806400, EPI_ISL_1806403, EPI_ISL_1806404, EPI_ISL_1806408, EPI_ISL_1806409, EPI_ISL_1806411, EPI_ISL_1806413, EPI_ISL_1806414, EPI_ISL_1806419, EPI_ISL_1806421, EPI_ISL_1806422, EPI_ISL_1806423, EPI_ISL_1806427, EPI_ISL_1806428, EPI_ISL_1806429, EPI_ISL_1806435, EPI_ISL_1806436, EPI_ISL_1806437, EPI_ISL_1806438, EPI_ISL_1806439, EPI_ISL_1806440, EPI_ISL_1806441, EPI_ISL_1806442, EPI_ISL_1806444, EPI_ISL_1806445, EPI_ISL_1806452, EPI_ISL_1806453, EPI_ISL_1806457, EPI_ISL_1806460, EPI_ISL_1806461, EPI_ISL_1806465, EPI_ISL_1806471, EPI_ISL_1806474, EPI_ISL_1806477, EPI_ISL_1806479, EPI_ISL_1806482, EPI_ISL_1806483, EPI_ISL_1806484, EPI_ISL_1806486, EPI_ISL_1806487, EPI_ISL_1806489, EPI_ISL_1806490, EPI_ISL_1806492, EPI_ISL_1806493, EPI_ISL_1806495, EPI_ISL_1806497, EPI_ISL_1806498, EPI_ISL_1806499, EPI_ISL_1806502, EPI_ISL_1806854, EPI_ISL_1806855, EPI_ISL_1806856, EPI_ISL_1806857, EPI_ISL_1806858, EPI_ISL_1806859, EPI_ISL_1806860, EPI_ISL_1806861, EPI_ISL_1806862, EPI_ISL_1806863, EPI_ISL_1806864, EPI_ISL_1806865, EPI_ISL_1806866, EPI_ISL_1806867, EPI_ISL_1806868, EPI_ISL_1806869, EPI_ISL_1806870, EPI_ISL_1806871, EPI_ISL_1806872, EPI_ISL_1806873, EPI_ISL_1806874, EPI_ISL_1806875, EPI_ISL_1806876, EPI_ISL_1806877, EPI_ISL_1806878, EPI_ISL_1806879, EPI_ISL_1806880, EPI_ISL_1806881, EPI_ISL_1806882, EPI_ISL_1806883, EPI_ISL_1806884, EPI_ISL_1806885, EPI_ISL_1806886, EPI_ISL_1806887, EPI_ISL_1806888, EPI_ISL_1806889, EPI_ISL_1806890, EPI_ISL_1806891, EPI_ISL_1806892, EPI_ISL_1806893, EPI_ISL_1806894, EPI_ISL_1806895, EPI_ISL_1806896, EPI_ISL_1806906, EPI_ISL_1806907, EPI_ISL_1806908, EPI_ISL_1806909, EPI_ISL_1806911, EPI_ISL_1806912, EPI_ISL_1806913, EPI_ISL_1806914, EPI_ISL_1806915, EPI_ISL_1806916, EPI_ISL_1806917, EPI_ISL_1806919, EPI_ISL_1806920, EPI_ISL_1806921, EPI_ISL_1806922, EPI_ISL_1806924, EPI_ISL_1806925, EPI_ISL_1806926, EPI_ISL_1806929, EPI_ISL_1806931, EPI_ISL_1806932, EPI_ISL_1806933, EPI_ISL_1806934, EPI_ISL_1806936, EPI_ISL_1806938, EPI_ISL_1806939, EPI_ISL_1806940, EPI_ISL_1806942, EPI_ISL_1806944, EPI_ISL_1806946, EPI_ISL_1806947, EPI_ISL_1806948, EPI_ISL_1806951, EPI_ISL_1806952, EPI_ISL_1806973, EPI_ISL_1806974, EPI_ISL_1806975, EPI_ISL_1806976, EPI_ISL_1806977, EPI_ISL_1806978, EPI_ISL_1806979, EPI_ISL_1806980, EPI_ISL_1806981, EPI_ISL_1806982, EPI_ISL_1806983, EPI_ISL_1806984, EPI_ISL_1806985, EPI_ISL_1806986, EPI_ISL_1806987, EPI_ISL_1806988, EPI_ISL_1806989, EPI_ISL_1806990, EPI_ISL_1806991, EPI_ISL_1806992, EPI_ISL_1806993, EPI_ISL_1806994, EPI_ISL_1806995, EPI_ISL_1806996, EPI_ISL_1806997, EPI_ISL_1806998, EPI_ISL_1806999, EPI_ISL_1807000, EPI_ISL_1807001, EPI_ISL_1807002, EPI_ISL_1807003, EPI_ISL_1807004, EPI_ISL_1807005, EPI_ISL_1807006, EPI_ISL_1807007, EPI_ISL_1807008, EPI_ISL_1807009, EPI_ISL_1807010, EPI_ISL_1807011, EPI_ISL_1807012, EPI_ISL_1807013, EPI_ISL_1807014, EPI_ISL_1807015, EPI_ISL_1807016, EPI_ISL_1807017, EPI_ISL_1807018, EPI_ISL_1807019, EPI_ISL_1807020, EPI_ISL_1807021, EPI_ISL_1807022, EPI_ISL_1807023, EPI_ISL_1807024, EPI_ISL_1807025, EPI_ISL_1807026, EPI_ISL_1807027, EPI_ISL_1807028, EPI_ISL_1807029, EPI_ISL_1807030, EPI_ISL_1807031, EPI_ISL_1807032, EPI_ISL_1807033, EPI_ISL_1807034, EPI_ISL_1807035, EPI_ISL_1807036, EPI_ISL_1807037, EPI_ISL_1807038, EPI_ISL_1807039, EPI_ISL_1807040, EPI_ISL_1807041, EPI_ISL_1807042, EPI_ISL_1807043, EPI_ISL_1807044, EPI_ISL_1807045, EPI_ISL_1807046, EPI_ISL_1807047, EPI_ISL_1807048, EPI_ISL_1807049, EPI_ISL_1807050, EPI_ISL_1807051, EPI_ISL_1807052, EPI_ISL_1807053, EPI_ISL_1807054, EPI_ISL_1807056, EPI_ISL_1807057, EPI_ISL_1807058, EPI_ISL_1807059, EPI_ISL_1807060, EPI_ISL_1807061, EPI_ISL_1807062, EPI_ISL_1807063, EPI_ISL_1807064, EPI_ISL_1807065, EPI_ISL_1807066, EPI_ISL_1807067, EPI_ISL_1807069, EPI_ISL_1807070, EPI_ISL_1807071, EPI_ISL_1807072, EPI_ISL_1807073, EPI_ISL_1807074, EPI_ISL_1807075, EPI_ISL_1807076, EPI_ISL_1807077, EPI_ISL_1807078, EPI_ISL_1807079, EPI_ISL_1807080, EPI_ISL_1807082, EPI_ISL_1807083, EPI_ISL_1807084, EPI_ISL_1807085, EPI_ISL_1807086, EPI_ISL_1807087, EPI_ISL_1807088, EPI_ISL_1807089, EPI_ISL_1807090, EPI_ISL_1807091, EPI_ISL_1807092, EPI_ISL_1807093, EPI_ISL_1807094, EPI_ISL_1807095, EPI_ISL_1807096, EPI_ISL_1807097, EPI_ISL_1807098, EPI_ISL_1807100, EPI_ISL_1807101, EPI_ISL_1807102, EPI_ISL_1807103, EPI_ISL_1807104, EPI_ISL_1807105, EPI_ISL_1807106, EPI_ISL_1807107, EPI_ISL_1807108, EPI_ISL_1807109, EPI_ISL_1807110, EPI_ISL_1807111, EPI_ISL_1807112, EPI_ISL_1807113, EPI_ISL_1807114, EPI_ISL_1807115, EPI_ISL_1807116, EPI_ISL_1807117, EPI_ISL_1807118, EPI_ISL_1807119, EPI_ISL_1807120, EPI_ISL_1807121, EPI_ISL_1807122, EPI_ISL_1807123, EPI_ISL_1807124, EPI_ISL_1807125, EPI_ISL_1807127, EPI_ISL_1807128, EPI_ISL_1807129, EPI_ISL_1807130, EPI_ISL_1807131, EPI_ISL_1807132, EPI_ISL_1807133, EPI_ISL_1807134, EPI_ISL_1807135, EPI_ISL_1807136, EPI_ISL_1807137, EPI_ISL_1807138, EPI_ISL_1807139, EPI_ISL_1807140, EPI_ISL_1807141, EPI_ISL_1807142, EPI_ISL_1807143, EPI_ISL_1807144, EPI_ISL_1807145, EPI_ISL_1807146, EPI_ISL_1807147, EPI_ISL_1807148, EPI_ISL_1807149, EPI_ISL_1807150, EPI_ISL_1807151, EPI_ISL_1807152, EPI_ISL_1807153, EPI_ISL_1807154, EPI_ISL_1807156, EPI_ISL_1807157, EPI_ISL_1807158, EPI_ISL_1807159, EPI_ISL_1807160, EPI_ISL_1807161, EPI_ISL_1807162, EPI_ISL_1807163, EPI_ISL_1807164, EPI_ISL_1807165, EPI_ISL_1807166, EPI_ISL_1807167, EPI_ISL_1807168, EPI_ISL_1807169, EPI_ISL_1807170, EPI_ISL_1807171, EPI_ISL_1807172, EPI_ISL_1807173, EPI_ISL_1807174, EPI_ISL_1807175, EPI_ISL_1807176, EPI_ISL_1807177, EPI_ISL_1807178, EPI_ISL_1829484, EPI_ISL_1829485, EPI_ISL_1829486, EPI_ISL_1829489, EPI_ISL_1829490, EPI_ISL_1829493, EPI_ISL_1829494, EPI_ISL_1829495, EPI_ISL_1829497, EPI_ISL_1829502, EPI_ISL_1829503, EPI_ISL_1829505, EPI_ISL_1829506, EPI_ISL_1829510, EPI_ISL_1829512, EPI_ISL_1829515, EPI_ISL_1829520, EPI_ISL_1829521, EPI_ISL_1829524, EPI_ISL_1829529, EPI_ISL_1829537, EPI_ISL_1829539, EPI_ISL_1829541, EPI_ISL_1829543, EPI_ISL_1829545, EPI_ISL_1829546, EPI_ISL_1829548, EPI_ISL_1829552, EPI_ISL_1829553, EPI_ISL_1829562, EPI_ISL_1829564, EPI_ISL_1829568, EPI_ISL_1829573, EPI_ISL_1829575, EPI_ISL_1829577, EPI_ISL_1829578, EPI_ISL_1829582, EPI_ISL_1829583, EPI_ISL_1829588, EPI_ISL_1829589, EPI_ISL_1829590, EPI_ISL_1829591, EPI_ISL_1829596, EPI_ISL_1829598, EPI_ISL_1829602, EPI_ISL_1829604, EPI_ISL_1829605, EPI_ISL_1829606, EPI_ISL_1829607, EPI_ISL_1829609, EPI_ISL_1829610, EPI_ISL_1829611, EPI_ISL_1829614, EPI_ISL_1829615, EPI_ISL_1829616, EPI_ISL_1829618, EPI_ISL_1829622, EPI_ISL_1829624, EPI_ISL_1829626, EPI_ISL_1829631, EPI_ISL_1829636, EPI_ISL_1829637, EPI_ISL_1829639, EPI_ISL_1829640, EPI_ISL_1829642, EPI_ISL_1829644, EPI_ISL_1829648, EPI_ISL_1829649, EPI_ISL_1829654, EPI_ISL_1829655, EPI_ISL_1829657, EPI_ISL_1829658, EPI_ISL_1829661, EPI_ISL_1829662, EPI_ISL_1829663, EPI_ISL_1829664, EPI_ISL_1829665, EPI_ISL_1829667, EPI_ISL_1829669, EPI_ISL_1829670, EPI_ISL_1829671, EPI_ISL_1829674, EPI_ISL_1829678, EPI_ISL_1829684, EPI_ISL_1829688, EPI_ISL_1829690, EPI_ISL_1829692, EPI_ISL_1829694, EPI_ISL_1829696, EPI_ISL_1829697, EPI_ISL_1829698, EPI_ISL_1829699, EPI_ISL_1829702, EPI_ISL_1829704, EPI_ISL_1829707, EPI_ISL_1829708, EPI_ISL_1829710, EPI_ISL_1829711, EPI_ISL_1829725, EPI_ISL_1829728, EPI_ISL_1829733, EPI_ISL_1829736, EPI_ISL_1829737, EPI_ISL_1829739, EPI_ISL_1829745, EPI_ISL_1829747, EPI_ISL_1829748, EPI_ISL_1829749, EPI_ISL_1829752, EPI_ISL_1829753, EPI_ISL_1829756, EPI_ISL_1829758, EPI_ISL_1829767, EPI_ISL_1829769, EPI_ISL_1829775, EPI_ISL_1829780, EPI_ISL_1829781, EPI_ISL_1829782, EPI_ISL_1829785, EPI_ISL_1829787, EPI_ISL_1829791, EPI_ISL_1829796, EPI_ISL_1829802, EPI_ISL_1829833, EPI_ISL_1829908, EPI_ISL_1829972, EPI_ISL_1829998, EPI_ISL_1830018, EPI_ISL_1830068, EPI_ISL_1830102, EPI_ISL_1830103, EPI_ISL_1830106, EPI_ISL_1830107, EPI_ISL_1830113, EPI_ISL_1830119, EPI_ISL_1830124, EPI_ISL_1830129, EPI_ISL_1830130, EPI_ISL_1830134, EPI_ISL_1830135, EPI_ISL_1830139, EPI_ISL_1830143, EPI_ISL_1830144, EPI_ISL_1830145, EPI_ISL_1830147, EPI_ISL_1830148, EPI_ISL_1830149, EPI_ISL_1830151, EPI_ISL_1830153, EPI_ISL_1830160, EPI_ISL_1830161, EPI_ISL_1830163, EPI_ISL_1830169, EPI_ISL_1830181, EPI_ISL_1830190, EPI_ISL_1830191, EPI_ISL_1830193, EPI_ISL_1830198, EPI_ISL_1830199, EPI_ISL_1830204, EPI_ISL_1830205, EPI_ISL_1830212, EPI_ISL_1830215, EPI_ISL_1830228, EPI_ISL_1830232, EPI_ISL_1830243, EPI_ISL_1830244, EPI_ISL_1830246, EPI_ISL_1830250, EPI_ISL_1830253, EPI_ISL_1830254, EPI_ISL_1830256, EPI_ISL_1830258, EPI_ISL_1830261, EPI_ISL_1830263, EPI_ISL_1830265, EPI_ISL_1830266, EPI_ISL_1830268, EPI_ISL_1830273, EPI_ISL_1830274, EPI_ISL_1830276, EPI_ISL_1830277, EPI_ISL_1830279, EPI_ISL_1830283, EPI_ISL_1830285, EPI_ISL_1830287, EPI_ISL_1830293, EPI_ISL_1830294, EPI_ISL_1830295, EPI_ISL_1830297, EPI_ISL_1830299, EPI_ISL_1830301, EPI_ISL_1830307, EPI_ISL_1830311, EPI_ISL_1830318, EPI_ISL_1830320, EPI_ISL_1830322, EPI_ISL_1830329, EPI_ISL_1830334, EPI_ISL_1830336, EPI_ISL_1830342, EPI_ISL_1830345, EPI_ISL_1830346, EPI_ISL_1830349, EPI_ISL_1830352, EPI_ISL_1830354, EPI_ISL_1830358, EPI_ISL_1830362, EPI_ISL_1830367, EPI_ISL_1830369, EPI_ISL_1830373, EPI_ISL_1830379, EPI_ISL_1830380, EPI_ISL_1830381, EPI_ISL_1830384, EPI_ISL_1830385, EPI_ISL_1830387, EPI_ISL_1830392, EPI_ISL_1830399, EPI_ISL_1830418, EPI_ISL_1830421, EPI_ISL_1830428, EPI_ISL_1830430, EPI_ISL_1830435, EPI_ISL_1830436, EPI_ISL_1830442, EPI_ISL_1830446, EPI_ISL_1830451, EPI_ISL_1830452, EPI_ISL_1830453, EPI_ISL_1830456, EPI_ISL_1830465, EPI_ISL_1830469, EPI_ISL_1830474, EPI_ISL_1830475, EPI_ISL_1830477, EPI_ISL_1830493, EPI_ISL_1830494, EPI_ISL_1830497, EPI_ISL_1830502, EPI_ISL_1830508, EPI_ISL_1830509, EPI_ISL_1830522, EPI_ISL_1830523, EPI_ISL_1830527, EPI_ISL_1830528, EPI_ISL_1830532, EPI_ISL_1830540, EPI_ISL_1830544, EPI_ISL_1830546, EPI_ISL_1830551, EPI_ISL_1830554, EPI_ISL_1830570, EPI_ISL_1830592, EPI_ISL_1830593, EPI_ISL_1830595, EPI_ISL_1830599, EPI_ISL_1830600, EPI_ISL_1830608, EPI_ISL_1830609, EPI_ISL_1830612, EPI_ISL_1830615, EPI_ISL_1830629, EPI_ISL_1830638, EPI_ISL_1830643, EPI_ISL_1830657, EPI_ISL_1830660, EPI_ISL_1830668, EPI_ISL_1830670, EPI_ISL_1830672, EPI_ISL_1830674, EPI_ISL_1830685, EPI_ISL_1830700, EPI_ISL_1830702, EPI_ISL_1830707, EPI_ISL_1830717, EPI_ISL_1830729, EPI_ISL_1830732, EPI_ISL_1830733, EPI_ISL_1830734, EPI_ISL_1830736, EPI_ISL_1830737, EPI_ISL_1830738, EPI_ISL_1830739, EPI_ISL_1830742, EPI_ISL_1830743, EPI_ISL_1830744, EPI_ISL_1830745, EPI_ISL_1830747, EPI_ISL_1830749, EPI_ISL_1830750, EPI_ISL_1830751, EPI_ISL_1830753, EPI_ISL_1830754, EPI_ISL_1830755, EPI_ISL_1830756, EPI_ISL_1830757, EPI_ISL_1830759, EPI_ISL_1830760, EPI_ISL_1830765, EPI_ISL_1830766, EPI_ISL_1830767, EPI_ISL_1830768, EPI_ISL_1830770, EPI_ISL_1830771, EPI_ISL_1830773, EPI_ISL_1830774, EPI_ISL_1830776, EPI_ISL_1830777, EPI_ISL_1830779, EPI_ISL_1830781, EPI_ISL_1830782, EPI_ISL_1830783, EPI_ISL_1830784, EPI_ISL_1830785, EPI_ISL_1830786, EPI_ISL_1830787, EPI_ISL_1830788, EPI_ISL_1830790, EPI_ISL_1830792, EPI_ISL_1830794, EPI_ISL_1830795, EPI_ISL_1830796, EPI_ISL_1830797, EPI_ISL_1830798, EPI_ISL_1830800, EPI_ISL_1830801, EPI_ISL_1830802, EPI_ISL_1830804, EPI_ISL_1830809, EPI_ISL_1830810, EPI_ISL_1830816, EPI_ISL_1830820, EPI_ISL_1830821, EPI_ISL_1830822, EPI_ISL_1830824, EPI_ISL_1830825, EPI_ISL_1830826, EPI_ISL_1830827, EPI_ISL_1830829, EPI_ISL_1830830, EPI_ISL_1830831, EPI_ISL_1830834, EPI_ISL_1830835, EPI_ISL_1830837, EPI_ISL_1830838, EPI_ISL_1830839, EPI_ISL_1830841, EPI_ISL_1830842, EPI_ISL_1830843, EPI_ISL_1830845, EPI_ISL_1830847, EPI_ISL_1830849, EPI_ISL_1830851, EPI_ISL_1830852, EPI_ISL_1830853, EPI_ISL_1830854, EPI_ISL_1830855, EPI_ISL_1830857, EPI_ISL_1830860, EPI_ISL_1830861, EPI_ISL_1830862, EPI_ISL_1830864, EPI_ISL_1830865, EPI_ISL_1830866, EPI_ISL_1830867, EPI_ISL_1830871, EPI_ISL_1830872, EPI_ISL_1830873, EPI_ISL_1830874, EPI_ISL_1830875, EPI_ISL_1830876, EPI_ISL_1830877, EPI_ISL_1830879, EPI_ISL_1830882, EPI_ISL_1830884, EPI_ISL_1830885, EPI_ISL_1830886, EPI_ISL_1830887, EPI_ISL_1830889, EPI_ISL_1830890, EPI_ISL_1830891, EPI_ISL_1830892, EPI_ISL_1830893, EPI_ISL_1830894, EPI_ISL_1830899, EPI_ISL_1830900, EPI_ISL_1830901, EPI_ISL_1830902, EPI_ISL_1830903, EPI_ISL_1830904, EPI_ISL_1830905, EPI_ISL_1830906, EPI_ISL_1830907, EPI_ISL_1830908, EPI_ISL_1830909, EPI_ISL_1830910, EPI_ISL_1830912, EPI_ISL_1830913, EPI_ISL_1830915, EPI_ISL_1830916, EPI_ISL_1830917, EPI_ISL_1830918, EPI_ISL_1830919, EPI_ISL_1830920, EPI_ISL_1830922, EPI_ISL_1830923, EPI_ISL_1830924, EPI_ISL_1830926, EPI_ISL_1830928, EPI_ISL_1830930, EPI_ISL_1830931, EPI_ISL_1830932, EPI_ISL_1830933, EPI_ISL_1830934, EPI_ISL_1830936, EPI_ISL_1830937, EPI_ISL_1830938, EPI_ISL_1830939, EPI_ISL_1830940, EPI_ISL_1830941, EPI_ISL_1830942, EPI_ISL_1830945, EPI_ISL_1830946, EPI_ISL_1830948, EPI_ISL_1830950, EPI_ISL_1830951, EPI_ISL_1830952, EPI_ISL_1830953, EPI_ISL_1830954, EPI_ISL_1830955, EPI_ISL_1830957, EPI_ISL_1830958, EPI_ISL_1830960, EPI_ISL_1830962, EPI_ISL_1830963, EPI_ISL_1830964, EPI_ISL_1830965, EPI_ISL_1830967, EPI_ISL_1830969, EPI_ISL_1830971, EPI_ISL_1830972, EPI_ISL_1830973, EPI_ISL_1830974, EPI_ISL_1830975, EPI_ISL_1830976, EPI_ISL_1830977, EPI_ISL_1830978, EPI_ISL_1830980, EPI_ISL_1830982, EPI_ISL_1830984, EPI_ISL_1830986, EPI_ISL_1830987, EPI_ISL_1830988, EPI_ISL_1830989, EPI_ISL_1830990, EPI_ISL_1830991, EPI_ISL_1830992, EPI_ISL_1830993, EPI_ISL_1830994, EPI_ISL_1830995, EPI_ISL_1830996, EPI_ISL_1830997, EPI_ISL_1830999, EPI_ISL_1831000, EPI_ISL_1831004, EPI_ISL_1831007, EPI_ISL_1831009, EPI_ISL_1831010, EPI_ISL_1831012, EPI_ISL_1831013, EPI_ISL_1831014, EPI_ISL_1831016, EPI_ISL_1831017, EPI_ISL_1831020, EPI_ISL_1831021, EPI_ISL_1831022, EPI_ISL_1831024, EPI_ISL_1831025, EPI_ISL_1831026, EPI_ISL_1831028, EPI_ISL_1831030, EPI_ISL_1831031, EPI_ISL_1831032, EPI_ISL_1831033, EPI_ISL_1831035, EPI_ISL_1831036, EPI_ISL_1831037, EPI_ISL_1831038, EPI_ISL_1831040, EPI_ISL_1831041, EPI_ISL_1831042, EPI_ISL_1831044, EPI_ISL_1831046, EPI_ISL_1831047, EPI_ISL_1831051, EPI_ISL_1831053, EPI_ISL_1831056, EPI_ISL_1831059, EPI_ISL_1831062, EPI_ISL_1831063, EPI_ISL_1831064, EPI_ISL_1831065, EPI_ISL_1831066, EPI_ISL_1831067, EPI_ISL_1831068, EPI_ISL_1831069, EPI_ISL_1831072, EPI_ISL_1831075, EPI_ISL_1831076, EPI_ISL_1831077, EPI_ISL_1831078, EPI_ISL_1831081, EPI_ISL_1831083, EPI_ISL_1831088, EPI_ISL_1831089, EPI_ISL_1831090, EPI_ISL_1831091, EPI_ISL_1831092, EPI_ISL_1831093, EPI_ISL_1831094, EPI_ISL_1831095, EPI_ISL_1831096, EPI_ISL_1831097, EPI_ISL_1831098, EPI_ISL_1831099, EPI_ISL_1831100, EPI_ISL_1831101, EPI_ISL_1831102, EPI_ISL_1831103, EPI_ISL_1831104, EPI_ISL_1831105, EPI_ISL_1831106, EPI_ISL_1831107, EPI_ISL_1831108, EPI_ISL_1831109, EPI_ISL_1831111, EPI_ISL_1831112, EPI_ISL_1831113, EPI_ISL_1831114, EPI_ISL_1831115, EPI_ISL_1831116, EPI_ISL_1831117, EPI_ISL_1831118, EPI_ISL_1831119, EPI_ISL_1831120, EPI_ISL_1831121, EPI_ISL_1831122, EPI_ISL_1831123, EPI_ISL_1831124, EPI_ISL_1831125, EPI_ISL_1831126, EPI_ISL_1831127, EPI_ISL_1831128, EPI_ISL_1831129, EPI_ISL_1831130, EPI_ISL_1831131, EPI_ISL_1831132, EPI_ISL_1831133, EPI_ISL_1831134, EPI_ISL_1831135, EPI_ISL_1831136, EPI_ISL_1831137, EPI_ISL_1831138, EPI_ISL_1831139, EPI_ISL_1831140, EPI_ISL_1831141, EPI_ISL_1831142, EPI_ISL_1831143, EPI_ISL_1831144, EPI_ISL_1831145, EPI_ISL_1831146, EPI_ISL_1831147, EPI_ISL_1831148, EPI_ISL_1831149, EPI_ISL_1831150, EPI_ISL_1831151, EPI_ISL_1831152, EPI_ISL_1831153, EPI_ISL_1831154, EPI_ISL_1831155, EPI_ISL_1831156, EPI_ISL_1831157, EPI_ISL_1831158, EPI_ISL_1831159, EPI_ISL_1831160, EPI_ISL_1831161, EPI_ISL_1831162, EPI_ISL_1831163, EPI_ISL_1831164, EPI_ISL_1831165, EPI_ISL_1831166, EPI_ISL_1831167, EPI_ISL_1831168, EPI_ISL_1831169, EPI_ISL_1831170, EPI_ISL_1831171, EPI_ISL_1831172, EPI_ISL_1831173, EPI_ISL_1831174, EPI_ISL_1831175, EPI_ISL_1831176, EPI_ISL_1831177, EPI_ISL_1831178, EPI_ISL_1831179, EPI_ISL_1831180, EPI_ISL_1831181, EPI_ISL_1831182, EPI_ISL_1831183, EPI_ISL_1831184, EPI_ISL_1831185, EPI_ISL_1831186, EPI_ISL_1831187, EPI_ISL_1831188, EPI_ISL_1831189, EPI_ISL_1831190, EPI_ISL_1831191, EPI_ISL_1831195, EPI_ISL_1831196, EPI_ISL_1831197, EPI_ISL_1831198, EPI_ISL_1831199, EPI_ISL_1831200, EPI_ISL_1831201, EPI_ISL_1831202, EPI_ISL_1831203, EPI_ISL_1831204, EPI_ISL_1831205, EPI_ISL_1831206, EPI_ISL_1831207, EPI_ISL_1831208, EPI_ISL_1831209, EPI_ISL_1831210, EPI_ISL_1831211, EPI_ISL_1831212, EPI_ISL_1831213, EPI_ISL_1831214, EPI_ISL_1831215, EPI_ISL_1831216, EPI_ISL_1831217, EPI_ISL_1831218, EPI_ISL_1831219, EPI_ISL_1831220, EPI_ISL_1831221, EPI_ISL_1831222, EPI_ISL_1831223, EPI_ISL_1831224, EPI_ISL_1831225, EPI_ISL_1831226, EPI_ISL_1831227, EPI_ISL_1831228, EPI_ISL_1831229, EPI_ISL_1831230, EPI_ISL_1831231, EPI_ISL_1831232, EPI_ISL_1831233, EPI_ISL_1831234, EPI_ISL_1831235, EPI_ISL_1831236, EPI_ISL_1831237, EPI_ISL_1831238, EPI_ISL_1831239, EPI_ISL_1831240, EPI_ISL_1831241, EPI_ISL_1831242, EPI_ISL_1831243, EPI_ISL_1831244, EPI_ISL_1831245, EPI_ISL_1831246, EPI_ISL_1831247, EPI_ISL_1831248, EPI_ISL_1831249, EPI_ISL_1831250, EPI_ISL_1831251, EPI_ISL_1831252, EPI_ISL_1831253, EPI_ISL_1831254, EPI_ISL_1831255, EPI_ISL_1831256, EPI_ISL_1831257, EPI_ISL_1831258, EPI_ISL_1831259, EPI_ISL_1831260, EPI_ISL_1912008, EPI_ISL_1912015, EPI_ISL_1912024, EPI_ISL_1912046, EPI_ISL_1912060, EPI_ISL_1912063, EPI_ISL_1912065, EPI_ISL_1912069, EPI_ISL_1912083, EPI_ISL_1912087, EPI_ISL_1912104, EPI_ISL_1912127, EPI_ISL_1912137, EPI_ISL_1912147, EPI_ISL_1912155, EPI_ISL_1912181, EPI_ISL_1912187, EPI_ISL_1912190, EPI_ISL_1912209, EPI_ISL_1912228, EPI_ISL_1912229, EPI_ISL_1912231, EPI_ISL_1912235, EPI_ISL_1912238, EPI_ISL_1912246, EPI_ISL_1912254, EPI_ISL_1912259, EPI_ISL_1912275, EPI_ISL_1912278, EPI_ISL_1912280, EPI_ISL_1912289, EPI_ISL_1912312, EPI_ISL_1912320, EPI_ISL_1912324, EPI_ISL_1912326, EPI_ISL_1912330, EPI_ISL_1912340, EPI_ISL_1985444, EPI_ISL_2005631, EPI_ISL_2005679, EPI_ISL_2005700, EPI_ISL_2005761, EPI_ISL_2006087, EPI_ISL_2006092, EPI_ISL_2006115, EPI_ISL_2006118, EPI_ISL_2006123, EPI_ISL_2006133, EPI_ISL_2006144, EPI_ISL_2006151, EPI_ISL_2006157, EPI_ISL_2006161, EPI_ISL_2006170, EPI_ISL_2006171, EPI_ISL_2006180, EPI_ISL_2006183, EPI_ISL_2006195, EPI_ISL_2006196, EPI_ISL_2006202, EPI_ISL_2006204, EPI_ISL_2006205, EPI_ISL_2006215, EPI_ISL_2006226, EPI_ISL_2006229, EPI_ISL_2006230, EPI_ISL_2006237, EPI_ISL_2006241, EPI_ISL_2006243, EPI_ISL_2006253, EPI_ISL_2006254, EPI_ISL_2006266, EPI_ISL_2006271, EPI_ISL_2006283, EPI_ISL_2006286, EPI_ISL_2006288, EPI_ISL_2006399, EPI_ISL_2022595, EPI_ISL_2022657, EPI_ISL_2022659, EPI_ISL_2022668, EPI_ISL_2022765

| | | | |
|---|---|---|---|
| see above | Lighthouse Lab in Glasgow | Wellcome Sanger Institute for the COVID-19 Genomics UK (COG-UK) Consortium | Anna Dominiczak and Alex Alderton; Carol Clugston; Cordelia Langford; David Gray; David K. Jackson; Dominic Kwiatkowski; Ewan Harrison; Harper VanSteenhouse; Ian Johnston; Jeffrey Barrett; John Sillitoe on behalf of the Wellcome Sanger Institute COVID-19 Surveillance Team; Roberto Amato; Sonia Goncalves; Yumi Kasai |

EPI_ISL_1697480, EPI_ISL_1697506, EPI_ISL_1697549, EPI_ISL_1697568, EPI_ISL_1697591, EPI_ISL_1697627, EPI_ISL_1697685, EPI_ISL_1697696, EPI_ISL_1697702, EPI_ISL_1697711, EPI_ISL_1697970, EPI_ISL_1697972, EPI_ISL_1697973, EPI_ISL_1697974, EPI_ISL_1697977, EPI_ISL_1697981, EPI_ISL_1697983, EPI_ISL_1697984, EPI_ISL_1697985, EPI_ISL_1697987, EPI_ISL_1697988, EPI_ISL_1697990, EPI_ISL_1697991, EPI_ISL_1697992, EPI_ISL_1697993, EPI_ISL_1697994, EPI_ISL_1697995, EPI_ISL_1697997, EPI_ISL_1697999, EPI_ISL_1698001, EPI_ISL_1698002, EPI_ISL_1698003, EPI_ISL_1698004, EPI_ISL_1698005, EPI_ISL_1698006, EPI_ISL_1698008, EPI_ISL_1698009, EPI_ISL_1698013, EPI_ISL_1698014, EPI_ISL_1698017, EPI_ISL_1698018, EPI_ISL_1698020, EPI_ISL_1698021, EPI_ISL_1698022, EPI_ISL_1698023, EPI_ISL_1698024, EPI_ISL_1698027, EPI_ISL_1698032, EPI_ISL_1698033, EPI_ISL_1698035, EPI_ISL_1698038, EPI_ISL_1698039, EPI_ISL_1698040, EPI_ISL_1698042, EPI_ISL_1698043, EPI_ISL_1698047, EPI_ISL_1698048, EPI_ISL_1698050, EPI_ISL_1698051, EPI_ISL_1698052, EPI_ISL_1698053, EPI_ISL_1698055, EPI_ISL_1698059, EPI_ISL_1698060, EPI_ISL_1698064,

EPI_ISL_1698065, EPI_ISL_1698068, EPI_ISL_1698070, EPI_ISL_1698072, EPI_ISL_1698074, EPI_ISL_1698076, EPI_ISL_1698079, EPI_ISL_1698081, EPI_ISL_1698083, EPI_ISL_1698084, EPI_ISL_1698085, EPI_ISL_1698086, EPI_ISL_1698088, EPI_ISL_1698089, EPI_ISL_1698092, EPI_ISL_1698093, EPI_ISL_1698094, EPI_ISL_1698095, EPI_ISL_1698096, EPI_ISL_1698097, EPI_ISL_1698098, EPI_ISL_1698101, EPI_ISL_1698104, EPI_ISL_1698105, EPI_ISL_1698106, EPI_ISL_1698108, EPI_ISL_1698110, EPI_ISL_1698111, EPI_ISL_1698112, EPI_ISL_1698119, EPI_ISL_1698120, EPI_ISL_1698121, EPI_ISL_1698122, EPI_ISL_1698123, EPI_ISL_1698124, EPI_ISL_1698125, EPI_ISL_1698126, EPI_ISL_1698127, EPI_ISL_1698128, EPI_ISL_1698133, EPI_ISL_1698134, EPI_ISL_1698135, EPI_ISL_1698137, EPI_ISL_1698138, EPI_ISL_1698139, EPI_ISL_1698140, EPI_ISL_1698141, EPI_ISL_1698142, EPI_ISL_1698143, EPI_ISL_1698144, EPI_ISL_1698145, EPI_ISL_1698147, EPI_ISL_1698148, EPI_ISL_1698149, EPI_ISL_1698151, EPI_ISL_1698154, EPI_ISL_1698155, EPI_ISL_1698158, EPI_ISL_1698159, EPI_ISL_1698160, EPI_ISL_1698161, EPI_ISL_1698162, EPI_ISL_1698163, EPI_ISL_1698167, EPI_ISL_1698168, EPI_ISL_1698170, EPI_ISL_1698171, EPI_ISL_1698174, EPI_ISL_1698175, EPI_ISL_1698177, EPI_ISL_1698178, EPI_ISL_1698179, EPI_ISL_1698181, EPI_ISL_1698184, EPI_ISL_1698187, EPI_ISL_1698189, EPI_ISL_1698191, EPI_ISL_1698192, EPI_ISL_1698196, EPI_ISL_1698198, EPI_ISL_1698200, EPI_ISL_1698201, EPI_ISL_1698206, EPI_ISL_1698208, EPI_ISL_1698210, EPI_ISL_1698211, EPI_ISL_1698212, EPI_ISL_1698216, EPI_ISL_1698217, EPI_ISL_1698218, EPI_ISL_1698220, EPI_ISL_1698222, EPI_ISL_1698224, EPI_ISL_1698230, EPI_ISL_1698232, EPI_ISL_1698233, EPI_ISL_1698235, EPI_ISL_1698236, EPI_ISL_1698237, EPI_ISL_1698238, EPI_ISL_1698239, EPI_ISL_1698240, EPI_ISL_1698243, EPI_ISL_1698244, EPI_ISL_1698245, EPI_ISL_1698246, EPI_ISL_1698248, EPI_ISL_1698249, EPI_ISL_1698250, EPI_ISL_1698251, EPI_ISL_1698253, EPI_ISL_1698322, EPI_ISL_1699297, EPI_ISL_1699302, EPI_ISL_1699321, EPI_ISL_1699335, EPI_ISL_1699336, EPI_ISL_1699355, EPI_ISL_1699363, EPI_ISL_1699420, EPI_ISL_1699555, EPI_ISL_1699559, EPI_ISL_1699566, EPI_ISL_1699578, EPI_ISL_1699580, EPI_ISL_1699582, EPI_ISL_1699598, EPI_ISL_1699605, EPI_ISL_1699609, EPI_ISL_1699620, EPI_ISL_1699622, EPI_ISL_1699628, EPI_ISL_1699635, EPI_ISL_1699636, EPI_ISL_1699638, EPI_ISL_1699659, EPI_ISL_1699661, EPI_ISL_1699666, EPI_ISL_1699667, EPI_ISL_1699675, EPI_ISL_1699676, EPI_ISL_1699680, EPI_ISL_1699691, EPI_ISL_1699692, EPI_ISL_1699698, EPI_ISL_1699710, EPI_ISL_1699715, EPI_ISL_1699722, EPI_ISL_1699724, EPI_ISL_1699729, EPI_ISL_1699730, EPI_ISL_1699735, EPI_ISL_1699736, EPI_ISL_1699740, EPI_ISL_1699759, EPI_ISL_1699767, EPI_ISL_1699770, EPI_ISL_1699772, EPI_ISL_1699788, EPI_ISL_1699794, EPI_ISL_1699796, EPI_ISL_1699800, EPI_ISL_1699803, EPI_ISL_1699805, EPI_ISL_1699811, EPI_ISL_1699815, EPI_ISL_1699822, EPI_ISL_1699840, EPI_ISL_1718323, EPI_ISL_1718328, EPI_ISL_1718333, EPI_ISL_1718334, EPI_ISL_1718337, EPI_ISL_1718344, EPI_ISL_1718347, EPI_ISL_1718349, EPI_ISL_1718350, EPI_ISL_1718353, EPI_ISL_1718355, EPI_ISL_1718358, EPI_ISL_1718359, EPI_ISL_1718371, EPI_ISL_1718374, EPI_ISL_1718377, EPI_ISL_1718378, EPI_ISL_1718382, EPI_ISL_1718386, EPI_ISL_1718391, EPI_ISL_1718395, EPI_ISL_1718397, EPI_ISL_1718402, EPI_ISL_1718403, EPI_ISL_1718404, EPI_ISL_1718405, EPI_ISL_1718407, EPI_ISL_1718410, EPI_ISL_1718412, EPI_ISL_1718415, EPI_ISL_1718418, EPI_ISL_1718420, EPI_ISL_1718424, EPI_ISL_1718427, EPI_ISL_1718432, EPI_ISL_1718435, EPI_ISL_1718440, EPI_ISL_1718441, EPI_ISL_1718443, EPI_ISL_1718445, EPI_ISL_1718447, EPI_ISL_1718448, EPI_ISL_1718449, EPI_ISL_1718451, EPI_ISL_1718456, EPI_ISL_1718457, EPI_ISL_1718461, EPI_ISL_1718462, EPI_ISL_1718463, EPI_ISL_1718467, EPI_ISL_1718468, EPI_ISL_1718472, EPI_ISL_1718473, EPI_ISL_1718475, EPI_ISL_1718478, EPI_ISL_1718481, EPI_ISL_1718485, EPI_ISL_1718487, EPI_ISL_1718488, EPI_ISL_1718489, EPI_ISL_1718491, EPI_ISL_1718494, EPI_ISL_1718495, EPI_ISL_1718502, EPI_ISL_1718503, EPI_ISL_1718506, EPI_ISL_1718508, EPI_ISL_1718509, EPI_ISL_1718513, EPI_ISL_1718514, EPI_ISL_1718515, EPI_ISL_1718516, EPI_ISL_1718520, EPI_ISL_1718523, EPI_ISL_1718524, EPI_ISL_1718525, EPI_ISL_1718526, EPI_ISL_1718529, EPI_ISL_1718530, EPI_ISL_1718533, EPI_ISL_1718534, EPI_ISL_1718535, EPI_ISL_1718537, EPI_ISL_1718539, EPI_ISL_1718546, EPI_ISL_1718547, EPI_ISL_1718550, EPI_ISL_1718551, EPI_ISL_1718553, EPI_ISL_1718554, EPI_ISL_1718556, EPI_ISL_1718558, EPI_ISL_1718559, EPI_ISL_1718562, EPI_ISL_1718564, EPI_ISL_1718566, EPI_ISL_1718568, EPI_ISL_1718572, EPI_ISL_1718573, EPI_ISL_1718574, EPI_ISL_1718575, EPI_ISL_1718577, EPI_ISL_1718581, EPI_ISL_1718586, EPI_ISL_1718587, EPI_ISL_1718590, EPI_ISL_1718594, EPI_ISL_1718596, EPI_ISL_1718600, EPI_ISL_1718602, EPI_ISL_1718606, EPI_ISL_1718610, EPI_ISL_1718611, EPI_ISL_1718612, EPI_ISL_1718613, EPI_ISL_1718617, EPI_ISL_1718618, EPI_ISL_1718622, EPI_ISL_1718640, EPI_ISL_1718642, EPI_ISL_1718645, EPI_ISL_1718663, EPI_ISL_1718668, EPI_ISL_1718685, EPI_ISL_1718686, EPI_ISL_1718688, EPI_ISL_1718691, EPI_ISL_1718692, EPI_ISL_1718695, EPI_ISL_1718697, EPI_ISL_1718701, EPI_ISL_1718707, EPI_ISL_1718742, EPI_ISL_1718744, EPI_ISL_1718746, EPI_ISL_1718750, EPI_ISL_1718760, EPI_ISL_1718783, EPI_ISL_1718789, EPI_ISL_1718791, EPI_ISL_1718810, EPI_ISL_1718818, EPI_ISL_1718822, EPI_ISL_1718838, EPI_ISL_1718846, EPI_ISL_1718857, EPI_ISL_1718858, EPI_ISL_1718860, EPI_ISL_1718869, EPI_ISL_1718875, EPI_ISL_1718885, EPI_ISL_1718886, EPI_ISL_1718887, EPI_ISL_1718888, EPI_ISL_1718889, EPI_ISL_1718890, EPI_ISL_1718891, EPI_ISL_1718892, EPI_ISL_1718893, EPI_ISL_1718894, EPI_ISL_1718895, EPI_ISL_1718896, EPI_ISL_1718898, EPI_ISL_1718899, EPI_ISL_1718900, EPI_ISL_1718901, EPI_ISL_1718902, EPI_ISL_1718903, EPI_ISL_1718905, EPI_ISL_1718906, EPI_ISL_1718907, EPI_ISL_1718908, EPI_ISL_1718909, EPI_ISL_1718910, EPI_ISL_1718911, EPI_ISL_1718914, EPI_ISL_1718915, EPI_ISL_1718916, EPI_ISL_1718917, EPI_ISL_1718918, EPI_ISL_1718919, EPI_ISL_1718920, EPI_ISL_1718922, EPI_ISL_1718923, EPI_ISL_1718924, EPI_ISL_1718925, EPI_ISL_1718926, EPI_ISL_1718933, EPI_ISL_1718934, EPI_ISL_1718935, EPI_ISL_1718936, EPI_ISL_1718937, EPI_ISL_1718938, EPI_ISL_1718939, EPI_ISL_1718940, EPI_ISL_1718941, EPI_ISL_1718944, EPI_ISL_1718945, EPI_ISL_1718946, EPI_ISL_1718948, EPI_ISL_1718949, EPI_ISL_1718950, EPI_ISL_1718951, EPI_ISL_1718952, EPI_ISL_1718953, EPI_ISL_1718954, EPI_ISL_1718955, EPI_ISL_1718956, EPI_ISL_1718957, EPI_ISL_1718958, EPI_ISL_1718960, EPI_ISL_1718961, EPI_ISL_1718962, EPI_ISL_1718963, EPI_ISL_1718964, EPI_ISL_1718965, EPI_ISL_1718966, EPI_ISL_1718967, EPI_ISL_1718969, EPI_ISL_1718970, EPI_ISL_1718972, EPI_ISL_1718974, EPI_ISL_1718975, EPI_ISL_1718976, EPI_ISL_1718977, EPI_ISL_1718978, EPI_ISL_1718982, EPI_ISL_1718983, EPI_ISL_1718984, EPI_ISL_1718985, EPI_ISL_1718986, EPI_ISL_1718987, EPI_ISL_1718988, EPI_ISL_1718990, EPI_ISL_1718993, EPI_ISL_1718994, EPI_ISL_1718995, EPI_ISL_1718996, EPI_ISL_1718998, EPI_ISL_1718999, EPI_ISL_1719000, EPI_ISL_1719001, EPI_ISL_1719002, EPI_ISL_1719003, EPI_ISL_1719005, EPI_ISL_1719006, EPI_ISL_1719008, EPI_ISL_1719012, EPI_ISL_1719014, EPI_ISL_1719015, EPI_ISL_1719016, EPI_ISL_1719018, EPI_ISL_1719019, EPI_ISL_1719020, EPI_ISL_1719021, EPI_ISL_1719023, EPI_ISL_1719024, EPI_ISL_1719025, EPI_ISL_1719028, EPI_ISL_1719029, EPI_ISL_1719030, EPI_ISL_1719031, EPI_ISL_1719032, EPI_ISL_1719034, EPI_ISL_1719035, EPI_ISL_1719037, EPI_ISL_1719038, EPI_ISL_1719039, EPI_ISL_1719040, EPI_ISL_1719041, EPI_ISL_1719043, EPI_ISL_1719044, EPI_ISL_1719046, EPI_ISL_1719047, EPI_ISL_1719049, EPI_ISL_1719050, EPI_ISL_1719052, EPI_ISL_1719053, EPI_ISL_1719054, EPI_ISL_1719055, EPI_ISL_1719056, EPI_ISL_1719057, EPI_ISL_1719058, EPI_ISL_1719059, EPI_ISL_1719060, EPI_ISL_1719061, EPI_ISL_1719063, EPI_ISL_1719064, EPI_ISL_1719067, EPI_ISL_1719068, EPI_ISL_1719069, EPI_ISL_1719070, EPI_ISL_1719072, EPI_ISL_1719073, EPI_ISL_1719074, EPI_ISL_1719075, EPI_ISL_1719076, EPI_ISL_1719077, EPI_ISL_1719078, EPI_ISL_1719079, EPI_ISL_1719080, EPI_ISL_1719081, EPI_ISL_1719082, EPI_ISL_1719083, EPI_ISL_1719085, EPI_ISL_1719086, EPI_ISL_1719087, EPI_ISL_1719090, EPI_ISL_1719091, EPI_ISL_1719092, EPI_ISL_1719093, EPI_ISL_1719094, EPI_ISL_1719095, EPI_ISL_1719096, EPI_ISL_1719097, EPI_ISL_1719098, EPI_ISL_1719100, EPI_ISL_1719101, EPI_ISL_1719102, EPI_ISL_1719103, EPI_ISL_1719104, EPI_ISL_1719105, EPI_ISL_1719106, EPI_ISL_1719108, EPI_ISL_1719109, EPI_ISL_1719110, EPI_ISL_1719111, EPI_ISL_1719112, EPI_ISL_1719113, EPI_ISL_1719114, EPI_ISL_1719116, EPI_ISL_1719117, EPI_ISL_1719119, EPI_ISL_1719121, EPI_ISL_1719122, EPI_ISL_1719123, EPI_ISL_1719124, EPI_ISL_1719125, EPI_ISL_1719126, EPI_ISL_1719129, EPI_ISL_1719130, EPI_ISL_1719131, EPI_ISL_1719132, EPI_ISL_1719133, EPI_ISL_1719135, EPI_ISL_1719136, EPI_ISL_1719137, EPI_ISL_1719138, EPI_ISL_1719139, EPI_ISL_1719140, EPI_ISL_1719141, EPI_ISL_1719143, EPI_ISL_1719146, EPI_ISL_1719147, EPI_ISL_1719148, EPI_ISL_1719149, EPI_ISL_1719151, EPI_ISL_1719152, EPI_ISL_1719153, EPI_ISL_1719154, EPI_ISL_1719155, EPI_ISL_1719156, EPI_ISL_1719158, EPI_ISL_1719160, EPI_ISL_1719161, EPI_ISL_1719162, EPI_ISL_1719163, EPI_ISL_1719164, EPI_ISL_1719165, EPI_ISL_1719166, EPI_ISL_1719169, EPI_ISL_1719170, EPI_ISL_1719171, EPI_ISL_1719172, EPI_ISL_1719173, EPI_ISL_1719174, EPI_ISL_1719176, EPI_ISL_1719178, EPI_ISL_1719179, EPI_ISL_1719180, EPI_ISL_1719181, EPI_ISL_1719182, EPI_ISL_1719183, EPI_ISL_1719184, EPI_ISL_1719185, EPI_ISL_1719186, EPI_ISL_1719187, EPI_ISL_1719188, EPI_ISL_1719189, EPI_ISL_1719190, EPI_ISL_1719192, EPI_ISL_1719193, EPI_ISL_1719194, EPI_ISL_1719198, EPI_ISL_1719199, EPI_ISL_1719200, EPI_ISL_1719201, EPI_ISL_1719202, EPI_ISL_1719203, EPI_ISL_1719204, EPI_ISL_1719205, EPI_ISL_1719206, EPI_ISL_1719207, EPI_ISL_1719208, EPI_ISL_1719209, EPI_ISL_1719210, EPI_ISL_1719211, EPI_ISL_1719212, EPI_ISL_1719213, EPI_ISL_1719215, EPI_ISL_1719217, EPI_ISL_1740785, EPI_ISL_1740793, EPI_ISL_1740799, EPI_ISL_1740802, EPI_ISL_1740807, EPI_ISL_1740813, EPI_ISL_1740814, EPI_ISL_1740821, EPI_ISL_1740822, EPI_ISL_1740824, EPI_ISL_1740842, EPI_ISL_1740843, EPI_ISL_1740844, EPI_ISL_1740851, EPI_ISL_1740856, EPI_ISL_1740858, EPI_ISL_1740860, EPI_ISL_1740862, EPI_ISL_1740864, EPI_ISL_1740874, EPI_ISL_1740875, EPI_ISL_1740882, EPI_ISL_1740887, EPI_ISL_1740888, EPI_ISL_1740891, EPI_ISL_1740893, EPI_ISL_1740898, EPI_ISL_1740900, EPI_ISL_1740909, EPI_ISL_1740915, EPI_ISL_1740922, EPI_ISL_1740931, EPI_ISL_1740935, EPI_ISL_1740936, EPI_ISL_1740945, EPI_ISL_1740947, EPI_ISL_1740953, EPI_ISL_1740954, EPI_ISL_1740963, EPI_ISL_1740965, EPI_ISL_1740970, EPI_ISL_1740971, EPI_ISL_1740972, EPI_ISL_1740976, EPI_ISL_1740989, EPI_ISL_1740999, EPI_ISL_1741008, EPI_ISL_1741016, EPI_ISL_1741020, EPI_ISL_1741021, EPI_ISL_1741024, EPI_ISL_1741036, EPI_ISL_1741040, EPI_ISL_1741042, EPI_ISL_1741044, EPI_ISL_1741048, EPI_ISL_1741054, EPI_ISL_1741057, EPI_ISL_1741276, EPI_ISL_1741277, EPI_ISL_1741278, EPI_ISL_1741279, EPI_ISL_1741280, EPI_ISL_1741281, EPI_ISL_1741282, EPI_ISL_1741284, EPI_ISL_1741285, EPI_ISL_1741286, EPI_ISL_1741288, EPI_ISL_1741289, EPI_ISL_1741290, EPI_ISL_1741291, EPI_ISL_1741292, EPI_ISL_1741293, EPI_ISL_1741295, EPI_ISL_1741296, EPI_ISL_1741297, EPI_ISL_1741300, EPI_ISL_1741302, EPI_ISL_1741307, EPI_ISL_1741313, EPI_ISL_1741315, EPI_ISL_1741316, EPI_ISL_1741317, EPI_ISL_1741318, EPI_ISL_1741319, EPI_ISL_1741320, EPI_ISL_1741321, EPI_ISL_1741322, EPI_ISL_1741324, EPI_ISL_1741327, EPI_ISL_1741328, EPI_ISL_1741329, EPI_ISL_1741330, EPI_ISL_1741332, EPI_ISL_1741333, EPI_ISL_1741335, EPI_ISL_1741337, EPI_ISL_1741339, EPI_ISL_1741340, EPI_ISL_1741341, EPI_ISL_1741342, EPI_ISL_1741343, EPI_ISL_1741344, EPI_ISL_1741345, EPI_ISL_1741346, EPI_ISL_1741347, EPI_ISL_1741348, EPI_ISL_1741349, EPI_ISL_1741350, EPI_ISL_1741353, EPI_ISL_1741354, EPI_ISL_1741355, EPI_ISL_1741356, EPI_ISL_1741357, EPI_ISL_1741359, EPI_ISL_1741360, EPI_ISL_1741361, EPI_ISL_1741364, EPI_ISL_1741365, EPI_ISL_1741367, EPI_ISL_1741369, EPI_ISL_1741377, EPI_ISL_1741379, EPI_ISL_1741380, EPI_ISL_1741381, EPI_ISL_1741383, EPI_ISL_1741385, EPI_ISL_1741386, EPI_ISL_1741387, EPI_ISL_1741388, EPI_ISL_1741389, EPI_ISL_1741395, EPI_ISL_1741397, EPI_ISL_1741398, EPI_ISL_1741400, EPI_ISL_1741401, EPI_ISL_1741402, EPI_ISL_1741403, EPI_ISL_1741408, EPI_ISL_1741409, EPI_ISL_1741411, EPI_ISL_1741413, EPI_ISL_1741415, EPI_ISL_1741416, EPI_ISL_1741418, EPI_ISL_1741419, EPI_ISL_1741422, EPI_ISL_1741423, EPI_ISL_1741424, EPI_ISL_1741425, EPI_ISL_1741426, EPI_ISL_1741428, EPI_ISL_1741430, EPI_ISL_1741431, EPI_ISL_1741434, EPI_ISL_1741436, EPI_ISL_1741437, EPI_ISL_1741441, EPI_ISL_1741442, EPI_ISL_1741443, EPI_ISL_1741444, EPI_ISL_1741445, EPI_ISL_1741447, EPI_ISL_1741448, EPI_ISL_1741449, EPI_ISL_1741450, EPI_ISL_1741454, EPI_ISL_1741455, EPI_ISL_1741456, EPI_ISL_1741457, EPI_ISL_1741458, EPI_ISL_1741459, EPI_ISL_1741460, EPI_ISL_1741461, EPI_ISL_1741462, EPI_ISL_1741464, EPI_ISL_1741465, EPI_ISL_1741467, EPI_ISL_1741470, EPI_ISL_1741471, EPI_ISL_1741472, EPI_ISL_1741473, EPI_ISL_1741474, EPI_ISL_1741476, EPI_ISL_1741477, EPI_ISL_1741478, EPI_ISL_1741479, EPI_ISL_1741480, EPI_ISL_1741483, EPI_ISL_1741484, EPI_ISL_1741485, EPI_ISL_1741489, EPI_ISL_1741490, EPI_ISL_1741491, EPI_ISL_1741492, EPI_ISL_1741493, EPI_ISL_1741533, EPI_ISL_1742274, EPI_ISL_1758456, EPI_ISL_1758466, EPI_ISL_1758472, EPI_ISL_1758476, EPI_ISL_1758484, EPI_ISL_1758485, EPI_ISL_1758492, EPI_ISL_1758495, EPI_ISL_1758497, EPI_ISL_1758503, EPI_ISL_1758523, EPI_ISL_1758526, EPI_ISL_1758533, EPI_ISL_1758540, EPI_ISL_1758549, EPI_ISL_1758550, EPI_ISL_1758559, EPI_ISL_1758569, EPI_ISL_1758570, EPI_ISL_1758587, EPI_ISL_1758598, EPI_ISL_1758608, EPI_ISL_1758616, EPI_ISL_1758619, EPI_ISL_1758628, EPI_ISL_1758632, EPI_ISL_1758636, EPI_ISL_1758662, EPI_ISL_1758664, EPI_ISL_1758886, EPI_ISL_1758904, EPI_ISL_1758910, EPI_ISL_1758911, EPI_ISL_1758919, EPI_ISL_1758932, EPI_ISL_1758946, EPI_ISL_1758964, EPI_ISL_1758980, EPI_ISL_1759000, EPI_ISL_1759041, EPI_ISL_1759046, EPI_ISL_1759055, EPI_ISL_1759059, EPI_ISL_1759065, EPI_ISL_1759084, EPI_ISL_1759100, EPI_ISL_1759103, EPI_ISL_1759106, EPI_ISL_1759108, EPI_ISL_1759114, EPI_ISL_1759117, EPI_ISL_1759121, EPI_ISL_1759123, EPI_ISL_1759151, EPI_ISL_1759158, EPI_ISL_1759164, EPI_ISL_1759170, EPI_ISL_1759175, EPI_ISL_1759176, EPI_ISL_1759177, EPI_ISL_1759178, EPI_ISL_1759179, EPI_ISL_1759181, EPI_ISL_1759182, EPI_ISL_1759183, EPI_ISL_1759185, EPI_ISL_1759188, EPI_ISL_1759189, EPI_ISL_1759191, EPI_ISL_1759193, EPI_ISL_1759195, EPI_ISL_1759196, EPI_ISL_1759197, EPI_ISL_1759198, EPI_ISL_1759200, EPI_ISL_1759201, EPI_ISL_1759203, EPI_ISL_1759205, EPI_ISL_1759206, EPI_ISL_1759207, EPI_ISL_1759210, EPI_ISL_1759211, EPI_ISL_1759212, EPI_ISL_1759213, EPI_ISL_1759214, EPI_ISL_1759217, EPI_ISL_1759220, EPI_ISL_1759223, EPI_ISL_1759224, EPI_ISL_1759225, EPI_ISL_1759226, EPI_ISL_1759228, EPI_ISL_1759230, EPI_ISL_1759232, EPI_ISL_1759233, EPI_ISL_1759235, EPI_ISL_1759236, EPI_ISL_1759238, EPI_ISL_1759239, EPI_ISL_1759240, EPI_ISL_1759241, EPI_ISL_1759244, EPI_ISL_1759247, EPI_ISL_1759249, EPI_ISL_1759250, EPI_ISL_1759252, EPI_ISL_1759253, EPI_ISL_1759254, EPI_ISL_1759255, EPI_ISL_1759256, EPI_ISL_1759258, EPI_ISL_1759259, EPI_ISL_1759260, EPI_ISL_1759262, EPI_ISL_1759263, EPI_ISL_1759264, EPI_ISL_1759271, EPI_ISL_1759272, EPI_ISL_1759273, EPI_ISL_1759276, EPI_ISL_1759278, EPI_ISL_1759279, EPI_ISL_1759281, EPI_ISL_1759283, EPI_ISL_1759284, EPI_ISL_1759285, EPI_ISL_1759291, EPI_ISL_1759292, EPI_ISL_1759296, EPI_ISL_1759298, EPI_ISL_1759299, EPI_ISL_1759303, EPI_ISL_1759304, EPI_ISL_1759310, EPI_ISL_1759311, EPI_ISL_1759312, EPI_ISL_1759313, EPI_ISL_1759316, EPI_ISL_1759318, EPI_ISL_1759319, EPI_ISL_1759321, EPI_ISL_1759323, EPI_ISL_1759324, EPI_ISL_1759326, EPI_ISL_1759328, EPI_ISL_1759330, EPI_ISL_1759332, EPI_ISL_1759334, EPI_ISL_1759335, EPI_ISL_1759336, EPI_ISL_1759338, EPI_ISL_1759342, EPI_ISL_1759343, EPI_ISL_1759344, EPI_ISL_1759345, EPI_ISL_1759348, EPI_ISL_1759349, EPI_ISL_1759816, EPI_ISL_1759818, EPI_ISL_1759819, EPI_ISL_1759820, EPI_ISL_1759821, EPI_ISL_1759822, EPI_ISL_1759825, EPI_ISL_1759826, EPI_ISL_1759830, EPI_ISL_1759834, EPI_ISL_1759835, EPI_ISL_1759836, EPI_ISL_1759837, EPI_ISL_1759838, EPI_ISL_1759839, EPI_ISL_1759841, EPI_ISL_1759843, EPI_ISL_1759845, EPI_ISL_1759846, EPI_ISL_1759847, EPI_ISL_1759849, EPI_ISL_1759851, EPI_ISL_1759852, EPI_ISL_1759853, EPI_ISL_1759855, EPI_ISL_1759856, EPI_ISL_1759857, EPI_ISL_1759858, EPI_ISL_1759860, EPI_ISL_1759862, EPI_ISL_1759868, EPI_ISL_1759870, EPI_ISL_1759871, EPI_ISL_1759872, EPI_ISL_1759874, EPI_ISL_1759875, EPI_ISL_1759876, EPI_ISL_1759880, EPI_ISL_1759882, EPI_ISL_1759885, EPI_ISL_1759886, EPI_ISL_1759887, EPI_ISL_1759889, EPI_ISL_1759893, EPI_ISL_1759895, EPI_ISL_1790431, EPI_ISL_1790433, EPI_ISL_1790434, EPI_ISL_1790435, EPI_ISL_1790436, EPI_ISL_1790437, EPI_ISL_1790440, EPI_ISL_1790441, EPI_ISL_1790443, EPI_ISL_1790444, EPI_ISL_1790446, EPI_ISL_1790449, EPI_ISL_1790450, EPI_ISL_1790452, EPI_ISL_1790453, EPI_ISL_1790454, EPI_ISL_1790455, EPI_ISL_1790458, EPI_ISL_1790460, EPI_ISL_1790466, EPI_ISL_1790468, EPI_ISL_1790469, EPI_ISL_1790470, EPI_ISL_1790471, EPI_ISL_1790473, EPI_ISL_1790475, EPI_ISL_1790476, EPI_ISL_1790477, EPI_ISL_1790478, EPI_ISL_1790479, EPI_ISL_1790481, EPI_ISL_1790485, EPI_ISL_1790486, EPI_ISL_1790487, EPI_ISL_1790488, EPI_ISL_1790489, EPI_ISL_1790490, EPI_ISL_1790492, EPI_ISL_1790494, EPI_ISL_1790495, EPI_ISL_1790496, EPI_ISL_1790498, EPI_ISL_1790499, EPI_ISL_1790501, EPI_ISL_1790503, EPI_ISL_1790507, EPI_ISL_1790508, EPI_ISL_1790509, EPI_ISL_1790511, EPI_ISL_1790512, EPI_ISL_1790513, EPI_ISL_1790514, EPI_ISL_1790516, EPI_ISL_1790517, EPI_ISL_1790518, EPI_ISL_1790519, EPI_ISL_1790520, EPI_ISL_1790521, EPI_ISL_1790522, EPI_ISL_1790523, EPI_ISL_1790526, EPI_ISL_1790527, EPI_ISL_1790528, EPI_ISL_1790529, EPI_ISL_1790530, EPI_ISL_1790532, EPI_ISL_1790535, EPI_ISL_1790537, EPI_ISL_1790539, EPI_ISL_1790540, EPI_ISL_1790541, EPI_ISL_1790545, EPI_ISL_1790550, EPI_ISL_1790551, EPI_ISL_1790556, EPI_ISL_1790559, EPI_ISL_1790560, EPI_ISL_1790561, EPI_ISL_1790562, EPI_ISL_1790564, EPI_ISL_1790565, EPI_ISL_1790566, EPI_ISL_1790568, EPI_ISL_1790570, EPI_ISL_1790572, EPI_ISL_1790573, EPI_ISL_1790575, EPI_ISL_1790578, EPI_ISL_1790580, EPI_ISL_1790582, EPI_ISL_1790583, EPI_ISL_1790586, EPI_ISL_1790587, EPI_ISL_1790588, EPI_ISL_1790590, EPI_ISL_1790592, EPI_ISL_1790593, EPI_ISL_1790594, EPI_ISL_1790595, EPI_ISL_1790597, EPI_ISL_1790598, EPI_ISL_1790599, EPI_ISL_1790600, EPI_ISL_1790602, EPI_ISL_1790604, EPI_ISL_1790605, EPI_ISL_1790607, EPI_ISL_1790608, EPI_ISL_1790609, EPI_ISL_1790610, EPI_ISL_1790611, EPI_ISL_1790612, EPI_ISL_1790613, EPI_ISL_1790614, EPI_ISL_1790619, EPI_ISL_1790620, EPI_ISL_1790622, EPI_ISL_1790623, EPI_ISL_1790624, EPI_ISL_1790625, EPI_ISL_1790629, EPI_ISL_1790631, EPI_ISL_1790633, EPI_ISL_1790637, EPI_ISL_1790638, EPI_ISL_1790640, EPI_ISL_1790641, EPI_ISL_1790642, EPI_ISL_1790643, EPI_ISL_1790645, EPI_ISL_1790646, EPI_ISL_1790648, EPI_ISL_1790649, EPI_ISL_1790654, EPI_ISL_1790655, EPI_ISL_1790656, EPI_ISL_1790657, EPI_ISL_1790658, EPI_ISL_1790661, EPI_ISL_1790664, EPI_ISL_1790668, EPI_ISL_1790669, EPI_ISL_1790671, EPI_ISL_1790672, EPI_ISL_1790673, EPI_ISL_1790674, EPI_ISL_1790675, EPI_ISL_1790677, EPI_ISL_1790678, EPI_ISL_1790679, EPI_ISL_1790680, EPI_ISL_1790681, EPI_ISL_1790683, EPI_ISL_1790686, EPI_ISL_1790687, EPI_ISL_1790688, EPI_ISL_1790689, EPI_ISL_1790690, EPI_ISL_1790695, EPI_ISL_1790696, EPI_ISL_1790697, EPI_ISL_1790698, EPI_ISL_1790699, EPI_ISL_1790700, EPI_ISL_1790701, EPI_ISL_1790702, EPI_ISL_1790703, EPI_ISL_1790706, EPI_ISL_1790707, EPI_ISL_1790708, EPI_ISL_1790709, EPI_ISL_1790711, EPI_ISL_1790712, EPI_ISL_1790714, EPI_ISL_1790715, EPI_ISL_1790716, EPI_ISL_1790719, EPI_ISL_1790720, EPI_ISL_1790724, EPI_ISL_1790725, EPI_ISL_1790727, EPI_ISL_1790728, EPI_ISL_1790729, EPI_ISL_1790730, EPI_ISL_1790732, EPI_ISL_1790733, EPI_ISL_1790734, EPI_ISL_1790735, EPI_ISL_1790737, EPI_ISL_1790738, EPI_ISL_1790740, EPI_ISL_1790741, EPI_ISL_1790742, EPI_ISL_1790743, EPI_ISL_1790744, EPI_ISL_1790746, EPI_ISL_1790749, EPI_ISL_1790751, EPI_ISL_1790752, EPI_ISL_1790753, EPI_ISL_1790754, EPI_ISL_1790757, EPI_ISL_1790758, EPI_ISL_1790761, EPI_ISL_1790763, EPI_ISL_1790764, EPI_ISL_1790766, EPI_ISL_1790768, EPI_ISL_1790769, EPI_ISL_1790770, EPI_ISL_1790774, EPI_ISL_1790776, EPI_ISL_1790778, EPI_ISL_1790779, EPI_ISL_1790780, EPI_ISL_1790781, EPI_ISL_1790783, EPI_ISL_1790784, EPI_ISL_1790785, EPI_ISL_1790786, EPI_ISL_1790789, EPI_ISL_1790792, EPI_ISL_1790796, EPI_ISL_1790798, EPI_ISL_1790799, EPI_ISL_1790800, EPI_ISL_1790801, EPI_ISL_1790802, EPI_ISL_1790803, EPI_ISL_1790804, EPI_ISL_1790810, EPI_ISL_1790812, EPI_ISL_1790814, EPI_ISL_1790815, EPI_ISL_1790816, EPI_ISL_1790817, EPI_ISL_1790818, EPI_ISL_1790819, EPI_ISL_1790820, EPI_ISL_1790821, EPI_ISL_1790822, EPI_ISL_1790824, EPI_ISL_1790825, EPI_ISL_1790826, EPI_ISL_1790827, EPI_ISL_1790829, EPI_ISL_1790832, EPI_ISL_1790833, EPI_ISL_1790834, EPI_ISL_1790836, EPI_ISL_1790837, EPI_ISL_1790839, EPI_ISL_1790840, EPI_ISL_1790841, EPI_ISL_1790845, EPI_ISL_1790847, EPI_ISL_1790848, EPI_ISL_1790850, EPI_ISL_1790852, EPI_ISL_1790853, EPI_ISL_1790856, EPI_ISL_1790857, EPI_ISL_1790861, EPI_ISL_1790862, EPI_ISL_1790864, EPI_ISL_1790865, EPI_ISL_1790866, EPI_ISL_1790867, EPI_ISL_1790868, EPI_ISL_1790869, EPI_ISL_1790870, EPI_ISL_1790877, EPI_ISL_1790878, EPI_ISL_1790879, EPI_ISL_1790883, EPI_ISL_1790885, EPI_ISL_1790886, EPI_ISL_1790888, EPI_ISL_1790893, EPI_ISL_1790894, EPI_ISL_1790896, EPI_ISL_1790898, EPI_ISL_1790899, EPI_ISL_1790900, EPI_ISL_1790901, EPI_ISL_1790902, EPI_ISL_1790903, EPI_ISL_1790904, EPI_ISL_1790905, EPI_ISL_1790907, EPI_ISL_1790908, EPI_ISL_1790909, EPI_ISL_1790910, EPI_ISL_1790911, EPI_ISL_1790914, EPI_ISL_1790915, EPI_ISL_1790917, EPI_ISL_1790923, EPI_ISL_1790924, EPI_ISL_1790925, EPI_ISL_1790927, EPI_ISL_1790928, EPI_ISL_1790931, EPI_ISL_1790935, EPI_ISL_1790940, EPI_ISL_1790942, EPI_ISL_1790944, EPI_ISL_1790948, EPI_ISL_1790949, EPI_ISL_1790954, EPI_ISL_1790957, EPI_ISL_1790959, EPI_ISL_1790960, EPI_ISL_1790962, EPI_ISL_1790964, EPI_ISL_1790967, EPI_ISL_1790972, EPI_ISL_1790976, EPI_ISL_1790977, EPI_ISL_1790979, EPI_ISL_1790980, EPI_ISL_1790984, EPI_ISL_1790985, EPI_ISL_1790989, EPI_ISL_1790991, EPI_ISL_1790993, EPI_ISL_1790997, EPI_ISL_1790999, EPI_ISL_1791000, EPI_ISL_1791002, EPI_ISL_1791003, EPI_ISL_1791005, EPI_ISL_1791006, EPI_ISL_1791007, EPI_ISL_1791009, EPI_ISL_1791012, EPI_ISL_1791013, EPI_ISL_1791018, EPI_ISL_1805963, EPI_ISL_1805966, EPI_ISL_1805967, EPI_ISL_1805971, EPI_ISL_1805972, EPI_ISL_1805975, EPI_ISL_1805976, EPI_ISL_1805977, EPI_ISL_1805980, EPI_ISL_1805984, EPI_ISL_1805989, EPI_ISL_1805990, EPI_ISL_1805996, EPI_ISL_1805998, EPI_ISL_1806000, EPI_ISL_1806003, EPI_ISL_1806006, EPI_ISL_1806009, EPI_ISL_1806011, EPI_ISL_1806012, EPI_ISL_1806014, EPI_ISL_1806018, EPI_ISL_1806022, EPI_ISL_1806024, EPI_ISL_1806028, EPI_ISL_1806031, EPI_ISL_1806032, EPI_ISL_1806036, EPI_ISL_1806037, EPI_ISL_1806040, EPI_ISL_1806041, EPI_ISL_1806042, EPI_ISL_1806054, EPI_ISL_1806057, EPI_ISL_1806059, EPI_ISL_1806060, EPI_ISL_1806066, EPI_ISL_1806068, EPI_ISL_1806070, EPI_ISL_1806072, EPI_ISL_1806079, EPI_ISL_1806081, EPI_ISL_1806084, EPI_ISL_1806097, EPI_ISL_1806098, EPI_ISL_1806101, EPI_ISL_1806103, EPI_ISL_1806105, EPI_ISL_1806107, EPI_ISL_1806110, EPI_ISL_1806113, EPI_ISL_1806114, EPI_ISL_1806115, EPI_ISL_1806116, EPI_ISL_1806117, EPI_ISL_1806118, EPI_ISL_1806120, EPI_ISL_1806127, EPI_ISL_1806129, EPI_ISL_1806130, EPI_ISL_1806132, EPI_ISL_1806133, EPI_ISL_1806137, EPI_ISL_1806138, EPI_ISL_1806148, EPI_ISL_1806151, EPI_ISL_1806160, EPI_ISL_1806164, EPI_ISL_1806165, EPI_ISL_1806173, EPI_ISL_1806177, EPI_ISL_1806179, EPI_ISL_1806187, EPI_ISL_1806190, EPI_ISL_1806192, EPI_ISL_1806193, EPI_ISL_1806196, EPI_ISL_1806198, EPI_ISL_1806216, EPI_ISL_1806221, EPI_ISL_1806222, EPI_ISL_1806230, EPI_ISL_1806240, EPI_ISL_1806243, EPI_ISL_1806251, EPI_ISL_1806255, EPI_ISL_1806264, EPI_ISL_1806266, EPI_ISL_1806271, EPI_ISL_1806272, EPI_ISL_1806276, EPI_ISL_1806290, EPI_ISL_1806291, EPI_ISL_1806294, EPI_ISL_1806306, EPI_ISL_1806314, EPI_ISL_1806315, EPI_ISL_1806327, EPI_ISL_1806342, EPI_ISL_1806343, EPI_ISL_1806344, EPI_ISL_1806352, EPI_ISL_1806363, EPI_ISL_1806366, EPI_ISL_1806367, EPI_ISL_1806377, EPI_ISL_1806379, EPI_ISL_1806380, EPI_ISL_1806390, EPI_ISL_1806392, EPI_ISL_1806393, EPI_ISL_1806394, EPI_ISL_1806397, EPI_ISL_1806402, EPI_ISL_1806405, EPI_ISL_1806406, EPI_ISL_1806407, EPI_ISL_1806410, EPI_ISL_1806412, EPI_ISL_1806417, EPI_ISL_1806420, EPI_ISL_1806426, EPI_ISL_1806430, EPI_ISL_1806443, EPI_ISL_1806446, EPI_ISL_1806447, EPI_ISL_1806450, EPI_ISL_1806454, EPI_ISL_1806455, EPI_ISL_1806468, EPI_ISL_1806469, EPI_ISL_1806470, EPI_ISL_1806473, EPI_ISL_1806475, EPI_ISL_1806476, EPI_ISL_1806480, EPI_ISL_1806481, EPI_ISL_1806491, EPI_ISL_1806494, EPI_ISL_1806503, EPI_ISL_1806506, EPI_ISL_1806508, EPI_ISL_1806524, EPI_ISL_1806526, EPI_ISL_1806536, EPI_ISL_1806558, EPI_ISL_1806566, EPI_ISL_1806569, EPI_ISL_1806587, EPI_ISL_1806596, EPI_ISL_1806601, EPI_ISL_1806606, EPI_ISL_1806611, EPI_ISL_1806621, EPI_ISL_1806631, EPI_ISL_1806632, EPI_ISL_1806639, EPI_ISL_1806642, EPI_ISL_1806643, EPI_ISL_1806646, EPI_ISL_1806652, EPI_ISL_1806653, EPI_ISL_1806667, EPI_ISL_1806673, EPI_ISL_1806674, EPI_ISL_1806675, EPI_ISL_1806681, EPI_ISL_1806687, EPI_ISL_1806709, EPI_ISL_1806720, EPI_ISL_1806723, EPI_ISL_1806725, EPI_ISL_1806735, EPI_ISL_1806741, EPI_ISL_1806748, EPI_ISL_1806753, EPI_ISL_1806761, EPI_ISL_1806767, EPI_ISL_1806772, EPI_ISL_1806777, EPI_ISL_1806798, EPI_ISL_1806802, EPI_ISL_1806806, EPI_ISL_1806808, EPI_ISL_1806821, EPI_ISL_1806830, EPI_ISL_1806832, EPI_ISL_1806835, EPI_ISL_1806840, EPI_ISL_1806847, EPI_ISL_1806848, EPI_ISL_1806897, EPI_ISL_1806899, EPI_ISL_1806900, EPI_ISL_1806901, EPI_ISL_1806902, EPI_ISL_1806903, EPI_ISL_1806904, EPI_ISL_1806905, EPI_ISL_1829206, EPI_ISL_1829208, EPI_ISL_1829239, EPI_ISL_1829264, EPI_ISL_1829287, EPI_ISL_1829315, EPI_ISL_1829316, EPI_ISL_1829372, EPI_ISL_1829391, EPI_ISL_1829421, EPI_ISL_1829519, EPI_ISL_1829534, EPI_ISL_1829635, EPI_ISL_1829651, EPI_ISL_1829773, EPI_ISL_1830104, EPI_ISL_1830111, EPI_ISL_1830112, EPI_ISL_1830116, EPI_ISL_1830123, EPI_ISL_1830132, EPI_ISL_1830136, EPI_ISL_1830156, EPI_ISL_1830162, EPI_ISL_1830168, EPI_ISL_1830172, EPI_ISL_1830177, EPI_ISL_1830178, EPI_ISL_1830179, EPI_ISL_1830180, EPI_ISL_1830186, EPI_ISL_1830196, EPI_ISL_1830197, EPI_ISL_1830200, EPI_ISL_1830202, EPI_ISL_1830213, EPI_ISL_1830222, EPI_ISL_1830223, EPI_ISL_1830229, EPI_ISL_1830239, EPI_ISL_1830257, EPI_ISL_1830264, EPI_ISL_1830269, EPI_ISL_1830272, EPI_ISL_1830278, EPI_ISL_1830281, EPI_ISL_1830282, EPI_ISL_1830288, EPI_ISL_1830305, EPI_ISL_1830310, EPI_ISL_1830317, EPI_ISL_1830323, EPI_ISL_1830324, EPI_ISL_1830327, EPI_ISL_1830333, EPI_ISL_1830335, EPI_ISL_1830337, EPI_ISL_1830341, EPI_ISL_1830355, EPI_ISL_1830359, EPI_ISL_1830372, EPI_ISL_1830374, EPI_ISL_1830376, EPI_ISL_1830390, EPI_ISL_1830417, EPI_ISL_1830457, EPI_ISL_1830462, EPI_ISL_1830511, EPI_ISL_1830515, EPI_ISL_1830525, EPI_ISL_1830529, EPI_ISL_1830565, EPI_ISL_1830569, EPI_ISL_1830603, EPI_ISL_1830631, EPI_ISL_1830659, EPI_ISL_1830676, EPI_ISL_1830704, EPI_ISL_1830706, EPI_ISL_1830708, EPI_ISL_1830719, EPI_ISL_1831087, EPI_ISL_1831269, EPI_ISL_1831270, EPI_ISL_1831271, EPI_ISL_1831272, EPI_ISL_1831273, EPI_ISL_1831274, EPI_ISL_1831275, EPI_ISL_1831276, EPI_ISL_1831277, EPI_ISL_1831278, EPI_ISL_1831279, EPI_ISL_1831280, EPI_ISL_1831281, EPI_ISL_1831282, EPI_ISL_1831283, EPI_ISL_1857827, EPI_ISL_1857846, EPI_ISL_1857848, EPI_ISL_1857849, EPI_ISL_1857851, EPI_ISL_1857858, EPI_ISL_1857861, EPI_ISL_1857870, EPI_ISL_1857871, EPI_ISL_1857873, EPI_ISL_1857895, EPI_ISL_1857915, EPI_ISL_1857924, EPI_ISL_1857931, EPI_ISL_1857940, EPI_ISL_1857944, EPI_ISL_1857953, EPI_ISL_1857956, EPI_ISL_1857958, EPI_ISL_1857959, EPI_ISL_1857965, EPI_ISL_1857966, EPI_ISL_1858004, EPI_ISL_1858008, EPI_ISL_1858013, EPI_ISL_1858017, EPI_ISL_1858024, EPI_ISL_1858043, EPI_ISL_1858118, EPI_ISL_1858129, EPI_ISL_1858131, EPI_ISL_1858141, EPI_ISL_1858215, EPI_ISL_1912003, EPI_ISL_1912004, EPI_ISL_1912005, EPI_ISL_1912009, EPI_ISL_1912017, EPI_ISL_1912018, EPI_ISL_1912021, EPI_ISL_1912030, EPI_ISL_1912033, EPI_ISL_1912034, EPI_ISL_1912035, EPI_ISL_1912037, EPI_ISL_1912039, EPI_ISL_1912044, EPI_ISL_1912052, EPI_ISL_1912058, EPI_ISL_1912061, EPI_ISL_1912070, EPI_ISL_1912071, EPI_ISL_1912073, EPI_ISL_1912075, EPI_ISL_1912081, EPI_ISL_1912082, EPI_ISL_1912094, EPI_ISL_1912096, EPI_ISL_1912113, EPI_ISL_1912117, EPI_ISL_1912125, EPI_ISL_1912128, EPI_ISL_1912130, EPI_ISL_1912144, EPI_ISL_1912151, EPI_ISL_1912153, EPI_ISL_1912154, EPI_ISL_1912158, EPI_ISL_1912165, EPI_ISL_1912168, EPI_ISL_1912171, EPI_ISL_1912179, EPI_ISL_1912186, EPI_ISL_1912189, EPI_ISL_1912196, EPI_ISL_1912199, EPI_ISL_1912201, EPI_ISL_1912207, EPI_ISL_1912211, EPI_ISL_1912215, EPI_ISL_1912217, EPI_ISL_1912221, EPI_ISL_1912240, EPI_ISL_1912242, EPI_ISL_1912249, EPI_ISL_1912256, EPI_ISL_1912264, EPI_ISL_1912265, EPI_ISL_1912270, EPI_ISL_1912276, EPI_ISL_1912284, EPI_ISL_1912291, EPI_ISL_1912298, EPI_ISL_1912301, EPI_ISL_1912304, EPI_ISL_1912316, EPI_ISL_1912323, EPI_ISL_1912331, EPI_ISL_1912334, EPI_ISL_1912335, EPI_ISL_1912341, EPI_ISL_1912345, EPI_ISL_1912773, EPI_ISL_1912778, EPI_ISL_1912779, EPI_ISL_1912780, EPI_ISL_1921514, EPI_ISL_1921520, EPI_ISL_1921734, EPI_ISL_1985193, EPI_ISL_1985194, EPI_ISL_1985196, EPI_ISL_1985198, EPI_ISL_1985200, EPI_ISL_1985201, EPI_ISL_1985202, EPI_ISL_1985203, EPI_ISL_1985204, EPI_ISL_1985206, EPI_ISL_1985207, EPI_ISL_1985210, EPI_ISL_1985211, EPI_ISL_1985213, EPI_ISL_1985215, EPI_ISL_1985216, EPI_ISL_1985220, EPI_ISL_1985223, EPI_ISL_1985225, EPI_ISL_1985226, EPI_ISL_1985227, EPI_ISL_1985228, EPI_ISL_1985230, EPI_ISL_1985231, EPI_ISL_1985232, EPI_ISL_1985234, EPI_ISL_1985235, EPI_ISL_1985238, EPI_ISL_1985239, EPI_ISL_1985241, EPI_ISL_1985242, EPI_ISL_1985243, EPI_ISL_1985245, EPI_ISL_1985247, EPI_ISL_1985248, EPI_ISL_1985251, EPI_ISL_1985254, EPI_ISL_1985256, EPI_ISL_1985260, EPI_ISL_1985261, EPI_ISL_1985264, EPI_ISL_1985265, EPI_ISL_1985267, EPI_ISL_1985268, EPI_ISL_1985270, EPI_ISL_1985273, EPI_ISL_1985274, EPI_ISL_1985275, EPI_ISL_1985277, EPI_ISL_1985278, EPI_ISL_1985281, EPI_ISL_1985282, EPI_ISL_1985283, EPI_ISL_1985284, EPI_ISL_1985285, EPI_ISL_1985287, EPI_ISL_1985288, EPI_ISL_1985289, EPI_ISL_1985290, EPI_ISL_1985292, EPI_ISL_1985294, EPI_ISL_1985295, EPI_ISL_1985296, EPI_ISL_1985298, EPI_ISL_1985299, EPI_ISL_1985300, EPI_ISL_1985302, EPI_ISL_1985303, EPI_ISL_1985305, EPI_ISL_1985308, EPI_ISL_1985312, EPI_ISL_1985313, EPI_ISL_1985315, EPI_ISL_1985316, EPI_ISL_1985317, EPI_ISL_1985321, EPI_ISL_1985322, EPI_ISL_1985325, EPI_ISL_1985327, EPI_ISL_1985328, EPI_ISL_1985329, EPI_ISL_1985330, EPI_ISL_1985331, EPI_ISL_1985332, EPI_ISL_1985334, EPI_ISL_1985335, EPI_ISL_1985339, EPI_ISL_1985343, EPI_ISL_1985344, EPI_ISL_1985345, EPI_ISL_1985350, EPI_ISL_1985351, EPI_ISL_1985352, EPI_ISL_1985353, EPI_ISL_1985356, EPI_ISL_1985358, EPI_ISL_1985361, EPI_ISL_1985363, EPI_ISL_1985365, EPI_ISL_1985367, EPI_ISL_1985368, EPI_ISL_1985369, EPI_ISL_1985371, EPI_ISL_1985372, EPI_ISL_1985375, EPI_ISL_1985376, EPI_ISL_1985378, EPI_ISL_1985379, EPI_ISL_1985380, EPI_ISL_1985386, EPI_ISL_1985392, EPI_ISL_1985393, EPI_ISL_1985395, EPI_ISL_1985397, EPI_ISL_1985399, EPI_ISL_1985403, EPI_ISL_1985404, EPI_ISL_1985405, EPI_ISL_1985406, EPI_ISL_1985408, EPI_ISL_1985409, EPI_ISL_1985410, EPI_ISL_1985412, EPI_ISL_1985415, EPI_ISL_1985416, EPI_ISL_1985418, EPI_ISL_1985422, EPI_ISL_1985423, EPI_ISL_1985424, EPI_ISL_1985425, EPI_ISL_1985428, EPI_ISL_1985430, EPI_ISL_1985433, EPI_ISL_1985434, EPI_ISL_1985435, EPI_ISL_1985438, EPI_ISL_1985439, EPI_ISL_1985441, EPI_ISL_1985442, EPI_ISL_1985443, EPI_ISL_1985446, EPI_ISL_1985447, EPI_ISL_1985449, EPI_ISL_1985450, EPI_ISL_1985452, EPI_ISL_1985453, EPI_ISL_1985455, EPI_ISL_1985456, EPI_ISL_1985457, EPI_ISL_1985459, EPI_ISL_1985460, EPI_ISL_1985463, EPI_ISL_1985464, EPI_ISL_1985465, EPI_ISL_1985467, EPI_ISL_1985468, EPI_ISL_1985469, EPI_ISL_1985471, EPI_ISL_1985475, EPI_ISL_1985476, EPI_ISL_1985478, EPI_ISL_1985481, EPI_ISL_1985483, EPI_ISL_1985484, EPI_ISL_1985485, EPI_ISL_1985486, EPI_ISL_1985487, EPI_ISL_1985490, EPI_ISL_1985492, EPI_ISL_1985498, EPI_ISL_1985500, EPI_ISL_1985502, EPI_ISL_1985506, EPI_ISL_1985507, EPI_ISL_1985508, EPI_ISL_1985510, EPI_ISL_1985512, EPI_ISL_1986732, EPI_ISL_1986782, EPI_ISL_1986801, EPI_ISL_1986869, EPI_ISL_1986877, EPI_ISL_1986901, EPI_ISL_1986941, EPI_ISL_1987019, EPI_ISL_1987257, EPI_ISL_2004625, EPI_ISL_2004639, EPI_ISL_2004757, EPI_ISL_2004804, EPI_ISL_2004834, EPI_ISL_2004840, EPI_ISL_2004841, EPI_ISL_2004869, EPI_ISL_2005650, EPI_ISL_2005685, EPI_ISL_2005725, EPI_ISL_2005754, EPI_ISL_2005757, EPI_ISL_2005795, EPI_ISL_2005799, EPI_ISL_2006463, EPI_ISL_2006522, EPI_ISL_2006523, EPI_ISL_2006524, EPI_ISL_2006525, EPI_ISL_2006526, EPI_ISL_2006527, EPI_ISL_2006528, EPI_ISL_2006529, EPI_ISL_2006530, EPI_ISL_2006531, EPI_ISL_2006532, EPI_ISL_2006533, EPI_ISL_2006534, EPI_ISL_2006535, EPI_ISL_2006537, EPI_ISL_2006538, EPI_ISL_2006539, EPI_ISL_2006540, EPI_ISL_2022501, EPI_ISL_2022504, EPI_ISL_2022513, EPI_ISL_2022514, EPI_ISL_2022523, EPI_ISL_2022526, EPI_ISL_2023027, EPI_ISL_2023029, EPI_ISL_2023030, EPI_ISL_2023031, EPI_ISL_2091472, EPI_ISL_2091489, EPI_ISL_2091745, EPI_ISL_2119908, EPI_ISL_2172107, EPI_ISL_2172316

| | | | |
|---|---|---|---|
| see above | Lighthouse Lab in Milton Keynes | Wellcome Sanger Institute for the COVID-19 Genomics UK (COG-UK) Consortium | Cordelia Langford; David K. Jackson; Dominic Kwiatkowski; Ewan Harrison; Ian Johnston; Jeffrey Barrett; John Sillitoe on behalf of the Wellcome Sanger Institute COVID-19 Surveillance Team; Roberto Amato; Sonia Goncalves; The Lighthouse Lab in Milton Keynes and Alex Alderton |
| EPI_ISL_1833013, EPI_ISL_1833021, EPI_ISL_1833022 | Lincolnshire Hospitals and DeepSeq Nottingham | COVID-19 Genomics UK (COG-UK) Consortium | Christopher Moore; Fei Sang; Joeseph Chappell; Johnny Debebe; Jonathan Ball; Matthew Carlisle; Matthew Loose; Nadine Holmes; Nichola Duckworth; Patrick McClure; Sarah Walsh; Tim Sloan; Victoria Wright |

EPI_ISL_1832436, EPI_ISL_1832438, EPI_ISL_1832441, EPI_ISL_1832443, EPI_ISL_1832445, EPI_ISL_1832447, EPI_ISL_1832449, EPI_ISL_1832451, EPI_ISL_1832453, EPI_ISL_1832455, EPI_ISL_1832457, EPI_ISL_1832461, EPI_ISL_1832463, EPI_ISL_1832471, EPI_ISL_1832474, EPI_ISL_1832476, EPI_ISL_1832514, EPI_ISL_1832516, EPI_ISL_1832518, EPI_ISL_1832522, EPI_ISL_1832524, EPI_ISL_1832526, EPI_ISL_1832528, EPI_ISL_1974928, EPI_ISL_1974932, EPI_ISL_1974944, EPI_ISL_1974945, EPI_ISL_1974951, EPI_ISL_1974952, EPI_ISL_1974954, EPI_ISL_1974955, EPI_ISL_1974957, EPI_ISL_1974958, EPI_ISL_1974966, EPI_ISL_1974967, EPI_ISL_1974969, EPI_ISL_1974973, EPI_ISL_1974974, EPI_ISL_1974978, EPI_ISL_1974980, EPI_ISL_1974982, EPI_ISL_1974986, EPI_ISL_1974988, EPI_ISL_1974990, EPI_ISL_1974991, EPI_ISL_1974993, EPI_ISL_1975011, EPI_ISL_1975012, EPI_ISL_1975014, EPI_ISL_1975017, EPI_ISL_1975018, EPI_ISL_1975020, EPI_ISL_2125954, EPI_ISL_2125958, EPI_ISL_2125971, EPI_ISL_2125973, EPI_ISL_2125975, EPI_ISL_2125980

| | | | |
|---|---|---|---|
| see above | Liverpool Clinical Laboratories | COVID-19 Genomics UK (COG-UK) Consortium | A Alrezaihi; Alessandro Gerada; Alistair Darby; Angela Cowell; Anita Lucaci; Anu Chawla; Cassie Olateju; Catherine Hartley; Charlotte Nelson; Ecaterina Vamos; Elaine O'Toole; Eleanor G Bentley; Ghada T Shawli; Hermione Webster; Isabel García-Dorival; James Johnson; James P Stewart; Jenifer Manson; Joanne Watts; Jones Benjamin; Jordan J Clark; Julian Hiscox; L Luu; Lucille Rainbow; M Almsaud; Margaret Hughes; Mark Whitehead; Matthew Gemmell; Miren Iturriza-Gomara; Muhannad Alruwaili; N.P Randle; Neil Swainston; PKF Gilmore; Parul Sharma; Rebekah Penrice-Randal; Richard Eccles; Richard Gregory; Sam Haldenby; Steve Paterson; Stuart D Armstrong; Trevor Ian Robinson; Ximeng Han |

EPI_ISL_1730402, EPI_ISL_1730403, EPI_ISL_1730404, EPI_ISL_1832710, EPI_ISL_1832714, EPI_ISL_1978358, EPI_ISL_1978364, EPI_ISL_1978366, EPI_ISL_1978367, EPI_ISL_1978369, EPI_ISL_1978371, EPI_ISL_1978372, EPI_ISL_1978374, EPI_ISL_1978375, EPI_ISL_1978376, EPI_ISL_1978377, EPI_ISL_1978378, EPI_ISL_1978379, EPI_ISL_1978380, EPI_ISL_1978381, EPI_ISL_1978382, EPI_ISL_1978383, EPI_ISL_1978384, EPI_ISL_1978385, EPI_ISL_1978387, EPI_ISL_1978388, EPI_ISL_1978389, EPI_ISL_1978390, EPI_ISL_1978391, EPI_ISL_1978392, EPI_ISL_1978393, EPI_ISL_1978394, EPI_ISL_1978395, EPI_ISL_1978397, EPI_ISL_1978402, EPI_ISL_1978403, EPI_ISL_1978405, EPI_ISL_1978406, EPI_ISL_1978408, EPI_ISL_1978412, EPI_ISL_1978413, EPI_ISL_1978415, EPI_ISL_1978417, EPI_ISL_1978419, EPI_ISL_1978420, EPI_ISL_1978424, EPI_ISL_1978425, EPI_ISL_1978434, EPI_ISL_1978435, EPI_ISL_1978437, EPI_ISL_1978438, EPI_ISL_1978439, EPI_ISL_1978440, EPI_ISL_1978441, EPI_ISL_1978442, EPI_ISL_1978444, EPI_ISL_1978445, EPI_ISL_1978446, EPI_ISL_1978447, EPI_ISL_1978448, EPI_ISL_1978449, EPI_ISL_1978450, EPI_ISL_1978451, EPI_ISL_1978452, EPI_ISL_1978455, EPI_ISL_1978456, EPI_ISL_1978457, EPI_ISL_1978459, EPI_ISL_1978461, EPI_ISL_1978462, EPI_ISL_1978466, EPI_ISL_1978470, EPI_ISL_1978471, EPI_ISL_1978473, EPI_ISL_1978475, EPI_ISL_1978476, EPI_ISL_1978478, EPI_ISL_1978480, EPI_ISL_1978481, EPI_ISL_1978483, EPI_ISL_1978485, EPI_ISL_1978487, EPI_ISL_1978488, EPI_ISL_1978489, EPI_ISL_1978491, EPI_ISL_1978493, EPI_ISL_1978494, EPI_ISL_1978496, EPI_ISL_1978497, EPI_ISL_1978499, EPI_ISL_1978500, EPI_ISL_1978502, EPI_ISL_1978503, EPI_ISL_1978505, EPI_ISL_1978506, EPI_ISL_1978507, EPI_ISL_1978509, EPI_ISL_1978510, EPI_ISL_1978512, EPI_ISL_1978514, EPI_ISL_1978515, EPI_ISL_1978517, EPI_ISL_1978518, EPI_ISL_1978520, EPI_ISL_1978525, EPI_ISL_1978526, EPI_ISL_1978528, EPI_ISL_1978529, EPI_ISL_1978531, EPI_ISL_1978533, EPI_ISL_1978556, EPI_ISL_1978561, EPI_ISL_1978563, EPI_ISL_1978564, EPI_ISL_1978565, EPI_ISL_1978566, EPI_ISL_1978567, EPI_ISL_1978570, EPI_ISL_1978572, EPI_ISL_1978574, EPI_ISL_1978577, EPI_ISL_1978579, EPI_ISL_1978581, EPI_ISL_1978583, EPI_ISL_1978585, EPI_ISL_1978586, EPI_ISL_1978588, EPI_ISL_1978590, EPI_ISL_1978591, EPI_ISL_1978593, EPI_ISL_1978595, EPI_ISL_1978597,

| | | | |
|---|---|---|---|
| EPI_ISL_1978598, EPI_ISL_1978600, EPI_ISL_1978602, EPI_ISL_1978604, EPI_ISL_1978606, EPI_ISL_1978607, EPI_ISL_1978609, EPI_ISL_1978611, EPI_ISL_1978613, EPI_ISL_1978614, EPI_ISL_1978616, EPI_ISL_1978617, EPI_ISL_1978619, EPI_ISL_1978621, EPI_ISL_1978623, EPI_ISL_1978626, EPI_ISL_1978660, EPI_ISL_1978662, EPI_ISL_1978663, EPI_ISL_1978665, EPI_ISL_1978669, EPI_ISL_1978670, EPI_ISL_1978682, EPI_ISL_1978686, EPI_ISL_1978736, EPI_ISL_1978741, EPI_ISL_1978743, EPI_ISL_1978891 | | | |
| see above | Northumbria University / South Tees Hospitals NHS Foundation Trust / North Cumbria Integrated Care NHS Foundation Trust / North Tees and Hartlepool NHS Foundation Trust / Newcastle Hospitals NHS Foundation Trust | COVID-19 Genomics UK (COG-UK) Consortium | Andrew Nelson; Brendan Payne; Clive Graham; Darren L Smith; Debra Padgett; Edward Barton; Emma Swindells; Garren Scott; Gary Black; Gary Eltringham; Giles S Holt; Greg R Young; Jane Greenaway; Jennifer Collins; John Allan; Joshua Loh; Lynn Dover; Matthew Bashton; Mohammad A Tariq; Paul Baker; Sarah Essex; Steve Liggett; Wen C Yew; Yusri Taha |
| EPI_ISL_1730607, EPI_ISL_1730608, EPI_ISL_1730609, EPI_ISL_1730610, EPI_ISL_1730611, EPI_ISL_1730612, EPI_ISL_1730613, EPI_ISL_1730614, EPI_ISL_1730615, EPI_ISL_1730616, EPI_ISL_1730617, EPI_ISL_1730618, EPI_ISL_1730619, EPI_ISL_1730620, EPI_ISL_1730621, EPI_ISL_1730622, EPI_ISL_1730623, EPI_ISL_1730625, EPI_ISL_1730626, EPI_ISL_1730627, EPI_ISL_1730628, EPI_ISL_1730629, EPI_ISL_1730630, EPI_ISL_1730633, EPI_ISL_1730634, EPI_ISL_1730635, EPI_ISL_1730637, EPI_ISL_1730640, EPI_ISL_1730641, EPI_ISL_1730642, EPI_ISL_1730643, EPI_ISL_1730644, EPI_ISL_1730645, EPI_ISL_1730646, EPI_ISL_1730647, EPI_ISL_1730648, EPI_ISL_1730649, EPI_ISL_1730650, EPI_ISL_1730651, EPI_ISL_1730652, EPI_ISL_1730653, EPI_ISL_1730654, EPI_ISL_1730655, EPI_ISL_1730656, EPI_ISL_1730657, EPI_ISL_1730658, EPI_ISL_1730659, EPI_ISL_1730660, EPI_ISL_1730661, EPI_ISL_1730662, EPI_ISL_1730663, EPI_ISL_1730664, EPI_ISL_1730665, EPI_ISL_1730666, EPI_ISL_1730690, EPI_ISL_1730691, EPI_ISL_1730692, EPI_ISL_1730693, EPI_ISL_1730694, EPI_ISL_1730695, EPI_ISL_1730696, EPI_ISL_1730697, EPI_ISL_1730698, EPI_ISL_1730699, EPI_ISL_1730700, EPI_ISL_1730701, EPI_ISL_1730702, EPI_ISL_1730703, EPI_ISL_1730704, EPI_ISL_1730705, EPI_ISL_1730706, EPI_ISL_1730707, EPI_ISL_1730708, EPI_ISL_1730709, EPI_ISL_1730710, EPI_ISL_1730711, EPI_ISL_1730712, EPI_ISL_1730713, EPI_ISL_1730714, EPI_ISL_1730715, EPI_ISL_1730716, EPI_ISL_1730717, EPI_ISL_1730718, EPI_ISL_1730719, EPI_ISL_1730720, EPI_ISL_1730721, EPI_ISL_1730722, EPI_ISL_1730723, EPI_ISL_1730724, EPI_ISL_1730725, EPI_ISL_1730726, EPI_ISL_1730727, EPI_ISL_1730728, EPI_ISL_1730729, EPI_ISL_1730730, EPI_ISL_1730731, EPI_ISL_1730732, EPI_ISL_1730733, EPI_ISL_1730734, EPI_ISL_1730735, EPI_ISL_1730736, EPI_ISL_1730737, EPI_ISL_1730738, EPI_ISL_1730739, EPI_ISL_1730740, EPI_ISL_1730741, EPI_ISL_1730742, EPI_ISL_1730743, EPI_ISL_1730744, EPI_ISL_1730745, EPI_ISL_1730746, EPI_ISL_1730747, EPI_ISL_1730748, EPI_ISL_1730749, EPI_ISL_1730750, EPI_ISL_1730751, EPI_ISL_1730752, EPI_ISL_1730753, EPI_ISL_1730754, EPI_ISL_1730755, EPI_ISL_1730756, EPI_ISL_1730757, EPI_ISL_1730758, EPI_ISL_1730759, EPI_ISL_1730760, EPI_ISL_1730761, EPI_ISL_1730762, EPI_ISL_1730763, EPI_ISL_1730764, EPI_ISL_1730767, EPI_ISL_1730768, EPI_ISL_1730769, EPI_ISL_1730770, EPI_ISL_1730771, EPI_ISL_1730772, EPI_ISL_1730774, EPI_ISL_1730775, EPI_ISL_1730776, EPI_ISL_1730777, EPI_ISL_1730778, EPI_ISL_1730779, EPI_ISL_1730780, EPI_ISL_1730781, EPI_ISL_1730782, EPI_ISL_1730783, EPI_ISL_1730785, EPI_ISL_1730787, EPI_ISL_1730788, EPI_ISL_1730790, EPI_ISL_1730791, EPI_ISL_1730792, EPI_ISL_1730793, EPI_ISL_1730794, EPI_ISL_1730795, EPI_ISL_1730796, EPI_ISL_1730798, EPI_ISL_1730799, EPI_ISL_1730800, EPI_ISL_1730801, EPI_ISL_1730802, EPI_ISL_1730803, EPI_ISL_1730804, EPI_ISL_1730805, EPI_ISL_1730806, EPI_ISL_1730807, EPI_ISL_1730808, EPI_ISL_1730809, EPI_ISL_1730810, EPI_ISL_1730811, EPI_ISL_1730812, EPI_ISL_1730813, EPI_ISL_1730814, EPI_ISL_1730815, EPI_ISL_1730816, EPI_ISL_1730817, EPI_ISL_1730818, EPI_ISL_1730819, EPI_ISL_1979275, EPI_ISL_1979283, EPI_ISL_1979305, EPI_ISL_1979306, EPI_ISL_1979308, EPI_ISL_1979341, EPI_ISL_1979343, EPI_ISL_1979344, EPI_ISL_1979346, EPI_ISL_1979347, EPI_ISL_1979349, EPI_ISL_1979351, EPI_ISL_1979353, EPI_ISL_1979354, EPI_ISL_1979357, EPI_ISL_1979361, EPI_ISL_1979364, EPI_ISL_1979381, EPI_ISL_1979410, EPI_ISL_1979415, EPI_ISL_1979447, EPI_ISL_1979461, EPI_ISL_1979463, EPI_ISL_1979464, EPI_ISL_1979466, EPI_ISL_1979470, EPI_ISL_1979472, EPI_ISL_1979475, EPI_ISL_1979478, EPI_ISL_1979481, EPI_ISL_1979488, EPI_ISL_1979656, EPI_ISL_1979659, EPI_ISL_1979661, EPI_ISL_1979662, EPI_ISL_1979665, EPI_ISL_1979669, EPI_ISL_1979672, EPI_ISL_1979674, EPI_ISL_1979677, EPI_ISL_1979685, EPI_ISL_1979693, EPI_ISL_1979698 | | | |
| see above | Originating lab: Wales Specialist Virology Centre Sequencing lab: Pathogen Genomics Unit | Public Health Wales Microbiology Cardiff Wales Specialist Virology Centre | Alec Birchley; Alexander Adams; Amy Gaskin; Angela Marchbank; Bree Gatica-Wilcox; Catherine Moore; Jason Coombes; Joanne Watkins; Joel Southgate; Johnathan Evans; Laura Gifford; Lauren Gilbert; Lee Graham; Malorie Perry; Matthew Bull; Nicole Pacchiarini; Sally Corden; Sara Kumziene-Summerhayes; Sara Rey; Sarah Taylor; Simon Cottrell; Sophie Jones; Tom Connor |
| EPI_ISL_1979038, EPI_ISL_1979050, EPI_ISL_1979051, EPI_ISL_1979058, EPI_ISL_1979060, EPI_ISL_1979066, EPI_ISL_1979068, EPI_ISL_1979069, EPI_ISL_1979082, EPI_ISL_1979090, EPI_ISL_1979095, EPI_ISL_1979097 | | | |
| see above | Oxford Viromics, NDM, University of Oxford; Oxford University Hospitals; Basingstoke and North Hampshire Hospital | COVID-19 Genomics UK (COG-UK) Consortium | Alex Mobbs; Amy Trebes; Anita Justice; Catrin Moore; Christophe Fraser; David Bonsall; David Buck; Emma Wise; George Macintyre; Jessica Lynch; John Todd; Mariateresa de Cesare; Matilde Mori; Monique Andersson; Nathan Moore; Nick Cortes; Robert Shaw; Stephen Kidd; Tanya Golubchik; Timothy Peto |
| EPI_ISL_1832723, EPI_ISL_1832742, EPI_ISL_1832771, EPI_ISL_1832773, EPI_ISL_1832775, EPI_ISL_1832778, EPI_ISL_1832780, EPI_ISL_1832782, EPI_ISL_1832784, EPI_ISL_1832786, EPI_ISL_1832788, EPI_ISL_1832791, EPI_ISL_1832793, EPI_ISL_1832794, EPI_ISL_1832796, EPI_ISL_1832797, EPI_ISL_1832798, EPI_ISL_1832799, EPI_ISL_1832801, EPI_ISL_1832804, EPI_ISL_1832805, EPI_ISL_1832812, EPI_ISL_1832814, EPI_ISL_1832816, EPI_ISL_1832818, EPI_ISL_1832820, EPI_ISL_1832822, EPI_ISL_1832824, EPI_ISL_1832826, EPI_ISL_1832828, EPI_ISL_1832830, EPI_ISL_1832832, EPI_ISL_1832834, EPI_ISL_1832836, EPI_ISL_1832839, EPI_ISL_1832840, EPI_ISL_1832850, EPI_ISL_1832867, EPI_ISL_1832871, EPI_ISL_1832872, EPI_ISL_1832873, EPI_ISL_1832874, EPI_ISL_1832876, EPI_ISL_1832877, EPI_ISL_1832878, EPI_ISL_1832879, EPI_ISL_1832880, EPI_ISL_1832881, EPI_ISL_1832882, EPI_ISL_1832883, EPI_ISL_1832884, EPI_ISL_1832885, EPI_ISL_1832886, EPI_ISL_1832887, EPI_ISL_1832888, EPI_ISL_1832889, EPI_ISL_1832890, EPI_ISL_1978900, EPI_ISL_1978944, EPI_ISL_1978946, EPI_ISL_1978948, EPI_ISL_1978949, EPI_ISL_1978951 | | | |
| see above | Quadram Institute Bioscience | COVID-19 Genomics UK (COG-UK) Consortium | Alexander J Trotter; Alison E. Mather; Alp Aydin; Ana P. Tedim; Anastasia Kolyva; Andrew Bell; Andrew J. Page; Claire Stuart; Dave J. Baker; Gemma L. Kay; John Wain; Justin O'Grady; Leonardo de Oliveira Martins; Lizzie Meadows; Maria Diaz; Mark Webber; Muhammed Yasir; Nabil-Fareed Alikhan; Ngozi Elumogo; Nicholas M. Thomson; Rachael Stanley; Rachel Gilroy; Reenesh Prakash; Samir Dervisevic; Samuel Bloomfield; Steven Rudder; Thanh Le-Viet |
| EPI_ISL_1730408, EPI_ISL_1730409, EPI_ISL_1730410, EPI_ISL_1730423, EPI_ISL_1730424, EPI_ISL_1730425, EPI_ISL_1833002, EPI_ISL_1833003, EPI_ISL_1833029, EPI_ISL_1833030, EPI_ISL_1833031, EPI_ISL_1833032 | | | |
| see above | Queens Medical Centre, Clinical Microbiology Department / DeepSeq Nottingham | COVID-19 Genomics UK (COG-UK) Consortium | Christopher Moore; Emily Park; Fei Sang; Gemma Clark; Hannah Howson-Wells; Jack Hill; Jayasree Dey; Johnny Debebe; Jonathan Ball; Joseph Chappell; Manjinder Khakh; Matthew Carlisle; Matthew Loose; Michelle M Lister; Nadine Holmes; Patrick McClure; Theocharis Tsoleridis; Timothy Byaruhanga; Vicki M Fleming; Victoria Wright; Wendy Smith |
| EPI_ISL_1697976, EPI_ISL_1697978, EPI_ISL_1697982, EPI_ISL_1697989, EPI_ISL_1697996, EPI_ISL_1698007, EPI_ISL_1698015, EPI_ISL_1698028, EPI_ISL_1698034, EPI_ISL_1698041, EPI_ISL_1698044, EPI_ISL_1698057, EPI_ISL_1698058, EPI_ISL_1698063, EPI_ISL_1698067, EPI_ISL_1698069, EPI_ISL_1698077, EPI_ISL_1698082, EPI_ISL_1698087, EPI_ISL_1698099, EPI_ISL_1698107, EPI_ISL_1698109, EPI_ISL_1698114, EPI_ISL_1698118, EPI_ISL_1698129, EPI_ISL_1698131, EPI_ISL_1698136, EPI_ISL_1698150, EPI_ISL_1698153, EPI_ISL_1698156, EPI_ISL_1698164, EPI_ISL_1698166, EPI_ISL_1698180, EPI_ISL_1698183, EPI_ISL_1698202, EPI_ISL_1698214, EPI_ISL_1698221, EPI_ISL_1698223, EPI_ISL_1698234, EPI_ISL_1698241, EPI_ISL_1718649, EPI_ISL_1718672, EPI_ISL_1718700, EPI_ISL_1718703, EPI_ISL_1718704, EPI_ISL_1718751, EPI_ISL_1718801, EPI_ISL_1718872, EPI_ISL_1718876, EPI_ISL_1740533, EPI_ISL_1740535, EPI_ISL_1740551, EPI_ISL_1740558, EPI_ISL_1740564, EPI_ISL_1740567, EPI_ISL_1740578, EPI_ISL_1740579, EPI_ISL_1740588, EPI_ISL_1740589, EPI_ISL_1740597, EPI_ISL_1740601, EPI_ISL_1740610, EPI_ISL_1740611, EPI_ISL_1740614, EPI_ISL_1740619, EPI_ISL_1740621, EPI_ISL_1740624, EPI_ISL_1740633, EPI_ISL_1740636, EPI_ISL_1740646, EPI_ISL_1740656, EPI_ISL_1740661, EPI_ISL_1740663, EPI_ISL_1740668, EPI_ISL_1740673, EPI_ISL_1740678, EPI_ISL_1740690, EPI_ISL_1740691, EPI_ISL_1740701, EPI_ISL_1740708, EPI_ISL_1740710, EPI_ISL_1740712, EPI_ISL_1740716, EPI_ISL_1740719, EPI_ISL_1740730, EPI_ISL_1740740, EPI_ISL_1740744, EPI_ISL_1740745, EPI_ISL_1740754, EPI_ISL_1740759, EPI_ISL_1740762, EPI_ISL_1740766, EPI_ISL_1740772, EPI_ISL_1740773, EPI_ISL_1740774, EPI_ISL_1740782, EPI_ISL_1740784, EPI_ISL_1741273, EPI_ISL_1741275, EPI_ISL_1741294, EPI_ISL_1741298, EPI_ISL_1741301, EPI_ISL_1741306, EPI_ISL_1741310, EPI_ISL_1741325, EPI_ISL_1741334, EPI_ISL_1741351, EPI_ISL_1741362, EPI_ISL_1741366, EPI_ISL_1741376, EPI_ISL_1741378, EPI_ISL_1741384, EPI_ISL_1741391, EPI_ISL_1741396, EPI_ISL_1741399, EPI_ISL_1741414, EPI_ISL_1741429, EPI_ISL_1741432, EPI_ISL_1741435, EPI_ISL_1741466, EPI_ISL_1741468, EPI_ISL_1741475, EPI_ISL_1741481, EPI_ISL_1741488, EPI_ISL_1741543, EPI_ISL_1741544, EPI_ISL_1758882, EPI_ISL_1758902, EPI_ISL_1758921, EPI_ISL_1758982, EPI_ISL_1759005, EPI_ISL_1759026, EPI_ISL_1759095, EPI_ISL_1759132, EPI_ISL_1790215, EPI_ISL_1790217, EPI_ISL_1790232, EPI_ISL_1790258, EPI_ISL_1790260, EPI_ISL_1790269, EPI_ISL_1790272, EPI_ISL_1790286, EPI_ISL_1790287, EPI_ISL_1790289, EPI_ISL_1790300, EPI_ISL_1790303, EPI_ISL_1790307, EPI_ISL_1790308, EPI_ISL_1790322, EPI_ISL_1790323, EPI_ISL_1790324, EPI_ISL_1790325, EPI_ISL_1790328, EPI_ISL_1790333, EPI_ISL_1790340, EPI_ISL_1790347, EPI_ISL_1790349, EPI_ISL_1790361, EPI_ISL_1790364, EPI_ISL_1790367, EPI_ISL_1790369, EPI_ISL_1790370, EPI_ISL_1790372, EPI_ISL_1790378, EPI_ISL_1790381, EPI_ISL_1790382, EPI_ISL_1790388, EPI_ISL_1790393, EPI_ISL_1790414, EPI_ISL_1790415, EPI_ISL_1790423, EPI_ISL_1790427, EPI_ISL_1790428, EPI_ISL_1806910, EPI_ISL_1806918, EPI_ISL_1806923, EPI_ISL_1806927, EPI_ISL_1806928, EPI_ISL_1806930, EPI_ISL_1806935, EPI_ISL_1806937, EPI_ISL_1806941, EPI_ISL_1806943, EPI_ISL_1806945, EPI_ISL_1806949, EPI_ISL_1806950, EPI_ISL_1806953, EPI_ISL_1806954, EPI_ISL_1806955, EPI_ISL_1806956, EPI_ISL_1806957, EPI_ISL_1806958, EPI_ISL_1806959, EPI_ISL_1806960, EPI_ISL_1806961, EPI_ISL_1806962, EPI_ISL_1806963, EPI_ISL_1806964, EPI_ISL_1806965, EPI_ISL_1806966, EPI_ISL_1806967, EPI_ISL_1806968, EPI_ISL_1806969, EPI_ISL_1806970, EPI_ISL_1806971, EPI_ISL_1806972, EPI_ISL_1807179, EPI_ISL_1807180, EPI_ISL_1807181, EPI_ISL_1807182, EPI_ISL_1807183, EPI_ISL_1829776, EPI_ISL_1829805, EPI_ISL_1829807, EPI_ISL_1829810, EPI_ISL_1829817, EPI_ISL_1829826, EPI_ISL_1829831, EPI_ISL_1829832, EPI_ISL_1829837, EPI_ISL_1829841, EPI_ISL_1829845, EPI_ISL_1829846, EPI_ISL_1829847, EPI_ISL_1829858, EPI_ISL_1829864, EPI_ISL_1829868, EPI_ISL_1829876, EPI_ISL_1829877, EPI_ISL_1829891, EPI_ISL_1829892, EPI_ISL_1829893, EPI_ISL_1829897, EPI_ISL_1829910, EPI_ISL_1829912, EPI_ISL_1829918, EPI_ISL_1829924, EPI_ISL_1829926, EPI_ISL_1829933, EPI_ISL_1829936, EPI_ISL_1829942, EPI_ISL_1829945, EPI_ISL_1829947, EPI_ISL_1829951, EPI_ISL_1829954, EPI_ISL_1829955, EPI_ISL_1829956, EPI_ISL_1829958, EPI_ISL_1829960, EPI_ISL_1829961, EPI_ISL_1829967, EPI_ISL_1829975, EPI_ISL_1829977, EPI_ISL_1829978, EPI_ISL_1829981, EPI_ISL_1829990, EPI_ISL_1829991, EPI_ISL_1829993, EPI_ISL_1829999, EPI_ISL_1830001, EPI_ISL_1830002, EPI_ISL_1830008, EPI_ISL_1830010, EPI_ISL_1830022, EPI_ISL_1830029, EPI_ISL_1830030, EPI_ISL_1830032, EPI_ISL_1830033, EPI_ISL_1830034, EPI_ISL_1830037, EPI_ISL_1830038, EPI_ISL_1830041, EPI_ISL_1830045, EPI_ISL_1830047, EPI_ISL_1830051, EPI_ISL_1830057, EPI_ISL_1830067, EPI_ISL_1830076, EPI_ISL_1830084, EPI_ISL_1830087, EPI_ISL_1830091, EPI_ISL_1830100, EPI_ISL_1830108, EPI_ISL_1830115, EPI_ISL_1830118, EPI_ISL_1830120, EPI_ISL_1830127, EPI_ISL_1830138, EPI_ISL_1830146, EPI_ISL_1830157, EPI_ISL_1830164, EPI_ISL_1830189, EPI_ISL_1830194, EPI_ISL_1830207, EPI_ISL_1830209, EPI_ISL_1830216, EPI_ISL_1830218, EPI_ISL_1830221, EPI_ISL_1830227, EPI_ISL_1830233, EPI_ISL_1830238, EPI_ISL_1830240, EPI_ISL_1830242, EPI_ISL_1830247, EPI_ISL_1830267, EPI_ISL_1830270, EPI_ISL_1830280, EPI_ISL_1830290, EPI_ISL_1830292, EPI_ISL_1830313, EPI_ISL_1830331, EPI_ISL_1830343, EPI_ISL_1830351, EPI_ISL_1830366, EPI_ISL_1830375, EPI_ISL_1830378, EPI_ISL_1857631, EPI_ISL_1912763, EPI_ISL_1912764, EPI_ISL_1912766, EPI_ISL_1912770, EPI_ISL_1912771, EPI_ISL_1912772, EPI_ISL_1912775, EPI_ISL_1912776, EPI_ISL_1912777, EPI_ISL_1912781, EPI_ISL_1912782, EPI_ISL_1912783, EPI_ISL_1921440, EPI_ISL_1921441, EPI_ISL_1921443, EPI_ISL_1921449, EPI_ISL_1921463, EPI_ISL_1921467, EPI_ISL_1921485, EPI_ISL_1921488, EPI_ISL_1921489, EPI_ISL_1921490, EPI_ISL_1921502, EPI_ISL_1921513, EPI_ISL_1921522, EPI_ISL_1921523, EPI_ISL_1921538, EPI_ISL_1921545, EPI_ISL_1921557, EPI_ISL_1921558, EPI_ISL_1921559, EPI_ISL_1921561, EPI_ISL_1921567, EPI_ISL_1921568, EPI_ISL_1921575, EPI_ISL_1921582, EPI_ISL_1921590, EPI_ISL_1921596, EPI_ISL_1921597, EPI_ISL_1921600, EPI_ISL_1921604, EPI_ISL_1921626, EPI_ISL_1921627, EPI_ISL_1921631, EPI_ISL_1921648, EPI_ISL_1921660, EPI_ISL_1921673, EPI_ISL_1921679, EPI_ISL_1921684, EPI_ISL_1921691, EPI_ISL_1921699, EPI_ISL_1921702, EPI_ISL_1921709, EPI_ISL_1921711, EPI_ISL_1921713, EPI_ISL_1921717, EPI_ISL_1921722, EPI_ISL_1921731, EPI_ISL_1921732, EPI_ISL_1985214, EPI_ISL_1985253, EPI_ISL_1985266, EPI_ISL_1985276, EPI_ISL_1985385, EPI_ISL_1985448, EPI_ISL_1985480, EPI_ISL_1985493, EPI_ISL_1985505, EPI_ISL_1985511, EPI_ISL_1987380, EPI_ISL_2005613, EPI_ISL_2005616, EPI_ISL_2005617, EPI_ISL_2005618, EPI_ISL_2005619, EPI_ISL_2005624, EPI_ISL_2005625, EPI_ISL_2005638, EPI_ISL_2005642, EPI_ISL_2005646, EPI_ISL_2005647, EPI_ISL_2005649, EPI_ISL_2005651, EPI_ISL_2005652, EPI_ISL_2005656, EPI_ISL_2005659, EPI_ISL_2005660, EPI_ISL_2005663, EPI_ISL_2005664, EPI_ISL_2005666, EPI_ISL_2005671, EPI_ISL_2005672, EPI_ISL_2005675, EPI_ISL_2005683, EPI_ISL_2005690, EPI_ISL_2005695, EPI_ISL_2005703, EPI_ISL_2005705, EPI_ISL_2005706, EPI_ISL_2005712, EPI_ISL_2005714, EPI_ISL_2005715, EPI_ISL_2005716, EPI_ISL_2005721, EPI_ISL_2005729, EPI_ISL_2005732, EPI_ISL_2005733, EPI_ISL_2005734, EPI_ISL_2005736, EPI_ISL_2005740, EPI_ISL_2005764, EPI_ISL_2005769, EPI_ISL_2005784, EPI_ISL_2005792, EPI_ISL_2005802, EPI_ISL_2005803, EPI_ISL_2005804, EPI_ISL_2005808, EPI_ISL_2022496, EPI_ISL_2022497, EPI_ISL_2022498, EPI_ISL_2022499, EPI_ISL_2022500, EPI_ISL_2022502, EPI_ISL_2022503, EPI_ISL_2022505, EPI_ISL_2022506, EPI_ISL_2022507, EPI_ISL_2022508, EPI_ISL_2022509, EPI_ISL_2022510, EPI_ISL_2022511, EPI_ISL_2022512, EPI_ISL_2022515, EPI_ISL_2022516, EPI_ISL_2022517, EPI_ISL_2022518, EPI_ISL_2022519, EPI_ISL_2022520, EPI_ISL_2022521, EPI_ISL_2022522, EPI_ISL_2022524, EPI_ISL_2022525, EPI_ISL_2022527, EPI_ISL_2022528, EPI_ISL_2022529, EPI_ISL_2023028 | | | |
| see above | Randox Laboratories | Wellcome Sanger Institute for the COVID-19 Genomics UK (COG-UK) Consortium | Cordelia Langford; David K. Jackson; Dominic Kwiatkowski; Ewan Harrison; Ian Johnston; Jeffrey Barrett; John Sillitoe on behalf of the Wellcome Sanger Institute COVID-19 Surveillance Team; Randox Laboratories and Alex Alderton; Roberto Amato; Sonia Goncalves |
| EPI_ISL_1978258, EPI_ISL_1978259, EPI_ISL_1978261, EPI_ISL_1978263, EPI_ISL_1978265, EPI_ISL_1978266, EPI_ISL_1978268, EPI_ISL_1978270, EPI_ISL_1978271, EPI_ISL_1978273, EPI_ISL_1978275, EPI_ISL_1978276, EPI_ISL_1978278, EPI_ISL_1978280, EPI_ISL_1978282, EPI_ISL_1978283, EPI_ISL_1978285, EPI_ISL_1978287, EPI_ISL_1978289, EPI_ISL_1978290, EPI_ISL_1978292, EPI_ISL_1978293, EPI_ISL_1978295, EPI_ISL_1978297, EPI_ISL_1978299, EPI_ISL_1978300, EPI_ISL_1978302, EPI_ISL_1978304, EPI_ISL_1978306, EPI_ISL_1978307, EPI_ISL_1978309, EPI_ISL_1978310, EPI_ISL_1978312, EPI_ISL_1978313, EPI_ISL_1978315, EPI_ISL_1978316, EPI_ISL_1978318, EPI_ISL_1978319, EPI_ISL_1978321, EPI_ISL_1978322, EPI_ISL_1978324, EPI_ISL_1978326, EPI_ISL_1978327, EPI_ISL_1978329, EPI_ISL_1978330, EPI_ISL_1978332, EPI_ISL_1978334, EPI_ISL_1978335, EPI_ISL_1978337, EPI_ISL_1978339, EPI_ISL_1978341, EPI_ISL_1978342, EPI_ISL_1978344, EPI_ISL_1978345, EPI_ISL_1978347, EPI_ISL_1978349, EPI_ISL_1978351, EPI_ISL_1978352, EPI_ISL_1978354, EPI_ISL_1978356 | | | |
| see above | Regional Virus Laboratory, Belfast Health and Social Care Trust | COVID-19 Genomics UK (COG-UK) Consortium | Alison Watt; Ciara Cox; Conall McCaughey; David Simpson; Derek Fairley; James McKenna; Mairead Connor; Susan Feeney; Tanya Curran; Zoltan Molnar |
| EPI_ISL_1833828, EPI_ISL_1833829 | St George's University Hospitals NHS Foundation Trust | COVID-19 Genomics UK (COG-UK) Consortium | Adam Witney; Cassie Pope; Irene Monahan; Joshua Taylor; Ken Laing; NgeeKeong Tan |
| EPI_ISL_1637102, EPI_ISL_1730327, EPI_ISL_1730330, EPI_ISL_1730331, EPI_ISL_1730332, EPI_ISL_1730333, EPI_ISL_1730335, EPI_ISL_1730340, EPI_ISL_1730346, EPI_ISL_1730347, EPI_ISL_1730348, EPI_ISL_1730349, EPI_ISL_1730350, EPI_ISL_1730351, EPI_ISL_1730352, EPI_ISL_1730353, EPI_ISL_1730354, EPI_ISL_1730363, EPI_ISL_1730364, EPI_ISL_1730365, EPI_ISL_1730366, EPI_ISL_1730374, EPI_ISL_1730376, EPI_ISL_1730378, EPI_ISL_1730385, EPI_ISL_1832580, EPI_ISL_1832582, EPI_ISL_1832584, EPI_ISL_1832585, EPI_ISL_1832590, EPI_ISL_1832591, EPI_ISL_1832598, EPI_ISL_1832602, EPI_ISL_1832604, EPI_ISL_1832606, EPI_ISL_1832611, EPI_ISL_1832614, EPI_ISL_1832615, EPI_ISL_1832617, EPI_ISL_1832623, EPI_ISL_1832633, EPI_ISL_1832640, EPI_ISL_1832641, EPI_ISL_1832643, EPI_ISL_1832646, EPI_ISL_1832649, EPI_ISL_1832651, EPI_ISL_1832653, EPI_ISL_1832655, EPI_ISL_1832657, EPI_ISL_1832661, EPI_ISL_1832663, EPI_ISL_1832665, EPI_ISL_1832667, EPI_ISL_1832669, EPI_ISL_1832671, EPI_ISL_1832672, EPI_ISL_1832674, EPI_ISL_1832676, EPI_ISL_1832679, EPI_ISL_1832682, EPI_ISL_1832701, EPI_ISL_2126134 | | | |
| see above | University College London, Great Ormond Street Hospital for Children NHS Foundation Trust, Imperial College Healthcare NHS Trust | COVID-19 Genomics UK (COG-UK) Consortium | Alison Holmes; Charlotte Williams; Helena Tutill; Jacqueline Findlay; James Price; Judith Breuer; Julianne Brown; Kathryn Harris; Leysa Forrest; Mark Kristiansen; Paola Niola; Paola Resende Silva; Patricia Dyal; Paul Randell; Rachel Williams; Samuel Weeks; Sergi Castellano; Sunando Roy; Tony Brooks; Yasmin Panchbhaya |
| EPI_ISL_1832336, EPI_ISL_1832337, EPI_ISL_1832338, EPI_ISL_1832339, EPI_ISL_1832340, EPI_ISL_2125909, EPI_ISL_2125911 | | | |
| see above | University Hospitals Of Leicester NHS Trust and DeepSeq Nottingham | COVID-19 Genomics UK (COG-UK) Consortium | Christopher Holmes; Christopher Moore; Fei Sang; Joeseph Chappell; Johnny Debebe; Jonathan Ball; Julian Tang; Karlie Fallon; Matthew Carlisle; Matthew Loose; Nadine Holmes; Patrick McClure; Paul Bird; Thomas Helmer; Victoria Wright |
| EPI_ISL_1729993, EPI_ISL_1730001, EPI_ISL_1730002, EPI_ISL_1730004, EPI_ISL_1730005, EPI_ISL_1730006, EPI_ISL_1730007, EPI_ISL_1730010, EPI_ISL_1730011, EPI_ISL_1730012, EPI_ISL_1730013, EPI_ISL_1730014, EPI_ISL_1730015, EPI_ISL_1832137, EPI_ISL_1832139, EPI_ISL_1832141, EPI_ISL_1832143, EPI_ISL_1832145, EPI_ISL_1832147, EPI_ISL_1832149, EPI_ISL_1832155, EPI_ISL_1974731, EPI_ISL_1974739 | | | |
| see above | University of Birmingham | COVID-19 Genomics UK (COG-UK) Consortium | Alex Richter; Andrew Bosworth; Andrew Bosworth. Queen Elizabeth Hospital: Anna Casey; Andrew D Beggs PHE Heartlands Lab: Husam Osman; Charlotte Poxon; Institute of Microbiology; Joanne Stockton; Josh Quick; Kasun Wanigasooriya; Mike Kidd; Nicholas Loman. University of Birmingham Testing Laboratory: Celina M Whalley; Oliver Pickles; Radoslaw Poplawski; Samuel Nicholls; University of Birmingham: Claire McMurray; Will Rowe |
| EPI_ISL_1730127, EPI_ISL_1730128, EPI_ISL_1730129, EPI_ISL_1730133, EPI_ISL_1730134, EPI_ISL_1730135, EPI_ISL_1832252, EPI_ISL_1832253, EPI_ISL_1832254, EPI_ISL_1832255, EPI_ISL_1832256, EPI_ISL_1832257, EPI_ISL_1832258, EPI_ISL_1832259, EPI_ISL_1832261, EPI_ISL_1832262, EPI_ISL_1832263, EPI_ISL_1832276, EPI_ISL_1974829, EPI_ISL_1974831, EPI_ISL_1974832, EPI_ISL_1974862, EPI_ISL_1974863, EPI_ISL_1974864, EPI_ISL_1974866, EPI_ISL_1974867, EPI_ISL_1974869, EPI_ISL_1974871, EPI_ISL_1974872, EPI_ISL_1974874, EPI_ISL_1974876, EPI_ISL_1974878, EPI_ISL_1974879, EPI_ISL_1974881, EPI_ISL_1974882, EPI_ISL_1974900, EPI_ISL_1974901, EPI_ISL_1974913, EPI_ISL_2125698, EPI_ISL_2125703, EPI_ISL_2125704, EPI_ISL_2125706, EPI_ISL_2125708, EPI_ISL_2125893, EPI_ISL_2125894 | | | |
| see above | Virology Department, Royal Infirmary of Edinburgh, NHS Lothian / School of Biological Sciences, University of Edinburgh | COVID-19 Genomics UK (COG-UK) Consortium | Colquhoun R; Cotton S; Dewar R; Gallagher A; Hill V; Jackson B; Maloney D; McCrone JT; McHugh M; O'Toole Á; Rambaut A; Rooke S; Scher E; Templeton K; Yu X |
| EPI_ISL_1730932, EPI_ISL_1730933, EPI_ISL_1730934, EPI_ISL_1730938, EPI_ISL_1730941, EPI_ISL_1730942, EPI_ISL_1730943, EPI_ISL_1730944, EPI_ISL_1730945, EPI_ISL_1730947, EPI_ISL_1730949, EPI_ISL_1730956, EPI_ISL_1730960, EPI_ISL_1730966, EPI_ISL_1730967, EPI_ISL_1730968, EPI_ISL_1730970, EPI_ISL_1730971, EPI_ISL_1730977, EPI_ISL_1730980, EPI_ISL_1730991, EPI_ISL_1731003, EPI_ISL_1731006, EPI_ISL_1731008, EPI_ISL_1731013, EPI_ISL_1731014, EPI_ISL_1731015, EPI_ISL_1731017, EPI_ISL_1731023, EPI_ISL_1731035, EPI_ISL_1731037, EPI_ISL_1731038, EPI_ISL_1731040, EPI_ISL_1731041, EPI_ISL_1833831, EPI_ISL_1833832, EPI_ISL_1833834, EPI_ISL_1833837, EPI_ISL_1833839, EPI_ISL_1833843, EPI_ISL_1833848, EPI_ISL_1833850, EPI_ISL_1833851, EPI_ISL_1833852, EPI_ISL_1833853, EPI_ISL_1833855, EPI_ISL_1833857, EPI_ISL_1833860, EPI_ISL_1833862, EPI_ISL_1833863, EPI_ISL_1833864, EPI_ISL_1833867, EPI_ISL_1833870, EPI_ISL_1833873, EPI_ISL_1833874, EPI_ISL_1833882, EPI_ISL_1833884, EPI_ISL_1833886, EPI_ISL_1833887, EPI_ISL_1833890, EPI_ISL_1833893, EPI_ISL_1833895, EPI_ISL_1833897, EPI_ISL_1833900, EPI_ISL_1833901, EPI_ISL_1833902, EPI_ISL_1833905, EPI_ISL_1833907, EPI_ISL_1833910, EPI_ISL_1833911, EPI_ISL_1833912, EPI_ISL_1833914, EPI_ISL_1833915, EPI_ISL_1833916, EPI_ISL_1980348, EPI_ISL_1980360, EPI_ISL_1980366, EPI_ISL_1980379, EPI_ISL_1980395, EPI_ISL_1980396, EPI_ISL_1980411, EPI_ISL_1980415, EPI_ISL_1980431, EPI_ISL_1980443, EPI_ISL_1980454 | | | |
| see above | Virology Department, Sheffield Teaching Hospitals NHS Foundation Trust/Department of Infection, Immunity and Cardiovascular Disease, The Medical School, University of Sheffield | COVID-19 Genomics UK (COG-UK) Consortium | Adri Angyal; Alex Keeley; Benjamin Lindsey; Cariad Evans; Danielle Groves; Dave Partridge; Katie Johnson; Laura Carrilero; Luke Green; Matthew Parker; Matthew Wyles; Mehmet Yavuz; Mohammad Raza; Nikki Smith; Paul Parsons; Rachel Tucker; Rebecca Brown; Thushan de Silva |
| EPI_ISL_1637487, EPI_ISL_1637500, EPI_ISL_1637502, EPI_ISL_1637503, EPI_ISL_1637512, EPI_ISL_1833137, EPI_ISL_1833148, EPI_ISL_1833179, EPI_ISL_1833180, EPI_ISL_1833181, EPI_ISL_1833182, EPI_ISL_1833183, EPI_ISL_1833184, EPI_ISL_1833185, EPI_ISL_1833187, EPI_ISL_1833188, EPI_ISL_1833191, EPI_ISL_1833192, EPI_ISL_1833193, EPI_ISL_1833194, EPI_ISL_1833195, EPI_ISL_1833197, EPI_ISL_1833198, EPI_ISL_1833199, EPI_ISL_1833201, EPI_ISL_1833202, EPI_ISL_1833203, EPI_ISL_1833204, EPI_ISL_1833206, EPI_ISL_1833207, EPI_ISL_1833208, EPI_ISL_1833209, EPI_ISL_1833210, EPI_ISL_1833211, EPI_ISL_1833212, EPI_ISL_1833213, EPI_ISL_1833214, EPI_ISL_1833215, EPI_ISL_1833216, EPI_ISL_1833217, EPI_ISL_1833218, EPI_ISL_1833219, EPI_ISL_1833220, EPI_ISL_1833221, EPI_ISL_1833222, EPI_ISL_1833223, EPI_ISL_1833224, EPI_ISL_1833225, EPI_ISL_1833226, EPI_ISL_1833227, EPI_ISL_1833228, EPI_ISL_1833229, EPI_ISL_1833230, EPI_ISL_1833231, EPI_ISL_1833232, EPI_ISL_1833233, EPI_ISL_1833234, EPI_ISL_1833235, EPI_ISL_1833236, EPI_ISL_1833237, EPI_ISL_1833238, EPI_ISL_1833239, EPI_ISL_1833240, EPI_ISL_1833241, EPI_ISL_1833242, EPI_ISL_1833243, EPI_ISL_1833244, EPI_ISL_1833245, EPI_ISL_1833246, EPI_ISL_1833247, EPI_ISL_1833248, EPI_ISL_1833249, EPI_ISL_1833250, EPI_ISL_1833251, EPI_ISL_1833252, EPI_ISL_1833253, EPI_ISL_1833254, EPI_ISL_1833255, EPI_ISL_1833256, EPI_ISL_1833257, EPI_ISL_1833258, EPI_ISL_1833259 | | | |
| see above | Wales Specialist Virology Centre Sequencing lab: Pathogen Genomics Unit | Public Health Wales Microbiology Cardiff Wales Specialist Virology Centre | Alec Birchley; Alexander Adams; Amy Gaskin; Angela Marchbank; Bree Gatica-Wilcox; Catherine Moore; Jason Coombes; Joanne Watkins; Joel Southgate; Johnathan Evans; Laura Gifford; Lauren Gilbert; Lee Graham; Malorie Perry; Matthew Bull; Nicole Pacchiarini; Sally Corden; Sara Kumziene-Summerhayes; Sara Rey; Sarah Taylor; Simon Cottrell; Sophie Jones; Tom Connor |
| EPI_ISL_1832301, EPI_ISL_1832302, EPI_ISL_1832303, EPI_ISL_1832304, EPI_ISL_1832306, EPI_ISL_1832307, EPI_ISL_1832309, EPI_ISL_1832310, EPI_ISL_1832311, EPI_ISL_1832312, EPI_ISL_2125498 | | | |
| see above | West of Scotland Specialist Virology Centre, NHSGGC / MRC-University of Glasgow Centre for Virus Research | COVID-19 Genomics UK (COG-UK) Consortium | Alasdair MacLean; Alice Broos; Ana da Silva Filipe; Antonia Ho; Daniel Mair; David L Robertson; Emma Thomson; Guy Mollett; Ioulia Tsatsani; James Shepherd; Jenna Nichols; Joseph Hughes; Kathy Li; Kathy Smollett; Kyriaki Nomikou; Lily Tong; Matthew Holden; Natasha Johnson; Rachel Blacow; Richard Orton; Rory Gunson; Sarah McDonald; Sharif Shaaban; Sreenu Vattipally; Stephen Carmichael |

We gratefully acknowledge the following Authors from the Originating laboratories responsible for obtaining the specimens, as well as the Submitting laboratories where the genome data were generated and shared via GISAID, on which this research is based.

All Submitters of data may be contacted directly via www.gisaid.org

Authors are sorted alphabetically.

| Accession ID | Originating Laboratory | Submitting Laboratory | Authors |
|---|---|---|---|
| EPI_ISL_1975057, EPI_ISL_1975059, EPI_ISL_1975060, EPI_ISL_1975062, EPI_ISL_1975064, EPI_ISL_1975066, EPI_ISL_1975067, EPI_ISL_1975077, EPI_ISL_1975104, EPI_ISL_1975106, EPI_ISL_1975107, EPI_ISL_2126006, EPI_ISL_2126007, EPI_ISL_2126018, EPI_ISL_2126019, EPI_ISL_2126023 | | | |
| see above | Barts Health NHS Trust | COVID-19 Genomics UK (COG-UK) Consortium | BROAD; Beatrix; CUTINO-MOGUEL; Claire; David; Dola; HARRINGTON; KELE; KULASEGARAN-SHYLINI; Maria-Teresa; OWOYEMI; Raghavendran |
| EPI_ISL_2128517 | Bioinformatics and Biostatistics Lab, Advanced Sequencing Facility | COVID-19 Genomics UK (COG-UK) Consortium | Aengus Stewart; Chelsea Sawyer; Harshil Patel; Jerome Nicod; Laura Cubitt; Margaret Crawford |
| EPI_ISL_1833278, EPI_ISL_1833282, EPI_ISL_1833283, EPI_ISL_1833286, EPI_ISL_1833288, EPI_ISL_1833341, EPI_ISL_1833345, EPI_ISL_1833346, EPI_ISL_1833351, EPI_ISL_1980109, EPI_ISL_1980111, EPI_ISL_1980112, EPI_ISL_1980210, EPI_ISL_1980212, EPI_ISL_1980213, EPI_ISL_2127897, EPI_ISL_2127899, EPI_ISL_2128176 | | | |
| see above | Centre for Enzyme Innovation, University of Portsmouth / Translational Research Laboratory, Portsmouth Hospitals NHS Trust | COVID-19 Genomics UK (COG-UK) Consortium | Angela Beckett; Christopher Fearn; Kate Cook; Katie Loveson; Salman Goudarzi; Samuel Robson; Scott Elliott; Sharon Glaysher |
| EPI_ISL_1832205, EPI_ISL_1832207, EPI_ISL_1832210, EPI_ISL_1832218, EPI_ISL_1832219, EPI_ISL_1832220, EPI_ISL_1832221, EPI_ISL_1832222, EPI_ISL_1832223, EPI_ISL_1832224, EPI_ISL_1832225, EPI_ISL_1832227, EPI_ISL_1832228, EPI_ISL_1832229, EPI_ISL_1832230, EPI_ISL_1832231, EPI_ISL_1832232, EPI_ISL_1832233, EPI_ISL_1974765, EPI_ISL_1974766, EPI_ISL_1974769, EPI_ISL_1974771, EPI_ISL_1974773, EPI_ISL_1974775, EPI_ISL_1974777, EPI_ISL_1974779, EPI_ISL_1974783, EPI_ISL_1974786 | | | |
| see above | Department of Pathology, University of Cambridge | COVID-19 Genomics UK (COG-UK) Consortium | Aminu S. Jahun; Ian Goodfellow; Iliana Georgana; Martin D. Curran; Myra Hosmillo; Rhys Izuagbe; Surendra Parmar; William L. Hamilton; Yasmin Chaudhry |
| EPI_ISL_2005342, EPI_ISL_2005344, EPI_ISL_2005345, EPI_ISL_2005350, EPI_ISL_2005354, EPI_ISL_2005356, EPI_ISL_2005368, EPI_ISL_2005376, EPI_ISL_2005383, EPI_ISL_2005399, EPI_ISL_2005402, EPI_ISL_2005403, EPI_ISL_2005407, EPI_ISL_2005409, EPI_ISL_2005415, EPI_ISL_2005416, EPI_ISL_2005417, EPI_ISL_2005419, EPI_ISL_2005420, EPI_ISL_2005424, EPI_ISL_2005428, EPI_ISL_2005432, EPI_ISL_2005440, EPI_ISL_2005453, EPI_ISL_2005455, EPI_ISL_2005465, EPI_ISL_2005470, EPI_ISL_2005471, EPI_ISL_2005473, EPI_ISL_2005475, EPI_ISL_2005480, EPI_ISL_2005483, EPI_ISL_2005487, EPI_ISL_2005490, EPI_ISL_2005494, EPI_ISL_2005497, EPI_ISL_2005498, EPI_ISL_2005501, EPI_ISL_2005524, EPI_ISL_2005527, EPI_ISL_2005534, EPI_ISL_2005535, EPI_ISL_2005536, EPI_ISL_2005542, EPI_ISL_2005544, EPI_ISL_2005545, EPI_ISL_2005546, EPI_ISL_2005554, EPI_ISL_2005559, EPI_ISL_2005562, EPI_ISL_2005563, EPI_ISL_2005566, EPI_ISL_2005569, EPI_ISL_2005572, EPI_ISL_2005575, EPI_ISL_2005583, EPI_ISL_2005587, EPI_ISL_2005591, EPI_ISL_2005592, EPI_ISL_2005594, EPI_ISL_2005604, EPI_ISL_2005607, EPI_ISL_2006543, EPI_ISL_2006550, EPI_ISL_2006552, EPI_ISL_2006577, EPI_ISL_2006578, EPI_ISL_2006582, EPI_ISL_2006592, EPI_ISL_2006595, EPI_ISL_2006597, EPI_ISL_2006602, EPI_ISL_2006603, EPI_ISL_2006608, EPI_ISL_2006613, EPI_ISL_2006616, EPI_ISL_2006618, EPI_ISL_2006621, EPI_ISL_2006623, EPI_ISL_2006626, EPI_ISL_2006632, EPI_ISL_2006653, EPI_ISL_2006656, EPI_ISL_2006664, EPI_ISL_2022535, EPI_ISL_2022545, EPI_ISL_2022549, EPI_ISL_2022554, EPI_ISL_2022557, EPI_ISL_2022558, EPI_ISL_2022560, EPI_ISL_2022561, EPI_ISL_2022565, EPI_ISL_2022568, EPI_ISL_2022570, EPI_ISL_2022571, EPI_ISL_2022572, EPI_ISL_2022573, EPI_ISL_2022574, EPI_ISL_2022575, EPI_ISL_2022576, EPI_ISL_2022581, EPI_ISL_2022583, EPI_ISL_2080361, EPI_ISL_2080364, EPI_ISL_2080365, EPI_ISL_2080366, EPI_ISL_2080368, EPI_ISL_2080369, EPI_ISL_2080370, EPI_ISL_2080372, EPI_ISL_2080373, EPI_ISL_2080374, EPI_ISL_2080379, EPI_ISL_2080385, EPI_ISL_2080387, EPI_ISL_2080388, EPI_ISL_2080391, EPI_ISL_2080393, EPI_ISL_2080394, EPI_ISL_2080396, EPI_ISL_2080398, EPI_ISL_2080399, EPI_ISL_2080400, EPI_ISL_2080402, EPI_ISL_2080403, EPI_ISL_2080404, EPI_ISL_2080407, EPI_ISL_2080409, EPI_ISL_2080412, EPI_ISL_2080413, EPI_ISL_2080417, EPI_ISL_2080418, EPI_ISL_2080419, EPI_ISL_2080420, EPI_ISL_2080424, EPI_ISL_2080425, EPI_ISL_2080429, EPI_ISL_2080431, EPI_ISL_2080432, EPI_ISL_2080433, EPI_ISL_2080434, EPI_ISL_2080438, EPI_ISL_2080440, EPI_ISL_2080441, EPI_ISL_2080442, EPI_ISL_2080446, EPI_ISL_2080451, EPI_ISL_2080452, EPI_ISL_2080453, EPI_ISL_2080454, EPI_ISL_2080455, EPI_ISL_2080457, EPI_ISL_2080458, EPI_ISL_2080459, EPI_ISL_2080461, EPI_ISL_2080462, EPI_ISL_2080464, EPI_ISL_2080465, EPI_ISL_2080468, EPI_ISL_2080470, EPI_ISL_2080474 | | | |
| see above | Health Services Laboratories | Wellcome Sanger Institute for the COVID-19 Genomics UK (COG-UK) Consortium | Cordelia Langford; David K. Jackson; Dominic Kwiatkowski; Ewan Harrison; Health Services Laboratories and Alex Alderton; Ian Johnston; Jeffrey Barrett; John Sillitoe on behalf of the Wellcome Sanger Institute COVID-19 Surveillance Team; Roberto Amato; Sonia Goncalves |
| EPI_ISL_1829209, EPI_ISL_1829211, EPI_ISL_1829212, EPI_ISL_1829213, EPI_ISL_1829214, EPI_ISL_1829216, EPI_ISL_1829220, EPI_ISL_1829222, EPI_ISL_1829223, EPI_ISL_1829227, EPI_ISL_1829229, EPI_ISL_1829232, EPI_ISL_1829233, EPI_ISL_1829237, EPI_ISL_1829242, EPI_ISL_1829243, EPI_ISL_1829245, EPI_ISL_1829252, EPI_ISL_1829254, EPI_ISL_1829255, EPI_ISL_1829258, EPI_ISL_1829260, EPI_ISL_1829262, EPI_ISL_1829271, EPI_ISL_1829272, EPI_ISL_1829273, EPI_ISL_1829274, EPI_ISL_1829276, EPI_ISL_1829278, EPI_ISL_1829279, EPI_ISL_1829281, EPI_ISL_1829282, EPI_ISL_1829285, EPI_ISL_1829289, EPI_ISL_1829291, EPI_ISL_1829292, EPI_ISL_1829296, EPI_ISL_1829297, EPI_ISL_1829299, EPI_ISL_1829300, EPI_ISL_1829302, EPI_ISL_1829304, EPI_ISL_1829307, EPI_ISL_1829310, EPI_ISL_1829311, EPI_ISL_1829318, EPI_ISL_1829319, EPI_ISL_1829320, EPI_ISL_1829325, EPI_ISL_1829333, EPI_ISL_1829337, EPI_ISL_1829342, EPI_ISL_1829344, EPI_ISL_1829345, EPI_ISL_1829352, EPI_ISL_1829353, EPI_ISL_1829356, EPI_ISL_1829357, EPI_ISL_1829359, EPI_ISL_1829360, EPI_ISL_1829362, EPI_ISL_1829373, EPI_ISL_1829374, EPI_ISL_1829375, EPI_ISL_1829376, EPI_ISL_1829382, EPI_ISL_1829400, EPI_ISL_1829403, EPI_ISL_1829406, EPI_ISL_1829408, EPI_ISL_1829411, EPI_ISL_1829412, EPI_ISL_1829413, EPI_ISL_1829419, EPI_ISL_1829420, EPI_ISL_1829427, EPI_ISL_1829430, EPI_ISL_1829433, EPI_ISL_1829434, EPI_ISL_1829435, EPI_ISL_1829437, EPI_ISL_1829438, EPI_ISL_1829440, EPI_ISL_1829444, EPI_ISL_1829452, EPI_ISL_1829456, EPI_ISL_1829457, EPI_ISL_1829458, EPI_ISL_1829460, EPI_ISL_1829462, EPI_ISL_1829474, EPI_ISL_1829804, EPI_ISL_1829806, EPI_ISL_1829808, EPI_ISL_1829809, EPI_ISL_1829811, EPI_ISL_1829813, EPI_ISL_1829815, EPI_ISL_1829816, EPI_ISL_1829818, EPI_ISL_1829819, EPI_ISL_1829821, EPI_ISL_1829822, EPI_ISL_1829823, EPI_ISL_1829825, EPI_ISL_1829830, EPI_ISL_1829838, EPI_ISL_1829839, EPI_ISL_1829840, EPI_ISL_1829842, EPI_ISL_1829843, EPI_ISL_1829848, EPI_ISL_1829850, EPI_ISL_1829851, EPI_ISL_1829853, EPI_ISL_1829854, EPI_ISL_1829855, EPI_ISL_1829859, EPI_ISL_1829860, EPI_ISL_1829862, EPI_ISL_1829863, EPI_ISL_1829866, EPI_ISL_1829867, EPI_ISL_1829869, EPI_ISL_1829871, EPI_ISL_1829874, EPI_ISL_1829875, EPI_ISL_1829878, EPI_ISL_1829879, EPI_ISL_1829880, EPI_ISL_1829881, EPI_ISL_1829882, EPI_ISL_1829883, EPI_ISL_1829884, EPI_ISL_1829885, EPI_ISL_1829886, EPI_ISL_1829887, EPI_ISL_1829889, EPI_ISL_1829896, EPI_ISL_1829898, EPI_ISL_1829900, EPI_ISL_1829902, EPI_ISL_1829904, EPI_ISL_1829905, EPI_ISL_1829909, EPI_ISL_1829913, EPI_ISL_1829914, EPI_ISL_1829915, EPI_ISL_1829917, EPI_ISL_1829920, EPI_ISL_1829921, EPI_ISL_1829922, EPI_ISL_1829925, EPI_ISL_1829929, EPI_ISL_1829930, EPI_ISL_1829935, EPI_ISL_1829937, EPI_ISL_1829938, EPI_ISL_1829939, EPI_ISL_1829940, EPI_ISL_1829943, EPI_ISL_1829944, EPI_ISL_1829950, EPI_ISL_1829952, EPI_ISL_1829953, EPI_ISL_1829962, EPI_ISL_1829963, EPI_ISL_1829964, EPI_ISL_1829965, EPI_ISL_1829966, EPI_ISL_1829969, EPI_ISL_1829970, EPI_ISL_1829973, EPI_ISL_1829974, EPI_ISL_1829976, EPI_ISL_1829979, EPI_ISL_1829982, EPI_ISL_1829983, EPI_ISL_1829984, EPI_ISL_1829985, EPI_ISL_1829986, EPI_ISL_1829987, EPI_ISL_1829988, EPI_ISL_1829994, EPI_ISL_1829995, EPI_ISL_1829996, EPI_ISL_1830003, EPI_ISL_1830004, EPI_ISL_1830006, EPI_ISL_1830007, EPI_ISL_1830011, EPI_ISL_1830012, EPI_ISL_1830013, EPI_ISL_1830014, EPI_ISL_1830019, EPI_ISL_1830020, EPI_ISL_1830021, EPI_ISL_1830023, EPI_ISL_1830024, EPI_ISL_1830025, EPI_ISL_1830031, EPI_ISL_1830035, EPI_ISL_1830036, EPI_ISL_1830039, EPI_ISL_1830040, EPI_ISL_1830042, EPI_ISL_1830044, EPI_ISL_1830046, EPI_ISL_1830049, EPI_ISL_1830052, EPI_ISL_1830053, EPI_ISL_1830054, EPI_ISL_1830055, EPI_ISL_1830058, EPI_ISL_1830061, EPI_ISL_1830062, EPI_ISL_1830064, EPI_ISL_1830065, EPI_ISL_1830066, EPI_ISL_1830069, EPI_ISL_1830071, EPI_ISL_1830072, EPI_ISL_1830074, EPI_ISL_1830075, EPI_ISL_1830077, EPI_ISL_1830078, EPI_ISL_1830080, EPI_ISL_1830082, EPI_ISL_1830083, EPI_ISL_1830085, EPI_ISL_1830089, EPI_ISL_1830090, EPI_ISL_1830093, EPI_ISL_1830094, EPI_ISL_1830095, EPI_ISL_1830098, EPI_ISL_1830105, EPI_ISL_1830201, EPI_ISL_1830211, EPI_ISL_1830219, EPI_ISL_1830220, EPI_ISL_1830309, EPI_ISL_1830314, EPI_ISL_1830321, EPI_ISL_1830348, EPI_ISL_1830363, EPI_ISL_1830371, EPI_ISL_1830377, EPI_ISL_1830393, EPI_ISL_1830395, EPI_ISL_1830396, EPI_ISL_1830407, EPI_ISL_1830415, EPI_ISL_1830420, EPI_ISL_1830425, EPI_ISL_1830426, EPI_ISL_1830427, EPI_ISL_1830433, EPI_ISL_1830434, EPI_ISL_1830439, EPI_ISL_1830443, EPI_ISL_1830444, EPI_ISL_1830445, EPI_ISL_1830455, EPI_ISL_1830458, EPI_ISL_1830460, EPI_ISL_1830463, EPI_ISL_1830466, EPI_ISL_1830467, EPI_ISL_1830468, EPI_ISL_1830471, EPI_ISL_1830478, EPI_ISL_1830479, EPI_ISL_1830480, EPI_ISL_1830484, EPI_ISL_1830486, EPI_ISL_1830487, EPI_ISL_1830491, EPI_ISL_1830500, EPI_ISL_1830504, EPI_ISL_1830507, EPI_ISL_1830513, EPI_ISL_1830516, EPI_ISL_1830519, EPI_ISL_1830520, EPI_ISL_1830521, EPI_ISL_1830530, EPI_ISL_1830533, EPI_ISL_1830534, EPI_ISL_1830535, EPI_ISL_1830536, EPI_ISL_1830538, EPI_ISL_1830541, EPI_ISL_1830543, EPI_ISL_1830547, EPI_ISL_1830548, EPI_ISL_1830550, EPI_ISL_1830553, EPI_ISL_1830558, EPI_ISL_1830561, EPI_ISL_1830566, EPI_ISL_1830567, EPI_ISL_1830571, EPI_ISL_1830573, EPI_ISL_1830575, EPI_ISL_1830577, EPI_ISL_1830582, EPI_ISL_1830586, EPI_ISL_1830587, EPI_ISL_1830588, EPI_ISL_1830591, EPI_ISL_1830602, EPI_ISL_1830610, EPI_ISL_1830611, EPI_ISL_1830616, EPI_ISL_1830617, EPI_ISL_1830618, EPI_ISL_1830620, EPI_ISL_1830622, EPI_ISL_1830625, EPI_ISL_1830627, EPI_ISL_1830628, EPI_ISL_1830636, EPI_ISL_1830639, EPI_ISL_1830640, EPI_ISL_1830641, EPI_ISL_1830642, EPI_ISL_1830645, EPI_ISL_1830646, EPI_ISL_1830647, EPI_ISL_1830653, EPI_ISL_1830655, EPI_ISL_1830662, EPI_ISL_1830663, EPI_ISL_1830666, EPI_ISL_1830678, EPI_ISL_1830680, EPI_ISL_1830683, EPI_ISL_1830684, EPI_ISL_1830686, EPI_ISL_1830691, EPI_ISL_1830696, EPI_ISL_1830697, EPI_ISL_1830698, EPI_ISL_1830699, EPI_ISL_1830705, EPI_ISL_1830711, EPI_ISL_1830721, EPI_ISL_1830723, EPI_ISL_1830724, EPI_ISL_1830728, EPI_ISL_1830730, EPI_ISL_1830740, EPI_ISL_1830741, EPI_ISL_1830746, EPI_ISL_1830752, EPI_ISL_1830761, EPI_ISL_1830762, EPI_ISL_1830764, EPI_ISL_1830775, EPI_ISL_1830778, EPI_ISL_1830780, EPI_ISL_1830789, EPI_ISL_1830793, EPI_ISL_1830799, EPI_ISL_1830803, EPI_ISL_1830808, EPI_ISL_1830812, EPI_ISL_1830814, EPI_ISL_1830815, EPI_ISL_1830818, EPI_ISL_1830832, EPI_ISL_1830833, EPI_ISL_1830858, EPI_ISL_1830859, EPI_ISL_1830863, EPI_ISL_1830880, EPI_ISL_1830881, EPI_ISL_1830888, EPI_ISL_1830895, EPI_ISL_1830897, EPI_ISL_1830898, EPI_ISL_1830921, EPI_ISL_1830925, EPI_ISL_1830927, EPI_ISL_1830929, EPI_ISL_1830935, EPI_ISL_1830943, EPI_ISL_1830961, EPI_ISL_1830981, EPI_ISL_1830985, EPI_ISL_1831003, EPI_ISL_1831018, EPI_ISL_1831019, EPI_ISL_1831034, EPI_ISL_1831043, EPI_ISL_1831050, EPI_ISL_1831058, EPI_ISL_1831071, EPI_ISL_1831073, EPI_ISL_1857806, EPI_ISL_1857807, EPI_ISL_1857808, EPI_ISL_1857809, EPI_ISL_1857811, EPI_ISL_1857812, EPI_ISL_1857815, EPI_ISL_1857816, EPI_ISL_1857817, EPI_ISL_1857819, EPI_ISL_1857821, EPI_ISL_1857822, EPI_ISL_1857824, EPI_ISL_1857825, EPI_ISL_1857828, EPI_ISL_1857829, EPI_ISL_1857830, EPI_ISL_1857832, EPI_ISL_1857833, EPI_ISL_1857834, EPI_ISL_1857835, EPI_ISL_1857836, EPI_ISL_1857837, EPI_ISL_1857838, EPI_ISL_1857839, EPI_ISL_1857840, EPI_ISL_1857841, EPI_ISL_1857843, EPI_ISL_1857850, EPI_ISL_1857854, EPI_ISL_1857856, EPI_ISL_1857857, EPI_ISL_1857859, EPI_ISL_1857860, EPI_ISL_1857863, EPI_ISL_1857864, EPI_ISL_1857868, EPI_ISL_1857869, EPI_ISL_1857872, EPI_ISL_1857875, EPI_ISL_1857876, EPI_ISL_1857877, EPI_ISL_1857878, EPI_ISL_1857879, EPI_ISL_1857880, EPI_ISL_1857883, EPI_ISL_1857885, EPI_ISL_1857886, EPI_ISL_1857887, EPI_ISL_1857890, EPI_ISL_1857891, EPI_ISL_1857892, EPI_ISL_1857893, EPI_ISL_1857896, EPI_ISL_1857904, EPI_ISL_1857908, EPI_ISL_1857912, EPI_ISL_1857913, EPI_ISL_1857914, EPI_ISL_1857917, EPI_ISL_1857920, EPI_ISL_1857921, EPI_ISL_1857922, EPI_ISL_1857923, EPI_ISL_1857926, EPI_ISL_1857927, EPI_ISL_1857928, EPI_ISL_1857932, EPI_ISL_1857933, EPI_ISL_1857934, EPI_ISL_1857937, EPI_ISL_1857941, EPI_ISL_1857942, EPI_ISL_1857943, EPI_ISL_1857945, EPI_ISL_1857948, EPI_ISL_1857952, EPI_ISL_1857954, EPI_ISL_1857955, EPI_ISL_1857957, EPI_ISL_1857960, EPI_ISL_1857961, EPI_ISL_1857963, EPI_ISL_1857964, EPI_ISL_1857967, EPI_ISL_1857969, EPI_ISL_1857970, EPI_ISL_1857972, EPI_ISL_1857974, EPI_ISL_1857977, EPI_ISL_1857980, EPI_ISL_1857982, EPI_ISL_1857986, EPI_ISL_1857988, EPI_ISL_1857989, EPI_ISL_1857993, EPI_ISL_1857994, EPI_ISL_1858001, EPI_ISL_1858006, EPI_ISL_1858009, EPI_ISL_1858010, EPI_ISL_1858012, EPI_ISL_1858015, EPI_ISL_1858018, EPI_ISL_1858020, EPI_ISL_1858021, EPI_ISL_1858023, EPI_ISL_1858026, EPI_ISL_1858031, EPI_ISL_1858032, EPI_ISL_1858034, EPI_ISL_1858035, EPI_ISL_1858037, EPI_ISL_1858041, EPI_ISL_1858048, EPI_ISL_1858049, EPI_ISL_1858051, EPI_ISL_1858054, EPI_ISL_1858057, EPI_ISL_1858065, EPI_ISL_1858066, EPI_ISL_1858070, EPI_ISL_1858071, EPI_ISL_1858073, EPI_ISL_1858075, EPI_ISL_1858076, EPI_ISL_1858079, EPI_ISL_1858081, EPI_ISL_1858084, EPI_ISL_1858086, EPI_ISL_1858087, EPI_ISL_1858089, EPI_ISL_1858092, EPI_ISL_1858094, EPI_ISL_1858097, EPI_ISL_1858099, EPI_ISL_1858101, EPI_ISL_1858104, EPI_ISL_1858107, EPI_ISL_1858108, EPI_ISL_1858110, EPI_ISL_1858113, EPI_ISL_1858114, EPI_ISL_1858119, EPI_ISL_1858121, EPI_ISL_1858124, EPI_ISL_1858127, EPI_ISL_1858132, EPI_ISL_1858134, EPI_ISL_1858136, EPI_ISL_1858138, EPI_ISL_1858139, EPI_ISL_1858143, EPI_ISL_1858147, EPI_ISL_1858152, EPI_ISL_1858155, EPI_ISL_1858156, EPI_ISL_1858158, EPI_ISL_1858207, EPI_ISL_1858208, EPI_ISL_1858211, EPI_ISL_1858213, EPI_ISL_1858217, EPI_ISL_1858221, EPI_ISL_1858228, EPI_ISL_1858237, EPI_ISL_1912007, EPI_ISL_1912014, EPI_ISL_1912025, EPI_ISL_1912040, EPI_ISL_1912042, EPI_ISL_1912043, EPI_ISL_1912054, EPI_ISL_1912062, EPI_ISL_1912067, EPI_ISL_1912077, EPI_ISL_1912079, EPI_ISL_1912080, EPI_ISL_1912089, EPI_ISL_1912090, EPI_ISL_1912092, EPI_ISL_1912095, EPI_ISL_1912097, EPI_ISL_1912098, EPI_ISL_1912100, EPI_ISL_1912101, EPI_ISL_1912106, EPI_ISL_1912112, EPI_ISL_1912115, EPI_ISL_1912120, EPI_ISL_1912121, EPI_ISL_1912122, EPI_ISL_1912123, EPI_ISL_1912133, EPI_ISL_1912134, EPI_ISL_1912135, EPI_ISL_1912136, EPI_ISL_1912138, EPI_ISL_1912139, EPI_ISL_1912146, EPI_ISL_1912148, EPI_ISL_1912156, EPI_ISL_1912160, EPI_ISL_1912161, EPI_ISL_1912162, EPI_ISL_1912163, EPI_ISL_1912166, EPI_ISL_1912169, EPI_ISL_1912170, EPI_ISL_1912176, EPI_ISL_1912178, EPI_ISL_1912180, EPI_ISL_1912182, EPI_ISL_1912183, EPI_ISL_1912184, EPI_ISL_1912185, EPI_ISL_1912188, EPI_ISL_1912191, EPI_ISL_1912193, EPI_ISL_1912194, EPI_ISL_1912197, EPI_ISL_1912198, EPI_ISL_1912202, EPI_ISL_1912204, EPI_ISL_1912206, EPI_ISL_1912214, EPI_ISL_1912223, EPI_ISL_1912225, EPI_ISL_1912226, EPI_ISL_1912248, EPI_ISL_1912261, EPI_ISL_1912262, EPI_ISL_1912268, EPI_ISL_1912283, EPI_ISL_1912286, EPI_ISL_1912288, EPI_ISL_1912294, EPI_ISL_1912297, EPI_ISL_1912307, EPI_ISL_1912309, EPI_ISL_1912314, EPI_ISL_1912318, EPI_ISL_1912321, EPI_ISL_1912325, EPI_ISL_1912328, EPI_ISL_1912329, EPI_ISL_1912332, EPI_ISL_1912333, EPI_ISL_1912342, EPI_ISL_1912344, EPI_ISL_1912347, EPI_ISL_1912348, EPI_ISL_1912351, EPI_ISL_1912352, EPI_ISL_1912353, EPI_ISL_1912354, EPI_ISL_1912355, EPI_ISL_1912358, EPI_ISL_1912359, EPI_ISL_1912360, EPI_ISL_1912361, EPI_ISL_1912362, EPI_ISL_1912364, EPI_ISL_1912365, EPI_ISL_1912369, EPI_ISL_1912371, EPI_ISL_1912373, EPI_ISL_1912374, EPI_ISL_1912375, EPI_ISL_1912376, EPI_ISL_1912377, EPI_ISL_1912379, EPI_ISL_1912381, EPI_ISL_1912382, EPI_ISL_1912383, EPI_ISL_1912384, EPI_ISL_1912385, EPI_ISL_1912387, EPI_ISL_1912389, EPI_ISL_1912390, EPI_ISL_1912393, EPI_ISL_1912394, EPI_ISL_1912396, EPI_ISL_1912397, EPI_ISL_1912398, EPI_ISL_1912399, EPI_ISL_1912401, EPI_ISL_1912402, EPI_ISL_1912403, EPI_ISL_1912405, EPI_ISL_1912407, EPI_ISL_1912408, EPI_ISL_1912409, EPI_ISL_1912413, EPI_ISL_1912414, EPI_ISL_1912415, EPI_ISL_1912416, EPI_ISL_1912418, EPI_ISL_1912422, EPI_ISL_1912425, EPI_ISL_1912426, EPI_ISL_1912427, EPI_ISL_1912428, EPI_ISL_1912429, EPI_ISL_1912430, EPI_ISL_1912431, EPI_ISL_1912432, EPI_ISL_1912434, EPI_ISL_1912435, EPI_ISL_1912436, EPI_ISL_1912437, EPI_ISL_1912438, EPI_ISL_1912439, EPI_ISL_1912442, EPI_ISL_1912443, EPI_ISL_1912444, EPI_ISL_1912446, EPI_ISL_1912447, EPI_ISL_1912448, EPI_ISL_1912449, EPI_ISL_1912452, EPI_ISL_1912453, EPI_ISL_1912454, EPI_ISL_1912455, EPI_ISL_1912456, EPI_ISL_1912458, EPI_ISL_1912459, EPI_ISL_1912460, EPI_ISL_1912461, EPI_ISL_1912463, EPI_ISL_1912464, EPI_ISL_1912465, EPI_ISL_1912466, EPI_ISL_1912467, EPI_ISL_1912468, EPI_ISL_1912469, EPI_ISL_1912471, EPI_ISL_1912474, EPI_ISL_1912475, EPI_ISL_1912479, EPI_ISL_1912480, EPI_ISL_1912482, EPI_ISL_1912483, EPI_ISL_1912484, EPI_ISL_1912486, EPI_ISL_1912487, EPI_ISL_1912488, EPI_ISL_1912490, EPI_ISL_1912492, EPI_ISL_1912493, EPI_ISL_1912494, EPI_ISL_1912495, EPI_ISL_1912496, EPI_ISL_1912498, EPI_ISL_1912499, EPI_ISL_1912503, EPI_ISL_1912504, EPI_ISL_1912505, EPI_ISL_1912507, EPI_ISL_1912508, EPI_ISL_1912509, EPI_ISL_1912510, EPI_ISL_1912514, EPI_ISL_1912515, EPI_ISL_1912516, EPI_ISL_1912517, EPI_ISL_1912521, EPI_ISL_1912522, EPI_ISL_1912523, EPI_ISL_1912524, EPI_ISL_1912526, EPI_ISL_1912528, EPI_ISL_1912529, EPI_ISL_1912530, EPI_ISL_1912531, EPI_ISL_1912532, EPI_ISL_1912533, EPI_ISL_1912534, EPI_ISL_1912535, EPI_ISL_1912536, EPI_ISL_1912537, EPI_ISL_1912538, EPI_ISL_1912539, EPI_ISL_1912540, EPI_ISL_1912541, EPI_ISL_1912542, EPI_ISL_1912543, EPI_ISL_1912544, EPI_ISL_1912548, EPI_ISL_1912551, EPI_ISL_1912552, EPI_ISL_1912553, EPI_ISL_1912554, EPI_ISL_1912555, EPI_ISL_1912556, EPI_ISL_1912557, EPI_ISL_1912559, EPI_ISL_1912560, EPI_ISL_1912561, EPI_ISL_1912562, EPI_ISL_1912563, EPI_ISL_1912564, EPI_ISL_1912566, EPI_ISL_1912567, EPI_ISL_1912568, EPI_ISL_1912570, EPI_ISL_1912571, EPI_ISL_1912572, EPI_ISL_1912574, EPI_ISL_1912575, EPI_ISL_1912577, EPI_ISL_1912578, EPI_ISL_1912579, EPI_ISL_1912580, EPI_ISL_1912583, EPI_ISL_1912584, EPI_ISL_1912585, EPI_ISL_1912586, EPI_ISL_1912587, EPI_ISL_1912590, EPI_ISL_1912592, EPI_ISL_1912593, EPI_ISL_1912729, EPI_ISL_1912737, EPI_ISL_1912740, EPI_ISL_1912751, EPI_ISL_1912767, EPI_ISL_1912784, EPI_ISL_1921166, EPI_ISL_1921167, EPI_ISL_1921168, EPI_ISL_1921169, EPI_ISL_1921170, EPI_ISL_1921171, EPI_ISL_1921172, EPI_ISL_1921173, EPI_ISL_1921174, EPI_ISL_1921175, EPI_ISL_1921176, EPI_ISL_1921177, EPI_ISL_1921178, EPI_ISL_1921179, EPI_ISL_1921180, EPI_ISL_1921181, EPI_ISL_1921182, EPI_ISL_1921183, EPI_ISL_1921184, EPI_ISL_1921185, EPI_ISL_1921186, EPI_ISL_1921187, EPI_ISL_1921188, EPI_ISL_1921189, EPI_ISL_1921190, EPI_ISL_1921191, EPI_ISL_1921192, EPI_ISL_1921193, EPI_ISL_1921194, EPI_ISL_1921195, EPI_ISL_1921196, EPI_ISL_1921197, EPI_ISL_1921198, EPI_ISL_1921199, EPI_ISL_1921200, EPI_ISL_1921201, EPI_ISL_1921202, EPI_ISL_1921203, EPI_ISL_1921204, EPI_ISL_1921205, EPI_ISL_1921206, EPI_ISL_1921207, EPI_ISL_1921208, EPI_ISL_1921209, EPI_ISL_1921210, EPI_ISL_1921211, EPI_ISL_1921212, EPI_ISL_1921213, EPI_ISL_1921214, EPI_ISL_1921215, EPI_ISL_1921216, EPI_ISL_1921217, EPI_ISL_1921218, EPI_ISL_1921219, EPI_ISL_1921220, EPI_ISL_1921221, EPI_ISL_1921222, EPI_ISL_1921223, EPI_ISL_1921224, EPI_ISL_1921225, EPI_ISL_1921226, EPI_ISL_1921227, EPI_ISL_1921228, EPI_ISL_1921229, EPI_ISL_1921230, EPI_ISL_1921231, EPI_ISL_1921232, EPI_ISL_1921233, EPI_ISL_1921234, EPI_ISL_1921235, EPI_ISL_1921236, EPI_ISL_1921237, EPI_ISL_1921238, EPI_ISL_1921239, EPI_ISL_1921240, EPI_ISL_1921241, EPI_ISL_1921242, EPI_ISL_1921243, EPI_ISL_1921244, EPI_ISL_1921245, EPI_ISL_1921246, EPI_ISL_1921247, EPI_ISL_1921248, EPI_ISL_1921249, EPI_ISL_1921250, EPI_ISL_1921251, EPI_ISL_1921252, EPI_ISL_1921253, EPI_ISL_1921254, EPI_ISL_1921255, EPI_ISL_1921256, EPI_ISL_1921257, EPI_ISL_1921258, EPI_ISL_1921259, EPI_ISL_1921261, EPI_ISL_1921262, EPI_ISL_1921263, EPI_ISL_1921264, EPI_ISL_1921265, EPI_ISL_1921266, EPI_ISL_1921267, EPI_ISL_1921268, EPI_ISL_1921269, EPI_ISL_1921270, EPI_ISL_1921271, EPI_ISL_1921272, EPI_ISL_1921273, EPI_ISL_1921274, EPI_ISL_1921275, EPI_ISL_1921276, EPI_ISL_1921277, EPI_ISL_1921278, EPI_ISL_1921279, EPI_ISL_1921280, EPI_ISL_1921281, EPI_ISL_1921282, EPI_ISL_1921283, EPI_ISL_1921284, EPI_ISL_1921285, EPI_ISL_1921286, EPI_ISL_1921287, EPI_ISL_1921289, EPI_ISL_1921290, EPI_ISL_1921291, EPI_ISL_1921292, EPI_ISL_1921293, EPI_ISL_1921294, EPI_ISL_1921295, EPI_ISL_1921296, EPI_ISL_1921297, EPI_ISL_1921298, EPI_ISL_1921299, EPI_ISL_1921300, EPI_ISL_1921301, EPI_ISL_1921302, EPI_ISL_1921303, EPI_ISL_1921304, EPI_ISL_1921305, EPI_ISL_1921306, EPI_ISL_1921307, EPI_ISL_1921308, EPI_ISL_1921309, EPI_ISL_1921310, EPI_ISL_1921311, EPI_ISL_1921312, EPI_ISL_1921313, EPI_ISL_1921314, EPI_ISL_1921315, EPI_ISL_1921316, EPI_ISL_1921317, EPI_ISL_1921318, EPI_ISL_1921319, EPI_ISL_1921320, EPI_ISL_1921321, EPI_ISL_1921322, EPI_ISL_1921323, EPI_ISL_1921324, EPI_ISL_1921325, EPI_ISL_1921326, EPI_ISL_1921327, EPI_ISL_1921328, EPI_ISL_1921329, EPI_ISL_1921330, EPI_ISL_1921331, EPI_ISL_1921332, EPI_ISL_1921333, EPI_ISL_1921334, EPI_ISL_1921335, EPI_ISL_1921336, EPI_ISL_1921337, EPI_ISL_1921338, EPI_ISL_1921339, EPI_ISL_1921340, EPI_ISL_1921341, EPI_ISL_1921342, EPI_ISL_1921343, EPI_ISL_1921344, EPI_ISL_1921345, EPI_ISL_1921346, EPI_ISL_1921347, EPI_ISL_1921348, EPI_ISL_1921349, EPI_ISL_1921350, EPI_ISL_1921351, EPI_ISL_1921352, EPI_ISL_1921353, EPI_ISL_1921354, EPI_ISL_1921355, EPI_ISL_1921356, EPI_ISL_1921357, EPI_ISL_1921358, EPI_ISL_1921359, EPI_ISL_1921360, EPI_ISL_1921361, EPI_ISL_1921362, EPI_ISL_1921363, EPI_ISL_1921364, EPI_ISL_1921365, EPI_ISL_1921366, EPI_ISL_1921367, EPI_ISL_1921368, EPI_ISL_1921369, EPI_ISL_1921370, EPI_ISL_1921371, EPI_ISL_1921372, EPI_ISL_1921373, EPI_ISL_1921374, EPI_ISL_1921375, EPI_ISL_1921376, EPI_ISL_1921377, EPI_ISL_1921378, EPI_ISL_1921379, EPI_ISL_1921380, EPI_ISL_1921381, EPI_ISL_1921382, EPI_ISL_1921383, EPI_ISL_1921384, EPI_ISL_1921385, EPI_ISL_1921386, EPI_ISL_1921387, EPI_ISL_1921388, EPI_ISL_1921389, EPI_ISL_1921390, EPI_ISL_1921391, EPI_ISL_1921392, EPI_ISL_1921393, EPI_ISL_1921394, EPI_ISL_1921395, EPI_ISL_1921396, EPI_ISL_1921397, EPI_ISL_1921398, EPI_ISL_1921399, EPI_ISL_1921400, EPI_ISL_1921401, EPI_ISL_1921402, EPI_ISL_1921403, EPI_ISL_1921404, EPI_ISL_1921405, EPI_ISL_1921406, EPI_ISL_1921407, EPI_ISL_1921408, EPI_ISL_1921409, EPI_ISL_1921410, EPI_ISL_1921411, EPI_ISL_1921412, EPI_ISL_1921413, EPI_ISL_1921414, EPI_ISL_1921415, EPI_ISL_1921416, EPI_ISL_1921417, EPI_ISL_1921418, EPI_ISL_1921419, EPI_ISL_1921420, EPI_ISL_1921421, EPI_ISL_1921422, EPI_ISL_1921423, EPI_ISL_1921424, EPI_ISL_1921426, EPI_ISL_1921427, EPI_ISL_1921428, EPI_ISL_1921429, EPI_ISL_1921430, EPI_ISL_1921431, EPI_ISL_1921432, EPI_ISL_1921433, EPI_ISL_1921434, EPI_ISL_1921435, EPI_ISL_1921436, EPI_ISL_1921437, EPI_ISL_1921438, EPI_ISL_1921444, EPI_ISL_1921446, EPI_ISL_1921459, EPI_ISL_1921465, EPI_ISL_1921486, EPI_ISL_1921494, EPI_ISL_1921495, EPI_ISL_1921496, EPI_ISL_1921506, EPI_ISL_1921507, EPI_ISL_1921509, EPI_ISL_1921515, EPI_ISL_1921525, EPI_ISL_1921533, EPI_ISL_1921552, EPI_ISL_1921553, EPI_ISL_1921555, EPI_ISL_1921572, EPI_ISL_1921588, EPI_ISL_1921598, EPI_ISL_1921613, EPI_ISL_1921618, EPI_ISL_1921619, EPI_ISL_1921634, EPI_ISL_1921637, EPI_ISL_1921645, EPI_ISL_1921656, EPI_ISL_1921662, EPI_ISL_1921666, EPI_ISL_1921667, EPI_ISL_1921669, EPI_ISL_1921676, EPI_ISL_1921685, EPI_ISL_1921686, EPI_ISL_1921698, EPI_ISL_1921700, EPI_ISL_1921703, EPI_ISL_1921725, EPI_ISL_1921735, EPI_ISL_1939677, EPI_ISL_1939690, EPI_ISL_1939719, EPI_ISL_1985497, EPI_ISL_1985685, EPI_ISL_1985688, EPI_ISL_1985689, EPI_ISL_1985694, EPI_ISL_1985696, EPI_ISL_1985716, EPI_ISL_1985748, EPI_ISL_1985762, EPI_ISL_1985811, EPI_ISL_1985820, EPI_ISL_1985822, EPI_ISL_1985826, EPI_ISL_1985841, EPI_ISL_1985864, EPI_ISL_1985871, EPI_ISL_1985872, EPI_ISL_1985876, EPI_ISL_1985886, EPI_ISL_1985892, EPI_ISL_1985896, EPI_ISL_1985901, EPI_ISL_1985910, EPI_ISL_1985957, EPI_ISL_1985971, EPI_ISL_1985979, EPI_ISL_1985980, EPI_ISL_1985983, EPI_ISL_1985990, EPI_ISL_1986006, EPI_ISL_1986030, EPI_ISL_1986032, EPI_ISL_1986036, EPI_ISL_1986046, EPI_ISL_1986062, EPI_ISL_1986068, EPI_ISL_1986075, EPI_ISL_1986076, EPI_ISL_1986092, EPI_ISL_1986096, EPI_ISL_1986099, EPI_ISL_1986107, EPI_ISL_1986125, EPI_ISL_1986128, EPI_ISL_1986130, EPI_ISL_1986136, EPI_ISL_1986139, EPI_ISL_1986166, EPI_ISL_1986167, EPI_ISL_1986171, EPI_ISL_1986180, EPI_ISL_1986185, EPI_ISL_1986186, EPI_ISL_1986208, EPI_ISL_1986218, EPI_ISL_1986224, EPI_ISL_1986237, EPI_ISL_1986252, EPI_ISL_1986267, EPI_ISL_1986290, EPI_ISL_1986311, EPI_ISL_1986315, EPI_ISL_1986318, EPI_ISL_1986339, EPI_ISL_1986365, EPI_ISL_1986502, EPI_ISL_1986516, EPI_ISL_1986523, EPI_ISL_1986528, EPI_ISL_1986539, EPI_ISL_1986542, EPI_ISL_1986563, EPI_ISL_1986567, EPI_ISL_1986572, EPI_ISL_1986573, EPI_ISL_1986574, EPI_ISL_1986579, EPI_ISL_1986580, EPI_ISL_1986582, EPI_ISL_1986585, EPI_ISL_1986589, EPI_ISL_1986595, EPI_ISL_1986601, EPI_ISL_1986607, EPI_ISL_1986611, EPI_ISL_1986612, EPI_ISL_1986617, EPI_ISL_1986618, EPI_ISL_1986626, EPI_ISL_1986628, EPI_ISL_1986632, EPI_ISL_1986634, EPI_ISL_1986637, EPI_ISL_1986639, EPI_ISL_1986649, EPI_ISL_1986658, EPI_ISL_1986660, EPI_ISL_1986663, EPI_ISL_1986666, EPI_ISL_1986668, EPI_ISL_1986671, EPI_ISL_1986677, EPI_ISL_1986678, EPI_ISL_1986679, EPI_ISL_1986682, EPI_ISL_1986684, EPI_ISL_1986689, EPI_ISL_1986690, EPI_ISL_1986692, EPI_ISL_1986693, EPI_ISL_1986699, EPI_ISL_1986701, EPI_ISL_1986702, EPI_ISL_1986703, EPI_ISL_1986707, EPI_ISL_1986712, EPI_ISL_1986716, EPI_ISL_1986718, EPI_ISL_1986720, EPI_ISL_1986721, EPI_ISL_1986722, EPI_ISL_1986730, EPI_ISL_1986733, EPI_ISL_1986739, EPI_ISL_1986740, EPI_ISL_1986741, EPI_ISL_1986743, EPI_ISL_1986748, EPI_ISL_1986749, EPI_ISL_1986756, EPI_ISL_1986761, EPI_ISL_1986763, EPI_ISL_1986768, EPI_ISL_1986771, EPI_ISL_1986773, EPI_ISL_1986775, EPI_ISL_1986780, EPI_ISL_1986781, EPI_ISL_1986790, EPI_ISL_1986792, EPI_ISL_1986793, EPI_ISL_1986803, EPI_ISL_1986810, EPI_ISL_1986812, EPI_ISL_1986813, EPI_ISL_1986820, EPI_ISL_1986822, EPI_ISL_1986829, EPI_ISL_1986840, EPI_ISL_1986845, EPI_ISL_1986853, EPI_ISL_1986854, EPI_ISL_1986858, EPI_ISL_1986860, EPI_ISL_1986861, EPI_ISL_1986864, EPI_ISL_1986865, EPI_ISL_1986866, EPI_ISL_1986871, EPI_ISL_1986873, EPI_ISL_1986876, EPI_ISL_1986878, EPI_ISL_1986879, EPI_ISL_1986886, EPI_ISL_1986887, EPI_ISL_1986891, EPI_ISL_1986893, EPI_ISL_1986895, EPI_ISL_1986896, EPI_ISL_1986900, EPI_ISL_1986904, EPI_ISL_1986921, EPI_ISL_1986930, EPI_ISL_1986933, EPI_ISL_1986944, EPI_ISL_1986957, EPI_ISL_1986958, EPI_ISL_1986961, EPI_ISL_1986962, EPI_ISL_1986963, EPI_ISL_1986964, EPI_ISL_1986970, EPI_ISL_1986972, EPI_ISL_1986974, EPI_ISL_1986976, EPI_ISL_1986980, EPI_ISL_1986986, EPI_ISL_1986992, EPI_ISL_1986993, EPI_ISL_1986995, EPI_ISL_1986996, EPI_ISL_1987000, EPI_ISL_1987001, EPI_ISL_1987006, EPI_ISL_1987007, EPI_ISL_1987009, EPI_ISL_1987011, EPI_ISL_1987012, EPI_ISL_1987016, EPI_ISL_1987018, EPI_ISL_1987020, EPI_ISL_1987024, EPI_ISL_1987025, EPI_ISL_1987028, EPI_ISL_1987030, EPI_ISL_1987032, EPI_ISL_1987034, EPI_ISL_1987040, EPI_ISL_1987041, EPI_ISL_1987046, EPI_ISL_1987048, EPI_ISL_1987049, EPI_ISL_1987050, EPI_ISL_1987054, EPI_ISL_1987055, EPI_ISL_1987056, EPI_ISL_1987061, EPI_ISL_1987062, EPI_ISL_1987063, EPI_ISL_1987068, EPI_ISL_1987070, EPI_ISL_1987073, EPI_ISL_1987074, EPI_ISL_1987123, EPI_ISL_1987126, EPI_ISL_1987128, EPI_ISL_1987129, EPI_ISL_1987130, EPI_ISL_1987136, EPI_ISL_1987140, EPI_ISL_1987146, EPI_ISL_1987149, EPI_ISL_1987151, EPI_ISL_1987152, EPI_ISL_1987153, EPI_ISL_1987154, EPI_ISL_1987156, EPI_ISL_1987157, EPI_ISL_1987161, EPI_ISL_1987162, EPI_ISL_1987165, EPI_ISL_1987167, EPI_ISL_1987168, EPI_ISL_1987172, EPI_ISL_1987173, EPI_ISL_1987175, EPI_ISL_1987179, EPI_ISL_1987180, EPI_ISL_1987188, EPI_ISL_1987189, EPI_ISL_1987192, EPI_ISL_1987196, EPI_ISL_1987197, EPI_ISL_1987198, EPI_ISL_1987203, EPI_ISL_1987204, EPI_ISL_1987206, EPI_ISL_1987207, EPI_ISL_1987211, EPI_ISL_1987212, EPI_ISL_1987213, EPI_ISL_1987214, EPI_ISL_1987218, EPI_ISL_1987221, EPI_ISL_1987223, EPI_ISL_1987224, EPI_ISL_1987225, EPI_ISL_1987227, EPI_ISL_1987230, EPI_ISL_1987233, EPI_ISL_1987234, EPI_ISL_1987235, EPI_ISL_1987236, EPI_ISL_1987239, EPI_ISL_1987241, EPI_ISL_1987250, EPI_ISL_1987253, EPI_ISL_1987255, EPI_ISL_1987258, EPI_ISL_1987264, EPI_ISL_1987265, EPI_ISL_1987267, EPI_ISL_1987269, EPI_ISL_1987275, EPI_ISL_1987276, EPI_ISL_1987278, EPI_ISL_1987287, EPI_ISL_1987289, EPI_ISL_1987294, EPI_ISL_1987295, EPI_ISL_1987297, EPI_ISL_1987298, EPI_ISL_1987299, EPI_ISL_1987303, EPI_ISL_1987311, EPI_ISL_1987321, EPI_ISL_1987325, EPI_ISL_1987326, EPI_ISL_1987333, EPI_ISL_1987334, EPI_ISL_1987337, EPI_ISL_1987341, EPI_ISL_1987348, EPI_ISL_1987354, EPI_ISL_1987364, EPI_ISL_1987366, EPI_ISL_1987369, EPI_ISL_1987375, EPI_ISL_1987376, EPI_ISL_1987377, EPI_ISL_2004359, EPI_ISL_2004369, EPI_ISL_2004385, EPI_ISL_2004392, EPI_ISL_2004414, EPI_ISL_2004449, EPI_ISL_2004452, EPI_ISL_2004457, EPI_ISL_2004468, EPI_ISL_2004490, EPI_ISL_2004499, EPI_ISL_2004518, EPI_ISL_2004528, EPI_ISL_2004537, EPI_ISL_2004539, EPI_ISL_2004560, EPI_ISL_2004563, EPI_ISL_2004579, EPI_ISL_2004594, EPI_ISL_2004884, EPI_ISL_2004887, EPI_ISL_2004888, EPI_ISL_2004889, EPI_ISL_2004891, EPI_ISL_2004892, EPI_ISL_2004904, EPI_ISL_2004905, EPI_ISL_2004906, EPI_ISL_2004912, EPI_ISL_2004915, EPI_ISL_2004918, EPI_ISL_2004923, EPI_ISL_2004924, EPI_ISL_2004932, EPI_ISL_2004934, EPI_ISL_2004938, EPI_ISL_2004939, EPI_ISL_2004941, EPI_ISL_2004945, EPI_ISL_2004952, EPI_ISL_2004953, EPI_ISL_2004955, EPI_ISL_2004958, EPI_ISL_2004962, EPI_ISL_2004963, EPI_ISL_2004967, EPI_ISL_2004969, EPI_ISL_2004970, EPI_ISL_2004974, EPI_ISL_2004980, EPI_ISL_2004987, EPI_ISL_2004995, EPI_ISL_2004999, EPI_ISL_2005004, EPI_ISL_2005007, EPI_ISL_2005008, EPI_ISL_2005009, EPI_ISL_2005010, EPI_ISL_2005012, EPI_ISL_2005013, EPI_ISL_2005014, EPI_ISL_2005017, EPI_ISL_2005020, EPI_ISL_2005022, EPI_ISL_2005023, EPI_ISL_2005025, EPI_ISL_2005031, EPI_ISL_2005034, EPI_ISL_2005040, EPI_ISL_2005043, EPI_ISL_2005048, EPI_ISL_2005049, EPI_ISL_2005050, EPI_ISL_2005060, EPI_ISL_2005063, EPI_ISL_2005065, EPI_ISL_2005066, EPI_ISL_2005068, EPI_ISL_2005072, EPI_ISL_2005073, EPI_ISL_2005075, EPI_ISL_2005079, EPI_ISL_2005081, EPI_ISL_2005087, EPI_ISL_2005088, EPI_ISL_2005093, EPI_ISL_2005096, EPI_ISL_2005105, EPI_ISL_2005107, EPI_ISL_2005110, EPI_ISL_2005111, EPI_ISL_2005112, EPI_ISL_2005114, EPI_ISL_2005116, EPI_ISL_2005117, EPI_ISL_2005119, EPI_ISL_2005120, EPI_ISL_2005122, EPI_ISL_2005123, EPI_ISL_2005126, EPI_ISL_2005129, EPI_ISL_2005136, EPI_ISL_2005139, EPI_ISL_2005140, EPI_ISL_2005141, EPI_ISL_2005145, EPI_ISL_2005146, EPI_ISL_2005151, EPI_ISL_2005152, EPI_ISL_2005160, EPI_ISL_2005163, EPI_ISL_2005166, EPI_ISL_2005167, EPI_ISL_2005172, EPI_ISL_2005173, EPI_ISL_2005174, EPI_ISL_2005176, EPI_ISL_2005178, EPI_ISL_2005180, EPI_ISL_2005181, EPI_ISL_2005184, EPI_ISL_2005185, EPI_ISL_2005188, EPI_ISL_2005189, EPI_ISL_2005197, EPI_ISL_2005199, EPI_ISL_2005200, EPI_ISL_2005201, EPI_ISL_2005202, EPI_ISL_2005204, EPI_ISL_2005207, EPI_ISL_2005209, EPI_ISL_2005212, EPI_ISL_2005213, | | | |

EPI_ISL_2005214, EPI_ISL_2005218, EPI_ISL_2005220, EPI_ISL_2005227, EPI_ISL_2005228, EPI_ISL_2005229, EPI_ISL_2005230, EPI_ISL_2005232, EPI_ISL_2005237, EPI_ISL_2005238, EPI_ISL_2005239, EPI_ISL_2005241, EPI_ISL_2005242, EPI_ISL_2005243, EPI_ISL_2005250, EPI_ISL_2005251, EPI_ISL_2005252, EPI_ISL_2005253, EPI_ISL_2005254, EPI_ISL_2005258, EPI_ISL_2005259, EPI_ISL_2005260, EPI_ISL_2005261, EPI_ISL_2005268, EPI_ISL_2005271, EPI_ISL_2005272, EPI_ISL_2005273, EPI_ISL_2005274, EPI_ISL_2005278, EPI_ISL_2005280, EPI_ISL_2005285, EPI_ISL_2005288, EPI_ISL_2005289, EPI_ISL_2005291, EPI_ISL_2005292, EPI_ISL_2005295, EPI_ISL_2005299, EPI_ISL_2005301, EPI_ISL_2005302, EPI_ISL_2005309, EPI_ISL_2005310, EPI_ISL_2005312, EPI_ISL_2005318, EPI_ISL_2005321, EPI_ISL_2005324, EPI_ISL_2005325, EPI_ISL_2005327, EPI_ISL_2005329, EPI_ISL_2005330, EPI_ISL_2005331, EPI_ISL_2005333, EPI_ISL_2005335, EPI_ISL_2005337, EPI_ISL_2005347, EPI_ISL_2005349, EPI_ISL_2005353, EPI_ISL_2005357, EPI_ISL_2005358, EPI_ISL_2005360, EPI_ISL_2005362, EPI_ISL_2005363, EPI_ISL_2005364, EPI_ISL_2005369, EPI_ISL_2005378, EPI_ISL_2005379, EPI_ISL_2005382, EPI_ISL_2005384, EPI_ISL_2005385, EPI_ISL_2005391, EPI_ISL_2005392, EPI_ISL_2005393, EPI_ISL_2005395, EPI_ISL_2005396, EPI_ISL_2005404, EPI_ISL_2005413, EPI_ISL_2005418, EPI_ISL_2005422, EPI_ISL_2005425, EPI_ISL_2005426, EPI_ISL_2005430, EPI_ISL_2005435, EPI_ISL_2005437, EPI_ISL_2005438, EPI_ISL_2005442, EPI_ISL_2005443, EPI_ISL_2005448, EPI_ISL_2005450, EPI_ISL_2005452, EPI_ISL_2005462, EPI_ISL_2005464, EPI_ISL_2005468, EPI_ISL_2005469, EPI_ISL_2005476, EPI_ISL_2005478, EPI_ISL_2005486, EPI_ISL_2005493, EPI_ISL_2005496, EPI_ISL_2005499, EPI_ISL_2005504, EPI_ISL_2005513, EPI_ISL_2005515, EPI_ISL_2005516, EPI_ISL_2005519, EPI_ISL_2005520, EPI_ISL_2005521, EPI_ISL_2005522, EPI_ISL_2005531, EPI_ISL_2005532, EPI_ISL_2005533, EPI_ISL_2005538, EPI_ISL_2005539, EPI_ISL_2005540, EPI_ISL_2005548, EPI_ISL_2005550, EPI_ISL_2005551, EPI_ISL_2005552, EPI_ISL_2005556, EPI_ISL_2005557, EPI_ISL_2005558, EPI_ISL_2005560, EPI_ISL_2005561, EPI_ISL_2005564, EPI_ISL_2005567, EPI_ISL_2005571, EPI_ISL_2005573, EPI_ISL_2005577, EPI_ISL_2005582, EPI_ISL_2005584, EPI_ISL_2005585, EPI_ISL_2005588, EPI_ISL_2005589, EPI_ISL_2005590, EPI_ISL_2005593, EPI_ISL_2005598, EPI_ISL_2005599, EPI_ISL_2005603, EPI_ISL_2005608, EPI_ISL_2005610, EPI_ISL_2005611, EPI_ISL_2005931, EPI_ISL_2006669, EPI_ISL_2006670, EPI_ISL_2006671, EPI_ISL_2006672, EPI_ISL_2021463, EPI_ISL_2021697, EPI_ISL_2021701, EPI_ISL_2021703, EPI_ISL_2021704, EPI_ISL_2021707, EPI_ISL_2021711, EPI_ISL_2021712, EPI_ISL_2021713, EPI_ISL_2021717, EPI_ISL_2021719, EPI_ISL_2021728, EPI_ISL_2021733, EPI_ISL_2021735, EPI_ISL_2021736, EPI_ISL_2021739, EPI_ISL_2021742, EPI_ISL_2021743, EPI_ISL_2021746, EPI_ISL_2021747, EPI_ISL_2021748, EPI_ISL_2021749, EPI_ISL_2021752, EPI_ISL_2021754, EPI_ISL_2021757, EPI_ISL_2021764, EPI_ISL_2021767, EPI_ISL_2021769, EPI_ISL_2021771, EPI_ISL_2021773, EPI_ISL_2021774, EPI_ISL_2021775, EPI_ISL_2021778, EPI_ISL_2021780, EPI_ISL_2021782, EPI_ISL_2021784, EPI_ISL_2021791, EPI_ISL_2021793, EPI_ISL_2021794, EPI_ISL_2021797, EPI_ISL_2021805, EPI_ISL_2021807, EPI_ISL_2021809, EPI_ISL_2021813, EPI_ISL_2021814, EPI_ISL_2021820, EPI_ISL_2021822, EPI_ISL_2021826, EPI_ISL_2021828, EPI_ISL_2021829, EPI_ISL_2021835, EPI_ISL_2021843, EPI_ISL_2021847, EPI_ISL_2021923, EPI_ISL_2021961, EPI_ISL_2021994, EPI_ISL_2022018, EPI_ISL_2022041, EPI_ISL_2022055, EPI_ISL_2022066, EPI_ISL_2022072, EPI_ISL_2022080, EPI_ISL_2022120, EPI_ISL_2022495, EPI_ISL_2022805, EPI_ISL_2022979, EPI_ISL_2080421, EPI_ISL_2080483, EPI_ISL_2080484, EPI_ISL_2091463, EPI_ISL_2091468, EPI_ISL_2091473, EPI_ISL_2091480, EPI_ISL_2091482, EPI_ISL_2091484, EPI_ISL_2091486, EPI_ISL_2091493, EPI_ISL_2091496, EPI_ISL_2091500, EPI_ISL_2091511, EPI_ISL_2091515, EPI_ISL_2091525, EPI_ISL_2091536, EPI_ISL_2091539, EPI_ISL_2091543, EPI_ISL_2091547, EPI_ISL_2091555, EPI_ISL_2091562, EPI_ISL_2091564, EPI_ISL_2091565, EPI_ISL_2091566, EPI_ISL_2091569, EPI_ISL_2091581, EPI_ISL_2091585, EPI_ISL_2091587, EPI_ISL_2091592, EPI_ISL_2091596, EPI_ISL_2091602, EPI_ISL_2091604, EPI_ISL_2091613, EPI_ISL_2091624, EPI_ISL_2091632, EPI_ISL_2091638, EPI_ISL_2091640, EPI_ISL_2091649, EPI_ISL_2091650, EPI_ISL_2091652, EPI_ISL_2091656, EPI_ISL_2091664, EPI_ISL_2091669, EPI_ISL_2091670, EPI_ISL_2091672, EPI_ISL_2091679, EPI_ISL_2091685, EPI_ISL_2091687, EPI_ISL_2091693, EPI_ISL_2091694, EPI_ISL_2091695, EPI_ISL_2091698, EPI_ISL_2091702, EPI_ISL_2091704, EPI_ISL_2091710, EPI_ISL_2091711, EPI_ISL_2091724, EPI_ISL_2091729, EPI_ISL_2091730, EPI_ISL_2091733, EPI_ISL_2091735, EPI_ISL_2091741, EPI_ISL_2091742, EPI_ISL_2091744, EPI_ISL_2091749, EPI_ISL_2091751, EPI_ISL_2091756, EPI_ISL_2091759, EPI_ISL_2091760, EPI_ISL_2091763, EPI_ISL_2091765, EPI_ISL_2091766, EPI_ISL_2091769, EPI_ISL_2091772, EPI_ISL_2091774, EPI_ISL_2091779, EPI_ISL_2091781, EPI_ISL_2091783, EPI_ISL_2091784, EPI_ISL_2091786, EPI_ISL_2091788, EPI_ISL_2091789, EPI_ISL_2091791, EPI_ISL_2092467, EPI_ISL_2092468, EPI_ISL_2092469, EPI_ISL_2092470, EPI_ISL_2092471, EPI_ISL_2092472, EPI_ISL_2092473, EPI_ISL_2092474, EPI_ISL_2092475, EPI_ISL_2092476, EPI_ISL_2092477, EPI_ISL_2092478, EPI_ISL_2092479, EPI_ISL_2092480, EPI_ISL_2092482, EPI_ISL_2092483, EPI_ISL_2092484, EPI_ISL_2092485, EPI_ISL_2092486, EPI_ISL_2092487, EPI_ISL_2092488, EPI_ISL_2092489, EPI_ISL_2092490, EPI_ISL_2092491, EPI_ISL_2092492, EPI_ISL_2092493, EPI_ISL_2092494, EPI_ISL_2092496, EPI_ISL_2092498, EPI_ISL_2092499, EPI_ISL_2092500, EPI_ISL_2092501, EPI_ISL_2092502, EPI_ISL_2092503, EPI_ISL_2092504, EPI_ISL_2092505, EPI_ISL_2092506, EPI_ISL_2092507, EPI_ISL_2092509, EPI_ISL_2092511, EPI_ISL_2092512, EPI_ISL_2092513, EPI_ISL_2092514, EPI_ISL_2092515, EPI_ISL_2092516, EPI_ISL_2092518, EPI_ISL_2092519, EPI_ISL_2092520, EPI_ISL_2092521, EPI_ISL_2092522, EPI_ISL_2092523, EPI_ISL_2092524, EPI_ISL_2092526, EPI_ISL_2092527, EPI_ISL_2092528, EPI_ISL_2092531, EPI_ISL_2092533, EPI_ISL_2092534, EPI_ISL_2092535, EPI_ISL_2092537, EPI_ISL_2092539, EPI_ISL_2092540, EPI_ISL_2092541, EPI_ISL_2092543, EPI_ISL_2092544, EPI_ISL_2092545, EPI_ISL_2092546, EPI_ISL_2092547, EPI_ISL_2092548, EPI_ISL_2092550, EPI_ISL_2092551, EPI_ISL_2092552, EPI_ISL_2092553, EPI_ISL_2092554, EPI_ISL_2092555, EPI_ISL_2092556, EPI_ISL_2092557, EPI_ISL_2092558, EPI_ISL_2092559, EPI_ISL_2092560, EPI_ISL_2092561, EPI_ISL_2092562, EPI_ISL_2092563, EPI_ISL_2092564, EPI_ISL_2092565, EPI_ISL_2092566, EPI_ISL_2092568, EPI_ISL_2092569, EPI_ISL_2092571, EPI_ISL_2092575, EPI_ISL_2092577, EPI_ISL_2092578, EPI_ISL_2092580, EPI_ISL_2092581, EPI_ISL_2092582, EPI_ISL_2092584, EPI_ISL_2092585, EPI_ISL_2092586, EPI_ISL_2092587, EPI_ISL_2092588, EPI_ISL_2092589, EPI_ISL_2092590, EPI_ISL_2092591, EPI_ISL_2092592, EPI_ISL_2092593, EPI_ISL_2092595, EPI_ISL_2092596, EPI_ISL_2092597, EPI_ISL_2092598, EPI_ISL_2092599, EPI_ISL_2092600, EPI_ISL_2092601, EPI_ISL_2092602, EPI_ISL_2092604, EPI_ISL_2092605, EPI_ISL_2092606, EPI_ISL_2092607, EPI_ISL_2092609, EPI_ISL_2092611, EPI_ISL_2092612, EPI_ISL_2092613, EPI_ISL_2092614, EPI_ISL_2092615, EPI_ISL_2092618, EPI_ISL_2092619, EPI_ISL_2092620, EPI_ISL_2092621, EPI_ISL_2092622, EPI_ISL_2092623, EPI_ISL_2092624, EPI_ISL_2092625, EPI_ISL_2092626, EPI_ISL_2092628, EPI_ISL_2092629, EPI_ISL_2092630, EPI_ISL_2092631, EPI_ISL_2092635, EPI_ISL_2092636, EPI_ISL_2092637, EPI_ISL_2092638, EPI_ISL_2092639, EPI_ISL_2092640, EPI_ISL_2092641, EPI_ISL_2092642, EPI_ISL_2092643, EPI_ISL_2092644, EPI_ISL_2092645, EPI_ISL_2092646, EPI_ISL_2138718, EPI_ISL_2172093, EPI_ISL_2172094, EPI_ISL_2172095, EPI_ISL_2172096, EPI_ISL_2172097, EPI_ISL_2172098, EPI_ISL_2172100, EPI_ISL_2172106, EPI_ISL_2172108, EPI_ISL_2172109, EPI_ISL_2172113, EPI_ISL_2172114, EPI_ISL_2172116, EPI_ISL_2172119, EPI_ISL_2172121, EPI_ISL_2172123, EPI_ISL_2172125, EPI_ISL_2172126, EPI_ISL_2172128, EPI_ISL_2172129, EPI_ISL_2172130, EPI_ISL_2172131, EPI_ISL_2172134, EPI_ISL_2172135, EPI_ISL_2172136, EPI_ISL_2172137, EPI_ISL_2172138, EPI_ISL_2172139, EPI_ISL_2172140, EPI_ISL_2172141, EPI_ISL_2172151, EPI_ISL_2172152, EPI_ISL_2172153, EPI_ISL_2172154, EPI_ISL_2172158, EPI_ISL_2172159, EPI_ISL_2172161, EPI_ISL_2172164, EPI_ISL_2172168, EPI_ISL_2172169, EPI_ISL_2172171, EPI_ISL_2172173, EPI_ISL_2172176, EPI_ISL_2172177, EPI_ISL_2172179, EPI_ISL_2172180, EPI_ISL_2172182, EPI_ISL_2172183, EPI_ISL_2172184, EPI_ISL_2172186, EPI_ISL_2172187, EPI_ISL_2172188, EPI_ISL_2172189, EPI_ISL_2172190, EPI_ISL_2172193, EPI_ISL_2172198, EPI_ISL_2172199, EPI_ISL_2172200, EPI_ISL_2172202, EPI_ISL_2172207, EPI_ISL_2172208, EPI_ISL_2172213, EPI_ISL_2172214, EPI_ISL_2172215, EPI_ISL_2172219, EPI_ISL_2172221, EPI_ISL_2172223, EPI_ISL_2172224, EPI_ISL_2172229, EPI_ISL_2172231, EPI_ISL_2172233, EPI_ISL_2172235, EPI_ISL_2172236, EPI_ISL_2172238, EPI_ISL_2172239, EPI_ISL_2172240, EPI_ISL_2172241, EPI_ISL_2172243, EPI_ISL_2172248, EPI_ISL_2172251, EPI_ISL_2172255, EPI_ISL_2172256, EPI_ISL_2172258, EPI_ISL_2172260, EPI_ISL_2172261, EPI_ISL_2172264, EPI_ISL_2172265, EPI_ISL_2172267, EPI_ISL_2172268, EPI_ISL_2172269, EPI_ISL_2172270, EPI_ISL_2172271, EPI_ISL_2172274, EPI_ISL_2172275, EPI_ISL_2172276, EPI_ISL_2172277, EPI_ISL_2172278, EPI_ISL_2172279, EPI_ISL_2172280, EPI_ISL_2172281, EPI_ISL_2172282, EPI_ISL_2172285, EPI_ISL_2172287, EPI_ISL_2172289, EPI_ISL_2172290, EPI_ISL_2172294, EPI_ISL_2172295, EPI_ISL_2172296, EPI_ISL_2172298, EPI_ISL_2172299, EPI_ISL_2172300, EPI_ISL_2172302, EPI_ISL_2172303, EPI_ISL_2172304, EPI_ISL_2172311, EPI_ISL_2172312, EPI_ISL_2172314, EPI_ISL_2172317, EPI_ISL_2172320, EPI_ISL_2172321, EPI_ISL_2172324, EPI_ISL_2172326, EPI_ISL_2172327, EPI_ISL_2172328, EPI_ISL_2172329, EPI_ISL_2172333, EPI_ISL_2172337, EPI_ISL_2172340, EPI_ISL_2172341, EPI_ISL_2172345, EPI_ISL_2172349, EPI_ISL_2172350, EPI_ISL_2172351, EPI_ISL_2172353, EPI_ISL_2172356, EPI_ISL_2172359, EPI_ISL_2172361, EPI_ISL_2172362, EPI_ISL_2172363, EPI_ISL_2172364, EPI_ISL_2172366, EPI_ISL_2172367, EPI_ISL_2172370, EPI_ISL_2172371, EPI_ISL_2172374, EPI_ISL_2172376, EPI_ISL_2172380, EPI_ISL_2172381, EPI_ISL_2172383, EPI_ISL_2172386, EPI_ISL_2172387, EPI_ISL_2172388, EPI_ISL_2172392, EPI_ISL_2172393, EPI_ISL_2172394, EPI_ISL_2172396, EPI_ISL_2172401, EPI_ISL_2172403, EPI_ISL_2172404

| | | | |
|---|---|---|---|
| see above | Lighthouse Lab in Alderley Park | Wellcome Sanger Institute for the COVID-19 Genomics UK (COG-UK) Consortium | Cordelia Langford; David K. Jackson; Dominic Kwiatkowski; Ewan Harrison; Ian Johnston; Jacquelyn Wynn; Jeffrey Barrett; John Sillitoe on behalf of the Wellcome Sanger Institute COVID-19 Surveillance Team; Mairead Hyland; Roberto Amato; Sonia Goncalves; The Lighthouse Lab in Alderley Park and Alex Alderton |
| EPI_ISL_1806592, EPI_ISL_1806792, EPI_ISL_1831086 | Lighthouse Lab in Cambridge | Wellcome Sanger Institute for the COVID-19 Genomics UK (COG-UK) Consortium | Cordelia Langford; David K. Jackson; Dominic Kwiatkowski; Ewan Harrison; Ian Johnston; Jeffrey Barrett; John Sillitoe on behalf of the Wellcome Sanger Institute COVID-19 Surveillance Team; Rob Howes; Roberto Amato; Sonia Goncalves; The Lighthouse Lab in Cambridge and Alex Alderton |

EPI_ISL_1829487, EPI_ISL_1829491, EPI_ISL_1829492, EPI_ISL_1829496, EPI_ISL_1829498, EPI_ISL_1829499, EPI_ISL_1829501, EPI_ISL_1829504, EPI_ISL_1829507, EPI_ISL_1829508, EPI_ISL_1829509, EPI_ISL_1829514, EPI_ISL_1829516, EPI_ISL_1829517, EPI_ISL_1829518, EPI_ISL_1829523, EPI_ISL_1829526, EPI_ISL_1829531, EPI_ISL_1829533, EPI_ISL_1829535, EPI_ISL_1829538, EPI_ISL_1829542, EPI_ISL_1829544, EPI_ISL_1829547, EPI_ISL_1829549, EPI_ISL_1829550, EPI_ISL_1829551, EPI_ISL_1829554, EPI_ISL_1829555, EPI_ISL_1829556, EPI_ISL_1829558, EPI_ISL_1829559, EPI_ISL_1829560, EPI_ISL_1829563, EPI_ISL_1829565, EPI_ISL_1829567, EPI_ISL_1829569, EPI_ISL_1829571, EPI_ISL_1829576, EPI_ISL_1829580, EPI_ISL_1829581, EPI_ISL_1829584, EPI_ISL_1829585, EPI_ISL_1829592, EPI_ISL_1829594, EPI_ISL_1829595, EPI_ISL_1829597, EPI_ISL_1829599, EPI_ISL_1829600, EPI_ISL_1829603, EPI_ISL_1829608, EPI_ISL_1829612, EPI_ISL_1829613, EPI_ISL_1829617, EPI_ISL_1829619, EPI_ISL_1829620, EPI_ISL_1829621, EPI_ISL_1829623, EPI_ISL_1829625, EPI_ISL_1829627, EPI_ISL_1829628, EPI_ISL_1829629, EPI_ISL_1829630, EPI_ISL_1829633, EPI_ISL_1829634, EPI_ISL_1829638, EPI_ISL_1829641, EPI_ISL_1829645, EPI_ISL_1829646, EPI_ISL_1829647, EPI_ISL_1829650, EPI_ISL_1829652, EPI_ISL_1829653, EPI_ISL_1829659, EPI_ISL_1829660, EPI_ISL_1829672, EPI_ISL_1829675, EPI_ISL_1829676, EPI_ISL_1829677, EPI_ISL_1829679, EPI_ISL_1829681, EPI_ISL_1829682, EPI_ISL_1829685, EPI_ISL_1829686, EPI_ISL_1829687, EPI_ISL_1829691, EPI_ISL_1829693, EPI_ISL_1829695, EPI_ISL_1829701, EPI_ISL_1829706, EPI_ISL_1829712, EPI_ISL_1829714, EPI_ISL_1829715, EPI_ISL_1829716, EPI_ISL_1829717, EPI_ISL_1829718, EPI_ISL_1829719, EPI_ISL_1829720, EPI_ISL_1829721, EPI_ISL_1829722, EPI_ISL_1829724, EPI_ISL_1829726, EPI_ISL_1829727, EPI_ISL_1829730, EPI_ISL_1829731, EPI_ISL_1829732, EPI_ISL_1829734, EPI_ISL_1829735, EPI_ISL_1829738, EPI_ISL_1829740, EPI_ISL_1829741, EPI_ISL_1829743, EPI_ISL_1829746, EPI_ISL_1829751, EPI_ISL_1829754, EPI_ISL_1829755, EPI_ISL_1829757, EPI_ISL_1829759, EPI_ISL_1829760, EPI_ISL_1829761, EPI_ISL_1829762, EPI_ISL_1829764, EPI_ISL_1829765, EPI_ISL_1829766, EPI_ISL_1829768, EPI_ISL_1829772, EPI_ISL_1829778, EPI_ISL_1829779, EPI_ISL_1829783, EPI_ISL_1829784, EPI_ISL_1829786, EPI_ISL_1829789, EPI_ISL_1829790, EPI_ISL_1829792, EPI_ISL_1829793, EPI_ISL_1829794, EPI_ISL_1829798, EPI_ISL_1829799, EPI_ISL_1829812, EPI_ISL_1829824, EPI_ISL_1829834, EPI_ISL_1829836, EPI_ISL_1829844, EPI_ISL_1829849, EPI_ISL_1829856, EPI_ISL_1829857, EPI_ISL_1829870, EPI_ISL_1829873, EPI_ISL_1829888, EPI_ISL_1829890, EPI_ISL_1829895, EPI_ISL_1829901, EPI_ISL_1829903, EPI_ISL_1829906, EPI_ISL_1829907, EPI_ISL_1829919, EPI_ISL_1829923, EPI_ISL_1829927, EPI_ISL_1829928, EPI_ISL_1829932, EPI_ISL_1829941, EPI_ISL_1829948, EPI_ISL_1829949, EPI_ISL_1829957, EPI_ISL_1829959, EPI_ISL_1829989, EPI_ISL_1829997, EPI_ISL_1830000, EPI_ISL_1830005, EPI_ISL_1830009, EPI_ISL_1830015, EPI_ISL_1830016, EPI_ISL_1830027, EPI_ISL_1830028, EPI_ISL_1830043, EPI_ISL_1830050, EPI_ISL_1830056, EPI_ISL_1830063, EPI_ISL_1830070, EPI_ISL_1830079, EPI_ISL_1830081, EPI_ISL_1830088, EPI_ISL_1830096, EPI_ISL_1830097, EPI_ISL_1830099, EPI_ISL_1831193, EPI_ISL_1857588, EPI_ISL_1857589, EPI_ISL_1857590, EPI_ISL_1857591, EPI_ISL_1857592, EPI_ISL_1857594, EPI_ISL_1857596, EPI_ISL_1857598, EPI_ISL_1857599, EPI_ISL_1857600, EPI_ISL_1857601, EPI_ISL_1857602, EPI_ISL_1857604, EPI_ISL_1857605, EPI_ISL_1857606, EPI_ISL_1857607, EPI_ISL_1857608, EPI_ISL_1857609, EPI_ISL_1857610, EPI_ISL_1857611, EPI_ISL_1857612, EPI_ISL_1857613, EPI_ISL_1857614, EPI_ISL_1857617, EPI_ISL_1857618, EPI_ISL_1857619, EPI_ISL_1857620, EPI_ISL_1857621, EPI_ISL_1857622, EPI_ISL_1857623, EPI_ISL_1857624, EPI_ISL_1857625, EPI_ISL_1857626, EPI_ISL_1857627, EPI_ISL_1857628, EPI_ISL_1857629, EPI_ISL_1857632, EPI_ISL_1857633, EPI_ISL_1857637, EPI_ISL_1857638, EPI_ISL_1857642, EPI_ISL_1857644, EPI_ISL_1857645, EPI_ISL_1857646, EPI_ISL_1857647, EPI_ISL_1857648, EPI_ISL_1857651, EPI_ISL_1857652, EPI_ISL_1857653, EPI_ISL_1857654, EPI_ISL_1857657, EPI_ISL_1857658, EPI_ISL_1857659, EPI_ISL_1857660, EPI_ISL_1857662, EPI_ISL_1857663, EPI_ISL_1857666, EPI_ISL_1857667, EPI_ISL_1857668, EPI_ISL_1857669, EPI_ISL_1857670, EPI_ISL_1857671, EPI_ISL_1857672, EPI_ISL_1857675, EPI_ISL_1857676, EPI_ISL_1857677, EPI_ISL_1857678, EPI_ISL_1857679, EPI_ISL_1857681, EPI_ISL_1857682, EPI_ISL_1857683, EPI_ISL_1857684, EPI_ISL_1857685, EPI_ISL_1857686, EPI_ISL_1857687, EPI_ISL_1857688, EPI_ISL_1857689, EPI_ISL_1857690, EPI_ISL_1857691, EPI_ISL_1857692, EPI_ISL_1857693, EPI_ISL_1857694, EPI_ISL_1857696, EPI_ISL_1857697, EPI_ISL_1857698, EPI_ISL_1857701, EPI_ISL_1857702, EPI_ISL_1857704, EPI_ISL_1857705, EPI_ISL_1857706, EPI_ISL_1857707, EPI_ISL_1857708, EPI_ISL_1857709, EPI_ISL_1857710, EPI_ISL_1857711, EPI_ISL_1857712, EPI_ISL_1857713, EPI_ISL_1857714, EPI_ISL_1857715, EPI_ISL_1857716, EPI_ISL_1857718, EPI_ISL_1857719, EPI_ISL_1857720, EPI_ISL_1857723, EPI_ISL_1857724, EPI_ISL_1857725, EPI_ISL_1857726, EPI_ISL_1857727, EPI_ISL_1857730, EPI_ISL_1857731, EPI_ISL_1857732, EPI_ISL_1857733, EPI_ISL_1857734, EPI_ISL_1857735, EPI_ISL_1857736, EPI_ISL_1857737, EPI_ISL_1857738, EPI_ISL_1857739, EPI_ISL_1857740, EPI_ISL_1857741, EPI_ISL_1857742, EPI_ISL_1857743, EPI_ISL_1857744, EPI_ISL_1857745, EPI_ISL_1857746, EPI_ISL_1857747, EPI_ISL_1857748, EPI_ISL_1857749, EPI_ISL_1857750, EPI_ISL_1857751, EPI_ISL_1857752, EPI_ISL_1857753, EPI_ISL_1857754, EPI_ISL_1857755, EPI_ISL_1857759, EPI_ISL_1857761, EPI_ISL_1857762, EPI_ISL_1857764, EPI_ISL_1857765, EPI_ISL_1857768, EPI_ISL_1857769, EPI_ISL_1857771, EPI_ISL_1857772, EPI_ISL_1857773, EPI_ISL_1857775, EPI_ISL_1857776, EPI_ISL_1857777, EPI_ISL_1857778, EPI_ISL_1857780, EPI_ISL_1857781, EPI_ISL_1857782, EPI_ISL_1857783, EPI_ISL_1857784, EPI_ISL_1857785, EPI_ISL_1857787, EPI_ISL_1857788, EPI_ISL_1857789, EPI_ISL_1857791, EPI_ISL_1857792, EPI_ISL_1857793, EPI_ISL_1857795, EPI_ISL_1857796, EPI_ISL_1857797, EPI_ISL_1857798, EPI_ISL_1857799, EPI_ISL_1857800, EPI_ISL_1857801, EPI_ISL_1857802, EPI_ISL_1857803, EPI_ISL_1857804, EPI_ISL_1857805, EPI_ISL_1857810, EPI_ISL_1857813, EPI_ISL_1857814, EPI_ISL_1857818, EPI_ISL_1857820, EPI_ISL_1857823, EPI_ISL_1857826, EPI_ISL_1857844, EPI_ISL_1857845, EPI_ISL_1857853, EPI_ISL_1857865, EPI_ISL_1857867, EPI_ISL_1857881, EPI_ISL_1857882, EPI_ISL_1857888, EPI_ISL_1857889, EPI_ISL_1857894, EPI_ISL_1857898, EPI_ISL_1857900, EPI_ISL_1857901, EPI_ISL_1857902, EPI_ISL_1857907, EPI_ISL_1857909, EPI_ISL_1857910, EPI_ISL_1857911, EPI_ISL_1857916, EPI_ISL_1857925, EPI_ISL_1857929, EPI_ISL_1857930, EPI_ISL_1857936, EPI_ISL_1857939, EPI_ISL_1857947, EPI_ISL_1857949, EPI_ISL_1857950, EPI_ISL_1857951, EPI_ISL_1857962, EPI_ISL_1857975, EPI_ISL_1857983, EPI_ISL_1857985, EPI_ISL_1857991, EPI_ISL_1857996, EPI_ISL_1857997, EPI_ISL_1858003, EPI_ISL_1858028, EPI_ISL_1858029, EPI_ISL_1858038, EPI_ISL_1858040, EPI_ISL_1858044, EPI_ISL_1858046, EPI_ISL_1858052, EPI_ISL_1858055, EPI_ISL_1858063, EPI_ISL_1858068, EPI_ISL_1858078, EPI_ISL_1858083, EPI_ISL_1858096, EPI_ISL_1858102, EPI_ISL_1858105, EPI_ISL_1858116, EPI_ISL_1858123, EPI_ISL_1858126, EPI_ISL_1858146, EPI_ISL_1858149, EPI_ISL_1858151, EPI_ISL_1858199, EPI_ISL_1858200, EPI_ISL_1858203, EPI_ISL_1858205, EPI_ISL_1858210, EPI_ISL_1858218, EPI_ISL_1858220, EPI_ISL_1858223, EPI_ISL_1858225, EPI_ISL_1858226, EPI_ISL_1858229, EPI_ISL_1858231, EPI_ISL_1858233, EPI_ISL_1858234, EPI_ISL_1912350, EPI_ISL_1912356, EPI_ISL_1912357, EPI_ISL_1912363, EPI_ISL_1912366, EPI_ISL_1912367, EPI_ISL_1912368, EPI_ISL_1912370, EPI_ISL_1912372, EPI_ISL_1912378, EPI_ISL_1912380, EPI_ISL_1912386, EPI_ISL_1912388, EPI_ISL_1912391, EPI_ISL_1912392, EPI_ISL_1912395, EPI_ISL_1912400, EPI_ISL_1912404, EPI_ISL_1912406, EPI_ISL_1912411, EPI_ISL_1912412, EPI_ISL_1912417, EPI_ISL_1912419, EPI_ISL_1912420, EPI_ISL_1912421, EPI_ISL_1912423, EPI_ISL_1912424, EPI_ISL_1912433, EPI_ISL_1912440, EPI_ISL_1912441, EPI_ISL_1912445, EPI_ISL_1912450, EPI_ISL_1912451, EPI_ISL_1912457, EPI_ISL_1912462, EPI_ISL_1912470, EPI_ISL_1912472, EPI_ISL_1912473, EPI_ISL_1912476, EPI_ISL_1912477, EPI_ISL_1912478, EPI_ISL_1912485, EPI_ISL_1912489, EPI_ISL_1912497, EPI_ISL_1912500, EPI_ISL_1912501, EPI_ISL_1912502, EPI_ISL_1912511, EPI_ISL_1912512, EPI_ISL_1912513, EPI_ISL_1912518, EPI_ISL_1912519, EPI_ISL_1912520, EPI_ISL_1912525, EPI_ISL_1912527, EPI_ISL_1912546, EPI_ISL_1912547, EPI_ISL_1912549, EPI_ISL_1912550, EPI_ISL_1912558, EPI_ISL_1912565, EPI_ISL_1912569, EPI_ISL_1912570, EPI_ISL_1912576, EPI_ISL_1912581, EPI_ISL_1912588, EPI_ISL_1912589, EPI_ISL_1912591, EPI_ISL_1912594, EPI_ISL_1912595, EPI_ISL_1912596, EPI_ISL_1912597, EPI_ISL_1912598, EPI_ISL_1912599, EPI_ISL_1912600, EPI_ISL_1912601, EPI_ISL_1912602, EPI_ISL_1912603, EPI_ISL_1912604, EPI_ISL_1912605, EPI_ISL_1912606, EPI_ISL_1912607, EPI_ISL_1912608, EPI_ISL_1912609, EPI_ISL_1912610, EPI_ISL_1912611, EPI_ISL_1912612, EPI_ISL_1912613, EPI_ISL_1912614, EPI_ISL_1912615, EPI_ISL_1912616, EPI_ISL_1912617, EPI_ISL_1912618, EPI_ISL_1912619, EPI_ISL_1912620, EPI_ISL_1912621, EPI_ISL_1912622, EPI_ISL_1912623, EPI_ISL_1912624, EPI_ISL_1912625, EPI_ISL_1912626, EPI_ISL_1912627, EPI_ISL_1912628, EPI_ISL_1912629, EPI_ISL_1912630, EPI_ISL_1912631, EPI_ISL_1912632, EPI_ISL_1912633, EPI_ISL_1912634, EPI_ISL_1912635, EPI_ISL_1912636, EPI_ISL_1912637, EPI_ISL_1912638, EPI_ISL_1912640, EPI_ISL_1912641, EPI_ISL_1912642, EPI_ISL_1912643, EPI_ISL_1912644, EPI_ISL_1912645, EPI_ISL_1912646, EPI_ISL_1912647, EPI_ISL_1912648, EPI_ISL_1912649, EPI_ISL_1912650, EPI_ISL_1912651, EPI_ISL_1912652, EPI_ISL_1912653, EPI_ISL_1912654, EPI_ISL_1912655, EPI_ISL_1912656, EPI_ISL_1912657, EPI_ISL_1912658, EPI_ISL_1912659, EPI_ISL_1912660, EPI_ISL_1912661, EPI_ISL_1912662, EPI_ISL_1912663, EPI_ISL_1912664, EPI_ISL_1912665, EPI_ISL_1912666, EPI_ISL_1912667, EPI_ISL_1912668, EPI_ISL_1912669, EPI_ISL_1912670, EPI_ISL_1912671, EPI_ISL_1912672, EPI_ISL_1912673, EPI_ISL_1912674, EPI_ISL_1912675, EPI_ISL_1912676, EPI_ISL_1912677, EPI_ISL_1912678, EPI_ISL_1912679, EPI_ISL_1912680, EPI_ISL_1912681, EPI_ISL_1912682, EPI_ISL_1912683, EPI_ISL_1912684, EPI_ISL_1912685, EPI_ISL_1912686, EPI_ISL_1912687, EPI_ISL_1912688, EPI_ISL_1912689, EPI_ISL_1912691, EPI_ISL_1912692, EPI_ISL_1912693, EPI_ISL_1912694, EPI_ISL_1912695, EPI_ISL_1912696, EPI_ISL_1912697, EPI_ISL_1912698, EPI_ISL_1912699, EPI_ISL_1912700, EPI_ISL_1912701, EPI_ISL_1912702, EPI_ISL_1912703, EPI_ISL_1912704, EPI_ISL_1912705, EPI_ISL_1912706, EPI_ISL_1912707, EPI_ISL_1912708, EPI_ISL_1912709, EPI_ISL_1912710, EPI_ISL_1912711, EPI_ISL_1912712, EPI_ISL_1912713, EPI_ISL_1912714, EPI_ISL_1912715, EPI_ISL_1912716, EPI_ISL_1912717, EPI_ISL_1912718, EPI_ISL_1912719, EPI_ISL_1912720, EPI_ISL_1912721, EPI_ISL_1912722, EPI_ISL_1912723, EPI_ISL_1912724, EPI_ISL_1912725, EPI_ISL_1912726, EPI_ISL_1912727, EPI_ISL_1912728, EPI_ISL_1912730, EPI_ISL_1912731, EPI_ISL_1912732, EPI_ISL_1912733, EPI_ISL_1912734, EPI_ISL_1912735, EPI_ISL_1912736, EPI_ISL_1912738, EPI_ISL_1912739, EPI_ISL_1912741, EPI_ISL_1912742, EPI_ISL_1912743, EPI_ISL_1912744, EPI_ISL_1912745, EPI_ISL_1912746, EPI_ISL_1912747, EPI_ISL_1912748, EPI_ISL_1912749, EPI_ISL_1912750, EPI_ISL_1921260, EPI_ISL_1921425, EPI_ISL_1921737, EPI_ISL_1939618, EPI_ISL_1939619, EPI_ISL_1939620, EPI_ISL_1939621, EPI_ISL_1939622, EPI_ISL_1939623, EPI_ISL_1939624, EPI_ISL_1939625, EPI_ISL_1939626, EPI_ISL_1939627, EPI_ISL_1939628, EPI_ISL_1939629, EPI_ISL_1939630, EPI_ISL_1939631, EPI_ISL_1939632, EPI_ISL_1939634, EPI_ISL_1939635, EPI_ISL_1939636, EPI_ISL_1939637, EPI_ISL_1939638, EPI_ISL_1939639, EPI_ISL_1939640, EPI_ISL_1939641, EPI_ISL_1939642, EPI_ISL_1939643, EPI_ISL_1939644, EPI_ISL_1939645, EPI_ISL_1939646, EPI_ISL_1939647, EPI_ISL_1939648, EPI_ISL_1939649, EPI_ISL_1939650, EPI_ISL_1939651, EPI_ISL_1939652, EPI_ISL_1939653, EPI_ISL_1939654, EPI_ISL_1939655, EPI_ISL_1939656, EPI_ISL_1939657, EPI_ISL_1939658, EPI_ISL_1939659, EPI_ISL_1939660, EPI_ISL_1939661, EPI_ISL_1939662, EPI_ISL_1939663, EPI_ISL_1939664, EPI_ISL_1939665, EPI_ISL_1939666, EPI_ISL_1939667, EPI_ISL_1939668, EPI_ISL_1939669, EPI_ISL_1939670, EPI_ISL_1939671, EPI_ISL_1939672, EPI_ISL_1939673, EPI_ISL_1939674, EPI_ISL_1939675, EPI_ISL_1939676, EPI_ISL_1939678, EPI_ISL_1939679, EPI_ISL_1939680, EPI_ISL_1939682, EPI_ISL_1939683, EPI_ISL_1939684, EPI_ISL_1939685, EPI_ISL_1939686, EPI_ISL_1939687, EPI_ISL_1939688, EPI_ISL_1939689, EPI_ISL_1939691, EPI_ISL_1939692, EPI_ISL_1939693, EPI_ISL_1939694, EPI_ISL_1939695, EPI_ISL_1939696, EPI_ISL_1939697, EPI_ISL_1939698, EPI_ISL_1939699, EPI_ISL_1939700, EPI_ISL_1939701, EPI_ISL_1939702, EPI_ISL_1939703, EPI_ISL_1939704, EPI_ISL_1939705, EPI_ISL_1939706, EPI_ISL_1939707, EPI_ISL_1939721, EPI_ISL_1939722, EPI_ISL_1939723, EPI_ISL_1939724, EPI_ISL_1939725, EPI_ISL_1939726, EPI_ISL_1939727, EPI_ISL_1939728, EPI_ISL_1939729, EPI_ISL_1939730, EPI_ISL_1939731, EPI_ISL_1939732, EPI_ISL_1939733, EPI_ISL_1939734, EPI_ISL_1939735, EPI_ISL_1939736, EPI_ISL_1939737, EPI_ISL_1939738, EPI_ISL_1939739, EPI_ISL_1939740, EPI_ISL_1939741, EPI_ISL_1939742, EPI_ISL_1939743, EPI_ISL_1939744, EPI_ISL_1939745, EPI_ISL_1939746, EPI_ISL_1939747, EPI_ISL_1939748, EPI_ISL_1939749, EPI_ISL_1939750, EPI_ISL_1939751, EPI_ISL_1939752, EPI_ISL_1939753, EPI_ISL_1939754, EPI_ISL_1939755, EPI_ISL_1939756, EPI_ISL_1939757, EPI_ISL_1939758, EPI_ISL_1939759, EPI_ISL_1939760, EPI_ISL_1939761, EPI_ISL_1939762, EPI_ISL_1939763, EPI_ISL_1939764, EPI_ISL_1939765, EPI_ISL_1939766, EPI_ISL_1939767, EPI_ISL_1939768, EPI_ISL_1939769, EPI_ISL_1939770, EPI_ISL_1939771, EPI_ISL_1939772, EPI_ISL_1939773, EPI_ISL_1939774, EPI_ISL_1939775, EPI_ISL_1939776, EPI_ISL_1939777, EPI_ISL_1939778, EPI_ISL_1939779, EPI_ISL_1939780, EPI_ISL_1939781, EPI_ISL_1939782, EPI_ISL_1939783, EPI_ISL_1939784, EPI_ISL_1939785, EPI_ISL_1939786, EPI_ISL_1939787, EPI_ISL_1939788, EPI_ISL_1939789, EPI_ISL_1939790, EPI_ISL_1939791, EPI_ISL_1939792, EPI_ISL_1939793, EPI_ISL_1939794, EPI_ISL_1939795, EPI_ISL_1939796, EPI_ISL_1939797, EPI_ISL_1939798, EPI_ISL_1939799, EPI_ISL_1939800, EPI_ISL_1939801, EPI_ISL_1939802, EPI_ISL_1939803, EPI_ISL_1939804, EPI_ISL_1939805, EPI_ISL_1939806, EPI_ISL_1939807, EPI_ISL_1939808, EPI_ISL_1939809, EPI_ISL_1939810, EPI_ISL_1939811, EPI_ISL_1939812, EPI_ISL_1939813, EPI_ISL_1939814, EPI_ISL_1939815, EPI_ISL_1939816, EPI_ISL_1939817, EPI_ISL_1939818, EPI_ISL_1939819, EPI_ISL_1939820, EPI_ISL_1939821, EPI_ISL_1939822, EPI_ISL_1939823, EPI_ISL_1939824, EPI_ISL_1939825, EPI_ISL_1939826, EPI_ISL_1986151, EPI_ISL_1986152, EPI_ISL_1986160, EPI_ISL_1986192, EPI_ISL_1986209, EPI_ISL_1986256, EPI_ISL_1986266, EPI_ISL_1986272, EPI_ISL_1986374, EPI_ISL_1986386, EPI_ISL_1986389, EPI_ISL_1986508, EPI_ISL_1986512, EPI_ISL_1986526, EPI_ISL_1986536, EPI_ISL_1986859, EPI_ISL_1986880, EPI_ISL_1986889, EPI_ISL_1986899, EPI_ISL_1986924, EPI_ISL_1986935, EPI_ISL_1986939, EPI_ISL_1986954, EPI_ISL_1986967, EPI_ISL_1986971, EPI_ISL_1986977, EPI_ISL_1986981, EPI_ISL_1986987, EPI_ISL_1986988, EPI_ISL_1986989, EPI_ISL_1987002, EPI_ISL_1987004, EPI_ISL_1987005, EPI_ISL_1987013, EPI_ISL_1987023, EPI_ISL_1987031, EPI_ISL_1987033, EPI_ISL_1987036, EPI_ISL_1987042, EPI_ISL_1987051, EPI_ISL_1987058, EPI_ISL_1987059, EPI_ISL_1987072, EPI_ISL_1987132, EPI_ISL_1987135, EPI_ISL_1987138, EPI_ISL_1987139, EPI_ISL_1987142, EPI_ISL_1987145, EPI_ISL_1987166, EPI_ISL_1987169, EPI_ISL_1987191, EPI_ISL_1987194, EPI_ISL_1987205, EPI_ISL_1987209, EPI_ISL_1987219, EPI_ISL_1987231, EPI_ISL_1987232, EPI_ISL_1987244, EPI_ISL_1987246, EPI_ISL_1987251, EPI_ISL_1987262, EPI_ISL_1987268, EPI_ISL_1987271, EPI_ISL_1987273, EPI_ISL_1987274, EPI_ISL_1987279, EPI_ISL_1987280, EPI_ISL_1987281, EPI_ISL_1987285, EPI_ISL_1987292, EPI_ISL_1987300, EPI_ISL_1987302, EPI_ISL_1987307, EPI_ISL_1987309, EPI_ISL_1987312, EPI_ISL_1987314, EPI_ISL_1987315, EPI_ISL_1987323, EPI_ISL_1987330, EPI_ISL_1987339, EPI_ISL_1987340, EPI_ISL_1987345, EPI_ISL_1987355, EPI_ISL_1987356, EPI_ISL_1987362, EPI_ISL_1987363, EPI_ISL_1987373, EPI_ISL_2004641, EPI_ISL_2004880, EPI_ISL_2004881, EPI_ISL_2004882, EPI_ISL_2004883, EPI_ISL_2004885, EPI_ISL_2004886, EPI_ISL_2004890, EPI_ISL_2004893, EPI_ISL_2004894, EPI_ISL_2004895, EPI_ISL_2004896, EPI_ISL_2004897, EPI_ISL_2004898, EPI_ISL_2004899, EPI_ISL_2004900, EPI_ISL_2004901, EPI_ISL_2004902, EPI_ISL_2004903, EPI_ISL_2004907, EPI_ISL_2004908, EPI_ISL_2004909, EPI_ISL_2004910, EPI_ISL_2004911, EPI_ISL_2004913, EPI_ISL_2004914, EPI_ISL_2004916, EPI_ISL_2004917, EPI_ISL_2004919, EPI_ISL_2004920, EPI_ISL_2004921, EPI_ISL_2004922, EPI_ISL_2004925, EPI_ISL_2004926, EPI_ISL_2004927, EPI_ISL_2004928, EPI_ISL_2004929, EPI_ISL_2004930, EPI_ISL_2004931, EPI_ISL_2004933, EPI_ISL_2004935, EPI_ISL_2004936, EPI_ISL_2004937, EPI_ISL_2004940, EPI_ISL_2004942, EPI_ISL_2004943, EPI_ISL_2004944, EPI_ISL_2004946, EPI_ISL_2004947, EPI_ISL_2004948, EPI_ISL_2004949, EPI_ISL_2004950, EPI_ISL_2004951, EPI_ISL_2004954, EPI_ISL_2004956, EPI_ISL_2004957, EPI_ISL_2004959, EPI_ISL_2004960, EPI_ISL_2004961, EPI_ISL_2004964, EPI_ISL_2004965, EPI_ISL_2004966, EPI_ISL_2004968, EPI_ISL_2004971, EPI_ISL_2004972, EPI_ISL_2004973, EPI_ISL_2004975, EPI_ISL_2004976, EPI_ISL_2004977, EPI_ISL_2004978, EPI_ISL_2004979, EPI_ISL_2004981, EPI_ISL_2004982, EPI_ISL_2004983, EPI_ISL_2004984, EPI_ISL_2004985, EPI_ISL_2004986, EPI_ISL_2004988, EPI_ISL_2004989, EPI_ISL_2004990, EPI_ISL_2004991, EPI_ISL_2004993, EPI_ISL_2004994, EPI_ISL_2004996, EPI_ISL_2004997, EPI_ISL_2004998, EPI_ISL_2005000, EPI_ISL_2005001, EPI_ISL_2005002, EPI_ISL_2005003, EPI_ISL_2005005, EPI_ISL_2005006, EPI_ISL_2005011, EPI_ISL_2005015, EPI_ISL_2005016, EPI_ISL_2005018, EPI_ISL_2005019, EPI_ISL_2005021, EPI_ISL_2005024, EPI_ISL_2005026, EPI_ISL_2005027, EPI_ISL_2005028, EPI_ISL_2005029, EPI_ISL_2005030, EPI_ISL_2005032, EPI_ISL_2005033, EPI_ISL_2005035, EPI_ISL_2005036, EPI_ISL_2005037, EPI_ISL_2005038, EPI_ISL_2005039, EPI_ISL_2005041, EPI_ISL_2005042, EPI_ISL_2005044, EPI_ISL_2005045, EPI_ISL_2005046, EPI_ISL_2005047, EPI_ISL_2005051, EPI_ISL_2005052, EPI_ISL_2005053, EPI_ISL_2005054, EPI_ISL_2005055, EPI_ISL_2005056, EPI_ISL_2005057, EPI_ISL_2005058, EPI_ISL_2005059, EPI_ISL_2005061, EPI_ISL_2005062, EPI_ISL_2005064, EPI_ISL_2005067, EPI_ISL_2005069, EPI_ISL_2005070, EPI_ISL_2005071, EPI_ISL_2005074, EPI_ISL_2005076, EPI_ISL_2005077, EPI_ISL_2005078, EPI_ISL_2005080, EPI_ISL_2005082, EPI_ISL_2005083, EPI_ISL_2005084, EPI_ISL_2005085, EPI_ISL_2005086, EPI_ISL_2005089, EPI_ISL_2005090, EPI_ISL_2005091, EPI_ISL_2005092, EPI_ISL_2005094, EPI_ISL_2005095, EPI_ISL_2005097, EPI_ISL_2005098, EPI_ISL_2005099, EPI_ISL_2005100, EPI_ISL_2005101, EPI_ISL_2005102, EPI_ISL_2005103, EPI_ISL_2005104, EPI_ISL_2005106, EPI_ISL_2005108, EPI_ISL_2005109, EPI_ISL_2005113, EPI_ISL_2005115, EPI_ISL_2005118, EPI_ISL_2005121, EPI_ISL_2005124, EPI_ISL_2005125, EPI_ISL_2005127, EPI_ISL_2005128, EPI_ISL_2005130, EPI_ISL_2005131, EPI_ISL_2005132, EPI_ISL_2005133, EPI_ISL_2005134, EPI_ISL_2005135, EPI_ISL_2005137, EPI_ISL_2005138, EPI_ISL_2005142, EPI_ISL_2005143, EPI_ISL_2005144, EPI_ISL_2005147, EPI_ISL_2005148, EPI_ISL_2005149, EPI_ISL_2005150, EPI_ISL_2005153, EPI_ISL_2005154, EPI_ISL_2005155, EPI_ISL_2005156, EPI_ISL_2005157, EPI_ISL_2005158, EPI_ISL_2005159, EPI_ISL_2005161, EPI_ISL_2005164, EPI_ISL_2005165, EPI_ISL_2005168, EPI_ISL_2005169, EPI_ISL_2005170, EPI_ISL_2005171, EPI_ISL_2005175, EPI_ISL_2005177, EPI_ISL_2005179, EPI_ISL_2005182, EPI_ISL_2005183, EPI_ISL_2005186, EPI_ISL_2005190, EPI_ISL_2005191, EPI_ISL_2005193, EPI_ISL_2005194, EPI_ISL_2005195, EPI_ISL_2005196, EPI_ISL_2005198, EPI_ISL_2005203, EPI_ISL_2005205, EPI_ISL_2005206, EPI_ISL_2005208, EPI_ISL_2005210, EPI_ISL_2005211, EPI_ISL_2005215, EPI_ISL_2005217, EPI_ISL_2005219, EPI_ISL_2005221, EPI_ISL_2005222, EPI_ISL_2005223, EPI_ISL_2005224, EPI_ISL_2005225, EPI_ISL_2005226, EPI_ISL_2005231, EPI_ISL_2005233, EPI_ISL_2005234, EPI_ISL_2005235, EPI_ISL_2005236, EPI_ISL_2005240, EPI_ISL_2005244, EPI_ISL_2005245, EPI_ISL_2005246, EPI_ISL_2005247, EPI_ISL_2005248, EPI_ISL_2005249, EPI_ISL_2005255, EPI_ISL_2005256, EPI_ISL_2005257, EPI_ISL_2005262, EPI_ISL_2005263, EPI_ISL_2005264, EPI_ISL_2005265, EPI_ISL_2005266, EPI_ISL_2005267, EPI_ISL_2005270, EPI_ISL_2005275, EPI_ISL_2005276, EPI_ISL_2005277, EPI_ISL_2005279, EPI_ISL_2005281, EPI_ISL_2005282, EPI_ISL_2005283, EPI_ISL_2005284, EPI_ISL_2005286, EPI_ISL_2005287, EPI_ISL_2005290, EPI_ISL_2005293, EPI_ISL_2005294, EPI_ISL_2005296, EPI_ISL_2005297, EPI_ISL_2005298, EPI_ISL_2005300, EPI_ISL_2005303, EPI_ISL_2005304, EPI_ISL_2005305, EPI_ISL_2005306, EPI_ISL_2005307, EPI_ISL_2005308, EPI_ISL_2005311, EPI_ISL_2005313, EPI_ISL_2005314, EPI_ISL_2005315, EPI_ISL_2005316, EPI_ISL_2005317, EPI_ISL_2005319, EPI_ISL_2005320, EPI_ISL_2005322, EPI_ISL_2005323, EPI_ISL_2005326, EPI_ISL_2005328, EPI_ISL_2005332, EPI_ISL_2005334, EPI_ISL_2005336, EPI_ISL_2005338, EPI_ISL_2005339, EPI_ISL_2005340, EPI_ISL_2005814, EPI_ISL_2005818, EPI_ISL_2005820, EPI_ISL_2005834, EPI_ISL_2005841, EPI_ISL_2005867, EPI_ISL_2005884, EPI_ISL_2005893, EPI_ISL_2005895, EPI_ISL_2005900, EPI_ISL_2005914, EPI_ISL_2005918, EPI_ISL_2005923, EPI_ISL_2005927, EPI_ISL_2005930, EPI_ISL_2005932, EPI_ISL_2005940, EPI_ISL_2005945, EPI_ISL_2005957, EPI_ISL_2005965, EPI_ISL_2005967, EPI_ISL_2005968, EPI_ISL_2005976, EPI_ISL_2005979, EPI_ISL_2005982, EPI_ISL_2005998, EPI_ISL_2006000, EPI_ISL_2006006, EPI_ISL_2006013, EPI_ISL_2006043, EPI_ISL_2006047, EPI_ISL_2006050, EPI_ISL_2006054, EPI_ISL_2006058, EPI_ISL_2006059, EPI_ISL_2006068, EPI_ISL_2006071, EPI_ISL_2006073, EPI_ISL_2006076, EPI_ISL_2006077, EPI_ISL_2006079, EPI_ISL_2006081, EPI_ISL_2006082, EPI_ISL_2006083, EPI_ISL_2006084, EPI_ISL_2006086, EPI_ISL_2006088, EPI_ISL_2006089, EPI_ISL_2006091, EPI_ISL_2006093, EPI_ISL_2006094, EPI_ISL_2006095, EPI_ISL_2006097, EPI_ISL_2006098, EPI_ISL_2006099, EPI_ISL_2006100, EPI_ISL_2006101, EPI_ISL_2006103, EPI_ISL_2006104, EPI_ISL_2006106, EPI_ISL_2006107, EPI_ISL_2006108, EPI_ISL_2006109, EPI_ISL_2006110, EPI_ISL_2006111, EPI_ISL_2006116, EPI_ISL_2006117, EPI_ISL_2006119, EPI_ISL_2006122, EPI_ISL_2006125, EPI_ISL_2006127, EPI_ISL_2006129, EPI_ISL_2006130, EPI_ISL_2006131, EPI_ISL_2006136, EPI_ISL_2006137, EPI_ISL_2006140, EPI_ISL_2006141, EPI_ISL_2006142, EPI_ISL_2006143, EPI_ISL_2006145, EPI_ISL_2006146, EPI_ISL_2006147, EPI_ISL_2006148, EPI_ISL_2006152, EPI_ISL_2006154, EPI_ISL_2006155, EPI_ISL_2006156, EPI_ISL_2006160, EPI_ISL_2006162, EPI_ISL_2006164, EPI_ISL_2006165, EPI_ISL_2006168, EPI_ISL_2006172, EPI_ISL_2006173, EPI_ISL_2006174, EPI_ISL_2006175, EPI_ISL_2006177, EPI_ISL_2006178, EPI_ISL_2006179, EPI_ISL_2006181, EPI_ISL_2006182, EPI_ISL_2006186, EPI_ISL_2006189, EPI_ISL_2006190, EPI_ISL_2006193, EPI_ISL_2006194, EPI_ISL_2006198, EPI_ISL_2006199, EPI_ISL_2006201, EPI_ISL_2006207, EPI_ISL_2006211, EPI_ISL_2006212, EPI_ISL_2006213, EPI_ISL_2006217, EPI_ISL_2006218, EPI_ISL_2006219, EPI_ISL_2006222, EPI_ISL_2006223, EPI_ISL_2006224, EPI_ISL_2006225, EPI_ISL_2006227, EPI_ISL_2006231, EPI_ISL_2006232, EPI_ISL_2006233, EPI_ISL_2006235, EPI_ISL_2006238, EPI_ISL_2006239, EPI_ISL_2006240, EPI_ISL_2006244, EPI_ISL_2006245, EPI_ISL_2006246, EPI_ISL_2006249, EPI_ISL_2006250, EPI_ISL_2006256, EPI_ISL_2006258, EPI_ISL_2006262, EPI_ISL_2006264, EPI_ISL_2006265, EPI_ISL_2006267, EPI_ISL_2006273, EPI_ISL_2006275, EPI_ISL_2006276, EPI_ISL_2006277, EPI_ISL_2006279, EPI_ISL_2006280, EPI_ISL_2006281, EPI_ISL_2006284, EPI_ISL_2006285, EPI_ISL_2006287, EPI_ISL_2006290, EPI_ISL_2006292, EPI_ISL_2006293, EPI_ISL_2006295, EPI_ISL_2006296, EPI_ISL_2006297, EPI_ISL_2006298, EPI_ISL_2006299, EPI_ISL_2006300, EPI_ISL_2006301, EPI_ISL_2006303, EPI_ISL_2006304, EPI_ISL_2006305, EPI_ISL_2006306, EPI_ISL_2006307, EPI_ISL_2006308, EPI_ISL_2006309, EPI_ISL_2006310, EPI_ISL_2006311, EPI_ISL_2006313, EPI_ISL_2006314, EPI_ISL_2006315, EPI_ISL_2006316, EPI_ISL_2006317, EPI_ISL_2006318, EPI_ISL_2006319, EPI_ISL_2006320, EPI_ISL_2006321, EPI_ISL_2006322, EPI_ISL_2006323, EPI_ISL_2006324, EPI_ISL_2006325, EPI_ISL_2006326, EPI_ISL_2006327, EPI_ISL_2006329, EPI_ISL_2006330, EPI_ISL_2006331, EPI_ISL_2006332, EPI_ISL_2006335, EPI_ISL_2006336, EPI_ISL_2006337, EPI_ISL_2006338, EPI_ISL_2006339, EPI_ISL_2006340, EPI_ISL_2006341, EPI_ISL_2006342, EPI_ISL_2006343, EPI_ISL_2006346, EPI_ISL_2006347, EPI_ISL_2006348, EPI_ISL_2006349, EPI_ISL_2006350, EPI_ISL_2006351, EPI_ISL_2006352, EPI_ISL_2006353, EPI_ISL_2006354, EPI_ISL_2006355, EPI_ISL_2006356, EPI_ISL_2006357, EPI_ISL_2006359, EPI_ISL_2006360, EPI_ISL_2006361, EPI_ISL_2006362, EPI_ISL_2006363, EPI_ISL_2006364, EPI_ISL_2006365, EPI_ISL_2006366, EPI_ISL_2006367, EPI_ISL_2006368, EPI_ISL_2006370, EPI_ISL_2006371, EPI_ISL_2006372, EPI_ISL_2006373, EPI_ISL_2006374, EPI_ISL_2006375, EPI_ISL_2006376, EPI_ISL_2006377, EPI_ISL_2006378, EPI_ISL_2006379, EPI_ISL_2006380, EPI_ISL_2006381, EPI_ISL_2006382, EPI_ISL_2006383, EPI_ISL_2006385, EPI_ISL_2006386, EPI_ISL_2006387, EPI_ISL_2006388, EPI_ISL_2006389, EPI_ISL_2006390, EPI_ISL_2006391, EPI_ISL_2006393, EPI_ISL_2006394, EPI_ISL_2006395, EPI_ISL_2006396, EPI_ISL_2006397, EPI_ISL_2006398, EPI_ISL_2006400, EPI_ISL_2006401, EPI_ISL_2006402, EPI_ISL_2006403, EPI_ISL_2006404, EPI_ISL_2006406, EPI_ISL_2006407, EPI_ISL_2006408, EPI_ISL_2006409, EPI_ISL_2006410, EPI_ISL_2006411, EPI_ISL_2006412, EPI_ISL_2006413, EPI_ISL_2006414, EPI_ISL_2006415, EPI_ISL_2006416, EPI_ISL_2006417, EPI_ISL_2006418, EPI_ISL_2006419, EPI_ISL_2006420, EPI_ISL_2006421, EPI_ISL_2006422, EPI_ISL_2006423, EPI_ISL_2006424, EPI_ISL_2006425, EPI_ISL_2006426, EPI_ISL_2006427, EPI_ISL_2006428, EPI_ISL_2006429, EPI_ISL_2006430, EPI_ISL_2006432, EPI_ISL_2006433, EPI_ISL_2006434, EPI_ISL_2006435, EPI_ISL_2006436, EPI_ISL_2006437, EPI_ISL_2006439, EPI_ISL_2006440, EPI_ISL_2006441, EPI_ISL_2006443, EPI_ISL_2006444, EPI_ISL_2006445, EPI_ISL_2006446, EPI_ISL_2006447, EPI_ISL_2006448, EPI_ISL_2006449, EPI_ISL_2006450, EPI_ISL_2006451, EPI_ISL_2006453, EPI_ISL_2006454, EPI_ISL_2006455, EPI_ISL_2006456, EPI_ISL_2006457, EPI_ISL_2006459, EPI_ISL_2006460, EPI_ISL_2006461, EPI_ISL_2006464, EPI_ISL_2006465, EPI_ISL_2006466, EPI_ISL_2006467, EPI_ISL_2006468, EPI_ISL_2006469, EPI_ISL_2006470, EPI_ISL_2006472, EPI_ISL_2006473, EPI_ISL_2006474, EPI_ISL_2006475, EPI_ISL_2006476, EPI_ISL_2006477, EPI_ISL_2006478, EPI_ISL_2006479, EPI_ISL_2006480, EPI_ISL_2006483, EPI_ISL_2006484, EPI_ISL_2006486, EPI_ISL_2006488, EPI_ISL_2006489, EPI_ISL_2006490, EPI_ISL_2006491, EPI_ISL_2006492, EPI_ISL_2006494, EPI_ISL_2006495, EPI_ISL_2006496, EPI_ISL_2006497, EPI_ISL_2006498, EPI_ISL_2006499, EPI_ISL_2006500, EPI_ISL_2006501, EPI_ISL_2006502, EPI_ISL_2006503, EPI_ISL_2006504, EPI_ISL_2006505, EPI_ISL_2006506, EPI_ISL_2006507, EPI_ISL_2006509, EPI_ISL_2006510, EPI_ISL_2006511, EPI_ISL_2006512, EPI_ISL_2006513, EPI_ISL_2006514, EPI_ISL_2006515, EPI_ISL_2006516, EPI_ISL_2006517, EPI_ISL_2006518, EPI_ISL_2006519, EPI_ISL_2006520, EPI_ISL_2006521, EPI_ISL_2021698, EPI_ISL_2021699, EPI_ISL_2021725, EPI_ISL_2021726, EPI_ISL_2021727, EPI_ISL_2021729, EPI_ISL_2021734, EPI_ISL_2021737, EPI_ISL_2021756, EPI_ISL_2021758, EPI_ISL_2021760, EPI_ISL_2021770, EPI_ISL_2021777, EPI_ISL_2021779, EPI_ISL_2021785, EPI_ISL_2021789, EPI_ISL_2021792, EPI_ISL_2021798, EPI_ISL_2021808, EPI_ISL_2021811, EPI_ISL_2021812, EPI_ISL_2021816, EPI_ISL_2021818, EPI_ISL_2021819, EPI_ISL_2021831, EPI_ISL_2021837, EPI_ISL_2021842, EPI_ISL_2022439, EPI_ISL_2022440, EPI_ISL_2022441, EPI_ISL_2022442, EPI_ISL_2022443, EPI_ISL_2022444, EPI_ISL_2022446, EPI_ISL_2022447, EPI_ISL_2022448, EPI_ISL_2022449, EPI_ISL_2022450, EPI_ISL_2022451, EPI_ISL_2022453, EPI_ISL_2022454, EPI_ISL_2022455, EPI_ISL_2022456, EPI_ISL_2022457, EPI_ISL_2022458, EPI_ISL_2022460, EPI_ISL_2022461, EPI_ISL_2022462, EPI_ISL_2022463, EPI_ISL_2022465, EPI_ISL_2022466, EPI_ISL_2022467, EPI_ISL_2022468, EPI_ISL_2022469, EPI_ISL_2022470,

EPI_ISL_2022471, EPI_ISL_2022472, EPI_ISL_2022473, EPI_ISL_2022474, EPI_ISL_2022475, EPI_ISL_2022476, EPI_ISL_2022477, EPI_ISL_2022478, EPI_ISL_2022479, EPI_ISL_2022480, EPI_ISL_2022481, EPI_ISL_2022482, EPI_ISL_2022483, EPI_ISL_2022484, EPI_ISL_2022485, EPI_ISL_2022486, EPI_ISL_2022487, EPI_ISL_2022488, EPI_ISL_2022489, EPI_ISL_2022490, EPI_ISL_2022491, EPI_ISL_2022492, EPI_ISL_2022493, EPI_ISL_2022540, EPI_ISL_2022544, EPI_ISL_2022551, EPI_ISL_2022553, EPI_ISL_2022555, EPI_ISL_2022582, EPI_ISL_2022586, EPI_ISL_2022591, EPI_ISL_2022592, EPI_ISL_2022593, EPI_ISL_2022594, EPI_ISL_2022596, EPI_ISL_2022597, EPI_ISL_2022598, EPI_ISL_2022599, EPI_ISL_2022600, EPI_ISL_2022601, EPI_ISL_2022602, EPI_ISL_2022603, EPI_ISL_2022604, EPI_ISL_2022605, EPI_ISL_2022606, EPI_ISL_2022607, EPI_ISL_2022608, EPI_ISL_2022609, EPI_ISL_2022610, EPI_ISL_2022611, EPI_ISL_2022612, EPI_ISL_2022613, EPI_ISL_2022614, EPI_ISL_2022615, EPI_ISL_2022616, EPI_ISL_2022617, EPI_ISL_2022618, EPI_ISL_2022619, EPI_ISL_2022620, EPI_ISL_2022621, EPI_ISL_2022622, EPI_ISL_2022623, EPI_ISL_2022624, EPI_ISL_2022625, EPI_ISL_2022626, EPI_ISL_2022627, EPI_ISL_2022628, EPI_ISL_2022629, EPI_ISL_2022630, EPI_ISL_2022631, EPI_ISL_2022632, EPI_ISL_2022633, EPI_ISL_2022634, EPI_ISL_2022635, EPI_ISL_2022636, EPI_ISL_2022637, EPI_ISL_2022638, EPI_ISL_2022639, EPI_ISL_2022640, EPI_ISL_2022641, EPI_ISL_2022642, EPI_ISL_2022643, EPI_ISL_2022644, EPI_ISL_2022646, EPI_ISL_2022647, EPI_ISL_2022648, EPI_ISL_2022649, EPI_ISL_2022650, EPI_ISL_2022651, EPI_ISL_2022652, EPI_ISL_2022653, EPI_ISL_2022654, EPI_ISL_2022655, EPI_ISL_2022656, EPI_ISL_2022658, EPI_ISL_2022661, EPI_ISL_2022662, EPI_ISL_2022663, EPI_ISL_2022664, EPI_ISL_2022665, EPI_ISL_2022666, EPI_ISL_2022669, EPI_ISL_2022670, EPI_ISL_2022671, EPI_ISL_2022672, EPI_ISL_2022673, EPI_ISL_2022674, EPI_ISL_2022675, EPI_ISL_2022676, EPI_ISL_2022677, EPI_ISL_2022678, EPI_ISL_2022679, EPI_ISL_2022680, EPI_ISL_2022681, EPI_ISL_2022682, EPI_ISL_2022683, EPI_ISL_2022684, EPI_ISL_2022685, EPI_ISL_2022686, EPI_ISL_2022687, EPI_ISL_2022688, EPI_ISL_2022689, EPI_ISL_2022690, EPI_ISL_2022691, EPI_ISL_2022692, EPI_ISL_2022693, EPI_ISL_2022694, EPI_ISL_2022695, EPI_ISL_2022696, EPI_ISL_2022697, EPI_ISL_2022698, EPI_ISL_2022699, EPI_ISL_2022700, EPI_ISL_2022701, EPI_ISL_2022702, EPI_ISL_2022703, EPI_ISL_2022704, EPI_ISL_2022705, EPI_ISL_2022706, EPI_ISL_2022707, EPI_ISL_2022708, EPI_ISL_2022709, EPI_ISL_2022710, EPI_ISL_2022711, EPI_ISL_2022712, EPI_ISL_2022713, EPI_ISL_2022714, EPI_ISL_2022715, EPI_ISL_2022716, EPI_ISL_2022717, EPI_ISL_2022719, EPI_ISL_2022720, EPI_ISL_2022721, EPI_ISL_2022722, EPI_ISL_2022723, EPI_ISL_2022724, EPI_ISL_2022725, EPI_ISL_2022726, EPI_ISL_2022727, EPI_ISL_2022728, EPI_ISL_2022729, EPI_ISL_2022730, EPI_ISL_2022731, EPI_ISL_2022732, EPI_ISL_2022733, EPI_ISL_2022734, EPI_ISL_2022735, EPI_ISL_2022736, EPI_ISL_2022737, EPI_ISL_2022738, EPI_ISL_2022739, EPI_ISL_2022740, EPI_ISL_2022741, EPI_ISL_2022742, EPI_ISL_2022743, EPI_ISL_2022744, EPI_ISL_2022745, EPI_ISL_2022746, EPI_ISL_2022747, EPI_ISL_2022748, EPI_ISL_2022749, EPI_ISL_2022750, EPI_ISL_2022751, EPI_ISL_2022752, EPI_ISL_2022753, EPI_ISL_2022754, EPI_ISL_2022755, EPI_ISL_2022756, EPI_ISL_2022757, EPI_ISL_2022758, EPI_ISL_2022759, EPI_ISL_2022760, EPI_ISL_2022761, EPI_ISL_2022762, EPI_ISL_2022763, EPI_ISL_2022764, EPI_ISL_2022766, EPI_ISL_2022767, EPI_ISL_2022768, EPI_ISL_2022769, EPI_ISL_2022770, EPI_ISL_2022771, EPI_ISL_2022772, EPI_ISL_2022773, EPI_ISL_2022774, EPI_ISL_2022775, EPI_ISL_2022776, EPI_ISL_2022777, EPI_ISL_2022778, EPI_ISL_2022779, EPI_ISL_2022780, EPI_ISL_2022817, EPI_ISL_2022842, EPI_ISL_2022948, EPI_ISL_2022949, EPI_ISL_2022950, EPI_ISL_2022951, EPI_ISL_2022952, EPI_ISL_2022953, EPI_ISL_2022954, EPI_ISL_2022955, EPI_ISL_2022956, EPI_ISL_2022957, EPI_ISL_2022958, EPI_ISL_2022959, EPI_ISL_2022960, EPI_ISL_2022961, EPI_ISL_2022962, EPI_ISL_2022963, EPI_ISL_2022964, EPI_ISL_2022965, EPI_ISL_2022966, EPI_ISL_2022967, EPI_ISL_2022968, EPI_ISL_2022969, EPI_ISL_2022970, EPI_ISL_2022971, EPI_ISL_2022972, EPI_ISL_2022973, EPI_ISL_2022974, EPI_ISL_2022975, EPI_ISL_2022976, EPI_ISL_2022977, EPI_ISL_2022978, EPI_ISL_2022980, EPI_ISL_2022981, EPI_ISL_2022982, EPI_ISL_2022983, EPI_ISL_2022984, EPI_ISL_2022985, EPI_ISL_2022986, EPI_ISL_2022987, EPI_ISL_2022988, EPI_ISL_2022989, EPI_ISL_2022990, EPI_ISL_2022991, EPI_ISL_2022992, EPI_ISL_2022993, EPI_ISL_2022994, EPI_ISL_2022995, EPI_ISL_2022996, EPI_ISL_2022997, EPI_ISL_2022998, EPI_ISL_2022999, EPI_ISL_2023000, EPI_ISL_2023001, EPI_ISL_2023002, EPI_ISL_2023003, EPI_ISL_2023004, EPI_ISL_2023005, EPI_ISL_2023006, EPI_ISL_2023007, EPI_ISL_2023008, EPI_ISL_2023009, EPI_ISL_2023010, EPI_ISL_2023011, EPI_ISL_2023012, EPI_ISL_2023013, EPI_ISL_2023014, EPI_ISL_2023015, EPI_ISL_2023016, EPI_ISL_2023017, EPI_ISL_2023018, EPI_ISL_2023019, EPI_ISL_2023020, EPI_ISL_2023021, EPI_ISL_2023022, EPI_ISL_2023023, EPI_ISL_2023024, EPI_ISL_2023025, EPI_ISL_2023026, EPI_ISL_2092481, EPI_ISL_2092495, EPI_ISL_2092497, EPI_ISL_2092508, EPI_ISL_2092510, EPI_ISL_2092517, EPI_ISL_2092525, EPI_ISL_2092529, EPI_ISL_2092530, EPI_ISL_2092532, EPI_ISL_2092536, EPI_ISL_2092538, EPI_ISL_2092542, EPI_ISL_2092549, EPI_ISL_2092567, EPI_ISL_2092570, EPI_ISL_2092572, EPI_ISL_2092573, EPI_ISL_2092576, EPI_ISL_2092579, EPI_ISL_2092583, EPI_ISL_2092594, EPI_ISL_2092603, EPI_ISL_2092608, EPI_ISL_2092610, EPI_ISL_2092616, EPI_ISL_2092617, EPI_ISL_2092627, EPI_ISL_2092632, EPI_ISL_2092633, EPI_ISL_2092634, EPI_ISL_2092647, EPI_ISL_2092648, EPI_ISL_2092649, EPI_ISL_2092650, EPI_ISL_2092651, EPI_ISL_2092652, EPI_ISL_2092653, EPI_ISL_2092654, EPI_ISL_2092655, EPI_ISL_2092656, EPI_ISL_2092657, EPI_ISL_2092658, EPI_ISL_2092659, EPI_ISL_2092660, EPI_ISL_2092661, EPI_ISL_2092662, EPI_ISL_2092663, EPI_ISL_2092664, EPI_ISL_2092665, EPI_ISL_2092666, EPI_ISL_2092667, EPI_ISL_2092668, EPI_ISL_2092669, EPI_ISL_2092670, EPI_ISL_2092671, EPI_ISL_2092672, EPI_ISL_2092673, EPI_ISL_2092674, EPI_ISL_2122707, EPI_ISL_2122708, EPI_ISL_2122710, EPI_ISL_2122712, EPI_ISL_2122713, EPI_ISL_2122714, EPI_ISL_2122716, EPI_ISL_2122718, EPI_ISL_2122719, EPI_ISL_2122721, EPI_ISL_2122722, EPI_ISL_2122724, EPI_ISL_2122725, EPI_ISL_2122727, EPI_ISL_2122728, EPI_ISL_2122730, EPI_ISL_2122731, EPI_ISL_2122733, EPI_ISL_2122735, EPI_ISL_2122736, EPI_ISL_2122738, EPI_ISL_2122739, EPI_ISL_2122741, EPI_ISL_2122742, EPI_ISL_2122744, EPI_ISL_2122745, EPI_ISL_2122747, EPI_ISL_2122748, EPI_ISL_2122750, EPI_ISL_2122751, EPI_ISL_2122752, EPI_ISL_2122754, EPI_ISL_2122756, EPI_ISL_2122758, EPI_ISL_2122760, EPI_ISL_2122761, EPI_ISL_2122763, EPI_ISL_2122764, EPI_ISL_2122766, EPI_ISL_2122767, EPI_ISL_2122769, EPI_ISL_2122771, EPI_ISL_2122772, EPI_ISL_2122773, EPI_ISL_2122774, EPI_ISL_2122775, EPI_ISL_2122776, EPI_ISL_2122779, EPI_ISL_2122781, EPI_ISL_2122782, EPI_ISL_2122784, EPI_ISL_2122789, EPI_ISL_2122792, EPI_ISL_2122794, EPI_ISL_2122796, EPI_ISL_2122797, EPI_ISL_2122798, EPI_ISL_2122800, EPI_ISL_2122801, EPI_ISL_2122803, EPI_ISL_2122804, EPI_ISL_2122806, EPI_ISL_2122807, EPI_ISL_2122809, EPI_ISL_2122810, EPI_ISL_2122811, EPI_ISL_2122813, EPI_ISL_2122814, EPI_ISL_2122817, EPI_ISL_2122819, EPI_ISL_2122820, EPI_ISL_2122822, EPI_ISL_2122823, EPI_ISL_2122825, EPI_ISL_2122826, EPI_ISL_2122827, EPI_ISL_2122829, EPI_ISL_2122830, EPI_ISL_2122833, EPI_ISL_2122835, EPI_ISL_2122836, EPI_ISL_2122837, EPI_ISL_2122839, EPI_ISL_2122840, EPI_ISL_2122841, EPI_ISL_2122842, EPI_ISL_2122844, EPI_ISL_2122846, EPI_ISL_2122847, EPI_ISL_2122849, EPI_ISL_2122850, EPI_ISL_2122851, EPI_ISL_2122853, EPI_ISL_2122854, EPI_ISL_2122856, EPI_ISL_2122857, EPI_ISL_2122859, EPI_ISL_2122860, EPI_ISL_2122862, EPI_ISL_2122864, EPI_ISL_2122865, EPI_ISL_2122866, EPI_ISL_2122868, EPI_ISL_2122869, EPI_ISL_2122872, EPI_ISL_2122874, EPI_ISL_2122875, EPI_ISL_2122876, EPI_ISL_2122878, EPI_ISL_2122879, EPI_ISL_2122881, EPI_ISL_2122882, EPI_ISL_2122884, EPI_ISL_2122885, EPI_ISL_2122887, EPI_ISL_2122888, EPI_ISL_2122891, EPI_ISL_2122893, EPI_ISL_2122894, EPI_ISL_2122896, EPI_ISL_2122897, EPI_ISL_2122899, EPI_ISL_2122900, EPI_ISL_2122902, EPI_ISL_2122903, EPI_ISL_2122905, EPI_ISL_2122906, EPI_ISL_2122908, EPI_ISL_2122909, EPI_ISL_2122910, EPI_ISL_2122912, EPI_ISL_2122913, EPI_ISL_2122915, EPI_ISL_2122916, EPI_ISL_2122917, EPI_ISL_2122919, EPI_ISL_2122920, EPI_ISL_2122922, EPI_ISL_2122923, EPI_ISL_2122925, EPI_ISL_2122926, EPI_ISL_2122928, EPI_ISL_2122930, EPI_ISL_2122931, EPI_ISL_2122933

| | | | |
|---|---|---|---|
| see above | Lighthouse Lab in Glasgow | Wellcome Sanger Institute for the COVID-19 Genomics UK (COG-UK) Consortium | Anna Dominiczak and Alex Alderton; Carol Clugston; Cordelia Langford; David Gray; David K. Jackson; Dominic Kwiatkowski; Ewan Harrison; Harper VanSteenhouse; Ian Johnston; Jeffrey Barrett; John Sillitoe on behalf of the Wellcome Sanger Institute COVID-19 Surveillance Team; Roberto Amato; Sonia Goncalves; Yumi Kasai |

EPI_ISL_1806184, EPI_ISL_1806636, EPI_ISL_1806828, EPI_ISL_1829207, EPI_ISL_1829210, EPI_ISL_1829217, EPI_ISL_1829218, EPI_ISL_1829219, EPI_ISL_1829221, EPI_ISL_1829224, EPI_ISL_1829225, EPI_ISL_1829226, EPI_ISL_1829228, EPI_ISL_1829230, EPI_ISL_1829231, EPI_ISL_1829234, EPI_ISL_1829235, EPI_ISL_1829236, EPI_ISL_1829238, EPI_ISL_1829240, EPI_ISL_1829241, EPI_ISL_1829244, EPI_ISL_1829246, EPI_ISL_1829247, EPI_ISL_1829248, EPI_ISL_1829249, EPI_ISL_1829250, EPI_ISL_1829251, EPI_ISL_1829253, EPI_ISL_1829256, EPI_ISL_1829257, EPI_ISL_1829259, EPI_ISL_1829261, EPI_ISL_1829263, EPI_ISL_1829265, EPI_ISL_1829266, EPI_ISL_1829267, EPI_ISL_1829268, EPI_ISL_1829270, EPI_ISL_1829275, EPI_ISL_1829277, EPI_ISL_1829280, EPI_ISL_1829283, EPI_ISL_1829284, EPI_ISL_1829286, EPI_ISL_1829288, EPI_ISL_1829290, EPI_ISL_1829293, EPI_ISL_1829295, EPI_ISL_1829298, EPI_ISL_1829301, EPI_ISL_1829303, EPI_ISL_1829305, EPI_ISL_1829306, EPI_ISL_1829308, EPI_ISL_1829309, EPI_ISL_1829312, EPI_ISL_1829313, EPI_ISL_1829314, EPI_ISL_1829317, EPI_ISL_1829321, EPI_ISL_1829322, EPI_ISL_1829323, EPI_ISL_1829324, EPI_ISL_1829326, EPI_ISL_1829328, EPI_ISL_1829329, EPI_ISL_1829330, EPI_ISL_1829331, EPI_ISL_1829332, EPI_ISL_1829334, EPI_ISL_1829335, EPI_ISL_1829336, EPI_ISL_1829338, EPI_ISL_1829339, EPI_ISL_1829340, EPI_ISL_1829341, EPI_ISL_1829343, EPI_ISL_1829346, EPI_ISL_1829347, EPI_ISL_1829348, EPI_ISL_1829349, EPI_ISL_1829350, EPI_ISL_1829351, EPI_ISL_1829354, EPI_ISL_1829355, EPI_ISL_1829358, EPI_ISL_1829361, EPI_ISL_1829364, EPI_ISL_1829365, EPI_ISL_1829366, EPI_ISL_1829367, EPI_ISL_1829368, EPI_ISL_1829369, EPI_ISL_1829370, EPI_ISL_1829371, EPI_ISL_1829377, EPI_ISL_1829378, EPI_ISL_1829379, EPI_ISL_1829380, EPI_ISL_1829381, EPI_ISL_1829383, EPI_ISL_1829384, EPI_ISL_1829385, EPI_ISL_1829386, EPI_ISL_1829387, EPI_ISL_1829388, EPI_ISL_1829389, EPI_ISL_1829390, EPI_ISL_1829392, EPI_ISL_1829393, EPI_ISL_1829394, EPI_ISL_1829395, EPI_ISL_1829396, EPI_ISL_1829397, EPI_ISL_1829398, EPI_ISL_1829399, EPI_ISL_1829401, EPI_ISL_1829402, EPI_ISL_1829404, EPI_ISL_1829405, EPI_ISL_1829407, EPI_ISL_1829409, EPI_ISL_1829410, EPI_ISL_1829414, EPI_ISL_1829415, EPI_ISL_1829416, EPI_ISL_1829417, EPI_ISL_1829418, EPI_ISL_1829422, EPI_ISL_1829423, EPI_ISL_1829424, EPI_ISL_1829425, EPI_ISL_1829426, EPI_ISL_1829428, EPI_ISL_1829429, EPI_ISL_1829431, EPI_ISL_1829436, EPI_ISL_1829439, EPI_ISL_1829441, EPI_ISL_1829443, EPI_ISL_1829445, EPI_ISL_1829446, EPI_ISL_1829447, EPI_ISL_1829448, EPI_ISL_1829449, EPI_ISL_1829450, EPI_ISL_1829451, EPI_ISL_1829453, EPI_ISL_1829454, EPI_ISL_1829455, EPI_ISL_1829459, EPI_ISL_1829463, EPI_ISL_1829464, EPI_ISL_1829465, EPI_ISL_1829466, EPI_ISL_1829467, EPI_ISL_1829468, EPI_ISL_1829469, EPI_ISL_1829470, EPI_ISL_1829471, EPI_ISL_1829472, EPI_ISL_1829473, EPI_ISL_1829475, EPI_ISL_1829476, EPI_ISL_1829477, EPI_ISL_1829478, EPI_ISL_1829479, EPI_ISL_1829480, EPI_ISL_1829481, EPI_ISL_1829482, EPI_ISL_1829483, EPI_ISL_1829488, EPI_ISL_1829511, EPI_ISL_1829513, EPI_ISL_1829522, EPI_ISL_1829527, EPI_ISL_1829528, EPI_ISL_1829530, EPI_ISL_1829536, EPI_ISL_1829557, EPI_ISL_1829566, EPI_ISL_1829572, EPI_ISL_1829574, EPI_ISL_1829579, EPI_ISL_1829586, EPI_ISL_1829587, EPI_ISL_1829632, EPI_ISL_1829656, EPI_ISL_1829668, EPI_ISL_1829680, EPI_ISL_1829683, EPI_ISL_1829689, EPI_ISL_1829705, EPI_ISL_1829709, EPI_ISL_1829713, EPI_ISL_1829729, EPI_ISL_1829742, EPI_ISL_1829744, EPI_ISL_1829750, EPI_ISL_1829763, EPI_ISL_1829770, EPI_ISL_1829771, EPI_ISL_1829774, EPI_ISL_1829777, EPI_ISL_1829788, EPI_ISL_1829800, EPI_ISL_1830389, EPI_ISL_1830394, EPI_ISL_1830400, EPI_ISL_1830401, EPI_ISL_1830403, EPI_ISL_1830404, EPI_ISL_1830406, EPI_ISL_1830408, EPI_ISL_1830410, EPI_ISL_1830411, EPI_ISL_1830412, EPI_ISL_1830413, EPI_ISL_1830419, EPI_ISL_1830423, EPI_ISL_1830424, EPI_ISL_1830431, EPI_ISL_1830432, EPI_ISL_1830437, EPI_ISL_1830438, EPI_ISL_1830447, EPI_ISL_1830449, EPI_ISL_1830450, EPI_ISL_1830454, EPI_ISL_1830459, EPI_ISL_1830472, EPI_ISL_1830473, EPI_ISL_1830481, EPI_ISL_1830482, EPI_ISL_1830483, EPI_ISL_1830485, EPI_ISL_1830489, EPI_ISL_1830492, EPI_ISL_1830495, EPI_ISL_1830496, EPI_ISL_1830498, EPI_ISL_1830499, EPI_ISL_1830501, EPI_ISL_1830503, EPI_ISL_1830506, EPI_ISL_1830510, EPI_ISL_1830512, EPI_ISL_1830514, EPI_ISL_1830518, EPI_ISL_1830524, EPI_ISL_1830526, EPI_ISL_1830531, EPI_ISL_1830537, EPI_ISL_1830542, EPI_ISL_1830545, EPI_ISL_1830555, EPI_ISL_1830559, EPI_ISL_1830562, EPI_ISL_1830563, EPI_ISL_1830564, EPI_ISL_1830568, EPI_ISL_1830572, EPI_ISL_1830574, EPI_ISL_1830576, EPI_ISL_1830578, EPI_ISL_1830579, EPI_ISL_1830585, EPI_ISL_1830589, EPI_ISL_1830594, EPI_ISL_1830597, EPI_ISL_1830601, EPI_ISL_1830604, EPI_ISL_1830606, EPI_ISL_1830607, EPI_ISL_1830614, EPI_ISL_1830619, EPI_ISL_1830623, EPI_ISL_1830624, EPI_ISL_1830626, EPI_ISL_1830630, EPI_ISL_1830633, EPI_ISL_1830635, EPI_ISL_1830637, EPI_ISL_1830648, EPI_ISL_1830649, EPI_ISL_1830650, EPI_ISL_1830651, EPI_ISL_1830652, EPI_ISL_1830654, EPI_ISL_1830656, EPI_ISL_1830661, EPI_ISL_1830665, EPI_ISL_1830667, EPI_ISL_1830671, EPI_ISL_1830673, EPI_ISL_1830675, EPI_ISL_1830677, EPI_ISL_1830679, EPI_ISL_1830681, EPI_ISL_1830682, EPI_ISL_1830688, EPI_ISL_1830689, EPI_ISL_1830692, EPI_ISL_1830693, EPI_ISL_1830695, EPI_ISL_1830710, EPI_ISL_1830712, EPI_ISL_1830713, EPI_ISL_1830714, EPI_ISL_1830715, EPI_ISL_1830716, EPI_ISL_1830718, EPI_ISL_1830720, EPI_ISL_1830722, EPI_ISL_1830725, EPI_ISL_1830726, EPI_ISL_1830727, EPI_ISL_1830813, EPI_ISL_1857831, EPI_ISL_1857842, EPI_ISL_1857847, EPI_ISL_1857852, EPI_ISL_1857855, EPI_ISL_1857862, EPI_ISL_1857866, EPI_ISL_1857874, EPI_ISL_1857884, EPI_ISL_1857897, EPI_ISL_1857899, EPI_ISL_1857903, EPI_ISL_1857905, EPI_ISL_1857906, EPI_ISL_1857918, EPI_ISL_1857919, EPI_ISL_1857935, EPI_ISL_1857938, EPI_ISL_1857946, EPI_ISL_1857979, EPI_ISL_1857999, EPI_ISL_1858059, EPI_ISL_1858062, EPI_ISL_1858091, EPI_ISL_1858111, EPI_ISL_1858144, EPI_ISL_1858154, EPI_ISL_1858202, EPI_ISL_1858239, EPI_ISL_1912006, EPI_ISL_1912010, EPI_ISL_1912011, EPI_ISL_1912013, EPI_ISL_1912020, EPI_ISL_1912022, EPI_ISL_1912031, EPI_ISL_1912032, EPI_ISL_1912045, EPI_ISL_1912047, EPI_ISL_1912048, EPI_ISL_1912050, EPI_ISL_1912051, EPI_ISL_1912053, EPI_ISL_1912055, EPI_ISL_1912059, EPI_ISL_1912072, EPI_ISL_1912084, EPI_ISL_1912103, EPI_ISL_1912105, EPI_ISL_1912108, EPI_ISL_1912109, EPI_ISL_1912110, EPI_ISL_1912116, EPI_ISL_1912118, EPI_ISL_1912126, EPI_ISL_1912129, EPI_ISL_1912131, EPI_ISL_1912132, EPI_ISL_1912140, EPI_ISL_1912141, EPI_ISL_1912143, EPI_ISL_1912145, EPI_ISL_1912149, EPI_ISL_1912150, EPI_ISL_1912152, EPI_ISL_1912159, EPI_ISL_1912164, EPI_ISL_1912172, EPI_ISL_1912177, EPI_ISL_1912192, EPI_ISL_1912195, EPI_ISL_1912212, EPI_ISL_1912232, EPI_ISL_1912234, EPI_ISL_1912243, EPI_ISL_1912245, EPI_ISL_1912257, EPI_ISL_1912272, EPI_ISL_1912281, EPI_ISL_1912295, EPI_ISL_1912311, EPI_ISL_1912319, EPI_ISL_1912327, EPI_ISL_1912336, EPI_ISL_1912339, EPI_ISL_1912343, EPI_ISL_1912349, EPI_ISL_1912752, EPI_ISL_1912753, EPI_ISL_1912754, EPI_ISL_1912755, EPI_ISL_1912756, EPI_ISL_1912757, EPI_ISL_1912758, EPI_ISL_1921439, EPI_ISL_1921442, EPI_ISL_1921445, EPI_ISL_1921447, EPI_ISL_1921448, EPI_ISL_1921450, EPI_ISL_1921451, EPI_ISL_1921452, EPI_ISL_1921454, EPI_ISL_1921455, EPI_ISL_1921456, EPI_ISL_1921457, EPI_ISL_1921458, EPI_ISL_1921460, EPI_ISL_1921461, EPI_ISL_1921462, EPI_ISL_1921464, EPI_ISL_1921466, EPI_ISL_1921468, EPI_ISL_1921469, EPI_ISL_1921470, EPI_ISL_1921471, EPI_ISL_1921472, EPI_ISL_1921473, EPI_ISL_1921474, EPI_ISL_1921475, EPI_ISL_1921476, EPI_ISL_1921477, EPI_ISL_1921478, EPI_ISL_1921479, EPI_ISL_1921480, EPI_ISL_1921481, EPI_ISL_1921482, EPI_ISL_1921483, EPI_ISL_1921484, EPI_ISL_1921487, EPI_ISL_1921491, EPI_ISL_1921492, EPI_ISL_1921493, EPI_ISL_1921497, EPI_ISL_1921498, EPI_ISL_1921499, EPI_ISL_1921500, EPI_ISL_1921501, EPI_ISL_1921503, EPI_ISL_1921504, EPI_ISL_1921505, EPI_ISL_1921508, EPI_ISL_1921510, EPI_ISL_1921511, EPI_ISL_1921512, EPI_ISL_1921516, EPI_ISL_1921517, EPI_ISL_1921518, EPI_ISL_1921519, EPI_ISL_1921521, EPI_ISL_1921524, EPI_ISL_1921526, EPI_ISL_1921527, EPI_ISL_1921528, EPI_ISL_1921529, EPI_ISL_1921530, EPI_ISL_1921531, EPI_ISL_1921532, EPI_ISL_1921534, EPI_ISL_1921535, EPI_ISL_1921536, EPI_ISL_1921537, EPI_ISL_1921539, EPI_ISL_1921540, EPI_ISL_1921541, EPI_ISL_1921542, EPI_ISL_1921543, EPI_ISL_1921544, EPI_ISL_1921546, EPI_ISL_1921547, EPI_ISL_1921548, EPI_ISL_1921549, EPI_ISL_1921550, EPI_ISL_1921551, EPI_ISL_1921554, EPI_ISL_1921556, EPI_ISL_1921560, EPI_ISL_1921562, EPI_ISL_1921563, EPI_ISL_1921564, EPI_ISL_1921565, EPI_ISL_1921566, EPI_ISL_1921569, EPI_ISL_1921570, EPI_ISL_1921571, EPI_ISL_1921573, EPI_ISL_1921574, EPI_ISL_1921576, EPI_ISL_1921577, EPI_ISL_1921578, EPI_ISL_1921579, EPI_ISL_1921580, EPI_ISL_1921581, EPI_ISL_1921583, EPI_ISL_1921584, EPI_ISL_1921585, EPI_ISL_1921586, EPI_ISL_1921587, EPI_ISL_1921589, EPI_ISL_1921591, EPI_ISL_1921592, EPI_ISL_1921593, EPI_ISL_1921594, EPI_ISL_1921595, EPI_ISL_1921599, EPI_ISL_1921601, EPI_ISL_1921602, EPI_ISL_1921603, EPI_ISL_1921605, EPI_ISL_1921606, EPI_ISL_1921607, EPI_ISL_1921608, EPI_ISL_1921609, EPI_ISL_1921610, EPI_ISL_1921611, EPI_ISL_1921612, EPI_ISL_1921614, EPI_ISL_1921615, EPI_ISL_1921616, EPI_ISL_1921617, EPI_ISL_1921620, EPI_ISL_1921621, EPI_ISL_1921622, EPI_ISL_1921623, EPI_ISL_1921624, EPI_ISL_1921625, EPI_ISL_1921628, EPI_ISL_1921629, EPI_ISL_1921630, EPI_ISL_1921632, EPI_ISL_1921633, EPI_ISL_1921635, EPI_ISL_1921636, EPI_ISL_1921638, EPI_ISL_1921639, EPI_ISL_1921640, EPI_ISL_1921641, EPI_ISL_1921642, EPI_ISL_1921643, EPI_ISL_1921644, EPI_ISL_1921646, EPI_ISL_1921647, EPI_ISL_1921649, EPI_ISL_1921650, EPI_ISL_1921651, EPI_ISL_1921652, EPI_ISL_1921653, EPI_ISL_1921654, EPI_ISL_1921655, EPI_ISL_1921657, EPI_ISL_1921658, EPI_ISL_1921659, EPI_ISL_1921661, EPI_ISL_1921663, EPI_ISL_1921664, EPI_ISL_1921665, EPI_ISL_1921668, EPI_ISL_1921670, EPI_ISL_1921671, EPI_ISL_1921672, EPI_ISL_1921674, EPI_ISL_1921675, EPI_ISL_1921677, EPI_ISL_1921678, EPI_ISL_1921680, EPI_ISL_1921681, EPI_ISL_1921682, EPI_ISL_1921683, EPI_ISL_1921687, EPI_ISL_1921688, EPI_ISL_1921689, EPI_ISL_1921690, EPI_ISL_1921692, EPI_ISL_1921693, EPI_ISL_1921694, EPI_ISL_1921695, EPI_ISL_1921696, EPI_ISL_1921697, EPI_ISL_1921701, EPI_ISL_1921704, EPI_ISL_1921705, EPI_ISL_1921706, EPI_ISL_1921707, EPI_ISL_1921708, EPI_ISL_1921710, EPI_ISL_1921712, EPI_ISL_1921714, EPI_ISL_1921715, EPI_ISL_1921716, EPI_ISL_1921718, EPI_ISL_1921719, EPI_ISL_1921720, EPI_ISL_1921721, EPI_ISL_1921723, EPI_ISL_1921724, EPI_ISL_1921726, EPI_ISL_1921727, EPI_ISL_1921728, EPI_ISL_1921729, EPI_ISL_1921730, EPI_ISL_1921733, EPI_ISL_1921736, EPI_ISL_1939708, EPI_ISL_1939709, EPI_ISL_1939710, EPI_ISL_1939711, EPI_ISL_1939712, EPI_ISL_1939713, EPI_ISL_1939714, EPI_ISL_1939715, EPI_ISL_1939716, EPI_ISL_1939717, EPI_ISL_1939718, EPI_ISL_1985513, EPI_ISL_1985514, EPI_ISL_1985515, EPI_ISL_1985516, EPI_ISL_1985518, EPI_ISL_1985519, EPI_ISL_1985520, EPI_ISL_1985521, EPI_ISL_1985523, EPI_ISL_1985525, EPI_ISL_1985526, EPI_ISL_1985527, EPI_ISL_1985528, EPI_ISL_1985529, EPI_ISL_1985531, EPI_ISL_1985532, EPI_ISL_1985533, EPI_ISL_1985534, EPI_ISL_1985535, EPI_ISL_1985536, EPI_ISL_1985539, EPI_ISL_1985540, EPI_ISL_1985541, EPI_ISL_1985543, EPI_ISL_1985544, EPI_ISL_1985545, EPI_ISL_1985546, EPI_ISL_1985547, EPI_ISL_1985548, EPI_ISL_1985549, EPI_ISL_1985551, EPI_ISL_1985552, EPI_ISL_1985553, EPI_ISL_1985554, EPI_ISL_1985555, EPI_ISL_1985558, EPI_ISL_1985559, EPI_ISL_1985560, EPI_ISL_1985561, EPI_ISL_1985562, EPI_ISL_1985563, EPI_ISL_1985565, EPI_ISL_1985566, EPI_ISL_1985567, EPI_ISL_1985568, EPI_ISL_1985570, EPI_ISL_1985572, EPI_ISL_1985573, EPI_ISL_1985574, EPI_ISL_1985575, EPI_ISL_1985578, EPI_ISL_1985579, EPI_ISL_1985580, EPI_ISL_1985581, EPI_ISL_1985582, EPI_ISL_1985583, EPI_ISL_1985584, EPI_ISL_1985586, EPI_ISL_1985587, EPI_ISL_1985588, EPI_ISL_1985589, EPI_ISL_1985590, EPI_ISL_1985592, EPI_ISL_1985593, EPI_ISL_1985594, EPI_ISL_1985595, EPI_ISL_1985596, EPI_ISL_1985597, EPI_ISL_1985599, EPI_ISL_1985600, EPI_ISL_1985601, EPI_ISL_1985602, EPI_ISL_1985603, EPI_ISL_1985604, EPI_ISL_1985605, EPI_ISL_1985607, EPI_ISL_1985608, EPI_ISL_1985609, EPI_ISL_1985610, EPI_ISL_1985611, EPI_ISL_1985612, EPI_ISL_1985613, EPI_ISL_1985614, EPI_ISL_1985616, EPI_ISL_1985617, EPI_ISL_1985618, EPI_ISL_1985619, EPI_ISL_1985620, EPI_ISL_1985622, EPI_ISL_1985623, EPI_ISL_1985624, EPI_ISL_1985625, EPI_ISL_1985626, EPI_ISL_1985627, EPI_ISL_1985629, EPI_ISL_1985630, EPI_ISL_1985631, EPI_ISL_1985632, EPI_ISL_1985633, EPI_ISL_1985635, EPI_ISL_1985636, EPI_ISL_1985637, EPI_ISL_1985638, EPI_ISL_1985639, EPI_ISL_1985640, EPI_ISL_1985642, EPI_ISL_1985643, EPI_ISL_1985644, EPI_ISL_1985645, EPI_ISL_1985646, EPI_ISL_1985647, EPI_ISL_1985648, EPI_ISL_1985650, EPI_ISL_1985651, EPI_ISL_1985652, EPI_ISL_1985653, EPI_ISL_1985654, EPI_ISL_1985656, EPI_ISL_1985657, EPI_ISL_1985658, EPI_ISL_1985659, EPI_ISL_1985660, EPI_ISL_1985661, EPI_ISL_1985663, EPI_ISL_1985664, EPI_ISL_1985665, EPI_ISL_1985666, EPI_ISL_1985667, EPI_ISL_1985668, EPI_ISL_1985669, EPI_ISL_1985670, EPI_ISL_1985671, EPI_ISL_1985673, EPI_ISL_1985674, EPI_ISL_1985675, EPI_ISL_1985676, EPI_ISL_1985677, EPI_ISL_1985678, EPI_ISL_1985680, EPI_ISL_1985681, EPI_ISL_1985682, EPI_ISL_1985683, EPI_ISL_1985684, EPI_ISL_1985687, EPI_ISL_1985690, EPI_ISL_1985691, EPI_ISL_1985697, EPI_ISL_1985698, EPI_ISL_1985699, EPI_ISL_1985701, EPI_ISL_1985702, EPI_ISL_1985703, EPI_ISL_1985704, EPI_ISL_1985705, EPI_ISL_1985706, EPI_ISL_1985708, EPI_ISL_1985710, EPI_ISL_1985713, EPI_ISL_1985715, EPI_ISL_1985717, EPI_ISL_1985718, EPI_ISL_1985720, EPI_ISL_1985722, EPI_ISL_1985723, EPI_ISL_1985724, EPI_ISL_1985725, EPI_ISL_1985726, EPI_ISL_1985728, EPI_ISL_1985730, EPI_ISL_1985731, EPI_ISL_1985732, EPI_ISL_1985733, EPI_ISL_1985735, EPI_ISL_1985736, EPI_ISL_1985738, EPI_ISL_1985739, EPI_ISL_1985740, EPI_ISL_1985741, EPI_ISL_1985742, EPI_ISL_1985743, EPI_ISL_1985747, EPI_ISL_1985749, EPI_ISL_1985750, EPI_ISL_1985752, EPI_ISL_1985753, EPI_ISL_1985754, EPI_ISL_1985758, EPI_ISL_1985760, EPI_ISL_1985763, EPI_ISL_1985764, EPI_ISL_1985766, EPI_ISL_1985769, EPI_ISL_1985804, EPI_ISL_1985806, EPI_ISL_1985807, EPI_ISL_1985808, EPI_ISL_1985809, EPI_ISL_1985812, EPI_ISL_1985813, EPI_ISL_1985814, EPI_ISL_1985815, EPI_ISL_1985816, EPI_ISL_1985817, EPI_ISL_1985819, EPI_ISL_1985821, EPI_ISL_1985823, EPI_ISL_1985827, EPI_ISL_1985829, EPI_ISL_1985831, EPI_ISL_1985832, EPI_ISL_1985833, EPI_ISL_1985834, EPI_ISL_1985836, EPI_ISL_1985837, EPI_ISL_1985839, EPI_ISL_1985840, EPI_ISL_1985842, EPI_ISL_1985843, EPI_ISL_1985844, EPI_ISL_1985846, EPI_ISL_1985847, EPI_ISL_1985850, EPI_ISL_1985851, EPI_ISL_1985853, EPI_ISL_1985854, EPI_ISL_1985855, EPI_ISL_1985856, EPI_ISL_1985857, EPI_ISL_1985858, EPI_ISL_1985860, EPI_ISL_1985863, EPI_ISL_1985865, EPI_ISL_1985867, EPI_ISL_1985868, EPI_ISL_1985869, EPI_ISL_1985870, EPI_ISL_1985875, EPI_ISL_1985877, EPI_ISL_1985878, EPI_ISL_1985879, EPI_ISL_1985880, EPI_ISL_1985882, EPI_ISL_1985883, EPI_ISL_1985884, EPI_ISL_1985885, EPI_ISL_1985887, EPI_ISL_1985888, EPI_ISL_1985891, EPI_ISL_1985893, EPI_ISL_1985894, EPI_ISL_1985895, EPI_ISL_1985898, EPI_ISL_1985899, EPI_ISL_1985900, EPI_ISL_1985903, EPI_ISL_1985905, EPI_ISL_1985906, EPI_ISL_1985907, EPI_ISL_1985908, EPI_ISL_1985911, EPI_ISL_1985913, EPI_ISL_1985917, EPI_ISL_1985918, EPI_ISL_1985922, EPI_ISL_1985923, EPI_ISL_1985924, EPI_ISL_1985926, EPI_ISL_1985928, EPI_ISL_1985930, EPI_ISL_1985931, EPI_ISL_1985932, EPI_ISL_1985933, EPI_ISL_1985935, EPI_ISL_1985936, EPI_ISL_1985937, EPI_ISL_1985938, EPI_ISL_1985939, EPI_ISL_1985940, EPI_ISL_1985942, EPI_ISL_1985943, EPI_ISL_1985944, EPI_ISL_1985945, EPI_ISL_1985946, EPI_ISL_1985947, EPI_ISL_1985949, EPI_ISL_1985951, EPI_ISL_1985952, EPI_ISL_1985955, EPI_ISL_1985958, EPI_ISL_1985960, EPI_ISL_1985961, EPI_ISL_1985962, EPI_ISL_1985963, EPI_ISL_1985964, EPI_ISL_1985968, EPI_ISL_1985969, EPI_ISL_1985970, EPI_ISL_1985973, EPI_ISL_1985974, EPI_ISL_1985975, EPI_ISL_1985978, EPI_ISL_1985981, EPI_ISL_1985985, EPI_ISL_1985987, EPI_ISL_1985988, EPI_ISL_1985991, EPI_ISL_1985994, EPI_ISL_1985995, EPI_ISL_1985996, EPI_ISL_1985997, EPI_ISL_1985998, EPI_ISL_1986000, EPI_ISL_1986001, EPI_ISL_1986002, EPI_ISL_1986003, EPI_ISL_1986004, EPI_ISL_1986007, EPI_ISL_1986008, EPI_ISL_1986011, EPI_ISL_1986012, EPI_ISL_1986014, EPI_ISL_1986015, EPI_ISL_1986016, EPI_ISL_1986017, EPI_ISL_1986019, EPI_ISL_1986022, EPI_ISL_1986023, EPI_ISL_1986024, EPI_ISL_1986025, EPI_ISL_1986026, EPI_ISL_1986029, EPI_ISL_1986031, EPI_ISL_1986033, EPI_ISL_1986035, EPI_ISL_1986037, EPI_ISL_1986038, EPI_ISL_1986039, EPI_ISL_1986040, EPI_ISL_1986042, EPI_ISL_1986044, EPI_ISL_1986048, EPI_ISL_1986049, EPI_ISL_1986050, EPI_ISL_1986052, EPI_ISL_1986053, EPI_ISL_1986055, EPI_ISL_1986056, EPI_ISL_1986057, EPI_ISL_1986059, EPI_ISL_1986061, EPI_ISL_1986063, EPI_ISL_1986065, EPI_ISL_1986066, EPI_ISL_1986072, EPI_ISL_1986073, EPI_ISL_1986077, EPI_ISL_1986078, EPI_ISL_1986080, EPI_ISL_1986081, EPI_ISL_1986085, EPI_ISL_1986086, EPI_ISL_1986088, EPI_ISL_1986089, EPI_ISL_1986090, EPI_ISL_1986091, EPI_ISL_1986093, EPI_ISL_1986095, EPI_ISL_1986098, EPI_ISL_1986100, EPI_ISL_1986101, EPI_ISL_1986103, EPI_ISL_1986104, EPI_ISL_1986106, EPI_ISL_1986108, EPI_ISL_1986109, EPI_ISL_1986119, EPI_ISL_1986120, EPI_ISL_1986121, EPI_ISL_1986122, EPI_ISL_1986127, EPI_ISL_1986135, EPI_ISL_1986137, EPI_ISL_1986140, EPI_ISL_1986142, EPI_ISL_1986143, EPI_ISL_1986145, EPI_ISL_1986146, EPI_ISL_1986150, EPI_ISL_1986153, EPI_ISL_1986158, EPI_ISL_1986161, EPI_ISL_1986164, EPI_ISL_1986165, EPI_ISL_1986170, EPI_ISL_1986173, EPI_ISL_1986177, EPI_ISL_1986178, EPI_ISL_1986183, EPI_ISL_1986187, EPI_ISL_1986188, EPI_ISL_1986190, EPI_ISL_1986193, EPI_ISL_1986195, EPI_ISL_1986196, EPI_ISL_1986198, EPI_ISL_1986199, EPI_ISL_1986202, EPI_ISL_1986207, EPI_ISL_1986211, EPI_ISL_1986214, EPI_ISL_1986217, EPI_ISL_1986222, EPI_ISL_1986223, EPI_ISL_1986225, EPI_ISL_1986226, EPI_ISL_1986229, EPI_ISL_1986232, EPI_ISL_1986234, EPI_ISL_1986236, EPI_ISL_1986239, EPI_ISL_1986240, EPI_ISL_1986243, EPI_ISL_1986244, EPI_ISL_1986245, EPI_ISL_1986246, EPI_ISL_1986247, EPI_ISL_1986248, EPI_ISL_1986249, EPI_ISL_1986253, EPI_ISL_1986258, EPI_ISL_1986259, EPI_ISL_1986260, EPI_ISL_1986262, EPI_ISL_1986271, EPI_ISL_1986276, EPI_ISL_1986280, EPI_ISL_1986286, EPI_ISL_1986287, EPI_ISL_1986288, EPI_ISL_1986289, EPI_ISL_1986291, EPI_ISL_1986293, EPI_ISL_1986295, EPI_ISL_1986300, EPI_ISL_1986301, EPI_ISL_1986303, EPI_ISL_1986306, EPI_ISL_1986308, EPI_ISL_1986309, EPI_ISL_1986313, EPI_ISL_1986316, EPI_ISL_1986321, EPI_ISL_1986322, EPI_ISL_1986329, EPI_ISL_1986330, EPI_ISL_1986333, EPI_ISL_1986340, EPI_ISL_1986341, EPI_ISL_1986344, EPI_ISL_1986345, EPI_ISL_1986346, EPI_ISL_1986347, EPI_ISL_1986348, EPI_ISL_1986352, EPI_ISL_1986356, EPI_ISL_1986358, EPI_ISL_1986359, EPI_ISL_1986363, EPI_ISL_1986367, EPI_ISL_1986368, EPI_ISL_1986375, EPI_ISL_1986380, EPI_ISL_1986382, EPI_ISL_1986383, EPI_ISL_1986384, EPI_ISL_1986388, EPI_ISL_1986391, EPI_ISL_1986496, EPI_ISL_1986498, EPI_ISL_1986499, EPI_ISL_1986500, EPI_ISL_1986503, EPI_ISL_1986504, EPI_ISL_1986506, EPI_ISL_1986513, EPI_ISL_1986514, EPI_ISL_1986525, EPI_ISL_1986527, EPI_ISL_1986530, EPI_ISL_1986531, EPI_ISL_1986532, EPI_ISL_1986537, EPI_ISL_1986540, EPI_ISL_1986543, EPI_ISL_1986544, EPI_ISL_1986545, EPI_ISL_1986546, EPI_ISL_1986547, EPI_ISL_1986549, EPI_ISL_1986551, EPI_ISL_1986554, EPI_ISL_1986555, EPI_ISL_1986557, EPI_ISL_1986561, EPI_ISL_1986564, EPI_ISL_1986565, EPI_ISL_1986566, EPI_ISL_1986568, EPI_ISL_1986578, EPI_ISL_1986581, EPI_ISL_1986584, EPI_ISL_1986587, EPI_ISL_1986591, EPI_ISL_1986598, EPI_ISL_1986599, EPI_ISL_1986603, EPI_ISL_1986606, EPI_ISL_1986614, EPI_ISL_1986620, EPI_ISL_1986629, EPI_ISL_1986630, EPI_ISL_1986633, EPI_ISL_1986651, EPI_ISL_1986652, EPI_ISL_1986653, EPI_ISL_1986656, EPI_ISL_1986662, EPI_ISL_1986674, EPI_ISL_1986676, EPI_ISL_1986680, EPI_ISL_1986697, EPI_ISL_1986704, EPI_ISL_1986705, EPI_ISL_1986714, EPI_ISL_1986715, EPI_ISL_1986725, EPI_ISL_1986727, EPI_ISL_1986729, EPI_ISL_1986735, EPI_ISL_1986744, EPI_ISL_1986747, EPI_ISL_1986752, EPI_ISL_1986754, EPI_ISL_1986755, EPI_ISL_1986760, EPI_ISL_1986769, EPI_ISL_1986774, EPI_ISL_1986787, EPI_ISL_1986794, EPI_ISL_1986798, EPI_ISL_1986809, EPI_ISL_1986814, EPI_ISL_1986819, EPI_ISL_1986821, EPI_ISL_1986825, EPI_ISL_1986827, EPI_ISL_1986830, EPI_ISL_1986834, EPI_ISL_1986837, EPI_ISL_1986846, EPI_ISL_1986848, EPI_ISL_1986851, EPI_ISL_1986856, EPI_ISL_1986857, EPI_ISL_1986885, EPI_ISL_1986897, EPI_ISL_1986903, EPI_ISL_1986906, EPI_ISL_1986909, EPI_ISL_1986914, EPI_ISL_1986920, EPI_ISL_1986931, EPI_ISL_1986932, EPI_ISL_1986937, EPI_ISL_1986948, EPI_ISL_1986950, EPI_ISL_1986951, EPI_ISL_1986955, EPI_ISL_1986965, EPI_ISL_1986966, EPI_ISL_1986969, EPI_ISL_1986973, EPI_ISL_1986978, EPI_ISL_1986982, EPI_ISL_1986985, EPI_ISL_1986991, EPI_ISL_1986994, EPI_ISL_1986997, EPI_ISL_1986999, EPI_ISL_1987010, EPI_ISL_1987014, EPI_ISL_1987017, EPI_ISL_1987027, EPI_ISL_1987038, EPI_ISL_1987039, EPI_ISL_1987043, EPI_ISL_1987052, EPI_ISL_1987064, EPI_ISL_1987066, EPI_ISL_1987067, EPI_ISL_1987075, EPI_ISL_1987076, EPI_ISL_1987122, EPI_ISL_1987124, EPI_ISL_1987141, EPI_ISL_1987144, EPI_ISL_1987158, EPI_ISL_1987164, EPI_ISL_1987176, EPI_ISL_1987181, EPI_ISL_1987184, EPI_ISL_1987186, EPI_ISL_1987187, EPI_ISL_1987199, EPI_ISL_1987201, EPI_ISL_1987202, EPI_ISL_1987217, EPI_ISL_1987226, EPI_ISL_1987228, EPI_ISL_1987237, EPI_ISL_1987240, EPI_ISL_1987247, EPI_ISL_1987248, EPI_ISL_1987252, EPI_ISL_1987263, EPI_ISL_1987266, EPI_ISL_1987272, EPI_ISL_1987288, EPI_ISL_1987290, EPI_ISL_1987304, EPI_ISL_1987305, EPI_ISL_1987306, EPI_ISL_1987310, EPI_ISL_1987313, EPI_ISL_1987316, EPI_ISL_1987318, EPI_ISL_1987319, EPI_ISL_1987320, EPI_ISL_1987327, EPI_ISL_1987328, EPI_ISL_1987332, EPI_ISL_1987342, EPI_ISL_1987346, EPI_ISL_1987347, EPI_ISL_1987349, EPI_ISL_1987350, EPI_ISL_1987357, EPI_ISL_1987360, EPI_ISL_1987361, EPI_ISL_1987368, EPI_ISL_1987370, EPI_ISL_1987372, EPI_ISL_1987374, EPI_ISL_1987379, EPI_ISL_1987381, EPI_ISL_1987382, EPI_ISL_2004372, EPI_ISL_2004398, EPI_ISL_2004627, EPI_ISL_2004632, EPI_ISL_2004635, EPI_ISL_2004637, EPI_ISL_2004643, EPI_ISL_2004646, EPI_ISL_2004648, EPI_ISL_2004651, EPI_ISL_2004653, EPI_ISL_2004660, EPI_ISL_2004667, EPI_ISL_2004668, EPI_ISL_2004670, EPI_ISL_2004674, EPI_ISL_2004675, EPI_ISL_2004676, EPI_ISL_2004677, EPI_ISL_2004679, EPI_ISL_2004680, EPI_ISL_2004682, EPI_ISL_2004685, EPI_ISL_2004687, EPI_ISL_2004691, EPI_ISL_2004693, EPI_ISL_2004695, EPI_ISL_2004697, EPI_ISL_2004701, EPI_ISL_2004702, EPI_ISL_2004709, EPI_ISL_2004713, EPI_ISL_2004715, EPI_ISL_2004717, EPI_ISL_2004724, EPI_ISL_2004726, EPI_ISL_2004728, EPI_ISL_2004729, EPI_ISL_2004734, EPI_ISL_2004738, EPI_ISL_2004744, EPI_ISL_2004745, EPI_ISL_2004748, EPI_ISL_2004751, EPI_ISL_2004752, EPI_ISL_2004753, EPI_ISL_2004755, EPI_ISL_2004758, EPI_ISL_2004762, EPI_ISL_2004767, EPI_ISL_2004768, EPI_ISL_2004769, EPI_ISL_2004773, EPI_ISL_2004775, EPI_ISL_2004787, EPI_ISL_2004788, EPI_ISL_2004789, EPI_ISL_2004790, EPI_ISL_2004794, EPI_ISL_2004795, EPI_ISL_2004796, EPI_ISL_2004798, EPI_ISL_2004799, EPI_ISL_2004805, EPI_ISL_2004806, EPI_ISL_2004807, EPI_ISL_2004812, EPI_ISL_2004815, EPI_ISL_2004816, EPI_ISL_2004817, EPI_ISL_2004818, EPI_ISL_2004833, EPI_ISL_2004836, EPI_ISL_2004837, EPI_ISL_2004846, EPI_ISL_2004847, EPI_ISL_2004849, EPI_ISL_2004850, EPI_ISL_2004852, EPI_ISL_2004860, EPI_ISL_2004862, EPI_ISL_2004864, EPI_ISL_2004868, EPI_ISL_2004871, EPI_ISL_2004876, EPI_ISL_2004878, EPI_ISL_2005269, EPI_ISL_2005341, EPI_ISL_2005343, EPI_ISL_2005355, EPI_ISL_2005359, EPI_ISL_2005361, EPI_ISL_2005365, EPI_ISL_2005366, EPI_ISL_2005367, EPI_ISL_2005370, EPI_ISL_2005372, EPI_ISL_2005373, EPI_ISL_2005375, EPI_ISL_2005377, EPI_ISL_2005380, EPI_ISL_2005381, EPI_ISL_2005386, EPI_ISL_2005387, EPI_ISL_2005388, EPI_ISL_2005389, EPI_ISL_2005394, EPI_ISL_2005400, EPI_ISL_2005406, EPI_ISL_2005412, EPI_ISL_2005414, EPI_ISL_2005421, EPI_ISL_2005423, EPI_ISL_2005429, EPI_ISL_2005434, EPI_ISL_2005439, EPI_ISL_2005441, EPI_ISL_2005444, EPI_ISL_2005445, EPI_ISL_2005446, EPI_ISL_2005447, EPI_ISL_2005449, EPI_ISL_2005451, EPI_ISL_2005457, EPI_ISL_2005458, EPI_ISL_2005459, EPI_ISL_2005460, EPI_ISL_2005461, EPI_ISL_2005463, EPI_ISL_2005466, EPI_ISL_2005467, EPI_ISL_2005472, EPI_ISL_2005477, EPI_ISL_2005479, EPI_ISL_2005481, EPI_ISL_2005484, EPI_ISL_2005489, EPI_ISL_2005495, EPI_ISL_2005502, EPI_ISL_2005503, EPI_ISL_2005506, EPI_ISL_2005507, EPI_ISL_2005509, EPI_ISL_2005510, EPI_ISL_2005511, EPI_ISL_2005512, EPI_ISL_2005514, EPI_ISL_2005517, EPI_ISL_2005518, EPI_ISL_2005523, EPI_ISL_2005525, EPI_ISL_2005529, EPI_ISL_2005537, EPI_ISL_2005553, EPI_ISL_2005555, EPI_ISL_2005568, EPI_ISL_2005579, EPI_ISL_2005581, EPI_ISL_2005597, EPI_ISL_2005600, EPI_ISL_2005601, EPI_ISL_2005605, EPI_ISL_2005606, EPI_ISL_2005609, EPI_ISL_2005612, EPI_ISL_2005676, EPI_ISL_2005796, EPI_ISL_2005810, EPI_ISL_2005835, EPI_ISL_2005847, EPI_ISL_2005862, EPI_ISL_2005869, EPI_ISL_2005878, EPI_ISL_2005881, EPI_ISL_2005896, EPI_ISL_2005959, EPI_ISL_2005961, EPI_ISL_2005970, EPI_ISL_2006003, EPI_ISL_2006015, EPI_ISL_2006016, EPI_ISL_2006027, EPI_ISL_2006029, EPI_ISL_2006036, EPI_ISL_2006051, EPI_ISL_2006344, EPI_ISL_2006458, EPI_ISL_2006667, EPI_ISL_2006668, EPI_ISL_2021041, EPI_ISL_2021046, EPI_ISL_2021047, EPI_ISL_2021062, EPI_ISL_2021063, EPI_ISL_2021065, EPI_ISL_2021067, EPI_ISL_2021068, EPI_ISL_2021074, EPI_ISL_2021081, EPI_ISL_2021086, EPI_ISL_2021087, EPI_ISL_2021097, EPI_ISL_2021104, EPI_ISL_2021107, EPI_ISL_2021109, EPI_ISL_2021115, EPI_ISL_2021123, EPI_ISL_2021128, EPI_ISL_2021130, EPI_ISL_2021132, EPI_ISL_2021133, EPI_ISL_2021136, EPI_ISL_2021139, EPI_ISL_2021149, EPI_ISL_2021157, EPI_ISL_2021167, EPI_ISL_2021185, EPI_ISL_2021191, EPI_ISL_2021219, EPI_ISL_2021221, EPI_ISL_2021227, EPI_ISL_2021263, EPI_ISL_2021267, EPI_ISL_2021268, EPI_ISL_2021270, EPI_ISL_2021278, EPI_ISL_2021294, EPI_ISL_2021301, EPI_ISL_2021334, EPI_ISL_2021351, EPI_ISL_2021352, EPI_ISL_2021356, EPI_ISL_2021371, EPI_ISL_2021381, EPI_ISL_2021384, EPI_ISL_2021391, EPI_ISL_2021406, EPI_ISL_2021407, EPI_ISL_2021409, EPI_ISL_2021412, EPI_ISL_2021416, EPI_ISL_2021417, EPI_ISL_2021424, EPI_ISL_2021429, EPI_ISL_2021433, EPI_ISL_2021435, EPI_ISL_2021436, EPI_ISL_2021438, EPI_ISL_2021440, EPI_ISL_2021441, EPI_ISL_2021442, EPI_ISL_2021445, EPI_ISL_2021447, EPI_ISL_2021452, EPI_ISL_2021458, EPI_ISL_2021479, EPI_ISL_2021480, EPI_ISL_2021481, EPI_ISL_2021495, EPI_ISL_2021501, EPI_ISL_2021502, EPI_ISL_2021504, EPI_ISL_2021506, EPI_ISL_2021507, EPI_ISL_2021510, EPI_ISL_2021518, EPI_ISL_2021522, EPI_ISL_2021534, EPI_ISL_2021535, EPI_ISL_2021538, EPI_ISL_2021539, EPI_ISL_2021545, EPI_ISL_2021554, EPI_ISL_2021555, EPI_ISL_2021556, EPI_ISL_2021557, EPI_ISL_2021566, EPI_ISL_2021567, EPI_ISL_2021571, EPI_ISL_2021577, EPI_ISL_2021578, EPI_ISL_2021580, EPI_ISL_2021584, EPI_ISL_2021586, EPI_ISL_2021587, EPI_ISL_2021588, EPI_ISL_2021590, EPI_ISL_2021595, EPI_ISL_2021597, EPI_ISL_2021611, EPI_ISL_2021626, EPI_ISL_2021630, EPI_ISL_2021632, EPI_ISL_2021642, EPI_ISL_2021645, EPI_ISL_2021646, EPI_ISL_2021655, EPI_ISL_2021666, EPI_ISL_2021668, EPI_ISL_2021673, EPI_ISL_2021676, EPI_ISL_2021677, EPI_ISL_2021678, EPI_ISL_2021681, EPI_ISL_2021685, EPI_ISL_2021686, EPI_ISL_2021687, EPI_ISL_2021688, EPI_ISL_2021691, EPI_ISL_2021695, EPI_ISL_2021696, EPI_ISL_2021700, EPI_ISL_2021702, EPI_ISL_2021705, EPI_ISL_2021706, EPI_ISL_2021709, EPI_ISL_2021716, EPI_ISL_2021718, EPI_ISL_2021720, EPI_ISL_2021723, EPI_ISL_2021724, EPI_ISL_2021730, EPI_ISL_2021731, EPI_ISL_2021732, EPI_ISL_2021738, EPI_ISL_2021741, EPI_ISL_2021744, EPI_ISL_2021753, EPI_ISL_2021761, EPI_ISL_2021762, EPI_ISL_2021766, EPI_ISL_2021772, EPI_ISL_2021781, EPI_ISL_2021787, EPI_ISL_2021790, EPI_ISL_2021795, EPI_ISL_2021796, EPI_ISL_2021799, EPI_ISL_2021800, EPI_ISL_2021804, EPI_ISL_2021810, EPI_ISL_2021817, EPI_ISL_2021821, EPI_ISL_2021824, EPI_ISL_2021827, EPI_ISL_2021832, EPI_ISL_2021833, EPI_ISL_2021834, EPI_ISL_2021836, EPI_ISL_2021838, EPI_ISL_2021839, EPI_ISL_2021840, EPI_ISL_2021841, EPI_ISL_2021844, EPI_ISL_2021845, EPI_ISL_2021846, EPI_ISL_2021848, EPI_ISL_2021860, EPI_ISL_2021863, EPI_ISL_2021899, EPI_ISL_2021906, EPI_ISL_2021920, EPI_ISL_2021936, EPI_ISL_2021942, EPI_ISL_2021945, EPI_ISL_2021964, EPI_ISL_2021968, EPI_ISL_2021987, EPI_ISL_2021997, EPI_ISL_2022001, EPI_ISL_2022004, EPI_ISL_2022007, EPI_ISL_2022056, EPI_ISL_2022057, EPI_ISL_2022058, EPI_ISL_2022067, EPI_ISL_2022088, EPI_ISL_2022117, EPI_ISL_2022118, EPI_ISL_2022136, EPI_ISL_2022144, EPI_ISL_2022445, EPI_ISL_2022452, EPI_ISL_2022494, EPI_ISL_2080477, EPI_ISL_2091469, EPI_ISL_2091477, EPI_ISL_2091481, EPI_ISL_2091487, EPI_ISL_2091492, EPI_ISL_2091494, EPI_ISL_2091495, EPI_ISL_2091497, EPI_ISL_2091502, EPI_ISL_2091510, EPI_ISL_2091514, EPI_ISL_2091518, EPI_ISL_2091521, EPI_ISL_2091522, EPI_ISL_2091524, EPI_ISL_2091526, EPI_ISL_2091528, EPI_ISL_2091532, EPI_ISL_2091533, EPI_ISL_2091534, EPI_ISL_2091540, EPI_ISL_2091542, EPI_ISL_2091549, EPI_ISL_2091553, EPI_ISL_2091554, EPI_ISL_2091557, EPI_ISL_2091558, EPI_ISL_2091582, EPI_ISL_2091583, EPI_ISL_2091586, EPI_ISL_2091590, EPI_ISL_2091603, EPI_ISL_2091605, EPI_ISL_2091614, EPI_ISL_2091622, EPI_ISL_2091625, EPI_ISL_2091627, EPI_ISL_2091630, EPI_ISL_2091633, EPI_ISL_2091636, EPI_ISL_2091639, EPI_ISL_2091642, EPI_ISL_2091643, EPI_ISL_2091651, EPI_ISL_2091659, EPI_ISL_2091660, EPI_ISL_2091665, EPI_ISL_2091666, EPI_ISL_2091671, EPI_ISL_2091678, EPI_ISL_2091681, EPI_ISL_2091691, EPI_ISL_2091699, EPI_ISL_2091703, EPI_ISL_2091713, EPI_ISL_2091719, EPI_ISL_2091728, EPI_ISL_2091736, EPI_ISL_2091738, EPI_ISL_2091740, EPI_ISL_2091743, EPI_ISL_2091746, EPI_ISL_2091750, EPI_ISL_2091757, EPI_ISL_2091761, EPI_ISL_2091764, EPI_ISL_2091785, EPI_ISL_2091793, EPI_ISL_2119009, EPI_ISL_2121029, EPI_ISL_2121052, EPI_ISL_2121153, EPI_ISL_2121516, EPI_ISL_2138175, EPI_ISL_2138192, EPI_ISL_2138280, EPI_ISL_2138301, EPI_ISL_2172099, EPI_ISL_2172101, EPI_ISL_2172102, EPI_ISL_2172103, EPI_ISL_2172104, EPI_ISL_2172105, EPI_ISL_2172110, EPI_ISL_2172111, EPI_ISL_2172115, EPI_ISL_2172117, EPI_ISL_2172118, EPI_ISL_2172120, EPI_ISL_2172121, EPI_ISL_2172127, EPI_ISL_2172132, EPI_ISL_2172133, EPI_ISL_2172142, EPI_ISL_2172143, EPI_ISL_2172144, EPI_ISL_2172145, EPI_ISL_2172146, EPI_ISL_2172147, EPI_ISL_2172149, EPI_ISL_2172150, EPI_ISL_2172155, EPI_ISL_2172156, EPI_ISL_2172157, EPI_ISL_2172160, EPI_ISL_2172162, EPI_ISL_2172163, EPI_ISL_2172165, EPI_ISL_2172166, EPI_ISL_2172167, EPI_ISL_2172174, EPI_ISL_2172175, EPI_ISL_2172178, EPI_ISL_2172181, EPI_ISL_2172185, EPI_ISL_2172191, EPI_ISL_2172192, EPI_ISL_2172194, EPI_ISL_2172195, EPI_ISL_2172196, EPI_ISL_2172197, EPI_ISL_2172201, EPI_ISL_2172203, EPI_ISL_2172204, EPI_ISL_2172206, EPI_ISL_2172209, EPI_ISL_2172210, EPI_ISL_2172211, EPI_ISL_2172212, EPI_ISL_2172217, EPI_ISL_2172218, EPI_ISL_2172220, EPI_ISL_2172222, EPI_ISL_2172225, EPI_ISL_2172226, EPI_ISL_2172227, EPI_ISL_2172228,

| | | | |
|---|---|---|---|
| EPI_ISL_2172230, EPI_ISL_2172232, EPI_ISL_2172237, EPI_ISL_2172244, EPI_ISL_2172245, EPI_ISL_2172246, EPI_ISL_2172247, EPI_ISL_2172249, EPI_ISL_2172250, EPI_ISL_2172252, EPI_ISL_2172253, EPI_ISL_2172254, EPI_ISL_2172257, EPI_ISL_2172259, EPI_ISL_2172262, EPI_ISL_2172263, EPI_ISL_2172266, EPI_ISL_2172273, EPI_ISL_2172283, EPI_ISL_2172284, EPI_ISL_2172288, EPI_ISL_2172291, EPI_ISL_2172292, EPI_ISL_2172297, EPI_ISL_2172305, EPI_ISL_2172306, EPI_ISL_2172307, EPI_ISL_2172308, EPI_ISL_2172309, EPI_ISL_2172310, EPI_ISL_2172313, EPI_ISL_2172315, EPI_ISL_2172318, EPI_ISL_2172319, EPI_ISL_2172322, EPI_ISL_2172323, EPI_ISL_2172325, EPI_ISL_2172330, EPI_ISL_2172331, EPI_ISL_2172332, EPI_ISL_2172334, EPI_ISL_2172336, EPI_ISL_2172338, EPI_ISL_2172339, EPI_ISL_2172342, EPI_ISL_2172343, EPI_ISL_2172346, EPI_ISL_2172347, EPI_ISL_2172348, EPI_ISL_2172352, EPI_ISL_2172354, EPI_ISL_2172355, EPI_ISL_2172357, EPI_ISL_2172358, EPI_ISL_2172360, EPI_ISL_2172365, EPI_ISL_2172368, EPI_ISL_2172369, EPI_ISL_2172372, EPI_ISL_2172373, EPI_ISL_2172375, EPI_ISL_2172377, EPI_ISL_2172378, EPI_ISL_2172379, EPI_ISL_2172382, EPI_ISL_2172384, EPI_ISL_2172385, EPI_ISL_2172390, EPI_ISL_2172391, EPI_ISL_2172395, EPI_ISL_2172397, EPI_ISL_2172398, EPI_ISL_2172399, EPI_ISL_2172400, EPI_ISL_2172402 | | | |
| see above | Lighthouse Lab in Milton Keynes | Wellcome Sanger Institute for the COVID-19 Genomics UK (COG-UK) Consortium | Cordelia Langford; David K. Jackson; Dominic Kwiatkowski; Ewan Harrison; Ian Johnston; Jeffrey Barrett; John Sillitoe on behalf of the Wellcome Sanger Institute COVID-19 Surveillance Team; Roberto Amato; Sonia Goncalves; The Lighthouse Lab in Milton Keynes and Alex Alderton |
| EPI_ISL_1974983, EPI_ISL_1974984, EPI_ISL_1974994, EPI_ISL_1974996, EPI_ISL_1974998, EPI_ISL_1974999, EPI_ISL_1975001, EPI_ISL_1975003, EPI_ISL_1975004, EPI_ISL_1975006, EPI_ISL_1975008, EPI_ISL_1975009, EPI_ISL_1975015, EPI_ISL_1975022, EPI_ISL_1975023, EPI_ISL_1975025, EPI_ISL_1975026, EPI_ISL_1975028, EPI_ISL_1975030, EPI_ISL_1975031, EPI_ISL_1975033, EPI_ISL_1975034, EPI_ISL_1975036, EPI_ISL_1975038, EPI_ISL_1975039, EPI_ISL_2125944, EPI_ISL_2125946, EPI_ISL_2125948, EPI_ISL_2125950, EPI_ISL_2125952, EPI_ISL_2125957, EPI_ISL_2125960, EPI_ISL_2125962, EPI_ISL_2125964, EPI_ISL_2125966, EPI_ISL_2125968, EPI_ISL_2125969, EPI_ISL_2125977, EPI_ISL_2125978, EPI_ISL_2125982, EPI_ISL_2125984, EPI_ISL_2125986 | | | |
| see above | Liverpool Clinical Laboratories | COVID-19 Genomics UK (COG-UK) Consortium | A Alrezaihi; Alessandro Gerada; Alistair Darby; Angela Cowell; Anita Lucaci; Anu Chawla; Cassie Olateju; Catherine Hartley; Charlotte Nelson; Ecaterina Vamos; Elaine O'Toole; Eleanor G Bentley; Ghada T Shawli; Hermione Webster; Isabel García-Dorival; James Johnson; James P Stewart; Jenifer Manson; Joanne Watts; Jones Benjamin; Jordan J Clark; Julian Hiscox; L Luu; Lucille Rainbow; M Almsaud; Margaret Hughes; Mark Whitehead; Matthew Gemmell; Miren Iturriza-Gomara; Muhannad Alruwaili; N.P Randle; Neil Swainston; PKF Gilmore; Parul Sharma; Rebekah Penrice-Randal; Richard Eccles; Richard Gregory; Sam Haldenby; Steve Paterson; Stuart D Armstrong; Trevor Ian Robinson; Ximeng Han |
| EPI_ISL_1832716, EPI_ISL_1832718, EPI_ISL_1832720, EPI_ISL_1832722, EPI_ISL_1978360, EPI_ISL_1978363, EPI_ISL_1978431, EPI_ISL_1978432, EPI_ISL_1978628, EPI_ISL_1978631, EPI_ISL_1978672, EPI_ISL_1978674, EPI_ISL_1978675, EPI_ISL_1978677, EPI_ISL_1978679, EPI_ISL_1978680, EPI_ISL_1978684, EPI_ISL_1978687, EPI_ISL_1978688, EPI_ISL_1978690, EPI_ISL_1978692, EPI_ISL_1978693, EPI_ISL_1978695, EPI_ISL_1978697, EPI_ISL_1978698, EPI_ISL_1978700, EPI_ISL_1978702, EPI_ISL_1978703, EPI_ISL_1978705, EPI_ISL_1978708, EPI_ISL_1978710, EPI_ISL_1978711, EPI_ISL_1978713, EPI_ISL_1978715, EPI_ISL_1978717, EPI_ISL_1978718, EPI_ISL_1978719, EPI_ISL_1978720, EPI_ISL_1978721, EPI_ISL_1978723, EPI_ISL_1978724, EPI_ISL_1978726, EPI_ISL_1978728, EPI_ISL_1978729, EPI_ISL_1978733, EPI_ISL_1978734, EPI_ISL_1978889, EPI_ISL_2126188, EPI_ISL_2126190, EPI_ISL_2126191, EPI_ISL_2126193, EPI_ISL_2126195, EPI_ISL_2126197, EPI_ISL_2126199, EPI_ISL_2126201, EPI_ISL_2126203, EPI_ISL_2126205, EPI_ISL_2126207, EPI_ISL_2126209, EPI_ISL_2126211, EPI_ISL_2126212, EPI_ISL_2126214, EPI_ISL_2126216, EPI_ISL_2126218, EPI_ISL_2126220, EPI_ISL_2126222, EPI_ISL_2126224, EPI_ISL_2126226, EPI_ISL_2126228, EPI_ISL_2126230, EPI_ISL_2126232, EPI_ISL_2126233, EPI_ISL_2126235, EPI_ISL_2126237, EPI_ISL_2126240, EPI_ISL_2126242, EPI_ISL_2126244, EPI_ISL_2126246, EPI_ISL_2126248, EPI_ISL_2126250, EPI_ISL_2126251, EPI_ISL_2126255, EPI_ISL_2126258, EPI_ISL_2126260, EPI_ISL_2126262, EPI_ISL_2126263, EPI_ISL_2126265, EPI_ISL_2126267, EPI_ISL_2126269, EPI_ISL_2126270, EPI_ISL_2126272, EPI_ISL_2126274, EPI_ISL_2126276, EPI_ISL_2126278, EPI_ISL_2126280, EPI_ISL_2126282, EPI_ISL_2126283, EPI_ISL_2126285, EPI_ISL_2126287, EPI_ISL_2126289, EPI_ISL_2126291, EPI_ISL_2126292, EPI_ISL_2126294, EPI_ISL_2126296, EPI_ISL_2126298, EPI_ISL_2126300, EPI_ISL_2126302, EPI_ISL_2126303, EPI_ISL_2126305, EPI_ISL_2126307, EPI_ISL_2126309, EPI_ISL_2126311, EPI_ISL_2126314, EPI_ISL_2126316, EPI_ISL_2126317, EPI_ISL_2126319, EPI_ISL_2126321, EPI_ISL_2126323, EPI_ISL_2126325, EPI_ISL_2126326, EPI_ISL_2126327, EPI_ISL_2126329, EPI_ISL_2126331, EPI_ISL_2126333, EPI_ISL_2126334, EPI_ISL_2126336, EPI_ISL_2126338, EPI_ISL_2126340, EPI_ISL_2126342, EPI_ISL_2126344, EPI_ISL_2126345, EPI_ISL_2126347, EPI_ISL_2126348, EPI_ISL_2126349, EPI_ISL_2126350, EPI_ISL_2126351, EPI_ISL_2126353, EPI_ISL_2126355, EPI_ISL_2126356, EPI_ISL_2126358, EPI_ISL_2126360, EPI_ISL_2126361, EPI_ISL_2126363, EPI_ISL_2126365, EPI_ISL_2126367, EPI_ISL_2126369, EPI_ISL_2126370, EPI_ISL_2126372, EPI_ISL_2126374, EPI_ISL_2126376, EPI_ISL_2126380, EPI_ISL_2126381, EPI_ISL_2126383, EPI_ISL_2126384, EPI_ISL_2126389, EPI_ISL_2126391, EPI_ISL_2126393, EPI_ISL_2126395, EPI_ISL_2126397, EPI_ISL_2126398, EPI_ISL_2126400, EPI_ISL_2126402, EPI_ISL_2126403, EPI_ISL_2126405, EPI_ISL_2126407, EPI_ISL_2126409, EPI_ISL_2126411, EPI_ISL_2126412, EPI_ISL_2126414, EPI_ISL_2126416, EPI_ISL_2126418, EPI_ISL_2126419, EPI_ISL_2126421, EPI_ISL_2126423, EPI_ISL_2126425, EPI_ISL_2126427, EPI_ISL_2126429, EPI_ISL_2126431, EPI_ISL_2126433, EPI_ISL_2126435, EPI_ISL_2126437, EPI_ISL_2126439, EPI_ISL_2126440, EPI_ISL_2126442, EPI_ISL_2126444, EPI_ISL_2126446, EPI_ISL_2126448, EPI_ISL_2126450, EPI_ISL_2126452, EPI_ISL_2126454, EPI_ISL_2126456, EPI_ISL_2126458, EPI_ISL_2126460, EPI_ISL_2126462, EPI_ISL_2126464, EPI_ISL_2126466, EPI_ISL_2126467, EPI_ISL_2126469, EPI_ISL_2126471, EPI_ISL_2126473, EPI_ISL_2126474, EPI_ISL_2126477, EPI_ISL_2126478, EPI_ISL_2126480, EPI_ISL_2126482, EPI_ISL_2126484, EPI_ISL_2126486, EPI_ISL_2126488, EPI_ISL_2126490, EPI_ISL_2126491, EPI_ISL_2126493, EPI_ISL_2126495, EPI_ISL_2126497, EPI_ISL_2126499, EPI_ISL_2126501, EPI_ISL_2126502, EPI_ISL_2126504, EPI_ISL_2126508, EPI_ISL_2126510, EPI_ISL_2126511, EPI_ISL_2126513, EPI_ISL_2126515, EPI_ISL_2126517, EPI_ISL_2126519, EPI_ISL_2126520, EPI_ISL_2126522, EPI_ISL_2126524, EPI_ISL_2126525, EPI_ISL_2126527, EPI_ISL_2126530, EPI_ISL_2126532, EPI_ISL_2126534, EPI_ISL_2126535, EPI_ISL_2126537, EPI_ISL_2126539, EPI_ISL_2126541, EPI_ISL_2126544, EPI_ISL_2126546, EPI_ISL_2126548, EPI_ISL_2126550, EPI_ISL_2126551, EPI_ISL_2126553, EPI_ISL_2126555, EPI_ISL_2126557, EPI_ISL_2126559, EPI_ISL_2126560, EPI_ISL_2126562, EPI_ISL_2126564, EPI_ISL_2126567, EPI_ISL_2126569, EPI_ISL_2126570, EPI_ISL_2126572, EPI_ISL_2126574, EPI_ISL_2126576, EPI_ISL_2126578, EPI_ISL_2126580, EPI_ISL_2126581, EPI_ISL_2126582, EPI_ISL_2126584, EPI_ISL_2126586, EPI_ISL_2126588, EPI_ISL_2126590, EPI_ISL_2126591, EPI_ISL_2126592, EPI_ISL_2126594, EPI_ISL_2126596, EPI_ISL_2126598, EPI_ISL_2126600, EPI_ISL_2126602, EPI_ISL_2126603, EPI_ISL_2126605, EPI_ISL_2126607, EPI_ISL_2126609, EPI_ISL_2126610, EPI_ISL_2126611, EPI_ISL_2126613, EPI_ISL_2126615, EPI_ISL_2126617, EPI_ISL_2126620, EPI_ISL_2126622, EPI_ISL_2126624, EPI_ISL_2126626, EPI_ISL_2126628, EPI_ISL_2126630, EPI_ISL_2126631, EPI_ISL_2126634, EPI_ISL_2126635, EPI_ISL_2126637, EPI_ISL_2126641, EPI_ISL_2126643, EPI_ISL_2126646, EPI_ISL_2126648, EPI_ISL_2126650, EPI_ISL_2126652, EPI_ISL_2126654, EPI_ISL_2126656, EPI_ISL_2126657, EPI_ISL_2126659, EPI_ISL_2126660, EPI_ISL_2126662, EPI_ISL_2126664, EPI_ISL_2126722, EPI_ISL_2126724, EPI_ISL_2126732, EPI_ISL_2126733, EPI_ISL_2126735, EPI_ISL_2126737, EPI_ISL_2126739, EPI_ISL_2126742, EPI_ISL_2126744, EPI_ISL_2126745, EPI_ISL_2126747, EPI_ISL_2126749, EPI_ISL_2126751, EPI_ISL_2126753, EPI_ISL_2126757, EPI_ISL_2126759, EPI_ISL_2126760, EPI_ISL_2126762, EPI_ISL_2126764, EPI_ISL_2126766, EPI_ISL_2126768, EPI_ISL_2126769, EPI_ISL_2126771, EPI_ISL_2126773, EPI_ISL_2126775, EPI_ISL_2126777, EPI_ISL_2126779, EPI_ISL_2126781, EPI_ISL_2126783, EPI_ISL_2126784, EPI_ISL_2126786, EPI_ISL_2126788, EPI_ISL_2126790, EPI_ISL_2126792, EPI_ISL_2126798, EPI_ISL_2127594, EPI_ISL_2127595, EPI_ISL_2127601, EPI_ISL_2127602 | | | |
| see above | Northumbria University / South Tees Hospitals NHS Foundation Trust / North Cumbria Integrated Care NHS Foundation Trust / North Tees and Hartlepool NHS Foundation Trust / Newcastle Hospitals NHS Foundation Trust | COVID-19 Genomics UK (COG-UK) Consortium | Andrew Nelson; Brendan Payne; Clive Graham; Darren L Smith; Debra Padgett; Edward Barton; Emma Swindells; Garren Scott; Gary Black; Gary Eltringham; Giles S Holt; Greg R Young; Jane Greenaway; Jennifer Collins; John Allan; Joshua Loh; Lynn Dover; Matthew Bashton; Mohammad A Tariq; Paul Baker; Sarah Essex; Steve Liggett; Wen C Yew; Yusri Taha |
| EPI_ISL_1979228, EPI_ISL_1979230, EPI_ISL_1979231, EPI_ISL_1979233, EPI_ISL_1979234, EPI_ISL_1979236, EPI_ISL_1979237, EPI_ISL_1979239, EPI_ISL_1979241, EPI_ISL_1979242, EPI_ISL_1979244, EPI_ISL_1979246, EPI_ISL_1979247, EPI_ISL_1979249, EPI_ISL_1979251, EPI_ISL_1979252, EPI_ISL_1979254, EPI_ISL_1979255, EPI_ISL_1979257, EPI_ISL_1979258, EPI_ISL_1979259, EPI_ISL_1979261, EPI_ISL_1979263, EPI_ISL_1979264, EPI_ISL_1979266, EPI_ISL_1979267, EPI_ISL_1979269, EPI_ISL_1979270, EPI_ISL_1979274, EPI_ISL_1979276, EPI_ISL_1979278, EPI_ISL_1979280, EPI_ISL_1979281, EPI_ISL_1979284, EPI_ISL_1979285, EPI_ISL_1979287, EPI_ISL_1979289, EPI_ISL_1979290, EPI_ISL_1979292, EPI_ISL_1979293, EPI_ISL_1979295, EPI_ISL_1979296, EPI_ISL_1979298, EPI_ISL_1979300, EPI_ISL_1979301, EPI_ISL_1979303, EPI_ISL_1979309, EPI_ISL_1979312, EPI_ISL_1979320, EPI_ISL_1979322, EPI_ISL_1979325, EPI_ISL_1979328, EPI_ISL_1979329, EPI_ISL_1979331, EPI_ISL_1979333, EPI_ISL_1979336, EPI_ISL_1979339, EPI_ISL_1979356, EPI_ISL_1979359, EPI_ISL_1979362, EPI_ISL_1979366, EPI_ISL_1979368, EPI_ISL_1979369, EPI_ISL_1979371, EPI_ISL_1979372, EPI_ISL_1979374, EPI_ISL_1979375, EPI_ISL_1979377, EPI_ISL_1979378, EPI_ISL_1979380, EPI_ISL_1979383, EPI_ISL_1979384, EPI_ISL_1979386, EPI_ISL_1979387, EPI_ISL_1979389, EPI_ISL_1979390, EPI_ISL_1979391, EPI_ISL_1979392, EPI_ISL_1979393, EPI_ISL_1979394, EPI_ISL_1979395, EPI_ISL_1979396, EPI_ISL_1979398, EPI_ISL_1979400, EPI_ISL_1979401, EPI_ISL_1979403, EPI_ISL_1979405, EPI_ISL_1979406, EPI_ISL_1979408, EPI_ISL_1979412, EPI_ISL_1979413, EPI_ISL_1979416, EPI_ISL_1979418, EPI_ISL_1979420, EPI_ISL_1979421, EPI_ISL_1979423, EPI_ISL_1979424, EPI_ISL_1979426, EPI_ISL_1979428, EPI_ISL_1979430, EPI_ISL_1979431, EPI_ISL_1979433, EPI_ISL_1979434, EPI_ISL_1979436, EPI_ISL_1979437, EPI_ISL_1979439, EPI_ISL_1979441, EPI_ISL_1979442, EPI_ISL_1979444, EPI_ISL_1979445, EPI_ISL_1979448, EPI_ISL_1979449, EPI_ISL_1979450, EPI_ISL_1979452, EPI_ISL_1979453, EPI_ISL_1979455, EPI_ISL_1979456, EPI_ISL_1979458, EPI_ISL_1979460, EPI_ISL_1979467, EPI_ISL_1979469, EPI_ISL_1979473, EPI_ISL_1979477, EPI_ISL_1979480, EPI_ISL_1979483, EPI_ISL_1979484, EPI_ISL_1979485, EPI_ISL_1979487, EPI_ISL_1979490, EPI_ISL_1979492, EPI_ISL_1979495, EPI_ISL_1979506, EPI_ISL_1979507, EPI_ISL_1979509, EPI_ISL_1979510, EPI_ISL_1979525, EPI_ISL_1979526, EPI_ISL_1979528, EPI_ISL_1979529, EPI_ISL_1979531, EPI_ISL_1979532, EPI_ISL_1979534, EPI_ISL_1979536, EPI_ISL_1979537, EPI_ISL_1979539, EPI_ISL_1979541, EPI_ISL_1979542, EPI_ISL_1979544, EPI_ISL_1979545, EPI_ISL_1979547, EPI_ISL_1979548, EPI_ISL_1979550, EPI_ISL_1979552, EPI_ISL_1979553, EPI_ISL_1979555, EPI_ISL_1979556, EPI_ISL_1979558, EPI_ISL_1979559, EPI_ISL_1979561, EPI_ISL_1979562, EPI_ISL_1979564, EPI_ISL_1979565, EPI_ISL_1979567, EPI_ISL_1979570, EPI_ISL_1979573, EPI_ISL_1979575, EPI_ISL_1979577, EPI_ISL_1979578, EPI_ISL_1979580, EPI_ISL_1979581, EPI_ISL_1979583, EPI_ISL_1979585, EPI_ISL_1979586, EPI_ISL_1979588, EPI_ISL_1979590, EPI_ISL_1979591, EPI_ISL_1979593, EPI_ISL_1979594, EPI_ISL_1979596, EPI_ISL_1979597, EPI_ISL_1979599, EPI_ISL_1979604, EPI_ISL_1979607, EPI_ISL_1979609, EPI_ISL_1979613, EPI_ISL_1979615, EPI_ISL_1979621, EPI_ISL_1979627, EPI_ISL_1979629, EPI_ISL_1979630, EPI_ISL_1979638, EPI_ISL_1979642, EPI_ISL_1979645, EPI_ISL_1979651, EPI_ISL_1979654, EPI_ISL_1979657, EPI_ISL_1979664, EPI_ISL_1979667, EPI_ISL_1979671, EPI_ISL_1979675, EPI_ISL_1979678, EPI_ISL_1979680, EPI_ISL_1979682, EPI_ISL_1979683, EPI_ISL_1979686, EPI_ISL_1979688, EPI_ISL_1979690, EPI_ISL_1979691, EPI_ISL_1979695, EPI_ISL_1979696, EPI_ISL_1979703, EPI_ISL_1979704, EPI_ISL_1979708, EPI_ISL_1979711, EPI_ISL_1979717, EPI_ISL_1979718, EPI_ISL_1979722, EPI_ISL_1979730, EPI_ISL_1979731, EPI_ISL_1979733, EPI_ISL_1979738, EPI_ISL_1979739 | | | |
| see above | Originating lab: Wales Specialist Virology Centre Sequencing lab: Pathogen Genomics Unit | Public Health Wales Microbiology Cardiff Wales Specialist Virology Centre | Alec Birchley; Alexander Adams; Amy Gaskin; Angela Marchbank; Bree Gatica-Wilcox; Catherine Moore; Jason Coombes; Joanne Watkins; Joel Southgate; Johnathan Evans; Laura Gifford; Lauren Gilbert; Lee Graham; Malorie Perry; Matthew Bull; Nicole Pacchiarini; Sally Corden; Sara Kumziene-Summerhayes; Sara Rey; Sarah Taylor; Simon Cottrell; Sophie Jones; Tom Connor |
| EPI_ISL_1832726, EPI_ISL_1974718, EPI_ISL_1978901, EPI_ISL_1978903, EPI_ISL_1978923, EPI_ISL_1978947, EPI_ISL_1978950 | | | |
| see above | Quadram Institute Bioscience | COVID-19 Genomics UK (COG-UK) Consortium | Alexander J Trotter; Alison E. Mather; Alp Aydin; Ana P. Tedim; Anastasia Kolyva; Andrew Bell; Andrew J. Page; Claire Stuart; Dave J. Baker; Gemma L. Kay; John Wain; Justin O'Grady; Leonardo de Oliveira Martins; Lizzie Meadows; Maria Diaz; Mark Webber; Muhammed Yasir; Nabil-Fareed Alikhan; Ngozi Elumogo; Nicholas M. Thomson; Rachael Stanley; Rachel Gilroy; Reenesh Prakash; Samir Dervisevic; Samuel Bloomfield; Steven Rudder; Thanh Le-Viet |
| EPI_ISL_1833023, EPI_ISL_1833024, EPI_ISL_1833033, EPI_ISL_1833034, EPI_ISL_1978961, EPI_ISL_1978962, EPI_ISL_1978963 | | | |
| see above | Queens Medical Centre, Clinical Microbiology Department / DeepSeq Nottingham | COVID-19 Genomics UK (COG-UK) Consortium | Christopher Moore; Emily Park; Fei Sang; Gemma Clark; Hannah Howson-Wells; Jack Hill; Jayasree Dey; Johnny Debebe; Jonathan Ball; Joseph Chappell; Manjinder Khakh; Matthew Carlisle; Matthew Loose; Michelle M Lister; Nadine Holmes; Patrick McClure; Theocharis Tsoleridis; Timothy Byaruhanga; Vicki M Fleming; Victoria Wright; Wendy Smith |
| EPI_ISL_1829803, EPI_ISL_1829814, EPI_ISL_1829820, EPI_ISL_1829827, EPI_ISL_1829829, EPI_ISL_1829852, EPI_ISL_1829861, EPI_ISL_1829894, EPI_ISL_1829911, EPI_ISL_1829931, EPI_ISL_1829968, EPI_ISL_1829971, EPI_ISL_1829980, EPI_ISL_1829992, EPI_ISL_1830017, EPI_ISL_1830059, EPI_ISL_1830060, EPI_ISL_1830073, EPI_ISL_1830086, EPI_ISL_1857593, EPI_ISL_1857595, EPI_ISL_1857603, EPI_ISL_1857615, EPI_ISL_1857616, EPI_ISL_1857630, EPI_ISL_1857634, EPI_ISL_1857635, EPI_ISL_1857636, EPI_ISL_1857639, EPI_ISL_1857640, EPI_ISL_1857641, EPI_ISL_1857643, EPI_ISL_1857649, EPI_ISL_1857650, EPI_ISL_1857655, EPI_ISL_1857661, EPI_ISL_1857664, EPI_ISL_1857665, EPI_ISL_1857673, EPI_ISL_1857674, EPI_ISL_1857680, EPI_ISL_1857695, EPI_ISL_1857699, EPI_ISL_1857700, EPI_ISL_1857703, EPI_ISL_1857717, EPI_ISL_1857721, EPI_ISL_1857722, EPI_ISL_1857728, EPI_ISL_1857729, EPI_ISL_1857756, EPI_ISL_1857757, EPI_ISL_1857758, EPI_ISL_1857760, EPI_ISL_1857766, EPI_ISL_1857767, EPI_ISL_1857770, EPI_ISL_1857774, EPI_ISL_1857779, EPI_ISL_1857786, EPI_ISL_1857790, EPI_ISL_1857794, EPI_ISL_1912639, EPI_ISL_1912690, EPI_ISL_1912759, EPI_ISL_1912760, EPI_ISL_1912761, EPI_ISL_1912762, EPI_ISL_1912765, EPI_ISL_1912768, EPI_ISL_1912769, EPI_ISL_1986592, EPI_ISL_1986613, EPI_ISL_1986655, EPI_ISL_1986661, EPI_ISL_1986688, EPI_ISL_1986695, EPI_ISL_1986757, EPI_ISL_1986759, EPI_ISL_1986832, EPI_ISL_1986867, EPI_ISL_1986870, EPI_ISL_1986943, EPI_ISL_1986947, EPI_ISL_1986949, EPI_ISL_1986956, EPI_ISL_1986959, EPI_ISL_1986979, EPI_ISL_1986983, EPI_ISL_1987003, EPI_ISL_1987022, EPI_ISL_1987026, EPI_ISL_1987035, EPI_ISL_1987044, EPI_ISL_1987047, EPI_ISL_1987057, EPI_ISL_1987127, EPI_ISL_1987133, EPI_ISL_1987134, EPI_ISL_1987143, EPI_ISL_1987147, EPI_ISL_1987150, EPI_ISL_1987160, EPI_ISL_1987170, EPI_ISL_1987174, EPI_ISL_1987178, EPI_ISL_1987182, EPI_ISL_1987183, EPI_ISL_1987190, EPI_ISL_1987195, EPI_ISL_1987210, EPI_ISL_1987216, EPI_ISL_1987220, EPI_ISL_1987242, EPI_ISL_1987243, EPI_ISL_1987249, EPI_ISL_1987256, EPI_ISL_1987259, EPI_ISL_1987260, EPI_ISL_1987282, EPI_ISL_1987283, EPI_ISL_1987284, EPI_ISL_1987291, EPI_ISL_1987296, EPI_ISL_1987322, EPI_ISL_1987329, EPI_ISL_1987335, EPI_ISL_1987336, EPI_ISL_1987343, EPI_ISL_1987351, EPI_ISL_1987353, EPI_ISL_1987358, EPI_ISL_1987365, EPI_ISL_1987367, EPI_ISL_2004350, EPI_ISL_2004448, EPI_ISL_2004546, EPI_ISL_2004595, EPI_ISL_2005162, EPI_ISL_2005187, EPI_ISL_2005192, EPI_ISL_2005816, EPI_ISL_2005823, EPI_ISL_2005828, EPI_ISL_2005837, EPI_ISL_2005838, EPI_ISL_2005839, EPI_ISL_2005845, EPI_ISL_2005850, EPI_ISL_2005852, EPI_ISL_2005858, EPI_ISL_2005863, EPI_ISL_2005866, EPI_ISL_2005868, EPI_ISL_2005877, EPI_ISL_2005886, EPI_ISL_2005890, EPI_ISL_2005898, EPI_ISL_2005901, EPI_ISL_2005902, EPI_ISL_2005905, EPI_ISL_2005906, EPI_ISL_2005911, EPI_ISL_2005913, EPI_ISL_2005919, EPI_ISL_2005924, EPI_ISL_2005926, EPI_ISL_2005939, EPI_ISL_2005941, EPI_ISL_2005946, EPI_ISL_2005947, EPI_ISL_2005953, EPI_ISL_2005955, EPI_ISL_2005958, EPI_ISL_2005962, EPI_ISL_2005964, EPI_ISL_2005984, EPI_ISL_2005985, EPI_ISL_2005988, EPI_ISL_2005989, EPI_ISL_2005992, EPI_ISL_2005993, EPI_ISL_2005996, EPI_ISL_2006002, EPI_ISL_2006007, EPI_ISL_2006008, EPI_ISL_2006010, EPI_ISL_2006012, EPI_ISL_2006018, EPI_ISL_2006021, EPI_ISL_2006024, EPI_ISL_2006034, EPI_ISL_2006035, EPI_ISL_2006039, EPI_ISL_2006040, EPI_ISL_2006044, EPI_ISL_2006053, EPI_ISL_2006062, EPI_ISL_2022459, EPI_ISL_2022464, EPI_ISL_2022543, EPI_ISL_2022546, EPI_ISL_2022578, EPI_ISL_2022585 | | | |
| see above | Randox Laboratories | Wellcome Sanger Institute for the COVID-19 Genomics UK (COG-UK) Consortium | Cordelia Langford; David K. Jackson; Dominic Kwiatkowski; Ewan Harrison; Ian Johnston; Jeffrey Barrett; John Sillitoe on behalf of the Wellcome Sanger Institute COVID-19 Surveillance Team; Randox Laboratories and Alex Alderton; Roberto Amato; Sonia Goncalves |
| EPI_ISL_1832588, EPI_ISL_1832596, EPI_ISL_1832599, EPI_ISL_1832608, EPI_ISL_1832610, EPI_ISL_1832619, EPI_ISL_1832625, EPI_ISL_1832627, EPI_ISL_1832629, EPI_ISL_1832631, EPI_ISL_1832636, EPI_ISL_1832638, EPI_ISL_1832681, EPI_ISL_1832684, EPI_ISL_1832686, EPI_ISL_1832688, EPI_ISL_1832690, EPI_ISL_1832692, EPI_ISL_1832694, EPI_ISL_1832696, EPI_ISL_1975116, EPI_ISL_1975120, EPI_ISL_1975121, EPI_ISL_1975124, EPI_ISL_1975126, EPI_ISL_1975127, EPI_ISL_1975128, EPI_ISL_1975136, EPI_ISL_1975138, EPI_ISL_1975139, EPI_ISL_1975141, EPI_ISL_1975142, EPI_ISL_1975144, EPI_ISL_1975145, EPI_ISL_1975147, EPI_ISL_1975149, EPI_ISL_1975151, EPI_ISL_1975152, EPI_ISL_1975153, EPI_ISL_1975155, EPI_ISL_1975157, EPI_ISL_1975158, EPI_ISL_1975160, EPI_ISL_1975162, EPI_ISL_1975163, EPI_ISL_1975165, EPI_ISL_1975166, EPI_ISL_1975168, EPI_ISL_1975170, EPI_ISL_1975172, EPI_ISL_1975174, EPI_ISL_1975175, EPI_ISL_1975177, EPI_ISL_2126144 | | | |
| see above | University College London, Great Ormond Street Hospital for Children NHS Foundation Trust, Imperial College Healthcare NHS Trust | COVID-19 Genomics UK (COG-UK) Consortium | Alison Holmes; Charlotte Williams; Helena Tutill; Jacqueline Findlay; James Price; Judith Breuer; Julianne Brown; Kathryn Harris; Leysa Forrest; Mark Kristiansen; Paola Niola; Paola Resende Silva; Patricia Dyal; Paul Randell; Rachel Williams; Samuel Weeks; Sergi Castellano; Sunando Roy; Tony Brooks; Yasmin Panchbhaya |
| EPI_ISL_2125896, EPI_ISL_2125898, EPI_ISL_2125899, EPI_ISL_2125901, EPI_ISL_2125903, EPI_ISL_2125905, EPI_ISL_2125921, EPI_ISL_2125927 | | | |
| see above | University Hospitals Of Leicester NHS Trust and DeepSeq Nottingham | COVID-19 Genomics UK (COG-UK) Consortium | Christopher Holmes; Christopher Moore; Fei Sang; Joeseph Chappell; Johnny Debebe; Jonathan Ball; Julian Tang; Karlie Fallon; Matthew Carlisle; Matthew Loose; Nadine Holmes; Patrick McClure; Paul Bird; Thomas Helmer; Victoria Wright |
| EPI_ISL_1832151, EPI_ISL_1832153, EPI_ISL_1832157, EPI_ISL_1832159, EPI_ISL_1974723, EPI_ISL_1974724, EPI_ISL_1974726, EPI_ISL_1974728, EPI_ISL_1974729, EPI_ISL_1974733, EPI_ISL_1974734, EPI_ISL_1974736, EPI_ISL_1974737, EPI_ISL_1974741, EPI_ISL_1974742, EPI_ISL_1974744, EPI_ISL_1974745, EPI_ISL_1974747, EPI_ISL_1974749, EPI_ISL_1974750, EPI_ISL_1974752, EPI_ISL_1974753, EPI_ISL_1974755, EPI_ISL_2125412, EPI_ISL_2125414, EPI_ISL_2125416, EPI_ISL_2125417, EPI_ISL_2125420, EPI_ISL_2125421, EPI_ISL_2125423, EPI_ISL_2125425 | | | |
| see above | University of Birmingham | COVID-19 Genomics UK (COG-UK) Consortium | Alex Richter; Andrew Bosworth; Andrew Bosworth. Queen Elizabeth Hospital: Anna Casey; Andrew D Beggs PHE Heartlands Lab: Husam Osman; Charlotte Poxon; Institute of Microbiology; Joanne Stockton; Josh Quick; Kasun Wanigasooriya; Mike Kidd; Nicholas Loman. University of Birmingham Testing Laboratory: Celina M Whalley; Oliver Pickles; Radoslaw Poplawski; Samuel Nicholls; University of Birmingham: Claire McMurray; Will Rowe |
| EPI_ISL_1832274, EPI_ISL_1832275, EPI_ISL_1974834, EPI_ISL_1974836, EPI_ISL_1974837, EPI_ISL_1974840, EPI_ISL_1974841, EPI_ISL_1974842, EPI_ISL_1974844, EPI_ISL_1974846, EPI_ISL_1974847, EPI_ISL_1974849, EPI_ISL_1974850, EPI_ISL_1974852, EPI_ISL_1974853, EPI_ISL_1974855, EPI_ISL_1974856, EPI_ISL_1974857, EPI_ISL_1974859, EPI_ISL_1974860, EPI_ISL_1974884, EPI_ISL_1974885, EPI_ISL_1974887, EPI_ISL_1974889, EPI_ISL_1974903, EPI_ISL_1974905, EPI_ISL_2125635, EPI_ISL_2125637, EPI_ISL_2125638, EPI_ISL_2125640, EPI_ISL_2125642, EPI_ISL_2125644, EPI_ISL_2125646, EPI_ISL_2125647, EPI_ISL_2125649, EPI_ISL_2125654, EPI_ISL_2125655, EPI_ISL_2125657, EPI_ISL_2125659, EPI_ISL_2125661, EPI_ISL_2125662, EPI_ISL_2125664, EPI_ISL_2125666, EPI_ISL_2125668, EPI_ISL_2125689, EPI_ISL_2125710, EPI_ISL_2125711, EPI_ISL_2125713, EPI_ISL_2125715, EPI_ISL_2125717, EPI_ISL_2125751, EPI_ISL_2125865, EPI_ISL_2125866, EPI_ISL_2125874, EPI_ISL_2125875, EPI_ISL_2125876, EPI_ISL_2125883 | | | |
| see above | Virology Department, Royal Infirmary of Edinburgh, NHS Lothian / School of Biological Sciences, University of Edinburgh | COVID-19 Genomics UK (COG-UK) Consortium | Colquhoun R; Cotton S; Dewar R; Gallagher A; Hill V; Jackson B; Maloney D; McCrone JT; McHugh M; O'Toole Á; Rambaut A; Rooke S; Scher E; Templeton K; Yu X |
| EPI_ISL_1833833, EPI_ISL_1833836, EPI_ISL_1833838, EPI_ISL_1833845, EPI_ISL_1833861, EPI_ISL_1833868, EPI_ISL_1833876, EPI_ISL_1833883, EPI_ISL_1833891, EPI_ISL_1833899, EPI_ISL_1833906, EPI_ISL_1980340, EPI_ISL_1980342, EPI_ISL_1980344, EPI_ISL_1980345, EPI_ISL_1980349, EPI_ISL_1980351, EPI_ISL_1980358, EPI_ISL_1980362, EPI_ISL_1980363, EPI_ISL_1980372, EPI_ISL_1980374, EPI_ISL_1980377, EPI_ISL_1980382, EPI_ISL_1980384, EPI_ISL_1980385, EPI_ISL_1980387, EPI_ISL_1980389, EPI_ISL_1980390, EPI_ISL_1980393, EPI_ISL_1980398, EPI_ISL_1980399, EPI_ISL_1980401, EPI_ISL_1980406, EPI_ISL_1980409, EPI_ISL_1980418, EPI_ISL_1980420, EPI_ISL_1980425, EPI_ISL_1980426, EPI_ISL_1980428, EPI_ISL_1980430, EPI_ISL_1980435, EPI_ISL_1980437, EPI_ISL_1980438, EPI_ISL_1980440, EPI_ISL_1980445, EPI_ISL_1980449, EPI_ISL_1980451, EPI_ISL_1980456, EPI_ISL_1980457, EPI_ISL_2128465, EPI_ISL_2128473, EPI_ISL_2128478, EPI_ISL_2128479, EPI_ISL_2128483, EPI_ISL_2128485, EPI_ISL_2128487, EPI_ISL_2128492, EPI_ISL_2128494, EPI_ISL_2128497, EPI_ISL_2128499, EPI_ISL_2128503 | | | |
| see above | Virology Department, Sheffield Teaching Hospitals NHS Foundation Trust/Department of Infection, Immunity and Cardiovascular Disease, The Medical School, University of Sheffield | COVID-19 Genomics UK (COG-UK) Consortium | Adri Angyal; Alex Keeley; Benjamin Lindsey; Cariad Evans; Danielle Groves; Dave Partridge; Katie Johnson; Laura Carrilero; Luke Green; Matthew Parker; Matthew Wyles; Mehmet Yavuz; Mohammad Raza; Nikki Smith; Paul Parsons; Rachel Tucker; Rebecca Brown; Thushan de Silva |
| EPI_ISL_2127876, EPI_ISL_2127878, EPI_ISL_2127882, EPI_ISL_2127885, EPI_ISL_2127888, EPI_ISL_2127889, EPI_ISL_2127892, EPI_ISL_2127893 | | | |
| see above | Wales Specialist Virology Centre Sequencing lab: Pathogen Genomics Unit | Public Health Wales Microbiology Cardiff Wales Specialist Virology Centre | Alec Birchley; Alexander Adams; Amy Gaskin; Angela Marchbank; Bree Gatica-Wilcox; Catherine Moore; Jason Coombes; Joanne Watkins; Joel Southgate; Johnathan Evans; Laura Gifford; Lauren Gilbert; Lee Graham; Malorie Perry; Matthew Bull; Nicole Pacchiarini; Sally Corden; Sara Kumziene-Summerhayes; Sara Rey; Sarah Taylor; Simon Cottrell; Sophie Jones; Tom Connor |

We gratefully acknowledge the following Authors from the Originating laboratories responsible for obtaining the specimens, as well as the Submitting laboratories where the genome data were generated and shared via GISAID, on which this research is based.

All Submitters of data may be contacted directly via www.gisaid.org

Authors are sorted alphabetically.

| Accession ID | Originating Laboratory | Submitting Laboratory | Authors |
|---|---|---|---|
| EPI_ISL_2686088, EPI_ISL_2686089, EPI_ISL_2686090, EPI_ISL_2686091, EPI_ISL_2686092, EPI_ISL_2686093 | Amazon Dx, AUV2 Laboratory | Amazon Dx, AUV2 Laboratory | Luke Meredith |
| EPI_ISL_2126010, EPI_ISL_2126011, EPI_ISL_2126013, EPI_ISL_2126024, EPI_ISL_2126026, EPI_ISL_2126029, EPI_ISL_2126031, EPI_ISL_2126033, EPI_ISL_2126035, EPI_ISL_2126062, EPI_ISL_2126115, EPI_ISL_2126117, EPI_ISL_2126119, EPI_ISL_2126121, EPI_ISL_2126123, EPI_ISL_2126125, EPI_ISL_2238912, EPI_ISL_2238913 | | | |
| see above | Barts Health NHS Trust | COVID-19 Genomics UK (COG-UK) Consortium | BROAD; Beatrix; CUTINO-MOGUEL; Claire; David; Dola; HARRINGTON; KELE; KULASEGARAN-SHYLINI; Maria-Teresa; OWOYEMI; Raghavendran |
| EPI_ISL_2128554, EPI_ISL_2128558, EPI_ISL_2128565, EPI_ISL_2128582, EPI_ISL_2128595, EPI_ISL_2128599 | Bioinformatics and Biostatistics Lab, Advanced Sequencing Facility | COVID-19 Genomics UK (COG-UK) Consortium | Aengus Stewart; Chelsea Sawyer; Harshil Patel; Jerome Nicod; Laura Cubitt; Margaret Crawford |
| EPI_ISL_1980114, EPI_ISL_1980116, EPI_ISL_1980118, EPI_ISL_1980119, EPI_ISL_1980121, EPI_ISL_1980122, EPI_ISL_1980215, EPI_ISL_2128178, EPI_ISL_2128180, EPI_ISL_2128182, EPI_ISL_2128225, EPI_ISL_2128227, EPI_ISL_2128229, EPI_ISL_2128231, EPI_ISL_2128233, EPI_ISL_2128234, EPI_ISL_2128370, EPI_ISL_2128372 | | | |
| see above | Centre for Enzyme Innovation, University of Portsmouth / Translational Research Laboratory, Portsmouth Hospitals NHS Trust | COVID-19 Genomics UK (COG-UK) Consortium | Angela Beckett; Christopher Fearn; Kate Cook; Katie Loveson; Salman Goudarzi; Samuel Robson; Scott Elliott; Sharon Glaysher |
| EPI_ISL_2240661, EPI_ISL_2240662, EPI_ISL_2240665, EPI_ISL_2240675 | Centre for Enzyme Innovation, University of Portsmouth / Translational Research Laboratory, Portsmouth Hospitals NHS Trust / Hampshire Hospitals NHS Foundation Trust / Bournemouth University / University Hospitals Dorset NHS Foundation Trust / Brighton and Sussex University Hospitals NHS Trust / Dartford and Gravesham NHS Trust / Isle of Wight NHS Trust / Maidstone and Tunbridge Wells NHS Trust / University Hospital Southampton NHS Foundation Trust | COVID-19 Genomics UK (COG-UK) Consortium | Adhyana Mahanama; Allyson Lloyd; Andrew Butt; Angela Beckett; Anibolina Castigador; Anna Mantzaouratou; Anoop Chauhan; Ben Macklin; Benjamin Cogger; Buddhini Samaraweera; Cassandra Malone; Christopher Fearn; Claire Thomas; Dorian Crudgington; Eleri Wilson-Davies; Emanuela Pelosi; Emily Macnaughton; Helen Umpleby; Helen Wheeler; Jacqui Prieto; Jonnie Seymour; Karen Withell; Kate Cook; Katie Loveson; Kelly Bicknell; Kevin Tucker; Kordo Saeed; Liz Sheridan; Magdalena Barrow; Matthew Harvey; Mohammed Hassan-Ibrahim; Nathan Moore; Nick Cortes; Rachael Jeremiah; Robert Impey; Salman Goudarzi; Samirakhon Raupova; Samuel Robson; Sarah Buchan; Sarah Jeremiah; Sarah Wyllie; Scott Elliott; Sharon Glaysher; Siona Silvieri; Stephen Aplin; Stephen Kidd; Thea Sass |
| EPI_ISL_1974763, EPI_ISL_1974768, EPI_ISL_1974778, EPI_ISL_1974781, EPI_ISL_1974784, EPI_ISL_1974796, EPI_ISL_2125456, EPI_ISL_2125458, EPI_ISL_2125459, EPI_ISL_2125461, EPI_ISL_2125463, EPI_ISL_2125465, EPI_ISL_2125467, EPI_ISL_2125469, EPI_ISL_2125471, EPI_ISL_2125472, EPI_ISL_2125474, EPI_ISL_2125476, EPI_ISL_2125484, EPI_ISL_2125486 | | | |
| see above | Department of Pathology, University of Cambridge | COVID-19 Genomics UK (COG-UK) Consortium | Aminu S. Jahun; Ian Goodfellow; Iliana Georgana; Martin D. Curran; Myra Hosmillo; Rhys Izuagbe; Surendra Parmar; William L. Hamilton; Yasmin Chaudhry |
| EPI_ISL_2022532, EPI_ISL_2022533, EPI_ISL_2022534, EPI_ISL_2022536, EPI_ISL_2022537, EPI_ISL_2022538, EPI_ISL_2022539, EPI_ISL_2022541, EPI_ISL_2022547, EPI_ISL_2022548, EPI_ISL_2022550, EPI_ISL_2022552, EPI_ISL_2022556, EPI_ISL_2022559, EPI_ISL_2022562, EPI_ISL_2022564, EPI_ISL_2022566, EPI_ISL_2022567, EPI_ISL_2022569, EPI_ISL_2022580, EPI_ISL_2022587, EPI_ISL_2022588, EPI_ISL_2022589, EPI_ISL_2022590, EPI_ISL_2080362, EPI_ISL_2080363, EPI_ISL_2080367, EPI_ISL_2080371, EPI_ISL_2080375, EPI_ISL_2080376, EPI_ISL_2080377, EPI_ISL_2080378, EPI_ISL_2080380, EPI_ISL_2080381, EPI_ISL_2080382, EPI_ISL_2080383, EPI_ISL_2080384, EPI_ISL_2080386, EPI_ISL_2080389, EPI_ISL_2080390, EPI_ISL_2080392, EPI_ISL_2080395, EPI_ISL_2080397, EPI_ISL_2080401, EPI_ISL_2080405, EPI_ISL_2080406, EPI_ISL_2080408, EPI_ISL_2080410, EPI_ISL_2080411, EPI_ISL_2080414, EPI_ISL_2080415, EPI_ISL_2080416, EPI_ISL_2080422, EPI_ISL_2080423, EPI_ISL_2080426, EPI_ISL_2080427, EPI_ISL_2080428, EPI_ISL_2080430, EPI_ISL_2080435, EPI_ISL_2080436, EPI_ISL_2080437, EPI_ISL_2080439, EPI_ISL_2080443, EPI_ISL_2080444, EPI_ISL_2080445, EPI_ISL_2080447, EPI_ISL_2080448, EPI_ISL_2080449, EPI_ISL_2080450, EPI_ISL_2080456, EPI_ISL_2080460, EPI_ISL_2080463, EPI_ISL_2080466, EPI_ISL_2080467, EPI_ISL_2080469, EPI_ISL_2080471, EPI_ISL_2080472, EPI_ISL_2080473, EPI_ISL_2138370, EPI_ISL_2138382, EPI_ISL_2138385, EPI_ISL_2138388, EPI_ISL_2138408, EPI_ISL_2138416, EPI_ISL_2138425, EPI_ISL_2138429, EPI_ISL_2138434, EPI_ISL_2138442, EPI_ISL_2138455, EPI_ISL_2138459, EPI_ISL_2138469, EPI_ISL_2138475, EPI_ISL_2138485, EPI_ISL_2138543, EPI_ISL_2138549, EPI_ISL_2138561, EPI_ISL_2138572, EPI_ISL_2138617, EPI_ISL_2138622, EPI_ISL_2138649, EPI_ISL_2138678, EPI_ISL_2138681, EPI_ISL_2138684, EPI_ISL_2138688, EPI_ISL_2138691 | | | |
| see above | Health Services Laboratories | Wellcome Sanger Institute for the COVID-19 Genomics UK (COG-UK) Consortium | Cordelia Langford; David K. Jackson; Dominic Kwiatkowski; Ewan Harrison; Health Services Laboratories and Alex Alderton; Ian Johnston; Jeffrey Barrett; John Sillitoe on behalf of the Wellcome Sanger Institute COVID-19 Surveillance Team; Roberto Amato; Sonia Goncalves |
| EPI_ISL_1921738, EPI_ISL_1985692, EPI_ISL_1985695, EPI_ISL_1985709, EPI_ISL_1985711, EPI_ISL_1985712, EPI_ISL_1985719, EPI_ISL_1985734, EPI_ISL_1985744, EPI_ISL_1985755, EPI_ISL_1985757, EPI_ISL_1985765, EPI_ISL_1985767, EPI_ISL_1985803, EPI_ISL_1985825, EPI_ISL_1985828, EPI_ISL_1985835, EPI_ISL_1985848, EPI_ISL_1985861, EPI_ISL_1985862, EPI_ISL_1985873, EPI_ISL_1985890, EPI_ISL_1985902, EPI_ISL_1985909, EPI_ISL_1985914, EPI_ISL_1985916, EPI_ISL_1985920, EPI_ISL_1985921, EPI_ISL_1985925, EPI_ISL_1985929, EPI_ISL_1985948, EPI_ISL_1985953, EPI_ISL_1985954, EPI_ISL_1985965, EPI_ISL_1985967, EPI_ISL_1985976, EPI_ISL_1985984, EPI_ISL_1985989, EPI_ISL_1986009, EPI_ISL_1986013, EPI_ISL_1986018, EPI_ISL_1986021, EPI_ISL_1986028, EPI_ISL_1986041, EPI_ISL_1986045, EPI_ISL_1986058, EPI_ISL_1986064, EPI_ISL_1986069, EPI_ISL_1986071, EPI_ISL_1986074, EPI_ISL_1986082, EPI_ISL_1986097, EPI_ISL_1986105, EPI_ISL_1986113, EPI_ISL_1986114, EPI_ISL_1986115, EPI_ISL_1986118, EPI_ISL_1986124, EPI_ISL_1986129, EPI_ISL_1986144, EPI_ISL_1986155, EPI_ISL_1986159, EPI_ISL_1986163, EPI_ISL_1986168, EPI_ISL_1986172, EPI_ISL_1986184, EPI_ISL_1986203, EPI_ISL_1986206, EPI_ISL_1986210, EPI_ISL_1986213, EPI_ISL_1986220, EPI_ISL_1986227, EPI_ISL_1986233, EPI_ISL_1986255, EPI_ISL_1986275, EPI_ISL_1986278, EPI_ISL_1986281, EPI_ISL_1986283, EPI_ISL_1986284, EPI_ISL_1986297, EPI_ISL_1986304, EPI_ISL_1986305, EPI_ISL_1986310, EPI_ISL_1986314, EPI_ISL_1986317, EPI_ISL_1986323, EPI_ISL_1986331, EPI_ISL_1986334, EPI_ISL_1986336, EPI_ISL_1986338, EPI_ISL_1986360, EPI_ISL_1986362, EPI_ISL_1986372, EPI_ISL_1986376, EPI_ISL_1986390, EPI_ISL_1986494, EPI_ISL_1986511, EPI_ISL_1986519, EPI_ISL_1986550, EPI_ISL_1986576, EPI_ISL_1986577, EPI_ISL_1986593, EPI_ISL_1986669, EPI_ISL_1986746, EPI_ISL_1986783, EPI_ISL_2004345, EPI_ISL_2004346, EPI_ISL_2004347, EPI_ISL_2004351, EPI_ISL_2004352, EPI_ISL_2004353, EPI_ISL_2004354, EPI_ISL_2004356, EPI_ISL_2004357, EPI_ISL_2004358, EPI_ISL_2004360, EPI_ISL_2004361, EPI_ISL_2004362, EPI_ISL_2004363, EPI_ISL_2004364, EPI_ISL_2004365, EPI_ISL_2004366, EPI_ISL_2004367, EPI_ISL_2004368, EPI_ISL_2004370, EPI_ISL_2004371, EPI_ISL_2004373, EPI_ISL_2004375, EPI_ISL_2004376, EPI_ISL_2004377, EPI_ISL_2004378, EPI_ISL_2004379, EPI_ISL_2004380, EPI_ISL_2004381, EPI_ISL_2004382, EPI_ISL_2004383, EPI_ISL_2004384, EPI_ISL_2004386, EPI_ISL_2004387, EPI_ISL_2004388, EPI_ISL_2004389, EPI_ISL_2004390, EPI_ISL_2004391, EPI_ISL_2004393, EPI_ISL_2004394, EPI_ISL_2004395, EPI_ISL_2004397, EPI_ISL_2004399, EPI_ISL_2004400, EPI_ISL_2004401, EPI_ISL_2004403, EPI_ISL_2004404, EPI_ISL_2004405, EPI_ISL_2004406, EPI_ISL_2004407, EPI_ISL_2004410, EPI_ISL_2004411, EPI_ISL_2004412, EPI_ISL_2004415, EPI_ISL_2004416, EPI_ISL_2004417, EPI_ISL_2004419, EPI_ISL_2004420, EPI_ISL_2004421, EPI_ISL_2004422, EPI_ISL_2004424, EPI_ISL_2004425, EPI_ISL_2004426, EPI_ISL_2004427, EPI_ISL_2004428, EPI_ISL_2004429, EPI_ISL_2004430, EPI_ISL_2004431, EPI_ISL_2004432, EPI_ISL_2004433, EPI_ISL_2004435, EPI_ISL_2004437, EPI_ISL_2004439, EPI_ISL_2004440, EPI_ISL_2004441, EPI_ISL_2004442, EPI_ISL_2004443, EPI_ISL_2004444, EPI_ISL_2004445, EPI_ISL_2004446, EPI_ISL_2004450, EPI_ISL_2004451, EPI_ISL_2004454, EPI_ISL_2004456, EPI_ISL_2004458, EPI_ISL_2004459, EPI_ISL_2004460, EPI_ISL_2004461, EPI_ISL_2004462, EPI_ISL_2004463, EPI_ISL_2004464, EPI_ISL_2004465, EPI_ISL_2004466, EPI_ISL_2004467, EPI_ISL_2004469, EPI_ISL_2004470, EPI_ISL_2004471, EPI_ISL_2004474, EPI_ISL_2004475, EPI_ISL_2004479, EPI_ISL_2004480, EPI_ISL_2004481, EPI_ISL_2004482, EPI_ISL_2004483, EPI_ISL_2004486, EPI_ISL_2004487, EPI_ISL_2004488, EPI_ISL_2004489, EPI_ISL_2004492, EPI_ISL_2004493, EPI_ISL_2004495, EPI_ISL_2004496, EPI_ISL_2004497, EPI_ISL_2004498, EPI_ISL_2004500, EPI_ISL_2004501, EPI_ISL_2004502, EPI_ISL_2004503, EPI_ISL_2004504, EPI_ISL_2004505, EPI_ISL_2004506, EPI_ISL_2004507, EPI_ISL_2004509, EPI_ISL_2004510, EPI_ISL_2004511, EPI_ISL_2004512, EPI_ISL_2004513, EPI_ISL_2004514, EPI_ISL_2004515, EPI_ISL_2004516, EPI_ISL_2004517, EPI_ISL_2004519, EPI_ISL_2004520, EPI_ISL_2004521, EPI_ISL_2004522, EPI_ISL_2004523, EPI_ISL_2004524, EPI_ISL_2004525, EPI_ISL_2004526, EPI_ISL_2004529, EPI_ISL_2004530, EPI_ISL_2004531, EPI_ISL_2004532, EPI_ISL_2004533, EPI_ISL_2004534, EPI_ISL_2004535, EPI_ISL_2004536, EPI_ISL_2004538, EPI_ISL_2004540, EPI_ISL_2004541, EPI_ISL_2004542, EPI_ISL_2004543, EPI_ISL_2004544, EPI_ISL_2004547, EPI_ISL_2004550, EPI_ISL_2004551, EPI_ISL_2004552, EPI_ISL_2004553, EPI_ISL_2004554, EPI_ISL_2004555, EPI_ISL_2004556, EPI_ISL_2004557, EPI_ISL_2004558, EPI_ISL_2004561, EPI_ISL_2004562, EPI_ISL_2004564, EPI_ISL_2004565, EPI_ISL_2004566, EPI_ISL_2004567, EPI_ISL_2004568, EPI_ISL_2004569, EPI_ISL_2004570, EPI_ISL_2004571, EPI_ISL_2004572, EPI_ISL_2004573, EPI_ISL_2004575, EPI_ISL_2004576, EPI_ISL_2004577, EPI_ISL_2004578, EPI_ISL_2004580, EPI_ISL_2004581, EPI_ISL_2004582, EPI_ISL_2004583, EPI_ISL_2004585, EPI_ISL_2004586, EPI_ISL_2004587, EPI_ISL_2004588, EPI_ISL_2004589, EPI_ISL_2004590, EPI_ISL_2004591, EPI_ISL_2004592, EPI_ISL_2004593, EPI_ISL_2004596, EPI_ISL_2004597, EPI_ISL_2004598, EPI_ISL_2004599, EPI_ISL_2004600, EPI_ISL_2004601, EPI_ISL_2004602, EPI_ISL_2004603, EPI_ISL_2004606, EPI_ISL_2004607, EPI_ISL_2004608, EPI_ISL_2004609, EPI_ISL_2004610, EPI_ISL_2004612, EPI_ISL_2004613, EPI_ISL_2004614, EPI_ISL_2004615, EPI_ISL_2004616, EPI_ISL_2004617, EPI_ISL_2004618, EPI_ISL_2004619, EPI_ISL_2004658, EPI_ISL_2004671, EPI_ISL_2004673, EPI_ISL_2004688, EPI_ISL_2004704, EPI_ISL_2004725, EPI_ISL_2004732, EPI_ISL_2004747, EPI_ISL_2004754, EPI_ISL_2004774, EPI_ISL_2004779, EPI_ISL_2004811, EPI_ISL_2004821, EPI_ISL_2004845, EPI_ISL_2005851, EPI_ISL_2021043, EPI_ISL_2021044, EPI_ISL_2021052, EPI_ISL_2021053, EPI_ISL_2021054, EPI_ISL_2021056, EPI_ISL_2021059, EPI_ISL_2021061, EPI_ISL_2021064, EPI_ISL_2021077, EPI_ISL_2021082, EPI_ISL_2021083, EPI_ISL_2021090, EPI_ISL_2021092, EPI_ISL_2021110, EPI_ISL_2021111, EPI_ISL_2021119, EPI_ISL_2021124, EPI_ISL_2021127, EPI_ISL_2021131, EPI_ISL_2021144, EPI_ISL_2021145, EPI_ISL_2021147, EPI_ISL_2021151, EPI_ISL_2021152, EPI_ISL_2021154, EPI_ISL_2021158, EPI_ISL_2021159, EPI_ISL_2021170, EPI_ISL_2021181, EPI_ISL_2021183, EPI_ISL_2021186, EPI_ISL_2021190, EPI_ISL_2021193, EPI_ISL_2021194, EPI_ISL_2021195, EPI_ISL_2021197, EPI_ISL_2021198, EPI_ISL_2021200, EPI_ISL_2021205, EPI_ISL_2021209, EPI_ISL_2021215, EPI_ISL_2021218, EPI_ISL_2021222, EPI_ISL_2021225, EPI_ISL_2021228, EPI_ISL_2021231, EPI_ISL_2021235, EPI_ISL_2021236, EPI_ISL_2021241, EPI_ISL_2021242, EPI_ISL_2021248, EPI_ISL_2021249, EPI_ISL_2021252, EPI_ISL_2021254, EPI_ISL_2021255, EPI_ISL_2021258, EPI_ISL_2021262, EPI_ISL_2021271, EPI_ISL_2021274, EPI_ISL_2021275, EPI_ISL_2021277, EPI_ISL_2021280, EPI_ISL_2021281, EPI_ISL_2021282, EPI_ISL_2021287, EPI_ISL_2021291, EPI_ISL_2021292, EPI_ISL_2021296, EPI_ISL_2021298, EPI_ISL_2021310, EPI_ISL_2021311, EPI_ISL_2021315, EPI_ISL_2021316, EPI_ISL_2021317, EPI_ISL_2021318, EPI_ISL_2021319, EPI_ISL_2021326, EPI_ISL_2021327, EPI_ISL_2021329, EPI_ISL_2021331, EPI_ISL_2021332, EPI_ISL_2021339, EPI_ISL_2021343, EPI_ISL_2021348, EPI_ISL_2021349, EPI_ISL_2021358, EPI_ISL_2021362, EPI_ISL_2021363, EPI_ISL_2021364, EPI_ISL_2021365, EPI_ISL_2021366, EPI_ISL_2021367, EPI_ISL_2021369, EPI_ISL_2021370, EPI_ISL_2021373, EPI_ISL_2021374, EPI_ISL_2021375, EPI_ISL_2021376, EPI_ISL_2021377, EPI_ISL_2021379, EPI_ISL_2021380, EPI_ISL_2021385, EPI_ISL_2021386, EPI_ISL_2021388, EPI_ISL_2021389, EPI_ISL_2021390, EPI_ISL_2021393, EPI_ISL_2021394, EPI_ISL_2021395, EPI_ISL_2021396, EPI_ISL_2021397, EPI_ISL_2021400, EPI_ISL_2021402, EPI_ISL_2021403, EPI_ISL_2021408, EPI_ISL_2021410, EPI_ISL_2021411, EPI_ISL_2021413, EPI_ISL_2021414, EPI_ISL_2021418, EPI_ISL_2021420, EPI_ISL_2021423, EPI_ISL_2021426, EPI_ISL_2021428, EPI_ISL_2021431, EPI_ISL_2021432, EPI_ISL_2021437, EPI_ISL_2021443, EPI_ISL_2021446, EPI_ISL_2021449, EPI_ISL_2021450, EPI_ISL_2021451, EPI_ISL_2021455, EPI_ISL_2021456, EPI_ISL_2021457, EPI_ISL_2021459, EPI_ISL_2021460, EPI_ISL_2021464, EPI_ISL_2021466, EPI_ISL_2021468, EPI_ISL_2021469, EPI_ISL_2021470, EPI_ISL_2021472, EPI_ISL_2021473, EPI_ISL_2021474, EPI_ISL_2021475, EPI_ISL_2021476, EPI_ISL_2021482, EPI_ISL_2021484, EPI_ISL_2021485, EPI_ISL_2021486, EPI_ISL_2021487, EPI_ISL_2021488, EPI_ISL_2021489, EPI_ISL_2021490, EPI_ISL_2021491, EPI_ISL_2021493, EPI_ISL_2021497, EPI_ISL_2021498, EPI_ISL_2021499, EPI_ISL_2021500, EPI_ISL_2021503, EPI_ISL_2021505, EPI_ISL_2021512, EPI_ISL_2021513, EPI_ISL_2021514, EPI_ISL_2021515, EPI_ISL_2021516, EPI_ISL_2021519, EPI_ISL_2021520, EPI_ISL_2021521, EPI_ISL_2021524, EPI_ISL_2021525, EPI_ISL_2021526, EPI_ISL_2021527, EPI_ISL_2021528, EPI_ISL_2021529, EPI_ISL_2021530, EPI_ISL_2021531, EPI_ISL_2021532, EPI_ISL_2021533, EPI_ISL_2021537, EPI_ISL_2021540, EPI_ISL_2021542, EPI_ISL_2021543, EPI_ISL_2021547, EPI_ISL_2021550, EPI_ISL_2021551, EPI_ISL_2021552, EPI_ISL_2021553, EPI_ISL_2021559, EPI_ISL_2021560, EPI_ISL_2021561, EPI_ISL_2021562, EPI_ISL_2021563, EPI_ISL_2021564, EPI_ISL_2021565, EPI_ISL_2021568, EPI_ISL_2021569, EPI_ISL_2021570, EPI_ISL_2021579, EPI_ISL_2021581, EPI_ISL_2021582, EPI_ISL_2021583, EPI_ISL_2021585, EPI_ISL_2021589, EPI_ISL_2021591, EPI_ISL_2021592, EPI_ISL_2021594, EPI_ISL_2021596, EPI_ISL_2021598, EPI_ISL_2021599, EPI_ISL_2021600, EPI_ISL_2021602, EPI_ISL_2021604, EPI_ISL_2021605, EPI_ISL_2021606, EPI_ISL_2021607, EPI_ISL_2021608, EPI_ISL_2021609, EPI_ISL_2021615, EPI_ISL_2021616, EPI_ISL_2021617, EPI_ISL_2021618, EPI_ISL_2021619, EPI_ISL_2021621, EPI_ISL_2021623, EPI_ISL_2021624, EPI_ISL_2021627, EPI_ISL_2021628, EPI_ISL_2021629, EPI_ISL_2021631, EPI_ISL_2021633, EPI_ISL_2021634, EPI_ISL_2021637, EPI_ISL_2021640, EPI_ISL_2021641, EPI_ISL_2021643, EPI_ISL_2021644, EPI_ISL_2021647, EPI_ISL_2021648, EPI_ISL_2021649, EPI_ISL_2021650, EPI_ISL_2021651, EPI_ISL_2021652, EPI_ISL_2021653, EPI_ISL_2021656, EPI_ISL_2021657, EPI_ISL_2021660, EPI_ISL_2021662, EPI_ISL_2021664, EPI_ISL_2021667, EPI_ISL_2021669, EPI_ISL_2021670, EPI_ISL_2021672, EPI_ISL_2021674, EPI_ISL_2021680, EPI_ISL_2021682, EPI_ISL_2021684, EPI_ISL_2021689, EPI_ISL_2021692, EPI_ISL_2021693, EPI_ISL_2021694, EPI_ISL_2021852, EPI_ISL_2021853, EPI_ISL_2021857, EPI_ISL_2021861, EPI_ISL_2021864, EPI_ISL_2021865, EPI_ISL_2021867, EPI_ISL_2021869, EPI_ISL_2021870, EPI_ISL_2021873, EPI_ISL_2021876, EPI_ISL_2021882, EPI_ISL_2021883, EPI_ISL_2021890, EPI_ISL_2021891, EPI_ISL_2021892, EPI_ISL_2021895, EPI_ISL_2021900, EPI_ISL_2021901, EPI_ISL_2021903, EPI_ISL_2021904, EPI_ISL_2021908, EPI_ISL_2021911, EPI_ISL_2021912, EPI_ISL_2021913, EPI_ISL_2021916, EPI_ISL_2021918, EPI_ISL_2021933, EPI_ISL_2021934, EPI_ISL_2021935, EPI_ISL_2021938, EPI_ISL_2021941, EPI_ISL_2021943, EPI_ISL_2021949, EPI_ISL_2021952, EPI_ISL_2021954, EPI_ISL_2021956, EPI_ISL_2021963, EPI_ISL_2021971, EPI_ISL_2021977, EPI_ISL_2021980, EPI_ISL_2021984, EPI_ISL_2021986, EPI_ISL_2021988, EPI_ISL_2021989, EPI_ISL_2021993, EPI_ISL_2022000, EPI_ISL_2022002, EPI_ISL_2022003, EPI_ISL_2022005, EPI_ISL_2022009, EPI_ISL_2022011, EPI_ISL_2022020, EPI_ISL_2022021, EPI_ISL_2022023, EPI_ISL_2022027, EPI_ISL_2022033, EPI_ISL_2022035, EPI_ISL_2022038, EPI_ISL_2022053, EPI_ISL_2022061, EPI_ISL_2022062, EPI_ISL_2022074, EPI_ISL_2022075, EPI_ISL_2022077, EPI_ISL_2022086, EPI_ISL_2022087, EPI_ISL_2022091, EPI_ISL_2022092, EPI_ISL_2022097, EPI_ISL_2022101, EPI_ISL_2022109, EPI_ISL_2022112, EPI_ISL_2022115, EPI_ISL_2022124, EPI_ISL_2022127, EPI_ISL_2022129, EPI_ISL_2022132, EPI_ISL_2022135, EPI_ISL_2022138, EPI_ISL_2022142, EPI_ISL_2022149, EPI_ISL_2022154, EPI_ISL_2022160, EPI_ISL_2022162, EPI_ISL_2022163, EPI_ISL_2022164, EPI_ISL_2022165, EPI_ISL_2022166, EPI_ISL_2022168, EPI_ISL_2022171, EPI_ISL_2022172, EPI_ISL_2022178, EPI_ISL_2022181, EPI_ISL_2022182, EPI_ISL_2022183, EPI_ISL_2022184, EPI_ISL_2022185, EPI_ISL_2022188, EPI_ISL_2022189, EPI_ISL_2022190, EPI_ISL_2022192, EPI_ISL_2022193, EPI_ISL_2022195, EPI_ISL_2022196, EPI_ISL_2022197, EPI_ISL_2022198, EPI_ISL_2022205, EPI_ISL_2022206, EPI_ISL_2022208, EPI_ISL_2022209, EPI_ISL_2022210, EPI_ISL_2022211, EPI_ISL_2022212, EPI_ISL_2022214, EPI_ISL_2022215, EPI_ISL_2022216, EPI_ISL_2022217, EPI_ISL_2022218, EPI_ISL_2022219, EPI_ISL_2022220, EPI_ISL_2022221, EPI_ISL_2022222, EPI_ISL_2022223, EPI_ISL_2022225, EPI_ISL_2022226, EPI_ISL_2022227, EPI_ISL_2022228, EPI_ISL_2022229, EPI_ISL_2022230, EPI_ISL_2022232, EPI_ISL_2022233, EPI_ISL_2022234, EPI_ISL_2022235, EPI_ISL_2022237, EPI_ISL_2022243, EPI_ISL_2022246, EPI_ISL_2022247, EPI_ISL_2022248, EPI_ISL_2022249, EPI_ISL_2022252, EPI_ISL_2022253, EPI_ISL_2022254, EPI_ISL_2022255, EPI_ISL_2022257, EPI_ISL_2022259, EPI_ISL_2022260, EPI_ISL_2022261, EPI_ISL_2022266, EPI_ISL_2022267, EPI_ISL_2022268, EPI_ISL_2022269, EPI_ISL_2022270, EPI_ISL_2022271, EPI_ISL_2022273, EPI_ISL_2022274, EPI_ISL_2022275, EPI_ISL_2022276, EPI_ISL_2022282, EPI_ISL_2022283, EPI_ISL_2022284, EPI_ISL_2022287, EPI_ISL_2022289, EPI_ISL_2022291, EPI_ISL_2022292, EPI_ISL_2022293, EPI_ISL_2022294, EPI_ISL_2022297, EPI_ISL_2022298, EPI_ISL_2022300, EPI_ISL_2022301, EPI_ISL_2022302, EPI_ISL_2022303, EPI_ISL_2022307, EPI_ISL_2022309, EPI_ISL_2022310, EPI_ISL_2022311, EPI_ISL_2022312, EPI_ISL_2022313, EPI_ISL_2022314, EPI_ISL_2022315, EPI_ISL_2022316, EPI_ISL_2022318, EPI_ISL_2022320, EPI_ISL_2022321, EPI_ISL_2022322, EPI_ISL_2022323, EPI_ISL_2022324, EPI_ISL_2022325, EPI_ISL_2022326, EPI_ISL_2022328, EPI_ISL_2022329, EPI_ISL_2022331, EPI_ISL_2022333, EPI_ISL_2022334, EPI_ISL_2022335, EPI_ISL_2022336, EPI_ISL_2022338, EPI_ISL_2022339, EPI_ISL_2022341, EPI_ISL_2022344, EPI_ISL_2022345, EPI_ISL_2022347, EPI_ISL_2022349, EPI_ISL_2022350, EPI_ISL_2022353, EPI_ISL_2022354, EPI_ISL_2022355, EPI_ISL_2022357, EPI_ISL_2022360, EPI_ISL_2022361, EPI_ISL_2022362, EPI_ISL_2022363, EPI_ISL_2022365, EPI_ISL_2022366, EPI_ISL_2022368, EPI_ISL_2022370, EPI_ISL_2022371, EPI_ISL_2022372, EPI_ISL_2022377, EPI_ISL_2022378, EPI_ISL_2022379, EPI_ISL_2022380, EPI_ISL_2022382, EPI_ISL_2022384, EPI_ISL_2022385, EPI_ISL_2022386, EPI_ISL_2022389, EPI_ISL_2022390, EPI_ISL_2022391, EPI_ISL_2022392, EPI_ISL_2022393, EPI_ISL_2022394, EPI_ISL_2022395, EPI_ISL_2022396, EPI_ISL_2022401, EPI_ISL_2022402, EPI_ISL_2022403, EPI_ISL_2022404, EPI_ISL_2022405, EPI_ISL_2022406, EPI_ISL_2022407, EPI_ISL_2022409, EPI_ISL_2022411, EPI_ISL_2022412, EPI_ISL_2022413, EPI_ISL_2022414, EPI_ISL_2022416, EPI_ISL_2022417, EPI_ISL_2022418, EPI_ISL_2022421, EPI_ISL_2022422, EPI_ISL_2022424, EPI_ISL_2022425, EPI_ISL_2022426, EPI_ISL_2022427, EPI_ISL_2022430, EPI_ISL_2022431, EPI_ISL_2022433, EPI_ISL_2022434, EPI_ISL_2022435, EPI_ISL_2022436, EPI_ISL_2022438, EPI_ISL_2022784, EPI_ISL_2022840, EPI_ISL_2022845, EPI_ISL_2080360, EPI_ISL_2080475, EPI_ISL_2080481, EPI_ISL_2080482, EPI_ISL_2091462, EPI_ISL_2091464, EPI_ISL_2091465, EPI_ISL_2091466, EPI_ISL_2091467, EPI_ISL_2091470, EPI_ISL_2091471, EPI_ISL_2091474, EPI_ISL_2091475, EPI_ISL_2091476, EPI_ISL_2091478, EPI_ISL_2091483, EPI_ISL_2091485, EPI_ISL_2091488, EPI_ISL_2091490, EPI_ISL_2091491, EPI_ISL_2091498, EPI_ISL_2091499, EPI_ISL_2091501, EPI_ISL_2091503, EPI_ISL_2091504, EPI_ISL_2091505, EPI_ISL_2091506, EPI_ISL_2091507, EPI_ISL_2091508, EPI_ISL_2091509, EPI_ISL_2091512, EPI_ISL_2091513, EPI_ISL_2091516, EPI_ISL_2091517, EPI_ISL_2091519, EPI_ISL_2091520, EPI_ISL_2091523, EPI_ISL_2091527, EPI_ISL_2091529, EPI_ISL_2091530, EPI_ISL_2091531, EPI_ISL_2091535, EPI_ISL_2091537, EPI_ISL_2091538, EPI_ISL_2091541, EPI_ISL_2091544, EPI_ISL_2091545, EPI_ISL_2091546, EPI_ISL_2091548, EPI_ISL_2091550, EPI_ISL_2091551, EPI_ISL_2091552, EPI_ISL_2091556, EPI_ISL_2091559, EPI_ISL_2091560, EPI_ISL_2091561, EPI_ISL_2091563, EPI_ISL_2091567, EPI_ISL_2091568, EPI_ISL_2091570, EPI_ISL_2091571, EPI_ISL_2091572, EPI_ISL_2091573, EPI_ISL_2091574, EPI_ISL_2091575, EPI_ISL_2091576, EPI_ISL_2091577, EPI_ISL_2091578, EPI_ISL_2091579, EPI_ISL_2091580, EPI_ISL_2091584, EPI_ISL_2091588, EPI_ISL_2091589, EPI_ISL_2091591, EPI_ISL_2091593, EPI_ISL_2091594, EPI_ISL_2091595, EPI_ISL_2091597, EPI_ISL_2091598, EPI_ISL_2091599, EPI_ISL_2091600, EPI_ISL_2091601, EPI_ISL_2091606, EPI_ISL_2091607, EPI_ISL_2091608, EPI_ISL_2091609, EPI_ISL_2091610, EPI_ISL_2091611, EPI_ISL_2091612, EPI_ISL_2091615, EPI_ISL_2091616, EPI_ISL_2091617, EPI_ISL_2091618, EPI_ISL_2091619, EPI_ISL_2091620, EPI_ISL_2091621, EPI_ISL_2091623, EPI_ISL_2091626, EPI_ISL_2091628, EPI_ISL_2091629, EPI_ISL_2091631, EPI_ISL_2091634, EPI_ISL_2091635, EPI_ISL_2091637, EPI_ISL_2091641, EPI_ISL_2091644, EPI_ISL_2091645, EPI_ISL_2091646, EPI_ISL_2091647, EPI_ISL_2091648, EPI_ISL_2091653, EPI_ISL_2091654, EPI_ISL_2091655, EPI_ISL_2091657, EPI_ISL_2091658, EPI_ISL_2091661, EPI_ISL_2091662, EPI_ISL_2091663, EPI_ISL_2091667, EPI_ISL_2091668, EPI_ISL_2091673, EPI_ISL_2091674, EPI_ISL_2091675, EPI_ISL_2091676, EPI_ISL_2091677, EPI_ISL_2091680, EPI_ISL_2091682, EPI_ISL_2091683, EPI_ISL_2091684, EPI_ISL_2091686, EPI_ISL_2091688, EPI_ISL_2091689, EPI_ISL_2091690, EPI_ISL_2091692, EPI_ISL_2091696, EPI_ISL_2091697, EPI_ISL_2091700, EPI_ISL_2091701, EPI_ISL_2091705, EPI_ISL_2091706, EPI_ISL_2091707, EPI_ISL_2091708, EPI_ISL_2091709, EPI_ISL_2091712, EPI_ISL_2091714, EPI_ISL_2091715, EPI_ISL_2091716, EPI_ISL_2091717, EPI_ISL_2091718, EPI_ISL_2091720, EPI_ISL_2091721, EPI_ISL_2091722, EPI_ISL_2091723, EPI_ISL_2091725, EPI_ISL_2091726, EPI_ISL_2091727, EPI_ISL_2091731, EPI_ISL_2091732, EPI_ISL_2091734, EPI_ISL_2091737, EPI_ISL_2091739, EPI_ISL_2091747, EPI_ISL_2091748, EPI_ISL_2091752, EPI_ISL_2091753, EPI_ISL_2091755, EPI_ISL_2091758, EPI_ISL_2091762, EPI_ISL_2091767, EPI_ISL_2091768, EPI_ISL_2091770, EPI_ISL_2091771, EPI_ISL_2091773, EPI_ISL_2091775, EPI_ISL_2091776, EPI_ISL_2091777, EPI_ISL_2091778, EPI_ISL_2091780, EPI_ISL_2091782, EPI_ISL_2091787, EPI_ISL_2091790, EPI_ISL_2091792, EPI_ISL_2091800, EPI_ISL_2091801, EPI_ISL_2091804, EPI_ISL_2091808, EPI_ISL_2091810, EPI_ISL_2091812, EPI_ISL_2091814, EPI_ISL_2091816, EPI_ISL_2091819, EPI_ISL_2091823, EPI_ISL_2091825, EPI_ISL_2091827, EPI_ISL_2091828, EPI_ISL_2091829, EPI_ISL_2091830, EPI_ISL_2091831, EPI_ISL_2091833, EPI_ISL_2091836, EPI_ISL_2091841, EPI_ISL_2091842, EPI_ISL_2091844, EPI_ISL_2091845, EPI_ISL_2091847, EPI_ISL_2091851, EPI_ISL_2091852, EPI_ISL_2091854, EPI_ISL_2091856, EPI_ISL_2091858, EPI_ISL_2091859, EPI_ISL_2091863, EPI_ISL_2091864, EPI_ISL_2091865, EPI_ISL_2091866, EPI_ISL_2091868, EPI_ISL_2091869, EPI_ISL_2091870, EPI_ISL_2091874, EPI_ISL_2091877, EPI_ISL_2091878, EPI_ISL_2091880, EPI_ISL_2091881, EPI_ISL_2091884, EPI_ISL_2091885, EPI_ISL_2091888, EPI_ISL_2091891, EPI_ISL_2091892, EPI_ISL_2091893, EPI_ISL_2091894, EPI_ISL_2091896, EPI_ISL_2091897, EPI_ISL_2091898, EPI_ISL_2091904, EPI_ISL_2091905, EPI_ISL_2091910, EPI_ISL_2091911, EPI_ISL_2091915, EPI_ISL_2091917, EPI_ISL_2091919, EPI_ISL_2091921, EPI_ISL_2091922, EPI_ISL_2091924, EPI_ISL_2091929, EPI_ISL_2091930, EPI_ISL_2091931, EPI_ISL_2091934, EPI_ISL_2091936, EPI_ISL_2091937, EPI_ISL_2091940, EPI_ISL_2091943, EPI_ISL_2091945, EPI_ISL_2091947, EPI_ISL_2091951, EPI_ISL_2091954, EPI_ISL_2091955, EPI_ISL_2091958, EPI_ISL_2091959, EPI_ISL_2091961, EPI_ISL_2091962, EPI_ISL_2091964, EPI_ISL_2091965, EPI_ISL_2091966, EPI_ISL_2091967, EPI_ISL_2091968, EPI_ISL_2091970, EPI_ISL_2091974, EPI_ISL_2091977, EPI_ISL_2091978, EPI_ISL_2091979, EPI_ISL_2091980, EPI_ISL_2091982, EPI_ISL_2091987, EPI_ISL_2091995, EPI_ISL_2091996, EPI_ISL_2091999, EPI_ISL_2092001, EPI_ISL_2092002, EPI_ISL_2092003, EPI_ISL_2092005, EPI_ISL_2092006, EPI_ISL_2092008, EPI_ISL_2092010, EPI_ISL_2092011, EPI_ISL_2092013, EPI_ISL_2092020, EPI_ISL_2092022, EPI_ISL_2092023, EPI_ISL_2092026, EPI_ISL_2092034, EPI_ISL_2092039, EPI_ISL_2092040, EPI_ISL_2092046, EPI_ISL_2092047, EPI_ISL_2092048, EPI_ISL_2092049, EPI_ISL_2092050, EPI_ISL_2092052, EPI_ISL_2092054, EPI_ISL_2092055, EPI_ISL_2092057, EPI_ISL_2092058, EPI_ISL_2092059, EPI_ISL_2092060, EPI_ISL_2092061, EPI_ISL_2092075, EPI_ISL_2092076, EPI_ISL_2092082, EPI_ISL_2092085, EPI_ISL_2092091, EPI_ISL_2092092, EPI_ISL_2092096, EPI_ISL_2092099, EPI_ISL_2092100, EPI_ISL_2092102, EPI_ISL_2092103, EPI_ISL_2092104, EPI_ISL_2092105, EPI_ISL_2092106, EPI_ISL_2092107, EPI_ISL_2092108, EPI_ISL_2092109, EPI_ISL_2092110, EPI_ISL_2092111, EPI_ISL_2092113, EPI_ISL_2092115, EPI_ISL_2092116, EPI_ISL_2092117, EPI_ISL_2092119, EPI_ISL_2092121, EPI_ISL_2092122, EPI_ISL_2092123, EPI_ISL_2092124, EPI_ISL_2092125, EPI_ISL_2092126, EPI_ISL_2092127, EPI_ISL_2092128, EPI_ISL_2092129, EPI_ISL_2092130, EPI_ISL_2092131, EPI_ISL_2092132, EPI_ISL_2092133, EPI_ISL_2092134, EPI_ISL_2092135, EPI_ISL_2092136, EPI_ISL_2092137, EPI_ISL_2092138, | | | |

EPI_ISL_2092139, EPI_ISL_2092142, EPI_ISL_2092144, EPI_ISL_2092145, EPI_ISL_2092146, EPI_ISL_2092147, EPI_ISL_2092148, EPI_ISL_2092149, EPI_ISL_2092150, EPI_ISL_2092151, EPI_ISL_2092153, EPI_ISL_2092154, EPI_ISL_2092155, EPI_ISL_2092157, EPI_ISL_2092158, EPI_ISL_2092159, EPI_ISL_2092160, EPI_ISL_2092161, EPI_ISL_2092164, EPI_ISL_2092165, EPI_ISL_2092166, EPI_ISL_2092168,
EPI_ISL_2092169, EPI_ISL_2092170, EPI_ISL_2092172, EPI_ISL_2092173, EPI_ISL_2092174, EPI_ISL_2092175, EPI_ISL_2092176, EPI_ISL_2092177, EPI_ISL_2092178, EPI_ISL_2092179, EPI_ISL_2092180, EPI_ISL_2092182, EPI_ISL_2092184, EPI_ISL_2092185, EPI_ISL_2092186, EPI_ISL_2092187, EPI_ISL_2092189, EPI_ISL_2092190, EPI_ISL_2092191, EPI_ISL_2092192, EPI_ISL_2092194, EPI_ISL_2092196,
EPI_ISL_2092197, EPI_ISL_2092198, EPI_ISL_2092199, EPI_ISL_2092201, EPI_ISL_2092204, EPI_ISL_2092205, EPI_ISL_2092206, EPI_ISL_2092207, EPI_ISL_2092208, EPI_ISL_2092209, EPI_ISL_2092210, EPI_ISL_2092211, EPI_ISL_2092212, EPI_ISL_2092213, EPI_ISL_2092215, EPI_ISL_2092216, EPI_ISL_2092217, EPI_ISL_2092218, EPI_ISL_2092219, EPI_ISL_2092220, EPI_ISL_2092221, EPI_ISL_2092224,
EPI_ISL_2092225, EPI_ISL_2092226, EPI_ISL_2092227, EPI_ISL_2092228, EPI_ISL_2092229, EPI_ISL_2092230, EPI_ISL_2092231, EPI_ISL_2092232, EPI_ISL_2092234, EPI_ISL_2092235, EPI_ISL_2092236, EPI_ISL_2092237, EPI_ISL_2092238, EPI_ISL_2092241, EPI_ISL_2092242, EPI_ISL_2092244, EPI_ISL_2092246, EPI_ISL_2092247, EPI_ISL_2092248, EPI_ISL_2092249, EPI_ISL_2092250, EPI_ISL_2092251,
EPI_ISL_2092252, EPI_ISL_2092253, EPI_ISL_2092254, EPI_ISL_2092255, EPI_ISL_2092257, EPI_ISL_2092258, EPI_ISL_2092260, EPI_ISL_2092261, EPI_ISL_2092262, EPI_ISL_2092263, EPI_ISL_2092267, EPI_ISL_2092268, EPI_ISL_2092269, EPI_ISL_2092271, EPI_ISL_2092274, EPI_ISL_2092275, EPI_ISL_2092276, EPI_ISL_2092277, EPI_ISL_2092279, EPI_ISL_2092281, EPI_ISL_2092282, EPI_ISL_2092283,
EPI_ISL_2092284, EPI_ISL_2092285, EPI_ISL_2092286, EPI_ISL_2092287, EPI_ISL_2092288, EPI_ISL_2092289, EPI_ISL_2092290, EPI_ISL_2092291, EPI_ISL_2092292, EPI_ISL_2092293, EPI_ISL_2092294, EPI_ISL_2092295, EPI_ISL_2092297, EPI_ISL_2092298, EPI_ISL_2092299, EPI_ISL_2092300, EPI_ISL_2092301, EPI_ISL_2092302, EPI_ISL_2092303, EPI_ISL_2092304, EPI_ISL_2092305, EPI_ISL_2092306,
EPI_ISL_2092307, EPI_ISL_2092308, EPI_ISL_2092309, EPI_ISL_2092310, EPI_ISL_2092311, EPI_ISL_2092313, EPI_ISL_2092314, EPI_ISL_2092315, EPI_ISL_2092316, EPI_ISL_2092317, EPI_ISL_2092318, EPI_ISL_2092319, EPI_ISL_2092320, EPI_ISL_2092321, EPI_ISL_2092322, EPI_ISL_2092323, EPI_ISL_2092324, EPI_ISL_2092325, EPI_ISL_2092326, EPI_ISL_2092327, EPI_ISL_2092328, EPI_ISL_2092329,
EPI_ISL_2092330, EPI_ISL_2092331, EPI_ISL_2092332, EPI_ISL_2092333, EPI_ISL_2092334, EPI_ISL_2092335, EPI_ISL_2092336, EPI_ISL_2092337, EPI_ISL_2092340, EPI_ISL_2092343, EPI_ISL_2092344, EPI_ISL_2092345, EPI_ISL_2092346, EPI_ISL_2092348, EPI_ISL_2092349, EPI_ISL_2092350, EPI_ISL_2092351, EPI_ISL_2092352, EPI_ISL_2092353, EPI_ISL_2092354, EPI_ISL_2092355, EPI_ISL_2092356,
EPI_ISL_2092357, EPI_ISL_2092358, EPI_ISL_2092359, EPI_ISL_2092360, EPI_ISL_2092361, EPI_ISL_2092363, EPI_ISL_2092364, EPI_ISL_2092366, EPI_ISL_2092367, EPI_ISL_2092368, EPI_ISL_2092369, EPI_ISL_2092370, EPI_ISL_2092371, EPI_ISL_2092372, EPI_ISL_2092374, EPI_ISL_2092375, EPI_ISL_2092376, EPI_ISL_2092377, EPI_ISL_2092378, EPI_ISL_2092379, EPI_ISL_2092381, EPI_ISL_2092382,
EPI_ISL_2092383, EPI_ISL_2092384, EPI_ISL_2092385, EPI_ISL_2092386, EPI_ISL_2092387, EPI_ISL_2092388, EPI_ISL_2092389, EPI_ISL_2092390, EPI_ISL_2092391, EPI_ISL_2092392, EPI_ISL_2092393, EPI_ISL_2092394, EPI_ISL_2092395, EPI_ISL_2092396, EPI_ISL_2092397, EPI_ISL_2092398, EPI_ISL_2092399, EPI_ISL_2092400, EPI_ISL_2092402, EPI_ISL_2092403, EPI_ISL_2092404, EPI_ISL_2092405,
EPI_ISL_2092407, EPI_ISL_2092408, EPI_ISL_2092409, EPI_ISL_2092410, EPI_ISL_2092412, EPI_ISL_2092413, EPI_ISL_2092415, EPI_ISL_2092418, EPI_ISL_2092420, EPI_ISL_2092421, EPI_ISL_2092422, EPI_ISL_2092423, EPI_ISL_2092424, EPI_ISL_2092426, EPI_ISL_2092427, EPI_ISL_2092429, EPI_ISL_2092430, EPI_ISL_2092431, EPI_ISL_2092432, EPI_ISL_2092433, EPI_ISL_2092434, EPI_ISL_2092435,
EPI_ISL_2092436, EPI_ISL_2092437, EPI_ISL_2092439, EPI_ISL_2092443, EPI_ISL_2092444, EPI_ISL_2092445, EPI_ISL_2092446, EPI_ISL_2092447, EPI_ISL_2092448, EPI_ISL_2092449, EPI_ISL_2092450, EPI_ISL_2092452, EPI_ISL_2092453, EPI_ISL_2092454, EPI_ISL_2092455, EPI_ISL_2092456, EPI_ISL_2092457, EPI_ISL_2092458, EPI_ISL_2092460, EPI_ISL_2092461, EPI_ISL_2092462, EPI_ISL_2092463,
EPI_ISL_2092464, EPI_ISL_2092465, EPI_ISL_2092466, EPI_ISL_2092741, EPI_ISL_2092742, EPI_ISL_2117512, EPI_ISL_2117525, EPI_ISL_2117528, EPI_ISL_2117533, EPI_ISL_2117539, EPI_ISL_2117546, EPI_ISL_2117552, EPI_ISL_2117553, EPI_ISL_2117557, EPI_ISL_2117560, EPI_ISL_2117565, EPI_ISL_2117568, EPI_ISL_2117569, EPI_ISL_2117573, EPI_ISL_2117577, EPI_ISL_2117580, EPI_ISL_2117588,
EPI_ISL_2117590, EPI_ISL_2117594, EPI_ISL_2117595, EPI_ISL_2117603, EPI_ISL_2117604, EPI_ISL_2117610, EPI_ISL_2117614, EPI_ISL_2117619, EPI_ISL_2117620, EPI_ISL_2117622, EPI_ISL_2117623, EPI_ISL_2117624, EPI_ISL_2117626, EPI_ISL_2117633, EPI_ISL_2117647, EPI_ISL_2117650, EPI_ISL_2117652, EPI_ISL_2117654, EPI_ISL_2117658, EPI_ISL_2117661, EPI_ISL_2117663, EPI_ISL_2117665,
EPI_ISL_2117670, EPI_ISL_2117672, EPI_ISL_2117673, EPI_ISL_2117674, EPI_ISL_2117684, EPI_ISL_2117685, EPI_ISL_2117687, EPI_ISL_2117688, EPI_ISL_2117690, EPI_ISL_2117691, EPI_ISL_2117693, EPI_ISL_2117694, EPI_ISL_2117696, EPI_ISL_2117699, EPI_ISL_2117702, EPI_ISL_2117708, EPI_ISL_2117717, EPI_ISL_2117719, EPI_ISL_2117720, EPI_ISL_2117723, EPI_ISL_2117729, EPI_ISL_2117742,
EPI_ISL_2117748, EPI_ISL_2117754, EPI_ISL_2117756, EPI_ISL_2117758, EPI_ISL_2117762, EPI_ISL_2117764, EPI_ISL_2117769, EPI_ISL_2117770, EPI_ISL_2117772, EPI_ISL_2117784, EPI_ISL_2117787, EPI_ISL_2117794, EPI_ISL_2117796, EPI_ISL_2117809, EPI_ISL_2117819, EPI_ISL_2117829, EPI_ISL_2117831, EPI_ISL_2117832, EPI_ISL_2117834, EPI_ISL_2117838, EPI_ISL_2117843, EPI_ISL_2117845,
EPI_ISL_2117851, EPI_ISL_2117860, EPI_ISL_2117865, EPI_ISL_2117871, EPI_ISL_2117877, EPI_ISL_2117879, EPI_ISL_2117880, EPI_ISL_2117895, EPI_ISL_2117903, EPI_ISL_2117908, EPI_ISL_2117919, EPI_ISL_2117931, EPI_ISL_2117933, EPI_ISL_2117934, EPI_ISL_2117935, EPI_ISL_2117938, EPI_ISL_2117939, EPI_ISL_2117941, EPI_ISL_2117949, EPI_ISL_2117954, EPI_ISL_2117957, EPI_ISL_2117966,
EPI_ISL_2117967, EPI_ISL_2117969, EPI_ISL_2117974, EPI_ISL_2117980, EPI_ISL_2117981, EPI_ISL_2117984, EPI_ISL_2117993, EPI_ISL_2117994, EPI_ISL_2117996, EPI_ISL_2118001, EPI_ISL_2118002, EPI_ISL_2118005, EPI_ISL_2118010, EPI_ISL_2118011, EPI_ISL_2118016, EPI_ISL_2118027, EPI_ISL_2118029, EPI_ISL_2118041, EPI_ISL_2118052, EPI_ISL_2118059, EPI_ISL_2118060, EPI_ISL_2118061,
EPI_ISL_2118064, EPI_ISL_2118065, EPI_ISL_2118068, EPI_ISL_2118070, EPI_ISL_2118072, EPI_ISL_2118076, EPI_ISL_2118083, EPI_ISL_2118099, EPI_ISL_2118100, EPI_ISL_2118102, EPI_ISL_2118106, EPI_ISL_2118108, EPI_ISL_2118111, EPI_ISL_2118114, EPI_ISL_2118120, EPI_ISL_2118129, EPI_ISL_2118132, EPI_ISL_2118142, EPI_ISL_2118148, EPI_ISL_2118158, EPI_ISL_2118162, EPI_ISL_2118170,
EPI_ISL_2118171, EPI_ISL_2118173, EPI_ISL_2118174, EPI_ISL_2118185, EPI_ISL_2118191, EPI_ISL_2118192, EPI_ISL_2118195, EPI_ISL_2118211, EPI_ISL_2118219, EPI_ISL_2118220, EPI_ISL_2118233, EPI_ISL_2118235, EPI_ISL_2118237, EPI_ISL_2118238, EPI_ISL_2118244, EPI_ISL_2118249, EPI_ISL_2118250, EPI_ISL_2118252, EPI_ISL_2118254, EPI_ISL_2118255, EPI_ISL_2118256, EPI_ISL_2118261,
EPI_ISL_2118262, EPI_ISL_2118264, EPI_ISL_2118265, EPI_ISL_2118267, EPI_ISL_2118268, EPI_ISL_2118270, EPI_ISL_2118271, EPI_ISL_2118272, EPI_ISL_2118274, EPI_ISL_2118310, EPI_ISL_2118312, EPI_ISL_2118314, EPI_ISL_2118317, EPI_ISL_2118318, EPI_ISL_2118320, EPI_ISL_2118321, EPI_ISL_2118322, EPI_ISL_2118327, EPI_ISL_2118329, EPI_ISL_2118330, EPI_ISL_2118331, EPI_ISL_2118333,
EPI_ISL_2118334, EPI_ISL_2118336, EPI_ISL_2118341, EPI_ISL_2118342, EPI_ISL_2118346, EPI_ISL_2118348, EPI_ISL_2118350, EPI_ISL_2118351, EPI_ISL_2118353, EPI_ISL_2118354, EPI_ISL_2118357, EPI_ISL_2118360, EPI_ISL_2118362, EPI_ISL_2118365, EPI_ISL_2118366, EPI_ISL_2118369, EPI_ISL_2118372, EPI_ISL_2118374, EPI_ISL_2118375, EPI_ISL_2118377, EPI_ISL_2118378, EPI_ISL_2118380,
EPI_ISL_2118381, EPI_ISL_2118384, EPI_ISL_2118385, EPI_ISL_2118387, EPI_ISL_2118388, EPI_ISL_2118391, EPI_ISL_2118393, EPI_ISL_2118394, EPI_ISL_2118396, EPI_ISL_2118397, EPI_ISL_2118404, EPI_ISL_2118408, EPI_ISL_2118411, EPI_ISL_2118414, EPI_ISL_2118416, EPI_ISL_2118419, EPI_ISL_2118421, EPI_ISL_2118422, EPI_ISL_2118424, EPI_ISL_2118427, EPI_ISL_2118428, EPI_ISL_2118430,
EPI_ISL_2118431, EPI_ISL_2118433, EPI_ISL_2118434, EPI_ISL_2118437, EPI_ISL_2118441, EPI_ISL_2118443, EPI_ISL_2118444, EPI_ISL_2118446, EPI_ISL_2118447, EPI_ISL_2118448, EPI_ISL_2118456, EPI_ISL_2118457, EPI_ISL_2118460, EPI_ISL_2118462, EPI_ISL_2118465, EPI_ISL_2118469, EPI_ISL_2118471, EPI_ISL_2118472, EPI_ISL_2118473, EPI_ISL_2118475, EPI_ISL_2118476, EPI_ISL_2118478,
EPI_ISL_2118479, EPI_ISL_2118481, EPI_ISL_2118484, EPI_ISL_2118485, EPI_ISL_2118487, EPI_ISL_2118488, EPI_ISL_2118489, EPI_ISL_2118491, EPI_ISL_2118492, EPI_ISL_2118494, EPI_ISL_2118496, EPI_ISL_2118497, EPI_ISL_2118500, EPI_ISL_2118502, EPI_ISL_2118504, EPI_ISL_2118506, EPI_ISL_2118509, EPI_ISL_2118510, EPI_ISL_2118512, EPI_ISL_2118513, EPI_ISL_2118515,
EPI_ISL_2118516, EPI_ISL_2118518, EPI_ISL_2118521, EPI_ISL_2118522, EPI_ISL_2118523, EPI_ISL_2118525, EPI_ISL_2118528, EPI_ISL_2118529, EPI_ISL_2118531, EPI_ISL_2118532, EPI_ISL_2118534, EPI_ISL_2118535, EPI_ISL_2118537, EPI_ISL_2118538, EPI_ISL_2118540, EPI_ISL_2118541, EPI_ISL_2118543, EPI_ISL_2118544, EPI_ISL_2118546, EPI_ISL_2118551, EPI_ISL_2118552, EPI_ISL_2118554,
EPI_ISL_2118556, EPI_ISL_2118558, EPI_ISL_2118560, EPI_ISL_2118561, EPI_ISL_2118564, EPI_ISL_2118566, EPI_ISL_2118567, EPI_ISL_2118569, EPI_ISL_2118573, EPI_ISL_2118575, EPI_ISL_2118579, EPI_ISL_2118581, EPI_ISL_2118582, EPI_ISL_2118585, EPI_ISL_2118586, EPI_ISL_2118589, EPI_ISL_2118590, EPI_ISL_2118593, EPI_ISL_2118595, EPI_ISL_2118596, EPI_ISL_2118598, EPI_ISL_2118599,
EPI_ISL_2118601, EPI_ISL_2118604, EPI_ISL_2118609, EPI_ISL_2118610, EPI_ISL_2118612, EPI_ISL_2118613, EPI_ISL_2118615, EPI_ISL_2118618, EPI_ISL_2118622, EPI_ISL_2118624, EPI_ISL_2118625, EPI_ISL_2118628, EPI_ISL_2118630, EPI_ISL_2118631, EPI_ISL_2118634, EPI_ISL_2118636, EPI_ISL_2118638, EPI_ISL_2118639, EPI_ISL_2118642, EPI_ISL_2118643, EPI_ISL_2118645, EPI_ISL_2118648,
EPI_ISL_2118649, EPI_ISL_2118652, EPI_ISL_2118654, EPI_ISL_2118655, EPI_ISL_2118657, EPI_ISL_2118658, EPI_ISL_2118659, EPI_ISL_2118661, EPI_ISL_2118662, EPI_ISL_2118667, EPI_ISL_2118668, EPI_ISL_2118670, EPI_ISL_2118671, EPI_ISL_2118673, EPI_ISL_2118675, EPI_ISL_2118676, EPI_ISL_2118678, EPI_ISL_2118679, EPI_ISL_2118684, EPI_ISL_2118685, EPI_ISL_2118687,
EPI_ISL_2118689, EPI_ISL_2118690, EPI_ISL_2118692, EPI_ISL_2118694, EPI_ISL_2118696, EPI_ISL_2118698, EPI_ISL_2118700, EPI_ISL_2118701, EPI_ISL_2118704, EPI_ISL_2118705, EPI_ISL_2118708, EPI_ISL_2118709, EPI_ISL_2118711, EPI_ISL_2118712, EPI_ISL_2118713, EPI_ISL_2118716, EPI_ISL_2118718, EPI_ISL_2118721, EPI_ISL_2118722, EPI_ISL_2118724, EPI_ISL_2118725, EPI_ISL_2118728,
EPI_ISL_2118730, EPI_ISL_2118732, EPI_ISL_2118733, EPI_ISL_2118735, EPI_ISL_2118738, EPI_ISL_2118739, EPI_ISL_2118740, EPI_ISL_2118742, EPI_ISL_2118744, EPI_ISL_2118745, EPI_ISL_2118748, EPI_ISL_2118750, EPI_ISL_2118751, EPI_ISL_2118756, EPI_ISL_2118758, EPI_ISL_2118762, EPI_ISL_2118763, EPI_ISL_2118765, EPI_ISL_2118766, EPI_ISL_2118770, EPI_ISL_2118773,
EPI_ISL_2118775, EPI_ISL_2118776, EPI_ISL_2118778, EPI_ISL_2118779, EPI_ISL_2118781, EPI_ISL_2118785, EPI_ISL_2118786, EPI_ISL_2118788, EPI_ISL_2118791, EPI_ISL_2118793, EPI_ISL_2118796, EPI_ISL_2118797, EPI_ISL_2118803, EPI_ISL_2118807, EPI_ISL_2118811, EPI_ISL_2118813, EPI_ISL_2118814, EPI_ISL_2118816, EPI_ISL_2118817, EPI_ISL_2118819, EPI_ISL_2118822, EPI_ISL_2118828,
EPI_ISL_2118829, EPI_ISL_2118830, EPI_ISL_2118831, EPI_ISL_2118832, EPI_ISL_2118835, EPI_ISL_2118843, EPI_ISL_2118844, EPI_ISL_2118849, EPI_ISL_2118851, EPI_ISL_2118852, EPI_ISL_2118855, EPI_ISL_2118856, EPI_ISL_2118864, EPI_ISL_2118865, EPI_ISL_2118868, EPI_ISL_2118876, EPI_ISL_2118882, EPI_ISL_2118885, EPI_ISL_2118886, EPI_ISL_2118890, EPI_ISL_2118891, EPI_ISL_2118897,
EPI_ISL_2118903, EPI_ISL_2118908, EPI_ISL_2118912, EPI_ISL_2118915, EPI_ISL_2118918, EPI_ISL_2118922, EPI_ISL_2118923, EPI_ISL_2118924, EPI_ISL_2118926, EPI_ISL_2118928, EPI_ISL_2118930, EPI_ISL_2118933, EPI_ISL_2118935, EPI_ISL_2118939, EPI_ISL_2118945, EPI_ISL_2118955, EPI_ISL_2118958, EPI_ISL_2118960, EPI_ISL_2118965, EPI_ISL_2118967, EPI_ISL_2118970, EPI_ISL_2118973,
EPI_ISL_2118979, EPI_ISL_2118983, EPI_ISL_2118985, EPI_ISL_2118988, EPI_ISL_2118989, EPI_ISL_2118991, EPI_ISL_2118992, EPI_ISL_2119006, EPI_ISL_2119013, EPI_ISL_2119015, EPI_ISL_2119017, EPI_ISL_2119020, EPI_ISL_2119022, EPI_ISL_2119024, EPI_ISL_2119027, EPI_ISL_2119039, EPI_ISL_2119040, EPI_ISL_2119041, EPI_ISL_2119043, EPI_ISL_2119045, EPI_ISL_2119048,
EPI_ISL_2119082, EPI_ISL_2119083, EPI_ISL_2119086, EPI_ISL_2119088, EPI_ISL_2119089, EPI_ISL_2119091, EPI_ISL_2119097, EPI_ISL_2119099, EPI_ISL_2119100, EPI_ISL_2119102, EPI_ISL_2119113, EPI_ISL_2119121, EPI_ISL_2119122, EPI_ISL_2119124, EPI_ISL_2119127, EPI_ISL_2119128, EPI_ISL_2119130, EPI_ISL_2119133, EPI_ISL_2119134, EPI_ISL_2119138, EPI_ISL_2119141, EPI_ISL_2119142,
EPI_ISL_2119146, EPI_ISL_2119149, EPI_ISL_2119150, EPI_ISL_2119152, EPI_ISL_2119155, EPI_ISL_2119158, EPI_ISL_2119160, EPI_ISL_2119163, EPI_ISL_2119166, EPI_ISL_2119169, EPI_ISL_2119172, EPI_ISL_2119183, EPI_ISL_2119185, EPI_ISL_2119192, EPI_ISL_2119196, EPI_ISL_2119199, EPI_ISL_2119204, EPI_ISL_2119206, EPI_ISL_2119210, EPI_ISL_2119211, EPI_ISL_2119216, EPI_ISL_2119218,
EPI_ISL_2119220, EPI_ISL_2119222, EPI_ISL_2119226, EPI_ISL_2119230, EPI_ISL_2119234, EPI_ISL_2119237, EPI_ISL_2119243, EPI_ISL_2119244, EPI_ISL_2119247, EPI_ISL_2119250, EPI_ISL_2119256, EPI_ISL_2119260, EPI_ISL_2119265, EPI_ISL_2119266, EPI_ISL_2119268, EPI_ISL_2119271, EPI_ISL_2119274, EPI_ISL_2119275, EPI_ISL_2119277, EPI_ISL_2119278, EPI_ISL_2119280, EPI_ISL_2119285,
EPI_ISL_2119289, EPI_ISL_2119294, EPI_ISL_2119301, EPI_ISL_2119309, EPI_ISL_2119311, EPI_ISL_2119320, EPI_ISL_2119324, EPI_ISL_2119327, EPI_ISL_2119330, EPI_ISL_2119336, EPI_ISL_2119338, EPI_ISL_2119343, EPI_ISL_2119349, EPI_ISL_2119370, EPI_ISL_2119395, EPI_ISL_2119406, EPI_ISL_2119410, EPI_ISL_2119416, EPI_ISL_2119428, EPI_ISL_2119432, EPI_ISL_2119439,
EPI_ISL_2119442, EPI_ISL_2119445, EPI_ISL_2119448, EPI_ISL_2119452, EPI_ISL_2119471, EPI_ISL_2119473, EPI_ISL_2119476, EPI_ISL_2119480, EPI_ISL_2119484, EPI_ISL_2119487, EPI_ISL_2119504, EPI_ISL_2119518, EPI_ISL_2119534, EPI_ISL_2119537, EPI_ISL_2119539, EPI_ISL_2119541, EPI_ISL_2119543, EPI_ISL_2119551, EPI_ISL_2119554, EPI_ISL_2119559, EPI_ISL_2119561, EPI_ISL_2119569,
EPI_ISL_2119571, EPI_ISL_2119574, EPI_ISL_2119584, EPI_ISL_2119587, EPI_ISL_2119591, EPI_ISL_2119595, EPI_ISL_2119599, EPI_ISL_2119609, EPI_ISL_2119620, EPI_ISL_2119622, EPI_ISL_2119623, EPI_ISL_2119626, EPI_ISL_2119628, EPI_ISL_2119631, EPI_ISL_2119633, EPI_ISL_2119638, EPI_ISL_2119639, EPI_ISL_2119646, EPI_ISL_2119651, EPI_ISL_2119679, EPI_ISL_2119691,
EPI_ISL_2119709, EPI_ISL_2119710, EPI_ISL_2119713, EPI_ISL_2119715, EPI_ISL_2119726, EPI_ISL_2119729, EPI_ISL_2119734, EPI_ISL_2119736, EPI_ISL_2119738, EPI_ISL_2119762, EPI_ISL_2119765, EPI_ISL_2119769, EPI_ISL_2119772, EPI_ISL_2119775, EPI_ISL_2119780, EPI_ISL_2119781, EPI_ISL_2119794, EPI_ISL_2119822, EPI_ISL_2119835, EPI_ISL_2119838, EPI_ISL_2119849, EPI_ISL_2119852,
EPI_ISL_2119855, EPI_ISL_2119859, EPI_ISL_2119867, EPI_ISL_2119872, EPI_ISL_2119873, EPI_ISL_2119902, EPI_ISL_2119915, EPI_ISL_2119919, EPI_ISL_2119943, EPI_ISL_2119951, EPI_ISL_2119958, EPI_ISL_2119961, EPI_ISL_2119988, EPI_ISL_2119994, EPI_ISL_2119999, EPI_ISL_2120012, EPI_ISL_2120015, EPI_ISL_2120017, EPI_ISL_2120022, EPI_ISL_2120031, EPI_ISL_2120043, EPI_ISL_2120051,
EPI_ISL_2120069, EPI_ISL_2120072, EPI_ISL_2120087, EPI_ISL_2120092, EPI_ISL_2120121, EPI_ISL_2120124, EPI_ISL_2120127, EPI_ISL_2120129, EPI_ISL_2120130, EPI_ISL_2120136, EPI_ISL_2120149, EPI_ISL_2120161, EPI_ISL_2120169, EPI_ISL_2120175, EPI_ISL_2120178, EPI_ISL_2120186, EPI_ISL_2120199, EPI_ISL_2120201, EPI_ISL_2120204, EPI_ISL_2120208, EPI_ISL_2120233, EPI_ISL_2120248,
EPI_ISL_2120261, EPI_ISL_2120265, EPI_ISL_2120304, EPI_ISL_2120336, EPI_ISL_2120347, EPI_ISL_2120364, EPI_ISL_2120370, EPI_ISL_2120374, EPI_ISL_2120378, EPI_ISL_2120382, EPI_ISL_2120385, EPI_ISL_2120387, EPI_ISL_2120415, EPI_ISL_2120425, EPI_ISL_2120434, EPI_ISL_2120445, EPI_ISL_2120453, EPI_ISL_2120455, EPI_ISL_2120458, EPI_ISL_2120460, EPI_ISL_2120461,
EPI_ISL_2120463, EPI_ISL_2120464, EPI_ISL_2120468, EPI_ISL_2120469, EPI_ISL_2120479, EPI_ISL_2120480, EPI_ISL_2120482, EPI_ISL_2120483, EPI_ISL_2120486, EPI_ISL_2120489, EPI_ISL_2120492, EPI_ISL_2120494, EPI_ISL_2120495, EPI_ISL_2120498, EPI_ISL_2120500, EPI_ISL_2120505, EPI_ISL_2120506, EPI_ISL_2120509, EPI_ISL_2120511, EPI_ISL_2120512, EPI_ISL_2120517, EPI_ISL_2120518,
EPI_ISL_2120523, EPI_ISL_2120525, EPI_ISL_2120529, EPI_ISL_2120530, EPI_ISL_2120533, EPI_ISL_2120536, EPI_ISL_2120539, EPI_ISL_2120542, EPI_ISL_2120544, EPI_ISL_2120547, EPI_ISL_2120551, EPI_ISL_2120552, EPI_ISL_2120554, EPI_ISL_2120556, EPI_ISL_2120559, EPI_ISL_2120562, EPI_ISL_2120563, EPI_ISL_2120567, EPI_ISL_2120569, EPI_ISL_2120572, EPI_ISL_2120579, EPI_ISL_2120582,
EPI_ISL_2120587, EPI_ISL_2120588, EPI_ISL_2120591, EPI_ISL_2120592, EPI_ISL_2120595, EPI_ISL_2120600, EPI_ISL_2120602, EPI_ISL_2120604, EPI_ISL_2120611, EPI_ISL_2120617, EPI_ISL_2120619, EPI_ISL_2120620, EPI_ISL_2120624, EPI_ISL_2120625, EPI_ISL_2120627, EPI_ISL_2120633, EPI_ISL_2120637, EPI_ISL_2120640, EPI_ISL_2120645, EPI_ISL_2120651, EPI_ISL_2120691, EPI_ISL_2120692,
EPI_ISL_2120694, EPI_ISL_2120702, EPI_ISL_2120705, EPI_ISL_2120706, EPI_ISL_2120709, EPI_ISL_2120710, EPI_ISL_2120712, EPI_ISL_2120715, EPI_ISL_2120721, EPI_ISL_2120727, EPI_ISL_2120728, EPI_ISL_2120730, EPI_ISL_2120732, EPI_ISL_2120737, EPI_ISL_2120742, EPI_ISL_2120743, EPI_ISL_2120748, EPI_ISL_2120750, EPI_ISL_2120757, EPI_ISL_2120759, EPI_ISL_2120763, EPI_ISL_2120764,
EPI_ISL_2120766, EPI_ISL_2120767, EPI_ISL_2120768, EPI_ISL_2120769, EPI_ISL_2120771, EPI_ISL_2120772, EPI_ISL_2120778, EPI_ISL_2120780, EPI_ISL_2120784, EPI_ISL_2120785, EPI_ISL_2120789, EPI_ISL_2120796, EPI_ISL_2120799, EPI_ISL_2120803, EPI_ISL_2120805, EPI_ISL_2120809, EPI_ISL_2120811, EPI_ISL_2120815, EPI_ISL_2120818, EPI_ISL_2120819, EPI_ISL_2120821, EPI_ISL_2120822,
EPI_ISL_2120824, EPI_ISL_2120825, EPI_ISL_2120831, EPI_ISL_2120834, EPI_ISL_2120843, EPI_ISL_2120845, EPI_ISL_2120851, EPI_ISL_2120854, EPI_ISL_2120856, EPI_ISL_2120857, EPI_ISL_2120860, EPI_ISL_2120865, EPI_ISL_2120869, EPI_ISL_2120872, EPI_ISL_2120879, EPI_ISL_2120885, EPI_ISL_2120888, EPI_ISL_2120889, EPI_ISL_2120894, EPI_ISL_2120895, EPI_ISL_2120897, EPI_ISL_2120901,
EPI_ISL_2120909, EPI_ISL_2120911, EPI_ISL_2120912, EPI_ISL_2120913, EPI_ISL_2120916, EPI_ISL_2120919, EPI_ISL_2120923, EPI_ISL_2120925, EPI_ISL_2120930, EPI_ISL_2120933, EPI_ISL_2120935, EPI_ISL_2120936, EPI_ISL_2120938, EPI_ISL_2120939, EPI_ISL_2120941, EPI_ISL_2120942, EPI_ISL_2120944, EPI_ISL_2120945, EPI_ISL_2120953, EPI_ISL_2120957, EPI_ISL_2120960, EPI_ISL_2120962,
EPI_ISL_2120963, EPI_ISL_2120972, EPI_ISL_2120976, EPI_ISL_2120977, EPI_ISL_2120981, EPI_ISL_2120984, EPI_ISL_2120989, EPI_ISL_2120990, EPI_ISL_2120993, EPI_ISL_2120995, EPI_ISL_2120998, EPI_ISL_2120999, EPI_ISL_2121001, EPI_ISL_2121008, EPI_ISL_2121017, EPI_ISL_2121019, EPI_ISL_2121095, EPI_ISL_2121188, EPI_ISL_2121370, EPI_ISL_2121546, EPI_ISL_2122128, EPI_ISL_2122130,
EPI_ISL_2122131, EPI_ISL_2122133, EPI_ISL_2122135, EPI_ISL_2122136, EPI_ISL_2122139, EPI_ISL_2122140, EPI_ISL_2122149, EPI_ISL_2122151, EPI_ISL_2122158, EPI_ISL_2122161, EPI_ISL_2122165, EPI_ISL_2122181, EPI_ISL_2122183, EPI_ISL_2122189, EPI_ISL_2122191, EPI_ISL_2122194, EPI_ISL_2122195, EPI_ISL_2122206, EPI_ISL_2122208, EPI_ISL_2122210, EPI_ISL_2122215, EPI_ISL_2122217,
EPI_ISL_2122218, EPI_ISL_2122221, EPI_ISL_2122229, EPI_ISL_2122235, EPI_ISL_2122240, EPI_ISL_2122241, EPI_ISL_2122245, EPI_ISL_2122248, EPI_ISL_2122251, EPI_ISL_2122254, EPI_ISL_2122275, EPI_ISL_2122276, EPI_ISL_2122282, EPI_ISL_2122286, EPI_ISL_2122289, EPI_ISL_2122298, EPI_ISL_2122301, EPI_ISL_2122310, EPI_ISL_2122322, EPI_ISL_2122325, EPI_ISL_2122328,
EPI_ISL_2122334, EPI_ISL_2122335, EPI_ISL_2122341, EPI_ISL_2122343, EPI_ISL_2122345, EPI_ISL_2122348, EPI_ISL_2122349, EPI_ISL_2122352, EPI_ISL_2122354, EPI_ISL_2122355, EPI_ISL_2122358, EPI_ISL_2122360, EPI_ISL_2122363, EPI_ISL_2122365, EPI_ISL_2122375, EPI_ISL_2122378, EPI_ISL_2122381, EPI_ISL_2122383, EPI_ISL_2122384, EPI_ISL_2122386, EPI_ISL_2122390, EPI_ISL_2122397,
EPI_ISL_2122398, EPI_ISL_2122400, EPI_ISL_2122403, EPI_ISL_2122408, EPI_ISL_2122409, EPI_ISL_2122412, EPI_ISL_2122414, EPI_ISL_2122420, EPI_ISL_2122421, EPI_ISL_2122424, EPI_ISL_2122429, EPI_ISL_2122430, EPI_ISL_2122432, EPI_ISL_2122433, EPI_ISL_2122435, EPI_ISL_2122438, EPI_ISL_2122439, EPI_ISL_2122441, EPI_ISL_2122442, EPI_ISL_2122444, EPI_ISL_2122445, EPI_ISL_2122448,
EPI_ISL_2122455, EPI_ISL_2122456, EPI_ISL_2122462, EPI_ISL_2122463, EPI_ISL_2122478, EPI_ISL_2122490, EPI_ISL_2122499, EPI_ISL_2122500, EPI_ISL_2122504, EPI_ISL_2122508, EPI_ISL_2122510, EPI_ISL_2122516, EPI_ISL_2122523, EPI_ISL_2122524, EPI_ISL_2122529, EPI_ISL_2122540, EPI_ISL_2122541, EPI_ISL_2122547, EPI_ISL_2122548,
EPI_ISL_2122553, EPI_ISL_2122557, EPI_ISL_2122560, EPI_ISL_2122563, EPI_ISL_2122565, EPI_ISL_2122583, EPI_ISL_2122587, EPI_ISL_2122593, EPI_ISL_2122597, EPI_ISL_2122602, EPI_ISL_2122604, EPI_ISL_2122606, EPI_ISL_2122609, EPI_ISL_2122623, EPI_ISL_2122629, EPI_ISL_2122630, EPI_ISL_2122638, EPI_ISL_2122644, EPI_ISL_2122646, EPI_ISL_2122647, EPI_ISL_2122654, EPI_ISL_2122655,
EPI_ISL_2122662, EPI_ISL_2122664, EPI_ISL_2122668, EPI_ISL_2122671, EPI_ISL_2122676, EPI_ISL_2122677, EPI_ISL_2122679, EPI_ISL_2122680, EPI_ISL_2122683, EPI_ISL_2122685, EPI_ISL_2122687, EPI_ISL_2122689, EPI_ISL_2122698, EPI_ISL_2122699, EPI_ISL_2122705, EPI_ISL_2137852, EPI_ISL_2137854, EPI_ISL_2137876, EPI_ISL_2137879, EPI_ISL_2137885, EPI_ISL_2137891,
EPI_ISL_2137893, EPI_ISL_2137898, EPI_ISL_2137899, EPI_ISL_2137900, EPI_ISL_2137903, EPI_ISL_2137904, EPI_ISL_2137907, EPI_ISL_2137910, EPI_ISL_2137950, EPI_ISL_2137952, EPI_ISL_2137968, EPI_ISL_2137972, EPI_ISL_2137979, EPI_ISL_2137990, EPI_ISL_2137993, EPI_ISL_2138000, EPI_ISL_2138003, EPI_ISL_2138004, EPI_ISL_2138006, EPI_ISL_2138020, EPI_ISL_2138022, EPI_ISL_2138024,
EPI_ISL_2138028, EPI_ISL_2138034, EPI_ISL_2138043, EPI_ISL_2138044, EPI_ISL_2138047, EPI_ISL_2138048, EPI_ISL_2138063, EPI_ISL_2138076, EPI_ISL_2138093, EPI_ISL_2138094, EPI_ISL_2138096, EPI_ISL_2138098, EPI_ISL_2138106, EPI_ISL_2138109, EPI_ISL_2138111, EPI_ISL_2138122, EPI_ISL_2138129, EPI_ISL_2138137, EPI_ISL_2138142, EPI_ISL_2138143, EPI_ISL_2138147,
EPI_ISL_2138155, EPI_ISL_2138158, EPI_ISL_2138159, EPI_ISL_2138163, EPI_ISL_2138164, EPI_ISL_2138167, EPI_ISL_2138173, EPI_ISL_2138176, EPI_ISL_2138178, EPI_ISL_2138181, EPI_ISL_2138183, EPI_ISL_2138184, EPI_ISL_2138198, EPI_ISL_2138205, EPI_ISL_2138208, EPI_ISL_2138210, EPI_ISL_2138215, EPI_ISL_2138224, EPI_ISL_2138232, EPI_ISL_2138238, EPI_ISL_2138242, EPI_ISL_2138255,
EPI_ISL_2138272, EPI_ISL_2138289, EPI_ISL_2138293, EPI_ISL_2138296, EPI_ISL_2138307, EPI_ISL_2138311, EPI_ISL_2138315, EPI_ISL_2138318, EPI_ISL_2138321, EPI_ISL_2138322, EPI_ISL_2138325, EPI_ISL_2138337, EPI_ISL_2138339, EPI_ISL_2138353, EPI_ISL_2138355, EPI_ISL_2138356, EPI_ISL_2138359, EPI_ISL_2138360, EPI_ISL_2138363, EPI_ISL_2138372, EPI_ISL_2138379,
EPI_ISL_2138386, EPI_ISL_2138390, EPI_ISL_2138391, EPI_ISL_2138398, EPI_ISL_2138406, EPI_ISL_2138407, EPI_ISL_2138409, EPI_ISL_2138414, EPI_ISL_2138415, EPI_ISL_2138420, EPI_ISL_2138428, EPI_ISL_2138430, EPI_ISL_2138431, EPI_ISL_2138440, EPI_ISL_2138441, EPI_ISL_2138446, EPI_ISL_2138448, EPI_ISL_2138451, EPI_ISL_2138454, EPI_ISL_2138456, EPI_ISL_2138458, EPI_ISL_2138460,
EPI_ISL_2138461, EPI_ISL_2138467, EPI_ISL_2138470, EPI_ISL_2138472, EPI_ISL_2138476, EPI_ISL_2138477, EPI_ISL_2138479, EPI_ISL_2138480, EPI_ISL_2138481, EPI_ISL_2138483, EPI_ISL_2138484, EPI_ISL_2138487, EPI_ISL_2138488, EPI_ISL_2138489, EPI_ISL_2138491, EPI_ISL_2138492, EPI_ISL_2138493, EPI_ISL_2138494, EPI_ISL_2138501, EPI_ISL_2138505, EPI_ISL_2138508, EPI_ISL_2138511,
EPI_ISL_2138512, EPI_ISL_2138520, EPI_ISL_2138522, EPI_ISL_2138524, EPI_ISL_2138528, EPI_ISL_2138532, EPI_ISL_2138533, EPI_ISL_2138538, EPI_ISL_2138544, EPI_ISL_2138553, EPI_ISL_2138558, EPI_ISL_2138559, EPI_ISL_2138560, EPI_ISL_2138562, EPI_ISL_2138563, EPI_ISL_2138565, EPI_ISL_2138568, EPI_ISL_2138569, EPI_ISL_2138571, EPI_ISL_2138577, EPI_ISL_2138578, EPI_ISL_2138579,
EPI_ISL_2138583, EPI_ISL_2138584, EPI_ISL_2138586, EPI_ISL_2138587, EPI_ISL_2138589, EPI_ISL_2138594, EPI_ISL_2138596, EPI_ISL_2138601, EPI_ISL_2138602, EPI_ISL_2138603, EPI_ISL_2138604, EPI_ISL_2138606, EPI_ISL_2138607, EPI_ISL_2138608, EPI_ISL_2138609, EPI_ISL_2138610, EPI_ISL_2138612, EPI_ISL_2138616, EPI_ISL_2138619, EPI_ISL_2138620, EPI_ISL_2138624, EPI_ISL_2138633,
EPI_ISL_2138635, EPI_ISL_2138636, EPI_ISL_2138641, EPI_ISL_2138644, EPI_ISL_2138647, EPI_ISL_2138650, EPI_ISL_2138653, EPI_ISL_2138663, EPI_ISL_2138665, EPI_ISL_2138670, EPI_ISL_2138674, EPI_ISL_2138675, EPI_ISL_2138683, EPI_ISL_2138686, EPI_ISL_2138689, EPI_ISL_2138692, EPI_ISL_2138693, EPI_ISL_2138697, EPI_ISL_2138698, EPI_ISL_2138705, EPI_ISL_2138710, EPI_ISL_2138713,
EPI_ISL_2138715, EPI_ISL_2138717, EPI_ISL_2152656, EPI_ISL_2152657, EPI_ISL_2152659, EPI_ISL_2152664, EPI_ISL_2153047, EPI_ISL_2171675, EPI_ISL_2171676, EPI_ISL_2171677, EPI_ISL_2171678, EPI_ISL_2171679, EPI_ISL_2171682, EPI_ISL_2171684, EPI_ISL_2171685, EPI_ISL_2171686, EPI_ISL_2171687, EPI_ISL_2171688, EPI_ISL_2171689, EPI_ISL_2171690, EPI_ISL_2171691, EPI_ISL_2171692,
EPI_ISL_2171693, EPI_ISL_2171694, EPI_ISL_2171696, EPI_ISL_2171697, EPI_ISL_2171698, EPI_ISL_2171699, EPI_ISL_2171700, EPI_ISL_2171701, EPI_ISL_2171702, EPI_ISL_2171704, EPI_ISL_2171705, EPI_ISL_2171709, EPI_ISL_2171710, EPI_ISL_2171711, EPI_ISL_2171712, EPI_ISL_2171713, EPI_ISL_2171714, EPI_ISL_2171716, EPI_ISL_2171717, EPI_ISL_2171718, EPI_ISL_2171719, EPI_ISL_2171724,
EPI_ISL_2171725, EPI_ISL_2171727, EPI_ISL_2171728, EPI_ISL_2171729, EPI_ISL_2171730, EPI_ISL_2171731, EPI_ISL_2171732, EPI_ISL_2171733, EPI_ISL_2171735, EPI_ISL_2171736, EPI_ISL_2171737, EPI_ISL_2171739, EPI_ISL_2171740, EPI_ISL_2171741, EPI_ISL_2171742, EPI_ISL_2171743, EPI_ISL_2171744, EPI_ISL_2171745, EPI_ISL_2171747, EPI_ISL_2171749, EPI_ISL_2171750, EPI_ISL_2171751,
EPI_ISL_2171752, EPI_ISL_2171753, EPI_ISL_2171754, EPI_ISL_2171755, EPI_ISL_2171757, EPI_ISL_2171758, EPI_ISL_2171759, EPI_ISL_2171760, EPI_ISL_2171761, EPI_ISL_2171765, EPI_ISL_2171766, EPI_ISL_2171767, EPI_ISL_2171768, EPI_ISL_2171769, EPI_ISL_2171775, EPI_ISL_2171776, EPI_ISL_2171777, EPI_ISL_2171778, EPI_ISL_2171779, EPI_ISL_2171783, EPI_ISL_2171785, EPI_ISL_2171787,
EPI_ISL_2171788, EPI_ISL_2171789, EPI_ISL_2171790, EPI_ISL_2171792, EPI_ISL_2171793, EPI_ISL_2171794, EPI_ISL_2171796, EPI_ISL_2171797, EPI_ISL_2171798, EPI_ISL_2171799, EPI_ISL_2171802, EPI_ISL_2171803, EPI_ISL_2171805, EPI_ISL_2171808, EPI_ISL_2171809, EPI_ISL_2171810, EPI_ISL_2171811, EPI_ISL_2171812, EPI_ISL_2171813, EPI_ISL_2171816, EPI_ISL_2171817,
EPI_ISL_2171829, EPI_ISL_2171857, EPI_ISL_2171954, EPI_ISL_2171967, EPI_ISL_2172033, EPI_ISL_2198709, EPI_ISL_2198722, EPI_ISL_2198741, EPI_ISL_2198781, EPI_ISL_2198803, EPI_ISL_2198812, EPI_ISL_2198848, EPI_ISL_2198899, EPI_ISL_2198901, EPI_ISL_2198905, EPI_ISL_2198936, EPI_ISL_2198939, EPI_ISL_2198943, EPI_ISL_2198963, EPI_ISL_2198965, EPI_ISL_2199357, EPI_ISL_2199409,
EPI_ISL_2199439, EPI_ISL_2199488, EPI_ISL_2199521, EPI_ISL_2199535, EPI_ISL_2199586, EPI_ISL_2199629, EPI_ISL_2199643, EPI_ISL_2199653, EPI_ISL_2199654, EPI_ISL_2199659, EPI_ISL_2199832, EPI_ISL_2236820, EPI_ISL_2236822, EPI_ISL_2236823, EPI_ISL_2236824, EPI_ISL_2236827, EPI_ISL_2236829, EPI_ISL_2236836, EPI_ISL_2236840, EPI_ISL_2236841, EPI_ISL_2236843,
EPI_ISL_2236844, EPI_ISL_2236845, EPI_ISL_2236847, EPI_ISL_2236849, EPI_ISL_2236852, EPI_ISL_2236857, EPI_ISL_2236866, EPI_ISL_2236867, EPI_ISL_2236868, EPI_ISL_2236870, EPI_ISL_2236872, EPI_ISL_2236873, EPI_ISL_2236875, EPI_ISL_2236879, EPI_ISL_2236885, EPI_ISL_2236887, EPI_ISL_2236890, EPI_ISL_2236892, EPI_ISL_2236894, EPI_ISL_2236895, EPI_ISL_2236896, EPI_ISL_2236898,
EPI_ISL_2236899, EPI_ISL_2236900, EPI_ISL_2236903, EPI_ISL_2236905, EPI_ISL_2236907, EPI_ISL_2236908, EPI_ISL_2236910, EPI_ISL_2236911, EPI_ISL_2236913, EPI_ISL_2236915, EPI_ISL_2236917, EPI_ISL_2236919, EPI_ISL_2236920, EPI_ISL_2236922, EPI_ISL_2236924, EPI_ISL_2236926, EPI_ISL_2236928, EPI_ISL_2236929, EPI_ISL_2236930, EPI_ISL_2236934, EPI_ISL_2236936,
EPI_ISL_2236937, EPI_ISL_2236939, EPI_ISL_2236941, EPI_ISL_2236942, EPI_ISL_2236945, EPI_ISL_2236946, EPI_ISL_2236952, EPI_ISL_2236953, EPI_ISL_2236954, EPI_ISL_2236958, EPI_ISL_2236960, EPI_ISL_2236962, EPI_ISL_2236963, EPI_ISL_2236964, EPI_ISL_2236966, EPI_ISL_2236967, EPI_ISL_2236968, EPI_ISL_2236969, EPI_ISL_2236971, EPI_ISL_2236974, EPI_ISL_2236981, EPI_ISL_2236982,
EPI_ISL_2236983, EPI_ISL_2236984, EPI_ISL_2236987, EPI_ISL_2236990, EPI_ISL_2236992, EPI_ISL_2236995, EPI_ISL_2236996, EPI_ISL_2236998, EPI_ISL_2236999, EPI_ISL_2237000, EPI_ISL_2237003, EPI_ISL_2237006, EPI_ISL_2237007, EPI_ISL_2237008, EPI_ISL_2237011, EPI_ISL_2237012, EPI_ISL_2237013, EPI_ISL_2237014, EPI_ISL_2237015, EPI_ISL_2237020,
EPI_ISL_2237023, EPI_ISL_2237024, EPI_ISL_2237025, EPI_ISL_2237026, EPI_ISL_2237027, EPI_ISL_2237028, EPI_ISL_2237030, EPI_ISL_2237032, EPI_ISL_2237035, EPI_ISL_2237036, EPI_ISL_2237037, EPI_ISL_2237038, EPI_ISL_2237040, EPI_ISL_2237041, EPI_ISL_2237045, EPI_ISL_2237048, EPI_ISL_2237051, EPI_ISL_2237054, EPI_ISL_2237057, EPI_ISL_2237059, EPI_ISL_2237062, EPI_ISL_2237063,
EPI_ISL_2237064, EPI_ISL_2237065, EPI_ISL_2237070, EPI_ISL_2237071, EPI_ISL_2237079, EPI_ISL_2237080, EPI_ISL_2237081, EPI_ISL_2237082, EPI_ISL_2237083, EPI_ISL_2237085, EPI_ISL_2237086, EPI_ISL_2237087, EPI_ISL_2237088, EPI_ISL_2237090, EPI_ISL_2237091, EPI_ISL_2237096, EPI_ISL_2237098, EPI_ISL_2237100, EPI_ISL_2237102, EPI_ISL_2237103, EPI_ISL_2237106, EPI_ISL_2237107,
EPI_ISL_2237109, EPI_ISL_2237110, EPI_ISL_2237112, EPI_ISL_2237117, EPI_ISL_2237118, EPI_ISL_2237119, EPI_ISL_2237121, EPI_ISL_2237124, EPI_ISL_2237125, EPI_ISL_2237127, EPI_ISL_2237129, EPI_ISL_2237130, EPI_ISL_2237132, EPI_ISL_2237133, EPI_ISL_2237141, EPI_ISL_2237145, EPI_ISL_2237147, EPI_ISL_2237148, EPI_ISL_2237152, EPI_ISL_2237153, EPI_ISL_2237154, EPI_ISL_2237155,
EPI_ISL_2237156, EPI_ISL_2237157, EPI_ISL_2238181, EPI_ISL_2238189, EPI_ISL_2238207, EPI_ISL_2238251, EPI_ISL_2238256, EPI_ISL_2238261, EPI_ISL_2238295, EPI_ISL_2238334, EPI_ISL_2238356, EPI_ISL_2238363, EPI_ISL_2238364, EPI_ISL_2238366, EPI_ISL_2238371, EPI_ISL_2238373, EPI_ISL_2238374, EPI_ISL_2238375, EPI_ISL_2238376, EPI_ISL_2238380, EPI_ISL_2238381, EPI_ISL_2238384,
EPI_ISL_2238394, EPI_ISL_2238395, EPI_ISL_2238397, EPI_ISL_2238398, EPI_ISL_2238401, EPI_ISL_2238409, EPI_ISL_2238410, EPI_ISL_2238411, EPI_ISL_2238414, EPI_ISL_2238415, EPI_ISL_2238416, EPI_ISL_2238417, EPI_ISL_2238418, EPI_ISL_2238419, EPI_ISL_2238421, EPI_ISL_2238422, EPI_ISL_2238423, EPI_ISL_2238426, EPI_ISL_2238427, EPI_ISL_2238431, EPI_ISL_2238433, EPI_ISL_2238438,
EPI_ISL_2238441, EPI_ISL_2238448, EPI_ISL_2238452, EPI_ISL_2238456, EPI_ISL_2238461, EPI_ISL_2238466, EPI_ISL_2238469, EPI_ISL_2238470, EPI_ISL_2238471, EPI_ISL_2238480, EPI_ISL_2238482, EPI_ISL_2238485, EPI_ISL_2238486, EPI_ISL_2238489, EPI_ISL_2238490, EPI_ISL_2238496, EPI_ISL_2238498, EPI_ISL_2238501, EPI_ISL_2238513, EPI_ISL_2238515, EPI_ISL_2238516, EPI_ISL_2238518,
EPI_ISL_2238521, EPI_ISL_2238522, EPI_ISL_2238523, EPI_ISL_2238525, EPI_ISL_2238526, EPI_ISL_2238530, EPI_ISL_2238531, EPI_ISL_2238533, EPI_ISL_2238537, EPI_ISL_2238542, EPI_ISL_2238545, EPI_ISL_2238548, EPI_ISL_2238549, EPI_ISL_2238551, EPI_ISL_2238553, EPI_ISL_2238556, EPI_ISL_2238561, EPI_ISL_2238564, EPI_ISL_2238566, EPI_ISL_2238567, EPI_ISL_2238581,
EPI_ISL_2238584, EPI_ISL_2238585, EPI_ISL_2238587, EPI_ISL_2238590, EPI_ISL_2238593, EPI_ISL_2238603, EPI_ISL_2238606, EPI_ISL_2238609, EPI_ISL_2238617, EPI_ISL_2238618, EPI_ISL_2238619, EPI_ISL_2238620, EPI_ISL_2238621, EPI_ISL_2238631, EPI_ISL_2238633, EPI_ISL_2238635, EPI_ISL_2238638, EPI_ISL_2238640, EPI_ISL_2238641, EPI_ISL_2238646,
EPI_ISL_2238649, EPI_ISL_2238653, EPI_ISL_2238656, EPI_ISL_2238659, EPI_ISL_2238661, EPI_ISL_2238663, EPI_ISL_2238669, EPI_ISL_2238670, EPI_ISL_2238675, EPI_ISL_2238676, EPI_ISL_2238677, EPI_ISL_2238678

| | | | |
|---|---|---|---|
| see above | Lighthouse Lab in Alderley Park | Wellcome Sanger Institute for the COVID-19 Genomics UK (COG-UK) Consortium | Cordelia Langford; David K. Jackson; Dominic Kwiatkowski; Ewan Harrison; Ian Johnston; Jacquelyn Wynn; Jeffrey Barrett; John Sillitoe on behalf of the Wellcome Sanger Institute COVID-19 Surveillance Team; Mairead Hyland; Roberto Amato; Sonia Goncalves; The Lighthouse Lab in Alderley Park and Alex Alderton |

EPI_ISL_1939681, EPI_ISL_1986111, EPI_ISL_1986112, EPI_ISL_1986116, EPI_ISL_1986126, EPI_ISL_1986131, EPI_ISL_1986132, EPI_ISL_1986134, EPI_ISL_1986138, EPI_ISL_1986148, EPI_ISL_1986149, EPI_ISL_1986156, EPI_ISL_1986157, EPI_ISL_1986175, EPI_ISL_1986176, EPI_ISL_1986179, EPI_ISL_1986182, EPI_ISL_1986189, EPI_ISL_1986194, EPI_ISL_1986200, EPI_ISL_1986201, EPI_ISL_1986204,
EPI_ISL_1986215, EPI_ISL_1986216, EPI_ISL_1986219, EPI_ISL_1986228, EPI_ISL_1986231, EPI_ISL_1986238, EPI_ISL_1986241, EPI_ISL_1986251, EPI_ISL_1986254, EPI_ISL_1986264, EPI_ISL_1986265, EPI_ISL_1986268, EPI_ISL_1986270, EPI_ISL_1986273, EPI_ISL_1986276, EPI_ISL_1986279, EPI_ISL_1986282, EPI_ISL_1986294, EPI_ISL_1986296, EPI_ISL_1986298, EPI_ISL_1986324, EPI_ISL_1986325,
EPI_ISL_1986326, EPI_ISL_1986332, EPI_ISL_1986349, EPI_ISL_1986353, EPI_ISL_1986354, EPI_ISL_1986355, EPI_ISL_1986366, EPI_ISL_1986370, EPI_ISL_1986373, EPI_ISL_1986378, EPI_ISL_1986381, EPI_ISL_1986493, EPI_ISL_1986497, EPI_ISL_1986505, EPI_ISL_1986509, EPI_ISL_1986510, EPI_ISL_1986517, EPI_ISL_1986518, EPI_ISL_1986520, EPI_ISL_1986522, EPI_ISL_1986524, EPI_ISL_1986533,
EPI_ISL_1986534, EPI_ISL_1986538, EPI_ISL_1986552, EPI_ISL_1986553, EPI_ISL_1986558, EPI_ISL_1986559, EPI_ISL_1986560, EPI_ISL_1986569, EPI_ISL_1986571, EPI_ISL_1986594, EPI_ISL_1986605, EPI_ISL_1986609, EPI_ISL_1986619, EPI_ISL_1986623, EPI_ISL_1986627, EPI_ISL_1986648, EPI_ISL_1986664, EPI_ISL_1986665, EPI_ISL_1986686, EPI_ISL_1986710, EPI_ISL_1986726, EPI_ISL_1986736,
EPI_ISL_1986753, EPI_ISL_1986762, EPI_ISL_1986770, EPI_ISL_1986778, EPI_ISL_1986785, EPI_ISL_1986797, EPI_ISL_1986804, EPI_ISL_1986805, EPI_ISL_1986806, EPI_ISL_1986807, EPI_ISL_1986816, EPI_ISL_1986823, EPI_ISL_1986831, EPI_ISL_1986842, EPI_ISL_1986874, EPI_ISL_1986882, EPI_ISL_1986890, EPI_ISL_1986907, EPI_ISL_1986912, EPI_ISL_1986923, EPI_ISL_1986925, EPI_ISL_1986927,
EPI_ISL_1986928, EPI_ISL_1986946, EPI_ISL_1986952, EPI_ISL_2004349, EPI_ISL_2004402, EPI_ISL_2004408, EPI_ISL_2004418, EPI_ISL_2004438, EPI_ISL_2004484, EPI_ISL_2004508, EPI_ISL_2004549, EPI_ISL_2004559, EPI_ISL_2004605, EPI_ISL_2004644, EPI_ISL_2004711, EPI_ISL_2004716, EPI_ISL_2004721, EPI_ISL_2004814, EPI_ISL_2004844, EPI_ISL_2004866, EPI_ISL_2004873, EPI_ISL_2004877,
EPI_ISL_2021040, EPI_ISL_2021042, EPI_ISL_2021045, EPI_ISL_2021048, EPI_ISL_2021049, EPI_ISL_2021050, EPI_ISL_2021051, EPI_ISL_2021057, EPI_ISL_2021058, EPI_ISL_2021060, EPI_ISL_2021066, EPI_ISL_2021070, EPI_ISL_2021072, EPI_ISL_2021075, EPI_ISL_2021076, EPI_ISL_2021078, EPI_ISL_2021079, EPI_ISL_2021080, EPI_ISL_2021084, EPI_ISL_2021085, EPI_ISL_2021088, EPI_ISL_2021089,
EPI_ISL_2021091, EPI_ISL_2021093, EPI_ISL_2021094, EPI_ISL_2021095, EPI_ISL_2021096, EPI_ISL_2021098, EPI_ISL_2021100, EPI_ISL_2021101, EPI_ISL_2021102, EPI_ISL_2021103, EPI_ISL_2021105, EPI_ISL_2021106, EPI_ISL_2021108, EPI_ISL_2021112, EPI_ISL_2021113, EPI_ISL_2021116, EPI_ISL_2021117, EPI_ISL_2021118, EPI_ISL_2021121, EPI_ISL_2021122, EPI_ISL_2021125, EPI_ISL_2021126,
EPI_ISL_2021129, EPI_ISL_2021134, EPI_ISL_2021135, EPI_ISL_2021138, EPI_ISL_2021140, EPI_ISL_2021142, EPI_ISL_2021146, EPI_ISL_2021150, EPI_ISL_2021153, EPI_ISL_2021155, EPI_ISL_2021156, EPI_ISL_2021160, EPI_ISL_2021162, EPI_ISL_2021163, EPI_ISL_2021164, EPI_ISL_2021165, EPI_ISL_2021166, EPI_ISL_2021168, EPI_ISL_2021169, EPI_ISL_2021172, EPI_ISL_2021173,
EPI_ISL_2021174, EPI_ISL_2021175, EPI_ISL_2021176, EPI_ISL_2021177, EPI_ISL_2021178, EPI_ISL_2021179, EPI_ISL_2021180, EPI_ISL_2021184, EPI_ISL_2021187, EPI_ISL_2021188, EPI_ISL_2021192, EPI_ISL_2021196, EPI_ISL_2021199, EPI_ISL_2021201, EPI_ISL_2021202, EPI_ISL_2021203, EPI_ISL_2021204, EPI_ISL_2021206, EPI_ISL_2021207, EPI_ISL_2021208, EPI_ISL_2021210, EPI_ISL_2021211,
EPI_ISL_2021212, EPI_ISL_2021213, EPI_ISL_2021214, EPI_ISL_2021216, EPI_ISL_2021217, EPI_ISL_2021220, EPI_ISL_2021223, EPI_ISL_2021226, EPI_ISL_2021230, EPI_ISL_2021232, EPI_ISL_2021233, EPI_ISL_2021234, EPI_ISL_2021237, EPI_ISL_2021238, EPI_ISL_2021239, EPI_ISL_2021240, EPI_ISL_2021243, EPI_ISL_2021244, EPI_ISL_2021245, EPI_ISL_2021246, EPI_ISL_2021247, EPI_ISL_2021250,
EPI_ISL_2021251, EPI_ISL_2021256, EPI_ISL_2021257, EPI_ISL_2021259, EPI_ISL_2021260, EPI_ISL_2021261, EPI_ISL_2021264, EPI_ISL_2021265, EPI_ISL_2021266, EPI_ISL_2021269, EPI_ISL_2021272, EPI_ISL_2021273, EPI_ISL_2021276, EPI_ISL_2021279, EPI_ISL_2021283, EPI_ISL_2021284, EPI_ISL_2021286, EPI_ISL_2021288, EPI_ISL_2021289, EPI_ISL_2021290, EPI_ISL_2021295, EPI_ISL_2021297,
EPI_ISL_2021299, EPI_ISL_2021300, EPI_ISL_2021302, EPI_ISL_2021303, EPI_ISL_2021304, EPI_ISL_2021305, EPI_ISL_2021306, EPI_ISL_2021307, EPI_ISL_2021308, EPI_ISL_2021312, EPI_ISL_2021313, EPI_ISL_2021314, EPI_ISL_2021320, EPI_ISL_2021321, EPI_ISL_2021322, EPI_ISL_2021323, EPI_ISL_2021324, EPI_ISL_2021325, EPI_ISL_2021328, EPI_ISL_2021333, EPI_ISL_2021335,
EPI_ISL_2021336, EPI_ISL_2021337, EPI_ISL_2021338, EPI_ISL_2021340, EPI_ISL_2021341, EPI_ISL_2021342, EPI_ISL_2021344, EPI_ISL_2021345, EPI_ISL_2021346, EPI_ISL_2021347, EPI_ISL_2021350, EPI_ISL_2021353, EPI_ISL_2021354, EPI_ISL_2021355, EPI_ISL_2021357, EPI_ISL_2021359, EPI_ISL_2021360, EPI_ISL_2021361, EPI_ISL_2021765, EPI_ISL_2021806, EPI_ISL_2022340, EPI_ISL_2022781,
EPI_ISL_2022782, EPI_ISL_2022783, EPI_ISL_2022785, EPI_ISL_2022786, EPI_ISL_2022787, EPI_ISL_2022788, EPI_ISL_2022789, EPI_ISL_2022790, EPI_ISL_2022791, EPI_ISL_2022792, EPI_ISL_2022793, EPI_ISL_2022794, EPI_ISL_2022795, EPI_ISL_2022796, EPI_ISL_2022797, EPI_ISL_2022798, EPI_ISL_2022799, EPI_ISL_2022800, EPI_ISL_2022801, EPI_ISL_2022802, EPI_ISL_2022803, EPI_ISL_2022804,

EPI_ISL_2022806, EPI_ISL_2022807, EPI_ISL_2022808, EPI_ISL_2022809, EPI_ISL_2022810, EPI_ISL_2022811, EPI_ISL_2022812, EPI_ISL_2022813, EPI_ISL_2022814, EPI_ISL_2022815, EPI_ISL_2022816, EPI_ISL_2022818, EPI_ISL_2022819, EPI_ISL_2022820, EPI_ISL_2022821, EPI_ISL_2022822, EPI_ISL_2022823, EPI_ISL_2022824, EPI_ISL_2022825, EPI_ISL_2022826, EPI_ISL_2022827, EPI_ISL_2022828,
EPI_ISL_2022829, EPI_ISL_2022830, EPI_ISL_2022831, EPI_ISL_2022832, EPI_ISL_2022833, EPI_ISL_2022834, EPI_ISL_2022835, EPI_ISL_2022836, EPI_ISL_2022837, EPI_ISL_2022838, EPI_ISL_2022839, EPI_ISL_2022841, EPI_ISL_2022843, EPI_ISL_2022844, EPI_ISL_2022846, EPI_ISL_2022847, EPI_ISL_2022848, EPI_ISL_2022849, EPI_ISL_2022850, EPI_ISL_2022851, EPI_ISL_2022852, EPI_ISL_2022853,
EPI_ISL_2022854, EPI_ISL_2022855, EPI_ISL_2022856, EPI_ISL_2022857, EPI_ISL_2022858, EPI_ISL_2022859, EPI_ISL_2022860, EPI_ISL_2022861, EPI_ISL_2022862, EPI_ISL_2022863, EPI_ISL_2022864, EPI_ISL_2022865, EPI_ISL_2022866, EPI_ISL_2022867, EPI_ISL_2022869, EPI_ISL_2022870, EPI_ISL_2022871, EPI_ISL_2022872, EPI_ISL_2022873, EPI_ISL_2022874, EPI_ISL_2022875,
EPI_ISL_2022876, EPI_ISL_2022877, EPI_ISL_2022878, EPI_ISL_2022879, EPI_ISL_2022880, EPI_ISL_2022881, EPI_ISL_2022882, EPI_ISL_2022883, EPI_ISL_2022884, EPI_ISL_2022885, EPI_ISL_2022886, EPI_ISL_2022887, EPI_ISL_2022888, EPI_ISL_2022889, EPI_ISL_2022890, EPI_ISL_2022891, EPI_ISL_2022892, EPI_ISL_2022893, EPI_ISL_2022894, EPI_ISL_2022895, EPI_ISL_2022896, EPI_ISL_2022897,
EPI_ISL_2022898, EPI_ISL_2022899, EPI_ISL_2022900, EPI_ISL_2022901, EPI_ISL_2022903, EPI_ISL_2022904, EPI_ISL_2022905, EPI_ISL_2022906, EPI_ISL_2022907, EPI_ISL_2022908, EPI_ISL_2022909, EPI_ISL_2022910, EPI_ISL_2022911, EPI_ISL_2022914, EPI_ISL_2022915, EPI_ISL_2022916, EPI_ISL_2022917, EPI_ISL_2022918, EPI_ISL_2022919, EPI_ISL_2022920, EPI_ISL_2022921, EPI_ISL_2022923,
EPI_ISL_2022924, EPI_ISL_2022925, EPI_ISL_2022926, EPI_ISL_2022927, EPI_ISL_2022928, EPI_ISL_2022929, EPI_ISL_2022930, EPI_ISL_2022931, EPI_ISL_2022932, EPI_ISL_2022933, EPI_ISL_2022934, EPI_ISL_2022935, EPI_ISL_2022936, EPI_ISL_2022937, EPI_ISL_2022938, EPI_ISL_2022939, EPI_ISL_2022940, EPI_ISL_2022941, EPI_ISL_2022942, EPI_ISL_2022943, EPI_ISL_2022944, EPI_ISL_2022945,
EPI_ISL_2022946, EPI_ISL_2022947, EPI_ISL_2092676, EPI_ISL_2092677, EPI_ISL_2092678, EPI_ISL_2092679, EPI_ISL_2092680, EPI_ISL_2092681, EPI_ISL_2092682, EPI_ISL_2092683, EPI_ISL_2092684, EPI_ISL_2092685, EPI_ISL_2092686, EPI_ISL_2092687, EPI_ISL_2092688, EPI_ISL_2092689, EPI_ISL_2092690, EPI_ISL_2092691, EPI_ISL_2092692, EPI_ISL_2092693, EPI_ISL_2092694, EPI_ISL_2092695,
EPI_ISL_2092696, EPI_ISL_2092697, EPI_ISL_2092698, EPI_ISL_2092699, EPI_ISL_2092700, EPI_ISL_2092701, EPI_ISL_2092702, EPI_ISL_2092703, EPI_ISL_2092704, EPI_ISL_2092705, EPI_ISL_2092706, EPI_ISL_2092707, EPI_ISL_2092708, EPI_ISL_2092709, EPI_ISL_2092710, EPI_ISL_2092711, EPI_ISL_2092712, EPI_ISL_2092713, EPI_ISL_2092714, EPI_ISL_2092715, EPI_ISL_2092716, EPI_ISL_2092717,
EPI_ISL_2092718, EPI_ISL_2092719, EPI_ISL_2092720, EPI_ISL_2092721, EPI_ISL_2092722, EPI_ISL_2092723, EPI_ISL_2092724, EPI_ISL_2092725, EPI_ISL_2092726, EPI_ISL_2092727, EPI_ISL_2092728, EPI_ISL_2092729, EPI_ISL_2092730, EPI_ISL_2092731, EPI_ISL_2092732, EPI_ISL_2092733, EPI_ISL_2092734, EPI_ISL_2092735, EPI_ISL_2092736, EPI_ISL_2092737, EPI_ISL_2092738, EPI_ISL_2092739,
EPI_ISL_2092740, EPI_ISL_2092743, EPI_ISL_2092744, EPI_ISL_2092745, EPI_ISL_2092746, EPI_ISL_2092747, EPI_ISL_2092748, EPI_ISL_2092749, EPI_ISL_2092750, EPI_ISL_2092751, EPI_ISL_2092752, EPI_ISL_2092753, EPI_ISL_2092754, EPI_ISL_2092755, EPI_ISL_2092756, EPI_ISL_2117519, EPI_ISL_2117521, EPI_ISL_2117532, EPI_ISL_2117536, EPI_ISL_2117554, EPI_ISL_2117563, EPI_ISL_2117571,
EPI_ISL_2117578, EPI_ISL_2117579, EPI_ISL_2117581, EPI_ISL_2117597, EPI_ISL_2117600, EPI_ISL_2117621, EPI_ISL_2117627, EPI_ISL_2117632, EPI_ISL_2117639, EPI_ISL_2117643, EPI_ISL_2117644, EPI_ISL_2117645, EPI_ISL_2117648, EPI_ISL_2117649, EPI_ISL_2117651, EPI_ISL_2117657, EPI_ISL_2117660, EPI_ISL_2117667, EPI_ISL_2117668, EPI_ISL_2117681, EPI_ISL_2117682, EPI_ISL_2117697,
EPI_ISL_2117703, EPI_ISL_2117713, EPI_ISL_2117714, EPI_ISL_2117716, EPI_ISL_2117726, EPI_ISL_2117727, EPI_ISL_2117731, EPI_ISL_2117734, EPI_ISL_2117736, EPI_ISL_2117745, EPI_ISL_2117749, EPI_ISL_2117751, EPI_ISL_2117752, EPI_ISL_2117760, EPI_ISL_2117761, EPI_ISL_2117765, EPI_ISL_2117767, EPI_ISL_2117773, EPI_ISL_2117779, EPI_ISL_2117781, EPI_ISL_2117799, EPI_ISL_2117804,
EPI_ISL_2117806, EPI_ISL_2117808, EPI_ISL_2117811, EPI_ISL_2117814, EPI_ISL_2117817, EPI_ISL_2117820, EPI_ISL_2117823, EPI_ISL_2117828, EPI_ISL_2117837, EPI_ISL_2117842, EPI_ISL_2117854, EPI_ISL_2117855, EPI_ISL_2117857, EPI_ISL_2117864, EPI_ISL_2117870, EPI_ISL_2117873, EPI_ISL_2117883, EPI_ISL_2117885, EPI_ISL_2117893, EPI_ISL_2117900, EPI_ISL_2117904, EPI_ISL_2117906,
EPI_ISL_2117907, EPI_ISL_2117909, EPI_ISL_2117912, EPI_ISL_2117915, EPI_ISL_2117921, EPI_ISL_2117922, EPI_ISL_2117925, EPI_ISL_2117927, EPI_ISL_2117930, EPI_ISL_2117945, EPI_ISL_2117946, EPI_ISL_2117948, EPI_ISL_2117951, EPI_ISL_2117958, EPI_ISL_2117963, EPI_ISL_2117970, EPI_ISL_2117977, EPI_ISL_2117978, EPI_ISL_2117985, EPI_ISL_2117988, EPI_ISL_2117990, EPI_ISL_2117997,
EPI_ISL_2117998, EPI_ISL_2118004, EPI_ISL_2118008, EPI_ISL_2118013, EPI_ISL_2118017, EPI_ISL_2118020, EPI_ISL_2118022, EPI_ISL_2118025, EPI_ISL_2118030, EPI_ISL_2118033, EPI_ISL_2118038, EPI_ISL_2118042, EPI_ISL_2118046, EPI_ISL_2118048, EPI_ISL_2118053, EPI_ISL_2118054, EPI_ISL_2118056, EPI_ISL_2118062, EPI_ISL_2118067, EPI_ISL_2118073, EPI_ISL_2118078, EPI_ISL_2118080,
EPI_ISL_2118081, EPI_ISL_2118086, EPI_ISL_2118087, EPI_ISL_2118088, EPI_ISL_2118090, EPI_ISL_2118091, EPI_ISL_2118094, EPI_ISL_2118103, EPI_ISL_2118105, EPI_ISL_2118109, EPI_ISL_2118125, EPI_ISL_2118126, EPI_ISL_2118127, EPI_ISL_2118130, EPI_ISL_2118133, EPI_ISL_2118135, EPI_ISL_2118137, EPI_ISL_2118139, EPI_ISL_2118143, EPI_ISL_2118145, EPI_ISL_2118146, EPI_ISL_2118149,
EPI_ISL_2118150, EPI_ISL_2118153, EPI_ISL_2118159, EPI_ISL_2118167, EPI_ISL_2118176, EPI_ISL_2118177, EPI_ISL_2118180, EPI_ISL_2118193, EPI_ISL_2118199, EPI_ISL_2118200, EPI_ISL_2118212, EPI_ISL_2118222, EPI_ISL_2118232, EPI_ISL_2118240, EPI_ISL_2118241, EPI_ISL_2118243, EPI_ISL_2118246, EPI_ISL_2118247, EPI_ISL_2118806, EPI_ISL_2119351, EPI_ISL_2119354, EPI_ISL_2119359,
EPI_ISL_2119364, EPI_ISL_2119375, EPI_ISL_2119378, EPI_ISL_2119386, EPI_ISL_2119387, EPI_ISL_2119389, EPI_ISL_2119392, EPI_ISL_2119398, EPI_ISL_2119403, EPI_ISL_2119420, EPI_ISL_2119424, EPI_ISL_2119425, EPI_ISL_2119430, EPI_ISL_2119431, EPI_ISL_2119435, EPI_ISL_2119438, EPI_ISL_2119441, EPI_ISL_2119444, EPI_ISL_2119449, EPI_ISL_2119460, EPI_ISL_2119462, EPI_ISL_2119463,
EPI_ISL_2119470, EPI_ISL_2119474, EPI_ISL_2119482, EPI_ISL_2119485, EPI_ISL_2119491, EPI_ISL_2119498, EPI_ISL_2119502, EPI_ISL_2119512, EPI_ISL_2119513, EPI_ISL_2119522, EPI_ISL_2119525, EPI_ISL_2119528, EPI_ISL_2119546, EPI_ISL_2119556, EPI_ISL_2119562, EPI_ISL_2119564, EPI_ISL_2119567, EPI_ISL_2119576, EPI_ISL_2119577, EPI_ISL_2119589, EPI_ISL_2119593, EPI_ISL_2119597,
EPI_ISL_2119610, EPI_ISL_2119625, EPI_ISL_2119635, EPI_ISL_2119642, EPI_ISL_2119652, EPI_ISL_2119656, EPI_ISL_2119658, EPI_ISL_2119661, EPI_ISL_2119662, EPI_ISL_2119664, EPI_ISL_2119671, EPI_ISL_2119680, EPI_ISL_2119682, EPI_ISL_2119683, EPI_ISL_2119692, EPI_ISL_2119699, EPI_ISL_2119712, EPI_ISL_2119717, EPI_ISL_2119724, EPI_ISL_2119741, EPI_ISL_2119745, EPI_ISL_2119748,
EPI_ISL_2119749, EPI_ISL_2119753, EPI_ISL_2119754, EPI_ISL_2119778, EPI_ISL_2119782, EPI_ISL_2119792, EPI_ISL_2119803, EPI_ISL_2119805, EPI_ISL_2119819, EPI_ISL_2119825, EPI_ISL_2119828, EPI_ISL_2119832, EPI_ISL_2119836, EPI_ISL_2119841, EPI_ISL_2119848, EPI_ISL_2119858, EPI_ISL_2119863, EPI_ISL_2119864, EPI_ISL_2119870, EPI_ISL_2119912, EPI_ISL_2119918, EPI_ISL_2119921,
EPI_ISL_2119924, EPI_ISL_2119925, EPI_ISL_2119928, EPI_ISL_2119930, EPI_ISL_2119933, EPI_ISL_2119934, EPI_ISL_2119938, EPI_ISL_2119939, EPI_ISL_2119941, EPI_ISL_2119950, EPI_ISL_2119964, EPI_ISL_2119967, EPI_ISL_2119968, EPI_ISL_2119981, EPI_ISL_2119984, EPI_ISL_2120005, EPI_ISL_2120010, EPI_ISL_2120016, EPI_ISL_2120019, EPI_ISL_2120021, EPI_ISL_2120024, EPI_ISL_2120025,
EPI_ISL_2120030, EPI_ISL_2120033, EPI_ISL_2120035, EPI_ISL_2120039, EPI_ISL_2120046, EPI_ISL_2120049, EPI_ISL_2120066, EPI_ISL_2120067, EPI_ISL_2120075, EPI_ISL_2120078, EPI_ISL_2120098, EPI_ISL_2120116, EPI_ISL_2120120, EPI_ISL_2120126, EPI_ISL_2120133, EPI_ISL_2120140, EPI_ISL_2120148, EPI_ISL_2120154, EPI_ISL_2120165, EPI_ISL_2120181, EPI_ISL_2120209, EPI_ISL_2120210,
EPI_ISL_2120218, EPI_ISL_2120222, EPI_ISL_2120224, EPI_ISL_2120226, EPI_ISL_2120229, EPI_ISL_2120230, EPI_ISL_2120238, EPI_ISL_2120245, EPI_ISL_2120247, EPI_ISL_2120250, EPI_ISL_2120253, EPI_ISL_2120259, EPI_ISL_2120262, EPI_ISL_2120267, EPI_ISL_2120280, EPI_ISL_2120282, EPI_ISL_2120290, EPI_ISL_2120295, EPI_ISL_2120296, EPI_ISL_2120307, EPI_ISL_2120319, EPI_ISL_2120322,
EPI_ISL_2120324, EPI_ISL_2120326, EPI_ISL_2120339, EPI_ISL_2120343, EPI_ISL_2120345, EPI_ISL_2120362, EPI_ISL_2120365, EPI_ISL_2120373, EPI_ISL_2120381, EPI_ISL_2120394, EPI_ISL_2120395, EPI_ISL_2120406, EPI_ISL_2120407, EPI_ISL_2120418, EPI_ISL_2120422, EPI_ISL_2120431, EPI_ISL_2120436, EPI_ISL_2120439, EPI_ISL_2120440, EPI_ISL_2120443, EPI_ISL_2120448,
EPI_ISL_2120466, EPI_ISL_2120472, EPI_ISL_2120474, EPI_ISL_2120484, EPI_ISL_2120497, EPI_ISL_2120508, EPI_ISL_2120515, EPI_ISL_2120520, EPI_ISL_2120528, EPI_ISL_2120532, EPI_ISL_2120535, EPI_ISL_2120549, EPI_ISL_2120557, EPI_ISL_2120564, EPI_ISL_2120566, EPI_ISL_2120570, EPI_ISL_2120597, EPI_ISL_2120598, EPI_ISL_2120601, EPI_ISL_2120616, EPI_ISL_2120622, EPI_ISL_2120631,
EPI_ISL_2120634, EPI_ISL_2120636, EPI_ISL_2120638, EPI_ISL_2120646, EPI_ISL_2120649, EPI_ISL_2120654, EPI_ISL_2120657, EPI_ISL_2120690, EPI_ISL_2120703, EPI_ISL_2120708, EPI_ISL_2120717, EPI_ISL_2120718, EPI_ISL_2120720, EPI_ISL_2120723, EPI_ISL_2120733, EPI_ISL_2120735, EPI_ISL_2120736, EPI_ISL_2120738, EPI_ISL_2120739, EPI_ISL_2120740, EPI_ISL_2120753, EPI_ISL_2120770,
EPI_ISL_2120773, EPI_ISL_2120774, EPI_ISL_2120775, EPI_ISL_2120781, EPI_ISL_2120787, EPI_ISL_2120790, EPI_ISL_2120798, EPI_ISL_2120806, EPI_ISL_2120808, EPI_ISL_2120814, EPI_ISL_2120816, EPI_ISL_2120827, EPI_ISL_2120833, EPI_ISL_2120840, EPI_ISL_2120842, EPI_ISL_2120846, EPI_ISL_2120848, EPI_ISL_2120849, EPI_ISL_2120858, EPI_ISL_2120871, EPI_ISL_2120873, EPI_ISL_2120874,
EPI_ISL_2120876, EPI_ISL_2120880, EPI_ISL_2120883, EPI_ISL_2120886, EPI_ISL_2120892, EPI_ISL_2120898, EPI_ISL_2120899, EPI_ISL_2120904, EPI_ISL_2120907, EPI_ISL_2120908, EPI_ISL_2120922, EPI_ISL_2120928, EPI_ISL_2120949, EPI_ISL_2120954, EPI_ISL_2120966, EPI_ISL_2120969, EPI_ISL_2120971, EPI_ISL_2120980, EPI_ISL_2120983, EPI_ISL_2120986, EPI_ISL_2120987, EPI_ISL_2120996,
EPI_ISL_2121004, EPI_ISL_2121014, EPI_ISL_2121016, EPI_ISL_2121020, EPI_ISL_2121028, EPI_ISL_2121034, EPI_ISL_2121035, EPI_ISL_2121038, EPI_ISL_2121039, EPI_ISL_2121042, EPI_ISL_2121049, EPI_ISL_2121053, EPI_ISL_2121055, EPI_ISL_2121058, EPI_ISL_2121061, EPI_ISL_2121063, EPI_ISL_2121064, EPI_ISL_2121067, EPI_ISL_2121070, EPI_ISL_2121073, EPI_ISL_2121075, EPI_ISL_2121081,
EPI_ISL_2121084, EPI_ISL_2121085, EPI_ISL_2121086, EPI_ISL_2121088, EPI_ISL_2121089, EPI_ISL_2121091, EPI_ISL_2121094, EPI_ISL_2121099, EPI_ISL_2121102, EPI_ISL_2121105, EPI_ISL_2121110, EPI_ISL_2121113, EPI_ISL_2121119, EPI_ISL_2121120, EPI_ISL_2121125, EPI_ISL_2121126, EPI_ISL_2121128, EPI_ISL_2121129, EPI_ISL_2121131, EPI_ISL_2121132, EPI_ISL_2121134, EPI_ISL_2121135,
EPI_ISL_2121139, EPI_ISL_2121143, EPI_ISL_2121144, EPI_ISL_2121146, EPI_ISL_2121147, EPI_ISL_2121149, EPI_ISL_2121154, EPI_ISL_2121160, EPI_ISL_2121163, EPI_ISL_2121169, EPI_ISL_2121171, EPI_ISL_2121172, EPI_ISL_2121174, EPI_ISL_2121175, EPI_ISL_2121179, EPI_ISL_2121180, EPI_ISL_2121185, EPI_ISL_2121186, EPI_ISL_2121190, EPI_ISL_2121192, EPI_ISL_2121193, EPI_ISL_2121199,
EPI_ISL_2121203, EPI_ISL_2121206, EPI_ISL_2121207, EPI_ISL_2121213, EPI_ISL_2121216, EPI_ISL_2121218, EPI_ISL_2121219, EPI_ISL_2121221, EPI_ISL_2121224, EPI_ISL_2121225, EPI_ISL_2121227, EPI_ISL_2121228, EPI_ISL_2121231, EPI_ISL_2121234, EPI_ISL_2121235, EPI_ISL_2121244, EPI_ISL_2121249, EPI_ISL_2121250, EPI_ISL_2121252, EPI_ISL_2121258, EPI_ISL_2121261, EPI_ISL_2121262,
EPI_ISL_2121268, EPI_ISL_2121269, EPI_ISL_2121271, EPI_ISL_2121274, EPI_ISL_2121279, EPI_ISL_2121280, EPI_ISL_2121281, EPI_ISL_2121283, EPI_ISL_2121285, EPI_ISL_2121288, EPI_ISL_2121290, EPI_ISL_2121297, EPI_ISL_2121299, EPI_ISL_2121302, EPI_ISL_2121303, EPI_ISL_2121310, EPI_ISL_2121319, EPI_ISL_2121321, EPI_ISL_2121322, EPI_ISL_2121324, EPI_ISL_2121325, EPI_ISL_2121327,
EPI_ISL_2121330, EPI_ISL_2121331, EPI_ISL_2121334, EPI_ISL_2121337, EPI_ISL_2121341, EPI_ISL_2121343, EPI_ISL_2121344, EPI_ISL_2121349, EPI_ISL_2121352, EPI_ISL_2121353, EPI_ISL_2121359, EPI_ISL_2121361, EPI_ISL_2121362, EPI_ISL_2121365, EPI_ISL_2121367, EPI_ISL_2121368, EPI_ISL_2121378, EPI_ISL_2121380, EPI_ISL_2121381, EPI_ISL_2121383, EPI_ISL_2121386, EPI_ISL_2121388,
EPI_ISL_2121390, EPI_ISL_2121391, EPI_ISL_2121394, EPI_ISL_2121396, EPI_ISL_2121405, EPI_ISL_2121407, EPI_ISL_2121408, EPI_ISL_2121410, EPI_ISL_2121414, EPI_ISL_2121419, EPI_ISL_2121428, EPI_ISL_2121431, EPI_ISL_2121434, EPI_ISL_2121470, EPI_ISL_2121478, EPI_ISL_2121487, EPI_ISL_2121490, EPI_ISL_2121492, EPI_ISL_2121496, EPI_ISL_2121503, EPI_ISL_2121504,
EPI_ISL_2121511, EPI_ISL_2121528, EPI_ISL_2121530, EPI_ISL_2121532, EPI_ISL_2121543, EPI_ISL_2121545, EPI_ISL_2121548, EPI_ISL_2121551, EPI_ISL_2121552, EPI_ISL_2121554, EPI_ISL_2121555, EPI_ISL_2121559, EPI_ISL_2121561, EPI_ISL_2121563, EPI_ISL_2121564, EPI_ISL_2121566, EPI_ISL_2121569, EPI_ISL_2121571, EPI_ISL_2121579, EPI_ISL_2121585, EPI_ISL_2121604,
EPI_ISL_2121606, EPI_ISL_2121620, EPI_ISL_2121623, EPI_ISL_2121637, EPI_ISL_2121640, EPI_ISL_2121642, EPI_ISL_2121643, EPI_ISL_2121654, EPI_ISL_2121656, EPI_ISL_2121658, EPI_ISL_2121663, EPI_ISL_2121673, EPI_ISL_2121674, EPI_ISL_2121682, EPI_ISL_2121694, EPI_ISL_2121699, EPI_ISL_2121702, EPI_ISL_2121705, EPI_ISL_2121713, EPI_ISL_2121721, EPI_ISL_2121722, EPI_ISL_2121724,
EPI_ISL_2121727, EPI_ISL_2121729, EPI_ISL_2121732, EPI_ISL_2121737, EPI_ISL_2121738, EPI_ISL_2121740, EPI_ISL_2121741, EPI_ISL_2121745, EPI_ISL_2121747, EPI_ISL_2121751, EPI_ISL_2121765, EPI_ISL_2121777, EPI_ISL_2121780, EPI_ISL_2121787, EPI_ISL_2121788, EPI_ISL_2121791, EPI_ISL_2121795, EPI_ISL_2121799, EPI_ISL_2121809, EPI_ISL_2121813, EPI_ISL_2121816, EPI_ISL_2121817,
EPI_ISL_2121825, EPI_ISL_2121830, EPI_ISL_2121831, EPI_ISL_2121840, EPI_ISL_2121843, EPI_ISL_2121844, EPI_ISL_2121846, EPI_ISL_2121849, EPI_ISL_2121869, EPI_ISL_2121878, EPI_ISL_2121881, EPI_ISL_2121885, EPI_ISL_2121894, EPI_ISL_2121896, EPI_ISL_2121917, EPI_ISL_2121923, EPI_ISL_2121925, EPI_ISL_2121928, EPI_ISL_2121950, EPI_ISL_2121954, EPI_ISL_2121965, EPI_ISL_2121968,
EPI_ISL_2121970, EPI_ISL_2121971, EPI_ISL_2121973, EPI_ISL_2121979, EPI_ISL_2121982, EPI_ISL_2121985, EPI_ISL_2121986, EPI_ISL_2121994, EPI_ISL_2121999, EPI_ISL_2122013, EPI_ISL_2122016, EPI_ISL_2122021, EPI_ISL_2122027, EPI_ISL_2122044, EPI_ISL_2122048, EPI_ISL_2122055, EPI_ISL_2122073, EPI_ISL_2122078, EPI_ISL_2122092, EPI_ISL_2122097, EPI_ISL_2122098, EPI_ISL_2122120,
EPI_ISL_2122123, EPI_ISL_2122143, EPI_ISL_2122146, EPI_ISL_2122152, EPI_ISL_2122153, EPI_ISL_2122156, EPI_ISL_2122169, EPI_ISL_2122170, EPI_ISL_2122174, EPI_ISL_2122178, EPI_ISL_2122186, EPI_ISL_2122192, EPI_ISL_2122213, EPI_ISL_2122220, EPI_ISL_2122223, EPI_ISL_2122224, EPI_ISL_2122226, EPI_ISL_2122231, EPI_ISL_2122238, EPI_ISL_2122243, EPI_ISL_2122250, EPI_ISL_2122253,
EPI_ISL_2122256, EPI_ISL_2122278, EPI_ISL_2122281, EPI_ISL_2122290, EPI_ISL_2122292, EPI_ISL_2122293, EPI_ISL_2122295, EPI_ISL_2122300, EPI_ISL_2122306, EPI_ISL_2122314, EPI_ISL_2122316, EPI_ISL_2122317, EPI_ISL_2122320, EPI_ISL_2122337, EPI_ISL_2122338, EPI_ISL_2122340, EPI_ISL_2122346, EPI_ISL_2122357, EPI_ISL_2122359, EPI_ISL_2122362, EPI_ISL_2122371, EPI_ISL_2122372,
EPI_ISL_2122374, EPI_ISL_2122377, EPI_ISL_2122392, EPI_ISL_2122394, EPI_ISL_2122395, EPI_ISL_2122406, EPI_ISL_2122410, EPI_ISL_2122415, EPI_ISL_2122418, EPI_ISL_2122423, EPI_ISL_2122426, EPI_ISL_2122428, EPI_ISL_2122437, EPI_ISL_2122447, EPI_ISL_2122450, EPI_ISL_2122452, EPI_ISL_2122453, EPI_ISL_2122459, EPI_ISL_2122465, EPI_ISL_2122467, EPI_ISL_2122476, EPI_ISL_2122479,
EPI_ISL_2122480, EPI_ISL_2122482, EPI_ISL_2122485, EPI_ISL_2122494, EPI_ISL_2122496, EPI_ISL_2122507, EPI_ISL_2122512, EPI_ISL_2122520, EPI_ISL_2122521, EPI_ISL_2122543, EPI_ISL_2122554, EPI_ISL_2122559, EPI_ISL_2122566, EPI_ISL_2122569, EPI_ISL_2122571, EPI_ISL_2122573, EPI_ISL_2122576, EPI_ISL_2122578, EPI_ISL_2122579, EPI_ISL_2122581, EPI_ISL_2122584, EPI_ISL_2122588,
EPI_ISL_2122594, EPI_ISL_2122596, EPI_ISL_2122603, EPI_ISL_2122612, EPI_ISL_2122619, EPI_ISL_2122626, EPI_ISL_2122627, EPI_ISL_2122633, EPI_ISL_2122635, EPI_ISL_2122641, EPI_ISL_2122642, EPI_ISL_2122649, EPI_ISL_2122652, EPI_ISL_2122656, EPI_ISL_2122659, EPI_ISL_2122669, EPI_ISL_2122674, EPI_ISL_2122693, EPI_ISL_2122695, EPI_ISL_2137923, EPI_ISL_2137953, EPI_ISL_2138005,
EPI_ISL_2138165, EPI_ISL_2138185, EPI_ISL_2138270, EPI_ISL_2138295, EPI_ISL_2138701, EPI_ISL_2152665, EPI_ISL_2152666, EPI_ISL_2152667, EPI_ISL_2152668, EPI_ISL_2152669, EPI_ISL_2152670, EPI_ISL_2152671, EPI_ISL_2152672, EPI_ISL_2152673, EPI_ISL_2152674, EPI_ISL_2152675, EPI_ISL_2152676, EPI_ISL_2152677, EPI_ISL_2152678, EPI_ISL_2152679, EPI_ISL_2152680, EPI_ISL_2152681,
EPI_ISL_2152682, EPI_ISL_2152683, EPI_ISL_2152684, EPI_ISL_2152685, EPI_ISL_2152686, EPI_ISL_2152687, EPI_ISL_2152688, EPI_ISL_2152689, EPI_ISL_2152690, EPI_ISL_2152691, EPI_ISL_2152692, EPI_ISL_2152693, EPI_ISL_2152694, EPI_ISL_2152695, EPI_ISL_2152696, EPI_ISL_2152697, EPI_ISL_2152698, EPI_ISL_2152699, EPI_ISL_2152700, EPI_ISL_2152701, EPI_ISL_2152702, EPI_ISL_2152703,
EPI_ISL_2152704, EPI_ISL_2152705, EPI_ISL_2152706, EPI_ISL_2152707, EPI_ISL_2152708, EPI_ISL_2152709, EPI_ISL_2152710, EPI_ISL_2152711, EPI_ISL_2152712, EPI_ISL_2152713, EPI_ISL_2152714, EPI_ISL_2152715, EPI_ISL_2152716, EPI_ISL_2152717, EPI_ISL_2152718, EPI_ISL_2152719, EPI_ISL_2152720, EPI_ISL_2152721, EPI_ISL_2152722, EPI_ISL_2152723, EPI_ISL_2152724, EPI_ISL_2152725,
EPI_ISL_2152726, EPI_ISL_2152727, EPI_ISL_2152728, EPI_ISL_2152729, EPI_ISL_2152730, EPI_ISL_2152731, EPI_ISL_2152732, EPI_ISL_2152733, EPI_ISL_2152734, EPI_ISL_2152735, EPI_ISL_2152736, EPI_ISL_2152737, EPI_ISL_2152738, EPI_ISL_2152739, EPI_ISL_2152740, EPI_ISL_2152741, EPI_ISL_2152742, EPI_ISL_2152743, EPI_ISL_2152744, EPI_ISL_2152745, EPI_ISL_2152746, EPI_ISL_2152747,
EPI_ISL_2152748, EPI_ISL_2152749, EPI_ISL_2152750, EPI_ISL_2152751, EPI_ISL_2152752, EPI_ISL_2152753, EPI_ISL_2152754, EPI_ISL_2152755, EPI_ISL_2152756, EPI_ISL_2152757, EPI_ISL_2152758, EPI_ISL_2152759, EPI_ISL_2152760, EPI_ISL_2152761, EPI_ISL_2152762, EPI_ISL_2152763, EPI_ISL_2152764, EPI_ISL_2152765, EPI_ISL_2152766, EPI_ISL_2152767, EPI_ISL_2152768, EPI_ISL_2152769,
EPI_ISL_2152770, EPI_ISL_2152771, EPI_ISL_2152772, EPI_ISL_2152773, EPI_ISL_2152774, EPI_ISL_2152775, EPI_ISL_2152776, EPI_ISL_2152777, EPI_ISL_2152778, EPI_ISL_2152779, EPI_ISL_2152780, EPI_ISL_2152781, EPI_ISL_2152782, EPI_ISL_2152783, EPI_ISL_2152784, EPI_ISL_2152785, EPI_ISL_2152786, EPI_ISL_2152787, EPI_ISL_2152788, EPI_ISL_2152789, EPI_ISL_2152790, EPI_ISL_2152791,
EPI_ISL_2152792, EPI_ISL_2152793, EPI_ISL_2152794, EPI_ISL_2152795, EPI_ISL_2152796, EPI_ISL_2152797, EPI_ISL_2152798, EPI_ISL_2152799, EPI_ISL_2152800, EPI_ISL_2152801, EPI_ISL_2152802, EPI_ISL_2152804, EPI_ISL_2152805, EPI_ISL_2152806, EPI_ISL_2152807, EPI_ISL_2152808, EPI_ISL_2152810, EPI_ISL_2152811, EPI_ISL_2152812, EPI_ISL_2152813, EPI_ISL_2152814, EPI_ISL_2152815,
EPI_ISL_2152816, EPI_ISL_2152817, EPI_ISL_2152818, EPI_ISL_2152819, EPI_ISL_2152820, EPI_ISL_2152821, EPI_ISL_2152822, EPI_ISL_2152823, EPI_ISL_2152824, EPI_ISL_2152825, EPI_ISL_2152826, EPI_ISL_2152827, EPI_ISL_2152828, EPI_ISL_2152829, EPI_ISL_2152830, EPI_ISL_2152831, EPI_ISL_2152832, EPI_ISL_2152833, EPI_ISL_2152834, EPI_ISL_2152835, EPI_ISL_2152836, EPI_ISL_2152837,
EPI_ISL_2152838, EPI_ISL_2152840, EPI_ISL_2152841, EPI_ISL_2152842, EPI_ISL_2152843, EPI_ISL_2152844, EPI_ISL_2152845, EPI_ISL_2152846, EPI_ISL_2152847, EPI_ISL_2152848, EPI_ISL_2152849, EPI_ISL_2152850, EPI_ISL_2152851, EPI_ISL_2152852, EPI_ISL_2152853, EPI_ISL_2152854, EPI_ISL_2152855, EPI_ISL_2152856, EPI_ISL_2152857, EPI_ISL_2152858, EPI_ISL_2152859, EPI_ISL_2152860,
EPI_ISL_2152862, EPI_ISL_2152863, EPI_ISL_2152864, EPI_ISL_2152866, EPI_ISL_2152958, EPI_ISL_2152960, EPI_ISL_2152962, EPI_ISL_2152964, EPI_ISL_2152965, EPI_ISL_2152967, EPI_ISL_2152970, EPI_ISL_2152971, EPI_ISL_2152972, EPI_ISL_2152976, EPI_ISL_2152977, EPI_ISL_2152979, EPI_ISL_2152984, EPI_ISL_2152986, EPI_ISL_2152987, EPI_ISL_2152988, EPI_ISL_2152989, EPI_ISL_2152991,
EPI_ISL_2152993, EPI_ISL_2152994, EPI_ISL_2152995, EPI_ISL_2152996, EPI_ISL_2152998, EPI_ISL_2153001, EPI_ISL_2153003, EPI_ISL_2153004, EPI_ISL_2153005, EPI_ISL_2153007, EPI_ISL_2153008, EPI_ISL_2153009, EPI_ISL_2153010, EPI_ISL_2153011, EPI_ISL_2153012, EPI_ISL_2153013, EPI_ISL_2153014, EPI_ISL_2153017, EPI_ISL_2153019, EPI_ISL_2153020, EPI_ISL_2153022, EPI_ISL_2153024,
EPI_ISL_2153026, EPI_ISL_2153029, EPI_ISL_2153030, EPI_ISL_2153033, EPI_ISL_2153034, EPI_ISL_2153035, EPI_ISL_2153036, EPI_ISL_2153037, EPI_ISL_2153039, EPI_ISL_2153040, EPI_ISL_2153041, EPI_ISL_2171707, EPI_ISL_2171721, EPI_ISL_2171726, EPI_ISL_2171746, EPI_ISL_2171763, EPI_ISL_2171773, EPI_ISL_2171784, EPI_ISL_2171807, EPI_ISL_2171815, EPI_ISL_2171832,
EPI_ISL_2171836, EPI_ISL_2171949, EPI_ISL_2199684, EPI_ISL_2199688, EPI_ISL_2199691, EPI_ISL_2199694, EPI_ISL_2199697, EPI_ISL_2199701, EPI_ISL_2199704, EPI_ISL_2199705, EPI_ISL_2199717, EPI_ISL_2199719, EPI_ISL_2199723, EPI_ISL_2199727, EPI_ISL_2199731, EPI_ISL_2199732, EPI_ISL_2199733, EPI_ISL_2199740, EPI_ISL_2199743, EPI_ISL_2199745, EPI_ISL_2199749, EPI_ISL_2199756,
EPI_ISL_2199757, EPI_ISL_2199763, EPI_ISL_2199770, EPI_ISL_2199774, EPI_ISL_2199776, EPI_ISL_2199780, EPI_ISL_2199783, EPI_ISL_2199784, EPI_ISL_2199785, EPI_ISL_2199794, EPI_ISL_2199800, EPI_ISL_2199823, EPI_ISL_2199836, EPI_ISL_2199840, EPI_ISL_2199842, EPI_ISL_2199853, EPI_ISL_2199858, EPI_ISL_2199862, EPI_ISL_2199866, EPI_ISL_2199867, EPI_ISL_2199869,
EPI_ISL_2199870, EPI_ISL_2199872, EPI_ISL_2199877, EPI_ISL_2199887, EPI_ISL_2199915, EPI_ISL_2199951, EPI_ISL_2199952, EPI_ISL_2236825, EPI_ISL_2236826, EPI_ISL_2236828, EPI_ISL_2236831, EPI_ISL_2236832, EPI_ISL_2236833, EPI_ISL_2236834, EPI_ISL_2236838, EPI_ISL_2236839, EPI_ISL_2236846, EPI_ISL_2236848, EPI_ISL_2236853, EPI_ISL_2236854, EPI_ISL_2236855, EPI_ISL_2236856,
EPI_ISL_2236859, EPI_ISL_2236876, EPI_ISL_2236878, EPI_ISL_2236880, EPI_ISL_2236881, EPI_ISL_2236882, EPI_ISL_2236883, EPI_ISL_2236888, EPI_ISL_2236893, EPI_ISL_2236897, EPI_ISL_2236906, EPI_ISL_2236909, EPI_ISL_2236912, EPI_ISL_2236916, EPI_ISL_2236918, EPI_ISL_2236933, EPI_ISL_2236935, EPI_ISL_2236940, EPI_ISL_2236943, EPI_ISL_2236944, EPI_ISL_2236947, EPI_ISL_2236948,
EPI_ISL_2236949, EPI_ISL_2236950, EPI_ISL_2236951, EPI_ISL_2236961, EPI_ISL_2236965, EPI_ISL_2236972, EPI_ISL_2236975, EPI_ISL_2236976, EPI_ISL_2236977, EPI_ISL_2236978, EPI_ISL_2236979, EPI_ISL_2236985, EPI_ISL_2236986, EPI_ISL_2236991, EPI_ISL_2237001, EPI_ISL_2237004, EPI_ISL_2237010, EPI_ISL_2237029, EPI_ISL_2237031, EPI_ISL_2237033, EPI_ISL_2237039, EPI_ISL_2237042,
EPI_ISL_2237043, EPI_ISL_2237049, EPI_ISL_2237050, EPI_ISL_2237052, EPI_ISL_2237056, EPI_ISL_2237060, EPI_ISL_2237068, EPI_ISL_2237077, EPI_ISL_2237078, EPI_ISL_2237089, EPI_ISL_2237092, EPI_ISL_2237093, EPI_ISL_2237097, EPI_ISL_2237101, EPI_ISL_2237105, EPI_ISL_2237116, EPI_ISL_2237122, EPI_ISL_2237123, EPI_ISL_2237126, EPI_ISL_2237134, EPI_ISL_2237135, EPI_ISL_2237137,
EPI_ISL_2237140, EPI_ISL_2237151, EPI_ISL_2237158, EPI_ISL_2238067, EPI_ISL_2238069, EPI_ISL_2238071, EPI_ISL_2238072, EPI_ISL_2238075, EPI_ISL_2238076, EPI_ISL_2238086, EPI_ISL_2238088, EPI_ISL_2238090, EPI_ISL_2238092, EPI_ISL_2238094, EPI_ISL_2238097, EPI_ISL_2238100, EPI_ISL_2238102, EPI_ISL_2238107, EPI_ISL_2238109, EPI_ISL_2238112, EPI_ISL_2238113, EPI_ISL_2238120,
EPI_ISL_2238123, EPI_ISL_2238125, EPI_ISL_2238126, EPI_ISL_2238128, EPI_ISL_2238130, EPI_ISL_2238132, EPI_ISL_2238133, EPI_ISL_2238135, EPI_ISL_2238136, EPI_ISL_2238139, EPI_ISL_2238143, EPI_ISL_2238144, EPI_ISL_2238145, EPI_ISL_2238147, EPI_ISL_2238148, EPI_ISL_2238149, EPI_ISL_2238150, EPI_ISL_2238153, EPI_ISL_2238155, EPI_ISL_2238156, EPI_ISL_2238157, EPI_ISL_2238158,
EPI_ISL_2238160, EPI_ISL_2238162, EPI_ISL_2238164, EPI_ISL_2238168, EPI_ISL_2238172, EPI_ISL_2238173, EPI_ISL_2238176, EPI_ISL_2238177, EPI_ISL_2238184, EPI_ISL_2238185, EPI_ISL_2238186, EPI_ISL_2238188, EPI_ISL_2238192, EPI_ISL_2238193, EPI_ISL_2238195, EPI_ISL_2238199, EPI_ISL_2238200, EPI_ISL_2238203, EPI_ISL_2238206, EPI_ISL_2238208, EPI_ISL_2238210, EPI_ISL_2238212,
EPI_ISL_2238213, EPI_ISL_2238214, EPI_ISL_2238215, EPI_ISL_2238216, EPI_ISL_2238217, EPI_ISL_2238219, EPI_ISL_2238221, EPI_ISL_2238222, EPI_ISL_2238223, EPI_ISL_2238224, EPI_ISL_2238225, EPI_ISL_2238227, EPI_ISL_2238231, EPI_ISL_2238233, EPI_ISL_2238234, EPI_ISL_2238237, EPI_ISL_2238241, EPI_ISL_2238242, EPI_ISL_2238244, EPI_ISL_2238245, EPI_ISL_2238247, EPI_ISL_2238250,
EPI_ISL_2238254, EPI_ISL_2238255, EPI_ISL_2238257, EPI_ISL_2238258, EPI_ISL_2238260, EPI_ISL_2238263, EPI_ISL_2238266, EPI_ISL_2238268, EPI_ISL_2238269, EPI_ISL_2238271, EPI_ISL_2238273, EPI_ISL_2238274, EPI_ISL_2238275, EPI_ISL_2238277, EPI_ISL_2238278, EPI_ISL_2238281, EPI_ISL_2238285, EPI_ISL_2238286, EPI_ISL_2238289, EPI_ISL_2238290, EPI_ISL_2238291, EPI_ISL_2238294,
EPI_ISL_2238296, EPI_ISL_2238297, EPI_ISL_2238298, EPI_ISL_2238299, EPI_ISL_2238301, EPI_ISL_2238303, EPI_ISL_2238307, EPI_ISL_2238308, EPI_ISL_2238311, EPI_ISL_2238313, EPI_ISL_2238314, EPI_ISL_2238318, EPI_ISL_2238321, EPI_ISL_2238322, EPI_ISL_2238323, EPI_ISL_2238336, EPI_ISL_2238337, EPI_ISL_2238339, EPI_ISL_2238340, EPI_ISL_2238343, EPI_ISL_2238344, EPI_ISL_2238347,
EPI_ISL_2238348, EPI_ISL_2238350, EPI_ISL_2238352, EPI_ISL_2238353, EPI_ISL_2238355, EPI_ISL_2238357, EPI_ISL_2238358, EPI_ISL_2238360, EPI_ISL_2238361, EPI_ISL_2238362, EPI_ISL_2238365, EPI_ISL_2238367, EPI_ISL_2238368, EPI_ISL_2238369, EPI_ISL_2238370, EPI_ISL_2238372, EPI_ISL_2238377, EPI_ISL_2238379, EPI_ISL_2238382, EPI_ISL_2238383, EPI_ISL_2238385,
EPI_ISL_2238386, EPI_ISL_2238387, EPI_ISL_2238388, EPI_ISL_2238389, EPI_ISL_2238390, EPI_ISL_2238392, EPI_ISL_2238393, EPI_ISL_2238396, EPI_ISL_2238399, EPI_ISL_2238400, EPI_ISL_2238402, EPI_ISL_2238403, EPI_ISL_2238404, EPI_ISL_2238405, EPI_ISL_2238406, EPI_ISL_2238407, EPI_ISL_2238408, EPI_ISL_2238412, EPI_ISL_2238413, EPI_ISL_2238420, EPI_ISL_2238424,
EPI_ISL_2238425, EPI_ISL_2238428, EPI_ISL_2238429, EPI_ISL_2238430, EPI_ISL_2238432, EPI_ISL_2238434, EPI_ISL_2238435, EPI_ISL_2238436, EPI_ISL_2238437, EPI_ISL_2238439, EPI_ISL_2238440, EPI_ISL_2238442, EPI_ISL_2238443, EPI_ISL_2238444, EPI_ISL_2238445, EPI_ISL_2238446, EPI_ISL_2238447, EPI_ISL_2238449, EPI_ISL_2238450, EPI_ISL_2238451, EPI_ISL_2238453, EPI_ISL_2238454,
EPI_ISL_2238455, EPI_ISL_2238457, EPI_ISL_2238458, EPI_ISL_2238459, EPI_ISL_2238460, EPI_ISL_2238462, EPI_ISL_2238463, EPI_ISL_2238464, EPI_ISL_2238465, EPI_ISL_2238467, EPI_ISL_2238468, EPI_ISL_2238472, EPI_ISL_2238473, EPI_ISL_2238474, EPI_ISL_2238476, EPI_ISL_2238477, EPI_ISL_2238478, EPI_ISL_2238479, EPI_ISL_2238483, EPI_ISL_2238484, EPI_ISL_2238487,
EPI_ISL_2238488, EPI_ISL_2238491, EPI_ISL_2238492, EPI_ISL_2238493, EPI_ISL_2238494, EPI_ISL_2238495, EPI_ISL_2238497, EPI_ISL_2238499, EPI_ISL_2238500, EPI_ISL_2238502, EPI_ISL_2238503, EPI_ISL_2238504, EPI_ISL_2238505, EPI_ISL_2238506, EPI_ISL_2238507, EPI_ISL_2238508, EPI_ISL_2238509, EPI_ISL_2238510, EPI_ISL_2238511, EPI_ISL_2238512, EPI_ISL_2238514, EPI_ISL_2238517,
EPI_ISL_2238519, EPI_ISL_2238520, EPI_ISL_2238524, EPI_ISL_2238527, EPI_ISL_2238528, EPI_ISL_2238529, EPI_ISL_2238532, EPI_ISL_2238534, EPI_ISL_2238535, EPI_ISL_2238536, EPI_ISL_2238538, EPI_ISL_2238539, EPI_ISL_2238541, EPI_ISL_2238544, EPI_ISL_2238546, EPI_ISL_2238547, EPI_ISL_2238550, EPI_ISL_2238552, EPI_ISL_2238555, EPI_ISL_2238557,
EPI_ISL_2238558, EPI_ISL_2238559, EPI_ISL_2238560, EPI_ISL_2238562, EPI_ISL_2238563, EPI_ISL_2238565, EPI_ISL_2238568, EPI_ISL_2238569, EPI_ISL_2238570, EPI_ISL_2238571, EPI_ISL_2238572, EPI_ISL_2238573, EPI_ISL_2238574, EPI_ISL_2238575, EPI_ISL_2238576, EPI_ISL_2238577, EPI_ISL_2238578, EPI_ISL_2238579, EPI_ISL_2238580, EPI_ISL_2238582, EPI_ISL_2238583, EPI_ISL_2238586,
EPI_ISL_2238588, EPI_ISL_2238589, EPI_ISL_2238591, EPI_ISL_2238592, EPI_ISL_2238595, EPI_ISL_2238596, EPI_ISL_2238597, EPI_ISL_2238598, EPI_ISL_2238599, EPI_ISL_2238600, EPI_ISL_2238601, EPI_ISL_2238602, EPI_ISL_2238605, EPI_ISL_2238607, EPI_ISL_2238608, EPI_ISL_2238612, EPI_ISL_2238613, EPI_ISL_2238614, EPI_ISL_2238615, EPI_ISL_2238616, EPI_ISL_2238622,
EPI_ISL_2238623, EPI_ISL_2238624, EPI_ISL_2238625, EPI_ISL_2238626, EPI_ISL_2238627, EPI_ISL_2238628, EPI_ISL_2238629, EPI_ISL_2238630, EPI_ISL_2238632, EPI_ISL_2238634, EPI_ISL_2238636, EPI_ISL_2238637, EPI_ISL_2238639, EPI_ISL_2238642, EPI_ISL_2238643, EPI_ISL_2238644, EPI_ISL_2238645, EPI_ISL_2238647, EPI_ISL_2238648, EPI_ISL_2238650, EPI_ISL_2238651, EPI_ISL_2238652,
EPI_ISL_2238654, EPI_ISL_2238655, EPI_ISL_2238657, EPI_ISL_2238658, EPI_ISL_2238660, EPI_ISL_2238662, EPI_ISL_2238664, EPI_ISL_2238666, EPI_ISL_2238667, EPI_ISL_2238668, EPI_ISL_2238671, EPI_ISL_2238672, EPI_ISL_2238673, EPI_ISL_2238679, EPI_ISL_2238680, EPI_ISL_2254089, EPI_ISL_2275746, EPI_ISL_2396345, EPI_ISL_2396348, EPI_ISL_2396349,
EPI_ISL_2396353, EPI_ISL_2396359, EPI_ISL_2396367, EPI_ISL_2396368, EPI_ISL_2396369, EPI_ISL_2396382, EPI_ISL_2396383, EPI_ISL_2396384, EPI_ISL_2396385, EPI_ISL_2396388, EPI_ISL_2396410, EPI_ISL_2396433, EPI_ISL_2396437, EPI_ISL_2396440, EPI_ISL_2396474, EPI_ISL_2457183, EPI_ISL_2457807, EPI_ISL_2457870, EPI_ISL_2457876, EPI_ISL_2486008

| | | | |
|---|---|---|---|
| see above | Lighthouse Lab in Glasgow | Wellcome Sanger Institute for the COVID-19 Genomics UK (COG-UK) Consortium | Anna Dominiczak and Alex Alderton; Carol Clugston; Cordelia Langford; David Gray; David K. Jackson; Dominic Kwiatkowski; Ewan Harrison; Harper VanSteenhouse; Ian Johnston; Jeffrey Barrett; John Sillitoe on behalf of the Wellcome Sanger Institute COVID-19 Surveillance Team; Roberto Amato; Sonia Goncalves; Yumi Kasai |

EPI_ISL_1985727, EPI_ISL_1985746, EPI_ISL_1985756, EPI_ISL_1985761, EPI_ISL_1985849, EPI_ISL_1985915, EPI_ISL_1985956, EPI_ISL_1985993, EPI_ISL_1986005, EPI_ISL_1986043, EPI_ISL_1986067, EPI_ISL_1986083, EPI_ISL_1986084, EPI_ISL_1986850, EPI_ISL_2004620, EPI_ISL_2004621, EPI_ISL_2004622, EPI_ISL_2004623, EPI_ISL_2004624, EPI_ISL_2004626, EPI_ISL_2004628, EPI_ISL_2004629,
EPI_ISL_2004630, EPI_ISL_2004631, EPI_ISL_2004633, EPI_ISL_2004634, EPI_ISL_2004636, EPI_ISL_2004638, EPI_ISL_2004640, EPI_ISL_2004642, EPI_ISL_2004645, EPI_ISL_2004647, EPI_ISL_2004649, EPI_ISL_2004650, EPI_ISL_2004652, EPI_ISL_2004654, EPI_ISL_2004655, EPI_ISL_2004656, EPI_ISL_2004657, EPI_ISL_2004659, EPI_ISL_2004661, EPI_ISL_2004662, EPI_ISL_2004663, EPI_ISL_2004664,
EPI_ISL_2004665, EPI_ISL_2004666, EPI_ISL_2004669, EPI_ISL_2004672, EPI_ISL_2004678, EPI_ISL_2004681, EPI_ISL_2004683, EPI_ISL_2004684, EPI_ISL_2004686, EPI_ISL_2004689, EPI_ISL_2004690, EPI_ISL_2004692, EPI_ISL_2004694, EPI_ISL_2004696, EPI_ISL_2004698, EPI_ISL_2004699, EPI_ISL_2004700, EPI_ISL_2004703, EPI_ISL_2004705, EPI_ISL_2004706, EPI_ISL_2004707, EPI_ISL_2004708,
EPI_ISL_2004710, EPI_ISL_2004712, EPI_ISL_2004714, EPI_ISL_2004718, EPI_ISL_2004719, EPI_ISL_2004720, EPI_ISL_2004722, EPI_ISL_2004723, EPI_ISL_2004727, EPI_ISL_2004730, EPI_ISL_2004731, EPI_ISL_2004733, EPI_ISL_2004735, EPI_ISL_2004736, EPI_ISL_2004737, EPI_ISL_2004739, EPI_ISL_2004740, EPI_ISL_2004741, EPI_ISL_2004742, EPI_ISL_2004743, EPI_ISL_2004746, EPI_ISL_2004749,
EPI_ISL_2004750, EPI_ISL_2004756, EPI_ISL_2004759, EPI_ISL_2004760, EPI_ISL_2004761, EPI_ISL_2004763, EPI_ISL_2004764, EPI_ISL_2004765, EPI_ISL_2004766, EPI_ISL_2004770, EPI_ISL_2004771, EPI_ISL_2004772, EPI_ISL_2004776, EPI_ISL_2004777, EPI_ISL_2004778, EPI_ISL_2004780, EPI_ISL_2004781, EPI_ISL_2004782, EPI_ISL_2004783, EPI_ISL_2004784, EPI_ISL_2004785, EPI_ISL_2004786,
EPI_ISL_2004791, EPI_ISL_2004792, EPI_ISL_2004793, EPI_ISL_2004797, EPI_ISL_2004800, EPI_ISL_2004801, EPI_ISL_2004802, EPI_ISL_2004803, EPI_ISL_2004808, EPI_ISL_2004809, EPI_ISL_2004810, EPI_ISL_2004813, EPI_ISL_2004819, EPI_ISL_2004820, EPI_ISL_2004822, EPI_ISL_2004823, EPI_ISL_2004824, EPI_ISL_2004825, EPI_ISL_2004826, EPI_ISL_2004827, EPI_ISL_2004828, EPI_ISL_2004829,
EPI_ISL_2004830, EPI_ISL_2004831, EPI_ISL_2004832, EPI_ISL_2004835, EPI_ISL_2004838, EPI_ISL_2004839, EPI_ISL_2004842, EPI_ISL_2004843, EPI_ISL_2004848, EPI_ISL_2004851, EPI_ISL_2004853, EPI_ISL_2004854, EPI_ISL_2004855, EPI_ISL_2004856, EPI_ISL_2004857, EPI_ISL_2004858, EPI_ISL_2004861, EPI_ISL_2004863, EPI_ISL_2004865, EPI_ISL_2004867, EPI_ISL_2004870,
EPI_ISL_2004872, EPI_ISL_2004874, EPI_ISL_2004875, EPI_ISL_2004879, EPI_ISL_2005811, EPI_ISL_2005812, EPI_ISL_2005813, EPI_ISL_2005815, EPI_ISL_2005817, EPI_ISL_2005819, EPI_ISL_2005821, EPI_ISL_2005822, EPI_ISL_2005824, EPI_ISL_2005825, EPI_ISL_2005826, EPI_ISL_2005827, EPI_ISL_2005829, EPI_ISL_2005830, EPI_ISL_2005831, EPI_ISL_2005832, EPI_ISL_2005833, EPI_ISL_2005836,
EPI_ISL_2005840, EPI_ISL_2005842, EPI_ISL_2005844, EPI_ISL_2005849, EPI_ISL_2005853, EPI_ISL_2005854, EPI_ISL_2005855, EPI_ISL_2005856, EPI_ISL_2005857, EPI_ISL_2005859, EPI_ISL_2005860, EPI_ISL_2005861, EPI_ISL_2005864, EPI_ISL_2005870, EPI_ISL_2005871, EPI_ISL_2005872, EPI_ISL_2005873, EPI_ISL_2005874, EPI_ISL_2005875, EPI_ISL_2005876,
EPI_ISL_2005879, EPI_ISL_2005880, EPI_ISL_2005882, EPI_ISL_2005883, EPI_ISL_2005885, EPI_ISL_2005887, EPI_ISL_2005888, EPI_ISL_2005889, EPI_ISL_2005891, EPI_ISL_2005892, EPI_ISL_2005894, EPI_ISL_2005897, EPI_ISL_2005899, EPI_ISL_2005903, EPI_ISL_2005904, EPI_ISL_2005907, EPI_ISL_2005908, EPI_ISL_2005909, EPI_ISL_2005910, EPI_ISL_2005912, EPI_ISL_2005915, EPI_ISL_2005916,
EPI_ISL_2005917, EPI_ISL_2005920, EPI_ISL_2005921, EPI_ISL_2005922, EPI_ISL_2005925, EPI_ISL_2005929, EPI_ISL_2005933, EPI_ISL_2005934, EPI_ISL_2005935, EPI_ISL_2005936, EPI_ISL_2005937, EPI_ISL_2005938, EPI_ISL_2005942, EPI_ISL_2005943, EPI_ISL_2005944, EPI_ISL_2005948, EPI_ISL_2005949, EPI_ISL_2005950, EPI_ISL_2005951, EPI_ISL_2005952, EPI_ISL_2005954, EPI_ISL_2005956,
EPI_ISL_2005960, EPI_ISL_2005963, EPI_ISL_2005966, EPI_ISL_2005969, EPI_ISL_2005971, EPI_ISL_2005972, EPI_ISL_2005973, EPI_ISL_2005974, EPI_ISL_2005975, EPI_ISL_2005977, EPI_ISL_2005978, EPI_ISL_2019425, EPI_ISL_2005981, EPI_ISL_2005983, EPI_ISL_2005986, EPI_ISL_2005987, EPI_ISL_2005990, EPI_ISL_2005991, EPI_ISL_2005994, EPI_ISL_2005995, EPI_ISL_2005997, EPI_ISL_2005999,
EPI_ISL_2006001, EPI_ISL_2006004, EPI_ISL_2006005, EPI_ISL_2006009, EPI_ISL_2006011, EPI_ISL_2006014, EPI_ISL_2006017, EPI_ISL_2006019, EPI_ISL_2006020, EPI_ISL_2006022, EPI_ISL_2006023, EPI_ISL_2006025, EPI_ISL_2006028, EPI_ISL_2006030, EPI_ISL_2006031, EPI_ISL_2006032, EPI_ISL_2006033, EPI_ISL_2006037, EPI_ISL_2006041, EPI_ISL_2006042, EPI_ISL_2006045,
EPI_ISL_2006046, EPI_ISL_2006048, EPI_ISL_2006049, EPI_ISL_2006052, EPI_ISL_2006055, EPI_ISL_2006056, EPI_ISL_2006057, EPI_ISL_2006060, EPI_ISL_2006061, EPI_ISL_2006063, EPI_ISL_2006064, EPI_ISL_2006065, EPI_ISL_2006066, EPI_ISL_2006067, EPI_ISL_2006069, EPI_ISL_2006070, EPI_ISL_2021055, EPI_ISL_2021069, EPI_ISL_2021071, EPI_ISL_2021073, EPI_ISL_2021099, EPI_ISL_2021114,
EPI_ISL_2021120, EPI_ISL_2021141, EPI_ISL_2021143, EPI_ISL_2021161, EPI_ISL_2021171, EPI_ISL_2021182, EPI_ISL_2021189, EPI_ISL_2021224, EPI_ISL_2021229, EPI_ISL_2021253, EPI_ISL_2021285, EPI_ISL_2021293, EPI_ISL_2021309, EPI_ISL_2021368, EPI_ISL_2021382, EPI_ISL_2021383, EPI_ISL_2021387, EPI_ISL_2021392, EPI_ISL_2021398, EPI_ISL_2021399, EPI_ISL_2021401, EPI_ISL_2021404,
EPI_ISL_2021405, EPI_ISL_2021415, EPI_ISL_2021419, EPI_ISL_2021421, EPI_ISL_2021422, EPI_ISL_2021425, EPI_ISL_2021427, EPI_ISL_2021430, EPI_ISL_2021434, EPI_ISL_2021439, EPI_ISL_2021444, EPI_ISL_2021448, EPI_ISL_2021453, EPI_ISL_2021454, EPI_ISL_2021461, EPI_ISL_2021462, EPI_ISL_2021465, EPI_ISL_2021467, EPI_ISL_2021471, EPI_ISL_2021477, EPI_ISL_2021478, EPI_ISL_2021483,
EPI_ISL_2021492, EPI_ISL_2021494, EPI_ISL_2021496, EPI_ISL_2021508, EPI_ISL_2021509, EPI_ISL_2021511, EPI_ISL_2021517, EPI_ISL_2021536, EPI_ISL_2021541, EPI_ISL_2021544, EPI_ISL_2021546, EPI_ISL_2021548, EPI_ISL_2021549, EPI_ISL_2021558, EPI_ISL_2021575, EPI_ISL_2021576, EPI_ISL_2021593, EPI_ISL_2021601, EPI_ISL_2021603, EPI_ISL_2021610, EPI_ISL_2021612, EPI_ISL_2021613,
EPI_ISL_2021614, EPI_ISL_2021620, EPI_ISL_2021622, EPI_ISL_2021625, EPI_ISL_2021635, EPI_ISL_2021636, EPI_ISL_2021638, EPI_ISL_2021654, EPI_ISL_2021658, EPI_ISL_2021659, EPI_ISL_2021661, EPI_ISL_2021663, EPI_ISL_2021665, EPI_ISL_2021671, EPI_ISL_2021675, EPI_ISL_2021679, EPI_ISL_2021683, EPI_ISL_2021690, EPI_ISL_2021708, EPI_ISL_2021755, EPI_ISL_2021783, EPI_ISL_2021823,
EPI_ISL_2021849, EPI_ISL_2021850, EPI_ISL_2021851, EPI_ISL_2021854, EPI_ISL_2021855, EPI_ISL_2021856, EPI_ISL_2021858, EPI_ISL_2021859, EPI_ISL_2021862, EPI_ISL_2021866, EPI_ISL_2021868, EPI_ISL_2021871, EPI_ISL_2021872, EPI_ISL_2021874, EPI_ISL_2021875, EPI_ISL_2021877, EPI_ISL_2021878, EPI_ISL_2021879, EPI_ISL_2021880, EPI_ISL_2021881, EPI_ISL_2021884, EPI_ISL_2021885,
EPI_ISL_2021886, EPI_ISL_2021887, EPI_ISL_2021888, EPI_ISL_2021889, EPI_ISL_2021893, EPI_ISL_2021894, EPI_ISL_2021896, EPI_ISL_2021897, EPI_ISL_2021898, EPI_ISL_2021902, EPI_ISL_2021905, EPI_ISL_2021907, EPI_ISL_2021909, EPI_ISL_2021910, EPI_ISL_2021914, EPI_ISL_2021915, EPI_ISL_2021917, EPI_ISL_2021919, EPI_ISL_2021921, EPI_ISL_2021922, EPI_ISL_2021924, EPI_ISL_2021925,
EPI_ISL_2021926, EPI_ISL_2021927, EPI_ISL_2021928, EPI_ISL_2021929, EPI_ISL_2021930, EPI_ISL_2021931, EPI_ISL_2021932, EPI_ISL_2021937, EPI_ISL_2021939, EPI_ISL_2021940, EPI_ISL_2021944, EPI_ISL_2021946, EPI_ISL_2021947, EPI_ISL_2021948, EPI_ISL_2021950, EPI_ISL_2021951, EPI_ISL_2021953, EPI_ISL_2021955, EPI_ISL_2021957, EPI_ISL_2021959, EPI_ISL_2021960,
EPI_ISL_2021962, EPI_ISL_2021965, EPI_ISL_2021966, EPI_ISL_2021967, EPI_ISL_2021969, EPI_ISL_2021970, EPI_ISL_2021972, EPI_ISL_2021973, EPI_ISL_2021974, EPI_ISL_2021975, EPI_ISL_2021976, EPI_ISL_2021978, EPI_ISL_2021979, EPI_ISL_2021981, EPI_ISL_2021982, EPI_ISL_2021983, EPI_ISL_2021985, EPI_ISL_2021990, EPI_ISL_2021991, EPI_ISL_2021992, EPI_ISL_2021995, EPI_ISL_2021996,
EPI_ISL_2021998, EPI_ISL_2021999, EPI_ISL_2022006, EPI_ISL_2022008, EPI_ISL_2022010, EPI_ISL_2022012, EPI_ISL_2022013, EPI_ISL_2022014, EPI_ISL_2022015, EPI_ISL_2022016, EPI_ISL_2022017, EPI_ISL_2022019, EPI_ISL_2022022, EPI_ISL_2022024, EPI_ISL_2022025, EPI_ISL_2022026, EPI_ISL_2022028, EPI_ISL_2022029, EPI_ISL_2022030, EPI_ISL_2022031, EPI_ISL_2022032, EPI_ISL_2022034,

EPI_ISL_2022036, EPI_ISL_2022037, EPI_ISL_2022039, EPI_ISL_2022040, EPI_ISL_2022042, EPI_ISL_2022043, EPI_ISL_2022044, EPI_ISL_2022045, EPI_ISL_2022046, EPI_ISL_2022047, EPI_ISL_2022048, EPI_ISL_2022049, EPI_ISL_2022050, EPI_ISL_2022051, EPI_ISL_2022052, EPI_ISL_2022054, EPI_ISL_2022059, EPI_ISL_2022060, EPI_ISL_2022063, EPI_ISL_2022064, EPI_ISL_2022065, EPI_ISL_2022068, EPI_ISL_2022069, EPI_ISL_2022070, EPI_ISL_2022071, EPI_ISL_2022073, EPI_ISL_2022076, EPI_ISL_2022078, EPI_ISL_2022079, EPI_ISL_2022081, EPI_ISL_2022082, EPI_ISL_2022083, EPI_ISL_2022084, EPI_ISL_2022085, EPI_ISL_2022089, EPI_ISL_2022090, EPI_ISL_2022093, EPI_ISL_2022094, EPI_ISL_2022095, EPI_ISL_2022096, EPI_ISL_2022098, EPI_ISL_2022099, EPI_ISL_2022100, EPI_ISL_2022102, EPI_ISL_2022103, EPI_ISL_2022104, EPI_ISL_2022105, EPI_ISL_2022106, EPI_ISL_2022107, EPI_ISL_2022108, EPI_ISL_2022110, EPI_ISL_2022111, EPI_ISL_2022113, EPI_ISL_2022114, EPI_ISL_2022116, EPI_ISL_2022119, EPI_ISL_2022121, EPI_ISL_2022122, EPI_ISL_2022123, EPI_ISL_2022125, EPI_ISL_2022126, EPI_ISL_2022128, EPI_ISL_2022130, EPI_ISL_2022131, EPI_ISL_2022133, EPI_ISL_2022134, EPI_ISL_2022137, EPI_ISL_2022139, EPI_ISL_2022140, EPI_ISL_2022141, EPI_ISL_2022143, EPI_ISL_2022145, EPI_ISL_2022146, EPI_ISL_2022147, EPI_ISL_2022148, EPI_ISL_2022150, EPI_ISL_2022151, EPI_ISL_2022152, EPI_ISL_2022153, EPI_ISL_2022155, EPI_ISL_2022156, EPI_ISL_2022157, EPI_ISL_2022158, EPI_ISL_2022159, EPI_ISL_2022161, EPI_ISL_2022167, EPI_ISL_2022169, EPI_ISL_2022170, EPI_ISL_2022173, EPI_ISL_2022174, EPI_ISL_2022175, EPI_ISL_2022176, EPI_ISL_2022177, EPI_ISL_2022180, EPI_ISL_2022186, EPI_ISL_2022187, EPI_ISL_2022191, EPI_ISL_2022194, EPI_ISL_2022199, EPI_ISL_2022200, EPI_ISL_2022201, EPI_ISL_2022202, EPI_ISL_2022203, EPI_ISL_2022204, EPI_ISL_2022207, EPI_ISL_2022213, EPI_ISL_2022224, EPI_ISL_2022231, EPI_ISL_2022236, EPI_ISL_2022238, EPI_ISL_2022239, EPI_ISL_2022240, EPI_ISL_2022241, EPI_ISL_2022242, EPI_ISL_2022244, EPI_ISL_2022245, EPI_ISL_2022250, EPI_ISL_2022251, EPI_ISL_2022256, EPI_ISL_2022258, EPI_ISL_2022262, EPI_ISL_2022263, EPI_ISL_2022264, EPI_ISL_2022265, EPI_ISL_2022272, EPI_ISL_2022277, EPI_ISL_2022279, EPI_ISL_2022280, EPI_ISL_2022281, EPI_ISL_2022285, EPI_ISL_2022286, EPI_ISL_2022288, EPI_ISL_2022290, EPI_ISL_2022295, EPI_ISL_2022296, EPI_ISL_2022299, EPI_ISL_2022304, EPI_ISL_2022305, EPI_ISL_2022306, EPI_ISL_2022308, EPI_ISL_2022317, EPI_ISL_2022319, EPI_ISL_2022327, EPI_ISL_2022330, EPI_ISL_2022332, EPI_ISL_2022337, EPI_ISL_2022342, EPI_ISL_2022343, EPI_ISL_2022348, EPI_ISL_2022351, EPI_ISL_2022352, EPI_ISL_2022356, EPI_ISL_2022358, EPI_ISL_2022359, EPI_ISL_2022364, EPI_ISL_2022367, EPI_ISL_2022369, EPI_ISL_2022373, EPI_ISL_2022374, EPI_ISL_2022375, EPI_ISL_2022376, EPI_ISL_2022381, EPI_ISL_2022383, EPI_ISL_2022387, EPI_ISL_2022388, EPI_ISL_2022397, EPI_ISL_2022398, EPI_ISL_2022399, EPI_ISL_2022400, EPI_ISL_2022408, EPI_ISL_2022410, EPI_ISL_2022415, EPI_ISL_2022419, EPI_ISL_2022420, EPI_ISL_2022423, EPI_ISL_2022428, EPI_ISL_2022429, EPI_ISL_2022437, EPI_ISL_2022531, EPI_ISL_2022542, EPI_ISL_2022577, EPI_ISL_2022579, EPI_ISL_2022584, EPI_ISL_2022902, EPI_ISL_2022912, EPI_ISL_2022913, EPI_ISL_2022922, EPI_ISL_2080476, EPI_ISL_2080478, EPI_ISL_2080480, EPI_ISL_2091794, EPI_ISL_2091795, EPI_ISL_2091796, EPI_ISL_2091797, EPI_ISL_2091798, EPI_ISL_2091799, EPI_ISL_2091802, EPI_ISL_2091803, EPI_ISL_2091805, EPI_ISL_2091806, EPI_ISL_2091807, EPI_ISL_2091809, EPI_ISL_2091811, EPI_ISL_2091813, EPI_ISL_2091815, EPI_ISL_2091817, EPI_ISL_2091818, EPI_ISL_2091820, EPI_ISL_2091821, EPI_ISL_2091822, EPI_ISL_2091826, EPI_ISL_2091832, EPI_ISL_2091834, EPI_ISL_2091835, EPI_ISL_2091837, EPI_ISL_2091838, EPI_ISL_2091839, EPI_ISL_2091840, EPI_ISL_2091843, EPI_ISL_2091848, EPI_ISL_2091849, EPI_ISL_2091850, EPI_ISL_2091853, EPI_ISL_2091855, EPI_ISL_2091857, EPI_ISL_2091860, EPI_ISL_2091861, EPI_ISL_2091862, EPI_ISL_2091867, EPI_ISL_2091871, EPI_ISL_2091872, EPI_ISL_2091873, EPI_ISL_2091875, EPI_ISL_2091876, EPI_ISL_2091879, EPI_ISL_2091882, EPI_ISL_2091883, EPI_ISL_2091886, EPI_ISL_2091887, EPI_ISL_2091890, EPI_ISL_2091895, EPI_ISL_2091899, EPI_ISL_2091900, EPI_ISL_2091901, EPI_ISL_2091902, EPI_ISL_2091903, EPI_ISL_2091906, EPI_ISL_2091908, EPI_ISL_2091909, EPI_ISL_2091912, EPI_ISL_2091913, EPI_ISL_2091914, EPI_ISL_2091916, EPI_ISL_2091918, EPI_ISL_2091920, EPI_ISL_2091923, EPI_ISL_2091925, EPI_ISL_2091926, EPI_ISL_2091927, EPI_ISL_2091928, EPI_ISL_2091932, EPI_ISL_2091933, EPI_ISL_2091935, EPI_ISL_2091938, EPI_ISL_2091939, EPI_ISL_2091941, EPI_ISL_2091942, EPI_ISL_2091944, EPI_ISL_2091946, EPI_ISL_2091948, EPI_ISL_2091949, EPI_ISL_2091950, EPI_ISL_2091952, EPI_ISL_2091953, EPI_ISL_2091956, EPI_ISL_2091957, EPI_ISL_2091960, EPI_ISL_2091963, EPI_ISL_2091969, EPI_ISL_2091971, EPI_ISL_2091972, EPI_ISL_2091973, EPI_ISL_2091976, EPI_ISL_2091981, EPI_ISL_2091983, EPI_ISL_2091984, EPI_ISL_2091986, EPI_ISL_2091988, EPI_ISL_2091989, EPI_ISL_2091990, EPI_ISL_2091991, EPI_ISL_2091993, EPI_ISL_2091994, EPI_ISL_2091997, EPI_ISL_2092000, EPI_ISL_2092004, EPI_ISL_2092007, EPI_ISL_2092009, EPI_ISL_2092012, EPI_ISL_2092014, EPI_ISL_2092015, EPI_ISL_2092017, EPI_ISL_2092018, EPI_ISL_2092021, EPI_ISL_2092024, EPI_ISL_2092025, EPI_ISL_2092027, EPI_ISL_2092028, EPI_ISL_2092029, EPI_ISL_2092030, EPI_ISL_2092031, EPI_ISL_2092032, EPI_ISL_2092033, EPI_ISL_2092035, EPI_ISL_2092036, EPI_ISL_2092037, EPI_ISL_2092041, EPI_ISL_2092043, EPI_ISL_2092044, EPI_ISL_2092045, EPI_ISL_2092051, EPI_ISL_2092053, EPI_ISL_2092056, EPI_ISL_2092062, EPI_ISL_2092063, EPI_ISL_2092064, EPI_ISL_2092065, EPI_ISL_2092066, EPI_ISL_2092067, EPI_ISL_2092068, EPI_ISL_2092069, EPI_ISL_2092070, EPI_ISL_2092071, EPI_ISL_2092072, EPI_ISL_2092073, EPI_ISL_2092074, EPI_ISL_2092077, EPI_ISL_2092078, EPI_ISL_2092079, EPI_ISL_2092080, EPI_ISL_2092081, EPI_ISL_2092084, EPI_ISL_2092086, EPI_ISL_2092087, EPI_ISL_2092088, EPI_ISL_2092089, EPI_ISL_2092090, EPI_ISL_2092093, EPI_ISL_2092094, EPI_ISL_2092095, EPI_ISL_2092097, EPI_ISL_2092098, EPI_ISL_2092101, EPI_ISL_2092675, EPI_ISL_2117515, EPI_ISL_2117516, EPI_ISL_2117523, EPI_ISL_2117529, EPI_ISL_2117538, EPI_ISL_2117542, EPI_ISL_2117544, EPI_ISL_2117549, EPI_ISL_2117556, EPI_ISL_2117559, EPI_ISL_2117562, EPI_ISL_2117566, EPI_ISL_2117572, EPI_ISL_2117582, EPI_ISL_2117584, EPI_ISL_2117585, EPI_ISL_2117586, EPI_ISL_2117591, EPI_ISL_2117592, EPI_ISL_2117598, EPI_ISL_2117601, EPI_ISL_2117606, EPI_ISL_2117607, EPI_ISL_2117608, EPI_ISL_2117611, EPI_ISL_2117613, EPI_ISL_2117615, EPI_ISL_2117617, EPI_ISL_2117629, EPI_ISL_2117630, EPI_ISL_2117635, EPI_ISL_2117636, EPI_ISL_2117638, EPI_ISL_2117641, EPI_ISL_2117656, EPI_ISL_2117664, EPI_ISL_2117676, EPI_ISL_2117678, EPI_ISL_2117689, EPI_ISL_2117700, EPI_ISL_2117705, EPI_ISL_2117706, EPI_ISL_2117709, EPI_ISL_2117711, EPI_ISL_2117722, EPI_ISL_2117725, EPI_ISL_2117728, EPI_ISL_2117730, EPI_ISL_2117733, EPI_ISL_2117737, EPI_ISL_2117739, EPI_ISL_2117740, EPI_ISL_2117743, EPI_ISL_2117747, EPI_ISL_2117755, EPI_ISL_2117783, EPI_ISL_2117785, EPI_ISL_2117788, EPI_ISL_2117790, EPI_ISL_2117791, EPI_ISL_2117793, EPI_ISL_2117797, EPI_ISL_2117800, EPI_ISL_2117801, EPI_ISL_2117803, EPI_ISL_2117812, EPI_ISL_2117816, EPI_ISL_2117822, EPI_ISL_2117825, EPI_ISL_2117835, EPI_ISL_2117840, EPI_ISL_2117846, EPI_ISL_2117848, EPI_ISL_2117850, EPI_ISL_2117859, EPI_ISL_2117862, EPI_ISL_2117867, EPI_ISL_2117868, EPI_ISL_2117874, EPI_ISL_2117876, EPI_ISL_2117882, EPI_ISL_2117886, EPI_ISL_2117888, EPI_ISL_2117894, EPI_ISL_2117897, EPI_ISL_2117898, EPI_ISL_2117901, EPI_ISL_2117905, EPI_ISL_2117910, EPI_ISL_2117913, EPI_ISL_2117916, EPI_ISL_2117918, EPI_ISL_2117924, EPI_ISL_2117928, EPI_ISL_2117936, EPI_ISL_2117942, EPI_ISL_2117943, EPI_ISL_2117952, EPI_ISL_2117955, EPI_ISL_2117960, EPI_ISL_2117961, EPI_ISL_2117964, EPI_ISL_2117972, EPI_ISL_2117975, EPI_ISL_2117981, EPI_ISL_2117987, EPI_ISL_2117991, EPI_ISL_2118000, EPI_ISL_2118007, EPI_ISL_2118014, EPI_ISL_2118019, EPI_ISL_2118026, EPI_ISL_2118032, EPI_ISL_2118035, EPI_ISL_2118036, EPI_ISL_2118040, EPI_ISL_2118045, EPI_ISL_2118050, EPI_ISL_2118058, EPI_ISL_2118084, EPI_ISL_2118089, EPI_ISL_2118093, EPI_ISL_2118096, EPI_ISL_2118097, EPI_ISL_2118113, EPI_ISL_2118116, EPI_ISL_2118118, EPI_ISL_2118119, EPI_ISL_2118122, EPI_ISL_2118123, EPI_ISL_2118140, EPI_ISL_2118152, EPI_ISL_2118155, EPI_ISL_2118156, EPI_ISL_2118161, EPI_ISL_2118164, EPI_ISL_2118168, EPI_ISL_2118179, EPI_ISL_2118182, EPI_ISL_2118183, EPI_ISL_2118186, EPI_ISL_2118188, EPI_ISL_2118189, EPI_ISL_2118197, EPI_ISL_2118201, EPI_ISL_2118205, EPI_ISL_2118206, EPI_ISL_2118208, EPI_ISL_2118210, EPI_ISL_2118214, EPI_ISL_2118215, EPI_ISL_2118217, EPI_ISL_2118223, EPI_ISL_2118225, EPI_ISL_2118226, EPI_ISL_2118228, EPI_ISL_2118229, EPI_ISL_2118231, EPI_ISL_2118234, EPI_ISL_2118258, EPI_ISL_2118259, EPI_ISL_2118311, EPI_ISL_2118316, EPI_ISL_2118324, EPI_ISL_2118325, EPI_ISL_2118338, EPI_ISL_2118339, EPI_ISL_2118343, EPI_ISL_2118345, EPI_ISL_2118356, EPI_ISL_2118359, EPI_ISL_2118363, EPI_ISL_2118368, EPI_ISL_2118370, EPI_ISL_2118382, EPI_ISL_2118390, EPI_ISL_2118401, EPI_ISL_2118402, EPI_ISL_2118406, EPI_ISL_2118407, EPI_ISL_2118410, EPI_ISL_2118413, EPI_ISL_2118425, EPI_ISL_2118435, EPI_ISL_2118436, EPI_ISL_2118438, EPI_ISL_2118440, EPI_ISL_2118450, EPI_ISL_2118452, EPI_ISL_2118453, EPI_ISL_2118459, EPI_ISL_2118463, EPI_ISL_2118466, EPI_ISL_2118468, EPI_ISL_2118499, EPI_ISL_2118503, EPI_ISL_2118507, EPI_ISL_2118519, EPI_ISL_2118526, EPI_ISL_2118549, EPI_ISL_2118557, EPI_ISL_2118562, EPI_ISL_2118570, EPI_ISL_2118572, EPI_ISL_2118576, EPI_ISL_2118578, EPI_ISL_2118583, EPI_ISL_2118588, EPI_ISL_2118592, EPI_ISL_2118603, EPI_ISL_2118605, EPI_ISL_2118607, EPI_ISL_2118616, EPI_ISL_2118619, EPI_ISL_2118621, EPI_ISL_2118627, EPI_ISL_2118633, EPI_ISL_2118640, EPI_ISL_2118647, EPI_ISL_2118651, EPI_ISL_2118664, EPI_ISL_2118665, EPI_ISL_2118681, EPI_ISL_2118695, EPI_ISL_2118702, EPI_ISL_2118706, EPI_ISL_2118707, EPI_ISL_2118715, EPI_ISL_2118719, EPI_ISL_2118727, EPI_ISL_2118736, EPI_ISL_2118747, EPI_ISL_2118753, EPI_ISL_2118754, EPI_ISL_2118759, EPI_ISL_2118760, EPI_ISL_2118767, EPI_ISL_2118772, EPI_ISL_2118782, EPI_ISL_2118783, EPI_ISL_2118789, EPI_ISL_2118794, EPI_ISL_2118798, EPI_ISL_2118800, EPI_ISL_2118801, EPI_ISL_2118804, EPI_ISL_2118809, EPI_ISL_2118810, EPI_ISL_2118820, EPI_ISL_2118823, EPI_ISL_2118825, EPI_ISL_2118826, EPI_ISL_2118834, EPI_ISL_2118837, EPI_ISL_2118838, EPI_ISL_2118840, EPI_ISL_2118841, EPI_ISL_2118846, EPI_ISL_2118847, EPI_ISL_2118854, EPI_ISL_2118858, EPI_ISL_2118859, EPI_ISL_2118861, EPI_ISL_2118862, EPI_ISL_2118866, EPI_ISL_2118870, EPI_ISL_2118871, EPI_ISL_2118874, EPI_ISL_2118877, EPI_ISL_2118879, EPI_ISL_2118880, EPI_ISL_2118883, EPI_ISL_2118888, EPI_ISL_2118892, EPI_ISL_2118894, EPI_ISL_2118896, EPI_ISL_2118899, EPI_ISL_2118900, EPI_ISL_2118902, EPI_ISL_2118905, EPI_ISL_2118906, EPI_ISL_2118909, EPI_ISL_2118911, EPI_ISL_2118914, EPI_ISL_2118917, EPI_ISL_2118919, EPI_ISL_2118920, EPI_ISL_2118921, EPI_ISL_2118925, EPI_ISL_2118927, EPI_ISL_2118929, EPI_ISL_2118931, EPI_ISL_2118932, EPI_ISL_2118934, EPI_ISL_2118936, EPI_ISL_2118938, EPI_ISL_2118941, EPI_ISL_2118942, EPI_ISL_2118944, EPI_ISL_2118947, EPI_ISL_2118949, EPI_ISL_2118950, EPI_ISL_2118952, EPI_ISL_2118953, EPI_ISL_2118956, EPI_ISL_2118959, EPI_ISL_2118962, EPI_ISL_2118964, EPI_ISL_2118966, EPI_ISL_2118969, EPI_ISL_2118972, EPI_ISL_2118974, EPI_ISL_2118975, EPI_ISL_2118976, EPI_ISL_2118977, EPI_ISL_2118980, EPI_ISL_2118982, EPI_ISL_2118986, EPI_ISL_2118994, EPI_ISL_2118995, EPI_ISL_2118996, EPI_ISL_2119000, EPI_ISL_2119001, EPI_ISL_2119003, EPI_ISL_2119007, EPI_ISL_2119011, EPI_ISL_2119012, EPI_ISL_2119018, EPI_ISL_2119021, EPI_ISL_2119025, EPI_ISL_2119028, EPI_ISL_2119030, EPI_ISL_2119031, EPI_ISL_2119033, EPI_ISL_2119034, EPI_ISL_2119036, EPI_ISL_2119037, EPI_ISL_2119046, EPI_ISL_2119080, EPI_ISL_2119085, EPI_ISL_2119092, EPI_ISL_2119094, EPI_ISL_2119095, EPI_ISL_2119103, EPI_ISL_2119105, EPI_ISL_2119106, EPI_ISL_2119107, EPI_ISL_2119109, EPI_ISL_2119110, EPI_ISL_2119112, EPI_ISL_2119115, EPI_ISL_2119116, EPI_ISL_2119118, EPI_ISL_2119119, EPI_ISL_2119125, EPI_ISL_2119131, EPI_ISL_2119136, EPI_ISL_2119139, EPI_ISL_2119144, EPI_ISL_2119148, EPI_ISL_2119154, EPI_ISL_2119157, EPI_ISL_2119161, EPI_ISL_2119168, EPI_ISL_2119171, EPI_ISL_2119174, EPI_ISL_2119175, EPI_ISL_2119177, EPI_ISL_2119178, EPI_ISL_2119179, EPI_ISL_2119181, EPI_ISL_2119186, EPI_ISL_2119188, EPI_ISL_2119189, EPI_ISL_2119191, EPI_ISL_2119193, EPI_ISL_2119194, EPI_ISL_2119197, EPI_ISL_2119200, EPI_ISL_2119202, EPI_ISL_2119203, EPI_ISL_2119208, EPI_ISL_2119213, EPI_ISL_2119214, EPI_ISL_2119219, EPI_ISL_2119223, EPI_ISL_2119225, EPI_ISL_2119228, EPI_ISL_2119231, EPI_ISL_2119233, EPI_ISL_2119235, EPI_ISL_2119238, EPI_ISL_2119240, EPI_ISL_2119242, EPI_ISL_2119246, EPI_ISL_2119252, EPI_ISL_2119254, EPI_ISL_2119255, EPI_ISL_2119258, EPI_ISL_2119261, EPI_ISL_2119263, EPI_ISL_2119270, EPI_ISL_2119273, EPI_ISL_2119281, EPI_ISL_2119283, EPI_ISL_2119292, EPI_ISL_2119293, EPI_ISL_2119296, EPI_ISL_2119298, EPI_ISL_2119303, EPI_ISL_2119304, EPI_ISL_2119306, EPI_ISL_2119307, EPI_ISL_2119312, EPI_ISL_2119314, EPI_ISL_2119317, EPI_ISL_2119319, EPI_ISL_2119322, EPI_ISL_2119325, EPI_ISL_2119328, EPI_ISL_2119332, EPI_ISL_2119333, EPI_ISL_2119335, EPI_ISL_2119339, EPI_ISL_2119341, EPI_ISL_2119344, EPI_ISL_2119348, EPI_ISL_2119352, EPI_ISL_2119356, EPI_ISL_2119358, EPI_ISL_2119361, EPI_ISL_2119363, EPI_ISL_2119367, EPI_ISL_2119368, EPI_ISL_2119371, EPI_ISL_2119373, EPI_ISL_2119376, EPI_ISL_2119379, EPI_ISL_2119381, EPI_ISL_2119383, EPI_ISL_2119385, EPI_ISL_2119390, EPI_ISL_2119393, EPI_ISL_2119397, EPI_ISL_2119400, EPI_ISL_2119402, EPI_ISL_2119405, EPI_ISL_2119408, EPI_ISL_2119411, EPI_ISL_2119412, EPI_ISL_2119413, EPI_ISL_2119414, EPI_ISL_2119417, EPI_ISL_2119419, EPI_ISL_2119422, EPI_ISL_2119427, EPI_ISL_2119434, EPI_ISL_2119437, EPI_ISL_2119440, EPI_ISL_2119447, EPI_ISL_2119450, EPI_ISL_2119453, EPI_ISL_2119454, EPI_ISL_2119456, EPI_ISL_2119457, EPI_ISL_2119458, EPI_ISL_2119465, EPI_ISL_2119467, EPI_ISL_2119468, EPI_ISL_2119477, EPI_ISL_2119479, EPI_ISL_2119489, EPI_ISL_2119492, EPI_ISL_2119495, EPI_ISL_2119497, EPI_ISL_2119500, EPI_ISL_2119503, EPI_ISL_2119506, EPI_ISL_2119507, EPI_ISL_2119509, EPI_ISL_2119510, EPI_ISL_2119515, EPI_ISL_2119517, EPI_ISL_2119520, EPI_ISL_2119523, EPI_ISL_2119526, EPI_ISL_2119529, EPI_ISL_2119531, EPI_ISL_2119532, EPI_ISL_2119535, EPI_ISL_2119545, EPI_ISL_2119548, EPI_ISL_2119549, EPI_ISL_2119552, EPI_ISL_2119557, EPI_ISL_2119565, EPI_ISL_2119572, EPI_ISL_2119579, EPI_ISL_2119580, EPI_ISL_2119590, EPI_ISL_2119596, EPI_ISL_2119601, EPI_ISL_2119602, EPI_ISL_2119604, EPI_ISL_2119606, EPI_ISL_2119607, EPI_ISL_2119612, EPI_ISL_2119613, EPI_ISL_2119615, EPI_ISL_2119617, EPI_ISL_2119618, EPI_ISL_2119630, EPI_ISL_2119641, EPI_ISL_2119644, EPI_ISL_2119647, EPI_ISL_2119649, EPI_ISL_2119655, EPI_ISL_2119657, EPI_ISL_2119666, EPI_ISL_2119667, EPI_ISL_2119669, EPI_ISL_2119672, EPI_ISL_2119674, EPI_ISL_2119676, EPI_ISL_2119677, EPI_ISL_2119685, EPI_ISL_2119686, EPI_ISL_2119688, EPI_ISL_2119689, EPI_ISL_2119694, EPI_ISL_2119696, EPI_ISL_2119697, EPI_ISL_2119701, EPI_ISL_2119702, EPI_ISL_2119704, EPI_ISL_2119706, EPI_ISL_2119707, EPI_ISL_2119718, EPI_ISL_2119720, EPI_ISL_2119723, EPI_ISL_2119727, EPI_ISL_2119731, EPI_ISL_2119732, EPI_ISL_2119735, EPI_ISL_2119740, EPI_ISL_2119743, EPI_ISL_2119746, EPI_ISL_2119751, EPI_ISL_2119756, EPI_ISL_2119757, EPI_ISL_2119759, EPI_ISL_2119761, EPI_ISL_2119764, EPI_ISL_2119767, EPI_ISL_2119770, EPI_ISL_2119774, EPI_ISL_2119777, EPI_ISL_2119784, EPI_ISL_2119786, EPI_ISL_2119787, EPI_ISL_2119789, EPI_ISL_2119790, EPI_ISL_2119795, EPI_ISL_2119797, EPI_ISL_2119798, EPI_ISL_2119800, EPI_ISL_2119801, EPI_ISL_2119806, EPI_ISL_2119808, EPI_ISL_2119809, EPI_ISL_2119811, EPI_ISL_2119812, EPI_ISL_2119814, EPI_ISL_2119816, EPI_ISL_2119817, EPI_ISL_2119820, EPI_ISL_2119823, EPI_ISL_2119826, EPI_ISL_2119830, EPI_ISL_2119833, EPI_ISL_2119839, EPI_ISL_2119843, EPI_ISL_2119844, EPI_ISL_2119846, EPI_ISL_2119851, EPI_ISL_2119854, EPI_ISL_2119857, EPI_ISL_2119861, EPI_ISL_2119865, EPI_ISL_2119869, EPI_ISL_2119875, EPI_ISL_2119904, EPI_ISL_2119906, EPI_ISL_2119907, EPI_ISL_2119910, EPI_ISL_2119911, EPI_ISL_2119914, EPI_ISL_2119917, EPI_ISL_2119922, EPI_ISL_2119927, EPI_ISL_2119931, EPI_ISL_2119936, EPI_ISL_2119942, EPI_ISL_2119945, EPI_ISL_2119946, EPI_ISL_2119948, EPI_ISL_2119952, EPI_ISL_2119954, EPI_ISL_2119955, EPI_ISL_2119957, EPI_ISL_2119960, EPI_ISL_2119963, EPI_ISL_2119965, EPI_ISL_2119970, EPI_ISL_2119972, EPI_ISL_2119973, EPI_ISL_2119975, EPI_ISL_2119976, EPI_ISL_2119977, EPI_ISL_2119979, EPI_ISL_2119980, EPI_ISL_2119982, EPI_ISL_2119985, EPI_ISL_2119986, EPI_ISL_2119987, EPI_ISL_2119990, EPI_ISL_2119991, EPI_ISL_2119993, EPI_ISL_2119996, EPI_ISL_2119997, EPI_ISL_2120000, EPI_ISL_2120002, EPI_ISL_2120003, EPI_ISL_2120006, EPI_ISL_2120008, EPI_ISL_2120009, EPI_ISL_2120013, EPI_ISL_2120027, EPI_ISL_2120028, EPI_ISL_2120036, EPI_ISL_2120037, EPI_ISL_2120040, EPI_ISL_2120042, EPI_ISL_2120045, EPI_ISL_2120048, EPI_ISL_2120052, EPI_ISL_2120053, EPI_ISL_2120054, EPI_ISL_2120057, EPI_ISL_2120059, EPI_ISL_2120061, EPI_ISL_2120062, EPI_ISL_2120064, EPI_ISL_2120068, EPI_ISL_2120071, EPI_ISL_2120074, EPI_ISL_2120077, EPI_ISL_2120080, EPI_ISL_2120082, EPI_ISL_2120084, EPI_ISL_2120085, EPI_ISL_2120088, EPI_ISL_2120089, EPI_ISL_2120091, EPI_ISL_2120094, EPI_ISL_2120095, EPI_ISL_2120096, EPI_ISL_2120100, EPI_ISL_2120101, EPI_ISL_2120103, EPI_ISL_2120104, EPI_ISL_2120106, EPI_ISL_2120107, EPI_ISL_2120109, EPI_ISL_2120110, EPI_ISL_2120112, EPI_ISL_2120113, EPI_ISL_2120114, EPI_ISL_2120115, EPI_ISL_2120117, EPI_ISL_2120119, EPI_ISL_2120123, EPI_ISL_2120132, EPI_ISL_2120135, EPI_ISL_2120138, EPI_ISL_2120141, EPI_ISL_2120143, EPI_ISL_2120146, EPI_ISL_2120150, EPI_ISL_2120152, EPI_ISL_2120155, EPI_ISL_2120157, EPI_ISL_2120158, EPI_ISL_2120160, EPI_ISL_2120163, EPI_ISL_2120166, EPI_ISL_2120168, EPI_ISL_2120171, EPI_ISL_2120172, EPI_ISL_2120173, EPI_ISL_2120176, EPI_ISL_2120179, EPI_ISL_2120182, EPI_ISL_2120184, EPI_ISL_2120185, EPI_ISL_2120188, EPI_ISL_2120191, EPI_ISL_2120192, EPI_ISL_2120193, EPI_ISL_2120195, EPI_ISL_2120196, EPI_ISL_2120198, EPI_ISL_2120202, EPI_ISL_2120206, EPI_ISL_2120207, EPI_ISL_2120211, EPI_ISL_2120213, EPI_ISL_2120214, EPI_ISL_2120216, EPI_ISL_2120217, EPI_ISL_2120220, EPI_ISL_2120221, EPI_ISL_2120227, EPI_ISL_2120232, EPI_ISL_2120236, EPI_ISL_2120239, EPI_ISL_2120240, EPI_ISL_2120242, EPI_ISL_2120244, EPI_ISL_2120252, EPI_ISL_2120254, EPI_ISL_2120256, EPI_ISL_2120257, EPI_ISL_2120264, EPI_ISL_2120268, EPI_ISL_2120270, EPI_ISL_2120271, EPI_ISL_2120273, EPI_ISL_2120274, EPI_ISL_2120276, EPI_ISL_2120277, EPI_ISL_2120279, EPI_ISL_2120283, EPI_ISL_2120285, EPI_ISL_2120286, EPI_ISL_2120287, EPI_ISL_2120289, EPI_ISL_2120292, EPI_ISL_2120293, EPI_ISL_2120298, EPI_ISL_2120299, EPI_ISL_2120301, EPI_ISL_2120305, EPI_ISL_2120310, EPI_ISL_2120311, EPI_ISL_2120313, EPI_ISL_2120314, EPI_ISL_2120316, EPI_ISL_2120317, EPI_ISL_2120320, EPI_ISL_2120323, EPI_ISL_2120328, EPI_ISL_2120329, EPI_ISL_2120331, EPI_ISL_2120334, EPI_ISL_2120337, EPI_ISL_2120340, EPI_ISL_2120342, EPI_ISL_2120346, EPI_ISL_2120349, EPI_ISL_2120350, EPI_ISL_2120352, EPI_ISL_2120353, EPI_ISL_2120355, EPI_ISL_2120357, EPI_ISL_2120358, EPI_ISL_2120360, EPI_ISL_2120367, EPI_ISL_2120369, EPI_ISL_2120371, EPI_ISL_2120376, EPI_ISL_2120379, EPI_ISL_2120384, EPI_ISL_2120388, EPI_ISL_2120390, EPI_ISL_2120391, EPI_ISL_2120393, EPI_ISL_2120397, EPI_ISL_2120400, EPI_ISL_2120402, EPI_ISL_2120405, EPI_ISL_2120409, EPI_ISL_2120410, EPI_ISL_2120412, EPI_ISL_2120413, EPI_ISL_2120417, EPI_ISL_2120419, EPI_ISL_2120420, EPI_ISL_2120423, EPI_ISL_2120426, EPI_ISL_2120428, EPI_ISL_2120430, EPI_ISL_2120433, EPI_ISL_2120437, EPI_ISL_2120442, EPI_ISL_2120449, EPI_ISL_2120450, EPI_ISL_2120452, EPI_ISL_2120457, EPI_ISL_2120471, EPI_ISL_2120476, EPI_ISL_2120477, EPI_ISL_2120487, EPI_ISL_2120491, EPI_ISL_2120503, EPI_ISL_2120514, EPI_ISL_2120522, EPI_ISL_2120526, EPI_ISL_2120538, EPI_ISL_2120541, EPI_ISL_2120545, EPI_ISL_2120560, EPI_ISL_2120578, EPI_ISL_2120581, EPI_ISL_2120584, EPI_ISL_2120585, EPI_ISL_2120590, EPI_ISL_2120594, EPI_ISL_2120605, EPI_ISL_2120607, EPI_ISL_2120608, EPI_ISL_2120610, EPI_ISL_2120613, EPI_ISL_2120614, EPI_ISL_2120628, EPI_ISL_2120630, EPI_ISL_2120642, EPI_ISL_2120643, EPI_ISL_2120648, EPI_ISL_2120652, EPI_ISL_2120655, EPI_ISL_2120695, EPI_ISL_2120697, EPI_ISL_2120700, EPI_ISL_2120713, EPI_ISL_2120724, EPI_ISL_2120726, EPI_ISL_2120745, EPI_ISL_2120746, EPI_ISL_2120751, EPI_ISL_2120754, EPI_ISL_2120756, EPI_ISL_2120760, EPI_ISL_2120761, EPI_ISL_2120776, EPI_ISL_2120782, EPI_ISL_2120791, EPI_ISL_2120793, EPI_ISL_2120795, EPI_ISL_2120800, EPI_ISL_2120802, EPI_ISL_2120812, EPI_ISL_2120828, EPI_ISL_2120830, EPI_ISL_2120836, EPI_ISL_2120837, EPI_ISL_2120839, EPI_ISL_2120852, EPI_ISL_2120861, EPI_ISL_2120866, EPI_ISL_2120868, EPI_ISL_2120877, EPI_ISL_2120882, EPI_ISL_2120891, EPI_ISL_2120902, EPI_ISL_2120905, EPI_ISL_2120910, EPI_ISL_2120915, EPI_ISL_2120918, EPI_ISL_2120921, EPI_ISL_2120926, EPI_ISL_2120929, EPI_ISL_2120932, EPI_ISL_2120943, EPI_ISL_2120947, EPI_ISL_2120948, EPI_ISL_2120951, EPI_ISL_2120956, EPI_ISL_2120959, EPI_ISL_2120965, EPI_ISL_2120968, EPI_ISL_2120979, EPI_ISL_2120992, EPI_ISL_2121005, EPI_ISL_2121007, EPI_ISL_2121010, EPI_ISL_2121011, EPI_ISL_2121013, EPI_ISL_2121021, EPI_ISL_2121023, EPI_ISL_2121025, EPI_ISL_2121026, EPI_ISL_2121031, EPI_ISL_2121032, EPI_ISL_2121037, EPI_ISL_2121041, EPI_ISL_2121044, EPI_ISL_2121046, EPI_ISL_2121047, EPI_ISL_2121050, EPI_ISL_2121056, EPI_ISL_2121057, EPI_ISL_2121060, EPI_ISL_2121066, EPI_ISL_2121069, EPI_ISL_2121072, EPI_ISL_2121076, EPI_ISL_2121078, EPI_ISL_2121079, EPI_ISL_2121082, EPI_ISL_2121092, EPI_ISL_2121097, EPI_ISL_2121098, EPI_ISL_2121101, EPI_ISL_2121104, EPI_ISL_2121107, EPI_ISL_2121108, EPI_ISL_2121111, EPI_ISL_2121114, EPI_ISL_2121116, EPI_ISL_2121117, EPI_ISL_2121122, EPI_ISL_2121123, EPI_ISL_2121137, EPI_ISL_2121138, EPI_ISL_2121141, EPI_ISL_2121150, EPI_ISL_2121151, EPI_ISL_2121156, EPI_ISL_2121157, EPI_ISL_2121159, EPI_ISL_2121162, EPI_ISL_2121165, EPI_ISL_2121166, EPI_ISL_2121168, EPI_ISL_2121177, EPI_ISL_2121182, EPI_ISL_2121183, EPI_ISL_2121195, EPI_ISL_2121196, EPI_ISL_2121198, EPI_ISL_2121201, EPI_ISL_2121204, EPI_ISL_2121209, EPI_ISL_2121210, EPI_ISL_2121215, EPI_ISL_2121222, EPI_ISL_2121230, EPI_ISL_2121232, EPI_ISL_2121237, EPI_ISL_2121239, EPI_ISL_2121240, EPI_ISL_2121242, EPI_ISL_2121243, EPI_ISL_2121246, EPI_ISL_2121247, EPI_ISL_2121253, EPI_ISL_2121255, EPI_ISL_2121256, EPI_ISL_2121259, EPI_ISL_2121263, EPI_ISL_2121265, EPI_ISL_2121266, EPI_ISL_2121273, EPI_ISL_2121275, EPI_ISL_2121277, EPI_ISL_2121286, EPI_ISL_2121291, EPI_ISL_2121293, EPI_ISL_2121294, EPI_ISL_2121296, EPI_ISL_2121300, EPI_ISL_2121305, EPI_ISL_2121306, EPI_ISL_2121308, EPI_ISL_2121309, EPI_ISL_2121312, EPI_ISL_2121313, EPI_ISL_2121315, EPI_ISL_2121316, EPI_ISL_2121318, EPI_ISL_2121333, EPI_ISL_2121335, EPI_ISL_2121338, EPI_ISL_2121340, EPI_ISL_2121346, EPI_ISL_2121347, EPI_ISL_2121350, EPI_ISL_2121355, EPI_ISL_2121356, EPI_ISL_2121358, EPI_ISL_2121364, EPI_ISL_2121371, EPI_ISL_2121373, EPI_ISL_2121374, EPI_ISL_2121376, EPI_ISL_2121382, EPI_ISL_2121385, EPI_ISL_2121393, EPI_ISL_2121397, EPI_ISL_2121399, EPI_ISL_2121401, EPI_ISL_2121402, EPI_ISL_2121404, EPI_ISL_2121413, EPI_ISL_2121416, EPI_ISL_2121417, EPI_ISL_2121420, EPI_ISL_2121422, EPI_ISL_2121425, EPI_ISL_2121426, EPI_ISL_2121429, EPI_ISL_2121432, EPI_ISL_2121436, EPI_ISL_2121437, EPI_ISL_2121469, EPI_ISL_2121472, EPI_ISL_2121473, EPI_ISL_2121475, EPI_ISL_2121479, EPI_ISL_2121483, EPI_ISL_2121484, EPI_ISL_2121489, EPI_ISL_2121493, EPI_ISL_2121495, EPI_ISL_2121498, EPI_ISL_2121500, EPI_ISL_2121501, EPI_ISL_2121506, EPI_ISL_2121508, EPI_ISL_2121509, EPI_ISL_2121513, EPI_ISL_2121514, EPI_ISL_2121517, EPI_ISL_2121519, EPI_ISL_2121521, EPI_ISL_2121522, EPI_ISL_2121524, EPI_ISL_2121525, EPI_ISL_2121527, EPI_ISL_2121533, EPI_ISL_2121536, EPI_ISL_2121538, EPI_ISL_2121540, EPI_ISL_2121542, EPI_ISL_2121547, EPI_ISL_2121549, EPI_ISL_2121557, EPI_ISL_2121568, EPI_ISL_2121572, EPI_ISL_2121574, EPI_ISL_2121576, EPI_ISL_2121577, EPI_ISL_2121580, EPI_ISL_2121582, EPI_ISL_2121584, EPI_ISL_2121587, EPI_ISL_2121588, EPI_ISL_2121590, EPI_ISL_2121591, EPI_ISL_2121593, EPI_ISL_2121594, EPI_ISL_2121596, EPI_ISL_2121598, EPI_ISL_2121600, EPI_ISL_2121601, EPI_ISL_2121603, EPI_ISL_2121607, EPI_ISL_2121609, EPI_ISL_2121611, EPI_ISL_2121613, EPI_ISL_2121614, EPI_ISL_2121615, EPI_ISL_2121617, EPI_ISL_2121618, EPI_ISL_2121622, EPI_ISL_2121625, EPI_ISL_2121626, EPI_ISL_2121628, EPI_ISL_2121629, EPI_ISL_2121631, EPI_ISL_2121632, EPI_ISL_2121634, EPI_ISL_2121636, EPI_ISL_2121639, EPI_ISL_2121645, EPI_ISL_2121647, EPI_ISL_2121648, EPI_ISL_2121650, EPI_ISL_2121651, EPI_ISL_2121653, EPI_ISL_2121657, EPI_ISL_2121660, EPI_ISL_2121662, EPI_ISL_2121665, EPI_ISL_2121666, EPI_ISL_2121668, EPI_ISL_2121670, EPI_ISL_2121671, EPI_ISL_2121676, EPI_ISL_2121677, EPI_ISL_2121679, EPI_ISL_2121680, EPI_ISL_2121684, EPI_ISL_2121685, EPI_ISL_2121687, EPI_ISL_2121688, EPI_ISL_2121690, EPI_ISL_2121691, EPI_ISL_2121693, EPI_ISL_2121696, EPI_ISL_2121697, EPI_ISL_2121700, EPI_ISL_2121704, EPI_ISL_2121707, EPI_ISL_2121708, EPI_ISL_2121710, EPI_ISL_2121711, EPI_ISL_2121714, EPI_ISL_2121716, EPI_ISL_2121718, EPI_ISL_2121719, EPI_ISL_2121725, EPI_ISL_2121731, EPI_ISL_2121733, EPI_ISL_2121735, EPI_ISL_2121743, EPI_ISL_2121748, EPI_ISL_2121750, EPI_ISL_2121753, EPI_ISL_2121755, EPI_ISL_2121756, EPI_ISL_2121758, EPI_ISL_2121759, EPI_ISL_2121761, EPI_ISL_2121762, EPI_ISL_2121764, EPI_ISL_2121767, EPI_ISL_2121769, EPI_ISL_2121770, EPI_ISL_2121772, EPI_ISL_2121773, EPI_ISL_2121775, EPI_ISL_2121778, EPI_ISL_2121781, EPI_ISL_2121782, EPI_ISL_2121784, EPI_ISL_2121785, EPI_ISL_2121790, EPI_ISL_2121793, EPI_ISL_2121796, EPI_ISL_2121798, EPI_ISL_2121801, EPI_ISL_2121802, EPI_ISL_2121804, EPI_ISL_2121806, EPI_ISL_2121807, EPI_ISL_2121810, EPI_ISL_2121812, EPI_ISL_2121819, EPI_ISL_2121820, EPI_ISL_2121822, EPI_ISL_2121823, EPI_ISL_2121826, EPI_ISL_2121828, EPI_ISL_2121833, EPI_ISL_2121835, EPI_ISL_2121837, EPI_ISL_2121838, EPI_ISL_2121841, EPI_ISL_2121847, EPI_ISL_2121850, EPI_ISL_2121852, EPI_ISL_2121853, EPI_ISL_2121855, EPI_ISL_2121856, EPI_ISL_2121858, EPI_ISL_2121859, EPI_ISL_2121861, EPI_ISL_2121863, EPI_ISL_2121864, EPI_ISL_2121866, EPI_ISL_2121867, EPI_ISL_2121870, EPI_ISL_2121872, EPI_ISL_2121874, EPI_ISL_2121875, EPI_ISL_2121877, EPI_ISL_2121880, EPI_ISL_2121883, EPI_ISL_2121886, EPI_ISL_2121888, EPI_ISL_2121889, EPI_ISL_2121891, EPI_ISL_2121893, EPI_ISL_2121897, EPI_ISL_2121899, EPI_ISL_2121901, EPI_ISL_2121902, EPI_ISL_2121904, EPI_ISL_2121906, EPI_ISL_2121907, EPI_ISL_2121909, EPI_ISL_2121911, EPI_ISL_2121912, EPI_ISL_2121914, EPI_ISL_2121915, EPI_ISL_2121918, EPI_ISL_2121920, EPI_ISL_2121922, EPI_ISL_2121927, EPI_ISL_2121930, EPI_ISL_2121931, EPI_ISL_2121933, EPI_ISL_2121934, EPI_ISL_2121936, EPI_ISL_2121938, EPI_ISL_2121939, EPI_ISL_2121941, EPI_ISL_2121942, EPI_ISL_2121944, EPI_ISL_2121946, EPI_ISL_2121947, EPI_ISL_2121949, EPI_ISL_2121952, EPI_ISL_2121955, EPI_ISL_2121957, EPI_ISL_2121958, EPI_ISL_2121960, EPI_ISL_2121962, EPI_ISL_2121963, EPI_ISL_2121966, EPI_ISL_2121974, EPI_ISL_2121976, EPI_ISL_2121977, EPI_ISL_2121980, EPI_ISL_2121983, EPI_ISL_2121988, EPI_ISL_2121989, EPI_ISL_2121991, EPI_ISL_2121995, EPI_ISL_2121997, EPI_ISL_2122000, EPI_ISL_2122002, EPI_ISL_2122004, EPI_ISL_2122005, EPI_ISL_2122007, EPI_ISL_2122009, EPI_ISL_2122011, EPI_ISL_2122014, EPI_ISL_2122018, EPI_ISL_2122019, EPI_ISL_2122023, EPI_ISL_2122024, EPI_ISL_2122026, EPI_ISL_2122029, EPI_ISL_2122031, EPI_ISL_2122032, EPI_ISL_2122034, EPI_ISL_2122036, EPI_ISL_2122037, EPI_ISL_2122039, EPI_ISL_2122040, EPI_ISL_2122042, EPI_ISL_2122043, EPI_ISL_2122045, EPI_ISL_2122046, EPI_ISL_2122050, EPI_ISL_2122051, EPI_ISL_2122053, EPI_ISL_2122056, EPI_ISL_2122058, EPI_ISL_2122059, EPI_ISL_2122060, EPI_ISL_2122062, EPI_ISL_2122063, EPI_ISL_2122065, EPI_ISL_2122066, EPI_ISL_2122068, EPI_ISL_2122069, EPI_ISL_2122071, EPI_ISL_2122074, EPI_ISL_2122076, EPI_ISL_2122080, EPI_ISL_2122082, EPI_ISL_2122084, EPI_ISL_2122085, EPI_ISL_2122086, EPI_ISL_2122088, EPI_ISL_2122089, EPI_ISL_2122091, EPI_ISL_2122094, EPI_ISL_2122096, EPI_ISL_2122100, EPI_ISL_2122102, EPI_ISL_2122103, EPI_ISL_2122105, EPI_ISL_2122106, EPI_ISL_2122108, EPI_ISL_2122109, EPI_ISL_2122111, EPI_ISL_2122113, EPI_ISL_2122114, EPI_ISL_2122116, EPI_ISL_2122117, EPI_ISL_2122119, EPI_ISL_2122122, EPI_ISL_2122147, EPI_ISL_2122148, EPI_ISL_2122150, EPI_ISL_2122155, EPI_ISL_2122167, EPI_ISL_2122176, EPI_ISL_2122180, EPI_ISL_2122184, EPI_ISL_2122187, EPI_ISL_2122200, EPI_ISL_2122202, EPI_ISL_2122203, EPI_ISL_2122205, EPI_ISL_2122211, EPI_ISL_2122234, EPI_ISL_2122279, EPI_ISL_2122284, EPI_ISL_2122287, EPI_ISL_2122303, EPI_ISL_2122308, EPI_ISL_2122309, EPI_ISL_2122312, EPI_ISL_2122319, EPI_ISL_2122323, EPI_ISL_2122326, EPI_ISL_2122331, EPI_ISL_2122332, EPI_ISL_2122351, EPI_ISL_2122369, EPI_ISL_2122388, EPI_ISL_2122402, EPI_ISL_2122468, EPI_ISL_2122470, EPI_ISL_2122473, EPI_ISL_2122484, EPI_ISL_2122497, EPI_ISL_2122515, EPI_ISL_2122518, EPI_ISL_2122525, EPI_ISL_2122532, EPI_ISL_2122533, EPI_ISL_2122535, EPI_ISL_2122537, EPI_ISL_2122538, EPI_ISL_2122551, EPI_ISL_2122562, EPI_ISL_2122568, EPI_ISL_2122586, EPI_ISL_2122590, EPI_ISL_2122591, EPI_ISL_2122598, EPI_ISL_2122600, EPI_ISL_2122611, EPI_ISL_2122613, EPI_ISL_2122616, EPI_ISL_2122621, EPI_ISL_2122632, EPI_ISL_2122639, EPI_ISL_2122661, EPI_ISL_2122665, EPI_ISL_2122667, EPI_ISL_2122681, EPI_ISL_2122701, EPI_ISL_2122702, EPI_ISL_2122704, EPI_ISL_2137847, EPI_ISL_2137856, EPI_ISL_2137864, EPI_ISL_2137868, EPI_ISL_2137886, EPI_ISL_2137901, EPI_ISL_2137902, EPI_ISL_2137930, EPI_ISL_2137938, EPI_ISL_2137951, EPI_ISL_2137970, EPI_ISL_2137974, EPI_ISL_2137985, EPI_ISL_2138009, EPI_ISL_2138010, EPI_ISL_2138011, EPI_ISL_2138025, EPI_ISL_2138026, EPI_ISL_2138030, EPI_ISL_2138038, EPI_ISL_2138056, EPI_ISL_2138070, EPI_ISL_2138085, EPI_ISL_2138088, EPI_ISL_2138100, EPI_ISL_2138101, EPI_ISL_2138108, EPI_ISL_2138110, EPI_ISL_2138113, EPI_ISL_2138114, EPI_ISL_2138115, EPI_ISL_2138116, EPI_ISL_2138117, EPI_ISL_2138118, EPI_ISL_2138119, EPI_ISL_2138121, EPI_ISL_2138123, EPI_ISL_2138124, EPI_ISL_2138125, EPI_ISL_2138126, EPI_ISL_2138127, EPI_ISL_2138132, EPI_ISL_2138133, EPI_ISL_2138134, EPI_ISL_2138135, EPI_ISL_2138138, EPI_ISL_2138141, EPI_ISL_2138146, EPI_ISL_2138149, EPI_ISL_2138150, EPI_ISL_2138151, EPI_ISL_2138152, EPI_ISL_2138153, EPI_ISL_2138154, EPI_ISL_2138162, EPI_ISL_2138166, EPI_ISL_2138169, EPI_ISL_2138170, EPI_ISL_2138172, EPI_ISL_2138174, EPI_ISL_2138179, EPI_ISL_2138186, EPI_ISL_2138187, EPI_ISL_2138188, EPI_ISL_2138190, EPI_ISL_2138191, EPI_ISL_2138195, EPI_ISL_2138196, EPI_ISL_2138197, EPI_ISL_2138199, EPI_ISL_2138200, EPI_ISL_2138201, EPI_ISL_2138202, EPI_ISL_2138203, EPI_ISL_2138204, EPI_ISL_2138207, EPI_ISL_2138211, EPI_ISL_2138212, EPI_ISL_2138213, EPI_ISL_2138216, EPI_ISL_2138218, EPI_ISL_2138220, EPI_ISL_2138221, EPI_ISL_2138229, EPI_ISL_2138228, EPI_ISL_2138229, EPI_ISL_2138230, EPI_ISL_2138233, EPI_ISL_2138235, EPI_ISL_2138239, EPI_ISL_2138241, EPI_ISL_2138243, EPI_ISL_2138244, EPI_ISL_2138245, EPI_ISL_2138250, EPI_ISL_2138251, EPI_ISL_2138252, EPI_ISL_2138253, EPI_ISL_2138254, EPI_ISL_2138261, EPI_ISL_2138263, EPI_ISL_2138267, EPI_ISL_2138269, EPI_ISL_2138275, EPI_ISL_2138279, EPI_ISL_2138281, EPI_ISL_2138282, EPI_ISL_2138284, EPI_ISL_2138290, EPI_ISL_2138299, EPI_ISL_2138303, EPI_ISL_2138304, EPI_ISL_2138308, EPI_ISL_2138309, EPI_ISL_2138310, EPI_ISL_2138313, EPI_ISL_2138316, EPI_ISL_2138323, EPI_ISL_2138324, EPI_ISL_2138326, EPI_ISL_2138327, EPI_ISL_2138329, EPI_ISL_2138330, EPI_ISL_2138332, EPI_ISL_2138334, EPI_ISL_2138335, EPI_ISL_2138341, EPI_ISL_2138342, EPI_ISL_2138345, EPI_ISL_2138346, EPI_ISL_2138347, EPI_ISL_2138349, EPI_ISL_2138351, EPI_ISL_2138352, EPI_ISL_2138358, EPI_ISL_2138361, EPI_ISL_2138362, EPI_ISL_2138364, EPI_ISL_2138365, EPI_ISL_2138368, EPI_ISL_2138376, EPI_ISL_2138383, EPI_ISL_2138392, EPI_ISL_2138394, EPI_ISL_2138395, EPI_ISL_2138403, EPI_ISL_2138405, EPI_ISL_2138411, EPI_ISL_2138413, EPI_ISL_2138417, EPI_ISL_2138419, EPI_ISL_2138432, EPI_ISL_2138437, EPI_ISL_2138443, EPI_ISL_2138444, EPI_ISL_2138445, EPI_ISL_2138453, EPI_ISL_2138457, EPI_ISL_2138462, EPI_ISL_2138463, EPI_ISL_2138464, EPI_ISL_2138466, EPI_ISL_2138490, EPI_ISL_2138495, EPI_ISL_2138496, EPI_ISL_2138497, EPI_ISL_2138498, EPI_ISL_2138502, EPI_ISL_2138504, EPI_ISL_2138506, EPI_ISL_2138509, EPI_ISL_2138510, EPI_ISL_2138525, EPI_ISL_2138529, EPI_ISL_2138530, EPI_ISL_2138531, EPI_ISL_2138537, EPI_ISL_2138539, EPI_ISL_2138546, EPI_ISL_2138550, EPI_ISL_2138555, EPI_ISL_2138557, EPI_ISL_2138566, EPI_ISL_2138570, EPI_ISL_2138573, EPI_ISL_2138575, EPI_ISL_2138581, EPI_ISL_2138585, EPI_ISL_2138588, EPI_ISL_2138590, EPI_ISL_2138593, EPI_ISL_2138599, EPI_ISL_2138613, EPI_ISL_2138638, EPI_ISL_2138639, EPI_ISL_2138643, EPI_ISL_2138645, EPI_ISL_2138651, EPI_ISL_2138654, EPI_ISL_2138660, EPI_ISL_2138666, EPI_ISL_2138667, EPI_ISL_2138671, EPI_ISL_2138677, EPI_ISL_2138679, EPI_ISL_2138699, EPI_ISL_2138700, EPI_ISL_2138702, EPI_ISL_2138703, EPI_ISL_2138704, EPI_ISL_2138706, EPI_ISL_2138707, EPI_ISL_2138708, EPI_ISL_2138709, EPI_ISL_2138711, EPI_ISL_2138712, EPI_ISL_2138716, EPI_ISL_2152661, EPI_ISL_2152663, EPI_ISL_2152803, EPI_ISL_2152809, EPI_ISL_2152839, EPI_ISL_2152861, EPI_ISL_2152865, EPI_ISL_2152950, EPI_ISL_2153016, EPI_ISL_2153042, EPI_ISL_2153043, EPI_ISL_2153044, EPI_ISL_2153045, EPI_ISL_2198660, EPI_ISL_2198670, EPI_ISL_2198685, EPI_ISL_2198825, EPI_ISL_2198846, EPI_ISL_2199018, EPI_ISL_2199422, EPI_ISL_2199606, EPI_ISL_2236821, EPI_ISL_2236830, EPI_ISL_2236835, EPI_ISL_2236842, EPI_ISL_2236850, EPI_ISL_2236851, EPI_ISL_2236858, EPI_ISL_2236860, EPI_ISL_2236861, EPI_ISL_2236862, EPI_ISL_2236863, EPI_ISL_2236864, EPI_ISL_2236865, EPI_ISL_2236869, EPI_ISL_2236871, EPI_ISL_2236874, EPI_ISL_2236877, EPI_ISL_2236884, EPI_ISL_2236886, EPI_ISL_2236889, EPI_ISL_2236891, EPI_ISL_2236901, EPI_ISL_2236904, EPI_ISL_2236914, EPI_ISL_2236921, EPI_ISL_2236923, EPI_ISL_2236925, EPI_ISL_2236927, EPI_ISL_2236931, EPI_ISL_2236932, EPI_ISL_2236938, EPI_ISL_2236955, EPI_ISL_2236956, EPI_ISL_2236957, EPI_ISL_2236959, EPI_ISL_2236970, EPI_ISL_2236973, EPI_ISL_2236980, EPI_ISL_2236988, EPI_ISL_2236989, EPI_ISL_2236993, EPI_ISL_2236994, EPI_ISL_2237002, EPI_ISL_2237009, EPI_ISL_2237016, EPI_ISL_2237017, EPI_ISL_2237018, EPI_ISL_2237019, EPI_ISL_2237021, EPI_ISL_2237022, EPI_ISL_2237034, EPI_ISL_2237044, EPI_ISL_2237046, EPI_ISL_2237047, EPI_ISL_2237053, EPI_ISL_2237055, EPI_ISL_2237058, EPI_ISL_2237061, EPI_ISL_2237066, EPI_ISL_2237067, EPI_ISL_2237069, EPI_ISL_2237072, EPI_ISL_2237073, EPI_ISL_2237074, EPI_ISL_2237075, EPI_ISL_2237076, EPI_ISL_2237084, EPI_ISL_2237094, EPI_ISL_2237099, EPI_ISL_2237104, EPI_ISL_2237108, EPI_ISL_2237111, EPI_ISL_2237113, EPI_ISL_2237114, EPI_ISL_2237120, EPI_ISL_2237128, EPI_ISL_2237131, EPI_ISL_2237136, EPI_ISL_2237138, EPI_ISL_2237139, EPI_ISL_2237142, EPI_ISL_2237143, EPI_ISL_2237144, EPI_ISL_2237146, EPI_ISL_2237149, EPI_ISL_2237150, EPI_ISL_2237801, EPI_ISL_2237871, EPI_ISL_2237907, EPI_ISL_2237916, EPI_ISL_2237942, EPI_ISL_2238030, EPI_ISL_2238058, EPI_ISL_2238068, EPI_ISL_2238070, EPI_ISL_2238073, EPI_ISL_2238074, EPI_ISL_2238077, EPI_ISL_2238078, EPI_ISL_2238079, EPI_ISL_2238080, EPI_ISL_2238081, EPI_ISL_2238082, EPI_ISL_2238083, EPI_ISL_2238084, EPI_ISL_2238085, EPI_ISL_2238087, EPI_ISL_2238089, EPI_ISL_2238091, EPI_ISL_2238093, EPI_ISL_2238095, EPI_ISL_2238098, EPI_ISL_2238099, EPI_ISL_2238101, EPI_ISL_2238103, EPI_ISL_2238104, EPI_ISL_2238105, EPI_ISL_2238106, EPI_ISL_2238108, EPI_ISL_2238110, EPI_ISL_2238111, EPI_ISL_2238114, EPI_ISL_2238115, EPI_ISL_2238116, EPI_ISL_2238117, EPI_ISL_2238118, EPI_ISL_2238119, EPI_ISL_2238121, EPI_ISL_2238122, EPI_ISL_2238124, EPI_ISL_2238127, EPI_ISL_2238129, EPI_ISL_2238131, EPI_ISL_2238134, EPI_ISL_2238137, EPI_ISL_2238138, EPI_ISL_2238140, EPI_ISL_2238141, EPI_ISL_2238142, EPI_ISL_2238146, EPI_ISL_2238151, EPI_ISL_2238152, EPI_ISL_2238154, EPI_ISL_2238159, EPI_ISL_2238161, EPI_ISL_2238163, EPI_ISL_2238165, EPI_ISL_2238166, EPI_ISL_2238167, EPI_ISL_2238169, EPI_ISL_2238170, EPI_ISL_2238171, EPI_ISL_2238174, EPI_ISL_2238175, EPI_ISL_2238178, EPI_ISL_2238179, EPI_ISL_2238180, EPI_ISL_2238182, EPI_ISL_2238183, EPI_ISL_2238187, EPI_ISL_2238190, EPI_ISL_2238191, EPI_ISL_2238194, EPI_ISL_2238196, EPI_ISL_2238197, EPI_ISL_2238198, EPI_ISL_2238201, EPI_ISL_2238202, EPI_ISL_2238204, EPI_ISL_2238205, EPI_ISL_2238209, EPI_ISL_2238211, EPI_ISL_2238218, EPI_ISL_2238220, EPI_ISL_2238226, EPI_ISL_2238228, EPI_ISL_2238229, EPI_ISL_2238230, EPI_ISL_2238232, EPI_ISL_2238235, EPI_ISL_2238236, EPI_ISL_2238238, EPI_ISL_2238239, EPI_ISL_2238240, EPI_ISL_2238243, EPI_ISL_2238246, EPI_ISL_2238248, EPI_ISL_2238249, EPI_ISL_2238252, EPI_ISL_2238253, EPI_ISL_2238259, EPI_ISL_2238262, EPI_ISL_2238264, EPI_ISL_2238265, EPI_ISL_2238267, EPI_ISL_2238270, EPI_ISL_2238272, EPI_ISL_2238276, EPI_ISL_2238279, EPI_ISL_2238280, EPI_ISL_2238282, EPI_ISL_2238283, EPI_ISL_2238284, EPI_ISL_2238287, EPI_ISL_2238288, EPI_ISL_2238292, EPI_ISL_2238293, EPI_ISL_2238300, EPI_ISL_2238302, EPI_ISL_2238304, EPI_ISL_2238305, EPI_ISL_2238306, EPI_ISL_2238309, EPI_ISL_2238310, EPI_ISL_2238312, EPI_ISL_2238315, EPI_ISL_2238316, EPI_ISL_2238317, EPI_ISL_2238319, EPI_ISL_2238320, EPI_ISL_2238324, EPI_ISL_2238325, EPI_ISL_2238326, EPI_ISL_2238327, EPI_ISL_2238328, EPI_ISL_2238329, EPI_ISL_2238330, EPI_ISL_2238331, EPI_ISL_2238332, EPI_ISL_2238333, EPI_ISL_2238335, EPI_ISL_2238338, EPI_ISL_2238341, EPI_ISL_2238342, EPI_ISL_2238345, EPI_ISL_2238346, EPI_ISL_2238349, EPI_ISL_2238351, EPI_ISL_2238354, EPI_ISL_2238694, EPI_ISL_2238696, EPI_ISL_2253691, EPI_ISL_2253715, EPI_ISL_2253749, EPI_ISL_2253763, EPI_ISL_2253820, EPI_ISL_2253860, EPI_ISL_2253912, EPI_ISL_2253929, EPI_ISL_2253930, EPI_ISL_2254106, EPI_ISL_2346713, EPI_ISL_2393592, EPI_ISL_2393600, EPI_ISL_2393724, EPI_ISL_2435475, EPI_ISL_2435482, EPI_ISL_2435485, EPI_ISL_2446297, EPI_ISL_2446461, EPI_ISL_2446506, EPI_ISL_2446673, EPI_ISL_2456794, EPI_ISL_2457503, EPI_ISL_2554620, EPI_ISL_2570290, EPI_ISL_2665389

| | | | |
|---|---|---|---|
| see above | Lighthouse Lab in Milton Keynes | Wellcome Sanger Institute for the COVID-19 Genomics UK (COG-UK) Consortium | Cordelia Langford; David K. Jackson; Dominic Kwiatkowski; Ewan Harrison; Ian Johnston; Jeffrey Barrett; John Sillitoe on behalf of the Wellcome Sanger Institute COVID-19 Surveillance Team; Roberto Amato; Sonia Goncalves; The Lighthouse Lab in Milton Keynes and Alex Alderton |
| EPI_ISL_2240448 | Lincolnshire Hospitals and DeepSeq Nottingham | COVID-19 Genomics | Christopher Moore; Fei Sang; Joeseph Chappell; Johnny Debebe; Jonathan Ball; Matthew Carlisle; Matthew Loose; Nadine Holmes; Nichola Duckworth; Patrick McClure; Sarah Walsh; Tim Sloan; Victoria Wright |

| | | | |
|---|---|---|---|
| | | UK (COG-UK) Consortium | |
| EPI_ISL_1975041, EPI_ISL_1975042, EPI_ISL_1975044, EPI_ISL_1975045, EPI_ISL_1975047, EPI_ISL_1975049, EPI_ISL_2125988, EPI_ISL_2125989, EPI_ISL_2125990, EPI_ISL_2125992, EPI_ISL_2125993, EPI_ISL_2125995, EPI_ISL_2125997, EPI_ISL_2238871, EPI_ISL_2238872, EPI_ISL_2238873, EPI_ISL_2238877, EPI_ISL_2238878, EPI_ISL_2238887, EPI_ISL_2355362, EPI_ISL_2355363, EPI_ISL_2355364, EPI_ISL_2355365, EPI_ISL_2434241, EPI_ISL_2434242, EPI_ISL_2434243 | | | |
| see above | Liverpool Clinical Laboratories | COVID-19 Genomics UK (COG-UK) Consortium | A Alrezaihi; Alessandro Gerada; Alistair Darby; Angela Cowell; Anita Lucaci; Anu Chawla; Cassie Olateju; Catherine Hartley; Charlotte Nelson; Ecaterina Vamos; Elaine O'Toole; Eleanor G Bentley; Ghada T Shawli; Hermione Webster; Isabel García-Dorival; James Johnson; James P Stewart; Jenifer Manson; Joanne Watts; Jones Benjamin; Jordan J Clark; Julian Hiscox; L Luu; Lucille Rainbow; M Almsaud; Margaret Hughes; Mark Whitehead; Matthew Gemmell; Miren Iturriza-Gomara; Muhannad Alruwaili; N.P Randle; Neil Swainston; PKF Gilmore; Parul Sharma; Rebekah Penrice-Randal; Richard Eccles; Richard Gregory; Sam Haldenby; Steve Paterson; Stuart D Armstrong; Trevor Ian Robinson; Ximeng Han |
| EPI_ISL_2126181, EPI_ISL_2126184, EPI_ISL_2126186, EPI_ISL_2126666, EPI_ISL_2126668, EPI_ISL_2126670, EPI_ISL_2126672, EPI_ISL_2126673, EPI_ISL_2126677, EPI_ISL_2126679, EPI_ISL_2126681, EPI_ISL_2126683, EPI_ISL_2126685, EPI_ISL_2126689, EPI_ISL_2126691, EPI_ISL_2126692, EPI_ISL_2126696, EPI_ISL_2126698, EPI_ISL_2126699, EPI_ISL_2126702, EPI_ISL_2126704, EPI_ISL_2126706, EPI_ISL_2126707, EPI_ISL_2126709, EPI_ISL_2126712, EPI_ISL_2126714, EPI_ISL_2126715, EPI_ISL_2126720, EPI_ISL_2126794, EPI_ISL_2126796, EPI_ISL_2127047, EPI_ISL_2127048, EPI_ISL_2127049, EPI_ISL_2127050, EPI_ISL_2127052, EPI_ISL_2127054, EPI_ISL_2127299, EPI_ISL_2127301, EPI_ISL_2127303, EPI_ISL_2127305, EPI_ISL_2127534, EPI_ISL_2127536, EPI_ISL_2127538, EPI_ISL_2127540, EPI_ISL_2127543, EPI_ISL_2127546, EPI_ISL_2127548, EPI_ISL_2127550, EPI_ISL_2127552, EPI_ISL_2127553, EPI_ISL_2127556, EPI_ISL_2127557, EPI_ISL_2127559, EPI_ISL_2127561, EPI_ISL_2127563, EPI_ISL_2127565, EPI_ISL_2127567, EPI_ISL_2127568, EPI_ISL_2127570, EPI_ISL_2127572, EPI_ISL_2127574, EPI_ISL_2127576, EPI_ISL_2127578, EPI_ISL_2127580, EPI_ISL_2127581, EPI_ISL_2127583, EPI_ISL_2127585, EPI_ISL_2127586, EPI_ISL_2127588, EPI_ISL_2127590, EPI_ISL_2127592, EPI_ISL_2239320, EPI_ISL_2239321, EPI_ISL_2239322, EPI_ISL_2239323, EPI_ISL_2239325, EPI_ISL_2239326, EPI_ISL_2239327, EPI_ISL_2239328, EPI_ISL_2239329, EPI_ISL_2239330, EPI_ISL_2239331, EPI_ISL_2239332, EPI_ISL_2239333, EPI_ISL_2239334, EPI_ISL_2239335, EPI_ISL_2239336, EPI_ISL_2239337, EPI_ISL_2239338, EPI_ISL_2239339, EPI_ISL_2239340, EPI_ISL_2239342, EPI_ISL_2239343, EPI_ISL_2239344, EPI_ISL_2239345, EPI_ISL_2239346, EPI_ISL_2239347, EPI_ISL_2239348, EPI_ISL_2239349, EPI_ISL_2239350, EPI_ISL_2239351, EPI_ISL_2239352, EPI_ISL_2239353, EPI_ISL_2239354, EPI_ISL_2239355, EPI_ISL_2239356, EPI_ISL_2239357, EPI_ISL_2239358, EPI_ISL_2239360, EPI_ISL_2239361, EPI_ISL_2239362, EPI_ISL_2239363, EPI_ISL_2239364, EPI_ISL_2239366, EPI_ISL_2239367, EPI_ISL_2239368, EPI_ISL_2239369, EPI_ISL_2239370, EPI_ISL_2239371, EPI_ISL_2239372, EPI_ISL_2239373, EPI_ISL_2239374, EPI_ISL_2239375, EPI_ISL_2239376, EPI_ISL_2239377, EPI_ISL_2239378, EPI_ISL_2239379, EPI_ISL_2239380, EPI_ISL_2239381, EPI_ISL_2239382, EPI_ISL_2239383, EPI_ISL_2239384, EPI_ISL_2239385, EPI_ISL_2239386, EPI_ISL_2239387, EPI_ISL_2239388, EPI_ISL_2239389, EPI_ISL_2239390, EPI_ISL_2239391, EPI_ISL_2239392, EPI_ISL_2239393, EPI_ISL_2239394, EPI_ISL_2239395, EPI_ISL_2239396, EPI_ISL_2239397, EPI_ISL_2239398, EPI_ISL_2239399, EPI_ISL_2239400, EPI_ISL_2239401, EPI_ISL_2239402, EPI_ISL_2239403, EPI_ISL_2239404, EPI_ISL_2239405, EPI_ISL_2239406, EPI_ISL_2239407, EPI_ISL_2239408, EPI_ISL_2239409, EPI_ISL_2239410, EPI_ISL_2239411, EPI_ISL_2239412, EPI_ISL_2239413, EPI_ISL_2239414, EPI_ISL_2239415, EPI_ISL_2239416, EPI_ISL_2239417, EPI_ISL_2239418, EPI_ISL_2239419, EPI_ISL_2239420, EPI_ISL_2239421, EPI_ISL_2239422, EPI_ISL_2239423, EPI_ISL_2239424, EPI_ISL_2239425, EPI_ISL_2239426, EPI_ISL_2239427, EPI_ISL_2239428, EPI_ISL_2239429, EPI_ISL_2239430, EPI_ISL_2239431, EPI_ISL_2239432, EPI_ISL_2239433, EPI_ISL_2239434, EPI_ISL_2239435, EPI_ISL_2239436, EPI_ISL_2239437, EPI_ISL_2239438, EPI_ISL_2239440, EPI_ISL_2239441, EPI_ISL_2239442, EPI_ISL_2239443, EPI_ISL_2239444, EPI_ISL_2239446, EPI_ISL_2239447, EPI_ISL_2239448, EPI_ISL_2239449, EPI_ISL_2239450, EPI_ISL_2239451, EPI_ISL_2239452, EPI_ISL_2239453, EPI_ISL_2239454, EPI_ISL_2239455, EPI_ISL_2239456, EPI_ISL_2239457, EPI_ISL_2239458, EPI_ISL_2239459, EPI_ISL_2239460, EPI_ISL_2239461, EPI_ISL_2239462, EPI_ISL_2239463, EPI_ISL_2239464, EPI_ISL_2239465, EPI_ISL_2239466, EPI_ISL_2239467, EPI_ISL_2239469, EPI_ISL_2239470, EPI_ISL_2239471, EPI_ISL_2239472, EPI_ISL_2239473, EPI_ISL_2239474, EPI_ISL_2239475, EPI_ISL_2239476, EPI_ISL_2239477, EPI_ISL_2239478, EPI_ISL_2239479, EPI_ISL_2239480, EPI_ISL_2239481, EPI_ISL_2239482, EPI_ISL_2239483, EPI_ISL_2239484, EPI_ISL_2239485, EPI_ISL_2239486, EPI_ISL_2239487, EPI_ISL_2239488, EPI_ISL_2239489, EPI_ISL_2239490, EPI_ISL_2239491, EPI_ISL_2239492, EPI_ISL_2239493, EPI_ISL_2239494, EPI_ISL_2239495, EPI_ISL_2239496, EPI_ISL_2239497, EPI_ISL_2239498, EPI_ISL_2239500, EPI_ISL_2239501, EPI_ISL_2239502, EPI_ISL_2239503, EPI_ISL_2239504, EPI_ISL_2239505, EPI_ISL_2239506, EPI_ISL_2239508, EPI_ISL_2239509, EPI_ISL_2239510, EPI_ISL_2239511, EPI_ISL_2239512, EPI_ISL_2239513, EPI_ISL_2239514, EPI_ISL_2239515, EPI_ISL_2239516, EPI_ISL_2239517, EPI_ISL_2239518, EPI_ISL_2239519, EPI_ISL_2239520, EPI_ISL_2239521, EPI_ISL_2239522, EPI_ISL_2239523, EPI_ISL_2239524, EPI_ISL_2239525, EPI_ISL_2239526, EPI_ISL_2239527, EPI_ISL_2239528, EPI_ISL_2239529, EPI_ISL_2239530, EPI_ISL_2239531, EPI_ISL_2239532, EPI_ISL_2239533, EPI_ISL_2239534, EPI_ISL_2239535, EPI_ISL_2239536, EPI_ISL_2239538, EPI_ISL_2239539, EPI_ISL_2239540, EPI_ISL_2239541, EPI_ISL_2239542, EPI_ISL_2239543, EPI_ISL_2239544, EPI_ISL_2239545, EPI_ISL_2239546, EPI_ISL_2239547, EPI_ISL_2239548, EPI_ISL_2239549, EPI_ISL_2239551, EPI_ISL_2239552, EPI_ISL_2239553, EPI_ISL_2239554, EPI_ISL_2239555, EPI_ISL_2239556, EPI_ISL_2239557, EPI_ISL_2239558, EPI_ISL_2239559, EPI_ISL_2239561, EPI_ISL_2239562, EPI_ISL_2239563, EPI_ISL_2239564, EPI_ISL_2239565, EPI_ISL_2239566, EPI_ISL_2239567, EPI_ISL_2239568, EPI_ISL_2239569, EPI_ISL_2239570, EPI_ISL_2239571, EPI_ISL_2239572, EPI_ISL_2239573, EPI_ISL_2239574, EPI_ISL_2239575, EPI_ISL_2239576, EPI_ISL_2239577, EPI_ISL_2239578, EPI_ISL_2239579, EPI_ISL_2239580, EPI_ISL_2239581, EPI_ISL_2239583, EPI_ISL_2239584, EPI_ISL_2239585, EPI_ISL_2239586, EPI_ISL_2239587, EPI_ISL_2239588, EPI_ISL_2239589, EPI_ISL_2239590, EPI_ISL_2239591, EPI_ISL_2239592, EPI_ISL_2239593, EPI_ISL_2239594, EPI_ISL_2239595, EPI_ISL_2239596, EPI_ISL_2239597, EPI_ISL_2239598, EPI_ISL_2239599, EPI_ISL_2239600, EPI_ISL_2239601, EPI_ISL_2239602, EPI_ISL_2239603, EPI_ISL_2239604, EPI_ISL_2239605, EPI_ISL_2239606, EPI_ISL_2239607, EPI_ISL_2239608, EPI_ISL_2239609, EPI_ISL_2239610, EPI_ISL_2239612, EPI_ISL_2239613, EPI_ISL_2239614, EPI_ISL_2239615, EPI_ISL_2239616, EPI_ISL_2239618, EPI_ISL_2239619, EPI_ISL_2239620, EPI_ISL_2239621, EPI_ISL_2239622, EPI_ISL_2239623, EPI_ISL_2239624, EPI_ISL_2239625, EPI_ISL_2239626, EPI_ISL_2239627, EPI_ISL_2239628, EPI_ISL_2239629, EPI_ISL_2239630, EPI_ISL_2239631, EPI_ISL_2239632, EPI_ISL_2239634, EPI_ISL_2239635, EPI_ISL_2239636, EPI_ISL_2239637, EPI_ISL_2239638, EPI_ISL_2239639, EPI_ISL_2239640, EPI_ISL_2239641, EPI_ISL_2239642, EPI_ISL_2239643, EPI_ISL_2239644, EPI_ISL_2239645, EPI_ISL_2239646, EPI_ISL_2239647, EPI_ISL_2239648, EPI_ISL_2239649, EPI_ISL_2239650, EPI_ISL_2239652, EPI_ISL_2239653, EPI_ISL_2239654, EPI_ISL_2239655, EPI_ISL_2239656, EPI_ISL_2239657, EPI_ISL_2239658, EPI_ISL_2239659, EPI_ISL_2239660, EPI_ISL_2239749, EPI_ISL_2239753, EPI_ISL_2239763, EPI_ISL_2239769, EPI_ISL_2239776, EPI_ISL_2239785, EPI_ISL_2239792, EPI_ISL_2239798, EPI_ISL_2239805, EPI_ISL_2239810, EPI_ISL_2239819, EPI_ISL_2239821, EPI_ISL_2239830, EPI_ISL_2239843, EPI_ISL_2239844, EPI_ISL_2239852, EPI_ISL_2239857, EPI_ISL_2239865, EPI_ISL_2239881, EPI_ISL_2239910, EPI_ISL_2239911, EPI_ISL_2239915, EPI_ISL_2239916, EPI_ISL_2239917, EPI_ISL_2239919, EPI_ISL_2239920, EPI_ISL_2239921, EPI_ISL_2239926, EPI_ISL_2239934, EPI_ISL_2239946, EPI_ISL_2239950, EPI_ISL_2239952, EPI_ISL_2239955, EPI_ISL_2239956, EPI_ISL_2239957, EPI_ISL_2239959, EPI_ISL_2239964, EPI_ISL_2239965, EPI_ISL_2239968, EPI_ISL_2239969, EPI_ISL_2239970, EPI_ISL_2239972, EPI_ISL_2239973, EPI_ISL_2239978, EPI_ISL_2239979, EPI_ISL_2239980, EPI_ISL_2239983, EPI_ISL_2239985, EPI_ISL_2240023, EPI_ISL_2240025, EPI_ISL_2240027, EPI_ISL_2240030, EPI_ISL_2240059, EPI_ISL_2240060, EPI_ISL_2240061, EPI_ISL_2240063, EPI_ISL_2240064, EPI_ISL_2240065, EPI_ISL_2240066, EPI_ISL_2240067, EPI_ISL_2240068, EPI_ISL_2240069, EPI_ISL_2240070, EPI_ISL_2240071, EPI_ISL_2240072, EPI_ISL_2240073, EPI_ISL_2240074, EPI_ISL_2240075, EPI_ISL_2240077, EPI_ISL_2240078, EPI_ISL_2240079, EPI_ISL_2240080, EPI_ISL_2240081, EPI_ISL_2240082, EPI_ISL_2240083, EPI_ISL_2240085, EPI_ISL_2240086, EPI_ISL_2240087, EPI_ISL_2240088, EPI_ISL_2240089, EPI_ISL_2240090, EPI_ISL_2240091, EPI_ISL_2240092, EPI_ISL_2240093, EPI_ISL_2240094, EPI_ISL_2240095, EPI_ISL_2240097, EPI_ISL_2240098, EPI_ISL_2240099, EPI_ISL_2240100, EPI_ISL_2240102, EPI_ISL_2240104, EPI_ISL_2240105, EPI_ISL_2240106, EPI_ISL_2240107, EPI_ISL_2240108, EPI_ISL_2240109, EPI_ISL_2240110, EPI_ISL_2240111, EPI_ISL_2240112, EPI_ISL_2240113, EPI_ISL_2240114, EPI_ISL_2240115, EPI_ISL_2240116, EPI_ISL_2240117, EPI_ISL_2240118, EPI_ISL_2240120, EPI_ISL_2240121, EPI_ISL_2240122, EPI_ISL_2240123, EPI_ISL_2240124, EPI_ISL_2240125, EPI_ISL_2240127, EPI_ISL_2240128, EPI_ISL_2240145, EPI_ISL_2240265, EPI_ISL_2240267, EPI_ISL_2240269, EPI_ISL_2240284, EPI_ISL_2240289, EPI_ISL_2240416, EPI_ISL_2240417, EPI_ISL_2356097, EPI_ISL_2356098, EPI_ISL_2356099, EPI_ISL_2356104, EPI_ISL_2356105, EPI_ISL_2356110, EPI_ISL_2356111, EPI_ISL_2356117, EPI_ISL_2356118, EPI_ISL_2356122, EPI_ISL_2356123, EPI_ISL_2356129, EPI_ISL_2356130, EPI_ISL_2356131, EPI_ISL_2356136, EPI_ISL_2356137, EPI_ISL_2356138, EPI_ISL_2356144, EPI_ISL_2356145, EPI_ISL_2356146, EPI_ISL_2356151, EPI_ISL_2356152, EPI_ISL_2356153, EPI_ISL_2356154, EPI_ISL_2356157, EPI_ISL_2356158, EPI_ISL_2356160, EPI_ISL_2356163, EPI_ISL_2356164, EPI_ISL_2356165, EPI_ISL_2356166, EPI_ISL_2356171, EPI_ISL_2356172, EPI_ISL_2356173, EPI_ISL_2356174, EPI_ISL_2356179, EPI_ISL_2356188, EPI_ISL_2356194, EPI_ISL_2356200, EPI_ISL_2356201, EPI_ISL_2356207, EPI_ISL_2356209, EPI_ISL_2356216, EPI_ISL_2356221, EPI_ISL_2356228, EPI_ISL_2356234, EPI_ISL_2356235, EPI_ISL_2356240, EPI_ISL_2356245, EPI_ISL_2356250, EPI_ISL_2356252, EPI_ISL_2356569, EPI_ISL_2356573, EPI_ISL_2356574, EPI_ISL_2356577, EPI_ISL_2356578, EPI_ISL_2356579, EPI_ISL_2356580, EPI_ISL_2356582, EPI_ISL_2356585, EPI_ISL_2356587, EPI_ISL_2356591, EPI_ISL_2356593, EPI_ISL_2356596, EPI_ISL_2356601, EPI_ISL_2356604, EPI_ISL_2356606, EPI_ISL_2356610, EPI_ISL_2356612, EPI_ISL_2356617, EPI_ISL_2356623, EPI_ISL_2356624, EPI_ISL_2356629, EPI_ISL_2356634, EPI_ISL_2356635, EPI_ISL_2356636, EPI_ISL_2356640, EPI_ISL_2356645, EPI_ISL_2356651, EPI_ISL_2356656, EPI_ISL_2356661, EPI_ISL_2356662, EPI_ISL_2356667, EPI_ISL_2356669, EPI_ISL_2356675, EPI_ISL_2356680, EPI_ISL_2434353, EPI_ISL_2434354, EPI_ISL_2434356, EPI_ISL_2434357, EPI_ISL_2434358, EPI_ISL_2434361, EPI_ISL_2434363, EPI_ISL_2434364, EPI_ISL_2434365, EPI_ISL_2434367, EPI_ISL_2434370, EPI_ISL_2434371, EPI_ISL_2434372, EPI_ISL_2434373, EPI_ISL_2434375, EPI_ISL_2434376, EPI_ISL_2434377, EPI_ISL_2434379, EPI_ISL_2434382, EPI_ISL_2434383, EPI_ISL_2434384, EPI_ISL_2434386, EPI_ISL_2434390, EPI_ISL_2434391, EPI_ISL_2434395, EPI_ISL_2434397, EPI_ISL_2434398, EPI_ISL_2434401, EPI_ISL_2434403, EPI_ISL_2434404, EPI_ISL_2434405, EPI_ISL_2434407, EPI_ISL_2434409, EPI_ISL_2434410, EPI_ISL_2434411, EPI_ISL_2434414, EPI_ISL_2434416, EPI_ISL_2434417, EPI_ISL_2434420, EPI_ISL_2434421, EPI_ISL_2434425, EPI_ISL_2434427, EPI_ISL_2434429, EPI_ISL_2434431, EPI_ISL_2434436, EPI_ISL_2434437, EPI_ISL_2434438, EPI_ISL_2434440, EPI_ISL_2434442, EPI_ISL_2434443, EPI_ISL_2434445, EPI_ISL_2434448, EPI_ISL_2434451, EPI_ISL_2434452, EPI_ISL_2434453, EPI_ISL_2434459, EPI_ISL_2434460, EPI_ISL_2434465, EPI_ISL_2434467, EPI_ISL_2434468, EPI_ISL_2434469, EPI_ISL_2434470, EPI_ISL_2434472, EPI_ISL_2434473, EPI_ISL_2434475, EPI_ISL_2434476, EPI_ISL_2434480, EPI_ISL_2434482, EPI_ISL_2434483, EPI_ISL_2434488, EPI_ISL_2434490, EPI_ISL_2434493, EPI_ISL_2434497, EPI_ISL_2434500, EPI_ISL_2434501, EPI_ISL_2434502, EPI_ISL_2434504, EPI_ISL_2434507, EPI_ISL_2434509, EPI_ISL_2434510, EPI_ISL_2434511, EPI_ISL_2434513, EPI_ISL_2434514, EPI_ISL_2434515, EPI_ISL_2434517, EPI_ISL_2434519, EPI_ISL_2434520, EPI_ISL_2434521, EPI_ISL_2434523, EPI_ISL_2434524, EPI_ISL_2434526, EPI_ISL_2434527, EPI_ISL_2434528, EPI_ISL_2434529, EPI_ISL_2434531, EPI_ISL_2434533, EPI_ISL_2434534, EPI_ISL_2434536, EPI_ISL_2434537, EPI_ISL_2434538, EPI_ISL_2434540, EPI_ISL_2434541, EPI_ISL_2434544, EPI_ISL_2434545, EPI_ISL_2434547, EPI_ISL_2434548, EPI_ISL_2434550, EPI_ISL_2434551, EPI_ISL_2434552, EPI_ISL_2434554, EPI_ISL_2434555, EPI_ISL_2434556, EPI_ISL_2434558, EPI_ISL_2434559, EPI_ISL_2434560, EPI_ISL_2434562, EPI_ISL_2434563, EPI_ISL_2434564, EPI_ISL_2434566, EPI_ISL_2434567, EPI_ISL_2434568, EPI_ISL_2434570, EPI_ISL_2434571, EPI_ISL_2434573, EPI_ISL_2434574, EPI_ISL_2434575, EPI_ISL_2434576, EPI_ISL_2434577, EPI_ISL_2434578, EPI_ISL_2434580, EPI_ISL_2434581, EPI_ISL_2434583, EPI_ISL_2434584, EPI_ISL_2434585, EPI_ISL_2434587, EPI_ISL_2434588, EPI_ISL_2434590, EPI_ISL_2434591, EPI_ISL_2434592, EPI_ISL_2434593, EPI_ISL_2434595, EPI_ISL_2434599, EPI_ISL_2434601, EPI_ISL_2434604, EPI_ISL_2434606, EPI_ISL_2434612, EPI_ISL_2434614, EPI_ISL_2434618, EPI_ISL_2434619, EPI_ISL_2434621, EPI_ISL_2434626, EPI_ISL_2434627, EPI_ISL_2434629, EPI_ISL_2434633, EPI_ISL_2434634, EPI_ISL_2434635, EPI_ISL_2434637, EPI_ISL_2434641, EPI_ISL_2434642, EPI_ISL_2434643, EPI_ISL_2434644, EPI_ISL_2434645, EPI_ISL_2434650, EPI_ISL_2434651, EPI_ISL_2434652, EPI_ISL_2434656, EPI_ISL_2434658, EPI_ISL_2434659, EPI_ISL_2434662, EPI_ISL_2434664, EPI_ISL_2434665, EPI_ISL_2434671, EPI_ISL_2434682, EPI_ISL_2434683, EPI_ISL_2434686, EPI_ISL_2434688, EPI_ISL_2434689, EPI_ISL_2434694, EPI_ISL_2434695, EPI_ISL_2434697, EPI_ISL_2434701, EPI_ISL_2434702, EPI_ISL_2434705, EPI_ISL_2434708, EPI_ISL_2434709, EPI_ISL_2434712, EPI_ISL_2434715, EPI_ISL_2434716, EPI_ISL_2434718, EPI_ISL_2434722, EPI_ISL_2434730, EPI_ISL_2434734, EPI_ISL_2434740, EPI_ISL_2434748, EPI_ISL_2434752, EPI_ISL_2434753, EPI_ISL_2434756, EPI_ISL_2434757 | | | |
| see above | Northumbria University / South Tees Hospitals NHS Foundation Trust / North Cumbria Integrated Care NHS Foundation Trust / North Tees and Hartlepool NHS Foundation Trust / Newcastle Hospitals NHS Foundation Trust | COVID-19 Genomics UK (COG-UK) Consortium | Andrew Nelson; Brendan Payne; Clive Graham; Darren L Smith; Debra Padgett; Edward Barton; Emma Swindells; Garren Scott; Gary Black; Gary Eltringham; Giles S Holt; Greg R Young; Jane Greenaway; Jennifer Collins; John Allan; Joshua Loh; Lynn Dover; Matthew Bashton; Mohammad A Tariq; Paul Baker; Sarah Essex; Steve Liggett; Wen C Yew; Yusri Taha |
| EPI_ISL_1979272, EPI_ISL_1979311, EPI_ISL_1979313, EPI_ISL_1979315, EPI_ISL_1979317, EPI_ISL_1979318, EPI_ISL_1979323, EPI_ISL_1979326, EPI_ISL_1979334, EPI_ISL_1979338, EPI_ISL_1979493, EPI_ISL_1979497, EPI_ISL_1979498, EPI_ISL_1979500, EPI_ISL_1979501, EPI_ISL_1979503, EPI_ISL_1979504, EPI_ISL_1979512, EPI_ISL_1979514, EPI_ISL_1979515, EPI_ISL_1979517, EPI_ISL_1979518, EPI_ISL_1979520, EPI_ISL_1979522, EPI_ISL_1979523, EPI_ISL_1979568, EPI_ISL_1979572, EPI_ISL_1979601, EPI_ISL_1979602, EPI_ISL_1979605, EPI_ISL_1979610, EPI_ISL_1979612, EPI_ISL_1979616, EPI_ISL_1979618, EPI_ISL_1979620, EPI_ISL_1979623, EPI_ISL_1979624, EPI_ISL_1979626, EPI_ISL_1979634, EPI_ISL_1979635, EPI_ISL_1979637, EPI_ISL_1979640, EPI_ISL_1979643, EPI_ISL_1979646, EPI_ISL_1979648, EPI_ISL_1979650, EPI_ISL_1979652, EPI_ISL_1979700, EPI_ISL_1979701, EPI_ISL_1979706, EPI_ISL_1979709, EPI_ISL_1979713, EPI_ISL_1979714, EPI_ISL_1979715, EPI_ISL_1979720, EPI_ISL_1979723, EPI_ISL_1979725, EPI_ISL_1979726, EPI_ISL_1979728, EPI_ISL_1979735, EPI_ISL_1979736, EPI_ISL_1979741, EPI_ISL_2240511, EPI_ISL_2240518, EPI_ISL_2240536, EPI_ISL_2240538, EPI_ISL_2240539, EPI_ISL_2240568, EPI_ISL_2240569, EPI_ISL_2240578, EPI_ISL_2240603, EPI_ISL_2240605, EPI_ISL_2240607, EPI_ISL_2240608, EPI_ISL_2240609, EPI_ISL_2240614, EPI_ISL_2240616, EPI_ISL_2240635, EPI_ISL_2240636, EPI_ISL_2240641, EPI_ISL_2240644, EPI_ISL_2240645, EPI_ISL_2240648, EPI_ISL_2240655, EPI_ISL_2240657, EPI_ISL_2240659 | | | |
| see above | Originating lab: Wales Specialist Virology Centre Sequencing lab: Pathogen Genomics Unit | Public Health Wales Microbiology Cardiff Wales Specialist Virology Centre | Alec Birchley; Alexander Adams; Amy Gaskin; Angela Marchbank; Bree Gatica-Wilcox; Catherine Moore; Jason Coombes; Joanne Watkins; Joel Southgate; Johnathan Evans; Laura Gifford; Lauren Gilbert; Lee Graham; Malorie Perry; Matthew Bull; Nicole Pacchiarini; Sally Corden; Sara Kumziene-Summerhayes; Sara Rey; Sarah Taylor; Simon Cottrell; Sophie Jones; Tom Connor |
| EPI_ISL_2240498, EPI_ISL_2240501, EPI_ISL_2240502, EPI_ISL_2240503, EPI_ISL_2240504, EPI_ISL_2240505, EPI_ISL_2240507, EPI_ISL_2240508, EPI_ISL_2240509, EPI_ISL_2356709, EPI_ISL_2356713, EPI_ISL_2356715 | | | |
| see above | Oxford Viromics, NDM, University of Oxford; Oxford University Hospitals; Basingstoke and North Hampshire Hospital | COVID-19 Genomics UK (COG-UK) Consortium | Alex Mobbs; Amy Trebes; Anita Justice; Catrin Moore; Christophe Fraser; David Bonsall; David Buck; Emma Wise; George Macintyre; Jessica Lynch; John Todd; Mariateresa de Cesare; Matilde Mori; Monique Andersson; Nathan Moore; Nick Cortes; Robert Shaw; Stephen Kidd; Tanya Golubchik; Timothy Peto |
| EPI_ISL_2240418, EPI_ISL_2240419, EPI_ISL_2240420, EPI_ISL_2240422, EPI_ISL_2240423, EPI_ISL_2240428, EPI_ISL_2240430, EPI_ISL_2240431, EPI_ISL_2240436, EPI_ISL_2356693 | | | |
| see above | Quadram Institute Bioscience | COVID-19 Genomics UK (COG-UK) Consortium | Alexander J Trotter; Alison E. Mather; Alp Aydin; Ana P. Tedim; Anastasia Kolyva; Andrew Bell; Andrew J. Page; Claire Stuart; Dave J. Baker; Gemma L. Kay; John Wain; Justin O'Grady; Leonardo de Oliveira Martins; Lizzie Meadows; Maria Diaz; Mark Webber; Muhammed Yasir; Nabil-Fareed Alikhan; Ngozi Elumogo; Nicholas M. Thomson; Rachael Stanley; Rachel Gilroy; Reenesh Prakash; Samir Dervisevic; Samuel Bloomfield; Steven Rudder; Thanh Le-Viet |
| EPI_ISL_1978958, EPI_ISL_1978959, EPI_ISL_1978960, EPI_ISL_2127604, EPI_ISL_2127606, EPI_ISL_2127608, EPI_ISL_2127610, EPI_ISL_2127611, EPI_ISL_2127613, EPI_ISL_2127615, EPI_ISL_2127617, EPI_ISL_2127619, EPI_ISL_2127621 | | | |
| see above | Queens Medical Centre, Clinical Microbiology Department / DeepSeq Nottingham | COVID-19 Genomics UK (COG-UK) Consortium | Christopher Moore; Emily Park; Fei Sang; Gemma Clark; Hannah Howson-Wells; Jack Hill; Jayasree Dey; Johnny Debebe; Jonathan Ball; Joseph Chappell; Manjinder Khakh; Matthew Carlisle; Matthew Loose; Michelle M Lister; Nadine Holmes; Patrick McClure; Theocharis Tsoleridis; Timothy Byaruhanga; Vicki M Fleming; Victoria Wright; Wendy Smith |
| EPI_ISL_1986673, EPI_ISL_1986751, EPI_ISL_1986863, EPI_ISL_2004344, EPI_ISL_2004348, EPI_ISL_2004355, EPI_ISL_2004374, EPI_ISL_2004396, EPI_ISL_2004409, EPI_ISL_2004413, EPI_ISL_2004423, EPI_ISL_2004434, EPI_ISL_2004436, EPI_ISL_2004447, EPI_ISL_2004455, EPI_ISL_2004472, EPI_ISL_2004473, EPI_ISL_2004476, EPI_ISL_2004477, EPI_ISL_2004478, EPI_ISL_2004485, EPI_ISL_2004491, EPI_ISL_2004494, EPI_ISL_2004527, EPI_ISL_2004545, EPI_ISL_2004548, EPI_ISL_2004574, EPI_ISL_2004584, EPI_ISL_2004604, EPI_ISL_2004611, EPI_ISL_2005846, EPI_ISL_2005865, EPI_ISL_2005928, EPI_ISL_2006026, EPI_ISL_2021372, EPI_ISL_2021378, EPI_ISL_2021523, EPI_ISL_2021572, EPI_ISL_2021573, EPI_ISL_2021574, EPI_ISL_2021639, EPI_ISL_2021714, EPI_ISL_2021715, EPI_ISL_2021721, EPI_ISL_2021722, EPI_ISL_2021740, EPI_ISL_2021745, EPI_ISL_2021750, EPI_ISL_2021751, EPI_ISL_2021759, EPI_ISL_2021763, EPI_ISL_2021768, EPI_ISL_2021776, EPI_ISL_2021786, EPI_ISL_2021788, EPI_ISL_2021801, EPI_ISL_2021802, EPI_ISL_2021803, EPI_ISL_2021815, EPI_ISL_2021825, EPI_ISL_2022530, EPI_ISL_2022563, EPI_ISL_2091824, EPI_ISL_2091846, EPI_ISL_2091889, EPI_ISL_2091907, EPI_ISL_2091975, EPI_ISL_2091985, EPI_ISL_2091992, EPI_ISL_2091998, EPI_ISL_2092016, EPI_ISL_2092019, EPI_ISL_2092038, EPI_ISL_2092042, EPI_ISL_2092083, EPI_ISL_2092112, EPI_ISL_2092114, EPI_ISL_2092118, EPI_ISL_2092120, EPI_ISL_2092140, EPI_ISL_2092141, EPI_ISL_2092143, EPI_ISL_2092152, EPI_ISL_2092156, EPI_ISL_2092162, EPI_ISL_2092163, EPI_ISL_2092167, EPI_ISL_2092171, EPI_ISL_2092181, EPI_ISL_2092183, EPI_ISL_2092188, EPI_ISL_2092193, EPI_ISL_2092195, EPI_ISL_2092200, EPI_ISL_2092202, EPI_ISL_2092203, EPI_ISL_2092214, EPI_ISL_2092222, EPI_ISL_2092223, EPI_ISL_2092233, EPI_ISL_2092239, EPI_ISL_2092240, EPI_ISL_2092243, EPI_ISL_2092245, EPI_ISL_2092256, EPI_ISL_2092259, EPI_ISL_2092264, EPI_ISL_2092265, EPI_ISL_2092266, EPI_ISL_2092270, EPI_ISL_2092272, EPI_ISL_2092273, EPI_ISL_2092278, EPI_ISL_2092280, EPI_ISL_2092296, EPI_ISL_2092312, EPI_ISL_2092338, EPI_ISL_2092339, EPI_ISL_2092341, EPI_ISL_2092342, EPI_ISL_2092347, EPI_ISL_2092362, EPI_ISL_2092365, EPI_ISL_2092373, EPI_ISL_2092380, EPI_ISL_2092401, EPI_ISL_2092406, EPI_ISL_2092411, EPI_ISL_2092414, EPI_ISL_2092416, EPI_ISL_2092417, EPI_ISL_2092419, EPI_ISL_2092425, EPI_ISL_2092428, EPI_ISL_2092438, EPI_ISL_2092440, EPI_ISL_2092441, EPI_ISL_2092442, EPI_ISL_2092451, EPI_ISL_2092459, EPI_ISL_2122125, EPI_ISL_2122126, EPI_ISL_2122142, EPI_ISL_2122144, EPI_ISL_2122159, EPI_ISL_2122162, EPI_ISL_2122164, EPI_ISL_2122171, EPI_ISL_2122197, EPI_ISL_2122198, EPI_ISL_2122232, EPI_ISL_2122237, EPI_ISL_2122297, EPI_ISL_2122304, EPI_ISL_2122366, EPI_ISL_2122380, EPI_ISL_2122405, EPI_ISL_2122416, EPI_ISL_2122471, EPI_ISL_2122474, EPI_ISL_2122488, EPI_ISL_2122491, EPI_ISL_2122502, EPI_ISL_2122503, EPI_ISL_2122505, EPI_ISL_2122513, EPI_ISL_2122550, EPI_ISL_2122556, EPI_ISL_2122572, EPI_ISL_2122575, EPI_ISL_2122607, EPI_ISL_2122618, EPI_ISL_2122625, EPI_ISL_2122636, EPI_ISL_2122658, EPI_ISL_2122691, EPI_ISL_2122692, EPI_ISL_2122696, EPI_ISL_2153046, EPI_ISL_2153048, EPI_ISL_2153049, EPI_ISL_2153050, EPI_ISL_2153051, EPI_ISL_2153052, EPI_ISL_2153053, EPI_ISL_2153054, EPI_ISL_2153055, EPI_ISL_2153056, EPI_ISL_2153057, EPI_ISL_2153058, EPI_ISL_2153059, EPI_ISL_2153060, EPI_ISL_2153061, EPI_ISL_2153062, EPI_ISL_2153063, EPI_ISL_2153064, EPI_ISL_2238359, EPI_ISL_2238475, EPI_ISL_2238611, EPI_ISL_2253818, EPI_ISL_2253894, EPI_ISL_2254107, EPI_ISL_2254108, EPI_ISL_2254109, EPI_ISL_2254110, EPI_ISL_2254111, EPI_ISL_2254112, EPI_ISL_2254113, EPI_ISL_2254114, EPI_ISL_2254115, EPI_ISL_2254116, EPI_ISL_2254117, EPI_ISL_2254118, EPI_ISL_2396845, EPI_ISL_2396849, EPI_ISL_2396855, EPI_ISL_2396862, EPI_ISL_2396863 | | | |
| see above | Randox Laboratories | Wellcome Sanger Institute for the COVID-19 Genomics UK (COG-UK) Consortium | Cordelia Langford; David K. Jackson; Dominic Kwiatkowski; Ewan Harrison; Ian Johnston; Jeffrey Barrett; John Sillitoe on behalf of the Wellcome Sanger Institute COVID-19 Surveillance Team; Randox Laboratories and Alex Alderton; Roberto Amato; Sonia Goncalves |
| EPI_ISL_2734939, EPI_ISL_2742304, EPI_ISL_2742306, EPI_ISL_2742309, EPI_ISL_2742313, EPI_ISL_2742315, EPI_ISL_2742319, EPI_ISL_2742320, EPI_ISL_2742321, EPI_ISL_2742322, EPI_ISL_2742323, EPI_ISL_2742325, EPI_ISL_2742326, EPI_ISL_2742328, EPI_ISL_2742329, EPI_ISL_2742330, EPI_ISL_2742331, EPI_ISL_2742332, EPI_ISL_2742334, EPI_ISL_2742336, EPI_ISL_2742340, EPI_ISL_2742342, EPI_ISL_2742343, EPI_ISL_2742348, EPI_ISL_2742351, EPI_ISL_2742354, EPI_ISL_2742355, EPI_ISL_2742356, EPI_ISL_2742358, EPI_ISL_2742359, EPI_ISL_2742360, EPI_ISL_2742361, EPI_ISL_2742362, EPI_ISL_2742363, EPI_ISL_2742364, EPI_ISL_2742365, EPI_ISL_2742366, EPI_ISL_2742368, EPI_ISL_2742369, EPI_ISL_2742370, EPI_ISL_2742371, EPI_ISL_2742372, EPI_ISL_2742373, EPI_ISL_2742374, EPI_ISL_2742375, EPI_ISL_2742376, EPI_ISL_2742380, EPI_ISL_2742381, EPI_ISL_2742425, EPI_ISL_2742426, EPI_ISL_2742428, EPI_ISL_2742432, EPI_ISL_2742436, EPI_ISL_2742438, EPI_ISL_2742446, EPI_ISL_2742452, EPI_ISL_2742470, EPI_ISL_2742479, EPI_ISL_2742488, EPI_ISL_2742934, EPI_ISL_2742935, EPI_ISL_2742940, EPI_ISL_2742954, EPI_ISL_2742973, EPI_ISL_2748742, EPI_ISL_2748764, EPI_ISL_2748959, EPI_ISL_2748979, EPI_ISL_2749405, EPI_ISL_2749444, EPI_ISL_2749447, EPI_ISL_2749462, EPI_ISL_2749530, EPI_ISL_2753576, EPI_ISL_2753589 | | | |
| see above | Respiratory Virus Unit, Microbiology Services Colindale, Public Health England | COVID-19 Genomics UK (COG-UK) Consortium | PHE Covid Sequencing Team |
| EPI_ISL_1975115, EPI_ISL_1975118, EPI_ISL_1975122, EPI_ISL_1975130, EPI_ISL_1975131, EPI_ISL_1975133, EPI_ISL_1975134, EPI_ISL_1975171, EPI_ISL_1975179, EPI_ISL_1975180, EPI_ISL_2126130, EPI_ISL_2126132, EPI_ISL_2126136, EPI_ISL_2126138, EPI_ISL_2126140, EPI_ISL_2126142, EPI_ISL_2126145, EPI_ISL_2126147, EPI_ISL_2126149, EPI_ISL_2126151, EPI_ISL_2126152, EPI_ISL_2126154, EPI_ISL_2126155, EPI_ISL_2126157, EPI_ISL_2126159, EPI_ISL_2126161, EPI_ISL_2126163, EPI_ISL_2126164, EPI_ISL_2126166, EPI_ISL_2126168, EPI_ISL_2126170, EPI_ISL_2126171, EPI_ISL_2126173, EPI_ISL_2126175, EPI_ISL_2126177, EPI_ISL_2238950, EPI_ISL_2238973, EPI_ISL_2238977, EPI_ISL_2238978, EPI_ISL_2238979, EPI_ISL_2238980, EPI_ISL_2238981, EPI_ISL_2238983 | | | |
| see above | University College London, Great Ormond Street Hospital for Children NHS Foundation Trust, Imperial College Healthcare NHS Trust | COVID-19 Genomics UK (COG-UK) Consortium | Alison Holmes; Charlotte Williams; Helena Tutill; Jacqueline Findlay; James Price; Judith Breuer; Julianne Brown; Kathryn Harris; Leysa Forrest; Mark Kristiansen; Paola Niola; Paola Resende Silva; Patricia Dyal; Paul Randell; Rachel Williams; Samuel Weeks; Sergi Castellano; Sunando Roy; Tony Brooks; Yasmin Panchbhaya |
| EPI_ISL_2238862, EPI_ISL_2238863 | University Hospitals Of Leicester NHS Trust and DeepSeq Nottingham | COVID-19 Genomics UK (COG-UK) Consortium | Christopher Holmes; Christopher Moore; Fei Sang; Joeseph Chappell; Johnny Debebe; Jonathan Ball; Julian Tang; Karlie Fallon; Matthew Carlisle; Matthew Loose; Nadine Holmes; Patrick McClure; Paul Bird; Thomas Helmer; Victoria Wright |
| EPI_ISL_1974757, EPI_ISL_2125426, EPI_ISL_2125428, EPI_ISL_2125430, EPI_ISL_2125432, EPI_ISL_2125434, EPI_ISL_2125436, EPI_ISL_2125438, EPI_ISL_2125439, EPI_ISL_2125441, EPI_ISL_2125443, EPI_ISL_2125444, EPI_ISL_2125446, EPI_ISL_2125447, EPI_ISL_2125449, EPI_ISL_2238731, EPI_ISL_2238732, EPI_ISL_2238733, EPI_ISL_2238734, EPI_ISL_2238735, EPI_ISL_2238736, EPI_ISL_2354503, EPI_ISL_2354524 | | | |
| see above | University of Birmingham | COVID-19 Genomics | Alex Richter; Andrew Bosworth; Andrew Bosworth. Queen Elizabeth Hospital: Anna Casey; Andrew D Beggs PHE Heartlands Lab: Husam Osman; Charlotte Poxon; Institute of Microbiology; Joanne Stockton; Josh Quick; |

| | | | |
|---|---|---|---|
| | | UK (COG-UK) Consortium | Kasun Wanigasooriya; Mike Kidd; Nicholas Loman. University of Birmingham Testing Laboratory: Celina M Whalley; Oliver Pickles; Radoslaw Poplawski; Samuel Nicholls; University of Birmingham: Claire McMurray; Will Rowe |
| EPI_ISL_2238830 | University of Exeter | COVID-19 Genomics UK (COG-UK) Consortium | Aaron Jeffries; Audrey Farbos; Ben Temperton; Jane Masoli; Joanna Warwick-Dugdale; Michelle Michelsen; Robyn Manley; Stephen Michell |
| EPI_ISL_2125651, EPI_ISL_2125652, EPI_ISL_2125669, EPI_ISL_2125671, EPI_ISL_2125673, EPI_ISL_2125675, EPI_ISL_2125677, EPI_ISL_2125678, EPI_ISL_2125680, EPI_ISL_2125682, EPI_ISL_2125684, EPI_ISL_2125686, EPI_ISL_2125687, EPI_ISL_2125719, EPI_ISL_2125720, EPI_ISL_2125722, EPI_ISL_2125724, EPI_ISL_2125726, EPI_ISL_2125728, EPI_ISL_2125740, EPI_ISL_2125742, EPI_ISL_2125744, EPI_ISL_2125745, EPI_ISL_2125748, EPI_ISL_2125749, EPI_ISL_2125753, EPI_ISL_2125755, EPI_ISL_2125756, EPI_ISL_2125758, EPI_ISL_2125888, EPI_ISL_2125890, EPI_ISL_2125891, EPI_ISL_2238751, EPI_ISL_2238752, EPI_ISL_2238756, EPI_ISL_2238770, EPI_ISL_2238771, EPI_ISL_2238809, EPI_ISL_2238810, EPI_ISL_2354558, EPI_ISL_2354563, EPI_ISL_2354579, EPI_ISL_2354582, EPI_ISL_2354584 | | | |
| see above | Virology Department, Royal Infirmary of Edinburgh, NHS Lothian / School of Biological Sciences, University of Edinburgh | COVID-19 Genomics UK (COG-UK) Consortium | Colquhoun R; Cotton S; Dewar R; Gallagher A; Hill V; Jackson B; Maloney D; McCrone JT; McHugh M; O’Toole Á; Rambaut A; Rooke S; Scher E; Templeton K; Yu X |
| EPI_ISL_1980347, EPI_ISL_1980356, EPI_ISL_1980359, EPI_ISL_1980368, EPI_ISL_1980375, EPI_ISL_1980403, EPI_ISL_1980404, EPI_ISL_1980407, EPI_ISL_1980417, EPI_ISL_1980422, EPI_ISL_1980423, EPI_ISL_1980441, EPI_ISL_1980448, EPI_ISL_2128460, EPI_ISL_2128463, EPI_ISL_2128467, EPI_ISL_2128469, EPI_ISL_2128471, EPI_ISL_2128474, EPI_ISL_2128476, EPI_ISL_2128481, EPI_ISL_2128491, EPI_ISL_2128495, EPI_ISL_2128502, EPI_ISL_2240682, EPI_ISL_2240687, EPI_ISL_2240705, EPI_ISL_2240708, EPI_ISL_2240717, EPI_ISL_2240721, EPI_ISL_2240727, EPI_ISL_2240728, EPI_ISL_2356884, EPI_ISL_2356885, EPI_ISL_2356886, EPI_ISL_2356890, EPI_ISL_2356892, EPI_ISL_2356894, EPI_ISL_2356895, EPI_ISL_2356900, EPI_ISL_2356904, EPI_ISL_2356909, EPI_ISL_2520940, EPI_ISL_2521001 | | | |
| see above | Virology Department, Sheffield Teaching Hospitals NHS Foundation Trust/Department of Infection, Immunity and Cardiovascular Disease, The Medical School, University of Sheffield | COVID-19 Genomics UK (COG-UK) Consortium | Adri Angyal; Alex Keeley; Benjamin Lindsey; Cariad Evans; Danielle Groves; Dave Partridge; Katie Johnson; Laura Carrilero; Luke Green; Matthew Parker; Matthew Wyles; Mehmet Yavuz; Mohammad Raza; Nikki Smith; Paul Parsons; Rachel Tucker; Rebecca Brown; Thushan de Silva |
| EPI_ISL_2127728, EPI_ISL_2127729, EPI_ISL_2127730, EPI_ISL_2127731, EPI_ISL_2127732, EPI_ISL_2127733, EPI_ISL_2127734, EPI_ISL_2127735, EPI_ISL_2127736, EPI_ISL_2127738, EPI_ISL_2127739, EPI_ISL_2127741, EPI_ISL_2127743, EPI_ISL_2127745, EPI_ISL_2127746, EPI_ISL_2127748, EPI_ISL_2127750, EPI_ISL_2127752, EPI_ISL_2127753, EPI_ISL_2127755, EPI_ISL_2127757, EPI_ISL_2127759, EPI_ISL_2127761, EPI_ISL_2127763, EPI_ISL_2127764, EPI_ISL_2127765, EPI_ISL_2127767, EPI_ISL_2127768, EPI_ISL_2127770, EPI_ISL_2127772, EPI_ISL_2127773, EPI_ISL_2127774, EPI_ISL_2127776, EPI_ISL_2127777, EPI_ISL_2127778, EPI_ISL_2127780, EPI_ISL_2127782, EPI_ISL_2127784, EPI_ISL_2127785, EPI_ISL_2127787, EPI_ISL_2127789, EPI_ISL_2127791, EPI_ISL_2127792, EPI_ISL_2127795, EPI_ISL_2127797, EPI_ISL_2127798, EPI_ISL_2127800, EPI_ISL_2127802, EPI_ISL_2127804, EPI_ISL_2127805, EPI_ISL_2127807, EPI_ISL_2127809, EPI_ISL_2127811, EPI_ISL_2127812, EPI_ISL_2127814, EPI_ISL_2127815, EPI_ISL_2127816, EPI_ISL_2127818, EPI_ISL_2127820, EPI_ISL_2127822, EPI_ISL_2127823, EPI_ISL_2127825, EPI_ISL_2127827, EPI_ISL_2127829, EPI_ISL_2127831, EPI_ISL_2127833, EPI_ISL_2127835, EPI_ISL_2127837, EPI_ISL_2127839, EPI_ISL_2127841, EPI_ISL_2127842, EPI_ISL_2127845, EPI_ISL_2127847, EPI_ISL_2127849, EPI_ISL_2127851, EPI_ISL_2127852, EPI_ISL_2127854, EPI_ISL_2127858, EPI_ISL_2127860, EPI_ISL_2127861, EPI_ISL_2127863, EPI_ISL_2127865, EPI_ISL_2127867, EPI_ISL_2127869, EPI_ISL_2127871, EPI_ISL_2127873, EPI_ISL_2127874, EPI_ISL_2127880, EPI_ISL_2127883, EPI_ISL_2356877 | | | |
| see above | Wales Specialist Virology Centre Sequencing lab: Pathogen Genomics Unit | Public Health Wales Microbiology Cardiff Wales Specialist Virology Centre | Alec Birchley; Alexander Adams; Amy Gaskin; Angela Marchbank; Bree Gatica-Wilcox; Catherine Moore; Jason Coombes; Joanne Watkins; Joel Southgate; Johnathan Evans; Laura Gifford; Lauren Gilbert; Lee Graham; Malorie Perry; Matthew Bull; Nicole Pacchiarini; Sally Corden; Sara Kumziene-Summerhayes; Sara Rey; Sarah Taylor; Simon Cottrell; Sophie Jones; Tom Connor |
| EPI_ISL_2125502, EPI_ISL_2434191, EPI_ISL_2434192, EPI_ISL_2434199, EPI_ISL_2434204 | West of Scotland Specialist Virology Centre, NHSGGC / MRC-University of Glasgow Centre for Virus Research | COVID-19 Genomics UK (COG-UK) Consortium | Alasdair MacLean; Alice Broos; Ana da Silva Filipe; Antonia Ho; Daniel Mair; David L Robertson; Emma Thomson; Guy Mollett; Ioulia Tsatsani; James Shepherd; Jenna Nichols; Joseph Hughes; Kathy Li; Kathy Smollett; Kyriaki Nomikou; Lily Tong; Matthew Holden; Natasha Johnson; Rachel Blacow; Richard Orton; Rory Gunson; Sarah McDonald; Sharif Shaaban; Sreenu Vattipally; Stephen Carmichael |

We gratefully acknowledge the following Authors from the Originating laboratories responsible for obtaining the specimens, as well as the Submitting laboratories where the genome data were generated and shared via GISAID, on which this research is based.

All Submitters of data may be contacted directly via www.gisaid.org

Authors are sorted alphabetically.

| Accession ID | Originating Laboratory | Submitting Laboratory | Authors |
|---|---|---|---|
| EPI_ISL_2686094, EPI_ISL_2686095, EPI_ISL_2686096, EPI_ISL_2686097, EPI_ISL_2686098, EPI_ISL_2686099, EPI_ISL_2686100, EPI_ISL_2686101, EPI_ISL_2686102, EPI_ISL_2686103 | | | |
| see above | Amazon Dx, AUV2 Laboratory | Amazon Dx, AUV2 Laboratory | Luke Meredith |
| EPI_ISL_2238944, EPI_ISL_2238945, EPI_ISL_2238946 | Barts Health NHS Trust | COVID-19 Genomics UK (COG-UK) Consortium | BROAD; Beatrix; CUTINO-MOGUEL; Claire; David; Dola; HARRINGTON; KELE; KULASEGARAN-SHYLINI; Maria-Teresa; OWOYEMI; Raghavendran |
| EPI_ISL_2128236, EPI_ISL_2128238 | Centre for Enzyme Innovation, University of Portsmouth / Translational Research Laboratory, Portsmouth Hospitals NHS Trust | COVID-19 Genomics UK (COG-UK) Consortium | Angela Beckett; Christopher Fearn; Kate Cook; Katie Loveson; Salman Goudarzi; Samuel Robson; Scott Elliott; Sharon Glaysher |
| EPI_ISL_2240663, EPI_ISL_2240667, EPI_ISL_2240668, EPI_ISL_2240669, EPI_ISL_2240670, EPI_ISL_2240671, EPI_ISL_2240673, EPI_ISL_2240674 | | | |
| see above | Centre for Enzyme Innovation, University of Portsmouth / Translational Research Laboratory, Portsmouth Hospitals NHS Trust / Hampshire Hospitals NHS Foundation Trust / Bournemouth University / University Hospitals Dorset NHS Foundation Trust / Brighton and Sussex University Hospitals NHS Trust / Dartford and Gravesham NHS Trust / Isle of Wight NHS Trust / Maidstone and Tunbridge Wells NHS Trust / University Hospital Southampton NHS Foundation Trust | COVID-19 Genomics UK (COG-UK) Consortium | Adhyana Mahanama; Allyson Lloyd; Andrew Butt; Angela Beckett; Anibolina Castigador; Anna Mantzaouratou; Anoop Chauhan; Ben Macklin; Benjamin Cogger; Buddhini Samaraweera; Cassandra Malone; Christopher Fearn; Claire Thomas; Dorian Crudgington; Eleri Wilson-Davies; Emanuela Pelosi; Emily Macnaughton; Helen Umpleby; Helen Wheeler; Jacqui Prieto; Jonnie Seymour; Karen Withell; Kate Cook; Katie Loveson; Kelly Bicknell; Kevin Tucker; Kordo Saeed; Liz Sheridan; Magdalena Barrow; Matthew Harvey; Mohammed Hassan-Ibrahim; Nathan Moore; Nick Cortes; Rachael Jeremiah; Robert Impey; Salman Goudarzi; Samirakhon Raupova; Samuel Robson; Sarah Buchan; Sarah Jeremiah; Sarah Wyllie; Scott Elliott; Sharon Glaysher; Siona Silvieri; Stephen Aplin; Stephen Kidd; Thea Sass |
| EPI_ISL_2125479, EPI_ISL_2125480, EPI_ISL_2125482, EPI_ISL_2238738, EPI_ISL_2238739, EPI_ISL_2238740, EPI_ISL_2238741, EPI_ISL_2238742, EPI_ISL_2238743, EPI_ISL_2238744, EPI_ISL_2238745, EPI_ISL_2238746, EPI_ISL_2238747, EPI_ISL_2238748, EPI_ISL_2238749, EPI_ISL_2238750 | | | |
| see above | Department of Pathology, University of Cambridge | COVID-19 Genomics UK (COG-UK) Consortium | Aminu S. Jahun; Ian Goodfellow; Iliana Georgana; Martin D. Curran; Myra Hosmillo; Rhys Izuagbe; Surendra Parmar; William L. Hamilton; Yasmin Chaudhry |
| EPI_ISL_2138418, EPI_ISL_2138436, EPI_ISL_2138471, EPI_ISL_2138503, EPI_ISL_2138516, EPI_ISL_2138517, EPI_ISL_2138521, EPI_ISL_2138523, EPI_ISL_2138542, EPI_ISL_2138580, EPI_ISL_2138582, EPI_ISL_2138597, EPI_ISL_2138600, EPI_ISL_2138694, EPI_ISL_2235545, EPI_ISL_2235570, EPI_ISL_2235571, EPI_ISL_2235575, EPI_ISL_2235580, EPI_ISL_2235599, EPI_ISL_2235603, EPI_ISL_2235604, EPI_ISL_2235616, EPI_ISL_2235622, EPI_ISL_2235633, EPI_ISL_2235638, EPI_ISL_2235639, EPI_ISL_2235644, EPI_ISL_2235652, EPI_ISL_2235675, EPI_ISL_2235694, EPI_ISL_2235696, EPI_ISL_2235707, EPI_ISL_2235710, EPI_ISL_2235722, EPI_ISL_2235723, EPI_ISL_2235728, EPI_ISL_2235730, EPI_ISL_2235743, EPI_ISL_2235749, EPI_ISL_2235754, EPI_ISL_2235784, EPI_ISL_2235786, EPI_ISL_2235787, EPI_ISL_2235792, EPI_ISL_2235803, EPI_ISL_2235821, EPI_ISL_2235851, EPI_ISL_2253222, EPI_ISL_2253223, EPI_ISL_2253224, EPI_ISL_2253225, EPI_ISL_2253231, EPI_ISL_2253236, EPI_ISL_2253237, EPI_ISL_2253240, EPI_ISL_2253244, EPI_ISL_2253246, EPI_ISL_2253251, EPI_ISL_2253254, EPI_ISL_2253257, EPI_ISL_2253260, EPI_ISL_2253261, EPI_ISL_2253262, EPI_ISL_2253265, EPI_ISL_2253267, EPI_ISL_2253268, EPI_ISL_2253269, EPI_ISL_2253275, EPI_ISL_2253276, EPI_ISL_2253278, EPI_ISL_2253281, EPI_ISL_2253283, EPI_ISL_2253285, EPI_ISL_2253286, EPI_ISL_2253288, EPI_ISL_2253289, EPI_ISL_2253290, EPI_ISL_2253291, EPI_ISL_2253292, EPI_ISL_2253293, EPI_ISL_2253297, EPI_ISL_2253298, EPI_ISL_2253299, EPI_ISL_2253301, EPI_ISL_2253302, EPI_ISL_2253303, EPI_ISL_2253306, EPI_ISL_2253308, EPI_ISL_2253310, EPI_ISL_2253311, EPI_ISL_2253312, EPI_ISL_2253315, EPI_ISL_2253316, EPI_ISL_2253318, EPI_ISL_2253320, EPI_ISL_2253321, EPI_ISL_2253325, EPI_ISL_2253327, EPI_ISL_2253330, EPI_ISL_2253334, EPI_ISL_2253335, EPI_ISL_2253338, EPI_ISL_2253340, EPI_ISL_2253341, EPI_ISL_2253343, EPI_ISL_2253346, EPI_ISL_2253347, EPI_ISL_2253349, EPI_ISL_2253350, EPI_ISL_2253352, EPI_ISL_2253354, EPI_ISL_2253355, EPI_ISL_2253360, EPI_ISL_2253362, EPI_ISL_2253365, EPI_ISL_2253367, EPI_ISL_2253375, EPI_ISL_2253379, EPI_ISL_2253381, EPI_ISL_2253382, EPI_ISL_2253385, EPI_ISL_2253409, EPI_ISL_2316281, EPI_ISL_2316293, EPI_ISL_2316297, EPI_ISL_2316306, EPI_ISL_2316336, EPI_ISL_2316359, EPI_ISL_2316365, EPI_ISL_2316384, EPI_ISL_2316386, EPI_ISL_2316411, EPI_ISL_2316413, EPI_ISL_2316430, EPI_ISL_2316459, EPI_ISL_2316502, EPI_ISL_2316505, EPI_ISL_2316507, EPI_ISL_2316517, EPI_ISL_2316533, EPI_ISL_2316535, EPI_ISL_2316592, EPI_ISL_2316636 | | | |
| see above | Health Services Laboratories | Wellcome Sanger Institute for the COVID-19 Genomics UK (COG-UK) Consortium | Cordelia Langford; David K. Jackson; Dominic Kwiatkowski; Ewan Harrison; Health Services Laboratories and Alex Alderton; Ian Johnston; Jeffrey Barrett; John Sillitoe on behalf of the Wellcome Sanger Institute COVID-19 Surveillance Team; Roberto Amato; Sonia Goncalves |
| EPI_ISL_2137844, EPI_ISL_2137845, EPI_ISL_2137846, EPI_ISL_2137848, EPI_ISL_2137850, EPI_ISL_2137875, EPI_ISL_2137877, EPI_ISL_2137882, EPI_ISL_2137887, EPI_ISL_2137888, EPI_ISL_2137890, EPI_ISL_2137894, EPI_ISL_2137896, EPI_ISL_2137911, EPI_ISL_2137919, EPI_ISL_2137920, EPI_ISL_2137925, EPI_ISL_2137928, EPI_ISL_2137929, EPI_ISL_2137933, EPI_ISL_2137947, EPI_ISL_2137954, EPI_ISL_2137956, EPI_ISL_2137963, EPI_ISL_2137967, EPI_ISL_2137976, EPI_ISL_2137978, EPI_ISL_2137983, EPI_ISL_2137997, EPI_ISL_2137998, EPI_ISL_2138001, EPI_ISL_2138002, EPI_ISL_2138012, EPI_ISL_2138019, EPI_ISL_2138023, EPI_ISL_2138031, EPI_ISL_2138032, EPI_ISL_2138046, EPI_ISL_2138049, EPI_ISL_2138054, EPI_ISL_2138062, EPI_ISL_2138066, EPI_ISL_2138071, EPI_ISL_2138073, EPI_ISL_2138075, EPI_ISL_2138078, EPI_ISL_2138082, EPI_ISL_2138087, EPI_ISL_2138092, EPI_ISL_2138107, EPI_ISL_2138112, EPI_ISL_2138120, EPI_ISL_2138128, EPI_ISL_2138136, EPI_ISL_2138139, EPI_ISL_2138144, EPI_ISL_2138171, EPI_ISL_2138193, EPI_ISL_2138206, EPI_ISL_2138214, EPI_ISL_2138219, EPI_ISL_2138226, EPI_ISL_2138237, EPI_ISL_2138246, EPI_ISL_2138247, EPI_ISL_2138248, EPI_ISL_2138260, EPI_ISL_2138262, EPI_ISL_2138264, EPI_ISL_2138268, EPI_ISL_2138273, EPI_ISL_2138274, EPI_ISL_2138276, EPI_ISL_2138283, EPI_ISL_2138285, EPI_ISL_2138287, EPI_ISL_2138288, EPI_ISL_2138294, EPI_ISL_2138300, EPI_ISL_2138306, EPI_ISL_2138319, EPI_ISL_2138328, EPI_ISL_2138331, EPI_ISL_2138344, EPI_ISL_2138354, EPI_ISL_2171680, EPI_ISL_2171683, EPI_ISL_2171695, EPI_ISL_2171703, EPI_ISL_2171708, EPI_ISL_2171715, EPI_ISL_2171722, EPI_ISL_2171723, EPI_ISL_2171734, EPI_ISL_2171738, EPI_ISL_2171756, EPI_ISL_2171770, EPI_ISL_2171771, EPI_ISL_2171772, EPI_ISL_2171780, EPI_ISL_2171781, EPI_ISL_2171782, EPI_ISL_2171786, EPI_ISL_2171791, EPI_ISL_2171800, EPI_ISL_2171801, EPI_ISL_2171806, EPI_ISL_2171814, EPI_ISL_2171818, EPI_ISL_2171819, EPI_ISL_2171823, EPI_ISL_2171824, EPI_ISL_2171825, EPI_ISL_2171826, EPI_ISL_2171828, EPI_ISL_2171830, EPI_ISL_2171831, EPI_ISL_2171833, EPI_ISL_2171834, EPI_ISL_2171835, EPI_ISL_2171838, EPI_ISL_2171839, EPI_ISL_2171842, EPI_ISL_2171843, EPI_ISL_2171844, EPI_ISL_2171845, EPI_ISL_2171846, EPI_ISL_2171847, EPI_ISL_2171848, EPI_ISL_2171849, EPI_ISL_2171851, EPI_ISL_2171852, EPI_ISL_2171855, EPI_ISL_2171856, EPI_ISL_2171858, EPI_ISL_2171859, EPI_ISL_2171860, EPI_ISL_2171862, EPI_ISL_2171864, EPI_ISL_2171865, EPI_ISL_2171866, EPI_ISL_2171868, EPI_ISL_2171869, EPI_ISL_2171870, EPI_ISL_2171874, EPI_ISL_2171877, EPI_ISL_2171878, EPI_ISL_2171879, EPI_ISL_2171880, EPI_ISL_2171882, EPI_ISL_2171884, EPI_ISL_2171888, EPI_ISL_2171891, EPI_ISL_2171894, EPI_ISL_2171896, EPI_ISL_2171897, EPI_ISL_2171898, EPI_ISL_2171900, EPI_ISL_2171901, EPI_ISL_2171902, EPI_ISL_2171903, EPI_ISL_2171904, EPI_ISL_2171905, EPI_ISL_2171907, EPI_ISL_2171908, EPI_ISL_2171913, EPI_ISL_2171915, EPI_ISL_2171917, EPI_ISL_2171919, EPI_ISL_2171921, EPI_ISL_2171922, EPI_ISL_2171923, EPI_ISL_2171924, EPI_ISL_2171925, EPI_ISL_2171926, EPI_ISL_2171927, EPI_ISL_2171929, EPI_ISL_2171930, EPI_ISL_2171931, EPI_ISL_2171932, EPI_ISL_2171933, EPI_ISL_2171934, EPI_ISL_2171935, EPI_ISL_2171938, EPI_ISL_2171942, EPI_ISL_2171944, EPI_ISL_2171945, EPI_ISL_2171946, EPI_ISL_2171947, EPI_ISL_2171948, EPI_ISL_2171950, EPI_ISL_2171952, EPI_ISL_2171953, EPI_ISL_2171955, EPI_ISL_2171956, EPI_ISL_2171958, EPI_ISL_2171960, EPI_ISL_2171962, EPI_ISL_2171964, EPI_ISL_2171965, EPI_ISL_2171969, EPI_ISL_2171972, EPI_ISL_2171973, EPI_ISL_2171974, EPI_ISL_2171975, EPI_ISL_2171977, EPI_ISL_2171979, EPI_ISL_2171980, EPI_ISL_2171981, EPI_ISL_2171983, EPI_ISL_2171984, EPI_ISL_2171985, EPI_ISL_2171987, EPI_ISL_2171988, EPI_ISL_2171989, EPI_ISL_2171990, EPI_ISL_2171991, EPI_ISL_2171992, EPI_ISL_2171993, EPI_ISL_2171996, EPI_ISL_2171997, EPI_ISL_2171998, EPI_ISL_2171999, EPI_ISL_2172000, EPI_ISL_2172002, EPI_ISL_2172003, EPI_ISL_2172004, EPI_ISL_2172005, EPI_ISL_2172007, EPI_ISL_2172010, EPI_ISL_2172013, EPI_ISL_2172016, EPI_ISL_2172017, EPI_ISL_2172018, EPI_ISL_2172019, EPI_ISL_2172020, EPI_ISL_2172021, EPI_ISL_2172022, EPI_ISL_2172023, EPI_ISL_2172024, EPI_ISL_2172025, EPI_ISL_2172026, EPI_ISL_2172029, EPI_ISL_2172030, EPI_ISL_2172031, EPI_ISL_2172032, EPI_ISL_2172036, EPI_ISL_2172039, EPI_ISL_2172040, EPI_ISL_2172041, EPI_ISL_2172042, EPI_ISL_2172043, EPI_ISL_2172044, EPI_ISL_2172046, EPI_ISL_2172047, EPI_ISL_2172049, EPI_ISL_2172050, EPI_ISL_2172051, EPI_ISL_2172052, EPI_ISL_2172053, EPI_ISL_2172055, EPI_ISL_2172057, EPI_ISL_2172058, EPI_ISL_2172059, EPI_ISL_2172062, EPI_ISL_2172063, EPI_ISL_2172065, EPI_ISL_2172066, EPI_ISL_2172067, EPI_ISL_2172068, EPI_ISL_2172073, EPI_ISL_2172075, EPI_ISL_2172077, EPI_ISL_2172079, EPI_ISL_2172080, EPI_ISL_2172082, EPI_ISL_2172083, EPI_ISL_2172084, EPI_ISL_2172086, EPI_ISL_2172087, EPI_ISL_2172090, EPI_ISL_2172091, EPI_ISL_2198346, EPI_ISL_2198348, EPI_ISL_2198350, EPI_ISL_2198351, EPI_ISL_2198357, EPI_ISL_2198359, EPI_ISL_2198362, EPI_ISL_2198363, EPI_ISL_2198365, EPI_ISL_2198366, EPI_ISL_2198370, EPI_ISL_2198372, EPI_ISL_2198380, EPI_ISL_2198381, EPI_ISL_2198382, EPI_ISL_2198386, EPI_ISL_2198387, EPI_ISL_2198388, EPI_ISL_2198396, EPI_ISL_2198397, EPI_ISL_2198400, EPI_ISL_2198401, EPI_ISL_2198406, EPI_ISL_2198412, EPI_ISL_2198413, EPI_ISL_2198414, EPI_ISL_2198417, EPI_ISL_2198420, EPI_ISL_2198423, EPI_ISL_2198424, EPI_ISL_2198425, EPI_ISL_2198430, EPI_ISL_2198431, EPI_ISL_2198435, EPI_ISL_2198437, EPI_ISL_2198438, EPI_ISL_2198439, EPI_ISL_2198440, EPI_ISL_2198446, EPI_ISL_2198450, EPI_ISL_2198454, EPI_ISL_2198461, EPI_ISL_2198462, EPI_ISL_2198466, EPI_ISL_2198476, EPI_ISL_2198479, EPI_ISL_2198482, EPI_ISL_2198489, EPI_ISL_2198490, EPI_ISL_2198491, EPI_ISL_2198494, EPI_ISL_2198496, EPI_ISL_2198501, EPI_ISL_2198503, EPI_ISL_2198504, EPI_ISL_2198505, EPI_ISL_2198512, EPI_ISL_2198515, EPI_ISL_2198517, EPI_ISL_2198518, EPI_ISL_2198520, EPI_ISL_2198525, EPI_ISL_2198527, EPI_ISL_2198529, EPI_ISL_2198531, EPI_ISL_2198532, EPI_ISL_2198533, EPI_ISL_2198536, EPI_ISL_2198537, EPI_ISL_2198538, EPI_ISL_2198539, EPI_ISL_2198547, EPI_ISL_2198554, EPI_ISL_2198557, EPI_ISL_2198558, EPI_ISL_2198559, EPI_ISL_2198562, EPI_ISL_2198563, EPI_ISL_2198565, EPI_ISL_2198567, EPI_ISL_2198568, EPI_ISL_2198569, EPI_ISL_2198571, EPI_ISL_2198580, EPI_ISL_2198582, EPI_ISL_2198584, EPI_ISL_2198588, EPI_ISL_2198590, EPI_ISL_2198592, EPI_ISL_2198593, EPI_ISL_2198596, EPI_ISL_2198598, EPI_ISL_2198602, EPI_ISL_2198603, EPI_ISL_2198604, EPI_ISL_2198605, EPI_ISL_2198606, EPI_ISL_2198611, EPI_ISL_2198613, EPI_ISL_2198614, EPI_ISL_2198616, EPI_ISL_2198617, EPI_ISL_2198622, EPI_ISL_2198626, EPI_ISL_2198630, EPI_ISL_2198635, EPI_ISL_2198640, EPI_ISL_2198645, EPI_ISL_2198648, EPI_ISL_2198650, EPI_ISL_2198651, EPI_ISL_2198656, EPI_ISL_2198658, EPI_ISL_2198662, EPI_ISL_2198664, EPI_ISL_2198666, EPI_ISL_2198668, EPI_ISL_2198672, EPI_ISL_2198673, EPI_ISL_2198676, EPI_ISL_2198677, EPI_ISL_2198678, EPI_ISL_2198680, EPI_ISL_2198681, EPI_ISL_2198683, EPI_ISL_2198684, EPI_ISL_2198686, EPI_ISL_2198689, EPI_ISL_2198690, EPI_ISL_2198692, EPI_ISL_2198695, EPI_ISL_2198701, EPI_ISL_2198702, EPI_ISL_2198703, EPI_ISL_2198711, EPI_ISL_2198713, EPI_ISL_2198714, EPI_ISL_2198715, EPI_ISL_2198719, EPI_ISL_2198720, EPI_ISL_2198725, EPI_ISL_2198727, EPI_ISL_2198729, EPI_ISL_2198731, EPI_ISL_2198732, EPI_ISL_2198733, EPI_ISL_2198736, EPI_ISL_2198739, EPI_ISL_2198740, EPI_ISL_2198745, EPI_ISL_2198749, EPI_ISL_2198751, EPI_ISL_2198754, EPI_ISL_2198756, EPI_ISL_2198760, EPI_ISL_2198762, EPI_ISL_2198763, EPI_ISL_2198767, EPI_ISL_2198773, EPI_ISL_2198775, EPI_ISL_2198776, EPI_ISL_2198782, EPI_ISL_2198784, EPI_ISL_2198785, EPI_ISL_2198786, EPI_ISL_2198796, EPI_ISL_2198801, EPI_ISL_2198802, EPI_ISL_2198806, EPI_ISL_2198808, EPI_ISL_2198811, EPI_ISL_2198813, EPI_ISL_2198815, EPI_ISL_2198820, EPI_ISL_2198822, EPI_ISL_2198827, EPI_ISL_2198831, EPI_ISL_2198833, EPI_ISL_2198835, EPI_ISL_2198838, EPI_ISL_2198839, EPI_ISL_2198844, EPI_ISL_2198845, EPI_ISL_2198850, EPI_ISL_2198852, EPI_ISL_2198856, EPI_ISL_2198858, EPI_ISL_2198860, EPI_ISL_2198865, EPI_ISL_2198867, EPI_ISL_2198868, EPI_ISL_2198870, EPI_ISL_2198871, EPI_ISL_2198872, EPI_ISL_2198876, EPI_ISL_2198877, EPI_ISL_2198878, EPI_ISL_2198879, EPI_ISL_2198882, EPI_ISL_2198886, EPI_ISL_2198892, EPI_ISL_2198894, EPI_ISL_2198895, EPI_ISL_2198896, EPI_ISL_2198898, EPI_ISL_2198900, EPI_ISL_2198904, EPI_ISL_2198907, EPI_ISL_2198908, EPI_ISL_2198909, EPI_ISL_2198915, EPI_ISL_2198916, EPI_ISL_2198917, EPI_ISL_2198919, EPI_ISL_2198920, EPI_ISL_2198921, EPI_ISL_2198922, EPI_ISL_2198925, EPI_ISL_2198926, EPI_ISL_2198927, EPI_ISL_2198932, EPI_ISL_2198938, EPI_ISL_2198940, EPI_ISL_2198948, EPI_ISL_2198952, EPI_ISL_2198953, EPI_ISL_2198954, EPI_ISL_2198956, EPI_ISL_2198957, EPI_ISL_2198961, EPI_ISL_2198964, EPI_ISL_2198968, EPI_ISL_2198970, EPI_ISL_2198972, EPI_ISL_2198975, EPI_ISL_2198980, EPI_ISL_2198986, EPI_ISL_2198987, EPI_ISL_2198988, EPI_ISL_2198990, EPI_ISL_2198991, EPI_ISL_2198995, EPI_ISL_2198996, EPI_ISL_2199003, EPI_ISL_2199024, EPI_ISL_2199025, EPI_ISL_2199032, EPI_ISL_2199034, EPI_ISL_2199035, EPI_ISL_2199039, EPI_ISL_2199048, EPI_ISL_2199049, EPI_ISL_2199053, EPI_ISL_2199057, EPI_ISL_2199060, EPI_ISL_2199064, EPI_ISL_2199069, EPI_ISL_2199072, EPI_ISL_2199077, EPI_ISL_2199079, EPI_ISL_2199083, EPI_ISL_2199084, EPI_ISL_2199085, EPI_ISL_2199088, EPI_ISL_2199090, EPI_ISL_2199100, EPI_ISL_2199102, EPI_ISL_2199103, EPI_ISL_2199108, EPI_ISL_2199111, EPI_ISL_2199112, EPI_ISL_2199113, EPI_ISL_2199114, EPI_ISL_2199119, EPI_ISL_2199121, EPI_ISL_2199122, EPI_ISL_2199123, EPI_ISL_2199130, EPI_ISL_2199134, EPI_ISL_2199140, EPI_ISL_2199149, EPI_ISL_2199156, EPI_ISL_2199161, EPI_ISL_2199164, EPI_ISL_2199173, EPI_ISL_2199176, EPI_ISL_2199179, EPI_ISL_2199180, EPI_ISL_2199190, EPI_ISL_2199191, EPI_ISL_2199192, EPI_ISL_2199195, EPI_ISL_2199198, EPI_ISL_2199201, EPI_ISL_2199205, EPI_ISL_2199207, EPI_ISL_2199211, EPI_ISL_2199215, EPI_ISL_2199216, EPI_ISL_2199218, EPI_ISL_2199219, EPI_ISL_2199224, EPI_ISL_2199236, EPI_ISL_2199241, EPI_ISL_2199242, EPI_ISL_2199247, EPI_ISL_2199250, EPI_ISL_2199251, EPI_ISL_2199253, EPI_ISL_2199254, EPI_ISL_2199257, EPI_ISL_2199258, EPI_ISL_2199270, EPI_ISL_2199271, EPI_ISL_2199280, EPI_ISL_2199283, EPI_ISL_2199284, EPI_ISL_2199285, EPI_ISL_2199286, EPI_ISL_2199295, EPI_ISL_2199302, EPI_ISL_2199311, EPI_ISL_2199313, EPI_ISL_2199318, EPI_ISL_2199320, EPI_ISL_2199321, EPI_ISL_2199324, EPI_ISL_2199329, EPI_ISL_2199330, EPI_ISL_2199332, EPI_ISL_2199342, EPI_ISL_2199343, EPI_ISL_2199344, EPI_ISL_2199345, EPI_ISL_2199346, EPI_ISL_2199347, EPI_ISL_2199348, EPI_ISL_2199349, EPI_ISL_2199350, EPI_ISL_2199351, EPI_ISL_2199352, EPI_ISL_2199353, EPI_ISL_2199355, EPI_ISL_2199356, EPI_ISL_2199359, EPI_ISL_2199360, EPI_ISL_2199361, EPI_ISL_2199363, EPI_ISL_2199364, EPI_ISL_2199365, EPI_ISL_2199366, EPI_ISL_2199367, EPI_ISL_2199368, EPI_ISL_2199370, EPI_ISL_2199371, EPI_ISL_2199372, EPI_ISL_2199373, EPI_ISL_2199374, EPI_ISL_2199375, EPI_ISL_2199376, EPI_ISL_2199377, EPI_ISL_2199378, EPI_ISL_2199379, EPI_ISL_2199380, EPI_ISL_2199381, EPI_ISL_2199382, EPI_ISL_2199384, EPI_ISL_2199385, EPI_ISL_2199386, EPI_ISL_2199387, EPI_ISL_2199388, EPI_ISL_2199389, EPI_ISL_2199390, EPI_ISL_2199393, EPI_ISL_2199394, EPI_ISL_2199395, EPI_ISL_2199396, EPI_ISL_2199397, EPI_ISL_2199398, EPI_ISL_2199399, EPI_ISL_2199400, EPI_ISL_2199401, EPI_ISL_2199402, EPI_ISL_2199403, EPI_ISL_2199404, EPI_ISL_2199405, EPI_ISL_2199407, EPI_ISL_2199408, EPI_ISL_2199410, EPI_ISL_2199411, EPI_ISL_2199412, EPI_ISL_2199414, EPI_ISL_2199415, EPI_ISL_2199416, EPI_ISL_2199417, EPI_ISL_2199418, EPI_ISL_2199420, EPI_ISL_2199421, EPI_ISL_2199423, EPI_ISL_2199424, EPI_ISL_2199425, EPI_ISL_2199426, EPI_ISL_2199427, EPI_ISL_2199428, EPI_ISL_2199431, EPI_ISL_2199432, EPI_ISL_2199433, EPI_ISL_2199434, EPI_ISL_2199435, EPI_ISL_2199436, EPI_ISL_2199437, EPI_ISL_2199438, EPI_ISL_2199440, EPI_ISL_2199441, EPI_ISL_2199442, EPI_ISL_2199443, EPI_ISL_2199444, EPI_ISL_2199445, EPI_ISL_2199446, EPI_ISL_2199448, EPI_ISL_2199449, EPI_ISL_2199450, EPI_ISL_2199451, EPI_ISL_2199452, EPI_ISL_2199453, EPI_ISL_2199454, EPI_ISL_2199455, EPI_ISL_2199456, EPI_ISL_2199457, EPI_ISL_2199458, EPI_ISL_2199460, EPI_ISL_2199461, EPI_ISL_2199462, EPI_ISL_2199464, EPI_ISL_2199465, EPI_ISL_2199467, EPI_ISL_2199468, EPI_ISL_2199469, EPI_ISL_2199471, EPI_ISL_2199472, EPI_ISL_2199473, EPI_ISL_2199474, EPI_ISL_2199475, EPI_ISL_2199476, EPI_ISL_2199477, EPI_ISL_2199478, EPI_ISL_2199479, EPI_ISL_2199480, EPI_ISL_2199481, EPI_ISL_2199482, EPI_ISL_2199483, EPI_ISL_2199484, EPI_ISL_2199485, EPI_ISL_2199486, EPI_ISL_2199487, EPI_ISL_2199489, EPI_ISL_2199490, EPI_ISL_2199491, EPI_ISL_2199492, EPI_ISL_2199493, EPI_ISL_2199494, EPI_ISL_2199495, EPI_ISL_2199496, EPI_ISL_2199497, EPI_ISL_2199498, EPI_ISL_2199499, EPI_ISL_2199500, EPI_ISL_2199501, EPI_ISL_2199503, EPI_ISL_2199505, EPI_ISL_2199506, EPI_ISL_2199507, EPI_ISL_2199508, EPI_ISL_2199509, EPI_ISL_2199510, EPI_ISL_2199511, EPI_ISL_2199512, EPI_ISL_2199513, EPI_ISL_2199514, EPI_ISL_2199515, EPI_ISL_2199516, EPI_ISL_2199517, EPI_ISL_2199518, EPI_ISL_2199522, EPI_ISL_2199523, EPI_ISL_2199524, EPI_ISL_2199525, EPI_ISL_2199526, EPI_ISL_2199527, EPI_ISL_2199528, EPI_ISL_2199529, EPI_ISL_2199530, EPI_ISL_2199531, EPI_ISL_2199532, EPI_ISL_2199533, EPI_ISL_2199534, EPI_ISL_2199536, EPI_ISL_2199537, EPI_ISL_2199538, EPI_ISL_2199539, EPI_ISL_2199540, EPI_ISL_2199541, EPI_ISL_2199542, EPI_ISL_2199543, EPI_ISL_2199544, EPI_ISL_2199545, EPI_ISL_2199546, EPI_ISL_2199547, EPI_ISL_2199548, EPI_ISL_2199550, EPI_ISL_2199551, EPI_ISL_2199552, EPI_ISL_2199553, EPI_ISL_2199554, EPI_ISL_2199555, EPI_ISL_2199556, EPI_ISL_2199557, EPI_ISL_2199558, EPI_ISL_2199559, EPI_ISL_2199560, EPI_ISL_2199561, EPI_ISL_2199562, EPI_ISL_2199563, EPI_ISL_2199564, EPI_ISL_2199565, EPI_ISL_2199566, EPI_ISL_2199567, EPI_ISL_2199568, EPI_ISL_2199569, EPI_ISL_2199570, EPI_ISL_2199572, EPI_ISL_2199573, EPI_ISL_2199574, EPI_ISL_2199575, EPI_ISL_2199576, EPI_ISL_2199577, EPI_ISL_2199578, EPI_ISL_2199579, EPI_ISL_2199580, EPI_ISL_2199581, EPI_ISL_2199582, EPI_ISL_2199583, EPI_ISL_2199584, EPI_ISL_2199585, EPI_ISL_2199587, EPI_ISL_2199588, EPI_ISL_2199589, EPI_ISL_2199590, EPI_ISL_2199591, EPI_ISL_2199592, EPI_ISL_2199593, EPI_ISL_2199594, EPI_ISL_2199595, EPI_ISL_2199596, EPI_ISL_2199597, EPI_ISL_2199598, EPI_ISL_2199599, EPI_ISL_2199600, EPI_ISL_2199601, EPI_ISL_2199602, EPI_ISL_2199603, EPI_ISL_2199604, EPI_ISL_2199605, EPI_ISL_2199607, EPI_ISL_2199608, EPI_ISL_2199609, EPI_ISL_2199610, EPI_ISL_2199611, EPI_ISL_2199612, EPI_ISL_2199613, EPI_ISL_2199615, EPI_ISL_2199616, EPI_ISL_2199617, EPI_ISL_2199618, EPI_ISL_2199619, EPI_ISL_2199620, EPI_ISL_2199621, EPI_ISL_2199622, EPI_ISL_2199623, EPI_ISL_2199625, EPI_ISL_2199626, EPI_ISL_2199627, EPI_ISL_2199628, EPI_ISL_2199631, EPI_ISL_2199632, EPI_ISL_2199633, EPI_ISL_2199634, EPI_ISL_2199635, EPI_ISL_2199636, EPI_ISL_2199637, EPI_ISL_2199638, EPI_ISL_2199639, EPI_ISL_2199640, EPI_ISL_2199641, EPI_ISL_2199642, EPI_ISL_2199644, EPI_ISL_2199645, EPI_ISL_2199646, EPI_ISL_2199647, EPI_ISL_2199648, EPI_ISL_2199649, EPI_ISL_2199650, EPI_ISL_2199651, EPI_ISL_2199652, EPI_ISL_2199656, EPI_ISL_2199657, EPI_ISL_2199658, EPI_ISL_2199660, EPI_ISL_2199661, EPI_ISL_2199662, EPI_ISL_2199663, EPI_ISL_2199664, EPI_ISL_2199665, EPI_ISL_2199666, EPI_ISL_2199667, EPI_ISL_2199668, EPI_ISL_2199671, EPI_ISL_2199672, EPI_ISL_2199673, EPI_ISL_2199674, EPI_ISL_2199675, EPI_ISL_2199676, EPI_ISL_2199677, EPI_ISL_2199678, EPI_ISL_2199679, EPI_ISL_2199680, EPI_ISL_2199681, EPI_ISL_2199682, EPI_ISL_2199683, EPI_ISL_2235528, EPI_ISL_2235530, EPI_ISL_2235535, EPI_ISL_2235537, EPI_ISL_2235539, EPI_ISL_2235540, EPI_ISL_2235560, EPI_ISL_2235562, EPI_ISL_2235568, EPI_ISL_2235573, EPI_ISL_2235577, EPI_ISL_2235578, EPI_ISL_2235583, EPI_ISL_2235586, EPI_ISL_2235592, EPI_ISL_2235596, EPI_ISL_2235600, EPI_ISL_2235620, EPI_ISL_2235626, EPI_ISL_2235628, EPI_ISL_2235629, EPI_ISL_2235635, EPI_ISL_2235636, EPI_ISL_2235645, EPI_ISL_2235651, EPI_ISL_2235653, EPI_ISL_2235671, EPI_ISL_2235674, EPI_ISL_2235677, EPI_ISL_2235685, EPI_ISL_2235700, EPI_ISL_2235702, EPI_ISL_2235712, EPI_ISL_2235714, EPI_ISL_2235721, EPI_ISL_2235736, EPI_ISL_2235742, EPI_ISL_2235746, EPI_ISL_2235757, EPI_ISL_2235759, EPI_ISL_2235760, EPI_ISL_2235762, EPI_ISL_2235763, EPI_ISL_2235773, EPI_ISL_2235775, EPI_ISL_2235777, EPI_ISL_2235782, EPI_ISL_2235794, EPI_ISL_2235795, EPI_ISL_2235802, EPI_ISL_2235804, EPI_ISL_2235806, EPI_ISL_2235812, EPI_ISL_2235816, EPI_ISL_2235823, EPI_ISL_2235827, EPI_ISL_2235832, EPI_ISL_2235837, EPI_ISL_2235843, EPI_ISL_2235847, EPI_ISL_2235849, EPI_ISL_2235858, EPI_ISL_2235863, EPI_ISL_2235864, EPI_ISL_2235865, EPI_ISL_2235866, EPI_ISL_2235868, EPI_ISL_2235870, EPI_ISL_2235871, EPI_ISL_2235875, EPI_ISL_2235876, EPI_ISL_2235877, EPI_ISL_2235878, EPI_ISL_2235879, EPI_ISL_2235880, EPI_ISL_2235881, EPI_ISL_2235882, EPI_ISL_2235883, EPI_ISL_2235884, EPI_ISL_2235885, EPI_ISL_2235887, EPI_ISL_2235888, EPI_ISL_2235889, EPI_ISL_2235890, EPI_ISL_2235891, EPI_ISL_2235895, EPI_ISL_2235903, EPI_ISL_2235904, EPI_ISL_2235905, EPI_ISL_2235909, EPI_ISL_2235911, EPI_ISL_2235912, EPI_ISL_2235914, EPI_ISL_2235916, EPI_ISL_2235917, EPI_ISL_2235918, EPI_ISL_2235919, EPI_ISL_2235920, EPI_ISL_2235923, EPI_ISL_2235924, EPI_ISL_2235925, EPI_ISL_2235926, EPI_ISL_2235927, EPI_ISL_2235928, EPI_ISL_2235929, EPI_ISL_2235930, EPI_ISL_2235932, EPI_ISL_2235933, EPI_ISL_2235934, EPI_ISL_2235936, EPI_ISL_2235937, EPI_ISL_2235938, EPI_ISL_2235939, EPI_ISL_2235940, EPI_ISL_2235941, EPI_ISL_2235943, EPI_ISL_2235944, EPI_ISL_2235945, EPI_ISL_2235946, EPI_ISL_2235947, EPI_ISL_2235948, EPI_ISL_2235949, EPI_ISL_2235951, EPI_ISL_2235956, EPI_ISL_2235960, EPI_ISL_2235961, EPI_ISL_2235963, EPI_ISL_2235964, EPI_ISL_2235966, EPI_ISL_2235967, EPI_ISL_2235968, EPI_ISL_2235969, EPI_ISL_2235973, EPI_ISL_2235974, EPI_ISL_2235977, EPI_ISL_2235978, EPI_ISL_2235979, EPI_ISL_2235984, EPI_ISL_2235985, EPI_ISL_2235988, EPI_ISL_2235998, EPI_ISL_2236000, EPI_ISL_2236001, EPI_ISL_2236004, EPI_ISL_2236005, EPI_ISL_2236007, EPI_ISL_2236012, EPI_ISL_2236018, EPI_ISL_2236019, EPI_ISL_2236021, EPI_ISL_2236022, EPI_ISL_2236027, EPI_ISL_2236028, EPI_ISL_2236030, EPI_ISL_2236031, EPI_ISL_2236033, EPI_ISL_2236035, EPI_ISL_2236036, EPI_ISL_2236039, EPI_ISL_2236042, EPI_ISL_2236043, EPI_ISL_2236044, EPI_ISL_2236045, EPI_ISL_2236047, EPI_ISL_2236048, EPI_ISL_2236050, EPI_ISL_2236051, EPI_ISL_2236056, EPI_ISL_2236061, EPI_ISL_2236062, EPI_ISL_2236066, EPI_ISL_2236068, EPI_ISL_2236069, EPI_ISL_2236070, EPI_ISL_2236072, EPI_ISL_2236073, EPI_ISL_2236074, EPI_ISL_2236075, EPI_ISL_2236076, EPI_ISL_2236077, EPI_ISL_2236081, EPI_ISL_2236082, EPI_ISL_2236085, EPI_ISL_2236087, EPI_ISL_2236089, EPI_ISL_2236090, EPI_ISL_2236093, EPI_ISL_2236094, EPI_ISL_2236095, EPI_ISL_2236096, EPI_ISL_2236098, EPI_ISL_2236099, EPI_ISL_2236102, EPI_ISL_2236103, EPI_ISL_2236106, EPI_ISL_2236107, EPI_ISL_2236109, EPI_ISL_2236114, EPI_ISL_2236116, EPI_ISL_2236119, EPI_ISL_2236122, EPI_ISL_2236124, EPI_ISL_2236125, EPI_ISL_2236126, EPI_ISL_2236129, EPI_ISL_2236131, EPI_ISL_2236132, EPI_ISL_2236135, EPI_ISL_2236136, EPI_ISL_2236138, EPI_ISL_2236139, EPI_ISL_2236140, EPI_ISL_2236142, EPI_ISL_2236143, EPI_ISL_2236144, EPI_ISL_2236147, EPI_ISL_2236148, EPI_ISL_2236154, EPI_ISL_2236161, EPI_ISL_2236166, EPI_ISL_2236169, EPI_ISL_2236170, EPI_ISL_2236171, EPI_ISL_2236174, EPI_ISL_2236176, EPI_ISL_2236177, EPI_ISL_2236180, EPI_ISL_2236181, EPI_ISL_2236184, EPI_ISL_2236187, EPI_ISL_2236189, EPI_ISL_2236190, EPI_ISL_2236192, EPI_ISL_2236195, EPI_ISL_2236196, EPI_ISL_2236197, EPI_ISL_2236198, EPI_ISL_2236200, EPI_ISL_2236201, EPI_ISL_2236202, EPI_ISL_2236205, EPI_ISL_2236208, EPI_ISL_2236209, EPI_ISL_2236212, EPI_ISL_2236213, EPI_ISL_2236214, EPI_ISL_2236215, EPI_ISL_2236217, EPI_ISL_2236218, EPI_ISL_2236219, EPI_ISL_2236220, EPI_ISL_2236222, EPI_ISL_2236223, EPI_ISL_2236224, EPI_ISL_2236225, EPI_ISL_2236226, EPI_ISL_2236227, EPI_ISL_2236228, EPI_ISL_2236229, EPI_ISL_2236230, EPI_ISL_2236231, EPI_ISL_2236232, EPI_ISL_2236233, EPI_ISL_2236234, EPI_ISL_2236235, EPI_ISL_2236237, EPI_ISL_2236238, EPI_ISL_2236239, EPI_ISL_2236240, EPI_ISL_2236241, EPI_ISL_2236242, EPI_ISL_2236243, EPI_ISL_2236244, EPI_ISL_2236245, EPI_ISL_2236247, EPI_ISL_2236248, EPI_ISL_2236249, EPI_ISL_2236250, EPI_ISL_2236251, EPI_ISL_2236252, EPI_ISL_2236253, EPI_ISL_2236254, EPI_ISL_2236255, EPI_ISL_2236259, EPI_ISL_2236261, EPI_ISL_2236262, EPI_ISL_2236263, EPI_ISL_2236264, EPI_ISL_2236265, EPI_ISL_2236266, EPI_ISL_2236267, EPI_ISL_2236268, EPI_ISL_2236269, EPI_ISL_2236270, EPI_ISL_2236271, EPI_ISL_2236272, EPI_ISL_2236273, EPI_ISL_2236274, EPI_ISL_2236275, EPI_ISL_2236276, EPI_ISL_2236277, EPI_ISL_2236279, EPI_ISL_2236280, EPI_ISL_2236281, EPI_ISL_2236282, EPI_ISL_2236283, EPI_ISL_2236284, EPI_ISL_2236286, EPI_ISL_2236288, EPI_ISL_2236289, EPI_ISL_2236290, EPI_ISL_2236291, EPI_ISL_2236293, EPI_ISL_2236294, EPI_ISL_2236295, EPI_ISL_2236296, EPI_ISL_2236297, EPI_ISL_2236298, EPI_ISL_2236299, EPI_ISL_2236300, EPI_ISL_2236301, EPI_ISL_2236302, EPI_ISL_2236306, EPI_ISL_2236307, EPI_ISL_2236308, EPI_ISL_2236309, EPI_ISL_2236310, EPI_ISL_2236311, EPI_ISL_2236312, EPI_ISL_2236313, EPI_ISL_2236314, EPI_ISL_2236315, EPI_ISL_2236316, EPI_ISL_2236317, EPI_ISL_2236318, EPI_ISL_2236319, EPI_ISL_2236320, EPI_ISL_2236321, EPI_ISL_2236322, EPI_ISL_2236323, EPI_ISL_2236324, EPI_ISL_2236325, EPI_ISL_2236326, EPI_ISL_2236327, EPI_ISL_2236328, EPI_ISL_2236329, EPI_ISL_2236330, EPI_ISL_2236331, EPI_ISL_2236332, EPI_ISL_2236333, EPI_ISL_2236334, EPI_ISL_2236335, EPI_ISL_2236337, EPI_ISL_2236338, EPI_ISL_2236339, EPI_ISL_2236341, EPI_ISL_2236345, EPI_ISL_2236347, EPI_ISL_2236348, EPI_ISL_2236349, EPI_ISL_2236350, EPI_ISL_2236351, EPI_ISL_2236352, EPI_ISL_2236354, EPI_ISL_2236355, EPI_ISL_2236356, EPI_ISL_2236358, EPI_ISL_2236359, EPI_ISL_2236361, EPI_ISL_2236362, EPI_ISL_2236363, EPI_ISL_2236364, EPI_ISL_2236365, EPI_ISL_2236366, EPI_ISL_2236367, EPI_ISL_2236368, EPI_ISL_2236369, EPI_ISL_2236370, EPI_ISL_2236371, EPI_ISL_2236372, EPI_ISL_2236373, EPI_ISL_2236374, EPI_ISL_2236375, EPI_ISL_2236376, EPI_ISL_2236377, EPI_ISL_2236378, EPI_ISL_2236379, EPI_ISL_2236380, EPI_ISL_2236382, EPI_ISL_2236383, EPI_ISL_2236384, EPI_ISL_2236385, EPI_ISL_2236386, EPI_ISL_2236387, EPI_ISL_2236388, EPI_ISL_2236389, EPI_ISL_2236391, EPI_ISL_2236393, EPI_ISL_2236395, EPI_ISL_2236396, EPI_ISL_2236397, EPI_ISL_2236398, EPI_ISL_2236399, EPI_ISL_2236400, EPI_ISL_2236401, EPI_ISL_2236402, EPI_ISL_2236403, EPI_ISL_2236404, EPI_ISL_2236405, EPI_ISL_2236406, EPI_ISL_2236407, EPI_ISL_2236408, EPI_ISL_2236410, EPI_ISL_2236412, EPI_ISL_2236413, EPI_ISL_2236414, EPI_ISL_2236415, EPI_ISL_2236416, EPI_ISL_2236417, EPI_ISL_2236418, EPI_ISL_2236419, EPI_ISL_2236421, EPI_ISL_2236422, EPI_ISL_2236423, EPI_ISL_2236424, EPI_ISL_2236425, EPI_ISL_2236426, EPI_ISL_2236427, EPI_ISL_2236428, EPI_ISL_2236429, EPI_ISL_2236430, EPI_ISL_2236431, EPI_ISL_2236432, EPI_ISL_2236434, EPI_ISL_2236435, EPI_ISL_2236436, EPI_ISL_2236437, EPI_ISL_2236438, EPI_ISL_2236439, EPI_ISL_2236440, EPI_ISL_2236441, EPI_ISL_2236442, EPI_ISL_2236444, EPI_ISL_2236446, EPI_ISL_2236447, EPI_ISL_2236448, EPI_ISL_2236449, EPI_ISL_2236451, EPI_ISL_2236452, EPI_ISL_2236453, EPI_ISL_2236454, EPI_ISL_2236455, EPI_ISL_2236456, EPI_ISL_2236457, EPI_ISL_2236458, EPI_ISL_2236459, EPI_ISL_2236460, EPI_ISL_2236461, EPI_ISL_2236462, EPI_ISL_2236464, | | | |

EPI_ISL_2236465, EPI_ISL_2236466, EPI_ISL_2236467, EPI_ISL_2236468, EPI_ISL_2236469, EPI_ISL_2236471, EPI_ISL_2236472, EPI_ISL_2236473, EPI_ISL_2237159, EPI_ISL_2237163, EPI_ISL_2237164, EPI_ISL_2237166, EPI_ISL_2237170, EPI_ISL_2237173, EPI_ISL_2237176, EPI_ISL_2237178, EPI_ISL_2237179, EPI_ISL_2237181, EPI_ISL_2237183, EPI_ISL_2237184, EPI_ISL_2237185, EPI_ISL_2237186,
EPI_ISL_2237187, EPI_ISL_2237188, EPI_ISL_2237190, EPI_ISL_2237191, EPI_ISL_2237193, EPI_ISL_2237195, EPI_ISL_2237197, EPI_ISL_2237200, EPI_ISL_2237201, EPI_ISL_2237202, EPI_ISL_2237203, EPI_ISL_2237204, EPI_ISL_2237205, EPI_ISL_2237207, EPI_ISL_2237209, EPI_ISL_2237210, EPI_ISL_2237211, EPI_ISL_2237212, EPI_ISL_2237214, EPI_ISL_2237215, EPI_ISL_2237216, EPI_ISL_2237217,
EPI_ISL_2237218, EPI_ISL_2237219, EPI_ISL_2237220, EPI_ISL_2237221, EPI_ISL_2237222, EPI_ISL_2237224, EPI_ISL_2237227, EPI_ISL_2237228, EPI_ISL_2237232, EPI_ISL_2237233, EPI_ISL_2237236, EPI_ISL_2237238, EPI_ISL_2237239, EPI_ISL_2237240, EPI_ISL_2237241, EPI_ISL_2237243, EPI_ISL_2237246, EPI_ISL_2237248, EPI_ISL_2237250, EPI_ISL_2237251, EPI_ISL_2237254, EPI_ISL_2237255,
EPI_ISL_2237256, EPI_ISL_2237258, EPI_ISL_2237259, EPI_ISL_2237260, EPI_ISL_2237261, EPI_ISL_2237263, EPI_ISL_2237264, EPI_ISL_2237267, EPI_ISL_2237268, EPI_ISL_2237270, EPI_ISL_2237271, EPI_ISL_2237272, EPI_ISL_2237273, EPI_ISL_2237275, EPI_ISL_2237277, EPI_ISL_2237278, EPI_ISL_2237279, EPI_ISL_2237280, EPI_ISL_2237287, EPI_ISL_2237291, EPI_ISL_2237292, EPI_ISL_2237294,
EPI_ISL_2237295, EPI_ISL_2237296, EPI_ISL_2237297, EPI_ISL_2237298, EPI_ISL_2237299, EPI_ISL_2237300, EPI_ISL_2237303, EPI_ISL_2237304, EPI_ISL_2237305, EPI_ISL_2237306, EPI_ISL_2237307, EPI_ISL_2237308, EPI_ISL_2237310, EPI_ISL_2237312, EPI_ISL_2237313, EPI_ISL_2237316, EPI_ISL_2237317, EPI_ISL_2237320, EPI_ISL_2237322, EPI_ISL_2237323, EPI_ISL_2237324, EPI_ISL_2237325,
EPI_ISL_2237327, EPI_ISL_2237328, EPI_ISL_2237331, EPI_ISL_2237332, EPI_ISL_2237333, EPI_ISL_2237334, EPI_ISL_2237336, EPI_ISL_2237337, EPI_ISL_2237339, EPI_ISL_2237340, EPI_ISL_2237341, EPI_ISL_2237342, EPI_ISL_2237347, EPI_ISL_2237351, EPI_ISL_2237353, EPI_ISL_2237354, EPI_ISL_2237357, EPI_ISL_2237358, EPI_ISL_2237360, EPI_ISL_2237361, EPI_ISL_2237362, EPI_ISL_2237363,
EPI_ISL_2237364, EPI_ISL_2237368, EPI_ISL_2237371, EPI_ISL_2237373, EPI_ISL_2237374, EPI_ISL_2237375, EPI_ISL_2237377, EPI_ISL_2237379, EPI_ISL_2237381, EPI_ISL_2237387, EPI_ISL_2237388, EPI_ISL_2237389, EPI_ISL_2237390, EPI_ISL_2237392, EPI_ISL_2237393, EPI_ISL_2237394, EPI_ISL_2237395, EPI_ISL_2237396, EPI_ISL_2237397, EPI_ISL_2237401, EPI_ISL_2237403,
EPI_ISL_2237404, EPI_ISL_2237407, EPI_ISL_2237408, EPI_ISL_2237409, EPI_ISL_2237412, EPI_ISL_2237418, EPI_ISL_2237422, EPI_ISL_2237435, EPI_ISL_2237438, EPI_ISL_2237439, EPI_ISL_2237443, EPI_ISL_2237446, EPI_ISL_2237448, EPI_ISL_2237454, EPI_ISL_2237461, EPI_ISL_2237465, EPI_ISL_2237469, EPI_ISL_2237472, EPI_ISL_2237473, EPI_ISL_2237486, EPI_ISL_2237487, EPI_ISL_2237488,
EPI_ISL_2237495, EPI_ISL_2237498, EPI_ISL_2237503, EPI_ISL_2237504, EPI_ISL_2237515, EPI_ISL_2237518, EPI_ISL_2237519, EPI_ISL_2237520, EPI_ISL_2237521, EPI_ISL_2237530, EPI_ISL_2237533, EPI_ISL_2237537, EPI_ISL_2237544, EPI_ISL_2237547, EPI_ISL_2237548, EPI_ISL_2237549, EPI_ISL_2237553, EPI_ISL_2237556, EPI_ISL_2237557, EPI_ISL_2237559, EPI_ISL_2237560, EPI_ISL_2237567,
EPI_ISL_2237570, EPI_ISL_2237571, EPI_ISL_2237572, EPI_ISL_2237574, EPI_ISL_2237584, EPI_ISL_2237585, EPI_ISL_2237586, EPI_ISL_2237599, EPI_ISL_2237600, EPI_ISL_2237601, EPI_ISL_2237605, EPI_ISL_2237606, EPI_ISL_2237611, EPI_ISL_2237612, EPI_ISL_2237618, EPI_ISL_2237620, EPI_ISL_2237625, EPI_ISL_2237627, EPI_ISL_2237628, EPI_ISL_2237629, EPI_ISL_2237649, EPI_ISL_2237656,
EPI_ISL_2237657, EPI_ISL_2237661, EPI_ISL_2237662, EPI_ISL_2237663, EPI_ISL_2237665, EPI_ISL_2237678, EPI_ISL_2237684, EPI_ISL_2237692, EPI_ISL_2237694, EPI_ISL_2237703, EPI_ISL_2237719, EPI_ISL_2237720, EPI_ISL_2237721, EPI_ISL_2237723, EPI_ISL_2237724, EPI_ISL_2237731, EPI_ISL_2237734, EPI_ISL_2237735, EPI_ISL_2237739, EPI_ISL_2237742, EPI_ISL_2237743, EPI_ISL_2237756,
EPI_ISL_2237764, EPI_ISL_2237770, EPI_ISL_2237777, EPI_ISL_2237779, EPI_ISL_2237781, EPI_ISL_2237800, EPI_ISL_2237804, EPI_ISL_2237818, EPI_ISL_2237823, EPI_ISL_2237826, EPI_ISL_2237827, EPI_ISL_2237828, EPI_ISL_2237831, EPI_ISL_2237833, EPI_ISL_2237835, EPI_ISL_2237839, EPI_ISL_2237843, EPI_ISL_2237849, EPI_ISL_2237851, EPI_ISL_2237857, EPI_ISL_2237858, EPI_ISL_2237862,
EPI_ISL_2237864, EPI_ISL_2237868, EPI_ISL_2237870, EPI_ISL_2237874, EPI_ISL_2237878, EPI_ISL_2237882, EPI_ISL_2237883, EPI_ISL_2237884, EPI_ISL_2237885, EPI_ISL_2237894, EPI_ISL_2237897, EPI_ISL_2237908, EPI_ISL_2237917, EPI_ISL_2237918, EPI_ISL_2237920, EPI_ISL_2237923, EPI_ISL_2237925, EPI_ISL_2237928, EPI_ISL_2237931, EPI_ISL_2237934, EPI_ISL_2237945, EPI_ISL_2237955,
EPI_ISL_2237958, EPI_ISL_2237964, EPI_ISL_2237968, EPI_ISL_2237993, EPI_ISL_2237996, EPI_ISL_2237997, EPI_ISL_2238000, EPI_ISL_2238006, EPI_ISL_2238009, EPI_ISL_2238011, EPI_ISL_2238012, EPI_ISL_2238017, EPI_ISL_2238021, EPI_ISL_2238023, EPI_ISL_2238029, EPI_ISL_2238031, EPI_ISL_2238032, EPI_ISL_2238034, EPI_ISL_2238038, EPI_ISL_2238042, EPI_ISL_2238043, EPI_ISL_2238051,
EPI_ISL_2238057, EPI_ISL_2238060, EPI_ISL_2238062, EPI_ISL_2238687, EPI_ISL_2238688, EPI_ISL_2238689, EPI_ISL_2238690, EPI_ISL_2238691, EPI_ISL_2238692, EPI_ISL_2253386, EPI_ISL_2253388, EPI_ISL_2253389, EPI_ISL_2253392, EPI_ISL_2253393, EPI_ISL_2253394, EPI_ISL_2253398, EPI_ISL_2253399, EPI_ISL_2253401, EPI_ISL_2253403, EPI_ISL_2253404, EPI_ISL_2253407, EPI_ISL_2253408,
EPI_ISL_2253410, EPI_ISL_2253412, EPI_ISL_2253414, EPI_ISL_2253415, EPI_ISL_2253417, EPI_ISL_2253418, EPI_ISL_2253420, EPI_ISL_2253421, EPI_ISL_2253422, EPI_ISL_2253423, EPI_ISL_2253424, EPI_ISL_2253425, EPI_ISL_2253427, EPI_ISL_2253433, EPI_ISL_2253435, EPI_ISL_2253436, EPI_ISL_2253437, EPI_ISL_2253438, EPI_ISL_2253440, EPI_ISL_2253441, EPI_ISL_2253442, EPI_ISL_2253444,
EPI_ISL_2253445, EPI_ISL_2253446, EPI_ISL_2253447, EPI_ISL_2253449, EPI_ISL_2253450, EPI_ISL_2253452, EPI_ISL_2253453, EPI_ISL_2253454, EPI_ISL_2253455, EPI_ISL_2253459, EPI_ISL_2253463, EPI_ISL_2253464, EPI_ISL_2253466, EPI_ISL_2253469, EPI_ISL_2253470, EPI_ISL_2253471, EPI_ISL_2253472, EPI_ISL_2253473, EPI_ISL_2253474, EPI_ISL_2253475, EPI_ISL_2253476, EPI_ISL_2253479,
EPI_ISL_2253483, EPI_ISL_2253484, EPI_ISL_2253485, EPI_ISL_2253488, EPI_ISL_2253492, EPI_ISL_2253495, EPI_ISL_2253497, EPI_ISL_2253498, EPI_ISL_2253500, EPI_ISL_2253501, EPI_ISL_2253502, EPI_ISL_2253503, EPI_ISL_2253504, EPI_ISL_2253505, EPI_ISL_2253506, EPI_ISL_2253509, EPI_ISL_2253510, EPI_ISL_2253514, EPI_ISL_2253515, EPI_ISL_2253516, EPI_ISL_2253517, EPI_ISL_2253518,
EPI_ISL_2253520, EPI_ISL_2253521, EPI_ISL_2253523, EPI_ISL_2253526, EPI_ISL_2253527, EPI_ISL_2253529, EPI_ISL_2253530, EPI_ISL_2253531, EPI_ISL_2253532, EPI_ISL_2253534, EPI_ISL_2253535, EPI_ISL_2253536, EPI_ISL_2253538, EPI_ISL_2253541, EPI_ISL_2253544, EPI_ISL_2253545, EPI_ISL_2253546, EPI_ISL_2253547, EPI_ISL_2253548, EPI_ISL_2253549, EPI_ISL_2253551, EPI_ISL_2253552,
EPI_ISL_2253554, EPI_ISL_2253555, EPI_ISL_2253556, EPI_ISL_2253558, EPI_ISL_2253559, EPI_ISL_2253560, EPI_ISL_2253561, EPI_ISL_2253562, EPI_ISL_2253566, EPI_ISL_2253568, EPI_ISL_2253569, EPI_ISL_2253570, EPI_ISL_2253571, EPI_ISL_2253575, EPI_ISL_2253581, EPI_ISL_2253582, EPI_ISL_2253584, EPI_ISL_2253585, EPI_ISL_2253586, EPI_ISL_2253587, EPI_ISL_2253588, EPI_ISL_2253589,
EPI_ISL_2253590, EPI_ISL_2253591, EPI_ISL_2253592, EPI_ISL_2253593, EPI_ISL_2253594, EPI_ISL_2253595, EPI_ISL_2253596, EPI_ISL_2253597, EPI_ISL_2253598, EPI_ISL_2253601, EPI_ISL_2253602, EPI_ISL_2253603, EPI_ISL_2253605, EPI_ISL_2253607, EPI_ISL_2253608, EPI_ISL_2253610, EPI_ISL_2253613, EPI_ISL_2253615, EPI_ISL_2253616, EPI_ISL_2253617, EPI_ISL_2253619, EPI_ISL_2253620,
EPI_ISL_2253622, EPI_ISL_2253623, EPI_ISL_2253624, EPI_ISL_2253625, EPI_ISL_2253626, EPI_ISL_2253628, EPI_ISL_2253630, EPI_ISL_2253631, EPI_ISL_2253633, EPI_ISL_2253636, EPI_ISL_2253637, EPI_ISL_2253639, EPI_ISL_2253641, EPI_ISL_2253643, EPI_ISL_2253644, EPI_ISL_2253645, EPI_ISL_2253646, EPI_ISL_2253647, EPI_ISL_2253648, EPI_ISL_2253649, EPI_ISL_2253650, EPI_ISL_2253651,
EPI_ISL_2253652, EPI_ISL_2253653, EPI_ISL_2253654, EPI_ISL_2253657, EPI_ISL_2253658, EPI_ISL_2253659, EPI_ISL_2253661, EPI_ISL_2253662, EPI_ISL_2253663, EPI_ISL_2253664, EPI_ISL_2253665, EPI_ISL_2253666, EPI_ISL_2253667, EPI_ISL_2253668, EPI_ISL_2253669, EPI_ISL_2253671, EPI_ISL_2253672, EPI_ISL_2253674, EPI_ISL_2253675, EPI_ISL_2253676, EPI_ISL_2253677, EPI_ISL_2253851,
EPI_ISL_2253939, EPI_ISL_2253940, EPI_ISL_2253942, EPI_ISL_2253944, EPI_ISL_2253945, EPI_ISL_2253946, EPI_ISL_2253947, EPI_ISL_2253948, EPI_ISL_2253950, EPI_ISL_2253951, EPI_ISL_2253954, EPI_ISL_2253955, EPI_ISL_2253956, EPI_ISL_2253958, EPI_ISL_2253959, EPI_ISL_2253960, EPI_ISL_2253961, EPI_ISL_2253963, EPI_ISL_2253964, EPI_ISL_2253965, EPI_ISL_2253966, EPI_ISL_2253967,
EPI_ISL_2253968, EPI_ISL_2253969, EPI_ISL_2253970, EPI_ISL_2253971, EPI_ISL_2253972, EPI_ISL_2253973, EPI_ISL_2253974, EPI_ISL_2253978, EPI_ISL_2253979, EPI_ISL_2253983, EPI_ISL_2253985, EPI_ISL_2253986, EPI_ISL_2253987, EPI_ISL_2253988, EPI_ISL_2253989, EPI_ISL_2253990, EPI_ISL_2253991, EPI_ISL_2253993, EPI_ISL_2253994, EPI_ISL_2253996, EPI_ISL_2253998, EPI_ISL_2253999,
EPI_ISL_2254001, EPI_ISL_2254002, EPI_ISL_2254003, EPI_ISL_2254004, EPI_ISL_2254006, EPI_ISL_2254007, EPI_ISL_2254008, EPI_ISL_2254009, EPI_ISL_2254011, EPI_ISL_2254013, EPI_ISL_2254017, EPI_ISL_2254018, EPI_ISL_2254019, EPI_ISL_2254021, EPI_ISL_2254022, EPI_ISL_2254023, EPI_ISL_2254024, EPI_ISL_2254025, EPI_ISL_2254026, EPI_ISL_2254027, EPI_ISL_2254030, EPI_ISL_2254031,
EPI_ISL_2254032, EPI_ISL_2254033, EPI_ISL_2254034, EPI_ISL_2254035, EPI_ISL_2254037, EPI_ISL_2254038, EPI_ISL_2254040, EPI_ISL_2254041, EPI_ISL_2254042, EPI_ISL_2254043, EPI_ISL_2254044, EPI_ISL_2254045, EPI_ISL_2254046, EPI_ISL_2254047, EPI_ISL_2254048, EPI_ISL_2254049, EPI_ISL_2254050, EPI_ISL_2254051, EPI_ISL_2254052, EPI_ISL_2254055, EPI_ISL_2254056, EPI_ISL_2254058,
EPI_ISL_2254059, EPI_ISL_2254062, EPI_ISL_2254063, EPI_ISL_2254064, EPI_ISL_2254065, EPI_ISL_2254066, EPI_ISL_2254067, EPI_ISL_2254068, EPI_ISL_2254069, EPI_ISL_2254070, EPI_ISL_2254072, EPI_ISL_2254085, EPI_ISL_2254088, EPI_ISL_2254090, EPI_ISL_2254091, EPI_ISL_2254092, EPI_ISL_2254093, EPI_ISL_2254104, EPI_ISL_2254119, EPI_ISL_2274983, EPI_ISL_2274995, EPI_ISL_2275034,
EPI_ISL_2275052, EPI_ISL_2275087, EPI_ISL_2275109, EPI_ISL_2275148, EPI_ISL_2275171, EPI_ISL_2275172, EPI_ISL_2275185, EPI_ISL_2275188, EPI_ISL_2275189, EPI_ISL_2275190, EPI_ISL_2275193, EPI_ISL_2275195, EPI_ISL_2275198, EPI_ISL_2275201, EPI_ISL_2275202, EPI_ISL_2275204, EPI_ISL_2275205, EPI_ISL_2275206, EPI_ISL_2275210, EPI_ISL_2275211, EPI_ISL_2275215, EPI_ISL_2275217,
EPI_ISL_2275219, EPI_ISL_2275221, EPI_ISL_2275229, EPI_ISL_2275231, EPI_ISL_2275232, EPI_ISL_2275236, EPI_ISL_2275237, EPI_ISL_2275238, EPI_ISL_2275239, EPI_ISL_2275242, EPI_ISL_2275243, EPI_ISL_2275244, EPI_ISL_2275245, EPI_ISL_2275247, EPI_ISL_2275252, EPI_ISL_2275256, EPI_ISL_2275261, EPI_ISL_2275262, EPI_ISL_2275264, EPI_ISL_2275267, EPI_ISL_2275268, EPI_ISL_2275269,
EPI_ISL_2275271, EPI_ISL_2275272, EPI_ISL_2275279, EPI_ISL_2275283, EPI_ISL_2275288, EPI_ISL_2275290, EPI_ISL_2275292, EPI_ISL_2275304, EPI_ISL_2275305, EPI_ISL_2275307, EPI_ISL_2275308, EPI_ISL_2275309, EPI_ISL_2275312, EPI_ISL_2275315, EPI_ISL_2275318, EPI_ISL_2275321, EPI_ISL_2275323, EPI_ISL_2275325, EPI_ISL_2275326, EPI_ISL_2275327, EPI_ISL_2275328, EPI_ISL_2275330,
EPI_ISL_2275337, EPI_ISL_2275339, EPI_ISL_2275341, EPI_ISL_2275343, EPI_ISL_2275345, EPI_ISL_2275346, EPI_ISL_2275347, EPI_ISL_2275348, EPI_ISL_2275349, EPI_ISL_2275354, EPI_ISL_2275356, EPI_ISL_2275358, EPI_ISL_2275360, EPI_ISL_2275362, EPI_ISL_2275363, EPI_ISL_2275364, EPI_ISL_2275366, EPI_ISL_2275375, EPI_ISL_2275376, EPI_ISL_2275377, EPI_ISL_2275380, EPI_ISL_2275383,
EPI_ISL_2275385, EPI_ISL_2275386, EPI_ISL_2275388, EPI_ISL_2275394, EPI_ISL_2275395, EPI_ISL_2275398, EPI_ISL_2275402, EPI_ISL_2296692, EPI_ISL_2296694, EPI_ISL_2316279, EPI_ISL_2316290, EPI_ISL_2316291, EPI_ISL_2316294, EPI_ISL_2316295, EPI_ISL_2316298, EPI_ISL_2316300, EPI_ISL_2316303, EPI_ISL_2316304, EPI_ISL_2316307, EPI_ISL_2316309, EPI_ISL_2316310, EPI_ISL_2316318,
EPI_ISL_2316323, EPI_ISL_2316326, EPI_ISL_2316327, EPI_ISL_2316330, EPI_ISL_2316333, EPI_ISL_2316335, EPI_ISL_2316337, EPI_ISL_2316339, EPI_ISL_2316342, EPI_ISL_2316345, EPI_ISL_2316347, EPI_ISL_2316349, EPI_ISL_2316350, EPI_ISL_2316353, EPI_ISL_2316356, EPI_ISL_2316361, EPI_ISL_2316362, EPI_ISL_2316364, EPI_ISL_2316366, EPI_ISL_2316369, EPI_ISL_2316371, EPI_ISL_2316372,
EPI_ISL_2316373, EPI_ISL_2316375, EPI_ISL_2316376, EPI_ISL_2316378, EPI_ISL_2316381, EPI_ISL_2316387, EPI_ISL_2316389, EPI_ISL_2316392, EPI_ISL_2316393, EPI_ISL_2316401, EPI_ISL_2316404, EPI_ISL_2316405, EPI_ISL_2316407, EPI_ISL_2316410, EPI_ISL_2316414, EPI_ISL_2316416, EPI_ISL_2316419, EPI_ISL_2316422, EPI_ISL_2316424, EPI_ISL_2316427, EPI_ISL_2316434, EPI_ISL_2316435,
EPI_ISL_2316437, EPI_ISL_2316441, EPI_ISL_2316445, EPI_ISL_2316448, EPI_ISL_2316452, EPI_ISL_2316453, EPI_ISL_2316456, EPI_ISL_2316458, EPI_ISL_2316461, EPI_ISL_2316463, EPI_ISL_2316464, EPI_ISL_2316468, EPI_ISL_2316469, EPI_ISL_2316471, EPI_ISL_2316473, EPI_ISL_2316476, EPI_ISL_2316485, EPI_ISL_2316487, EPI_ISL_2316491, EPI_ISL_2316496, EPI_ISL_2316498, EPI_ISL_2316500,
EPI_ISL_2316501, EPI_ISL_2316504, EPI_ISL_2316509, EPI_ISL_2316511, EPI_ISL_2316512, EPI_ISL_2316514, EPI_ISL_2316515, EPI_ISL_2316516, EPI_ISL_2316519, EPI_ISL_2316524, EPI_ISL_2316527, EPI_ISL_2316532, EPI_ISL_2316536, EPI_ISL_2316538, EPI_ISL_2316539, EPI_ISL_2316541, EPI_ISL_2316542, EPI_ISL_2316545, EPI_ISL_2316548, EPI_ISL_2316550, EPI_ISL_2316551,
EPI_ISL_2316553, EPI_ISL_2316554, EPI_ISL_2316556, EPI_ISL_2316560, EPI_ISL_2316562, EPI_ISL_2316563, EPI_ISL_2316566, EPI_ISL_2316568, EPI_ISL_2316569, EPI_ISL_2316575, EPI_ISL_2316576, EPI_ISL_2316578, EPI_ISL_2316579, EPI_ISL_2316582, EPI_ISL_2316584, EPI_ISL_2316586, EPI_ISL_2316588, EPI_ISL_2316590, EPI_ISL_2316594, EPI_ISL_2316596, EPI_ISL_2316598, EPI_ISL_2316601,
EPI_ISL_2316603, EPI_ISL_2316604, EPI_ISL_2316606, EPI_ISL_2316607, EPI_ISL_2316610, EPI_ISL_2316611, EPI_ISL_2316613, EPI_ISL_2316614, EPI_ISL_2316618, EPI_ISL_2316632, EPI_ISL_2316633, EPI_ISL_2316634, EPI_ISL_2316637, EPI_ISL_2316641, EPI_ISL_2316649, EPI_ISL_2316651, EPI_ISL_2316656, EPI_ISL_2316660, EPI_ISL_2316663, EPI_ISL_2317165, EPI_ISL_2317173,
EPI_ISL_2317174, EPI_ISL_2317176, EPI_ISL_2317178, EPI_ISL_2317179, EPI_ISL_2317180, EPI_ISL_2317181, EPI_ISL_2317185, EPI_ISL_2317191, EPI_ISL_2317195, EPI_ISL_2317198, EPI_ISL_2317203, EPI_ISL_2317206, EPI_ISL_2317213, EPI_ISL_2317214, EPI_ISL_2317219, EPI_ISL_2317224, EPI_ISL_2317227, EPI_ISL_2317232, EPI_ISL_2317233, EPI_ISL_2317235, EPI_ISL_2317237, EPI_ISL_2317238,
EPI_ISL_2317240, EPI_ISL_2317244, EPI_ISL_2317246, EPI_ISL_2317247, EPI_ISL_2317250, EPI_ISL_2317251, EPI_ISL_2317256, EPI_ISL_2317263, EPI_ISL_2317265, EPI_ISL_2317267, EPI_ISL_2317270, EPI_ISL_2317285, EPI_ISL_2317287, EPI_ISL_2317289, EPI_ISL_2317292, EPI_ISL_2317294, EPI_ISL_2317295, EPI_ISL_2317300, EPI_ISL_2317304, EPI_ISL_2317305, EPI_ISL_2317307, EPI_ISL_2317310,
EPI_ISL_2317312, EPI_ISL_2317321, EPI_ISL_2317322, EPI_ISL_2317328, EPI_ISL_2317331, EPI_ISL_2317332, EPI_ISL_2317338, EPI_ISL_2317339, EPI_ISL_2317345, EPI_ISL_2317346, EPI_ISL_2317350, EPI_ISL_2317354, EPI_ISL_2317355, EPI_ISL_2317358, EPI_ISL_2317359, EPI_ISL_2317368, EPI_ISL_2317370, EPI_ISL_2317371, EPI_ISL_2317376, EPI_ISL_2317381, EPI_ISL_2317382, EPI_ISL_2317383,
EPI_ISL_2317384, EPI_ISL_2317385, EPI_ISL_2317386, EPI_ISL_2317390, EPI_ISL_2317393, EPI_ISL_2317394, EPI_ISL_2317398, EPI_ISL_2317399, EPI_ISL_2317404, EPI_ISL_2317408, EPI_ISL_2317409, EPI_ISL_2317410, EPI_ISL_2317419, EPI_ISL_2317426, EPI_ISL_2317428, EPI_ISL_2317430, EPI_ISL_2317431, EPI_ISL_2317432, EPI_ISL_2317442, EPI_ISL_2317444, EPI_ISL_2317451, EPI_ISL_2317455,
EPI_ISL_2317457, EPI_ISL_2317473, EPI_ISL_2317483, EPI_ISL_2317486, EPI_ISL_2317488, EPI_ISL_2317491, EPI_ISL_2317493, EPI_ISL_2317494, EPI_ISL_2317495, EPI_ISL_2317507, EPI_ISL_2317511, EPI_ISL_2317519, EPI_ISL_2317524, EPI_ISL_2317526, EPI_ISL_2317530, EPI_ISL_2317534, EPI_ISL_2317537, EPI_ISL_2317538, EPI_ISL_2317539, EPI_ISL_2317540, EPI_ISL_2317547, EPI_ISL_2317556,
EPI_ISL_2317561, EPI_ISL_2317563, EPI_ISL_2317574, EPI_ISL_2317575, EPI_ISL_2317576, EPI_ISL_2317581, EPI_ISL_2317585, EPI_ISL_2317591, EPI_ISL_2317592, EPI_ISL_2317595, EPI_ISL_2317598, EPI_ISL_2317601, EPI_ISL_2317603, EPI_ISL_2317614, EPI_ISL_2317618, EPI_ISL_2317620, EPI_ISL_2317622, EPI_ISL_2317623, EPI_ISL_2317624, EPI_ISL_2317625, EPI_ISL_2317629, EPI_ISL_2317630,
EPI_ISL_2317637, EPI_ISL_2317646, EPI_ISL_2317647, EPI_ISL_2317649, EPI_ISL_2317654, EPI_ISL_2317655, EPI_ISL_2317656, EPI_ISL_2317661, EPI_ISL_2317662, EPI_ISL_2317663, EPI_ISL_2317669, EPI_ISL_2317670, EPI_ISL_2317672, EPI_ISL_2317674, EPI_ISL_2317676, EPI_ISL_2317677, EPI_ISL_2317678, EPI_ISL_2317681, EPI_ISL_2317690, EPI_ISL_2317701, EPI_ISL_2317707, EPI_ISL_2317712,
EPI_ISL_2317714, EPI_ISL_2317725, EPI_ISL_2317727, EPI_ISL_2317730, EPI_ISL_2317731, EPI_ISL_2317733, EPI_ISL_2317735, EPI_ISL_2317738, EPI_ISL_2317743, EPI_ISL_2317744, EPI_ISL_2317749, EPI_ISL_2317753, EPI_ISL_2317754, EPI_ISL_2317759, EPI_ISL_2317760, EPI_ISL_2317762, EPI_ISL_2317766, EPI_ISL_2317772, EPI_ISL_2317775, EPI_ISL_2317778, EPI_ISL_2317782, EPI_ISL_2317785,
EPI_ISL_2317788, EPI_ISL_2317789, EPI_ISL_2317790, EPI_ISL_2317791, EPI_ISL_2317792, EPI_ISL_2317795, EPI_ISL_2317796, EPI_ISL_2317799, EPI_ISL_2317800, EPI_ISL_2317804, EPI_ISL_2317806, EPI_ISL_2317807, EPI_ISL_2317813, EPI_ISL_2317814, EPI_ISL_2317816, EPI_ISL_2317817, EPI_ISL_2317819, EPI_ISL_2317823, EPI_ISL_2317832, EPI_ISL_2317833, EPI_ISL_2317835, EPI_ISL_2317836,
EPI_ISL_2317839, EPI_ISL_2317840, EPI_ISL_2317842, EPI_ISL_2317843, EPI_ISL_2317853, EPI_ISL_2317854, EPI_ISL_2317856, EPI_ISL_2317857, EPI_ISL_2317859, EPI_ISL_2317860, EPI_ISL_2317867, EPI_ISL_2317868, EPI_ISL_2317872, EPI_ISL_2317874, EPI_ISL_2317878, EPI_ISL_2317880, EPI_ISL_2317884, EPI_ISL_2317885, EPI_ISL_2317888,
EPI_ISL_2317891, EPI_ISL_2317893, EPI_ISL_2317894, EPI_ISL_2317896, EPI_ISL_2317897, EPI_ISL_2317898, EPI_ISL_2317900, EPI_ISL_2317901, EPI_ISL_2317903, EPI_ISL_2317905, EPI_ISL_2317908, EPI_ISL_2317910, EPI_ISL_2317912, EPI_ISL_2317913, EPI_ISL_2317914, EPI_ISL_2317917, EPI_ISL_2317918, EPI_ISL_2317921, EPI_ISL_2317924, EPI_ISL_2317925, EPI_ISL_2317926, EPI_ISL_2317931,
EPI_ISL_2317933, EPI_ISL_2317934, EPI_ISL_2317936, EPI_ISL_2317939, EPI_ISL_2317941, EPI_ISL_2317944, EPI_ISL_2317946, EPI_ISL_2317948, EPI_ISL_2317951, EPI_ISL_2317956, EPI_ISL_2317957, EPI_ISL_2317958, EPI_ISL_2317963, EPI_ISL_2317964, EPI_ISL_2317967, EPI_ISL_2317968, EPI_ISL_2317970, EPI_ISL_2317974, EPI_ISL_2317977, EPI_ISL_2317987, EPI_ISL_2317988, EPI_ISL_2317990,
EPI_ISL_2317991, EPI_ISL_2317992, EPI_ISL_2317993, EPI_ISL_2317996, EPI_ISL_2317999, EPI_ISL_2318001, EPI_ISL_2318002, EPI_ISL_2318005, EPI_ISL_2318007, EPI_ISL_2318008, EPI_ISL_2318010, EPI_ISL_2318014, EPI_ISL_2318018, EPI_ISL_2318019, EPI_ISL_2318025, EPI_ISL_2318035, EPI_ISL_2318038, EPI_ISL_2318045, EPI_ISL_2318046, EPI_ISL_2318047, EPI_ISL_2318048, EPI_ISL_2318053,
EPI_ISL_2318056, EPI_ISL_2318061, EPI_ISL_2318084, EPI_ISL_2318086, EPI_ISL_2318087, EPI_ISL_2318091, EPI_ISL_2318092, EPI_ISL_2318100, EPI_ISL_2318113, EPI_ISL_2318115, EPI_ISL_2318119, EPI_ISL_2318125, EPI_ISL_2318126, EPI_ISL_2318129, EPI_ISL_2318134, EPI_ISL_2318140, EPI_ISL_2318142, EPI_ISL_2318148, EPI_ISL_2318159, EPI_ISL_2318171,
EPI_ISL_2318179, EPI_ISL_2318182, EPI_ISL_2318185, EPI_ISL_2318186, EPI_ISL_2318192, EPI_ISL_2318193, EPI_ISL_2318195, EPI_ISL_2318197, EPI_ISL_2318202, EPI_ISL_2318205, EPI_ISL_2318211, EPI_ISL_2318214, EPI_ISL_2318215, EPI_ISL_2318217, EPI_ISL_2318225, EPI_ISL_2318228, EPI_ISL_2318229, EPI_ISL_2318233, EPI_ISL_2318235, EPI_ISL_2318242, EPI_ISL_2318246, EPI_ISL_2318252,
EPI_ISL_2318253, EPI_ISL_2318254, EPI_ISL_2318259, EPI_ISL_2318267, EPI_ISL_2318271, EPI_ISL_2318275, EPI_ISL_2318277, EPI_ISL_2318289, EPI_ISL_2318293, EPI_ISL_2318298, EPI_ISL_2318300, EPI_ISL_2318302, EPI_ISL_2318306, EPI_ISL_2318311, EPI_ISL_2318322, EPI_ISL_2318329, EPI_ISL_2318330, EPI_ISL_2318333, EPI_ISL_2318339, EPI_ISL_2318349, EPI_ISL_2318351, EPI_ISL_2318352,
EPI_ISL_2318353, EPI_ISL_2318360, EPI_ISL_2318368, EPI_ISL_2318370, EPI_ISL_2318371, EPI_ISL_2318372, EPI_ISL_2318374, EPI_ISL_2318378, EPI_ISL_2318379, EPI_ISL_2318385, EPI_ISL_2318393, EPI_ISL_2318398, EPI_ISL_2318402, EPI_ISL_2318409, EPI_ISL_2318414, EPI_ISL_2318423, EPI_ISL_2318426, EPI_ISL_2318429, EPI_ISL_2318433, EPI_ISL_2318438, EPI_ISL_2318439, EPI_ISL_2318444,
EPI_ISL_2318446, EPI_ISL_2318454, EPI_ISL_2318456, EPI_ISL_2318457, EPI_ISL_2318462, EPI_ISL_2318465, EPI_ISL_2318467, EPI_ISL_2318477, EPI_ISL_2318482, EPI_ISL_2318484, EPI_ISL_2318490, EPI_ISL_2318491, EPI_ISL_2318492, EPI_ISL_2318493, EPI_ISL_2318506, EPI_ISL_2318511, EPI_ISL_2318512, EPI_ISL_2318514, EPI_ISL_2318516, EPI_ISL_2318530, EPI_ISL_2318545, EPI_ISL_2318547,
EPI_ISL_2318550, EPI_ISL_2318551, EPI_ISL_2318553, EPI_ISL_2318560, EPI_ISL_2318562, EPI_ISL_2318564, EPI_ISL_2318569, EPI_ISL_2318578, EPI_ISL_2318589, EPI_ISL_2318590, EPI_ISL_2318593, EPI_ISL_2318601, EPI_ISL_2318605, EPI_ISL_2318611, EPI_ISL_2318614, EPI_ISL_2318618, EPI_ISL_2318621, EPI_ISL_2318627, EPI_ISL_2318630, EPI_ISL_2318642, EPI_ISL_2318647,
EPI_ISL_2318648, EPI_ISL_2318653, EPI_ISL_2318663, EPI_ISL_2318668, EPI_ISL_2318674, EPI_ISL_2318676, EPI_ISL_2318679, EPI_ISL_2318681, EPI_ISL_2318683, EPI_ISL_2318690, EPI_ISL_2318693, EPI_ISL_2318697, EPI_ISL_2318707, EPI_ISL_2318709, EPI_ISL_2318711, EPI_ISL_2346738, EPI_ISL_2346776, EPI_ISL_2346800, EPI_ISL_2346839, EPI_ISL_2346843, EPI_ISL_2346853, EPI_ISL_2346855,
EPI_ISL_2346877, EPI_ISL_2346951, EPI_ISL_2346975, EPI_ISL_2346977, EPI_ISL_2347020, EPI_ISL_2347063, EPI_ISL_2347072, EPI_ISL_2347085, EPI_ISL_2347551, EPI_ISL_2347635, EPI_ISL_2347639, EPI_ISL_2347651, EPI_ISL_2347676, EPI_ISL_2347681, EPI_ISL_2347745, EPI_ISL_2347779, EPI_ISL_2347805, EPI_ISL_2347828, EPI_ISL_2347840, EPI_ISL_2348186, EPI_ISL_2348189, EPI_ISL_2348203,
EPI_ISL_2348233, EPI_ISL_2348307, EPI_ISL_2348422, EPI_ISL_2348423, EPI_ISL_2348478, EPI_ISL_2348479, EPI_ISL_2396016, EPI_ISL_2396017, EPI_ISL_2396019, EPI_ISL_2396020, EPI_ISL_2396021, EPI_ISL_2396022, EPI_ISL_2396025, EPI_ISL_2396027, EPI_ISL_2396028, EPI_ISL_2396030, EPI_ISL_2396031, EPI_ISL_2396032, EPI_ISL_2396033, EPI_ISL_2396034, EPI_ISL_2396036, EPI_ISL_2396037,
EPI_ISL_2396039, EPI_ISL_2396040, EPI_ISL_2396041, EPI_ISL_2396043, EPI_ISL_2396044, EPI_ISL_2396045, EPI_ISL_2396046, EPI_ISL_2396047, EPI_ISL_2396048, EPI_ISL_2396049, EPI_ISL_2396050, EPI_ISL_2396052, EPI_ISL_2396055, EPI_ISL_2396056, EPI_ISL_2396057, EPI_ISL_2396058, EPI_ISL_2396059, EPI_ISL_2396060, EPI_ISL_2396061, EPI_ISL_2396065, EPI_ISL_2396066,
EPI_ISL_2396067, EPI_ISL_2396070, EPI_ISL_2396071, EPI_ISL_2396072, EPI_ISL_2396073, EPI_ISL_2396076, EPI_ISL_2396077, EPI_ISL_2396078, EPI_ISL_2396079, EPI_ISL_2396080, EPI_ISL_2396081, EPI_ISL_2396082, EPI_ISL_2396083, EPI_ISL_2396084, EPI_ISL_2396085, EPI_ISL_2396086, EPI_ISL_2396087, EPI_ISL_2396088, EPI_ISL_2396090, EPI_ISL_2396092, EPI_ISL_2396093, EPI_ISL_2396094,
EPI_ISL_2396096, EPI_ISL_2396097, EPI_ISL_2396098, EPI_ISL_2396099, EPI_ISL_2396100, EPI_ISL_2396101, EPI_ISL_2396102, EPI_ISL_2396103, EPI_ISL_2396104, EPI_ISL_2396105, EPI_ISL_2396106, EPI_ISL_2396107, EPI_ISL_2396108, EPI_ISL_2396109, EPI_ISL_2396110, EPI_ISL_2396112, EPI_ISL_2396114, EPI_ISL_2396115, EPI_ISL_2396116, EPI_ISL_2396117, EPI_ISL_2396118, EPI_ISL_2396119,
EPI_ISL_2396120, EPI_ISL_2396121, EPI_ISL_2396123, EPI_ISL_2396124, EPI_ISL_2396125, EPI_ISL_2396126, EPI_ISL_2396127, EPI_ISL_2396128, EPI_ISL_2396129, EPI_ISL_2396130, EPI_ISL_2396131, EPI_ISL_2396132, EPI_ISL_2396133, EPI_ISL_2396134, EPI_ISL_2396137, EPI_ISL_2396140, EPI_ISL_2396141, EPI_ISL_2396143, EPI_ISL_2396144, EPI_ISL_2396145, EPI_ISL_2396146, EPI_ISL_2396149,
EPI_ISL_2396150, EPI_ISL_2396151, EPI_ISL_2396152, EPI_ISL_2396153, EPI_ISL_2396154, EPI_ISL_2396155, EPI_ISL_2396156, EPI_ISL_2396157, EPI_ISL_2396158, EPI_ISL_2396159, EPI_ISL_2396161, EPI_ISL_2396162, EPI_ISL_2396163, EPI_ISL_2396165, EPI_ISL_2396167, EPI_ISL_2396168, EPI_ISL_2396170, EPI_ISL_2396171, EPI_ISL_2396173, EPI_ISL_2396174, EPI_ISL_2396176, EPI_ISL_2396177,
EPI_ISL_2396179, EPI_ISL_2396180, EPI_ISL_2396181, EPI_ISL_2396182, EPI_ISL_2396183, EPI_ISL_2396184, EPI_ISL_2396185, EPI_ISL_2396186, EPI_ISL_2396188, EPI_ISL_2396189, EPI_ISL_2396190, EPI_ISL_2396191, EPI_ISL_2396192, EPI_ISL_2396195, EPI_ISL_2396196, EPI_ISL_2396197, EPI_ISL_2396201, EPI_ISL_2396202, EPI_ISL_2396203, EPI_ISL_2396204, EPI_ISL_2396206, EPI_ISL_2396207,
EPI_ISL_2396208, EPI_ISL_2396209, EPI_ISL_2396210, EPI_ISL_2396211, EPI_ISL_2396212, EPI_ISL_2396214, EPI_ISL_2396216, EPI_ISL_2396217, EPI_ISL_2396218, EPI_ISL_2396219, EPI_ISL_2396220, EPI_ISL_2396221, EPI_ISL_2396223, EPI_ISL_2396224, EPI_ISL_2396226, EPI_ISL_2396230, EPI_ISL_2396232, EPI_ISL_2396233, EPI_ISL_2396234, EPI_ISL_2396237, EPI_ISL_2396238, EPI_ISL_2396239,
EPI_ISL_2396240, EPI_ISL_2396241, EPI_ISL_2396242, EPI_ISL_2396243, EPI_ISL_2396244, EPI_ISL_2396245, EPI_ISL_2396247, EPI_ISL_2396248, EPI_ISL_2396249, EPI_ISL_2396251, EPI_ISL_2396252, EPI_ISL_2396254, EPI_ISL_2396255, EPI_ISL_2396256, EPI_ISL_2396257, EPI_ISL_2396258, EPI_ISL_2396259, EPI_ISL_2396261, EPI_ISL_2396262, EPI_ISL_2396263, EPI_ISL_2396264, EPI_ISL_2396266,
EPI_ISL_2396267, EPI_ISL_2396268, EPI_ISL_2396269, EPI_ISL_2396270, EPI_ISL_2396271, EPI_ISL_2396274, EPI_ISL_2396275, EPI_ISL_2396276, EPI_ISL_2396277, EPI_ISL_2396279, EPI_ISL_2396280, EPI_ISL_2396284, EPI_ISL_2396285, EPI_ISL_2396286, EPI_ISL_2396287, EPI_ISL_2396288, EPI_ISL_2396290, EPI_ISL_2396291, EPI_ISL_2396292, EPI_ISL_2396293, EPI_ISL_2396294, EPI_ISL_2396295,
EPI_ISL_2396297, EPI_ISL_2396299, EPI_ISL_2396300, EPI_ISL_2396301, EPI_ISL_2396302, EPI_ISL_2396303, EPI_ISL_2396304, EPI_ISL_2396305, EPI_ISL_2396307, EPI_ISL_2396309, EPI_ISL_2396310, EPI_ISL_2396311, EPI_ISL_2396313, EPI_ISL_2396314, EPI_ISL_2396315, EPI_ISL_2396316, EPI_ISL_2396317, EPI_ISL_2396318, EPI_ISL_2396320, EPI_ISL_2396321, EPI_ISL_2396322, EPI_ISL_2396326,
EPI_ISL_2396328, EPI_ISL_2396329, EPI_ISL_2396330, EPI_ISL_2396331, EPI_ISL_2396332, EPI_ISL_2396333, EPI_ISL_2396335, EPI_ISL_2396337, EPI_ISL_2396338, EPI_ISL_2396339, EPI_ISL_2396340, EPI_ISL_2396341, EPI_ISL_2396342, EPI_ISL_2396343, EPI_ISL_2704268, EPI_ISL_2767582

| | | | |
|---|---|---|---|
| see above | Lighthouse Lab in Alderley Park | Wellcome Sanger Institute for the COVID-19 Genomics UK (COG-UK) Consortium | Cordelia Langford; David K. Jackson; Dominic Kwiatkowski; Ewan Harrison; Ian Johnston; Jacquelyn Wynn; Jeffrey Barrett; John Sillitoe on behalf of the Wellcome Sanger Institute COVID-19 Surveillance Team; Mairead Hyland; Roberto Amato; Sonia Goncalves; The Lighthouse Lab in Alderley Park and Alex Alderton |

EPI_ISL_2117679, EPI_ISL_2138033, EPI_ISL_2138050, EPI_ISL_2152867, EPI_ISL_2152868, EPI_ISL_2152869, EPI_ISL_2152870, EPI_ISL_2152871, EPI_ISL_2152872, EPI_ISL_2152873, EPI_ISL_2152874, EPI_ISL_2152875, EPI_ISL_2152876, EPI_ISL_2152877, EPI_ISL_2152878, EPI_ISL_2152879, EPI_ISL_2152880, EPI_ISL_2152881, EPI_ISL_2152882, EPI_ISL_2152883, EPI_ISL_2152884, EPI_ISL_2152885,
EPI_ISL_2152886, EPI_ISL_2152887, EPI_ISL_2152888, EPI_ISL_2152890, EPI_ISL_2152891, EPI_ISL_2152892, EPI_ISL_2152893, EPI_ISL_2152895, EPI_ISL_2152897, EPI_ISL_2152898, EPI_ISL_2152899, EPI_ISL_2152900, EPI_ISL_2152901, EPI_ISL_2152902, EPI_ISL_2152903, EPI_ISL_2152904, EPI_ISL_2152906, EPI_ISL_2152907, EPI_ISL_2152908, EPI_ISL_2152909, EPI_ISL_2152910,
EPI_ISL_2152911, EPI_ISL_2152912, EPI_ISL_2152913, EPI_ISL_2152914, EPI_ISL_2152915, EPI_ISL_2152916, EPI_ISL_2152917, EPI_ISL_2152918, EPI_ISL_2152919, EPI_ISL_2152920, EPI_ISL_2152921, EPI_ISL_2152922, EPI_ISL_2152924, EPI_ISL_2152925, EPI_ISL_2152926, EPI_ISL_2152927, EPI_ISL_2152928, EPI_ISL_2152929, EPI_ISL_2152930, EPI_ISL_2152931, EPI_ISL_2152932, EPI_ISL_2152933,
EPI_ISL_2152934, EPI_ISL_2152935, EPI_ISL_2152936, EPI_ISL_2152937, EPI_ISL_2152938, EPI_ISL_2152939, EPI_ISL_2152940, EPI_ISL_2152941, EPI_ISL_2152942, EPI_ISL_2152943, EPI_ISL_2152944, EPI_ISL_2152945, EPI_ISL_2152946, EPI_ISL_2152947, EPI_ISL_2152949, EPI_ISL_2152951, EPI_ISL_2152952, EPI_ISL_2152953, EPI_ISL_2152954, EPI_ISL_2152955, EPI_ISL_2152956,
EPI_ISL_2152957, EPI_ISL_2152959, EPI_ISL_2152961, EPI_ISL_2152963, EPI_ISL_2152966, EPI_ISL_2152968, EPI_ISL_2152969, EPI_ISL_2152973, EPI_ISL_2152974, EPI_ISL_2152975, EPI_ISL_2152978, EPI_ISL_2152980, EPI_ISL_2152981, EPI_ISL_2152982, EPI_ISL_2152983, EPI_ISL_2152985, EPI_ISL_2152990, EPI_ISL_2152992, EPI_ISL_2152997, EPI_ISL_2152999, EPI_ISL_2153000, EPI_ISL_2153002,
EPI_ISL_2153006, EPI_ISL_2153015, EPI_ISL_2153018, EPI_ISL_2153021, EPI_ISL_2153023, EPI_ISL_2153025, EPI_ISL_2153027, EPI_ISL_2153028, EPI_ISL_2153031, EPI_ISL_2153032, EPI_ISL_2171681, EPI_ISL_2171706, EPI_ISL_2171720, EPI_ISL_2171748, EPI_ISL_2171762, EPI_ISL_2171764, EPI_ISL_2171774, EPI_ISL_2171804, EPI_ISL_2171821, EPI_ISL_2171822, EPI_ISL_2171827,
EPI_ISL_2171837, EPI_ISL_2171840, EPI_ISL_2171841, EPI_ISL_2171850, EPI_ISL_2171853, EPI_ISL_2171854, EPI_ISL_2171861, EPI_ISL_2171867, EPI_ISL_2171871, EPI_ISL_2171872, EPI_ISL_2171873, EPI_ISL_2171875, EPI_ISL_2171876, EPI_ISL_2171881, EPI_ISL_2171883, EPI_ISL_2171885, EPI_ISL_2171886, EPI_ISL_2171889, EPI_ISL_2171890, EPI_ISL_2171893, EPI_ISL_2171906, EPI_ISL_2171909,
EPI_ISL_2171911, EPI_ISL_2171912, EPI_ISL_2171914, EPI_ISL_2171916, EPI_ISL_2171918, EPI_ISL_2171920, EPI_ISL_2171928, EPI_ISL_2171936, EPI_ISL_2171937, EPI_ISL_2171939, EPI_ISL_2171940, EPI_ISL_2171951, EPI_ISL_2171957, EPI_ISL_2171959, EPI_ISL_2171961, EPI_ISL_2171963, EPI_ISL_2171966, EPI_ISL_2171970, EPI_ISL_2171971, EPI_ISL_2171976, EPI_ISL_2171978, EPI_ISL_2171982,
EPI_ISL_2171994, EPI_ISL_2171995, EPI_ISL_2172001, EPI_ISL_2172006, EPI_ISL_2172008, EPI_ISL_2172009, EPI_ISL_2172011, EPI_ISL_2172012, EPI_ISL_2172015, EPI_ISL_2172027, EPI_ISL_2172035, EPI_ISL_2172037, EPI_ISL_2172038, EPI_ISL_2172045, EPI_ISL_2172048, EPI_ISL_2172056, EPI_ISL_2172060, EPI_ISL_2172061, EPI_ISL_2172064, EPI_ISL_2172069, EPI_ISL_2172070, EPI_ISL_2172071,
EPI_ISL_2172074, EPI_ISL_2172076, EPI_ISL_2172078, EPI_ISL_2172085, EPI_ISL_2172088, EPI_ISL_2172089, EPI_ISL_2198671, EPI_ISL_2198708, EPI_ISL_2198735, EPI_ISL_2198738, EPI_ISL_2198753, EPI_ISL_2198755, EPI_ISL_2198789, EPI_ISL_2198804, EPI_ISL_2198807, EPI_ISL_2198817, EPI_ISL_2198855, EPI_ISL_2198880, EPI_ISL_2198947, EPI_ISL_2198966, EPI_ISL_2198998, EPI_ISL_2198999,
EPI_ISL_2199000, EPI_ISL_2199004, EPI_ISL_2199005, EPI_ISL_2199007, EPI_ISL_2199008, EPI_ISL_2199009, EPI_ISL_2199010, EPI_ISL_2199011, EPI_ISL_2199014, EPI_ISL_2199015, EPI_ISL_2199016, EPI_ISL_2199017, EPI_ISL_2199019, EPI_ISL_2199022, EPI_ISL_2199023, EPI_ISL_2199026, EPI_ISL_2199029, EPI_ISL_2199030, EPI_ISL_2199031, EPI_ISL_2199033, EPI_ISL_2199038, EPI_ISL_2199040,
EPI_ISL_2199041, EPI_ISL_2199042, EPI_ISL_2199043, EPI_ISL_2199044, EPI_ISL_2199046, EPI_ISL_2199047, EPI_ISL_2199050, EPI_ISL_2199052, EPI_ISL_2199054, EPI_ISL_2199058, EPI_ISL_2199059, EPI_ISL_2199062, EPI_ISL_2199063, EPI_ISL_2199065, EPI_ISL_2199067, EPI_ISL_2199070, EPI_ISL_2199073, EPI_ISL_2199074, EPI_ISL_2199076, EPI_ISL_2199078, EPI_ISL_2199080, EPI_ISL_2199081,
EPI_ISL_2199082, EPI_ISL_2199087, EPI_ISL_2199089, EPI_ISL_2199091, EPI_ISL_2199092, EPI_ISL_2199094, EPI_ISL_2199095, EPI_ISL_2199096, EPI_ISL_2199099, EPI_ISL_2199104, EPI_ISL_2199105, EPI_ISL_2199106, EPI_ISL_2199107, EPI_ISL_2199109, EPI_ISL_2199115, EPI_ISL_2199116, EPI_ISL_2199117, EPI_ISL_2199118, EPI_ISL_2199120, EPI_ISL_2199125, EPI_ISL_2199127, EPI_ISL_2199129,
EPI_ISL_2199132, EPI_ISL_2199133, EPI_ISL_2199137, EPI_ISL_2199138, EPI_ISL_2199139, EPI_ISL_2199141, EPI_ISL_2199142, EPI_ISL_2199143, EPI_ISL_2199145, EPI_ISL_2199148, EPI_ISL_2199150, EPI_ISL_2199151, EPI_ISL_2199153, EPI_ISL_2199154, EPI_ISL_2199155, EPI_ISL_2199157, EPI_ISL_2199159, EPI_ISL_2199162, EPI_ISL_2199163, EPI_ISL_2199165, EPI_ISL_2199168, EPI_ISL_2199169,
EPI_ISL_2199170, EPI_ISL_2199171, EPI_ISL_2199172, EPI_ISL_2199174, EPI_ISL_2199175, EPI_ISL_2199178, EPI_ISL_2199181, EPI_ISL_2199183, EPI_ISL_2199185, EPI_ISL_2199186, EPI_ISL_2199187, EPI_ISL_2199189, EPI_ISL_2199193, EPI_ISL_2199194, EPI_ISL_2199197, EPI_ISL_2199199, EPI_ISL_2199200, EPI_ISL_2199203, EPI_ISL_2199204, EPI_ISL_2199208, EPI_ISL_2199210, EPI_ISL_2199212,
EPI_ISL_2199213, EPI_ISL_2199217, EPI_ISL_2199220, EPI_ISL_2199221, EPI_ISL_2199222, EPI_ISL_2199223, EPI_ISL_2199225, EPI_ISL_2199227, EPI_ISL_2199229, EPI_ISL_2199230, EPI_ISL_2199231, EPI_ISL_2199234, EPI_ISL_2199237, EPI_ISL_2199238, EPI_ISL_2199244, EPI_ISL_2199245, EPI_ISL_2199246, EPI_ISL_2199248, EPI_ISL_2199249, EPI_ISL_2199252, EPI_ISL_2199255,
EPI_ISL_2199256, EPI_ISL_2199259, EPI_ISL_2199261, EPI_ISL_2199262, EPI_ISL_2199266, EPI_ISL_2199267, EPI_ISL_2199272, EPI_ISL_2199273, EPI_ISL_2199274, EPI_ISL_2199275, EPI_ISL_2199276, EPI_ISL_2199277, EPI_ISL_2199278, EPI_ISL_2199279, EPI_ISL_2199282, EPI_ISL_2199287, EPI_ISL_2199288, EPI_ISL_2199290, EPI_ISL_2199293, EPI_ISL_2199294, EPI_ISL_2199296, EPI_ISL_2199298,
EPI_ISL_2199299, EPI_ISL_2199300, EPI_ISL_2199304, EPI_ISL_2199305, EPI_ISL_2199310, EPI_ISL_2199312, EPI_ISL_2199315, EPI_ISL_2199316, EPI_ISL_2199319, EPI_ISL_2199326, EPI_ISL_2199327, EPI_ISL_2199328, EPI_ISL_2199331, EPI_ISL_2199334, EPI_ISL_2199335, EPI_ISL_2199336, EPI_ISL_2199337, EPI_ISL_2199338, EPI_ISL_2199339, EPI_ISL_2199341, EPI_ISL_2199685,
EPI_ISL_2199686, EPI_ISL_2199687, EPI_ISL_2199689, EPI_ISL_2199690, EPI_ISL_2199692, EPI_ISL_2199693, EPI_ISL_2199695, EPI_ISL_2199696, EPI_ISL_2199698, EPI_ISL_2199699, EPI_ISL_2199700, EPI_ISL_2199702, EPI_ISL_2199703, EPI_ISL_2199706, EPI_ISL_2199707, EPI_ISL_2199708, EPI_ISL_2199709, EPI_ISL_2199710, EPI_ISL_2199711, EPI_ISL_2199712, EPI_ISL_2199713, EPI_ISL_2199714,
EPI_ISL_2199715, EPI_ISL_2199716, EPI_ISL_2199718, EPI_ISL_2199720, EPI_ISL_2199721, EPI_ISL_2199722, EPI_ISL_2199724, EPI_ISL_2199725, EPI_ISL_2199726, EPI_ISL_2199728, EPI_ISL_2199729, EPI_ISL_2199730, EPI_ISL_2199734, EPI_ISL_2199735, EPI_ISL_2199736, EPI_ISL_2199737, EPI_ISL_2199738, EPI_ISL_2199739, EPI_ISL_2199741, EPI_ISL_2199742, EPI_ISL_2199744, EPI_ISL_2199746,
EPI_ISL_2199747, EPI_ISL_2199748, EPI_ISL_2199750, EPI_ISL_2199751, EPI_ISL_2199752, EPI_ISL_2199753, EPI_ISL_2199754, EPI_ISL_2199755, EPI_ISL_2199758, EPI_ISL_2199759, EPI_ISL_2199760, EPI_ISL_2199761, EPI_ISL_2199762, EPI_ISL_2199764, EPI_ISL_2199765, EPI_ISL_2199766, EPI_ISL_2199767, EPI_ISL_2199768, EPI_ISL_2199769, EPI_ISL_2199771, EPI_ISL_2199772, EPI_ISL_2199773,
EPI_ISL_2199775, EPI_ISL_2199777, EPI_ISL_2199778, EPI_ISL_2199781, EPI_ISL_2199782, EPI_ISL_2199786, EPI_ISL_2199787, EPI_ISL_2199788, EPI_ISL_2199789, EPI_ISL_2199790, EPI_ISL_2199791, EPI_ISL_2199792, EPI_ISL_2199793, EPI_ISL_2199795, EPI_ISL_2199796, EPI_ISL_2199798, EPI_ISL_2199799, EPI_ISL_2199801, EPI_ISL_2199802, EPI_ISL_2199803, EPI_ISL_2199804,
EPI_ISL_2199805, EPI_ISL_2199806, EPI_ISL_2199807, EPI_ISL_2199808, EPI_ISL_2199809, EPI_ISL_2199810, EPI_ISL_2199811, EPI_ISL_2199812, EPI_ISL_2199813, EPI_ISL_2199814, EPI_ISL_2199815, EPI_ISL_2199816, EPI_ISL_2199817, EPI_ISL_2199818, EPI_ISL_2199819, EPI_ISL_2199820, EPI_ISL_2199821, EPI_ISL_2199822, EPI_ISL_2199824, EPI_ISL_2199825, EPI_ISL_2199826, EPI_ISL_2199827,
EPI_ISL_2199828, EPI_ISL_2199829, EPI_ISL_2199830, EPI_ISL_2199831, EPI_ISL_2199833, EPI_ISL_2199834, EPI_ISL_2199835, EPI_ISL_2199837, EPI_ISL_2199838, EPI_ISL_2199839, EPI_ISL_2199841, EPI_ISL_2199843, EPI_ISL_2199844, EPI_ISL_2199845, EPI_ISL_2199846, EPI_ISL_2199847, EPI_ISL_2199848, EPI_ISL_2199849, EPI_ISL_2199850, EPI_ISL_2199851, EPI_ISL_2199852, EPI_ISL_2199854,
EPI_ISL_2199855, EPI_ISL_2199856, EPI_ISL_2199857, EPI_ISL_2199859, EPI_ISL_2199860, EPI_ISL_2199861, EPI_ISL_2199863, EPI_ISL_2199864, EPI_ISL_2199865, EPI_ISL_2199868, EPI_ISL_2199871, EPI_ISL_2199873, EPI_ISL_2199874, EPI_ISL_2199875, EPI_ISL_2199876, EPI_ISL_2199878, EPI_ISL_2199879, EPI_ISL_2199880, EPI_ISL_2199881, EPI_ISL_2199882, EPI_ISL_2199883, EPI_ISL_2199884,
EPI_ISL_2199885, EPI_ISL_2199886, EPI_ISL_2199888, EPI_ISL_2199889, EPI_ISL_2199890, EPI_ISL_2199891, EPI_ISL_2199892, EPI_ISL_2199893, EPI_ISL_2199894, EPI_ISL_2199895, EPI_ISL_2199896, EPI_ISL_2199897, EPI_ISL_2199898, EPI_ISL_2199899, EPI_ISL_2199900, EPI_ISL_2199901, EPI_ISL_2199902, EPI_ISL_2199903, EPI_ISL_2199904, EPI_ISL_2199905, EPI_ISL_2199906, EPI_ISL_2199907,
EPI_ISL_2199908, EPI_ISL_2199909, EPI_ISL_2199910, EPI_ISL_2199911, EPI_ISL_2199912, EPI_ISL_2199913, EPI_ISL_2199914, EPI_ISL_2199916, EPI_ISL_2199917, EPI_ISL_2199918, EPI_ISL_2199919, EPI_ISL_2199920, EPI_ISL_2199921, EPI_ISL_2199922, EPI_ISL_2199923, EPI_ISL_2199924, EPI_ISL_2199925, EPI_ISL_2199926, EPI_ISL_2199927, EPI_ISL_2199928, EPI_ISL_2199929, EPI_ISL_2199930,
EPI_ISL_2199931, EPI_ISL_2199932, EPI_ISL_2199933, EPI_ISL_2199934, EPI_ISL_2199935, EPI_ISL_2199936, EPI_ISL_2199937, EPI_ISL_2199938, EPI_ISL_2199939, EPI_ISL_2199940, EPI_ISL_2199941, EPI_ISL_2199942, EPI_ISL_2199943, EPI_ISL_2199944, EPI_ISL_2199946, EPI_ISL_2199947, EPI_ISL_2199948, EPI_ISL_2199949, EPI_ISL_2199950, EPI_ISL_2199953, EPI_ISL_2199954,
EPI_ISL_2199955, EPI_ISL_2199956, EPI_ISL_2199957, EPI_ISL_2199958, EPI_ISL_2199959, EPI_ISL_2199960, EPI_ISL_2199961, EPI_ISL_2199962, EPI_ISL_2199963, EPI_ISL_2199964, EPI_ISL_2199965, EPI_ISL_2199966, EPI_ISL_2199967, EPI_ISL_2199968, EPI_ISL_2199969, EPI_ISL_2199970, EPI_ISL_2199971, EPI_ISL_2199972, EPI_ISL_2199973, EPI_ISL_2199974, EPI_ISL_2199975, EPI_ISL_2199976,
EPI_ISL_2199977, EPI_ISL_2235529, EPI_ISL_2235532, EPI_ISL_2235533, EPI_ISL_2235534, EPI_ISL_2235536, EPI_ISL_2235538, EPI_ISL_2235541, EPI_ISL_2235542, EPI_ISL_2235543, EPI_ISL_2235544, EPI_ISL_2235546, EPI_ISL_2235552, EPI_ISL_2235554, EPI_ISL_2235555, EPI_ISL_2235556, EPI_ISL_2235558, EPI_ISL_2235561, EPI_ISL_2235563, EPI_ISL_2235565, EPI_ISL_2235566, EPI_ISL_2235572,
EPI_ISL_2235574, EPI_ISL_2235576, EPI_ISL_2235579, EPI_ISL_2235582, EPI_ISL_2235585, EPI_ISL_2235587, EPI_ISL_2235589, EPI_ISL_2235590, EPI_ISL_2235593, EPI_ISL_2235595, EPI_ISL_2235601, EPI_ISL_2235602, EPI_ISL_2235605, EPI_ISL_2235606, EPI_ISL_2235607, EPI_ISL_2235608, EPI_ISL_2235612, EPI_ISL_2235614, EPI_ISL_2235617, EPI_ISL_2235619, EPI_ISL_2235623, EPI_ISL_2235625,
EPI_ISL_2235631, EPI_ISL_2235632, EPI_ISL_2235640, EPI_ISL_2235642, EPI_ISL_2235643, EPI_ISL_2235646, EPI_ISL_2235647, EPI_ISL_2235648, EPI_ISL_2235653, EPI_ISL_2235654, EPI_ISL_2235655, EPI_ISL_2235658, EPI_ISL_2235661, EPI_ISL_2235662, EPI_ISL_2235664, EPI_ISL_2235665, EPI_ISL_2235667, EPI_ISL_2235668, EPI_ISL_2235670, EPI_ISL_2235672, EPI_ISL_2235676, EPI_ISL_2235678,

EPI_ISL_2235680, EPI_ISL_2235681, EPI_ISL_2235682, EPI_ISL_2235683, EPI_ISL_2235684, EPI_ISL_2235686, EPI_ISL_2235687, EPI_ISL_2235688, EPI_ISL_2235689, EPI_ISL_2235690, EPI_ISL_2235691, EPI_ISL_2235692, EPI_ISL_2235693, EPI_ISL_2235697, EPI_ISL_2235699, EPI_ISL_2235701, EPI_ISL_2235703, EPI_ISL_2235704, EPI_ISL_2235705, EPI_ISL_2235708, EPI_ISL_2235709, EPI_ISL_2235711, EPI_ISL_2235715, EPI_ISL_2235716, EPI_ISL_2235717, EPI_ISL_2235718, EPI_ISL_2235719, EPI_ISL_2235724, EPI_ISL_2235727, EPI_ISL_2235733, EPI_ISL_2235735, EPI_ISL_2235738, EPI_ISL_2235739, EPI_ISL_2235740, EPI_ISL_2235745, EPI_ISL_2235747, EPI_ISL_2235750, EPI_ISL_2235753, EPI_ISL_2235755, EPI_ISL_2235758, EPI_ISL_2235761, EPI_ISL_2235764, EPI_ISL_2235766, EPI_ISL_2235767, EPI_ISL_2235771, EPI_ISL_2235772, EPI_ISL_2235774, EPI_ISL_2235776, EPI_ISL_2235778, EPI_ISL_2235779, EPI_ISL_2235781, EPI_ISL_2235783, EPI_ISL_2235785, EPI_ISL_2235788, EPI_ISL_2235789, EPI_ISL_2235790, EPI_ISL_2235796, EPI_ISL_2235797, EPI_ISL_2235799, EPI_ISL_2235807, EPI_ISL_2235808, EPI_ISL_2235809, EPI_ISL_2235813, EPI_ISL_2235814, EPI_ISL_2235815, EPI_ISL_2235819, EPI_ISL_2235820, EPI_ISL_2235822, EPI_ISL_2235824, EPI_ISL_2235825, EPI_ISL_2235826, EPI_ISL_2235829, EPI_ISL_2235830, EPI_ISL_2235836, EPI_ISL_2235838, EPI_ISL_2235841, EPI_ISL_2235842, EPI_ISL_2235844, EPI_ISL_2235848, EPI_ISL_2235850, EPI_ISL_2235852, EPI_ISL_2235853, EPI_ISL_2235854, EPI_ISL_2235855, EPI_ISL_2235860, EPI_ISL_2235861, EPI_ISL_2235862, EPI_ISL_2235869, EPI_ISL_2235872, EPI_ISL_2235874, EPI_ISL_2235892, EPI_ISL_2235898, EPI_ISL_2235900, EPI_ISL_2235902, EPI_ISL_2235913, EPI_ISL_2235922, EPI_ISL_2235935, EPI_ISL_2235952, EPI_ISL_2235953, EPI_ISL_2235954, EPI_ISL_2235957, EPI_ISL_2235958, EPI_ISL_2235959, EPI_ISL_2235962, EPI_ISL_2235965, EPI_ISL_2235975, EPI_ISL_2235980, EPI_ISL_2235981, EPI_ISL_2235982, EPI_ISL_2235986, EPI_ISL_2235987, EPI_ISL_2235990, EPI_ISL_2235991, EPI_ISL_2235993, EPI_ISL_2235995, EPI_ISL_2235997, EPI_ISL_2236002, EPI_ISL_2236008, EPI_ISL_2236014, EPI_ISL_2236015, EPI_ISL_2236017, EPI_ISL_2236020, EPI_ISL_2236025, EPI_ISL_2236029, EPI_ISL_2236032, EPI_ISL_2236037, EPI_ISL_2236038, EPI_ISL_2236040, EPI_ISL_2236041, EPI_ISL_2236046, EPI_ISL_2236054, EPI_ISL_2236057, EPI_ISL_2236058, EPI_ISL_2236060, EPI_ISL_2236064, EPI_ISL_2236078, EPI_ISL_2236079, EPI_ISL_2236080, EPI_ISL_2236091, EPI_ISL_2236092, EPI_ISL_2236100, EPI_ISL_2236105, EPI_ISL_2236110, EPI_ISL_2236111, EPI_ISL_2236112, EPI_ISL_2236115, EPI_ISL_2236117, EPI_ISL_2236123, EPI_ISL_2236127, EPI_ISL_2236128, EPI_ISL_2236130, EPI_ISL_2236133, EPI_ISL_2236146, EPI_ISL_2236150, EPI_ISL_2236151, EPI_ISL_2236152, EPI_ISL_2236153, EPI_ISL_2236156, EPI_ISL_2236157, EPI_ISL_2236160, EPI_ISL_2236162, EPI_ISL_2236164, EPI_ISL_2236165, EPI_ISL_2236167, EPI_ISL_2236168, EPI_ISL_2236182, EPI_ISL_2236185, EPI_ISL_2236186, EPI_ISL_2236191, EPI_ISL_2236204, EPI_ISL_2236211, EPI_ISL_2236256, EPI_ISL_2236260, EPI_ISL_2236278, EPI_ISL_2236285, EPI_ISL_2236287, EPI_ISL_2236304, EPI_ISL_2236305, EPI_ISL_2236340, EPI_ISL_2236342, EPI_ISL_2236343, EPI_ISL_2236346, EPI_ISL_2236360, EPI_ISL_2236390, EPI_ISL_2236409, EPI_ISL_2236411, EPI_ISL_2236420, EPI_ISL_2236450, EPI_ISL_2236474, EPI_ISL_2236477, EPI_ISL_2236482, EPI_ISL_2236489, EPI_ISL_2236490, EPI_ISL_2236493, EPI_ISL_2236498, EPI_ISL_2236500, EPI_ISL_2236502, EPI_ISL_2236503, EPI_ISL_2236507, EPI_ISL_2236510, EPI_ISL_2236515, EPI_ISL_2236518, EPI_ISL_2236521, EPI_ISL_2236522, EPI_ISL_2236523, EPI_ISL_2236525, EPI_ISL_2236533, EPI_ISL_2236538, EPI_ISL_2236539, EPI_ISL_2236540, EPI_ISL_2236541, EPI_ISL_2236542, EPI_ISL_2236549, EPI_ISL_2236558, EPI_ISL_2236559, EPI_ISL_2236564, EPI_ISL_2236569, EPI_ISL_2236572, EPI_ISL_2236573, EPI_ISL_2236574, EPI_ISL_2236577, EPI_ISL_2236584, EPI_ISL_2236586, EPI_ISL_2236593, EPI_ISL_2236595, EPI_ISL_2236597, EPI_ISL_2236617, EPI_ISL_2236626, EPI_ISL_2236633, EPI_ISL_2236634, EPI_ISL_2236635, EPI_ISL_2236639, EPI_ISL_2236650, EPI_ISL_2236654, EPI_ISL_2236667, EPI_ISL_2236674, EPI_ISL_2236678, EPI_ISL_2236679, EPI_ISL_2236680, EPI_ISL_2236682, EPI_ISL_2236683, EPI_ISL_2236686, EPI_ISL_2236687, EPI_ISL_2236693, EPI_ISL_2236697, EPI_ISL_2236702, EPI_ISL_2236706, EPI_ISL_2236709, EPI_ISL_2236710, EPI_ISL_2236714, EPI_ISL_2236737, EPI_ISL_2236744, EPI_ISL_2236748, EPI_ISL_2236749, EPI_ISL_2236750, EPI_ISL_2236752, EPI_ISL_2236754, EPI_ISL_2236755, EPI_ISL_2236756, EPI_ISL_2236759, EPI_ISL_2236765, EPI_ISL_2236768, EPI_ISL_2236770, EPI_ISL_2236771, EPI_ISL_2236773, EPI_ISL_2236776, EPI_ISL_2236781, EPI_ISL_2236785, EPI_ISL_2236793, EPI_ISL_2236796, EPI_ISL_2236799, EPI_ISL_2236800, EPI_ISL_2236801, EPI_ISL_2236804, EPI_ISL_2236806, EPI_ISL_2236808, EPI_ISL_2236817, EPI_ISL_2237160, EPI_ISL_2237161, EPI_ISL_2237162, EPI_ISL_2237165, EPI_ISL_2237167, EPI_ISL_2237168, EPI_ISL_2237169, EPI_ISL_2237171, EPI_ISL_2237172, EPI_ISL_2237174, EPI_ISL_2237175, EPI_ISL_2237177, EPI_ISL_2237180, EPI_ISL_2237182, EPI_ISL_2237189, EPI_ISL_2237192, EPI_ISL_2237196, EPI_ISL_2237198, EPI_ISL_2237199, EPI_ISL_2237206, EPI_ISL_2237208, EPI_ISL_2237213, EPI_ISL_2237223, EPI_ISL_2237225, EPI_ISL_2237226, EPI_ISL_2237229, EPI_ISL_2237230, EPI_ISL_2237231, EPI_ISL_2237234, EPI_ISL_2237235, EPI_ISL_2237237, EPI_ISL_2237242, EPI_ISL_2237245, EPI_ISL_2237247, EPI_ISL_2237249, EPI_ISL_2237252, EPI_ISL_2237253, EPI_ISL_2237257, EPI_ISL_2237262, EPI_ISL_2237265, EPI_ISL_2237266, EPI_ISL_2237269, EPI_ISL_2237274, EPI_ISL_2237276, EPI_ISL_2237281, EPI_ISL_2237282, EPI_ISL_2237283, EPI_ISL_2237284, EPI_ISL_2237285, EPI_ISL_2237286, EPI_ISL_2237288, EPI_ISL_2237289, EPI_ISL_2237290, EPI_ISL_2237293, EPI_ISL_2237301, EPI_ISL_2237302, EPI_ISL_2237309, EPI_ISL_2237311, EPI_ISL_2237314, EPI_ISL_2237315, EPI_ISL_2237318, EPI_ISL_2237319, EPI_ISL_2237321, EPI_ISL_2237326, EPI_ISL_2237329, EPI_ISL_2237330, EPI_ISL_2237335, EPI_ISL_2237338, EPI_ISL_2237343, EPI_ISL_2237344, EPI_ISL_2237345, EPI_ISL_2237346, EPI_ISL_2237348, EPI_ISL_2237349, EPI_ISL_2237350, EPI_ISL_2237352, EPI_ISL_2237355, EPI_ISL_2237356, EPI_ISL_2237359, EPI_ISL_2237365, EPI_ISL_2237366, EPI_ISL_2237367, EPI_ISL_2237369, EPI_ISL_2237370, EPI_ISL_2237372, EPI_ISL_2237376, EPI_ISL_2237378, EPI_ISL_2237380, EPI_ISL_2237382, EPI_ISL_2237383, EPI_ISL_2237384, EPI_ISL_2237385, EPI_ISL_2237386, EPI_ISL_2237391, EPI_ISL_2237399, EPI_ISL_2237400, EPI_ISL_2237402, EPI_ISL_2237405, EPI_ISL_2237406, EPI_ISL_2237410, EPI_ISL_2237411, EPI_ISL_2237413, EPI_ISL_2237414, EPI_ISL_2237427, EPI_ISL_2237429, EPI_ISL_2237431, EPI_ISL_2237436, EPI_ISL_2237437, EPI_ISL_2237440, EPI_ISL_2237441, EPI_ISL_2237445, EPI_ISL_2237447, EPI_ISL_2237451, EPI_ISL_2237452, EPI_ISL_2237453, EPI_ISL_2237455, EPI_ISL_2237458, EPI_ISL_2237463, EPI_ISL_2237464, EPI_ISL_2237466, EPI_ISL_2237468, EPI_ISL_2237475, EPI_ISL_2237480, EPI_ISL_2237483, EPI_ISL_2237494, EPI_ISL_2237496, EPI_ISL_2237500, EPI_ISL_2237509, EPI_ISL_2237512, EPI_ISL_2237529, EPI_ISL_2237532, EPI_ISL_2237539, EPI_ISL_2237545, EPI_ISL_2237546, EPI_ISL_2237551, EPI_ISL_2237558, EPI_ISL_2237575, EPI_ISL_2237577, EPI_ISL_2237580, EPI_ISL_2237587, EPI_ISL_2237589, EPI_ISL_2237595, EPI_ISL_2237610, EPI_ISL_2237614, EPI_ISL_2237616, EPI_ISL_2237622, EPI_ISL_2237623, EPI_ISL_2237624, EPI_ISL_2237626, EPI_ISL_2237635, EPI_ISL_2237641, EPI_ISL_2237647, EPI_ISL_2237654, EPI_ISL_2237655, EPI_ISL_2237664, EPI_ISL_2237667, EPI_ISL_2237670, EPI_ISL_2237675, EPI_ISL_2237685, EPI_ISL_2237686, EPI_ISL_2237687, EPI_ISL_2237697, EPI_ISL_2237698, EPI_ISL_2237699, EPI_ISL_2237702, EPI_ISL_2237704, EPI_ISL_2237708, EPI_ISL_2237713, EPI_ISL_2237716, EPI_ISL_2237726, EPI_ISL_2237732, EPI_ISL_2237733, EPI_ISL_2237744, EPI_ISL_2237754, EPI_ISL_2237755, EPI_ISL_2237757, EPI_ISL_2237758, EPI_ISL_2237759, EPI_ISL_2237761, EPI_ISL_2237763, EPI_ISL_2237765, EPI_ISL_2237769, EPI_ISL_2237771, EPI_ISL_2237772, EPI_ISL_2237773, EPI_ISL_2237775, EPI_ISL_2237776, EPI_ISL_2237782, EPI_ISL_2237783, EPI_ISL_2237784, EPI_ISL_2237788, EPI_ISL_2237793, EPI_ISL_2237795, EPI_ISL_2237796, EPI_ISL_2237797, EPI_ISL_2237798, EPI_ISL_2237799, EPI_ISL_2237802, EPI_ISL_2237803, EPI_ISL_2237806, EPI_ISL_2237809, EPI_ISL_2237813, EPI_ISL_2237814, EPI_ISL_2237815, EPI_ISL_2237816, EPI_ISL_2237819, EPI_ISL_2237820, EPI_ISL_2237821, EPI_ISL_2237830, EPI_ISL_2237832, EPI_ISL_2237834, EPI_ISL_2237838, EPI_ISL_2237840, EPI_ISL_2237841, EPI_ISL_2237842, EPI_ISL_2237844, EPI_ISL_2237845, EPI_ISL_2237852, EPI_ISL_2237854, EPI_ISL_2237856, EPI_ISL_2237860, EPI_ISL_2237861, EPI_ISL_2237863, EPI_ISL_2237865, EPI_ISL_2237867, EPI_ISL_2237872, EPI_ISL_2237877, EPI_ISL_2237879, EPI_ISL_2237880, EPI_ISL_2237881, EPI_ISL_2237889, EPI_ISL_2237891, EPI_ISL_2237892, EPI_ISL_2237893, EPI_ISL_2237898, EPI_ISL_2237899, EPI_ISL_2237900, EPI_ISL_2237904, EPI_ISL_2237906, EPI_ISL_2237909, EPI_ISL_2237910, EPI_ISL_2237911, EPI_ISL_2237913, EPI_ISL_2237914, EPI_ISL_2237922, EPI_ISL_2237926, EPI_ISL_2237930, EPI_ISL_2237932, EPI_ISL_2237937, EPI_ISL_2237938, EPI_ISL_2237939, EPI_ISL_2237940, EPI_ISL_2237941, EPI_ISL_2237948, EPI_ISL_2237952, EPI_ISL_2237956, EPI_ISL_2237957, EPI_ISL_2237960, EPI_ISL_2237961, EPI_ISL_2237965, EPI_ISL_2237966, EPI_ISL_2237970, EPI_ISL_2237971, EPI_ISL_2237972, EPI_ISL_2237979, EPI_ISL_2237980, EPI_ISL_2237981, EPI_ISL_2237984, EPI_ISL_2237985, EPI_ISL_2237987, EPI_ISL_2237990, EPI_ISL_2237992, EPI_ISL_2237995, EPI_ISL_2237998, EPI_ISL_2237999, EPI_ISL_2238004, EPI_ISL_2238010, EPI_ISL_2238019, EPI_ISL_2238020, EPI_ISL_2238024, EPI_ISL_2238025, EPI_ISL_2238026, EPI_ISL_2238028, EPI_ISL_2238037, EPI_ISL_2238039, EPI_ISL_2238040, EPI_ISL_2238041, EPI_ISL_2238045, EPI_ISL_2238046, EPI_ISL_2238048, EPI_ISL_2238049, EPI_ISL_2238054, EPI_ISL_2238061, EPI_ISL_2238064, EPI_ISL_2253264, EPI_ISL_2253430, EPI_ISL_2253451, EPI_ISL_2253465, EPI_ISL_2253494, EPI_ISL_2253539, EPI_ISL_2253609, EPI_ISL_2253611, EPI_ISL_2253621, EPI_ISL_2253627, EPI_ISL_2253941, EPI_ISL_2253949, EPI_ISL_2253952, EPI_ISL_2253957, EPI_ISL_2253975, EPI_ISL_2253981, EPI_ISL_2253992, EPI_ISL_2253997, EPI_ISL_2254016, EPI_ISL_2254036, EPI_ISL_2254054, EPI_ISL_2254061, EPI_ISL_2254086, EPI_ISL_2254120, EPI_ISL_2275000, EPI_ISL_2275006, EPI_ISL_2275030, EPI_ISL_2275088, EPI_ISL_2275102, EPI_ISL_2275131, EPI_ISL_2275158, EPI_ISL_2275186, EPI_ISL_2275192, EPI_ISL_2275194, EPI_ISL_2275207, EPI_ISL_2275209, EPI_ISL_2275213, EPI_ISL_2275214, EPI_ISL_2275218, EPI_ISL_2275225, EPI_ISL_2275227, EPI_ISL_2275234, EPI_ISL_2275240, EPI_ISL_2275241, EPI_ISL_2275248, EPI_ISL_2275250, EPI_ISL_2275251, EPI_ISL_2275254, EPI_ISL_2275255, EPI_ISL_2275257, EPI_ISL_2275265, EPI_ISL_2275266, EPI_ISL_2275274, EPI_ISL_2275276, EPI_ISL_2275278, EPI_ISL_2275282, EPI_ISL_2275286, EPI_ISL_2275289, EPI_ISL_2275291, EPI_ISL_2275295, EPI_ISL_2275298, EPI_ISL_2275301, EPI_ISL_2275302, EPI_ISL_2275306, EPI_ISL_2275316, EPI_ISL_2275319, EPI_ISL_2275320, EPI_ISL_2275324, EPI_ISL_2275329, EPI_ISL_2275332, EPI_ISL_2275335, EPI_ISL_2275336, EPI_ISL_2275338, EPI_ISL_2275340, EPI_ISL_2275351, EPI_ISL_2275359, EPI_ISL_2275369, EPI_ISL_2275370, EPI_ISL_2275371, EPI_ISL_2275381, EPI_ISL_2275382, EPI_ISL_2275384, EPI_ISL_2275387, EPI_ISL_2275389, EPI_ISL_2275390, EPI_ISL_2275393, EPI_ISL_2275396, EPI_ISL_2275399, EPI_ISL_2275406, EPI_ISL_2275407, EPI_ISL_2275408, EPI_ISL_2275409, EPI_ISL_2275410, EPI_ISL_2275411, EPI_ISL_2275412, EPI_ISL_2275413, EPI_ISL_2275414, EPI_ISL_2275415, EPI_ISL_2275416, EPI_ISL_2275417, EPI_ISL_2275418, EPI_ISL_2275419, EPI_ISL_2275420, EPI_ISL_2275421, EPI_ISL_2275422, EPI_ISL_2275423, EPI_ISL_2275424, EPI_ISL_2275425, EPI_ISL_2275426, EPI_ISL_2275427, EPI_ISL_2275428, EPI_ISL_2275429, EPI_ISL_2275430, EPI_ISL_2275431, EPI_ISL_2275432, EPI_ISL_2275433, EPI_ISL_2275434, EPI_ISL_2275435, EPI_ISL_2275436, EPI_ISL_2275437, EPI_ISL_2275438, EPI_ISL_2275439, EPI_ISL_2275440, EPI_ISL_2275441, EPI_ISL_2275442, EPI_ISL_2275443, EPI_ISL_2275444, EPI_ISL_2275445, EPI_ISL_2275446, EPI_ISL_2275447, EPI_ISL_2275448, EPI_ISL_2275449, EPI_ISL_2275450, EPI_ISL_2275451, EPI_ISL_2275452, EPI_ISL_2275453, EPI_ISL_2275454, EPI_ISL_2275455, EPI_ISL_2275456, EPI_ISL_2275457, EPI_ISL_2275458, EPI_ISL_2275459, EPI_ISL_2275460, EPI_ISL_2275461, EPI_ISL_2275462, EPI_ISL_2275463, EPI_ISL_2275464, EPI_ISL_2275465, EPI_ISL_2275466, EPI_ISL_2275467, EPI_ISL_2275468, EPI_ISL_2275469, EPI_ISL_2275470, EPI_ISL_2275471, EPI_ISL_2275472, EPI_ISL_2275473, EPI_ISL_2275474, EPI_ISL_2275475, EPI_ISL_2275476, EPI_ISL_2275477, EPI_ISL_2275478, EPI_ISL_2275479, EPI_ISL_2275480, EPI_ISL_2275481, EPI_ISL_2275482, EPI_ISL_2275483, EPI_ISL_2275484, EPI_ISL_2275485, EPI_ISL_2275486, EPI_ISL_2275487, EPI_ISL_2275488, EPI_ISL_2275489, EPI_ISL_2275490, EPI_ISL_2275491, EPI_ISL_2275492, EPI_ISL_2275493, EPI_ISL_2275494, EPI_ISL_2275495, EPI_ISL_2275496, EPI_ISL_2275497, EPI_ISL_2275498, EPI_ISL_2275499, EPI_ISL_2275500, EPI_ISL_2275501, EPI_ISL_2275502, EPI_ISL_2275503, EPI_ISL_2275504, EPI_ISL_2275505, EPI_ISL_2275506, EPI_ISL_2275507, EPI_ISL_2275508, EPI_ISL_2275509, EPI_ISL_2275510, EPI_ISL_2275511, EPI_ISL_2275512, EPI_ISL_2275513, EPI_ISL_2275514, EPI_ISL_2275515, EPI_ISL_2275516, EPI_ISL_2275517, EPI_ISL_2275518, EPI_ISL_2275519, EPI_ISL_2275520, EPI_ISL_2275521, EPI_ISL_2275522, EPI_ISL_2275523, EPI_ISL_2275524, EPI_ISL_2275525, EPI_ISL_2275526, EPI_ISL_2275527, EPI_ISL_2275528, EPI_ISL_2275529, EPI_ISL_2275530, EPI_ISL_2275531, EPI_ISL_2275532, EPI_ISL_2275533, EPI_ISL_2275534, EPI_ISL_2275535, EPI_ISL_2275536, EPI_ISL_2275537, EPI_ISL_2275538, EPI_ISL_2275539, EPI_ISL_2275540, EPI_ISL_2275541, EPI_ISL_2275542, EPI_ISL_2275543, EPI_ISL_2275544, EPI_ISL_2275545, EPI_ISL_2275546, EPI_ISL_2275547, EPI_ISL_2275548, EPI_ISL_2275549, EPI_ISL_2275550, EPI_ISL_2275551, EPI_ISL_2275552, EPI_ISL_2275553, EPI_ISL_2275554, EPI_ISL_2275555, EPI_ISL_2275556, EPI_ISL_2275557, EPI_ISL_2275558, EPI_ISL_2275559, EPI_ISL_2275560, EPI_ISL_2275561, EPI_ISL_2275562, EPI_ISL_2275563, EPI_ISL_2275564, EPI_ISL_2275565, EPI_ISL_2275566, EPI_ISL_2275567, EPI_ISL_2275568, EPI_ISL_2275569, EPI_ISL_2275570, EPI_ISL_2275571, EPI_ISL_2275572, EPI_ISL_2275573, EPI_ISL_2275574, EPI_ISL_2275575, EPI_ISL_2275577, EPI_ISL_2275578, EPI_ISL_2275579, EPI_ISL_2275580, EPI_ISL_2275581, EPI_ISL_2275582, EPI_ISL_2275583, EPI_ISL_2275584, EPI_ISL_2275585, EPI_ISL_2275586, EPI_ISL_2275587, EPI_ISL_2275588, EPI_ISL_2275589, EPI_ISL_2275590, EPI_ISL_2275591, EPI_ISL_2275592, EPI_ISL_2275593, EPI_ISL_2275594, EPI_ISL_2275596, EPI_ISL_2275597, EPI_ISL_2275598, EPI_ISL_2275599, EPI_ISL_2275600, EPI_ISL_2275601, EPI_ISL_2275602, EPI_ISL_2275603, EPI_ISL_2275604, EPI_ISL_2275605, EPI_ISL_2275606, EPI_ISL_2275607, EPI_ISL_2275608, EPI_ISL_2275609, EPI_ISL_2275610, EPI_ISL_2275611, EPI_ISL_2275612, EPI_ISL_2275613, EPI_ISL_2275614, EPI_ISL_2275615, EPI_ISL_2275616, EPI_ISL_2275617, EPI_ISL_2275618, EPI_ISL_2275619, EPI_ISL_2275620, EPI_ISL_2275621, EPI_ISL_2275622, EPI_ISL_2275623, EPI_ISL_2275624, EPI_ISL_2275625, EPI_ISL_2275626, EPI_ISL_2275627, EPI_ISL_2275628, EPI_ISL_2275629, EPI_ISL_2275630, EPI_ISL_2275631, EPI_ISL_2275632, EPI_ISL_2275633, EPI_ISL_2275634, EPI_ISL_2275635, EPI_ISL_2275636, EPI_ISL_2275637, EPI_ISL_2275638, EPI_ISL_2275639, EPI_ISL_2275640, EPI_ISL_2275641, EPI_ISL_2275642, EPI_ISL_2275643, EPI_ISL_2275644, EPI_ISL_2275645, EPI_ISL_2275646, EPI_ISL_2275647, EPI_ISL_2275648, EPI_ISL_2275649, EPI_ISL_2275650, EPI_ISL_2275651, EPI_ISL_2275652, EPI_ISL_2275653, EPI_ISL_2275654, EPI_ISL_2275655, EPI_ISL_2275656, EPI_ISL_2275657, EPI_ISL_2275658, EPI_ISL_2275659, EPI_ISL_2275660, EPI_ISL_2275661, EPI_ISL_2275662, EPI_ISL_2275663, EPI_ISL_2275664, EPI_ISL_2275665, EPI_ISL_2275666, EPI_ISL_2275667, EPI_ISL_2275668, EPI_ISL_2275669, EPI_ISL_2275670, EPI_ISL_2275671, EPI_ISL_2275672, EPI_ISL_2275673, EPI_ISL_2275674, EPI_ISL_2275675, EPI_ISL_2275676, EPI_ISL_2275677, EPI_ISL_2275678, EPI_ISL_2275679, EPI_ISL_2275680, EPI_ISL_2275681, EPI_ISL_2275682, EPI_ISL_2275683, EPI_ISL_2275684, EPI_ISL_2275685, EPI_ISL_2275686, EPI_ISL_2275687, EPI_ISL_2275688, EPI_ISL_2275689, EPI_ISL_2275690, EPI_ISL_2275691, EPI_ISL_2275692, EPI_ISL_2275693, EPI_ISL_2275694, EPI_ISL_2275695, EPI_ISL_2275696, EPI_ISL_2275697, EPI_ISL_2275698, EPI_ISL_2275699, EPI_ISL_2275700, EPI_ISL_2275701, EPI_ISL_2275702, EPI_ISL_2275703, EPI_ISL_2275704, EPI_ISL_2275705, EPI_ISL_2275706, EPI_ISL_2275707, EPI_ISL_2275708, EPI_ISL_2275709, EPI_ISL_2275710, EPI_ISL_2275711, EPI_ISL_2275712, EPI_ISL_2275713, EPI_ISL_2275714, EPI_ISL_2275715, EPI_ISL_2275716, EPI_ISL_2275717, EPI_ISL_2275718, EPI_ISL_2275719, EPI_ISL_2275720, EPI_ISL_2275721, EPI_ISL_2275722, EPI_ISL_2275723, EPI_ISL_2275724, EPI_ISL_2275725, EPI_ISL_2275726, EPI_ISL_2275727, EPI_ISL_2275728, EPI_ISL_2275729, EPI_ISL_2275730, EPI_ISL_2275731, EPI_ISL_2275732, EPI_ISL_2275733, EPI_ISL_2275734, EPI_ISL_2275735, EPI_ISL_2275736, EPI_ISL_2275737, EPI_ISL_2275738, EPI_ISL_2275739, EPI_ISL_2275740, EPI_ISL_2275741, EPI_ISL_2275742, EPI_ISL_2275743, EPI_ISL_2275744, EPI_ISL_2275745, EPI_ISL_2316283, EPI_ISL_2316477, EPI_ISL_2316523, EPI_ISL_2316640, EPI_ISL_2316752, EPI_ISL_2316756, EPI_ISL_2316786, EPI_ISL_2316867, EPI_ISL_2317002, EPI_ISL_2317114, EPI_ISL_2317821, EPI_ISL_2318865, EPI_ISL_2318866, EPI_ISL_2318867, EPI_ISL_2318868, EPI_ISL_2318869, EPI_ISL_2318870, EPI_ISL_2318871, EPI_ISL_2318872, EPI_ISL_2318873, EPI_ISL_2318874, EPI_ISL_2318875, EPI_ISL_2318876, EPI_ISL_2318877, EPI_ISL_2318878, EPI_ISL_2318879, EPI_ISL_2318880, EPI_ISL_2318881, EPI_ISL_2318882, EPI_ISL_2318883, EPI_ISL_2318884, EPI_ISL_2318885, EPI_ISL_2318886, EPI_ISL_2318887, EPI_ISL_2318888, EPI_ISL_2318889, EPI_ISL_2318890, EPI_ISL_2318891, EPI_ISL_2318892, EPI_ISL_2318893, EPI_ISL_2318894, EPI_ISL_2318895, EPI_ISL_2318896, EPI_ISL_2354217, EPI_ISL_2354218, EPI_ISL_2354219, EPI_ISL_2354220, EPI_ISL_2354221, EPI_ISL_2354222, EPI_ISL_2354223, EPI_ISL_2354224, EPI_ISL_2354225, EPI_ISL_2354226, EPI_ISL_2354227, EPI_ISL_2354228, EPI_ISL_2354229, EPI_ISL_2354230, EPI_ISL_2354231, EPI_ISL_2354232, EPI_ISL_2354233, EPI_ISL_2354234, EPI_ISL_2354235, EPI_ISL_2354236, EPI_ISL_2354237, EPI_ISL_2354238, EPI_ISL_2354239, EPI_ISL_2354240, EPI_ISL_2354241, EPI_ISL_2354242, EPI_ISL_2354243, EPI_ISL_2354244, EPI_ISL_2354245, EPI_ISL_2354246, EPI_ISL_2354247, EPI_ISL_2354248, EPI_ISL_2354249, EPI_ISL_2354250, EPI_ISL_2354251, EPI_ISL_2354252, EPI_ISL_2354253, EPI_ISL_2354254, EPI_ISL_2354255, EPI_ISL_2354256, EPI_ISL_2354257, EPI_ISL_2354258, EPI_ISL_2354259, EPI_ISL_2354260, EPI_ISL_2354261, EPI_ISL_2354262, EPI_ISL_2354263, EPI_ISL_2354264, EPI_ISL_2354265, EPI_ISL_2354266, EPI_ISL_2354267, EPI_ISL_2354268, EPI_ISL_2354269, EPI_ISL_2354270, EPI_ISL_2354271, EPI_ISL_2354272, EPI_ISL_2354273, EPI_ISL_2354274, EPI_ISL_2354275, EPI_ISL_2354276, EPI_ISL_2354277, EPI_ISL_2354278, EPI_ISL_2354279, EPI_ISL_2354280, EPI_ISL_2354281, EPI_ISL_2354282, EPI_ISL_2354283, EPI_ISL_2354284, EPI_ISL_2354285, EPI_ISL_2354286, EPI_ISL_2354287, EPI_ISL_2354288, EPI_ISL_2354289, EPI_ISL_2354290, EPI_ISL_2354291, EPI_ISL_2354314, EPI_ISL_2354334, EPI_ISL_2354345, EPI_ISL_2354355, EPI_ISL_2354379, EPI_ISL_2354391, EPI_ISL_2354392, EPI_ISL_2354394, EPI_ISL_2354400, EPI_ISL_2354404, EPI_ISL_2354405, EPI_ISL_2354415, EPI_ISL_2354416, EPI_ISL_2354417, EPI_ISL_2354425, EPI_ISL_2354435, EPI_ISL_2354442, EPI_ISL_2354443, EPI_ISL_2354449, EPI_ISL_2354450, EPI_ISL_2354455, EPI_ISL_2354457, EPI_ISL_2354463, EPI_ISL_2354470, EPI_ISL_2354472, EPI_ISL_2447332

| | | | |
|---|---|---|---|
| see above | Lighthouse Lab in Glasgow | Wellcome Sanger Institute for the COVID-19 Genomics UK (COG-UK) Consortium | Anna Dominiczak and Alex Alderton; Carol Clugston; Cordelia Langford; David Gray; David K. Jackson; Dominic Kwiatkowski; Ewan Harrison; Harper VanSteenhouse; Ian Johnston; Jeffrey Barrett; John Sillitoe on behalf of the Wellcome Sanger Institute COVID-19 Surveillance Team; Roberto Amato; Sonia Goncalves; Yumi Kasai |

EPI_ISL_2118023, EPI_ISL_2137849, EPI_ISL_2137851, EPI_ISL_2137853, EPI_ISL_2137855, EPI_ISL_2137857, EPI_ISL_2137858, EPI_ISL_2137859, EPI_ISL_2137860, EPI_ISL_2137861, EPI_ISL_2137862, EPI_ISL_2137863, EPI_ISL_2137865, EPI_ISL_2137866, EPI_ISL_2137867, EPI_ISL_2137869, EPI_ISL_2137870, EPI_ISL_2137871, EPI_ISL_2137872, EPI_ISL_2137873, EPI_ISL_2137874, EPI_ISL_2137878, EPI_ISL_2137880, EPI_ISL_2137881, EPI_ISL_2137883, EPI_ISL_2137884, EPI_ISL_2137889, EPI_ISL_2137895, EPI_ISL_2137897, EPI_ISL_2137905, EPI_ISL_2137906, EPI_ISL_2137908, EPI_ISL_2137909, EPI_ISL_2137912, EPI_ISL_2137913, EPI_ISL_2137914, EPI_ISL_2137915, EPI_ISL_2137916, EPI_ISL_2137917, EPI_ISL_2137918, EPI_ISL_2137921, EPI_ISL_2137922, EPI_ISL_2137924, EPI_ISL_2137926, EPI_ISL_2137927, EPI_ISL_2137931, EPI_ISL_2137932, EPI_ISL_2137934, EPI_ISL_2137935, EPI_ISL_2137936, EPI_ISL_2137937, EPI_ISL_2137939, EPI_ISL_2137940, EPI_ISL_2137941, EPI_ISL_2137942, EPI_ISL_2137943, EPI_ISL_2137944, EPI_ISL_2137945, EPI_ISL_2137946, EPI_ISL_2137948, EPI_ISL_2137949, EPI_ISL_2137955, EPI_ISL_2137957, EPI_ISL_2137958, EPI_ISL_2137959, EPI_ISL_2137960, EPI_ISL_2137961, EPI_ISL_2137962, EPI_ISL_2137964, EPI_ISL_2137969, EPI_ISL_2137971, EPI_ISL_2137973, EPI_ISL_2137975, EPI_ISL_2137977, EPI_ISL_2137980, EPI_ISL_2137981, EPI_ISL_2137982, EPI_ISL_2137984, EPI_ISL_2137986, EPI_ISL_2137988, EPI_ISL_2137991, EPI_ISL_2137992, EPI_ISL_2137994, EPI_ISL_2137995, EPI_ISL_2137996, EPI_ISL_2137999, EPI_ISL_2138007, EPI_ISL_2138008, EPI_ISL_2138013, EPI_ISL_2138014, EPI_ISL_2138015, EPI_ISL_2138016, EPI_ISL_2138017, EPI_ISL_2138018, EPI_ISL_2138021, EPI_ISL_2138027, EPI_ISL_2138029, EPI_ISL_2138035, EPI_ISL_2138036, EPI_ISL_2138037, EPI_ISL_2138039, EPI_ISL_2138040, EPI_ISL_2138042, EPI_ISL_2138045, EPI_ISL_2138051, EPI_ISL_2138052, EPI_ISL_2138053, EPI_ISL_2138055, EPI_ISL_2138057, EPI_ISL_2138058, EPI_ISL_2138059, EPI_ISL_2138060, EPI_ISL_2138061, EPI_ISL_2138064, EPI_ISL_2138065, EPI_ISL_2138067, EPI_ISL_2138068, EPI_ISL_2138069, EPI_ISL_2138072, EPI_ISL_2138074, EPI_ISL_2138077, EPI_ISL_2138079, EPI_ISL_2138080, EPI_ISL_2138081, EPI_ISL_2138083, EPI_ISL_2138084, EPI_ISL_2138086, EPI_ISL_2138089, EPI_ISL_2138090, EPI_ISL_2138091, EPI_ISL_2138095, EPI_ISL_2138097, EPI_ISL_2138099, EPI_ISL_2138102, EPI_ISL_2138103, EPI_ISL_2138104, EPI_ISL_2138105, EPI_ISL_2138130, EPI_ISL_2138131, EPI_ISL_2138140, EPI_ISL_2138145, EPI_ISL_2138148, EPI_ISL_2138156, EPI_ISL_2138157, EPI_ISL_2138160, EPI_ISL_2138161, EPI_ISL_2138168, EPI_ISL_2138177, EPI_ISL_2138180, EPI_ISL_2138182, EPI_ISL_2138189, EPI_ISL_2138194, EPI_ISL_2138209, EPI_ISL_2138217, EPI_ISL_2138222, EPI_ISL_2138223, EPI_ISL_2138225, EPI_ISL_2138231, EPI_ISL_2138234, EPI_ISL_2138236, EPI_ISL_2138240, EPI_ISL_2138249, EPI_ISL_2138256, EPI_ISL_2138257, EPI_ISL_2138258, EPI_ISL_2138259, EPI_ISL_2138265, EPI_ISL_2138266, EPI_ISL_2138271, EPI_ISL_2138277, EPI_ISL_2138278, EPI_ISL_2138286, EPI_ISL_2138292, EPI_ISL_2138297, EPI_ISL_2138298, EPI_ISL_2138302, EPI_ISL_2138305, EPI_ISL_2138312, EPI_ISL_2138314, EPI_ISL_2138317, EPI_ISL_2138320, EPI_ISL_2138333, EPI_ISL_2138336, EPI_ISL_2138338, EPI_ISL_2138340, EPI_ISL_2138343, EPI_ISL_2138348, EPI_ISL_2138350, EPI_ISL_2138357, EPI_ISL_2138366, EPI_ISL_2138367, EPI_ISL_2138369, EPI_ISL_2138371, EPI_ISL_2138373, EPI_ISL_2138374, EPI_ISL_2138377, EPI_ISL_2138378, EPI_ISL_2138380, EPI_ISL_2138381, EPI_ISL_2138384, EPI_ISL_2138387, EPI_ISL_2138389, EPI_ISL_2138393, EPI_ISL_2138396, EPI_ISL_2138397, EPI_ISL_2138399, EPI_ISL_2138400, EPI_ISL_2138401, EPI_ISL_2138402, EPI_ISL_2138404, EPI_ISL_2138410, EPI_ISL_2138412, EPI_ISL_2138421, EPI_ISL_2138422, EPI_ISL_2138423, EPI_ISL_2138424, EPI_ISL_2138426, EPI_ISL_2138427, EPI_ISL_2138433, EPI_ISL_2138435, EPI_ISL_2138438, EPI_ISL_2138439, EPI_ISL_2138447, EPI_ISL_2138449, EPI_ISL_2138450, EPI_ISL_2138452, EPI_ISL_2138465, EPI_ISL_2138468, EPI_ISL_2138473, EPI_ISL_2138474, EPI_ISL_2138478, EPI_ISL_2138482, EPI_ISL_2138486, EPI_ISL_2138499, EPI_ISL_2138500, EPI_ISL_2138507, EPI_ISL_2138513, EPI_ISL_2138514, EPI_ISL_2138515, EPI_ISL_2138518, EPI_ISL_2138519, EPI_ISL_2138526, EPI_ISL_2138527, EPI_ISL_2138534, EPI_ISL_2138535, EPI_ISL_2138536, EPI_ISL_2138540, EPI_ISL_2138545, EPI_ISL_2138547, EPI_ISL_2138548, EPI_ISL_2138551, EPI_ISL_2138552, EPI_ISL_2138554, EPI_ISL_2138564, EPI_ISL_2138567, EPI_ISL_2138574, EPI_ISL_2138576, EPI_ISL_2138592, EPI_ISL_2138595, EPI_ISL_2138598, EPI_ISL_2138605, EPI_ISL_2138611, EPI_ISL_2138614, EPI_ISL_2138615, EPI_ISL_2138618, EPI_ISL_2138621, EPI_ISL_2138623, EPI_ISL_2138625, EPI_ISL_2138626, EPI_ISL_2138627, EPI_ISL_2138628, EPI_ISL_2138629, EPI_ISL_2138630, EPI_ISL_2138631, EPI_ISL_2138632, EPI_ISL_2138634, EPI_ISL_2138637, EPI_ISL_2138640, EPI_ISL_2138642, EPI_ISL_2138646, EPI_ISL_2138648, EPI_ISL_2138652, EPI_ISL_2138655, EPI_ISL_2138656, EPI_ISL_2138657, EPI_ISL_2138658, EPI_ISL_2138659, EPI_ISL_2138661, EPI_ISL_2138662, EPI_ISL_2138664, EPI_ISL_2138668, EPI_ISL_2138669, EPI_ISL_2138672, EPI_ISL_2138673, EPI_ISL_2138676, EPI_ISL_2138680, EPI_ISL_2138682, EPI_ISL_2138686, EPI_ISL_2138687, EPI_ISL_2138690, EPI_ISL_2138695, EPI_ISL_2138696, EPI_ISL_2138714, EPI_ISL_2152658, EPI_ISL_2152660, EPI_ISL_2152662, EPI_ISL_2152889, EPI_ISL_2152896, EPI_ISL_2152905, EPI_ISL_2152923, EPI_ISL_2171863, EPI_ISL_2171887, EPI_ISL_2171892, EPI_ISL_2171895, EPI_ISL_2171899, EPI_ISL_2171910, EPI_ISL_2171941, EPI_ISL_2171943, EPI_ISL_2171986, EPI_ISL_2172014, EPI_ISL_2172028, EPI_ISL_2172034, EPI_ISL_2172054, EPI_ISL_2172072, EPI_ISL_2172081, EPI_ISL_2198344, EPI_ISL_2198345, EPI_ISL_2198347, EPI_ISL_2198349, EPI_ISL_2198352, EPI_ISL_2198353, EPI_ISL_2198354, EPI_ISL_2198355, EPI_ISL_2198356, EPI_ISL_2198358, EPI_ISL_2198360, EPI_ISL_2198361, EPI_ISL_2198364, EPI_ISL_2198367, EPI_ISL_2198368, EPI_ISL_2198369, EPI_ISL_2198371, EPI_ISL_2198373, EPI_ISL_2198374, EPI_ISL_2198375, EPI_ISL_2198376, EPI_ISL_2198377, EPI_ISL_2198378, EPI_ISL_2198379, EPI_ISL_2198383, EPI_ISL_2198384, EPI_ISL_2198385, EPI_ISL_2198389, EPI_ISL_2198390, EPI_ISL_2198391, EPI_ISL_2198392, EPI_ISL_2198393, EPI_ISL_2198394, EPI_ISL_2198395, EPI_ISL_2198398, EPI_ISL_2198399, EPI_ISL_2198402, EPI_ISL_2198403, EPI_ISL_2198404, EPI_ISL_2198405, EPI_ISL_2198407, EPI_ISL_2198408, EPI_ISL_2198409, EPI_ISL_2198410, EPI_ISL_2198411, EPI_ISL_2198415, EPI_ISL_2198416, EPI_ISL_2198418, EPI_ISL_2198419, EPI_ISL_2198421, EPI_ISL_2198422, EPI_ISL_2198426, EPI_ISL_2198427, EPI_ISL_2198428, EPI_ISL_2198429, EPI_ISL_2198432, EPI_ISL_2198433, EPI_ISL_2198434, EPI_ISL_2198436, EPI_ISL_2198437, EPI_ISL_2198443, EPI_ISL_2198444, EPI_ISL_2198445, EPI_ISL_2198447, EPI_ISL_2198448, EPI_ISL_2198449, EPI_ISL_2198451, EPI_ISL_2198452, EPI_ISL_2198453, EPI_ISL_2198455, EPI_ISL_2198456, EPI_ISL_2198457, EPI_ISL_2198458, EPI_ISL_2198459, EPI_ISL_2198460, EPI_ISL_2198463, EPI_ISL_2198464, EPI_ISL_2198465, EPI_ISL_2198467, EPI_ISL_2198468, EPI_ISL_2198469, EPI_ISL_2198470, EPI_ISL_2198471, EPI_ISL_2198472, EPI_ISL_2198473, EPI_ISL_2198474, EPI_ISL_2198475, EPI_ISL_2198477, EPI_ISL_2198478, EPI_ISL_2198480, EPI_ISL_2198481, EPI_ISL_2198483, EPI_ISL_2198484, EPI_ISL_2198485, EPI_ISL_2198486, EPI_ISL_2198487, EPI_ISL_2198488, EPI_ISL_2198492, EPI_ISL_2198493, EPI_ISL_2198495, EPI_ISL_2198497, EPI_ISL_2198498, EPI_ISL_2198499, EPI_ISL_2198500, EPI_ISL_2198502, EPI_ISL_2198506, EPI_ISL_2198507, EPI_ISL_2198508, EPI_ISL_2198509, EPI_ISL_2198510, EPI_ISL_2198511, EPI_ISL_2198513, EPI_ISL_2198514, EPI_ISL_2198516, EPI_ISL_2198519, EPI_ISL_2198521, EPI_ISL_2198522, EPI_ISL_2198523, EPI_ISL_2198524, EPI_ISL_2198526, EPI_ISL_2198528, EPI_ISL_2198530, EPI_ISL_2198534, EPI_ISL_2198535, EPI_ISL_2198540, EPI_ISL_2198541, EPI_ISL_2198542, EPI_ISL_2198543, EPI_ISL_2198544, EPI_ISL_2198545, EPI_ISL_2198546, EPI_ISL_2198548, EPI_ISL_2198549, EPI_ISL_2198550, EPI_ISL_2198551, EPI_ISL_2198553, EPI_ISL_2198555, EPI_ISL_2198556, EPI_ISL_2198560, EPI_ISL_2198561, EPI_ISL_2198564, EPI_ISL_2198566, EPI_ISL_2198570, EPI_ISL_2198572, EPI_ISL_2198573, EPI_ISL_2198576, EPI_ISL_2198577, EPI_ISL_2198578, EPI_ISL_2198579, EPI_ISL_2198581, EPI_ISL_2198583, EPI_ISL_2198585, EPI_ISL_2198586, EPI_ISL_2198587, EPI_ISL_2198589, EPI_ISL_2198591, EPI_ISL_2198594, EPI_ISL_2198595, EPI_ISL_2198597, EPI_ISL_2198599, EPI_ISL_2198600, EPI_ISL_2198601, EPI_ISL_2198607, EPI_ISL_2198608, EPI_ISL_2198609, EPI_ISL_2198610, EPI_ISL_2198612, EPI_ISL_2198615, EPI_ISL_2198618, EPI_ISL_2198619, EPI_ISL_2198620, EPI_ISL_2198621, EPI_ISL_2198623, EPI_ISL_2198624, EPI_ISL_2198625, EPI_ISL_2198627, EPI_ISL_2198628, EPI_ISL_2198629, EPI_ISL_2198631, EPI_ISL_2198632, EPI_ISL_2198633, EPI_ISL_2198634, EPI_ISL_2198636, EPI_ISL_2198637, EPI_ISL_2198638, EPI_ISL_2198639, EPI_ISL_2198641, EPI_ISL_2198642, EPI_ISL_2198644, EPI_ISL_2198646, EPI_ISL_2198647, EPI_ISL_2198649, EPI_ISL_2198652, EPI_ISL_2198653, EPI_ISL_2198654, EPI_ISL_2198655, EPI_ISL_2198657, EPI_ISL_2198659, EPI_ISL_2198661, EPI_ISL_2198663, EPI_ISL_2198665, EPI_ISL_2198667, EPI_ISL_2198669, EPI_ISL_2198674, EPI_ISL_2198675, EPI_ISL_2198679, EPI_ISL_2198682, EPI_ISL_2198687, EPI_ISL_2198688, EPI_ISL_2198691, EPI_ISL_2198693, EPI_ISL_2198694, EPI_ISL_2198696, EPI_ISL_2198697, EPI_ISL_2198698, EPI_ISL_2198699, EPI_ISL_2198700, EPI_ISL_2198704, EPI_ISL_2198705, EPI_ISL_2198706, EPI_ISL_2198707, EPI_ISL_2198710, EPI_ISL_2198712, EPI_ISL_2198716, EPI_ISL_2198717, EPI_ISL_2198718, EPI_ISL_2198721, EPI_ISL_2198723, EPI_ISL_2198724, EPI_ISL_2198726, EPI_ISL_2198728, EPI_ISL_2198730, EPI_ISL_2198734, EPI_ISL_2198737, EPI_ISL_2198742, EPI_ISL_2198743, EPI_ISL_2198744, EPI_ISL_2198746, EPI_ISL_2198747, EPI_ISL_2198748, EPI_ISL_2198750, EPI_ISL_2198752, EPI_ISL_2198757, EPI_ISL_2198758, EPI_ISL_2198759, EPI_ISL_2198761, EPI_ISL_2198764, EPI_ISL_2198765, EPI_ISL_2198766, EPI_ISL_2198768, EPI_ISL_2198769, EPI_ISL_2198770, EPI_ISL_2198771, EPI_ISL_2198772, EPI_ISL_2198774, EPI_ISL_2198777, EPI_ISL_2198778, EPI_ISL_2198779, EPI_ISL_2198780, EPI_ISL_2198783, EPI_ISL_2198787, EPI_ISL_2198788, EPI_ISL_2198790, EPI_ISL_2198791, EPI_ISL_2198792, EPI_ISL_2198793, EPI_ISL_2198794, EPI_ISL_2198795, EPI_ISL_2198797, EPI_ISL_2198798, EPI_ISL_2198799, EPI_ISL_2198800, EPI_ISL_2198805, EPI_ISL_2198809, EPI_ISL_2198810, EPI_ISL_2198814, EPI_ISL_2198816, EPI_ISL_2198818, EPI_ISL_2198819, EPI_ISL_2198821, EPI_ISL_2198823, EPI_ISL_2198824, EPI_ISL_2198826, EPI_ISL_2198828, EPI_ISL_2198830, EPI_ISL_2198832, EPI_ISL_2198834, EPI_ISL_2198836, EPI_ISL_2198837, EPI_ISL_2198840, EPI_ISL_2198841, EPI_ISL_2198842, EPI_ISL_2198843, EPI_ISL_2198847, EPI_ISL_2198849, EPI_ISL_2198851, EPI_ISL_2198853, EPI_ISL_2198854, EPI_ISL_2198857, EPI_ISL_2198859, EPI_ISL_2198861, EPI_ISL_2198862, EPI_ISL_2198863, EPI_ISL_2198864, EPI_ISL_2198866, EPI_ISL_2198869, EPI_ISL_2198873, EPI_ISL_2198874, EPI_ISL_2198875, EPI_ISL_2198881, EPI_ISL_2198883, EPI_ISL_2198884, EPI_ISL_2198885, EPI_ISL_2198887, EPI_ISL_2198888, EPI_ISL_2198889, EPI_ISL_2198890, EPI_ISL_2198891, EPI_ISL_2198893, EPI_ISL_2198897, EPI_ISL_2198902, EPI_ISL_2198903, EPI_ISL_2198906, EPI_ISL_2198910, EPI_ISL_2198911, EPI_ISL_2198912, EPI_ISL_2198913, EPI_ISL_2198914, EPI_ISL_2198918, EPI_ISL_2198923, EPI_ISL_2198924, EPI_ISL_2198928, EPI_ISL_2198929, EPI_ISL_2198930, EPI_ISL_2198931, EPI_ISL_2198933, EPI_ISL_2198934, EPI_ISL_2198935, EPI_ISL_2198937, EPI_ISL_2198941, EPI_ISL_2198942, EPI_ISL_2198944, EPI_ISL_2198945, EPI_ISL_2198946, EPI_ISL_2198949, EPI_ISL_2198950, EPI_ISL_2198951, EPI_ISL_2198955, EPI_ISL_2198958, EPI_ISL_2198959, EPI_ISL_2198960, EPI_ISL_2198962, EPI_ISL_2198967, EPI_ISL_2198969, EPI_ISL_2198971, EPI_ISL_2198973, EPI_ISL_2198974, EPI_ISL_2198976, EPI_ISL_2198977, EPI_ISL_2198978, EPI_ISL_2198979, EPI_ISL_2198981, EPI_ISL_2198983, EPI_ISL_2198984, EPI_ISL_2198985, EPI_ISL_2198989, EPI_ISL_2198992, EPI_ISL_2198993, EPI_ISL_2198994, EPI_ISL_2198997, EPI_ISL_2199001, EPI_ISL_2199002, EPI_ISL_2199006, EPI_ISL_2199012, EPI_ISL_2199013, EPI_ISL_2199020, EPI_ISL_2199021, EPI_ISL_2199027, EPI_ISL_2199028, EPI_ISL_2199036, EPI_ISL_2199037, EPI_ISL_2199045, EPI_ISL_2199051, EPI_ISL_2199055, EPI_ISL_2199056, EPI_ISL_2199061, EPI_ISL_2199066, EPI_ISL_2199068, EPI_ISL_2199071, EPI_ISL_2199075, EPI_ISL_2199086, EPI_ISL_2199093, EPI_ISL_2199097, EPI_ISL_2199098, EPI_ISL_2199101, EPI_ISL_2199110, EPI_ISL_2199124, EPI_ISL_2199126, EPI_ISL_2199128, EPI_ISL_2199131, EPI_ISL_2199135, EPI_ISL_2199136, EPI_ISL_2199144, EPI_ISL_2199146, EPI_ISL_2199147, EPI_ISL_2199152, EPI_ISL_2199158, EPI_ISL_2199160, EPI_ISL_2199166, EPI_ISL_2199167, EPI_ISL_2199177, EPI_ISL_2199182, EPI_ISL_2199184, EPI_ISL_2199188, EPI_ISL_2199202, EPI_ISL_2199206, EPI_ISL_2199209, EPI_ISL_2199214, EPI_ISL_2199226, EPI_ISL_2199228, EPI_ISL_2199232, EPI_ISL_2199233, EPI_ISL_2199239, EPI_ISL_2199240, EPI_ISL_2199243, EPI_ISL_2199260, EPI_ISL_2199263, EPI_ISL_2199264, EPI_ISL_2199265, EPI_ISL_2199268, EPI_ISL_2199269, EPI_ISL_2199281, EPI_ISL_2199289, EPI_ISL_2199291, EPI_ISL_2199292, EPI_ISL_2199297, EPI_ISL_2199301, EPI_ISL_2199306, EPI_ISL_2199307, EPI_ISL_2199308, EPI_ISL_2199309, EPI_ISL_2199314, EPI_ISL_2199317, EPI_ISL_2199322, EPI_ISL_2199323, EPI_ISL_2199325, EPI_ISL_2199333, EPI_ISL_2199340, EPI_ISL_2199362, EPI_ISL_2199369, EPI_ISL_2199383, EPI_ISL_2199391, EPI_ISL_2199406, EPI_ISL_2199413, EPI_ISL_2199419, EPI_ISL_2199429, EPI_ISL_2199430, EPI_ISL_2199459, EPI_ISL_2199463, EPI_ISL_2199466, EPI_ISL_2199470, EPI_ISL_2199504, EPI_ISL_2199519, EPI_ISL_2199520, EPI_ISL_2199549, EPI_ISL_2199571, EPI_ISL_2199614, EPI_ISL_2199624, EPI_ISL_2199630, EPI_ISL_2199669, EPI_ISL_2199670, EPI_ISL_2199978, EPI_ISL_2235531, EPI_ISL_2235547, EPI_ISL_2235548, EPI_ISL_2235549, EPI_ISL_2235550, EPI_ISL_2235551, EPI_ISL_2235553, EPI_ISL_2235557, EPI_ISL_2235559, EPI_ISL_2235564, EPI_ISL_2235567, EPI_ISL_2235569, EPI_ISL_2235581, EPI_ISL_2235584, EPI_ISL_2235588, EPI_ISL_2235591, EPI_ISL_2235594, EPI_ISL_2235597, EPI_ISL_2235598, EPI_ISL_2235609, EPI_ISL_2235610, EPI_ISL_2235611, EPI_ISL_2235613, EPI_ISL_2235615, EPI_ISL_2235618, EPI_ISL_2235621, EPI_ISL_2235625, EPI_ISL_2235627, EPI_ISL_2235630, EPI_ISL_2235634, EPI_ISL_2235641, EPI_ISL_2235649, EPI_ISL_2235650, EPI_ISL_2235656, EPI_ISL_2235657, EPI_ISL_2235660, EPI_ISL_2235663, EPI_ISL_2235666, EPI_ISL_2235669, EPI_ISL_2235673, EPI_ISL_2235679, EPI_ISL_2235695, EPI_ISL_2235698, EPI_ISL_2235706, EPI_ISL_2235713, EPI_ISL_2235720, EPI_ISL_2235725, EPI_ISL_2235726, EPI_ISL_2235729, EPI_ISL_2235731, EPI_ISL_2235732, EPI_ISL_2235734, EPI_ISL_2235737, EPI_ISL_2235741, EPI_ISL_2235744, EPI_ISL_2235748, EPI_ISL_2235751, EPI_ISL_2235752, EPI_ISL_2235756, EPI_ISL_2235765, EPI_ISL_2235768, EPI_ISL_2235769, EPI_ISL_2235770, EPI_ISL_2235780, EPI_ISL_2235791, EPI_ISL_2235793, EPI_ISL_2235798, EPI_ISL_2235800, EPI_ISL_2235801, EPI_ISL_2235805, EPI_ISL_2235810, EPI_ISL_2235811, EPI_ISL_2235817, EPI_ISL_2235818, EPI_ISL_2235828, EPI_ISL_2235831, EPI_ISL_2235833, EPI_ISL_2235834, EPI_ISL_2235835, EPI_ISL_2235839, EPI_ISL_2235840, EPI_ISL_2235845, EPI_ISL_2235846, EPI_ISL_2235856, EPI_ISL_2235857, EPI_ISL_2235859, EPI_ISL_2235873, EPI_ISL_2235886, EPI_ISL_2235893, EPI_ISL_2235894, EPI_ISL_2235896, EPI_ISL_2235897, EPI_ISL_2235899, EPI_ISL_2235901, EPI_ISL_2235906, EPI_ISL_2235907, EPI_ISL_2235910, EPI_ISL_2235915, EPI_ISL_2235921, EPI_ISL_2235931, EPI_ISL_2235942, EPI_ISL_2235950, EPI_ISL_2235955, EPI_ISL_2235970, EPI_ISL_2235971, EPI_ISL_2235972, EPI_ISL_2235976, EPI_ISL_2235983, EPI_ISL_2235992, EPI_ISL_2235994, EPI_ISL_2235996, EPI_ISL_2235999, EPI_ISL_2236003, EPI_ISL_2236006, EPI_ISL_2236009, EPI_ISL_2236010, EPI_ISL_2236011, EPI_ISL_2236013, EPI_ISL_2236016, EPI_ISL_2236023, EPI_ISL_2236024, EPI_ISL_2236026, EPI_ISL_2236034, EPI_ISL_2236049, EPI_ISL_2236052, EPI_ISL_2236053, EPI_ISL_2236055, EPI_ISL_2236059, EPI_ISL_2236063, EPI_ISL_2236065, EPI_ISL_2236067, EPI_ISL_2236071, EPI_ISL_2236083, EPI_ISL_2236084, EPI_ISL_2236086, EPI_ISL_2236088, EPI_ISL_2236097, EPI_ISL_2236101, EPI_ISL_2236104, EPI_ISL_2236108, EPI_ISL_2236118, EPI_ISL_2236120, EPI_ISL_2236121, EPI_ISL_2236134, EPI_ISL_2236137, EPI_ISL_2236141, EPI_ISL_2236145, EPI_ISL_2236149, EPI_ISL_2236155, EPI_ISL_2236158, EPI_ISL_2236159, EPI_ISL_2236163, EPI_ISL_2236172, EPI_ISL_2236173, EPI_ISL_2236175, EPI_ISL_2236178, EPI_ISL_2236179, EPI_ISL_2236183, EPI_ISL_2236188, EPI_ISL_2236193, EPI_ISL_2236194, EPI_ISL_2236199, EPI_ISL_2236206, EPI_ISL_2236208, EPI_ISL_2236210, EPI_ISL_2236216, EPI_ISL_2236221, EPI_ISL_2236236, EPI_ISL_2236246, EPI_ISL_2236257, EPI_ISL_2236292, EPI_ISL_2236303, EPI_ISL_2236336, EPI_ISL_2236344, EPI_ISL_2236357, EPI_ISL_2236381, EPI_ISL_2236392, EPI_ISL_2236394, EPI_ISL_2236433, EPI_ISL_2236443, EPI_ISL_2236445, EPI_ISL_2236463, EPI_ISL_2236470, EPI_ISL_2236475, EPI_ISL_2236476, EPI_ISL_2236478, EPI_ISL_2236479, EPI_ISL_2236480, EPI_ISL_2236481, EPI_ISL_2236483, EPI_ISL_2236484, EPI_ISL_2236485, EPI_ISL_2236486, EPI_ISL_2236487, EPI_ISL_2236488, EPI_ISL_2236491, EPI_ISL_2236492, EPI_ISL_2236494, EPI_ISL_2236495, EPI_ISL_2236496, EPI_ISL_2236497, EPI_ISL_2236499, EPI_ISL_2236501, EPI_ISL_2236504, EPI_ISL_2236505, EPI_ISL_2236506, EPI_ISL_2236508, EPI_ISL_2236509, EPI_ISL_2236511, EPI_ISL_2236512, EPI_ISL_2236513, EPI_ISL_2236514, EPI_ISL_2236516, EPI_ISL_2236517, EPI_ISL_2236519, EPI_ISL_2236520, EPI_ISL_2236524, EPI_ISL_2236526, EPI_ISL_2236527, EPI_ISL_2236528, EPI_ISL_2236529, EPI_ISL_2236530, EPI_ISL_2236531, EPI_ISL_2236532, EPI_ISL_2236534, EPI_ISL_2236535, EPI_ISL_2236536, EPI_ISL_2236537, EPI_ISL_2236543, EPI_ISL_2236544, EPI_ISL_2236545, EPI_ISL_2236546, EPI_ISL_2236547, EPI_ISL_2236548, EPI_ISL_2236550, EPI_ISL_2236551, EPI_ISL_2236552, EPI_ISL_2236553, EPI_ISL_2236554, EPI_ISL_2236555, EPI_ISL_2236556, EPI_ISL_2236557, EPI_ISL_2236560, EPI_ISL_2236561, EPI_ISL_2236562, EPI_ISL_2236563, EPI_ISL_2236565, EPI_ISL_2236566, EPI_ISL_2236567, EPI_ISL_2236568, EPI_ISL_2236570, EPI_ISL_2236571, EPI_ISL_2236575, EPI_ISL_2236576, EPI_ISL_2236578, EPI_ISL_2236579, EPI_ISL_2236580, EPI_ISL_2236581, EPI_ISL_2236582, EPI_ISL_2236583, EPI_ISL_2236585, EPI_ISL_2236587, EPI_ISL_2236588, EPI_ISL_2236589, EPI_ISL_2236590, EPI_ISL_2236591, EPI_ISL_2236592, EPI_ISL_2236594, EPI_ISL_2236596, EPI_ISL_2236598, EPI_ISL_2236599, EPI_ISL_2236600, EPI_ISL_2236601, EPI_ISL_2236602, EPI_ISL_2236603, EPI_ISL_2236604, EPI_ISL_2236605, EPI_ISL_2236606, EPI_ISL_2236607, EPI_ISL_2236608, EPI_ISL_2236609, EPI_ISL_2236610, EPI_ISL_2236611, EPI_ISL_2236612, EPI_ISL_2236613, EPI_ISL_2236614, EPI_ISL_2236615, EPI_ISL_2236616, EPI_ISL_2236618, EPI_ISL_2236619, EPI_ISL_2236620, EPI_ISL_2236621, EPI_ISL_2236622, EPI_ISL_2236623, EPI_ISL_2236624, EPI_ISL_2236625, EPI_ISL_2236627, EPI_ISL_2236628, EPI_ISL_2236629, EPI_ISL_2236630, EPI_ISL_2236631, EPI_ISL_2236632, EPI_ISL_2236636, EPI_ISL_2236637, EPI_ISL_2236638, EPI_ISL_2236640, EPI_ISL_2236641, EPI_ISL_2236642, EPI_ISL_2236643, EPI_ISL_2236644, EPI_ISL_2236645, EPI_ISL_2236646, EPI_ISL_2236647, EPI_ISL_2236648, EPI_ISL_2236649, EPI_ISL_2236651, EPI_ISL_2236652, EPI_ISL_2236653, EPI_ISL_2236655, EPI_ISL_2236656, EPI_ISL_2236657, EPI_ISL_2236658, EPI_ISL_2236659, EPI_ISL_2236660, EPI_ISL_2236661, EPI_ISL_2236662, EPI_ISL_2236663, EPI_ISL_2236664, EPI_ISL_2236665, EPI_ISL_2236666, EPI_ISL_2236668, EPI_ISL_2236669,

EPI_ISL_2236670, EPI_ISL_2236671, EPI_ISL_2236672, EPI_ISL_2236673, EPI_ISL_2236675, EPI_ISL_2236676, EPI_ISL_2236677, EPI_ISL_2236681, EPI_ISL_2236684, EPI_ISL_2236685, EPI_ISL_2236688, EPI_ISL_2236689, EPI_ISL_2236690, EPI_ISL_2236691, EPI_ISL_2236692, EPI_ISL_2236694, EPI_ISL_2236695, EPI_ISL_2236696, EPI_ISL_2236698, EPI_ISL_2236699, EPI_ISL_2236700, EPI_ISL_2236701, EPI_ISL_2236703, EPI_ISL_2236704, EPI_ISL_2236705, EPI_ISL_2236707, EPI_ISL_2236708, EPI_ISL_2236711, EPI_ISL_2236712, EPI_ISL_2236713, EPI_ISL_2236715, EPI_ISL_2236716, EPI_ISL_2236717, EPI_ISL_2236718, EPI_ISL_2236719, EPI_ISL_2236720, EPI_ISL_2236721, EPI_ISL_2236722, EPI_ISL_2236723, EPI_ISL_2236724, EPI_ISL_2236725, EPI_ISL_2236726, EPI_ISL_2236727, EPI_ISL_2236728, EPI_ISL_2236729, EPI_ISL_2236730, EPI_ISL_2236731, EPI_ISL_2236732, EPI_ISL_2236733, EPI_ISL_2236734, EPI_ISL_2236735, EPI_ISL_2236736, EPI_ISL_2236738, EPI_ISL_2236739, EPI_ISL_2236740, EPI_ISL_2236741, EPI_ISL_2236742, EPI_ISL_2236743, EPI_ISL_2236745, EPI_ISL_2236746, EPI_ISL_2236747, EPI_ISL_2236751, EPI_ISL_2236753, EPI_ISL_2236757, EPI_ISL_2236758, EPI_ISL_2236760, EPI_ISL_2236761, EPI_ISL_2236762, EPI_ISL_2236763, EPI_ISL_2236764, EPI_ISL_2236766, EPI_ISL_2236767, EPI_ISL_2236769, EPI_ISL_2236772, EPI_ISL_2236774, EPI_ISL_2236775, EPI_ISL_2236777, EPI_ISL_2236778, EPI_ISL_2236779, EPI_ISL_2236780, EPI_ISL_2236782, EPI_ISL_2236783, EPI_ISL_2236784, EPI_ISL_2236786, EPI_ISL_2236787, EPI_ISL_2236788, EPI_ISL_2236789, EPI_ISL_2236790, EPI_ISL_2236791, EPI_ISL_2236792, EPI_ISL_2236794, EPI_ISL_2236795, EPI_ISL_2236797, EPI_ISL_2236798, EPI_ISL_2236802, EPI_ISL_2236803, EPI_ISL_2236805, EPI_ISL_2236807, EPI_ISL_2236809, EPI_ISL_2236810, EPI_ISL_2236811, EPI_ISL_2236812, EPI_ISL_2236813, EPI_ISL_2236814, EPI_ISL_2236815, EPI_ISL_2236816, EPI_ISL_2236818, EPI_ISL_2236819, EPI_ISL_2237115, EPI_ISL_2237415, EPI_ISL_2237416, EPI_ISL_2237417, EPI_ISL_2237419, EPI_ISL_2237420, EPI_ISL_2237421, EPI_ISL_2237423, EPI_ISL_2237424, EPI_ISL_2237425, EPI_ISL_2237426, EPI_ISL_2237428, EPI_ISL_2237430, EPI_ISL_2237432, EPI_ISL_2237433, EPI_ISL_2237434, EPI_ISL_2237442, EPI_ISL_2237444, EPI_ISL_2237449, EPI_ISL_2237450, EPI_ISL_2237456, EPI_ISL_2237457, EPI_ISL_2237459, EPI_ISL_2237460, EPI_ISL_2237462, EPI_ISL_2237467, EPI_ISL_2237470, EPI_ISL_2237471, EPI_ISL_2237474, EPI_ISL_2237476, EPI_ISL_2237477, EPI_ISL_2237478, EPI_ISL_2237479, EPI_ISL_2237481, EPI_ISL_2237482, EPI_ISL_2237484, EPI_ISL_2237485, EPI_ISL_2237489, EPI_ISL_2237490, EPI_ISL_2237491, EPI_ISL_2237492, EPI_ISL_2237493, EPI_ISL_2237497, EPI_ISL_2237499, EPI_ISL_2237501, EPI_ISL_2237502, EPI_ISL_2237505, EPI_ISL_2237506, EPI_ISL_2237507, EPI_ISL_2237508, EPI_ISL_2237510, EPI_ISL_2237511, EPI_ISL_2237513, EPI_ISL_2237514, EPI_ISL_2237516, EPI_ISL_2237517, EPI_ISL_2237522, EPI_ISL_2237523, EPI_ISL_2237524, EPI_ISL_2237525, EPI_ISL_2237526, EPI_ISL_2237527, EPI_ISL_2237528, EPI_ISL_2237531, EPI_ISL_2237534, EPI_ISL_2237535, EPI_ISL_2237536, EPI_ISL_2237538, EPI_ISL_2237540, EPI_ISL_2237541, EPI_ISL_2237542, EPI_ISL_2237543, EPI_ISL_2237550, EPI_ISL_2237552, EPI_ISL_2237554, EPI_ISL_2237555, EPI_ISL_2237561, EPI_ISL_2237562, EPI_ISL_2237563, EPI_ISL_2237564, EPI_ISL_2237565, EPI_ISL_2237566, EPI_ISL_2237568, EPI_ISL_2237569, EPI_ISL_2237573, EPI_ISL_2237576, EPI_ISL_2237578, EPI_ISL_2237579, EPI_ISL_2237581, EPI_ISL_2237582, EPI_ISL_2237583, EPI_ISL_2237588, EPI_ISL_2237590, EPI_ISL_2237591, EPI_ISL_2237592, EPI_ISL_2237593, EPI_ISL_2237594, EPI_ISL_2237596, EPI_ISL_2237597, EPI_ISL_2237598, EPI_ISL_2237602, EPI_ISL_2237603, EPI_ISL_2237604, EPI_ISL_2237607, EPI_ISL_2237608, EPI_ISL_2237609, EPI_ISL_2237613, EPI_ISL_2237615, EPI_ISL_2237617, EPI_ISL_2237619, EPI_ISL_2237621, EPI_ISL_2237630, EPI_ISL_2237631, EPI_ISL_2237632, EPI_ISL_2237633, EPI_ISL_2237634, EPI_ISL_2237636, EPI_ISL_2237637, EPI_ISL_2237638, EPI_ISL_2237639, EPI_ISL_2237640, EPI_ISL_2237642, EPI_ISL_2237643, EPI_ISL_2237644, EPI_ISL_2237645, EPI_ISL_2237646, EPI_ISL_2237648, EPI_ISL_2237650, EPI_ISL_2237651, EPI_ISL_2237652, EPI_ISL_2237653, EPI_ISL_2237658, EPI_ISL_2237659, EPI_ISL_2237660, EPI_ISL_2237666, EPI_ISL_2237668, EPI_ISL_2237669, EPI_ISL_2237671, EPI_ISL_2237672, EPI_ISL_2237673, EPI_ISL_2237674, EPI_ISL_2237676, EPI_ISL_2237677, EPI_ISL_2237679, EPI_ISL_2237680, EPI_ISL_2237681, EPI_ISL_2237682, EPI_ISL_2237683, EPI_ISL_2237688, EPI_ISL_2237689, EPI_ISL_2237690, EPI_ISL_2237691, EPI_ISL_2237693, EPI_ISL_2237695, EPI_ISL_2237696, EPI_ISL_2237700, EPI_ISL_2237701, EPI_ISL_2237705, EPI_ISL_2237706, EPI_ISL_2237707, EPI_ISL_2237709, EPI_ISL_2237710, EPI_ISL_2237711, EPI_ISL_2237712, EPI_ISL_2237714, EPI_ISL_2237715, EPI_ISL_2237717, EPI_ISL_2237718, EPI_ISL_2237722, EPI_ISL_2237725, EPI_ISL_2237727, EPI_ISL_2237728, EPI_ISL_2237729, EPI_ISL_2237730, EPI_ISL_2237736, EPI_ISL_2237737, EPI_ISL_2237738, EPI_ISL_2237740, EPI_ISL_2237741, EPI_ISL_2237745, EPI_ISL_2237746, EPI_ISL_2237747, EPI_ISL_2237748, EPI_ISL_2237749, EPI_ISL_2237750, EPI_ISL_2237751, EPI_ISL_2237752, EPI_ISL_2237753, EPI_ISL_2237760, EPI_ISL_2237762, EPI_ISL_2237766, EPI_ISL_2237767, EPI_ISL_2237768, EPI_ISL_2237774, EPI_ISL_2237778, EPI_ISL_2237780, EPI_ISL_2237785, EPI_ISL_2237786, EPI_ISL_2237787, EPI_ISL_2237789, EPI_ISL_2237790, EPI_ISL_2237791, EPI_ISL_2237792, EPI_ISL_2237794, EPI_ISL_2237805, EPI_ISL_2237807, EPI_ISL_2237808, EPI_ISL_2237810, EPI_ISL_2237811, EPI_ISL_2237812, EPI_ISL_2237817, EPI_ISL_2237822, EPI_ISL_2237824, EPI_ISL_2237825, EPI_ISL_2237829, EPI_ISL_2237836, EPI_ISL_2237837, EPI_ISL_2237846, EPI_ISL_2237847, EPI_ISL_2237848, EPI_ISL_2237850, EPI_ISL_2237853, EPI_ISL_2237855, EPI_ISL_2237859, EPI_ISL_2237866, EPI_ISL_2237869, EPI_ISL_2237873, EPI_ISL_2237875, EPI_ISL_2237876, EPI_ISL_2237886, EPI_ISL_2237887, EPI_ISL_2237888, EPI_ISL_2237890, EPI_ISL_2237895, EPI_ISL_2237896, EPI_ISL_2237901, EPI_ISL_2237902, EPI_ISL_2237903, EPI_ISL_2237905, EPI_ISL_2237912, EPI_ISL_2237915, EPI_ISL_2237919, EPI_ISL_2237921, EPI_ISL_2237924, EPI_ISL_2237927, EPI_ISL_2237929, EPI_ISL_2237933, EPI_ISL_2237935, EPI_ISL_2237936, EPI_ISL_2237943, EPI_ISL_2237944, EPI_ISL_2237946, EPI_ISL_2237947, EPI_ISL_2237949, EPI_ISL_2237950, EPI_ISL_2237951, EPI_ISL_2237953, EPI_ISL_2237954, EPI_ISL_2237959, EPI_ISL_2237962, EPI_ISL_2237963, EPI_ISL_2237967, EPI_ISL_2237969, EPI_ISL_2237973, EPI_ISL_2237974, EPI_ISL_2237975, EPI_ISL_2237976, EPI_ISL_2237977, EPI_ISL_2237978, EPI_ISL_2237982, EPI_ISL_2237983, EPI_ISL_2237988, EPI_ISL_2237989, EPI_ISL_2237991, EPI_ISL_2237994, EPI_ISL_2238001, EPI_ISL_2238002, EPI_ISL_2238003, EPI_ISL_2238005, EPI_ISL_2238007, EPI_ISL_2238008, EPI_ISL_2238013, EPI_ISL_2238014, EPI_ISL_2238015, EPI_ISL_2238016, EPI_ISL_2238018, EPI_ISL_2238022, EPI_ISL_2238027, EPI_ISL_2238033, EPI_ISL_2238035, EPI_ISL_2238036, EPI_ISL_2238044, EPI_ISL_2238047, EPI_ISL_2238050, EPI_ISL_2238052, EPI_ISL_2238053, EPI_ISL_2238055, EPI_ISL_2238056, EPI_ISL_2238059, EPI_ISL_2238063, EPI_ISL_2238065, EPI_ISL_2238066, EPI_ISL_2238681, EPI_ISL_2238682, EPI_ISL_2238683, EPI_ISL_2238684, EPI_ISL_2238685, EPI_ISL_2238686, EPI_ISL_2238693, EPI_ISL_2253227, EPI_ISL_2253230, EPI_ISL_2253235, EPI_ISL_2253238, EPI_ISL_2253245, EPI_ISL_2253247, EPI_ISL_2253248, EPI_ISL_2253255, EPI_ISL_2253256, EPI_ISL_2253271, EPI_ISL_2253272, EPI_ISL_2253273, EPI_ISL_2253294, EPI_ISL_2253300, EPI_ISL_2253305, EPI_ISL_2253307, EPI_ISL_2253314, EPI_ISL_2253329, EPI_ISL_2253331, EPI_ISL_2253342, EPI_ISL_2253344, EPI_ISL_2253353, EPI_ISL_2253356, EPI_ISL_2253357, EPI_ISL_2253358, EPI_ISL_2253359, EPI_ISL_2253377, EPI_ISL_2253380, EPI_ISL_2253390, EPI_ISL_2253391, EPI_ISL_2253395, EPI_ISL_2253397, EPI_ISL_2253402, EPI_ISL_2253406, EPI_ISL_2253411, EPI_ISL_2253416, EPI_ISL_2253419, EPI_ISL_2253429, EPI_ISL_2253434, EPI_ISL_2253443, EPI_ISL_2253448, EPI_ISL_2253456, EPI_ISL_2253458, EPI_ISL_2253461, EPI_ISL_2253462, EPI_ISL_2253468, EPI_ISL_2253477, EPI_ISL_2253478, EPI_ISL_2253480, EPI_ISL_2253481, EPI_ISL_2253482, EPI_ISL_2253487, EPI_ISL_2253489, EPI_ISL_2253493, EPI_ISL_2253496, EPI_ISL_2253512, EPI_ISL_2253513, EPI_ISL_2253522, EPI_ISL_2253524, EPI_ISL_2253525, EPI_ISL_2253533, EPI_ISL_2253542, EPI_ISL_2253553, EPI_ISL_2253557, EPI_ISL_2253565, EPI_ISL_2253567, EPI_ISL_2253572, EPI_ISL_2253573, EPI_ISL_2253574, EPI_ISL_2253576, EPI_ISL_2253577, EPI_ISL_2253580, EPI_ISL_2253583, EPI_ISL_2253599, EPI_ISL_2253600, EPI_ISL_2253612, EPI_ISL_2253618, EPI_ISL_2253629, EPI_ISL_2253632, EPI_ISL_2253634, EPI_ISL_2253635, EPI_ISL_2253638, EPI_ISL_2253640, EPI_ISL_2253642, EPI_ISL_2253655, EPI_ISL_2253656, EPI_ISL_2253670, EPI_ISL_2253678, EPI_ISL_2253679, EPI_ISL_2253680, EPI_ISL_2253681, EPI_ISL_2253682, EPI_ISL_2253683, EPI_ISL_2253684, EPI_ISL_2253685, EPI_ISL_2253686, EPI_ISL_2253687, EPI_ISL_2253688, EPI_ISL_2253689, EPI_ISL_2253690, EPI_ISL_2253692, EPI_ISL_2253693, EPI_ISL_2253694, EPI_ISL_2253695, EPI_ISL_2253697, EPI_ISL_2253698, EPI_ISL_2253699, EPI_ISL_2253700, EPI_ISL_2253701, EPI_ISL_2253702, EPI_ISL_2253703, EPI_ISL_2253704, EPI_ISL_2253706, EPI_ISL_2253707, EPI_ISL_2253708, EPI_ISL_2253709, EPI_ISL_2253710, EPI_ISL_2253711, EPI_ISL_2253712, EPI_ISL_2253713, EPI_ISL_2253714, EPI_ISL_2253716, EPI_ISL_2253717, EPI_ISL_2253718, EPI_ISL_2253719, EPI_ISL_2253720, EPI_ISL_2253721, EPI_ISL_2253722, EPI_ISL_2253723, EPI_ISL_2253724, EPI_ISL_2253725, EPI_ISL_2253726, EPI_ISL_2253729, EPI_ISL_2253730, EPI_ISL_2253731, EPI_ISL_2253732, EPI_ISL_2253733, EPI_ISL_2253734, EPI_ISL_2253735, EPI_ISL_2253736, EPI_ISL_2253737, EPI_ISL_2253738, EPI_ISL_2253739, EPI_ISL_2253740, EPI_ISL_2253741, EPI_ISL_2253742, EPI_ISL_2253743, EPI_ISL_2253744, EPI_ISL_2253745, EPI_ISL_2253746, EPI_ISL_2253747, EPI_ISL_2253748, EPI_ISL_2253750, EPI_ISL_2253751, EPI_ISL_2253752, EPI_ISL_2253753, EPI_ISL_2253754, EPI_ISL_2253755, EPI_ISL_2253756, EPI_ISL_2253757, EPI_ISL_2253758, EPI_ISL_2253759, EPI_ISL_2253760, EPI_ISL_2253761, EPI_ISL_2253762, EPI_ISL_2253764, EPI_ISL_2253765, EPI_ISL_2253767, EPI_ISL_2253768, EPI_ISL_2253769, EPI_ISL_2253770, EPI_ISL_2253771, EPI_ISL_2253772, EPI_ISL_2253773, EPI_ISL_2253775, EPI_ISL_2253776, EPI_ISL_2253778, EPI_ISL_2253779, EPI_ISL_2253780, EPI_ISL_2253781, EPI_ISL_2253782, EPI_ISL_2253783, EPI_ISL_2253784, EPI_ISL_2253785, EPI_ISL_2253786, EPI_ISL_2253787, EPI_ISL_2253788, EPI_ISL_2253789, EPI_ISL_2253790, EPI_ISL_2253791, EPI_ISL_2253792, EPI_ISL_2253793, EPI_ISL_2253795, EPI_ISL_2253796, EPI_ISL_2253797, EPI_ISL_2253798, EPI_ISL_2253799, EPI_ISL_2253800, EPI_ISL_2253801, EPI_ISL_2253803, EPI_ISL_2253804, EPI_ISL_2253805, EPI_ISL_2253806, EPI_ISL_2253807, EPI_ISL_2253808, EPI_ISL_2253809, EPI_ISL_2253810, EPI_ISL_2253811, EPI_ISL_2253812, EPI_ISL_2253813, EPI_ISL_2253814, EPI_ISL_2253815, EPI_ISL_2253816, EPI_ISL_2253817, EPI_ISL_2253819, EPI_ISL_2253821, EPI_ISL_2253822, EPI_ISL_2253823, EPI_ISL_2253824, EPI_ISL_2253825, EPI_ISL_2253826, EPI_ISL_2253827, EPI_ISL_2253828, EPI_ISL_2253829, EPI_ISL_2253830, EPI_ISL_2253831, EPI_ISL_2253832, EPI_ISL_2253833, EPI_ISL_2253834, EPI_ISL_2253835, EPI_ISL_2253836, EPI_ISL_2253837, EPI_ISL_2253838, EPI_ISL_2253839, EPI_ISL_2253840, EPI_ISL_2253841, EPI_ISL_2253842, EPI_ISL_2253843, EPI_ISL_2253844, EPI_ISL_2253845, EPI_ISL_2253846, EPI_ISL_2253847, EPI_ISL_2253848, EPI_ISL_2253849, EPI_ISL_2253850, EPI_ISL_2253852, EPI_ISL_2253853, EPI_ISL_2253854, EPI_ISL_2253855, EPI_ISL_2253856, EPI_ISL_2253858, EPI_ISL_2253859, EPI_ISL_2253861, EPI_ISL_2253863, EPI_ISL_2253864, EPI_ISL_2253865, EPI_ISL_2253866, EPI_ISL_2253867, EPI_ISL_2253868, EPI_ISL_2253869, EPI_ISL_2253870, EPI_ISL_2253871, EPI_ISL_2253872, EPI_ISL_2253873, EPI_ISL_2253874, EPI_ISL_2253876, EPI_ISL_2253877, EPI_ISL_2253879, EPI_ISL_2253880, EPI_ISL_2253881, EPI_ISL_2253883, EPI_ISL_2253884, EPI_ISL_2253885, EPI_ISL_2253886, EPI_ISL_2253887, EPI_ISL_2253888, EPI_ISL_2253889, EPI_ISL_2253890, EPI_ISL_2253891, EPI_ISL_2253892, EPI_ISL_2253893, EPI_ISL_2253895, EPI_ISL_2253896, EPI_ISL_2253897, EPI_ISL_2253899, EPI_ISL_2253900, EPI_ISL_2253901, EPI_ISL_2253902, EPI_ISL_2253903, EPI_ISL_2253904, EPI_ISL_2253905, EPI_ISL_2253906, EPI_ISL_2253907, EPI_ISL_2253908, EPI_ISL_2253909, EPI_ISL_2253910, EPI_ISL_2253911, EPI_ISL_2253913, EPI_ISL_2253914, EPI_ISL_2253915, EPI_ISL_2253916, EPI_ISL_2253917, EPI_ISL_2253918, EPI_ISL_2253919, EPI_ISL_2253920, EPI_ISL_2253921, EPI_ISL_2253922, EPI_ISL_2253923, EPI_ISL_2253925, EPI_ISL_2253926, EPI_ISL_2253927, EPI_ISL_2253928, EPI_ISL_2253932, EPI_ISL_2253933, EPI_ISL_2253934, EPI_ISL_2253935, EPI_ISL_2253936, EPI_ISL_2253937, EPI_ISL_2253943, EPI_ISL_2253953, EPI_ISL_2253962, EPI_ISL_2253976, EPI_ISL_2253977, EPI_ISL_2253980, EPI_ISL_2253982, EPI_ISL_2253984, EPI_ISL_2253995, EPI_ISL_2254000, EPI_ISL_2254005, EPI_ISL_2254010, EPI_ISL_2254012, EPI_ISL_2254014, EPI_ISL_2254015, EPI_ISL_2254020, EPI_ISL_2254028, EPI_ISL_2254029, EPI_ISL_2254039, EPI_ISL_2254053, EPI_ISL_2254057, EPI_ISL_2254060, EPI_ISL_2254071, EPI_ISL_2254087, EPI_ISL_2254121, EPI_ISL_2275014, EPI_ISL_2275029, EPI_ISL_2275038, EPI_ISL_2275053, EPI_ISL_2275064, EPI_ISL_2275066, EPI_ISL_2275071, EPI_ISL_2275080, EPI_ISL_2275085, EPI_ISL_2275094, EPI_ISL_2275097, EPI_ISL_2275100, EPI_ISL_2275175, EPI_ISL_2275184, EPI_ISL_2275187, EPI_ISL_2275191, EPI_ISL_2275197, EPI_ISL_2275199, EPI_ISL_2275200, EPI_ISL_2275203, EPI_ISL_2275212, EPI_ISL_2275216, EPI_ISL_2275220, EPI_ISL_2275222, EPI_ISL_2275223, EPI_ISL_2275228, EPI_ISL_2275230, EPI_ISL_2275235, EPI_ISL_2275246, EPI_ISL_2275249, EPI_ISL_2275253, EPI_ISL_2275258, EPI_ISL_2275259, EPI_ISL_2275260, EPI_ISL_2275263, EPI_ISL_2275270, EPI_ISL_2275273, EPI_ISL_2275275, EPI_ISL_2275280, EPI_ISL_2275281, EPI_ISL_2275284, EPI_ISL_2275285, EPI_ISL_2275293, EPI_ISL_2275294, EPI_ISL_2275296, EPI_ISL_2275297, EPI_ISL_2275299, EPI_ISL_2275300, EPI_ISL_2275303, EPI_ISL_2275310, EPI_ISL_2275311, EPI_ISL_2275313, EPI_ISL_2275314, EPI_ISL_2275317, EPI_ISL_2275322, EPI_ISL_2275331, EPI_ISL_2275333, EPI_ISL_2275334, EPI_ISL_2275342, EPI_ISL_2275344, EPI_ISL_2275350, EPI_ISL_2275352, EPI_ISL_2275353, EPI_ISL_2275355, EPI_ISL_2275357, EPI_ISL_2275361, EPI_ISL_2275365, EPI_ISL_2275367, EPI_ISL_2275368, EPI_ISL_2275372, EPI_ISL_2275373, EPI_ISL_2275374, EPI_ISL_2275378, EPI_ISL_2275379, EPI_ISL_2275391, EPI_ISL_2275392, EPI_ISL_2275397, EPI_ISL_2275400, EPI_ISL_2275401, EPI_ISL_2275403, EPI_ISL_2275404, EPI_ISL_2275405, EPI_ISL_2275576, EPI_ISL_2296691, EPI_ISL_2296693, EPI_ISL_2296695, EPI_ISL_2296696, EPI_ISL_2296697, EPI_ISL_2296698, EPI_ISL_2316284, EPI_ISL_2316285, EPI_ISL_2316287, EPI_ISL_2316288, EPI_ISL_2316302, EPI_ISL_2316311, EPI_ISL_2316313, EPI_ISL_2316314, EPI_ISL_2316316, EPI_ISL_2316317, EPI_ISL_2316320, EPI_ISL_2316322, EPI_ISL_2316325, EPI_ISL_2316329, EPI_ISL_2316332, EPI_ISL_2316340, EPI_ISL_2316343, EPI_ISL_2316346, EPI_ISL_2316351, EPI_ISL_2316354, EPI_ISL_2316358, EPI_ISL_2316368, EPI_ISL_2316379, EPI_ISL_2316390, EPI_ISL_2316395, EPI_ISL_2316398, EPI_ISL_2316400, EPI_ISL_2316403, EPI_ISL_2316408, EPI_ISL_2316417, EPI_ISL_2316420, EPI_ISL_2316423, EPI_ISL_2316426, EPI_ISL_2316428, EPI_ISL_2316431, EPI_ISL_2316432, EPI_ISL_2316438, EPI_ISL_2316440, EPI_ISL_2316442, EPI_ISL_2316444, EPI_ISL_2316447, EPI_ISL_2316450, EPI_ISL_2316451, EPI_ISL_2316454, EPI_ISL_2316457, EPI_ISL_2316460, EPI_ISL_2316462, EPI_ISL_2316465, EPI_ISL_2316467, EPI_ISL_2316472, EPI_ISL_2316475, EPI_ISL_2316479, EPI_ISL_2316480, EPI_ISL_2316482, EPI_ISL_2316483, EPI_ISL_2316484, EPI_ISL_2316486, EPI_ISL_2316488, EPI_ISL_2316489, EPI_ISL_2316492, EPI_ISL_2316494, EPI_ISL_2316495, EPI_ISL_2316508, EPI_ISL_2316510, EPI_ISL_2316520, EPI_ISL_2316522, EPI_ISL_2316526, EPI_ISL_2316529, EPI_ISL_2316530, EPI_ISL_2316544, EPI_ISL_2316557, EPI_ISL_2316559, EPI_ISL_2316564, EPI_ISL_2316570, EPI_ISL_2316572, EPI_ISL_2316573, EPI_ISL_2316581, EPI_ISL_2316587, EPI_ISL_2316593, EPI_ISL_2316608, EPI_ISL_2316616, EPI_ISL_2316619, EPI_ISL_2316621, EPI_ISL_2316623, EPI_ISL_2316624, EPI_ISL_2316626, EPI_ISL_2316627, EPI_ISL_2316629, EPI_ISL_2316630, EPI_ISL_2316639, EPI_ISL_2316643, EPI_ISL_2316645, EPI_ISL_2316646, EPI_ISL_2316652, EPI_ISL_2316653, EPI_ISL_2316655, EPI_ISL_2316658, EPI_ISL_2316659, EPI_ISL_2316662, EPI_ISL_2317243, EPI_ISL_2317253, EPI_ISL_2317363, EPI_ISL_2317464, EPI_ISL_2317468, EPI_ISL_2317477, EPI_ISL_2317492, EPI_ISL_2317502, EPI_ISL_2317522, EPI_ISL_2317528, EPI_ISL_2317533, EPI_ISL_2317543, EPI_ISL_2317583, EPI_ISL_2317602, EPI_ISL_2317606, EPI_ISL_2317628, EPI_ISL_2317631, EPI_ISL_2317634, EPI_ISL_2317635, EPI_ISL_2317636, EPI_ISL_2317706, EPI_ISL_2317709, EPI_ISL_2317739, EPI_ISL_2317740, EPI_ISL_2317742, EPI_ISL_2317755, EPI_ISL_2317761, EPI_ISL_2317765, EPI_ISL_2317770, EPI_ISL_2317773, EPI_ISL_2317781, EPI_ISL_2317793, EPI_ISL_2317797, EPI_ISL_2317798, EPI_ISL_2317801, EPI_ISL_2317805, EPI_ISL_2317809, EPI_ISL_2317815, EPI_ISL_2317818, EPI_ISL_2317825, EPI_ISL_2317831, EPI_ISL_2317834, EPI_ISL_2317837, EPI_ISL_2317848, EPI_ISL_2317852, EPI_ISL_2317855, EPI_ISL_2317864, EPI_ISL_2317865, EPI_ISL_2317870, EPI_ISL_2317873, EPI_ISL_2317875, EPI_ISL_2317883, EPI_ISL_2317887, EPI_ISL_2317890, EPI_ISL_2317899, EPI_ISL_2317911, EPI_ISL_2317919, EPI_ISL_2317922, EPI_ISL_2317929, EPI_ISL_2317935, EPI_ISL_2317937, EPI_ISL_2317942, EPI_ISL_2317945, EPI_ISL_2317949, EPI_ISL_2317955, EPI_ISL_2317960, EPI_ISL_2317971, EPI_ISL_2317975, EPI_ISL_2317982, EPI_ISL_2317984, EPI_ISL_2317985, EPI_ISL_2318003, EPI_ISL_2318006, EPI_ISL_2346568, EPI_ISL_2346998, EPI_ISL_2348480, EPI_ISL_2348484, EPI_ISL_2348485, EPI_ISL_2352812, EPI_ISL_2393503, EPI_ISL_2393618, EPI_ISL_2393716, EPI_ISL_2396820, EPI_ISL_2435397, EPI_ISL_2435423, EPI_ISL_2435503, EPI_ISL_2438544, EPI_ISL_2446425, EPI_ISL_2446531, EPI_ISL_2446620

| | | | |
|---|---|---|---|
| see above | Lighthouse Lab in Milton Keynes | Wellcome Sanger Institute for the COVID-19 Genomics UK (COG-UK) Consortium | Cordelia Langford; David K. Jackson; Dominic Kwiatkowski; Ewan Harrison; Ian Johnston; Jeffrey Barrett; John Sillitoe on behalf of the Wellcome Sanger Institute COVID-19 Surveillance Team; Roberto Amato; Sonia Goncalves; The Lighthouse Lab in Milton Keynes and Alex Alderton |

EPI_ISL_2278042, EPI_ISL_2278043, EPI_ISL_2278044, EPI_ISL_2278045, EPI_ISL_2278046, EPI_ISL_2278047, EPI_ISL_2278048, EPI_ISL_2278049, EPI_ISL_2278050, EPI_ISL_2278051, EPI_ISL_2278052, EPI_ISL_2278053, EPI_ISL_2278054, EPI_ISL_2278055, EPI_ISL_2278056, EPI_ISL_2278057, EPI_ISL_2278058, EPI_ISL_2278059, EPI_ISL_2278060, EPI_ISL_2278061, EPI_ISL_2278062, EPI_ISL_2278063, EPI_ISL_2278064, EPI_ISL_2278065, EPI_ISL_2278066, EPI_ISL_2278067, EPI_ISL_2278068, EPI_ISL_2278069, EPI_ISL_2278070, EPI_ISL_2278071, EPI_ISL_2278072, EPI_ISL_2278073, EPI_ISL_2278074, EPI_ISL_2278075, EPI_ISL_2278076, EPI_ISL_2278077, EPI_ISL_2278078, EPI_ISL_2278079, EPI_ISL_2278080, EPI_ISL_2278081, EPI_ISL_2278082, EPI_ISL_2278083, EPI_ISL_2278084, EPI_ISL_2278085, EPI_ISL_2278086, EPI_ISL_2278088, EPI_ISL_2278089, EPI_ISL_2278091, EPI_ISL_2278093, EPI_ISL_2278094, EPI_ISL_2278096, EPI_ISL_2278098, EPI_ISL_2278099, EPI_ISL_2278101, EPI_ISL_2278102, EPI_ISL_2278103, EPI_ISL_2278105, EPI_ISL_2278107, EPI_ISL_2278109, EPI_ISL_2278110, EPI_ISL_2278112, EPI_ISL_2278113, EPI_ISL_2278115, EPI_ISL_2278116, EPI_ISL_2278118, EPI_ISL_2278120, EPI_ISL_2278121, EPI_ISL_2278123, EPI_ISL_2278125, EPI_ISL_2278127, EPI_ISL_2278128, EPI_ISL_2278130, EPI_ISL_2278131, EPI_ISL_2278133, EPI_ISL_2278134, EPI_ISL_2278136, EPI_ISL_2278138, EPI_ISL_2278139, EPI_ISL_2278141, EPI_ISL_2278142, EPI_ISL_2278144, EPI_ISL_2278146, EPI_ISL_2278147, EPI_ISL_2278149, EPI_ISL_2278151, EPI_ISL_2278152, EPI_ISL_2278154, EPI_ISL_2278156, EPI_ISL_2278157, EPI_ISL_2278159, EPI_ISL_2278161, EPI_ISL_2278162, EPI_ISL_2278163, EPI_ISL_2278165, EPI_ISL_2278166, EPI_ISL_2278168, EPI_ISL_2278170, EPI_ISL_2278171, EPI_ISL_2278173

| | | | |
|---|---|---|---|
| see above | Lighthouse Laboratory Plymouth | Wellcome Sanger Institute for the COVID-19 Genomics UK (COG-UK) Consortium | Cordelia Langford; David K. Jackson; Dominic Kwiatkowski; Ewan Harrison; Ian Johnston; Jeffrey Barrett; John Sillitoe on behalf of the Wellcome Sanger Institute COVID-19 Surveillance Team; Lighthouse Laboratory Plymouth and Alex Alderton; Roberto Amato; Sonia Goncalves |

EPI_ISL_2238879, EPI_ISL_2238880, EPI_ISL_2238881, EPI_ISL_2238885, EPI_ISL_2238886, EPI_ISL_2238889, EPI_ISL_2238890, EPI_ISL_2238891, EPI_ISL_2238892, EPI_ISL_2238893, EPI_ISL_2238894, EPI_ISL_2355350, EPI_ISL_2355351, EPI_ISL_2355352, EPI_ISL_2355353, EPI_ISL_2355354, EPI_ISL_2355355, EPI_ISL_2355356, EPI_ISL_2355357, EPI_ISL_2355358, EPI_ISL_2355359, EPI_ISL_2355360, EPI_ISL_2355361, EPI_ISL_2355366, EPI_ISL_2355367, EPI_ISL_2355369, EPI_ISL_2355370, EPI_ISL_2355371, EPI_ISL_2434250, EPI_ISL_2434252

| | | | |
|---|---|---|---|
| see above | Liverpool Clinical Laboratories | COVID-19 Genomics UK (COG-UK) Consortium | A Alrezaihi; Alessandro Gerada; Alistair Darby; Angela Cowell; Anita Lucaci; Anu Chawla; Cassie Olateju; Catherine Hartley; Charlotte Nelson; Ecaterina Vamos; Elaine O'Toole; Eleanor G Bentley; Ghada T Shawli; Hermione Webster; Isabel García-Dorival; James Johnson; James P Stewart; Jenifer Manson; Joanne Watts; Jones Benjamin; Jordan J Clark; Julian Hiscox; L Luu; Lucille Rainbow; M Almsaud; Margaret Hughes; Mark Whitehead; Matthew Gemmell; Miren Iturriza-Gomara; Muhannad Alruwaili; N.P Randle; Neil Swainston; PKF Gilmore; Parul Sharma; Rebekah Penrice-Randal; Richard Eccles; Richard Gregory; Sam Haldenby; Steve Paterson; Stuart D Armstrong; Trevor Ian Robinson; Ximeng Han |

EPI_ISL_2239661, EPI_ISL_2239662, EPI_ISL_2239663, EPI_ISL_2239664, EPI_ISL_2239665, EPI_ISL_2239667, EPI_ISL_2239668, EPI_ISL_2239669, EPI_ISL_2239670, EPI_ISL_2239671, EPI_ISL_2239672, EPI_ISL_2239673, EPI_ISL_2239674, EPI_ISL_2239676, EPI_ISL_2239677, EPI_ISL_2239678, EPI_ISL_2239679, EPI_ISL_2239680, EPI_ISL_2239681, EPI_ISL_2239682, EPI_ISL_2239683, EPI_ISL_2239684, EPI_ISL_2239685, EPI_ISL_2239686, EPI_ISL_2239687, EPI_ISL_2239688, EPI_ISL_2239689, EPI_ISL_2239690, EPI_ISL_2239691, EPI_ISL_2239692, EPI_ISL_2239693, EPI_ISL_2239694, EPI_ISL_2239696, EPI_ISL_2239697, EPI_ISL_2239699, EPI_ISL_2239700, EPI_ISL_2239701, EPI_ISL_2239702, EPI_ISL_2239703, EPI_ISL_2239704, EPI_ISL_2239705, EPI_ISL_2239706, EPI_ISL_2239707, EPI_ISL_2239708, EPI_ISL_2239709, EPI_ISL_2239710, EPI_ISL_2239711, EPI_ISL_2239712, EPI_ISL_2239713, EPI_ISL_2239714, EPI_ISL_2239715, EPI_ISL_2239716, EPI_ISL_2239717, EPI_ISL_2239718, EPI_ISL_2239719, EPI_ISL_2239720, EPI_ISL_2239721, EPI_ISL_2239722, EPI_ISL_2239724, EPI_ISL_2239725, EPI_ISL_2239726, EPI_ISL_2239727, EPI_ISL_2239728, EPI_ISL_2239729, EPI_ISL_2239730, EPI_ISL_2239731, EPI_ISL_2239732, EPI_ISL_2239733, EPI_ISL_2239734, EPI_ISL_2239735, EPI_ISL_2239736, EPI_ISL_2239737, EPI_ISL_2239738, EPI_ISL_2239739, EPI_ISL_2239740, EPI_ISL_2239741, EPI_ISL_2239742, EPI_ISL_2239743, EPI_ISL_2239744, EPI_ISL_2239745, EPI_ISL_2239746, EPI_ISL_2239747, EPI_ISL_2239748, EPI_ISL_2239750, EPI_ISL_2239751, EPI_ISL_2239752, EPI_ISL_2239754, EPI_ISL_2239755, EPI_ISL_2239756, EPI_ISL_2239757, EPI_ISL_2239758, EPI_ISL_2239759, EPI_ISL_2239760, EPI_ISL_2239761, EPI_ISL_2239762, EPI_ISL_2239764, EPI_ISL_2239765, EPI_ISL_2239766, EPI_ISL_2239767, EPI_ISL_2239768, EPI_ISL_2239770, EPI_ISL_2239771, EPI_ISL_2239772, EPI_ISL_2239773, EPI_ISL_2239774, EPI_ISL_2239775, EPI_ISL_2239777, EPI_ISL_2239778, EPI_ISL_2239779, EPI_ISL_2239780, EPI_ISL_2239781, EPI_ISL_2239782, EPI_ISL_2239783, EPI_ISL_2239784, EPI_ISL_2239786, EPI_ISL_2239787, EPI_ISL_2239788, EPI_ISL_2239789, EPI_ISL_2239790, EPI_ISL_2239791, EPI_ISL_2239793, EPI_ISL_2239794, EPI_ISL_2239795, EPI_ISL_2239796, EPI_ISL_2239797, EPI_ISL_2239799, EPI_ISL_2239800, EPI_ISL_2239801, EPI_ISL_2239802, EPI_ISL_2239803, EPI_ISL_2239804, EPI_ISL_2239806, EPI_ISL_2239807, EPI_ISL_2239808, EPI_ISL_2239809, EPI_ISL_2239811, EPI_ISL_2239812, EPI_ISL_2239813, EPI_ISL_2239814, EPI_ISL_2239815, EPI_ISL_2239816, EPI_ISL_2239817, EPI_ISL_2239818, EPI_ISL_2239820, EPI_ISL_2239822, EPI_ISL_2239823, EPI_ISL_2239837, EPI_ISL_2239838, EPI_ISL_2239839, EPI_ISL_2239846, EPI_ISL_2239847, EPI_ISL_2239848, EPI_ISL_2239850, EPI_ISL_2239851, EPI_ISL_2239853, EPI_ISL_2239859, EPI_ISL_2239866, EPI_ISL_2239867, EPI_ISL_2239871, EPI_ISL_2239872, EPI_ISL_2239876, EPI_ISL_2239877, EPI_ISL_2239878, EPI_ISL_2239879, EPI_ISL_2239880, EPI_ISL_2239882, EPI_ISL_2239883, EPI_ISL_2239884, EPI_ISL_2239886, EPI_ISL_2239888, EPI_ISL_2239889, EPI_ISL_2239890, EPI_ISL_2239891, EPI_ISL_2239892, EPI_ISL_2239893, EPI_ISL_2239894, EPI_ISL_2239895, EPI_ISL_2239896, EPI_ISL_2239897, EPI_ISL_2239899, EPI_ISL_2239900, EPI_ISL_2239901, EPI_ISL_2239902, EPI_ISL_2239903, EPI_ISL_2239904, EPI_ISL_2239905, EPI_ISL_2239906, EPI_ISL_2239913, EPI_ISL_2239914, EPI_ISL_2239918, EPI_ISL_2239922, EPI_ISL_2239923, EPI_ISL_2239927, EPI_ISL_2239928, EPI_ISL_2239929, EPI_ISL_2239931, EPI_ISL_2239935, EPI_ISL_2239940, EPI_ISL_2239941, EPI_ISL_2239942, EPI_ISL_2239943, EPI_ISL_2239947, EPI_ISL_2239948, EPI_ISL_2239949, EPI_ISL_2239962, EPI_ISL_2239967, EPI_ISL_2239974, EPI_ISL_2239976, EPI_ISL_2239977, EPI_ISL_2239981, EPI_ISL_2239982, EPI_ISL_2239984, EPI_ISL_2239986, EPI_ISL_2239988, EPI_ISL_2239990, EPI_ISL_2239991, EPI_ISL_2239992, EPI_ISL_2239993, EPI_ISL_2239994, EPI_ISL_2239995, EPI_ISL_2239996, EPI_ISL_2239997, EPI_ISL_2239998, EPI_ISL_2239999, EPI_ISL_2240000, EPI_ISL_2240001, EPI_ISL_2240002, EPI_ISL_2240003, EPI_ISL_2240004, EPI_ISL_2240005, EPI_ISL_2240006, EPI_ISL_2240007, EPI_ISL_2240008, EPI_ISL_2240009, EPI_ISL_2240010, EPI_ISL_2240011, EPI_ISL_2240013, EPI_ISL_2240014, EPI_ISL_2240015, EPI_ISL_2240017, EPI_ISL_2240018, EPI_ISL_2240019, EPI_ISL_2240020, EPI_ISL_2240022, EPI_ISL_2240028, EPI_ISL_2240033, EPI_ISL_2240038, EPI_ISL_2240039, EPI_ISL_2240040, EPI_ISL_2240041, EPI_ISL_2240042, EPI_ISL_2240043, EPI_ISL_2240044, EPI_ISL_2240046, EPI_ISL_2240047, EPI_ISL_2240049, EPI_ISL_2240051, EPI_ISL_2240052, EPI_ISL_2240053, EPI_ISL_2240054, EPI_ISL_2240055, EPI_ISL_2240056, EPI_ISL_2240057, EPI_ISL_2240058, EPI_ISL_2240076, EPI_ISL_2240084, EPI_ISL_2240129, EPI_ISL_2240130, EPI_ISL_2240131, EPI_ISL_2240132, EPI_ISL_2240133, EPI_ISL_2240134, EPI_ISL_2240135, EPI_ISL_2240136, EPI_ISL_2240137, EPI_ISL_2240138, EPI_ISL_2240139, EPI_ISL_2240140, EPI_ISL_2240141, EPI_ISL_2240142, EPI_ISL_2240143, EPI_ISL_2240144, EPI_ISL_2240146, EPI_ISL_2240147, EPI_ISL_2240148, EPI_ISL_2240149, EPI_ISL_2240150, EPI_ISL_2240151, EPI_ISL_2240152, EPI_ISL_2240153, EPI_ISL_2240154, EPI_ISL_2240155, EPI_ISL_2240156, EPI_ISL_2240157, EPI_ISL_2240158, EPI_ISL_2240159, EPI_ISL_2240160, EPI_ISL_2240161, EPI_ISL_2240162, EPI_ISL_2240163, EPI_ISL_2240164, EPI_ISL_2240165, EPI_ISL_2240166, EPI_ISL_2240167, EPI_ISL_2240168, EPI_ISL_2240169, EPI_ISL_2240170, EPI_ISL_2240171, EPI_ISL_2240172, EPI_ISL_2240173, EPI_ISL_2240174, EPI_ISL_2240175, EPI_ISL_2240177, EPI_ISL_2240178, EPI_ISL_2240179, EPI_ISL_2240180, EPI_ISL_2240181, EPI_ISL_2240182, EPI_ISL_2240183, EPI_ISL_2240184, EPI_ISL_2240185, EPI_ISL_2240186, EPI_ISL_2240187, EPI_ISL_2240188, EPI_ISL_2240189, EPI_ISL_2240190, EPI_ISL_2240191, EPI_ISL_2240192, EPI_ISL_2240193, EPI_ISL_2240194, EPI_ISL_2240195, EPI_ISL_2240196, EPI_ISL_2240197, EPI_ISL_2240198, EPI_ISL_2240199, EPI_ISL_2240200, EPI_ISL_2240201, EPI_ISL_2240202, EPI_ISL_2240203, EPI_ISL_2240204, EPI_ISL_2240205, EPI_ISL_2240206, EPI_ISL_2240207, EPI_ISL_2240208, EPI_ISL_2240209, EPI_ISL_2240210, EPI_ISL_2240272, EPI_ISL_2355440, EPI_ISL_2355443, EPI_ISL_2355444, EPI_ISL_2355446, EPI_ISL_2355447, EPI_ISL_2355448, EPI_ISL_2355449, EPI_ISL_2355450, EPI_ISL_2355451, EPI_ISL_2355452, EPI_ISL_2355919, EPI_ISL_2355920, EPI_ISL_2355921, EPI_ISL_2355923, EPI_ISL_2355924, EPI_ISL_2355925, EPI_ISL_2355926, EPI_ISL_2355927, EPI_ISL_2355928, EPI_ISL_2355929, EPI_ISL_2355931, EPI_ISL_2355932, EPI_ISL_2355933, EPI_ISL_2355934, EPI_ISL_2355935, EPI_ISL_2355936, EPI_ISL_2355937, EPI_ISL_2355939, EPI_ISL_2355940, EPI_ISL_2355941, EPI_ISL_2355942, EPI_ISL_2355943, EPI_ISL_2355944, EPI_ISL_2355945, EPI_ISL_2355947, EPI_ISL_2355948, EPI_ISL_2355949, EPI_ISL_2355950, EPI_ISL_2355951, EPI_ISL_2355952, EPI_ISL_2355953, EPI_ISL_2355955, EPI_ISL_2355956, EPI_ISL_2355957, EPI_ISL_2355958, EPI_ISL_2355959, EPI_ISL_2355960, EPI_ISL_2355961, EPI_ISL_2355963, EPI_ISL_2355964, EPI_ISL_2355965, EPI_ISL_2355966, EPI_ISL_2355967, EPI_ISL_2355968, EPI_ISL_2355969, EPI_ISL_2355971, EPI_ISL_2355972, EPI_ISL_2355973, EPI_ISL_2355974, EPI_ISL_2355975, EPI_ISL_2355976, EPI_ISL_2355977, EPI_ISL_2355978, EPI_ISL_2355979, EPI_ISL_2355980, EPI_ISL_2355981, EPI_ISL_2355982, EPI_ISL_2355983, EPI_ISL_2355984, EPI_ISL_2355985, EPI_ISL_2355986, EPI_ISL_2355987, EPI_ISL_2355988, EPI_ISL_2355989, EPI_ISL_2355990, EPI_ISL_2355991, EPI_ISL_2355992, EPI_ISL_2355993, EPI_ISL_2355994, EPI_ISL_2355995, EPI_ISL_2355996, EPI_ISL_2355997, EPI_ISL_2355998, EPI_ISL_2355999, EPI_ISL_2356000, EPI_ISL_2356001, EPI_ISL_2356002, EPI_ISL_2356003, EPI_ISL_2356004, EPI_ISL_2356005, EPI_ISL_2356006, EPI_ISL_2356007, EPI_ISL_2356008, EPI_ISL_2356009, EPI_ISL_2356010, EPI_ISL_2356011, EPI_ISL_2356012, EPI_ISL_2356013, EPI_ISL_2356014, EPI_ISL_2356015, EPI_ISL_2356016, EPI_ISL_2356017, EPI_ISL_2356018, EPI_ISL_2356020, EPI_ISL_2356021, EPI_ISL_2356022, EPI_ISL_2356023, EPI_ISL_2356024, EPI_ISL_2356025, EPI_ISL_2356026, EPI_ISL_2356028, EPI_ISL_2356029, EPI_ISL_2356030, EPI_ISL_2356031, EPI_ISL_2356032, EPI_ISL_2356033, EPI_ISL_2356034, EPI_ISL_2356035, EPI_ISL_2356036, EPI_ISL_2356037, EPI_ISL_2356038, EPI_ISL_2356039, EPI_ISL_2356040, EPI_ISL_2356041, EPI_ISL_2356042, EPI_ISL_2356043, EPI_ISL_2356044, EPI_ISL_2356045, EPI_ISL_2356046, EPI_ISL_2356047, EPI_ISL_2356048, EPI_ISL_2356049, EPI_ISL_2356052, EPI_ISL_2356053, EPI_ISL_2356054, EPI_ISL_2356055, EPI_ISL_2356056, EPI_ISL_2356059, EPI_ISL_2356060, EPI_ISL_2356061, EPI_ISL_2356062, EPI_ISL_2356063, EPI_ISL_2356066, EPI_ISL_2356067, EPI_ISL_2356068, EPI_ISL_2356069, EPI_ISL_2356070, EPI_ISL_2356071, EPI_ISL_2356074, EPI_ISL_2356075, EPI_ISL_2356076, EPI_ISL_2356077, EPI_ISL_2356080, EPI_ISL_2356081, EPI_ISL_2356082, EPI_ISL_2356083, EPI_ISL_2356084, EPI_ISL_2356085, EPI_ISL_2356087, EPI_ISL_2356088, EPI_ISL_2356089, EPI_ISL_2356090, EPI_ISL_2356091, EPI_ISL_2356093, EPI_ISL_2356176, EPI_ISL_2356177, EPI_ISL_2356178, EPI_ISL_2356182, EPI_ISL_2356183, EPI_ISL_2356184, EPI_ISL_2356185, EPI_ISL_2356190, EPI_ISL_2356191, EPI_ISL_2356192, EPI_ISL_2356193, EPI_ISL_2356197, EPI_ISL_2356198, EPI_ISL_2356199, EPI_ISL_2356203, EPI_ISL_2356204, EPI_ISL_2356205, EPI_ISL_2356206, EPI_ISL_2356211, EPI_ISL_2356212, EPI_ISL_2356213, EPI_ISL_2356214, EPI_ISL_2356217, EPI_ISL_2356218, EPI_ISL_2356219, EPI_ISL_2356220, EPI_ISL_2356224, EPI_ISL_2356225, EPI_ISL_2356226, EPI_ISL_2356227, EPI_ISL_2356231, EPI_ISL_2356232, EPI_ISL_2356233, EPI_ISL_2356236, EPI_ISL_2356237, EPI_ISL_2356238, EPI_ISL_2356239, EPI_ISL_2356243, EPI_ISL_2356244, EPI_ISL_2356247, EPI_ISL_2356248, EPI_ISL_2356249, EPI_ISL_2356275, EPI_ISL_2356681, EPI_ISL_2356682, EPI_ISL_2434368, EPI_ISL_2434378, EPI_ISL_2434380, EPI_ISL_2434385, EPI_ISL_2434387, EPI_ISL_2434393, EPI_ISL_2434394, EPI_ISL_2434400, EPI_ISL_2434406, EPI_ISL_2434408, EPI_ISL_2434413, EPI_ISL_2434415, EPI_ISL_2434419, EPI_ISL_2434423, EPI_ISL_2434426, EPI_ISL_2434428, EPI_ISL_2434430, EPI_ISL_2434432, EPI_ISL_2434434, EPI_ISL_2434435, EPI_ISL_2434439, EPI_ISL_2434441, EPI_ISL_2434444, EPI_ISL_2434446, EPI_ISL_2434447, EPI_ISL_2434449, EPI_ISL_2434450, EPI_ISL_2434454, EPI_ISL_2434456, EPI_ISL_2434457, EPI_ISL_2434458, EPI_ISL_2434461, EPI_ISL_2434462, EPI_ISL_2434463, EPI_ISL_2434464, EPI_ISL_2434466, EPI_ISL_2434471, EPI_ISL_2434474, EPI_ISL_2434478, EPI_ISL_2434479, EPI_ISL_2434481, EPI_ISL_2434485, EPI_ISL_2434486, EPI_ISL_2434487, EPI_ISL_2434491, EPI_ISL_2434492, EPI_ISL_2434494, EPI_ISL_2434495, EPI_ISL_2434496, EPI_ISL_2434498, EPI_ISL_2434499, EPI_ISL_2434503, EPI_ISL_2434505, EPI_ISL_2434506, EPI_ISL_2434508, EPI_ISL_2434516, EPI_ISL_2434522, EPI_ISL_2434542, EPI_ISL_2434543, EPI_ISL_2434549, EPI_ISL_2434557, EPI_ISL_2434572, EPI_ISL_2434579, EPI_ISL_2434586, EPI_ISL_2434594, EPI_ISL_2434596, EPI_ISL_2434597, EPI_ISL_2434598, EPI_ISL_2434602, EPI_ISL_2434603, EPI_ISL_2434607, EPI_ISL_2434608, EPI_ISL_2434609, EPI_ISL_2434610, EPI_ISL_2434615, EPI_ISL_2434616, EPI_ISL_2434617, EPI_ISL_2434622, EPI_ISL_2434623, EPI_ISL_2434624, EPI_ISL_2434625, EPI_ISL_2434630, EPI_ISL_2434631, EPI_ISL_2434632, EPI_ISL_2434638, EPI_ISL_2434639, EPI_ISL_2434640, EPI_ISL_2434646, EPI_ISL_2434647, EPI_ISL_2434648, EPI_ISL_2434649, EPI_ISL_2434653, EPI_ISL_2434654, EPI_ISL_2434655, EPI_ISL_2434657, EPI_ISL_2434660, EPI_ISL_2434661, EPI_ISL_2434663, EPI_ISL_2434666, EPI_ISL_2434667, EPI_ISL_2434668, EPI_ISL_2434670, EPI_ISL_2434674, EPI_ISL_2434675, EPI_ISL_2434676, EPI_ISL_2434677, EPI_ISL_2434678

| | | | |
|---|---|---|---|
| see above | Northumbria University / South Tees Hospitals NHS Foundation Trust / North Cumbria Integrated Care NHS Foundation Trust / North Tees and Hartlepool NHS Foundation Trust / Newcastle Hospitals NHS Foundation Trust | COVID-19 Genomics UK (COG-UK) Consortium | Andrew Nelson; Brendan Payne; Clive Graham; Darren L Smith; Debra Padgett; Edward Barton; Emma Swindells; Garren Scott; Gary Black; Gary Eltringham; Giles S Holt; Greg R Young; Jane Greenaway; Jennifer Collins; John Allan; Joshua Loh; Lynn Dover; Matthew Bashton; Mohammad A Tariq; Paul Baker; Sarah Essex; Steve Liggett; Wen C Yew; Yusri Taha |

EPI_ISL_2240512, EPI_ISL_2240513, EPI_ISL_2240514, EPI_ISL_2240515, EPI_ISL_2240516, EPI_ISL_2240517, EPI_ISL_2240519, EPI_ISL_2240520, EPI_ISL_2240521, EPI_ISL_2240522, EPI_ISL_2240523, EPI_ISL_2240524, EPI_ISL_2240525, EPI_ISL_2240526, EPI_ISL_2240527, EPI_ISL_2240528, EPI_ISL_2240529, EPI_ISL_2240530, EPI_ISL_2240531, EPI_ISL_2240532, EPI_ISL_2240533, EPI_ISL_2240534, EPI_ISL_2240535, EPI_ISL_2240537, EPI_ISL_2240540, EPI_ISL_2240541, EPI_ISL_2240542, EPI_ISL_2240543, EPI_ISL_2240544, EPI_ISL_2240545, EPI_ISL_2240546, EPI_ISL_2240547, EPI_ISL_2240548, EPI_ISL_2240549, EPI_ISL_2240550, EPI_ISL_2240551, EPI_ISL_2240552, EPI_ISL_2240553, EPI_ISL_2240554, EPI_ISL_2240555, EPI_ISL_2240556, EPI_ISL_2240557, EPI_ISL_2240558, EPI_ISL_2240559, EPI_ISL_2240560, EPI_ISL_2240561, EPI_ISL_2240562, EPI_ISL_2240563, EPI_ISL_2240564, EPI_ISL_2240565, EPI_ISL_2240566, EPI_ISL_2240567, EPI_ISL_2240570, EPI_ISL_2240571, EPI_ISL_2240572, EPI_ISL_2240573, EPI_ISL_2240574, EPI_ISL_2240575, EPI_ISL_2240576, EPI_ISL_2240579, EPI_ISL_2240580, EPI_ISL_2240581, EPI_ISL_2240582, EPI_ISL_2240583, EPI_ISL_2240584, EPI_ISL_2240585, EPI_ISL_2240587, EPI_ISL_2240588, EPI_ISL_2240589, EPI_ISL_2240590, EPI_ISL_2240591, EPI_ISL_2240592, EPI_ISL_2240593, EPI_ISL_2240594, EPI_ISL_2240595, EPI_ISL_2240596, EPI_ISL_2240597, EPI_ISL_2240598, EPI_ISL_2240599, EPI_ISL_2240600, EPI_ISL_2240601, EPI_ISL_2240602, EPI_ISL_2240604, EPI_ISL_2240606, EPI_ISL_2240610, EPI_ISL_2240611, EPI_ISL_2240612, EPI_ISL_2240613,

| | | | |
|---|---|---|---|
| EPI_ISL_2240615, EPI_ISL_2240617, EPI_ISL_2240618, EPI_ISL_2240619, EPI_ISL_2240620, EPI_ISL_2240621, EPI_ISL_2240622, EPI_ISL_2240623, EPI_ISL_2240624, EPI_ISL_2240625, EPI_ISL_2240626, EPI_ISL_2240627, EPI_ISL_2240628, EPI_ISL_2240629, EPI_ISL_2240630, EPI_ISL_2240632, EPI_ISL_2240633, EPI_ISL_2240634, EPI_ISL_2240637, EPI_ISL_2240638, EPI_ISL_2240639, EPI_ISL_2240640, EPI_ISL_2240642, EPI_ISL_2240643, EPI_ISL_2240646, EPI_ISL_2240647, EPI_ISL_2240649, EPI_ISL_2240650, EPI_ISL_2240651, EPI_ISL_2240652, EPI_ISL_2240653, EPI_ISL_2240654, EPI_ISL_2240656, EPI_ISL_2240658, EPI_ISL_2240660, EPI_ISL_2434782, EPI_ISL_2434804 | | | |
| see above | Originating lab: Wales Specialist Virology Centre Sequencing lab: Pathogen Genomics Unit | Public Health Wales Microbiology Cardiff Wales Specialist Virology Centre | Alec Birchley; Alexander Adams; Amy Gaskin; Angela Marchbank; Bree Gatica-Wilcox; Catherine Moore; Jason Coombes; Joanne Watkins; Joel Southgate; Johnathan Evans; Laura Gifford; Lauren Gilbert; Lee Graham; Malorie Perry; Matthew Bull; Nicole Pacchiarini; Sally Corden; Sara Kumziene-Summerhayes; Sara Rey; Sarah Taylor; Simon Cottrell; Sophie Jones; Tom Connor |
| EPI_ISL_2356708, EPI_ISL_2356710, EPI_ISL_2356711, EPI_ISL_2356712, EPI_ISL_2356714, EPI_ISL_2356716, EPI_ISL_2356717, EPI_ISL_2434761, EPI_ISL_2434762, EPI_ISL_2434766, EPI_ISL_2434767, EPI_ISL_2434769, EPI_ISL_2434770, EPI_ISL_2434771, EPI_ISL_2434774, EPI_ISL_2434776, EPI_ISL_2434777, EPI_ISL_2434778, EPI_ISL_2434779 | | | |
| see above | Oxford Viromics, NDM, University of Oxford; Oxford University Hospitals; Basingstoke and North Hampshire Hospital | COVID-19 Genomics UK (COG-UK) Consortium | Alex Mobbs; Amy Trebes; Anita Justice; Catrin Moore; Christophe Fraser; David Bonsall; David Buck; Emma Wise; George Macintyre; Jessica Lynch; John Todd; Mariateresa de Cesare; Matilde Mori; Monique Andersson; Nathan Moore; Nick Cortes; Robert Shaw; Stephen Kidd; Tanya Golubchik; Timothy Peto |
| EPI_ISL_2356686, EPI_ISL_2356687, EPI_ISL_2356694 | Quadram Institute Bioscience | COVID-19 Genomics UK (COG-UK) Consortium | Alexander J Trotter; Alison E. Mather; Alp Aydin; Ana P. Tedim; Anastasia Kolyva; Andrew Bell; Andrew J. Page; Claire Stuart; Dave J. Baker; Gemma L. Kay; John Wain; Justin O'Grady; Leonardo de Oliveira Martins; Lizzie Meadows; Maria Diaz; Mark Webber; Muhammed Yasir; Nabil-Fareed Alikhan; Ngozi Elumogo; Nicholas M. Thomson; Rachael Stanley; Rachel Gilroy; Reenesh Prakash; Samir Dervisevic; Samuel Bloomfield; Steven Rudder; Thanh Le-Viet |
| EPI_ISL_2127623, EPI_ISL_2127624, EPI_ISL_2240449, EPI_ISL_2240450, EPI_ISL_2240451, EPI_ISL_2240452, EPI_ISL_2240453, EPI_ISL_2240454, EPI_ISL_2240461, EPI_ISL_2240462, EPI_ISL_2240463, EPI_ISL_2355343, EPI_ISL_2434758 | | | |
| see above | Queens Medical Centre, Clinical Microbiology Department / DeepSeq Nottingham | COVID-19 Genomics UK (COG-UK) Consortium | Christopher Moore; Emily Park; Fei Sang; Gemma Clark; Hannah Howson-Wells; Jack Hill; Jayasree Dey; Johnny Debebe; Jonathan Ball; Joseph Chappell; Manjinder Khakh; Matthew Carlisle; Matthew Loose; Michelle M Lister; Nadine Holmes; Patrick McClure; Theocharis Tsoleridis; Timothy Byaruhanga; Vicki M Fleming; Victoria Wright; Wendy Smith |
| EPI_ISL_2237194, EPI_ISL_2237244, EPI_ISL_2253696, EPI_ISL_2253705, EPI_ISL_2253727, EPI_ISL_2253728, EPI_ISL_2253766, EPI_ISL_2253774, EPI_ISL_2253777, EPI_ISL_2253794, EPI_ISL_2253802, EPI_ISL_2253857, EPI_ISL_2253862, EPI_ISL_2253875, EPI_ISL_2253878, EPI_ISL_2253882, EPI_ISL_2253898, EPI_ISL_2253924, EPI_ISL_2253931, EPI_ISL_2253938, EPI_ISL_2254073, EPI_ISL_2254074, EPI_ISL_2254075, EPI_ISL_2254076, EPI_ISL_2254077, EPI_ISL_2254078, EPI_ISL_2254079, EPI_ISL_2254080, EPI_ISL_2254081, EPI_ISL_2254082, EPI_ISL_2254083, EPI_ISL_2254084, EPI_ISL_2254094, EPI_ISL_2254095, EPI_ISL_2254096, EPI_ISL_2254097, EPI_ISL_2254098, EPI_ISL_2254099, EPI_ISL_2254100, EPI_ISL_2254101, EPI_ISL_2254102, EPI_ISL_2254103, EPI_ISL_2254105, EPI_ISL_2317192, EPI_ISL_2317756, EPI_ISL_2317769, EPI_ISL_2317774, EPI_ISL_2317776, EPI_ISL_2317784, EPI_ISL_2317787, EPI_ISL_2317808, EPI_ISL_2317810, EPI_ISL_2317811, EPI_ISL_2317846, EPI_ISL_2317847, EPI_ISL_2317851, EPI_ISL_2317886, EPI_ISL_2317906, EPI_ISL_2317916, EPI_ISL_2317930, EPI_ISL_2317947, EPI_ISL_2317954, EPI_ISL_2317966, EPI_ISL_2317978, EPI_ISL_2317979, EPI_ISL_2317980, EPI_ISL_2317994, EPI_ISL_2318000, EPI_ISL_2396819, EPI_ISL_2396821, EPI_ISL_2396822, EPI_ISL_2396823, EPI_ISL_2396824, EPI_ISL_2396825, EPI_ISL_2396826, EPI_ISL_2396827, EPI_ISL_2396828, EPI_ISL_2396829, EPI_ISL_2396830, EPI_ISL_2396831, EPI_ISL_2396839, EPI_ISL_2396840, EPI_ISL_2396841, EPI_ISL_2396842, EPI_ISL_2396843, EPI_ISL_2396844, EPI_ISL_2396846, EPI_ISL_2396847, EPI_ISL_2396848, EPI_ISL_2396850, EPI_ISL_2396851, EPI_ISL_2396852, EPI_ISL_2396853, EPI_ISL_2396854, EPI_ISL_2396856, EPI_ISL_2396857, EPI_ISL_2396858, EPI_ISL_2396859, EPI_ISL_2396860, EPI_ISL_2396861 | | | |
| see above | Randox Laboratories | Wellcome Sanger Institute for the COVID-19 Genomics UK (COG-UK) Consortium | Cordelia Langford; David K. Jackson; Dominic Kwiatkowski; Ewan Harrison; Ian Johnston; Jeffrey Barrett; John Sillitoe on behalf of the Wellcome Sanger Institute COVID-19 Surveillance Team; Randox Laboratories and Alex Alderton; Roberto Amato; Sonia Goncalves |
| EPI_ISL_2734942, EPI_ISL_2734943, EPI_ISL_2737725, EPI_ISL_2742429, EPI_ISL_2742430, EPI_ISL_2742433, EPI_ISL_2742435, EPI_ISL_2742440, EPI_ISL_2742441, EPI_ISL_2742443, EPI_ISL_2742444, EPI_ISL_2742447, EPI_ISL_2742448, EPI_ISL_2742449, EPI_ISL_2742450, EPI_ISL_2742451, EPI_ISL_2742458, EPI_ISL_2742459, EPI_ISL_2742463, EPI_ISL_2742464, EPI_ISL_2742465, EPI_ISL_2742466, EPI_ISL_2742467, EPI_ISL_2742469, EPI_ISL_2742471, EPI_ISL_2742473, EPI_ISL_2742474, EPI_ISL_2742476, EPI_ISL_2742477, EPI_ISL_2742478, EPI_ISL_2742482, EPI_ISL_2742486, EPI_ISL_2742489, EPI_ISL_2742490, EPI_ISL_2742491, EPI_ISL_2742492, EPI_ISL_2742493, EPI_ISL_2742495, EPI_ISL_2742496, EPI_ISL_2742499, EPI_ISL_2742500, EPI_ISL_2742501, EPI_ISL_2742502, EPI_ISL_2742503, EPI_ISL_2742504, EPI_ISL_2742505, EPI_ISL_2742506, EPI_ISL_2742507, EPI_ISL_2742508, EPI_ISL_2742509, EPI_ISL_2742510, EPI_ISL_2742511, EPI_ISL_2742512, EPI_ISL_2742517, EPI_ISL_2742518, EPI_ISL_2742521, EPI_ISL_2742522, EPI_ISL_2742524, EPI_ISL_2742525, EPI_ISL_2742526, EPI_ISL_2742528, EPI_ISL_2742529, EPI_ISL_2742530, EPI_ISL_2742531, EPI_ISL_2742533, EPI_ISL_2742534, EPI_ISL_2742536, EPI_ISL_2742537, EPI_ISL_2742538, EPI_ISL_2742539, EPI_ISL_2742540, EPI_ISL_2742541, EPI_ISL_2742542, EPI_ISL_2742543, EPI_ISL_2742544, EPI_ISL_2742545, EPI_ISL_2742546, EPI_ISL_2742547, EPI_ISL_2742548, EPI_ISL_2742549, EPI_ISL_2742550, EPI_ISL_2742551, EPI_ISL_2742552, EPI_ISL_2742553, EPI_ISL_2742554, EPI_ISL_2742555, EPI_ISL_2742556, EPI_ISL_2742557, EPI_ISL_2742558, EPI_ISL_2742559, EPI_ISL_2742560, EPI_ISL_2742561, EPI_ISL_2742562, EPI_ISL_2742563, EPI_ISL_2742564, EPI_ISL_2742565, EPI_ISL_2742566, EPI_ISL_2742567, EPI_ISL_2742568, EPI_ISL_2742569, EPI_ISL_2742570, EPI_ISL_2742571, EPI_ISL_2742572, EPI_ISL_2742573, EPI_ISL_2742574, EPI_ISL_2742575, EPI_ISL_2742576, EPI_ISL_2742577, EPI_ISL_2742578, EPI_ISL_2742579, EPI_ISL_2742580, EPI_ISL_2742581, EPI_ISL_2742582, EPI_ISL_2742583, EPI_ISL_2742584, EPI_ISL_2742585, EPI_ISL_2742586, EPI_ISL_2742587, EPI_ISL_2742589, EPI_ISL_2742590, EPI_ISL_2742591, EPI_ISL_2742592, EPI_ISL_2742593, EPI_ISL_2742594, EPI_ISL_2742595, EPI_ISL_2742596, EPI_ISL_2742597, EPI_ISL_2742598, EPI_ISL_2742599, EPI_ISL_2742600, EPI_ISL_2742601, EPI_ISL_2742602, EPI_ISL_2742603, EPI_ISL_2742604, EPI_ISL_2742605, EPI_ISL_2742606, EPI_ISL_2742607, EPI_ISL_2742608, EPI_ISL_2742609, EPI_ISL_2742610, EPI_ISL_2742611, EPI_ISL_2742612, EPI_ISL_2742613, EPI_ISL_2742614, EPI_ISL_2742615, EPI_ISL_2742616, EPI_ISL_2742617, EPI_ISL_2742618, EPI_ISL_2742620, EPI_ISL_2742621, EPI_ISL_2742622, EPI_ISL_2742623, EPI_ISL_2742625, EPI_ISL_2742626, EPI_ISL_2742627, EPI_ISL_2742629, EPI_ISL_2742636, EPI_ISL_2742637, EPI_ISL_2742638, EPI_ISL_2742639, EPI_ISL_2742640, EPI_ISL_2742641, EPI_ISL_2742642, EPI_ISL_2742644, EPI_ISL_2742646, EPI_ISL_2742647, EPI_ISL_2742648, EPI_ISL_2742649, EPI_ISL_2742650, EPI_ISL_2742651, EPI_ISL_2742653, EPI_ISL_2742654, EPI_ISL_2742655, EPI_ISL_2742656, EPI_ISL_2742657, EPI_ISL_2742658, EPI_ISL_2742659, EPI_ISL_2742660, EPI_ISL_2742661, EPI_ISL_2742663, EPI_ISL_2742664, EPI_ISL_2742665, EPI_ISL_2742666, EPI_ISL_2742667, EPI_ISL_2742668, EPI_ISL_2742669, EPI_ISL_2742670, EPI_ISL_2742671, EPI_ISL_2742672, EPI_ISL_2742676, EPI_ISL_2742677, EPI_ISL_2742845, EPI_ISL_2742895, EPI_ISL_2742922, EPI_ISL_2742923, EPI_ISL_2742924, EPI_ISL_2742925, EPI_ISL_2742926, EPI_ISL_2742927, EPI_ISL_2742928, EPI_ISL_2742929, EPI_ISL_2742930, EPI_ISL_2742931, EPI_ISL_2742932, EPI_ISL_2742933, EPI_ISL_2742936, EPI_ISL_2742937, EPI_ISL_2742938, EPI_ISL_2742939, EPI_ISL_2742941, EPI_ISL_2742942, EPI_ISL_2742943, EPI_ISL_2742944, EPI_ISL_2742945, EPI_ISL_2742946, EPI_ISL_2742947, EPI_ISL_2742948, EPI_ISL_2742949, EPI_ISL_2742951, EPI_ISL_2742952, EPI_ISL_2742953, EPI_ISL_2742955, EPI_ISL_2742956, EPI_ISL_2742957, EPI_ISL_2742958, EPI_ISL_2742959, EPI_ISL_2742960, EPI_ISL_2742961, EPI_ISL_2742963, EPI_ISL_2742964, EPI_ISL_2742965, EPI_ISL_2742966, EPI_ISL_2742967, EPI_ISL_2742968, EPI_ISL_2742969, EPI_ISL_2742970, EPI_ISL_2742971, EPI_ISL_2742972, EPI_ISL_2742974, EPI_ISL_2742975, EPI_ISL_2742976, EPI_ISL_2742977, EPI_ISL_2742978, EPI_ISL_2743208, EPI_ISL_2749589, EPI_ISL_2749592, EPI_ISL_2753581, EPI_ISL_2753582, EPI_ISL_2753585, EPI_ISL_2753586, EPI_ISL_2753590, EPI_ISL_2817378 | | | |
| see above | Respiratory Virus Unit, Microbiology Services Colindale, Public Health England | COVID-19 Genomics UK (COG-UK) Consortium | PHE Covid Sequencing Team |
| EPI_ISL_2238947, EPI_ISL_2238948, EPI_ISL_2238949, EPI_ISL_2238951, EPI_ISL_2238952, EPI_ISL_2238953, EPI_ISL_2238954, EPI_ISL_2238955, EPI_ISL_2238956, EPI_ISL_2238957, EPI_ISL_2238958, EPI_ISL_2238959, EPI_ISL_2238960, EPI_ISL_2238961, EPI_ISL_2238962, EPI_ISL_2238963, EPI_ISL_2238964, EPI_ISL_2238965, EPI_ISL_2238966, EPI_ISL_2238967, EPI_ISL_2238968, EPI_ISL_2238969, EPI_ISL_2238970, EPI_ISL_2238971, EPI_ISL_2238972, EPI_ISL_2238974, EPI_ISL_2238975, EPI_ISL_2238976, EPI_ISL_2238982, EPI_ISL_2355385, EPI_ISL_2355392, EPI_ISL_2355396, EPI_ISL_2355398, EPI_ISL_2355399, EPI_ISL_2355404, EPI_ISL_2355405, EPI_ISL_2355406, EPI_ISL_2355407, EPI_ISL_2355408, EPI_ISL_2355410, EPI_ISL_2355413, EPI_ISL_2355414, EPI_ISL_2355418, EPI_ISL_2355419, EPI_ISL_2355420, EPI_ISL_2355421, EPI_ISL_2355422, EPI_ISL_2355423, EPI_ISL_2355424 | | | |
| see above | University College London, Great Ormond Street Hospital for Children NHS Foundation Trust, Imperial College Healthcare NHS Trust | COVID-19 Genomics UK (COG-UK) Consortium | Alison Holmes; Charlotte Williams; Helena Tutill; Jacqueline Findlay; James Price; Judith Breuer; Julianne Brown; Kathryn Harris; Leysa Forrest; Mark Kristiansen; Paola Niola; Paola Resende Silva; Patricia Dyal; Paul Randell; Rachel Williams; Samuel Weeks; Sergi Castellano; Sunando Roy; Tony Brooks; Yasmin Panchbhaya |
| EPI_ISL_2238864, EPI_ISL_2238865, EPI_ISL_2238866, EPI_ISL_2238867, EPI_ISL_2238868 | University Hospitals Of Leicester NHS Trust and DeepSeq Nottingham | COVID-19 Genomics UK (COG-UK) Consortium | Christopher Holmes; Christopher Moore; Fei Sang; Joeseph Chappell; Johnny Debebe; Jonathan Ball; Julian Tang; Karlie Fallon; Matthew Carlisle; Matthew Loose; Nadine Holmes; Patrick McClure; Paul Bird; Thomas Helmer; Victoria Wright |
| EPI_ISL_2354505, EPI_ISL_2354506, EPI_ISL_2354507, EPI_ISL_2354508, EPI_ISL_2354509, EPI_ISL_2354523, EPI_ISL_2354526, EPI_ISL_2354527, EPI_ISL_2354528, EPI_ISL_2354529, EPI_ISL_2354530, EPI_ISL_2354531 | | | |
| see above | University of Birmingham | COVID-19 Genomics UK (COG-UK) Consortium | Alex Richter; Andrew Bosworth; Andrew Bosworth. Queen Elizabeth Hospital: Anna Casey; Andrew D Beggs PHE Heartlands Lab: Husam Osman; Charlotte Poxon; Institute of Microbiology; Joanne Stockton; Josh Quick; Kasun Wanigasooriya; Mike Kidd; Nicholas Loman. University of Birmingham Testing Laboratory: Celina M Whalley; Oliver Pickles; Radoslaw Poplawski; Samuel Nicholls; University of Birmingham: Claire McMurray; Will Rowe |
| EPI_ISL_2238753, EPI_ISL_2238754, EPI_ISL_2238757, EPI_ISL_2238759, EPI_ISL_2238760, EPI_ISL_2238762, EPI_ISL_2238763, EPI_ISL_2238764, EPI_ISL_2238765, EPI_ISL_2238788, EPI_ISL_2238789, EPI_ISL_2238790, EPI_ISL_2238791, EPI_ISL_2238792, EPI_ISL_2238793, EPI_ISL_2238794, EPI_ISL_2238795, EPI_ISL_2238796, EPI_ISL_2238797, EPI_ISL_2238798, EPI_ISL_2238799, EPI_ISL_2238800, EPI_ISL_2238801, EPI_ISL_2238802, EPI_ISL_2238803, EPI_ISL_2238804, EPI_ISL_2238805, EPI_ISL_2238806, EPI_ISL_2238807, EPI_ISL_2238808, EPI_ISL_2238811, EPI_ISL_2238812, EPI_ISL_2238813, EPI_ISL_2238814, EPI_ISL_2238815, EPI_ISL_2238816, EPI_ISL_2238817, EPI_ISL_2238818, EPI_ISL_2238819, EPI_ISL_2354550, EPI_ISL_2354551, EPI_ISL_2354552, EPI_ISL_2354553, EPI_ISL_2354554, EPI_ISL_2354555, EPI_ISL_2354556, EPI_ISL_2354557, EPI_ISL_2354560, EPI_ISL_2354562, EPI_ISL_2354564, EPI_ISL_2354567, EPI_ISL_2354568, EPI_ISL_2354581, EPI_ISL_2354583, EPI_ISL_2354585, EPI_ISL_2354591, EPI_ISL_2354593, EPI_ISL_2354595, EPI_ISL_2354596, EPI_ISL_2354611, EPI_ISL_2354612, EPI_ISL_2354617, EPI_ISL_2354618, EPI_ISL_2354624, EPI_ISL_2354625, EPI_ISL_2354626, EPI_ISL_2354627, EPI_ISL_2354628, EPI_ISL_2354629, EPI_ISL_2354633, EPI_ISL_2622335, EPI_ISL_2622336, EPI_ISL_2622337, EPI_ISL_2622338, EPI_ISL_2622339 | | | |
| see above | Virology Department, Royal Infirmary of Edinburgh, NHS Lothian / School of Biological Sciences, University of Edinburgh | COVID-19 Genomics UK (COG-UK) Consortium | Colquhoun R; Cotton S; Dewar R; Gallagher A; Hill V; Jackson B; Maloney D; McCrone JT; McHugh M; O'Toole Á; Rambaut A; Rooke S; Scher E; Templeton K; Yu X |
| EPI_ISL_2240679, EPI_ISL_2240684, EPI_ISL_2240685, EPI_ISL_2240686, EPI_ISL_2240688, EPI_ISL_2240689, EPI_ISL_2240692, EPI_ISL_2240693, EPI_ISL_2240695, EPI_ISL_2240696, EPI_ISL_2240698, EPI_ISL_2240699, EPI_ISL_2240700, EPI_ISL_2240701, EPI_ISL_2240702, EPI_ISL_2240703, EPI_ISL_2240706, EPI_ISL_2240710, EPI_ISL_2240711, EPI_ISL_2240713, EPI_ISL_2240716, EPI_ISL_2240718, EPI_ISL_2240719, EPI_ISL_2240723, EPI_ISL_2240724, EPI_ISL_2240725, EPI_ISL_2240726, EPI_ISL_2356888, EPI_ISL_2356898, EPI_ISL_2356902, EPI_ISL_2356908, EPI_ISL_2434968, EPI_ISL_2520946, EPI_ISL_2521054, EPI_ISL_2521082, EPI_ISL_2521099, EPI_ISL_2521139 | | | |
| see above | Virology Department, Sheffield Teaching Hospitals NHS Foundation Trust/Department of Infection, Immunity and Cardiovascular Disease, The Medical School, University of Sheffield | COVID-19 Genomics UK (COG-UK) Consortium | Adri Angyal; Alex Keeley; Benjamin Lindsey; Cariad Evans; Danielle Groves; Dave Partridge; Katie Johnson; Laura Carrilero; Luke Green; Matthew Parker; Matthew Wyles; Mehmet Yavuz; Mohammad Raza; Nikki Smith; Paul Parsons; Rachel Tucker; Rebecca Brown; Thushan de Silva |
| EPI_ISL_2356741, EPI_ISL_2356742, EPI_ISL_2356743, EPI_ISL_2356744, EPI_ISL_2356745, EPI_ISL_2356746, EPI_ISL_2356748, EPI_ISL_2356749, EPI_ISL_2356750, EPI_ISL_2356751, EPI_ISL_2356755, EPI_ISL_2356756, EPI_ISL_2356757, EPI_ISL_2356758, EPI_ISL_2356759, EPI_ISL_2356760, EPI_ISL_2356761, EPI_ISL_2356762, EPI_ISL_2356764, EPI_ISL_2356765, EPI_ISL_2356766, EPI_ISL_2356767, EPI_ISL_2356768, EPI_ISL_2356769, EPI_ISL_2356770, EPI_ISL_2356771, EPI_ISL_2356772, EPI_ISL_2356773, EPI_ISL_2356774, EPI_ISL_2356775, EPI_ISL_2356776, EPI_ISL_2356777, EPI_ISL_2356778, EPI_ISL_2356779, EPI_ISL_2356780, EPI_ISL_2356781, EPI_ISL_2356782, EPI_ISL_2356783, EPI_ISL_2356784, EPI_ISL_2356785, EPI_ISL_2356786, EPI_ISL_2356787, EPI_ISL_2356788, EPI_ISL_2356789, EPI_ISL_2356794, EPI_ISL_2356797, EPI_ISL_2356798, EPI_ISL_2356799, EPI_ISL_2356802, EPI_ISL_2356803, EPI_ISL_2356804, EPI_ISL_2356805, EPI_ISL_2356807, EPI_ISL_2356808, EPI_ISL_2356810, EPI_ISL_2356812, EPI_ISL_2356813, EPI_ISL_2356867, EPI_ISL_2356868, EPI_ISL_2356869, EPI_ISL_2356870, EPI_ISL_2356871, EPI_ISL_2356872, EPI_ISL_2356873, EPI_ISL_2356874, EPI_ISL_2356876, EPI_ISL_2520663 | | | |
| see above | Wales Specialist Virology Centre Sequencing lab: Pathogen Genomics Unit | Public Health Wales Microbiology Cardiff Wales Specialist Virology Centre | Alec Birchley; Alexander Adams; Amy Gaskin; Angela Marchbank; Bree Gatica-Wilcox; Catherine Moore; Jason Coombes; Joanne Watkins; Joel Southgate; Johnathan Evans; Laura Gifford; Lauren Gilbert; Lee Graham; Malorie Perry; Matthew Bull; Nicole Pacchiarini; Sally Corden; Sara Kumziene-Summerhayes; Sara Rey; Sarah Taylor; Simon Cottrell; Sophie Jones; Tom Connor |
| EPI_ISL_2434193, EPI_ISL_2434194, EPI_ISL_2434195, EPI_ISL_2434196, EPI_ISL_2434197, EPI_ISL_2434198, EPI_ISL_2434200, EPI_ISL_2434201, EPI_ISL_2517289, EPI_ISL_2815896, EPI_ISL_2815914, EPI_ISL_2815915 | | | |
| see above | West of Scotland Specialist Virology Centre, NHSGGC / MRC-University of Glasgow Centre for Virus Research | COVID-19 Genomics UK (COG-UK) Consortium | Alasdair MacLean; Alice Broos; Ana da Silva Filipe; Antonia Ho; Daniel Mair; David L Robertson; Emma Thomson; Guy Mollett; Ioulia Tsatsani; James Shepherd; Jenna Nichols; Joseph Hughes; Kathy Li; Kathy Smollett; Kyriaki Nomikou; Lily Tong; Matthew Holden; Natasha Johnson; Rachel Blacow; Richard Orton; Rory Gunson; Sarah McDonald; Sharif Shaaban; Sreenu Vattipally; Stephen Carmichael |

We gratefully acknowledge the following Authors from the Originating laboratories responsible for obtaining the specimens, as well as the Submitting laboratories where the genome data were generated and shared via GISAID, on which this research is based.

All Submitters of data may be contacted directly via www.gisaid.org

Authors are sorted alphabetically.

| Accession ID | Originating Laboratory | Submitting Laboratory | Authors |
|---|---|---|---|
| EPI_ISL_2686104, EPI_ISL_2686105, EPI_ISL_2686106, EPI_ISL_2686107, EPI_ISL_2686108, EPI_ISL_2686109, EPI_ISL_2686110, EPI_ISL_2686111 | | | |
| see above | Amazon Dx, AUV2 Laboratory | Amazon Dx, AUV2 Laboratory | Luke Meredith |
| EPI_ISL_2355373, EPI_ISL_2355374, EPI_ISL_2355375, EPI_ISL_2434259, EPI_ISL_2434265, EPI_ISL_2434266, EPI_ISL_2434267 | | | |
| see above | Barts Health NHS Trust | COVID-19 Genomics UK (COG-UK) Consortium | BROAD; Beatrix; CUTINO-MOGUEL; Claire; David; Dola; HARRINGTON; KELE; KULASEGARAN-SHYLINI; Maria-Teresa; OWOYEMI; Raghavendran |
| EPI_ISL_2354542, EPI_ISL_2354544, EPI_ISL_2354545, EPI_ISL_2354546 | Department of Pathology, University of Cambridge | COVID-19 Genomics UK (COG-UK) Consortium | Aminu S. Jahun; Ian Goodfellow; Iliana Georgana; Martin D. Curran; Myra Hosmillo; Rhys Izuagbe; Surendra Parmar; William L. Hamilton; Yasmin Chaudhry |
| EPI_ISL_2253228, EPI_ISL_2253232, EPI_ISL_2253304, EPI_ISL_2253319, EPI_ISL_2253326, EPI_ISL_2253332, EPI_ISL_2253364, EPI_ISL_2253366, EPI_ISL_2253383, EPI_ISL_2253396, EPI_ISL_2253431, EPI_ISL_2253457, EPI_ISL_2253460, EPI_ISL_2253486, EPI_ISL_2253491, EPI_ISL_2253499, EPI_ISL_2253519, EPI_ISL_2253578, EPI_ISL_2253614, EPI_ISL_2253660, EPI_ISL_2317170, EPI_ISL_2317171, EPI_ISL_2317187, EPI_ISL_2317188, EPI_ISL_2317193, EPI_ISL_2317199, EPI_ISL_2317204, EPI_ISL_2317218, EPI_ISL_2317223, EPI_ISL_2317236, EPI_ISL_2317241, EPI_ISL_2317245, EPI_ISL_2317248, EPI_ISL_2317252, EPI_ISL_2317255, EPI_ISL_2317257, EPI_ISL_2317262, EPI_ISL_2317279, EPI_ISL_2317282, EPI_ISL_2317286, EPI_ISL_2317296, EPI_ISL_2317297, EPI_ISL_2317319, EPI_ISL_2317333, EPI_ISL_2317337, EPI_ISL_2317343, EPI_ISL_2317351, EPI_ISL_2317357, EPI_ISL_2317364, EPI_ISL_2317372, EPI_ISL_2317378, EPI_ISL_2317380, EPI_ISL_2317387, EPI_ISL_2317391, EPI_ISL_2317402, EPI_ISL_2317411, EPI_ISL_2318033, EPI_ISL_2318042, EPI_ISL_2318052, EPI_ISL_2318066, EPI_ISL_2318103, EPI_ISL_2318109, EPI_ISL_2318112, EPI_ISL_2318114, EPI_ISL_2318124, EPI_ISL_2318152, EPI_ISL_2318183, EPI_ISL_2318206, EPI_ISL_2318216, EPI_ISL_2318221, EPI_ISL_2318248, EPI_ISL_2318265, EPI_ISL_2318341, EPI_ISL_2347353, EPI_ISL_2347373, EPI_ISL_2347382, EPI_ISL_2347386, EPI_ISL_2347391, EPI_ISL_2347403, EPI_ISL_2347408, EPI_ISL_2347410, EPI_ISL_2347436, EPI_ISL_2347445, EPI_ISL_2347452, EPI_ISL_2347469, EPI_ISL_2347473, EPI_ISL_2347480, EPI_ISL_2347488, EPI_ISL_2347506, EPI_ISL_2347512, EPI_ISL_2347523, EPI_ISL_2347531, EPI_ISL_2347536, EPI_ISL_2347541, EPI_ISL_2347557, EPI_ISL_2347558, EPI_ISL_2347564, EPI_ISL_2347568, EPI_ISL_2347569, EPI_ISL_2347577, EPI_ISL_2347580, EPI_ISL_2347612, EPI_ISL_2347636, EPI_ISL_2347652, EPI_ISL_2347683, EPI_ISL_2347708, EPI_ISL_2347770, EPI_ISL_2347773, EPI_ISL_2347776, EPI_ISL_2347804, EPI_ISL_2347807, EPI_ISL_2347861, EPI_ISL_2347872, EPI_ISL_2347893, EPI_ISL_2347901, EPI_ISL_2347912, EPI_ISL_2347913, EPI_ISL_2347919, EPI_ISL_2347922, EPI_ISL_2347930, EPI_ISL_2347941, EPI_ISL_2347956, EPI_ISL_2347967, EPI_ISL_2347968, EPI_ISL_2347999, EPI_ISL_2348004, EPI_ISL_2348007, EPI_ISL_2348012, EPI_ISL_2348016, EPI_ISL_2348029, EPI_ISL_2348038, EPI_ISL_2348046, EPI_ISL_2348057, EPI_ISL_2348058, EPI_ISL_2348065, EPI_ISL_2348084, EPI_ISL_2348090, EPI_ISL_2348094, EPI_ISL_2348098, EPI_ISL_2348111, EPI_ISL_2348126, EPI_ISL_2348136, EPI_ISL_2348162, EPI_ISL_2436937, EPI_ISL_2436940, EPI_ISL_2436949, EPI_ISL_2436970, EPI_ISL_2436979, EPI_ISL_2436989, EPI_ISL_2436995, EPI_ISL_2436999, EPI_ISL_2437003, EPI_ISL_2437004, EPI_ISL_2437006, EPI_ISL_2437009, EPI_ISL_2437017, EPI_ISL_2437022, EPI_ISL_2437023, EPI_ISL_2437025, EPI_ISL_2437039, EPI_ISL_2437044, EPI_ISL_2437075, EPI_ISL_2437095, EPI_ISL_2437104, EPI_ISL_2437109, EPI_ISL_2437114, EPI_ISL_2437117, EPI_ISL_2437122, EPI_ISL_2437123, EPI_ISL_2437131, EPI_ISL_2437136, EPI_ISL_2437137, EPI_ISL_2437142, EPI_ISL_2437147, EPI_ISL_2437151, EPI_ISL_2437168, EPI_ISL_2437170, EPI_ISL_2437174, EPI_ISL_2437176, EPI_ISL_2437181, EPI_ISL_2437196, EPI_ISL_2437214, EPI_ISL_2437223, EPI_ISL_2437247, EPI_ISL_2437255, EPI_ISL_2437263, EPI_ISL_2437264, EPI_ISL_2447266, EPI_ISL_2447304, EPI_ISL_2447349 | | | |
| see above | Health Services Laboratories | Wellcome Sanger Institute for the COVID-19 Genomics UK (COG-UK) Consortium | Cordelia Langford; David K. Jackson; Dominic Kwiatkowski; Ewan Harrison; Health Services Laboratories and Alex Alderton; Ian Johnston; Jeffrey Barrett; John Sillitoe on behalf of the Wellcome Sanger Institute COVID-19 Surveillance Team; Roberto Amato; Sonia Goncalves |
| EPI_ISL_2274987, EPI_ISL_2275002, EPI_ISL_2275012, EPI_ISL_2275055, EPI_ISL_2275057, EPI_ISL_2275062, EPI_ISL_2275103, EPI_ISL_2275111, EPI_ISL_2275128, EPI_ISL_2275137, EPI_ISL_2275139, EPI_ISL_2275155, EPI_ISL_2275179, EPI_ISL_2317750, EPI_ISL_2317752, EPI_ISL_2317758, EPI_ISL_2317763, EPI_ISL_2317764, EPI_ISL_2317767, EPI_ISL_2317768, EPI_ISL_2317771, EPI_ISL_2317777, EPI_ISL_2317783, EPI_ISL_2317786, EPI_ISL_2317794, EPI_ISL_2317812, EPI_ISL_2317820, EPI_ISL_2317822, EPI_ISL_2317827, EPI_ISL_2317828, EPI_ISL_2317845, EPI_ISL_2317849, EPI_ISL_2317850, EPI_ISL_2317858, EPI_ISL_2317861, EPI_ISL_2317863, EPI_ISL_2317866, EPI_ISL_2317869, EPI_ISL_2317871, EPI_ISL_2317876, EPI_ISL_2317877, EPI_ISL_2317882, EPI_ISL_2317889, EPI_ISL_2317895, EPI_ISL_2317902, EPI_ISL_2317904, EPI_ISL_2317915, EPI_ISL_2317920, EPI_ISL_2317923, EPI_ISL_2317927, EPI_ISL_2317928, EPI_ISL_2317932, EPI_ISL_2317938, EPI_ISL_2317940, EPI_ISL_2317950, EPI_ISL_2317952, EPI_ISL_2317953, EPI_ISL_2317959, EPI_ISL_2317961, EPI_ISL_2317962, EPI_ISL_2317965, EPI_ISL_2317972, EPI_ISL_2317976, EPI_ISL_2317981, EPI_ISL_2317983, EPI_ISL_2317986, EPI_ISL_2317989, EPI_ISL_2318004, EPI_ISL_2318012, EPI_ISL_2318013, EPI_ISL_2318015, EPI_ISL_2318022, EPI_ISL_2318023, EPI_ISL_2318026, EPI_ISL_2318027, EPI_ISL_2318028, EPI_ISL_2318029, EPI_ISL_2318030, EPI_ISL_2318031, EPI_ISL_2318034, EPI_ISL_2318036, EPI_ISL_2318039, EPI_ISL_2318040, EPI_ISL_2318041, EPI_ISL_2318049, EPI_ISL_2318050, EPI_ISL_2318051, EPI_ISL_2318055, EPI_ISL_2318058, EPI_ISL_2318059, EPI_ISL_2318062, EPI_ISL_2318068, EPI_ISL_2318071, EPI_ISL_2318072, EPI_ISL_2318073, EPI_ISL_2318074, EPI_ISL_2318075, EPI_ISL_2318077, EPI_ISL_2318078, EPI_ISL_2318079, EPI_ISL_2318080, EPI_ISL_2318088, EPI_ISL_2318089, EPI_ISL_2318094, EPI_ISL_2318095, EPI_ISL_2318096, EPI_ISL_2318097, EPI_ISL_2318098, EPI_ISL_2318099, EPI_ISL_2318101, EPI_ISL_2318104, EPI_ISL_2318105, EPI_ISL_2318106, EPI_ISL_2318107, EPI_ISL_2318108, EPI_ISL_2318116, EPI_ISL_2318121, EPI_ISL_2318122, EPI_ISL_2318127, EPI_ISL_2318128, EPI_ISL_2318131, EPI_ISL_2318132, EPI_ISL_2318133, EPI_ISL_2318135, EPI_ISL_2318136, EPI_ISL_2318137, EPI_ISL_2318138, EPI_ISL_2318139, EPI_ISL_2318144, EPI_ISL_2318147, EPI_ISL_2318149, EPI_ISL_2318151, EPI_ISL_2318154, EPI_ISL_2318155, EPI_ISL_2318157, EPI_ISL_2318160, EPI_ISL_2318161, EPI_ISL_2318162, EPI_ISL_2318163, EPI_ISL_2318165, EPI_ISL_2318166, EPI_ISL_2318168, EPI_ISL_2318169, EPI_ISL_2318170, EPI_ISL_2318172, EPI_ISL_2318176, EPI_ISL_2318177, EPI_ISL_2318178, EPI_ISL_2318180, EPI_ISL_2318184, EPI_ISL_2318187, EPI_ISL_2318188, EPI_ISL_2318189, EPI_ISL_2318190, EPI_ISL_2318191, EPI_ISL_2318194, EPI_ISL_2318198, EPI_ISL_2318199, EPI_ISL_2318201, EPI_ISL_2318203, EPI_ISL_2318204, EPI_ISL_2318207, EPI_ISL_2318209, EPI_ISL_2318210, EPI_ISL_2318212, EPI_ISL_2318213, EPI_ISL_2318219, EPI_ISL_2318220, EPI_ISL_2318222, EPI_ISL_2318223, EPI_ISL_2318227, EPI_ISL_2318230, EPI_ISL_2318232, EPI_ISL_2318234, EPI_ISL_2318236, EPI_ISL_2318237, EPI_ISL_2318238, EPI_ISL_2318239, EPI_ISL_2318240, EPI_ISL_2318244, EPI_ISL_2318247, EPI_ISL_2318250, EPI_ISL_2318256, EPI_ISL_2318257, EPI_ISL_2318258, EPI_ISL_2318260, EPI_ISL_2318261, EPI_ISL_2318262, EPI_ISL_2318263, EPI_ISL_2318266, EPI_ISL_2318268, EPI_ISL_2318270, EPI_ISL_2318272, EPI_ISL_2318273, EPI_ISL_2318274, EPI_ISL_2318276, EPI_ISL_2318281, EPI_ISL_2318282, EPI_ISL_2318283, EPI_ISL_2318284, EPI_ISL_2318285, EPI_ISL_2318286, EPI_ISL_2318287, EPI_ISL_2318288, EPI_ISL_2318291, EPI_ISL_2318294, EPI_ISL_2318295, EPI_ISL_2318296, EPI_ISL_2318297, EPI_ISL_2318299, EPI_ISL_2318303, EPI_ISL_2318304, EPI_ISL_2318305, EPI_ISL_2318307, EPI_ISL_2318308, EPI_ISL_2318310, EPI_ISL_2318313, EPI_ISL_2318314, EPI_ISL_2318315, EPI_ISL_2318318, EPI_ISL_2318319, EPI_ISL_2318321, EPI_ISL_2318323, EPI_ISL_2318324, EPI_ISL_2318325, EPI_ISL_2318326, EPI_ISL_2318327, EPI_ISL_2318328, EPI_ISL_2318332, EPI_ISL_2318335, EPI_ISL_2318336, EPI_ISL_2318337, EPI_ISL_2318338, EPI_ISL_2318342, EPI_ISL_2318344, EPI_ISL_2318345, EPI_ISL_2318346, EPI_ISL_2318347, EPI_ISL_2318348, EPI_ISL_2318354, EPI_ISL_2318355, EPI_ISL_2318356, EPI_ISL_2318357, EPI_ISL_2318359, EPI_ISL_2318362, EPI_ISL_2318363, EPI_ISL_2318364, EPI_ISL_2318365, EPI_ISL_2318369, EPI_ISL_2318375, EPI_ISL_2318381, EPI_ISL_2318382, EPI_ISL_2318384, EPI_ISL_2318387, EPI_ISL_2318391, EPI_ISL_2318394, EPI_ISL_2318395, EPI_ISL_2318396, EPI_ISL_2318397, EPI_ISL_2318401, EPI_ISL_2318404, EPI_ISL_2318405, EPI_ISL_2318406, EPI_ISL_2318407, EPI_ISL_2318408, EPI_ISL_2318410, EPI_ISL_2318411, EPI_ISL_2318413, EPI_ISL_2318416, EPI_ISL_2318417, EPI_ISL_2318418, EPI_ISL_2318420, EPI_ISL_2318421, EPI_ISL_2318422, EPI_ISL_2318424, EPI_ISL_2318425, EPI_ISL_2318427, EPI_ISL_2318430, EPI_ISL_2318431, EPI_ISL_2318432, EPI_ISL_2318435, EPI_ISL_2318436, EPI_ISL_2318440, EPI_ISL_2318441, EPI_ISL_2318442, EPI_ISL_2318443, EPI_ISL_2318447, EPI_ISL_2318448, EPI_ISL_2318449, EPI_ISL_2318451, EPI_ISL_2318452, EPI_ISL_2318455, EPI_ISL_2318458, EPI_ISL_2318459, EPI_ISL_2318460, EPI_ISL_2318461, EPI_ISL_2318466, EPI_ISL_2318476, EPI_ISL_2318478, EPI_ISL_2318479, EPI_ISL_2318480, EPI_ISL_2318483, EPI_ISL_2318485, EPI_ISL_2318494, EPI_ISL_2318495, EPI_ISL_2318497, EPI_ISL_2318498, EPI_ISL_2318499, EPI_ISL_2318502, EPI_ISL_2318503, EPI_ISL_2318505, EPI_ISL_2318508, EPI_ISL_2318517, EPI_ISL_2318518, EPI_ISL_2318520, EPI_ISL_2318522, EPI_ISL_2318523, EPI_ISL_2318524, EPI_ISL_2318525, EPI_ISL_2318526, EPI_ISL_2318527, EPI_ISL_2318528, EPI_ISL_2318529, EPI_ISL_2318531, EPI_ISL_2318533, EPI_ISL_2318534, EPI_ISL_2318535, EPI_ISL_2318538, EPI_ISL_2318539, EPI_ISL_2318540, EPI_ISL_2318541, EPI_ISL_2318542, EPI_ISL_2318543, EPI_ISL_2318546, EPI_ISL_2318549, EPI_ISL_2318552, EPI_ISL_2318554, EPI_ISL_2318556, EPI_ISL_2318557, EPI_ISL_2318558, EPI_ISL_2318559, EPI_ISL_2318561, EPI_ISL_2318566, EPI_ISL_2318567, EPI_ISL_2318568, EPI_ISL_2318570, EPI_ISL_2318571, EPI_ISL_2318574, EPI_ISL_2318580, EPI_ISL_2318582, EPI_ISL_2318583, EPI_ISL_2318584, EPI_ISL_2318586, EPI_ISL_2318587, EPI_ISL_2318588, EPI_ISL_2318591, EPI_ISL_2318592, EPI_ISL_2318595, EPI_ISL_2318596, EPI_ISL_2318597, EPI_ISL_2318598, EPI_ISL_2318599, EPI_ISL_2318600, EPI_ISL_2318602, EPI_ISL_2318603, EPI_ISL_2318606, EPI_ISL_2318608, EPI_ISL_2318609, EPI_ISL_2318610, EPI_ISL_2318612, EPI_ISL_2318613, EPI_ISL_2318616, EPI_ISL_2318617, EPI_ISL_2318619, EPI_ISL_2318620, EPI_ISL_2318622, EPI_ISL_2318623, EPI_ISL_2318626, EPI_ISL_2318628, EPI_ISL_2318631, EPI_ISL_2318632, EPI_ISL_2318633, EPI_ISL_2318634, EPI_ISL_2318635, EPI_ISL_2318638, EPI_ISL_2318639, EPI_ISL_2318640, EPI_ISL_2318643, EPI_ISL_2318644, EPI_ISL_2318645, EPI_ISL_2318646, EPI_ISL_2318651, EPI_ISL_2318652, EPI_ISL_2318654, EPI_ISL_2318655, EPI_ISL_2318656, EPI_ISL_2318657, EPI_ISL_2318660, EPI_ISL_2318661, EPI_ISL_2318662, EPI_ISL_2318664, EPI_ISL_2318667, EPI_ISL_2318669, EPI_ISL_2318670, EPI_ISL_2318671, EPI_ISL_2318672, EPI_ISL_2318673, EPI_ISL_2318675, EPI_ISL_2318680, EPI_ISL_2318682, EPI_ISL_2318684, EPI_ISL_2318685, EPI_ISL_2318689, EPI_ISL_2318691, EPI_ISL_2318692, EPI_ISL_2318694, EPI_ISL_2318696, EPI_ISL_2318698, EPI_ISL_2318699, EPI_ISL_2318701, EPI_ISL_2318703, EPI_ISL_2318706, EPI_ISL_2318708, EPI_ISL_2318713, EPI_ISL_2318714, EPI_ISL_2346474, EPI_ISL_2346475, EPI_ISL_2346478, EPI_ISL_2346482, EPI_ISL_2346485, EPI_ISL_2346487, EPI_ISL_2346492, EPI_ISL_2346495, EPI_ISL_2346497, EPI_ISL_2346498, EPI_ISL_2346499, EPI_ISL_2346501, EPI_ISL_2346502, EPI_ISL_2346504, EPI_ISL_2346508, EPI_ISL_2346509, EPI_ISL_2346514, EPI_ISL_2346515, EPI_ISL_2346516, EPI_ISL_2346517, EPI_ISL_2346518, EPI_ISL_2346520, EPI_ISL_2346522, EPI_ISL_2346526, EPI_ISL_2346528, EPI_ISL_2346537, EPI_ISL_2346541, EPI_ISL_2346543, EPI_ISL_2346544, EPI_ISL_2346545, EPI_ISL_2346554, EPI_ISL_2346555, EPI_ISL_2346556, EPI_ISL_2346557, EPI_ISL_2346559, EPI_ISL_2346564, EPI_ISL_2346567, EPI_ISL_2346569, EPI_ISL_2346571, EPI_ISL_2346572, EPI_ISL_2346588, EPI_ISL_2346591, EPI_ISL_2346592, EPI_ISL_2346593, EPI_ISL_2346597, EPI_ISL_2346602, EPI_ISL_2346603, EPI_ISL_2346605, EPI_ISL_2346606, EPI_ISL_2346609, EPI_ISL_2346613, EPI_ISL_2346619, EPI_ISL_2346620, EPI_ISL_2346624, EPI_ISL_2346627, EPI_ISL_2346629, EPI_ISL_2346633, EPI_ISL_2346634, EPI_ISL_2346635, EPI_ISL_2346636, EPI_ISL_2346637, EPI_ISL_2346639, EPI_ISL_2346643, EPI_ISL_2346644, EPI_ISL_2346650, EPI_ISL_2346651, EPI_ISL_2346652, EPI_ISL_2346655, EPI_ISL_2346656, EPI_ISL_2346659, EPI_ISL_2346660, EPI_ISL_2346661, EPI_ISL_2346663, EPI_ISL_2346666, EPI_ISL_2346668, EPI_ISL_2346669, EPI_ISL_2346672, EPI_ISL_2346675, EPI_ISL_2346676, EPI_ISL_2346684, EPI_ISL_2346690, EPI_ISL_2346692, EPI_ISL_2346698, EPI_ISL_2346703, EPI_ISL_2346705, EPI_ISL_2346709, EPI_ISL_2346711, EPI_ISL_2346715, EPI_ISL_2346719, EPI_ISL_2346721, EPI_ISL_2346722, EPI_ISL_2346723, EPI_ISL_2346725, EPI_ISL_2346728, EPI_ISL_2346729, EPI_ISL_2346730, EPI_ISL_2346732, EPI_ISL_2346733, EPI_ISL_2346734, EPI_ISL_2346735, EPI_ISL_2346737, EPI_ISL_2346740, EPI_ISL_2346741, EPI_ISL_2346742, EPI_ISL_2346744, EPI_ISL_2346745, EPI_ISL_2346747, EPI_ISL_2346748, EPI_ISL_2346749, EPI_ISL_2346750, EPI_ISL_2346751, EPI_ISL_2346752, EPI_ISL_2346753, EPI_ISL_2346754, EPI_ISL_2346755, EPI_ISL_2346756, EPI_ISL_2346757, EPI_ISL_2346759, EPI_ISL_2346761, EPI_ISL_2346762, EPI_ISL_2346763, EPI_ISL_2346764, EPI_ISL_2346765, EPI_ISL_2346766, EPI_ISL_2346767, EPI_ISL_2346768, EPI_ISL_2346771, EPI_ISL_2346772, EPI_ISL_2346773, EPI_ISL_2346775, EPI_ISL_2346777, EPI_ISL_2346780, EPI_ISL_2346781, EPI_ISL_2346782, EPI_ISL_2346783, EPI_ISL_2346784, EPI_ISL_2346786, EPI_ISL_2346788, EPI_ISL_2346789, EPI_ISL_2346790, EPI_ISL_2346792, EPI_ISL_2346793, EPI_ISL_2346795, EPI_ISL_2346796, EPI_ISL_2346798, EPI_ISL_2346799, EPI_ISL_2346802, EPI_ISL_2346803, EPI_ISL_2346804, EPI_ISL_2346805, EPI_ISL_2346808, EPI_ISL_2346811, EPI_ISL_2346812, EPI_ISL_2346813, EPI_ISL_2346814, EPI_ISL_2346815, EPI_ISL_2346816, EPI_ISL_2346817, EPI_ISL_2346818, EPI_ISL_2346821, EPI_ISL_2346823, EPI_ISL_2346824, EPI_ISL_2346825, EPI_ISL_2346826, EPI_ISL_2346827, EPI_ISL_2346829, EPI_ISL_2346832, EPI_ISL_2346834, EPI_ISL_2346835, EPI_ISL_2346836, EPI_ISL_2346841, EPI_ISL_2346842, EPI_ISL_2346844, EPI_ISL_2346845, EPI_ISL_2346846, EPI_ISL_2346848, EPI_ISL_2346850, EPI_ISL_2346851, EPI_ISL_2346852, EPI_ISL_2346854, EPI_ISL_2346856, EPI_ISL_2346857, EPI_ISL_2346859, EPI_ISL_2346860, EPI_ISL_2346861, EPI_ISL_2346865, EPI_ISL_2346866, EPI_ISL_2346867, EPI_ISL_2346868, EPI_ISL_2346869, EPI_ISL_2346870, EPI_ISL_2346871, EPI_ISL_2346873, EPI_ISL_2346874, EPI_ISL_2346875, EPI_ISL_2346878, EPI_ISL_2346879, EPI_ISL_2346880, EPI_ISL_2346881, EPI_ISL_2346883, EPI_ISL_2346884, EPI_ISL_2346885, EPI_ISL_2346886, EPI_ISL_2346887, EPI_ISL_2346889, EPI_ISL_2346891, EPI_ISL_2346892, EPI_ISL_2346894, EPI_ISL_2346895, EPI_ISL_2346896, EPI_ISL_2346897, EPI_ISL_2346898, EPI_ISL_2346899, EPI_ISL_2346900, EPI_ISL_2346901, EPI_ISL_2346902, EPI_ISL_2346905, EPI_ISL_2346906, EPI_ISL_2346908, EPI_ISL_2346909, EPI_ISL_2346911, EPI_ISL_2346912, EPI_ISL_2346913, EPI_ISL_2346914, EPI_ISL_2346915, EPI_ISL_2346916, EPI_ISL_2346917, EPI_ISL_2346918, EPI_ISL_2346923, EPI_ISL_2346924, EPI_ISL_2346925, EPI_ISL_2346926, EPI_ISL_2346927, EPI_ISL_2346929, EPI_ISL_2346930, EPI_ISL_2346931, EPI_ISL_2346932, EPI_ISL_2346933, EPI_ISL_2346934, EPI_ISL_2346937, EPI_ISL_2346938, EPI_ISL_2346939, EPI_ISL_2346940, EPI_ISL_2346942, EPI_ISL_2346943, EPI_ISL_2346945, EPI_ISL_2346947, EPI_ISL_2346948, EPI_ISL_2346949, EPI_ISL_2346950, EPI_ISL_2346952, EPI_ISL_2346954, EPI_ISL_2346956, EPI_ISL_2346958, EPI_ISL_2346959, EPI_ISL_2346960, EPI_ISL_2346961, EPI_ISL_2346962, EPI_ISL_2346963, EPI_ISL_2346965, EPI_ISL_2346966, EPI_ISL_2346967, EPI_ISL_2346968, EPI_ISL_2346970, EPI_ISL_2346972, EPI_ISL_2346974, EPI_ISL_2346976, EPI_ISL_2346979, EPI_ISL_2346981, EPI_ISL_2346982, EPI_ISL_2346983, EPI_ISL_2346984, EPI_ISL_2346985, EPI_ISL_2346986, EPI_ISL_2346988, EPI_ISL_2346989, EPI_ISL_2346990, EPI_ISL_2346991, EPI_ISL_2346992, EPI_ISL_2346993, EPI_ISL_2346994, EPI_ISL_2346995, EPI_ISL_2346996, EPI_ISL_2346997, EPI_ISL_2346999, EPI_ISL_2347001, EPI_ISL_2347002, EPI_ISL_2347003, EPI_ISL_2347004, EPI_ISL_2347005, EPI_ISL_2347006, EPI_ISL_2347008, EPI_ISL_2347011, EPI_ISL_2347015, EPI_ISL_2347018, EPI_ISL_2347019, EPI_ISL_2347021, EPI_ISL_2347024, EPI_ISL_2347025, EPI_ISL_2347026, EPI_ISL_2347027, EPI_ISL_2347029, EPI_ISL_2347030, EPI_ISL_2347031, EPI_ISL_2347032, EPI_ISL_2347033, EPI_ISL_2347035, EPI_ISL_2347036, EPI_ISL_2347037, EPI_ISL_2347038, EPI_ISL_2347039, EPI_ISL_2347040, EPI_ISL_2347041, EPI_ISL_2347044, EPI_ISL_2347045, EPI_ISL_2347046, EPI_ISL_2347047, EPI_ISL_2347048, EPI_ISL_2347049, EPI_ISL_2347050, EPI_ISL_2347052, EPI_ISL_2347054, EPI_ISL_2347055, EPI_ISL_2347056, EPI_ISL_2347057, EPI_ISL_2347058, EPI_ISL_2347060, EPI_ISL_2347061, EPI_ISL_2347062, EPI_ISL_2347064, EPI_ISL_2347066, EPI_ISL_2347068, EPI_ISL_2347069, EPI_ISL_2347071, EPI_ISL_2347073, EPI_ISL_2347074, EPI_ISL_2347075, EPI_ISL_2347076, EPI_ISL_2347077, EPI_ISL_2347078, EPI_ISL_2347079, EPI_ISL_2347080, EPI_ISL_2347086, EPI_ISL_2347087, EPI_ISL_2347334, EPI_ISL_2347335, EPI_ISL_2347336, EPI_ISL_2347337, EPI_ISL_2347338, EPI_ISL_2347339, EPI_ISL_2347340, EPI_ISL_2347341, EPI_ISL_2347342, EPI_ISL_2347343, EPI_ISL_2347344, EPI_ISL_2347345, EPI_ISL_2347346, EPI_ISL_2347348, EPI_ISL_2347349, EPI_ISL_2347350, EPI_ISL_2347351, EPI_ISL_2347352, EPI_ISL_2347354, EPI_ISL_2347355, EPI_ISL_2347356, EPI_ISL_2347357, EPI_ISL_2347358, EPI_ISL_2347359, EPI_ISL_2347360, EPI_ISL_2347361, EPI_ISL_2347362, EPI_ISL_2347363, EPI_ISL_2347364, EPI_ISL_2347365, EPI_ISL_2347366, EPI_ISL_2347367, EPI_ISL_2347368, EPI_ISL_2347369, EPI_ISL_2347370, EPI_ISL_2347371, EPI_ISL_2347372, EPI_ISL_2347374, EPI_ISL_2347375, EPI_ISL_2347376, EPI_ISL_2347377, EPI_ISL_2347378, EPI_ISL_2347379, EPI_ISL_2347380, EPI_ISL_2347381, EPI_ISL_2347383, EPI_ISL_2347384, EPI_ISL_2347385, EPI_ISL_2347387, EPI_ISL_2347389, EPI_ISL_2347390, EPI_ISL_2347392, EPI_ISL_2347393, EPI_ISL_2347394, EPI_ISL_2347395, EPI_ISL_2347396, EPI_ISL_2347397, EPI_ISL_2347398, EPI_ISL_2347399, EPI_ISL_2347400, EPI_ISL_2347401, EPI_ISL_2347402, EPI_ISL_2347404, EPI_ISL_2347405, EPI_ISL_2347406, EPI_ISL_2347407, EPI_ISL_2347409, EPI_ISL_2347413, EPI_ISL_2347414, EPI_ISL_2347415, EPI_ISL_2347416, EPI_ISL_2347417, EPI_ISL_2347419, EPI_ISL_2347420, EPI_ISL_2347421, EPI_ISL_2347422, EPI_ISL_2347423, EPI_ISL_2347424, EPI_ISL_2347425, EPI_ISL_2347427, EPI_ISL_2347428, EPI_ISL_2347429, EPI_ISL_2347430, EPI_ISL_2347431, EPI_ISL_2347432, EPI_ISL_2347433, EPI_ISL_2347434, EPI_ISL_2347435, EPI_ISL_2347438, EPI_ISL_2347439, EPI_ISL_2347440, EPI_ISL_2347442, EPI_ISL_2347443, EPI_ISL_2347444, EPI_ISL_2347446, EPI_ISL_2347447, EPI_ISL_2347448, EPI_ISL_2347449, EPI_ISL_2347450, EPI_ISL_2347451, EPI_ISL_2347453, EPI_ISL_2347454, EPI_ISL_2347455, EPI_ISL_2347456, EPI_ISL_2347457, EPI_ISL_2347458, EPI_ISL_2347459, EPI_ISL_2347460, EPI_ISL_2347461, EPI_ISL_2347463, EPI_ISL_2347464, EPI_ISL_2347465, EPI_ISL_2347466, EPI_ISL_2347467, EPI_ISL_2347468, EPI_ISL_2347470, EPI_ISL_2347471, EPI_ISL_2347472, EPI_ISL_2347474, EPI_ISL_2347475, EPI_ISL_2347476, EPI_ISL_2347477, EPI_ISL_2347478, EPI_ISL_2347479, EPI_ISL_2347481, EPI_ISL_2347482, EPI_ISL_2347483, EPI_ISL_2347484, EPI_ISL_2347485, EPI_ISL_2347486, EPI_ISL_2347487, EPI_ISL_2347489, EPI_ISL_2347490, EPI_ISL_2347491, EPI_ISL_2347492, EPI_ISL_2347493, EPI_ISL_2347494, EPI_ISL_2347495, EPI_ISL_2347496, EPI_ISL_2347497, EPI_ISL_2347498, EPI_ISL_2347499, EPI_ISL_2347500, EPI_ISL_2347501, EPI_ISL_2347502, EPI_ISL_2347503, EPI_ISL_2347504, EPI_ISL_2347505, EPI_ISL_2347507, EPI_ISL_2347508, EPI_ISL_2347509, EPI_ISL_2347510, EPI_ISL_2347511, EPI_ISL_2347513, EPI_ISL_2347514, EPI_ISL_2347515, EPI_ISL_2347516, EPI_ISL_2347517, EPI_ISL_2347518, EPI_ISL_2347519, EPI_ISL_2347520, EPI_ISL_2347521, EPI_ISL_2347522, EPI_ISL_2347524, EPI_ISL_2347525, EPI_ISL_2347526, EPI_ISL_2347527, EPI_ISL_2347528, EPI_ISL_2347529, EPI_ISL_2347530, EPI_ISL_2347532, EPI_ISL_2347533, EPI_ISL_2347534, EPI_ISL_2347535, EPI_ISL_2347537, EPI_ISL_2347538, EPI_ISL_2347539, EPI_ISL_2347540, EPI_ISL_2347542, EPI_ISL_2347543, EPI_ISL_2347544, EPI_ISL_2347545, EPI_ISL_2347546, EPI_ISL_2347547, EPI_ISL_2347548, EPI_ISL_2347549, EPI_ISL_2347550, EPI_ISL_2347552, EPI_ISL_2347553, EPI_ISL_2347555, EPI_ISL_2347556, EPI_ISL_2347559, EPI_ISL_2347560, EPI_ISL_2347561, EPI_ISL_2347562, EPI_ISL_2347563, EPI_ISL_2347565, EPI_ISL_2347566, EPI_ISL_2347567, EPI_ISL_2347570, EPI_ISL_2347572, EPI_ISL_2347573, EPI_ISL_2347574, EPI_ISL_2347575, EPI_ISL_2347576, EPI_ISL_2347578, EPI_ISL_2347579, EPI_ISL_2347581, EPI_ISL_2347582, EPI_ISL_2347583, EPI_ISL_2347584, EPI_ISL_2347585, EPI_ISL_2347586, EPI_ISL_2347587, EPI_ISL_2347588, EPI_ISL_2347589, EPI_ISL_2347590, EPI_ISL_2347591, EPI_ISL_2347592, EPI_ISL_2347593, EPI_ISL_2347594, EPI_ISL_2347595, EPI_ISL_2347596, EPI_ISL_2347597, EPI_ISL_2347598, EPI_ISL_2347599, EPI_ISL_2347600, EPI_ISL_2347601, EPI_ISL_2347602, EPI_ISL_2347603, EPI_ISL_2347604, EPI_ISL_2347605, EPI_ISL_2347606, EPI_ISL_2347607, EPI_ISL_2347608, EPI_ISL_2347609, EPI_ISL_2347610, EPI_ISL_2347611, EPI_ISL_2347613, EPI_ISL_2347614, EPI_ISL_2347615, EPI_ISL_2347616, EPI_ISL_2347617, EPI_ISL_2347619, EPI_ISL_2347620, EPI_ISL_2347621, EPI_ISL_2347622, EPI_ISL_2347623, EPI_ISL_2347624, EPI_ISL_2347625, EPI_ISL_2347626, EPI_ISL_2347627, EPI_ISL_2347628, EPI_ISL_2347629, EPI_ISL_2347630, EPI_ISL_2347631, EPI_ISL_2347632, EPI_ISL_2347633, EPI_ISL_2347634, EPI_ISL_2347637, EPI_ISL_2347640, EPI_ISL_2347641, EPI_ISL_2347642, EPI_ISL_2347644, EPI_ISL_2347645, EPI_ISL_2347646, EPI_ISL_2347647, EPI_ISL_2347648, EPI_ISL_2347649, EPI_ISL_2347650, EPI_ISL_2347654, EPI_ISL_2347656, EPI_ISL_2347657, EPI_ISL_2347659, EPI_ISL_2347660, EPI_ISL_2347661, EPI_ISL_2347664, EPI_ISL_2347665, EPI_ISL_2347666, EPI_ISL_2347667, EPI_ISL_2347668, EPI_ISL_2347670, EPI_ISL_2347671, EPI_ISL_2347672, EPI_ISL_2347674, EPI_ISL_2347675, EPI_ISL_2347677, EPI_ISL_2347678, EPI_ISL_2347679, EPI_ISL_2347680, EPI_ISL_2347682, EPI_ISL_2347685, EPI_ISL_2347686, EPI_ISL_2347687, EPI_ISL_2347688, EPI_ISL_2347689, EPI_ISL_2347690, EPI_ISL_2347691, EPI_ISL_2347692, EPI_ISL_2347693, EPI_ISL_2347695, EPI_ISL_2347696, EPI_ISL_2347697, EPI_ISL_2347698, EPI_ISL_2347699, EPI_ISL_2347700, EPI_ISL_2347701, EPI_ISL_2347702, EPI_ISL_2347703, EPI_ISL_2347705, EPI_ISL_2347706, EPI_ISL_2347709, EPI_ISL_2347711, EPI_ISL_2347713, EPI_ISL_2347714, EPI_ISL_2347715, EPI_ISL_2347716, EPI_ISL_2347717, EPI_ISL_2347718, EPI_ISL_2347719, EPI_ISL_2347720, EPI_ISL_2347722, EPI_ISL_2347723, EPI_ISL_2347724, EPI_ISL_2347725, EPI_ISL_2347726, EPI_ISL_2347727, EPI_ISL_2347729, EPI_ISL_2347731, EPI_ISL_2347732, EPI_ISL_2347733, EPI_ISL_2347734, EPI_ISL_2347735, EPI_ISL_2347736, EPI_ISL_2347737, EPI_ISL_2347738, EPI_ISL_2347739, EPI_ISL_2347740, EPI_ISL_2347742, EPI_ISL_2347743, EPI_ISL_2347746, EPI_ISL_2347747, EPI_ISL_2347748, EPI_ISL_2347750, EPI_ISL_2347751, EPI_ISL_2347752, EPI_ISL_2347754, EPI_ISL_2347755, EPI_ISL_2347757, EPI_ISL_2347758, EPI_ISL_2347759, EPI_ISL_2347760, EPI_ISL_2347761, EPI_ISL_2347762, EPI_ISL_2347763, EPI_ISL_2347764, EPI_ISL_2347766, EPI_ISL_2347767, EPI_ISL_2347768, EPI_ISL_2347769, EPI_ISL_2347771, EPI_ISL_2347772, EPI_ISL_2347774, EPI_ISL_2347775, EPI_ISL_2347777, EPI_ISL_2347778, EPI_ISL_2347780, EPI_ISL_2347781, EPI_ISL_2347782, EPI_ISL_2347783, EPI_ISL_2347785, EPI_ISL_2347786, EPI_ISL_2347787, EPI_ISL_2347788, EPI_ISL_2347789, EPI_ISL_2347790, EPI_ISL_2347794, EPI_ISL_2347795, EPI_ISL_2347796, EPI_ISL_2347797, EPI_ISL_2347798, EPI_ISL_2347799, EPI_ISL_2347800, EPI_ISL_2347801, EPI_ISL_2347802, EPI_ISL_2347803, EPI_ISL_2347806, EPI_ISL_2347809, EPI_ISL_2347810, EPI_ISL_2347811, EPI_ISL_2347812, EPI_ISL_2347813, EPI_ISL_2347814, EPI_ISL_2347815, EPI_ISL_2347816, EPI_ISL_2347817, EPI_ISL_2347818, EPI_ISL_2347820, EPI_ISL_2347821, EPI_ISL_2347822, EPI_ISL_2347823, EPI_ISL_2347824, EPI_ISL_2347826, EPI_ISL_2347827, EPI_ISL_2347829, EPI_ISL_2347830, EPI_ISL_2347831, EPI_ISL_2347833, EPI_ISL_2347834, EPI_ISL_2347835, EPI_ISL_2347836, EPI_ISL_2347837, EPI_ISL_2347838, EPI_ISL_2347839, EPI_ISL_2347841, EPI_ISL_2347842, EPI_ISL_2347844, EPI_ISL_2347845, EPI_ISL_2347846, EPI_ISL_2347847, EPI_ISL_2347848, EPI_ISL_2347849, EPI_ISL_2347850, EPI_ISL_2347852, EPI_ISL_2347853, EPI_ISL_2347854, EPI_ISL_2347855, EPI_ISL_2347856, EPI_ISL_2347857, EPI_ISL_2347858, EPI_ISL_2347859, EPI_ISL_2347860, EPI_ISL_2347862, EPI_ISL_2347864, EPI_ISL_2347867, EPI_ISL_2347870, EPI_ISL_2347873, EPI_ISL_2347875, EPI_ISL_2347876, EPI_ISL_2347877, EPI_ISL_2347881, EPI_ISL_2347884, EPI_ISL_2347885, EPI_ISL_2347887, EPI_ISL_2347888, EPI_ISL_2347889, EPI_ISL_2347890, EPI_ISL_2347891, EPI_ISL_2347892, EPI_ISL_2347896, EPI_ISL_2347897, EPI_ISL_2347899, EPI_ISL_2347900, EPI_ISL_2347902, EPI_ISL_2347904, EPI_ISL_2347906, EPI_ISL_2347907, EPI_ISL_2347908, EPI_ISL_2347910, EPI_ISL_2347914, EPI_ISL_2347915, EPI_ISL_2347916, EPI_ISL_2347917, EPI_ISL_2347918, EPI_ISL_2347921, EPI_ISL_2347925, EPI_ISL_2347926, EPI_ISL_2347927, EPI_ISL_2347928, EPI_ISL_2347929, EPI_ISL_2347931, EPI_ISL_2347932, EPI_ISL_2347938, EPI_ISL_2347940, EPI_ISL_2347942, EPI_ISL_2347945, EPI_ISL_2347947, EPI_ISL_2347949, EPI_ISL_2347950, EPI_ISL_2347952, EPI_ISL_2347954, EPI_ISL_2347957, EPI_ISL_2347958, EPI_ISL_2347959, EPI_ISL_2347960, EPI_ISL_2347962, EPI_ISL_2347963, EPI_ISL_2347964, EPI_ISL_2347965, EPI_ISL_2347969, EPI_ISL_2347970, EPI_ISL_2347972, EPI_ISL_2347973, EPI_ISL_2347977, EPI_ISL_2347978, EPI_ISL_2347979, EPI_ISL_2347980, EPI_ISL_2347981, EPI_ISL_2347982, EPI_ISL_2347984, EPI_ISL_2347986, EPI_ISL_2347988, EPI_ISL_2347989, EPI_ISL_2347990, EPI_ISL_2347991, EPI_ISL_2347993, EPI_ISL_2347994, EPI_ISL_2347995, EPI_ISL_2348000, EPI_ISL_2348001, EPI_ISL_2348002, EPI_ISL_2348003, EPI_ISL_2348005, EPI_ISL_2348008, EPI_ISL_2348009, EPI_ISL_2348018, EPI_ISL_2348021, EPI_ISL_2348023, EPI_ISL_2348024, EPI_ISL_2348025, EPI_ISL_2348026, EPI_ISL_2348027, EPI_ISL_2348031, EPI_ISL_2348032, EPI_ISL_2348034, EPI_ISL_2348036, EPI_ISL_2348043, EPI_ISL_2348044, EPI_ISL_2348045, EPI_ISL_2348047, EPI_ISL_2348048, EPI_ISL_2348049, EPI_ISL_2348052, EPI_ISL_2348054, EPI_ISL_2348055, EPI_ISL_2348059, EPI_ISL_2348060, EPI_ISL_2348063, EPI_ISL_2348066, EPI_ISL_2348067, EPI_ISL_2348069, EPI_ISL_2348070, EPI_ISL_2348072, EPI_ISL_2348075, EPI_ISL_2348076, EPI_ISL_2348077, EPI_ISL_2348078, EPI_ISL_2348079, EPI_ISL_2348080, EPI_ISL_2348083, EPI_ISL_2348085, EPI_ISL_2348086, EPI_ISL_2348087, EPI_ISL_2348089, EPI_ISL_2348091, EPI_ISL_2348095, EPI_ISL_2348097, EPI_ISL_2348099, EPI_ISL_2348100, EPI_ISL_2348101, EPI_ISL_2348102, EPI_ISL_2348104, EPI_ISL_2348106, EPI_ISL_2348107, EPI_ISL_2348108, EPI_ISL_2348110, EPI_ISL_2348113, EPI_ISL_2348114, EPI_ISL_2348115, EPI_ISL_2348116, EPI_ISL_2348117, EPI_ISL_2348119, EPI_ISL_2348120, EPI_ISL_2348123, EPI_ISL_2348124, EPI_ISL_2348125, EPI_ISL_2348128, EPI_ISL_2348131, EPI_ISL_2348133, EPI_ISL_2348135, EPI_ISL_2348137, EPI_ISL_2348138, EPI_ISL_2348139, EPI_ISL_2348140, EPI_ISL_2348141, EPI_ISL_2348142, EPI_ISL_2348143, EPI_ISL_2348145, EPI_ISL_2348146, EPI_ISL_2348147, EPI_ISL_2348148, EPI_ISL_2348154, EPI_ISL_2348155, EPI_ISL_2348156, EPI_ISL_2348158, EPI_ISL_2348160, EPI_ISL_2348161, EPI_ISL_2348165, EPI_ISL_2348166, EPI_ISL_2348167, EPI_ISL_2348169, EPI_ISL_2348170, EPI_ISL_2348172, EPI_ISL_2348175, EPI_ISL_2348176, EPI_ISL_2348177, EPI_ISL_2348178, EPI_ISL_2348179, EPI_ISL_2348180, EPI_ISL_2348182, EPI_ISL_2348183, EPI_ISL_2348184, EPI_ISL_2348185, EPI_ISL_2348188, EPI_ISL_2348194, EPI_ISL_2348195, EPI_ISL_2348199, EPI_ISL_2348200, EPI_ISL_2348201, EPI_ISL_2348202, EPI_ISL_2348204, EPI_ISL_2348211, EPI_ISL_2348212, EPI_ISL_2348216, EPI_ISL_2348218, EPI_ISL_2348219, EPI_ISL_2348222, EPI_ISL_2348224, EPI_ISL_2348229, EPI_ISL_2348231, EPI_ISL_2348232, EPI_ISL_2348239, EPI_ISL_2348240, EPI_ISL_2348241, EPI_ISL_2348245, EPI_ISL_2348252, EPI_ISL_2348253, EPI_ISL_2348254, EPI_ISL_2348257, EPI_ISL_2348258, EPI_ISL_2348261, EPI_ISL_2348262, EPI_ISL_2348264, EPI_ISL_2348265, EPI_ISL_2348268, EPI_ISL_2348271, EPI_ISL_2348272, EPI_ISL_2348273, EPI_ISL_2348274, EPI_ISL_2348275, EPI_ISL_2348277, EPI_ISL_2348279, EPI_ISL_2348280, EPI_ISL_2348284, EPI_ISL_2348285, EPI_ISL_2348286, EPI_ISL_2348287, EPI_ISL_2348288, EPI_ISL_2348290, EPI_ISL_2348292, EPI_ISL_2348296, EPI_ISL_2348297, EPI_ISL_2348301, EPI_ISL_2348302, EPI_ISL_2348303, EPI_ISL_2348304, EPI_ISL_2348306, EPI_ISL_2348308, EPI_ISL_2348309, EPI_ISL_2348311, EPI_ISL_2348315, EPI_ISL_2348316, EPI_ISL_2348317, EPI_ISL_2348323, EPI_ISL_2348326, EPI_ISL_2348327, EPI_ISL_2348328, EPI_ISL_2348331, EPI_ISL_2348336, EPI_ISL_2348338, EPI_ISL_2348342, EPI_ISL_2348344, EPI_ISL_2348345, EPI_ISL_2348346, EPI_ISL_2348347, EPI_ISL_2348348, EPI_ISL_2348349, EPI_ISL_2348354, EPI_ISL_2348356, EPI_ISL_2348361, EPI_ISL_2348362, EPI_ISL_2348363, EPI_ISL_2348364, EPI_ISL_2348365, EPI_ISL_2348367, EPI_ISL_2348369, EPI_ISL_2348370, EPI_ISL_2348371, EPI_ISL_2348372, EPI_ISL_2348373, EPI_ISL_2348376, EPI_ISL_2348377, EPI_ISL_2348378, EPI_ISL_2348379, EPI_ISL_2348381, EPI_ISL_2348382, EPI_ISL_2348383, EPI_ISL_2348384, EPI_ISL_2348386, EPI_ISL_2348391, EPI_ISL_2348392, EPI_ISL_2348400, EPI_ISL_2348401, EPI_ISL_2348407, EPI_ISL_2348409, EPI_ISL_2348411, EPI_ISL_2348414, EPI_ISL_2348418, EPI_ISL_2348420, EPI_ISL_2348421, EPI_ISL_2348424, EPI_ISL_2348425, EPI_ISL_2348426, EPI_ISL_2348427, EPI_ISL_2348428, EPI_ISL_2348429, EPI_ISL_2348433, EPI_ISL_2348435, EPI_ISL_2348436, EPI_ISL_2348437, EPI_ISL_2348438, EPI_ISL_2348442, EPI_ISL_2348446, EPI_ISL_2348447, EPI_ISL_2348448, EPI_ISL_2348450, EPI_ISL_2348451, EPI_ISL_2348452, EPI_ISL_2348454, EPI_ISL_2348457, EPI_ISL_2348459, EPI_ISL_2348461, EPI_ISL_2348462, EPI_ISL_2348464, EPI_ISL_2348465, EPI_ISL_2348467, EPI_ISL_2348469, EPI_ISL_2348474, EPI_ISL_2348475, EPI_ISL_2348476, EPI_ISL_2352695, EPI_ISL_2352698, EPI_ISL_2352701, EPI_ISL_2352702, EPI_ISL_2352705, EPI_ISL_2352706, EPI_ISL_2352707, EPI_ISL_2352709, EPI_ISL_2352711, EPI_ISL_2352712, EPI_ISL_2352713, EPI_ISL_2352717, EPI_ISL_2352724, EPI_ISL_2352725, EPI_ISL_2352728, EPI_ISL_2352729, EPI_ISL_2352730, EPI_ISL_2352732, EPI_ISL_2352734, EPI_ISL_2352735, EPI_ISL_2352740, EPI_ISL_2352741, EPI_ISL_2352743, EPI_ISL_2352751, EPI_ISL_2352752, EPI_ISL_2352756, EPI_ISL_2352757, EPI_ISL_2352761, EPI_ISL_2352764, EPI_ISL_2352765, EPI_ISL_2352766, EPI_ISL_2352769, EPI_ISL_2352772, EPI_ISL_2352774, EPI_ISL_2352775, EPI_ISL_2352780, EPI_ISL_2352781, EPI_ISL_2352784, EPI_ISL_2352785, EPI_ISL_2352786, EPI_ISL_2352791, EPI_ISL_2352799, EPI_ISL_2352801, EPI_ISL_2352805, EPI_ISL_2352808, EPI_ISL_2352809, EPI_ISL_2352813, EPI_ISL_2352815, EPI_ISL_2352818, EPI_ISL_2352826, EPI_ISL_2352827, EPI_ISL_2352833, EPI_ISL_2352836, EPI_ISL_2352840, EPI_ISL_2352843, EPI_ISL_2352844, EPI_ISL_2352848, EPI_ISL_2352850, EPI_ISL_2352851, EPI_ISL_2352853, EPI_ISL_2352861, EPI_ISL_2352866, EPI_ISL_2352870, EPI_ISL_2352872, EPI_ISL_2352873, EPI_ISL_2352876, EPI_ISL_2352885, EPI_ISL_2352894, EPI_ISL_2352898, EPI_ISL_2352903, EPI_ISL_2352907, EPI_ISL_2352913, EPI_ISL_2352915, EPI_ISL_2352916, EPI_ISL_2352918, EPI_ISL_2352922, EPI_ISL_2352930, EPI_ISL_2352932, EPI_ISL_2352934, EPI_ISL_2352935, EPI_ISL_2352936, EPI_ISL_2352937, EPI_ISL_2352941, EPI_ISL_2352942, EPI_ISL_2352947, EPI_ISL_2352950, EPI_ISL_2352951, EPI_ISL_2352953, EPI_ISL_2352958, EPI_ISL_2352960, | | | |

EPI_ISL_2352963, EPI_ISL_2352965, EPI_ISL_2352968, EPI_ISL_2352970, EPI_ISL_2352974, EPI_ISL_2352976, EPI_ISL_2352980, EPI_ISL_2352981, EPI_ISL_2352983, EPI_ISL_2352984, EPI_ISL_2352990, EPI_ISL_2352991, EPI_ISL_2352994, EPI_ISL_2352997, EPI_ISL_2352998, EPI_ISL_2352999, EPI_ISL_2353000, EPI_ISL_2353001, EPI_ISL_2353004, EPI_ISL_2353005, EPI_ISL_2353006, EPI_ISL_2353007, EPI_ISL_2353008, EPI_ISL_2353010, EPI_ISL_2353011, EPI_ISL_2353012, EPI_ISL_2353015, EPI_ISL_2353016, EPI_ISL_2353017, EPI_ISL_2353018, EPI_ISL_2353019, EPI_ISL_2353020, EPI_ISL_2353021, EPI_ISL_2353022, EPI_ISL_2353023, EPI_ISL_2353025, EPI_ISL_2353026, EPI_ISL_2353028, EPI_ISL_2353029, EPI_ISL_2353031, EPI_ISL_2353032, EPI_ISL_2353034, EPI_ISL_2353035, EPI_ISL_2353036, EPI_ISL_2353041, EPI_ISL_2353042, EPI_ISL_2353043, EPI_ISL_2353044, EPI_ISL_2353045, EPI_ISL_2353046, EPI_ISL_2353047, EPI_ISL_2353049, EPI_ISL_2353050, EPI_ISL_2353055, EPI_ISL_2353056, EPI_ISL_2353057, EPI_ISL_2353059, EPI_ISL_2353060, EPI_ISL_2353061, EPI_ISL_2353063, EPI_ISL_2353069, EPI_ISL_2353070, EPI_ISL_2353072, EPI_ISL_2353075, EPI_ISL_2353076, EPI_ISL_2353077, EPI_ISL_2353078, EPI_ISL_2353079, EPI_ISL_2353080, EPI_ISL_2353081, EPI_ISL_2353082, EPI_ISL_2353083, EPI_ISL_2353092, EPI_ISL_2353093, EPI_ISL_2353094, EPI_ISL_2353095, EPI_ISL_2353096, EPI_ISL_2353098, EPI_ISL_2353099, EPI_ISL_2353100, EPI_ISL_2353101, EPI_ISL_2353102, EPI_ISL_2353104, EPI_ISL_2353105, EPI_ISL_2353107, EPI_ISL_2353108, EPI_ISL_2353109, EPI_ISL_2353110, EPI_ISL_2353111, EPI_ISL_2353112, EPI_ISL_2353113, EPI_ISL_2353116, EPI_ISL_2353117, EPI_ISL_2353118, EPI_ISL_2353119, EPI_ISL_2353121, EPI_ISL_2353122, EPI_ISL_2353124, EPI_ISL_2353125, EPI_ISL_2353128, EPI_ISL_2353129, EPI_ISL_2353131, EPI_ISL_2353132, EPI_ISL_2353135, EPI_ISL_2353139, EPI_ISL_2353141, EPI_ISL_2353142, EPI_ISL_2353144, EPI_ISL_2353146, EPI_ISL_2353147, EPI_ISL_2353148, EPI_ISL_2353149, EPI_ISL_2353150, EPI_ISL_2353151, EPI_ISL_2353152, EPI_ISL_2353153, EPI_ISL_2353154, EPI_ISL_2353155, EPI_ISL_2353156, EPI_ISL_2353158, EPI_ISL_2353159, EPI_ISL_2353161, EPI_ISL_2353162, EPI_ISL_2353163, EPI_ISL_2353164, EPI_ISL_2353165, EPI_ISL_2353166, EPI_ISL_2353167, EPI_ISL_2353168, EPI_ISL_2353170, EPI_ISL_2353171, EPI_ISL_2353172, EPI_ISL_2353173, EPI_ISL_2353174, EPI_ISL_2353176, EPI_ISL_2353179, EPI_ISL_2353181, EPI_ISL_2353182, EPI_ISL_2353186, EPI_ISL_2353187, EPI_ISL_2353190, EPI_ISL_2353192, EPI_ISL_2353195, EPI_ISL_2353202, EPI_ISL_2353203, EPI_ISL_2353206, EPI_ISL_2353210, EPI_ISL_2353214, EPI_ISL_2353215, EPI_ISL_2353216, EPI_ISL_2353218, EPI_ISL_2353220, EPI_ISL_2353221, EPI_ISL_2353222, EPI_ISL_2353224, EPI_ISL_2353225, EPI_ISL_2353226, EPI_ISL_2353227, EPI_ISL_2353228, EPI_ISL_2353230, EPI_ISL_2353231, EPI_ISL_2353235, EPI_ISL_2353236, EPI_ISL_2353237, EPI_ISL_2353238, EPI_ISL_2353239, EPI_ISL_2353241, EPI_ISL_2353242, EPI_ISL_2353243, EPI_ISL_2353245, EPI_ISL_2353248, EPI_ISL_2353249, EPI_ISL_2353250, EPI_ISL_2353251, EPI_ISL_2353257, EPI_ISL_2353265, EPI_ISL_2353266, EPI_ISL_2353270, EPI_ISL_2353272, EPI_ISL_2353273, EPI_ISL_2353275, EPI_ISL_2353276, EPI_ISL_2353281, EPI_ISL_2353284, EPI_ISL_2353285, EPI_ISL_2353286, EPI_ISL_2353291, EPI_ISL_2353297, EPI_ISL_2353301, EPI_ISL_2353303, EPI_ISL_2353306, EPI_ISL_2353307, EPI_ISL_2353308, EPI_ISL_2353309, EPI_ISL_2353316, EPI_ISL_2353321, EPI_ISL_2353323, EPI_ISL_2353327, EPI_ISL_2353332, EPI_ISL_2353336, EPI_ISL_2353337, EPI_ISL_2353339, EPI_ISL_2353350, EPI_ISL_2353354, EPI_ISL_2353360, EPI_ISL_2353364, EPI_ISL_2353368, EPI_ISL_2353372, EPI_ISL_2353374, EPI_ISL_2353377, EPI_ISL_2353379, EPI_ISL_2353381, EPI_ISL_2353386, EPI_ISL_2353387, EPI_ISL_2353392, EPI_ISL_2353394, EPI_ISL_2353396, EPI_ISL_2353398, EPI_ISL_2353399, EPI_ISL_2353405, EPI_ISL_2353406, EPI_ISL_2353410, EPI_ISL_2353412, EPI_ISL_2353418, EPI_ISL_2353428, EPI_ISL_2353431, EPI_ISL_2353433, EPI_ISL_2353436, EPI_ISL_2353437, EPI_ISL_2353440, EPI_ISL_2353444, EPI_ISL_2353450, EPI_ISL_2353458, EPI_ISL_2353461, EPI_ISL_2353472, EPI_ISL_2353474, EPI_ISL_2353478, EPI_ISL_2353483, EPI_ISL_2353486, EPI_ISL_2353488, EPI_ISL_2353494, EPI_ISL_2353496, EPI_ISL_2353497, EPI_ISL_2353499, EPI_ISL_2353508, EPI_ISL_2353514, EPI_ISL_2353520, EPI_ISL_2353523, EPI_ISL_2353524, EPI_ISL_2353530, EPI_ISL_2353532, EPI_ISL_2353533, EPI_ISL_2353535, EPI_ISL_2353538, EPI_ISL_2353540, EPI_ISL_2353542, EPI_ISL_2353549, EPI_ISL_2353551, EPI_ISL_2353554, EPI_ISL_2353556, EPI_ISL_2353562, EPI_ISL_2353566, EPI_ISL_2353569, EPI_ISL_2353579, EPI_ISL_2353582, EPI_ISL_2353584, EPI_ISL_2353586, EPI_ISL_2353587, EPI_ISL_2353588, EPI_ISL_2353590, EPI_ISL_2353593, EPI_ISL_2353594, EPI_ISL_2353595, EPI_ISL_2353596, EPI_ISL_2353597, EPI_ISL_2353599, EPI_ISL_2353602, EPI_ISL_2353603, EPI_ISL_2353605, EPI_ISL_2353606, EPI_ISL_2353607, EPI_ISL_2353608, EPI_ISL_2353609, EPI_ISL_2353610, EPI_ISL_2353611, EPI_ISL_2353612, EPI_ISL_2353618, EPI_ISL_2353619, EPI_ISL_2353621, EPI_ISL_2353624, EPI_ISL_2353625, EPI_ISL_2353626, EPI_ISL_2353627, EPI_ISL_2353628, EPI_ISL_2353630, EPI_ISL_2353631, EPI_ISL_2353632, EPI_ISL_2353635, EPI_ISL_2353636, EPI_ISL_2353637, EPI_ISL_2353641, EPI_ISL_2353642, EPI_ISL_2353646, EPI_ISL_2353647, EPI_ISL_2353649, EPI_ISL_2353650, EPI_ISL_2353651, EPI_ISL_2353653, EPI_ISL_2353654, EPI_ISL_2353656, EPI_ISL_2353657, EPI_ISL_2353659, EPI_ISL_2353660, EPI_ISL_2353663, EPI_ISL_2353664, EPI_ISL_2353665, EPI_ISL_2353667, EPI_ISL_2353672, EPI_ISL_2353673, EPI_ISL_2353675, EPI_ISL_2353676, EPI_ISL_2353679, EPI_ISL_2353680, EPI_ISL_2353682, EPI_ISL_2353683, EPI_ISL_2353687, EPI_ISL_2353689, EPI_ISL_2353690, EPI_ISL_2353691, EPI_ISL_2353693, EPI_ISL_2353694, EPI_ISL_2353695, EPI_ISL_2353696, EPI_ISL_2353697, EPI_ISL_2353698, EPI_ISL_2353699, EPI_ISL_2353707, EPI_ISL_2353708, EPI_ISL_2353709, EPI_ISL_2353713, EPI_ISL_2353714, EPI_ISL_2353716, EPI_ISL_2353717, EPI_ISL_2353721, EPI_ISL_2353723, EPI_ISL_2353724, EPI_ISL_2353725, EPI_ISL_2353729, EPI_ISL_2353730, EPI_ISL_2353731, EPI_ISL_2353732, EPI_ISL_2353733, EPI_ISL_2353734, EPI_ISL_2353738, EPI_ISL_2353741, EPI_ISL_2353744, EPI_ISL_2353745, EPI_ISL_2353747, EPI_ISL_2353749, EPI_ISL_2353751, EPI_ISL_2353752, EPI_ISL_2353753, EPI_ISL_2353754, EPI_ISL_2353755, EPI_ISL_2353757, EPI_ISL_2353759, EPI_ISL_2353760, EPI_ISL_2353761, EPI_ISL_2353762, EPI_ISL_2353764, EPI_ISL_2353765, EPI_ISL_2353766, EPI_ISL_2353768, EPI_ISL_2353770, EPI_ISL_2353772, EPI_ISL_2353773, EPI_ISL_2353774, EPI_ISL_2353775, EPI_ISL_2353777, EPI_ISL_2353778, EPI_ISL_2353780, EPI_ISL_2353781, EPI_ISL_2353782, EPI_ISL_2353783, EPI_ISL_2353784, EPI_ISL_2353785, EPI_ISL_2353786, EPI_ISL_2353787, EPI_ISL_2353788, EPI_ISL_2353789, EPI_ISL_2353790, EPI_ISL_2353791, EPI_ISL_2353792, EPI_ISL_2353793, EPI_ISL_2353794, EPI_ISL_2353795, EPI_ISL_2353797, EPI_ISL_2353798, EPI_ISL_2353799, EPI_ISL_2353800, EPI_ISL_2353802, EPI_ISL_2353806, EPI_ISL_2353809, EPI_ISL_2353811, EPI_ISL_2353812, EPI_ISL_2353815, EPI_ISL_2353816, EPI_ISL_2353817, EPI_ISL_2353818, EPI_ISL_2353822, EPI_ISL_2353823, EPI_ISL_2353824, EPI_ISL_2353826, EPI_ISL_2353828, EPI_ISL_2353829, EPI_ISL_2353830, EPI_ISL_2353831, EPI_ISL_2353832, EPI_ISL_2353833, EPI_ISL_2353836, EPI_ISL_2353838, EPI_ISL_2353839, EPI_ISL_2353840, EPI_ISL_2353841, EPI_ISL_2353842, EPI_ISL_2353843, EPI_ISL_2353846, EPI_ISL_2353847, EPI_ISL_2353848, EPI_ISL_2353850, EPI_ISL_2353851, EPI_ISL_2353854, EPI_ISL_2353857, EPI_ISL_2353860, EPI_ISL_2353861, EPI_ISL_2353862, EPI_ISL_2353863, EPI_ISL_2365972, EPI_ISL_2365973, EPI_ISL_2365974, EPI_ISL_2365975, EPI_ISL_2365976, EPI_ISL_2365979, EPI_ISL_2365980, EPI_ISL_2365981, EPI_ISL_2365983, EPI_ISL_2365985, EPI_ISL_2365986, EPI_ISL_2365988, EPI_ISL_2365989, EPI_ISL_2365990, EPI_ISL_2365991, EPI_ISL_2365992, EPI_ISL_2365993, EPI_ISL_2365994, EPI_ISL_2365996, EPI_ISL_2365997, EPI_ISL_2365998, EPI_ISL_2365999, EPI_ISL_2366000, EPI_ISL_2366001, EPI_ISL_2366003, EPI_ISL_2366005, EPI_ISL_2366008, EPI_ISL_2366009, EPI_ISL_2366010, EPI_ISL_2366013, EPI_ISL_2366014, EPI_ISL_2366016, EPI_ISL_2366018, EPI_ISL_2366019, EPI_ISL_2366020, EPI_ISL_2366021, EPI_ISL_2366024, EPI_ISL_2366025, EPI_ISL_2366026, EPI_ISL_2366027, EPI_ISL_2366029, EPI_ISL_2366031, EPI_ISL_2366033, EPI_ISL_2366034, EPI_ISL_2366035, EPI_ISL_2366036, EPI_ISL_2366043, EPI_ISL_2366044, EPI_ISL_2366045, EPI_ISL_2366046, EPI_ISL_2366048, EPI_ISL_2366052, EPI_ISL_2366053, EPI_ISL_2366054, EPI_ISL_2366056, EPI_ISL_2366057, EPI_ISL_2366061, EPI_ISL_2366062, EPI_ISL_2366064, EPI_ISL_2366067, EPI_ISL_2366069, EPI_ISL_2366070, EPI_ISL_2366071, EPI_ISL_2366073, EPI_ISL_2366074, EPI_ISL_2366075, EPI_ISL_2366076, EPI_ISL_2366077, EPI_ISL_2366078, EPI_ISL_2366079, EPI_ISL_2366080, EPI_ISL_2366082, EPI_ISL_2366083, EPI_ISL_2366084, EPI_ISL_2366085, EPI_ISL_2366089, EPI_ISL_2366092, EPI_ISL_2366093, EPI_ISL_2366095, EPI_ISL_2366096, EPI_ISL_2366097, EPI_ISL_2366098, EPI_ISL_2366102, EPI_ISL_2366103, EPI_ISL_2366104, EPI_ISL_2366107, EPI_ISL_2366108, EPI_ISL_2366109, EPI_ISL_2366114, EPI_ISL_2366115, EPI_ISL_2366116, EPI_ISL_2366119, EPI_ISL_2366121, EPI_ISL_2366123, EPI_ISL_2366124, EPI_ISL_2366126, EPI_ISL_2366127, EPI_ISL_2366128, EPI_ISL_2366129, EPI_ISL_2366130, EPI_ISL_2366131, EPI_ISL_2366132, EPI_ISL_2366135, EPI_ISL_2366136, EPI_ISL_2366137, EPI_ISL_2366138, EPI_ISL_2366139, EPI_ISL_2366141, EPI_ISL_2366142, EPI_ISL_2366144, EPI_ISL_2366145, EPI_ISL_2366146, EPI_ISL_2366149, EPI_ISL_2366150, EPI_ISL_2366151, EPI_ISL_2366154, EPI_ISL_2366155, EPI_ISL_2366156, EPI_ISL_2366158, EPI_ISL_2366159, EPI_ISL_2366160, EPI_ISL_2366161, EPI_ISL_2366170, EPI_ISL_2366171, EPI_ISL_2366172, EPI_ISL_2366173, EPI_ISL_2366174, EPI_ISL_2366177, EPI_ISL_2366178, EPI_ISL_2366181, EPI_ISL_2366184, EPI_ISL_2366185, EPI_ISL_2366186, EPI_ISL_2366187, EPI_ISL_2366188, EPI_ISL_2366191, EPI_ISL_2366192, EPI_ISL_2366195, EPI_ISL_2366197, EPI_ISL_2366198, EPI_ISL_2366199, EPI_ISL_2366200, EPI_ISL_2366201, EPI_ISL_2366202, EPI_ISL_2366205, EPI_ISL_2366206, EPI_ISL_2366207, EPI_ISL_2366208, EPI_ISL_2366209, EPI_ISL_2366210, EPI_ISL_2366211, EPI_ISL_2366213, EPI_ISL_2366214, EPI_ISL_2366219, EPI_ISL_2366220, EPI_ISL_2366222, EPI_ISL_2366224, EPI_ISL_2366225, EPI_ISL_2366227, EPI_ISL_2366230, EPI_ISL_2366231, EPI_ISL_2366232, EPI_ISL_2366236, EPI_ISL_2366237, EPI_ISL_2366241, EPI_ISL_2366245, EPI_ISL_2366246, EPI_ISL_2366247, EPI_ISL_2366250, EPI_ISL_2366251, EPI_ISL_2366252, EPI_ISL_2366253, EPI_ISL_2366255, EPI_ISL_2366256, EPI_ISL_2366257, EPI_ISL_2366258, EPI_ISL_2366259, EPI_ISL_2366262, EPI_ISL_2366265, EPI_ISL_2366266, EPI_ISL_2366268, EPI_ISL_2366302, EPI_ISL_2366305, EPI_ISL_2366308, EPI_ISL_2366311, EPI_ISL_2366313, EPI_ISL_2366316, EPI_ISL_2366318, EPI_ISL_2366325, EPI_ISL_2366326, EPI_ISL_2366330, EPI_ISL_2366331, EPI_ISL_2366339, EPI_ISL_2366343, EPI_ISL_2366352, EPI_ISL_2366356, EPI_ISL_2366357, EPI_ISL_2366359, EPI_ISL_2366361, EPI_ISL_2366362, EPI_ISL_2366366, EPI_ISL_2366375, EPI_ISL_2366376, EPI_ISL_2366377, EPI_ISL_2366384, EPI_ISL_2366389, EPI_ISL_2366392, EPI_ISL_2366395, EPI_ISL_2366397, EPI_ISL_2366398, EPI_ISL_2366403, EPI_ISL_2366404, EPI_ISL_2366411, EPI_ISL_2366416, EPI_ISL_2366417, EPI_ISL_2366426, EPI_ISL_2366427, EPI_ISL_2366433, EPI_ISL_2366439, EPI_ISL_2366441, EPI_ISL_2366443, EPI_ISL_2366446, EPI_ISL_2366447, EPI_ISL_2366457, EPI_ISL_2366460, EPI_ISL_2366465, EPI_ISL_2366468, EPI_ISL_2366469, EPI_ISL_2366472, EPI_ISL_2366477, EPI_ISL_2366484, EPI_ISL_2366485, EPI_ISL_2366493, EPI_ISL_2366499, EPI_ISL_2366500, EPI_ISL_2366504, EPI_ISL_2366513, EPI_ISL_2366516, EPI_ISL_2366522, EPI_ISL_2366523, EPI_ISL_2366525, EPI_ISL_2366528, EPI_ISL_2366535, EPI_ISL_2366541, EPI_ISL_2366543, EPI_ISL_2366548, EPI_ISL_2366561, EPI_ISL_2366562, EPI_ISL_2366569, EPI_ISL_2366581, EPI_ISL_2366584, EPI_ISL_2366593, EPI_ISL_2366595, EPI_ISL_2366596, EPI_ISL_2366597, EPI_ISL_2366603, EPI_ISL_2366607, EPI_ISL_2366608, EPI_ISL_2366609, EPI_ISL_2366610, EPI_ISL_2366611, EPI_ISL_2366612, EPI_ISL_2366613, EPI_ISL_2366614, EPI_ISL_2366615, EPI_ISL_2366616, EPI_ISL_2366617, EPI_ISL_2366618, EPI_ISL_2366619, EPI_ISL_2366620, EPI_ISL_2392269, EPI_ISL_2392288, EPI_ISL_2392408, EPI_ISL_2392945, EPI_ISL_2392947, EPI_ISL_2392949, EPI_ISL_2392950, EPI_ISL_2392956, EPI_ISL_2392960, EPI_ISL_2392961, EPI_ISL_2392963, EPI_ISL_2392971, EPI_ISL_2392973, EPI_ISL_2392979, EPI_ISL_2392985, EPI_ISL_2392986, EPI_ISL_2392988, EPI_ISL_2392989, EPI_ISL_2392991, EPI_ISL_2392996, EPI_ISL_2393008, EPI_ISL_2393009, EPI_ISL_2393013, EPI_ISL_2393014, EPI_ISL_2393016, EPI_ISL_2393018, EPI_ISL_2393019, EPI_ISL_2393022, EPI_ISL_2393028, EPI_ISL_2393033, EPI_ISL_2393035, EPI_ISL_2393037, EPI_ISL_2393039, EPI_ISL_2393041, EPI_ISL_2393042, EPI_ISL_2393046, EPI_ISL_2393048, EPI_ISL_2393049, EPI_ISL_2393052, EPI_ISL_2393054, EPI_ISL_2393056, EPI_ISL_2393057, EPI_ISL_2393059, EPI_ISL_2393060, EPI_ISL_2393064, EPI_ISL_2393066, EPI_ISL_2393067, EPI_ISL_2393068, EPI_ISL_2393070, EPI_ISL_2393073, EPI_ISL_2393074, EPI_ISL_2393080, EPI_ISL_2393083, EPI_ISL_2393084, EPI_ISL_2393086, EPI_ISL_2393094, EPI_ISL_2393095, EPI_ISL_2393098, EPI_ISL_2393099, EPI_ISL_2393102, EPI_ISL_2393108, EPI_ISL_2393110, EPI_ISL_2393113, EPI_ISL_2393115, EPI_ISL_2393116, EPI_ISL_2393124, EPI_ISL_2393127, EPI_ISL_2393143, EPI_ISL_2393147, EPI_ISL_2393151, EPI_ISL_2393160, EPI_ISL_2393162, EPI_ISL_2393163, EPI_ISL_2393165, EPI_ISL_2393166, EPI_ISL_2393167, EPI_ISL_2393196, EPI_ISL_2393207, EPI_ISL_2393212, EPI_ISL_2393225, EPI_ISL_2393231, EPI_ISL_2393237, EPI_ISL_2393247, EPI_ISL_2393253, EPI_ISL_2393262, EPI_ISL_2393270, EPI_ISL_2393271, EPI_ISL_2393272, EPI_ISL_2393274, EPI_ISL_2393280, EPI_ISL_2393290, EPI_ISL_2393301, EPI_ISL_2393307, EPI_ISL_2393314, EPI_ISL_2393316, EPI_ISL_2393322, EPI_ISL_2393325, EPI_ISL_2393339, EPI_ISL_2393340, EPI_ISL_2393362, EPI_ISL_2393373, EPI_ISL_2393377, EPI_ISL_2393380, EPI_ISL_2393385, EPI_ISL_2393388, EPI_ISL_2393391, EPI_ISL_2393401, EPI_ISL_2393408, EPI_ISL_2393417, EPI_ISL_2393418, EPI_ISL_2393426, EPI_ISL_2393429, EPI_ISL_2393443, EPI_ISL_2393444, EPI_ISL_2393455, EPI_ISL_2393482, EPI_ISL_2393487, EPI_ISL_2393488, EPI_ISL_2393493, EPI_ISL_2393501, EPI_ISL_2393509, EPI_ISL_2393510, EPI_ISL_2393511, EPI_ISL_2393515, EPI_ISL_2393516, EPI_ISL_2393517, EPI_ISL_2393518, EPI_ISL_2393521, EPI_ISL_2393522, EPI_ISL_2393528, EPI_ISL_2393535, EPI_ISL_2393542, EPI_ISL_2393547, EPI_ISL_2393551, EPI_ISL_2393554, EPI_ISL_2393556, EPI_ISL_2393561, EPI_ISL_2393571, EPI_ISL_2393574, EPI_ISL_2393577, EPI_ISL_2393581, EPI_ISL_2393582, EPI_ISL_2393591, EPI_ISL_2393593, EPI_ISL_2393595, EPI_ISL_2393596, EPI_ISL_2393601, EPI_ISL_2393603, EPI_ISL_2393604, EPI_ISL_2393605, EPI_ISL_2393607, EPI_ISL_2393608, EPI_ISL_2393609, EPI_ISL_2393612, EPI_ISL_2393615, EPI_ISL_2393623, EPI_ISL_2393629, EPI_ISL_2393631, EPI_ISL_2393632, EPI_ISL_2393633, EPI_ISL_2393634, EPI_ISL_2393636, EPI_ISL_2393639, EPI_ISL_2393640, EPI_ISL_2393641, EPI_ISL_2393645, EPI_ISL_2393649, EPI_ISL_2393659, EPI_ISL_2393664, EPI_ISL_2393669, EPI_ISL_2393671, EPI_ISL_2393672, EPI_ISL_2393676, EPI_ISL_2393681, EPI_ISL_2393684, EPI_ISL_2393685, EPI_ISL_2393689, EPI_ISL_2393691, EPI_ISL_2393697, EPI_ISL_2393698, EPI_ISL_2393699, EPI_ISL_2393704, EPI_ISL_2393705, EPI_ISL_2393713, EPI_ISL_2393721, EPI_ISL_2393722, EPI_ISL_2393725, EPI_ISL_2393732, EPI_ISL_2393734, EPI_ISL_2393736, EPI_ISL_2393737, EPI_ISL_2393741, EPI_ISL_2393743, EPI_ISL_2393744, EPI_ISL_2393745, EPI_ISL_2393751, EPI_ISL_2393752, EPI_ISL_2393762, EPI_ISL_2393770, EPI_ISL_2393774, EPI_ISL_2393775, EPI_ISL_2393782, EPI_ISL_2393788, EPI_ISL_2393793, EPI_ISL_2393800, EPI_ISL_2393802, EPI_ISL_2393803, EPI_ISL_2393806, EPI_ISL_2393808, EPI_ISL_2393811, EPI_ISL_2393813, EPI_ISL_2393833, EPI_ISL_2393844, EPI_ISL_2393845, EPI_ISL_2393852, EPI_ISL_2393855, EPI_ISL_2393862, EPI_ISL_2393863, EPI_ISL_2393872, EPI_ISL_2393875, EPI_ISL_2393887, EPI_ISL_2393890, EPI_ISL_2393892, EPI_ISL_2393895, EPI_ISL_2393905, EPI_ISL_2393906, EPI_ISL_2393909, EPI_ISL_2393922, EPI_ISL_2393927, EPI_ISL_2393930, EPI_ISL_2393932, EPI_ISL_2393947, EPI_ISL_2393950, EPI_ISL_2393957, EPI_ISL_2393959, EPI_ISL_2393963, EPI_ISL_2393966, EPI_ISL_2393968, EPI_ISL_2393973, EPI_ISL_2393980, EPI_ISL_2393981, EPI_ISL_2393984, EPI_ISL_2393985, EPI_ISL_2393994, EPI_ISL_2394000, EPI_ISL_2394002, EPI_ISL_2394005, EPI_ISL_2394007, EPI_ISL_2394011, EPI_ISL_2394014, EPI_ISL_2394018, EPI_ISL_2394021, EPI_ISL_2394022, EPI_ISL_2394023, EPI_ISL_2394028, EPI_ISL_2394039, EPI_ISL_2394125, EPI_ISL_2394187, EPI_ISL_2394215, EPI_ISL_2394320, EPI_ISL_2394340, EPI_ISL_2394348, EPI_ISL_2394355, EPI_ISL_2394358, EPI_ISL_2394359, EPI_ISL_2394362, EPI_ISL_2394364, EPI_ISL_2394366, EPI_ISL_2394368, EPI_ISL_2394373, EPI_ISL_2394376, EPI_ISL_2394379, EPI_ISL_2394380, EPI_ISL_2394381, EPI_ISL_2394385, EPI_ISL_2394386, EPI_ISL_2394387, EPI_ISL_2394388, EPI_ISL_2394390, EPI_ISL_2394392, EPI_ISL_2394394, EPI_ISL_2394398, EPI_ISL_2394400, EPI_ISL_2394401, EPI_ISL_2394404, EPI_ISL_2394405, EPI_ISL_2394407, EPI_ISL_2394410, EPI_ISL_2394412, EPI_ISL_2394420, EPI_ISL_2394424, EPI_ISL_2394426, EPI_ISL_2394429, EPI_ISL_2394430, EPI_ISL_2394435, EPI_ISL_2394436, EPI_ISL_2394437, EPI_ISL_2394441, EPI_ISL_2394444, EPI_ISL_2394445, EPI_ISL_2394446, EPI_ISL_2394448, EPI_ISL_2394451, EPI_ISL_2394453, EPI_ISL_2394455, EPI_ISL_2394456, EPI_ISL_2394466, EPI_ISL_2394471, EPI_ISL_2394472, EPI_ISL_2394478, EPI_ISL_2394479, EPI_ISL_2394480, EPI_ISL_2394486, EPI_ISL_2394500, EPI_ISL_2394501, EPI_ISL_2394502, EPI_ISL_2394504, EPI_ISL_2394508, EPI_ISL_2394516, EPI_ISL_2394524, EPI_ISL_2394525, EPI_ISL_2394527, EPI_ISL_2394532, EPI_ISL_2394536, EPI_ISL_2394538, EPI_ISL_2394539, EPI_ISL_2394541, EPI_ISL_2394544, EPI_ISL_2394552, EPI_ISL_2394556, EPI_ISL_2394560, EPI_ISL_2394564, EPI_ISL_2394570, EPI_ISL_2394574, EPI_ISL_2394575, EPI_ISL_2394579, EPI_ISL_2394591, EPI_ISL_2394596, EPI_ISL_2394613, EPI_ISL_2394622, EPI_ISL_2394625, EPI_ISL_2394632, EPI_ISL_2394633, EPI_ISL_2394648, EPI_ISL_2394650, EPI_ISL_2394659, EPI_ISL_2394667, EPI_ISL_2394673, EPI_ISL_2394674, EPI_ISL_2394683, EPI_ISL_2394693, EPI_ISL_2394704, EPI_ISL_2394706, EPI_ISL_2394708, EPI_ISL_2394709, EPI_ISL_2394712, EPI_ISL_2394714, EPI_ISL_2394724, EPI_ISL_2394737, EPI_ISL_2394746, EPI_ISL_2394764, EPI_ISL_2394779, EPI_ISL_2394785, EPI_ISL_2394791, EPI_ISL_2394826, EPI_ISL_2394829, EPI_ISL_2394832, EPI_ISL_2394836, EPI_ISL_2394843, EPI_ISL_2394847, EPI_ISL_2394848, EPI_ISL_2394853, EPI_ISL_2394861, EPI_ISL_2394889, EPI_ISL_2394897, EPI_ISL_2394951, EPI_ISL_2394997, EPI_ISL_2395042, EPI_ISL_2395047, EPI_ISL_2395053, EPI_ISL_2395055, EPI_ISL_2395056, EPI_ISL_2395057, EPI_ISL_2395060, EPI_ISL_2395061, EPI_ISL_2395062, EPI_ISL_2395063, EPI_ISL_2395065, EPI_ISL_2395068, EPI_ISL_2395070, EPI_ISL_2395074, EPI_ISL_2395076, EPI_ISL_2395079, EPI_ISL_2395081, EPI_ISL_2395083, EPI_ISL_2395084, EPI_ISL_2395085, EPI_ISL_2395086, EPI_ISL_2395089, EPI_ISL_2395090, EPI_ISL_2395093, EPI_ISL_2395095, EPI_ISL_2395096, EPI_ISL_2395098, EPI_ISL_2395099, EPI_ISL_2395100, EPI_ISL_2395102, EPI_ISL_2395103, EPI_ISL_2395104, EPI_ISL_2395105, EPI_ISL_2395106, EPI_ISL_2395107, EPI_ISL_2395110, EPI_ISL_2395115, EPI_ISL_2395117, EPI_ISL_2395119, EPI_ISL_2395124, EPI_ISL_2395125, EPI_ISL_2395126, EPI_ISL_2395127, EPI_ISL_2395128, EPI_ISL_2395130, EPI_ISL_2395131, EPI_ISL_2395133, EPI_ISL_2395137, EPI_ISL_2395140, EPI_ISL_2395141, EPI_ISL_2395144, EPI_ISL_2395145, EPI_ISL_2395146, EPI_ISL_2395148, EPI_ISL_2395149, EPI_ISL_2395151, EPI_ISL_2395152, EPI_ISL_2395154, EPI_ISL_2395156, EPI_ISL_2395158, EPI_ISL_2395161, EPI_ISL_2395166, EPI_ISL_2395170, EPI_ISL_2395171, EPI_ISL_2395173, EPI_ISL_2395174, EPI_ISL_2395178, EPI_ISL_2395183, EPI_ISL_2395185, EPI_ISL_2395187, EPI_ISL_2395190, EPI_ISL_2395191, EPI_ISL_2395196, EPI_ISL_2395198, EPI_ISL_2395200, EPI_ISL_2395203, EPI_ISL_2395204, EPI_ISL_2395205, EPI_ISL_2395207, EPI_ISL_2395208, EPI_ISL_2395210, EPI_ISL_2395211, EPI_ISL_2395215, EPI_ISL_2395216, EPI_ISL_2395217, EPI_ISL_2395219, EPI_ISL_2395221, EPI_ISL_2395224, EPI_ISL_2395225, EPI_ISL_2395226, EPI_ISL_2395227, EPI_ISL_2395229, EPI_ISL_2395230, EPI_ISL_2395231, EPI_ISL_2395232, EPI_ISL_2395233, EPI_ISL_2395237, EPI_ISL_2395240, EPI_ISL_2395241, EPI_ISL_2395242, EPI_ISL_2395244, EPI_ISL_2395246, EPI_ISL_2395247, EPI_ISL_2395255, EPI_ISL_2395256, EPI_ISL_2395257, EPI_ISL_2395258, EPI_ISL_2395259, EPI_ISL_2395261, EPI_ISL_2395263, EPI_ISL_2395265, EPI_ISL_2395267, EPI_ISL_2395268, EPI_ISL_2395271, EPI_ISL_2395272, EPI_ISL_2395275, EPI_ISL_2395278, EPI_ISL_2395281, EPI_ISL_2395287, EPI_ISL_2395288, EPI_ISL_2395294, EPI_ISL_2395297, EPI_ISL_2395300, EPI_ISL_2395301, EPI_ISL_2395303, EPI_ISL_2395304, EPI_ISL_2395306, EPI_ISL_2395311, EPI_ISL_2395312, EPI_ISL_2395313, EPI_ISL_2395314, EPI_ISL_2395315, EPI_ISL_2395324, EPI_ISL_2395325, EPI_ISL_2395326, EPI_ISL_2395328, EPI_ISL_2395329, EPI_ISL_2395330, EPI_ISL_2395332, EPI_ISL_2395333, EPI_ISL_2395336, EPI_ISL_2395342, EPI_ISL_2395343, EPI_ISL_2395360, EPI_ISL_2395361, EPI_ISL_2395363, EPI_ISL_2395364, EPI_ISL_2395367, EPI_ISL_2395368, EPI_ISL_2395379, EPI_ISL_2395380, EPI_ISL_2395381, EPI_ISL_2395387, EPI_ISL_2395390, EPI_ISL_2395391, EPI_ISL_2395400, EPI_ISL_2395411, EPI_ISL_2395422, EPI_ISL_2395426, EPI_ISL_2395437, EPI_ISL_2395456, EPI_ISL_2395460, EPI_ISL_2395461, EPI_ISL_2395462, EPI_ISL_2395467, EPI_ISL_2395469, EPI_ISL_2395478, EPI_ISL_2395484, EPI_ISL_2395485, EPI_ISL_2395486, EPI_ISL_2395491, EPI_ISL_2395494, EPI_ISL_2395497, EPI_ISL_2395505, EPI_ISL_2395521, EPI_ISL_2395524, EPI_ISL_2395530, EPI_ISL_2395535, EPI_ISL_2395551, EPI_ISL_2395567, EPI_ISL_2395568, EPI_ISL_2395569, EPI_ISL_2395575, EPI_ISL_2395576, EPI_ISL_2395584, EPI_ISL_2395587, EPI_ISL_2395588, EPI_ISL_2395589, EPI_ISL_2395595, EPI_ISL_2395596, EPI_ISL_2395599, EPI_ISL_2395600, EPI_ISL_2395603, EPI_ISL_2395604, EPI_ISL_2395618, EPI_ISL_2395622, EPI_ISL_2395624, EPI_ISL_2395629, EPI_ISL_2395634, EPI_ISL_2395640, EPI_ISL_2395643, EPI_ISL_2395646, EPI_ISL_2395648, EPI_ISL_2395658, EPI_ISL_2395659, EPI_ISL_2395664, EPI_ISL_2395674, EPI_ISL_2395675, EPI_ISL_2395679, EPI_ISL_2395682, EPI_ISL_2395683, EPI_ISL_2396018, EPI_ISL_2396023, EPI_ISL_2396024, EPI_ISL_2396026, EPI_ISL_2396029, EPI_ISL_2396035, EPI_ISL_2396038, EPI_ISL_2396042, EPI_ISL_2396051, EPI_ISL_2396053, EPI_ISL_2396062, EPI_ISL_2396063, EPI_ISL_2396064, EPI_ISL_2396068, EPI_ISL_2396069, EPI_ISL_2396074, EPI_ISL_2396075, EPI_ISL_2396089, EPI_ISL_2396091, EPI_ISL_2396095, EPI_ISL_2396111, EPI_ISL_2396113, EPI_ISL_2396122, EPI_ISL_2396135, EPI_ISL_2396136, EPI_ISL_2396138, EPI_ISL_2396139, EPI_ISL_2396142, EPI_ISL_2396147, EPI_ISL_2396148, EPI_ISL_2396160, EPI_ISL_2396164, EPI_ISL_2396166, EPI_ISL_2396169, EPI_ISL_2396172, EPI_ISL_2396175, EPI_ISL_2396178, EPI_ISL_2396187, EPI_ISL_2396193, EPI_ISL_2396194, EPI_ISL_2396198, EPI_ISL_2396199, EPI_ISL_2396205, EPI_ISL_2396213, EPI_ISL_2396215, EPI_ISL_2396222, EPI_ISL_2396225, EPI_ISL_2396227, EPI_ISL_2396228, EPI_ISL_2396229, EPI_ISL_2396231, EPI_ISL_2396235, EPI_ISL_2396236, EPI_ISL_2396246, EPI_ISL_2396250, EPI_ISL_2396253, EPI_ISL_2396260, EPI_ISL_2396265, EPI_ISL_2396272, EPI_ISL_2396273, EPI_ISL_2396278, EPI_ISL_2396281, EPI_ISL_2396282, EPI_ISL_2396283, EPI_ISL_2396289, EPI_ISL_2396296, EPI_ISL_2396298, EPI_ISL_2396306, EPI_ISL_2396308, EPI_ISL_2396312, EPI_ISL_2396319, EPI_ISL_2396323, EPI_ISL_2396324, EPI_ISL_2396325, EPI_ISL_2396327, EPI_ISL_2396334, EPI_ISL_2396336, EPI_ISL_2396389, EPI_ISL_2396798, EPI_ISL_2396799, EPI_ISL_2396809, EPI_ISL_2396817, EPI_ISL_2407094, EPI_ISL_2408049, EPI_ISL_2408057, EPI_ISL_2408058, EPI_ISL_2435271, EPI_ISL_2436929, EPI_ISL_2436931, EPI_ISL_2436935, EPI_ISL_2436936, EPI_ISL_2436941, EPI_ISL_2436942, EPI_ISL_2436943, EPI_ISL_2436944, EPI_ISL_2436945, EPI_ISL_2436946, EPI_ISL_2436948, EPI_ISL_2436950, EPI_ISL_2436951, EPI_ISL_2436952, EPI_ISL_2436954, EPI_ISL_2436956, EPI_ISL_2436957, EPI_ISL_2436958, EPI_ISL_2436962, EPI_ISL_2436963, EPI_ISL_2436964, EPI_ISL_2436965, EPI_ISL_2436966, EPI_ISL_2436969, EPI_ISL_2436971, EPI_ISL_2436973, EPI_ISL_2436976, EPI_ISL_2436977, EPI_ISL_2436978, EPI_ISL_2436980, EPI_ISL_2436982, EPI_ISL_2436984, EPI_ISL_2436986, EPI_ISL_2436987, EPI_ISL_2436990, EPI_ISL_2436991, EPI_ISL_2436993, EPI_ISL_2436994, EPI_ISL_2436996, EPI_ISL_2437001, EPI_ISL_2437002, EPI_ISL_2437011, EPI_ISL_2437012, EPI_ISL_2437015, EPI_ISL_2437018, EPI_ISL_2437019, EPI_ISL_2437020, EPI_ISL_2437027, EPI_ISL_2437028, EPI_ISL_2437031, EPI_ISL_2437032, EPI_ISL_2437033, EPI_ISL_2437035, EPI_ISL_2437037, EPI_ISL_2437038, EPI_ISL_2437040, EPI_ISL_2437041, EPI_ISL_2437042, EPI_ISL_2437043, EPI_ISL_2437045, EPI_ISL_2437046, EPI_ISL_2437047, EPI_ISL_2437048, EPI_ISL_2437051, EPI_ISL_2437052, EPI_ISL_2437053, EPI_ISL_2437054, EPI_ISL_2437055, EPI_ISL_2437056, EPI_ISL_2437057, EPI_ISL_2437059, EPI_ISL_2437060, EPI_ISL_2437061, EPI_ISL_2437062, EPI_ISL_2437063, EPI_ISL_2437065, EPI_ISL_2437069, EPI_ISL_2437071, EPI_ISL_2437072, EPI_ISL_2437073, EPI_ISL_2437076, EPI_ISL_2437077, EPI_ISL_2437079, EPI_ISL_2437080, EPI_ISL_2437081, EPI_ISL_2437082, EPI_ISL_2437083, EPI_ISL_2437084, EPI_ISL_2437085, EPI_ISL_2437086, EPI_ISL_2437087, EPI_ISL_2437091, EPI_ISL_2437096, EPI_ISL_2437097, EPI_ISL_2437099, EPI_ISL_2437103, EPI_ISL_2437105, EPI_ISL_2437106, EPI_ISL_2437107, EPI_ISL_2437110, EPI_ISL_2437111, EPI_ISL_2437112, EPI_ISL_2437113, EPI_ISL_2437115, EPI_ISL_2437118, EPI_ISL_2437119, EPI_ISL_2437120, EPI_ISL_2437121, EPI_ISL_2437124, EPI_ISL_2437125, EPI_ISL_2437126, EPI_ISL_2437129, EPI_ISL_2437130, EPI_ISL_2437133, EPI_ISL_2437134, EPI_ISL_2437135, EPI_ISL_2437140, EPI_ISL_2437145, EPI_ISL_2437148, EPI_ISL_2437152, EPI_ISL_2437153, EPI_ISL_2437154, EPI_ISL_2437155, EPI_ISL_2437158, EPI_ISL_2437159, EPI_ISL_2437161, EPI_ISL_2437162, EPI_ISL_2437163, EPI_ISL_2437167, EPI_ISL_2437169, EPI_ISL_2437171, EPI_ISL_2437172, EPI_ISL_2437173, EPI_ISL_2437177, EPI_ISL_2437178, EPI_ISL_2437179, EPI_ISL_2437180, EPI_ISL_2437182, EPI_ISL_2437183, EPI_ISL_2437184, EPI_ISL_2437186, EPI_ISL_2437187, EPI_ISL_2437189, EPI_ISL_2437190, EPI_ISL_2437191, EPI_ISL_2437193, EPI_ISL_2437194, EPI_ISL_2437198, EPI_ISL_2437200, EPI_ISL_2437202, EPI_ISL_2437204, EPI_ISL_2437205, EPI_ISL_2437207, EPI_ISL_2437208, EPI_ISL_2437210, EPI_ISL_2437211, EPI_ISL_2437212, EPI_ISL_2437213, EPI_ISL_2437215, EPI_ISL_2437218, EPI_ISL_2437219, EPI_ISL_2437220, EPI_ISL_2437221, EPI_ISL_2437222, EPI_ISL_2437224, EPI_ISL_2437225, EPI_ISL_2437226, EPI_ISL_2437227, EPI_ISL_2437228, EPI_ISL_2437229, EPI_ISL_2437230, EPI_ISL_2437232, EPI_ISL_2437238, EPI_ISL_2437239, EPI_ISL_2437241, EPI_ISL_2437242, EPI_ISL_2437243, EPI_ISL_2437244, EPI_ISL_2437246, EPI_ISL_2437248, EPI_ISL_2437249, EPI_ISL_2437250, EPI_ISL_2437251, EPI_ISL_2437252, EPI_ISL_2437254, EPI_ISL_2437256, EPI_ISL_2437257, EPI_ISL_2437258, EPI_ISL_2437259, EPI_ISL_2437262, EPI_ISL_2437265, EPI_ISL_2437266, EPI_ISL_2437267, EPI_ISL_2437268, EPI_ISL_2437269, EPI_ISL_2437270, EPI_ISL_2437271, EPI_ISL_2437275, EPI_ISL_2437276, EPI_ISL_2437576, EPI_ISL_2446414, EPI_ISL_2446416, EPI_ISL_2446436, EPI_ISL_2446450, EPI_ISL_2446451, EPI_ISL_2446459, EPI_ISL_2446463, EPI_ISL_2446465, EPI_ISL_2446472, EPI_ISL_2446478, EPI_ISL_2446491, EPI_ISL_2446499, EPI_ISL_2446503, EPI_ISL_2446505, EPI_ISL_2446512, EPI_ISL_2446513, EPI_ISL_2446518, EPI_ISL_2446524, EPI_ISL_2446527, EPI_ISL_2446534, EPI_ISL_2446538, EPI_ISL_2446542, EPI_ISL_2446550, EPI_ISL_2446555, EPI_ISL_2446556, EPI_ISL_2446557, EPI_ISL_2446558, EPI_ISL_2446561, EPI_ISL_2446562, EPI_ISL_2446563, EPI_ISL_2446572, EPI_ISL_2446583, EPI_ISL_2446588, EPI_ISL_2446590, EPI_ISL_2446600, EPI_ISL_2446603, EPI_ISL_2446607, EPI_ISL_2446608, EPI_ISL_2446610, EPI_ISL_2446616, EPI_ISL_2446632, EPI_ISL_2446642, EPI_ISL_2446648, EPI_ISL_2446653, EPI_ISL_2446657, EPI_ISL_2446658, EPI_ISL_2446665, EPI_ISL_2446667, EPI_ISL_2446670, EPI_ISL_2446677, EPI_ISL_2446694, EPI_ISL_2446706, EPI_ISL_2446707, EPI_ISL_2446709, EPI_ISL_2446719, EPI_ISL_2446722, EPI_ISL_2446725, EPI_ISL_2446727, EPI_ISL_2446729, EPI_ISL_2446731, EPI_ISL_2446733, EPI_ISL_2446734, EPI_ISL_2446739, EPI_ISL_2446742, EPI_ISL_2446746, EPI_ISL_2447758, EPI_ISL_2511983, EPI_ISL_2512167, EPI_ISL_2512221, EPI_ISL_2830830

| | | | |
|---|---|---|---|
| see above | Lighthouse Lab in Alderley Park | Wellcome Sanger Institute for the COVID-19 Genomics UK (COG-UK) Consortium | Cordelia Langford; David K. Jackson; Dominic Kwiatkowski; Ewan Harrison; Ian Johnston; Jacquelyn Wynn; Jeffrey Barrett; John Sillitoe on behalf of the Wellcome Sanger Institute COVID-19 Surveillance Team; Mairead Hyland; Roberto Amato; Sonia Goncalves; The Lighthouse Lab in Alderley Park and Alex Alderton |

EPI_ISL_2316665, EPI_ISL_2316666, EPI_ISL_2316668, EPI_ISL_2316673, EPI_ISL_2316674, EPI_ISL_2316679, EPI_ISL_2316680, EPI_ISL_2316682, EPI_ISL_2316683, EPI_ISL_2316684, EPI_ISL_2316686, EPI_ISL_2316687, EPI_ISL_2316689, EPI_ISL_2316690, EPI_ISL_2316693, EPI_ISL_2316696, EPI_ISL_2316697, EPI_ISL_2316699, EPI_ISL_2316700, EPI_ISL_2316701, EPI_ISL_2316703, EPI_ISL_2316704, EPI_ISL_2316707, EPI_ISL_2316710, EPI_ISL_2316713, EPI_ISL_2316716, EPI_ISL_2316719, EPI_ISL_2316720, EPI_ISL_2316721, EPI_ISL_2316724, EPI_ISL_2316726, EPI_ISL_2316727, EPI_ISL_2316729, EPI_ISL_2316732, EPI_ISL_2316736, EPI_ISL_2316737, EPI_ISL_2316739, EPI_ISL_2316740, EPI_ISL_2316743, EPI_ISL_2316745, EPI_ISL_2316746, EPI_ISL_2316747, EPI_ISL_2316750, EPI_ISL_2316758, EPI_ISL_2316761, EPI_ISL_2316762, EPI_ISL_2316765, EPI_ISL_2316767, EPI_ISL_2316771, EPI_ISL_2316773, EPI_ISL_2316777, EPI_ISL_2316778, EPI_ISL_2316780, EPI_ISL_2316781, EPI_ISL_2316783, EPI_ISL_2316784, EPI_ISL_2316787, EPI_ISL_2316789, EPI_ISL_2316790, EPI_ISL_2316791, EPI_ISL_2316793, EPI_ISL_2316796, EPI_ISL_2316797, EPI_ISL_2316800, EPI_ISL_2316802, EPI_ISL_2316803, EPI_ISL_2316805, EPI_ISL_2316806, EPI_ISL_2316808, EPI_ISL_2316813, EPI_ISL_2316815, EPI_ISL_2316816, EPI_ISL_2316818, EPI_ISL_2316819, EPI_ISL_2316821, EPI_ISL_2316828, EPI_ISL_2316830, EPI_ISL_2316831, EPI_ISL_2316832, EPI_ISL_2316835, EPI_ISL_2316838, EPI_ISL_2316839, EPI_ISL_2316841, EPI_ISL_2316842, EPI_ISL_2316844, EPI_ISL_2316845, EPI_ISL_2316850, EPI_ISL_2316851, EPI_ISL_2316852, EPI_ISL_2316855, EPI_ISL_2316857, EPI_ISL_2316859, EPI_ISL_2316862, EPI_ISL_2316865, EPI_ISL_2316868, EPI_ISL_2316870, EPI_ISL_2316872, EPI_ISL_2316873, EPI_ISL_2316876, EPI_ISL_2316878, EPI_ISL_2316879, EPI_ISL_2316882, EPI_ISL_2316885, EPI_ISL_2316889, EPI_ISL_2316890, EPI_ISL_2316892, EPI_ISL_2316893, EPI_ISL_2316895, EPI_ISL_2316896, EPI_ISL_2316899, EPI_ISL_2316900, EPI_ISL_2316903, EPI_ISL_2316904, EPI_ISL_2316907, EPI_ISL_2316908, EPI_ISL_2316910, EPI_ISL_2316913, EPI_ISL_2316914, EPI_ISL_2316916, EPI_ISL_2316917, EPI_ISL_2316919, EPI_ISL_2316922, EPI_ISL_2316926, EPI_ISL_2316927, EPI_ISL_2316929, EPI_ISL_2316930, EPI_ISL_2316932, EPI_ISL_2316935, EPI_ISL_2316941, EPI_ISL_2316944, EPI_ISL_2316947, EPI_ISL_2316949, EPI_ISL_2316951, EPI_ISL_2316953, EPI_ISL_2316954, EPI_ISL_2316957, EPI_ISL_2316958, EPI_ISL_2316961, EPI_ISL_2316963, EPI_ISL_2316964, EPI_ISL_2316965, EPI_ISL_2316968, EPI_ISL_2316970, EPI_ISL_2316971, EPI_ISL_2316973, EPI_ISL_2316976, EPI_ISL_2316978, EPI_ISL_2316979, EPI_ISL_2316980, EPI_ISL_2316983, EPI_ISL_2316985, EPI_ISL_2316986, EPI_ISL_2316987, EPI_ISL_2316989, EPI_ISL_2316992, EPI_ISL_2316993, EPI_ISL_2316995, EPI_ISL_2316998, EPI_ISL_2316999, EPI_ISL_2317038, EPI_ISL_2317039, EPI_ISL_2317042, EPI_ISL_2317043, EPI_ISL_2317045, EPI_ISL_2317046, EPI_ISL_2317047, EPI_ISL_2317049, EPI_ISL_2317050, EPI_ISL_2317051, EPI_ISL_2317052, EPI_ISL_2317053, EPI_ISL_2317054, EPI_ISL_2317056, EPI_ISL_2317057, EPI_ISL_2317058, EPI_ISL_2317061, EPI_ISL_2317062, EPI_ISL_2317063, EPI_ISL_2317064, EPI_ISL_2317065, EPI_ISL_2317066, EPI_ISL_2317068, EPI_ISL_2317069, EPI_ISL_2317070, EPI_ISL_2317071, EPI_ISL_2317072, EPI_ISL_2317074, EPI_ISL_2317077, EPI_ISL_2317078, EPI_ISL_2317081, EPI_ISL_2317083, EPI_ISL_2317084, EPI_ISL_2317086, EPI_ISL_2317087, EPI_ISL_2317088, EPI_ISL_2317089, EPI_ISL_2317091, EPI_ISL_2317093, EPI_ISL_2317096, EPI_ISL_2317097, EPI_ISL_2317098, EPI_ISL_2317101, EPI_ISL_2317103, EPI_ISL_2317106, EPI_ISL_2317109, EPI_ISL_2317111, EPI_ISL_2317112, EPI_ISL_2317113, EPI_ISL_2317116, EPI_ISL_2317118, EPI_ISL_2317119, EPI_ISL_2317120, EPI_ISL_2317123, EPI_ISL_2317124, EPI_ISL_2317125, EPI_ISL_2317127, EPI_ISL_2317129, EPI_ISL_2317130, EPI_ISL_2317131, EPI_ISL_2317132, EPI_ISL_2317135, EPI_ISL_2317136, EPI_ISL_2317137, EPI_ISL_2317138, EPI_ISL_2317139, EPI_ISL_2317140, EPI_ISL_2317141, EPI_ISL_2317142, EPI_ISL_2317144, EPI_ISL_2317145, EPI_ISL_2317148, EPI_ISL_2317150, EPI_ISL_2317153, EPI_ISL_2317155, EPI_ISL_2317157, EPI_ISL_2317159, EPI_ISL_2317160, EPI_ISL_2317161, EPI_ISL_2318016, EPI_ISL_2318017, EPI_ISL_2318020, EPI_ISL_2318021, EPI_ISL_2318024, EPI_ISL_2318032, EPI_ISL_2318044, EPI_ISL_2318054, EPI_ISL_2318057, EPI_ISL_2318060, EPI_ISL_2318063, EPI_ISL_2318064, EPI_ISL_2318065, EPI_ISL_2318067, EPI_ISL_2318069, EPI_ISL_2318070, EPI_ISL_2318076, EPI_ISL_2318081, EPI_ISL_2318083, EPI_ISL_2318085, EPI_ISL_2318090, EPI_ISL_2318093, EPI_ISL_2318102, EPI_ISL_2318110, EPI_ISL_2318111, EPI_ISL_2318117, EPI_ISL_2318118, EPI_ISL_2318120, EPI_ISL_2318123, EPI_ISL_2318130, EPI_ISL_2318141, EPI_ISL_2318143, EPI_ISL_2318145, EPI_ISL_2318150, EPI_ISL_2318153, EPI_ISL_2318156, EPI_ISL_2318158, EPI_ISL_2318164, EPI_ISL_2318167, EPI_ISL_2318173, EPI_ISL_2318174, EPI_ISL_2318175, EPI_ISL_2318181, EPI_ISL_2318196, EPI_ISL_2318200, EPI_ISL_2318208, EPI_ISL_2318218, EPI_ISL_2318224, EPI_ISL_2318226, EPI_ISL_2318231, EPI_ISL_2318241, EPI_ISL_2318243, EPI_ISL_2318245, EPI_ISL_2318249, EPI_ISL_2318251, EPI_ISL_2318255, EPI_ISL_2318264, EPI_ISL_2318269, EPI_ISL_2318278, EPI_ISL_2318279, EPI_ISL_2318280, EPI_ISL_2318290, EPI_ISL_2318292, EPI_ISL_2318301, EPI_ISL_2318309, EPI_ISL_2318312, EPI_ISL_2318316, EPI_ISL_2318317, EPI_ISL_2318320, EPI_ISL_2318331, EPI_ISL_2318334, EPI_ISL_2318340, EPI_ISL_2318343, EPI_ISL_2318350, EPI_ISL_2318358, EPI_ISL_2318361, EPI_ISL_2318366, EPI_ISL_2318367, EPI_ISL_2318373, EPI_ISL_2318376, EPI_ISL_2318377, EPI_ISL_2318380, EPI_ISL_2318383, EPI_ISL_2318386, EPI_ISL_2318388, EPI_ISL_2318389, EPI_ISL_2318390, EPI_ISL_2318392, EPI_ISL_2318399, EPI_ISL_2318400, EPI_ISL_2318403, EPI_ISL_2318412, EPI_ISL_2318415, EPI_ISL_2318419, EPI_ISL_2318428, EPI_ISL_2318434, EPI_ISL_2318437, EPI_ISL_2318445, EPI_ISL_2318450, EPI_ISL_2318453, EPI_ISL_2318463, EPI_ISL_2318464, EPI_ISL_2318468, EPI_ISL_2318469, EPI_ISL_2318470, EPI_ISL_2318471, EPI_ISL_2318472, EPI_ISL_2318473, EPI_ISL_2318474, EPI_ISL_2318475, EPI_ISL_2318481, EPI_ISL_2318486, EPI_ISL_2318487, EPI_ISL_2318488, EPI_ISL_2318489, EPI_ISL_2318496, EPI_ISL_2318500, EPI_ISL_2318501, EPI_ISL_2318504, EPI_ISL_2318507, EPI_ISL_2318509, EPI_ISL_2318510, EPI_ISL_2318513, EPI_ISL_2318515, EPI_ISL_2318519, EPI_ISL_2318521, EPI_ISL_2318532, EPI_ISL_2318536, EPI_ISL_2318537, EPI_ISL_2318544, EPI_ISL_2318548, EPI_ISL_2318553, EPI_ISL_2318555, EPI_ISL_2318565, EPI_ISL_2318573, EPI_ISL_2318575, EPI_ISL_2318576, EPI_ISL_2318577, EPI_ISL_2318579, EPI_ISL_2318581, EPI_ISL_2318585, EPI_ISL_2318594, EPI_ISL_2318604, EPI_ISL_2318607, EPI_ISL_2318615, EPI_ISL_2318624, EPI_ISL_2318625, EPI_ISL_2318629, EPI_ISL_2318637, EPI_ISL_2318641, EPI_ISL_2318649, EPI_ISL_2318650, EPI_ISL_2318658, EPI_ISL_2318659, EPI_ISL_2318665, EPI_ISL_2318666, EPI_ISL_2318677, EPI_ISL_2318678, EPI_ISL_2318686, EPI_ISL_2318687, EPI_ISL_2318688, EPI_ISL_2318695, EPI_ISL_2318700, EPI_ISL_2318702, EPI_ISL_2318704, EPI_ISL_2318705, EPI_ISL_2318710, EPI_ISL_2318712, EPI_ISL_2318715, EPI_ISL_2318716, EPI_ISL_2318717, EPI_ISL_2318718, EPI_ISL_2318719, EPI_ISL_2318720, EPI_ISL_2318721, EPI_ISL_2318722, EPI_ISL_2318723, EPI_ISL_2318724, EPI_ISL_2318725, EPI_ISL_2318726, EPI_ISL_2318727, EPI_ISL_2318728, EPI_ISL_2318729, EPI_ISL_2318730, EPI_ISL_2318731, EPI_ISL_2318732, EPI_ISL_2318733, EPI_ISL_2318734, EPI_ISL_2318735, EPI_ISL_2318737, EPI_ISL_2318738, EPI_ISL_2318739, EPI_ISL_2318740, EPI_ISL_2318741, EPI_ISL_2318742, EPI_ISL_2318743, EPI_ISL_2318744, EPI_ISL_2318745, EPI_ISL_2318746, EPI_ISL_2318747, EPI_ISL_2318748, EPI_ISL_2318749, EPI_ISL_2318750, EPI_ISL_2318751, EPI_ISL_2318752, EPI_ISL_2318753, EPI_ISL_2318754, EPI_ISL_2318755, EPI_ISL_2318756, EPI_ISL_2318757, EPI_ISL_2318758, EPI_ISL_2318759, EPI_ISL_2318760, EPI_ISL_2318761, EPI_ISL_2318762, EPI_ISL_2318763, EPI_ISL_2318764, EPI_ISL_2318765, EPI_ISL_2318766, EPI_ISL_2318767, EPI_ISL_2318768, EPI_ISL_2318769, EPI_ISL_2318770, EPI_ISL_2318771, EPI_ISL_2318772, EPI_ISL_2318773, EPI_ISL_2318774, EPI_ISL_2318775, EPI_ISL_2318776, EPI_ISL_2318777, EPI_ISL_2318778, EPI_ISL_2318779, EPI_ISL_2318780, EPI_ISL_2318781, EPI_ISL_2318782, EPI_ISL_2318783, EPI_ISL_2318784, EPI_ISL_2318785, EPI_ISL_2318786, EPI_ISL_2318787, EPI_ISL_2318788, EPI_ISL_2318789, EPI_ISL_2318790, EPI_ISL_2318791, EPI_ISL_2318792, EPI_ISL_2318793, EPI_ISL_2318794, EPI_ISL_2318795, EPI_ISL_2318796, EPI_ISL_2318797, EPI_ISL_2318798, EPI_ISL_2318799, EPI_ISL_2318800, EPI_ISL_2318801, EPI_ISL_2318802, EPI_ISL_2318803, EPI_ISL_2318804, EPI_ISL_2318805, EPI_ISL_2318806, EPI_ISL_2318807, EPI_ISL_2318808, EPI_ISL_2318809, EPI_ISL_2318810, EPI_ISL_2318811, EPI_ISL_2318812, EPI_ISL_2318813, EPI_ISL_2318814, EPI_ISL_2318815, EPI_ISL_2318816, EPI_ISL_2318817, EPI_ISL_2318818, EPI_ISL_2318819, EPI_ISL_2318820, EPI_ISL_2318821, EPI_ISL_2318822, EPI_ISL_2318823, EPI_ISL_2318825, EPI_ISL_2318826, EPI_ISL_2318827, EPI_ISL_2318828, EPI_ISL_2318829, EPI_ISL_2318830, EPI_ISL_2318831, EPI_ISL_2318832, EPI_ISL_2318833, EPI_ISL_2318834, EPI_ISL_2318835, EPI_ISL_2318836, EPI_ISL_2318837, EPI_ISL_2318838, EPI_ISL_2318839, EPI_ISL_2318840, EPI_ISL_2318841, EPI_ISL_2318842, EPI_ISL_2318843, EPI_ISL_2318844, EPI_ISL_2318845, EPI_ISL_2318846, EPI_ISL_2318847, EPI_ISL_2318848, EPI_ISL_2318849, EPI_ISL_2318850, EPI_ISL_2318851, EPI_ISL_2318852, EPI_ISL_2318853, EPI_ISL_2318854, EPI_ISL_2318855, EPI_ISL_2318856, EPI_ISL_2318857, EPI_ISL_2318858, EPI_ISL_2318859, EPI_ISL_2318860, EPI_ISL_2318861, EPI_ISL_2318862, EPI_ISL_2318863, EPI_ISL_2318864, EPI_ISL_2346483, EPI_ISL_2346494, EPI_ISL_2346523, EPI_ISL_2346524, EPI_ISL_2346570, EPI_ISL_2346618, EPI_ISL_2346646, EPI_ISL_2346687, EPI_ISL_2346714, EPI_ISL_2347179, EPI_ISL_2347192, EPI_ISL_2347200, EPI_ISL_2347267, EPI_ISL_2347268, EPI_ISL_2347347, EPI_ISL_2347418, EPI_ISL_2347426, EPI_ISL_2347441, EPI_ISL_2347655, EPI_ISL_2347663, EPI_ISL_2347673, EPI_ISL_2347749, EPI_ISL_2347784, EPI_ISL_2347825, EPI_ISL_2347832, EPI_ISL_2348226, EPI_ISL_2348251, EPI_ISL_2348276, EPI_ISL_2348282, EPI_ISL_2348374, EPI_ISL_2348388, EPI_ISL_2348441, EPI_ISL_2352720, EPI_ISL_2352737, EPI_ISL_2352739, EPI_ISL_2352742, EPI_ISL_2352800, EPI_ISL_2352804, EPI_ISL_2352807, EPI_ISL_2352814, EPI_ISL_2352821, EPI_ISL_2352834, EPI_ISL_2352858, EPI_ISL_2352863, EPI_ISL_2352865, EPI_ISL_2352871, EPI_ISL_2352889, EPI_ISL_2352892, EPI_ISL_2352901, EPI_ISL_2352920, EPI_ISL_2352926, EPI_ISL_2352931, EPI_ISL_2352938, EPI_ISL_2352944, EPI_ISL_2352948, EPI_ISL_2352952, EPI_ISL_2352954, EPI_ISL_2352989, EPI_ISL_2352993, EPI_ISL_2353592, EPI_ISL_2353614, EPI_ISL_2353622, EPI_ISL_2353668, EPI_ISL_2353677, EPI_ISL_2353678, EPI_ISL_2353688, EPI_ISL_2353700, EPI_ISL_2353715, EPI_ISL_2353727, EPI_ISL_2353825, EPI_ISL_2353827, EPI_ISL_2353845, EPI_ISL_2353859, EPI_ISL_2353864, EPI_ISL_2353865, EPI_ISL_2353866, EPI_ISL_2353867, EPI_ISL_2353868, EPI_ISL_2353869, EPI_ISL_2353870, EPI_ISL_2353871, EPI_ISL_2353872, EPI_ISL_2353873, EPI_ISL_2353874, EPI_ISL_2353875, EPI_ISL_2353876, EPI_ISL_2353877, EPI_ISL_2353878, EPI_ISL_2353879, EPI_ISL_2353880, EPI_ISL_2353881, EPI_ISL_2353882, EPI_ISL_2353883, EPI_ISL_2353884, EPI_ISL_2353885, EPI_ISL_2353886, EPI_ISL_2353887, EPI_ISL_2353888, EPI_ISL_2353889, EPI_ISL_2353890, EPI_ISL_2353891,

EPI_ISL_2353892, EPI_ISL_2353893, EPI_ISL_2353894, EPI_ISL_2353895, EPI_ISL_2353896, EPI_ISL_2353897, EPI_ISL_2353898, EPI_ISL_2353899, EPI_ISL_2353900, EPI_ISL_2353901, EPI_ISL_2353902, EPI_ISL_2353903, EPI_ISL_2353904, EPI_ISL_2353905, EPI_ISL_2353906, EPI_ISL_2353907, EPI_ISL_2353908, EPI_ISL_2353909, EPI_ISL_2353910, EPI_ISL_2353911, EPI_ISL_2353912, EPI_ISL_2353913,
EPI_ISL_2353914, EPI_ISL_2353915, EPI_ISL_2353916, EPI_ISL_2353917, EPI_ISL_2353918, EPI_ISL_2353919, EPI_ISL_2353920, EPI_ISL_2353921, EPI_ISL_2353922, EPI_ISL_2353923, EPI_ISL_2353924, EPI_ISL_2353925, EPI_ISL_2353926, EPI_ISL_2353927, EPI_ISL_2353928, EPI_ISL_2353929, EPI_ISL_2353930, EPI_ISL_2353931, EPI_ISL_2353932, EPI_ISL_2353933, EPI_ISL_2353934, EPI_ISL_2353935,
EPI_ISL_2353936, EPI_ISL_2353937, EPI_ISL_2353938, EPI_ISL_2353939, EPI_ISL_2353941, EPI_ISL_2353942, EPI_ISL_2353943, EPI_ISL_2353944, EPI_ISL_2353945, EPI_ISL_2353946, EPI_ISL_2353947, EPI_ISL_2353948, EPI_ISL_2353949, EPI_ISL_2353950, EPI_ISL_2353951, EPI_ISL_2353952, EPI_ISL_2353953, EPI_ISL_2353954, EPI_ISL_2353955, EPI_ISL_2353956, EPI_ISL_2353957, EPI_ISL_2353958,
EPI_ISL_2353959, EPI_ISL_2353960, EPI_ISL_2353961, EPI_ISL_2353962, EPI_ISL_2353963, EPI_ISL_2353964, EPI_ISL_2353965, EPI_ISL_2353966, EPI_ISL_2353967, EPI_ISL_2353968, EPI_ISL_2353969, EPI_ISL_2353970, EPI_ISL_2353971, EPI_ISL_2353972, EPI_ISL_2353973, EPI_ISL_2353974, EPI_ISL_2353975, EPI_ISL_2353976, EPI_ISL_2353977, EPI_ISL_2353978, EPI_ISL_2353979, EPI_ISL_2353980,
EPI_ISL_2353981, EPI_ISL_2353982, EPI_ISL_2353983, EPI_ISL_2353984, EPI_ISL_2353985, EPI_ISL_2353986, EPI_ISL_2353987, EPI_ISL_2353988, EPI_ISL_2353989, EPI_ISL_2353990, EPI_ISL_2353991, EPI_ISL_2353992, EPI_ISL_2353993, EPI_ISL_2353994, EPI_ISL_2353995, EPI_ISL_2353996, EPI_ISL_2353997, EPI_ISL_2353998, EPI_ISL_2353999, EPI_ISL_2354000, EPI_ISL_2354001, EPI_ISL_2354002,
EPI_ISL_2354003, EPI_ISL_2354004, EPI_ISL_2354005, EPI_ISL_2354006, EPI_ISL_2354007, EPI_ISL_2354008, EPI_ISL_2354009, EPI_ISL_2354010, EPI_ISL_2354011, EPI_ISL_2354012, EPI_ISL_2354013, EPI_ISL_2354014, EPI_ISL_2354015, EPI_ISL_2354016, EPI_ISL_2354017, EPI_ISL_2354019, EPI_ISL_2354020, EPI_ISL_2354021, EPI_ISL_2354022, EPI_ISL_2354023, EPI_ISL_2354024, EPI_ISL_2354025,
EPI_ISL_2354026, EPI_ISL_2354027, EPI_ISL_2354028, EPI_ISL_2354029, EPI_ISL_2354030, EPI_ISL_2354031, EPI_ISL_2354032, EPI_ISL_2354033, EPI_ISL_2354034, EPI_ISL_2354035, EPI_ISL_2354036, EPI_ISL_2354037, EPI_ISL_2354038, EPI_ISL_2354039, EPI_ISL_2354040, EPI_ISL_2354041, EPI_ISL_2354042, EPI_ISL_2354043, EPI_ISL_2354044, EPI_ISL_2354045, EPI_ISL_2354046, EPI_ISL_2354047,
EPI_ISL_2354048, EPI_ISL_2354049, EPI_ISL_2354050, EPI_ISL_2354051, EPI_ISL_2354052, EPI_ISL_2354053, EPI_ISL_2354054, EPI_ISL_2354055, EPI_ISL_2354056, EPI_ISL_2354057, EPI_ISL_2354058, EPI_ISL_2354059, EPI_ISL_2354060, EPI_ISL_2354061, EPI_ISL_2354062, EPI_ISL_2354064, EPI_ISL_2354065, EPI_ISL_2354066, EPI_ISL_2354067, EPI_ISL_2354068, EPI_ISL_2354069, EPI_ISL_2354070,
EPI_ISL_2354071, EPI_ISL_2354072, EPI_ISL_2354073, EPI_ISL_2354074, EPI_ISL_2354076, EPI_ISL_2354077, EPI_ISL_2354078, EPI_ISL_2354079, EPI_ISL_2354080, EPI_ISL_2354081, EPI_ISL_2354082, EPI_ISL_2354083, EPI_ISL_2354084, EPI_ISL_2354085, EPI_ISL_2354086, EPI_ISL_2354087, EPI_ISL_2354088, EPI_ISL_2354089, EPI_ISL_2354090, EPI_ISL_2354091, EPI_ISL_2354092, EPI_ISL_2354093,
EPI_ISL_2354094, EPI_ISL_2354095, EPI_ISL_2354096, EPI_ISL_2354097, EPI_ISL_2354098, EPI_ISL_2354099, EPI_ISL_2354100, EPI_ISL_2354101, EPI_ISL_2354102, EPI_ISL_2354103, EPI_ISL_2354104, EPI_ISL_2354105, EPI_ISL_2354106, EPI_ISL_2354107, EPI_ISL_2354108, EPI_ISL_2354109, EPI_ISL_2354110, EPI_ISL_2354111, EPI_ISL_2354112, EPI_ISL_2354113, EPI_ISL_2354114, EPI_ISL_2354115,
EPI_ISL_2354116, EPI_ISL_2354117, EPI_ISL_2354118, EPI_ISL_2354119, EPI_ISL_2354120, EPI_ISL_2354121, EPI_ISL_2354122, EPI_ISL_2354123, EPI_ISL_2354124, EPI_ISL_2354125, EPI_ISL_2354126, EPI_ISL_2354127, EPI_ISL_2354128, EPI_ISL_2354129, EPI_ISL_2354130, EPI_ISL_2354131, EPI_ISL_2354132, EPI_ISL_2354133, EPI_ISL_2354134, EPI_ISL_2354135, EPI_ISL_2354136, EPI_ISL_2354137,
EPI_ISL_2354138, EPI_ISL_2354139, EPI_ISL_2354140, EPI_ISL_2354141, EPI_ISL_2354142, EPI_ISL_2354143, EPI_ISL_2354144, EPI_ISL_2354145, EPI_ISL_2354146, EPI_ISL_2354147, EPI_ISL_2354148, EPI_ISL_2354149, EPI_ISL_2354150, EPI_ISL_2354151, EPI_ISL_2354152, EPI_ISL_2354153, EPI_ISL_2354154, EPI_ISL_2354155, EPI_ISL_2354156, EPI_ISL_2354157, EPI_ISL_2354158, EPI_ISL_2354159,
EPI_ISL_2354160, EPI_ISL_2354161, EPI_ISL_2354162, EPI_ISL_2354163, EPI_ISL_2354164, EPI_ISL_2354165, EPI_ISL_2354166, EPI_ISL_2354167, EPI_ISL_2354168, EPI_ISL_2354169, EPI_ISL_2354170, EPI_ISL_2354171, EPI_ISL_2354172, EPI_ISL_2354173, EPI_ISL_2354174, EPI_ISL_2354175, EPI_ISL_2354176, EPI_ISL_2354177, EPI_ISL_2354178, EPI_ISL_2354179, EPI_ISL_2354180, EPI_ISL_2354181,
EPI_ISL_2354182, EPI_ISL_2354183, EPI_ISL_2354184, EPI_ISL_2354185, EPI_ISL_2354186, EPI_ISL_2354187, EPI_ISL_2354188, EPI_ISL_2354189, EPI_ISL_2354190, EPI_ISL_2354191, EPI_ISL_2354192, EPI_ISL_2354193, EPI_ISL_2354194, EPI_ISL_2354195, EPI_ISL_2354196, EPI_ISL_2354197, EPI_ISL_2354198, EPI_ISL_2354199, EPI_ISL_2354201, EPI_ISL_2354202, EPI_ISL_2354203, EPI_ISL_2354204,
EPI_ISL_2354205, EPI_ISL_2354206, EPI_ISL_2354207, EPI_ISL_2354208, EPI_ISL_2354209, EPI_ISL_2354210, EPI_ISL_2354211, EPI_ISL_2354212, EPI_ISL_2354213, EPI_ISL_2354214, EPI_ISL_2354215, EPI_ISL_2354216, EPI_ISL_2354292, EPI_ISL_2354293, EPI_ISL_2354294, EPI_ISL_2354295, EPI_ISL_2354296, EPI_ISL_2354297, EPI_ISL_2354298, EPI_ISL_2354299, EPI_ISL_2354300, EPI_ISL_2354301,
EPI_ISL_2354302, EPI_ISL_2354303, EPI_ISL_2354304, EPI_ISL_2354305, EPI_ISL_2354306, EPI_ISL_2354307, EPI_ISL_2354308, EPI_ISL_2354309, EPI_ISL_2354310, EPI_ISL_2354311, EPI_ISL_2354312, EPI_ISL_2354313, EPI_ISL_2354315, EPI_ISL_2354316, EPI_ISL_2354317, EPI_ISL_2354318, EPI_ISL_2354319, EPI_ISL_2354320, EPI_ISL_2354321, EPI_ISL_2354322, EPI_ISL_2354323, EPI_ISL_2354324,
EPI_ISL_2354325, EPI_ISL_2354326, EPI_ISL_2354327, EPI_ISL_2354328, EPI_ISL_2354329, EPI_ISL_2354330, EPI_ISL_2354331, EPI_ISL_2354332, EPI_ISL_2354333, EPI_ISL_2354335, EPI_ISL_2354336, EPI_ISL_2354337, EPI_ISL_2354338, EPI_ISL_2354339, EPI_ISL_2354340, EPI_ISL_2354341, EPI_ISL_2354342, EPI_ISL_2354343, EPI_ISL_2354344, EPI_ISL_2354346, EPI_ISL_2354347, EPI_ISL_2354348,
EPI_ISL_2354349, EPI_ISL_2354350, EPI_ISL_2354351, EPI_ISL_2354352, EPI_ISL_2354353, EPI_ISL_2354354, EPI_ISL_2354356, EPI_ISL_2354357, EPI_ISL_2354358, EPI_ISL_2354359, EPI_ISL_2354360, EPI_ISL_2354361, EPI_ISL_2354362, EPI_ISL_2354363, EPI_ISL_2354364, EPI_ISL_2354365, EPI_ISL_2354366, EPI_ISL_2354367, EPI_ISL_2354368, EPI_ISL_2354369, EPI_ISL_2354370, EPI_ISL_2354371,
EPI_ISL_2354372, EPI_ISL_2354373, EPI_ISL_2354374, EPI_ISL_2354375, EPI_ISL_2354376, EPI_ISL_2354377, EPI_ISL_2354378, EPI_ISL_2354380, EPI_ISL_2354381, EPI_ISL_2354382, EPI_ISL_2354383, EPI_ISL_2354384, EPI_ISL_2354385, EPI_ISL_2354386, EPI_ISL_2354387, EPI_ISL_2354388, EPI_ISL_2354389, EPI_ISL_2354390, EPI_ISL_2354393, EPI_ISL_2354395, EPI_ISL_2354396, EPI_ISL_2354397,
EPI_ISL_2354398, EPI_ISL_2354399, EPI_ISL_2354401, EPI_ISL_2354402, EPI_ISL_2354403, EPI_ISL_2354406, EPI_ISL_2354407, EPI_ISL_2354408, EPI_ISL_2354409, EPI_ISL_2354410, EPI_ISL_2354411, EPI_ISL_2354412, EPI_ISL_2354413, EPI_ISL_2354414, EPI_ISL_2354418, EPI_ISL_2354419, EPI_ISL_2354420, EPI_ISL_2354421, EPI_ISL_2354422, EPI_ISL_2354423, EPI_ISL_2354424, EPI_ISL_2354426,
EPI_ISL_2354427, EPI_ISL_2354428, EPI_ISL_2354429, EPI_ISL_2354430, EPI_ISL_2354431, EPI_ISL_2354432, EPI_ISL_2354433, EPI_ISL_2354434, EPI_ISL_2354436, EPI_ISL_2354437, EPI_ISL_2354438, EPI_ISL_2354439, EPI_ISL_2354440, EPI_ISL_2354441, EPI_ISL_2354444, EPI_ISL_2354445, EPI_ISL_2354446, EPI_ISL_2354447, EPI_ISL_2354448, EPI_ISL_2354451, EPI_ISL_2354452, EPI_ISL_2354453,
EPI_ISL_2354454, EPI_ISL_2354456, EPI_ISL_2354458, EPI_ISL_2354459, EPI_ISL_2354460, EPI_ISL_2354461, EPI_ISL_2354462, EPI_ISL_2354464, EPI_ISL_2354465, EPI_ISL_2354466, EPI_ISL_2354467, EPI_ISL_2354468, EPI_ISL_2354469, EPI_ISL_2354471, EPI_ISL_2354473, EPI_ISL_2354474, EPI_ISL_2354475, EPI_ISL_2354476, EPI_ISL_2354477, EPI_ISL_2366270, EPI_ISL_2366296, EPI_ISL_2366298,
EPI_ISL_2366303, EPI_ISL_2366310, EPI_ISL_2366314, EPI_ISL_2366321, EPI_ISL_2366322, EPI_ISL_2366340, EPI_ISL_2366344, EPI_ISL_2366353, EPI_ISL_2366381, EPI_ISL_2366405, EPI_ISL_2366451, EPI_ISL_2366479, EPI_ISL_2366486, EPI_ISL_2366491, EPI_ISL_2366503, EPI_ISL_2366527, EPI_ISL_2366529, EPI_ISL_2366530, EPI_ISL_2366538, EPI_ISL_2366552, EPI_ISL_2366564, EPI_ISL_2366578,
EPI_ISL_2366588, EPI_ISL_2394467, EPI_ISL_2395454, EPI_ISL_2395471, EPI_ISL_2395483, EPI_ISL_2395557, EPI_ISL_2395685, EPI_ISL_2395687, EPI_ISL_2395688, EPI_ISL_2395690, EPI_ISL_2395692, EPI_ISL_2395693, EPI_ISL_2395694, EPI_ISL_2395695, EPI_ISL_2395700, EPI_ISL_2395701, EPI_ISL_2395704, EPI_ISL_2395707, EPI_ISL_2395708, EPI_ISL_2395709, EPI_ISL_2395711, EPI_ISL_2395715,
EPI_ISL_2395716, EPI_ISL_2395718, EPI_ISL_2395722, EPI_ISL_2395723, EPI_ISL_2395724, EPI_ISL_2395726, EPI_ISL_2395728, EPI_ISL_2395729, EPI_ISL_2395730, EPI_ISL_2395732, EPI_ISL_2395733, EPI_ISL_2395734, EPI_ISL_2395737, EPI_ISL_2395738, EPI_ISL_2395739, EPI_ISL_2395740, EPI_ISL_2395742, EPI_ISL_2395745, EPI_ISL_2395747, EPI_ISL_2395753, EPI_ISL_2395755, EPI_ISL_2395756,
EPI_ISL_2395758, EPI_ISL_2395760, EPI_ISL_2395761, EPI_ISL_2395763, EPI_ISL_2395764, EPI_ISL_2395766, EPI_ISL_2395767, EPI_ISL_2395769, EPI_ISL_2395770, EPI_ISL_2395771, EPI_ISL_2395773, EPI_ISL_2395774, EPI_ISL_2395775, EPI_ISL_2395779, EPI_ISL_2395784, EPI_ISL_2395786, EPI_ISL_2395787, EPI_ISL_2395788, EPI_ISL_2395791, EPI_ISL_2395792, EPI_ISL_2395793, EPI_ISL_2395794,
EPI_ISL_2395795, EPI_ISL_2395796, EPI_ISL_2395799, EPI_ISL_2395800, EPI_ISL_2395801, EPI_ISL_2395804, EPI_ISL_2395806, EPI_ISL_2395808, EPI_ISL_2395810, EPI_ISL_2395812, EPI_ISL_2395813, EPI_ISL_2395815, EPI_ISL_2395816, EPI_ISL_2395817, EPI_ISL_2395818, EPI_ISL_2395821, EPI_ISL_2395823, EPI_ISL_2395824, EPI_ISL_2395826, EPI_ISL_2395829, EPI_ISL_2395830, EPI_ISL_2395833,
EPI_ISL_2395835, EPI_ISL_2395838, EPI_ISL_2395841, EPI_ISL_2395842, EPI_ISL_2395843, EPI_ISL_2395846, EPI_ISL_2395848, EPI_ISL_2395850, EPI_ISL_2395853, EPI_ISL_2395854, EPI_ISL_2395855, EPI_ISL_2395859, EPI_ISL_2395862, EPI_ISL_2395863, EPI_ISL_2395865, EPI_ISL_2395866, EPI_ISL_2395868, EPI_ISL_2395870, EPI_ISL_2395871, EPI_ISL_2395872, EPI_ISL_2395873, EPI_ISL_2395877,
EPI_ISL_2395879, EPI_ISL_2395885, EPI_ISL_2395886, EPI_ISL_2395888, EPI_ISL_2395894, EPI_ISL_2395895, EPI_ISL_2395896, EPI_ISL_2395898, EPI_ISL_2395899, EPI_ISL_2395900, EPI_ISL_2395903, EPI_ISL_2395905, EPI_ISL_2395906, EPI_ISL_2395908, EPI_ISL_2395911, EPI_ISL_2395913, EPI_ISL_2395914, EPI_ISL_2395915, EPI_ISL_2395916, EPI_ISL_2395918, EPI_ISL_2395919, EPI_ISL_2395921,
EPI_ISL_2395922, EPI_ISL_2395925, EPI_ISL_2395926, EPI_ISL_2395930, EPI_ISL_2395931, EPI_ISL_2395932, EPI_ISL_2395933, EPI_ISL_2395935, EPI_ISL_2395937, EPI_ISL_2395938, EPI_ISL_2395939, EPI_ISL_2395940, EPI_ISL_2395946, EPI_ISL_2395950, EPI_ISL_2395952, EPI_ISL_2395960, EPI_ISL_2395962, EPI_ISL_2395968, EPI_ISL_2395969, EPI_ISL_2395970, EPI_ISL_2395975, EPI_ISL_2395976,
EPI_ISL_2395980, EPI_ISL_2395981, EPI_ISL_2395982, EPI_ISL_2395983, EPI_ISL_2395984, EPI_ISL_2395985, EPI_ISL_2395989, EPI_ISL_2395993, EPI_ISL_2395997, EPI_ISL_2395999, EPI_ISL_2396000, EPI_ISL_2396001, EPI_ISL_2396002, EPI_ISL_2396003, EPI_ISL_2396004, EPI_ISL_2396008, EPI_ISL_2396010, EPI_ISL_2396015, EPI_ISL_2396350, EPI_ISL_2396355, EPI_ISL_2396357, EPI_ISL_2396358,
EPI_ISL_2396360, EPI_ISL_2396365, EPI_ISL_2396366, EPI_ISL_2396373, EPI_ISL_2396375, EPI_ISL_2396377, EPI_ISL_2396380, EPI_ISL_2396381, EPI_ISL_2396390, EPI_ISL_2396392, EPI_ISL_2396395, EPI_ISL_2396397, EPI_ISL_2396398, EPI_ISL_2396399, EPI_ISL_2396401, EPI_ISL_2396402, EPI_ISL_2396404, EPI_ISL_2396406, EPI_ISL_2396407, EPI_ISL_2396408, EPI_ISL_2396409, EPI_ISL_2396411,
EPI_ISL_2396412, EPI_ISL_2396413, EPI_ISL_2396414, EPI_ISL_2396415, EPI_ISL_2396418, EPI_ISL_2396422, EPI_ISL_2396423, EPI_ISL_2396424, EPI_ISL_2396425, EPI_ISL_2396427, EPI_ISL_2396428, EPI_ISL_2396430, EPI_ISL_2396431, EPI_ISL_2396432, EPI_ISL_2396434, EPI_ISL_2396435, EPI_ISL_2396436, EPI_ISL_2396439, EPI_ISL_2396445, EPI_ISL_2396446, EPI_ISL_2396452, EPI_ISL_2396454,
EPI_ISL_2396456, EPI_ISL_2396457, EPI_ISL_2396460, EPI_ISL_2396462, EPI_ISL_2396464, EPI_ISL_2396466, EPI_ISL_2396468, EPI_ISL_2396470, EPI_ISL_2396471, EPI_ISL_2396472, EPI_ISL_2396476, EPI_ISL_2396477, EPI_ISL_2396478, EPI_ISL_2396480, EPI_ISL_2396481, EPI_ISL_2396482, EPI_ISL_2396484, EPI_ISL_2396487, EPI_ISL_2396489, EPI_ISL_2396490, EPI_ISL_2396491, EPI_ISL_2396492,
EPI_ISL_2396493, EPI_ISL_2396495, EPI_ISL_2396496, EPI_ISL_2396498, EPI_ISL_2396499, EPI_ISL_2396501, EPI_ISL_2396502, EPI_ISL_2396503, EPI_ISL_2396506, EPI_ISL_2396507, EPI_ISL_2396508, EPI_ISL_2396509, EPI_ISL_2396512, EPI_ISL_2396513, EPI_ISL_2396514, EPI_ISL_2396515, EPI_ISL_2396516, EPI_ISL_2396517, EPI_ISL_2396518, EPI_ISL_2396519, EPI_ISL_2396520, EPI_ISL_2396521,
EPI_ISL_2396522, EPI_ISL_2396523, EPI_ISL_2396524, EPI_ISL_2396525, EPI_ISL_2396526, EPI_ISL_2396527, EPI_ISL_2396528, EPI_ISL_2396529, EPI_ISL_2396530, EPI_ISL_2396531, EPI_ISL_2396532, EPI_ISL_2396533, EPI_ISL_2396534, EPI_ISL_2396535, EPI_ISL_2396536, EPI_ISL_2396537, EPI_ISL_2396538, EPI_ISL_2396539, EPI_ISL_2396540, EPI_ISL_2396541, EPI_ISL_2396542, EPI_ISL_2396543,
EPI_ISL_2396544, EPI_ISL_2396545, EPI_ISL_2396546, EPI_ISL_2396547, EPI_ISL_2396548, EPI_ISL_2396549, EPI_ISL_2396550, EPI_ISL_2396551, EPI_ISL_2396552, EPI_ISL_2396553, EPI_ISL_2396554, EPI_ISL_2396555, EPI_ISL_2396556, EPI_ISL_2396557, EPI_ISL_2396558, EPI_ISL_2396559, EPI_ISL_2396560, EPI_ISL_2396561, EPI_ISL_2396562, EPI_ISL_2396563, EPI_ISL_2396564, EPI_ISL_2396565,
EPI_ISL_2396566, EPI_ISL_2396567, EPI_ISL_2396568, EPI_ISL_2396569, EPI_ISL_2396570, EPI_ISL_2396571, EPI_ISL_2396572, EPI_ISL_2396573, EPI_ISL_2396574, EPI_ISL_2396575, EPI_ISL_2396576, EPI_ISL_2396577, EPI_ISL_2396578, EPI_ISL_2396579, EPI_ISL_2396580, EPI_ISL_2396581, EPI_ISL_2396582, EPI_ISL_2396583, EPI_ISL_2396584, EPI_ISL_2396585, EPI_ISL_2396587, EPI_ISL_2396588,
EPI_ISL_2396589, EPI_ISL_2396590, EPI_ISL_2396591, EPI_ISL_2396592, EPI_ISL_2396593, EPI_ISL_2396594, EPI_ISL_2396595, EPI_ISL_2396596, EPI_ISL_2396597, EPI_ISL_2396598, EPI_ISL_2396599, EPI_ISL_2396600, EPI_ISL_2396601, EPI_ISL_2396602, EPI_ISL_2396603, EPI_ISL_2396604, EPI_ISL_2396605, EPI_ISL_2396606, EPI_ISL_2396607, EPI_ISL_2396608, EPI_ISL_2396609, EPI_ISL_2396610,
EPI_ISL_2396611, EPI_ISL_2396612, EPI_ISL_2396613, EPI_ISL_2396614, EPI_ISL_2396615, EPI_ISL_2396616, EPI_ISL_2396617, EPI_ISL_2396618, EPI_ISL_2396619, EPI_ISL_2396620, EPI_ISL_2396621, EPI_ISL_2396622, EPI_ISL_2396623, EPI_ISL_2396624, EPI_ISL_2396625, EPI_ISL_2396626, EPI_ISL_2396627, EPI_ISL_2396628, EPI_ISL_2396629, EPI_ISL_2396630, EPI_ISL_2396631, EPI_ISL_2396632,
EPI_ISL_2396633, EPI_ISL_2396634, EPI_ISL_2396635, EPI_ISL_2396636, EPI_ISL_2396637, EPI_ISL_2396638, EPI_ISL_2396639, EPI_ISL_2396640, EPI_ISL_2396641, EPI_ISL_2396642, EPI_ISL_2396643, EPI_ISL_2396644, EPI_ISL_2396645, EPI_ISL_2396646, EPI_ISL_2396647, EPI_ISL_2396648, EPI_ISL_2396649, EPI_ISL_2396650, EPI_ISL_2396651, EPI_ISL_2396652, EPI_ISL_2396653, EPI_ISL_2396654,
EPI_ISL_2396655, EPI_ISL_2396656, EPI_ISL_2396657, EPI_ISL_2396658, EPI_ISL_2396659, EPI_ISL_2396660, EPI_ISL_2396661, EPI_ISL_2396662, EPI_ISL_2396663, EPI_ISL_2396664, EPI_ISL_2396665, EPI_ISL_2396666, EPI_ISL_2396667, EPI_ISL_2396668, EPI_ISL_2396669, EPI_ISL_2396670, EPI_ISL_2396671, EPI_ISL_2396672, EPI_ISL_2396673, EPI_ISL_2396674, EPI_ISL_2396675, EPI_ISL_2396676,
EPI_ISL_2396677, EPI_ISL_2396678, EPI_ISL_2396679, EPI_ISL_2396680, EPI_ISL_2396681, EPI_ISL_2396682, EPI_ISL_2396683, EPI_ISL_2396684, EPI_ISL_2396685, EPI_ISL_2396686, EPI_ISL_2396687, EPI_ISL_2396688, EPI_ISL_2396689, EPI_ISL_2396690, EPI_ISL_2396691, EPI_ISL_2396692, EPI_ISL_2396693, EPI_ISL_2396694, EPI_ISL_2396695, EPI_ISL_2396696, EPI_ISL_2396697, EPI_ISL_2396698,
EPI_ISL_2396699, EPI_ISL_2396700, EPI_ISL_2396701, EPI_ISL_2396702, EPI_ISL_2396703, EPI_ISL_2396704, EPI_ISL_2396705, EPI_ISL_2396706, EPI_ISL_2396708, EPI_ISL_2396709, EPI_ISL_2396710, EPI_ISL_2396713, EPI_ISL_2396714, EPI_ISL_2396715, EPI_ISL_2396716, EPI_ISL_2396717, EPI_ISL_2396719, EPI_ISL_2396720, EPI_ISL_2396721, EPI_ISL_2396722, EPI_ISL_2396723, EPI_ISL_2396724,
EPI_ISL_2396725, EPI_ISL_2396726, EPI_ISL_2396727, EPI_ISL_2396728, EPI_ISL_2396729, EPI_ISL_2396730, EPI_ISL_2396731, EPI_ISL_2396732, EPI_ISL_2396733, EPI_ISL_2396734, EPI_ISL_2396735, EPI_ISL_2396736, EPI_ISL_2396737, EPI_ISL_2396738, EPI_ISL_2396739, EPI_ISL_2396740, EPI_ISL_2396741, EPI_ISL_2396742, EPI_ISL_2396743, EPI_ISL_2396744, EPI_ISL_2396745, EPI_ISL_2396746,
EPI_ISL_2396747, EPI_ISL_2396748, EPI_ISL_2396749, EPI_ISL_2396750, EPI_ISL_2396751, EPI_ISL_2396752, EPI_ISL_2396754, EPI_ISL_2396755, EPI_ISL_2396756, EPI_ISL_2396757, EPI_ISL_2396758, EPI_ISL_2396759, EPI_ISL_2396760, EPI_ISL_2396761, EPI_ISL_2396762, EPI_ISL_2396763, EPI_ISL_2396764, EPI_ISL_2396765, EPI_ISL_2396766, EPI_ISL_2396767, EPI_ISL_2396768, EPI_ISL_2396769,
EPI_ISL_2396770, EPI_ISL_2396771, EPI_ISL_2396772, EPI_ISL_2396773, EPI_ISL_2396774, EPI_ISL_2396775, EPI_ISL_2396776, EPI_ISL_2396777, EPI_ISL_2396778, EPI_ISL_2396779, EPI_ISL_2396780, EPI_ISL_2396781, EPI_ISL_2396782, EPI_ISL_2396783, EPI_ISL_2396784, EPI_ISL_2396785, EPI_ISL_2396786, EPI_ISL_2396787, EPI_ISL_2396788, EPI_ISL_2396789, EPI_ISL_2396790, EPI_ISL_2396791,
EPI_ISL_2396792, EPI_ISL_2396793, EPI_ISL_2396794, EPI_ISL_2396795, EPI_ISL_2396796, EPI_ISL_2396797, EPI_ISL_2407302, EPI_ISL_2407317, EPI_ISL_2407320, EPI_ISL_2407327, EPI_ISL_2407343, EPI_ISL_2407347, EPI_ISL_2407348, EPI_ISL_2407354, EPI_ISL_2407360, EPI_ISL_2407391, EPI_ISL_2407412, EPI_ISL_2407413, EPI_ISL_2407420, EPI_ISL_2407422, EPI_ISL_2407425, EPI_ISL_2407445,
EPI_ISL_2407461, EPI_ISL_2407525, EPI_ISL_2407548, EPI_ISL_2407573, EPI_ISL_2407592, EPI_ISL_2407909, EPI_ISL_2407922, EPI_ISL_2407946, EPI_ISL_2407954, EPI_ISL_2407997, EPI_ISL_2407998, EPI_ISL_2408000, EPI_ISL_2408001, EPI_ISL_2408002, EPI_ISL_2408004, EPI_ISL_2408006, EPI_ISL_2408007, EPI_ISL_2408009, EPI_ISL_2408011, EPI_ISL_2408015, EPI_ISL_2408017, EPI_ISL_2408018,
EPI_ISL_2408020, EPI_ISL_2408022, EPI_ISL_2408023, EPI_ISL_2408024, EPI_ISL_2408025, EPI_ISL_2408031, EPI_ISL_2408032, EPI_ISL_2408033, EPI_ISL_2408034, EPI_ISL_2408035, EPI_ISL_2408036, EPI_ISL_2408038, EPI_ISL_2408039, EPI_ISL_2408040, EPI_ISL_2408041, EPI_ISL_2408043, EPI_ISL_2408044, EPI_ISL_2408045, EPI_ISL_2408046, EPI_ISL_2408047, EPI_ISL_2408048, EPI_ISL_2408050,
EPI_ISL_2408051, EPI_ISL_2408052, EPI_ISL_2408053, EPI_ISL_2408054, EPI_ISL_2408056, EPI_ISL_2408059, EPI_ISL_2408060, EPI_ISL_2447286, EPI_ISL_2447288, EPI_ISL_2447299, EPI_ISL_2447323, EPI_ISL_2447327, EPI_ISL_2447336, EPI_ISL_2457690, EPI_ISL_2457692, EPI_ISL_2457694, EPI_ISL_2457695, EPI_ISL_2457696, EPI_ISL_2457698, EPI_ISL_2457699, EPI_ISL_2457702, EPI_ISL_2457704,
EPI_ISL_2457705, EPI_ISL_2457707, EPI_ISL_2457708, EPI_ISL_2457710, EPI_ISL_2457713, EPI_ISL_2457716, EPI_ISL_2457718, EPI_ISL_2457719, EPI_ISL_2457721, EPI_ISL_2457726, EPI_ISL_2457727, EPI_ISL_2457728, EPI_ISL_2457729, EPI_ISL_2457730, EPI_ISL_2457732, EPI_ISL_2457733, EPI_ISL_2457735, EPI_ISL_2457737, EPI_ISL_2457738, EPI_ISL_2457741, EPI_ISL_2457743, EPI_ISL_2457744,
EPI_ISL_2457745, EPI_ISL_2457746, EPI_ISL_2457747, EPI_ISL_2457748, EPI_ISL_2457749, EPI_ISL_2457750, EPI_ISL_2457752, EPI_ISL_2457753, EPI_ISL_2457754, EPI_ISL_2457755, EPI_ISL_2457756, EPI_ISL_2457760, EPI_ISL_2457763, EPI_ISL_2457764, EPI_ISL_2457767, EPI_ISL_2457768, EPI_ISL_2457769, EPI_ISL_2457770, EPI_ISL_2457771, EPI_ISL_2457772, EPI_ISL_2457773, EPI_ISL_2457775,
EPI_ISL_2457776, EPI_ISL_2457777, EPI_ISL_2457778, EPI_ISL_2457782, EPI_ISL_2457785, EPI_ISL_2457787, EPI_ISL_2457788, EPI_ISL_2457789, EPI_ISL_2457790, EPI_ISL_2457792, EPI_ISL_2457793, EPI_ISL_2457794, EPI_ISL_2457796, EPI_ISL_2457797, EPI_ISL_2457799, EPI_ISL_2457801, EPI_ISL_2457802, EPI_ISL_2457804, EPI_ISL_2457805, EPI_ISL_2457806, EPI_ISL_2457808, EPI_ISL_2457811,
EPI_ISL_2457812, EPI_ISL_2457813, EPI_ISL_2457815, EPI_ISL_2457817, EPI_ISL_2457818, EPI_ISL_2457819, EPI_ISL_2457820, EPI_ISL_2457823, EPI_ISL_2457824, EPI_ISL_2457825, EPI_ISL_2457826, EPI_ISL_2457828, EPI_ISL_2457829, EPI_ISL_2457831, EPI_ISL_2457837, EPI_ISL_2457838, EPI_ISL_2457839, EPI_ISL_2457840, EPI_ISL_2457844, EPI_ISL_2457845, EPI_ISL_2457849, EPI_ISL_2457850,
EPI_ISL_2457851, EPI_ISL_2457852, EPI_ISL_2457854, EPI_ISL_2457855, EPI_ISL_2457856, EPI_ISL_2457859, EPI_ISL_2457862, EPI_ISL_2457865, EPI_ISL_2457866, EPI_ISL_2457867, EPI_ISL_2457868, EPI_ISL_2457871, EPI_ISL_2457874, EPI_ISL_2457877, EPI_ISL_2457878, EPI_ISL_2457880, EPI_ISL_2457882

| | | | |
|---|---|---|---|
| see above | Lighthouse Lab in Glasgow | Wellcome Sanger Institute for the COVID-19 Genomics UK (COG-UK) Consortium | Anna Dominiczak and Alex Alderton; Carol Clugston; Cordelia Langford; David Gray; David K. Jackson; Dominic Kwiatkowski; Ewan Harrison; Harper VanSteenhouse; Ian Johnston; Jeffrey Barrett; John Sillitoe on behalf of the Wellcome Sanger Institute COVID-19 Surveillance Team; Roberto Amato; Sonia Goncalves; Yumi Kasai |

EPI_ISL_2235908, EPI_ISL_2253226, EPI_ISL_2253229, EPI_ISL_2253233, EPI_ISL_2253239, EPI_ISL_2253241, EPI_ISL_2253242, EPI_ISL_2253243, EPI_ISL_2253249, EPI_ISL_2253250, EPI_ISL_2253252, EPI_ISL_2253253, EPI_ISL_2253258, EPI_ISL_2253259, EPI_ISL_2253263, EPI_ISL_2253266, EPI_ISL_2253270, EPI_ISL_2253274, EPI_ISL_2253277, EPI_ISL_2253279, EPI_ISL_2253280, EPI_ISL_2253282,
EPI_ISL_2253284, EPI_ISL_2253287, EPI_ISL_2253295, EPI_ISL_2253296, EPI_ISL_2253309, EPI_ISL_2253313, EPI_ISL_2253317, EPI_ISL_2253322, EPI_ISL_2253323, EPI_ISL_2253324, EPI_ISL_2253328, EPI_ISL_2253333, EPI_ISL_2253336, EPI_ISL_2253337, EPI_ISL_2253339, EPI_ISL_2253345, EPI_ISL_2253348, EPI_ISL_2253351, EPI_ISL_2253361, EPI_ISL_2253363, EPI_ISL_2253368, EPI_ISL_2253369,
EPI_ISL_2253370, EPI_ISL_2253371, EPI_ISL_2253372, EPI_ISL_2253373, EPI_ISL_2253374, EPI_ISL_2253376, EPI_ISL_2253378, EPI_ISL_2253384, EPI_ISL_2253387, EPI_ISL_2253400, EPI_ISL_2253405, EPI_ISL_2253413, EPI_ISL_2253426, EPI_ISL_2253428, EPI_ISL_2253432, EPI_ISL_2253439, EPI_ISL_2253467, EPI_ISL_2253490, EPI_ISL_2253507, EPI_ISL_2253508, EPI_ISL_2253511, EPI_ISL_2253528,
EPI_ISL_2253537, EPI_ISL_2253540, EPI_ISL_2253543, EPI_ISL_2253550, EPI_ISL_2253563, EPI_ISL_2253564, EPI_ISL_2253579, EPI_ISL_2253604, EPI_ISL_2253606, EPI_ISL_2253673, EPI_ISL_2274980, EPI_ISL_2274981, EPI_ISL_2274982, EPI_ISL_2274984, EPI_ISL_2274985, EPI_ISL_2274986, EPI_ISL_2274988, EPI_ISL_2274989, EPI_ISL_2274990, EPI_ISL_2274991, EPI_ISL_2274992, EPI_ISL_2274993,
EPI_ISL_2274994, EPI_ISL_2274996, EPI_ISL_2274997, EPI_ISL_2274998, EPI_ISL_2274999, EPI_ISL_2275001, EPI_ISL_2275003, EPI_ISL_2275004, EPI_ISL_2275005, EPI_ISL_2275007, EPI_ISL_2275008, EPI_ISL_2275009, EPI_ISL_2275010, EPI_ISL_2275011, EPI_ISL_2275013, EPI_ISL_2275015, EPI_ISL_2275016, EPI_ISL_2275017, EPI_ISL_2275018, EPI_ISL_2275019, EPI_ISL_2275020, EPI_ISL_2275021,
EPI_ISL_2275022, EPI_ISL_2275023, EPI_ISL_2275024, EPI_ISL_2275025, EPI_ISL_2275026, EPI_ISL_2275027, EPI_ISL_2275028, EPI_ISL_2275031, EPI_ISL_2275032, EPI_ISL_2275033, EPI_ISL_2275035, EPI_ISL_2275036, EPI_ISL_2275037, EPI_ISL_2275039, EPI_ISL_2275040, EPI_ISL_2275041, EPI_ISL_2275042, EPI_ISL_2275043, EPI_ISL_2275044, EPI_ISL_2275045, EPI_ISL_2275046, EPI_ISL_2275047,
EPI_ISL_2275048, EPI_ISL_2275049, EPI_ISL_2275050, EPI_ISL_2275051, EPI_ISL_2275054, EPI_ISL_2275056, EPI_ISL_2275058, EPI_ISL_2275059, EPI_ISL_2275060, EPI_ISL_2275061, EPI_ISL_2275063, EPI_ISL_2275065, EPI_ISL_2275067, EPI_ISL_2275068, EPI_ISL_2275069, EPI_ISL_2275070, EPI_ISL_2275072, EPI_ISL_2275073, EPI_ISL_2275074, EPI_ISL_2275075, EPI_ISL_2275076, EPI_ISL_2275077,
EPI_ISL_2275078, EPI_ISL_2275079, EPI_ISL_2275081, EPI_ISL_2275082, EPI_ISL_2275083, EPI_ISL_2275084, EPI_ISL_2275086, EPI_ISL_2275089, EPI_ISL_2275090, EPI_ISL_2275091, EPI_ISL_2275092, EPI_ISL_2275093, EPI_ISL_2275095, EPI_ISL_2275096, EPI_ISL_2275098, EPI_ISL_2275099, EPI_ISL_2275101, EPI_ISL_2275104, EPI_ISL_2275105, EPI_ISL_2275106, EPI_ISL_2275107, EPI_ISL_2275108,
EPI_ISL_2275110, EPI_ISL_2275112, EPI_ISL_2275113, EPI_ISL_2275114, EPI_ISL_2275115, EPI_ISL_2275116, EPI_ISL_2275117, EPI_ISL_2275118, EPI_ISL_2275119, EPI_ISL_2275120, EPI_ISL_2275121, EPI_ISL_2275122, EPI_ISL_2275123, EPI_ISL_2275124, EPI_ISL_2275125, EPI_ISL_2275126, EPI_ISL_2275127, EPI_ISL_2275129, EPI_ISL_2275130, EPI_ISL_2275132, EPI_ISL_2275133, EPI_ISL_2275134,
EPI_ISL_2275135, EPI_ISL_2275136, EPI_ISL_2275138, EPI_ISL_2275140, EPI_ISL_2275141, EPI_ISL_2275142, EPI_ISL_2275143, EPI_ISL_2275144, EPI_ISL_2275145, EPI_ISL_2275146, EPI_ISL_2275147, EPI_ISL_2275149, EPI_ISL_2275150, EPI_ISL_2275151, EPI_ISL_2275152, EPI_ISL_2275153, EPI_ISL_2275154, EPI_ISL_2275156, EPI_ISL_2275157, EPI_ISL_2275159, EPI_ISL_2275160, EPI_ISL_2275161,
EPI_ISL_2275162, EPI_ISL_2275163, EPI_ISL_2275164, EPI_ISL_2275165, EPI_ISL_2275166, EPI_ISL_2275167, EPI_ISL_2275168, EPI_ISL_2275169, EPI_ISL_2275170, EPI_ISL_2275173, EPI_ISL_2275174, EPI_ISL_2275176, EPI_ISL_2275177, EPI_ISL_2275178, EPI_ISL_2275180, EPI_ISL_2275181, EPI_ISL_2275182, EPI_ISL_2275183, EPI_ISL_2275196, EPI_ISL_2275208, EPI_ISL_2275224, EPI_ISL_2275226,
EPI_ISL_2275233, EPI_ISL_2275277, EPI_ISL_2275287, EPI_ISL_2296699, EPI_ISL_2316669, EPI_ISL_2316671, EPI_ISL_2316676, EPI_ISL_2316677, EPI_ISL_2316692, EPI_ISL_2316695, EPI_ISL_2316706, EPI_ISL_2316709, EPI_ISL_2316712, EPI_ISL_2316715, EPI_ISL_2316723, EPI_ISL_2316730, EPI_ISL_2316734, EPI_ISL_2316742, EPI_ISL_2316749, EPI_ISL_2316753, EPI_ISL_2316755, EPI_ISL_2316759,
EPI_ISL_2316763, EPI_ISL_2316766, EPI_ISL_2316769, EPI_ISL_2316774, EPI_ISL_2316776, EPI_ISL_2316794, EPI_ISL_2316801, EPI_ISL_2316809, EPI_ISL_2316810, EPI_ISL_2316812, EPI_ISL_2316822, EPI_ISL_2316824, EPI_ISL_2316825, EPI_ISL_2316827, EPI_ISL_2316834, EPI_ISL_2316837, EPI_ISL_2316846, EPI_ISL_2316848, EPI_ISL_2316854, EPI_ISL_2316860, EPI_ISL_2316864, EPI_ISL_2316866,
EPI_ISL_2316869, EPI_ISL_2316875, EPI_ISL_2316881, EPI_ISL_2316883, EPI_ISL_2316886, EPI_ISL_2316887, EPI_ISL_2316897, EPI_ISL_2316902, EPI_ISL_2316906, EPI_ISL_2316912, EPI_ISL_2316920, EPI_ISL_2316923, EPI_ISL_2316924, EPI_ISL_2316933, EPI_ISL_2316936, EPI_ISL_2316940, EPI_ISL_2316943, EPI_ISL_2316946, EPI_ISL_2316948, EPI_ISL_2316956, EPI_ISL_2316960, EPI_ISL_2316967,
EPI_ISL_2316972, EPI_ISL_2316975, EPI_ISL_2316982, EPI_ISL_2316991, EPI_ISL_2316996, EPI_ISL_2317000, EPI_ISL_2317037, EPI_ISL_2317040, EPI_ISL_2317041, EPI_ISL_2317044, EPI_ISL_2317048, EPI_ISL_2317055, EPI_ISL_2317059, EPI_ISL_2317060, EPI_ISL_2317067, EPI_ISL_2317073, EPI_ISL_2317075, EPI_ISL_2317076, EPI_ISL_2317079, EPI_ISL_2317080, EPI_ISL_2317082, EPI_ISL_2317085,
EPI_ISL_2317090, EPI_ISL_2317092, EPI_ISL_2317094, EPI_ISL_2317095, EPI_ISL_2317099, EPI_ISL_2317100, EPI_ISL_2317102, EPI_ISL_2317104, EPI_ISL_2317105, EPI_ISL_2317108, EPI_ISL_2317110, EPI_ISL_2317115, EPI_ISL_2317117, EPI_ISL_2317121, EPI_ISL_2317122, EPI_ISL_2317126, EPI_ISL_2317128, EPI_ISL_2317133, EPI_ISL_2317134, EPI_ISL_2317143, EPI_ISL_2317146, EPI_ISL_2317147,
EPI_ISL_2317149, EPI_ISL_2317151, EPI_ISL_2317152, EPI_ISL_2317154, EPI_ISL_2317156, EPI_ISL_2317163, EPI_ISL_2317164, EPI_ISL_2317166, EPI_ISL_2317167, EPI_ISL_2317168, EPI_ISL_2317169, EPI_ISL_2317172, EPI_ISL_2317175, EPI_ISL_2317177, EPI_ISL_2317182, EPI_ISL_2317183, EPI_ISL_2317184, EPI_ISL_2317186, EPI_ISL_2317194, EPI_ISL_2317196, EPI_ISL_2317197, EPI_ISL_2317201,
EPI_ISL_2317205, EPI_ISL_2317207, EPI_ISL_2317209, EPI_ISL_2317210, EPI_ISL_2317212, EPI_ISL_2317216, EPI_ISL_2317220, EPI_ISL_2317222, EPI_ISL_2317225, EPI_ISL_2317226, EPI_ISL_2317229, EPI_ISL_2317230, EPI_ISL_2317231, EPI_ISL_2317234, EPI_ISL_2317239, EPI_ISL_2317242, EPI_ISL_2317249, EPI_ISL_2317254, EPI_ISL_2317259, EPI_ISL_2317260, EPI_ISL_2317264, EPI_ISL_2317268,
EPI_ISL_2317269, EPI_ISL_2317271, EPI_ISL_2317273, EPI_ISL_2317274, EPI_ISL_2317276, EPI_ISL_2317277, EPI_ISL_2317280, EPI_ISL_2317281, EPI_ISL_2317284, EPI_ISL_2317288, EPI_ISL_2317290, EPI_ISL_2317291, EPI_ISL_2317298, EPI_ISL_2317299, EPI_ISL_2317301, EPI_ISL_2317306, EPI_ISL_2317308, EPI_ISL_2317311, EPI_ISL_2317315, EPI_ISL_2317316, EPI_ISL_2317320, EPI_ISL_2317323,
EPI_ISL_2317324, EPI_ISL_2317325, EPI_ISL_2317326, EPI_ISL_2317327, EPI_ISL_2317329, EPI_ISL_2317330, EPI_ISL_2317334, EPI_ISL_2317336, EPI_ISL_2317340, EPI_ISL_2317341, EPI_ISL_2317342, EPI_ISL_2317347, EPI_ISL_2317349, EPI_ISL_2317352, EPI_ISL_2317353, EPI_ISL_2317360, EPI_ISL_2317361, EPI_ISL_2317365, EPI_ISL_2317367, EPI_ISL_2317373, EPI_ISL_2317374, EPI_ISL_2317375,
EPI_ISL_2317377, EPI_ISL_2317379, EPI_ISL_2317389, EPI_ISL_2317392, EPI_ISL_2317395, EPI_ISL_2317396, EPI_ISL_2317400, EPI_ISL_2317401, EPI_ISL_2317403, EPI_ISL_2317406, EPI_ISL_2317407, EPI_ISL_2317412, EPI_ISL_2317413, EPI_ISL_2317414, EPI_ISL_2317415, EPI_ISL_2317416, EPI_ISL_2317417, EPI_ISL_2317418, EPI_ISL_2317420, EPI_ISL_2317421, EPI_ISL_2317422, EPI_ISL_2317423,
EPI_ISL_2317424, EPI_ISL_2317425, EPI_ISL_2317427, EPI_ISL_2317429, EPI_ISL_2317433, EPI_ISL_2317434, EPI_ISL_2317435, EPI_ISL_2317436, EPI_ISL_2317437, EPI_ISL_2317438, EPI_ISL_2317439, EPI_ISL_2317440, EPI_ISL_2317441, EPI_ISL_2317443, EPI_ISL_2317445, EPI_ISL_2317446, EPI_ISL_2317447, EPI_ISL_2317448, EPI_ISL_2317449, EPI_ISL_2317450, EPI_ISL_2317452, EPI_ISL_2317453,
EPI_ISL_2317454, EPI_ISL_2317456, EPI_ISL_2317458, EPI_ISL_2317459, EPI_ISL_2317460, EPI_ISL_2317461, EPI_ISL_2317462, EPI_ISL_2317463, EPI_ISL_2317465, EPI_ISL_2317466, EPI_ISL_2317467, EPI_ISL_2317469, EPI_ISL_2317470, EPI_ISL_2317471, EPI_ISL_2317472, EPI_ISL_2317474, EPI_ISL_2317475, EPI_ISL_2317476, EPI_ISL_2317478, EPI_ISL_2317479, EPI_ISL_2317480, EPI_ISL_2317481,
EPI_ISL_2317482, EPI_ISL_2317484, EPI_ISL_2317485, EPI_ISL_2317487, EPI_ISL_2317489, EPI_ISL_2317490, EPI_ISL_2317496, EPI_ISL_2317497, EPI_ISL_2317498, EPI_ISL_2317499, EPI_ISL_2317500, EPI_ISL_2317501, EPI_ISL_2317503, EPI_ISL_2317504, EPI_ISL_2317505, EPI_ISL_2317506, EPI_ISL_2317508, EPI_ISL_2317509, EPI_ISL_2317510, EPI_ISL_2317512, EPI_ISL_2317513, EPI_ISL_2317514,
EPI_ISL_2317515, EPI_ISL_2317516, EPI_ISL_2317517, EPI_ISL_2317518, EPI_ISL_2317520, EPI_ISL_2317521, EPI_ISL_2317523, EPI_ISL_2317525, EPI_ISL_2317527, EPI_ISL_2317529, EPI_ISL_2317531, EPI_ISL_2317532, EPI_ISL_2317535, EPI_ISL_2317536, EPI_ISL_2317541, EPI_ISL_2317542, EPI_ISL_2317544, EPI_ISL_2317545, EPI_ISL_2317546, EPI_ISL_2317548, EPI_ISL_2317549, EPI_ISL_2317550,
EPI_ISL_2317551, EPI_ISL_2317552, EPI_ISL_2317553, EPI_ISL_2317554, EPI_ISL_2317555, EPI_ISL_2317557, EPI_ISL_2317558, EPI_ISL_2317559, EPI_ISL_2317560, EPI_ISL_2317562, EPI_ISL_2317564, EPI_ISL_2317565, EPI_ISL_2317566, EPI_ISL_2317567, EPI_ISL_2317568, EPI_ISL_2317569, EPI_ISL_2317570, EPI_ISL_2317571, EPI_ISL_2317572, EPI_ISL_2317573, EPI_ISL_2317577, EPI_ISL_2317578,
EPI_ISL_2317579, EPI_ISL_2317580, EPI_ISL_2317582, EPI_ISL_2317584, EPI_ISL_2317586, EPI_ISL_2317587, EPI_ISL_2317588, EPI_ISL_2317589, EPI_ISL_2317590, EPI_ISL_2317593, EPI_ISL_2317594, EPI_ISL_2317596, EPI_ISL_2317597, EPI_ISL_2317599, EPI_ISL_2317600, EPI_ISL_2317604, EPI_ISL_2317605, EPI_ISL_2317607, EPI_ISL_2317608, EPI_ISL_2317609, EPI_ISL_2317610, EPI_ISL_2317611,
EPI_ISL_2317612, EPI_ISL_2317613, EPI_ISL_2317615, EPI_ISL_2317616, EPI_ISL_2317617, EPI_ISL_2317619, EPI_ISL_2317621, EPI_ISL_2317626, EPI_ISL_2317627, EPI_ISL_2317632, EPI_ISL_2317633, EPI_ISL_2317638, EPI_ISL_2317639, EPI_ISL_2317640, EPI_ISL_2317641, EPI_ISL_2317642, EPI_ISL_2317643, EPI_ISL_2317644, EPI_ISL_2317645, EPI_ISL_2317648, EPI_ISL_2317650, EPI_ISL_2317651,
EPI_ISL_2317652, EPI_ISL_2317653, EPI_ISL_2317657, EPI_ISL_2317658, EPI_ISL_2317659, EPI_ISL_2317660, EPI_ISL_2317664, EPI_ISL_2317665, EPI_ISL_2317666, EPI_ISL_2317667, EPI_ISL_2317668, EPI_ISL_2317671, EPI_ISL_2317673, EPI_ISL_2317675, EPI_ISL_2317679, EPI_ISL_2317680, EPI_ISL_2317682, EPI_ISL_2317683, EPI_ISL_2317684, EPI_ISL_2317685, EPI_ISL_2317686, EPI_ISL_2317687,
EPI_ISL_2317688, EPI_ISL_2317689, EPI_ISL_2317691, EPI_ISL_2317692, EPI_ISL_2317693, EPI_ISL_2317694, EPI_ISL_2317695, EPI_ISL_2317696, EPI_ISL_2317697, EPI_ISL_2317698, EPI_ISL_2317699, EPI_ISL_2317700, EPI_ISL_2317702, EPI_ISL_2317703, EPI_ISL_2317704, EPI_ISL_2317705, EPI_ISL_2317708, EPI_ISL_2317710, EPI_ISL_2317711, EPI_ISL_2317713, EPI_ISL_2317715, EPI_ISL_2317716,
EPI_ISL_2317717, EPI_ISL_2317718, EPI_ISL_2317719, EPI_ISL_2317720, EPI_ISL_2317721, EPI_ISL_2317722, EPI_ISL_2317723, EPI_ISL_2317724, EPI_ISL_2317726, EPI_ISL_2317728, EPI_ISL_2317729, EPI_ISL_2317732, EPI_ISL_2317734, EPI_ISL_2317736, EPI_ISL_2317737, EPI_ISL_2317741, EPI_ISL_2317745, EPI_ISL_2317746, EPI_ISL_2317747, EPI_ISL_2317748, EPI_ISL_2317751, EPI_ISL_2317757,
EPI_ISL_2317779, EPI_ISL_2317780, EPI_ISL_2317802, EPI_ISL_2317803, EPI_ISL_2317824, EPI_ISL_2317826, EPI_ISL_2317829, EPI_ISL_2317830, EPI_ISL_2317838, EPI_ISL_2317862, EPI_ISL_2317879, EPI_ISL_2317892, EPI_ISL_2317907, EPI_ISL_2317909, EPI_ISL_2317943, EPI_ISL_2317969, EPI_ISL_2317973, EPI_ISL_2317995, EPI_ISL_2317997, EPI_ISL_2317998, EPI_ISL_2318009, EPI_ISL_2318011,
EPI_ISL_2346472, EPI_ISL_2346473, EPI_ISL_2346476, EPI_ISL_2346477, EPI_ISL_2346479, EPI_ISL_2346480, EPI_ISL_2346481, EPI_ISL_2346484, EPI_ISL_2346486, EPI_ISL_2346488, EPI_ISL_2346489, EPI_ISL_2346490, EPI_ISL_2346491, EPI_ISL_2346493, EPI_ISL_2346496, EPI_ISL_2346500, EPI_ISL_2346503, EPI_ISL_2346505, EPI_ISL_2346506, EPI_ISL_2346507, EPI_ISL_2346510, EPI_ISL_2346511,
EPI_ISL_2346512, EPI_ISL_2346513, EPI_ISL_2346519, EPI_ISL_2346521, EPI_ISL_2346525, EPI_ISL_2346527, EPI_ISL_2346529, EPI_ISL_2346530, EPI_ISL_2346531, EPI_ISL_2346532, EPI_ISL_2346533, EPI_ISL_2346534, EPI_ISL_2346535, EPI_ISL_2346536, EPI_ISL_2346538, EPI_ISL_2346539, EPI_ISL_2346540, EPI_ISL_2346542, EPI_ISL_2346546, EPI_ISL_2346547, EPI_ISL_2346548, EPI_ISL_2346549,
EPI_ISL_2346550, EPI_ISL_2346551, EPI_ISL_2346552, EPI_ISL_2346553, EPI_ISL_2346558, EPI_ISL_2346560, EPI_ISL_2346561, EPI_ISL_2346562, EPI_ISL_2346563, EPI_ISL_2346565, EPI_ISL_2346566, EPI_ISL_2346573, EPI_ISL_2346574, EPI_ISL_2346575, EPI_ISL_2346576, EPI_ISL_2346577, EPI_ISL_2346578, EPI_ISL_2346579, EPI_ISL_2346580, EPI_ISL_2346581, EPI_ISL_2346582, EPI_ISL_2346583,
EPI_ISL_2346584, EPI_ISL_2346585, EPI_ISL_2346586, EPI_ISL_2346587, EPI_ISL_2346589, EPI_ISL_2346590, EPI_ISL_2346594, EPI_ISL_2346595, EPI_ISL_2346596, EPI_ISL_2346598, EPI_ISL_2346599, EPI_ISL_2346600, EPI_ISL_2346601, EPI_ISL_2346604, EPI_ISL_2346607, EPI_ISL_2346608, EPI_ISL_2346610, EPI_ISL_2346611, EPI_ISL_2346612, EPI_ISL_2346614, EPI_ISL_2346615, EPI_ISL_2346616,
EPI_ISL_2346617, EPI_ISL_2346621, EPI_ISL_2346623, EPI_ISL_2346625, EPI_ISL_2346626, EPI_ISL_2346628, EPI_ISL_2346630, EPI_ISL_2346631, EPI_ISL_2346632, EPI_ISL_2346638, EPI_ISL_2346640, EPI_ISL_2346641, EPI_ISL_2346642, EPI_ISL_2346645, EPI_ISL_2346647, EPI_ISL_2346648, EPI_ISL_2346649, EPI_ISL_2346653, EPI_ISL_2346654, EPI_ISL_2346657, EPI_ISL_2346658, EPI_ISL_2346662,
EPI_ISL_2346664, EPI_ISL_2346665, EPI_ISL_2346667, EPI_ISL_2346670, EPI_ISL_2346671, EPI_ISL_2346673, EPI_ISL_2346677, EPI_ISL_2346678, EPI_ISL_2346679, EPI_ISL_2346680, EPI_ISL_2346681, EPI_ISL_2346682, EPI_ISL_2346683, EPI_ISL_2346685, EPI_ISL_2346686, EPI_ISL_2346688, EPI_ISL_2346689, EPI_ISL_2346691, EPI_ISL_2346693, EPI_ISL_2346694, EPI_ISL_2346695, EPI_ISL_2346696,
EPI_ISL_2346697, EPI_ISL_2346699, EPI_ISL_2346701, EPI_ISL_2346702, EPI_ISL_2346704, EPI_ISL_2346706, EPI_ISL_2346707, EPI_ISL_2346708, EPI_ISL_2346710, EPI_ISL_2346712, EPI_ISL_2346716, EPI_ISL_2346717, EPI_ISL_2346718, EPI_ISL_2346720, EPI_ISL_2346724, EPI_ISL_2346726, EPI_ISL_2346727, EPI_ISL_2346731, EPI_ISL_2346736, EPI_ISL_2346739, EPI_ISL_2346743, EPI_ISL_2346746,
EPI_ISL_2346758, EPI_ISL_2346760, EPI_ISL_2346769, EPI_ISL_2346770, EPI_ISL_2346774, EPI_ISL_2346778, EPI_ISL_2346779, EPI_ISL_2346785, EPI_ISL_2346791, EPI_ISL_2346794, EPI_ISL_2346797, EPI_ISL_2346801, EPI_ISL_2346806, EPI_ISL_2346807, EPI_ISL_2346809, EPI_ISL_2346810, EPI_ISL_2346819, EPI_ISL_2346820, EPI_ISL_2346822, EPI_ISL_2346828, EPI_ISL_2346830, EPI_ISL_2346831,
EPI_ISL_2346833, EPI_ISL_2346837, EPI_ISL_2346838, EPI_ISL_2346840, EPI_ISL_2346847, EPI_ISL_2346849, EPI_ISL_2346858, EPI_ISL_2346862, EPI_ISL_2346864, EPI_ISL_2346872, EPI_ISL_2346876, EPI_ISL_2346882, EPI_ISL_2346888, EPI_ISL_2346890, EPI_ISL_2346893, EPI_ISL_2346904, EPI_ISL_2346907, EPI_ISL_2346910, EPI_ISL_2346919, EPI_ISL_2346920, EPI_ISL_2346921, EPI_ISL_2346922,
EPI_ISL_2346928, EPI_ISL_2346935, EPI_ISL_2346936, EPI_ISL_2346941, EPI_ISL_2346944, EPI_ISL_2346946, EPI_ISL_2346953, EPI_ISL_2346955, EPI_ISL_2346964, EPI_ISL_2346969, EPI_ISL_2346971, EPI_ISL_2346973, EPI_ISL_2346978, EPI_ISL_2346980, EPI_ISL_2346987, EPI_ISL_2347000, EPI_ISL_2347007, EPI_ISL_2347009, EPI_ISL_2347010, EPI_ISL_2347012, EPI_ISL_2347013, EPI_ISL_2347014,
EPI_ISL_2347016, EPI_ISL_2347017, EPI_ISL_2347022, EPI_ISL_2347023, EPI_ISL_2347028, EPI_ISL_2347034, EPI_ISL_2347043, EPI_ISL_2347051, EPI_ISL_2347053, EPI_ISL_2347059, EPI_ISL_2347065, EPI_ISL_2347067, EPI_ISL_2347070, EPI_ISL_2347081, EPI_ISL_2347082, EPI_ISL_2347083, EPI_ISL_2347084, EPI_ISL_2347088, EPI_ISL_2347089, EPI_ISL_2347090, EPI_ISL_2347091, EPI_ISL_2347092,
EPI_ISL_2347093, EPI_ISL_2347094, EPI_ISL_2347095, EPI_ISL_2347096, EPI_ISL_2347097, EPI_ISL_2347098, EPI_ISL_2347099, EPI_ISL_2347100, EPI_ISL_2347101, EPI_ISL_2347102, EPI_ISL_2347103, EPI_ISL_2347104, EPI_ISL_2347105, EPI_ISL_2347106, EPI_ISL_2347107, EPI_ISL_2347108, EPI_ISL_2347109, EPI_ISL_2347110, EPI_ISL_2347111, EPI_ISL_2347112, EPI_ISL_2347113, EPI_ISL_2347114,
EPI_ISL_2347115, EPI_ISL_2347116, EPI_ISL_2347117, EPI_ISL_2347118, EPI_ISL_2347119, EPI_ISL_2347120, EPI_ISL_2347121, EPI_ISL_2347122, EPI_ISL_2347123, EPI_ISL_2347124, EPI_ISL_2347125, EPI_ISL_2347126, EPI_ISL_2347127, EPI_ISL_2347128, EPI_ISL_2347129, EPI_ISL_2347130, EPI_ISL_2347131, EPI_ISL_2347132, EPI_ISL_2347133, EPI_ISL_2347134, EPI_ISL_2347135, EPI_ISL_2347136,
EPI_ISL_2347137, EPI_ISL_2347138, EPI_ISL_2347139, EPI_ISL_2347140, EPI_ISL_2347141, EPI_ISL_2347142, EPI_ISL_2347143, EPI_ISL_2347144, EPI_ISL_2347145, EPI_ISL_2347146, EPI_ISL_2347147, EPI_ISL_2347148, EPI_ISL_2347149, EPI_ISL_2347150, EPI_ISL_2347151, EPI_ISL_2347152, EPI_ISL_2347153, EPI_ISL_2347154, EPI_ISL_2347155, EPI_ISL_2347156, EPI_ISL_2347157, EPI_ISL_2347158,
EPI_ISL_2347159, EPI_ISL_2347160, EPI_ISL_2347161, EPI_ISL_2347162, EPI_ISL_2347163, EPI_ISL_2347164, EPI_ISL_2347165, EPI_ISL_2347166, EPI_ISL_2347167, EPI_ISL_2347168, EPI_ISL_2347169, EPI_ISL_2347170, EPI_ISL_2347171, EPI_ISL_2347172, EPI_ISL_2347173, EPI_ISL_2347174, EPI_ISL_2347175, EPI_ISL_2347176, EPI_ISL_2347177, EPI_ISL_2347178, EPI_ISL_2347180, EPI_ISL_2347181,
EPI_ISL_2347182, EPI_ISL_2347183, EPI_ISL_2347184, EPI_ISL_2347185, EPI_ISL_2347186, EPI_ISL_2347187, EPI_ISL_2347188, EPI_ISL_2347189, EPI_ISL_2347190, EPI_ISL_2347191, EPI_ISL_2347193, EPI_ISL_2347194, EPI_ISL_2347195, EPI_ISL_2347196, EPI_ISL_2347197, EPI_ISL_2347198, EPI_ISL_2347199, EPI_ISL_2347201, EPI_ISL_2347202, EPI_ISL_2347203, EPI_ISL_2347204, EPI_ISL_2347205,
EPI_ISL_2347206, EPI_ISL_2347207, EPI_ISL_2347208, EPI_ISL_2347209, EPI_ISL_2347210, EPI_ISL_2347211, EPI_ISL_2347212, EPI_ISL_2347213, EPI_ISL_2347214, EPI_ISL_2347215, EPI_ISL_2347216, EPI_ISL_2347217, EPI_ISL_2347218, EPI_ISL_2347219, EPI_ISL_2347220, EPI_ISL_2347221, EPI_ISL_2347222, EPI_ISL_2347223, EPI_ISL_2347224, EPI_ISL_2347225, EPI_ISL_2347226, EPI_ISL_2347227,
EPI_ISL_2347228, EPI_ISL_2347229, EPI_ISL_2347230, EPI_ISL_2347231, EPI_ISL_2347232, EPI_ISL_2347233, EPI_ISL_2347234, EPI_ISL_2347235, EPI_ISL_2347236, EPI_ISL_2347237, EPI_ISL_2347238, EPI_ISL_2347239, EPI_ISL_2347240, EPI_ISL_2347241, EPI_ISL_2347242, EPI_ISL_2347243, EPI_ISL_2347244, EPI_ISL_2347245, EPI_ISL_2347246, EPI_ISL_2347247, EPI_ISL_2347248, EPI_ISL_2347249,

| | | | |
|---|---|---|---|
| EPI_ISL_2347250, EPI_ISL_2347251, EPI_ISL_2347252, EPI_ISL_2347253, EPI_ISL_2347254, EPI_ISL_2347255, EPI_ISL_2347256, EPI_ISL_2347257, EPI_ISL_2347258, EPI_ISL_2347259, EPI_ISL_2347260, EPI_ISL_2347261, EPI_ISL_2347262, EPI_ISL_2347263, EPI_ISL_2347264, EPI_ISL_2347265, EPI_ISL_2347266, EPI_ISL_2347269, EPI_ISL_2347270, EPI_ISL_2347271, EPI_ISL_2347272, EPI_ISL_2347273, EPI_ISL_2347274, EPI_ISL_2347275, EPI_ISL_2347276, EPI_ISL_2347277, EPI_ISL_2347278, EPI_ISL_2347279, EPI_ISL_2347280, EPI_ISL_2347281, EPI_ISL_2347282, EPI_ISL_2347283, EPI_ISL_2347284, EPI_ISL_2347285, EPI_ISL_2347286, EPI_ISL_2347287, EPI_ISL_2347288, EPI_ISL_2347289, EPI_ISL_2347290, EPI_ISL_2347291, EPI_ISL_2347292, EPI_ISL_2347293, EPI_ISL_2347294, EPI_ISL_2347295, EPI_ISL_2347296, EPI_ISL_2347297, EPI_ISL_2347298, EPI_ISL_2347299, EPI_ISL_2347300, EPI_ISL_2347301, EPI_ISL_2347302, EPI_ISL_2347303, EPI_ISL_2347304, EPI_ISL_2347305, EPI_ISL_2347306, EPI_ISL_2347307, EPI_ISL_2347308, EPI_ISL_2347309, EPI_ISL_2347310, EPI_ISL_2347311, EPI_ISL_2347312, EPI_ISL_2347313, EPI_ISL_2347314, EPI_ISL_2347315, EPI_ISL_2347316, EPI_ISL_2347317, EPI_ISL_2347318, EPI_ISL_2347319, EPI_ISL_2347320, EPI_ISL_2347321, EPI_ISL_2347322, EPI_ISL_2347323, EPI_ISL_2347324, EPI_ISL_2347325, EPI_ISL_2347326, EPI_ISL_2347327, EPI_ISL_2347328, EPI_ISL_2347329, EPI_ISL_2347330, EPI_ISL_2347331, EPI_ISL_2347332, EPI_ISL_2347333, EPI_ISL_2347388, EPI_ISL_2347411, EPI_ISL_2347412, EPI_ISL_2347437, EPI_ISL_2347462, EPI_ISL_2347571, EPI_ISL_2347865, EPI_ISL_2347874, EPI_ISL_2347878, EPI_ISL_2347879, EPI_ISL_2347882, EPI_ISL_2347883, EPI_ISL_2347886, EPI_ISL_2347895, EPI_ISL_2347898, EPI_ISL_2347903, EPI_ISL_2347909, EPI_ISL_2347920, EPI_ISL_2347923, EPI_ISL_2347933, EPI_ISL_2347934, EPI_ISL_2347935, EPI_ISL_2347936, EPI_ISL_2347937, EPI_ISL_2347939, EPI_ISL_2347943, EPI_ISL_2347944, EPI_ISL_2347946, EPI_ISL_2347948, EPI_ISL_2347951, EPI_ISL_2347953, EPI_ISL_2347955, EPI_ISL_2347961, EPI_ISL_2347966, EPI_ISL_2347971, EPI_ISL_2347975, EPI_ISL_2347976, EPI_ISL_2347987, EPI_ISL_2347992, EPI_ISL_2347996, EPI_ISL_2348010, EPI_ISL_2348011, EPI_ISL_2348015, EPI_ISL_2348017, EPI_ISL_2348019, EPI_ISL_2348020, EPI_ISL_2348022, EPI_ISL_2348028, EPI_ISL_2348033, EPI_ISL_2348035, EPI_ISL_2348039, EPI_ISL_2348040, EPI_ISL_2348041, EPI_ISL_2348042, EPI_ISL_2348050, EPI_ISL_2348051, EPI_ISL_2348062, EPI_ISL_2348063, EPI_ISL_2348068, EPI_ISL_2348071, EPI_ISL_2348074, EPI_ISL_2348081, EPI_ISL_2348082, EPI_ISL_2348088, EPI_ISL_2348093, EPI_ISL_2348096, EPI_ISL_2348103, EPI_ISL_2348105, EPI_ISL_2348112, EPI_ISL_2348122, EPI_ISL_2348127, EPI_ISL_2348129, EPI_ISL_2348130, EPI_ISL_2348132, EPI_ISL_2348134, EPI_ISL_2348149, EPI_ISL_2348151, EPI_ISL_2348152, EPI_ISL_2348157, EPI_ISL_2348163, EPI_ISL_2348168, EPI_ISL_2348171, EPI_ISL_2348173, EPI_ISL_2348181, EPI_ISL_2348187, EPI_ISL_2348190, EPI_ISL_2348191, EPI_ISL_2348192, EPI_ISL_2348193, EPI_ISL_2348196, EPI_ISL_2348197, EPI_ISL_2348198, EPI_ISL_2348205, EPI_ISL_2348206, EPI_ISL_2348207, EPI_ISL_2348208, EPI_ISL_2348209, EPI_ISL_2348210, EPI_ISL_2348213, EPI_ISL_2348214, EPI_ISL_2348215, EPI_ISL_2348217, EPI_ISL_2348220, EPI_ISL_2348221, EPI_ISL_2348223, EPI_ISL_2348225, EPI_ISL_2348227, EPI_ISL_2348228, EPI_ISL_2348230, EPI_ISL_2348234, EPI_ISL_2348235, EPI_ISL_2348236, EPI_ISL_2348237, EPI_ISL_2348238, EPI_ISL_2348242, EPI_ISL_2348243, EPI_ISL_2348244, EPI_ISL_2348246, EPI_ISL_2348247, EPI_ISL_2348248, EPI_ISL_2348249, EPI_ISL_2348250, EPI_ISL_2348255, EPI_ISL_2348256, EPI_ISL_2348259, EPI_ISL_2348260, EPI_ISL_2348263, EPI_ISL_2348266, EPI_ISL_2348267, EPI_ISL_2348270, EPI_ISL_2348278, EPI_ISL_2348281, EPI_ISL_2348283, EPI_ISL_2348289, EPI_ISL_2348291, EPI_ISL_2348293, EPI_ISL_2348294, EPI_ISL_2348295, EPI_ISL_2348298, EPI_ISL_2348299, EPI_ISL_2348300, EPI_ISL_2348305, EPI_ISL_2348310, EPI_ISL_2348312, EPI_ISL_2348313, EPI_ISL_2348314, EPI_ISL_2348318, EPI_ISL_2348319, EPI_ISL_2348320, EPI_ISL_2348321, EPI_ISL_2348322, EPI_ISL_2348324, EPI_ISL_2348325, EPI_ISL_2348329, EPI_ISL_2348330, EPI_ISL_2348332, EPI_ISL_2348333, EPI_ISL_2348334, EPI_ISL_2348335, EPI_ISL_2348337, EPI_ISL_2348339, EPI_ISL_2348340, EPI_ISL_2348341, EPI_ISL_2348343, EPI_ISL_2348350, EPI_ISL_2348351, EPI_ISL_2348352, EPI_ISL_2348353, EPI_ISL_2348355, EPI_ISL_2348357, EPI_ISL_2348358, EPI_ISL_2348359, EPI_ISL_2348360, EPI_ISL_2348366, EPI_ISL_2348368, EPI_ISL_2348375, EPI_ISL_2348380, EPI_ISL_2348385, EPI_ISL_2348387, EPI_ISL_2348389, EPI_ISL_2348390, EPI_ISL_2348393, EPI_ISL_2348394, EPI_ISL_2348395, EPI_ISL_2348396, EPI_ISL_2348397, EPI_ISL_2348398, EPI_ISL_2348399, EPI_ISL_2348402, EPI_ISL_2348403, EPI_ISL_2348404, EPI_ISL_2348405, EPI_ISL_2348406, EPI_ISL_2348408, EPI_ISL_2348410, EPI_ISL_2348412, EPI_ISL_2348413, EPI_ISL_2348415, EPI_ISL_2348416, EPI_ISL_2348417, EPI_ISL_2348419, EPI_ISL_2348430, EPI_ISL_2348431, EPI_ISL_2348432, EPI_ISL_2348434, EPI_ISL_2348439, EPI_ISL_2348440, EPI_ISL_2348443, EPI_ISL_2348444, EPI_ISL_2348445, EPI_ISL_2348449, EPI_ISL_2348453, EPI_ISL_2348455, EPI_ISL_2348456, EPI_ISL_2348458, EPI_ISL_2348460, EPI_ISL_2348463, EPI_ISL_2348466, EPI_ISL_2348468, EPI_ISL_2348470, EPI_ISL_2348471, EPI_ISL_2348472, EPI_ISL_2348473, EPI_ISL_2348477, EPI_ISL_2348481, EPI_ISL_2348482, EPI_ISL_2348483, EPI_ISL_2352689, EPI_ISL_2352693, EPI_ISL_2352694, EPI_ISL_2352700, EPI_ISL_2352704, EPI_ISL_2352708, EPI_ISL_2352714, EPI_ISL_2352718, EPI_ISL_2352719, EPI_ISL_2352722, EPI_ISL_2352726, EPI_ISL_2352736, EPI_ISL_2352744, EPI_ISL_2352759, EPI_ISL_2352760, EPI_ISL_2352763, EPI_ISL_2352767, EPI_ISL_2352768, EPI_ISL_2352771, EPI_ISL_2352777, EPI_ISL_2352779, EPI_ISL_2352782, EPI_ISL_2352787, EPI_ISL_2352789, EPI_ISL_2352792, EPI_ISL_2352794, EPI_ISL_2352797, EPI_ISL_2352803, EPI_ISL_2352817, EPI_ISL_2352819, EPI_ISL_2352820, EPI_ISL_2352824, EPI_ISL_2352828, EPI_ISL_2352829, EPI_ISL_2352830, EPI_ISL_2352832, EPI_ISL_2352835, EPI_ISL_2352837, EPI_ISL_2352838, EPI_ISL_2352841, EPI_ISL_2352842, EPI_ISL_2352849, EPI_ISL_2352852, EPI_ISL_2352857, EPI_ISL_2352860, EPI_ISL_2352867, EPI_ISL_2352869, EPI_ISL_2352874, EPI_ISL_2352877, EPI_ISL_2352878, EPI_ISL_2352879, EPI_ISL_2352880, EPI_ISL_2352881, EPI_ISL_2352887, EPI_ISL_2352888, EPI_ISL_2352899, EPI_ISL_2352902, EPI_ISL_2352908, EPI_ISL_2352909, EPI_ISL_2352912, EPI_ISL_2352917, EPI_ISL_2352923, EPI_ISL_2352925, EPI_ISL_2352933, EPI_ISL_2352939, EPI_ISL_2352949, EPI_ISL_2352955, EPI_ISL_2352956, EPI_ISL_2352959, EPI_ISL_2352961, EPI_ISL_2352962, EPI_ISL_2352966, EPI_ISL_2352969, EPI_ISL_2352973, EPI_ISL_2352975, EPI_ISL_2352977, EPI_ISL_2352985, EPI_ISL_2352986, EPI_ISL_2352987, EPI_ISL_2352988, EPI_ISL_2352992, EPI_ISL_2352995, EPI_ISL_2352996, EPI_ISL_2353002, EPI_ISL_2353003, EPI_ISL_2353009, EPI_ISL_2353013, EPI_ISL_2353014, EPI_ISL_2353024, EPI_ISL_2353027, EPI_ISL_2353030, EPI_ISL_2353033, EPI_ISL_2353037, EPI_ISL_2353038, EPI_ISL_2353039, EPI_ISL_2353040, EPI_ISL_2353048, EPI_ISL_2353051, EPI_ISL_2353053, EPI_ISL_2353054, EPI_ISL_2353058, EPI_ISL_2353064, EPI_ISL_2353065, EPI_ISL_2353066, EPI_ISL_2353067, EPI_ISL_2353068, EPI_ISL_2353071, EPI_ISL_2353073, EPI_ISL_2353074, EPI_ISL_2353084, EPI_ISL_2353085, EPI_ISL_2353086, EPI_ISL_2353087, EPI_ISL_2353088, EPI_ISL_2353089, EPI_ISL_2353090, EPI_ISL_2353091, EPI_ISL_2353097, EPI_ISL_2353103, EPI_ISL_2353106, EPI_ISL_2353114, EPI_ISL_2353115, EPI_ISL_2353120, EPI_ISL_2353123, EPI_ISL_2353126, EPI_ISL_2353127, EPI_ISL_2353130, EPI_ISL_2353133, EPI_ISL_2353134, EPI_ISL_2353136, EPI_ISL_2353138, EPI_ISL_2353140, EPI_ISL_2353143, EPI_ISL_2353145, EPI_ISL_2353157, EPI_ISL_2353160, EPI_ISL_2353169, EPI_ISL_2353175, EPI_ISL_2353177, EPI_ISL_2353178, EPI_ISL_2353180, EPI_ISL_2353184, EPI_ISL_2353185, EPI_ISL_2353188, EPI_ISL_2353189, EPI_ISL_2353191, EPI_ISL_2353193, EPI_ISL_2353194, EPI_ISL_2353196, EPI_ISL_2353197, EPI_ISL_2353198, EPI_ISL_2353199, EPI_ISL_2353200, EPI_ISL_2353201, EPI_ISL_2353204, EPI_ISL_2353205, EPI_ISL_2353207, EPI_ISL_2353208, EPI_ISL_2353209, EPI_ISL_2353211, EPI_ISL_2353212, EPI_ISL_2353213, EPI_ISL_2353217, EPI_ISL_2353219, EPI_ISL_2353223, EPI_ISL_2353229, EPI_ISL_2353232, EPI_ISL_2353233, EPI_ISL_2353234, EPI_ISL_2353240, EPI_ISL_2353244, EPI_ISL_2353246, EPI_ISL_2353247, EPI_ISL_2353252, EPI_ISL_2353253, EPI_ISL_2353255, EPI_ISL_2353256, EPI_ISL_2353259, EPI_ISL_2353260, EPI_ISL_2353261, EPI_ISL_2353262, EPI_ISL_2353263, EPI_ISL_2353267, EPI_ISL_2353268, EPI_ISL_2353269, EPI_ISL_2353274, EPI_ISL_2353278, EPI_ISL_2353279, EPI_ISL_2353282, EPI_ISL_2353287, EPI_ISL_2353288, EPI_ISL_2353289, EPI_ISL_2353290, EPI_ISL_2353292, EPI_ISL_2353293, EPI_ISL_2353295, EPI_ISL_2353296, EPI_ISL_2353298, EPI_ISL_2353299, EPI_ISL_2353300, EPI_ISL_2353302, EPI_ISL_2353304, EPI_ISL_2353305, EPI_ISL_2353310, EPI_ISL_2353311, EPI_ISL_2353313, EPI_ISL_2353314, EPI_ISL_2353317, EPI_ISL_2353319, EPI_ISL_2353320, EPI_ISL_2353322, EPI_ISL_2353324, EPI_ISL_2353325, EPI_ISL_2353326, EPI_ISL_2353328, EPI_ISL_2353329, EPI_ISL_2353330, EPI_ISL_2353334, EPI_ISL_2353340, EPI_ISL_2353341, EPI_ISL_2353342, EPI_ISL_2353344, EPI_ISL_2353345, EPI_ISL_2353351, EPI_ISL_2353352, EPI_ISL_2353353, EPI_ISL_2353356, EPI_ISL_2353358, EPI_ISL_2353359, EPI_ISL_2353362, EPI_ISL_2353363, EPI_ISL_2353365, EPI_ISL_2353366, EPI_ISL_2353367, EPI_ISL_2353369, EPI_ISL_2353371, EPI_ISL_2353373, EPI_ISL_2353375, EPI_ISL_2353378, EPI_ISL_2353383, EPI_ISL_2353384, EPI_ISL_2353385, EPI_ISL_2353388, EPI_ISL_2353389, EPI_ISL_2353390, EPI_ISL_2353391, EPI_ISL_2353393, EPI_ISL_2353395, EPI_ISL_2353397, EPI_ISL_2353400, EPI_ISL_2353401, EPI_ISL_2353403, EPI_ISL_2353408, EPI_ISL_2353411, EPI_ISL_2353414, EPI_ISL_2353415, EPI_ISL_2353416, EPI_ISL_2353419, EPI_ISL_2353421, EPI_ISL_2353422, EPI_ISL_2353423, EPI_ISL_2353424, EPI_ISL_2353425, EPI_ISL_2353426, EPI_ISL_2353427, EPI_ISL_2353430, EPI_ISL_2353432, EPI_ISL_2353434, EPI_ISL_2353435, EPI_ISL_2353439, EPI_ISL_2353441, EPI_ISL_2353442, EPI_ISL_2353443, EPI_ISL_2353445, EPI_ISL_2353446, EPI_ISL_2353449, EPI_ISL_2353451, EPI_ISL_2353452, EPI_ISL_2353453, EPI_ISL_2353456, EPI_ISL_2353460, EPI_ISL_2353462, EPI_ISL_2353463, EPI_ISL_2353464, EPI_ISL_2353465, EPI_ISL_2353466, EPI_ISL_2353469, EPI_ISL_2353470, EPI_ISL_2353473, EPI_ISL_2353475, EPI_ISL_2353476, EPI_ISL_2353477, EPI_ISL_2353481, EPI_ISL_2353484, EPI_ISL_2353485, EPI_ISL_2353487, EPI_ISL_2353490, EPI_ISL_2353492, EPI_ISL_2353493, EPI_ISL_2353495, EPI_ISL_2353498, EPI_ISL_2353500, EPI_ISL_2353501, EPI_ISL_2353502, EPI_ISL_2353503, EPI_ISL_2353505, EPI_ISL_2353506, EPI_ISL_2353507, EPI_ISL_2353510, EPI_ISL_2353511, EPI_ISL_2353512, EPI_ISL_2353513, EPI_ISL_2353515, EPI_ISL_2353516, EPI_ISL_2353517, EPI_ISL_2353519, EPI_ISL_2353521, EPI_ISL_2353522, EPI_ISL_2353525, EPI_ISL_2353527, EPI_ISL_2353528, EPI_ISL_2353529, EPI_ISL_2353531, EPI_ISL_2353534, EPI_ISL_2353536, EPI_ISL_2353537, EPI_ISL_2353539, EPI_ISL_2353544, EPI_ISL_2353546, EPI_ISL_2353550, EPI_ISL_2353552, EPI_ISL_2353553, EPI_ISL_2353555, EPI_ISL_2353559, EPI_ISL_2353560, EPI_ISL_2353563, EPI_ISL_2353564, EPI_ISL_2353568, EPI_ISL_2353570, EPI_ISL_2353571, EPI_ISL_2353572, EPI_ISL_2353573, EPI_ISL_2353574, EPI_ISL_2353575, EPI_ISL_2353577, EPI_ISL_2353580, EPI_ISL_2353581, EPI_ISL_2353585, EPI_ISL_2353589, EPI_ISL_2353591, EPI_ISL_2353600, EPI_ISL_2353601, EPI_ISL_2353604, EPI_ISL_2353613, EPI_ISL_2353615, EPI_ISL_2353616, EPI_ISL_2353617, EPI_ISL_2353620, EPI_ISL_2353623, EPI_ISL_2353629, EPI_ISL_2353633, EPI_ISL_2353634, EPI_ISL_2353638, EPI_ISL_2353639, EPI_ISL_2353640, EPI_ISL_2353643, EPI_ISL_2353644, EPI_ISL_2353645, EPI_ISL_2353648, EPI_ISL_2353652, EPI_ISL_2353655, EPI_ISL_2353658, EPI_ISL_2353661, EPI_ISL_2353662, EPI_ISL_2353666, EPI_ISL_2353669, EPI_ISL_2353670, EPI_ISL_2353671, EPI_ISL_2353674, EPI_ISL_2353681, EPI_ISL_2353684, EPI_ISL_2353685, EPI_ISL_2353686, EPI_ISL_2353692, EPI_ISL_2353701, EPI_ISL_2353702, EPI_ISL_2353703, EPI_ISL_2353704, EPI_ISL_2353705, EPI_ISL_2353706, EPI_ISL_2353710, EPI_ISL_2353711, EPI_ISL_2353712, EPI_ISL_2353718, EPI_ISL_2353719, EPI_ISL_2353720, EPI_ISL_2353722, EPI_ISL_2353726, EPI_ISL_2353728, EPI_ISL_2353735, EPI_ISL_2353736, EPI_ISL_2353737, EPI_ISL_2353739, EPI_ISL_2353740, EPI_ISL_2353742, EPI_ISL_2353743, EPI_ISL_2353746, EPI_ISL_2353748, EPI_ISL_2353750, EPI_ISL_2353756, EPI_ISL_2353758, EPI_ISL_2353763, EPI_ISL_2353767, EPI_ISL_2353769, EPI_ISL_2353771, EPI_ISL_2353776, EPI_ISL_2353779, EPI_ISL_2353796, EPI_ISL_2353801, EPI_ISL_2353803, EPI_ISL_2353804, EPI_ISL_2353805, EPI_ISL_2353807, EPI_ISL_2353808, EPI_ISL_2353810, EPI_ISL_2353813, EPI_ISL_2353814, EPI_ISL_2353819, EPI_ISL_2353820, EPI_ISL_2353821, EPI_ISL_2353834, EPI_ISL_2353835, EPI_ISL_2353837, EPI_ISL_2353844, EPI_ISL_2353852, EPI_ISL_2353853, EPI_ISL_2353855, EPI_ISL_2353856, EPI_ISL_2353858, EPI_ISL_2353940, EPI_ISL_2354018, EPI_ISL_2354063, EPI_ISL_2365995, EPI_ISL_2366002, EPI_ISL_2366040, EPI_ISL_2366041, EPI_ISL_2366059, EPI_ISL_2366068, EPI_ISL_2366081, EPI_ISL_2366100, EPI_ISL_2366125, EPI_ISL_2366152, EPI_ISL_2366153, EPI_ISL_2366175, EPI_ISL_2366176, EPI_ISL_2366190, EPI_ISL_2366216, EPI_ISL_2366218, EPI_ISL_2366260, EPI_ISL_2366263, EPI_ISL_2366267, EPI_ISL_2366271, EPI_ISL_2366286, EPI_ISL_2366294, EPI_ISL_2366323, EPI_ISL_2366354, EPI_ISL_2366450, EPI_ISL_2366481, EPI_ISL_2366488, EPI_ISL_2366490, EPI_ISL_2366492, EPI_ISL_2366494, EPI_ISL_2366495, EPI_ISL_2366496, EPI_ISL_2366497, EPI_ISL_2366498, EPI_ISL_2366501, EPI_ISL_2366502, EPI_ISL_2366505, EPI_ISL_2366506, EPI_ISL_2366507, EPI_ISL_2366508, EPI_ISL_2366509, EPI_ISL_2366510, EPI_ISL_2366511, EPI_ISL_2366512, EPI_ISL_2366514, EPI_ISL_2366515, EPI_ISL_2366517, EPI_ISL_2366518, EPI_ISL_2366519, EPI_ISL_2366520, EPI_ISL_2366521, EPI_ISL_2366524, EPI_ISL_2366526, EPI_ISL_2366531, EPI_ISL_2366532, EPI_ISL_2366533, EPI_ISL_2366534, EPI_ISL_2366536, EPI_ISL_2366537, EPI_ISL_2366539, EPI_ISL_2366540, EPI_ISL_2366542, EPI_ISL_2366544, EPI_ISL_2366545, EPI_ISL_2366546, EPI_ISL_2366547, EPI_ISL_2366549, EPI_ISL_2366550, EPI_ISL_2366551, EPI_ISL_2366553, EPI_ISL_2366554, EPI_ISL_2366555, EPI_ISL_2366556, EPI_ISL_2366557, EPI_ISL_2366558, EPI_ISL_2366559, EPI_ISL_2366560, EPI_ISL_2366563, EPI_ISL_2366565, EPI_ISL_2366566, EPI_ISL_2366567, EPI_ISL_2366568, EPI_ISL_2366570, EPI_ISL_2366571, EPI_ISL_2366572, EPI_ISL_2366573, EPI_ISL_2366574, EPI_ISL_2366575, EPI_ISL_2366576, EPI_ISL_2366577, EPI_ISL_2366579, EPI_ISL_2366580, EPI_ISL_2366582, EPI_ISL_2366583, EPI_ISL_2366585, EPI_ISL_2366586, EPI_ISL_2366587, EPI_ISL_2366589, EPI_ISL_2366590, EPI_ISL_2366591, EPI_ISL_2366592, EPI_ISL_2366594, EPI_ISL_2366598, EPI_ISL_2366599, EPI_ISL_2366600, EPI_ISL_2366601, EPI_ISL_2366602, EPI_ISL_2366604, EPI_ISL_2366605, EPI_ISL_2366606, EPI_ISL_2393537, EPI_ISL_2393562, EPI_ISL_2393594, EPI_ISL_2393714, EPI_ISL_2393766, EPI_ISL_2393818, EPI_ISL_2395389, EPI_ISL_2395452, EPI_ISL_2395500, EPI_ISL_2395642, EPI_ISL_2395647, EPI_ISL_2395686, EPI_ISL_2395697, EPI_ISL_2395781, EPI_ISL_2396586, EPI_ISL_2396753, EPI_ISL_2396810, EPI_ISL_2396812, EPI_ISL_2396813, EPI_ISL_2396814, EPI_ISL_2396815, EPI_ISL_2396816, EPI_ISL_2396818, EPI_ISL_2436932, EPI_ISL_2436939, EPI_ISL_2436968, EPI_ISL_2436975, EPI_ISL_2436988, EPI_ISL_2436992, EPI_ISL_2437000, EPI_ISL_2437007, EPI_ISL_2437008, EPI_ISL_2437014, EPI_ISL_2437016, EPI_ISL_2437021, EPI_ISL_2437024, EPI_ISL_2437026, EPI_ISL_2437030, EPI_ISL_2437036, EPI_ISL_2437050, EPI_ISL_2437058, EPI_ISL_2437070, EPI_ISL_2437074, EPI_ISL_2437089, EPI_ISL_2437098, EPI_ISL_2437102, EPI_ISL_2437108, EPI_ISL_2437116, EPI_ISL_2437127, EPI_ISL_2437132, EPI_ISL_2437138, EPI_ISL_2437139, EPI_ISL_2437143, EPI_ISL_2437144, EPI_ISL_2437156, EPI_ISL_2437157, EPI_ISL_2437164, EPI_ISL_2437165, EPI_ISL_2437175, EPI_ISL_2437185, EPI_ISL_2437195, EPI_ISL_2437197, EPI_ISL_2437201, EPI_ISL_2437206, EPI_ISL_2437217, EPI_ISL_2437231, EPI_ISL_2437233, EPI_ISL_2437234, EPI_ISL_2437245, EPI_ISL_2437273, EPI_ISL_2438543, EPI_ISL_2539373, EPI_ISL_2539394, EPI_ISL_2540197, EPI_ISL_2540361, EPI_ISL_2540833, EPI_ISL_2540899 | | | |
| see above | Lighthouse Lab in Milton Keynes | Wellcome Sanger Institute for the COVID-19 Genomics UK (COG-UK) Consortium | Cordelia Langford; David K. Jackson; Dominic Kwiatkowski; Ewan Harrison; Ian Johnston; Jeffrey Barrett; John Sillitoe on behalf of the Wellcome Sanger Institute COVID-19 Surveillance Team; Roberto Amato; Sonia Goncalves; The Lighthouse Lab in Milton Keynes and Alex Alderton |
| EPI_ISL_2278011, EPI_ISL_2278012, EPI_ISL_2278013, EPI_ISL_2278014, EPI_ISL_2278015, EPI_ISL_2278016, EPI_ISL_2278017, EPI_ISL_2278018, EPI_ISL_2278019, EPI_ISL_2278020, EPI_ISL_2278021, EPI_ISL_2278022, EPI_ISL_2278023, EPI_ISL_2278024, EPI_ISL_2278025, EPI_ISL_2278026, EPI_ISL_2278027, EPI_ISL_2278028, EPI_ISL_2278029, EPI_ISL_2278030, EPI_ISL_2278031, EPI_ISL_2278032, EPI_ISL_2278033, EPI_ISL_2278034, EPI_ISL_2278035, EPI_ISL_2278036, EPI_ISL_2278037, EPI_ISL_2278038, EPI_ISL_2278039, EPI_ISL_2278040, EPI_ISL_2278041, EPI_ISL_2317162, EPI_ISL_2317189, EPI_ISL_2317190, EPI_ISL_2317200, EPI_ISL_2317202, EPI_ISL_2317208, EPI_ISL_2317211, EPI_ISL_2317215, EPI_ISL_2317221, EPI_ISL_2317258, EPI_ISL_2317261, EPI_ISL_2317266, EPI_ISL_2317272, EPI_ISL_2317275, EPI_ISL_2317278, EPI_ISL_2317283, EPI_ISL_2317293, EPI_ISL_2317302, EPI_ISL_2317303, EPI_ISL_2317309, EPI_ISL_2317313, EPI_ISL_2317317, EPI_ISL_2317318, EPI_ISL_2317344, EPI_ISL_2317348, EPI_ISL_2317356, EPI_ISL_2317362, EPI_ISL_2317366, EPI_ISL_2317369, EPI_ISL_2317388, EPI_ISL_2317397, EPI_ISL_2317405, EPI_ISL_2347618, EPI_ISL_2347638, EPI_ISL_2347643, EPI_ISL_2347658, EPI_ISL_2347669, EPI_ISL_2347684, EPI_ISL_2347704, EPI_ISL_2347710, EPI_ISL_2347712, EPI_ISL_2347721, EPI_ISL_2347728, EPI_ISL_2347730, EPI_ISL_2347753, EPI_ISL_2347756, EPI_ISL_2347791, EPI_ISL_2347792, EPI_ISL_2347793, EPI_ISL_2347819, EPI_ISL_2347843, EPI_ISL_2347863, EPI_ISL_2347866, EPI_ISL_2347868, EPI_ISL_2347869, EPI_ISL_2347871, EPI_ISL_2347894, EPI_ISL_2347905, EPI_ISL_2347911, EPI_ISL_2347924, EPI_ISL_2347983, EPI_ISL_2347985, EPI_ISL_2347997, EPI_ISL_2347998, EPI_ISL_2348006, EPI_ISL_2348013, EPI_ISL_2348030, EPI_ISL_2348037, EPI_ISL_2348053, EPI_ISL_2348056, EPI_ISL_2348061, EPI_ISL_2348073, EPI_ISL_2348092, EPI_ISL_2348109, EPI_ISL_2348118, EPI_ISL_2348121, EPI_ISL_2348148, EPI_ISL_2348150, EPI_ISL_2348159, EPI_ISL_2353254, EPI_ISL_2353258, EPI_ISL_2353264, EPI_ISL_2353277, EPI_ISL_2353283, EPI_ISL_2353294, EPI_ISL_2353312, EPI_ISL_2353315, EPI_ISL_2353318, EPI_ISL_2353333, EPI_ISL_2353335, EPI_ISL_2353346, EPI_ISL_2353347, EPI_ISL_2353348, EPI_ISL_2353349, EPI_ISL_2353355, EPI_ISL_2353361, EPI_ISL_2353370, EPI_ISL_2353376, EPI_ISL_2353380, EPI_ISL_2353382, EPI_ISL_2353402, EPI_ISL_2353407, EPI_ISL_2353409, EPI_ISL_2353413, EPI_ISL_2353417, EPI_ISL_2353420, EPI_ISL_2353429, EPI_ISL_2353438, EPI_ISL_2353447, EPI_ISL_2353448, EPI_ISL_2353454, EPI_ISL_2353457, EPI_ISL_2353459, EPI_ISL_2353467, EPI_ISL_2353468, EPI_ISL_2353471, EPI_ISL_2353479, EPI_ISL_2353480, EPI_ISL_2353482, EPI_ISL_2353489, EPI_ISL_2353491, EPI_ISL_2353504, EPI_ISL_2353509, EPI_ISL_2353526, EPI_ISL_2353541, EPI_ISL_2353543, EPI_ISL_2353545, EPI_ISL_2353547, EPI_ISL_2353548, EPI_ISL_2353557, EPI_ISL_2353558, EPI_ISL_2353561, EPI_ISL_2353565, EPI_ISL_2353567, EPI_ISL_2353576, EPI_ISL_2353578, EPI_ISL_2353583, EPI_ISL_2396712, EPI_ISL_2447233, EPI_ISL_2447242, EPI_ISL_2447259, EPI_ISL_2447272, EPI_ISL_2447298, EPI_ISL_2447317, EPI_ISL_2447347 | | | |
| see above | Lighthouse Laboratory Plymouth | Wellcome Sanger Institute for the COVID-19 Genomics UK (COG-UK) Consortium | Cordelia Langford; David K. Jackson; Dominic Kwiatkowski; Ewan Harrison; Ian Johnston; Jeffrey Barrett; John Sillitoe on behalf of the Wellcome Sanger Institute COVID-19 Surveillance Team; Lighthouse Laboratory Plymouth and Alex Alderton; Roberto Amato; Sonia Goncalves |
| EPI_ISL_2355368, EPI_ISL_2355372, EPI_ISL_2434246, EPI_ISL_2434248, EPI_ISL_2434249 | Liverpool Clinical Laboratories | COVID-19 Genomics UK (COG-UK) Consortium | A Alrezaihi; Alessandro Gerada; Alistair Darby; Angela Cowell; Anita Lucaci; Anu Chawla; Cassie Olateju; Catherine Hartley; Charlotte Nelson; Ecaterina Vamos; Elaine O'Toole; Eleanor G Bentley; Ghada T Shawli; Hermione Webster; Isabel García-Dorival; James Johnson; James P Stewart; Jenifer Manson; Joanne Watts; Jones Benjamin; Jordan J Clark; Julian Hiscox; L Luu; Lucille Rainbow; M Almsaud; Margaret Hughes; Mark Whitehead; Matthew Gemmell; Miren Iturriza-Gomara; Muhannad Alruwaili; N.P Randle; Neil Swainston; PKF Gilmore; Parul Sharma; Rebekah Penrice-Randal; Richard Eccles; Richard Gregory; Sam Haldenby; Steve Paterson; Stuart D Armstrong; Trevor Ian Robinson; Ximeng Han |
| EPI_ISL_2355922, EPI_ISL_2355930, EPI_ISL_2355938, EPI_ISL_2355946, EPI_ISL_2355954, EPI_ISL_2355962, EPI_ISL_2355970, EPI_ISL_2356050, EPI_ISL_2356051, EPI_ISL_2356057, EPI_ISL_2356058, EPI_ISL_2356064, EPI_ISL_2356065, EPI_ISL_2356072, EPI_ISL_2356073, EPI_ISL_2356078, EPI_ISL_2356079, EPI_ISL_2356086, EPI_ISL_2356092, EPI_ISL_2356094, EPI_ISL_2356095, EPI_ISL_2356096, EPI_ISL_2356100, EPI_ISL_2356101, EPI_ISL_2356102, EPI_ISL_2356103, EPI_ISL_2356106, EPI_ISL_2356107, EPI_ISL_2356108, EPI_ISL_2356109, EPI_ISL_2356113, EPI_ISL_2356114, EPI_ISL_2356115, EPI_ISL_2356116, EPI_ISL_2356119, EPI_ISL_2356120, EPI_ISL_2356121, EPI_ISL_2356124, EPI_ISL_2356125, EPI_ISL_2356126, EPI_ISL_2356127, EPI_ISL_2356128, EPI_ISL_2356132, EPI_ISL_2356133, EPI_ISL_2356134, EPI_ISL_2356135, EPI_ISL_2356139, EPI_ISL_2356140, EPI_ISL_2356141, EPI_ISL_2356142, EPI_ISL_2356143, EPI_ISL_2356147, EPI_ISL_2356148, EPI_ISL_2356149, EPI_ISL_2356150, EPI_ISL_2356155, EPI_ISL_2356156, EPI_ISL_2356161, EPI_ISL_2356162, EPI_ISL_2356168, EPI_ISL_2356169, EPI_ISL_2356170, EPI_ISL_2356175, EPI_ISL_2356181, EPI_ISL_2356189, EPI_ISL_2356196, EPI_ISL_2356210, EPI_ISL_2356242, EPI_ISL_2356246, EPI_ISL_2356568, EPI_ISL_2356571, EPI_ISL_2356572, EPI_ISL_2356575, EPI_ISL_2356576, EPI_ISL_2356581, EPI_ISL_2356583, EPI_ISL_2356584, EPI_ISL_2356588, EPI_ISL_2356589, EPI_ISL_2356590, EPI_ISL_2356594, EPI_ISL_2356597, EPI_ISL_2356598, EPI_ISL_2356599, EPI_ISL_2356600, EPI_ISL_2356602, EPI_ISL_2356605, EPI_ISL_2356608, EPI_ISL_2356609, EPI_ISL_2356611, EPI_ISL_2356613, EPI_ISL_2356614, EPI_ISL_2356615, EPI_ISL_2356616, EPI_ISL_2356618, EPI_ISL_2356619, EPI_ISL_2356620, EPI_ISL_2356621, EPI_ISL_2356622, EPI_ISL_2356625, EPI_ISL_2356626, EPI_ISL_2356627, EPI_ISL_2356628, EPI_ISL_2356630, EPI_ISL_2356631, EPI_ISL_2356632, EPI_ISL_2356633, EPI_ISL_2356637, EPI_ISL_2356638, EPI_ISL_2356639, EPI_ISL_2356643, EPI_ISL_2356644, EPI_ISL_2356647, EPI_ISL_2356648, EPI_ISL_2356649, EPI_ISL_2356650, EPI_ISL_2356652, EPI_ISL_2356653, EPI_ISL_2356654, EPI_ISL_2356655, EPI_ISL_2356658, EPI_ISL_2356659, EPI_ISL_2356660, EPI_ISL_2356663, EPI_ISL_2356664, EPI_ISL_2356665, EPI_ISL_2356666, EPI_ISL_2356670, EPI_ISL_2356671, EPI_ISL_2356672, EPI_ISL_2356673, EPI_ISL_2356674, EPI_ISL_2356676, EPI_ISL_2356677, EPI_ISL_2356678, EPI_ISL_2356679, EPI_ISL_2356683, EPI_ISL_2356684, EPI_ISL_2434272, EPI_ISL_2434273, EPI_ISL_2434274, EPI_ISL_2434275, EPI_ISL_2434276, EPI_ISL_2434277, EPI_ISL_2434280, EPI_ISL_2434281, EPI_ISL_2434282, EPI_ISL_2434283, EPI_ISL_2434284, EPI_ISL_2434285, EPI_ISL_2434286, EPI_ISL_2434288, EPI_ISL_2434289, EPI_ISL_2434290, EPI_ISL_2434291, EPI_ISL_2434292, EPI_ISL_2434294, EPI_ISL_2434295, EPI_ISL_2434296, EPI_ISL_2434297, EPI_ISL_2434298, EPI_ISL_2434299, EPI_ISL_2434302, EPI_ISL_2434303, EPI_ISL_2434304, EPI_ISL_2434305, EPI_ISL_2434306, EPI_ISL_2434307, EPI_ISL_2434310, EPI_ISL_2434311, EPI_ISL_2434312, EPI_ISL_2434313, EPI_ISL_2434314, EPI_ISL_2434315, EPI_ISL_2434318, EPI_ISL_2434319, EPI_ISL_2434320, EPI_ISL_2434322, EPI_ISL_2434326, EPI_ISL_2434327, EPI_ISL_2434328, EPI_ISL_2434329, EPI_ISL_2434332, EPI_ISL_2434333, EPI_ISL_2434334, EPI_ISL_2434338, EPI_ISL_2434339, EPI_ISL_2434340, EPI_ISL_2434344, EPI_ISL_2434346, EPI_ISL_2434348, EPI_ISL_2434349, EPI_ISL_2434369, EPI_ISL_2434691, EPI_ISL_2434706 | | | |
| see above | Northumbria University / South Tees Hospitals NHS Foundation Trust / North Cumbria Integrated Care NHS Foundation Trust / North Tees and Hartlepool NHS Foundation Trust / Newcastle Hospitals NHS Foundation Trust | COVID-19 Genomics UK (COG-UK) Consortium | Andrew Nelson; Brendan Payne; Clive Graham; Darren L Smith; Debra Padgett; Edward Barton; Emma Swindells; Garren Scott; Gary Black; Gary Eltringham; Giles S Holt; Greg R Young; Jane Greenaway; Jennifer Collins; John Allan; Joshua Loh; Lynn Dover; Matthew Bashton; Mohammad A Tariq; Paul Baker; Sarah Essex; Steve Liggett; Wen C Yew; Yusri Taha |
| EPI_ISL_2434785, EPI_ISL_2434786, EPI_ISL_2434787, EPI_ISL_2434789, EPI_ISL_2434790, EPI_ISL_2434791, EPI_ISL_2434792, EPI_ISL_2434795, EPI_ISL_2434798, EPI_ISL_2434800, EPI_ISL_2434801, EPI_ISL_2434802, EPI_ISL_2434803, EPI_ISL_2434805, EPI_ISL_2434806, EPI_ISL_2434807, EPI_ISL_2434808, EPI_ISL_2434809, EPI_ISL_2434811, EPI_ISL_2434812, EPI_ISL_2434813, EPI_ISL_2434817, EPI_ISL_2434818, EPI_ISL_2434819, EPI_ISL_2434820, EPI_ISL_2434823, EPI_ISL_2434825, EPI_ISL_2434826, EPI_ISL_2434827, EPI_ISL_2434828, EPI_ISL_2434829, EPI_ISL_2434831, EPI_ISL_2434836, EPI_ISL_2434842, EPI_ISL_2434879, EPI_ISL_2434880, EPI_ISL_2434881, EPI_ISL_2434882, EPI_ISL_2434884, EPI_ISL_2434885, EPI_ISL_2434886, EPI_ISL_2434887, EPI_ISL_2434888, EPI_ISL_2434889, EPI_ISL_2434890, EPI_ISL_2434893 | | | |
| see above | Originating lab: Wales Specialist Virology Centre Sequencing lab: Pathogen Genomics Unit | Public Health Wales Microbiology Cardiff Wales Specialist Virology Centre | Alec Birchley; Alexander Adams; Amy Gaskin; Angela Marchbank; Bree Gatica-Wilcox; Catherine Moore; Jason Coombes; Joanne Watkins; Joel Southgate; Johnathan Evans; Laura Gifford; Lauren Gilbert; Lee Graham; Malorie Perry; Matthew Bull; Nicole Pacchiarini; Sally Corden; Sara Kumziene-Summerhayes; Sara Rey; Sarah Taylor; Simon Cottrell; Sophie Jones; Tom Connor |
| EPI_ISL_2434763, EPI_ISL_2434764, EPI_ISL_2434765, EPI_ISL_2434768, EPI_ISL_2434772, EPI_ISL_2434773, EPI_ISL_2434775, EPI_ISL_2624574, EPI_ISL_2624575, EPI_ISL_2624578, EPI_ISL_2624580, EPI_ISL_2624581 | | | |
| see above | Oxford Viromics, NDM, University of Oxford; Oxford University Hospitals; Basingstoke and North Hampshire Hospital | COVID-19 Genomics UK (COG-UK) Consortium | Alex Mobbs; Amy Trebes; Anita Justice; Catrin Moore; Christophe Fraser; David Bonsall; David Buck; Emma Wise; George Macintyre; Jessica Lynch; John Todd; Mariateresa de Cesare; Matilde Mori; Monique Andersson; Nathan Moore; Nick Cortes; Robert Shaw; Stephen Kidd; Tanya Golubchik; Timothy Peto |
| EPI_ISL_2356695, EPI_ISL_2356696, EPI_ISL_2434188, EPI_ISL_2623767, EPI_ISL_2623768, EPI_ISL_2624009, EPI_ISL_2624010, EPI_ISL_2624011, EPI_ISL_2624012, EPI_ISL_2624013, EPI_ISL_2624015, EPI_ISL_2624023, EPI_ISL_2624024, EPI_ISL_2624025, EPI_ISL_2624045, EPI_ISL_2624046, EPI_ISL_2624061, EPI_ISL_2624062, EPI_ISL_2624063, EPI_ISL_2624064, EPI_ISL_2624066, EPI_ISL_2624069, EPI_ISL_2624070, EPI_ISL_2624071, EPI_ISL_2624072, EPI_ISL_2624073, EPI_ISL_2624075, EPI_ISL_2624076, EPI_ISL_2624077, EPI_ISL_2624078, EPI_ISL_2624079, EPI_ISL_2624080, EPI_ISL_2624081, EPI_ISL_2624082, EPI_ISL_2624083, EPI_ISL_2624084, EPI_ISL_2624085, EPI_ISL_2624086, EPI_ISL_2624087, EPI_ISL_2624088, EPI_ISL_2624089, EPI_ISL_2624090, EPI_ISL_2624091, EPI_ISL_2624092, EPI_ISL_2624093, EPI_ISL_2624094, EPI_ISL_2624095, EPI_ISL_2624096, EPI_ISL_2624097, EPI_ISL_2624098, EPI_ISL_2624099, EPI_ISL_2624100, EPI_ISL_2624101, EPI_ISL_2624102, EPI_ISL_2624103, EPI_ISL_2624104, EPI_ISL_2624105, EPI_ISL_2624110, EPI_ISL_2624127, EPI_ISL_2624147, EPI_ISL_2624170, EPI_ISL_2624171, EPI_ISL_2624172, EPI_ISL_2624173, EPI_ISL_2624174, EPI_ISL_2624175, EPI_ISL_2624176, EPI_ISL_2624177, EPI_ISL_2624178, EPI_ISL_2624179, EPI_ISL_2624180, EPI_ISL_2624181, EPI_ISL_2624182, EPI_ISL_2624183, EPI_ISL_2624184, EPI_ISL_2624185, EPI_ISL_2624186, EPI_ISL_2624187, EPI_ISL_2624188, EPI_ISL_2624189, EPI_ISL_2624190, EPI_ISL_2624191, EPI_ISL_2624192, EPI_ISL_2624193, EPI_ISL_2624194, EPI_ISL_2624195, EPI_ISL_2624196, EPI_ISL_2624197, EPI_ISL_2624198, EPI_ISL_2624199, EPI_ISL_2624200, EPI_ISL_2624201, EPI_ISL_2624202, EPI_ISL_2624203, EPI_ISL_2624204, EPI_ISL_2624205, EPI_ISL_2624206, EPI_ISL_2624207, EPI_ISL_2624208, EPI_ISL_2624209, EPI_ISL_2624210, EPI_ISL_2624211, EPI_ISL_2624212, EPI_ISL_2624213, EPI_ISL_2624214, EPI_ISL_2624215, EPI_ISL_2624216, EPI_ISL_2624217, EPI_ISL_2624218, EPI_ISL_2624219, EPI_ISL_2624220, EPI_ISL_2624221, EPI_ISL_2624223, EPI_ISL_2624224, EPI_ISL_2624225, EPI_ISL_2624226, EPI_ISL_2624227, EPI_ISL_2624228, EPI_ISL_2624229, EPI_ISL_2624230, EPI_ISL_2624231, EPI_ISL_2624232, EPI_ISL_2624233, EPI_ISL_2624234, EPI_ISL_2624235, EPI_ISL_2624236, EPI_ISL_2624237, EPI_ISL_2624238, EPI_ISL_2624239, EPI_ISL_2624240, EPI_ISL_2624241, EPI_ISL_2624242, EPI_ISL_2624243, EPI_ISL_2624244, EPI_ISL_2624245, EPI_ISL_2624246, EPI_ISL_2624247, EPI_ISL_2624248, EPI_ISL_2624249, EPI_ISL_2624250, EPI_ISL_2624251, EPI_ISL_2624252, EPI_ISL_2624253, EPI_ISL_2624254, EPI_ISL_2624255, EPI_ISL_2624256, EPI_ISL_2624257, EPI_ISL_2624258, EPI_ISL_2624259, EPI_ISL_2624260, EPI_ISL_2624261, EPI_ISL_2624262, EPI_ISL_2624263, EPI_ISL_2624264, EPI_ISL_2624265, EPI_ISL_2624266, EPI_ISL_2624267, EPI_ISL_2624268, EPI_ISL_2624269, EPI_ISL_2624270, EPI_ISL_2624271, EPI_ISL_2624272, EPI_ISL_2624273, EPI_ISL_2624274, EPI_ISL_2624275, EPI_ISL_2624276, EPI_ISL_2624277, EPI_ISL_2624278, EPI_ISL_2624279, EPI_ISL_2624280, EPI_ISL_2624281, EPI_ISL_2624282, EPI_ISL_2624283, EPI_ISL_2624284, EPI_ISL_2624285, EPI_ISL_2624286, EPI_ISL_2624287, EPI_ISL_2624288, EPI_ISL_2624356, EPI_ISL_2624357, EPI_ISL_2624358, EPI_ISL_2624359, EPI_ISL_2624360, EPI_ISL_2624361, EPI_ISL_2624362, EPI_ISL_2624363, EPI_ISL_2624364, EPI_ISL_2624365, EPI_ISL_2624366, EPI_ISL_2624367, EPI_ISL_2624368, EPI_ISL_2624369, EPI_ISL_2624370, EPI_ISL_2624371, EPI_ISL_2624372, EPI_ISL_2624373, EPI_ISL_2624374, EPI_ISL_2624375, EPI_ISL_2624376, EPI_ISL_2624377, EPI_ISL_2624378, EPI_ISL_2624379, EPI_ISL_2624380, EPI_ISL_2624381, EPI_ISL_2624382, EPI_ISL_2624383, EPI_ISL_2624384, EPI_ISL_2624385, EPI_ISL_2624386, EPI_ISL_2624387, EPI_ISL_2624388, EPI_ISL_2624389, EPI_ISL_2624390, EPI_ISL_2624391, EPI_ISL_2624392, EPI_ISL_2624393, EPI_ISL_2624394, EPI_ISL_2624395, EPI_ISL_2624396, EPI_ISL_2624397, EPI_ISL_2624398, EPI_ISL_2624399, EPI_ISL_2624400, EPI_ISL_2624401, EPI_ISL_2624402, EPI_ISL_2624403, EPI_ISL_2624404, EPI_ISL_2624405, EPI_ISL_2624406, EPI_ISL_2706884 | | | |
| see above | Quadram Institute Bioscience | COVID-19 Genomics UK (COG-UK) Consortium | Alexander J Trotter; Alison E. Mather; Alp Aydin; Ana P. Tedim; Anastasia Kolyva; Andrew Bell; Andrew J. Page; Claire Stuart; Dave J. Baker; Gemma L. Kay; John Wain; Justin O'Grady; Leonardo de Oliveira Martins; Lizzie Meadows; Maria Diaz; Mark Webber; Muhammed Yasir; Nabil-Fareed Alikhan; Ngozi Elumogo; Nicholas M. Thomson; Rachael Stanley; Rachel Gilroy; Reenesh Prakash; Samir Dervisevic; Samuel Bloomfield; Steven Rudder; Thanh Le-Viet |
| EPI_ISL_2240458, EPI_ISL_2240459, EPI_ISL_2240460, EPI_ISL_2355344, EPI_ISL_2355345, EPI_ISL_2356699, EPI_ISL_2356700, EPI_ISL_2356701, EPI_ISL_2356702, EPI_ISL_2356703, EPI_ISL_2356704, EPI_ISL_2356705, EPI_ISL_2356706 | | | |
| see above | Queens Medical Centre, Clinical Microbiology Department / DeepSeq Nottingham | COVID-19 Genomics UK (COG-UK) Consortium | Christopher Moore; Emily Park; Fei Sang; Gemma Clark; Hannah Howson-Wells; Jack Hill; Jayasree Dey; Johnny Debebe; Jonathan Ball; Joseph Chappell; Manjinder Khakh; Matthew Carlisle; Matthew Loose; Michelle M Lister; Nadine Holmes; Patrick McClure; Theocharis Tsoleridis; Timothy Byaruhanga; Vicki M Fleming; Victoria Wright; Wendy Smith |
| EPI_ISL_2317217, EPI_ISL_2317228, EPI_ISL_2317314, EPI_ISL_2317335, EPI_ISL_2346622, EPI_ISL_2346674, EPI_ISL_2346700, EPI_ISL_2347653, EPI_ISL_2347694, EPI_ISL_2347707, EPI_ISL_2347741, EPI_ISL_2347744, EPI_ISL_2347808, EPI_ISL_2347851, EPI_ISL_2347880, EPI_ISL_2348014, EPI_ISL_2348144, EPI_ISL_2348153, EPI_ISL_2353271, EPI_ISL_2353280, EPI_ISL_2353331, EPI_ISL_2353338, EPI_ISL_2353404, EPI_ISL_2353455, EPI_ISL_2353518, EPI_ISL_2396707, EPI_ISL_2396711, EPI_ISL_2396718, EPI_ISL_2396800, EPI_ISL_2396801, EPI_ISL_2396802, EPI_ISL_2396803, EPI_ISL_2396804, EPI_ISL_2396805, EPI_ISL_2396806, EPI_ISL_2396807, EPI_ISL_2396808, EPI_ISL_2396811, EPI_ISL_2396832, EPI_ISL_2396833, EPI_ISL_2396834, EPI_ISL_2396835, EPI_ISL_2396836, EPI_ISL_2396837, EPI_ISL_2396838, EPI_ISL_2447270, EPI_ISL_2447289, EPI_ISL_2447291, EPI_ISL_2447314, EPI_ISL_2447321 | | | |
| see above | Randox Laboratories | Wellcome Sanger Institute | Cordelia Langford; David K. Jackson; Dominic Kwiatkowski; Ewan Harrison; Ian Johnston; Jeffrey Barrett; John Sillitoe on behalf of the Wellcome Sanger Institute COVID-19 Surveillance Team; Randox Laboratories and Alex Alderton; Roberto Amato; Sonia |

| | | | |
|---|---|---|---|
| | | for the COVID-19 Genomics UK (COG-UK) Consortium | Goncalves |
| EPI_ISL_2742827, EPI_ISL_2742832, EPI_ISL_2742856, EPI_ISL_2742857, EPI_ISL_2742860, EPI_ISL_2742865, EPI_ISL_2742866, EPI_ISL_2742867, EPI_ISL_2742868, EPI_ISL_2742869, EPI_ISL_2742870, EPI_ISL_2742871, EPI_ISL_2742872, EPI_ISL_2742873, EPI_ISL_2742874, EPI_ISL_2742875, EPI_ISL_2742876, EPI_ISL_2742883, EPI_ISL_2742887, EPI_ISL_2742888, EPI_ISL_2742889, EPI_ISL_2742898, EPI_ISL_2742900, EPI_ISL_2742904, EPI_ISL_2742906, EPI_ISL_2742908, EPI_ISL_2742909, EPI_ISL_2742917, EPI_ISL_2742918, EPI_ISL_2742920, EPI_ISL_2742980, EPI_ISL_2742981, EPI_ISL_2742983, EPI_ISL_2742984, EPI_ISL_2742985, EPI_ISL_2742988, EPI_ISL_2742989, EPI_ISL_2742993, EPI_ISL_2742996, EPI_ISL_2742997, EPI_ISL_2742998, EPI_ISL_2743000, EPI_ISL_2743008, EPI_ISL_2743037, EPI_ISL_2743049, EPI_ISL_2743057, EPI_ISL_2743067, EPI_ISL_2743068, EPI_ISL_2743069, EPI_ISL_2743070, EPI_ISL_2743071, EPI_ISL_2743072, EPI_ISL_2743073, EPI_ISL_2743074, EPI_ISL_2743075, EPI_ISL_2743076, EPI_ISL_2743077, EPI_ISL_2743087, EPI_ISL_2743088, EPI_ISL_2743102, EPI_ISL_2743109, EPI_ISL_2743166, EPI_ISL_2743172, EPI_ISL_2743173, EPI_ISL_2743193, EPI_ISL_2743195, EPI_ISL_2743205, EPI_ISL_2743253, EPI_ISL_2744245, EPI_ISL_2744246, EPI_ISL_2744507, EPI_ISL_2744512, EPI_ISL_2744519, EPI_ISL_2744710, EPI_ISL_2744711, EPI_ISL_2744713, EPI_ISL_2749140, EPI_ISL_2749170, EPI_ISL_2749590 | | | |
| see above | Respiratory Virus Unit, Microbiology Services Colindale, Public Health England | COVID-19 Genomics UK (COG-UK) Consortium | PHE Covid Sequencing Team |
| EPI_ISL_2355376, EPI_ISL_2355377, EPI_ISL_2355378, EPI_ISL_2355379, EPI_ISL_2355380, EPI_ISL_2355381, EPI_ISL_2355382, EPI_ISL_2355383, EPI_ISL_2355384, EPI_ISL_2355386, EPI_ISL_2355387, EPI_ISL_2355388, EPI_ISL_2355389, EPI_ISL_2355390, EPI_ISL_2355391, EPI_ISL_2355393, EPI_ISL_2355394, EPI_ISL_2355395, EPI_ISL_2355397, EPI_ISL_2355400, EPI_ISL_2355401, EPI_ISL_2355402, EPI_ISL_2355403, EPI_ISL_2355409, EPI_ISL_2355411, EPI_ISL_2355412, EPI_ISL_2355415, EPI_ISL_2355416, EPI_ISL_2355417, EPI_ISL_2355425, EPI_ISL_2355426, EPI_ISL_2355427, EPI_ISL_2355430, EPI_ISL_2355431, EPI_ISL_2355432, EPI_ISL_2355433, EPI_ISL_2355434, EPI_ISL_2355435, EPI_ISL_2355436, EPI_ISL_2355437, EPI_ISL_2355438, EPI_ISL_2355439 | | | |
| see above | University College London, Great Ormond Street Hospital for Children NHS Foundation Trust, Imperial College Healthcare NHS Trust | COVID-19 Genomics UK (COG-UK) Consortium | Alison Holmes; Charlotte Williams; Helena Tutill; Jacqueline Findlay; James Price; Judith Breuer; Julianne Brown; Kathryn Harris; Leysa Forrest; Mark Kristiansen; Paola Niola; Paola Resende Silva; Patricia Dyal; Paul Randell; Rachel Williams; Samuel Weeks; Sergi Castellano; Sunando Roy; Tony Brooks; Yasmin Panchbhaya |
| EPI_ISL_2238869, EPI_ISL_2238870, EPI_ISL_2355346 | University Hospitals Of Leicester NHS Trust and DeepSeq Nottingham | COVID-19 Genomics UK (COG-UK) Consortium | Christopher Holmes; Christopher Moore; Fei Sang; Joeseph Chappell; Johnny Debebe; Jonathan Ball; Julian Tang; Karlie Fallon; Matthew Carlisle; Matthew Loose; Nadine Holmes; Patrick McClure; Paul Bird; Thomas Helmer; Victoria Wright |
| EPI_ISL_2354510, EPI_ISL_2354511, EPI_ISL_2354513, EPI_ISL_2354514, EPI_ISL_2354516, EPI_ISL_2354517, EPI_ISL_2354518, EPI_ISL_2354519, EPI_ISL_2354520, EPI_ISL_2354532, EPI_ISL_2354534 | | | |
| see above | University of Birmingham | COVID-19 Genomics UK (COG-UK) Consortium | Alex Richter; Andrew Bosworth; Andrew Bosworth. Queen Elizabeth Hospital: Anna Casey; Andrew D Beggs PHE Heartlands Lab: Husam Osman; Charlotte Poxon; Institute of Microbiology; Joanne Stockton; Josh Quick; Kasun Wanigasooriya; Mike Kidd; Nicholas Loman. University of Birmingham Testing Laboratory: Celina M Whalley; Oliver Pickles; Radoslaw Poplawski; Samuel Nicholls; University of Birmingham: Claire McMurray; Will Rowe |
| EPI_ISL_2354569, EPI_ISL_2354570, EPI_ISL_2354571, EPI_ISL_2354572, EPI_ISL_2354573, EPI_ISL_2354574, EPI_ISL_2354575, EPI_ISL_2354576, EPI_ISL_2354577, EPI_ISL_2354578, EPI_ISL_2354580, EPI_ISL_2354586, EPI_ISL_2354587, EPI_ISL_2354588, EPI_ISL_2354589, EPI_ISL_2354590, EPI_ISL_2354592, EPI_ISL_2354594, EPI_ISL_2354597, EPI_ISL_2354598, EPI_ISL_2354599, EPI_ISL_2354600, EPI_ISL_2354601, EPI_ISL_2354602, EPI_ISL_2354603, EPI_ISL_2354604, EPI_ISL_2354605, EPI_ISL_2354606, EPI_ISL_2354607, EPI_ISL_2354608, EPI_ISL_2354609, EPI_ISL_2354610, EPI_ISL_2354614, EPI_ISL_2354615, EPI_ISL_2354616, EPI_ISL_2354619, EPI_ISL_2354620, EPI_ISL_2354621, EPI_ISL_2354622, EPI_ISL_2354623, EPI_ISL_2354630, EPI_ISL_2354631, EPI_ISL_2354632, EPI_ISL_2434215, EPI_ISL_2434216, EPI_ISL_2434235, EPI_ISL_2517302, EPI_ISL_2517303 | | | |
| see above | Virology Department, Royal Infirmary of Edinburgh, NHS Lothian / School of Biological Sciences, University of Edinburgh | COVID-19 Genomics UK (COG-UK) Consortium | Colquhoun R; Cotton S; Dewar R; Gallagher A; Hill V; Jackson B; Maloney D; McCrone JT; McHugh M; O'Toole Á; Rambaut A; Rooke S; Scher E; Templeton K; Yu X |
| EPI_ISL_2356887, EPI_ISL_2356889, EPI_ISL_2356891, EPI_ISL_2356893, EPI_ISL_2356896, EPI_ISL_2356897, EPI_ISL_2356899, EPI_ISL_2356901, EPI_ISL_2356903, EPI_ISL_2356905, EPI_ISL_2356906, EPI_ISL_2356907, EPI_ISL_2520941, EPI_ISL_2520959, EPI_ISL_2521057, EPI_ISL_2521069, EPI_ISL_2521080, EPI_ISL_2521131 | | | |
| see above | Virology Department, Sheffield Teaching Hospitals NHS Foundation Trust/Department of Infection, Immunity and Cardiovascular Disease, The Medical School, University of Sheffield | COVID-19 Genomics UK (COG-UK) Consortium | Adri Angyal; Alex Keeley; Benjamin Lindsey; Cariad Evans; Danielle Groves; Dave Partridge; Katie Johnson; Laura Carrilero; Luke Green; Matthew Parker; Matthew Wyles; Mehmet Yavuz; Mohammad Raza; Nikki Smith; Paul Parsons; Rachel Tucker; Rebecca Brown; Thushan de Silva |
| EPI_ISL_2356719, EPI_ISL_2356720, EPI_ISL_2356721, EPI_ISL_2356722, EPI_ISL_2356723, EPI_ISL_2356724, EPI_ISL_2356725, EPI_ISL_2356726, EPI_ISL_2356727, EPI_ISL_2356728, EPI_ISL_2356729, EPI_ISL_2356730, EPI_ISL_2356731, EPI_ISL_2356732, EPI_ISL_2356733, EPI_ISL_2356734, EPI_ISL_2356735, EPI_ISL_2356736, EPI_ISL_2356737, EPI_ISL_2356738, EPI_ISL_2356739, EPI_ISL_2356740, EPI_ISL_2356747, EPI_ISL_2356752, EPI_ISL_2356753, EPI_ISL_2356754, EPI_ISL_2356763, EPI_ISL_2356790, EPI_ISL_2356791, EPI_ISL_2356792, EPI_ISL_2356793, EPI_ISL_2356795, EPI_ISL_2356796, EPI_ISL_2356800, EPI_ISL_2356801, EPI_ISL_2356809, EPI_ISL_2356811, EPI_ISL_2356814, EPI_ISL_2356815, EPI_ISL_2356816, EPI_ISL_2356817, EPI_ISL_2356818, EPI_ISL_2356819, EPI_ISL_2356820, EPI_ISL_2356821, EPI_ISL_2356822, EPI_ISL_2356823, EPI_ISL_2356824, EPI_ISL_2356825, EPI_ISL_2356826, EPI_ISL_2356827, EPI_ISL_2356828, EPI_ISL_2356829, EPI_ISL_2356830, EPI_ISL_2356831, EPI_ISL_2356832, EPI_ISL_2356833, EPI_ISL_2356834, EPI_ISL_2356835, EPI_ISL_2356836, EPI_ISL_2356837, EPI_ISL_2356838, EPI_ISL_2356839, EPI_ISL_2356840, EPI_ISL_2356841, EPI_ISL_2356842, EPI_ISL_2356843, EPI_ISL_2356844, EPI_ISL_2356845, EPI_ISL_2356846, EPI_ISL_2356847, EPI_ISL_2356848, EPI_ISL_2356849, EPI_ISL_2356850, EPI_ISL_2356851, EPI_ISL_2356852, EPI_ISL_2356853, EPI_ISL_2356854, EPI_ISL_2356855, EPI_ISL_2356856, EPI_ISL_2356857, EPI_ISL_2356858, EPI_ISL_2356859, EPI_ISL_2356860, EPI_ISL_2356861, EPI_ISL_2356862, EPI_ISL_2356863, EPI_ISL_2356864, EPI_ISL_2356865, EPI_ISL_2356866, EPI_ISL_2520734 | | | |
| see above | Wales Specialist Virology Centre Sequencing lab: Pathogen Genomics Unit | Public Health Wales Microbiology Cardiff Wales Specialist Virology Centre | Alec Birchley; Alexander Adams; Amy Gaskin; Angela Marchbank; Bree Gatica-Wilcox; Catherine Moore; Jason Coombes; Joanne Watkins; Joel Southgate; Johnathan Evans; Laura Gifford; Lauren Gilbert; Lee Graham; Malorie Perry; Matthew Bull; Nicole Pacchiarini; Sally Corden; Sara Kumziene-Summerhayes; Sara Rey; Sarah Taylor; Simon Cottrell; Sophie Jones; Tom Connor |
| EPI_ISL_2517230, EPI_ISL_2517231, EPI_ISL_2517232, EPI_ISL_2517233, EPI_ISL_2517234, EPI_ISL_2517235, EPI_ISL_2517236, EPI_ISL_2517237, EPI_ISL_2517254, EPI_ISL_2517255, EPI_ISL_2517256, EPI_ISL_2517257, EPI_ISL_2517259, EPI_ISL_2517260, EPI_ISL_2517261, EPI_ISL_2517278, EPI_ISL_2517279, EPI_ISL_2517280, EPI_ISL_2704755 | | | |
| see above | West of Scotland Specialist Virology Centre, NHSGGC / MRC-University of Glasgow Centre for Virus Research | COVID-19 Genomics UK (COG-UK) Consortium | Alasdair MacLean; Alice Broos; Ana da Silva Filipe; Antonia Ho; Daniel Mair; David L Robertson; Emma Thomson; Guy Mollett; Ioulia Tsatsani; James Shepherd; Jenna Nichols; Joseph Hughes; Kathy Li; Kathy Smollett; Kyriaki Nomikou; Lily Tong; Matthew Holden; Natasha Johnson; Rachel Blacow; Richard Orton; Rory Gunson; Sarah McDonald; Sharif Shaaban; Sreenu Vattipally; Stephen Carmichael |

We gratefully acknowledge the following Authors from the Originating laboratories responsible for obtaining the specimens, as well as the Submitting laboratories where the genome data were generated and shared via GISAID, on which this research is based.

All Submitters of data may be contacted directly via www.gisaid.org

Authors are sorted alphabetically.

| Accession ID | Originating Laboratory | Submitting Laboratory | Authors |
|---|---|---|---|
| EPI_ISL_2686112, EPI_ISL_2686113, EPI_ISL_2686114, EPI_ISL_2686115, EPI_ISL_2686116, EPI_ISL_2686117, EPI_ISL_2686118, EPI_ISL_2686119, EPI_ISL_2686120, EPI_ISL_2686121 | | | |
| see above | Amazon Dx, AUV2 Laboratory | Amazon Dx, AUV2 Laboratory | Luke Meredith |
| EPI_ISL_2434260, EPI_ISL_2434261, EPI_ISL_2434262, EPI_ISL_2434263, EPI_ISL_2434264 | Barts Health NHS Trust | COVID-19 Genomics UK (COG-UK) Consortium | BROAD; Beatrix; CUTINO-MOGUEL; Claire; David; Dola; HARRINGTON; KELE; KULASEGARAN-SHYLINI; Maria-Teresa; OWOYEMI; Raghavendran |
| EPI_ISL_2394586, EPI_ISL_2394601, EPI_ISL_2394607, EPI_ISL_2394612, EPI_ISL_2394614, EPI_ISL_2394624, EPI_ISL_2394638, EPI_ISL_2394640, EPI_ISL_2394642, EPI_ISL_2394668, EPI_ISL_2394687, EPI_ISL_2394694, EPI_ISL_2394703, EPI_ISL_2394717, EPI_ISL_2394719, EPI_ISL_2394738, EPI_ISL_2394747, EPI_ISL_2394755, EPI_ISL_2394760, EPI_ISL_2394784, EPI_ISL_2394797, EPI_ISL_2394804, EPI_ISL_2394807, EPI_ISL_2394821, EPI_ISL_2394825, EPI_ISL_2394840, EPI_ISL_2394844, EPI_ISL_2394850, EPI_ISL_2394858, EPI_ISL_2436236, EPI_ISL_2436953, EPI_ISL_2436959, EPI_ISL_2436961, EPI_ISL_2436967, EPI_ISL_2436974, EPI_ISL_2436981, EPI_ISL_2436983, EPI_ISL_2436985, EPI_ISL_2436998, EPI_ISL_2437010, EPI_ISL_2437049, EPI_ISL_2437064, EPI_ISL_2437067, EPI_ISL_2437078, EPI_ISL_2437088, EPI_ISL_2437090, EPI_ISL_2437092, EPI_ISL_2437100, EPI_ISL_2437101, EPI_ISL_2437128, EPI_ISL_2437146, EPI_ISL_2437160, EPI_ISL_2437166, EPI_ISL_2437188, EPI_ISL_2437192, EPI_ISL_2437236, EPI_ISL_2437272, EPI_ISL_2437274, EPI_ISL_2437347, EPI_ISL_2437494, EPI_ISL_2447237, EPI_ISL_2447238, EPI_ISL_2447239, EPI_ISL_2447244, EPI_ISL_2447249, EPI_ISL_2447253, EPI_ISL_2447256, EPI_ISL_2447258, EPI_ISL_2447264, EPI_ISL_2447268, EPI_ISL_2447271, EPI_ISL_2447278, EPI_ISL_2447282, EPI_ISL_2447285, EPI_ISL_2447297, EPI_ISL_2447308, EPI_ISL_2447322, EPI_ISL_2447335, EPI_ISL_2447340, EPI_ISL_2447343, EPI_ISL_2447358, EPI_ISL_2447361, EPI_ISL_2447362, EPI_ISL_2456863, EPI_ISL_2456878, EPI_ISL_2456969, EPI_ISL_2456998, EPI_ISL_2457055, EPI_ISL_2457123, EPI_ISL_2457125, EPI_ISL_2457127 | | | |
| see above | Health Services Laboratories | Wellcome Sanger Institute for the COVID-19 Genomics UK (COG-UK) Consortium | Cordelia Langford; David K. Jackson; Dominic Kwiatkowski; Ewan Harrison; Health Services Laboratories and Alex Alderton; Ian Johnston; Jeffrey Barrett; John Sillitoe on behalf of the Wellcome Sanger Institute COVID-19 Surveillance Team; Roberto Amato; Sonia Goncalves |
| EPI_ISL_2318898, EPI_ISL_2392197, EPI_ISL_2392200, EPI_ISL_2392201, EPI_ISL_2392202, EPI_ISL_2392203, EPI_ISL_2392204, EPI_ISL_2392205, EPI_ISL_2392206, EPI_ISL_2392207, EPI_ISL_2392208, EPI_ISL_2392209, EPI_ISL_2392212, EPI_ISL_2392213, EPI_ISL_2392214, EPI_ISL_2392215, EPI_ISL_2392216, EPI_ISL_2392217, EPI_ISL_2392218, EPI_ISL_2392219, EPI_ISL_2392221, EPI_ISL_2392222, EPI_ISL_2392226, EPI_ISL_2392228, EPI_ISL_2392229, EPI_ISL_2392231, EPI_ISL_2392232, EPI_ISL_2392233, EPI_ISL_2392234, EPI_ISL_2392237, EPI_ISL_2392238, EPI_ISL_2392239, EPI_ISL_2392240, EPI_ISL_2392241, EPI_ISL_2392244, EPI_ISL_2392245, EPI_ISL_2392246, EPI_ISL_2392247, EPI_ISL_2392248, EPI_ISL_2392250, EPI_ISL_2392251, EPI_ISL_2392252, EPI_ISL_2392253, EPI_ISL_2392254, EPI_ISL_2392255, EPI_ISL_2392256, EPI_ISL_2392257, EPI_ISL_2392258, EPI_ISL_2392259, EPI_ISL_2392260, EPI_ISL_2392261, EPI_ISL_2392262, EPI_ISL_2392264, EPI_ISL_2392267, EPI_ISL_2392268, EPI_ISL_2392270, EPI_ISL_2392271, EPI_ISL_2392272, EPI_ISL_2392273, EPI_ISL_2392274, EPI_ISL_2392275, EPI_ISL_2392276, EPI_ISL_2392278, EPI_ISL_2392280, EPI_ISL_2392281, EPI_ISL_2392282, EPI_ISL_2392283, EPI_ISL_2392284, EPI_ISL_2392285, EPI_ISL_2392286, EPI_ISL_2392287, EPI_ISL_2392289, EPI_ISL_2392290, EPI_ISL_2392291, EPI_ISL_2392293, EPI_ISL_2392295, EPI_ISL_2392296, EPI_ISL_2392297, EPI_ISL_2392298, EPI_ISL_2392299, EPI_ISL_2392300, EPI_ISL_2392301, EPI_ISL_2392302, EPI_ISL_2392303, EPI_ISL_2392305, EPI_ISL_2392306, EPI_ISL_2392307, EPI_ISL_2392308, EPI_ISL_2392309, EPI_ISL_2392311, EPI_ISL_2392312, EPI_ISL_2392313, EPI_ISL_2392314, EPI_ISL_2392315, EPI_ISL_2392317, EPI_ISL_2392318, EPI_ISL_2392319, EPI_ISL_2392320, EPI_ISL_2392321, EPI_ISL_2392322, EPI_ISL_2392324, EPI_ISL_2392325, EPI_ISL_2392326, EPI_ISL_2392327, EPI_ISL_2392328, EPI_ISL_2392329, EPI_ISL_2392330, EPI_ISL_2392331, EPI_ISL_2392332, EPI_ISL_2392334, EPI_ISL_2392335, EPI_ISL_2392337, EPI_ISL_2392339, EPI_ISL_2392340, EPI_ISL_2392341, EPI_ISL_2392342, EPI_ISL_2392343, EPI_ISL_2392344, EPI_ISL_2392346, EPI_ISL_2392347, EPI_ISL_2392348, EPI_ISL_2392350, EPI_ISL_2392351, EPI_ISL_2392352, EPI_ISL_2392354, EPI_ISL_2392357, EPI_ISL_2392358, EPI_ISL_2392359, EPI_ISL_2392360, EPI_ISL_2392361, EPI_ISL_2392363, EPI_ISL_2392364, EPI_ISL_2392365, EPI_ISL_2392366, EPI_ISL_2392367, EPI_ISL_2392369, EPI_ISL_2392370, EPI_ISL_2392371, EPI_ISL_2392372, EPI_ISL_2392373, EPI_ISL_2392374, EPI_ISL_2392375, EPI_ISL_2392376, EPI_ISL_2392377, EPI_ISL_2392378, EPI_ISL_2392379, EPI_ISL_2392380, EPI_ISL_2392382, EPI_ISL_2392383, EPI_ISL_2392384, EPI_ISL_2392385, EPI_ISL_2392386, EPI_ISL_2392388, EPI_ISL_2392392, EPI_ISL_2392394, EPI_ISL_2392395, EPI_ISL_2392396, EPI_ISL_2392399, EPI_ISL_2392400, EPI_ISL_2392401, EPI_ISL_2392402, EPI_ISL_2392403, EPI_ISL_2392406, EPI_ISL_2392407, EPI_ISL_2392409, EPI_ISL_2392410, EPI_ISL_2392411, EPI_ISL_2392412, EPI_ISL_2392413, EPI_ISL_2392414, EPI_ISL_2392415, EPI_ISL_2392416, EPI_ISL_2392417, EPI_ISL_2392418, EPI_ISL_2392419, EPI_ISL_2392422, EPI_ISL_2392423, EPI_ISL_2392424, EPI_ISL_2392425, EPI_ISL_2392426, EPI_ISL_2392427, EPI_ISL_2392428, EPI_ISL_2392429, EPI_ISL_2392431, EPI_ISL_2392432, EPI_ISL_2392434, EPI_ISL_2392435, EPI_ISL_2392437, EPI_ISL_2392439, EPI_ISL_2392440, EPI_ISL_2392443, EPI_ISL_2392444, EPI_ISL_2392446, EPI_ISL_2392447, EPI_ISL_2392449, EPI_ISL_2392450, EPI_ISL_2392451, EPI_ISL_2392452, EPI_ISL_2392453, EPI_ISL_2392455, EPI_ISL_2392458, EPI_ISL_2392459, EPI_ISL_2392460, EPI_ISL_2392461, EPI_ISL_2392463, EPI_ISL_2392464, EPI_ISL_2392465, EPI_ISL_2392466, EPI_ISL_2392467, EPI_ISL_2392469, EPI_ISL_2392470, EPI_ISL_2392471, EPI_ISL_2392472, EPI_ISL_2392473, EPI_ISL_2392474, EPI_ISL_2392475, EPI_ISL_2392476, EPI_ISL_2392477, EPI_ISL_2392478, EPI_ISL_2392479, EPI_ISL_2392480, EPI_ISL_2392482, EPI_ISL_2392483, EPI_ISL_2392484, EPI_ISL_2392485, EPI_ISL_2392486, EPI_ISL_2392487, EPI_ISL_2392488, EPI_ISL_2392489, EPI_ISL_2392490, EPI_ISL_2392491, EPI_ISL_2392492, EPI_ISL_2392493, EPI_ISL_2392494, EPI_ISL_2392495, EPI_ISL_2392496, EPI_ISL_2392497, EPI_ISL_2392498, EPI_ISL_2392499, EPI_ISL_2392500, EPI_ISL_2392502, EPI_ISL_2392503, EPI_ISL_2392505, EPI_ISL_2392506, EPI_ISL_2392507, EPI_ISL_2392508, EPI_ISL_2392509, EPI_ISL_2392511, EPI_ISL_2392512, EPI_ISL_2392514, EPI_ISL_2392515, EPI_ISL_2392516, EPI_ISL_2392517, EPI_ISL_2392518, EPI_ISL_2392519, EPI_ISL_2392520, EPI_ISL_2392521, EPI_ISL_2392526, EPI_ISL_2392527, EPI_ISL_2392528, EPI_ISL_2392529, EPI_ISL_2392530, EPI_ISL_2392534, EPI_ISL_2392536, EPI_ISL_2392537, EPI_ISL_2392538, EPI_ISL_2392539, EPI_ISL_2392541, EPI_ISL_2392542, EPI_ISL_2392544, EPI_ISL_2392545, EPI_ISL_2392546, EPI_ISL_2392547, EPI_ISL_2392548, EPI_ISL_2392549, EPI_ISL_2392556, EPI_ISL_2392557, EPI_ISL_2392562, EPI_ISL_2392564, EPI_ISL_2392565, EPI_ISL_2392566, EPI_ISL_2392569, EPI_ISL_2392572, EPI_ISL_2392573, EPI_ISL_2392574, EPI_ISL_2392575, EPI_ISL_2392580, EPI_ISL_2392584, EPI_ISL_2392585, EPI_ISL_2392587, EPI_ISL_2392594, EPI_ISL_2392596, EPI_ISL_2392597, EPI_ISL_2392598, EPI_ISL_2392600, EPI_ISL_2392601, EPI_ISL_2392603, EPI_ISL_2392605, EPI_ISL_2392607, EPI_ISL_2392608, EPI_ISL_2392611, EPI_ISL_2392613, EPI_ISL_2392614, EPI_ISL_2392617, EPI_ISL_2392619, EPI_ISL_2392620, EPI_ISL_2392621, EPI_ISL_2392622, EPI_ISL_2392625, EPI_ISL_2392628, EPI_ISL_2392630, EPI_ISL_2392632, EPI_ISL_2392633, EPI_ISL_2392634, EPI_ISL_2392635, EPI_ISL_2392636, EPI_ISL_2392638, EPI_ISL_2392642, EPI_ISL_2392644, EPI_ISL_2392645, EPI_ISL_2392646, EPI_ISL_2392647, EPI_ISL_2392648, EPI_ISL_2392650, EPI_ISL_2392651, EPI_ISL_2392652, EPI_ISL_2392653, EPI_ISL_2392654, EPI_ISL_2392655, EPI_ISL_2392656, EPI_ISL_2392657, EPI_ISL_2392658, EPI_ISL_2392659, EPI_ISL_2392661, EPI_ISL_2392662, EPI_ISL_2392663, EPI_ISL_2392665, EPI_ISL_2392667, EPI_ISL_2392668, EPI_ISL_2392669, EPI_ISL_2392670, EPI_ISL_2392671, EPI_ISL_2392674, EPI_ISL_2392675, EPI_ISL_2392676, EPI_ISL_2392680, EPI_ISL_2392682, EPI_ISL_2392684, EPI_ISL_2392688, EPI_ISL_2392689, EPI_ISL_2392690, EPI_ISL_2392691, EPI_ISL_2392692, EPI_ISL_2392693, EPI_ISL_2392694, EPI_ISL_2392696, EPI_ISL_2392698, EPI_ISL_2392699, EPI_ISL_2392701, EPI_ISL_2392702, EPI_ISL_2392706, EPI_ISL_2392707, EPI_ISL_2392708, EPI_ISL_2392712, EPI_ISL_2392713, EPI_ISL_2392714, EPI_ISL_2392715, EPI_ISL_2392718, EPI_ISL_2392720, EPI_ISL_2392721, EPI_ISL_2392723, EPI_ISL_2392725, EPI_ISL_2392726, EPI_ISL_2392728, EPI_ISL_2392729, EPI_ISL_2392730, EPI_ISL_2392731, EPI_ISL_2392735, EPI_ISL_2392736, EPI_ISL_2392738, EPI_ISL_2392742, EPI_ISL_2392745, EPI_ISL_2392750, EPI_ISL_2392751, EPI_ISL_2392753, EPI_ISL_2392754, EPI_ISL_2392755, EPI_ISL_2392756, EPI_ISL_2392760, EPI_ISL_2392761, EPI_ISL_2392763, EPI_ISL_2392944, EPI_ISL_2392946, EPI_ISL_2392951, EPI_ISL_2392952, EPI_ISL_2392953, EPI_ISL_2392954, EPI_ISL_2392955, EPI_ISL_2392957, EPI_ISL_2392959, EPI_ISL_2392962, EPI_ISL_2392964, EPI_ISL_2392965, EPI_ISL_2392966, EPI_ISL_2392967, EPI_ISL_2392968, EPI_ISL_2392969, EPI_ISL_2392970, EPI_ISL_2392974, EPI_ISL_2392975, EPI_ISL_2392976, EPI_ISL_2392977, EPI_ISL_2392978, EPI_ISL_2392980, EPI_ISL_2392981, EPI_ISL_2392982, EPI_ISL_2392983, EPI_ISL_2392984, EPI_ISL_2392987, EPI_ISL_2392990, EPI_ISL_2392992, EPI_ISL_2392993, EPI_ISL_2392994, EPI_ISL_2392997, EPI_ISL_2392998, EPI_ISL_2392999, EPI_ISL_2393000, EPI_ISL_2393001, EPI_ISL_2393002, EPI_ISL_2393003, EPI_ISL_2393005, EPI_ISL_2393006, EPI_ISL_2393007, EPI_ISL_2393010, EPI_ISL_2393011, EPI_ISL_2393012, EPI_ISL_2393015, EPI_ISL_2393017, EPI_ISL_2393020, EPI_ISL_2393023, EPI_ISL_2393024, EPI_ISL_2393025, EPI_ISL_2393026, EPI_ISL_2393027, EPI_ISL_2393029, EPI_ISL_2393030, EPI_ISL_2393031, EPI_ISL_2393032, EPI_ISL_2393034, EPI_ISL_2393036, EPI_ISL_2393038, EPI_ISL_2393040, EPI_ISL_2393043, EPI_ISL_2393044, EPI_ISL_2393045, EPI_ISL_2393047, EPI_ISL_2393050, EPI_ISL_2393051, EPI_ISL_2393053, EPI_ISL_2393055, EPI_ISL_2393058, EPI_ISL_2393061, EPI_ISL_2393062, EPI_ISL_2393063, EPI_ISL_2393065, EPI_ISL_2393069, EPI_ISL_2393071, EPI_ISL_2393072, EPI_ISL_2393075, EPI_ISL_2393076, EPI_ISL_2393077, EPI_ISL_2393078, EPI_ISL_2393079, EPI_ISL_2393081, EPI_ISL_2393082, EPI_ISL_2393087, EPI_ISL_2393088, EPI_ISL_2393089, EPI_ISL_2393090, EPI_ISL_2393091, EPI_ISL_2393092, EPI_ISL_2393093, EPI_ISL_2393096, EPI_ISL_2393097, EPI_ISL_2393101, EPI_ISL_2393103, EPI_ISL_2393104, EPI_ISL_2393105, EPI_ISL_2393106, EPI_ISL_2393107, EPI_ISL_2393109, EPI_ISL_2393111, EPI_ISL_2393112, EPI_ISL_2393117, EPI_ISL_2393118, EPI_ISL_2393119, EPI_ISL_2393120, EPI_ISL_2393121, EPI_ISL_2393122, EPI_ISL_2393123, EPI_ISL_2393125, EPI_ISL_2393126, EPI_ISL_2393128, EPI_ISL_2393129, EPI_ISL_2393130, EPI_ISL_2393132, EPI_ISL_2393133, EPI_ISL_2393134, EPI_ISL_2393136, EPI_ISL_2393149, EPI_ISL_2393153, EPI_ISL_2393161, EPI_ISL_2393164, EPI_ISL_2393169, EPI_ISL_2393170, EPI_ISL_2393173, EPI_ISL_2393174, EPI_ISL_2393175, EPI_ISL_2393176, EPI_ISL_2393178, EPI_ISL_2393179, EPI_ISL_2393181, EPI_ISL_2393182, EPI_ISL_2393183, EPI_ISL_2393184, EPI_ISL_2393186, EPI_ISL_2393190, EPI_ISL_2393192, EPI_ISL_2393193, EPI_ISL_2393194, EPI_ISL_2393197, EPI_ISL_2393198, EPI_ISL_2393201, EPI_ISL_2393202, EPI_ISL_2393203, EPI_ISL_2393204, EPI_ISL_2393209, EPI_ISL_2393210, EPI_ISL_2393213, EPI_ISL_2393216, EPI_ISL_2393217, EPI_ISL_2393220, EPI_ISL_2393222, EPI_ISL_2393223, EPI_ISL_2393224, EPI_ISL_2393227, EPI_ISL_2393230, EPI_ISL_2393232, EPI_ISL_2393234, EPI_ISL_2393235, EPI_ISL_2393238, EPI_ISL_2393239, EPI_ISL_2393241, EPI_ISL_2393242, EPI_ISL_2393246, EPI_ISL_2393248, EPI_ISL_2393252, EPI_ISL_2393254, EPI_ISL_2393255, EPI_ISL_2393256, EPI_ISL_2393257, EPI_ISL_2393258, EPI_ISL_2393260, EPI_ISL_2393261, EPI_ISL_2393263, EPI_ISL_2393265, EPI_ISL_2393266, EPI_ISL_2393268, EPI_ISL_2393269, EPI_ISL_2393275, EPI_ISL_2393277, EPI_ISL_2393282, EPI_ISL_2393283, EPI_ISL_2393285, EPI_ISL_2393286, EPI_ISL_2393289, EPI_ISL_2393291, EPI_ISL_2393292, EPI_ISL_2393297, EPI_ISL_2393299, EPI_ISL_2393300, EPI_ISL_2393302, EPI_ISL_2393306, EPI_ISL_2393310, EPI_ISL_2393312, EPI_ISL_2393313, EPI_ISL_2393326, EPI_ISL_2393329, EPI_ISL_2393330, EPI_ISL_2393331, EPI_ISL_2393333, EPI_ISL_2393334, EPI_ISL_2393335, EPI_ISL_2393341, EPI_ISL_2393344, EPI_ISL_2393346, EPI_ISL_2393348, EPI_ISL_2393354, EPI_ISL_2393355, EPI_ISL_2393356, EPI_ISL_2393361, EPI_ISL_2393363, EPI_ISL_2393364, EPI_ISL_2393365, EPI_ISL_2393366, EPI_ISL_2393369, EPI_ISL_2393371, EPI_ISL_2393378, EPI_ISL_2393379, EPI_ISL_2393381, EPI_ISL_2393382, EPI_ISL_2393386, EPI_ISL_2393390, EPI_ISL_2393398, EPI_ISL_2393399, EPI_ISL_2393400, EPI_ISL_2393402, EPI_ISL_2393403, EPI_ISL_2393404, EPI_ISL_2393409, EPI_ISL_2393411, EPI_ISL_2393412, EPI_ISL_2393415, EPI_ISL_2393416, EPI_ISL_2393421, EPI_ISL_2393422, EPI_ISL_2393423, EPI_ISL_2393427, EPI_ISL_2393432, EPI_ISL_2393433, EPI_ISL_2393436, EPI_ISL_2393438, EPI_ISL_2393447, EPI_ISL_2393456, EPI_ISL_2393457, EPI_ISL_2393459, EPI_ISL_2393461, EPI_ISL_2393463, EPI_ISL_2393464, EPI_ISL_2393465, EPI_ISL_2393466, EPI_ISL_2393468, EPI_ISL_2393472, EPI_ISL_2393474, EPI_ISL_2393476, EPI_ISL_2393478, EPI_ISL_2393479, EPI_ISL_2393480, EPI_ISL_2393485, EPI_ISL_2393489, EPI_ISL_2393494, EPI_ISL_2393495, EPI_ISL_2393496, EPI_ISL_2393497, EPI_ISL_2393499, EPI_ISL_2393502, EPI_ISL_2393504, EPI_ISL_2393505, EPI_ISL_2393506, EPI_ISL_2393507, EPI_ISL_2393508, EPI_ISL_2393512, EPI_ISL_2393513, EPI_ISL_2393525, EPI_ISL_2393526, EPI_ISL_2393527, EPI_ISL_2393529, EPI_ISL_2393530, EPI_ISL_2393531, EPI_ISL_2393532, EPI_ISL_2393534, EPI_ISL_2393538, EPI_ISL_2393539, EPI_ISL_2393540, EPI_ISL_2393541, EPI_ISL_2393544, EPI_ISL_2393545, EPI_ISL_2393546, EPI_ISL_2393548, EPI_ISL_2393549, EPI_ISL_2393550, EPI_ISL_2393557, EPI_ISL_2393558, EPI_ISL_2393563, EPI_ISL_2393564, EPI_ISL_2393567, EPI_ISL_2393568, EPI_ISL_2393570, EPI_ISL_2393575, EPI_ISL_2393578, EPI_ISL_2393579, EPI_ISL_2393580, EPI_ISL_2393583, EPI_ISL_2393585, EPI_ISL_2393587, EPI_ISL_2393588, EPI_ISL_2393590, EPI_ISL_2393602, EPI_ISL_2393606, EPI_ISL_2393610, EPI_ISL_2393614, EPI_ISL_2393617, EPI_ISL_2393624, EPI_ISL_2393627, EPI_ISL_2393628, EPI_ISL_2393630, EPI_ISL_2393635, EPI_ISL_2393637, EPI_ISL_2393638, EPI_ISL_2393642, EPI_ISL_2393643, EPI_ISL_2393646, EPI_ISL_2393648, EPI_ISL_2393651, EPI_ISL_2393657, EPI_ISL_2393658, EPI_ISL_2393661, EPI_ISL_2393662, EPI_ISL_2393667, EPI_ISL_2393670, EPI_ISL_2393673, EPI_ISL_2393675, EPI_ISL_2393682, EPI_ISL_2393688, EPI_ISL_2393690, EPI_ISL_2393692, EPI_ISL_2393694, EPI_ISL_2393696, EPI_ISL_2393702, EPI_ISL_2393706, EPI_ISL_2393707, EPI_ISL_2393709, EPI_ISL_2393715, EPI_ISL_2393717, EPI_ISL_2393720, EPI_ISL_2393730, EPI_ISL_2393731, EPI_ISL_2393735, EPI_ISL_2393738, EPI_ISL_2393739, EPI_ISL_2393750, EPI_ISL_2393753, EPI_ISL_2393755, EPI_ISL_2393756, EPI_ISL_2393757, EPI_ISL_2393758, EPI_ISL_2393759, EPI_ISL_2393760, EPI_ISL_2393761, EPI_ISL_2393764, EPI_ISL_2393768, EPI_ISL_2393769, EPI_ISL_2393777, EPI_ISL_2393780, EPI_ISL_2393781, EPI_ISL_2393783, EPI_ISL_2393789, EPI_ISL_2393791, EPI_ISL_2393792, EPI_ISL_2393794, EPI_ISL_2393795, EPI_ISL_2393797, EPI_ISL_2393801, EPI_ISL_2393805, EPI_ISL_2393807, EPI_ISL_2393809, EPI_ISL_2393812, EPI_ISL_2393815, EPI_ISL_2393822, EPI_ISL_2393823, EPI_ISL_2393825, EPI_ISL_2393832, EPI_ISL_2393835, EPI_ISL_2393840, EPI_ISL_2393848, EPI_ISL_2393850, EPI_ISL_2393851, EPI_ISL_2393853, EPI_ISL_2393856, EPI_ISL_2393857, EPI_ISL_2393866, EPI_ISL_2393867, EPI_ISL_2393868, EPI_ISL_2393870, EPI_ISL_2393876, EPI_ISL_2393878, EPI_ISL_2393880, EPI_ISL_2393884, EPI_ISL_2393885, EPI_ISL_2393900, EPI_ISL_2393901, EPI_ISL_2393908, EPI_ISL_2393910, EPI_ISL_2393914, EPI_ISL_2393915, EPI_ISL_2393918, EPI_ISL_2393920, EPI_ISL_2393923, EPI_ISL_2393924, EPI_ISL_2393934, EPI_ISL_2393936, EPI_ISL_2393938, EPI_ISL_2393943, EPI_ISL_2393948, EPI_ISL_2393952, EPI_ISL_2393960, EPI_ISL_2393961, EPI_ISL_2393964, EPI_ISL_2393971, EPI_ISL_2393974, EPI_ISL_2393979, EPI_ISL_2393986, EPI_ISL_2393987, EPI_ISL_2393989, EPI_ISL_2393990, EPI_ISL_2393993, EPI_ISL_2393995, EPI_ISL_2394008, EPI_ISL_2394009, EPI_ISL_2394010, EPI_ISL_2394012, EPI_ISL_2394013, EPI_ISL_2394019, EPI_ISL_2394029, EPI_ISL_2394030, EPI_ISL_2394035, EPI_ISL_2394036, EPI_ISL_2394037, EPI_ISL_2394038, EPI_ISL_2394040, EPI_ISL_2394041, EPI_ISL_2394042, EPI_ISL_2394043, EPI_ISL_2394044, EPI_ISL_2394046, EPI_ISL_2394047, EPI_ISL_2394048, EPI_ISL_2394049, EPI_ISL_2394051, EPI_ISL_2394053, EPI_ISL_2394054, EPI_ISL_2394055, EPI_ISL_2394056, EPI_ISL_2394057, EPI_ISL_2394058, EPI_ISL_2394059, EPI_ISL_2394060, EPI_ISL_2394061, EPI_ISL_2394062, EPI_ISL_2394063, EPI_ISL_2394064, EPI_ISL_2394065, EPI_ISL_2394066, EPI_ISL_2394067, EPI_ISL_2394068, EPI_ISL_2394070, EPI_ISL_2394071, EPI_ISL_2394072, EPI_ISL_2394073, EPI_ISL_2394074, EPI_ISL_2394075, EPI_ISL_2394076, EPI_ISL_2394077, EPI_ISL_2394078, EPI_ISL_2394079, EPI_ISL_2394080, EPI_ISL_2394081, EPI_ISL_2394082, EPI_ISL_2394083, EPI_ISL_2394084, EPI_ISL_2394085, EPI_ISL_2394086, EPI_ISL_2394087, EPI_ISL_2394088, EPI_ISL_2394089, EPI_ISL_2394091, EPI_ISL_2394092, EPI_ISL_2394093, EPI_ISL_2394095, EPI_ISL_2394096, EPI_ISL_2394098, EPI_ISL_2394099, EPI_ISL_2394100, EPI_ISL_2394101, EPI_ISL_2394102, EPI_ISL_2394103, EPI_ISL_2394104, EPI_ISL_2394105, EPI_ISL_2394106, EPI_ISL_2394107, EPI_ISL_2394108, EPI_ISL_2394109, EPI_ISL_2394110, EPI_ISL_2394111, EPI_ISL_2394112, EPI_ISL_2394113, EPI_ISL_2394114, EPI_ISL_2394116, EPI_ISL_2394117, EPI_ISL_2394118, EPI_ISL_2394119, EPI_ISL_2394120, EPI_ISL_2394121, EPI_ISL_2394122, EPI_ISL_2394123, EPI_ISL_2394124, EPI_ISL_2394126, EPI_ISL_2394127, EPI_ISL_2394128, EPI_ISL_2394129, EPI_ISL_2394130, EPI_ISL_2394131, EPI_ISL_2394132, EPI_ISL_2394133, EPI_ISL_2394134, EPI_ISL_2394135, EPI_ISL_2394136, EPI_ISL_2394138, EPI_ISL_2394139, EPI_ISL_2394140, EPI_ISL_2394141, EPI_ISL_2394142, EPI_ISL_2394143, EPI_ISL_2394144, EPI_ISL_2394145, EPI_ISL_2394147, EPI_ISL_2394148, EPI_ISL_2394149, EPI_ISL_2394150, EPI_ISL_2394151, EPI_ISL_2394152, EPI_ISL_2394153, EPI_ISL_2394154, EPI_ISL_2394155, EPI_ISL_2394156, EPI_ISL_2394157, EPI_ISL_2394158, EPI_ISL_2394159, EPI_ISL_2394160, EPI_ISL_2394161, EPI_ISL_2394162, EPI_ISL_2394163, EPI_ISL_2394164, EPI_ISL_2394165, EPI_ISL_2394166, EPI_ISL_2394167, EPI_ISL_2394168, EPI_ISL_2394169, EPI_ISL_2394171, EPI_ISL_2394172, EPI_ISL_2394173, EPI_ISL_2394174, EPI_ISL_2394175, EPI_ISL_2394177, EPI_ISL_2394178, EPI_ISL_2394179, EPI_ISL_2394181, EPI_ISL_2394182, EPI_ISL_2394183, EPI_ISL_2394184, EPI_ISL_2394185, EPI_ISL_2394186, EPI_ISL_2394188, EPI_ISL_2394189, EPI_ISL_2394190, EPI_ISL_2394191, EPI_ISL_2394192, EPI_ISL_2394193, EPI_ISL_2394194, EPI_ISL_2394195, EPI_ISL_2394196, EPI_ISL_2394197, EPI_ISL_2394198, EPI_ISL_2394199, EPI_ISL_2394200, EPI_ISL_2394201, EPI_ISL_2394202, EPI_ISL_2394203, EPI_ISL_2394204, EPI_ISL_2394205, EPI_ISL_2394206, EPI_ISL_2394207, EPI_ISL_2394208, EPI_ISL_2394209, EPI_ISL_2394210, EPI_ISL_2394211, EPI_ISL_2394212, EPI_ISL_2394213, EPI_ISL_2394214, EPI_ISL_2394217, EPI_ISL_2394218, EPI_ISL_2394219, EPI_ISL_2394220, EPI_ISL_2394221, EPI_ISL_2394222, EPI_ISL_2394223, EPI_ISL_2394224, EPI_ISL_2394225, EPI_ISL_2394226, EPI_ISL_2394227, EPI_ISL_2394228, EPI_ISL_2394229, EPI_ISL_2394230, EPI_ISL_2394231, EPI_ISL_2394232, EPI_ISL_2394233, EPI_ISL_2394234, EPI_ISL_2394235, EPI_ISL_2394236, EPI_ISL_2394237, EPI_ISL_2394238, EPI_ISL_2394239, EPI_ISL_2394240, EPI_ISL_2394241, EPI_ISL_2394243, EPI_ISL_2394244, EPI_ISL_2394245, EPI_ISL_2394248, EPI_ISL_2394249, EPI_ISL_2394250, EPI_ISL_2394251, EPI_ISL_2394252, EPI_ISL_2394253, EPI_ISL_2394254, EPI_ISL_2394255, EPI_ISL_2394257, EPI_ISL_2394258, EPI_ISL_2394259, EPI_ISL_2394261, EPI_ISL_2394262, EPI_ISL_2394263, EPI_ISL_2394264, EPI_ISL_2394265, EPI_ISL_2394266, EPI_ISL_2394267, EPI_ISL_2394268, EPI_ISL_2394269, EPI_ISL_2394270, EPI_ISL_2394271, EPI_ISL_2394273, EPI_ISL_2394274, EPI_ISL_2394275, EPI_ISL_2394276, EPI_ISL_2394277, EPI_ISL_2394278, EPI_ISL_2394279, EPI_ISL_2394280, EPI_ISL_2394281, EPI_ISL_2394283, EPI_ISL_2394284, EPI_ISL_2394285, EPI_ISL_2394286, EPI_ISL_2394287, EPI_ISL_2394288, EPI_ISL_2394289, EPI_ISL_2394290, EPI_ISL_2394291, EPI_ISL_2394292, EPI_ISL_2394293, EPI_ISL_2394294, EPI_ISL_2394295, EPI_ISL_2394296, EPI_ISL_2394297, EPI_ISL_2394298, EPI_ISL_2394299, EPI_ISL_2394300, EPI_ISL_2394301, EPI_ISL_2394302, EPI_ISL_2394304, EPI_ISL_2394305, EPI_ISL_2394306, EPI_ISL_2394307, EPI_ISL_2394308, EPI_ISL_2394309, EPI_ISL_2394310, EPI_ISL_2394311, EPI_ISL_2394313, EPI_ISL_2394314, EPI_ISL_2394315, EPI_ISL_2394317, EPI_ISL_2394318, EPI_ISL_2394319, EPI_ISL_2394324, EPI_ISL_2394325, EPI_ISL_2394326, EPI_ISL_2394329, EPI_ISL_2394330, EPI_ISL_2394331, EPI_ISL_2394332, EPI_ISL_2394333, EPI_ISL_2394335, EPI_ISL_2394337, EPI_ISL_2394338, EPI_ISL_2394339, EPI_ISL_2394343, EPI_ISL_2394344, EPI_ISL_2394345, EPI_ISL_2394347, EPI_ISL_2394349, EPI_ISL_2394351, EPI_ISL_2394352, EPI_ISL_2394357, EPI_ISL_2394361, EPI_ISL_2394365, EPI_ISL_2394367, EPI_ISL_2394370, EPI_ISL_2394371, EPI_ISL_2394375, EPI_ISL_2394377, EPI_ISL_2394378, EPI_ISL_2394382, EPI_ISL_2394389, EPI_ISL_2394391, EPI_ISL_2394393, EPI_ISL_2394395, EPI_ISL_2394396, EPI_ISL_2394397, EPI_ISL_2394399, EPI_ISL_2394402, EPI_ISL_2394403, EPI_ISL_2394406, EPI_ISL_2394409, EPI_ISL_2394411, EPI_ISL_2394413, EPI_ISL_2394416, EPI_ISL_2394418, EPI_ISL_2394422, EPI_ISL_2394423, EPI_ISL_2394432, EPI_ISL_2394433, EPI_ISL_2394434, EPI_ISL_2394438, EPI_ISL_2394439, EPI_ISL_2394440, EPI_ISL_2394443, EPI_ISL_2394457, EPI_ISL_2394458, EPI_ISL_2394460, EPI_ISL_2394461, EPI_ISL_2394464, EPI_ISL_2394468, EPI_ISL_2394475, EPI_ISL_2394476, EPI_ISL_2394477, EPI_ISL_2394482, EPI_ISL_2394483, EPI_ISL_2394485, EPI_ISL_2394487, EPI_ISL_2394491, EPI_ISL_2394494, EPI_ISL_2394495, EPI_ISL_2394496, EPI_ISL_2394497, EPI_ISL_2394498, EPI_ISL_2394503, EPI_ISL_2394505, EPI_ISL_2394506, EPI_ISL_2394507, EPI_ISL_2394510, EPI_ISL_2394512, EPI_ISL_2394513, EPI_ISL_2394514, EPI_ISL_2394517, EPI_ISL_2394518, EPI_ISL_2394521, EPI_ISL_2394522, EPI_ISL_2394523, EPI_ISL_2394526, EPI_ISL_2394529, EPI_ISL_2394530, EPI_ISL_2394531, EPI_ISL_2394535, EPI_ISL_2394543, EPI_ISL_2394545, EPI_ISL_2394546, EPI_ISL_2394553, EPI_ISL_2394554, EPI_ISL_2394555, EPI_ISL_2394558, EPI_ISL_2394562, EPI_ISL_2394563, EPI_ISL_2394565, EPI_ISL_2394567, EPI_ISL_2394568, EPI_ISL_2394573, EPI_ISL_2394577, EPI_ISL_2394578, EPI_ISL_2394580, EPI_ISL_2394581, EPI_ISL_2394582, EPI_ISL_2394583, EPI_ISL_2394584, EPI_ISL_2394587, EPI_ISL_2394588, EPI_ISL_2394589, EPI_ISL_2394590, EPI_ISL_2394592, EPI_ISL_2394593, EPI_ISL_2394594, EPI_ISL_2394595, EPI_ISL_2394597, EPI_ISL_2394598, EPI_ISL_2394599, EPI_ISL_2394600, EPI_ISL_2394602, EPI_ISL_2394603, EPI_ISL_2394604, EPI_ISL_2394605, EPI_ISL_2394606, EPI_ISL_2394608, EPI_ISL_2394609, EPI_ISL_2394610, EPI_ISL_2394611, EPI_ISL_2394615, EPI_ISL_2394616, EPI_ISL_2394617, EPI_ISL_2394619, EPI_ISL_2394620, EPI_ISL_2394621, EPI_ISL_2394623, EPI_ISL_2394626, EPI_ISL_2394627, EPI_ISL_2394628, EPI_ISL_2394629, EPI_ISL_2394630, EPI_ISL_2394631, EPI_ISL_2394634, EPI_ISL_2394635, EPI_ISL_2394636, EPI_ISL_2394637, EPI_ISL_2394639, EPI_ISL_2394641, EPI_ISL_2394643, EPI_ISL_2394644, EPI_ISL_2394645, EPI_ISL_2394646, EPI_ISL_2394647, EPI_ISL_2394649, EPI_ISL_2394651, EPI_ISL_2394652, EPI_ISL_2394653, EPI_ISL_2394654, EPI_ISL_2394655, EPI_ISL_2394656, EPI_ISL_2394657, EPI_ISL_2394658, EPI_ISL_2394660, EPI_ISL_2394661, EPI_ISL_2394662, EPI_ISL_2394663, EPI_ISL_2394664, EPI_ISL_2394665, EPI_ISL_2394666, EPI_ISL_2394669, EPI_ISL_2394670, EPI_ISL_2394671, EPI_ISL_2394672, EPI_ISL_2394675, EPI_ISL_2394676, EPI_ISL_2394677, EPI_ISL_2394678, EPI_ISL_2394679, EPI_ISL_2394680, EPI_ISL_2394681, EPI_ISL_2394682, EPI_ISL_2394684, EPI_ISL_2394685, EPI_ISL_2394686, EPI_ISL_2394688, EPI_ISL_2394690, EPI_ISL_2394691, EPI_ISL_2394692, EPI_ISL_2394695, EPI_ISL_2394696, EPI_ISL_2394697, EPI_ISL_2394698, EPI_ISL_2394699, EPI_ISL_2394701, EPI_ISL_2394702, EPI_ISL_2394705, EPI_ISL_2394711, EPI_ISL_2394713, EPI_ISL_2394715, EPI_ISL_2394716, EPI_ISL_2394718, EPI_ISL_2394720, EPI_ISL_2394721, EPI_ISL_2394722, EPI_ISL_2394723, EPI_ISL_2394725, EPI_ISL_2394726, EPI_ISL_2394727, EPI_ISL_2394728, EPI_ISL_2394729, EPI_ISL_2394730, EPI_ISL_2394731, EPI_ISL_2394732, EPI_ISL_2394733, EPI_ISL_2394734, EPI_ISL_2394735, EPI_ISL_2394736, EPI_ISL_2394739, EPI_ISL_2394740, EPI_ISL_2394741, EPI_ISL_2394743, EPI_ISL_2394744, EPI_ISL_2394745, EPI_ISL_2394748, EPI_ISL_2394749, EPI_ISL_2394750, EPI_ISL_2394751, EPI_ISL_2394752, EPI_ISL_2394753, EPI_ISL_2394754, EPI_ISL_2394756, EPI_ISL_2394757, EPI_ISL_2394758, EPI_ISL_2394759, EPI_ISL_2394761, EPI_ISL_2394763, EPI_ISL_2394765, EPI_ISL_2394766, EPI_ISL_2394768, EPI_ISL_2394769, EPI_ISL_2394770, EPI_ISL_2394771, EPI_ISL_2394772, EPI_ISL_2394773, EPI_ISL_2394774, EPI_ISL_2394775, EPI_ISL_2394776, EPI_ISL_2394777, EPI_ISL_2394778, EPI_ISL_2394781, EPI_ISL_2394782, EPI_ISL_2394783, EPI_ISL_2394786, EPI_ISL_2394787, EPI_ISL_2394788, EPI_ISL_2394789, EPI_ISL_2394790, EPI_ISL_2394792, EPI_ISL_2394793, EPI_ISL_2394794, EPI_ISL_2394795, EPI_ISL_2394796, EPI_ISL_2394798, EPI_ISL_2394799, EPI_ISL_2394800, EPI_ISL_2394801, EPI_ISL_2394802, EPI_ISL_2394805, EPI_ISL_2394806, EPI_ISL_2394808, EPI_ISL_2394809, EPI_ISL_2394810, EPI_ISL_2394811, EPI_ISL_2394812, EPI_ISL_2394813, EPI_ISL_2394814, EPI_ISL_2394815, EPI_ISL_2394816, EPI_ISL_2394817, EPI_ISL_2394818, EPI_ISL_2394819, EPI_ISL_2394820, EPI_ISL_2394822, EPI_ISL_2394823, EPI_ISL_2394824, EPI_ISL_2394827, EPI_ISL_2394828, EPI_ISL_2394830, EPI_ISL_2394831, EPI_ISL_2394833, EPI_ISL_2394834, EPI_ISL_2394835, EPI_ISL_2394837, EPI_ISL_2394838, EPI_ISL_2394839, EPI_ISL_2394841, EPI_ISL_2394842, EPI_ISL_2394845, EPI_ISL_2394849, EPI_ISL_2394851, EPI_ISL_2394852, EPI_ISL_2394854, EPI_ISL_2394855, EPI_ISL_2394857, EPI_ISL_2394859, EPI_ISL_2394860, EPI_ISL_2394862, EPI_ISL_2394863, EPI_ISL_2394864, EPI_ISL_2394866, EPI_ISL_2394871, EPI_ISL_2394873, EPI_ISL_2394879, EPI_ISL_2394881, EPI_ISL_2394884, EPI_ISL_2394885, EPI_ISL_2394887, EPI_ISL_2394888, EPI_ISL_2394890, EPI_ISL_2394891, EPI_ISL_2394898, EPI_ISL_2394899, EPI_ISL_2394900, EPI_ISL_2394906, EPI_ISL_2394909, EPI_ISL_2394912, EPI_ISL_2394914, EPI_ISL_2394915, EPI_ISL_2394916, EPI_ISL_2394921, EPI_ISL_2394922, EPI_ISL_2394924, EPI_ISL_2394928, EPI_ISL_2394932, EPI_ISL_2394935, EPI_ISL_2394938, EPI_ISL_2394939, EPI_ISL_2394940, EPI_ISL_2394941, EPI_ISL_2394942, EPI_ISL_2394945, EPI_ISL_2394946, EPI_ISL_2394947, EPI_ISL_2394948, EPI_ISL_2394949, EPI_ISL_2394952, EPI_ISL_2394954, EPI_ISL_2394955, EPI_ISL_2394958, EPI_ISL_2394959, EPI_ISL_2394960, EPI_ISL_2394962, EPI_ISL_2394964, EPI_ISL_2394967, EPI_ISL_2394968, EPI_ISL_2394969, EPI_ISL_2394970, EPI_ISL_2394971, EPI_ISL_2394973, EPI_ISL_2394976, EPI_ISL_2394977, EPI_ISL_2394979, EPI_ISL_2394980, EPI_ISL_2394985, EPI_ISL_2394986, EPI_ISL_2394988, EPI_ISL_2394989, EPI_ISL_2394994, EPI_ISL_2394995, EPI_ISL_2394998, EPI_ISL_2395000, EPI_ISL_2395002, EPI_ISL_2395003, EPI_ISL_2395004, EPI_ISL_2395006, EPI_ISL_2395009, EPI_ISL_2395011, EPI_ISL_2395014, EPI_ISL_2395016, EPI_ISL_2395018, EPI_ISL_2395019, EPI_ISL_2395021, EPI_ISL_2395023, EPI_ISL_2395024, EPI_ISL_2395025, EPI_ISL_2395027, EPI_ISL_2395030, EPI_ISL_2395032, EPI_ISL_2395036, EPI_ISL_2395038, EPI_ISL_2395039, EPI_ISL_2395041, EPI_ISL_2395045, EPI_ISL_2395046, EPI_ISL_2395048, EPI_ISL_2395049, EPI_ISL_2395050, EPI_ISL_2395051, EPI_ISL_2395052, EPI_ISL_2395054, EPI_ISL_2395058, EPI_ISL_2395059, EPI_ISL_2395064, EPI_ISL_2395066, EPI_ISL_2395067, EPI_ISL_2395069, EPI_ISL_2395071, EPI_ISL_2395072, EPI_ISL_2395075, EPI_ISL_2395077, EPI_ISL_2395078, EPI_ISL_2395080, EPI_ISL_2395087, EPI_ISL_2395088, EPI_ISL_2395091, EPI_ISL_2395092, EPI_ISL_2395094, EPI_ISL_2395097, EPI_ISL_2395101, EPI_ISL_2395108, EPI_ISL_2395111, EPI_ISL_2395113, EPI_ISL_2395114, EPI_ISL_2395116, EPI_ISL_2395118, EPI_ISL_2395120, EPI_ISL_2395121, EPI_ISL_2395122, EPI_ISL_2395129, EPI_ISL_2395134, EPI_ISL_2395135, EPI_ISL_2395136, EPI_ISL_2395138, EPI_ISL_2395139, EPI_ISL_2395142, EPI_ISL_2395143, EPI_ISL_2395147, EPI_ISL_2395155, EPI_ISL_2395159, EPI_ISL_2395160, EPI_ISL_2395162, EPI_ISL_2395163, EPI_ISL_2395164, EPI_ISL_2395165, EPI_ISL_2395167, EPI_ISL_2395168, EPI_ISL_2395169, EPI_ISL_2395175, EPI_ISL_2395177, EPI_ISL_2395179, EPI_ISL_2395180, EPI_ISL_2395181, EPI_ISL_2395182, EPI_ISL_2395186, EPI_ISL_2395188, EPI_ISL_2395189, EPI_ISL_2395192, EPI_ISL_2395193, EPI_ISL_2395194, EPI_ISL_2395197, EPI_ISL_2395199, EPI_ISL_2395201, EPI_ISL_2395202, EPI_ISL_2395206, EPI_ISL_2395214, EPI_ISL_2395218, EPI_ISL_2395220, EPI_ISL_2395222, EPI_ISL_2395234, EPI_ISL_2395235, EPI_ISL_2395236, EPI_ISL_2395239, EPI_ISL_2395243, EPI_ISL_2395245, EPI_ISL_2395250, EPI_ISL_2395252, EPI_ISL_2395253, EPI_ISL_2395254, EPI_ISL_2395260, EPI_ISL_2395262, EPI_ISL_2395264, EPI_ISL_2395266, EPI_ISL_2395269, EPI_ISL_2395270, EPI_ISL_2395274, EPI_ISL_2395276, EPI_ISL_2395277, EPI_ISL_2395279, EPI_ISL_2395282, EPI_ISL_2395283, EPI_ISL_2395284, EPI_ISL_2395285, EPI_ISL_2395286, EPI_ISL_2395289, EPI_ISL_2395290, EPI_ISL_2395291, EPI_ISL_2395292, EPI_ISL_2395293, EPI_ISL_2395295, EPI_ISL_2395296, EPI_ISL_2395298, EPI_ISL_2395299, EPI_ISL_2395302, EPI_ISL_2395305, EPI_ISL_2395310, EPI_ISL_2395316, EPI_ISL_2395317, EPI_ISL_2395320, EPI_ISL_2395321, EPI_ISL_2395322, EPI_ISL_2395323, EPI_ISL_2395327, EPI_ISL_2395331, EPI_ISL_2395334, EPI_ISL_2395335, EPI_ISL_2406895, EPI_ISL_2406897, EPI_ISL_2406901, EPI_ISL_2406902, EPI_ISL_2406903, EPI_ISL_2406906, EPI_ISL_2406908, EPI_ISL_2406911, EPI_ISL_2406914, EPI_ISL_2406915, EPI_ISL_2406916, EPI_ISL_2406919, EPI_ISL_2406920, EPI_ISL_2406923, EPI_ISL_2406925, EPI_ISL_2406927, EPI_ISL_2406928, EPI_ISL_2406932, EPI_ISL_2406933, EPI_ISL_2406934, EPI_ISL_2406939, EPI_ISL_2406943, EPI_ISL_2406945, EPI_ISL_2406946, EPI_ISL_2406949, EPI_ISL_2406950, EPI_ISL_2406951, EPI_ISL_2406953, EPI_ISL_2406954, EPI_ISL_2406956, EPI_ISL_2406961, EPI_ISL_2406962, EPI_ISL_2406963, EPI_ISL_2406965, EPI_ISL_2406967, EPI_ISL_2406973, EPI_ISL_2406976, EPI_ISL_2406977, EPI_ISL_2406979, EPI_ISL_2406980, EPI_ISL_2406981, EPI_ISL_2406983, EPI_ISL_2406985, EPI_ISL_2406986, EPI_ISL_2406988, EPI_ISL_2406989, EPI_ISL_2406992, EPI_ISL_2406994, EPI_ISL_2406996, EPI_ISL_2406998, EPI_ISL_2407000, EPI_ISL_2407005, EPI_ISL_2407007, EPI_ISL_2407010, EPI_ISL_2407013, EPI_ISL_2407014, EPI_ISL_2407015, EPI_ISL_2407020, EPI_ISL_2407025, EPI_ISL_2407029, EPI_ISL_2407030, EPI_ISL_2407032, EPI_ISL_2407033, EPI_ISL_2407037, EPI_ISL_2407038, EPI_ISL_2407040, EPI_ISL_2407042, EPI_ISL_2407046, EPI_ISL_2407051, EPI_ISL_2407053, EPI_ISL_2407055, EPI_ISL_2407062, EPI_ISL_2407064, EPI_ISL_2407066, EPI_ISL_2407067, EPI_ISL_2407083, EPI_ISL_2407084, EPI_ISL_2407086, EPI_ISL_2407087, EPI_ISL_2407091, EPI_ISL_2407095, EPI_ISL_2407096, EPI_ISL_2407101, EPI_ISL_2407103, EPI_ISL_2407105, EPI_ISL_2407106, EPI_ISL_2407108, EPI_ISL_2407111, EPI_ISL_2407115, EPI_ISL_2407116, EPI_ISL_2407117, EPI_ISL_2407118, EPI_ISL_2407119, EPI_ISL_2407120, EPI_ISL_2407121, EPI_ISL_2407122, EPI_ISL_2407123, EPI_ISL_2407124, EPI_ISL_2407125, EPI_ISL_2407126, EPI_ISL_2407127, EPI_ISL_2407128, EPI_ISL_2407130, EPI_ISL_2407131, EPI_ISL_2407132, EPI_ISL_2407135, EPI_ISL_2407136, EPI_ISL_2407137, EPI_ISL_2407138, EPI_ISL_2407139, EPI_ISL_2407140, EPI_ISL_2407141, EPI_ISL_2407142, EPI_ISL_2407143, EPI_ISL_2407144, EPI_ISL_2407145, EPI_ISL_2407146, EPI_ISL_2407147, EPI_ISL_2407148, EPI_ISL_2407151, EPI_ISL_2407152, EPI_ISL_2407153, EPI_ISL_2407154, EPI_ISL_2407155, EPI_ISL_2407156, EPI_ISL_2407157, EPI_ISL_2407158, EPI_ISL_2407159, EPI_ISL_2407160, EPI_ISL_2407162, EPI_ISL_2407163, EPI_ISL_2407164, EPI_ISL_2407165, EPI_ISL_2407166, EPI_ISL_2407167, EPI_ISL_2407168, EPI_ISL_2407169, EPI_ISL_2407170, EPI_ISL_2407171, EPI_ISL_2407172, EPI_ISL_2407173, EPI_ISL_2407175, EPI_ISL_2407177, EPI_ISL_2407178, EPI_ISL_2407179, EPI_ISL_2407180, EPI_ISL_2407181, EPI_ISL_2407182, EPI_ISL_2407183, EPI_ISL_2407184, EPI_ISL_2407185, EPI_ISL_2407186, EPI_ISL_2407187, EPI_ISL_2407188, EPI_ISL_2407189, EPI_ISL_2407190, EPI_ISL_2407191, EPI_ISL_2407192, EPI_ISL_2407193, EPI_ISL_2407194, EPI_ISL_2407195, EPI_ISL_2407196, EPI_ISL_2407197, EPI_ISL_2407198, EPI_ISL_2407199, EPI_ISL_2407200, EPI_ISL_2407201, EPI_ISL_2407202, EPI_ISL_2407203, EPI_ISL_2407204, EPI_ISL_2407205, EPI_ISL_2407206, EPI_ISL_2407207, EPI_ISL_2407209, EPI_ISL_2407210, EPI_ISL_2407211, EPI_ISL_2407212, EPI_ISL_2407213, EPI_ISL_2407214, EPI_ISL_2407215, EPI_ISL_2407216, EPI_ISL_2407217, | | | |

EPI_ISL_2407218, EPI_ISL_2407220, EPI_ISL_2407221, EPI_ISL_2407222, EPI_ISL_2407223, EPI_ISL_2407224, EPI_ISL_2407225, EPI_ISL_2407226, EPI_ISL_2407227, EPI_ISL_2407228, EPI_ISL_2407229, EPI_ISL_2407230, EPI_ISL_2407231, EPI_ISL_2407232, EPI_ISL_2407233, EPI_ISL_2407234, EPI_ISL_2407235, EPI_ISL_2407236, EPI_ISL_2407237, EPI_ISL_2407238, EPI_ISL_2407239, EPI_ISL_2407242,
EPI_ISL_2407244, EPI_ISL_2407245, EPI_ISL_2407246, EPI_ISL_2407247, EPI_ISL_2407248, EPI_ISL_2407250, EPI_ISL_2407251, EPI_ISL_2407252, EPI_ISL_2407253, EPI_ISL_2407254, EPI_ISL_2407256, EPI_ISL_2407257, EPI_ISL_2407258, EPI_ISL_2407260, EPI_ISL_2407262, EPI_ISL_2407263, EPI_ISL_2407264, EPI_ISL_2407265, EPI_ISL_2407266, EPI_ISL_2407267, EPI_ISL_2407268, EPI_ISL_2407269,
EPI_ISL_2407270, EPI_ISL_2407271, EPI_ISL_2407272, EPI_ISL_2407273, EPI_ISL_2407274, EPI_ISL_2407275, EPI_ISL_2407276, EPI_ISL_2407277, EPI_ISL_2407278, EPI_ISL_2407279, EPI_ISL_2407280, EPI_ISL_2407281, EPI_ISL_2407282, EPI_ISL_2407283, EPI_ISL_2407284, EPI_ISL_2407467, EPI_ISL_2407610, EPI_ISL_2407613, EPI_ISL_2407644, EPI_ISL_2407647, EPI_ISL_2434996, EPI_ISL_2435000,
EPI_ISL_2435005, EPI_ISL_2435024, EPI_ISL_2435047, EPI_ISL_2435070, EPI_ISL_2435080, EPI_ISL_2435084, EPI_ISL_2435085, EPI_ISL_2435098, EPI_ISL_2435103, EPI_ISL_2435104, EPI_ISL_2435111, EPI_ISL_2435123, EPI_ISL_2435127, EPI_ISL_2435133, EPI_ISL_2435143, EPI_ISL_2435157, EPI_ISL_2435174, EPI_ISL_2435195, EPI_ISL_2435205, EPI_ISL_2435211, EPI_ISL_2435228, EPI_ISL_2435244,
EPI_ISL_2435253, EPI_ISL_2435266, EPI_ISL_2435285, EPI_ISL_2435290, EPI_ISL_2435303, EPI_ISL_2435314, EPI_ISL_2435316, EPI_ISL_2435497, EPI_ISL_2435512, EPI_ISL_2435530, EPI_ISL_2435531, EPI_ISL_2435533, EPI_ISL_2435534, EPI_ISL_2435536, EPI_ISL_2435538, EPI_ISL_2435539, EPI_ISL_2435540, EPI_ISL_2435541, EPI_ISL_2435543, EPI_ISL_2435545, EPI_ISL_2435546, EPI_ISL_2435548,
EPI_ISL_2435550, EPI_ISL_2435551, EPI_ISL_2435554, EPI_ISL_2435555, EPI_ISL_2435557, EPI_ISL_2435561, EPI_ISL_2435562, EPI_ISL_2435564, EPI_ISL_2435567, EPI_ISL_2435568, EPI_ISL_2435569, EPI_ISL_2435570, EPI_ISL_2435571, EPI_ISL_2435572, EPI_ISL_2435573, EPI_ISL_2435574, EPI_ISL_2435575, EPI_ISL_2435576, EPI_ISL_2435577, EPI_ISL_2435578, EPI_ISL_2435579, EPI_ISL_2435581,
EPI_ISL_2435582, EPI_ISL_2435583, EPI_ISL_2435584, EPI_ISL_2435585, EPI_ISL_2435588, EPI_ISL_2435591, EPI_ISL_2435594, EPI_ISL_2435595, EPI_ISL_2435596, EPI_ISL_2435599, EPI_ISL_2435602, EPI_ISL_2435603, EPI_ISL_2435604, EPI_ISL_2435606, EPI_ISL_2435607, EPI_ISL_2435608, EPI_ISL_2435609, EPI_ISL_2435610, EPI_ISL_2435611, EPI_ISL_2435612, EPI_ISL_2435613, EPI_ISL_2435614,
EPI_ISL_2435615, EPI_ISL_2435616, EPI_ISL_2435618, EPI_ISL_2435619, EPI_ISL_2435620, EPI_ISL_2435622, EPI_ISL_2435623, EPI_ISL_2435624, EPI_ISL_2435625, EPI_ISL_2435626, EPI_ISL_2435627, EPI_ISL_2435629, EPI_ISL_2435630, EPI_ISL_2435631, EPI_ISL_2435632, EPI_ISL_2435633, EPI_ISL_2435634, EPI_ISL_2435636, EPI_ISL_2435637, EPI_ISL_2435638, EPI_ISL_2435639, EPI_ISL_2435640,
EPI_ISL_2435642, EPI_ISL_2435643, EPI_ISL_2435646, EPI_ISL_2435647, EPI_ISL_2435648, EPI_ISL_2435649, EPI_ISL_2435650, EPI_ISL_2435652, EPI_ISL_2435653, EPI_ISL_2435654, EPI_ISL_2435655, EPI_ISL_2435656, EPI_ISL_2435657, EPI_ISL_2435658, EPI_ISL_2435659, EPI_ISL_2435660, EPI_ISL_2435661, EPI_ISL_2435663, EPI_ISL_2435664, EPI_ISL_2435665, EPI_ISL_2435668, EPI_ISL_2435669,
EPI_ISL_2435670, EPI_ISL_2435672, EPI_ISL_2435673, EPI_ISL_2435674, EPI_ISL_2435676, EPI_ISL_2435679, EPI_ISL_2435680, EPI_ISL_2435681, EPI_ISL_2435682, EPI_ISL_2435683, EPI_ISL_2435684, EPI_ISL_2435685, EPI_ISL_2435686, EPI_ISL_2435687, EPI_ISL_2435688, EPI_ISL_2435689, EPI_ISL_2435691, EPI_ISL_2435694, EPI_ISL_2435695, EPI_ISL_2435696, EPI_ISL_2435698, EPI_ISL_2435699,
EPI_ISL_2435700, EPI_ISL_2435702, EPI_ISL_2435704, EPI_ISL_2435705, EPI_ISL_2435706, EPI_ISL_2435707, EPI_ISL_2435708, EPI_ISL_2435709, EPI_ISL_2435710, EPI_ISL_2435711, EPI_ISL_2435713, EPI_ISL_2435715, EPI_ISL_2435717, EPI_ISL_2435718, EPI_ISL_2435725, EPI_ISL_2435726, EPI_ISL_2435727, EPI_ISL_2435728, EPI_ISL_2435729, EPI_ISL_2435730, EPI_ISL_2435731, EPI_ISL_2435732,
EPI_ISL_2435734, EPI_ISL_2435735, EPI_ISL_2435737, EPI_ISL_2435738, EPI_ISL_2435739, EPI_ISL_2435741, EPI_ISL_2435742, EPI_ISL_2435743, EPI_ISL_2435744, EPI_ISL_2435745, EPI_ISL_2435746, EPI_ISL_2435751, EPI_ISL_2435752, EPI_ISL_2435754, EPI_ISL_2435755, EPI_ISL_2435756, EPI_ISL_2435757, EPI_ISL_2435760, EPI_ISL_2435761, EPI_ISL_2435763, EPI_ISL_2435764, EPI_ISL_2435765,
EPI_ISL_2435768, EPI_ISL_2435769, EPI_ISL_2435770, EPI_ISL_2435771, EPI_ISL_2435772, EPI_ISL_2435773, EPI_ISL_2435775, EPI_ISL_2435776, EPI_ISL_2435777, EPI_ISL_2435778, EPI_ISL_2435780, EPI_ISL_2435781, EPI_ISL_2435782, EPI_ISL_2435783, EPI_ISL_2435784, EPI_ISL_2435785, EPI_ISL_2435786, EPI_ISL_2435787, EPI_ISL_2435788, EPI_ISL_2435790, EPI_ISL_2435792, EPI_ISL_2435794,
EPI_ISL_2435795, EPI_ISL_2435796, EPI_ISL_2435797, EPI_ISL_2435799, EPI_ISL_2435802, EPI_ISL_2435803, EPI_ISL_2435804, EPI_ISL_2435806, EPI_ISL_2435808, EPI_ISL_2435809, EPI_ISL_2435810, EPI_ISL_2435811, EPI_ISL_2435813, EPI_ISL_2435816, EPI_ISL_2435817, EPI_ISL_2435820, EPI_ISL_2435821, EPI_ISL_2435822, EPI_ISL_2435823, EPI_ISL_2435824, EPI_ISL_2435825, EPI_ISL_2435826,
EPI_ISL_2435827, EPI_ISL_2435828, EPI_ISL_2435829, EPI_ISL_2435832, EPI_ISL_2435833, EPI_ISL_2435834, EPI_ISL_2435835, EPI_ISL_2435838, EPI_ISL_2435839, EPI_ISL_2435840, EPI_ISL_2435841, EPI_ISL_2435843, EPI_ISL_2435844, EPI_ISL_2435845, EPI_ISL_2435846, EPI_ISL_2435847, EPI_ISL_2435848, EPI_ISL_2435849, EPI_ISL_2435850, EPI_ISL_2435851, EPI_ISL_2435852, EPI_ISL_2435853,
EPI_ISL_2435854, EPI_ISL_2435855, EPI_ISL_2435857, EPI_ISL_2435858, EPI_ISL_2435860, EPI_ISL_2435861, EPI_ISL_2435864, EPI_ISL_2435865, EPI_ISL_2435867, EPI_ISL_2435869, EPI_ISL_2435870, EPI_ISL_2435871, EPI_ISL_2435872, EPI_ISL_2435873, EPI_ISL_2435874, EPI_ISL_2435876, EPI_ISL_2435877, EPI_ISL_2435878, EPI_ISL_2435879, EPI_ISL_2435881, EPI_ISL_2435883, EPI_ISL_2435884,
EPI_ISL_2435885, EPI_ISL_2435887, EPI_ISL_2435889, EPI_ISL_2435891, EPI_ISL_2435892, EPI_ISL_2435893, EPI_ISL_2435894, EPI_ISL_2435895, EPI_ISL_2435897, EPI_ISL_2435898, EPI_ISL_2435901, EPI_ISL_2435902, EPI_ISL_2435903, EPI_ISL_2435904, EPI_ISL_2435906, EPI_ISL_2435907, EPI_ISL_2435910, EPI_ISL_2435911, EPI_ISL_2435912, EPI_ISL_2435913, EPI_ISL_2435914, EPI_ISL_2435915,
EPI_ISL_2435916, EPI_ISL_2435917, EPI_ISL_2435919, EPI_ISL_2435920, EPI_ISL_2435921, EPI_ISL_2435922, EPI_ISL_2435924, EPI_ISL_2435925, EPI_ISL_2435927, EPI_ISL_2435931, EPI_ISL_2435932, EPI_ISL_2435933, EPI_ISL_2435934, EPI_ISL_2435939, EPI_ISL_2435942, EPI_ISL_2435943, EPI_ISL_2435945, EPI_ISL_2435946, EPI_ISL_2435947, EPI_ISL_2435948, EPI_ISL_2435950, EPI_ISL_2435951,
EPI_ISL_2435953, EPI_ISL_2435957, EPI_ISL_2435958, EPI_ISL_2435959, EPI_ISL_2435960, EPI_ISL_2435961, EPI_ISL_2435962, EPI_ISL_2435963, EPI_ISL_2435964, EPI_ISL_2435965, EPI_ISL_2435968, EPI_ISL_2435970, EPI_ISL_2435972, EPI_ISL_2435974, EPI_ISL_2435975, EPI_ISL_2435976, EPI_ISL_2435977, EPI_ISL_2435979, EPI_ISL_2435980, EPI_ISL_2435982, EPI_ISL_2435983, EPI_ISL_2435985,
EPI_ISL_2435987, EPI_ISL_2435988, EPI_ISL_2435992, EPI_ISL_2435993, EPI_ISL_2435994, EPI_ISL_2435995, EPI_ISL_2435996, EPI_ISL_2435997, EPI_ISL_2435998, EPI_ISL_2436000, EPI_ISL_2436001, EPI_ISL_2436004, EPI_ISL_2436006, EPI_ISL_2436007, EPI_ISL_2436008, EPI_ISL_2436010, EPI_ISL_2436011, EPI_ISL_2436012, EPI_ISL_2436013, EPI_ISL_2436014, EPI_ISL_2436015, EPI_ISL_2436016,
EPI_ISL_2436017, EPI_ISL_2436023, EPI_ISL_2436024, EPI_ISL_2436028, EPI_ISL_2436029, EPI_ISL_2436030, EPI_ISL_2436031, EPI_ISL_2436032, EPI_ISL_2436033, EPI_ISL_2436035, EPI_ISL_2436036, EPI_ISL_2436037, EPI_ISL_2436038, EPI_ISL_2436039, EPI_ISL_2436040, EPI_ISL_2436041, EPI_ISL_2436042, EPI_ISL_2436043, EPI_ISL_2436044, EPI_ISL_2436045, EPI_ISL_2436047, EPI_ISL_2436048,
EPI_ISL_2436049, EPI_ISL_2436050, EPI_ISL_2436051, EPI_ISL_2436052, EPI_ISL_2436055, EPI_ISL_2436056, EPI_ISL_2436057, EPI_ISL_2436058, EPI_ISL_2436059, EPI_ISL_2436064, EPI_ISL_2436065, EPI_ISL_2436066, EPI_ISL_2436067, EPI_ISL_2436068, EPI_ISL_2436069, EPI_ISL_2436070, EPI_ISL_2436071, EPI_ISL_2436073, EPI_ISL_2436075, EPI_ISL_2436076, EPI_ISL_2436077, EPI_ISL_2436078,
EPI_ISL_2436080, EPI_ISL_2436082, EPI_ISL_2436083, EPI_ISL_2436084, EPI_ISL_2436085, EPI_ISL_2436087, EPI_ISL_2436088, EPI_ISL_2436090, EPI_ISL_2436093, EPI_ISL_2436094, EPI_ISL_2436095, EPI_ISL_2436096, EPI_ISL_2436098, EPI_ISL_2436099, EPI_ISL_2436100, EPI_ISL_2436103, EPI_ISL_2436104, EPI_ISL_2436105, EPI_ISL_2436106, EPI_ISL_2436108, EPI_ISL_2436109, EPI_ISL_2436110,
EPI_ISL_2436111, EPI_ISL_2436112, EPI_ISL_2436115, EPI_ISL_2436117, EPI_ISL_2436118, EPI_ISL_2436120, EPI_ISL_2436121, EPI_ISL_2436122, EPI_ISL_2436125, EPI_ISL_2436127, EPI_ISL_2436128, EPI_ISL_2436131, EPI_ISL_2436133, EPI_ISL_2436134, EPI_ISL_2436135, EPI_ISL_2436138, EPI_ISL_2436141, EPI_ISL_2436142, EPI_ISL_2436143, EPI_ISL_2436144, EPI_ISL_2436148, EPI_ISL_2436149,
EPI_ISL_2436150, EPI_ISL_2436151, EPI_ISL_2436153, EPI_ISL_2436155, EPI_ISL_2436156, EPI_ISL_2436157, EPI_ISL_2436159, EPI_ISL_2436160, EPI_ISL_2436162, EPI_ISL_2436163, EPI_ISL_2436164, EPI_ISL_2436166, EPI_ISL_2436167, EPI_ISL_2436168, EPI_ISL_2436169, EPI_ISL_2436170, EPI_ISL_2436172, EPI_ISL_2436173, EPI_ISL_2436174, EPI_ISL_2436177, EPI_ISL_2436178, EPI_ISL_2436180,
EPI_ISL_2436183, EPI_ISL_2436185, EPI_ISL_2436186, EPI_ISL_2436187, EPI_ISL_2436188, EPI_ISL_2436190, EPI_ISL_2436191, EPI_ISL_2436192, EPI_ISL_2436193, EPI_ISL_2436194, EPI_ISL_2436195, EPI_ISL_2436197, EPI_ISL_2436200, EPI_ISL_2436202, EPI_ISL_2436207, EPI_ISL_2436208, EPI_ISL_2436210, EPI_ISL_2436211, EPI_ISL_2436212, EPI_ISL_2436213, EPI_ISL_2436214, EPI_ISL_2436217,
EPI_ISL_2436218, EPI_ISL_2436219, EPI_ISL_2436220, EPI_ISL_2436224, EPI_ISL_2436225, EPI_ISL_2436243, EPI_ISL_2436252, EPI_ISL_2436254, EPI_ISL_2436265, EPI_ISL_2436292, EPI_ISL_2436314, EPI_ISL_2436329, EPI_ISL_2436332, EPI_ISL_2436362, EPI_ISL_2436364, EPI_ISL_2436371, EPI_ISL_2436379, EPI_ISL_2436380, EPI_ISL_2436399, EPI_ISL_2436426, EPI_ISL_2436437, EPI_ISL_2436446,
EPI_ISL_2436451, EPI_ISL_2436453, EPI_ISL_2436476, EPI_ISL_2436479, EPI_ISL_2436504, EPI_ISL_2436505, EPI_ISL_2436516, EPI_ISL_2436524, EPI_ISL_2436533, EPI_ISL_2436542, EPI_ISL_2436543, EPI_ISL_2436555, EPI_ISL_2436562, EPI_ISL_2436566, EPI_ISL_2436573, EPI_ISL_2436575, EPI_ISL_2436579, EPI_ISL_2436581, EPI_ISL_2436585, EPI_ISL_2436596, EPI_ISL_2436603, EPI_ISL_2436604,
EPI_ISL_2436605, EPI_ISL_2436611, EPI_ISL_2436614, EPI_ISL_2436619, EPI_ISL_2436621, EPI_ISL_2436627, EPI_ISL_2436629, EPI_ISL_2436632, EPI_ISL_2436633, EPI_ISL_2436661, EPI_ISL_2436666, EPI_ISL_2436681, EPI_ISL_2436685, EPI_ISL_2436687, EPI_ISL_2436690, EPI_ISL_2436693, EPI_ISL_2436701, EPI_ISL_2436705, EPI_ISL_2436718, EPI_ISL_2436719, EPI_ISL_2436742, EPI_ISL_2436749,
EPI_ISL_2436754, EPI_ISL_2436755, EPI_ISL_2436756, EPI_ISL_2436763, EPI_ISL_2436774, EPI_ISL_2436775, EPI_ISL_2436780, EPI_ISL_2436782, EPI_ISL_2436786, EPI_ISL_2436788, EPI_ISL_2436801, EPI_ISL_2436803, EPI_ISL_2436807, EPI_ISL_2436812, EPI_ISL_2436815, EPI_ISL_2436821, EPI_ISL_2436822, EPI_ISL_2436825, EPI_ISL_2436826, EPI_ISL_2436831, EPI_ISL_2436841, EPI_ISL_2436842,
EPI_ISL_2436847, EPI_ISL_2436849, EPI_ISL_2436861, EPI_ISL_2436863, EPI_ISL_2436866, EPI_ISL_2436870, EPI_ISL_2436884, EPI_ISL_2436892, EPI_ISL_2436895, EPI_ISL_2436898, EPI_ISL_2436900, EPI_ISL_2436911, EPI_ISL_2436912, EPI_ISL_2436916, EPI_ISL_2436918, EPI_ISL_2436919, EPI_ISL_2436920, EPI_ISL_2436923, EPI_ISL_2437277, EPI_ISL_2437279, EPI_ISL_2437281, EPI_ISL_2437284,
EPI_ISL_2437285, EPI_ISL_2437286, EPI_ISL_2437287, EPI_ISL_2437288, EPI_ISL_2437289, EPI_ISL_2437292, EPI_ISL_2437293, EPI_ISL_2437297, EPI_ISL_2437302, EPI_ISL_2437306, EPI_ISL_2437307, EPI_ISL_2437309, EPI_ISL_2437310, EPI_ISL_2437311, EPI_ISL_2437314, EPI_ISL_2437318, EPI_ISL_2437319, EPI_ISL_2437320, EPI_ISL_2437321, EPI_ISL_2437324, EPI_ISL_2437327, EPI_ISL_2437328,
EPI_ISL_2437333, EPI_ISL_2437334, EPI_ISL_2437336, EPI_ISL_2437338, EPI_ISL_2437339, EPI_ISL_2437340, EPI_ISL_2437343, EPI_ISL_2437346, EPI_ISL_2437348, EPI_ISL_2437349, EPI_ISL_2437350, EPI_ISL_2437357, EPI_ISL_2437358, EPI_ISL_2437359, EPI_ISL_2437361, EPI_ISL_2437363, EPI_ISL_2437364, EPI_ISL_2437365, EPI_ISL_2437366, EPI_ISL_2437367, EPI_ISL_2437368, EPI_ISL_2437369,
EPI_ISL_2437372, EPI_ISL_2437373, EPI_ISL_2437374, EPI_ISL_2437376, EPI_ISL_2437377, EPI_ISL_2437380, EPI_ISL_2437381, EPI_ISL_2437383, EPI_ISL_2437384, EPI_ISL_2437385, EPI_ISL_2437386, EPI_ISL_2437387, EPI_ISL_2437390, EPI_ISL_2437391, EPI_ISL_2437392, EPI_ISL_2437393, EPI_ISL_2437394, EPI_ISL_2437395, EPI_ISL_2437396, EPI_ISL_2437397, EPI_ISL_2437401, EPI_ISL_2437406,
EPI_ISL_2437410, EPI_ISL_2437411, EPI_ISL_2437413, EPI_ISL_2437414, EPI_ISL_2437416, EPI_ISL_2437417, EPI_ISL_2437418, EPI_ISL_2437419, EPI_ISL_2437420, EPI_ISL_2437421, EPI_ISL_2437425, EPI_ISL_2437428, EPI_ISL_2437429, EPI_ISL_2437434, EPI_ISL_2437438, EPI_ISL_2437441, EPI_ISL_2437444, EPI_ISL_2437445, EPI_ISL_2437446, EPI_ISL_2437449, EPI_ISL_2437450, EPI_ISL_2437451,
EPI_ISL_2437454, EPI_ISL_2437460, EPI_ISL_2437461, EPI_ISL_2437464, EPI_ISL_2437465, EPI_ISL_2437469, EPI_ISL_2437476, EPI_ISL_2437478, EPI_ISL_2437480, EPI_ISL_2437482, EPI_ISL_2437483, EPI_ISL_2437486, EPI_ISL_2437487, EPI_ISL_2437492, EPI_ISL_2437493, EPI_ISL_2437495, EPI_ISL_2437496, EPI_ISL_2437499, EPI_ISL_2437500, EPI_ISL_2437501, EPI_ISL_2437502, EPI_ISL_2437504,
EPI_ISL_2437505, EPI_ISL_2437506, EPI_ISL_2437507, EPI_ISL_2437508, EPI_ISL_2437509, EPI_ISL_2437510, EPI_ISL_2437511, EPI_ISL_2437512, EPI_ISL_2437514, EPI_ISL_2437515, EPI_ISL_2437517, EPI_ISL_2437518, EPI_ISL_2437520, EPI_ISL_2437522, EPI_ISL_2437523, EPI_ISL_2437526, EPI_ISL_2437527, EPI_ISL_2437528, EPI_ISL_2437529, EPI_ISL_2437530, EPI_ISL_2437531, EPI_ISL_2437534,
EPI_ISL_2437535, EPI_ISL_2437537, EPI_ISL_2437538, EPI_ISL_2437541, EPI_ISL_2437542, EPI_ISL_2437543, EPI_ISL_2437544, EPI_ISL_2437550, EPI_ISL_2437551, EPI_ISL_2437554, EPI_ISL_2437555, EPI_ISL_2437558, EPI_ISL_2437560, EPI_ISL_2437562, EPI_ISL_2437567, EPI_ISL_2437568, EPI_ISL_2437569, EPI_ISL_2437570, EPI_ISL_2437574, EPI_ISL_2437577, EPI_ISL_2437579, EPI_ISL_2437580,
EPI_ISL_2437581, EPI_ISL_2437582, EPI_ISL_2437587, EPI_ISL_2437591, EPI_ISL_2437592, EPI_ISL_2437593, EPI_ISL_2437595, EPI_ISL_2437597, EPI_ISL_2437598, EPI_ISL_2437599, EPI_ISL_2437601, EPI_ISL_2437602, EPI_ISL_2437605, EPI_ISL_2437606, EPI_ISL_2437608, EPI_ISL_2437609, EPI_ISL_2437612, EPI_ISL_2437613, EPI_ISL_2437614, EPI_ISL_2437617, EPI_ISL_2437621, EPI_ISL_2437622,
EPI_ISL_2437630, EPI_ISL_2437632, EPI_ISL_2437637, EPI_ISL_2437639, EPI_ISL_2437641, EPI_ISL_2437642, EPI_ISL_2437644, EPI_ISL_2437649, EPI_ISL_2437651, EPI_ISL_2437654, EPI_ISL_2437668, EPI_ISL_2437671, EPI_ISL_2437675, EPI_ISL_2437676, EPI_ISL_2437682, EPI_ISL_2437686, EPI_ISL_2437703, EPI_ISL_2437714, EPI_ISL_2437718, EPI_ISL_2437720, EPI_ISL_2437721, EPI_ISL_2437722,
EPI_ISL_2437727, EPI_ISL_2437731, EPI_ISL_2437735, EPI_ISL_2437738, EPI_ISL_2437743, EPI_ISL_2437756, EPI_ISL_2437758, EPI_ISL_2437759, EPI_ISL_2437769, EPI_ISL_2437774, EPI_ISL_2437776, EPI_ISL_2437779, EPI_ISL_2437786, EPI_ISL_2437787, EPI_ISL_2437793, EPI_ISL_2437800, EPI_ISL_2437807, EPI_ISL_2437810, EPI_ISL_2437817, EPI_ISL_2437820, EPI_ISL_2437824, EPI_ISL_2437826,
EPI_ISL_2437827, EPI_ISL_2437828, EPI_ISL_2437829, EPI_ISL_2437831, EPI_ISL_2437833, EPI_ISL_2437834, EPI_ISL_2437837, EPI_ISL_2437839, EPI_ISL_2437841, EPI_ISL_2437847, EPI_ISL_2437851, EPI_ISL_2437858, EPI_ISL_2437859, EPI_ISL_2437866, EPI_ISL_2437867, EPI_ISL_2437871, EPI_ISL_2437875, EPI_ISL_2437877, EPI_ISL_2437882, EPI_ISL_2437889, EPI_ISL_2437895, EPI_ISL_2437896,
EPI_ISL_2437898, EPI_ISL_2437899, EPI_ISL_2437900, EPI_ISL_2437902, EPI_ISL_2437906, EPI_ISL_2437912, EPI_ISL_2437916, EPI_ISL_2437918, EPI_ISL_2437919, EPI_ISL_2437921, EPI_ISL_2437925, EPI_ISL_2437927, EPI_ISL_2437929, EPI_ISL_2437932, EPI_ISL_2437935, EPI_ISL_2437936, EPI_ISL_2437938, EPI_ISL_2437941, EPI_ISL_2437942, EPI_ISL_2437944, EPI_ISL_2437961, EPI_ISL_2437976,
EPI_ISL_2437978, EPI_ISL_2437983, EPI_ISL_2437987, EPI_ISL_2437989, EPI_ISL_2438051, EPI_ISL_2438053, EPI_ISL_2438056, EPI_ISL_2438058, EPI_ISL_2438085, EPI_ISL_2438095, EPI_ISL_2438096, EPI_ISL_2438099, EPI_ISL_2438108, EPI_ISL_2438109, EPI_ISL_2438113, EPI_ISL_2438116, EPI_ISL_2438118, EPI_ISL_2438131, EPI_ISL_2438156, EPI_ISL_2438164, EPI_ISL_2438171, EPI_ISL_2438182,
EPI_ISL_2438198, EPI_ISL_2438205, EPI_ISL_2438207, EPI_ISL_2438217, EPI_ISL_2438218, EPI_ISL_2438230, EPI_ISL_2438235, EPI_ISL_2438246, EPI_ISL_2438269, EPI_ISL_2438282, EPI_ISL_2438289, EPI_ISL_2438293, EPI_ISL_2438298, EPI_ISL_2438301, EPI_ISL_2438305, EPI_ISL_2438310, EPI_ISL_2438324, EPI_ISL_2438338, EPI_ISL_2438346, EPI_ISL_2438352, EPI_ISL_2438363, EPI_ISL_2438372,
EPI_ISL_2438374, EPI_ISL_2438519, EPI_ISL_2446137, EPI_ISL_2446231, EPI_ISL_2446306, EPI_ISL_2446318, EPI_ISL_2446362, EPI_ISL_2446412, EPI_ISL_2446413, EPI_ISL_2446415, EPI_ISL_2446419, EPI_ISL_2446420, EPI_ISL_2446421, EPI_ISL_2446422, EPI_ISL_2446423, EPI_ISL_2446424, EPI_ISL_2446426, EPI_ISL_2446427, EPI_ISL_2446429, EPI_ISL_2446430, EPI_ISL_2446431, EPI_ISL_2446432,
EPI_ISL_2446434, EPI_ISL_2446435, EPI_ISL_2446439, EPI_ISL_2446440, EPI_ISL_2446441, EPI_ISL_2446443, EPI_ISL_2446444, EPI_ISL_2446445, EPI_ISL_2446446, EPI_ISL_2446447, EPI_ISL_2446448, EPI_ISL_2446449, EPI_ISL_2446452, EPI_ISL_2446454, EPI_ISL_2446455, EPI_ISL_2446457, EPI_ISL_2446462, EPI_ISL_2446464, EPI_ISL_2446467, EPI_ISL_2446468, EPI_ISL_2446469, EPI_ISL_2446470,
EPI_ISL_2446471, EPI_ISL_2446474, EPI_ISL_2446476, EPI_ISL_2446479, EPI_ISL_2446482, EPI_ISL_2446483, EPI_ISL_2446484, EPI_ISL_2446485, EPI_ISL_2446486, EPI_ISL_2446487, EPI_ISL_2446488, EPI_ISL_2446489, EPI_ISL_2446490, EPI_ISL_2446492, EPI_ISL_2446493, EPI_ISL_2446494, EPI_ISL_2446496, EPI_ISL_2446497, EPI_ISL_2446501, EPI_ISL_2446502, EPI_ISL_2446504, EPI_ISL_2446508,
EPI_ISL_2446509, EPI_ISL_2446510, EPI_ISL_2446511, EPI_ISL_2446515, EPI_ISL_2446516, EPI_ISL_2446517, EPI_ISL_2446522, EPI_ISL_2446523, EPI_ISL_2446525, EPI_ISL_2446530, EPI_ISL_2446532, EPI_ISL_2446535, EPI_ISL_2446536, EPI_ISL_2446540, EPI_ISL_2446541, EPI_ISL_2446543, EPI_ISL_2446544, EPI_ISL_2446545, EPI_ISL_2446547, EPI_ISL_2446548, EPI_ISL_2446549, EPI_ISL_2446551,
EPI_ISL_2446552, EPI_ISL_2446553, EPI_ISL_2446554, EPI_ISL_2446560, EPI_ISL_2446565, EPI_ISL_2446566, EPI_ISL_2446567, EPI_ISL_2446568, EPI_ISL_2446571, EPI_ISL_2446575, EPI_ISL_2446576, EPI_ISL_2446577, EPI_ISL_2446579, EPI_ISL_2446580, EPI_ISL_2446581, EPI_ISL_2446582, EPI_ISL_2446584, EPI_ISL_2446585, EPI_ISL_2446586, EPI_ISL_2446587, EPI_ISL_2446591, EPI_ISL_2446592,
EPI_ISL_2446593, EPI_ISL_2446594, EPI_ISL_2446595, EPI_ISL_2446597, EPI_ISL_2446599, EPI_ISL_2446602, EPI_ISL_2446604, EPI_ISL_2446606, EPI_ISL_2446609, EPI_ISL_2446611, EPI_ISL_2446612, EPI_ISL_2446613, EPI_ISL_2446614, EPI_ISL_2446615, EPI_ISL_2446617, EPI_ISL_2446618, EPI_ISL_2446621, EPI_ISL_2446622, EPI_ISL_2446623, EPI_ISL_2446624, EPI_ISL_2446626, EPI_ISL_2446627,
EPI_ISL_2446631, EPI_ISL_2446633, EPI_ISL_2446634, EPI_ISL_2446635, EPI_ISL_2446636, EPI_ISL_2446637, EPI_ISL_2446639, EPI_ISL_2446641, EPI_ISL_2446644, EPI_ISL_2446647, EPI_ISL_2446649, EPI_ISL_2446650, EPI_ISL_2446651, EPI_ISL_2446652, EPI_ISL_2446654, EPI_ISL_2446655, EPI_ISL_2446656, EPI_ISL_2446661, EPI_ISL_2446663, EPI_ISL_2446664, EPI_ISL_2446666, EPI_ISL_2446668,
EPI_ISL_2446669, EPI_ISL_2446671, EPI_ISL_2446672, EPI_ISL_2446674, EPI_ISL_2446675, EPI_ISL_2446679, EPI_ISL_2446682, EPI_ISL_2446683, EPI_ISL_2446684, EPI_ISL_2446685, EPI_ISL_2446688, EPI_ISL_2446691, EPI_ISL_2446693, EPI_ISL_2446695, EPI_ISL_2446696, EPI_ISL_2446697, EPI_ISL_2446698, EPI_ISL_2446699, EPI_ISL_2446701, EPI_ISL_2446702, EPI_ISL_2446703, EPI_ISL_2446704,
EPI_ISL_2446705, EPI_ISL_2446708, EPI_ISL_2446710, EPI_ISL_2446711, EPI_ISL_2446712, EPI_ISL_2446714, EPI_ISL_2446715, EPI_ISL_2446716, EPI_ISL_2446717, EPI_ISL_2446718, EPI_ISL_2446720, EPI_ISL_2446721, EPI_ISL_2446726, EPI_ISL_2446728, EPI_ISL_2446730, EPI_ISL_2446732, EPI_ISL_2446735, EPI_ISL_2446737, EPI_ISL_2446738, EPI_ISL_2446740, EPI_ISL_2446741, EPI_ISL_2446743,
EPI_ISL_2446746, EPI_ISL_2446748, EPI_ISL_2446758, EPI_ISL_2446759, EPI_ISL_2446761, EPI_ISL_2446777, EPI_ISL_2446785, EPI_ISL_2446787, EPI_ISL_2446812, EPI_ISL_2446822, EPI_ISL_2446832, EPI_ISL_2446837, EPI_ISL_2446841, EPI_ISL_2446850, EPI_ISL_2446851, EPI_ISL_2446859, EPI_ISL_2446875, EPI_ISL_2446882, EPI_ISL_2446904, EPI_ISL_2446912, EPI_ISL_2446913, EPI_ISL_2446929,
EPI_ISL_2446941, EPI_ISL_2446950, EPI_ISL_2446957, EPI_ISL_2446958, EPI_ISL_2446961, EPI_ISL_2446974, EPI_ISL_2446977, EPI_ISL_2447005, EPI_ISL_2447008, EPI_ISL_2447377, EPI_ISL_2447378, EPI_ISL_2447379, EPI_ISL_2447391, EPI_ISL_2447427, EPI_ISL_2447429, EPI_ISL_2447451, EPI_ISL_2447454, EPI_ISL_2447516, EPI_ISL_2447518, EPI_ISL_2447525, EPI_ISL_2447581, EPI_ISL_2447595,
EPI_ISL_2447637, EPI_ISL_2447725, EPI_ISL_2447752, EPI_ISL_2447756, EPI_ISL_2447760, EPI_ISL_2447761, EPI_ISL_2447762, EPI_ISL_2456833, EPI_ISL_2456834, EPI_ISL_2456837, EPI_ISL_2456839, EPI_ISL_2456847, EPI_ISL_2456854, EPI_ISL_2456855, EPI_ISL_2456856, EPI_ISL_2456858, EPI_ISL_2456864, EPI_ISL_2456865, EPI_ISL_2456867, EPI_ISL_2456868, EPI_ISL_2456872, EPI_ISL_2456874,
EPI_ISL_2456875, EPI_ISL_2456880, EPI_ISL_2456881, EPI_ISL_2456884, EPI_ISL_2456885, EPI_ISL_2456887, EPI_ISL_2456890, EPI_ISL_2456891, EPI_ISL_2456892, EPI_ISL_2456897, EPI_ISL_2456900, EPI_ISL_2456902, EPI_ISL_2456903, EPI_ISL_2456904, EPI_ISL_2456905, EPI_ISL_2456907, EPI_ISL_2456910, EPI_ISL_2456912, EPI_ISL_2456913, EPI_ISL_2456923, EPI_ISL_2456929, EPI_ISL_2456931,
EPI_ISL_2456933, EPI_ISL_2456935, EPI_ISL_2456936, EPI_ISL_2456937, EPI_ISL_2456940, EPI_ISL_2456943, EPI_ISL_2456944, EPI_ISL_2456945, EPI_ISL_2456946, EPI_ISL_2456949, EPI_ISL_2456950, EPI_ISL_2456953, EPI_ISL_2456954, EPI_ISL_2456957, EPI_ISL_2456958, EPI_ISL_2456959, EPI_ISL_2456961, EPI_ISL_2456963, EPI_ISL_2456966, EPI_ISL_2456972, EPI_ISL_2456974, EPI_ISL_2456977,
EPI_ISL_2456978, EPI_ISL_2456984, EPI_ISL_2456987, EPI_ISL_2456989, EPI_ISL_2456993, EPI_ISL_2456995, EPI_ISL_2456999, EPI_ISL_2457000, EPI_ISL_2457004, EPI_ISL_2457005, EPI_ISL_2457006, EPI_ISL_2457008, EPI_ISL_2457010, EPI_ISL_2457012, EPI_ISL_2457013, EPI_ISL_2457015, EPI_ISL_2457018, EPI_ISL_2457025, EPI_ISL_2457026, EPI_ISL_2457027, EPI_ISL_2457028, EPI_ISL_2457031,
EPI_ISL_2457032, EPI_ISL_2457038, EPI_ISL_2457039, EPI_ISL_2457042, EPI_ISL_2457049, EPI_ISL_2457051, EPI_ISL_2457052, EPI_ISL_2457056, EPI_ISL_2457066, EPI_ISL_2457067, EPI_ISL_2457068, EPI_ISL_2457073, EPI_ISL_2457074, EPI_ISL_2457075, EPI_ISL_2457076, EPI_ISL_2457081, EPI_ISL_2457083, EPI_ISL_2457086, EPI_ISL_2457088, EPI_ISL_2457093, EPI_ISL_2457095, EPI_ISL_2457098,
EPI_ISL_2457101, EPI_ISL_2457111, EPI_ISL_2457113, EPI_ISL_2457116, EPI_ISL_2457117, EPI_ISL_2457119, EPI_ISL_2457120, EPI_ISL_2457121, EPI_ISL_2457504, EPI_ISL_2457506, EPI_ISL_2469465, EPI_ISL_2469468, EPI_ISL_2485285, EPI_ISL_2485469, EPI_ISL_2485489, EPI_ISL_2513612, EPI_ISL_2513615, EPI_ISL_2513618, EPI_ISL_2513621, EPI_ISL_2513622, EPI_ISL_2513623, EPI_ISL_2513625,
EPI_ISL_2513626, EPI_ISL_2513628, EPI_ISL_2513629, EPI_ISL_2513631, EPI_ISL_2513635, EPI_ISL_2513646, EPI_ISL_2513648, EPI_ISL_2513649, EPI_ISL_2513653, EPI_ISL_2513656, EPI_ISL_2513658, EPI_ISL_2513659, EPI_ISL_2513663, EPI_ISL_2513664, EPI_ISL_2513667, EPI_ISL_2513668, EPI_ISL_2513670, EPI_ISL_2513671, EPI_ISL_2513675, EPI_ISL_2513677, EPI_ISL_2513679, EPI_ISL_2513681,
EPI_ISL_2513683, EPI_ISL_2513685, EPI_ISL_2513687, EPI_ISL_2513688, EPI_ISL_2513690, EPI_ISL_2513693, EPI_ISL_2513697, EPI_ISL_2513699, EPI_ISL_2513700, EPI_ISL_2513701, EPI_ISL_2513703, EPI_ISL_2513704, EPI_ISL_2513705, EPI_ISL_2513709, EPI_ISL_2513714, EPI_ISL_2513716, EPI_ISL_2513718, EPI_ISL_2513720, EPI_ISL_2513722, EPI_ISL_2513723, EPI_ISL_2513726,
EPI_ISL_2513727, EPI_ISL_2513736, EPI_ISL_2513738, EPI_ISL_2513739, EPI_ISL_2513740, EPI_ISL_2513741, EPI_ISL_2513746, EPI_ISL_2513747, EPI_ISL_2513750, EPI_ISL_2513752, EPI_ISL_2513754, EPI_ISL_2513757, EPI_ISL_2513758, EPI_ISL_2513759, EPI_ISL_2513762, EPI_ISL_2513763, EPI_ISL_2513764, EPI_ISL_2513765, EPI_ISL_2513766, EPI_ISL_2513768, EPI_ISL_2513770, EPI_ISL_2513774,
EPI_ISL_2513776, EPI_ISL_2513779, EPI_ISL_2513780, EPI_ISL_2513781, EPI_ISL_2513783, EPI_ISL_2513785, EPI_ISL_2513787, EPI_ISL_2513788, EPI_ISL_2513790, EPI_ISL_2513793, EPI_ISL_2513795, EPI_ISL_2513796, EPI_ISL_2513797, EPI_ISL_2513798, EPI_ISL_2513799, EPI_ISL_2515169, EPI_ISL_2515182, EPI_ISL_2515186, EPI_ISL_2515252, EPI_ISL_2515263, EPI_ISL_2515293,
EPI_ISL_2515309, EPI_ISL_2515317, EPI_ISL_2515332, EPI_ISL_2515338, EPI_ISL_2516044, EPI_ISL_2516201

| | | | |
|---|---|---|---|
| see above | Lighthouse Lab in Alderley Park | Wellcome Sanger Institute for the COVID-19 Genomics UK (COG-UK) Consortium | Cordelia Langford; David K. Jackson; Dominic Kwiatkowski; Ewan Harrison; Ian Johnston; Jacquelyn Wynn; Jeffrey Barrett; John Sillitoe on behalf of the Wellcome Sanger Institute COVID-19 Surveillance Team; Mairead Hyland; Roberto Amato; Sonia Goncalves; The Lighthouse Lab in Alderley Park and Alex Alderton |

EPI_ISL_2392389, EPI_ISL_2392579, EPI_ISL_2392582, EPI_ISL_2392593, EPI_ISL_2392610, EPI_ISL_2392626, EPI_ISL_2392639, EPI_ISL_2392640, EPI_ISL_2392664, EPI_ISL_2392666, EPI_ISL_2392672, EPI_ISL_2392673, EPI_ISL_2392678, EPI_ISL_2392685, EPI_ISL_2392695, EPI_ISL_2392710, EPI_ISL_2392716, EPI_ISL_2392724, EPI_ISL_2392732, EPI_ISL_2392772, EPI_ISL_2392785, EPI_ISL_2392798,
EPI_ISL_2392840, EPI_ISL_2392844, EPI_ISL_2392889, EPI_ISL_2392904, EPI_ISL_2392919, EPI_ISL_2392928, EPI_ISL_2392948, EPI_ISL_2392958, EPI_ISL_2392972, EPI_ISL_2392995, EPI_ISL_2393004, EPI_ISL_2393021, EPI_ISL_2393085, EPI_ISL_2393100, EPI_ISL_2393114, EPI_ISL_2393131, EPI_ISL_2393491, EPI_ISL_2393492, EPI_ISL_2393553, EPI_ISL_2393555, EPI_ISL_2393586, EPI_ISL_2393589,
EPI_ISL_2393597, EPI_ISL_2393611, EPI_ISL_2393655, EPI_ISL_2393663, EPI_ISL_2393678, EPI_ISL_2393683, EPI_ISL_2393693, EPI_ISL_2393701, EPI_ISL_2393729, EPI_ISL_2393773, EPI_ISL_2393798, EPI_ISL_2393824, EPI_ISL_2393831, EPI_ISL_2393838, EPI_ISL_2393847, EPI_ISL_2393899, EPI_ISL_2393902, EPI_ISL_2393926, EPI_ISL_2393942, EPI_ISL_2393945, EPI_ISL_2393958, EPI_ISL_2393965,
EPI_ISL_2393972, EPI_ISL_2393988, EPI_ISL_2393996, EPI_ISL_2393997, EPI_ISL_2394001, EPI_ISL_2394031, EPI_ISL_2394033, EPI_ISL_2394045, EPI_ISL_2394050, EPI_ISL_2394052, EPI_ISL_2394069, EPI_ISL_2394090, EPI_ISL_2394094, EPI_ISL_2394097, EPI_ISL_2394115, EPI_ISL_2394137, EPI_ISL_2394146, EPI_ISL_2394170, EPI_ISL_2394176, EPI_ISL_2394180, EPI_ISL_2394216, EPI_ISL_2394242,
EPI_ISL_2394246, EPI_ISL_2394247, EPI_ISL_2394256, EPI_ISL_2394260, EPI_ISL_2394272, EPI_ISL_2394282, EPI_ISL_2394303, EPI_ISL_2394312, EPI_ISL_2394316, EPI_ISL_2394321, EPI_ISL_2394322, EPI_ISL_2394323, EPI_ISL_2394327, EPI_ISL_2394328, EPI_ISL_2394334, EPI_ISL_2394336, EPI_ISL_2394341, EPI_ISL_2394342, EPI_ISL_2394346, EPI_ISL_2394350, EPI_ISL_2394353, EPI_ISL_2394354,
EPI_ISL_2394356, EPI_ISL_2394360, EPI_ISL_2394363, EPI_ISL_2394369, EPI_ISL_2394372, EPI_ISL_2394374, EPI_ISL_2394383, EPI_ISL_2394384, EPI_ISL_2394408, EPI_ISL_2394414, EPI_ISL_2394415, EPI_ISL_2394417, EPI_ISL_2394419, EPI_ISL_2394421, EPI_ISL_2394425, EPI_ISL_2394427, EPI_ISL_2394428, EPI_ISL_2394431, EPI_ISL_2394442, EPI_ISL_2394447, EPI_ISL_2394449, EPI_ISL_2394450,
EPI_ISL_2394452, EPI_ISL_2394454, EPI_ISL_2394459, EPI_ISL_2394462, EPI_ISL_2394463, EPI_ISL_2394465, EPI_ISL_2394469, EPI_ISL_2394470, EPI_ISL_2394473, EPI_ISL_2394474, EPI_ISL_2394481, EPI_ISL_2394484, EPI_ISL_2394488, EPI_ISL_2394489, EPI_ISL_2394490, EPI_ISL_2394492, EPI_ISL_2394493, EPI_ISL_2394499, EPI_ISL_2394509, EPI_ISL_2394511, EPI_ISL_2394519, EPI_ISL_2394520,
EPI_ISL_2394528, EPI_ISL_2394533, EPI_ISL_2394534, EPI_ISL_2394537, EPI_ISL_2394540, EPI_ISL_2394542, EPI_ISL_2394547, EPI_ISL_2394548, EPI_ISL_2394549, EPI_ISL_2394550, EPI_ISL_2394551, EPI_ISL_2394557, EPI_ISL_2394559, EPI_ISL_2394561, EPI_ISL_2394566, EPI_ISL_2394569, EPI_ISL_2394571, EPI_ISL_2394572, EPI_ISL_2394576, EPI_ISL_2394585, EPI_ISL_2394618, EPI_ISL_2394689,
EPI_ISL_2394700, EPI_ISL_2394707, EPI_ISL_2394710, EPI_ISL_2394742, EPI_ISL_2394762, EPI_ISL_2394767, EPI_ISL_2394780, EPI_ISL_2394803, EPI_ISL_2394846, EPI_ISL_2394856, EPI_ISL_2394878, EPI_ISL_2394892, EPI_ISL_2394893, EPI_ISL_2394919, EPI_ISL_2394975, EPI_ISL_2394991, EPI_ISL_2395015, EPI_ISL_2395043, EPI_ISL_2395044, EPI_ISL_2395073, EPI_ISL_2395082, EPI_ISL_2395109,
EPI_ISL_2395112, EPI_ISL_2395123, EPI_ISL_2395132, EPI_ISL_2395150, EPI_ISL_2395153, EPI_ISL_2395172, EPI_ISL_2395176, EPI_ISL_2395184, EPI_ISL_2395195, EPI_ISL_2395209, EPI_ISL_2395212, EPI_ISL_2395213, EPI_ISL_2395223, EPI_ISL_2395228, EPI_ISL_2395238, EPI_ISL_2395248, EPI_ISL_2395249, EPI_ISL_2395251, EPI_ISL_2395273, EPI_ISL_2395280, EPI_ISL_2395307, EPI_ISL_2395308,
EPI_ISL_2395309, EPI_ISL_2395318, EPI_ISL_2395319, EPI_ISL_2395337, EPI_ISL_2395340, EPI_ISL_2395344, EPI_ISL_2395348, EPI_ISL_2395350, EPI_ISL_2395362, EPI_ISL_2395365, EPI_ISL_2395374, EPI_ISL_2395377, EPI_ISL_2395378, EPI_ISL_2395385, EPI_ISL_2395386, EPI_ISL_2395388, EPI_ISL_2395393, EPI_ISL_2395394, EPI_ISL_2395398, EPI_ISL_2395399, EPI_ISL_2395401, EPI_ISL_2395403,
EPI_ISL_2395406, EPI_ISL_2395413, EPI_ISL_2395416, EPI_ISL_2395423, EPI_ISL_2395424, EPI_ISL_2395429, EPI_ISL_2395430, EPI_ISL_2395431, EPI_ISL_2395432, EPI_ISL_2395435, EPI_ISL_2395440, EPI_ISL_2395442, EPI_ISL_2395448, EPI_ISL_2395449, EPI_ISL_2395450, EPI_ISL_2395455, EPI_ISL_2395458, EPI_ISL_2395459, EPI_ISL_2395465, EPI_ISL_2395472, EPI_ISL_2395473, EPI_ISL_2395474,
EPI_ISL_2395481, EPI_ISL_2395492, EPI_ISL_2395504, EPI_ISL_2395509, EPI_ISL_2395510, EPI_ISL_2395515, EPI_ISL_2395517, EPI_ISL_2395526, EPI_ISL_2395527, EPI_ISL_2395528, EPI_ISL_2395531, EPI_ISL_2395537, EPI_ISL_2395538, EPI_ISL_2395539, EPI_ISL_2395544, EPI_ISL_2395547, EPI_ISL_2395555, EPI_ISL_2395558, EPI_ISL_2395562, EPI_ISL_2395566, EPI_ISL_2395580, EPI_ISL_2395586,
EPI_ISL_2395597, EPI_ISL_2395601, EPI_ISL_2395602, EPI_ISL_2395606, EPI_ISL_2395610, EPI_ISL_2395621, EPI_ISL_2395626, EPI_ISL_2395628, EPI_ISL_2395632, EPI_ISL_2395635, EPI_ISL_2395636, EPI_ISL_2395637, EPI_ISL_2395650, EPI_ISL_2395654, EPI_ISL_2395656, EPI_ISL_2395660, EPI_ISL_2395661, EPI_ISL_2395666, EPI_ISL_2395667, EPI_ISL_2395670, EPI_ISL_2395671, EPI_ISL_2395676,
EPI_ISL_2395678, EPI_ISL_2395680, EPI_ISL_2395681, EPI_ISL_2395712, EPI_ISL_2395743, EPI_ISL_2395752, EPI_ISL_2395827, EPI_ISL_2395860, EPI_ISL_2395867, EPI_ISL_2395912, EPI_ISL_2406590, EPI_ISL_2406591, EPI_ISL_2406592, EPI_ISL_2406593, EPI_ISL_2406594, EPI_ISL_2406595, EPI_ISL_2406596, EPI_ISL_2406597, EPI_ISL_2406598, EPI_ISL_2406599, EPI_ISL_2406600, EPI_ISL_2406601,
EPI_ISL_2406602, EPI_ISL_2406603, EPI_ISL_2406604, EPI_ISL_2406605, EPI_ISL_2406606, EPI_ISL_2406607, EPI_ISL_2406608, EPI_ISL_2406609, EPI_ISL_2406610, EPI_ISL_2406611, EPI_ISL_2406612, EPI_ISL_2406613, EPI_ISL_2406614, EPI_ISL_2406615, EPI_ISL_2406616, EPI_ISL_2406617, EPI_ISL_2406618, EPI_ISL_2406619, EPI_ISL_2406620, EPI_ISL_2406621, EPI_ISL_2406623, EPI_ISL_2406624,
EPI_ISL_2406625, EPI_ISL_2406626, EPI_ISL_2406627, EPI_ISL_2406628, EPI_ISL_2406629, EPI_ISL_2406630, EPI_ISL_2406631, EPI_ISL_2406632, EPI_ISL_2406633, EPI_ISL_2406634, EPI_ISL_2406635, EPI_ISL_2406636, EPI_ISL_2406637, EPI_ISL_2406638, EPI_ISL_2406639, EPI_ISL_2406640, EPI_ISL_2406641, EPI_ISL_2406642, EPI_ISL_2406643, EPI_ISL_2406644, EPI_ISL_2406645, EPI_ISL_2406646,
EPI_ISL_2406647, EPI_ISL_2406648, EPI_ISL_2406649, EPI_ISL_2406650, EPI_ISL_2406651, EPI_ISL_2406652, EPI_ISL_2406653, EPI_ISL_2406654, EPI_ISL_2406655, EPI_ISL_2406656, EPI_ISL_2406657, EPI_ISL_2406658, EPI_ISL_2406659, EPI_ISL_2406660, EPI_ISL_2406661, EPI_ISL_2406662, EPI_ISL_2406663, EPI_ISL_2406664, EPI_ISL_2406665, EPI_ISL_2406666, EPI_ISL_2406667, EPI_ISL_2406668,
EPI_ISL_2406669, EPI_ISL_2406670, EPI_ISL_2406671, EPI_ISL_2406672, EPI_ISL_2406673, EPI_ISL_2406674, EPI_ISL_2406675, EPI_ISL_2406676, EPI_ISL_2406677, EPI_ISL_2406678, EPI_ISL_2406679, EPI_ISL_2406680, EPI_ISL_2406681, EPI_ISL_2406682, EPI_ISL_2406683, EPI_ISL_2406684, EPI_ISL_2406685, EPI_ISL_2406686, EPI_ISL_2406687, EPI_ISL_2406688, EPI_ISL_2406689, EPI_ISL_2406690,
EPI_ISL_2406691, EPI_ISL_2406692, EPI_ISL_2406693, EPI_ISL_2406694, EPI_ISL_2406695, EPI_ISL_2406696, EPI_ISL_2406697, EPI_ISL_2406698, EPI_ISL_2406699, EPI_ISL_2406700, EPI_ISL_2406701, EPI_ISL_2406702, EPI_ISL_2406703, EPI_ISL_2406704, EPI_ISL_2406705, EPI_ISL_2406706, EPI_ISL_2406707, EPI_ISL_2406708, EPI_ISL_2406709, EPI_ISL_2406710, EPI_ISL_2406711, EPI_ISL_2406712,
EPI_ISL_2406713, EPI_ISL_2406714, EPI_ISL_2406715, EPI_ISL_2406716, EPI_ISL_2406717, EPI_ISL_2406718, EPI_ISL_2406719, EPI_ISL_2406720, EPI_ISL_2406721, EPI_ISL_2406722, EPI_ISL_2406723, EPI_ISL_2406724, EPI_ISL_2406725, EPI_ISL_2406726, EPI_ISL_2406727, EPI_ISL_2406728, EPI_ISL_2406729, EPI_ISL_2406730, EPI_ISL_2406731, EPI_ISL_2406732, EPI_ISL_2406733, EPI_ISL_2406734,
EPI_ISL_2406735, EPI_ISL_2406736, EPI_ISL_2406738, EPI_ISL_2406739, EPI_ISL_2406740, EPI_ISL_2406741, EPI_ISL_2406742, EPI_ISL_2406743, EPI_ISL_2406744, EPI_ISL_2406745, EPI_ISL_2406747, EPI_ISL_2406748, EPI_ISL_2406749, EPI_ISL_2406750, EPI_ISL_2406751, EPI_ISL_2406752, EPI_ISL_2406753, EPI_ISL_2406754, EPI_ISL_2406755, EPI_ISL_2406756, EPI_ISL_2406757,
EPI_ISL_2406758, EPI_ISL_2406759, EPI_ISL_2406760, EPI_ISL_2406761, EPI_ISL_2406762, EPI_ISL_2406763, EPI_ISL_2406764, EPI_ISL_2406765, EPI_ISL_2406766, EPI_ISL_2406767, EPI_ISL_2406768, EPI_ISL_2406769, EPI_ISL_2406770, EPI_ISL_2406771, EPI_ISL_2406772, EPI_ISL_2406773, EPI_ISL_2406774, EPI_ISL_2406775, EPI_ISL_2406776, EPI_ISL_2406777, EPI_ISL_2406778, EPI_ISL_2406779,
EPI_ISL_2406780, EPI_ISL_2406781, EPI_ISL_2406782, EPI_ISL_2406783, EPI_ISL_2406784, EPI_ISL_2406785, EPI_ISL_2406786, EPI_ISL_2406787, EPI_ISL_2406788, EPI_ISL_2406789, EPI_ISL_2406790, EPI_ISL_2406791, EPI_ISL_2406792, EPI_ISL_2406793, EPI_ISL_2406794, EPI_ISL_2406795, EPI_ISL_2406796, EPI_ISL_2406797, EPI_ISL_2406798, EPI_ISL_2406799, EPI_ISL_2406800, EPI_ISL_2406801,
EPI_ISL_2406802, EPI_ISL_2406803, EPI_ISL_2406804, EPI_ISL_2406805, EPI_ISL_2406806, EPI_ISL_2406807, EPI_ISL_2406808, EPI_ISL_2406809, EPI_ISL_2406810, EPI_ISL_2406811, EPI_ISL_2406812, EPI_ISL_2406813, EPI_ISL_2406814, EPI_ISL_2406815, EPI_ISL_2406816, EPI_ISL_2406817, EPI_ISL_2406818, EPI_ISL_2406819, EPI_ISL_2406820, EPI_ISL_2406821, EPI_ISL_2406822, EPI_ISL_2406823,
EPI_ISL_2406824, EPI_ISL_2406825, EPI_ISL_2406826, EPI_ISL_2406827, EPI_ISL_2406828, EPI_ISL_2406829, EPI_ISL_2406830, EPI_ISL_2406831, EPI_ISL_2406832, EPI_ISL_2406833, EPI_ISL_2406834, EPI_ISL_2406835, EPI_ISL_2406836, EPI_ISL_2406837, EPI_ISL_2406838, EPI_ISL_2406839, EPI_ISL_2406840, EPI_ISL_2406841, EPI_ISL_2406842, EPI_ISL_2406843, EPI_ISL_2406844, EPI_ISL_2406845,
EPI_ISL_2406846, EPI_ISL_2406847, EPI_ISL_2406848, EPI_ISL_2406849, EPI_ISL_2406850, EPI_ISL_2406851, EPI_ISL_2406852, EPI_ISL_2406853, EPI_ISL_2406854, EPI_ISL_2406855, EPI_ISL_2406856, EPI_ISL_2406858, EPI_ISL_2406859, EPI_ISL_2406860, EPI_ISL_2406861, EPI_ISL_2406862, EPI_ISL_2406863, EPI_ISL_2406864, EPI_ISL_2406865, EPI_ISL_2406866, EPI_ISL_2406867, EPI_ISL_2406868,
EPI_ISL_2406869, EPI_ISL_2406870, EPI_ISL_2406871, EPI_ISL_2406872, EPI_ISL_2406873, EPI_ISL_2406874, EPI_ISL_2406875, EPI_ISL_2406876, EPI_ISL_2406877, EPI_ISL_2406878, EPI_ISL_2406879, EPI_ISL_2406880, EPI_ISL_2406881, EPI_ISL_2406882, EPI_ISL_2406883, EPI_ISL_2406884, EPI_ISL_2406885, EPI_ISL_2406886, EPI_ISL_2406887, EPI_ISL_2406889, EPI_ISL_2406890,
EPI_ISL_2406891, EPI_ISL_2406892, EPI_ISL_2406893, EPI_ISL_2406894, EPI_ISL_2406898, EPI_ISL_2406909, EPI_ISL_2406912, EPI_ISL_2406916, EPI_ISL_2406922, EPI_ISL_2406938, EPI_ISL_2406940, EPI_ISL_2406942, EPI_ISL_2406948, EPI_ISL_2406952, EPI_ISL_2406957, EPI_ISL_2406959, EPI_ISL_2406966, EPI_ISL_2406974, EPI_ISL_2406978, EPI_ISL_2406993, EPI_ISL_2406995, EPI_ISL_2406997, EPI_ISL_2406999,
EPI_ISL_2407001, EPI_ISL_2407017, EPI_ISL_2407019, EPI_ISL_2407022, EPI_ISL_2407023, EPI_ISL_2407026, EPI_ISL_2407027, EPI_ISL_2407028, EPI_ISL_2407034, EPI_ISL_2407039, EPI_ISL_2407044, EPI_ISL_2407050, EPI_ISL_2407056, EPI_ISL_2407057, EPI_ISL_2407061, EPI_ISL_2407063, EPI_ISL_2407072, EPI_ISL_2407076, EPI_ISL_2407085, EPI_ISL_2407088, EPI_ISL_2407089,
EPI_ISL_2407093, EPI_ISL_2407112, EPI_ISL_2407113, EPI_ISL_2407114, EPI_ISL_2407129, EPI_ISL_2407133, EPI_ISL_2407134, EPI_ISL_2407149, EPI_ISL_2407150, EPI_ISL_2407161, EPI_ISL_2407174, EPI_ISL_2407176, EPI_ISL_2407208, EPI_ISL_2407219, EPI_ISL_2407240, EPI_ISL_2407241, EPI_ISL_2407243, EPI_ISL_2407249, EPI_ISL_2407255, EPI_ISL_2407259, EPI_ISL_2407261, EPI_ISL_2407285,
EPI_ISL_2407286, EPI_ISL_2407287, EPI_ISL_2407288, EPI_ISL_2407289, EPI_ISL_2407290, EPI_ISL_2407291, EPI_ISL_2407292, EPI_ISL_2407293, EPI_ISL_2407294, EPI_ISL_2407295, EPI_ISL_2407296, EPI_ISL_2407297, EPI_ISL_2407298, EPI_ISL_2407299, EPI_ISL_2407300, EPI_ISL_2407301, EPI_ISL_2407303, EPI_ISL_2407304, EPI_ISL_2407305, EPI_ISL_2407306, EPI_ISL_2407307, EPI_ISL_2407308,
EPI_ISL_2407309, EPI_ISL_2407310, EPI_ISL_2407311, EPI_ISL_2407312, EPI_ISL_2407313, EPI_ISL_2407314, EPI_ISL_2407315, EPI_ISL_2407316, EPI_ISL_2407318, EPI_ISL_2407319, EPI_ISL_2407321, EPI_ISL_2407322, EPI_ISL_2407323, EPI_ISL_2407324, EPI_ISL_2407325, EPI_ISL_2407326, EPI_ISL_2407328, EPI_ISL_2407329, EPI_ISL_2407330, EPI_ISL_2407331, EPI_ISL_2407332, EPI_ISL_2407333,
EPI_ISL_2407334, EPI_ISL_2407335, EPI_ISL_2407336, EPI_ISL_2407337, EPI_ISL_2407338, EPI_ISL_2407339, EPI_ISL_2407340, EPI_ISL_2407341, EPI_ISL_2407342, EPI_ISL_2407344, EPI_ISL_2407345, EPI_ISL_2407346, EPI_ISL_2407349, EPI_ISL_2407350, EPI_ISL_2407351, EPI_ISL_2407352, EPI_ISL_2407353, EPI_ISL_2407355, EPI_ISL_2407356, EPI_ISL_2407357, EPI_ISL_2407358, EPI_ISL_2407359,
EPI_ISL_2407361, EPI_ISL_2407362, EPI_ISL_2407363, EPI_ISL_2407364, EPI_ISL_2407365, EPI_ISL_2407366, EPI_ISL_2407367, EPI_ISL_2407368, EPI_ISL_2407369, EPI_ISL_2407370, EPI_ISL_2407371, EPI_ISL_2407372, EPI_ISL_2407373, EPI_ISL_2407374, EPI_ISL_2407375, EPI_ISL_2407376, EPI_ISL_2407377, EPI_ISL_2407378, EPI_ISL_2407379, EPI_ISL_2407380, EPI_ISL_2407381, EPI_ISL_2407382,
EPI_ISL_2407383, EPI_ISL_2407384, EPI_ISL_2407386, EPI_ISL_2407387, EPI_ISL_2407388, EPI_ISL_2407389, EPI_ISL_2407390, EPI_ISL_2407392, EPI_ISL_2407393, EPI_ISL_2407394, EPI_ISL_2407395, EPI_ISL_2407396, EPI_ISL_2407397, EPI_ISL_2407398, EPI_ISL_2407399, EPI_ISL_2407400, EPI_ISL_2407401, EPI_ISL_2407402, EPI_ISL_2407403, EPI_ISL_2407404, EPI_ISL_2407405,
EPI_ISL_2407406, EPI_ISL_2407407, EPI_ISL_2407408, EPI_ISL_2407409, EPI_ISL_2407410, EPI_ISL_2407411, EPI_ISL_2407414, EPI_ISL_2407415, EPI_ISL_2407416, EPI_ISL_2407417, EPI_ISL_2407418, EPI_ISL_2407419, EPI_ISL_2407421, EPI_ISL_2407423, EPI_ISL_2407424, EPI_ISL_2407426, EPI_ISL_2407427, EPI_ISL_2407428, EPI_ISL_2407429, EPI_ISL_2407430, EPI_ISL_2407431, EPI_ISL_2407432,
EPI_ISL_2407433, EPI_ISL_2407434, EPI_ISL_2407435, EPI_ISL_2407436, EPI_ISL_2407437, EPI_ISL_2407438, EPI_ISL_2407439, EPI_ISL_2407440, EPI_ISL_2407441, EPI_ISL_2407442, EPI_ISL_2407443, EPI_ISL_2407444, EPI_ISL_2407446, EPI_ISL_2407447, EPI_ISL_2407448, EPI_ISL_2407449, EPI_ISL_2407450, EPI_ISL_2407451, EPI_ISL_2407452, EPI_ISL_2407453, EPI_ISL_2407454, EPI_ISL_2407455,
EPI_ISL_2407456, EPI_ISL_2407457, EPI_ISL_2407458, EPI_ISL_2407459, EPI_ISL_2407460, EPI_ISL_2407462, EPI_ISL_2407463, EPI_ISL_2407464, EPI_ISL_2407465, EPI_ISL_2407466, EPI_ISL_2407468, EPI_ISL_2407469, EPI_ISL_2407470, EPI_ISL_2407471, EPI_ISL_2407472, EPI_ISL_2407473, EPI_ISL_2407474, EPI_ISL_2407475, EPI_ISL_2407476, EPI_ISL_2407477, EPI_ISL_2407478, EPI_ISL_2407479,

EPI_ISL_2407480, EPI_ISL_2407481, EPI_ISL_2407482, EPI_ISL_2407483, EPI_ISL_2407484, EPI_ISL_2407485, EPI_ISL_2407486, EPI_ISL_2407487, EPI_ISL_2407488, EPI_ISL_2407489, EPI_ISL_2407490, EPI_ISL_2407491, EPI_ISL_2407492, EPI_ISL_2407493, EPI_ISL_2407494, EPI_ISL_2407495, EPI_ISL_2407496, EPI_ISL_2407497, EPI_ISL_2407498, EPI_ISL_2407499, EPI_ISL_2407500, EPI_ISL_2407501, EPI_ISL_2407502, EPI_ISL_2407503, EPI_ISL_2407504, EPI_ISL_2407505, EPI_ISL_2407506, EPI_ISL_2407507, EPI_ISL_2407508, EPI_ISL_2407509, EPI_ISL_2407510, EPI_ISL_2407511, EPI_ISL_2407512, EPI_ISL_2407513, EPI_ISL_2407514, EPI_ISL_2407515, EPI_ISL_2407516, EPI_ISL_2407517, EPI_ISL_2407518, EPI_ISL_2407519, EPI_ISL_2407520, EPI_ISL_2407521, EPI_ISL_2407522, EPI_ISL_2407523, EPI_ISL_2407524, EPI_ISL_2407526, EPI_ISL_2407527, EPI_ISL_2407528, EPI_ISL_2407529, EPI_ISL_2407530, EPI_ISL_2407531, EPI_ISL_2407532, EPI_ISL_2407533, EPI_ISL_2407534, EPI_ISL_2407535, EPI_ISL_2407536, EPI_ISL_2407537, EPI_ISL_2407538, EPI_ISL_2407539, EPI_ISL_2407540, EPI_ISL_2407541, EPI_ISL_2407542, EPI_ISL_2407543, EPI_ISL_2407544, EPI_ISL_2407545, EPI_ISL_2407546, EPI_ISL_2407547, EPI_ISL_2407549, EPI_ISL_2407550, EPI_ISL_2407551, EPI_ISL_2407552, EPI_ISL_2407553, EPI_ISL_2407554, EPI_ISL_2407555, EPI_ISL_2407556, EPI_ISL_2407557, EPI_ISL_2407558, EPI_ISL_2407559, EPI_ISL_2407560, EPI_ISL_2407561, EPI_ISL_2407562, EPI_ISL_2407563, EPI_ISL_2407564, EPI_ISL_2407565, EPI_ISL_2407566, EPI_ISL_2407567, EPI_ISL_2407568, EPI_ISL_2407569, EPI_ISL_2407570, EPI_ISL_2407571, EPI_ISL_2407572, EPI_ISL_2407574, EPI_ISL_2407575, EPI_ISL_2407576, EPI_ISL_2407577, EPI_ISL_2407578, EPI_ISL_2407579, EPI_ISL_2407580, EPI_ISL_2407581, EPI_ISL_2407582, EPI_ISL_2407583, EPI_ISL_2407584, EPI_ISL_2407585, EPI_ISL_2407586, EPI_ISL_2407587, EPI_ISL_2407588, EPI_ISL_2407589, EPI_ISL_2407590, EPI_ISL_2407591, EPI_ISL_2407593, EPI_ISL_2407594, EPI_ISL_2407595, EPI_ISL_2407597, EPI_ISL_2407598, EPI_ISL_2407599, EPI_ISL_2407600, EPI_ISL_2407601, EPI_ISL_2407602, EPI_ISL_2407603, EPI_ISL_2407604, EPI_ISL_2407605, EPI_ISL_2407606, EPI_ISL_2407607, EPI_ISL_2407608, EPI_ISL_2407609, EPI_ISL_2407611, EPI_ISL_2407612, EPI_ISL_2407614, EPI_ISL_2407615, EPI_ISL_2407616, EPI_ISL_2407617, EPI_ISL_2407618, EPI_ISL_2407619, EPI_ISL_2407620, EPI_ISL_2407621, EPI_ISL_2407622, EPI_ISL_2407623, EPI_ISL_2407624, EPI_ISL_2407625, EPI_ISL_2407626, EPI_ISL_2407627, EPI_ISL_2407628, EPI_ISL_2407629, EPI_ISL_2407630, EPI_ISL_2407631, EPI_ISL_2407632, EPI_ISL_2407633, EPI_ISL_2407634, EPI_ISL_2407635, EPI_ISL_2407636, EPI_ISL_2407638, EPI_ISL_2407639, EPI_ISL_2407640, EPI_ISL_2407641, EPI_ISL_2407642, EPI_ISL_2407643, EPI_ISL_2407645, EPI_ISL_2407646, EPI_ISL_2407648, EPI_ISL_2407649, EPI_ISL_2407650, EPI_ISL_2407651, EPI_ISL_2407653, EPI_ISL_2407654, EPI_ISL_2407655, EPI_ISL_2407656, EPI_ISL_2407657, EPI_ISL_2407658, EPI_ISL_2407659, EPI_ISL_2407660, EPI_ISL_2407661, EPI_ISL_2407662, EPI_ISL_2407663, EPI_ISL_2407664, EPI_ISL_2407665, EPI_ISL_2407666, EPI_ISL_2407667, EPI_ISL_2407668, EPI_ISL_2407669, EPI_ISL_2407670, EPI_ISL_2407671, EPI_ISL_2407672, EPI_ISL_2407673, EPI_ISL_2407674, EPI_ISL_2407675, EPI_ISL_2407676, EPI_ISL_2407677, EPI_ISL_2407678, EPI_ISL_2407679, EPI_ISL_2407680, EPI_ISL_2407681, EPI_ISL_2407682, EPI_ISL_2407683, EPI_ISL_2407684, EPI_ISL_2407685, EPI_ISL_2407686, EPI_ISL_2407687, EPI_ISL_2407688, EPI_ISL_2407689, EPI_ISL_2407690, EPI_ISL_2407691, EPI_ISL_2407692, EPI_ISL_2407693, EPI_ISL_2407694, EPI_ISL_2407695, EPI_ISL_2407696, EPI_ISL_2407697, EPI_ISL_2407698, EPI_ISL_2407699, EPI_ISL_2407700, EPI_ISL_2407701, EPI_ISL_2407702, EPI_ISL_2407703, EPI_ISL_2407704, EPI_ISL_2407705, EPI_ISL_2407706, EPI_ISL_2407707, EPI_ISL_2407708, EPI_ISL_2407709, EPI_ISL_2407710, EPI_ISL_2407711, EPI_ISL_2407712, EPI_ISL_2407713, EPI_ISL_2407714, EPI_ISL_2407715, EPI_ISL_2407716, EPI_ISL_2407717, EPI_ISL_2407718, EPI_ISL_2407719, EPI_ISL_2407720, EPI_ISL_2407721, EPI_ISL_2407722, EPI_ISL_2407723, EPI_ISL_2407724, EPI_ISL_2407725, EPI_ISL_2407726, EPI_ISL_2407727, EPI_ISL_2407728, EPI_ISL_2407729, EPI_ISL_2407730, EPI_ISL_2407731, EPI_ISL_2407732, EPI_ISL_2407733, EPI_ISL_2407734, EPI_ISL_2407735, EPI_ISL_2407736, EPI_ISL_2407737, EPI_ISL_2407738, EPI_ISL_2407739, EPI_ISL_2407740, EPI_ISL_2407741, EPI_ISL_2407742, EPI_ISL_2407743, EPI_ISL_2407744, EPI_ISL_2407745, EPI_ISL_2407746, EPI_ISL_2407747, EPI_ISL_2407748, EPI_ISL_2407749, EPI_ISL_2407750, EPI_ISL_2407751, EPI_ISL_2407752, EPI_ISL_2407753, EPI_ISL_2407754, EPI_ISL_2407755, EPI_ISL_2407756, EPI_ISL_2407757, EPI_ISL_2407758, EPI_ISL_2407759, EPI_ISL_2407760, EPI_ISL_2407761, EPI_ISL_2407762, EPI_ISL_2407763, EPI_ISL_2407764, EPI_ISL_2407765, EPI_ISL_2407766, EPI_ISL_2407767, EPI_ISL_2407768, EPI_ISL_2407769, EPI_ISL_2407770, EPI_ISL_2407771, EPI_ISL_2407772, EPI_ISL_2407773, EPI_ISL_2407774, EPI_ISL_2407775, EPI_ISL_2407776, EPI_ISL_2407777, EPI_ISL_2407778, EPI_ISL_2407779, EPI_ISL_2407780, EPI_ISL_2407781, EPI_ISL_2407782, EPI_ISL_2407783, EPI_ISL_2407784, EPI_ISL_2407785, EPI_ISL_2407786, EPI_ISL_2407787, EPI_ISL_2407788, EPI_ISL_2407789, EPI_ISL_2407790, EPI_ISL_2407791, EPI_ISL_2407792, EPI_ISL_2407793, EPI_ISL_2407794, EPI_ISL_2407795, EPI_ISL_2407796, EPI_ISL_2407797, EPI_ISL_2407798, EPI_ISL_2407799, EPI_ISL_2407800, EPI_ISL_2407801, EPI_ISL_2407802, EPI_ISL_2407803, EPI_ISL_2407804, EPI_ISL_2407805, EPI_ISL_2407806, EPI_ISL_2407807, EPI_ISL_2407808, EPI_ISL_2407809, EPI_ISL_2407810, EPI_ISL_2407812, EPI_ISL_2407813, EPI_ISL_2407814, EPI_ISL_2407815, EPI_ISL_2407816, EPI_ISL_2407817, EPI_ISL_2407818, EPI_ISL_2407819, EPI_ISL_2407820, EPI_ISL_2407821, EPI_ISL_2407822, EPI_ISL_2407823, EPI_ISL_2407824, EPI_ISL_2407825, EPI_ISL_2407826, EPI_ISL_2407827, EPI_ISL_2407828, EPI_ISL_2407829, EPI_ISL_2407830, EPI_ISL_2407831, EPI_ISL_2407832, EPI_ISL_2407833, EPI_ISL_2407834, EPI_ISL_2407835, EPI_ISL_2407836, EPI_ISL_2407837, EPI_ISL_2407838, EPI_ISL_2407839, EPI_ISL_2407840, EPI_ISL_2407841, EPI_ISL_2407842, EPI_ISL_2407843, EPI_ISL_2407844, EPI_ISL_2407845, EPI_ISL_2407846, EPI_ISL_2407847, EPI_ISL_2407848, EPI_ISL_2407849, EPI_ISL_2407850, EPI_ISL_2407851, EPI_ISL_2407852, EPI_ISL_2407853, EPI_ISL_2407854, EPI_ISL_2407855, EPI_ISL_2407856, EPI_ISL_2407857, EPI_ISL_2407858, EPI_ISL_2407859, EPI_ISL_2407860, EPI_ISL_2407861, EPI_ISL_2407862, EPI_ISL_2407863, EPI_ISL_2407864, EPI_ISL_2407865, EPI_ISL_2407866, EPI_ISL_2407867, EPI_ISL_2407868, EPI_ISL_2407869, EPI_ISL_2407870, EPI_ISL_2407871, EPI_ISL_2407872, EPI_ISL_2407873, EPI_ISL_2407874, EPI_ISL_2407875, EPI_ISL_2407876, EPI_ISL_2407877, EPI_ISL_2407878, EPI_ISL_2407879, EPI_ISL_2407880, EPI_ISL_2407881, EPI_ISL_2407882, EPI_ISL_2407883, EPI_ISL_2407884, EPI_ISL_2407885, EPI_ISL_2407886, EPI_ISL_2407887, EPI_ISL_2407888, EPI_ISL_2407889, EPI_ISL_2407890, EPI_ISL_2407891, EPI_ISL_2407892, EPI_ISL_2407893, EPI_ISL_2407894, EPI_ISL_2407895, EPI_ISL_2407896, EPI_ISL_2407897, EPI_ISL_2407898, EPI_ISL_2407899, EPI_ISL_2407900, EPI_ISL_2407901, EPI_ISL_2407902, EPI_ISL_2407903, EPI_ISL_2407904, EPI_ISL_2407905, EPI_ISL_2407906, EPI_ISL_2407907, EPI_ISL_2407908, EPI_ISL_2407910, EPI_ISL_2407911, EPI_ISL_2407912, EPI_ISL_2407913, EPI_ISL_2407914, EPI_ISL_2407915, EPI_ISL_2407916, EPI_ISL_2407917, EPI_ISL_2407918, EPI_ISL_2407919, EPI_ISL_2407920, EPI_ISL_2407921, EPI_ISL_2407923, EPI_ISL_2407924, EPI_ISL_2407925, EPI_ISL_2407926, EPI_ISL_2407927, EPI_ISL_2407928, EPI_ISL_2407929, EPI_ISL_2407930, EPI_ISL_2407931, EPI_ISL_2407932, EPI_ISL_2407933, EPI_ISL_2407934, EPI_ISL_2407935, EPI_ISL_2407936, EPI_ISL_2407937, EPI_ISL_2407938, EPI_ISL_2407939, EPI_ISL_2407940, EPI_ISL_2407941, EPI_ISL_2407942, EPI_ISL_2407943, EPI_ISL_2407944, EPI_ISL_2407945, EPI_ISL_2407947, EPI_ISL_2407948, EPI_ISL_2407949, EPI_ISL_2407950, EPI_ISL_2407951, EPI_ISL_2407952, EPI_ISL_2407953, EPI_ISL_2407955, EPI_ISL_2407956, EPI_ISL_2407957, EPI_ISL_2407958, EPI_ISL_2407959, EPI_ISL_2407960, EPI_ISL_2407961, EPI_ISL_2407962, EPI_ISL_2407963, EPI_ISL_2407964, EPI_ISL_2407965, EPI_ISL_2407966, EPI_ISL_2407967, EPI_ISL_2407968, EPI_ISL_2407969, EPI_ISL_2407970, EPI_ISL_2407971, EPI_ISL_2407972, EPI_ISL_2407973, EPI_ISL_2407974, EPI_ISL_2407975, EPI_ISL_2407976, EPI_ISL_2407977, EPI_ISL_2407978, EPI_ISL_2407979, EPI_ISL_2407980, EPI_ISL_2407981, EPI_ISL_2407982, EPI_ISL_2407983, EPI_ISL_2407984, EPI_ISL_2407985, EPI_ISL_2407986, EPI_ISL_2407987, EPI_ISL_2407988, EPI_ISL_2407989, EPI_ISL_2407990, EPI_ISL_2407991, EPI_ISL_2407992, EPI_ISL_2407993, EPI_ISL_2407994, EPI_ISL_2407995, EPI_ISL_2407996, EPI_ISL_2407999, EPI_ISL_2408003, EPI_ISL_2408005, EPI_ISL_2408008, EPI_ISL_2408010, EPI_ISL_2408012, EPI_ISL_2408013, EPI_ISL_2408014, EPI_ISL_2408016, EPI_ISL_2408019, EPI_ISL_2408021, EPI_ISL_2408026, EPI_ISL_2408027, EPI_ISL_2408028, EPI_ISL_2408029, EPI_ISL_2408030, EPI_ISL_2408037, EPI_ISL_2408042, EPI_ISL_2408055, EPI_ISL_2435095, EPI_ISL_2435558, EPI_ISL_2436267, EPI_ISL_2436275, EPI_ISL_2436315, EPI_ISL_2436442, EPI_ISL_2436547, EPI_ISL_2436595, EPI_ISL_2436620, EPI_ISL_2436635, EPI_ISL_2436699, EPI_ISL_2436713, EPI_ISL_2436752, EPI_ISL_2436868, EPI_ISL_2437278, EPI_ISL_2437280, EPI_ISL_2437290, EPI_ISL_2437295, EPI_ISL_2437299, EPI_ISL_2437301, EPI_ISL_2437308, EPI_ISL_2437313, EPI_ISL_2437315, EPI_ISL_2437316, EPI_ISL_2437330, EPI_ISL_2437331, EPI_ISL_2437345, EPI_ISL_2437354, EPI_ISL_2437355, EPI_ISL_2437375, EPI_ISL_2437382, EPI_ISL_2437388, EPI_ISL_2437398, EPI_ISL_2437400, EPI_ISL_2437402, EPI_ISL_2437403, EPI_ISL_2437405, EPI_ISL_2437407, EPI_ISL_2437412, EPI_ISL_2437415, EPI_ISL_2437423, EPI_ISL_2437427, EPI_ISL_2437430, EPI_ISL_2437437, EPI_ISL_2437448, EPI_ISL_2437453, EPI_ISL_2437457, EPI_ISL_2437463, EPI_ISL_2437466, EPI_ISL_2437467, EPI_ISL_2437468, EPI_ISL_2437470, EPI_ISL_2437474, EPI_ISL_2437479, EPI_ISL_2437481, EPI_ISL_2437484, EPI_ISL_2437485, EPI_ISL_2437490, EPI_ISL_2437491, EPI_ISL_2437519, EPI_ISL_2437521, EPI_ISL_2437524, EPI_ISL_2437525, EPI_ISL_2437548, EPI_ISL_2437552, EPI_ISL_2437556, EPI_ISL_2437557, EPI_ISL_2437563, EPI_ISL_2437566, EPI_ISL_2437571, EPI_ISL_2437572, EPI_ISL_2437573, EPI_ISL_2437583, EPI_ISL_2437586, EPI_ISL_2437600, EPI_ISL_2437603, EPI_ISL_2437610, EPI_ISL_2437672, EPI_ISL_2437689, EPI_ISL_2437838, EPI_ISL_2437973, EPI_ISL_2438047, EPI_ISL_2438064, EPI_ISL_2438067, EPI_ISL_2438106, EPI_ISL_2438117, EPI_ISL_2438121, EPI_ISL_2438127, EPI_ISL_2438130, EPI_ISL_2438150, EPI_ISL_2438186, EPI_ISL_2438197, EPI_ISL_2438211, EPI_ISL_2438212, EPI_ISL_2438248, EPI_ISL_2438260, EPI_ISL_2438267, EPI_ISL_2438273, EPI_ISL_2438299, EPI_ISL_2438344, EPI_ISL_2438345, EPI_ISL_2438362, EPI_ISL_2438364, EPI_ISL_2438378, EPI_ISL_2438379, EPI_ISL_2438380, EPI_ISL_2438381, EPI_ISL_2438382, EPI_ISL_2438383, EPI_ISL_2438384, EPI_ISL_2438385, EPI_ISL_2438386, EPI_ISL_2438387, EPI_ISL_2438388, EPI_ISL_2438389, EPI_ISL_2438390, EPI_ISL_2438391, EPI_ISL_2438392, EPI_ISL_2438393, EPI_ISL_2438394, EPI_ISL_2438395, EPI_ISL_2438396, EPI_ISL_2438397, EPI_ISL_2438398, EPI_ISL_2438399, EPI_ISL_2438400, EPI_ISL_2438403, EPI_ISL_2438404, EPI_ISL_2438405, EPI_ISL_2438406, EPI_ISL_2438407, EPI_ISL_2438408, EPI_ISL_2438409, EPI_ISL_2438410, EPI_ISL_2438411, EPI_ISL_2438412, EPI_ISL_2438413, EPI_ISL_2438414, EPI_ISL_2438415, EPI_ISL_2438416, EPI_ISL_2438417, EPI_ISL_2438418, EPI_ISL_2438419, EPI_ISL_2438420, EPI_ISL_2438421, EPI_ISL_2438422, EPI_ISL_2438423, EPI_ISL_2438424, EPI_ISL_2438425, EPI_ISL_2438426, EPI_ISL_2438428, EPI_ISL_2438429, EPI_ISL_2438430, EPI_ISL_2438431, EPI_ISL_2438432, EPI_ISL_2438433, EPI_ISL_2438434, EPI_ISL_2438435, EPI_ISL_2438436, EPI_ISL_2438437, EPI_ISL_2438438, EPI_ISL_2438439, EPI_ISL_2438440, EPI_ISL_2438441, EPI_ISL_2438442, EPI_ISL_2438443, EPI_ISL_2438444, EPI_ISL_2438445, EPI_ISL_2438446, EPI_ISL_2438447, EPI_ISL_2438448, EPI_ISL_2438449, EPI_ISL_2438450, EPI_ISL_2438451, EPI_ISL_2438452, EPI_ISL_2438453, EPI_ISL_2438454, EPI_ISL_2438455, EPI_ISL_2438456, EPI_ISL_2438457, EPI_ISL_2438458, EPI_ISL_2438459, EPI_ISL_2438460, EPI_ISL_2438461, EPI_ISL_2438462, EPI_ISL_2438463, EPI_ISL_2438464, EPI_ISL_2438465, EPI_ISL_2438466, EPI_ISL_2438467, EPI_ISL_2438468, EPI_ISL_2438469, EPI_ISL_2438470, EPI_ISL_2438471, EPI_ISL_2438472, EPI_ISL_2438473, EPI_ISL_2438474, EPI_ISL_2438475, EPI_ISL_2438476, EPI_ISL_2438477, EPI_ISL_2438478, EPI_ISL_2438479, EPI_ISL_2438480, EPI_ISL_2438481, EPI_ISL_2438482, EPI_ISL_2438483, EPI_ISL_2438484, EPI_ISL_2438485, EPI_ISL_2438486, EPI_ISL_2438487, EPI_ISL_2438488, EPI_ISL_2438489, EPI_ISL_2438490, EPI_ISL_2438491, EPI_ISL_2438492, EPI_ISL_2438493, EPI_ISL_2438494, EPI_ISL_2438495, EPI_ISL_2438496, EPI_ISL_2438497, EPI_ISL_2438498, EPI_ISL_2438499, EPI_ISL_2438500, EPI_ISL_2438501, EPI_ISL_2438502, EPI_ISL_2438503, EPI_ISL_2438504, EPI_ISL_2438505, EPI_ISL_2438506, EPI_ISL_2438507, EPI_ISL_2438508, EPI_ISL_2438509, EPI_ISL_2438510, EPI_ISL_2438511, EPI_ISL_2438512, EPI_ISL_2438513, EPI_ISL_2438514, EPI_ISL_2438515, EPI_ISL_2438516, EPI_ISL_2438517, EPI_ISL_2438518, EPI_ISL_2438520, EPI_ISL_2438521, EPI_ISL_2438522, EPI_ISL_2438523, EPI_ISL_2438524, EPI_ISL_2438525, EPI_ISL_2438526, EPI_ISL_2438527, EPI_ISL_2438528, EPI_ISL_2438529, EPI_ISL_2438530, EPI_ISL_2438531, EPI_ISL_2438532, EPI_ISL_2438533, EPI_ISL_2438534, EPI_ISL_2438535, EPI_ISL_2438536, EPI_ISL_2438537, EPI_ISL_2438538, EPI_ISL_2438539, EPI_ISL_2438540, EPI_ISL_2438541, EPI_ISL_2438542, EPI_ISL_2457193, EPI_ISL_2457687, EPI_ISL_2457689, EPI_ISL_2457691, EPI_ISL_2457693, EPI_ISL_2457700, EPI_ISL_2457701, EPI_ISL_2457703, EPI_ISL_2457709, EPI_ISL_2457714, EPI_ISL_2457723, EPI_ISL_2457731, EPI_ISL_2457758, EPI_ISL_2457761, EPI_ISL_2457774, EPI_ISL_2457781, EPI_ISL_2457783, EPI_ISL_2457784, EPI_ISL_2457791, EPI_ISL_2457795, EPI_ISL_2457798, EPI_ISL_2457800, EPI_ISL_2457809, EPI_ISL_2457810, EPI_ISL_2457822, EPI_ISL_2457832, EPI_ISL_2457834, EPI_ISL_2457836, EPI_ISL_2457846, EPI_ISL_2457847, EPI_ISL_2457857, EPI_ISL_2457861, EPI_ISL_2457864, EPI_ISL_2457883, EPI_ISL_2488186, EPI_ISL_2511359, EPI_ISL_2511489, EPI_ISL_2513390, EPI_ISL_2513613, EPI_ISL_2513614, EPI_ISL_2513616, EPI_ISL_2513617, EPI_ISL_2513619, EPI_ISL_2513620, EPI_ISL_2513624, EPI_ISL_2513627, EPI_ISL_2513630, EPI_ISL_2513632, EPI_ISL_2513633, EPI_ISL_2513634, EPI_ISL_2513636, EPI_ISL_2513637, EPI_ISL_2513638, EPI_ISL_2513639, EPI_ISL_2513640, EPI_ISL_2513641, EPI_ISL_2513642, EPI_ISL_2513643, EPI_ISL_2513644, EPI_ISL_2513645, EPI_ISL_2513647, EPI_ISL_2513650, EPI_ISL_2513651, EPI_ISL_2513652, EPI_ISL_2513654, EPI_ISL_2513655, EPI_ISL_2513657, EPI_ISL_2513660, EPI_ISL_2513661, EPI_ISL_2513662, EPI_ISL_2513665, EPI_ISL_2513666, EPI_ISL_2513669, EPI_ISL_2513672, EPI_ISL_2513673, EPI_ISL_2513674, EPI_ISL_2513676, EPI_ISL_2513678, EPI_ISL_2513680, EPI_ISL_2513682, EPI_ISL_2513684, EPI_ISL_2513689, EPI_ISL_2513691, EPI_ISL_2513692, EPI_ISL_2513694, EPI_ISL_2513695, EPI_ISL_2513696, EPI_ISL_2513698, EPI_ISL_2513702, EPI_ISL_2513706, EPI_ISL_2513707, EPI_ISL_2513708, EPI_ISL_2513710, EPI_ISL_2513711, EPI_ISL_2513712, EPI_ISL_2513713, EPI_ISL_2513715, EPI_ISL_2513717, EPI_ISL_2513719, EPI_ISL_2513721, EPI_ISL_2513724, EPI_ISL_2513725, EPI_ISL_2513728, EPI_ISL_2513729, EPI_ISL_2513730, EPI_ISL_2513731, EPI_ISL_2513732, EPI_ISL_2513733, EPI_ISL_2513734, EPI_ISL_2513735, EPI_ISL_2513737, EPI_ISL_2513742, EPI_ISL_2513743, EPI_ISL_2513744, EPI_ISL_2513745, EPI_ISL_2513748, EPI_ISL_2513749, EPI_ISL_2513751, EPI_ISL_2513753, EPI_ISL_2513755, EPI_ISL_2513756, EPI_ISL_2513760, EPI_ISL_2513761, EPI_ISL_2513767, EPI_ISL_2513769, EPI_ISL_2513771, EPI_ISL_2513772, EPI_ISL_2513773, EPI_ISL_2513775, EPI_ISL_2513777, EPI_ISL_2513778, EPI_ISL_2513782, EPI_ISL_2513786, EPI_ISL_2513789, EPI_ISL_2513791, EPI_ISL_2513792, EPI_ISL_2513794, EPI_ISL_2513800, EPI_ISL_2542022, EPI_ISL_2542543, EPI_ISL_2542544, EPI_ISL_2542545, EPI_ISL_2542546, EPI_ISL_2542547, EPI_ISL_2542548, EPI_ISL_2542549, EPI_ISL_2542550, EPI_ISL_2542551, EPI_ISL_2542552, EPI_ISL_2542553, EPI_ISL_2542554, EPI_ISL_2542555, EPI_ISL_2542556, EPI_ISL_2542557, EPI_ISL_2542558, EPI_ISL_2542559, EPI_ISL_2542560, EPI_ISL_2542561, EPI_ISL_2542562, EPI_ISL_2542563, EPI_ISL_2542564, EPI_ISL_2542565, EPI_ISL_2542566, EPI_ISL_2542567, EPI_ISL_2542568, EPI_ISL_2542569, EPI_ISL_2542570, EPI_ISL_2542571, EPI_ISL_2542572, EPI_ISL_2542573, EPI_ISL_2542574, EPI_ISL_2542575, EPI_ISL_2542576, EPI_ISL_2542577, EPI_ISL_2542578, EPI_ISL_2542579, EPI_ISL_2542580, EPI_ISL_2542581, EPI_ISL_2542582, EPI_ISL_2542583, EPI_ISL_2542584, EPI_ISL_2542585, EPI_ISL_2542586, EPI_ISL_2542587, EPI_ISL_2542588, EPI_ISL_2542589, EPI_ISL_2542590, EPI_ISL_2542591, EPI_ISL_2542592, EPI_ISL_2542593, EPI_ISL_2542594, EPI_ISL_2542595, EPI_ISL_2542596, EPI_ISL_2542597, EPI_ISL_2542598, EPI_ISL_2542599, EPI_ISL_2542600, EPI_ISL_2542601, EPI_ISL_2542602, EPI_ISL_2542603, EPI_ISL_2542604, EPI_ISL_2542605, EPI_ISL_2542606, EPI_ISL_2542607, EPI_ISL_2542608, EPI_ISL_2542609, EPI_ISL_2542610, EPI_ISL_2542611, EPI_ISL_2542612, EPI_ISL_2542613, EPI_ISL_2542614, EPI_ISL_2542615, EPI_ISL_2542616, EPI_ISL_2542617, EPI_ISL_2542618, EPI_ISL_2542619, EPI_ISL_2542620, EPI_ISL_2542621, EPI_ISL_2542622, EPI_ISL_2542623, EPI_ISL_2542624, EPI_ISL_2542625, EPI_ISL_2542626, EPI_ISL_2542627, EPI_ISL_2542628, EPI_ISL_2542629, EPI_ISL_2542630, EPI_ISL_2542631, EPI_ISL_2542632, EPI_ISL_2542633, EPI_ISL_2542634, EPI_ISL_2542635, EPI_ISL_2542636, EPI_ISL_2542637, EPI_ISL_2542638, EPI_ISL_2542639, EPI_ISL_2542640, EPI_ISL_2542641, EPI_ISL_2542642, EPI_ISL_2542643, EPI_ISL_2542644, EPI_ISL_2542645, EPI_ISL_2542646, EPI_ISL_2542647, EPI_ISL_2542648, EPI_ISL_2542649, EPI_ISL_2542650, EPI_ISL_2542651, EPI_ISL_2542652, EPI_ISL_2542653, EPI_ISL_2542654, EPI_ISL_2542655, EPI_ISL_2542656, EPI_ISL_2542657, EPI_ISL_2542658, EPI_ISL_2542659, EPI_ISL_2542660, EPI_ISL_2542661, EPI_ISL_2542662, EPI_ISL_2542663, EPI_ISL_2542664, EPI_ISL_2542665, EPI_ISL_2542666, EPI_ISL_2542667, EPI_ISL_2542668, EPI_ISL_2542669, EPI_ISL_2542670, EPI_ISL_2542671, EPI_ISL_2542672, EPI_ISL_2542673, EPI_ISL_2542674, EPI_ISL_2542675, EPI_ISL_2542676, EPI_ISL_2542677, EPI_ISL_2542678, EPI_ISL_2542679, EPI_ISL_2542680, EPI_ISL_2542681, EPI_ISL_2542682, EPI_ISL_2542683, EPI_ISL_2542684, EPI_ISL_2542685, EPI_ISL_2542686, EPI_ISL_2542687, EPI_ISL_2542688, EPI_ISL_2542689, EPI_ISL_2542690, EPI_ISL_2542691, EPI_ISL_2542692, EPI_ISL_2542693, EPI_ISL_2542694, EPI_ISL_2814659

| | | | |
|---|---|---|---|
| see above | Lighthouse Lab in Glasgow | Wellcome Sanger Institute for the COVID-19 Genomics UK (COG-UK) Consortium | Anna Dominiczak and Alex Alderton; Carol Clugston; Cordelia Langford; David Gray; David K. Jackson; Dominic Kwiatkowski; Ewan Harrison; Harper VanSteenhouse; Ian Johnston; Jeffrey Barrett; John Sillitoe on behalf of the Wellcome Sanger Institute COVID-19 Surveillance Team; Roberto Amato; Sonia Goncalves; Yumi Kasai |

EPI_ISL_2352690, EPI_ISL_2352691, EPI_ISL_2352692, EPI_ISL_2352696, EPI_ISL_2352697, EPI_ISL_2352699, EPI_ISL_2352703, EPI_ISL_2352710, EPI_ISL_2352715, EPI_ISL_2352716, EPI_ISL_2352721, EPI_ISL_2352723, EPI_ISL_2352727, EPI_ISL_2352733, EPI_ISL_2352738, EPI_ISL_2352745, EPI_ISL_2352746, EPI_ISL_2352747, EPI_ISL_2352748, EPI_ISL_2352749, EPI_ISL_2352750, EPI_ISL_2352754, EPI_ISL_2352755, EPI_ISL_2352758, EPI_ISL_2352762, EPI_ISL_2352770, EPI_ISL_2352773, EPI_ISL_2352776, EPI_ISL_2352778, EPI_ISL_2352783, EPI_ISL_2352788, EPI_ISL_2352790, EPI_ISL_2352793, EPI_ISL_2352795, EPI_ISL_2352796, EPI_ISL_2352798, EPI_ISL_2352802, EPI_ISL_2352806, EPI_ISL_2352810, EPI_ISL_2352811, EPI_ISL_2352816, EPI_ISL_2352822, EPI_ISL_2352823, EPI_ISL_2352825, EPI_ISL_2352831, EPI_ISL_2352839, EPI_ISL_2352846, EPI_ISL_2352847, EPI_ISL_2352854, EPI_ISL_2352855, EPI_ISL_2352856, EPI_ISL_2352859, EPI_ISL_2352862, EPI_ISL_2352864, EPI_ISL_2352868, EPI_ISL_2352883, EPI_ISL_2352886, EPI_ISL_2352890, EPI_ISL_2352891, EPI_ISL_2352893, EPI_ISL_2352895, EPI_ISL_2352896, EPI_ISL_2352897, EPI_ISL_2352900, EPI_ISL_2352904, EPI_ISL_2352905, EPI_ISL_2352906, EPI_ISL_2352910, EPI_ISL_2352911, EPI_ISL_2352914, EPI_ISL_2352919, EPI_ISL_2352921, EPI_ISL_2352924, EPI_ISL_2352927, EPI_ISL_2352928, EPI_ISL_2352929, EPI_ISL_2352940, EPI_ISL_2352943, EPI_ISL_2352945, EPI_ISL_2352946, EPI_ISL_2352957, EPI_ISL_2352964, EPI_ISL_2352967, EPI_ISL_2352971, EPI_ISL_2352972, EPI_ISL_2352978, EPI_ISL_2352979, EPI_ISL_2352982, EPI_ISL_2365971, EPI_ISL_2365977, EPI_ISL_2365984, EPI_ISL_2365987, EPI_ISL_2366004, EPI_ISL_2366006, EPI_ISL_2366007, EPI_ISL_2366011, EPI_ISL_2366012, EPI_ISL_2366015, EPI_ISL_2366023, EPI_ISL_2366038, EPI_ISL_2366042, EPI_ISL_2366047, EPI_ISL_2366049, EPI_ISL_2366050, EPI_ISL_2366051, EPI_ISL_2366072, EPI_ISL_2366094, EPI_ISL_2366105, EPI_ISL_2366110, EPI_ISL_2366112, EPI_ISL_2366113, EPI_ISL_2366117, EPI_ISL_2366120, EPI_ISL_2366122, EPI_ISL_2366133, EPI_ISL_2366134, EPI_ISL_2366140, EPI_ISL_2366143, EPI_ISL_2366147, EPI_ISL_2366148, EPI_ISL_2366157, EPI_ISL_2366162, EPI_ISL_2366163, EPI_ISL_2366166, EPI_ISL_2366169, EPI_ISL_2366182, EPI_ISL_2366189, EPI_ISL_2366193, EPI_ISL_2366194, EPI_ISL_2366196, EPI_ISL_2366203, EPI_ISL_2366215, EPI_ISL_2366221, EPI_ISL_2366226, EPI_ISL_2366228, EPI_ISL_2366229, EPI_ISL_2366233, EPI_ISL_2366234, EPI_ISL_2366235, EPI_ISL_2366239, EPI_ISL_2366240, EPI_ISL_2366243, EPI_ISL_2366244, EPI_ISL_2366248, EPI_ISL_2366249, EPI_ISL_2366254, EPI_ISL_2366261, EPI_ISL_2366264, EPI_ISL_2366269, EPI_ISL_2366273, EPI_ISL_2366275, EPI_ISL_2366276, EPI_ISL_2366278, EPI_ISL_2366279, EPI_ISL_2366280, EPI_ISL_2366281, EPI_ISL_2366282, EPI_ISL_2366283, EPI_ISL_2366284, EPI_ISL_2366285, EPI_ISL_2366288, EPI_ISL_2366289, EPI_ISL_2366290, EPI_ISL_2366291, EPI_ISL_2366292, EPI_ISL_2366293, EPI_ISL_2366295, EPI_ISL_2366297, EPI_ISL_2366301, EPI_ISL_2366306, EPI_ISL_2366307, EPI_ISL_2366309, EPI_ISL_2366315, EPI_ISL_2366317, EPI_ISL_2366319, EPI_ISL_2366320, EPI_ISL_2366324, EPI_ISL_2366327, EPI_ISL_2366328, EPI_ISL_2366329, EPI_ISL_2366332, EPI_ISL_2366333, EPI_ISL_2366334, EPI_ISL_2366336, EPI_ISL_2366337, EPI_ISL_2366338, EPI_ISL_2366341, EPI_ISL_2366342, EPI_ISL_2366345, EPI_ISL_2366346, EPI_ISL_2366347, EPI_ISL_2366348, EPI_ISL_2366349, EPI_ISL_2366350, EPI_ISL_2366355, EPI_ISL_2366358, EPI_ISL_2366360, EPI_ISL_2366363, EPI_ISL_2366364, EPI_ISL_2366365, EPI_ISL_2366368, EPI_ISL_2366369, EPI_ISL_2366370, EPI_ISL_2366371, EPI_ISL_2366372, EPI_ISL_2366373, EPI_ISL_2366374, EPI_ISL_2366378, EPI_ISL_2366379, EPI_ISL_2366380, EPI_ISL_2366383, EPI_ISL_2366386, EPI_ISL_2366387, EPI_ISL_2366393, EPI_ISL_2366394, EPI_ISL_2366396, EPI_ISL_2366399, EPI_ISL_2366400, EPI_ISL_2366402, EPI_ISL_2366406, EPI_ISL_2366408, EPI_ISL_2366409, EPI_ISL_2366414, EPI_ISL_2366415, EPI_ISL_2366418, EPI_ISL_2366421, EPI_ISL_2366422, EPI_ISL_2366423, EPI_ISL_2366424, EPI_ISL_2366425, EPI_ISL_2366428, EPI_ISL_2366429, EPI_ISL_2366430, EPI_ISL_2366431, EPI_ISL_2366432, EPI_ISL_2366435, EPI_ISL_2366436, EPI_ISL_2366437, EPI_ISL_2366440, EPI_ISL_2366442, EPI_ISL_2366444, EPI_ISL_2366445, EPI_ISL_2366449, EPI_ISL_2366452, EPI_ISL_2366453, EPI_ISL_2366454, EPI_ISL_2366455, EPI_ISL_2366456, EPI_ISL_2366458, EPI_ISL_2366459, EPI_ISL_2366462, EPI_ISL_2366463, EPI_ISL_2366466, EPI_ISL_2366471, EPI_ISL_2366473, EPI_ISL_2366474, EPI_ISL_2366475, EPI_ISL_2366476, EPI_ISL_2366478, EPI_ISL_2366482, EPI_ISL_2366483, EPI_ISL_2366487, EPI_ISL_2366489, EPI_ISL_2392196, EPI_ISL_2392198, EPI_ISL_2392199, EPI_ISL_2392210, EPI_ISL_2392211, EPI_ISL_2392220, EPI_ISL_2392223, EPI_ISL_2392224, EPI_ISL_2392225, EPI_ISL_2392227, EPI_ISL_2392230, EPI_ISL_2392235, EPI_ISL_2392236, EPI_ISL_2392242, EPI_ISL_2392243, EPI_ISL_2392249, EPI_ISL_2392263, EPI_ISL_2392265, EPI_ISL_2392266, EPI_ISL_2392277, EPI_ISL_2392279, EPI_ISL_2392292, EPI_ISL_2392294, EPI_ISL_2392304, EPI_ISL_2392310, EPI_ISL_2392316, EPI_ISL_2392323, EPI_ISL_2392333, EPI_ISL_2392336, EPI_ISL_2392338, EPI_ISL_2392345, EPI_ISL_2392349, EPI_ISL_2392353, EPI_ISL_2392355, EPI_ISL_2392356, EPI_ISL_2392362, EPI_ISL_2392368, EPI_ISL_2392381, EPI_ISL_2392387, EPI_ISL_2392390, EPI_ISL_2392391, EPI_ISL_2392393, EPI_ISL_2392397, EPI_ISL_2392398, EPI_ISL_2392404, EPI_ISL_2392405, EPI_ISL_2392420, EPI_ISL_2392421, EPI_ISL_2392430, EPI_ISL_2392433, EPI_ISL_2392436, EPI_ISL_2392438, EPI_ISL_2392441, EPI_ISL_2392442, EPI_ISL_2392445, EPI_ISL_2392448, EPI_ISL_2392454, EPI_ISL_2392456, EPI_ISL_2392457, EPI_ISL_2392462, EPI_ISL_2392468, EPI_ISL_2392481, EPI_ISL_2392501, EPI_ISL_2392504, EPI_ISL_2392510, EPI_ISL_2392513, EPI_ISL_2392522, EPI_ISL_2392523, EPI_ISL_2392524, EPI_ISL_2392525, EPI_ISL_2392531, EPI_ISL_2392533, EPI_ISL_2392540, EPI_ISL_2392552, EPI_ISL_2392553, EPI_ISL_2392555, EPI_ISL_2392558, EPI_ISL_2392560, EPI_ISL_2392561, EPI_ISL_2392568, EPI_ISL_2392576, EPI_ISL_2392581, EPI_ISL_2392588, EPI_ISL_2392604, EPI_ISL_2392615, EPI_ISL_2392641, EPI_ISL_2392660, EPI_ISL_2392679, EPI_ISL_2392700, EPI_ISL_2392705, EPI_ISL_2392709, EPI_ISL_2392717, EPI_ISL_2392733, EPI_ISL_2392737, EPI_ISL_2392739, EPI_ISL_2392740, EPI_ISL_2392747, EPI_ISL_2392757, EPI_ISL_2392778, EPI_ISL_2392793, EPI_ISL_2392802, EPI_ISL_2392807, EPI_ISL_2392808, EPI_ISL_2392813, EPI_ISL_2392815, EPI_ISL_2392816, EPI_ISL_2392817, EPI_ISL_2392818, EPI_ISL_2392821, EPI_ISL_2392824, EPI_ISL_2392827, EPI_ISL_2392839, EPI_ISL_2392859, EPI_ISL_2392866, EPI_ISL_2392903, EPI_ISL_2392916, EPI_ISL_2392927, EPI_ISL_2392936, EPI_ISL_2392942, EPI_ISL_2392943, EPI_ISL_2393135, EPI_ISL_2393138, EPI_ISL_2393139, EPI_ISL_2393141, EPI_ISL_2393142, EPI_ISL_2393144, EPI_ISL_2393145, EPI_ISL_2393146, EPI_ISL_2393148, EPI_ISL_2393152, EPI_ISL_2393154, EPI_ISL_2393155, EPI_ISL_2393156, EPI_ISL_2393168, EPI_ISL_2393171, EPI_ISL_2393172, EPI_ISL_2393187, EPI_ISL_2393188, EPI_ISL_2393191, EPI_ISL_2393200, EPI_ISL_2393205, EPI_ISL_2393206, EPI_ISL_2393208, EPI_ISL_2393214, EPI_ISL_2393218, EPI_ISL_2393221, EPI_ISL_2393236, EPI_ISL_2393249, EPI_ISL_2393259, EPI_ISL_2393267, EPI_ISL_2393276, EPI_ISL_2393288, EPI_ISL_2393294, EPI_ISL_2393296, EPI_ISL_2393303, EPI_ISL_2393305, EPI_ISL_2393308, EPI_ISL_2393309, EPI_ISL_2393315, EPI_ISL_2393317, EPI_ISL_2393319, EPI_ISL_2393320, EPI_ISL_2393321, EPI_ISL_2393324, EPI_ISL_2393327, EPI_ISL_2393337, EPI_ISL_2393338, EPI_ISL_2393345, EPI_ISL_2393349, EPI_ISL_2393350, EPI_ISL_2393352, EPI_ISL_2393357, EPI_ISL_2393359, EPI_ISL_2393367, EPI_ISL_2393370, EPI_ISL_2393375, EPI_ISL_2393383, EPI_ISL_2393384, EPI_ISL_2393389, EPI_ISL_2393392, EPI_ISL_2393393, EPI_ISL_2393394, EPI_ISL_2393395, EPI_ISL_2393396, EPI_ISL_2393397, EPI_ISL_2393407, EPI_ISL_2393410, EPI_ISL_2393413, EPI_ISL_2393414, EPI_ISL_2393419, EPI_ISL_2393425, EPI_ISL_2393430, EPI_ISL_2393434, EPI_ISL_2393437, EPI_ISL_2393439, EPI_ISL_2393440, EPI_ISL_2393442, EPI_ISL_2393448, EPI_ISL_2393450, EPI_ISL_2393451, EPI_ISL_2393452, EPI_ISL_2393454, EPI_ISL_2393458, EPI_ISL_2393460, EPI_ISL_2393462, EPI_ISL_2393467, EPI_ISL_2393469, EPI_ISL_2393473, EPI_ISL_2393475, EPI_ISL_2393477, EPI_ISL_2393481, EPI_ISL_2393486, EPI_ISL_2393490, EPI_ISL_2393498, EPI_ISL_2393500, EPI_ISL_2393514, EPI_ISL_2393519, EPI_ISL_2393520, EPI_ISL_2393523, EPI_ISL_2393524, EPI_ISL_2393533, EPI_ISL_2393536, EPI_ISL_2393543, EPI_ISL_2393552, EPI_ISL_2393559, EPI_ISL_2393560, EPI_ISL_2393565, EPI_ISL_2393566, EPI_ISL_2393569, EPI_ISL_2393572, EPI_ISL_2393573, EPI_ISL_2393576, EPI_ISL_2393584, EPI_ISL_2393598, EPI_ISL_2393599, EPI_ISL_2393613, EPI_ISL_2393616, EPI_ISL_2393619, EPI_ISL_2393620, EPI_ISL_2393621, EPI_ISL_2393622, EPI_ISL_2393625, EPI_ISL_2393626, EPI_ISL_2393644, EPI_ISL_2393647, EPI_ISL_2393650, EPI_ISL_2393652, EPI_ISL_2393653, EPI_ISL_2393654, EPI_ISL_2393656, EPI_ISL_2393660, EPI_ISL_2393665, EPI_ISL_2393666, EPI_ISL_2393668, EPI_ISL_2393674, EPI_ISL_2393677, EPI_ISL_2393679, EPI_ISL_2393680, EPI_ISL_2393686, EPI_ISL_2393695, EPI_ISL_2393700, EPI_ISL_2393708, EPI_ISL_2393710, EPI_ISL_2393711, EPI_ISL_2393712, EPI_ISL_2393718, EPI_ISL_2393719, EPI_ISL_2393723, EPI_ISL_2393726, EPI_ISL_2393727, EPI_ISL_2393733, EPI_ISL_2393740, EPI_ISL_2393742, EPI_ISL_2393746, EPI_ISL_2393747, EPI_ISL_2393748, EPI_ISL_2393749, EPI_ISL_2393754, EPI_ISL_2393763, EPI_ISL_2393765, EPI_ISL_2393767, EPI_ISL_2393771, EPI_ISL_2393772, EPI_ISL_2393776, EPI_ISL_2393778, EPI_ISL_2393779, EPI_ISL_2393784, EPI_ISL_2393785, EPI_ISL_2393786, EPI_ISL_2393787, EPI_ISL_2393790, EPI_ISL_2393796, EPI_ISL_2393799, EPI_ISL_2393804, EPI_ISL_2393810, EPI_ISL_2393814, EPI_ISL_2393816, EPI_ISL_2393817, EPI_ISL_2393819, EPI_ISL_2393820, EPI_ISL_2393821, EPI_ISL_2393826, EPI_ISL_2393827, EPI_ISL_2393828, EPI_ISL_2393829, EPI_ISL_2393830, EPI_ISL_2393834, EPI_ISL_2393836, EPI_ISL_2393837, EPI_ISL_2393839, EPI_ISL_2393841, EPI_ISL_2393842, EPI_ISL_2393843, EPI_ISL_2393846, EPI_ISL_2393849, EPI_ISL_2393854, EPI_ISL_2393858, EPI_ISL_2393859, EPI_ISL_2393860, EPI_ISL_2393861, EPI_ISL_2393864, EPI_ISL_2393865, EPI_ISL_2393869, EPI_ISL_2393871, EPI_ISL_2393873, EPI_ISL_2393874, EPI_ISL_2393877, EPI_ISL_2393879, EPI_ISL_2393881, EPI_ISL_2393882, EPI_ISL_2393883, EPI_ISL_2393886, EPI_ISL_2393888, EPI_ISL_2393889, EPI_ISL_2393891, EPI_ISL_2393893, EPI_ISL_2393894, EPI_ISL_2393896, EPI_ISL_2393897, EPI_ISL_2393898, EPI_ISL_2393903, EPI_ISL_2393904, EPI_ISL_2393907, EPI_ISL_2393911, EPI_ISL_2393912, EPI_ISL_2393913, EPI_ISL_2393916, EPI_ISL_2393917, EPI_ISL_2393919, EPI_ISL_2393921, EPI_ISL_2393925, EPI_ISL_2393928, EPI_ISL_2393929, EPI_ISL_2393931, EPI_ISL_2393933, EPI_ISL_2393935, EPI_ISL_2393937, EPI_ISL_2393939, EPI_ISL_2393940, EPI_ISL_2393941, EPI_ISL_2393944, EPI_ISL_2393946, EPI_ISL_2393949, EPI_ISL_2393951, EPI_ISL_2393953, EPI_ISL_2393954, EPI_ISL_2393955, EPI_ISL_2393956, EPI_ISL_2393962, EPI_ISL_2393967, EPI_ISL_2393969, EPI_ISL_2393970, EPI_ISL_2393975, EPI_ISL_2393976, EPI_ISL_2393977, EPI_ISL_2393978, EPI_ISL_2393982, EPI_ISL_2393983, EPI_ISL_2393991, EPI_ISL_2393992, EPI_ISL_2393998, EPI_ISL_2393999, EPI_ISL_2394003, EPI_ISL_2394004, EPI_ISL_2394006, EPI_ISL_2394015, EPI_ISL_2394016, EPI_ISL_2394017, EPI_ISL_2394020, EPI_ISL_2394024, EPI_ISL_2394025, EPI_ISL_2394026, EPI_ISL_2394027, EPI_ISL_2394032, EPI_ISL_2394034, EPI_ISL_2394865, EPI_ISL_2394867, EPI_ISL_2394868, EPI_ISL_2394869, EPI_ISL_2394870, EPI_ISL_2394872, EPI_ISL_2394874, EPI_ISL_2394875, EPI_ISL_2394876, EPI_ISL_2394877, EPI_ISL_2394880, EPI_ISL_2394882, EPI_ISL_2394883, EPI_ISL_2394886, EPI_ISL_2394894, EPI_ISL_2394895, EPI_ISL_2394896, EPI_ISL_2394901, EPI_ISL_2394902, EPI_ISL_2394903, EPI_ISL_2394904, EPI_ISL_2394905, EPI_ISL_2394907, EPI_ISL_2394908, EPI_ISL_2394910, EPI_ISL_2394911, EPI_ISL_2394913, EPI_ISL_2394917, EPI_ISL_2394918, EPI_ISL_2394920, EPI_ISL_2394923, EPI_ISL_2394925, EPI_ISL_2394926, EPI_ISL_2394927, EPI_ISL_2394929, EPI_ISL_2394930, EPI_ISL_2394931, EPI_ISL_2394933, EPI_ISL_2394934, EPI_ISL_2394936, EPI_ISL_2394937, EPI_ISL_2394943, EPI_ISL_2394944, EPI_ISL_2394950, EPI_ISL_2394953, EPI_ISL_2394956, EPI_ISL_2394957, EPI_ISL_2394961, EPI_ISL_2394963, EPI_ISL_2394965, EPI_ISL_2394966, EPI_ISL_2394972, EPI_ISL_2394974, EPI_ISL_2394978, EPI_ISL_2394981, EPI_ISL_2394982, EPI_ISL_2394983, EPI_ISL_2394984, EPI_ISL_2394987, EPI_ISL_2394990, EPI_ISL_2394992, EPI_ISL_2394993, EPI_ISL_2394996, EPI_ISL_2394999, EPI_ISL_2395001, EPI_ISL_2395005, EPI_ISL_2395007, EPI_ISL_2395008, EPI_ISL_2395010, EPI_ISL_2395012, EPI_ISL_2395013, EPI_ISL_2395017, EPI_ISL_2395020, EPI_ISL_2395022, EPI_ISL_2395026, EPI_ISL_2395028, EPI_ISL_2395029, EPI_ISL_2395031, EPI_ISL_2395033, EPI_ISL_2395034, EPI_ISL_2395035, EPI_ISL_2395037, EPI_ISL_2395040, EPI_ISL_2395338, EPI_ISL_2395339, EPI_ISL_2395341, EPI_ISL_2395345, EPI_ISL_2395346, EPI_ISL_2395347, EPI_ISL_2395349, EPI_ISL_2395351, EPI_ISL_2395352, EPI_ISL_2395353, EPI_ISL_2395354, EPI_ISL_2395355, EPI_ISL_2395356, EPI_ISL_2395357, EPI_ISL_2395358, EPI_ISL_2395359, EPI_ISL_2395366, EPI_ISL_2395369, EPI_ISL_2395370, EPI_ISL_2395371, EPI_ISL_2395372, EPI_ISL_2395373, EPI_ISL_2395375, EPI_ISL_2395376, EPI_ISL_2395382, EPI_ISL_2395383, EPI_ISL_2395384, EPI_ISL_2395392, EPI_ISL_2395395, EPI_ISL_2395396, EPI_ISL_2395397, EPI_ISL_2395402, EPI_ISL_2395404, EPI_ISL_2395405, EPI_ISL_2395407, EPI_ISL_2395408, EPI_ISL_2395409, EPI_ISL_2395410, EPI_ISL_2395412, EPI_ISL_2395414, EPI_ISL_2395415, EPI_ISL_2395417, EPI_ISL_2395418, EPI_ISL_2395419, EPI_ISL_2395420, EPI_ISL_2395421, EPI_ISL_2395425, EPI_ISL_2395427, EPI_ISL_2395428, EPI_ISL_2395433, EPI_ISL_2395434, EPI_ISL_2395436, EPI_ISL_2395438, EPI_ISL_2395439, EPI_ISL_2395441, EPI_ISL_2395443, EPI_ISL_2395444, EPI_ISL_2395445, EPI_ISL_2395446, EPI_ISL_2395447, EPI_ISL_2395451, EPI_ISL_2395453, EPI_ISL_2395457, EPI_ISL_2395463, EPI_ISL_2395464, EPI_ISL_2395466, EPI_ISL_2395468, EPI_ISL_2395470, EPI_ISL_2395475, EPI_ISL_2395476, EPI_ISL_2395477, EPI_ISL_2395479, EPI_ISL_2395480, EPI_ISL_2395482, EPI_ISL_2395487, EPI_ISL_2395488, EPI_ISL_2395489, EPI_ISL_2395490, EPI_ISL_2395493, EPI_ISL_2395495, EPI_ISL_2395496, EPI_ISL_2395498, EPI_ISL_2395499, EPI_ISL_2395501, EPI_ISL_2395503, EPI_ISL_2395506, EPI_ISL_2395507, EPI_ISL_2395508, EPI_ISL_2395511, EPI_ISL_2395512, EPI_ISL_2395513, EPI_ISL_2395514, EPI_ISL_2395516, EPI_ISL_2395518, EPI_ISL_2395519, EPI_ISL_2395520, EPI_ISL_2395522, EPI_ISL_2395523, EPI_ISL_2395525, EPI_ISL_2395529, EPI_ISL_2395532, EPI_ISL_2395533, EPI_ISL_2395534, EPI_ISL_2395536, EPI_ISL_2395540, EPI_ISL_2395541, EPI_ISL_2395542, EPI_ISL_2395543, EPI_ISL_2395545, EPI_ISL_2395546, EPI_ISL_2395548, EPI_ISL_2395549, EPI_ISL_2395550, EPI_ISL_2395552, EPI_ISL_2395553, EPI_ISL_2395554, EPI_ISL_2395556, EPI_ISL_2395559, EPI_ISL_2395560, EPI_ISL_2395561, EPI_ISL_2395563, EPI_ISL_2395564, EPI_ISL_2395565, EPI_ISL_2395570, EPI_ISL_2395571, EPI_ISL_2395572, EPI_ISL_2395573, EPI_ISL_2395574, EPI_ISL_2395577, EPI_ISL_2395578, EPI_ISL_2395579, EPI_ISL_2395581, EPI_ISL_2395582, EPI_ISL_2395583, EPI_ISL_2395585, EPI_ISL_2395590, EPI_ISL_2395591, EPI_ISL_2395592, EPI_ISL_2395593, EPI_ISL_2395594, EPI_ISL_2395598, EPI_ISL_2395605, EPI_ISL_2395607, EPI_ISL_2395608, EPI_ISL_2395609, EPI_ISL_2395611, EPI_ISL_2395612, EPI_ISL_2395613, EPI_ISL_2395614, EPI_ISL_2395615, EPI_ISL_2395616, EPI_ISL_2395617, EPI_ISL_2395619, EPI_ISL_2395620, EPI_ISL_2395623, EPI_ISL_2395625, EPI_ISL_2395627, EPI_ISL_2395630, EPI_ISL_2395631, EPI_ISL_2395633, EPI_ISL_2395638, EPI_ISL_2395639, EPI_ISL_2395641, EPI_ISL_2395644, EPI_ISL_2395645, EPI_ISL_2395649, EPI_ISL_2395651, EPI_ISL_2395652, EPI_ISL_2395653, EPI_ISL_2395655, EPI_ISL_2395657, EPI_ISL_2395662, EPI_ISL_2395663, EPI_ISL_2395665, EPI_ISL_2395668, EPI_ISL_2395669, EPI_ISL_2395672, EPI_ISL_2395673, EPI_ISL_2395677, EPI_ISL_2395684, EPI_ISL_2395689, EPI_ISL_2395691, EPI_ISL_2395696, EPI_ISL_2395698, EPI_ISL_2395699, EPI_ISL_2395702, EPI_ISL_2395703, EPI_ISL_2395705, EPI_ISL_2395706, EPI_ISL_2395710, EPI_ISL_2395713, EPI_ISL_2395714, EPI_ISL_2395717, EPI_ISL_2395719, EPI_ISL_2395720, EPI_ISL_2395721, EPI_ISL_2395725, EPI_ISL_2395727, EPI_ISL_2395731, EPI_ISL_2395735, EPI_ISL_2395736, EPI_ISL_2395741, EPI_ISL_2395744, EPI_ISL_2395746, EPI_ISL_2395748, EPI_ISL_2395749, EPI_ISL_2395750, EPI_ISL_2395751, EPI_ISL_2395754, EPI_ISL_2395757, EPI_ISL_2395759, EPI_ISL_2395762, EPI_ISL_2395765, EPI_ISL_2395768, EPI_ISL_2395772, EPI_ISL_2395776, EPI_ISL_2395777, EPI_ISL_2395778, EPI_ISL_2395780, EPI_ISL_2395782, EPI_ISL_2395783, EPI_ISL_2395785, EPI_ISL_2395789, EPI_ISL_2395790, EPI_ISL_2395797, EPI_ISL_2395798, EPI_ISL_2395802, EPI_ISL_2395803, EPI_ISL_2395805, EPI_ISL_2395807, EPI_ISL_2395809, EPI_ISL_2395811, EPI_ISL_2395814, EPI_ISL_2395819, EPI_ISL_2395820, EPI_ISL_2395822, EPI_ISL_2395825, EPI_ISL_2395828, EPI_ISL_2395831, EPI_ISL_2395832, EPI_ISL_2395834, EPI_ISL_2395836, EPI_ISL_2395837, EPI_ISL_2395839, EPI_ISL_2395840, EPI_ISL_2395844, EPI_ISL_2395845, EPI_ISL_2395847, EPI_ISL_2395849, EPI_ISL_2395851, EPI_ISL_2395852, EPI_ISL_2395856, EPI_ISL_2395857, EPI_ISL_2395858, EPI_ISL_2395861, EPI_ISL_2395864, EPI_ISL_2395869, EPI_ISL_2395874, EPI_ISL_2395875, EPI_ISL_2395876, EPI_ISL_2395878, EPI_ISL_2395880, EPI_ISL_2395881, EPI_ISL_2395882, EPI_ISL_2395883, EPI_ISL_2395884, EPI_ISL_2395887, EPI_ISL_2395889, EPI_ISL_2395890, EPI_ISL_2395891, EPI_ISL_2395892, EPI_ISL_2395893, EPI_ISL_2395897, EPI_ISL_2395901, EPI_ISL_2395902, EPI_ISL_2395904, EPI_ISL_2395907, EPI_ISL_2395909, EPI_ISL_2395910, EPI_ISL_2395917, EPI_ISL_2395920, EPI_ISL_2395923, EPI_ISL_2395924, EPI_ISL_2395927, EPI_ISL_2395928, EPI_ISL_2395929, EPI_ISL_2395934, EPI_ISL_2395936, EPI_ISL_2395941, EPI_ISL_2395942, EPI_ISL_2395943, EPI_ISL_2395944, EPI_ISL_2395945, EPI_ISL_2395947, EPI_ISL_2395948, EPI_ISL_2395949, EPI_ISL_2395951, EPI_ISL_2395953, EPI_ISL_2395954, EPI_ISL_2395955, EPI_ISL_2395956, EPI_ISL_2395957, EPI_ISL_2395958, EPI_ISL_2395959, EPI_ISL_2395961, EPI_ISL_2395963, EPI_ISL_2395964, EPI_ISL_2395965, EPI_ISL_2395966, EPI_ISL_2395967, EPI_ISL_2395971, EPI_ISL_2395972, EPI_ISL_2395973, EPI_ISL_2395974, EPI_ISL_2395977, EPI_ISL_2395979, EPI_ISL_2395986, EPI_ISL_2395987, EPI_ISL_2395988, EPI_ISL_2395990, EPI_ISL_2395991, EPI_ISL_2395992, EPI_ISL_2395994, EPI_ISL_2395995, EPI_ISL_2395996, EPI_ISL_2395998, EPI_ISL_2396005, EPI_ISL_2396006, EPI_ISL_2396007, EPI_ISL_2396009, EPI_ISL_2396011, EPI_ISL_2396012, EPI_ISL_2396013, EPI_ISL_2396014, EPI_ISL_2396370, EPI_ISL_2406622, EPI_ISL_2406746, EPI_ISL_2406896, EPI_ISL_2406899, EPI_ISL_2406900, EPI_ISL_2406904, EPI_ISL_2406905, EPI_ISL_2406907, EPI_ISL_2406910, EPI_ISL_2406913, EPI_ISL_2406917, EPI_ISL_2406918, EPI_ISL_2406921, EPI_ISL_2406924, EPI_ISL_2406926, EPI_ISL_2406929, EPI_ISL_2406930, EPI_ISL_2406931, EPI_ISL_2406935, EPI_ISL_2406936, EPI_ISL_2406937, EPI_ISL_2406941, EPI_ISL_2406944, EPI_ISL_2406947, EPI_ISL_2406955, EPI_ISL_2406958, EPI_ISL_2406960, EPI_ISL_2406964, EPI_ISL_2406968, EPI_ISL_2406969, EPI_ISL_2406970, EPI_ISL_2406971, EPI_ISL_2406972, EPI_ISL_2406975, EPI_ISL_2406982, EPI_ISL_2406984, EPI_ISL_2406987, EPI_ISL_2406990, EPI_ISL_2406991, EPI_ISL_2407002, EPI_ISL_2407003, EPI_ISL_2407004, EPI_ISL_2407006, EPI_ISL_2407008, EPI_ISL_2407009, EPI_ISL_2407011, EPI_ISL_2407012, EPI_ISL_2407016, EPI_ISL_2407018, EPI_ISL_2407021, EPI_ISL_2407024, EPI_ISL_2407031, EPI_ISL_2407035, EPI_ISL_2407036, EPI_ISL_2407041, EPI_ISL_2407043, EPI_ISL_2407045, EPI_ISL_2407047, EPI_ISL_2407048, EPI_ISL_2407049, EPI_ISL_2407052, EPI_ISL_2407054, EPI_ISL_2407058, EPI_ISL_2407059, EPI_ISL_2407060, EPI_ISL_2407065, EPI_ISL_2407068, EPI_ISL_2407069, EPI_ISL_2407070, EPI_ISL_2407071, EPI_ISL_2407073, EPI_ISL_2407074, EPI_ISL_2407075, EPI_ISL_2407077, EPI_ISL_2407078, EPI_ISL_2407079, EPI_ISL_2407080, EPI_ISL_2407081, EPI_ISL_2407090, EPI_ISL_2407092, EPI_ISL_2407097, EPI_ISL_2407098, EPI_ISL_2407099, EPI_ISL_2407100, EPI_ISL_2407102, EPI_ISL_2407104, EPI_ISL_2407107, EPI_ISL_2407109, EPI_ISL_2407110, EPI_ISL_2408061, EPI_ISL_2435382, EPI_ISL_2435446, EPI_ISL_2435454, EPI_ISL_2435526, EPI_ISL_2435560, EPI_ISL_2435651, EPI_ISL_2435690, EPI_ISL_2435807, EPI_ISL_2436429, EPI_ISL_2436565, EPI_ISL_2436642, EPI_ISL_2436648, EPI_ISL_2436715, EPI_ISL_2436731, EPI_ISL_2436930, EPI_ISL_2436933, EPI_ISL_2436934, EPI_ISL_2436938, EPI_ISL_2436947, EPI_ISL_2436955, EPI_ISL_2436960, EPI_ISL_2436972, EPI_ISL_2436997, EPI_ISL_2437005, EPI_ISL_2437013, EPI_ISL_2437029, EPI_ISL_2437034, EPI_ISL_2437066, EPI_ISL_2437068, EPI_ISL_2437094, EPI_ISL_2437141, EPI_ISL_2437149, EPI_ISL_2437150, EPI_ISL_2437199, EPI_ISL_2437203, EPI_ISL_2437209, EPI_ISL_2437216, EPI_ISL_2437235, EPI_ISL_2437237, EPI_ISL_2437240, EPI_ISL_2437253, EPI_ISL_2437260, EPI_ISL_2437261, EPI_ISL_2437298, EPI_ISL_2437304, EPI_ISL_2437322, EPI_ISL_2437325, EPI_ISL_2437341, EPI_ISL_2437342, EPI_ISL_2437370, EPI_ISL_2437408, EPI_ISL_2437433, EPI_ISL_2437456, EPI_ISL_2437458, EPI_ISL_2437459, EPI_ISL_2437489, EPI_ISL_2437561, EPI_ISL_2437607, EPI_ISL_2437697, EPI_ISL_2437730, EPI_ISL_2437752, EPI_ISL_2437757, EPI_ISL_2437794, EPI_ISL_2437842, EPI_ISL_2437931, EPI_ISL_2437947, EPI_ISL_2438076, EPI_ISL_2438081, EPI_ISL_2438183, EPI_ISL_2438276, EPI_ISL_2438370, EPI_ISL_2438401, EPI_ISL_2438427, EPI_ISL_2446220, EPI_ISL_2446417, EPI_ISL_2446418, EPI_ISL_2446428, EPI_ISL_2446433, EPI_ISL_2446437, EPI_ISL_2446438, EPI_ISL_2446442, EPI_ISL_2446453, EPI_ISL_2446456, EPI_ISL_2446458, EPI_ISL_2446460, EPI_ISL_2446466, EPI_ISL_2446473, EPI_ISL_2446475,

| | | | |
|---|---|---|---|
| EPI_ISL_2446477, EPI_ISL_2446480, EPI_ISL_2446481, EPI_ISL_2446495, EPI_ISL_2446498, EPI_ISL_2446500, EPI_ISL_2446507, EPI_ISL_2446514, EPI_ISL_2446519, EPI_ISL_2446520, EPI_ISL_2446521, EPI_ISL_2446526, EPI_ISL_2446528, EPI_ISL_2446529, EPI_ISL_2446533, EPI_ISL_2446537, EPI_ISL_2446539, EPI_ISL_2446546, EPI_ISL_2446559, EPI_ISL_2446564, EPI_ISL_2446569, EPI_ISL_2446570, EPI_ISL_2446573, EPI_ISL_2446574, EPI_ISL_2446578, EPI_ISL_2446589, EPI_ISL_2446598, EPI_ISL_2446601, EPI_ISL_2446605, EPI_ISL_2446619, EPI_ISL_2446625, EPI_ISL_2446628, EPI_ISL_2446629, EPI_ISL_2446630, EPI_ISL_2446638, EPI_ISL_2446640, EPI_ISL_2446643, EPI_ISL_2446645, EPI_ISL_2446646, EPI_ISL_2446659, EPI_ISL_2446660, EPI_ISL_2446662, EPI_ISL_2446676, EPI_ISL_2446678, EPI_ISL_2446680, EPI_ISL_2446681, EPI_ISL_2446686, EPI_ISL_2446687, EPI_ISL_2446689, EPI_ISL_2446690, EPI_ISL_2446692, EPI_ISL_2446700, EPI_ISL_2446713, EPI_ISL_2446723, EPI_ISL_2446724, EPI_ISL_2446736, EPI_ISL_2446744, EPI_ISL_2446745, EPI_ISL_2446749, EPI_ISL_2446750, EPI_ISL_2456251, EPI_ISL_2456829, EPI_ISL_2456830, EPI_ISL_2456835, EPI_ISL_2456836, EPI_ISL_2456838, EPI_ISL_2456840, EPI_ISL_2456841, EPI_ISL_2456843, EPI_ISL_2456845, EPI_ISL_2456848, EPI_ISL_2456849, EPI_ISL_2456850, EPI_ISL_2456851, EPI_ISL_2456852, EPI_ISL_2456853, EPI_ISL_2456857, EPI_ISL_2456859, EPI_ISL_2456861, EPI_ISL_2456862, EPI_ISL_2456866, EPI_ISL_2456869, EPI_ISL_2456871, EPI_ISL_2456873, EPI_ISL_2456876, EPI_ISL_2456879, EPI_ISL_2456888, EPI_ISL_2456889, EPI_ISL_2456893, EPI_ISL_2456894, EPI_ISL_2456895, EPI_ISL_2456896, EPI_ISL_2456901, EPI_ISL_2456909, EPI_ISL_2456911, EPI_ISL_2456916, EPI_ISL_2456917, EPI_ISL_2456922, EPI_ISL_2456926, EPI_ISL_2456930, EPI_ISL_2456932, EPI_ISL_2456934, EPI_ISL_2456938, EPI_ISL_2456939, EPI_ISL_2456941, EPI_ISL_2456942, EPI_ISL_2456948, EPI_ISL_2456952, EPI_ISL_2456955, EPI_ISL_2456956, EPI_ISL_2456967, EPI_ISL_2456970, EPI_ISL_2456971, EPI_ISL_2456973, EPI_ISL_2456980, EPI_ISL_2456983, EPI_ISL_2456985, EPI_ISL_2456986, EPI_ISL_2456991, EPI_ISL_2456992, EPI_ISL_2456994, EPI_ISL_2456997, EPI_ISL_2457007, EPI_ISL_2457014, EPI_ISL_2457016, EPI_ISL_2457020, EPI_ISL_2457021, EPI_ISL_2457022, EPI_ISL_2457033, EPI_ISL_2457035, EPI_ISL_2457037, EPI_ISL_2457043, EPI_ISL_2457044, EPI_ISL_2457045, EPI_ISL_2457047, EPI_ISL_2457048, EPI_ISL_2457050, EPI_ISL_2457053, EPI_ISL_2457054, EPI_ISL_2457057, EPI_ISL_2457060, EPI_ISL_2457063, EPI_ISL_2457064, EPI_ISL_2457069, EPI_ISL_2457070, EPI_ISL_2457071, EPI_ISL_2457072, EPI_ISL_2457077, EPI_ISL_2457078, EPI_ISL_2457079, EPI_ISL_2457080, EPI_ISL_2457082, EPI_ISL_2457084, EPI_ISL_2457085, EPI_ISL_2457089, EPI_ISL_2457091, EPI_ISL_2457094, EPI_ISL_2457096, EPI_ISL_2457097, EPI_ISL_2457099, EPI_ISL_2457102, EPI_ISL_2457104, EPI_ISL_2457109, EPI_ISL_2457110, EPI_ISL_2457112, EPI_ISL_2457118, EPI_ISL_2457124, EPI_ISL_2457126, EPI_ISL_2457128, EPI_ISL_2457130, EPI_ISL_2457502, EPI_ISL_2457505, EPI_ISL_2540945 | | | |
| see above | Lighthouse Lab in Milton Keynes | Wellcome Sanger Institute for the COVID-19 Genomics UK (COG-UK) Consortium | Cordelia Langford; David K. Jackson; Dominic Kwiatkowski; Ewan Harrison; Ian Johnston; Jeffrey Barrett; John Sillitoe on behalf of the Wellcome Sanger Institute COVID-19 Surveillance Team; Roberto Amato; Sonia Goncalves; The Lighthouse Lab in Milton Keynes and Alex Alderton |
| EPI_ISL_2393137, EPI_ISL_2393140, EPI_ISL_2393150, EPI_ISL_2393157, EPI_ISL_2393159, EPI_ISL_2393177, EPI_ISL_2393180, EPI_ISL_2393189, EPI_ISL_2393195, EPI_ISL_2393199, EPI_ISL_2393211, EPI_ISL_2393215, EPI_ISL_2393219, EPI_ISL_2393226, EPI_ISL_2393228, EPI_ISL_2393233, EPI_ISL_2393240, EPI_ISL_2393243, EPI_ISL_2393244, EPI_ISL_2393245, EPI_ISL_2393250, EPI_ISL_2393251, EPI_ISL_2393264, EPI_ISL_2393278, EPI_ISL_2393279, EPI_ISL_2393281, EPI_ISL_2393284, EPI_ISL_2393293, EPI_ISL_2393295, EPI_ISL_2393304, EPI_ISL_2393311, EPI_ISL_2393318, EPI_ISL_2393323, EPI_ISL_2393328, EPI_ISL_2393332, EPI_ISL_2393342, EPI_ISL_2393343, EPI_ISL_2393347, EPI_ISL_2393351, EPI_ISL_2393353, EPI_ISL_2393358, EPI_ISL_2393360, EPI_ISL_2393368, EPI_ISL_2393372, EPI_ISL_2393374, EPI_ISL_2393376, EPI_ISL_2393387, EPI_ISL_2393405, EPI_ISL_2393406, EPI_ISL_2393420, EPI_ISL_2393424, EPI_ISL_2393428, EPI_ISL_2393431, EPI_ISL_2393435, EPI_ISL_2393441, EPI_ISL_2393445, EPI_ISL_2393446, EPI_ISL_2393453, EPI_ISL_2393470, EPI_ISL_2393483, EPI_ISL_2393484, EPI_ISL_2447234, EPI_ISL_2447236, EPI_ISL_2447241, EPI_ISL_2447243, EPI_ISL_2447245, EPI_ISL_2447247, EPI_ISL_2447248, EPI_ISL_2447251, EPI_ISL_2447252, EPI_ISL_2447254, EPI_ISL_2447257, EPI_ISL_2447262, EPI_ISL_2447265, EPI_ISL_2447267, EPI_ISL_2447276, EPI_ISL_2447279, EPI_ISL_2447283, EPI_ISL_2447293, EPI_ISL_2447294, EPI_ISL_2447296, EPI_ISL_2447302, EPI_ISL_2447306, EPI_ISL_2447307, EPI_ISL_2447315, EPI_ISL_2447318, EPI_ISL_2447319, EPI_ISL_2447325, EPI_ISL_2447326, EPI_ISL_2447330, EPI_ISL_2447334, EPI_ISL_2447337, EPI_ISL_2447338, EPI_ISL_2447339, EPI_ISL_2447341, EPI_ISL_2447346, EPI_ISL_2447348, EPI_ISL_2447350, EPI_ISL_2447355, EPI_ISL_2447356, EPI_ISL_2447357, EPI_ISL_2447359, EPI_ISL_2514019 | | | |
| see above | Lighthouse Laboratory Plymouth | Wellcome Sanger Institute for the COVID-19 Genomics UK (COG-UK) Consortium | Cordelia Langford; David K. Jackson; Dominic Kwiatkowski; Ewan Harrison; Ian Johnston; Jeffrey Barrett; John Sillitoe on behalf of the Wellcome Sanger Institute COVID-19 Surveillance Team; Lighthouse Laboratory Plymouth and Alex Alderton; Roberto Amato; Sonia Goncalves |
| EPI_ISL_2434251, EPI_ISL_2434253, EPI_ISL_2434254, EPI_ISL_2434255, EPI_ISL_2434256, EPI_ISL_2434257, EPI_ISL_2434258 | | | |
| see above | Liverpool Clinical Laboratories | COVID-19 Genomics UK (COG-UK) Consortium | A Alrezaihi; Alessandro Gerada; Alistair Darby; Angela Cowell; Anita Lucaci; Anu Chawla; Cassie Olateju; Catherine Hartley; Charlotte Nelson; Ecaterina Vamos; Elaine O'Toole; Eleanor G Bentley; Ghada T Shawli; Hermione Webster; Isabel García-Dorival; James Johnson; James P Stewart; Jenifer Manson; Joanne Watts; Jones Benjamin; Jordan J Clark; Julian Hiscox; L Luu; Lucille Rainbow; M Almsaud; Margaret Hughes; Mark Whitehead; Matthew Gemmell; Miren Iturriza-Gomara; Muhannad Alruwaili; N.P Randle; Neil Swainston; PKF Gilmore; Parul Sharma; Rebekah Penrice-Randal; Richard Eccles; Richard Gregory; Sam Haldenby; Steve Paterson; Stuart D Armstrong; Trevor Ian Robinson; Ximeng Han |
| EPI_ISL_2434278, EPI_ISL_2434279, EPI_ISL_2434287, EPI_ISL_2434293, EPI_ISL_2434300, EPI_ISL_2434301, EPI_ISL_2434308, EPI_ISL_2434309, EPI_ISL_2434316, EPI_ISL_2434317, EPI_ISL_2434321, EPI_ISL_2434323, EPI_ISL_2434324, EPI_ISL_2434330, EPI_ISL_2434331, EPI_ISL_2434335, EPI_ISL_2434336, EPI_ISL_2434337, EPI_ISL_2434341, EPI_ISL_2434343, EPI_ISL_2434347, EPI_ISL_2434351, EPI_ISL_2434352, EPI_ISL_2434362, EPI_ISL_2434374, EPI_ISL_2434381, EPI_ISL_2434396, EPI_ISL_2434680, EPI_ISL_2434681, EPI_ISL_2434685, EPI_ISL_2434687, EPI_ISL_2434692, EPI_ISL_2434693, EPI_ISL_2434698, EPI_ISL_2434699, EPI_ISL_2434700, EPI_ISL_2434707, EPI_ISL_2434713, EPI_ISL_2434714, EPI_ISL_2434719, EPI_ISL_2434720, EPI_ISL_2434721, EPI_ISL_2434726, EPI_ISL_2434727, EPI_ISL_2434731, EPI_ISL_2434732, EPI_ISL_2434733, EPI_ISL_2434737, EPI_ISL_2434738, EPI_ISL_2434739, EPI_ISL_2434743, EPI_ISL_2434744, EPI_ISL_2434745, EPI_ISL_2434750, EPI_ISL_2434751, EPI_ISL_2434755, EPI_ISL_2517859, EPI_ISL_2517860, EPI_ISL_2517861, EPI_ISL_2517862, EPI_ISL_2517863, EPI_ISL_2517864, EPI_ISL_2517865, EPI_ISL_2517866, EPI_ISL_2517867, EPI_ISL_2517868, EPI_ISL_2517869, EPI_ISL_2517870, EPI_ISL_2517871, EPI_ISL_2517872, EPI_ISL_2517873, EPI_ISL_2517874, EPI_ISL_2517875, EPI_ISL_2517876, EPI_ISL_2517877, EPI_ISL_2517878, EPI_ISL_2517879, EPI_ISL_2517880, EPI_ISL_2517881, EPI_ISL_2517882, EPI_ISL_2517883, EPI_ISL_2517884, EPI_ISL_2517885, EPI_ISL_2517886, EPI_ISL_2517887, EPI_ISL_2517888, EPI_ISL_2517889, EPI_ISL_2517890, EPI_ISL_2517891, EPI_ISL_2517892, EPI_ISL_2517893, EPI_ISL_2517894, EPI_ISL_2517895, EPI_ISL_2517896, EPI_ISL_2517897, EPI_ISL_2517898, EPI_ISL_2517899, EPI_ISL_2517900, EPI_ISL_2517901, EPI_ISL_2517902, EPI_ISL_2517903, EPI_ISL_2517904, EPI_ISL_2517905, EPI_ISL_2517906, EPI_ISL_2517907, EPI_ISL_2517908, EPI_ISL_2517910, EPI_ISL_2517911, EPI_ISL_2517912, EPI_ISL_2517913, EPI_ISL_2517914, EPI_ISL_2517915, EPI_ISL_2517916, EPI_ISL_2517917, EPI_ISL_2517918, EPI_ISL_2517919, EPI_ISL_2517920, EPI_ISL_2517921, EPI_ISL_2517922, EPI_ISL_2517923, EPI_ISL_2517924, EPI_ISL_2517925, EPI_ISL_2517926, EPI_ISL_2517927, EPI_ISL_2517928, EPI_ISL_2517929, EPI_ISL_2517930, EPI_ISL_2517931, EPI_ISL_2517932, EPI_ISL_2517933, EPI_ISL_2517934, EPI_ISL_2517935, EPI_ISL_2517936, EPI_ISL_2517937, EPI_ISL_2517938, EPI_ISL_2517939, EPI_ISL_2517940, EPI_ISL_2517941, EPI_ISL_2517942, EPI_ISL_2517943, EPI_ISL_2517944, EPI_ISL_2517945, EPI_ISL_2517946, EPI_ISL_2517947, EPI_ISL_2517948, EPI_ISL_2517949, EPI_ISL_2517950, EPI_ISL_2517951, EPI_ISL_2517952, EPI_ISL_2517953, EPI_ISL_2517954, EPI_ISL_2517955, EPI_ISL_2517956, EPI_ISL_2517957, EPI_ISL_2517958, EPI_ISL_2517959, EPI_ISL_2517960, EPI_ISL_2517961, EPI_ISL_2517962, EPI_ISL_2517963, EPI_ISL_2517964, EPI_ISL_2517965, EPI_ISL_2517966, EPI_ISL_2517967, EPI_ISL_2517968, EPI_ISL_2517969, EPI_ISL_2517970, EPI_ISL_2517971, EPI_ISL_2517972, EPI_ISL_2517973, EPI_ISL_2517974, EPI_ISL_2517975, EPI_ISL_2517976, EPI_ISL_2517977, EPI_ISL_2517978, EPI_ISL_2517979, EPI_ISL_2517980, EPI_ISL_2517981, EPI_ISL_2517982, EPI_ISL_2517983, EPI_ISL_2517984, EPI_ISL_2517985, EPI_ISL_2517986, EPI_ISL_2517987, EPI_ISL_2517988, EPI_ISL_2517989, EPI_ISL_2517990, EPI_ISL_2517991, EPI_ISL_2517992, EPI_ISL_2517993, EPI_ISL_2517994, EPI_ISL_2517995, EPI_ISL_2517996, EPI_ISL_2517997, EPI_ISL_2517998, EPI_ISL_2517999, EPI_ISL_2518000, EPI_ISL_2518001, EPI_ISL_2518002, EPI_ISL_2518003, EPI_ISL_2518004, EPI_ISL_2518005, EPI_ISL_2518007, EPI_ISL_2518008, EPI_ISL_2518009, EPI_ISL_2518010, EPI_ISL_2518011, EPI_ISL_2518012, EPI_ISL_2518013, EPI_ISL_2518014, EPI_ISL_2518015, EPI_ISL_2518016, EPI_ISL_2518017, EPI_ISL_2518018, EPI_ISL_2518019, EPI_ISL_2518020, EPI_ISL_2518021, EPI_ISL_2518022, EPI_ISL_2518023, EPI_ISL_2518024, EPI_ISL_2518025, EPI_ISL_2518026, EPI_ISL_2518027, EPI_ISL_2518028, EPI_ISL_2518029, EPI_ISL_2518030, EPI_ISL_2518031, EPI_ISL_2518032, EPI_ISL_2518033, EPI_ISL_2518034, EPI_ISL_2518035, EPI_ISL_2518036, EPI_ISL_2518037, EPI_ISL_2518038, EPI_ISL_2518039, EPI_ISL_2518040, EPI_ISL_2518041, EPI_ISL_2518042, EPI_ISL_2518043, EPI_ISL_2518044, EPI_ISL_2518045, EPI_ISL_2518046, EPI_ISL_2518047, EPI_ISL_2518048, EPI_ISL_2518049, EPI_ISL_2518050, EPI_ISL_2518051, EPI_ISL_2518052, EPI_ISL_2518053, EPI_ISL_2518054, EPI_ISL_2518055, EPI_ISL_2518056, EPI_ISL_2518057, EPI_ISL_2518058, EPI_ISL_2518059, EPI_ISL_2518060, EPI_ISL_2518061, EPI_ISL_2518062, EPI_ISL_2518063, EPI_ISL_2518064, EPI_ISL_2518065, EPI_ISL_2518066, EPI_ISL_2518067, EPI_ISL_2518068, EPI_ISL_2518069, EPI_ISL_2518070, EPI_ISL_2518071, EPI_ISL_2518072, EPI_ISL_2518073, EPI_ISL_2518074, EPI_ISL_2518075, EPI_ISL_2518076, EPI_ISL_2518077, EPI_ISL_2518078, EPI_ISL_2518079, EPI_ISL_2518080, EPI_ISL_2518081, EPI_ISL_2518082, EPI_ISL_2518083, EPI_ISL_2518084, EPI_ISL_2518085, EPI_ISL_2518086, EPI_ISL_2518087, EPI_ISL_2518088, EPI_ISL_2518089, EPI_ISL_2518090, EPI_ISL_2518091, EPI_ISL_2518092, EPI_ISL_2518093, EPI_ISL_2518094, EPI_ISL_2518095, EPI_ISL_2518096, EPI_ISL_2518097, EPI_ISL_2518098, EPI_ISL_2518099, EPI_ISL_2518100, EPI_ISL_2518101, EPI_ISL_2518102, EPI_ISL_2518103, EPI_ISL_2518104, EPI_ISL_2518105, EPI_ISL_2518106, EPI_ISL_2518107, EPI_ISL_2518108, EPI_ISL_2518109, EPI_ISL_2518110, EPI_ISL_2518111, EPI_ISL_2518112, EPI_ISL_2518113, EPI_ISL_2518115, EPI_ISL_2518116, EPI_ISL_2518117, EPI_ISL_2518119, EPI_ISL_2518120, EPI_ISL_2518121, EPI_ISL_2518122, EPI_ISL_2518123, EPI_ISL_2518124, EPI_ISL_2518125, EPI_ISL_2518126, EPI_ISL_2518127, EPI_ISL_2518128, EPI_ISL_2518129, EPI_ISL_2518130, EPI_ISL_2518131, EPI_ISL_2518133, EPI_ISL_2518135, EPI_ISL_2518136, EPI_ISL_2518137, EPI_ISL_2518139, EPI_ISL_2518140, EPI_ISL_2518141, EPI_ISL_2518143, EPI_ISL_2518144, EPI_ISL_2518145, EPI_ISL_2518146, EPI_ISL_2518147, EPI_ISL_2518148, EPI_ISL_2518149, EPI_ISL_2518150, EPI_ISL_2518151, EPI_ISL_2518152, EPI_ISL_2518153, EPI_ISL_2518154, EPI_ISL_2518155, EPI_ISL_2518156, EPI_ISL_2518157, EPI_ISL_2518158, EPI_ISL_2518159, EPI_ISL_2518160, EPI_ISL_2518161, EPI_ISL_2518162, EPI_ISL_2518163, EPI_ISL_2518165, EPI_ISL_2518166, EPI_ISL_2518167, EPI_ISL_2518168, EPI_ISL_2518169, EPI_ISL_2518170, EPI_ISL_2518171, EPI_ISL_2518172, EPI_ISL_2518173, EPI_ISL_2518174, EPI_ISL_2518175, EPI_ISL_2518176, EPI_ISL_2518178, EPI_ISL_2518179, EPI_ISL_2518180, EPI_ISL_2518181, EPI_ISL_2518182, EPI_ISL_2518185, EPI_ISL_2518186, EPI_ISL_2518187, EPI_ISL_2518188, EPI_ISL_2518189, EPI_ISL_2518190, EPI_ISL_2518191, EPI_ISL_2518192, EPI_ISL_2518193, EPI_ISL_2518194, EPI_ISL_2518195, EPI_ISL_2518196, EPI_ISL_2518197, EPI_ISL_2518198, EPI_ISL_2518199, EPI_ISL_2518200, EPI_ISL_2518201, EPI_ISL_2518202, EPI_ISL_2518204, EPI_ISL_2518205, EPI_ISL_2518207, EPI_ISL_2518208, EPI_ISL_2518209, EPI_ISL_2518210, EPI_ISL_2518211, EPI_ISL_2518212, EPI_ISL_2518213, EPI_ISL_2518214, EPI_ISL_2518215, EPI_ISL_2518216, EPI_ISL_2518217, EPI_ISL_2518218, EPI_ISL_2518219, EPI_ISL_2518220, EPI_ISL_2518221, EPI_ISL_2518222, EPI_ISL_2518223, EPI_ISL_2518224, EPI_ISL_2518225, EPI_ISL_2518226, EPI_ISL_2518227, EPI_ISL_2518228, EPI_ISL_2518230, EPI_ISL_2518231, EPI_ISL_2518232, EPI_ISL_2518233, EPI_ISL_2518234, EPI_ISL_2518235, EPI_ISL_2518236, EPI_ISL_2518237, EPI_ISL_2518238, EPI_ISL_2518239, EPI_ISL_2518240, EPI_ISL_2518241, EPI_ISL_2518242, EPI_ISL_2518243, EPI_ISL_2518244, EPI_ISL_2518245, EPI_ISL_2518246, EPI_ISL_2518247, EPI_ISL_2518248, EPI_ISL_2518249, EPI_ISL_2518250, EPI_ISL_2518251, EPI_ISL_2518252, EPI_ISL_2518253, EPI_ISL_2518254, EPI_ISL_2518255, EPI_ISL_2518256, EPI_ISL_2518257, EPI_ISL_2518258, EPI_ISL_2518259, EPI_ISL_2518260, EPI_ISL_2518261, EPI_ISL_2518262, EPI_ISL_2518263, EPI_ISL_2518264, EPI_ISL_2518265, EPI_ISL_2518267, EPI_ISL_2518268, EPI_ISL_2518269, EPI_ISL_2518270, EPI_ISL_2518272, EPI_ISL_2518273, EPI_ISL_2518274, EPI_ISL_2518275, EPI_ISL_2518276, EPI_ISL_2518277, EPI_ISL_2518278, EPI_ISL_2518279, EPI_ISL_2518280, EPI_ISL_2518281, EPI_ISL_2518282, EPI_ISL_2518283, EPI_ISL_2518284, EPI_ISL_2518285, EPI_ISL_2518287, EPI_ISL_2518288, EPI_ISL_2518289, EPI_ISL_2518290, EPI_ISL_2518291, EPI_ISL_2518292, EPI_ISL_2518293, EPI_ISL_2518294, EPI_ISL_2518295, EPI_ISL_2518296, EPI_ISL_2518297, EPI_ISL_2518298, EPI_ISL_2518299, EPI_ISL_2518300, EPI_ISL_2518301, EPI_ISL_2518302, EPI_ISL_2518303, EPI_ISL_2518304, EPI_ISL_2518305, EPI_ISL_2518306, EPI_ISL_2518307, EPI_ISL_2518308, EPI_ISL_2518309, EPI_ISL_2518460, EPI_ISL_2518463, EPI_ISL_2518677 | | | |
| see above | Northumbria University / South Tees Hospitals NHS Foundation Trust / North Cumbria Integrated Care NHS Foundation Trust / North Tees and Hartlepool NHS Foundation Trust / Newcastle Hospitals NHS Foundation Trust | COVID-19 Genomics UK (COG-UK) Consortium | Andrew Nelson; Brendan Payne; Clive Graham; Darren L Smith; Debra Padgett; Edward Barton; Emma Swindells; Garren Scott; Gary Black; Gary Eltringham; Giles S Holt; Greg R Young; Jane Greenaway; Jennifer Collins; John Allan; Joshua Loh; Lynn Dover; Matthew Bashton; Mohammad A Tariq; Paul Baker; Sarah Essex; Steve Liggett; Wen C Yew; Yusri Taha |
| EPI_ISL_2434783, EPI_ISL_2434784, EPI_ISL_2434788, EPI_ISL_2434793, EPI_ISL_2434794, EPI_ISL_2434796, EPI_ISL_2434797, EPI_ISL_2434799, EPI_ISL_2434810, EPI_ISL_2434814, EPI_ISL_2434815, EPI_ISL_2434816, EPI_ISL_2434821, EPI_ISL_2434822, EPI_ISL_2434824, EPI_ISL_2434830, EPI_ISL_2434832, EPI_ISL_2434833, EPI_ISL_2434834, EPI_ISL_2434835, EPI_ISL_2434837, EPI_ISL_2434838, EPI_ISL_2434839, EPI_ISL_2434840, EPI_ISL_2434841, EPI_ISL_2434843, EPI_ISL_2434844, EPI_ISL_2434849, EPI_ISL_2434850, EPI_ISL_2434853, EPI_ISL_2434856, EPI_ISL_2434857, EPI_ISL_2434860, EPI_ISL_2434861, EPI_ISL_2434862, EPI_ISL_2434863, EPI_ISL_2434864, EPI_ISL_2434868, EPI_ISL_2434872, EPI_ISL_2434873, EPI_ISL_2434874, EPI_ISL_2434875, EPI_ISL_2434876, EPI_ISL_2434877, EPI_ISL_2434878, EPI_ISL_2434883, EPI_ISL_2434891, EPI_ISL_2434892, EPI_ISL_2434895, EPI_ISL_2434897, EPI_ISL_2434899, EPI_ISL_2434905, EPI_ISL_2434907, EPI_ISL_2434908, EPI_ISL_2434909, EPI_ISL_2434911, EPI_ISL_2434912, EPI_ISL_2434914, EPI_ISL_2434915, EPI_ISL_2434917, EPI_ISL_2434923, EPI_ISL_2434928, EPI_ISL_2434930, EPI_ISL_2434932, EPI_ISL_2434933, EPI_ISL_2434934, EPI_ISL_2434936, EPI_ISL_2434938, EPI_ISL_2434941, EPI_ISL_2434943, EPI_ISL_2434944, EPI_ISL_2434947, EPI_ISL_2434949, EPI_ISL_2434950, EPI_ISL_2434953, EPI_ISL_2434954, EPI_ISL_2434955, EPI_ISL_2434963 | | | |
| see above | Originating lab: Wales Specialist Virology Centre Sequencing lab: Pathogen Genomics Unit | Public Health Wales Microbiology Cardiff Wales Specialist Virology Centre | Alec Birchley; Alexander Adams; Amy Gaskin; Angela Marchbank; Bree Gatica-Wilcox; Catherine Moore; Jason Coombes; Joanne Watkins; Joel Southgate; Johnathan Evans; Laura Gifford; Lauren Gilbert; Lee Graham; Malorie Perry; Matthew Bull; Nicole Pacchiarini; Sally Corden; Sara Kumziene-Summerhayes; Sara Rey; Sarah Taylor; Simon Cottrell; Sophie Jones; Tom Connor |
| EPI_ISL_2624537, EPI_ISL_2624543, EPI_ISL_2624549, EPI_ISL_2624551, EPI_ISL_2624552, EPI_ISL_2624553, EPI_ISL_2624558, EPI_ISL_2624559, EPI_ISL_2624572 | | | |
| see above | Oxford Viromics, NDM, University of Oxford; Oxford University Hospitals; Basingstoke and North Hampshire Hospital | COVID-19 Genomics UK (COG-UK) Consortium | Alex Mobbs; Amy Trebes; Anita Justice; Catrin Moore; Christophe Fraser; David Bonsall; David Buck; Emma Wise; George Macintyre; Jessica Lynch; John Todd; Mariateresa de Cesare; Matilde Mori; Monique Andersson; Nathan Moore; Nick Cortes; Robert Shaw; Stephen Kidd; Tanya Golubchik; Timothy Peto |
| EPI_ISL_2434177, EPI_ISL_2434179, EPI_ISL_2434181, EPI_ISL_2434189, EPI_ISL_2519015, EPI_ISL_2519125, EPI_ISL_2519131, EPI_ISL_2519167, EPI_ISL_2519172, EPI_ISL_2519186, EPI_ISL_2519760, EPI_ISL_2519790, EPI_ISL_2519972, EPI_ISL_2519973, EPI_ISL_2519975, EPI_ISL_2519979, EPI_ISL_2519987, EPI_ISL_2519988, EPI_ISL_2519989, EPI_ISL_2519992, EPI_ISL_2519994, EPI_ISL_2519996, EPI_ISL_2519997, EPI_ISL_2520001, EPI_ISL_2520003, EPI_ISL_2520005, EPI_ISL_2520018, EPI_ISL_2520019, EPI_ISL_2520022, EPI_ISL_2520023, EPI_ISL_2520024, EPI_ISL_2520028, EPI_ISL_2520030, EPI_ISL_2520031, EPI_ISL_2520032, EPI_ISL_2520050, EPI_ISL_2520055, EPI_ISL_2520056, EPI_ISL_2520058, EPI_ISL_2520063, EPI_ISL_2520064, EPI_ISL_2520067, EPI_ISL_2520071, EPI_ISL_2520072, EPI_ISL_2520073, EPI_ISL_2520074, EPI_ISL_2520076, EPI_ISL_2520077, EPI_ISL_2520078, EPI_ISL_2520079, EPI_ISL_2520081, EPI_ISL_2520082, EPI_ISL_2520083, EPI_ISL_2520089, EPI_ISL_2520090, EPI_ISL_2520097, EPI_ISL_2520098, EPI_ISL_2520099, EPI_ISL_2520100, EPI_ISL_2520102, EPI_ISL_2520105, EPI_ISL_2623648, EPI_ISL_2623649, EPI_ISL_2623650, EPI_ISL_2623651, EPI_ISL_2623652, EPI_ISL_2623653, EPI_ISL_2623654, EPI_ISL_2623655, EPI_ISL_2623656, EPI_ISL_2623657, EPI_ISL_2623659, EPI_ISL_2623660, EPI_ISL_2623661, EPI_ISL_2623662, EPI_ISL_2623663, EPI_ISL_2623664, EPI_ISL_2623666, EPI_ISL_2623667, EPI_ISL_2623668, EPI_ISL_2623669, EPI_ISL_2623670, EPI_ISL_2623671, EPI_ISL_2623672, EPI_ISL_2623673, EPI_ISL_2623674, EPI_ISL_2623675, EPI_ISL_2623676, EPI_ISL_2623677, EPI_ISL_2623678, EPI_ISL_2623679, EPI_ISL_2623680, EPI_ISL_2623684, EPI_ISL_2623685, EPI_ISL_2623686, EPI_ISL_2623687, EPI_ISL_2623688, EPI_ISL_2623689, EPI_ISL_2623690, EPI_ISL_2623691, EPI_ISL_2623692, EPI_ISL_2623693, EPI_ISL_2623694, EPI_ISL_2623695, EPI_ISL_2623696, EPI_ISL_2623697, EPI_ISL_2623698, EPI_ISL_2623699, EPI_ISL_2623700, EPI_ISL_2623701, EPI_ISL_2623702, EPI_ISL_2623703, EPI_ISL_2623704, EPI_ISL_2623705, EPI_ISL_2623706, EPI_ISL_2623707, EPI_ISL_2623708, EPI_ISL_2623709, EPI_ISL_2623710, EPI_ISL_2623711, EPI_ISL_2623712, EPI_ISL_2623713, EPI_ISL_2623714, EPI_ISL_2623715, EPI_ISL_2623716, EPI_ISL_2623717, EPI_ISL_2623718, EPI_ISL_2623719, EPI_ISL_2623720, EPI_ISL_2623721, EPI_ISL_2623722, EPI_ISL_2623723, EPI_ISL_2623724, EPI_ISL_2623725, EPI_ISL_2623726, EPI_ISL_2623727, EPI_ISL_2623728, EPI_ISL_2623729, EPI_ISL_2623730, EPI_ISL_2623731, EPI_ISL_2623732, EPI_ISL_2623733, EPI_ISL_2623734, EPI_ISL_2623735, EPI_ISL_2623736, EPI_ISL_2623737, EPI_ISL_2623738, EPI_ISL_2623739, EPI_ISL_2623740, EPI_ISL_2623741, EPI_ISL_2623742, EPI_ISL_2623743, EPI_ISL_2623744, EPI_ISL_2623745, EPI_ISL_2623746, EPI_ISL_2623747, EPI_ISL_2623748, EPI_ISL_2623749, EPI_ISL_2623750, EPI_ISL_2623751, EPI_ISL_2623752, EPI_ISL_2623753, EPI_ISL_2623754, EPI_ISL_2623756, EPI_ISL_2623757, EPI_ISL_2623758, EPI_ISL_2623759, EPI_ISL_2623760, EPI_ISL_2623761, EPI_ISL_2623762, EPI_ISL_2623763, EPI_ISL_2623764, EPI_ISL_2623765, EPI_ISL_2623766, EPI_ISL_2623769, EPI_ISL_2623770, EPI_ISL_2623771, EPI_ISL_2623772, EPI_ISL_2623773, EPI_ISL_2623774, EPI_ISL_2623775, EPI_ISL_2623776, EPI_ISL_2623777, EPI_ISL_2623778, EPI_ISL_2623779, EPI_ISL_2623780, EPI_ISL_2623781, EPI_ISL_2623782, EPI_ISL_2623783, EPI_ISL_2623784, EPI_ISL_2623785, EPI_ISL_2623786, EPI_ISL_2623787, EPI_ISL_2623788, EPI_ISL_2623789, EPI_ISL_2623790, EPI_ISL_2623791, EPI_ISL_2623792, EPI_ISL_2623793, EPI_ISL_2623794, EPI_ISL_2623797, EPI_ISL_2623801, EPI_ISL_2623803, EPI_ISL_2623825, EPI_ISL_2623826, EPI_ISL_2623827, EPI_ISL_2623832, EPI_ISL_2623835, EPI_ISL_2623836, EPI_ISL_2623843, EPI_ISL_2623850, EPI_ISL_2623852, EPI_ISL_2623853, EPI_ISL_2623869, EPI_ISL_2623873, EPI_ISL_2623874, EPI_ISL_2623875, EPI_ISL_2623876, EPI_ISL_2623877, EPI_ISL_2623878, EPI_ISL_2623898, EPI_ISL_2623899, EPI_ISL_2623900, EPI_ISL_2623901, EPI_ISL_2623902, EPI_ISL_2623903, EPI_ISL_2623904, EPI_ISL_2623905, EPI_ISL_2623906, EPI_ISL_2623907, EPI_ISL_2623912, EPI_ISL_2623913, EPI_ISL_2623914, EPI_ISL_2623915, EPI_ISL_2623916, EPI_ISL_2623917, EPI_ISL_2623918, EPI_ISL_2623919, EPI_ISL_2623920, EPI_ISL_2623921, EPI_ISL_2623922, EPI_ISL_2623923, EPI_ISL_2623924, EPI_ISL_2623925, EPI_ISL_2623926, EPI_ISL_2623927, EPI_ISL_2623928, EPI_ISL_2623929, EPI_ISL_2623930, EPI_ISL_2623931, EPI_ISL_2623932, EPI_ISL_2623933, EPI_ISL_2623934, EPI_ISL_2623935, EPI_ISL_2623936, EPI_ISL_2623937, EPI_ISL_2623938, EPI_ISL_2623939, EPI_ISL_2623942, EPI_ISL_2623954, EPI_ISL_2623955, EPI_ISL_2623960, EPI_ISL_2623968, EPI_ISL_2623969, EPI_ISL_2623971, EPI_ISL_2623972, EPI_ISL_2623975, EPI_ISL_2623998, EPI_ISL_2624001, EPI_ISL_2624002, EPI_ISL_2624003, EPI_ISL_2624004, EPI_ISL_2624005, EPI_ISL_2624006, EPI_ISL_2624007, EPI_ISL_2624008, EPI_ISL_2624014, EPI_ISL_2624016, EPI_ISL_2624017, EPI_ISL_2624018, EPI_ISL_2624019, EPI_ISL_2624020, EPI_ISL_2624021, EPI_ISL_2624022, EPI_ISL_2624026, EPI_ISL_2624027, EPI_ISL_2624028, EPI_ISL_2624029, EPI_ISL_2624030, EPI_ISL_2624031, EPI_ISL_2624032, EPI_ISL_2624033, EPI_ISL_2624034, EPI_ISL_2624035, EPI_ISL_2624036, EPI_ISL_2624037, EPI_ISL_2624038, EPI_ISL_2624039, EPI_ISL_2624040, EPI_ISL_2624041, EPI_ISL_2624042, EPI_ISL_2624043, EPI_ISL_2624044, EPI_ISL_2624047, EPI_ISL_2624048, EPI_ISL_2624049, EPI_ISL_2624050, EPI_ISL_2624051, EPI_ISL_2624052, EPI_ISL_2624053, EPI_ISL_2624054, EPI_ISL_2624055, EPI_ISL_2624056, EPI_ISL_2624057, EPI_ISL_2624058, EPI_ISL_2624059, EPI_ISL_2624060, EPI_ISL_2624065, EPI_ISL_2624067, EPI_ISL_2624068, EPI_ISL_2624074, EPI_ISL_2624090, EPI_ISL_2624106, EPI_ISL_2624107, EPI_ISL_2624108, EPI_ISL_2624109, EPI_ISL_2624111, EPI_ISL_2624112, EPI_ISL_2624113, EPI_ISL_2624114, EPI_ISL_2624115, EPI_ISL_2624116, EPI_ISL_2624117, EPI_ISL_2624118, EPI_ISL_2624119, EPI_ISL_2624120, EPI_ISL_2624121, EPI_ISL_2624122, EPI_ISL_2624123, EPI_ISL_2624124, EPI_ISL_2624125, EPI_ISL_2624126, EPI_ISL_2624128, EPI_ISL_2624129, EPI_ISL_2624130, EPI_ISL_2624131, EPI_ISL_2624132, EPI_ISL_2624133, EPI_ISL_2624134, EPI_ISL_2624135, EPI_ISL_2624136, EPI_ISL_2624137, EPI_ISL_2624138, EPI_ISL_2624139, EPI_ISL_2624140, EPI_ISL_2624141, EPI_ISL_2624142, EPI_ISL_2624143, EPI_ISL_2624144, EPI_ISL_2624145, EPI_ISL_2624146, EPI_ISL_2624148, EPI_ISL_2624149, EPI_ISL_2624150, EPI_ISL_2624151, EPI_ISL_2624152, EPI_ISL_2624153, EPI_ISL_2624154, EPI_ISL_2624155, EPI_ISL_2624156, EPI_ISL_2624157, EPI_ISL_2624158, EPI_ISL_2624159, EPI_ISL_2624160, EPI_ISL_2624161, EPI_ISL_2624162, EPI_ISL_2624163, EPI_ISL_2624164, EPI_ISL_2624165, EPI_ISL_2624166, EPI_ISL_2624167, EPI_ISL_2624168, EPI_ISL_2624169, EPI_ISL_2624222, EPI_ISL_2624290, EPI_ISL_2624296, EPI_ISL_2624320, EPI_ISL_2624346, EPI_ISL_2624347, EPI_ISL_2624348, EPI_ISL_2706644, EPI_ISL_2706645, EPI_ISL_2706646, EPI_ISL_2706647, EPI_ISL_2706648, EPI_ISL_2706649, EPI_ISL_2706650, EPI_ISL_2706651, EPI_ISL_2706652, EPI_ISL_2706653, EPI_ISL_2706654, EPI_ISL_2706655, EPI_ISL_2706656, EPI_ISL_2706657, EPI_ISL_2706658, EPI_ISL_2706659, EPI_ISL_2706660, EPI_ISL_2706661, EPI_ISL_2706662, EPI_ISL_2706663, EPI_ISL_2706664, EPI_ISL_2706665, EPI_ISL_2706666, EPI_ISL_2706667, EPI_ISL_2706668, EPI_ISL_2706669, EPI_ISL_2706670, EPI_ISL_2706671, EPI_ISL_2706672, EPI_ISL_2706673, EPI_ISL_2706674, EPI_ISL_2706675, EPI_ISL_2706676, EPI_ISL_2706677, EPI_ISL_2706678, EPI_ISL_2706679, EPI_ISL_2706680, EPI_ISL_2706681, EPI_ISL_2706682, EPI_ISL_2706683, EPI_ISL_2706684, EPI_ISL_2706685, EPI_ISL_2706686, EPI_ISL_2706687, EPI_ISL_2706688, EPI_ISL_2706689, EPI_ISL_2706690, EPI_ISL_2706691, EPI_ISL_2706692, EPI_ISL_2706693, EPI_ISL_2706694, EPI_ISL_2706695, EPI_ISL_2706696, EPI_ISL_2706697, EPI_ISL_2706698, EPI_ISL_2706699, EPI_ISL_2706700, EPI_ISL_2706701, EPI_ISL_2706702, EPI_ISL_2706703, EPI_ISL_2706704, EPI_ISL_2706705, EPI_ISL_2706706, EPI_ISL_2706707, EPI_ISL_2706708, EPI_ISL_2706709, EPI_ISL_2706710, EPI_ISL_2706711, EPI_ISL_2706712, EPI_ISL_2706713, EPI_ISL_2706714, EPI_ISL_2706769, EPI_ISL_2706770, EPI_ISL_2706771, EPI_ISL_2706772, EPI_ISL_2706773, EPI_ISL_2706774, EPI_ISL_2706775, EPI_ISL_2706776, EPI_ISL_2706777, EPI_ISL_2706778, EPI_ISL_2706779, EPI_ISL_2706780, EPI_ISL_2706781, EPI_ISL_2706782, EPI_ISL_2706783, EPI_ISL_2706784, EPI_ISL_2706785, EPI_ISL_2706786, EPI_ISL_2706787, EPI_ISL_2706788, EPI_ISL_2706789, EPI_ISL_2706790, EPI_ISL_2706791, EPI_ISL_2706792, EPI_ISL_2706793, EPI_ISL_2706794, EPI_ISL_2706795, EPI_ISL_2706796, EPI_ISL_2706797, EPI_ISL_2706798, EPI_ISL_2706799, EPI_ISL_2706800, EPI_ISL_2706801, EPI_ISL_2706802, EPI_ISL_2706803, EPI_ISL_2706804, EPI_ISL_2706805, EPI_ISL_2706806, EPI_ISL_2706807, EPI_ISL_2706808, EPI_ISL_2706810, EPI_ISL_2706811, EPI_ISL_2706812, EPI_ISL_2706813, EPI_ISL_2706814, EPI_ISL_2706815, EPI_ISL_2706816, EPI_ISL_2706817, EPI_ISL_2706818, EPI_ISL_2706819, EPI_ISL_2706820, EPI_ISL_2706821, EPI_ISL_2706822, EPI_ISL_2706823, EPI_ISL_2706824, EPI_ISL_2706825, EPI_ISL_2706826, EPI_ISL_2706827, EPI_ISL_2706828, EPI_ISL_2706829, EPI_ISL_2706831, EPI_ISL_2706832, EPI_ISL_2706833, EPI_ISL_2706834, EPI_ISL_2706836, EPI_ISL_2706837, EPI_ISL_2706838, EPI_ISL_2706839, EPI_ISL_2706840, EPI_ISL_2706841, EPI_ISL_2706842, EPI_ISL_2706843, EPI_ISL_2706844, EPI_ISL_2706845, EPI_ISL_2706846, EPI_ISL_2706847, EPI_ISL_2706848, EPI_ISL_2706849, EPI_ISL_2706850, EPI_ISL_2706851, EPI_ISL_2706852, EPI_ISL_2706853, EPI_ISL_2706854, EPI_ISL_2706855, EPI_ISL_2706856, EPI_ISL_2706857, EPI_ISL_2706858, EPI_ISL_2706859, EPI_ISL_2706860, EPI_ISL_2706861, EPI_ISL_2706862, EPI_ISL_2706863, EPI_ISL_2706864, EPI_ISL_2706865, EPI_ISL_2706866, EPI_ISL_2706867, EPI_ISL_2706868, EPI_ISL_2706869, EPI_ISL_2706870, EPI_ISL_2706871, EPI_ISL_2706872, EPI_ISL_2706873, EPI_ISL_2706874, EPI_ISL_2706875, EPI_ISL_2706876, EPI_ISL_2706877, EPI_ISL_2706878, EPI_ISL_2706879, EPI_ISL_2706880, EPI_ISL_2706881, EPI_ISL_2706882, EPI_ISL_2706883, EPI_ISL_2706885, EPI_ISL_2706886, EPI_ISL_2706887, EPI_ISL_2706888, EPI_ISL_2706889, EPI_ISL_2706890, EPI_ISL_2706891, EPI_ISL_2706892, EPI_ISL_2706893, EPI_ISL_2706894, EPI_ISL_2706895, EPI_ISL_2706896, EPI_ISL_2706897, EPI_ISL_2706898, EPI_ISL_2706899, EPI_ISL_2706900, EPI_ISL_2706901, EPI_ISL_2706902, EPI_ISL_2706903, EPI_ISL_2706904, EPI_ISL_2706905, EPI_ISL_2706906, EPI_ISL_2706907, EPI_ISL_2706908, EPI_ISL_2706909, EPI_ISL_2706910, EPI_ISL_2706911, EPI_ISL_2706912, EPI_ISL_2706913, EPI_ISL_2706914, EPI_ISL_2706915, EPI_ISL_2706916, EPI_ISL_2706917 | | | |
| see above | Quadram Institute Bioscience | COVID-19 Genomics UK (COG-UK) Consortium | Alexander J Trotter; Alison E. Mather; Alp Aydin; Ana P. Tedim; Anastasia Kolyva; Andrew Bell; Andrew J. Page; Claire Stuart; Dave J. Baker; Gemma L. Kay; John Wain; Justin O'Grady; Leonardo de Oliveira Martins; Lizzie Meadows; Maria Diaz; Mark Webber; Muhammed Yasir; Nabil-Fareed Alikhan; Ngozi Elumogo; Nicholas M. Thomson; Rachael Stanley; Rachel Gilroy; Reenesh Prakash; Samir Dervisevic; Samuel Bloomfield; Steven Rudder; Thanh Le-Viet |
| EPI_ISL_2434759, EPI_ISL_2434760 | Queens Medical Centre, Clinical Microbiology Department / DeepSeq Nottingham | COVID-19 Genomics UK (COG-UK) Consortium | Christopher Moore; Emily Park; Fei Sang; Gemma Clark; Hannah Howson-Wells; Jack Hill; Jayasree Dey; Johnny Debebe; Jonathan Ball; Joseph Chappell; Manjinder Khakh; Matthew Carlisle; Matthew Loose; Michelle M Lister; Nadine Holmes; Patrick McClure; Theocharis Tsoleridis; Timothy Byaruhanga; Vicki M Fleming; Victoria Wright; Wendy Smith |
| EPI_ISL_2393158, EPI_ISL_2393185, EPI_ISL_2393229, EPI_ISL_2393273, EPI_ISL_2393287, EPI_ISL_2393298, EPI_ISL_2393336, EPI_ISL_2393449, EPI_ISL_2393471, EPI_ISL_2447763, EPI_ISL_2447764, EPI_ISL_2447771, EPI_ISL_2447772, EPI_ISL_2447773, EPI_ISL_2447774, EPI_ISL_2447775, EPI_ISL_2447776 | | | |
| see above | Randox Laboratories | Wellcome Sanger Institute for the COVID-19 Genomics UK (COG-UK) Consortium | Cordelia Langford; David K. Jackson; Dominic Kwiatkowski; Ewan Harrison; Ian Johnston; Jeffrey Barrett; John Sillitoe on behalf of the Wellcome Sanger Institute COVID-19 Surveillance Team; Randox Laboratories and Alex Alderton; Roberto Amato; Sonia Goncalves |
| EPI_ISL_2742910, EPI_ISL_2743005, EPI_ISL_2743010, EPI_ISL_2743016, EPI_ISL_2743018, EPI_ISL_2743024, EPI_ISL_2743034, EPI_ISL_2743043, EPI_ISL_2743047, EPI_ISL_2743060, EPI_ISL_2743080, EPI_ISL_2743085, EPI_ISL_2743089, EPI_ISL_2743093, EPI_ISL_2743103, EPI_ISL_2743111, EPI_ISL_2743112, EPI_ISL_2743118, EPI_ISL_2743124, EPI_ISL_2743125, EPI_ISL_2743126, EPI_ISL_2743135, EPI_ISL_2743138, EPI_ISL_2743139, EPI_ISL_2743140, EPI_ISL_2743147, EPI_ISL_2743176, EPI_ISL_2743204, EPI_ISL_2743206, EPI_ISL_2743211, EPI_ISL_2743213, EPI_ISL_2743220, EPI_ISL_2743221, EPI_ISL_2743224, EPI_ISL_2743232, EPI_ISL_2743236, EPI_ISL_2743237, EPI_ISL_2743241, EPI_ISL_2743245, EPI_ISL_2743255, EPI_ISL_2743256, EPI_ISL_2743257, EPI_ISL_2743258, EPI_ISL_2743269, EPI_ISL_2743273, EPI_ISL_2743275, EPI_ISL_2743279, EPI_ISL_2743288, EPI_ISL_2743291, EPI_ISL_2743292, EPI_ISL_2743302, EPI_ISL_2743307, EPI_ISL_2743309, EPI_ISL_2743310, EPI_ISL_2743316, EPI_ISL_2743323, EPI_ISL_2743327, EPI_ISL_2743328, EPI_ISL_2743329, EPI_ISL_2743337, EPI_ISL_2743343, EPI_ISL_2743347, EPI_ISL_2743357, EPI_ISL_2743359, EPI_ISL_2743364, EPI_ISL_2743382, EPI_ISL_2743390, EPI_ISL_2743393, EPI_ISL_2743394, EPI_ISL_2743395, EPI_ISL_2743400, EPI_ISL_2743402, EPI_ISL_2743404, EPI_ISL_2743412, EPI_ISL_2743413, EPI_ISL_2743416, EPI_ISL_2743428, EPI_ISL_2743439, EPI_ISL_2743440, EPI_ISL_2743441, EPI_ISL_2743442, EPI_ISL_2743444, EPI_ISL_2743445, EPI_ISL_2743446, EPI_ISL_2743447, EPI_ISL_2743449, EPI_ISL_2743451, EPI_ISL_2743453, EPI_ISL_2743468, EPI_ISL_2743481, EPI_ISL_2743509, EPI_ISL_2743512, EPI_ISL_2743513, EPI_ISL_2743515, EPI_ISL_2743516, EPI_ISL_2743519, EPI_ISL_2743523, EPI_ISL_2743524, EPI_ISL_2743526, EPI_ISL_2743531, EPI_ISL_2743585, EPI_ISL_2743586, EPI_ISL_2743593, EPI_ISL_2743605, EPI_ISL_2743662, EPI_ISL_2743727, EPI_ISL_2743729, EPI_ISL_2743732, EPI_ISL_2743733, EPI_ISL_2743971, EPI_ISL_2743996, EPI_ISL_2744005, EPI_ISL_2744017, EPI_ISL_2744072, EPI_ISL_2744235, EPI_ISL_2744237, EPI_ISL_2744242, EPI_ISL_2744244, EPI_ISL_2744248, EPI_ISL_2744250, EPI_ISL_2744251, EPI_ISL_2744253, EPI_ISL_2744254, EPI_ISL_2744255, EPI_ISL_2744257, EPI_ISL_2744258, EPI_ISL_2744259, EPI_ISL_2744260, EPI_ISL_2744261, EPI_ISL_2744262, EPI_ISL_2744263, EPI_ISL_2744264, EPI_ISL_2744270, EPI_ISL_2744502, EPI_ISL_2744719, EPI_ISL_2744720, EPI_ISL_2744725, EPI_ISL_2744726, EPI_ISL_2745218, EPI_ISL_2745225, EPI_ISL_2746707, EPI_ISL_2746793, EPI_ISL_2817839 | | | |

| | | | |
|---|---|---|---|
| see above | Respiratory Virus Unit, Microbiology Services Colindale, Public Health England | COVID-19 Genomics UK (COG-UK) Consortium | PHE Covid Sequencing Team |
| EPI_ISL_2355428, EPI_ISL_2355429 | University College London, Great Ormond Street Hospital for Children NHS Foundation Trust, Imperial College Healthcare NHS Trust | COVID-19 Genomics UK (COG-UK) Consortium | Alison Holmes; Charlotte Williams; Helena Tutill; Jacqueline Findlay; James Price; Judith Breuer; Julianne Brown; Kathryn Harris; Leysa Forrest; Mark Kristiansen; Paola Niola; Paola Resende Silva; Patricia Dyal; Paul Randell; Rachel Williams; Samuel Weeks; Sergi Castellano; Sunando Roy; Tony Brooks; Yasmin Panchbhaya |
| EPI_ISL_2354515 | University of Birmingham | COVID-19 Genomics UK (COG-UK) Consortium | Alex Richter; Andrew Bosworth; Andrew Bosworth. Queen Elizabeth Hospital: Anna Casey; Andrew D Beggs PHE Heartlands Lab: Husam Osman; Charlotte Poxon; Institute of Microbiology; Joanne Stockton; Josh Quick; Kasun Wanigasooriya; Mike Kidd; Nicholas Loman. University of Birmingham Testing Laboratory: Celina M Whalley; Oliver Pickles; Radoslaw Poplawski; Samuel Nicholls; University of Birmingham: Claire McMurray; Will Rowe |
| EPI_ISL_2434212, EPI_ISL_2434213, EPI_ISL_2434214, EPI_ISL_2434217, EPI_ISL_2434218, EPI_ISL_2434219, EPI_ISL_2434220, EPI_ISL_2434221, EPI_ISL_2434222, EPI_ISL_2434225, EPI_ISL_2434226, EPI_ISL_2434227, EPI_ISL_2517301 | | | |
| see above | Virology Department, Royal Infirmary of Edinburgh, NHS Lothian / School of Biological Sciences, University of Edinburgh | COVID-19 Genomics UK (COG-UK) Consortium | Colquhoun R; Cotton S; Dewar R; Gallagher A; Hill V; Jackson B; Maloney D; McCrone JT; McHugh M; O'Toole Á; Rambaut A; Rooke S; Scher E; Templeton K; Yu X |
| EPI_ISL_2521092 | Virology Department, Sheffield Teaching Hospitals NHS Foundation Trust/Department of Infection, Immunity and Cardiovascular Disease, The Medical School, University of Sheffield | COVID-19 Genomics UK (COG-UK) Consortium | Adri Angyal; Alex Keeley; Benjamin Lindsey; Cariad Evans; Danielle Groves; Dave Partridge; Katie Johnson; Laura Carrilero; Luke Green; Matthew Parker; Matthew Wyles; Mehmet Yavuz; Mohammad Raza; Nikki Smith; Paul Parsons; Rachel Tucker; Rebecca Brown; Thushan de Silva |
| EPI_ISL_2520732, EPI_ISL_2520733 | Wales Specialist Virology Centre Sequencing lab: Pathogen Genomics Unit | Public Health Wales Microbiology Cardiff Wales Specialist Virology Centre | Alec Birchley; Alexander Adams; Amy Gaskin; Angela Marchbank; Bree Gatica-Wilcox; Catherine Moore; Jason Coombes; Joanne Watkins; Joel Southgate; Johnathan Evans; Laura Gifford; Lauren Gilbert; Lee Graham; Malorie Perry; Matthew Bull; Nicole Pacchiarini; Sally Corden; Sara Kumziene-Summerhayes; Sara Rey; Sarah Taylor; Simon Cottrell; Sophie Jones; Tom Connor |
| EPI_ISL_2517238, EPI_ISL_2517239, EPI_ISL_2517240, EPI_ISL_2517241, EPI_ISL_2517242, EPI_ISL_2517243, EPI_ISL_2517244, EPI_ISL_2517245, EPI_ISL_2517246, EPI_ISL_2517247, EPI_ISL_2517252, EPI_ISL_2517258, EPI_ISL_2517262, EPI_ISL_2517263, EPI_ISL_2517264, EPI_ISL_2517265, EPI_ISL_2517266, EPI_ISL_2517267, EPI_ISL_2517268, EPI_ISL_2517269, EPI_ISL_2517270, EPI_ISL_2517271, EPI_ISL_2517281, EPI_ISL_2517282, EPI_ISL_2517283, EPI_ISL_2517284, EPI_ISL_2517285, EPI_ISL_2517286, EPI_ISL_2517287, EPI_ISL_2517288, EPI_ISL_2517290, EPI_ISL_2722632, EPI_ISL_2722664 | | | |
| see above | West of Scotland Specialist Virology Centre, NHSGGC / MRC-University of Glasgow Centre for Virus Research | COVID-19 Genomics UK (COG-UK) Consortium | Alasdair MacLean; Alice Broos; Ana da Silva Filipe; Antonia Ho; Daniel Mair; David L Robertson; Emma Thomson; Guy Mollett; Ioulia Tsatsani; James Shepherd; Jenna Nichols; Joseph Hughes; Kathy Li; Kathy Smollett; Kyriaki Nomikou; Lily Tong; Matthew Holden; Natasha Johnson; Rachel Blacow; Richard Orton; Rory Gunson; Sarah McDonald; Sharif Shaaban; Sreenu Vattipally; Stephen Carmichael |

We gratefully acknowledge the following Authors from the Originating laboratories responsible for obtaining the specimens, as well as the Submitting laboratories where the genome data were generated and shared via GISAID, on which this research is based.

All Submitters of data may be contacted directly via www.gisaid.org

Authors are sorted alphabetically.

| Accession ID | Originating Laboratory | Submitting Laboratory | Authors |
|---|---|---|---|
| EPI_ISL_2686122, EPI_ISL_2686123, EPI_ISL_2686124, EPI_ISL_2686125, EPI_ISL_2686126, EPI_ISL_2686127, EPI_ISL_2686128, EPI_ISL_2686129 | | | |
| see above | Amazon Dx, AUV2 Laboratory | Amazon Dx, AUV2 Laboratory | Luke Meredith |
| EPI_ISL_2434268, EPI_ISL_2434269, EPI_ISL_2434270, EPI_ISL_2434271, EPI_ISL_2517305, EPI_ISL_2517306, EPI_ISL_2517307 | | | |
| see above | Barts Health NHS Trust | COVID-19 Genomics UK (COG-UK) Consortium | BROAD; Beatrix; CUTINO-MOGUEL; Claire; David; Dola; HARRINGTON; KELE; KULASEGARAN-SHYLINI; Maria-Teresa; OWOYEMI; Raghavendran |
| EPI_ISL_2436237, EPI_ISL_2436238, EPI_ISL_2436239, EPI_ISL_2436240, EPI_ISL_2436250, EPI_ISL_2436261, EPI_ISL_2436264, EPI_ISL_2436270, EPI_ISL_2436274, EPI_ISL_2436278, EPI_ISL_2436285, EPI_ISL_2436286, EPI_ISL_2436289, EPI_ISL_2436294, EPI_ISL_2436298, EPI_ISL_2436317, EPI_ISL_2436319, EPI_ISL_2436320, EPI_ISL_2436338, EPI_ISL_2436339, EPI_ISL_2436349, EPI_ISL_2436350, EPI_ISL_2436358, EPI_ISL_2436372, EPI_ISL_2436378, EPI_ISL_2436381, EPI_ISL_2436397, EPI_ISL_2436398, EPI_ISL_2436404, EPI_ISL_2436410, EPI_ISL_2436415, EPI_ISL_2436419, EPI_ISL_2436422, EPI_ISL_2436430, EPI_ISL_2436435, EPI_ISL_2436439, EPI_ISL_2436440, EPI_ISL_2436443, EPI_ISL_2436448, EPI_ISL_2436463, EPI_ISL_2436472, EPI_ISL_2436475, EPI_ISL_2436477, EPI_ISL_2436478, EPI_ISL_2436482, EPI_ISL_2436484, EPI_ISL_2436485, EPI_ISL_2436486, EPI_ISL_2436487, EPI_ISL_2436489, EPI_ISL_2436493, EPI_ISL_2436500, EPI_ISL_2436501, EPI_ISL_2436508, EPI_ISL_2436515, EPI_ISL_2436525, EPI_ISL_2436535, EPI_ISL_2436550, EPI_ISL_2436553, EPI_ISL_2436556, EPI_ISL_2436561, EPI_ISL_2436563, EPI_ISL_2437326, EPI_ISL_2437329, EPI_ISL_2437362, EPI_ISL_2437399, EPI_ISL_2437404, EPI_ISL_2437426, EPI_ISL_2437432, EPI_ISL_2437472, EPI_ISL_2437497, EPI_ISL_2437513, EPI_ISL_2437536, EPI_ISL_2437547, EPI_ISL_2437596, EPI_ISL_2446158, EPI_ISL_2446184, EPI_ISL_2446187, EPI_ISL_2446214, EPI_ISL_2446219, EPI_ISL_2446230, EPI_ISL_2446245, EPI_ISL_2446250, EPI_ISL_2446258, EPI_ISL_2446287, EPI_ISL_2446402, EPI_ISL_2447364, EPI_ISL_2447368, EPI_ISL_2447384, EPI_ISL_2447408, EPI_ISL_2447444, EPI_ISL_2447445, EPI_ISL_2447466, EPI_ISL_2447472, EPI_ISL_2447493, EPI_ISL_2447495, EPI_ISL_2447496, EPI_ISL_2447502, EPI_ISL_2447507, EPI_ISL_2447513, EPI_ISL_2447519, EPI_ISL_2447529, EPI_ISL_2447541, EPI_ISL_2447545, EPI_ISL_2447550, EPI_ISL_2447551, EPI_ISL_2447561, EPI_ISL_2447563, EPI_ISL_2447564, EPI_ISL_2447568, EPI_ISL_2447576, EPI_ISL_2447580, EPI_ISL_2447600, EPI_ISL_2447616, EPI_ISL_2447632, EPI_ISL_2447651, EPI_ISL_2447669, EPI_ISL_2447675, EPI_ISL_2447705, EPI_ISL_2447708, EPI_ISL_2447709, EPI_ISL_2447727, EPI_ISL_2447729, EPI_ISL_2447747, EPI_ISL_2447749, EPI_ISL_2447759, EPI_ISL_2456826, EPI_ISL_2456842, EPI_ISL_2456844, EPI_ISL_2456882, EPI_ISL_2456886, EPI_ISL_2456906, EPI_ISL_2456908, EPI_ISL_2456918, EPI_ISL_2456919, EPI_ISL_2456920, EPI_ISL_2456921, EPI_ISL_2456924, EPI_ISL_2456927, EPI_ISL_2456928, EPI_ISL_2456951, EPI_ISL_2456960, EPI_ISL_2456962, EPI_ISL_2456964, EPI_ISL_2456968, EPI_ISL_2456975, EPI_ISL_2456979, EPI_ISL_2456982, EPI_ISL_2456988, EPI_ISL_2456996, EPI_ISL_2457003, EPI_ISL_2457011, EPI_ISL_2457017, EPI_ISL_2457019, EPI_ISL_2457023, EPI_ISL_2457030, EPI_ISL_2457034, EPI_ISL_2457040, EPI_ISL_2457041, EPI_ISL_2457046, EPI_ISL_2457062, EPI_ISL_2457087, EPI_ISL_2457090, EPI_ISL_2457092, EPI_ISL_2457100, EPI_ISL_2457103, EPI_ISL_2457105, EPI_ISL_2457114, EPI_ISL_2457122, EPI_ISL_2469241, EPI_ISL_2469466, EPI_ISL_2513803, EPI_ISL_2513805, EPI_ISL_2513807, EPI_ISL_2513808, EPI_ISL_2513809, EPI_ISL_2513810, EPI_ISL_2513811, EPI_ISL_2513812, EPI_ISL_2513813, EPI_ISL_2513814, EPI_ISL_2513816, EPI_ISL_2513817, EPI_ISL_2513820, EPI_ISL_2513822, EPI_ISL_2513825, EPI_ISL_2513832, EPI_ISL_2513833, EPI_ISL_2513834, EPI_ISL_2513835, EPI_ISL_2513836, EPI_ISL_2513837, EPI_ISL_2513838, EPI_ISL_2513840, EPI_ISL_2513844, EPI_ISL_2513846, EPI_ISL_2513847, EPI_ISL_2513848, EPI_ISL_2513849, EPI_ISL_2513850, EPI_ISL_2513851, EPI_ISL_2513852, EPI_ISL_2513854, EPI_ISL_2513855, EPI_ISL_2513856, EPI_ISL_2513857, EPI_ISL_2513858, EPI_ISL_2513859, EPI_ISL_2513860, EPI_ISL_2513862, EPI_ISL_2513865, EPI_ISL_2513868, EPI_ISL_2513875, EPI_ISL_2513878, EPI_ISL_2513879, EPI_ISL_2513881, EPI_ISL_2513882, EPI_ISL_2513886, EPI_ISL_2513888, EPI_ISL_2513889, EPI_ISL_2513891, EPI_ISL_2513893, EPI_ISL_2513894, EPI_ISL_2513895, EPI_ISL_2513896, EPI_ISL_2513898, EPI_ISL_2513899, EPI_ISL_2513901, EPI_ISL_2513902, EPI_ISL_2513907, EPI_ISL_2513908, EPI_ISL_2513909, EPI_ISL_2513911, EPI_ISL_2513912, EPI_ISL_2513913, EPI_ISL_2513914, EPI_ISL_2513915, EPI_ISL_2513919, EPI_ISL_2513920, EPI_ISL_2513921, EPI_ISL_2513922, EPI_ISL_2513923, EPI_ISL_2513926, EPI_ISL_2513927, EPI_ISL_2513931, EPI_ISL_2513933, EPI_ISL_2513937, EPI_ISL_2513938, EPI_ISL_2513942, EPI_ISL_2513943, EPI_ISL_2513948, EPI_ISL_2513949, EPI_ISL_2513950, EPI_ISL_2513951, EPI_ISL_2513955, EPI_ISL_2513956, EPI_ISL_2513958, EPI_ISL_2513965, EPI_ISL_2513967, EPI_ISL_2513968, EPI_ISL_2513971, EPI_ISL_2513975, EPI_ISL_2513976, EPI_ISL_2513977, EPI_ISL_2513978, EPI_ISL_2513979, EPI_ISL_2513981, EPI_ISL_2513982, EPI_ISL_2513984, EPI_ISL_2513987, EPI_ISL_2513989, EPI_ISL_2513992, EPI_ISL_2513997, EPI_ISL_2513999, EPI_ISL_2514000, EPI_ISL_2514002, EPI_ISL_2514003, EPI_ISL_2514004, EPI_ISL_2514005, EPI_ISL_2514007, EPI_ISL_2514009, EPI_ISL_2514011, EPI_ISL_2514014, EPI_ISL_2514021, EPI_ISL_2514022, EPI_ISL_2514024, EPI_ISL_2514025, EPI_ISL_2514026, EPI_ISL_2514028, EPI_ISL_2514029, EPI_ISL_2514030, EPI_ISL_2514031, EPI_ISL_2514032, EPI_ISL_2514033, EPI_ISL_2514035, EPI_ISL_2514036, EPI_ISL_2514037, EPI_ISL_2514038, EPI_ISL_2514039, EPI_ISL_2514041, EPI_ISL_2514048, EPI_ISL_2514050, EPI_ISL_2514053, EPI_ISL_2514056, EPI_ISL_2514057, EPI_ISL_2514060, EPI_ISL_2514062, EPI_ISL_2514063, EPI_ISL_2514071, EPI_ISL_2514072 | | | |
| see above | Health Services Laboratories | Wellcome Sanger Institute for the COVID-19 Genomics UK (COG-UK) Consortium | Cordelia Langford; David K. Jackson; Dominic Kwiatkowski; Ewan Harrison; Health Services Laboratories and Alex Alderton; Ian Johnston; Jeffrey Barrett; John Sillitoe on behalf of the Wellcome Sanger Institute COVID-19 Surveillance Team; Roberto Amato; Sonia Goncalves |
| EPI_ISL_2434992, EPI_ISL_2434993, EPI_ISL_2434994, EPI_ISL_2434995, EPI_ISL_2434997, EPI_ISL_2434998, EPI_ISL_2434999, EPI_ISL_2435002, EPI_ISL_2435003, EPI_ISL_2435007, EPI_ISL_2435008, EPI_ISL_2435009, EPI_ISL_2435010, EPI_ISL_2435011, EPI_ISL_2435012, EPI_ISL_2435013, EPI_ISL_2435014, EPI_ISL_2435015, EPI_ISL_2435016, EPI_ISL_2435017, EPI_ISL_2435018, EPI_ISL_2435019, EPI_ISL_2435020, EPI_ISL_2435021, EPI_ISL_2435022, EPI_ISL_2435023, EPI_ISL_2435025, EPI_ISL_2435026, EPI_ISL_2435027, EPI_ISL_2435031, EPI_ISL_2435033, EPI_ISL_2435035, EPI_ISL_2435037, EPI_ISL_2435038, EPI_ISL_2435039, EPI_ISL_2435040, EPI_ISL_2435043, EPI_ISL_2435046, EPI_ISL_2435048, EPI_ISL_2435049, EPI_ISL_2435050, EPI_ISL_2435051, EPI_ISL_2435053, EPI_ISL_2435054, EPI_ISL_2435056, EPI_ISL_2435057, EPI_ISL_2435058, EPI_ISL_2435059, EPI_ISL_2435060, EPI_ISL_2435063, EPI_ISL_2435064, EPI_ISL_2435065, EPI_ISL_2435066, EPI_ISL_2435067, EPI_ISL_2435068, EPI_ISL_2435069, EPI_ISL_2435071, EPI_ISL_2435072, EPI_ISL_2435073, EPI_ISL_2435076, EPI_ISL_2435078, EPI_ISL_2435079, EPI_ISL_2435081, EPI_ISL_2435082, EPI_ISL_2435083, EPI_ISL_2435087, EPI_ISL_2435089, EPI_ISL_2435090, EPI_ISL_2435092, EPI_ISL_2435093, EPI_ISL_2435094, EPI_ISL_2435097, EPI_ISL_2435100, EPI_ISL_2435101, EPI_ISL_2435102, EPI_ISL_2435105, EPI_ISL_2435106, EPI_ISL_2435109, EPI_ISL_2435112, EPI_ISL_2435113, EPI_ISL_2435116, EPI_ISL_2435118, EPI_ISL_2435119, EPI_ISL_2435120, EPI_ISL_2435121, EPI_ISL_2435122, EPI_ISL_2435125, EPI_ISL_2435128, EPI_ISL_2435130, EPI_ISL_2435131, EPI_ISL_2435134, EPI_ISL_2435135, EPI_ISL_2435137, EPI_ISL_2435140, EPI_ISL_2435142, EPI_ISL_2435144, EPI_ISL_2435146, EPI_ISL_2435147, EPI_ISL_2435148, EPI_ISL_2435149, EPI_ISL_2435150, EPI_ISL_2435151, EPI_ISL_2435153, EPI_ISL_2435154, EPI_ISL_2435155, EPI_ISL_2435156, EPI_ISL_2435158, EPI_ISL_2435159, EPI_ISL_2435160, EPI_ISL_2435161, EPI_ISL_2435162, EPI_ISL_2435165, EPI_ISL_2435166, EPI_ISL_2435167, EPI_ISL_2435169, EPI_ISL_2435170, EPI_ISL_2435171, EPI_ISL_2435172, EPI_ISL_2435176, EPI_ISL_2435177, EPI_ISL_2435180, EPI_ISL_2435181, EPI_ISL_2435182, EPI_ISL_2435183, EPI_ISL_2435184, EPI_ISL_2435185, EPI_ISL_2435186, EPI_ISL_2435187, EPI_ISL_2435188, EPI_ISL_2435189, EPI_ISL_2435190, EPI_ISL_2435191, EPI_ISL_2435192, EPI_ISL_2435193, EPI_ISL_2435194, EPI_ISL_2435196, EPI_ISL_2435197, EPI_ISL_2435199, EPI_ISL_2435200, EPI_ISL_2435201, EPI_ISL_2435202, EPI_ISL_2435203, EPI_ISL_2435204, EPI_ISL_2435206, EPI_ISL_2435207, EPI_ISL_2435208, EPI_ISL_2435209, EPI_ISL_2435210, EPI_ISL_2435212, EPI_ISL_2435213, EPI_ISL_2435215, EPI_ISL_2435217, EPI_ISL_2435218, EPI_ISL_2435219, EPI_ISL_2435221, EPI_ISL_2435222, EPI_ISL_2435223, EPI_ISL_2435224, EPI_ISL_2435229, EPI_ISL_2435230, EPI_ISL_2435231, EPI_ISL_2435232, EPI_ISL_2435233, EPI_ISL_2435234, EPI_ISL_2435235, EPI_ISL_2435236, EPI_ISL_2435237, EPI_ISL_2435238, EPI_ISL_2435239, EPI_ISL_2435240, EPI_ISL_2435241, EPI_ISL_2435242, EPI_ISL_2435243, EPI_ISL_2435245, EPI_ISL_2435246, EPI_ISL_2435248, EPI_ISL_2435249, EPI_ISL_2435250, EPI_ISL_2435251, EPI_ISL_2435256, EPI_ISL_2435257, EPI_ISL_2435259, EPI_ISL_2435260, EPI_ISL_2435261, EPI_ISL_2435262, EPI_ISL_2435264, EPI_ISL_2435265, EPI_ISL_2435267, EPI_ISL_2435268, EPI_ISL_2435269, EPI_ISL_2435270, EPI_ISL_2435272, EPI_ISL_2435273, EPI_ISL_2435274, EPI_ISL_2435276, EPI_ISL_2435277, EPI_ISL_2435279, EPI_ISL_2435280, EPI_ISL_2435283, EPI_ISL_2435284, EPI_ISL_2435286, EPI_ISL_2435287, EPI_ISL_2435288, EPI_ISL_2435289, EPI_ISL_2435291, EPI_ISL_2435293, EPI_ISL_2435294, EPI_ISL_2435297, EPI_ISL_2435298, EPI_ISL_2435301, EPI_ISL_2435302, EPI_ISL_2435304, EPI_ISL_2435307, EPI_ISL_2435308, EPI_ISL_2435309, EPI_ISL_2435310, EPI_ISL_2435312, EPI_ISL_2435317, EPI_ISL_2435320, EPI_ISL_2435321, EPI_ISL_2435323, EPI_ISL_2435324, EPI_ISL_2435326, EPI_ISL_2435327, EPI_ISL_2435328, EPI_ISL_2435331, EPI_ISL_2435332, EPI_ISL_2435334, EPI_ISL_2435336, EPI_ISL_2435338, EPI_ISL_2435339, EPI_ISL_2435340, EPI_ISL_2435341, EPI_ISL_2435342, EPI_ISL_2435344, EPI_ISL_2435345, EPI_ISL_2435348, EPI_ISL_2435350, EPI_ISL_2435351, EPI_ISL_2435355, EPI_ISL_2435357, EPI_ISL_2435359, EPI_ISL_2435360, EPI_ISL_2435362, EPI_ISL_2435363, EPI_ISL_2435365, EPI_ISL_2435369, EPI_ISL_2435370, EPI_ISL_2435371, EPI_ISL_2435372, EPI_ISL_2435373, EPI_ISL_2435374, EPI_ISL_2435375, EPI_ISL_2435376, EPI_ISL_2435377, EPI_ISL_2435380, EPI_ISL_2435381, EPI_ISL_2435383, EPI_ISL_2435387, EPI_ISL_2435390, EPI_ISL_2435393, EPI_ISL_2435394, EPI_ISL_2435401, EPI_ISL_2435402, EPI_ISL_2435403, EPI_ISL_2435404, EPI_ISL_2435406, EPI_ISL_2435407, EPI_ISL_2435408, EPI_ISL_2435410, EPI_ISL_2435412, EPI_ISL_2435414, EPI_ISL_2435416, EPI_ISL_2435417, EPI_ISL_2435418, EPI_ISL_2435420, EPI_ISL_2435421, EPI_ISL_2435422, EPI_ISL_2435425, EPI_ISL_2435426, EPI_ISL_2435427, EPI_ISL_2435429, EPI_ISL_2435431, EPI_ISL_2435434, EPI_ISL_2435435, EPI_ISL_2435437, EPI_ISL_2435440, EPI_ISL_2435451, EPI_ISL_2435452, EPI_ISL_2435456, EPI_ISL_2435458, EPI_ISL_2435466, EPI_ISL_2435467, EPI_ISL_2435468, EPI_ISL_2435469, EPI_ISL_2435474, EPI_ISL_2435476, EPI_ISL_2435478, EPI_ISL_2435479, EPI_ISL_2435480, EPI_ISL_2435481, EPI_ISL_2435484, EPI_ISL_2435486, EPI_ISL_2435488, EPI_ISL_2435489, EPI_ISL_2435491, EPI_ISL_2435498, EPI_ISL_2435500, EPI_ISL_2435508, EPI_ISL_2435509, EPI_ISL_2435510, EPI_ISL_2435514, EPI_ISL_2435515, EPI_ISL_2435516, EPI_ISL_2435517, EPI_ISL_2435518, EPI_ISL_2435519, EPI_ISL_2435522, EPI_ISL_2435524, EPI_ISL_2435527, EPI_ISL_2435868, EPI_ISL_2435875, EPI_ISL_2435880, EPI_ISL_2435882, EPI_ISL_2435886, EPI_ISL_2435888, EPI_ISL_2435890, EPI_ISL_2435896, EPI_ISL_2435899, EPI_ISL_2435900, EPI_ISL_2435905, EPI_ISL_2435908, EPI_ISL_2435909, EPI_ISL_2435918, EPI_ISL_2435923, EPI_ISL_2435926, EPI_ISL_2435928, EPI_ISL_2435929, EPI_ISL_2435930, EPI_ISL_2435935, EPI_ISL_2435936, EPI_ISL_2435937, EPI_ISL_2435938, EPI_ISL_2435940, EPI_ISL_2435941, EPI_ISL_2435944, EPI_ISL_2435949, EPI_ISL_2435952, EPI_ISL_2435954, EPI_ISL_2435955, EPI_ISL_2435956, EPI_ISL_2435966, EPI_ISL_2435967, EPI_ISL_2435969, EPI_ISL_2435971, EPI_ISL_2435973, EPI_ISL_2435978, EPI_ISL_2435981, EPI_ISL_2435984, EPI_ISL_2435986, EPI_ISL_2435989, EPI_ISL_2435990, EPI_ISL_2435991, EPI_ISL_2435999, EPI_ISL_2436002, EPI_ISL_2436003, EPI_ISL_2436005, EPI_ISL_2436009, EPI_ISL_2436018, EPI_ISL_2436019, EPI_ISL_2436020, EPI_ISL_2436021, EPI_ISL_2436022, EPI_ISL_2436025, EPI_ISL_2436026, EPI_ISL_2436027, EPI_ISL_2436034, EPI_ISL_2436046, EPI_ISL_2436053, EPI_ISL_2436054, EPI_ISL_2436060, EPI_ISL_2436061, EPI_ISL_2436062, EPI_ISL_2436063, EPI_ISL_2436072, EPI_ISL_2436074, EPI_ISL_2436079, EPI_ISL_2436081, EPI_ISL_2436086, EPI_ISL_2436089, EPI_ISL_2436091, EPI_ISL_2436092, EPI_ISL_2436097, EPI_ISL_2436101, EPI_ISL_2436102, EPI_ISL_2436107, EPI_ISL_2436113, EPI_ISL_2436114, EPI_ISL_2436116, EPI_ISL_2436119, EPI_ISL_2436123, EPI_ISL_2436124, EPI_ISL_2436126, EPI_ISL_2436129, EPI_ISL_2436130, EPI_ISL_2436132, EPI_ISL_2436136, EPI_ISL_2436137, EPI_ISL_2436139, EPI_ISL_2436140, EPI_ISL_2436145, EPI_ISL_2436146, EPI_ISL_2436147, EPI_ISL_2436152, EPI_ISL_2436154, EPI_ISL_2436158, EPI_ISL_2436161, EPI_ISL_2436165, EPI_ISL_2436171, EPI_ISL_2436175, EPI_ISL_2436176, EPI_ISL_2436179, EPI_ISL_2436181, EPI_ISL_2436182, EPI_ISL_2436184, EPI_ISL_2436189, EPI_ISL_2436196, EPI_ISL_2436198, EPI_ISL_2436199, EPI_ISL_2436201, EPI_ISL_2436203, EPI_ISL_2436204, EPI_ISL_2436205, EPI_ISL_2436206, EPI_ISL_2436209, EPI_ISL_2436215, EPI_ISL_2436216, EPI_ISL_2436228, EPI_ISL_2436248, EPI_ISL_2436251, EPI_ISL_2436260, EPI_ISL_2436263, EPI_ISL_2436282, EPI_ISL_2436284, EPI_ISL_2436291, EPI_ISL_2436297, EPI_ISL_2436300, EPI_ISL_2436302, EPI_ISL_2436306, EPI_ISL_2436324, EPI_ISL_2436326, EPI_ISL_2436327, EPI_ISL_2436330, EPI_ISL_2436331, EPI_ISL_2436333, EPI_ISL_2436337, EPI_ISL_2436342, EPI_ISL_2436347, EPI_ISL_2436357, EPI_ISL_2436360, EPI_ISL_2436367, EPI_ISL_2436374, EPI_ISL_2436375, EPI_ISL_2436383, EPI_ISL_2436392, EPI_ISL_2436402, EPI_ISL_2436403, EPI_ISL_2436409, EPI_ISL_2436412, EPI_ISL_2436414, EPI_ISL_2436421, EPI_ISL_2436425, EPI_ISL_2436431, EPI_ISL_2436438, EPI_ISL_2436450, EPI_ISL_2436452, EPI_ISL_2436456, EPI_ISL_2436457, EPI_ISL_2436461, EPI_ISL_2436468, EPI_ISL_2436473, EPI_ISL_2436474, EPI_ISL_2436491, EPI_ISL_2436502, EPI_ISL_2436517, EPI_ISL_2436522, EPI_ISL_2436532, EPI_ISL_2436536, EPI_ISL_2436538, EPI_ISL_2436541, EPI_ISL_2436551, EPI_ISL_2436557, EPI_ISL_2436569, EPI_ISL_2436570, EPI_ISL_2436576, EPI_ISL_2436578, EPI_ISL_2436580, EPI_ISL_2436590, EPI_ISL_2436591, EPI_ISL_2436599, EPI_ISL_2436602, EPI_ISL_2436606, EPI_ISL_2436609, EPI_ISL_2436610, EPI_ISL_2436612, EPI_ISL_2436613, EPI_ISL_2436615, EPI_ISL_2436616, EPI_ISL_2436618, EPI_ISL_2436622, EPI_ISL_2436624, EPI_ISL_2436626, EPI_ISL_2436637, EPI_ISL_2436638, EPI_ISL_2436640, EPI_ISL_2436643, EPI_ISL_2436645, EPI_ISL_2436646, EPI_ISL_2436647, EPI_ISL_2436652, EPI_ISL_2436653, EPI_ISL_2436654, EPI_ISL_2436655, EPI_ISL_2436658, EPI_ISL_2436659, EPI_ISL_2436660, EPI_ISL_2436663, EPI_ISL_2436667, EPI_ISL_2436671, EPI_ISL_2436673, EPI_ISL_2436675, EPI_ISL_2436677, EPI_ISL_2436679, EPI_ISL_2436688, EPI_ISL_2436691, EPI_ISL_2436694, EPI_ISL_2436695, EPI_ISL_2436700, EPI_ISL_2436702, EPI_ISL_2436703, EPI_ISL_2436704, EPI_ISL_2436706, EPI_ISL_2436707, EPI_ISL_2436711, EPI_ISL_2436716, EPI_ISL_2436717, EPI_ISL_2436722, EPI_ISL_2436724, EPI_ISL_2436725, EPI_ISL_2436730, EPI_ISL_2436734, EPI_ISL_2436741, EPI_ISL_2436745, EPI_ISL_2436747, EPI_ISL_2436750, EPI_ISL_2436751, EPI_ISL_2436758, EPI_ISL_2436772, EPI_ISL_2436777, EPI_ISL_2436784, EPI_ISL_2436785, EPI_ISL_2436794, EPI_ISL_2436799, EPI_ISL_2436800, EPI_ISL_2436810, EPI_ISL_2436811, EPI_ISL_2436814, EPI_ISL_2436817, EPI_ISL_2436820, EPI_ISL_2436829, EPI_ISL_2436832, EPI_ISL_2436836, EPI_ISL_2436837, EPI_ISL_2436838, EPI_ISL_2436839, EPI_ISL_2436840, EPI_ISL_2436844, EPI_ISL_2436846, EPI_ISL_2436853, EPI_ISL_2436862, EPI_ISL_2436869, EPI_ISL_2436871, EPI_ISL_2436872, EPI_ISL_2436874, EPI_ISL_2436875, EPI_ISL_2436876, EPI_ISL_2436878, EPI_ISL_2436880, EPI_ISL_2436882, EPI_ISL_2436888, EPI_ISL_2436891, EPI_ISL_2436893, EPI_ISL_2436903, EPI_ISL_2436906, EPI_ISL_2436908, EPI_ISL_2436909, EPI_ISL_2436910, EPI_ISL_2436913, EPI_ISL_2436914, EPI_ISL_2436915, EPI_ISL_2436917, EPI_ISL_2436921, EPI_ISL_2436924, EPI_ISL_2436925, EPI_ISL_2436928, EPI_ISL_2437958, EPI_ISL_2437969, EPI_ISL_2437975, EPI_ISL_2437979, EPI_ISL_2437984, EPI_ISL_2437991, EPI_ISL_2438046, EPI_ISL_2438107, EPI_ISL_2438111, EPI_ISL_2438119, EPI_ISL_2438120, EPI_ISL_2438124, EPI_ISL_2438132, EPI_ISL_2438136, EPI_ISL_2438148, EPI_ISL_2438166, EPI_ISL_2438167, EPI_ISL_2438175, EPI_ISL_2438178, EPI_ISL_2438189, EPI_ISL_2438203, EPI_ISL_2438213, EPI_ISL_2438221, EPI_ISL_2438226, EPI_ISL_2438229, EPI_ISL_2438237, EPI_ISL_2438238, EPI_ISL_2438245, EPI_ISL_2438247, EPI_ISL_2438251, EPI_ISL_2438287, EPI_ISL_2438294, EPI_ISL_2438295, EPI_ISL_2438309, EPI_ISL_2438316, EPI_ISL_2438323, EPI_ISL_2438329, EPI_ISL_2438335, EPI_ISL_2438358, EPI_ISL_2438368, EPI_ISL_2438376, EPI_ISL_2438402, EPI_ISL_2446095, EPI_ISL_2446101, EPI_ISL_2446103, EPI_ISL_2446104, EPI_ISL_2446118, EPI_ISL_2446121, EPI_ISL_2446122, EPI_ISL_2446124, EPI_ISL_2446127, EPI_ISL_2446128, EPI_ISL_2446130, EPI_ISL_2446135, EPI_ISL_2446142, EPI_ISL_2446143, EPI_ISL_2446144, EPI_ISL_2446149, EPI_ISL_2446152, EPI_ISL_2446153, EPI_ISL_2446154, EPI_ISL_2446159, EPI_ISL_2446162, EPI_ISL_2446167, EPI_ISL_2446169, EPI_ISL_2446170, EPI_ISL_2446171, EPI_ISL_2446175, EPI_ISL_2446185, EPI_ISL_2446186, EPI_ISL_2446190, EPI_ISL_2446191, EPI_ISL_2446195, EPI_ISL_2446199, EPI_ISL_2446202, EPI_ISL_2446204, EPI_ISL_2446205, EPI_ISL_2446206, EPI_ISL_2446208, EPI_ISL_2446209, EPI_ISL_2446212, EPI_ISL_2446215, EPI_ISL_2446216, EPI_ISL_2446218, EPI_ISL_2446221, EPI_ISL_2446222, EPI_ISL_2446241, EPI_ISL_2446242, EPI_ISL_2446244, EPI_ISL_2446247, EPI_ISL_2446251, EPI_ISL_2446253, EPI_ISL_2446256, EPI_ISL_2446259, EPI_ISL_2446260, EPI_ISL_2446262, EPI_ISL_2446263, EPI_ISL_2446265, EPI_ISL_2446267, EPI_ISL_2446268, EPI_ISL_2446271, EPI_ISL_2446274, EPI_ISL_2446275, EPI_ISL_2446279, EPI_ISL_2446280, EPI_ISL_2446282, EPI_ISL_2446284, EPI_ISL_2446285, EPI_ISL_2446288, EPI_ISL_2446290, EPI_ISL_2446291, EPI_ISL_2446292, EPI_ISL_2446294, EPI_ISL_2446299, EPI_ISL_2446301, EPI_ISL_2446303, EPI_ISL_2446304, EPI_ISL_2446305, EPI_ISL_2446308, EPI_ISL_2446309, EPI_ISL_2446310, EPI_ISL_2446313, EPI_ISL_2446317, EPI_ISL_2446321, EPI_ISL_2446326, EPI_ISL_2446327, EPI_ISL_2446330, EPI_ISL_2446334, EPI_ISL_2446335, EPI_ISL_2446340, EPI_ISL_2446344, EPI_ISL_2446346, EPI_ISL_2446349, EPI_ISL_2446351, EPI_ISL_2446352, EPI_ISL_2446355, EPI_ISL_2446357, EPI_ISL_2446359, EPI_ISL_2446360, EPI_ISL_2446370, EPI_ISL_2446372, EPI_ISL_2446373, EPI_ISL_2446374, EPI_ISL_2446378, EPI_ISL_2446379, EPI_ISL_2446381, EPI_ISL_2446382, EPI_ISL_2446383, EPI_ISL_2446386, EPI_ISL_2446387, EPI_ISL_2446388, EPI_ISL_2446392, EPI_ISL_2446393, EPI_ISL_2446396, EPI_ISL_2446397, EPI_ISL_2446398, EPI_ISL_2446399, EPI_ISL_2446406, EPI_ISL_2446409, EPI_ISL_2446410, EPI_ISL_2446411, EPI_ISL_2446751, EPI_ISL_2446757, EPI_ISL_2446763, EPI_ISL_2446781, EPI_ISL_2446782, EPI_ISL_2446784, EPI_ISL_2446789, EPI_ISL_2446791, EPI_ISL_2446794, EPI_ISL_2446797, EPI_ISL_2446798, EPI_ISL_2446799, EPI_ISL_2446810, EPI_ISL_2446818, EPI_ISL_2446819, EPI_ISL_2446824, EPI_ISL_2446826, EPI_ISL_2446830, EPI_ISL_2446834, EPI_ISL_2446839, EPI_ISL_2446842, EPI_ISL_2446848, EPI_ISL_2446852, EPI_ISL_2446856, EPI_ISL_2446857, EPI_ISL_2446860, EPI_ISL_2446864, EPI_ISL_2446869, EPI_ISL_2446871, EPI_ISL_2446872, EPI_ISL_2446877, EPI_ISL_2446881, EPI_ISL_2446886, EPI_ISL_2446893, EPI_ISL_2446915, EPI_ISL_2446920, EPI_ISL_2446924, EPI_ISL_2446933, EPI_ISL_2446934, EPI_ISL_2446935, EPI_ISL_2446936, EPI_ISL_2446947, EPI_ISL_2446953, EPI_ISL_2446962, EPI_ISL_2446968, EPI_ISL_2446970, EPI_ISL_2446971, EPI_ISL_2446981, EPI_ISL_2446982, EPI_ISL_2446983, EPI_ISL_2446984, EPI_ISL_2446985, EPI_ISL_2446988, EPI_ISL_2446990, EPI_ISL_2446993, EPI_ISL_2446994, EPI_ISL_2446997, EPI_ISL_2446999, EPI_ISL_2447004, EPI_ISL_2447007, EPI_ISL_2447011, EPI_ISL_2447017, EPI_ISL_2447019, EPI_ISL_2447020, EPI_ISL_2447022, EPI_ISL_2447023, EPI_ISL_2447025, EPI_ISL_2447029, EPI_ISL_2447030, EPI_ISL_2447031, EPI_ISL_2447035, EPI_ISL_2447036, EPI_ISL_2447039, EPI_ISL_2447045, EPI_ISL_2447049, EPI_ISL_2447051, EPI_ISL_2447052, EPI_ISL_2447057, EPI_ISL_2447063, EPI_ISL_2447066, EPI_ISL_2447067, EPI_ISL_2447071, EPI_ISL_2447074, EPI_ISL_2447075, EPI_ISL_2447076, EPI_ISL_2447080, EPI_ISL_2447084, EPI_ISL_2447088, EPI_ISL_2447090, EPI_ISL_2447094, EPI_ISL_2447100, EPI_ISL_2447102, EPI_ISL_2447103, EPI_ISL_2447104, EPI_ISL_2447106, EPI_ISL_2447107, EPI_ISL_2447108, EPI_ISL_2447110, EPI_ISL_2447112, EPI_ISL_2447118, EPI_ISL_2447121, EPI_ISL_2447126, EPI_ISL_2447127, EPI_ISL_2447128, EPI_ISL_2447129, EPI_ISL_2447136, EPI_ISL_2447141, EPI_ISL_2447142, EPI_ISL_2447143, EPI_ISL_2447146, EPI_ISL_2447147, EPI_ISL_2447148, EPI_ISL_2447152, EPI_ISL_2447153, EPI_ISL_2447154, EPI_ISL_2447155, EPI_ISL_2447157, EPI_ISL_2447160, EPI_ISL_2447164, EPI_ISL_2447168, EPI_ISL_2447169, EPI_ISL_2447172, EPI_ISL_2447180, EPI_ISL_2447183, EPI_ISL_2447189, EPI_ISL_2447191, EPI_ISL_2447192, EPI_ISL_2447193, EPI_ISL_2447195, EPI_ISL_2447198, EPI_ISL_2447199, EPI_ISL_2447201, EPI_ISL_2447202, EPI_ISL_2447204, EPI_ISL_2447205, EPI_ISL_2447209, EPI_ISL_2447211, EPI_ISL_2447213, EPI_ISL_2447214, EPI_ISL_2447218, EPI_ISL_2447219, EPI_ISL_2447220, EPI_ISL_2447221, EPI_ISL_2447222, EPI_ISL_2447223, EPI_ISL_2447225, EPI_ISL_2447227, EPI_ISL_2447228, EPI_ISL_2447232, EPI_ISL_2447365, EPI_ISL_2447366, EPI_ISL_2447370, EPI_ISL_2447373, EPI_ISL_2447375, EPI_ISL_2447380, EPI_ISL_2447381, EPI_ISL_2447383, EPI_ISL_2447385, EPI_ISL_2447386, EPI_ISL_2447387, EPI_ISL_2447388, EPI_ISL_2447393, EPI_ISL_2447394, EPI_ISL_2447396, EPI_ISL_2447397, EPI_ISL_2447401, EPI_ISL_2447402, EPI_ISL_2447405, EPI_ISL_2447407, EPI_ISL_2447409, EPI_ISL_2447412, EPI_ISL_2447416, EPI_ISL_2447420, EPI_ISL_2447421, EPI_ISL_2447422, EPI_ISL_2447425, EPI_ISL_2447426, EPI_ISL_2447428, EPI_ISL_2447431, EPI_ISL_2447438, EPI_ISL_2447439, EPI_ISL_2447442, EPI_ISL_2447443, EPI_ISL_2447446, EPI_ISL_2447447, EPI_ISL_2447449, EPI_ISL_2447452, EPI_ISL_2447460, EPI_ISL_2447467, EPI_ISL_2447470, EPI_ISL_2447475, EPI_ISL_2447480, EPI_ISL_2447481, EPI_ISL_2447482, EPI_ISL_2447483, EPI_ISL_2447484, EPI_ISL_2447486, EPI_ISL_2447487, EPI_ISL_2447488, EPI_ISL_2447490, EPI_ISL_2447491, EPI_ISL_2447498, EPI_ISL_2447500, EPI_ISL_2447504, EPI_ISL_2447505, EPI_ISL_2447506, EPI_ISL_2447509, EPI_ISL_2447510, EPI_ISL_2447514, EPI_ISL_2447515, EPI_ISL_2447517, EPI_ISL_2447520, EPI_ISL_2447526, EPI_ISL_2447527, EPI_ISL_2447530, EPI_ISL_2447531, EPI_ISL_2447532, EPI_ISL_2447534, EPI_ISL_2447535, EPI_ISL_2447538, EPI_ISL_2447539, EPI_ISL_2447540, EPI_ISL_2447543, EPI_ISL_2447546, EPI_ISL_2447547, EPI_ISL_2447548, EPI_ISL_2447549, EPI_ISL_2447553, EPI_ISL_2447555, EPI_ISL_2447556, EPI_ISL_2447559, EPI_ISL_2447560, EPI_ISL_2447565, EPI_ISL_2447567, EPI_ISL_2447569, EPI_ISL_2447570, EPI_ISL_2447571, EPI_ISL_2447573, EPI_ISL_2447577, EPI_ISL_2447578, EPI_ISL_2447579, EPI_ISL_2447584, EPI_ISL_2447585, EPI_ISL_2447586, EPI_ISL_2447588, EPI_ISL_2447589, EPI_ISL_2447590, EPI_ISL_2447591, EPI_ISL_2447593, EPI_ISL_2447594, EPI_ISL_2447596, EPI_ISL_2447597, EPI_ISL_2447601, EPI_ISL_2447602, EPI_ISL_2447603, EPI_ISL_2447619, EPI_ISL_2447622, EPI_ISL_2447626, EPI_ISL_2447627, EPI_ISL_2447630, EPI_ISL_2447633, EPI_ISL_2447635, EPI_ISL_2447638, EPI_ISL_2447640, EPI_ISL_2447641, EPI_ISL_2447645, EPI_ISL_2447646, EPI_ISL_2447649, EPI_ISL_2447652, EPI_ISL_2447655, EPI_ISL_2447658, EPI_ISL_2447660, EPI_ISL_2447666, EPI_ISL_2447668, EPI_ISL_2447671, EPI_ISL_2447674, EPI_ISL_2447677, EPI_ISL_2447682, EPI_ISL_2447684, EPI_ISL_2447685, EPI_ISL_2447687, EPI_ISL_2447688, EPI_ISL_2447690, EPI_ISL_2447696, EPI_ISL_2447701, EPI_ISL_2447703, EPI_ISL_2447706, EPI_ISL_2447707, EPI_ISL_2447710, EPI_ISL_2447713, EPI_ISL_2447714, EPI_ISL_2447717, EPI_ISL_2447719, EPI_ISL_2447722, EPI_ISL_2447724, EPI_ISL_2447726, EPI_ISL_2447728, EPI_ISL_2447730, EPI_ISL_2447731, EPI_ISL_2447736, EPI_ISL_2447738, EPI_ISL_2447740, EPI_ISL_2447745, EPI_ISL_2447746, EPI_ISL_2447748, EPI_ISL_2447751, EPI_ISL_2447753, EPI_ISL_2447755, EPI_ISL_2447757, EPI_ISL_2456008, EPI_ISL_2456009, EPI_ISL_2456010, EPI_ISL_2456011, EPI_ISL_2456018, EPI_ISL_2456031, EPI_ISL_2456035, EPI_ISL_2456039, EPI_ISL_2456045, EPI_ISL_2456049, EPI_ISL_2456054, EPI_ISL_2456055, EPI_ISL_2456057, EPI_ISL_2456060, EPI_ISL_2456061, EPI_ISL_2456067, EPI_ISL_2456070, EPI_ISL_2456071, EPI_ISL_2456072, EPI_ISL_2456079, EPI_ISL_2456083, EPI_ISL_2456090, EPI_ISL_2456092, EPI_ISL_2456094, EPI_ISL_2456095, EPI_ISL_2456096, EPI_ISL_2456097, EPI_ISL_2456109, EPI_ISL_2456112, EPI_ISL_2456125, EPI_ISL_2456128, EPI_ISL_2456130, EPI_ISL_2456134, EPI_ISL_2456139, EPI_ISL_2456142, EPI_ISL_2456144, EPI_ISL_2456145, EPI_ISL_2456151, EPI_ISL_2456162, EPI_ISL_2456164, EPI_ISL_2456165, EPI_ISL_2456170, EPI_ISL_2456173, EPI_ISL_2456192, EPI_ISL_2456197, EPI_ISL_2456201, EPI_ISL_2456204, EPI_ISL_2456205, EPI_ISL_2456208, EPI_ISL_2456210, EPI_ISL_2456224, EPI_ISL_2456227, EPI_ISL_2456233, EPI_ISL_2456235, EPI_ISL_2456236, EPI_ISL_2456239, EPI_ISL_2456242, EPI_ISL_2456249, EPI_ISL_2456254, EPI_ISL_2456263, EPI_ISL_2456270, EPI_ISL_2456272, EPI_ISL_2456275, EPI_ISL_2456278, EPI_ISL_2456287, EPI_ISL_2456292, EPI_ISL_2456299, EPI_ISL_2456322, EPI_ISL_2456325, EPI_ISL_2456326, EPI_ISL_2456328, EPI_ISL_2456335, EPI_ISL_2456348, EPI_ISL_2456361, EPI_ISL_2456363, EPI_ISL_2456369, EPI_ISL_2456378, EPI_ISL_2456385, EPI_ISL_2456387, EPI_ISL_2456394, EPI_ISL_2456396, EPI_ISL_2456397, EPI_ISL_2456399, EPI_ISL_2456406, EPI_ISL_2456415, EPI_ISL_2456424, EPI_ISL_2456426, EPI_ISL_2456429, EPI_ISL_2456450, EPI_ISL_2456497, EPI_ISL_2456515, EPI_ISL_2456546, EPI_ISL_2456581, EPI_ISL_2456588, EPI_ISL_2456603, EPI_ISL_2456667, EPI_ISL_2456674, EPI_ISL_2456746, EPI_ISL_2456758, EPI_ISL_2456768, EPI_ISL_2456779, EPI_ISL_2456817, EPI_ISL_2456822, EPI_ISL_2457132, EPI_ISL_2457135, EPI_ISL_2457136, EPI_ISL_2457137, EPI_ISL_2457138, EPI_ISL_2457139, EPI_ISL_2457140, EPI_ISL_2457142, EPI_ISL_2457143, EPI_ISL_2457144, EPI_ISL_2457146, EPI_ISL_2457147, EPI_ISL_2457148, EPI_ISL_2457155, EPI_ISL_2457156, EPI_ISL_2457157, EPI_ISL_2457159, EPI_ISL_2457162, EPI_ISL_2457163, EPI_ISL_2457164, EPI_ISL_2457174, EPI_ISL_2457179, EPI_ISL_2457180, EPI_ISL_2457181, EPI_ISL_2457182, EPI_ISL_2457185, EPI_ISL_2457188, EPI_ISL_2457189, EPI_ISL_2457196, EPI_ISL_2457197, EPI_ISL_2457198, EPI_ISL_2457200, EPI_ISL_2457201, EPI_ISL_2457206, EPI_ISL_2457208, EPI_ISL_2457209, EPI_ISL_2457211, EPI_ISL_2457212, EPI_ISL_2457213, EPI_ISL_2457214, EPI_ISL_2457215, EPI_ISL_2457217, EPI_ISL_2457218, EPI_ISL_2457220, EPI_ISL_2457223, EPI_ISL_2457225, EPI_ISL_2457226, EPI_ISL_2457230, EPI_ISL_2457232, EPI_ISL_2457233, EPI_ISL_2457234, EPI_ISL_2457238, EPI_ISL_2457239, EPI_ISL_2457242, EPI_ISL_2457243, EPI_ISL_2457244, EPI_ISL_2457245, EPI_ISL_2457247, EPI_ISL_2457249, EPI_ISL_2457252, EPI_ISL_2457253, EPI_ISL_2457254, EPI_ISL_2457255, EPI_ISL_2457256, EPI_ISL_2457258, EPI_ISL_2457260, EPI_ISL_2457261, EPI_ISL_2457263, EPI_ISL_2457265, EPI_ISL_2457267, EPI_ISL_2457268, EPI_ISL_2457270, EPI_ISL_2457274, EPI_ISL_2457275, EPI_ISL_2457276, EPI_ISL_2457277, EPI_ISL_2457279, EPI_ISL_2457282, EPI_ISL_2457283, EPI_ISL_2457285, EPI_ISL_2457287, EPI_ISL_2457289, EPI_ISL_2457290, EPI_ISL_2457293, EPI_ISL_2457294, EPI_ISL_2457300, EPI_ISL_2457301, EPI_ISL_2457302, EPI_ISL_2457304, EPI_ISL_2457306, EPI_ISL_2457311, EPI_ISL_2457313, EPI_ISL_2457314, EPI_ISL_2457315, EPI_ISL_2457318, EPI_ISL_2457322, EPI_ISL_2457323, EPI_ISL_2457324, EPI_ISL_2457326, EPI_ISL_2457327, EPI_ISL_2457331, EPI_ISL_2457332, EPI_ISL_2457334, EPI_ISL_2457336, EPI_ISL_2457337, EPI_ISL_2457338, EPI_ISL_2457344, EPI_ISL_2457345, EPI_ISL_2457347, EPI_ISL_2457350, EPI_ISL_2457353, EPI_ISL_2457354, EPI_ISL_2457355, EPI_ISL_2457356, EPI_ISL_2457357, EPI_ISL_2457358, EPI_ISL_2457360, EPI_ISL_2457364, EPI_ISL_2457365, EPI_ISL_2457367, EPI_ISL_2457368, EPI_ISL_2457373, EPI_ISL_2457375, EPI_ISL_2457377, EPI_ISL_2457378, EPI_ISL_2457379, EPI_ISL_2457380, EPI_ISL_2457382, EPI_ISL_2457383, EPI_ISL_2457384, EPI_ISL_2457386, EPI_ISL_2457387, EPI_ISL_2457388, EPI_ISL_2457391, EPI_ISL_2457394, EPI_ISL_2457395, EPI_ISL_2457397, EPI_ISL_2457400, EPI_ISL_2457401, EPI_ISL_2457404, EPI_ISL_2457405, EPI_ISL_2457406, EPI_ISL_2457407, EPI_ISL_2457409, EPI_ISL_2457410, EPI_ISL_2457413, EPI_ISL_2457414, EPI_ISL_2457415, EPI_ISL_2457418, EPI_ISL_2457421, EPI_ISL_2457422, EPI_ISL_2457423, EPI_ISL_2457424, EPI_ISL_2457426, EPI_ISL_2457428, EPI_ISL_2457429, EPI_ISL_2457433, EPI_ISL_2457434, EPI_ISL_2457436, EPI_ISL_2457437, EPI_ISL_2457438, EPI_ISL_2457439, EPI_ISL_2457444, EPI_ISL_2457445, EPI_ISL_2457448, EPI_ISL_2457450, EPI_ISL_2457454, EPI_ISL_2457455, EPI_ISL_2457456, EPI_ISL_2457458, EPI_ISL_2457459, EPI_ISL_2457460, EPI_ISL_2457461, EPI_ISL_2457463, EPI_ISL_2457465, EPI_ISL_2457466, EPI_ISL_2457467, EPI_ISL_2457468, EPI_ISL_2457472, EPI_ISL_2457884, EPI_ISL_2467945, EPI_ISL_2467947, EPI_ISL_2467953, EPI_ISL_2467954, EPI_ISL_2467955, EPI_ISL_2467957, EPI_ISL_2467961, EPI_ISL_2467963, EPI_ISL_2467968, EPI_ISL_2467970, EPI_ISL_2467971, EPI_ISL_2467975, EPI_ISL_2467978, EPI_ISL_2467979, EPI_ISL_2467980, EPI_ISL_2467982, EPI_ISL_2467986, EPI_ISL_2467987, EPI_ISL_2467988, EPI_ISL_2467992, EPI_ISL_2467994, EPI_ISL_2467997, EPI_ISL_2467998, EPI_ISL_2467999, EPI_ISL_2468002, EPI_ISL_2468003, EPI_ISL_2468005, EPI_ISL_2468007, EPI_ISL_2468009, EPI_ISL_2468010, EPI_ISL_2468012, EPI_ISL_2468015, EPI_ISL_2468016, EPI_ISL_2468019, EPI_ISL_2468022, EPI_ISL_2468023, EPI_ISL_2468024, EPI_ISL_2468026, EPI_ISL_2468027, EPI_ISL_2468028, EPI_ISL_2468029, EPI_ISL_2468030, EPI_ISL_2468031, EPI_ISL_2468033, EPI_ISL_2468039, EPI_ISL_2468040, EPI_ISL_2468042, EPI_ISL_2468043, EPI_ISL_2468045, EPI_ISL_2468048, EPI_ISL_2468049, EPI_ISL_2468052, EPI_ISL_2468053, EPI_ISL_2468054, EPI_ISL_2468057, EPI_ISL_2468060, EPI_ISL_2468062, EPI_ISL_2468065, EPI_ISL_2468066, EPI_ISL_2468067, EPI_ISL_2468069, EPI_ISL_2468071, EPI_ISL_2468073, EPI_ISL_2468074, EPI_ISL_2468077, EPI_ISL_2468078, EPI_ISL_2468079, EPI_ISL_2468081, EPI_ISL_2468083, EPI_ISL_2468084, EPI_ISL_2468089, EPI_ISL_2468090, EPI_ISL_2468092, EPI_ISL_2468097, EPI_ISL_2468098, EPI_ISL_2468100, EPI_ISL_2468104, EPI_ISL_2468105, EPI_ISL_2468106, EPI_ISL_2468109, EPI_ISL_2468110, EPI_ISL_2468112, EPI_ISL_2468113, EPI_ISL_2468114, EPI_ISL_2468117, EPI_ISL_2468118, EPI_ISL_2468120, EPI_ISL_2468122, EPI_ISL_2468123, EPI_ISL_2468125, EPI_ISL_2468126, EPI_ISL_2468127, EPI_ISL_2468129, EPI_ISL_2468131, EPI_ISL_2468132, EPI_ISL_2468135, EPI_ISL_2468137, EPI_ISL_2468139, EPI_ISL_2468140, EPI_ISL_2468141, EPI_ISL_2468142, EPI_ISL_2468145, EPI_ISL_2468146, EPI_ISL_2468147, EPI_ISL_2468148, EPI_ISL_2468152, EPI_ISL_2468153, EPI_ISL_2468155, EPI_ISL_2468156, EPI_ISL_2468157, EPI_ISL_2468161, EPI_ISL_2468162, EPI_ISL_2468163, EPI_ISL_2468164, EPI_ISL_2468166, EPI_ISL_2468167, EPI_ISL_2468168, EPI_ISL_2468171, EPI_ISL_2468173, EPI_ISL_2468177, EPI_ISL_2468178, EPI_ISL_2468180, EPI_ISL_2468181, EPI_ISL_2468182, EPI_ISL_2468184, EPI_ISL_2468185, EPI_ISL_2468186, EPI_ISL_2468188, EPI_ISL_2468193, EPI_ISL_2468196, EPI_ISL_2468197, EPI_ISL_2468198, EPI_ISL_2468200, EPI_ISL_2468204, EPI_ISL_2468205, EPI_ISL_2468207, EPI_ISL_2468212, EPI_ISL_2468213, EPI_ISL_2468214, EPI_ISL_2468217, EPI_ISL_2468218, EPI_ISL_2468219, EPI_ISL_2468220, EPI_ISL_2468227, EPI_ISL_2468228, EPI_ISL_2468231, EPI_ISL_2468234, EPI_ISL_2468238, EPI_ISL_2468239, EPI_ISL_2468240, EPI_ISL_2468242, EPI_ISL_2468249, EPI_ISL_2468251, EPI_ISL_2468252, EPI_ISL_2468255, EPI_ISL_2468256, EPI_ISL_2468257, EPI_ISL_2468261, EPI_ISL_2468262, EPI_ISL_2468263, EPI_ISL_2468267, EPI_ISL_2468268, EPI_ISL_2468269, EPI_ISL_2468271, EPI_ISL_2468272, EPI_ISL_2468274, EPI_ISL_2468277, EPI_ISL_2468279, EPI_ISL_2468280, EPI_ISL_2468281, EPI_ISL_2468283, EPI_ISL_2468287, EPI_ISL_2468291, EPI_ISL_2468295, EPI_ISL_2468303, EPI_ISL_2468310, EPI_ISL_2468314, EPI_ISL_2468322, EPI_ISL_2468329, EPI_ISL_2468332, EPI_ISL_2468336, EPI_ISL_2468340, EPI_ISL_2468347, EPI_ISL_2468353, EPI_ISL_2468354, EPI_ISL_2468358, EPI_ISL_2468363, EPI_ISL_2468364, EPI_ISL_2468368, EPI_ISL_2468375, EPI_ISL_2468377, EPI_ISL_2468381, EPI_ISL_2468389, EPI_ISL_2468398, EPI_ISL_2468402, EPI_ISL_2468404, EPI_ISL_2468417, EPI_ISL_2468422, EPI_ISL_2468440, EPI_ISL_2468451, EPI_ISL_2468453, EPI_ISL_2468457, EPI_ISL_2468460, EPI_ISL_2468465, EPI_ISL_2468471, EPI_ISL_2468472, EPI_ISL_2468474, EPI_ISL_2468478, EPI_ISL_2468483, EPI_ISL_2468487, EPI_ISL_2468495, EPI_ISL_2468496, EPI_ISL_2468504, EPI_ISL_2468506, EPI_ISL_2468519, EPI_ISL_2468525, EPI_ISL_2468528, EPI_ISL_2468531, EPI_ISL_2468532, EPI_ISL_2468542, EPI_ISL_2468550, EPI_ISL_2468551, EPI_ISL_2468558, EPI_ISL_2468566, EPI_ISL_2468568, EPI_ISL_2468570, EPI_ISL_2468586, EPI_ISL_2468589, EPI_ISL_2468590, EPI_ISL_2468592, EPI_ISL_2468594, EPI_ISL_2468598, EPI_ISL_2468599, | | | |

EPI_ISL_2468600, EPI_ISL_2468602, EPI_ISL_2468603, EPI_ISL_2468604, EPI_ISL_2468605, EPI_ISL_2468609, EPI_ISL_2468611, EPI_ISL_2468613, EPI_ISL_2468614, EPI_ISL_2468617, EPI_ISL_2468618, EPI_ISL_2468619, EPI_ISL_2468622, EPI_ISL_2468625, EPI_ISL_2468626, EPI_ISL_2468628, EPI_ISL_2468629, EPI_ISL_2468631, EPI_ISL_2468633, EPI_ISL_2468634, EPI_ISL_2468635, EPI_ISL_2468637,
EPI_ISL_2468639, EPI_ISL_2468641, EPI_ISL_2468643, EPI_ISL_2468645, EPI_ISL_2468649, EPI_ISL_2468650, EPI_ISL_2468653, EPI_ISL_2468655, EPI_ISL_2468658, EPI_ISL_2468660, EPI_ISL_2468662, EPI_ISL_2468663, EPI_ISL_2468664, EPI_ISL_2468665, EPI_ISL_2468667, EPI_ISL_2468670, EPI_ISL_2468672, EPI_ISL_2468675, EPI_ISL_2468676, EPI_ISL_2468677, EPI_ISL_2468678, EPI_ISL_2468679,
EPI_ISL_2468683, EPI_ISL_2468684, EPI_ISL_2468690, EPI_ISL_2468692, EPI_ISL_2468693, EPI_ISL_2468694, EPI_ISL_2468696, EPI_ISL_2468698, EPI_ISL_2468699, EPI_ISL_2468701, EPI_ISL_2468702, EPI_ISL_2468703, EPI_ISL_2468704, EPI_ISL_2468706, EPI_ISL_2468707, EPI_ISL_2468710, EPI_ISL_2468711, EPI_ISL_2468714, EPI_ISL_2468716, EPI_ISL_2468717, EPI_ISL_2468719, EPI_ISL_2468720,
EPI_ISL_2468723, EPI_ISL_2468726, EPI_ISL_2468729, EPI_ISL_2468730, EPI_ISL_2468733, EPI_ISL_2468735, EPI_ISL_2468737, EPI_ISL_2468738, EPI_ISL_2468740, EPI_ISL_2468741, EPI_ISL_2468742, EPI_ISL_2468743, EPI_ISL_2468747, EPI_ISL_2468749, EPI_ISL_2468751, EPI_ISL_2468752, EPI_ISL_2468753, EPI_ISL_2468754, EPI_ISL_2468756, EPI_ISL_2468757, EPI_ISL_2468758, EPI_ISL_2468760,
EPI_ISL_2468761, EPI_ISL_2468762, EPI_ISL_2468764, EPI_ISL_2468766, EPI_ISL_2468768, EPI_ISL_2468769, EPI_ISL_2468771, EPI_ISL_2468772, EPI_ISL_2468773, EPI_ISL_2468774, EPI_ISL_2468775, EPI_ISL_2468776, EPI_ISL_2468778, EPI_ISL_2468779, EPI_ISL_2468780, EPI_ISL_2468781, EPI_ISL_2468782, EPI_ISL_2468783, EPI_ISL_2468785, EPI_ISL_2468787, EPI_ISL_2468788, EPI_ISL_2468789,
EPI_ISL_2468793, EPI_ISL_2468797, EPI_ISL_2468798, EPI_ISL_2468799, EPI_ISL_2468801, EPI_ISL_2468802, EPI_ISL_2468804, EPI_ISL_2468807, EPI_ISL_2468811, EPI_ISL_2468814, EPI_ISL_2468815, EPI_ISL_2468818, EPI_ISL_2468819, EPI_ISL_2468820, EPI_ISL_2468822, EPI_ISL_2468823, EPI_ISL_2468824, EPI_ISL_2468827, EPI_ISL_2468828, EPI_ISL_2468831, EPI_ISL_2468833, EPI_ISL_2468834,
EPI_ISL_2468835, EPI_ISL_2468837, EPI_ISL_2468839, EPI_ISL_2468843, EPI_ISL_2468844, EPI_ISL_2468845, EPI_ISL_2468846, EPI_ISL_2468848, EPI_ISL_2468851, EPI_ISL_2468853, EPI_ISL_2468854, EPI_ISL_2468855, EPI_ISL_2468857, EPI_ISL_2468859, EPI_ISL_2468860, EPI_ISL_2468862, EPI_ISL_2468864, EPI_ISL_2468865, EPI_ISL_2468870, EPI_ISL_2468871, EPI_ISL_2468874,
EPI_ISL_2468877, EPI_ISL_2468878, EPI_ISL_2468880, EPI_ISL_2468882, EPI_ISL_2468883, EPI_ISL_2468884, EPI_ISL_2468888, EPI_ISL_2468889, EPI_ISL_2468891, EPI_ISL_2468892, EPI_ISL_2468893, EPI_ISL_2468896, EPI_ISL_2468898, EPI_ISL_2468900, EPI_ISL_2468901, EPI_ISL_2468902, EPI_ISL_2468903, EPI_ISL_2468906, EPI_ISL_2468907, EPI_ISL_2468912, EPI_ISL_2468913, EPI_ISL_2468914,
EPI_ISL_2468924, EPI_ISL_2468925, EPI_ISL_2468926, EPI_ISL_2468927, EPI_ISL_2468928, EPI_ISL_2468929, EPI_ISL_2468932, EPI_ISL_2468934, EPI_ISL_2468938, EPI_ISL_2468939, EPI_ISL_2468940, EPI_ISL_2468941, EPI_ISL_2468942, EPI_ISL_2468943, EPI_ISL_2468945, EPI_ISL_2468946, EPI_ISL_2468947, EPI_ISL_2468949, EPI_ISL_2468950, EPI_ISL_2468951, EPI_ISL_2468952, EPI_ISL_2468953,
EPI_ISL_2468954, EPI_ISL_2468955, EPI_ISL_2468957, EPI_ISL_2468958, EPI_ISL_2468959, EPI_ISL_2468960, EPI_ISL_2468962, EPI_ISL_2468963, EPI_ISL_2468964, EPI_ISL_2468965, EPI_ISL_2468966, EPI_ISL_2468967, EPI_ISL_2468968, EPI_ISL_2468969, EPI_ISL_2468970, EPI_ISL_2468971, EPI_ISL_2468972, EPI_ISL_2468973, EPI_ISL_2468974, EPI_ISL_2468975, EPI_ISL_2468976, EPI_ISL_2468977,
EPI_ISL_2468978, EPI_ISL_2468979, EPI_ISL_2468980, EPI_ISL_2468981, EPI_ISL_2468982, EPI_ISL_2468983, EPI_ISL_2468985, EPI_ISL_2468986, EPI_ISL_2468987, EPI_ISL_2468988, EPI_ISL_2468989, EPI_ISL_2468990, EPI_ISL_2468991, EPI_ISL_2468992, EPI_ISL_2468993, EPI_ISL_2468994, EPI_ISL_2468995, EPI_ISL_2468996, EPI_ISL_2468997, EPI_ISL_2468998, EPI_ISL_2469000, EPI_ISL_2469001,
EPI_ISL_2469002, EPI_ISL_2469003, EPI_ISL_2469004, EPI_ISL_2469005, EPI_ISL_2469006, EPI_ISL_2469007, EPI_ISL_2469008, EPI_ISL_2469009, EPI_ISL_2469010, EPI_ISL_2469011, EPI_ISL_2469012, EPI_ISL_2469013, EPI_ISL_2469014, EPI_ISL_2469015, EPI_ISL_2469016, EPI_ISL_2469017, EPI_ISL_2469018, EPI_ISL_2469019, EPI_ISL_2469020, EPI_ISL_2469021, EPI_ISL_2469022, EPI_ISL_2469023,
EPI_ISL_2469024, EPI_ISL_2469025, EPI_ISL_2469026, EPI_ISL_2469027, EPI_ISL_2469028, EPI_ISL_2469029, EPI_ISL_2469030, EPI_ISL_2469031, EPI_ISL_2469032, EPI_ISL_2469033, EPI_ISL_2469034, EPI_ISL_2469035, EPI_ISL_2469036, EPI_ISL_2469037, EPI_ISL_2469038, EPI_ISL_2469039, EPI_ISL_2469040, EPI_ISL_2469041, EPI_ISL_2469042, EPI_ISL_2469043, EPI_ISL_2469044, EPI_ISL_2469045,
EPI_ISL_2469046, EPI_ISL_2469047, EPI_ISL_2469048, EPI_ISL_2469049, EPI_ISL_2469050, EPI_ISL_2469051, EPI_ISL_2469052, EPI_ISL_2469053, EPI_ISL_2469054, EPI_ISL_2469055, EPI_ISL_2469056, EPI_ISL_2469057, EPI_ISL_2469058, EPI_ISL_2469059, EPI_ISL_2469060, EPI_ISL_2469061, EPI_ISL_2469062, EPI_ISL_2469063, EPI_ISL_2469064, EPI_ISL_2469065, EPI_ISL_2469066, EPI_ISL_2469067,
EPI_ISL_2469068, EPI_ISL_2469069, EPI_ISL_2469070, EPI_ISL_2469071, EPI_ISL_2469073, EPI_ISL_2469074, EPI_ISL_2469075, EPI_ISL_2469076, EPI_ISL_2469077, EPI_ISL_2469078, EPI_ISL_2469079, EPI_ISL_2469080, EPI_ISL_2469081, EPI_ISL_2469082, EPI_ISL_2469083, EPI_ISL_2469084, EPI_ISL_2469085, EPI_ISL_2469086, EPI_ISL_2469087, EPI_ISL_2469088, EPI_ISL_2469089, EPI_ISL_2469090,
EPI_ISL_2469091, EPI_ISL_2469092, EPI_ISL_2469093, EPI_ISL_2469094, EPI_ISL_2469095, EPI_ISL_2469096, EPI_ISL_2469097, EPI_ISL_2469098, EPI_ISL_2469099, EPI_ISL_2469100, EPI_ISL_2469101, EPI_ISL_2469102, EPI_ISL_2469103, EPI_ISL_2469104, EPI_ISL_2469105, EPI_ISL_2469106, EPI_ISL_2469107, EPI_ISL_2469108, EPI_ISL_2469109, EPI_ISL_2469110, EPI_ISL_2469111, EPI_ISL_2469112,
EPI_ISL_2469113, EPI_ISL_2469114, EPI_ISL_2469115, EPI_ISL_2469116, EPI_ISL_2469117, EPI_ISL_2469118, EPI_ISL_2469119, EPI_ISL_2469120, EPI_ISL_2469121, EPI_ISL_2469122, EPI_ISL_2469123, EPI_ISL_2469124, EPI_ISL_2469125, EPI_ISL_2469126, EPI_ISL_2469127, EPI_ISL_2469128, EPI_ISL_2469129, EPI_ISL_2469130, EPI_ISL_2469131, EPI_ISL_2469132, EPI_ISL_2469133, EPI_ISL_2469134,
EPI_ISL_2469135, EPI_ISL_2469136, EPI_ISL_2469137, EPI_ISL_2469138, EPI_ISL_2469139, EPI_ISL_2469140, EPI_ISL_2469141, EPI_ISL_2469142, EPI_ISL_2469143, EPI_ISL_2469144, EPI_ISL_2469145, EPI_ISL_2469146, EPI_ISL_2469147, EPI_ISL_2469148, EPI_ISL_2469149, EPI_ISL_2469150, EPI_ISL_2469151, EPI_ISL_2469152, EPI_ISL_2469153, EPI_ISL_2469154, EPI_ISL_2469155, EPI_ISL_2469156,
EPI_ISL_2469157, EPI_ISL_2469158, EPI_ISL_2469159, EPI_ISL_2469160, EPI_ISL_2469161, EPI_ISL_2469162, EPI_ISL_2469163, EPI_ISL_2469164, EPI_ISL_2469165, EPI_ISL_2469166, EPI_ISL_2469167, EPI_ISL_2469168, EPI_ISL_2469169, EPI_ISL_2469170, EPI_ISL_2469171, EPI_ISL_2469172, EPI_ISL_2469173, EPI_ISL_2469174, EPI_ISL_2469175, EPI_ISL_2469176, EPI_ISL_2469177, EPI_ISL_2469178,
EPI_ISL_2469179, EPI_ISL_2469180, EPI_ISL_2469181, EPI_ISL_2469182, EPI_ISL_2469183, EPI_ISL_2469184, EPI_ISL_2469185, EPI_ISL_2469186, EPI_ISL_2469187, EPI_ISL_2469188, EPI_ISL_2469190, EPI_ISL_2469191, EPI_ISL_2469192, EPI_ISL_2469193, EPI_ISL_2469194, EPI_ISL_2469195, EPI_ISL_2469196, EPI_ISL_2469197, EPI_ISL_2469198, EPI_ISL_2469199, EPI_ISL_2469200, EPI_ISL_2469201,
EPI_ISL_2469202, EPI_ISL_2469203, EPI_ISL_2469204, EPI_ISL_2469205, EPI_ISL_2469206, EPI_ISL_2469207, EPI_ISL_2469208, EPI_ISL_2469209, EPI_ISL_2469210, EPI_ISL_2469211, EPI_ISL_2469213, EPI_ISL_2469214, EPI_ISL_2469215, EPI_ISL_2469216, EPI_ISL_2469217, EPI_ISL_2469218, EPI_ISL_2469219, EPI_ISL_2469220, EPI_ISL_2469221, EPI_ISL_2469223, EPI_ISL_2469224, EPI_ISL_2469225,
EPI_ISL_2469226, EPI_ISL_2469229, EPI_ISL_2469232, EPI_ISL_2469238, EPI_ISL_2469242, EPI_ISL_2469243, EPI_ISL_2469244, EPI_ISL_2469247, EPI_ISL_2469250, EPI_ISL_2469254, EPI_ISL_2469255, EPI_ISL_2469256, EPI_ISL_2469257, EPI_ISL_2469264, EPI_ISL_2469266, EPI_ISL_2469269, EPI_ISL_2469270, EPI_ISL_2469271, EPI_ISL_2469280, EPI_ISL_2469283, EPI_ISL_2469285, EPI_ISL_2469288,
EPI_ISL_2469290, EPI_ISL_2469292, EPI_ISL_2469294, EPI_ISL_2469296, EPI_ISL_2469306, EPI_ISL_2469307, EPI_ISL_2469308, EPI_ISL_2469310, EPI_ISL_2469311, EPI_ISL_2469312, EPI_ISL_2469313, EPI_ISL_2469319, EPI_ISL_2469324, EPI_ISL_2469325, EPI_ISL_2469328, EPI_ISL_2469329, EPI_ISL_2469333, EPI_ISL_2469334, EPI_ISL_2469335, EPI_ISL_2469342, EPI_ISL_2469343, EPI_ISL_2469346,
EPI_ISL_2469348, EPI_ISL_2469350, EPI_ISL_2469351, EPI_ISL_2469352, EPI_ISL_2469358, EPI_ISL_2469359, EPI_ISL_2469361, EPI_ISL_2469367, EPI_ISL_2469368, EPI_ISL_2469375, EPI_ISL_2469377, EPI_ISL_2469378, EPI_ISL_2469379, EPI_ISL_2469383, EPI_ISL_2469384, EPI_ISL_2469387, EPI_ISL_2469391, EPI_ISL_2469393, EPI_ISL_2469395, EPI_ISL_2469398, EPI_ISL_2469401, EPI_ISL_2469408,
EPI_ISL_2469409, EPI_ISL_2469411, EPI_ISL_2469413, EPI_ISL_2469419, EPI_ISL_2469420, EPI_ISL_2469421, EPI_ISL_2469423, EPI_ISL_2469425, EPI_ISL_2469429, EPI_ISL_2469433, EPI_ISL_2469437, EPI_ISL_2469438, EPI_ISL_2469441, EPI_ISL_2469443, EPI_ISL_2469445, EPI_ISL_2469448, EPI_ISL_2469449, EPI_ISL_2469451, EPI_ISL_2469467, EPI_ISL_2485259, EPI_ISL_2485262, EPI_ISL_2485264,
EPI_ISL_2485266, EPI_ISL_2485269, EPI_ISL_2485277, EPI_ISL_2485280, EPI_ISL_2485281, EPI_ISL_2485293, EPI_ISL_2485303, EPI_ISL_2485305, EPI_ISL_2485310, EPI_ISL_2485311, EPI_ISL_2485313, EPI_ISL_2485314, EPI_ISL_2485319, EPI_ISL_2485331, EPI_ISL_2485340, EPI_ISL_2485342, EPI_ISL_2485354, EPI_ISL_2485373, EPI_ISL_2485377, EPI_ISL_2485397, EPI_ISL_2485398, EPI_ISL_2485399,
EPI_ISL_2485402, EPI_ISL_2485404, EPI_ISL_2485407, EPI_ISL_2485408, EPI_ISL_2485412, EPI_ISL_2485417, EPI_ISL_2485426, EPI_ISL_2485430, EPI_ISL_2485437, EPI_ISL_2485439, EPI_ISL_2485441, EPI_ISL_2485446, EPI_ISL_2485447, EPI_ISL_2485460, EPI_ISL_2485463, EPI_ISL_2485470, EPI_ISL_2485488, EPI_ISL_2485498, EPI_ISL_2485502, EPI_ISL_2485503, EPI_ISL_2485508, EPI_ISL_2485515,
EPI_ISL_2485524, EPI_ISL_2485534, EPI_ISL_2485565, EPI_ISL_2485568, EPI_ISL_2485617, EPI_ISL_2485626, EPI_ISL_2485656, EPI_ISL_2485658, EPI_ISL_2485683, EPI_ISL_2485705, EPI_ISL_2485780, EPI_ISL_2485802, EPI_ISL_2485836, EPI_ISL_2486531, EPI_ISL_2486535, EPI_ISL_2486536, EPI_ISL_2486537, EPI_ISL_2486538, EPI_ISL_2486548, EPI_ISL_2486550, EPI_ISL_2486552, EPI_ISL_2486554,
EPI_ISL_2486555, EPI_ISL_2486556, EPI_ISL_2486557, EPI_ISL_2486558, EPI_ISL_2486561, EPI_ISL_2486563, EPI_ISL_2486566, EPI_ISL_2486567, EPI_ISL_2486568, EPI_ISL_2486571, EPI_ISL_2486581, EPI_ISL_2486585, EPI_ISL_2486597, EPI_ISL_2486600, EPI_ISL_2486601, EPI_ISL_2486603, EPI_ISL_2486604, EPI_ISL_2486606, EPI_ISL_2486609, EPI_ISL_2486617, EPI_ISL_2486622, EPI_ISL_2486627,
EPI_ISL_2486628, EPI_ISL_2486630, EPI_ISL_2486632, EPI_ISL_2486633, EPI_ISL_2486635, EPI_ISL_2486638, EPI_ISL_2486640, EPI_ISL_2486641, EPI_ISL_2486642, EPI_ISL_2486644, EPI_ISL_2486645, EPI_ISL_2486647, EPI_ISL_2486648, EPI_ISL_2486651, EPI_ISL_2486654, EPI_ISL_2486657, EPI_ISL_2486661, EPI_ISL_2486663, EPI_ISL_2486664, EPI_ISL_2486668, EPI_ISL_2486670, EPI_ISL_2486672,
EPI_ISL_2486673, EPI_ISL_2486674, EPI_ISL_2486675, EPI_ISL_2486676, EPI_ISL_2486678, EPI_ISL_2486681, EPI_ISL_2486682, EPI_ISL_2486685, EPI_ISL_2486687, EPI_ISL_2486699, EPI_ISL_2486701, EPI_ISL_2486704, EPI_ISL_2486707, EPI_ISL_2486709, EPI_ISL_2486711, EPI_ISL_2486713, EPI_ISL_2486714, EPI_ISL_2486718, EPI_ISL_2486723, EPI_ISL_2486724, EPI_ISL_2486727, EPI_ISL_2486733,
EPI_ISL_2486735, EPI_ISL_2486736, EPI_ISL_2486737, EPI_ISL_2486738, EPI_ISL_2486742, EPI_ISL_2486743, EPI_ISL_2486748, EPI_ISL_2486749, EPI_ISL_2486751, EPI_ISL_2486755, EPI_ISL_2486760, EPI_ISL_2486761, EPI_ISL_2486766, EPI_ISL_2486768, EPI_ISL_2486769, EPI_ISL_2486771, EPI_ISL_2486774, EPI_ISL_2486777, EPI_ISL_2486780, EPI_ISL_2486782, EPI_ISL_2486785, EPI_ISL_2486786,
EPI_ISL_2486787, EPI_ISL_2486790, EPI_ISL_2486791, EPI_ISL_2486793, EPI_ISL_2486794, EPI_ISL_2486795, EPI_ISL_2486797, EPI_ISL_2486800, EPI_ISL_2486802, EPI_ISL_2486809, EPI_ISL_2486812, EPI_ISL_2486814, EPI_ISL_2486815, EPI_ISL_2486816, EPI_ISL_2486821, EPI_ISL_2486822, EPI_ISL_2486823, EPI_ISL_2486826, EPI_ISL_2486827, EPI_ISL_2486829, EPI_ISL_2486831, EPI_ISL_2486832,
EPI_ISL_2486835, EPI_ISL_2486838, EPI_ISL_2486845, EPI_ISL_2486849, EPI_ISL_2486851, EPI_ISL_2486855, EPI_ISL_2486860, EPI_ISL_2486862, EPI_ISL_2486863, EPI_ISL_2486864, EPI_ISL_2486865, EPI_ISL_2486868, EPI_ISL_2486872, EPI_ISL_2486873, EPI_ISL_2486875, EPI_ISL_2486878, EPI_ISL_2486882, EPI_ISL_2486883, EPI_ISL_2486886, EPI_ISL_2486888, EPI_ISL_2486890, EPI_ISL_2486894,
EPI_ISL_2486912, EPI_ISL_2486923, EPI_ISL_2486998, EPI_ISL_2487008, EPI_ISL_2487019, EPI_ISL_2487027, EPI_ISL_2487031, EPI_ISL_2487032, EPI_ISL_2487047, EPI_ISL_2487069, EPI_ISL_2487084, EPI_ISL_2487091, EPI_ISL_2487104, EPI_ISL_2487132, EPI_ISL_2487133, EPI_ISL_2487176, EPI_ISL_2487197, EPI_ISL_2487585, EPI_ISL_2487586, EPI_ISL_2487591, EPI_ISL_2487596, EPI_ISL_2487601,
EPI_ISL_2487611, EPI_ISL_2487623, EPI_ISL_2487626, EPI_ISL_2487630, EPI_ISL_2487631, EPI_ISL_2487635, EPI_ISL_2487640, EPI_ISL_2487649, EPI_ISL_2487650, EPI_ISL_2487662, EPI_ISL_2487664, EPI_ISL_2487670, EPI_ISL_2487687, EPI_ISL_2487689, EPI_ISL_2487693, EPI_ISL_2487700, EPI_ISL_2487706, EPI_ISL_2487712, EPI_ISL_2487716, EPI_ISL_2487724, EPI_ISL_2487725, EPI_ISL_2487726,
EPI_ISL_2487729, EPI_ISL_2487769, EPI_ISL_2487783, EPI_ISL_2487805, EPI_ISL_2487809, EPI_ISL_2487813, EPI_ISL_2487814, EPI_ISL_2487818, EPI_ISL_2487823, EPI_ISL_2487828, EPI_ISL_2487831, EPI_ISL_2487836, EPI_ISL_2487848, EPI_ISL_2487851, EPI_ISL_2487858, EPI_ISL_2487861, EPI_ISL_2487867, EPI_ISL_2487881, EPI_ISL_2487884, EPI_ISL_2487895, EPI_ISL_2487896, EPI_ISL_2487897,
EPI_ISL_2487898, EPI_ISL_2487902, EPI_ISL_2487913, EPI_ISL_2487914, EPI_ISL_2487917, EPI_ISL_2487929, EPI_ISL_2488080, EPI_ISL_2488081, EPI_ISL_2488094, EPI_ISL_2488238, EPI_ISL_2488240, EPI_ISL_2488241, EPI_ISL_2488438, EPI_ISL_2488505, EPI_ISL_2511385, EPI_ISL_2511565, EPI_ISL_2511580, EPI_ISL_2511862, EPI_ISL_2511965, EPI_ISL_2512108, EPI_ISL_2512986, EPI_ISL_2513013,
EPI_ISL_2513064, EPI_ISL_2513128, EPI_ISL_2513184, EPI_ISL_2513200, EPI_ISL_2513212, EPI_ISL_2513226, EPI_ISL_2513227, EPI_ISL_2513245, EPI_ISL_2513251, EPI_ISL_2513291, EPI_ISL_2513292, EPI_ISL_2513293, EPI_ISL_2513295, EPI_ISL_2513296, EPI_ISL_2513297, EPI_ISL_2513298, EPI_ISL_2513299, EPI_ISL_2513302, EPI_ISL_2513304, EPI_ISL_2513305, EPI_ISL_2513307, EPI_ISL_2513308,
EPI_ISL_2513310, EPI_ISL_2513312, EPI_ISL_2513315, EPI_ISL_2513316, EPI_ISL_2513317, EPI_ISL_2513319, EPI_ISL_2513321, EPI_ISL_2513323, EPI_ISL_2513324, EPI_ISL_2513325, EPI_ISL_2513327, EPI_ISL_2513328, EPI_ISL_2513329, EPI_ISL_2513330, EPI_ISL_2513331, EPI_ISL_2513332, EPI_ISL_2513333, EPI_ISL_2513336, EPI_ISL_2513337, EPI_ISL_2513338, EPI_ISL_2513340, EPI_ISL_2513343,
EPI_ISL_2513344, EPI_ISL_2513345, EPI_ISL_2513347, EPI_ISL_2513349, EPI_ISL_2513350, EPI_ISL_2513352, EPI_ISL_2513353, EPI_ISL_2513354, EPI_ISL_2513355, EPI_ISL_2513356, EPI_ISL_2513357, EPI_ISL_2513358, EPI_ISL_2513359, EPI_ISL_2513360, EPI_ISL_2513361, EPI_ISL_2513363, EPI_ISL_2513364, EPI_ISL_2513365, EPI_ISL_2513367, EPI_ISL_2513369, EPI_ISL_2513371, EPI_ISL_2513372,
EPI_ISL_2513374, EPI_ISL_2513378, EPI_ISL_2513380, EPI_ISL_2513381, EPI_ISL_2513382, EPI_ISL_2513383, EPI_ISL_2513384, EPI_ISL_2513385, EPI_ISL_2513386, EPI_ISL_2513387, EPI_ISL_2513388, EPI_ISL_2513389, EPI_ISL_2513391, EPI_ISL_2513392, EPI_ISL_2513393, EPI_ISL_2513395, EPI_ISL_2513397, EPI_ISL_2513398, EPI_ISL_2513399, EPI_ISL_2513400, EPI_ISL_2513402, EPI_ISL_2513403,
EPI_ISL_2513405, EPI_ISL_2513407, EPI_ISL_2513408, EPI_ISL_2513409, EPI_ISL_2513410, EPI_ISL_2513412, EPI_ISL_2513414, EPI_ISL_2513415, EPI_ISL_2513417, EPI_ISL_2513418, EPI_ISL_2513419, EPI_ISL_2513420, EPI_ISL_2513423, EPI_ISL_2513424, EPI_ISL_2513425, EPI_ISL_2513426, EPI_ISL_2513427, EPI_ISL_2513428, EPI_ISL_2513429, EPI_ISL_2513430, EPI_ISL_2513431, EPI_ISL_2513432,
EPI_ISL_2513433, EPI_ISL_2513435, EPI_ISL_2513436, EPI_ISL_2513437, EPI_ISL_2513439, EPI_ISL_2513440, EPI_ISL_2513441, EPI_ISL_2513443, EPI_ISL_2513444, EPI_ISL_2513446, EPI_ISL_2513448, EPI_ISL_2513449, EPI_ISL_2513451, EPI_ISL_2513452, EPI_ISL_2513453, EPI_ISL_2513454, EPI_ISL_2513456, EPI_ISL_2513458, EPI_ISL_2513459, EPI_ISL_2513460, EPI_ISL_2513461, EPI_ISL_2513463,
EPI_ISL_2513464, EPI_ISL_2513466, EPI_ISL_2513467, EPI_ISL_2513468, EPI_ISL_2513471, EPI_ISL_2513472, EPI_ISL_2513474, EPI_ISL_2513475, EPI_ISL_2513477, EPI_ISL_2513478, EPI_ISL_2513479, EPI_ISL_2513480, EPI_ISL_2513481, EPI_ISL_2513482, EPI_ISL_2513483, EPI_ISL_2513484, EPI_ISL_2513485, EPI_ISL_2513486, EPI_ISL_2513487, EPI_ISL_2513488, EPI_ISL_2513489, EPI_ISL_2513491,
EPI_ISL_2513492, EPI_ISL_2513493, EPI_ISL_2513494, EPI_ISL_2513495, EPI_ISL_2513500, EPI_ISL_2513501, EPI_ISL_2513503, EPI_ISL_2513504, EPI_ISL_2513506, EPI_ISL_2513507, EPI_ISL_2513508, EPI_ISL_2513509, EPI_ISL_2513510, EPI_ISL_2513511, EPI_ISL_2513512, EPI_ISL_2513513, EPI_ISL_2513514, EPI_ISL_2513516, EPI_ISL_2513519, EPI_ISL_2513520, EPI_ISL_2513521, EPI_ISL_2513522,
EPI_ISL_2513523, EPI_ISL_2513527, EPI_ISL_2513528, EPI_ISL_2513529, EPI_ISL_2513530, EPI_ISL_2513531, EPI_ISL_2513534, EPI_ISL_2513535, EPI_ISL_2513537, EPI_ISL_2513539, EPI_ISL_2513540, EPI_ISL_2513543, EPI_ISL_2513545, EPI_ISL_2513546, EPI_ISL_2513547, EPI_ISL_2513548, EPI_ISL_2513549, EPI_ISL_2513551, EPI_ISL_2513552, EPI_ISL_2513553, EPI_ISL_2513554, EPI_ISL_2513555,
EPI_ISL_2513556, EPI_ISL_2513559, EPI_ISL_2513560, EPI_ISL_2513561, EPI_ISL_2513562, EPI_ISL_2513565, EPI_ISL_2513567, EPI_ISL_2513569, EPI_ISL_2513570, EPI_ISL_2513571, EPI_ISL_2513572, EPI_ISL_2513573, EPI_ISL_2513574, EPI_ISL_2513576, EPI_ISL_2513577, EPI_ISL_2513578, EPI_ISL_2513580, EPI_ISL_2513581, EPI_ISL_2513582, EPI_ISL_2513583, EPI_ISL_2513584, EPI_ISL_2513585,
EPI_ISL_2513586, EPI_ISL_2513589, EPI_ISL_2513590, EPI_ISL_2513592, EPI_ISL_2513593, EPI_ISL_2513594, EPI_ISL_2513595, EPI_ISL_2513597, EPI_ISL_2513598, EPI_ISL_2513599, EPI_ISL_2513600, EPI_ISL_2513601, EPI_ISL_2513602, EPI_ISL_2513603, EPI_ISL_2513604, EPI_ISL_2513605, EPI_ISL_2513606, EPI_ISL_2513608, EPI_ISL_2513610, EPI_ISL_2513611, EPI_ISL_2513801, EPI_ISL_2513802,
EPI_ISL_2513804, EPI_ISL_2513821, EPI_ISL_2513823, EPI_ISL_2513826, EPI_ISL_2513827, EPI_ISL_2513828, EPI_ISL_2513829, EPI_ISL_2513830, EPI_ISL_2513839, EPI_ISL_2513841, EPI_ISL_2513842, EPI_ISL_2513861, EPI_ISL_2513864, EPI_ISL_2513869, EPI_ISL_2513871, EPI_ISL_2513873, EPI_ISL_2513874, EPI_ISL_2513880, EPI_ISL_2513883, EPI_ISL_2513884, EPI_ISL_2513892, EPI_ISL_2513897,
EPI_ISL_2513900, EPI_ISL_2513903, EPI_ISL_2513904, EPI_ISL_2513905, EPI_ISL_2513916, EPI_ISL_2513918, EPI_ISL_2513924, EPI_ISL_2513925, EPI_ISL_2513928, EPI_ISL_2513929, EPI_ISL_2513930, EPI_ISL_2513932, EPI_ISL_2513935, EPI_ISL_2513936, EPI_ISL_2513939, EPI_ISL_2513940, EPI_ISL_2513945, EPI_ISL_2513946, EPI_ISL_2513949, EPI_ISL_2513953, EPI_ISL_2513954, EPI_ISL_2513957,
EPI_ISL_2513959, EPI_ISL_2513960, EPI_ISL_2513961, EPI_ISL_2513962, EPI_ISL_2513963, EPI_ISL_2513966, EPI_ISL_2513970, EPI_ISL_2513972, EPI_ISL_2513973, EPI_ISL_2513974, EPI_ISL_2513980, EPI_ISL_2513983, EPI_ISL_2513985, EPI_ISL_2513986, EPI_ISL_2513990, EPI_ISL_2513994, EPI_ISL_2513995, EPI_ISL_2513998, EPI_ISL_2514001, EPI_ISL_2514006, EPI_ISL_2514008, EPI_ISL_2514010,
EPI_ISL_2514015, EPI_ISL_2514016, EPI_ISL_2514018, EPI_ISL_2514023, EPI_ISL_2514027, EPI_ISL_2514045, EPI_ISL_2514046, EPI_ISL_2514047, EPI_ISL_2514049, EPI_ISL_2514052, EPI_ISL_2514055, EPI_ISL_2514058, EPI_ISL_2514059, EPI_ISL_2514061, EPI_ISL_2514065, EPI_ISL_2514066, EPI_ISL_2514067, EPI_ISL_2514068, EPI_ISL_2514070, EPI_ISL_2514073, EPI_ISL_2514074,
EPI_ISL_2514148, EPI_ISL_2514177, EPI_ISL_2514208, EPI_ISL_2514226, EPI_ISL_2514238, EPI_ISL_2514245, EPI_ISL_2514286, EPI_ISL_2514302, EPI_ISL_2514348, EPI_ISL_2514409, EPI_ISL_2514411, EPI_ISL_2514413, EPI_ISL_2514414, EPI_ISL_2514417, EPI_ISL_2514420, EPI_ISL_2514423, EPI_ISL_2514424, EPI_ISL_2514425, EPI_ISL_2514428, EPI_ISL_2514429, EPI_ISL_2514430, EPI_ISL_2514431,
EPI_ISL_2514432, EPI_ISL_2514433, EPI_ISL_2514434, EPI_ISL_2514435, EPI_ISL_2514436, EPI_ISL_2514438, EPI_ISL_2514440, EPI_ISL_2514443, EPI_ISL_2514444, EPI_ISL_2514445, EPI_ISL_2514447, EPI_ISL_2514449, EPI_ISL_2514454, EPI_ISL_2514456, EPI_ISL_2514457, EPI_ISL_2514461, EPI_ISL_2514462, EPI_ISL_2514464, EPI_ISL_2514466, EPI_ISL_2514468, EPI_ISL_2514470, EPI_ISL_2514471,
EPI_ISL_2514472, EPI_ISL_2514473, EPI_ISL_2514474, EPI_ISL_2514477, EPI_ISL_2514480, EPI_ISL_2514481, EPI_ISL_2514484, EPI_ISL_2514485, EPI_ISL_2514487, EPI_ISL_2514488, EPI_ISL_2514490, EPI_ISL_2514495, EPI_ISL_2514498, EPI_ISL_2514499, EPI_ISL_2514501, EPI_ISL_2514503, EPI_ISL_2514504, EPI_ISL_2514506, EPI_ISL_2514508,
EPI_ISL_2514511, EPI_ISL_2514513, EPI_ISL_2514514, EPI_ISL_2514516, EPI_ISL_2514519, EPI_ISL_2514522, EPI_ISL_2514524, EPI_ISL_2514527, EPI_ISL_2514528, EPI_ISL_2514533, EPI_ISL_2514534, EPI_ISL_2514535, EPI_ISL_2514538, EPI_ISL_2514540, EPI_ISL_2514541, EPI_ISL_2514542, EPI_ISL_2514543, EPI_ISL_2514545, EPI_ISL_2514546, EPI_ISL_2514547, EPI_ISL_2514549, EPI_ISL_2514551,
EPI_ISL_2514555, EPI_ISL_2514557, EPI_ISL_2514558, EPI_ISL_2514564, EPI_ISL_2514566, EPI_ISL_2514568, EPI_ISL_2514569, EPI_ISL_2514572, EPI_ISL_2514573, EPI_ISL_2514576, EPI_ISL_2514579, EPI_ISL_2514581, EPI_ISL_2514583, EPI_ISL_2514584, EPI_ISL_2514585, EPI_ISL_2514586, EPI_ISL_2514587, EPI_ISL_2514588, EPI_ISL_2514589, EPI_ISL_2514593,
EPI_ISL_2514594, EPI_ISL_2514595, EPI_ISL_2514596, EPI_ISL_2514597, EPI_ISL_2514600, EPI_ISL_2514602, EPI_ISL_2514603, EPI_ISL_2514604, EPI_ISL_2514605, EPI_ISL_2514607, EPI_ISL_2514614, EPI_ISL_2514618, EPI_ISL_2514619, EPI_ISL_2514620, EPI_ISL_2514621, EPI_ISL_2514624, EPI_ISL_2514625, EPI_ISL_2514627, EPI_ISL_2514628, EPI_ISL_2514629, EPI_ISL_2514632, EPI_ISL_2514635,
EPI_ISL_2514636, EPI_ISL_2514637, EPI_ISL_2514641, EPI_ISL_2514642, EPI_ISL_2514644, EPI_ISL_2514647, EPI_ISL_2514648, EPI_ISL_2514649, EPI_ISL_2514650, EPI_ISL_2514651, EPI_ISL_2514652, EPI_ISL_2514653, EPI_ISL_2514656, EPI_ISL_2514657, EPI_ISL_2514658, EPI_ISL_2514659, EPI_ISL_2514661, EPI_ISL_2514662, EPI_ISL_2514663, EPI_ISL_2514664, EPI_ISL_2514665,
EPI_ISL_2514670, EPI_ISL_2514672, EPI_ISL_2514673, EPI_ISL_2514674, EPI_ISL_2514675, EPI_ISL_2514676, EPI_ISL_2514677, EPI_ISL_2514679, EPI_ISL_2514681, EPI_ISL_2514682, EPI_ISL_2514686, EPI_ISL_2514689, EPI_ISL_2514690, EPI_ISL_2514691, EPI_ISL_2514695, EPI_ISL_2514696, EPI_ISL_2514700, EPI_ISL_2514701, EPI_ISL_2514704, EPI_ISL_2514705, EPI_ISL_2514706, EPI_ISL_2514707,
EPI_ISL_2514708, EPI_ISL_2514709, EPI_ISL_2514713, EPI_ISL_2514714, EPI_ISL_2514720, EPI_ISL_2514721, EPI_ISL_2514722, EPI_ISL_2514723, EPI_ISL_2514724, EPI_ISL_2514727, EPI_ISL_2514928, EPI_ISL_2514929, EPI_ISL_2514935, EPI_ISL_2514937, EPI_ISL_2514938, EPI_ISL_2514939, EPI_ISL_2514941, EPI_ISL_2514942, EPI_ISL_2514945, EPI_ISL_2514946, EPI_ISL_2514947, EPI_ISL_2514950,
EPI_ISL_2514951, EPI_ISL_2514954, EPI_ISL_2514956, EPI_ISL_2514958, EPI_ISL_2514959, EPI_ISL_2514960, EPI_ISL_2514961, EPI_ISL_2514962, EPI_ISL_2514965, EPI_ISL_2514966, EPI_ISL_2514967, EPI_ISL_2514969, EPI_ISL_2514971, EPI_ISL_2514972, EPI_ISL_2514976, EPI_ISL_2514977, EPI_ISL_2514979, EPI_ISL_2514984, EPI_ISL_2514986, EPI_ISL_2514992, EPI_ISL_2514993, EPI_ISL_2514995,
EPI_ISL_2514996, EPI_ISL_2514997, EPI_ISL_2515000, EPI_ISL_2515002, EPI_ISL_2515005, EPI_ISL_2515012, EPI_ISL_2515013, EPI_ISL_2515014, EPI_ISL_2515015, EPI_ISL_2515016, EPI_ISL_2515019, EPI_ISL_2515024, EPI_ISL_2515025, EPI_ISL_2515026, EPI_ISL_2515027, EPI_ISL_2515029, EPI_ISL_2515030, EPI_ISL_2515031, EPI_ISL_2515033, EPI_ISL_2515034,
EPI_ISL_2515035, EPI_ISL_2515037, EPI_ISL_2515038, EPI_ISL_2515039, EPI_ISL_2515040, EPI_ISL_2515045, EPI_ISL_2515047, EPI_ISL_2515048, EPI_ISL_2515051, EPI_ISL_2515054, EPI_ISL_2515059, EPI_ISL_2515060, EPI_ISL_2515062, EPI_ISL_2515063, EPI_ISL_2515066, EPI_ISL_2515071, EPI_ISL_2515074, EPI_ISL_2515075, EPI_ISL_2515076, EPI_ISL_2515081, EPI_ISL_2515084, EPI_ISL_2515085,
EPI_ISL_2515088, EPI_ISL_2515091, EPI_ISL_2515094, EPI_ISL_2515100, EPI_ISL_2515103, EPI_ISL_2515104, EPI_ISL_2515105, EPI_ISL_2515106, EPI_ISL_2515108, EPI_ISL_2515109, EPI_ISL_2515111, EPI_ISL_2515112, EPI_ISL_2515117, EPI_ISL_2515118, EPI_ISL_2515119, EPI_ISL_2515120, EPI_ISL_2515121, EPI_ISL_2515122, EPI_ISL_2515123, EPI_ISL_2515124, EPI_ISL_2515126, EPI_ISL_2515127,
EPI_ISL_2515128, EPI_ISL_2515129, EPI_ISL_2515130, EPI_ISL_2515131, EPI_ISL_2515132, EPI_ISL_2515133, EPI_ISL_2515134, EPI_ISL_2515135, EPI_ISL_2515136, EPI_ISL_2515137, EPI_ISL_2515138, EPI_ISL_2515139, EPI_ISL_2515140, EPI_ISL_2515141, EPI_ISL_2515142, EPI_ISL_2515143, EPI_ISL_2515144, EPI_ISL_2515146, EPI_ISL_2515147, EPI_ISL_2515148, EPI_ISL_2515149, EPI_ISL_2515150,
EPI_ISL_2515151, EPI_ISL_2515153, EPI_ISL_2515154, EPI_ISL_2515156, EPI_ISL_2515157, EPI_ISL_2515158, EPI_ISL_2515159, EPI_ISL_2515160, EPI_ISL_2515161, EPI_ISL_2515162, EPI_ISL_2515163, EPI_ISL_2515164, EPI_ISL_2515165, EPI_ISL_2515166, EPI_ISL_2515167, EPI_ISL_2515168, EPI_ISL_2515170, EPI_ISL_2515171, EPI_ISL_2515172, EPI_ISL_2515173, EPI_ISL_2515174, EPI_ISL_2515177,
EPI_ISL_2515178, EPI_ISL_2515179, EPI_ISL_2515180, EPI_ISL_2515181, EPI_ISL_2515183, EPI_ISL_2515184, EPI_ISL_2515185, EPI_ISL_2515187, EPI_ISL_2515188, EPI_ISL_2515189, EPI_ISL_2515190, EPI_ISL_2515191, EPI_ISL_2515192, EPI_ISL_2515193, EPI_ISL_2515195, EPI_ISL_2515196, EPI_ISL_2515197, EPI_ISL_2515198, EPI_ISL_2515199, EPI_ISL_2515200, EPI_ISL_2515201, EPI_ISL_2515202,
EPI_ISL_2515204, EPI_ISL_2515205, EPI_ISL_2515206, EPI_ISL_2515207, EPI_ISL_2515208, EPI_ISL_2515209, EPI_ISL_2515210, EPI_ISL_2515212, EPI_ISL_2515213, EPI_ISL_2515214, EPI_ISL_2515215, EPI_ISL_2515216, EPI_ISL_2515217, EPI_ISL_2515218, EPI_ISL_2515219, EPI_ISL_2515220, EPI_ISL_2515221, EPI_ISL_2515222, EPI_ISL_2515223, EPI_ISL_2515224, EPI_ISL_2515225,
EPI_ISL_2515226, EPI_ISL_2515227, EPI_ISL_2515228, EPI_ISL_2515229, EPI_ISL_2515230, EPI_ISL_2515232, EPI_ISL_2515233, EPI_ISL_2515234, EPI_ISL_2515235, EPI_ISL_2515236, EPI_ISL_2515237, EPI_ISL_2515238, EPI_ISL_2515239, EPI_ISL_2515240, EPI_ISL_2515241, EPI_ISL_2515242, EPI_ISL_2515243, EPI_ISL_2515244, EPI_ISL_2515245, EPI_ISL_2515246, EPI_ISL_2515247, EPI_ISL_2515248,
EPI_ISL_2515250, EPI_ISL_2515251, EPI_ISL_2515253, EPI_ISL_2515254, EPI_ISL_2515255, EPI_ISL_2515256, EPI_ISL_2515257, EPI_ISL_2515258, EPI_ISL_2515259, EPI_ISL_2515260, EPI_ISL_2515261, EPI_ISL_2515262, EPI_ISL_2515266, EPI_ISL_2515267, EPI_ISL_2515268, EPI_ISL_2515269, EPI_ISL_2515271, EPI_ISL_2515272, EPI_ISL_2515273, EPI_ISL_2515274, EPI_ISL_2515276, EPI_ISL_2515277,
EPI_ISL_2515278, EPI_ISL_2515279, EPI_ISL_2515280, EPI_ISL_2515281, EPI_ISL_2515282, EPI_ISL_2515283, EPI_ISL_2515284, EPI_ISL_2515285, EPI_ISL_2515286, EPI_ISL_2515287, EPI_ISL_2515288, EPI_ISL_2515289, EPI_ISL_2515290, EPI_ISL_2515291, EPI_ISL_2515292, EPI_ISL_2515294, EPI_ISL_2515296, EPI_ISL_2515297, EPI_ISL_2515298, EPI_ISL_2515299, EPI_ISL_2515300, EPI_ISL_2515301,
EPI_ISL_2515302, EPI_ISL_2515303, EPI_ISL_2515304, EPI_ISL_2515305, EPI_ISL_2515306, EPI_ISL_2515307, EPI_ISL_2515308, EPI_ISL_2515310, EPI_ISL_2515311, EPI_ISL_2515312, EPI_ISL_2515313, EPI_ISL_2515315, EPI_ISL_2515316, EPI_ISL_2515318, EPI_ISL_2515319, EPI_ISL_2515320, EPI_ISL_2515321, EPI_ISL_2515322, EPI_ISL_2515323, EPI_ISL_2515325, EPI_ISL_2515326,
EPI_ISL_2515327, EPI_ISL_2515328, EPI_ISL_2515329, EPI_ISL_2515330, EPI_ISL_2515333, EPI_ISL_2515335, EPI_ISL_2515336, EPI_ISL_2515337, EPI_ISL_2515339, EPI_ISL_2515340, EPI_ISL_2515341, EPI_ISL_2515342, EPI_ISL_2515343, EPI_ISL_2515344, EPI_ISL_2515345, EPI_ISL_2515346, EPI_ISL_2515347, EPI_ISL_2515348, EPI_ISL_2515349, EPI_ISL_2515350, EPI_ISL_2515351, EPI_ISL_2515352,
EPI_ISL_2515353, EPI_ISL_2515354, EPI_ISL_2515355, EPI_ISL_2515356, EPI_ISL_2515357, EPI_ISL_2515358, EPI_ISL_2515360, EPI_ISL_2515361, EPI_ISL_2515362, EPI_ISL_2515363, EPI_ISL_2515364, EPI_ISL_2515365, EPI_ISL_2515366, EPI_ISL_2515367, EPI_ISL_2515368, EPI_ISL_2515369, EPI_ISL_2515370, EPI_ISL_2515371, EPI_ISL_2515372, EPI_ISL_2515373, EPI_ISL_2515374, EPI_ISL_2515375,
EPI_ISL_2515376, EPI_ISL_2515377, EPI_ISL_2515378, EPI_ISL_2515379, EPI_ISL_2515380, EPI_ISL_2515381, EPI_ISL_2515382, EPI_ISL_2515384, EPI_ISL_2515385, EPI_ISL_2515386, EPI_ISL_2515387, EPI_ISL_2515388, EPI_ISL_2515389, EPI_ISL_2515390, EPI_ISL_2515391, EPI_ISL_2515392, EPI_ISL_2515393, EPI_ISL_2515394, EPI_ISL_2515395, EPI_ISL_2515396, EPI_ISL_2515397, EPI_ISL_2515645,
EPI_ISL_2515661, EPI_ISL_2515699, EPI_ISL_2515709, EPI_ISL_2515713, EPI_ISL_2515734, EPI_ISL_2515766, EPI_ISL_2515780, EPI_ISL_2515796, EPI_ISL_2515805, EPI_ISL_2515829, EPI_ISL_2515912, EPI_ISL_2515952, EPI_ISL_2516003, EPI_ISL_2516020, EPI_ISL_2516023, EPI_ISL_2516025, EPI_ISL_2516031, EPI_ISL_2516033, EPI_ISL_2516039, EPI_ISL_2516042, EPI_ISL_2516051, EPI_ISL_2516055,
EPI_ISL_2516056, EPI_ISL_2516058, EPI_ISL_2516061, EPI_ISL_2516073, EPI_ISL_2516076, EPI_ISL_2516081, EPI_ISL_2516085, EPI_ISL_2516093, EPI_ISL_2516097, EPI_ISL_2516100, EPI_ISL_2516106, EPI_ISL_2516107, EPI_ISL_2516113, EPI_ISL_2516123, EPI_ISL_2516128, EPI_ISL_2516133, EPI_ISL_2516134, EPI_ISL_2516135, EPI_ISL_2516148, EPI_ISL_2516164, EPI_ISL_2516165, EPI_ISL_2516167,
EPI_ISL_2516173, EPI_ISL_2516188, EPI_ISL_2516190, EPI_ISL_2516192, EPI_ISL_2516198, EPI_ISL_2516200, EPI_ISL_2516223, EPI_ISL_2516227, EPI_ISL_2516228, EPI_ISL_2516231, EPI_ISL_2516233, EPI_ISL_2516240, EPI_ISL_2516245, EPI_ISL_2516251, EPI_ISL_2516261, EPI_ISL_2516265, EPI_ISL_2516267, EPI_ISL_2516273, EPI_ISL_2516279, EPI_ISL_2516295,
EPI_ISL_2516298, EPI_ISL_2516300, EPI_ISL_2516305, EPI_ISL_2516307, EPI_ISL_2516308, EPI_ISL_2516312, EPI_ISL_2516313, EPI_ISL_2516314, EPI_ISL_2516315, EPI_ISL_2537942, EPI_ISL_2537944, EPI_ISL_2537959, EPI_ISL_2537962, EPI_ISL_2537982, EPI_ISL_2538008, EPI_ISL_2538013, EPI_ISL_2538028, EPI_ISL_2538057, EPI_ISL_2538060, EPI_ISL_2538061, EPI_ISL_2538070, EPI_ISL_2538136,
EPI_ISL_2538138, EPI_ISL_2538164, EPI_ISL_2538395, EPI_ISL_2538801, EPI_ISL_2539174, EPI_ISL_2539177, EPI_ISL_2539181, EPI_ISL_2539188, EPI_ISL_2539190, EPI_ISL_2539191, EPI_ISL_2539197, EPI_ISL_2539198, EPI_ISL_2539199, EPI_ISL_2539201, EPI_ISL_2539202, EPI_ISL_2539204, EPI_ISL_2539205, EPI_ISL_2539206, EPI_ISL_2539208, EPI_ISL_2539210, EPI_ISL_2539216,
EPI_ISL_2539218, EPI_ISL_2539219, EPI_ISL_2539220, EPI_ISL_2539224, EPI_ISL_2539227, EPI_ISL_2539228, EPI_ISL_2539230, EPI_ISL_2539236, EPI_ISL_2539238, EPI_ISL_2539239, EPI_ISL_2539242, EPI_ISL_2539248, EPI_ISL_2539254, EPI_ISL_2539255, EPI_ISL_2539256, EPI_ISL_2539258, EPI_ISL_2539259, EPI_ISL_2539260, EPI_ISL_2539267, EPI_ISL_2539269, EPI_ISL_2539270, EPI_ISL_2539274,
EPI_ISL_2539275, EPI_ISL_2539283, EPI_ISL_2539284, EPI_ISL_2539286, EPI_ISL_2539289, EPI_ISL_2539291, EPI_ISL_2539292, EPI_ISL_2539294, EPI_ISL_2539296, EPI_ISL_2539301, EPI_ISL_2539302, EPI_ISL_2539304, EPI_ISL_2539305, EPI_ISL_2539306, EPI_ISL_2539308, EPI_ISL_2539311, EPI_ISL_2539314, EPI_ISL_2539315, EPI_ISL_2539317, EPI_ISL_2539324, EPI_ISL_2539326,
EPI_ISL_2539328, EPI_ISL_2539333, EPI_ISL_2539337, EPI_ISL_2539346, EPI_ISL_2539349, EPI_ISL_2539351, EPI_ISL_2539353, EPI_ISL_2539355, EPI_ISL_2539362, EPI_ISL_2539365, EPI_ISL_2539370, EPI_ISL_2539371, EPI_ISL_2539372, EPI_ISL_2539376, EPI_ISL_2539382, EPI_ISL_2539386, EPI_ISL_2539387, EPI_ISL_2539389, EPI_ISL_2539391, EPI_ISL_2539393, EPI_ISL_2539398, EPI_ISL_2539401,
EPI_ISL_2539402, EPI_ISL_2539407, EPI_ISL_2539409, EPI_ISL_2539410, EPI_ISL_2539412, EPI_ISL_2539413, EPI_ISL_2539414, EPI_ISL_2539416, EPI_ISL_2539419, EPI_ISL_2539423, EPI_ISL_2539425, EPI_ISL_2539426, EPI_ISL_2539432, EPI_ISL_2539433, EPI_ISL_2539434, EPI_ISL_2539436, EPI_ISL_2539442, EPI_ISL_2539445, EPI_ISL_2539450, EPI_ISL_2539452, EPI_ISL_2539454, EPI_ISL_2539456,
EPI_ISL_2539457, EPI_ISL_2539462, EPI_ISL_2539464, EPI_ISL_2539465, EPI_ISL_2539466, EPI_ISL_2539467, EPI_ISL_2539468, EPI_ISL_2539469, EPI_ISL_2539470, EPI_ISL_2539471, EPI_ISL_2539472, EPI_ISL_2539474, EPI_ISL_2539480, EPI_ISL_2539482, EPI_ISL_2539485, EPI_ISL_2539487, EPI_ISL_2539488, EPI_ISL_2539489, EPI_ISL_2539494, EPI_ISL_2539498, EPI_ISL_2539503, EPI_ISL_2539504,
EPI_ISL_2539507, EPI_ISL_2539511, EPI_ISL_2539515, EPI_ISL_2539517, EPI_ISL_2539519, EPI_ISL_2539520, EPI_ISL_2539521, EPI_ISL_2539522, EPI_ISL_2539530, EPI_ISL_2539532, EPI_ISL_2539534, EPI_ISL_2539535, EPI_ISL_2539544, EPI_ISL_2539552, EPI_ISL_2539553, EPI_ISL_2539557, EPI_ISL_2539558, EPI_ISL_2539559, EPI_ISL_2539560, EPI_ISL_2539562, EPI_ISL_2539565, EPI_ISL_2539566,
EPI_ISL_2539567, EPI_ISL_2539570, EPI_ISL_2539572, EPI_ISL_2539577, EPI_ISL_2539582, EPI_ISL_2539590, EPI_ISL_2539594, EPI_ISL_2539595, EPI_ISL_2539596, EPI_ISL_2539597, EPI_ISL_2539598, EPI_ISL_2539601, EPI_ISL_2539603, EPI_ISL_2539605, EPI_ISL_2539606, EPI_ISL_2539609, EPI_ISL_2539610, EPI_ISL_2539612, EPI_ISL_2539613, EPI_ISL_2539615, EPI_ISL_2539616, EPI_ISL_2539622,
EPI_ISL_2539623, EPI_ISL_2539625, EPI_ISL_2539626, EPI_ISL_2539628, EPI_ISL_2539632, EPI_ISL_2539635, EPI_ISL_2539637, EPI_ISL_2539653, EPI_ISL_2539654, EPI_ISL_2539655, EPI_ISL_2539657, EPI_ISL_2539660, EPI_ISL_2539662, EPI_ISL_2539666, EPI_ISL_2539667, EPI_ISL_2539669, EPI_ISL_2539670, EPI_ISL_2539671, EPI_ISL_2539675, EPI_ISL_2539679, EPI_ISL_2539682, EPI_ISL_2539683,
EPI_ISL_2539684, EPI_ISL_2539687, EPI_ISL_2539688, EPI_ISL_2539689, EPI_ISL_2539694, EPI_ISL_2539696, EPI_ISL_2539698, EPI_ISL_2539699, EPI_ISL_2539706, EPI_ISL_2539707, EPI_ISL_2539708, EPI_ISL_2539710, EPI_ISL_2539713, EPI_ISL_2539717, EPI_ISL_2539719, EPI_ISL_2539720, EPI_ISL_2539722, EPI_ISL_2539724, EPI_ISL_2539730, EPI_ISL_2539735, EPI_ISL_2541815, EPI_ISL_2542785,
EPI_ISL_2542786, EPI_ISL_2542787, EPI_ISL_2542788, EPI_ISL_2543048, EPI_ISL_2543148, EPI_ISL_2543407, EPI_ISL_2553438, EPI_ISL_2553505, EPI_ISL_2553510, EPI_ISL_2553638, EPI_ISL_2555002, EPI_ISL_2569441, EPI_ISL_2640510

| | | | |
|---|---|---|---|
| see above | Lighthouse Lab in Alderley Park | Wellcome Sanger Institute for the COVID-19 Genomics UK (COG-UK) Consortium | Cordelia Langford; David K. Jackson; Dominic Kwiatkowski; Ewan Harrison; Ian Johnston; Jacquelyn Wynn; Jeffrey Barrett; John Sillitoe on behalf of the Wellcome Sanger Institute COVID-19 Surveillance Team; Mairead Hyland; Roberto Amato; Sonia Goncalves; The Lighthouse Lab in Alderley Park and Alex Alderton |

EPI_ISL_2407596, EPI_ISL_2407637, EPI_ISL_2435001, EPI_ISL_2435004, EPI_ISL_2435006, EPI_ISL_2435028, EPI_ISL_2435029, EPI_ISL_2435030, EPI_ISL_2435032, EPI_ISL_2435034, EPI_ISL_2435036, EPI_ISL_2435041, EPI_ISL_2435042, EPI_ISL_2435044, EPI_ISL_2435045, EPI_ISL_2435052, EPI_ISL_2435055, EPI_ISL_2435061, EPI_ISL_2435062, EPI_ISL_2435074, EPI_ISL_2435075, EPI_ISL_2435077,
EPI_ISL_2435086, EPI_ISL_2435088, EPI_ISL_2435091, EPI_ISL_2435096, EPI_ISL_2435099, EPI_ISL_2435107, EPI_ISL_2435108, EPI_ISL_2435110, EPI_ISL_2435114, EPI_ISL_2435115, EPI_ISL_2435117, EPI_ISL_2435124, EPI_ISL_2435129, EPI_ISL_2435132, EPI_ISL_2435136, EPI_ISL_2435138, EPI_ISL_2435139, EPI_ISL_2435141, EPI_ISL_2435145, EPI_ISL_2435152, EPI_ISL_2435163,
EPI_ISL_2435164, EPI_ISL_2435168, EPI_ISL_2435173, EPI_ISL_2435175, EPI_ISL_2435178, EPI_ISL_2435179, EPI_ISL_2435198, EPI_ISL_2435214, EPI_ISL_2435216, EPI_ISL_2435220, EPI_ISL_2435225, EPI_ISL_2435226, EPI_ISL_2435227, EPI_ISL_2435247, EPI_ISL_2435252, EPI_ISL_2435254, EPI_ISL_2435255, EPI_ISL_2435258, EPI_ISL_2435263, EPI_ISL_2435275, EPI_ISL_2435278, EPI_ISL_2435281,
EPI_ISL_2435282, EPI_ISL_2435292, EPI_ISL_2435295, EPI_ISL_2435296, EPI_ISL_2435299, EPI_ISL_2435300, EPI_ISL_2435305, EPI_ISL_2435306, EPI_ISL_2435311, EPI_ISL_2435313, EPI_ISL_2435315, EPI_ISL_2435318, EPI_ISL_2435319, EPI_ISL_2435322, EPI_ISL_2435325, EPI_ISL_2435329, EPI_ISL_2435330, EPI_ISL_2435333, EPI_ISL_2435335, EPI_ISL_2435337, EPI_ISL_2435354, EPI_ISL_2435379,
EPI_ISL_2435385, EPI_ISL_2435392, EPI_ISL_2435396, EPI_ISL_2435439, EPI_ISL_2435442, EPI_ISL_2435447, EPI_ISL_2435460, EPI_ISL_2435470, EPI_ISL_2435471, EPI_ISL_2435477, EPI_ISL_2435493, EPI_ISL_2435532, EPI_ISL_2435556, EPI_ISL_2435587, EPI_ISL_2435589, EPI_ISL_2435598, EPI_ISL_2435601, EPI_ISL_2435605, EPI_ISL_2435628, EPI_ISL_2435667, EPI_ISL_2435677, EPI_ISL_2435678,
EPI_ISL_2435693, EPI_ISL_2435701, EPI_ISL_2435703, EPI_ISL_2435714, EPI_ISL_2435733, EPI_ISL_2435736, EPI_ISL_2435740, EPI_ISL_2435747, EPI_ISL_2435750, EPI_ISL_2435753, EPI_ISL_2435767, EPI_ISL_2435774, EPI_ISL_2435791, EPI_ISL_2435801, EPI_ISL_2435805, EPI_ISL_2435812, EPI_ISL_2435819, EPI_ISL_2435836, EPI_ISL_2435837, EPI_ISL_2435842, EPI_ISL_2435862, EPI_ISL_2435865,
EPI_ISL_2436222, EPI_ISL_2436227, EPI_ISL_2436229, EPI_ISL_2436231, EPI_ISL_2436233, EPI_ISL_2436234, EPI_ISL_2436241, EPI_ISL_2436242, EPI_ISL_2436253, EPI_ISL_2436257, EPI_ISL_2436258, EPI_ISL_2436262, EPI_ISL_2436269, EPI_ISL_2436271, EPI_ISL_2436272, EPI_ISL_2436273, EPI_ISL_2436277, EPI_ISL_2436280, EPI_ISL_2436287, EPI_ISL_2436296, EPI_ISL_2436303, EPI_ISL_2436304,
EPI_ISL_2436307, EPI_ISL_2436308, EPI_ISL_2436309, EPI_ISL_2436311, EPI_ISL_2436316, EPI_ISL_2436318, EPI_ISL_2436321, EPI_ISL_2436322, EPI_ISL_2436325, EPI_ISL_2436334, EPI_ISL_2436336, EPI_ISL_2436340, EPI_ISL_2436343, EPI_ISL_2436344, EPI_ISL_2436346, EPI_ISL_2436348, EPI_ISL_2436351, EPI_ISL_2436352, EPI_ISL_2436354, EPI_ISL_2436355, EPI_ISL_2436356, EPI_ISL_2436361,
EPI_ISL_2436368, EPI_ISL_2436369, EPI_ISL_2436370, EPI_ISL_2436376, EPI_ISL_2436377, EPI_ISL_2436388, EPI_ISL_2436389, EPI_ISL_2436391, EPI_ISL_2436400, EPI_ISL_2436406, EPI_ISL_2436407, EPI_ISL_2436408, EPI_ISL_2436411, EPI_ISL_2436413, EPI_ISL_2436417, EPI_ISL_2436423, EPI_ISL_2436428, EPI_ISL_2436434, EPI_ISL_2436436, EPI_ISL_2436445, EPI_ISL_2436447, EPI_ISL_2436449,
EPI_ISL_2436454, EPI_ISL_2436462, EPI_ISL_2436464, EPI_ISL_2436465, EPI_ISL_2436470, EPI_ISL_2436471, EPI_ISL_2436483, EPI_ISL_2436511, EPI_ISL_2436514, EPI_ISL_2436521, EPI_ISL_2436528, EPI_ISL_2436529, EPI_ISL_2436534, EPI_ISL_2436552, EPI_ISL_2436560, EPI_ISL_2436568, EPI_ISL_2436574, EPI_ISL_2436577, EPI_ISL_2436582, EPI_ISL_2436583, EPI_ISL_2436586, EPI_ISL_2436588,
EPI_ISL_2436597, EPI_ISL_2436598, EPI_ISL_2436607, EPI_ISL_2436623, EPI_ISL_2436630, EPI_ISL_2436634, EPI_ISL_2436636, EPI_ISL_2436641, EPI_ISL_2436650, EPI_ISL_2436656, EPI_ISL_2436662, EPI_ISL_2436664, EPI_ISL_2436668, EPI_ISL_2436682, EPI_ISL_2436686, EPI_ISL_2436689, EPI_ISL_2436697, EPI_ISL_2436708, EPI_ISL_2436709, EPI_ISL_2436712, EPI_ISL_2436721, EPI_ISL_2436723,
EPI_ISL_2436727, EPI_ISL_2436729, EPI_ISL_2436733, EPI_ISL_2436737, EPI_ISL_2436743, EPI_ISL_2436748, EPI_ISL_2436753, EPI_ISL_2436759, EPI_ISL_2436760, EPI_ISL_2436762, EPI_ISL_2436767, EPI_ISL_2436769, EPI_ISL_2436771, EPI_ISL_2436773, EPI_ISL_2436781, EPI_ISL_2436787, EPI_ISL_2436789, EPI_ISL_2436790, EPI_ISL_2436791, EPI_ISL_2436796, EPI_ISL_2436797, EPI_ISL_2436798,
EPI_ISL_2436804, EPI_ISL_2436805, EPI_ISL_2436806, EPI_ISL_2436808, EPI_ISL_2436813, EPI_ISL_2436816, EPI_ISL_2436819, EPI_ISL_2436827, EPI_ISL_2436828, EPI_ISL_2436830, EPI_ISL_2436835, EPI_ISL_2436843, EPI_ISL_2436848, EPI_ISL_2436850, EPI_ISL_2436856, EPI_ISL_2436859, EPI_ISL_2436860, EPI_ISL_2436864, EPI_ISL_2436873, EPI_ISL_2436877, EPI_ISL_2436885, EPI_ISL_2436886,
EPI_ISL_2436887, EPI_ISL_2436889, EPI_ISL_2436897, EPI_ISL_2436899, EPI_ISL_2436901, EPI_ISL_2436902, EPI_ISL_2436904, EPI_ISL_2436922, EPI_ISL_2436927, EPI_ISL_2437620, EPI_ISL_2437634, EPI_ISL_2437635, EPI_ISL_2437638, EPI_ISL_2437640, EPI_ISL_2437645, EPI_ISL_2437648, EPI_ISL_2437650, EPI_ISL_2437652, EPI_ISL_2437656, EPI_ISL_2437657, EPI_ISL_2437659, EPI_ISL_2437660,

EPI_ISL_2437661, EPI_ISL_2437664, EPI_ISL_2437667, EPI_ISL_2437669, EPI_ISL_2437670, EPI_ISL_2437687, EPI_ISL_2437692, EPI_ISL_2437694, EPI_ISL_2437698, EPI_ISL_2437700, EPI_ISL_2437702, EPI_ISL_2437705, EPI_ISL_2437706, EPI_ISL_2437709, EPI_ISL_2437715, EPI_ISL_2437719, EPI_ISL_2437723, EPI_ISL_2437737, EPI_ISL_2437741, EPI_ISL_2437746, EPI_ISL_2437749, EPI_ISL_2437750,
EPI_ISL_2437761, EPI_ISL_2437763, EPI_ISL_2437775, EPI_ISL_2437778, EPI_ISL_2437780, EPI_ISL_2437788, EPI_ISL_2437789, EPI_ISL_2437791, EPI_ISL_2437792, EPI_ISL_2437798, EPI_ISL_2437802, EPI_ISL_2437803, EPI_ISL_2437806, EPI_ISL_2437812, EPI_ISL_2437815, EPI_ISL_2437816, EPI_ISL_2437818, EPI_ISL_2437819, EPI_ISL_2437830, EPI_ISL_2437848, EPI_ISL_2437852, EPI_ISL_2437853,
EPI_ISL_2437860, EPI_ISL_2437861, EPI_ISL_2437862, EPI_ISL_2437865, EPI_ISL_2437872, EPI_ISL_2437880, EPI_ISL_2437883, EPI_ISL_2437885, EPI_ISL_2437886, EPI_ISL_2437890, EPI_ISL_2437892, EPI_ISL_2437893, EPI_ISL_2437897, EPI_ISL_2437907, EPI_ISL_2437910, EPI_ISL_2437914, EPI_ISL_2437915, EPI_ISL_2437917, EPI_ISL_2437920, EPI_ISL_2437924, EPI_ISL_2437926, EPI_ISL_2437928,
EPI_ISL_2437934, EPI_ISL_2437937, EPI_ISL_2437945, EPI_ISL_2437946, EPI_ISL_2437955, EPI_ISL_2437956, EPI_ISL_2437957, EPI_ISL_2437963, EPI_ISL_2437964, EPI_ISL_2437966, EPI_ISL_2437968, EPI_ISL_2437970, EPI_ISL_2437972, EPI_ISL_2437974, EPI_ISL_2437981, EPI_ISL_2437982, EPI_ISL_2437985, EPI_ISL_2437986, EPI_ISL_2437990, EPI_ISL_2437992, EPI_ISL_2437993, EPI_ISL_2437994,
EPI_ISL_2438037, EPI_ISL_2438038, EPI_ISL_2438040, EPI_ISL_2438042, EPI_ISL_2438043, EPI_ISL_2438044, EPI_ISL_2438049, EPI_ISL_2438063, EPI_ISL_2438069, EPI_ISL_2438074, EPI_ISL_2438075, EPI_ISL_2438078, EPI_ISL_2438079, EPI_ISL_2438082, EPI_ISL_2438084, EPI_ISL_2438087, EPI_ISL_2438091, EPI_ISL_2438093, EPI_ISL_2438097, EPI_ISL_2438100, EPI_ISL_2438102, EPI_ISL_2438103,
EPI_ISL_2438104, EPI_ISL_2438105, EPI_ISL_2438110, EPI_ISL_2438112, EPI_ISL_2438114, EPI_ISL_2438115, EPI_ISL_2438122, EPI_ISL_2438125, EPI_ISL_2438129, EPI_ISL_2438133, EPI_ISL_2438134, EPI_ISL_2438135, EPI_ISL_2438139, EPI_ISL_2438143, EPI_ISL_2438144, EPI_ISL_2438147, EPI_ISL_2438149, EPI_ISL_2438151, EPI_ISL_2438152, EPI_ISL_2438153, EPI_ISL_2438154, EPI_ISL_2438157,
EPI_ISL_2438158, EPI_ISL_2438160, EPI_ISL_2438161, EPI_ISL_2438163, EPI_ISL_2438168, EPI_ISL_2438169, EPI_ISL_2438170, EPI_ISL_2438172, EPI_ISL_2438174, EPI_ISL_2438179, EPI_ISL_2438181, EPI_ISL_2438184, EPI_ISL_2438185, EPI_ISL_2438187, EPI_ISL_2438190, EPI_ISL_2438192, EPI_ISL_2438195, EPI_ISL_2438200, EPI_ISL_2438201, EPI_ISL_2438202, EPI_ISL_2438204, EPI_ISL_2438209,
EPI_ISL_2438210, EPI_ISL_2438214, EPI_ISL_2438219, EPI_ISL_2438222, EPI_ISL_2438227, EPI_ISL_2438228, EPI_ISL_2438232, EPI_ISL_2438234, EPI_ISL_2438236, EPI_ISL_2438239, EPI_ISL_2438240, EPI_ISL_2438242, EPI_ISL_2438243, EPI_ISL_2438249, EPI_ISL_2438250, EPI_ISL_2438253, EPI_ISL_2438256, EPI_ISL_2438257, EPI_ISL_2438258, EPI_ISL_2438259, EPI_ISL_2438261, EPI_ISL_2438262,
EPI_ISL_2438263, EPI_ISL_2438265, EPI_ISL_2438268, EPI_ISL_2438272, EPI_ISL_2438275, EPI_ISL_2438277, EPI_ISL_2438278, EPI_ISL_2438280, EPI_ISL_2438284, EPI_ISL_2438285, EPI_ISL_2438286, EPI_ISL_2438291, EPI_ISL_2438296, EPI_ISL_2438297, EPI_ISL_2438300, EPI_ISL_2438303, EPI_ISL_2438304, EPI_ISL_2438306, EPI_ISL_2438307, EPI_ISL_2438308, EPI_ISL_2438311, EPI_ISL_2438315,
EPI_ISL_2438317, EPI_ISL_2438318, EPI_ISL_2438319, EPI_ISL_2438320, EPI_ISL_2438321, EPI_ISL_2438322, EPI_ISL_2438325, EPI_ISL_2438327, EPI_ISL_2438328, EPI_ISL_2438330, EPI_ISL_2438331, EPI_ISL_2438332, EPI_ISL_2438333, EPI_ISL_2438334, EPI_ISL_2438336, EPI_ISL_2438337, EPI_ISL_2438339, EPI_ISL_2438347, EPI_ISL_2438348, EPI_ISL_2438349, EPI_ISL_2438350, EPI_ISL_2438351,
EPI_ISL_2438353, EPI_ISL_2438354, EPI_ISL_2438355, EPI_ISL_2438356, EPI_ISL_2438357, EPI_ISL_2438359, EPI_ISL_2438366, EPI_ISL_2438369, EPI_ISL_2438373, EPI_ISL_2438375, EPI_ISL_2446106, EPI_ISL_2446110, EPI_ISL_2446112, EPI_ISL_2446115, EPI_ISL_2446116, EPI_ISL_2446123, EPI_ISL_2446125, EPI_ISL_2446131, EPI_ISL_2446134, EPI_ISL_2446141, EPI_ISL_2446147, EPI_ISL_2446157,
EPI_ISL_2446168, EPI_ISL_2446179, EPI_ISL_2446181, EPI_ISL_2446194, EPI_ISL_2446196, EPI_ISL_2446200, EPI_ISL_2446211, EPI_ISL_2446228, EPI_ISL_2446229, EPI_ISL_2446236, EPI_ISL_2446240, EPI_ISL_2446248, EPI_ISL_2446254, EPI_ISL_2446257, EPI_ISL_2446278, EPI_ISL_2446289, EPI_ISL_2446298, EPI_ISL_2446307, EPI_ISL_2446314, EPI_ISL_2446333, EPI_ISL_2446341, EPI_ISL_2446348,
EPI_ISL_2446361, EPI_ISL_2446363, EPI_ISL_2446365, EPI_ISL_2446367, EPI_ISL_2446368, EPI_ISL_2446376, EPI_ISL_2446380, EPI_ISL_2446385, EPI_ISL_2446389, EPI_ISL_2446390, EPI_ISL_2446394, EPI_ISL_2446395, EPI_ISL_2446404, EPI_ISL_2446407, EPI_ISL_2446843, EPI_ISL_2446844, EPI_ISL_2446854, EPI_ISL_2446858, EPI_ISL_2446900, EPI_ISL_2446917, EPI_ISL_2446952, EPI_ISL_2446978,
EPI_ISL_2446995, EPI_ISL_2447026, EPI_ISL_2447028, EPI_ISL_2447032, EPI_ISL_2447038, EPI_ISL_2447040, EPI_ISL_2447042, EPI_ISL_2447048, EPI_ISL_2447053, EPI_ISL_2447058, EPI_ISL_2447064, EPI_ISL_2447065, EPI_ISL_2447070, EPI_ISL_2447077, EPI_ISL_2447079, EPI_ISL_2447082, EPI_ISL_2447085, EPI_ISL_2447089, EPI_ISL_2447097, EPI_ISL_2447098, EPI_ISL_2447099, EPI_ISL_2447114,
EPI_ISL_2447116, EPI_ISL_2447120, EPI_ISL_2447123, EPI_ISL_2447125, EPI_ISL_2447130, EPI_ISL_2447132, EPI_ISL_2447151, EPI_ISL_2447162, EPI_ISL_2447165, EPI_ISL_2447167, EPI_ISL_2447173, EPI_ISL_2447176, EPI_ISL_2447177, EPI_ISL_2447178, EPI_ISL_2447184, EPI_ISL_2447197, EPI_ISL_2447200, EPI_ISL_2447208, EPI_ISL_2447212, EPI_ISL_2447224, EPI_ISL_2456752, EPI_ISL_2457131,
EPI_ISL_2457133, EPI_ISL_2457134, EPI_ISL_2457141, EPI_ISL_2457145, EPI_ISL_2457149, EPI_ISL_2457150, EPI_ISL_2457151, EPI_ISL_2457152, EPI_ISL_2457153, EPI_ISL_2457154, EPI_ISL_2457158, EPI_ISL_2457160, EPI_ISL_2457161, EPI_ISL_2457165, EPI_ISL_2457166, EPI_ISL_2457167, EPI_ISL_2457168, EPI_ISL_2457169, EPI_ISL_2457170, EPI_ISL_2457171, EPI_ISL_2457172, EPI_ISL_2457175,
EPI_ISL_2457176, EPI_ISL_2457177, EPI_ISL_2457178, EPI_ISL_2457184, EPI_ISL_2457186, EPI_ISL_2457187, EPI_ISL_2457190, EPI_ISL_2457191, EPI_ISL_2457192, EPI_ISL_2457194, EPI_ISL_2457195, EPI_ISL_2457199, EPI_ISL_2457202, EPI_ISL_2457203, EPI_ISL_2457204, EPI_ISL_2457205, EPI_ISL_2457207, EPI_ISL_2457210, EPI_ISL_2457216, EPI_ISL_2457219, EPI_ISL_2457221, EPI_ISL_2457222,
EPI_ISL_2457224, EPI_ISL_2457227, EPI_ISL_2457228, EPI_ISL_2457229, EPI_ISL_2457231, EPI_ISL_2457235, EPI_ISL_2457236, EPI_ISL_2457237, EPI_ISL_2457240, EPI_ISL_2457241, EPI_ISL_2457246, EPI_ISL_2457248, EPI_ISL_2457250, EPI_ISL_2457251, EPI_ISL_2457257, EPI_ISL_2457259, EPI_ISL_2457262, EPI_ISL_2457264, EPI_ISL_2457266, EPI_ISL_2457269, EPI_ISL_2457271, EPI_ISL_2457272,
EPI_ISL_2457273, EPI_ISL_2457278, EPI_ISL_2457280, EPI_ISL_2457281, EPI_ISL_2457284, EPI_ISL_2457286, EPI_ISL_2457288, EPI_ISL_2457291, EPI_ISL_2457292, EPI_ISL_2457295, EPI_ISL_2457296, EPI_ISL_2457297, EPI_ISL_2457298, EPI_ISL_2457299, EPI_ISL_2457303, EPI_ISL_2457305, EPI_ISL_2457307, EPI_ISL_2457308, EPI_ISL_2457309, EPI_ISL_2457310, EPI_ISL_2457312, EPI_ISL_2457316,
EPI_ISL_2457319, EPI_ISL_2457320, EPI_ISL_2457321, EPI_ISL_2457328, EPI_ISL_2457329, EPI_ISL_2457330, EPI_ISL_2457333, EPI_ISL_2457335, EPI_ISL_2457339, EPI_ISL_2457340, EPI_ISL_2457341, EPI_ISL_2457342, EPI_ISL_2457348, EPI_ISL_2457349, EPI_ISL_2457351, EPI_ISL_2457352, EPI_ISL_2457359, EPI_ISL_2457361, EPI_ISL_2457362, EPI_ISL_2457363, EPI_ISL_2457366, EPI_ISL_2457369,
EPI_ISL_2457370, EPI_ISL_2457371, EPI_ISL_2457372, EPI_ISL_2457374, EPI_ISL_2457376, EPI_ISL_2457381, EPI_ISL_2457385, EPI_ISL_2457389, EPI_ISL_2457390, EPI_ISL_2457392, EPI_ISL_2457393, EPI_ISL_2457396, EPI_ISL_2457398, EPI_ISL_2457399, EPI_ISL_2457402, EPI_ISL_2457403, EPI_ISL_2457408, EPI_ISL_2457411, EPI_ISL_2457412, EPI_ISL_2457416, EPI_ISL_2457417, EPI_ISL_2457419,
EPI_ISL_2457420, EPI_ISL_2457425, EPI_ISL_2457427, EPI_ISL_2457430, EPI_ISL_2457431, EPI_ISL_2457432, EPI_ISL_2457435, EPI_ISL_2457440, EPI_ISL_2457441, EPI_ISL_2457442, EPI_ISL_2457443, EPI_ISL_2457446, EPI_ISL_2457447, EPI_ISL_2457449, EPI_ISL_2457451, EPI_ISL_2457452, EPI_ISL_2457453, EPI_ISL_2457457, EPI_ISL_2457462, EPI_ISL_2457464, EPI_ISL_2457469, EPI_ISL_2457470,
EPI_ISL_2457471, EPI_ISL_2457473, EPI_ISL_2457474, EPI_ISL_2457475, EPI_ISL_2457476, EPI_ISL_2457477, EPI_ISL_2457478, EPI_ISL_2457479, EPI_ISL_2457480, EPI_ISL_2457481, EPI_ISL_2457482, EPI_ISL_2457483, EPI_ISL_2457484, EPI_ISL_2457485, EPI_ISL_2457486, EPI_ISL_2457487, EPI_ISL_2457488, EPI_ISL_2457489, EPI_ISL_2457490, EPI_ISL_2457491, EPI_ISL_2457492, EPI_ISL_2457493,
EPI_ISL_2457494, EPI_ISL_2457495, EPI_ISL_2457496, EPI_ISL_2457497, EPI_ISL_2457498, EPI_ISL_2457499, EPI_ISL_2457500, EPI_ISL_2457501, EPI_ISL_2457507, EPI_ISL_2457508, EPI_ISL_2457509, EPI_ISL_2457510, EPI_ISL_2457511, EPI_ISL_2457512, EPI_ISL_2457513, EPI_ISL_2457514, EPI_ISL_2457515, EPI_ISL_2457516, EPI_ISL_2457517, EPI_ISL_2457518, EPI_ISL_2457519, EPI_ISL_2457520,
EPI_ISL_2457521, EPI_ISL_2457522, EPI_ISL_2457523, EPI_ISL_2457524, EPI_ISL_2457525, EPI_ISL_2457526, EPI_ISL_2457527, EPI_ISL_2457528, EPI_ISL_2457529, EPI_ISL_2457530, EPI_ISL_2457531, EPI_ISL_2457532, EPI_ISL_2457533, EPI_ISL_2457534, EPI_ISL_2457535, EPI_ISL_2457536, EPI_ISL_2457537, EPI_ISL_2457538, EPI_ISL_2457539, EPI_ISL_2457540, EPI_ISL_2457541, EPI_ISL_2457542,
EPI_ISL_2457543, EPI_ISL_2457544, EPI_ISL_2457545, EPI_ISL_2457546, EPI_ISL_2457547, EPI_ISL_2457548, EPI_ISL_2457549, EPI_ISL_2457550, EPI_ISL_2457551, EPI_ISL_2457552, EPI_ISL_2457553, EPI_ISL_2457554, EPI_ISL_2457555, EPI_ISL_2457556, EPI_ISL_2457557, EPI_ISL_2457558, EPI_ISL_2457559, EPI_ISL_2457560, EPI_ISL_2457561, EPI_ISL_2457562, EPI_ISL_2457563, EPI_ISL_2457564,
EPI_ISL_2457565, EPI_ISL_2457566, EPI_ISL_2457567, EPI_ISL_2457568, EPI_ISL_2457569, EPI_ISL_2457570, EPI_ISL_2457571, EPI_ISL_2457572, EPI_ISL_2457573, EPI_ISL_2457574, EPI_ISL_2457575, EPI_ISL_2457576, EPI_ISL_2457577, EPI_ISL_2457578, EPI_ISL_2457579, EPI_ISL_2457580, EPI_ISL_2457581, EPI_ISL_2457582, EPI_ISL_2457583, EPI_ISL_2457584, EPI_ISL_2457585, EPI_ISL_2457586,
EPI_ISL_2457587, EPI_ISL_2457588, EPI_ISL_2457589, EPI_ISL_2457590, EPI_ISL_2457591, EPI_ISL_2457592, EPI_ISL_2457593, EPI_ISL_2457594, EPI_ISL_2457595, EPI_ISL_2457596, EPI_ISL_2457597, EPI_ISL_2457598, EPI_ISL_2457599, EPI_ISL_2457600, EPI_ISL_2457601, EPI_ISL_2457602, EPI_ISL_2457603, EPI_ISL_2457604, EPI_ISL_2457605, EPI_ISL_2457606, EPI_ISL_2457607, EPI_ISL_2457608,
EPI_ISL_2457609, EPI_ISL_2457610, EPI_ISL_2457611, EPI_ISL_2457612, EPI_ISL_2457613, EPI_ISL_2457614, EPI_ISL_2457615, EPI_ISL_2457616, EPI_ISL_2457617, EPI_ISL_2457618, EPI_ISL_2457619, EPI_ISL_2457620, EPI_ISL_2457621, EPI_ISL_2457622, EPI_ISL_2457623, EPI_ISL_2457624, EPI_ISL_2457625, EPI_ISL_2457626, EPI_ISL_2457627, EPI_ISL_2457628, EPI_ISL_2457629, EPI_ISL_2457630,
EPI_ISL_2457631, EPI_ISL_2457632, EPI_ISL_2457633, EPI_ISL_2457634, EPI_ISL_2457635, EPI_ISL_2457636, EPI_ISL_2457637, EPI_ISL_2457638, EPI_ISL_2457639, EPI_ISL_2457640, EPI_ISL_2457641, EPI_ISL_2457642, EPI_ISL_2457643, EPI_ISL_2457644, EPI_ISL_2457645, EPI_ISL_2457646, EPI_ISL_2457647, EPI_ISL_2457648, EPI_ISL_2457649, EPI_ISL_2457650, EPI_ISL_2457651, EPI_ISL_2457652,
EPI_ISL_2457653, EPI_ISL_2457654, EPI_ISL_2457655, EPI_ISL_2457656, EPI_ISL_2457657, EPI_ISL_2457658, EPI_ISL_2457659, EPI_ISL_2457660, EPI_ISL_2457661, EPI_ISL_2457662, EPI_ISL_2457663, EPI_ISL_2457664, EPI_ISL_2457665, EPI_ISL_2457666, EPI_ISL_2457667, EPI_ISL_2457668, EPI_ISL_2457669, EPI_ISL_2457670, EPI_ISL_2457671, EPI_ISL_2457672, EPI_ISL_2457673, EPI_ISL_2457674,
EPI_ISL_2457675, EPI_ISL_2457676, EPI_ISL_2457677, EPI_ISL_2457678, EPI_ISL_2457679, EPI_ISL_2457680, EPI_ISL_2457681, EPI_ISL_2457682, EPI_ISL_2457683, EPI_ISL_2457684, EPI_ISL_2457685, EPI_ISL_2457686, EPI_ISL_2457886, EPI_ISL_2457888, EPI_ISL_2457889, EPI_ISL_2457890, EPI_ISL_2457891, EPI_ISL_2457892, EPI_ISL_2457893, EPI_ISL_2457894, EPI_ISL_2457895, EPI_ISL_2457896,
EPI_ISL_2457897, EPI_ISL_2457898, EPI_ISL_2457899, EPI_ISL_2457900, EPI_ISL_2457901, EPI_ISL_2457903, EPI_ISL_2457904, EPI_ISL_2457905, EPI_ISL_2457906, EPI_ISL_2457907, EPI_ISL_2457908, EPI_ISL_2457909, EPI_ISL_2457910, EPI_ISL_2457911, EPI_ISL_2457912, EPI_ISL_2457913, EPI_ISL_2457914, EPI_ISL_2457915, EPI_ISL_2457916, EPI_ISL_2457917, EPI_ISL_2457918,
EPI_ISL_2457919, EPI_ISL_2457920, EPI_ISL_2457921, EPI_ISL_2457922, EPI_ISL_2457923, EPI_ISL_2457924, EPI_ISL_2457925, EPI_ISL_2457926, EPI_ISL_2457927, EPI_ISL_2457928, EPI_ISL_2457929, EPI_ISL_2457930, EPI_ISL_2457931, EPI_ISL_2457932, EPI_ISL_2457933, EPI_ISL_2457934, EPI_ISL_2457935, EPI_ISL_2457936, EPI_ISL_2457937, EPI_ISL_2457938, EPI_ISL_2457939, EPI_ISL_2457940,
EPI_ISL_2457941, EPI_ISL_2457942, EPI_ISL_2457943, EPI_ISL_2457944, EPI_ISL_2457945, EPI_ISL_2457946, EPI_ISL_2457947, EPI_ISL_2457948, EPI_ISL_2457949, EPI_ISL_2457950, EPI_ISL_2457951, EPI_ISL_2457952, EPI_ISL_2457953, EPI_ISL_2457954, EPI_ISL_2457955, EPI_ISL_2457956, EPI_ISL_2457957, EPI_ISL_2457958, EPI_ISL_2457959, EPI_ISL_2457960, EPI_ISL_2457961, EPI_ISL_2457962,
EPI_ISL_2457963, EPI_ISL_2457964, EPI_ISL_2457965, EPI_ISL_2457966, EPI_ISL_2457967, EPI_ISL_2457968, EPI_ISL_2457969, EPI_ISL_2457970, EPI_ISL_2457971, EPI_ISL_2457972, EPI_ISL_2457973, EPI_ISL_2457974, EPI_ISL_2457975, EPI_ISL_2457976, EPI_ISL_2457977, EPI_ISL_2457978, EPI_ISL_2457979, EPI_ISL_2457980, EPI_ISL_2457981, EPI_ISL_2457982, EPI_ISL_2457983, EPI_ISL_2457984,
EPI_ISL_2457985, EPI_ISL_2457986, EPI_ISL_2457987, EPI_ISL_2457988, EPI_ISL_2457989, EPI_ISL_2457990, EPI_ISL_2457991, EPI_ISL_2457992, EPI_ISL_2457993, EPI_ISL_2457994, EPI_ISL_2457995, EPI_ISL_2457996, EPI_ISL_2457997, EPI_ISL_2457998, EPI_ISL_2457999, EPI_ISL_2458000, EPI_ISL_2458001, EPI_ISL_2458002, EPI_ISL_2458003, EPI_ISL_2458004, EPI_ISL_2458005, EPI_ISL_2458006,
EPI_ISL_2458007, EPI_ISL_2458008, EPI_ISL_2458009, EPI_ISL_2458010, EPI_ISL_2458011, EPI_ISL_2458012, EPI_ISL_2458013, EPI_ISL_2458014, EPI_ISL_2458015, EPI_ISL_2458016, EPI_ISL_2458017, EPI_ISL_2458018, EPI_ISL_2458019, EPI_ISL_2458020, EPI_ISL_2458021, EPI_ISL_2458022, EPI_ISL_2458023, EPI_ISL_2458024, EPI_ISL_2458025, EPI_ISL_2458026, EPI_ISL_2458027, EPI_ISL_2458028,
EPI_ISL_2458029, EPI_ISL_2458030, EPI_ISL_2458031, EPI_ISL_2458032, EPI_ISL_2458033, EPI_ISL_2458034, EPI_ISL_2458035, EPI_ISL_2458036, EPI_ISL_2458037, EPI_ISL_2458038, EPI_ISL_2458039, EPI_ISL_2458040, EPI_ISL_2458041, EPI_ISL_2458042, EPI_ISL_2458043, EPI_ISL_2458044, EPI_ISL_2458045, EPI_ISL_2458046, EPI_ISL_2458047, EPI_ISL_2458048, EPI_ISL_2458049, EPI_ISL_2458050,
EPI_ISL_2458051, EPI_ISL_2458052, EPI_ISL_2458053, EPI_ISL_2458054, EPI_ISL_2458055, EPI_ISL_2458056, EPI_ISL_2469212, EPI_ISL_2469268, EPI_ISL_2469318, EPI_ISL_2469382, EPI_ISL_2469434, EPI_ISL_2486458, EPI_ISL_2486504, EPI_ISL_2487339, EPI_ISL_2487439, EPI_ISL_2487549, EPI_ISL_2487834, EPI_ISL_2487997, EPI_ISL_2488101, EPI_ISL_2488102, EPI_ISL_2488103, EPI_ISL_2488104,
EPI_ISL_2488105, EPI_ISL_2488106, EPI_ISL_2488107, EPI_ISL_2488108, EPI_ISL_2488109, EPI_ISL_2488110, EPI_ISL_2488111, EPI_ISL_2488112, EPI_ISL_2488113, EPI_ISL_2488114, EPI_ISL_2488115, EPI_ISL_2488116, EPI_ISL_2488117, EPI_ISL_2488118, EPI_ISL_2488119, EPI_ISL_2488120, EPI_ISL_2488121, EPI_ISL_2488122, EPI_ISL_2488123, EPI_ISL_2488124, EPI_ISL_2488125, EPI_ISL_2488126,
EPI_ISL_2488127, EPI_ISL_2488128, EPI_ISL_2488129, EPI_ISL_2488130, EPI_ISL_2488131, EPI_ISL_2488132, EPI_ISL_2488133, EPI_ISL_2488134, EPI_ISL_2488135, EPI_ISL_2488136, EPI_ISL_2488137, EPI_ISL_2488138, EPI_ISL_2488139, EPI_ISL_2488140, EPI_ISL_2488141, EPI_ISL_2488142, EPI_ISL_2488143, EPI_ISL_2488144, EPI_ISL_2488145, EPI_ISL_2488146, EPI_ISL_2488147, EPI_ISL_2488148,
EPI_ISL_2488149, EPI_ISL_2488150, EPI_ISL_2488151, EPI_ISL_2488152, EPI_ISL_2488153, EPI_ISL_2488154, EPI_ISL_2488155, EPI_ISL_2488156, EPI_ISL_2488157, EPI_ISL_2488158, EPI_ISL_2488159, EPI_ISL_2488160, EPI_ISL_2488161, EPI_ISL_2488162, EPI_ISL_2488163, EPI_ISL_2488164, EPI_ISL_2488165, EPI_ISL_2488166, EPI_ISL_2488167, EPI_ISL_2488168, EPI_ISL_2488169, EPI_ISL_2488170,
EPI_ISL_2488171, EPI_ISL_2488172, EPI_ISL_2488173, EPI_ISL_2488174, EPI_ISL_2488175, EPI_ISL_2488176, EPI_ISL_2488177, EPI_ISL_2488178, EPI_ISL_2488179, EPI_ISL_2488180, EPI_ISL_2488181, EPI_ISL_2488182, EPI_ISL_2488183, EPI_ISL_2488184, EPI_ISL_2488185, EPI_ISL_2488187, EPI_ISL_2488188, EPI_ISL_2488189, EPI_ISL_2488190, EPI_ISL_2488191, EPI_ISL_2488192, EPI_ISL_2488193,
EPI_ISL_2488194, EPI_ISL_2488195, EPI_ISL_2488196, EPI_ISL_2488197, EPI_ISL_2488198, EPI_ISL_2488199, EPI_ISL_2488200, EPI_ISL_2488201, EPI_ISL_2488202, EPI_ISL_2488203, EPI_ISL_2488204, EPI_ISL_2488205, EPI_ISL_2488206, EPI_ISL_2488207, EPI_ISL_2488208, EPI_ISL_2488209, EPI_ISL_2488210, EPI_ISL_2488211, EPI_ISL_2488212, EPI_ISL_2488213, EPI_ISL_2488214, EPI_ISL_2488215,
EPI_ISL_2488216, EPI_ISL_2488217, EPI_ISL_2488218, EPI_ISL_2488219, EPI_ISL_2488220, EPI_ISL_2488221, EPI_ISL_2488222, EPI_ISL_2488223, EPI_ISL_2488224, EPI_ISL_2488225, EPI_ISL_2488226, EPI_ISL_2488227, EPI_ISL_2488228, EPI_ISL_2488229, EPI_ISL_2488230, EPI_ISL_2488231, EPI_ISL_2488232, EPI_ISL_2488233, EPI_ISL_2488234, EPI_ISL_2488235, EPI_ISL_2488243, EPI_ISL_2488244,
EPI_ISL_2488245, EPI_ISL_2488246, EPI_ISL_2488247, EPI_ISL_2488248, EPI_ISL_2488249, EPI_ISL_2488250, EPI_ISL_2488251, EPI_ISL_2488252, EPI_ISL_2488253, EPI_ISL_2488254, EPI_ISL_2488255, EPI_ISL_2488256, EPI_ISL_2488257, EPI_ISL_2488258, EPI_ISL_2488259, EPI_ISL_2488260, EPI_ISL_2488261, EPI_ISL_2488262, EPI_ISL_2488263, EPI_ISL_2488264, EPI_ISL_2488265, EPI_ISL_2488266,
EPI_ISL_2488267, EPI_ISL_2488268, EPI_ISL_2488269, EPI_ISL_2488270, EPI_ISL_2488271, EPI_ISL_2488272, EPI_ISL_2488273, EPI_ISL_2488274, EPI_ISL_2488275, EPI_ISL_2488276, EPI_ISL_2488277, EPI_ISL_2488278, EPI_ISL_2488279, EPI_ISL_2488280, EPI_ISL_2488281, EPI_ISL_2488283, EPI_ISL_2488284, EPI_ISL_2488285, EPI_ISL_2488286, EPI_ISL_2488287, EPI_ISL_2488288, EPI_ISL_2488289,
EPI_ISL_2488290, EPI_ISL_2488291, EPI_ISL_2488292, EPI_ISL_2488293, EPI_ISL_2488294, EPI_ISL_2488295, EPI_ISL_2488296, EPI_ISL_2488297, EPI_ISL_2488298, EPI_ISL_2488299, EPI_ISL_2488300, EPI_ISL_2488301, EPI_ISL_2488302, EPI_ISL_2488303, EPI_ISL_2488304, EPI_ISL_2488305, EPI_ISL_2488306, EPI_ISL_2488307, EPI_ISL_2488308, EPI_ISL_2488309, EPI_ISL_2488310, EPI_ISL_2488311,
EPI_ISL_2488312, EPI_ISL_2488313, EPI_ISL_2488314, EPI_ISL_2488316, EPI_ISL_2488317, EPI_ISL_2488318, EPI_ISL_2488319, EPI_ISL_2488320, EPI_ISL_2488321, EPI_ISL_2488322, EPI_ISL_2488323, EPI_ISL_2488324, EPI_ISL_2488325, EPI_ISL_2488326, EPI_ISL_2488327, EPI_ISL_2488328, EPI_ISL_2488329, EPI_ISL_2488330, EPI_ISL_2488331, EPI_ISL_2488332, EPI_ISL_2488333, EPI_ISL_2488334,
EPI_ISL_2488335, EPI_ISL_2488336, EPI_ISL_2488337, EPI_ISL_2488338, EPI_ISL_2488339, EPI_ISL_2488340, EPI_ISL_2488341, EPI_ISL_2488342, EPI_ISL_2488343, EPI_ISL_2488344, EPI_ISL_2488345, EPI_ISL_2488346, EPI_ISL_2488347, EPI_ISL_2488348, EPI_ISL_2488349, EPI_ISL_2488350, EPI_ISL_2488351, EPI_ISL_2488352, EPI_ISL_2488353, EPI_ISL_2488354, EPI_ISL_2488355, EPI_ISL_2488356,
EPI_ISL_2488357, EPI_ISL_2488358, EPI_ISL_2488359, EPI_ISL_2488360, EPI_ISL_2488361, EPI_ISL_2488362, EPI_ISL_2488363, EPI_ISL_2488364, EPI_ISL_2488365, EPI_ISL_2488366, EPI_ISL_2488367, EPI_ISL_2488368, EPI_ISL_2488369, EPI_ISL_2488370, EPI_ISL_2488371, EPI_ISL_2488372, EPI_ISL_2488373, EPI_ISL_2488374, EPI_ISL_2488375, EPI_ISL_2488376, EPI_ISL_2488377, EPI_ISL_2488378,
EPI_ISL_2488379, EPI_ISL_2488380, EPI_ISL_2488381, EPI_ISL_2488382, EPI_ISL_2488383, EPI_ISL_2488384, EPI_ISL_2488385, EPI_ISL_2488386, EPI_ISL_2488387, EPI_ISL_2488388, EPI_ISL_2488389, EPI_ISL_2488390, EPI_ISL_2488391, EPI_ISL_2488392, EPI_ISL_2488393, EPI_ISL_2488394, EPI_ISL_2488395, EPI_ISL_2488396, EPI_ISL_2488397, EPI_ISL_2488398, EPI_ISL_2488399, EPI_ISL_2488400,
EPI_ISL_2488401, EPI_ISL_2488402, EPI_ISL_2488403, EPI_ISL_2488404, EPI_ISL_2488405, EPI_ISL_2488406, EPI_ISL_2488407, EPI_ISL_2488408, EPI_ISL_2488409, EPI_ISL_2488410, EPI_ISL_2488411, EPI_ISL_2488413, EPI_ISL_2488414, EPI_ISL_2488415, EPI_ISL_2488416, EPI_ISL_2488417, EPI_ISL_2488418, EPI_ISL_2488419, EPI_ISL_2488420, EPI_ISL_2488421, EPI_ISL_2488422, EPI_ISL_2488423,
EPI_ISL_2488424, EPI_ISL_2488425, EPI_ISL_2488426, EPI_ISL_2488427, EPI_ISL_2488428, EPI_ISL_2488429, EPI_ISL_2488430, EPI_ISL_2488431, EPI_ISL_2488432, EPI_ISL_2488433, EPI_ISL_2488434, EPI_ISL_2488435, EPI_ISL_2488436, EPI_ISL_2488437, EPI_ISL_2488439, EPI_ISL_2488440, EPI_ISL_2488441, EPI_ISL_2488442, EPI_ISL_2488443, EPI_ISL_2488444, EPI_ISL_2488445, EPI_ISL_2488446,
EPI_ISL_2488447, EPI_ISL_2488448, EPI_ISL_2488449, EPI_ISL_2488450, EPI_ISL_2488451, EPI_ISL_2488452, EPI_ISL_2488453, EPI_ISL_2488454, EPI_ISL_2488455, EPI_ISL_2488456, EPI_ISL_2488457, EPI_ISL_2488458, EPI_ISL_2488459, EPI_ISL_2488460, EPI_ISL_2488461, EPI_ISL_2488462, EPI_ISL_2488463, EPI_ISL_2488464, EPI_ISL_2488465, EPI_ISL_2488466, EPI_ISL_2488467, EPI_ISL_2488468,
EPI_ISL_2488469, EPI_ISL_2488470, EPI_ISL_2488471, EPI_ISL_2488472, EPI_ISL_2488473, EPI_ISL_2488474, EPI_ISL_2488475, EPI_ISL_2488476, EPI_ISL_2488477, EPI_ISL_2488478, EPI_ISL_2488479, EPI_ISL_2488480, EPI_ISL_2488481, EPI_ISL_2488482, EPI_ISL_2488483, EPI_ISL_2488484, EPI_ISL_2488485, EPI_ISL_2488486, EPI_ISL_2488487, EPI_ISL_2488488, EPI_ISL_2488489, EPI_ISL_2488490,
EPI_ISL_2488491, EPI_ISL_2488492, EPI_ISL_2488493, EPI_ISL_2488494, EPI_ISL_2488496, EPI_ISL_2488497, EPI_ISL_2488498, EPI_ISL_2488499, EPI_ISL_2488500, EPI_ISL_2488501, EPI_ISL_2488502, EPI_ISL_2488503, EPI_ISL_2488504, EPI_ISL_2488506, EPI_ISL_2488507, EPI_ISL_2488508, EPI_ISL_2488511, EPI_ISL_2488513, EPI_ISL_2488515, EPI_ISL_2488518,
EPI_ISL_2488519, EPI_ISL_2488521, EPI_ISL_2488522, EPI_ISL_2488524, EPI_ISL_2488525, EPI_ISL_2488527, EPI_ISL_2488528, EPI_ISL_2488531, EPI_ISL_2488532, EPI_ISL_2488533, EPI_ISL_2488534, EPI_ISL_2488535, EPI_ISL_2488536, EPI_ISL_2488537, EPI_ISL_2488538, EPI_ISL_2488539, EPI_ISL_2488540, EPI_ISL_2488541, EPI_ISL_2488542, EPI_ISL_2488544, EPI_ISL_2488545, EPI_ISL_2488546,
EPI_ISL_2488547, EPI_ISL_2488548, EPI_ISL_2488549, EPI_ISL_2488550, EPI_ISL_2488552, EPI_ISL_2488554, EPI_ISL_2488555, EPI_ISL_2488556, EPI_ISL_2488558, EPI_ISL_2488559, EPI_ISL_2488560, EPI_ISL_2488561, EPI_ISL_2488562, EPI_ISL_2488564, EPI_ISL_2488565, EPI_ISL_2488566, EPI_ISL_2488568, EPI_ISL_2488570, EPI_ISL_2488572, EPI_ISL_2488573, EPI_ISL_2488574, EPI_ISL_2488575,
EPI_ISL_2488576, EPI_ISL_2488577, EPI_ISL_2488579, EPI_ISL_2488580, EPI_ISL_2488581, EPI_ISL_2488582, EPI_ISL_2488585, EPI_ISL_2488586, EPI_ISL_2488587, EPI_ISL_2488588, EPI_ISL_2488589, EPI_ISL_2488590, EPI_ISL_2488591, EPI_ISL_2488592, EPI_ISL_2488593, EPI_ISL_2488594, EPI_ISL_2488595, EPI_ISL_2488597, EPI_ISL_2488600, EPI_ISL_2488601, EPI_ISL_2488602, EPI_ISL_2488603,
EPI_ISL_2488604, EPI_ISL_2488605, EPI_ISL_2488609, EPI_ISL_2488610, EPI_ISL_2488611, EPI_ISL_2488613, EPI_ISL_2488614, EPI_ISL_2488615, EPI_ISL_2488616, EPI_ISL_2488617, EPI_ISL_2488618, EPI_ISL_2488619, EPI_ISL_2488620, EPI_ISL_2488621, EPI_ISL_2488622, EPI_ISL_2488623, EPI_ISL_2488624, EPI_ISL_2488627, EPI_ISL_2488628, EPI_ISL_2488629, EPI_ISL_2488631, EPI_ISL_2488632,
EPI_ISL_2488633, EPI_ISL_2488634, EPI_ISL_2488636, EPI_ISL_2488637, EPI_ISL_2488638, EPI_ISL_2488639, EPI_ISL_2488640, EPI_ISL_2488641, EPI_ISL_2488642, EPI_ISL_2488644, EPI_ISL_2488645, EPI_ISL_2488647, EPI_ISL_2488648, EPI_ISL_2488649, EPI_ISL_2488650, EPI_ISL_2488651, EPI_ISL_2488652, EPI_ISL_2488654, EPI_ISL_2488655, EPI_ISL_2488657, EPI_ISL_2488658,
EPI_ISL_2488660, EPI_ISL_2488661, EPI_ISL_2488662, EPI_ISL_2488664, EPI_ISL_2488665, EPI_ISL_2488666, EPI_ISL_2488668, EPI_ISL_2488669, EPI_ISL_2488671, EPI_ISL_2488672, EPI_ISL_2488673, EPI_ISL_2488674, EPI_ISL_2488675, EPI_ISL_2488676, EPI_ISL_2488677, EPI_ISL_2488679, EPI_ISL_2488680, EPI_ISL_2488681, EPI_ISL_2488682, EPI_ISL_2488683, EPI_ISL_2488684, EPI_ISL_2488685,
EPI_ISL_2488686, EPI_ISL_2488687, EPI_ISL_2488688, EPI_ISL_2488689, EPI_ISL_2488691, EPI_ISL_2488692, EPI_ISL_2488695, EPI_ISL_2488697, EPI_ISL_2488698, EPI_ISL_2488699, EPI_ISL_2488700, EPI_ISL_2488701, EPI_ISL_2488702, EPI_ISL_2488703, EPI_ISL_2488704, EPI_ISL_2488705, EPI_ISL_2511439, EPI_ISL_2511528, EPI_ISL_2513294, EPI_ISL_2513300, EPI_ISL_2513301, EPI_ISL_2513303,
EPI_ISL_2513306, EPI_ISL_2513309, EPI_ISL_2513311, EPI_ISL_2513313, EPI_ISL_2513314, EPI_ISL_2513318, EPI_ISL_2513320, EPI_ISL_2513322, EPI_ISL_2513326, EPI_ISL_2513334, EPI_ISL_2513335, EPI_ISL_2513339, EPI_ISL_2513341, EPI_ISL_2513342, EPI_ISL_2513346, EPI_ISL_2513348, EPI_ISL_2513351, EPI_ISL_2513362, EPI_ISL_2513366, EPI_ISL_2513368, EPI_ISL_2513370, EPI_ISL_2513373,
EPI_ISL_2513375, EPI_ISL_2513376, EPI_ISL_2513377, EPI_ISL_2513379, EPI_ISL_2513394, EPI_ISL_2513396, EPI_ISL_2513401, EPI_ISL_2513404, EPI_ISL_2513406, EPI_ISL_2513411, EPI_ISL_2513413, EPI_ISL_2513416, EPI_ISL_2513421, EPI_ISL_2513422, EPI_ISL_2513434, EPI_ISL_2513438, EPI_ISL_2513442, EPI_ISL_2513445, EPI_ISL_2513447, EPI_ISL_2513450, EPI_ISL_2513455, EPI_ISL_2513457,
EPI_ISL_2513462, EPI_ISL_2513465, EPI_ISL_2513469, EPI_ISL_2513470, EPI_ISL_2513473, EPI_ISL_2513476, EPI_ISL_2513490, EPI_ISL_2513496, EPI_ISL_2513497, EPI_ISL_2513498, EPI_ISL_2513499, EPI_ISL_2513502, EPI_ISL_2513505, EPI_ISL_2513515, EPI_ISL_2513517, EPI_ISL_2513518, EPI_ISL_2513524, EPI_ISL_2513525, EPI_ISL_2513526, EPI_ISL_2513532, EPI_ISL_2513533, EPI_ISL_2513536,
EPI_ISL_2513538, EPI_ISL_2513541, EPI_ISL_2513542, EPI_ISL_2513544, EPI_ISL_2513550, EPI_ISL_2513557, EPI_ISL_2513558, EPI_ISL_2513563, EPI_ISL_2513564, EPI_ISL_2513566, EPI_ISL_2513568, EPI_ISL_2513575, EPI_ISL_2513579, EPI_ISL_2513587, EPI_ISL_2513588, EPI_ISL_2513591, EPI_ISL_2513596, EPI_ISL_2513607, EPI_ISL_2513609, EPI_ISL_2513910, EPI_ISL_2513964, EPI_ISL_2514927,
EPI_ISL_2514933, EPI_ISL_2514934, EPI_ISL_2514948, EPI_ISL_2514955, EPI_ISL_2514964, EPI_ISL_2514968, EPI_ISL_2514973, EPI_ISL_2514974, EPI_ISL_2514975, EPI_ISL_2514978, EPI_ISL_2514982, EPI_ISL_2514983, EPI_ISL_2514985, EPI_ISL_2514988, EPI_ISL_2514989, EPI_ISL_2514991, EPI_ISL_2514998, EPI_ISL_2515004, EPI_ISL_2515009, EPI_ISL_2515020, EPI_ISL_2515041, EPI_ISL_2515042,
EPI_ISL_2515044, EPI_ISL_2515046, EPI_ISL_2515049, EPI_ISL_2515052, EPI_ISL_2515057, EPI_ISL_2515058, EPI_ISL_2515061, EPI_ISL_2515065, EPI_ISL_2515072, EPI_ISL_2515073, EPI_ISL_2515079, EPI_ISL_2515082, EPI_ISL_2515087, EPI_ISL_2515090, EPI_ISL_2515096, EPI_ISL_2515098, EPI_ISL_2515099, EPI_ISL_2515107, EPI_ISL_2515114, EPI_ISL_2515125, EPI_ISL_2515145, EPI_ISL_2515152,
EPI_ISL_2515155, EPI_ISL_2515175, EPI_ISL_2515176, EPI_ISL_2515194, EPI_ISL_2515203, EPI_ISL_2515231, EPI_ISL_2515249, EPI_ISL_2515264, EPI_ISL_2515265, EPI_ISL_2515270, EPI_ISL_2515275, EPI_ISL_2515295, EPI_ISL_2515324, EPI_ISL_2515331, EPI_ISL_2515334, EPI_ISL_2515359, EPI_ISL_2515383, EPI_ISL_2539225, EPI_ISL_2539352, EPI_ISL_2539415, EPI_ISL_2539424, EPI_ISL_2539427,
EPI_ISL_2539439, EPI_ISL_2539440, EPI_ISL_2539446, EPI_ISL_2539447, EPI_ISL_2539455, EPI_ISL_2539458, EPI_ISL_2539460, EPI_ISL_2539463, EPI_ISL_2539473, EPI_ISL_2539475, EPI_ISL_2539476, EPI_ISL_2539478, EPI_ISL_2539490, EPI_ISL_2539491, EPI_ISL_2539492, EPI_ISL_2539493, EPI_ISL_2539496, EPI_ISL_2539499, EPI_ISL_2539500, EPI_ISL_2539501, EPI_ISL_2539502, EPI_ISL_2539506,
EPI_ISL_2539514, EPI_ISL_2539526, EPI_ISL_2539533, EPI_ISL_2539539, EPI_ISL_2539546, EPI_ISL_2539548, EPI_ISL_2539550, EPI_ISL_2539551, EPI_ISL_2539554, EPI_ISL_2539556, EPI_ISL_2539564, EPI_ISL_2539568, EPI_ISL_2539569, EPI_ISL_2539571, EPI_ISL_2539575, EPI_ISL_2539576, EPI_ISL_2539578, EPI_ISL_2539581, EPI_ISL_2539585, EPI_ISL_2539586, EPI_ISL_2539599,
EPI_ISL_2539600, EPI_ISL_2539604, EPI_ISL_2539608, EPI_ISL_2539614, EPI_ISL_2539617, EPI_ISL_2539621, EPI_ISL_2539627, EPI_ISL_2539629, EPI_ISL_2539631, EPI_ISL_2539633, EPI_ISL_2539634, EPI_ISL_2539639, EPI_ISL_2539641, EPI_ISL_2539643, EPI_ISL_2539644, EPI_ISL_2539651, EPI_ISL_2539656, EPI_ISL_2539658, EPI_ISL_2539659, EPI_ISL_2539668, EPI_ISL_2539673, EPI_ISL_2539674,
EPI_ISL_2539676, EPI_ISL_2539680, EPI_ISL_2539685, EPI_ISL_2539686, EPI_ISL_2539691, EPI_ISL_2539700, EPI_ISL_2539701, EPI_ISL_2539703, EPI_ISL_2539709, EPI_ISL_2539716, EPI_ISL_2539718, EPI_ISL_2539726, EPI_ISL_2539729, EPI_ISL_2539731, EPI_ISL_2539734, EPI_ISL_2542695, EPI_ISL_2542696, EPI_ISL_2542697, EPI_ISL_2542698, EPI_ISL_2542699,
EPI_ISL_2542700, EPI_ISL_2542701, EPI_ISL_2542702, EPI_ISL_2542703, EPI_ISL_2542704, EPI_ISL_2542705, EPI_ISL_2542706, EPI_ISL_2542707, EPI_ISL_2542708, EPI_ISL_2542709, EPI_ISL_2542710, EPI_ISL_2542711, EPI_ISL_2542712, EPI_ISL_2542713, EPI_ISL_2542714, EPI_ISL_2542715, EPI_ISL_2542716, EPI_ISL_2542717, EPI_ISL_2542718, EPI_ISL_2542719, EPI_ISL_2542720, EPI_ISL_2542721,
EPI_ISL_2542722, EPI_ISL_2542723, EPI_ISL_2542724, EPI_ISL_2542725, EPI_ISL_2542726, EPI_ISL_2542727, EPI_ISL_2542728, EPI_ISL_2542729, EPI_ISL_2542730, EPI_ISL_2542731, EPI_ISL_2542732, EPI_ISL_2542733, EPI_ISL_2542734, EPI_ISL_2542735, EPI_ISL_2542736, EPI_ISL_2542737, EPI_ISL_2542738, EPI_ISL_2542739, EPI_ISL_2542740, EPI_ISL_2542741, EPI_ISL_2542742, EPI_ISL_2542743,
EPI_ISL_2542744, EPI_ISL_2542745, EPI_ISL_2542746, EPI_ISL_2542747, EPI_ISL_2542748, EPI_ISL_2542749, EPI_ISL_2542750, EPI_ISL_2542751, EPI_ISL_2542752, EPI_ISL_2542753, EPI_ISL_2542754, EPI_ISL_2542755, EPI_ISL_2542756, EPI_ISL_2542757, EPI_ISL_2542758, EPI_ISL_2542759, EPI_ISL_2542760, EPI_ISL_2542761, EPI_ISL_2542762, EPI_ISL_2542763, EPI_ISL_2542764, EPI_ISL_2542765,
EPI_ISL_2542766, EPI_ISL_2542767, EPI_ISL_2542768, EPI_ISL_2542769, EPI_ISL_2542770, EPI_ISL_2542771, EPI_ISL_2542772, EPI_ISL_2542773, EPI_ISL_2542774, EPI_ISL_2542775, EPI_ISL_2542776, EPI_ISL_2542777, EPI_ISL_2542778, EPI_ISL_2542779, EPI_ISL_2542780, EPI_ISL_2542781, EPI_ISL_2542782, EPI_ISL_2542783, EPI_ISL_2542784, EPI_ISL_2542953, EPI_ISL_2542954, EPI_ISL_2542955,
EPI_ISL_2542956, EPI_ISL_2542957, EPI_ISL_2542958, EPI_ISL_2542959, EPI_ISL_2542960, EPI_ISL_2542961, EPI_ISL_2542962, EPI_ISL_2542963, EPI_ISL_2542964, EPI_ISL_2542965, EPI_ISL_2542966, EPI_ISL_2542967, EPI_ISL_2542968, EPI_ISL_2542969, EPI_ISL_2542970, EPI_ISL_2542971, EPI_ISL_2542972, EPI_ISL_2542973, EPI_ISL_2542974, EPI_ISL_2542975, EPI_ISL_2542976, EPI_ISL_2542977,
EPI_ISL_2542978, EPI_ISL_2542979, EPI_ISL_2542980, EPI_ISL_2542981, EPI_ISL_2542982, EPI_ISL_2542983, EPI_ISL_2542984, EPI_ISL_2542985, EPI_ISL_2542986, EPI_ISL_2542987, EPI_ISL_2542988, EPI_ISL_2542989, EPI_ISL_2542990, EPI_ISL_2542991, EPI_ISL_2542992, EPI_ISL_2542993, EPI_ISL_2542994, EPI_ISL_2542995, EPI_ISL_2542996, EPI_ISL_2542997, EPI_ISL_2542998, EPI_ISL_2542999,
EPI_ISL_2543000, EPI_ISL_2543001, EPI_ISL_2543002, EPI_ISL_2543003, EPI_ISL_2543004, EPI_ISL_2543005, EPI_ISL_2543006, EPI_ISL_2543007, EPI_ISL_2543008, EPI_ISL_2543009, EPI_ISL_2543010, EPI_ISL_2543011, EPI_ISL_2543012, EPI_ISL_2543013, EPI_ISL_2543014, EPI_ISL_2543015, EPI_ISL_2543016, EPI_ISL_2543017, EPI_ISL_2543018, EPI_ISL_2543019, EPI_ISL_2543020, EPI_ISL_2543021,
EPI_ISL_2543022, EPI_ISL_2543023, EPI_ISL_2543024, EPI_ISL_2543025, EPI_ISL_2543026, EPI_ISL_2543027, EPI_ISL_2543028, EPI_ISL_2543029, EPI_ISL_2543030, EPI_ISL_2543031, EPI_ISL_2543032, EPI_ISL_2543033, EPI_ISL_2543034, EPI_ISL_2543035, EPI_ISL_2543036, EPI_ISL_2543037, EPI_ISL_2543038, EPI_ISL_2543039, EPI_ISL_2543040, EPI_ISL_2543041, EPI_ISL_2543042, EPI_ISL_2543043,
EPI_ISL_2543044, EPI_ISL_2543045, EPI_ISL_2543046, EPI_ISL_2543047, EPI_ISL_2543049, EPI_ISL_2543050, EPI_ISL_2543051, EPI_ISL_2543052, EPI_ISL_2543053, EPI_ISL_2543054, EPI_ISL_2543055, EPI_ISL_2543056, EPI_ISL_2543057, EPI_ISL_2543058, EPI_ISL_2543059, EPI_ISL_2543060, EPI_ISL_2543061, EPI_ISL_2543062, EPI_ISL_2543063, EPI_ISL_2543064, EPI_ISL_2543065, EPI_ISL_2543066,
EPI_ISL_2543067, EPI_ISL_2543068, EPI_ISL_2543069, EPI_ISL_2543070, EPI_ISL_2543071, EPI_ISL_2543072, EPI_ISL_2543073, EPI_ISL_2543074, EPI_ISL_2543075, EPI_ISL_2543076, EPI_ISL_2543077, EPI_ISL_2543078, EPI_ISL_2543079, EPI_ISL_2543080, EPI_ISL_2543081, EPI_ISL_2543082, EPI_ISL_2543083, EPI_ISL_2543084, EPI_ISL_2543085, EPI_ISL_2543086, EPI_ISL_2543087, EPI_ISL_2543088,
EPI_ISL_2543089, EPI_ISL_2543090, EPI_ISL_2543091, EPI_ISL_2543092, EPI_ISL_2543093, EPI_ISL_2543094, EPI_ISL_2543095, EPI_ISL_2543096, EPI_ISL_2543097, EPI_ISL_2543098, EPI_ISL_2543099, EPI_ISL_2543100, EPI_ISL_2543101, EPI_ISL_2543102, EPI_ISL_2543103, EPI_ISL_2543104, EPI_ISL_2543105, EPI_ISL_2543106, EPI_ISL_2543107, EPI_ISL_2543108, EPI_ISL_2543109, EPI_ISL_2543110,
EPI_ISL_2543111, EPI_ISL_2543112, EPI_ISL_2543113, EPI_ISL_2543114, EPI_ISL_2543115, EPI_ISL_2543116, EPI_ISL_2543117, EPI_ISL_2543118, EPI_ISL_2543119, EPI_ISL_2543120, EPI_ISL_2543121, EPI_ISL_2543122, EPI_ISL_2543124, EPI_ISL_2543125, EPI_ISL_2543126, EPI_ISL_2543127, EPI_ISL_2543128, EPI_ISL_2543129, EPI_ISL_2543130, EPI_ISL_2543131, EPI_ISL_2543132,
EPI_ISL_2543133, EPI_ISL_2543134, EPI_ISL_2543135, EPI_ISL_2543136, EPI_ISL_2543137, EPI_ISL_2543138, EPI_ISL_2543139, EPI_ISL_2543140, EPI_ISL_2543141, EPI_ISL_2543142, EPI_ISL_2543143, EPI_ISL_2543144, EPI_ISL_2543145, EPI_ISL_2543146, EPI_ISL_2543147, EPI_ISL_2543149, EPI_ISL_2543150, EPI_ISL_2543151, EPI_ISL_2543152, EPI_ISL_2543153, EPI_ISL_2543154, EPI_ISL_2543155,
EPI_ISL_2543156, EPI_ISL_2543157, EPI_ISL_2543158, EPI_ISL_2543159, EPI_ISL_2543160, EPI_ISL_2543161, EPI_ISL_2543162, EPI_ISL_2543164, EPI_ISL_2543165, EPI_ISL_2543166, EPI_ISL_2543167, EPI_ISL_2543168, EPI_ISL_2543169, EPI_ISL_2543170, EPI_ISL_2543171, EPI_ISL_2543172, EPI_ISL_2543173, EPI_ISL_2543174, EPI_ISL_2543175, EPI_ISL_2543176, EPI_ISL_2543177,
EPI_ISL_2543178, EPI_ISL_2543179, EPI_ISL_2543180, EPI_ISL_2543181, EPI_ISL_2543182, EPI_ISL_2543183, EPI_ISL_2543184, EPI_ISL_2543185, EPI_ISL_2543186, EPI_ISL_2543187, EPI_ISL_2543188, EPI_ISL_2543189, EPI_ISL_2543190, EPI_ISL_2543191, EPI_ISL_2543192, EPI_ISL_2543305, EPI_ISL_2543306, EPI_ISL_2543307, EPI_ISL_2543308, EPI_ISL_2543309, EPI_ISL_2543310, EPI_ISL_2543311,
EPI_ISL_2543312, EPI_ISL_2543313, EPI_ISL_2543314, EPI_ISL_2543315, EPI_ISL_2543316, EPI_ISL_2543317, EPI_ISL_2543318, EPI_ISL_2543319, EPI_ISL_2543320, EPI_ISL_2543321, EPI_ISL_2543322, EPI_ISL_2543323, EPI_ISL_2543324, EPI_ISL_2543325, EPI_ISL_2543326, EPI_ISL_2543327, EPI_ISL_2543328, EPI_ISL_2543329, EPI_ISL_2543330, EPI_ISL_2543331, EPI_ISL_2543332, EPI_ISL_2543333,
EPI_ISL_2543340, EPI_ISL_2543354, EPI_ISL_2543375, EPI_ISL_2543381, EPI_ISL_2554915, EPI_ISL_2554916, EPI_ISL_2554917, EPI_ISL_2554918, EPI_ISL_2554919, EPI_ISL_2554920, EPI_ISL_2554921, EPI_ISL_2554922, EPI_ISL_2554923, EPI_ISL_2554924, EPI_ISL_2554925, EPI_ISL_2554926, EPI_ISL_2554927, EPI_ISL_2554928, EPI_ISL_2554929, EPI_ISL_2554930, EPI_ISL_2554931, EPI_ISL_2554932,
EPI_ISL_2554933, EPI_ISL_2554934, EPI_ISL_2554935, EPI_ISL_2554936, EPI_ISL_2554937, EPI_ISL_2554938, EPI_ISL_2554939, EPI_ISL_2554940, EPI_ISL_2554941, EPI_ISL_2554942, EPI_ISL_2554943, EPI_ISL_2554944, EPI_ISL_2554945, EPI_ISL_2554946, EPI_ISL_2554947, EPI_ISL_2554948, EPI_ISL_2554949, EPI_ISL_2554950, EPI_ISL_2554951, EPI_ISL_2554952, EPI_ISL_2554953, EPI_ISL_2554954,
EPI_ISL_2554955, EPI_ISL_2554956, EPI_ISL_2554957, EPI_ISL_2554958, EPI_ISL_2554959, EPI_ISL_2554960, EPI_ISL_2554961, EPI_ISL_2554962, EPI_ISL_2554963, EPI_ISL_2554964, EPI_ISL_2554965, EPI_ISL_2554966, EPI_ISL_2554967, EPI_ISL_2554968, EPI_ISL_2554969, EPI_ISL_2554970, EPI_ISL_2554971, EPI_ISL_2554972, EPI_ISL_2554973, EPI_ISL_2554974, EPI_ISL_2554975, EPI_ISL_2554976,
EPI_ISL_2554977, EPI_ISL_2554978, EPI_ISL_2554979, EPI_ISL_2554980, EPI_ISL_2554981, EPI_ISL_2554982, EPI_ISL_2554983, EPI_ISL_2554984, EPI_ISL_2554985, EPI_ISL_2554986, EPI_ISL_2554987, EPI_ISL_2554988, EPI_ISL_2554989, EPI_ISL_2554990, EPI_ISL_2554992, EPI_ISL_2554994, EPI_ISL_2554995, EPI_ISL_2554996, EPI_ISL_2554997, EPI_ISL_2554998,
EPI_ISL_2554999, EPI_ISL_2555000, EPI_ISL_2555001, EPI_ISL_2555003, EPI_ISL_2555013, EPI_ISL_2555014, EPI_ISL_2555019, EPI_ISL_2555035, EPI_ISL_2555039, EPI_ISL_2555052, EPI_ISL_2555525

| | | | |
|---|---|---|---|
| see above | Lighthouse Lab in Glasgow | Wellcome Sanger Institute for the COVID-19 Genomics UK (COG-UK) Consortium | Anna Dominiczak and Alex Alderton; Carol Clugston; Cordelia Langford; David Gray; David K. Jackson; Dominic Kwiatkowski; Ewan Harrison; Harper VanSteenhouse; Ian Johnston; Jeffrey Barrett; John Sillitoe on behalf of the Wellcome Sanger Institute COVID-19 Surveillance Team; Roberto Amato; Sonia Goncalves; Yumi Kasai |

EPI_ISL_2354200, EPI_ISL_2392532, EPI_ISL_2392535, EPI_ISL_2392543, EPI_ISL_2392550, EPI_ISL_2392551, EPI_ISL_2392554, EPI_ISL_2392559, EPI_ISL_2392563, EPI_ISL_2392567, EPI_ISL_2392570, EPI_ISL_2392571, EPI_ISL_2392577, EPI_ISL_2392578, EPI_ISL_2392583, EPI_ISL_2392586, EPI_ISL_2392589, EPI_ISL_2392590, EPI_ISL_2392591, EPI_ISL_2392592, EPI_ISL_2392595, EPI_ISL_2392599,
EPI_ISL_2392602, EPI_ISL_2392606, EPI_ISL_2392609, EPI_ISL_2392612, EPI_ISL_2392616, EPI_ISL_2392618, EPI_ISL_2392623, EPI_ISL_2392624, EPI_ISL_2392627, EPI_ISL_2392629, EPI_ISL_2392631, EPI_ISL_2392637, EPI_ISL_2392643, EPI_ISL_2392649, EPI_ISL_2392677, EPI_ISL_2392681, EPI_ISL_2392683, EPI_ISL_2392686, EPI_ISL_2392687, EPI_ISL_2392697, EPI_ISL_2392703, EPI_ISL_2392704,
EPI_ISL_2392711, EPI_ISL_2392719, EPI_ISL_2392722, EPI_ISL_2392727, EPI_ISL_2392734, EPI_ISL_2392741, EPI_ISL_2392743, EPI_ISL_2392746, EPI_ISL_2392748, EPI_ISL_2392749, EPI_ISL_2392752, EPI_ISL_2392758, EPI_ISL_2392759, EPI_ISL_2392762, EPI_ISL_2392764, EPI_ISL_2392765, EPI_ISL_2392766, EPI_ISL_2392767, EPI_ISL_2392768, EPI_ISL_2392769, EPI_ISL_2392770,
EPI_ISL_2392771, EPI_ISL_2392773, EPI_ISL_2392774, EPI_ISL_2392775, EPI_ISL_2392776, EPI_ISL_2392777, EPI_ISL_2392779, EPI_ISL_2392780, EPI_ISL_2392781, EPI_ISL_2392782, EPI_ISL_2392783, EPI_ISL_2392784, EPI_ISL_2392786, EPI_ISL_2392787, EPI_ISL_2392788, EPI_ISL_2392789, EPI_ISL_2392790, EPI_ISL_2392791, EPI_ISL_2392792, EPI_ISL_2392794, EPI_ISL_2392795, EPI_ISL_2392796,
EPI_ISL_2392797, EPI_ISL_2392799, EPI_ISL_2392800, EPI_ISL_2392801, EPI_ISL_2392803, EPI_ISL_2392804, EPI_ISL_2392805, EPI_ISL_2392806, EPI_ISL_2392809, EPI_ISL_2392810, EPI_ISL_2392811, EPI_ISL_2392812, EPI_ISL_2392814, EPI_ISL_2392819, EPI_ISL_2392820, EPI_ISL_2392822, EPI_ISL_2392823, EPI_ISL_2392825, EPI_ISL_2392826, EPI_ISL_2392828, EPI_ISL_2392829, EPI_ISL_2392830,
EPI_ISL_2392831, EPI_ISL_2392832, EPI_ISL_2392833, EPI_ISL_2392834, EPI_ISL_2392835, EPI_ISL_2392836, EPI_ISL_2392837, EPI_ISL_2392838, EPI_ISL_2392841, EPI_ISL_2392842, EPI_ISL_2392843, EPI_ISL_2392845, EPI_ISL_2392846, EPI_ISL_2392847, EPI_ISL_2392848, EPI_ISL_2392849, EPI_ISL_2392850, EPI_ISL_2392851, EPI_ISL_2392852, EPI_ISL_2392853, EPI_ISL_2392854, EPI_ISL_2392855,
EPI_ISL_2392856, EPI_ISL_2392857, EPI_ISL_2392858, EPI_ISL_2392860, EPI_ISL_2392861, EPI_ISL_2392862, EPI_ISL_2392863, EPI_ISL_2392864, EPI_ISL_2392865, EPI_ISL_2392867, EPI_ISL_2392868, EPI_ISL_2392869, EPI_ISL_2392870, EPI_ISL_2392871, EPI_ISL_2392872, EPI_ISL_2392873, EPI_ISL_2392874, EPI_ISL_2392875, EPI_ISL_2392876, EPI_ISL_2392877, EPI_ISL_2392878, EPI_ISL_2392879,
EPI_ISL_2392880, EPI_ISL_2392881, EPI_ISL_2392882, EPI_ISL_2392883, EPI_ISL_2392884, EPI_ISL_2392885, EPI_ISL_2392886, EPI_ISL_2392887, EPI_ISL_2392888, EPI_ISL_2392890, EPI_ISL_2392891, EPI_ISL_2392892, EPI_ISL_2392893, EPI_ISL_2392894, EPI_ISL_2392895, EPI_ISL_2392896, EPI_ISL_2392897, EPI_ISL_2392898, EPI_ISL_2392899, EPI_ISL_2392900, EPI_ISL_2392901, EPI_ISL_2392902,
EPI_ISL_2392905, EPI_ISL_2392906, EPI_ISL_2392907, EPI_ISL_2392908, EPI_ISL_2392909, EPI_ISL_2392910, EPI_ISL_2392911, EPI_ISL_2392912, EPI_ISL_2392913, EPI_ISL_2392914, EPI_ISL_2392915, EPI_ISL_2392917, EPI_ISL_2392918, EPI_ISL_2392920, EPI_ISL_2392921, EPI_ISL_2392922, EPI_ISL_2392923, EPI_ISL_2392924, EPI_ISL_2392925, EPI_ISL_2392926, EPI_ISL_2392929, EPI_ISL_2392930,

EPI_ISL_2392931, EPI_ISL_2392932, EPI_ISL_2392933, EPI_ISL_2392934, EPI_ISL_2392935, EPI_ISL_2392937, EPI_ISL_2392938, EPI_ISL_2392939, EPI_ISL_2392940, EPI_ISL_2392941, EPI_ISL_2393703, EPI_ISL_2406857, EPI_ISL_2435343, EPI_ISL_2435346, EPI_ISL_2435347, EPI_ISL_2435349, EPI_ISL_2435353, EPI_ISL_2435356, EPI_ISL_2435358, EPI_ISL_2435361, EPI_ISL_2435364, EPI_ISL_2435366, EPI_ISL_2435367, EPI_ISL_2435368, EPI_ISL_2435378, EPI_ISL_2435384, EPI_ISL_2435386, EPI_ISL_2435388, EPI_ISL_2435389, EPI_ISL_2435391, EPI_ISL_2435395, EPI_ISL_2435398, EPI_ISL_2435399, EPI_ISL_2435400, EPI_ISL_2435405, EPI_ISL_2435409, EPI_ISL_2435413, EPI_ISL_2435415, EPI_ISL_2435419, EPI_ISL_2435424, EPI_ISL_2435428, EPI_ISL_2435430, EPI_ISL_2435432, EPI_ISL_2435433, EPI_ISL_2435436, EPI_ISL_2435438, EPI_ISL_2435441, EPI_ISL_2435443, EPI_ISL_2435444, EPI_ISL_2435445, EPI_ISL_2435446, EPI_ISL_2435449, EPI_ISL_2435450, EPI_ISL_2435453, EPI_ISL_2435455, EPI_ISL_2435457, EPI_ISL_2435459, EPI_ISL_2435461, EPI_ISL_2435462, EPI_ISL_2435463, EPI_ISL_2435464, EPI_ISL_2435465, EPI_ISL_2435472, EPI_ISL_2435473, EPI_ISL_2435483, EPI_ISL_2435487, EPI_ISL_2435490, EPI_ISL_2435492, EPI_ISL_2435494, EPI_ISL_2435495, EPI_ISL_2435496, EPI_ISL_2435499, EPI_ISL_2435501, EPI_ISL_2435502, EPI_ISL_2435505, EPI_ISL_2435506, EPI_ISL_2435507, EPI_ISL_2435511, EPI_ISL_2435513, EPI_ISL_2435520, EPI_ISL_2435521, EPI_ISL_2435523, EPI_ISL_2435525, EPI_ISL_2435528, EPI_ISL_2435529, EPI_ISL_2435535, EPI_ISL_2435537, EPI_ISL_2435542, EPI_ISL_2435544, EPI_ISL_2435547, EPI_ISL_2435549, EPI_ISL_2435552, EPI_ISL_2435553, EPI_ISL_2435559, EPI_ISL_2435563, EPI_ISL_2435565, EPI_ISL_2435566, EPI_ISL_2435580, EPI_ISL_2435586, EPI_ISL_2435590, EPI_ISL_2435592, EPI_ISL_2435593, EPI_ISL_2435597, EPI_ISL_2435600, EPI_ISL_2435617, EPI_ISL_2435621, EPI_ISL_2435635, EPI_ISL_2435641, EPI_ISL_2435644, EPI_ISL_2435645, EPI_ISL_2435662, EPI_ISL_2435666, EPI_ISL_2435671, EPI_ISL_2435675, EPI_ISL_2435692, EPI_ISL_2435697, EPI_ISL_2435712, EPI_ISL_2435716, EPI_ISL_2435719, EPI_ISL_2435720, EPI_ISL_2435721, EPI_ISL_2435722, EPI_ISL_2435723, EPI_ISL_2435724, EPI_ISL_2435748, EPI_ISL_2435749, EPI_ISL_2435758, EPI_ISL_2435759, EPI_ISL_2435762, EPI_ISL_2435766, EPI_ISL_2435779, EPI_ISL_2435789, EPI_ISL_2435793, EPI_ISL_2435798, EPI_ISL_2435800, EPI_ISL_2435814, EPI_ISL_2435815, EPI_ISL_2435818, EPI_ISL_2435830, EPI_ISL_2435831, EPI_ISL_2435856, EPI_ISL_2435859, EPI_ISL_2435866, EPI_ISL_2436232, EPI_ISL_2436235, EPI_ISL_2436245, EPI_ISL_2436246, EPI_ISL_2436249, EPI_ISL_2436255, EPI_ISL_2436256, EPI_ISL_2436279, EPI_ISL_2436281, EPI_ISL_2436283, EPI_ISL_2436293, EPI_ISL_2436295, EPI_ISL_2436299, EPI_ISL_2436301, EPI_ISL_2436305, EPI_ISL_2436310, EPI_ISL_2436313, EPI_ISL_2436335, EPI_ISL_2436353, EPI_ISL_2436359, EPI_ISL_2436365, EPI_ISL_2436373, EPI_ISL_2436382, EPI_ISL_2436384, EPI_ISL_2436385, EPI_ISL_2436386, EPI_ISL_2436387, EPI_ISL_2436390, EPI_ISL_2436393, EPI_ISL_2436394, EPI_ISL_2436395, EPI_ISL_2436396, EPI_ISL_2436401, EPI_ISL_2436405, EPI_ISL_2436420, EPI_ISL_2436424, EPI_ISL_2436427, EPI_ISL_2436432, EPI_ISL_2436433, EPI_ISL_2436441, EPI_ISL_2436444, EPI_ISL_2436455, EPI_ISL_2436458, EPI_ISL_2436460, EPI_ISL_2436466, EPI_ISL_2436480, EPI_ISL_2436481, EPI_ISL_2436488, EPI_ISL_2436492, EPI_ISL_2436494, EPI_ISL_2436495, EPI_ISL_2436496, EPI_ISL_2436497, EPI_ISL_2436498, EPI_ISL_2436503, EPI_ISL_2436506, EPI_ISL_2436507, EPI_ISL_2436509, EPI_ISL_2436510, EPI_ISL_2436512, EPI_ISL_2436513, EPI_ISL_2436518, EPI_ISL_2436520, EPI_ISL_2436523, EPI_ISL_2436527, EPI_ISL_2436530, EPI_ISL_2436531, EPI_ISL_2436540, EPI_ISL_2436545, EPI_ISL_2436546, EPI_ISL_2436554, EPI_ISL_2436558, EPI_ISL_2436571, EPI_ISL_2436572, EPI_ISL_2436584, EPI_ISL_2436587, EPI_ISL_2436589, EPI_ISL_2436592, EPI_ISL_2436593, EPI_ISL_2436600, EPI_ISL_2436601, EPI_ISL_2436608, EPI_ISL_2436617, EPI_ISL_2436625, EPI_ISL_2436628, EPI_ISL_2436631, EPI_ISL_2436639, EPI_ISL_2436644, EPI_ISL_2436649, EPI_ISL_2436651, EPI_ISL_2436657, EPI_ISL_2436665, EPI_ISL_2436669, EPI_ISL_2436670, EPI_ISL_2436672, EPI_ISL_2436674, EPI_ISL_2436676, EPI_ISL_2436678, EPI_ISL_2436680, EPI_ISL_2436683, EPI_ISL_2436684, EPI_ISL_2436692, EPI_ISL_2436696, EPI_ISL_2436698, EPI_ISL_2436710, EPI_ISL_2436714, EPI_ISL_2436720, EPI_ISL_2436726, EPI_ISL_2436728, EPI_ISL_2436732, EPI_ISL_2436735, EPI_ISL_2436736, EPI_ISL_2436738, EPI_ISL_2436739, EPI_ISL_2436740, EPI_ISL_2436744, EPI_ISL_2436746, EPI_ISL_2436757, EPI_ISL_2436761, EPI_ISL_2436764, EPI_ISL_2436765, EPI_ISL_2436766, EPI_ISL_2436768, EPI_ISL_2436770, EPI_ISL_2436776, EPI_ISL_2436778, EPI_ISL_2436779, EPI_ISL_2436783, EPI_ISL_2436791, EPI_ISL_2436793, EPI_ISL_2436795, EPI_ISL_2436802, EPI_ISL_2436809, EPI_ISL_2436818, EPI_ISL_2436823, EPI_ISL_2436824, EPI_ISL_2436833, EPI_ISL_2436834, EPI_ISL_2436845, EPI_ISL_2436851, EPI_ISL_2436852, EPI_ISL_2436854, EPI_ISL_2436855, EPI_ISL_2436857, EPI_ISL_2436858, EPI_ISL_2436865, EPI_ISL_2436867, EPI_ISL_2436879, EPI_ISL_2436881, EPI_ISL_2436883, EPI_ISL_2436890, EPI_ISL_2436894, EPI_ISL_2436896, EPI_ISL_2436905, EPI_ISL_2436907, EPI_ISL_2436926, EPI_ISL_2437303, EPI_ISL_2437317, EPI_ISL_2437332, EPI_ISL_2437335, EPI_ISL_2437352, EPI_ISL_2437356, EPI_ISL_2437360, EPI_ISL_2437371, EPI_ISL_2437431, EPI_ISL_2437435, EPI_ISL_2437436, EPI_ISL_2437439, EPI_ISL_2437447, EPI_ISL_2437452, EPI_ISL_2437455, EPI_ISL_2437471, EPI_ISL_2437475, EPI_ISL_2437477, EPI_ISL_2437503, EPI_ISL_2437516, EPI_ISL_2437532, EPI_ISL_2437533, EPI_ISL_2437539, EPI_ISL_2437546, EPI_ISL_2437549, EPI_ISL_2437553, EPI_ISL_2437559, EPI_ISL_2437564, EPI_ISL_2437575, EPI_ISL_2437584, EPI_ISL_2437585, EPI_ISL_2437604, EPI_ISL_2437615, EPI_ISL_2437616, EPI_ISL_2437618, EPI_ISL_2437619, EPI_ISL_2437623, EPI_ISL_2437624, EPI_ISL_2437625, EPI_ISL_2437626, EPI_ISL_2437627, EPI_ISL_2437628, EPI_ISL_2437629, EPI_ISL_2437631, EPI_ISL_2437633, EPI_ISL_2437636, EPI_ISL_2437643, EPI_ISL_2437646, EPI_ISL_2437647, EPI_ISL_2437653, EPI_ISL_2437655, EPI_ISL_2437658, EPI_ISL_2437662, EPI_ISL_2437663, EPI_ISL_2437665, EPI_ISL_2437666, EPI_ISL_2437673, EPI_ISL_2437674, EPI_ISL_2437677, EPI_ISL_2437678, EPI_ISL_2437679, EPI_ISL_2437680, EPI_ISL_2437681, EPI_ISL_2437683, EPI_ISL_2437684, EPI_ISL_2437685, EPI_ISL_2437688, EPI_ISL_2437690, EPI_ISL_2437691, EPI_ISL_2437693, EPI_ISL_2437695, EPI_ISL_2437696, EPI_ISL_2437699, EPI_ISL_2437701, EPI_ISL_2437704, EPI_ISL_2437707, EPI_ISL_2437708, EPI_ISL_2437710, EPI_ISL_2437711, EPI_ISL_2437712, EPI_ISL_2437713, EPI_ISL_2437716, EPI_ISL_2437717, EPI_ISL_2437724, EPI_ISL_2437725, EPI_ISL_2437726, EPI_ISL_2437728, EPI_ISL_2437729, EPI_ISL_2437732, EPI_ISL_2437733, EPI_ISL_2437734, EPI_ISL_2437736, EPI_ISL_2437739, EPI_ISL_2437740, EPI_ISL_2437742, EPI_ISL_2437744, EPI_ISL_2437745, EPI_ISL_2437747, EPI_ISL_2437748, EPI_ISL_2437751, EPI_ISL_2437753, EPI_ISL_2437754, EPI_ISL_2437755, EPI_ISL_2437760, EPI_ISL_2437762, EPI_ISL_2437764, EPI_ISL_2437765, EPI_ISL_2437766, EPI_ISL_2437767, EPI_ISL_2437768, EPI_ISL_2437770, EPI_ISL_2437771, EPI_ISL_2437772, EPI_ISL_2437773, EPI_ISL_2437777, EPI_ISL_2437781, EPI_ISL_2437782, EPI_ISL_2437783, EPI_ISL_2437784, EPI_ISL_2437785, EPI_ISL_2437790, EPI_ISL_2437795, EPI_ISL_2437796, EPI_ISL_2437797, EPI_ISL_2437799, EPI_ISL_2437801, EPI_ISL_2437804, EPI_ISL_2437805, EPI_ISL_2437808, EPI_ISL_2437809, EPI_ISL_2437811, EPI_ISL_2437813, EPI_ISL_2437814, EPI_ISL_2437821, EPI_ISL_2437822, EPI_ISL_2437823, EPI_ISL_2437825, EPI_ISL_2437832, EPI_ISL_2437835, EPI_ISL_2437836, EPI_ISL_2437840, EPI_ISL_2437843, EPI_ISL_2437844, EPI_ISL_2437845, EPI_ISL_2437846, EPI_ISL_2437849, EPI_ISL_2437850, EPI_ISL_2437854, EPI_ISL_2437855, EPI_ISL_2437856, EPI_ISL_2437857, EPI_ISL_2437863, EPI_ISL_2437864, EPI_ISL_2437868, EPI_ISL_2437869, EPI_ISL_2437870, EPI_ISL_2437873, EPI_ISL_2437874, EPI_ISL_2437876, EPI_ISL_2437878, EPI_ISL_2437879, EPI_ISL_2437881, EPI_ISL_2437884, EPI_ISL_2437887, EPI_ISL_2437888, EPI_ISL_2437891, EPI_ISL_2437894, EPI_ISL_2437901, EPI_ISL_2437903, EPI_ISL_2437904, EPI_ISL_2437905, EPI_ISL_2437908, EPI_ISL_2437909, EPI_ISL_2437911, EPI_ISL_2437913, EPI_ISL_2437922, EPI_ISL_2437923, EPI_ISL_2437930, EPI_ISL_2437933, EPI_ISL_2437939, EPI_ISL_2437940, EPI_ISL_2437943, EPI_ISL_2437949, EPI_ISL_2437950, EPI_ISL_2437951, EPI_ISL_2437952, EPI_ISL_2437953, EPI_ISL_2437954, EPI_ISL_2437959, EPI_ISL_2437960, EPI_ISL_2437962, EPI_ISL_2437965, EPI_ISL_2437967, EPI_ISL_2437971, EPI_ISL_2437977, EPI_ISL_2437980, EPI_ISL_2437988, EPI_ISL_2438054, EPI_ISL_2438060, EPI_ISL_2438061, EPI_ISL_2438066, EPI_ISL_2438070, EPI_ISL_2438072, EPI_ISL_2438088, EPI_ISL_2438123, EPI_ISL_2438126, EPI_ISL_2438128, EPI_ISL_2438137, EPI_ISL_2438138, EPI_ISL_2438140, EPI_ISL_2438141, EPI_ISL_2438142, EPI_ISL_2438145, EPI_ISL_2438146, EPI_ISL_2438155, EPI_ISL_2438159, EPI_ISL_2438162, EPI_ISL_2438165, EPI_ISL_2438173, EPI_ISL_2438177, EPI_ISL_2438180, EPI_ISL_2438188, EPI_ISL_2438191, EPI_ISL_2438193, EPI_ISL_2438194, EPI_ISL_2438196, EPI_ISL_2438199, EPI_ISL_2438206, EPI_ISL_2438208, EPI_ISL_2438215, EPI_ISL_2438216, EPI_ISL_2438220, EPI_ISL_2438223, EPI_ISL_2438224, EPI_ISL_2438225, EPI_ISL_2438231, EPI_ISL_2438233, EPI_ISL_2438241, EPI_ISL_2438244, EPI_ISL_2438252, EPI_ISL_2438254, EPI_ISL_2438255, EPI_ISL_2438264, EPI_ISL_2438266, EPI_ISL_2438270, EPI_ISL_2438271, EPI_ISL_2438274, EPI_ISL_2438279, EPI_ISL_2438281, EPI_ISL_2438283, EPI_ISL_2438288, EPI_ISL_2438290, EPI_ISL_2438292, EPI_ISL_2438302, EPI_ISL_2438312, EPI_ISL_2438313, EPI_ISL_2438314, EPI_ISL_2438326, EPI_ISL_2438340, EPI_ISL_2438341, EPI_ISL_2438342, EPI_ISL_2438343, EPI_ISL_2438360, EPI_ISL_2438361, EPI_ISL_2438365, EPI_ISL_2438367, EPI_ISL_2438371, EPI_ISL_2438377, EPI_ISL_2446097, EPI_ISL_2446098, EPI_ISL_2446100, EPI_ISL_2446108, EPI_ISL_2446109, EPI_ISL_2446113, EPI_ISL_2446119, EPI_ISL_2446126, EPI_ISL_2446129, EPI_ISL_2446132, EPI_ISL_2446133, EPI_ISL_2446136, EPI_ISL_2446138, EPI_ISL_2446139, EPI_ISL_2446140, EPI_ISL_2446145, EPI_ISL_2446146, EPI_ISL_2446148, EPI_ISL_2446150, EPI_ISL_2446151, EPI_ISL_2446155, EPI_ISL_2446156, EPI_ISL_2446160, EPI_ISL_2446161, EPI_ISL_2446163, EPI_ISL_2446164, EPI_ISL_2446165, EPI_ISL_2446166, EPI_ISL_2446172, EPI_ISL_2446173, EPI_ISL_2446174, EPI_ISL_2446176, EPI_ISL_2446177, EPI_ISL_2446178, EPI_ISL_2446180, EPI_ISL_2446182, EPI_ISL_2446183, EPI_ISL_2446188, EPI_ISL_2446189, EPI_ISL_2446191, EPI_ISL_2446192, EPI_ISL_2446197, EPI_ISL_2446198, EPI_ISL_2446201, EPI_ISL_2446203, EPI_ISL_2446207, EPI_ISL_2446210, EPI_ISL_2446213, EPI_ISL_2446217, EPI_ISL_2446223, EPI_ISL_2446224, EPI_ISL_2446225, EPI_ISL_2446226, EPI_ISL_2446227, EPI_ISL_2446232, EPI_ISL_2446233, EPI_ISL_2446234, EPI_ISL_2446235, EPI_ISL_2446237, EPI_ISL_2446238, EPI_ISL_2446239, EPI_ISL_2446243, EPI_ISL_2446246, EPI_ISL_2446249, EPI_ISL_2446252, EPI_ISL_2446255, EPI_ISL_2446261, EPI_ISL_2446264, EPI_ISL_2446269, EPI_ISL_2446270, EPI_ISL_2446272, EPI_ISL_2446273, EPI_ISL_2446276, EPI_ISL_2446277, EPI_ISL_2446281, EPI_ISL_2446283, EPI_ISL_2446286, EPI_ISL_2446293, EPI_ISL_2446295, EPI_ISL_2446296, EPI_ISL_2446300, EPI_ISL_2446302, EPI_ISL_2446311, EPI_ISL_2446312, EPI_ISL_2446315, EPI_ISL_2446316, EPI_ISL_2446319, EPI_ISL_2446320, EPI_ISL_2446322, EPI_ISL_2446323, EPI_ISL_2446324, EPI_ISL_2446325, EPI_ISL_2446328, EPI_ISL_2446329, EPI_ISL_2446331, EPI_ISL_2446332, EPI_ISL_2446336, EPI_ISL_2446337, EPI_ISL_2446338, EPI_ISL_2446339, EPI_ISL_2446342, EPI_ISL_2446343, EPI_ISL_2446345, EPI_ISL_2446347, EPI_ISL_2446350, EPI_ISL_2446353, EPI_ISL_2446354, EPI_ISL_2446356, EPI_ISL_2446358, EPI_ISL_2446364, EPI_ISL_2446366, EPI_ISL_2446369, EPI_ISL_2446371, EPI_ISL_2446375, EPI_ISL_2446377, EPI_ISL_2446384, EPI_ISL_2446391, EPI_ISL_2446400, EPI_ISL_2446401, EPI_ISL_2446403, EPI_ISL_2446405, EPI_ISL_2446408, EPI_ISL_2446752, EPI_ISL_2446753, EPI_ISL_2446754, EPI_ISL_2446755, EPI_ISL_2446756, EPI_ISL_2446760, EPI_ISL_2446761, EPI_ISL_2446762, EPI_ISL_2446764, EPI_ISL_2446765, EPI_ISL_2446766, EPI_ISL_2446767, EPI_ISL_2446768, EPI_ISL_2446769, EPI_ISL_2446770, EPI_ISL_2446772, EPI_ISL_2446773, EPI_ISL_2446774, EPI_ISL_2446775, EPI_ISL_2446776, EPI_ISL_2446778, EPI_ISL_2446779, EPI_ISL_2446780, EPI_ISL_2446783, EPI_ISL_2446786, EPI_ISL_2446788, EPI_ISL_2446790, EPI_ISL_2446792, EPI_ISL_2446793, EPI_ISL_2446795, EPI_ISL_2446796, EPI_ISL_2446800, EPI_ISL_2446801, EPI_ISL_2446802, EPI_ISL_2446803, EPI_ISL_2446804, EPI_ISL_2446805, EPI_ISL_2446806, EPI_ISL_2446807, EPI_ISL_2446808, EPI_ISL_2446809, EPI_ISL_2446811, EPI_ISL_2446813, EPI_ISL_2446814, EPI_ISL_2446815, EPI_ISL_2446816, EPI_ISL_2446817, EPI_ISL_2446820, EPI_ISL_2446821, EPI_ISL_2446823, EPI_ISL_2446825, EPI_ISL_2446827, EPI_ISL_2446828, EPI_ISL_2446829, EPI_ISL_2446831, EPI_ISL_2446833, EPI_ISL_2446835, EPI_ISL_2446836, EPI_ISL_2446838, EPI_ISL_2446840, EPI_ISL_2446845, EPI_ISL_2446846, EPI_ISL_2446847, EPI_ISL_2446849, EPI_ISL_2446853, EPI_ISL_2446855, EPI_ISL_2446861, EPI_ISL_2446863, EPI_ISL_2446865, EPI_ISL_2446866, EPI_ISL_2446867, EPI_ISL_2446868, EPI_ISL_2446870, EPI_ISL_2446873, EPI_ISL_2446874, EPI_ISL_2446876, EPI_ISL_2446878, EPI_ISL_2446879, EPI_ISL_2446880, EPI_ISL_2446883, EPI_ISL_2446884, EPI_ISL_2446885, EPI_ISL_2446887, EPI_ISL_2446888, EPI_ISL_2446889, EPI_ISL_2446890, EPI_ISL_2446891, EPI_ISL_2446892, EPI_ISL_2446894, EPI_ISL_2446895, EPI_ISL_2446896, EPI_ISL_2446897, EPI_ISL_2446898, EPI_ISL_2446899, EPI_ISL_2446901, EPI_ISL_2446902, EPI_ISL_2446903, EPI_ISL_2446905, EPI_ISL_2446906, EPI_ISL_2446907, EPI_ISL_2446908, EPI_ISL_2446909, EPI_ISL_2446910, EPI_ISL_2446911, EPI_ISL_2446914, EPI_ISL_2446916, EPI_ISL_2446918, EPI_ISL_2446919, EPI_ISL_2446921, EPI_ISL_2446922, EPI_ISL_2446923, EPI_ISL_2446925, EPI_ISL_2446926, EPI_ISL_2446927, EPI_ISL_2446928, EPI_ISL_2446930, EPI_ISL_2446931, EPI_ISL_2446932, EPI_ISL_2446937, EPI_ISL_2446938, EPI_ISL_2446939, EPI_ISL_2446940, EPI_ISL_2446942, EPI_ISL_2446943, EPI_ISL_2446944, EPI_ISL_2446945, EPI_ISL_2446946, EPI_ISL_2446948, EPI_ISL_2446949, EPI_ISL_2446951, EPI_ISL_2446954, EPI_ISL_2446955, EPI_ISL_2446956, EPI_ISL_2446959, EPI_ISL_2446960, EPI_ISL_2446963, EPI_ISL_2446964, EPI_ISL_2446965, EPI_ISL_2446966, EPI_ISL_2446967, EPI_ISL_2446969, EPI_ISL_2446972, EPI_ISL_2446973, EPI_ISL_2446975, EPI_ISL_2446976, EPI_ISL_2446979, EPI_ISL_2446980, EPI_ISL_2446986, EPI_ISL_2446987, EPI_ISL_2446989, EPI_ISL_2446991, EPI_ISL_2446992, EPI_ISL_2446996, EPI_ISL_2446998, EPI_ISL_2447000, EPI_ISL_2447001, EPI_ISL_2447002, EPI_ISL_2447003, EPI_ISL_2447006, EPI_ISL_2447009, EPI_ISL_2447010, EPI_ISL_2447012, EPI_ISL_2447013, EPI_ISL_2447014, EPI_ISL_2447015, EPI_ISL_2447016, EPI_ISL_2447018, EPI_ISL_2447021, EPI_ISL_2447024, EPI_ISL_2447027, EPI_ISL_2447033, EPI_ISL_2447034, EPI_ISL_2447037, EPI_ISL_2447041, EPI_ISL_2447043, EPI_ISL_2447044, EPI_ISL_2447046, EPI_ISL_2447047, EPI_ISL_2447050, EPI_ISL_2447054, EPI_ISL_2447055, EPI_ISL_2447056, EPI_ISL_2447059, EPI_ISL_2447060, EPI_ISL_2447061, EPI_ISL_2447062, EPI_ISL_2447068, EPI_ISL_2447072, EPI_ISL_2447073, EPI_ISL_2447078, EPI_ISL_2447081, EPI_ISL_2447083, EPI_ISL_2447086, EPI_ISL_2447087, EPI_ISL_2447091, EPI_ISL_2447092, EPI_ISL_2447093, EPI_ISL_2447095, EPI_ISL_2447096, EPI_ISL_2447101, EPI_ISL_2447105, EPI_ISL_2447109, EPI_ISL_2447111, EPI_ISL_2447113, EPI_ISL_2447115, EPI_ISL_2447117, EPI_ISL_2447119, EPI_ISL_2447122, EPI_ISL_2447124, EPI_ISL_2447131, EPI_ISL_2447133, EPI_ISL_2447134, EPI_ISL_2447135, EPI_ISL_2447137, EPI_ISL_2447138, EPI_ISL_2447139, EPI_ISL_2447140, EPI_ISL_2447144, EPI_ISL_2447145, EPI_ISL_2447149, EPI_ISL_2447150, EPI_ISL_2447156, EPI_ISL_2447158, EPI_ISL_2447159, EPI_ISL_2447161, EPI_ISL_2447163, EPI_ISL_2447166, EPI_ISL_2447170, EPI_ISL_2447171, EPI_ISL_2447174, EPI_ISL_2447175, EPI_ISL_2447179, EPI_ISL_2447181, EPI_ISL_2447182, EPI_ISL_2447185, EPI_ISL_2447186, EPI_ISL_2447187, EPI_ISL_2447190, EPI_ISL_2447194, EPI_ISL_2447196, EPI_ISL_2447203, EPI_ISL_2447206, EPI_ISL_2447207, EPI_ISL_2447210, EPI_ISL_2447215, EPI_ISL_2447216, EPI_ISL_2447217, EPI_ISL_2447226, EPI_ISL_2447229, EPI_ISL_2447230, EPI_ISL_2447231, EPI_ISL_2447363, EPI_ISL_2447369, EPI_ISL_2447371, EPI_ISL_2447372, EPI_ISL_2447374, EPI_ISL_2447376, EPI_ISL_2447389, EPI_ISL_2447390, EPI_ISL_2447392, EPI_ISL_2447395, EPI_ISL_2447398, EPI_ISL_2447399, EPI_ISL_2447400, EPI_ISL_2447411, EPI_ISL_2447413, EPI_ISL_2447414, EPI_ISL_2447415, EPI_ISL_2447417, EPI_ISL_2447418, EPI_ISL_2447423, EPI_ISL_2447424, EPI_ISL_2447430, EPI_ISL_2447432, EPI_ISL_2447434, EPI_ISL_2447436, EPI_ISL_2447437, EPI_ISL_2447448, EPI_ISL_2447450, EPI_ISL_2447456, EPI_ISL_2447457, EPI_ISL_2447458, EPI_ISL_2447459, EPI_ISL_2447461, EPI_ISL_2447462, EPI_ISL_2447464, EPI_ISL_2447468, EPI_ISL_2447473, EPI_ISL_2447474, EPI_ISL_2447476, EPI_ISL_2447477, EPI_ISL_2447478, EPI_ISL_2447479, EPI_ISL_2447485, EPI_ISL_2447492, EPI_ISL_2447497, EPI_ISL_2447499, EPI_ISL_2447501, EPI_ISL_2447508, EPI_ISL_2447511, EPI_ISL_2447523, EPI_ISL_2447524, EPI_ISL_2447528, EPI_ISL_2447533, EPI_ISL_2447536, EPI_ISL_2447542, EPI_ISL_2447544, EPI_ISL_2447552, EPI_ISL_2447557, EPI_ISL_2447562, EPI_ISL_2447566, EPI_ISL_2447572, EPI_ISL_2447575, EPI_ISL_2447582, EPI_ISL_2447587, EPI_ISL_2447598, EPI_ISL_2447599, EPI_ISL_2447614, EPI_ISL_2447621, EPI_ISL_2447643, EPI_ISL_2447647, EPI_ISL_2447657, EPI_ISL_2447665, EPI_ISL_2447691, EPI_ISL_2447698, EPI_ISL_2447699, EPI_ISL_2447702, EPI_ISL_2447711, EPI_ISL_2447732, EPI_ISL_2447733, EPI_ISL_2447735, EPI_ISL_2447737, EPI_ISL_2447741, EPI_ISL_2447742, EPI_ISL_2447743, EPI_ISL_2447750, EPI_ISL_2447754, EPI_ISL_2447770, EPI_ISL_2456006, EPI_ISL_2456014, EPI_ISL_2456015, EPI_ISL_2456016, EPI_ISL_2456019, EPI_ISL_2456020, EPI_ISL_2456022, EPI_ISL_2456023, EPI_ISL_2456024, EPI_ISL_2456026, EPI_ISL_2456028, EPI_ISL_2456029, EPI_ISL_2456030, EPI_ISL_2456033, EPI_ISL_2456036, EPI_ISL_2456037, EPI_ISL_2456038, EPI_ISL_2456040, EPI_ISL_2456044, EPI_ISL_2456048, EPI_ISL_2456051, EPI_ISL_2456053, EPI_ISL_2456059, EPI_ISL_2456062, EPI_ISL_2456064, EPI_ISL_2456066, EPI_ISL_2456069, EPI_ISL_2456077, EPI_ISL_2456078, EPI_ISL_2456084, EPI_ISL_2456085, EPI_ISL_2456086, EPI_ISL_2456087, EPI_ISL_2456088, EPI_ISL_2456093, EPI_ISL_2456098, EPI_ISL_2456102, EPI_ISL_2456104, EPI_ISL_2456106, EPI_ISL_2456107, EPI_ISL_2456108, EPI_ISL_2456116, EPI_ISL_2456121, EPI_ISL_2456122, EPI_ISL_2456124, EPI_ISL_2456127, EPI_ISL_2456133, EPI_ISL_2456137, EPI_ISL_2456140, EPI_ISL_2456154, EPI_ISL_2456167, EPI_ISL_2456171, EPI_ISL_2456183, EPI_ISL_2456186, EPI_ISL_2456189, EPI_ISL_2456190, EPI_ISL_2456195, EPI_ISL_2456207, EPI_ISL_2456213, EPI_ISL_2456220, EPI_ISL_2456221, EPI_ISL_2456226, EPI_ISL_2456230, EPI_ISL_2456246, EPI_ISL_2456248, EPI_ISL_2456256, EPI_ISL_2456259, EPI_ISL_2456260, EPI_ISL_2456265, EPI_ISL_2456266, EPI_ISL_2456271, EPI_ISL_2456281, EPI_ISL_2456286, EPI_ISL_2456290, EPI_ISL_2456293, EPI_ISL_2456295, EPI_ISL_2456296, EPI_ISL_2456298, EPI_ISL_2456301, EPI_ISL_2456302, EPI_ISL_2456305, EPI_ISL_2456308, EPI_ISL_2456309, EPI_ISL_2456312, EPI_ISL_2456315, EPI_ISL_2456317, EPI_ISL_2456319, EPI_ISL_2456320, EPI_ISL_2456333, EPI_ISL_2456339, EPI_ISL_2456341, EPI_ISL_2456342, EPI_ISL_2456344, EPI_ISL_2456345, EPI_ISL_2456347, EPI_ISL_2456349, EPI_ISL_2456354, EPI_ISL_2456364, EPI_ISL_2456366, EPI_ISL_2456371, EPI_ISL_2456375, EPI_ISL_2456381, EPI_ISL_2456390, EPI_ISL_2456402, EPI_ISL_2456408, EPI_ISL_2456409, EPI_ISL_2456411, EPI_ISL_2456414, EPI_ISL_2456421, EPI_ISL_2456431, EPI_ISL_2456432, EPI_ISL_2456433, EPI_ISL_2456437, EPI_ISL_2456452, EPI_ISL_2456454, EPI_ISL_2456459, EPI_ISL_2456503, EPI_ISL_2456504, EPI_ISL_2456518, EPI_ISL_2456520, EPI_ISL_2456536, EPI_ISL_2456548, EPI_ISL_2456552, EPI_ISL_2456554, EPI_ISL_2456555, EPI_ISL_2456564, EPI_ISL_2456566, EPI_ISL_2456573, EPI_ISL_2456582, EPI_ISL_2456585, EPI_ISL_2456594, EPI_ISL_2456610, EPI_ISL_2456619, EPI_ISL_2456621, EPI_ISL_2456626, EPI_ISL_2456637, EPI_ISL_2456664, EPI_ISL_2456677, EPI_ISL_2456682, EPI_ISL_2456747, EPI_ISL_2456749, EPI_ISL_2456755, EPI_ISL_2456763, EPI_ISL_2456778, EPI_ISL_2456793, EPI_ISL_2456799, EPI_ISL_2456814, EPI_ISL_2456823, EPI_ISL_2456827, EPI_ISL_2456828, EPI_ISL_2456831, EPI_ISL_2456832, EPI_ISL_2456846, EPI_ISL_2456860, EPI_ISL_2456870, EPI_ISL_2456877, EPI_ISL_2456883, EPI_ISL_2456898, EPI_ISL_2456899, EPI_ISL_2456914, EPI_ISL_2456915, EPI_ISL_2456925, EPI_ISL_2456947, EPI_ISL_2456965, EPI_ISL_2456976, EPI_ISL_2456981, EPI_ISL_2456990, EPI_ISL_2457001, EPI_ISL_2457002, EPI_ISL_2457009, EPI_ISL_2457024, EPI_ISL_2457029, EPI_ISL_2457036, EPI_ISL_2457059, EPI_ISL_2457065, EPI_ISL_2457106, EPI_ISL_2457107, EPI_ISL_2457108, EPI_ISL_2457115, EPI_ISL_2457129, EPI_ISL_2457887, EPI_ISL_2468099, EPI_ISL_2468362, EPI_ISL_2468458, EPI_ISL_2468480, EPI_ISL_2468517, EPI_ISL_2468615, EPI_ISL_2468732, EPI_ISL_2469259, EPI_ISL_2469272, EPI_ISL_2469341, EPI_ISL_2469388, EPI_ISL_2469410, EPI_ISL_2469416, EPI_ISL_2469431, EPI_ISL_2469435, EPI_ISL_2469439, EPI_ISL_2469442, EPI_ISL_2469463, EPI_ISL_2485299, EPI_ISL_2485351, EPI_ISL_2485370, EPI_ISL_2485372, EPI_ISL_2485380, EPI_ISL_2485396, EPI_ISL_2485435, EPI_ISL_2485442, EPI_ISL_2485540, EPI_ISL_2485564, EPI_ISL_2485732, EPI_ISL_2485741, EPI_ISL_2485801, EPI_ISL_2485814, EPI_ISL_2485819, EPI_ISL_2485869, EPI_ISL_2486023, EPI_ISL_2486908, EPI_ISL_2486925, EPI_ISL_2486976, EPI_ISL_2487036, EPI_ISL_2487077, EPI_ISL_2487113, EPI_ISL_2487121, EPI_ISL_2487153, EPI_ISL_2487173, EPI_ISL_2487210, EPI_ISL_2488315, EPI_ISL_2511705, EPI_ISL_2511716, EPI_ISL_2511890, EPI_ISL_2514410, EPI_ISL_2514412, EPI_ISL_2514415, EPI_ISL_2514416, EPI_ISL_2514418, EPI_ISL_2514419, EPI_ISL_2514421, EPI_ISL_2514422, EPI_ISL_2514426, EPI_ISL_2514427, EPI_ISL_2514437, EPI_ISL_2514439, EPI_ISL_2514441, EPI_ISL_2514442, EPI_ISL_2514448, EPI_ISL_2514450, EPI_ISL_2514451, EPI_ISL_2514452, EPI_ISL_2514453, EPI_ISL_2514455, EPI_ISL_2514458, EPI_ISL_2514459, EPI_ISL_2514460, EPI_ISL_2514463, EPI_ISL_2514465, EPI_ISL_2514467, EPI_ISL_2514469, EPI_ISL_2514475, EPI_ISL_2514476, EPI_ISL_2514478, EPI_ISL_2514479, EPI_ISL_2514483, EPI_ISL_2514486, EPI_ISL_2514489, EPI_ISL_2514492, EPI_ISL_2514493, EPI_ISL_2514494, EPI_ISL_2514497, EPI_ISL_2514500, EPI_ISL_2514502, EPI_ISL_2514505, EPI_ISL_2514507, EPI_ISL_2514509, EPI_ISL_2514510, EPI_ISL_2514512, EPI_ISL_2514515, EPI_ISL_2514517, EPI_ISL_2514518, EPI_ISL_2514520, EPI_ISL_2514521, EPI_ISL_2514523, EPI_ISL_2514525, EPI_ISL_2514526, EPI_ISL_2514529, EPI_ISL_2514530, EPI_ISL_2514531, EPI_ISL_2514532, EPI_ISL_2514536, EPI_ISL_2514537, EPI_ISL_2514539, EPI_ISL_2514544, EPI_ISL_2514548, EPI_ISL_2514550, EPI_ISL_2514552, EPI_ISL_2514553, EPI_ISL_2514554, EPI_ISL_2514556, EPI_ISL_2514559, EPI_ISL_2514560, EPI_ISL_2514561, EPI_ISL_2514562, EPI_ISL_2514563, EPI_ISL_2514567, EPI_ISL_2514570, EPI_ISL_2514571, EPI_ISL_2514574, EPI_ISL_2514575, EPI_ISL_2514577, EPI_ISL_2514580, EPI_ISL_2514582, EPI_ISL_2514590, EPI_ISL_2514591, EPI_ISL_2514592, EPI_ISL_2514598, EPI_ISL_2514599, EPI_ISL_2514601, EPI_ISL_2514606, EPI_ISL_2514608, EPI_ISL_2514609, EPI_ISL_2514610, EPI_ISL_2514611, EPI_ISL_2514612, EPI_ISL_2514613, EPI_ISL_2514615, EPI_ISL_2514616, EPI_ISL_2514617, EPI_ISL_2514622, EPI_ISL_2514623, EPI_ISL_2514626, EPI_ISL_2514630, EPI_ISL_2514631, EPI_ISL_2514633, EPI_ISL_2514634, EPI_ISL_2514638, EPI_ISL_2514639, EPI_ISL_2514640, EPI_ISL_2514645, EPI_ISL_2514646, EPI_ISL_2514654, EPI_ISL_2514655, EPI_ISL_2514660, EPI_ISL_2514666, EPI_ISL_2514667, EPI_ISL_2514668, EPI_ISL_2514669, EPI_ISL_2514671, EPI_ISL_2514678, EPI_ISL_2514680, EPI_ISL_2514683, EPI_ISL_2514684, EPI_ISL_2514685, EPI_ISL_2514687, EPI_ISL_2514688, EPI_ISL_2514692, EPI_ISL_2514693, EPI_ISL_2514694, EPI_ISL_2514697, EPI_ISL_2514698, EPI_ISL_2514702, EPI_ISL_2514703, EPI_ISL_2514711, EPI_ISL_2514712, EPI_ISL_2514715, EPI_ISL_2514716, EPI_ISL_2514717, EPI_ISL_2514718, EPI_ISL_2514719, EPI_ISL_2514725, EPI_ISL_2514726, EPI_ISL_2539178, EPI_ISL_2539179, EPI_ISL_2539182, EPI_ISL_2539193, EPI_ISL_2539194, EPI_ISL_2539195, EPI_ISL_2539200, EPI_ISL_2539203, EPI_ISL_2539207, EPI_ISL_2539209, EPI_ISL_2539211, EPI_ISL_2539214, EPI_ISL_2539215, EPI_ISL_2539223, EPI_ISL_2539226, EPI_ISL_2539231, EPI_ISL_2539243, EPI_ISL_2539244, EPI_ISL_2539246, EPI_ISL_2539247, EPI_ISL_2539249, EPI_ISL_2539250, EPI_ISL_2539251, EPI_ISL_2539257, EPI_ISL_2539262, EPI_ISL_2539263, EPI_ISL_2539265, EPI_ISL_2539271, EPI_ISL_2539272, EPI_ISL_2539277, EPI_ISL_2539279, EPI_ISL_2539280, EPI_ISL_2539282, EPI_ISL_2539287, EPI_ISL_2539297, EPI_ISL_2539298, EPI_ISL_2539300, EPI_ISL_2539303, EPI_ISL_2539307, EPI_ISL_2539309, EPI_ISL_2539316, EPI_ISL_2539320, EPI_ISL_2539321, EPI_ISL_2539322, EPI_ISL_2539325, EPI_ISL_2539327, EPI_ISL_2539329, EPI_ISL_2539331, EPI_ISL_2539336, EPI_ISL_2539338, EPI_ISL_2539343, EPI_ISL_2539344, EPI_ISL_2539346, EPI_ISL_2539350, EPI_ISL_2539356, EPI_ISL_2539357, EPI_ISL_2539358, EPI_ISL_2539359, EPI_ISL_2539363, EPI_ISL_2539366, EPI_ISL_2539369, EPI_ISL_2539374, EPI_ISL_2539379, EPI_ISL_2539380, EPI_ISL_2539383, EPI_ISL_2539384, EPI_ISL_2539390, EPI_ISL_2539392, EPI_ISL_2539395, EPI_ISL_2539396, EPI_ISL_2539403, EPI_ISL_2539404, EPI_ISL_2539405, EPI_ISL_2539406, EPI_ISL_2539408, EPI_ISL_2539420, EPI_ISL_2539428, EPI_ISL_2539431, EPI_ISL_2539435, EPI_ISL_2539438, EPI_ISL_2539441, EPI_ISL_2539448, EPI_ISL_2539451, EPI_ISL_2539453, EPI_ISL_2539459, EPI_ISL_2539461, EPI_ISL_2539481, EPI_ISL_2539483, EPI_ISL_2539484, EPI_ISL_2539495, EPI_ISL_2539505, EPI_ISL_2539509, EPI_ISL_2539512, EPI_ISL_2539516, EPI_ISL_2539518, EPI_ISL_2539524, EPI_ISL_2539525, EPI_ISL_2539531, EPI_ISL_2539536, EPI_ISL_2539538, EPI_ISL_2539540, EPI_ISL_2539543, EPI_ISL_2539545, EPI_ISL_2539547, EPI_ISL_2539549, EPI_ISL_2539555, EPI_ISL_2539561, EPI_ISL_2539563, EPI_ISL_2539574, EPI_ISL_2539579, EPI_ISL_2539583, EPI_ISL_2539584, EPI_ISL_2539588, EPI_ISL_2539591, EPI_ISL_2539611, EPI_ISL_2539624, EPI_ISL_2539630, EPI_ISL_2539636, EPI_ISL_2539638, EPI_ISL_2539640, EPI_ISL_2539642, EPI_ISL_2539645, EPI_ISL_2539647, EPI_ISL_2539661, EPI_ISL_2539664, EPI_ISL_2539665, EPI_ISL_2539677, EPI_ISL_2539690, EPI_ISL_2539693, EPI_ISL_2539697, EPI_ISL_2539702, EPI_ISL_2539712, EPI_ISL_2539715, EPI_ISL_2539721, EPI_ISL_2539723, EPI_ISL_2539725, EPI_ISL_2539728, EPI_ISL_2540704, EPI_ISL_2540766, EPI_ISL_2641715

| | | | |
|---|---|---|---|
| see above | Lighthouse Lab in Milton Keynes | Wellcome Sanger Institute for the COVID-19 Genomics UK (COG-UK) Consortium | Cordelia Langford; David K. Jackson; Dominic Kwiatkowski; Ewan Harrison; Ian Johnston; Jeffrey Barrett; John Sillitoe on behalf of the Wellcome Sanger Institute COVID-19 Surveillance Team; Roberto Amato; Sonia Goncalves; The Lighthouse Lab in Milton Keynes and Alex Alderton |

EPI_ISL_2436221, EPI_ISL_2436223, EPI_ISL_2436226, EPI_ISL_2436230, EPI_ISL_2436244, EPI_ISL_2436247, EPI_ISL_2436259, EPI_ISL_2436266, EPI_ISL_2436268, EPI_ISL_2436276, EPI_ISL_2436288, EPI_ISL_2436290, EPI_ISL_2436312, EPI_ISL_2436323, EPI_ISL_2436328, EPI_ISL_2436341, EPI_ISL_2436345, EPI_ISL_2436363, EPI_ISL_2436366, EPI_ISL_2436416, EPI_ISL_2436418, EPI_ISL_2436459, EPI_ISL_2436467, EPI_ISL_2436490, EPI_ISL_2436499, EPI_ISL_2436519, EPI_ISL_2436526, EPI_ISL_2436537, EPI_ISL_2436539, EPI_ISL_2436544, EPI_ISL_2436548, EPI_ISL_2436549, EPI_ISL_2436559, EPI_ISL_2436564, EPI_ISL_2436567, EPI_ISL_2437282, EPI_ISL_2437283, EPI_ISL_2437291, EPI_ISL_2437294, EPI_ISL_2437296, EPI_ISL_2437305, EPI_ISL_2437312, EPI_ISL_2437323, EPI_ISL_2437337, EPI_ISL_2437344, EPI_ISL_2437351, EPI_ISL_2437353, EPI_ISL_2437378, EPI_ISL_2437389, EPI_ISL_2437409, EPI_ISL_2437422, EPI_ISL_2437424, EPI_ISL_2437440, EPI_ISL_2437442, EPI_ISL_2437443, EPI_ISL_2437462, EPI_ISL_2437473, EPI_ISL_2437488, EPI_ISL_2437498, EPI_ISL_2437540, EPI_ISL_2437545, EPI_ISL_2437565, EPI_ISL_2437578, EPI_ISL_2437588, EPI_ISL_2437589, EPI_ISL_2437590, EPI_ISL_2437594, EPI_ISL_2437611, EPI_ISL_2447367, EPI_ISL_2447382, EPI_ISL_2447403, EPI_ISL_2447404, EPI_ISL_2447406, EPI_ISL_2447410, EPI_ISL_2447419, EPI_ISL_2447433, EPI_ISL_2447435, EPI_ISL_2447440, EPI_ISL_2447441, EPI_ISL_2447453, EPI_ISL_2447455, EPI_ISL_2447463, EPI_ISL_2447465, EPI_ISL_2447469, EPI_ISL_2447471, EPI_ISL_2447489, EPI_ISL_2447494, EPI_ISL_2447503, EPI_ISL_2447512, EPI_ISL_2447521, EPI_ISL_2447522, EPI_ISL_2447537, EPI_ISL_2447554, EPI_ISL_2447558, EPI_ISL_2447574, EPI_ISL_2447583, EPI_ISL_2447592, EPI_ISL_2447624, EPI_ISL_2447629, EPI_ISL_2447654, EPI_ISL_2447661, EPI_ISL_2447663, EPI_ISL_2447672, EPI_ISL_2447679, EPI_ISL_2447681, EPI_ISL_2447704, EPI_ISL_2447712, EPI_ISL_2447715, EPI_ISL_2447716, EPI_ISL_2447718, EPI_ISL_2447720, EPI_ISL_2447721, EPI_ISL_2447723, EPI_ISL_2447734, EPI_ISL_2447739, EPI_ISL_2447744, EPI_ISL_2457885, EPI_ISL_2513806, EPI_ISL_2513815, EPI_ISL_2513818, EPI_ISL_2513819, EPI_ISL_2513824, EPI_ISL_2513831, EPI_ISL_2513843, EPI_ISL_2513845, EPI_ISL_2513853, EPI_ISL_2513863, EPI_ISL_2513866, EPI_ISL_2513867, EPI_ISL_2513870, EPI_ISL_2513872, EPI_ISL_2513876, EPI_ISL_2513877, EPI_ISL_2513885, EPI_ISL_2513887, EPI_ISL_2513890, EPI_ISL_2513906, EPI_ISL_2513917, EPI_ISL_2513934, EPI_ISL_2513941, EPI_ISL_2513944, EPI_ISL_2513947, EPI_ISL_2513969, EPI_ISL_2513988, EPI_ISL_2513993, EPI_ISL_2513996, EPI_ISL_2514012, EPI_ISL_2514013, EPI_ISL_2514017, EPI_ISL_2514020, EPI_ISL_2514034, EPI_ISL_2514040, EPI_ISL_2514043, EPI_ISL_2514044, EPI_ISL_2514051, EPI_ISL_2514054, EPI_ISL_2514064, EPI_ISL_2514069, EPI_ISL_2514930, EPI_ISL_2514931, EPI_ISL_2514932, EPI_ISL_2514936, EPI_ISL_2514940, EPI_ISL_2514943, EPI_ISL_2514944, EPI_ISL_2514949, EPI_ISL_2514952, EPI_ISL_2514953, EPI_ISL_2514957, EPI_ISL_2514963, EPI_ISL_2514970, EPI_ISL_2514980, EPI_ISL_2514981, EPI_ISL_2514987, EPI_ISL_2514990, EPI_ISL_2514994, EPI_ISL_2514999, EPI_ISL_2515001, EPI_ISL_2515006, EPI_ISL_2515007, EPI_ISL_2515008, EPI_ISL_2515010, EPI_ISL_2515011, EPI_ISL_2515017, EPI_ISL_2515018, EPI_ISL_2515021, EPI_ISL_2515022, EPI_ISL_2515028, EPI_ISL_2515032, EPI_ISL_2515036, EPI_ISL_2515043, EPI_ISL_2515053, EPI_ISL_2515055, EPI_ISL_2515056, EPI_ISL_2515064, EPI_ISL_2515068, EPI_ISL_2515069, EPI_ISL_2515070, EPI_ISL_2515077, EPI_ISL_2515078, EPI_ISL_2515080, EPI_ISL_2515083, EPI_ISL_2515086, EPI_ISL_2515089, EPI_ISL_2515092, EPI_ISL_2515093, EPI_ISL_2515095, EPI_ISL_2515097, EPI_ISL_2515101, EPI_ISL_2515102, EPI_ISL_2515110, EPI_ISL_2515113, EPI_ISL_2515115, EPI_ISL_2515116, EPI_ISL_2913942

| | | | |
|---|---|---|---|
| see above | Lighthouse Laboratory Plymouth | Wellcome Sanger Institute for the COVID-19 Genomics UK (COG-UK) Consortium | Cordelia Langford; David K. Jackson; Dominic Kwiatkowski; Ewan Harrison; Ian Johnston; Jeffrey Barrett; John Sillitoe on behalf of the Wellcome Sanger Institute COVID-19 Surveillance Team; Lighthouse Laboratory Plymouth and Alex Alderton; Roberto Amato; Sonia Goncalves |

EPI_ISL_2518310, EPI_ISL_2518311, EPI_ISL_2518312, EPI_ISL_2518313, EPI_ISL_2518314, EPI_ISL_2518315, EPI_ISL_2518316, EPI_ISL_2518317, EPI_ISL_2518318, EPI_ISL_2518319, EPI_ISL_2518320, EPI_ISL_2518321, EPI_ISL_2518322, EPI_ISL_2518323, EPI_ISL_2518324, EPI_ISL_2518325, EPI_ISL_2518326, EPI_ISL_2518327, EPI_ISL_2518328, EPI_ISL_2518329, EPI_ISL_2518330, EPI_ISL_2518331, EPI_ISL_2518332, EPI_ISL_2518334, EPI_ISL_2518335, EPI_ISL_2518336, EPI_ISL_2518337, EPI_ISL_2518338, EPI_ISL_2518339, EPI_ISL_2518340, EPI_ISL_2518341, EPI_ISL_2518342, EPI_ISL_2518343, EPI_ISL_2518344, EPI_ISL_2518345, EPI_ISL_2518346, EPI_ISL_2518347, EPI_ISL_2518348, EPI_ISL_2518349, EPI_ISL_2518350, EPI_ISL_2518351, EPI_ISL_2518352, EPI_ISL_2518353, EPI_ISL_2518354, EPI_ISL_2518355, EPI_ISL_2518356, EPI_ISL_2518357, EPI_ISL_2518358, EPI_ISL_2518359, EPI_ISL_2518360, EPI_ISL_2518361, EPI_ISL_2518362, EPI_ISL_2518363, EPI_ISL_2518364, EPI_ISL_2518365, EPI_ISL_2518366, EPI_ISL_2518367, EPI_ISL_2518369, EPI_ISL_2518370, EPI_ISL_2518371, EPI_ISL_2518372, EPI_ISL_2518373, EPI_ISL_2518374, EPI_ISL_2518375, EPI_ISL_2518376, EPI_ISL_2518377, EPI_ISL_2518378, EPI_ISL_2518379, EPI_ISL_2518380, EPI_ISL_2518381, EPI_ISL_2518382, EPI_ISL_2518383, EPI_ISL_2518384, EPI_ISL_2518385, EPI_ISL_2518386, EPI_ISL_2518387, EPI_ISL_2518388, EPI_ISL_2518389, EPI_ISL_2518390, EPI_ISL_2518391, EPI_ISL_2518392, EPI_ISL_2518393, EPI_ISL_2518394, EPI_ISL_2518395, EPI_ISL_2518396, EPI_ISL_2518397, EPI_ISL_2518398, EPI_ISL_2518399, EPI_ISL_2518400, EPI_ISL_2518401, EPI_ISL_2518402, EPI_ISL_2518403, EPI_ISL_2518404, EPI_ISL_2518405, EPI_ISL_2518406, EPI_ISL_2518407, EPI_ISL_2518408, EPI_ISL_2518409, EPI_ISL_2518410, EPI_ISL_2518411, EPI_ISL_2518412, EPI_ISL_2518413, EPI_ISL_2518414, EPI_ISL_2518415, EPI_ISL_2518416, EPI_ISL_2518417, EPI_ISL_2518418, EPI_ISL_2518419, EPI_ISL_2518420, EPI_ISL_2518421, EPI_ISL_2518422, EPI_ISL_2518423, EPI_ISL_2518424, EPI_ISL_2518425, EPI_ISL_2518426, EPI_ISL_2518427, EPI_ISL_2518428, EPI_ISL_2518429, EPI_ISL_2518430, EPI_ISL_2518431, EPI_ISL_2518432, EPI_ISL_2518433, EPI_ISL_2518434, EPI_ISL_2518435, EPI_ISL_2518436, EPI_ISL_2518437, EPI_ISL_2518438, EPI_ISL_2518439, EPI_ISL_2518440, EPI_ISL_2518441, EPI_ISL_2518442, EPI_ISL_2518443, EPI_ISL_2518444, EPI_ISL_2518445, EPI_ISL_2518446, EPI_ISL_2518447, EPI_ISL_2518448, EPI_ISL_2518449, EPI_ISL_2518450, EPI_ISL_2518451, EPI_ISL_2518452, EPI_ISL_2518453, EPI_ISL_2518454, EPI_ISL_2518455, EPI_ISL_2518457, EPI_ISL_2518458, EPI_ISL_2518459, EPI_ISL_2518461, EPI_ISL_2518462, EPI_ISL_2518464, EPI_ISL_2518465, EPI_ISL_2518466, EPI_ISL_2518467, EPI_ISL_2518468, EPI_ISL_2518469, EPI_ISL_2518470, EPI_ISL_2518471, EPI_ISL_2518472, EPI_ISL_2518473, EPI_ISL_2518474, EPI_ISL_2518475, EPI_ISL_2518476, EPI_ISL_2518477, EPI_ISL_2518478, EPI_ISL_2518479, EPI_ISL_2518480, EPI_ISL_2518481, EPI_ISL_2518483, EPI_ISL_2518484, EPI_ISL_2518485, EPI_ISL_2518486, EPI_ISL_2518487, EPI_ISL_2518488, EPI_ISL_2518489, EPI_ISL_2518490, EPI_ISL_2518491, EPI_ISL_2518492, EPI_ISL_2518493, EPI_ISL_2518494, EPI_ISL_2518495, EPI_ISL_2518496, EPI_ISL_2518497, EPI_ISL_2518498, EPI_ISL_2518499, EPI_ISL_2518500, EPI_ISL_2518501, EPI_ISL_2518502, EPI_ISL_2518503, EPI_ISL_2518504, EPI_ISL_2518506, EPI_ISL_2518507, EPI_ISL_2518508, EPI_ISL_2518509, EPI_ISL_2518510, EPI_ISL_2518511, EPI_ISL_2518512, EPI_ISL_2518513, EPI_ISL_2518514, EPI_ISL_2518515, EPI_ISL_2518516, EPI_ISL_2518517, EPI_ISL_2518518, EPI_ISL_2518519, EPI_ISL_2518520, EPI_ISL_2518522, EPI_ISL_2518523, EPI_ISL_2518524, EPI_ISL_2518525, EPI_ISL_2518526, EPI_ISL_2518528, EPI_ISL_2518529, EPI_ISL_2518530, EPI_ISL_2518531, EPI_ISL_2518532, EPI_ISL_2518533, EPI_ISL_2518534, EPI_ISL_2518535, EPI_ISL_2518536, EPI_ISL_2518537, EPI_ISL_2518538, EPI_ISL_2518539, EPI_ISL_2518540, EPI_ISL_2518541, EPI_ISL_2518542, EPI_ISL_2518543, EPI_ISL_2518544, EPI_ISL_2518545, EPI_ISL_2518546, EPI_ISL_2518548, EPI_ISL_2518549, EPI_ISL_2518550, EPI_ISL_2518551, EPI_ISL_2518552, EPI_ISL_2518553, EPI_ISL_2518554, EPI_ISL_2518555, EPI_ISL_2518556, EPI_ISL_2518557, EPI_ISL_2518558, EPI_ISL_2518559, EPI_ISL_2518560, EPI_ISL_2518561, EPI_ISL_2518562, EPI_ISL_2518563, EPI_ISL_2518564, EPI_ISL_2518565, EPI_ISL_2518566, EPI_ISL_2518567, EPI_ISL_2518568, EPI_ISL_2518569, EPI_ISL_2518570, EPI_ISL_2518571, EPI_ISL_2518572, EPI_ISL_2518573, EPI_ISL_2518574, EPI_ISL_2518575, EPI_ISL_2518576, EPI_ISL_2518577, EPI_ISL_2518578, EPI_ISL_2518579, EPI_ISL_2518580, EPI_ISL_2518581, EPI_ISL_2518582, EPI_ISL_2518583, EPI_ISL_2518584, EPI_ISL_2518585, EPI_ISL_2518586, EPI_ISL_2518587, EPI_ISL_2518588, EPI_ISL_2518589, EPI_ISL_2518590, EPI_ISL_2518591, EPI_ISL_2518592, EPI_ISL_2518593, EPI_ISL_2518594, EPI_ISL_2518595, EPI_ISL_2518597, EPI_ISL_2518598, EPI_ISL_2518599, EPI_ISL_2518600, EPI_ISL_2518601, EPI_ISL_2518602, EPI_ISL_2518603, EPI_ISL_2518604, EPI_ISL_2518605, EPI_ISL_2518606, EPI_ISL_2518607, EPI_ISL_2518608, EPI_ISL_2518610, EPI_ISL_2518611, EPI_ISL_2518612, EPI_ISL_2518614, EPI_ISL_2518615, EPI_ISL_2518616, EPI_ISL_2518617, EPI_ISL_2518618, EPI_ISL_2518619, EPI_ISL_2518620, EPI_ISL_2518621, EPI_ISL_2518622, EPI_ISL_2518623, EPI_ISL_2518624, EPI_ISL_2518626, EPI_ISL_2518627, EPI_ISL_2518628, EPI_ISL_2518629, EPI_ISL_2518630, EPI_ISL_2518631, EPI_ISL_2518632, EPI_ISL_2518633, EPI_ISL_2518634, EPI_ISL_2518635, EPI_ISL_2518636, EPI_ISL_2518637, EPI_ISL_2518638, EPI_ISL_2518639, EPI_ISL_2518640, EPI_ISL_2518641, EPI_ISL_2518642, EPI_ISL_2518643, EPI_ISL_2518644, EPI_ISL_2518645, EPI_ISL_2518646, EPI_ISL_2518647, EPI_ISL_2518648, EPI_ISL_2518649, EPI_ISL_2518650, EPI_ISL_2518651, EPI_ISL_2518652, EPI_ISL_2518653, EPI_ISL_2518654, EPI_ISL_2518655, EPI_ISL_2518656, EPI_ISL_2518657, EPI_ISL_2518658, EPI_ISL_2518659, EPI_ISL_2518660, EPI_ISL_2518662, EPI_ISL_2518663, EPI_ISL_2518664, EPI_ISL_2518665, EPI_ISL_2518666, EPI_ISL_2518667, EPI_ISL_2518668, EPI_ISL_2518669, EPI_ISL_2518670, EPI_ISL_2518671, EPI_ISL_2518672, EPI_ISL_2518673, EPI_ISL_2518674, EPI_ISL_2518675, EPI_ISL_2518676, EPI_ISL_2518678, EPI_ISL_2518679, EPI_ISL_2518680, EPI_ISL_2518681, EPI_ISL_2518682, EPI_ISL_2518683, EPI_ISL_2518684, EPI_ISL_2518685, EPI_ISL_2518686, EPI_ISL_2518687, EPI_ISL_2518688, EPI_ISL_2518689, EPI_ISL_2518690, EPI_ISL_2518691, EPI_ISL_2518692, EPI_ISL_2518693, EPI_ISL_2518695, EPI_ISL_2518696, EPI_ISL_2518697, EPI_ISL_2518698, EPI_ISL_2518699, EPI_ISL_2518700, EPI_ISL_2518701, EPI_ISL_2518702, EPI_ISL_2518703, EPI_ISL_2518704, EPI_ISL_2518705, EPI_ISL_2518706, EPI_ISL_2518707, EPI_ISL_2518708, EPI_ISL_2518709, EPI_ISL_2518710, EPI_ISL_2518711, EPI_ISL_2518712, EPI_ISL_2518713, EPI_ISL_2518714, EPI_ISL_2518715, EPI_ISL_2518716, EPI_ISL_2518717, EPI_ISL_2518718, EPI_ISL_2518719, EPI_ISL_2518720, EPI_ISL_2518721, EPI_ISL_2518722, EPI_ISL_2518723, EPI_ISL_2518724, EPI_ISL_2518725, EPI_ISL_2518726, EPI_ISL_2518728, EPI_ISL_2518729, EPI_ISL_2518730, EPI_ISL_2518732, EPI_ISL_2518734, EPI_ISL_2518735, EPI_ISL_2518736, EPI_ISL_2518737, EPI_ISL_2518738, EPI_ISL_2518739, EPI_ISL_2518740, EPI_ISL_2518741, EPI_ISL_2518742, EPI_ISL_2518743, EPI_ISL_2518744, EPI_ISL_2518745, EPI_ISL_2518746, EPI_ISL_2518747, EPI_ISL_2518749, EPI_ISL_2518751, EPI_ISL_2518752, EPI_ISL_2518753, EPI_ISL_2518755, EPI_ISL_2518756, EPI_ISL_2518757, EPI_ISL_2518758, EPI_ISL_2518759, EPI_ISL_2518760, EPI_ISL_2518761, EPI_ISL_2518763, EPI_ISL_2518764, EPI_ISL_2518765, EPI_ISL_2518766, EPI_ISL_2518767, EPI_ISL_2518768, EPI_ISL_2518769, EPI_ISL_2518770, EPI_ISL_2518771, EPI_ISL_2518772, EPI_ISL_2518773, EPI_ISL_2518774, EPI_ISL_2518775, EPI_ISL_2518776, EPI_ISL_2518777, EPI_ISL_2518778, EPI_ISL_2518779, EPI_ISL_2518780, EPI_ISL_2518782, EPI_ISL_2518783, EPI_ISL_2518784, EPI_ISL_2518786, EPI_ISL_2518787, EPI_ISL_2518788, EPI_ISL_2518790, EPI_ISL_2518791, EPI_ISL_2518792, EPI_ISL_2518793, EPI_ISL_2518794, EPI_ISL_2518795, EPI_ISL_2518796, EPI_ISL_2518797, EPI_ISL_2518798, EPI_ISL_2518799, EPI_ISL_2518800, EPI_ISL_2518801, EPI_ISL_2518802, EPI_ISL_2518803, EPI_ISL_2518804, EPI_ISL_2518805, EPI_ISL_2518806, EPI_ISL_2518807, EPI_ISL_2518808, EPI_ISL_2518809, EPI_ISL_2518810, EPI_ISL_2518811, EPI_ISL_2518813, EPI_ISL_2518814,

| | | | |
|---|---|---|---|
| EPI_ISL_2518815, EPI_ISL_2518816, EPI_ISL_2518817, EPI_ISL_2518818, EPI_ISL_2518819, EPI_ISL_2518820, EPI_ISL_2518821, EPI_ISL_2518822, EPI_ISL_2518823, EPI_ISL_2518824, EPI_ISL_2518825, EPI_ISL_2518826, EPI_ISL_2518827, EPI_ISL_2518828, EPI_ISL_2518829, EPI_ISL_2518830, EPI_ISL_2518831, EPI_ISL_2518832, EPI_ISL_2518833, EPI_ISL_2518834, EPI_ISL_2518835, EPI_ISL_2518836, EPI_ISL_2518837, EPI_ISL_2518838, EPI_ISL_2518839, EPI_ISL_2518840, EPI_ISL_2518841, EPI_ISL_2518842, EPI_ISL_2518843, EPI_ISL_2518844, EPI_ISL_2518845, EPI_ISL_2518846, EPI_ISL_2518847, EPI_ISL_2518848, EPI_ISL_2518849, EPI_ISL_2518850, EPI_ISL_2518851, EPI_ISL_2518852, EPI_ISL_2518853, EPI_ISL_2518854, EPI_ISL_2518855, EPI_ISL_2518856, EPI_ISL_2518857, EPI_ISL_2518858, EPI_ISL_2518859, EPI_ISL_2518860, EPI_ISL_2518861, EPI_ISL_2518862, EPI_ISL_2518863, EPI_ISL_2518864, EPI_ISL_2518865, EPI_ISL_2518866, EPI_ISL_2518867, EPI_ISL_2518868, EPI_ISL_2518869, EPI_ISL_2518870, EPI_ISL_2518871, EPI_ISL_2518872, EPI_ISL_2518873, EPI_ISL_2518874, EPI_ISL_2518875, EPI_ISL_2518876, EPI_ISL_2518878, EPI_ISL_2518879, EPI_ISL_2518880, EPI_ISL_2518881, EPI_ISL_2518882, EPI_ISL_2518883, EPI_ISL_2518884, EPI_ISL_2518885, EPI_ISL_2518886, EPI_ISL_2518887, EPI_ISL_2518888, EPI_ISL_2518889, EPI_ISL_2518890, EPI_ISL_2518891, EPI_ISL_2518892, EPI_ISL_2518893, EPI_ISL_2518894, EPI_ISL_2518895, EPI_ISL_2518896, EPI_ISL_2518897, EPI_ISL_2518898, EPI_ISL_2518899, EPI_ISL_2518900, EPI_ISL_2518901, EPI_ISL_2518902, EPI_ISL_2518903, EPI_ISL_2518904, EPI_ISL_2518905, EPI_ISL_2518906, EPI_ISL_2518907, EPI_ISL_2518908, EPI_ISL_2518909, EPI_ISL_2518910, EPI_ISL_2518911, EPI_ISL_2518912, EPI_ISL_2518913, EPI_ISL_2518914, EPI_ISL_2518921, EPI_ISL_2518929, EPI_ISL_2518934, EPI_ISL_2518937, EPI_ISL_2518940, EPI_ISL_2518941, EPI_ISL_2518946, EPI_ISL_2518949, EPI_ISL_2518951, EPI_ISL_2518960, EPI_ISL_2518964, EPI_ISL_2518965, EPI_ISL_2518969, EPI_ISL_2518970 | | | |
| see above | Northumbria University / South Tees Hospitals NHS Foundation Trust / North Cumbria Integrated Care NHS Foundation Trust / North Tees and Hartlepool NHS Foundation Trust / Newcastle Hospitals NHS Foundation Trust | COVID-19 Genomics UK (COG-UK) Consortium | Andrew Nelson; Brendan Payne; Clive Graham; Darren L Smith; Debra Padgett; Edward Barton; Emma Swindells; Garren Scott; Gary Black; Gary Eltringham; Giles S Holt; Greg R Young; Jane Greenaway; Jennifer Collins; John Allan; Joshua Loh; Lynn Dover; Matthew Bashton; Mohammad A Tariq; Paul Baker; Sarah Essex; Steve Liggett; Wen C Yew; Yusri Taha |
| EPI_ISL_2434845, EPI_ISL_2434846, EPI_ISL_2434847, EPI_ISL_2434848, EPI_ISL_2434851, EPI_ISL_2434852, EPI_ISL_2434854, EPI_ISL_2434855, EPI_ISL_2434858, EPI_ISL_2434859, EPI_ISL_2434865, EPI_ISL_2434866, EPI_ISL_2434867, EPI_ISL_2434869, EPI_ISL_2434870, EPI_ISL_2434871, EPI_ISL_2434894, EPI_ISL_2434896, EPI_ISL_2434898, EPI_ISL_2434900, EPI_ISL_2434901, EPI_ISL_2434902, EPI_ISL_2434903, EPI_ISL_2434904, EPI_ISL_2434906, EPI_ISL_2434910, EPI_ISL_2434913, EPI_ISL_2434916, EPI_ISL_2434918, EPI_ISL_2434919, EPI_ISL_2434920, EPI_ISL_2434921, EPI_ISL_2434922, EPI_ISL_2434924, EPI_ISL_2434925, EPI_ISL_2434926, EPI_ISL_2434927, EPI_ISL_2434929, EPI_ISL_2434931, EPI_ISL_2434935, EPI_ISL_2434937, EPI_ISL_2434939, EPI_ISL_2434940, EPI_ISL_2434942, EPI_ISL_2434945, EPI_ISL_2434946, EPI_ISL_2434951, EPI_ISL_2434952, EPI_ISL_2434956, EPI_ISL_2434957, EPI_ISL_2434958, EPI_ISL_2434959, EPI_ISL_2434960, EPI_ISL_2434962, EPI_ISL_2434964 | | | |
| see above | Originating lab: Wales Specialist Virology Centre Sequencing lab: Pathogen Genomics Unit | Public Health Wales Microbiology Cardiff Wales Specialist Virology Centre | Alec Birchley; Alexander Adams; Amy Gaskin; Angela Marchbank; Bree Gatica-Wilcox; Catherine Moore; Jason Coombes; Joanne Watkins; Joel Southgate; Johnathan Evans; Laura Gifford; Lauren Gilbert; Lee Graham; Malorie Perry; Matthew Bull; Nicole Pacchiarini; Sally Corden; Sara Kumziene-Summerhayes; Sara Rey; Sarah Taylor; Simon Cottrell; Sophie Jones; Tom Connor |
| EPI_ISL_2624544, EPI_ISL_2624547, EPI_ISL_2624554, EPI_ISL_2624555, EPI_ISL_2624556, EPI_ISL_2624557, EPI_ISL_2624560, EPI_ISL_2624561, EPI_ISL_2624564, EPI_ISL_2624565, EPI_ISL_2624567, EPI_ISL_2624569, EPI_ISL_2624570, EPI_ISL_2624571, EPI_ISL_2624573, EPI_ISL_2624579, EPI_ISL_2624585, EPI_ISL_2624586 | | | |
| see above | Oxford Viromics, NDM, University of Oxford; Oxford University Hospitals; Basingstoke and North Hampshire Hospital | COVID-19 Genomics UK (COG-UK) Consortium | Alex Mobbs; Amy Trebes; Anita Justice; Catrin Moore; Christophe Fraser; David Bonsall; David Buck; Emma Wise; George Macintyre; Jessica Lynch; John Todd; Mariateresa de Cesare; Matilde Mori; Monique Andersson; Nathan Moore; Nick Cortes; Robert Shaw; Stephen Kidd; Tanya Golubchik; Timothy Peto |
| EPI_ISL_2518971, EPI_ISL_2518972, EPI_ISL_2518973, EPI_ISL_2518974, EPI_ISL_2518975, EPI_ISL_2518976, EPI_ISL_2518977, EPI_ISL_2518978, EPI_ISL_2518979, EPI_ISL_2518980, EPI_ISL_2518981, EPI_ISL_2518982, EPI_ISL_2518983, EPI_ISL_2518984, EPI_ISL_2518985, EPI_ISL_2518986, EPI_ISL_2518987, EPI_ISL_2518988, EPI_ISL_2518989, EPI_ISL_2518990, EPI_ISL_2518991, EPI_ISL_2518992, EPI_ISL_2518993, EPI_ISL_2518994, EPI_ISL_2518995, EPI_ISL_2518996, EPI_ISL_2518997, EPI_ISL_2518998, EPI_ISL_2518999, EPI_ISL_2519000, EPI_ISL_2519001, EPI_ISL_2519002, EPI_ISL_2519003, EPI_ISL_2519004, EPI_ISL_2519005, EPI_ISL_2519006, EPI_ISL_2519007, EPI_ISL_2519008, EPI_ISL_2519009, EPI_ISL_2519010, EPI_ISL_2519011, EPI_ISL_2519012, EPI_ISL_2519013, EPI_ISL_2519014, EPI_ISL_2519016, EPI_ISL_2519017, EPI_ISL_2519018, EPI_ISL_2519019, EPI_ISL_2519020, EPI_ISL_2519021, EPI_ISL_2519022, EPI_ISL_2519023, EPI_ISL_2519024, EPI_ISL_2519025, EPI_ISL_2519026, EPI_ISL_2519027, EPI_ISL_2519028, EPI_ISL_2519029, EPI_ISL_2519030, EPI_ISL_2519031, EPI_ISL_2519032, EPI_ISL_2519033, EPI_ISL_2519034, EPI_ISL_2519035, EPI_ISL_2519036, EPI_ISL_2519037, EPI_ISL_2519038, EPI_ISL_2519039, EPI_ISL_2519040, EPI_ISL_2519041, EPI_ISL_2519042, EPI_ISL_2519043, EPI_ISL_2519044, EPI_ISL_2519045, EPI_ISL_2519046, EPI_ISL_2519047, EPI_ISL_2519048, EPI_ISL_2519049, EPI_ISL_2519050, EPI_ISL_2519051, EPI_ISL_2519052, EPI_ISL_2519053, EPI_ISL_2519054, EPI_ISL_2519055, EPI_ISL_2519056, EPI_ISL_2519057, EPI_ISL_2519058, EPI_ISL_2519059, EPI_ISL_2519060, EPI_ISL_2519061, EPI_ISL_2519062, EPI_ISL_2519063, EPI_ISL_2519064, EPI_ISL_2519065, EPI_ISL_2519066, EPI_ISL_2519067, EPI_ISL_2519068, EPI_ISL_2519069, EPI_ISL_2519070, EPI_ISL_2519071, EPI_ISL_2519072, EPI_ISL_2519073, EPI_ISL_2519074, EPI_ISL_2519075, EPI_ISL_2519076, EPI_ISL_2519077, EPI_ISL_2519078, EPI_ISL_2519079, EPI_ISL_2519080, EPI_ISL_2519081, EPI_ISL_2519082, EPI_ISL_2519083, EPI_ISL_2519084, EPI_ISL_2519085, EPI_ISL_2519086, EPI_ISL_2519087, EPI_ISL_2519088, EPI_ISL_2519089, EPI_ISL_2519090, EPI_ISL_2519091, EPI_ISL_2519092, EPI_ISL_2519093, EPI_ISL_2519094, EPI_ISL_2519095, EPI_ISL_2519096, EPI_ISL_2519097, EPI_ISL_2519098, EPI_ISL_2519099, EPI_ISL_2519100, EPI_ISL_2519101, EPI_ISL_2519102, EPI_ISL_2519103, EPI_ISL_2519104, EPI_ISL_2519105, EPI_ISL_2519106, EPI_ISL_2519107, EPI_ISL_2519108, EPI_ISL_2519109, EPI_ISL_2519110, EPI_ISL_2519111, EPI_ISL_2519112, EPI_ISL_2519113, EPI_ISL_2519114, EPI_ISL_2519115, EPI_ISL_2519116, EPI_ISL_2519117, EPI_ISL_2519118, EPI_ISL_2519119, EPI_ISL_2519120, EPI_ISL_2519121, EPI_ISL_2519122, EPI_ISL_2519123, EPI_ISL_2519124, EPI_ISL_2519126, EPI_ISL_2519127, EPI_ISL_2519128, EPI_ISL_2519129, EPI_ISL_2519130, EPI_ISL_2519132, EPI_ISL_2519133, EPI_ISL_2519134, EPI_ISL_2519135, EPI_ISL_2519136, EPI_ISL_2519137, EPI_ISL_2519138, EPI_ISL_2519139, EPI_ISL_2519140, EPI_ISL_2519141, EPI_ISL_2519142, EPI_ISL_2519143, EPI_ISL_2519144, EPI_ISL_2519145, EPI_ISL_2519146, EPI_ISL_2519147, EPI_ISL_2519148, EPI_ISL_2519149, EPI_ISL_2519150, EPI_ISL_2519151, EPI_ISL_2519152, EPI_ISL_2519153, EPI_ISL_2519154, EPI_ISL_2519155, EPI_ISL_2519156, EPI_ISL_2519157, EPI_ISL_2519158, EPI_ISL_2519159, EPI_ISL_2519160, EPI_ISL_2519161, EPI_ISL_2519162, EPI_ISL_2519163, EPI_ISL_2519164, EPI_ISL_2519165, EPI_ISL_2519166, EPI_ISL_2519168, EPI_ISL_2519169, EPI_ISL_2519170, EPI_ISL_2519171, EPI_ISL_2519173, EPI_ISL_2519174, EPI_ISL_2519175, EPI_ISL_2519176, EPI_ISL_2519177, EPI_ISL_2519178, EPI_ISL_2519179, EPI_ISL_2519180, EPI_ISL_2519181, EPI_ISL_2519182, EPI_ISL_2519183, EPI_ISL_2519184, EPI_ISL_2519185, EPI_ISL_2519187, EPI_ISL_2519188, EPI_ISL_2519189, EPI_ISL_2519190, EPI_ISL_2519191, EPI_ISL_2519192, EPI_ISL_2519193, EPI_ISL_2519194, EPI_ISL_2519195, EPI_ISL_2519196, EPI_ISL_2519197, EPI_ISL_2519504, EPI_ISL_2519524, EPI_ISL_2519541, EPI_ISL_2519544, EPI_ISL_2519545, EPI_ISL_2519546, EPI_ISL_2519623, EPI_ISL_2519674, EPI_ISL_2519686, EPI_ISL_2519727, EPI_ISL_2519728, EPI_ISL_2519729, EPI_ISL_2519730, EPI_ISL_2519731, EPI_ISL_2519732, EPI_ISL_2519733, EPI_ISL_2519734, EPI_ISL_2519735, EPI_ISL_2519736, EPI_ISL_2519737, EPI_ISL_2519738, EPI_ISL_2519739, EPI_ISL_2519740, EPI_ISL_2519741, EPI_ISL_2519751, EPI_ISL_2519755, EPI_ISL_2519756, EPI_ISL_2519757, EPI_ISL_2519758, EPI_ISL_2519759, EPI_ISL_2519761, EPI_ISL_2519762, EPI_ISL_2519763, EPI_ISL_2519765, EPI_ISL_2519766, EPI_ISL_2519767, EPI_ISL_2519768, EPI_ISL_2519769, EPI_ISL_2519770, EPI_ISL_2519771, EPI_ISL_2519772, EPI_ISL_2519773, EPI_ISL_2519775, EPI_ISL_2519776, EPI_ISL_2519777, EPI_ISL_2519778, EPI_ISL_2519779, EPI_ISL_2519780, EPI_ISL_2519781, EPI_ISL_2519782, EPI_ISL_2519783, EPI_ISL_2519784, EPI_ISL_2519785, EPI_ISL_2519786, EPI_ISL_2519787, EPI_ISL_2519788, EPI_ISL_2519789, EPI_ISL_2519791, EPI_ISL_2519793, EPI_ISL_2519794, EPI_ISL_2519795, EPI_ISL_2519796, EPI_ISL_2519797, EPI_ISL_2519798, EPI_ISL_2519799, EPI_ISL_2519800, EPI_ISL_2519801, EPI_ISL_2519802, EPI_ISL_2519803, EPI_ISL_2519804, EPI_ISL_2519806, EPI_ISL_2519807, EPI_ISL_2519808, EPI_ISL_2519809, EPI_ISL_2519811, EPI_ISL_2519812, EPI_ISL_2519813, EPI_ISL_2519814, EPI_ISL_2519815, EPI_ISL_2519816, EPI_ISL_2519817, EPI_ISL_2519818, EPI_ISL_2519819, EPI_ISL_2519820, EPI_ISL_2519821, EPI_ISL_2519822, EPI_ISL_2519823, EPI_ISL_2519824, EPI_ISL_2519825, EPI_ISL_2519827, EPI_ISL_2519829, EPI_ISL_2519830, EPI_ISL_2519832, EPI_ISL_2519833, EPI_ISL_2519834, EPI_ISL_2519835, EPI_ISL_2519836, EPI_ISL_2519837, EPI_ISL_2519838, EPI_ISL_2519839, EPI_ISL_2519840, EPI_ISL_2519841, EPI_ISL_2519842, EPI_ISL_2519843, EPI_ISL_2519844, EPI_ISL_2519846, EPI_ISL_2519847, EPI_ISL_2519848, EPI_ISL_2519849, EPI_ISL_2519850, EPI_ISL_2519851, EPI_ISL_2519852, EPI_ISL_2519853, EPI_ISL_2519854, EPI_ISL_2519855, EPI_ISL_2519856, EPI_ISL_2519857, EPI_ISL_2519858, EPI_ISL_2519859, EPI_ISL_2519860, EPI_ISL_2519862, EPI_ISL_2519863, EPI_ISL_2519865, EPI_ISL_2519866, EPI_ISL_2519867, EPI_ISL_2519868, EPI_ISL_2519869, EPI_ISL_2519870, EPI_ISL_2519871, EPI_ISL_2519872, EPI_ISL_2519874, EPI_ISL_2519875, EPI_ISL_2519876, EPI_ISL_2519877, EPI_ISL_2519878, EPI_ISL_2519879, EPI_ISL_2519880, EPI_ISL_2519881, EPI_ISL_2519882, EPI_ISL_2519883, EPI_ISL_2519884, EPI_ISL_2519885, EPI_ISL_2519886, EPI_ISL_2519887, EPI_ISL_2519888, EPI_ISL_2519889, EPI_ISL_2519890, EPI_ISL_2519891, EPI_ISL_2519892, EPI_ISL_2519893, EPI_ISL_2519894, EPI_ISL_2519895, EPI_ISL_2519896, EPI_ISL_2519897, EPI_ISL_2519899, EPI_ISL_2519900, EPI_ISL_2519901, EPI_ISL_2519902, EPI_ISL_2519903, EPI_ISL_2519904, EPI_ISL_2519905, EPI_ISL_2519906, EPI_ISL_2519907, EPI_ISL_2519908, EPI_ISL_2519909, EPI_ISL_2519910, EPI_ISL_2519911, EPI_ISL_2519912, EPI_ISL_2519913, EPI_ISL_2519914, EPI_ISL_2519915, EPI_ISL_2519916, EPI_ISL_2519917, EPI_ISL_2519918, EPI_ISL_2519919, EPI_ISL_2519920, EPI_ISL_2519921, EPI_ISL_2519922, EPI_ISL_2519923, EPI_ISL_2519924, EPI_ISL_2519925, EPI_ISL_2519926, EPI_ISL_2519927, EPI_ISL_2519928, EPI_ISL_2519929, EPI_ISL_2519930, EPI_ISL_2519931, EPI_ISL_2519932, EPI_ISL_2519933, EPI_ISL_2519934, EPI_ISL_2519935, EPI_ISL_2519936, EPI_ISL_2519937, EPI_ISL_2519938, EPI_ISL_2519939, EPI_ISL_2519940, EPI_ISL_2519941, EPI_ISL_2519942, EPI_ISL_2519943, EPI_ISL_2519944, EPI_ISL_2519946, EPI_ISL_2519947, EPI_ISL_2519948, EPI_ISL_2519949, EPI_ISL_2519950, EPI_ISL_2519951, EPI_ISL_2519952, EPI_ISL_2519953, EPI_ISL_2519954, EPI_ISL_2519955, EPI_ISL_2519956, EPI_ISL_2519957, EPI_ISL_2519958, EPI_ISL_2519959, EPI_ISL_2519960, EPI_ISL_2519961, EPI_ISL_2519962, EPI_ISL_2519963, EPI_ISL_2519964, EPI_ISL_2519965, EPI_ISL_2519966, EPI_ISL_2519967, EPI_ISL_2519968, EPI_ISL_2519969, EPI_ISL_2519970, EPI_ISL_2519971, EPI_ISL_2519974, EPI_ISL_2519976, EPI_ISL_2519977, EPI_ISL_2519978, EPI_ISL_2519980, EPI_ISL_2519981, EPI_ISL_2519982, EPI_ISL_2519983, EPI_ISL_2519984, EPI_ISL_2519985, EPI_ISL_2519986, EPI_ISL_2519990, EPI_ISL_2519991, EPI_ISL_2519993, EPI_ISL_2519995, EPI_ISL_2519998, EPI_ISL_2519999, EPI_ISL_2520000, EPI_ISL_2520002, EPI_ISL_2520004, EPI_ISL_2520006, EPI_ISL_2520007, EPI_ISL_2520008, EPI_ISL_2520009, EPI_ISL_2520010, EPI_ISL_2520011, EPI_ISL_2520012, EPI_ISL_2520013, EPI_ISL_2520014, EPI_ISL_2520015, EPI_ISL_2520016, EPI_ISL_2520017, EPI_ISL_2520020, EPI_ISL_2520021, EPI_ISL_2520025, EPI_ISL_2520026, EPI_ISL_2520027, EPI_ISL_2520029, EPI_ISL_2520033, EPI_ISL_2520034, EPI_ISL_2520035, EPI_ISL_2520036, EPI_ISL_2520037, EPI_ISL_2520038, EPI_ISL_2520039, EPI_ISL_2520040, EPI_ISL_2520041, EPI_ISL_2520042, EPI_ISL_2520043, EPI_ISL_2520044, EPI_ISL_2520045, EPI_ISL_2520046, EPI_ISL_2520047, EPI_ISL_2520048, EPI_ISL_2520049, EPI_ISL_2520051, EPI_ISL_2520052, EPI_ISL_2520053, EPI_ISL_2520054, EPI_ISL_2520059, EPI_ISL_2520060, EPI_ISL_2520061, EPI_ISL_2520062, EPI_ISL_2520065, EPI_ISL_2520066, EPI_ISL_2520068, EPI_ISL_2520069, EPI_ISL_2520070, EPI_ISL_2520075, EPI_ISL_2520080, EPI_ISL_2520084, EPI_ISL_2520085, EPI_ISL_2520086, EPI_ISL_2520087, EPI_ISL_2520088, EPI_ISL_2520091, EPI_ISL_2520092, EPI_ISL_2520093, EPI_ISL_2520094, EPI_ISL_2520095, EPI_ISL_2520096, EPI_ISL_2520101, EPI_ISL_2520103, EPI_ISL_2520104, EPI_ISL_2520176, EPI_ISL_2623658, EPI_ISL_2623665, EPI_ISL_2623681, EPI_ISL_2623683, EPI_ISL_2623795, EPI_ISL_2623796, EPI_ISL_2623798, EPI_ISL_2623799, EPI_ISL_2623800, EPI_ISL_2623802, EPI_ISL_2623804, EPI_ISL_2623805, EPI_ISL_2623806, EPI_ISL_2623807, EPI_ISL_2623808, EPI_ISL_2623809, EPI_ISL_2623810, EPI_ISL_2623811, EPI_ISL_2623812, EPI_ISL_2623813, EPI_ISL_2623814, EPI_ISL_2623815, EPI_ISL_2623816, EPI_ISL_2623817, EPI_ISL_2623818, EPI_ISL_2623819, EPI_ISL_2623820, EPI_ISL_2623821, EPI_ISL_2623822, EPI_ISL_2623823, EPI_ISL_2623824, EPI_ISL_2623828, EPI_ISL_2623829, EPI_ISL_2623830, EPI_ISL_2623831, EPI_ISL_2623833, EPI_ISL_2623834, EPI_ISL_2623837, EPI_ISL_2623838, EPI_ISL_2623839, EPI_ISL_2623840, EPI_ISL_2623841, EPI_ISL_2623842, EPI_ISL_2623844, EPI_ISL_2623845, EPI_ISL_2623846, EPI_ISL_2623847, EPI_ISL_2623848, EPI_ISL_2623849, EPI_ISL_2623851, EPI_ISL_2623854, EPI_ISL_2623855, EPI_ISL_2623856, EPI_ISL_2623857, EPI_ISL_2623858, EPI_ISL_2623859, EPI_ISL_2623860, EPI_ISL_2623861, EPI_ISL_2623862, EPI_ISL_2623863, EPI_ISL_2623864, EPI_ISL_2623865, EPI_ISL_2623866, EPI_ISL_2623867, EPI_ISL_2623868, EPI_ISL_2623870, EPI_ISL_2623871, EPI_ISL_2623872, EPI_ISL_2623879, EPI_ISL_2623880, EPI_ISL_2623881, EPI_ISL_2623882, EPI_ISL_2623883, EPI_ISL_2623884, EPI_ISL_2623885, EPI_ISL_2623886, EPI_ISL_2623887, EPI_ISL_2623888, EPI_ISL_2623889, EPI_ISL_2623890, EPI_ISL_2623891, EPI_ISL_2623892, EPI_ISL_2623893, EPI_ISL_2623894, EPI_ISL_2623895, EPI_ISL_2623896, EPI_ISL_2623897, EPI_ISL_2623908, EPI_ISL_2623909, EPI_ISL_2623910, EPI_ISL_2623911, EPI_ISL_2623940, EPI_ISL_2623944, EPI_ISL_2623945, EPI_ISL_2623946, EPI_ISL_2623947, EPI_ISL_2623948, EPI_ISL_2623949, EPI_ISL_2623950, EPI_ISL_2623951, EPI_ISL_2623952, EPI_ISL_2623953, EPI_ISL_2623954, EPI_ISL_2623956, EPI_ISL_2623957, EPI_ISL_2623958, EPI_ISL_2623959, EPI_ISL_2623961, EPI_ISL_2623962, EPI_ISL_2623963, EPI_ISL_2623964, EPI_ISL_2623965, EPI_ISL_2623966, EPI_ISL_2623967, EPI_ISL_2623970, EPI_ISL_2623973, EPI_ISL_2623974, EPI_ISL_2623976, EPI_ISL_2623977, EPI_ISL_2623978, EPI_ISL_2623979, EPI_ISL_2623980, EPI_ISL_2623981, EPI_ISL_2623982, EPI_ISL_2623983, EPI_ISL_2623984, EPI_ISL_2623985, EPI_ISL_2623986, EPI_ISL_2623987, EPI_ISL_2623988, EPI_ISL_2623989, EPI_ISL_2623990, EPI_ISL_2623991, EPI_ISL_2623992, EPI_ISL_2623993, EPI_ISL_2623994, EPI_ISL_2623995, EPI_ISL_2623996, EPI_ISL_2623997, EPI_ISL_2623999, EPI_ISL_2624000, EPI_ISL_2624291, EPI_ISL_2624292, EPI_ISL_2624293, EPI_ISL_2624294, EPI_ISL_2624295, EPI_ISL_2624297, EPI_ISL_2624298, EPI_ISL_2624299, EPI_ISL_2624300, EPI_ISL_2624301, EPI_ISL_2624302, EPI_ISL_2624303, EPI_ISL_2624304, EPI_ISL_2624305, EPI_ISL_2624306, EPI_ISL_2624307, EPI_ISL_2624308, EPI_ISL_2624309, EPI_ISL_2624310, EPI_ISL_2624311, EPI_ISL_2624312, EPI_ISL_2624313, EPI_ISL_2624314, EPI_ISL_2624315, EPI_ISL_2624316, EPI_ISL_2624317, EPI_ISL_2624318, EPI_ISL_2624319, EPI_ISL_2624321, EPI_ISL_2624322, EPI_ISL_2624323, EPI_ISL_2624324, EPI_ISL_2624325, EPI_ISL_2624326, EPI_ISL_2624327, EPI_ISL_2624328, EPI_ISL_2624329, EPI_ISL_2624330, EPI_ISL_2624331, EPI_ISL_2624332, EPI_ISL_2624333, EPI_ISL_2624334, EPI_ISL_2624335, EPI_ISL_2624336, EPI_ISL_2624337, EPI_ISL_2624338, EPI_ISL_2624339, EPI_ISL_2624340, EPI_ISL_2624341, EPI_ISL_2624342, EPI_ISL_2624343, EPI_ISL_2624344, EPI_ISL_2624345, EPI_ISL_2624349, EPI_ISL_2624350, EPI_ISL_2624351, EPI_ISL_2624352, EPI_ISL_2624353, EPI_ISL_2624354, EPI_ISL_2624355, EPI_ISL_2706809, EPI_ISL_2706830 | | | |
| see above | Quadram Institute Bioscience | COVID-19 Genomics UK (COG-UK) Consortium | Alexander J Trotter; Alison E. Mather; Alp Aydin; Ana P. Tedim; Anastasia Kolyva; Andrew Bell; Andrew J. Page; Claire Stuart; Dave J. Baker; Gemma L. Kay; John Wain; Justin O'Grady; Leonardo de Oliveira Martins; Lizzie Meadows; Maria Diaz; Mark Webber; Muhammed Yasir; Nabil-Fareed Alikhan; Ngozi Elumogo; Nicholas M. Thomson; Rachael Stanley; Rachel Gilroy; Reenesh Prakash; Samir Dervisevic; Samuel Bloomfield; Steven Rudder; Thanh Le-Viet |
| EPI_ISL_2447765, EPI_ISL_2447766, EPI_ISL_2447767, EPI_ISL_2447768, EPI_ISL_2447769 | Randox Laboratories | Wellcome Sanger Institute for the COVID-19 Genomics UK (COG-UK) Consortium | Cordelia Langford; David K. Jackson; Dominic Kwiatkowski; Ewan Harrison; Ian Johnston; Jeffrey Barrett; John Sillitoe on behalf of the Wellcome Sanger Institute COVID-19 Surveillance Team; Randox Laboratories and Alex Alderton; Roberto Amato; Sonia Goncalves |
| EPI_ISL_2743338, EPI_ISL_2743350, EPI_ISL_2743351, EPI_ISL_2743361, EPI_ISL_2743363, EPI_ISL_2743379, EPI_ISL_2743387, EPI_ISL_2743460, EPI_ISL_2743467, EPI_ISL_2743469, EPI_ISL_2743479, EPI_ISL_2743480, EPI_ISL_2743491, EPI_ISL_2743493, EPI_ISL_2743495, EPI_ISL_2743496, EPI_ISL_2743497, EPI_ISL_2743500, EPI_ISL_2743540, EPI_ISL_2743541, EPI_ISL_2743557, EPI_ISL_2743576, EPI_ISL_2743580, EPI_ISL_2743601, EPI_ISL_2743602, EPI_ISL_2743614, EPI_ISL_2743615, EPI_ISL_2743622, EPI_ISL_2743625, EPI_ISL_2743634, EPI_ISL_2743637, EPI_ISL_2743641, EPI_ISL_2743654, EPI_ISL_2743661, EPI_ISL_2743680, EPI_ISL_2743682, EPI_ISL_2743685, EPI_ISL_2743690, EPI_ISL_2743693, EPI_ISL_2743696, EPI_ISL_2743710, EPI_ISL_2743717, EPI_ISL_2743722, EPI_ISL_2743736, EPI_ISL_2743742, EPI_ISL_2743745, EPI_ISL_2743755, EPI_ISL_2743760, EPI_ISL_2743761, EPI_ISL_2743762, EPI_ISL_2743767, EPI_ISL_2743774, EPI_ISL_2743777, EPI_ISL_2743779, EPI_ISL_2743783, EPI_ISL_2743786, EPI_ISL_2743787, EPI_ISL_2743794, EPI_ISL_2743799, EPI_ISL_2743809, EPI_ISL_2743810, EPI_ISL_2743832, EPI_ISL_2743834, EPI_ISL_2743931, EPI_ISL_2743951, EPI_ISL_2743952, EPI_ISL_2743964, EPI_ISL_2743967, EPI_ISL_2743978, EPI_ISL_2743980, EPI_ISL_2743981, EPI_ISL_2743988, EPI_ISL_2744001, EPI_ISL_2744002, EPI_ISL_2744003, EPI_ISL_2744004, EPI_ISL_2744006, EPI_ISL_2744008, EPI_ISL_2744009, EPI_ISL_2744011, EPI_ISL_2744012, EPI_ISL_2744013, EPI_ISL_2744015, EPI_ISL_2744019, EPI_ISL_2744025, EPI_ISL_2744026, EPI_ISL_2744032, EPI_ISL_2744034, EPI_ISL_2744036, EPI_ISL_2744037, EPI_ISL_2744038, EPI_ISL_2744055, EPI_ISL_2744060, EPI_ISL_2744132, EPI_ISL_2744230, EPI_ISL_2744297, EPI_ISL_2744634, EPI_ISL_2744709, EPI_ISL_2744712, EPI_ISL_2744714, EPI_ISL_2744715, EPI_ISL_2744716, EPI_ISL_2744717, EPI_ISL_2744718, EPI_ISL_2744721, EPI_ISL_2744723, EPI_ISL_2744724, EPI_ISL_2744727, EPI_ISL_2745228, EPI_ISL_2745417, EPI_ISL_2749193 | | | |
| see above | Respiratory Virus Unit, Microbiology Services Colindale, Public Health England | COVID-19 Genomics UK (COG-UK) Consortium | PHE Covid Sequencing Team |
| EPI_ISL_2434223, EPI_ISL_2434224, EPI_ISL_2434228, EPI_ISL_2434229, EPI_ISL_2434230, EPI_ISL_2434231, EPI_ISL_2434232, EPI_ISL_2434233, EPI_ISL_2434234, EPI_ISL_2434236, EPI_ISL_2517291, EPI_ISL_2517292, EPI_ISL_2517293, EPI_ISL_2517294, EPI_ISL_2517295, EPI_ISL_2517297, EPI_ISL_2517298, EPI_ISL_2517299, EPI_ISL_2517300, EPI_ISL_2622340, EPI_ISL_2704831, EPI_ISL_2704832, EPI_ISL_2734360 | | | |
| see above | Virology Department, Royal Infirmary of Edinburgh, NHS Lothian / School of Biological Sciences, University of Edinburgh | COVID-19 Genomics UK (COG-UK) Consortium | Colquhoun R; Cotton S; Dewar R; Gallagher A; Hill V; Jackson B; Maloney D; McCrone JT; McHugh M; O'Toole Á; Rambaut A; Rooke S; Scher E; Templeton K; Yu X |
| EPI_ISL_2520945, EPI_ISL_2520986, EPI_ISL_2521005, EPI_ISL_2521043, EPI_ISL_2521085, EPI_ISL_2521110, EPI_ISL_2521146 | | | |
| see above | Virology Department, Sheffield Teaching Hospitals NHS Foundation Trust/Department of Infection, Immunity and Cardiovascular Disease, The Medical School, University of Sheffield | COVID-19 Genomics UK (COG-UK) Consortium | Adri Angyal; Alex Keeley; Benjamin Lindsey; Cariad Evans; Danielle Groves; Dave Partridge; Katie Johnson; Laura Carrilero; Luke Green; Matthew Parker; Matthew Wyles; Mehmet Yavuz; Mohammad Raza; Nikki Smith; Paul Parsons; Rachel Tucker; Rebecca Brown; Thushan de Silva |
| EPI_ISL_2520534, EPI_ISL_2520535, EPI_ISL_2520536, EPI_ISL_2520537, EPI_ISL_2520538, EPI_ISL_2520539, EPI_ISL_2520540, EPI_ISL_2520542, EPI_ISL_2520543, EPI_ISL_2520544, EPI_ISL_2520545, EPI_ISL_2520546, EPI_ISL_2520547, EPI_ISL_2520548, EPI_ISL_2520549, EPI_ISL_2520550, EPI_ISL_2520551, EPI_ISL_2520552, EPI_ISL_2520553, EPI_ISL_2520554, EPI_ISL_2520555, EPI_ISL_2520556, EPI_ISL_2520723, EPI_ISL_2520724, EPI_ISL_2520725, EPI_ISL_2520726, EPI_ISL_2520727, EPI_ISL_2520728, EPI_ISL_2520729, EPI_ISL_2520730, EPI_ISL_2520731, EPI_ISL_2520735, EPI_ISL_2520736, EPI_ISL_2520737, EPI_ISL_2520738, EPI_ISL_2520739, EPI_ISL_2520740, EPI_ISL_2520741, EPI_ISL_2520742, EPI_ISL_2520743, EPI_ISL_2520744, EPI_ISL_2520745, EPI_ISL_2520746, EPI_ISL_2520747, EPI_ISL_2520748, EPI_ISL_2520749, EPI_ISL_2520750, EPI_ISL_2520751, EPI_ISL_2520752, EPI_ISL_2520753, EPI_ISL_2520754, EPI_ISL_2520755, EPI_ISL_2520756, EPI_ISL_2520757, EPI_ISL_2520758, EPI_ISL_2520759, EPI_ISL_2520760, EPI_ISL_2520766, EPI_ISL_2520767, EPI_ISL_2520768, EPI_ISL_2520769, EPI_ISL_2520770, EPI_ISL_2520773, EPI_ISL_2520774, EPI_ISL_2520775, EPI_ISL_2520779, EPI_ISL_2520782 | | | |
| see above | Wales Specialist Virology Centre Sequencing lab: Pathogen Genomics Unit | Public Health Wales Microbiology Cardiff Wales Specialist Virology Centre | Alec Birchley; Alexander Adams; Amy Gaskin; Angela Marchbank; Bree Gatica-Wilcox; Catherine Moore; Jason Coombes; Joanne Watkins; Joel Southgate; Johnathan Evans; Laura Gifford; Lauren Gilbert; Lee Graham; Malorie Perry; Matthew Bull; Nicole Pacchiarini; Sally Corden; Sara Kumziene-Summerhayes; Sara Rey; Sarah Taylor; Simon Cottrell; Sophie Jones; Tom Connor |
| EPI_ISL_2517248, EPI_ISL_2517249, EPI_ISL_2517250, EPI_ISL_2517251, EPI_ISL_2517253, EPI_ISL_2517272, EPI_ISL_2517273, EPI_ISL_2517274, EPI_ISL_2517275, EPI_ISL_2517276, EPI_ISL_2517277, EPI_ISL_2704807, EPI_ISL_2722633, EPI_ISL_2722634, EPI_ISL_2722635, EPI_ISL_2722636, EPI_ISL_2722637, EPI_ISL_2722639, EPI_ISL_2722640, EPI_ISL_2722641, EPI_ISL_2722642, EPI_ISL_2722643, EPI_ISL_2722644, EPI_ISL_2722645, EPI_ISL_2722648, EPI_ISL_2722649, EPI_ISL_2722650, EPI_ISL_2722674, EPI_ISL_2722675, EPI_ISL_2722676 | | | |
| see above | West of Scotland Specialist Virology Centre, NHSGGC / MRC-University of Glasgow Centre for Virus Research | COVID-19 Genomics UK (COG-UK) Consortium | Alasdair MacLean; Alice Broos; Ana da Silva Filipe; Antonia Ho; Daniel Mair; David L Robertson; Emma Thomson; Guy Mollett; Ioulia Tsatsani; James Shepherd; Jenna Nichols; Joseph Hughes; Kathy Li; Kathy Smollett; Kyriaki Nomikou; Lily Tong; Matthew Holden; Natasha Johnson; Rachel Blacow; Richard Orton; Rory Gunson; Sarah McDonald; Sharif Shaaban; Sreenu Vattipally; Stephen Carmichael |

We gratefully acknowledge the following Authors from the Originating laboratories responsible for obtaining the specimens, as well as the Submitting laboratories where the genome data were generated and shared via GISAID, on which this research is based.

All Submitters of data may be contacted directly via www.gisaid.org

Authors are sorted alphabetically.

| Accession ID | Originating Laboratory | Submitting Laboratory | Authors |
|---|---|---|---|
| EPI_ISL_2686130, EPI_ISL_2686131, EPI_ISL_2686132, EPI_ISL_2686133, EPI_ISL_2686134, EPI_ISL_2686135, EPI_ISL_2686136, EPI_ISL_2686137, EPI_ISL_2686138, EPI_ISL_2686139, EPI_ISL_2686140, EPI_ISL_2686141 | | | |
| see above | Amazon Dx, AUV2 Laboratory | Amazon Dx, AUV2 Laboratory | Luke Meredith |
| EPI_ISL_2517308, EPI_ISL_2517309, EPI_ISL_2517310, EPI_ISL_2517312, EPI_ISL_2517313, EPI_ISL_2517314, EPI_ISL_2517315, EPI_ISL_2517316, EPI_ISL_2517317, EPI_ISL_2517318, EPI_ISL_2517319, EPI_ISL_2517320, EPI_ISL_2517321 | | | |
| see above | Barts Health NHS Trust | COVID-19 Genomics UK (COG-UK) Consortium | BROAD; Beatrix; CUTINO-MOGUEL; Claire; David; Dola; HARRINGTON; KELE; KULASEGARAN-SHYLINI; Maria-Teresa; OWOYEMI; Raghavendran |
| EPI_ISL_2935022 | Berkshire and Surrey Pathology Services Lighthouse Laboratory | Wellcome Sanger Institute for the COVID-19 Genomics UK (COG-UK) Consortium | Berkshire and Surrey Pathology Services Lighthouse Laboratory and Alex Alderton; Cordelia Langford; David K. Jackson; Dominic Kwiatkowski; Ewan Harrison; Ian Johnston; Jeffrey Barrett; John Sillitoe on behalf of the Wellcome Sanger Institute COVID-19 Surveillance Team; Roberto Amato; Sonia Goncalves |
| EPI_ISL_2467962, EPI_ISL_2467969, EPI_ISL_2467984, EPI_ISL_2467985, EPI_ISL_2467989, EPI_ISL_2468014, EPI_ISL_2468021, EPI_ISL_2468036, EPI_ISL_2468085, EPI_ISL_2468115, EPI_ISL_2468116, EPI_ISL_2468119, EPI_ISL_2468136, EPI_ISL_2468154, EPI_ISL_2468158, EPI_ISL_2468176, EPI_ISL_2468187, EPI_ISL_2468194, EPI_ISL_2468203, EPI_ISL_2468206, EPI_ISL_2468210, EPI_ISL_2468241, EPI_ISL_2468259, EPI_ISL_2468264, EPI_ISL_2468276, EPI_ISL_2468290, EPI_ISL_2468296, EPI_ISL_2468297, EPI_ISL_2468302, EPI_ISL_2468306, EPI_ISL_2468309, EPI_ISL_2468311, EPI_ISL_2468313, EPI_ISL_2468315, EPI_ISL_2468317, EPI_ISL_2468319, EPI_ISL_2468320, EPI_ISL_2468331, EPI_ISL_2468337, EPI_ISL_2468342, EPI_ISL_2468348, EPI_ISL_2468350, EPI_ISL_2468361, EPI_ISL_2468365, EPI_ISL_2468373, EPI_ISL_2468383, EPI_ISL_2468385, EPI_ISL_2468386, EPI_ISL_2468399, EPI_ISL_2468401, EPI_ISL_2468415, EPI_ISL_2468421, EPI_ISL_2468423, EPI_ISL_2468426, EPI_ISL_2468427, EPI_ISL_2468430, EPI_ISL_2468431, EPI_ISL_2468432, EPI_ISL_2468434, EPI_ISL_2468438, EPI_ISL_2468439, EPI_ISL_2468445, EPI_ISL_2468450, EPI_ISL_2468456, EPI_ISL_2468459, EPI_ISL_2468463, EPI_ISL_2468473, EPI_ISL_2468476, EPI_ISL_2468484, EPI_ISL_2468486, EPI_ISL_2468489, EPI_ISL_2468493, EPI_ISL_2468499, EPI_ISL_2468501, EPI_ISL_2468520, EPI_ISL_2468526, EPI_ISL_2468536, EPI_ISL_2468537, EPI_ISL_2468538, EPI_ISL_2468547, EPI_ISL_2468549, EPI_ISL_2468554, EPI_ISL_2468555, EPI_ISL_2468560, EPI_ISL_2468562, EPI_ISL_2468567, EPI_ISL_2468575, EPI_ISL_2468576, EPI_ISL_2468577, EPI_ISL_2468579, EPI_ISL_2469227, EPI_ISL_2469228, EPI_ISL_2469230, EPI_ISL_2469233, EPI_ISL_2469234, EPI_ISL_2469235, EPI_ISL_2469236, EPI_ISL_2469237, EPI_ISL_2469239, EPI_ISL_2469240, EPI_ISL_2469245, EPI_ISL_2469246, EPI_ISL_2469248, EPI_ISL_2469251, EPI_ISL_2469252, EPI_ISL_2469258, EPI_ISL_2469261, EPI_ISL_2469262, EPI_ISL_2469263, EPI_ISL_2469265, EPI_ISL_2469267, EPI_ISL_2469273, EPI_ISL_2469274, EPI_ISL_2469276, EPI_ISL_2469277, EPI_ISL_2469278, EPI_ISL_2469279, EPI_ISL_2469284, EPI_ISL_2469286, EPI_ISL_2469287, EPI_ISL_2469289, EPI_ISL_2469293, EPI_ISL_2469295, EPI_ISL_2469298, EPI_ISL_2469300, EPI_ISL_2469303, EPI_ISL_2469304, EPI_ISL_2469305, EPI_ISL_2469309, EPI_ISL_2469315, EPI_ISL_2469317, EPI_ISL_2469320, EPI_ISL_2469321, EPI_ISL_2469322, EPI_ISL_2469330, EPI_ISL_2469336, EPI_ISL_2469338, EPI_ISL_2469340, EPI_ISL_2469344, EPI_ISL_2469345, EPI_ISL_2469347, EPI_ISL_2469349, EPI_ISL_2469353, EPI_ISL_2469354, EPI_ISL_2469356, EPI_ISL_2469362, EPI_ISL_2469363, EPI_ISL_2469364, EPI_ISL_2469365, EPI_ISL_2469369, EPI_ISL_2469370, EPI_ISL_2469371, EPI_ISL_2469373, EPI_ISL_2469376, EPI_ISL_2469380, EPI_ISL_2469381, EPI_ISL_2469385, EPI_ISL_2469386, EPI_ISL_2469389, EPI_ISL_2469390, EPI_ISL_2469392, EPI_ISL_2469394, EPI_ISL_2469397, EPI_ISL_2469399, EPI_ISL_2469400, EPI_ISL_2469402, EPI_ISL_2469404, EPI_ISL_2469405, EPI_ISL_2469406, EPI_ISL_2469407, EPI_ISL_2469414, EPI_ISL_2469417, EPI_ISL_2469418, EPI_ISL_2469427, EPI_ISL_2469428, EPI_ISL_2469430, EPI_ISL_2469432, EPI_ISL_2469436, EPI_ISL_2469446, EPI_ISL_2469450, EPI_ISL_2485328, EPI_ISL_2485345, EPI_ISL_2485367, EPI_ISL_2485371, EPI_ISL_2485379, EPI_ISL_2485392, EPI_ISL_2485393, EPI_ISL_2485403, EPI_ISL_2485432, EPI_ISL_2485445, EPI_ISL_2485467, EPI_ISL_2485471, EPI_ISL_2485473, EPI_ISL_2485486, EPI_ISL_2485492, EPI_ISL_2485497, EPI_ISL_2485499, EPI_ISL_2485507, EPI_ISL_2485509, EPI_ISL_2485517, EPI_ISL_2485521, EPI_ISL_2485654, EPI_ISL_2485681, EPI_ISL_2485698, EPI_ISL_2486895, EPI_ISL_2486916, EPI_ISL_2486918, EPI_ISL_2486929, EPI_ISL_2486934, EPI_ISL_2486944, EPI_ISL_2486951, EPI_ISL_2487035, EPI_ISL_2487088, EPI_ISL_2487096, EPI_ISL_2487116, EPI_ISL_2487118, EPI_ISL_2487148, EPI_ISL_2487172, EPI_ISL_2487175, EPI_ISL_2487192, EPI_ISL_2487209, EPI_ISL_2487213, EPI_ISL_2487949, EPI_ISL_2515644, EPI_ISL_2516010, EPI_ISL_2516011, EPI_ISL_2516017, EPI_ISL_2516022, EPI_ISL_2516027, EPI_ISL_2516028, EPI_ISL_2516032, EPI_ISL_2516046, EPI_ISL_2516047, EPI_ISL_2516050, EPI_ISL_2516052, EPI_ISL_2516053, EPI_ISL_2516060, EPI_ISL_2516063, EPI_ISL_2516065, EPI_ISL_2516067, EPI_ISL_2516069, EPI_ISL_2516071, EPI_ISL_2516074, EPI_ISL_2516083, EPI_ISL_2516091, EPI_ISL_2516095, EPI_ISL_2516099, EPI_ISL_2516101, EPI_ISL_2516102, EPI_ISL_2516108, EPI_ISL_2516110, EPI_ISL_2516115, EPI_ISL_2516116, EPI_ISL_2516119, EPI_ISL_2516122, EPI_ISL_2516126, EPI_ISL_2516136, EPI_ISL_2516137, EPI_ISL_2516138, EPI_ISL_2516139, EPI_ISL_2516144, EPI_ISL_2516153, EPI_ISL_2516155, EPI_ISL_2516160, EPI_ISL_2516162, EPI_ISL_2516163, EPI_ISL_2516168, EPI_ISL_2516170, EPI_ISL_2516176, EPI_ISL_2516177, EPI_ISL_2516185, EPI_ISL_2516195, EPI_ISL_2516199, EPI_ISL_2516202, EPI_ISL_2516203, EPI_ISL_2516204, EPI_ISL_2516208, EPI_ISL_2516211, EPI_ISL_2516212, EPI_ISL_2516216, EPI_ISL_2516217, EPI_ISL_2516222, EPI_ISL_2516226, EPI_ISL_2516230, EPI_ISL_2516235, EPI_ISL_2516236, EPI_ISL_2516241, EPI_ISL_2516248, EPI_ISL_2516252, EPI_ISL_2516254, EPI_ISL_2516257, EPI_ISL_2516259, EPI_ISL_2516264, EPI_ISL_2516266, EPI_ISL_2516268, EPI_ISL_2516270, EPI_ISL_2516274, EPI_ISL_2516280, EPI_ISL_2516281, EPI_ISL_2516282, EPI_ISL_2516284, EPI_ISL_2516289, EPI_ISL_2516290, EPI_ISL_2516292, EPI_ISL_2516297, EPI_ISL_2516299, EPI_ISL_2516301, EPI_ISL_2516302, EPI_ISL_2516306 | | | |
| see above | Health Services Laboratories | Wellcome Sanger Institute for the COVID-19 Genomics UK (COG-UK) Consortium | Cordelia Langford; David K. Jackson; Dominic Kwiatkowski; Ewan Harrison; Health Services Laboratories and Alex Alderton; Ian Johnston; Jeffrey Barrett; John Sillitoe on behalf of the Wellcome Sanger Institute COVID-19 Surveillance Team; Roberto Amato; Sonia Goncalves |
| EPI_ISL_2468300, EPI_ISL_2468301, EPI_ISL_2468308, EPI_ISL_2468312, EPI_ISL_2468318, EPI_ISL_2468323, EPI_ISL_2468338, EPI_ISL_2468343, EPI_ISL_2468359, EPI_ISL_2468374, EPI_ISL_2468378, EPI_ISL_2468382, EPI_ISL_2468384, EPI_ISL_2468388, EPI_ISL_2468392, EPI_ISL_2468396, EPI_ISL_2468405, EPI_ISL_2468408, EPI_ISL_2468447, EPI_ISL_2468469, EPI_ISL_2468470, EPI_ISL_2468475, EPI_ISL_2468490, EPI_ISL_2468491, EPI_ISL_2468497, EPI_ISL_2468512, EPI_ISL_2468513, EPI_ISL_2468516, EPI_ISL_2468521, EPI_ISL_2468522, EPI_ISL_2468527, EPI_ISL_2468530, EPI_ISL_2468540, EPI_ISL_2468552, EPI_ISL_2468553, EPI_ISL_2468574, EPI_ISL_2468582, EPI_ISL_2468584, EPI_ISL_2468585, EPI_ISL_2468591, EPI_ISL_2468596, EPI_ISL_2468606, EPI_ISL_2468607, EPI_ISL_2468608, EPI_ISL_2468610, EPI_ISL_2468616, EPI_ISL_2468621, EPI_ISL_2468624, EPI_ISL_2468630, EPI_ISL_2468632, EPI_ISL_2468640, EPI_ISL_2468646, EPI_ISL_2468651, EPI_ISL_2468652, EPI_ISL_2468654, EPI_ISL_2468657, EPI_ISL_2468668, EPI_ISL_2468671, EPI_ISL_2468680, EPI_ISL_2468681, EPI_ISL_2468685, EPI_ISL_2468687, EPI_ISL_2468697, EPI_ISL_2468700, EPI_ISL_2468705, EPI_ISL_2468712, EPI_ISL_2468713, EPI_ISL_2468718, EPI_ISL_2468721, EPI_ISL_2468727, EPI_ISL_2468728, EPI_ISL_2468739, EPI_ISL_2468745, EPI_ISL_2468746, EPI_ISL_2468748, EPI_ISL_2468767, EPI_ISL_2468770, EPI_ISL_2468795, EPI_ISL_2468796, EPI_ISL_2468800, EPI_ISL_2468803, EPI_ISL_2468805, EPI_ISL_2468806, EPI_ISL_2468808, EPI_ISL_2468810, EPI_ISL_2468812, EPI_ISL_2468817, EPI_ISL_2468821, EPI_ISL_2468825, EPI_ISL_2468829, EPI_ISL_2468832, EPI_ISL_2468841, EPI_ISL_2468842, EPI_ISL_2468847, EPI_ISL_2468849, EPI_ISL_2468850, EPI_ISL_2468861, EPI_ISL_2468863, EPI_ISL_2468867, EPI_ISL_2468869, EPI_ISL_2468872, EPI_ISL_2468873, EPI_ISL_2468879, EPI_ISL_2468886, EPI_ISL_2468897, EPI_ISL_2468904, EPI_ISL_2468905, EPI_ISL_2468908, EPI_ISL_2468909, EPI_ISL_2468911, EPI_ISL_2468915, EPI_ISL_2468917, EPI_ISL_2468921, EPI_ISL_2468923, EPI_ISL_2468936, EPI_ISL_2468937, EPI_ISL_2468956, EPI_ISL_2485265, EPI_ISL_2485267, EPI_ISL_2485268, EPI_ISL_2485270, EPI_ISL_2485272, EPI_ISL_2485274, EPI_ISL_2485275, EPI_ISL_2485283, EPI_ISL_2485284, EPI_ISL_2485287, EPI_ISL_2485295, EPI_ISL_2485296, EPI_ISL_2485298, EPI_ISL_2485302, EPI_ISL_2485307, EPI_ISL_2485309, EPI_ISL_2485315, EPI_ISL_2485320, EPI_ISL_2485322, EPI_ISL_2485323, EPI_ISL_2485324, EPI_ISL_2485326, EPI_ISL_2485327, EPI_ISL_2485329, EPI_ISL_2485334, EPI_ISL_2485338, EPI_ISL_2485339, EPI_ISL_2485341, EPI_ISL_2485344, EPI_ISL_2485350, EPI_ISL_2485356, EPI_ISL_2485357, EPI_ISL_2485358, EPI_ISL_2485359, EPI_ISL_2485360, EPI_ISL_2485362, EPI_ISL_2485363, EPI_ISL_2485365, EPI_ISL_2485366, EPI_ISL_2485368, EPI_ISL_2485369, EPI_ISL_2485375, EPI_ISL_2485376, EPI_ISL_2485378, EPI_ISL_2485386, EPI_ISL_2485388, EPI_ISL_2485389, EPI_ISL_2485390, EPI_ISL_2485391, EPI_ISL_2485394, EPI_ISL_2485395, EPI_ISL_2485405, EPI_ISL_2485409, EPI_ISL_2485411, EPI_ISL_2485419, EPI_ISL_2485420, EPI_ISL_2485423, EPI_ISL_2485424, EPI_ISL_2485425, EPI_ISL_2485427, EPI_ISL_2485428, EPI_ISL_2485429, EPI_ISL_2485433, EPI_ISL_2485436, EPI_ISL_2485438, EPI_ISL_2485448, EPI_ISL_2485449, EPI_ISL_2485452, EPI_ISL_2485454, EPI_ISL_2485455, EPI_ISL_2485458, EPI_ISL_2485459, EPI_ISL_2485461, EPI_ISL_2485464, EPI_ISL_2485466, EPI_ISL_2485478, EPI_ISL_2485480, EPI_ISL_2485481, EPI_ISL_2485484, EPI_ISL_2485491, EPI_ISL_2485493, EPI_ISL_2485495, EPI_ISL_2485496, EPI_ISL_2485500, EPI_ISL_2485501, EPI_ISL_2485510, EPI_ISL_2485511, EPI_ISL_2485516, EPI_ISL_2485518, EPI_ISL_2485519, EPI_ISL_2485520, EPI_ISL_2485523, EPI_ISL_2485526, EPI_ISL_2485527, EPI_ISL_2485530, EPI_ISL_2485533, EPI_ISL_2485535, EPI_ISL_2485536, EPI_ISL_2485538, EPI_ISL_2485542, EPI_ISL_2485543, EPI_ISL_2485545, EPI_ISL_2485547, EPI_ISL_2485548, EPI_ISL_2485549, EPI_ISL_2485551, EPI_ISL_2485552, EPI_ISL_2485553, EPI_ISL_2485555, EPI_ISL_2485556, EPI_ISL_2485557, EPI_ISL_2485558, EPI_ISL_2485561, EPI_ISL_2485562, EPI_ISL_2485563, EPI_ISL_2485566, EPI_ISL_2485570, EPI_ISL_2485572, EPI_ISL_2485573, EPI_ISL_2485574, EPI_ISL_2485575, EPI_ISL_2485576, EPI_ISL_2485577, EPI_ISL_2485578, EPI_ISL_2485583, EPI_ISL_2485585, EPI_ISL_2485587, EPI_ISL_2485591, EPI_ISL_2485593, EPI_ISL_2485594, EPI_ISL_2485598, EPI_ISL_2485599, EPI_ISL_2485601, EPI_ISL_2485603, EPI_ISL_2485604, EPI_ISL_2485607, EPI_ISL_2485609, EPI_ISL_2485611, EPI_ISL_2485613, EPI_ISL_2485614, EPI_ISL_2485615, EPI_ISL_2485620, EPI_ISL_2485621, EPI_ISL_2485624, EPI_ISL_2485625, EPI_ISL_2485627, EPI_ISL_2485629, EPI_ISL_2485630, EPI_ISL_2485631, EPI_ISL_2485632, EPI_ISL_2485634, EPI_ISL_2485635, EPI_ISL_2485636, EPI_ISL_2485637, EPI_ISL_2485638, EPI_ISL_2485639, EPI_ISL_2485640, EPI_ISL_2485644, EPI_ISL_2485645, EPI_ISL_2485649, EPI_ISL_2485650, EPI_ISL_2485653, EPI_ISL_2485655, EPI_ISL_2485657, EPI_ISL_2485659, EPI_ISL_2485661, EPI_ISL_2485662, EPI_ISL_2485663, EPI_ISL_2485668, EPI_ISL_2485669, EPI_ISL_2485670, EPI_ISL_2485671, EPI_ISL_2485674, EPI_ISL_2485675, EPI_ISL_2485678, EPI_ISL_2485682, EPI_ISL_2485686, EPI_ISL_2485687, EPI_ISL_2485691, EPI_ISL_2485695, EPI_ISL_2485696, EPI_ISL_2485699, EPI_ISL_2485700, EPI_ISL_2485702, EPI_ISL_2485709, EPI_ISL_2485713, EPI_ISL_2485718, EPI_ISL_2485719, EPI_ISL_2485720, EPI_ISL_2485721, EPI_ISL_2485722, EPI_ISL_2485724, EPI_ISL_2485728, EPI_ISL_2485731, EPI_ISL_2485733, EPI_ISL_2485734, EPI_ISL_2485736, EPI_ISL_2485737, EPI_ISL_2485738, EPI_ISL_2485740, EPI_ISL_2485747, EPI_ISL_2485748, EPI_ISL_2485753, EPI_ISL_2485754, EPI_ISL_2485759, EPI_ISL_2485761, EPI_ISL_2485766, EPI_ISL_2485769, EPI_ISL_2485770, EPI_ISL_2485772, EPI_ISL_2485774, EPI_ISL_2485775, EPI_ISL_2485776, EPI_ISL_2485778, EPI_ISL_2485781, EPI_ISL_2485782, EPI_ISL_2485783, EPI_ISL_2485784, EPI_ISL_2485786, EPI_ISL_2485787, EPI_ISL_2485788, EPI_ISL_2485789, EPI_ISL_2485790, EPI_ISL_2485793, EPI_ISL_2485797, EPI_ISL_2485798, EPI_ISL_2485799, EPI_ISL_2485803, EPI_ISL_2485805, EPI_ISL_2485808, EPI_ISL_2485810, EPI_ISL_2485813, EPI_ISL_2485816, EPI_ISL_2485817, EPI_ISL_2485818, EPI_ISL_2485822, EPI_ISL_2485823, EPI_ISL_2485824, EPI_ISL_2485826, EPI_ISL_2485827, EPI_ISL_2485828, EPI_ISL_2485831, EPI_ISL_2485832, EPI_ISL_2485833, EPI_ISL_2485834, EPI_ISL_2485837, EPI_ISL_2485838, EPI_ISL_2485840, EPI_ISL_2485841, EPI_ISL_2485842, EPI_ISL_2485843, EPI_ISL_2485847, EPI_ISL_2485848, EPI_ISL_2485852, EPI_ISL_2485853, EPI_ISL_2485857, EPI_ISL_2485861, EPI_ISL_2485866, EPI_ISL_2485870, EPI_ISL_2485876, EPI_ISL_2485880, EPI_ISL_2485881, EPI_ISL_2485896, EPI_ISL_2485899, EPI_ISL_2485900, EPI_ISL_2485910, EPI_ISL_2485916, EPI_ISL_2485922, EPI_ISL_2485923, EPI_ISL_2485934, EPI_ISL_2485937, EPI_ISL_2485941, EPI_ISL_2485942, EPI_ISL_2485945, EPI_ISL_2485946, EPI_ISL_2485953, EPI_ISL_2485954, EPI_ISL_2485955, EPI_ISL_2485956, EPI_ISL_2485957, EPI_ISL_2485970, EPI_ISL_2485976, EPI_ISL_2485978, EPI_ISL_2485983, EPI_ISL_2485991, EPI_ISL_2485997, EPI_ISL_2485998, EPI_ISL_2486002, EPI_ISL_2486003, EPI_ISL_2486007, EPI_ISL_2486011, EPI_ISL_2486013, EPI_ISL_2486028, EPI_ISL_2486029, EPI_ISL_2486034, EPI_ISL_2486035, EPI_ISL_2486037, EPI_ISL_2486039, EPI_ISL_2486044, EPI_ISL_2486047, EPI_ISL_2486050, EPI_ISL_2486054, EPI_ISL_2486055, EPI_ISL_2486060, EPI_ISL_2486065, EPI_ISL_2486068, EPI_ISL_2486077, EPI_ISL_2486080, EPI_ISL_2486084, EPI_ISL_2486085, EPI_ISL_2486089, EPI_ISL_2486090, EPI_ISL_2486091, EPI_ISL_2486092, EPI_ISL_2486095, EPI_ISL_2486096, EPI_ISL_2486097, EPI_ISL_2486098, EPI_ISL_2486099, EPI_ISL_2486100, EPI_ISL_2486101, EPI_ISL_2486102, EPI_ISL_2486105, EPI_ISL_2486110, EPI_ISL_2486120, EPI_ISL_2486121, EPI_ISL_2486126, EPI_ISL_2486130, EPI_ISL_2486140, EPI_ISL_2486144, EPI_ISL_2486146, EPI_ISL_2486150, EPI_ISL_2486154, EPI_ISL_2486157, EPI_ISL_2486161, EPI_ISL_2486162, EPI_ISL_2486163, EPI_ISL_2486168, EPI_ISL_2486171, EPI_ISL_2486174, EPI_ISL_2486180, EPI_ISL_2486183, EPI_ISL_2486530, EPI_ISL_2486532, EPI_ISL_2486533, EPI_ISL_2486534, EPI_ISL_2486539, EPI_ISL_2486540, EPI_ISL_2486541, EPI_ISL_2486542, EPI_ISL_2486543, EPI_ISL_2486544, EPI_ISL_2486545, EPI_ISL_2486546, EPI_ISL_2486547, EPI_ISL_2486549, EPI_ISL_2486551, EPI_ISL_2486553, EPI_ISL_2486559, EPI_ISL_2486560, EPI_ISL_2486562, EPI_ISL_2486564, EPI_ISL_2486565, EPI_ISL_2486569, EPI_ISL_2486570, EPI_ISL_2486572, EPI_ISL_2486573, EPI_ISL_2486574, EPI_ISL_2486575, EPI_ISL_2486576, EPI_ISL_2486577, EPI_ISL_2486578, EPI_ISL_2486579, EPI_ISL_2486580, EPI_ISL_2486582, EPI_ISL_2486583, EPI_ISL_2486584, EPI_ISL_2486586, EPI_ISL_2486587, EPI_ISL_2486588, EPI_ISL_2486589, EPI_ISL_2486590, EPI_ISL_2486591, EPI_ISL_2486592, EPI_ISL_2486593, EPI_ISL_2486594, EPI_ISL_2486595, EPI_ISL_2486596, EPI_ISL_2486598, EPI_ISL_2486599, EPI_ISL_2486602, EPI_ISL_2486605, EPI_ISL_2486607, EPI_ISL_2486608, EPI_ISL_2486610, EPI_ISL_2486611, EPI_ISL_2486612, EPI_ISL_2486613, EPI_ISL_2486614, EPI_ISL_2486615, EPI_ISL_2486616, EPI_ISL_2486618, EPI_ISL_2486619, EPI_ISL_2486620, EPI_ISL_2486621, EPI_ISL_2486623, EPI_ISL_2486624, EPI_ISL_2486625, EPI_ISL_2486626, EPI_ISL_2486629, EPI_ISL_2486631, EPI_ISL_2486634, EPI_ISL_2486636, EPI_ISL_2486637, EPI_ISL_2486639, EPI_ISL_2486643, EPI_ISL_2486646, EPI_ISL_2486649, EPI_ISL_2486650, EPI_ISL_2486652, EPI_ISL_2486653, EPI_ISL_2486655, EPI_ISL_2486656, EPI_ISL_2486658, EPI_ISL_2486659, EPI_ISL_2486660, EPI_ISL_2486662, EPI_ISL_2486665, EPI_ISL_2486666, EPI_ISL_2486667, EPI_ISL_2486669, EPI_ISL_2486671, EPI_ISL_2486677, EPI_ISL_2486679, EPI_ISL_2486680, EPI_ISL_2486683, EPI_ISL_2486684, EPI_ISL_2486686, EPI_ISL_2486688, EPI_ISL_2486689, EPI_ISL_2486691, EPI_ISL_2486692, EPI_ISL_2486693, EPI_ISL_2486694, EPI_ISL_2486695, EPI_ISL_2486696, EPI_ISL_2486697, EPI_ISL_2486698, EPI_ISL_2486700, EPI_ISL_2486702, EPI_ISL_2486703, EPI_ISL_2486705, EPI_ISL_2486706, EPI_ISL_2486708, EPI_ISL_2486710, EPI_ISL_2486712, EPI_ISL_2486715, EPI_ISL_2486716, EPI_ISL_2486717, EPI_ISL_2486719, EPI_ISL_2486720, EPI_ISL_2486721, EPI_ISL_2486722, EPI_ISL_2486725, EPI_ISL_2486726, EPI_ISL_2486728, EPI_ISL_2486729, EPI_ISL_2486730, EPI_ISL_2486731, EPI_ISL_2486732, EPI_ISL_2486734, EPI_ISL_2486739, EPI_ISL_2486740, EPI_ISL_2486741, EPI_ISL_2486744, EPI_ISL_2486745, EPI_ISL_2486746, EPI_ISL_2486747, EPI_ISL_2486750, EPI_ISL_2486752, EPI_ISL_2486753, EPI_ISL_2486754, EPI_ISL_2486756, EPI_ISL_2486757, EPI_ISL_2486758, EPI_ISL_2486759, EPI_ISL_2486762, EPI_ISL_2486763, EPI_ISL_2486764, EPI_ISL_2486765, EPI_ISL_2486767, EPI_ISL_2486770, EPI_ISL_2486772, EPI_ISL_2486773, EPI_ISL_2486775, EPI_ISL_2486776, EPI_ISL_2486778, EPI_ISL_2486779, EPI_ISL_2486781, EPI_ISL_2486783, EPI_ISL_2486784, EPI_ISL_2486788, EPI_ISL_2486789, EPI_ISL_2486792, EPI_ISL_2486796, EPI_ISL_2486798, EPI_ISL_2486799, EPI_ISL_2486801, EPI_ISL_2486803, EPI_ISL_2486804, EPI_ISL_2486805, EPI_ISL_2486806, EPI_ISL_2486807, EPI_ISL_2486808, EPI_ISL_2486810, EPI_ISL_2486811, EPI_ISL_2486813, EPI_ISL_2486817, EPI_ISL_2486818, EPI_ISL_2486819, EPI_ISL_2486820, EPI_ISL_2486824, EPI_ISL_2486825, EPI_ISL_2486828, EPI_ISL_2486830, EPI_ISL_2486833, EPI_ISL_2486834, EPI_ISL_2486836, EPI_ISL_2486837, EPI_ISL_2486839, EPI_ISL_2486840, EPI_ISL_2486841, EPI_ISL_2486842, EPI_ISL_2486843, EPI_ISL_2486844, EPI_ISL_2486846, EPI_ISL_2486847, EPI_ISL_2486848, EPI_ISL_2486850, EPI_ISL_2486852, EPI_ISL_2486853, EPI_ISL_2486854, EPI_ISL_2486856, EPI_ISL_2486857, EPI_ISL_2486858, EPI_ISL_2486859, EPI_ISL_2486861, EPI_ISL_2486866, EPI_ISL_2486867, EPI_ISL_2486869, EPI_ISL_2486870, EPI_ISL_2486871, EPI_ISL_2486874, EPI_ISL_2486876, EPI_ISL_2486877, EPI_ISL_2486879, EPI_ISL_2486880, EPI_ISL_2486881, EPI_ISL_2486884, EPI_ISL_2486885, EPI_ISL_2486887, EPI_ISL_2486889, EPI_ISL_2486891, EPI_ISL_2486892, EPI_ISL_2486893, EPI_ISL_2486897, EPI_ISL_2486901, EPI_ISL_2486902, EPI_ISL_2486903, EPI_ISL_2486905, EPI_ISL_2486906, EPI_ISL_2486907, EPI_ISL_2486909, EPI_ISL_2486910, EPI_ISL_2486911, EPI_ISL_2486913, EPI_ISL_2486914, EPI_ISL_2486915, EPI_ISL_2486917, EPI_ISL_2486919, EPI_ISL_2486920, EPI_ISL_2486926, EPI_ISL_2486927, EPI_ISL_2486928, EPI_ISL_2486935, EPI_ISL_2486936, EPI_ISL_2486937, EPI_ISL_2486939, EPI_ISL_2486940, EPI_ISL_2486943, EPI_ISL_2486946, EPI_ISL_2486948, EPI_ISL_2486954, EPI_ISL_2486955, EPI_ISL_2486957, EPI_ISL_2486958, EPI_ISL_2486959, EPI_ISL_2486960, EPI_ISL_2486962, EPI_ISL_2486964, EPI_ISL_2486965, EPI_ISL_2486966, EPI_ISL_2486967, EPI_ISL_2486968, EPI_ISL_2486970, EPI_ISL_2486972, EPI_ISL_2486973, EPI_ISL_2486974, EPI_ISL_2486975, EPI_ISL_2486980, EPI_ISL_2486985, EPI_ISL_2486986, EPI_ISL_2486988, EPI_ISL_2486990, EPI_ISL_2486991, EPI_ISL_2486992, EPI_ISL_2486993, EPI_ISL_2486994, EPI_ISL_2486997, EPI_ISL_2486999, EPI_ISL_2487000, EPI_ISL_2487002, EPI_ISL_2487003, EPI_ISL_2487004, EPI_ISL_2487007, EPI_ISL_2487009, EPI_ISL_2487010, EPI_ISL_2487012, EPI_ISL_2487016, EPI_ISL_2487018, EPI_ISL_2487023, EPI_ISL_2487024, EPI_ISL_2487028, EPI_ISL_2487029, EPI_ISL_2487033, EPI_ISL_2487037, EPI_ISL_2487039, EPI_ISL_2487040, EPI_ISL_2487042, EPI_ISL_2487045, EPI_ISL_2487046, EPI_ISL_2487049, EPI_ISL_2487051, EPI_ISL_2487053, EPI_ISL_2487054, EPI_ISL_2487056, EPI_ISL_2487059, EPI_ISL_2487060, EPI_ISL_2487063, EPI_ISL_2487064, EPI_ISL_2487066, EPI_ISL_2487068, EPI_ISL_2487072, EPI_ISL_2487074, EPI_ISL_2487075, EPI_ISL_2487076, EPI_ISL_2487078, EPI_ISL_2487081, EPI_ISL_2487085, EPI_ISL_2487086, EPI_ISL_2487087, EPI_ISL_2487093, EPI_ISL_2487094, EPI_ISL_2487095, EPI_ISL_2487098, EPI_ISL_2487099, EPI_ISL_2487100, EPI_ISL_2487101, EPI_ISL_2487102, EPI_ISL_2487103, EPI_ISL_2487105, EPI_ISL_2487108, EPI_ISL_2487109, EPI_ISL_2487110, EPI_ISL_2487112, EPI_ISL_2487114, EPI_ISL_2487115, EPI_ISL_2487119, EPI_ISL_2487120, EPI_ISL_2487122, EPI_ISL_2487123, EPI_ISL_2487125, EPI_ISL_2487127, EPI_ISL_2487128, EPI_ISL_2487131, EPI_ISL_2487134, EPI_ISL_2487135, EPI_ISL_2487138, EPI_ISL_2487139, EPI_ISL_2487140, EPI_ISL_2487142, EPI_ISL_2487146, EPI_ISL_2487149, EPI_ISL_2487152, EPI_ISL_2487154, EPI_ISL_2487155, EPI_ISL_2487156, EPI_ISL_2487157, EPI_ISL_2487160, EPI_ISL_2487162, EPI_ISL_2487163, EPI_ISL_2487165, EPI_ISL_2487166, EPI_ISL_2487167, EPI_ISL_2487168, EPI_ISL_2487178, EPI_ISL_2487179, EPI_ISL_2487180, EPI_ISL_2487182, EPI_ISL_2487184, EPI_ISL_2487185, EPI_ISL_2487186, EPI_ISL_2487190, EPI_ISL_2487191, EPI_ISL_2487193, EPI_ISL_2487198, EPI_ISL_2487199, EPI_ISL_2487204, EPI_ISL_2487205, EPI_ISL_2487208, EPI_ISL_2487211, EPI_ISL_2487214, EPI_ISL_2487217, EPI_ISL_2511360, EPI_ISL_2511362, EPI_ISL_2511366, EPI_ISL_2511367, EPI_ISL_2511370, EPI_ISL_2511373, EPI_ISL_2511377, EPI_ISL_2511378, EPI_ISL_2511379, EPI_ISL_2511383, EPI_ISL_2511386, EPI_ISL_2511389, EPI_ISL_2511393, EPI_ISL_2511394, EPI_ISL_2511397, EPI_ISL_2511405, EPI_ISL_2511408, EPI_ISL_2511409, EPI_ISL_2511410, EPI_ISL_2511416, EPI_ISL_2511419, EPI_ISL_2511421, EPI_ISL_2511423, EPI_ISL_2511424, EPI_ISL_2511425, EPI_ISL_2511427, EPI_ISL_2511430, EPI_ISL_2511434, EPI_ISL_2511437, EPI_ISL_2511442, EPI_ISL_2511446, EPI_ISL_2511449, EPI_ISL_2511450, EPI_ISL_2511451, EPI_ISL_2511453, EPI_ISL_2511456, EPI_ISL_2511479, EPI_ISL_2511481, EPI_ISL_2511482, EPI_ISL_2511484, EPI_ISL_2511493, EPI_ISL_2511497, EPI_ISL_2511499, EPI_ISL_2511505, EPI_ISL_2511510, EPI_ISL_2511511, EPI_ISL_2511518, EPI_ISL_2511523, EPI_ISL_2511525, EPI_ISL_2511526, EPI_ISL_2511529, EPI_ISL_2511532, EPI_ISL_2511535, EPI_ISL_2511537, EPI_ISL_2511541, EPI_ISL_2511558, EPI_ISL_2511561, EPI_ISL_2511562, EPI_ISL_2511563, EPI_ISL_2511566, EPI_ISL_2511567, EPI_ISL_2511570, EPI_ISL_2511574, EPI_ISL_2511575, EPI_ISL_2511582, EPI_ISL_2511585, EPI_ISL_2511587, EPI_ISL_2511593, EPI_ISL_2511595, EPI_ISL_2511599, EPI_ISL_2511602, EPI_ISL_2511606, EPI_ISL_2511610, EPI_ISL_2511612, EPI_ISL_2511614, EPI_ISL_2511615, EPI_ISL_2511616, EPI_ISL_2511618, EPI_ISL_2511626, EPI_ISL_2511628, EPI_ISL_2511629, EPI_ISL_2511634, EPI_ISL_2511638, EPI_ISL_2511651, EPI_ISL_2511652, EPI_ISL_2511657, EPI_ISL_2511658, EPI_ISL_2511659, EPI_ISL_2511685, EPI_ISL_2511687, EPI_ISL_2511694, EPI_ISL_2511696, EPI_ISL_2511697, EPI_ISL_2511699, EPI_ISL_2511701, EPI_ISL_2511708, EPI_ISL_2511709, EPI_ISL_2511713, EPI_ISL_2511717, EPI_ISL_2511718, EPI_ISL_2511720, EPI_ISL_2511731, EPI_ISL_2511732, EPI_ISL_2511735, EPI_ISL_2511742, EPI_ISL_2511746, EPI_ISL_2511757, EPI_ISL_2511761, EPI_ISL_2511762, EPI_ISL_2511765, EPI_ISL_2511769, EPI_ISL_2511773, EPI_ISL_2511776, EPI_ISL_2511779, EPI_ISL_2511781, EPI_ISL_2511784, EPI_ISL_2511787, EPI_ISL_2511790, EPI_ISL_2511793, EPI_ISL_2511799, EPI_ISL_2511804, EPI_ISL_2511811, EPI_ISL_2511813, EPI_ISL_2511814, EPI_ISL_2511816, EPI_ISL_2511819, EPI_ISL_2511824, EPI_ISL_2511825, EPI_ISL_2511828, EPI_ISL_2511832, EPI_ISL_2511834, EPI_ISL_2511848, EPI_ISL_2511853, EPI_ISL_2511867, EPI_ISL_2511869, EPI_ISL_2511870, EPI_ISL_2511874, EPI_ISL_2511875, EPI_ISL_2511880, EPI_ISL_2511889, EPI_ISL_2511900, EPI_ISL_2511902, EPI_ISL_2511904, EPI_ISL_2511905, EPI_ISL_2511909, EPI_ISL_2511920, EPI_ISL_2511934, EPI_ISL_2511936, EPI_ISL_2511940, EPI_ISL_2511944, EPI_ISL_2511946, EPI_ISL_2511948, EPI_ISL_2511956, EPI_ISL_2511961, EPI_ISL_2511973, EPI_ISL_2511976, EPI_ISL_2511978, EPI_ISL_2511981, EPI_ISL_2511982, EPI_ISL_2511995, EPI_ISL_2512006, EPI_ISL_2512013, EPI_ISL_2512014, EPI_ISL_2512022, EPI_ISL_2512033, EPI_ISL_2512037, EPI_ISL_2512040, EPI_ISL_2512043, EPI_ISL_2512044, EPI_ISL_2512045, EPI_ISL_2512046, EPI_ISL_2512048, EPI_ISL_2512049, EPI_ISL_2512050, EPI_ISL_2512062, EPI_ISL_2512066, EPI_ISL_2512071, EPI_ISL_2512074, EPI_ISL_2512078, EPI_ISL_2512080, EPI_ISL_2512081, EPI_ISL_2512082, EPI_ISL_2512084, EPI_ISL_2512098, EPI_ISL_2512107, EPI_ISL_2512116, EPI_ISL_2512120, EPI_ISL_2512125, EPI_ISL_2512126, EPI_ISL_2512127, EPI_ISL_2512138, EPI_ISL_2512140, EPI_ISL_2512160, EPI_ISL_2512165, EPI_ISL_2512168, EPI_ISL_2512172, EPI_ISL_2512173, EPI_ISL_2512177, EPI_ISL_2512182, EPI_ISL_2512187, EPI_ISL_2512196, EPI_ISL_2512198, EPI_ISL_2512200, EPI_ISL_2512202, EPI_ISL_2512204, EPI_ISL_2512207, EPI_ISL_2512208, EPI_ISL_2512219, EPI_ISL_2512225, EPI_ISL_2512232, EPI_ISL_2512243, EPI_ISL_2512249, EPI_ISL_2512254, EPI_ISL_2512262, EPI_ISL_2512267, EPI_ISL_2512269, EPI_ISL_2512277, EPI_ISL_2512279, EPI_ISL_2512282, EPI_ISL_2512285, EPI_ISL_2512928, EPI_ISL_2512937, EPI_ISL_2512938, EPI_ISL_2512941, EPI_ISL_2512947, EPI_ISL_2512949, EPI_ISL_2512953, EPI_ISL_2512954, EPI_ISL_2512957, EPI_ISL_2512958, EPI_ISL_2512960, EPI_ISL_2512965, EPI_ISL_2512967, EPI_ISL_2512968, EPI_ISL_2512972, EPI_ISL_2512975, EPI_ISL_2512976, EPI_ISL_2512980, EPI_ISL_2512983, EPI_ISL_2512985, EPI_ISL_2512987, EPI_ISL_2512991, EPI_ISL_2512993, EPI_ISL_2512994, EPI_ISL_2512996, EPI_ISL_2513002, EPI_ISL_2513003, EPI_ISL_2513004, EPI_ISL_2513009, EPI_ISL_2513010, EPI_ISL_2513011, EPI_ISL_2513012, EPI_ISL_2513015, EPI_ISL_2513016, EPI_ISL_2513017, EPI_ISL_2513019, EPI_ISL_2513020, EPI_ISL_2513024, EPI_ISL_2513025, EPI_ISL_2513027, EPI_ISL_2513033, EPI_ISL_2513038, EPI_ISL_2513042, EPI_ISL_2513045, EPI_ISL_2513046, EPI_ISL_2513048, EPI_ISL_2513051, EPI_ISL_2513052, EPI_ISL_2513055, EPI_ISL_2513058, EPI_ISL_2513061, EPI_ISL_2513062, EPI_ISL_2513065, EPI_ISL_2513068, EPI_ISL_2513069, EPI_ISL_2513070, EPI_ISL_2513081, EPI_ISL_2513085, EPI_ISL_2513086, EPI_ISL_2513087, EPI_ISL_2513088, EPI_ISL_2513092, EPI_ISL_2513098, EPI_ISL_2513100, EPI_ISL_2513101, EPI_ISL_2513102, EPI_ISL_2513104, EPI_ISL_2513106, EPI_ISL_2513108, EPI_ISL_2513110, EPI_ISL_2513113, EPI_ISL_2513118, EPI_ISL_2513127, EPI_ISL_2513129, EPI_ISL_2513130, EPI_ISL_2513137, EPI_ISL_2513139, EPI_ISL_2513141, EPI_ISL_2513148, EPI_ISL_2513153, EPI_ISL_2513154, EPI_ISL_2513161, EPI_ISL_2513162, EPI_ISL_2513163, EPI_ISL_2513166, EPI_ISL_2513170, EPI_ISL_2513172, EPI_ISL_2513175, EPI_ISL_2513176, EPI_ISL_2513177, EPI_ISL_2513180, EPI_ISL_2513182, EPI_ISL_2513183, EPI_ISL_2513188, EPI_ISL_2513190, EPI_ISL_2513191, EPI_ISL_2513192, EPI_ISL_2513193, EPI_ISL_2513194, EPI_ISL_2513196, EPI_ISL_2513197, EPI_ISL_2513199, EPI_ISL_2513204, EPI_ISL_2513206, EPI_ISL_2513207, EPI_ISL_2513210, EPI_ISL_2513217, EPI_ISL_2513218, EPI_ISL_2513223, EPI_ISL_2513228, EPI_ISL_2513229, EPI_ISL_2513231, EPI_ISL_2513233, EPI_ISL_2513236, EPI_ISL_2513237, EPI_ISL_2513240, EPI_ISL_2513244, EPI_ISL_2513249, EPI_ISL_2513254, EPI_ISL_2513255, EPI_ISL_2513259, EPI_ISL_2513261, EPI_ISL_2513266, EPI_ISL_2513269, EPI_ISL_2513273, EPI_ISL_2513274, EPI_ISL_2513276, EPI_ISL_2513278, EPI_ISL_2513281, EPI_ISL_2513282, EPI_ISL_2513284, EPI_ISL_2513285, EPI_ISL_2513287, EPI_ISL_2513288, EPI_ISL_2513290, EPI_ISL_2514075, EPI_ISL_2514077, EPI_ISL_2514080, EPI_ISL_2514081, EPI_ISL_2514082, EPI_ISL_2514083, EPI_ISL_2514084, EPI_ISL_2514085, EPI_ISL_2514086, EPI_ISL_2514088, EPI_ISL_2514089, EPI_ISL_2514090, EPI_ISL_2514091, EPI_ISL_2514092, EPI_ISL_2514094, EPI_ISL_2514095, EPI_ISL_2514096, EPI_ISL_2514097, EPI_ISL_2514098, EPI_ISL_2514100, EPI_ISL_2514101, EPI_ISL_2514104, EPI_ISL_2514105, EPI_ISL_2514110, EPI_ISL_2514111, EPI_ISL_2514112, EPI_ISL_2514114, EPI_ISL_2514115, EPI_ISL_2514117, EPI_ISL_2514119, EPI_ISL_2514122, EPI_ISL_2514123, EPI_ISL_2514124, EPI_ISL_2514127, EPI_ISL_2514133, EPI_ISL_2514137, EPI_ISL_2514138, EPI_ISL_2514141, EPI_ISL_2514142, EPI_ISL_2514143, EPI_ISL_2514145, EPI_ISL_2514146, EPI_ISL_2514147, EPI_ISL_2514150, EPI_ISL_2514151, EPI_ISL_2514153, EPI_ISL_2514154, EPI_ISL_2514155, EPI_ISL_2514157, EPI_ISL_2514158, EPI_ISL_2514159, EPI_ISL_2514160, EPI_ISL_2514163, EPI_ISL_2514166, EPI_ISL_2514167, EPI_ISL_2514169, EPI_ISL_2514170, EPI_ISL_2514171, EPI_ISL_2514172, EPI_ISL_2514175, EPI_ISL_2514176, EPI_ISL_2514178, EPI_ISL_2514179, EPI_ISL_2514182, EPI_ISL_2514186, EPI_ISL_2514188, EPI_ISL_2514191, EPI_ISL_2514192, EPI_ISL_2514194, EPI_ISL_2514195, EPI_ISL_2514197, EPI_ISL_2514199, EPI_ISL_2514200, EPI_ISL_2514201, EPI_ISL_2514202, EPI_ISL_2514204, EPI_ISL_2514205, EPI_ISL_2514207, EPI_ISL_2514210, EPI_ISL_2514212, EPI_ISL_2514213, EPI_ISL_2514214, EPI_ISL_2514216, EPI_ISL_2514217, EPI_ISL_2514219, EPI_ISL_2514220, EPI_ISL_2514221, EPI_ISL_2514224, EPI_ISL_2514225, EPI_ISL_2514227, EPI_ISL_2514228, EPI_ISL_2514229, EPI_ISL_2514230, EPI_ISL_2514231, EPI_ISL_2514232, EPI_ISL_2514237, EPI_ISL_2514240, EPI_ISL_2514241, EPI_ISL_2514243, EPI_ISL_2514246, EPI_ISL_2514247, EPI_ISL_2514248, EPI_ISL_2514249, EPI_ISL_2514251, EPI_ISL_2514252, EPI_ISL_2514255, EPI_ISL_2514257, EPI_ISL_2514258, EPI_ISL_2514259, EPI_ISL_2514260, EPI_ISL_2514261, EPI_ISL_2514262, EPI_ISL_2514265, EPI_ISL_2514266, EPI_ISL_2514267, EPI_ISL_2514270, EPI_ISL_2514272, EPI_ISL_2514273, EPI_ISL_2514274, EPI_ISL_2514276, EPI_ISL_2514277, EPI_ISL_2514279, EPI_ISL_2514280, EPI_ISL_2514281, EPI_ISL_2514283, EPI_ISL_2514287, EPI_ISL_2514288, EPI_ISL_2514289, EPI_ISL_2514290, EPI_ISL_2514293, EPI_ISL_2514294, EPI_ISL_2514301, EPI_ISL_2514303, EPI_ISL_2514309, EPI_ISL_2514312, EPI_ISL_2514316, EPI_ISL_2514317, EPI_ISL_2514318, EPI_ISL_2514320, EPI_ISL_2514321, EPI_ISL_2514324, EPI_ISL_2514325, EPI_ISL_2514326, EPI_ISL_2514328, EPI_ISL_2514329, EPI_ISL_2514332, EPI_ISL_2514333, EPI_ISL_2514334, EPI_ISL_2514335, EPI_ISL_2514336, EPI_ISL_2514337, EPI_ISL_2514339, EPI_ISL_2514341, EPI_ISL_2514342, EPI_ISL_2514345, EPI_ISL_2514346, EPI_ISL_2514352, EPI_ISL_2514353, EPI_ISL_2514355, EPI_ISL_2514358, EPI_ISL_2514360, EPI_ISL_2514364, EPI_ISL_2514367, EPI_ISL_2514368, EPI_ISL_2514369, EPI_ISL_2514371, EPI_ISL_2514372, EPI_ISL_2514373, EPI_ISL_2514374, EPI_ISL_2514377, EPI_ISL_2514378, EPI_ISL_2514380, EPI_ISL_2514381, EPI_ISL_2514382, EPI_ISL_2514384, EPI_ISL_2514385, EPI_ISL_2514390, EPI_ISL_2514391, EPI_ISL_2514392, EPI_ISL_2514393, EPI_ISL_2514394, EPI_ISL_2514398, EPI_ISL_2514399, EPI_ISL_2514400, EPI_ISL_2514402, EPI_ISL_2514404, EPI_ISL_2514405, EPI_ISL_2514406, EPI_ISL_2514407, EPI_ISL_2514408, EPI_ISL_2515399, EPI_ISL_2515406, EPI_ISL_2515409, EPI_ISL_2515421, EPI_ISL_2515424, EPI_ISL_2515425, EPI_ISL_2515428, EPI_ISL_2515438, EPI_ISL_2515440, EPI_ISL_2515443, EPI_ISL_2515445, EPI_ISL_2515447, EPI_ISL_2515454, EPI_ISL_2515462, EPI_ISL_2515465, EPI_ISL_2515468, EPI_ISL_2515478, EPI_ISL_2515483, EPI_ISL_2515487, EPI_ISL_2515491, EPI_ISL_2515492, EPI_ISL_2515495, EPI_ISL_2515497, EPI_ISL_2515501, EPI_ISL_2515507, EPI_ISL_2515512, EPI_ISL_2515517, EPI_ISL_2515522, EPI_ISL_2515524, EPI_ISL_2515527, EPI_ISL_2515529, EPI_ISL_2515538, EPI_ISL_2515540, EPI_ISL_2515542, EPI_ISL_2515544, EPI_ISL_2515546, EPI_ISL_2515549, EPI_ISL_2515550, EPI_ISL_2515551, EPI_ISL_2515557, EPI_ISL_2515558, EPI_ISL_2515564, EPI_ISL_2515566, EPI_ISL_2515569, EPI_ISL_2515571, EPI_ISL_2515572, EPI_ISL_2515573, EPI_ISL_2515579, EPI_ISL_2515580, EPI_ISL_2515581, EPI_ISL_2515583, EPI_ISL_2515584, EPI_ISL_2515588, EPI_ISL_2515591, EPI_ISL_2515598, EPI_ISL_2515599, EPI_ISL_2515601, EPI_ISL_2515602, EPI_ISL_2515605, EPI_ISL_2515617, EPI_ISL_2515622, EPI_ISL_2515623, EPI_ISL_2515625, EPI_ISL_2515629, EPI_ISL_2515640, EPI_ISL_2515648, EPI_ISL_2515649, EPI_ISL_2515650, EPI_ISL_2515651, EPI_ISL_2515652, EPI_ISL_2515653, EPI_ISL_2515654, EPI_ISL_2515655, EPI_ISL_2515656, EPI_ISL_2515657, EPI_ISL_2515658, EPI_ISL_2515659, EPI_ISL_2515660, EPI_ISL_2515662, EPI_ISL_2515663, EPI_ISL_2515664, EPI_ISL_2515665, EPI_ISL_2515669, EPI_ISL_2515670, EPI_ISL_2515671, EPI_ISL_2515673, EPI_ISL_2515674, EPI_ISL_2515675, EPI_ISL_2515676, EPI_ISL_2515677, EPI_ISL_2515678, EPI_ISL_2515680, EPI_ISL_2515682, EPI_ISL_2515683, EPI_ISL_2515684, EPI_ISL_2515687, EPI_ISL_2515688, EPI_ISL_2515689, EPI_ISL_2515690, EPI_ISL_2515691, EPI_ISL_2515692, EPI_ISL_2515693, EPI_ISL_2515694, EPI_ISL_2515695, EPI_ISL_2515696, EPI_ISL_2515697, EPI_ISL_2515702, EPI_ISL_2515703, EPI_ISL_2515705, EPI_ISL_2515706, EPI_ISL_2515707, EPI_ISL_2515708, EPI_ISL_2515710, EPI_ISL_2515711, EPI_ISL_2515714, EPI_ISL_2515717, EPI_ISL_2515718, EPI_ISL_2515720, EPI_ISL_2515722, EPI_ISL_2515726, EPI_ISL_2515728, EPI_ISL_2515729, EPI_ISL_2515731, EPI_ISL_2515733, EPI_ISL_2515735, EPI_ISL_2515736, EPI_ISL_2515737, EPI_ISL_2515738, EPI_ISL_2515739, EPI_ISL_2515740, EPI_ISL_2515741, EPI_ISL_2515742, EPI_ISL_2515743, EPI_ISL_2515744, EPI_ISL_2515745, EPI_ISL_2515746, EPI_ISL_2515747, EPI_ISL_2515749, EPI_ISL_2515750, EPI_ISL_2515752, EPI_ISL_2515753, EPI_ISL_2515754, EPI_ISL_2515755, EPI_ISL_2515758, EPI_ISL_2515759, EPI_ISL_2515760, EPI_ISL_2515761, EPI_ISL_2515762, EPI_ISL_2515764, EPI_ISL_2515768, EPI_ISL_2515769, EPI_ISL_2515770, EPI_ISL_2515771, EPI_ISL_2515774, EPI_ISL_2515775, | | | |

EPI_ISL_2515776, EPI_ISL_2515777, EPI_ISL_2515779, EPI_ISL_2515781, EPI_ISL_2515782, EPI_ISL_2515783, EPI_ISL_2515784, EPI_ISL_2515785, EPI_ISL_2515786, EPI_ISL_2515787, EPI_ISL_2515788, EPI_ISL_2515790, EPI_ISL_2515791, EPI_ISL_2515792, EPI_ISL_2515795, EPI_ISL_2515798, EPI_ISL_2515799, EPI_ISL_2515801, EPI_ISL_2515802, EPI_ISL_2515803, EPI_ISL_2515804, EPI_ISL_2515806,
EPI_ISL_2515807, EPI_ISL_2515809, EPI_ISL_2515810, EPI_ISL_2515811, EPI_ISL_2515812, EPI_ISL_2515813, EPI_ISL_2515814, EPI_ISL_2515815, EPI_ISL_2515817, EPI_ISL_2515818, EPI_ISL_2515819, EPI_ISL_2515820, EPI_ISL_2515821, EPI_ISL_2515822, EPI_ISL_2515824, EPI_ISL_2515825, EPI_ISL_2515827, EPI_ISL_2515828, EPI_ISL_2515830, EPI_ISL_2515831, EPI_ISL_2515832, EPI_ISL_2515833,
EPI_ISL_2515834, EPI_ISL_2515835, EPI_ISL_2515836, EPI_ISL_2515837, EPI_ISL_2515838, EPI_ISL_2515839, EPI_ISL_2515840, EPI_ISL_2515841, EPI_ISL_2515842, EPI_ISL_2515843, EPI_ISL_2515844, EPI_ISL_2515845, EPI_ISL_2515847, EPI_ISL_2515848, EPI_ISL_2515850, EPI_ISL_2515852, EPI_ISL_2515853, EPI_ISL_2515854, EPI_ISL_2515855, EPI_ISL_2515856, EPI_ISL_2515857, EPI_ISL_2515858,
EPI_ISL_2515859, EPI_ISL_2515861, EPI_ISL_2515862, EPI_ISL_2515863, EPI_ISL_2515864, EPI_ISL_2515865, EPI_ISL_2515867, EPI_ISL_2515869, EPI_ISL_2515870, EPI_ISL_2515871, EPI_ISL_2515873, EPI_ISL_2515875, EPI_ISL_2515876, EPI_ISL_2515879, EPI_ISL_2515881, EPI_ISL_2515882, EPI_ISL_2515883, EPI_ISL_2515887, EPI_ISL_2515889, EPI_ISL_2515890, EPI_ISL_2515891, EPI_ISL_2515892,
EPI_ISL_2515893, EPI_ISL_2515895, EPI_ISL_2515897, EPI_ISL_2515899, EPI_ISL_2515900, EPI_ISL_2515901, EPI_ISL_2515903, EPI_ISL_2515906, EPI_ISL_2515908, EPI_ISL_2515911, EPI_ISL_2515913, EPI_ISL_2515914, EPI_ISL_2515916, EPI_ISL_2515917, EPI_ISL_2515919, EPI_ISL_2515920, EPI_ISL_2515921, EPI_ISL_2515922, EPI_ISL_2515923, EPI_ISL_2515924, EPI_ISL_2515925, EPI_ISL_2515926,
EPI_ISL_2515927, EPI_ISL_2515928, EPI_ISL_2515929, EPI_ISL_2515930, EPI_ISL_2515931, EPI_ISL_2515932, EPI_ISL_2515934, EPI_ISL_2515935, EPI_ISL_2515936, EPI_ISL_2515939, EPI_ISL_2515940, EPI_ISL_2515941, EPI_ISL_2515942, EPI_ISL_2515943, EPI_ISL_2515944, EPI_ISL_2515945, EPI_ISL_2515946, EPI_ISL_2515947, EPI_ISL_2515950, EPI_ISL_2515951, EPI_ISL_2515953, EPI_ISL_2515954,
EPI_ISL_2515955, EPI_ISL_2515956, EPI_ISL_2515957, EPI_ISL_2515958, EPI_ISL_2515959, EPI_ISL_2515960, EPI_ISL_2515961, EPI_ISL_2515962, EPI_ISL_2515965, EPI_ISL_2515966, EPI_ISL_2515967, EPI_ISL_2515968, EPI_ISL_2515969, EPI_ISL_2515970, EPI_ISL_2515972, EPI_ISL_2515974, EPI_ISL_2515975, EPI_ISL_2515976, EPI_ISL_2515977, EPI_ISL_2515979, EPI_ISL_2515980, EPI_ISL_2515981,
EPI_ISL_2515983, EPI_ISL_2515984, EPI_ISL_2515985, EPI_ISL_2515987, EPI_ISL_2515990, EPI_ISL_2515991, EPI_ISL_2515992, EPI_ISL_2515993, EPI_ISL_2515994, EPI_ISL_2515995, EPI_ISL_2515996, EPI_ISL_2515998, EPI_ISL_2515999, EPI_ISL_2516001, EPI_ISL_2516002, EPI_ISL_2516004, EPI_ISL_2516005, EPI_ISL_2516006, EPI_ISL_2516008, EPI_ISL_2516009, EPI_ISL_2516012, EPI_ISL_2516013,
EPI_ISL_2516015, EPI_ISL_2516016, EPI_ISL_2516018, EPI_ISL_2516019, EPI_ISL_2516026, EPI_ISL_2516029, EPI_ISL_2516030, EPI_ISL_2516034, EPI_ISL_2516036, EPI_ISL_2516037, EPI_ISL_2516038, EPI_ISL_2516040, EPI_ISL_2516043, EPI_ISL_2516045, EPI_ISL_2516048, EPI_ISL_2516049, EPI_ISL_2516054, EPI_ISL_2516057, EPI_ISL_2516059, EPI_ISL_2516062, EPI_ISL_2516064, EPI_ISL_2516066,
EPI_ISL_2516068, EPI_ISL_2516070, EPI_ISL_2516072, EPI_ISL_2516075, EPI_ISL_2516077, EPI_ISL_2516078, EPI_ISL_2516079, EPI_ISL_2516080, EPI_ISL_2516082, EPI_ISL_2516084, EPI_ISL_2516087, EPI_ISL_2516088, EPI_ISL_2516089, EPI_ISL_2516092, EPI_ISL_2516094, EPI_ISL_2516096, EPI_ISL_2516098, EPI_ISL_2516104, EPI_ISL_2516105, EPI_ISL_2516111, EPI_ISL_2516114, EPI_ISL_2516117,
EPI_ISL_2516118, EPI_ISL_2516120, EPI_ISL_2516121, EPI_ISL_2516124, EPI_ISL_2516129, EPI_ISL_2516130, EPI_ISL_2516132, EPI_ISL_2516140, EPI_ISL_2516141, EPI_ISL_2516142, EPI_ISL_2516146, EPI_ISL_2516150, EPI_ISL_2516151, EPI_ISL_2516152, EPI_ISL_2516154, EPI_ISL_2516156, EPI_ISL_2516157, EPI_ISL_2516159, EPI_ISL_2516161, EPI_ISL_2516166, EPI_ISL_2516171, EPI_ISL_2516172,
EPI_ISL_2516174, EPI_ISL_2516175, EPI_ISL_2516178, EPI_ISL_2516179, EPI_ISL_2516180, EPI_ISL_2516181, EPI_ISL_2516182, EPI_ISL_2516184, EPI_ISL_2516186, EPI_ISL_2516187, EPI_ISL_2516189, EPI_ISL_2516191, EPI_ISL_2516193, EPI_ISL_2516194, EPI_ISL_2516196, EPI_ISL_2516197, EPI_ISL_2516205, EPI_ISL_2516206, EPI_ISL_2516207, EPI_ISL_2516209, EPI_ISL_2516210, EPI_ISL_2516213,
EPI_ISL_2516214, EPI_ISL_2516215, EPI_ISL_2516218, EPI_ISL_2516219, EPI_ISL_2516220, EPI_ISL_2516221, EPI_ISL_2516229, EPI_ISL_2516232, EPI_ISL_2516234, EPI_ISL_2516237, EPI_ISL_2516238, EPI_ISL_2516242, EPI_ISL_2516244, EPI_ISL_2516247, EPI_ISL_2516249, EPI_ISL_2516250, EPI_ISL_2516253, EPI_ISL_2516255, EPI_ISL_2516256, EPI_ISL_2516258, EPI_ISL_2516262, EPI_ISL_2516263,
EPI_ISL_2516269, EPI_ISL_2516271, EPI_ISL_2516272, EPI_ISL_2516276, EPI_ISL_2516277, EPI_ISL_2516278, EPI_ISL_2516283, EPI_ISL_2516285, EPI_ISL_2516287, EPI_ISL_2516291, EPI_ISL_2516304, EPI_ISL_2516309, EPI_ISL_2516311, EPI_ISL_2537809, EPI_ISL_2537812, EPI_ISL_2537813, EPI_ISL_2537814, EPI_ISL_2537815, EPI_ISL_2537816, EPI_ISL_2537817, EPI_ISL_2537819, EPI_ISL_2537820,
EPI_ISL_2537821, EPI_ISL_2537822, EPI_ISL_2537824, EPI_ISL_2537825, EPI_ISL_2537827, EPI_ISL_2537828, EPI_ISL_2537830, EPI_ISL_2537831, EPI_ISL_2537832, EPI_ISL_2537833, EPI_ISL_2537835, EPI_ISL_2537837, EPI_ISL_2537839, EPI_ISL_2537840, EPI_ISL_2537842, EPI_ISL_2537843, EPI_ISL_2537844, EPI_ISL_2537845, EPI_ISL_2537847, EPI_ISL_2537848, EPI_ISL_2537850,
EPI_ISL_2537851, EPI_ISL_2537853, EPI_ISL_2537854, EPI_ISL_2537855, EPI_ISL_2537857, EPI_ISL_2537858, EPI_ISL_2537859, EPI_ISL_2537860, EPI_ISL_2537861, EPI_ISL_2537863, EPI_ISL_2537866, EPI_ISL_2537867, EPI_ISL_2537868, EPI_ISL_2537870, EPI_ISL_2537871, EPI_ISL_2537875, EPI_ISL_2537876, EPI_ISL_2537878, EPI_ISL_2537883, EPI_ISL_2537884, EPI_ISL_2537885, EPI_ISL_2537888,
EPI_ISL_2537889, EPI_ISL_2537890, EPI_ISL_2537891, EPI_ISL_2537893, EPI_ISL_2537894, EPI_ISL_2537900, EPI_ISL_2537901, EPI_ISL_2537902, EPI_ISL_2537905, EPI_ISL_2537909, EPI_ISL_2537910, EPI_ISL_2537912, EPI_ISL_2537913, EPI_ISL_2537914, EPI_ISL_2537915, EPI_ISL_2537916, EPI_ISL_2537917, EPI_ISL_2537918, EPI_ISL_2537919, EPI_ISL_2537920, EPI_ISL_2537924, EPI_ISL_2537925,
EPI_ISL_2537926, EPI_ISL_2537927, EPI_ISL_2537928, EPI_ISL_2537930, EPI_ISL_2537933, EPI_ISL_2537934, EPI_ISL_2537935, EPI_ISL_2537936, EPI_ISL_2537938, EPI_ISL_2537939, EPI_ISL_2537943, EPI_ISL_2537945, EPI_ISL_2537946, EPI_ISL_2537947, EPI_ISL_2537949, EPI_ISL_2537951, EPI_ISL_2537953, EPI_ISL_2537955, EPI_ISL_2537956, EPI_ISL_2537957, EPI_ISL_2537960, EPI_ISL_2537961,
EPI_ISL_2537963, EPI_ISL_2537964, EPI_ISL_2537965, EPI_ISL_2537966, EPI_ISL_2537967, EPI_ISL_2537968, EPI_ISL_2537970, EPI_ISL_2537971, EPI_ISL_2537972, EPI_ISL_2537973, EPI_ISL_2537974, EPI_ISL_2537975, EPI_ISL_2537976, EPI_ISL_2537977, EPI_ISL_2537978, EPI_ISL_2537980, EPI_ISL_2537981, EPI_ISL_2537983, EPI_ISL_2537987, EPI_ISL_2537988, EPI_ISL_2537989,
EPI_ISL_2537991, EPI_ISL_2537992, EPI_ISL_2537993, EPI_ISL_2537994, EPI_ISL_2537995, EPI_ISL_2537997, EPI_ISL_2537999, EPI_ISL_2538000, EPI_ISL_2538001, EPI_ISL_2538002, EPI_ISL_2538003, EPI_ISL_2538004, EPI_ISL_2538006, EPI_ISL_2538007, EPI_ISL_2538009, EPI_ISL_2538010, EPI_ISL_2538012, EPI_ISL_2538014, EPI_ISL_2538015, EPI_ISL_2538016, EPI_ISL_2538018, EPI_ISL_2538019,
EPI_ISL_2538020, EPI_ISL_2538021, EPI_ISL_2538022, EPI_ISL_2538023, EPI_ISL_2538024, EPI_ISL_2538025, EPI_ISL_2538027, EPI_ISL_2538029, EPI_ISL_2538030, EPI_ISL_2538031, EPI_ISL_2538033, EPI_ISL_2538034, EPI_ISL_2538035, EPI_ISL_2538036, EPI_ISL_2538037, EPI_ISL_2538039, EPI_ISL_2538040, EPI_ISL_2538042, EPI_ISL_2538043, EPI_ISL_2538044, EPI_ISL_2538045, EPI_ISL_2538046,
EPI_ISL_2538047, EPI_ISL_2538049, EPI_ISL_2538050, EPI_ISL_2538051, EPI_ISL_2538052, EPI_ISL_2538053, EPI_ISL_2538054, EPI_ISL_2538055, EPI_ISL_2538056, EPI_ISL_2538058, EPI_ISL_2538059, EPI_ISL_2538062, EPI_ISL_2538063, EPI_ISL_2538064, EPI_ISL_2538065, EPI_ISL_2538066, EPI_ISL_2538067, EPI_ISL_2538068, EPI_ISL_2538069, EPI_ISL_2538071, EPI_ISL_2538073, EPI_ISL_2538074,
EPI_ISL_2538077, EPI_ISL_2538078, EPI_ISL_2538079, EPI_ISL_2538080, EPI_ISL_2538081, EPI_ISL_2538082, EPI_ISL_2538083, EPI_ISL_2538084, EPI_ISL_2538085, EPI_ISL_2538087, EPI_ISL_2538088, EPI_ISL_2538089, EPI_ISL_2538091, EPI_ISL_2538092, EPI_ISL_2538094, EPI_ISL_2538095, EPI_ISL_2538096, EPI_ISL_2538099, EPI_ISL_2538100, EPI_ISL_2538101, EPI_ISL_2538102, EPI_ISL_2538103,
EPI_ISL_2538104, EPI_ISL_2538105, EPI_ISL_2538106, EPI_ISL_2538107, EPI_ISL_2538108, EPI_ISL_2538109, EPI_ISL_2538110, EPI_ISL_2538112, EPI_ISL_2538113, EPI_ISL_2538114, EPI_ISL_2538115, EPI_ISL_2538116, EPI_ISL_2538117, EPI_ISL_2538118, EPI_ISL_2538119, EPI_ISL_2538120, EPI_ISL_2538121, EPI_ISL_2538122, EPI_ISL_2538123, EPI_ISL_2538125, EPI_ISL_2538127, EPI_ISL_2538128,
EPI_ISL_2538129, EPI_ISL_2538130, EPI_ISL_2538132, EPI_ISL_2538133, EPI_ISL_2538134, EPI_ISL_2538135, EPI_ISL_2538137, EPI_ISL_2538139, EPI_ISL_2538140, EPI_ISL_2538141, EPI_ISL_2538142, EPI_ISL_2538143, EPI_ISL_2538145, EPI_ISL_2538146, EPI_ISL_2538147, EPI_ISL_2538148, EPI_ISL_2538150, EPI_ISL_2538151, EPI_ISL_2538152, EPI_ISL_2538153, EPI_ISL_2538154,
EPI_ISL_2538156, EPI_ISL_2538157, EPI_ISL_2538159, EPI_ISL_2538160, EPI_ISL_2538161, EPI_ISL_2538162, EPI_ISL_2538163, EPI_ISL_2538169, EPI_ISL_2538174, EPI_ISL_2538183, EPI_ISL_2538187, EPI_ISL_2538210, EPI_ISL_2538213, EPI_ISL_2538219, EPI_ISL_2538220, EPI_ISL_2538235, EPI_ISL_2538244, EPI_ISL_2538245, EPI_ISL_2538256, EPI_ISL_2538258, EPI_ISL_2538261, EPI_ISL_2538269,
EPI_ISL_2538270, EPI_ISL_2538274, EPI_ISL_2538278, EPI_ISL_2538281, EPI_ISL_2538286, EPI_ISL_2538301, EPI_ISL_2538314, EPI_ISL_2538319, EPI_ISL_2538324, EPI_ISL_2538351, EPI_ISL_2538354, EPI_ISL_2538414, EPI_ISL_2538415, EPI_ISL_2538423, EPI_ISL_2538446, EPI_ISL_2538460, EPI_ISL_2538466, EPI_ISL_2538496, EPI_ISL_2538497, EPI_ISL_2538505,
EPI_ISL_2538517, EPI_ISL_2538545, EPI_ISL_2538576, EPI_ISL_2538579, EPI_ISL_2538592, EPI_ISL_2538603, EPI_ISL_2538606, EPI_ISL_2538621, EPI_ISL_2538629, EPI_ISL_2538633, EPI_ISL_2538638, EPI_ISL_2538648, EPI_ISL_2538652, EPI_ISL_2538653, EPI_ISL_2538660, EPI_ISL_2538663, EPI_ISL_2538675, EPI_ISL_2538680, EPI_ISL_2538683, EPI_ISL_2538684, EPI_ISL_2538687, EPI_ISL_2538724,
EPI_ISL_2538728, EPI_ISL_2538730, EPI_ISL_2538741, EPI_ISL_2538749, EPI_ISL_2538751, EPI_ISL_2538771, EPI_ISL_2538781, EPI_ISL_2538800, EPI_ISL_2538817, EPI_ISL_2538821, EPI_ISL_2538826, EPI_ISL_2538832, EPI_ISL_2538836, EPI_ISL_2538846, EPI_ISL_2538848, EPI_ISL_2538855, EPI_ISL_2538857, EPI_ISL_2538858, EPI_ISL_2538869, EPI_ISL_2538871, EPI_ISL_2538922, EPI_ISL_2538926,
EPI_ISL_2538927, EPI_ISL_2538929, EPI_ISL_2538957, EPI_ISL_2538960, EPI_ISL_2538961, EPI_ISL_2538963, EPI_ISL_2538978, EPI_ISL_2538981, EPI_ISL_2538983, EPI_ISL_2538994, EPI_ISL_2539008, EPI_ISL_2539025, EPI_ISL_2539042, EPI_ISL_2539058, EPI_ISL_2539061, EPI_ISL_2539068, EPI_ISL_2539072, EPI_ISL_2539073, EPI_ISL_2539108, EPI_ISL_2539123, EPI_ISL_2539131, EPI_ISL_2540373,
EPI_ISL_2540376, EPI_ISL_2540378, EPI_ISL_2540379, EPI_ISL_2540381, EPI_ISL_2540383, EPI_ISL_2540384, EPI_ISL_2540391, EPI_ISL_2540392, EPI_ISL_2540395, EPI_ISL_2540396, EPI_ISL_2540397, EPI_ISL_2540411, EPI_ISL_2540419, EPI_ISL_2540420, EPI_ISL_2540436, EPI_ISL_2540441, EPI_ISL_2540451, EPI_ISL_2540454, EPI_ISL_2540461, EPI_ISL_2540482, EPI_ISL_2540487, EPI_ISL_2540488,
EPI_ISL_2540489, EPI_ISL_2540490, EPI_ISL_2540501, EPI_ISL_2540502, EPI_ISL_2540505, EPI_ISL_2540517, EPI_ISL_2540520, EPI_ISL_2540524, EPI_ISL_2540526, EPI_ISL_2540536, EPI_ISL_2540540, EPI_ISL_2540551, EPI_ISL_2540553, EPI_ISL_2540554, EPI_ISL_2540560, EPI_ISL_2540562, EPI_ISL_2540564, EPI_ISL_2540566, EPI_ISL_2540567, EPI_ISL_2540569, EPI_ISL_2540598, EPI_ISL_2540624,
EPI_ISL_2540639, EPI_ISL_2540647, EPI_ISL_2540758, EPI_ISL_2540794, EPI_ISL_2540836, EPI_ISL_2540846, EPI_ISL_2540913, EPI_ISL_2540937, EPI_ISL_2541640, EPI_ISL_2541650, EPI_ISL_2541660, EPI_ISL_2541710, EPI_ISL_2541719, EPI_ISL_2541728, EPI_ISL_2541729, EPI_ISL_2541735, EPI_ISL_2541736, EPI_ISL_2541739, EPI_ISL_2541761, EPI_ISL_2541776, EPI_ISL_2541781,
EPI_ISL_2541783, EPI_ISL_2541792, EPI_ISL_2541827, EPI_ISL_2541847, EPI_ISL_2541848, EPI_ISL_2541852, EPI_ISL_2541857, EPI_ISL_2541891, EPI_ISL_2541901, EPI_ISL_2541920, EPI_ISL_2541923, EPI_ISL_2541924, EPI_ISL_2541955, EPI_ISL_2541965, EPI_ISL_2541969, EPI_ISL_2541970, EPI_ISL_2541976, EPI_ISL_2541984, EPI_ISL_2541988, EPI_ISL_2552326, EPI_ISL_2552603, EPI_ISL_2553098,
EPI_ISL_2553108, EPI_ISL_2553145, EPI_ISL_2553188, EPI_ISL_2553223, EPI_ISL_2553290, EPI_ISL_2553309, EPI_ISL_2553314, EPI_ISL_2553350, EPI_ISL_2553368, EPI_ISL_2553403, EPI_ISL_2553460, EPI_ISL_2553464, EPI_ISL_2553478, EPI_ISL_2553519, EPI_ISL_2553613, EPI_ISL_2553622, EPI_ISL_2553628, EPI_ISL_2553631, EPI_ISL_2553684, EPI_ISL_2553696, EPI_ISL_2553713, EPI_ISL_2553760,
EPI_ISL_2554242, EPI_ISL_2554846, EPI_ISL_2555526, EPI_ISL_2567656, EPI_ISL_2567659, EPI_ISL_2567664, EPI_ISL_2567666, EPI_ISL_2567668, EPI_ISL_2567669, EPI_ISL_2567680, EPI_ISL_2567684, EPI_ISL_2567688, EPI_ISL_2567692, EPI_ISL_2567694, EPI_ISL_2567705, EPI_ISL_2567732, EPI_ISL_2567734, EPI_ISL_2567735, EPI_ISL_2567741, EPI_ISL_2567745, EPI_ISL_2567757, EPI_ISL_2567766,
EPI_ISL_2567775, EPI_ISL_2567785, EPI_ISL_2567788, EPI_ISL_2567791, EPI_ISL_2567801, EPI_ISL_2567813, EPI_ISL_2567815, EPI_ISL_2567831, EPI_ISL_2567835, EPI_ISL_2567836, EPI_ISL_2567851, EPI_ISL_2567854, EPI_ISL_2567857, EPI_ISL_2567861, EPI_ISL_2567862, EPI_ISL_2567863, EPI_ISL_2567868, EPI_ISL_2567871, EPI_ISL_2567876, EPI_ISL_2567889, EPI_ISL_2567890,
EPI_ISL_2567906, EPI_ISL_2567908, EPI_ISL_2567910, EPI_ISL_2567911, EPI_ISL_2567914, EPI_ISL_2567915, EPI_ISL_2567919, EPI_ISL_2567935, EPI_ISL_2568208, EPI_ISL_2569592, EPI_ISL_2569641, EPI_ISL_2570524, EPI_ISL_2570528, EPI_ISL_2570566, EPI_ISL_2570604, EPI_ISL_2570615, EPI_ISL_2570630, EPI_ISL_2570634, EPI_ISL_2570664, EPI_ISL_2570711, EPI_ISL_2570746, EPI_ISL_2570769,
EPI_ISL_2640357, EPI_ISL_2640391, EPI_ISL_2640419, EPI_ISL_2640427, EPI_ISL_2640458, EPI_ISL_2640490, EPI_ISL_2640523, EPI_ISL_2640530, EPI_ISL_2640552, EPI_ISL_2640559, EPI_ISL_2640621, EPI_ISL_2641571

| | | | |
|---|---|---|---|
| see above | Lighthouse Lab in Alderley Park | Wellcome Sanger Institute for the COVID-19 Genomics UK (COG-UK) Consortium | Cordelia Langford; David K. Jackson; Dominic Kwiatkowski; Ewan Harrison; Ian Johnston; Jacquelyn Wynn; Jeffrey Barrett; John Sillitoe on behalf of the Wellcome Sanger Institute COVID-19 Surveillance Team; Mairead Hyland; Roberto Amato; Sonia Goncalves; The Lighthouse Lab in Alderley Park and Alex Alderton |

EPI_ISL_2437948, EPI_ISL_2457317, EPI_ISL_2457325, EPI_ISL_2457343, EPI_ISL_2457346, EPI_ISL_2468298, EPI_ISL_2468307, EPI_ISL_2468324, EPI_ISL_2468330, EPI_ISL_2468400, EPI_ISL_2468406, EPI_ISL_2468425, EPI_ISL_2468444, EPI_ISL_2468449, EPI_ISL_2468494, EPI_ISL_2468508, EPI_ISL_2468511, EPI_ISL_2468514, EPI_ISL_2468539, EPI_ISL_2468557, EPI_ISL_2468581, EPI_ISL_2468948,
EPI_ISL_2468961, EPI_ISL_2468984, EPI_ISL_2468999, EPI_ISL_2469072, EPI_ISL_2469189, EPI_ISL_2469222, EPI_ISL_2485559, EPI_ISL_2485571, EPI_ISL_2485595, EPI_ISL_2485622, EPI_ISL_2485660, EPI_ISL_2485676, EPI_ISL_2485679, EPI_ISL_2485684, EPI_ISL_2485688, EPI_ISL_2485716, EPI_ISL_2485744, EPI_ISL_2485763, EPI_ISL_2485811, EPI_ISL_2485845, EPI_ISL_2485849, EPI_ISL_2485851,
EPI_ISL_2485854, EPI_ISL_2485859, EPI_ISL_2485860, EPI_ISL_2485862, EPI_ISL_2485863, EPI_ISL_2485865, EPI_ISL_2485871, EPI_ISL_2485872, EPI_ISL_2485873, EPI_ISL_2485874, EPI_ISL_2485879, EPI_ISL_2485882, EPI_ISL_2485884, EPI_ISL_2485885, EPI_ISL_2485886, EPI_ISL_2485887, EPI_ISL_2485888, EPI_ISL_2485889, EPI_ISL_2485891, EPI_ISL_2485895, EPI_ISL_2485897, EPI_ISL_2485902,
EPI_ISL_2485903, EPI_ISL_2485904, EPI_ISL_2485905, EPI_ISL_2485906, EPI_ISL_2485907, EPI_ISL_2485909, EPI_ISL_2485911, EPI_ISL_2485912, EPI_ISL_2485914, EPI_ISL_2485915, EPI_ISL_2485917, EPI_ISL_2485919, EPI_ISL_2485920, EPI_ISL_2485921, EPI_ISL_2485924, EPI_ISL_2485925, EPI_ISL_2485926, EPI_ISL_2485928, EPI_ISL_2485929, EPI_ISL_2485931, EPI_ISL_2485933, EPI_ISL_2485935,
EPI_ISL_2485936, EPI_ISL_2485938, EPI_ISL_2485939, EPI_ISL_2485943, EPI_ISL_2485944, EPI_ISL_2485947, EPI_ISL_2485948, EPI_ISL_2485949, EPI_ISL_2485951, EPI_ISL_2485952, EPI_ISL_2485958, EPI_ISL_2485959, EPI_ISL_2485960, EPI_ISL_2485961, EPI_ISL_2485963, EPI_ISL_2485964, EPI_ISL_2485966, EPI_ISL_2485967, EPI_ISL_2485968, EPI_ISL_2485969, EPI_ISL_2485971, EPI_ISL_2485972,
EPI_ISL_2485973, EPI_ISL_2485974, EPI_ISL_2485975, EPI_ISL_2485980, EPI_ISL_2485981, EPI_ISL_2485982, EPI_ISL_2485984, EPI_ISL_2485985, EPI_ISL_2485987, EPI_ISL_2485988, EPI_ISL_2485989, EPI_ISL_2485990, EPI_ISL_2485992, EPI_ISL_2485993, EPI_ISL_2485994, EPI_ISL_2485996, EPI_ISL_2485999, EPI_ISL_2486001, EPI_ISL_2486004, EPI_ISL_2486009, EPI_ISL_2486010, EPI_ISL_2486012,
EPI_ISL_2486015, EPI_ISL_2486018, EPI_ISL_2486019, EPI_ISL_2486020, EPI_ISL_2486021, EPI_ISL_2486022, EPI_ISL_2486025, EPI_ISL_2486026, EPI_ISL_2486027, EPI_ISL_2486040, EPI_ISL_2486041, EPI_ISL_2486043, EPI_ISL_2486046, EPI_ISL_2486048, EPI_ISL_2486049, EPI_ISL_2486052, EPI_ISL_2486053, EPI_ISL_2486057, EPI_ISL_2486059, EPI_ISL_2486061, EPI_ISL_2486062, EPI_ISL_2486063,
EPI_ISL_2486066, EPI_ISL_2486067, EPI_ISL_2486069, EPI_ISL_2486070, EPI_ISL_2486072, EPI_ISL_2486073, EPI_ISL_2486074, EPI_ISL_2486075, EPI_ISL_2486079, EPI_ISL_2486081, EPI_ISL_2486082, EPI_ISL_2486086, EPI_ISL_2486088, EPI_ISL_2486094, EPI_ISL_2486103, EPI_ISL_2486106, EPI_ISL_2486108, EPI_ISL_2486111, EPI_ISL_2486112, EPI_ISL_2486114, EPI_ISL_2486115, EPI_ISL_2486116,
EPI_ISL_2486117, EPI_ISL_2486118, EPI_ISL_2486122, EPI_ISL_2486123, EPI_ISL_2486125, EPI_ISL_2486129, EPI_ISL_2486131, EPI_ISL_2486133, EPI_ISL_2486134, EPI_ISL_2486138, EPI_ISL_2486139, EPI_ISL_2486143, EPI_ISL_2486145, EPI_ISL_2486148, EPI_ISL_2486149, EPI_ISL_2486151, EPI_ISL_2486152, EPI_ISL_2486153, EPI_ISL_2486155, EPI_ISL_2486156, EPI_ISL_2486158, EPI_ISL_2486160,
EPI_ISL_2486164, EPI_ISL_2486165, EPI_ISL_2486166, EPI_ISL_2486167, EPI_ISL_2486169, EPI_ISL_2486172, EPI_ISL_2486175, EPI_ISL_2486177, EPI_ISL_2486178, EPI_ISL_2486179, EPI_ISL_2486182, EPI_ISL_2486184, EPI_ISL_2486187, EPI_ISL_2486188, EPI_ISL_2486189, EPI_ISL_2486190, EPI_ISL_2486192, EPI_ISL_2486193, EPI_ISL_2486195, EPI_ISL_2486196, EPI_ISL_2486197, EPI_ISL_2486198,
EPI_ISL_2486199, EPI_ISL_2486200, EPI_ISL_2486201, EPI_ISL_2486202, EPI_ISL_2486203, EPI_ISL_2486205, EPI_ISL_2486206, EPI_ISL_2486207, EPI_ISL_2486208, EPI_ISL_2486209, EPI_ISL_2486210, EPI_ISL_2486211, EPI_ISL_2486212, EPI_ISL_2486213, EPI_ISL_2486214, EPI_ISL_2486215, EPI_ISL_2486216, EPI_ISL_2486217, EPI_ISL_2486218, EPI_ISL_2486219, EPI_ISL_2486220, EPI_ISL_2486222,
EPI_ISL_2486223, EPI_ISL_2486224, EPI_ISL_2486225, EPI_ISL_2486226, EPI_ISL_2486227, EPI_ISL_2486228, EPI_ISL_2486229, EPI_ISL_2486230, EPI_ISL_2486231, EPI_ISL_2486232, EPI_ISL_2486233, EPI_ISL_2486234, EPI_ISL_2486235, EPI_ISL_2486236, EPI_ISL_2486237, EPI_ISL_2486238, EPI_ISL_2486239, EPI_ISL_2486240, EPI_ISL_2486242, EPI_ISL_2486243, EPI_ISL_2486244, EPI_ISL_2486245,
EPI_ISL_2486246, EPI_ISL_2486247, EPI_ISL_2486249, EPI_ISL_2486250, EPI_ISL_2486251, EPI_ISL_2486252, EPI_ISL_2486253, EPI_ISL_2486254, EPI_ISL_2486256, EPI_ISL_2486257, EPI_ISL_2486258, EPI_ISL_2486259, EPI_ISL_2486260, EPI_ISL_2486261, EPI_ISL_2486262, EPI_ISL_2486263, EPI_ISL_2486264, EPI_ISL_2486265, EPI_ISL_2486266, EPI_ISL_2486267, EPI_ISL_2486268, EPI_ISL_2486269,
EPI_ISL_2486270, EPI_ISL_2486271, EPI_ISL_2486272, EPI_ISL_2486273, EPI_ISL_2486274, EPI_ISL_2486275, EPI_ISL_2486276, EPI_ISL_2486277, EPI_ISL_2486278, EPI_ISL_2486279, EPI_ISL_2486280, EPI_ISL_2486281, EPI_ISL_2486282, EPI_ISL_2486283, EPI_ISL_2486285, EPI_ISL_2486287, EPI_ISL_2486288, EPI_ISL_2486289, EPI_ISL_2486290, EPI_ISL_2486291, EPI_ISL_2486292, EPI_ISL_2486293,
EPI_ISL_2486294, EPI_ISL_2486295, EPI_ISL_2486296, EPI_ISL_2486297, EPI_ISL_2486298, EPI_ISL_2486299, EPI_ISL_2486300, EPI_ISL_2486301, EPI_ISL_2486302, EPI_ISL_2486303, EPI_ISL_2486304, EPI_ISL_2486306, EPI_ISL_2486307, EPI_ISL_2486308, EPI_ISL_2486309, EPI_ISL_2486311, EPI_ISL_2486312, EPI_ISL_2486313, EPI_ISL_2486314, EPI_ISL_2486315, EPI_ISL_2486316, EPI_ISL_2486317,
EPI_ISL_2486318, EPI_ISL_2486319, EPI_ISL_2486321, EPI_ISL_2486322, EPI_ISL_2486323, EPI_ISL_2486324, EPI_ISL_2486325, EPI_ISL_2486326, EPI_ISL_2486327, EPI_ISL_2486328, EPI_ISL_2486329, EPI_ISL_2486330, EPI_ISL_2486331, EPI_ISL_2486332, EPI_ISL_2486333, EPI_ISL_2486334, EPI_ISL_2486335, EPI_ISL_2486336, EPI_ISL_2486337, EPI_ISL_2486338, EPI_ISL_2486339, EPI_ISL_2486341,
EPI_ISL_2486344, EPI_ISL_2486345, EPI_ISL_2486346, EPI_ISL_2486347, EPI_ISL_2486348, EPI_ISL_2486349, EPI_ISL_2486350, EPI_ISL_2486351, EPI_ISL_2486352, EPI_ISL_2486353, EPI_ISL_2486354, EPI_ISL_2486355, EPI_ISL_2486356, EPI_ISL_2486357, EPI_ISL_2486359, EPI_ISL_2486360, EPI_ISL_2486361, EPI_ISL_2486362, EPI_ISL_2486363, EPI_ISL_2486365, EPI_ISL_2486366, EPI_ISL_2486367,
EPI_ISL_2486368, EPI_ISL_2486369, EPI_ISL_2486370, EPI_ISL_2486371, EPI_ISL_2486373, EPI_ISL_2486374, EPI_ISL_2486375, EPI_ISL_2486376, EPI_ISL_2486377, EPI_ISL_2486378, EPI_ISL_2486380, EPI_ISL_2486381, EPI_ISL_2486382, EPI_ISL_2486383, EPI_ISL_2486384, EPI_ISL_2486385, EPI_ISL_2486386, EPI_ISL_2486387, EPI_ISL_2486388, EPI_ISL_2486389, EPI_ISL_2486390, EPI_ISL_2486391,
EPI_ISL_2486392, EPI_ISL_2486393, EPI_ISL_2486394, EPI_ISL_2486395, EPI_ISL_2486396, EPI_ISL_2486397, EPI_ISL_2486398, EPI_ISL_2486399, EPI_ISL_2486400, EPI_ISL_2486401, EPI_ISL_2486402, EPI_ISL_2486403, EPI_ISL_2486404, EPI_ISL_2486405, EPI_ISL_2486406, EPI_ISL_2486407, EPI_ISL_2486408, EPI_ISL_2486409, EPI_ISL_2486410, EPI_ISL_2486411, EPI_ISL_2486412, EPI_ISL_2486413,
EPI_ISL_2486414, EPI_ISL_2486415, EPI_ISL_2486416, EPI_ISL_2486417, EPI_ISL_2486418, EPI_ISL_2486419, EPI_ISL_2486420, EPI_ISL_2486421, EPI_ISL_2486422, EPI_ISL_2486423, EPI_ISL_2486424, EPI_ISL_2486425, EPI_ISL_2486426, EPI_ISL_2486427, EPI_ISL_2486428, EPI_ISL_2486429, EPI_ISL_2486430, EPI_ISL_2486431, EPI_ISL_2486432, EPI_ISL_2486433, EPI_ISL_2486434, EPI_ISL_2486435,
EPI_ISL_2486436, EPI_ISL_2486437, EPI_ISL_2486438, EPI_ISL_2486439, EPI_ISL_2486440, EPI_ISL_2486441, EPI_ISL_2486442, EPI_ISL_2486443, EPI_ISL_2486444, EPI_ISL_2486445, EPI_ISL_2486447, EPI_ISL_2486448, EPI_ISL_2486450, EPI_ISL_2486451, EPI_ISL_2486452, EPI_ISL_2486453, EPI_ISL_2486454, EPI_ISL_2486455, EPI_ISL_2486456, EPI_ISL_2486457, EPI_ISL_2486459, EPI_ISL_2486460,
EPI_ISL_2486461, EPI_ISL_2486462, EPI_ISL_2486463, EPI_ISL_2486464, EPI_ISL_2486467, EPI_ISL_2486468, EPI_ISL_2486471, EPI_ISL_2486472, EPI_ISL_2486473, EPI_ISL_2486474, EPI_ISL_2486475, EPI_ISL_2486476, EPI_ISL_2486478, EPI_ISL_2486479, EPI_ISL_2486480, EPI_ISL_2486481, EPI_ISL_2486482, EPI_ISL_2486483, EPI_ISL_2486484, EPI_ISL_2486485, EPI_ISL_2486486, EPI_ISL_2486487,
EPI_ISL_2486488, EPI_ISL_2486489, EPI_ISL_2486491, EPI_ISL_2486492, EPI_ISL_2486493, EPI_ISL_2486494, EPI_ISL_2486495, EPI_ISL_2486496, EPI_ISL_2486497, EPI_ISL_2486498, EPI_ISL_2486500, EPI_ISL_2486501, EPI_ISL_2486502, EPI_ISL_2486503, EPI_ISL_2486505, EPI_ISL_2486506, EPI_ISL_2486507, EPI_ISL_2486508, EPI_ISL_2486509, EPI_ISL_2486510, EPI_ISL_2486512, EPI_ISL_2486513,
EPI_ISL_2486514, EPI_ISL_2486515, EPI_ISL_2486517, EPI_ISL_2486518, EPI_ISL_2486519, EPI_ISL_2486521, EPI_ISL_2486522, EPI_ISL_2486523, EPI_ISL_2486524, EPI_ISL_2486525, EPI_ISL_2486526, EPI_ISL_2486527, EPI_ISL_2486528, EPI_ISL_2486529, EPI_ISL_2487207, EPI_ISL_2487218, EPI_ISL_2487219, EPI_ISL_2487220, EPI_ISL_2487221, EPI_ISL_2487222, EPI_ISL_2487223, EPI_ISL_2487224,
EPI_ISL_2487225, EPI_ISL_2487226, EPI_ISL_2487227, EPI_ISL_2487228, EPI_ISL_2487229, EPI_ISL_2487230, EPI_ISL_2487231, EPI_ISL_2487232, EPI_ISL_2487233, EPI_ISL_2487234, EPI_ISL_2487235, EPI_ISL_2487236, EPI_ISL_2487237, EPI_ISL_2487238, EPI_ISL_2487239, EPI_ISL_2487240, EPI_ISL_2487241, EPI_ISL_2487242, EPI_ISL_2487243, EPI_ISL_2487244, EPI_ISL_2487245, EPI_ISL_2487246,
EPI_ISL_2487247, EPI_ISL_2487248, EPI_ISL_2487249, EPI_ISL_2487250, EPI_ISL_2487251, EPI_ISL_2487252, EPI_ISL_2487253, EPI_ISL_2487254, EPI_ISL_2487255, EPI_ISL_2487256, EPI_ISL_2487257, EPI_ISL_2487258, EPI_ISL_2487259, EPI_ISL_2487261, EPI_ISL_2487262, EPI_ISL_2487263, EPI_ISL_2487264, EPI_ISL_2487265, EPI_ISL_2487266, EPI_ISL_2487267, EPI_ISL_2487268, EPI_ISL_2487269,
EPI_ISL_2487270, EPI_ISL_2487271, EPI_ISL_2487272, EPI_ISL_2487273, EPI_ISL_2487274, EPI_ISL_2487275, EPI_ISL_2487276, EPI_ISL_2487277, EPI_ISL_2487278, EPI_ISL_2487279, EPI_ISL_2487280, EPI_ISL_2487281, EPI_ISL_2487284, EPI_ISL_2487285, EPI_ISL_2487286, EPI_ISL_2487287, EPI_ISL_2487288, EPI_ISL_2487289, EPI_ISL_2487290, EPI_ISL_2487291, EPI_ISL_2487292,
EPI_ISL_2487293, EPI_ISL_2487294, EPI_ISL_2487295, EPI_ISL_2487296, EPI_ISL_2487297, EPI_ISL_2487298, EPI_ISL_2487299, EPI_ISL_2487300, EPI_ISL_2487301, EPI_ISL_2487302, EPI_ISL_2487303, EPI_ISL_2487304, EPI_ISL_2487305, EPI_ISL_2487306, EPI_ISL_2487307, EPI_ISL_2487308, EPI_ISL_2487309, EPI_ISL_2487310, EPI_ISL_2487311, EPI_ISL_2487312, EPI_ISL_2487313, EPI_ISL_2487314,
EPI_ISL_2487315, EPI_ISL_2487316, EPI_ISL_2487317, EPI_ISL_2487318, EPI_ISL_2487319, EPI_ISL_2487321, EPI_ISL_2487322, EPI_ISL_2487323, EPI_ISL_2487324, EPI_ISL_2487325, EPI_ISL_2487326, EPI_ISL_2487327, EPI_ISL_2487328, EPI_ISL_2487329, EPI_ISL_2487330, EPI_ISL_2487331, EPI_ISL_2487332, EPI_ISL_2487333, EPI_ISL_2487334, EPI_ISL_2487335, EPI_ISL_2487336,
EPI_ISL_2487337, EPI_ISL_2487338, EPI_ISL_2487340, EPI_ISL_2487341, EPI_ISL_2487342, EPI_ISL_2487343, EPI_ISL_2487344, EPI_ISL_2487345, EPI_ISL_2487346, EPI_ISL_2487347, EPI_ISL_2487348, EPI_ISL_2487349, EPI_ISL_2487351, EPI_ISL_2487352, EPI_ISL_2487353, EPI_ISL_2487354, EPI_ISL_2487355, EPI_ISL_2487356, EPI_ISL_2487357, EPI_ISL_2487358, EPI_ISL_2487359, EPI_ISL_2487360,
EPI_ISL_2487361, EPI_ISL_2487362, EPI_ISL_2487363, EPI_ISL_2487364, EPI_ISL_2487365, EPI_ISL_2487366, EPI_ISL_2487367, EPI_ISL_2487368, EPI_ISL_2487369, EPI_ISL_2487370, EPI_ISL_2487371, EPI_ISL_2487372, EPI_ISL_2487373, EPI_ISL_2487374, EPI_ISL_2487375, EPI_ISL_2487376, EPI_ISL_2487377, EPI_ISL_2487378, EPI_ISL_2487379, EPI_ISL_2487380, EPI_ISL_2487381, EPI_ISL_2487382,
EPI_ISL_2487383, EPI_ISL_2487384, EPI_ISL_2487385, EPI_ISL_2487386, EPI_ISL_2487387, EPI_ISL_2487388, EPI_ISL_2487389, EPI_ISL_2487390, EPI_ISL_2487391, EPI_ISL_2487392, EPI_ISL_2487393, EPI_ISL_2487394, EPI_ISL_2487395, EPI_ISL_2487396, EPI_ISL_2487397, EPI_ISL_2487398, EPI_ISL_2487399, EPI_ISL_2487400, EPI_ISL_2487401, EPI_ISL_2487402, EPI_ISL_2487403, EPI_ISL_2487404,
EPI_ISL_2487405, EPI_ISL_2487406, EPI_ISL_2487407, EPI_ISL_2487408, EPI_ISL_2487409, EPI_ISL_2487411, EPI_ISL_2487412, EPI_ISL_2487413, EPI_ISL_2487414, EPI_ISL_2487415, EPI_ISL_2487416, EPI_ISL_2487417, EPI_ISL_2487418, EPI_ISL_2487419, EPI_ISL_2487420, EPI_ISL_2487421, EPI_ISL_2487422, EPI_ISL_2487423, EPI_ISL_2487424, EPI_ISL_2487426, EPI_ISL_2487427,
EPI_ISL_2487428, EPI_ISL_2487429, EPI_ISL_2487430, EPI_ISL_2487431, EPI_ISL_2487432, EPI_ISL_2487433, EPI_ISL_2487434, EPI_ISL_2487435, EPI_ISL_2487436, EPI_ISL_2487437, EPI_ISL_2487438, EPI_ISL_2487440, EPI_ISL_2487441, EPI_ISL_2487442, EPI_ISL_2487443, EPI_ISL_2487444, EPI_ISL_2487445, EPI_ISL_2487446, EPI_ISL_2487448, EPI_ISL_2487449, EPI_ISL_2487450, EPI_ISL_2487451,
EPI_ISL_2487452, EPI_ISL_2487453, EPI_ISL_2487454, EPI_ISL_2487455, EPI_ISL_2487456, EPI_ISL_2487457, EPI_ISL_2487458, EPI_ISL_2487459, EPI_ISL_2487460, EPI_ISL_2487461, EPI_ISL_2487463, EPI_ISL_2487464, EPI_ISL_2487465, EPI_ISL_2487466, EPI_ISL_2487467, EPI_ISL_2487468, EPI_ISL_2487469, EPI_ISL_2487470, EPI_ISL_2487471, EPI_ISL_2487472, EPI_ISL_2487473, EPI_ISL_2487475,
EPI_ISL_2487476, EPI_ISL_2487477, EPI_ISL_2487478, EPI_ISL_2487479, EPI_ISL_2487480, EPI_ISL_2487481, EPI_ISL_2487482, EPI_ISL_2487483, EPI_ISL_2487484, EPI_ISL_2487485, EPI_ISL_2487486, EPI_ISL_2487487, EPI_ISL_2487488, EPI_ISL_2487489, EPI_ISL_2487490, EPI_ISL_2487491, EPI_ISL_2487492, EPI_ISL_2487493, EPI_ISL_2487494, EPI_ISL_2487495, EPI_ISL_2487496, EPI_ISL_2487497,
EPI_ISL_2487498, EPI_ISL_2487499, EPI_ISL_2487500, EPI_ISL_2487501, EPI_ISL_2487502, EPI_ISL_2487504, EPI_ISL_2487505, EPI_ISL_2487506, EPI_ISL_2487507, EPI_ISL_2487508, EPI_ISL_2487509, EPI_ISL_2487511, EPI_ISL_2487512, EPI_ISL_2487513, EPI_ISL_2487514, EPI_ISL_2487515, EPI_ISL_2487516, EPI_ISL_2487517, EPI_ISL_2487518, EPI_ISL_2487519,
EPI_ISL_2487520, EPI_ISL_2487521, EPI_ISL_2487522, EPI_ISL_2487523, EPI_ISL_2487524, EPI_ISL_2487525, EPI_ISL_2487526, EPI_ISL_2487528, EPI_ISL_2487529, EPI_ISL_2487530, EPI_ISL_2487531, EPI_ISL_2487532, EPI_ISL_2487533, EPI_ISL_2487534, EPI_ISL_2487535, EPI_ISL_2487536, EPI_ISL_2487537, EPI_ISL_2487538, EPI_ISL_2487539, EPI_ISL_2487540, EPI_ISL_2487541, EPI_ISL_2487542,
EPI_ISL_2487543, EPI_ISL_2487544, EPI_ISL_2487545, EPI_ISL_2487546, EPI_ISL_2487547, EPI_ISL_2487548, EPI_ISL_2487550, EPI_ISL_2487551, EPI_ISL_2487552, EPI_ISL_2487553, EPI_ISL_2487554, EPI_ISL_2487555, EPI_ISL_2487556, EPI_ISL_2487557, EPI_ISL_2487560, EPI_ISL_2487561, EPI_ISL_2487562, EPI_ISL_2487563, EPI_ISL_2487564, EPI_ISL_2487565, EPI_ISL_2487566,
EPI_ISL_2487567, EPI_ISL_2487568, EPI_ISL_2487569, EPI_ISL_2487570, EPI_ISL_2487571, EPI_ISL_2487572, EPI_ISL_2487573, EPI_ISL_2487574, EPI_ISL_2487575, EPI_ISL_2487576, EPI_ISL_2487577, EPI_ISL_2487578, EPI_ISL_2487579, EPI_ISL_2487580, EPI_ISL_2487581, EPI_ISL_2487582, EPI_ISL_2487583, EPI_ISL_2487584, EPI_ISL_2487587, EPI_ISL_2487588, EPI_ISL_2487589, EPI_ISL_2487590,
EPI_ISL_2487592, EPI_ISL_2487593, EPI_ISL_2487594, EPI_ISL_2487595, EPI_ISL_2487597, EPI_ISL_2487598, EPI_ISL_2487599, EPI_ISL_2487600, EPI_ISL_2487602, EPI_ISL_2487603, EPI_ISL_2487604, EPI_ISL_2487605, EPI_ISL_2487606, EPI_ISL_2487607, EPI_ISL_2487608, EPI_ISL_2487609, EPI_ISL_2487610, EPI_ISL_2487612, EPI_ISL_2487613, EPI_ISL_2487614, EPI_ISL_2487615, EPI_ISL_2487616,
EPI_ISL_2487617, EPI_ISL_2487618, EPI_ISL_2487619, EPI_ISL_2487620, EPI_ISL_2487621, EPI_ISL_2487622, EPI_ISL_2487624, EPI_ISL_2487626, EPI_ISL_2487627, EPI_ISL_2487628, EPI_ISL_2487629, EPI_ISL_2487632, EPI_ISL_2487633, EPI_ISL_2487634, EPI_ISL_2487636, EPI_ISL_2487637, EPI_ISL_2487638, EPI_ISL_2487639, EPI_ISL_2487641, EPI_ISL_2487642, EPI_ISL_2487643, EPI_ISL_2487644,
EPI_ISL_2487645, EPI_ISL_2487646, EPI_ISL_2487647, EPI_ISL_2487648, EPI_ISL_2487651, EPI_ISL_2487652, EPI_ISL_2487653, EPI_ISL_2487654, EPI_ISL_2487655, EPI_ISL_2487656, EPI_ISL_2487657, EPI_ISL_2487658, EPI_ISL_2487659, EPI_ISL_2487660, EPI_ISL_2487661, EPI_ISL_2487663, EPI_ISL_2487665, EPI_ISL_2487666, EPI_ISL_2487667, EPI_ISL_2487668, EPI_ISL_2487669, EPI_ISL_2487671,
EPI_ISL_2487672, EPI_ISL_2487673, EPI_ISL_2487674, EPI_ISL_2487675, EPI_ISL_2487676, EPI_ISL_2487677, EPI_ISL_2487678, EPI_ISL_2487679, EPI_ISL_2487680, EPI_ISL_2487681, EPI_ISL_2487682, EPI_ISL_2487683, EPI_ISL_2487684, EPI_ISL_2487685, EPI_ISL_2487686, EPI_ISL_2487688, EPI_ISL_2487690, EPI_ISL_2487691, EPI_ISL_2487692, EPI_ISL_2487694, EPI_ISL_2487695, EPI_ISL_2487696,
EPI_ISL_2487697, EPI_ISL_2487698, EPI_ISL_2487699, EPI_ISL_2487701, EPI_ISL_2487702, EPI_ISL_2487703, EPI_ISL_2487704, EPI_ISL_2487705, EPI_ISL_2487707, EPI_ISL_2487708, EPI_ISL_2487709, EPI_ISL_2487710, EPI_ISL_2487711, EPI_ISL_2487713, EPI_ISL_2487714, EPI_ISL_2487715, EPI_ISL_2487717, EPI_ISL_2487718, EPI_ISL_2487719, EPI_ISL_2487720, EPI_ISL_2487721, EPI_ISL_2487722,
EPI_ISL_2487723, EPI_ISL_2487727, EPI_ISL_2487728, EPI_ISL_2487730, EPI_ISL_2487731, EPI_ISL_2487732, EPI_ISL_2487733, EPI_ISL_2487734, EPI_ISL_2487735, EPI_ISL_2487736, EPI_ISL_2487737, EPI_ISL_2487738, EPI_ISL_2487739, EPI_ISL_2487740, EPI_ISL_2487741, EPI_ISL_2487742, EPI_ISL_2487743, EPI_ISL_2487744, EPI_ISL_2487745, EPI_ISL_2487746, EPI_ISL_2487747, EPI_ISL_2487748,
EPI_ISL_2487749, EPI_ISL_2487750, EPI_ISL_2487751, EPI_ISL_2487752, EPI_ISL_2487753, EPI_ISL_2487754, EPI_ISL_2487755, EPI_ISL_2487756, EPI_ISL_2487757, EPI_ISL_2487758, EPI_ISL_2487759, EPI_ISL_2487760, EPI_ISL_2487761, EPI_ISL_2487762, EPI_ISL_2487763, EPI_ISL_2487764, EPI_ISL_2487765, EPI_ISL_2487766, EPI_ISL_2487767, EPI_ISL_2487768, EPI_ISL_2487770, EPI_ISL_2487771,
EPI_ISL_2487772, EPI_ISL_2487773, EPI_ISL_2487774, EPI_ISL_2487775, EPI_ISL_2487776, EPI_ISL_2487777, EPI_ISL_2487778, EPI_ISL_2487779, EPI_ISL_2487780, EPI_ISL_2487781, EPI_ISL_2487782, EPI_ISL_2487784, EPI_ISL_2487785, EPI_ISL_2487786, EPI_ISL_2487787, EPI_ISL_2487788, EPI_ISL_2487789, EPI_ISL_2487790, EPI_ISL_2487791, EPI_ISL_2487792, EPI_ISL_2487793, EPI_ISL_2487794,
EPI_ISL_2487795, EPI_ISL_2487796, EPI_ISL_2487797, EPI_ISL_2487798, EPI_ISL_2487799, EPI_ISL_2487800, EPI_ISL_2487801, EPI_ISL_2487802, EPI_ISL_2487803, EPI_ISL_2487804, EPI_ISL_2487806, EPI_ISL_2487807, EPI_ISL_2487808, EPI_ISL_2487810, EPI_ISL_2487811, EPI_ISL_2487812, EPI_ISL_2487815, EPI_ISL_2487816, EPI_ISL_2487817, EPI_ISL_2487819, EPI_ISL_2487820, EPI_ISL_2487821,
EPI_ISL_2487822, EPI_ISL_2487824, EPI_ISL_2487825, EPI_ISL_2487826, EPI_ISL_2487827, EPI_ISL_2487829, EPI_ISL_2487830, EPI_ISL_2487832, EPI_ISL_2487833, EPI_ISL_2487835, EPI_ISL_2487837, EPI_ISL_2487838, EPI_ISL_2487839, EPI_ISL_2487840, EPI_ISL_2487841, EPI_ISL_2487842, EPI_ISL_2487843, EPI_ISL_2487844, EPI_ISL_2487845, EPI_ISL_2487846, EPI_ISL_2487847, EPI_ISL_2487849,
EPI_ISL_2487850, EPI_ISL_2487852, EPI_ISL_2487853, EPI_ISL_2487854, EPI_ISL_2487855, EPI_ISL_2487856, EPI_ISL_2487857, EPI_ISL_2487859, EPI_ISL_2487860, EPI_ISL_2487862, EPI_ISL_2487863, EPI_ISL_2487864, EPI_ISL_2487865, EPI_ISL_2487866, EPI_ISL_2487868, EPI_ISL_2487869, EPI_ISL_2487870, EPI_ISL_2487871, EPI_ISL_2487872, EPI_ISL_2487873, EPI_ISL_2487874, EPI_ISL_2487875,
EPI_ISL_2487876, EPI_ISL_2487877, EPI_ISL_2487878, EPI_ISL_2487879, EPI_ISL_2487880, EPI_ISL_2487882, EPI_ISL_2487883, EPI_ISL_2487885, EPI_ISL_2487886, EPI_ISL_2487887, EPI_ISL_2487888, EPI_ISL_2487889, EPI_ISL_2487890, EPI_ISL_2487891, EPI_ISL_2487892, EPI_ISL_2487893, EPI_ISL_2487894, EPI_ISL_2487899, EPI_ISL_2487900, EPI_ISL_2487901, EPI_ISL_2487903, EPI_ISL_2487904,
EPI_ISL_2487905, EPI_ISL_2487906, EPI_ISL_2487907, EPI_ISL_2487908, EPI_ISL_2487909, EPI_ISL_2487910, EPI_ISL_2487911, EPI_ISL_2487912, EPI_ISL_2487915, EPI_ISL_2487916, EPI_ISL_2487918, EPI_ISL_2487919, EPI_ISL_2487920, EPI_ISL_2487921, EPI_ISL_2487922, EPI_ISL_2487923, EPI_ISL_2487924, EPI_ISL_2487925, EPI_ISL_2487926, EPI_ISL_2487927, EPI_ISL_2487928, EPI_ISL_2487930,
EPI_ISL_2487931, EPI_ISL_2487932, EPI_ISL_2487933, EPI_ISL_2487934, EPI_ISL_2487935, EPI_ISL_2487936, EPI_ISL_2487937, EPI_ISL_2487938, EPI_ISL_2487939, EPI_ISL_2487940, EPI_ISL_2487941, EPI_ISL_2487942, EPI_ISL_2487943, EPI_ISL_2487944, EPI_ISL_2487945, EPI_ISL_2487946, EPI_ISL_2487947, EPI_ISL_2487948, EPI_ISL_2487950, EPI_ISL_2487951, EPI_ISL_2487952, EPI_ISL_2487953,
EPI_ISL_2487954, EPI_ISL_2487955, EPI_ISL_2487956, EPI_ISL_2487957, EPI_ISL_2487958, EPI_ISL_2487959, EPI_ISL_2487960, EPI_ISL_2487961, EPI_ISL_2487962, EPI_ISL_2487963, EPI_ISL_2487964, EPI_ISL_2487965, EPI_ISL_2487967, EPI_ISL_2487968, EPI_ISL_2487969, EPI_ISL_2487970, EPI_ISL_2487971, EPI_ISL_2487972, EPI_ISL_2487973, EPI_ISL_2487974, EPI_ISL_2487975,
EPI_ISL_2487976, EPI_ISL_2487977, EPI_ISL_2487978, EPI_ISL_2487979, EPI_ISL_2487980, EPI_ISL_2487981, EPI_ISL_2487982, EPI_ISL_2487983, EPI_ISL_2487984, EPI_ISL_2487985, EPI_ISL_2487986, EPI_ISL_2487987, EPI_ISL_2487988, EPI_ISL_2487990, EPI_ISL_2487991, EPI_ISL_2487992, EPI_ISL_2487993, EPI_ISL_2487994, EPI_ISL_2487995, EPI_ISL_2487996, EPI_ISL_2487998, EPI_ISL_2487999,
EPI_ISL_2488000, EPI_ISL_2488001, EPI_ISL_2488002, EPI_ISL_2488003, EPI_ISL_2488004, EPI_ISL_2488005, EPI_ISL_2488006, EPI_ISL_2488007, EPI_ISL_2488008, EPI_ISL_2488009, EPI_ISL_2488010, EPI_ISL_2488011, EPI_ISL_2488012, EPI_ISL_2488013, EPI_ISL_2488014, EPI_ISL_2488015, EPI_ISL_2488016, EPI_ISL_2488017, EPI_ISL_2488018, EPI_ISL_2488019, EPI_ISL_2488020, EPI_ISL_2488021,
EPI_ISL_2488022, EPI_ISL_2488023, EPI_ISL_2488024, EPI_ISL_2488025, EPI_ISL_2488026, EPI_ISL_2488027, EPI_ISL_2488028, EPI_ISL_2488029, EPI_ISL_2488030, EPI_ISL_2488031, EPI_ISL_2488032, EPI_ISL_2488033, EPI_ISL_2488034, EPI_ISL_2488035, EPI_ISL_2488036, EPI_ISL_2488037, EPI_ISL_2488038, EPI_ISL_2488039, EPI_ISL_2488040, EPI_ISL_2488041, EPI_ISL_2488042, EPI_ISL_2488043,
EPI_ISL_2488044, EPI_ISL_2488045, EPI_ISL_2488046, EPI_ISL_2488047, EPI_ISL_2488048, EPI_ISL_2488049, EPI_ISL_2488050, EPI_ISL_2488051, EPI_ISL_2488052, EPI_ISL_2488053, EPI_ISL_2488054, EPI_ISL_2488055, EPI_ISL_2488056, EPI_ISL_2488057, EPI_ISL_2488058, EPI_ISL_2488059, EPI_ISL_2488060, EPI_ISL_2488061, EPI_ISL_2488062, EPI_ISL_2488063, EPI_ISL_2488064, EPI_ISL_2488065,
EPI_ISL_2488066, EPI_ISL_2488067, EPI_ISL_2488068, EPI_ISL_2488069, EPI_ISL_2488070, EPI_ISL_2488071, EPI_ISL_2488072, EPI_ISL_2488073, EPI_ISL_2488074, EPI_ISL_2488075, EPI_ISL_2488076, EPI_ISL_2488077, EPI_ISL_2488078, EPI_ISL_2488079, EPI_ISL_2488082, EPI_ISL_2488083, EPI_ISL_2488084, EPI_ISL_2488085, EPI_ISL_2488086, EPI_ISL_2488087, EPI_ISL_2488088, EPI_ISL_2488089,
EPI_ISL_2488090, EPI_ISL_2488091, EPI_ISL_2488092, EPI_ISL_2488093, EPI_ISL_2488095, EPI_ISL_2488096, EPI_ISL_2488097, EPI_ISL_2488098, EPI_ISL_2488099, EPI_ISL_2488100, EPI_ISL_2488509, EPI_ISL_2488510, EPI_ISL_2488514, EPI_ISL_2488516, EPI_ISL_2488517, EPI_ISL_2488520, EPI_ISL_2488523, EPI_ISL_2488526, EPI_ISL_2488529, EPI_ISL_2488530, EPI_ISL_2488543, EPI_ISL_2488551,
EPI_ISL_2488553, EPI_ISL_2488557, EPI_ISL_2488563, EPI_ISL_2488567, EPI_ISL_2488569, EPI_ISL_2488571, EPI_ISL_2488578, EPI_ISL_2488583, EPI_ISL_2488584, EPI_ISL_2488596, EPI_ISL_2488598, EPI_ISL_2488599, EPI_ISL_2488606, EPI_ISL_2488607, EPI_ISL_2488608, EPI_ISL_2488612, EPI_ISL_2488625, EPI_ISL_2488626, EPI_ISL_2488630, EPI_ISL_2488635, EPI_ISL_2488643, EPI_ISL_2488646,
EPI_ISL_2488656, EPI_ISL_2488659, EPI_ISL_2488667, EPI_ISL_2488670, EPI_ISL_2488678, EPI_ISL_2488690, EPI_ISL_2488693, EPI_ISL_2488694, EPI_ISL_2488696, EPI_ISL_2488706, EPI_ISL_2488707, EPI_ISL_2510875, EPI_ISL_2512453, EPI_ISL_2514733, EPI_ISL_2514734, EPI_ISL_2514735, EPI_ISL_2514736, EPI_ISL_2514738, EPI_ISL_2514740, EPI_ISL_2514748, EPI_ISL_2514749,
EPI_ISL_2514751, EPI_ISL_2514752, EPI_ISL_2514753, EPI_ISL_2514758, EPI_ISL_2514761, EPI_ISL_2514765, EPI_ISL_2514769, EPI_ISL_2514771, EPI_ISL_2514773, EPI_ISL_2514774, EPI_ISL_2514775, EPI_ISL_2514776, EPI_ISL_2514777, EPI_ISL_2514779, EPI_ISL_2514780, EPI_ISL_2514782, EPI_ISL_2514787, EPI_ISL_2514789, EPI_ISL_2514790, EPI_ISL_2514793, EPI_ISL_2514794, EPI_ISL_2514799,
EPI_ISL_2514802, EPI_ISL_2514807, EPI_ISL_2514808, EPI_ISL_2514814, EPI_ISL_2514818, EPI_ISL_2514820, EPI_ISL_2514822, EPI_ISL_2514825, EPI_ISL_2514827, EPI_ISL_2514831, EPI_ISL_2514836, EPI_ISL_2514840, EPI_ISL_2514841, EPI_ISL_2514843, EPI_ISL_2514847, EPI_ISL_2514849, EPI_ISL_2514852, EPI_ISL_2514855, EPI_ISL_2514856, EPI_ISL_2514859, EPI_ISL_2514861, EPI_ISL_2514862,
EPI_ISL_2514863, EPI_ISL_2514865, EPI_ISL_2514879, EPI_ISL_2514884, EPI_ISL_2514886, EPI_ISL_2514888, EPI_ISL_2514899, EPI_ISL_2514903, EPI_ISL_2514912, EPI_ISL_2514917, EPI_ISL_2514918, EPI_ISL_2514924, EPI_ISL_2515463, EPI_ISL_2516014, EPI_ISL_2516021, EPI_ISL_2516041, EPI_ISL_2516086, EPI_ISL_2516090, EPI_ISL_2516109, EPI_ISL_2516112, EPI_ISL_2516125, EPI_ISL_2516127,
EPI_ISL_2516131, EPI_ISL_2516143, EPI_ISL_2516145, EPI_ISL_2516149, EPI_ISL_2516158, EPI_ISL_2516169, EPI_ISL_2516183, EPI_ISL_2516224, EPI_ISL_2516243, EPI_ISL_2516246, EPI_ISL_2516260, EPI_ISL_2516286, EPI_ISL_2516288, EPI_ISL_2516293, EPI_ISL_2516303, EPI_ISL_2542112, EPI_ISL_2542333, EPI_ISL_2542369, EPI_ISL_2542379, EPI_ISL_2542381, EPI_ISL_2542434, EPI_ISL_2542790,

| | | | |
|---|---|---|---|
| EPI_ISL_2542791, EPI_ISL_2542792, EPI_ISL_2542793, EPI_ISL_2542794, EPI_ISL_2542795, EPI_ISL_2542796, EPI_ISL_2542797, EPI_ISL_2542798, EPI_ISL_2542799, EPI_ISL_2542801, EPI_ISL_2542802, EPI_ISL_2542804, EPI_ISL_2542805, EPI_ISL_2542806, EPI_ISL_2542809, EPI_ISL_2542810, EPI_ISL_2542811, EPI_ISL_2542812, EPI_ISL_2542813, EPI_ISL_2542814, EPI_ISL_2542816, EPI_ISL_2542817, EPI_ISL_2542818, EPI_ISL_2542819, EPI_ISL_2542820, EPI_ISL_2542821, EPI_ISL_2542822, EPI_ISL_2542823, EPI_ISL_2542824, EPI_ISL_2542825, EPI_ISL_2542826, EPI_ISL_2542828, EPI_ISL_2542829, EPI_ISL_2542830, EPI_ISL_2542831, EPI_ISL_2542832, EPI_ISL_2542834, EPI_ISL_2542835, EPI_ISL_2542836, EPI_ISL_2542837, EPI_ISL_2542838, EPI_ISL_2542840, EPI_ISL_2542841, EPI_ISL_2542842, EPI_ISL_2542844, EPI_ISL_2542847, EPI_ISL_2542848, EPI_ISL_2542849, EPI_ISL_2542850, EPI_ISL_2542851, EPI_ISL_2542853, EPI_ISL_2542855, EPI_ISL_2542856, EPI_ISL_2542858, EPI_ISL_2542859, EPI_ISL_2542860, EPI_ISL_2542862, EPI_ISL_2542863, EPI_ISL_2542864, EPI_ISL_2542865, EPI_ISL_2542866, EPI_ISL_2542867, EPI_ISL_2542868, EPI_ISL_2542869, EPI_ISL_2542870, EPI_ISL_2542871, EPI_ISL_2542872, EPI_ISL_2542873, EPI_ISL_2542874, EPI_ISL_2542875, EPI_ISL_2542879, EPI_ISL_2542880, EPI_ISL_2542882, EPI_ISL_2542883, EPI_ISL_2542885, EPI_ISL_2542887, EPI_ISL_2542889, EPI_ISL_2542890, EPI_ISL_2542891, EPI_ISL_2542892, EPI_ISL_2542894, EPI_ISL_2542897, EPI_ISL_2542898, EPI_ISL_2542900, EPI_ISL_2542901, EPI_ISL_2542902, EPI_ISL_2542903, EPI_ISL_2542904, EPI_ISL_2542906, EPI_ISL_2542909, EPI_ISL_2542912, EPI_ISL_2542914, EPI_ISL_2542915, EPI_ISL_2542916, EPI_ISL_2542917, EPI_ISL_2542918, EPI_ISL_2542921, EPI_ISL_2542924, EPI_ISL_2542925, EPI_ISL_2542926, EPI_ISL_2542929, EPI_ISL_2542930, EPI_ISL_2542931, EPI_ISL_2542933, EPI_ISL_2542934, EPI_ISL_2542935, EPI_ISL_2542936, EPI_ISL_2542938, EPI_ISL_2542939, EPI_ISL_2542940, EPI_ISL_2542942, EPI_ISL_2542943, EPI_ISL_2542944, EPI_ISL_2542948, EPI_ISL_2542950, EPI_ISL_2542951, EPI_ISL_2542952, EPI_ISL_2543231, EPI_ISL_2543257, EPI_ISL_2543336, EPI_ISL_2543337, EPI_ISL_2543338, EPI_ISL_2543339, EPI_ISL_2543341, EPI_ISL_2543342, EPI_ISL_2543343, EPI_ISL_2543344, EPI_ISL_2543345, EPI_ISL_2543346, EPI_ISL_2543347, EPI_ISL_2543348, EPI_ISL_2543349, EPI_ISL_2543350, EPI_ISL_2543351, EPI_ISL_2543352, EPI_ISL_2543353, EPI_ISL_2543355, EPI_ISL_2543356, EPI_ISL_2543357, EPI_ISL_2543358, EPI_ISL_2543359, EPI_ISL_2543360, EPI_ISL_2543361, EPI_ISL_2543362, EPI_ISL_2543363, EPI_ISL_2543364, EPI_ISL_2543366, EPI_ISL_2543367, EPI_ISL_2543368, EPI_ISL_2543369, EPI_ISL_2543370, EPI_ISL_2543371, EPI_ISL_2543372, EPI_ISL_2543373, EPI_ISL_2543374, EPI_ISL_2543376, EPI_ISL_2543377, EPI_ISL_2543378, EPI_ISL_2543379, EPI_ISL_2543380, EPI_ISL_2543382, EPI_ISL_2543383, EPI_ISL_2543384, EPI_ISL_2543385, EPI_ISL_2543386, EPI_ISL_2543387, EPI_ISL_2543388, EPI_ISL_2543389, EPI_ISL_2543390, EPI_ISL_2543391, EPI_ISL_2543392, EPI_ISL_2543393, EPI_ISL_2543394, EPI_ISL_2543395, EPI_ISL_2543396, EPI_ISL_2543397, EPI_ISL_2543398, EPI_ISL_2543399, EPI_ISL_2543400, EPI_ISL_2543401, EPI_ISL_2543402, EPI_ISL_2543403, EPI_ISL_2543404, EPI_ISL_2543405, EPI_ISL_2543406, EPI_ISL_2543408, EPI_ISL_2543409, EPI_ISL_2543410, EPI_ISL_2543411, EPI_ISL_2543412, EPI_ISL_2543413, EPI_ISL_2543414, EPI_ISL_2543415, EPI_ISL_2543416, EPI_ISL_2543417, EPI_ISL_2543418, EPI_ISL_2543419, EPI_ISL_2543420, EPI_ISL_2543421, EPI_ISL_2543422, EPI_ISL_2543423, EPI_ISL_2543424, EPI_ISL_2543425, EPI_ISL_2543426, EPI_ISL_2543427, EPI_ISL_2543428, EPI_ISL_2543429, EPI_ISL_2543430, EPI_ISL_2543431, EPI_ISL_2543432, EPI_ISL_2555008, EPI_ISL_2555009, EPI_ISL_2555010, EPI_ISL_2555011, EPI_ISL_2555012, EPI_ISL_2555015, EPI_ISL_2555016, EPI_ISL_2555017, EPI_ISL_2555018, EPI_ISL_2555020, EPI_ISL_2555021, EPI_ISL_2555022, EPI_ISL_2555023, EPI_ISL_2555024, EPI_ISL_2555025, EPI_ISL_2555026, EPI_ISL_2555027, EPI_ISL_2555028, EPI_ISL_2555029, EPI_ISL_2555030, EPI_ISL_2555031, EPI_ISL_2555032, EPI_ISL_2555033, EPI_ISL_2555034, EPI_ISL_2555036, EPI_ISL_2555037, EPI_ISL_2555038, EPI_ISL_2555040, EPI_ISL_2555041, EPI_ISL_2555042, EPI_ISL_2555043, EPI_ISL_2555044, EPI_ISL_2555045, EPI_ISL_2555046, EPI_ISL_2555047, EPI_ISL_2555048, EPI_ISL_2555049, EPI_ISL_2555050, EPI_ISL_2555051, EPI_ISL_2555053, EPI_ISL_2555149, EPI_ISL_2555159, EPI_ISL_2555174, EPI_ISL_2555187, EPI_ISL_2555191, EPI_ISL_2555196, EPI_ISL_2555200, EPI_ISL_2555205, EPI_ISL_2555267, EPI_ISL_2605435, EPI_ISL_2936960 | | | |
| see above | Lighthouse Lab in Glasgow | Wellcome Sanger Institute for the COVID-19 Genomics UK (COG-UK) Consortium | Anna Dominiczak and Alex Alderton; Carol Clugston; Cordelia Langford; David Gray; David K. Jackson; Dominic Kwiatkowski; Ewan Harrison; Harper VanSteenhouse; Ian Johnston; Jeffrey Barrett; John Sillitoe on behalf of the Wellcome Sanger Institute COVID-19 Surveillance Team; Roberto Amato; Sonia Goncalves; Yumi Kasai |
| EPI_ISL_2456005, EPI_ISL_2456012, EPI_ISL_2456013, EPI_ISL_2456017, EPI_ISL_2456021, EPI_ISL_2456025, EPI_ISL_2456027, EPI_ISL_2456032, EPI_ISL_2456034, EPI_ISL_2456041, EPI_ISL_2456042, EPI_ISL_2456043, EPI_ISL_2456046, EPI_ISL_2456047, EPI_ISL_2456050, EPI_ISL_2456052, EPI_ISL_2456056, EPI_ISL_2456058, EPI_ISL_2456063, EPI_ISL_2456065, EPI_ISL_2456068, EPI_ISL_2456073, EPI_ISL_2456074, EPI_ISL_2456075, EPI_ISL_2456076, EPI_ISL_2456080, EPI_ISL_2456081, EPI_ISL_2456082, EPI_ISL_2456089, EPI_ISL_2456091, EPI_ISL_2456099, EPI_ISL_2456100, EPI_ISL_2456101, EPI_ISL_2456103, EPI_ISL_2456105, EPI_ISL_2456110, EPI_ISL_2456113, EPI_ISL_2456118, EPI_ISL_2456119, EPI_ISL_2456131, EPI_ISL_2456148, EPI_ISL_2456150, EPI_ISL_2456152, EPI_ISL_2456155, EPI_ISL_2456157, EPI_ISL_2456158, EPI_ISL_2456159, EPI_ISL_2456161, EPI_ISL_2456168, EPI_ISL_2456174, EPI_ISL_2456176, EPI_ISL_2456177, EPI_ISL_2456178, EPI_ISL_2456180, EPI_ISL_2456181, EPI_ISL_2456184, EPI_ISL_2456187, EPI_ISL_2456194, EPI_ISL_2456198, EPI_ISL_2456199, EPI_ISL_2456202, EPI_ISL_2456211, EPI_ISL_2456214, EPI_ISL_2456216, EPI_ISL_2456217, EPI_ISL_2456219, EPI_ISL_2456223, EPI_ISL_2456229, EPI_ISL_2456232, EPI_ISL_2456238, EPI_ISL_2456241, EPI_ISL_2456243, EPI_ISL_2456245, EPI_ISL_2456252, EPI_ISL_2456257, EPI_ISL_2456262, EPI_ISL_2456268, EPI_ISL_2456269, EPI_ISL_2456274, EPI_ISL_2456277, EPI_ISL_2456280, EPI_ISL_2456283, EPI_ISL_2456284, EPI_ISL_2456289, EPI_ISL_2456304, EPI_ISL_2456306, EPI_ISL_2456311, EPI_ISL_2456314, EPI_ISL_2456323, EPI_ISL_2456332, EPI_ISL_2456336, EPI_ISL_2456351, EPI_ISL_2456352, EPI_ISL_2456355, EPI_ISL_2456357, EPI_ISL_2456358, EPI_ISL_2456360, EPI_ISL_2456368, EPI_ISL_2456372, EPI_ISL_2456373, EPI_ISL_2456376, EPI_ISL_2456379, EPI_ISL_2456382, EPI_ISL_2456384, EPI_ISL_2456388, EPI_ISL_2456391, EPI_ISL_2456398, EPI_ISL_2456400, EPI_ISL_2456403, EPI_ISL_2456405, EPI_ISL_2456412, EPI_ISL_2456416, EPI_ISL_2456417, EPI_ISL_2456420, EPI_ISL_2456423, EPI_ISL_2456427, EPI_ISL_2456435, EPI_ISL_2456436, EPI_ISL_2456440, EPI_ISL_2456442, EPI_ISL_2456443, EPI_ISL_2456445, EPI_ISL_2456446, EPI_ISL_2456448, EPI_ISL_2456451, EPI_ISL_2456456, EPI_ISL_2456457, EPI_ISL_2456460, EPI_ISL_2456461, EPI_ISL_2456463, EPI_ISL_2456464, EPI_ISL_2456466, EPI_ISL_2456467, EPI_ISL_2456469, EPI_ISL_2456471, EPI_ISL_2456472, EPI_ISL_2456473, EPI_ISL_2456475, EPI_ISL_2456476, EPI_ISL_2456478, EPI_ISL_2456479, EPI_ISL_2456481, EPI_ISL_2456482, EPI_ISL_2456484, EPI_ISL_2456485, EPI_ISL_2456487, EPI_ISL_2456488, EPI_ISL_2456490, EPI_ISL_2456491, EPI_ISL_2456493, EPI_ISL_2456494, EPI_ISL_2456495, EPI_ISL_2456498, EPI_ISL_2456500, EPI_ISL_2456501, EPI_ISL_2456506, EPI_ISL_2456507, EPI_ISL_2456509, EPI_ISL_2456511, EPI_ISL_2456512, EPI_ISL_2456514, EPI_ISL_2456517, EPI_ISL_2456521, EPI_ISL_2456523, EPI_ISL_2456524, EPI_ISL_2456526, EPI_ISL_2456527, EPI_ISL_2456529, EPI_ISL_2456530, EPI_ISL_2456532, EPI_ISL_2456533, EPI_ISL_2456535, EPI_ISL_2456538, EPI_ISL_2456539, EPI_ISL_2456540, EPI_ISL_2456542, EPI_ISL_2456543, EPI_ISL_2456545, EPI_ISL_2456549, EPI_ISL_2456551, EPI_ISL_2456557, EPI_ISL_2456559, EPI_ISL_2456560, EPI_ISL_2456562, EPI_ISL_2456563, EPI_ISL_2456567, EPI_ISL_2456569, EPI_ISL_2456570, EPI_ISL_2456572, EPI_ISL_2456575, EPI_ISL_2456576, EPI_ISL_2456578, EPI_ISL_2456579, EPI_ISL_2456584, EPI_ISL_2456588, EPI_ISL_2456590, EPI_ISL_2456591, EPI_ISL_2456593, EPI_ISL_2456596, EPI_ISL_2456597, EPI_ISL_2456599, EPI_ISL_2456600, EPI_ISL_2456602, EPI_ISL_2456605, EPI_ISL_2456606, EPI_ISL_2456608, EPI_ISL_2456611, EPI_ISL_2456613, EPI_ISL_2456614, EPI_ISL_2456615, EPI_ISL_2456617, EPI_ISL_2456620, EPI_ISL_2456623, EPI_ISL_2456624, EPI_ISL_2456627, EPI_ISL_2456628, EPI_ISL_2456630, EPI_ISL_2456631, EPI_ISL_2456633, EPI_ISL_2456634, EPI_ISL_2456636, EPI_ISL_2456639, EPI_ISL_2456640, EPI_ISL_2456642, EPI_ISL_2456643, EPI_ISL_2456644, EPI_ISL_2456646, EPI_ISL_2456647, EPI_ISL_2456648, EPI_ISL_2456650, EPI_ISL_2456651, EPI_ISL_2456653, EPI_ISL_2456655, EPI_ISL_2456656, EPI_ISL_2456658, EPI_ISL_2456659, EPI_ISL_2456661, EPI_ISL_2456662, EPI_ISL_2456665, EPI_ISL_2456668, EPI_ISL_2456669, EPI_ISL_2456671, EPI_ISL_2456672, EPI_ISL_2456675, EPI_ISL_2456679, EPI_ISL_2456680, EPI_ISL_2456683, EPI_ISL_2456684, EPI_ISL_2456686, EPI_ISL_2456687, EPI_ISL_2456689, EPI_ISL_2456690, EPI_ISL_2456692, EPI_ISL_2456693, EPI_ISL_2456695, EPI_ISL_2456696, EPI_ISL_2456748, EPI_ISL_2456750, EPI_ISL_2456751, EPI_ISL_2456753, EPI_ISL_2456754, EPI_ISL_2456756, EPI_ISL_2456757, EPI_ISL_2456759, EPI_ISL_2456760, EPI_ISL_2456761, EPI_ISL_2456762, EPI_ISL_2456764, EPI_ISL_2456765, EPI_ISL_2456766, EPI_ISL_2456767, EPI_ISL_2456769, EPI_ISL_2456770, EPI_ISL_2456771, EPI_ISL_2456772, EPI_ISL_2456773, EPI_ISL_2456774, EPI_ISL_2456775, EPI_ISL_2456776, EPI_ISL_2456777, EPI_ISL_2456780, EPI_ISL_2456781, EPI_ISL_2456782, EPI_ISL_2456783, EPI_ISL_2456784, EPI_ISL_2456785, EPI_ISL_2456786, EPI_ISL_2456787, EPI_ISL_2456788, EPI_ISL_2456789, EPI_ISL_2456790, EPI_ISL_2456791, EPI_ISL_2456792, EPI_ISL_2456795, EPI_ISL_2456796, EPI_ISL_2456797, EPI_ISL_2456798, EPI_ISL_2456800, EPI_ISL_2456801, EPI_ISL_2456802, EPI_ISL_2456803, EPI_ISL_2456804, EPI_ISL_2456805, EPI_ISL_2456806, EPI_ISL_2456807, EPI_ISL_2456808, EPI_ISL_2456809, EPI_ISL_2456810, EPI_ISL_2456811, EPI_ISL_2456812, EPI_ISL_2456813, EPI_ISL_2456815, EPI_ISL_2456816, EPI_ISL_2456818, EPI_ISL_2456819, EPI_ISL_2456820, EPI_ISL_2456821, EPI_ISL_2456824, EPI_ISL_2456825, EPI_ISL_2467946, EPI_ISL_2467951, EPI_ISL_2467956, EPI_ISL_2467959, EPI_ISL_2467960, EPI_ISL_2467965, EPI_ISL_2467966, EPI_ISL_2467967, EPI_ISL_2467972, EPI_ISL_2467974, EPI_ISL_2467977, EPI_ISL_2467981, EPI_ISL_2467983, EPI_ISL_2467995, EPI_ISL_2468000, EPI_ISL_2468001, EPI_ISL_2468004, EPI_ISL_2468006, EPI_ISL_2468008, EPI_ISL_2468018, EPI_ISL_2468025, EPI_ISL_2468034, EPI_ISL_2468035, EPI_ISL_2468038, EPI_ISL_2468044, EPI_ISL_2468050, EPI_ISL_2468055, EPI_ISL_2468058, EPI_ISL_2468059, EPI_ISL_2468061, EPI_ISL_2468063, EPI_ISL_2468070, EPI_ISL_2468076, EPI_ISL_2468082, EPI_ISL_2468086, EPI_ISL_2468091, EPI_ISL_2468094, EPI_ISL_2468095, EPI_ISL_2468096, EPI_ISL_2468102, EPI_ISL_2468107, EPI_ISL_2468111, EPI_ISL_2468121, EPI_ISL_2468124, EPI_ISL_2468128, EPI_ISL_2468133, EPI_ISL_2468138, EPI_ISL_2468143, EPI_ISL_2468149, EPI_ISL_2468150, EPI_ISL_2468151, EPI_ISL_2468159, EPI_ISL_2468160, EPI_ISL_2468169, EPI_ISL_2468172, EPI_ISL_2468174, EPI_ISL_2468175, EPI_ISL_2468179, EPI_ISL_2468183, EPI_ISL_2468189, EPI_ISL_2468192, EPI_ISL_2468195, EPI_ISL_2468201, EPI_ISL_2468202, EPI_ISL_2468208, EPI_ISL_2468211, EPI_ISL_2468215, EPI_ISL_2468222, EPI_ISL_2468223, EPI_ISL_2468225, EPI_ISL_2468226, EPI_ISL_2468229, EPI_ISL_2468230, EPI_ISL_2468232, EPI_ISL_2468233, EPI_ISL_2468237, EPI_ISL_2468243, EPI_ISL_2468244, EPI_ISL_2468245, EPI_ISL_2468250, EPI_ISL_2468258, EPI_ISL_2468260, EPI_ISL_2468265, EPI_ISL_2468266, EPI_ISL_2468270, EPI_ISL_2468273, EPI_ISL_2468278, EPI_ISL_2468282, EPI_ISL_2468284, EPI_ISL_2468286, EPI_ISL_2468293, EPI_ISL_2468299, EPI_ISL_2468305, EPI_ISL_2468316, EPI_ISL_2468321, EPI_ISL_2468325, EPI_ISL_2468326, EPI_ISL_2468327, EPI_ISL_2468328, EPI_ISL_2468333, EPI_ISL_2468334, EPI_ISL_2468335, EPI_ISL_2468339, EPI_ISL_2468341, EPI_ISL_2468345, EPI_ISL_2468346, EPI_ISL_2468353, EPI_ISL_2468355, EPI_ISL_2468356, EPI_ISL_2468360, EPI_ISL_2468366, EPI_ISL_2468369, EPI_ISL_2468371, EPI_ISL_2468376, EPI_ISL_2468379, EPI_ISL_2468380, EPI_ISL_2468387, EPI_ISL_2468390, EPI_ISL_2468391, EPI_ISL_2468394, EPI_ISL_2468403, EPI_ISL_2468407, EPI_ISL_2468409, EPI_ISL_2468410, EPI_ISL_2468411, EPI_ISL_2468412, EPI_ISL_2468414, EPI_ISL_2468416, EPI_ISL_2468418, EPI_ISL_2468419, EPI_ISL_2468420, EPI_ISL_2468424, EPI_ISL_2468428, EPI_ISL_2468429, EPI_ISL_2468433, EPI_ISL_2468436, EPI_ISL_2468437, EPI_ISL_2468441, EPI_ISL_2468442, EPI_ISL_2468448, EPI_ISL_2468452, EPI_ISL_2468454, EPI_ISL_2468455, EPI_ISL_2468461, EPI_ISL_2468462, EPI_ISL_2468467, EPI_ISL_2468468, EPI_ISL_2468479, EPI_ISL_2468481, EPI_ISL_2468482, EPI_ISL_2468485, EPI_ISL_2468488, EPI_ISL_2468492, EPI_ISL_2468500, EPI_ISL_2468502, EPI_ISL_2468503, EPI_ISL_2468505, EPI_ISL_2468507, EPI_ISL_2468509, EPI_ISL_2468510, EPI_ISL_2468515, EPI_ISL_2468518, EPI_ISL_2468533, EPI_ISL_2468534, EPI_ISL_2468541, EPI_ISL_2468543, EPI_ISL_2468544, EPI_ISL_2468548, EPI_ISL_2468559, EPI_ISL_2468563, EPI_ISL_2468564, EPI_ISL_2468569, EPI_ISL_2468571, EPI_ISL_2468572, EPI_ISL_2468578, EPI_ISL_2468580, EPI_ISL_2468583, EPI_ISL_2468587, EPI_ISL_2468588, EPI_ISL_2468593, EPI_ISL_2468595, EPI_ISL_2468597, EPI_ISL_2468601, EPI_ISL_2468612, EPI_ISL_2468620, EPI_ISL_2468623, EPI_ISL_2468627, EPI_ISL_2468636, EPI_ISL_2468638, EPI_ISL_2468642, EPI_ISL_2468644, EPI_ISL_2468647, EPI_ISL_2468648, EPI_ISL_2468656, EPI_ISL_2468659, EPI_ISL_2468661, EPI_ISL_2468666, EPI_ISL_2468669, EPI_ISL_2468673, EPI_ISL_2468674, EPI_ISL_2468682, EPI_ISL_2468686, EPI_ISL_2468688, EPI_ISL_2468689, EPI_ISL_2468691, EPI_ISL_2468695, EPI_ISL_2468708, EPI_ISL_2468709, EPI_ISL_2468715, EPI_ISL_2468722, EPI_ISL_2468724, EPI_ISL_2468725, EPI_ISL_2468731, EPI_ISL_2468734, EPI_ISL_2468736, EPI_ISL_2468744, EPI_ISL_2468750, EPI_ISL_2468755, EPI_ISL_2468759, EPI_ISL_2468763, EPI_ISL_2468765, EPI_ISL_2468777, EPI_ISL_2468784, EPI_ISL_2468786, EPI_ISL_2468790, EPI_ISL_2468791, EPI_ISL_2468792, EPI_ISL_2468794, EPI_ISL_2468809, EPI_ISL_2468813, EPI_ISL_2468816, EPI_ISL_2468826, EPI_ISL_2468830, EPI_ISL_2468836, EPI_ISL_2468838, EPI_ISL_2468840, EPI_ISL_2468856, EPI_ISL_2468858, EPI_ISL_2468866, EPI_ISL_2468868, EPI_ISL_2468875, EPI_ISL_2468876, EPI_ISL_2468881, EPI_ISL_2468885, EPI_ISL_2468887, EPI_ISL_2468890, EPI_ISL_2468894, EPI_ISL_2468895, EPI_ISL_2468899, EPI_ISL_2468910, EPI_ISL_2468916, EPI_ISL_2468918, EPI_ISL_2468919, EPI_ISL_2468920, EPI_ISL_2468922, EPI_ISL_2468930, EPI_ISL_2468931, EPI_ISL_2468933, EPI_ISL_2468935, EPI_ISL_2469231, EPI_ISL_2469249, EPI_ISL_2469253, EPI_ISL_2469260, EPI_ISL_2469275, EPI_ISL_2469281, EPI_ISL_2469282, EPI_ISL_2469291, EPI_ISL_2469297, EPI_ISL_2469299, EPI_ISL_2469301, EPI_ISL_2469302, EPI_ISL_2469314, EPI_ISL_2469316, EPI_ISL_2469323, EPI_ISL_2469326, EPI_ISL_2469327, EPI_ISL_2469331, EPI_ISL_2469332, EPI_ISL_2469337, EPI_ISL_2469339, EPI_ISL_2469355, EPI_ISL_2469357, EPI_ISL_2469360, EPI_ISL_2469366, EPI_ISL_2469372, EPI_ISL_2469374, EPI_ISL_2469396, EPI_ISL_2469403, EPI_ISL_2469412, EPI_ISL_2469415, EPI_ISL_2469422, EPI_ISL_2469424, EPI_ISL_2469426, EPI_ISL_2469440, EPI_ISL_2469444, EPI_ISL_2469447, EPI_ISL_2469452, EPI_ISL_2469453, EPI_ISL_2469454, EPI_ISL_2469455, EPI_ISL_2469456, EPI_ISL_2469457, EPI_ISL_2469458, EPI_ISL_2469459, EPI_ISL_2469460, EPI_ISL_2469461, EPI_ISL_2469462, EPI_ISL_2469464, EPI_ISL_2485258, EPI_ISL_2485260, EPI_ISL_2485261, EPI_ISL_2485271, EPI_ISL_2485276, EPI_ISL_2485278, EPI_ISL_2485279, EPI_ISL_2485282, EPI_ISL_2485286, EPI_ISL_2485290, EPI_ISL_2485291, EPI_ISL_2485292, EPI_ISL_2485300, EPI_ISL_2485301, EPI_ISL_2485304, EPI_ISL_2485306, EPI_ISL_2485308, EPI_ISL_2485312, EPI_ISL_2485316, EPI_ISL_2485325, EPI_ISL_2485332, EPI_ISL_2485335, EPI_ISL_2485336, EPI_ISL_2485337, EPI_ISL_2485343, EPI_ISL_2485347, EPI_ISL_2485348, EPI_ISL_2485353, EPI_ISL_2485355, EPI_ISL_2485361, EPI_ISL_2485364, EPI_ISL_2485374, EPI_ISL_2485381, EPI_ISL_2485383, EPI_ISL_2485384, EPI_ISL_2485410, EPI_ISL_2485413, EPI_ISL_2485416, EPI_ISL_2485421, EPI_ISL_2485431, EPI_ISL_2485434, EPI_ISL_2485440, EPI_ISL_2485443, EPI_ISL_2485450, EPI_ISL_2485451, EPI_ISL_2485453, EPI_ISL_2485456, EPI_ISL_2485457, EPI_ISL_2485462, EPI_ISL_2485465, EPI_ISL_2485468, EPI_ISL_2485472, EPI_ISL_2485476, EPI_ISL_2485477, EPI_ISL_2485485, EPI_ISL_2485490, EPI_ISL_2485494, EPI_ISL_2485504, EPI_ISL_2485505, EPI_ISL_2485506, EPI_ISL_2485513, EPI_ISL_2485522, EPI_ISL_2485525, EPI_ISL_2485528, EPI_ISL_2485529, EPI_ISL_2485531, EPI_ISL_2485532, EPI_ISL_2485537, EPI_ISL_2485539, EPI_ISL_2485546, EPI_ISL_2485554, EPI_ISL_2485560, EPI_ISL_2485569, EPI_ISL_2485579, EPI_ISL_2485580, EPI_ISL_2485581, EPI_ISL_2485582, EPI_ISL_2485586, EPI_ISL_2485588, EPI_ISL_2485589, EPI_ISL_2485590, EPI_ISL_2485592, EPI_ISL_2485596, EPI_ISL_2485597, EPI_ISL_2485600, EPI_ISL_2485605, EPI_ISL_2485606, EPI_ISL_2485610, EPI_ISL_2485616, EPI_ISL_2485623, EPI_ISL_2485628, EPI_ISL_2485633, EPI_ISL_2485641, EPI_ISL_2485642, EPI_ISL_2485643, EPI_ISL_2485646, EPI_ISL_2485648, EPI_ISL_2485651, EPI_ISL_2485652, EPI_ISL_2485664, EPI_ISL_2485665, EPI_ISL_2485666, EPI_ISL_2485667, EPI_ISL_2485672, EPI_ISL_2485673, EPI_ISL_2485680, EPI_ISL_2485685, EPI_ISL_2485689, EPI_ISL_2485694, EPI_ISL_2485701, EPI_ISL_2485703, EPI_ISL_2485704, EPI_ISL_2485706, EPI_ISL_2485707, EPI_ISL_2485708, EPI_ISL_2485710, EPI_ISL_2485715, EPI_ISL_2485717, EPI_ISL_2485723, EPI_ISL_2485725, EPI_ISL_2485726, EPI_ISL_2485727, EPI_ISL_2485729, EPI_ISL_2485730, EPI_ISL_2485735, EPI_ISL_2485739, EPI_ISL_2485742, EPI_ISL_2485743, EPI_ISL_2485745, EPI_ISL_2485746, EPI_ISL_2485750, EPI_ISL_2485751, EPI_ISL_2485752, EPI_ISL_2485755, EPI_ISL_2485758, EPI_ISL_2485760, EPI_ISL_2485764, EPI_ISL_2485768, EPI_ISL_2485771, EPI_ISL_2485773, EPI_ISL_2485777, EPI_ISL_2485779, EPI_ISL_2485785, EPI_ISL_2485791, EPI_ISL_2485792, EPI_ISL_2485794, EPI_ISL_2485796, EPI_ISL_2485800, EPI_ISL_2485804, EPI_ISL_2485806, EPI_ISL_2485809, EPI_ISL_2485812, EPI_ISL_2485815, EPI_ISL_2485821, EPI_ISL_2485825, EPI_ISL_2485829, EPI_ISL_2485839, EPI_ISL_2485844, EPI_ISL_2485846, EPI_ISL_2485850, EPI_ISL_2485855, EPI_ISL_2485856, EPI_ISL_2485858, EPI_ISL_2485864, EPI_ISL_2485867, EPI_ISL_2485868, EPI_ISL_2485875, EPI_ISL_2485877, EPI_ISL_2485878, EPI_ISL_2485883, EPI_ISL_2485890, EPI_ISL_2485892, EPI_ISL_2485893, EPI_ISL_2485894, EPI_ISL_2485898, EPI_ISL_2485901, EPI_ISL_2485908, EPI_ISL_2485913, EPI_ISL_2485918, EPI_ISL_2485927, EPI_ISL_2485930, EPI_ISL_2485932, EPI_ISL_2485940, EPI_ISL_2485950, EPI_ISL_2485962, EPI_ISL_2485965, EPI_ISL_2485977, EPI_ISL_2485979, EPI_ISL_2485986, EPI_ISL_2485995, EPI_ISL_2486000, EPI_ISL_2486005, EPI_ISL_2486006, EPI_ISL_2486014, EPI_ISL_2486016, EPI_ISL_2486017, EPI_ISL_2486024, EPI_ISL_2486030, EPI_ISL_2486031, EPI_ISL_2486032, EPI_ISL_2486033, EPI_ISL_2486036, EPI_ISL_2486038, EPI_ISL_2486042, EPI_ISL_2486045, EPI_ISL_2486051, EPI_ISL_2486056, EPI_ISL_2486058, EPI_ISL_2486064, EPI_ISL_2486071, EPI_ISL_2486076, EPI_ISL_2486078, EPI_ISL_2486083, EPI_ISL_2486087, EPI_ISL_2486093, EPI_ISL_2486104, EPI_ISL_2486107, EPI_ISL_2486109, EPI_ISL_2486113, EPI_ISL_2486119, EPI_ISL_2486124, EPI_ISL_2486127, EPI_ISL_2486128, EPI_ISL_2486132, EPI_ISL_2486135, EPI_ISL_2486136, EPI_ISL_2486137, EPI_ISL_2486141, EPI_ISL_2486142, EPI_ISL_2486147, EPI_ISL_2486159, EPI_ISL_2486170, EPI_ISL_2486173, EPI_ISL_2486176, EPI_ISL_2486181, EPI_ISL_2486185, EPI_ISL_2486186, EPI_ISL_2486191, EPI_ISL_2486896, EPI_ISL_2486898, EPI_ISL_2486899, EPI_ISL_2486900, EPI_ISL_2486904, EPI_ISL_2486921, EPI_ISL_2486922, EPI_ISL_2486924, EPI_ISL_2486930, EPI_ISL_2486931, EPI_ISL_2486932, EPI_ISL_2486933, EPI_ISL_2486938, EPI_ISL_2486941, EPI_ISL_2486945, EPI_ISL_2486947, EPI_ISL_2486949, EPI_ISL_2486950, EPI_ISL_2486952, EPI_ISL_2486953, EPI_ISL_2486956, EPI_ISL_2486961, EPI_ISL_2486963, EPI_ISL_2486969, EPI_ISL_2486971, EPI_ISL_2486977, EPI_ISL_2486978, EPI_ISL_2486979, EPI_ISL_2486981, EPI_ISL_2486982, EPI_ISL_2486983, EPI_ISL_2486984, EPI_ISL_2486987, EPI_ISL_2486989, EPI_ISL_2486995, EPI_ISL_2486996, EPI_ISL_2487001, EPI_ISL_2487005, EPI_ISL_2487006, EPI_ISL_2487011, EPI_ISL_2487013, EPI_ISL_2487014, EPI_ISL_2487015, EPI_ISL_2487017, EPI_ISL_2487020, EPI_ISL_2487021, EPI_ISL_2487022, EPI_ISL_2487025, EPI_ISL_2487026, EPI_ISL_2487030, EPI_ISL_2487034, EPI_ISL_2487038, EPI_ISL_2487041, EPI_ISL_2487043, EPI_ISL_2487044, EPI_ISL_2487048, EPI_ISL_2487050, EPI_ISL_2487052, EPI_ISL_2487055, EPI_ISL_2487057, EPI_ISL_2487058, EPI_ISL_2487061, EPI_ISL_2487062, EPI_ISL_2487065, EPI_ISL_2487067, EPI_ISL_2487070, EPI_ISL_2487071, EPI_ISL_2487073, EPI_ISL_2487079, EPI_ISL_2487080, EPI_ISL_2487082, EPI_ISL_2487083, EPI_ISL_2487089, EPI_ISL_2487090, EPI_ISL_2487092, EPI_ISL_2487097, EPI_ISL_2487106, EPI_ISL_2487107, EPI_ISL_2487111, EPI_ISL_2487117, EPI_ISL_2487124, EPI_ISL_2487126, EPI_ISL_2487129, EPI_ISL_2487130, EPI_ISL_2487136, EPI_ISL_2487137, EPI_ISL_2487141, EPI_ISL_2487143, EPI_ISL_2487144, EPI_ISL_2487145, EPI_ISL_2487147, EPI_ISL_2487150, EPI_ISL_2487151, EPI_ISL_2487158, EPI_ISL_2487159, EPI_ISL_2487161, EPI_ISL_2487164, EPI_ISL_2487169, EPI_ISL_2487170, EPI_ISL_2487171, EPI_ISL_2487174, EPI_ISL_2487177, EPI_ISL_2487181, EPI_ISL_2487183, EPI_ISL_2487187, EPI_ISL_2487188, EPI_ISL_2487189, EPI_ISL_2487194, EPI_ISL_2487195, EPI_ISL_2487196, EPI_ISL_2487200, EPI_ISL_2487201, EPI_ISL_2487202, EPI_ISL_2487203, EPI_ISL_2487206, EPI_ISL_2487212, EPI_ISL_2487215, EPI_ISL_2487216, EPI_ISL_2488239, EPI_ISL_2488242, EPI_ISL_2488282, EPI_ISL_2510881, EPI_ISL_2510911, EPI_ISL_2510938, EPI_ISL_2510944, EPI_ISL_2511042, EPI_ISL_2511044, EPI_ISL_2511067, EPI_ISL_2511083, EPI_ISL_2511398, EPI_ISL_2511498, EPI_ISL_2511503, EPI_ISL_2511627, EPI_ISL_2511680, EPI_ISL_2511681, EPI_ISL_2511786, EPI_ISL_2511789, EPI_ISL_2511792, EPI_ISL_2511927, EPI_ISL_2511937, EPI_ISL_2512385, EPI_ISL_2512386, EPI_ISL_2512393, EPI_ISL_2512404, EPI_ISL_2512410, EPI_ISL_2512412, EPI_ISL_2512430, EPI_ISL_2512441, EPI_ISL_2512454, EPI_ISL_2512467, EPI_ISL_2512486, EPI_ISL_2512540, EPI_ISL_2512550, EPI_ISL_2512587, EPI_ISL_2512600, EPI_ISL_2512641, EPI_ISL_2512648, EPI_ISL_2512649, EPI_ISL_2512668, EPI_ISL_2512931, EPI_ISL_2512956, EPI_ISL_2513000, EPI_ISL_2513005, EPI_ISL_2513021, EPI_ISL_2513043, EPI_ISL_2513125, EPI_ISL_2513157, EPI_ISL_2514107, EPI_ISL_2514131, EPI_ISL_2514156, EPI_ISL_2514185, EPI_ISL_2514211, EPI_ISL_2514344, EPI_ISL_2515543, EPI_ISL_2515646, EPI_ISL_2515647, EPI_ISL_2515666, EPI_ISL_2515668, EPI_ISL_2515700, EPI_ISL_2515724, EPI_ISL_2515725, EPI_ISL_2515727, EPI_ISL_2515846, EPI_ISL_2515874, EPI_ISL_2515880, EPI_ISL_2515915, EPI_ISL_2515938, EPI_ISL_2515963, EPI_ISL_2515964, EPI_ISL_2515973, EPI_ISL_2515978, EPI_ISL_2516000, EPI_ISL_2516310, EPI_ISL_2539172, EPI_ISL_2539173, EPI_ISL_2539175, EPI_ISL_2539180, EPI_ISL_2539183, EPI_ISL_2539184, EPI_ISL_2539185, EPI_ISL_2539186, EPI_ISL_2539189, EPI_ISL_2539192, EPI_ISL_2539196, EPI_ISL_2539212, EPI_ISL_2539213, EPI_ISL_2539217, EPI_ISL_2539221, EPI_ISL_2539222, EPI_ISL_2539229, EPI_ISL_2539232, EPI_ISL_2539233, EPI_ISL_2539235, EPI_ISL_2539237, EPI_ISL_2539240, EPI_ISL_2539241, EPI_ISL_2539245, EPI_ISL_2539252, EPI_ISL_2539253, EPI_ISL_2539261, EPI_ISL_2539264, EPI_ISL_2539266, EPI_ISL_2539268, EPI_ISL_2539273, EPI_ISL_2539276, EPI_ISL_2539278, EPI_ISL_2539281, EPI_ISL_2539285, EPI_ISL_2539288, EPI_ISL_2539290, EPI_ISL_2539293, EPI_ISL_2539295, EPI_ISL_2539299, EPI_ISL_2539310, EPI_ISL_2539312, EPI_ISL_2539313, EPI_ISL_2539319, EPI_ISL_2539323, EPI_ISL_2539330, EPI_ISL_2539332, EPI_ISL_2539334, EPI_ISL_2539339, EPI_ISL_2539340, EPI_ISL_2539341, EPI_ISL_2539342, EPI_ISL_2539345, EPI_ISL_2539347, EPI_ISL_2539354, EPI_ISL_2539360, EPI_ISL_2539361, EPI_ISL_2539364, EPI_ISL_2539367, EPI_ISL_2539368, EPI_ISL_2539375, EPI_ISL_2539377, EPI_ISL_2539378, EPI_ISL_2539385, EPI_ISL_2539388, EPI_ISL_2539397, EPI_ISL_2539399, EPI_ISL_2539400, EPI_ISL_2539411, EPI_ISL_2539417, EPI_ISL_2539418, EPI_ISL_2539421, EPI_ISL_2539422, EPI_ISL_2539429, EPI_ISL_2539433, EPI_ISL_2539437, EPI_ISL_2539443, EPI_ISL_2539444, EPI_ISL_2539449, EPI_ISL_2539477, EPI_ISL_2539479, EPI_ISL_2539486, EPI_ISL_2539497, EPI_ISL_2539508, EPI_ISL_2539510, EPI_ISL_2539513, EPI_ISL_2539523, EPI_ISL_2539527, EPI_ISL_2539528, EPI_ISL_2539529, EPI_ISL_2539537, EPI_ISL_2539541, EPI_ISL_2539573, EPI_ISL_2539580, EPI_ISL_2539587, EPI_ISL_2539589, EPI_ISL_2539592, EPI_ISL_2539593, EPI_ISL_2539602, EPI_ISL_2539607, EPI_ISL_2539618, EPI_ISL_2539619, EPI_ISL_2539620, EPI_ISL_2539646, EPI_ISL_2539648, EPI_ISL_2539649, EPI_ISL_2539650, EPI_ISL_2539652, EPI_ISL_2539663, EPI_ISL_2539672, EPI_ISL_2539692, EPI_ISL_2539695, EPI_ISL_2539705, EPI_ISL_2539711, EPI_ISL_2539727, EPI_ISL_2539732, EPI_ISL_2539733, EPI_ISL_2540139, EPI_ISL_2541052, EPI_ISL_2541166, EPI_ISL_2640511 | | | |
| see above | Lighthouse Lab in Milton Keynes | Wellcome Sanger Institute for the COVID-19 Genomics UK (COG-UK) Consortium | Cordelia Langford; David K. Jackson; Dominic Kwiatkowski; Ewan Harrison; Ian Johnston; Jeffrey Barrett; John Sillitoe on behalf of the Wellcome Sanger Institute COVID-19 Surveillance Team; Roberto Amato; Sonia Goncalves; The Lighthouse Lab in Milton Keynes and Alex Alderton |
| EPI_ISL_2467944, EPI_ISL_2467948, EPI_ISL_2467949, EPI_ISL_2467950, EPI_ISL_2467952, EPI_ISL_2467958, EPI_ISL_2467964, EPI_ISL_2467973, EPI_ISL_2467976, EPI_ISL_2467990, EPI_ISL_2467991, EPI_ISL_2467993, EPI_ISL_2467996, EPI_ISL_2468011, EPI_ISL_2468013, EPI_ISL_2468017, EPI_ISL_2468020, EPI_ISL_2468032, EPI_ISL_2468037, EPI_ISL_2468041, EPI_ISL_2468046, EPI_ISL_2468047, EPI_ISL_2468051, EPI_ISL_2468056, EPI_ISL_2468064, EPI_ISL_2468068, EPI_ISL_2468072, EPI_ISL_2468075, EPI_ISL_2468080, EPI_ISL_2468087, EPI_ISL_2468088, EPI_ISL_2468093, EPI_ISL_2468101, EPI_ISL_2468103, EPI_ISL_2468108, EPI_ISL_2468130, EPI_ISL_2468134, EPI_ISL_2468165, EPI_ISL_2468170, EPI_ISL_2468190, EPI_ISL_2468199, EPI_ISL_2468209, EPI_ISL_2468216, EPI_ISL_2468221, EPI_ISL_2468235, EPI_ISL_2468236, EPI_ISL_2468246, EPI_ISL_2468247, EPI_ISL_2468248, EPI_ISL_2468253, EPI_ISL_2468254, EPI_ISL_2468275, EPI_ISL_2468285, EPI_ISL_2468288, EPI_ISL_2468289, EPI_ISL_2468292, EPI_ISL_2468294, EPI_ISL_2468304, EPI_ISL_2468344, EPI_ISL_2468349, EPI_ISL_2468352, EPI_ISL_2468357, EPI_ISL_2468367, EPI_ISL_2468370, EPI_ISL_2468372, EPI_ISL_2468393, EPI_ISL_2468395, EPI_ISL_2468397, EPI_ISL_2468413, EPI_ISL_2468443, EPI_ISL_2468446, EPI_ISL_2468464, EPI_ISL_2468466, EPI_ISL_2468477, EPI_ISL_2468498, EPI_ISL_2468523, EPI_ISL_2468524, EPI_ISL_2468529, EPI_ISL_2468535, EPI_ISL_2468545, EPI_ISL_2468546, EPI_ISL_2468556, EPI_ISL_2468561, EPI_ISL_2468565, EPI_ISL_2468573, EPI_ISL_2485263, EPI_ISL_2485422, EPI_ISL_2487989, EPI_ISL_2488236, EPI_ISL_2488237, EPI_ISL_2516316, EPI_ISL_2639219 | | | |
| see above | Lighthouse Laboratory Plymouth | Wellcome Sanger Institute for the COVID-19 Genomics UK (COG-UK) Consortium | Cordelia Langford; David K. Jackson; Dominic Kwiatkowski; Ewan Harrison; Ian Johnston; Jeffrey Barrett; John Sillitoe on behalf of the Wellcome Sanger Institute COVID-19 Surveillance Team; Lighthouse Laboratory Plymouth and Alex Alderton; Roberto Amato; Sonia Goncalves |
| EPI_ISL_2518915, EPI_ISL_2518916, EPI_ISL_2518917, EPI_ISL_2518918, EPI_ISL_2518919, EPI_ISL_2518920, EPI_ISL_2518922, EPI_ISL_2518923, EPI_ISL_2518924, EPI_ISL_2518925, EPI_ISL_2518926, EPI_ISL_2518927, EPI_ISL_2518928, EPI_ISL_2518930, EPI_ISL_2518931, EPI_ISL_2518932, EPI_ISL_2518933, EPI_ISL_2518935, EPI_ISL_2518936, EPI_ISL_2518938, EPI_ISL_2518939, EPI_ISL_2518942, EPI_ISL_2518943, EPI_ISL_2518944, EPI_ISL_2518945, EPI_ISL_2518947, EPI_ISL_2518948, EPI_ISL_2518950, EPI_ISL_2518952, EPI_ISL_2518953, EPI_ISL_2518954, EPI_ISL_2518955, EPI_ISL_2518956, EPI_ISL_2518957, EPI_ISL_2518958, EPI_ISL_2518959, EPI_ISL_2518961, EPI_ISL_2518962, EPI_ISL_2518963, EPI_ISL_2518966, EPI_ISL_2518967, EPI_ISL_2518968, EPI_ISL_2623258 | | | |
| see above | Northumbria University / South Tees Hospitals NHS Foundation Trust / North Cumbria Integrated Care NHS Foundation Trust / North Tees and Hartlepool NHS Foundation Trust / Newcastle Hospitals NHS Foundation Trust | COVID-19 Genomics UK (COG-UK) Consortium | Andrew Nelson; Brendan Payne; Clive Graham; Darren L Smith; Debra Padgett; Edward Barton; Emma Swindells; Garren Scott; Gary Black; Gary Eltringham; Giles S Holt; Greg R Young; Jane Greenaway; Jennifer Collins; John Allan; Joshua Loh; Lynn Dover; Matthew Bashton; Mohammad A Tariq; Paul Baker; Sarah Essex; Steve Liggett; Wen C Yew; Yusri Taha |
| EPI_ISL_2624526, EPI_ISL_2624539, EPI_ISL_2624540, EPI_ISL_2624545, EPI_ISL_2624577, EPI_ISL_2624584 | Oxford Viromics, NDM, University of Oxford; Oxford University Hospitals; Basingstoke and North Hampshire Hospital | COVID-19 Genomics UK (COG-UK) Consortium | Alex Mobbs; Amy Trebes; Anita Justice; Catrin Moore; Christophe Fraser; David Bonsall; David Buck; Emma Wise; George Macintyre; Jessica Lynch; John Todd; Mariateresa de Cesare; Matilde Mori; Monique Andersson; Nathan Moore; Nick Cortes; Robert Shaw; Stephen Kidd; Tanya Golubchik; Timothy Peto |
| EPI_ISL_2519198, EPI_ISL_2519200, EPI_ISL_2519201, EPI_ISL_2519202, EPI_ISL_2519203, EPI_ISL_2519204, EPI_ISL_2519205, EPI_ISL_2519206, EPI_ISL_2519207, EPI_ISL_2519208, EPI_ISL_2519209, EPI_ISL_2519210, EPI_ISL_2519211, EPI_ISL_2519212, EPI_ISL_2519213, EPI_ISL_2519214, EPI_ISL_2519215, EPI_ISL_2519216, EPI_ISL_2519217, EPI_ISL_2519218, EPI_ISL_2519219, EPI_ISL_2519220, EPI_ISL_2519221, EPI_ISL_2519222, EPI_ISL_2519223, EPI_ISL_2519224, EPI_ISL_2519225, EPI_ISL_2519226, EPI_ISL_2519227, EPI_ISL_2519228, EPI_ISL_2519229, EPI_ISL_2519230, EPI_ISL_2519231, EPI_ISL_2519232, EPI_ISL_2519233, EPI_ISL_2519234, EPI_ISL_2519235, EPI_ISL_2519236, EPI_ISL_2519237, EPI_ISL_2519238, EPI_ISL_2519239, EPI_ISL_2519240, EPI_ISL_2519241, EPI_ISL_2519242, EPI_ISL_2519243, EPI_ISL_2519244, EPI_ISL_2519245, EPI_ISL_2519246, EPI_ISL_2519247, EPI_ISL_2519248, EPI_ISL_2519249, EPI_ISL_2519250, EPI_ISL_2519251, EPI_ISL_2519252, EPI_ISL_2519253, EPI_ISL_2519254, EPI_ISL_2519255, EPI_ISL_2519256, EPI_ISL_2519257, EPI_ISL_2519258, EPI_ISL_2519259, EPI_ISL_2519260, EPI_ISL_2519261, EPI_ISL_2519262, EPI_ISL_2519263, EPI_ISL_2519264, EPI_ISL_2519334, EPI_ISL_2519349, EPI_ISL_2519368, EPI_ISL_2519369, EPI_ISL_2519468, EPI_ISL_2519470, EPI_ISL_2519471, EPI_ISL_2519473, EPI_ISL_2519478, EPI_ISL_2519505, EPI_ISL_2519506, EPI_ISL_2519507, EPI_ISL_2519508, EPI_ISL_2519509, EPI_ISL_2519510, EPI_ISL_2519511, EPI_ISL_2519512, EPI_ISL_2519513, EPI_ISL_2519514, EPI_ISL_2519515, EPI_ISL_2519516, EPI_ISL_2519517, EPI_ISL_2519518, EPI_ISL_2519519, EPI_ISL_2519520, EPI_ISL_2519521, EPI_ISL_2519522, EPI_ISL_2519523, EPI_ISL_2519527, EPI_ISL_2519528, EPI_ISL_2519529, EPI_ISL_2519530, EPI_ISL_2519531, EPI_ISL_2519532, EPI_ISL_2519533, EPI_ISL_2519535, EPI_ISL_2519536, EPI_ISL_2519537, EPI_ISL_2519538, EPI_ISL_2519540, EPI_ISL_2519542, EPI_ISL_2519543, EPI_ISL_2519547, EPI_ISL_2519548, EPI_ISL_2519549, EPI_ISL_2519551, EPI_ISL_2519552, EPI_ISL_2519553, EPI_ISL_2519554, EPI_ISL_2519555, EPI_ISL_2519556, EPI_ISL_2519557, EPI_ISL_2519558, EPI_ISL_2519559, EPI_ISL_2519560, EPI_ISL_2519561, EPI_ISL_2519562, EPI_ISL_2519564, EPI_ISL_2519565, EPI_ISL_2519566, EPI_ISL_2519567, EPI_ISL_2519568, EPI_ISL_2519569, EPI_ISL_2519570, EPI_ISL_2519571, EPI_ISL_2519572, EPI_ISL_2519574, EPI_ISL_2519575, EPI_ISL_2519576, EPI_ISL_2519577, EPI_ISL_2519578, EPI_ISL_2519579, EPI_ISL_2519580, EPI_ISL_2519581, EPI_ISL_2519582, EPI_ISL_2519583, EPI_ISL_2519584, EPI_ISL_2519585, EPI_ISL_2519586, EPI_ISL_2519587, EPI_ISL_2519588, EPI_ISL_2519589, EPI_ISL_2519590, EPI_ISL_2519591, EPI_ISL_2519592, EPI_ISL_2519593, EPI_ISL_2519594, EPI_ISL_2519595, EPI_ISL_2519596, EPI_ISL_2519597, EPI_ISL_2519598, EPI_ISL_2519599, EPI_ISL_2519600, EPI_ISL_2519601, EPI_ISL_2519602, EPI_ISL_2519603, EPI_ISL_2519604, EPI_ISL_2519605, EPI_ISL_2519606, EPI_ISL_2519607, EPI_ISL_2519608, EPI_ISL_2519609, EPI_ISL_2519610, EPI_ISL_2519611, EPI_ISL_2519612, EPI_ISL_2519613, EPI_ISL_2519614, EPI_ISL_2519615, EPI_ISL_2519616, EPI_ISL_2519617, EPI_ISL_2519618, EPI_ISL_2519619, EPI_ISL_2519620, EPI_ISL_2519621, EPI_ISL_2519622, EPI_ISL_2519624, EPI_ISL_2519625, EPI_ISL_2519626, EPI_ISL_2519627, EPI_ISL_2519628, EPI_ISL_2519629, EPI_ISL_2519630, EPI_ISL_2519631, EPI_ISL_2519632, EPI_ISL_2519633, EPI_ISL_2519634, EPI_ISL_2519635, EPI_ISL_2519636, EPI_ISL_2519637, EPI_ISL_2519638, EPI_ISL_2519639, EPI_ISL_2519640, EPI_ISL_2519641, EPI_ISL_2519642, EPI_ISL_2519643, EPI_ISL_2519644, EPI_ISL_2519645, EPI_ISL_2519647, EPI_ISL_2519648, EPI_ISL_2519649, EPI_ISL_2519650, EPI_ISL_2519651, EPI_ISL_2519652, EPI_ISL_2519653, EPI_ISL_2519654, EPI_ISL_2519655, EPI_ISL_2519656, EPI_ISL_2519657, EPI_ISL_2519658, EPI_ISL_2519659, EPI_ISL_2519660, EPI_ISL_2519661, EPI_ISL_2519662, EPI_ISL_2519663, EPI_ISL_2519664, EPI_ISL_2519665, EPI_ISL_2519666, EPI_ISL_2519667, EPI_ISL_2519668, EPI_ISL_2519669, EPI_ISL_2519670, EPI_ISL_2519671, EPI_ISL_2519672, EPI_ISL_2519673, EPI_ISL_2519675, EPI_ISL_2519676, EPI_ISL_2519677, EPI_ISL_2519678, EPI_ISL_2519679, EPI_ISL_2519680, EPI_ISL_2519681, EPI_ISL_2519682, EPI_ISL_2519683, EPI_ISL_2519684, EPI_ISL_2519685, EPI_ISL_2519687, EPI_ISL_2519688, EPI_ISL_2519690, EPI_ISL_2519691, EPI_ISL_2519692, EPI_ISL_2519693, EPI_ISL_2519694, EPI_ISL_2519695, EPI_ISL_2519696, EPI_ISL_2519697, EPI_ISL_2519698, EPI_ISL_2519699, EPI_ISL_2519700, EPI_ISL_2519701, EPI_ISL_2519702, EPI_ISL_2519703, EPI_ISL_2519704, EPI_ISL_2519705, EPI_ISL_2519706, EPI_ISL_2519707, EPI_ISL_2519708, EPI_ISL_2519709, EPI_ISL_2519710, EPI_ISL_2519711, EPI_ISL_2519712, EPI_ISL_2519713, EPI_ISL_2519714, EPI_ISL_2519715, EPI_ISL_2519716, EPI_ISL_2519717, EPI_ISL_2519718, EPI_ISL_2519719, EPI_ISL_2519720, EPI_ISL_2519721, EPI_ISL_2519722, EPI_ISL_2519723, EPI_ISL_2519724, EPI_ISL_2519725, EPI_ISL_2519726, EPI_ISL_2519742, EPI_ISL_2519743, EPI_ISL_2519744, EPI_ISL_2519745, EPI_ISL_2519746, EPI_ISL_2519747, EPI_ISL_2519748, EPI_ISL_2519749, EPI_ISL_2519750, EPI_ISL_2519753, EPI_ISL_2519754, EPI_ISL_2519764, EPI_ISL_2519805, EPI_ISL_2519826, EPI_ISL_2519831, EPI_ISL_2519873, EPI_ISL_2519898, EPI_ISL_2519945, EPI_ISL_2520106, EPI_ISL_2520107, EPI_ISL_2520108, EPI_ISL_2520109, EPI_ISL_2520110, EPI_ISL_2520111, EPI_ISL_2520112, EPI_ISL_2520113, EPI_ISL_2520114, EPI_ISL_2520115, EPI_ISL_2520116, EPI_ISL_2520117, EPI_ISL_2520119, EPI_ISL_2520120, EPI_ISL_2520121, EPI_ISL_2520122, EPI_ISL_2520123, EPI_ISL_2520124, EPI_ISL_2520125, EPI_ISL_2520126, EPI_ISL_2520127, EPI_ISL_2520128, EPI_ISL_2520129, EPI_ISL_2520130, EPI_ISL_2520131, EPI_ISL_2520132, EPI_ISL_2520133, EPI_ISL_2520134, EPI_ISL_2520135, EPI_ISL_2520136, EPI_ISL_2520137, EPI_ISL_2520138, EPI_ISL_2520139, EPI_ISL_2520140, EPI_ISL_2520141, EPI_ISL_2520142, EPI_ISL_2520143, EPI_ISL_2520144, EPI_ISL_2520145, EPI_ISL_2520146, EPI_ISL_2520147, EPI_ISL_2520148, EPI_ISL_2520149, EPI_ISL_2520150, EPI_ISL_2520151, EPI_ISL_2520152, EPI_ISL_2520153, EPI_ISL_2520154, EPI_ISL_2520155, EPI_ISL_2520156, EPI_ISL_2520157, EPI_ISL_2520158, EPI_ISL_2520159, EPI_ISL_2520160, EPI_ISL_2520161, EPI_ISL_2520162, EPI_ISL_2520164, EPI_ISL_2520165, EPI_ISL_2520166, EPI_ISL_2520167, EPI_ISL_2520168, EPI_ISL_2520169, EPI_ISL_2520170, EPI_ISL_2520171, EPI_ISL_2520172, EPI_ISL_2520173, EPI_ISL_2520174, EPI_ISL_2520175, EPI_ISL_2520177, EPI_ISL_2520178, EPI_ISL_2520179, EPI_ISL_2520180, EPI_ISL_2520181, EPI_ISL_2520182, EPI_ISL_2520183, EPI_ISL_2520184, EPI_ISL_2520185, EPI_ISL_2520186, EPI_ISL_2520187, EPI_ISL_2520188, EPI_ISL_2520189, EPI_ISL_2520190, EPI_ISL_2520191, EPI_ISL_2520192, EPI_ISL_2520193, EPI_ISL_2520194, EPI_ISL_2520195, EPI_ISL_2520196, EPI_ISL_2520197, EPI_ISL_2520198, EPI_ISL_2520199, EPI_ISL_2520200, EPI_ISL_2520201, EPI_ISL_2520202, | | | |

| | | | |
|---|---|---|---|
| EPI_ISL_2520203, EPI_ISL_2520204, EPI_ISL_2520205, EPI_ISL_2520206, EPI_ISL_2520207, EPI_ISL_2520208, EPI_ISL_2520209, EPI_ISL_2520210, EPI_ISL_2520211, EPI_ISL_2520212, EPI_ISL_2520213, EPI_ISL_2520214, EPI_ISL_2520215, EPI_ISL_2520216, EPI_ISL_2520217, EPI_ISL_2520218, EPI_ISL_2520219, EPI_ISL_2520220, EPI_ISL_2520221, EPI_ISL_2520222, EPI_ISL_2520223, EPI_ISL_2520224, EPI_ISL_2520225, EPI_ISL_2520226, EPI_ISL_2520227, EPI_ISL_2520228, EPI_ISL_2520229, EPI_ISL_2520230, EPI_ISL_2520231, EPI_ISL_2520232, EPI_ISL_2520233, EPI_ISL_2520235, EPI_ISL_2520236, EPI_ISL_2520237, EPI_ISL_2520238, EPI_ISL_2520239, EPI_ISL_2520240, EPI_ISL_2520243, EPI_ISL_2520244, EPI_ISL_2520245, EPI_ISL_2520246, EPI_ISL_2520247, EPI_ISL_2520248, EPI_ISL_2520249, EPI_ISL_2520250, EPI_ISL_2520251, EPI_ISL_2520252, EPI_ISL_2520253, EPI_ISL_2520254, EPI_ISL_2520255, EPI_ISL_2520257, EPI_ISL_2520258, EPI_ISL_2520259, EPI_ISL_2520260, EPI_ISL_2520261, EPI_ISL_2520262, EPI_ISL_2520264, EPI_ISL_2520266, EPI_ISL_2520267, EPI_ISL_2520268, EPI_ISL_2520269, EPI_ISL_2520270, EPI_ISL_2520271, EPI_ISL_2520272, EPI_ISL_2520273, EPI_ISL_2520274, EPI_ISL_2520275, EPI_ISL_2520276, EPI_ISL_2520277, EPI_ISL_2520278, EPI_ISL_2520279, EPI_ISL_2520280, EPI_ISL_2520281, EPI_ISL_2520282, EPI_ISL_2520283, EPI_ISL_2520284, EPI_ISL_2520285, EPI_ISL_2520286, EPI_ISL_2520287, EPI_ISL_2520295, EPI_ISL_2520297, EPI_ISL_2520298, EPI_ISL_2520299, EPI_ISL_2520302, EPI_ISL_2520303, EPI_ISL_2520304, EPI_ISL_2520305, EPI_ISL_2520306, EPI_ISL_2520307, EPI_ISL_2520308, EPI_ISL_2520310, EPI_ISL_2520311, EPI_ISL_2520337, EPI_ISL_2520358, EPI_ISL_2520362, EPI_ISL_2520377, EPI_ISL_2706718, EPI_ISL_2706725, EPI_ISL_2706727, EPI_ISL_2706731, EPI_ISL_2706744, EPI_ISL_2706745, EPI_ISL_2706755, EPI_ISL_2706758, EPI_ISL_2706766, EPI_ISL_2706928, EPI_ISL_2706929, EPI_ISL_2706932, EPI_ISL_2706933, EPI_ISL_2706934, EPI_ISL_2706935, EPI_ISL_2706936, EPI_ISL_2706937, EPI_ISL_2706938, EPI_ISL_2706940, EPI_ISL_2706941, EPI_ISL_2706942, EPI_ISL_2706955, EPI_ISL_2706972, EPI_ISL_2706974 | | | |
| see above | Quadram Institute Bioscience | COVID-19 Genomics UK (COG-UK) Consortium | Alexander J Trotter; Alison E. Mather; Alp Aydin; Ana P. Tedim; Anastasia Kolyva; Andrew Bell; Andrew J. Page; Claire Stuart; Dave J. Baker; Gemma L. Kay; John Wain; Justin O’Grady; Leonardo de Oliveira Martins; Lizzie Meadows; Maria Diaz; Mark Webber; Muhammed Yasir; Nabil-Fareed Alikhan; Ngozi Elumogo; Nicholas M. Thomson; Rachael Stanley; Rachel Gilroy; Reenesh Prakash; Samir Dervisevic; Samuel Bloomfield; Steven Rudder; Thanh Le-Viet |
| EPI_ISL_2520422, EPI_ISL_2520426, EPI_ISL_2520452, EPI_ISL_2520457, EPI_ISL_2520465, EPI_ISL_2520466, EPI_ISL_2520467, EPI_ISL_2520472 | | | |
| see above | Queens Medical Centre, Clinical Microbiology Department / DeepSeq Nottingham | COVID-19 Genomics UK (COG-UK) Consortium | Christopher Moore; Emily Park; Fei Sang; Gemma Clark; Hannah Howson-Wells; Jack Hill; Jayasree Dey; Johnny Debebe; Jonathan Ball; Joseph Chappell; Manjinder Khakh; Matthew Carlisle; Matthew Loose; Michelle M Lister; Nadine Holmes; Patrick McClure; Theocharis Tsoleridis; Timothy Byaruhanga; Vicki M Fleming; Victoria Wright; Wendy Smith |
| EPI_ISL_2743608, EPI_ISL_2743768, EPI_ISL_2743771, EPI_ISL_2743801, EPI_ISL_2743842, EPI_ISL_2743844, EPI_ISL_2743850, EPI_ISL_2743851, EPI_ISL_2743854, EPI_ISL_2743855, EPI_ISL_2743856, EPI_ISL_2743867, EPI_ISL_2743868, EPI_ISL_2743876, EPI_ISL_2743879, EPI_ISL_2743880, EPI_ISL_2743885, EPI_ISL_2743887, EPI_ISL_2743899, EPI_ISL_2743905, EPI_ISL_2743914, EPI_ISL_2743916, EPI_ISL_2743917, EPI_ISL_2743918, EPI_ISL_2743920, EPI_ISL_2743936, EPI_ISL_2743939, EPI_ISL_2744028, EPI_ISL_2744076, EPI_ISL_2744095, EPI_ISL_2744121, EPI_ISL_2744123, EPI_ISL_2744127, EPI_ISL_2744128, EPI_ISL_2744129, EPI_ISL_2744131, EPI_ISL_2744134, EPI_ISL_2744150, EPI_ISL_2744155, EPI_ISL_2744174, EPI_ISL_2744181, EPI_ISL_2744195, EPI_ISL_2744229, EPI_ISL_2744273, EPI_ISL_2744275, EPI_ISL_2744298, EPI_ISL_2744340, EPI_ISL_2744349, EPI_ISL_2744399, EPI_ISL_2744419, EPI_ISL_2744423, EPI_ISL_2744425, EPI_ISL_2744428, EPI_ISL_2744914, EPI_ISL_2744919, EPI_ISL_2744920, EPI_ISL_2744922, EPI_ISL_2744925, EPI_ISL_2744932, EPI_ISL_2744938, EPI_ISL_2744939, EPI_ISL_2744942, EPI_ISL_2744949, EPI_ISL_2744951, EPI_ISL_2744957, EPI_ISL_2744963, EPI_ISL_2744970, EPI_ISL_2744971, EPI_ISL_2744975, EPI_ISL_2744979, EPI_ISL_2745230, EPI_ISL_2745231, EPI_ISL_2745233, EPI_ISL_2745234, EPI_ISL_2745235, EPI_ISL_2745236, EPI_ISL_2745425, EPI_ISL_2746574, EPI_ISL_2904265, EPI_ISL_2904273, EPI_ISL_2904277, EPI_ISL_2904280, EPI_ISL_2904284, EPI_ISL_2904287, EPI_ISL_2904288, EPI_ISL_2904291, EPI_ISL_2904298, EPI_ISL_2904299, EPI_ISL_2904304, EPI_ISL_2904314, EPI_ISL_2904316 | | | |
| see above | Respiratory Virus Unit, Microbiology Services Colindale, Public Health England | COVID-19 Genomics UK (COG-UK) Consortium | PHE Covid Sequencing Team |
| EPI_ISL_2517296, EPI_ISL_2622341, EPI_ISL_2622342, EPI_ISL_2622343, EPI_ISL_2622344, EPI_ISL_2622345, EPI_ISL_2622346, EPI_ISL_2622348, EPI_ISL_2622349, EPI_ISL_2622350, EPI_ISL_2622351, EPI_ISL_2622352, EPI_ISL_2622353, EPI_ISL_2622354, EPI_ISL_2622355, EPI_ISL_2622360, EPI_ISL_2622361, EPI_ISL_2622372, EPI_ISL_2704833, EPI_ISL_2704834, EPI_ISL_2704835, EPI_ISL_2704836, EPI_ISL_2704837, EPI_ISL_2704838, EPI_ISL_2704839, EPI_ISL_2704840, EPI_ISL_2704841 | | | |
| see above | Virology Department, Royal Infirmary of Edinburgh, NHS Lothian / School of Biological Sciences, University of Edinburgh | COVID-19 Genomics UK (COG-UK) Consortium | Colquhoun R; Cotton S; Dewar R; Gallagher A; Hill V; Jackson B; Maloney D; McCrone JT; McHugh M; O’Toole Á; Rambaut A; Rooke S; Scher E; Templeton K; Yu X |
| EPI_ISL_2434966, EPI_ISL_2434967, EPI_ISL_2434969, EPI_ISL_2520929, EPI_ISL_2520930, EPI_ISL_2520932, EPI_ISL_2520933, EPI_ISL_2520935, EPI_ISL_2520937, EPI_ISL_2520939, EPI_ISL_2520943, EPI_ISL_2520947, EPI_ISL_2520948, EPI_ISL_2520949, EPI_ISL_2520951, EPI_ISL_2520952, EPI_ISL_2520953, EPI_ISL_2520955, EPI_ISL_2520956, EPI_ISL_2520957, EPI_ISL_2520958, EPI_ISL_2520960, EPI_ISL_2520963, EPI_ISL_2520964, EPI_ISL_2520965, EPI_ISL_2520966, EPI_ISL_2520967, EPI_ISL_2520971, EPI_ISL_2520974, EPI_ISL_2520977, EPI_ISL_2520978, EPI_ISL_2520979, EPI_ISL_2520980, EPI_ISL_2520981, EPI_ISL_2520982, EPI_ISL_2520988, EPI_ISL_2520989, EPI_ISL_2520991, EPI_ISL_2520992, EPI_ISL_2520993, EPI_ISL_2520995, EPI_ISL_2520996, EPI_ISL_2520997, EPI_ISL_2520999, EPI_ISL_2521000, EPI_ISL_2521004, EPI_ISL_2521008, EPI_ISL_2521013, EPI_ISL_2521014, EPI_ISL_2521016, EPI_ISL_2521018, EPI_ISL_2521019, EPI_ISL_2521021, EPI_ISL_2521023, EPI_ISL_2521026, EPI_ISL_2521027, EPI_ISL_2521029, EPI_ISL_2521032, EPI_ISL_2521034, EPI_ISL_2521035, EPI_ISL_2521036, EPI_ISL_2521037, EPI_ISL_2521040, EPI_ISL_2521041, EPI_ISL_2521042, EPI_ISL_2521044, EPI_ISL_2521045, EPI_ISL_2521046, EPI_ISL_2521047, EPI_ISL_2521053, EPI_ISL_2521055, EPI_ISL_2521059, EPI_ISL_2521060, EPI_ISL_2521064, EPI_ISL_2521065, EPI_ISL_2521066, EPI_ISL_2521067, EPI_ISL_2521072, EPI_ISL_2521073, EPI_ISL_2521074, EPI_ISL_2521076, EPI_ISL_2521078, EPI_ISL_2521083, EPI_ISL_2521084, EPI_ISL_2521086, EPI_ISL_2521087, EPI_ISL_2521088, EPI_ISL_2521089, EPI_ISL_2521091, EPI_ISL_2521094, EPI_ISL_2521095, EPI_ISL_2521096, EPI_ISL_2521097, EPI_ISL_2521100, EPI_ISL_2521101, EPI_ISL_2521102, EPI_ISL_2521103, EPI_ISL_2521104, EPI_ISL_2521105, EPI_ISL_2521107, EPI_ISL_2521108, EPI_ISL_2521109, EPI_ISL_2521111, EPI_ISL_2521112, EPI_ISL_2521113, EPI_ISL_2521115, EPI_ISL_2521116, EPI_ISL_2521117, EPI_ISL_2521120, EPI_ISL_2521121, EPI_ISL_2521122, EPI_ISL_2521123, EPI_ISL_2521124, EPI_ISL_2521126, EPI_ISL_2521127, EPI_ISL_2521129, EPI_ISL_2521134, EPI_ISL_2521136, EPI_ISL_2521137, EPI_ISL_2521138, EPI_ISL_2521140, EPI_ISL_2521141, EPI_ISL_2521142, EPI_ISL_2521147, EPI_ISL_2521148, EPI_ISL_2521149, EPI_ISL_2625252, EPI_ISL_2625260, EPI_ISL_2625270, EPI_ISL_2625288, EPI_ISL_2625354, EPI_ISL_2625413, EPI_ISL_2625479, EPI_ISL_2625483, EPI_ISL_2625529, EPI_ISL_2625555, EPI_ISL_2625556, EPI_ISL_2625564, EPI_ISL_2625615, EPI_ISL_2625617 | | | |
| see above | Virology Department, Sheffield Teaching Hospitals NHS Foundation Trust/Department of Infection, Immunity and Cardiovascular Disease, The Medical School, University of Sheffield | COVID-19 Genomics UK (COG-UK) Consortium | Adri Angyal; Alex Keeley; Benjamin Lindsey; Cariad Evans; Danielle Groves; Dave Partridge; Katie Johnson; Laura Carrilero; Luke Green; Matthew Parker; Matthew Wyles; Mehmet Yavuz; Mohammad Raza; Nikki Smith; Paul Parsons; Rachel Tucker; Rebecca Brown; Thushan de Silva |
| EPI_ISL_2520541, EPI_ISL_2520557, EPI_ISL_2520559, EPI_ISL_2520560, EPI_ISL_2520565, EPI_ISL_2520566, EPI_ISL_2520569, EPI_ISL_2520571, EPI_ISL_2520573, EPI_ISL_2520574, EPI_ISL_2520576, EPI_ISL_2520578, EPI_ISL_2520580, EPI_ISL_2520581, EPI_ISL_2520582, EPI_ISL_2520583, EPI_ISL_2520586, EPI_ISL_2520587, EPI_ISL_2520588, EPI_ISL_2520593, EPI_ISL_2520594, EPI_ISL_2520595, EPI_ISL_2520596, EPI_ISL_2520597, EPI_ISL_2520599, EPI_ISL_2520600, EPI_ISL_2520603, EPI_ISL_2520604, EPI_ISL_2520605, EPI_ISL_2520606, EPI_ISL_2520615, EPI_ISL_2520616, EPI_ISL_2520617, EPI_ISL_2520618, EPI_ISL_2520619, EPI_ISL_2520621, EPI_ISL_2520622, EPI_ISL_2520625, EPI_ISL_2520628, EPI_ISL_2520629, EPI_ISL_2520630, EPI_ISL_2520631, EPI_ISL_2520632, EPI_ISL_2520633, EPI_ISL_2520635, EPI_ISL_2520636, EPI_ISL_2520637, EPI_ISL_2520639, EPI_ISL_2520641, EPI_ISL_2520642, EPI_ISL_2520643, EPI_ISL_2520651, EPI_ISL_2520652, EPI_ISL_2520653, EPI_ISL_2520654, EPI_ISL_2520655, EPI_ISL_2520657, EPI_ISL_2520659, EPI_ISL_2520660, EPI_ISL_2520661, EPI_ISL_2520665, EPI_ISL_2520667, EPI_ISL_2520668, EPI_ISL_2520669, EPI_ISL_2520670, EPI_ISL_2520671, EPI_ISL_2520672, EPI_ISL_2520673, EPI_ISL_2520674, EPI_ISL_2520676, EPI_ISL_2520678, EPI_ISL_2520683, EPI_ISL_2520686, EPI_ISL_2520688, EPI_ISL_2520691, EPI_ISL_2520693, EPI_ISL_2520694, EPI_ISL_2520695, EPI_ISL_2520696, EPI_ISL_2520697, EPI_ISL_2520698, EPI_ISL_2520700, EPI_ISL_2520701, EPI_ISL_2520707, EPI_ISL_2520710, EPI_ISL_2520716, EPI_ISL_2520717, EPI_ISL_2520719, EPI_ISL_2520721, EPI_ISL_2520761, EPI_ISL_2520762, EPI_ISL_2520763, EPI_ISL_2520764, EPI_ISL_2520765, EPI_ISL_2520771, EPI_ISL_2520772, EPI_ISL_2520776, EPI_ISL_2520777, EPI_ISL_2520778, EPI_ISL_2520780, EPI_ISL_2520781, EPI_ISL_2520783, EPI_ISL_2520785, EPI_ISL_2520790, EPI_ISL_2520791, EPI_ISL_2520792, EPI_ISL_2520793, EPI_ISL_2520794, EPI_ISL_2520795, EPI_ISL_2520798, EPI_ISL_2520799, EPI_ISL_2520800, EPI_ISL_2520801, EPI_ISL_2520802, EPI_ISL_2520804, EPI_ISL_2520807, EPI_ISL_2520808, EPI_ISL_2520811 | | | |
| see above | Wales Specialist Virology Centre Sequencing lab: Pathogen Genomics Unit | Public Health Wales Microbiology Cardiff Wales Specialist Virology Centre | Alec Birchley; Alexander Adams; Amy Gaskin; Angela Marchbank; Bree Gatica-Wilcox; Catherine Moore; Jason Coombes; Joanne Watkins; Joel Southgate; Johnathan Evans; Laura Gifford; Lauren Gilbert; Lee Graham; Malorie Perry; Matthew Bull; Nicole Pacchiarini; Sally Corden; Sara Kumziene-Summerhayes; Sara Rey; Sarah Taylor; Simon Cottrell; Sophie Jones; Tom Connor |
| EPI_ISL_2704802, EPI_ISL_2704809, EPI_ISL_2704815, EPI_ISL_2722638, EPI_ISL_2722646, EPI_ISL_2722647, EPI_ISL_2722651, EPI_ISL_2722652, EPI_ISL_2722653, EPI_ISL_2722654, EPI_ISL_2722655, EPI_ISL_2722656, EPI_ISL_2722657, EPI_ISL_2722658, EPI_ISL_2722659, EPI_ISL_2722660, EPI_ISL_2722661, EPI_ISL_2722670, EPI_ISL_2722678, EPI_ISL_2734271, EPI_ISL_2734282, EPI_ISL_2734283, EPI_ISL_2734284, EPI_ISL_2734285, EPI_ISL_2734288, EPI_ISL_2734289, EPI_ISL_2734290, EPI_ISL_2815916, EPI_ISL_2900728 | | | |
| see above | West of Scotland Specialist Virology Centre, NHSGGC / MRC-University of Glasgow Centre for Virus Research | COVID-19 Genomics UK (COG-UK) Consortium | Alasdair MacLean; Alice Broos; Ana da Silva Filipe; Antonia Ho; Daniel Mair; David L Robertson; Emma Thomson; Guy Mollett; Ioulia Tsatsani; James Shepherd; Jenna Nichols; Joseph Hughes; Kathy Li; Kathy Smollett; Kyriaki Nomikou; Lily Tong; Matthew Holden; Natasha Johnson; Rachel Blacow; Richard Orton; Rory Gunson; Sarah McDonald; Sharif Shaaban; Sreenu Vattipally; Stephen Carmichael |

We gratefully acknowledge the following Authors from the Originating laboratories responsible for obtaining the specimens, as well as the Submitting laboratories where the genome data were generated and shared via GISAID, on which this research is based.

All Submitters of data may be contacted directly via www.gisaid.org

Authors are sorted alphabetically.

| Accession ID | Originating Laboratory | Submitting Laboratory | Authors |
|---|---|---|---|
| EPI_ISL_2686142, EPI_ISL_2686143, EPI_ISL_2686144, EPI_ISL_2686145, EPI_ISL_2686151 | Amazon Dx, AUV2 Laboratory | Amazon Dx, AUV2 Laboratory | Luke Meredith |
| EPI_ISL_2622435, EPI_ISL_2622436, EPI_ISL_2622437, EPI_ISL_2622438, EPI_ISL_2622439, EPI_ISL_2622440, EPI_ISL_2622441, EPI_ISL_2622442 | | | |
| see above | Barts Health NHS Trust | COVID-19 Genomics UK (COG-UK) Consortium | BROAD; Beatrix; CUTINO-MOGUEL; Claire; David; Dola; HARRINGTON; KELE; KULASEGARAN-SHYLINI; Maria-Teresa; OWOYEMI; Raghavendran |
| EPI_ISL_2520824, EPI_ISL_2520876, EPI_ISL_2520878, EPI_ISL_2520880, EPI_ISL_2520886, EPI_ISL_2520888, EPI_ISL_2520889, EPI_ISL_2520891, EPI_ISL_2520894, EPI_ISL_2520895, EPI_ISL_2520900, EPI_ISL_2520903, EPI_ISL_2520913 | | | |
| see above | Centre for Enzyme Innovation, University of Portsmouth / Translational Research Laboratory, Portsmouth Hospitals NHS Trust / Hampshire Hospitals NHS Foundation Trust / Bournemouth University / University Hospitals Dorset NHS Foundation Trust / Brighton and Sussex University Hospitals NHS Trust / Dartford and Gravesham NHS Trust / Isle of Wight NHS Trust / Maidstone and Tunbridge Wells NHS Trust / University Hospital Southampton NHS Foundation Trust | COVID-19 Genomics UK (COG-UK) Consortium | Adhyana Mahanama; Allyson Lloyd; Andrew Butt; Angela Beckett; Anibolina Castigador; Anna Mantzaouratou; Anoop Chauhan; Ben Macklin; Benjamin Cogger; Buddhini Samaraweera; Cassandra Malone; Christopher Fearn; Claire Thomas; Dorian Crudgington; Eleri Wilson-Davies; Emanuela Pelosi; Emily Macnaughton; Helen Umpleby; Helen Wheeler; Jacqui Prieto; Jonnie Seymour; Karen Withell; Kate Cook; Katie Loveson; Kelly Bicknell; Kevin Tucker; Kordo Saeed; Liz Sheridan; Magdalena Barrow; Matthew Harvey; Mohammed Hassan-Ibrahim; Nathan Moore; Nick Cortes; Rachael Jeremiah; Robert Impey; Salman Goudarzi; Samirakhon Raupova; Samuel Robson; Sarah Buchan; Sarah Jeremiah; Sarah Wyllie; Scott Elliott; Sharon Glaysher; Siona Silvieri; Stephen Aplin; Stephen Kidd; Thea Sass |
| EPI_ISL_2722214 | Department of Pathology, University of Cambridge | COVID-19 Genomics UK (COG-UK) Consortium | Aminu S. Jahun; Ian Goodfellow; Iliana Georgana; Martin D. Curran; Myra Hosmillo; Rhys Izuagbe; Surendra Parmar; William L. Hamilton; Yasmin Chaudhry |
| EPI_ISL_2511097, EPI_ISL_2511098, EPI_ISL_2511102, EPI_ISL_2511107, EPI_ISL_2511110, EPI_ISL_2511112, EPI_ISL_2511114, EPI_ISL_2511115, EPI_ISL_2511116, EPI_ISL_2511117, EPI_ISL_2511118, EPI_ISL_2511128, EPI_ISL_2511129, EPI_ISL_2511132, EPI_ISL_2511141, EPI_ISL_2511144, EPI_ISL_2511146, EPI_ISL_2511149, EPI_ISL_2511154, EPI_ISL_2511159, EPI_ISL_2511160, EPI_ISL_2511162, EPI_ISL_2511163, EPI_ISL_2511167, EPI_ISL_2511169, EPI_ISL_2511171, EPI_ISL_2511174, EPI_ISL_2511177, EPI_ISL_2511179, EPI_ISL_2511180, EPI_ISL_2511182, EPI_ISL_2511187, EPI_ISL_2511189, EPI_ISL_2511190, EPI_ISL_2511191, EPI_ISL_2511192, EPI_ISL_2511193, EPI_ISL_2511194, EPI_ISL_2511201, EPI_ISL_2511205, EPI_ISL_2511212, EPI_ISL_2511213, EPI_ISL_2511218, EPI_ISL_2511220, EPI_ISL_2511222, EPI_ISL_2511230, EPI_ISL_2511236, EPI_ISL_2511241, EPI_ISL_2511242, EPI_ISL_2511243, EPI_ISL_2511245, EPI_ISL_2511246, EPI_ISL_2511249, EPI_ISL_2511250, EPI_ISL_2511256, EPI_ISL_2511260, EPI_ISL_2511269, EPI_ISL_2511270, EPI_ISL_2511272, EPI_ISL_2511274, EPI_ISL_2511275, EPI_ISL_2511280, EPI_ISL_2511281, EPI_ISL_2511282, EPI_ISL_2511290, EPI_ISL_2511294, EPI_ISL_2511295, EPI_ISL_2511296, EPI_ISL_2511297, EPI_ISL_2511306, EPI_ISL_2511308, EPI_ISL_2511310, EPI_ISL_2511311, EPI_ISL_2511319, EPI_ISL_2511321, EPI_ISL_2511323, EPI_ISL_2511324, EPI_ISL_2511326, EPI_ISL_2511329, EPI_ISL_2511330, EPI_ISL_2511332, EPI_ISL_2511334, EPI_ISL_2511335, EPI_ISL_2511336, EPI_ISL_2511338, EPI_ISL_2511340, EPI_ISL_2511341, EPI_ISL_2511343, EPI_ISL_2511344, EPI_ISL_2511349, EPI_ISL_2511351, EPI_ISL_2511938, EPI_ISL_2511939, EPI_ISL_2511941, EPI_ISL_2511942, EPI_ISL_2511943, EPI_ISL_2511945, EPI_ISL_2511947, EPI_ISL_2511949, EPI_ISL_2511950, EPI_ISL_2511951, EPI_ISL_2511952, EPI_ISL_2511953, EPI_ISL_2511954, EPI_ISL_2511955, EPI_ISL_2511957, EPI_ISL_2511958, EPI_ISL_2511959, EPI_ISL_2511960, EPI_ISL_2511962, EPI_ISL_2511963, EPI_ISL_2511964, EPI_ISL_2511966, EPI_ISL_2511967, EPI_ISL_2511968, EPI_ISL_2511969, EPI_ISL_2511970, EPI_ISL_2511971, EPI_ISL_2511972, EPI_ISL_2511974, EPI_ISL_2511975, EPI_ISL_2511977, EPI_ISL_2511979, EPI_ISL_2511980, EPI_ISL_2511984, EPI_ISL_2511985, EPI_ISL_2511986, EPI_ISL_2511987, EPI_ISL_2511988, EPI_ISL_2511989, EPI_ISL_2511990, EPI_ISL_2511991, EPI_ISL_2511992, EPI_ISL_2511993, EPI_ISL_2511994, EPI_ISL_2511996, EPI_ISL_2511997, EPI_ISL_2511998, EPI_ISL_2511999, EPI_ISL_2512000, EPI_ISL_2512001, EPI_ISL_2512002, EPI_ISL_2512003, EPI_ISL_2512004, EPI_ISL_2512005, EPI_ISL_2512007, EPI_ISL_2512008, EPI_ISL_2512009, EPI_ISL_2512010, EPI_ISL_2512011, EPI_ISL_2512012, EPI_ISL_2512015, EPI_ISL_2512016, EPI_ISL_2512017, EPI_ISL_2512018, EPI_ISL_2512019, EPI_ISL_2512020, EPI_ISL_2512021, EPI_ISL_2512023, EPI_ISL_2512024, EPI_ISL_2512025, EPI_ISL_2512026, EPI_ISL_2512027, EPI_ISL_2512028, EPI_ISL_2512029, EPI_ISL_2512030, EPI_ISL_2512031, EPI_ISL_2512032, EPI_ISL_2512034, EPI_ISL_2512035, EPI_ISL_2512036, EPI_ISL_2512038, EPI_ISL_2512039, EPI_ISL_2512041, EPI_ISL_2512042, EPI_ISL_2512047, EPI_ISL_2512051, EPI_ISL_2512052, EPI_ISL_2512053, EPI_ISL_2512054, EPI_ISL_2512055, EPI_ISL_2512057, EPI_ISL_2512058, EPI_ISL_2512059, EPI_ISL_2512060, EPI_ISL_2512061, EPI_ISL_2512063, EPI_ISL_2512065, EPI_ISL_2512067, EPI_ISL_2512068, EPI_ISL_2512069, EPI_ISL_2512070, EPI_ISL_2512072, EPI_ISL_2512073, EPI_ISL_2512075, EPI_ISL_2512076, EPI_ISL_2512077, EPI_ISL_2512079, EPI_ISL_2512085, EPI_ISL_2512086, EPI_ISL_2512087, EPI_ISL_2512088, EPI_ISL_2512089, EPI_ISL_2512090, EPI_ISL_2512091, EPI_ISL_2512092, EPI_ISL_2512093, EPI_ISL_2512094, EPI_ISL_2512095, EPI_ISL_2512096, EPI_ISL_2512097, EPI_ISL_2512099, EPI_ISL_2512100, EPI_ISL_2512101, EPI_ISL_2512102, EPI_ISL_2512103, EPI_ISL_2512104, EPI_ISL_2512105, EPI_ISL_2512106, EPI_ISL_2512109, EPI_ISL_2512110, EPI_ISL_2512112, EPI_ISL_2512113, EPI_ISL_2512114, EPI_ISL_2512115, EPI_ISL_2512117, EPI_ISL_2512118, EPI_ISL_2512119, EPI_ISL_2512121, EPI_ISL_2512122, EPI_ISL_2512123, EPI_ISL_2512124, EPI_ISL_2512128, EPI_ISL_2512129, EPI_ISL_2512130, EPI_ISL_2512131, EPI_ISL_2512132, EPI_ISL_2512133, EPI_ISL_2512134, EPI_ISL_2512135, EPI_ISL_2512136, EPI_ISL_2512137, EPI_ISL_2512139, EPI_ISL_2512142, EPI_ISL_2512143, EPI_ISL_2512144, EPI_ISL_2512145, EPI_ISL_2512146, EPI_ISL_2512147, EPI_ISL_2512148, EPI_ISL_2512149, EPI_ISL_2512150, EPI_ISL_2512151, EPI_ISL_2512152, EPI_ISL_2512153, EPI_ISL_2512154, EPI_ISL_2512155, EPI_ISL_2512156, EPI_ISL_2512157, EPI_ISL_2512158, EPI_ISL_2512159, EPI_ISL_2512161, EPI_ISL_2512162, EPI_ISL_2512163, EPI_ISL_2512164, EPI_ISL_2512166, EPI_ISL_2512169, EPI_ISL_2512170, EPI_ISL_2512171, EPI_ISL_2512174, EPI_ISL_2512175, EPI_ISL_2512176, EPI_ISL_2512178, EPI_ISL_2512179, EPI_ISL_2512180, EPI_ISL_2512181, EPI_ISL_2512183, EPI_ISL_2512184, EPI_ISL_2512185, EPI_ISL_2512186, EPI_ISL_2512188, EPI_ISL_2512189, EPI_ISL_2512190, EPI_ISL_2512191, EPI_ISL_2512192, EPI_ISL_2512193, EPI_ISL_2512194, EPI_ISL_2512195, EPI_ISL_2512197, EPI_ISL_2512199, EPI_ISL_2512201, EPI_ISL_2512203, EPI_ISL_2512205, EPI_ISL_2512206, EPI_ISL_2512209, EPI_ISL_2512210, EPI_ISL_2512211, EPI_ISL_2512212, EPI_ISL_2512213, EPI_ISL_2512214, EPI_ISL_2512215, EPI_ISL_2512216, EPI_ISL_2512218, EPI_ISL_2512220, EPI_ISL_2512222, EPI_ISL_2512223, EPI_ISL_2512224, EPI_ISL_2512226, EPI_ISL_2512227, EPI_ISL_2512228, EPI_ISL_2512229, EPI_ISL_2512230, EPI_ISL_2512231, EPI_ISL_2512233, EPI_ISL_2512234, EPI_ISL_2512236, EPI_ISL_2512237, EPI_ISL_2512238, EPI_ISL_2512239, EPI_ISL_2512240, EPI_ISL_2512241, EPI_ISL_2512242, EPI_ISL_2512245, EPI_ISL_2512246, EPI_ISL_2512247, EPI_ISL_2512248, EPI_ISL_2512250, EPI_ISL_2512251, EPI_ISL_2512252, EPI_ISL_2512253, EPI_ISL_2512255, EPI_ISL_2512256, EPI_ISL_2512257, EPI_ISL_2512258, EPI_ISL_2512259, EPI_ISL_2512260, EPI_ISL_2512261, EPI_ISL_2512263, EPI_ISL_2512264, EPI_ISL_2512265, EPI_ISL_2512266, EPI_ISL_2512268, EPI_ISL_2512270, EPI_ISL_2512271, EPI_ISL_2512272, EPI_ISL_2512273, EPI_ISL_2512274, EPI_ISL_2512275, EPI_ISL_2512276, EPI_ISL_2512278, EPI_ISL_2512280, EPI_ISL_2512281, EPI_ISL_2512283, EPI_ISL_2512284, EPI_ISL_2512286, EPI_ISL_2512287, EPI_ISL_2512288, EPI_ISL_2512289, EPI_ISL_2512290, EPI_ISL_2512291, EPI_ISL_2512292, EPI_ISL_2512293, EPI_ISL_2512296, EPI_ISL_2512305, EPI_ISL_2512307, EPI_ISL_2512309, EPI_ISL_2512311, EPI_ISL_2512313, EPI_ISL_2512318, EPI_ISL_2512320, EPI_ISL_2512325, EPI_ISL_2512330, EPI_ISL_2512332, EPI_ISL_2512334, EPI_ISL_2512344, EPI_ISL_2512348, EPI_ISL_2512359, EPI_ISL_2512363, EPI_ISL_2512364, EPI_ISL_2512367, EPI_ISL_2512369, EPI_ISL_2512924, EPI_ISL_2512930, EPI_ISL_2512934, EPI_ISL_2512935, EPI_ISL_2512942, EPI_ISL_2512943, EPI_ISL_2512944, EPI_ISL_2512945, EPI_ISL_2512946, EPI_ISL_2512951, EPI_ISL_2512955, EPI_ISL_2512961, EPI_ISL_2512963, EPI_ISL_2512964, EPI_ISL_2512970, EPI_ISL_2512971, EPI_ISL_2512977, EPI_ISL_2512978, EPI_ISL_2512981, EPI_ISL_2512982, EPI_ISL_2512992, EPI_ISL_2512999, EPI_ISL_2513001, EPI_ISL_2513008, EPI_ISL_2513023, EPI_ISL_2513029, EPI_ISL_2513037, EPI_ISL_2513040, EPI_ISL_2513041, EPI_ISL_2513044, EPI_ISL_2513053, EPI_ISL_2513054, EPI_ISL_2513056, EPI_ISL_2513057, EPI_ISL_2513060, EPI_ISL_2513071, EPI_ISL_2513072, EPI_ISL_2513073, EPI_ISL_2513079, EPI_ISL_2513082, EPI_ISL_2513084, EPI_ISL_2513089, EPI_ISL_2513097, EPI_ISL_2513099, EPI_ISL_2513103, EPI_ISL_2513116, EPI_ISL_2513120, EPI_ISL_2513121, EPI_ISL_2513122, EPI_ISL_2513124, EPI_ISL_2513126, EPI_ISL_2513131, EPI_ISL_2513133, EPI_ISL_2513136, EPI_ISL_2513138, EPI_ISL_2513140, EPI_ISL_2513142, EPI_ISL_2513143, EPI_ISL_2513145, EPI_ISL_2513149, EPI_ISL_2513151, EPI_ISL_2513164, EPI_ISL_2513165, EPI_ISL_2513169, EPI_ISL_2513173, EPI_ISL_2513178, EPI_ISL_2513179, EPI_ISL_2513185, EPI_ISL_2513187, EPI_ISL_2513189, EPI_ISL_2513198, EPI_ISL_2513203, EPI_ISL_2513208, EPI_ISL_2513211, EPI_ISL_2513214, EPI_ISL_2513216, EPI_ISL_2513219, EPI_ISL_2513220, EPI_ISL_2513222, EPI_ISL_2513224, EPI_ISL_2513230, EPI_ISL_2513234, EPI_ISL_2513239, EPI_ISL_2513247, EPI_ISL_2513253, EPI_ISL_2513256, EPI_ISL_2513262, EPI_ISL_2513268, EPI_ISL_2513270, EPI_ISL_2513271, EPI_ISL_2513280, EPI_ISL_2516024, EPI_ISL_2516035, EPI_ISL_2516147, EPI_ISL_2516239, EPI_ISL_2516294, EPI_ISL_2539746, EPI_ISL_2539761, EPI_ISL_2539763, EPI_ISL_2539771, EPI_ISL_2539790, EPI_ISL_2539795, EPI_ISL_2539806, EPI_ISL_2539812, EPI_ISL_2539817, EPI_ISL_2539839, EPI_ISL_2539841, EPI_ISL_2539852, EPI_ISL_2539853, EPI_ISL_2539861, EPI_ISL_2539863, EPI_ISL_2539864, EPI_ISL_2539879, EPI_ISL_2539886, EPI_ISL_2539890, EPI_ISL_2539895, EPI_ISL_2539898, EPI_ISL_2539900, EPI_ISL_2539902, EPI_ISL_2539907, EPI_ISL_2539914, EPI_ISL_2539921, EPI_ISL_2539924, EPI_ISL_2539927, EPI_ISL_2539931, EPI_ISL_2539939, EPI_ISL_2539941, EPI_ISL_2539943, EPI_ISL_2539950, EPI_ISL_2539963, EPI_ISL_2539969, EPI_ISL_2539971, EPI_ISL_2539977, EPI_ISL_2539979, EPI_ISL_2539988, EPI_ISL_2539990, EPI_ISL_2539995, EPI_ISL_2540005, EPI_ISL_2540016, EPI_ISL_2540951, EPI_ISL_2540954, EPI_ISL_2540971, EPI_ISL_2540974, EPI_ISL_2540977, EPI_ISL_2540979, EPI_ISL_2540984, EPI_ISL_2540992, EPI_ISL_2540997, EPI_ISL_2541000, EPI_ISL_2541003, EPI_ISL_2541005, EPI_ISL_2541009, EPI_ISL_2541014, EPI_ISL_2541022, EPI_ISL_2541027, EPI_ISL_2541028, EPI_ISL_2541035, EPI_ISL_2541055, EPI_ISL_2541060, EPI_ISL_2541061, EPI_ISL_2541072, EPI_ISL_2541074, EPI_ISL_2541076, EPI_ISL_2541077, EPI_ISL_2541078, EPI_ISL_2541079, EPI_ISL_2541084, EPI_ISL_2541094, EPI_ISL_2541103, EPI_ISL_2541107, EPI_ISL_2541112, EPI_ISL_2541116, EPI_ISL_2541120, EPI_ISL_2541142, EPI_ISL_2541143, EPI_ISL_2541147, EPI_ISL_2541157, EPI_ISL_2541163, EPI_ISL_2541170, EPI_ISL_2541177, EPI_ISL_2541181, EPI_ISL_2541184, EPI_ISL_2541185, EPI_ISL_2541194, EPI_ISL_2541201, EPI_ISL_2541212, EPI_ISL_2541224, EPI_ISL_2541225, EPI_ISL_2541228, EPI_ISL_2541229, EPI_ISL_2541231, EPI_ISL_2541241, EPI_ISL_2541244, EPI_ISL_2541251, EPI_ISL_2541258, EPI_ISL_2541280, EPI_ISL_2541287, EPI_ISL_2541298, EPI_ISL_2541300, EPI_ISL_2541312, EPI_ISL_2541317, EPI_ISL_2541319, EPI_ISL_2541321, EPI_ISL_2541364, EPI_ISL_2541380, EPI_ISL_2541408, EPI_ISL_2541410, EPI_ISL_2541414, EPI_ISL_2541452, EPI_ISL_2541459, EPI_ISL_2541471, EPI_ISL_2541476, EPI_ISL_2541510, EPI_ISL_2541511, EPI_ISL_2541522, EPI_ISL_2541523, EPI_ISL_2541528, EPI_ISL_2541544, EPI_ISL_2541551, EPI_ISL_2541581, EPI_ISL_2541582, EPI_ISL_2541597, EPI_ISL_2541602, EPI_ISL_2541609, EPI_ISL_2541612, EPI_ISL_2541616, EPI_ISL_2541622, EPI_ISL_2541630, EPI_ISL_2541635, EPI_ISL_2542104, EPI_ISL_2542105, EPI_ISL_2552614, EPI_ISL_2552624, EPI_ISL_2552636, EPI_ISL_2552637, EPI_ISL_2552640, EPI_ISL_2552646, EPI_ISL_2552648, EPI_ISL_2552651, EPI_ISL_2552662, EPI_ISL_2552664, EPI_ISL_2552666, EPI_ISL_2552670, EPI_ISL_2552685, EPI_ISL_2552696, EPI_ISL_2552697, EPI_ISL_2552698, EPI_ISL_2552700, EPI_ISL_2552712, EPI_ISL_2552719, EPI_ISL_2552720, EPI_ISL_2552723, EPI_ISL_2552725, EPI_ISL_2552731, EPI_ISL_2552734, EPI_ISL_2552744, EPI_ISL_2552745, EPI_ISL_2552746, EPI_ISL_2552748, EPI_ISL_2552749, EPI_ISL_2552751, EPI_ISL_2552756, EPI_ISL_2552767, EPI_ISL_2552768, EPI_ISL_2552772, EPI_ISL_2552773, EPI_ISL_2552775, EPI_ISL_2552777, EPI_ISL_2552782, EPI_ISL_2552783, EPI_ISL_2552790, EPI_ISL_2552794, EPI_ISL_2552795, EPI_ISL_2552801, EPI_ISL_2552803, EPI_ISL_2552805, EPI_ISL_2552808, EPI_ISL_2552810, EPI_ISL_2552814, EPI_ISL_2552816, EPI_ISL_2552819, EPI_ISL_2552820, EPI_ISL_2552823, EPI_ISL_2552838, EPI_ISL_2552839, EPI_ISL_2552844, EPI_ISL_2552852, EPI_ISL_2552859, EPI_ISL_2552860, EPI_ISL_2552875, EPI_ISL_2552882, EPI_ISL_2552887, EPI_ISL_2552888, EPI_ISL_2552889, EPI_ISL_2552890, EPI_ISL_2552901, EPI_ISL_2552905, EPI_ISL_2552914, EPI_ISL_2552919, EPI_ISL_2552923, EPI_ISL_2555057, EPI_ISL_2555064, EPI_ISL_2641358, EPI_ISL_2641419, EPI_ISL_2641449, EPI_ISL_2641450, EPI_ISL_2641495, EPI_ISL_2641535, EPI_ISL_2641556, EPI_ISL_2641583, EPI_ISL_2641590, EPI_ISL_2641688, EPI_ISL_2641697 | | | |
| see above | Health Services Laboratories | Wellcome Sanger Institute for the COVID-19 Genomics UK (COG-UK) Consortium | Cordelia Langford; David K. Jackson; Dominic Kwiatkowski; Ewan Harrison; Health Services Laboratories and Alex Alderton; Ian Johnston; Jeffrey Barrett; John Sillitoe on behalf of the Wellcome Sanger Institute COVID-19 Surveillance Team; Roberto Amato; Sonia Goncalves |
| EPI_ISL_2511361, EPI_ISL_2511376, EPI_ISL_2511391, EPI_ISL_2511395, EPI_ISL_2511415, EPI_ISL_2511417, EPI_ISL_2511443, EPI_ISL_2511468, EPI_ISL_2511524, EPI_ISL_2511527, EPI_ISL_2511539, EPI_ISL_2511545, EPI_ISL_2511551, EPI_ISL_2511598, EPI_ISL_2511631, EPI_ISL_2511646, EPI_ISL_2511648, EPI_ISL_2511650, EPI_ISL_2511653, EPI_ISL_2511665, EPI_ISL_2511677, EPI_ISL_2511684, EPI_ISL_2511689, EPI_ISL_2511711, EPI_ISL_2511721, EPI_ISL_2511726, EPI_ISL_2511744, EPI_ISL_2511754, EPI_ISL_2511764, EPI_ISL_2511820, EPI_ISL_2511833, EPI_ISL_2511842, EPI_ISL_2511847, EPI_ISL_2511852, EPI_ISL_2511863, EPI_ISL_2511864, EPI_ISL_2511871, EPI_ISL_2511873, EPI_ISL_2511897, EPI_ISL_2511898, EPI_ISL_2511916, EPI_ISL_2511928, EPI_ISL_2511935, EPI_ISL_2512064, EPI_ISL_2512111, EPI_ISL_2512141, EPI_ISL_2512217, EPI_ISL_2512235, EPI_ISL_2512244, EPI_ISL_2512923, EPI_ISL_2512926, EPI_ISL_2512939, EPI_ISL_2512950, EPI_ISL_2512959, EPI_ISL_2512973, EPI_ISL_2512974, EPI_ISL_2512979, EPI_ISL_2512988, EPI_ISL_2512989, EPI_ISL_2512995, EPI_ISL_2513028, EPI_ISL_2513035, EPI_ISL_2513059, EPI_ISL_2513066, EPI_ISL_2513067, EPI_ISL_2513074, EPI_ISL_2513076, EPI_ISL_2513077, EPI_ISL_2513078, EPI_ISL_2513096, EPI_ISL_2513111, EPI_ISL_2513112, EPI_ISL_2513117, EPI_ISL_2513135, EPI_ISL_2513144, EPI_ISL_2513146, EPI_ISL_2513150, EPI_ISL_2513155, EPI_ISL_2513159, EPI_ISL_2513160, EPI_ISL_2513168, EPI_ISL_2513171, EPI_ISL_2513174, EPI_ISL_2513181, EPI_ISL_2513195, EPI_ISL_2513205, EPI_ISL_2513213, EPI_ISL_2513232, EPI_ISL_2513235, EPI_ISL_2513238, EPI_ISL_2513242, EPI_ISL_2513243, EPI_ISL_2513252, EPI_ISL_2513260, EPI_ISL_2513265, EPI_ISL_2513272, EPI_ISL_2513286, EPI_ISL_2514076, EPI_ISL_2514078, EPI_ISL_2514079, EPI_ISL_2514087, EPI_ISL_2514093, EPI_ISL_2514099, EPI_ISL_2514106, EPI_ISL_2514108, EPI_ISL_2514109, EPI_ISL_2514113, EPI_ISL_2514120, EPI_ISL_2514128, EPI_ISL_2514130, EPI_ISL_2514132, EPI_ISL_2514139, EPI_ISL_2514140, EPI_ISL_2514149, EPI_ISL_2514161, EPI_ISL_2514164, EPI_ISL_2514165, EPI_ISL_2514173, EPI_ISL_2514180, EPI_ISL_2514184, EPI_ISL_2514187, EPI_ISL_2514189, EPI_ISL_2514196, EPI_ISL_2514198, EPI_ISL_2514206, EPI_ISL_2514209, EPI_ISL_2514215, EPI_ISL_2514223, EPI_ISL_2514233, EPI_ISL_2514235, EPI_ISL_2514239, EPI_ISL_2514242, EPI_ISL_2514244, EPI_ISL_2514256, EPI_ISL_2514263, EPI_ISL_2514264, EPI_ISL_2514271, EPI_ISL_2514275, EPI_ISL_2514278, EPI_ISL_2514282, EPI_ISL_2514285, EPI_ISL_2514295, EPI_ISL_2514296, EPI_ISL_2514297, EPI_ISL_2514299, EPI_ISL_2514305, EPI_ISL_2514306, EPI_ISL_2514307, EPI_ISL_2514308, EPI_ISL_2514313, EPI_ISL_2514315, EPI_ISL_2514319, EPI_ISL_2514322, EPI_ISL_2514331, EPI_ISL_2514338, EPI_ISL_2514340, EPI_ISL_2514343, EPI_ISL_2514349, EPI_ISL_2514350, EPI_ISL_2514351, EPI_ISL_2514361, EPI_ISL_2514362, EPI_ISL_2514363, EPI_ISL_2514387, EPI_ISL_2514388, EPI_ISL_2514397, EPI_ISL_2515401, EPI_ISL_2515403, EPI_ISL_2515407, EPI_ISL_2515427, EPI_ISL_2515429, EPI_ISL_2515435, EPI_ISL_2515446, EPI_ISL_2515452, EPI_ISL_2515475, EPI_ISL_2515482, EPI_ISL_2515484, EPI_ISL_2515493, EPI_ISL_2515494, EPI_ISL_2515509, EPI_ISL_2515514, EPI_ISL_2515515, EPI_ISL_2515541, EPI_ISL_2515547, EPI_ISL_2515554, EPI_ISL_2515555, EPI_ISL_2515562, EPI_ISL_2515563, EPI_ISL_2515585, EPI_ISL_2515606, EPI_ISL_2515626, EPI_ISL_2515630, EPI_ISL_2515632, EPI_ISL_2537500, EPI_ISL_2537502, EPI_ISL_2537509, EPI_ISL_2537519, EPI_ISL_2537527, EPI_ISL_2537559, EPI_ISL_2537564, EPI_ISL_2537568, EPI_ISL_2537579, EPI_ISL_2537591, EPI_ISL_2537602, EPI_ISL_2537605, EPI_ISL_2537623, EPI_ISL_2537630, EPI_ISL_2537634, EPI_ISL_2537648, EPI_ISL_2537652, EPI_ISL_2537655, EPI_ISL_2537656, EPI_ISL_2537671, EPI_ISL_2537675, EPI_ISL_2537678, EPI_ISL_2537682, EPI_ISL_2537698, EPI_ISL_2537699, EPI_ISL_2537707, EPI_ISL_2537709, EPI_ISL_2537723, EPI_ISL_2537730, EPI_ISL_2537738, EPI_ISL_2537742, EPI_ISL_2537749, EPI_ISL_2537763, EPI_ISL_2537764, EPI_ISL_2537768, EPI_ISL_2537772, EPI_ISL_2537775, EPI_ISL_2537785, EPI_ISL_2537786, EPI_ISL_2537787, EPI_ISL_2537793, EPI_ISL_2537795, EPI_ISL_2537796, EPI_ISL_2537808, EPI_ISL_2537810, EPI_ISL_2537811, EPI_ISL_2537818, EPI_ISL_2537823, EPI_ISL_2537826, EPI_ISL_2537829, EPI_ISL_2537834, EPI_ISL_2537836, EPI_ISL_2537838, EPI_ISL_2537846, EPI_ISL_2537849, EPI_ISL_2537852, EPI_ISL_2537856, EPI_ISL_2537862, EPI_ISL_2537864, EPI_ISL_2537865, EPI_ISL_2537869, EPI_ISL_2537872, EPI_ISL_2537873, EPI_ISL_2537874, EPI_ISL_2537877, EPI_ISL_2537879, EPI_ISL_2537880, EPI_ISL_2537881, EPI_ISL_2537882, EPI_ISL_2537886, EPI_ISL_2537887, EPI_ISL_2537892, EPI_ISL_2537895, EPI_ISL_2537896, EPI_ISL_2537897, EPI_ISL_2537898, EPI_ISL_2537899, EPI_ISL_2537903, EPI_ISL_2537904, EPI_ISL_2537906, EPI_ISL_2537907, EPI_ISL_2537908, EPI_ISL_2537911, EPI_ISL_2537921, EPI_ISL_2537922, EPI_ISL_2537923, EPI_ISL_2537929, EPI_ISL_2537931, EPI_ISL_2537932, EPI_ISL_2537937, EPI_ISL_2537940, EPI_ISL_2537941, EPI_ISL_2537948, EPI_ISL_2537950, EPI_ISL_2537952, EPI_ISL_2537954, EPI_ISL_2537958, EPI_ISL_2537969, EPI_ISL_2537979, EPI_ISL_2537984, EPI_ISL_2537986, EPI_ISL_2537990, EPI_ISL_2537996, EPI_ISL_2537998, EPI_ISL_2538005, EPI_ISL_2538011, EPI_ISL_2538017, EPI_ISL_2538026, EPI_ISL_2538032, EPI_ISL_2538038, EPI_ISL_2538041, EPI_ISL_2538048, EPI_ISL_2538072, EPI_ISL_2538075, EPI_ISL_2538076, EPI_ISL_2538086, EPI_ISL_2538090, EPI_ISL_2538093, EPI_ISL_2538097, EPI_ISL_2538098, EPI_ISL_2538111, EPI_ISL_2538124, EPI_ISL_2538131, EPI_ISL_2538149, EPI_ISL_2538155, EPI_ISL_2538158, EPI_ISL_2538165, EPI_ISL_2538166, EPI_ISL_2538167, EPI_ISL_2538168, EPI_ISL_2538170, EPI_ISL_2538171, EPI_ISL_2538172, EPI_ISL_2538173, EPI_ISL_2538175, EPI_ISL_2538176, EPI_ISL_2538177, EPI_ISL_2538178, EPI_ISL_2538179, EPI_ISL_2538180, EPI_ISL_2538181, EPI_ISL_2538182, EPI_ISL_2538184, EPI_ISL_2538185, EPI_ISL_2538186, EPI_ISL_2538188, EPI_ISL_2538189, EPI_ISL_2538190, EPI_ISL_2538191, EPI_ISL_2538192, EPI_ISL_2538193, EPI_ISL_2538194, EPI_ISL_2538195, EPI_ISL_2538196, EPI_ISL_2538197, EPI_ISL_2538198, EPI_ISL_2538199, EPI_ISL_2538200, EPI_ISL_2538201, EPI_ISL_2538202, EPI_ISL_2538203, EPI_ISL_2538204, EPI_ISL_2538205, EPI_ISL_2538206, EPI_ISL_2538207, EPI_ISL_2538208, EPI_ISL_2538209, EPI_ISL_2538211, EPI_ISL_2538212, EPI_ISL_2538214, EPI_ISL_2538215, EPI_ISL_2538216, EPI_ISL_2538217, EPI_ISL_2538218, EPI_ISL_2538221, EPI_ISL_2538222, EPI_ISL_2538223, EPI_ISL_2538224, EPI_ISL_2538225, EPI_ISL_2538226, EPI_ISL_2538227, EPI_ISL_2538228, EPI_ISL_2538229, EPI_ISL_2538230, EPI_ISL_2538231, EPI_ISL_2538232, EPI_ISL_2538233, EPI_ISL_2538234, EPI_ISL_2538236, EPI_ISL_2538237, EPI_ISL_2538238, EPI_ISL_2538239, EPI_ISL_2538240, EPI_ISL_2538241, EPI_ISL_2538242, EPI_ISL_2538243, EPI_ISL_2538246, EPI_ISL_2538247, EPI_ISL_2538248, EPI_ISL_2538249, EPI_ISL_2538250, EPI_ISL_2538251, EPI_ISL_2538252, EPI_ISL_2538253, EPI_ISL_2538254, EPI_ISL_2538255, EPI_ISL_2538257, EPI_ISL_2538259, EPI_ISL_2538260, EPI_ISL_2538262, EPI_ISL_2538263, EPI_ISL_2538264, EPI_ISL_2538265, EPI_ISL_2538266, EPI_ISL_2538267, EPI_ISL_2538268, EPI_ISL_2538271, EPI_ISL_2538272, EPI_ISL_2538273, EPI_ISL_2538275, EPI_ISL_2538276, EPI_ISL_2538277, EPI_ISL_2538279, EPI_ISL_2538280, EPI_ISL_2538282, EPI_ISL_2538283, EPI_ISL_2538284, EPI_ISL_2538285, EPI_ISL_2538287, EPI_ISL_2538288, EPI_ISL_2538289, EPI_ISL_2538290, EPI_ISL_2538291, EPI_ISL_2538292, EPI_ISL_2538293, EPI_ISL_2538294, EPI_ISL_2538295, EPI_ISL_2538296, EPI_ISL_2538297, EPI_ISL_2538298, EPI_ISL_2538299, EPI_ISL_2538300, EPI_ISL_2538302, EPI_ISL_2538303, EPI_ISL_2538304, EPI_ISL_2538305, EPI_ISL_2538306, EPI_ISL_2538307, EPI_ISL_2538308, EPI_ISL_2538309, EPI_ISL_2538310, EPI_ISL_2538311, EPI_ISL_2538312, EPI_ISL_2538313, EPI_ISL_2538315, EPI_ISL_2538316, EPI_ISL_2538317, EPI_ISL_2538318, EPI_ISL_2538320, EPI_ISL_2538321, EPI_ISL_2538322, EPI_ISL_2538323, EPI_ISL_2538325, EPI_ISL_2538326, EPI_ISL_2538327, EPI_ISL_2538328, EPI_ISL_2538329, EPI_ISL_2538330, EPI_ISL_2538331, EPI_ISL_2538332, EPI_ISL_2538333, EPI_ISL_2538334, EPI_ISL_2538335, EPI_ISL_2538336, EPI_ISL_2538337, EPI_ISL_2538338, EPI_ISL_2538339, EPI_ISL_2538340, EPI_ISL_2538341, EPI_ISL_2538342, EPI_ISL_2538343, EPI_ISL_2538345, EPI_ISL_2538346, EPI_ISL_2538347, EPI_ISL_2538348, EPI_ISL_2538349, EPI_ISL_2538350, EPI_ISL_2538353, EPI_ISL_2538355, EPI_ISL_2538356, EPI_ISL_2538357, EPI_ISL_2538358, EPI_ISL_2538359, EPI_ISL_2538360, EPI_ISL_2538361, EPI_ISL_2538362, EPI_ISL_2538363, EPI_ISL_2538364, EPI_ISL_2538365, EPI_ISL_2538366, EPI_ISL_2538367, EPI_ISL_2538368, EPI_ISL_2538369, EPI_ISL_2538370, EPI_ISL_2538371, EPI_ISL_2538372, EPI_ISL_2538373, EPI_ISL_2538374, EPI_ISL_2538375, EPI_ISL_2538376, EPI_ISL_2538377, EPI_ISL_2538378, EPI_ISL_2538379, EPI_ISL_2538380, EPI_ISL_2538381, EPI_ISL_2538382, EPI_ISL_2538383, EPI_ISL_2538384, EPI_ISL_2538385, EPI_ISL_2538386, EPI_ISL_2538387, EPI_ISL_2538388, EPI_ISL_2538389, EPI_ISL_2538390, EPI_ISL_2538391, EPI_ISL_2538392, EPI_ISL_2538393, EPI_ISL_2538394, EPI_ISL_2538396, EPI_ISL_2538398, EPI_ISL_2538399, EPI_ISL_2538400, EPI_ISL_2538401, EPI_ISL_2538402, EPI_ISL_2538403, EPI_ISL_2538404, EPI_ISL_2538405, EPI_ISL_2538406, EPI_ISL_2538407, EPI_ISL_2538408, EPI_ISL_2538409, EPI_ISL_2538410, EPI_ISL_2538411, EPI_ISL_2538412, EPI_ISL_2538413, EPI_ISL_2538416, EPI_ISL_2538417, EPI_ISL_2538418, EPI_ISL_2538419, EPI_ISL_2538420, EPI_ISL_2538421, EPI_ISL_2538422, EPI_ISL_2538424, EPI_ISL_2538425, EPI_ISL_2538426, EPI_ISL_2538427, EPI_ISL_2538428, EPI_ISL_2538429, EPI_ISL_2538430, EPI_ISL_2538431, EPI_ISL_2538432, EPI_ISL_2538433, EPI_ISL_2538435, EPI_ISL_2538436, EPI_ISL_2538437, EPI_ISL_2538438, EPI_ISL_2538439, EPI_ISL_2538440, EPI_ISL_2538441, EPI_ISL_2538442, EPI_ISL_2538443, EPI_ISL_2538444, EPI_ISL_2538447, EPI_ISL_2538448, EPI_ISL_2538449, EPI_ISL_2538450, EPI_ISL_2538451, EPI_ISL_2538452, EPI_ISL_2538453, EPI_ISL_2538454, EPI_ISL_2538455, EPI_ISL_2538456, EPI_ISL_2538457, EPI_ISL_2538458, EPI_ISL_2538459, EPI_ISL_2538461, EPI_ISL_2538462, EPI_ISL_2538463, EPI_ISL_2538464, EPI_ISL_2538465, EPI_ISL_2538467, EPI_ISL_2538468, EPI_ISL_2538469, EPI_ISL_2538470, EPI_ISL_2538471, EPI_ISL_2538472, EPI_ISL_2538474, EPI_ISL_2538475, EPI_ISL_2538476, EPI_ISL_2538477, EPI_ISL_2538478, EPI_ISL_2538479, EPI_ISL_2538480, EPI_ISL_2538481, EPI_ISL_2538482, EPI_ISL_2538483, EPI_ISL_2538484, EPI_ISL_2538485, EPI_ISL_2538486, EPI_ISL_2538487, EPI_ISL_2538488, EPI_ISL_2538489, EPI_ISL_2538490, EPI_ISL_2538491, EPI_ISL_2538492, EPI_ISL_2538493, EPI_ISL_2538494, EPI_ISL_2538495, EPI_ISL_2538496, EPI_ISL_2538499, EPI_ISL_2538500, EPI_ISL_2538501, EPI_ISL_2538502, EPI_ISL_2538503, EPI_ISL_2538504, EPI_ISL_2538506, EPI_ISL_2538507, EPI_ISL_2538508, EPI_ISL_2538509, EPI_ISL_2538510, EPI_ISL_2538511, EPI_ISL_2538512, EPI_ISL_2538513, EPI_ISL_2538514, EPI_ISL_2538515, EPI_ISL_2538516, EPI_ISL_2538518, EPI_ISL_2538519, EPI_ISL_2538520, EPI_ISL_2538521, EPI_ISL_2538522, EPI_ISL_2538523, EPI_ISL_2538524, EPI_ISL_2538525, EPI_ISL_2538526, EPI_ISL_2538527, EPI_ISL_2538528, EPI_ISL_2538529, EPI_ISL_2538530, EPI_ISL_2538531, EPI_ISL_2538532, EPI_ISL_2538533, EPI_ISL_2538536, EPI_ISL_2538537, EPI_ISL_2538541, EPI_ISL_2538543, EPI_ISL_2538544, EPI_ISL_2538546, EPI_ISL_2538547, EPI_ISL_2538548, EPI_ISL_2538549, EPI_ISL_2538550, EPI_ISL_2538551, EPI_ISL_2538552, EPI_ISL_2538553, EPI_ISL_2538554, EPI_ISL_2538555, EPI_ISL_2538557, EPI_ISL_2538558, EPI_ISL_2538559, EPI_ISL_2538562, EPI_ISL_2538564, EPI_ISL_2538565, EPI_ISL_2538566, EPI_ISL_2538569, EPI_ISL_2538571, EPI_ISL_2538572, EPI_ISL_2538573, EPI_ISL_2538574, EPI_ISL_2538575, EPI_ISL_2538578, EPI_ISL_2538580, EPI_ISL_2538581, EPI_ISL_2538583, EPI_ISL_2538584, EPI_ISL_2538585, EPI_ISL_2538586, EPI_ISL_2538588, EPI_ISL_2538589, EPI_ISL_2538590, EPI_ISL_2538591, EPI_ISL_2538596, EPI_ISL_2538597, EPI_ISL_2538599, EPI_ISL_2538600, EPI_ISL_2538602, EPI_ISL_2538604, EPI_ISL_2538605, EPI_ISL_2538610, EPI_ISL_2538612, EPI_ISL_2538614, EPI_ISL_2538615, EPI_ISL_2538616, EPI_ISL_2538617, EPI_ISL_2538618, EPI_ISL_2538619, EPI_ISL_2538622, EPI_ISL_2538624, EPI_ISL_2538625, EPI_ISL_2538627, EPI_ISL_2538628, EPI_ISL_2538630, EPI_ISL_2538632, EPI_ISL_2538634, EPI_ISL_2538635, EPI_ISL_2538636, EPI_ISL_2538637, EPI_ISL_2538639, EPI_ISL_2538640, EPI_ISL_2538643, EPI_ISL_2538644, EPI_ISL_2538645, EPI_ISL_2538646, EPI_ISL_2538647, EPI_ISL_2538649, EPI_ISL_2538650, EPI_ISL_2538651, EPI_ISL_2538654, EPI_ISL_2538655, EPI_ISL_2538656, EPI_ISL_2538657, EPI_ISL_2538658, EPI_ISL_2538659, EPI_ISL_2538661, EPI_ISL_2538662, EPI_ISL_2538664, EPI_ISL_2538665, EPI_ISL_2538666, EPI_ISL_2538667, EPI_ISL_2538669, EPI_ISL_2538671, EPI_ISL_2538673, EPI_ISL_2538674, EPI_ISL_2538676, EPI_ISL_2538677, EPI_ISL_2538679, EPI_ISL_2538681, EPI_ISL_2538682, EPI_ISL_2538686, EPI_ISL_2538688, EPI_ISL_2538689, EPI_ISL_2538690, EPI_ISL_2538691, EPI_ISL_2538692, EPI_ISL_2538693, EPI_ISL_2538695, EPI_ISL_2538697, EPI_ISL_2538698, EPI_ISL_2538701, EPI_ISL_2538704, EPI_ISL_2538705, EPI_ISL_2538706, EPI_ISL_2538707, EPI_ISL_2538709, EPI_ISL_2538714, EPI_ISL_2538715, EPI_ISL_2538716, EPI_ISL_2538718, EPI_ISL_2538720, EPI_ISL_2538721, EPI_ISL_2538722, EPI_ISL_2538725, EPI_ISL_2538726, EPI_ISL_2538727, EPI_ISL_2538729, EPI_ISL_2538731, EPI_ISL_2538734, EPI_ISL_2538735, EPI_ISL_2538736, EPI_ISL_2538737, EPI_ISL_2538738, EPI_ISL_2538739, EPI_ISL_2538742, EPI_ISL_2538743, EPI_ISL_2538744, EPI_ISL_2538746, EPI_ISL_2538747, EPI_ISL_2538750, EPI_ISL_2538752, EPI_ISL_2538753, EPI_ISL_2538756, EPI_ISL_2538759, EPI_ISL_2538760, EPI_ISL_2538761, EPI_ISL_2538763, EPI_ISL_2538765, EPI_ISL_2538766, EPI_ISL_2538767, EPI_ISL_2538769, EPI_ISL_2538770, EPI_ISL_2538772, EPI_ISL_2538773, EPI_ISL_2538774, EPI_ISL_2538776, EPI_ISL_2538778, EPI_ISL_2538780, EPI_ISL_2538782, EPI_ISL_2538784, EPI_ISL_2538786, EPI_ISL_2538788, EPI_ISL_2538789, EPI_ISL_2538790, EPI_ISL_2538791, EPI_ISL_2538792, EPI_ISL_2538793, EPI_ISL_2538794, EPI_ISL_2538795, EPI_ISL_2538796, EPI_ISL_2538797, EPI_ISL_2538798, EPI_ISL_2538803, EPI_ISL_2538805, EPI_ISL_2538806, EPI_ISL_2538807, EPI_ISL_2538808, EPI_ISL_2538809, EPI_ISL_2538812, EPI_ISL_2538814, EPI_ISL_2538815, EPI_ISL_2538819, EPI_ISL_2538820, EPI_ISL_2538822, EPI_ISL_2538823, EPI_ISL_2538824, EPI_ISL_2538825, EPI_ISL_2538827, EPI_ISL_2538828, EPI_ISL_2538829, EPI_ISL_2538831, EPI_ISL_2538833, EPI_ISL_2538834, EPI_ISL_2538835, EPI_ISL_2538837, EPI_ISL_2538838, EPI_ISL_2538839, EPI_ISL_2538840, EPI_ISL_2538841, EPI_ISL_2538842, EPI_ISL_2538843, EPI_ISL_2538844, EPI_ISL_2538845, EPI_ISL_2538847, EPI_ISL_2538849, EPI_ISL_2538850, EPI_ISL_2538851, EPI_ISL_2538852, EPI_ISL_2538853, EPI_ISL_2538854, EPI_ISL_2538856, EPI_ISL_2538859, EPI_ISL_2538860, EPI_ISL_2538861, EPI_ISL_2538863, EPI_ISL_2538864, EPI_ISL_2538865, EPI_ISL_2538866, EPI_ISL_2538867, EPI_ISL_2538868, EPI_ISL_2538870, EPI_ISL_2538872, EPI_ISL_2538873, EPI_ISL_2538874, EPI_ISL_2538875, EPI_ISL_2538876, EPI_ISL_2538877, EPI_ISL_2538878, EPI_ISL_2538879, EPI_ISL_2538880, EPI_ISL_2538881, | | | |

EPI_ISL_2538882, EPI_ISL_2538883, EPI_ISL_2538884, EPI_ISL_2538885, EPI_ISL_2538886, EPI_ISL_2538887, EPI_ISL_2538888, EPI_ISL_2538889, EPI_ISL_2538890, EPI_ISL_2538891, EPI_ISL_2538892, EPI_ISL_2538893, EPI_ISL_2538894, EPI_ISL_2538895, EPI_ISL_2538896, EPI_ISL_2538897, EPI_ISL_2538898, EPI_ISL_2538899, EPI_ISL_2538900, EPI_ISL_2538901, EPI_ISL_2538902, EPI_ISL_2538903,
EPI_ISL_2538904, EPI_ISL_2538905, EPI_ISL_2538906, EPI_ISL_2538907, EPI_ISL_2538908, EPI_ISL_2538909, EPI_ISL_2538910, EPI_ISL_2538911, EPI_ISL_2538912, EPI_ISL_2538913, EPI_ISL_2538914, EPI_ISL_2538916, EPI_ISL_2538917, EPI_ISL_2538918, EPI_ISL_2538919, EPI_ISL_2538920, EPI_ISL_2538921, EPI_ISL_2538923, EPI_ISL_2538924, EPI_ISL_2538925, EPI_ISL_2538928, EPI_ISL_2538931,
EPI_ISL_2538932, EPI_ISL_2538933, EPI_ISL_2538934, EPI_ISL_2538935, EPI_ISL_2538936, EPI_ISL_2538937, EPI_ISL_2538938, EPI_ISL_2538939, EPI_ISL_2538940, EPI_ISL_2538941, EPI_ISL_2538942, EPI_ISL_2538943, EPI_ISL_2538944, EPI_ISL_2538945, EPI_ISL_2538946, EPI_ISL_2538947, EPI_ISL_2538948, EPI_ISL_2538949, EPI_ISL_2538950, EPI_ISL_2538951, EPI_ISL_2538952, EPI_ISL_2538953,
EPI_ISL_2538954, EPI_ISL_2538955, EPI_ISL_2538956, EPI_ISL_2538958, EPI_ISL_2538959, EPI_ISL_2538962, EPI_ISL_2538964, EPI_ISL_2538965, EPI_ISL_2538966, EPI_ISL_2538967, EPI_ISL_2538968, EPI_ISL_2538969, EPI_ISL_2538970, EPI_ISL_2538971, EPI_ISL_2538972, EPI_ISL_2538973, EPI_ISL_2538974, EPI_ISL_2538975, EPI_ISL_2538976, EPI_ISL_2538977, EPI_ISL_2538979, EPI_ISL_2538980,
EPI_ISL_2538982, EPI_ISL_2538984, EPI_ISL_2538985, EPI_ISL_2538986, EPI_ISL_2538987, EPI_ISL_2538988, EPI_ISL_2538989, EPI_ISL_2538990, EPI_ISL_2538991, EPI_ISL_2538992, EPI_ISL_2538993, EPI_ISL_2538995, EPI_ISL_2538996, EPI_ISL_2538997, EPI_ISL_2538998, EPI_ISL_2538999, EPI_ISL_2539000, EPI_ISL_2539001, EPI_ISL_2539002, EPI_ISL_2539003, EPI_ISL_2539004, EPI_ISL_2539005,
EPI_ISL_2539006, EPI_ISL_2539007, EPI_ISL_2539009, EPI_ISL_2539010, EPI_ISL_2539011, EPI_ISL_2539012, EPI_ISL_2539013, EPI_ISL_2539014, EPI_ISL_2539015, EPI_ISL_2539016, EPI_ISL_2539017, EPI_ISL_2539018, EPI_ISL_2539019, EPI_ISL_2539020, EPI_ISL_2539021, EPI_ISL_2539022, EPI_ISL_2539023, EPI_ISL_2539024, EPI_ISL_2539026, EPI_ISL_2539027, EPI_ISL_2539028, EPI_ISL_2539029,
EPI_ISL_2539030, EPI_ISL_2539031, EPI_ISL_2539032, EPI_ISL_2539033, EPI_ISL_2539034, EPI_ISL_2539035, EPI_ISL_2539036, EPI_ISL_2539037, EPI_ISL_2539038, EPI_ISL_2539039, EPI_ISL_2539040, EPI_ISL_2539041, EPI_ISL_2539043, EPI_ISL_2539044, EPI_ISL_2539045, EPI_ISL_2539046, EPI_ISL_2539047, EPI_ISL_2539048, EPI_ISL_2539049, EPI_ISL_2539050, EPI_ISL_2539051, EPI_ISL_2539052,
EPI_ISL_2539053, EPI_ISL_2539054, EPI_ISL_2539055, EPI_ISL_2539056, EPI_ISL_2540730, EPI_ISL_2539057, EPI_ISL_2539059, EPI_ISL_2539060, EPI_ISL_2539062, EPI_ISL_2539063, EPI_ISL_2539064, EPI_ISL_2539065, EPI_ISL_2539066, EPI_ISL_2539067, EPI_ISL_2539069, EPI_ISL_2539070, EPI_ISL_2539071, EPI_ISL_2539074, EPI_ISL_2539075, EPI_ISL_2539076, EPI_ISL_2539077, EPI_ISL_2539078, EPI_ISL_2539079,
EPI_ISL_2539080, EPI_ISL_2539081, EPI_ISL_2539082, EPI_ISL_2539083, EPI_ISL_2539084, EPI_ISL_2539085, EPI_ISL_2539086, EPI_ISL_2539087, EPI_ISL_2539088, EPI_ISL_2539089, EPI_ISL_2539090, EPI_ISL_2539091, EPI_ISL_2539092, EPI_ISL_2539093, EPI_ISL_2539094, EPI_ISL_2539095, EPI_ISL_2539096, EPI_ISL_2539097, EPI_ISL_2539098, EPI_ISL_2539099, EPI_ISL_2539100, EPI_ISL_2539101,
EPI_ISL_2539102, EPI_ISL_2539103, EPI_ISL_2539104, EPI_ISL_2539105, EPI_ISL_2539106, EPI_ISL_2539107, EPI_ISL_2539109, EPI_ISL_2539110, EPI_ISL_2539111, EPI_ISL_2539112, EPI_ISL_2539113, EPI_ISL_2539114, EPI_ISL_2539115, EPI_ISL_2539116, EPI_ISL_2539117, EPI_ISL_2539118, EPI_ISL_2539119, EPI_ISL_2539120, EPI_ISL_2539121, EPI_ISL_2539122, EPI_ISL_2539124, EPI_ISL_2539125,
EPI_ISL_2539126, EPI_ISL_2539127, EPI_ISL_2539128, EPI_ISL_2539129, EPI_ISL_2539130, EPI_ISL_2539132, EPI_ISL_2539133, EPI_ISL_2539134, EPI_ISL_2539135, EPI_ISL_2539136, EPI_ISL_2539137, EPI_ISL_2539138, EPI_ISL_2539139, EPI_ISL_2539140, EPI_ISL_2539141, EPI_ISL_2539142, EPI_ISL_2539143, EPI_ISL_2539144, EPI_ISL_2539145, EPI_ISL_2539146, EPI_ISL_2539147, EPI_ISL_2539148,
EPI_ISL_2539149, EPI_ISL_2539150, EPI_ISL_2539151, EPI_ISL_2539152, EPI_ISL_2539153, EPI_ISL_2539154, EPI_ISL_2539155, EPI_ISL_2539156, EPI_ISL_2539157, EPI_ISL_2539158, EPI_ISL_2539159, EPI_ISL_2539160, EPI_ISL_2539161, EPI_ISL_2539162, EPI_ISL_2539163, EPI_ISL_2539164, EPI_ISL_2539165, EPI_ISL_2539166, EPI_ISL_2539167, EPI_ISL_2539168, EPI_ISL_2539169, EPI_ISL_2539170,
EPI_ISL_2539171, EPI_ISL_2540377, EPI_ISL_2540385, EPI_ISL_2540386, EPI_ISL_2540387, EPI_ISL_2540390, EPI_ISL_2540394, EPI_ISL_2540399, EPI_ISL_2540401, EPI_ISL_2540403, EPI_ISL_2540404, EPI_ISL_2540407, EPI_ISL_2540408, EPI_ISL_2540409, EPI_ISL_2540410, EPI_ISL_2540412, EPI_ISL_2540413, EPI_ISL_2540414, EPI_ISL_2540417, EPI_ISL_2540418, EPI_ISL_2540422, EPI_ISL_2540423,
EPI_ISL_2540424, EPI_ISL_2540425, EPI_ISL_2540426, EPI_ISL_2540427, EPI_ISL_2540428, EPI_ISL_2540429, EPI_ISL_2540430, EPI_ISL_2540431, EPI_ISL_2540432, EPI_ISL_2540433, EPI_ISL_2540434, EPI_ISL_2540437, EPI_ISL_2540439, EPI_ISL_2540443, EPI_ISL_2540444, EPI_ISL_2540445, EPI_ISL_2540447, EPI_ISL_2540449, EPI_ISL_2540450, EPI_ISL_2540452, EPI_ISL_2540453, EPI_ISL_2540456,
EPI_ISL_2540458, EPI_ISL_2540460, EPI_ISL_2540462, EPI_ISL_2540463, EPI_ISL_2540464, EPI_ISL_2540467, EPI_ISL_2540468, EPI_ISL_2540469, EPI_ISL_2540470, EPI_ISL_2540472, EPI_ISL_2540474, EPI_ISL_2540475, EPI_ISL_2540477, EPI_ISL_2540478, EPI_ISL_2540479, EPI_ISL_2540480, EPI_ISL_2540481, EPI_ISL_2540483, EPI_ISL_2540484, EPI_ISL_2540492, EPI_ISL_2540493, EPI_ISL_2540494,
EPI_ISL_2540496, EPI_ISL_2540497, EPI_ISL_2540498, EPI_ISL_2540499, EPI_ISL_2540500, EPI_ISL_2540506, EPI_ISL_2540508, EPI_ISL_2540509, EPI_ISL_2540510, EPI_ISL_2540513, EPI_ISL_2540514, EPI_ISL_2540516, EPI_ISL_2540522, EPI_ISL_2540523, EPI_ISL_2540525, EPI_ISL_2540527, EPI_ISL_2540528, EPI_ISL_2540529, EPI_ISL_2540530, EPI_ISL_2540531, EPI_ISL_2540532, EPI_ISL_2540533,
EPI_ISL_2540534, EPI_ISL_2540535, EPI_ISL_2540537, EPI_ISL_2540538, EPI_ISL_2540539, EPI_ISL_2540541, EPI_ISL_2540543, EPI_ISL_2540545, EPI_ISL_2540546, EPI_ISL_2540547, EPI_ISL_2540548, EPI_ISL_2540550, EPI_ISL_2540552, EPI_ISL_2540555, EPI_ISL_2540556, EPI_ISL_2540557, EPI_ISL_2540559, EPI_ISL_2540561, EPI_ISL_2540563, EPI_ISL_2540565, EPI_ISL_2540570, EPI_ISL_2540571,
EPI_ISL_2540572, EPI_ISL_2540574, EPI_ISL_2540576, EPI_ISL_2540577, EPI_ISL_2540579, EPI_ISL_2540580, EPI_ISL_2540581, EPI_ISL_2540582, EPI_ISL_2540583, EPI_ISL_2540584, EPI_ISL_2540587, EPI_ISL_2540588, EPI_ISL_2540589, EPI_ISL_2540590, EPI_ISL_2540592, EPI_ISL_2540593, EPI_ISL_2540595, EPI_ISL_2540596, EPI_ISL_2540597, EPI_ISL_2540602, EPI_ISL_2540605, EPI_ISL_2540610,
EPI_ISL_2540627, EPI_ISL_2540632, EPI_ISL_2540636, EPI_ISL_2540637, EPI_ISL_2540643, EPI_ISL_2540644, EPI_ISL_2540646, EPI_ISL_2540648, EPI_ISL_2540649, EPI_ISL_2540654, EPI_ISL_2540658, EPI_ISL_2540662, EPI_ISL_2540672, EPI_ISL_2540673, EPI_ISL_2540679, EPI_ISL_2540689, EPI_ISL_2540693, EPI_ISL_2540695, EPI_ISL_2540696, EPI_ISL_2540701, EPI_ISL_2540703, EPI_ISL_2540709,
EPI_ISL_2540715, EPI_ISL_2540719, EPI_ISL_2540722, EPI_ISL_2540727, EPI_ISL_2540730, EPI_ISL_2540739, EPI_ISL_2540740, EPI_ISL_2540741, EPI_ISL_2540742, EPI_ISL_2540745, EPI_ISL_2540747, EPI_ISL_2540761, EPI_ISL_2540763, EPI_ISL_2540767, EPI_ISL_2540768, EPI_ISL_2540769, EPI_ISL_2540782, EPI_ISL_2540790, EPI_ISL_2540798, EPI_ISL_2540799, EPI_ISL_2540800, EPI_ISL_2540802,
EPI_ISL_2540803, EPI_ISL_2540804, EPI_ISL_2540805, EPI_ISL_2540806, EPI_ISL_2540809, EPI_ISL_2540816, EPI_ISL_2540831, EPI_ISL_2540832, EPI_ISL_2540840, EPI_ISL_2540841, EPI_ISL_2540853, EPI_ISL_2540863, EPI_ISL_2540867, EPI_ISL_2540868, EPI_ISL_2540883, EPI_ISL_2540892, EPI_ISL_2540893, EPI_ISL_2540900, EPI_ISL_2540904, EPI_ISL_2540909, EPI_ISL_2540917, EPI_ISL_2540921,
EPI_ISL_2540923, EPI_ISL_2540929, EPI_ISL_2540930, EPI_ISL_2540936, EPI_ISL_2541636, EPI_ISL_2541637, EPI_ISL_2541638, EPI_ISL_2541639, EPI_ISL_2541641, EPI_ISL_2541642, EPI_ISL_2541643, EPI_ISL_2541644, EPI_ISL_2541645, EPI_ISL_2541646, EPI_ISL_2541647, EPI_ISL_2541648, EPI_ISL_2541649, EPI_ISL_2541651, EPI_ISL_2541652, EPI_ISL_2541653, EPI_ISL_2541654, EPI_ISL_2541655,
EPI_ISL_2541656, EPI_ISL_2541657, EPI_ISL_2541658, EPI_ISL_2541659, EPI_ISL_2541661, EPI_ISL_2541662, EPI_ISL_2541663, EPI_ISL_2541664, EPI_ISL_2541665, EPI_ISL_2541666, EPI_ISL_2541667, EPI_ISL_2541668, EPI_ISL_2541669, EPI_ISL_2541670, EPI_ISL_2541671, EPI_ISL_2541672, EPI_ISL_2541673, EPI_ISL_2541674, EPI_ISL_2541675, EPI_ISL_2541676, EPI_ISL_2541677, EPI_ISL_2541678,
EPI_ISL_2541679, EPI_ISL_2541680, EPI_ISL_2541681, EPI_ISL_2541682, EPI_ISL_2541683, EPI_ISL_2541684, EPI_ISL_2541685, EPI_ISL_2541686, EPI_ISL_2541687, EPI_ISL_2541688, EPI_ISL_2541689, EPI_ISL_2541690, EPI_ISL_2541691, EPI_ISL_2541692, EPI_ISL_2541693, EPI_ISL_2541694, EPI_ISL_2541695, EPI_ISL_2541696, EPI_ISL_2541697, EPI_ISL_2541698, EPI_ISL_2541699, EPI_ISL_2541700,
EPI_ISL_2541701, EPI_ISL_2541702, EPI_ISL_2541703, EPI_ISL_2541704, EPI_ISL_2541705, EPI_ISL_2541706, EPI_ISL_2541707, EPI_ISL_2541708, EPI_ISL_2541709, EPI_ISL_2541711, EPI_ISL_2541712, EPI_ISL_2541713, EPI_ISL_2541714, EPI_ISL_2541715, EPI_ISL_2541716, EPI_ISL_2541717, EPI_ISL_2541718, EPI_ISL_2541720, EPI_ISL_2541721, EPI_ISL_2541722, EPI_ISL_2541723, EPI_ISL_2541724,
EPI_ISL_2541725, EPI_ISL_2541726, EPI_ISL_2541727, EPI_ISL_2541730, EPI_ISL_2541731, EPI_ISL_2541732, EPI_ISL_2541733, EPI_ISL_2541734, EPI_ISL_2541737, EPI_ISL_2541738, EPI_ISL_2541740, EPI_ISL_2541741, EPI_ISL_2541742, EPI_ISL_2541743, EPI_ISL_2541744, EPI_ISL_2541745, EPI_ISL_2541746, EPI_ISL_2541747, EPI_ISL_2541748, EPI_ISL_2541749, EPI_ISL_2541750, EPI_ISL_2541751,
EPI_ISL_2541752, EPI_ISL_2541753, EPI_ISL_2541754, EPI_ISL_2541755, EPI_ISL_2541756, EPI_ISL_2541757, EPI_ISL_2541758, EPI_ISL_2541759, EPI_ISL_2541760, EPI_ISL_2541763, EPI_ISL_2541764, EPI_ISL_2541765, EPI_ISL_2541766, EPI_ISL_2541767, EPI_ISL_2541768, EPI_ISL_2541769, EPI_ISL_2541770, EPI_ISL_2541771, EPI_ISL_2541772, EPI_ISL_2541773, EPI_ISL_2541774, EPI_ISL_2541775,
EPI_ISL_2541777, EPI_ISL_2541778, EPI_ISL_2541779, EPI_ISL_2541780, EPI_ISL_2541782, EPI_ISL_2541784, EPI_ISL_2541785, EPI_ISL_2541786, EPI_ISL_2541787, EPI_ISL_2541788, EPI_ISL_2541789, EPI_ISL_2541790, EPI_ISL_2541791, EPI_ISL_2541793, EPI_ISL_2541794, EPI_ISL_2541795, EPI_ISL_2541796, EPI_ISL_2541797, EPI_ISL_2541798, EPI_ISL_2541799, EPI_ISL_2541800, EPI_ISL_2541801,
EPI_ISL_2541802, EPI_ISL_2541803, EPI_ISL_2541804, EPI_ISL_2541805, EPI_ISL_2541806, EPI_ISL_2541807, EPI_ISL_2541808, EPI_ISL_2541809, EPI_ISL_2541810, EPI_ISL_2541811, EPI_ISL_2541812, EPI_ISL_2541813, EPI_ISL_2541814, EPI_ISL_2541816, EPI_ISL_2541817, EPI_ISL_2541818, EPI_ISL_2541819, EPI_ISL_2541820, EPI_ISL_2541821, EPI_ISL_2541822, EPI_ISL_2541823, EPI_ISL_2541824,
EPI_ISL_2541825, EPI_ISL_2541826, EPI_ISL_2541828, EPI_ISL_2541829, EPI_ISL_2541830, EPI_ISL_2541831, EPI_ISL_2541832, EPI_ISL_2541833, EPI_ISL_2541834, EPI_ISL_2541835, EPI_ISL_2541836, EPI_ISL_2541837, EPI_ISL_2541838, EPI_ISL_2541839, EPI_ISL_2541840, EPI_ISL_2541841, EPI_ISL_2541842, EPI_ISL_2541843, EPI_ISL_2541844, EPI_ISL_2541845, EPI_ISL_2541846, EPI_ISL_2541849,
EPI_ISL_2541850, EPI_ISL_2541851, EPI_ISL_2541853, EPI_ISL_2541854, EPI_ISL_2541855, EPI_ISL_2541856, EPI_ISL_2541858, EPI_ISL_2541859, EPI_ISL_2541860, EPI_ISL_2541861, EPI_ISL_2541862, EPI_ISL_2541863, EPI_ISL_2541864, EPI_ISL_2541865, EPI_ISL_2541866, EPI_ISL_2541867, EPI_ISL_2541868, EPI_ISL_2541869, EPI_ISL_2541870, EPI_ISL_2541871, EPI_ISL_2541872, EPI_ISL_2541873,
EPI_ISL_2541874, EPI_ISL_2541875, EPI_ISL_2541876, EPI_ISL_2541877, EPI_ISL_2541878, EPI_ISL_2541879, EPI_ISL_2541880, EPI_ISL_2541881, EPI_ISL_2541882, EPI_ISL_2541883, EPI_ISL_2541884, EPI_ISL_2541885, EPI_ISL_2541886, EPI_ISL_2541887, EPI_ISL_2541888, EPI_ISL_2541889, EPI_ISL_2541890, EPI_ISL_2541892, EPI_ISL_2541893, EPI_ISL_2541894, EPI_ISL_2541895, EPI_ISL_2541896,
EPI_ISL_2541897, EPI_ISL_2541898, EPI_ISL_2541899, EPI_ISL_2541900, EPI_ISL_2541902, EPI_ISL_2541903, EPI_ISL_2541904, EPI_ISL_2541905, EPI_ISL_2541906, EPI_ISL_2541907, EPI_ISL_2541908, EPI_ISL_2541909, EPI_ISL_2541910, EPI_ISL_2541911, EPI_ISL_2541912, EPI_ISL_2541913, EPI_ISL_2541914, EPI_ISL_2541915, EPI_ISL_2541916, EPI_ISL_2541917, EPI_ISL_2541918, EPI_ISL_2541919,
EPI_ISL_2541921, EPI_ISL_2541922, EPI_ISL_2541925, EPI_ISL_2541926, EPI_ISL_2541927, EPI_ISL_2541928, EPI_ISL_2541929, EPI_ISL_2541930, EPI_ISL_2541931, EPI_ISL_2541932, EPI_ISL_2541933, EPI_ISL_2541934, EPI_ISL_2541935, EPI_ISL_2541936, EPI_ISL_2541937, EPI_ISL_2541938, EPI_ISL_2541939, EPI_ISL_2541940, EPI_ISL_2541941, EPI_ISL_2541942, EPI_ISL_2541943, EPI_ISL_2541944,
EPI_ISL_2541945, EPI_ISL_2541946, EPI_ISL_2541947, EPI_ISL_2541948, EPI_ISL_2541949, EPI_ISL_2541950, EPI_ISL_2541951, EPI_ISL_2541952, EPI_ISL_2541953, EPI_ISL_2541954, EPI_ISL_2541956, EPI_ISL_2541957, EPI_ISL_2541958, EPI_ISL_2541959, EPI_ISL_2541960, EPI_ISL_2541961, EPI_ISL_2541962, EPI_ISL_2541963, EPI_ISL_2541964, EPI_ISL_2541966, EPI_ISL_2541967, EPI_ISL_2541968,
EPI_ISL_2541971, EPI_ISL_2541972, EPI_ISL_2541973, EPI_ISL_2541974, EPI_ISL_2541975, EPI_ISL_2541977, EPI_ISL_2541978, EPI_ISL_2541979, EPI_ISL_2541980, EPI_ISL_2541981, EPI_ISL_2541983, EPI_ISL_2541985, EPI_ISL_2541986, EPI_ISL_2541987, EPI_ISL_2543222, EPI_ISL_2551590, EPI_ISL_2551591, EPI_ISL_2551599, EPI_ISL_2551600, EPI_ISL_2551602, EPI_ISL_2551603,
EPI_ISL_2551606, EPI_ISL_2551607, EPI_ISL_2551608, EPI_ISL_2551613, EPI_ISL_2551620, EPI_ISL_2551622, EPI_ISL_2551627, EPI_ISL_2551629, EPI_ISL_2551633, EPI_ISL_2551634, EPI_ISL_2551635, EPI_ISL_2551638, EPI_ISL_2551642, EPI_ISL_2551643, EPI_ISL_2551645, EPI_ISL_2551649, EPI_ISL_2551651, EPI_ISL_2551652, EPI_ISL_2551654, EPI_ISL_2551655, EPI_ISL_2551658, EPI_ISL_2551661,
EPI_ISL_2551662, EPI_ISL_2551669, EPI_ISL_2551670, EPI_ISL_2551675, EPI_ISL_2551681, EPI_ISL_2551686, EPI_ISL_2551688, EPI_ISL_2551689, EPI_ISL_2551693, EPI_ISL_2551696, EPI_ISL_2551697, EPI_ISL_2551702, EPI_ISL_2551704, EPI_ISL_2551710, EPI_ISL_2551714, EPI_ISL_2551716, EPI_ISL_2551718, EPI_ISL_2551721, EPI_ISL_2551723, EPI_ISL_2551727, EPI_ISL_2551730, EPI_ISL_2551736,
EPI_ISL_2551738, EPI_ISL_2551742, EPI_ISL_2551743, EPI_ISL_2551744, EPI_ISL_2551745, EPI_ISL_2551748, EPI_ISL_2551753, EPI_ISL_2551755, EPI_ISL_2551756, EPI_ISL_2551761, EPI_ISL_2551765, EPI_ISL_2551768, EPI_ISL_2551769, EPI_ISL_2551773, EPI_ISL_2551778, EPI_ISL_2551780, EPI_ISL_2551781, EPI_ISL_2551795, EPI_ISL_2551797, EPI_ISL_2551799, EPI_ISL_2551800,
EPI_ISL_2551802, EPI_ISL_2551807, EPI_ISL_2551809, EPI_ISL_2551810, EPI_ISL_2551815, EPI_ISL_2551816, EPI_ISL_2551822, EPI_ISL_2551830, EPI_ISL_2551834, EPI_ISL_2551835, EPI_ISL_2551840, EPI_ISL_2551842, EPI_ISL_2551844, EPI_ISL_2551845, EPI_ISL_2551846, EPI_ISL_2551847, EPI_ISL_2551849, EPI_ISL_2551856, EPI_ISL_2551859, EPI_ISL_2551863, EPI_ISL_2551865, EPI_ISL_2551866,
EPI_ISL_2551867, EPI_ISL_2551874, EPI_ISL_2551875, EPI_ISL_2551876, EPI_ISL_2551877, EPI_ISL_2551879, EPI_ISL_2551882, EPI_ISL_2551883, EPI_ISL_2551885, EPI_ISL_2551889, EPI_ISL_2551890, EPI_ISL_2551892, EPI_ISL_2551893, EPI_ISL_2551899, EPI_ISL_2551902, EPI_ISL_2551905, EPI_ISL_2551908, EPI_ISL_2551911, EPI_ISL_2551916, EPI_ISL_2551917,
EPI_ISL_2551923, EPI_ISL_2551925, EPI_ISL_2552293, EPI_ISL_2552314, EPI_ISL_2552330, EPI_ISL_2552384, EPI_ISL_2552425, EPI_ISL_2552459, EPI_ISL_2552461, EPI_ISL_2552477, EPI_ISL_2552489, EPI_ISL_2552491, EPI_ISL_2552499, EPI_ISL_2552512, EPI_ISL_2552573, EPI_ISL_2552575, EPI_ISL_2552593, EPI_ISL_2552602, EPI_ISL_2552609, EPI_ISL_2552611, EPI_ISL_2552616, EPI_ISL_2552623,
EPI_ISL_2552627, EPI_ISL_2552635, EPI_ISL_2552639, EPI_ISL_2552642, EPI_ISL_2552655, EPI_ISL_2552660, EPI_ISL_2552661, EPI_ISL_2552663, EPI_ISL_2552680, EPI_ISL_2552681, EPI_ISL_2552690, EPI_ISL_2552691, EPI_ISL_2552692, EPI_ISL_2552695, EPI_ISL_2552703, EPI_ISL_2552706, EPI_ISL_2552713, EPI_ISL_2552715, EPI_ISL_2552718, EPI_ISL_2552738, EPI_ISL_2552741, EPI_ISL_2552742,
EPI_ISL_2552754, EPI_ISL_2552759, EPI_ISL_2552765, EPI_ISL_2552812, EPI_ISL_2552834, EPI_ISL_2552836, EPI_ISL_2552837, EPI_ISL_2552841, EPI_ISL_2552851, EPI_ISL_2552854, EPI_ISL_2552857, EPI_ISL_2552866, EPI_ISL_2552878, EPI_ISL_2552892, EPI_ISL_2552898, EPI_ISL_2552924, EPI_ISL_2552942, EPI_ISL_2552951, EPI_ISL_2552969, EPI_ISL_2552980, EPI_ISL_2552991, EPI_ISL_2553003,
EPI_ISL_2553022, EPI_ISL_2553027, EPI_ISL_2553044, EPI_ISL_2553047, EPI_ISL_2553052, EPI_ISL_2553055, EPI_ISL_2553062, EPI_ISL_2553068, EPI_ISL_2553079, EPI_ISL_2553080, EPI_ISL_2553089, EPI_ISL_2553091, EPI_ISL_2553092, EPI_ISL_2553093, EPI_ISL_2553109, EPI_ISL_2553112, EPI_ISL_2553113, EPI_ISL_2553117, EPI_ISL_2553119, EPI_ISL_2553124,
EPI_ISL_2553125, EPI_ISL_2553131, EPI_ISL_2553132, EPI_ISL_2553133, EPI_ISL_2553134, EPI_ISL_2553137, EPI_ISL_2553143, EPI_ISL_2553146, EPI_ISL_2553153, EPI_ISL_2553169, EPI_ISL_2553170, EPI_ISL_2553172, EPI_ISL_2553174, EPI_ISL_2553177, EPI_ISL_2553183, EPI_ISL_2553189, EPI_ISL_2553190, EPI_ISL_2553202, EPI_ISL_2553216, EPI_ISL_2553219, EPI_ISL_2553229, EPI_ISL_2553231,
EPI_ISL_2553239, EPI_ISL_2553242, EPI_ISL_2553247, EPI_ISL_2553255, EPI_ISL_2553263, EPI_ISL_2553264, EPI_ISL_2553268, EPI_ISL_2553270, EPI_ISL_2553272, EPI_ISL_2553274, EPI_ISL_2553278, EPI_ISL_2553282, EPI_ISL_2553303, EPI_ISL_2553306, EPI_ISL_2553318, EPI_ISL_2553321, EPI_ISL_2553329, EPI_ISL_2553332, EPI_ISL_2553333, EPI_ISL_2553337, EPI_ISL_2553340, EPI_ISL_2553345,
EPI_ISL_2553346, EPI_ISL_2553352, EPI_ISL_2553355, EPI_ISL_2553371, EPI_ISL_2553373, EPI_ISL_2553375, EPI_ISL_2553381, EPI_ISL_2553386, EPI_ISL_2553390, EPI_ISL_2553391, EPI_ISL_2553405, EPI_ISL_2553408, EPI_ISL_2553410, EPI_ISL_2553414, EPI_ISL_2553418, EPI_ISL_2553419, EPI_ISL_2553420, EPI_ISL_2553422, EPI_ISL_2553425, EPI_ISL_2553427, EPI_ISL_2553436, EPI_ISL_2553440,
EPI_ISL_2553442, EPI_ISL_2553443, EPI_ISL_2553444, EPI_ISL_2553445, EPI_ISL_2553446, EPI_ISL_2553447, EPI_ISL_2553448, EPI_ISL_2553449, EPI_ISL_2553451, EPI_ISL_2553452, EPI_ISL_2553453, EPI_ISL_2553454, EPI_ISL_2553455, EPI_ISL_2553456, EPI_ISL_2553457, EPI_ISL_2553461, EPI_ISL_2553463, EPI_ISL_2553468, EPI_ISL_2553469, EPI_ISL_2553470, EPI_ISL_2553471, EPI_ISL_2553472,
EPI_ISL_2553473, EPI_ISL_2553474, EPI_ISL_2553475, EPI_ISL_2553476, EPI_ISL_2553477, EPI_ISL_2553479, EPI_ISL_2553480, EPI_ISL_2553481, EPI_ISL_2553482, EPI_ISL_2553483, EPI_ISL_2553484, EPI_ISL_2553485, EPI_ISL_2553486, EPI_ISL_2553487, EPI_ISL_2553488, EPI_ISL_2553489, EPI_ISL_2553490, EPI_ISL_2553492, EPI_ISL_2553494, EPI_ISL_2553496, EPI_ISL_2553497, EPI_ISL_2553500,
EPI_ISL_2553503, EPI_ISL_2553504, EPI_ISL_2553506, EPI_ISL_2553509, EPI_ISL_2553511, EPI_ISL_2553512, EPI_ISL_2553513, EPI_ISL_2553514, EPI_ISL_2553515, EPI_ISL_2553516, EPI_ISL_2553517, EPI_ISL_2553518, EPI_ISL_2553520, EPI_ISL_2553521, EPI_ISL_2553522, EPI_ISL_2553523, EPI_ISL_2553525, EPI_ISL_2553526, EPI_ISL_2553527, EPI_ISL_2553532, EPI_ISL_2553533, EPI_ISL_2553534,
EPI_ISL_2553538, EPI_ISL_2553540, EPI_ISL_2553542, EPI_ISL_2553543, EPI_ISL_2553544, EPI_ISL_2553545, EPI_ISL_2553547, EPI_ISL_2553548, EPI_ISL_2553550, EPI_ISL_2553552, EPI_ISL_2553553, EPI_ISL_2553554, EPI_ISL_2553555, EPI_ISL_2553556, EPI_ISL_2553557, EPI_ISL_2553558, EPI_ISL_2553559, EPI_ISL_2553560, EPI_ISL_2553561, EPI_ISL_2553562, EPI_ISL_2553564, EPI_ISL_2553567,
EPI_ISL_2553568, EPI_ISL_2553570, EPI_ISL_2553572, EPI_ISL_2553574, EPI_ISL_2553575, EPI_ISL_2553576, EPI_ISL_2553577, EPI_ISL_2553578, EPI_ISL_2553579, EPI_ISL_2553580, EPI_ISL_2553581, EPI_ISL_2553582, EPI_ISL_2553584, EPI_ISL_2553585, EPI_ISL_2553586, EPI_ISL_2553587, EPI_ISL_2553588, EPI_ISL_2553589, EPI_ISL_2553590, EPI_ISL_2553591, EPI_ISL_2553594, EPI_ISL_2553597,
EPI_ISL_2553598, EPI_ISL_2553599, EPI_ISL_2553601, EPI_ISL_2553602, EPI_ISL_2553603, EPI_ISL_2553604, EPI_ISL_2553605, EPI_ISL_2553606, EPI_ISL_2553607, EPI_ISL_2553608, EPI_ISL_2553609, EPI_ISL_2553610, EPI_ISL_2553611, EPI_ISL_2553612, EPI_ISL_2553615, EPI_ISL_2553616, EPI_ISL_2553618, EPI_ISL_2553619, EPI_ISL_2553620, EPI_ISL_2553621, EPI_ISL_2553624, EPI_ISL_2553625,
EPI_ISL_2553629, EPI_ISL_2553633, EPI_ISL_2553634, EPI_ISL_2553636, EPI_ISL_2553639, EPI_ISL_2553640, EPI_ISL_2553642, EPI_ISL_2553644, EPI_ISL_2553647, EPI_ISL_2553648, EPI_ISL_2553650, EPI_ISL_2553651, EPI_ISL_2553652, EPI_ISL_2553653, EPI_ISL_2553654, EPI_ISL_2553655, EPI_ISL_2553656, EPI_ISL_2553658, EPI_ISL_2553659, EPI_ISL_2553660, EPI_ISL_2553661, EPI_ISL_2553662,
EPI_ISL_2553663, EPI_ISL_2553664, EPI_ISL_2553667, EPI_ISL_2553668, EPI_ISL_2553669, EPI_ISL_2553670, EPI_ISL_2553672, EPI_ISL_2553674, EPI_ISL_2553676, EPI_ISL_2553677, EPI_ISL_2553678, EPI_ISL_2553679, EPI_ISL_2553681, EPI_ISL_2553683, EPI_ISL_2553686, EPI_ISL_2553688, EPI_ISL_2553689, EPI_ISL_2553693, EPI_ISL_2553694, EPI_ISL_2553695, EPI_ISL_2553697, EPI_ISL_2553698,
EPI_ISL_2553700, EPI_ISL_2553701, EPI_ISL_2553702, EPI_ISL_2553703, EPI_ISL_2553704, EPI_ISL_2553705, EPI_ISL_2553707, EPI_ISL_2553708, EPI_ISL_2553709, EPI_ISL_2553711, EPI_ISL_2553712, EPI_ISL_2553714, EPI_ISL_2553718, EPI_ISL_2553719, EPI_ISL_2553720, EPI_ISL_2553721, EPI_ISL_2553722, EPI_ISL_2553723, EPI_ISL_2553724, EPI_ISL_2553726, EPI_ISL_2553727, EPI_ISL_2553729,
EPI_ISL_2553730, EPI_ISL_2553732, EPI_ISL_2553734, EPI_ISL_2553735, EPI_ISL_2553736, EPI_ISL_2553737, EPI_ISL_2553738, EPI_ISL_2553739, EPI_ISL_2553740, EPI_ISL_2553744, EPI_ISL_2553745, EPI_ISL_2553746, EPI_ISL_2553748, EPI_ISL_2553751, EPI_ISL_2553752, EPI_ISL_2553753, EPI_ISL_2553754, EPI_ISL_2553755, EPI_ISL_2553756, EPI_ISL_2553757, EPI_ISL_2553758,
EPI_ISL_2553761, EPI_ISL_2553763, EPI_ISL_2553765, EPI_ISL_2553766, EPI_ISL_2553767, EPI_ISL_2553768, EPI_ISL_2553769, EPI_ISL_2553770, EPI_ISL_2553771, EPI_ISL_2553773, EPI_ISL_2553777, EPI_ISL_2553778, EPI_ISL_2553781, EPI_ISL_2553782, EPI_ISL_2553783, EPI_ISL_2553784, EPI_ISL_2553786, EPI_ISL_2554142, EPI_ISL_2554147, EPI_ISL_2554156, EPI_ISL_2554158, EPI_ISL_2554162,
EPI_ISL_2554166, EPI_ISL_2554168, EPI_ISL_2554170, EPI_ISL_2554172, EPI_ISL_2554177, EPI_ISL_2554178, EPI_ISL_2554181, EPI_ISL_2554184, EPI_ISL_2554189, EPI_ISL_2554190, EPI_ISL_2554197, EPI_ISL_2554198, EPI_ISL_2554201, EPI_ISL_2554202, EPI_ISL_2554205, EPI_ISL_2554207, EPI_ISL_2554215, EPI_ISL_2554218, EPI_ISL_2554223, EPI_ISL_2554225, EPI_ISL_2554227, EPI_ISL_2554229,
EPI_ISL_2554238, EPI_ISL_2554239, EPI_ISL_2554247, EPI_ISL_2554252, EPI_ISL_2554253, EPI_ISL_2554265, EPI_ISL_2554267, EPI_ISL_2554272, EPI_ISL_2554278, EPI_ISL_2554286, EPI_ISL_2554292, EPI_ISL_2554293, EPI_ISL_2554299, EPI_ISL_2554309, EPI_ISL_2554325, EPI_ISL_2554327, EPI_ISL_2554328, EPI_ISL_2554339, EPI_ISL_2554342, EPI_ISL_2554344, EPI_ISL_2554358, EPI_ISL_2554363,
EPI_ISL_2554369, EPI_ISL_2554373, EPI_ISL_2554377, EPI_ISL_2554383, EPI_ISL_2554389, EPI_ISL_2554390, EPI_ISL_2554402, EPI_ISL_2554405, EPI_ISL_2554410, EPI_ISL_2554414, EPI_ISL_2554417, EPI_ISL_2554420, EPI_ISL_2554423, EPI_ISL_2554426, EPI_ISL_2554429, EPI_ISL_2554432, EPI_ISL_2554434, EPI_ISL_2554442, EPI_ISL_2554443, EPI_ISL_2554444, EPI_ISL_2554457, EPI_ISL_2554459,
EPI_ISL_2554469, EPI_ISL_2554471, EPI_ISL_2554477, EPI_ISL_2554479, EPI_ISL_2554482, EPI_ISL_2554484, EPI_ISL_2554490, EPI_ISL_2554847, EPI_ISL_2554885, EPI_ISL_2567635, EPI_ISL_2567636, EPI_ISL_2567637, EPI_ISL_2567638, EPI_ISL_2567639, EPI_ISL_2567640, EPI_ISL_2567641, EPI_ISL_2567642, EPI_ISL_2567643, EPI_ISL_2567644, EPI_ISL_2567645, EPI_ISL_2567646, EPI_ISL_2567647,
EPI_ISL_2567648, EPI_ISL_2567649, EPI_ISL_2567650, EPI_ISL_2567651, EPI_ISL_2567652, EPI_ISL_2567653, EPI_ISL_2567654, EPI_ISL_2567655, EPI_ISL_2567657, EPI_ISL_2567658, EPI_ISL_2567660, EPI_ISL_2567661, EPI_ISL_2567662, EPI_ISL_2567663, EPI_ISL_2567665, EPI_ISL_2567667, EPI_ISL_2567670, EPI_ISL_2567671, EPI_ISL_2567672, EPI_ISL_2567673, EPI_ISL_2567674, EPI_ISL_2567675,
EPI_ISL_2567676, EPI_ISL_2567677, EPI_ISL_2567678, EPI_ISL_2567679, EPI_ISL_2567681, EPI_ISL_2567682, EPI_ISL_2567683, EPI_ISL_2567685, EPI_ISL_2567686, EPI_ISL_2567687, EPI_ISL_2567689, EPI_ISL_2567690, EPI_ISL_2567691, EPI_ISL_2567693, EPI_ISL_2567695, EPI_ISL_2567696, EPI_ISL_2567697, EPI_ISL_2567698, EPI_ISL_2567699, EPI_ISL_2567700, EPI_ISL_2567701, EPI_ISL_2567702,
EPI_ISL_2567703, EPI_ISL_2567704, EPI_ISL_2567706, EPI_ISL_2567707, EPI_ISL_2567708, EPI_ISL_2567709, EPI_ISL_2567710, EPI_ISL_2567711, EPI_ISL_2567712, EPI_ISL_2567713, EPI_ISL_2567714, EPI_ISL_2567715, EPI_ISL_2567716, EPI_ISL_2567717, EPI_ISL_2567719, EPI_ISL_2567720, EPI_ISL_2567721, EPI_ISL_2567722, EPI_ISL_2567723, EPI_ISL_2567724, EPI_ISL_2567725,
EPI_ISL_2567726, EPI_ISL_2567727, EPI_ISL_2567728, EPI_ISL_2567729, EPI_ISL_2567730, EPI_ISL_2567731, EPI_ISL_2567733, EPI_ISL_2567736, EPI_ISL_2567737, EPI_ISL_2567738, EPI_ISL_2567739, EPI_ISL_2567740, EPI_ISL_2567742, EPI_ISL_2567743, EPI_ISL_2567744, EPI_ISL_2567746, EPI_ISL_2567747, EPI_ISL_2567748, EPI_ISL_2567749, EPI_ISL_2567750, EPI_ISL_2567751, EPI_ISL_2567752,
EPI_ISL_2567753, EPI_ISL_2567754, EPI_ISL_2567755, EPI_ISL_2567756, EPI_ISL_2567758, EPI_ISL_2567759, EPI_ISL_2567760, EPI_ISL_2567761, EPI_ISL_2567762, EPI_ISL_2567763, EPI_ISL_2567764, EPI_ISL_2567765, EPI_ISL_2567767, EPI_ISL_2567768, EPI_ISL_2567769, EPI_ISL_2567770, EPI_ISL_2567771, EPI_ISL_2567772, EPI_ISL_2567773, EPI_ISL_2567774, EPI_ISL_2567776, EPI_ISL_2567777,
EPI_ISL_2567778, EPI_ISL_2567779, EPI_ISL_2567780, EPI_ISL_2567781, EPI_ISL_2567782, EPI_ISL_2567783, EPI_ISL_2567784, EPI_ISL_2567786, EPI_ISL_2567787, EPI_ISL_2567789, EPI_ISL_2567790, EPI_ISL_2567792, EPI_ISL_2567793, EPI_ISL_2567794, EPI_ISL_2567795, EPI_ISL_2567796, EPI_ISL_2567797, EPI_ISL_2567798, EPI_ISL_2567799, EPI_ISL_2567800, EPI_ISL_2567802, EPI_ISL_2567803,
EPI_ISL_2567804, EPI_ISL_2567805, EPI_ISL_2567806, EPI_ISL_2567807, EPI_ISL_2567808, EPI_ISL_2567809, EPI_ISL_2567810, EPI_ISL_2567811, EPI_ISL_2567812, EPI_ISL_2567814, EPI_ISL_2567816, EPI_ISL_2567817, EPI_ISL_2567818, EPI_ISL_2567819, EPI_ISL_2567820, EPI_ISL_2567821, EPI_ISL_2567822, EPI_ISL_2567823, EPI_ISL_2567825, EPI_ISL_2567826, EPI_ISL_2567827,
EPI_ISL_2567828, EPI_ISL_2567829, EPI_ISL_2567830, EPI_ISL_2567832, EPI_ISL_2567833, EPI_ISL_2567834, EPI_ISL_2567837, EPI_ISL_2567838, EPI_ISL_2567839, EPI_ISL_2567840, EPI_ISL_2567841, EPI_ISL_2567842, EPI_ISL_2567843, EPI_ISL_2567844, EPI_ISL_2567845, EPI_ISL_2567846, EPI_ISL_2567847, EPI_ISL_2567848, EPI_ISL_2567849, EPI_ISL_2567850, EPI_ISL_2567852, EPI_ISL_2567853,
EPI_ISL_2567855, EPI_ISL_2567856, EPI_ISL_2567858, EPI_ISL_2567859, EPI_ISL_2567860, EPI_ISL_2567864, EPI_ISL_2567865, EPI_ISL_2567866, EPI_ISL_2567867, EPI_ISL_2567869, EPI_ISL_2567870, EPI_ISL_2567872, EPI_ISL_2567874, EPI_ISL_2567875, EPI_ISL_2567877, EPI_ISL_2567878, EPI_ISL_2567879, EPI_ISL_2567880, EPI_ISL_2567881, EPI_ISL_2567883, EPI_ISL_2567884,
EPI_ISL_2567885, EPI_ISL_2567886, EPI_ISL_2567887, EPI_ISL_2567888, EPI_ISL_2567891, EPI_ISL_2567892, EPI_ISL_2567893, EPI_ISL_2567894, EPI_ISL_2567895, EPI_ISL_2567896, EPI_ISL_2567897, EPI_ISL_2567898, EPI_ISL_2567899, EPI_ISL_2567900, EPI_ISL_2567901, EPI_ISL_2567902, EPI_ISL_2567903, EPI_ISL_2567904, EPI_ISL_2567905, EPI_ISL_2567907, EPI_ISL_2567909, EPI_ISL_2567912,
EPI_ISL_2567913, EPI_ISL_2567916, EPI_ISL_2567917, EPI_ISL_2567918, EPI_ISL_2567920, EPI_ISL_2567922, EPI_ISL_2567923, EPI_ISL_2567924, EPI_ISL_2567928, EPI_ISL_2567929, EPI_ISL_2567930, EPI_ISL_2567931, EPI_ISL_2567932, EPI_ISL_2567933, EPI_ISL_2567934, EPI_ISL_2567936, EPI_ISL_2567937, EPI_ISL_2567938, EPI_ISL_2567939, EPI_ISL_2567940, EPI_ISL_2567941,
EPI_ISL_2567942, EPI_ISL_2567943, EPI_ISL_2567944, EPI_ISL_2567945, EPI_ISL_2567946, EPI_ISL_2567948, EPI_ISL_2567949, EPI_ISL_2567951, EPI_ISL_2567953, EPI_ISL_2567956, EPI_ISL_2567957, EPI_ISL_2567959, EPI_ISL_2567965, EPI_ISL_2567968, EPI_ISL_2567972, EPI_ISL_2567976, EPI_ISL_2567977, EPI_ISL_2567978, EPI_ISL_2567984, EPI_ISL_2567986, EPI_ISL_2567987, EPI_ISL_2567989,
EPI_ISL_2567990, EPI_ISL_2567992, EPI_ISL_2567998, EPI_ISL_2568000, EPI_ISL_2568003, EPI_ISL_2568005, EPI_ISL_2568006, EPI_ISL_2568011, EPI_ISL_2568012, EPI_ISL_2568014, EPI_ISL_2568019, EPI_ISL_2568020, EPI_ISL_2568021, EPI_ISL_2568026, EPI_ISL_2568027, EPI_ISL_2568031, EPI_ISL_2568034, EPI_ISL_2568036, EPI_ISL_2568037, EPI_ISL_2568047, EPI_ISL_2568051, EPI_ISL_2568052,
EPI_ISL_2568055, EPI_ISL_2568056, EPI_ISL_2568058, EPI_ISL_2568061, EPI_ISL_2568063, EPI_ISL_2568070, EPI_ISL_2568071, EPI_ISL_2568072, EPI_ISL_2568081, EPI_ISL_2568085, EPI_ISL_2568087, EPI_ISL_2568090, EPI_ISL_2568092, EPI_ISL_2568100, EPI_ISL_2568101, EPI_ISL_2568115, EPI_ISL_2568141, EPI_ISL_2568177, EPI_ISL_2568198, EPI_ISL_2568215, EPI_ISL_2568238, EPI_ISL_2568267,
EPI_ISL_2568294, EPI_ISL_2568338, EPI_ISL_2568339, EPI_ISL_2568378, EPI_ISL_2568381, EPI_ISL_2568383, EPI_ISL_2568584, EPI_ISL_2568630, EPI_ISL_2568654, EPI_ISL_2568664, EPI_ISL_2568780, EPI_ISL_2568782, EPI_ISL_2568804, EPI_ISL_2568851, EPI_ISL_2568917, EPI_ISL_2568967, EPI_ISL_2568987, EPI_ISL_2569122, EPI_ISL_2569151, EPI_ISL_2569168, EPI_ISL_2569169, EPI_ISL_2569180,
EPI_ISL_2569184, EPI_ISL_2569186, EPI_ISL_2569201, EPI_ISL_2569205, EPI_ISL_2569220, EPI_ISL_2569241, EPI_ISL_2569249, EPI_ISL_2569273, EPI_ISL_2569274, EPI_ISL_2569281, EPI_ISL_2569309, EPI_ISL_2569316, EPI_ISL_2569324, EPI_ISL_2569325, EPI_ISL_2569332, EPI_ISL_2569338, EPI_ISL_2569365, EPI_ISL_2569379, EPI_ISL_2569386, EPI_ISL_2569395, EPI_ISL_2569418, EPI_ISL_2569419,
EPI_ISL_2569432, EPI_ISL_2569437, EPI_ISL_2569438, EPI_ISL_2569446, EPI_ISL_2569454, EPI_ISL_2569456, EPI_ISL_2569460, EPI_ISL_2569461, EPI_ISL_2569470, EPI_ISL_2569471, EPI_ISL_2569489, EPI_ISL_2569494, EPI_ISL_2569498, EPI_ISL_2569500, EPI_ISL_2569505, EPI_ISL_2569512, EPI_ISL_2569521, EPI_ISL_2569523, EPI_ISL_2569524, EPI_ISL_2569529, EPI_ISL_2569573, EPI_ISL_2569589,
EPI_ISL_2569599, EPI_ISL_2569621, EPI_ISL_2569622, EPI_ISL_2569660, EPI_ISL_2569698, EPI_ISL_2569778, EPI_ISL_2569878, EPI_ISL_2569931, EPI_ISL_2569934, EPI_ISL_2569936, EPI_ISL_2569937, EPI_ISL_2569938, EPI_ISL_2569939, EPI_ISL_2569940, EPI_ISL_2569941, EPI_ISL_2569942, EPI_ISL_2569944, EPI_ISL_2569948, EPI_ISL_2569949, EPI_ISL_2569951, EPI_ISL_2569952, EPI_ISL_2569954,
EPI_ISL_2569955, EPI_ISL_2569958, EPI_ISL_2569959, EPI_ISL_2569960, EPI_ISL_2569961, EPI_ISL_2569963, EPI_ISL_2569965, EPI_ISL_2569966, EPI_ISL_2569967, EPI_ISL_2569968, EPI_ISL_2569971, EPI_ISL_2569977, EPI_ISL_2569980, EPI_ISL_2569981, EPI_ISL_2569983, EPI_ISL_2569984, EPI_ISL_2569986, EPI_ISL_2569989, EPI_ISL_2569991, EPI_ISL_2569993, EPI_ISL_2569996, EPI_ISL_2569998,
EPI_ISL_2570001, EPI_ISL_2570002, EPI_ISL_2570008, EPI_ISL_2570010, EPI_ISL_2570011, EPI_ISL_2570015, EPI_ISL_2570018, EPI_ISL_2570019, EPI_ISL_2570020, EPI_ISL_2570021, EPI_ISL_2570023, EPI_ISL_2570024, EPI_ISL_2570027, EPI_ISL_2570038, EPI_ISL_2570040, EPI_ISL_2570042, EPI_ISL_2570043, EPI_ISL_2570046, EPI_ISL_2570048, EPI_ISL_2570050, EPI_ISL_2570052, EPI_ISL_2570054,
EPI_ISL_2570055, EPI_ISL_2570056, EPI_ISL_2570057, EPI_ISL_2570058, EPI_ISL_2570060, EPI_ISL_2570062, EPI_ISL_2570065, EPI_ISL_2570066, EPI_ISL_2570067, EPI_ISL_2570069, EPI_ISL_2570076, EPI_ISL_2570077, EPI_ISL_2570078, EPI_ISL_2570080, EPI_ISL_2570088, EPI_ISL_2570089, EPI_ISL_2570091, EPI_ISL_2570092, EPI_ISL_2570094, EPI_ISL_2570102, EPI_ISL_2570103, EPI_ISL_2570105,
EPI_ISL_2570107, EPI_ISL_2570108, EPI_ISL_2570110, EPI_ISL_2570111, EPI_ISL_2570112, EPI_ISL_2570114, EPI_ISL_2570117, EPI_ISL_2570118, EPI_ISL_2570119, EPI_ISL_2570120, EPI_ISL_2570121, EPI_ISL_2570125, EPI_ISL_2570126, EPI_ISL_2570127, EPI_ISL_2570128, EPI_ISL_2570132, EPI_ISL_2570134, EPI_ISL_2570138, EPI_ISL_2570139, EPI_ISL_2570141, EPI_ISL_2570143, EPI_ISL_2570145,
EPI_ISL_2570148, EPI_ISL_2570149, EPI_ISL_2570150, EPI_ISL_2570154, EPI_ISL_2570155, EPI_ISL_2570157, EPI_ISL_2570159, EPI_ISL_2570161, EPI_ISL_2570162, EPI_ISL_2570163, EPI_ISL_2570172, EPI_ISL_2570177, EPI_ISL_2570178, EPI_ISL_2570181, EPI_ISL_2570191, EPI_ISL_2570193, EPI_ISL_2570194, EPI_ISL_2570195, EPI_ISL_2570196, EPI_ISL_2570198, EPI_ISL_2570199, EPI_ISL_2570200,
EPI_ISL_2570203, EPI_ISL_2570204, EPI_ISL_2570206, EPI_ISL_2570207, EPI_ISL_2570211, EPI_ISL_2570213, EPI_ISL_2570215, EPI_ISL_2570217, EPI_ISL_2570219, EPI_ISL_2570222, EPI_ISL_2570223, EPI_ISL_2570224, EPI_ISL_2570226, EPI_ISL_2570230, EPI_ISL_2570231, EPI_ISL_2570233, EPI_ISL_2570234, EPI_ISL_2570235, EPI_ISL_2570240, EPI_ISL_2570241, EPI_ISL_2570243, EPI_ISL_2570244,
EPI_ISL_2570249, EPI_ISL_2570251, EPI_ISL_2570254, EPI_ISL_2570256, EPI_ISL_2570259, EPI_ISL_2570264, EPI_ISL_2570268, EPI_ISL_2570273, EPI_ISL_2570274, EPI_ISL_2570276, EPI_ISL_2570293, EPI_ISL_2570298, EPI_ISL_2570300, EPI_ISL_2570301, EPI_ISL_2570308, EPI_ISL_2570310, EPI_ISL_2570314, EPI_ISL_2570317, EPI_ISL_2570324, EPI_ISL_2570331, EPI_ISL_2570337, EPI_ISL_2570347,
EPI_ISL_2570352, EPI_ISL_2570353, EPI_ISL_2570367, EPI_ISL_2570372, EPI_ISL_2570386, EPI_ISL_2570389, EPI_ISL_2570400, EPI_ISL_2570402, EPI_ISL_2570406, EPI_ISL_2570417, EPI_ISL_2570439, EPI_ISL_2570442, EPI_ISL_2570459, EPI_ISL_2570465, EPI_ISL_2570473, EPI_ISL_2570475, EPI_ISL_2570477, EPI_ISL_2570478, EPI_ISL_2570480, EPI_ISL_2570481, EPI_ISL_2570484, EPI_ISL_2570486,
EPI_ISL_2570487, EPI_ISL_2570491, EPI_ISL_2570492, EPI_ISL_2570493, EPI_ISL_2570494, EPI_ISL_2570495, EPI_ISL_2570496, EPI_ISL_2570497, EPI_ISL_2570499, EPI_ISL_2570503, EPI_ISL_2570504, EPI_ISL_2570506, EPI_ISL_2570507, EPI_ISL_2570508, EPI_ISL_2570510, EPI_ISL_2570511, EPI_ISL_2570512, EPI_ISL_2570515, EPI_ISL_2570516, EPI_ISL_2570518, EPI_ISL_2570519, EPI_ISL_2570520,
EPI_ISL_2570522, EPI_ISL_2570523, EPI_ISL_2570525, EPI_ISL_2570526, EPI_ISL_2570527, EPI_ISL_2570530, EPI_ISL_2570531, EPI_ISL_2570532, EPI_ISL_2570533, EPI_ISL_2570535, EPI_ISL_2570536, EPI_ISL_2570538, EPI_ISL_2570539, EPI_ISL_2570540, EPI_ISL_2570541, EPI_ISL_2570544, EPI_ISL_2570546, EPI_ISL_2570547, EPI_ISL_2570548, EPI_ISL_2570549, EPI_ISL_2570551, EPI_ISL_2570553,
EPI_ISL_2570554, EPI_ISL_2570556, EPI_ISL_2570558, EPI_ISL_2570560, EPI_ISL_2570561, EPI_ISL_2570562, EPI_ISL_2570563, EPI_ISL_2570568, EPI_ISL_2570569, EPI_ISL_2570570, EPI_ISL_2570571, EPI_ISL_2570572, EPI_ISL_2570573, EPI_ISL_2570577, EPI_ISL_2570578, EPI_ISL_2570579, EPI_ISL_2570580, EPI_ISL_2570581, EPI_ISL_2570584, EPI_ISL_2570586, EPI_ISL_2570587,
EPI_ISL_2570588, EPI_ISL_2570592, EPI_ISL_2570594, EPI_ISL_2570595, EPI_ISL_2570596, EPI_ISL_2570597, EPI_ISL_2570603, EPI_ISL_2570605, EPI_ISL_2570606, EPI_ISL_2570607, EPI_ISL_2570612, EPI_ISL_2570613, EPI_ISL_2570614, EPI_ISL_2570619, EPI_ISL_2570623, EPI_ISL_2570624, EPI_ISL_2570625, EPI_ISL_2570626, EPI_ISL_2570627, EPI_ISL_2570628, EPI_ISL_2570629, EPI_ISL_2570631,
EPI_ISL_2570632, EPI_ISL_2570633, EPI_ISL_2570636, EPI_ISL_2570637, EPI_ISL_2570639, EPI_ISL_2570640, EPI_ISL_2570642, EPI_ISL_2570643, EPI_ISL_2570645, EPI_ISL_2570649, EPI_ISL_2570651, EPI_ISL_2570652, EPI_ISL_2570653, EPI_ISL_2570654, EPI_ISL_2570655, EPI_ISL_2570656, EPI_ISL_2570658, EPI_ISL_2570660, EPI_ISL_2570661, EPI_ISL_2570662, EPI_ISL_2570665, EPI_ISL_2570666,
EPI_ISL_2570668, EPI_ISL_2570669, EPI_ISL_2570670, EPI_ISL_2570671, EPI_ISL_2570672, EPI_ISL_2570676, EPI_ISL_2570677, EPI_ISL_2570678, EPI_ISL_2570679, EPI_ISL_2570680, EPI_ISL_2570682, EPI_ISL_2570685, EPI_ISL_2570686, EPI_ISL_2570687, EPI_ISL_2570688, EPI_ISL_2570689, EPI_ISL_2570690, EPI_ISL_2570693, EPI_ISL_2570694, EPI_ISL_2570695, EPI_ISL_2570697, EPI_ISL_2570698,
EPI_ISL_2570699, EPI_ISL_2570700, EPI_ISL_2570701, EPI_ISL_2570702, EPI_ISL_2570705, EPI_ISL_2570706, EPI_ISL_2570707, EPI_ISL_2570708, EPI_ISL_2570710, EPI_ISL_2570712, EPI_ISL_2570714, EPI_ISL_2570715, EPI_ISL_2570716, EPI_ISL_2570719, EPI_ISL_2570721, EPI_ISL_2570722, EPI_ISL_2570723, EPI_ISL_2570724, EPI_ISL_2570726, EPI_ISL_2570727, EPI_ISL_2570728, EPI_ISL_2570731,
EPI_ISL_2570732, EPI_ISL_2570733, EPI_ISL_2570736, EPI_ISL_2570737, EPI_ISL_2570740, EPI_ISL_2570741, EPI_ISL_2570742, EPI_ISL_2570743, EPI_ISL_2570744, EPI_ISL_2570745, EPI_ISL_2570747, EPI_ISL_2570749, EPI_ISL_2570750, EPI_ISL_2570752, EPI_ISL_2570753, EPI_ISL_2570756, EPI_ISL_2570758, EPI_ISL_2570759, EPI_ISL_2570760, EPI_ISL_2570762, EPI_ISL_2570763, EPI_ISL_2570765,
EPI_ISL_2570766, EPI_ISL_2570768, EPI_ISL_2570772, EPI_ISL_2570774, EPI_ISL_2570775, EPI_ISL_2570777, EPI_ISL_2570779, EPI_ISL_2570781, EPI_ISL_2570782, EPI_ISL_2570784, EPI_ISL_2570786, EPI_ISL_2570787, EPI_ISL_2570788, EPI_ISL_2570790, EPI_ISL_2570792, EPI_ISL_2570794, EPI_ISL_2570795, EPI_ISL_2570796, EPI_ISL_2570797, EPI_ISL_2570798, EPI_ISL_2570799, EPI_ISL_2570801,
EPI_ISL_2570802, EPI_ISL_2570803, EPI_ISL_2605128, EPI_ISL_2605261, EPI_ISL_2605353, EPI_ISL_2605418, EPI_ISL_2605463, EPI_ISL_2605523, EPI_ISL_2605556, EPI_ISL_2605572, EPI_ISL_2605612, EPI_ISL_2605696, EPI_ISL_2605756, EPI_ISL_2605767, EPI_ISL_2605780, EPI_ISL_2605781, EPI_ISL_2605791, EPI_ISL_2605800, EPI_ISL_2605811, EPI_ISL_2605821, EPI_ISL_2605837,
EPI_ISL_2605840, EPI_ISL_2605853, EPI_ISL_2605856, EPI_ISL_2605881, EPI_ISL_2605951, EPI_ISL_2605977, EPI_ISL_2606003, EPI_ISL_2606062, EPI_ISL_2606065, EPI_ISL_2606082, EPI_ISL_2606105, EPI_ISL_2606117, EPI_ISL_2606129, EPI_ISL_2606147, EPI_ISL_2606161, EPI_ISL_2606180, EPI_ISL_2606191, EPI_ISL_2606208, EPI_ISL_2606210, EPI_ISL_2606245, EPI_ISL_2606280,
EPI_ISL_2606286, EPI_ISL_2606288, EPI_ISL_2606298, EPI_ISL_2606306, EPI_ISL_2606313, EPI_ISL_2606350, EPI_ISL_2606360, EPI_ISL_2606372, EPI_ISL_2606389, EPI_ISL_2606436, EPI_ISL_2606729, EPI_ISL_2606757, EPI_ISL_2606820, EPI_ISL_2607014, EPI_ISL_2607252, EPI_ISL_2607254, EPI_ISL_2607255, EPI_ISL_2607258, EPI_ISL_2607259, EPI_ISL_2607636, EPI_ISL_2640327, EPI_ISL_2640333,
EPI_ISL_2640337, EPI_ISL_2640338, EPI_ISL_2640339, EPI_ISL_2640340, EPI_ISL_2640342, EPI_ISL_2640343, EPI_ISL_2640346, EPI_ISL_2640349, EPI_ISL_2640351, EPI_ISL_2640352, EPI_ISL_2640353, EPI_ISL_2640358, EPI_ISL_2640359, EPI_ISL_2640364, EPI_ISL_2640366, EPI_ISL_2640374, EPI_ISL_2640375, EPI_ISL_2640376, EPI_ISL_2640377, EPI_ISL_2640378, EPI_ISL_2640381,
EPI_ISL_2640386, EPI_ISL_2640389, EPI_ISL_2640392, EPI_ISL_2640393, EPI_ISL_2640395, EPI_ISL_2640396, EPI_ISL_2640397, EPI_ISL_2640401, EPI_ISL_2640409, EPI_ISL_2640410, EPI_ISL_2640414, EPI_ISL_2640415, EPI_ISL_2640418, EPI_ISL_2640420, EPI_ISL_2640423, EPI_ISL_2640424, EPI_ISL_2640426, EPI_ISL_2640431, EPI_ISL_2640437, EPI_ISL_2640438, EPI_ISL_2640439, EPI_ISL_2640441,
EPI_ISL_2640442, EPI_ISL_2640443, EPI_ISL_2640444, EPI_ISL_2640445, EPI_ISL_2640447, EPI_ISL_2640450, EPI_ISL_2640452, EPI_ISL_2640453, EPI_ISL_2640455, EPI_ISL_2640460, EPI_ISL_2640461, EPI_ISL_2640462, EPI_ISL_2640463, EPI_ISL_2640466, EPI_ISL_2640468, EPI_ISL_2640471, EPI_ISL_2640473, EPI_ISL_2640474, EPI_ISL_2640481, EPI_ISL_2640484, EPI_ISL_2640485, EPI_ISL_2640491,
EPI_ISL_2640493, EPI_ISL_2640494, EPI_ISL_2640499, EPI_ISL_2640500, EPI_ISL_2640502, EPI_ISL_2640512, EPI_ISL_2640513, EPI_ISL_2640515, EPI_ISL_2640517, EPI_ISL_2640520, EPI_ISL_2640521, EPI_ISL_2640526, EPI_ISL_2640527, EPI_ISL_2640528, EPI_ISL_2640531, EPI_ISL_2640533, EPI_ISL_2640534, EPI_ISL_2640535, EPI_ISL_2640536, EPI_ISL_2640541, EPI_ISL_2640542, EPI_ISL_2640544,
EPI_ISL_2640546, EPI_ISL_2640554, EPI_ISL_2640556, EPI_ISL_2640557, EPI_ISL_2640560, EPI_ISL_2640562, EPI_ISL_2640563, EPI_ISL_2640566, EPI_ISL_2640569, EPI_ISL_2640570, EPI_ISL_2640572, EPI_ISL_2640573, EPI_ISL_2640574, EPI_ISL_2640575, EPI_ISL_2640577, EPI_ISL_2640578, EPI_ISL_2640580, EPI_ISL_2640581, EPI_ISL_2640583, EPI_ISL_2640584, EPI_ISL_2640585, EPI_ISL_2640587,
EPI_ISL_2640590, EPI_ISL_2640591, EPI_ISL_2640592, EPI_ISL_2640593, EPI_ISL_2640594, EPI_ISL_2640595, EPI_ISL_2640596, EPI_ISL_2640597, EPI_ISL_2640598, EPI_ISL_2640599, EPI_ISL_2640602, EPI_ISL_2640603, EPI_ISL_2640605, EPI_ISL_2640610, EPI_ISL_2640611, EPI_ISL_2640612, EPI_ISL_2640613, EPI_ISL_2640614, EPI_ISL_2640615, EPI_ISL_2640618, EPI_ISL_2640620, EPI_ISL_2640622,
EPI_ISL_2640630, EPI_ISL_2640631, EPI_ISL_2640632, EPI_ISL_2640633, EPI_ISL_2640636, EPI_ISL_2640637, EPI_ISL_2640640, EPI_ISL_2640642, EPI_ISL_2640645, EPI_ISL_2640646, EPI_ISL_2640648, EPI_ISL_2640649, EPI_ISL_2640651, EPI_ISL_2640652, EPI_ISL_2640662, EPI_ISL_2640663, EPI_ISL_2640666, EPI_ISL_2640667, EPI_ISL_2640668, EPI_ISL_2640671, EPI_ISL_2640676, EPI_ISL_2640686,
EPI_ISL_2640687, EPI_ISL_2640688, EPI_ISL_2640691, EPI_ISL_2640693, EPI_ISL_2640694, EPI_ISL_2640698, EPI_ISL_2640699, EPI_ISL_2640701, EPI_ISL_2640702, EPI_ISL_2640703, EPI_ISL_2640704, EPI_ISL_2640710, EPI_ISL_2640717, EPI_ISL_2640719, EPI_ISL_2640720, EPI_ISL_2640721, EPI_ISL_2640722, EPI_ISL_2640728, EPI_ISL_2640732, EPI_ISL_2640735, EPI_ISL_2640737, EPI_ISL_2640738,
EPI_ISL_2640742, EPI_ISL_2640743, EPI_ISL_2640746, EPI_ISL_2640747, EPI_ISL_2640748, EPI_ISL_2640752, EPI_ISL_2640754, EPI_ISL_2640756, EPI_ISL_2640758, EPI_ISL_2640759, EPI_ISL_2640761, EPI_ISL_2640768, EPI_ISL_2640774, EPI_ISL_2640775, EPI_ISL_2640776, EPI_ISL_2640781, EPI_ISL_2640784, EPI_ISL_2640794, EPI_ISL_2640795, EPI_ISL_2640799, EPI_ISL_2640801, EPI_ISL_2640804,

EPI_ISL_2640808, EPI_ISL_2640815, EPI_ISL_2640817, EPI_ISL_2640818, EPI_ISL_2640819, EPI_ISL_2640821, EPI_ISL_2640822, EPI_ISL_2640826, EPI_ISL_2640833, EPI_ISL_2640838, EPI_ISL_2640848, EPI_ISL_2640853, EPI_ISL_2640859, EPI_ISL_2640864, EPI_ISL_2640866, EPI_ISL_2640870, EPI_ISL_2640871, EPI_ISL_2640872, EPI_ISL_2640876, EPI_ISL_2640879, EPI_ISL_2640886, EPI_ISL_2640889,
EPI_ISL_2640894, EPI_ISL_2640896, EPI_ISL_2640898, EPI_ISL_2640902, EPI_ISL_2640906, EPI_ISL_2640917, EPI_ISL_2640918, EPI_ISL_2640927, EPI_ISL_2640928, EPI_ISL_2640930, EPI_ISL_2640934, EPI_ISL_2640937, EPI_ISL_2640943, EPI_ISL_2640945, EPI_ISL_2640958, EPI_ISL_2640961, EPI_ISL_2640967, EPI_ISL_2640972, EPI_ISL_2640973, EPI_ISL_2640976, EPI_ISL_2640978, EPI_ISL_2640980,
EPI_ISL_2640984, EPI_ISL_2640985, EPI_ISL_2640986, EPI_ISL_2640987, EPI_ISL_2640990, EPI_ISL_2640995, EPI_ISL_2640999, EPI_ISL_2641002, EPI_ISL_2641005, EPI_ISL_2641007, EPI_ISL_2641008, EPI_ISL_2641009, EPI_ISL_2641010, EPI_ISL_2641013, EPI_ISL_2641014, EPI_ISL_2641016, EPI_ISL_2641017, EPI_ISL_2641020, EPI_ISL_2641021, EPI_ISL_2641023, EPI_ISL_2641024, EPI_ISL_2641026,
EPI_ISL_2641028, EPI_ISL_2641029, EPI_ISL_2641034, EPI_ISL_2641036, EPI_ISL_2641038, EPI_ISL_2641042, EPI_ISL_2641044, EPI_ISL_2641045, EPI_ISL_2641048, EPI_ISL_2641049, EPI_ISL_2641050, EPI_ISL_2641054, EPI_ISL_2641055, EPI_ISL_2641056, EPI_ISL_2641057, EPI_ISL_2641058, EPI_ISL_2641059, EPI_ISL_2641063, EPI_ISL_2641066, EPI_ISL_2641069, EPI_ISL_2641071, EPI_ISL_2641073,
EPI_ISL_2641076, EPI_ISL_2641077, EPI_ISL_2641079, EPI_ISL_2641082, EPI_ISL_2641086, EPI_ISL_2641087, EPI_ISL_2641090, EPI_ISL_2641101, EPI_ISL_2641102, EPI_ISL_2641103, EPI_ISL_2641105, EPI_ISL_2641106, EPI_ISL_2641108, EPI_ISL_2641109, EPI_ISL_2641115, EPI_ISL_2641120, EPI_ISL_2641121, EPI_ISL_2641123, EPI_ISL_2641129, EPI_ISL_2641131, EPI_ISL_2641133, EPI_ISL_2641134,
EPI_ISL_2641135, EPI_ISL_2641136, EPI_ISL_2641139, EPI_ISL_2641146, EPI_ISL_2641147, EPI_ISL_2641151, EPI_ISL_2641155, EPI_ISL_2641157, EPI_ISL_2641158, EPI_ISL_2641161, EPI_ISL_2641166, EPI_ISL_2641170, EPI_ISL_2641175, EPI_ISL_2641182, EPI_ISL_2641184, EPI_ISL_2641186, EPI_ISL_2641189, EPI_ISL_2641191, EPI_ISL_2641193, EPI_ISL_2641201, EPI_ISL_2641202, EPI_ISL_2641204,
EPI_ISL_2641205, EPI_ISL_2641206, EPI_ISL_2641207, EPI_ISL_2641209, EPI_ISL_2641210, EPI_ISL_2641211, EPI_ISL_2641212, EPI_ISL_2641216, EPI_ISL_2641219, EPI_ISL_2641220, EPI_ISL_2641221, EPI_ISL_2641223, EPI_ISL_2641225, EPI_ISL_2641232, EPI_ISL_2641233, EPI_ISL_2641234, EPI_ISL_2641239, EPI_ISL_2641240, EPI_ISL_2641241, EPI_ISL_2641243, EPI_ISL_2641244, EPI_ISL_2641248,
EPI_ISL_2641249, EPI_ISL_2641250, EPI_ISL_2641251, EPI_ISL_2641253, EPI_ISL_2641255, EPI_ISL_2641258, EPI_ISL_2641259, EPI_ISL_2641261, EPI_ISL_2641267, EPI_ISL_2641268, EPI_ISL_2641270, EPI_ISL_2641271, EPI_ISL_2641273, EPI_ISL_2641277, EPI_ISL_2641278, EPI_ISL_2641279, EPI_ISL_2641284, EPI_ISL_2641285, EPI_ISL_2641290, EPI_ISL_2641291, EPI_ISL_2641292, EPI_ISL_2641295,
EPI_ISL_2641296, EPI_ISL_2641297, EPI_ISL_2641303, EPI_ISL_2641304, EPI_ISL_2641306, EPI_ISL_2641310, EPI_ISL_2641311, EPI_ISL_2641312, EPI_ISL_2641321, EPI_ISL_2641323, EPI_ISL_2641326, EPI_ISL_2641328, EPI_ISL_2641330, EPI_ISL_2641333, EPI_ISL_2641334, EPI_ISL_2641337, EPI_ISL_2641338, EPI_ISL_2641343, EPI_ISL_2641346, EPI_ISL_2641347, EPI_ISL_2641348, EPI_ISL_2641349,
EPI_ISL_2641351, EPI_ISL_2641352, EPI_ISL_2641354, EPI_ISL_2641355, EPI_ISL_2641357, EPI_ISL_2641362, EPI_ISL_2641363, EPI_ISL_2641367, EPI_ISL_2641368, EPI_ISL_2641372, EPI_ISL_2641377, EPI_ISL_2641378, EPI_ISL_2641379, EPI_ISL_2641381, EPI_ISL_2641382, EPI_ISL_2641383, EPI_ISL_2641386, EPI_ISL_2641387, EPI_ISL_2641391, EPI_ISL_2641402, EPI_ISL_2641406, EPI_ISL_2641413,
EPI_ISL_2641414, EPI_ISL_2641415, EPI_ISL_2641442, EPI_ISL_2641460, EPI_ISL_2641469, EPI_ISL_2641472, EPI_ISL_2641480, EPI_ISL_2641484, EPI_ISL_2641492, EPI_ISL_2641494, EPI_ISL_2641500, EPI_ISL_2641505, EPI_ISL_2641510, EPI_ISL_2641514, EPI_ISL_2641515, EPI_ISL_2641518, EPI_ISL_2641525, EPI_ISL_2641532, EPI_ISL_2641534, EPI_ISL_2641539, EPI_ISL_2641544, EPI_ISL_2641545,
EPI_ISL_2641547, EPI_ISL_2641549, EPI_ISL_2641566, EPI_ISL_2641577, EPI_ISL_2641585, EPI_ISL_2641588, EPI_ISL_2641592, EPI_ISL_2641594, EPI_ISL_2641603, EPI_ISL_2641610, EPI_ISL_2641612, EPI_ISL_2641615, EPI_ISL_2641617, EPI_ISL_2641620, EPI_ISL_2641621, EPI_ISL_2641624, EPI_ISL_2641626, EPI_ISL_2641627, EPI_ISL_2641633, EPI_ISL_2641634, EPI_ISL_2641640, EPI_ISL_2641644,
EPI_ISL_2641652, EPI_ISL_2641653, EPI_ISL_2641655, EPI_ISL_2641657, EPI_ISL_2641665, EPI_ISL_2641669, EPI_ISL_2641670, EPI_ISL_2641679, EPI_ISL_2641682, EPI_ISL_2641687, EPI_ISL_2641713, EPI_ISL_2641714, EPI_ISL_2651999, EPI_ISL_2652002

| | | | |
|---|---|---|---|
| see above | Lighthouse Lab in Alderley Park | Wellcome Sanger Institute for the COVID-19 Genomics UK (COG-UK) Consortium | Cordelia Langford; David K. Jackson; Dominic Kwiatkowski; Ewan Harrison; Ian Johnston; Jacquelyn Wynn; Jeffrey Barrett; John Sillitoe on behalf of the Wellcome Sanger Institute COVID-19 Surveillance Team; Mairead Hyland; Roberto Amato; Sonia Goncalves; The Lighthouse Lab in Alderley Park and Alex Alderton |

EPI_ISL_2486221, EPI_ISL_2486241, EPI_ISL_2486248, EPI_ISL_2486255, EPI_ISL_2486284, EPI_ISL_2486286, EPI_ISL_2486305, EPI_ISL_2486310, EPI_ISL_2486320, EPI_ISL_2486340, EPI_ISL_2486343, EPI_ISL_2486358, EPI_ISL_2486364, EPI_ISL_2486372, EPI_ISL_2486379, EPI_ISL_2486446, EPI_ISL_2486449, EPI_ISL_2486465, EPI_ISL_2486466, EPI_ISL_2486469, EPI_ISL_2486470, EPI_ISL_2486477,
EPI_ISL_2486490, EPI_ISL_2486499, EPI_ISL_2486511, EPI_ISL_2486520, EPI_ISL_2488412, EPI_ISL_2510882, EPI_ISL_2510886, EPI_ISL_2510893, EPI_ISL_2510895, EPI_ISL_2510902, EPI_ISL_2510904, EPI_ISL_2510910, EPI_ISL_2510930, EPI_ISL_2510932, EPI_ISL_2510933, EPI_ISL_2510953, EPI_ISL_2510956, EPI_ISL_2510962, EPI_ISL_2510965, EPI_ISL_2510997, EPI_ISL_2511011, EPI_ISL_2511014,
EPI_ISL_2511019, EPI_ISL_2511021, EPI_ISL_2511028, EPI_ISL_2511037, EPI_ISL_2511051, EPI_ISL_2511061, EPI_ISL_2511062, EPI_ISL_2511070, EPI_ISL_2511080, EPI_ISL_2511081, EPI_ISL_2511087, EPI_ISL_2511088, EPI_ISL_2511093, EPI_ISL_2511094, EPI_ISL_2511096, EPI_ISL_2511099, EPI_ISL_2511100, EPI_ISL_2511101, EPI_ISL_2511103, EPI_ISL_2511104, EPI_ISL_2511105, EPI_ISL_2511106,
EPI_ISL_2511108, EPI_ISL_2511109, EPI_ISL_2511111, EPI_ISL_2511113, EPI_ISL_2511119, EPI_ISL_2511120, EPI_ISL_2511121, EPI_ISL_2511122, EPI_ISL_2511123, EPI_ISL_2511124, EPI_ISL_2511125, EPI_ISL_2511126, EPI_ISL_2511127, EPI_ISL_2511130, EPI_ISL_2511131, EPI_ISL_2511133, EPI_ISL_2511135, EPI_ISL_2511136, EPI_ISL_2511137, EPI_ISL_2511138, EPI_ISL_2511139, EPI_ISL_2511140,
EPI_ISL_2511142, EPI_ISL_2511143, EPI_ISL_2511145, EPI_ISL_2511147, EPI_ISL_2511148, EPI_ISL_2511150, EPI_ISL_2511151, EPI_ISL_2511152, EPI_ISL_2511153, EPI_ISL_2511155, EPI_ISL_2511156, EPI_ISL_2511157, EPI_ISL_2511158, EPI_ISL_2511161, EPI_ISL_2511164, EPI_ISL_2511165, EPI_ISL_2511166, EPI_ISL_2511168, EPI_ISL_2511170, EPI_ISL_2511172, EPI_ISL_2511173, EPI_ISL_2511175,
EPI_ISL_2511176, EPI_ISL_2511178, EPI_ISL_2511181, EPI_ISL_2511183, EPI_ISL_2511184, EPI_ISL_2511185, EPI_ISL_2511186, EPI_ISL_2511188, EPI_ISL_2511195, EPI_ISL_2511196, EPI_ISL_2511197, EPI_ISL_2511198, EPI_ISL_2511199, EPI_ISL_2511200, EPI_ISL_2511202, EPI_ISL_2511203, EPI_ISL_2511204, EPI_ISL_2511206, EPI_ISL_2511207, EPI_ISL_2511208, EPI_ISL_2511209, EPI_ISL_2511210,
EPI_ISL_2511211, EPI_ISL_2511214, EPI_ISL_2511215, EPI_ISL_2511216, EPI_ISL_2511217, EPI_ISL_2511219, EPI_ISL_2511221, EPI_ISL_2511223, EPI_ISL_2511224, EPI_ISL_2511225, EPI_ISL_2511226, EPI_ISL_2511227, EPI_ISL_2511228, EPI_ISL_2511229, EPI_ISL_2511231, EPI_ISL_2511232, EPI_ISL_2511233, EPI_ISL_2511234, EPI_ISL_2511235, EPI_ISL_2511237, EPI_ISL_2511238, EPI_ISL_2511239,
EPI_ISL_2511240, EPI_ISL_2511244, EPI_ISL_2511247, EPI_ISL_2511248, EPI_ISL_2511251, EPI_ISL_2511252, EPI_ISL_2511253, EPI_ISL_2511254, EPI_ISL_2511255, EPI_ISL_2511257, EPI_ISL_2511258, EPI_ISL_2511259, EPI_ISL_2511261, EPI_ISL_2511262, EPI_ISL_2511263, EPI_ISL_2511264, EPI_ISL_2511265, EPI_ISL_2511266, EPI_ISL_2511267, EPI_ISL_2511268, EPI_ISL_2511271, EPI_ISL_2511273,
EPI_ISL_2511276, EPI_ISL_2511277, EPI_ISL_2511278, EPI_ISL_2511279, EPI_ISL_2511283, EPI_ISL_2511284, EPI_ISL_2511285, EPI_ISL_2511286, EPI_ISL_2511287, EPI_ISL_2511288, EPI_ISL_2511289, EPI_ISL_2511291, EPI_ISL_2511292, EPI_ISL_2511293, EPI_ISL_2511298, EPI_ISL_2511299, EPI_ISL_2511300, EPI_ISL_2511301, EPI_ISL_2511302, EPI_ISL_2511303, EPI_ISL_2511304, EPI_ISL_2511305,
EPI_ISL_2511307, EPI_ISL_2511309, EPI_ISL_2511312, EPI_ISL_2511313, EPI_ISL_2511314, EPI_ISL_2511315, EPI_ISL_2511316, EPI_ISL_2511317, EPI_ISL_2511318, EPI_ISL_2511320, EPI_ISL_2511322, EPI_ISL_2511325, EPI_ISL_2511328, EPI_ISL_2511331, EPI_ISL_2511333, EPI_ISL_2511337, EPI_ISL_2511339, EPI_ISL_2511342, EPI_ISL_2511345, EPI_ISL_2511346, EPI_ISL_2511347, EPI_ISL_2511348,
EPI_ISL_2511350, EPI_ISL_2511404, EPI_ISL_2511441, EPI_ISL_2511492, EPI_ISL_2511502, EPI_ISL_2511504, EPI_ISL_2511571, EPI_ISL_2511573, EPI_ISL_2511596, EPI_ISL_2511643, EPI_ISL_2511644, EPI_ISL_2511645, EPI_ISL_2511679, EPI_ISL_2511683, EPI_ISL_2511690, EPI_ISL_2511704, EPI_ISL_2511706, EPI_ISL_2511707, EPI_ISL_2511714, EPI_ISL_2511723, EPI_ISL_2511728, EPI_ISL_2511729,
EPI_ISL_2511743, EPI_ISL_2511750, EPI_ISL_2511755, EPI_ISL_2511763, EPI_ISL_2511778, EPI_ISL_2511810, EPI_ISL_2511821, EPI_ISL_2511829, EPI_ISL_2511836, EPI_ISL_2511838, EPI_ISL_2511844, EPI_ISL_2511846, EPI_ISL_2511851, EPI_ISL_2511854, EPI_ISL_2511856, EPI_ISL_2511858, EPI_ISL_2511886, EPI_ISL_2511887, EPI_ISL_2511893, EPI_ISL_2511894, EPI_ISL_2511895, EPI_ISL_2511906,
EPI_ISL_2511907, EPI_ISL_2511910, EPI_ISL_2511915, EPI_ISL_2511921, EPI_ISL_2511923, EPI_ISL_2512294, EPI_ISL_2512295, EPI_ISL_2512297, EPI_ISL_2512298, EPI_ISL_2512299, EPI_ISL_2512300, EPI_ISL_2512301, EPI_ISL_2512302, EPI_ISL_2512303, EPI_ISL_2512304, EPI_ISL_2512306, EPI_ISL_2512308, EPI_ISL_2512310, EPI_ISL_2512312, EPI_ISL_2512314, EPI_ISL_2512315, EPI_ISL_2512316,
EPI_ISL_2512317, EPI_ISL_2512319, EPI_ISL_2512321, EPI_ISL_2512322, EPI_ISL_2512323, EPI_ISL_2512324, EPI_ISL_2512326, EPI_ISL_2512327, EPI_ISL_2512328, EPI_ISL_2512329, EPI_ISL_2512331, EPI_ISL_2512333, EPI_ISL_2512335, EPI_ISL_2512336, EPI_ISL_2512337, EPI_ISL_2512338, EPI_ISL_2512339, EPI_ISL_2512340, EPI_ISL_2512341, EPI_ISL_2512342, EPI_ISL_2512343, EPI_ISL_2512345,
EPI_ISL_2512346, EPI_ISL_2512347, EPI_ISL_2512349, EPI_ISL_2512350, EPI_ISL_2512351, EPI_ISL_2512352, EPI_ISL_2512353, EPI_ISL_2512354, EPI_ISL_2512355, EPI_ISL_2512356, EPI_ISL_2512357, EPI_ISL_2512358, EPI_ISL_2512360, EPI_ISL_2512361, EPI_ISL_2512362, EPI_ISL_2512365, EPI_ISL_2512366, EPI_ISL_2512368, EPI_ISL_2512370, EPI_ISL_2512371, EPI_ISL_2512372, EPI_ISL_2512373,
EPI_ISL_2512374, EPI_ISL_2512384, EPI_ISL_2512389, EPI_ISL_2512395, EPI_ISL_2512397, EPI_ISL_2512425, EPI_ISL_2512435, EPI_ISL_2512436, EPI_ISL_2512439, EPI_ISL_2512444, EPI_ISL_2512456, EPI_ISL_2512457, EPI_ISL_2512463, EPI_ISL_2512469, EPI_ISL_2512470, EPI_ISL_2512480, EPI_ISL_2512492, EPI_ISL_2512497, EPI_ISL_2512506, EPI_ISL_2512507, EPI_ISL_2512514, EPI_ISL_2512520,
EPI_ISL_2512525, EPI_ISL_2512528, EPI_ISL_2512532, EPI_ISL_2512547, EPI_ISL_2512575, EPI_ISL_2512589, EPI_ISL_2512601, EPI_ISL_2512606, EPI_ISL_2512646, EPI_ISL_2512666, EPI_ISL_2512684, EPI_ISL_2512687, EPI_ISL_2512688, EPI_ISL_2512689, EPI_ISL_2512691, EPI_ISL_2512692, EPI_ISL_2512694, EPI_ISL_2512698, EPI_ISL_2512699, EPI_ISL_2512705, EPI_ISL_2512707, EPI_ISL_2512710,
EPI_ISL_2512711, EPI_ISL_2512713, EPI_ISL_2512714, EPI_ISL_2512721, EPI_ISL_2512730, EPI_ISL_2512732, EPI_ISL_2512737, EPI_ISL_2512748, EPI_ISL_2512754, EPI_ISL_2512755, EPI_ISL_2512757, EPI_ISL_2512760, EPI_ISL_2512764, EPI_ISL_2512766, EPI_ISL_2512767, EPI_ISL_2512770, EPI_ISL_2512772, EPI_ISL_2512775, EPI_ISL_2512778, EPI_ISL_2512781, EPI_ISL_2512786, EPI_ISL_2512789,
EPI_ISL_2512791, EPI_ISL_2512792, EPI_ISL_2512794, EPI_ISL_2512802, EPI_ISL_2512804, EPI_ISL_2512807, EPI_ISL_2512809, EPI_ISL_2512810, EPI_ISL_2512815, EPI_ISL_2512829, EPI_ISL_2512844, EPI_ISL_2512851, EPI_ISL_2512854, EPI_ISL_2512856, EPI_ISL_2512859, EPI_ISL_2512861, EPI_ISL_2512862, EPI_ISL_2512863, EPI_ISL_2512865, EPI_ISL_2512868, EPI_ISL_2512869, EPI_ISL_2512870,
EPI_ISL_2512876, EPI_ISL_2512882, EPI_ISL_2512884, EPI_ISL_2512885, EPI_ISL_2512887, EPI_ISL_2512889, EPI_ISL_2512890, EPI_ISL_2512897, EPI_ISL_2512898, EPI_ISL_2512901, EPI_ISL_2512902, EPI_ISL_2512904, EPI_ISL_2512907, EPI_ISL_2512908, EPI_ISL_2512914, EPI_ISL_2512915, EPI_ISL_2512919, EPI_ISL_2514729, EPI_ISL_2514731, EPI_ISL_2514732, EPI_ISL_2514737, EPI_ISL_2514739,
EPI_ISL_2514743, EPI_ISL_2514745, EPI_ISL_2514766, EPI_ISL_2514770, EPI_ISL_2514772, EPI_ISL_2514788, EPI_ISL_2514801, EPI_ISL_2514806, EPI_ISL_2514811, EPI_ISL_2514821, EPI_ISL_2514823, EPI_ISL_2514824, EPI_ISL_2514828, EPI_ISL_2514832, EPI_ISL_2514835, EPI_ISL_2514838, EPI_ISL_2514839, EPI_ISL_2514842, EPI_ISL_2514846, EPI_ISL_2514848, EPI_ISL_2514853, EPI_ISL_2514858,
EPI_ISL_2514870, EPI_ISL_2514872, EPI_ISL_2514875, EPI_ISL_2514877, EPI_ISL_2514878, EPI_ISL_2514887, EPI_ISL_2514889, EPI_ISL_2514890, EPI_ISL_2514894, EPI_ISL_2514898, EPI_ISL_2514902, EPI_ISL_2514904, EPI_ISL_2514910, EPI_ISL_2514911, EPI_ISL_2514913, EPI_ISL_2514920, EPI_ISL_2514923, EPI_ISL_2515398, EPI_ISL_2515411, EPI_ISL_2515412, EPI_ISL_2515414, EPI_ISL_2515417,
EPI_ISL_2515422, EPI_ISL_2515423, EPI_ISL_2515434, EPI_ISL_2515436, EPI_ISL_2515441, EPI_ISL_2515449, EPI_ISL_2515450, EPI_ISL_2515453, EPI_ISL_2515455, EPI_ISL_2515456, EPI_ISL_2515460, EPI_ISL_2515464, EPI_ISL_2515477, EPI_ISL_2515479, EPI_ISL_2515496, EPI_ISL_2515502, EPI_ISL_2515503, EPI_ISL_2515504, EPI_ISL_2515506, EPI_ISL_2515516, EPI_ISL_2515525, EPI_ISL_2515526,
EPI_ISL_2515530, EPI_ISL_2515533, EPI_ISL_2515548, EPI_ISL_2515552, EPI_ISL_2515553, EPI_ISL_2515560, EPI_ISL_2515574, EPI_ISL_2515586, EPI_ISL_2515589, EPI_ISL_2515594, EPI_ISL_2515596, EPI_ISL_2515608, EPI_ISL_2515609, EPI_ISL_2515614, EPI_ISL_2515618, EPI_ISL_2515621, EPI_ISL_2515627, EPI_ISL_2515633, EPI_ISL_2515636, EPI_ISL_2538534, EPI_ISL_2538535, EPI_ISL_2538539,
EPI_ISL_2538556, EPI_ISL_2538563, EPI_ISL_2538567, EPI_ISL_2538570, EPI_ISL_2538577, EPI_ISL_2538587, EPI_ISL_2538593, EPI_ISL_2538594, EPI_ISL_2538601, EPI_ISL_2538607, EPI_ISL_2538608, EPI_ISL_2538609, EPI_ISL_2538613, EPI_ISL_2538631, EPI_ISL_2538641, EPI_ISL_2538668, EPI_ISL_2538670, EPI_ISL_2538678, EPI_ISL_2538694, EPI_ISL_2538696, EPI_ISL_2538699, EPI_ISL_2538700,
EPI_ISL_2538708, EPI_ISL_2538712, EPI_ISL_2538713, EPI_ISL_2538717, EPI_ISL_2538719, EPI_ISL_2538733, EPI_ISL_2538740, EPI_ISL_2538748, EPI_ISL_2538762, EPI_ISL_2538764, EPI_ISL_2538775, EPI_ISL_2538785, EPI_ISL_2538787, EPI_ISL_2538810, EPI_ISL_2538811, EPI_ISL_2538813, EPI_ISL_2538862, EPI_ISL_2538915, EPI_ISL_2538930, EPI_ISL_2540374, EPI_ISL_2540375, EPI_ISL_2540380,
EPI_ISL_2540382, EPI_ISL_2540388, EPI_ISL_2540389, EPI_ISL_2540393, EPI_ISL_2540398, EPI_ISL_2540400, EPI_ISL_2540402, EPI_ISL_2540405, EPI_ISL_2540406, EPI_ISL_2540415, EPI_ISL_2540416, EPI_ISL_2540421, EPI_ISL_2540435, EPI_ISL_2540438, EPI_ISL_2540440, EPI_ISL_2540442, EPI_ISL_2540446, EPI_ISL_2540448, EPI_ISL_2540455, EPI_ISL_2540457, EPI_ISL_2540459, EPI_ISL_2540465,
EPI_ISL_2540466, EPI_ISL_2540471, EPI_ISL_2540473, EPI_ISL_2540476, EPI_ISL_2540485, EPI_ISL_2540486, EPI_ISL_2540491, EPI_ISL_2540495, EPI_ISL_2540503, EPI_ISL_2540504, EPI_ISL_2540507, EPI_ISL_2540511, EPI_ISL_2540512, EPI_ISL_2540515, EPI_ISL_2540518, EPI_ISL_2540519, EPI_ISL_2540521, EPI_ISL_2540542, EPI_ISL_2540544, EPI_ISL_2540549, EPI_ISL_2540558, EPI_ISL_2540568,
EPI_ISL_2540573, EPI_ISL_2540575, EPI_ISL_2540578, EPI_ISL_2540585, EPI_ISL_2540586, EPI_ISL_2540978, EPI_ISL_2540999, EPI_ISL_2541031, EPI_ISL_2541050, EPI_ISL_2541075, EPI_ISL_2541989, EPI_ISL_2541990, EPI_ISL_2541991, EPI_ISL_2541992, EPI_ISL_2541993, EPI_ISL_2541994, EPI_ISL_2541995, EPI_ISL_2541996, EPI_ISL_2541997, EPI_ISL_2541998, EPI_ISL_2541999, EPI_ISL_2542000,
EPI_ISL_2542001, EPI_ISL_2542002, EPI_ISL_2542003, EPI_ISL_2542004, EPI_ISL_2542005, EPI_ISL_2542006, EPI_ISL_2542007, EPI_ISL_2542008, EPI_ISL_2542009, EPI_ISL_2542010, EPI_ISL_2542011, EPI_ISL_2542012, EPI_ISL_2542013, EPI_ISL_2542014, EPI_ISL_2542015, EPI_ISL_2542016, EPI_ISL_2542017, EPI_ISL_2542018, EPI_ISL_2542019, EPI_ISL_2542020, EPI_ISL_2542021, EPI_ISL_2542024,
EPI_ISL_2542025, EPI_ISL_2542026, EPI_ISL_2542027, EPI_ISL_2542029, EPI_ISL_2542030, EPI_ISL_2542031, EPI_ISL_2542032, EPI_ISL_2542033, EPI_ISL_2542034, EPI_ISL_2542035, EPI_ISL_2542036, EPI_ISL_2542037, EPI_ISL_2542038, EPI_ISL_2542039, EPI_ISL_2542040, EPI_ISL_2542041, EPI_ISL_2542042, EPI_ISL_2542043, EPI_ISL_2542044, EPI_ISL_2542045, EPI_ISL_2542046, EPI_ISL_2542047,
EPI_ISL_2542048, EPI_ISL_2542049, EPI_ISL_2542050, EPI_ISL_2542051, EPI_ISL_2542052, EPI_ISL_2542053, EPI_ISL_2542054, EPI_ISL_2542055, EPI_ISL_2542056, EPI_ISL_2542057, EPI_ISL_2542058, EPI_ISL_2542059, EPI_ISL_2542060, EPI_ISL_2542061, EPI_ISL_2542062, EPI_ISL_2542063, EPI_ISL_2542064, EPI_ISL_2542065, EPI_ISL_2542066, EPI_ISL_2542067, EPI_ISL_2542068, EPI_ISL_2542069,
EPI_ISL_2542070, EPI_ISL_2542071, EPI_ISL_2542072, EPI_ISL_2542073, EPI_ISL_2542074, EPI_ISL_2542075, EPI_ISL_2542076, EPI_ISL_2542077, EPI_ISL_2542078, EPI_ISL_2542079, EPI_ISL_2542080, EPI_ISL_2542081, EPI_ISL_2542082, EPI_ISL_2542083, EPI_ISL_2542084, EPI_ISL_2542085, EPI_ISL_2542086, EPI_ISL_2542087, EPI_ISL_2542088, EPI_ISL_2542089, EPI_ISL_2542090, EPI_ISL_2542091,
EPI_ISL_2542092, EPI_ISL_2542093, EPI_ISL_2542094, EPI_ISL_2542095, EPI_ISL_2542096, EPI_ISL_2542097, EPI_ISL_2542098, EPI_ISL_2542099, EPI_ISL_2542100, EPI_ISL_2542101, EPI_ISL_2542102, EPI_ISL_2542103, EPI_ISL_2542106, EPI_ISL_2542107, EPI_ISL_2542108, EPI_ISL_2542109, EPI_ISL_2542110, EPI_ISL_2542111, EPI_ISL_2542113, EPI_ISL_2542114, EPI_ISL_2542115, EPI_ISL_2542116,
EPI_ISL_2542117, EPI_ISL_2542118, EPI_ISL_2542119, EPI_ISL_2542120, EPI_ISL_2542121, EPI_ISL_2542122, EPI_ISL_2542123, EPI_ISL_2542124, EPI_ISL_2542125, EPI_ISL_2542126, EPI_ISL_2542127, EPI_ISL_2542128, EPI_ISL_2542129, EPI_ISL_2542130, EPI_ISL_2542131, EPI_ISL_2542132, EPI_ISL_2542133, EPI_ISL_2542134, EPI_ISL_2542135, EPI_ISL_2542136, EPI_ISL_2542137, EPI_ISL_2542138,
EPI_ISL_2542139, EPI_ISL_2542140, EPI_ISL_2542141, EPI_ISL_2542142, EPI_ISL_2542143, EPI_ISL_2542144, EPI_ISL_2542145, EPI_ISL_2542146, EPI_ISL_2542147, EPI_ISL_2542148, EPI_ISL_2542149, EPI_ISL_2542150, EPI_ISL_2542151, EPI_ISL_2542152, EPI_ISL_2542153, EPI_ISL_2542154, EPI_ISL_2542155, EPI_ISL_2542156, EPI_ISL_2542157, EPI_ISL_2542158, EPI_ISL_2542159, EPI_ISL_2542160,
EPI_ISL_2542161, EPI_ISL_2542162, EPI_ISL_2542163, EPI_ISL_2542164, EPI_ISL_2542165, EPI_ISL_2542166, EPI_ISL_2542167, EPI_ISL_2542168, EPI_ISL_2542169, EPI_ISL_2542170, EPI_ISL_2542171, EPI_ISL_2542172, EPI_ISL_2542173, EPI_ISL_2542174, EPI_ISL_2542175, EPI_ISL_2542176, EPI_ISL_2542177, EPI_ISL_2542178, EPI_ISL_2542179, EPI_ISL_2542180, EPI_ISL_2542181, EPI_ISL_2542182,
EPI_ISL_2542183, EPI_ISL_2542184, EPI_ISL_2542185, EPI_ISL_2542186, EPI_ISL_2542187, EPI_ISL_2542188, EPI_ISL_2542189, EPI_ISL_2542190, EPI_ISL_2542191, EPI_ISL_2542192, EPI_ISL_2542193, EPI_ISL_2542194, EPI_ISL_2542195, EPI_ISL_2542196, EPI_ISL_2542197, EPI_ISL_2542198, EPI_ISL_2542199, EPI_ISL_2542200, EPI_ISL_2542201, EPI_ISL_2542202, EPI_ISL_2542203, EPI_ISL_2542204,
EPI_ISL_2542205, EPI_ISL_2542206, EPI_ISL_2542207, EPI_ISL_2542208, EPI_ISL_2542209, EPI_ISL_2542210, EPI_ISL_2542211, EPI_ISL_2542212, EPI_ISL_2542213, EPI_ISL_2542214, EPI_ISL_2542215, EPI_ISL_2542216, EPI_ISL_2542217, EPI_ISL_2542218, EPI_ISL_2542219, EPI_ISL_2542220, EPI_ISL_2542221, EPI_ISL_2542222, EPI_ISL_2542223, EPI_ISL_2542224, EPI_ISL_2542225, EPI_ISL_2542226,
EPI_ISL_2542227, EPI_ISL_2542228, EPI_ISL_2542229, EPI_ISL_2542230, EPI_ISL_2542231, EPI_ISL_2542232, EPI_ISL_2542233, EPI_ISL_2542234, EPI_ISL_2542235, EPI_ISL_2542236, EPI_ISL_2542237, EPI_ISL_2542238, EPI_ISL_2542239, EPI_ISL_2542240, EPI_ISL_2542241, EPI_ISL_2542242, EPI_ISL_2542243, EPI_ISL_2542244, EPI_ISL_2542245, EPI_ISL_2542246, EPI_ISL_2542247, EPI_ISL_2542248,
EPI_ISL_2542249, EPI_ISL_2542250, EPI_ISL_2542251, EPI_ISL_2542252, EPI_ISL_2542253, EPI_ISL_2542254, EPI_ISL_2542255, EPI_ISL_2542256, EPI_ISL_2542257, EPI_ISL_2542258, EPI_ISL_2542259, EPI_ISL_2542260, EPI_ISL_2542261, EPI_ISL_2542262, EPI_ISL_2542263, EPI_ISL_2542264, EPI_ISL_2542265, EPI_ISL_2542266, EPI_ISL_2542267, EPI_ISL_2542268, EPI_ISL_2542269, EPI_ISL_2542270,
EPI_ISL_2542271, EPI_ISL_2542272, EPI_ISL_2542273, EPI_ISL_2542274, EPI_ISL_2542275, EPI_ISL_2542276, EPI_ISL_2542277, EPI_ISL_2542278, EPI_ISL_2542279, EPI_ISL_2542280, EPI_ISL_2542281, EPI_ISL_2542282, EPI_ISL_2542283, EPI_ISL_2542284, EPI_ISL_2542285, EPI_ISL_2542286, EPI_ISL_2542287, EPI_ISL_2542288, EPI_ISL_2542289, EPI_ISL_2542290, EPI_ISL_2542291, EPI_ISL_2542292,
EPI_ISL_2542293, EPI_ISL_2542294, EPI_ISL_2542295, EPI_ISL_2542296, EPI_ISL_2542297, EPI_ISL_2542298, EPI_ISL_2542299, EPI_ISL_2542300, EPI_ISL_2542301, EPI_ISL_2542302, EPI_ISL_2542303, EPI_ISL_2542304, EPI_ISL_2542305, EPI_ISL_2542306, EPI_ISL_2542307, EPI_ISL_2542308, EPI_ISL_2542309, EPI_ISL_2542310, EPI_ISL_2542311, EPI_ISL_2542312, EPI_ISL_2542313, EPI_ISL_2542314,
EPI_ISL_2542315, EPI_ISL_2542316, EPI_ISL_2542317, EPI_ISL_2542318, EPI_ISL_2542319, EPI_ISL_2542320, EPI_ISL_2542321, EPI_ISL_2542322, EPI_ISL_2542323, EPI_ISL_2542324, EPI_ISL_2542325, EPI_ISL_2542326, EPI_ISL_2542327, EPI_ISL_2542328, EPI_ISL_2542329, EPI_ISL_2542330, EPI_ISL_2542331, EPI_ISL_2542332, EPI_ISL_2542334, EPI_ISL_2542335, EPI_ISL_2542336, EPI_ISL_2542337,
EPI_ISL_2542338, EPI_ISL_2542339, EPI_ISL_2542340, EPI_ISL_2542341, EPI_ISL_2542342, EPI_ISL_2542343, EPI_ISL_2542344, EPI_ISL_2542345, EPI_ISL_2542346, EPI_ISL_2542347, EPI_ISL_2542348, EPI_ISL_2542349, EPI_ISL_2542350, EPI_ISL_2542351, EPI_ISL_2542352, EPI_ISL_2542353, EPI_ISL_2542354, EPI_ISL_2542355, EPI_ISL_2542356, EPI_ISL_2542357, EPI_ISL_2542358, EPI_ISL_2542359,
EPI_ISL_2542360, EPI_ISL_2542361, EPI_ISL_2542362, EPI_ISL_2542363, EPI_ISL_2542364, EPI_ISL_2542365, EPI_ISL_2542366, EPI_ISL_2542367, EPI_ISL_2542368, EPI_ISL_2542370, EPI_ISL_2542371, EPI_ISL_2542372, EPI_ISL_2542373, EPI_ISL_2542374, EPI_ISL_2542375, EPI_ISL_2542376, EPI_ISL_2542377, EPI_ISL_2542378, EPI_ISL_2542380, EPI_ISL_2542382, EPI_ISL_2542383, EPI_ISL_2542384,
EPI_ISL_2542385, EPI_ISL_2542386, EPI_ISL_2542387, EPI_ISL_2542388, EPI_ISL_2542389, EPI_ISL_2542390, EPI_ISL_2542391, EPI_ISL_2542392, EPI_ISL_2542393, EPI_ISL_2542394, EPI_ISL_2542395, EPI_ISL_2542396, EPI_ISL_2542397, EPI_ISL_2542398, EPI_ISL_2542399, EPI_ISL_2542400, EPI_ISL_2542401, EPI_ISL_2542402, EPI_ISL_2542403, EPI_ISL_2542404, EPI_ISL_2542405, EPI_ISL_2542406,
EPI_ISL_2542407, EPI_ISL_2542408, EPI_ISL_2542409, EPI_ISL_2542410, EPI_ISL_2542411, EPI_ISL_2542412, EPI_ISL_2542413, EPI_ISL_2542414, EPI_ISL_2542415, EPI_ISL_2542416, EPI_ISL_2542417, EPI_ISL_2542418, EPI_ISL_2542419, EPI_ISL_2542420, EPI_ISL_2542421, EPI_ISL_2542422, EPI_ISL_2542423, EPI_ISL_2542424, EPI_ISL_2542425, EPI_ISL_2542426, EPI_ISL_2542427, EPI_ISL_2542428,
EPI_ISL_2542429, EPI_ISL_2542430, EPI_ISL_2542431, EPI_ISL_2542432, EPI_ISL_2542433, EPI_ISL_2542435, EPI_ISL_2542436, EPI_ISL_2542437, EPI_ISL_2542438, EPI_ISL_2542439, EPI_ISL_2542440, EPI_ISL_2542441, EPI_ISL_2542442, EPI_ISL_2542443, EPI_ISL_2542444, EPI_ISL_2542445, EPI_ISL_2542446, EPI_ISL_2542447, EPI_ISL_2542448, EPI_ISL_2542449, EPI_ISL_2542450, EPI_ISL_2542451,
EPI_ISL_2542452, EPI_ISL_2542453, EPI_ISL_2542454, EPI_ISL_2542455, EPI_ISL_2542456, EPI_ISL_2542457, EPI_ISL_2542458, EPI_ISL_2542459, EPI_ISL_2542460, EPI_ISL_2542461, EPI_ISL_2542462, EPI_ISL_2542463, EPI_ISL_2542464, EPI_ISL_2542465, EPI_ISL_2542466, EPI_ISL_2542467, EPI_ISL_2542468, EPI_ISL_2542469, EPI_ISL_2542470, EPI_ISL_2542471, EPI_ISL_2542472, EPI_ISL_2542473,
EPI_ISL_2542474, EPI_ISL_2542475, EPI_ISL_2542476, EPI_ISL_2542477, EPI_ISL_2542478, EPI_ISL_2542479, EPI_ISL_2542480, EPI_ISL_2542481, EPI_ISL_2542482, EPI_ISL_2542483, EPI_ISL_2542484, EPI_ISL_2542485, EPI_ISL_2542486, EPI_ISL_2542487, EPI_ISL_2542488, EPI_ISL_2542489, EPI_ISL_2542490, EPI_ISL_2542491, EPI_ISL_2542492, EPI_ISL_2542493, EPI_ISL_2542494, EPI_ISL_2542495,
EPI_ISL_2542496, EPI_ISL_2542497, EPI_ISL_2542498, EPI_ISL_2542499, EPI_ISL_2542500, EPI_ISL_2542501, EPI_ISL_2542502, EPI_ISL_2542503, EPI_ISL_2542504, EPI_ISL_2542505, EPI_ISL_2542506, EPI_ISL_2542507, EPI_ISL_2542508, EPI_ISL_2542509, EPI_ISL_2542510, EPI_ISL_2542511, EPI_ISL_2542512, EPI_ISL_2542513, EPI_ISL_2542514, EPI_ISL_2542515, EPI_ISL_2542516, EPI_ISL_2542517,
EPI_ISL_2542518, EPI_ISL_2542519, EPI_ISL_2542520, EPI_ISL_2542521, EPI_ISL_2542522, EPI_ISL_2542523, EPI_ISL_2542524, EPI_ISL_2542525, EPI_ISL_2542526, EPI_ISL_2542527, EPI_ISL_2542528, EPI_ISL_2542529, EPI_ISL_2542530, EPI_ISL_2542531, EPI_ISL_2542532, EPI_ISL_2542533, EPI_ISL_2542534, EPI_ISL_2542535, EPI_ISL_2542536, EPI_ISL_2542537, EPI_ISL_2542538, EPI_ISL_2542539,
EPI_ISL_2542540, EPI_ISL_2542541, EPI_ISL_2542542, EPI_ISL_2542789, EPI_ISL_2542800, EPI_ISL_2542803, EPI_ISL_2542807, EPI_ISL_2542808, EPI_ISL_2542815, EPI_ISL_2542827, EPI_ISL_2542833, EPI_ISL_2542839, EPI_ISL_2542843, EPI_ISL_2542845, EPI_ISL_2542846, EPI_ISL_2542852, EPI_ISL_2542854, EPI_ISL_2542857, EPI_ISL_2542861, EPI_ISL_2542876, EPI_ISL_2542877, EPI_ISL_2542878,
EPI_ISL_2542881, EPI_ISL_2542884, EPI_ISL_2542886, EPI_ISL_2542888, EPI_ISL_2542893, EPI_ISL_2542895, EPI_ISL_2542896, EPI_ISL_2542899, EPI_ISL_2542905, EPI_ISL_2542907, EPI_ISL_2542908, EPI_ISL_2542910, EPI_ISL_2542911, EPI_ISL_2542913, EPI_ISL_2542919, EPI_ISL_2542920, EPI_ISL_2542922, EPI_ISL_2542923, EPI_ISL_2542927, EPI_ISL_2542928, EPI_ISL_2542932, EPI_ISL_2542937,
EPI_ISL_2542941, EPI_ISL_2542945, EPI_ISL_2542946, EPI_ISL_2542947, EPI_ISL_2542949, EPI_ISL_2543193, EPI_ISL_2543194, EPI_ISL_2543195, EPI_ISL_2543196, EPI_ISL_2543197, EPI_ISL_2543198, EPI_ISL_2543199, EPI_ISL_2543200, EPI_ISL_2543201, EPI_ISL_2543202, EPI_ISL_2543203, EPI_ISL_2543204, EPI_ISL_2543205, EPI_ISL_2543206, EPI_ISL_2543207, EPI_ISL_2543208, EPI_ISL_2543209,
EPI_ISL_2543210, EPI_ISL_2543211, EPI_ISL_2543212, EPI_ISL_2543213, EPI_ISL_2543214, EPI_ISL_2543215, EPI_ISL_2543216, EPI_ISL_2543217, EPI_ISL_2543218, EPI_ISL_2543219, EPI_ISL_2543220, EPI_ISL_2543221, EPI_ISL_2543223, EPI_ISL_2543224, EPI_ISL_2543225, EPI_ISL_2543226, EPI_ISL_2543227, EPI_ISL_2543228, EPI_ISL_2543229, EPI_ISL_2543230, EPI_ISL_2543232, EPI_ISL_2543233,
EPI_ISL_2543234, EPI_ISL_2543235, EPI_ISL_2543236, EPI_ISL_2543237, EPI_ISL_2543238, EPI_ISL_2543239, EPI_ISL_2543240, EPI_ISL_2543241, EPI_ISL_2543242, EPI_ISL_2543243, EPI_ISL_2543244, EPI_ISL_2543245, EPI_ISL_2543246, EPI_ISL_2543247, EPI_ISL_2543248, EPI_ISL_2543249, EPI_ISL_2543250, EPI_ISL_2543251, EPI_ISL_2543252, EPI_ISL_2543253, EPI_ISL_2543254, EPI_ISL_2543255,
EPI_ISL_2543256, EPI_ISL_2543258, EPI_ISL_2543259, EPI_ISL_2543260, EPI_ISL_2543261, EPI_ISL_2543262, EPI_ISL_2543263, EPI_ISL_2543264, EPI_ISL_2543265, EPI_ISL_2543266, EPI_ISL_2543267, EPI_ISL_2543268, EPI_ISL_2543269, EPI_ISL_2543270, EPI_ISL_2543271, EPI_ISL_2543272, EPI_ISL_2543273, EPI_ISL_2543274, EPI_ISL_2543275, EPI_ISL_2543276, EPI_ISL_2543277, EPI_ISL_2543278,
EPI_ISL_2543279, EPI_ISL_2543280, EPI_ISL_2543281, EPI_ISL_2543282, EPI_ISL_2543283, EPI_ISL_2543284, EPI_ISL_2543285, EPI_ISL_2543286, EPI_ISL_2543287, EPI_ISL_2543288, EPI_ISL_2543289, EPI_ISL_2543290, EPI_ISL_2543291, EPI_ISL_2543292, EPI_ISL_2543293, EPI_ISL_2543294, EPI_ISL_2543295, EPI_ISL_2543296, EPI_ISL_2543297, EPI_ISL_2543298, EPI_ISL_2543299, EPI_ISL_2543300,
EPI_ISL_2543301, EPI_ISL_2543302, EPI_ISL_2543303, EPI_ISL_2543304, EPI_ISL_2551977, EPI_ISL_2552007, EPI_ISL_2553066, EPI_ISL_2553149, EPI_ISL_2553175, EPI_ISL_2553187, EPI_ISL_2553191, EPI_ISL_2553209, EPI_ISL_2553218, EPI_ISL_2553280, EPI_ISL_2553311, EPI_ISL_2553364, EPI_ISL_2553383, EPI_ISL_2553387, EPI_ISL_2553430, EPI_ISL_2553524, EPI_ISL_2553530, EPI_ISL_2553549,
EPI_ISL_2553571, EPI_ISL_2553595, EPI_ISL_2553632, EPI_ISL_2553706, EPI_ISL_2553717, EPI_ISL_2553725, EPI_ISL_2553742, EPI_ISL_2553749, EPI_ISL_2553774, EPI_ISL_2553779, EPI_ISL_2554219, EPI_ISL_2554262, EPI_ISL_2554288, EPI_ISL_2554314, EPI_ISL_2554374, EPI_ISL_2554848, EPI_ISL_2554849, EPI_ISL_2554850, EPI_ISL_2554851, EPI_ISL_2554852, EPI_ISL_2554853, EPI_ISL_2554854,
EPI_ISL_2554855, EPI_ISL_2554856, EPI_ISL_2554857, EPI_ISL_2554858, EPI_ISL_2554859, EPI_ISL_2554860, EPI_ISL_2554861, EPI_ISL_2554862, EPI_ISL_2554863, EPI_ISL_2554864, EPI_ISL_2554865, EPI_ISL_2554866, EPI_ISL_2554867, EPI_ISL_2554868, EPI_ISL_2554869, EPI_ISL_2554870, EPI_ISL_2554871, EPI_ISL_2554872, EPI_ISL_2554873, EPI_ISL_2554874, EPI_ISL_2554875, EPI_ISL_2554876,
EPI_ISL_2554877, EPI_ISL_2554878, EPI_ISL_2554879, EPI_ISL_2554880, EPI_ISL_2554881, EPI_ISL_2554882, EPI_ISL_2554883, EPI_ISL_2554884, EPI_ISL_2554886, EPI_ISL_2554887, EPI_ISL_2554888, EPI_ISL_2554889, EPI_ISL_2554890, EPI_ISL_2554891, EPI_ISL_2554892, EPI_ISL_2554893, EPI_ISL_2554894, EPI_ISL_2554895, EPI_ISL_2554896, EPI_ISL_2554897, EPI_ISL_2554898, EPI_ISL_2554899,
EPI_ISL_2554900, EPI_ISL_2554901, EPI_ISL_2554902, EPI_ISL_2554903, EPI_ISL_2554904, EPI_ISL_2554905, EPI_ISL_2554906, EPI_ISL_2554907, EPI_ISL_2554908, EPI_ISL_2554909, EPI_ISL_2554910, EPI_ISL_2554911, EPI_ISL_2554912, EPI_ISL_2554913, EPI_ISL_2554914, EPI_ISL_2555056, EPI_ISL_2555058, EPI_ISL_2555059, EPI_ISL_2555060, EPI_ISL_2555061, EPI_ISL_2555062, EPI_ISL_2555063,
EPI_ISL_2555065, EPI_ISL_2555066, EPI_ISL_2555067, EPI_ISL_2555068, EPI_ISL_2555069, EPI_ISL_2555070, EPI_ISL_2555071, EPI_ISL_2555072, EPI_ISL_2555073, EPI_ISL_2555074, EPI_ISL_2555075, EPI_ISL_2555076, EPI_ISL_2555077, EPI_ISL_2555078, EPI_ISL_2555079, EPI_ISL_2555080, EPI_ISL_2555081, EPI_ISL_2555082, EPI_ISL_2555083, EPI_ISL_2555084, EPI_ISL_2555085, EPI_ISL_2555086,
EPI_ISL_2555087, EPI_ISL_2555088, EPI_ISL_2555089, EPI_ISL_2555090, EPI_ISL_2555091, EPI_ISL_2555092, EPI_ISL_2555093, EPI_ISL_2555094, EPI_ISL_2555095, EPI_ISL_2555096, EPI_ISL_2555097, EPI_ISL_2555098, EPI_ISL_2555099, EPI_ISL_2555100, EPI_ISL_2555101, EPI_ISL_2555102, EPI_ISL_2555103, EPI_ISL_2555104, EPI_ISL_2555105, EPI_ISL_2555106, EPI_ISL_2555107, EPI_ISL_2555108,
EPI_ISL_2555109, EPI_ISL_2555110, EPI_ISL_2555111, EPI_ISL_2555112, EPI_ISL_2555113, EPI_ISL_2555114, EPI_ISL_2555115, EPI_ISL_2555116, EPI_ISL_2555117, EPI_ISL_2555118, EPI_ISL_2555119, EPI_ISL_2555120, EPI_ISL_2555121, EPI_ISL_2555122, EPI_ISL_2555123, EPI_ISL_2555124, EPI_ISL_2555125, EPI_ISL_2555126, EPI_ISL_2555127, EPI_ISL_2555128, EPI_ISL_2555129, EPI_ISL_2555130,
EPI_ISL_2555131, EPI_ISL_2555132, EPI_ISL_2555134, EPI_ISL_2555135, EPI_ISL_2555136, EPI_ISL_2555137, EPI_ISL_2555138, EPI_ISL_2555139, EPI_ISL_2555140, EPI_ISL_2555141, EPI_ISL_2555142, EPI_ISL_2555143, EPI_ISL_2555144, EPI_ISL_2555145, EPI_ISL_2555146, EPI_ISL_2555147, EPI_ISL_2555148, EPI_ISL_2555150, EPI_ISL_2555151, EPI_ISL_2555152, EPI_ISL_2555153, EPI_ISL_2555154,
EPI_ISL_2555154, EPI_ISL_2555155, EPI_ISL_2555156, EPI_ISL_2555157, EPI_ISL_2555158, EPI_ISL_2555160, EPI_ISL_2555161, EPI_ISL_2555162, EPI_ISL_2555163, EPI_ISL_2555164, EPI_ISL_2555165, EPI_ISL_2555166, EPI_ISL_2555167, EPI_ISL_2555168, EPI_ISL_2555169, EPI_ISL_2555170, EPI_ISL_2555171, EPI_ISL_2555172, EPI_ISL_2555173, EPI_ISL_2555175, EPI_ISL_2555176, EPI_ISL_2555177,
EPI_ISL_2555178, EPI_ISL_2555179, EPI_ISL_2555180, EPI_ISL_2555181, EPI_ISL_2555182, EPI_ISL_2555183, EPI_ISL_2555184, EPI_ISL_2555185, EPI_ISL_2555186, EPI_ISL_2555188, EPI_ISL_2555189, EPI_ISL_2555190, EPI_ISL_2555192, EPI_ISL_2555193, EPI_ISL_2555194, EPI_ISL_2555195, EPI_ISL_2555197, EPI_ISL_2555198, EPI_ISL_2555199, EPI_ISL_2555201, EPI_ISL_2555202, EPI_ISL_2555203,
EPI_ISL_2555204, EPI_ISL_2555206, EPI_ISL_2555207, EPI_ISL_2555208, EPI_ISL_2555209, EPI_ISL_2555210, EPI_ISL_2555211, EPI_ISL_2555212, EPI_ISL_2555213, EPI_ISL_2555214, EPI_ISL_2555215, EPI_ISL_2555216, EPI_ISL_2555217, EPI_ISL_2555218, EPI_ISL_2555219, EPI_ISL_2555220, EPI_ISL_2555221, EPI_ISL_2555222, EPI_ISL_2555223, EPI_ISL_2555224, EPI_ISL_2555225, EPI_ISL_2555226,
EPI_ISL_2555227, EPI_ISL_2555228, EPI_ISL_2555229, EPI_ISL_2555230, EPI_ISL_2555231, EPI_ISL_2555232, EPI_ISL_2555233, EPI_ISL_2555234, EPI_ISL_2555235, EPI_ISL_2555236, EPI_ISL_2555237, EPI_ISL_2555238, EPI_ISL_2555239, EPI_ISL_2555240, EPI_ISL_2555241, EPI_ISL_2555242, EPI_ISL_2555243, EPI_ISL_2555244, EPI_ISL_2555245, EPI_ISL_2555246, EPI_ISL_2555247, EPI_ISL_2555248,
EPI_ISL_2555249, EPI_ISL_2555250, EPI_ISL_2555251, EPI_ISL_2555252, EPI_ISL_2555253, EPI_ISL_2555254, EPI_ISL_2555255, EPI_ISL_2555256, EPI_ISL_2555257, EPI_ISL_2555258, EPI_ISL_2555259, EPI_ISL_2555260, EPI_ISL_2555261, EPI_ISL_2555262, EPI_ISL_2555263, EPI_ISL_2555264, EPI_ISL_2555265, EPI_ISL_2555266, EPI_ISL_2555268, EPI_ISL_2555269, EPI_ISL_2555270, EPI_ISL_2555271,
EPI_ISL_2555272, EPI_ISL_2555273, EPI_ISL_2555274, EPI_ISL_2555275, EPI_ISL_2555276, EPI_ISL_2555277, EPI_ISL_2555278, EPI_ISL_2555279, EPI_ISL_2555280, EPI_ISL_2555281, EPI_ISL_2555282, EPI_ISL_2555283, EPI_ISL_2555284, EPI_ISL_2555285, EPI_ISL_2555286, EPI_ISL_2555287, EPI_ISL_2555288, EPI_ISL_2555289, EPI_ISL_2555290, EPI_ISL_2555291, EPI_ISL_2555292, EPI_ISL_2555293,
EPI_ISL_2555294, EPI_ISL_2555295, EPI_ISL_2555296, EPI_ISL_2555297, EPI_ISL_2555298, EPI_ISL_2555299, EPI_ISL_2555300, EPI_ISL_2555301, EPI_ISL_2555302, EPI_ISL_2555303, EPI_ISL_2555304, EPI_ISL_2555305, EPI_ISL_2555306, EPI_ISL_2555307, EPI_ISL_2555308, EPI_ISL_2555309, EPI_ISL_2555310, EPI_ISL_2555311, EPI_ISL_2555312, EPI_ISL_2555313, EPI_ISL_2555314, EPI_ISL_2555315,
EPI_ISL_2555316, EPI_ISL_2555317, EPI_ISL_2555318, EPI_ISL_2555319, EPI_ISL_2555320, EPI_ISL_2555321, EPI_ISL_2555322, EPI_ISL_2555323, EPI_ISL_2555324, EPI_ISL_2555325, EPI_ISL_2555326, EPI_ISL_2555327, EPI_ISL_2555328, EPI_ISL_2555329, EPI_ISL_2555330, EPI_ISL_2555331, EPI_ISL_2555332, EPI_ISL_2555333, EPI_ISL_2555334, EPI_ISL_2555335, EPI_ISL_2555336, EPI_ISL_2555337,
EPI_ISL_2555338, EPI_ISL_2555339, EPI_ISL_2555340, EPI_ISL_2555341, EPI_ISL_2555342, EPI_ISL_2555343, EPI_ISL_2555344, EPI_ISL_2555345, EPI_ISL_2555346, EPI_ISL_2555347, EPI_ISL_2555348, EPI_ISL_2555349, EPI_ISL_2555350, EPI_ISL_2555351, EPI_ISL_2555352, EPI_ISL_2555353, EPI_ISL_2555354, EPI_ISL_2555355, EPI_ISL_2555356, EPI_ISL_2555357, EPI_ISL_2555358, EPI_ISL_2555359,
EPI_ISL_2555360, EPI_ISL_2555361, EPI_ISL_2555362, EPI_ISL_2555363, EPI_ISL_2555364, EPI_ISL_2555365, EPI_ISL_2555366, EPI_ISL_2555367, EPI_ISL_2555368, EPI_ISL_2555369, EPI_ISL_2555370, EPI_ISL_2555371, EPI_ISL_2555372, EPI_ISL_2555373, EPI_ISL_2555374, EPI_ISL_2555375, EPI_ISL_2555376, EPI_ISL_2555377, EPI_ISL_2555378, EPI_ISL_2555379, EPI_ISL_2555380, EPI_ISL_2555381,
EPI_ISL_2555382, EPI_ISL_2555383, EPI_ISL_2555384, EPI_ISL_2555385, EPI_ISL_2555386, EPI_ISL_2555387, EPI_ISL_2555388, EPI_ISL_2555389, EPI_ISL_2555390, EPI_ISL_2555391, EPI_ISL_2555392, EPI_ISL_2555393, EPI_ISL_2555394, EPI_ISL_2555395, EPI_ISL_2555396, EPI_ISL_2555397, EPI_ISL_2555398, EPI_ISL_2555399, EPI_ISL_2555400, EPI_ISL_2555401, EPI_ISL_2555402, EPI_ISL_2555403,
EPI_ISL_2555404, EPI_ISL_2555405, EPI_ISL_2555406, EPI_ISL_2555407, EPI_ISL_2555408, EPI_ISL_2555409, EPI_ISL_2555410, EPI_ISL_2555411, EPI_ISL_2555412, EPI_ISL_2555413, EPI_ISL_2555414, EPI_ISL_2555415, EPI_ISL_2555416, EPI_ISL_2555417, EPI_ISL_2555418, EPI_ISL_2555419, EPI_ISL_2555420, EPI_ISL_2555421, EPI_ISL_2555422, EPI_ISL_2555423, EPI_ISL_2555424, EPI_ISL_2555425,
EPI_ISL_2555426, EPI_ISL_2555427, EPI_ISL_2555428, EPI_ISL_2555429, EPI_ISL_2555430, EPI_ISL_2555431, EPI_ISL_2555432, EPI_ISL_2555433, EPI_ISL_2555434, EPI_ISL_2555435, EPI_ISL_2555436, EPI_ISL_2555437, EPI_ISL_2555438, EPI_ISL_2555439, EPI_ISL_2555440, EPI_ISL_2555441, EPI_ISL_2555442, EPI_ISL_2555443, EPI_ISL_2555444, EPI_ISL_2555445, EPI_ISL_2555446, EPI_ISL_2555447,
EPI_ISL_2555448, EPI_ISL_2555449, EPI_ISL_2555450, EPI_ISL_2555451, EPI_ISL_2555452, EPI_ISL_2555453, EPI_ISL_2555454, EPI_ISL_2555455, EPI_ISL_2555456, EPI_ISL_2555457, EPI_ISL_2555458, EPI_ISL_2555459, EPI_ISL_2555460, EPI_ISL_2555461, EPI_ISL_2555462, EPI_ISL_2555463, EPI_ISL_2555464, EPI_ISL_2555465, EPI_ISL_2555466, EPI_ISL_2555467, EPI_ISL_2555468, EPI_ISL_2555469,
EPI_ISL_2555470, EPI_ISL_2555471, EPI_ISL_2555472, EPI_ISL_2555473, EPI_ISL_2555474, EPI_ISL_2555475, EPI_ISL_2555476, EPI_ISL_2555477, EPI_ISL_2555478, EPI_ISL_2555479, EPI_ISL_2555480, EPI_ISL_2555481, EPI_ISL_2555482, EPI_ISL_2555483, EPI_ISL_2555484, EPI_ISL_2555485, EPI_ISL_2555486, EPI_ISL_2555487, EPI_ISL_2555488, EPI_ISL_2555489, EPI_ISL_2555490, EPI_ISL_2555491,
EPI_ISL_2555492, EPI_ISL_2555493, EPI_ISL_2555494, EPI_ISL_2555495, EPI_ISL_2555496, EPI_ISL_2555497, EPI_ISL_2555498, EPI_ISL_2555499, EPI_ISL_2555500, EPI_ISL_2555501, EPI_ISL_2555502, EPI_ISL_2555503, EPI_ISL_2555504, EPI_ISL_2555505, EPI_ISL_2555506, EPI_ISL_2555507, EPI_ISL_2555508, EPI_ISL_2555509, EPI_ISL_2555510, EPI_ISL_2555511, EPI_ISL_2555512, EPI_ISL_2555513,
EPI_ISL_2555514, EPI_ISL_2555515, EPI_ISL_2555516, EPI_ISL_2555517, EPI_ISL_2555518, EPI_ISL_2555519, EPI_ISL_2555520, EPI_ISL_2555521, EPI_ISL_2555522, EPI_ISL_2555523, EPI_ISL_2569092, EPI_ISL_2570277, EPI_ISL_2570291, EPI_ISL_2570292, EPI_ISL_2570373, EPI_ISL_2570436, EPI_ISL_2570441, EPI_ISL_2570443, EPI_ISL_2570445, EPI_ISL_2607229, EPI_ISL_2607274, EPI_ISL_2607280,
EPI_ISL_2607312, EPI_ISL_2607329, EPI_ISL_2607337, EPI_ISL_2607356, EPI_ISL_2607953, EPI_ISL_2607956, EPI_ISL_2607959, EPI_ISL_2607960, EPI_ISL_2607961, EPI_ISL_2607962, EPI_ISL_2607963, EPI_ISL_2607964, EPI_ISL_2607966, EPI_ISL_2607968, EPI_ISL_2607969, EPI_ISL_2607973, EPI_ISL_2607974, EPI_ISL_2607975, EPI_ISL_2607976, EPI_ISL_2607977, EPI_ISL_2607978, EPI_ISL_2607980,
EPI_ISL_2607982, EPI_ISL_2607983, EPI_ISL_2607984, EPI_ISL_2607986, EPI_ISL_2607988, EPI_ISL_2607990, EPI_ISL_2607991, EPI_ISL_2607992, EPI_ISL_2607993, EPI_ISL_2607994, EPI_ISL_2607995, EPI_ISL_2607996, EPI_ISL_2607997, EPI_ISL_2607998, EPI_ISL_2607999, EPI_ISL_2608000, EPI_ISL_2608003, EPI_ISL_2608006, EPI_ISL_2608009, EPI_ISL_2608010, EPI_ISL_2608011, EPI_ISL_2608013,
EPI_ISL_2608015, EPI_ISL_2608016, EPI_ISL_2608017, EPI_ISL_2608018, EPI_ISL_2608019, EPI_ISL_2608021, EPI_ISL_2608022, EPI_ISL_2608024, EPI_ISL_2608026, EPI_ISL_2608028, EPI_ISL_2608029, EPI_ISL_2608036, EPI_ISL_2608037, EPI_ISL_2608038, EPI_ISL_2608039, EPI_ISL_2608040, EPI_ISL_2608041, EPI_ISL_2608042, EPI_ISL_2608043, EPI_ISL_2608045, EPI_ISL_2608047, EPI_ISL_2608048,
EPI_ISL_2608049, EPI_ISL_2608050, EPI_ISL_2608051, EPI_ISL_2608053, EPI_ISL_2608054, EPI_ISL_2608058, EPI_ISL_2608059, EPI_ISL_2608060, EPI_ISL_2608061, EPI_ISL_2608064, EPI_ISL_2608065, EPI_ISL_2608066, EPI_ISL_2608067, EPI_ISL_2608072, EPI_ISL_2608074, EPI_ISL_2608075, EPI_ISL_2608077, EPI_ISL_2608078, EPI_ISL_2608079, EPI_ISL_2608084, EPI_ISL_2608085, EPI_ISL_2608088,
EPI_ISL_2608090, EPI_ISL_2608091, EPI_ISL_2608092, EPI_ISL_2608093, EPI_ISL_2608095, EPI_ISL_2608096, EPI_ISL_2608098, EPI_ISL_2608101, EPI_ISL_2608103, EPI_ISL_2608104, EPI_ISL_2608107, EPI_ISL_2608108, EPI_ISL_2608117, EPI_ISL_2640678, EPI_ISL_2640713, EPI_ISL_2640726, EPI_ISL_2640740, EPI_ISL_2640744, EPI_ISL_2640765, EPI_ISL_2640823, EPI_ISL_2640830, EPI_ISL_2640931,

EPI_ISL_2640933, EPI_ISL_2640935, EPI_ISL_2640955, EPI_ISL_2640977, EPI_ISL_2640992

| | | | |
|---|---|---|---|
| see above | Lighthouse Lab in Glasgow | Wellcome Sanger Institute for the COVID-19 Genomics UK (COG-UK) Consortium | Anna Dominiczak and Alex Alderton; Carol Clugston; Cordelia Langford; David Gray; David K. Jackson; Dominic Kwiatkowski; Ewan Harrison; Harper VanSteenhouse; Ian Johnston; Jeffrey Barrett; John Sillitoe on behalf of the Wellcome Sanger Institute COVID-19 Surveillance Team; Roberto Amato; Sonia Goncalves; Yumi Kasai |

EPI_ISL_2485541, EPI_ISL_2485544, EPI_ISL_2485550, EPI_ISL_2485567, EPI_ISL_2485584, EPI_ISL_2485602, EPI_ISL_2485608, EPI_ISL_2485612, EPI_ISL_2485618, EPI_ISL_2485619, EPI_ISL_2485647, EPI_ISL_2485677, EPI_ISL_2485690, EPI_ISL_2485692, EPI_ISL_2485693, EPI_ISL_2485697, EPI_ISL_2485711, EPI_ISL_2485712, EPI_ISL_2485714, EPI_ISL_2485749, EPI_ISL_2485756, EPI_ISL_2485762, EPI_ISL_2485765, EPI_ISL_2485767, EPI_ISL_2485795, EPI_ISL_2485807, EPI_ISL_2485820, EPI_ISL_2485830, EPI_ISL_2485835, EPI_ISL_2510872, EPI_ISL_2510873, EPI_ISL_2510876, EPI_ISL_2510878, EPI_ISL_2510879, EPI_ISL_2510884, EPI_ISL_2510887, EPI_ISL_2510889, EPI_ISL_2510890, EPI_ISL_2510892, EPI_ISL_2510896, EPI_ISL_2510898, EPI_ISL_2510899, EPI_ISL_2510901, EPI_ISL_2510906, EPI_ISL_2510907, EPI_ISL_2510909, EPI_ISL_2510913, EPI_ISL_2510914, EPI_ISL_2510916, EPI_ISL_2510917, EPI_ISL_2510919, EPI_ISL_2510920, EPI_ISL_2510922, EPI_ISL_2510923, EPI_ISL_2510925, EPI_ISL_2510926, EPI_ISL_2510927, EPI_ISL_2510929, EPI_ISL_2510935, EPI_ISL_2510937, EPI_ISL_2510940, EPI_ISL_2510941, EPI_ISL_2510942, EPI_ISL_2510946, EPI_ISL_2510947, EPI_ISL_2510949, EPI_ISL_2510950, EPI_ISL_2510952, EPI_ISL_2510955, EPI_ISL_2510958, EPI_ISL_2510959, EPI_ISL_2510961, EPI_ISL_2510963, EPI_ISL_2510966, EPI_ISL_2510968, EPI_ISL_2510969, EPI_ISL_2510971, EPI_ISL_2510972, EPI_ISL_2510974, EPI_ISL_2510975, EPI_ISL_2510977, EPI_ISL_2510978, EPI_ISL_2510980, EPI_ISL_2510981, EPI_ISL_2510983, EPI_ISL_2510984, EPI_ISL_2510986, EPI_ISL_2510987, EPI_ISL_2510989, EPI_ISL_2510990, EPI_ISL_2510993, EPI_ISL_2510994, EPI_ISL_2510995, EPI_ISL_2510998, EPI_ISL_2511000, EPI_ISL_2511001, EPI_ISL_2511003, EPI_ISL_2511004, EPI_ISL_2511006, EPI_ISL_2511007, EPI_ISL_2511009, EPI_ISL_2511010, EPI_ISL_2511013, EPI_ISL_2511016, EPI_ISL_2511018, EPI_ISL_2511020, EPI_ISL_2511022, EPI_ISL_2511023, EPI_ISL_2511024, EPI_ISL_2511025, EPI_ISL_2511026, EPI_ISL_2511027, EPI_ISL_2511029, EPI_ISL_2511030, EPI_ISL_2511031, EPI_ISL_2511032, EPI_ISL_2511033, EPI_ISL_2511034, EPI_ISL_2511035, EPI_ISL_2511036, EPI_ISL_2511038, EPI_ISL_2511039, EPI_ISL_2511040, EPI_ISL_2511041, EPI_ISL_2511043, EPI_ISL_2511045, EPI_ISL_2511046, EPI_ISL_2511047, EPI_ISL_2511048, EPI_ISL_2511049, EPI_ISL_2511050, EPI_ISL_2511052, EPI_ISL_2511053, EPI_ISL_2511054, EPI_ISL_2511055, EPI_ISL_2511056, EPI_ISL_2511057, EPI_ISL_2511058, EPI_ISL_2511059, EPI_ISL_2511060, EPI_ISL_2511063, EPI_ISL_2511064, EPI_ISL_2511065, EPI_ISL_2511066, EPI_ISL_2511068, EPI_ISL_2511069, EPI_ISL_2511071, EPI_ISL_2511072, EPI_ISL_2511073, EPI_ISL_2511074, EPI_ISL_2511075, EPI_ISL_2511076, EPI_ISL_2511077, EPI_ISL_2511078, EPI_ISL_2511079, EPI_ISL_2511082, EPI_ISL_2511084, EPI_ISL_2511085, EPI_ISL_2511086, EPI_ISL_2511089, EPI_ISL_2511090, EPI_ISL_2511091, EPI_ISL_2511092, EPI_ISL_2511095, EPI_ISL_2511352, EPI_ISL_2511353, EPI_ISL_2511354, EPI_ISL_2511355, EPI_ISL_2511357, EPI_ISL_2511358, EPI_ISL_2511363, EPI_ISL_2511364, EPI_ISL_2511365, EPI_ISL_2511368, EPI_ISL_2511369, EPI_ISL_2511371, EPI_ISL_2511372, EPI_ISL_2511375, EPI_ISL_2511380, EPI_ISL_2511381, EPI_ISL_2511382, EPI_ISL_2511387, EPI_ISL_2511388, EPI_ISL_2511392, EPI_ISL_2511396, EPI_ISL_2511399, EPI_ISL_2511401, EPI_ISL_2511402, EPI_ISL_2511412, EPI_ISL_2511413, EPI_ISL_2511420, EPI_ISL_2511422, EPI_ISL_2511428, EPI_ISL_2511429, EPI_ISL_2511431, EPI_ISL_2511432, EPI_ISL_2511435, EPI_ISL_2511438, EPI_ISL_2511440, EPI_ISL_2511444, EPI_ISL_2511445, EPI_ISL_2511447, EPI_ISL_2511448, EPI_ISL_2511452, EPI_ISL_2511454, EPI_ISL_2511455, EPI_ISL_2511457, EPI_ISL_2511458, EPI_ISL_2511460, EPI_ISL_2511464, EPI_ISL_2511465, EPI_ISL_2511466, EPI_ISL_2511470, EPI_ISL_2511472, EPI_ISL_2511473, EPI_ISL_2511474, EPI_ISL_2511475, EPI_ISL_2511476, EPI_ISL_2511477, EPI_ISL_2511478, EPI_ISL_2511485, EPI_ISL_2511486, EPI_ISL_2511487, EPI_ISL_2511488, EPI_ISL_2511490, EPI_ISL_2511491, EPI_ISL_2511494, EPI_ISL_2511495, EPI_ISL_2511496, EPI_ISL_2511506, EPI_ISL_2511507, EPI_ISL_2511508, EPI_ISL_2511509, EPI_ISL_2511512, EPI_ISL_2511513, EPI_ISL_2511515, EPI_ISL_2511517, EPI_ISL_2511519, EPI_ISL_2511520, EPI_ISL_2511521, EPI_ISL_2511538, EPI_ISL_2511542, EPI_ISL_2511544, EPI_ISL_2511549, EPI_ISL_2511550, EPI_ISL_2511555, EPI_ISL_2511556, EPI_ISL_2511557, EPI_ISL_2511559, EPI_ISL_2511560, EPI_ISL_2511568, EPI_ISL_2511572, EPI_ISL_2511576, EPI_ISL_2511577, EPI_ISL_2511578, EPI_ISL_2511579, EPI_ISL_2511584, EPI_ISL_2511588, EPI_ISL_2511589, EPI_ISL_2511594, EPI_ISL_2511597, EPI_ISL_2511600, EPI_ISL_2511601, EPI_ISL_2511603, EPI_ISL_2511605, EPI_ISL_2511607, EPI_ISL_2511608, EPI_ISL_2511611, EPI_ISL_2511617, EPI_ISL_2511619, EPI_ISL_2511620, EPI_ISL_2511622, EPI_ISL_2511624, EPI_ISL_2511625, EPI_ISL_2511630, EPI_ISL_2511633, EPI_ISL_2511636, EPI_ISL_2511637, EPI_ISL_2511640, EPI_ISL_2511641, EPI_ISL_2511647, EPI_ISL_2511649, EPI_ISL_2511654, EPI_ISL_2511655, EPI_ISL_2511656, EPI_ISL_2511660, EPI_ISL_2511661, EPI_ISL_2511662, EPI_ISL_2511663, EPI_ISL_2511664, EPI_ISL_2511666, EPI_ISL_2511667, EPI_ISL_2511668, EPI_ISL_2511669, EPI_ISL_2511670, EPI_ISL_2511671, EPI_ISL_2511672, EPI_ISL_2511673, EPI_ISL_2511674, EPI_ISL_2511675, EPI_ISL_2511676, EPI_ISL_2511678, EPI_ISL_2511682, EPI_ISL_2511686, EPI_ISL_2511688, EPI_ISL_2511691, EPI_ISL_2511692, EPI_ISL_2511693, EPI_ISL_2511695, EPI_ISL_2511698, EPI_ISL_2511700, EPI_ISL_2511702, EPI_ISL_2511703, EPI_ISL_2511710, EPI_ISL_2511712, EPI_ISL_2511715, EPI_ISL_2511719, EPI_ISL_2511722, EPI_ISL_2511724, EPI_ISL_2511725, EPI_ISL_2511727, EPI_ISL_2511730, EPI_ISL_2511733, EPI_ISL_2511734, EPI_ISL_2511736, EPI_ISL_2511737, EPI_ISL_2511738, EPI_ISL_2511739, EPI_ISL_2511740, EPI_ISL_2511741, EPI_ISL_2511745, EPI_ISL_2511747, EPI_ISL_2511748, EPI_ISL_2511749, EPI_ISL_2511751, EPI_ISL_2511752, EPI_ISL_2511753, EPI_ISL_2511756, EPI_ISL_2511758, EPI_ISL_2511759, EPI_ISL_2511760, EPI_ISL_2511766, EPI_ISL_2511767, EPI_ISL_2511768, EPI_ISL_2511770, EPI_ISL_2511771, EPI_ISL_2511772, EPI_ISL_2511774, EPI_ISL_2511775, EPI_ISL_2511777, EPI_ISL_2511780, EPI_ISL_2511782, EPI_ISL_2511783, EPI_ISL_2511785, EPI_ISL_2511788, EPI_ISL_2511791, EPI_ISL_2511794, EPI_ISL_2511795, EPI_ISL_2511796, EPI_ISL_2511797, EPI_ISL_2511798, EPI_ISL_2511800, EPI_ISL_2511801, EPI_ISL_2511802, EPI_ISL_2511803, EPI_ISL_2511805, EPI_ISL_2511806, EPI_ISL_2511807, EPI_ISL_2511808, EPI_ISL_2511809, EPI_ISL_2511812, EPI_ISL_2511815, EPI_ISL_2511817, EPI_ISL_2511818, EPI_ISL_2511822, EPI_ISL_2511823, EPI_ISL_2511826, EPI_ISL_2511827, EPI_ISL_2511830, EPI_ISL_2511831, EPI_ISL_2511835, EPI_ISL_2511837, EPI_ISL_2511839, EPI_ISL_2511840, EPI_ISL_2511841, EPI_ISL_2511843, EPI_ISL_2511845, EPI_ISL_2511849, EPI_ISL_2511850, EPI_ISL_2511855, EPI_ISL_2511857, EPI_ISL_2511859, EPI_ISL_2511860, EPI_ISL_2511861, EPI_ISL_2511865, EPI_ISL_2511866, EPI_ISL_2511868, EPI_ISL_2511872, EPI_ISL_2511876, EPI_ISL_2511877, EPI_ISL_2511878, EPI_ISL_2511879, EPI_ISL_2511881, EPI_ISL_2511882, EPI_ISL_2511883, EPI_ISL_2511884, EPI_ISL_2511885, EPI_ISL_2511888, EPI_ISL_2511891, EPI_ISL_2511892, EPI_ISL_2511896, EPI_ISL_2511899, EPI_ISL_2511901, EPI_ISL_2511903, EPI_ISL_2511908, EPI_ISL_2511911, EPI_ISL_2511912, EPI_ISL_2511913, EPI_ISL_2511914, EPI_ISL_2511917, EPI_ISL_2511918, EPI_ISL_2511919, EPI_ISL_2511922, EPI_ISL_2511924, EPI_ISL_2511925, EPI_ISL_2511926, EPI_ISL_2511929, EPI_ISL_2511930, EPI_ISL_2511931, EPI_ISL_2511932, EPI_ISL_2511933, EPI_ISL_2512375, EPI_ISL_2512376, EPI_ISL_2512377, EPI_ISL_2512378, EPI_ISL_2512379, EPI_ISL_2512380, EPI_ISL_2512381, EPI_ISL_2512382, EPI_ISL_2512383, EPI_ISL_2512387, EPI_ISL_2512388, EPI_ISL_2512390, EPI_ISL_2512391, EPI_ISL_2512392, EPI_ISL_2512394, EPI_ISL_2512396, EPI_ISL_2512398, EPI_ISL_2512399, EPI_ISL_2512400, EPI_ISL_2512401, EPI_ISL_2512402, EPI_ISL_2512403, EPI_ISL_2512405, EPI_ISL_2512406, EPI_ISL_2512407, EPI_ISL_2512408, EPI_ISL_2512409, EPI_ISL_2512411, EPI_ISL_2512413, EPI_ISL_2512414, EPI_ISL_2512415, EPI_ISL_2512416, EPI_ISL_2512417, EPI_ISL_2512418, EPI_ISL_2512419, EPI_ISL_2512420, EPI_ISL_2512421, EPI_ISL_2512422, EPI_ISL_2512423, EPI_ISL_2512424, EPI_ISL_2512426, EPI_ISL_2512427, EPI_ISL_2512428, EPI_ISL_2512429, EPI_ISL_2512431, EPI_ISL_2512432, EPI_ISL_2512433, EPI_ISL_2512434, EPI_ISL_2512437, EPI_ISL_2512438, EPI_ISL_2512440, EPI_ISL_2512442, EPI_ISL_2512443, EPI_ISL_2512445, EPI_ISL_2512446, EPI_ISL_2512447, EPI_ISL_2512448, EPI_ISL_2512449, EPI_ISL_2512450, EPI_ISL_2512451, EPI_ISL_2512452, EPI_ISL_2512455, EPI_ISL_2512458, EPI_ISL_2512459, EPI_ISL_2512460, EPI_ISL_2512461, EPI_ISL_2512462, EPI_ISL_2512464, EPI_ISL_2512465, EPI_ISL_2512466, EPI_ISL_2512468, EPI_ISL_2512471, EPI_ISL_2512472, EPI_ISL_2512473, EPI_ISL_2512474, EPI_ISL_2512475, EPI_ISL_2512476, EPI_ISL_2512477, EPI_ISL_2512478, EPI_ISL_2512479, EPI_ISL_2512481, EPI_ISL_2512482, EPI_ISL_2512483, EPI_ISL_2512484, EPI_ISL_2512485, EPI_ISL_2512487, EPI_ISL_2512488, EPI_ISL_2512489, EPI_ISL_2512490, EPI_ISL_2512491, EPI_ISL_2512493, EPI_ISL_2512494, EPI_ISL_2512495, EPI_ISL_2512496, EPI_ISL_2512498, EPI_ISL_2512499, EPI_ISL_2512500, EPI_ISL_2512501, EPI_ISL_2512502, EPI_ISL_2512503, EPI_ISL_2512504, EPI_ISL_2512505, EPI_ISL_2512508, EPI_ISL_2512509, EPI_ISL_2512510, EPI_ISL_2512511, EPI_ISL_2512512, EPI_ISL_2512513, EPI_ISL_2512515, EPI_ISL_2512516, EPI_ISL_2512517, EPI_ISL_2512518, EPI_ISL_2512519, EPI_ISL_2512521, EPI_ISL_2512522, EPI_ISL_2512523, EPI_ISL_2512524, EPI_ISL_2512526, EPI_ISL_2512527, EPI_ISL_2512529, EPI_ISL_2512530, EPI_ISL_2512531, EPI_ISL_2512533, EPI_ISL_2512534, EPI_ISL_2512535, EPI_ISL_2512536, EPI_ISL_2512537, EPI_ISL_2512538, EPI_ISL_2512539, EPI_ISL_2512541, EPI_ISL_2512542, EPI_ISL_2512543, EPI_ISL_2512544, EPI_ISL_2512545, EPI_ISL_2512546, EPI_ISL_2512548, EPI_ISL_2512549, EPI_ISL_2512551, EPI_ISL_2512552, EPI_ISL_2512553, EPI_ISL_2512554, EPI_ISL_2512555, EPI_ISL_2512556, EPI_ISL_2512557, EPI_ISL_2512558, EPI_ISL_2512559, EPI_ISL_2512560, EPI_ISL_2512561, EPI_ISL_2512562, EPI_ISL_2512563, EPI_ISL_2512564, EPI_ISL_2512565, EPI_ISL_2512566, EPI_ISL_2512567, EPI_ISL_2512568, EPI_ISL_2512569, EPI_ISL_2512570, EPI_ISL_2512571, EPI_ISL_2512572, EPI_ISL_2512573, EPI_ISL_2512574, EPI_ISL_2512576, EPI_ISL_2512577, EPI_ISL_2512578, EPI_ISL_2512579, EPI_ISL_2512580, EPI_ISL_2512581, EPI_ISL_2512582, EPI_ISL_2512583, EPI_ISL_2512584, EPI_ISL_2512585, EPI_ISL_2512586, EPI_ISL_2512588, EPI_ISL_2512590, EPI_ISL_2512591, EPI_ISL_2512592, EPI_ISL_2512593, EPI_ISL_2512594, EPI_ISL_2512595, EPI_ISL_2512596, EPI_ISL_2512597, EPI_ISL_2512598, EPI_ISL_2512599, EPI_ISL_2512602, EPI_ISL_2512603, EPI_ISL_2512604, EPI_ISL_2512605, EPI_ISL_2512607, EPI_ISL_2512608, EPI_ISL_2512609, EPI_ISL_2512610, EPI_ISL_2512611, EPI_ISL_2512612, EPI_ISL_2512613, EPI_ISL_2512614, EPI_ISL_2512615, EPI_ISL_2512616, EPI_ISL_2512617, EPI_ISL_2512618, EPI_ISL_2512619, EPI_ISL_2512620, EPI_ISL_2512621, EPI_ISL_2512622, EPI_ISL_2512623, EPI_ISL_2512624, EPI_ISL_2512625, EPI_ISL_2512626, EPI_ISL_2512627, EPI_ISL_2512628, EPI_ISL_2512629, EPI_ISL_2512630, EPI_ISL_2512631, EPI_ISL_2512633, EPI_ISL_2512634, EPI_ISL_2512635, EPI_ISL_2512636, EPI_ISL_2512637, EPI_ISL_2512638, EPI_ISL_2512639, EPI_ISL_2512640, EPI_ISL_2512642, EPI_ISL_2512643, EPI_ISL_2512644, EPI_ISL_2512645, EPI_ISL_2512647, EPI_ISL_2512650, EPI_ISL_2512651, EPI_ISL_2512652, EPI_ISL_2512653, EPI_ISL_2512654, EPI_ISL_2512655, EPI_ISL_2512656, EPI_ISL_2512657, EPI_ISL_2512658, EPI_ISL_2512659, EPI_ISL_2512660, EPI_ISL_2512661, EPI_ISL_2512662, EPI_ISL_2512663, EPI_ISL_2512664, EPI_ISL_2512665, EPI_ISL_2512667, EPI_ISL_2512669, EPI_ISL_2512670, EPI_ISL_2512671, EPI_ISL_2512672, EPI_ISL_2512673, EPI_ISL_2512674, EPI_ISL_2512675, EPI_ISL_2512676, EPI_ISL_2512677, EPI_ISL_2512678, EPI_ISL_2512679, EPI_ISL_2512680, EPI_ISL_2512681, EPI_ISL_2512682, EPI_ISL_2512683, EPI_ISL_2512685, EPI_ISL_2512686, EPI_ISL_2512690, EPI_ISL_2512693, EPI_ISL_2512695, EPI_ISL_2512696, EPI_ISL_2512697, EPI_ISL_2512700, EPI_ISL_2512701, EPI_ISL_2512702, EPI_ISL_2512703, EPI_ISL_2512704, EPI_ISL_2512706, EPI_ISL_2512708, EPI_ISL_2512709, EPI_ISL_2512712, EPI_ISL_2512715, EPI_ISL_2512716, EPI_ISL_2512717, EPI_ISL_2512718, EPI_ISL_2512719, EPI_ISL_2512720, EPI_ISL_2512722, EPI_ISL_2512723, EPI_ISL_2512724, EPI_ISL_2512725, EPI_ISL_2512726, EPI_ISL_2512727, EPI_ISL_2512728, EPI_ISL_2512729, EPI_ISL_2512731, EPI_ISL_2512733, EPI_ISL_2512734, EPI_ISL_2512735, EPI_ISL_2512736, EPI_ISL_2512738, EPI_ISL_2512739, EPI_ISL_2512740, EPI_ISL_2512741, EPI_ISL_2512742, EPI_ISL_2512743, EPI_ISL_2512744, EPI_ISL_2512745, EPI_ISL_2512746, EPI_ISL_2512747, EPI_ISL_2512749, EPI_ISL_2512750, EPI_ISL_2512751, EPI_ISL_2512752, EPI_ISL_2512753, EPI_ISL_2512756, EPI_ISL_2512758, EPI_ISL_2512759, EPI_ISL_2512761, EPI_ISL_2512762, EPI_ISL_2512763, EPI_ISL_2512765, EPI_ISL_2512768, EPI_ISL_2512769, EPI_ISL_2512771, EPI_ISL_2512773, EPI_ISL_2512774, EPI_ISL_2512776, EPI_ISL_2512777, EPI_ISL_2512779, EPI_ISL_2512780, EPI_ISL_2512782, EPI_ISL_2512783, EPI_ISL_2512784, EPI_ISL_2512785, EPI_ISL_2512787, EPI_ISL_2512788, EPI_ISL_2512790, EPI_ISL_2512793, EPI_ISL_2512795, EPI_ISL_2512796, EPI_ISL_2512797, EPI_ISL_2512798, EPI_ISL_2512799, EPI_ISL_2512800, EPI_ISL_2512801, EPI_ISL_2512803, EPI_ISL_2512805, EPI_ISL_2512806, EPI_ISL_2512808, EPI_ISL_2512811, EPI_ISL_2512813, EPI_ISL_2512814, EPI_ISL_2512816, EPI_ISL_2512817, EPI_ISL_2512818, EPI_ISL_2512819, EPI_ISL_2512820, EPI_ISL_2512821, EPI_ISL_2512822, EPI_ISL_2512823, EPI_ISL_2512824, EPI_ISL_2512825, EPI_ISL_2512826, EPI_ISL_2512827, EPI_ISL_2512828, EPI_ISL_2512830, EPI_ISL_2512831, EPI_ISL_2512832, EPI_ISL_2512833, EPI_ISL_2512834, EPI_ISL_2512835, EPI_ISL_2512836, EPI_ISL_2512837, EPI_ISL_2512838, EPI_ISL_2512839, EPI_ISL_2512840, EPI_ISL_2512841, EPI_ISL_2512842, EPI_ISL_2512843, EPI_ISL_2512845, EPI_ISL_2512846, EPI_ISL_2512847, EPI_ISL_2512848, EPI_ISL_2512849, EPI_ISL_2512850, EPI_ISL_2512852, EPI_ISL_2512853, EPI_ISL_2512855, EPI_ISL_2512857, EPI_ISL_2512858, EPI_ISL_2512860, EPI_ISL_2512864, EPI_ISL_2512866, EPI_ISL_2512867, EPI_ISL_2512871, EPI_ISL_2512872, EPI_ISL_2512873, EPI_ISL_2512874, EPI_ISL_2512875, EPI_ISL_2512877, EPI_ISL_2512878, EPI_ISL_2512879, EPI_ISL_2512880, EPI_ISL_2512881, EPI_ISL_2512883, EPI_ISL_2512886, EPI_ISL_2512888, EPI_ISL_2512891, EPI_ISL_2512892, EPI_ISL_2512893, EPI_ISL_2512894, EPI_ISL_2512895, EPI_ISL_2512896, EPI_ISL_2512899, EPI_ISL_2512900, EPI_ISL_2512903, EPI_ISL_2512905, EPI_ISL_2512906, EPI_ISL_2512909, EPI_ISL_2512910, EPI_ISL_2512911, EPI_ISL_2512912, EPI_ISL_2512913, EPI_ISL_2512916, EPI_ISL_2512917, EPI_ISL_2512918, EPI_ISL_2512920, EPI_ISL_2512921, EPI_ISL_2512922, EPI_ISL_2512925, EPI_ISL_2512927, EPI_ISL_2512929, EPI_ISL_2512932, EPI_ISL_2512933, EPI_ISL_2512936, EPI_ISL_2512940, EPI_ISL_2512948, EPI_ISL_2512952, EPI_ISL_2512962, EPI_ISL_2512966, EPI_ISL_2512969, EPI_ISL_2512984, EPI_ISL_2512990, EPI_ISL_2512997, EPI_ISL_2512998, EPI_ISL_2513006, EPI_ISL_2513007, EPI_ISL_2513014, EPI_ISL_2513018, EPI_ISL_2513022, EPI_ISL_2513026, EPI_ISL_2513030, EPI_ISL_2513031, EPI_ISL_2513032, EPI_ISL_2513034, EPI_ISL_2513036, EPI_ISL_2513039, EPI_ISL_2513047, EPI_ISL_2513049, EPI_ISL_2513050, EPI_ISL_2513063, EPI_ISL_2513075, EPI_ISL_2513080, EPI_ISL_2513083, EPI_ISL_2513090, EPI_ISL_2513091, EPI_ISL_2513093, EPI_ISL_2513094, EPI_ISL_2513095, EPI_ISL_2513105, EPI_ISL_2513107, EPI_ISL_2513109, EPI_ISL_2513114, EPI_ISL_2513115, EPI_ISL_2513119, EPI_ISL_2513123, EPI_ISL_2513132, EPI_ISL_2513134, EPI_ISL_2513147, EPI_ISL_2513152, EPI_ISL_2513156, EPI_ISL_2513158, EPI_ISL_2513167, EPI_ISL_2513186, EPI_ISL_2513201, EPI_ISL_2513202, EPI_ISL_2513209, EPI_ISL_2513215, EPI_ISL_2513221, EPI_ISL_2513225, EPI_ISL_2513241, EPI_ISL_2513246, EPI_ISL_2513248, EPI_ISL_2513250, EPI_ISL_2513257, EPI_ISL_2513258, EPI_ISL_2513263, EPI_ISL_2513264, EPI_ISL_2513267, EPI_ISL_2513275, EPI_ISL_2513277, EPI_ISL_2513279, EPI_ISL_2513283, EPI_ISL_2513289, EPI_ISL_2514102, EPI_ISL_2514103, EPI_ISL_2514116, EPI_ISL_2514118, EPI_ISL_2514121, EPI_ISL_2514125, EPI_ISL_2514126, EPI_ISL_2514129, EPI_ISL_2514134, EPI_ISL_2514135, EPI_ISL_2514136, EPI_ISL_2514144, EPI_ISL_2514152, EPI_ISL_2514162, EPI_ISL_2514168, EPI_ISL_2514174, EPI_ISL_2514181, EPI_ISL_2514183, EPI_ISL_2514190, EPI_ISL_2514193, EPI_ISL_2514203, EPI_ISL_2514218, EPI_ISL_2514222, EPI_ISL_2514236, EPI_ISL_2514250, EPI_ISL_2514253, EPI_ISL_2514254, EPI_ISL_2514268, EPI_ISL_2514269, EPI_ISL_2514284, EPI_ISL_2514291, EPI_ISL_2514292, EPI_ISL_2514298, EPI_ISL_2514300, EPI_ISL_2514304, EPI_ISL_2514310, EPI_ISL_2514311, EPI_ISL_2514314, EPI_ISL_2514323, EPI_ISL_2514327, EPI_ISL_2514330, EPI_ISL_2514347, EPI_ISL_2514354, EPI_ISL_2514356, EPI_ISL_2514357, EPI_ISL_2514359, EPI_ISL_2514365, EPI_ISL_2514366, EPI_ISL_2514370, EPI_ISL_2514375, EPI_ISL_2514376, EPI_ISL_2514379, EPI_ISL_2514383, EPI_ISL_2514386, EPI_ISL_2514389, EPI_ISL_2514395, EPI_ISL_2514396, EPI_ISL_2514401, EPI_ISL_2514403, EPI_ISL_2514728, EPI_ISL_2514730, EPI_ISL_2514742, EPI_ISL_2514744, EPI_ISL_2514746, EPI_ISL_2514747, EPI_ISL_2514750, EPI_ISL_2514754, EPI_ISL_2514755, EPI_ISL_2514756, EPI_ISL_2514757, EPI_ISL_2514759, EPI_ISL_2514760, EPI_ISL_2514762, EPI_ISL_2514763, EPI_ISL_2514764, EPI_ISL_2514767, EPI_ISL_2514768, EPI_ISL_2514778, EPI_ISL_2514781, EPI_ISL_2514783, EPI_ISL_2514784, EPI_ISL_2514785, EPI_ISL_2514786, EPI_ISL_2514791, EPI_ISL_2514792, EPI_ISL_2514795, EPI_ISL_2514796, EPI_ISL_2514797, EPI_ISL_2514798, EPI_ISL_2514800, EPI_ISL_2514803, EPI_ISL_2514804, EPI_ISL_2514805, EPI_ISL_2514809, EPI_ISL_2514810, EPI_ISL_2514812, EPI_ISL_2514813, EPI_ISL_2514815, EPI_ISL_2514816, EPI_ISL_2514817, EPI_ISL_2514819, EPI_ISL_2514826, EPI_ISL_2514829, EPI_ISL_2514830, EPI_ISL_2514834, EPI_ISL_2514837, EPI_ISL_2514844, EPI_ISL_2514845, EPI_ISL_2514850, EPI_ISL_2514854, EPI_ISL_2514857, EPI_ISL_2514860, EPI_ISL_2514861, EPI_ISL_2514864, EPI_ISL_2514866, EPI_ISL_2514867, EPI_ISL_2514868, EPI_ISL_2514869, EPI_ISL_2514871, EPI_ISL_2514873, EPI_ISL_2514874, EPI_ISL_2514876, EPI_ISL_2514880, EPI_ISL_2514881, EPI_ISL_2514882, EPI_ISL_2514883, EPI_ISL_2514885, EPI_ISL_2514891, EPI_ISL_2514892, EPI_ISL_2514893, EPI_ISL_2514895, EPI_ISL_2514896, EPI_ISL_2514897, EPI_ISL_2514900, EPI_ISL_2514901, EPI_ISL_2514905, EPI_ISL_2514906, EPI_ISL_2514907, EPI_ISL_2514908, EPI_ISL_2514909, EPI_ISL_2514914, EPI_ISL_2514915, EPI_ISL_2514916, EPI_ISL_2514919, EPI_ISL_2514922, EPI_ISL_2514925, EPI_ISL_2514926, EPI_ISL_2515400, EPI_ISL_2515404, EPI_ISL_2515405, EPI_ISL_2515408, EPI_ISL_2515410, EPI_ISL_2515413, EPI_ISL_2515415, EPI_ISL_2515416, EPI_ISL_2515420, EPI_ISL_2515426, EPI_ISL_2515431, EPI_ISL_2515433, EPI_ISL_2515437, EPI_ISL_2515439, EPI_ISL_2515442, EPI_ISL_2515444, EPI_ISL_2515451, EPI_ISL_2515457, EPI_ISL_2515458, EPI_ISL_2515459, EPI_ISL_2515461, EPI_ISL_2515466, EPI_ISL_2515467, EPI_ISL_2515470, EPI_ISL_2515471, EPI_ISL_2515473, EPI_ISL_2515480, EPI_ISL_2515481, EPI_ISL_2515485, EPI_ISL_2515486, EPI_ISL_2515489, EPI_ISL_2515490, EPI_ISL_2515498, EPI_ISL_2515499, EPI_ISL_2515505, EPI_ISL_2515508, EPI_ISL_2515510, EPI_ISL_2515513, EPI_ISL_2515518, EPI_ISL_2515519, EPI_ISL_2515520, EPI_ISL_2515521, EPI_ISL_2515523, EPI_ISL_2515531, EPI_ISL_2515532, EPI_ISL_2515534, EPI_ISL_2515536, EPI_ISL_2515537, EPI_ISL_2515539, EPI_ISL_2515559, EPI_ISL_2515561, EPI_ISL_2515567, EPI_ISL_2515568, EPI_ISL_2515570, EPI_ISL_2515575, EPI_ISL_2515576, EPI_ISL_2515578, EPI_ISL_2515582, EPI_ISL_2515587, EPI_ISL_2515590, EPI_ISL_2515592, EPI_ISL_2515595, EPI_ISL_2515597, EPI_ISL_2515604, EPI_ISL_2515610, EPI_ISL_2515611, EPI_ISL_2515612, EPI_ISL_2515613, EPI_ISL_2515615, EPI_ISL_2515616, EPI_ISL_2515619, EPI_ISL_2515624, EPI_ISL_2515631, EPI_ISL_2515634, EPI_ISL_2515635, EPI_ISL_2515637, EPI_ISL_2515638, EPI_ISL_2515641, EPI_ISL_2515642, EPI_ISL_2515643, EPI_ISL_2537507, EPI_ISL_2537520, EPI_ISL_2537524, EPI_ISL_2537540, EPI_ISL_2537553, EPI_ISL_2537586, EPI_ISL_2537592, EPI_ISL_2537601, EPI_ISL_2537618, EPI_ISL_2537640, EPI_ISL_2537661, EPI_ISL_2537666, EPI_ISL_2537676, EPI_ISL_2537680, EPI_ISL_2537683, EPI_ISL_2537761, EPI_ISL_2537769, EPI_ISL_2538642, EPI_ISL_2538672, EPI_ISL_2538745, EPI_ISL_2538777, EPI_ISL_2538816, EPI_ISL_2539736, EPI_ISL_2539737, EPI_ISL_2539738, EPI_ISL_2539739, EPI_ISL_2539742, EPI_ISL_2539743, EPI_ISL_2539745, EPI_ISL_2539748, EPI_ISL_2539749, EPI_ISL_2539752, EPI_ISL_2539754, EPI_ISL_2539757, EPI_ISL_2539758, EPI_ISL_2539759, EPI_ISL_2539760, EPI_ISL_2539762, EPI_ISL_2539766, EPI_ISL_2539767, EPI_ISL_2539768, EPI_ISL_2539769, EPI_ISL_2539770, EPI_ISL_2539772, EPI_ISL_2539773, EPI_ISL_2539774, EPI_ISL_2539776, EPI_ISL_2539777, EPI_ISL_2539778, EPI_ISL_2539780, EPI_ISL_2539787, EPI_ISL_2539788, EPI_ISL_2539789, EPI_ISL_2539791, EPI_ISL_2539793, EPI_ISL_2539794, EPI_ISL_2539797, EPI_ISL_2539798, EPI_ISL_2539799, EPI_ISL_2539801, EPI_ISL_2539803, EPI_ISL_2539804, EPI_ISL_2539805, EPI_ISL_2539807, EPI_ISL_2539808, EPI_ISL_2539809, EPI_ISL_2539810, EPI_ISL_2539815, EPI_ISL_2539819, EPI_ISL_2539820, EPI_ISL_2539821, EPI_ISL_2539824, EPI_ISL_2539825, EPI_ISL_2539826, EPI_ISL_2539827, EPI_ISL_2539829, EPI_ISL_2539830, EPI_ISL_2539831, EPI_ISL_2539832, EPI_ISL_2539835, EPI_ISL_2539836, EPI_ISL_2539838, EPI_ISL_2539840, EPI_ISL_2539843, EPI_ISL_2539845, EPI_ISL_2539846, EPI_ISL_2539847, EPI_ISL_2539848, EPI_ISL_2539849, EPI_ISL_2539850, EPI_ISL_2539851, EPI_ISL_2539855, EPI_ISL_2539856, EPI_ISL_2539857, EPI_ISL_2539860, EPI_ISL_2539865, EPI_ISL_2539866, EPI_ISL_2539867, EPI_ISL_2539868, EPI_ISL_2539869, EPI_ISL_2539870, EPI_ISL_2539871, EPI_ISL_2539872, EPI_ISL_2539875, EPI_ISL_2539876, EPI_ISL_2539880, EPI_ISL_2539881, EPI_ISL_2539885, EPI_ISL_2539887, EPI_ISL_2539889, EPI_ISL_2539891, EPI_ISL_2539892, EPI_ISL_2539894, EPI_ISL_2539897, EPI_ISL_2539899, EPI_ISL_2539903, EPI_ISL_2539904, EPI_ISL_2539906, EPI_ISL_2539909, EPI_ISL_2539912, EPI_ISL_2539913, EPI_ISL_2539915, EPI_ISL_2539916, EPI_ISL_2539917, EPI_ISL_2539918, EPI_ISL_2539919, EPI_ISL_2539920, EPI_ISL_2539922, EPI_ISL_2539923, EPI_ISL_2539925, EPI_ISL_2539926, EPI_ISL_2539928, EPI_ISL_2539930, EPI_ISL_2539933, EPI_ISL_2539934, EPI_ISL_2539936, EPI_ISL_2539938, EPI_ISL_2539942, EPI_ISL_2539945, EPI_ISL_2539946, EPI_ISL_2539947, EPI_ISL_2539949, EPI_ISL_2539953, EPI_ISL_2539954, EPI_ISL_2539955, EPI_ISL_2539956, EPI_ISL_2539958, EPI_ISL_2539959, EPI_ISL_2539960, EPI_ISL_2539961, EPI_ISL_2539964, EPI_ISL_2539966, EPI_ISL_2539968, EPI_ISL_2539970, EPI_ISL_2539972, EPI_ISL_2539973, EPI_ISL_2539975, EPI_ISL_2539976, EPI_ISL_2539978, EPI_ISL_2539980, EPI_ISL_2539981, EPI_ISL_2539983, EPI_ISL_2539984, EPI_ISL_2539985, EPI_ISL_2539992, EPI_ISL_2539993, EPI_ISL_2539994, EPI_ISL_2539996, EPI_ISL_2539997, EPI_ISL_2539998, EPI_ISL_2539999, EPI_ISL_2540000, EPI_ISL_2540001, EPI_ISL_2540006, EPI_ISL_2540007, EPI_ISL_2540008, EPI_ISL_2540009, EPI_ISL_2540010, EPI_ISL_2540011, EPI_ISL_2540012, EPI_ISL_2540013, EPI_ISL_2540014, EPI_ISL_2540017, EPI_ISL_2540018, EPI_ISL_2540019, EPI_ISL_2540020, EPI_ISL_2540021, EPI_ISL_2540022, EPI_ISL_2540023, EPI_ISL_2540025, EPI_ISL_2540026, EPI_ISL_2540027, EPI_ISL_2540028, EPI_ISL_2540029, EPI_ISL_2540036, EPI_ISL_2540038, EPI_ISL_2540039, EPI_ISL_2540040, EPI_ISL_2540042, EPI_ISL_2540051, EPI_ISL_2540053, EPI_ISL_2540054, EPI_ISL_2540056, EPI_ISL_2540058, EPI_ISL_2540069, EPI_ISL_2540074, EPI_ISL_2540076, EPI_ISL_2540078, EPI_ISL_2540082, EPI_ISL_2540083, EPI_ISL_2540092, EPI_ISL_2540093, EPI_ISL_2540095, EPI_ISL_2540096, EPI_ISL_2540097, EPI_ISL_2540099, EPI_ISL_2540101, EPI_ISL_2540108, EPI_ISL_2540113, EPI_ISL_2540116, EPI_ISL_2540118, EPI_ISL_2540119, EPI_ISL_2540129, EPI_ISL_2540132, EPI_ISL_2540142, EPI_ISL_2540150, EPI_ISL_2540156, EPI_ISL_2540159, EPI_ISL_2540160, EPI_ISL_2540165, EPI_ISL_2540166, EPI_ISL_2540169, EPI_ISL_2540173, EPI_ISL_2540174, EPI_ISL_2540177, EPI_ISL_2540180, EPI_ISL_2540186, EPI_ISL_2540187, EPI_ISL_2540188, EPI_ISL_2540189, EPI_ISL_2540191, EPI_ISL_2540192, EPI_ISL_2540194, EPI_ISL_2540195, EPI_ISL_2540201, EPI_ISL_2540204, EPI_ISL_2540220, EPI_ISL_2540223, EPI_ISL_2540224, EPI_ISL_2540227, EPI_ISL_2540232, EPI_ISL_2540235, EPI_ISL_2540236, EPI_ISL_2540237, EPI_ISL_2540239, EPI_ISL_2540246, EPI_ISL_2540248, EPI_ISL_2540249, EPI_ISL_2540254, EPI_ISL_2540256, EPI_ISL_2540257, EPI_ISL_2540258, EPI_ISL_2540260, EPI_ISL_2540269, EPI_ISL_2540273, EPI_ISL_2540276, EPI_ISL_2540283, EPI_ISL_2540284, EPI_ISL_2540286, EPI_ISL_2540287, EPI_ISL_2540302, EPI_ISL_2540304, EPI_ISL_2540309, EPI_ISL_2540318, EPI_ISL_2540323, EPI_ISL_2540329, EPI_ISL_2540330, EPI_ISL_2540331, EPI_ISL_2540333, EPI_ISL_2540334, EPI_ISL_2540338, EPI_ISL_2540344, EPI_ISL_2540347, EPI_ISL_2540354, EPI_ISL_2540357, EPI_ISL_2540358, EPI_ISL_2540363, EPI_ISL_2540368, EPI_ISL_2540369, EPI_ISL_2540370, EPI_ISL_2540599, EPI_ISL_2540606, EPI_ISL_2540608, EPI_ISL_2540609, EPI_ISL_2540612, EPI_ISL_2540614, EPI_ISL_2540617, EPI_ISL_2540625, EPI_ISL_2540628, EPI_ISL_2540633, EPI_ISL_2540645, EPI_ISL_2540652, EPI_ISL_2540653, EPI_ISL_2540657, EPI_ISL_2540665, EPI_ISL_2540671, EPI_ISL_2540681, EPI_ISL_2540687, EPI_ISL_2540692, EPI_ISL_2540707, EPI_ISL_2540713, EPI_ISL_2540728, EPI_ISL_2540737, EPI_ISL_2540746, EPI_ISL_2540749, EPI_ISL_2540783, EPI_ISL_2540784, EPI_ISL_2540801, EPI_ISL_2540823, EPI_ISL_2540824, EPI_ISL_2540834, EPI_ISL_2540835, EPI_ISL_2540857, EPI_ISL_2540861, EPI_ISL_2540877, EPI_ISL_2540878, EPI_ISL_2540885, EPI_ISL_2540886, EPI_ISL_2540891, EPI_ISL_2540912, EPI_ISL_2540914, EPI_ISL_2540925, EPI_ISL_2540931, EPI_ISL_2540946, EPI_ISL_2540947, EPI_ISL_2540949, EPI_ISL_2540953, EPI_ISL_2540955, EPI_ISL_2540956, EPI_ISL_2540960, EPI_ISL_2540964, EPI_ISL_2540965, EPI_ISL_2540966, EPI_ISL_2540967, EPI_ISL_2540969, EPI_ISL_2540973, EPI_ISL_2540981, EPI_ISL_2540983, EPI_ISL_2540985, EPI_ISL_2540989, EPI_ISL_2540996, EPI_ISL_2541010, EPI_ISL_2541011, EPI_ISL_2541012, EPI_ISL_2541015, EPI_ISL_2541016, EPI_ISL_2541020, EPI_ISL_2541023, EPI_ISL_2541026, EPI_ISL_2541030, EPI_ISL_2541033, EPI_ISL_2541037, EPI_ISL_2541040, EPI_ISL_2541042, EPI_ISL_2541045, EPI_ISL_2541046, EPI_ISL_2541048, EPI_ISL_2541051, EPI_ISL_2541054, EPI_ISL_2541059, EPI_ISL_2541063, EPI_ISL_2541064, EPI_ISL_2541070, EPI_ISL_2541080, EPI_ISL_2541087, EPI_ISL_2541091, EPI_ISL_2541093, EPI_ISL_2541096, EPI_ISL_2541099, EPI_ISL_2541100, EPI_ISL_2541102, EPI_ISL_2541104, EPI_ISL_2541109, EPI_ISL_2541110, EPI_ISL_2541114, EPI_ISL_2541125, EPI_ISL_2541127, EPI_ISL_2541135, EPI_ISL_2541136, EPI_ISL_2541137, EPI_ISL_2541148, EPI_ISL_2541153, EPI_ISL_2541154, EPI_ISL_2541159, EPI_ISL_2541160, EPI_ISL_2541162, EPI_ISL_2541164, EPI_ISL_2541172, EPI_ISL_2541175, EPI_ISL_2541176, EPI_ISL_2541180, EPI_ISL_2541183, EPI_ISL_2541187, EPI_ISL_2541188, EPI_ISL_2541191, EPI_ISL_2541193, EPI_ISL_2541198, EPI_ISL_2541199, EPI_ISL_2541200, EPI_ISL_2541202, EPI_ISL_2541204, EPI_ISL_2541206, EPI_ISL_2541207, EPI_ISL_2541209, EPI_ISL_2541213, EPI_ISL_2541215, EPI_ISL_2541223, EPI_ISL_2541239, EPI_ISL_2541242, EPI_ISL_2541247, EPI_ISL_2541248, EPI_ISL_2541253, EPI_ISL_2541260, EPI_ISL_2541261, EPI_ISL_2541262, EPI_ISL_2541263, EPI_ISL_2541268, EPI_ISL_2541269, EPI_ISL_2541271, EPI_ISL_2541274, EPI_ISL_2541282, EPI_ISL_2541283, EPI_ISL_2541286, EPI_ISL_2541288, EPI_ISL_2541295, EPI_ISL_2541296, EPI_ISL_2541297, EPI_ISL_2541306, EPI_ISL_2541309, EPI_ISL_2541320, EPI_ISL_2541322, EPI_ISL_2541323, EPI_ISL_2541324, EPI_ISL_2541330, EPI_ISL_2541334, EPI_ISL_2541338, EPI_ISL_2541341, EPI_ISL_2541342, EPI_ISL_2541345, EPI_ISL_2541347, EPI_ISL_2541365, EPI_ISL_2541366, EPI_ISL_2541369, EPI_ISL_2541375, EPI_ISL_2541377, EPI_ISL_2541396, EPI_ISL_2541398, EPI_ISL_2541405, EPI_ISL_2541407, EPI_ISL_2541421, EPI_ISL_2541425, EPI_ISL_2541432, EPI_ISL_2541433, EPI_ISL_2541440, EPI_ISL_2541443, EPI_ISL_2541447, EPI_ISL_2541450, EPI_ISL_2541464, EPI_ISL_2541474, EPI_ISL_2541475, EPI_ISL_2541478, EPI_ISL_2541481, EPI_ISL_2541484, EPI_ISL_2541489, EPI_ISL_2541491, EPI_ISL_2541496, EPI_ISL_2541500, EPI_ISL_2541501, EPI_ISL_2541509, EPI_ISL_2541519, EPI_ISL_2541520, EPI_ISL_2541521, EPI_ISL_2541524, EPI_ISL_2541529, EPI_ISL_2541532, EPI_ISL_2541535, EPI_ISL_2541536, EPI_ISL_2541538, EPI_ISL_2541542, EPI_ISL_2541545, EPI_ISL_2541548, EPI_ISL_2541554, EPI_ISL_2541556, EPI_ISL_2541558, EPI_ISL_2541561, EPI_ISL_2541562, EPI_ISL_2541564, EPI_ISL_2541567, EPI_ISL_2541569, EPI_ISL_2541578, EPI_ISL_2541584, EPI_ISL_2541591, EPI_ISL_2541593, EPI_ISL_2541598, EPI_ISL_2541600, EPI_ISL_2541603, EPI_ISL_2541608, EPI_ISL_2541618, EPI_ISL_2541619, EPI_ISL_2541624, EPI_ISL_2541628, EPI_ISL_2541631, EPI_ISL_2541634, EPI_ISL_2551713, EPI_ISL_2551734, EPI_ISL_2552625, EPI_ISL_2552629, EPI_ISL_2552753, EPI_ISL_2552903, EPI_ISL_2553197, EPI_ISL_2553384, EPI_ISL_2553413, EPI_ISL_2553999, EPI_ISL_2554070, EPI_ISL_2555055, EPI_ISL_2570225, EPI_ISL_2640329, EPI_ISL_2640363, EPI_ISL_2640385, EPI_ISL_2640390, EPI_ISL_2640394, EPI_ISL_2640399, EPI_ISL_2640400, EPI_ISL_2640402, EPI_ISL_2640404, EPI_ISL_2640411, EPI_ISL_2640429, EPI_ISL_2640440, EPI_ISL_2640506, EPI_ISL_2640550, EPI_ISL_2640564, EPI_ISL_2640589, EPI_ISL_2640617, EPI_ISL_2640655, EPI_ISL_2641097, EPI_ISL_2641099, EPI_ISL_2641119, EPI_ISL_2765598, EPI_ISL_2765666, EPI_ISL_2765678, EPI_ISL_2765693, EPI_ISL_2765826, EPI_ISL_2765859, EPI_ISL_2781486, EPI_ISL_2925467

| | | | |
|---|---|---|---|
| see above | Lighthouse Lab in Milton Keynes | Wellcome Sanger Institute for the COVID-19 Genomics UK (COG-UK) Consortium | Cordelia Langford; David K. Jackson; Dominic Kwiatkowski; Ewan Harrison; Ian Johnston; Jeffrey Barrett; John Sillitoe on behalf of the Wellcome Sanger Institute COVID-19 Surveillance Team; Roberto Amato; Sonia Goncalves; The Lighthouse Lab in Milton Keynes and Alex Alderton |

EPI_ISL_2485257, EPI_ISL_2485273, EPI_ISL_2485288, EPI_ISL_2485289, EPI_ISL_2485294, EPI_ISL_2485297, EPI_ISL_2485317, EPI_ISL_2485318, EPI_ISL_2485321, EPI_ISL_2485330, EPI_ISL_2485333, EPI_ISL_2485346, EPI_ISL_2485349, EPI_ISL_2485352, EPI_ISL_2485382, EPI_ISL_2485385, EPI_ISL_2485387, EPI_ISL_2485400, EPI_ISL_2485401, EPI_ISL_2485406, EPI_ISL_2485415, EPI_ISL_2485418, EPI_ISL_2485474, EPI_ISL_2485475, EPI_ISL_2485479, EPI_ISL_2485482, EPI_ISL_2485483, EPI_ISL_2485487, EPI_ISL_2485512, EPI_ISL_2485514, EPI_ISL_2487282, EPI_ISL_2487425, EPI_ISL_2487447, EPI_ISL_2487462, EPI_ISL_2487474, EPI_ISL_2487527, EPI_ISL_2487558, EPI_ISL_2511356, EPI_ISL_2511374, EPI_ISL_2511384, EPI_ISL_2511390, EPI_ISL_2511400, EPI_ISL_2511403, EPI_ISL_2511406, EPI_ISL_2511407, EPI_ISL_2511411, EPI_ISL_2511414, EPI_ISL_2511418, EPI_ISL_2511426, EPI_ISL_2511433, EPI_ISL_2511436, EPI_ISL_2511459, EPI_ISL_2511461, EPI_ISL_2511462, EPI_ISL_2511463, EPI_ISL_2511467, EPI_ISL_2511469, EPI_ISL_2511471, EPI_ISL_2511480, EPI_ISL_2511483, EPI_ISL_2511500, EPI_ISL_2511501, EPI_ISL_2511514, EPI_ISL_2511516, EPI_ISL_2511522, EPI_ISL_2511530, EPI_ISL_2511531, EPI_ISL_2511533, EPI_ISL_2511534, EPI_ISL_2511536, EPI_ISL_2511540, EPI_ISL_2511543, EPI_ISL_2511546, EPI_ISL_2511547, EPI_ISL_2511548, EPI_ISL_2511552, EPI_ISL_2511553, EPI_ISL_2511554, EPI_ISL_2511564, EPI_ISL_2511569, EPI_ISL_2511581, EPI_ISL_2511583, EPI_ISL_2511586, EPI_ISL_2511590, EPI_ISL_2511591, EPI_ISL_2511592, EPI_ISL_2511604, EPI_ISL_2511609, EPI_ISL_2511613, EPI_ISL_2511621, EPI_ISL_2511623, EPI_ISL_2511632, EPI_ISL_2511635, EPI_ISL_2511639, EPI_ISL_2511642, EPI_ISL_2512056, EPI_ISL_2512083, EPI_ISL_2515402, EPI_ISL_2515418, EPI_ISL_2515419, EPI_ISL_2515432, EPI_ISL_2515448, EPI_ISL_2515469, EPI_ISL_2515472, EPI_ISL_2515474, EPI_ISL_2515488, EPI_ISL_2515500, EPI_ISL_2515511, EPI_ISL_2515528, EPI_ISL_2515535, EPI_ISL_2515545, EPI_ISL_2515556, EPI_ISL_2515565, EPI_ISL_2515577, EPI_ISL_2515593, EPI_ISL_2515600, EPI_ISL_2515603, EPI_ISL_2515607, EPI_ISL_2515620, EPI_ISL_2515628, EPI_ISL_2515639, EPI_ISL_2515667, EPI_ISL_2515672, EPI_ISL_2515679, EPI_ISL_2515681, EPI_ISL_2515685, EPI_ISL_2515686, EPI_ISL_2515698, EPI_ISL_2515701, EPI_ISL_2515704, EPI_ISL_2515712, EPI_ISL_2515715, EPI_ISL_2515716, EPI_ISL_2515719, EPI_ISL_2515721, EPI_ISL_2515723, EPI_ISL_2515730, EPI_ISL_2515732, EPI_ISL_2515748, EPI_ISL_2515751, EPI_ISL_2515756, EPI_ISL_2515757, EPI_ISL_2515763, EPI_ISL_2515765, EPI_ISL_2515767, EPI_ISL_2515772, EPI_ISL_2515773, EPI_ISL_2515778, EPI_ISL_2515789, EPI_ISL_2515793, EPI_ISL_2515794, EPI_ISL_2515797, EPI_ISL_2515800, EPI_ISL_2515808, EPI_ISL_2515816, EPI_ISL_2515823, EPI_ISL_2515849, EPI_ISL_2515851, EPI_ISL_2515860, EPI_ISL_2515866, EPI_ISL_2515868, EPI_ISL_2515872, EPI_ISL_2515877, EPI_ISL_2515884, EPI_ISL_2515885, EPI_ISL_2515886, EPI_ISL_2515888, EPI_ISL_2515894, EPI_ISL_2515896, EPI_ISL_2515898, EPI_ISL_2515902, EPI_ISL_2515904, EPI_ISL_2515905, EPI_ISL_2515907, EPI_ISL_2515909, EPI_ISL_2515910, EPI_ISL_2515918, EPI_ISL_2515933, EPI_ISL_2515937, EPI_ISL_2515948, EPI_ISL_2515949, EPI_ISL_2515971, EPI_ISL_2515982, EPI_ISL_2515986, EPI_ISL_2515988, EPI_ISL_2515989, EPI_ISL_2515997, EPI_ISL_2516007, EPI_ISL_2516103, EPI_ISL_2516296, EPI_ISL_2539740, EPI_ISL_2539741, EPI_ISL_2539744, EPI_ISL_2539747, EPI_ISL_2539750, EPI_ISL_2539751, EPI_ISL_2539753, EPI_ISL_2539755, EPI_ISL_2539756, EPI_ISL_2539764, EPI_ISL_2539765, EPI_ISL_2539775, EPI_ISL_2539779, EPI_ISL_2539781, EPI_ISL_2539782, EPI_ISL_2539783, EPI_ISL_2539784, EPI_ISL_2539785, EPI_ISL_2539786, EPI_ISL_2539792, EPI_ISL_2539796, EPI_ISL_2539800, EPI_ISL_2539802, EPI_ISL_2539811, EPI_ISL_2539813, EPI_ISL_2539816, EPI_ISL_2539818, EPI_ISL_2539822, EPI_ISL_2539823, EPI_ISL_2539828, EPI_ISL_2539833, EPI_ISL_2539834, EPI_ISL_2539837, EPI_ISL_2539842, EPI_ISL_2539844, EPI_ISL_2539854, EPI_ISL_2539858, EPI_ISL_2539859, EPI_ISL_2539862, EPI_ISL_2539873, EPI_ISL_2539874, EPI_ISL_2539877, EPI_ISL_2539878, EPI_ISL_2539882, EPI_ISL_2539883, EPI_ISL_2539884, EPI_ISL_2539888, EPI_ISL_2539893, EPI_ISL_2539896, EPI_ISL_2539901, EPI_ISL_2539905, EPI_ISL_2539908, EPI_ISL_2539910, EPI_ISL_2539911, EPI_ISL_2539929, EPI_ISL_2539932, EPI_ISL_2539935, EPI_ISL_2539937, EPI_ISL_2539940, EPI_ISL_2539944, EPI_ISL_2539948, EPI_ISL_2539951, EPI_ISL_2539952, EPI_ISL_2539957, EPI_ISL_2539962, EPI_ISL_2539965, EPI_ISL_2539967, EPI_ISL_2539974, EPI_ISL_2539982, EPI_ISL_2539986, EPI_ISL_2539989, EPI_ISL_2539991, EPI_ISL_2540002, EPI_ISL_2540003, EPI_ISL_2540004, EPI_ISL_2540015, EPI_ISL_2540024, EPI_ISL_2540030, EPI_ISL_2540031, EPI_ISL_2540796, EPI_ISL_2541249, EPI_ISL_2541477, EPI_ISL_2541589, EPI_ISL_2542023, EPI_ISL_2542028, EPI_ISL_2543334, EPI_ISL_2543335

| | | | |
|---|---|---|---|
| see above | Lighthouse Laboratory Plymouth | Wellcome Sanger Institute for the COVID-19 Genomics UK (COG-UK) Consortium | Cordelia Langford; David K. Jackson; Dominic Kwiatkowski; Ewan Harrison; Ian Johnston; Jeffrey Barrett; John Sillitoe on behalf of the Wellcome Sanger Institute COVID-19 Surveillance Team; Lighthouse Laboratory Plymouth and Alex Alderton; Roberto Amato; Sonia Goncalves |

| | | | |
|---|---|---|---|
| EPI_ISL_2623125, EPI_ISL_2623126, EPI_ISL_2623127, EPI_ISL_2623128, EPI_ISL_2623129, EPI_ISL_2623130, EPI_ISL_2623131, EPI_ISL_2623132, EPI_ISL_2623133, EPI_ISL_2623134, EPI_ISL_2623135, EPI_ISL_2623136, EPI_ISL_2623137, EPI_ISL_2623138, EPI_ISL_2623139, EPI_ISL_2623140, EPI_ISL_2623141, EPI_ISL_2623142, EPI_ISL_2623143, EPI_ISL_2623144, EPI_ISL_2623145, EPI_ISL_2623146, EPI_ISL_2623147, EPI_ISL_2623148, EPI_ISL_2623149, EPI_ISL_2623150, EPI_ISL_2623151, EPI_ISL_2623152, EPI_ISL_2623153, EPI_ISL_2623154, EPI_ISL_2623155, EPI_ISL_2623156, EPI_ISL_2623157, EPI_ISL_2623158, EPI_ISL_2623159, EPI_ISL_2623160, EPI_ISL_2623161, EPI_ISL_2623162, EPI_ISL_2623163, EPI_ISL_2623164, EPI_ISL_2623165, EPI_ISL_2623166, EPI_ISL_2623167, EPI_ISL_2623168, EPI_ISL_2623169, EPI_ISL_2623170, EPI_ISL_2623171, EPI_ISL_2623172, EPI_ISL_2623173, EPI_ISL_2623174, EPI_ISL_2623175, EPI_ISL_2623176, EPI_ISL_2623177, EPI_ISL_2623178, EPI_ISL_2623179, EPI_ISL_2623180, EPI_ISL_2623181, EPI_ISL_2623182, EPI_ISL_2623183, EPI_ISL_2623184, EPI_ISL_2623185, EPI_ISL_2623186, EPI_ISL_2623187, EPI_ISL_2623188, EPI_ISL_2623189, EPI_ISL_2623190, EPI_ISL_2623191, EPI_ISL_2623192, EPI_ISL_2623193, EPI_ISL_2623194, EPI_ISL_2623195, EPI_ISL_2623196, EPI_ISL_2623197, EPI_ISL_2623198, EPI_ISL_2623199, EPI_ISL_2623200, EPI_ISL_2623201, EPI_ISL_2623202, EPI_ISL_2623203, EPI_ISL_2623204, EPI_ISL_2623205, EPI_ISL_2623206, EPI_ISL_2623207, EPI_ISL_2623208, EPI_ISL_2623209, EPI_ISL_2623210, EPI_ISL_2623211, EPI_ISL_2623212, EPI_ISL_2623213, EPI_ISL_2623214, EPI_ISL_2623215, EPI_ISL_2623216, EPI_ISL_2623217, EPI_ISL_2623218, EPI_ISL_2623219, EPI_ISL_2623220, EPI_ISL_2623221, EPI_ISL_2623222, EPI_ISL_2623223, EPI_ISL_2623224, EPI_ISL_2623225, EPI_ISL_2623226, EPI_ISL_2623228, EPI_ISL_2623229, EPI_ISL_2623230, EPI_ISL_2623231, EPI_ISL_2623232, EPI_ISL_2623233, EPI_ISL_2623234, EPI_ISL_2623235, EPI_ISL_2623236, EPI_ISL_2623238, EPI_ISL_2623239, EPI_ISL_2623240, EPI_ISL_2623241, EPI_ISL_2623242, EPI_ISL_2623243, EPI_ISL_2623244, EPI_ISL_2623245, EPI_ISL_2623246, EPI_ISL_2623247, EPI_ISL_2623248, EPI_ISL_2623249, EPI_ISL_2623250, EPI_ISL_2623251, EPI_ISL_2623252, EPI_ISL_2623253, EPI_ISL_2623254, EPI_ISL_2623255, EPI_ISL_2623256, EPI_ISL_2623257, EPI_ISL_2623260, EPI_ISL_2623261, EPI_ISL_2623262, EPI_ISL_2623263, EPI_ISL_2623264, EPI_ISL_2623265, EPI_ISL_2623266, EPI_ISL_2623267, EPI_ISL_2623268, EPI_ISL_2623270, EPI_ISL_2623271, EPI_ISL_2623275, EPI_ISL_2623276, EPI_ISL_2623279, EPI_ISL_2623287, EPI_ISL_2623291, EPI_ISL_2623300, EPI_ISL_2623303, EPI_ISL_2623305, EPI_ISL_2623308, EPI_ISL_2623312, EPI_ISL_2623317, EPI_ISL_2623322, EPI_ISL_2623323, EPI_ISL_2623324, EPI_ISL_2623334, EPI_ISL_2623340, EPI_ISL_2623344, EPI_ISL_2623349, EPI_ISL_2623354, EPI_ISL_2623361, EPI_ISL_2623363, EPI_ISL_2623367, EPI_ISL_2623368, EPI_ISL_2623382, EPI_ISL_2623483 | | | |
| see above | Northumbria University / South Tees Hospitals NHS Foundation Trust / North Cumbria Integrated Care NHS Foundation Trust / North Tees and Hartlepool NHS Foundation Trust / Newcastle Hospitals NHS Foundation Trust | COVID-19 Genomics UK (COG-UK) Consortium | Andrew Nelson; Brendan Payne; Clive Graham; Darren L Smith; Debra Padgett; Edward Barton; Emma Swindells; Garren Scott; Gary Black; Gary Eltringham; Giles S Holt; Greg R Young; Jane Greenaway; Jennifer Collins; John Allan; Joshua Loh; Lynn Dover; Matthew Bashton; Mohammad A Tariq; Paul Baker; Sarah Essex; Steve Liggett; Wen C Yew; Yusri Taha |
| EPI_ISL_2624587, EPI_ISL_2624601, EPI_ISL_2624609, EPI_ISL_2624610, EPI_ISL_2624625, EPI_ISL_2624637, EPI_ISL_2624643, EPI_ISL_2624648, EPI_ISL_2624651, EPI_ISL_2624652, EPI_ISL_2624653, EPI_ISL_2624654, EPI_ISL_2624657, EPI_ISL_2624658, EPI_ISL_2624659, EPI_ISL_2624662, EPI_ISL_2624664, EPI_ISL_2624696, EPI_ISL_2624715, EPI_ISL_2624717, EPI_ISL_2624720, EPI_ISL_2624724, EPI_ISL_2624726, EPI_ISL_2624727, EPI_ISL_2624728, EPI_ISL_2624730, EPI_ISL_2624732, EPI_ISL_2624739, EPI_ISL_2624741, EPI_ISL_2624742, EPI_ISL_2624746, EPI_ISL_2624789, EPI_ISL_2624796, EPI_ISL_2624797, EPI_ISL_2624800, EPI_ISL_2624803, EPI_ISL_2624809, EPI_ISL_2624813, EPI_ISL_2624827, EPI_ISL_2624854, EPI_ISL_2624866, EPI_ISL_2624878, EPI_ISL_2624879, EPI_ISL_2624880, EPI_ISL_2624882, EPI_ISL_2624886, EPI_ISL_2624888, EPI_ISL_2624897, EPI_ISL_2624907, EPI_ISL_2624913, EPI_ISL_2624914, EPI_ISL_2624917, EPI_ISL_2624924, EPI_ISL_2624927, EPI_ISL_2624929 | | | |
| see above | Originating lab: Wales Specialist Virology Centre Sequencing lab: Pathogen Genomics Unit | Public Health Wales Microbiology Cardiff Wales Specialist Virology Centre | Alec Birchley; Alexander Adams; Amy Gaskin; Angela Marchbank; Bree Gatica-Wilcox; Catherine Moore; Jason Coombes; Joanne Watkins; Joel Southgate; Johnathan Evans; Laura Gifford; Lauren Gilbert; Lee Graham; Malorie Perry; Matthew Bull; Nicole Pacchiarini; Sally Corden; Sara Kumziene-Summerhayes; Sara Rey; Sarah Taylor; Simon Cottrell; Sophie Jones; Tom Connor |
| EPI_ISL_2624525, EPI_ISL_2624528, EPI_ISL_2624530, EPI_ISL_2624531, EPI_ISL_2624532, EPI_ISL_2624533, EPI_ISL_2624534, EPI_ISL_2624535, EPI_ISL_2624536, EPI_ISL_2624538, EPI_ISL_2624541, EPI_ISL_2624542, EPI_ISL_2624546, EPI_ISL_2707322, EPI_ISL_2707323, EPI_ISL_2707329, EPI_ISL_2707358 | | | |
| see above | Oxford Viromics, NDM, University of Oxford; Oxford University Hospitals; Basingstoke and North Hampshire Hospital | COVID-19 Genomics UK (COG-UK) Consortium | Alex Mobbs; Amy Trebes; Anita Justice; Catrin Moore; Christophe Fraser; David Bonsall; David Buck; Emma Wise; George Macintyre; Jessica Lynch; John Todd; Mariateresa de Cesare; Matilde Mori; Monique Andersson; Nathan Moore; Nick Cortes; Robert Shaw; Stephen Kidd; Tanya Golubchik; Timothy Peto |
| EPI_ISL_2519199, EPI_ISL_2519265, EPI_ISL_2519266, EPI_ISL_2519267, EPI_ISL_2519268, EPI_ISL_2519269, EPI_ISL_2519270, EPI_ISL_2519271, EPI_ISL_2519272, EPI_ISL_2519273, EPI_ISL_2519274, EPI_ISL_2519275, EPI_ISL_2519276, EPI_ISL_2519277, EPI_ISL_2519278, EPI_ISL_2519279, EPI_ISL_2519280, EPI_ISL_2519281, EPI_ISL_2519282, EPI_ISL_2519283, EPI_ISL_2519284, EPI_ISL_2519285, EPI_ISL_2519286, EPI_ISL_2519287, EPI_ISL_2519288, EPI_ISL_2519289, EPI_ISL_2519290, EPI_ISL_2519291, EPI_ISL_2519292, EPI_ISL_2519293, EPI_ISL_2519294, EPI_ISL_2519295, EPI_ISL_2519296, EPI_ISL_2519297, EPI_ISL_2519298, EPI_ISL_2519299, EPI_ISL_2519300, EPI_ISL_2519301, EPI_ISL_2519302, EPI_ISL_2519303, EPI_ISL_2519304, EPI_ISL_2519305, EPI_ISL_2519306, EPI_ISL_2519307, EPI_ISL_2519308, EPI_ISL_2519309, EPI_ISL_2519310, EPI_ISL_2519311, EPI_ISL_2519312, EPI_ISL_2519313, EPI_ISL_2519314, EPI_ISL_2519315, EPI_ISL_2519316, EPI_ISL_2519317, EPI_ISL_2519318, EPI_ISL_2519319, EPI_ISL_2519320, EPI_ISL_2519321, EPI_ISL_2519322, EPI_ISL_2519323, EPI_ISL_2519324, EPI_ISL_2519325, EPI_ISL_2519326, EPI_ISL_2519327, EPI_ISL_2519328, EPI_ISL_2519329, EPI_ISL_2519330, EPI_ISL_2519331, EPI_ISL_2519332, EPI_ISL_2519333, EPI_ISL_2519335, EPI_ISL_2519336, EPI_ISL_2519337, EPI_ISL_2519338, EPI_ISL_2519339, EPI_ISL_2519340, EPI_ISL_2519341, EPI_ISL_2519342, EPI_ISL_2519343, EPI_ISL_2519344, EPI_ISL_2519345, EPI_ISL_2519346, EPI_ISL_2519347, EPI_ISL_2519348, EPI_ISL_2519350, EPI_ISL_2519351, EPI_ISL_2519352, EPI_ISL_2519353, EPI_ISL_2519354, EPI_ISL_2519355, EPI_ISL_2519356, EPI_ISL_2519357, EPI_ISL_2519358, EPI_ISL_2519359, EPI_ISL_2519360, EPI_ISL_2519361, EPI_ISL_2519362, EPI_ISL_2519363, EPI_ISL_2519364, EPI_ISL_2519365, EPI_ISL_2519366, EPI_ISL_2519367, EPI_ISL_2519370, EPI_ISL_2519371, EPI_ISL_2519372, EPI_ISL_2519373, EPI_ISL_2519374, EPI_ISL_2519375, EPI_ISL_2519376, EPI_ISL_2519377, EPI_ISL_2519378, EPI_ISL_2519379, EPI_ISL_2519380, EPI_ISL_2519381, EPI_ISL_2519382, EPI_ISL_2519383, EPI_ISL_2519384, EPI_ISL_2519385, EPI_ISL_2519386, EPI_ISL_2519387, EPI_ISL_2519388, EPI_ISL_2519389, EPI_ISL_2519390, EPI_ISL_2519391, EPI_ISL_2519392, EPI_ISL_2519393, EPI_ISL_2519394, EPI_ISL_2519395, EPI_ISL_2519396, EPI_ISL_2519397, EPI_ISL_2519398, EPI_ISL_2519399, EPI_ISL_2519400, EPI_ISL_2519401, EPI_ISL_2519402, EPI_ISL_2519403, EPI_ISL_2519404, EPI_ISL_2519405, EPI_ISL_2519406, EPI_ISL_2519407, EPI_ISL_2519408, EPI_ISL_2519409, EPI_ISL_2519410, EPI_ISL_2519411, EPI_ISL_2519412, EPI_ISL_2519413, EPI_ISL_2519414, EPI_ISL_2519415, EPI_ISL_2519416, EPI_ISL_2519417, EPI_ISL_2519418, EPI_ISL_2519419, EPI_ISL_2519420, EPI_ISL_2519421, EPI_ISL_2519422, EPI_ISL_2519423, EPI_ISL_2519424, EPI_ISL_2519425, EPI_ISL_2519426, EPI_ISL_2519427, EPI_ISL_2519428, EPI_ISL_2519429, EPI_ISL_2519430, EPI_ISL_2519431, EPI_ISL_2519432, EPI_ISL_2519433, EPI_ISL_2519434, EPI_ISL_2519435, EPI_ISL_2519436, EPI_ISL_2519437, EPI_ISL_2519438, EPI_ISL_2519439, EPI_ISL_2519440, EPI_ISL_2519441, EPI_ISL_2519442, EPI_ISL_2519443, EPI_ISL_2519444, EPI_ISL_2519445, EPI_ISL_2519446, EPI_ISL_2519447, EPI_ISL_2519448, EPI_ISL_2519449, EPI_ISL_2519450, EPI_ISL_2519451, EPI_ISL_2519452, EPI_ISL_2519453, EPI_ISL_2519454, EPI_ISL_2519455, EPI_ISL_2519456, EPI_ISL_2519457, EPI_ISL_2519458, EPI_ISL_2519459, EPI_ISL_2519460, EPI_ISL_2519461, EPI_ISL_2519462, EPI_ISL_2519463, EPI_ISL_2519464, EPI_ISL_2519465, EPI_ISL_2519466, EPI_ISL_2519467, EPI_ISL_2519469, EPI_ISL_2519472, EPI_ISL_2519474, EPI_ISL_2519475, EPI_ISL_2519476, EPI_ISL_2519477, EPI_ISL_2519479, EPI_ISL_2519480, EPI_ISL_2519481, EPI_ISL_2519482, EPI_ISL_2519483, EPI_ISL_2519484, EPI_ISL_2519485, EPI_ISL_2519486, EPI_ISL_2519487, EPI_ISL_2519488, EPI_ISL_2519489, EPI_ISL_2519490, EPI_ISL_2519491, EPI_ISL_2519492, EPI_ISL_2519493, EPI_ISL_2519494, EPI_ISL_2519495, EPI_ISL_2519496, EPI_ISL_2519497, EPI_ISL_2519498, EPI_ISL_2519499, EPI_ISL_2519500, EPI_ISL_2519501, EPI_ISL_2519502, EPI_ISL_2519503, EPI_ISL_2519525, EPI_ISL_2519526, EPI_ISL_2519534, EPI_ISL_2519539, EPI_ISL_2519550, EPI_ISL_2519563, EPI_ISL_2519573, EPI_ISL_2520118, EPI_ISL_2520163, EPI_ISL_2520234, EPI_ISL_2520241, EPI_ISL_2520242, EPI_ISL_2520256, EPI_ISL_2520263, EPI_ISL_2520265, EPI_ISL_2520288, EPI_ISL_2520289, EPI_ISL_2520290, EPI_ISL_2520291, EPI_ISL_2520292, EPI_ISL_2520293, EPI_ISL_2520294, EPI_ISL_2520296, EPI_ISL_2520300, EPI_ISL_2520301, EPI_ISL_2520309, EPI_ISL_2520312, EPI_ISL_2520313, EPI_ISL_2520314, EPI_ISL_2520315, EPI_ISL_2520316, EPI_ISL_2520317, EPI_ISL_2520318, EPI_ISL_2520319, EPI_ISL_2520320, EPI_ISL_2520321, EPI_ISL_2520322, EPI_ISL_2520323, EPI_ISL_2520324, EPI_ISL_2520325, EPI_ISL_2520326, EPI_ISL_2520327, EPI_ISL_2520328, EPI_ISL_2520329, EPI_ISL_2520330, EPI_ISL_2520331, EPI_ISL_2520332, EPI_ISL_2520333, EPI_ISL_2520334, EPI_ISL_2520335, EPI_ISL_2520336, EPI_ISL_2520338, EPI_ISL_2520339, EPI_ISL_2520340, EPI_ISL_2520341, EPI_ISL_2520342, EPI_ISL_2520343, EPI_ISL_2520344, EPI_ISL_2520345, EPI_ISL_2520346, EPI_ISL_2520347, EPI_ISL_2520348, EPI_ISL_2520349, EPI_ISL_2520350, EPI_ISL_2520351, EPI_ISL_2520352, EPI_ISL_2520353, EPI_ISL_2520354, EPI_ISL_2520355, EPI_ISL_2520356, EPI_ISL_2520357, EPI_ISL_2520359, EPI_ISL_2520360, EPI_ISL_2520361, EPI_ISL_2520363, EPI_ISL_2520364, EPI_ISL_2520365, EPI_ISL_2520366, EPI_ISL_2520367, EPI_ISL_2520368, EPI_ISL_2520369, EPI_ISL_2520370, EPI_ISL_2520371, EPI_ISL_2520372, EPI_ISL_2520373, EPI_ISL_2520374, EPI_ISL_2520375, EPI_ISL_2520376, EPI_ISL_2520378, EPI_ISL_2520379, EPI_ISL_2520380, EPI_ISL_2520381, EPI_ISL_2520382, EPI_ISL_2706715, EPI_ISL_2706716, EPI_ISL_2706717, EPI_ISL_2706719, EPI_ISL_2706720, EPI_ISL_2706721, EPI_ISL_2706722, EPI_ISL_2706723, EPI_ISL_2706724, EPI_ISL_2706726, EPI_ISL_2706728, EPI_ISL_2706729, EPI_ISL_2706730, EPI_ISL_2706732, EPI_ISL_2706733, EPI_ISL_2706734, EPI_ISL_2706735, EPI_ISL_2706736, EPI_ISL_2706737, EPI_ISL_2706738, EPI_ISL_2706739, EPI_ISL_2706740, EPI_ISL_2706741, EPI_ISL_2706742, EPI_ISL_2706743, EPI_ISL_2706746, EPI_ISL_2706747, EPI_ISL_2706748, EPI_ISL_2706749, EPI_ISL_2706750, EPI_ISL_2706751, EPI_ISL_2706752, EPI_ISL_2706753, EPI_ISL_2706754, EPI_ISL_2706756, EPI_ISL_2706757, EPI_ISL_2706759, EPI_ISL_2706760, EPI_ISL_2706761, EPI_ISL_2706762, EPI_ISL_2706763, EPI_ISL_2706764, EPI_ISL_2706765, EPI_ISL_2706767, EPI_ISL_2706768, EPI_ISL_2706918, EPI_ISL_2706919, EPI_ISL_2706920, EPI_ISL_2706921, EPI_ISL_2706922, EPI_ISL_2706923, EPI_ISL_2706924, EPI_ISL_2706925, EPI_ISL_2706926, EPI_ISL_2706927, EPI_ISL_2706930, EPI_ISL_2706931, EPI_ISL_2706939, EPI_ISL_2706943, EPI_ISL_2706944, EPI_ISL_2706945, EPI_ISL_2706946, EPI_ISL_2706947, EPI_ISL_2706948, EPI_ISL_2706949, EPI_ISL_2706950, EPI_ISL_2706951, EPI_ISL_2706952, EPI_ISL_2706953, EPI_ISL_2706954, EPI_ISL_2706956, EPI_ISL_2706957, EPI_ISL_2706958, EPI_ISL_2706959, EPI_ISL_2706960, EPI_ISL_2706961, EPI_ISL_2706962, EPI_ISL_2706963, EPI_ISL_2706964, EPI_ISL_2706965, EPI_ISL_2706966, EPI_ISL_2706967, EPI_ISL_2706968, EPI_ISL_2706969, EPI_ISL_2706970, EPI_ISL_2706971, EPI_ISL_2706973, EPI_ISL_2706975, EPI_ISL_2706976, EPI_ISL_2706977, EPI_ISL_2706978, EPI_ISL_2706979, EPI_ISL_2706980, EPI_ISL_2706981, EPI_ISL_2706982, EPI_ISL_2706983, EPI_ISL_2706984, EPI_ISL_2706985, EPI_ISL_2706986, EPI_ISL_2706987, EPI_ISL_2706988, EPI_ISL_2706989, EPI_ISL_2706990, EPI_ISL_2706991 | | | |
| see above | Quadram Institute Bioscience | COVID-19 Genomics UK (COG-UK) Consortium | Alexander J Trotter; Alison E. Mather; Alp Aydin; Ana P. Tedim; Anastasia Kolyva; Andrew Bell; Andrew J. Page; Claire Stuart; Dave J. Baker; Gemma L. Kay; John Wain; Justin O'Grady; Leonardo de Oliveira Martins; Lizzie Meadows; Maria Diaz; Mark Webber; Muhammed Yasir; Nabil-Fareed Alikhan; Ngozi Elumogo; Nicholas M. Thomson; Rachael Stanley; Rachel Gilroy; Reenesh Prakash; Samir Dervisevic; Samuel Bloomfield; Steven Rudder; Thanh Le-Viet |
| EPI_ISL_2520383, EPI_ISL_2520384, EPI_ISL_2520385, EPI_ISL_2520386, EPI_ISL_2520387, EPI_ISL_2520388, EPI_ISL_2520389, EPI_ISL_2520390, EPI_ISL_2520391, EPI_ISL_2520392, EPI_ISL_2520393, EPI_ISL_2520394, EPI_ISL_2520395, EPI_ISL_2520396, EPI_ISL_2520397, EPI_ISL_2520398, EPI_ISL_2520399, EPI_ISL_2520400, EPI_ISL_2520401, EPI_ISL_2520402, EPI_ISL_2520403, EPI_ISL_2520404, EPI_ISL_2520405, EPI_ISL_2520406, EPI_ISL_2520407, EPI_ISL_2520408, EPI_ISL_2520409, EPI_ISL_2520410, EPI_ISL_2520411, EPI_ISL_2520412, EPI_ISL_2520413, EPI_ISL_2520414, EPI_ISL_2520415, EPI_ISL_2520416, EPI_ISL_2520417, EPI_ISL_2520418, EPI_ISL_2520419, EPI_ISL_2520420, EPI_ISL_2520421, EPI_ISL_2520423, EPI_ISL_2520424, EPI_ISL_2520425, EPI_ISL_2520427, EPI_ISL_2520428, EPI_ISL_2520429, EPI_ISL_2520430, EPI_ISL_2520431, EPI_ISL_2520432, EPI_ISL_2520433, EPI_ISL_2520434, EPI_ISL_2520435, EPI_ISL_2520436, EPI_ISL_2520437, EPI_ISL_2520438, EPI_ISL_2520439, EPI_ISL_2520440, EPI_ISL_2520441, EPI_ISL_2520442, EPI_ISL_2520443, EPI_ISL_2520444, EPI_ISL_2520445, EPI_ISL_2520446, EPI_ISL_2520447, EPI_ISL_2520448, EPI_ISL_2520449, EPI_ISL_2520450, EPI_ISL_2520451, EPI_ISL_2520453, EPI_ISL_2520454, EPI_ISL_2520455, EPI_ISL_2520456, EPI_ISL_2520458, EPI_ISL_2520459, EPI_ISL_2520460, EPI_ISL_2520461, EPI_ISL_2520462, EPI_ISL_2520463, EPI_ISL_2520464, EPI_ISL_2520468, EPI_ISL_2520469, EPI_ISL_2520470, EPI_ISL_2520471, EPI_ISL_2520473, EPI_ISL_2520474, EPI_ISL_2520475, EPI_ISL_2520476, EPI_ISL_2520477, EPI_ISL_2520478, EPI_ISL_2520479, EPI_ISL_2520480, EPI_ISL_2520481, EPI_ISL_2520482, EPI_ISL_2520483, EPI_ISL_2520484, EPI_ISL_2520485, EPI_ISL_2520486, EPI_ISL_2520487, EPI_ISL_2520488, EPI_ISL_2520489, EPI_ISL_2520490, EPI_ISL_2520491, EPI_ISL_2520492, EPI_ISL_2520493, EPI_ISL_2520494, EPI_ISL_2520495, EPI_ISL_2520496, EPI_ISL_2520497, EPI_ISL_2520498, EPI_ISL_2520499, EPI_ISL_2520500, EPI_ISL_2520501, EPI_ISL_2520502, EPI_ISL_2520503, EPI_ISL_2520504, EPI_ISL_2520505, EPI_ISL_2520506, EPI_ISL_2520507, EPI_ISL_2520508, EPI_ISL_2520509, EPI_ISL_2520510, EPI_ISL_2520511, EPI_ISL_2520512, EPI_ISL_2520513, EPI_ISL_2520514, EPI_ISL_2520515, EPI_ISL_2520516, EPI_ISL_2520517, EPI_ISL_2520518, EPI_ISL_2520519, EPI_ISL_2520520, EPI_ISL_2520521, EPI_ISL_2520522, EPI_ISL_2520523, EPI_ISL_2520524, EPI_ISL_2520525, EPI_ISL_2520526, EPI_ISL_2520527, EPI_ISL_2520528, EPI_ISL_2520529, EPI_ISL_2520530, EPI_ISL_2520531, EPI_ISL_2520532, EPI_ISL_2520533 | | | |
| see above | Queens Medical Centre, Clinical Microbiology Department / DeepSeq Nottingham | COVID-19 Genomics UK (COG-UK) Consortium | Christopher Moore; Emily Park; Fei Sang; Gemma Clark; Hannah Howson-Wells; Jack Hill; Jayasree Dey; Johnny Debebe; Jonathan Ball; Joseph Chappell; Manjinder Khakh; Matthew Carlisle; Matthew Loose; Michelle M Lister; Nadine Holmes; Patrick McClure; Theocharis Tsoleridis; Timothy Byaruhanga; Vicki M Fleming; Victoria Wright; Wendy Smith |
| EPI_ISL_2743947, EPI_ISL_2744022, EPI_ISL_2744098, EPI_ISL_2744139, EPI_ISL_2744141, EPI_ISL_2744142, EPI_ISL_2744148, EPI_ISL_2744153, EPI_ISL_2744158, EPI_ISL_2744166, EPI_ISL_2744167, EPI_ISL_2744172, EPI_ISL_2744175, EPI_ISL_2744179, EPI_ISL_2744187, EPI_ISL_2744197, EPI_ISL_2744200, EPI_ISL_2744206, EPI_ISL_2744217, EPI_ISL_2744225, EPI_ISL_2744308, EPI_ISL_2744309, EPI_ISL_2744315, EPI_ISL_2744318, EPI_ISL_2744321, EPI_ISL_2744324, EPI_ISL_2744331, EPI_ISL_2744335, EPI_ISL_2744347, EPI_ISL_2744348, EPI_ISL_2744356, EPI_ISL_2744357, EPI_ISL_2744362, EPI_ISL_2744365, EPI_ISL_2744367, EPI_ISL_2744371, EPI_ISL_2744375, EPI_ISL_2744384, EPI_ISL_2744389, EPI_ISL_2744391, EPI_ISL_2744395, EPI_ISL_2744404, EPI_ISL_2744414, EPI_ISL_2744420, EPI_ISL_2744434, EPI_ISL_2744451, EPI_ISL_2744457, EPI_ISL_2744460, EPI_ISL_2744479, EPI_ISL_2744490, EPI_ISL_2744492, EPI_ISL_2744495, EPI_ISL_2744500, EPI_ISL_2744528, EPI_ISL_2744529, EPI_ISL_2744530, EPI_ISL_2744535, EPI_ISL_2744537, EPI_ISL_2744538, EPI_ISL_2744539, EPI_ISL_2744540, EPI_ISL_2744541, EPI_ISL_2744561, EPI_ISL_2744562, EPI_ISL_2744565, EPI_ISL_2744566, EPI_ISL_2744573, EPI_ISL_2744583, EPI_ISL_2744588, EPI_ISL_2744601, EPI_ISL_2744605, EPI_ISL_2744610, EPI_ISL_2744611, EPI_ISL_2744613, EPI_ISL_2744614, EPI_ISL_2744618, EPI_ISL_2744625, EPI_ISL_2744638, EPI_ISL_2744658, EPI_ISL_2744659, EPI_ISL_2744661, EPI_ISL_2744706, EPI_ISL_2744845, EPI_ISL_2744933, EPI_ISL_2745011, EPI_ISL_2745012, EPI_ISL_2745212, EPI_ISL_2745282, EPI_ISL_2745306, EPI_ISL_2745309, EPI_ISL_2745313, EPI_ISL_2745331, EPI_ISL_2745419, EPI_ISL_2745422, EPI_ISL_2745423, EPI_ISL_2745424, EPI_ISL_2745431, EPI_ISL_2745435, EPI_ISL_2745436, EPI_ISL_2745446, EPI_ISL_2745449, EPI_ISL_2745469, EPI_ISL_2745504, EPI_ISL_2746318, EPI_ISL_2746708, EPI_ISL_2817358, EPI_ISL_2817359 | | | |
| see above | Respiratory Virus Unit, Microbiology Services Colindale, Public Health England | COVID-19 Genomics UK (COG-UK) Consortium | PHE Covid Sequencing Team |
| EPI_ISL_2517322, EPI_ISL_2517323, EPI_ISL_2517324, EPI_ISL_2517325, EPI_ISL_2517326, EPI_ISL_2517327, EPI_ISL_2517328, EPI_ISL_2517329, EPI_ISL_2517330, EPI_ISL_2517331, EPI_ISL_2517332, EPI_ISL_2517333, EPI_ISL_2517334, EPI_ISL_2517335, EPI_ISL_2517337, EPI_ISL_2517338, EPI_ISL_2517339, EPI_ISL_2517340, EPI_ISL_2517341, EPI_ISL_2517342, EPI_ISL_2517343, EPI_ISL_2517344, EPI_ISL_2517345, EPI_ISL_2517346, EPI_ISL_2517347, EPI_ISL_2517348, EPI_ISL_2517349, EPI_ISL_2517350, EPI_ISL_2517351, EPI_ISL_2517352, EPI_ISL_2517353, EPI_ISL_2517354, EPI_ISL_2517355, EPI_ISL_2517356, EPI_ISL_2517357, EPI_ISL_2517358, EPI_ISL_2517359, EPI_ISL_2517360, EPI_ISL_2517361, EPI_ISL_2517362, EPI_ISL_2517363, EPI_ISL_2517364, EPI_ISL_2517365, EPI_ISL_2517366, EPI_ISL_2517367, EPI_ISL_2517368, EPI_ISL_2517369, EPI_ISL_2517370, EPI_ISL_2517371, EPI_ISL_2517372, EPI_ISL_2517373, EPI_ISL_2517374, EPI_ISL_2517375, EPI_ISL_2517376, EPI_ISL_2517377, EPI_ISL_2517378, EPI_ISL_2517379, EPI_ISL_2517380, EPI_ISL_2517381, EPI_ISL_2517382, EPI_ISL_2517383, EPI_ISL_2517384, EPI_ISL_2517385, EPI_ISL_2517386, EPI_ISL_2517387, EPI_ISL_2517388, EPI_ISL_2517389, EPI_ISL_2517390, EPI_ISL_2517391, EPI_ISL_2517392, EPI_ISL_2517393, EPI_ISL_2517394, EPI_ISL_2517395, EPI_ISL_2517396, EPI_ISL_2517397, EPI_ISL_2517398, EPI_ISL_2517399, EPI_ISL_2517400, EPI_ISL_2517401, EPI_ISL_2517402, EPI_ISL_2517403, EPI_ISL_2517404, EPI_ISL_2517405, EPI_ISL_2517406, EPI_ISL_2517407, EPI_ISL_2517409, EPI_ISL_2517410, EPI_ISL_2517411, EPI_ISL_2517412, EPI_ISL_2517413, EPI_ISL_2517414, EPI_ISL_2517415, EPI_ISL_2517416, EPI_ISL_2517417, EPI_ISL_2517418, EPI_ISL_2517419, EPI_ISL_2517420, EPI_ISL_2517421, EPI_ISL_2517422, EPI_ISL_2517423, EPI_ISL_2517425, EPI_ISL_2517426, EPI_ISL_2517427, EPI_ISL_2517428, EPI_ISL_2517429, EPI_ISL_2517430, EPI_ISL_2517431, EPI_ISL_2517432, EPI_ISL_2517433, EPI_ISL_2517434, EPI_ISL_2517435, EPI_ISL_2517436, EPI_ISL_2517437, EPI_ISL_2517438, EPI_ISL_2517439, EPI_ISL_2517440, EPI_ISL_2517441, EPI_ISL_2517442, EPI_ISL_2517443, EPI_ISL_2517444, EPI_ISL_2517445, EPI_ISL_2517446, EPI_ISL_2517447, EPI_ISL_2517449, EPI_ISL_2517450, EPI_ISL_2517451, EPI_ISL_2517452, EPI_ISL_2517453, EPI_ISL_2517454, EPI_ISL_2517455, EPI_ISL_2517456, EPI_ISL_2517457, EPI_ISL_2517458, EPI_ISL_2517459, EPI_ISL_2517460, EPI_ISL_2517461, EPI_ISL_2517462, EPI_ISL_2517463, EPI_ISL_2517464, EPI_ISL_2517465, EPI_ISL_2517466, EPI_ISL_2517468, EPI_ISL_2517469, EPI_ISL_2517470, EPI_ISL_2517471, EPI_ISL_2517472, EPI_ISL_2517473, EPI_ISL_2517474, EPI_ISL_2517475, EPI_ISL_2517476, EPI_ISL_2517478, EPI_ISL_2517479, EPI_ISL_2517480, EPI_ISL_2517481, EPI_ISL_2517482, EPI_ISL_2517490, EPI_ISL_2517491, EPI_ISL_2517496, EPI_ISL_2517498, EPI_ISL_2517505, EPI_ISL_2517519, EPI_ISL_2517524, EPI_ISL_2517529, EPI_ISL_2517535, EPI_ISL_2517544, EPI_ISL_2517560, EPI_ISL_2517668, EPI_ISL_2517681, EPI_ISL_2517730, EPI_ISL_2517731, EPI_ISL_2517733, EPI_ISL_2517742, EPI_ISL_2517752, EPI_ISL_2517763, EPI_ISL_2517771, EPI_ISL_2517776, EPI_ISL_2517777, EPI_ISL_2517780, EPI_ISL_2517781, EPI_ISL_2517785, EPI_ISL_2517791, EPI_ISL_2517807, EPI_ISL_2517808, EPI_ISL_2517813, EPI_ISL_2517815, EPI_ISL_2517817, EPI_ISL_2517818, EPI_ISL_2517822, EPI_ISL_2517825, EPI_ISL_2517826, EPI_ISL_2517841, EPI_ISL_2517844, EPI_ISL_2517857, EPI_ISL_2622477, EPI_ISL_2622485, EPI_ISL_2622492, EPI_ISL_2622499, EPI_ISL_2622507, EPI_ISL_2622508, EPI_ISL_2622509, EPI_ISL_2622512, EPI_ISL_2622514, EPI_ISL_2622516, EPI_ISL_2622518, EPI_ISL_2622519, EPI_ISL_2622522, EPI_ISL_2622532, EPI_ISL_2622534, EPI_ISL_2622536, EPI_ISL_2622538, EPI_ISL_2622553, EPI_ISL_2622592, EPI_ISL_2622733 | | | |
| see above | University College London, Great Ormond Street Hospital for Children NHS Foundation Trust, Imperial College Healthcare NHS Trust | COVID-19 Genomics UK (COG-UK) Consortium | Alison Holmes; Charlotte Williams; Helena Tutill; Jacqueline Findlay; James Price; Judith Breuer; Julianne Brown; Kathryn Harris; Leysa Forrest; Mark Kristiansen; Paola Niola; Paola Resende Silva; Patricia Dyal; Paul Randell; Rachel Williams; Samuel Weeks; Sergi Castellano; Sunando Roy; Tony Brooks; Yasmin Panchbhaya |
| EPI_ISL_2622356, EPI_ISL_2622357, EPI_ISL_2622358, EPI_ISL_2622362, EPI_ISL_2622363, EPI_ISL_2622364, EPI_ISL_2622365, EPI_ISL_2622366, EPI_ISL_2622367, EPI_ISL_2622368, EPI_ISL_2622369, EPI_ISL_2622370, EPI_ISL_2704826, EPI_ISL_2704842, EPI_ISL_2704843, EPI_ISL_2704844, EPI_ISL_2704845, EPI_ISL_2704846, EPI_ISL_2704863, EPI_ISL_2704885, EPI_ISL_2704928, EPI_ISL_2734364, EPI_ISL_2734365, EPI_ISL_2734366, EPI_ISL_2734370, EPI_ISL_2734371 | | | |
| see above | Virology Department, Royal Infirmary of Edinburgh, NHS Lothian / School of Biological Sciences, University of Edinburgh | COVID-19 Genomics UK (COG-UK) Consortium | Colquhoun R; Cotton S; Dewar R; Gallagher A; Hill V; Jackson B; Maloney D; McCrone JT; McHugh M; O'Toole Á; Rambaut A; Rooke S; Scher E; Templeton K; Yu X |
| EPI_ISL_2520928, EPI_ISL_2520931, EPI_ISL_2520934, EPI_ISL_2520936, EPI_ISL_2520938, EPI_ISL_2520942, EPI_ISL_2520944, EPI_ISL_2520962, EPI_ISL_2520968, EPI_ISL_2520970, EPI_ISL_2520972, EPI_ISL_2520973, EPI_ISL_2520975, EPI_ISL_2520983, EPI_ISL_2520984, EPI_ISL_2520985, EPI_ISL_2520987, EPI_ISL_2520990, EPI_ISL_2520994, EPI_ISL_2520998, EPI_ISL_2521002, EPI_ISL_2521003, EPI_ISL_2521006, EPI_ISL_2521007, EPI_ISL_2521009, EPI_ISL_2521010, EPI_ISL_2521011, EPI_ISL_2521012, EPI_ISL_2521015, EPI_ISL_2521017, EPI_ISL_2521022, EPI_ISL_2521025, EPI_ISL_2521028, EPI_ISL_2521030, EPI_ISL_2521031, EPI_ISL_2521033, EPI_ISL_2521038, EPI_ISL_2521039, EPI_ISL_2521048, EPI_ISL_2521049, EPI_ISL_2521050, EPI_ISL_2521051, EPI_ISL_2521052, EPI_ISL_2521056, EPI_ISL_2521058, EPI_ISL_2521061, EPI_ISL_2521062, EPI_ISL_2521063, EPI_ISL_2521068, EPI_ISL_2521070, EPI_ISL_2521071, EPI_ISL_2521075, EPI_ISL_2521077, EPI_ISL_2521079, EPI_ISL_2521081, EPI_ISL_2521090, EPI_ISL_2521093, EPI_ISL_2521098, EPI_ISL_2521106, EPI_ISL_2521114, EPI_ISL_2521119, EPI_ISL_2521125, EPI_ISL_2521128, EPI_ISL_2521130, EPI_ISL_2521132, EPI_ISL_2521133, EPI_ISL_2521135, EPI_ISL_2521143, EPI_ISL_2521144, EPI_ISL_2521145, EPI_ISL_2625213, EPI_ISL_2625215, EPI_ISL_2625216, EPI_ISL_2625218, EPI_ISL_2625219, EPI_ISL_2625222, EPI_ISL_2625223, EPI_ISL_2625226, EPI_ISL_2625227, EPI_ISL_2625228, EPI_ISL_2625229, EPI_ISL_2625231, EPI_ISL_2625233, EPI_ISL_2625235, EPI_ISL_2625239, EPI_ISL_2625240, EPI_ISL_2625241, EPI_ISL_2625244, EPI_ISL_2625246, EPI_ISL_2625250, EPI_ISL_2625254, EPI_ISL_2625256, EPI_ISL_2625257, EPI_ISL_2625264, EPI_ISL_2625267, EPI_ISL_2625269, EPI_ISL_2625273, EPI_ISL_2625276, EPI_ISL_2625278, EPI_ISL_2625284, EPI_ISL_2625295, EPI_ISL_2625298, EPI_ISL_2625302, EPI_ISL_2625305, EPI_ISL_2625309, EPI_ISL_2625312, EPI_ISL_2625315, EPI_ISL_2625319, EPI_ISL_2625320, EPI_ISL_2625328, EPI_ISL_2625333, EPI_ISL_2625334, EPI_ISL_2625335, EPI_ISL_2625338, EPI_ISL_2625339, EPI_ISL_2625344, EPI_ISL_2625352, EPI_ISL_2625356, EPI_ISL_2625357, EPI_ISL_2625359, EPI_ISL_2625360, EPI_ISL_2625363, EPI_ISL_2625367, EPI_ISL_2625369, EPI_ISL_2625370, EPI_ISL_2625382, EPI_ISL_2625383, EPI_ISL_2625385, EPI_ISL_2625390, EPI_ISL_2625392, EPI_ISL_2625396, EPI_ISL_2625397, EPI_ISL_2625400, EPI_ISL_2625401, EPI_ISL_2625402, EPI_ISL_2625403, EPI_ISL_2625404, EPI_ISL_2625405, EPI_ISL_2625406, EPI_ISL_2625407, EPI_ISL_2625411, EPI_ISL_2625416, EPI_ISL_2625418, EPI_ISL_2625419, EPI_ISL_2625422, EPI_ISL_2625426, EPI_ISL_2625428, EPI_ISL_2625429, EPI_ISL_2625430, EPI_ISL_2625434, EPI_ISL_2625440, EPI_ISL_2625441, EPI_ISL_2625443, EPI_ISL_2625444, EPI_ISL_2625446, EPI_ISL_2625447, EPI_ISL_2625448, EPI_ISL_2625450, EPI_ISL_2625453, EPI_ISL_2625455, EPI_ISL_2625456, EPI_ISL_2625457, EPI_ISL_2625458, EPI_ISL_2625464, EPI_ISL_2625465, EPI_ISL_2625466, EPI_ISL_2625469, EPI_ISL_2625471, EPI_ISL_2625474, EPI_ISL_2625478, EPI_ISL_2625481, EPI_ISL_2625482, EPI_ISL_2625484, EPI_ISL_2625485, EPI_ISL_2625486, EPI_ISL_2625489, EPI_ISL_2625500, EPI_ISL_2625504, EPI_ISL_2625505, EPI_ISL_2625511, EPI_ISL_2625512, EPI_ISL_2625518, EPI_ISL_2625519, EPI_ISL_2625520, EPI_ISL_2625522, EPI_ISL_2625523, EPI_ISL_2625524, EPI_ISL_2625525, EPI_ISL_2625526, EPI_ISL_2625531, EPI_ISL_2625532, EPI_ISL_2625533, EPI_ISL_2625534, EPI_ISL_2625535, EPI_ISL_2625537, EPI_ISL_2625546, EPI_ISL_2625549, EPI_ISL_2625551, EPI_ISL_2625552, EPI_ISL_2625553, EPI_ISL_2625557, EPI_ISL_2625558, EPI_ISL_2625567, EPI_ISL_2625568, EPI_ISL_2625577, EPI_ISL_2625578, EPI_ISL_2625583, EPI_ISL_2625587, EPI_ISL_2625588, EPI_ISL_2625589, EPI_ISL_2625591, EPI_ISL_2625592, EPI_ISL_2625594, EPI_ISL_2625603, EPI_ISL_2625607, EPI_ISL_2625608, EPI_ISL_2625611, EPI_ISL_2625612, EPI_ISL_2625613 | | | |
| see above | Virology Department, Sheffield Teaching Hospitals NHS Foundation Trust/Department of Infection, Immunity and Cardiovascular Disease, The Medical School, University of Sheffield | COVID-19 Genomics UK (COG-UK) Consortium | Adri Angyal; Alex Keeley; Benjamin Lindsey; Cariad Evans; Danielle Groves; Dave Partridge; Katie Johnson; Laura Carrilero; Luke Green; Matthew Parker; Matthew Wyles; Mehmet Yavuz; Mohammad Raza; Nikki Smith; Paul Parsons; Rachel Tucker; Rebecca Brown; Thushan de Silva |
| EPI_ISL_2520558, EPI_ISL_2520561, EPI_ISL_2520562, EPI_ISL_2520563, EPI_ISL_2520564, EPI_ISL_2520567, EPI_ISL_2520568, EPI_ISL_2520570, EPI_ISL_2520572, EPI_ISL_2520575, EPI_ISL_2520577, EPI_ISL_2520579, EPI_ISL_2520584, EPI_ISL_2520585, EPI_ISL_2520589, EPI_ISL_2520590, EPI_ISL_2520591, EPI_ISL_2520592, EPI_ISL_2520598, EPI_ISL_2520601, EPI_ISL_2520602, EPI_ISL_2520607, EPI_ISL_2520608, EPI_ISL_2520609, EPI_ISL_2520610, EPI_ISL_2520611, EPI_ISL_2520612, EPI_ISL_2520613, EPI_ISL_2520614, EPI_ISL_2520620, EPI_ISL_2520623, EPI_ISL_2520624, EPI_ISL_2520626, EPI_ISL_2520627, EPI_ISL_2520634, EPI_ISL_2520638, EPI_ISL_2520640, EPI_ISL_2520644, EPI_ISL_2520645, EPI_ISL_2520646, EPI_ISL_2520647, EPI_ISL_2520648, EPI_ISL_2520649, EPI_ISL_2520650, EPI_ISL_2520656, EPI_ISL_2520658, EPI_ISL_2520662, EPI_ISL_2520664, EPI_ISL_2520666, EPI_ISL_2520675, EPI_ISL_2520677, EPI_ISL_2520679, EPI_ISL_2520680, EPI_ISL_2520681, EPI_ISL_2520682, EPI_ISL_2520684, EPI_ISL_2520685, EPI_ISL_2520687, EPI_ISL_2520689, EPI_ISL_2520690, EPI_ISL_2520692, EPI_ISL_2520699, EPI_ISL_2520702, EPI_ISL_2520703, EPI_ISL_2520704, EPI_ISL_2520705, EPI_ISL_2520706, EPI_ISL_2520708, EPI_ISL_2520709, EPI_ISL_2520711, EPI_ISL_2520712, EPI_ISL_2520713, EPI_ISL_2520714, EPI_ISL_2520715, EPI_ISL_2520718, EPI_ISL_2520720, EPI_ISL_2520722, EPI_ISL_2520784, EPI_ISL_2520786, EPI_ISL_2520787, EPI_ISL_2520788, EPI_ISL_2520789, EPI_ISL_2520796, EPI_ISL_2520797, EPI_ISL_2520803, EPI_ISL_2520805, EPI_ISL_2520806, EPI_ISL_2520809, EPI_ISL_2520810, EPI_ISL_2520812 | | | |
| see above | Wales Specialist Virology Centre Sequencing lab: Pathogen Genomics Unit | Public Health Wales Microbiology Cardiff | Alec Birchley; Alexander Adams; Amy Gaskin; Angela Marchbank; Bree Gatica-Wilcox; Catherine Moore; Jason Coombes; Joanne Watkins; Joel Southgate; Johnathan Evans; Laura Gifford; Lauren Gilbert; Lee Graham; Malorie Perry; Matthew Bull; Nicole Pacchiarini; Sally Corden; Sara Kumziene-Summerhayes; Sara Rey; Sarah Taylor; Simon Cottrell; Sophie Jones; Tom Connor |

| | | | |
|---|---|---|---|
| | | Wales Specialist Virology Centre | |
| EPI_ISL_2722662, EPI_ISL_2722663, EPI_ISL_2722665, EPI_ISL_2722666, EPI_ISL_2722667, EPI_ISL_2722668, EPI_ISL_2722669, EPI_ISL_2722671, EPI_ISL_2722672, EPI_ISL_2722673, EPI_ISL_2722677, EPI_ISL_2722679, EPI_ISL_2734270, EPI_ISL_2734275, EPI_ISL_2734280, EPI_ISL_2734286, EPI_ISL_2734287, EPI_ISL_2734318 | | | |
| see above | West of Scotland Specialist Virology Centre, NHSGGC / MRC-University of Glasgow Centre for Virus Research | COVID-19 Genomics UK (COG-UK) Consortium | Alasdair MacLean; Alice Broos; Ana da Silva Filipe; Antonia Ho; Daniel Mair; David L Robertson; Emma Thomson; Guy Mollett; Ioulia Tsatsani; James Shepherd; Jenna Nichols; Joseph Hughes; Kathy Li; Kathy Smollett; Kyriaki Nomikou; Lily Tong; Matthew Holden; Natasha Johnson; Rachel Blacow; Richard Orton; Rory Gunson; Sarah McDonald; Sharif Shaaban; Sreenu Vattipally; Stephen Carmichael |

We gratefully acknowledge the following Authors from the Originating laboratories responsible for obtaining the specimens, as well as the Submitting laboratories where the genome data were generated and shared via GISAID, on which this research is based.

All Submitters of data may be contacted directly via www.gisaid.org

Authors are sorted alphabetically.

| Accession ID | Originating Laboratory | Submitting Laboratory | Authors |
|---|---|---|---|
| EPI_ISL_2686146, EPI_ISL_2686147, EPI_ISL_2686148, EPI_ISL_2686149, EPI_ISL_2686150, EPI_ISL_2686152, EPI_ISL_2686153, EPI_ISL_2686154, EPI_ISL_2686155, EPI_ISL_2686156, EPI_ISL_2686157, EPI_ISL_2686158, EPI_ISL_2686164, EPI_ISL_2686168 | | | |
| see above | Amazon Dx, AUV2 Laboratory | Amazon Dx, AUV2 Laboratory | Luke Meredith |
| EPI_ISL_2622443, EPI_ISL_2622444, EPI_ISL_2622445, EPI_ISL_2622446, EPI_ISL_2622467, EPI_ISL_2622468, EPI_ISL_2622469, EPI_ISL_2622470 | | | |
| see above | Barts Health NHS Trust | COVID-19 Genomics UK (COG-UK) Consortium | BROAD; Beatrix; CUTINO-MOGUEL; Claire; David; Dola; HARRINGTON; KELE; KULASEGARAN-SHYLINI; Maria-Teresa; OWOYEMI; Raghavendran |
| EPI_ISL_2520813, EPI_ISL_2520815, EPI_ISL_2520816, EPI_ISL_2520817, EPI_ISL_2520818, EPI_ISL_2520819, EPI_ISL_2520820, EPI_ISL_2520821, EPI_ISL_2520822, EPI_ISL_2520823, EPI_ISL_2520825, EPI_ISL_2520826, EPI_ISL_2520827, EPI_ISL_2520828, EPI_ISL_2520829, EPI_ISL_2520830, EPI_ISL_2520831, EPI_ISL_2520832, EPI_ISL_2520833, EPI_ISL_2520834, EPI_ISL_2520835, EPI_ISL_2520836, EPI_ISL_2520837, EPI_ISL_2520838, EPI_ISL_2520839, EPI_ISL_2520840, EPI_ISL_2520842, EPI_ISL_2520843, EPI_ISL_2520844, EPI_ISL_2520845, EPI_ISL_2520846, EPI_ISL_2520847, EPI_ISL_2520848, EPI_ISL_2520849, EPI_ISL_2520850, EPI_ISL_2520851, EPI_ISL_2520852, EPI_ISL_2520853, EPI_ISL_2520854, EPI_ISL_2520855, EPI_ISL_2520856, EPI_ISL_2520857, EPI_ISL_2520858, EPI_ISL_2520859, EPI_ISL_2520860, EPI_ISL_2520861, EPI_ISL_2520862, EPI_ISL_2520863, EPI_ISL_2520864, EPI_ISL_2520865, EPI_ISL_2520866, EPI_ISL_2520867, EPI_ISL_2520868, EPI_ISL_2520869, EPI_ISL_2520870, EPI_ISL_2520871, EPI_ISL_2520872, EPI_ISL_2520873, EPI_ISL_2520874, EPI_ISL_2520875, EPI_ISL_2520877, EPI_ISL_2520879, EPI_ISL_2520881, EPI_ISL_2520882, EPI_ISL_2520883, EPI_ISL_2520884, EPI_ISL_2520885, EPI_ISL_2520887, EPI_ISL_2520890, EPI_ISL_2520892, EPI_ISL_2520893, EPI_ISL_2520896, EPI_ISL_2520897, EPI_ISL_2520898, EPI_ISL_2520899, EPI_ISL_2520901, EPI_ISL_2520902, EPI_ISL_2520904, EPI_ISL_2520905, EPI_ISL_2520906, EPI_ISL_2520907, EPI_ISL_2520908, EPI_ISL_2520909, EPI_ISL_2520910, EPI_ISL_2520911, EPI_ISL_2520912, EPI_ISL_2520914, EPI_ISL_2520915, EPI_ISL_2520917, EPI_ISL_2520918, EPI_ISL_2520919, EPI_ISL_2520920, EPI_ISL_2520921, EPI_ISL_2520923, EPI_ISL_2520927, EPI_ISL_2624977, EPI_ISL_2624991, EPI_ISL_2625073 | | | |
| see above | Centre for Enzyme Innovation, University of Portsmouth / Translational Research Laboratory, Portsmouth Hospitals NHS Trust / Hampshire Hospitals NHS Foundation Trust / Bournemouth University / University Hospitals Dorset NHS Foundation Trust / Brighton and Sussex University Hospitals NHS Trust / Dartford and Gravesham NHS Trust / Isle of Wight NHS Trust / Maidstone and Tunbridge Wells NHS Trust / University Hospital Southampton NHS Foundation Trust | COVID-19 Genomics UK (COG-UK) Consortium | Adhyana Mahanama; Allyson Lloyd; Andrew Butt; Angela Beckett; Anibolina Castigador; Anna Mantzaouratou; Anoop Chauhan; Ben Macklin; Benjamin Cogger; Buddhini Samaraweera; Cassandra Malone; Christopher Fearn; Claire Thomas; Dorian Crudgington; Eleri Wilson-Davies; Emanuela Pelosi; Emily Macnaughton; Helen Umpleby; Helen Wheeler; Jacqui Prieto; Jonnie Seymour; Karen Withell; Kate Cook; Katie Loveson; Kelly Bicknell; Kevin Tucker; Kordo Saeed; Liz Sheridan; Magdalena Barrow; Matthew Harvey; Mohammed Hassan-Ibrahim; Nathan Moore; Nick Cortes; Rachael Jeremiah; Robert Impey; Salman Goudarzi; Samirakhon Raupova; Samuel Robson; Sarah Buchan; Sarah Jeremiah; Sarah Wyllie; Scott Elliott; Sharon Glaysher; Siona Silvieri; Stephen Aplin; Stephen Kidd; Thea Sass |
| EPI_ISL_2722124, EPI_ISL_2722126, EPI_ISL_2722136, EPI_ISL_2722144, EPI_ISL_2722146, EPI_ISL_2722153, EPI_ISL_2722159, EPI_ISL_2722160, EPI_ISL_2722161, EPI_ISL_2722162, EPI_ISL_2722163, EPI_ISL_2722164, EPI_ISL_2722165, EPI_ISL_2722166, EPI_ISL_2722167, EPI_ISL_2722168, EPI_ISL_2722169, EPI_ISL_2722170, EPI_ISL_2722171, EPI_ISL_2722173, EPI_ISL_2722174, EPI_ISL_2722175, EPI_ISL_2722176, EPI_ISL_2722177, EPI_ISL_2722178, EPI_ISL_2722179, EPI_ISL_2722180, EPI_ISL_2722181, EPI_ISL_2722182, EPI_ISL_2722183, EPI_ISL_2722184, EPI_ISL_2722185, EPI_ISL_2722186, EPI_ISL_2722187, EPI_ISL_2722188, EPI_ISL_2722189, EPI_ISL_2722190, EPI_ISL_2722191, EPI_ISL_2722192, EPI_ISL_2722193, EPI_ISL_2722194, EPI_ISL_2722195, EPI_ISL_2722196, EPI_ISL_2722197, EPI_ISL_2722198, EPI_ISL_2722199, EPI_ISL_2722200, EPI_ISL_2722201, EPI_ISL_2722202, EPI_ISL_2722203, EPI_ISL_2722204, EPI_ISL_2722205, EPI_ISL_2722206, EPI_ISL_2722207, EPI_ISL_2722208, EPI_ISL_2722209, EPI_ISL_2722210, EPI_ISL_2722211, EPI_ISL_2722212, EPI_ISL_2722213, EPI_ISL_2722215, EPI_ISL_2722216, EPI_ISL_2722217, EPI_ISL_2722218, EPI_ISL_2722219, EPI_ISL_2722220, EPI_ISL_2722221, EPI_ISL_2722222, EPI_ISL_2722223, EPI_ISL_2722224, EPI_ISL_2722225, EPI_ISL_2722226, EPI_ISL_2722227, EPI_ISL_2722228, EPI_ISL_2722229, EPI_ISL_2722230, EPI_ISL_2722231, EPI_ISL_2722232, EPI_ISL_2722233, EPI_ISL_2722234, EPI_ISL_2722235, EPI_ISL_2722236, EPI_ISL_2722237, EPI_ISL_2722238, EPI_ISL_2722239, EPI_ISL_2722240, EPI_ISL_2722241, EPI_ISL_2722242, EPI_ISL_2722243, EPI_ISL_2722244, EPI_ISL_2722245, EPI_ISL_2722246, EPI_ISL_2722247, EPI_ISL_2722248, EPI_ISL_2722249, EPI_ISL_2722250, EPI_ISL_2722251, EPI_ISL_2722252, EPI_ISL_2722253, EPI_ISL_2722254, EPI_ISL_2722255, EPI_ISL_2722256, EPI_ISL_2722257, EPI_ISL_2722258, EPI_ISL_2722259, EPI_ISL_2722260, EPI_ISL_2722261, EPI_ISL_2722262, EPI_ISL_2722263, EPI_ISL_2722264, EPI_ISL_2722265, EPI_ISL_2722266, EPI_ISL_2722267, EPI_ISL_2722268, EPI_ISL_2722269, EPI_ISL_2722270, EPI_ISL_2722271, EPI_ISL_2722272, EPI_ISL_2722273, EPI_ISL_2722274, EPI_ISL_2722275, EPI_ISL_2722276, EPI_ISL_2722277, EPI_ISL_2722278, EPI_ISL_2722279, EPI_ISL_2722280, EPI_ISL_2722281, EPI_ISL_2722282, EPI_ISL_2722283, EPI_ISL_2722284, EPI_ISL_2722285, EPI_ISL_2722286, EPI_ISL_2722287, EPI_ISL_2722288, EPI_ISL_2722289, EPI_ISL_2722290 | | | |
| see above | Department of Pathology, University of Cambridge | COVID-19 Genomics UK (COG-UK) Consortium | Aminu S. Jahun; Ian Goodfellow; Iliana Georgana; Martin D. Curran; Myra Hosmillo; Rhys Izuagbe; Surendra Parmar; William L. Hamilton; Yasmin Chaudhry |
| EPI_ISL_2537494, EPI_ISL_2537510, EPI_ISL_2537515, EPI_ISL_2537522, EPI_ISL_2537525, EPI_ISL_2537532, EPI_ISL_2537555, EPI_ISL_2537556, EPI_ISL_2537557, EPI_ISL_2537561, EPI_ISL_2537567, EPI_ISL_2537570, EPI_ISL_2537576, EPI_ISL_2537578, EPI_ISL_2537584, EPI_ISL_2537589, EPI_ISL_2537594, EPI_ISL_2537604, EPI_ISL_2537615, EPI_ISL_2537616, EPI_ISL_2537617, EPI_ISL_2537620, EPI_ISL_2537621, EPI_ISL_2537624, EPI_ISL_2537649, EPI_ISL_2537650, EPI_ISL_2537659, EPI_ISL_2537694, EPI_ISL_2537697, EPI_ISL_2537703, EPI_ISL_2537704, EPI_ISL_2537718, EPI_ISL_2537735, EPI_ISL_2537755, EPI_ISL_2537760, EPI_ISL_2537765, EPI_ISL_2537783, EPI_ISL_2537784, EPI_ISL_2537798, EPI_ISL_2537799, EPI_ISL_2537805, EPI_ISL_2537806, EPI_ISL_2552612, EPI_ISL_2552613, EPI_ISL_2552619, EPI_ISL_2552633, EPI_ISL_2552634, EPI_ISL_2552653, EPI_ISL_2552668, EPI_ISL_2552674, EPI_ISL_2552675, EPI_ISL_2552677, EPI_ISL_2552699, EPI_ISL_2552709, EPI_ISL_2552717, EPI_ISL_2552730, EPI_ISL_2552737, EPI_ISL_2552781, EPI_ISL_2552807, EPI_ISL_2552822, EPI_ISL_2552848, EPI_ISL_2552849, EPI_ISL_2552864, EPI_ISL_2552871, EPI_ISL_2552877, EPI_ISL_2552883, EPI_ISL_2552912, EPI_ISL_2553917, EPI_ISL_2553924, EPI_ISL_2553939, EPI_ISL_2554080, EPI_ISL_2555004, EPI_ISL_2555006, EPI_ISL_2641353, EPI_ISL_2641356, EPI_ISL_2641360, EPI_ISL_2641369, EPI_ISL_2641370, EPI_ISL_2641384, EPI_ISL_2641390, EPI_ISL_2641394, EPI_ISL_2641398, EPI_ISL_2641401, EPI_ISL_2641407, EPI_ISL_2641409, EPI_ISL_2641412, EPI_ISL_2641420, EPI_ISL_2641421, EPI_ISL_2641422, EPI_ISL_2641426, EPI_ISL_2641429, EPI_ISL_2641431, EPI_ISL_2641432, EPI_ISL_2641437, EPI_ISL_2641443, EPI_ISL_2641446, EPI_ISL_2641458, EPI_ISL_2641462, EPI_ISL_2641463, EPI_ISL_2641466, EPI_ISL_2641468, EPI_ISL_2641470, EPI_ISL_2641488, EPI_ISL_2641489, EPI_ISL_2641490, EPI_ISL_2641491, EPI_ISL_2641504, EPI_ISL_2641506, EPI_ISL_2641507, EPI_ISL_2641512, EPI_ISL_2641517, EPI_ISL_2641523, EPI_ISL_2641524, EPI_ISL_2641528, EPI_ISL_2641530, EPI_ISL_2641531, EPI_ISL_2641533, EPI_ISL_2641538, EPI_ISL_2641540, EPI_ISL_2641552, EPI_ISL_2641554, EPI_ISL_2641558, EPI_ISL_2641562, EPI_ISL_2641563, EPI_ISL_2641567, EPI_ISL_2641570, EPI_ISL_2641572, EPI_ISL_2641573, EPI_ISL_2641574, EPI_ISL_2641575, EPI_ISL_2641582, EPI_ISL_2641586, EPI_ISL_2641587, EPI_ISL_2641593, EPI_ISL_2641599, EPI_ISL_2641606, EPI_ISL_2641616, EPI_ISL_2641618, EPI_ISL_2641622, EPI_ISL_2641628, EPI_ISL_2641629, EPI_ISL_2641631, EPI_ISL_2641632, EPI_ISL_2641639, EPI_ISL_2641648, EPI_ISL_2641650, EPI_ISL_2641658, EPI_ISL_2641660, EPI_ISL_2641664, EPI_ISL_2641671, EPI_ISL_2641673, EPI_ISL_2641677, EPI_ISL_2641684, EPI_ISL_2641703, EPI_ISL_2641706, EPI_ISL_2641709 | | | |
| see above | Health Services Laboratories | Wellcome Sanger Institute for the COVID-19 Genomics UK (COG-UK) Consortium | Cordelia Langford; David K. Jackson; Dominic Kwiatkowski; Ewan Harrison; Health Services Laboratories and Alex Alderton; Ian Johnston; Jeffrey Barrett; John Sillitoe on behalf of the Wellcome Sanger Institute COVID-19 Surveillance Team; Roberto Amato; Sonia Goncalves |
| EPI_ISL_2537496, EPI_ISL_2537503, EPI_ISL_2537505, EPI_ISL_2537506, EPI_ISL_2537508, EPI_ISL_2537512, EPI_ISL_2537523, EPI_ISL_2537533, EPI_ISL_2537536, EPI_ISL_2537538, EPI_ISL_2537542, EPI_ISL_2537543, EPI_ISL_2537549, EPI_ISL_2537550, EPI_ISL_2537558, EPI_ISL_2537560, EPI_ISL_2537583, EPI_ISL_2537588, EPI_ISL_2537600, EPI_ISL_2537606, EPI_ISL_2537607, EPI_ISL_2537609, EPI_ISL_2537625, EPI_ISL_2537628, EPI_ISL_2537629, EPI_ISL_2537642, EPI_ISL_2537644, EPI_ISL_2537645, EPI_ISL_2537646, EPI_ISL_2537660, EPI_ISL_2537662, EPI_ISL_2537663, EPI_ISL_2537684, EPI_ISL_2537685, EPI_ISL_2537691, EPI_ISL_2537710, EPI_ISL_2537711, EPI_ISL_2537712, EPI_ISL_2537715, EPI_ISL_2537716, EPI_ISL_2537719, EPI_ISL_2537721, EPI_ISL_2537722, EPI_ISL_2537728, EPI_ISL_2537731, EPI_ISL_2537733, EPI_ISL_2537737, EPI_ISL_2537741, EPI_ISL_2537744, EPI_ISL_2537745, EPI_ISL_2537746, EPI_ISL_2537747, EPI_ISL_2537751, EPI_ISL_2537752, EPI_ISL_2537754, EPI_ISL_2537756, EPI_ISL_2537762, EPI_ISL_2537766, EPI_ISL_2537780, EPI_ISL_2537782, EPI_ISL_2537789, EPI_ISL_2537792, EPI_ISL_2537800, EPI_ISL_2537801, EPI_ISL_2551583, EPI_ISL_2551584, EPI_ISL_2551585, EPI_ISL_2551586, EPI_ISL_2551588, EPI_ISL_2551589, EPI_ISL_2551592, EPI_ISL_2551593, EPI_ISL_2551594, EPI_ISL_2551595, EPI_ISL_2551596, EPI_ISL_2551597, EPI_ISL_2551598, EPI_ISL_2551605, EPI_ISL_2551609, EPI_ISL_2551610, EPI_ISL_2551611, EPI_ISL_2551612, EPI_ISL_2551616, EPI_ISL_2551617, EPI_ISL_2551618, EPI_ISL_2551623, EPI_ISL_2551625, EPI_ISL_2551626, EPI_ISL_2551631, EPI_ISL_2551636, EPI_ISL_2551637, EPI_ISL_2551639, EPI_ISL_2551640, EPI_ISL_2551641, EPI_ISL_2551644, EPI_ISL_2551646, EPI_ISL_2551647, EPI_ISL_2551656, EPI_ISL_2551657, EPI_ISL_2551659, EPI_ISL_2551660, EPI_ISL_2551663, EPI_ISL_2551672, EPI_ISL_2551673, EPI_ISL_2551674, EPI_ISL_2551677, EPI_ISL_2551678, EPI_ISL_2551679, EPI_ISL_2551680, EPI_ISL_2551684, EPI_ISL_2551687, EPI_ISL_2551690, EPI_ISL_2551691, EPI_ISL_2551692, EPI_ISL_2551695, EPI_ISL_2551703, EPI_ISL_2551706, EPI_ISL_2551708, EPI_ISL_2551709, EPI_ISL_2551715, EPI_ISL_2551719, EPI_ISL_2551725, EPI_ISL_2551726, EPI_ISL_2551729, EPI_ISL_2551731, EPI_ISL_2551733, EPI_ISL_2551735, EPI_ISL_2551737, EPI_ISL_2551739, EPI_ISL_2551741, EPI_ISL_2551749, EPI_ISL_2551750, EPI_ISL_2551752, EPI_ISL_2551754, EPI_ISL_2551759, EPI_ISL_2551760, EPI_ISL_2551762, EPI_ISL_2551763, EPI_ISL_2551767, EPI_ISL_2551771, EPI_ISL_2551772, EPI_ISL_2551777, EPI_ISL_2551779, EPI_ISL_2551782, EPI_ISL_2551783, EPI_ISL_2551786, EPI_ISL_2551787, EPI_ISL_2551789, EPI_ISL_2551790, EPI_ISL_2551791, EPI_ISL_2551792, EPI_ISL_2551793, EPI_ISL_2551794, EPI_ISL_2551798, EPI_ISL_2551801, EPI_ISL_2551803, EPI_ISL_2551804, EPI_ISL_2551812, EPI_ISL_2551814, EPI_ISL_2551817, EPI_ISL_2551820, EPI_ISL_2551823, EPI_ISL_2551824, EPI_ISL_2551825, EPI_ISL_2551827, EPI_ISL_2551831, EPI_ISL_2551832, EPI_ISL_2551836, EPI_ISL_2551837, EPI_ISL_2551839, EPI_ISL_2551848, EPI_ISL_2551851, EPI_ISL_2551852, EPI_ISL_2551853, EPI_ISL_2551854, EPI_ISL_2551855, EPI_ISL_2551857, EPI_ISL_2551858, EPI_ISL_2551861, EPI_ISL_2551880, EPI_ISL_2551881, EPI_ISL_2551884, EPI_ISL_2551886, EPI_ISL_2551887, EPI_ISL_2551888, EPI_ISL_2551896, EPI_ISL_2551897, EPI_ISL_2551900, EPI_ISL_2551903, EPI_ISL_2551904, EPI_ISL_2551906, EPI_ISL_2551907, EPI_ISL_2551910, EPI_ISL_2551912, EPI_ISL_2551913, EPI_ISL_2551914, EPI_ISL_2551915, EPI_ISL_2551918, EPI_ISL_2551920, EPI_ISL_2551922, EPI_ISL_2551926, EPI_ISL_2552260, EPI_ISL_2552261, EPI_ISL_2552262, EPI_ISL_2552263, EPI_ISL_2552264, EPI_ISL_2552265, EPI_ISL_2552266, EPI_ISL_2552267, EPI_ISL_2552268, EPI_ISL_2552269, EPI_ISL_2552270, EPI_ISL_2552271, EPI_ISL_2552272, EPI_ISL_2552273, EPI_ISL_2552274, EPI_ISL_2552275, EPI_ISL_2552276, EPI_ISL_2552277, EPI_ISL_2552278, EPI_ISL_2552279, EPI_ISL_2552280, EPI_ISL_2552281, EPI_ISL_2552282, EPI_ISL_2552283, EPI_ISL_2552284, EPI_ISL_2552285, EPI_ISL_2552286, EPI_ISL_2552287, EPI_ISL_2552288, EPI_ISL_2552289, EPI_ISL_2552290, EPI_ISL_2552291, EPI_ISL_2552292, EPI_ISL_2552294, EPI_ISL_2552295, EPI_ISL_2552296, EPI_ISL_2552297, EPI_ISL_2552298, EPI_ISL_2552299, EPI_ISL_2552300, EPI_ISL_2552301, EPI_ISL_2552302, EPI_ISL_2552303, EPI_ISL_2552304, EPI_ISL_2552305, EPI_ISL_2552306, EPI_ISL_2552307, EPI_ISL_2552308, EPI_ISL_2552309, EPI_ISL_2552310, EPI_ISL_2552311, EPI_ISL_2552312, EPI_ISL_2552313, EPI_ISL_2552315, EPI_ISL_2552316, EPI_ISL_2552317, EPI_ISL_2552318, EPI_ISL_2552319, EPI_ISL_2552320, EPI_ISL_2552321, EPI_ISL_2552322, EPI_ISL_2552323, EPI_ISL_2552324, EPI_ISL_2552325, EPI_ISL_2552327, EPI_ISL_2552328, EPI_ISL_2552329, EPI_ISL_2552331, EPI_ISL_2552332, EPI_ISL_2552333, EPI_ISL_2552334, EPI_ISL_2552335, EPI_ISL_2552337, EPI_ISL_2552338, EPI_ISL_2552339, EPI_ISL_2552340, EPI_ISL_2552341, EPI_ISL_2552342, EPI_ISL_2552343, EPI_ISL_2552344, EPI_ISL_2552345, EPI_ISL_2552346, EPI_ISL_2552347, EPI_ISL_2552348, EPI_ISL_2552349, EPI_ISL_2552350, EPI_ISL_2552351, EPI_ISL_2552352, EPI_ISL_2552353, EPI_ISL_2552354, EPI_ISL_2552355, EPI_ISL_2552356, EPI_ISL_2552357, EPI_ISL_2552358, EPI_ISL_2552359, EPI_ISL_2552360, EPI_ISL_2552361, EPI_ISL_2552362, EPI_ISL_2552363, EPI_ISL_2552364, EPI_ISL_2552365, EPI_ISL_2552366, EPI_ISL_2552367, EPI_ISL_2552368, EPI_ISL_2552369, EPI_ISL_2552370, EPI_ISL_2552371, EPI_ISL_2552372, EPI_ISL_2552373, EPI_ISL_2552374, EPI_ISL_2552375, EPI_ISL_2552376, EPI_ISL_2552377, EPI_ISL_2552378, EPI_ISL_2552379, EPI_ISL_2552380, EPI_ISL_2552381, EPI_ISL_2552382, EPI_ISL_2552383, EPI_ISL_2552385, EPI_ISL_2552386, EPI_ISL_2552387, EPI_ISL_2552388, EPI_ISL_2552389, EPI_ISL_2552390, EPI_ISL_2552391, EPI_ISL_2552392, EPI_ISL_2552393, EPI_ISL_2552394, EPI_ISL_2552395, EPI_ISL_2552396, EPI_ISL_2552397, EPI_ISL_2552398, EPI_ISL_2552399, EPI_ISL_2552400, EPI_ISL_2552401, EPI_ISL_2552402, EPI_ISL_2552403, EPI_ISL_2552404, EPI_ISL_2552405, EPI_ISL_2552406, EPI_ISL_2552407, EPI_ISL_2552408, EPI_ISL_2552409, EPI_ISL_2552410, EPI_ISL_2552411, EPI_ISL_2552412, EPI_ISL_2552413, EPI_ISL_2552414, EPI_ISL_2552415, EPI_ISL_2552416, EPI_ISL_2552417, EPI_ISL_2552418, EPI_ISL_2552419, EPI_ISL_2552420, EPI_ISL_2552421, EPI_ISL_2552422, EPI_ISL_2552423, EPI_ISL_2552424, EPI_ISL_2552426, EPI_ISL_2552427, EPI_ISL_2552428, EPI_ISL_2552429, EPI_ISL_2552430, EPI_ISL_2552431, EPI_ISL_2552432, EPI_ISL_2552433, EPI_ISL_2552434, EPI_ISL_2552435, EPI_ISL_2552436, EPI_ISL_2552437, EPI_ISL_2552438, EPI_ISL_2552439, EPI_ISL_2552440, EPI_ISL_2552441, EPI_ISL_2552442, EPI_ISL_2552443, EPI_ISL_2552444, EPI_ISL_2552445, EPI_ISL_2552446, EPI_ISL_2552447, EPI_ISL_2552448, EPI_ISL_2552449, EPI_ISL_2552450, EPI_ISL_2552451, EPI_ISL_2552452, EPI_ISL_2552453, EPI_ISL_2552454, EPI_ISL_2552455, EPI_ISL_2552456, EPI_ISL_2552457, EPI_ISL_2552458, EPI_ISL_2552460, EPI_ISL_2552462, EPI_ISL_2552463, EPI_ISL_2552465, EPI_ISL_2552466, EPI_ISL_2552467, EPI_ISL_2552468, EPI_ISL_2552469, EPI_ISL_2552470, EPI_ISL_2552471, EPI_ISL_2552472, EPI_ISL_2552473, EPI_ISL_2552474, EPI_ISL_2552475, EPI_ISL_2552476, EPI_ISL_2552478, EPI_ISL_2552479, EPI_ISL_2552480, EPI_ISL_2552481, EPI_ISL_2552482, EPI_ISL_2552483, EPI_ISL_2552484, EPI_ISL_2552485, EPI_ISL_2552486, EPI_ISL_2552487, EPI_ISL_2552488, EPI_ISL_2552490, EPI_ISL_2552492, EPI_ISL_2552493, EPI_ISL_2552494, EPI_ISL_2552495, EPI_ISL_2552496, EPI_ISL_2552497, EPI_ISL_2552498, EPI_ISL_2552500, EPI_ISL_2552501, EPI_ISL_2552502, EPI_ISL_2552503, EPI_ISL_2552504, EPI_ISL_2552505, EPI_ISL_2552506, EPI_ISL_2552507, EPI_ISL_2552508, EPI_ISL_2552509, EPI_ISL_2552510, EPI_ISL_2552511, EPI_ISL_2552513, EPI_ISL_2552514, EPI_ISL_2552515, EPI_ISL_2552516, EPI_ISL_2552517, EPI_ISL_2552518, EPI_ISL_2552519, EPI_ISL_2552520, EPI_ISL_2552521, EPI_ISL_2552522, EPI_ISL_2552523, EPI_ISL_2552525, EPI_ISL_2552526, EPI_ISL_2552527, EPI_ISL_2552528, EPI_ISL_2552529, EPI_ISL_2552530, EPI_ISL_2552531, EPI_ISL_2552532, EPI_ISL_2552533, EPI_ISL_2552534, EPI_ISL_2552535, EPI_ISL_2552536, EPI_ISL_2552537, EPI_ISL_2552538, EPI_ISL_2552539, EPI_ISL_2552540, EPI_ISL_2552541, EPI_ISL_2552542, EPI_ISL_2552543, EPI_ISL_2552544, EPI_ISL_2552545, EPI_ISL_2552546, EPI_ISL_2552547, EPI_ISL_2552548, EPI_ISL_2552549, EPI_ISL_2552550, EPI_ISL_2552551, EPI_ISL_2552552, EPI_ISL_2552553, EPI_ISL_2552554, EPI_ISL_2552555, EPI_ISL_2552556, EPI_ISL_2552558, EPI_ISL_2552559, EPI_ISL_2552560, EPI_ISL_2552561, EPI_ISL_2552562, EPI_ISL_2552563, EPI_ISL_2552564, EPI_ISL_2552565, EPI_ISL_2552566, EPI_ISL_2552567, EPI_ISL_2552568, EPI_ISL_2552569, EPI_ISL_2552570, EPI_ISL_2552571, EPI_ISL_2552572, EPI_ISL_2552576, EPI_ISL_2552577, EPI_ISL_2552578, EPI_ISL_2552579, EPI_ISL_2552580, EPI_ISL_2552581, EPI_ISL_2552582, EPI_ISL_2552583, EPI_ISL_2552584, EPI_ISL_2552585, EPI_ISL_2552586, EPI_ISL_2552587, EPI_ISL_2552588, EPI_ISL_2552589, EPI_ISL_2552590, EPI_ISL_2552591, EPI_ISL_2552592, EPI_ISL_2552594, EPI_ISL_2552595, EPI_ISL_2552596, EPI_ISL_2552597, EPI_ISL_2552598, EPI_ISL_2552599, EPI_ISL_2552600, EPI_ISL_2552601, EPI_ISL_2552604, EPI_ISL_2552605, EPI_ISL_2552606, EPI_ISL_2552622, EPI_ISL_2552626, EPI_ISL_2552632, EPI_ISL_2552649, EPI_ISL_2552650, EPI_ISL_2552658, EPI_ISL_2552684, EPI_ISL_2552689, EPI_ISL_2552710, EPI_ISL_2552750, EPI_ISL_2552763, EPI_ISL_2552770, EPI_ISL_2552779, EPI_ISL_2552787, EPI_ISL_2552817, EPI_ISL_2552824, EPI_ISL_2552831, EPI_ISL_2552833, EPI_ISL_2552840, EPI_ISL_2552845, EPI_ISL_2552861, EPI_ISL_2552865, EPI_ISL_2552870, EPI_ISL_2552879, EPI_ISL_2552881, EPI_ISL_2552885, EPI_ISL_2552893, EPI_ISL_2552925, EPI_ISL_2552926, EPI_ISL_2552927, EPI_ISL_2552928, EPI_ISL_2552929, EPI_ISL_2552930, EPI_ISL_2552931, EPI_ISL_2552932, EPI_ISL_2552933, EPI_ISL_2552936, EPI_ISL_2552937, EPI_ISL_2552938, EPI_ISL_2552941, EPI_ISL_2552943, EPI_ISL_2552945, EPI_ISL_2552948, EPI_ISL_2552949, EPI_ISL_2552950, EPI_ISL_2552952, EPI_ISL_2552954, EPI_ISL_2552955, EPI_ISL_2552958, EPI_ISL_2552959, EPI_ISL_2552960, EPI_ISL_2552961, EPI_ISL_2552962, EPI_ISL_2552963, EPI_ISL_2552964, EPI_ISL_2552966, EPI_ISL_2552967, EPI_ISL_2552968, EPI_ISL_2552970, EPI_ISL_2552972, EPI_ISL_2552975, EPI_ISL_2552976, EPI_ISL_2552977, EPI_ISL_2552978, EPI_ISL_2552981, EPI_ISL_2552983, EPI_ISL_2552984, EPI_ISL_2552985, EPI_ISL_2552986, EPI_ISL_2552987, EPI_ISL_2552988, EPI_ISL_2552989, EPI_ISL_2552990, EPI_ISL_2552992, EPI_ISL_2552994, EPI_ISL_2552995, EPI_ISL_2552996, EPI_ISL_2552997, EPI_ISL_2552998, EPI_ISL_2552999, EPI_ISL_2553000, EPI_ISL_2553001, EPI_ISL_2553002, EPI_ISL_2553004, EPI_ISL_2553005, EPI_ISL_2553006, EPI_ISL_2553007, EPI_ISL_2553011, EPI_ISL_2553014, EPI_ISL_2553016, EPI_ISL_2553017, EPI_ISL_2553018, EPI_ISL_2553019, EPI_ISL_2553020, EPI_ISL_2553021, EPI_ISL_2553023, EPI_ISL_2553024, EPI_ISL_2553025, EPI_ISL_2553026, EPI_ISL_2553029, EPI_ISL_2553030, EPI_ISL_2553032, EPI_ISL_2553033, EPI_ISL_2553034, EPI_ISL_2553035, EPI_ISL_2553037, EPI_ISL_2553038, EPI_ISL_2553039, EPI_ISL_2553040, EPI_ISL_2553042, EPI_ISL_2553043, EPI_ISL_2553045, EPI_ISL_2553048, EPI_ISL_2553050, EPI_ISL_2553053, EPI_ISL_2553054, EPI_ISL_2553057, EPI_ISL_2553058, EPI_ISL_2553059, EPI_ISL_2553060, EPI_ISL_2553063, EPI_ISL_2553064, EPI_ISL_2553065, EPI_ISL_2553067, EPI_ISL_2553069, EPI_ISL_2553071, EPI_ISL_2553072, EPI_ISL_2553074, EPI_ISL_2553075, EPI_ISL_2553076, EPI_ISL_2553077, EPI_ISL_2553078, EPI_ISL_2553082, EPI_ISL_2553083, EPI_ISL_2553084, EPI_ISL_2553086, EPI_ISL_2553087, EPI_ISL_2553088, EPI_ISL_2553437, EPI_ISL_2553439, EPI_ISL_2553459, EPI_ISL_2553499, EPI_ISL_2553502, EPI_ISL_2553541, EPI_ISL_2553617, EPI_ISL_2553731, EPI_ISL_2553750, EPI_ISL_2553785, EPI_ISL_2553787, EPI_ISL_2553788, EPI_ISL_2553789, EPI_ISL_2553792, EPI_ISL_2553794, EPI_ISL_2553799, EPI_ISL_2553801, EPI_ISL_2553802, EPI_ISL_2553806, EPI_ISL_2553812, EPI_ISL_2553819, EPI_ISL_2553825, EPI_ISL_2553830, EPI_ISL_2553836, EPI_ISL_2553842, EPI_ISL_2553845, EPI_ISL_2553850, EPI_ISL_2553855, EPI_ISL_2553867, EPI_ISL_2553870, EPI_ISL_2553871, EPI_ISL_2553878, EPI_ISL_2553881, EPI_ISL_2553887, EPI_ISL_2553889, EPI_ISL_2553896, EPI_ISL_2553900, EPI_ISL_2553903, EPI_ISL_2553904, EPI_ISL_2553914, EPI_ISL_2553916, EPI_ISL_2553921, EPI_ISL_2553927, EPI_ISL_2553929, EPI_ISL_2553932, EPI_ISL_2553938, EPI_ISL_2553942, EPI_ISL_2553949, EPI_ISL_2553950, EPI_ISL_2553952, EPI_ISL_2553963, EPI_ISL_2553966, EPI_ISL_2553971, EPI_ISL_2553976, EPI_ISL_2553977, EPI_ISL_2553978, EPI_ISL_2553980, EPI_ISL_2553984, EPI_ISL_2553992, EPI_ISL_2554001, EPI_ISL_2554007, EPI_ISL_2554015, EPI_ISL_2554019, EPI_ISL_2554021, EPI_ISL_2554023, EPI_ISL_2554026, EPI_ISL_2554032, EPI_ISL_2554039, EPI_ISL_2554040, EPI_ISL_2554041, EPI_ISL_2554042, EPI_ISL_2554043, EPI_ISL_2554047, EPI_ISL_2554052, EPI_ISL_2554053, EPI_ISL_2554055, EPI_ISL_2554056, EPI_ISL_2554065, EPI_ISL_2554068, EPI_ISL_2554112, EPI_ISL_2554113, EPI_ISL_2554118, EPI_ISL_2554125, EPI_ISL_2554126, EPI_ISL_2554129, EPI_ISL_2554132, EPI_ISL_2554143, EPI_ISL_2554148, EPI_ISL_2554149, EPI_ISL_2554150, EPI_ISL_2554154, EPI_ISL_2554157, EPI_ISL_2554164, EPI_ISL_2554167, EPI_ISL_2554169, EPI_ISL_2554179, EPI_ISL_2554182, EPI_ISL_2554188, EPI_ISL_2554191, EPI_ISL_2554192, EPI_ISL_2554195, EPI_ISL_2554199, EPI_ISL_2554200, EPI_ISL_2554203, EPI_ISL_2554210, EPI_ISL_2554211, EPI_ISL_2554216, EPI_ISL_2554221, EPI_ISL_2554224, EPI_ISL_2554230, EPI_ISL_2554233, EPI_ISL_2554236, EPI_ISL_2554240, EPI_ISL_2554241, EPI_ISL_2554245, EPI_ISL_2554246, EPI_ISL_2554249, EPI_ISL_2554250, EPI_ISL_2554251, EPI_ISL_2554254, EPI_ISL_2554255, EPI_ISL_2554256, EPI_ISL_2554258, EPI_ISL_2554259, EPI_ISL_2554261, EPI_ISL_2554264, EPI_ISL_2554275, EPI_ISL_2554282, EPI_ISL_2554284, EPI_ISL_2554287, EPI_ISL_2554289, EPI_ISL_2554290, EPI_ISL_2554291, EPI_ISL_2554294, EPI_ISL_2554304, EPI_ISL_2554313, EPI_ISL_2554326, EPI_ISL_2554329, EPI_ISL_2554333, EPI_ISL_2554334, EPI_ISL_2554335, EPI_ISL_2554337, EPI_ISL_2554341, EPI_ISL_2554345, EPI_ISL_2554346, EPI_ISL_2554349, EPI_ISL_2554351, EPI_ISL_2554357, EPI_ISL_2554364, EPI_ISL_2554368, EPI_ISL_2554370, EPI_ISL_2554378, EPI_ISL_2554379, EPI_ISL_2554380, EPI_ISL_2554384, EPI_ISL_2554388, EPI_ISL_2554397, EPI_ISL_2554400, EPI_ISL_2554403, EPI_ISL_2554408, EPI_ISL_2554418, EPI_ISL_2554419, EPI_ISL_2554424, EPI_ISL_2554427, EPI_ISL_2554428, EPI_ISL_2554430, EPI_ISL_2554439, EPI_ISL_2554445, EPI_ISL_2554446, EPI_ISL_2554448, EPI_ISL_2554451, EPI_ISL_2554452, EPI_ISL_2554454, EPI_ISL_2554460, EPI_ISL_2554466, EPI_ISL_2554468, EPI_ISL_2554472, EPI_ISL_2554473, EPI_ISL_2554480, EPI_ISL_2554486, EPI_ISL_2554488, EPI_ISL_2554489, EPI_ISL_2555005, EPI_ISL_2555007, EPI_ISL_2567954, EPI_ISL_2567961, EPI_ISL_2567964, EPI_ISL_2567969, EPI_ISL_2567994, EPI_ISL_2568016, EPI_ISL_2568018, EPI_ISL_2568022, EPI_ISL_2568023, EPI_ISL_2568040, EPI_ISL_2568046, EPI_ISL_2568050, EPI_ISL_2568064, EPI_ISL_2568069, EPI_ISL_2568073, EPI_ISL_2568076, EPI_ISL_2568084, EPI_ISL_2568089, EPI_ISL_2568095, EPI_ISL_2568097, EPI_ISL_2568104, EPI_ISL_2568105, EPI_ISL_2568110, EPI_ISL_2568112, EPI_ISL_2568117, EPI_ISL_2568118, EPI_ISL_2568119, EPI_ISL_2568122, EPI_ISL_2568124, EPI_ISL_2568126, EPI_ISL_2568128, EPI_ISL_2568133, EPI_ISL_2568135, EPI_ISL_2568136, EPI_ISL_2568137, EPI_ISL_2568139, EPI_ISL_2568140, EPI_ISL_2568142, EPI_ISL_2568143, EPI_ISL_2568145, EPI_ISL_2568147, EPI_ISL_2568150, EPI_ISL_2568151, EPI_ISL_2568152, EPI_ISL_2568153, EPI_ISL_2568154, EPI_ISL_2568155, EPI_ISL_2568158, EPI_ISL_2568159, EPI_ISL_2568160, EPI_ISL_2568161, EPI_ISL_2568163, EPI_ISL_2568164, EPI_ISL_2568165, EPI_ISL_2568166, EPI_ISL_2568168, EPI_ISL_2568169, EPI_ISL_2568170, EPI_ISL_2568172, EPI_ISL_2568173, EPI_ISL_2568174, EPI_ISL_2568179, EPI_ISL_2568180, EPI_ISL_2568181, EPI_ISL_2568183, EPI_ISL_2568184, EPI_ISL_2568185, EPI_ISL_2568186, EPI_ISL_2568187, EPI_ISL_2568189, EPI_ISL_2568193, EPI_ISL_2568196, EPI_ISL_2568197, EPI_ISL_2568199, EPI_ISL_2568200, EPI_ISL_2568204, EPI_ISL_2568205, EPI_ISL_2568206, EPI_ISL_2568209, EPI_ISL_2568211, EPI_ISL_2568212, EPI_ISL_2568213, EPI_ISL_2568214, EPI_ISL_2568216, EPI_ISL_2568220, EPI_ISL_2568221, EPI_ISL_2568223, EPI_ISL_2568224, EPI_ISL_2568225, EPI_ISL_2568226, EPI_ISL_2568231, EPI_ISL_2568232, EPI_ISL_2568234, EPI_ISL_2568237, EPI_ISL_2568239, EPI_ISL_2568240, EPI_ISL_2568241, EPI_ISL_2568242, EPI_ISL_2568245, EPI_ISL_2568246, EPI_ISL_2568248, EPI_ISL_2568250, EPI_ISL_2568252, EPI_ISL_2568254, EPI_ISL_2568259, EPI_ISL_2568260, EPI_ISL_2568261, EPI_ISL_2568262, EPI_ISL_2568268, EPI_ISL_2568269, EPI_ISL_2568270, EPI_ISL_2568272, EPI_ISL_2568273, EPI_ISL_2568276, EPI_ISL_2568282, EPI_ISL_2568283, EPI_ISL_2568285, EPI_ISL_2568288, EPI_ISL_2568289, EPI_ISL_2568291, EPI_ISL_2568292, EPI_ISL_2568293, EPI_ISL_2568295, EPI_ISL_2568298, EPI_ISL_2568300, EPI_ISL_2568303, EPI_ISL_2568304, EPI_ISL_2568305, EPI_ISL_2568306, EPI_ISL_2568308, EPI_ISL_2568310, EPI_ISL_2568313, EPI_ISL_2568314, EPI_ISL_2568315, EPI_ISL_2568316, EPI_ISL_2568317, EPI_ISL_2568319, EPI_ISL_2568320, EPI_ISL_2568321, EPI_ISL_2568323, EPI_ISL_2568327, EPI_ISL_2568328, EPI_ISL_2568330, EPI_ISL_2568331, EPI_ISL_2568332, EPI_ISL_2568333, EPI_ISL_2568334, EPI_ISL_2568335, EPI_ISL_2568336, EPI_ISL_2568340, EPI_ISL_2568341, EPI_ISL_2568342, EPI_ISL_2568343, EPI_ISL_2568344, EPI_ISL_2568345, EPI_ISL_2568347, EPI_ISL_2568348, EPI_ISL_2568349, EPI_ISL_2568350, EPI_ISL_2568351, EPI_ISL_2568354, EPI_ISL_2568355, EPI_ISL_2568356, EPI_ISL_2568357, EPI_ISL_2568358, EPI_ISL_2568359, EPI_ISL_2568360, EPI_ISL_2568361, EPI_ISL_2568363, EPI_ISL_2568364, EPI_ISL_2568365, EPI_ISL_2568368, EPI_ISL_2568369, EPI_ISL_2568371, EPI_ISL_2568373, EPI_ISL_2568374, EPI_ISL_2568376, EPI_ISL_2568377, EPI_ISL_2568379, EPI_ISL_2568380, EPI_ISL_2568382, EPI_ISL_2568384, EPI_ISL_2568385, EPI_ISL_2568389, EPI_ISL_2568392, EPI_ISL_2568394, EPI_ISL_2568398, EPI_ISL_2568399, EPI_ISL_2568400, EPI_ISL_2568401, EPI_ISL_2568402, EPI_ISL_2568404, EPI_ISL_2568406, EPI_ISL_2568407, EPI_ISL_2568408, EPI_ISL_2568409, EPI_ISL_2568410, EPI_ISL_2568411, EPI_ISL_2568412, EPI_ISL_2568416, EPI_ISL_2568419, EPI_ISL_2568421, EPI_ISL_2568423, EPI_ISL_2568426, EPI_ISL_2568428, EPI_ISL_2568429, EPI_ISL_2568430, EPI_ISL_2568432, EPI_ISL_2568433, EPI_ISL_2568434, EPI_ISL_2568435, EPI_ISL_2568438, EPI_ISL_2568439, EPI_ISL_2568440, EPI_ISL_2568442, EPI_ISL_2568443, EPI_ISL_2568444, EPI_ISL_2568445, EPI_ISL_2568446, EPI_ISL_2568447, EPI_ISL_2568449, EPI_ISL_2568451, EPI_ISL_2568452, EPI_ISL_2568453, EPI_ISL_2568454, EPI_ISL_2568455, EPI_ISL_2568457, EPI_ISL_2568458, EPI_ISL_2568459, EPI_ISL_2568460, EPI_ISL_2568461, EPI_ISL_2568464, EPI_ISL_2568466, EPI_ISL_2568468, EPI_ISL_2568469, EPI_ISL_2568470, EPI_ISL_2568471, EPI_ISL_2568473, EPI_ISL_2568475, EPI_ISL_2568477, EPI_ISL_2568480, EPI_ISL_2568481, EPI_ISL_2568482, EPI_ISL_2568483, EPI_ISL_2568484, EPI_ISL_2568485, EPI_ISL_2568486, EPI_ISL_2568487, EPI_ISL_2568488, EPI_ISL_2568489, EPI_ISL_2568490, EPI_ISL_2568491, EPI_ISL_2568492, EPI_ISL_2568493, EPI_ISL_2568494, EPI_ISL_2568495, EPI_ISL_2568496, EPI_ISL_2568498, EPI_ISL_2568499, EPI_ISL_2568500, EPI_ISL_2568501, EPI_ISL_2568503, EPI_ISL_2568504, EPI_ISL_2568505, EPI_ISL_2568506, EPI_ISL_2568507, EPI_ISL_2568508, EPI_ISL_2568510, EPI_ISL_2568511, EPI_ISL_2568513, EPI_ISL_2568514, EPI_ISL_2568516, EPI_ISL_2568517, EPI_ISL_2568518, EPI_ISL_2568520, EPI_ISL_2568521, EPI_ISL_2568523, EPI_ISL_2568524, EPI_ISL_2568525, EPI_ISL_2568526, EPI_ISL_2568527, EPI_ISL_2568528, EPI_ISL_2568529, EPI_ISL_2568530, EPI_ISL_2568531, EPI_ISL_2568532, EPI_ISL_2568533, EPI_ISL_2568535, EPI_ISL_2568536, EPI_ISL_2568537, EPI_ISL_2568539, EPI_ISL_2568540, EPI_ISL_2568541, EPI_ISL_2568542, EPI_ISL_2568543, EPI_ISL_2568545, EPI_ISL_2568547, EPI_ISL_2568548, EPI_ISL_2568549, EPI_ISL_2568550, EPI_ISL_2568551, EPI_ISL_2568552, EPI_ISL_2568553, EPI_ISL_2568555, EPI_ISL_2568556, EPI_ISL_2568557, EPI_ISL_2568558, EPI_ISL_2568560, EPI_ISL_2568561, EPI_ISL_2568562, EPI_ISL_2568564, EPI_ISL_2568565, EPI_ISL_2568569, EPI_ISL_2568570, EPI_ISL_2568571, EPI_ISL_2568572, EPI_ISL_2568573, EPI_ISL_2568574, EPI_ISL_2568575, EPI_ISL_2568576, EPI_ISL_2568577, EPI_ISL_2568578, EPI_ISL_2568582, EPI_ISL_2568585, EPI_ISL_2568589, EPI_ISL_2568590, EPI_ISL_2568591, EPI_ISL_2568592, EPI_ISL_2568594, EPI_ISL_2568596, EPI_ISL_2568597, EPI_ISL_2568598, EPI_ISL_2568599, EPI_ISL_2568600, EPI_ISL_2568601, EPI_ISL_2568602, EPI_ISL_2568604, EPI_ISL_2568605, EPI_ISL_2568607, EPI_ISL_2568608, EPI_ISL_2568609, | | | |

EPI_ISL_2568610, EPI_ISL_2568612, EPI_ISL_2568613, EPI_ISL_2568614, EPI_ISL_2568615, EPI_ISL_2568616, EPI_ISL_2568617, EPI_ISL_2568618, EPI_ISL_2568620, EPI_ISL_2568621, EPI_ISL_2568622, EPI_ISL_2568624, EPI_ISL_2568625, EPI_ISL_2568626, EPI_ISL_2568627, EPI_ISL_2568629, EPI_ISL_2568631, EPI_ISL_2568632, EPI_ISL_2568635, EPI_ISL_2568637, EPI_ISL_2568638, EPI_ISL_2568639,
EPI_ISL_2568640, EPI_ISL_2568642, EPI_ISL_2568643, EPI_ISL_2568644, EPI_ISL_2568646, EPI_ISL_2568648, EPI_ISL_2568649, EPI_ISL_2568652, EPI_ISL_2568653, EPI_ISL_2568655, EPI_ISL_2568656, EPI_ISL_2568657, EPI_ISL_2568660, EPI_ISL_2568661, EPI_ISL_2568665, EPI_ISL_2568672, EPI_ISL_2568675, EPI_ISL_2568678, EPI_ISL_2568684, EPI_ISL_2568688, EPI_ISL_2568693, EPI_ISL_2568698,
EPI_ISL_2568711, EPI_ISL_2568719, EPI_ISL_2568730, EPI_ISL_2568740, EPI_ISL_2568746, EPI_ISL_2568752, EPI_ISL_2568753, EPI_ISL_2568756, EPI_ISL_2568758, EPI_ISL_2568760, EPI_ISL_2568764, EPI_ISL_2568775, EPI_ISL_2568776, EPI_ISL_2568784, EPI_ISL_2568787, EPI_ISL_2568810, EPI_ISL_2568812, EPI_ISL_2568823, EPI_ISL_2568829, EPI_ISL_2568841, EPI_ISL_2568842,
EPI_ISL_2568845, EPI_ISL_2568846, EPI_ISL_2568849, EPI_ISL_2568850, EPI_ISL_2568854, EPI_ISL_2568856, EPI_ISL_2568864, EPI_ISL_2568870, EPI_ISL_2568872, EPI_ISL_2568874, EPI_ISL_2568877, EPI_ISL_2568878, EPI_ISL_2568879, EPI_ISL_2568883, EPI_ISL_2568884, EPI_ISL_2568890, EPI_ISL_2568891, EPI_ISL_2568894, EPI_ISL_2568895, EPI_ISL_2568896, EPI_ISL_2568901, EPI_ISL_2568903,
EPI_ISL_2568904, EPI_ISL_2568905, EPI_ISL_2568906, EPI_ISL_2568907, EPI_ISL_2568911, EPI_ISL_2568913, EPI_ISL_2568915, EPI_ISL_2568924, EPI_ISL_2568928, EPI_ISL_2568931, EPI_ISL_2568932, EPI_ISL_2568934, EPI_ISL_2568937, EPI_ISL_2568939, EPI_ISL_2568940, EPI_ISL_2568941, EPI_ISL_2568945, EPI_ISL_2568948, EPI_ISL_2568949, EPI_ISL_2568952, EPI_ISL_2568959, EPI_ISL_2568962,
EPI_ISL_2568963, EPI_ISL_2568965, EPI_ISL_2568970, EPI_ISL_2568976, EPI_ISL_2568978, EPI_ISL_2568981, EPI_ISL_2568982, EPI_ISL_2568986, EPI_ISL_2568989, EPI_ISL_2568999, EPI_ISL_2569000, EPI_ISL_2569001, EPI_ISL_2569003, EPI_ISL_2569006, EPI_ISL_2569010, EPI_ISL_2569012, EPI_ISL_2569015, EPI_ISL_2569017, EPI_ISL_2569026, EPI_ISL_2569029, EPI_ISL_2569031, EPI_ISL_2569038,
EPI_ISL_2569039, EPI_ISL_2569041, EPI_ISL_2569043, EPI_ISL_2569044, EPI_ISL_2569048, EPI_ISL_2569049, EPI_ISL_2569050, EPI_ISL_2569052, EPI_ISL_2569057, EPI_ISL_2569061, EPI_ISL_2569065, EPI_ISL_2569075, EPI_ISL_2569085, EPI_ISL_2569086, EPI_ISL_2569089, EPI_ISL_2569096, EPI_ISL_2569113, EPI_ISL_2569117, EPI_ISL_2569124, EPI_ISL_2569127, EPI_ISL_2569128, EPI_ISL_2569129,
EPI_ISL_2569130, EPI_ISL_2569133, EPI_ISL_2569139, EPI_ISL_2569152, EPI_ISL_2569161, EPI_ISL_2569171, EPI_ISL_2569181, EPI_ISL_2569185, EPI_ISL_2569188, EPI_ISL_2569189, EPI_ISL_2569191, EPI_ISL_2569192, EPI_ISL_2569203, EPI_ISL_2569211, EPI_ISL_2569222, EPI_ISL_2569226, EPI_ISL_2569230, EPI_ISL_2569235, EPI_ISL_2569244, EPI_ISL_2569251, EPI_ISL_2569252, EPI_ISL_2569257,
EPI_ISL_2569258, EPI_ISL_2569260, EPI_ISL_2569276, EPI_ISL_2569277, EPI_ISL_2569282, EPI_ISL_2569285, EPI_ISL_2569292, EPI_ISL_2569293, EPI_ISL_2569294, EPI_ISL_2569299, EPI_ISL_2569303, EPI_ISL_2569307, EPI_ISL_2569311, EPI_ISL_2569318, EPI_ISL_2569319, EPI_ISL_2569320, EPI_ISL_2569321, EPI_ISL_2569322, EPI_ISL_2569323, EPI_ISL_2569326, EPI_ISL_2569327, EPI_ISL_2569328,
EPI_ISL_2569329, EPI_ISL_2569330, EPI_ISL_2569331, EPI_ISL_2569333, EPI_ISL_2569334, EPI_ISL_2569335, EPI_ISL_2569336, EPI_ISL_2569339, EPI_ISL_2569340, EPI_ISL_2569341, EPI_ISL_2569342, EPI_ISL_2569344, EPI_ISL_2569345, EPI_ISL_2569346, EPI_ISL_2569347, EPI_ISL_2569348, EPI_ISL_2569349, EPI_ISL_2569350, EPI_ISL_2569351, EPI_ISL_2569352, EPI_ISL_2569353, EPI_ISL_2569354,
EPI_ISL_2569355, EPI_ISL_2569356, EPI_ISL_2569357, EPI_ISL_2569358, EPI_ISL_2569360, EPI_ISL_2569361, EPI_ISL_2569362, EPI_ISL_2569363, EPI_ISL_2569364, EPI_ISL_2569366, EPI_ISL_2569367, EPI_ISL_2569368, EPI_ISL_2569369, EPI_ISL_2569370, EPI_ISL_2569371, EPI_ISL_2569372, EPI_ISL_2569373, EPI_ISL_2569374, EPI_ISL_2569375, EPI_ISL_2569376, EPI_ISL_2569377, EPI_ISL_2569380,
EPI_ISL_2569381, EPI_ISL_2569382, EPI_ISL_2569383, EPI_ISL_2569384, EPI_ISL_2569385, EPI_ISL_2569387, EPI_ISL_2569388, EPI_ISL_2569389, EPI_ISL_2569390, EPI_ISL_2569391, EPI_ISL_2569392, EPI_ISL_2569393, EPI_ISL_2569394, EPI_ISL_2569396, EPI_ISL_2569397, EPI_ISL_2569398, EPI_ISL_2569399, EPI_ISL_2569400, EPI_ISL_2569401, EPI_ISL_2569402, EPI_ISL_2569403, EPI_ISL_2569405,
EPI_ISL_2569406, EPI_ISL_2569407, EPI_ISL_2569408, EPI_ISL_2569409, EPI_ISL_2569410, EPI_ISL_2569411, EPI_ISL_2569412, EPI_ISL_2569413, EPI_ISL_2569414, EPI_ISL_2569415, EPI_ISL_2569416, EPI_ISL_2569417, EPI_ISL_2569420, EPI_ISL_2569421, EPI_ISL_2569422, EPI_ISL_2569423, EPI_ISL_2569424, EPI_ISL_2569425, EPI_ISL_2569426, EPI_ISL_2569427, EPI_ISL_2569428, EPI_ISL_2569429,
EPI_ISL_2569430, EPI_ISL_2569431, EPI_ISL_2569433, EPI_ISL_2569434, EPI_ISL_2569436, EPI_ISL_2569439, EPI_ISL_2569440, EPI_ISL_2569442, EPI_ISL_2569443, EPI_ISL_2569444, EPI_ISL_2569445, EPI_ISL_2569447, EPI_ISL_2569448, EPI_ISL_2569449, EPI_ISL_2569450, EPI_ISL_2569451, EPI_ISL_2569452, EPI_ISL_2569453, EPI_ISL_2569455, EPI_ISL_2569457, EPI_ISL_2569458, EPI_ISL_2569459,
EPI_ISL_2569462, EPI_ISL_2569463, EPI_ISL_2569464, EPI_ISL_2569465, EPI_ISL_2569466, EPI_ISL_2569467, EPI_ISL_2569468, EPI_ISL_2569469, EPI_ISL_2569472, EPI_ISL_2569473, EPI_ISL_2569475, EPI_ISL_2569476, EPI_ISL_2569477, EPI_ISL_2569478, EPI_ISL_2569479, EPI_ISL_2569481, EPI_ISL_2569482, EPI_ISL_2569483, EPI_ISL_2569484, EPI_ISL_2569485, EPI_ISL_2569486, EPI_ISL_2569487,
EPI_ISL_2569488, EPI_ISL_2569490, EPI_ISL_2569491, EPI_ISL_2569492, EPI_ISL_2569493, EPI_ISL_2569495, EPI_ISL_2569496, EPI_ISL_2569497, EPI_ISL_2569499, EPI_ISL_2569501, EPI_ISL_2569502, EPI_ISL_2569503, EPI_ISL_2569504, EPI_ISL_2569506, EPI_ISL_2569507, EPI_ISL_2569508, EPI_ISL_2569509, EPI_ISL_2569510, EPI_ISL_2569511, EPI_ISL_2569513, EPI_ISL_2569514, EPI_ISL_2569515,
EPI_ISL_2569516, EPI_ISL_2569517, EPI_ISL_2569519, EPI_ISL_2569520, EPI_ISL_2569522, EPI_ISL_2569525, EPI_ISL_2569526, EPI_ISL_2569527, EPI_ISL_2569528, EPI_ISL_2569530, EPI_ISL_2569531, EPI_ISL_2569532, EPI_ISL_2569533, EPI_ISL_2569534, EPI_ISL_2569535, EPI_ISL_2569536, EPI_ISL_2569537, EPI_ISL_2569538, EPI_ISL_2569539, EPI_ISL_2569540, EPI_ISL_2569541, EPI_ISL_2569542,
EPI_ISL_2569543, EPI_ISL_2569544, EPI_ISL_2569545, EPI_ISL_2569546, EPI_ISL_2569547, EPI_ISL_2569548, EPI_ISL_2569549, EPI_ISL_2569550, EPI_ISL_2569552, EPI_ISL_2569553, EPI_ISL_2569554, EPI_ISL_2569555, EPI_ISL_2569556, EPI_ISL_2569557, EPI_ISL_2569558, EPI_ISL_2569559, EPI_ISL_2569560, EPI_ISL_2569561, EPI_ISL_2569562, EPI_ISL_2569563, EPI_ISL_2569564, EPI_ISL_2569565,
EPI_ISL_2569566, EPI_ISL_2569567, EPI_ISL_2569568, EPI_ISL_2569569, EPI_ISL_2569570, EPI_ISL_2569571, EPI_ISL_2569572, EPI_ISL_2569574, EPI_ISL_2569575, EPI_ISL_2569576, EPI_ISL_2569577, EPI_ISL_2569578, EPI_ISL_2569579, EPI_ISL_2569581, EPI_ISL_2569582, EPI_ISL_2569583, EPI_ISL_2569584, EPI_ISL_2569585, EPI_ISL_2569586, EPI_ISL_2569587, EPI_ISL_2569588, EPI_ISL_2569590,
EPI_ISL_2569591, EPI_ISL_2569593, EPI_ISL_2569594, EPI_ISL_2569595, EPI_ISL_2569596, EPI_ISL_2569597, EPI_ISL_2569598, EPI_ISL_2569600, EPI_ISL_2569601, EPI_ISL_2569602, EPI_ISL_2569603, EPI_ISL_2569604, EPI_ISL_2569605, EPI_ISL_2569606, EPI_ISL_2569607, EPI_ISL_2569608, EPI_ISL_2569609, EPI_ISL_2569610, EPI_ISL_2569612, EPI_ISL_2569613, EPI_ISL_2569614, EPI_ISL_2569615,
EPI_ISL_2569616, EPI_ISL_2569617, EPI_ISL_2569618, EPI_ISL_2569619, EPI_ISL_2569620, EPI_ISL_2569623, EPI_ISL_2569624, EPI_ISL_2569625, EPI_ISL_2569626, EPI_ISL_2569628, EPI_ISL_2569629, EPI_ISL_2569630, EPI_ISL_2569631, EPI_ISL_2569632, EPI_ISL_2569633, EPI_ISL_2569634, EPI_ISL_2569635, EPI_ISL_2569636, EPI_ISL_2569637, EPI_ISL_2569638, EPI_ISL_2569639, EPI_ISL_2569640,
EPI_ISL_2569642, EPI_ISL_2569643, EPI_ISL_2569644, EPI_ISL_2569645, EPI_ISL_2569646, EPI_ISL_2569647, EPI_ISL_2569648, EPI_ISL_2569649, EPI_ISL_2569650, EPI_ISL_2569651, EPI_ISL_2569652, EPI_ISL_2569653, EPI_ISL_2569654, EPI_ISL_2569655, EPI_ISL_2569656, EPI_ISL_2569657, EPI_ISL_2569658, EPI_ISL_2569659, EPI_ISL_2569661, EPI_ISL_2569662, EPI_ISL_2569664, EPI_ISL_2569673,
EPI_ISL_2569675, EPI_ISL_2569677, EPI_ISL_2569678, EPI_ISL_2569689, EPI_ISL_2569690, EPI_ISL_2569692, EPI_ISL_2569695, EPI_ISL_2569697, EPI_ISL_2569705, EPI_ISL_2569706, EPI_ISL_2569707, EPI_ISL_2569708, EPI_ISL_2569710, EPI_ISL_2569711, EPI_ISL_2569714, EPI_ISL_2569716, EPI_ISL_2569723, EPI_ISL_2569726, EPI_ISL_2569728, EPI_ISL_2569729, EPI_ISL_2569731, EPI_ISL_2569732,
EPI_ISL_2569733, EPI_ISL_2569736, EPI_ISL_2569740, EPI_ISL_2569744, EPI_ISL_2569747, EPI_ISL_2569753, EPI_ISL_2569757, EPI_ISL_2569759, EPI_ISL_2569764, EPI_ISL_2569766, EPI_ISL_2569771, EPI_ISL_2569773, EPI_ISL_2569776, EPI_ISL_2569780, EPI_ISL_2569781, EPI_ISL_2569783, EPI_ISL_2569786, EPI_ISL_2569792, EPI_ISL_2569795, EPI_ISL_2569797, EPI_ISL_2569801, EPI_ISL_2569802,
EPI_ISL_2569803, EPI_ISL_2569811, EPI_ISL_2569813, EPI_ISL_2569814, EPI_ISL_2569820, EPI_ISL_2569822, EPI_ISL_2569823, EPI_ISL_2569827, EPI_ISL_2569829, EPI_ISL_2569831, EPI_ISL_2569834, EPI_ISL_2569837, EPI_ISL_2569838, EPI_ISL_2569839, EPI_ISL_2569847, EPI_ISL_2569850, EPI_ISL_2569854, EPI_ISL_2569856, EPI_ISL_2569857, EPI_ISL_2569864, EPI_ISL_2569865,
EPI_ISL_2569868, EPI_ISL_2569870, EPI_ISL_2569873, EPI_ISL_2569883, EPI_ISL_2569888, EPI_ISL_2569889, EPI_ISL_2569894, EPI_ISL_2569895, EPI_ISL_2569896, EPI_ISL_2569900, EPI_ISL_2569904, EPI_ISL_2569905, EPI_ISL_2569907, EPI_ISL_2569908, EPI_ISL_2569915, EPI_ISL_2569916, EPI_ISL_2569918, EPI_ISL_2569920, EPI_ISL_2569927, EPI_ISL_2569928, EPI_ISL_2569929, EPI_ISL_2569930,
EPI_ISL_2569932, EPI_ISL_2569933, EPI_ISL_2569945, EPI_ISL_2569946, EPI_ISL_2569947, EPI_ISL_2569953, EPI_ISL_2569957, EPI_ISL_2569964, EPI_ISL_2569970, EPI_ISL_2569972, EPI_ISL_2569974, EPI_ISL_2569975, EPI_ISL_2569978, EPI_ISL_2569985, EPI_ISL_2569987, EPI_ISL_2569988, EPI_ISL_2569990, EPI_ISL_2569992, EPI_ISL_2570003, EPI_ISL_2570005, EPI_ISL_2570006,
EPI_ISL_2570007, EPI_ISL_2570013, EPI_ISL_2570014, EPI_ISL_2570025, EPI_ISL_2570029, EPI_ISL_2570030, EPI_ISL_2570034, EPI_ISL_2570035, EPI_ISL_2570036, EPI_ISL_2570037, EPI_ISL_2570045, EPI_ISL_2570053, EPI_ISL_2570059, EPI_ISL_2570063, EPI_ISL_2570068, EPI_ISL_2570070, EPI_ISL_2570081, EPI_ISL_2570082, EPI_ISL_2570084, EPI_ISL_2570086, EPI_ISL_2570087, EPI_ISL_2570090,
EPI_ISL_2570095, EPI_ISL_2570098, EPI_ISL_2570100, EPI_ISL_2570101, EPI_ISL_2570113, EPI_ISL_2570115, EPI_ISL_2570116, EPI_ISL_2570123, EPI_ISL_2570129, EPI_ISL_2570130, EPI_ISL_2570133, EPI_ISL_2570135, EPI_ISL_2570136, EPI_ISL_2570137, EPI_ISL_2570142, EPI_ISL_2570146, EPI_ISL_2570147, EPI_ISL_2570151, EPI_ISL_2570158, EPI_ISL_2570160, EPI_ISL_2570164, EPI_ISL_2570165,
EPI_ISL_2570168, EPI_ISL_2570170, EPI_ISL_2570171, EPI_ISL_2570175, EPI_ISL_2570188, EPI_ISL_2570189, EPI_ISL_2570201, EPI_ISL_2570202, EPI_ISL_2570209, EPI_ISL_2570210, EPI_ISL_2570218, EPI_ISL_2570220, EPI_ISL_2570221, EPI_ISL_2570228, EPI_ISL_2570229, EPI_ISL_2570232, EPI_ISL_2570238, EPI_ISL_2570245, EPI_ISL_2570247, EPI_ISL_2570248, EPI_ISL_2570250, EPI_ISL_2570253,
EPI_ISL_2570255, EPI_ISL_2570257, EPI_ISL_2570258, EPI_ISL_2570261, EPI_ISL_2570262, EPI_ISL_2570263, EPI_ISL_2570265, EPI_ISL_2570266, EPI_ISL_2570267, EPI_ISL_2570269, EPI_ISL_2570270, EPI_ISL_2570272, EPI_ISL_2570474, EPI_ISL_2570489, EPI_ISL_2570514, EPI_ISL_2570534, EPI_ISL_2570550, EPI_ISL_2570557, EPI_ISL_2570564, EPI_ISL_2570567, EPI_ISL_2570583, EPI_ISL_2570591,
EPI_ISL_2570616, EPI_ISL_2570622, EPI_ISL_2570674, EPI_ISL_2570681, EPI_ISL_2570703, EPI_ISL_2570730, EPI_ISL_2570734, EPI_ISL_2570735, EPI_ISL_2570773, EPI_ISL_2570780, EPI_ISL_2570783, EPI_ISL_2570785, EPI_ISL_2570793, EPI_ISL_2605129, EPI_ISL_2605133, EPI_ISL_2605138, EPI_ISL_2605150, EPI_ISL_2605152, EPI_ISL_2605156, EPI_ISL_2605162, EPI_ISL_2605165, EPI_ISL_2605169,
EPI_ISL_2605176, EPI_ISL_2605177, EPI_ISL_2605180, EPI_ISL_2605188, EPI_ISL_2605189, EPI_ISL_2605192, EPI_ISL_2605200, EPI_ISL_2605203, EPI_ISL_2605214, EPI_ISL_2605216, EPI_ISL_2605223, EPI_ISL_2605227, EPI_ISL_2605233, EPI_ISL_2605235, EPI_ISL_2605241, EPI_ISL_2605248, EPI_ISL_2605255, EPI_ISL_2605258, EPI_ISL_2605275, EPI_ISL_2605280, EPI_ISL_2605282, EPI_ISL_2605283,
EPI_ISL_2605286, EPI_ISL_2605288, EPI_ISL_2605289, EPI_ISL_2605296, EPI_ISL_2605298, EPI_ISL_2605302, EPI_ISL_2605305, EPI_ISL_2605314, EPI_ISL_2605322, EPI_ISL_2605323, EPI_ISL_2605324, EPI_ISL_2605325, EPI_ISL_2605334, EPI_ISL_2605344, EPI_ISL_2605345, EPI_ISL_2605347, EPI_ISL_2605348, EPI_ISL_2605350, EPI_ISL_2605361, EPI_ISL_2605364, EPI_ISL_2605366, EPI_ISL_2605373,
EPI_ISL_2605375, EPI_ISL_2605376, EPI_ISL_2605415, EPI_ISL_2605427, EPI_ISL_2605430, EPI_ISL_2605440, EPI_ISL_2605442, EPI_ISL_2605445, EPI_ISL_2605447, EPI_ISL_2605448, EPI_ISL_2605452, EPI_ISL_2605455, EPI_ISL_2605470, EPI_ISL_2605471, EPI_ISL_2605475, EPI_ISL_2605476, EPI_ISL_2605479, EPI_ISL_2605481, EPI_ISL_2605484, EPI_ISL_2605485, EPI_ISL_2605487, EPI_ISL_2605488,
EPI_ISL_2605490, EPI_ISL_2605493, EPI_ISL_2605494, EPI_ISL_2605498, EPI_ISL_2605501, EPI_ISL_2605504, EPI_ISL_2605508, EPI_ISL_2605510, EPI_ISL_2605515, EPI_ISL_2605516, EPI_ISL_2605519, EPI_ISL_2605520, EPI_ISL_2605526, EPI_ISL_2605529, EPI_ISL_2605534, EPI_ISL_2605536, EPI_ISL_2605539, EPI_ISL_2605548, EPI_ISL_2605549, EPI_ISL_2605552, EPI_ISL_2605560, EPI_ISL_2605561,
EPI_ISL_2605564, EPI_ISL_2605566, EPI_ISL_2605569, EPI_ISL_2605570, EPI_ISL_2605573, EPI_ISL_2605578, EPI_ISL_2605580, EPI_ISL_2605583, EPI_ISL_2605584, EPI_ISL_2605589, EPI_ISL_2605592, EPI_ISL_2605597, EPI_ISL_2605598, EPI_ISL_2605601, EPI_ISL_2605603, EPI_ISL_2605606, EPI_ISL_2605609, EPI_ISL_2605618, EPI_ISL_2605623, EPI_ISL_2605630, EPI_ISL_2605632, EPI_ISL_2605637,
EPI_ISL_2605643, EPI_ISL_2605645, EPI_ISL_2605646, EPI_ISL_2605648, EPI_ISL_2605654, EPI_ISL_2605659, EPI_ISL_2605661, EPI_ISL_2605662, EPI_ISL_2605665, EPI_ISL_2605667, EPI_ISL_2605669, EPI_ISL_2605670, EPI_ISL_2605671, EPI_ISL_2605673, EPI_ISL_2605675, EPI_ISL_2605676, EPI_ISL_2605680, EPI_ISL_2605683, EPI_ISL_2605684, EPI_ISL_2605685, EPI_ISL_2605686, EPI_ISL_2605687,
EPI_ISL_2605688, EPI_ISL_2605690, EPI_ISL_2605692, EPI_ISL_2605694, EPI_ISL_2605695, EPI_ISL_2605697, EPI_ISL_2605701, EPI_ISL_2605705, EPI_ISL_2605706, EPI_ISL_2605708, EPI_ISL_2605711, EPI_ISL_2605712, EPI_ISL_2605717, EPI_ISL_2605720, EPI_ISL_2605721, EPI_ISL_2605723, EPI_ISL_2605725, EPI_ISL_2605726, EPI_ISL_2605727, EPI_ISL_2605728, EPI_ISL_2605731, EPI_ISL_2605732,
EPI_ISL_2605733, EPI_ISL_2605734, EPI_ISL_2605736, EPI_ISL_2605739, EPI_ISL_2605740, EPI_ISL_2605741, EPI_ISL_2605742, EPI_ISL_2605743, EPI_ISL_2605744, EPI_ISL_2605747, EPI_ISL_2605749, EPI_ISL_2605750, EPI_ISL_2605751, EPI_ISL_2605752, EPI_ISL_2605753, EPI_ISL_2605754, EPI_ISL_2605755, EPI_ISL_2605757, EPI_ISL_2605758, EPI_ISL_2605762, EPI_ISL_2605763, EPI_ISL_2605764,
EPI_ISL_2605768, EPI_ISL_2605769, EPI_ISL_2605770, EPI_ISL_2605773, EPI_ISL_2605774, EPI_ISL_2605775, EPI_ISL_2605776, EPI_ISL_2605777, EPI_ISL_2605778, EPI_ISL_2605779, EPI_ISL_2605783, EPI_ISL_2605786, EPI_ISL_2605788, EPI_ISL_2605790, EPI_ISL_2605793, EPI_ISL_2605794, EPI_ISL_2605795, EPI_ISL_2605797, EPI_ISL_2605799, EPI_ISL_2605801, EPI_ISL_2605802, EPI_ISL_2605803,
EPI_ISL_2605804, EPI_ISL_2605805, EPI_ISL_2605806, EPI_ISL_2605807, EPI_ISL_2605809, EPI_ISL_2605810, EPI_ISL_2605812, EPI_ISL_2605813, EPI_ISL_2605814, EPI_ISL_2605816, EPI_ISL_2605817, EPI_ISL_2605818, EPI_ISL_2605820, EPI_ISL_2605822, EPI_ISL_2605824, EPI_ISL_2605825, EPI_ISL_2605827, EPI_ISL_2605829, EPI_ISL_2605830, EPI_ISL_2605831, EPI_ISL_2605833, EPI_ISL_2605834,
EPI_ISL_2605835, EPI_ISL_2605838, EPI_ISL_2605841, EPI_ISL_2605842, EPI_ISL_2605843, EPI_ISL_2605844, EPI_ISL_2605845, EPI_ISL_2605846, EPI_ISL_2605850, EPI_ISL_2605852, EPI_ISL_2605854, EPI_ISL_2605855, EPI_ISL_2605857, EPI_ISL_2605859, EPI_ISL_2605860, EPI_ISL_2605861, EPI_ISL_2605862, EPI_ISL_2605863, EPI_ISL_2605865, EPI_ISL_2605866, EPI_ISL_2605867, EPI_ISL_2605868,
EPI_ISL_2605869, EPI_ISL_2605870, EPI_ISL_2605871, EPI_ISL_2605872, EPI_ISL_2605874, EPI_ISL_2605875, EPI_ISL_2605876, EPI_ISL_2605877, EPI_ISL_2605878, EPI_ISL_2605879, EPI_ISL_2605880, EPI_ISL_2605882, EPI_ISL_2605884, EPI_ISL_2605885, EPI_ISL_2605887, EPI_ISL_2605888, EPI_ISL_2605889, EPI_ISL_2605890, EPI_ISL_2605891, EPI_ISL_2605892, EPI_ISL_2605893, EPI_ISL_2605894,
EPI_ISL_2605895, EPI_ISL_2605896, EPI_ISL_2605898, EPI_ISL_2605899, EPI_ISL_2605901, EPI_ISL_2605903, EPI_ISL_2605904, EPI_ISL_2605906, EPI_ISL_2605907, EPI_ISL_2605909, EPI_ISL_2605910, EPI_ISL_2605912, EPI_ISL_2605913, EPI_ISL_2605914, EPI_ISL_2605915, EPI_ISL_2605916, EPI_ISL_2605917, EPI_ISL_2605918, EPI_ISL_2605919, EPI_ISL_2605920, EPI_ISL_2605921, EPI_ISL_2605922,
EPI_ISL_2605923, EPI_ISL_2605924, EPI_ISL_2605926, EPI_ISL_2605927, EPI_ISL_2605928, EPI_ISL_2605929, EPI_ISL_2605930, EPI_ISL_2605931, EPI_ISL_2605932, EPI_ISL_2605933, EPI_ISL_2605934, EPI_ISL_2605935, EPI_ISL_2605937, EPI_ISL_2605939, EPI_ISL_2605941, EPI_ISL_2605942, EPI_ISL_2605943, EPI_ISL_2605944, EPI_ISL_2605945, EPI_ISL_2605947, EPI_ISL_2605949, EPI_ISL_2605950,
EPI_ISL_2605953, EPI_ISL_2605954, EPI_ISL_2605955, EPI_ISL_2605956, EPI_ISL_2605958, EPI_ISL_2605959, EPI_ISL_2605960, EPI_ISL_2605961, EPI_ISL_2605963, EPI_ISL_2605964, EPI_ISL_2605965, EPI_ISL_2605968, EPI_ISL_2605969, EPI_ISL_2605970, EPI_ISL_2605971, EPI_ISL_2605973, EPI_ISL_2605975, EPI_ISL_2605976, EPI_ISL_2605978, EPI_ISL_2605980, EPI_ISL_2605981, EPI_ISL_2605982,
EPI_ISL_2605983, EPI_ISL_2605984, EPI_ISL_2605985, EPI_ISL_2605987, EPI_ISL_2605988, EPI_ISL_2605989, EPI_ISL_2605991, EPI_ISL_2605993, EPI_ISL_2605995, EPI_ISL_2605997, EPI_ISL_2605998, EPI_ISL_2605999, EPI_ISL_2606000, EPI_ISL_2606002, EPI_ISL_2606004, EPI_ISL_2606005, EPI_ISL_2606006, EPI_ISL_2606007, EPI_ISL_2606008, EPI_ISL_2606009, EPI_ISL_2606010, EPI_ISL_2606011,
EPI_ISL_2606014, EPI_ISL_2606015, EPI_ISL_2606016, EPI_ISL_2606017, EPI_ISL_2606019, EPI_ISL_2606020, EPI_ISL_2606021, EPI_ISL_2606022, EPI_ISL_2606024, EPI_ISL_2606025, EPI_ISL_2606026, EPI_ISL_2606029, EPI_ISL_2606030, EPI_ISL_2606031, EPI_ISL_2606032, EPI_ISL_2606033, EPI_ISL_2606034, EPI_ISL_2606035, EPI_ISL_2606036, EPI_ISL_2606038, EPI_ISL_2606039, EPI_ISL_2606041,
EPI_ISL_2606042, EPI_ISL_2606043, EPI_ISL_2606044, EPI_ISL_2606045, EPI_ISL_2606046, EPI_ISL_2606047, EPI_ISL_2606048, EPI_ISL_2606051, EPI_ISL_2606052, EPI_ISL_2606053, EPI_ISL_2606054, EPI_ISL_2606057, EPI_ISL_2606058, EPI_ISL_2606059, EPI_ISL_2606060, EPI_ISL_2606061, EPI_ISL_2606063, EPI_ISL_2606064, EPI_ISL_2606066, EPI_ISL_2606067, EPI_ISL_2606068, EPI_ISL_2606071,
EPI_ISL_2606073, EPI_ISL_2606074, EPI_ISL_2606075, EPI_ISL_2606077, EPI_ISL_2606078, EPI_ISL_2606079, EPI_ISL_2606080, EPI_ISL_2606081, EPI_ISL_2606084, EPI_ISL_2606085, EPI_ISL_2606086, EPI_ISL_2606087, EPI_ISL_2606089, EPI_ISL_2606091, EPI_ISL_2606092, EPI_ISL_2606093, EPI_ISL_2606097, EPI_ISL_2606098, EPI_ISL_2606099, EPI_ISL_2606100, EPI_ISL_2606101, EPI_ISL_2606102,
EPI_ISL_2606103, EPI_ISL_2606107, EPI_ISL_2606108, EPI_ISL_2606109, EPI_ISL_2606110, EPI_ISL_2606112, EPI_ISL_2606114, EPI_ISL_2606115, EPI_ISL_2606116, EPI_ISL_2606120, EPI_ISL_2606121, EPI_ISL_2606122, EPI_ISL_2606123, EPI_ISL_2606124, EPI_ISL_2606126, EPI_ISL_2606127, EPI_ISL_2606128, EPI_ISL_2606131, EPI_ISL_2606132, EPI_ISL_2606134, EPI_ISL_2606135, EPI_ISL_2606137,
EPI_ISL_2606139, EPI_ISL_2606144, EPI_ISL_2606146, EPI_ISL_2606148, EPI_ISL_2606150, EPI_ISL_2606151, EPI_ISL_2606154, EPI_ISL_2606155, EPI_ISL_2606156, EPI_ISL_2606157, EPI_ISL_2606158, EPI_ISL_2606159, EPI_ISL_2606160, EPI_ISL_2606162, EPI_ISL_2606163, EPI_ISL_2606164, EPI_ISL_2606165, EPI_ISL_2606166, EPI_ISL_2606167, EPI_ISL_2606170, EPI_ISL_2606171, EPI_ISL_2606172,
EPI_ISL_2606173, EPI_ISL_2606174, EPI_ISL_2606175, EPI_ISL_2606176, EPI_ISL_2606178, EPI_ISL_2606179, EPI_ISL_2606181, EPI_ISL_2606182, EPI_ISL_2606183, EPI_ISL_2606185, EPI_ISL_2606186, EPI_ISL_2606187, EPI_ISL_2606188, EPI_ISL_2606189, EPI_ISL_2606190, EPI_ISL_2606194, EPI_ISL_2606196, EPI_ISL_2606197, EPI_ISL_2606198, EPI_ISL_2606199, EPI_ISL_2606200, EPI_ISL_2606201,
EPI_ISL_2606202, EPI_ISL_2606204, EPI_ISL_2606205, EPI_ISL_2606206, EPI_ISL_2606207, EPI_ISL_2606209, EPI_ISL_2606211, EPI_ISL_2606213, EPI_ISL_2606214, EPI_ISL_2606215, EPI_ISL_2606218, EPI_ISL_2606219, EPI_ISL_2606220, EPI_ISL_2606221, EPI_ISL_2606222, EPI_ISL_2606223, EPI_ISL_2606226, EPI_ISL_2606227, EPI_ISL_2606228, EPI_ISL_2606229, EPI_ISL_2606230,
EPI_ISL_2606232, EPI_ISL_2606233, EPI_ISL_2606235, EPI_ISL_2606236, EPI_ISL_2606237, EPI_ISL_2606238, EPI_ISL_2606239, EPI_ISL_2606240, EPI_ISL_2606241, EPI_ISL_2606242, EPI_ISL_2606243, EPI_ISL_2606244, EPI_ISL_2606247, EPI_ISL_2606249, EPI_ISL_2606250, EPI_ISL_2606251, EPI_ISL_2606255, EPI_ISL_2606256, EPI_ISL_2606257, EPI_ISL_2606258, EPI_ISL_2606259, EPI_ISL_2606260,
EPI_ISL_2606262, EPI_ISL_2606263, EPI_ISL_2606264, EPI_ISL_2606265, EPI_ISL_2606266, EPI_ISL_2606267, EPI_ISL_2606268, EPI_ISL_2606269, EPI_ISL_2606270, EPI_ISL_2606271, EPI_ISL_2606272, EPI_ISL_2606273, EPI_ISL_2606274, EPI_ISL_2606275, EPI_ISL_2606276, EPI_ISL_2606277, EPI_ISL_2606278, EPI_ISL_2606279, EPI_ISL_2606282, EPI_ISL_2606284, EPI_ISL_2606285, EPI_ISL_2606287,
EPI_ISL_2606290, EPI_ISL_2606291, EPI_ISL_2606293, EPI_ISL_2606294, EPI_ISL_2606295, EPI_ISL_2606296, EPI_ISL_2606297, EPI_ISL_2606299, EPI_ISL_2606300, EPI_ISL_2606302, EPI_ISL_2606303, EPI_ISL_2606304, EPI_ISL_2606305, EPI_ISL_2606307, EPI_ISL_2606308, EPI_ISL_2606310, EPI_ISL_2606311, EPI_ISL_2606312, EPI_ISL_2606314, EPI_ISL_2606315, EPI_ISL_2606316,
EPI_ISL_2606317, EPI_ISL_2606318, EPI_ISL_2606319, EPI_ISL_2606322, EPI_ISL_2606324, EPI_ISL_2606326, EPI_ISL_2606327, EPI_ISL_2606328, EPI_ISL_2606330, EPI_ISL_2606331, EPI_ISL_2606332, EPI_ISL_2606333, EPI_ISL_2606335, EPI_ISL_2606336, EPI_ISL_2606338, EPI_ISL_2606340, EPI_ISL_2606341, EPI_ISL_2606342, EPI_ISL_2606343, EPI_ISL_2606344, EPI_ISL_2606345, EPI_ISL_2606346,
EPI_ISL_2606347, EPI_ISL_2606348, EPI_ISL_2606349, EPI_ISL_2606351, EPI_ISL_2606352, EPI_ISL_2606353, EPI_ISL_2606354, EPI_ISL_2606355, EPI_ISL_2606357, EPI_ISL_2606359, EPI_ISL_2606361, EPI_ISL_2606362, EPI_ISL_2606365, EPI_ISL_2606366, EPI_ISL_2606367, EPI_ISL_2606368, EPI_ISL_2606369, EPI_ISL_2606370, EPI_ISL_2606371, EPI_ISL_2606373, EPI_ISL_2606374, EPI_ISL_2606375,
EPI_ISL_2606376, EPI_ISL_2606377, EPI_ISL_2606379, EPI_ISL_2606380, EPI_ISL_2606381, EPI_ISL_2606382, EPI_ISL_2606383, EPI_ISL_2606384, EPI_ISL_2606386, EPI_ISL_2606387, EPI_ISL_2606388, EPI_ISL_2606390, EPI_ISL_2606391, EPI_ISL_2606393, EPI_ISL_2606394, EPI_ISL_2606395, EPI_ISL_2606397, EPI_ISL_2606398, EPI_ISL_2606399, EPI_ISL_2606400, EPI_ISL_2606401, EPI_ISL_2606402,
EPI_ISL_2606403, EPI_ISL_2606404, EPI_ISL_2606405, EPI_ISL_2606407, EPI_ISL_2606408, EPI_ISL_2606409, EPI_ISL_2606411, EPI_ISL_2606412, EPI_ISL_2606413, EPI_ISL_2606414, EPI_ISL_2606415, EPI_ISL_2606416, EPI_ISL_2606417, EPI_ISL_2606418, EPI_ISL_2606419, EPI_ISL_2606420, EPI_ISL_2606421, EPI_ISL_2606422, EPI_ISL_2606423, EPI_ISL_2606424, EPI_ISL_2606427, EPI_ISL_2606428,
EPI_ISL_2606429, EPI_ISL_2606430, EPI_ISL_2606431, EPI_ISL_2606432, EPI_ISL_2606433, EPI_ISL_2606434, EPI_ISL_2606435, EPI_ISL_2606438, EPI_ISL_2606439, EPI_ISL_2606440, EPI_ISL_2606441, EPI_ISL_2606442, EPI_ISL_2606443, EPI_ISL_2606444, EPI_ISL_2606445, EPI_ISL_2606446, EPI_ISL_2606447, EPI_ISL_2606448, EPI_ISL_2606449, EPI_ISL_2606451, EPI_ISL_2606452, EPI_ISL_2606730,
EPI_ISL_2606734, EPI_ISL_2606735, EPI_ISL_2606738, EPI_ISL_2606746, EPI_ISL_2606752, EPI_ISL_2606755, EPI_ISL_2606759, EPI_ISL_2606761, EPI_ISL_2606762, EPI_ISL_2606764, EPI_ISL_2606766, EPI_ISL_2606769, EPI_ISL_2606772, EPI_ISL_2606773, EPI_ISL_2606776, EPI_ISL_2606777, EPI_ISL_2606779, EPI_ISL_2606780, EPI_ISL_2606782, EPI_ISL_2606783, EPI_ISL_2606792, EPI_ISL_2606796,
EPI_ISL_2606798, EPI_ISL_2606801, EPI_ISL_2606802, EPI_ISL_2606808, EPI_ISL_2606809, EPI_ISL_2606810, EPI_ISL_2606811, EPI_ISL_2606812, EPI_ISL_2606822, EPI_ISL_2606825, EPI_ISL_2606826, EPI_ISL_2606828, EPI_ISL_2606831, EPI_ISL_2606832, EPI_ISL_2606836, EPI_ISL_2606837, EPI_ISL_2606838, EPI_ISL_2606857, EPI_ISL_2606858, EPI_ISL_2606861, EPI_ISL_2606862, EPI_ISL_2606866,
EPI_ISL_2606867, EPI_ISL_2606868, EPI_ISL_2606876, EPI_ISL_2606880, EPI_ISL_2606881, EPI_ISL_2606883, EPI_ISL_2606886, EPI_ISL_2606888, EPI_ISL_2606889, EPI_ISL_2606891, EPI_ISL_2606895, EPI_ISL_2606896, EPI_ISL_2606897, EPI_ISL_2606900, EPI_ISL_2606903, EPI_ISL_2606906, EPI_ISL_2606907, EPI_ISL_2606911, EPI_ISL_2606914, EPI_ISL_2606915, EPI_ISL_2606917, EPI_ISL_2606919,
EPI_ISL_2606923, EPI_ISL_2606924, EPI_ISL_2606934, EPI_ISL_2606940, EPI_ISL_2606948, EPI_ISL_2606956, EPI_ISL_2606957, EPI_ISL_2606958, EPI_ISL_2606961, EPI_ISL_2606965, EPI_ISL_2606967, EPI_ISL_2606968, EPI_ISL_2606972, EPI_ISL_2606973, EPI_ISL_2606975, EPI_ISL_2606976, EPI_ISL_2606977, EPI_ISL_2606979, EPI_ISL_2606981, EPI_ISL_2606988, EPI_ISL_2606992, EPI_ISL_2606999,
EPI_ISL_2607006, EPI_ISL_2607011, EPI_ISL_2607013, EPI_ISL_2607021, EPI_ISL_2607025, EPI_ISL_2607028, EPI_ISL_2607030, EPI_ISL_2607031, EPI_ISL_2607033, EPI_ISL_2607035, EPI_ISL_2607036, EPI_ISL_2607038, EPI_ISL_2607040, EPI_ISL_2607251, EPI_ISL_2607253, EPI_ISL_2607516, EPI_ISL_2607546, EPI_ISL_2607901, EPI_ISL_2607902, EPI_ISL_2607912, EPI_ISL_2607914, EPI_ISL_2607917,
EPI_ISL_2607929, EPI_ISL_2607932, EPI_ISL_2607934, EPI_ISL_2607946, EPI_ISL_2607947, EPI_ISL_2607950, EPI_ISL_2607951, EPI_ISL_2640078, EPI_ISL_2640079, EPI_ISL_2640080, EPI_ISL_2640081, EPI_ISL_2640082, EPI_ISL_2640083, EPI_ISL_2640161, EPI_ISL_2640328, EPI_ISL_2640406, EPI_ISL_2640436, EPI_ISL_2640549, EPI_ISL_2640586, EPI_ISL_2640600, EPI_ISL_2640609, EPI_ISL_2640627,
EPI_ISL_2640684, EPI_ISL_2640705, EPI_ISL_2640707, EPI_ISL_2640708, EPI_ISL_2640714, EPI_ISL_2640715, EPI_ISL_2640724, EPI_ISL_2640729, EPI_ISL_2640734, EPI_ISL_2640745, EPI_ISL_2640749, EPI_ISL_2640750, EPI_ISL_2640753, EPI_ISL_2640764, EPI_ISL_2640766, EPI_ISL_2640767, EPI_ISL_2640772, EPI_ISL_2640778, EPI_ISL_2640787, EPI_ISL_2640788, EPI_ISL_2640789, EPI_ISL_2640816,
EPI_ISL_2640827, EPI_ISL_2640831, EPI_ISL_2640832, EPI_ISL_2640835, EPI_ISL_2640837, EPI_ISL_2640842, EPI_ISL_2640843, EPI_ISL_2640854, EPI_ISL_2640860, EPI_ISL_2640861, EPI_ISL_2640862, EPI_ISL_2640863, EPI_ISL_2640869, EPI_ISL_2640877, EPI_ISL_2640881, EPI_ISL_2640882, EPI_ISL_2640883, EPI_ISL_2640884, EPI_ISL_2640891, EPI_ISL_2640892, EPI_ISL_2640897, EPI_ISL_2640904,
EPI_ISL_2640914, EPI_ISL_2640920, EPI_ISL_2640925, EPI_ISL_2640926, EPI_ISL_2640938, EPI_ISL_2640941, EPI_ISL_2640942, EPI_ISL_2640950, EPI_ISL_2640953, EPI_ISL_2640954, EPI_ISL_2640962, EPI_ISL_2640969, EPI_ISL_2640970, EPI_ISL_2640975, EPI_ISL_2640982, EPI_ISL_2640988, EPI_ISL_2641004, EPI_ISL_2641018, EPI_ISL_2641022, EPI_ISL_2641032, EPI_ISL_2641070, EPI_ISL_2641088,
EPI_ISL_2641098, EPI_ISL_2641114, EPI_ISL_2641116, EPI_ISL_2641143, EPI_ISL_2641180, EPI_ISL_2641197, EPI_ISL_2641218, EPI_ISL_2641231, EPI_ISL_2641245, EPI_ISL_2641247, EPI_ISL_2641252, EPI_ISL_2641260, EPI_ISL_2641287, EPI_ISL_2641307, EPI_ISL_2641340, EPI_ISL_2641344, EPI_ISL_2641350, EPI_ISL_2641395, EPI_ISL_2641423, EPI_ISL_2641493, EPI_ISL_2641498, EPI_ISL_2641511,
EPI_ISL_2641513, EPI_ISL_2641520, EPI_ISL_2641568, EPI_ISL_2641569, EPI_ISL_2641625, EPI_ISL_2641630, EPI_ISL_2641638, EPI_ISL_2651998

| | | | |
|---|---|---|---|
| see above | Lighthouse Lab in Alderley Park | Wellcome Sanger Institute for the COVID-19 Genomics UK (COG-UK) Consortium | Cordelia Langford; David K. Jackson; Dominic Kwiatkowski; Ewan Harrison; Ian Johnston; Jacquelyn Wynn; Jeffrey Barrett; John Sillitoe on behalf of the Wellcome Sanger Institute COVID-19 Surveillance Team; Mairead Hyland; Roberto Amato; Sonia Goncalves; The Lighthouse Lab in Alderley Park and Alex Alderton |

EPI_ISL_2486204, EPI_ISL_2486342, EPI_ISL_2486516, EPI_ISL_2487350, EPI_ISL_2551933, EPI_ISL_2551936, EPI_ISL_2551942, EPI_ISL_2551943, EPI_ISL_2551945, EPI_ISL_2551946, EPI_ISL_2551948, EPI_ISL_2551950, EPI_ISL_2551963, EPI_ISL_2551964, EPI_ISL_2551966, EPI_ISL_2551968, EPI_ISL_2551969, EPI_ISL_2551970, EPI_ISL_2551971, EPI_ISL_2551978, EPI_ISL_2551983, EPI_ISL_2551986,
EPI_ISL_2551990, EPI_ISL_2551992, EPI_ISL_2551994, EPI_ISL_2551995, EPI_ISL_2551997, EPI_ISL_2552008, EPI_ISL_2552013, EPI_ISL_2552067, EPI_ISL_2552069, EPI_ISL_2552072, EPI_ISL_2552111, EPI_ISL_2552114, EPI_ISL_2552123, EPI_ISL_2552132, EPI_ISL_2552139, EPI_ISL_2552161, EPI_ISL_2552221, EPI_ISL_2552238, EPI_ISL_2552242, EPI_ISL_2552607, EPI_ISL_2552630,
EPI_ISL_2552631, EPI_ISL_2552638, EPI_ISL_2552641, EPI_ISL_2552643, EPI_ISL_2552644, EPI_ISL_2552645, EPI_ISL_2552647, EPI_ISL_2552652, EPI_ISL_2552654, EPI_ISL_2552659, EPI_ISL_2552665, EPI_ISL_2552671, EPI_ISL_2552673, EPI_ISL_2552686, EPI_ISL_2552687, EPI_ISL_2552693, EPI_ISL_2552702, EPI_ISL_2552707, EPI_ISL_2552708, EPI_ISL_2552714, EPI_ISL_2552721, EPI_ISL_2552724,
EPI_ISL_2552726, EPI_ISL_2552729, EPI_ISL_2552732, EPI_ISL_2552735, EPI_ISL_2552736, EPI_ISL_2552743, EPI_ISL_2552747, EPI_ISL_2552761, EPI_ISL_2552762, EPI_ISL_2552766, EPI_ISL_2552771, EPI_ISL_2552774, EPI_ISL_2552778, EPI_ISL_2552786, EPI_ISL_2552788, EPI_ISL_2552791, EPI_ISL_2552792, EPI_ISL_2552796, EPI_ISL_2552804, EPI_ISL_2552806, EPI_ISL_2552809, EPI_ISL_2552818,
EPI_ISL_2552821, EPI_ISL_2552826, EPI_ISL_2552827, EPI_ISL_2552828, EPI_ISL_2552829, EPI_ISL_2552832, EPI_ISL_2552842, EPI_ISL_2552843, EPI_ISL_2552847, EPI_ISL_2552850, EPI_ISL_2552853, EPI_ISL_2552855, EPI_ISL_2552858, EPI_ISL_2552862, EPI_ISL_2552863, EPI_ISL_2552867, EPI_ISL_2552869, EPI_ISL_2552873, EPI_ISL_2552874, EPI_ISL_2552876, EPI_ISL_2552894, EPI_ISL_2552895,
EPI_ISL_2552896, EPI_ISL_2552900, EPI_ISL_2552902, EPI_ISL_2552910, EPI_ISL_2552911, EPI_ISL_2552915, EPI_ISL_2552920, EPI_ISL_2552921, EPI_ISL_2552934, EPI_ISL_2552935, EPI_ISL_2552939, EPI_ISL_2552940, EPI_ISL_2552944, EPI_ISL_2552946, EPI_ISL_2552947, EPI_ISL_2552953, EPI_ISL_2552956, EPI_ISL_2552957, EPI_ISL_2552965, EPI_ISL_2552971, EPI_ISL_2552973,
EPI_ISL_2552974, EPI_ISL_2552979, EPI_ISL_2552982, EPI_ISL_2552993, EPI_ISL_2553008, EPI_ISL_2553009, EPI_ISL_2553010, EPI_ISL_2553012, EPI_ISL_2553013, EPI_ISL_2553015, EPI_ISL_2553031, EPI_ISL_2553036, EPI_ISL_2553041, EPI_ISL_2553046, EPI_ISL_2553049, EPI_ISL_2553051, EPI_ISL_2553056, EPI_ISL_2553061, EPI_ISL_2553070, EPI_ISL_2553073, EPI_ISL_2553081, EPI_ISL_2553085,
EPI_ISL_2553094, EPI_ISL_2553095, EPI_ISL_2553096, EPI_ISL_2553097, EPI_ISL_2553099, EPI_ISL_2553100, EPI_ISL_2553101, EPI_ISL_2553103, EPI_ISL_2553104, EPI_ISL_2553105, EPI_ISL_2553106, EPI_ISL_2553107, EPI_ISL_2553115, EPI_ISL_2553118, EPI_ISL_2553120, EPI_ISL_2553121, EPI_ISL_2553122, EPI_ISL_2553123, EPI_ISL_2553129, EPI_ISL_2553130, EPI_ISL_2553135, EPI_ISL_2553136,
EPI_ISL_2553139, EPI_ISL_2553140, EPI_ISL_2553141, EPI_ISL_2553144, EPI_ISL_2553148, EPI_ISL_2553150, EPI_ISL_2553151, EPI_ISL_2553152, EPI_ISL_2553155, EPI_ISL_2553156, EPI_ISL_2553158, EPI_ISL_2553159, EPI_ISL_2553160, EPI_ISL_2553162, EPI_ISL_2553165, EPI_ISL_2553167, EPI_ISL_2553171, EPI_ISL_2553173, EPI_ISL_2553176, EPI_ISL_2553178, EPI_ISL_2553180, EPI_ISL_2553182,
EPI_ISL_2553184, EPI_ISL_2553185, EPI_ISL_2553186, EPI_ISL_2553192, EPI_ISL_2553196, EPI_ISL_2553198, EPI_ISL_2553200, EPI_ISL_2553201, EPI_ISL_2553205, EPI_ISL_2553206, EPI_ISL_2553207, EPI_ISL_2553208, EPI_ISL_2553211, EPI_ISL_2553212, EPI_ISL_2553213, EPI_ISL_2553214, EPI_ISL_2553215, EPI_ISL_2553217, EPI_ISL_2553220, EPI_ISL_2553221, EPI_ISL_2553222, EPI_ISL_2553225,
EPI_ISL_2553226, EPI_ISL_2553232, EPI_ISL_2553233, EPI_ISL_2553234, EPI_ISL_2553236, EPI_ISL_2553238, EPI_ISL_2553243, EPI_ISL_2553248, EPI_ISL_2553249, EPI_ISL_2553251, EPI_ISL_2553252, EPI_ISL_2553253, EPI_ISL_2553254, EPI_ISL_2553257, EPI_ISL_2553258, EPI_ISL_2553259, EPI_ISL_2553260, EPI_ISL_2553261, EPI_ISL_2553266, EPI_ISL_2553267, EPI_ISL_2553276, EPI_ISL_2553277,
EPI_ISL_2553279, EPI_ISL_2553281, EPI_ISL_2553284, EPI_ISL_2553285, EPI_ISL_2553287, EPI_ISL_2553288, EPI_ISL_2553291, EPI_ISL_2553293, EPI_ISL_2553295, EPI_ISL_2553297, EPI_ISL_2553299, EPI_ISL_2553300, EPI_ISL_2553301, EPI_ISL_2553305, EPI_ISL_2553307, EPI_ISL_2553308, EPI_ISL_2553310, EPI_ISL_2553316, EPI_ISL_2553317, EPI_ISL_2553319, EPI_ISL_2553323, EPI_ISL_2553324,
EPI_ISL_2553326, EPI_ISL_2553327, EPI_ISL_2553328, EPI_ISL_2553330, EPI_ISL_2553331, EPI_ISL_2553334, EPI_ISL_2553335, EPI_ISL_2553336, EPI_ISL_2553341, EPI_ISL_2553342, EPI_ISL_2553343, EPI_ISL_2553344, EPI_ISL_2553348, EPI_ISL_2553349, EPI_ISL_2553353, EPI_ISL_2553354, EPI_ISL_2553357, EPI_ISL_2553358, EPI_ISL_2553363, EPI_ISL_2553365, EPI_ISL_2553366, EPI_ISL_2553367,
EPI_ISL_2553369, EPI_ISL_2553370, EPI_ISL_2553372, EPI_ISL_2553374, EPI_ISL_2553376, EPI_ISL_2553377, EPI_ISL_2553378, EPI_ISL_2553382, EPI_ISL_2553385, EPI_ISL_2553392, EPI_ISL_2553393, EPI_ISL_2553394, EPI_ISL_2553395, EPI_ISL_2553400, EPI_ISL_2553401, EPI_ISL_2553402, EPI_ISL_2553404, EPI_ISL_2553406, EPI_ISL_2553407, EPI_ISL_2553409, EPI_ISL_2553415, EPI_ISL_2553417,
EPI_ISL_2553423, EPI_ISL_2553428, EPI_ISL_2553431, EPI_ISL_2553432, EPI_ISL_2553433, EPI_ISL_2553435, EPI_ISL_2553441, EPI_ISL_2553450, EPI_ISL_2553458, EPI_ISL_2553462, EPI_ISL_2553465, EPI_ISL_2553466, EPI_ISL_2553467, EPI_ISL_2553491, EPI_ISL_2553493, EPI_ISL_2553495, EPI_ISL_2553498, EPI_ISL_2553501, EPI_ISL_2553507, EPI_ISL_2553508, EPI_ISL_2553528, EPI_ISL_2553529,
EPI_ISL_2553531, EPI_ISL_2553535, EPI_ISL_2553536, EPI_ISL_2553537, EPI_ISL_2553539, EPI_ISL_2553546, EPI_ISL_2553551, EPI_ISL_2553563, EPI_ISL_2553565, EPI_ISL_2553566, EPI_ISL_2553569, EPI_ISL_2553573, EPI_ISL_2553583, EPI_ISL_2553592, EPI_ISL_2553593, EPI_ISL_2553596, EPI_ISL_2553600, EPI_ISL_2553614, EPI_ISL_2553623, EPI_ISL_2553627, EPI_ISL_2553630,
EPI_ISL_2553635, EPI_ISL_2553637, EPI_ISL_2553641, EPI_ISL_2553643, EPI_ISL_2553645, EPI_ISL_2553646, EPI_ISL_2553649, EPI_ISL_2553657, EPI_ISL_2553665, EPI_ISL_2553666, EPI_ISL_2553671, EPI_ISL_2553673, EPI_ISL_2553675, EPI_ISL_2553680, EPI_ISL_2553682, EPI_ISL_2553685, EPI_ISL_2553687, EPI_ISL_2553690, EPI_ISL_2553691, EPI_ISL_2553692, EPI_ISL_2553699, EPI_ISL_2553710,
EPI_ISL_2553715, EPI_ISL_2553716, EPI_ISL_2553728, EPI_ISL_2553741, EPI_ISL_2553743, EPI_ISL_2553747, EPI_ISL_2553759, EPI_ISL_2553762, EPI_ISL_2553764, EPI_ISL_2553772, EPI_ISL_2553775, EPI_ISL_2553776, EPI_ISL_2553780, EPI_ISL_2554144, EPI_ISL_2554146, EPI_ISL_2554152, EPI_ISL_2554153, EPI_ISL_2554160, EPI_ISL_2554163, EPI_ISL_2554165, EPI_ISL_2554171, EPI_ISL_2554175,
EPI_ISL_2554176, EPI_ISL_2554180, EPI_ISL_2554183, EPI_ISL_2554186, EPI_ISL_2554187, EPI_ISL_2554193, EPI_ISL_2554196, EPI_ISL_2554214, EPI_ISL_2554220, EPI_ISL_2554222, EPI_ISL_2554226, EPI_ISL_2554228, EPI_ISL_2554237, EPI_ISL_2554243, EPI_ISL_2554244, EPI_ISL_2554248, EPI_ISL_2554257, EPI_ISL_2554260, EPI_ISL_2554269, EPI_ISL_2554270, EPI_ISL_2554271, EPI_ISL_2554273,
EPI_ISL_2554279, EPI_ISL_2554281, EPI_ISL_2554300, EPI_ISL_2554301, EPI_ISL_2554312, EPI_ISL_2554318, EPI_ISL_2554320, EPI_ISL_2554321, EPI_ISL_2554322, EPI_ISL_2554323, EPI_ISL_2554330, EPI_ISL_2554347, EPI_ISL_2554348, EPI_ISL_2554355, EPI_ISL_2554356, EPI_ISL_2554360, EPI_ISL_2554361, EPI_ISL_2554362, EPI_ISL_2554365, EPI_ISL_2554366, EPI_ISL_2554372, EPI_ISL_2554381,
EPI_ISL_2554382, EPI_ISL_2554387, EPI_ISL_2554392, EPI_ISL_2554395, EPI_ISL_2554396, EPI_ISL_2554398, EPI_ISL_2554401, EPI_ISL_2554404, EPI_ISL_2554411, EPI_ISL_2554412, EPI_ISL_2554416, EPI_ISL_2554421, EPI_ISL_2554422, EPI_ISL_2554425, EPI_ISL_2554431, EPI_ISL_2554433, EPI_ISL_2554438, EPI_ISL_2554440, EPI_ISL_2554441, EPI_ISL_2554447, EPI_ISL_2554449, EPI_ISL_2554456,
EPI_ISL_2554464, EPI_ISL_2554470, EPI_ISL_2554474, EPI_ISL_2554475, EPI_ISL_2554476, EPI_ISL_2554478, EPI_ISL_2554481, EPI_ISL_2554483, EPI_ISL_2554485, EPI_ISL_2554487, EPI_ISL_2554491, EPI_ISL_2554496, EPI_ISL_2554515, EPI_ISL_2554548, EPI_ISL_2554613, EPI_ISL_2554646, EPI_ISL_2554667, EPI_ISL_2554677, EPI_ISL_2554679, EPI_ISL_2554686, EPI_ISL_2554697, EPI_ISL_2554703,
EPI_ISL_2554706, EPI_ISL_2554737, EPI_ISL_2554745, EPI_ISL_2554751, EPI_ISL_2554756, EPI_ISL_2554782, EPI_ISL_2554784, EPI_ISL_2554797, EPI_ISL_2554803, EPI_ISL_2554812, EPI_ISL_2554824, EPI_ISL_2567947, EPI_ISL_2567950, EPI_ISL_2567952, EPI_ISL_2567955, EPI_ISL_2567958, EPI_ISL_2567960, EPI_ISL_2567962, EPI_ISL_2567963, EPI_ISL_2567966, EPI_ISL_2567967, EPI_ISL_2567970,
EPI_ISL_2567971, EPI_ISL_2567973, EPI_ISL_2567974, EPI_ISL_2567975, EPI_ISL_2567979, EPI_ISL_2567980, EPI_ISL_2567981, EPI_ISL_2567982, EPI_ISL_2567983, EPI_ISL_2567985, EPI_ISL_2567988, EPI_ISL_2567991, EPI_ISL_2567993, EPI_ISL_2567995, EPI_ISL_2567996, EPI_ISL_2567997, EPI_ISL_2567999, EPI_ISL_2568001, EPI_ISL_2568002, EPI_ISL_2568004, EPI_ISL_2568007, EPI_ISL_2568008,
EPI_ISL_2568009, EPI_ISL_2568010, EPI_ISL_2568013, EPI_ISL_2568015, EPI_ISL_2568017, EPI_ISL_2568024, EPI_ISL_2568025, EPI_ISL_2568028, EPI_ISL_2568029, EPI_ISL_2568030, EPI_ISL_2568032, EPI_ISL_2568033, EPI_ISL_2568035, EPI_ISL_2568038, EPI_ISL_2568039, EPI_ISL_2568041, EPI_ISL_2568042, EPI_ISL_2568043, EPI_ISL_2568044, EPI_ISL_2568045, EPI_ISL_2568048, EPI_ISL_2568049,
EPI_ISL_2568053, EPI_ISL_2568054, EPI_ISL_2568057, EPI_ISL_2568059, EPI_ISL_2568060, EPI_ISL_2568062, EPI_ISL_2568065, EPI_ISL_2568066, EPI_ISL_2568067, EPI_ISL_2568068, EPI_ISL_2568074, EPI_ISL_2568075, EPI_ISL_2568077, EPI_ISL_2568078, EPI_ISL_2568079, EPI_ISL_2568080, EPI_ISL_2568082, EPI_ISL_2568083, EPI_ISL_2568088, EPI_ISL_2568091, EPI_ISL_2568093,
EPI_ISL_2568094, EPI_ISL_2568096, EPI_ISL_2568098, EPI_ISL_2568099, EPI_ISL_2568102, EPI_ISL_2568103, EPI_ISL_2568106, EPI_ISL_2568107, EPI_ISL_2568108, EPI_ISL_2568111, EPI_ISL_2568129, EPI_ISL_2568138, EPI_ISL_2568162, EPI_ISL_2568217, EPI_ISL_2568219, EPI_ISL_2568255, EPI_ISL_2568280, EPI_ISL_2568307, EPI_ISL_2568346, EPI_ISL_2568375, EPI_ISL_2568395, EPI_ISL_2568405,
EPI_ISL_2568413, EPI_ISL_2568417, EPI_ISL_2568431, EPI_ISL_2568437, EPI_ISL_2568474, EPI_ISL_2568544, EPI_ISL_2568546, EPI_ISL_2568559, EPI_ISL_2568579, EPI_ISL_2568587, EPI_ISL_2568588, EPI_ISL_2568593, EPI_ISL_2568606, EPI_ISL_2568636, EPI_ISL_2568641, EPI_ISL_2568645, EPI_ISL_2569016, EPI_ISL_2569020, EPI_ISL_2569023, EPI_ISL_2569027, EPI_ISL_2569032,
EPI_ISL_2569035, EPI_ISL_2569046, EPI_ISL_2569054, EPI_ISL_2569066, EPI_ISL_2569070, EPI_ISL_2569072, EPI_ISL_2569078, EPI_ISL_2569080, EPI_ISL_2569090, EPI_ISL_2569094, EPI_ISL_2569098, EPI_ISL_2569100, EPI_ISL_2569103, EPI_ISL_2569111, EPI_ISL_2569123, EPI_ISL_2569125, EPI_ISL_2569134, EPI_ISL_2569135, EPI_ISL_2569137, EPI_ISL_2569143, EPI_ISL_2569147, EPI_ISL_2569166,
EPI_ISL_2569167, EPI_ISL_2569170, EPI_ISL_2569174, EPI_ISL_2569177, EPI_ISL_2569179, EPI_ISL_2569206, EPI_ISL_2569214, EPI_ISL_2569216, EPI_ISL_2569219, EPI_ISL_2569225, EPI_ISL_2569228, EPI_ISL_2569243, EPI_ISL_2569248, EPI_ISL_2569253, EPI_ISL_2569255, EPI_ISL_2569261, EPI_ISL_2569262, EPI_ISL_2569267, EPI_ISL_2569278, EPI_ISL_2569280, EPI_ISL_2569283, EPI_ISL_2569288,

EPI_ISL_2569295, EPI_ISL_2569301, EPI_ISL_2569312, EPI_ISL_2569314, EPI_ISL_2570287, EPI_ISL_2570297, EPI_ISL_2570336, EPI_ISL_2570350, EPI_ISL_2570376, EPI_ISL_2570378, EPI_ISL_2570392, EPI_ISL_2570394, EPI_ISL_2570396, EPI_ISL_2570397, EPI_ISL_2570401, EPI_ISL_2570407, EPI_ISL_2570427, EPI_ISL_2570450, EPI_ISL_2605716, EPI_ISL_2607042, EPI_ISL_2607044, EPI_ISL_2607046, EPI_ISL_2607049, EPI_ISL_2607052, EPI_ISL_2607053, EPI_ISL_2607055, EPI_ISL_2607060, EPI_ISL_2607062, EPI_ISL_2607063, EPI_ISL_2607065, EPI_ISL_2607068, EPI_ISL_2607069, EPI_ISL_2607070, EPI_ISL_2607073, EPI_ISL_2607074, EPI_ISL_2607079, EPI_ISL_2607083, EPI_ISL_2607085, EPI_ISL_2607088, EPI_ISL_2607089, EPI_ISL_2607090, EPI_ISL_2607092, EPI_ISL_2607095, EPI_ISL_2607096, EPI_ISL_2607099, EPI_ISL_2607101, EPI_ISL_2607102, EPI_ISL_2607104, EPI_ISL_2607106, EPI_ISL_2607108, EPI_ISL_2607115, EPI_ISL_2607116, EPI_ISL_2607117, EPI_ISL_2607118, EPI_ISL_2607119, EPI_ISL_2607122, EPI_ISL_2607123, EPI_ISL_2607124, EPI_ISL_2607128, EPI_ISL_2607129, EPI_ISL_2607174, EPI_ISL_2607182, EPI_ISL_2607260, EPI_ISL_2607261, EPI_ISL_2607262, EPI_ISL_2607263, EPI_ISL_2607264, EPI_ISL_2607265, EPI_ISL_2607266, EPI_ISL_2607267, EPI_ISL_2607268, EPI_ISL_2607269, EPI_ISL_2607270, EPI_ISL_2607271, EPI_ISL_2607272, EPI_ISL_2607273, EPI_ISL_2607275, EPI_ISL_2607276, EPI_ISL_2607277, EPI_ISL_2607278, EPI_ISL_2607279, EPI_ISL_2607281, EPI_ISL_2607282, EPI_ISL_2607283, EPI_ISL_2607284, EPI_ISL_2607285, EPI_ISL_2607286, EPI_ISL_2607287, EPI_ISL_2607288, EPI_ISL_2607289, EPI_ISL_2607290, EPI_ISL_2607291, EPI_ISL_2607292, EPI_ISL_2607293, EPI_ISL_2607294, EPI_ISL_2607295, EPI_ISL_2607296, EPI_ISL_2607297, EPI_ISL_2607298, EPI_ISL_2607299, EPI_ISL_2607300, EPI_ISL_2607301, EPI_ISL_2607302, EPI_ISL_2607303, EPI_ISL_2607304, EPI_ISL_2607305, EPI_ISL_2607306, EPI_ISL_2607307, EPI_ISL_2607308, EPI_ISL_2607309, EPI_ISL_2607310, EPI_ISL_2607311, EPI_ISL_2607313, EPI_ISL_2607314, EPI_ISL_2607315, EPI_ISL_2607316, EPI_ISL_2607317, EPI_ISL_2607318, EPI_ISL_2607319, EPI_ISL_2607320, EPI_ISL_2607321, EPI_ISL_2607322, EPI_ISL_2607323, EPI_ISL_2607324, EPI_ISL_2607325, EPI_ISL_2607326, EPI_ISL_2607327, EPI_ISL_2607328, EPI_ISL_2607330, EPI_ISL_2607331, EPI_ISL_2607332, EPI_ISL_2607333, EPI_ISL_2607334, EPI_ISL_2607335, EPI_ISL_2607336, EPI_ISL_2607338, EPI_ISL_2607339, EPI_ISL_2607340, EPI_ISL_2607341, EPI_ISL_2607342, EPI_ISL_2607343, EPI_ISL_2607344, EPI_ISL_2607345, EPI_ISL_2607346, EPI_ISL_2607347, EPI_ISL_2607348, EPI_ISL_2607349, EPI_ISL_2607350, EPI_ISL_2607351, EPI_ISL_2607352, EPI_ISL_2607353, EPI_ISL_2607354, EPI_ISL_2607355, EPI_ISL_2607357, EPI_ISL_2607358, EPI_ISL_2607359, EPI_ISL_2607360, EPI_ISL_2607361, EPI_ISL_2607362, EPI_ISL_2607363, EPI_ISL_2607364, EPI_ISL_2607365, EPI_ISL_2607366, EPI_ISL_2607367, EPI_ISL_2607368, EPI_ISL_2607369, EPI_ISL_2607370, EPI_ISL_2607371, EPI_ISL_2607372, EPI_ISL_2607373, EPI_ISL_2607374, EPI_ISL_2607375, EPI_ISL_2607376, EPI_ISL_2607377, EPI_ISL_2607378, EPI_ISL_2607379, EPI_ISL_2607380, EPI_ISL_2607381, EPI_ISL_2607382, EPI_ISL_2607383, EPI_ISL_2607384, EPI_ISL_2607385, EPI_ISL_2607386, EPI_ISL_2607387, EPI_ISL_2607388, EPI_ISL_2607389, EPI_ISL_2607390, EPI_ISL_2607391, EPI_ISL_2607392, EPI_ISL_2607393, EPI_ISL_2607394, EPI_ISL_2607395, EPI_ISL_2607396, EPI_ISL_2607397, EPI_ISL_2607398, EPI_ISL_2607399, EPI_ISL_2607400, EPI_ISL_2607401, EPI_ISL_2607402, EPI_ISL_2607403, EPI_ISL_2607404, EPI_ISL_2607405, EPI_ISL_2607406, EPI_ISL_2607407, EPI_ISL_2607408, EPI_ISL_2607409, EPI_ISL_2607410, EPI_ISL_2607411, EPI_ISL_2607412, EPI_ISL_2607413, EPI_ISL_2607414, EPI_ISL_2607415, EPI_ISL_2607416, EPI_ISL_2607417, EPI_ISL_2607418, EPI_ISL_2607419, EPI_ISL_2607420, EPI_ISL_2607421, EPI_ISL_2607422, EPI_ISL_2607423, EPI_ISL_2607424, EPI_ISL_2607425, EPI_ISL_2607426, EPI_ISL_2607427, EPI_ISL_2607428, EPI_ISL_2607429, EPI_ISL_2607430, EPI_ISL_2607431, EPI_ISL_2607432, EPI_ISL_2607433, EPI_ISL_2607434, EPI_ISL_2607435, EPI_ISL_2607436, EPI_ISL_2607437, EPI_ISL_2607438, EPI_ISL_2607439, EPI_ISL_2607440, EPI_ISL_2607441, EPI_ISL_2607444, EPI_ISL_2607449, EPI_ISL_2607450, EPI_ISL_2607451, EPI_ISL_2607452, EPI_ISL_2607454, EPI_ISL_2607455, EPI_ISL_2607460, EPI_ISL_2607462, EPI_ISL_2607463, EPI_ISL_2607467, EPI_ISL_2607468, EPI_ISL_2607471, EPI_ISL_2607472, EPI_ISL_2607477, EPI_ISL_2607479, EPI_ISL_2607485, EPI_ISL_2607486, EPI_ISL_2607488, EPI_ISL_2607494, EPI_ISL_2607500, EPI_ISL_2607502, EPI_ISL_2607503, EPI_ISL_2607506, EPI_ISL_2607513, EPI_ISL_2607515, EPI_ISL_2607639, EPI_ISL_2607645, EPI_ISL_2607648, EPI_ISL_2607651, EPI_ISL_2607653, EPI_ISL_2607654, EPI_ISL_2607655, EPI_ISL_2607664, EPI_ISL_2607665, EPI_ISL_2607676, EPI_ISL_2607682, EPI_ISL_2607692, EPI_ISL_2607693, EPI_ISL_2607697, EPI_ISL_2607700, EPI_ISL_2607703, EPI_ISL_2607704, EPI_ISL_2607708, EPI_ISL_2607710, EPI_ISL_2607712, EPI_ISL_2607715, EPI_ISL_2607718, EPI_ISL_2607722, EPI_ISL_2607723, EPI_ISL_2607730, EPI_ISL_2607736, EPI_ISL_2607746, EPI_ISL_2607749, EPI_ISL_2607760, EPI_ISL_2607762, EPI_ISL_2607770, EPI_ISL_2607771, EPI_ISL_2607773, EPI_ISL_2607783, EPI_ISL_2607788, EPI_ISL_2607789, EPI_ISL_2607801, EPI_ISL_2607805, EPI_ISL_2607809, EPI_ISL_2607811, EPI_ISL_2607812, EPI_ISL_2607815, EPI_ISL_2607817, EPI_ISL_2607821, EPI_ISL_2607826, EPI_ISL_2607827, EPI_ISL_2607829, EPI_ISL_2607831, EPI_ISL_2607832, EPI_ISL_2607834, EPI_ISL_2607841, EPI_ISL_2607842, EPI_ISL_2607843, EPI_ISL_2607846, EPI_ISL_2607849, EPI_ISL_2607852, EPI_ISL_2607856, EPI_ISL_2607859, EPI_ISL_2607863, EPI_ISL_2607864, EPI_ISL_2607865, EPI_ISL_2607869, EPI_ISL_2607873, EPI_ISL_2607874, EPI_ISL_2607875, EPI_ISL_2607878, EPI_ISL_2607883, EPI_ISL_2607884, EPI_ISL_2607889, EPI_ISL_2607893, EPI_ISL_2607897, EPI_ISL_2607903, EPI_ISL_2607904, EPI_ISL_2607905, EPI_ISL_2607906, EPI_ISL_2607907, EPI_ISL_2607908, EPI_ISL_2607909, EPI_ISL_2607911, EPI_ISL_2607913, EPI_ISL_2607916, EPI_ISL_2607918, EPI_ISL_2607919, EPI_ISL_2607920, EPI_ISL_2607921, EPI_ISL_2607922, EPI_ISL_2607923, EPI_ISL_2607924, EPI_ISL_2607925, EPI_ISL_2607926, EPI_ISL_2607927, EPI_ISL_2607928, EPI_ISL_2607930, EPI_ISL_2607931, EPI_ISL_2607933, EPI_ISL_2607935, EPI_ISL_2607936, EPI_ISL_2607937, EPI_ISL_2607938, EPI_ISL_2607939, EPI_ISL_2607940, EPI_ISL_2607941, EPI_ISL_2607942, EPI_ISL_2607943, EPI_ISL_2607944, EPI_ISL_2607945, EPI_ISL_2607949, EPI_ISL_2607952, EPI_ISL_2607954, EPI_ISL_2607955, EPI_ISL_2607957, EPI_ISL_2607958, EPI_ISL_2607965, EPI_ISL_2607967, EPI_ISL_2607970, EPI_ISL_2607971, EPI_ISL_2607972, EPI_ISL_2607979, EPI_ISL_2607981, EPI_ISL_2607985, EPI_ISL_2607987, EPI_ISL_2607989, EPI_ISL_2608001, EPI_ISL_2608002, EPI_ISL_2608004, EPI_ISL_2608005, EPI_ISL_2608008, EPI_ISL_2608012, EPI_ISL_2608014, EPI_ISL_2608020, EPI_ISL_2608023, EPI_ISL_2608025, EPI_ISL_2608027, EPI_ISL_2608030, EPI_ISL_2608031, EPI_ISL_2608032, EPI_ISL_2608033, EPI_ISL_2608035, EPI_ISL_2608044, EPI_ISL_2608046, EPI_ISL_2608052, EPI_ISL_2608055, EPI_ISL_2608056, EPI_ISL_2608057, EPI_ISL_2608062, EPI_ISL_2608063, EPI_ISL_2608068, EPI_ISL_2608069, EPI_ISL_2608070, EPI_ISL_2608071, EPI_ISL_2608073, EPI_ISL_2608076, EPI_ISL_2608080, EPI_ISL_2608081, EPI_ISL_2608082, EPI_ISL_2608083, EPI_ISL_2608086, EPI_ISL_2608087, EPI_ISL_2608089, EPI_ISL_2608094, EPI_ISL_2608097, EPI_ISL_2608099, EPI_ISL_2608100, EPI_ISL_2608102, EPI_ISL_2608105, EPI_ISL_2608106, EPI_ISL_2608110, EPI_ISL_2608111, EPI_ISL_2608112, EPI_ISL_2608113, EPI_ISL_2608114, EPI_ISL_2608115, EPI_ISL_2608116, EPI_ISL_2608118, EPI_ISL_2608119, EPI_ISL_2608120, EPI_ISL_2608121, EPI_ISL_2608122, EPI_ISL_2608123, EPI_ISL_2608125, EPI_ISL_2608126, EPI_ISL_2608127, EPI_ISL_2608128, EPI_ISL_2608129, EPI_ISL_2608130, EPI_ISL_2608131, EPI_ISL_2608132, EPI_ISL_2608133, EPI_ISL_2608134, EPI_ISL_2608135, EPI_ISL_2608136, EPI_ISL_2608137, EPI_ISL_2608138, EPI_ISL_2608139, EPI_ISL_2608140, EPI_ISL_2608141, EPI_ISL_2608142, EPI_ISL_2608143, EPI_ISL_2608144, EPI_ISL_2608145, EPI_ISL_2608146, EPI_ISL_2608147, EPI_ISL_2608148, EPI_ISL_2608149, EPI_ISL_2608150, EPI_ISL_2608151, EPI_ISL_2608152, EPI_ISL_2608153, EPI_ISL_2608154, EPI_ISL_2608155, EPI_ISL_2608156, EPI_ISL_2608157, EPI_ISL_2608158, EPI_ISL_2608159, EPI_ISL_2608160, EPI_ISL_2608161, EPI_ISL_2608162, EPI_ISL_2608163, EPI_ISL_2608164, EPI_ISL_2608165, EPI_ISL_2608166, EPI_ISL_2608167, EPI_ISL_2608168, EPI_ISL_2608169, EPI_ISL_2608170, EPI_ISL_2608171, EPI_ISL_2608172, EPI_ISL_2608173, EPI_ISL_2608174, EPI_ISL_2608175, EPI_ISL_2608176, EPI_ISL_2608177, EPI_ISL_2608178, EPI_ISL_2608179, EPI_ISL_2608180, EPI_ISL_2608181, EPI_ISL_2608182, EPI_ISL_2608183, EPI_ISL_2608184, EPI_ISL_2608185, EPI_ISL_2608186, EPI_ISL_2608187, EPI_ISL_2608188, EPI_ISL_2608189, EPI_ISL_2608190, EPI_ISL_2608191, EPI_ISL_2608192, EPI_ISL_2608193, EPI_ISL_2608194, EPI_ISL_2608195, EPI_ISL_2608196, EPI_ISL_2608197, EPI_ISL_2608198, EPI_ISL_2608199, EPI_ISL_2608200, EPI_ISL_2608201, EPI_ISL_2608202, EPI_ISL_2608203, EPI_ISL_2608204, EPI_ISL_2608205, EPI_ISL_2608206, EPI_ISL_2608207, EPI_ISL_2608209, EPI_ISL_2608210, EPI_ISL_2608211, EPI_ISL_2608212, EPI_ISL_2608213, EPI_ISL_2608214, EPI_ISL_2608215, EPI_ISL_2608216, EPI_ISL_2608217, EPI_ISL_2608218, EPI_ISL_2608219, EPI_ISL_2608220, EPI_ISL_2608221, EPI_ISL_2608222, EPI_ISL_2608223, EPI_ISL_2608224, EPI_ISL_2608225, EPI_ISL_2608226, EPI_ISL_2608227, EPI_ISL_2608228, EPI_ISL_2608229, EPI_ISL_2608230, EPI_ISL_2608231, EPI_ISL_2608232, EPI_ISL_2608233, EPI_ISL_2608234, EPI_ISL_2608235, EPI_ISL_2608236, EPI_ISL_2608237, EPI_ISL_2608238, EPI_ISL_2608239, EPI_ISL_2608240, EPI_ISL_2608241, EPI_ISL_2608242, EPI_ISL_2608243, EPI_ISL_2608244, EPI_ISL_2608245, EPI_ISL_2608246, EPI_ISL_2608247, EPI_ISL_2608248, EPI_ISL_2608249, EPI_ISL_2608250, EPI_ISL_2608251, EPI_ISL_2608252, EPI_ISL_2608253, EPI_ISL_2608254, EPI_ISL_2608255, EPI_ISL_2608256, EPI_ISL_2608257, EPI_ISL_2608258, EPI_ISL_2608259, EPI_ISL_2608260, EPI_ISL_2608261, EPI_ISL_2608262, EPI_ISL_2608263, EPI_ISL_2608264, EPI_ISL_2608265, EPI_ISL_2608266, EPI_ISL_2608267, EPI_ISL_2608268, EPI_ISL_2608269, EPI_ISL_2608270, EPI_ISL_2608271, EPI_ISL_2608272, EPI_ISL_2608273, EPI_ISL_2608274, EPI_ISL_2608275, EPI_ISL_2608276, EPI_ISL_2608277, EPI_ISL_2608278, EPI_ISL_2608279, EPI_ISL_2608280, EPI_ISL_2608281, EPI_ISL_2608282, EPI_ISL_2608283, EPI_ISL_2640092, EPI_ISL_2640105, EPI_ISL_2640116, EPI_ISL_2640147, EPI_ISL_2640153, EPI_ISL_2640219, EPI_ISL_2640272, EPI_ISL_2640306, EPI_ISL_2640313, EPI_ISL_2640677, EPI_ISL_2640680, EPI_ISL_2640682, EPI_ISL_2640685, EPI_ISL_2640690, EPI_ISL_2640695, EPI_ISL_2640696, EPI_ISL_2640700, EPI_ISL_2640706, EPI_ISL_2640716, EPI_ISL_2640718, EPI_ISL_2640725, EPI_ISL_2640727, EPI_ISL_2640736, EPI_ISL_2640739, EPI_ISL_2640741, EPI_ISL_2640751, EPI_ISL_2640760, EPI_ISL_2640762, EPI_ISL_2640763, EPI_ISL_2640770, EPI_ISL_2640773, EPI_ISL_2640777, EPI_ISL_2640783, EPI_ISL_2640797, EPI_ISL_2640798, EPI_ISL_2640802, EPI_ISL_2640803, EPI_ISL_2640805, EPI_ISL_2640807, EPI_ISL_2640813, EPI_ISL_2640814, EPI_ISL_2640825, EPI_ISL_2640829, EPI_ISL_2640836, EPI_ISL_2640840, EPI_ISL_2640844, EPI_ISL_2640846, EPI_ISL_2640847, EPI_ISL_2640849, EPI_ISL_2640850, EPI_ISL_2640852, EPI_ISL_2640856, EPI_ISL_2640857, EPI_ISL_2640875, EPI_ISL_2640878, EPI_ISL_2640880, EPI_ISL_2640895, EPI_ISL_2640900, EPI_ISL_2640901, EPI_ISL_2640905, EPI_ISL_2640908, EPI_ISL_2640909, EPI_ISL_2640910, EPI_ISL_2640913, EPI_ISL_2640915, EPI_ISL_2640919, EPI_ISL_2640921, EPI_ISL_2640922, EPI_ISL_2640923, EPI_ISL_2640940, EPI_ISL_2640951, EPI_ISL_2640960, EPI_ISL_2640963, EPI_ISL_2640965, EPI_ISL_2640966, EPI_ISL_2640968, EPI_ISL_2640974, EPI_ISL_2640996, EPI_ISL_2640997, EPI_ISL_2640998, EPI_ISL_2641000, EPI_ISL_2641006, EPI_ISL_2641012, EPI_ISL_2641019, EPI_ISL_2641027, EPI_ISL_2641037, EPI_ISL_2641039, EPI_ISL_2641041, EPI_ISL_2641046, EPI_ISL_2641060, EPI_ISL_2641065, EPI_ISL_2641067, EPI_ISL_2641068, EPI_ISL_2641081, EPI_ISL_2641085, EPI_ISL_2641089, EPI_ISL_2641091, EPI_ISL_2641092, EPI_ISL_2641093, EPI_ISL_2641095, EPI_ISL_2641096, EPI_ISL_2641104, EPI_ISL_2641107, EPI_ISL_2641117, EPI_ISL_2641122, EPI_ISL_2641124, EPI_ISL_2641125, EPI_ISL_2641130, EPI_ISL_2641132, EPI_ISL_2641138, EPI_ISL_2641140, EPI_ISL_2641141, EPI_ISL_2641142, EPI_ISL_2641144, EPI_ISL_2641145, EPI_ISL_2641153, EPI_ISL_2641154, EPI_ISL_2641156, EPI_ISL_2641164, EPI_ISL_2641165, EPI_ISL_2641169, EPI_ISL_2641176, EPI_ISL_2641177, EPI_ISL_2641178, EPI_ISL_2641179, EPI_ISL_2641181, EPI_ISL_2641183, EPI_ISL_2641185, EPI_ISL_2641188, EPI_ISL_2641195, EPI_ISL_2641196, EPI_ISL_2641198, EPI_ISL_2641200, EPI_ISL_2641203, EPI_ISL_2641208, EPI_ISL_2641215, EPI_ISL_2641217, EPI_ISL_2641224, EPI_ISL_2641226, EPI_ISL_2641228, EPI_ISL_2641229, EPI_ISL_2641230, EPI_ISL_2641238, EPI_ISL_2641242, EPI_ISL_2641246, EPI_ISL_2641257, EPI_ISL_2641262, EPI_ISL_2641265, EPI_ISL_2641269, EPI_ISL_2641276, EPI_ISL_2641281, EPI_ISL_2641293, EPI_ISL_2641294, EPI_ISL_2641300, EPI_ISL_2641301, EPI_ISL_2641302, EPI_ISL_2641305, EPI_ISL_2641309, EPI_ISL_2641314, EPI_ISL_2641315, EPI_ISL_2641316, EPI_ISL_2641317, EPI_ISL_2641319, EPI_ISL_2641320, EPI_ISL_2641322, EPI_ISL_2641324, EPI_ISL_2641327, EPI_ISL_2641332, EPI_ISL_2641339, EPI_ISL_2641341, EPI_ISL_2641345, EPI_ISL_2641359, EPI_ISL_2641361, EPI_ISL_2641364, EPI_ISL_2641365, EPI_ISL_2641366, EPI_ISL_2641371, EPI_ISL_2641373, EPI_ISL_2641374, EPI_ISL_2641375, EPI_ISL_2641376, EPI_ISL_2641380, EPI_ISL_2641385, EPI_ISL_2641388, EPI_ISL_2641389, EPI_ISL_2641392, EPI_ISL_2641393, EPI_ISL_2641396, EPI_ISL_2641397, EPI_ISL_2641399, EPI_ISL_2641400, EPI_ISL_2641403, EPI_ISL_2641404, EPI_ISL_2641405, EPI_ISL_2641408, EPI_ISL_2641410, EPI_ISL_2641416, EPI_ISL_2641417, EPI_ISL_2641418, EPI_ISL_2641424, EPI_ISL_2641425, EPI_ISL_2641427, EPI_ISL_2641428, EPI_ISL_2641430, EPI_ISL_2641433, EPI_ISL_2641434, EPI_ISL_2641435, EPI_ISL_2641436, EPI_ISL_2641438, EPI_ISL_2641439, EPI_ISL_2641440, EPI_ISL_2641441, EPI_ISL_2641444, EPI_ISL_2641445, EPI_ISL_2641447, EPI_ISL_2641448, EPI_ISL_2641451, EPI_ISL_2641452, EPI_ISL_2641453, EPI_ISL_2641454, EPI_ISL_2641455, EPI_ISL_2641456, EPI_ISL_2641457, EPI_ISL_2641459, EPI_ISL_2641461, EPI_ISL_2641464, EPI_ISL_2641465, EPI_ISL_2641467, EPI_ISL_2641471, EPI_ISL_2641473, EPI_ISL_2641474, EPI_ISL_2641475, EPI_ISL_2641476, EPI_ISL_2641477, EPI_ISL_2641478, EPI_ISL_2641479, EPI_ISL_2641481, EPI_ISL_2641482, EPI_ISL_2641483, EPI_ISL_2641485, EPI_ISL_2641486, EPI_ISL_2641487, EPI_ISL_2641496, EPI_ISL_2641497, EPI_ISL_2641499, EPI_ISL_2641501, EPI_ISL_2641502, EPI_ISL_2641503, EPI_ISL_2641508, EPI_ISL_2641509, EPI_ISL_2641516, EPI_ISL_2641519, EPI_ISL_2641521, EPI_ISL_2641522, EPI_ISL_2641526, EPI_ISL_2641527, EPI_ISL_2641529, EPI_ISL_2641536, EPI_ISL_2641537, EPI_ISL_2641541, EPI_ISL_2641542, EPI_ISL_2641543, EPI_ISL_2641546, EPI_ISL_2641548, EPI_ISL_2641550, EPI_ISL_2641551, EPI_ISL_2641553, EPI_ISL_2641555, EPI_ISL_2641557, EPI_ISL_2641559, EPI_ISL_2641560, EPI_ISL_2641561, EPI_ISL_2641564, EPI_ISL_2641565, EPI_ISL_2641576, EPI_ISL_2641578, EPI_ISL_2641579, EPI_ISL_2641580, EPI_ISL_2641581, EPI_ISL_2641584, EPI_ISL_2641589, EPI_ISL_2641591, EPI_ISL_2641595, EPI_ISL_2641596, EPI_ISL_2641597, EPI_ISL_2641598, EPI_ISL_2641600, EPI_ISL_2641601, EPI_ISL_2641602, EPI_ISL_2641604, EPI_ISL_2641605, EPI_ISL_2641607, EPI_ISL_2641608, EPI_ISL_2641609, EPI_ISL_2641611, EPI_ISL_2641613, EPI_ISL_2641614, EPI_ISL_2641619, EPI_ISL_2641623, EPI_ISL_2641635, EPI_ISL_2641636, EPI_ISL_2641637, EPI_ISL_2641641, EPI_ISL_2641642, EPI_ISL_2641643, EPI_ISL_2641645, EPI_ISL_2641646, EPI_ISL_2641647, EPI_ISL_2641649, EPI_ISL_2641651, EPI_ISL_2641654, EPI_ISL_2641656, EPI_ISL_2641659, EPI_ISL_2641663, EPI_ISL_2641666, EPI_ISL_2641667, EPI_ISL_2641668, EPI_ISL_2641672, EPI_ISL_2641674, EPI_ISL_2641675, EPI_ISL_2641676, EPI_ISL_2641678, EPI_ISL_2641680, EPI_ISL_2641681, EPI_ISL_2641683, EPI_ISL_2641685, EPI_ISL_2641686, EPI_ISL_2641689, EPI_ISL_2641690, EPI_ISL_2641691, EPI_ISL_2641692, EPI_ISL_2641693, EPI_ISL_2641694, EPI_ISL_2641695, EPI_ISL_2641696, EPI_ISL_2641698, EPI_ISL_2641699, EPI_ISL_2641700, EPI_ISL_2641701, EPI_ISL_2641702, EPI_ISL_2641704, EPI_ISL_2641705, EPI_ISL_2641707, EPI_ISL_2641708, EPI_ISL_2641710, EPI_ISL_2641711, EPI_ISL_2652001

| | | | |
|---|---|---|---|
| see above | Lighthouse Lab in Glasgow | Wellcome Sanger Institute for the COVID-19 Genomics UK (COG-UK) Consortium | Anna Dominiczak and Alex Alderton; Carol Clugston; Cordelia Langford; David Gray; David K. Jackson; Dominic Kwiatkowski; Ewan Harrison; Harper VanSteenhouse; Ian Johnston; Jeffrey Barrett; John Sillitoe on behalf of the Wellcome Sanger Institute COVID-19 Surveillance Team; Roberto Amato; Sonia Goncalves; Yumi Kasai |

EPI_ISL_2511134, EPI_ISL_2511327, EPI_ISL_2537493, EPI_ISL_2537495, EPI_ISL_2537497, EPI_ISL_2537498, EPI_ISL_2537499, EPI_ISL_2537501, EPI_ISL_2537504, EPI_ISL_2537511, EPI_ISL_2537513, EPI_ISL_2537514, EPI_ISL_2537516, EPI_ISL_2537517, EPI_ISL_2537518, EPI_ISL_2537521, EPI_ISL_2537526, EPI_ISL_2537528, EPI_ISL_2537529, EPI_ISL_2537530, EPI_ISL_2537531, EPI_ISL_2537534, EPI_ISL_2537535, EPI_ISL_2537537, EPI_ISL_2537539, EPI_ISL_2537541, EPI_ISL_2537544, EPI_ISL_2537545, EPI_ISL_2537546, EPI_ISL_2537547, EPI_ISL_2537548, EPI_ISL_2537551, EPI_ISL_2537552, EPI_ISL_2537554, EPI_ISL_2537562, EPI_ISL_2537563, EPI_ISL_2537565, EPI_ISL_2537566, EPI_ISL_2537569, EPI_ISL_2537571, EPI_ISL_2537572, EPI_ISL_2537573, EPI_ISL_2537574, EPI_ISL_2537575, EPI_ISL_2537577, EPI_ISL_2537580, EPI_ISL_2537582, EPI_ISL_2537585, EPI_ISL_2537587, EPI_ISL_2537590, EPI_ISL_2537593, EPI_ISL_2537595, EPI_ISL_2537596, EPI_ISL_2537597, EPI_ISL_2537598, EPI_ISL_2537599, EPI_ISL_2537603, EPI_ISL_2537608, EPI_ISL_2537610, EPI_ISL_2537611, EPI_ISL_2537612, EPI_ISL_2537613, EPI_ISL_2537614, EPI_ISL_2537619, EPI_ISL_2537622, EPI_ISL_2537626, EPI_ISL_2537627, EPI_ISL_2537631, EPI_ISL_2537632, EPI_ISL_2537633, EPI_ISL_2537635, EPI_ISL_2537636, EPI_ISL_2537637, EPI_ISL_2537638, EPI_ISL_2537639, EPI_ISL_2537641, EPI_ISL_2537643, EPI_ISL_2537647, EPI_ISL_2537651, EPI_ISL_2537653, EPI_ISL_2537654, EPI_ISL_2537657, EPI_ISL_2537658, EPI_ISL_2537664, EPI_ISL_2537665, EPI_ISL_2537667, EPI_ISL_2537668, EPI_ISL_2537669, EPI_ISL_2537670, EPI_ISL_2537672, EPI_ISL_2537673, EPI_ISL_2537674, EPI_ISL_2537677, EPI_ISL_2537679, EPI_ISL_2537681, EPI_ISL_2537686, EPI_ISL_2537687, EPI_ISL_2537688, EPI_ISL_2537689, EPI_ISL_2537690, EPI_ISL_2537692, EPI_ISL_2537693, EPI_ISL_2537695, EPI_ISL_2537696, EPI_ISL_2537700, EPI_ISL_2537701, EPI_ISL_2537702, EPI_ISL_2537705, EPI_ISL_2537706, EPI_ISL_2537708, EPI_ISL_2537713, EPI_ISL_2537714, EPI_ISL_2537717, EPI_ISL_2537720, EPI_ISL_2537724, EPI_ISL_2537725, EPI_ISL_2537726, EPI_ISL_2537727, EPI_ISL_2537729, EPI_ISL_2537732, EPI_ISL_2537734, EPI_ISL_2537736, EPI_ISL_2537739, EPI_ISL_2537740, EPI_ISL_2537743, EPI_ISL_2537748, EPI_ISL_2537750, EPI_ISL_2537753, EPI_ISL_2537757, EPI_ISL_2537758, EPI_ISL_2537759, EPI_ISL_2537767, EPI_ISL_2537770, EPI_ISL_2537771, EPI_ISL_2537773, EPI_ISL_2537774, EPI_ISL_2537776, EPI_ISL_2537777, EPI_ISL_2537778, EPI_ISL_2537779, EPI_ISL_2537781, EPI_ISL_2537788, EPI_ISL_2537790, EPI_ISL_2537791, EPI_ISL_2537794, EPI_ISL_2537797, EPI_ISL_2537802, EPI_ISL_2537803, EPI_ISL_2537804, EPI_ISL_2537807, EPI_ISL_2538538, EPI_ISL_2538540, EPI_ISL_2538542, EPI_ISL_2538560, EPI_ISL_2538561, EPI_ISL_2538568, EPI_ISL_2538582, EPI_ISL_2538595, EPI_ISL_2538598, EPI_ISL_2538611, EPI_ISL_2538620, EPI_ISL_2538623, EPI_ISL_2538626, EPI_ISL_2538702, EPI_ISL_2538703, EPI_ISL_2538710, EPI_ISL_2538711, EPI_ISL_2538723, EPI_ISL_2538732, EPI_ISL_2538757, EPI_ISL_2538758, EPI_ISL_2538768, EPI_ISL_2538779, EPI_ISL_2538783, EPI_ISL_2538799, EPI_ISL_2538802, EPI_ISL_2538818, EPI_ISL_2538830, EPI_ISL_2540032, EPI_ISL_2540033, EPI_ISL_2540035, EPI_ISL_2540037, EPI_ISL_2540041, EPI_ISL_2540043, EPI_ISL_2540044, EPI_ISL_2540045, EPI_ISL_2540046, EPI_ISL_2540047, EPI_ISL_2540048, EPI_ISL_2540049, EPI_ISL_2540050, EPI_ISL_2540052, EPI_ISL_2540055, EPI_ISL_2540057, EPI_ISL_2540060, EPI_ISL_2540061, EPI_ISL_2540062, EPI_ISL_2540063, EPI_ISL_2540064, EPI_ISL_2540065, EPI_ISL_2540066, EPI_ISL_2540067, EPI_ISL_2540068, EPI_ISL_2540070, EPI_ISL_2540071, EPI_ISL_2540072, EPI_ISL_2540073, EPI_ISL_2540075, EPI_ISL_2540077, EPI_ISL_2540079, EPI_ISL_2540080, EPI_ISL_2540081, EPI_ISL_2540084, EPI_ISL_2540085, EPI_ISL_2540086, EPI_ISL_2540087, EPI_ISL_2540088, EPI_ISL_2540089, EPI_ISL_2540090, EPI_ISL_2540091, EPI_ISL_2540094, EPI_ISL_2540098, EPI_ISL_2540100, EPI_ISL_2540102, EPI_ISL_2540103, EPI_ISL_2540104, EPI_ISL_2540105, EPI_ISL_2540106, EPI_ISL_2540107, EPI_ISL_2540109, EPI_ISL_2540110, EPI_ISL_2540111, EPI_ISL_2540114, EPI_ISL_2540115, EPI_ISL_2540117, EPI_ISL_2540120, EPI_ISL_2540121, EPI_ISL_2540122, EPI_ISL_2540123, EPI_ISL_2540124, EPI_ISL_2540125, EPI_ISL_2540126, EPI_ISL_2540127, EPI_ISL_2540128, EPI_ISL_2540130, EPI_ISL_2540131, EPI_ISL_2540133, EPI_ISL_2540134, EPI_ISL_2540135, EPI_ISL_2540136, EPI_ISL_2540137, EPI_ISL_2540138, EPI_ISL_2540140, EPI_ISL_2540141, EPI_ISL_2540143, EPI_ISL_2540144, EPI_ISL_2540145, EPI_ISL_2540146, EPI_ISL_2540147, EPI_ISL_2540148, EPI_ISL_2540149, EPI_ISL_2540151, EPI_ISL_2540152, EPI_ISL_2540153, EPI_ISL_2540154, EPI_ISL_2540155, EPI_ISL_2540157, EPI_ISL_2540158, EPI_ISL_2540161, EPI_ISL_2540162, EPI_ISL_2540163, EPI_ISL_2540164, EPI_ISL_2540167, EPI_ISL_2540168, EPI_ISL_2540170, EPI_ISL_2540171, EPI_ISL_2540172, EPI_ISL_2540175, EPI_ISL_2540176, EPI_ISL_2540178, EPI_ISL_2540181, EPI_ISL_2540182, EPI_ISL_2540183, EPI_ISL_2540184, EPI_ISL_2540185, EPI_ISL_2540190, EPI_ISL_2540193, EPI_ISL_2540196, EPI_ISL_2540198, EPI_ISL_2540199, EPI_ISL_2540200, EPI_ISL_2540202, EPI_ISL_2540203, EPI_ISL_2540205, EPI_ISL_2540206, EPI_ISL_2540207, EPI_ISL_2540208, EPI_ISL_2540209, EPI_ISL_2540210, EPI_ISL_2540211, EPI_ISL_2540212, EPI_ISL_2540213, EPI_ISL_2540214, EPI_ISL_2540215, EPI_ISL_2540216, EPI_ISL_2540217, EPI_ISL_2540218, EPI_ISL_2540219, EPI_ISL_2540221, EPI_ISL_2540222, EPI_ISL_2540225, EPI_ISL_2540226, EPI_ISL_2540228, EPI_ISL_2540229, EPI_ISL_2540230, EPI_ISL_2540231, EPI_ISL_2540233, EPI_ISL_2540234, EPI_ISL_2540238, EPI_ISL_2540240, EPI_ISL_2540241, EPI_ISL_2540242, EPI_ISL_2540243, EPI_ISL_2540244, EPI_ISL_2540245, EPI_ISL_2540247, EPI_ISL_2540250, EPI_ISL_2540251, EPI_ISL_2540252, EPI_ISL_2540253, EPI_ISL_2540255, EPI_ISL_2540259, EPI_ISL_2540261, EPI_ISL_2540262, EPI_ISL_2540263, EPI_ISL_2540264, EPI_ISL_2540265, EPI_ISL_2540266, EPI_ISL_2540267, EPI_ISL_2540268, EPI_ISL_2540270, EPI_ISL_2540271, EPI_ISL_2540272, EPI_ISL_2540274, EPI_ISL_2540275, EPI_ISL_2540277, EPI_ISL_2540278, EPI_ISL_2540279, EPI_ISL_2540280, EPI_ISL_2540281, EPI_ISL_2540282, EPI_ISL_2540285, EPI_ISL_2540288, EPI_ISL_2540289, EPI_ISL_2540290, EPI_ISL_2540291, EPI_ISL_2540292, EPI_ISL_2540293, EPI_ISL_2540294, EPI_ISL_2540295, EPI_ISL_2540296, EPI_ISL_2540297, EPI_ISL_2540298, EPI_ISL_2540299, EPI_ISL_2540300, EPI_ISL_2540301, EPI_ISL_2540303, EPI_ISL_2540305, EPI_ISL_2540306, EPI_ISL_2540307, EPI_ISL_2540308, EPI_ISL_2540310, EPI_ISL_2540311, EPI_ISL_2540312, EPI_ISL_2540313, EPI_ISL_2540315, EPI_ISL_2540316, EPI_ISL_2540317, EPI_ISL_2540319, EPI_ISL_2540320, EPI_ISL_2540321, EPI_ISL_2540322, EPI_ISL_2540324, EPI_ISL_2540325, EPI_ISL_2540326, EPI_ISL_2540327, EPI_ISL_2540328, EPI_ISL_2540332, EPI_ISL_2540335, EPI_ISL_2540336, EPI_ISL_2540337, EPI_ISL_2540339, EPI_ISL_2540340, EPI_ISL_2540341, EPI_ISL_2540342, EPI_ISL_2540343, EPI_ISL_2540345, EPI_ISL_2540346, EPI_ISL_2540348, EPI_ISL_2540349, EPI_ISL_2540350, EPI_ISL_2540351, EPI_ISL_2540352, EPI_ISL_2540353, EPI_ISL_2540355, EPI_ISL_2540356, EPI_ISL_2540359, EPI_ISL_2540360, EPI_ISL_2540362, EPI_ISL_2540364, EPI_ISL_2540365, EPI_ISL_2540366, EPI_ISL_2540367, EPI_ISL_2540371, EPI_ISL_2540372, EPI_ISL_2540594, EPI_ISL_2540600, EPI_ISL_2540601, EPI_ISL_2540603, EPI_ISL_2540611, EPI_ISL_2540616, EPI_ISL_2540618, EPI_ISL_2540619, EPI_ISL_2540623, EPI_ISL_2540626, EPI_ISL_2540630, EPI_ISL_2540631, EPI_ISL_2540634, EPI_ISL_2540635, EPI_ISL_2540640, EPI_ISL_2540642, EPI_ISL_2540650, EPI_ISL_2540655, EPI_ISL_2540656, EPI_ISL_2540659, EPI_ISL_2540661, EPI_ISL_2540666, EPI_ISL_2540667, EPI_ISL_2540668, EPI_ISL_2540676, EPI_ISL_2540678, EPI_ISL_2540680, EPI_ISL_2540682, EPI_ISL_2540684, EPI_ISL_2540685, EPI_ISL_2540686, EPI_ISL_2540688, EPI_ISL_2540694, EPI_ISL_2540697, EPI_ISL_2540698, EPI_ISL_2540702, EPI_ISL_2540705, EPI_ISL_2540708, EPI_ISL_2540710, EPI_ISL_2540711, EPI_ISL_2540712, EPI_ISL_2540714, EPI_ISL_2540716, EPI_ISL_2540724, EPI_ISL_2540725, EPI_ISL_2540726, EPI_ISL_2540729, EPI_ISL_2540731, EPI_ISL_2540732, EPI_ISL_2540734, EPI_ISL_2540736, EPI_ISL_2540738, EPI_ISL_2540752, EPI_ISL_2540754, EPI_ISL_2540755, EPI_ISL_2540756, EPI_ISL_2540757, EPI_ISL_2540760, EPI_ISL_2540764, EPI_ISL_2540765, EPI_ISL_2540771, EPI_ISL_2540775, EPI_ISL_2540777, EPI_ISL_2540780, EPI_ISL_2540781, EPI_ISL_2540786, EPI_ISL_2540787, EPI_ISL_2540788, EPI_ISL_2540789, EPI_ISL_2540791, EPI_ISL_2540792, EPI_ISL_2540795, EPI_ISL_2540797, EPI_ISL_2540808, EPI_ISL_2540810, EPI_ISL_2540811, EPI_ISL_2540812, EPI_ISL_2540813, EPI_ISL_2540814, EPI_ISL_2540815, EPI_ISL_2540817, EPI_ISL_2540818, EPI_ISL_2540819, EPI_ISL_2540822, EPI_ISL_2540828, EPI_ISL_2540830, EPI_ISL_2540837, EPI_ISL_2540838, EPI_ISL_2540844, EPI_ISL_2540845, EPI_ISL_2540847, EPI_ISL_2540848, EPI_ISL_2540849, EPI_ISL_2540850, EPI_ISL_2540852, EPI_ISL_2540856, EPI_ISL_2540858, EPI_ISL_2540859, EPI_ISL_2540864, EPI_ISL_2540865, EPI_ISL_2540866, EPI_ISL_2540870, EPI_ISL_2540871, EPI_ISL_2540873, EPI_ISL_2540874, EPI_ISL_2540886, EPI_ISL_2540887, EPI_ISL_2540888, EPI_ISL_2540890, EPI_ISL_2540895, EPI_ISL_2540901, EPI_ISL_2540905, EPI_ISL_2540907, EPI_ISL_2540908, EPI_ISL_2540910, EPI_ISL_2540911, EPI_ISL_2540915, EPI_ISL_2540918, EPI_ISL_2540922, EPI_ISL_2540927, EPI_ISL_2540928, EPI_ISL_2540932, EPI_ISL_2540938, EPI_ISL_2540940, EPI_ISL_2540941, EPI_ISL_2540942, EPI_ISL_2540943, EPI_ISL_2540948, EPI_ISL_2540950, EPI_ISL_2540952, EPI_ISL_2540957, EPI_ISL_2540958, EPI_ISL_2540959, EPI_ISL_2540961, EPI_ISL_2540962, EPI_ISL_2540963, EPI_ISL_2540968, EPI_ISL_2540970, EPI_ISL_2540972, EPI_ISL_2540975, EPI_ISL_2540976, EPI_ISL_2540980, EPI_ISL_2540982, EPI_ISL_2540986, EPI_ISL_2540987, EPI_ISL_2540988, EPI_ISL_2540990, EPI_ISL_2540993, EPI_ISL_2540994, EPI_ISL_2540995, EPI_ISL_2541001, EPI_ISL_2541002, EPI_ISL_2541004, EPI_ISL_2541006, EPI_ISL_2541007, EPI_ISL_2541008, EPI_ISL_2541013, EPI_ISL_2541017, EPI_ISL_2541018, EPI_ISL_2541019, EPI_ISL_2541021, EPI_ISL_2541024, EPI_ISL_2541025, EPI_ISL_2541029, EPI_ISL_2541032, EPI_ISL_2541034, EPI_ISL_2541036, EPI_ISL_2541038, EPI_ISL_2541039, EPI_ISL_2541041, EPI_ISL_2541043, EPI_ISL_2541044, EPI_ISL_2541047, EPI_ISL_2541049, EPI_ISL_2541053, EPI_ISL_2541056, EPI_ISL_2541057, EPI_ISL_2541058, EPI_ISL_2541062, EPI_ISL_2541065, EPI_ISL_2541066, EPI_ISL_2541067, EPI_ISL_2541068, EPI_ISL_2541069, EPI_ISL_2541071, EPI_ISL_2541073, EPI_ISL_2541081, EPI_ISL_2541082, EPI_ISL_2541083, EPI_ISL_2541085, EPI_ISL_2541086, EPI_ISL_2541088, EPI_ISL_2541089, EPI_ISL_2541090, EPI_ISL_2541092, EPI_ISL_2541095, EPI_ISL_2541097, EPI_ISL_2541098, EPI_ISL_2541101, EPI_ISL_2541105, EPI_ISL_2541106, EPI_ISL_2541108, EPI_ISL_2541111, EPI_ISL_2541113, EPI_ISL_2541115, EPI_ISL_2541117, EPI_ISL_2541118, EPI_ISL_2541119, EPI_ISL_2541121, EPI_ISL_2541122, EPI_ISL_2541123, EPI_ISL_2541124, EPI_ISL_2541126, EPI_ISL_2541128, EPI_ISL_2541129, EPI_ISL_2541130, EPI_ISL_2541131, EPI_ISL_2541133, EPI_ISL_2541134, EPI_ISL_2541138, EPI_ISL_2541139, EPI_ISL_2541140, EPI_ISL_2541141, EPI_ISL_2541145, EPI_ISL_2541146, EPI_ISL_2541149, EPI_ISL_2541150, EPI_ISL_2541152, EPI_ISL_2541155, EPI_ISL_2541156, EPI_ISL_2541158, EPI_ISL_2541161, EPI_ISL_2541165, EPI_ISL_2541167, EPI_ISL_2541168, EPI_ISL_2541169, EPI_ISL_2541171, EPI_ISL_2541173, EPI_ISL_2541174, EPI_ISL_2541178, EPI_ISL_2541179, EPI_ISL_2541182, EPI_ISL_2541186, EPI_ISL_2541189, EPI_ISL_2541190, EPI_ISL_2541192, EPI_ISL_2541195, EPI_ISL_2541196, EPI_ISL_2541197, EPI_ISL_2541203, EPI_ISL_2541205, EPI_ISL_2541208, EPI_ISL_2541210, EPI_ISL_2541211, EPI_ISL_2541214, EPI_ISL_2541216, EPI_ISL_2541217, EPI_ISL_2541218, EPI_ISL_2541219, EPI_ISL_2541220, EPI_ISL_2541221, EPI_ISL_2541222, EPI_ISL_2541226, EPI_ISL_2541230, EPI_ISL_2541232, EPI_ISL_2541236, EPI_ISL_2541237, EPI_ISL_2541238, EPI_ISL_2541240, EPI_ISL_2541245, EPI_ISL_2541246, EPI_ISL_2541252, EPI_ISL_2541254, EPI_ISL_2541255, EPI_ISL_2541256, EPI_ISL_2541259, EPI_ISL_2541264, EPI_ISL_2541265, EPI_ISL_2541266, EPI_ISL_2541267, EPI_ISL_2541270, EPI_ISL_2541272, EPI_ISL_2541273, EPI_ISL_2541275, EPI_ISL_2541276, EPI_ISL_2541277, EPI_ISL_2541278, EPI_ISL_2541279, EPI_ISL_2541281, EPI_ISL_2541284, EPI_ISL_2541285, EPI_ISL_2541289, EPI_ISL_2541290, EPI_ISL_2541292, EPI_ISL_2541293, EPI_ISL_2541294, EPI_ISL_2541299, EPI_ISL_2541301, EPI_ISL_2541302, EPI_ISL_2541303, EPI_ISL_2541304, EPI_ISL_2541308, EPI_ISL_2541310, EPI_ISL_2541313, EPI_ISL_2541314, EPI_ISL_2541318, EPI_ISL_2541325, EPI_ISL_2541326, EPI_ISL_2541327, EPI_ISL_2541328, EPI_ISL_2541329, EPI_ISL_2541331, EPI_ISL_2541333, EPI_ISL_2541335, EPI_ISL_2541336, EPI_ISL_2541343, EPI_ISL_2541344, EPI_ISL_2541349, EPI_ISL_2541350, EPI_ISL_2541353, EPI_ISL_2541356, EPI_ISL_2541357, EPI_ISL_2541359, EPI_ISL_2541360, EPI_ISL_2541362, EPI_ISL_2541368, EPI_ISL_2541371, EPI_ISL_2541372, EPI_ISL_2541374, EPI_ISL_2541378, EPI_ISL_2541382, EPI_ISL_2541383, EPI_ISL_2541385, EPI_ISL_2541388, EPI_ISL_2541389, EPI_ISL_2541392, EPI_ISL_2541394, EPI_ISL_2541395, EPI_ISL_2541399, EPI_ISL_2541401, EPI_ISL_2541402, EPI_ISL_2541404, EPI_ISL_2541411, EPI_ISL_2541413, EPI_ISL_2541416, EPI_ISL_2541419, EPI_ISL_2541422, EPI_ISL_2541427, EPI_ISL_2541429, EPI_ISL_2541430, EPI_ISL_2541435, EPI_ISL_2541436, EPI_ISL_2541438, EPI_ISL_2541441, EPI_ISL_2541444, EPI_ISL_2541446, EPI_ISL_2541449, EPI_ISL_2541454, EPI_ISL_2541455, EPI_ISL_2541457, EPI_ISL_2541460, EPI_ISL_2541462, EPI_ISL_2541467, EPI_ISL_2541469, EPI_ISL_2541472, EPI_ISL_2541479, EPI_ISL_2541482, EPI_ISL_2541483, EPI_ISL_2541485, EPI_ISL_2541487, EPI_ISL_2541488, EPI_ISL_2541490, EPI_ISL_2541492, EPI_ISL_2541493, EPI_ISL_2541494, EPI_ISL_2541497, EPI_ISL_2541498, EPI_ISL_2541499, EPI_ISL_2541504, EPI_ISL_2541505, EPI_ISL_2541506, EPI_ISL_2541508, EPI_ISL_2541513, EPI_ISL_2541515, EPI_ISL_2541516, EPI_ISL_2541517, EPI_ISL_2541518, EPI_ISL_2541525, EPI_ISL_2541526, EPI_ISL_2541527, EPI_ISL_2541530, EPI_ISL_2541531, EPI_ISL_2541533, EPI_ISL_2541534, EPI_ISL_2541537, EPI_ISL_2541540, EPI_ISL_2541541, EPI_ISL_2541543, EPI_ISL_2541546, EPI_ISL_2541547, EPI_ISL_2541549, EPI_ISL_2541552, EPI_ISL_2541553, EPI_ISL_2541555, EPI_ISL_2541559, EPI_ISL_2541560, EPI_ISL_2541565, EPI_ISL_2541566, EPI_ISL_2541570, EPI_ISL_2541571, EPI_ISL_2541573, EPI_ISL_2541574, EPI_ISL_2541576, EPI_ISL_2541577, EPI_ISL_2541579, EPI_ISL_2541580, EPI_ISL_2541583, EPI_ISL_2541585, EPI_ISL_2541587, EPI_ISL_2541588, EPI_ISL_2541594, EPI_ISL_2541595, EPI_ISL_2541596, EPI_ISL_2541599, EPI_ISL_2541601, EPI_ISL_2541606, EPI_ISL_2541607, EPI_ISL_2541613, EPI_ISL_2541617, EPI_ISL_2541621, EPI_ISL_2541623, EPI_ISL_2541625, EPI_ISL_2541627, EPI_ISL_2541629, EPI_ISL_2551614, EPI_ISL_2551615, EPI_ISL_2551619, EPI_ISL_2551630, EPI_ISL_2551648, EPI_ISL_2551653, EPI_ISL_2551664, EPI_ISL_2551665, EPI_ISL_2551666, EPI_ISL_2551668, EPI_ISL_2551683, EPI_ISL_2551685, EPI_ISL_2551694, EPI_ISL_2551700, EPI_ISL_2551701, EPI_ISL_2551705, EPI_ISL_2551717, EPI_ISL_2551720, EPI_ISL_2551722, EPI_ISL_2551724, EPI_ISL_2551728, EPI_ISL_2551732, EPI_ISL_2551746, EPI_ISL_2551751, EPI_ISL_2551764, EPI_ISL_2551766, EPI_ISL_2551770, EPI_ISL_2551775, EPI_ISL_2551776, EPI_ISL_2551784, EPI_ISL_2551785, EPI_ISL_2551788, EPI_ISL_2551796, EPI_ISL_2551805, EPI_ISL_2551808, EPI_ISL_2551811, EPI_ISL_2551813, EPI_ISL_2551818, EPI_ISL_2551826, EPI_ISL_2551838, EPI_ISL_2551843, EPI_ISL_2551860, EPI_ISL_2551864, EPI_ISL_2551868, EPI_ISL_2551870, EPI_ISL_2551871, EPI_ISL_2551873, EPI_ISL_2551891, EPI_ISL_2551894, EPI_ISL_2551895, EPI_ISL_2551919, EPI_ISL_2551924, EPI_ISL_2552021, EPI_ISL_2552039, EPI_ISL_2552060, EPI_ISL_2552066, EPI_ISL_2552085, EPI_ISL_2552146, EPI_ISL_2552171, EPI_ISL_2552197, EPI_ISL_2552214, EPI_ISL_2552220, EPI_ISL_2552241, EPI_ISL_2552608, EPI_ISL_2552610, EPI_ISL_2552615, EPI_ISL_2552617, EPI_ISL_2552618, EPI_ISL_2552620, EPI_ISL_2552621, EPI_ISL_2552628, EPI_ISL_2552656, EPI_ISL_2552657, EPI_ISL_2552667, EPI_ISL_2552669, EPI_ISL_2552672, EPI_ISL_2552676, EPI_ISL_2552678, EPI_ISL_2552679, EPI_ISL_2552682, EPI_ISL_2552683, EPI_ISL_2552688, EPI_ISL_2552694, EPI_ISL_2552701, EPI_ISL_2552704, EPI_ISL_2552711, EPI_ISL_2552716, EPI_ISL_2552722, EPI_ISL_2552727, EPI_ISL_2552728, EPI_ISL_2552733, EPI_ISL_2552739, EPI_ISL_2552740, EPI_ISL_2552752, EPI_ISL_2552755, EPI_ISL_2552757, EPI_ISL_2552758, EPI_ISL_2552760, EPI_ISL_2552764, EPI_ISL_2552769, EPI_ISL_2552776, EPI_ISL_2552780, EPI_ISL_2552784, EPI_ISL_2552785, EPI_ISL_2552789, EPI_ISL_2552793, EPI_ISL_2552797, EPI_ISL_2552798, EPI_ISL_2552799, EPI_ISL_2552800, EPI_ISL_2552802, EPI_ISL_2552811, EPI_ISL_2552813, EPI_ISL_2552815, EPI_ISL_2552825, EPI_ISL_2552830, EPI_ISL_2552835, EPI_ISL_2552846, EPI_ISL_2552856, EPI_ISL_2552868, EPI_ISL_2552872, EPI_ISL_2552880, EPI_ISL_2552884, EPI_ISL_2552886, EPI_ISL_2552891, EPI_ISL_2552897, EPI_ISL_2552899, EPI_ISL_2552904, EPI_ISL_2552906, EPI_ISL_2552907, EPI_ISL_2552908, EPI_ISL_2552909, EPI_ISL_2552916, EPI_ISL_2552917, EPI_ISL_2552918, EPI_ISL_2552922, EPI_ISL_2553090, EPI_ISL_2553102, EPI_ISL_2553110, EPI_ISL_2553111, EPI_ISL_2553114, EPI_ISL_2553126, EPI_ISL_2553127, EPI_ISL_2553138, EPI_ISL_2553142, EPI_ISL_2553147, EPI_ISL_2553154, EPI_ISL_2553157, EPI_ISL_2553161, EPI_ISL_2553163, EPI_ISL_2553164, EPI_ISL_2553166, EPI_ISL_2553168, EPI_ISL_2553179, EPI_ISL_2553181, EPI_ISL_2553193, EPI_ISL_2553194, EPI_ISL_2553195, EPI_ISL_2553199, EPI_ISL_2553203, EPI_ISL_2553204, EPI_ISL_2553210, EPI_ISL_2553224, EPI_ISL_2553227, EPI_ISL_2553228, EPI_ISL_2553230, EPI_ISL_2553235, EPI_ISL_2553237, EPI_ISL_2553240, EPI_ISL_2553241, EPI_ISL_2553244, EPI_ISL_2553245, EPI_ISL_2553246, EPI_ISL_2553250, EPI_ISL_2553256, EPI_ISL_2553262, EPI_ISL_2553265, EPI_ISL_2553269, EPI_ISL_2553271, EPI_ISL_2553273, EPI_ISL_2553275, EPI_ISL_2553283, EPI_ISL_2553286, EPI_ISL_2553289, EPI_ISL_2553292, EPI_ISL_2553294, EPI_ISL_2553298, EPI_ISL_2553302, EPI_ISL_2553304, EPI_ISL_2553312, EPI_ISL_2553313, EPI_ISL_2553315, EPI_ISL_2553320, EPI_ISL_2553322, EPI_ISL_2553325, EPI_ISL_2553338, EPI_ISL_2553339, EPI_ISL_2553347, EPI_ISL_2553351, EPI_ISL_2553356, EPI_ISL_2553359, EPI_ISL_2553360, EPI_ISL_2553361, EPI_ISL_2553362, EPI_ISL_2553379, EPI_ISL_2553380, EPI_ISL_2553388, EPI_ISL_2553389, EPI_ISL_2553396, EPI_ISL_2553397, EPI_ISL_2553398, EPI_ISL_2553399, EPI_ISL_2553412, EPI_ISL_2553416, EPI_ISL_2553421, EPI_ISL_2553424, EPI_ISL_2553426, EPI_ISL_2553429, EPI_ISL_2553434, EPI_ISL_2553793, EPI_ISL_2553809, EPI_ISL_2553814, EPI_ISL_2553815, EPI_ISL_2553816, EPI_ISL_2553827, EPI_ISL_2553831, EPI_ISL_2553832, EPI_ISL_2553837, EPI_ISL_2553854, EPI_ISL_2553861, EPI_ISL_2553864, EPI_ISL_2553868, EPI_ISL_2553892, EPI_ISL_2553893, EPI_ISL_2553909, EPI_ISL_2553915, EPI_ISL_2553918, EPI_ISL_2553933, EPI_ISL_2553955, EPI_ISL_2553960, EPI_ISL_2553968, EPI_ISL_2553969, EPI_ISL_2553970, EPI_ISL_2553974, EPI_ISL_2553993, EPI_ISL_2553996, EPI_ISL_2554006, EPI_ISL_2554013, EPI_ISL_2554034, EPI_ISL_2554058, EPI_ISL_2554060, EPI_ISL_2554063, EPI_ISL_2554064, EPI_ISL_2554066, EPI_ISL_2554071, EPI_ISL_2554076, EPI_ISL_2554111, EPI_ISL_2554119, EPI_ISL_2554121, EPI_ISL_2554135, EPI_ISL_2554138, EPI_ISL_2554509, EPI_ISL_2554518, EPI_ISL_2554541, EPI_ISL_2554575, EPI_ISL_2554604, EPI_ISL_2554636, EPI_ISL_2554656, EPI_ISL_2554665, EPI_ISL_2554705, EPI_ISL_2554722, EPI_ISL_2554774, EPI_ISL_2554788, EPI_ISL_2554804, EPI_ISL_2554811, EPI_ISL_2554838, EPI_ISL_2555054, EPI_ISL_2555524, EPI_ISL_2568651, EPI_ISL_2569019, EPI_ISL_2569025, EPI_ISL_2569028, EPI_ISL_2569033, EPI_ISL_2569036, EPI_ISL_2569045, EPI_ISL_2569058, EPI_ISL_2569059, EPI_ISL_2569062, EPI_ISL_2569064, EPI_ISL_2569069, EPI_ISL_2569084, EPI_ISL_2569088, EPI_ISL_2569091, EPI_ISL_2569102, EPI_ISL_2569105, EPI_ISL_2569115, EPI_ISL_2569118, EPI_ISL_2569138, EPI_ISL_2569142, EPI_ISL_2569144, EPI_ISL_2569150, EPI_ISL_2569155, EPI_ISL_2569156, EPI_ISL_2569160, EPI_ISL_2569165, EPI_ISL_2569172, EPI_ISL_2569193, EPI_ISL_2569196, EPI_ISL_2569199, EPI_ISL_2569200, EPI_ISL_2569202, EPI_ISL_2569204, EPI_ISL_2569209, EPI_ISL_2569212, EPI_ISL_2569213, EPI_ISL_2569224, EPI_ISL_2569236, EPI_ISL_2569237, EPI_ISL_2569247, EPI_ISL_2569264, EPI_ISL_2569265, EPI_ISL_2569266, EPI_ISL_2569269, EPI_ISL_2569284, EPI_ISL_2569286, EPI_ISL_2569308, EPI_ISL_2569317, EPI_ISL_2569962, EPI_ISL_2569973, EPI_ISL_2569979, EPI_ISL_2570000, EPI_ISL_2570012, EPI_ISL_2570017, EPI_ISL_2570031, EPI_ISL_2570041, EPI_ISL_2570049, EPI_ISL_2570061, EPI_ISL_2570071, EPI_ISL_2570074, EPI_ISL_2570079, EPI_ISL_2570083, EPI_ISL_2570097, EPI_ISL_2570106, EPI_ISL_2570173, EPI_ISL_2570179, EPI_ISL_2570183, EPI_ISL_2570190, EPI_ISL_2570192, EPI_ISL_2570214, EPI_ISL_2570216, EPI_ISL_2570227, EPI_ISL_2570239, EPI_ISL_2570252, EPI_ISL_2570260, EPI_ISL_2570278, EPI_ISL_2570280, EPI_ISL_2570281, EPI_ISL_2570282, EPI_ISL_2570283, EPI_ISL_2570285, EPI_ISL_2570286, EPI_ISL_2570289, EPI_ISL_2570294, EPI_ISL_2570303, EPI_ISL_2570305, EPI_ISL_2570309, EPI_ISL_2570312, EPI_ISL_2570313, EPI_ISL_2570316, EPI_ISL_2570323, EPI_ISL_2570325, EPI_ISL_2570326, EPI_ISL_2570327, EPI_ISL_2570329, EPI_ISL_2570330, EPI_ISL_2570332, EPI_ISL_2570333, EPI_ISL_2570338, EPI_ISL_2570339, EPI_ISL_2570340, EPI_ISL_2570341, EPI_ISL_2570342, EPI_ISL_2570343, EPI_ISL_2570345, EPI_ISL_2570346, EPI_ISL_2570348, EPI_ISL_2570349, EPI_ISL_2570351, EPI_ISL_2570356, EPI_ISL_2570357, EPI_ISL_2570358, EPI_ISL_2570359, EPI_ISL_2570360, EPI_ISL_2570361, EPI_ISL_2570362, EPI_ISL_2570363, EPI_ISL_2570368, EPI_ISL_2570369, EPI_ISL_2570370, EPI_ISL_2570375, EPI_ISL_2570379, EPI_ISL_2570380, EPI_ISL_2570384, EPI_ISL_2570385, EPI_ISL_2570387, EPI_ISL_2570388, EPI_ISL_2570390, EPI_ISL_2570391, EPI_ISL_2570393, EPI_ISL_2570395, EPI_ISL_2570403, EPI_ISL_2570405, EPI_ISL_2570408, EPI_ISL_2570410, EPI_ISL_2570412, EPI_ISL_2570413, EPI_ISL_2570415, EPI_ISL_2570416, EPI_ISL_2570418, EPI_ISL_2570423, EPI_ISL_2570424, EPI_ISL_2570428, EPI_ISL_2570430, EPI_ISL_2570431, EPI_ISL_2570433, EPI_ISL_2570434, EPI_ISL_2570437, EPI_ISL_2570440, EPI_ISL_2570449, EPI_ISL_2570451, EPI_ISL_2570452, EPI_ISL_2570455, EPI_ISL_2570457, EPI_ISL_2570458, EPI_ISL_2570460, EPI_ISL_2570462, EPI_ISL_2570463, EPI_ISL_2570464, EPI_ISL_2570467, EPI_ISL_2570468, EPI_ISL_2570472, EPI_ISL_2606558, EPI_ISL_2606708, EPI_ISL_2607257, EPI_ISL_2608007, EPI_ISL_2608034, EPI_ISL_2640336, EPI_ISL_2640341,

EPI_ISL_2640344, EPI_ISL_2640345, EPI_ISL_2640347, EPI_ISL_2640348, EPI_ISL_2640350, EPI_ISL_2640354, EPI_ISL_2640356, EPI_ISL_2640360, EPI_ISL_2640361, EPI_ISL_2640362, EPI_ISL_2640365, EPI_ISL_2640367, EPI_ISL_2640368, EPI_ISL_2640369, EPI_ISL_2640370, EPI_ISL_2640371, EPI_ISL_2640372, EPI_ISL_2640373, EPI_ISL_2640379, EPI_ISL_2640380, EPI_ISL_2640382, EPI_ISL_2640383, EPI_ISL_2640384, EPI_ISL_2640387, EPI_ISL_2640388, EPI_ISL_2640398, EPI_ISL_2640403, EPI_ISL_2640405, EPI_ISL_2640407, EPI_ISL_2640408, EPI_ISL_2640412, EPI_ISL_2640413, EPI_ISL_2640416, EPI_ISL_2640417, EPI_ISL_2640421, EPI_ISL_2640422, EPI_ISL_2640425, EPI_ISL_2640428, EPI_ISL_2640430, EPI_ISL_2640432, EPI_ISL_2640433, EPI_ISL_2640434, EPI_ISL_2640435, EPI_ISL_2640446, EPI_ISL_2640448, EPI_ISL_2640449, EPI_ISL_2640451, EPI_ISL_2640454, EPI_ISL_2640456, EPI_ISL_2640457, EPI_ISL_2640459, EPI_ISL_2640464, EPI_ISL_2640465, EPI_ISL_2640467, EPI_ISL_2640469, EPI_ISL_2640470, EPI_ISL_2640472, EPI_ISL_2640475, EPI_ISL_2640476, EPI_ISL_2640477, EPI_ISL_2640478, EPI_ISL_2640479, EPI_ISL_2640480, EPI_ISL_2640482, EPI_ISL_2640483, EPI_ISL_2640486, EPI_ISL_2640487, EPI_ISL_2640488, EPI_ISL_2640489, EPI_ISL_2640492, EPI_ISL_2640495, EPI_ISL_2640496, EPI_ISL_2640497, EPI_ISL_2640498, EPI_ISL_2640501, EPI_ISL_2640503, EPI_ISL_2640504, EPI_ISL_2640505, EPI_ISL_2640507, EPI_ISL_2640508, EPI_ISL_2640509, EPI_ISL_2640514, EPI_ISL_2640516, EPI_ISL_2640518, EPI_ISL_2640519, EPI_ISL_2640522, EPI_ISL_2640524, EPI_ISL_2640525, EPI_ISL_2640529, EPI_ISL_2640532, EPI_ISL_2640537, EPI_ISL_2640538, EPI_ISL_2640539, EPI_ISL_2640540, EPI_ISL_2640543, EPI_ISL_2640545, EPI_ISL_2640547, EPI_ISL_2640548, EPI_ISL_2640551, EPI_ISL_2640553, EPI_ISL_2640555, EPI_ISL_2640558, EPI_ISL_2640561, EPI_ISL_2640565, EPI_ISL_2640567, EPI_ISL_2640568, EPI_ISL_2640571, EPI_ISL_2640576, EPI_ISL_2640579, EPI_ISL_2640582, EPI_ISL_2640588, EPI_ISL_2640601, EPI_ISL_2640604, EPI_ISL_2640606, EPI_ISL_2640607, EPI_ISL_2640608, EPI_ISL_2640616, EPI_ISL_2640619, EPI_ISL_2640623, EPI_ISL_2640624, EPI_ISL_2640625, EPI_ISL_2640626, EPI_ISL_2640628, EPI_ISL_2640629, EPI_ISL_2640634, EPI_ISL_2640635, EPI_ISL_2640638, EPI_ISL_2640639, EPI_ISL_2640641, EPI_ISL_2640643, EPI_ISL_2640644, EPI_ISL_2640647, EPI_ISL_2640650, EPI_ISL_2640653, EPI_ISL_2640654, EPI_ISL_2640656, EPI_ISL_2640657, EPI_ISL_2640658, EPI_ISL_2640659, EPI_ISL_2640660, EPI_ISL_2640661, EPI_ISL_2640664, EPI_ISL_2640665, EPI_ISL_2640669, EPI_ISL_2640670, EPI_ISL_2640672, EPI_ISL_2640673, EPI_ISL_2640674, EPI_ISL_2640675, EPI_ISL_2640679, EPI_ISL_2640681, EPI_ISL_2640683, EPI_ISL_2640689, EPI_ISL_2640692, EPI_ISL_2640697, EPI_ISL_2640709, EPI_ISL_2640711, EPI_ISL_2640712, EPI_ISL_2640723, EPI_ISL_2640730, EPI_ISL_2640731, EPI_ISL_2640733, EPI_ISL_2640755, EPI_ISL_2640757, EPI_ISL_2640769, EPI_ISL_2640771, EPI_ISL_2640779, EPI_ISL_2640780, EPI_ISL_2640782, EPI_ISL_2640785, EPI_ISL_2640786, EPI_ISL_2640790, EPI_ISL_2640791, EPI_ISL_2640793, EPI_ISL_2640796, EPI_ISL_2640800, EPI_ISL_2640806, EPI_ISL_2640809, EPI_ISL_2640810, EPI_ISL_2640811, EPI_ISL_2640812, EPI_ISL_2640820, EPI_ISL_2640824, EPI_ISL_2640828, EPI_ISL_2640834, EPI_ISL_2640839, EPI_ISL_2640841, EPI_ISL_2640845, EPI_ISL_2640851, EPI_ISL_2640855, EPI_ISL_2640858, EPI_ISL_2640865, EPI_ISL_2640867, EPI_ISL_2640868, EPI_ISL_2640873, EPI_ISL_2640874, EPI_ISL_2640885, EPI_ISL_2640887, EPI_ISL_2640888, EPI_ISL_2640890, EPI_ISL_2640893, EPI_ISL_2640899, EPI_ISL_2640903, EPI_ISL_2640907, EPI_ISL_2640911, EPI_ISL_2640912, EPI_ISL_2640916, EPI_ISL_2640924, EPI_ISL_2640929, EPI_ISL_2640932, EPI_ISL_2640936, EPI_ISL_2640939, EPI_ISL_2640944, EPI_ISL_2640946, EPI_ISL_2640947, EPI_ISL_2640948, EPI_ISL_2640949, EPI_ISL_2640952, EPI_ISL_2640956, EPI_ISL_2640957, EPI_ISL_2640959, EPI_ISL_2640964, EPI_ISL_2640971, EPI_ISL_2640979, EPI_ISL_2640981, EPI_ISL_2640983, EPI_ISL_2640989, EPI_ISL_2640991, EPI_ISL_2640993, EPI_ISL_2640994, EPI_ISL_2641001, EPI_ISL_2641003, EPI_ISL_2641011, EPI_ISL_2641015, EPI_ISL_2641025, EPI_ISL_2641030, EPI_ISL_2641031, EPI_ISL_2641033, EPI_ISL_2641035, EPI_ISL_2641040, EPI_ISL_2641043, EPI_ISL_2641047, EPI_ISL_2641051, EPI_ISL_2641052, EPI_ISL_2641053, EPI_ISL_2641061, EPI_ISL_2641062, EPI_ISL_2641064, EPI_ISL_2641072, EPI_ISL_2641074, EPI_ISL_2641075, EPI_ISL_2641078, EPI_ISL_2641080, EPI_ISL_2641083, EPI_ISL_2641084, EPI_ISL_2641094, EPI_ISL_2641100, EPI_ISL_2641110, EPI_ISL_2641111, EPI_ISL_2641112, EPI_ISL_2641113, EPI_ISL_2641118, EPI_ISL_2641126, EPI_ISL_2641127, EPI_ISL_2641128, EPI_ISL_2641148, EPI_ISL_2641149, EPI_ISL_2641150, EPI_ISL_2641152, EPI_ISL_2641159, EPI_ISL_2641160, EPI_ISL_2641162, EPI_ISL_2641163, EPI_ISL_2641167, EPI_ISL_2641168, EPI_ISL_2641171, EPI_ISL_2641172, EPI_ISL_2641173, EPI_ISL_2641174, EPI_ISL_2641187, EPI_ISL_2641190, EPI_ISL_2641192, EPI_ISL_2641194, EPI_ISL_2641199, EPI_ISL_2641213, EPI_ISL_2641214, EPI_ISL_2641222, EPI_ISL_2641227, EPI_ISL_2641235, EPI_ISL_2641236, EPI_ISL_2641237, EPI_ISL_2641254, EPI_ISL_2641256, EPI_ISL_2641263, EPI_ISL_2641264, EPI_ISL_2641266, EPI_ISL_2641272, EPI_ISL_2641274, EPI_ISL_2641275, EPI_ISL_2641280, EPI_ISL_2641282, EPI_ISL_2641283, EPI_ISL_2641286, EPI_ISL_2641288, EPI_ISL_2641289, EPI_ISL_2641298, EPI_ISL_2641299, EPI_ISL_2641308, EPI_ISL_2641313, EPI_ISL_2641318, EPI_ISL_2641325, EPI_ISL_2641329, EPI_ISL_2641331, EPI_ISL_2641335, EPI_ISL_2641336, EPI_ISL_2641342, EPI_ISL_2652000, EPI_ISL_2663843

| | | | |
|---|---|---|---|
| see above | Lighthouse Lab in Milton Keynes | Wellcome Sanger Institute for the COVID-19 Genomics UK (COG-UK) Consortium | Cordelia Langford; David K. Jackson; Dominic Kwiatkowski; Ewan Harrison; Ian Johnston; Jeffrey Barrett; John Sillitoe on behalf of the Wellcome Sanger Institute COVID-19 Surveillance Team; Roberto Amato; Sonia Goncalves; The Lighthouse Lab in Milton Keynes and Alex Alderton |

EPI_ISL_2515476, EPI_ISL_2540591, EPI_ISL_2540604, EPI_ISL_2540607, EPI_ISL_2540615, EPI_ISL_2540620, EPI_ISL_2540621, EPI_ISL_2540622, EPI_ISL_2540629, EPI_ISL_2540638, EPI_ISL_2540641, EPI_ISL_2540651, EPI_ISL_2540660, EPI_ISL_2540663, EPI_ISL_2540664, EPI_ISL_2540669, EPI_ISL_2540670, EPI_ISL_2540674, EPI_ISL_2540675, EPI_ISL_2540677, EPI_ISL_2540683, EPI_ISL_2540690, EPI_ISL_2540691, EPI_ISL_2540699, EPI_ISL_2540700, EPI_ISL_2540706, EPI_ISL_2540717, EPI_ISL_2540720, EPI_ISL_2540721, EPI_ISL_2540723, EPI_ISL_2540733, EPI_ISL_2540735, EPI_ISL_2540743, EPI_ISL_2540744, EPI_ISL_2540748, EPI_ISL_2540750, EPI_ISL_2540751, EPI_ISL_2540753, EPI_ISL_2540759, EPI_ISL_2540762, EPI_ISL_2540770, EPI_ISL_2540772, EPI_ISL_2540773, EPI_ISL_2540774, EPI_ISL_2540776, EPI_ISL_2540778, EPI_ISL_2540779, EPI_ISL_2540785, EPI_ISL_2540793, EPI_ISL_2540807, EPI_ISL_2540820, EPI_ISL_2540821, EPI_ISL_2540825, EPI_ISL_2540826, EPI_ISL_2540827, EPI_ISL_2540829, EPI_ISL_2540839, EPI_ISL_2540842, EPI_ISL_2540843, EPI_ISL_2540851, EPI_ISL_2540854, EPI_ISL_2540855, EPI_ISL_2540860, EPI_ISL_2540862, EPI_ISL_2540869, EPI_ISL_2540872, EPI_ISL_2540875, EPI_ISL_2540876, EPI_ISL_2540879, EPI_ISL_2540881, EPI_ISL_2540882, EPI_ISL_2540884, EPI_ISL_2540889, EPI_ISL_2540894, EPI_ISL_2540896, EPI_ISL_2540897, EPI_ISL_2540898, EPI_ISL_2540902, EPI_ISL_2540903, EPI_ISL_2540906, EPI_ISL_2540916, EPI_ISL_2540919, EPI_ISL_2540920, EPI_ISL_2540924, EPI_ISL_2540926, EPI_ISL_2540933, EPI_ISL_2540934, EPI_ISL_2540935, EPI_ISL_2540939, EPI_ISL_2540944, EPI_ISL_2541227, EPI_ISL_2541233, EPI_ISL_2541234, EPI_ISL_2541235, EPI_ISL_2541243, EPI_ISL_2541250, EPI_ISL_2541291, EPI_ISL_2541305, EPI_ISL_2541307, EPI_ISL_2541311, EPI_ISL_2541315, EPI_ISL_2541316, EPI_ISL_2541332, EPI_ISL_2541337, EPI_ISL_2541339, EPI_ISL_2541340, EPI_ISL_2541354, EPI_ISL_2541386, EPI_ISL_2541391, EPI_ISL_2541424, EPI_ISL_2541466, EPI_ISL_2541480, EPI_ISL_2541486, EPI_ISL_2541495, EPI_ISL_2541502, EPI_ISL_2541503, EPI_ISL_2541507, EPI_ISL_2541512, EPI_ISL_2541514, EPI_ISL_2541539, EPI_ISL_2541550, EPI_ISL_2541557, EPI_ISL_2541563, EPI_ISL_2541568, EPI_ISL_2541572, EPI_ISL_2541575, EPI_ISL_2541586, EPI_ISL_2541590, EPI_ISL_2541592, EPI_ISL_2541604, EPI_ISL_2541605, EPI_ISL_2541610, EPI_ISL_2541614, EPI_ISL_2541615, EPI_ISL_2541620, EPI_ISL_2541632, EPI_ISL_2541633, EPI_ISL_2568716, EPI_ISL_2568869, EPI_ISL_2569095, EPI_ISL_2569149, EPI_ISL_2569163, EPI_ISL_2569194, EPI_ISL_2570275, EPI_ISL_2570279, EPI_ISL_2570284, EPI_ISL_2570288, EPI_ISL_2570295, EPI_ISL_2570296, EPI_ISL_2570299, EPI_ISL_2570302, EPI_ISL_2570304, EPI_ISL_2570306, EPI_ISL_2570307, EPI_ISL_2570311, EPI_ISL_2570315, EPI_ISL_2570318, EPI_ISL_2570319, EPI_ISL_2570320, EPI_ISL_2570321, EPI_ISL_2570322, EPI_ISL_2570328, EPI_ISL_2570334, EPI_ISL_2570335, EPI_ISL_2570344, EPI_ISL_2570354, EPI_ISL_2570355, EPI_ISL_2570365, EPI_ISL_2570366, EPI_ISL_2570371, EPI_ISL_2570374, EPI_ISL_2570377, EPI_ISL_2570382, EPI_ISL_2570383, EPI_ISL_2570398, EPI_ISL_2570399, EPI_ISL_2570404, EPI_ISL_2570409, EPI_ISL_2570411, EPI_ISL_2570414, EPI_ISL_2570419, EPI_ISL_2570420, EPI_ISL_2570421, EPI_ISL_2570422, EPI_ISL_2570425, EPI_ISL_2570426, EPI_ISL_2570429, EPI_ISL_2570432, EPI_ISL_2570435, EPI_ISL_2570438, EPI_ISL_2570444, EPI_ISL_2570446, EPI_ISL_2570447, EPI_ISL_2570448, EPI_ISL_2570453, EPI_ISL_2570454, EPI_ISL_2570456, EPI_ISL_2570461, EPI_ISL_2570466, EPI_ISL_2570469, EPI_ISL_2570470, EPI_ISL_2570471, EPI_ISL_2570476, EPI_ISL_2570479, EPI_ISL_2570482, EPI_ISL_2570483, EPI_ISL_2570485, EPI_ISL_2570488, EPI_ISL_2570490, EPI_ISL_2570498, EPI_ISL_2570500, EPI_ISL_2570501, EPI_ISL_2570502, EPI_ISL_2570505, EPI_ISL_2570509, EPI_ISL_2570513, EPI_ISL_2570517, EPI_ISL_2570521, EPI_ISL_2570529, EPI_ISL_2570537, EPI_ISL_2570542, EPI_ISL_2570543, EPI_ISL_2570545, EPI_ISL_2570552, EPI_ISL_2570555, EPI_ISL_2570559, EPI_ISL_2570565, EPI_ISL_2570574, EPI_ISL_2570575, EPI_ISL_2570576, EPI_ISL_2570585, EPI_ISL_2570589, EPI_ISL_2570590, EPI_ISL_2570593, EPI_ISL_2570598, EPI_ISL_2570599, EPI_ISL_2570600, EPI_ISL_2570601, EPI_ISL_2570602, EPI_ISL_2570608, EPI_ISL_2570609, EPI_ISL_2570610, EPI_ISL_2570611, EPI_ISL_2570620, EPI_ISL_2570621, EPI_ISL_2570635, EPI_ISL_2570638, EPI_ISL_2570641, EPI_ISL_2570644, EPI_ISL_2570646, EPI_ISL_2570647, EPI_ISL_2570648, EPI_ISL_2570650, EPI_ISL_2570657, EPI_ISL_2570659, EPI_ISL_2570663, EPI_ISL_2570667, EPI_ISL_2570673, EPI_ISL_2570675, EPI_ISL_2570683, EPI_ISL_2570684, EPI_ISL_2570691, EPI_ISL_2570692, EPI_ISL_2570696, EPI_ISL_2570704, EPI_ISL_2570709, EPI_ISL_2570713, EPI_ISL_2570717, EPI_ISL_2570718, EPI_ISL_2570720, EPI_ISL_2570725, EPI_ISL_2570729, EPI_ISL_2570738, EPI_ISL_2570739, EPI_ISL_2570748, EPI_ISL_2570751, EPI_ISL_2570754, EPI_ISL_2570755, EPI_ISL_2570757, EPI_ISL_2570761, EPI_ISL_2570764, EPI_ISL_2570767, EPI_ISL_2570770, EPI_ISL_2570771, EPI_ISL_2570776, EPI_ISL_2570778, EPI_ISL_2570789, EPI_ISL_2570791, EPI_ISL_2570800, EPI_ISL_2607910, EPI_ISL_2608109, EPI_ISL_2641712

| | | | |
|---|---|---|---|
| see above | Lighthouse Laboratory Plymouth | Wellcome Sanger Institute for the COVID-19 Genomics UK (COG-UK) Consortium | Cordelia Langford; David K. Jackson; Dominic Kwiatkowski; Ewan Harrison; Ian Johnston; Jeffrey Barrett; John Sillitoe on behalf of the Wellcome Sanger Institute COVID-19 Surveillance Team; Lighthouse Laboratory Plymouth and Alex Alderton; Roberto Amato; Sonia Goncalves |

EPI_ISL_2623269, EPI_ISL_2623272, EPI_ISL_2623273, EPI_ISL_2623277, EPI_ISL_2623278, EPI_ISL_2623280, EPI_ISL_2623281, EPI_ISL_2623282, EPI_ISL_2623283, EPI_ISL_2623284, EPI_ISL_2623285, EPI_ISL_2623286, EPI_ISL_2623288, EPI_ISL_2623289, EPI_ISL_2623290, EPI_ISL_2623292, EPI_ISL_2623293, EPI_ISL_2623294, EPI_ISL_2623295, EPI_ISL_2623296, EPI_ISL_2623297, EPI_ISL_2623298, EPI_ISL_2623299, EPI_ISL_2623301, EPI_ISL_2623302, EPI_ISL_2623304, EPI_ISL_2623306, EPI_ISL_2623307, EPI_ISL_2623309, EPI_ISL_2623310, EPI_ISL_2623311, EPI_ISL_2623313, EPI_ISL_2623314, EPI_ISL_2623315, EPI_ISL_2623316, EPI_ISL_2623318, EPI_ISL_2623320, EPI_ISL_2623321, EPI_ISL_2623326, EPI_ISL_2623327, EPI_ISL_2623328, EPI_ISL_2623329, EPI_ISL_2623330, EPI_ISL_2623331, EPI_ISL_2623332, EPI_ISL_2623333, EPI_ISL_2623335, EPI_ISL_2623336, EPI_ISL_2623337, EPI_ISL_2623338, EPI_ISL_2623339, EPI_ISL_2623341, EPI_ISL_2623342, EPI_ISL_2623343, EPI_ISL_2623345, EPI_ISL_2623346, EPI_ISL_2623347, EPI_ISL_2623350, EPI_ISL_2623351, EPI_ISL_2623352, EPI_ISL_2623353, EPI_ISL_2623355, EPI_ISL_2623357, EPI_ISL_2623359, EPI_ISL_2623360, EPI_ISL_2623362, EPI_ISL_2623364, EPI_ISL_2623365, EPI_ISL_2623366, EPI_ISL_2623369, EPI_ISL_2623370, EPI_ISL_2623371, EPI_ISL_2623372, EPI_ISL_2623373, EPI_ISL_2623374, EPI_ISL_2623375, EPI_ISL_2623376, EPI_ISL_2623377, EPI_ISL_2623378, EPI_ISL_2623379, EPI_ISL_2623380, EPI_ISL_2623381, EPI_ISL_2623384, EPI_ISL_2623385, EPI_ISL_2623386, EPI_ISL_2623387, EPI_ISL_2623388, EPI_ISL_2623389, EPI_ISL_2623390, EPI_ISL_2623391, EPI_ISL_2623392, EPI_ISL_2623393, EPI_ISL_2623394, EPI_ISL_2623395, EPI_ISL_2623396, EPI_ISL_2623397, EPI_ISL_2623398, EPI_ISL_2623399, EPI_ISL_2623400, EPI_ISL_2623401, EPI_ISL_2623402, EPI_ISL_2623403, EPI_ISL_2623404, EPI_ISL_2623405, EPI_ISL_2623406, EPI_ISL_2623407, EPI_ISL_2623408, EPI_ISL_2623409, EPI_ISL_2623410, EPI_ISL_2623412, EPI_ISL_2623413, EPI_ISL_2623414, EPI_ISL_2623415, EPI_ISL_2623416, EPI_ISL_2623417, EPI_ISL_2623418, EPI_ISL_2623419, EPI_ISL_2623420, EPI_ISL_2623421, EPI_ISL_2623422, EPI_ISL_2623423, EPI_ISL_2623424, EPI_ISL_2623425, EPI_ISL_2623426, EPI_ISL_2623427, EPI_ISL_2623428, EPI_ISL_2623429, EPI_ISL_2623430, EPI_ISL_2623431, EPI_ISL_2623432, EPI_ISL_2623433, EPI_ISL_2623434, EPI_ISL_2623435, EPI_ISL_2623436, EPI_ISL_2623437, EPI_ISL_2623438, EPI_ISL_2623439, EPI_ISL_2623440, EPI_ISL_2623441, EPI_ISL_2623442, EPI_ISL_2623443, EPI_ISL_2623444, EPI_ISL_2623445, EPI_ISL_2623446, EPI_ISL_2623447, EPI_ISL_2623448, EPI_ISL_2623449, EPI_ISL_2623450, EPI_ISL_2623451, EPI_ISL_2623452, EPI_ISL_2623453, EPI_ISL_2623454, EPI_ISL_2623455, EPI_ISL_2623456, EPI_ISL_2623457, EPI_ISL_2623458, EPI_ISL_2623459, EPI_ISL_2623461, EPI_ISL_2623462, EPI_ISL_2623463, EPI_ISL_2623464, EPI_ISL_2623465, EPI_ISL_2623466, EPI_ISL_2623467, EPI_ISL_2623468, EPI_ISL_2623469, EPI_ISL_2623470, EPI_ISL_2623471, EPI_ISL_2623472, EPI_ISL_2623473, EPI_ISL_2623474, EPI_ISL_2623475, EPI_ISL_2623476, EPI_ISL_2623477, EPI_ISL_2623478, EPI_ISL_2623479, EPI_ISL_2623480, EPI_ISL_2623481, EPI_ISL_2623482, EPI_ISL_2623484, EPI_ISL_2623485, EPI_ISL_2623486, EPI_ISL_2623487, EPI_ISL_2623488, EPI_ISL_2623489, EPI_ISL_2623490, EPI_ISL_2623491, EPI_ISL_2623492, EPI_ISL_2623493, EPI_ISL_2623494, EPI_ISL_2623495, EPI_ISL_2623496, EPI_ISL_2623497, EPI_ISL_2623498, EPI_ISL_2623499, EPI_ISL_2623501, EPI_ISL_2623502, EPI_ISL_2623503, EPI_ISL_2623504, EPI_ISL_2623505, EPI_ISL_2623506, EPI_ISL_2623507, EPI_ISL_2623508, EPI_ISL_2623509, EPI_ISL_2623510, EPI_ISL_2623511, EPI_ISL_2623512, EPI_ISL_2623513, EPI_ISL_2623514, EPI_ISL_2623515, EPI_ISL_2623516, EPI_ISL_2623517, EPI_ISL_2623518, EPI_ISL_2623519, EPI_ISL_2623520, EPI_ISL_2623521, EPI_ISL_2623522, EPI_ISL_2623523, EPI_ISL_2623524, EPI_ISL_2623525, EPI_ISL_2623526, EPI_ISL_2623527, EPI_ISL_2623528, EPI_ISL_2623529, EPI_ISL_2623530, EPI_ISL_2623531, EPI_ISL_2623532, EPI_ISL_2623533, EPI_ISL_2623534, EPI_ISL_2623535, EPI_ISL_2623536, EPI_ISL_2623537, EPI_ISL_2623538, EPI_ISL_2623539, EPI_ISL_2623540, EPI_ISL_2623541, EPI_ISL_2623542, EPI_ISL_2623543, EPI_ISL_2623544, EPI_ISL_2623545, EPI_ISL_2623546, EPI_ISL_2623547, EPI_ISL_2623548, EPI_ISL_2623549, EPI_ISL_2623550, EPI_ISL_2623551, EPI_ISL_2623552, EPI_ISL_2623553, EPI_ISL_2623554

| | | | |
|---|---|---|---|
| see above | Northumbria University / South Tees Hospitals NHS Foundation Trust / North Cumbria Integrated Care NHS Foundation Trust / North Tees and Hartlepool NHS Foundation Trust / Newcastle Hospitals NHS Foundation Trust | COVID-19 Genomics UK (COG-UK) Consortium | Andrew Nelson; Brendan Payne; Clive Graham; Darren L Smith; Debra Padgett; Edward Barton; Emma Swindells; Garren Scott; Gary Black; Gary Eltringham; Giles S Holt; Greg R Young; Jane Greenaway; Jennifer Collins; John Allan; Joshua Loh; Lynn Dover; Matthew Bashton; Mohammad A Tariq; Paul Baker; Sarah Essex; Steve Liggett; Wen C Yew; Yusri Taha |

EPI_ISL_2624588, EPI_ISL_2624589, EPI_ISL_2624590, EPI_ISL_2624591, EPI_ISL_2624592, EPI_ISL_2624593, EPI_ISL_2624596, EPI_ISL_2624598, EPI_ISL_2624599, EPI_ISL_2624600, EPI_ISL_2624604, EPI_ISL_2624606, EPI_ISL_2624608, EPI_ISL_2624611, EPI_ISL_2624612, EPI_ISL_2624613, EPI_ISL_2624614, EPI_ISL_2624615, EPI_ISL_2624616, EPI_ISL_2624617, EPI_ISL_2624618, EPI_ISL_2624619, EPI_ISL_2624620, EPI_ISL_2624621, EPI_ISL_2624622, EPI_ISL_2624623, EPI_ISL_2624624, EPI_ISL_2624626, EPI_ISL_2624627, EPI_ISL_2624628, EPI_ISL_2624629, EPI_ISL_2624630, EPI_ISL_2624631, EPI_ISL_2624632, EPI_ISL_2624633, EPI_ISL_2624634, EPI_ISL_2624635, EPI_ISL_2624636, EPI_ISL_2624638, EPI_ISL_2624639, EPI_ISL_2624640, EPI_ISL_2624641, EPI_ISL_2624642, EPI_ISL_2624644, EPI_ISL_2624645, EPI_ISL_2624646, EPI_ISL_2624647, EPI_ISL_2624649, EPI_ISL_2624650, EPI_ISL_2624655, EPI_ISL_2624656, EPI_ISL_2624661, EPI_ISL_2624663, EPI_ISL_2624665, EPI_ISL_2624666, EPI_ISL_2624667, EPI_ISL_2624668, EPI_ISL_2624669, EPI_ISL_2624670, EPI_ISL_2624697, EPI_ISL_2624698, EPI_ISL_2624699, EPI_ISL_2624700, EPI_ISL_2624701, EPI_ISL_2624702, EPI_ISL_2624703, EPI_ISL_2624704, EPI_ISL_2624705, EPI_ISL_2624706, EPI_ISL_2624707, EPI_ISL_2624708, EPI_ISL_2624709, EPI_ISL_2624710, EPI_ISL_2624711, EPI_ISL_2624712, EPI_ISL_2624713, EPI_ISL_2624714, EPI_ISL_2624716, EPI_ISL_2624719, EPI_ISL_2624721, EPI_ISL_2624722, EPI_ISL_2624723, EPI_ISL_2624725, EPI_ISL_2624729, EPI_ISL_2624731, EPI_ISL_2624733, EPI_ISL_2624734, EPI_ISL_2624735, EPI_ISL_2624736, EPI_ISL_2624737, EPI_ISL_2624738, EPI_ISL_2624743, EPI_ISL_2624744, EPI_ISL_2624745, EPI_ISL_2624747, EPI_ISL_2624748, EPI_ISL_2624749, EPI_ISL_2624750, EPI_ISL_2624779, EPI_ISL_2624780, EPI_ISL_2624781, EPI_ISL_2624782, EPI_ISL_2624783, EPI_ISL_2624784, EPI_ISL_2624786, EPI_ISL_2624787, EPI_ISL_2624790, EPI_ISL_2624791, EPI_ISL_2624792, EPI_ISL_2624793, EPI_ISL_2624794, EPI_ISL_2624795, EPI_ISL_2624798, EPI_ISL_2624799, EPI_ISL_2624801, EPI_ISL_2624802, EPI_ISL_2624804, EPI_ISL_2624812, EPI_ISL_2624820, EPI_ISL_2624822, EPI_ISL_2624852, EPI_ISL_2624853, EPI_ISL_2624855, EPI_ISL_2624856, EPI_ISL_2624857, EPI_ISL_2624859, EPI_ISL_2624860, EPI_ISL_2624862, EPI_ISL_2624864, EPI_ISL_2624867, EPI_ISL_2624869, EPI_ISL_2624870, EPI_ISL_2624871, EPI_ISL_2624872, EPI_ISL_2624873, EPI_ISL_2624875, EPI_ISL_2624876, EPI_ISL_2624877, EPI_ISL_2624881, EPI_ISL_2624883, EPI_ISL_2624884, EPI_ISL_2624885, EPI_ISL_2624887, EPI_ISL_2624889, EPI_ISL_2624890, EPI_ISL_2624891, EPI_ISL_2624893, EPI_ISL_2624894, EPI_ISL_2624895, EPI_ISL_2624896, EPI_ISL_2624898, EPI_ISL_2624899, EPI_ISL_2624900, EPI_ISL_2624902, EPI_ISL_2624903, EPI_ISL_2624904, EPI_ISL_2624905, EPI_ISL_2624906, EPI_ISL_2624908, EPI_ISL_2624909, EPI_ISL_2624910, EPI_ISL_2624911, EPI_ISL_2624912, EPI_ISL_2624915, EPI_ISL_2624916, EPI_ISL_2624918, EPI_ISL_2624919, EPI_ISL_2624920, EPI_ISL_2624921, EPI_ISL_2624922, EPI_ISL_2624923, EPI_ISL_2624925, EPI_ISL_2624926, EPI_ISL_2624928

| | | | |
|---|---|---|---|
| see above | Originating lab: Wales Specialist Virology Centre Sequencing lab: Pathogen Genomics Unit | Public Health Wales Microbiology Cardiff Wales Specialist Virology Centre | Alec Birchley; Alexander Adams; Amy Gaskin; Angela Marchbank; Bree Gatica-Wilcox; Catherine Moore; Jason Coombes; Joanne Watkins; Joel Southgate; Johnathan Evans; Laura Gifford; Lauren Gilbert; Lee Graham; Malorie Perry; Matthew Bull; Nicole Pacchiarini; Sally Corden; Sara Kumziene-Summerhayes; Sara Rey; Sarah Taylor; Simon Cottrell; Sophie Jones; Tom Connor |

EPI_ISL_2707312, EPI_ISL_2707315, EPI_ISL_2707317, EPI_ISL_2707319, EPI_ISL_2707324, EPI_ISL_2707326, EPI_ISL_2707328, EPI_ISL_2707331, EPI_ISL_2707333, EPI_ISL_2707341, EPI_ISL_2707343, EPI_ISL_2707346, EPI_ISL_2707351, EPI_ISL_2707352, EPI_ISL_2707357

| | | | |
|---|---|---|---|
| see above | Oxford Viromics, NDM, University of Oxford; Oxford University Hospitals; Basingstoke and North Hampshire Hospital | COVID-19 Genomics UK (COG-UK) Consortium | Alex Mobbs; Amy Trebes; Anita Justice; Catrin Moore; Christophe Fraser; David Bonsall; David Buck; Emma Wise; George Macintyre; Jessica Lynch; John Todd; Mariateresa de Cesare; Matilde Mori; Monique Andersson; Nathan Moore; Nick Cortes; Robert Shaw; Stephen Kidd; Tanya Golubchik; Timothy Peto |

EPI_ISL_2624420, EPI_ISL_2624421, EPI_ISL_2624422, EPI_ISL_2624423, EPI_ISL_2624441, EPI_ISL_2624442, EPI_ISL_2624447, EPI_ISL_2624448, EPI_ISL_2624450, EPI_ISL_2624453, EPI_ISL_2624464

| | | | |
|---|---|---|---|
| see above | Queens Medical Centre, Clinical Microbiology Department / DeepSeq Nottingham | COVID-19 Genomics UK (COG-UK) Consortium | Christopher Moore; Emily Park; Fei Sang; Gemma Clark; Hannah Howson-Wells; Jack Hill; Jayasree Dey; Johnny Debebe; Jonathan Ball; Joseph Chappell; Manjinder Khakh; Matthew Carlisle; Matthew Loose; Michelle M Lister; Nadine Holmes; Patrick McClure; Theocharis Tsoleridis; Timothy Byaruhanga; Vicki M Fleming; Victoria Wright; Wendy Smith |

EPI_ISL_2744136, EPI_ISL_2744415, EPI_ISL_2744447, EPI_ISL_2744452, EPI_ISL_2744489, EPI_ISL_2744571, EPI_ISL_2744584, EPI_ISL_2744640, EPI_ISL_2744649, EPI_ISL_2744654, EPI_ISL_2744668, EPI_ISL_2744685, EPI_ISL_2744690, EPI_ISL_2744694, EPI_ISL_2744696, EPI_ISL_2744698, EPI_ISL_2744701, EPI_ISL_2744707, EPI_ISL_2744732, EPI_ISL_2744755, EPI_ISL_2744765, EPI_ISL_2744766, EPI_ISL_2744771, EPI_ISL_2744779, EPI_ISL_2744789, EPI_ISL_2744791, EPI_ISL_2744793, EPI_ISL_2744808, EPI_ISL_2744809, EPI_ISL_2744813, EPI_ISL_2744822, EPI_ISL_2744825, EPI_ISL_2744838, EPI_ISL_2744849, EPI_ISL_2744852, EPI_ISL_2744859, EPI_ISL_2744860, EPI_ISL_2744877, EPI_ISL_2744880, EPI_ISL_2744882, EPI_ISL_2744884, EPI_ISL_2744890, EPI_ISL_2744891, EPI_ISL_2744895, EPI_ISL_2744896, EPI_ISL_2744994, EPI_ISL_2744995, EPI_ISL_2745013, EPI_ISL_2745014, EPI_ISL_2745015, EPI_ISL_2745018, EPI_ISL_2745025, EPI_ISL_2745052, EPI_ISL_2745055, EPI_ISL_2745064, EPI_ISL_2745239, EPI_ISL_2745257, EPI_ISL_2745258, EPI_ISL_2745287, EPI_ISL_2745293, EPI_ISL_2745295, EPI_ISL_2745297, EPI_ISL_2745333, EPI_ISL_2745432, EPI_ISL_2745433, EPI_ISL_2745442, EPI_ISL_2745454, EPI_ISL_2745456, EPI_ISL_2745459, EPI_ISL_2745467, EPI_ISL_2745474, EPI_ISL_2745484, EPI_ISL_2746576, EPI_ISL_2749280, EPI_ISL_2817361, EPI_ISL_2817362, EPI_ISL_2817363, EPI_ISL_2817364, EPI_ISL_2817370

| | | | |
|---|---|---|---|
| see above | Respiratory Virus Unit, Microbiology Services Colindale, Public Health England | COVID-19 Genomics UK (COG-UK) Consortium | PHE Covid Sequencing Team |

EPI_ISL_2517336, EPI_ISL_2517408, EPI_ISL_2517424, EPI_ISL_2517448, EPI_ISL_2517467, EPI_ISL_2517477, EPI_ISL_2517483, EPI_ISL_2517484, EPI_ISL_2517485, EPI_ISL_2517486, EPI_ISL_2517487, EPI_ISL_2517488, EPI_ISL_2517489, EPI_ISL_2517492, EPI_ISL_2517493, EPI_ISL_2517494, EPI_ISL_2517495, EPI_ISL_2517497, EPI_ISL_2517499, EPI_ISL_2517500, EPI_ISL_2517501, EPI_ISL_2517502, EPI_ISL_2517503, EPI_ISL_2517504, EPI_ISL_2517506, EPI_ISL_2517507, EPI_ISL_2517508, EPI_ISL_2517509, EPI_ISL_2517510, EPI_ISL_2517511, EPI_ISL_2517512, EPI_ISL_2517513, EPI_ISL_2517514, EPI_ISL_2517515, EPI_ISL_2517516, EPI_ISL_2517517, EPI_ISL_2517518, EPI_ISL_2517520, EPI_ISL_2517521, EPI_ISL_2517522, EPI_ISL_2517523, EPI_ISL_2517525, EPI_ISL_2517526, EPI_ISL_2517527, EPI_ISL_2517528, EPI_ISL_2517530, EPI_ISL_2517531, EPI_ISL_2517532, EPI_ISL_2517533, EPI_ISL_2517534, EPI_ISL_2517536, EPI_ISL_2517537, EPI_ISL_2517538, EPI_ISL_2517539, EPI_ISL_2517540, EPI_ISL_2517541, EPI_ISL_2517542, EPI_ISL_2517543, EPI_ISL_2517545, EPI_ISL_2517546, EPI_ISL_2517547, EPI_ISL_2517548, EPI_ISL_2517549, EPI_ISL_2517550, EPI_ISL_2517551, EPI_ISL_2517552, EPI_ISL_2517553, EPI_ISL_2517554, EPI_ISL_2517555, EPI_ISL_2517556, EPI_ISL_2517557, EPI_ISL_2517558, EPI_ISL_2517559, EPI_ISL_2517561, EPI_ISL_2517562, EPI_ISL_2517563, EPI_ISL_2517564, EPI_ISL_2517565, EPI_ISL_2517566, EPI_ISL_2517567, EPI_ISL_2517568, EPI_ISL_2517569, EPI_ISL_2517570, EPI_ISL_2517571, EPI_ISL_2517572, EPI_ISL_2517573, EPI_ISL_2517574, EPI_ISL_2517575, EPI_ISL_2517576, EPI_ISL_2517577, EPI_ISL_2517578, EPI_ISL_2517579, EPI_ISL_2517580, EPI_ISL_2517581, EPI_ISL_2517582, EPI_ISL_2517583, EPI_ISL_2517584, EPI_ISL_2517585, EPI_ISL_2517586, EPI_ISL_2517587, EPI_ISL_2517588, EPI_ISL_2517589, EPI_ISL_2517590, EPI_ISL_2517591, EPI_ISL_2517592, EPI_ISL_2517593, EPI_ISL_2517594, EPI_ISL_2517595, EPI_ISL_2517596, EPI_ISL_2517597, EPI_ISL_2517598, EPI_ISL_2517599, EPI_ISL_2517600, EPI_ISL_2517601, EPI_ISL_2517602, EPI_ISL_2517603, EPI_ISL_2517604, EPI_ISL_2517605, EPI_ISL_2517606, EPI_ISL_2517607, EPI_ISL_2517608, EPI_ISL_2517609, EPI_ISL_2517610, EPI_ISL_2517611, EPI_ISL_2517612, EPI_ISL_2517613, EPI_ISL_2517614, EPI_ISL_2517615, EPI_ISL_2517616, EPI_ISL_2517617, EPI_ISL_2517618, EPI_ISL_2517619, EPI_ISL_2517620, EPI_ISL_2517621, EPI_ISL_2517622, EPI_ISL_2517623, EPI_ISL_2517624, EPI_ISL_2517625, EPI_ISL_2517626, EPI_ISL_2517627, EPI_ISL_2517628, EPI_ISL_2517629, EPI_ISL_2517630, EPI_ISL_2517631, EPI_ISL_2517632, EPI_ISL_2517633, EPI_ISL_2517634, EPI_ISL_2517635, EPI_ISL_2517636, EPI_ISL_2517637, EPI_ISL_2517638, EPI_ISL_2517639, EPI_ISL_2517640, EPI_ISL_2517641, EPI_ISL_2517642, EPI_ISL_2517643, EPI_ISL_2517644, EPI_ISL_2517645, EPI_ISL_2517646, EPI_ISL_2517647, EPI_ISL_2517648, EPI_ISL_2517649, EPI_ISL_2517650, EPI_ISL_2517651, EPI_ISL_2517652, EPI_ISL_2517653, EPI_ISL_2517654, EPI_ISL_2517655, EPI_ISL_2517656, EPI_ISL_2517657, EPI_ISL_2517658, EPI_ISL_2517659, EPI_ISL_2517660, EPI_ISL_2517661, EPI_ISL_2517662, EPI_ISL_2517663, EPI_ISL_2517664, EPI_ISL_2517665, EPI_ISL_2517666, EPI_ISL_2517667, EPI_ISL_2517669, EPI_ISL_2517670, EPI_ISL_2517671, EPI_ISL_2517672, EPI_ISL_2517673, EPI_ISL_2517674, EPI_ISL_2517675, EPI_ISL_2517676, EPI_ISL_2517677, EPI_ISL_2517678, EPI_ISL_2517679, EPI_ISL_2517680, EPI_ISL_2517682, EPI_ISL_2517683, EPI_ISL_2517684, EPI_ISL_2517685, EPI_ISL_2517686, EPI_ISL_2517687, EPI_ISL_2517688, EPI_ISL_2517689, EPI_ISL_2517690, EPI_ISL_2517691, EPI_ISL_2517692, EPI_ISL_2517693, EPI_ISL_2517694, EPI_ISL_2517695, EPI_ISL_2517696, EPI_ISL_2517697, EPI_ISL_2517698, EPI_ISL_2517699, EPI_ISL_2517700, EPI_ISL_2517701, EPI_ISL_2517702, EPI_ISL_2517703, EPI_ISL_2517704, EPI_ISL_2517705, EPI_ISL_2517706, EPI_ISL_2517707, EPI_ISL_2517708, EPI_ISL_2517709, EPI_ISL_2517710, EPI_ISL_2517711, EPI_ISL_2517712, EPI_ISL_2517713, EPI_ISL_2517714, EPI_ISL_2517715, EPI_ISL_2517716, EPI_ISL_2517717, EPI_ISL_2517718, EPI_ISL_2517719, EPI_ISL_2517720, EPI_ISL_2517721, EPI_ISL_2517722, EPI_ISL_2517723, EPI_ISL_2517724, EPI_ISL_2517725, EPI_ISL_2517726, EPI_ISL_2517727, EPI_ISL_2517728, EPI_ISL_2517729, EPI_ISL_2517732, EPI_ISL_2517734, EPI_ISL_2517735, EPI_ISL_2517736, EPI_ISL_2517737, EPI_ISL_2517738, EPI_ISL_2517739, EPI_ISL_2517740, EPI_ISL_2517741, EPI_ISL_2517743, EPI_ISL_2517744, EPI_ISL_2517745, EPI_ISL_2517746, EPI_ISL_2517747, EPI_ISL_2517748, EPI_ISL_2517749, EPI_ISL_2517750, EPI_ISL_2517751, EPI_ISL_2517753, EPI_ISL_2517754, EPI_ISL_2517755, EPI_ISL_2517756, EPI_ISL_2517757, EPI_ISL_2517758, EPI_ISL_2517759, EPI_ISL_2517760, EPI_ISL_2517761, EPI_ISL_2517762, EPI_ISL_2517764, EPI_ISL_2517765, EPI_ISL_2517766, EPI_ISL_2517767, EPI_ISL_2517768, EPI_ISL_2517769, EPI_ISL_2517770, EPI_ISL_2517772, EPI_ISL_2517773, EPI_ISL_2517774, EPI_ISL_2517775, EPI_ISL_2517778, EPI_ISL_2517779, EPI_ISL_2517782, EPI_ISL_2517783, EPI_ISL_2517784, EPI_ISL_2517786, EPI_ISL_2517788, EPI_ISL_2517789, EPI_ISL_2517790, EPI_ISL_2517792, EPI_ISL_2517793, EPI_ISL_2517794, EPI_ISL_2517795, EPI_ISL_2517796, EPI_ISL_2517797, EPI_ISL_2517798, EPI_ISL_2517799, EPI_ISL_2517800, EPI_ISL_2517801, EPI_ISL_2517802, EPI_ISL_2517803, EPI_ISL_2517804, EPI_ISL_2517805, EPI_ISL_2517806, EPI_ISL_2517809, EPI_ISL_2517810, EPI_ISL_2517811, EPI_ISL_2517812, EPI_ISL_2517814, EPI_ISL_2517816, EPI_ISL_2517819, EPI_ISL_2517820, EPI_ISL_2517821, EPI_ISL_2517823, EPI_ISL_2517824, EPI_ISL_2517827, EPI_ISL_2517828, EPI_ISL_2517829, EPI_ISL_2517830, EPI_ISL_2517831, EPI_ISL_2517832, EPI_ISL_2517833, EPI_ISL_2517834, EPI_ISL_2517835, EPI_ISL_2517836, EPI_ISL_2517837, EPI_ISL_2517838, EPI_ISL_2517839, EPI_ISL_2517840, EPI_ISL_2517843, EPI_ISL_2517845, EPI_ISL_2517846, EPI_ISL_2517848, EPI_ISL_2517849, EPI_ISL_2517850, EPI_ISL_2517852, EPI_ISL_2517853, EPI_ISL_2517854, EPI_ISL_2517855, EPI_ISL_2517858, EPI_ISL_2622478, EPI_ISL_2622479, EPI_ISL_2622480, EPI_ISL_2622481, EPI_ISL_2622482, EPI_ISL_2622486, EPI_ISL_2622487, EPI_ISL_2622488, EPI_ISL_2622489, EPI_ISL_2622490, EPI_ISL_2622491, EPI_ISL_2622494, EPI_ISL_2622495, EPI_ISL_2622496, EPI_ISL_2622497, EPI_ISL_2622498, EPI_ISL_2622501, EPI_ISL_2622502, EPI_ISL_2622503, EPI_ISL_2622504, EPI_ISL_2622505, EPI_ISL_2622506, EPI_ISL_2622510, EPI_ISL_2622511, EPI_ISL_2622513, EPI_ISL_2622515, EPI_ISL_2622517, EPI_ISL_2622520, EPI_ISL_2622521, EPI_ISL_2622523, EPI_ISL_2622524, EPI_ISL_2622525, EPI_ISL_2622526, EPI_ISL_2622527, EPI_ISL_2622528, EPI_ISL_2622530, EPI_ISL_2622531, EPI_ISL_2622533, EPI_ISL_2622535, EPI_ISL_2622537, EPI_ISL_2622539, EPI_ISL_2622540, EPI_ISL_2622541, EPI_ISL_2622542, EPI_ISL_2622543, EPI_ISL_2622544, EPI_ISL_2622545, EPI_ISL_2622546, EPI_ISL_2622547, EPI_ISL_2622548, EPI_ISL_2622549, EPI_ISL_2622550, EPI_ISL_2622551, EPI_ISL_2622552, EPI_ISL_2622554, EPI_ISL_2622555, EPI_ISL_2622559, EPI_ISL_2622560, EPI_ISL_2622561, EPI_ISL_2622562, EPI_ISL_2622563, EPI_ISL_2622564, EPI_ISL_2622566, EPI_ISL_2622567, EPI_ISL_2622569, EPI_ISL_2622570, EPI_ISL_2622571, EPI_ISL_2622572, EPI_ISL_2622573, EPI_ISL_2622574, EPI_ISL_2622576, EPI_ISL_2622578, EPI_ISL_2622579, EPI_ISL_2622580, EPI_ISL_2622581, EPI_ISL_2622582, EPI_ISL_2622583, EPI_ISL_2622584, EPI_ISL_2622587, EPI_ISL_2622588, EPI_ISL_2622590, EPI_ISL_2622591, EPI_ISL_2622593, EPI_ISL_2622594, EPI_ISL_2622595, EPI_ISL_2622596, EPI_ISL_2622597, EPI_ISL_2622598, EPI_ISL_2622600, EPI_ISL_2622601, EPI_ISL_2622602, EPI_ISL_2622603, EPI_ISL_2622604, EPI_ISL_2622605, EPI_ISL_2622606, EPI_ISL_2622607, EPI_ISL_2622608, EPI_ISL_2622609, EPI_ISL_2622610, EPI_ISL_2622611, EPI_ISL_2622612, EPI_ISL_2622613, EPI_ISL_2622614, EPI_ISL_2622615, EPI_ISL_2622616, EPI_ISL_2622617, EPI_ISL_2622618, EPI_ISL_2622619, EPI_ISL_2622620, EPI_ISL_2622621, EPI_ISL_2622622, EPI_ISL_2622623, EPI_ISL_2622624, EPI_ISL_2622626, EPI_ISL_2622627, EPI_ISL_2622628, EPI_ISL_2622629, EPI_ISL_2622630, EPI_ISL_2622631, EPI_ISL_2622632, EPI_ISL_2622634, EPI_ISL_2622635, EPI_ISL_2622636, EPI_ISL_2622637, EPI_ISL_2622638, EPI_ISL_2622639, EPI_ISL_2622640, EPI_ISL_2622641, EPI_ISL_2622642, EPI_ISL_2622643, EPI_ISL_2622644, EPI_ISL_2622645, EPI_ISL_2622646, EPI_ISL_2622647, EPI_ISL_2622648, EPI_ISL_2622649, EPI_ISL_2622650, EPI_ISL_2622651, EPI_ISL_2622652, EPI_ISL_2622653, EPI_ISL_2622654, EPI_ISL_2622655, EPI_ISL_2622656, EPI_ISL_2622657, EPI_ISL_2622658, EPI_ISL_2622659, EPI_ISL_2622661, EPI_ISL_2622662, EPI_ISL_2622663, EPI_ISL_2622664, EPI_ISL_2622666, EPI_ISL_2622667, EPI_ISL_2622668, EPI_ISL_2622669, EPI_ISL_2622670, EPI_ISL_2622671, EPI_ISL_2622672, EPI_ISL_2622673, EPI_ISL_2622674, EPI_ISL_2622675, EPI_ISL_2622676, EPI_ISL_2622677, EPI_ISL_2622678, EPI_ISL_2622679, EPI_ISL_2622680, EPI_ISL_2622681, EPI_ISL_2622682,

| | | | |
|---|---|---|---|
| EPI_ISL_2622683, EPI_ISL_2622684, EPI_ISL_2622685, EPI_ISL_2622686, EPI_ISL_2622687, EPI_ISL_2622688, EPI_ISL_2622689, EPI_ISL_2622690, EPI_ISL_2622691, EPI_ISL_2622692, EPI_ISL_2622693, EPI_ISL_2622694, EPI_ISL_2622695, EPI_ISL_2622696, EPI_ISL_2622697, EPI_ISL_2622698, EPI_ISL_2622699, EPI_ISL_2622700, EPI_ISL_2622701, EPI_ISL_2622702, EPI_ISL_2622703, EPI_ISL_2622704, EPI_ISL_2622705, EPI_ISL_2622706, EPI_ISL_2622707, EPI_ISL_2622708, EPI_ISL_2622709, EPI_ISL_2622710, EPI_ISL_2622711, EPI_ISL_2622712, EPI_ISL_2622713, EPI_ISL_2622714, EPI_ISL_2622715, EPI_ISL_2622716, EPI_ISL_2622717, EPI_ISL_2622718, EPI_ISL_2622719, EPI_ISL_2622720, EPI_ISL_2622721, EPI_ISL_2622722, EPI_ISL_2622723, EPI_ISL_2622724, EPI_ISL_2622725, EPI_ISL_2622726, EPI_ISL_2622727, EPI_ISL_2622728, EPI_ISL_2622729, EPI_ISL_2622730, EPI_ISL_2622731, EPI_ISL_2622732, EPI_ISL_2622734, EPI_ISL_2622735, EPI_ISL_2622736, EPI_ISL_2622738, EPI_ISL_2622739, EPI_ISL_2622740, EPI_ISL_2622741, EPI_ISL_2622742, EPI_ISL_2622743, EPI_ISL_2622744, EPI_ISL_2622745, EPI_ISL_2622746, EPI_ISL_2622747, EPI_ISL_2622748, EPI_ISL_2622749, EPI_ISL_2622750, EPI_ISL_2622751, EPI_ISL_2622752, EPI_ISL_2622753, EPI_ISL_2622754, EPI_ISL_2622755, EPI_ISL_2622756, EPI_ISL_2622757, EPI_ISL_2622758, EPI_ISL_2622759, EPI_ISL_2622760, EPI_ISL_2622761 | | | |
| see above | University College London, Great Ormond Street Hospital for Children NHS Foundation Trust, Imperial College Healthcare NHS Trust | COVID-19 Genomics UK (COG-UK) Consortium | Alison Holmes; Charlotte Williams; Helena Tutill; Jacqueline Findlay; James Price; Judith Breuer; Julianne Brown; Kathryn Harris; Leysa Forrest; Mark Kristiansen; Paola Niola; Paola Resende Silva; Patricia Dyal; Paul Randell; Rachel Williams; Samuel Weeks; Sergi Castellano; Sunando Roy; Tony Brooks; Yasmin Panchbhaya |
| EPI_ISL_2721908, EPI_ISL_2721921, EPI_ISL_2721937, EPI_ISL_2721985, EPI_ISL_2721987, EPI_ISL_2721989, EPI_ISL_2721990, EPI_ISL_2721992, EPI_ISL_2721993, EPI_ISL_2721995, EPI_ISL_2721996, EPI_ISL_2721998, EPI_ISL_2721999, EPI_ISL_2722001, EPI_ISL_2722003, EPI_ISL_2722005, EPI_ISL_2722008, EPI_ISL_2722009, EPI_ISL_2722011, EPI_ISL_2722013, EPI_ISL_2722015, EPI_ISL_2722016, EPI_ISL_2722018, EPI_ISL_2722020, EPI_ISL_2722021, EPI_ISL_2722023, EPI_ISL_2722025, EPI_ISL_2722026, EPI_ISL_2722069, EPI_ISL_2722102 | | | |
| see above | University of Birmingham | COVID-19 Genomics UK (COG-UK) Consortium | Alex Richter; Andrew Bosworth; Andrew Bosworth. Queen Elizabeth Hospital: Anna Casey; Andrew D Beggs PHE Heartlands Lab: Husam Osman; Charlotte Poxon; Institute of Microbiology; Joanne Stockton; Josh Quick; Kasun Wanigasooriya; Mike Kidd; Nicholas Loman. University of Birmingham Testing Laboratory: Celina M Whalley; Oliver Pickles; Radoslaw Poplawski; Samuel Nicholls; University of Birmingham: Claire McMurray; Will Rowe |
| EPI_ISL_2622359, EPI_ISL_2622371, EPI_ISL_2622373, EPI_ISL_2622374, EPI_ISL_2622375, EPI_ISL_2622376, EPI_ISL_2704823, EPI_ISL_2704824, EPI_ISL_2704825, EPI_ISL_2704827, EPI_ISL_2704828, EPI_ISL_2704829, EPI_ISL_2704847, EPI_ISL_2704854, EPI_ISL_2704874, EPI_ISL_2704886, EPI_ISL_2704887, EPI_ISL_2704888, EPI_ISL_2704889, EPI_ISL_2704890, EPI_ISL_2704891, EPI_ISL_2704892, EPI_ISL_2704893, EPI_ISL_2704894, EPI_ISL_2704895, EPI_ISL_2704931, EPI_ISL_2734362, EPI_ISL_2734363, EPI_ISL_2734367, EPI_ISL_2734368, EPI_ISL_2734369 | | | |
| see above | Virology Department, Royal Infirmary of Edinburgh, NHS Lothian / School of Biological Sciences, University of Edinburgh | COVID-19 Genomics UK (COG-UK) Consortium | Colquhoun R; Cotton S; Dewar R; Gallagher A; Hill V; Jackson B; Maloney D; McCrone JT; McHugh M; O'Toole Á; Rambaut A; Rooke S; Scher E; Templeton K; Yu X |
| EPI_ISL_2625293 | Virology Department, Sheffield Teaching Hospitals NHS Foundation Trust/Department of Infection, Immunity and Cardiovascular Disease, The Medical School, University of Sheffield | COVID-19 Genomics UK (COG-UK) Consortium | Adri Angyal; Alex Keeley; Benjamin Lindsey; Cariad Evans; Danielle Groves; Dave Partridge; Katie Johnson; Laura Carrilero; Luke Green; Matthew Parker; Matthew Wyles; Mehmet Yavuz; Mohammad Raza; Nikki Smith; Paul Parsons; Rachel Tucker; Rebecca Brown; Thushan de Silva |
| EPI_ISL_2704790, EPI_ISL_2734269, EPI_ISL_2734272, EPI_ISL_2734273, EPI_ISL_2734274, EPI_ISL_2734278, EPI_ISL_2734279, EPI_ISL_2734308, EPI_ISL_2734311, EPI_ISL_2734312, EPI_ISL_2734314, EPI_ISL_2734315, EPI_ISL_2734317, EPI_ISL_2900691 | | | |
| see above | West of Scotland Specialist Virology Centre, NHSGGC / MRC-University of Glasgow Centre for Virus Research | COVID-19 Genomics UK (COG-UK) Consortium | Alasdair MacLean; Alice Broos; Ana da Silva Filipe; Antonia Ho; Daniel Mair; David L Robertson; Emma Thomson; Guy Mollett; Ioulia Tsatsani; James Shepherd; Jenna Nichols; Joseph Hughes; Kathy Li; Kathy Smollett; Kyriaki Nomikou; Lily Tong; Matthew Holden; Natasha Johnson; Rachel Blacow; Richard Orton; Rory Gunson; Sarah McDonald; Sharif Shaaban; Sreenu Vattipally; Stephen Carmichael |

We gratefully acknowledge the following Authors from the Originating laboratories responsible for obtaining the specimens, as well as the Submitting laboratories where the genome data were generated and shared via GISAID, on which this research is based.

All Submitters of data may be contacted directly via www.gisaid.org

Authors are sorted alphabetically.

| Accession ID | Originating Laboratory | Submitting Laboratory | Authors |
|---|---|---|---|
| EPI_ISL_2686159, EPI_ISL_2686160, EPI_ISL_2686161, EPI_ISL_2686162, EPI_ISL_2686163, EPI_ISL_2686165, EPI_ISL_2686166, EPI_ISL_2686167, EPI_ISL_2686169, EPI_ISL_2686170, EPI_ISL_2686171, EPI_ISL_2686178 | | | |
| see above | Amazon Dx, AUV2 Laboratory | Amazon Dx, AUV2 Laboratory | Luke Meredith |
| EPI_ISL_2622447, EPI_ISL_2622448, EPI_ISL_2622449, EPI_ISL_2622450, EPI_ISL_2622451, EPI_ISL_2622452, EPI_ISL_2622453, EPI_ISL_2622454, EPI_ISL_2622455, EPI_ISL_2622456, EPI_ISL_2622457, EPI_ISL_2622458, EPI_ISL_2622459, EPI_ISL_2622460, EPI_ISL_2622461, EPI_ISL_2622462, EPI_ISL_2622463, EPI_ISL_2622464, EPI_ISL_2622465, EPI_ISL_2622466, EPI_ISL_2622471, EPI_ISL_2622472, EPI_ISL_2622474, EPI_ISL_2622475, EPI_ISL_2705483, EPI_ISL_2705485, EPI_ISL_2705486, EPI_ISL_2705487 | | | |
| see above | Barts Health NHS Trust | COVID-19 Genomics UK (COG-UK) Consortium | BROAD; Beatrix; CUTINO-MOGUEL; Claire; David; Dola; HARRINGTON; KELE; KULASEGARAN-SHYLINI; Maria-Teresa; OWOYEMI; Raghavendran |
| EPI_ISL_2624966, EPI_ISL_2624967, EPI_ISL_2624968, EPI_ISL_2624969, EPI_ISL_2624970, EPI_ISL_2624971, EPI_ISL_2624972, EPI_ISL_2624973, EPI_ISL_2624974, EPI_ISL_2624975, EPI_ISL_2624976, EPI_ISL_2624978, EPI_ISL_2624979, EPI_ISL_2624980, EPI_ISL_2624981, EPI_ISL_2624982, EPI_ISL_2624983, EPI_ISL_2624984, EPI_ISL_2624985, EPI_ISL_2624986, EPI_ISL_2624987, EPI_ISL_2624988, EPI_ISL_2624989, EPI_ISL_2624990, EPI_ISL_2624992, EPI_ISL_2624993, EPI_ISL_2624994, EPI_ISL_2624995, EPI_ISL_2624996, EPI_ISL_2624997, EPI_ISL_2624998, EPI_ISL_2624999, EPI_ISL_2625000, EPI_ISL_2625001, EPI_ISL_2625002, EPI_ISL_2625003, EPI_ISL_2625004, EPI_ISL_2625005, EPI_ISL_2625006, EPI_ISL_2625007, EPI_ISL_2625008, EPI_ISL_2625009, EPI_ISL_2625010, EPI_ISL_2625011, EPI_ISL_2625012, EPI_ISL_2625013, EPI_ISL_2625014, EPI_ISL_2625015, EPI_ISL_2625016, EPI_ISL_2625017, EPI_ISL_2625021, EPI_ISL_2625022, EPI_ISL_2625023, EPI_ISL_2625024, EPI_ISL_2625026, EPI_ISL_2625027, EPI_ISL_2625028, EPI_ISL_2625029, EPI_ISL_2625030, EPI_ISL_2625031, EPI_ISL_2625032, EPI_ISL_2625033, EPI_ISL_2625034, EPI_ISL_2625035, EPI_ISL_2625036, EPI_ISL_2625037, EPI_ISL_2625038, EPI_ISL_2625039, EPI_ISL_2625040, EPI_ISL_2625041, EPI_ISL_2625042, EPI_ISL_2625043, EPI_ISL_2625044, EPI_ISL_2625045, EPI_ISL_2625046, EPI_ISL_2625047, EPI_ISL_2625048, EPI_ISL_2625049, EPI_ISL_2625050, EPI_ISL_2625051, EPI_ISL_2625052, EPI_ISL_2625053, EPI_ISL_2625054, EPI_ISL_2625056, EPI_ISL_2625057, EPI_ISL_2625058, EPI_ISL_2625059, EPI_ISL_2625060, EPI_ISL_2625061, EPI_ISL_2625062, EPI_ISL_2625063, EPI_ISL_2625064, EPI_ISL_2625065, EPI_ISL_2625066, EPI_ISL_2625067, EPI_ISL_2625068, EPI_ISL_2625069, EPI_ISL_2625070, EPI_ISL_2625071, EPI_ISL_2625072, EPI_ISL_2625074, EPI_ISL_2625075, EPI_ISL_2625086, EPI_ISL_2625087, EPI_ISL_2625088, EPI_ISL_2625089, EPI_ISL_2625090, EPI_ISL_2625091, EPI_ISL_2625092, EPI_ISL_2625093, EPI_ISL_2625094, EPI_ISL_2625095, EPI_ISL_2625097, EPI_ISL_2625098, EPI_ISL_2625099, EPI_ISL_2625100, EPI_ISL_2625101, EPI_ISL_2625102, EPI_ISL_2625103, EPI_ISL_2625104, EPI_ISL_2625105, EPI_ISL_2625106, EPI_ISL_2625107, EPI_ISL_2625108, EPI_ISL_2625109, EPI_ISL_2625110, EPI_ISL_2625112, EPI_ISL_2625113, EPI_ISL_2625114, EPI_ISL_2625115, EPI_ISL_2625116, EPI_ISL_2625117, EPI_ISL_2625118, EPI_ISL_2625119, EPI_ISL_2625120, EPI_ISL_2625121, EPI_ISL_2625122, EPI_ISL_2625123, EPI_ISL_2625124, EPI_ISL_2625125, EPI_ISL_2625126, EPI_ISL_2625127, EPI_ISL_2625128, EPI_ISL_2625129, EPI_ISL_2625130, EPI_ISL_2625131, EPI_ISL_2625132, EPI_ISL_2625133, EPI_ISL_2625134, EPI_ISL_2625135, EPI_ISL_2625136, EPI_ISL_2625137, EPI_ISL_2625139, EPI_ISL_2625140, EPI_ISL_2625141, EPI_ISL_2625142, EPI_ISL_2625143, EPI_ISL_2625144, EPI_ISL_2625145, EPI_ISL_2625146, EPI_ISL_2625147, EPI_ISL_2625148, EPI_ISL_2625149, EPI_ISL_2625150, EPI_ISL_2625151, EPI_ISL_2625152, EPI_ISL_2625153, EPI_ISL_2625154, EPI_ISL_2625155, EPI_ISL_2625156, EPI_ISL_2625157, EPI_ISL_2625158, EPI_ISL_2625159, EPI_ISL_2625160, EPI_ISL_2625161, EPI_ISL_2625162, EPI_ISL_2625163, EPI_ISL_2625164, EPI_ISL_2625165, EPI_ISL_2625166, EPI_ISL_2625167, EPI_ISL_2625168, EPI_ISL_2625169, EPI_ISL_2625170, EPI_ISL_2625171, EPI_ISL_2625172, EPI_ISL_2625173, EPI_ISL_2625174, EPI_ISL_2625175, EPI_ISL_2625176, EPI_ISL_2625177, EPI_ISL_2625178, EPI_ISL_2625180, EPI_ISL_2625181, EPI_ISL_2625182, EPI_ISL_2625183, EPI_ISL_2625184, EPI_ISL_2625185, EPI_ISL_2625186, EPI_ISL_2625187, EPI_ISL_2625188, EPI_ISL_2625189, EPI_ISL_2625190, EPI_ISL_2625191, EPI_ISL_2625192, EPI_ISL_2625193, EPI_ISL_2625194, EPI_ISL_2625195, EPI_ISL_2625196, EPI_ISL_2625197, EPI_ISL_2625198, EPI_ISL_2625199, EPI_ISL_2625200, EPI_ISL_2625201, EPI_ISL_2625202, EPI_ISL_2625203, EPI_ISL_2625204, EPI_ISL_2625205, EPI_ISL_2625206, EPI_ISL_2625207, EPI_ISL_2625208, EPI_ISL_2708145, EPI_ISL_2708146, EPI_ISL_2708148, EPI_ISL_2708149, EPI_ISL_2708150, EPI_ISL_2708151, EPI_ISL_2708152, EPI_ISL_2708153, EPI_ISL_2708154, EPI_ISL_2708155, EPI_ISL_2708156, EPI_ISL_2708157, EPI_ISL_2708158, EPI_ISL_2708159, EPI_ISL_2708160, EPI_ISL_2708161, EPI_ISL_2708162, EPI_ISL_2708163, EPI_ISL_2708164, EPI_ISL_2708165, EPI_ISL_2708166 | | | |
| see above | Centre for Enzyme Innovation, University of Portsmouth / Translational Research Laboratory, Portsmouth Hospitals NHS Trust / Hampshire Hospitals NHS Foundation Trust / Bournemouth University / University Hospitals Dorset NHS Foundation Trust / Brighton and Sussex University Hospitals NHS Trust / Dartford and Gravesham NHS Trust / Isle of Wight NHS Trust / Maidstone and Tunbridge Wells NHS Trust / University Hospital Southampton NHS Foundation Trust | COVID-19 Genomics UK (COG-UK) Consortium | Adhyana Mahanama; Allyson Lloyd; Andrew Butt; Angela Beckett; Anibolina Castigador; Anna Mantzaouratou; Anoop Chauhan; Ben Macklin; Benjamin Cogger; Buddhini Samaraweera; Cassandra Malone; Christopher Fearn; Claire Thomas; Dorian Crudgington; Eleri Wilson-Davies; Emanuela Pelosi; Emily Macnaughton; Helen Umpleby; Helen Wheeler; Jacqui Prieto; Jonnie Seymour; Karen Withell; Kate Cook; Katie Loveson; Kelly Bicknell; Kevin Tucker; Kordo Saeed; Liz Sheridan; Magdalena Barrow; Matthew Harvey; Mohammed Hassan-Ibrahim; Nathan Moore; Nick Cortes; Rachael Jeremiah; Robert Impey; Salman Goudarzi; Samirakhon Raupova; Samuel Robson; Sarah Buchan; Sarah Jeremiah; Sarah Wyllie; Scott Elliott; Sharon Glaysher; Siona Silvieri; Stephen Aplin; Stephen Kidd; Thea Sass |
| EPI_ISL_2622176, EPI_ISL_2622177, EPI_ISL_2622178, EPI_ISL_2622179, EPI_ISL_2622180, EPI_ISL_2622181, EPI_ISL_2622182, EPI_ISL_2622183, EPI_ISL_2622184, EPI_ISL_2622185, EPI_ISL_2622186, EPI_ISL_2622187, EPI_ISL_2622188, EPI_ISL_2622189, EPI_ISL_2622190, EPI_ISL_2622191, EPI_ISL_2622192, EPI_ISL_2622193, EPI_ISL_2622194, EPI_ISL_2622195, EPI_ISL_2622196, EPI_ISL_2622197, EPI_ISL_2622198, EPI_ISL_2622199, EPI_ISL_2622200, EPI_ISL_2622201, EPI_ISL_2622202, EPI_ISL_2622203, EPI_ISL_2622204, EPI_ISL_2622205, EPI_ISL_2622206, EPI_ISL_2622207, EPI_ISL_2622208, EPI_ISL_2622209, EPI_ISL_2622210, EPI_ISL_2622211, EPI_ISL_2622212, EPI_ISL_2622213, EPI_ISL_2622214, EPI_ISL_2622215, EPI_ISL_2622216, EPI_ISL_2622217, EPI_ISL_2622218, EPI_ISL_2622219, EPI_ISL_2622220, EPI_ISL_2622221, EPI_ISL_2622222, EPI_ISL_2622223, EPI_ISL_2622224, EPI_ISL_2622225, EPI_ISL_2622226, EPI_ISL_2622227, EPI_ISL_2622228, EPI_ISL_2622229, EPI_ISL_2622230, EPI_ISL_2622231, EPI_ISL_2622232, EPI_ISL_2622233, EPI_ISL_2622234, EPI_ISL_2622235, EPI_ISL_2622236, EPI_ISL_2622237, EPI_ISL_2622238, EPI_ISL_2622239, EPI_ISL_2622240, EPI_ISL_2622241, EPI_ISL_2622242, EPI_ISL_2622243, EPI_ISL_2622244, EPI_ISL_2622245, EPI_ISL_2622246, EPI_ISL_2622247, EPI_ISL_2622248, EPI_ISL_2622249, EPI_ISL_2622250, EPI_ISL_2622251, EPI_ISL_2622252, EPI_ISL_2622253, EPI_ISL_2622254, EPI_ISL_2622255, EPI_ISL_2622256, EPI_ISL_2622257, EPI_ISL_2622258, EPI_ISL_2622259, EPI_ISL_2622260, EPI_ISL_2622261, EPI_ISL_2622262, EPI_ISL_2622263, EPI_ISL_2622264, EPI_ISL_2622265, EPI_ISL_2622266, EPI_ISL_2622267, EPI_ISL_2622268, EPI_ISL_2622269, EPI_ISL_2704678, EPI_ISL_2704679, EPI_ISL_2704680, EPI_ISL_2704681, EPI_ISL_2704682, EPI_ISL_2704683, EPI_ISL_2704684, EPI_ISL_2704685, EPI_ISL_2704686, EPI_ISL_2704687, EPI_ISL_2704688, EPI_ISL_2704689, EPI_ISL_2704690, EPI_ISL_2704691, EPI_ISL_2704692, EPI_ISL_2704693, EPI_ISL_2704694, EPI_ISL_2704695, EPI_ISL_2704696, EPI_ISL_2704697, EPI_ISL_2704698, EPI_ISL_2704699, EPI_ISL_2704700, EPI_ISL_2704701, EPI_ISL_2704702, EPI_ISL_2704703, EPI_ISL_2704704, EPI_ISL_2704705, EPI_ISL_2704706, EPI_ISL_2704707, EPI_ISL_2704708, EPI_ISL_2704709, EPI_ISL_2704710, EPI_ISL_2704711, EPI_ISL_2704712, EPI_ISL_2704713, EPI_ISL_2704714, EPI_ISL_2704715, EPI_ISL_2704716, EPI_ISL_2704717, EPI_ISL_2704718, EPI_ISL_2704719, EPI_ISL_2704720, EPI_ISL_2704721, EPI_ISL_2704722, EPI_ISL_2704723, EPI_ISL_2704724, EPI_ISL_2704725, EPI_ISL_2704726, EPI_ISL_2704727, EPI_ISL_2704728, EPI_ISL_2704729, EPI_ISL_2704730, EPI_ISL_2704731, EPI_ISL_2704732, EPI_ISL_2704733, EPI_ISL_2704734, EPI_ISL_2704735, EPI_ISL_2704736, EPI_ISL_2704737, EPI_ISL_2704738, EPI_ISL_2704739, EPI_ISL_2704740, EPI_ISL_2704741, EPI_ISL_2704742, EPI_ISL_2704743, EPI_ISL_2722108, EPI_ISL_2722109, EPI_ISL_2722110, EPI_ISL_2722111, EPI_ISL_2722112, EPI_ISL_2722113, EPI_ISL_2722114, EPI_ISL_2722115, EPI_ISL_2722116, EPI_ISL_2722117, EPI_ISL_2722118, EPI_ISL_2722119, EPI_ISL_2722120, EPI_ISL_2722121, EPI_ISL_2722122, EPI_ISL_2722123, EPI_ISL_2722125, EPI_ISL_2722127, EPI_ISL_2722128, EPI_ISL_2722129, EPI_ISL_2722130, EPI_ISL_2722131, EPI_ISL_2722132, EPI_ISL_2722133, EPI_ISL_2722134, EPI_ISL_2722135, EPI_ISL_2722137, EPI_ISL_2722138, EPI_ISL_2722139, EPI_ISL_2722140, EPI_ISL_2722141, EPI_ISL_2722142, EPI_ISL_2722143, EPI_ISL_2722145, EPI_ISL_2722147, EPI_ISL_2722148, EPI_ISL_2722149, EPI_ISL_2722150, EPI_ISL_2722151, EPI_ISL_2722152, EPI_ISL_2722154, EPI_ISL_2722155, EPI_ISL_2722156, EPI_ISL_2722157, EPI_ISL_2722158, EPI_ISL_2722172, EPI_ISL_2722364, EPI_ISL_2722365, EPI_ISL_2722366, EPI_ISL_2722367, EPI_ISL_2722368, EPI_ISL_2722369, EPI_ISL_2722370, EPI_ISL_2722371, EPI_ISL_2722372, EPI_ISL_2722373, EPI_ISL_2722374, EPI_ISL_2722375, EPI_ISL_2722376, EPI_ISL_2722377, EPI_ISL_2722378, EPI_ISL_2722379, EPI_ISL_2722380, EPI_ISL_2722381, EPI_ISL_2722382, EPI_ISL_2722383, EPI_ISL_2722384, EPI_ISL_2722385, EPI_ISL_2722386, EPI_ISL_2722387, EPI_ISL_2722388, EPI_ISL_2722389, EPI_ISL_2722390, EPI_ISL_2722391, EPI_ISL_2722392, EPI_ISL_2722393, EPI_ISL_2722394, EPI_ISL_2722395, EPI_ISL_2722396, EPI_ISL_2722397, EPI_ISL_2722398, EPI_ISL_2722399, EPI_ISL_2733919, EPI_ISL_2733920, EPI_ISL_2900365 | | | |
| see above | Department of Pathology, University of Cambridge | COVID-19 Genomics UK (COG-UK) Consortium | Aminu S. Jahun; Ian Goodfellow; Iliana Georgana; Martin D. Curran; Myra Hosmillo; Rhys Izuagbe; Surendra Parmar; William L. Hamilton; Yasmin Chaudhry |
| EPI_ISL_2541611, EPI_ISL_2553790, EPI_ISL_2553791, EPI_ISL_2553797, EPI_ISL_2553800, EPI_ISL_2553803, EPI_ISL_2553804, EPI_ISL_2553805, EPI_ISL_2553807, EPI_ISL_2553810, EPI_ISL_2553811, EPI_ISL_2553813, EPI_ISL_2553817, EPI_ISL_2553820, EPI_ISL_2553821, EPI_ISL_2553822, EPI_ISL_2553824, EPI_ISL_2553826, EPI_ISL_2553833, EPI_ISL_2553834, EPI_ISL_2553838, EPI_ISL_2553839, EPI_ISL_2553841, EPI_ISL_2553843, EPI_ISL_2553844, EPI_ISL_2553847, EPI_ISL_2553849, EPI_ISL_2553851, EPI_ISL_2553856, EPI_ISL_2553857, EPI_ISL_2553858, EPI_ISL_2553860, EPI_ISL_2553862, EPI_ISL_2553865, EPI_ISL_2553866, EPI_ISL_2553869, EPI_ISL_2553872, EPI_ISL_2553874, EPI_ISL_2553875, EPI_ISL_2553876, EPI_ISL_2553877, EPI_ISL_2553879, EPI_ISL_2553882, EPI_ISL_2553883, EPI_ISL_2553884, EPI_ISL_2553888, EPI_ISL_2553890, EPI_ISL_2553894, EPI_ISL_2553895, EPI_ISL_2553897, EPI_ISL_2553898, EPI_ISL_2553899, EPI_ISL_2553901, EPI_ISL_2553902, EPI_ISL_2553906, EPI_ISL_2553907, EPI_ISL_2553908, EPI_ISL_2553910, EPI_ISL_2553911, EPI_ISL_2553912, EPI_ISL_2553913, EPI_ISL_2553920, EPI_ISL_2553922, EPI_ISL_2553925, EPI_ISL_2553926, EPI_ISL_2553928, EPI_ISL_2553930, EPI_ISL_2553931, EPI_ISL_2553934, EPI_ISL_2553935, EPI_ISL_2553936, EPI_ISL_2553937, EPI_ISL_2553940, EPI_ISL_2553941, EPI_ISL_2553943, EPI_ISL_2553945, EPI_ISL_2553946, EPI_ISL_2553948, EPI_ISL_2553951, EPI_ISL_2553953, EPI_ISL_2553954, EPI_ISL_2553956, EPI_ISL_2553957, EPI_ISL_2553959, EPI_ISL_2553961, EPI_ISL_2553962, EPI_ISL_2553965, EPI_ISL_2553967, EPI_ISL_2553979, EPI_ISL_2553981, EPI_ISL_2553982, EPI_ISL_2553983, EPI_ISL_2553985, EPI_ISL_2553986, EPI_ISL_2553987, EPI_ISL_2553989, EPI_ISL_2553990, EPI_ISL_2553991, EPI_ISL_2553994, EPI_ISL_2553995, EPI_ISL_2553997, EPI_ISL_2554000, EPI_ISL_2554002, EPI_ISL_2554003, EPI_ISL_2554005, EPI_ISL_2554009, EPI_ISL_2554010, EPI_ISL_2554011, EPI_ISL_2554012, EPI_ISL_2554014, EPI_ISL_2554016, EPI_ISL_2554018, EPI_ISL_2554020, EPI_ISL_2554022, EPI_ISL_2554028, EPI_ISL_2554029, EPI_ISL_2554031, EPI_ISL_2554033, EPI_ISL_2554035, EPI_ISL_2554037, EPI_ISL_2554044, EPI_ISL_2554045, EPI_ISL_2554046, EPI_ISL_2554048, EPI_ISL_2554054, EPI_ISL_2554057, EPI_ISL_2554061, EPI_ISL_2554062, EPI_ISL_2554067, EPI_ISL_2554069, EPI_ISL_2554072, EPI_ISL_2554073, EPI_ISL_2554074, EPI_ISL_2554075, EPI_ISL_2554079, EPI_ISL_2554081, EPI_ISL_2554082, EPI_ISL_2554116, EPI_ISL_2554117, EPI_ISL_2554120, EPI_ISL_2554122, EPI_ISL_2554123, EPI_ISL_2554124, EPI_ISL_2554127, EPI_ISL_2554128, EPI_ISL_2554130, EPI_ISL_2554131, EPI_ISL_2554134, EPI_ISL_2554136, EPI_ISL_2554140, EPI_ISL_2570271, EPI_ISL_2608124, EPI_ISL_2638934, EPI_ISL_2638956, EPI_ISL_2638961, EPI_ISL_2638972, EPI_ISL_2639001, EPI_ISL_2639053, EPI_ISL_2639073, EPI_ISL_2639098, EPI_ISL_2639113, EPI_ISL_2639130, EPI_ISL_2639144, EPI_ISL_2639157, EPI_ISL_2639168, EPI_ISL_2639182, EPI_ISL_2639183, EPI_ISL_2639203, EPI_ISL_2639204, EPI_ISL_2639230, EPI_ISL_2639233 | | | |
| see above | Health Services Laboratories | Wellcome Sanger Institute for the COVID-19 Genomics UK (COG-UK) Consortium | Cordelia Langford; David K. Jackson; Dominic Kwiatkowski; Ewan Harrison; Health Services Laboratories and Alex Alderton; Ian Johnston; Jeffrey Barrett; John Sillitoe on behalf of the Wellcome Sanger Institute COVID-19 Surveillance Team; Roberto Amato; Sonia Goncalves |
| EPI_ISL_2552336, EPI_ISL_2552464, EPI_ISL_2552524, EPI_ISL_2552557, EPI_ISL_2552574, EPI_ISL_2568203, EPI_ISL_2568312, EPI_ISL_2569337, EPI_ISL_2569343, EPI_ISL_2569359, EPI_ISL_2569378, EPI_ISL_2569404, EPI_ISL_2569435, EPI_ISL_2569474, EPI_ISL_2569480, EPI_ISL_2569518, EPI_ISL_2569551, EPI_ISL_2569580, EPI_ISL_2569611, EPI_ISL_2569627, EPI_ISL_2605126, EPI_ISL_2605130, EPI_ISL_2605132, EPI_ISL_2605137, EPI_ISL_2605140, EPI_ISL_2605141, EPI_ISL_2605144, EPI_ISL_2605149, EPI_ISL_2605158, EPI_ISL_2605159, EPI_ISL_2605161, EPI_ISL_2605164, EPI_ISL_2605167, EPI_ISL_2605168, EPI_ISL_2605171, EPI_ISL_2605173, EPI_ISL_2605174, EPI_ISL_2605179, EPI_ISL_2605182, EPI_ISL_2605186, EPI_ISL_2605191, EPI_ISL_2605194, EPI_ISL_2605197, EPI_ISL_2605205, EPI_ISL_2605208, EPI_ISL_2605211, EPI_ISL_2605213, EPI_ISL_2605217, EPI_ISL_2605219, EPI_ISL_2605225, EPI_ISL_2605226, EPI_ISL_2605229, EPI_ISL_2605230, EPI_ISL_2605232, EPI_ISL_2605236, EPI_ISL_2605238, EPI_ISL_2605239, EPI_ISL_2605242, EPI_ISL_2605243, EPI_ISL_2605245, EPI_ISL_2605246, EPI_ISL_2605251, EPI_ISL_2605252, EPI_ISL_2605254, EPI_ISL_2605257, EPI_ISL_2605264, EPI_ISL_2605266, EPI_ISL_2605267, EPI_ISL_2605269, EPI_ISL_2605272, EPI_ISL_2605278, EPI_ISL_2605281, EPI_ISL_2605285, EPI_ISL_2605292, EPI_ISL_2605301, EPI_ISL_2605304, EPI_ISL_2605307, EPI_ISL_2605308, EPI_ISL_2605313, EPI_ISL_2605316, EPI_ISL_2605317, EPI_ISL_2605319, EPI_ISL_2605320, EPI_ISL_2605321, EPI_ISL_2605336, EPI_ISL_2605337, EPI_ISL_2605338, EPI_ISL_2605340, EPI_ISL_2605351, EPI_ISL_2605354, EPI_ISL_2605357, EPI_ISL_2605360, EPI_ISL_2605367, EPI_ISL_2605372, EPI_ISL_2605378, EPI_ISL_2605379, EPI_ISL_2605382, EPI_ISL_2605387, EPI_ISL_2605388, EPI_ISL_2605391, EPI_ISL_2605394, EPI_ISL_2605395, EPI_ISL_2605397, EPI_ISL_2605398, EPI_ISL_2605401, EPI_ISL_2605403, EPI_ISL_2605406, EPI_ISL_2605410, EPI_ISL_2605412, EPI_ISL_2605414, EPI_ISL_2605417, EPI_ISL_2605420, EPI_ISL_2605422, EPI_ISL_2605424, EPI_ISL_2605425, EPI_ISL_2605426, EPI_ISL_2605428, EPI_ISL_2605429, EPI_ISL_2605432, EPI_ISL_2605433, EPI_ISL_2605436, EPI_ISL_2605438, EPI_ISL_2605441, EPI_ISL_2605450, EPI_ISL_2605453, EPI_ISL_2605458, EPI_ISL_2605461, EPI_ISL_2605464, EPI_ISL_2605465, EPI_ISL_2605473, EPI_ISL_2605478, EPI_ISL_2605491, EPI_ISL_2605496, EPI_ISL_2605507, EPI_ISL_2605511, EPI_ISL_2605513, EPI_ISL_2605517, EPI_ISL_2605522, EPI_ISL_2605525, EPI_ISL_2605528, EPI_ISL_2605531, EPI_ISL_2605533, EPI_ISL_2605537, EPI_ISL_2605540, EPI_ISL_2605542, EPI_ISL_2605543, EPI_ISL_2605546, EPI_ISL_2605551, EPI_ISL_2605558, EPI_ISL_2605567, EPI_ISL_2605576, EPI_ISL_2605581, EPI_ISL_2605586, EPI_ISL_2605588, EPI_ISL_2605591, EPI_ISL_2605600, EPI_ISL_2605604, EPI_ISL_2605607, EPI_ISL_2605611, EPI_ISL_2605614, EPI_ISL_2605615, EPI_ISL_2605617, EPI_ISL_2605620, EPI_ISL_2605621, EPI_ISL_2605624, EPI_ISL_2605626, EPI_ISL_2605627, EPI_ISL_2605634, EPI_ISL_2605635, EPI_ISL_2605639, EPI_ISL_2605640, EPI_ISL_2605649, EPI_ISL_2605651, EPI_ISL_2605652, EPI_ISL_2605655, EPI_ISL_2605657, EPI_ISL_2605658, EPI_ISL_2605660, EPI_ISL_2605663, EPI_ISL_2605664, EPI_ISL_2605666, EPI_ISL_2605668, EPI_ISL_2605672, EPI_ISL_2605674, EPI_ISL_2605677, EPI_ISL_2605678, EPI_ISL_2605679, EPI_ISL_2605681, EPI_ISL_2605682, EPI_ISL_2605689, EPI_ISL_2605691, EPI_ISL_2605693, EPI_ISL_2605698, EPI_ISL_2605699, EPI_ISL_2605700, EPI_ISL_2605702, EPI_ISL_2605703, EPI_ISL_2605704, EPI_ISL_2605707, EPI_ISL_2605709, EPI_ISL_2605713, EPI_ISL_2605714, EPI_ISL_2605715, EPI_ISL_2605719, EPI_ISL_2605729, EPI_ISL_2605730, EPI_ISL_2605735, EPI_ISL_2605737, EPI_ISL_2605745, EPI_ISL_2605746, EPI_ISL_2605748, EPI_ISL_2605759, EPI_ISL_2605760, EPI_ISL_2605761, EPI_ISL_2605765, EPI_ISL_2605771, EPI_ISL_2605772, EPI_ISL_2605782, EPI_ISL_2605784, EPI_ISL_2605785, EPI_ISL_2605787, EPI_ISL_2605789, EPI_ISL_2605792, EPI_ISL_2605796, EPI_ISL_2605798, EPI_ISL_2605808, EPI_ISL_2605815, EPI_ISL_2605819, EPI_ISL_2605823, EPI_ISL_2605826, EPI_ISL_2605828, EPI_ISL_2605832, EPI_ISL_2605836, EPI_ISL_2605839, EPI_ISL_2605847, EPI_ISL_2605848, EPI_ISL_2605849, EPI_ISL_2605851, EPI_ISL_2605858, EPI_ISL_2605864, EPI_ISL_2605873, EPI_ISL_2605883, EPI_ISL_2605886, EPI_ISL_2605897, EPI_ISL_2605900, EPI_ISL_2605902, EPI_ISL_2605905, EPI_ISL_2605925, EPI_ISL_2605936, EPI_ISL_2605938, EPI_ISL_2605940, EPI_ISL_2605946, EPI_ISL_2605948, EPI_ISL_2605952, EPI_ISL_2605957, EPI_ISL_2605962, EPI_ISL_2605966, EPI_ISL_2605967, EPI_ISL_2605972, EPI_ISL_2605974, EPI_ISL_2605979, EPI_ISL_2605986, EPI_ISL_2605990, EPI_ISL_2605992, EPI_ISL_2605994, EPI_ISL_2605996, EPI_ISL_2606001, EPI_ISL_2606013, EPI_ISL_2606018, EPI_ISL_2606027, EPI_ISL_2606028, EPI_ISL_2606037, EPI_ISL_2606040, EPI_ISL_2606049, EPI_ISL_2606050, EPI_ISL_2606056, EPI_ISL_2606069, EPI_ISL_2606070, EPI_ISL_2606072, EPI_ISL_2606076, EPI_ISL_2606083, EPI_ISL_2606090, EPI_ISL_2606094, EPI_ISL_2606095, EPI_ISL_2606096, EPI_ISL_2606104, EPI_ISL_2606106, EPI_ISL_2606111, EPI_ISL_2606113, EPI_ISL_2606118, EPI_ISL_2606119, EPI_ISL_2606125, EPI_ISL_2606130, EPI_ISL_2606133, EPI_ISL_2606136, EPI_ISL_2606138, EPI_ISL_2606140, EPI_ISL_2606141, EPI_ISL_2606142, EPI_ISL_2606143, EPI_ISL_2606145, EPI_ISL_2606149, EPI_ISL_2606152, EPI_ISL_2606153, EPI_ISL_2606168, EPI_ISL_2606169, EPI_ISL_2606177, EPI_ISL_2606184, EPI_ISL_2606192, EPI_ISL_2606193, EPI_ISL_2606195, EPI_ISL_2606203, EPI_ISL_2606216, EPI_ISL_2606217, EPI_ISL_2606224, EPI_ISL_2606225, EPI_ISL_2606231, EPI_ISL_2606234, EPI_ISL_2606246, EPI_ISL_2606248, EPI_ISL_2606252, EPI_ISL_2606253, EPI_ISL_2606254, EPI_ISL_2606261, EPI_ISL_2606281, EPI_ISL_2606283, EPI_ISL_2606289, EPI_ISL_2606292, EPI_ISL_2606309, EPI_ISL_2606320, EPI_ISL_2606321, EPI_ISL_2606323, EPI_ISL_2606325, EPI_ISL_2606329, EPI_ISL_2606334, EPI_ISL_2606337, EPI_ISL_2606339, EPI_ISL_2606356, EPI_ISL_2606358, EPI_ISL_2606363, EPI_ISL_2606364, EPI_ISL_2606378, EPI_ISL_2606385, EPI_ISL_2606392, EPI_ISL_2606396, EPI_ISL_2606406, EPI_ISL_2606410, EPI_ISL_2606425, EPI_ISL_2606426, EPI_ISL_2606437, EPI_ISL_2606450, EPI_ISL_2606727, EPI_ISL_2606739, EPI_ISL_2606784, EPI_ISL_2606814, EPI_ISL_2606841, EPI_ISL_2606843, EPI_ISL_2606853, EPI_ISL_2606854, EPI_ISL_2606873, EPI_ISL_2606877, EPI_ISL_2606918, EPI_ISL_2606921, EPI_ISL_2606922, EPI_ISL_2606978, EPI_ISL_2606984, EPI_ISL_2606986, EPI_ISL_2606993, EPI_ISL_2607001, EPI_ISL_2607008, EPI_ISL_2607029, EPI_ISL_2607034, EPI_ISL_2638163, EPI_ISL_2638166, EPI_ISL_2638169, EPI_ISL_2638171, EPI_ISL_2638173, EPI_ISL_2638178, EPI_ISL_2638182, EPI_ISL_2638184, EPI_ISL_2638187, EPI_ISL_2638192, EPI_ISL_2638203, EPI_ISL_2638205, EPI_ISL_2638210, EPI_ISL_2638216, EPI_ISL_2638217, EPI_ISL_2638220, EPI_ISL_2638222, EPI_ISL_2638224, EPI_ISL_2638228, EPI_ISL_2638231, EPI_ISL_2638234, EPI_ISL_2638235, EPI_ISL_2638238, EPI_ISL_2638246, EPI_ISL_2638247, EPI_ISL_2638248, EPI_ISL_2638259, EPI_ISL_2638269, EPI_ISL_2638272, EPI_ISL_2638275, EPI_ISL_2638278, EPI_ISL_2638279, EPI_ISL_2638283, EPI_ISL_2638290, EPI_ISL_2638293, EPI_ISL_2638300, EPI_ISL_2638321, EPI_ISL_2638325, EPI_ISL_2638332, EPI_ISL_2638353, EPI_ISL_2638361, EPI_ISL_2638370, EPI_ISL_2638375, EPI_ISL_2638381, EPI_ISL_2638384, EPI_ISL_2638388, EPI_ISL_2638615, EPI_ISL_2638616, EPI_ISL_2638617, EPI_ISL_2638618, EPI_ISL_2638619, EPI_ISL_2638622, EPI_ISL_2638623, EPI_ISL_2638626, EPI_ISL_2638627, EPI_ISL_2638628, EPI_ISL_2638629, EPI_ISL_2638632, EPI_ISL_2638633, EPI_ISL_2638634, EPI_ISL_2638635, EPI_ISL_2638636, EPI_ISL_2638637, EPI_ISL_2638638, EPI_ISL_2638641, EPI_ISL_2638642, EPI_ISL_2638643, EPI_ISL_2638644, EPI_ISL_2638646, EPI_ISL_2638647, EPI_ISL_2638648, EPI_ISL_2638649, EPI_ISL_2638650, EPI_ISL_2638652, EPI_ISL_2638653, EPI_ISL_2638654, EPI_ISL_2638655, EPI_ISL_2638656, EPI_ISL_2638658, EPI_ISL_2638659, EPI_ISL_2638661, EPI_ISL_2638663, EPI_ISL_2638664, EPI_ISL_2638668, EPI_ISL_2638669, EPI_ISL_2638671, EPI_ISL_2638672, EPI_ISL_2638673, EPI_ISL_2638677, EPI_ISL_2638679, EPI_ISL_2638680, EPI_ISL_2638684, EPI_ISL_2638685, EPI_ISL_2638686, EPI_ISL_2638688, EPI_ISL_2638689, EPI_ISL_2638690, EPI_ISL_2638693, EPI_ISL_2638694, EPI_ISL_2638697, EPI_ISL_2638698, EPI_ISL_2638699, EPI_ISL_2638700, EPI_ISL_2638701, EPI_ISL_2638702, EPI_ISL_2638703, EPI_ISL_2638704, EPI_ISL_2638707, EPI_ISL_2638708, EPI_ISL_2638709, EPI_ISL_2638710, EPI_ISL_2638711, EPI_ISL_2638715, EPI_ISL_2638716, EPI_ISL_2638717, EPI_ISL_2638718, EPI_ISL_2638719, EPI_ISL_2638720, EPI_ISL_2638721, EPI_ISL_2638724, EPI_ISL_2638725, EPI_ISL_2638726, EPI_ISL_2638727, EPI_ISL_2638733, EPI_ISL_2638735, EPI_ISL_2638736, EPI_ISL_2638738, EPI_ISL_2638739, EPI_ISL_2638740, EPI_ISL_2638742, EPI_ISL_2638746, EPI_ISL_2638747, EPI_ISL_2638748, EPI_ISL_2638749, EPI_ISL_2638750, EPI_ISL_2638752, EPI_ISL_2638753, EPI_ISL_2638755, EPI_ISL_2638756, EPI_ISL_2638757, EPI_ISL_2638758, EPI_ISL_2638761, EPI_ISL_2638762, EPI_ISL_2638763, EPI_ISL_2638764, EPI_ISL_2638765, EPI_ISL_2638766, EPI_ISL_2638768, EPI_ISL_2638770, EPI_ISL_2638772, EPI_ISL_2638773, EPI_ISL_2638774, EPI_ISL_2638776, EPI_ISL_2638777, EPI_ISL_2638778, EPI_ISL_2638779, EPI_ISL_2638780, EPI_ISL_2638781, EPI_ISL_2638782, EPI_ISL_2638783, EPI_ISL_2638785, EPI_ISL_2638786, EPI_ISL_2638787, EPI_ISL_2638788, EPI_ISL_2638790, EPI_ISL_2638791, EPI_ISL_2638792, EPI_ISL_2638793, EPI_ISL_2638796, EPI_ISL_2638797, EPI_ISL_2638799, EPI_ISL_2638800, EPI_ISL_2638801, EPI_ISL_2638803, EPI_ISL_2638808, EPI_ISL_2638811, EPI_ISL_2638812, EPI_ISL_2638814, EPI_ISL_2638815, EPI_ISL_2638816, EPI_ISL_2638817, EPI_ISL_2638820, EPI_ISL_2638821, EPI_ISL_2638827, EPI_ISL_2638828, EPI_ISL_2638831, EPI_ISL_2638832, EPI_ISL_2638838, EPI_ISL_2638839, EPI_ISL_2638840, EPI_ISL_2638841, EPI_ISL_2638845, EPI_ISL_2638846, EPI_ISL_2638847, EPI_ISL_2638849, EPI_ISL_2638850, EPI_ISL_2638855, EPI_ISL_2638856, EPI_ISL_2638857, EPI_ISL_2638863, EPI_ISL_2638864, EPI_ISL_2638866, EPI_ISL_2638867, EPI_ISL_2638868, EPI_ISL_2638872, EPI_ISL_2638874, EPI_ISL_2638875, EPI_ISL_2638877, EPI_ISL_2638879, EPI_ISL_2638881, EPI_ISL_2638884, EPI_ISL_2638889, EPI_ISL_2638890, EPI_ISL_2638891, EPI_ISL_2638895, EPI_ISL_2638896, EPI_ISL_2638897, EPI_ISL_2638898, EPI_ISL_2638900, EPI_ISL_2638903, EPI_ISL_2638905, EPI_ISL_2638906, EPI_ISL_2638908, EPI_ISL_2638909, EPI_ISL_2638910, EPI_ISL_2638911, EPI_ISL_2638912, EPI_ISL_2638913, EPI_ISL_2638914, EPI_ISL_2638915, EPI_ISL_2638925, EPI_ISL_2638962, EPI_ISL_2639012, EPI_ISL_2639019, EPI_ISL_2639038, EPI_ISL_2639045, EPI_ISL_2639061, EPI_ISL_2639064, EPI_ISL_2639086, EPI_ISL_2639129, EPI_ISL_2639133, EPI_ISL_2639146, EPI_ISL_2639150, EPI_ISL_2639166, EPI_ISL_2639175, EPI_ISL_2639238, EPI_ISL_2639249, EPI_ISL_2639257, EPI_ISL_2639258, EPI_ISL_2639259, EPI_ISL_2639260, EPI_ISL_2639261, EPI_ISL_2639262, EPI_ISL_2639263, EPI_ISL_2639264, EPI_ISL_2639267, EPI_ISL_2639268, EPI_ISL_2639269, EPI_ISL_2639270, EPI_ISL_2639271, EPI_ISL_2639272, EPI_ISL_2639273, EPI_ISL_2639274, EPI_ISL_2639275, EPI_ISL_2639276, EPI_ISL_2639277, EPI_ISL_2639278, EPI_ISL_2639280, EPI_ISL_2639281, EPI_ISL_2639282, EPI_ISL_2639283, EPI_ISL_2639286, EPI_ISL_2639287, EPI_ISL_2639288, EPI_ISL_2639289, EPI_ISL_2639290, EPI_ISL_2639293, EPI_ISL_2639294, EPI_ISL_2639295, EPI_ISL_2639297, EPI_ISL_2639298, EPI_ISL_2639299, EPI_ISL_2639300, EPI_ISL_2639302, EPI_ISL_2639303, EPI_ISL_2639306, EPI_ISL_2639307, EPI_ISL_2639308, EPI_ISL_2639309, EPI_ISL_2639310, EPI_ISL_2639311, EPI_ISL_2639312, EPI_ISL_2639316, EPI_ISL_2639317, EPI_ISL_2639319, EPI_ISL_2639320, EPI_ISL_2639321, EPI_ISL_2639322, EPI_ISL_2639323, EPI_ISL_2639324, EPI_ISL_2639325, EPI_ISL_2639326, EPI_ISL_2639327, EPI_ISL_2639328, EPI_ISL_2639329, EPI_ISL_2639330, EPI_ISL_2639331, EPI_ISL_2639332, EPI_ISL_2639333, EPI_ISL_2639334, EPI_ISL_2639335, EPI_ISL_2639336, EPI_ISL_2639337, EPI_ISL_2639338, EPI_ISL_2639339, EPI_ISL_2639340, EPI_ISL_2639341, EPI_ISL_2639342, EPI_ISL_2639343, EPI_ISL_2639344, EPI_ISL_2639345, EPI_ISL_2639347, EPI_ISL_2639348, EPI_ISL_2639350, EPI_ISL_2639352, EPI_ISL_2639353, EPI_ISL_2639354, EPI_ISL_2639355, EPI_ISL_2639356, EPI_ISL_2639357, EPI_ISL_2639358, EPI_ISL_2639359, EPI_ISL_2639361, EPI_ISL_2639362, EPI_ISL_2639363, EPI_ISL_2639364, EPI_ISL_2639365, EPI_ISL_2639366, EPI_ISL_2639367, EPI_ISL_2639368, EPI_ISL_2639369, EPI_ISL_2639370, EPI_ISL_2639371, EPI_ISL_2639372, EPI_ISL_2639373, EPI_ISL_2639374, EPI_ISL_2639375, EPI_ISL_2639376, EPI_ISL_2639377, EPI_ISL_2639378, EPI_ISL_2639379, EPI_ISL_2639380, EPI_ISL_2639381, EPI_ISL_2639382, EPI_ISL_2639383, EPI_ISL_2639384, EPI_ISL_2639385, EPI_ISL_2639386, EPI_ISL_2639387, EPI_ISL_2639388, EPI_ISL_2639389, EPI_ISL_2639390, EPI_ISL_2639391, EPI_ISL_2639392, EPI_ISL_2639393, EPI_ISL_2639394, EPI_ISL_2639395, EPI_ISL_2639396, EPI_ISL_2639397, EPI_ISL_2639398, EPI_ISL_2639400, EPI_ISL_2639401, EPI_ISL_2639403, EPI_ISL_2639404, EPI_ISL_2639406, EPI_ISL_2639407, EPI_ISL_2639408, EPI_ISL_2639411, EPI_ISL_2639412, EPI_ISL_2639413, EPI_ISL_2639414, EPI_ISL_2639415, EPI_ISL_2639416, EPI_ISL_2639417, EPI_ISL_2639419, EPI_ISL_2639420, EPI_ISL_2639423, EPI_ISL_2639424, EPI_ISL_2639425, EPI_ISL_2639426, EPI_ISL_2639427, EPI_ISL_2639428, EPI_ISL_2639429, EPI_ISL_2639430, EPI_ISL_2639431, EPI_ISL_2639433, EPI_ISL_2639435, EPI_ISL_2639436, EPI_ISL_2639438, EPI_ISL_2639439, EPI_ISL_2639441, EPI_ISL_2639442, EPI_ISL_2639443, EPI_ISL_2639444, EPI_ISL_2639446, EPI_ISL_2639448, EPI_ISL_2639449, EPI_ISL_2639451, EPI_ISL_2639453, EPI_ISL_2639454, EPI_ISL_2639456, EPI_ISL_2639457, EPI_ISL_2639459, EPI_ISL_2639460, EPI_ISL_2639461, EPI_ISL_2639462, EPI_ISL_2639463, EPI_ISL_2639464, EPI_ISL_2639465, EPI_ISL_2639466, EPI_ISL_2639468, EPI_ISL_2639469, EPI_ISL_2639470, EPI_ISL_2639471, EPI_ISL_2639473, EPI_ISL_2639474, EPI_ISL_2639476, EPI_ISL_2639477, EPI_ISL_2639478, EPI_ISL_2639479, EPI_ISL_2639480, EPI_ISL_2639481, EPI_ISL_2639482, EPI_ISL_2639483, EPI_ISL_2639484, EPI_ISL_2639485, EPI_ISL_2639486, EPI_ISL_2639487, EPI_ISL_2639488, EPI_ISL_2639489, EPI_ISL_2639490, EPI_ISL_2639491, EPI_ISL_2639492, EPI_ISL_2639494, EPI_ISL_2639495, EPI_ISL_2639497, EPI_ISL_2639498, EPI_ISL_2639501, EPI_ISL_2639503, EPI_ISL_2639505, EPI_ISL_2639506, EPI_ISL_2639507, EPI_ISL_2639508, EPI_ISL_2639509, EPI_ISL_2639510, EPI_ISL_2639513, EPI_ISL_2639514, EPI_ISL_2639515, EPI_ISL_2639517, EPI_ISL_2639519, EPI_ISL_2639520, EPI_ISL_2639521, EPI_ISL_2639522, EPI_ISL_2639523, EPI_ISL_2639524, EPI_ISL_2639525, EPI_ISL_2639527, EPI_ISL_2639528, EPI_ISL_2639529, EPI_ISL_2639530, EPI_ISL_2639532, EPI_ISL_2639533, EPI_ISL_2639534, EPI_ISL_2639536, EPI_ISL_2639537, EPI_ISL_2639538, EPI_ISL_2639540, EPI_ISL_2639541, EPI_ISL_2639542, EPI_ISL_2639543, EPI_ISL_2639544, EPI_ISL_2639545, EPI_ISL_2639546, EPI_ISL_2639549, EPI_ISL_2639550, EPI_ISL_2639551, EPI_ISL_2639554, EPI_ISL_2639555, EPI_ISL_2639556, EPI_ISL_2639557, EPI_ISL_2639558, EPI_ISL_2639559, EPI_ISL_2639560, EPI_ISL_2639561, EPI_ISL_2639562, EPI_ISL_2639563, EPI_ISL_2639564, EPI_ISL_2639565, EPI_ISL_2639567, EPI_ISL_2639568, EPI_ISL_2639569, EPI_ISL_2639572, EPI_ISL_2639573, EPI_ISL_2639574, EPI_ISL_2639577, EPI_ISL_2639580, EPI_ISL_2639581, EPI_ISL_2639582, EPI_ISL_2639584, EPI_ISL_2639585, EPI_ISL_2639587, EPI_ISL_2639590, EPI_ISL_2639591, EPI_ISL_2639593, EPI_ISL_2639594, EPI_ISL_2639595, EPI_ISL_2639597, EPI_ISL_2639598, EPI_ISL_2639600, EPI_ISL_2639601, EPI_ISL_2639602, EPI_ISL_2639603, EPI_ISL_2639604, EPI_ISL_2639607, EPI_ISL_2639608, EPI_ISL_2639609, EPI_ISL_2639610, EPI_ISL_2639611, EPI_ISL_2639612, EPI_ISL_2639614, EPI_ISL_2639615, EPI_ISL_2639616, EPI_ISL_2639618, EPI_ISL_2639619, EPI_ISL_2639622, EPI_ISL_2639623, EPI_ISL_2639624, EPI_ISL_2639627, EPI_ISL_2639630, EPI_ISL_2639632, EPI_ISL_2639634, EPI_ISL_2639635, EPI_ISL_2639637, EPI_ISL_2639638, EPI_ISL_2639639, EPI_ISL_2639641, EPI_ISL_2639642, EPI_ISL_2639643, EPI_ISL_2639644, EPI_ISL_2639645, EPI_ISL_2639646, EPI_ISL_2639647, | | | |

EPI_ISL_2639649, EPI_ISL_2639650, EPI_ISL_2639651, EPI_ISL_2639653, EPI_ISL_2639654, EPI_ISL_2639660, EPI_ISL_2639662, EPI_ISL_2639663, EPI_ISL_2639664, EPI_ISL_2639665, EPI_ISL_2639666, EPI_ISL_2639670, EPI_ISL_2639672, EPI_ISL_2639673, EPI_ISL_2639675, EPI_ISL_2639676, EPI_ISL_2639678, EPI_ISL_2639679, EPI_ISL_2639680, EPI_ISL_2639681, EPI_ISL_2639683, EPI_ISL_2639685, EPI_ISL_2639687, EPI_ISL_2639688, EPI_ISL_2639691, EPI_ISL_2639692, EPI_ISL_2639700, EPI_ISL_2639701, EPI_ISL_2639705, EPI_ISL_2639707, EPI_ISL_2639708, EPI_ISL_2639709, EPI_ISL_2639711, EPI_ISL_2639712, EPI_ISL_2639714, EPI_ISL_2639715, EPI_ISL_2639718, EPI_ISL_2639719, EPI_ISL_2639722, EPI_ISL_2639724, EPI_ISL_2639725, EPI_ISL_2639726, EPI_ISL_2639729, EPI_ISL_2639730, EPI_ISL_2639734, EPI_ISL_2639735, EPI_ISL_2639736, EPI_ISL_2639737, EPI_ISL_2639739, EPI_ISL_2639742, EPI_ISL_2639744, EPI_ISL_2639745, EPI_ISL_2639747, EPI_ISL_2639749, EPI_ISL_2639751, EPI_ISL_2639752, EPI_ISL_2639753, EPI_ISL_2639754, EPI_ISL_2639756, EPI_ISL_2639758, EPI_ISL_2639761, EPI_ISL_2639763, EPI_ISL_2639766, EPI_ISL_2639767, EPI_ISL_2639768, EPI_ISL_2639770, EPI_ISL_2639771, EPI_ISL_2639772, EPI_ISL_2639776, EPI_ISL_2639778, EPI_ISL_2639779, EPI_ISL_2639780, EPI_ISL_2639783, EPI_ISL_2639785, EPI_ISL_2639786, EPI_ISL_2639787, EPI_ISL_2639789, EPI_ISL_2639792, EPI_ISL_2639793, EPI_ISL_2639794, EPI_ISL_2639795, EPI_ISL_2639800, EPI_ISL_2639801, EPI_ISL_2639802, EPI_ISL_2639803, EPI_ISL_2639807, EPI_ISL_2639808, EPI_ISL_2639809, EPI_ISL_2639810, EPI_ISL_2639814, EPI_ISL_2639815, EPI_ISL_2639816, EPI_ISL_2639817, EPI_ISL_2639819, EPI_ISL_2639821, EPI_ISL_2639823, EPI_ISL_2639824, EPI_ISL_2639825, EPI_ISL_2639826, EPI_ISL_2639827, EPI_ISL_2639830, EPI_ISL_2639831, EPI_ISL_2639834, EPI_ISL_2639837, EPI_ISL_2639842, EPI_ISL_2639843, EPI_ISL_2639846, EPI_ISL_2639848, EPI_ISL_2639849, EPI_ISL_2639850, EPI_ISL_2639851, EPI_ISL_2639853, EPI_ISL_2639854, EPI_ISL_2639855, EPI_ISL_2639856, EPI_ISL_2639857, EPI_ISL_2639859, EPI_ISL_2639861, EPI_ISL_2639865, EPI_ISL_2639866, EPI_ISL_2639868, EPI_ISL_2639869, EPI_ISL_2639870, EPI_ISL_2639871, EPI_ISL_2639873, EPI_ISL_2639874, EPI_ISL_2639875, EPI_ISL_2639876, EPI_ISL_2639877, EPI_ISL_2639879, EPI_ISL_2639881, EPI_ISL_2639882, EPI_ISL_2639883, EPI_ISL_2639884, EPI_ISL_2639886, EPI_ISL_2639891, EPI_ISL_2639892, EPI_ISL_2639893, EPI_ISL_2639895, EPI_ISL_2639899, EPI_ISL_2639900, EPI_ISL_2639903, EPI_ISL_2639904, EPI_ISL_2639905, EPI_ISL_2639908, EPI_ISL_2639909, EPI_ISL_2639910, EPI_ISL_2639911, EPI_ISL_2639912, EPI_ISL_2639913, EPI_ISL_2639914, EPI_ISL_2639915, EPI_ISL_2639916, EPI_ISL_2639918, EPI_ISL_2639919, EPI_ISL_2639923, EPI_ISL_2639925, EPI_ISL_2639926, EPI_ISL_2639929, EPI_ISL_2639930, EPI_ISL_2640069, EPI_ISL_2640332, EPI_ISL_2640334, EPI_ISL_2651641, EPI_ISL_2651644, EPI_ISL_2651645, EPI_ISL_2651646, EPI_ISL_2651647, EPI_ISL_2651648, EPI_ISL_2651649, EPI_ISL_2651654, EPI_ISL_2651659, EPI_ISL_2651662, EPI_ISL_2651665, EPI_ISL_2651666, EPI_ISL_2651669, EPI_ISL_2651671, EPI_ISL_2651675, EPI_ISL_2651676, EPI_ISL_2651678, EPI_ISL_2651680, EPI_ISL_2651681, EPI_ISL_2651683, EPI_ISL_2651688, EPI_ISL_2651690, EPI_ISL_2651691, EPI_ISL_2651696, EPI_ISL_2651698, EPI_ISL_2651699, EPI_ISL_2651702, EPI_ISL_2651704, EPI_ISL_2651706, EPI_ISL_2651715, EPI_ISL_2651719, EPI_ISL_2651720, EPI_ISL_2651726, EPI_ISL_2651727, EPI_ISL_2651730, EPI_ISL_2651732, EPI_ISL_2651742, EPI_ISL_2651746, EPI_ISL_2651753, EPI_ISL_2651754, EPI_ISL_2651755, EPI_ISL_2651756, EPI_ISL_2651759, EPI_ISL_2651760, EPI_ISL_2651762, EPI_ISL_2651763, EPI_ISL_2651946, EPI_ISL_2651947, EPI_ISL_2664015, EPI_ISL_2664017, EPI_ISL_2664021, EPI_ISL_2664022, EPI_ISL_2664024, EPI_ISL_2664031, EPI_ISL_2664038, EPI_ISL_2664042, EPI_ISL_2664043, EPI_ISL_2664049, EPI_ISL_2664069, EPI_ISL_2664070, EPI_ISL_2664074, EPI_ISL_2664078, EPI_ISL_2664085, EPI_ISL_2664088, EPI_ISL_2664096, EPI_ISL_2664098, EPI_ISL_2664104, EPI_ISL_2664123, EPI_ISL_2664125, EPI_ISL_2664126, EPI_ISL_2664137, EPI_ISL_2664140, EPI_ISL_2664163, EPI_ISL_2664164, EPI_ISL_2664166, EPI_ISL_2664168, EPI_ISL_2664170, EPI_ISL_2664171, EPI_ISL_2664188, EPI_ISL_2664191, EPI_ISL_2664194, EPI_ISL_2664198, EPI_ISL_2664201, EPI_ISL_2664205, EPI_ISL_2664207, EPI_ISL_2664209, EPI_ISL_2664210, EPI_ISL_2664211, EPI_ISL_2664212, EPI_ISL_2664225, EPI_ISL_2664226, EPI_ISL_2664229, EPI_ISL_2664253, EPI_ISL_2664258, EPI_ISL_2664259, EPI_ISL_2664260, EPI_ISL_2664264, EPI_ISL_2664294, EPI_ISL_2664296, EPI_ISL_2664301, EPI_ISL_2664302, EPI_ISL_2664304, EPI_ISL_2664319, EPI_ISL_2664346, EPI_ISL_2664365, EPI_ISL_2664366, EPI_ISL_2664369, EPI_ISL_2664373, EPI_ISL_2664389, EPI_ISL_2664390, EPI_ISL_2664392, EPI_ISL_2664397, EPI_ISL_2664399, EPI_ISL_2664401, EPI_ISL_2664408, EPI_ISL_2664422, EPI_ISL_2664440, EPI_ISL_2664488, EPI_ISL_2664516, EPI_ISL_2664549, EPI_ISL_2664556, EPI_ISL_2664566, EPI_ISL_2664584, EPI_ISL_2664610, EPI_ISL_2664680, EPI_ISL_2664706, EPI_ISL_2664709, EPI_ISL_2664953, EPI_ISL_2664976, EPI_ISL_2665042, EPI_ISL_2665106, EPI_ISL_2665116, EPI_ISL_2665120, EPI_ISL_2665121, EPI_ISL_2665160, EPI_ISL_2665184, EPI_ISL_2665225, EPI_ISL_2665260, EPI_ISL_2665296, EPI_ISL_2665363, EPI_ISL_2681759, EPI_ISL_2681767, EPI_ISL_2681814, EPI_ISL_2681859, EPI_ISL_2682307, EPI_ISL_2682348, EPI_ISL_2682388, EPI_ISL_2682402, EPI_ISL_2682409, EPI_ISL_2682433, EPI_ISL_2682449, EPI_ISL_2682474, EPI_ISL_2682475, EPI_ISL_2682479, EPI_ISL_2682488, EPI_ISL_2682489, EPI_ISL_2682512, EPI_ISL_2682562, EPI_ISL_2682662, EPI_ISL_2682666, EPI_ISL_2682680, EPI_ISL_2682690, EPI_ISL_2682695, EPI_ISL_2682719, EPI_ISL_2682725, EPI_ISL_2682732, EPI_ISL_2682733, EPI_ISL_2682734, EPI_ISL_2682736, EPI_ISL_2682738, EPI_ISL_2682739, EPI_ISL_2682740, EPI_ISL_2682746, EPI_ISL_2682753, EPI_ISL_2682757, EPI_ISL_2682762, EPI_ISL_2682763, EPI_ISL_2682765, EPI_ISL_2682773, EPI_ISL_2682776, EPI_ISL_2682781, EPI_ISL_2682784, EPI_ISL_2682788, EPI_ISL_2682790, EPI_ISL_2682794, EPI_ISL_2682801, EPI_ISL_2682802, EPI_ISL_2682804, EPI_ISL_2682805, EPI_ISL_2682812, EPI_ISL_2682816, EPI_ISL_2682817, EPI_ISL_2682818, EPI_ISL_2682821, EPI_ISL_2682822, EPI_ISL_2682823, EPI_ISL_2682825, EPI_ISL_2682830, EPI_ISL_2682832, EPI_ISL_2682838, EPI_ISL_2682843, EPI_ISL_2682844, EPI_ISL_2682854, EPI_ISL_2682856, EPI_ISL_2682857, EPI_ISL_2682862, EPI_ISL_2682864, EPI_ISL_2682865, EPI_ISL_2682866, EPI_ISL_2682875, EPI_ISL_2682876, EPI_ISL_2682878, EPI_ISL_2682882, EPI_ISL_2682884, EPI_ISL_2682890, EPI_ISL_2682893, EPI_ISL_2682897, EPI_ISL_2682898, EPI_ISL_2682907, EPI_ISL_2682910, EPI_ISL_2682913, EPI_ISL_2682919, EPI_ISL_2682920, EPI_ISL_2682928, EPI_ISL_2682936, EPI_ISL_2682937, EPI_ISL_2682939, EPI_ISL_2682949, EPI_ISL_2682952, EPI_ISL_2682957, EPI_ISL_2682958, EPI_ISL_2682960, EPI_ISL_2682966, EPI_ISL_2682971, EPI_ISL_2700097, EPI_ISL_2700999, EPI_ISL_2701035, EPI_ISL_2701062, EPI_ISL_2701064, EPI_ISL_2701075, EPI_ISL_2701079, EPI_ISL_2701131, EPI_ISL_2701193, EPI_ISL_2701213, EPI_ISL_2701243, EPI_ISL_2701285, EPI_ISL_2701287, EPI_ISL_2701291, EPI_ISL_2701293, EPI_ISL_2701344, EPI_ISL_2701352, EPI_ISL_2701353, EPI_ISL_2701377, EPI_ISL_2701392, EPI_ISL_2701412, EPI_ISL_2701419, EPI_ISL_2701421, EPI_ISL_2701447, EPI_ISL_2701472, EPI_ISL_2701484, EPI_ISL_2701503, EPI_ISL_2701510, EPI_ISL_2701524, EPI_ISL_2701566, EPI_ISL_2701573, EPI_ISL_2701598, EPI_ISL_2701603, EPI_ISL_2701607, EPI_ISL_2701611, EPI_ISL_2701660, EPI_ISL_2701695, EPI_ISL_2701696, EPI_ISL_2701717, EPI_ISL_2701720, EPI_ISL_2701749, EPI_ISL_2701777, EPI_ISL_2701782, EPI_ISL_2701795, EPI_ISL_2701808, EPI_ISL_2701809, EPI_ISL_2701815, EPI_ISL_2701820, EPI_ISL_2701821, EPI_ISL_2701831, EPI_ISL_2701853, EPI_ISL_2701854, EPI_ISL_2701862, EPI_ISL_2701883, EPI_ISL_2701888, EPI_ISL_2701913, EPI_ISL_2701937, EPI_ISL_2701939, EPI_ISL_2701941, EPI_ISL_2701979, EPI_ISL_2701987, EPI_ISL_2701993, EPI_ISL_2701996, EPI_ISL_2702022, EPI_ISL_2702024, EPI_ISL_2702035, EPI_ISL_2702384, EPI_ISL_2702469, EPI_ISL_2702471, EPI_ISL_2702482, EPI_ISL_2702487, EPI_ISL_2702519, EPI_ISL_2702522, EPI_ISL_2702538, EPI_ISL_2702539, EPI_ISL_2702540, EPI_ISL_2702544, EPI_ISL_2702560, EPI_ISL_2702568, EPI_ISL_2702570, EPI_ISL_2702590, EPI_ISL_2702594, EPI_ISL_2702600, EPI_ISL_2702605, EPI_ISL_2702607, EPI_ISL_2702610, EPI_ISL_2702612, EPI_ISL_2702630, EPI_ISL_2702631, EPI_ISL_2702657, EPI_ISL_2702661, EPI_ISL_2702665, EPI_ISL_2702674, EPI_ISL_2702689, EPI_ISL_2702701, EPI_ISL_2702706, EPI_ISL_2702708, EPI_ISL_2702710, EPI_ISL_2702713, EPI_ISL_2702717, EPI_ISL_2702722, EPI_ISL_2702727, EPI_ISL_2702728, EPI_ISL_2702738, EPI_ISL_2702757, EPI_ISL_2702769, EPI_ISL_2702779, EPI_ISL_2702784, EPI_ISL_2702812, EPI_ISL_2703186, EPI_ISL_2703199, EPI_ISL_2703217, EPI_ISL_2703225, EPI_ISL_2703284, EPI_ISL_2703300, EPI_ISL_2703333, EPI_ISL_2703353, EPI_ISL_2703358, EPI_ISL_2703387, EPI_ISL_2703388, EPI_ISL_2703390, EPI_ISL_2703395, EPI_ISL_2703411, EPI_ISL_2703497, EPI_ISL_2703755, EPI_ISL_2703785, EPI_ISL_2703793, EPI_ISL_2704283, EPI_ISL_2704287, EPI_ISL_2704351, EPI_ISL_2704373, EPI_ISL_2704424, EPI_ISL_2704491, EPI_ISL_2704668, EPI_ISL_2704670, EPI_ISL_2717791, EPI_ISL_2718283, EPI_ISL_2718296, EPI_ISL_2718303, EPI_ISL_2718304, EPI_ISL_2718308, EPI_ISL_2718311, EPI_ISL_2718312, EPI_ISL_2718314, EPI_ISL_2718316, EPI_ISL_2718320, EPI_ISL_2718322, EPI_ISL_2718326, EPI_ISL_2718330, EPI_ISL_2718335, EPI_ISL_2718337, EPI_ISL_2718344, EPI_ISL_2718346, EPI_ISL_2718347, EPI_ISL_2718348, EPI_ISL_2718351, EPI_ISL_2718352, EPI_ISL_2718354, EPI_ISL_2718356, EPI_ISL_2718362, EPI_ISL_2718363, EPI_ISL_2718365, EPI_ISL_2718370, EPI_ISL_2718371, EPI_ISL_2718372, EPI_ISL_2718373, EPI_ISL_2718378, EPI_ISL_2718379, EPI_ISL_2718381, EPI_ISL_2718382, EPI_ISL_2718383, EPI_ISL_2718385, EPI_ISL_2718388, EPI_ISL_2718390, EPI_ISL_2718391, EPI_ISL_2718393, EPI_ISL_2718400, EPI_ISL_2718403, EPI_ISL_2718404, EPI_ISL_2718406, EPI_ISL_2718412, EPI_ISL_2718414, EPI_ISL_2718415, EPI_ISL_2718417, EPI_ISL_2718418, EPI_ISL_2718423, EPI_ISL_2718424, EPI_ISL_2718426, EPI_ISL_2718434, EPI_ISL_2718436, EPI_ISL_2718442, EPI_ISL_2718443, EPI_ISL_2718447, EPI_ISL_2718448, EPI_ISL_2718449, EPI_ISL_2718454, EPI_ISL_2718455, EPI_ISL_2718456, EPI_ISL_2718461, EPI_ISL_2718463, EPI_ISL_2718465, EPI_ISL_2718469, EPI_ISL_2718471, EPI_ISL_2718472, EPI_ISL_2718474, EPI_ISL_2718475, EPI_ISL_2718476, EPI_ISL_2718479, EPI_ISL_2718484, EPI_ISL_2718487, EPI_ISL_2718489, EPI_ISL_2718490, EPI_ISL_2718493, EPI_ISL_2718494, EPI_ISL_2718495, EPI_ISL_2718499, EPI_ISL_2718502, EPI_ISL_2718509, EPI_ISL_2718512, EPI_ISL_2718518, EPI_ISL_2718519, EPI_ISL_2718520, EPI_ISL_2718521, EPI_ISL_2718522, EPI_ISL_2718527, EPI_ISL_2718528, EPI_ISL_2718530, EPI_ISL_2767449, EPI_ISL_2809211, EPI_ISL_2853260, EPI_ISL_2853261, EPI_ISL_2853262, EPI_ISL_2853266, EPI_ISL_2853267, EPI_ISL_2853269, EPI_ISL_2853270, EPI_ISL_2853271, EPI_ISL_2853272, EPI_ISL_2853273, EPI_ISL_2853274, EPI_ISL_2853276, EPI_ISL_2853277, EPI_ISL_2853278, EPI_ISL_2853279, EPI_ISL_2853280, EPI_ISL_2853282, EPI_ISL_2853283, EPI_ISL_2853284, EPI_ISL_2853286, EPI_ISL_2853287, EPI_ISL_2853289, EPI_ISL_2853291, EPI_ISL_2853292, EPI_ISL_2853293, EPI_ISL_2853294, EPI_ISL_2853298, EPI_ISL_2853302, EPI_ISL_2853303, EPI_ISL_2853304, EPI_ISL_2853305, EPI_ISL_2853306, EPI_ISL_2853308, EPI_ISL_2853309, EPI_ISL_2853310, EPI_ISL_2853313, EPI_ISL_2853316, EPI_ISL_2853317, EPI_ISL_2853318, EPI_ISL_2853319, EPI_ISL_2853320, EPI_ISL_2853321, EPI_ISL_2853323, EPI_ISL_2853324, EPI_ISL_2853325, EPI_ISL_2853326, EPI_ISL_2853328, EPI_ISL_2853329, EPI_ISL_2853330, EPI_ISL_2853332, EPI_ISL_2853333, EPI_ISL_2853334, EPI_ISL_2853337, EPI_ISL_2853338, EPI_ISL_2853339, EPI_ISL_2853340, EPI_ISL_2853341, EPI_ISL_2853342, EPI_ISL_2853343, EPI_ISL_2853344, EPI_ISL_2853345, EPI_ISL_2853349, EPI_ISL_2853350, EPI_ISL_2853351, EPI_ISL_2853352, EPI_ISL_2853353, EPI_ISL_2853354, EPI_ISL_2853355, EPI_ISL_2853357, EPI_ISL_2853358, EPI_ISL_2853359, EPI_ISL_2853360, EPI_ISL_2853362, EPI_ISL_2853366, EPI_ISL_2853367, EPI_ISL_2853369, EPI_ISL_2853370, EPI_ISL_2853373, EPI_ISL_2853375, EPI_ISL_2853376, EPI_ISL_2853378, EPI_ISL_2853379, EPI_ISL_2853380, EPI_ISL_2853381, EPI_ISL_2853382, EPI_ISL_2853384, EPI_ISL_2853385, EPI_ISL_2853386, EPI_ISL_2853387, EPI_ISL_2853388, EPI_ISL_2853389, EPI_ISL_2853390, EPI_ISL_2853391, EPI_ISL_2853392, EPI_ISL_2853393, EPI_ISL_2853395, EPI_ISL_2853396, EPI_ISL_2853397, EPI_ISL_2853398, EPI_ISL_2853402, EPI_ISL_2853404, EPI_ISL_2853405, EPI_ISL_2853406, EPI_ISL_2853407, EPI_ISL_2853408, EPI_ISL_2853409, EPI_ISL_2853410, EPI_ISL_2853412, EPI_ISL_2853413, EPI_ISL_2853414, EPI_ISL_2853415, EPI_ISL_2853416, EPI_ISL_2853418, EPI_ISL_2853419, EPI_ISL_2853421, EPI_ISL_2853422, EPI_ISL_2853423, EPI_ISL_2853425, EPI_ISL_2853426, EPI_ISL_2853427, EPI_ISL_2853430, EPI_ISL_2853431, EPI_ISL_2853432, EPI_ISL_2853434, EPI_ISL_2853435, EPI_ISL_2853436, EPI_ISL_2853437, EPI_ISL_2853439, EPI_ISL_2853440, EPI_ISL_2853441, EPI_ISL_2853442, EPI_ISL_2853443, EPI_ISL_2853444, EPI_ISL_2853445, EPI_ISL_2853446, EPI_ISL_2853448, EPI_ISL_2853449, EPI_ISL_2853450, EPI_ISL_2853451, EPI_ISL_2853452, EPI_ISL_2853453, EPI_ISL_2853455, EPI_ISL_2853462, EPI_ISL_2853463, EPI_ISL_2853464, EPI_ISL_2853465, EPI_ISL_2853466, EPI_ISL_2853468, EPI_ISL_2853469, EPI_ISL_2853470, EPI_ISL_2853471, EPI_ISL_2853472, EPI_ISL_2853474, EPI_ISL_2853475, EPI_ISL_2853477, EPI_ISL_2853478, EPI_ISL_2853480, EPI_ISL_2853481, EPI_ISL_2853482, EPI_ISL_2853484, EPI_ISL_2853485, EPI_ISL_2853486, EPI_ISL_2853488, EPI_ISL_2853489, EPI_ISL_2853492, EPI_ISL_2853494, EPI_ISL_2853496, EPI_ISL_2853497, EPI_ISL_2853498, EPI_ISL_2853499, EPI_ISL_2853500, EPI_ISL_2853503, EPI_ISL_2853504, EPI_ISL_2853505, EPI_ISL_2853506, EPI_ISL_2853507, EPI_ISL_2853509, EPI_ISL_2853510, EPI_ISL_2853511, EPI_ISL_2853512, EPI_ISL_2853513, EPI_ISL_2853515, EPI_ISL_2853517, EPI_ISL_2853518, EPI_ISL_2853519, EPI_ISL_2853520, EPI_ISL_2853521, EPI_ISL_2853522, EPI_ISL_2853523, EPI_ISL_2853524, EPI_ISL_2853525, EPI_ISL_2853527, EPI_ISL_2853528, EPI_ISL_2853529, EPI_ISL_2853530, EPI_ISL_2853531, EPI_ISL_2853532, EPI_ISL_2853535, EPI_ISL_2853537, EPI_ISL_2853538, EPI_ISL_2853539, EPI_ISL_2853541, EPI_ISL_2853542, EPI_ISL_2853546, EPI_ISL_2853548, EPI_ISL_2853549, EPI_ISL_2853550, EPI_ISL_2853551, EPI_ISL_2853552, EPI_ISL_2853553, EPI_ISL_2853554, EPI_ISL_2853555, EPI_ISL_2853556, EPI_ISL_2853558, EPI_ISL_2853559, EPI_ISL_2853560, EPI_ISL_2853561, EPI_ISL_2853562, EPI_ISL_2853563, EPI_ISL_2853564, EPI_ISL_2853565, EPI_ISL_2853566, EPI_ISL_2853568, EPI_ISL_2853569, EPI_ISL_2853570, EPI_ISL_2853571, EPI_ISL_2853572, EPI_ISL_2853573, EPI_ISL_2853575, EPI_ISL_2853576, EPI_ISL_2853577, EPI_ISL_2853578, EPI_ISL_2853579, EPI_ISL_2853580, EPI_ISL_2853581, EPI_ISL_2853583, EPI_ISL_2853584, EPI_ISL_2853585, EPI_ISL_2868229, EPI_ISL_2868230, EPI_ISL_2868231, EPI_ISL_2868232, EPI_ISL_2868233, EPI_ISL_2868235

| | | | |
|---|---|---|---|
| see above | Lighthouse Lab in Alderley Park | Wellcome Sanger Institute for the COVID-19 Genomics UK (COG-UK) Consortium | Cordelia Langford; David K. Jackson; Dominic Kwiatkowski; Ewan Harrison; Ian Johnston; Jacquelyn Wynn; Jeffrey Barrett; John Sillitoe on behalf of the Wellcome Sanger Institute COVID-19 Surveillance Team; Mairead Hyland; Roberto Amato; Sonia Goncalves; The Lighthouse Lab in Alderley Park and Alex Alderton |

EPI_ISL_2551927, EPI_ISL_2551928, EPI_ISL_2551929, EPI_ISL_2551930, EPI_ISL_2551931, EPI_ISL_2551932, EPI_ISL_2551934, EPI_ISL_2551935, EPI_ISL_2551937, EPI_ISL_2551938, EPI_ISL_2551939, EPI_ISL_2551940, EPI_ISL_2551941, EPI_ISL_2551944, EPI_ISL_2551947, EPI_ISL_2551949, EPI_ISL_2551951, EPI_ISL_2551952, EPI_ISL_2551953, EPI_ISL_2551954, EPI_ISL_2551955, EPI_ISL_2551956, EPI_ISL_2551957, EPI_ISL_2551958, EPI_ISL_2551959, EPI_ISL_2551960, EPI_ISL_2551961, EPI_ISL_2551962, EPI_ISL_2551965, EPI_ISL_2551967, EPI_ISL_2551972, EPI_ISL_2551973, EPI_ISL_2551974, EPI_ISL_2551975, EPI_ISL_2551976, EPI_ISL_2551979, EPI_ISL_2551980, EPI_ISL_2551981, EPI_ISL_2551982, EPI_ISL_2551984, EPI_ISL_2551985, EPI_ISL_2551987, EPI_ISL_2551988, EPI_ISL_2551989, EPI_ISL_2551991, EPI_ISL_2551993, EPI_ISL_2551996, EPI_ISL_2551998, EPI_ISL_2552000, EPI_ISL_2552001, EPI_ISL_2552002, EPI_ISL_2552003, EPI_ISL_2552004, EPI_ISL_2552005, EPI_ISL_2552006, EPI_ISL_2552009, EPI_ISL_2552010, EPI_ISL_2552011, EPI_ISL_2552012, EPI_ISL_2552014, EPI_ISL_2552015, EPI_ISL_2552016, EPI_ISL_2552017, EPI_ISL_2552027, EPI_ISL_2552031, EPI_ISL_2552032, EPI_ISL_2552051, EPI_ISL_2552058, EPI_ISL_2552064, EPI_ISL_2552081, EPI_ISL_2552090, EPI_ISL_2552094, EPI_ISL_2552095, EPI_ISL_2552098, EPI_ISL_2552106, EPI_ISL_2552112, EPI_ISL_2552116, EPI_ISL_2552124, EPI_ISL_2552125, EPI_ISL_2552133, EPI_ISL_2552135, EPI_ISL_2552140, EPI_ISL_2552143, EPI_ISL_2552145, EPI_ISL_2552149, EPI_ISL_2552154, EPI_ISL_2552157, EPI_ISL_2552166, EPI_ISL_2552168, EPI_ISL_2552170, EPI_ISL_2552174, EPI_ISL_2552175, EPI_ISL_2552179, EPI_ISL_2552185, EPI_ISL_2552192, EPI_ISL_2552193, EPI_ISL_2552208, EPI_ISL_2552216, EPI_ISL_2552230, EPI_ISL_2552231, EPI_ISL_2552234, EPI_ISL_2552252, EPI_ISL_2552255, EPI_ISL_2552256, EPI_ISL_2554495, EPI_ISL_2554499, EPI_ISL_2554503, EPI_ISL_2554507, EPI_ISL_2554508, EPI_ISL_2554511, EPI_ISL_2554512, EPI_ISL_2554516, EPI_ISL_2554517, EPI_ISL_2554522, EPI_ISL_2554525, EPI_ISL_2554527, EPI_ISL_2554528, EPI_ISL_2554529, EPI_ISL_2554531, EPI_ISL_2554537, EPI_ISL_2554538, EPI_ISL_2554542, EPI_ISL_2554546, EPI_ISL_2554547, EPI_ISL_2554549, EPI_ISL_2554554, EPI_ISL_2554559, EPI_ISL_2554560, EPI_ISL_2554561, EPI_ISL_2554562, EPI_ISL_2554564, EPI_ISL_2554565, EPI_ISL_2554566, EPI_ISL_2554569, EPI_ISL_2554571, EPI_ISL_2554572, EPI_ISL_2554573, EPI_ISL_2554574, EPI_ISL_2554576, EPI_ISL_2554578, EPI_ISL_2554581, EPI_ISL_2554584, EPI_ISL_2554588, EPI_ISL_2554590, EPI_ISL_2554592, EPI_ISL_2554593, EPI_ISL_2554597, EPI_ISL_2554598, EPI_ISL_2554600, EPI_ISL_2554601, EPI_ISL_2554602, EPI_ISL_2554603, EPI_ISL_2554608, EPI_ISL_2554611, EPI_ISL_2554615, EPI_ISL_2554616, EPI_ISL_2554617, EPI_ISL_2554618, EPI_ISL_2554619, EPI_ISL_2554622, EPI_ISL_2554625, EPI_ISL_2554626, EPI_ISL_2554627, EPI_ISL_2554630, EPI_ISL_2554632, EPI_ISL_2554634, EPI_ISL_2554642, EPI_ISL_2554643, EPI_ISL_2554645, EPI_ISL_2554649, EPI_ISL_2554650, EPI_ISL_2554651, EPI_ISL_2554652, EPI_ISL_2554654, EPI_ISL_2554657, EPI_ISL_2554658, EPI_ISL_2554661, EPI_ISL_2554662, EPI_ISL_2554663, EPI_ISL_2554664, EPI_ISL_2554666, EPI_ISL_2554671, EPI_ISL_2554672, EPI_ISL_2554673, EPI_ISL_2554674, EPI_ISL_2554681, EPI_ISL_2554682, EPI_ISL_2554683, EPI_ISL_2554688, EPI_ISL_2554690, EPI_ISL_2554691, EPI_ISL_2554692, EPI_ISL_2554695, EPI_ISL_2554698, EPI_ISL_2554699, EPI_ISL_2554702, EPI_ISL_2554707, EPI_ISL_2554713, EPI_ISL_2554714, EPI_ISL_2554716, EPI_ISL_2554717, EPI_ISL_2554718, EPI_ISL_2554719, EPI_ISL_2554721, EPI_ISL_2554723, EPI_ISL_2554729, EPI_ISL_2554732, EPI_ISL_2554736, EPI_ISL_2554741, EPI_ISL_2554743, EPI_ISL_2554744, EPI_ISL_2554748, EPI_ISL_2554749, EPI_ISL_2554750, EPI_ISL_2554754, EPI_ISL_2554758, EPI_ISL_2554763, EPI_ISL_2554770, EPI_ISL_2554777, EPI_ISL_2554779, EPI_ISL_2554785, EPI_ISL_2554786, EPI_ISL_2554787, EPI_ISL_2554789, EPI_ISL_2554792, EPI_ISL_2554796, EPI_ISL_2554798, EPI_ISL_2554801, EPI_ISL_2554802, EPI_ISL_2554807, EPI_ISL_2554809, EPI_ISL_2554810, EPI_ISL_2554813, EPI_ISL_2554814, EPI_ISL_2554819, EPI_ISL_2554820, EPI_ISL_2554822, EPI_ISL_2554823, EPI_ISL_2554825, EPI_ISL_2554826, EPI_ISL_2554827, EPI_ISL_2554830, EPI_ISL_2554834, EPI_ISL_2554836, EPI_ISL_2554839, EPI_ISL_2554840, EPI_ISL_2554841, EPI_ISL_2554842, EPI_ISL_2554843, EPI_ISL_2554845, EPI_ISL_2568125, EPI_ISL_2568134, EPI_ISL_2568266, EPI_ISL_2568279, EPI_ISL_2568391, EPI_ISL_2568403, EPI_ISL_2568414, EPI_ISL_2568418, EPI_ISL_2568420, EPI_ISL_2568422, EPI_ISL_2568424, EPI_ISL_2568425, EPI_ISL_2568427, EPI_ISL_2568436, EPI_ISL_2568441, EPI_ISL_2568448, EPI_ISL_2568450, EPI_ISL_2568456, EPI_ISL_2568462, EPI_ISL_2568463, EPI_ISL_2568465, EPI_ISL_2568467, EPI_ISL_2568472, EPI_ISL_2568476, EPI_ISL_2568478, EPI_ISL_2568479, EPI_ISL_2568497, EPI_ISL_2568502, EPI_ISL_2568509, EPI_ISL_2568512, EPI_ISL_2568515, EPI_ISL_2568519, EPI_ISL_2568522, EPI_ISL_2568534, EPI_ISL_2568538, EPI_ISL_2568554, EPI_ISL_2568563, EPI_ISL_2568566, EPI_ISL_2568567, EPI_ISL_2568568, EPI_ISL_2568580, EPI_ISL_2568581, EPI_ISL_2568583, EPI_ISL_2568586, EPI_ISL_2568595, EPI_ISL_2568603, EPI_ISL_2568611, EPI_ISL_2568619, EPI_ISL_2568623, EPI_ISL_2568628, EPI_ISL_2568633, EPI_ISL_2568634, EPI_ISL_2568647, EPI_ISL_2568650, EPI_ISL_2569688, EPI_ISL_2569696, EPI_ISL_2569749, EPI_ISL_2569752, EPI_ISL_2605135, EPI_ISL_2605142, EPI_ISL_2605146, EPI_ISL_2605147, EPI_ISL_2605153, EPI_ISL_2605155, EPI_ISL_2605183, EPI_ISL_2605185, EPI_ISL_2605195, EPI_ISL_2605199, EPI_ISL_2605202, EPI_ISL_2605206, EPI_ISL_2605210, EPI_ISL_2605220, EPI_ISL_2605222, EPI_ISL_2605249, EPI_ISL_2605260, EPI_ISL_2605263, EPI_ISL_2605270, EPI_ISL_2605274, EPI_ISL_2605291, EPI_ISL_2605294, EPI_ISL_2605295, EPI_ISL_2605299, EPI_ISL_2605310, EPI_ISL_2605311, EPI_ISL_2605318, EPI_ISL_2605326, EPI_ISL_2605330, EPI_ISL_2605333, EPI_ISL_2605342, EPI_ISL_2605355, EPI_ISL_2605358, EPI_ISL_2605363, EPI_ISL_2605369, EPI_ISL_2605370, EPI_ISL_2605380, EPI_ISL_2605384, EPI_ISL_2605385, EPI_ISL_2605390, EPI_ISL_2605392, EPI_ISL_2605400, EPI_ISL_2605404, EPI_ISL_2605407, EPI_ISL_2605409, EPI_ISL_2605416, EPI_ISL_2605419, EPI_ISL_2605421, EPI_ISL_2605423, EPI_ISL_2605444, EPI_ISL_2605456, EPI_ISL_2605460, EPI_ISL_2605467, EPI_ISL_2605468, EPI_ISL_2605482, EPI_ISL_2605499, EPI_ISL_2605505, EPI_ISL_2605545, EPI_ISL_2605554, EPI_ISL_2605555, EPI_ISL_2605563, EPI_ISL_2605574, EPI_ISL_2605594, EPI_ISL_2605595, EPI_ISL_2605629, EPI_ISL_2605638, EPI_ISL_2605642, EPI_ISL_2605656, EPI_ISL_2605710, EPI_ISL_2605718, EPI_ISL_2605722, EPI_ISL_2605724, EPI_ISL_2605738, EPI_ISL_2605908, EPI_ISL_2606012, EPI_ISL_2606055, EPI_ISL_2606457, EPI_ISL_2606470, EPI_ISL_2606479, EPI_ISL_2606484, EPI_ISL_2606535, EPI_ISL_2606557, EPI_ISL_2606593, EPI_ISL_2606624, EPI_ISL_2606686, EPI_ISL_2606690, EPI_ISL_2606706, EPI_ISL_2606709, EPI_ISL_2606712, EPI_ISL_2606721, EPI_ISL_2606736, EPI_ISL_2606737, EPI_ISL_2606740, EPI_ISL_2606754, EPI_ISL_2606758, EPI_ISL_2606760, EPI_ISL_2606768, EPI_ISL_2606770, EPI_ISL_2606771, EPI_ISL_2606774, EPI_ISL_2606775, EPI_ISL_2606778, EPI_ISL_2606786, EPI_ISL_2606788, EPI_ISL_2606790, EPI_ISL_2606793, EPI_ISL_2606803, EPI_ISL_2606806, EPI_ISL_2606807, EPI_ISL_2606819, EPI_ISL_2606829, EPI_ISL_2606839, EPI_ISL_2606840, EPI_ISL_2606845, EPI_ISL_2606846, EPI_ISL_2606848, EPI_ISL_2606864, EPI_ISL_2606871, EPI_ISL_2606875, EPI_ISL_2606882, EPI_ISL_2606885, EPI_ISL_2606893, EPI_ISL_2606894, EPI_ISL_2606899, EPI_ISL_2606905, EPI_ISL_2606909, EPI_ISL_2606920, EPI_ISL_2606925, EPI_ISL_2606930, EPI_ISL_2606931, EPI_ISL_2606937, EPI_ISL_2606938, EPI_ISL_2606944, EPI_ISL_2606954, EPI_ISL_2606959, EPI_ISL_2606960, EPI_ISL_2606971, EPI_ISL_2606982, EPI_ISL_2606983, EPI_ISL_2606995, EPI_ISL_2607005, EPI_ISL_2607007, EPI_ISL_2607016, EPI_ISL_2607043, EPI_ISL_2607045, EPI_ISL_2607047, EPI_ISL_2607048, EPI_ISL_2607050, EPI_ISL_2607051, EPI_ISL_2607054, EPI_ISL_2607056, EPI_ISL_2607057, EPI_ISL_2607058, EPI_ISL_2607059, EPI_ISL_2607061, EPI_ISL_2607064, EPI_ISL_2607066, EPI_ISL_2607067, EPI_ISL_2607071, EPI_ISL_2607072, EPI_ISL_2607075, EPI_ISL_2607076, EPI_ISL_2607077, EPI_ISL_2607078, EPI_ISL_2607080, EPI_ISL_2607081, EPI_ISL_2607082, EPI_ISL_2607084, EPI_ISL_2607086, EPI_ISL_2607087, EPI_ISL_2607091, EPI_ISL_2607093, EPI_ISL_2607094, EPI_ISL_2607097, EPI_ISL_2607098, EPI_ISL_2607103, EPI_ISL_2607105, EPI_ISL_2607107, EPI_ISL_2607109, EPI_ISL_2607110, EPI_ISL_2607111, EPI_ISL_2607112, EPI_ISL_2607113, EPI_ISL_2607114, EPI_ISL_2607120, EPI_ISL_2607121, EPI_ISL_2607125, EPI_ISL_2607126, EPI_ISL_2607127, EPI_ISL_2607130, EPI_ISL_2607131, EPI_ISL_2607132, EPI_ISL_2607133, EPI_ISL_2607134, EPI_ISL_2607135, EPI_ISL_2607136, EPI_ISL_2607137, EPI_ISL_2607138, EPI_ISL_2607139, EPI_ISL_2607140, EPI_ISL_2607141, EPI_ISL_2607142, EPI_ISL_2607143, EPI_ISL_2607144, EPI_ISL_2607145, EPI_ISL_2607146, EPI_ISL_2607147, EPI_ISL_2607148, EPI_ISL_2607149, EPI_ISL_2607150, EPI_ISL_2607151, EPI_ISL_2607152, EPI_ISL_2607153, EPI_ISL_2607154, EPI_ISL_2607156, EPI_ISL_2607157, EPI_ISL_2607158, EPI_ISL_2607159, EPI_ISL_2607160, EPI_ISL_2607161, EPI_ISL_2607162, EPI_ISL_2607163, EPI_ISL_2607164, EPI_ISL_2607165, EPI_ISL_2607166, EPI_ISL_2607167, EPI_ISL_2607168, EPI_ISL_2607169, EPI_ISL_2607170, EPI_ISL_2607171, EPI_ISL_2607172, EPI_ISL_2607173, EPI_ISL_2607175, EPI_ISL_2607176, EPI_ISL_2607177, EPI_ISL_2607178, EPI_ISL_2607179, EPI_ISL_2607180, EPI_ISL_2607181, EPI_ISL_2607183, EPI_ISL_2607184, EPI_ISL_2607185, EPI_ISL_2607186, EPI_ISL_2607187, EPI_ISL_2607188, EPI_ISL_2607189, EPI_ISL_2607190, EPI_ISL_2607191, EPI_ISL_2607192, EPI_ISL_2607193, EPI_ISL_2607194, EPI_ISL_2607195, EPI_ISL_2607196, EPI_ISL_2607197, EPI_ISL_2607198, EPI_ISL_2607199, EPI_ISL_2607200, EPI_ISL_2607201, EPI_ISL_2607202, EPI_ISL_2607203, EPI_ISL_2607204, EPI_ISL_2607205, EPI_ISL_2607206, EPI_ISL_2607207, EPI_ISL_2607208, EPI_ISL_2607209, EPI_ISL_2607210, EPI_ISL_2607211, EPI_ISL_2607212, EPI_ISL_2607213, EPI_ISL_2607214, EPI_ISL_2607215, EPI_ISL_2607216, EPI_ISL_2607217, EPI_ISL_2607218, EPI_ISL_2607219, EPI_ISL_2607220, EPI_ISL_2607221, EPI_ISL_2607222, EPI_ISL_2607223, EPI_ISL_2607224, EPI_ISL_2607225, EPI_ISL_2607226, EPI_ISL_2607227, EPI_ISL_2607228, EPI_ISL_2607230, EPI_ISL_2607231, EPI_ISL_2607232, EPI_ISL_2607233, EPI_ISL_2607234, EPI_ISL_2607235, EPI_ISL_2607236, EPI_ISL_2607237, EPI_ISL_2607238, EPI_ISL_2607239, EPI_ISL_2607240, EPI_ISL_2607241, EPI_ISL_2607242, EPI_ISL_2607243, EPI_ISL_2607244, EPI_ISL_2607245, EPI_ISL_2607246, EPI_ISL_2607247, EPI_ISL_2607248, EPI_ISL_2607249, EPI_ISL_2607250, EPI_ISL_2607442, EPI_ISL_2607443, EPI_ISL_2607445, EPI_ISL_2607446, EPI_ISL_2607447, EPI_ISL_2607448, EPI_ISL_2607453, EPI_ISL_2607456, EPI_ISL_2607457, EPI_ISL_2607458, EPI_ISL_2607459, EPI_ISL_2607461, EPI_ISL_2607464, EPI_ISL_2607465, EPI_ISL_2607466, EPI_ISL_2607469, EPI_ISL_2607470, EPI_ISL_2607473, EPI_ISL_2607474, EPI_ISL_2607475, EPI_ISL_2607476, EPI_ISL_2607478, EPI_ISL_2607480, EPI_ISL_2607481, EPI_ISL_2607482, EPI_ISL_2607484, EPI_ISL_2607487, EPI_ISL_2607489, EPI_ISL_2607490, EPI_ISL_2607491, EPI_ISL_2607492, EPI_ISL_2607493, EPI_ISL_2607495, EPI_ISL_2607496, EPI_ISL_2607498, EPI_ISL_2607499, EPI_ISL_2607501, EPI_ISL_2607505, EPI_ISL_2607508, EPI_ISL_2607509, EPI_ISL_2607510, EPI_ISL_2607511, EPI_ISL_2607512, EPI_ISL_2607514, EPI_ISL_2607517, EPI_ISL_2607518, EPI_ISL_2607519, EPI_ISL_2607520, EPI_ISL_2607521, EPI_ISL_2607522, EPI_ISL_2607523, EPI_ISL_2607524, EPI_ISL_2607525, EPI_ISL_2607526, EPI_ISL_2607527, EPI_ISL_2607528, EPI_ISL_2607529, EPI_ISL_2607530, EPI_ISL_2607531, EPI_ISL_2607532, EPI_ISL_2607533, EPI_ISL_2607534, EPI_ISL_2607535, EPI_ISL_2607536, EPI_ISL_2607537, EPI_ISL_2607538, EPI_ISL_2607539, EPI_ISL_2607540, EPI_ISL_2607541, EPI_ISL_2607542, EPI_ISL_2607543, EPI_ISL_2607544, EPI_ISL_2607545, EPI_ISL_2607547, EPI_ISL_2607548, EPI_ISL_2607549, EPI_ISL_2607550, EPI_ISL_2607551, EPI_ISL_2607552, EPI_ISL_2607553, EPI_ISL_2607554, EPI_ISL_2607555, EPI_ISL_2607556, EPI_ISL_2607557, EPI_ISL_2607558, EPI_ISL_2607559, EPI_ISL_2607560, EPI_ISL_2607561, EPI_ISL_2607562, EPI_ISL_2607563, EPI_ISL_2607564, EPI_ISL_2607565, EPI_ISL_2607566, EPI_ISL_2607567, EPI_ISL_2607568, EPI_ISL_2607569, EPI_ISL_2607570, EPI_ISL_2607571, EPI_ISL_2607572, EPI_ISL_2607573, EPI_ISL_2607574, EPI_ISL_2607575, EPI_ISL_2607576, EPI_ISL_2607577, EPI_ISL_2607578, EPI_ISL_2607579, EPI_ISL_2607580, EPI_ISL_2607581, EPI_ISL_2607582, EPI_ISL_2607583, EPI_ISL_2607584, EPI_ISL_2607585, EPI_ISL_2607586, EPI_ISL_2607587, EPI_ISL_2607588, EPI_ISL_2607589, EPI_ISL_2607590, EPI_ISL_2607591, EPI_ISL_2607592, EPI_ISL_2607593, EPI_ISL_2607594, EPI_ISL_2607595, EPI_ISL_2607596, EPI_ISL_2607597, EPI_ISL_2607598, EPI_ISL_2607599, EPI_ISL_2607600, EPI_ISL_2607601, EPI_ISL_2607602, EPI_ISL_2607603, EPI_ISL_2607604, EPI_ISL_2607605, EPI_ISL_2607606, EPI_ISL_2607607, EPI_ISL_2607608, EPI_ISL_2607609, EPI_ISL_2607610, EPI_ISL_2607611, EPI_ISL_2607612, EPI_ISL_2607613, EPI_ISL_2607614, EPI_ISL_2607615, EPI_ISL_2607616, EPI_ISL_2607617, EPI_ISL_2607618, EPI_ISL_2607619, EPI_ISL_2607620, EPI_ISL_2607621, EPI_ISL_2607622, EPI_ISL_2607623, EPI_ISL_2607624, EPI_ISL_2607625, EPI_ISL_2607626, EPI_ISL_2607627, EPI_ISL_2607628, EPI_ISL_2607629, EPI_ISL_2607630, EPI_ISL_2607631, EPI_ISL_2607632, EPI_ISL_2607633, EPI_ISL_2607634, EPI_ISL_2607635, EPI_ISL_2607638, EPI_ISL_2607640, EPI_ISL_2607641, EPI_ISL_2607642, EPI_ISL_2607643, EPI_ISL_2607644, EPI_ISL_2607646, EPI_ISL_2607647, EPI_ISL_2607649, EPI_ISL_2607650, EPI_ISL_2607652, EPI_ISL_2607656, EPI_ISL_2607657, EPI_ISL_2607658, EPI_ISL_2607659, EPI_ISL_2607660, EPI_ISL_2607661, EPI_ISL_2607662, EPI_ISL_2607663, EPI_ISL_2607666, EPI_ISL_2607667, EPI_ISL_2607668, EPI_ISL_2607669, EPI_ISL_2607670, EPI_ISL_2607671, EPI_ISL_2607672, EPI_ISL_2607673, EPI_ISL_2607674, EPI_ISL_2607675, EPI_ISL_2607677, EPI_ISL_2607679, EPI_ISL_2607680, EPI_ISL_2607681, EPI_ISL_2607683, EPI_ISL_2607684, EPI_ISL_2607685, EPI_ISL_2607686, EPI_ISL_2607687, EPI_ISL_2607688, EPI_ISL_2607689, EPI_ISL_2607690, EPI_ISL_2607691, EPI_ISL_2607694, EPI_ISL_2607695, EPI_ISL_2607696, EPI_ISL_2607698, EPI_ISL_2607699, EPI_ISL_2607701, EPI_ISL_2607702, EPI_ISL_2607705, EPI_ISL_2607707, EPI_ISL_2607709, EPI_ISL_2607711, EPI_ISL_2607713, EPI_ISL_2607714, EPI_ISL_2607716, EPI_ISL_2607717, EPI_ISL_2607719, EPI_ISL_2607720, EPI_ISL_2607721, EPI_ISL_2607724, EPI_ISL_2607725, EPI_ISL_2607726, EPI_ISL_2607727, EPI_ISL_2607728, EPI_ISL_2607729, EPI_ISL_2607731, EPI_ISL_2607732, EPI_ISL_2607733, EPI_ISL_2607734, EPI_ISL_2607735, EPI_ISL_2607737, EPI_ISL_2607738, EPI_ISL_2607739, EPI_ISL_2607740, EPI_ISL_2607741, EPI_ISL_2607742, EPI_ISL_2607743, EPI_ISL_2607744, EPI_ISL_2607745, EPI_ISL_2607747, EPI_ISL_2607748, EPI_ISL_2607750, EPI_ISL_2607751, EPI_ISL_2607752, EPI_ISL_2607753, EPI_ISL_2607754, EPI_ISL_2607755, EPI_ISL_2607756, EPI_ISL_2607757, EPI_ISL_2607758, EPI_ISL_2607759, EPI_ISL_2607761, EPI_ISL_2607764, EPI_ISL_2607765, EPI_ISL_2607766, EPI_ISL_2607767, EPI_ISL_2607768, EPI_ISL_2607769, EPI_ISL_2607772, EPI_ISL_2607774, EPI_ISL_2607775, EPI_ISL_2607776, EPI_ISL_2607777, EPI_ISL_2607778, EPI_ISL_2607779, EPI_ISL_2607780, EPI_ISL_2607781, EPI_ISL_2607782, EPI_ISL_2607784, EPI_ISL_2607785, EPI_ISL_2607786, EPI_ISL_2607787, EPI_ISL_2607790, EPI_ISL_2607791, EPI_ISL_2607792, EPI_ISL_2607793, EPI_ISL_2607794, EPI_ISL_2607795, EPI_ISL_2607796, EPI_ISL_2607797, EPI_ISL_2607798, EPI_ISL_2607799, EPI_ISL_2607800, EPI_ISL_2607802, EPI_ISL_2607803, EPI_ISL_2607804, EPI_ISL_2607806, EPI_ISL_2607807, EPI_ISL_2607808, EPI_ISL_2607810, EPI_ISL_2607813, EPI_ISL_2607814, EPI_ISL_2607816, EPI_ISL_2607818, EPI_ISL_2607819, EPI_ISL_2607820, EPI_ISL_2607822, EPI_ISL_2607823, EPI_ISL_2607824, EPI_ISL_2607825, EPI_ISL_2607828, EPI_ISL_2607830, EPI_ISL_2607833, EPI_ISL_2607835, EPI_ISL_2607836, EPI_ISL_2607837, EPI_ISL_2607838, EPI_ISL_2607839, EPI_ISL_2607840, EPI_ISL_2607844, EPI_ISL_2607845, EPI_ISL_2607847, EPI_ISL_2607848, EPI_ISL_2607850, EPI_ISL_2607851, EPI_ISL_2607853, EPI_ISL_2607854, EPI_ISL_2607855, EPI_ISL_2607857, EPI_ISL_2607858, EPI_ISL_2607860, EPI_ISL_2607861, EPI_ISL_2607862, EPI_ISL_2607866, EPI_ISL_2607867, EPI_ISL_2607868, EPI_ISL_2607870, EPI_ISL_2607871, EPI_ISL_2607872, EPI_ISL_2607876, EPI_ISL_2607877, EPI_ISL_2607879, EPI_ISL_2607880, EPI_ISL_2607881, EPI_ISL_2607882, EPI_ISL_2607885, EPI_ISL_2607886, EPI_ISL_2607887, EPI_ISL_2607888, EPI_ISL_2607890, EPI_ISL_2607891, EPI_ISL_2607892, EPI_ISL_2607894, EPI_ISL_2607895, EPI_ISL_2607896, EPI_ISL_2607898, EPI_ISL_2638165, EPI_ISL_2638223, EPI_ISL_2638307, EPI_ISL_2638377, EPI_ISL_2638419, EPI_ISL_2638434, EPI_ISL_2638435, EPI_ISL_2638453, EPI_ISL_2638480, EPI_ISL_2638512, EPI_ISL_2638572, EPI_ISL_2638574, EPI_ISL_2638595, EPI_ISL_2638625, EPI_ISL_2638674, EPI_ISL_2638675, EPI_ISL_2638676, EPI_ISL_2638681, EPI_ISL_2638683, EPI_ISL_2638691, EPI_ISL_2638692, EPI_ISL_2638695, EPI_ISL_2638696, EPI_ISL_2638728, EPI_ISL_2638729, EPI_ISL_2638730, EPI_ISL_2638731, EPI_ISL_2638741, EPI_ISL_2638745, EPI_ISL_2638751, EPI_ISL_2638754, EPI_ISL_2638759, EPI_ISL_2638760, EPI_ISL_2638767, EPI_ISL_2638769, EPI_ISL_2638771, EPI_ISL_2638775, EPI_ISL_2638789, EPI_ISL_2638805, EPI_ISL_2638807, EPI_ISL_2638813, EPI_ISL_2638822, EPI_ISL_2638829, EPI_ISL_2638833, EPI_ISL_2638834, EPI_ISL_2638844, EPI_ISL_2638848, EPI_ISL_2638852, EPI_ISL_2638853, EPI_ISL_2638854, EPI_ISL_2638858, EPI_ISL_2638859, EPI_ISL_2638865, EPI_ISL_2638869, EPI_ISL_2638887, EPI_ISL_2638902, EPI_ISL_2638904, EPI_ISL_2639659, EPI_ISL_2639677, EPI_ISL_2639702, EPI_ISL_2639710, EPI_ISL_2639731, EPI_ISL_2639733, EPI_ISL_2639765, EPI_ISL_2639777, EPI_ISL_2639932, EPI_ISL_2639933, EPI_ISL_2639934, EPI_ISL_2639935, EPI_ISL_2639936, EPI_ISL_2639937, EPI_ISL_2639938, EPI_ISL_2639939, EPI_ISL_2639940, EPI_ISL_2639941, EPI_ISL_2639942, EPI_ISL_2639943, EPI_ISL_2639944, EPI_ISL_2639945, EPI_ISL_2639946, EPI_ISL_2639947, EPI_ISL_2639948, EPI_ISL_2639949, EPI_ISL_2639950, EPI_ISL_2639951, EPI_ISL_2639952, EPI_ISL_2639953, EPI_ISL_2639954, EPI_ISL_2639955, EPI_ISL_2639956, EPI_ISL_2639957, EPI_ISL_2639958, EPI_ISL_2639959, EPI_ISL_2639960, EPI_ISL_2639961, EPI_ISL_2639962, EPI_ISL_2639963, EPI_ISL_2639964, EPI_ISL_2639965, EPI_ISL_2639966, EPI_ISL_2639967, EPI_ISL_2639968, EPI_ISL_2639969, EPI_ISL_2639970, EPI_ISL_2639971, EPI_ISL_2639972, EPI_ISL_2639973, EPI_ISL_2639974, EPI_ISL_2639975, EPI_ISL_2639976, EPI_ISL_2639977, EPI_ISL_2639978, EPI_ISL_2639979, EPI_ISL_2639980, EPI_ISL_2639981, EPI_ISL_2639982, EPI_ISL_2639983, EPI_ISL_2639984, EPI_ISL_2639985, EPI_ISL_2639986, EPI_ISL_2639987, EPI_ISL_2639988, EPI_ISL_2639989, EPI_ISL_2639990, EPI_ISL_2639991, EPI_ISL_2639992, EPI_ISL_2639993, EPI_ISL_2639994, EPI_ISL_2639995, EPI_ISL_2639996, EPI_ISL_2639997, EPI_ISL_2639998, EPI_ISL_2639999, EPI_ISL_2640000, EPI_ISL_2640001, EPI_ISL_2640002, EPI_ISL_2640003, EPI_ISL_2640004, EPI_ISL_2640005, EPI_ISL_2640006, EPI_ISL_2640007, EPI_ISL_2640008, EPI_ISL_2640009, EPI_ISL_2640010, EPI_ISL_2640011, EPI_ISL_2640012, EPI_ISL_2640013, EPI_ISL_2640014, EPI_ISL_2640015, EPI_ISL_2640016, EPI_ISL_2640017, EPI_ISL_2640018, EPI_ISL_2640019, EPI_ISL_2640020, EPI_ISL_2640021, EPI_ISL_2640022, EPI_ISL_2640023, EPI_ISL_2640024, EPI_ISL_2640025, EPI_ISL_2640026, EPI_ISL_2640027, EPI_ISL_2640028, EPI_ISL_2640029, EPI_ISL_2640030, EPI_ISL_2640031, EPI_ISL_2640032, EPI_ISL_2640033, EPI_ISL_2640034, EPI_ISL_2640035, EPI_ISL_2640036, EPI_ISL_2640037, EPI_ISL_2640038, EPI_ISL_2640039, EPI_ISL_2640040, EPI_ISL_2640041, EPI_ISL_2640042, EPI_ISL_2640043, EPI_ISL_2640044, EPI_ISL_2640045, EPI_ISL_2640046, EPI_ISL_2640047, EPI_ISL_2640048, EPI_ISL_2640049, EPI_ISL_2640050, EPI_ISL_2640051, EPI_ISL_2640052, EPI_ISL_2640053, EPI_ISL_2640054, EPI_ISL_2640055, EPI_ISL_2640056, EPI_ISL_2640057, EPI_ISL_2640058, EPI_ISL_2640059, EPI_ISL_2640060, EPI_ISL_2640061, EPI_ISL_2640062, EPI_ISL_2640063, EPI_ISL_2640064, EPI_ISL_2640065, EPI_ISL_2640066, EPI_ISL_2640067, EPI_ISL_2640068, EPI_ISL_2640070, EPI_ISL_2640071, EPI_ISL_2640072, EPI_ISL_2640073, EPI_ISL_2640075, EPI_ISL_2640076, EPI_ISL_2640077, EPI_ISL_2640084, EPI_ISL_2640085, EPI_ISL_2640086, EPI_ISL_2640087, EPI_ISL_2640088, EPI_ISL_2640089, EPI_ISL_2640090, EPI_ISL_2640091, EPI_ISL_2640093, EPI_ISL_2640094, EPI_ISL_2640095, EPI_ISL_2640096, EPI_ISL_2640097, EPI_ISL_2640098, EPI_ISL_2640099, EPI_ISL_2640100, EPI_ISL_2640101, EPI_ISL_2640102, EPI_ISL_2640103, EPI_ISL_2640104, EPI_ISL_2640106, EPI_ISL_2640107, EPI_ISL_2640109, EPI_ISL_2640110, EPI_ISL_2640111, EPI_ISL_2640112, EPI_ISL_2640113, EPI_ISL_2640114, EPI_ISL_2640115, EPI_ISL_2640117, EPI_ISL_2640118, EPI_ISL_2640119, EPI_ISL_2640120, EPI_ISL_2640121, EPI_ISL_2640124, EPI_ISL_2640125, EPI_ISL_2640126, EPI_ISL_2640127, EPI_ISL_2640128, EPI_ISL_2640129, EPI_ISL_2640130, EPI_ISL_2640131, EPI_ISL_2640132, EPI_ISL_2640133, EPI_ISL_2640134, EPI_ISL_2640135, EPI_ISL_2640136, EPI_ISL_2640137, EPI_ISL_2640138, EPI_ISL_2640139, EPI_ISL_2640140, EPI_ISL_2640141, EPI_ISL_2640142, EPI_ISL_2640143, EPI_ISL_2640144, EPI_ISL_2640145, EPI_ISL_2640146, EPI_ISL_2640148, EPI_ISL_2640149, EPI_ISL_2640150, EPI_ISL_2640151, EPI_ISL_2640152, EPI_ISL_2640154, EPI_ISL_2640155, EPI_ISL_2640156, EPI_ISL_2640157, EPI_ISL_2640158, EPI_ISL_2640159, EPI_ISL_2640160, EPI_ISL_2640162, EPI_ISL_2640163, EPI_ISL_2640164, EPI_ISL_2640165, EPI_ISL_2640166, EPI_ISL_2640167, EPI_ISL_2640168, EPI_ISL_2640169, EPI_ISL_2640170, EPI_ISL_2640171, EPI_ISL_2640172, EPI_ISL_2640173, EPI_ISL_2640174, EPI_ISL_2640175, EPI_ISL_2640176, EPI_ISL_2640177, EPI_ISL_2640178, EPI_ISL_2640179, EPI_ISL_2640180, EPI_ISL_2640181, EPI_ISL_2640182, EPI_ISL_2640183, EPI_ISL_2640184, EPI_ISL_2640185, EPI_ISL_2640186, EPI_ISL_2640187, EPI_ISL_2640188, EPI_ISL_2640189, EPI_ISL_2640190, EPI_ISL_2640191, EPI_ISL_2640192, EPI_ISL_2640193, EPI_ISL_2640194, EPI_ISL_2640195, EPI_ISL_2640196, EPI_ISL_2640197, EPI_ISL_2640198, EPI_ISL_2640199, EPI_ISL_2640200, EPI_ISL_2640201, EPI_ISL_2640202, EPI_ISL_2640203, EPI_ISL_2640204, EPI_ISL_2640205, EPI_ISL_2640206, EPI_ISL_2640207, EPI_ISL_2640208, EPI_ISL_2640209, EPI_ISL_2640210, EPI_ISL_2640211, EPI_ISL_2640212, EPI_ISL_2640213, EPI_ISL_2640214, EPI_ISL_2640215, EPI_ISL_2640216, EPI_ISL_2640217, EPI_ISL_2640218, EPI_ISL_2640220, EPI_ISL_2640221, EPI_ISL_2640222, EPI_ISL_2640223, EPI_ISL_2640224, EPI_ISL_2640225, EPI_ISL_2640226, EPI_ISL_2640227, EPI_ISL_2640228, EPI_ISL_2640229, EPI_ISL_2640230, EPI_ISL_2640231, EPI_ISL_2640232, EPI_ISL_2640233, EPI_ISL_2640234, EPI_ISL_2640235, EPI_ISL_2640236, EPI_ISL_2640237, EPI_ISL_2640238, EPI_ISL_2640239, EPI_ISL_2640240, EPI_ISL_2640241, EPI_ISL_2640242, EPI_ISL_2640243, EPI_ISL_2640244, EPI_ISL_2640245, EPI_ISL_2640246, EPI_ISL_2640247, EPI_ISL_2640248, EPI_ISL_2640249, EPI_ISL_2640250, EPI_ISL_2640251, EPI_ISL_2640252, EPI_ISL_2640253, EPI_ISL_2640254, EPI_ISL_2640255, EPI_ISL_2640256, EPI_ISL_2640257, EPI_ISL_2640258, EPI_ISL_2640259, EPI_ISL_2640260, EPI_ISL_2640261, EPI_ISL_2640262, EPI_ISL_2640263, EPI_ISL_2640264, EPI_ISL_2640265, EPI_ISL_2640266, EPI_ISL_2640267, EPI_ISL_2640268, EPI_ISL_2640269, EPI_ISL_2640270, EPI_ISL_2640271, EPI_ISL_2640272, EPI_ISL_2640273, EPI_ISL_2640274, EPI_ISL_2640275, EPI_ISL_2640276, EPI_ISL_2640277, EPI_ISL_2640278, EPI_ISL_2640279, EPI_ISL_2640280, EPI_ISL_2640281, EPI_ISL_2640282, EPI_ISL_2640283, EPI_ISL_2640284, EPI_ISL_2640285, EPI_ISL_2640286, EPI_ISL_2640287, EPI_ISL_2640288, EPI_ISL_2640289, EPI_ISL_2640290, EPI_ISL_2640291, EPI_ISL_2640292, EPI_ISL_2640293, EPI_ISL_2640294, EPI_ISL_2640295, EPI_ISL_2640296, EPI_ISL_2640297, EPI_ISL_2640298, EPI_ISL_2640299, EPI_ISL_2640300, EPI_ISL_2640301, EPI_ISL_2640302, EPI_ISL_2640303, EPI_ISL_2640304, EPI_ISL_2640305, EPI_ISL_2640307, EPI_ISL_2640308, EPI_ISL_2640309, EPI_ISL_2640310, EPI_ISL_2640311, EPI_ISL_2640312, EPI_ISL_2640314, EPI_ISL_2640315, EPI_ISL_2640316, EPI_ISL_2640317, EPI_ISL_2640318, EPI_ISL_2640319, EPI_ISL_2640320, EPI_ISL_2640321, EPI_ISL_2640322,

EPI_ISL_2640323, EPI_ISL_2640324, EPI_ISL_2640325, EPI_ISL_2651643, EPI_ISL_2651652, EPI_ISL_2651661, EPI_ISL_2651697, EPI_ISL_2651725, EPI_ISL_2651736, EPI_ISL_2651786, EPI_ISL_2651847, EPI_ISL_2651867, EPI_ISL_2651892, EPI_ISL_2651893, EPI_ISL_2651894, EPI_ISL_2651895, EPI_ISL_2651896, EPI_ISL_2651897, EPI_ISL_2651898, EPI_ISL_2651899, EPI_ISL_2651900, EPI_ISL_2651901, EPI_ISL_2651902, EPI_ISL_2651903, EPI_ISL_2651904, EPI_ISL_2651905, EPI_ISL_2651906, EPI_ISL_2651907, EPI_ISL_2651908, EPI_ISL_2651909, EPI_ISL_2651910, EPI_ISL_2651911, EPI_ISL_2651912, EPI_ISL_2651913, EPI_ISL_2651914, EPI_ISL_2651915, EPI_ISL_2651916, EPI_ISL_2651917, EPI_ISL_2651918, EPI_ISL_2651919, EPI_ISL_2651920, EPI_ISL_2651921, EPI_ISL_2651922, EPI_ISL_2651923, EPI_ISL_2651924, EPI_ISL_2651925, EPI_ISL_2651926, EPI_ISL_2651927, EPI_ISL_2651928, EPI_ISL_2651929, EPI_ISL_2651930, EPI_ISL_2651931, EPI_ISL_2651932, EPI_ISL_2651933, EPI_ISL_2651934, EPI_ISL_2651935, EPI_ISL_2651936, EPI_ISL_2651937, EPI_ISL_2651938, EPI_ISL_2651939, EPI_ISL_2651940, EPI_ISL_2651941, EPI_ISL_2651997, EPI_ISL_2663817, EPI_ISL_2663818, EPI_ISL_2663823, EPI_ISL_2663827, EPI_ISL_2663828, EPI_ISL_2663829, EPI_ISL_2663838, EPI_ISL_2663849, EPI_ISL_2663855, EPI_ISL_2663865, EPI_ISL_2663866, EPI_ISL_2663871, EPI_ISL_2663882, EPI_ISL_2663886, EPI_ISL_2663902, EPI_ISL_2663903, EPI_ISL_2663907, EPI_ISL_2663917, EPI_ISL_2663930, EPI_ISL_2663931, EPI_ISL_2663933, EPI_ISL_2663934, EPI_ISL_2663941, EPI_ISL_2663951, EPI_ISL_2663952, EPI_ISL_2663958, EPI_ISL_2663963, EPI_ISL_2663966, EPI_ISL_2663968, EPI_ISL_2663975, EPI_ISL_2663986, EPI_ISL_2663988, EPI_ISL_2664006, EPI_ISL_2664008, EPI_ISL_2664010, EPI_ISL_2681876, EPI_ISL_2681928, EPI_ISL_2681929, EPI_ISL_2681930, EPI_ISL_2681931, EPI_ISL_2681932, EPI_ISL_2681933, EPI_ISL_2681934, EPI_ISL_2681935, EPI_ISL_2681936, EPI_ISL_2681937, EPI_ISL_2681938, EPI_ISL_2681940, EPI_ISL_2681941, EPI_ISL_2681942, EPI_ISL_2681943, EPI_ISL_2681944, EPI_ISL_2681945, EPI_ISL_2681946, EPI_ISL_2681947, EPI_ISL_2681948, EPI_ISL_2681949, EPI_ISL_2681950, EPI_ISL_2681951, EPI_ISL_2681952, EPI_ISL_2681953, EPI_ISL_2681954, EPI_ISL_2681955, EPI_ISL_2681956, EPI_ISL_2681957, EPI_ISL_2681958, EPI_ISL_2681959, EPI_ISL_2681960, EPI_ISL_2681961, EPI_ISL_2681962, EPI_ISL_2681963, EPI_ISL_2681965, EPI_ISL_2681966, EPI_ISL_2681967, EPI_ISL_2681968, EPI_ISL_2681969, EPI_ISL_2681970, EPI_ISL_2681971, EPI_ISL_2681972, EPI_ISL_2681973, EPI_ISL_2681974, EPI_ISL_2681975, EPI_ISL_2681976, EPI_ISL_2681977, EPI_ISL_2681978, EPI_ISL_2681979, EPI_ISL_2681981, EPI_ISL_2681982, EPI_ISL_2681983, EPI_ISL_2681984, EPI_ISL_2681985, EPI_ISL_2681986, EPI_ISL_2681987, EPI_ISL_2681988, EPI_ISL_2681989, EPI_ISL_2681990, EPI_ISL_2681991, EPI_ISL_2681992, EPI_ISL_2681993, EPI_ISL_2681994, EPI_ISL_2681995, EPI_ISL_2681996, EPI_ISL_2681997, EPI_ISL_2681998, EPI_ISL_2681999, EPI_ISL_2682000, EPI_ISL_2682001, EPI_ISL_2682002, EPI_ISL_2682003, EPI_ISL_2682004, EPI_ISL_2682005, EPI_ISL_2682006, EPI_ISL_2682007, EPI_ISL_2682008, EPI_ISL_2682009, EPI_ISL_2682010, EPI_ISL_2682011, EPI_ISL_2682012, EPI_ISL_2682013, EPI_ISL_2682014, EPI_ISL_2682015, EPI_ISL_2682016, EPI_ISL_2682017, EPI_ISL_2682018, EPI_ISL_2682019, EPI_ISL_2682020, EPI_ISL_2682022, EPI_ISL_2682023, EPI_ISL_2682024, EPI_ISL_2682025, EPI_ISL_2682026, EPI_ISL_2682027, EPI_ISL_2682028, EPI_ISL_2682029, EPI_ISL_2682030, EPI_ISL_2682031, EPI_ISL_2682032, EPI_ISL_2682033, EPI_ISL_2682034, EPI_ISL_2682035, EPI_ISL_2682036, EPI_ISL_2682037, EPI_ISL_2682038, EPI_ISL_2682039, EPI_ISL_2682040, EPI_ISL_2682041, EPI_ISL_2682042, EPI_ISL_2682043, EPI_ISL_2682044, EPI_ISL_2682045, EPI_ISL_2682046, EPI_ISL_2682047, EPI_ISL_2682048, EPI_ISL_2682049, EPI_ISL_2682050, EPI_ISL_2682051, EPI_ISL_2682052, EPI_ISL_2682053, EPI_ISL_2682054, EPI_ISL_2682055, EPI_ISL_2682056, EPI_ISL_2682057, EPI_ISL_2682058, EPI_ISL_2682063, EPI_ISL_2682198, EPI_ISL_2683070, EPI_ISL_2683435, EPI_ISL_2683468, EPI_ISL_2683483, EPI_ISL_2701043, EPI_ISL_2701140, EPI_ISL_2701176, EPI_ISL_2701258, EPI_ISL_2718295, EPI_ISL_2718297, EPI_ISL_2718305, EPI_ISL_2718309, EPI_ISL_2718315, EPI_ISL_2718323, EPI_ISL_2718325, EPI_ISL_2718328, EPI_ISL_2718336, EPI_ISL_2718340, EPI_ISL_2718342, EPI_ISL_2718343, EPI_ISL_2718353, EPI_ISL_2718357, EPI_ISL_2718358, EPI_ISL_2718366, EPI_ISL_2718368, EPI_ISL_2718376, EPI_ISL_2718377, EPI_ISL_2718380, EPI_ISL_2718389, EPI_ISL_2718394, EPI_ISL_2718395, EPI_ISL_2718397, EPI_ISL_2718398, EPI_ISL_2718410, EPI_ISL_2718416, EPI_ISL_2718421, EPI_ISL_2718425, EPI_ISL_2718429, EPI_ISL_2718435, EPI_ISL_2718438, EPI_ISL_2718439, EPI_ISL_2718440, EPI_ISL_2718445, EPI_ISL_2718446, EPI_ISL_2718451, EPI_ISL_2718453, EPI_ISL_2718458, EPI_ISL_2718462, EPI_ISL_2718466, EPI_ISL_2718467, EPI_ISL_2718470, EPI_ISL_2718477, EPI_ISL_2718478, EPI_ISL_2718480, EPI_ISL_2718486, EPI_ISL_2718488, EPI_ISL_2718504, EPI_ISL_2718506, EPI_ISL_2718507, EPI_ISL_2718511, EPI_ISL_2718513, EPI_ISL_2718515, EPI_ISL_2718525, EPI_ISL_2731148, EPI_ISL_2731149, EPI_ISL_2731150, EPI_ISL_2731151, EPI_ISL_2731152, EPI_ISL_2731153, EPI_ISL_2731154, EPI_ISL_2731155, EPI_ISL_2731156, EPI_ISL_2731157, EPI_ISL_2731158, EPI_ISL_2731159, EPI_ISL_2731160, EPI_ISL_2731161, EPI_ISL_2731162, EPI_ISL_2731163, EPI_ISL_2731164, EPI_ISL_2731165, EPI_ISL_2731166, EPI_ISL_2731167, EPI_ISL_2731168, EPI_ISL_2731169, EPI_ISL_2731170, EPI_ISL_2731171, EPI_ISL_2731172, EPI_ISL_2731173, EPI_ISL_2731174, EPI_ISL_2731175, EPI_ISL_2731176, EPI_ISL_2731177, EPI_ISL_2731178, EPI_ISL_2731179, EPI_ISL_2731180, EPI_ISL_2731181, EPI_ISL_2731182, EPI_ISL_2731183, EPI_ISL_2731184, EPI_ISL_2731185, EPI_ISL_2731186, EPI_ISL_2731188, EPI_ISL_2731189, EPI_ISL_2731190, EPI_ISL_2731191, EPI_ISL_2731192, EPI_ISL_2731193, EPI_ISL_2731194, EPI_ISL_2731195, EPI_ISL_2731196, EPI_ISL_2731197, EPI_ISL_2731198, EPI_ISL_2731199, EPI_ISL_2731200, EPI_ISL_2731201, EPI_ISL_2731202, EPI_ISL_2731203, EPI_ISL_2731204, EPI_ISL_2731205, EPI_ISL_2731206, EPI_ISL_2731207, EPI_ISL_2731208, EPI_ISL_2731209, EPI_ISL_2731210, EPI_ISL_2731211, EPI_ISL_2731212, EPI_ISL_2731213, EPI_ISL_2731214, EPI_ISL_2731215, EPI_ISL_2731216, EPI_ISL_2731217, EPI_ISL_2731218, EPI_ISL_2731219, EPI_ISL_2731220, EPI_ISL_2731221, EPI_ISL_2731222, EPI_ISL_2731223, EPI_ISL_2731224, EPI_ISL_2731225, EPI_ISL_2731226, EPI_ISL_2731227, EPI_ISL_2731228, EPI_ISL_2731229, EPI_ISL_2731230, EPI_ISL_2731231, EPI_ISL_2731232, EPI_ISL_2731233, EPI_ISL_2731234, EPI_ISL_2731235, EPI_ISL_2731236, EPI_ISL_2731237, EPI_ISL_2731238, EPI_ISL_2731239, EPI_ISL_2731240, EPI_ISL_2731241, EPI_ISL_2731242, EPI_ISL_2731243, EPI_ISL_2731244, EPI_ISL_2731245, EPI_ISL_2731246, EPI_ISL_2731247, EPI_ISL_2731248, EPI_ISL_2731249, EPI_ISL_2731250, EPI_ISL_2731251, EPI_ISL_2731252, EPI_ISL_2731253, EPI_ISL_2731254, EPI_ISL_2731255, EPI_ISL_2731256, EPI_ISL_2731257, EPI_ISL_2731258, EPI_ISL_2731259, EPI_ISL_2731260, EPI_ISL_2731261, EPI_ISL_2731262, EPI_ISL_2731263, EPI_ISL_2731264, EPI_ISL_2731265, EPI_ISL_2731266, EPI_ISL_2731267, EPI_ISL_2731268, EPI_ISL_2731269

| | | | |
|---|---|---|---|
| see above | Lighthouse Lab in Glasgow | Wellcome Sanger Institute for the COVID-19 Genomics UK (COG-UK) Consortium | Anna Dominiczak and Alex Alderton; Carol Clugston; Cordelia Langford; David Gray; David K. Jackson; Dominic Kwiatkowski; Ewan Harrison; Harper VanSteenhouse; Ian Johnston; Jeffrey Barrett; John Sillitoe on behalf of the Wellcome Sanger Institute COVID-19 Surveillance Team; Roberto Amato; Sonia Goncalves; Yumi Kasai |

EPI_ISL_2551587, EPI_ISL_2551601, EPI_ISL_2551604, EPI_ISL_2551621, EPI_ISL_2551624, EPI_ISL_2551628, EPI_ISL_2551632, EPI_ISL_2551650, EPI_ISL_2551667, EPI_ISL_2551671, EPI_ISL_2551676, EPI_ISL_2551682, EPI_ISL_2551699, EPI_ISL_2551707, EPI_ISL_2551711, EPI_ISL_2551712, EPI_ISL_2551740, EPI_ISL_2551747, EPI_ISL_2551757, EPI_ISL_2551758, EPI_ISL_2551806, EPI_ISL_2551819, EPI_ISL_2551821, EPI_ISL_2551829, EPI_ISL_2551833, EPI_ISL_2551841, EPI_ISL_2551850, EPI_ISL_2551862, EPI_ISL_2551869, EPI_ISL_2551872, EPI_ISL_2551901, EPI_ISL_2551909, EPI_ISL_2551921, EPI_ISL_2552018, EPI_ISL_2552019, EPI_ISL_2552022, EPI_ISL_2552023, EPI_ISL_2552024, EPI_ISL_2552025, EPI_ISL_2552026, EPI_ISL_2552028, EPI_ISL_2552029, EPI_ISL_2552030, EPI_ISL_2552033, EPI_ISL_2552034, EPI_ISL_2552035, EPI_ISL_2552036, EPI_ISL_2552037, EPI_ISL_2552038, EPI_ISL_2552040, EPI_ISL_2552041, EPI_ISL_2552042, EPI_ISL_2552043, EPI_ISL_2552044, EPI_ISL_2552045, EPI_ISL_2552046, EPI_ISL_2552047, EPI_ISL_2552048, EPI_ISL_2552049, EPI_ISL_2552050, EPI_ISL_2552053, EPI_ISL_2552054, EPI_ISL_2552055, EPI_ISL_2552056, EPI_ISL_2552057, EPI_ISL_2552059, EPI_ISL_2552061, EPI_ISL_2552063, EPI_ISL_2552065, EPI_ISL_2552068, EPI_ISL_2552071, EPI_ISL_2552073, EPI_ISL_2552074, EPI_ISL_2552075, EPI_ISL_2552076, EPI_ISL_2552077, EPI_ISL_2552078, EPI_ISL_2552080, EPI_ISL_2552082, EPI_ISL_2552084, EPI_ISL_2552086, EPI_ISL_2552087, EPI_ISL_2552088, EPI_ISL_2552089, EPI_ISL_2552091, EPI_ISL_2552092, EPI_ISL_2552093, EPI_ISL_2552096, EPI_ISL_2552097, EPI_ISL_2552099, EPI_ISL_2552100, EPI_ISL_2552101, EPI_ISL_2552102, EPI_ISL_2552103, EPI_ISL_2552104, EPI_ISL_2552107, EPI_ISL_2552108, EPI_ISL_2552109, EPI_ISL_2552110, EPI_ISL_2552113, EPI_ISL_2552115, EPI_ISL_2552117, EPI_ISL_2552118, EPI_ISL_2552119, EPI_ISL_2552120, EPI_ISL_2552121, EPI_ISL_2552122, EPI_ISL_2552126, EPI_ISL_2552127, EPI_ISL_2552128, EPI_ISL_2552129, EPI_ISL_2552130, EPI_ISL_2552131, EPI_ISL_2552134, EPI_ISL_2552136, EPI_ISL_2552137, EPI_ISL_2552138, EPI_ISL_2552144, EPI_ISL_2552147, EPI_ISL_2552148, EPI_ISL_2552150, EPI_ISL_2552151, EPI_ISL_2552152, EPI_ISL_2552153, EPI_ISL_2552155, EPI_ISL_2552156, EPI_ISL_2552158, EPI_ISL_2552159, EPI_ISL_2552160, EPI_ISL_2552162, EPI_ISL_2552163, EPI_ISL_2552164, EPI_ISL_2552165, EPI_ISL_2552167, EPI_ISL_2552169, EPI_ISL_2552172, EPI_ISL_2552173, EPI_ISL_2552176, EPI_ISL_2552177, EPI_ISL_2552178, EPI_ISL_2552180, EPI_ISL_2552181, EPI_ISL_2552182, EPI_ISL_2552183, EPI_ISL_2552186, EPI_ISL_2552187, EPI_ISL_2552188, EPI_ISL_2552189, EPI_ISL_2552191, EPI_ISL_2552194, EPI_ISL_2552195, EPI_ISL_2552196, EPI_ISL_2552198, EPI_ISL_2552199, EPI_ISL_2552200, EPI_ISL_2552201, EPI_ISL_2552202, EPI_ISL_2552203, EPI_ISL_2552204, EPI_ISL_2552205, EPI_ISL_2552206, EPI_ISL_2552207, EPI_ISL_2552209, EPI_ISL_2552210, EPI_ISL_2552211, EPI_ISL_2552212, EPI_ISL_2552213, EPI_ISL_2552215, EPI_ISL_2552217, EPI_ISL_2552218, EPI_ISL_2552219, EPI_ISL_2552222, EPI_ISL_2552223, EPI_ISL_2552224, EPI_ISL_2552225, EPI_ISL_2552226, EPI_ISL_2552227, EPI_ISL_2552228, EPI_ISL_2552229, EPI_ISL_2552232, EPI_ISL_2552233, EPI_ISL_2552236, EPI_ISL_2552237, EPI_ISL_2552239, EPI_ISL_2552243, EPI_ISL_2552244, EPI_ISL_2552245, EPI_ISL_2552246, EPI_ISL_2552247, EPI_ISL_2552248, EPI_ISL_2552249, EPI_ISL_2552250, EPI_ISL_2552251, EPI_ISL_2552253, EPI_ISL_2552254, EPI_ISL_2552257, EPI_ISL_2552258, EPI_ISL_2552259, EPI_ISL_2553795, EPI_ISL_2553796, EPI_ISL_2553798, EPI_ISL_2553808, EPI_ISL_2553818, EPI_ISL_2553823, EPI_ISL_2553828, EPI_ISL_2553829, EPI_ISL_2553835, EPI_ISL_2553840, EPI_ISL_2553846, EPI_ISL_2553852, EPI_ISL_2553853, EPI_ISL_2553859, EPI_ISL_2553863, EPI_ISL_2553873, EPI_ISL_2553880, EPI_ISL_2553885, EPI_ISL_2553886, EPI_ISL_2553891, EPI_ISL_2553905, EPI_ISL_2553919, EPI_ISL_2553923, EPI_ISL_2553944, EPI_ISL_2553947, EPI_ISL_2553958, EPI_ISL_2553964, EPI_ISL_2553972, EPI_ISL_2553973, EPI_ISL_2553975, EPI_ISL_2553988, EPI_ISL_2553998, EPI_ISL_2554004, EPI_ISL_2554008, EPI_ISL_2554017, EPI_ISL_2554024, EPI_ISL_2554025, EPI_ISL_2554027, EPI_ISL_2554030, EPI_ISL_2554036, EPI_ISL_2554038, EPI_ISL_2554049, EPI_ISL_2554050, EPI_ISL_2554051, EPI_ISL_2554059, EPI_ISL_2554077, EPI_ISL_2554078, EPI_ISL_2554114, EPI_ISL_2554115, EPI_ISL_2554133, EPI_ISL_2554137, EPI_ISL_2554139, EPI_ISL_2554493, EPI_ISL_2554494, EPI_ISL_2554497, EPI_ISL_2554498, EPI_ISL_2554501, EPI_ISL_2554502, EPI_ISL_2554504, EPI_ISL_2554505, EPI_ISL_2554506, EPI_ISL_2554510, EPI_ISL_2554513, EPI_ISL_2554514, EPI_ISL_2554519, EPI_ISL_2554521, EPI_ISL_2554523, EPI_ISL_2554524, EPI_ISL_2554526, EPI_ISL_2554530, EPI_ISL_2554532, EPI_ISL_2554533, EPI_ISL_2554534, EPI_ISL_2554535, EPI_ISL_2554536, EPI_ISL_2554539, EPI_ISL_2554540, EPI_ISL_2554543, EPI_ISL_2554544, EPI_ISL_2554545, EPI_ISL_2554550, EPI_ISL_2554551, EPI_ISL_2554552, EPI_ISL_2554553, EPI_ISL_2554555, EPI_ISL_2554556, EPI_ISL_2554557, EPI_ISL_2554558, EPI_ISL_2554563, EPI_ISL_2554568, EPI_ISL_2554570, EPI_ISL_2554577, EPI_ISL_2554579, EPI_ISL_2554580, EPI_ISL_2554582, EPI_ISL_2554583, EPI_ISL_2554585, EPI_ISL_2554586, EPI_ISL_2554587, EPI_ISL_2554589, EPI_ISL_2554591, EPI_ISL_2554594, EPI_ISL_2554595, EPI_ISL_2554596, EPI_ISL_2554599, EPI_ISL_2554605, EPI_ISL_2554606, EPI_ISL_2554607, EPI_ISL_2554609, EPI_ISL_2554610, EPI_ISL_2554612, EPI_ISL_2554614, EPI_ISL_2554621, EPI_ISL_2554623, EPI_ISL_2554624, EPI_ISL_2554628, EPI_ISL_2554629, EPI_ISL_2554631, EPI_ISL_2554633, EPI_ISL_2554635, EPI_ISL_2554638, EPI_ISL_2554639, EPI_ISL_2554640, EPI_ISL_2554641, EPI_ISL_2554644, EPI_ISL_2554647, EPI_ISL_2554648, EPI_ISL_2554653, EPI_ISL_2554655, EPI_ISL_2554659, EPI_ISL_2554660, EPI_ISL_2554668, EPI_ISL_2554669, EPI_ISL_2554670, EPI_ISL_2554675, EPI_ISL_2554676, EPI_ISL_2554678, EPI_ISL_2554680, EPI_ISL_2554684, EPI_ISL_2554685, EPI_ISL_2554687, EPI_ISL_2554689, EPI_ISL_2554693, EPI_ISL_2554694, EPI_ISL_2554696, EPI_ISL_2554700, EPI_ISL_2554701, EPI_ISL_2554704, EPI_ISL_2554708, EPI_ISL_2554709, EPI_ISL_2554710, EPI_ISL_2554711, EPI_ISL_2554712, EPI_ISL_2554715, EPI_ISL_2554720, EPI_ISL_2554724, EPI_ISL_2554725, EPI_ISL_2554726, EPI_ISL_2554727, EPI_ISL_2554728, EPI_ISL_2554733, EPI_ISL_2554734, EPI_ISL_2554735, EPI_ISL_2554738, EPI_ISL_2554739, EPI_ISL_2554740, EPI_ISL_2554742, EPI_ISL_2554746, EPI_ISL_2554747, EPI_ISL_2554752, EPI_ISL_2554753, EPI_ISL_2554755, EPI_ISL_2554757, EPI_ISL_2554759, EPI_ISL_2554760, EPI_ISL_2554761, EPI_ISL_2554762, EPI_ISL_2554764, EPI_ISL_2554765, EPI_ISL_2554766, EPI_ISL_2554767, EPI_ISL_2554768, EPI_ISL_2554769, EPI_ISL_2554771, EPI_ISL_2554772, EPI_ISL_2554773, EPI_ISL_2554775, EPI_ISL_2554776, EPI_ISL_2554778, EPI_ISL_2554780, EPI_ISL_2554781, EPI_ISL_2554783, EPI_ISL_2554790, EPI_ISL_2554791, EPI_ISL_2554793, EPI_ISL_2554794, EPI_ISL_2554795, EPI_ISL_2554799, EPI_ISL_2554800, EPI_ISL_2554805, EPI_ISL_2554806, EPI_ISL_2554808, EPI_ISL_2554815, EPI_ISL_2554816, EPI_ISL_2554817, EPI_ISL_2554818, EPI_ISL_2554821, EPI_ISL_2554828, EPI_ISL_2554829, EPI_ISL_2554831, EPI_ISL_2554832, EPI_ISL_2554833, EPI_ISL_2554835, EPI_ISL_2554837, EPI_ISL_2554844, EPI_ISL_2568109, EPI_ISL_2568113, EPI_ISL_2568114, EPI_ISL_2568116, EPI_ISL_2568120, EPI_ISL_2568121, EPI_ISL_2568123, EPI_ISL_2568127, EPI_ISL_2568130, EPI_ISL_2568131, EPI_ISL_2568132, EPI_ISL_2568144, EPI_ISL_2568146, EPI_ISL_2568148, EPI_ISL_2568149, EPI_ISL_2568156, EPI_ISL_2568157, EPI_ISL_2568167, EPI_ISL_2568171, EPI_ISL_2568175, EPI_ISL_2568176, EPI_ISL_2568178, EPI_ISL_2568182, EPI_ISL_2568188, EPI_ISL_2568190, EPI_ISL_2568191, EPI_ISL_2568192, EPI_ISL_2568194, EPI_ISL_2568195, EPI_ISL_2568202, EPI_ISL_2568207, EPI_ISL_2568210, EPI_ISL_2568218, EPI_ISL_2568222, EPI_ISL_2568227, EPI_ISL_2568228, EPI_ISL_2568229, EPI_ISL_2568230, EPI_ISL_2568233, EPI_ISL_2568235, EPI_ISL_2568236, EPI_ISL_2568243, EPI_ISL_2568244, EPI_ISL_2568247, EPI_ISL_2568249, EPI_ISL_2568251, EPI_ISL_2568253, EPI_ISL_2568256, EPI_ISL_2568257, EPI_ISL_2568258, EPI_ISL_2568263, EPI_ISL_2568264, EPI_ISL_2568265, EPI_ISL_2568271, EPI_ISL_2568274, EPI_ISL_2568275, EPI_ISL_2568277, EPI_ISL_2568278, EPI_ISL_2568281, EPI_ISL_2568284, EPI_ISL_2568286, EPI_ISL_2568287, EPI_ISL_2568290, EPI_ISL_2568296, EPI_ISL_2568297, EPI_ISL_2568299, EPI_ISL_2568301, EPI_ISL_2568302, EPI_ISL_2568309, EPI_ISL_2568311, EPI_ISL_2568318, EPI_ISL_2568322, EPI_ISL_2568324, EPI_ISL_2568325, EPI_ISL_2568326, EPI_ISL_2568329, EPI_ISL_2568337, EPI_ISL_2568352, EPI_ISL_2568353, EPI_ISL_2568362, EPI_ISL_2568366, EPI_ISL_2568367, EPI_ISL_2568370, EPI_ISL_2568372, EPI_ISL_2568386, EPI_ISL_2568387, EPI_ISL_2568388, EPI_ISL_2568390, EPI_ISL_2568393, EPI_ISL_2568396, EPI_ISL_2568397, EPI_ISL_2568658, EPI_ISL_2568659, EPI_ISL_2568662, EPI_ISL_2568669, EPI_ISL_2568670, EPI_ISL_2568671, EPI_ISL_2568676, EPI_ISL_2568679, EPI_ISL_2568681, EPI_ISL_2568682, EPI_ISL_2568683, EPI_ISL_2568685, EPI_ISL_2568686, EPI_ISL_2568687, EPI_ISL_2568692, EPI_ISL_2568695, EPI_ISL_2568697, EPI_ISL_2568699, EPI_ISL_2568700, EPI_ISL_2568701, EPI_ISL_2568702, EPI_ISL_2568703, EPI_ISL_2568705, EPI_ISL_2568706, EPI_ISL_2568707, EPI_ISL_2568709, EPI_ISL_2568710, EPI_ISL_2568712, EPI_ISL_2568713, EPI_ISL_2568714, EPI_ISL_2568717, EPI_ISL_2568718, EPI_ISL_2568722, EPI_ISL_2568723, EPI_ISL_2568726, EPI_ISL_2568727, EPI_ISL_2568731, EPI_ISL_2568732, EPI_ISL_2568733, EPI_ISL_2568734, EPI_ISL_2568735, EPI_ISL_2568736, EPI_ISL_2568737, EPI_ISL_2568738, EPI_ISL_2568742, EPI_ISL_2568743, EPI_ISL_2568745, EPI_ISL_2568747, EPI_ISL_2568748, EPI_ISL_2568749, EPI_ISL_2568750, EPI_ISL_2568754, EPI_ISL_2568755, EPI_ISL_2568757, EPI_ISL_2568759, EPI_ISL_2568762, EPI_ISL_2568763, EPI_ISL_2568765, EPI_ISL_2568766, EPI_ISL_2568768, EPI_ISL_2568769, EPI_ISL_2568770, EPI_ISL_2568771, EPI_ISL_2568772, EPI_ISL_2568773, EPI_ISL_2568774, EPI_ISL_2568777, EPI_ISL_2568778, EPI_ISL_2568781, EPI_ISL_2568783, EPI_ISL_2568786, EPI_ISL_2568788, EPI_ISL_2568790, EPI_ISL_2568791, EPI_ISL_2568793, EPI_ISL_2568794, EPI_ISL_2568795, EPI_ISL_2568798, EPI_ISL_2568799, EPI_ISL_2568800, EPI_ISL_2568803, EPI_ISL_2568805, EPI_ISL_2568809, EPI_ISL_2568813, EPI_ISL_2568814, EPI_ISL_2568816, EPI_ISL_2568817, EPI_ISL_2568818, EPI_ISL_2568819, EPI_ISL_2568821, EPI_ISL_2568822, EPI_ISL_2568824, EPI_ISL_2568825, EPI_ISL_2568826, EPI_ISL_2568827, EPI_ISL_2568828, EPI_ISL_2568830, EPI_ISL_2568831, EPI_ISL_2568833, EPI_ISL_2568834, EPI_ISL_2568835, EPI_ISL_2568836, EPI_ISL_2568838, EPI_ISL_2568843, EPI_ISL_2568848, EPI_ISL_2568853, EPI_ISL_2568857, EPI_ISL_2568858, EPI_ISL_2568859, EPI_ISL_2568860, EPI_ISL_2568861, EPI_ISL_2568862, EPI_ISL_2568865, EPI_ISL_2568866, EPI_ISL_2568867, EPI_ISL_2568871, EPI_ISL_2568875, EPI_ISL_2568880, EPI_ISL_2568882, EPI_ISL_2568885, EPI_ISL_2568886, EPI_ISL_2568888, EPI_ISL_2568893, EPI_ISL_2568899, EPI_ISL_2568902, EPI_ISL_2568908, EPI_ISL_2568909, EPI_ISL_2568910, EPI_ISL_2568919, EPI_ISL_2568922, EPI_ISL_2568925, EPI_ISL_2568927, EPI_ISL_2568935, EPI_ISL_2568938, EPI_ISL_2568942, EPI_ISL_2568943, EPI_ISL_2568946, EPI_ISL_2568947, EPI_ISL_2568951, EPI_ISL_2568953, EPI_ISL_2568954, EPI_ISL_2568955, EPI_ISL_2568957, EPI_ISL_2568964, EPI_ISL_2568966, EPI_ISL_2568968, EPI_ISL_2568969, EPI_ISL_2568971, EPI_ISL_2568972, EPI_ISL_2568974, EPI_ISL_2568975, EPI_ISL_2568977, EPI_ISL_2568980, EPI_ISL_2568983, EPI_ISL_2568984, EPI_ISL_2568985, EPI_ISL_2568990, EPI_ISL_2568991, EPI_ISL_2568992, EPI_ISL_2568993, EPI_ISL_2568995, EPI_ISL_2568996, EPI_ISL_2568997, EPI_ISL_2568998, EPI_ISL_2569004, EPI_ISL_2569007, EPI_ISL_2569008, EPI_ISL_2569009, EPI_ISL_2569018, EPI_ISL_2569024, EPI_ISL_2569034, EPI_ISL_2569042, EPI_ISL_2569047, EPI_ISL_2569053, EPI_ISL_2569068, EPI_ISL_2569071, EPI_ISL_2569074, EPI_ISL_2569079, EPI_ISL_2569081, EPI_ISL_2569082, EPI_ISL_2569093, EPI_ISL_2569101, EPI_ISL_2569108, EPI_ISL_2569109, EPI_ISL_2569110, EPI_ISL_2569119, EPI_ISL_2569136, EPI_ISL_2569140, EPI_ISL_2569141, EPI_ISL_2569145, EPI_ISL_2569148, EPI_ISL_2569176, EPI_ISL_2569182, EPI_ISL_2569190, EPI_ISL_2569197, EPI_ISL_2569208, EPI_ISL_2569231, EPI_ISL_2569234, EPI_ISL_2569256, EPI_ISL_2569263, EPI_ISL_2569270, EPI_ISL_2569287, EPI_ISL_2569297, EPI_ISL_2569298, EPI_ISL_2569300, EPI_ISL_2569304, EPI_ISL_2569663, EPI_ISL_2569666, EPI_ISL_2569667, EPI_ISL_2569668, EPI_ISL_2569670, EPI_ISL_2569671, EPI_ISL_2569672, EPI_ISL_2569676, EPI_ISL_2569679, EPI_ISL_2569680, EPI_ISL_2569681, EPI_ISL_2569683, EPI_ISL_2569685, EPI_ISL_2569686, EPI_ISL_2569687, EPI_ISL_2569691, EPI_ISL_2569693, EPI_ISL_2569700, EPI_ISL_2569701, EPI_ISL_2569702, EPI_ISL_2569704, EPI_ISL_2569712, EPI_ISL_2569713, EPI_ISL_2569715, EPI_ISL_2569717, EPI_ISL_2569718, EPI_ISL_2569720, EPI_ISL_2569722, EPI_ISL_2569730, EPI_ISL_2569734, EPI_ISL_2569737, EPI_ISL_2569738, EPI_ISL_2569741, EPI_ISL_2569742, EPI_ISL_2569743, EPI_ISL_2569745, EPI_ISL_2569746, EPI_ISL_2569748, EPI_ISL_2569754, EPI_ISL_2569755, EPI_ISL_2569756, EPI_ISL_2569760, EPI_ISL_2569761, EPI_ISL_2569765, EPI_ISL_2569767, EPI_ISL_2569774, EPI_ISL_2569775, EPI_ISL_2569779, EPI_ISL_2569784, EPI_ISL_2569787, EPI_ISL_2569790, EPI_ISL_2569791, EPI_ISL_2569793, EPI_ISL_2569794, EPI_ISL_2569798, EPI_ISL_2569800, EPI_ISL_2569808, EPI_ISL_2569809, EPI_ISL_2569810, EPI_ISL_2569812, EPI_ISL_2569815, EPI_ISL_2569818, EPI_ISL_2569826, EPI_ISL_2569828, EPI_ISL_2569830, EPI_ISL_2569832, EPI_ISL_2569833, EPI_ISL_2569835, EPI_ISL_2569840, EPI_ISL_2569842, EPI_ISL_2569843, EPI_ISL_2569846, EPI_ISL_2569848, EPI_ISL_2569853, EPI_ISL_2569855, EPI_ISL_2569861, EPI_ISL_2569862, EPI_ISL_2569863, EPI_ISL_2569871, EPI_ISL_2569872, EPI_ISL_2569875, EPI_ISL_2569876, EPI_ISL_2569877, EPI_ISL_2569879, EPI_ISL_2569882, EPI_ISL_2569884, EPI_ISL_2569885, EPI_ISL_2569892, EPI_ISL_2569897, EPI_ISL_2569898, EPI_ISL_2569899, EPI_ISL_2569903, EPI_ISL_2569909, EPI_ISL_2569912, EPI_ISL_2569914, EPI_ISL_2569917, EPI_ISL_2569919, EPI_ISL_2569921, EPI_ISL_2569923, EPI_ISL_2569924, EPI_ISL_2569925, EPI_ISL_2569926, EPI_ISL_2569935, EPI_ISL_2569943, EPI_ISL_2569950, EPI_ISL_2569956, EPI_ISL_2569969, EPI_ISL_2569976, EPI_ISL_2569982, EPI_ISL_2569995, EPI_ISL_2569999, EPI_ISL_2570004, EPI_ISL_2570009, EPI_ISL_2570016, EPI_ISL_2570022, EPI_ISL_2570026, EPI_ISL_2570028, EPI_ISL_2570032, EPI_ISL_2570033, EPI_ISL_2570039, EPI_ISL_2570044, EPI_ISL_2570047, EPI_ISL_2570051, EPI_ISL_2570064, EPI_ISL_2570072, EPI_ISL_2570073, EPI_ISL_2570075, EPI_ISL_2570085, EPI_ISL_2570093, EPI_ISL_2570096, EPI_ISL_2570099, EPI_ISL_2570104, EPI_ISL_2570109, EPI_ISL_2570122, EPI_ISL_2570124, EPI_ISL_2570131, EPI_ISL_2570140, EPI_ISL_2570144, EPI_ISL_2570152, EPI_ISL_2570153, EPI_ISL_2570156, EPI_ISL_2570166, EPI_ISL_2570167, EPI_ISL_2570169, EPI_ISL_2570174, EPI_ISL_2570176, EPI_ISL_2570180, EPI_ISL_2570182, EPI_ISL_2570184, EPI_ISL_2570185, EPI_ISL_2570186, EPI_ISL_2570187, EPI_ISL_2570197, EPI_ISL_2570205, EPI_ISL_2570208, EPI_ISL_2570212, EPI_ISL_2570236, EPI_ISL_2570237, EPI_ISL_2570242, EPI_ISL_2570246, EPI_ISL_2605766, EPI_ISL_2606023, EPI_ISL_2606453, EPI_ISL_2606454, EPI_ISL_2606455, EPI_ISL_2606458, EPI_ISL_2606459, EPI_ISL_2606460, EPI_ISL_2606462, EPI_ISL_2606464, EPI_ISL_2606466, EPI_ISL_2606467, EPI_ISL_2606468, EPI_ISL_2606471, EPI_ISL_2606472, EPI_ISL_2606473, EPI_ISL_2606476, EPI_ISL_2606477, EPI_ISL_2606478, EPI_ISL_2606480, EPI_ISL_2606483, EPI_ISL_2606486, EPI_ISL_2606488, EPI_ISL_2606489, EPI_ISL_2606490, EPI_ISL_2606491, EPI_ISL_2606492, EPI_ISL_2606493, EPI_ISL_2606495, EPI_ISL_2606499, EPI_ISL_2606501, EPI_ISL_2606502, EPI_ISL_2606503, EPI_ISL_2606507, EPI_ISL_2606508, EPI_ISL_2606511, EPI_ISL_2606513, EPI_ISL_2606514, EPI_ISL_2606516, EPI_ISL_2606518, EPI_ISL_2606519, EPI_ISL_2606522, EPI_ISL_2606523, EPI_ISL_2606527, EPI_ISL_2606528, EPI_ISL_2606529, EPI_ISL_2606530, EPI_ISL_2606531, EPI_ISL_2606532, EPI_ISL_2606533, EPI_ISL_2606534, EPI_ISL_2606536, EPI_ISL_2606537, EPI_ISL_2606539, EPI_ISL_2606541, EPI_ISL_2606542, EPI_ISL_2606543, EPI_ISL_2606544, EPI_ISL_2606545, EPI_ISL_2606546, EPI_ISL_2606548, EPI_ISL_2606551, EPI_ISL_2606554, EPI_ISL_2606555, EPI_ISL_2606556, EPI_ISL_2606559, EPI_ISL_2606560, EPI_ISL_2606561, EPI_ISL_2606562, EPI_ISL_2606563, EPI_ISL_2606564, EPI_ISL_2606566, EPI_ISL_2606567, EPI_ISL_2606570, EPI_ISL_2606574, EPI_ISL_2606575, EPI_ISL_2606578, EPI_ISL_2606580, EPI_ISL_2606581, EPI_ISL_2606582, EPI_ISL_2606584, EPI_ISL_2606585, EPI_ISL_2606587, EPI_ISL_2606591, EPI_ISL_2606594, EPI_ISL_2606596, EPI_ISL_2606598, EPI_ISL_2606599, EPI_ISL_2606600, EPI_ISL_2606601, EPI_ISL_2606603, EPI_ISL_2606604, EPI_ISL_2606606, EPI_ISL_2606607, EPI_ISL_2606608, EPI_ISL_2606609, EPI_ISL_2606610, EPI_ISL_2606612, EPI_ISL_2606613, EPI_ISL_2606614, EPI_ISL_2606615, EPI_ISL_2606616, EPI_ISL_2606617, EPI_ISL_2606618, EPI_ISL_2606620, EPI_ISL_2606621, EPI_ISL_2606622, EPI_ISL_2606623, EPI_ISL_2606626, EPI_ISL_2606627, EPI_ISL_2606628, EPI_ISL_2606629, EPI_ISL_2606631, EPI_ISL_2606632, EPI_ISL_2606633, EPI_ISL_2606634, EPI_ISL_2606638, EPI_ISL_2606639, EPI_ISL_2606641, EPI_ISL_2606642, EPI_ISL_2606643, EPI_ISL_2606644, EPI_ISL_2606646, EPI_ISL_2606648, EPI_ISL_2606649, EPI_ISL_2606650, EPI_ISL_2606651, EPI_ISL_2606653, EPI_ISL_2606656, EPI_ISL_2606658, EPI_ISL_2606659, EPI_ISL_2606660, EPI_ISL_2606661, EPI_ISL_2606662, EPI_ISL_2606663, EPI_ISL_2606666, EPI_ISL_2606667, EPI_ISL_2606668, EPI_ISL_2606670, EPI_ISL_2606672, EPI_ISL_2606675, EPI_ISL_2606676, EPI_ISL_2606677, EPI_ISL_2606679, EPI_ISL_2606680, EPI_ISL_2606681, EPI_ISL_2606683, EPI_ISL_2606684, EPI_ISL_2606687, EPI_ISL_2606688, EPI_ISL_2606689, EPI_ISL_2606692, EPI_ISL_2606693, EPI_ISL_2606694, EPI_ISL_2606696, EPI_ISL_2606698, EPI_ISL_2606699, EPI_ISL_2606701, EPI_ISL_2606702, EPI_ISL_2606703, EPI_ISL_2606704, EPI_ISL_2606705, EPI_ISL_2606707, EPI_ISL_2606710, EPI_ISL_2606711, EPI_ISL_2606714, EPI_ISL_2606718, EPI_ISL_2606719, EPI_ISL_2606720, EPI_ISL_2606722, EPI_ISL_2606724, EPI_ISL_2606725, EPI_ISL_2606726, EPI_ISL_2606731, EPI_ISL_2606732, EPI_ISL_2606733, EPI_ISL_2606741, EPI_ISL_2606742, EPI_ISL_2606744, EPI_ISL_2606745, EPI_ISL_2606747, EPI_ISL_2606748, EPI_ISL_2606749, EPI_ISL_2606750, EPI_ISL_2606751, EPI_ISL_2606753, EPI_ISL_2606756, EPI_ISL_2606767, EPI_ISL_2606781, EPI_ISL_2606785, EPI_ISL_2606787, EPI_ISL_2606789, EPI_ISL_2606791, EPI_ISL_2606794, EPI_ISL_2606797, EPI_ISL_2606804, EPI_ISL_2606805, EPI_ISL_2606816, EPI_ISL_2606817, EPI_ISL_2606821, EPI_ISL_2606827, EPI_ISL_2606835, EPI_ISL_2606842, EPI_ISL_2606844, EPI_ISL_2606847, EPI_ISL_2606849, EPI_ISL_2606850, EPI_ISL_2606851, EPI_ISL_2606852, EPI_ISL_2606859, EPI_ISL_2606860, EPI_ISL_2606865, EPI_ISL_2606870, EPI_ISL_2606872, EPI_ISL_2606878, EPI_ISL_2606879, EPI_ISL_2606884, EPI_ISL_2606887, EPI_ISL_2606892, EPI_ISL_2606898, EPI_ISL_2606902, EPI_ISL_2606908, EPI_ISL_2606910, EPI_ISL_2606913, EPI_ISL_2606926, EPI_ISL_2606928, EPI_ISL_2606929, EPI_ISL_2606932, EPI_ISL_2606933, EPI_ISL_2606935, EPI_ISL_2606939, EPI_ISL_2606941, EPI_ISL_2606943, EPI_ISL_2606947, EPI_ISL_2606951, EPI_ISL_2606952, EPI_ISL_2606953, EPI_ISL_2606955, EPI_ISL_2606963, EPI_ISL_2606964, EPI_ISL_2606966, EPI_ISL_2606969, EPI_ISL_2606974, EPI_ISL_2606987, EPI_ISL_2606997, EPI_ISL_2607000, EPI_ISL_2607002, EPI_ISL_2607004, EPI_ISL_2607010, EPI_ISL_2607012, EPI_ISL_2607017, EPI_ISL_2607019, EPI_ISL_2607020, EPI_ISL_2607023, EPI_ISL_2607024, EPI_ISL_2607027, EPI_ISL_2607032, EPI_ISL_2607037, EPI_ISL_2607039, EPI_ISL_2607041, EPI_ISL_2607100, EPI_ISL_2607256, EPI_ISL_2607504, EPI_ISL_2607637, EPI_ISL_2607899, EPI_ISL_2607900, EPI_ISL_2607915, EPI_ISL_2638265, EPI_ISL_2638329, EPI_ISL_2638404, EPI_ISL_2638406, EPI_ISL_2638409, EPI_ISL_2638428, EPI_ISL_2638430, EPI_ISL_2638437, EPI_ISL_2638441, EPI_ISL_2638460, EPI_ISL_2638465, EPI_ISL_2638478, EPI_ISL_2638487, EPI_ISL_2638504, EPI_ISL_2638510, EPI_ISL_2638517, EPI_ISL_2638535, EPI_ISL_2638542, EPI_ISL_2638545, EPI_ISL_2638554, EPI_ISL_2638557, EPI_ISL_2638564, EPI_ISL_2638565, EPI_ISL_2638570, EPI_ISL_2638571, EPI_ISL_2638579, EPI_ISL_2638588, EPI_ISL_2638597, EPI_ISL_2638601, EPI_ISL_2638608, EPI_ISL_2638610, EPI_ISL_2638612, EPI_ISL_2638613, EPI_ISL_2638614, EPI_ISL_2638621, EPI_ISL_2638624, EPI_ISL_2638631, EPI_ISL_2638639, EPI_ISL_2638640, EPI_ISL_2638645, EPI_ISL_2638660, EPI_ISL_2638666, EPI_ISL_2638667, EPI_ISL_2638678, EPI_ISL_2638682, EPI_ISL_2638687, EPI_ISL_2638705, EPI_ISL_2638706, EPI_ISL_2638712, EPI_ISL_2638713, EPI_ISL_2638714, EPI_ISL_2638722, EPI_ISL_2638723, EPI_ISL_2638732, EPI_ISL_2638734, EPI_ISL_2638737, EPI_ISL_2638743, EPI_ISL_2638784, EPI_ISL_2638794, EPI_ISL_2638795, EPI_ISL_2638802, EPI_ISL_2638804, EPI_ISL_2638806, EPI_ISL_2638809, EPI_ISL_2638810, EPI_ISL_2638823, EPI_ISL_2638824, EPI_ISL_2638825, EPI_ISL_2638826, EPI_ISL_2638830, EPI_ISL_2638835, EPI_ISL_2638843, EPI_ISL_2638851, EPI_ISL_2638860, EPI_ISL_2638861, EPI_ISL_2638862, EPI_ISL_2638870, EPI_ISL_2638871, EPI_ISL_2638876, EPI_ISL_2638878, EPI_ISL_2638880, EPI_ISL_2638882, EPI_ISL_2638883, EPI_ISL_2638886, EPI_ISL_2638888, EPI_ISL_2638892, EPI_ISL_2638899, EPI_ISL_2638907, EPI_ISL_2638921, EPI_ISL_2638980, EPI_ISL_2638986, EPI_ISL_2638988, EPI_ISL_2638990, EPI_ISL_2638992, EPI_ISL_2638997, EPI_ISL_2639002, EPI_ISL_2639004, EPI_ISL_2639018, EPI_ISL_2639028, EPI_ISL_2639031, EPI_ISL_2639060, EPI_ISL_2639063, EPI_ISL_2639074, EPI_ISL_2639092, EPI_ISL_2639110, EPI_ISL_2639120, EPI_ISL_2639145, EPI_ISL_2639156, EPI_ISL_2639170, EPI_ISL_2639195, EPI_ISL_2639197, EPI_ISL_2639198, EPI_ISL_2639202, EPI_ISL_2639223, EPI_ISL_2639227, EPI_ISL_2639252, EPI_ISL_2640122, EPI_ISL_2640123, EPI_ISL_2640326, EPI_ISL_2651414, EPI_ISL_2651425, EPI_ISL_2651445, EPI_ISL_2651449, EPI_ISL_2651461, EPI_ISL_2651478, EPI_ISL_2651543, EPI_ISL_2651942, EPI_ISL_2651944, EPI_ISL_2663797, EPI_ISL_2663798, EPI_ISL_2663799, EPI_ISL_2663800, EPI_ISL_2663801, EPI_ISL_2663802, EPI_ISL_2663803, EPI_ISL_2663804, EPI_ISL_2663805, EPI_ISL_2663806, EPI_ISL_2663807, EPI_ISL_2663808, EPI_ISL_2663809, EPI_ISL_2663810, EPI_ISL_2663811, EPI_ISL_2663812, EPI_ISL_2663813, EPI_ISL_2663814, EPI_ISL_2663815, EPI_ISL_2663816, EPI_ISL_2663819, EPI_ISL_2663820, EPI_ISL_2663821, EPI_ISL_2663822, EPI_ISL_2663824, EPI_ISL_2663825, EPI_ISL_2663826, EPI_ISL_2663830, EPI_ISL_2663831, EPI_ISL_2663832, EPI_ISL_2663833, EPI_ISL_2663834, EPI_ISL_2663835, EPI_ISL_2663836, EPI_ISL_2663837, EPI_ISL_2663839, EPI_ISL_2663840, EPI_ISL_2663841, EPI_ISL_2663842, EPI_ISL_2663844, EPI_ISL_2663845, EPI_ISL_2663846, EPI_ISL_2663847, EPI_ISL_2663848, EPI_ISL_2663850, EPI_ISL_2663851, EPI_ISL_2663852, EPI_ISL_2663853, EPI_ISL_2663854, EPI_ISL_2663856, EPI_ISL_2663857, EPI_ISL_2663858, EPI_ISL_2663859, EPI_ISL_2663860, EPI_ISL_2663861, EPI_ISL_2663862, EPI_ISL_2663863, EPI_ISL_2663864, EPI_ISL_2663867, EPI_ISL_2663868, EPI_ISL_2663869, EPI_ISL_2663870, EPI_ISL_2663872, EPI_ISL_2663873, EPI_ISL_2663874, EPI_ISL_2663875, EPI_ISL_2663876, EPI_ISL_2663877, EPI_ISL_2663878, EPI_ISL_2663879, EPI_ISL_2663880, EPI_ISL_2663881, EPI_ISL_2663883, EPI_ISL_2663884, EPI_ISL_2663885, EPI_ISL_2663887, EPI_ISL_2663888, EPI_ISL_2663889, EPI_ISL_2663890, EPI_ISL_2663891, EPI_ISL_2663892, EPI_ISL_2663893, EPI_ISL_2663894, EPI_ISL_2663895, EPI_ISL_2663896, EPI_ISL_2663897, EPI_ISL_2663898, EPI_ISL_2663899, EPI_ISL_2663900, EPI_ISL_2663901, EPI_ISL_2663904, EPI_ISL_2663905, EPI_ISL_2663906, EPI_ISL_2663908, EPI_ISL_2663909, EPI_ISL_2663910, EPI_ISL_2663911, EPI_ISL_2663912, EPI_ISL_2663913, EPI_ISL_2663914, EPI_ISL_2663915, EPI_ISL_2663916, EPI_ISL_2663918, EPI_ISL_2663919, EPI_ISL_2663920, EPI_ISL_2663921, EPI_ISL_2663922, EPI_ISL_2663923, EPI_ISL_2663924, EPI_ISL_2663925, EPI_ISL_2663926, EPI_ISL_2663927, EPI_ISL_2663928, EPI_ISL_2663929, EPI_ISL_2663932, EPI_ISL_2663935, EPI_ISL_2663936, EPI_ISL_2663937, EPI_ISL_2663938, EPI_ISL_2663939, EPI_ISL_2663940, EPI_ISL_2663942, EPI_ISL_2663943, EPI_ISL_2663944, EPI_ISL_2663945, EPI_ISL_2663946, EPI_ISL_2663947, EPI_ISL_2663948, EPI_ISL_2663949, EPI_ISL_2663950, EPI_ISL_2663953, EPI_ISL_2663954, EPI_ISL_2663955, EPI_ISL_2663956, EPI_ISL_2663957, EPI_ISL_2663959, EPI_ISL_2663960, EPI_ISL_2663961, EPI_ISL_2663962, EPI_ISL_2663964, EPI_ISL_2663965, EPI_ISL_2663967, EPI_ISL_2663969, EPI_ISL_2663970, EPI_ISL_2663971, EPI_ISL_2663972, EPI_ISL_2663973, EPI_ISL_2663974, EPI_ISL_2663976, EPI_ISL_2663977, EPI_ISL_2663978, EPI_ISL_2663979, EPI_ISL_2663980, EPI_ISL_2663981, EPI_ISL_2663982, EPI_ISL_2663983, EPI_ISL_2663984, EPI_ISL_2663985, EPI_ISL_2663987, EPI_ISL_2663989, EPI_ISL_2663990, EPI_ISL_2663991, EPI_ISL_2663992, EPI_ISL_2663993, EPI_ISL_2663994, EPI_ISL_2663995, EPI_ISL_2663996, EPI_ISL_2663997, EPI_ISL_2663998, EPI_ISL_2663999, EPI_ISL_2664000, EPI_ISL_2664001, EPI_ISL_2664002, EPI_ISL_2664003, EPI_ISL_2664004, EPI_ISL_2664005, EPI_ISL_2664007, EPI_ISL_2664009, EPI_ISL_2664423, EPI_ISL_2664937, EPI_ISL_2665215, EPI_ISL_2665235, EPI_ISL_2665248, EPI_ISL_2665321, EPI_ISL_2665388, EPI_ISL_2681939, EPI_ISL_2681964, EPI_ISL_2681980, EPI_ISL_2682021, EPI_ISL_2682062, EPI_ISL_2682188, EPI_ISL_2682280, EPI_ISL_2699988, EPI_ISL_2703804

| | | | |
|---|---|---|---|
| see above | Lighthouse Lab in Milton Keynes | Wellcome Sanger Institute for the COVID-19 Genomics UK (COG-UK) Consortium | Cordelia Langford; David K. Jackson; Dominic Kwiatkowski; Ewan Harrison; Ian Johnston; Jeffrey Barrett; John Sillitoe on behalf of the Wellcome Sanger Institute COVID-19 Surveillance Team; Roberto Amato; Sonia Goncalves; The Lighthouse Lab in Milton Keynes and Alex Alderton |

EPI_ISL_2552020, EPI_ISL_2552052, EPI_ISL_2552062, EPI_ISL_2552070, EPI_ISL_2552079, EPI_ISL_2552083, EPI_ISL_2552105, EPI_ISL_2552141, EPI_ISL_2552142, EPI_ISL_2552184, EPI_ISL_2552190, EPI_ISL_2552235, EPI_ISL_2552240, EPI_ISL_2554141, EPI_ISL_2554145, EPI_ISL_2554151, EPI_ISL_2554155, EPI_ISL_2554159, EPI_ISL_2554161, EPI_ISL_2554173, EPI_ISL_2554174, EPI_ISL_2554185, EPI_ISL_2554194, EPI_ISL_2554204, EPI_ISL_2554206, EPI_ISL_2554208, EPI_ISL_2554209, EPI_ISL_2554212, EPI_ISL_2554213, EPI_ISL_2554217, EPI_ISL_2554231, EPI_ISL_2554232, EPI_ISL_2554234, EPI_ISL_2554235, EPI_ISL_2554263, EPI_ISL_2554266, EPI_ISL_2554268, EPI_ISL_2554274, EPI_ISL_2554276, EPI_ISL_2554277, EPI_ISL_2554280, EPI_ISL_2554283, EPI_ISL_2554285, EPI_ISL_2554295, EPI_ISL_2554296, EPI_ISL_2554297, EPI_ISL_2554298, EPI_ISL_2554302, EPI_ISL_2554303, EPI_ISL_2554305, EPI_ISL_2554306, EPI_ISL_2554307, EPI_ISL_2554308, EPI_ISL_2554310, EPI_ISL_2554311, EPI_ISL_2554315, EPI_ISL_2554316, EPI_ISL_2554317, EPI_ISL_2554319, EPI_ISL_2554324, EPI_ISL_2554331, EPI_ISL_2554332, EPI_ISL_2554336, EPI_ISL_2554338, EPI_ISL_2554340, EPI_ISL_2554343, EPI_ISL_2554350, EPI_ISL_2554352, EPI_ISL_2554353, EPI_ISL_2554354, EPI_ISL_2554359, EPI_ISL_2554366, EPI_ISL_2554371, EPI_ISL_2554375, EPI_ISL_2554376, EPI_ISL_2554385, EPI_ISL_2554386, EPI_ISL_2554391, EPI_ISL_2554393, EPI_ISL_2554394, EPI_ISL_2554399, EPI_ISL_2554406, EPI_ISL_2554407, EPI_ISL_2554409, EPI_ISL_2554413, EPI_ISL_2554415, EPI_ISL_2554435, EPI_ISL_2554436, EPI_ISL_2554437, EPI_ISL_2554450, EPI_ISL_2554453, EPI_ISL_2554455, EPI_ISL_2554458, EPI_ISL_2554461, EPI_ISL_2554462, EPI_ISL_2554463, EPI_ISL_2554465, EPI_ISL_2554467, EPI_ISL_2554492, EPI_ISL_2568663, EPI_ISL_2568666, EPI_ISL_2568667, EPI_ISL_2568668, EPI_ISL_2568673, EPI_ISL_2568674, EPI_ISL_2568677, EPI_ISL_2568680, EPI_ISL_2568689, EPI_ISL_2568690, EPI_ISL_2568691, EPI_ISL_2568694, EPI_ISL_2568696, EPI_ISL_2568704, EPI_ISL_2568708, EPI_ISL_2568715, EPI_ISL_2568720, EPI_ISL_2568721, EPI_ISL_2568724, EPI_ISL_2568725, EPI_ISL_2568728, EPI_ISL_2568729, EPI_ISL_2568739, EPI_ISL_2568741, EPI_ISL_2568744, EPI_ISL_2568751, EPI_ISL_2568761, EPI_ISL_2568767, EPI_ISL_2568779, EPI_ISL_2568785, EPI_ISL_2568789, EPI_ISL_2568792, EPI_ISL_2568796, EPI_ISL_2568797, EPI_ISL_2568801, EPI_ISL_2568802, EPI_ISL_2568806, EPI_ISL_2568807, EPI_ISL_2568808, EPI_ISL_2568815, EPI_ISL_2568820, EPI_ISL_2568832, EPI_ISL_2568837, EPI_ISL_2568839, EPI_ISL_2568840, EPI_ISL_2568844, EPI_ISL_2568847, EPI_ISL_2568852, EPI_ISL_2568855, EPI_ISL_2568863, EPI_ISL_2568868, EPI_ISL_2568873, EPI_ISL_2568876, EPI_ISL_2568881, EPI_ISL_2568887, EPI_ISL_2568889, EPI_ISL_2568892, EPI_ISL_2568897, EPI_ISL_2568898, EPI_ISL_2568900, EPI_ISL_2568912, EPI_ISL_2568914, EPI_ISL_2568916, EPI_ISL_2568918, EPI_ISL_2568920, EPI_ISL_2568921, EPI_ISL_2568923, EPI_ISL_2568926, EPI_ISL_2568929, EPI_ISL_2568930, EPI_ISL_2568933, EPI_ISL_2568936, EPI_ISL_2568944, EPI_ISL_2568950, EPI_ISL_2568956, EPI_ISL_2568958, EPI_ISL_2568960, EPI_ISL_2568961, EPI_ISL_2568973, EPI_ISL_2568979, EPI_ISL_2568988, EPI_ISL_2568994, EPI_ISL_2569002, EPI_ISL_2569005, EPI_ISL_2569011, EPI_ISL_2569013, EPI_ISL_2569014, EPI_ISL_2569021, EPI_ISL_2569022, EPI_ISL_2569030, EPI_ISL_2569037, EPI_ISL_2569040, EPI_ISL_2569051, EPI_ISL_2569055, EPI_ISL_2569056, EPI_ISL_2569060, EPI_ISL_2569063, EPI_ISL_2569067, EPI_ISL_2569073, EPI_ISL_2569076, EPI_ISL_2569077, EPI_ISL_2569083, EPI_ISL_2569087, EPI_ISL_2569097, EPI_ISL_2569099, EPI_ISL_2569104, EPI_ISL_2569106, EPI_ISL_2569107, EPI_ISL_2569112, EPI_ISL_2569114, EPI_ISL_2569116, EPI_ISL_2569120, EPI_ISL_2569121, EPI_ISL_2569126, EPI_ISL_2569131, EPI_ISL_2569132, EPI_ISL_2569146, EPI_ISL_2569153, EPI_ISL_2569154, EPI_ISL_2569157, EPI_ISL_2569158, EPI_ISL_2569159, EPI_ISL_2569162, EPI_ISL_2569164, EPI_ISL_2569173, EPI_ISL_2569175, EPI_ISL_2569178, EPI_ISL_2569183, EPI_ISL_2569187, EPI_ISL_2569195, EPI_ISL_2569198, EPI_ISL_2569207, EPI_ISL_2569210, EPI_ISL_2569215, EPI_ISL_2569217, EPI_ISL_2569218, EPI_ISL_2569221, EPI_ISL_2569223, EPI_ISL_2569227, EPI_ISL_2569232, EPI_ISL_2569233, EPI_ISL_2569238, EPI_ISL_2569239, EPI_ISL_2569240, EPI_ISL_2569242, EPI_ISL_2569245, EPI_ISL_2569246, EPI_ISL_2569250, EPI_ISL_2569254, EPI_ISL_2569259, EPI_ISL_2569268, EPI_ISL_2569271, EPI_ISL_2569272, EPI_ISL_2569275, EPI_ISL_2569279, EPI_ISL_2569289, EPI_ISL_2569290, EPI_ISL_2569291, EPI_ISL_2569296, EPI_ISL_2569302, EPI_ISL_2569305, EPI_ISL_2569306, EPI_ISL_2569310, EPI_ISL_2569313, EPI_ISL_2569315, EPI_ISL_2569665, EPI_ISL_2569669, EPI_ISL_2569674, EPI_ISL_2569682, EPI_ISL_2569684, EPI_ISL_2569694, EPI_ISL_2569699, EPI_ISL_2569703, EPI_ISL_2569709, EPI_ISL_2569719, EPI_ISL_2569721, EPI_ISL_2569724, EPI_ISL_2569725, EPI_ISL_2569727, EPI_ISL_2569735, EPI_ISL_2569739, EPI_ISL_2569750, EPI_ISL_2569751, EPI_ISL_2569758, EPI_ISL_2569762, EPI_ISL_2569763, EPI_ISL_2569768,

| | | | |
|---|---|---|---|
| EPI_ISL_2569769, EPI_ISL_2569770, EPI_ISL_2569772, EPI_ISL_2569777, EPI_ISL_2569782, EPI_ISL_2569785, EPI_ISL_2569788, EPI_ISL_2569789, EPI_ISL_2569796, EPI_ISL_2569799, EPI_ISL_2569804, EPI_ISL_2569805, EPI_ISL_2569806, EPI_ISL_2569807, EPI_ISL_2569816, EPI_ISL_2569817, EPI_ISL_2569819, EPI_ISL_2569821, EPI_ISL_2569825, EPI_ISL_2569836, EPI_ISL_2569841, EPI_ISL_2569844, EPI_ISL_2569845, EPI_ISL_2569849, EPI_ISL_2569851, EPI_ISL_2569852, EPI_ISL_2569858, EPI_ISL_2569859, EPI_ISL_2569860, EPI_ISL_2569866, EPI_ISL_2569867, EPI_ISL_2569869, EPI_ISL_2569874, EPI_ISL_2569880, EPI_ISL_2569881, EPI_ISL_2569886, EPI_ISL_2569887, EPI_ISL_2569890, EPI_ISL_2569891, EPI_ISL_2569893, EPI_ISL_2569901, EPI_ISL_2569902, EPI_ISL_2569906, EPI_ISL_2569910, EPI_ISL_2569911, EPI_ISL_2569913, EPI_ISL_2569922, EPI_ISL_2606456, EPI_ISL_2606461, EPI_ISL_2606463, EPI_ISL_2606465, EPI_ISL_2606469, EPI_ISL_2606474, EPI_ISL_2606475, EPI_ISL_2606481, EPI_ISL_2606482, EPI_ISL_2606485, EPI_ISL_2606487, EPI_ISL_2606494, EPI_ISL_2606496, EPI_ISL_2606497, EPI_ISL_2606498, EPI_ISL_2606500, EPI_ISL_2606504, EPI_ISL_2606505, EPI_ISL_2606506, EPI_ISL_2606509, EPI_ISL_2606510, EPI_ISL_2606512, EPI_ISL_2606515, EPI_ISL_2606517, EPI_ISL_2606520, EPI_ISL_2606521, EPI_ISL_2606524, EPI_ISL_2606525, EPI_ISL_2606526, EPI_ISL_2606538, EPI_ISL_2606540, EPI_ISL_2606547, EPI_ISL_2606549, EPI_ISL_2606550, EPI_ISL_2606552, EPI_ISL_2606553, EPI_ISL_2606565, EPI_ISL_2606568, EPI_ISL_2606569, EPI_ISL_2606571, EPI_ISL_2606572, EPI_ISL_2606573, EPI_ISL_2606576, EPI_ISL_2606577, EPI_ISL_2606579, EPI_ISL_2606583, EPI_ISL_2606586, EPI_ISL_2606588, EPI_ISL_2606589, EPI_ISL_2606590, EPI_ISL_2606592, EPI_ISL_2606595, EPI_ISL_2606597, EPI_ISL_2606602, EPI_ISL_2606605, EPI_ISL_2606611, EPI_ISL_2606619, EPI_ISL_2606625, EPI_ISL_2606630, EPI_ISL_2606635, EPI_ISL_2606636, EPI_ISL_2606637, EPI_ISL_2606640, EPI_ISL_2606645, EPI_ISL_2606647, EPI_ISL_2606652, EPI_ISL_2606654, EPI_ISL_2606655, EPI_ISL_2606657, EPI_ISL_2606664, EPI_ISL_2606665, EPI_ISL_2606669, EPI_ISL_2606671, EPI_ISL_2606673, EPI_ISL_2606674, EPI_ISL_2606678, EPI_ISL_2606682, EPI_ISL_2606685, EPI_ISL_2606691, EPI_ISL_2606695, EPI_ISL_2606697, EPI_ISL_2606700, EPI_ISL_2606713, EPI_ISL_2606715, EPI_ISL_2606716, EPI_ISL_2606717, EPI_ISL_2606723, EPI_ISL_2606728, EPI_ISL_2606743, EPI_ISL_2606763, EPI_ISL_2606765, EPI_ISL_2606795, EPI_ISL_2606799, EPI_ISL_2606800, EPI_ISL_2606813, EPI_ISL_2606815, EPI_ISL_2606818, EPI_ISL_2606823, EPI_ISL_2606824, EPI_ISL_2606830, EPI_ISL_2606833, EPI_ISL_2606834, EPI_ISL_2606855, EPI_ISL_2606856, EPI_ISL_2606863, EPI_ISL_2606869, EPI_ISL_2606874, EPI_ISL_2606890, EPI_ISL_2606901, EPI_ISL_2606904, EPI_ISL_2606912, EPI_ISL_2606916, EPI_ISL_2606927, EPI_ISL_2606936, EPI_ISL_2606942, EPI_ISL_2606945, EPI_ISL_2606946, EPI_ISL_2606949, EPI_ISL_2606950, EPI_ISL_2606962, EPI_ISL_2606970, EPI_ISL_2606980, EPI_ISL_2606985, EPI_ISL_2606989, EPI_ISL_2606990, EPI_ISL_2606991, EPI_ISL_2606994, EPI_ISL_2606996, EPI_ISL_2606998, EPI_ISL_2607003, EPI_ISL_2607009, EPI_ISL_2607015, EPI_ISL_2607018, EPI_ISL_2607022, EPI_ISL_2607026, EPI_ISL_2607948, EPI_ISL_2638620, EPI_ISL_2638630, EPI_ISL_2638651, EPI_ISL_2638665, EPI_ISL_2638744, EPI_ISL_2638818, EPI_ISL_2638819, EPI_ISL_2638836, EPI_ISL_2638842, EPI_ISL_2638873, EPI_ISL_2638894, EPI_ISL_2638901, EPI_ISL_2638923, EPI_ISL_2638975, EPI_ISL_2638984, EPI_ISL_2638998, EPI_ISL_2639023, EPI_ISL_2639029, EPI_ISL_2639048, EPI_ISL_2639062, EPI_ISL_2639104, EPI_ISL_2639119, EPI_ISL_2639161, EPI_ISL_2639169, EPI_ISL_2639200, EPI_ISL_2639652, EPI_ISL_2639669, EPI_ISL_2639706, EPI_ISL_2639727, EPI_ISL_2639784, EPI_ISL_2639835, EPI_ISL_2639887, EPI_ISL_2639890, EPI_ISL_2639897, EPI_ISL_2639906, EPI_ISL_2639907, EPI_ISL_2640108, EPI_ISL_2640330, EPI_ISL_2640331, EPI_ISL_2640335, EPI_ISL_2651804, EPI_ISL_2651943, EPI_ISL_2718321, EPI_ISL_2718341, EPI_ISL_2853259, EPI_ISL_2853297, EPI_ISL_2853364, EPI_ISL_2853365, EPI_ISL_2853400, EPI_ISL_2853483, EPI_ISL_2853526, EPI_ISL_2853534, EPI_ISL_2853574 | | | |
| see above | Lighthouse Laboratory Plymouth | Wellcome Sanger Institute for the COVID-19 Genomics UK (COG-UK) Consortium | Cordelia Langford; David K. Jackson; Dominic Kwiatkowski; Ewan Harrison; Ian Johnston; Jeffrey Barrett; John Sillitoe on behalf of the Wellcome Sanger Institute COVID-19 Surveillance Team; Lighthouse Laboratory Plymouth and Alex Alderton; Roberto Amato; Sonia Goncalves |
| EPI_ISL_2622377, EPI_ISL_2622378, EPI_ISL_2622379, EPI_ISL_2622380, EPI_ISL_2622381, EPI_ISL_2622382, EPI_ISL_2622383, EPI_ISL_2622384, EPI_ISL_2622385, EPI_ISL_2622386, EPI_ISL_2622387, EPI_ISL_2622388, EPI_ISL_2622389, EPI_ISL_2622390, EPI_ISL_2622391, EPI_ISL_2622392, EPI_ISL_2622393, EPI_ISL_2622395, EPI_ISL_2622396, EPI_ISL_2622397, EPI_ISL_2622398, EPI_ISL_2622399, EPI_ISL_2622400, EPI_ISL_2622402, EPI_ISL_2622403, EPI_ISL_2622404, EPI_ISL_2622405, EPI_ISL_2622406, EPI_ISL_2622407, EPI_ISL_2622408, EPI_ISL_2622409, EPI_ISL_2622410, EPI_ISL_2622411, EPI_ISL_2622412, EPI_ISL_2622413, EPI_ISL_2622414, EPI_ISL_2622415, EPI_ISL_2622416, EPI_ISL_2622417, EPI_ISL_2622418, EPI_ISL_2622419, EPI_ISL_2622421, EPI_ISL_2622422, EPI_ISL_2622423, EPI_ISL_2622424, EPI_ISL_2622425, EPI_ISL_2622426, EPI_ISL_2622427, EPI_ISL_2622428, EPI_ISL_2622429, EPI_ISL_2622430, EPI_ISL_2622431, EPI_ISL_2622432, EPI_ISL_2622434, EPI_ISL_2705472, EPI_ISL_2705473, EPI_ISL_2705474, EPI_ISL_2705475, EPI_ISL_2705476, EPI_ISL_2705477, EPI_ISL_2705478, EPI_ISL_2705479, EPI_ISL_2705480, EPI_ISL_2705481, EPI_ISL_2705482 | | | |
| see above | Liverpool Clinical Laboratories | COVID-19 Genomics UK (COG-UK) Consortium | A Alrezaihi; Alessandro Gerada; Alistair Darby; Angela Cowell; Anita Lucaci; Anu Chawla; Cassie Olateju; Catherine Hartley; Charlotte Nelson; Ecaterina Vamos; Elaine O'Toole; Eleanor G Bentley; Ghada T Shawli; Hermione Webster; Isabel García-Dorival; James Johnson; James P Stewart; Jenifer Manson; Joanne Watts; Jones Benjamin; Jordan J Clark; Julian Hiscox; L Luu; Lucille Rainbow; M Almsaud; Margaret Hughes; Mark Whitehead; Matthew Gemmell; Miren Iturriza-Gomara; Muhannad Alruwaili; N.P Randle; Neil Swainston; PKF Gilmore; Parul Sharma; Rebekah Penrice-Randal; Richard Eccles; Richard Gregory; Sam Haldenby; Steve Paterson; Stuart D Armstrong; Trevor Ian Robinson; Ximeng Han |
| EPI_ISL_2705930, EPI_ISL_2705931, EPI_ISL_2705932, EPI_ISL_2705933, EPI_ISL_2705934, EPI_ISL_2705935, EPI_ISL_2705936, EPI_ISL_2705937, EPI_ISL_2705939, EPI_ISL_2705940, EPI_ISL_2705941, EPI_ISL_2705942, EPI_ISL_2705943, EPI_ISL_2705944, EPI_ISL_2705945, EPI_ISL_2705946, EPI_ISL_2705947, EPI_ISL_2705948, EPI_ISL_2705949, EPI_ISL_2705950, EPI_ISL_2705951, EPI_ISL_2705952, EPI_ISL_2705953, EPI_ISL_2705954, EPI_ISL_2705955, EPI_ISL_2705956, EPI_ISL_2705957, EPI_ISL_2705958, EPI_ISL_2705959, EPI_ISL_2705960, EPI_ISL_2705961, EPI_ISL_2705962, EPI_ISL_2705963, EPI_ISL_2705964, EPI_ISL_2705965, EPI_ISL_2705966, EPI_ISL_2705967, EPI_ISL_2705968, EPI_ISL_2705969, EPI_ISL_2705970, EPI_ISL_2705971, EPI_ISL_2705972, EPI_ISL_2705973, EPI_ISL_2705974, EPI_ISL_2705975, EPI_ISL_2705976, EPI_ISL_2705977, EPI_ISL_2705978, EPI_ISL_2705979, EPI_ISL_2705980, EPI_ISL_2705981, EPI_ISL_2705982, EPI_ISL_2705983, EPI_ISL_2705984, EPI_ISL_2705985, EPI_ISL_2705986, EPI_ISL_2705987, EPI_ISL_2705988, EPI_ISL_2705989, EPI_ISL_2705990, EPI_ISL_2705991, EPI_ISL_2705992, EPI_ISL_2705993, EPI_ISL_2705994, EPI_ISL_2705995, EPI_ISL_2705996, EPI_ISL_2705997, EPI_ISL_2705998, EPI_ISL_2705999, EPI_ISL_2706000, EPI_ISL_2706001, EPI_ISL_2706002, EPI_ISL_2706003, EPI_ISL_2706004, EPI_ISL_2706005, EPI_ISL_2706006, EPI_ISL_2706007, EPI_ISL_2706008, EPI_ISL_2706009, EPI_ISL_2706010, EPI_ISL_2706011, EPI_ISL_2706012, EPI_ISL_2706013, EPI_ISL_2706014, EPI_ISL_2706015, EPI_ISL_2706016, EPI_ISL_2706017, EPI_ISL_2706018, EPI_ISL_2706019, EPI_ISL_2706020, EPI_ISL_2706022, EPI_ISL_2706023, EPI_ISL_2706024, EPI_ISL_2706025, EPI_ISL_2706026, EPI_ISL_2706027, EPI_ISL_2706028, EPI_ISL_2706029, EPI_ISL_2706030, EPI_ISL_2706031, EPI_ISL_2706032, EPI_ISL_2706033, EPI_ISL_2706034, EPI_ISL_2706035, EPI_ISL_2706036, EPI_ISL_2706037, EPI_ISL_2706038, EPI_ISL_2706039, EPI_ISL_2706040, EPI_ISL_2706041, EPI_ISL_2706042, EPI_ISL_2706043, EPI_ISL_2706044, EPI_ISL_2706045, EPI_ISL_2706046, EPI_ISL_2706047, EPI_ISL_2706048, EPI_ISL_2706049, EPI_ISL_2706050, EPI_ISL_2706051, EPI_ISL_2706052, EPI_ISL_2706053, EPI_ISL_2706054, EPI_ISL_2706055, EPI_ISL_2706056, EPI_ISL_2706057, EPI_ISL_2706058, EPI_ISL_2706059, EPI_ISL_2706060, EPI_ISL_2706061, EPI_ISL_2706062, EPI_ISL_2706063, EPI_ISL_2706064, EPI_ISL_2706065, EPI_ISL_2706066, EPI_ISL_2706067, EPI_ISL_2706068, EPI_ISL_2706069, EPI_ISL_2706070, EPI_ISL_2706072, EPI_ISL_2706073, EPI_ISL_2706074, EPI_ISL_2706075, EPI_ISL_2706076, EPI_ISL_2706077, EPI_ISL_2706078, EPI_ISL_2706079, EPI_ISL_2706080, EPI_ISL_2706081, EPI_ISL_2706082, EPI_ISL_2706083, EPI_ISL_2706084, EPI_ISL_2706085, EPI_ISL_2706086, EPI_ISL_2706087, EPI_ISL_2706088, EPI_ISL_2706089, EPI_ISL_2706090, EPI_ISL_2706091, EPI_ISL_2706092, EPI_ISL_2706093, EPI_ISL_2706094, EPI_ISL_2706095, EPI_ISL_2706096, EPI_ISL_2706097, EPI_ISL_2706098, EPI_ISL_2706099, EPI_ISL_2706100, EPI_ISL_2706101, EPI_ISL_2706102, EPI_ISL_2706103, EPI_ISL_2706104, EPI_ISL_2706105, EPI_ISL_2706106, EPI_ISL_2706107, EPI_ISL_2706108, EPI_ISL_2706109, EPI_ISL_2706110, EPI_ISL_2706111, EPI_ISL_2706112, EPI_ISL_2706113, EPI_ISL_2706114, EPI_ISL_2706115, EPI_ISL_2706116, EPI_ISL_2706117, EPI_ISL_2706118, EPI_ISL_2706119, EPI_ISL_2706120, EPI_ISL_2706121, EPI_ISL_2706122, EPI_ISL_2706123, EPI_ISL_2706124, EPI_ISL_2706125, EPI_ISL_2706126, EPI_ISL_2706127, EPI_ISL_2706128, EPI_ISL_2706129, EPI_ISL_2706130, EPI_ISL_2706131, EPI_ISL_2706132, EPI_ISL_2706133, EPI_ISL_2706135, EPI_ISL_2706136, EPI_ISL_2706137, EPI_ISL_2706138, EPI_ISL_2706139, EPI_ISL_2706140, EPI_ISL_2706141, EPI_ISL_2706142, EPI_ISL_2706143, EPI_ISL_2706144, EPI_ISL_2706145, EPI_ISL_2706146, EPI_ISL_2706147, EPI_ISL_2706148, EPI_ISL_2706149, EPI_ISL_2706150, EPI_ISL_2706151, EPI_ISL_2706152, EPI_ISL_2706153, EPI_ISL_2706154, EPI_ISL_2706155, EPI_ISL_2706159, EPI_ISL_2706162, EPI_ISL_2706163, EPI_ISL_2706170, EPI_ISL_2706176, EPI_ISL_2706177, EPI_ISL_2706181, EPI_ISL_2706182, EPI_ISL_2706183, EPI_ISL_2706187, EPI_ISL_2706203, EPI_ISL_2706213, EPI_ISL_2706218, EPI_ISL_2706250, EPI_ISL_2706257, EPI_ISL_2706263, EPI_ISL_2706285, EPI_ISL_2706302, EPI_ISL_2706316, EPI_ISL_2706323, EPI_ISL_2706326, EPI_ISL_2706331, EPI_ISL_2706338, EPI_ISL_2706343, EPI_ISL_2706345, EPI_ISL_2706350, EPI_ISL_2706372, EPI_ISL_2706380, EPI_ISL_2706381, EPI_ISL_2706387, EPI_ISL_2706394, EPI_ISL_2706399, EPI_ISL_2706415, EPI_ISL_2706418, EPI_ISL_2706430, EPI_ISL_2706433, EPI_ISL_2706444, EPI_ISL_2706450, EPI_ISL_2706460, EPI_ISL_2706468, EPI_ISL_2706475, EPI_ISL_2706487, EPI_ISL_2706489, EPI_ISL_2706491, EPI_ISL_2706494 | | | |
| see above | Northumbria University / South Tees Hospitals NHS Foundation Trust / North Cumbria Integrated Care NHS Foundation Trust / North Tees and Hartlepool NHS Foundation Trust / Newcastle Hospitals NHS Foundation Trust | COVID-19 Genomics UK (COG-UK) Consortium | Andrew Nelson; Brendan Payne; Clive Graham; Darren L Smith; Debra Padgett; Edward Barton; Emma Swindells; Garren Scott; Gary Black; Gary Eltringham; Giles S Holt; Greg R Young; Jane Greenaway; Jennifer Collins; John Allan; Joshua Loh; Lynn Dover; Matthew Bashton; Mohammad A Tariq; Paul Baker; Sarah Essex; Steve Liggett; Wen C Yew; Yusri Taha |
| EPI_ISL_2624594, EPI_ISL_2624595, EPI_ISL_2624597, EPI_ISL_2624602, EPI_ISL_2624603, EPI_ISL_2624605, EPI_ISL_2624607, EPI_ISL_2624660, EPI_ISL_2624671, EPI_ISL_2624672, EPI_ISL_2624673, EPI_ISL_2624674, EPI_ISL_2624675, EPI_ISL_2624676, EPI_ISL_2624677, EPI_ISL_2624678, EPI_ISL_2624679, EPI_ISL_2624680, EPI_ISL_2624681, EPI_ISL_2624682, EPI_ISL_2624683, EPI_ISL_2624684, EPI_ISL_2624685, EPI_ISL_2624686, EPI_ISL_2624687, EPI_ISL_2624688, EPI_ISL_2624689, EPI_ISL_2624690, EPI_ISL_2624691, EPI_ISL_2624692, EPI_ISL_2624693, EPI_ISL_2624694, EPI_ISL_2624695, EPI_ISL_2624718, EPI_ISL_2624740, EPI_ISL_2624751, EPI_ISL_2624752, EPI_ISL_2624753, EPI_ISL_2624754, EPI_ISL_2624755, EPI_ISL_2624756, EPI_ISL_2624757, EPI_ISL_2624758, EPI_ISL_2624759, EPI_ISL_2624760, EPI_ISL_2624761, EPI_ISL_2624762, EPI_ISL_2624763, EPI_ISL_2624764, EPI_ISL_2624765, EPI_ISL_2624766, EPI_ISL_2624767, EPI_ISL_2624768, EPI_ISL_2624769, EPI_ISL_2624770, EPI_ISL_2624771, EPI_ISL_2624772, EPI_ISL_2624773, EPI_ISL_2624774, EPI_ISL_2624775, EPI_ISL_2624776, EPI_ISL_2624777, EPI_ISL_2624778, EPI_ISL_2624785, EPI_ISL_2624788, EPI_ISL_2624805, EPI_ISL_2624806, EPI_ISL_2624807, EPI_ISL_2624808, EPI_ISL_2624810, EPI_ISL_2624811, EPI_ISL_2624814, EPI_ISL_2624815, EPI_ISL_2624816, EPI_ISL_2624817, EPI_ISL_2624818, EPI_ISL_2624819, EPI_ISL_2624821, EPI_ISL_2624823, EPI_ISL_2624824, EPI_ISL_2624825, EPI_ISL_2624826, EPI_ISL_2624828, EPI_ISL_2624829, EPI_ISL_2624830, EPI_ISL_2624831, EPI_ISL_2624832, EPI_ISL_2624833, EPI_ISL_2624834, EPI_ISL_2624835, EPI_ISL_2624836, EPI_ISL_2624837, EPI_ISL_2624838, EPI_ISL_2624839, EPI_ISL_2624840, EPI_ISL_2624841, EPI_ISL_2624842, EPI_ISL_2624843, EPI_ISL_2624844, EPI_ISL_2624845, EPI_ISL_2624846, EPI_ISL_2624847, EPI_ISL_2624848, EPI_ISL_2624849, EPI_ISL_2624850, EPI_ISL_2624851, EPI_ISL_2624858, EPI_ISL_2624861, EPI_ISL_2624863, EPI_ISL_2624865, EPI_ISL_2624868, EPI_ISL_2624874, EPI_ISL_2624892, EPI_ISL_2624901, EPI_ISL_2707366, EPI_ISL_2707368, EPI_ISL_2707376, EPI_ISL_2707380, EPI_ISL_2707382, EPI_ISL_2707386, EPI_ISL_2707392, EPI_ISL_2707397, EPI_ISL_2707401, EPI_ISL_2707402, EPI_ISL_2707403, EPI_ISL_2707405, EPI_ISL_2707406, EPI_ISL_2707407, EPI_ISL_2707408, EPI_ISL_2707409, EPI_ISL_2707411, EPI_ISL_2707412, EPI_ISL_2707413, EPI_ISL_2707417, EPI_ISL_2707418, EPI_ISL_2707419, EPI_ISL_2707422, EPI_ISL_2707425, EPI_ISL_2707427, EPI_ISL_2707430, EPI_ISL_2707432, EPI_ISL_2707434, EPI_ISL_2707440, EPI_ISL_2707449, EPI_ISL_2707460, EPI_ISL_2707468, EPI_ISL_2707471, EPI_ISL_2707472, EPI_ISL_2707475, EPI_ISL_2707476, EPI_ISL_2707477, EPI_ISL_2707478, EPI_ISL_2707488, EPI_ISL_2707491, EPI_ISL_2707492, EPI_ISL_2707494, EPI_ISL_2707495, EPI_ISL_2707498, EPI_ISL_2707499, EPI_ISL_2707500, EPI_ISL_2707501, EPI_ISL_2707503, EPI_ISL_2707506, EPI_ISL_2707507, EPI_ISL_2707508, EPI_ISL_2707509, EPI_ISL_2707511, EPI_ISL_2707512, EPI_ISL_2707515, EPI_ISL_2707517, EPI_ISL_2707520, EPI_ISL_2707521, EPI_ISL_2707522, EPI_ISL_2707526, EPI_ISL_2707527, EPI_ISL_2707528, EPI_ISL_2707529, EPI_ISL_2707530, EPI_ISL_2707534, EPI_ISL_2707535, EPI_ISL_2707537, EPI_ISL_2707538, EPI_ISL_2707539, EPI_ISL_2707540, EPI_ISL_2707543, EPI_ISL_2707544, EPI_ISL_2707551, EPI_ISL_2707552, EPI_ISL_2707553, EPI_ISL_2707556, EPI_ISL_2707557, EPI_ISL_2707562, EPI_ISL_2707587, EPI_ISL_2707593, EPI_ISL_2707596, EPI_ISL_2707604, EPI_ISL_2707627, EPI_ISL_2707629, EPI_ISL_2707632, EPI_ISL_2707633, EPI_ISL_2707634, EPI_ISL_2707635, EPI_ISL_2707641, EPI_ISL_2707642, EPI_ISL_2707646, EPI_ISL_2707647, EPI_ISL_2707651, EPI_ISL_2707652, EPI_ISL_2707655, EPI_ISL_2707656, EPI_ISL_2707657, EPI_ISL_2707658, EPI_ISL_2707659, EPI_ISL_2707660, EPI_ISL_2707661, EPI_ISL_2707662, EPI_ISL_2707663, EPI_ISL_2707664, EPI_ISL_2707665, EPI_ISL_2707666, EPI_ISL_2707667, EPI_ISL_2707668, EPI_ISL_2707669, EPI_ISL_2707670, EPI_ISL_2707671, EPI_ISL_2707672, EPI_ISL_2707673, EPI_ISL_2707674, EPI_ISL_2707675, EPI_ISL_2707676, EPI_ISL_2707677, EPI_ISL_2707678, EPI_ISL_2707679, EPI_ISL_2707680, EPI_ISL_2707681, EPI_ISL_2707682, EPI_ISL_2707683, EPI_ISL_2707684, EPI_ISL_2707687, EPI_ISL_2707688, EPI_ISL_2707689, EPI_ISL_2707690, EPI_ISL_2707691, EPI_ISL_2707692, EPI_ISL_2707693, EPI_ISL_2707695, EPI_ISL_2707696, EPI_ISL_2707697, EPI_ISL_2707698, EPI_ISL_2707699, EPI_ISL_2707700, EPI_ISL_2707701, EPI_ISL_2707702, EPI_ISL_2707703, EPI_ISL_2707704, EPI_ISL_2707705, EPI_ISL_2707706, EPI_ISL_2707707, EPI_ISL_2707791, EPI_ISL_2707792, EPI_ISL_2707793, EPI_ISL_2707794, EPI_ISL_2707795, EPI_ISL_2707796, EPI_ISL_2707797, EPI_ISL_2707798, EPI_ISL_2707799, EPI_ISL_2707800, EPI_ISL_2707801, EPI_ISL_2707802, EPI_ISL_2707803, EPI_ISL_2707804, EPI_ISL_2707805, EPI_ISL_2707806, EPI_ISL_2707807, EPI_ISL_2707808, EPI_ISL_2707809, EPI_ISL_2707810, EPI_ISL_2707811, EPI_ISL_2707812, EPI_ISL_2707813, EPI_ISL_2707814, EPI_ISL_2707815, EPI_ISL_2707816, EPI_ISL_2707817, EPI_ISL_2707845, EPI_ISL_2707846, EPI_ISL_2707847, EPI_ISL_2707848, EPI_ISL_2707849, EPI_ISL_2707850, EPI_ISL_2707851, EPI_ISL_2707852, EPI_ISL_2707853, EPI_ISL_2707854, EPI_ISL_2707855, EPI_ISL_2707856, EPI_ISL_2707857, EPI_ISL_2707858, EPI_ISL_2707859, EPI_ISL_2707860, EPI_ISL_2707861, EPI_ISL_2707862, EPI_ISL_2707863, EPI_ISL_2707864, EPI_ISL_2707865, EPI_ISL_2707866, EPI_ISL_2707867, EPI_ISL_2707868, EPI_ISL_2707869, EPI_ISL_2707870, EPI_ISL_2707871, EPI_ISL_2707872, EPI_ISL_2707873, EPI_ISL_2707874, EPI_ISL_2707875, EPI_ISL_2707876, EPI_ISL_2707877, EPI_ISL_2707878, EPI_ISL_2707879, EPI_ISL_2707880, EPI_ISL_2707881, EPI_ISL_2707883, EPI_ISL_2707884, EPI_ISL_2707886, EPI_ISL_2707887, EPI_ISL_2707888, EPI_ISL_2707898, EPI_ISL_2707899, EPI_ISL_2707901, EPI_ISL_2707904, EPI_ISL_2707907, EPI_ISL_2707908, EPI_ISL_2707910, EPI_ISL_2707911, EPI_ISL_2707912, EPI_ISL_2707913, EPI_ISL_2707915, EPI_ISL_2707917, EPI_ISL_2707918, EPI_ISL_2707919, EPI_ISL_2707920, EPI_ISL_2707921, EPI_ISL_2707922, EPI_ISL_2707923, EPI_ISL_2707925, EPI_ISL_2707927, EPI_ISL_2707928, EPI_ISL_2707929, EPI_ISL_2707932, EPI_ISL_2707933, EPI_ISL_2707937, EPI_ISL_2707940, EPI_ISL_2707943, EPI_ISL_2707946, EPI_ISL_2707952, EPI_ISL_2707953, EPI_ISL_2707954, EPI_ISL_2707957, EPI_ISL_2707958, EPI_ISL_2707959, EPI_ISL_2707960, EPI_ISL_2707964, EPI_ISL_2707969, EPI_ISL_2707970, EPI_ISL_2707981, EPI_ISL_2707983, EPI_ISL_2707984, EPI_ISL_2707985, EPI_ISL_2707986, EPI_ISL_2707987, EPI_ISL_2707988, EPI_ISL_2707989, EPI_ISL_2707990, EPI_ISL_2707991, EPI_ISL_2707992, EPI_ISL_2707993, EPI_ISL_2707994, EPI_ISL_2707995, EPI_ISL_2707996, EPI_ISL_2707997, EPI_ISL_2707998, EPI_ISL_2707999, EPI_ISL_2708000, EPI_ISL_2708001, EPI_ISL_2708002, EPI_ISL_2708003, EPI_ISL_2708004, EPI_ISL_2708005, EPI_ISL_2708006, EPI_ISL_2708007, EPI_ISL_2708008, EPI_ISL_2708010, EPI_ISL_2708022, EPI_ISL_2708023, EPI_ISL_2708024, EPI_ISL_2708025, EPI_ISL_2708026, EPI_ISL_2708028, EPI_ISL_2708032, EPI_ISL_2708034, EPI_ISL_2708035, EPI_ISL_2708036, EPI_ISL_2708037, EPI_ISL_2708038, EPI_ISL_2708039, EPI_ISL_2708040, EPI_ISL_2708044, EPI_ISL_2708045, EPI_ISL_2708046, EPI_ISL_2708047, EPI_ISL_2708048, EPI_ISL_2708050, EPI_ISL_2708052, EPI_ISL_2708053, EPI_ISL_2708056, EPI_ISL_2708057, EPI_ISL_2708059, EPI_ISL_2708060, EPI_ISL_2708061, EPI_ISL_2708062, EPI_ISL_2708063, EPI_ISL_2708064, EPI_ISL_2708065, EPI_ISL_2708066, EPI_ISL_2708067, EPI_ISL_2708068, EPI_ISL_2708069, EPI_ISL_2708070, EPI_ISL_2708071, EPI_ISL_2708072, EPI_ISL_2708073, EPI_ISL_2708074, EPI_ISL_2708075, EPI_ISL_2708076, EPI_ISL_2708077, EPI_ISL_2708078, EPI_ISL_2708079, EPI_ISL_2708080, EPI_ISL_2708081, EPI_ISL_2708082, EPI_ISL_2708083, EPI_ISL_2708084, EPI_ISL_2708085, EPI_ISL_2708086, EPI_ISL_2708087, EPI_ISL_2708091, EPI_ISL_2708092, EPI_ISL_2708096, EPI_ISL_2708098, EPI_ISL_2708099, EPI_ISL_2708104, EPI_ISL_2708105, EPI_ISL_2708106, EPI_ISL_2708108, EPI_ISL_2708113, EPI_ISL_2708114, EPI_ISL_2708116, EPI_ISL_2708118, EPI_ISL_2708119, EPI_ISL_2708120, EPI_ISL_2708121, EPI_ISL_2708122, EPI_ISL_2708123, EPI_ISL_2708124, EPI_ISL_2708125, EPI_ISL_2708126, EPI_ISL_2708127, EPI_ISL_2708128, EPI_ISL_2708129, EPI_ISL_2708130, EPI_ISL_2708131, EPI_ISL_2708132, EPI_ISL_2708133, EPI_ISL_2708134, EPI_ISL_2708135, EPI_ISL_2708136, EPI_ISL_2708137, EPI_ISL_2708138, EPI_ISL_2708139, EPI_ISL_2708140, EPI_ISL_2708141, EPI_ISL_2708142, EPI_ISL_2708143, EPI_ISL_2708144, EPI_ISL_2750032, EPI_ISL_2750033, EPI_ISL_2750034, EPI_ISL_2750036, EPI_ISL_2750039, EPI_ISL_2750042, EPI_ISL_2750043, EPI_ISL_2750061, EPI_ISL_2750488, EPI_ISL_2750493, EPI_ISL_2818330, EPI_ISL_2818518, EPI_ISL_2818519, EPI_ISL_2818521 | | | |
| see above | Originating lab: Wales Specialist Virology Centre Sequencing lab: Pathogen Genomics Unit | Public Health Wales Microbiology Cardiff Wales Specialist Virology Centre | Alec Birchley; Alexander Adams; Amy Gaskin; Angela Marchbank; Bree Gatica-Wilcox; Catherine Moore; Jason Coombes; Joanne Watkins; Joel Southgate; Johnathan Evans; Laura Gifford; Lauren Gilbert; Lee Graham; Malorie Perry; Matthew Bull; Nicole Pacchiarini; Sally Corden; Sara Kumziene-Summerhayes; Sara Rey; Sarah Taylor; Simon Cottrell; Sophie Jones; Tom Connor |
| EPI_ISL_2707311, EPI_ISL_2707313, EPI_ISL_2707314, EPI_ISL_2707318, EPI_ISL_2707320, EPI_ISL_2707321, EPI_ISL_2707325, EPI_ISL_2707327, EPI_ISL_2707332, EPI_ISL_2707335, EPI_ISL_2707336, EPI_ISL_2707337, EPI_ISL_2707338, EPI_ISL_2707339, EPI_ISL_2707340, EPI_ISL_2707342, EPI_ISL_2707345, EPI_ISL_2707347, EPI_ISL_2707348, EPI_ISL_2707349, EPI_ISL_2707350, EPI_ISL_2707353, EPI_ISL_2707354, EPI_ISL_2707355, EPI_ISL_2707356, EPI_ISL_2735686, EPI_ISL_2735693, EPI_ISL_2735699, EPI_ISL_2735700, EPI_ISL_2735701, EPI_ISL_2735705, EPI_ISL_2735706, EPI_ISL_2735707, EPI_ISL_2903123 | | | |
| see above | Oxford Viromics, NDM, University of Oxford; Oxford University Hospitals; Basingstoke and North Hampshire Hospital | COVID-19 Genomics UK (COG-UK) Consortium | Alex Mobbs; Amy Trebes; Anita Justice; Catrin Moore; Christophe Fraser; David Bonsall; David Buck; Emma Wise; George Macintyre; Jessica Lynch; John Todd; Mariateresa de Cesare; Matilde Mori; Monique Andersson; Nathan Moore; Nick Cortes; Robert Shaw; Stephen Kidd; Tanya Golubchik; Timothy Peto |
| EPI_ISL_2707080 | Quadram Institute Bioscience | COVID-19 Genomics UK (COG-UK) Consortium | Alexander J Trotter; Alison E. Mather; Alp Aydin; Ana P. Tedim; Anastasia Kolyva; Andrew Bell; Andrew J. Page; Claire Stuart; Dave J. Baker; Gemma L. Kay; John Wain; Justin O'Grady; Leonardo de Oliveira Martins; Lizzie Meadows; Maria Diaz; Mark Webber; Muhammed Yasir; Nabil-Fareed Alikhan; Ngozi Elumogo; Nicholas M. Thomson; Rachael Stanley; Rachel Gilroy; Reenesh Prakash; Samir Dervisevic; Samuel Bloomfield; Steven Rudder; Thanh Le-Viet |
| EPI_ISL_2624407, EPI_ISL_2624408, EPI_ISL_2624409, EPI_ISL_2624410, EPI_ISL_2624411, EPI_ISL_2624412, EPI_ISL_2624413, EPI_ISL_2624414, EPI_ISL_2624415, EPI_ISL_2624416, EPI_ISL_2624417, EPI_ISL_2624418, EPI_ISL_2624419, EPI_ISL_2624424, EPI_ISL_2624425, EPI_ISL_2624426, EPI_ISL_2624427, EPI_ISL_2624428, EPI_ISL_2624429, EPI_ISL_2624430, EPI_ISL_2624431, EPI_ISL_2624432, EPI_ISL_2624433, EPI_ISL_2624434, EPI_ISL_2624435, EPI_ISL_2624436, EPI_ISL_2624437, EPI_ISL_2624438, EPI_ISL_2624439, EPI_ISL_2624440, EPI_ISL_2624443, EPI_ISL_2624444, EPI_ISL_2624445, EPI_ISL_2624446, EPI_ISL_2624449, EPI_ISL_2624451, EPI_ISL_2624452, EPI_ISL_2624454, EPI_ISL_2624455, EPI_ISL_2624456, EPI_ISL_2624457, EPI_ISL_2624458, EPI_ISL_2624459, EPI_ISL_2624460, EPI_ISL_2624461, EPI_ISL_2624462, EPI_ISL_2624463, EPI_ISL_2624465, EPI_ISL_2624466, EPI_ISL_2624467, EPI_ISL_2624468, EPI_ISL_2624469, EPI_ISL_2624470, EPI_ISL_2624471, EPI_ISL_2624472, EPI_ISL_2624473, EPI_ISL_2624474, EPI_ISL_2624475, EPI_ISL_2624476, EPI_ISL_2624477, EPI_ISL_2624478, EPI_ISL_2624479, EPI_ISL_2624480, EPI_ISL_2624481, EPI_ISL_2624482, EPI_ISL_2624483, EPI_ISL_2624484, EPI_ISL_2624485, EPI_ISL_2624486, EPI_ISL_2624487, EPI_ISL_2624488, EPI_ISL_2624489, EPI_ISL_2624490, EPI_ISL_2624491, EPI_ISL_2624492, EPI_ISL_2624493, EPI_ISL_2624494, EPI_ISL_2624495, EPI_ISL_2624496, EPI_ISL_2624497, EPI_ISL_2624498, EPI_ISL_2624499, EPI_ISL_2624500, EPI_ISL_2624501, EPI_ISL_2624502, EPI_ISL_2624503, EPI_ISL_2624504, EPI_ISL_2624505, EPI_ISL_2624506, EPI_ISL_2624507, EPI_ISL_2624508, EPI_ISL_2624509, EPI_ISL_2624510, EPI_ISL_2624511, EPI_ISL_2624512, EPI_ISL_2624513, EPI_ISL_2624514, EPI_ISL_2624515, EPI_ISL_2624516, EPI_ISL_2624517, EPI_ISL_2624518, EPI_ISL_2624519, EPI_ISL_2624520, EPI_ISL_2624521, EPI_ISL_2624522, EPI_ISL_2624523, EPI_ISL_2624524 | | | |
| see above | Queens Medical Centre, Clinical Microbiology Department / DeepSeq Nottingham | COVID-19 Genomics UK (COG-UK) Consortium | Christopher Moore; Emily Park; Fei Sang; Gemma Clark; Hannah Howson-Wells; Jack Hill; Jayasree Dey; Johnny Debebe; Jonathan Ball; Joseph Chappell; Manjinder Khakh; Matthew Carlisle; Matthew Loose; Michelle M Lister; Nadine Holmes; Patrick McClure; Theocharis Tsoleridis; Timothy Byaruhanga; Vicki M Fleming; Victoria Wright; Wendy Smith |
| EPI_ISL_2744759, EPI_ISL_2744840, EPI_ISL_2744903, EPI_ISL_2744910, EPI_ISL_2744999, EPI_ISL_2745002, EPI_ISL_2745017, EPI_ISL_2745019, EPI_ISL_2745020, EPI_ISL_2745032, EPI_ISL_2745057, EPI_ISL_2745065, EPI_ISL_2745070, EPI_ISL_2745077, EPI_ISL_2745086, EPI_ISL_2745090, EPI_ISL_2745115, EPI_ISL_2745118, EPI_ISL_2745119, EPI_ISL_2745121, EPI_ISL_2745122, EPI_ISL_2745146, EPI_ISL_2745153, EPI_ISL_2745157, EPI_ISL_2745162, EPI_ISL_2745163, EPI_ISL_2745166, EPI_ISL_2745169, EPI_ISL_2745173, EPI_ISL_2745174, EPI_ISL_2745177, EPI_ISL_2745179, EPI_ISL_2745180, EPI_ISL_2745184, EPI_ISL_2745186, EPI_ISL_2745190, EPI_ISL_2745194, EPI_ISL_2745197, EPI_ISL_2745202, EPI_ISL_2745204, EPI_ISL_2745205, EPI_ISL_2745210, EPI_ISL_2745241, EPI_ISL_2745248, EPI_ISL_2745251, EPI_ISL_2745253, EPI_ISL_2745255, EPI_ISL_2745261, EPI_ISL_2745263, EPI_ISL_2745272, EPI_ISL_2745276, EPI_ISL_2745355, EPI_ISL_2745356, EPI_ISL_2745362, EPI_ISL_2745366, EPI_ISL_2745376, EPI_ISL_2745380, EPI_ISL_2745381, EPI_ISL_2745389, EPI_ISL_2745392, EPI_ISL_2745410, EPI_ISL_2745411, EPI_ISL_2745486, EPI_ISL_2745487, EPI_ISL_2745488, EPI_ISL_2745489, EPI_ISL_2745491, EPI_ISL_2745493, EPI_ISL_2745496, EPI_ISL_2745497, EPI_ISL_2745516, EPI_ISL_2745518, EPI_ISL_2745520, EPI_ISL_2745521, EPI_ISL_2745527, EPI_ISL_2745528, EPI_ISL_2745531, EPI_ISL_2745547, EPI_ISL_2745554, EPI_ISL_2745556, EPI_ISL_2745560, EPI_ISL_2745561, EPI_ISL_2745563, EPI_ISL_2745571, EPI_ISL_2745572, EPI_ISL_2745573, EPI_ISL_2745574, EPI_ISL_2745575, EPI_ISL_2745576, EPI_ISL_2745577, EPI_ISL_2745578, EPI_ISL_2745579, EPI_ISL_2745581, EPI_ISL_2745582, EPI_ISL_2745584, EPI_ISL_2745586, EPI_ISL_2745587, EPI_ISL_2745588, EPI_ISL_2745589, EPI_ISL_2745590, EPI_ISL_2745591, EPI_ISL_2745593, EPI_ISL_2745594, EPI_ISL_2745595, EPI_ISL_2745597, EPI_ISL_2745599, EPI_ISL_2745600, EPI_ISL_2745612, EPI_ISL_2745613, EPI_ISL_2745615, EPI_ISL_2745616, EPI_ISL_2745622, EPI_ISL_2745627, EPI_ISL_2745628, EPI_ISL_2745630, EPI_ISL_2745632, EPI_ISL_2745633, EPI_ISL_2745641, EPI_ISL_2745642, EPI_ISL_2745649, EPI_ISL_2745652, EPI_ISL_2745653, EPI_ISL_2745661, EPI_ISL_2745669, EPI_ISL_2745671, EPI_ISL_2745675, EPI_ISL_2745678, EPI_ISL_2745680, EPI_ISL_2745681, EPI_ISL_2745685, EPI_ISL_2745688, EPI_ISL_2745690, EPI_ISL_2745703, EPI_ISL_2745714, EPI_ISL_2745719, EPI_ISL_2745736, EPI_ISL_2745739, EPI_ISL_2745740, EPI_ISL_2745741, EPI_ISL_2745750, EPI_ISL_2745754, EPI_ISL_2745756, EPI_ISL_2745761, EPI_ISL_2745764, EPI_ISL_2745768, EPI_ISL_2745772, EPI_ISL_2745773, EPI_ISL_2745782, EPI_ISL_2745783, EPI_ISL_2745786, EPI_ISL_2745788, EPI_ISL_2745790, EPI_ISL_2745791, EPI_ISL_2745800, EPI_ISL_2745802, EPI_ISL_2745803, EPI_ISL_2745804, EPI_ISL_2745810, EPI_ISL_2745811, EPI_ISL_2745814, EPI_ISL_2745817, EPI_ISL_2745822, EPI_ISL_2745825, EPI_ISL_2745838, EPI_ISL_2745839, EPI_ISL_2745840, EPI_ISL_2745845, EPI_ISL_2745846, EPI_ISL_2745851, EPI_ISL_2745865, EPI_ISL_2745869, EPI_ISL_2745878, EPI_ISL_2745882, EPI_ISL_2745884, EPI_ISL_2745886, EPI_ISL_2745887, EPI_ISL_2745888, EPI_ISL_2745891, EPI_ISL_2745896, EPI_ISL_2745897, EPI_ISL_2745900, EPI_ISL_2745901, EPI_ISL_2745903, EPI_ISL_2745905, EPI_ISL_2745909, EPI_ISL_2745912, EPI_ISL_2745913, EPI_ISL_2745917, EPI_ISL_2745918, EPI_ISL_2745928, EPI_ISL_2745931, EPI_ISL_2745932, EPI_ISL_2745938, EPI_ISL_2745939, EPI_ISL_2745947, EPI_ISL_2745948, EPI_ISL_2745951, EPI_ISL_2745952, EPI_ISL_2745953, EPI_ISL_2745958, EPI_ISL_2745961, EPI_ISL_2745966, EPI_ISL_2745967, EPI_ISL_2745973, EPI_ISL_2745975, EPI_ISL_2745992, EPI_ISL_2745995, EPI_ISL_2746000, EPI_ISL_2746004, EPI_ISL_2746007, EPI_ISL_2746013, EPI_ISL_2746016, EPI_ISL_2746017, EPI_ISL_2746019, EPI_ISL_2746020, EPI_ISL_2746023, EPI_ISL_2746026, EPI_ISL_2746032, EPI_ISL_2746033, EPI_ISL_2746035, EPI_ISL_2746045, EPI_ISL_2746046, EPI_ISL_2746057, EPI_ISL_2746060, EPI_ISL_2746062, EPI_ISL_2746066, EPI_ISL_2746069, EPI_ISL_2746070, EPI_ISL_2746072, EPI_ISL_2746073, EPI_ISL_2746080, EPI_ISL_2746088, EPI_ISL_2746093, EPI_ISL_2746094, EPI_ISL_2746099, EPI_ISL_2746104, EPI_ISL_2746105, EPI_ISL_2746108, EPI_ISL_2746131, EPI_ISL_2746143, EPI_ISL_2746153, EPI_ISL_2746155, EPI_ISL_2746162, EPI_ISL_2746171, EPI_ISL_2746178, EPI_ISL_2746184, EPI_ISL_2746193, EPI_ISL_2746199, EPI_ISL_2746200, EPI_ISL_2746203, EPI_ISL_2746205, EPI_ISL_2746206, EPI_ISL_2746212, EPI_ISL_2746217, EPI_ISL_2746218, EPI_ISL_2746219, EPI_ISL_2746223, EPI_ISL_2746226, EPI_ISL_2746350, EPI_ISL_2746360, EPI_ISL_2746361, EPI_ISL_2746362, EPI_ISL_2746363, EPI_ISL_2746364, EPI_ISL_2746366, EPI_ISL_2746367, EPI_ISL_2746368, EPI_ISL_2746369, EPI_ISL_2746370, EPI_ISL_2746371, EPI_ISL_2746372, EPI_ISL_2746373, EPI_ISL_2746374, EPI_ISL_2746375, EPI_ISL_2746376, EPI_ISL_2746377, EPI_ISL_2746379, EPI_ISL_2746380, EPI_ISL_2746382, EPI_ISL_2746383, EPI_ISL_2746384, EPI_ISL_2746385, EPI_ISL_2746386, EPI_ISL_2746387, EPI_ISL_2746390, EPI_ISL_2746393, EPI_ISL_2746400, EPI_ISL_2746402, EPI_ISL_2746406, EPI_ISL_2746428, EPI_ISL_2746511, EPI_ISL_2746521, EPI_ISL_2746532, EPI_ISL_2746543, EPI_ISL_2746545, EPI_ISL_2746575, EPI_ISL_2746597, EPI_ISL_2746616, EPI_ISL_2746638, EPI_ISL_2746866, EPI_ISL_2748031, EPI_ISL_2748034, EPI_ISL_2749291, EPI_ISL_2749292, EPI_ISL_2749346, EPI_ISL_2749355, EPI_ISL_2749361, EPI_ISL_2749369, EPI_ISL_2749377, EPI_ISL_2817355, EPI_ISL_2817356, EPI_ISL_2817360, EPI_ISL_2817365, EPI_ISL_2817366, EPI_ISL_2817367, EPI_ISL_2817368, EPI_ISL_2817369, EPI_ISL_2817371, EPI_ISL_2817372, EPI_ISL_2817373, EPI_ISL_2817374, EPI_ISL_2817375, EPI_ISL_2817376, EPI_ISL_2817377, EPI_ISL_2817379, EPI_ISL_2817380, EPI_ISL_2817381, EPI_ISL_2817382, EPI_ISL_2817383, EPI_ISL_2817384, EPI_ISL_2817385, EPI_ISL_2817386, EPI_ISL_2817387, EPI_ISL_2817388, EPI_ISL_2817389, EPI_ISL_2817391, EPI_ISL_2903893, EPI_ISL_2904268, EPI_ISL_2904269, EPI_ISL_2904290, EPI_ISL_2904294, EPI_ISL_2904295, EPI_ISL_2904302, EPI_ISL_2904303, EPI_ISL_2904305, EPI_ISL_2904306, EPI_ISL_2904307, EPI_ISL_2904311, EPI_ISL_2904313, EPI_ISL_2904317, EPI_ISL_2904321, EPI_ISL_2904323, EPI_ISL_2904324, EPI_ISL_2904325, EPI_ISL_2904326 | | | |
| see above | Respiratory Virus Unit, Microbiology Services Colindale, Public Health England | COVID-19 Genomics UK (COG-UK) Consortium | PHE Covid Sequencing Team |
| EPI_ISL_2517787, EPI_ISL_2517842, EPI_ISL_2517847, EPI_ISL_2517851, EPI_ISL_2517856, EPI_ISL_2622476, EPI_ISL_2622483, EPI_ISL_2622484, EPI_ISL_2622493, EPI_ISL_2622500, EPI_ISL_2622529, EPI_ISL_2622556, EPI_ISL_2622557, EPI_ISL_2622558, EPI_ISL_2622565, EPI_ISL_2622568, EPI_ISL_2622575, EPI_ISL_2622577, EPI_ISL_2622585, EPI_ISL_2622586, EPI_ISL_2622589, EPI_ISL_2622599, | | | |

EPI_ISL_2622625, EPI_ISL_2622633, EPI_ISL_2622660, EPI_ISL_2622665, EPI_ISL_2622737, EPI_ISL_2622762, EPI_ISL_2622763, EPI_ISL_2622764, EPI_ISL_2622765, EPI_ISL_2622766, EPI_ISL_2622767, EPI_ISL_2622768, EPI_ISL_2622769, EPI_ISL_2622770, EPI_ISL_2622771, EPI_ISL_2622772, EPI_ISL_2622773, EPI_ISL_2622774, EPI_ISL_2622775, EPI_ISL_2622776, EPI_ISL_2622777, EPI_ISL_2622778, EPI_ISL_2622779, EPI_ISL_2622780, EPI_ISL_2622781, EPI_ISL_2622782, EPI_ISL_2622783, EPI_ISL_2622784, EPI_ISL_2622785, EPI_ISL_2622786, EPI_ISL_2622787, EPI_ISL_2622788, EPI_ISL_2622789, EPI_ISL_2622790, EPI_ISL_2622791, EPI_ISL_2622792, EPI_ISL_2622793, EPI_ISL_2622794, EPI_ISL_2622795, EPI_ISL_2622796, EPI_ISL_2622797, EPI_ISL_2622798, EPI_ISL_2622799, EPI_ISL_2622800, EPI_ISL_2622801, EPI_ISL_2622802, EPI_ISL_2622803, EPI_ISL_2622804, EPI_ISL_2622805, EPI_ISL_2622806, EPI_ISL_2622807, EPI_ISL_2622808, EPI_ISL_2622809, EPI_ISL_2622810, EPI_ISL_2622811, EPI_ISL_2622812, EPI_ISL_2622813, EPI_ISL_2622814, EPI_ISL_2622815, EPI_ISL_2622816, EPI_ISL_2622817, EPI_ISL_2622818, EPI_ISL_2622819, EPI_ISL_2622820, EPI_ISL_2622821, EPI_ISL_2622822, EPI_ISL_2622823, EPI_ISL_2622824, EPI_ISL_2622825, EPI_ISL_2622826, EPI_ISL_2622827, EPI_ISL_2622828, EPI_ISL_2622829, EPI_ISL_2622830, EPI_ISL_2622831, EPI_ISL_2622832, EPI_ISL_2622833, EPI_ISL_2622834, EPI_ISL_2622835, EPI_ISL_2622836, EPI_ISL_2622837, EPI_ISL_2622838, EPI_ISL_2622839, EPI_ISL_2622840, EPI_ISL_2622841, EPI_ISL_2622842, EPI_ISL_2622843, EPI_ISL_2622844, EPI_ISL_2622845, EPI_ISL_2622846, EPI_ISL_2622847, EPI_ISL_2622848, EPI_ISL_2622849, EPI_ISL_2622850, EPI_ISL_2622851, EPI_ISL_2622852, EPI_ISL_2622853, EPI_ISL_2622854, EPI_ISL_2622855, EPI_ISL_2622856, EPI_ISL_2622857, EPI_ISL_2622858, EPI_ISL_2622859, EPI_ISL_2622860, EPI_ISL_2622861, EPI_ISL_2622862, EPI_ISL_2622863, EPI_ISL_2622864, EPI_ISL_2622865, EPI_ISL_2622866, EPI_ISL_2622867, EPI_ISL_2622868, EPI_ISL_2622869, EPI_ISL_2622870, EPI_ISL_2622871, EPI_ISL_2622872, EPI_ISL_2622873, EPI_ISL_2622874, EPI_ISL_2622875, EPI_ISL_2622876, EPI_ISL_2622877, EPI_ISL_2622878, EPI_ISL_2622879, EPI_ISL_2622880, EPI_ISL_2622881, EPI_ISL_2622882, EPI_ISL_2622883, EPI_ISL_2622884, EPI_ISL_2622885, EPI_ISL_2622886, EPI_ISL_2622887, EPI_ISL_2622888, EPI_ISL_2622889, EPI_ISL_2622890, EPI_ISL_2622891, EPI_ISL_2622892, EPI_ISL_2622893, EPI_ISL_2622894, EPI_ISL_2622895, EPI_ISL_2622896, EPI_ISL_2622897, EPI_ISL_2622898, EPI_ISL_2622899, EPI_ISL_2622900, EPI_ISL_2622901, EPI_ISL_2622902, EPI_ISL_2622903, EPI_ISL_2622904, EPI_ISL_2622905, EPI_ISL_2622906, EPI_ISL_2622907, EPI_ISL_2622908, EPI_ISL_2622909, EPI_ISL_2622910, EPI_ISL_2622911, EPI_ISL_2622912, EPI_ISL_2622913, EPI_ISL_2622914, EPI_ISL_2622915, EPI_ISL_2622916, EPI_ISL_2622917, EPI_ISL_2622918, EPI_ISL_2622919, EPI_ISL_2622920, EPI_ISL_2622921, EPI_ISL_2622922, EPI_ISL_2622923, EPI_ISL_2622924, EPI_ISL_2622925, EPI_ISL_2622926, EPI_ISL_2622927, EPI_ISL_2622928, EPI_ISL_2622929, EPI_ISL_2622930, EPI_ISL_2622931, EPI_ISL_2622932, EPI_ISL_2622933, EPI_ISL_2622934, EPI_ISL_2622935, EPI_ISL_2622936, EPI_ISL_2622937, EPI_ISL_2622938, EPI_ISL_2622939, EPI_ISL_2622940, EPI_ISL_2622941, EPI_ISL_2622942, EPI_ISL_2622943, EPI_ISL_2622944, EPI_ISL_2622945, EPI_ISL_2622946, EPI_ISL_2622947, EPI_ISL_2622948, EPI_ISL_2622949, EPI_ISL_2622950, EPI_ISL_2622951, EPI_ISL_2622952, EPI_ISL_2622953, EPI_ISL_2622954, EPI_ISL_2622955, EPI_ISL_2622956, EPI_ISL_2622957, EPI_ISL_2622958, EPI_ISL_2622959, EPI_ISL_2622960, EPI_ISL_2622961, EPI_ISL_2622962, EPI_ISL_2622963, EPI_ISL_2622964, EPI_ISL_2622965, EPI_ISL_2622966, EPI_ISL_2622967, EPI_ISL_2622968, EPI_ISL_2622969, EPI_ISL_2622970, EPI_ISL_2622971, EPI_ISL_2622972, EPI_ISL_2622973, EPI_ISL_2622974, EPI_ISL_2622975, EPI_ISL_2622976, EPI_ISL_2622977, EPI_ISL_2622978, EPI_ISL_2622979, EPI_ISL_2622980, EPI_ISL_2622981, EPI_ISL_2622982, EPI_ISL_2622983, EPI_ISL_2622984, EPI_ISL_2622985, EPI_ISL_2622986, EPI_ISL_2622987, EPI_ISL_2622988, EPI_ISL_2622989, EPI_ISL_2622990, EPI_ISL_2622991, EPI_ISL_2622992, EPI_ISL_2622993, EPI_ISL_2622994, EPI_ISL_2622995, EPI_ISL_2622996, EPI_ISL_2622997, EPI_ISL_2622998, EPI_ISL_2622999, EPI_ISL_2623000, EPI_ISL_2623001, EPI_ISL_2623002, EPI_ISL_2623003, EPI_ISL_2623004, EPI_ISL_2623005, EPI_ISL_2623006, EPI_ISL_2623007, EPI_ISL_2623008, EPI_ISL_2623009, EPI_ISL_2623010, EPI_ISL_2623011, EPI_ISL_2623012, EPI_ISL_2623013, EPI_ISL_2623014, EPI_ISL_2623015, EPI_ISL_2623016, EPI_ISL_2623017, EPI_ISL_2623018, EPI_ISL_2623019, EPI_ISL_2623020, EPI_ISL_2623021, EPI_ISL_2623022, EPI_ISL_2623023, EPI_ISL_2623024, EPI_ISL_2623025, EPI_ISL_2623026, EPI_ISL_2623027, EPI_ISL_2623028, EPI_ISL_2623029, EPI_ISL_2623030, EPI_ISL_2623031, EPI_ISL_2623032, EPI_ISL_2623033, EPI_ISL_2623034, EPI_ISL_2623035, EPI_ISL_2623036, EPI_ISL_2623037, EPI_ISL_2623038, EPI_ISL_2623039, EPI_ISL_2623040, EPI_ISL_2623041, EPI_ISL_2623042, EPI_ISL_2623043, EPI_ISL_2623044, EPI_ISL_2623045, EPI_ISL_2623046, EPI_ISL_2623047, EPI_ISL_2623048, EPI_ISL_2623049, EPI_ISL_2623050, EPI_ISL_2623051, EPI_ISL_2623052, EPI_ISL_2623053, EPI_ISL_2623054, EPI_ISL_2623055, EPI_ISL_2623056, EPI_ISL_2623057, EPI_ISL_2623058, EPI_ISL_2623059, EPI_ISL_2623060, EPI_ISL_2623061, EPI_ISL_2623062, EPI_ISL_2623063, EPI_ISL_2623064, EPI_ISL_2623065, EPI_ISL_2623066, EPI_ISL_2623067, EPI_ISL_2623068, EPI_ISL_2623069, EPI_ISL_2623070, EPI_ISL_2623071, EPI_ISL_2623072, EPI_ISL_2623073, EPI_ISL_2623074, EPI_ISL_2623075, EPI_ISL_2623076, EPI_ISL_2623077, EPI_ISL_2623078, EPI_ISL_2623079, EPI_ISL_2623080, EPI_ISL_2623081, EPI_ISL_2623082, EPI_ISL_2623083, EPI_ISL_2623084, EPI_ISL_2623085, EPI_ISL_2623086, EPI_ISL_2623087, EPI_ISL_2623088, EPI_ISL_2623089, EPI_ISL_2623090, EPI_ISL_2623091, EPI_ISL_2623092, EPI_ISL_2623093, EPI_ISL_2623094, EPI_ISL_2623095, EPI_ISL_2623096, EPI_ISL_2623097, EPI_ISL_2623098, EPI_ISL_2623099, EPI_ISL_2623100, EPI_ISL_2623101, EPI_ISL_2623102, EPI_ISL_2623103, EPI_ISL_2623104, EPI_ISL_2623105, EPI_ISL_2623106, EPI_ISL_2623107, EPI_ISL_2623108, EPI_ISL_2623109, EPI_ISL_2623110, EPI_ISL_2623111, EPI_ISL_2623112, EPI_ISL_2623113, EPI_ISL_2623114, EPI_ISL_2623115, EPI_ISL_2623116, EPI_ISL_2623117, EPI_ISL_2623118, EPI_ISL_2623119, EPI_ISL_2623120, EPI_ISL_2623121, EPI_ISL_2623122, EPI_ISL_2623123, EPI_ISL_2623124, EPI_ISL_2705496, EPI_ISL_2705497, EPI_ISL_2705498, EPI_ISL_2705499, EPI_ISL_2705500, EPI_ISL_2705501, EPI_ISL_2705502, EPI_ISL_2705503, EPI_ISL_2705504, EPI_ISL_2705505, EPI_ISL_2705506, EPI_ISL_2705507, EPI_ISL_2705508, EPI_ISL_2705509, EPI_ISL_2705510, EPI_ISL_2705511, EPI_ISL_2705512, EPI_ISL_2705513, EPI_ISL_2705514, EPI_ISL_2705515, EPI_ISL_2705516, EPI_ISL_2705517, EPI_ISL_2705518, EPI_ISL_2705519, EPI_ISL_2705520, EPI_ISL_2705521, EPI_ISL_2705522, EPI_ISL_2705523, EPI_ISL_2705524, EPI_ISL_2705525, EPI_ISL_2705526, EPI_ISL_2705527, EPI_ISL_2705528, EPI_ISL_2705529, EPI_ISL_2705530, EPI_ISL_2705531, EPI_ISL_2705532, EPI_ISL_2705533, EPI_ISL_2705534, EPI_ISL_2705535, EPI_ISL_2705536, EPI_ISL_2705537, EPI_ISL_2705538, EPI_ISL_2705539, EPI_ISL_2705540, EPI_ISL_2705541, EPI_ISL_2705542, EPI_ISL_2705543, EPI_ISL_2705544, EPI_ISL_2705545, EPI_ISL_2705546, EPI_ISL_2705547, EPI_ISL_2705548, EPI_ISL_2705549, EPI_ISL_2705550, EPI_ISL_2705551, EPI_ISL_2705552, EPI_ISL_2705553, EPI_ISL_2705554, EPI_ISL_2705555, EPI_ISL_2705556, EPI_ISL_2705557, EPI_ISL_2705558, EPI_ISL_2705559, EPI_ISL_2705560, EPI_ISL_2705561, EPI_ISL_2705571, EPI_ISL_2705574, EPI_ISL_2705577, EPI_ISL_2705587, EPI_ISL_2705590, EPI_ISL_2705591, EPI_ISL_2705592, EPI_ISL_2705596, EPI_ISL_2705598, EPI_ISL_2705599, EPI_ISL_2705603, EPI_ISL_2705613, EPI_ISL_2705615, EPI_ISL_2705631, EPI_ISL_2705632, EPI_ISL_2705634, EPI_ISL_2705635, EPI_ISL_2705636, EPI_ISL_2705638, EPI_ISL_2705639, EPI_ISL_2705641, EPI_ISL_2705642, EPI_ISL_2705643, EPI_ISL_2705644, EPI_ISL_2705645, EPI_ISL_2705646, EPI_ISL_2705647, EPI_ISL_2705648, EPI_ISL_2705650, EPI_ISL_2705651, EPI_ISL_2705652, EPI_ISL_2705654, EPI_ISL_2705656, EPI_ISL_2705657, EPI_ISL_2705658, EPI_ISL_2705659, EPI_ISL_2705660, EPI_ISL_2705661, EPI_ISL_2705662, EPI_ISL_2705663, EPI_ISL_2705664, EPI_ISL_2705665, EPI_ISL_2705668, EPI_ISL_2705669, EPI_ISL_2705670, EPI_ISL_2705672, EPI_ISL_2705673, EPI_ISL_2705674, EPI_ISL_2705676, EPI_ISL_2705678, EPI_ISL_2705679, EPI_ISL_2705680, EPI_ISL_2705684, EPI_ISL_2705685, EPI_ISL_2705686, EPI_ISL_2705689, EPI_ISL_2705692, EPI_ISL_2705694, EPI_ISL_2705695, EPI_ISL_2705696, EPI_ISL_2705697, EPI_ISL_2705700, EPI_ISL_2705702, EPI_ISL_2705705, EPI_ISL_2705706, EPI_ISL_2705707, EPI_ISL_2705709, EPI_ISL_2705727, EPI_ISL_2705730, EPI_ISL_2705736, EPI_ISL_2705770, EPI_ISL_2705774, EPI_ISL_2705781, EPI_ISL_2705785, EPI_ISL_2705790, EPI_ISL_2705799, EPI_ISL_2705801, EPI_ISL_2705804, EPI_ISL_2705812, EPI_ISL_2705818, EPI_ISL_2705824, EPI_ISL_2705828, EPI_ISL_2705833, EPI_ISL_2705842, EPI_ISL_2705846, EPI_ISL_2705848, EPI_ISL_2705849, EPI_ISL_2901186, EPI_ISL_2901187, EPI_ISL_2901188, EPI_ISL_2901189, EPI_ISL_2901190, EPI_ISL_2901191, EPI_ISL_2901192, EPI_ISL_2901193, EPI_ISL_2901194, EPI_ISL_2901195, EPI_ISL_2901196, EPI_ISL_2901197, EPI_ISL_2901198, EPI_ISL_2901199, EPI_ISL_2901200, EPI_ISL_2901201, EPI_ISL_2901202, EPI_ISL_2901203, EPI_ISL_2901204, EPI_ISL_2901206, EPI_ISL_2901207, EPI_ISL_2901208, EPI_ISL_2901209, EPI_ISL_2901210, EPI_ISL_2901211, EPI_ISL_2901212, EPI_ISL_2901214, EPI_ISL_2901215, EPI_ISL_2901216, EPI_ISL_2901217, EPI_ISL_2901218, EPI_ISL_2901219, EPI_ISL_2901220, EPI_ISL_2901221, EPI_ISL_2901222, EPI_ISL_2901223, EPI_ISL_2901224, EPI_ISL_2901225, EPI_ISL_2901226, EPI_ISL_2901227, EPI_ISL_2901228, EPI_ISL_2901229, EPI_ISL_2901230, EPI_ISL_2901231, EPI_ISL_2901232, EPI_ISL_2901233, EPI_ISL_2901235, EPI_ISL_2901236, EPI_ISL_2901237, EPI_ISL_2901238, EPI_ISL_2901239, EPI_ISL_2901240, EPI_ISL_2901241, EPI_ISL_2901242, EPI_ISL_2901243, EPI_ISL_2901244, EPI_ISL_2901245, EPI_ISL_2901246, EPI_ISL_2901247, EPI_ISL_2901248, EPI_ISL_2901250, EPI_ISL_2901252, EPI_ISL_2901253, EPI_ISL_2901254, EPI_ISL_2901257, EPI_ISL_2901258, EPI_ISL_2901259, EPI_ISL_2901260, EPI_ISL_2901261, EPI_ISL_2901263, EPI_ISL_2901264, EPI_ISL_2901265, EPI_ISL_2901266, EPI_ISL_2901267, EPI_ISL_2901268, EPI_ISL_2901269, EPI_ISL_2901270, EPI_ISL_2901271, EPI_ISL_2901272, EPI_ISL_2901273, EPI_ISL_2901274, EPI_ISL_2901275, EPI_ISL_2901276, EPI_ISL_2901277, EPI_ISL_2901279, EPI_ISL_2901280, EPI_ISL_2901281, EPI_ISL_2901282, EPI_ISL_2901283, EPI_ISL_2901284, EPI_ISL_2901285, EPI_ISL_2901286, EPI_ISL_2901287, EPI_ISL_2901289, EPI_ISL_2901290, EPI_ISL_2901291, EPI_ISL_2901293, EPI_ISL_2901295, EPI_ISL_2901296, EPI_ISL_2901297, EPI_ISL_2901298, EPI_ISL_2901299, EPI_ISL_2901300, EPI_ISL_2901302, EPI_ISL_2901303, EPI_ISL_2901304, EPI_ISL_2901305, EPI_ISL_2901306, EPI_ISL_2901307, EPI_ISL_2901308, EPI_ISL_2901309, EPI_ISL_2901310, EPI_ISL_2901311, EPI_ISL_2901312, EPI_ISL_2901313, EPI_ISL_2901314, EPI_ISL_2901315, EPI_ISL_2901316, EPI_ISL_2901317, EPI_ISL_2901318, EPI_ISL_2901320, EPI_ISL_2901321, EPI_ISL_2901322, EPI_ISL_2901323, EPI_ISL_2901324, EPI_ISL_2901325, EPI_ISL_2901326, EPI_ISL_2901327, EPI_ISL_2901328

| | | | |
|---|---|---|---|
| see above | University College London, Great Ormond Street Hospital for Children NHS Foundation Trust, Imperial College Healthcare NHS Trust | COVID-19 Genomics UK (COG-UK) Consortium | Alison Holmes; Charlotte Williams; Helena Tutill; Jacqueline Findlay; James Price; Judith Breuer; Julianne Brown; Kathryn Harris; Leysa Forrest; Marius Cotic; Mark Kristiansen; Nadua Bayzid; Paola Niola; Paola Resende Silva; Patricia Dyal; Paul Randell; Rachel Williams; Samuel Weeks; Sergi Castellano; Sunando Roy; Tony Brooks; Yasmin Panchbhaya |

EPI_ISL_2721853, EPI_ISL_2721855, EPI_ISL_2721856, EPI_ISL_2721858, EPI_ISL_2721860, EPI_ISL_2721861, EPI_ISL_2721863, EPI_ISL_2721865, EPI_ISL_2721866, EPI_ISL_2721870, EPI_ISL_2721871, EPI_ISL_2721873, EPI_ISL_2721874, EPI_ISL_2721876, EPI_ISL_2721878, EPI_ISL_2721880, EPI_ISL_2721881, EPI_ISL_2721884, EPI_ISL_2721886, EPI_ISL_2721887, EPI_ISL_2721889, EPI_ISL_2721893, EPI_ISL_2721895, EPI_ISL_2721896, EPI_ISL_2721898, EPI_ISL_2721900, EPI_ISL_2721901, EPI_ISL_2721903, EPI_ISL_2721905, EPI_ISL_2721906, EPI_ISL_2721910, EPI_ISL_2721911, EPI_ISL_2721913, EPI_ISL_2721914, EPI_ISL_2721915, EPI_ISL_2721917, EPI_ISL_2721919, EPI_ISL_2721922, EPI_ISL_2721924, EPI_ISL_2721926, EPI_ISL_2721927, EPI_ISL_2721929, EPI_ISL_2721931, EPI_ISL_2721935, EPI_ISL_2721939, EPI_ISL_2721940, EPI_ISL_2721943, EPI_ISL_2721944, EPI_ISL_2721948, EPI_ISL_2721949, EPI_ISL_2721951, EPI_ISL_2721953, EPI_ISL_2721954, EPI_ISL_2721956, EPI_ISL_2721960, EPI_ISL_2721961, EPI_ISL_2721963, EPI_ISL_2721965, EPI_ISL_2721966, EPI_ISL_2721968, EPI_ISL_2721971, EPI_ISL_2721973, EPI_ISL_2721974, EPI_ISL_2721976, EPI_ISL_2721977, EPI_ISL_2721979, EPI_ISL_2721981, EPI_ISL_2721982, EPI_ISL_2721984

| | | | |
|---|---|---|---|
| see above | University of Birmingham | COVID-19 Genomics UK (COG-UK) Consortium | Alex Richter; Andrew Bosworth; Andrew Bosworth. Queen Elizabeth Hospital: Anna Casey; Andrew D Beggs PHE Heartlands Lab: Husam Osman; Charlotte Poxon; Institute of Microbiology; Joanne Stockton; Josh Quick; Kasun Wanigasooriya; Mike Kidd; Nicholas Loman. University of Birmingham Testing Laboratory: Celina M Whalley; Oliver Pickles; Radoslaw Poplawski; Samuel Nicholls; University of Birmingham: Claire McMurray; Will Rowe |

EPI_ISL_2704830, EPI_ISL_2704848, EPI_ISL_2704849, EPI_ISL_2704850, EPI_ISL_2704851, EPI_ISL_2704852, EPI_ISL_2704853, EPI_ISL_2704855, EPI_ISL_2704856, EPI_ISL_2704857, EPI_ISL_2704858, EPI_ISL_2704859, EPI_ISL_2704860, EPI_ISL_2704861, EPI_ISL_2704862, EPI_ISL_2704864, EPI_ISL_2704865, EPI_ISL_2704866, EPI_ISL_2704867, EPI_ISL_2704868, EPI_ISL_2704869, EPI_ISL_2704870, EPI_ISL_2704871, EPI_ISL_2704872, EPI_ISL_2704873, EPI_ISL_2704875, EPI_ISL_2704876, EPI_ISL_2704877, EPI_ISL_2704878, EPI_ISL_2704879, EPI_ISL_2704880, EPI_ISL_2704881, EPI_ISL_2704882, EPI_ISL_2704883, EPI_ISL_2704884, EPI_ISL_2704896, EPI_ISL_2704897, EPI_ISL_2704898, EPI_ISL_2704899, EPI_ISL_2704900, EPI_ISL_2704901, EPI_ISL_2704902, EPI_ISL_2704903, EPI_ISL_2704904, EPI_ISL_2704905, EPI_ISL_2704906, EPI_ISL_2704914, EPI_ISL_2704917, EPI_ISL_2704918, EPI_ISL_2704919, EPI_ISL_2704920, EPI_ISL_2704921, EPI_ISL_2704922, EPI_ISL_2704923, EPI_ISL_2704929, EPI_ISL_2704930, EPI_ISL_2704932, EPI_ISL_2734332, EPI_ISL_2734333, EPI_ISL_2734361, EPI_ISL_2734372, EPI_ISL_2734373, EPI_ISL_2734374, EPI_ISL_2734375, EPI_ISL_2734376, EPI_ISL_2734416, EPI_ISL_2734417, EPI_ISL_2734418, EPI_ISL_2734419, EPI_ISL_2734420, EPI_ISL_2734421, EPI_ISL_2734422, EPI_ISL_2734423, EPI_ISL_2734424, EPI_ISL_2734425, EPI_ISL_2734426, EPI_ISL_2734427, EPI_ISL_2734428, EPI_ISL_2734430

| | | | |
|---|---|---|---|
| see above | Virology Department, Royal Infirmary of Edinburgh, NHS Lothian / School of Biological Sciences, University of Edinburgh | COVID-19 Genomics UK (COG-UK) Consortium | Colquhoun R; Cotton S; Dewar R; Gallagher A; Hill V; Jackson B; Maloney D; McCrone JT; McHugh M; O'Toole Á; Rambaut A; Rooke S; Scher E; Templeton K; Yu X |

EPI_ISL_2625209, EPI_ISL_2625210, EPI_ISL_2625211, EPI_ISL_2625214, EPI_ISL_2625217, EPI_ISL_2625220, EPI_ISL_2625221, EPI_ISL_2625224, EPI_ISL_2625225, EPI_ISL_2625230, EPI_ISL_2625232, EPI_ISL_2625234, EPI_ISL_2625236, EPI_ISL_2625237, EPI_ISL_2625238, EPI_ISL_2625242, EPI_ISL_2625243, EPI_ISL_2625245, EPI_ISL_2625247, EPI_ISL_2625248, EPI_ISL_2625249, EPI_ISL_2625251, EPI_ISL_2625253, EPI_ISL_2625255, EPI_ISL_2625258, EPI_ISL_2625259, EPI_ISL_2625261, EPI_ISL_2625262, EPI_ISL_2625263, EPI_ISL_2625265, EPI_ISL_2625266, EPI_ISL_2625268, EPI_ISL_2625271, EPI_ISL_2625272, EPI_ISL_2625274, EPI_ISL_2625275, EPI_ISL_2625277, EPI_ISL_2625279, EPI_ISL_2625280, EPI_ISL_2625281, EPI_ISL_2625282, EPI_ISL_2625283, EPI_ISL_2625285, EPI_ISL_2625287, EPI_ISL_2625289, EPI_ISL_2625290, EPI_ISL_2625291, EPI_ISL_2625292, EPI_ISL_2625294, EPI_ISL_2625297, EPI_ISL_2625299, EPI_ISL_2625300, EPI_ISL_2625301, EPI_ISL_2625303, EPI_ISL_2625304, EPI_ISL_2625307, EPI_ISL_2625308, EPI_ISL_2625310, EPI_ISL_2625311, EPI_ISL_2625313, EPI_ISL_2625314, EPI_ISL_2625316, EPI_ISL_2625317, EPI_ISL_2625318, EPI_ISL_2625321, EPI_ISL_2625322, EPI_ISL_2625323, EPI_ISL_2625324, EPI_ISL_2625325, EPI_ISL_2625326, EPI_ISL_2625327, EPI_ISL_2625329, EPI_ISL_2625330, EPI_ISL_2625331, EPI_ISL_2625332, EPI_ISL_2625340, EPI_ISL_2625341, EPI_ISL_2625342, EPI_ISL_2625343, EPI_ISL_2625345, EPI_ISL_2625346, EPI_ISL_2625347, EPI_ISL_2625348, EPI_ISL_2625349, EPI_ISL_2625350, EPI_ISL_2625351, EPI_ISL_2625353, EPI_ISL_2625355, EPI_ISL_2625358, EPI_ISL_2625361, EPI_ISL_2625362, EPI_ISL_2625364, EPI_ISL_2625365, EPI_ISL_2625366, EPI_ISL_2625368, EPI_ISL_2625371, EPI_ISL_2625372, EPI_ISL_2625373, EPI_ISL_2625374, EPI_ISL_2625375, EPI_ISL_2625376, EPI_ISL_2625377, EPI_ISL_2625378, EPI_ISL_2625379, EPI_ISL_2625380, EPI_ISL_2625381, EPI_ISL_2625384, EPI_ISL_2625386, EPI_ISL_2625387, EPI_ISL_2625388, EPI_ISL_2625389, EPI_ISL_2625391, EPI_ISL_2625393, EPI_ISL_2625394, EPI_ISL_2625395, EPI_ISL_2625399, EPI_ISL_2625408, EPI_ISL_2625409, EPI_ISL_2625410, EPI_ISL_2625412, EPI_ISL_2625414, EPI_ISL_2625415, EPI_ISL_2625417, EPI_ISL_2625420, EPI_ISL_2625421, EPI_ISL_2625423, EPI_ISL_2625424, EPI_ISL_2625425, EPI_ISL_2625427, EPI_ISL_2625431, EPI_ISL_2625432, EPI_ISL_2625433, EPI_ISL_2625435, EPI_ISL_2625436, EPI_ISL_2625437, EPI_ISL_2625438, EPI_ISL_2625445, EPI_ISL_2625449, EPI_ISL_2625451, EPI_ISL_2625452, EPI_ISL_2625454, EPI_ISL_2625459, EPI_ISL_2625460, EPI_ISL_2625461, EPI_ISL_2625463, EPI_ISL_2625467, EPI_ISL_2625468, EPI_ISL_2625470, EPI_ISL_2625472, EPI_ISL_2625473, EPI_ISL_2625475, EPI_ISL_2625476, EPI_ISL_2625477, EPI_ISL_2625480, EPI_ISL_2625487, EPI_ISL_2625488, EPI_ISL_2625490, EPI_ISL_2625491, EPI_ISL_2625492, EPI_ISL_2625493, EPI_ISL_2625494, EPI_ISL_2625495, EPI_ISL_2625496, EPI_ISL_2625497, EPI_ISL_2625498, EPI_ISL_2625499, EPI_ISL_2625501, EPI_ISL_2625502, EPI_ISL_2625503, EPI_ISL_2625506, EPI_ISL_2625507, EPI_ISL_2625508, EPI_ISL_2625509, EPI_ISL_2625510, EPI_ISL_2625513, EPI_ISL_2625514, EPI_ISL_2625515, EPI_ISL_2625516, EPI_ISL_2625517, EPI_ISL_2625521, EPI_ISL_2625527, EPI_ISL_2625528, EPI_ISL_2625530, EPI_ISL_2625536, EPI_ISL_2625538, EPI_ISL_2625539, EPI_ISL_2625540, EPI_ISL_2625541, EPI_ISL_2625542, EPI_ISL_2625543, EPI_ISL_2625544, EPI_ISL_2625545, EPI_ISL_2625547, EPI_ISL_2625548, EPI_ISL_2625550, EPI_ISL_2625554, EPI_ISL_2625559, EPI_ISL_2625560, EPI_ISL_2625561, EPI_ISL_2625562, EPI_ISL_2625563, EPI_ISL_2625565, EPI_ISL_2625566, EPI_ISL_2625569, EPI_ISL_2625570, EPI_ISL_2625571, EPI_ISL_2625572, EPI_ISL_2625573, EPI_ISL_2625574, EPI_ISL_2625575, EPI_ISL_2625576, EPI_ISL_2625579, EPI_ISL_2625580, EPI_ISL_2625581, EPI_ISL_2625582, EPI_ISL_2625584, EPI_ISL_2625585, EPI_ISL_2625586, EPI_ISL_2625590, EPI_ISL_2625593, EPI_ISL_2625596, EPI_ISL_2625598, EPI_ISL_2625599, EPI_ISL_2625600, EPI_ISL_2625601, EPI_ISL_2625602, EPI_ISL_2625604, EPI_ISL_2625605, EPI_ISL_2625609, EPI_ISL_2625610, EPI_ISL_2625614, EPI_ISL_2625616, EPI_ISL_2625618, EPI_ISL_2625619, EPI_ISL_2625620

| | | | |
|---|---|---|---|
| see above | Virology Department, Sheffield Teaching Hospitals NHS Foundation Trust/Department of Infection, Immunity and Cardiovascular Disease, The Medical School, University of Sheffield | COVID-19 Genomics UK (COG-UK) Consortium | Adri Angyal; Alex Keeley; Benjamin Lindsey; Cariad Evans; Danielle Groves; Dave Partridge; Katie Johnson; Laura Carrilero; Luke Green; Matthew Parker; Matthew Wyles; Mehmet Yavuz; Mohammad Raza; Nikki Smith; Paul Parsons; Rachel Tucker; Rebecca Brown; Thushan de Silva |

EPI_ISL_2704744, EPI_ISL_2704745, EPI_ISL_2704746, EPI_ISL_2704747, EPI_ISL_2704749, EPI_ISL_2704752, EPI_ISL_2704754, EPI_ISL_2704756, EPI_ISL_2704762, EPI_ISL_2704766, EPI_ISL_2704770, EPI_ISL_2704777, EPI_ISL_2704778, EPI_ISL_2704784, EPI_ISL_2704788, EPI_ISL_2704793, EPI_ISL_2704794, EPI_ISL_2704796, EPI_ISL_2704799, EPI_ISL_2704800, EPI_ISL_2704803, EPI_ISL_2704806, EPI_ISL_2704808, EPI_ISL_2704810, EPI_ISL_2704813, EPI_ISL_2704816, EPI_ISL_2704817, EPI_ISL_2704820, EPI_ISL_2704822, EPI_ISL_2734268, EPI_ISL_2734276, EPI_ISL_2734277, EPI_ISL_2734281, EPI_ISL_2734291, EPI_ISL_2734292, EPI_ISL_2734293, EPI_ISL_2734294, EPI_ISL_2734295, EPI_ISL_2734296, EPI_ISL_2734297, EPI_ISL_2734298, EPI_ISL_2734299, EPI_ISL_2734300, EPI_ISL_2734301, EPI_ISL_2734302, EPI_ISL_2734303, EPI_ISL_2734304, EPI_ISL_2734305, EPI_ISL_2734306, EPI_ISL_2734307, EPI_ISL_2734309, EPI_ISL_2734310, EPI_ISL_2734313, EPI_ISL_2734316, EPI_ISL_2815913

| | | | |
|---|---|---|---|
| see above | West of Scotland Specialist Virology Centre, NHSGGC / MRC-University of Glasgow Centre for Virus Research | COVID-19 Genomics UK (COG-UK) Consortium | Alasdair MacLean; Alice Broos; Ana da Silva Filipe; Antonia Ho; Daniel Mair; David L Robertson; Emma Thomson; Guy Mollett; Ioulia Tsatsani; James Shepherd; Jenna Nichols; Joseph Hughes; Kathy Li; Kathy Smollett; Kyriaki Nomikou; Lily Tong; Matthew Holden; Natasha Johnson; Rachel Blacow; Richard Orton; Rory Gunson; Sarah McDonald; Sharif Shaaban; Sreenu Vattipally; Stephen Carmichael |

We gratefully acknowledge the following Authors from the Originating laboratories responsible for obtaining the specimens, as well as the Submitting laboratories where the genome data were generated and shared via GISAID, on which this research is based.

All Submitters of data may be contacted directly via www.gisaid.org

Authors are sorted alphabetically.

| Accession ID | Originating Laboratory | Submitting Laboratory | Authors |
|---|---|---|---|
| EPI_ISL_2686172, EPI_ISL_2686173, EPI_ISL_2686174, EPI_ISL_2686175, EPI_ISL_2686176, EPI_ISL_2686177, EPI_ISL_2686179, EPI_ISL_2686180, EPI_ISL_2686181, EPI_ISL_2686182, EPI_ISL_2686183, EPI_ISL_2686184, EPI_ISL_2686185, EPI_ISL_2686186, EPI_ISL_2686187, EPI_ISL_2686188, EPI_ISL_2686189, EPI_ISL_2686190, EPI_ISL_2686191, EPI_ISL_2686192, EPI_ISL_2686193, EPI_ISL_2686194, EPI_ISL_2686197, EPI_ISL_2686199, EPI_ISL_2686200 | | | |
| see above | Amazon Dx, AUV2 Laboratory | Amazon Dx, AUV2 Laboratory | Luke Meredith |
| EPI_ISL_2622473, EPI_ISL_2705488, EPI_ISL_2705489, EPI_ISL_2705490, EPI_ISL_2705491, EPI_ISL_2705492, EPI_ISL_2705493, EPI_ISL_2705494, EPI_ISL_2705495 | | | |
| see above | Barts Health NHS Trust | COVID-19 Genomics UK (COG-UK) Consortium | BROAD; Beatrix; CUTINO-MOGUEL; Claire; David; Dola; HARRINGTON; KELE; KULASEGARAN-SHYLINI; Maria-Teresa; OWOYEMI; Raghavendran |
| EPI_ISL_2708174, EPI_ISL_2708176, EPI_ISL_2708177, EPI_ISL_2708200 | Centre for Enzyme Innovation, University of Portsmouth / Translational Research Laboratory, Portsmouth Hospitals NHS Trust / Hampshire Hospitals NHS Foundation Trust / Bournemouth University / University Hospitals Dorset NHS Foundation Trust / Brighton and Sussex University Hospitals NHS Trust / Dartford and Gravesham NHS Trust / Isle of Wight NHS Trust / Maidstone and Tunbridge Wells NHS Trust / University Hospital Southampton NHS Foundation Trust | COVID-19 Genomics UK (COG-UK) Consortium | Adhyana Mahanama; Allyson Lloyd; Andrew Butt; Angela Beckett; Anibolina Castigador; Anna Mantzaouratou; Anoop Chauhan; Ben Macklin; Benjamin Cogger; Buddhini Samaraweera; Cassandra Malone; Christopher Fearn; Claire Thomas; Dorian Crudgington; Eleri Wilson-Davies; Emanuela Pelosi; Emily Macnaughton; Helen Umpleby; Helen Wheeler; Jacqui Prieto; Jonnie Seymour; Karen Withell; Kate Cook; Katie Loveson; Kelly Bicknell; Kevin Tucker; Kordo Saeed; Liz Sheridan; Magdalena Barrow; Matthew Harvey; Mohammed Hassan-Ibrahim; Nathan Moore; Nick Cortes; Rachael Jeremiah; Robert Impey; Salman Goudarzi; Samirakhon Raupova; Samuel Robson; Sarah Buchan; Sarah Jeremiah; Sarah Wyllie; Scott Elliott; Sharon Glaysher; Siona Silvieri; Stephen Aplin; Stephen Kidd; Thea Sass |
| EPI_ISL_2722291, EPI_ISL_2722292, EPI_ISL_2722293, EPI_ISL_2722295 | Department of Pathology, University of Cambridge | COVID-19 Genomics UK (COG-UK) Consortium | Aminu S. Jahun; Ian Goodfellow; Iliana Georgana; Martin D. Curran; Myra Hosmillo; Rhys Izuagbe; Surendra Parmar; William L. Hamilton; Yasmin Chaudhry |
| EPI_ISL_2638172, EPI_ISL_2638175, EPI_ISL_2638177, EPI_ISL_2638181, EPI_ISL_2638186, EPI_ISL_2638198, EPI_ISL_2638221, EPI_ISL_2638232, EPI_ISL_2638258, EPI_ISL_2638260, EPI_ISL_2638294, EPI_ISL_2638295, EPI_ISL_2638313, EPI_ISL_2638320, EPI_ISL_2638330, EPI_ISL_2638343, EPI_ISL_2638344, EPI_ISL_2638354, EPI_ISL_2638356, EPI_ISL_2638368, EPI_ISL_2638369, EPI_ISL_2638376, EPI_ISL_2638939, EPI_ISL_2638947, EPI_ISL_2638954, EPI_ISL_2638960, EPI_ISL_2638968, EPI_ISL_2638976, EPI_ISL_2638978, EPI_ISL_2638979, EPI_ISL_2638982, EPI_ISL_2638991, EPI_ISL_2638994, EPI_ISL_2638995, EPI_ISL_2639005, EPI_ISL_2639022, EPI_ISL_2639030, EPI_ISL_2639033, EPI_ISL_2639036, EPI_ISL_2639042, EPI_ISL_2639043, EPI_ISL_2639051, EPI_ISL_2639059, EPI_ISL_2639066, EPI_ISL_2639072, EPI_ISL_2639084, EPI_ISL_2639101, EPI_ISL_2639108, EPI_ISL_2639116, EPI_ISL_2639121, EPI_ISL_2639137, EPI_ISL_2639141, EPI_ISL_2639160, EPI_ISL_2639164, EPI_ISL_2639187, EPI_ISL_2639193, EPI_ISL_2639207, EPI_ISL_2639208, EPI_ISL_2639213, EPI_ISL_2639228, EPI_ISL_2639241, EPI_ISL_2639245, EPI_ISL_2664483, EPI_ISL_2664540, EPI_ISL_2664546, EPI_ISL_2664596, EPI_ISL_2664700, EPI_ISL_2664817, EPI_ISL_2664838, EPI_ISL_2664849, EPI_ISL_2664879, EPI_ISL_2664932, EPI_ISL_2664956, EPI_ISL_2664960, EPI_ISL_2665010, EPI_ISL_2682059, EPI_ISL_2682064, EPI_ISL_2682065, EPI_ISL_2682074, EPI_ISL_2682075, EPI_ISL_2682076, EPI_ISL_2682077, EPI_ISL_2682078, EPI_ISL_2682079, EPI_ISL_2682080, EPI_ISL_2682087, EPI_ISL_2682088, EPI_ISL_2682089, EPI_ISL_2682090, EPI_ISL_2682092, EPI_ISL_2682095, EPI_ISL_2682096, EPI_ISL_2682098, EPI_ISL_2682101, EPI_ISL_2682102, EPI_ISL_2682103, EPI_ISL_2682104, EPI_ISL_2682106, EPI_ISL_2682111, EPI_ISL_2682113, EPI_ISL_2682115, EPI_ISL_2682117, EPI_ISL_2682121, EPI_ISL_2682131, EPI_ISL_2682132, EPI_ISL_2682134, EPI_ISL_2682136, EPI_ISL_2682137, EPI_ISL_2682139, EPI_ISL_2682142, EPI_ISL_2682146, EPI_ISL_2682151, EPI_ISL_2682153, EPI_ISL_2682159, EPI_ISL_2682164, EPI_ISL_2682165, EPI_ISL_2682167, EPI_ISL_2682168, EPI_ISL_2682169, EPI_ISL_2682174, EPI_ISL_2682175, EPI_ISL_2682183, EPI_ISL_2682185, EPI_ISL_2682186, EPI_ISL_2682189, EPI_ISL_2682191, EPI_ISL_2682194, EPI_ISL_2682195, EPI_ISL_2682199, EPI_ISL_2682200, EPI_ISL_2682202, EPI_ISL_2682207, EPI_ISL_2682208, EPI_ISL_2682210, EPI_ISL_2682211, EPI_ISL_2682215, EPI_ISL_2682216, EPI_ISL_2682218, EPI_ISL_2682221, EPI_ISL_2682222, EPI_ISL_2682225, EPI_ISL_2682227, EPI_ISL_2682228, EPI_ISL_2682231, EPI_ISL_2682232, EPI_ISL_2682237, EPI_ISL_2682238, EPI_ISL_2682240, EPI_ISL_2682241, EPI_ISL_2682242, EPI_ISL_2682244, EPI_ISL_2682245, EPI_ISL_2682248, EPI_ISL_2682249, EPI_ISL_2682251, EPI_ISL_2682254, EPI_ISL_2682258, EPI_ISL_2682261, EPI_ISL_2682266, EPI_ISL_2682268, EPI_ISL_2682269, EPI_ISL_2682270, EPI_ISL_2682271, EPI_ISL_2682273, EPI_ISL_2682278, EPI_ISL_2682279, EPI_ISL_2682282, EPI_ISL_2682283, EPI_ISL_2682333, EPI_ISL_2704246 | | | |
| see above | Health Services Laboratories | Wellcome Sanger Institute for the COVID-19 Genomics UK (COG-UK) Consortium | Cordelia Langford; David K. Jackson; Dominic Kwiatkowski; Ewan Harrison; Health Services Laboratories and Alex Alderton; Ian Johnston; Jeffrey Barrett; John Sillitoe on behalf of the Wellcome Sanger Institute COVID-19 Surveillance Team; Roberto Amato; Sonia Goncalves |
| EPI_ISL_2638167, EPI_ISL_2638179, EPI_ISL_2638185, EPI_ISL_2638188, EPI_ISL_2638189, EPI_ISL_2638190, EPI_ISL_2638193, EPI_ISL_2638194, EPI_ISL_2638195, EPI_ISL_2638200, EPI_ISL_2638204, EPI_ISL_2638206, EPI_ISL_2638207, EPI_ISL_2638208, EPI_ISL_2638211, EPI_ISL_2638212, EPI_ISL_2638213, EPI_ISL_2638218, EPI_ISL_2638227, EPI_ISL_2638233, EPI_ISL_2638236, EPI_ISL_2638240, EPI_ISL_2638241, EPI_ISL_2638242, EPI_ISL_2638245, EPI_ISL_2638250, EPI_ISL_2638251, EPI_ISL_2638253, EPI_ISL_2638263, EPI_ISL_2638267, EPI_ISL_2638273, EPI_ISL_2638281, EPI_ISL_2638284, EPI_ISL_2638286, EPI_ISL_2638288, EPI_ISL_2638289, EPI_ISL_2638297, EPI_ISL_2638301, EPI_ISL_2638304, EPI_ISL_2638305, EPI_ISL_2638309, EPI_ISL_2638311, EPI_ISL_2638312, EPI_ISL_2638314, EPI_ISL_2638315, EPI_ISL_2638323, EPI_ISL_2638326, EPI_ISL_2638331, EPI_ISL_2638334, EPI_ISL_2638337, EPI_ISL_2638340, EPI_ISL_2638345, EPI_ISL_2638346, EPI_ISL_2638350, EPI_ISL_2638363, EPI_ISL_2638364, EPI_ISL_2638373, EPI_ISL_2638374, EPI_ISL_2638378, EPI_ISL_2638380, EPI_ISL_2638382, EPI_ISL_2638385, EPI_ISL_2638387, EPI_ISL_2638389, EPI_ISL_2638390, EPI_ISL_2639256, EPI_ISL_2639265, EPI_ISL_2639266, EPI_ISL_2639279, EPI_ISL_2639284, EPI_ISL_2639285, EPI_ISL_2639291, EPI_ISL_2639292, EPI_ISL_2639296, EPI_ISL_2639301, EPI_ISL_2639304, EPI_ISL_2639305, EPI_ISL_2639313, EPI_ISL_2639314, EPI_ISL_2639315, EPI_ISL_2639318, EPI_ISL_2639346, EPI_ISL_2639349, EPI_ISL_2639351, EPI_ISL_2639360, EPI_ISL_2639399, EPI_ISL_2639402, EPI_ISL_2639405, EPI_ISL_2639409, EPI_ISL_2639410, EPI_ISL_2639418, EPI_ISL_2639421, EPI_ISL_2639422, EPI_ISL_2639432, EPI_ISL_2639434, EPI_ISL_2639437, EPI_ISL_2639440, EPI_ISL_2639445, EPI_ISL_2639447, EPI_ISL_2639450, EPI_ISL_2639452, EPI_ISL_2639455, EPI_ISL_2639458, EPI_ISL_2639467, EPI_ISL_2639472, EPI_ISL_2639475, EPI_ISL_2639493, EPI_ISL_2639496, EPI_ISL_2639499, EPI_ISL_2639500, EPI_ISL_2639502, EPI_ISL_2639504, EPI_ISL_2639511, EPI_ISL_2639512, EPI_ISL_2639516, EPI_ISL_2639526, EPI_ISL_2639531, EPI_ISL_2639535, EPI_ISL_2639539, EPI_ISL_2639547, EPI_ISL_2639548, EPI_ISL_2639552, EPI_ISL_2639553, EPI_ISL_2639566, EPI_ISL_2639570, EPI_ISL_2639571, EPI_ISL_2639575, EPI_ISL_2639576, EPI_ISL_2639578, EPI_ISL_2639579, EPI_ISL_2639583, EPI_ISL_2639586, EPI_ISL_2639588, EPI_ISL_2639589, EPI_ISL_2639592, EPI_ISL_2639596, EPI_ISL_2639605, EPI_ISL_2639606, EPI_ISL_2639613, EPI_ISL_2651948, EPI_ISL_2664013, EPI_ISL_2664014, EPI_ISL_2664025, EPI_ISL_2664032, EPI_ISL_2664037, EPI_ISL_2664039, EPI_ISL_2664041, EPI_ISL_2664046, EPI_ISL_2664047, EPI_ISL_2664056, EPI_ISL_2664062, EPI_ISL_2664067, EPI_ISL_2664068, EPI_ISL_2664071, EPI_ISL_2664075, EPI_ISL_2664082, EPI_ISL_2664089, EPI_ISL_2664090, EPI_ISL_2664094, EPI_ISL_2664097, EPI_ISL_2664100, EPI_ISL_2664106, EPI_ISL_2664107, EPI_ISL_2664108, EPI_ISL_2664109, EPI_ISL_2664111, EPI_ISL_2664119, EPI_ISL_2664121, EPI_ISL_2664128, EPI_ISL_2664135, EPI_ISL_2664153, EPI_ISL_2664157, EPI_ISL_2664159, EPI_ISL_2664162, EPI_ISL_2664167, EPI_ISL_2664175, EPI_ISL_2664178, EPI_ISL_2664186, EPI_ISL_2664187, EPI_ISL_2664189, EPI_ISL_2664195, EPI_ISL_2664202, EPI_ISL_2664203, EPI_ISL_2664213, EPI_ISL_2664214, EPI_ISL_2664216, EPI_ISL_2664217, EPI_ISL_2664219, EPI_ISL_2664232, EPI_ISL_2664236, EPI_ISL_2664242, EPI_ISL_2664246, EPI_ISL_2664248, EPI_ISL_2664251, EPI_ISL_2664263, EPI_ISL_2664265, EPI_ISL_2664270, EPI_ISL_2664273, EPI_ISL_2664282, EPI_ISL_2664286, EPI_ISL_2664291, EPI_ISL_2664292, EPI_ISL_2664295, EPI_ISL_2664297, EPI_ISL_2664300, EPI_ISL_2664308, EPI_ISL_2664311, EPI_ISL_2664316, EPI_ISL_2664317, EPI_ISL_2664318, EPI_ISL_2664324, EPI_ISL_2664329, EPI_ISL_2664335, EPI_ISL_2664336, EPI_ISL_2664337, EPI_ISL_2664341, EPI_ISL_2664349, EPI_ISL_2664350, EPI_ISL_2664356, EPI_ISL_2664358, EPI_ISL_2664360, EPI_ISL_2664362, EPI_ISL_2664368, EPI_ISL_2664370, EPI_ISL_2664372, EPI_ISL_2664374, EPI_ISL_2664375, EPI_ISL_2664377, EPI_ISL_2664380, EPI_ISL_2664384, EPI_ISL_2664396, EPI_ISL_2664400, EPI_ISL_2664402, EPI_ISL_2664403, EPI_ISL_2664406, EPI_ISL_2664411, EPI_ISL_2664413, EPI_ISL_2664416, EPI_ISL_2664418, EPI_ISL_2664420, EPI_ISL_2664424, EPI_ISL_2664428, EPI_ISL_2664429, EPI_ISL_2664437, EPI_ISL_2664439, EPI_ISL_2664442, EPI_ISL_2664447, EPI_ISL_2664448, EPI_ISL_2664449, EPI_ISL_2664450, EPI_ISL_2664456, EPI_ISL_2664457, EPI_ISL_2664458, EPI_ISL_2664462, EPI_ISL_2664464, EPI_ISL_2664465, EPI_ISL_2664467, EPI_ISL_2664470, EPI_ISL_2664471, EPI_ISL_2664474, EPI_ISL_2664477, EPI_ISL_2664486, EPI_ISL_2664491, EPI_ISL_2664495, EPI_ISL_2664500, EPI_ISL_2664503, EPI_ISL_2664508, EPI_ISL_2664514, EPI_ISL_2664519, EPI_ISL_2664536, EPI_ISL_2664539, EPI_ISL_2664542, EPI_ISL_2664545, EPI_ISL_2664547, EPI_ISL_2664548, EPI_ISL_2664552, EPI_ISL_2664557, EPI_ISL_2664562, EPI_ISL_2664564, EPI_ISL_2664567, EPI_ISL_2664570, EPI_ISL_2664573, EPI_ISL_2664574, EPI_ISL_2664579, EPI_ISL_2664581, EPI_ISL_2664582, EPI_ISL_2664585, EPI_ISL_2664597, EPI_ISL_2664598, EPI_ISL_2664599, EPI_ISL_2664601, EPI_ISL_2664612, EPI_ISL_2664615, EPI_ISL_2664617, EPI_ISL_2664620, EPI_ISL_2664636, EPI_ISL_2664640, EPI_ISL_2664643, EPI_ISL_2664644, EPI_ISL_2664648, EPI_ISL_2664651, EPI_ISL_2664652, EPI_ISL_2664653, EPI_ISL_2664659, EPI_ISL_2664664, EPI_ISL_2664665, EPI_ISL_2664668, EPI_ISL_2664685, EPI_ISL_2664691, EPI_ISL_2664693, EPI_ISL_2664705, EPI_ISL_2664708, EPI_ISL_2664710, EPI_ISL_2664711, EPI_ISL_2664712, EPI_ISL_2664715, EPI_ISL_2664717, EPI_ISL_2664718, EPI_ISL_2664721, EPI_ISL_2664722, EPI_ISL_2664723, EPI_ISL_2664730, EPI_ISL_2664731, EPI_ISL_2664738, EPI_ISL_2664739, EPI_ISL_2664742, EPI_ISL_2664744, EPI_ISL_2664753, EPI_ISL_2664756, EPI_ISL_2664759, EPI_ISL_2664760, EPI_ISL_2664761, EPI_ISL_2664763, EPI_ISL_2664766, EPI_ISL_2664767, EPI_ISL_2664768, EPI_ISL_2664775, EPI_ISL_2664776, EPI_ISL_2664777, EPI_ISL_2664778, EPI_ISL_2664782, EPI_ISL_2664785, EPI_ISL_2664786, EPI_ISL_2664789, EPI_ISL_2664791, EPI_ISL_2664797, EPI_ISL_2664798, EPI_ISL_2664807, EPI_ISL_2664808, EPI_ISL_2664813, EPI_ISL_2664814, EPI_ISL_2664815, EPI_ISL_2664821, EPI_ISL_2664829, EPI_ISL_2664831, EPI_ISL_2664832, EPI_ISL_2664835, EPI_ISL_2664841, EPI_ISL_2664842, EPI_ISL_2664846, EPI_ISL_2664847, EPI_ISL_2664848, EPI_ISL_2664854, EPI_ISL_2664859, EPI_ISL_2664860, EPI_ISL_2664861, EPI_ISL_2664862, EPI_ISL_2664863, EPI_ISL_2664864, EPI_ISL_2664870, EPI_ISL_2664872, EPI_ISL_2664883, EPI_ISL_2664889, EPI_ISL_2664893, EPI_ISL_2664904, EPI_ISL_2664907, EPI_ISL_2664909, EPI_ISL_2664911, EPI_ISL_2664913, EPI_ISL_2664916, EPI_ISL_2664920, EPI_ISL_2664921, EPI_ISL_2664925, EPI_ISL_2664927, EPI_ISL_2664935, EPI_ISL_2664936, EPI_ISL_2664938, EPI_ISL_2664939, EPI_ISL_2664941, EPI_ISL_2664943, EPI_ISL_2664947, EPI_ISL_2664948, EPI_ISL_2664949, EPI_ISL_2664952, EPI_ISL_2664954, EPI_ISL_2664959, EPI_ISL_2664962, EPI_ISL_2664963, EPI_ISL_2664964, EPI_ISL_2664965, EPI_ISL_2664968, EPI_ISL_2664969, EPI_ISL_2664973, EPI_ISL_2664978, EPI_ISL_2664981, EPI_ISL_2664983, EPI_ISL_2664984, EPI_ISL_2664985, EPI_ISL_2664990, EPI_ISL_2664991, EPI_ISL_2664992, EPI_ISL_2664996, EPI_ISL_2665001, EPI_ISL_2665005, EPI_ISL_2665006, EPI_ISL_2665007, EPI_ISL_2665009, EPI_ISL_2665016, EPI_ISL_2665018, EPI_ISL_2665022, EPI_ISL_2665026, EPI_ISL_2665030, EPI_ISL_2665033, EPI_ISL_2665041, EPI_ISL_2665043, EPI_ISL_2665044, EPI_ISL_2665053, EPI_ISL_2665054, EPI_ISL_2665056, EPI_ISL_2665058, EPI_ISL_2665070, EPI_ISL_2665075, EPI_ISL_2665077, EPI_ISL_2665085, EPI_ISL_2665095, EPI_ISL_2665096, EPI_ISL_2665097, EPI_ISL_2665113, EPI_ISL_2665119, EPI_ISL_2665123, EPI_ISL_2665127, EPI_ISL_2665128, EPI_ISL_2665131, EPI_ISL_2665133, EPI_ISL_2665139, EPI_ISL_2665140, EPI_ISL_2665141, EPI_ISL_2665147, EPI_ISL_2665150, EPI_ISL_2665157, EPI_ISL_2665158, EPI_ISL_2665164, EPI_ISL_2665167, EPI_ISL_2665168, EPI_ISL_2665172, EPI_ISL_2665179, EPI_ISL_2665180, EPI_ISL_2665186, EPI_ISL_2665187, EPI_ISL_2665189, EPI_ISL_2665190, EPI_ISL_2665191, EPI_ISL_2665192, EPI_ISL_2665198, EPI_ISL_2665200, EPI_ISL_2665208, EPI_ISL_2665218, EPI_ISL_2665230, EPI_ISL_2665236, EPI_ISL_2665246, EPI_ISL_2665247, EPI_ISL_2665251, EPI_ISL_2665255, EPI_ISL_2665262, EPI_ISL_2665266, EPI_ISL_2665273, EPI_ISL_2665275, EPI_ISL_2665277, EPI_ISL_2665278, EPI_ISL_2665281, EPI_ISL_2665282, EPI_ISL_2665285, EPI_ISL_2665293, EPI_ISL_2665297, EPI_ISL_2665302, EPI_ISL_2665311, EPI_ISL_2665330, EPI_ISL_2665332, EPI_ISL_2665347, EPI_ISL_2665353, EPI_ISL_2665359, EPI_ISL_2665360, EPI_ISL_2665370, EPI_ISL_2665374, EPI_ISL_2665377, EPI_ISL_2665379, EPI_ISL_2665380, EPI_ISL_2681642, EPI_ISL_2681643, EPI_ISL_2681644, EPI_ISL_2681645, EPI_ISL_2681647, EPI_ISL_2681651, EPI_ISL_2681652, EPI_ISL_2681653, EPI_ISL_2681654, EPI_ISL_2681655, EPI_ISL_2681658, EPI_ISL_2681659, EPI_ISL_2681662, EPI_ISL_2681663, EPI_ISL_2681668, EPI_ISL_2681669, EPI_ISL_2681671, EPI_ISL_2681674, EPI_ISL_2681676, EPI_ISL_2681677, EPI_ISL_2681678, EPI_ISL_2681680, EPI_ISL_2681683, EPI_ISL_2681684, EPI_ISL_2681686, EPI_ISL_2681687, EPI_ISL_2681692, EPI_ISL_2681693, EPI_ISL_2681694, EPI_ISL_2681695, EPI_ISL_2681697, EPI_ISL_2681698, EPI_ISL_2681699, EPI_ISL_2681700, EPI_ISL_2681702, EPI_ISL_2681703, EPI_ISL_2681704, EPI_ISL_2681707, EPI_ISL_2681708, EPI_ISL_2681709, EPI_ISL_2681710, EPI_ISL_2681712, EPI_ISL_2681714, EPI_ISL_2681717, EPI_ISL_2681722, EPI_ISL_2681723, EPI_ISL_2681724, EPI_ISL_2681725, EPI_ISL_2681726, EPI_ISL_2681729, EPI_ISL_2681732, EPI_ISL_2681734, EPI_ISL_2681737, EPI_ISL_2681739, EPI_ISL_2681740, EPI_ISL_2681742, EPI_ISL_2681744, EPI_ISL_2681745, EPI_ISL_2681747, EPI_ISL_2681749, EPI_ISL_2681750, EPI_ISL_2681751, EPI_ISL_2681752, EPI_ISL_2681754, EPI_ISL_2681755, EPI_ISL_2681757, EPI_ISL_2681758, EPI_ISL_2681760, EPI_ISL_2681763, EPI_ISL_2681765, EPI_ISL_2681766, EPI_ISL_2681772, EPI_ISL_2681774, EPI_ISL_2681775, EPI_ISL_2681776, EPI_ISL_2681777, EPI_ISL_2681780, EPI_ISL_2681781, EPI_ISL_2681782, EPI_ISL_2681783, EPI_ISL_2681784, EPI_ISL_2681785, EPI_ISL_2681786, EPI_ISL_2681787, EPI_ISL_2681789, EPI_ISL_2681790, EPI_ISL_2681791, EPI_ISL_2681793, EPI_ISL_2681794, EPI_ISL_2681796, EPI_ISL_2681799, EPI_ISL_2681800, EPI_ISL_2681801, EPI_ISL_2681802, EPI_ISL_2681803, EPI_ISL_2681804, EPI_ISL_2681805, EPI_ISL_2681807, EPI_ISL_2681808, EPI_ISL_2681809, EPI_ISL_2681811, EPI_ISL_2681812, EPI_ISL_2681813, EPI_ISL_2681815, EPI_ISL_2681816, EPI_ISL_2681817, EPI_ISL_2681818, EPI_ISL_2681820, EPI_ISL_2681822, EPI_ISL_2681823, EPI_ISL_2681824, EPI_ISL_2681825, EPI_ISL_2681826, EPI_ISL_2681827, EPI_ISL_2681831, EPI_ISL_2681832, EPI_ISL_2681833, EPI_ISL_2681834, EPI_ISL_2681835, EPI_ISL_2681837, EPI_ISL_2681839, EPI_ISL_2681841, EPI_ISL_2681845, EPI_ISL_2681846, EPI_ISL_2681850, EPI_ISL_2681851, EPI_ISL_2681852, EPI_ISL_2681853, EPI_ISL_2681855, EPI_ISL_2681856, EPI_ISL_2681857, EPI_ISL_2681858, EPI_ISL_2681860, EPI_ISL_2681861, EPI_ISL_2681864, EPI_ISL_2681866, EPI_ISL_2681869, EPI_ISL_2681870, EPI_ISL_2681871, EPI_ISL_2681873, EPI_ISL_2681877, EPI_ISL_2681880, EPI_ISL_2681881, EPI_ISL_2681882, EPI_ISL_2681885, EPI_ISL_2681887, EPI_ISL_2681890, EPI_ISL_2681892, EPI_ISL_2681894, EPI_ISL_2681895, EPI_ISL_2681898, EPI_ISL_2681899, EPI_ISL_2681901, EPI_ISL_2681903, EPI_ISL_2681904, EPI_ISL_2681905, EPI_ISL_2681906, EPI_ISL_2681907, EPI_ISL_2681908, EPI_ISL_2681909, EPI_ISL_2681910, EPI_ISL_2681911, EPI_ISL_2681915, EPI_ISL_2681917, EPI_ISL_2681919, EPI_ISL_2681922, EPI_ISL_2681924, EPI_ISL_2681925, EPI_ISL_2681927, EPI_ISL_2682285, EPI_ISL_2682286, EPI_ISL_2682287, EPI_ISL_2682289, EPI_ISL_2682290, EPI_ISL_2682293, EPI_ISL_2682294, EPI_ISL_2682295, EPI_ISL_2682297, EPI_ISL_2682298, EPI_ISL_2682299, EPI_ISL_2682300, EPI_ISL_2682301, EPI_ISL_2682302, EPI_ISL_2682303, EPI_ISL_2682304, EPI_ISL_2682305, EPI_ISL_2682306, EPI_ISL_2682308, EPI_ISL_2682309, EPI_ISL_2682310, EPI_ISL_2682311, EPI_ISL_2682312, EPI_ISL_2682314, EPI_ISL_2682315, EPI_ISL_2682316, EPI_ISL_2682317, EPI_ISL_2682318, EPI_ISL_2682319, EPI_ISL_2682321, EPI_ISL_2682322, EPI_ISL_2682323, EPI_ISL_2682324, EPI_ISL_2682326, EPI_ISL_2682327, EPI_ISL_2682328, EPI_ISL_2682329, EPI_ISL_2682330, EPI_ISL_2682331, EPI_ISL_2682332, EPI_ISL_2682335, EPI_ISL_2682336, EPI_ISL_2682337, EPI_ISL_2682339, EPI_ISL_2682340, EPI_ISL_2682341, EPI_ISL_2682342, EPI_ISL_2682343, EPI_ISL_2682344, EPI_ISL_2682345, EPI_ISL_2682346, EPI_ISL_2682347, EPI_ISL_2682349, EPI_ISL_2682352, EPI_ISL_2682353, EPI_ISL_2682355, EPI_ISL_2682356, EPI_ISL_2682358, EPI_ISL_2682359, EPI_ISL_2682360, EPI_ISL_2682361, EPI_ISL_2682362, EPI_ISL_2682363, EPI_ISL_2682364, EPI_ISL_2682365, EPI_ISL_2682366, EPI_ISL_2682367, EPI_ISL_2682368, EPI_ISL_2682369, EPI_ISL_2682370, EPI_ISL_2682372, EPI_ISL_2682373, EPI_ISL_2682374, EPI_ISL_2682375, EPI_ISL_2682376, EPI_ISL_2682377, EPI_ISL_2682378, EPI_ISL_2682379, EPI_ISL_2682380, EPI_ISL_2682381, EPI_ISL_2682382, EPI_ISL_2682383, EPI_ISL_2682384, EPI_ISL_2682385, EPI_ISL_2682386, EPI_ISL_2682387, EPI_ISL_2682389, EPI_ISL_2682390, EPI_ISL_2682391, EPI_ISL_2682392, EPI_ISL_2682393, EPI_ISL_2682394, EPI_ISL_2682396, EPI_ISL_2682397, EPI_ISL_2682398, EPI_ISL_2682399, EPI_ISL_2682401, EPI_ISL_2682403, EPI_ISL_2682404, EPI_ISL_2682405, EPI_ISL_2682406, EPI_ISL_2682407, EPI_ISL_2682408, EPI_ISL_2682410, EPI_ISL_2682411, EPI_ISL_2682412, EPI_ISL_2682413, EPI_ISL_2682414, EPI_ISL_2682415, EPI_ISL_2682417, EPI_ISL_2682418, EPI_ISL_2682419, EPI_ISL_2682420, EPI_ISL_2682421, EPI_ISL_2682422, EPI_ISL_2682423, EPI_ISL_2682424, EPI_ISL_2682425, EPI_ISL_2682426, EPI_ISL_2682427, EPI_ISL_2682428, EPI_ISL_2682429, EPI_ISL_2682430, EPI_ISL_2682431, EPI_ISL_2682432, EPI_ISL_2682434, EPI_ISL_2682435, EPI_ISL_2682436, EPI_ISL_2682437, EPI_ISL_2682438, EPI_ISL_2682439, EPI_ISL_2682440, EPI_ISL_2682441, EPI_ISL_2682443, EPI_ISL_2682444, EPI_ISL_2682446, EPI_ISL_2682447, EPI_ISL_2682448, EPI_ISL_2682450, EPI_ISL_2682451, EPI_ISL_2682452, EPI_ISL_2682453, EPI_ISL_2682454, EPI_ISL_2682456, EPI_ISL_2682458, EPI_ISL_2682459, EPI_ISL_2682460, EPI_ISL_2682461, EPI_ISL_2682462, EPI_ISL_2682463, EPI_ISL_2682464, EPI_ISL_2682465, EPI_ISL_2682466, EPI_ISL_2682468, EPI_ISL_2682469, EPI_ISL_2682470, EPI_ISL_2682472, EPI_ISL_2682476, EPI_ISL_2682477, EPI_ISL_2682478, EPI_ISL_2682480, EPI_ISL_2682481, EPI_ISL_2682482, EPI_ISL_2682483, EPI_ISL_2682484, EPI_ISL_2682485, EPI_ISL_2682486, EPI_ISL_2682487, EPI_ISL_2682491, EPI_ISL_2682492, EPI_ISL_2682493, EPI_ISL_2682494, EPI_ISL_2682495, EPI_ISL_2682496, EPI_ISL_2682497, EPI_ISL_2682498, EPI_ISL_2682499, EPI_ISL_2682500, EPI_ISL_2682501, EPI_ISL_2682502, EPI_ISL_2682504, EPI_ISL_2682505, EPI_ISL_2682506, EPI_ISL_2682507, EPI_ISL_2682508, EPI_ISL_2682509, EPI_ISL_2682510, EPI_ISL_2682511, EPI_ISL_2682513, EPI_ISL_2682514, EPI_ISL_2682515, EPI_ISL_2682516, EPI_ISL_2682517, EPI_ISL_2682518, EPI_ISL_2682519, EPI_ISL_2682520, EPI_ISL_2682521, EPI_ISL_2682522, EPI_ISL_2682525, EPI_ISL_2682527, EPI_ISL_2682528, EPI_ISL_2682529, EPI_ISL_2682530, EPI_ISL_2682531, EPI_ISL_2682532, EPI_ISL_2682533, EPI_ISL_2682534, EPI_ISL_2682535, EPI_ISL_2682536, EPI_ISL_2682539, EPI_ISL_2682540, EPI_ISL_2682541, EPI_ISL_2682542, EPI_ISL_2682543, EPI_ISL_2682544, EPI_ISL_2682547, EPI_ISL_2682548, EPI_ISL_2682550, EPI_ISL_2682551, EPI_ISL_2682552, EPI_ISL_2682553, EPI_ISL_2682554, EPI_ISL_2682555, EPI_ISL_2682556, EPI_ISL_2682557, EPI_ISL_2682559, EPI_ISL_2682560, EPI_ISL_2682561, EPI_ISL_2682563, EPI_ISL_2682564, EPI_ISL_2682565, EPI_ISL_2682566, EPI_ISL_2682567, EPI_ISL_2682568, EPI_ISL_2682570, EPI_ISL_2682571, EPI_ISL_2682572, EPI_ISL_2682573, EPI_ISL_2682574, EPI_ISL_2682575, EPI_ISL_2682576, EPI_ISL_2682577, EPI_ISL_2682578, EPI_ISL_2682579, EPI_ISL_2682580, EPI_ISL_2682581, EPI_ISL_2682582, EPI_ISL_2682583, EPI_ISL_2682584, EPI_ISL_2682585, EPI_ISL_2682586, EPI_ISL_2682587, EPI_ISL_2682588, EPI_ISL_2682589, EPI_ISL_2682590, EPI_ISL_2682591, EPI_ISL_2682592, EPI_ISL_2682593, EPI_ISL_2682596, EPI_ISL_2682598, EPI_ISL_2682599, EPI_ISL_2682600, EPI_ISL_2682601, EPI_ISL_2682602, EPI_ISL_2682603, EPI_ISL_2682604, EPI_ISL_2682605, EPI_ISL_2682606, EPI_ISL_2682607, EPI_ISL_2682608, EPI_ISL_2682609, EPI_ISL_2682610, EPI_ISL_2682611, EPI_ISL_2682612, EPI_ISL_2682613, EPI_ISL_2682614, EPI_ISL_2682615, EPI_ISL_2682616, EPI_ISL_2682617, EPI_ISL_2682618, EPI_ISL_2682619, EPI_ISL_2682620, EPI_ISL_2682621, EPI_ISL_2682622, EPI_ISL_2682624, EPI_ISL_2682625, EPI_ISL_2682626, EPI_ISL_2682627, EPI_ISL_2682628, EPI_ISL_2682629, EPI_ISL_2682631, EPI_ISL_2682632, EPI_ISL_2682633, EPI_ISL_2682634, EPI_ISL_2682635, EPI_ISL_2682636, EPI_ISL_2682638, EPI_ISL_2682639, EPI_ISL_2682640, EPI_ISL_2682641, EPI_ISL_2682642, EPI_ISL_2682643, EPI_ISL_2682644, EPI_ISL_2682645, EPI_ISL_2682647, EPI_ISL_2682649, EPI_ISL_2682650, EPI_ISL_2682652, EPI_ISL_2682654, EPI_ISL_2682658, EPI_ISL_2682664, EPI_ISL_2682667, EPI_ISL_2682668, EPI_ISL_2682671, EPI_ISL_2682673, EPI_ISL_2682674, EPI_ISL_2682675, EPI_ISL_2682678, EPI_ISL_2682681, EPI_ISL_2682683, EPI_ISL_2682687, EPI_ISL_2682688, EPI_ISL_2682697, EPI_ISL_2682700, EPI_ISL_2682702, EPI_ISL_2682705, EPI_ISL_2682706, EPI_ISL_2682707, EPI_ISL_2682709, EPI_ISL_2682713, EPI_ISL_2682714, EPI_ISL_2682717, EPI_ISL_2682720, EPI_ISL_2682721, EPI_ISL_2682722, EPI_ISL_2682723, EPI_ISL_2682726, EPI_ISL_2682735, EPI_ISL_2682741, EPI_ISL_2682743, EPI_ISL_2682748, EPI_ISL_2682749, EPI_ISL_2682755, EPI_ISL_2682756, EPI_ISL_2682759, EPI_ISL_2682764, EPI_ISL_2682770, EPI_ISL_2682775, EPI_ISL_2682779, EPI_ISL_2682782, EPI_ISL_2682786, EPI_ISL_2682787, EPI_ISL_2682792, EPI_ISL_2682796, EPI_ISL_2682798, EPI_ISL_2682807, EPI_ISL_2682808, EPI_ISL_2682811, EPI_ISL_2682815, EPI_ISL_2682819, EPI_ISL_2682824, EPI_ISL_2682826, EPI_ISL_2682829, EPI_ISL_2682833, EPI_ISL_2682836, EPI_ISL_2682837, EPI_ISL_2682842, EPI_ISL_2682845, EPI_ISL_2682853, EPI_ISL_2682858, EPI_ISL_2682873, EPI_ISL_2682881, EPI_ISL_2682886, EPI_ISL_2682888, EPI_ISL_2682896, EPI_ISL_2682902, EPI_ISL_2682903, EPI_ISL_2682914, EPI_ISL_2682924, EPI_ISL_2682925, EPI_ISL_2682929, EPI_ISL_2682932, EPI_ISL_2682933, EPI_ISL_2682935, EPI_ISL_2682938, EPI_ISL_2682941, EPI_ISL_2682942, EPI_ISL_2682943, EPI_ISL_2682944, EPI_ISL_2682945, EPI_ISL_2682950, EPI_ISL_2682954, EPI_ISL_2682955, EPI_ISL_2682961, EPI_ISL_2682969, EPI_ISL_2683005, EPI_ISL_2683016, EPI_ISL_2683111, EPI_ISL_2683112, EPI_ISL_2683114, EPI_ISL_2699868, EPI_ISL_2699870, EPI_ISL_2699871, EPI_ISL_2699873, EPI_ISL_2699886, EPI_ISL_2699887, EPI_ISL_2699889, EPI_ISL_2699892, EPI_ISL_2699896, EPI_ISL_2699899, EPI_ISL_2699907, EPI_ISL_2699908, EPI_ISL_2699909, EPI_ISL_2699910, EPI_ISL_2699913, EPI_ISL_2699914, EPI_ISL_2699919, EPI_ISL_2699928, EPI_ISL_2699931, EPI_ISL_2699932, EPI_ISL_2699933, EPI_ISL_2699934, EPI_ISL_2699936, EPI_ISL_2699946, EPI_ISL_2699947, EPI_ISL_2699952, EPI_ISL_2699953, EPI_ISL_2699955, EPI_ISL_2699965, EPI_ISL_2699968, EPI_ISL_2699970, EPI_ISL_2699972, EPI_ISL_2699973, EPI_ISL_2699974, EPI_ISL_2699978, EPI_ISL_2699982, EPI_ISL_2699984, EPI_ISL_2699992, EPI_ISL_2699994, EPI_ISL_2700000, EPI_ISL_2700012, EPI_ISL_2700017, EPI_ISL_2700018, EPI_ISL_2700023, EPI_ISL_2700024, EPI_ISL_2700026, EPI_ISL_2700045, EPI_ISL_2700047, EPI_ISL_2700050, EPI_ISL_2700057, EPI_ISL_2700069, EPI_ISL_2700077, EPI_ISL_2700078, EPI_ISL_2700079, EPI_ISL_2700081, EPI_ISL_2700082, EPI_ISL_2700083, EPI_ISL_2700089, EPI_ISL_2700098, EPI_ISL_2700099, EPI_ISL_2700101, EPI_ISL_2700104, EPI_ISL_2700114, EPI_ISL_2700121, EPI_ISL_2700122, EPI_ISL_2700124, EPI_ISL_2700128, EPI_ISL_2700141, EPI_ISL_2700142, EPI_ISL_2700143, EPI_ISL_2700152, EPI_ISL_2700159, EPI_ISL_2700164, EPI_ISL_2700165, EPI_ISL_2700166, EPI_ISL_2700171, EPI_ISL_2700178, EPI_ISL_2700183, EPI_ISL_2700184, EPI_ISL_2700185, EPI_ISL_2700188, EPI_ISL_2700191, EPI_ISL_2700193, EPI_ISL_2700195, EPI_ISL_2700196, EPI_ISL_2700198, EPI_ISL_2700200, EPI_ISL_2700201, EPI_ISL_2700208, EPI_ISL_2700209, EPI_ISL_2700212, EPI_ISL_2700579, EPI_ISL_2700580, EPI_ISL_2700581, EPI_ISL_2700582, EPI_ISL_2700583, EPI_ISL_2700585, EPI_ISL_2700586, EPI_ISL_2700587, EPI_ISL_2700588, EPI_ISL_2700590, EPI_ISL_2700591, EPI_ISL_2700595, EPI_ISL_2700596, EPI_ISL_2700598, EPI_ISL_2700599, EPI_ISL_2700600, EPI_ISL_2700601, EPI_ISL_2700602, EPI_ISL_2700606, EPI_ISL_2700608, EPI_ISL_2700611, EPI_ISL_2700612, EPI_ISL_2700613, EPI_ISL_2700615, EPI_ISL_2700616, EPI_ISL_2700617, EPI_ISL_2700620, EPI_ISL_2700621, EPI_ISL_2700622, EPI_ISL_2700624, EPI_ISL_2700626, EPI_ISL_2700627, EPI_ISL_2700628, EPI_ISL_2700629, EPI_ISL_2700630, EPI_ISL_2700632, EPI_ISL_2700634, EPI_ISL_2700637, EPI_ISL_2700638, EPI_ISL_2700639, EPI_ISL_2700640, EPI_ISL_2700641, EPI_ISL_2700645, EPI_ISL_2700647, EPI_ISL_2700648, EPI_ISL_2700649, EPI_ISL_2700652, EPI_ISL_2700653, EPI_ISL_2700655, EPI_ISL_2700656, EPI_ISL_2700658, EPI_ISL_2700659, EPI_ISL_2700663, EPI_ISL_2700667, EPI_ISL_2700669, EPI_ISL_2700673, EPI_ISL_2700674, EPI_ISL_2700675, EPI_ISL_2700676, EPI_ISL_2700677, EPI_ISL_2700678, EPI_ISL_2700679, EPI_ISL_2700682, EPI_ISL_2700683, EPI_ISL_2700684, EPI_ISL_2700685, EPI_ISL_2700686, EPI_ISL_2700689, EPI_ISL_2700692, EPI_ISL_2700693, EPI_ISL_2700695, EPI_ISL_2700696, EPI_ISL_2700697, EPI_ISL_2700700, EPI_ISL_2700701, EPI_ISL_2700703, EPI_ISL_2700704, EPI_ISL_2700705, EPI_ISL_2700710, EPI_ISL_2700711, EPI_ISL_2700712, EPI_ISL_2700714, EPI_ISL_2700715, EPI_ISL_2700717, EPI_ISL_2700719, EPI_ISL_2700720, EPI_ISL_2700721, EPI_ISL_2700722, EPI_ISL_2700723, EPI_ISL_2700724, EPI_ISL_2700725, EPI_ISL_2700727, EPI_ISL_2700729, EPI_ISL_2700730, EPI_ISL_2700731, EPI_ISL_2700732, EPI_ISL_2700734, EPI_ISL_2700737, EPI_ISL_2700739, EPI_ISL_2700740, EPI_ISL_2700742, EPI_ISL_2700743, EPI_ISL_2700744, EPI_ISL_2700746, EPI_ISL_2700747, EPI_ISL_2700748, EPI_ISL_2700751, EPI_ISL_2700752, EPI_ISL_2700755, EPI_ISL_2700758, EPI_ISL_2700759, EPI_ISL_2700760, EPI_ISL_2700765, EPI_ISL_2700766, EPI_ISL_2700769, EPI_ISL_2700772, EPI_ISL_2700776, EPI_ISL_2700777, EPI_ISL_2700781, EPI_ISL_2700784, EPI_ISL_2700785, EPI_ISL_2700786, EPI_ISL_2700788, EPI_ISL_2700789, EPI_ISL_2700791, EPI_ISL_2700792, EPI_ISL_2700793, EPI_ISL_2700795, EPI_ISL_2700797, EPI_ISL_2700798, EPI_ISL_2700800, EPI_ISL_2700802, EPI_ISL_2700803, EPI_ISL_2700805, EPI_ISL_2700806, EPI_ISL_2700807, EPI_ISL_2700808, EPI_ISL_2700809, EPI_ISL_2700810, EPI_ISL_2700812, EPI_ISL_2700813, EPI_ISL_2700816, EPI_ISL_2700817, EPI_ISL_2700818, EPI_ISL_2700819, EPI_ISL_2700820, EPI_ISL_2700823, EPI_ISL_2700826, EPI_ISL_2700830, EPI_ISL_2700833, EPI_ISL_2700835, EPI_ISL_2700836, EPI_ISL_2700837, EPI_ISL_2700838, EPI_ISL_2700839, EPI_ISL_2700840, EPI_ISL_2700841, EPI_ISL_2700842, EPI_ISL_2700843, EPI_ISL_2700851, EPI_ISL_2700852, EPI_ISL_2700855, EPI_ISL_2700856, EPI_ISL_2700857, EPI_ISL_2700858, EPI_ISL_2700860, EPI_ISL_2700861, EPI_ISL_2700864, EPI_ISL_2700865, EPI_ISL_2700867, EPI_ISL_2700868, EPI_ISL_2700872, EPI_ISL_2700873, EPI_ISL_2700875, EPI_ISL_2700876, EPI_ISL_2700877, EPI_ISL_2700878, EPI_ISL_2700879, EPI_ISL_2700880, EPI_ISL_2700882, EPI_ISL_2700884, EPI_ISL_2700885, EPI_ISL_2700886, EPI_ISL_2700887, EPI_ISL_2700888, EPI_ISL_2700891, EPI_ISL_2700892, EPI_ISL_2700894, EPI_ISL_2700895, EPI_ISL_2700896, EPI_ISL_2700898, EPI_ISL_2700900, EPI_ISL_2700901, EPI_ISL_2700905, EPI_ISL_2700907, EPI_ISL_2700908, EPI_ISL_2700909, EPI_ISL_2700910, EPI_ISL_2700911, EPI_ISL_2700912, EPI_ISL_2700913, EPI_ISL_2700915, EPI_ISL_2700916, EPI_ISL_2700918, EPI_ISL_2700919, EPI_ISL_2700921, EPI_ISL_2700925, EPI_ISL_2700927, EPI_ISL_2700928, EPI_ISL_2700929, EPI_ISL_2700931, EPI_ISL_2700933, EPI_ISL_2700936, EPI_ISL_2700937, EPI_ISL_2700940, EPI_ISL_2700941, EPI_ISL_2700942, EPI_ISL_2700943, EPI_ISL_2700944, EPI_ISL_2700951, EPI_ISL_2700952, EPI_ISL_2700955, EPI_ISL_2700959, EPI_ISL_2700967, EPI_ISL_2700969, EPI_ISL_2700971, EPI_ISL_2700975, EPI_ISL_2700978, EPI_ISL_2700980, EPI_ISL_2700981, EPI_ISL_2700982, EPI_ISL_2700984, EPI_ISL_2700988, EPI_ISL_2700990, EPI_ISL_2700991, EPI_ISL_2700992, EPI_ISL_2700993, EPI_ISL_2700994, EPI_ISL_2700995, EPI_ISL_2700997, EPI_ISL_2700998, EPI_ISL_2701000, EPI_ISL_2701003, EPI_ISL_2701007, EPI_ISL_2701012, EPI_ISL_2701013, EPI_ISL_2701014, EPI_ISL_2701016, EPI_ISL_2701018, EPI_ISL_2701020, EPI_ISL_2701021, EPI_ISL_2701023, EPI_ISL_2701026, EPI_ISL_2701027, EPI_ISL_2701028, EPI_ISL_2701029, EPI_ISL_2701030, EPI_ISL_2701031, EPI_ISL_2701037, EPI_ISL_2701039, EPI_ISL_2701040, EPI_ISL_2701041, EPI_ISL_2701045, EPI_ISL_2701046, EPI_ISL_2701048, EPI_ISL_2701051, EPI_ISL_2701052, EPI_ISL_2701054, EPI_ISL_2701056, EPI_ISL_2701057, EPI_ISL_2701058, EPI_ISL_2701060, | | | |

EPI_ISL_2701063, EPI_ISL_2701066, EPI_ISL_2701067, EPI_ISL_2701069, EPI_ISL_2701071, EPI_ISL_2701074, EPI_ISL_2701076, EPI_ISL_2701077, EPI_ISL_2701078, EPI_ISL_2701085, EPI_ISL_2701087, EPI_ISL_2701096, EPI_ISL_2701097, EPI_ISL_2701098, EPI_ISL_2701100, EPI_ISL_2701101, EPI_ISL_2701102, EPI_ISL_2701104, EPI_ISL_2701106, EPI_ISL_2701108, EPI_ISL_2701109, EPI_ISL_2701110,
EPI_ISL_2701116, EPI_ISL_2701117, EPI_ISL_2701119, EPI_ISL_2701121, EPI_ISL_2701122, EPI_ISL_2701124, EPI_ISL_2701127, EPI_ISL_2701129, EPI_ISL_2701133, EPI_ISL_2701135, EPI_ISL_2701136, EPI_ISL_2701137, EPI_ISL_2701138, EPI_ISL_2701141, EPI_ISL_2701143, EPI_ISL_2701145, EPI_ISL_2701146, EPI_ISL_2701148, EPI_ISL_2701152, EPI_ISL_2701154, EPI_ISL_2701155, EPI_ISL_2701159,
EPI_ISL_2701160, EPI_ISL_2701161, EPI_ISL_2701162, EPI_ISL_2701163, EPI_ISL_2701164, EPI_ISL_2701166, EPI_ISL_2701167, EPI_ISL_2701168, EPI_ISL_2701169, EPI_ISL_2701170, EPI_ISL_2701175, EPI_ISL_2701177, EPI_ISL_2701180, EPI_ISL_2701182, EPI_ISL_2701184, EPI_ISL_2701185, EPI_ISL_2701188, EPI_ISL_2701189, EPI_ISL_2701190, EPI_ISL_2701195, EPI_ISL_2701196,
EPI_ISL_2701197, EPI_ISL_2701199, EPI_ISL_2701201, EPI_ISL_2701206, EPI_ISL_2701207, EPI_ISL_2701209, EPI_ISL_2701212, EPI_ISL_2701215, EPI_ISL_2701216, EPI_ISL_2701218, EPI_ISL_2701225, EPI_ISL_2701226, EPI_ISL_2701227, EPI_ISL_2701229, EPI_ISL_2701230, EPI_ISL_2701231, EPI_ISL_2701235, EPI_ISL_2701237, EPI_ISL_2701238, EPI_ISL_2701240, EPI_ISL_2701244, EPI_ISL_2701246,
EPI_ISL_2701247, EPI_ISL_2701248, EPI_ISL_2701249, EPI_ISL_2701250, EPI_ISL_2701253, EPI_ISL_2701255, EPI_ISL_2701262, EPI_ISL_2701263, EPI_ISL_2701265, EPI_ISL_2701267, EPI_ISL_2701268, EPI_ISL_2701270, EPI_ISL_2701271, EPI_ISL_2701272, EPI_ISL_2701281, EPI_ISL_2701284, EPI_ISL_2701286, EPI_ISL_2701289, EPI_ISL_2701290, EPI_ISL_2701292, EPI_ISL_2701294, EPI_ISL_2701301,
EPI_ISL_2701302, EPI_ISL_2701304, EPI_ISL_2701306, EPI_ISL_2701310, EPI_ISL_2701312, EPI_ISL_2701313, EPI_ISL_2701320, EPI_ISL_2701328, EPI_ISL_2701332, EPI_ISL_2701334, EPI_ISL_2701342, EPI_ISL_2701355, EPI_ISL_2701357, EPI_ISL_2701359, EPI_ISL_2701367, EPI_ISL_2701368, EPI_ISL_2701376, EPI_ISL_2701378, EPI_ISL_2701384, EPI_ISL_2701386, EPI_ISL_2701387, EPI_ISL_2701393,
EPI_ISL_2701395, EPI_ISL_2701396, EPI_ISL_2701398, EPI_ISL_2701406, EPI_ISL_2701410, EPI_ISL_2701413, EPI_ISL_2701416, EPI_ISL_2701418, EPI_ISL_2701429, EPI_ISL_2701433, EPI_ISL_2701435, EPI_ISL_2701440, EPI_ISL_2701442, EPI_ISL_2701446, EPI_ISL_2701450, EPI_ISL_2701451, EPI_ISL_2701461, EPI_ISL_2701463, EPI_ISL_2701467, EPI_ISL_2701477, EPI_ISL_2701483,
EPI_ISL_2701494, EPI_ISL_2701501, EPI_ISL_2701502, EPI_ISL_2701511, EPI_ISL_2701532, EPI_ISL_2701535, EPI_ISL_2701544, EPI_ISL_2701550, EPI_ISL_2701553, EPI_ISL_2701554, EPI_ISL_2701555, EPI_ISL_2701560, EPI_ISL_2701571, EPI_ISL_2701575, EPI_ISL_2701581, EPI_ISL_2701584, EPI_ISL_2701588, EPI_ISL_2701592, EPI_ISL_2701593, EPI_ISL_2701594, EPI_ISL_2701601, EPI_ISL_2701606,
EPI_ISL_2701608, EPI_ISL_2701613, EPI_ISL_2701616, EPI_ISL_2701617, EPI_ISL_2701618, EPI_ISL_2701619, EPI_ISL_2701622, EPI_ISL_2701625, EPI_ISL_2701628, EPI_ISL_2701631, EPI_ISL_2701632, EPI_ISL_2701634, EPI_ISL_2701639, EPI_ISL_2701644, EPI_ISL_2701645, EPI_ISL_2701649, EPI_ISL_2701664, EPI_ISL_2701665, EPI_ISL_2701666, EPI_ISL_2701669, EPI_ISL_2701673, EPI_ISL_2701675,
EPI_ISL_2701684, EPI_ISL_2701685, EPI_ISL_2701686, EPI_ISL_2701690, EPI_ISL_2701693, EPI_ISL_2701705, EPI_ISL_2701707, EPI_ISL_2701709, EPI_ISL_2701716, EPI_ISL_2701726, EPI_ISL_2701735, EPI_ISL_2701739, EPI_ISL_2701744, EPI_ISL_2701746, EPI_ISL_2701752, EPI_ISL_2701754, EPI_ISL_2701765, EPI_ISL_2701773, EPI_ISL_2701778, EPI_ISL_2701779, EPI_ISL_2701780, EPI_ISL_2701786,
EPI_ISL_2701796, EPI_ISL_2701801, EPI_ISL_2701802, EPI_ISL_2701812, EPI_ISL_2701813, EPI_ISL_2701817, EPI_ISL_2701819, EPI_ISL_2701826, EPI_ISL_2701827, EPI_ISL_2701829, EPI_ISL_2701834, EPI_ISL_2701836, EPI_ISL_2701837, EPI_ISL_2701846, EPI_ISL_2701856, EPI_ISL_2701860, EPI_ISL_2701868, EPI_ISL_2701870, EPI_ISL_2701880, EPI_ISL_2701884, EPI_ISL_2701890,
EPI_ISL_2701897, EPI_ISL_2701901, EPI_ISL_2701904, EPI_ISL_2701906, EPI_ISL_2701910, EPI_ISL_2701919, EPI_ISL_2701933, EPI_ISL_2701938, EPI_ISL_2701945, EPI_ISL_2701950, EPI_ISL_2701956, EPI_ISL_2701963, EPI_ISL_2701965, EPI_ISL_2701966, EPI_ISL_2701968, EPI_ISL_2701978, EPI_ISL_2701982, EPI_ISL_2701984, EPI_ISL_2701990, EPI_ISL_2701995, EPI_ISL_2701997, EPI_ISL_2702004,
EPI_ISL_2702011, EPI_ISL_2702014, EPI_ISL_2702020, EPI_ISL_2702030, EPI_ISL_2702034, EPI_ISL_2702037, EPI_ISL_2702039, EPI_ISL_2702040, EPI_ISL_2702045, EPI_ISL_2702048, EPI_ISL_2702055, EPI_ISL_2702058, EPI_ISL_2702067, EPI_ISL_2702071, EPI_ISL_2702072, EPI_ISL_2702078, EPI_ISL_2702082, EPI_ISL_2702084, EPI_ISL_2702085, EPI_ISL_2702089, EPI_ISL_2702095, EPI_ISL_2702098,
EPI_ISL_2702100, EPI_ISL_2702107, EPI_ISL_2702109, EPI_ISL_2702111, EPI_ISL_2702113, EPI_ISL_2702117, EPI_ISL_2702120, EPI_ISL_2702129, EPI_ISL_2702130, EPI_ISL_2702136, EPI_ISL_2702140, EPI_ISL_2702148, EPI_ISL_2702150, EPI_ISL_2702151, EPI_ISL_2702154, EPI_ISL_2702156, EPI_ISL_2702160, EPI_ISL_2702170, EPI_ISL_2702176, EPI_ISL_2702180, EPI_ISL_2702184, EPI_ISL_2702194,
EPI_ISL_2702197, EPI_ISL_2702198, EPI_ISL_2702201, EPI_ISL_2702213, EPI_ISL_2702216, EPI_ISL_2702222, EPI_ISL_2702224, EPI_ISL_2702226, EPI_ISL_2702229, EPI_ISL_2702231, EPI_ISL_2702237, EPI_ISL_2702241, EPI_ISL_2702242, EPI_ISL_2702256, EPI_ISL_2702257, EPI_ISL_2702258, EPI_ISL_2702274, EPI_ISL_2702281, EPI_ISL_2702288, EPI_ISL_2702300, EPI_ISL_2702301, EPI_ISL_2702317,
EPI_ISL_2702318, EPI_ISL_2702331, EPI_ISL_2702334, EPI_ISL_2702341, EPI_ISL_2702345, EPI_ISL_2702352, EPI_ISL_2702357, EPI_ISL_2702359, EPI_ISL_2702362, EPI_ISL_2702366, EPI_ISL_2702368, EPI_ISL_2702375, EPI_ISL_2702379, EPI_ISL_2702391, EPI_ISL_2702408, EPI_ISL_2702421, EPI_ISL_2702422, EPI_ISL_2702458, EPI_ISL_2702459, EPI_ISL_2702461, EPI_ISL_2702462, EPI_ISL_2702463,
EPI_ISL_2702464, EPI_ISL_2702465, EPI_ISL_2702466, EPI_ISL_2702468, EPI_ISL_2702473, EPI_ISL_2702476, EPI_ISL_2702477, EPI_ISL_2702478, EPI_ISL_2702480, EPI_ISL_2702481, EPI_ISL_2702484, EPI_ISL_2702485, EPI_ISL_2702486, EPI_ISL_2702488, EPI_ISL_2702490, EPI_ISL_2702491, EPI_ISL_2702492, EPI_ISL_2702493, EPI_ISL_2702494, EPI_ISL_2702495, EPI_ISL_2702496, EPI_ISL_2702497,
EPI_ISL_2702498, EPI_ISL_2702499, EPI_ISL_2702500, EPI_ISL_2702502, EPI_ISL_2702503, EPI_ISL_2702504, EPI_ISL_2702506, EPI_ISL_2702507, EPI_ISL_2702509, EPI_ISL_2702510, EPI_ISL_2702511, EPI_ISL_2702512, EPI_ISL_2702513, EPI_ISL_2702514, EPI_ISL_2702515, EPI_ISL_2702517, EPI_ISL_2702518, EPI_ISL_2702523, EPI_ISL_2702525, EPI_ISL_2702526, EPI_ISL_2702527, EPI_ISL_2702528,
EPI_ISL_2702529, EPI_ISL_2702530, EPI_ISL_2702531, EPI_ISL_2702532, EPI_ISL_2702533, EPI_ISL_2702535, EPI_ISL_2702536, EPI_ISL_2702537, EPI_ISL_2702541, EPI_ISL_2702542, EPI_ISL_2702543, EPI_ISL_2702545, EPI_ISL_2702546, EPI_ISL_2702547, EPI_ISL_2702548, EPI_ISL_2702549, EPI_ISL_2702550, EPI_ISL_2702551, EPI_ISL_2702552, EPI_ISL_2702554, EPI_ISL_2702555, EPI_ISL_2702556,
EPI_ISL_2702557, EPI_ISL_2702558, EPI_ISL_2702559, EPI_ISL_2702562, EPI_ISL_2702563, EPI_ISL_2702564, EPI_ISL_2702565, EPI_ISL_2702566, EPI_ISL_2702567, EPI_ISL_2702569, EPI_ISL_2702571, EPI_ISL_2702572, EPI_ISL_2702573, EPI_ISL_2702575, EPI_ISL_2702577, EPI_ISL_2702578, EPI_ISL_2702579, EPI_ISL_2702582, EPI_ISL_2702583, EPI_ISL_2702584, EPI_ISL_2702585, EPI_ISL_2702586,
EPI_ISL_2702588, EPI_ISL_2702589, EPI_ISL_2702591, EPI_ISL_2702595, EPI_ISL_2702597, EPI_ISL_2702598, EPI_ISL_2702599, EPI_ISL_2702601, EPI_ISL_2702602, EPI_ISL_2702603, EPI_ISL_2702612, EPI_ISL_2702613, EPI_ISL_2702615, EPI_ISL_2702616, EPI_ISL_2702617, EPI_ISL_2702618, EPI_ISL_2702620, EPI_ISL_2702621, EPI_ISL_2702623, EPI_ISL_2702624, EPI_ISL_2702625,
EPI_ISL_2702626, EPI_ISL_2702627, EPI_ISL_2702628, EPI_ISL_2702632, EPI_ISL_2702633, EPI_ISL_2702636, EPI_ISL_2702637, EPI_ISL_2702638, EPI_ISL_2702640, EPI_ISL_2702641, EPI_ISL_2702643, EPI_ISL_2702644, EPI_ISL_2702646, EPI_ISL_2702647, EPI_ISL_2702650, EPI_ISL_2702651, EPI_ISL_2702652, EPI_ISL_2702653, EPI_ISL_2702654, EPI_ISL_2702655, EPI_ISL_2702656, EPI_ISL_2702658,
EPI_ISL_2702660, EPI_ISL_2702664, EPI_ISL_2702667, EPI_ISL_2702668, EPI_ISL_2702669, EPI_ISL_2702671, EPI_ISL_2702673, EPI_ISL_2702675, EPI_ISL_2702677, EPI_ISL_2702678, EPI_ISL_2702679, EPI_ISL_2702680, EPI_ISL_2702681, EPI_ISL_2702682, EPI_ISL_2702684, EPI_ISL_2702685, EPI_ISL_2702686, EPI_ISL_2702692, EPI_ISL_2702693, EPI_ISL_2702695, EPI_ISL_2702697, EPI_ISL_2702699,
EPI_ISL_2702702, EPI_ISL_2702703, EPI_ISL_2702704, EPI_ISL_2702707, EPI_ISL_2702709, EPI_ISL_2702711, EPI_ISL_2702714, EPI_ISL_2702715, EPI_ISL_2702716, EPI_ISL_2702718, EPI_ISL_2702720, EPI_ISL_2702721, EPI_ISL_2702724, EPI_ISL_2702726, EPI_ISL_2702730, EPI_ISL_2702731, EPI_ISL_2702732, EPI_ISL_2702734, EPI_ISL_2702735, EPI_ISL_2702737, EPI_ISL_2702739, EPI_ISL_2702740,
EPI_ISL_2702742, EPI_ISL_2702743, EPI_ISL_2702744, EPI_ISL_2702745, EPI_ISL_2702747, EPI_ISL_2702749, EPI_ISL_2702750, EPI_ISL_2702751, EPI_ISL_2702752, EPI_ISL_2702756, EPI_ISL_2702758, EPI_ISL_2702759, EPI_ISL_2702760, EPI_ISL_2702761, EPI_ISL_2702762, EPI_ISL_2702763, EPI_ISL_2702764, EPI_ISL_2702766, EPI_ISL_2702767, EPI_ISL_2702768, EPI_ISL_2702771, EPI_ISL_2702772,
EPI_ISL_2702774, EPI_ISL_2702776, EPI_ISL_2702777, EPI_ISL_2702778, EPI_ISL_2702781, EPI_ISL_2702785, EPI_ISL_2702788, EPI_ISL_2702789, EPI_ISL_2702790, EPI_ISL_2702791, EPI_ISL_2702792, EPI_ISL_2702795, EPI_ISL_2702797, EPI_ISL_2702798, EPI_ISL_2702799, EPI_ISL_2702801, EPI_ISL_2702802, EPI_ISL_2702803, EPI_ISL_2702806, EPI_ISL_2702807, EPI_ISL_2702809, EPI_ISL_2702811,
EPI_ISL_2702815, EPI_ISL_2702817, EPI_ISL_2702822, EPI_ISL_2702823, EPI_ISL_2702829, EPI_ISL_2702840, EPI_ISL_2702843, EPI_ISL_2702847, EPI_ISL_2702850, EPI_ISL_2702854, EPI_ISL_2702857, EPI_ISL_2702858, EPI_ISL_2702859, EPI_ISL_2702860, EPI_ISL_2702864, EPI_ISL_2702865, EPI_ISL_2702868, EPI_ISL_2702876, EPI_ISL_2702884, EPI_ISL_2702897, EPI_ISL_2702905,
EPI_ISL_2702916, EPI_ISL_2702921, EPI_ISL_2702924, EPI_ISL_2702926, EPI_ISL_2702934, EPI_ISL_2702943, EPI_ISL_2702945, EPI_ISL_2702950, EPI_ISL_2702958, EPI_ISL_2702962, EPI_ISL_2702964, EPI_ISL_2702977, EPI_ISL_2702982, EPI_ISL_2702984, EPI_ISL_2702991, EPI_ISL_2702992, EPI_ISL_2702993, EPI_ISL_2702997, EPI_ISL_2703003, EPI_ISL_2703004, EPI_ISL_2703005, EPI_ISL_2703006,
EPI_ISL_2703007, EPI_ISL_2703008, EPI_ISL_2703017, EPI_ISL_2703022, EPI_ISL_2703024, EPI_ISL_2703028, EPI_ISL_2703029, EPI_ISL_2703030, EPI_ISL_2703033, EPI_ISL_2703035, EPI_ISL_2703045, EPI_ISL_2703050, EPI_ISL_2703063, EPI_ISL_2703071, EPI_ISL_2703084, EPI_ISL_2703086, EPI_ISL_2703090, EPI_ISL_2703093, EPI_ISL_2703095, EPI_ISL_2703096, EPI_ISL_2703101, EPI_ISL_2703103,
EPI_ISL_2703106, EPI_ISL_2703115, EPI_ISL_2703120, EPI_ISL_2703122, EPI_ISL_2703128, EPI_ISL_2703130, EPI_ISL_2703133, EPI_ISL_2703136, EPI_ISL_2703137, EPI_ISL_2703139, EPI_ISL_2703155, EPI_ISL_2703156, EPI_ISL_2703157, EPI_ISL_2703158, EPI_ISL_2703160, EPI_ISL_2703161, EPI_ISL_2703162, EPI_ISL_2703164, EPI_ISL_2703165, EPI_ISL_2703166, EPI_ISL_2703168, EPI_ISL_2703170,
EPI_ISL_2703171, EPI_ISL_2703172, EPI_ISL_2703173, EPI_ISL_2703174, EPI_ISL_2703175, EPI_ISL_2703176, EPI_ISL_2703177, EPI_ISL_2703178, EPI_ISL_2703179, EPI_ISL_2703183, EPI_ISL_2703188, EPI_ISL_2703190, EPI_ISL_2703191, EPI_ISL_2703192, EPI_ISL_2703193, EPI_ISL_2703194, EPI_ISL_2703196, EPI_ISL_2703197, EPI_ISL_2703198, EPI_ISL_2703201, EPI_ISL_2703202, EPI_ISL_2703203,
EPI_ISL_2703204, EPI_ISL_2703205, EPI_ISL_2703206, EPI_ISL_2703207, EPI_ISL_2703208, EPI_ISL_2703209, EPI_ISL_2703211, EPI_ISL_2703213, EPI_ISL_2703214, EPI_ISL_2703216, EPI_ISL_2703218, EPI_ISL_2703219, EPI_ISL_2703220, EPI_ISL_2703222, EPI_ISL_2703223, EPI_ISL_2703224, EPI_ISL_2703226, EPI_ISL_2703227, EPI_ISL_2703228, EPI_ISL_2703229, EPI_ISL_2703230, EPI_ISL_2703231,
EPI_ISL_2703232, EPI_ISL_2703233, EPI_ISL_2703234, EPI_ISL_2703235, EPI_ISL_2703236, EPI_ISL_2703238, EPI_ISL_2703239, EPI_ISL_2703240, EPI_ISL_2703241, EPI_ISL_2703242, EPI_ISL_2703244, EPI_ISL_2703245, EPI_ISL_2703246, EPI_ISL_2703247, EPI_ISL_2703249, EPI_ISL_2703250, EPI_ISL_2703251, EPI_ISL_2703252, EPI_ISL_2703253, EPI_ISL_2703254, EPI_ISL_2703256, EPI_ISL_2703257,
EPI_ISL_2703258, EPI_ISL_2703259, EPI_ISL_2703260, EPI_ISL_2703262, EPI_ISL_2703263, EPI_ISL_2703264, EPI_ISL_2703265, EPI_ISL_2703266, EPI_ISL_2703268, EPI_ISL_2703272, EPI_ISL_2703274, EPI_ISL_2703277, EPI_ISL_2703278, EPI_ISL_2703280, EPI_ISL_2703281, EPI_ISL_2703282, EPI_ISL_2703283, EPI_ISL_2703287, EPI_ISL_2703288, EPI_ISL_2703289, EPI_ISL_2703290, EPI_ISL_2703293,
EPI_ISL_2703294, EPI_ISL_2703295, EPI_ISL_2703296, EPI_ISL_2703297, EPI_ISL_2703298, EPI_ISL_2703299, EPI_ISL_2703301, EPI_ISL_2703302, EPI_ISL_2703303, EPI_ISL_2703304, EPI_ISL_2703305, EPI_ISL_2703307, EPI_ISL_2703309, EPI_ISL_2703310, EPI_ISL_2703311, EPI_ISL_2703313, EPI_ISL_2703314, EPI_ISL_2703315, EPI_ISL_2703316, EPI_ISL_2703319, EPI_ISL_2703321, EPI_ISL_2703324,
EPI_ISL_2703325, EPI_ISL_2703326, EPI_ISL_2703328, EPI_ISL_2703331, EPI_ISL_2703332, EPI_ISL_2703334, EPI_ISL_2703335, EPI_ISL_2703336, EPI_ISL_2703338, EPI_ISL_2703339, EPI_ISL_2703340, EPI_ISL_2703342, EPI_ISL_2703343, EPI_ISL_2703344, EPI_ISL_2703345, EPI_ISL_2703346, EPI_ISL_2703347, EPI_ISL_2703350, EPI_ISL_2703351, EPI_ISL_2703352, EPI_ISL_2703354, EPI_ISL_2703355,
EPI_ISL_2703356, EPI_ISL_2703357, EPI_ISL_2703359, EPI_ISL_2703360, EPI_ISL_2703361, EPI_ISL_2703363, EPI_ISL_2703364, EPI_ISL_2703365, EPI_ISL_2703367, EPI_ISL_2703369, EPI_ISL_2703370, EPI_ISL_2703371, EPI_ISL_2703372, EPI_ISL_2703375, EPI_ISL_2703376, EPI_ISL_2703377, EPI_ISL_2703378, EPI_ISL_2703380, EPI_ISL_2703385, EPI_ISL_2703386, EPI_ISL_2703389, EPI_ISL_2703391,
EPI_ISL_2703392, EPI_ISL_2703393, EPI_ISL_2703394, EPI_ISL_2703396, EPI_ISL_2703397, EPI_ISL_2703399, EPI_ISL_2703400, EPI_ISL_2703401, EPI_ISL_2703402, EPI_ISL_2703403, EPI_ISL_2703406, EPI_ISL_2703407, EPI_ISL_2703408, EPI_ISL_2703409, EPI_ISL_2703410, EPI_ISL_2703413, EPI_ISL_2703414, EPI_ISL_2703415, EPI_ISL_2703416, EPI_ISL_2703417, EPI_ISL_2703418, EPI_ISL_2703419,
EPI_ISL_2703421, EPI_ISL_2703422, EPI_ISL_2703425, EPI_ISL_2703426, EPI_ISL_2703427, EPI_ISL_2703428, EPI_ISL_2703429, EPI_ISL_2703430, EPI_ISL_2703431, EPI_ISL_2703432, EPI_ISL_2703433, EPI_ISL_2703434, EPI_ISL_2703435, EPI_ISL_2703436, EPI_ISL_2703437, EPI_ISL_2703439, EPI_ISL_2703440, EPI_ISL_2703442, EPI_ISL_2703444, EPI_ISL_2703447, EPI_ISL_2703448, EPI_ISL_2703450,
EPI_ISL_2703451, EPI_ISL_2703452, EPI_ISL_2703453, EPI_ISL_2703455, EPI_ISL_2703456, EPI_ISL_2703458, EPI_ISL_2703459, EPI_ISL_2703460, EPI_ISL_2703461, EPI_ISL_2703462, EPI_ISL_2703463, EPI_ISL_2703464, EPI_ISL_2703465, EPI_ISL_2703466, EPI_ISL_2703468, EPI_ISL_2703470, EPI_ISL_2703471, EPI_ISL_2703472, EPI_ISL_2703474, EPI_ISL_2703476, EPI_ISL_2703477, EPI_ISL_2703478,
EPI_ISL_2703479, EPI_ISL_2703480, EPI_ISL_2703481, EPI_ISL_2703482, EPI_ISL_2703483, EPI_ISL_2703485, EPI_ISL_2703486, EPI_ISL_2703487, EPI_ISL_2703488, EPI_ISL_2703490, EPI_ISL_2703491, EPI_ISL_2703492, EPI_ISL_2703494, EPI_ISL_2703495, EPI_ISL_2703496, EPI_ISL_2703498, EPI_ISL_2703499, EPI_ISL_2703500, EPI_ISL_2703501, EPI_ISL_2703503, EPI_ISL_2703510, EPI_ISL_2703511,
EPI_ISL_2703512, EPI_ISL_2703514, EPI_ISL_2703518, EPI_ISL_2703520, EPI_ISL_2703521, EPI_ISL_2703523, EPI_ISL_2703526, EPI_ISL_2703529, EPI_ISL_2703531, EPI_ISL_2703532, EPI_ISL_2703533, EPI_ISL_2703535, EPI_ISL_2703539, EPI_ISL_2703540, EPI_ISL_2703542, EPI_ISL_2703543, EPI_ISL_2703544, EPI_ISL_2703546, EPI_ISL_2703547, EPI_ISL_2703551, EPI_ISL_2703553, EPI_ISL_2703557,
EPI_ISL_2703563, EPI_ISL_2703565, EPI_ISL_2703566, EPI_ISL_2703567, EPI_ISL_2703568, EPI_ISL_2703569, EPI_ISL_2703574, EPI_ISL_2703575, EPI_ISL_2703584, EPI_ISL_2703585, EPI_ISL_2703589, EPI_ISL_2703590, EPI_ISL_2703593, EPI_ISL_2703595, EPI_ISL_2703596, EPI_ISL_2703597, EPI_ISL_2703602, EPI_ISL_2703603, EPI_ISL_2703608, EPI_ISL_2703609, EPI_ISL_2703610, EPI_ISL_2703611,
EPI_ISL_2703612, EPI_ISL_2703614, EPI_ISL_2703615, EPI_ISL_2703616, EPI_ISL_2703617, EPI_ISL_2703620, EPI_ISL_2703624, EPI_ISL_2703625, EPI_ISL_2703626, EPI_ISL_2703627, EPI_ISL_2703629, EPI_ISL_2703630, EPI_ISL_2703631, EPI_ISL_2703644, EPI_ISL_2703645, EPI_ISL_2703646, EPI_ISL_2703650, EPI_ISL_2703651, EPI_ISL_2703653, EPI_ISL_2703655, EPI_ISL_2703656, EPI_ISL_2703660,
EPI_ISL_2703661, EPI_ISL_2703664, EPI_ISL_2703668, EPI_ISL_2703669, EPI_ISL_2703670, EPI_ISL_2703671, EPI_ISL_2703672, EPI_ISL_2703674, EPI_ISL_2703676, EPI_ISL_2703678, EPI_ISL_2703681, EPI_ISL_2703683, EPI_ISL_2703686, EPI_ISL_2703687, EPI_ISL_2703688, EPI_ISL_2703689, EPI_ISL_2703693, EPI_ISL_2703698, EPI_ISL_2703699, EPI_ISL_2703702, EPI_ISL_2703704,
EPI_ISL_2703705, EPI_ISL_2703706, EPI_ISL_2703715, EPI_ISL_2703718, EPI_ISL_2703723, EPI_ISL_2703724, EPI_ISL_2703726, EPI_ISL_2703727, EPI_ISL_2703730, EPI_ISL_2703732, EPI_ISL_2703733, EPI_ISL_2703734, EPI_ISL_2703735, EPI_ISL_2703736, EPI_ISL_2703738, EPI_ISL_2703742, EPI_ISL_2703743, EPI_ISL_2703744, EPI_ISL_2703747, EPI_ISL_2703749, EPI_ISL_2703750, EPI_ISL_2703752,
EPI_ISL_2703754, EPI_ISL_2703756, EPI_ISL_2703760, EPI_ISL_2703761, EPI_ISL_2703763, EPI_ISL_2703765, EPI_ISL_2703771, EPI_ISL_2703772, EPI_ISL_2703773, EPI_ISL_2703774, EPI_ISL_2703775, EPI_ISL_2703776, EPI_ISL_2703778, EPI_ISL_2703781, EPI_ISL_2703784, EPI_ISL_2703786, EPI_ISL_2703787, EPI_ISL_2703792, EPI_ISL_2703794, EPI_ISL_2703795, EPI_ISL_2703803, EPI_ISL_2703810,
EPI_ISL_2703812, EPI_ISL_2703815, EPI_ISL_2703816, EPI_ISL_2703817, EPI_ISL_2703819, EPI_ISL_2703820, EPI_ISL_2703821, EPI_ISL_2703822, EPI_ISL_2703823, EPI_ISL_2703824, EPI_ISL_2703825, EPI_ISL_2703828, EPI_ISL_2704185, EPI_ISL_2704186, EPI_ISL_2704187, EPI_ISL_2704189, EPI_ISL_2704192, EPI_ISL_2704193, EPI_ISL_2704195, EPI_ISL_2704197, EPI_ISL_2704199, EPI_ISL_2704201,
EPI_ISL_2704202, EPI_ISL_2704203, EPI_ISL_2704205, EPI_ISL_2704206, EPI_ISL_2704207, EPI_ISL_2704208, EPI_ISL_2704209, EPI_ISL_2704210, EPI_ISL_2704211, EPI_ISL_2704212, EPI_ISL_2704213, EPI_ISL_2704216, EPI_ISL_2704217, EPI_ISL_2704219, EPI_ISL_2704220, EPI_ISL_2704221, EPI_ISL_2704223, EPI_ISL_2704226, EPI_ISL_2704228, EPI_ISL_2704230, EPI_ISL_2704231, EPI_ISL_2704233,
EPI_ISL_2704235, EPI_ISL_2704236, EPI_ISL_2704238, EPI_ISL_2704239, EPI_ISL_2704240, EPI_ISL_2704241, EPI_ISL_2704243, EPI_ISL_2704245, EPI_ISL_2704247, EPI_ISL_2704248, EPI_ISL_2704250, EPI_ISL_2704251, EPI_ISL_2704252, EPI_ISL_2704253, EPI_ISL_2704254, EPI_ISL_2704256, EPI_ISL_2704258, EPI_ISL_2704260, EPI_ISL_2704261, EPI_ISL_2704262, EPI_ISL_2704263, EPI_ISL_2704264,
EPI_ISL_2704265, EPI_ISL_2704267, EPI_ISL_2704269, EPI_ISL_2704272, EPI_ISL_2704273, EPI_ISL_2704276, EPI_ISL_2704278, EPI_ISL_2704279, EPI_ISL_2704280, EPI_ISL_2704281, EPI_ISL_2704282, EPI_ISL_2704284, EPI_ISL_2704285, EPI_ISL_2704286, EPI_ISL_2704290, EPI_ISL_2704293, EPI_ISL_2704294, EPI_ISL_2704295, EPI_ISL_2704296, EPI_ISL_2704297, EPI_ISL_2704298, EPI_ISL_2704299,
EPI_ISL_2704300, EPI_ISL_2704302, EPI_ISL_2704303, EPI_ISL_2704304, EPI_ISL_2704305, EPI_ISL_2704306, EPI_ISL_2704308, EPI_ISL_2704309, EPI_ISL_2704311, EPI_ISL_2704313, EPI_ISL_2704315, EPI_ISL_2704320, EPI_ISL_2704321, EPI_ISL_2704322, EPI_ISL_2704323, EPI_ISL_2704324, EPI_ISL_2704325, EPI_ISL_2704326, EPI_ISL_2704327, EPI_ISL_2704328, EPI_ISL_2704329, EPI_ISL_2704330,
EPI_ISL_2704331, EPI_ISL_2704332, EPI_ISL_2704334, EPI_ISL_2704335, EPI_ISL_2704339, EPI_ISL_2704341, EPI_ISL_2704342, EPI_ISL_2704343, EPI_ISL_2704344, EPI_ISL_2704345, EPI_ISL_2704347, EPI_ISL_2704348, EPI_ISL_2704350, EPI_ISL_2704352, EPI_ISL_2704353, EPI_ISL_2704354, EPI_ISL_2704355, EPI_ISL_2704357, EPI_ISL_2704358, EPI_ISL_2704360, EPI_ISL_2704362, EPI_ISL_2704363,
EPI_ISL_2704364, EPI_ISL_2704365, EPI_ISL_2704366, EPI_ISL_2704367, EPI_ISL_2704368, EPI_ISL_2704369, EPI_ISL_2704370, EPI_ISL_2704372, EPI_ISL_2704374, EPI_ISL_2704375, EPI_ISL_2704376, EPI_ISL_2704377, EPI_ISL_2704380, EPI_ISL_2704381, EPI_ISL_2704382, EPI_ISL_2704383, EPI_ISL_2704384, EPI_ISL_2704386, EPI_ISL_2704387, EPI_ISL_2704388, EPI_ISL_2704391, EPI_ISL_2704393,
EPI_ISL_2704394, EPI_ISL_2704395, EPI_ISL_2704397, EPI_ISL_2704398, EPI_ISL_2704399, EPI_ISL_2704401, EPI_ISL_2704403, EPI_ISL_2704405, EPI_ISL_2704406, EPI_ISL_2704407, EPI_ISL_2704408, EPI_ISL_2704409, EPI_ISL_2704410, EPI_ISL_2704411, EPI_ISL_2704416, EPI_ISL_2704417, EPI_ISL_2704418, EPI_ISL_2704420, EPI_ISL_2704422, EPI_ISL_2704423, EPI_ISL_2704425, EPI_ISL_2704426,
EPI_ISL_2704427, EPI_ISL_2704428, EPI_ISL_2704429, EPI_ISL_2704430, EPI_ISL_2704431, EPI_ISL_2704432, EPI_ISL_2704433, EPI_ISL_2704434, EPI_ISL_2704435, EPI_ISL_2704436, EPI_ISL_2704437, EPI_ISL_2704438, EPI_ISL_2704439, EPI_ISL_2704440, EPI_ISL_2704441, EPI_ISL_2704442, EPI_ISL_2704443, EPI_ISL_2704444, EPI_ISL_2704446, EPI_ISL_2704447, EPI_ISL_2704448, EPI_ISL_2704450,
EPI_ISL_2704452, EPI_ISL_2704453, EPI_ISL_2704454, EPI_ISL_2704455, EPI_ISL_2704456, EPI_ISL_2704458, EPI_ISL_2704459, EPI_ISL_2704460, EPI_ISL_2704461, EPI_ISL_2704462, EPI_ISL_2704463, EPI_ISL_2704464, EPI_ISL_2704465, EPI_ISL_2704468, EPI_ISL_2704471, EPI_ISL_2704472, EPI_ISL_2704473, EPI_ISL_2704474, EPI_ISL_2704475, EPI_ISL_2704476, EPI_ISL_2704477, EPI_ISL_2704479,
EPI_ISL_2704483, EPI_ISL_2704486, EPI_ISL_2704487, EPI_ISL_2704488, EPI_ISL_2704489, EPI_ISL_2704490, EPI_ISL_2704493, EPI_ISL_2704495, EPI_ISL_2704497, EPI_ISL_2704499, EPI_ISL_2704500, EPI_ISL_2704501, EPI_ISL_2704502, EPI_ISL_2704503, EPI_ISL_2704504, EPI_ISL_2704505, EPI_ISL_2704506, EPI_ISL_2704507, EPI_ISL_2704508, EPI_ISL_2704509, EPI_ISL_2704510,
EPI_ISL_2704512, EPI_ISL_2704514, EPI_ISL_2704515, EPI_ISL_2704516, EPI_ISL_2704517, EPI_ISL_2704518, EPI_ISL_2704519, EPI_ISL_2704520, EPI_ISL_2704521, EPI_ISL_2704522, EPI_ISL_2704523, EPI_ISL_2704524, EPI_ISL_2704525, EPI_ISL_2704526, EPI_ISL_2704528, EPI_ISL_2704529, EPI_ISL_2704530, EPI_ISL_2704531, EPI_ISL_2704669, EPI_ISL_2704672, EPI_ISL_2704673, EPI_ISL_2704674,
EPI_ISL_2716981, EPI_ISL_2716984, EPI_ISL_2716989, EPI_ISL_2716990, EPI_ISL_2716991, EPI_ISL_2716994, EPI_ISL_2717004, EPI_ISL_2717013, EPI_ISL_2717019, EPI_ISL_2717020, EPI_ISL_2717021, EPI_ISL_2717037, EPI_ISL_2717040, EPI_ISL_2717053, EPI_ISL_2717065, EPI_ISL_2717068, EPI_ISL_2717074, EPI_ISL_2717076, EPI_ISL_2717091, EPI_ISL_2717101, EPI_ISL_2717112, EPI_ISL_2717117,
EPI_ISL_2717118, EPI_ISL_2717135, EPI_ISL_2717136, EPI_ISL_2717141, EPI_ISL_2717162, EPI_ISL_2717163, EPI_ISL_2717165, EPI_ISL_2717167, EPI_ISL_2717168, EPI_ISL_2717169, EPI_ISL_2717172, EPI_ISL_2717174, EPI_ISL_2717175, EPI_ISL_2717212, EPI_ISL_2717218, EPI_ISL_2717220, EPI_ISL_2717223, EPI_ISL_2717228, EPI_ISL_2717245, EPI_ISL_2717256, EPI_ISL_2717278,
EPI_ISL_2717279, EPI_ISL_2717291, EPI_ISL_2717307, EPI_ISL_2717322, EPI_ISL_2717326, EPI_ISL_2717327, EPI_ISL_2717334, EPI_ISL_2717343, EPI_ISL_2717358, EPI_ISL_2717359, EPI_ISL_2717361, EPI_ISL_2717368, EPI_ISL_2717383, EPI_ISL_2717403, EPI_ISL_2717413, EPI_ISL_2717425, EPI_ISL_2717444, EPI_ISL_2717445, EPI_ISL_2717446, EPI_ISL_2717451, EPI_ISL_2717457,
EPI_ISL_2717458, EPI_ISL_2717462, EPI_ISL_2717494, EPI_ISL_2717502, EPI_ISL_2717510, EPI_ISL_2717517, EPI_ISL_2717527, EPI_ISL_2717533, EPI_ISL_2717541, EPI_ISL_2717550, EPI_ISL_2717569, EPI_ISL_2717580, EPI_ISL_2717582, EPI_ISL_2717596, EPI_ISL_2717601, EPI_ISL_2717603, EPI_ISL_2717606, EPI_ISL_2717622, EPI_ISL_2717627, EPI_ISL_2717629, EPI_ISL_2717643, EPI_ISL_2717644,
EPI_ISL_2717645, EPI_ISL_2717650, EPI_ISL_2717657, EPI_ISL_2717658, EPI_ISL_2717659, EPI_ISL_2717660, EPI_ISL_2717678, EPI_ISL_2717696, EPI_ISL_2717702, EPI_ISL_2717709, EPI_ISL_2717714, EPI_ISL_2717720, EPI_ISL_2717721, EPI_ISL_2717724, EPI_ISL_2717731, EPI_ISL_2717733, EPI_ISL_2717743, EPI_ISL_2717746, EPI_ISL_2717749, EPI_ISL_2717752, EPI_ISL_2717755, EPI_ISL_2717757,
EPI_ISL_2717764, EPI_ISL_2717769, EPI_ISL_2717770, EPI_ISL_2717772, EPI_ISL_2717807, EPI_ISL_2717837, EPI_ISL_2717839, EPI_ISL_2717863, EPI_ISL_2717866, EPI_ISL_2717895, EPI_ISL_2717897, EPI_ISL_2717956, EPI_ISL_2717968, EPI_ISL_2717976, EPI_ISL_2717981, EPI_ISL_2718036, EPI_ISL_2718047, EPI_ISL_2718088, EPI_ISL_2718128, EPI_ISL_2718136, EPI_ISL_2718281, EPI_ISL_2718282,
EPI_ISL_2718284, EPI_ISL_2718288, EPI_ISL_2718290, EPI_ISL_2718291, EPI_ISL_2718292, EPI_ISL_2718293, EPI_ISL_2729046, EPI_ISL_2729081, EPI_ISL_2729094, EPI_ISL_2729175, EPI_ISL_2729305, EPI_ISL_2729383, EPI_ISL_2729408, EPI_ISL_2729490, EPI_ISL_2729491, EPI_ISL_2729525, EPI_ISL_2729614, EPI_ISL_2729622, EPI_ISL_2729627, EPI_ISL_2729677, EPI_ISL_2729698,
EPI_ISL_2730127, EPI_ISL_2730412, EPI_ISL_2730447, EPI_ISL_2730527, EPI_ISL_2765778, EPI_ISL_2765824, EPI_ISL_2765910, EPI_ISL_2765914, EPI_ISL_2766079, EPI_ISL_2766107, EPI_ISL_2766605, EPI_ISL_2766617, EPI_ISL_2766767, EPI_ISL_2766837, EPI_ISL_2766864, EPI_ISL_2766910, EPI_ISL_2767038, EPI_ISL_2767042, EPI_ISL_2767071, EPI_ISL_2767078, EPI_ISL_2767175, EPI_ISL_2767182,
EPI_ISL_2767222, EPI_ISL_2767235, EPI_ISL_2767267, EPI_ISL_2767314, EPI_ISL_2767336, EPI_ISL_2767350, EPI_ISL_2767360, EPI_ISL_2767372, EPI_ISL_2767375, EPI_ISL_2767376, EPI_ISL_2767397, EPI_ISL_2767406, EPI_ISL_2767407, EPI_ISL_2767409, EPI_ISL_2767421, EPI_ISL_2767424, EPI_ISL_2767432, EPI_ISL_2767436, EPI_ISL_2767437, EPI_ISL_2767461, EPI_ISL_2767466, EPI_ISL_2767475,
EPI_ISL_2767484, EPI_ISL_2767489, EPI_ISL_2767494, EPI_ISL_2767499, EPI_ISL_2767517, EPI_ISL_2767530, EPI_ISL_2767537, EPI_ISL_2767542, EPI_ISL_2767558, EPI_ISL_2767572, EPI_ISL_2767573, EPI_ISL_2767574, EPI_ISL_2767581, EPI_ISL_2767596, EPI_ISL_2767597, EPI_ISL_2767600, EPI_ISL_2767605, EPI_ISL_2767609, EPI_ISL_2767610, EPI_ISL_2767626, EPI_ISL_2767633, EPI_ISL_2767634,
EPI_ISL_2767644, EPI_ISL_2767667, EPI_ISL_2767729, EPI_ISL_2767752, EPI_ISL_2767800, EPI_ISL_2767806, EPI_ISL_2767812, EPI_ISL_2767813, EPI_ISL_2767880, EPI_ISL_2767932, EPI_ISL_2767941, EPI_ISL_2767972, EPI_ISL_2768003, EPI_ISL_2780918, EPI_ISL_2780950, EPI_ISL_2781560, EPI_ISL_2781652, EPI_ISL_2781679, EPI_ISL_2781731, EPI_ISL_2781761, EPI_ISL_2781778, EPI_ISL_2781800,
EPI_ISL_2781801, EPI_ISL_2781804, EPI_ISL_2781805, EPI_ISL_2781806, EPI_ISL_2781811, EPI_ISL_2781823, EPI_ISL_2781828, EPI_ISL_2781829, EPI_ISL_2781835, EPI_ISL_2781838, EPI_ISL_2781842, EPI_ISL_2781843, EPI_ISL_2781848, EPI_ISL_2781857, EPI_ISL_2781871, EPI_ISL_2781872, EPI_ISL_2781881, EPI_ISL_2781885, EPI_ISL_2781897, EPI_ISL_2781900, EPI_ISL_2781902, EPI_ISL_2781904,
EPI_ISL_2781905, EPI_ISL_2781907, EPI_ISL_2781912, EPI_ISL_2781918, EPI_ISL_2781924, EPI_ISL_2781936, EPI_ISL_2781939, EPI_ISL_2781942, EPI_ISL_2781944, EPI_ISL_2781954, EPI_ISL_2781955, EPI_ISL_2781958, EPI_ISL_2781967, EPI_ISL_2781972, EPI_ISL_2781976, EPI_ISL_2781984, EPI_ISL_2781986, EPI_ISL_2781991, EPI_ISL_2781998, EPI_ISL_2782002, EPI_ISL_2782010, EPI_ISL_2782014,
EPI_ISL_2782015, EPI_ISL_2782018, EPI_ISL_2782019, EPI_ISL_2782023, EPI_ISL_2782026, EPI_ISL_2782031, EPI_ISL_2782038, EPI_ISL_2782042, EPI_ISL_2782066, EPI_ISL_2782096, EPI_ISL_2782147, EPI_ISL_2782187, EPI_ISL_2782196, EPI_ISL_2782257, EPI_ISL_2782259, EPI_ISL_2782287, EPI_ISL_2782291, EPI_ISL_2782300, EPI_ISL_2782302, EPI_ISL_2804785, EPI_ISL_2807364, EPI_ISL_2809149,
EPI_ISL_2809167, EPI_ISL_2809201, EPI_ISL_2809216, EPI_ISL_2809275, EPI_ISL_2809333, EPI_ISL_2809488, EPI_ISL_2831323, EPI_ISL_2831336, EPI_ISL_2831341, EPI_ISL_2831342, EPI_ISL_2831361, EPI_ISL_2831373, EPI_ISL_2831392, EPI_ISL_2831402, EPI_ISL_2831411, EPI_ISL_2831415, EPI_ISL_2831422, EPI_ISL_2831430, EPI_ISL_2831440, EPI_ISL_2831449, EPI_ISL_2831450, EPI_ISL_2831473,
EPI_ISL_2831487, EPI_ISL_2831495, EPI_ISL_2831497, EPI_ISL_2831500, EPI_ISL_2831504, EPI_ISL_2831514, EPI_ISL_2831523, EPI_ISL_2831538, EPI_ISL_2831554, EPI_ISL_2831556, EPI_ISL_2831561, EPI_ISL_2831563, EPI_ISL_2831571, EPI_ISL_2832097, EPI_ISL_2852576

| | | | |
|---|---|---|---|
| see above | Lighthouse Lab in Alderley Park | Wellcome Sanger Institute for the COVID-19 Genomics UK (COG-UK) Consortium | Cordelia Langford; David K. Jackson; Dominic Kwiatkowski; Ewan Harrison; Ian Johnston; Jacquelyn Wynn; Jeffrey Barrett; John Sillitoe on behalf of the Wellcome Sanger Institute COVID-19 Surveillance Team; Mairead Hyland; Roberto Amato; Sonia Goncalves; The Lighthouse Lab in Alderley Park and Alex Alderton |

EPI_ISL_2607497, EPI_ISL_2638249, EPI_ISL_2638271, EPI_ISL_2638287, EPI_ISL_2638324, EPI_ISL_2638349, EPI_ISL_2638397, EPI_ISL_2638399, EPI_ISL_2638403, EPI_ISL_2638410, EPI_ISL_2638414, EPI_ISL_2638416, EPI_ISL_2638420, EPI_ISL_2638423, EPI_ISL_2638427, EPI_ISL_2638432, EPI_ISL_2638440, EPI_ISL_2638443, EPI_ISL_2638449, EPI_ISL_2638454, EPI_ISL_2638456, EPI_ISL_2638458,
EPI_ISL_2638476, EPI_ISL_2638479, EPI_ISL_2638482, EPI_ISL_2638485, EPI_ISL_2638486, EPI_ISL_2638494, EPI_ISL_2638498, EPI_ISL_2638501, EPI_ISL_2638503, EPI_ISL_2638505, EPI_ISL_2638506, EPI_ISL_2638516, EPI_ISL_2638520, EPI_ISL_2638527, EPI_ISL_2638530, EPI_ISL_2638534, EPI_ISL_2638538, EPI_ISL_2638539, EPI_ISL_2638544, EPI_ISL_2638548, EPI_ISL_2638562, EPI_ISL_2638563,
EPI_ISL_2638568, EPI_ISL_2638577, EPI_ISL_2638581, EPI_ISL_2638586, EPI_ISL_2638589, EPI_ISL_2638590, EPI_ISL_2638591, EPI_ISL_2638592, EPI_ISL_2638600, EPI_ISL_2638605, EPI_ISL_2638606, EPI_ISL_2638609, EPI_ISL_2639617, EPI_ISL_2639621, EPI_ISL_2639625, EPI_ISL_2639640, EPI_ISL_2639657, EPI_ISL_2639667, EPI_ISL_2639674, EPI_ISL_2639689, EPI_ISL_2639697,
EPI_ISL_2639713, EPI_ISL_2639716, EPI_ISL_2639723, EPI_ISL_2639728, EPI_ISL_2639741, EPI_ISL_2639743, EPI_ISL_2639746, EPI_ISL_2639750, EPI_ISL_2639760, EPI_ISL_2639769, EPI_ISL_2639775, EPI_ISL_2639796, EPI_ISL_2639797, EPI_ISL_2639804, EPI_ISL_2639806, EPI_ISL_2639812, EPI_ISL_2639818, EPI_ISL_2639829, EPI_ISL_2639833, EPI_ISL_2639852, EPI_ISL_2639862, EPI_ISL_2639863,
EPI_ISL_2639864, EPI_ISL_2639878, EPI_ISL_2639896, EPI_ISL_2639902, EPI_ISL_2639921, EPI_ISL_2651410, EPI_ISL_2651412, EPI_ISL_2651413, EPI_ISL_2651418, EPI_ISL_2651427, EPI_ISL_2651430, EPI_ISL_2651431, EPI_ISL_2651432, EPI_ISL_2651433, EPI_ISL_2651434, EPI_ISL_2651437, EPI_ISL_2651438, EPI_ISL_2651451, EPI_ISL_2651453, EPI_ISL_2651456, EPI_ISL_2651457,
EPI_ISL_2651458, EPI_ISL_2651465, EPI_ISL_2651466, EPI_ISL_2651469, EPI_ISL_2651472, EPI_ISL_2651476, EPI_ISL_2651477, EPI_ISL_2651480, EPI_ISL_2651481, EPI_ISL_2651482, EPI_ISL_2651486, EPI_ISL_2651492, EPI_ISL_2651499, EPI_ISL_2651500, EPI_ISL_2651502, EPI_ISL_2651503, EPI_ISL_2651510, EPI_ISL_2651515, EPI_ISL_2651516, EPI_ISL_2651521, EPI_ISL_2651522, EPI_ISL_2651528,
EPI_ISL_2651531, EPI_ISL_2651532, EPI_ISL_2651533, EPI_ISL_2651538, EPI_ISL_2651539, EPI_ISL_2651547, EPI_ISL_2651551, EPI_ISL_2651552, EPI_ISL_2651556, EPI_ISL_2651557, EPI_ISL_2651561, EPI_ISL_2651565, EPI_ISL_2651566, EPI_ISL_2651571, EPI_ISL_2651572, EPI_ISL_2651574, EPI_ISL_2651576, EPI_ISL_2651577, EPI_ISL_2651582, EPI_ISL_2651588, EPI_ISL_2651592, EPI_ISL_2651594,
EPI_ISL_2651595, EPI_ISL_2651598, EPI_ISL_2651605, EPI_ISL_2651606, EPI_ISL_2651609, EPI_ISL_2651611, EPI_ISL_2651613, EPI_ISL_2651615, EPI_ISL_2651623, EPI_ISL_2651627, EPI_ISL_2651632, EPI_ISL_2651634, EPI_ISL_2651638, EPI_ISL_2651640, EPI_ISL_2651642, EPI_ISL_2651650, EPI_ISL_2651651, EPI_ISL_2651653, EPI_ISL_2651655, EPI_ISL_2651656, EPI_ISL_2651657, EPI_ISL_2651658,
EPI_ISL_2651663, EPI_ISL_2651664, EPI_ISL_2651667, EPI_ISL_2651668, EPI_ISL_2651670, EPI_ISL_2651672, EPI_ISL_2651673, EPI_ISL_2651674, EPI_ISL_2651677, EPI_ISL_2651679, EPI_ISL_2651682, EPI_ISL_2651684, EPI_ISL_2651685, EPI_ISL_2651686, EPI_ISL_2651687, EPI_ISL_2651689, EPI_ISL_2651692, EPI_ISL_2651693, EPI_ISL_2651694, EPI_ISL_2651695, EPI_ISL_2651700, EPI_ISL_2651701,
EPI_ISL_2651703, EPI_ISL_2651707, EPI_ISL_2651708, EPI_ISL_2651709, EPI_ISL_2651710, EPI_ISL_2651711, EPI_ISL_2651712, EPI_ISL_2651713, EPI_ISL_2651714, EPI_ISL_2651717, EPI_ISL_2651718, EPI_ISL_2651721, EPI_ISL_2651722, EPI_ISL_2651723, EPI_ISL_2651724, EPI_ISL_2651728, EPI_ISL_2651729, EPI_ISL_2651731, EPI_ISL_2651733, EPI_ISL_2651734, EPI_ISL_2651735, EPI_ISL_2651737,
EPI_ISL_2651738, EPI_ISL_2651739, EPI_ISL_2651740, EPI_ISL_2651741, EPI_ISL_2651743, EPI_ISL_2651744, EPI_ISL_2651745, EPI_ISL_2651747, EPI_ISL_2651748, EPI_ISL_2651749, EPI_ISL_2651750, EPI_ISL_2651751, EPI_ISL_2651752, EPI_ISL_2651757, EPI_ISL_2651758, EPI_ISL_2651761, EPI_ISL_2651764, EPI_ISL_2651765, EPI_ISL_2651766, EPI_ISL_2651767, EPI_ISL_2651768, EPI_ISL_2651769,
EPI_ISL_2651770, EPI_ISL_2651771, EPI_ISL_2651772, EPI_ISL_2651773, EPI_ISL_2651774, EPI_ISL_2651775, EPI_ISL_2651776, EPI_ISL_2651777, EPI_ISL_2651778, EPI_ISL_2651779, EPI_ISL_2651780, EPI_ISL_2651781, EPI_ISL_2651782, EPI_ISL_2651783, EPI_ISL_2651784, EPI_ISL_2651785, EPI_ISL_2651787, EPI_ISL_2651788, EPI_ISL_2651789, EPI_ISL_2651790, EPI_ISL_2651791, EPI_ISL_2651792,
EPI_ISL_2651793, EPI_ISL_2651794, EPI_ISL_2651795, EPI_ISL_2651796, EPI_ISL_2651797, EPI_ISL_2651798, EPI_ISL_2651799, EPI_ISL_2651800, EPI_ISL_2651801, EPI_ISL_2651802, EPI_ISL_2651803, EPI_ISL_2651805, EPI_ISL_2651806, EPI_ISL_2651807, EPI_ISL_2651808, EPI_ISL_2651809, EPI_ISL_2651810, EPI_ISL_2651811, EPI_ISL_2651812, EPI_ISL_2651813, EPI_ISL_2651814, EPI_ISL_2651815,
EPI_ISL_2651816, EPI_ISL_2651817, EPI_ISL_2651818, EPI_ISL_2651819, EPI_ISL_2651820, EPI_ISL_2651821, EPI_ISL_2651822, EPI_ISL_2651823, EPI_ISL_2651824, EPI_ISL_2651825, EPI_ISL_2651826, EPI_ISL_2651827, EPI_ISL_2651828, EPI_ISL_2651829, EPI_ISL_2651830, EPI_ISL_2651831, EPI_ISL_2651832, EPI_ISL_2651833, EPI_ISL_2651834, EPI_ISL_2651835, EPI_ISL_2651836, EPI_ISL_2651837,
EPI_ISL_2651838, EPI_ISL_2651839, EPI_ISL_2651840, EPI_ISL_2651841, EPI_ISL_2651842, EPI_ISL_2651843, EPI_ISL_2651844, EPI_ISL_2651845, EPI_ISL_2651846, EPI_ISL_2651849, EPI_ISL_2651850, EPI_ISL_2651851, EPI_ISL_2651852, EPI_ISL_2651853, EPI_ISL_2651854, EPI_ISL_2651855, EPI_ISL_2651856, EPI_ISL_2651857, EPI_ISL_2651858, EPI_ISL_2651859, EPI_ISL_2651860,
EPI_ISL_2651861, EPI_ISL_2651862, EPI_ISL_2651863, EPI_ISL_2651864, EPI_ISL_2651865, EPI_ISL_2651866, EPI_ISL_2651868, EPI_ISL_2651869, EPI_ISL_2651870, EPI_ISL_2651871, EPI_ISL_2651872, EPI_ISL_2651873, EPI_ISL_2651874, EPI_ISL_2651875, EPI_ISL_2651876, EPI_ISL_2651877, EPI_ISL_2651878, EPI_ISL_2651879, EPI_ISL_2651880, EPI_ISL_2651881, EPI_ISL_2651882, EPI_ISL_2651883,
EPI_ISL_2651884, EPI_ISL_2651885, EPI_ISL_2651886, EPI_ISL_2651887, EPI_ISL_2651888, EPI_ISL_2651889, EPI_ISL_2651890, EPI_ISL_2651891, EPI_ISL_2651949, EPI_ISL_2651950, EPI_ISL_2651951, EPI_ISL_2651952, EPI_ISL_2651953, EPI_ISL_2651954, EPI_ISL_2651955, EPI_ISL_2651956, EPI_ISL_2651957, EPI_ISL_2651958, EPI_ISL_2651959, EPI_ISL_2651960, EPI_ISL_2651961, EPI_ISL_2651962,
EPI_ISL_2651963, EPI_ISL_2651964, EPI_ISL_2651965, EPI_ISL_2651966, EPI_ISL_2651967, EPI_ISL_2651968, EPI_ISL_2651969, EPI_ISL_2651970, EPI_ISL_2651971, EPI_ISL_2651972, EPI_ISL_2651973, EPI_ISL_2651974, EPI_ISL_2651975, EPI_ISL_2651976, EPI_ISL_2651977, EPI_ISL_2651978, EPI_ISL_2651979, EPI_ISL_2651980, EPI_ISL_2651981, EPI_ISL_2651982, EPI_ISL_2651983, EPI_ISL_2651984,
EPI_ISL_2651985, EPI_ISL_2651986, EPI_ISL_2651987, EPI_ISL_2651988, EPI_ISL_2651989, EPI_ISL_2651990, EPI_ISL_2651991, EPI_ISL_2651992, EPI_ISL_2651993, EPI_ISL_2651994, EPI_ISL_2651995, EPI_ISL_2651996, EPI_ISL_2664011, EPI_ISL_2664012, EPI_ISL_2664018, EPI_ISL_2664027, EPI_ISL_2664035, EPI_ISL_2664036, EPI_ISL_2664040, EPI_ISL_2664054, EPI_ISL_2664060, EPI_ISL_2664063,
EPI_ISL_2664087, EPI_ISL_2664092, EPI_ISL_2664095, EPI_ISL_2664099, EPI_ISL_2664105, EPI_ISL_2664110, EPI_ISL_2664114, EPI_ISL_2664116, EPI_ISL_2664122, EPI_ISL_2664124, EPI_ISL_2664127, EPI_ISL_2664129, EPI_ISL_2664131, EPI_ISL_2664132, EPI_ISL_2664134, EPI_ISL_2664143, EPI_ISL_2664146, EPI_ISL_2664150, EPI_ISL_2664154, EPI_ISL_2664156, EPI_ISL_2664158,
EPI_ISL_2664160, EPI_ISL_2664172, EPI_ISL_2664176, EPI_ISL_2664180, EPI_ISL_2664182, EPI_ISL_2664184, EPI_ISL_2664190, EPI_ISL_2664193, EPI_ISL_2664199, EPI_ISL_2664200, EPI_ISL_2664206, EPI_ISL_2664208, EPI_ISL_2664223, EPI_ISL_2664224, EPI_ISL_2664234, EPI_ISL_2664237, EPI_ISL_2664240, EPI_ISL_2664250, EPI_ISL_2664254, EPI_ISL_2664266, EPI_ISL_2664267, EPI_ISL_2664269,
EPI_ISL_2664272, EPI_ISL_2664274, EPI_ISL_2664275, EPI_ISL_2664276, EPI_ISL_2664277, EPI_ISL_2664280, EPI_ISL_2664281, EPI_ISL_2664283, EPI_ISL_2664285, EPI_ISL_2664289, EPI_ISL_2664293, EPI_ISL_2664299, EPI_ISL_2664310, EPI_ISL_2664313, EPI_ISL_2664314, EPI_ISL_2664320, EPI_ISL_2664323, EPI_ISL_2664330, EPI_ISL_2664331, EPI_ISL_2664338, EPI_ISL_2664339, EPI_ISL_2664343,
EPI_ISL_2664353, EPI_ISL_2664355, EPI_ISL_2664361, EPI_ISL_2664363, EPI_ISL_2664364, EPI_ISL_2664376, EPI_ISL_2664381, EPI_ISL_2664382, EPI_ISL_2664383, EPI_ISL_2664386, EPI_ISL_2664387, EPI_ISL_2664404, EPI_ISL_2664407, EPI_ISL_2664410, EPI_ISL_2664419, EPI_ISL_2664427, EPI_ISL_2664434, EPI_ISL_2664435, EPI_ISL_2664436, EPI_ISL_2664441, EPI_ISL_2664445, EPI_ISL_2664446,
EPI_ISL_2664451, EPI_ISL_2664453, EPI_ISL_2664454, EPI_ISL_2664455, EPI_ISL_2664466, EPI_ISL_2664473, EPI_ISL_2664482, EPI_ISL_2664492, EPI_ISL_2664526, EPI_ISL_2664528, EPI_ISL_2664558, EPI_ISL_2664568, EPI_ISL_2664569, EPI_ISL_2664571, EPI_ISL_2664572, EPI_ISL_2664575, EPI_ISL_2664587, EPI_ISL_2664592, EPI_ISL_2664605, EPI_ISL_2664632, EPI_ISL_2664647,
EPI_ISL_2664650, EPI_ISL_2664662, EPI_ISL_2664669, EPI_ISL_2664681, EPI_ISL_2664683, EPI_ISL_2664684, EPI_ISL_2664686, EPI_ISL_2664689, EPI_ISL_2664694, EPI_ISL_2664719, EPI_ISL_2664724, EPI_ISL_2664733, EPI_ISL_2664752, EPI_ISL_2664804, EPI_ISL_2664806, EPI_ISL_2664828, EPI_ISL_2664871, EPI_ISL_2664881, EPI_ISL_2664906, EPI_ISL_2664910, EPI_ISL_2664924, EPI_ISL_2664929,
EPI_ISL_2664934, EPI_ISL_2664945, EPI_ISL_2664971, EPI_ISL_2664977, EPI_ISL_2664998, EPI_ISL_2665008, EPI_ISL_2681656, EPI_ISL_2681657, EPI_ISL_2681660, EPI_ISL_2681665, EPI_ISL_2681666, EPI_ISL_2681673, EPI_ISL_2681711, EPI_ISL_2681718, EPI_ISL_2681720, EPI_ISL_2681721, EPI_ISL_2681727, EPI_ISL_2681728, EPI_ISL_2681768, EPI_ISL_2681779, EPI_ISL_2681829, EPI_ISL_2681844,
EPI_ISL_2681847, EPI_ISL_2681862, EPI_ISL_2681865, EPI_ISL_2681867, EPI_ISL_2681878, EPI_ISL_2681889, EPI_ISL_2681902, EPI_ISL_2681914, EPI_ISL_2681923, EPI_ISL_2681926, EPI_ISL_2682068, EPI_ISL_2682069, EPI_ISL_2682071, EPI_ISL_2682073, EPI_ISL_2682082, EPI_ISL_2682083, EPI_ISL_2682085, EPI_ISL_2682093, EPI_ISL_2682097, EPI_ISL_2682100, EPI_ISL_2682105, EPI_ISL_2682107,
EPI_ISL_2682114, EPI_ISL_2682120, EPI_ISL_2682123, EPI_ISL_2682124, EPI_ISL_2682130, EPI_ISL_2682133, EPI_ISL_2682138, EPI_ISL_2682140, EPI_ISL_2682144, EPI_ISL_2682145, EPI_ISL_2682149, EPI_ISL_2682157, EPI_ISL_2682160, EPI_ISL_2682161, EPI_ISL_2682166, EPI_ISL_2682171, EPI_ISL_2682173, EPI_ISL_2682176, EPI_ISL_2682190, EPI_ISL_2682192, EPI_ISL_2682204,
EPI_ISL_2682212, EPI_ISL_2682213, EPI_ISL_2682226, EPI_ISL_2682229, EPI_ISL_2682230, EPI_ISL_2682233, EPI_ISL_2682252, EPI_ISL_2682256, EPI_ISL_2682260, EPI_ISL_2682262, EPI_ISL_2682265, EPI_ISL_2682274, EPI_ISL_2682276, EPI_ISL_2682284, EPI_ISL_2682653, EPI_ISL_2682655, EPI_ISL_2682656, EPI_ISL_2682657, EPI_ISL_2682659, EPI_ISL_2682660, EPI_ISL_2682661, EPI_ISL_2682663,

EPI_ISL_2682665, EPI_ISL_2682669, EPI_ISL_2682670, EPI_ISL_2682672, EPI_ISL_2682676, EPI_ISL_2682677, EPI_ISL_2682679, EPI_ISL_2682682, EPI_ISL_2682684, EPI_ISL_2682685, EPI_ISL_2682686, EPI_ISL_2682689, EPI_ISL_2682691, EPI_ISL_2682692, EPI_ISL_2682693, EPI_ISL_2682694, EPI_ISL_2682696, EPI_ISL_2682698, EPI_ISL_2682699, EPI_ISL_2682701, EPI_ISL_2682703, EPI_ISL_2682704,
EPI_ISL_2682708, EPI_ISL_2682710, EPI_ISL_2682711, EPI_ISL_2682712, EPI_ISL_2682715, EPI_ISL_2682716, EPI_ISL_2682718, EPI_ISL_2682724, EPI_ISL_2682727, EPI_ISL_2682728, EPI_ISL_2682729, EPI_ISL_2682730, EPI_ISL_2682731, EPI_ISL_2682737, EPI_ISL_2682742, EPI_ISL_2682744, EPI_ISL_2682745, EPI_ISL_2682747, EPI_ISL_2682750, EPI_ISL_2682751, EPI_ISL_2682752, EPI_ISL_2682754,
EPI_ISL_2682758, EPI_ISL_2682760, EPI_ISL_2682761, EPI_ISL_2682766, EPI_ISL_2682767, EPI_ISL_2682768, EPI_ISL_2682769, EPI_ISL_2682771, EPI_ISL_2682772, EPI_ISL_2682774, EPI_ISL_2682777, EPI_ISL_2682778, EPI_ISL_2682780, EPI_ISL_2682783, EPI_ISL_2682785, EPI_ISL_2682789, EPI_ISL_2682791, EPI_ISL_2682793, EPI_ISL_2682795, EPI_ISL_2682797, EPI_ISL_2682799, EPI_ISL_2682800,
EPI_ISL_2682803, EPI_ISL_2682806, EPI_ISL_2682809, EPI_ISL_2682810, EPI_ISL_2682813, EPI_ISL_2682814, EPI_ISL_2682820, EPI_ISL_2682827, EPI_ISL_2682828, EPI_ISL_2682831, EPI_ISL_2682834, EPI_ISL_2682835, EPI_ISL_2682839, EPI_ISL_2682840, EPI_ISL_2682841, EPI_ISL_2682846, EPI_ISL_2682847, EPI_ISL_2682848, EPI_ISL_2682849, EPI_ISL_2682850, EPI_ISL_2682851, EPI_ISL_2682852,
EPI_ISL_2682855, EPI_ISL_2682859, EPI_ISL_2682860, EPI_ISL_2682861, EPI_ISL_2682863, EPI_ISL_2682867, EPI_ISL_2682868, EPI_ISL_2682869, EPI_ISL_2682870, EPI_ISL_2682871, EPI_ISL_2682872, EPI_ISL_2682874, EPI_ISL_2682877, EPI_ISL_2682879, EPI_ISL_2682880, EPI_ISL_2682883, EPI_ISL_2682885, EPI_ISL_2682887, EPI_ISL_2682889, EPI_ISL_2682891, EPI_ISL_2682892, EPI_ISL_2682894,
EPI_ISL_2682895, EPI_ISL_2682899, EPI_ISL_2682900, EPI_ISL_2682901, EPI_ISL_2682904, EPI_ISL_2682905, EPI_ISL_2682906, EPI_ISL_2682908, EPI_ISL_2682909, EPI_ISL_2682911, EPI_ISL_2682912, EPI_ISL_2682915, EPI_ISL_2682916, EPI_ISL_2682917, EPI_ISL_2682918, EPI_ISL_2682921, EPI_ISL_2682922, EPI_ISL_2682923, EPI_ISL_2682926, EPI_ISL_2682927, EPI_ISL_2682930, EPI_ISL_2682931,
EPI_ISL_2682934, EPI_ISL_2682940, EPI_ISL_2682946, EPI_ISL_2682947, EPI_ISL_2682948, EPI_ISL_2682951, EPI_ISL_2682953, EPI_ISL_2682956, EPI_ISL_2682959, EPI_ISL_2682962, EPI_ISL_2682963, EPI_ISL_2682964, EPI_ISL_2682965, EPI_ISL_2682967, EPI_ISL_2682968, EPI_ISL_2682970, EPI_ISL_2682972, EPI_ISL_2682973, EPI_ISL_2682974, EPI_ISL_2682975, EPI_ISL_2682976, EPI_ISL_2682977,
EPI_ISL_2682978, EPI_ISL_2682979, EPI_ISL_2682980, EPI_ISL_2682981, EPI_ISL_2682982, EPI_ISL_2682983, EPI_ISL_2682984, EPI_ISL_2682985, EPI_ISL_2682986, EPI_ISL_2682987, EPI_ISL_2682988, EPI_ISL_2682989, EPI_ISL_2682990, EPI_ISL_2682991, EPI_ISL_2682992, EPI_ISL_2682993, EPI_ISL_2682994, EPI_ISL_2682995, EPI_ISL_2682996, EPI_ISL_2682997, EPI_ISL_2682998, EPI_ISL_2682999,
EPI_ISL_2683000, EPI_ISL_2683001, EPI_ISL_2683002, EPI_ISL_2683003, EPI_ISL_2683004, EPI_ISL_2683006, EPI_ISL_2683007, EPI_ISL_2683008, EPI_ISL_2683009, EPI_ISL_2683010, EPI_ISL_2683011, EPI_ISL_2683012, EPI_ISL_2683013, EPI_ISL_2683014, EPI_ISL_2683015, EPI_ISL_2683017, EPI_ISL_2683018, EPI_ISL_2683019, EPI_ISL_2683020, EPI_ISL_2683021, EPI_ISL_2683022, EPI_ISL_2683023,
EPI_ISL_2683024, EPI_ISL_2683025, EPI_ISL_2683026, EPI_ISL_2683027, EPI_ISL_2683028, EPI_ISL_2683029, EPI_ISL_2683030, EPI_ISL_2683031, EPI_ISL_2683032, EPI_ISL_2683033, EPI_ISL_2683034, EPI_ISL_2683035, EPI_ISL_2683036, EPI_ISL_2683037, EPI_ISL_2683038, EPI_ISL_2683039, EPI_ISL_2683040, EPI_ISL_2683041, EPI_ISL_2683042, EPI_ISL_2683043, EPI_ISL_2683044, EPI_ISL_2683045,
EPI_ISL_2683046, EPI_ISL_2683047, EPI_ISL_2683048, EPI_ISL_2683049, EPI_ISL_2683050, EPI_ISL_2683051, EPI_ISL_2683052, EPI_ISL_2683053, EPI_ISL_2683054, EPI_ISL_2683055, EPI_ISL_2683056, EPI_ISL_2683057, EPI_ISL_2683058, EPI_ISL_2683059, EPI_ISL_2683060, EPI_ISL_2683061, EPI_ISL_2683062, EPI_ISL_2683063, EPI_ISL_2683064, EPI_ISL_2683065, EPI_ISL_2683066, EPI_ISL_2683067,
EPI_ISL_2683068, EPI_ISL_2683069, EPI_ISL_2683071, EPI_ISL_2683072, EPI_ISL_2683073, EPI_ISL_2683074, EPI_ISL_2683075, EPI_ISL_2683076, EPI_ISL_2683077, EPI_ISL_2683078, EPI_ISL_2683079, EPI_ISL_2683080, EPI_ISL_2683081, EPI_ISL_2683082, EPI_ISL_2683083, EPI_ISL_2683084, EPI_ISL_2683085, EPI_ISL_2683086, EPI_ISL_2683087, EPI_ISL_2683088, EPI_ISL_2683089, EPI_ISL_2683090,
EPI_ISL_2683091, EPI_ISL_2683092, EPI_ISL_2683093, EPI_ISL_2683094, EPI_ISL_2683095, EPI_ISL_2683096, EPI_ISL_2683097, EPI_ISL_2683098, EPI_ISL_2683099, EPI_ISL_2683100, EPI_ISL_2683101, EPI_ISL_2683102, EPI_ISL_2683103, EPI_ISL_2683104, EPI_ISL_2683105, EPI_ISL_2683106, EPI_ISL_2683107, EPI_ISL_2683108, EPI_ISL_2683109, EPI_ISL_2683110, EPI_ISL_2683115, EPI_ISL_2683116,
EPI_ISL_2683117, EPI_ISL_2683118, EPI_ISL_2683119, EPI_ISL_2683120, EPI_ISL_2683121, EPI_ISL_2683122, EPI_ISL_2683123, EPI_ISL_2683124, EPI_ISL_2683125, EPI_ISL_2683126, EPI_ISL_2683127, EPI_ISL_2683128, EPI_ISL_2683129, EPI_ISL_2683130, EPI_ISL_2683131, EPI_ISL_2683132, EPI_ISL_2683133, EPI_ISL_2683134, EPI_ISL_2683135, EPI_ISL_2683136, EPI_ISL_2683137, EPI_ISL_2683138,
EPI_ISL_2683139, EPI_ISL_2683140, EPI_ISL_2683141, EPI_ISL_2683142, EPI_ISL_2683143, EPI_ISL_2683144, EPI_ISL_2683145, EPI_ISL_2683146, EPI_ISL_2683147, EPI_ISL_2683148, EPI_ISL_2683149, EPI_ISL_2683150, EPI_ISL_2683151, EPI_ISL_2683152, EPI_ISL_2683153, EPI_ISL_2683154, EPI_ISL_2683155, EPI_ISL_2683156, EPI_ISL_2683157, EPI_ISL_2683158, EPI_ISL_2683159, EPI_ISL_2683160,
EPI_ISL_2683161, EPI_ISL_2683162, EPI_ISL_2683163, EPI_ISL_2683164, EPI_ISL_2683165, EPI_ISL_2683166, EPI_ISL_2683167, EPI_ISL_2683168, EPI_ISL_2683169, EPI_ISL_2683170, EPI_ISL_2683171, EPI_ISL_2683172, EPI_ISL_2683173, EPI_ISL_2683174, EPI_ISL_2683175, EPI_ISL_2683176, EPI_ISL_2683177, EPI_ISL_2683178, EPI_ISL_2683179, EPI_ISL_2683180, EPI_ISL_2683181, EPI_ISL_2683182,
EPI_ISL_2683183, EPI_ISL_2683184, EPI_ISL_2683185, EPI_ISL_2683186, EPI_ISL_2683187, EPI_ISL_2683188, EPI_ISL_2683189, EPI_ISL_2683190, EPI_ISL_2683191, EPI_ISL_2683192, EPI_ISL_2683193, EPI_ISL_2683194, EPI_ISL_2683195, EPI_ISL_2683196, EPI_ISL_2683197, EPI_ISL_2683198, EPI_ISL_2683199, EPI_ISL_2683200, EPI_ISL_2683201, EPI_ISL_2683202, EPI_ISL_2683203, EPI_ISL_2683204,
EPI_ISL_2683205, EPI_ISL_2683206, EPI_ISL_2683207, EPI_ISL_2683208, EPI_ISL_2683209, EPI_ISL_2683210, EPI_ISL_2683211, EPI_ISL_2683212, EPI_ISL_2683213, EPI_ISL_2683214, EPI_ISL_2683215, EPI_ISL_2683216, EPI_ISL_2683217, EPI_ISL_2683218, EPI_ISL_2683219, EPI_ISL_2683220, EPI_ISL_2683221, EPI_ISL_2683222, EPI_ISL_2683223, EPI_ISL_2683224, EPI_ISL_2683225, EPI_ISL_2683226,
EPI_ISL_2683227, EPI_ISL_2683228, EPI_ISL_2683229, EPI_ISL_2683230, EPI_ISL_2683231, EPI_ISL_2683232, EPI_ISL_2683233, EPI_ISL_2683234, EPI_ISL_2683235, EPI_ISL_2683236, EPI_ISL_2683237, EPI_ISL_2683238, EPI_ISL_2683239, EPI_ISL_2683240, EPI_ISL_2683241, EPI_ISL_2683242, EPI_ISL_2683243, EPI_ISL_2683244, EPI_ISL_2683245, EPI_ISL_2683246, EPI_ISL_2683248, EPI_ISL_2683249,
EPI_ISL_2683250, EPI_ISL_2683251, EPI_ISL_2683252, EPI_ISL_2683253, EPI_ISL_2683254, EPI_ISL_2683255, EPI_ISL_2683256, EPI_ISL_2683257, EPI_ISL_2683258, EPI_ISL_2683259, EPI_ISL_2683260, EPI_ISL_2683261, EPI_ISL_2683262, EPI_ISL_2683263, EPI_ISL_2683264, EPI_ISL_2683265, EPI_ISL_2683266, EPI_ISL_2683267, EPI_ISL_2683268, EPI_ISL_2683269, EPI_ISL_2683270, EPI_ISL_2683271,
EPI_ISL_2683272, EPI_ISL_2683273, EPI_ISL_2683274, EPI_ISL_2683275, EPI_ISL_2683276, EPI_ISL_2683277, EPI_ISL_2683278, EPI_ISL_2683279, EPI_ISL_2683280, EPI_ISL_2683281, EPI_ISL_2683282, EPI_ISL_2683283, EPI_ISL_2683284, EPI_ISL_2683285, EPI_ISL_2683286, EPI_ISL_2683287, EPI_ISL_2683288, EPI_ISL_2683289, EPI_ISL_2683290, EPI_ISL_2683291, EPI_ISL_2683292, EPI_ISL_2683293,
EPI_ISL_2683294, EPI_ISL_2683295, EPI_ISL_2683296, EPI_ISL_2683297, EPI_ISL_2683298, EPI_ISL_2683299, EPI_ISL_2683300, EPI_ISL_2683301, EPI_ISL_2683302, EPI_ISL_2683303, EPI_ISL_2683304, EPI_ISL_2683305, EPI_ISL_2683306, EPI_ISL_2683307, EPI_ISL_2683308, EPI_ISL_2683309, EPI_ISL_2683310, EPI_ISL_2683311, EPI_ISL_2683312, EPI_ISL_2683313, EPI_ISL_2683314, EPI_ISL_2683315,
EPI_ISL_2683316, EPI_ISL_2683317, EPI_ISL_2683318, EPI_ISL_2683319, EPI_ISL_2683320, EPI_ISL_2683321, EPI_ISL_2683322, EPI_ISL_2683323, EPI_ISL_2683324, EPI_ISL_2683325, EPI_ISL_2683326, EPI_ISL_2683327, EPI_ISL_2683328, EPI_ISL_2683329, EPI_ISL_2683330, EPI_ISL_2683331, EPI_ISL_2683332, EPI_ISL_2683333, EPI_ISL_2683334, EPI_ISL_2683335, EPI_ISL_2683336, EPI_ISL_2683337,
EPI_ISL_2683338, EPI_ISL_2683339, EPI_ISL_2683340, EPI_ISL_2683341, EPI_ISL_2683342, EPI_ISL_2683343, EPI_ISL_2683344, EPI_ISL_2683345, EPI_ISL_2683346, EPI_ISL_2683347, EPI_ISL_2683348, EPI_ISL_2683349, EPI_ISL_2683350, EPI_ISL_2683351, EPI_ISL_2683352, EPI_ISL_2683353, EPI_ISL_2683354, EPI_ISL_2683355, EPI_ISL_2683356, EPI_ISL_2683357, EPI_ISL_2683358, EPI_ISL_2683359,
EPI_ISL_2683360, EPI_ISL_2683361, EPI_ISL_2683362, EPI_ISL_2683363, EPI_ISL_2683364, EPI_ISL_2683365, EPI_ISL_2683366, EPI_ISL_2683367, EPI_ISL_2683368, EPI_ISL_2683369, EPI_ISL_2683370, EPI_ISL_2683371, EPI_ISL_2683372, EPI_ISL_2683373, EPI_ISL_2683374, EPI_ISL_2683375, EPI_ISL_2683376, EPI_ISL_2683377, EPI_ISL_2683378, EPI_ISL_2683379, EPI_ISL_2683380, EPI_ISL_2683381,
EPI_ISL_2683382, EPI_ISL_2683383, EPI_ISL_2683384, EPI_ISL_2683385, EPI_ISL_2683386, EPI_ISL_2683387, EPI_ISL_2683388, EPI_ISL_2683389, EPI_ISL_2683390, EPI_ISL_2683391, EPI_ISL_2683392, EPI_ISL_2683393, EPI_ISL_2683394, EPI_ISL_2683395, EPI_ISL_2683396, EPI_ISL_2683397, EPI_ISL_2683398, EPI_ISL_2683399, EPI_ISL_2683400, EPI_ISL_2683401, EPI_ISL_2683402, EPI_ISL_2683403,
EPI_ISL_2683404, EPI_ISL_2683405, EPI_ISL_2683406, EPI_ISL_2683407, EPI_ISL_2683408, EPI_ISL_2683409, EPI_ISL_2683410, EPI_ISL_2683411, EPI_ISL_2683412, EPI_ISL_2683413, EPI_ISL_2683414, EPI_ISL_2683415, EPI_ISL_2683416, EPI_ISL_2683417, EPI_ISL_2683418, EPI_ISL_2683419, EPI_ISL_2683420, EPI_ISL_2683421, EPI_ISL_2683422, EPI_ISL_2683423, EPI_ISL_2683424, EPI_ISL_2683425,
EPI_ISL_2683426, EPI_ISL_2683427, EPI_ISL_2683429, EPI_ISL_2683430, EPI_ISL_2683431, EPI_ISL_2683432, EPI_ISL_2683433, EPI_ISL_2683434, EPI_ISL_2683436, EPI_ISL_2683437, EPI_ISL_2683438, EPI_ISL_2683439, EPI_ISL_2683440, EPI_ISL_2683441, EPI_ISL_2683442, EPI_ISL_2683443, EPI_ISL_2683444, EPI_ISL_2683445, EPI_ISL_2683446, EPI_ISL_2683447, EPI_ISL_2683448, EPI_ISL_2683449,
EPI_ISL_2683450, EPI_ISL_2683451, EPI_ISL_2683452, EPI_ISL_2683453, EPI_ISL_2683454, EPI_ISL_2683455, EPI_ISL_2683456, EPI_ISL_2683457, EPI_ISL_2683458, EPI_ISL_2683459, EPI_ISL_2683461, EPI_ISL_2683462, EPI_ISL_2683463, EPI_ISL_2683464, EPI_ISL_2683465, EPI_ISL_2683466, EPI_ISL_2683467, EPI_ISL_2683469, EPI_ISL_2683470, EPI_ISL_2683471, EPI_ISL_2683472, EPI_ISL_2683473,
EPI_ISL_2683474, EPI_ISL_2683475, EPI_ISL_2683476, EPI_ISL_2683477, EPI_ISL_2683478, EPI_ISL_2683479, EPI_ISL_2683480, EPI_ISL_2683481, EPI_ISL_2683482, EPI_ISL_2683484, EPI_ISL_2683485, EPI_ISL_2683486, EPI_ISL_2683487, EPI_ISL_2683488, EPI_ISL_2683489, EPI_ISL_2683490, EPI_ISL_2699534, EPI_ISL_2699540, EPI_ISL_2699544, EPI_ISL_2699563, EPI_ISL_2699577, EPI_ISL_2699583,
EPI_ISL_2699593, EPI_ISL_2699617, EPI_ISL_2699642, EPI_ISL_2699644, EPI_ISL_2699667, EPI_ISL_2699683, EPI_ISL_2699692, EPI_ISL_2699698, EPI_ISL_2699704, EPI_ISL_2699715, EPI_ISL_2699728, EPI_ISL_2699741, EPI_ISL_2699743, EPI_ISL_2699745, EPI_ISL_2699751, EPI_ISL_2699753, EPI_ISL_2699761, EPI_ISL_2699764, EPI_ISL_2699776, EPI_ISL_2699777, EPI_ISL_2699789, EPI_ISL_2699804,
EPI_ISL_2699811, EPI_ISL_2699812, EPI_ISL_2699829, EPI_ISL_2699832, EPI_ISL_2699838, EPI_ISL_2699854, EPI_ISL_2699860, EPI_ISL_2700294, EPI_ISL_2700319, EPI_ISL_2700485, EPI_ISL_2700939, EPI_ISL_2700945, EPI_ISL_2700946, EPI_ISL_2700947, EPI_ISL_2700948, EPI_ISL_2700949, EPI_ISL_2700953, EPI_ISL_2700954, EPI_ISL_2700957, EPI_ISL_2700958, EPI_ISL_2700960, EPI_ISL_2700961,
EPI_ISL_2700962, EPI_ISL_2700963, EPI_ISL_2700964, EPI_ISL_2700965, EPI_ISL_2700968, EPI_ISL_2700970, EPI_ISL_2700972, EPI_ISL_2700976, EPI_ISL_2700977, EPI_ISL_2700979, EPI_ISL_2700985, EPI_ISL_2700986, EPI_ISL_2701002, EPI_ISL_2701005, EPI_ISL_2701006, EPI_ISL_2701009, EPI_ISL_2701010, EPI_ISL_2701011, EPI_ISL_2701015, EPI_ISL_2701019, EPI_ISL_2701022, EPI_ISL_2701025,
EPI_ISL_2701032, EPI_ISL_2701033, EPI_ISL_2701036, EPI_ISL_2701044, EPI_ISL_2701049, EPI_ISL_2701050, EPI_ISL_2701053, EPI_ISL_2701065, EPI_ISL_2701073, EPI_ISL_2701080, EPI_ISL_2701081, EPI_ISL_2701082, EPI_ISL_2701083, EPI_ISL_2701088, EPI_ISL_2701090, EPI_ISL_2701094, EPI_ISL_2701095, EPI_ISL_2701103, EPI_ISL_2701105, EPI_ISL_2701107, EPI_ISL_2701111, EPI_ISL_2701112,
EPI_ISL_2701113, EPI_ISL_2701114, EPI_ISL_2701125, EPI_ISL_2701126, EPI_ISL_2701128, EPI_ISL_2701139, EPI_ISL_2701142, EPI_ISL_2701149, EPI_ISL_2701150, EPI_ISL_2701151, EPI_ISL_2701156, EPI_ISL_2701157, EPI_ISL_2701171, EPI_ISL_2701173, EPI_ISL_2701178, EPI_ISL_2701179, EPI_ISL_2701181, EPI_ISL_2701183, EPI_ISL_2701191, EPI_ISL_2701192, EPI_ISL_2701198, EPI_ISL_2701202,
EPI_ISL_2701205, EPI_ISL_2701210, EPI_ISL_2701222, EPI_ISL_2701223, EPI_ISL_2701228, EPI_ISL_2701242, EPI_ISL_2701251, EPI_ISL_2701256, EPI_ISL_2701257, EPI_ISL_2701264, EPI_ISL_2701266, EPI_ISL_2701269, EPI_ISL_2701276, EPI_ISL_2701277, EPI_ISL_2701278, EPI_ISL_2701279, EPI_ISL_2701280, EPI_ISL_2701283, EPI_ISL_2701296, EPI_ISL_2701299, EPI_ISL_2701322, EPI_ISL_2701381,
EPI_ISL_2701469, EPI_ISL_2701482, EPI_ISL_2701563, EPI_ISL_2701858, EPI_ISL_2702023, EPI_ISL_2702033, EPI_ISL_2702036, EPI_ISL_2702397, EPI_ISL_2702398, EPI_ISL_2702399, EPI_ISL_2702400, EPI_ISL_2702401, EPI_ISL_2702402, EPI_ISL_2702403, EPI_ISL_2702404, EPI_ISL_2702405, EPI_ISL_2702406, EPI_ISL_2702407, EPI_ISL_2702409, EPI_ISL_2702410, EPI_ISL_2702411, EPI_ISL_2702412,
EPI_ISL_2702413, EPI_ISL_2702414, EPI_ISL_2702415, EPI_ISL_2702416, EPI_ISL_2702417, EPI_ISL_2702418, EPI_ISL_2702419, EPI_ISL_2702420, EPI_ISL_2702423, EPI_ISL_2702424, EPI_ISL_2702425, EPI_ISL_2702426, EPI_ISL_2702427, EPI_ISL_2702428, EPI_ISL_2702429, EPI_ISL_2702430, EPI_ISL_2702431, EPI_ISL_2702432, EPI_ISL_2702433, EPI_ISL_2702434, EPI_ISL_2702435, EPI_ISL_2702436,
EPI_ISL_2702437, EPI_ISL_2702438, EPI_ISL_2702439, EPI_ISL_2702440, EPI_ISL_2702441, EPI_ISL_2702442, EPI_ISL_2702443, EPI_ISL_2702444, EPI_ISL_2702445, EPI_ISL_2702446, EPI_ISL_2702447, EPI_ISL_2702448, EPI_ISL_2702449, EPI_ISL_2702450, EPI_ISL_2702451, EPI_ISL_2702452, EPI_ISL_2702453, EPI_ISL_2702454, EPI_ISL_2702455, EPI_ISL_2702456, EPI_ISL_2702457, EPI_ISL_2702460,
EPI_ISL_2702467, EPI_ISL_2702470, EPI_ISL_2702472, EPI_ISL_2702474, EPI_ISL_2702475, EPI_ISL_2702479, EPI_ISL_2702483, EPI_ISL_2702489, EPI_ISL_2702501, EPI_ISL_2702505, EPI_ISL_2702508, EPI_ISL_2702516, EPI_ISL_2702520, EPI_ISL_2702521, EPI_ISL_2702524, EPI_ISL_2702534, EPI_ISL_2702553, EPI_ISL_2702561, EPI_ISL_2702574, EPI_ISL_2702576, EPI_ISL_2702580, EPI_ISL_2702581,
EPI_ISL_2702587, EPI_ISL_2702592, EPI_ISL_2702593, EPI_ISL_2702596, EPI_ISL_2702604, EPI_ISL_2702606, EPI_ISL_2702608, EPI_ISL_2702609, EPI_ISL_2702619, EPI_ISL_2702622, EPI_ISL_2702629, EPI_ISL_2702634, EPI_ISL_2702635, EPI_ISL_2702639, EPI_ISL_2702642, EPI_ISL_2702645, EPI_ISL_2702648, EPI_ISL_2702649, EPI_ISL_2702659, EPI_ISL_2702662, EPI_ISL_2702663, EPI_ISL_2702666,
EPI_ISL_2702670, EPI_ISL_2702672, EPI_ISL_2702676, EPI_ISL_2702683, EPI_ISL_2702687, EPI_ISL_2702688, EPI_ISL_2702690, EPI_ISL_2702691, EPI_ISL_2702694, EPI_ISL_2702696, EPI_ISL_2702698, EPI_ISL_2702700, EPI_ISL_2702705, EPI_ISL_2702712, EPI_ISL_2702719, EPI_ISL_2702723, EPI_ISL_2702725, EPI_ISL_2702729, EPI_ISL_2702733, EPI_ISL_2702736, EPI_ISL_2702741, EPI_ISL_2702746,
EPI_ISL_2702748, EPI_ISL_2702753, EPI_ISL_2702754, EPI_ISL_2702755, EPI_ISL_2702765, EPI_ISL_2702770, EPI_ISL_2702773, EPI_ISL_2702775, EPI_ISL_2702780, EPI_ISL_2702782, EPI_ISL_2702783, EPI_ISL_2702786, EPI_ISL_2702787, EPI_ISL_2702793, EPI_ISL_2702794, EPI_ISL_2702796, EPI_ISL_2702800, EPI_ISL_2702804, EPI_ISL_2702805, EPI_ISL_2702808, EPI_ISL_2702810, EPI_ISL_2703504,
EPI_ISL_2703506, EPI_ISL_2703509, EPI_ISL_2703513, EPI_ISL_2703517, EPI_ISL_2703524, EPI_ISL_2703525, EPI_ISL_2703528, EPI_ISL_2703530, EPI_ISL_2703536, EPI_ISL_2703537, EPI_ISL_2703550, EPI_ISL_2703554, EPI_ISL_2703561, EPI_ISL_2703562, EPI_ISL_2703578, EPI_ISL_2703581, EPI_ISL_2703582, EPI_ISL_2703588, EPI_ISL_2703591, EPI_ISL_2703594, EPI_ISL_2703598, EPI_ISL_2703604,
EPI_ISL_2703606, EPI_ISL_2703613, EPI_ISL_2703622, EPI_ISL_2703632, EPI_ISL_2703633, EPI_ISL_2703635, EPI_ISL_2703636, EPI_ISL_2703639, EPI_ISL_2703642, EPI_ISL_2703647, EPI_ISL_2703657, EPI_ISL_2703659, EPI_ISL_2703663, EPI_ISL_2703666, EPI_ISL_2703667, EPI_ISL_2703673, EPI_ISL_2703675, EPI_ISL_2703677, EPI_ISL_2703680, EPI_ISL_2703684, EPI_ISL_2703690, EPI_ISL_2703691,
EPI_ISL_2703692, EPI_ISL_2703696, EPI_ISL_2703697, EPI_ISL_2703708, EPI_ISL_2703713, EPI_ISL_2703714, EPI_ISL_2703720, EPI_ISL_2703722, EPI_ISL_2703725, EPI_ISL_2703729, EPI_ISL_2703731, EPI_ISL_2703737, EPI_ISL_2703739, EPI_ISL_2703740, EPI_ISL_2703748, EPI_ISL_2703751, EPI_ISL_2703757, EPI_ISL_2703758, EPI_ISL_2703767, EPI_ISL_2703769, EPI_ISL_2703770, EPI_ISL_2703777,
EPI_ISL_2703779, EPI_ISL_2703780, EPI_ISL_2703783, EPI_ISL_2703788, EPI_ISL_2703789, EPI_ISL_2703790, EPI_ISL_2703791, EPI_ISL_2703796, EPI_ISL_2703797, EPI_ISL_2703806, EPI_ISL_2703807, EPI_ISL_2703809, EPI_ISL_2703811, EPI_ISL_2703813, EPI_ISL_2703814, EPI_ISL_2703832, EPI_ISL_2703833, EPI_ISL_2704533, EPI_ISL_2704534, EPI_ISL_2704536, EPI_ISL_2704537, EPI_ISL_2704538,
EPI_ISL_2704539, EPI_ISL_2704540, EPI_ISL_2704541, EPI_ISL_2704542, EPI_ISL_2704543, EPI_ISL_2704544, EPI_ISL_2704545, EPI_ISL_2704546, EPI_ISL_2704547, EPI_ISL_2704548, EPI_ISL_2704549, EPI_ISL_2704550, EPI_ISL_2704551, EPI_ISL_2704552, EPI_ISL_2704553, EPI_ISL_2704554, EPI_ISL_2704555, EPI_ISL_2704556, EPI_ISL_2704557, EPI_ISL_2704558, EPI_ISL_2704559, EPI_ISL_2704560,
EPI_ISL_2704561, EPI_ISL_2704562, EPI_ISL_2704563, EPI_ISL_2704564, EPI_ISL_2704565, EPI_ISL_2704566, EPI_ISL_2704567, EPI_ISL_2704568, EPI_ISL_2704569, EPI_ISL_2704570, EPI_ISL_2704571, EPI_ISL_2704572, EPI_ISL_2704573, EPI_ISL_2704574, EPI_ISL_2704575, EPI_ISL_2704576, EPI_ISL_2704577, EPI_ISL_2704578, EPI_ISL_2704579, EPI_ISL_2704580, EPI_ISL_2704581, EPI_ISL_2704582,
EPI_ISL_2704583, EPI_ISL_2704584, EPI_ISL_2704585, EPI_ISL_2704586, EPI_ISL_2704587, EPI_ISL_2704588, EPI_ISL_2704589, EPI_ISL_2704590, EPI_ISL_2704591, EPI_ISL_2704592, EPI_ISL_2704593, EPI_ISL_2704594, EPI_ISL_2704595, EPI_ISL_2704596, EPI_ISL_2704597, EPI_ISL_2704598, EPI_ISL_2704599, EPI_ISL_2704600, EPI_ISL_2704601, EPI_ISL_2704602, EPI_ISL_2704603, EPI_ISL_2704604,
EPI_ISL_2704605, EPI_ISL_2704606, EPI_ISL_2704607, EPI_ISL_2704608, EPI_ISL_2704609, EPI_ISL_2704610, EPI_ISL_2704611, EPI_ISL_2704612, EPI_ISL_2704613, EPI_ISL_2704614, EPI_ISL_2704615, EPI_ISL_2704616, EPI_ISL_2704617, EPI_ISL_2704618, EPI_ISL_2704619, EPI_ISL_2704620, EPI_ISL_2704621, EPI_ISL_2704622, EPI_ISL_2704623, EPI_ISL_2704624, EPI_ISL_2704625, EPI_ISL_2704626,
EPI_ISL_2704627, EPI_ISL_2704628, EPI_ISL_2704629, EPI_ISL_2704630, EPI_ISL_2704631, EPI_ISL_2704632, EPI_ISL_2704633, EPI_ISL_2704634, EPI_ISL_2704635, EPI_ISL_2704636, EPI_ISL_2704637, EPI_ISL_2704638, EPI_ISL_2704639, EPI_ISL_2704640, EPI_ISL_2704641, EPI_ISL_2704642, EPI_ISL_2704643, EPI_ISL_2704644, EPI_ISL_2704645, EPI_ISL_2704646, EPI_ISL_2704647, EPI_ISL_2704648,
EPI_ISL_2704649, EPI_ISL_2704650, EPI_ISL_2704651, EPI_ISL_2704652, EPI_ISL_2704653, EPI_ISL_2704654, EPI_ISL_2704655, EPI_ISL_2704656, EPI_ISL_2704657, EPI_ISL_2704658, EPI_ISL_2704659, EPI_ISL_2704660, EPI_ISL_2704661, EPI_ISL_2704662, EPI_ISL_2704663, EPI_ISL_2704664, EPI_ISL_2704665, EPI_ISL_2704667, EPI_ISL_2704676, EPI_ISL_2809953

| | | | |
|---|---|---|---|
| see above | Lighthouse Lab in Glasgow | Wellcome Sanger Institute for the COVID-19 Genomics UK (COG-UK) Consortium | Anna Dominiczak and Alex Alderton; Carol Clugston; Cordelia Langford; David Gray; David K. Jackson; Dominic Kwiatkowski; Ewan Harrison; Harper VanSteenhouse; Ian Johnston; Jeffrey Barrett; John Sillitoe on behalf of the Wellcome Sanger Institute COVID-19 Surveillance Team; Roberto Amato; Sonia Goncalves; Yumi Kasai |

EPI_ISL_2638161, EPI_ISL_2638162, EPI_ISL_2638164, EPI_ISL_2638168, EPI_ISL_2638170, EPI_ISL_2638174, EPI_ISL_2638176, EPI_ISL_2638180, EPI_ISL_2638183, EPI_ISL_2638191, EPI_ISL_2638196, EPI_ISL_2638197, EPI_ISL_2638199, EPI_ISL_2638201, EPI_ISL_2638202, EPI_ISL_2638209, EPI_ISL_2638214, EPI_ISL_2638215, EPI_ISL_2638219, EPI_ISL_2638225, EPI_ISL_2638226, EPI_ISL_2638229,
EPI_ISL_2638230, EPI_ISL_2638237, EPI_ISL_2638239, EPI_ISL_2638243, EPI_ISL_2638244, EPI_ISL_2638252, EPI_ISL_2638254, EPI_ISL_2638255, EPI_ISL_2638256, EPI_ISL_2638257, EPI_ISL_2638261, EPI_ISL_2638262, EPI_ISL_2638264, EPI_ISL_2638266, EPI_ISL_2638268, EPI_ISL_2638270, EPI_ISL_2638274, EPI_ISL_2638276, EPI_ISL_2638277, EPI_ISL_2638280, EPI_ISL_2638282, EPI_ISL_2638285,
EPI_ISL_2638291, EPI_ISL_2638292, EPI_ISL_2638296, EPI_ISL_2638298, EPI_ISL_2638299, EPI_ISL_2638302, EPI_ISL_2638303, EPI_ISL_2638306, EPI_ISL_2638308, EPI_ISL_2638310, EPI_ISL_2638316, EPI_ISL_2638317, EPI_ISL_2638318, EPI_ISL_2638319, EPI_ISL_2638322, EPI_ISL_2638327, EPI_ISL_2638328, EPI_ISL_2638333, EPI_ISL_2638335, EPI_ISL_2638336, EPI_ISL_2638338, EPI_ISL_2638339,
EPI_ISL_2638341, EPI_ISL_2638342, EPI_ISL_2638347, EPI_ISL_2638348, EPI_ISL_2638351, EPI_ISL_2638352, EPI_ISL_2638355, EPI_ISL_2638357, EPI_ISL_2638358, EPI_ISL_2638359, EPI_ISL_2638360, EPI_ISL_2638362, EPI_ISL_2638365, EPI_ISL_2638366, EPI_ISL_2638367, EPI_ISL_2638371, EPI_ISL_2638372, EPI_ISL_2638379, EPI_ISL_2638383, EPI_ISL_2638386, EPI_ISL_2638391, EPI_ISL_2638392,
EPI_ISL_2638393, EPI_ISL_2638394, EPI_ISL_2638400, EPI_ISL_2638405, EPI_ISL_2638408, EPI_ISL_2638411, EPI_ISL_2638418, EPI_ISL_2638422, EPI_ISL_2638424, EPI_ISL_2638429, EPI_ISL_2638436, EPI_ISL_2638444, EPI_ISL_2638446, EPI_ISL_2638447, EPI_ISL_2638450, EPI_ISL_2638452, EPI_ISL_2638457, EPI_ISL_2638462, EPI_ISL_2638473, EPI_ISL_2638477, EPI_ISL_2638490, EPI_ISL_2638508,
EPI_ISL_2638509, EPI_ISL_2638513, EPI_ISL_2638514, EPI_ISL_2638515, EPI_ISL_2638521, EPI_ISL_2638523, EPI_ISL_2638528, EPI_ISL_2638536, EPI_ISL_2638537, EPI_ISL_2638540, EPI_ISL_2638560, EPI_ISL_2638561, EPI_ISL_2638566, EPI_ISL_2638567, EPI_ISL_2638573, EPI_ISL_2638576, EPI_ISL_2638578, EPI_ISL_2638580, EPI_ISL_2638582, EPI_ISL_2638585, EPI_ISL_2638593, EPI_ISL_2638603,
EPI_ISL_2638607, EPI_ISL_2638611, EPI_ISL_2638657, EPI_ISL_2638662, EPI_ISL_2638670, EPI_ISL_2638798, EPI_ISL_2638837, EPI_ISL_2638885, EPI_ISL_2638893, EPI_ISL_2638924, EPI_ISL_2638930, EPI_ISL_2638943, EPI_ISL_2638945, EPI_ISL_2638950, EPI_ISL_2638953, EPI_ISL_2638959, EPI_ISL_2638981, EPI_ISL_2639010, EPI_ISL_2639011, EPI_ISL_2639014, EPI_ISL_2639016, EPI_ISL_2639020,
EPI_ISL_2639034, EPI_ISL_2639037, EPI_ISL_2639054, EPI_ISL_2639057, EPI_ISL_2639069, EPI_ISL_2639088, EPI_ISL_2639099, EPI_ISL_2639118, EPI_ISL_2639140, EPI_ISL_2639143, EPI_ISL_2639162, EPI_ISL_2639176, EPI_ISL_2639215, EPI_ISL_2639225, EPI_ISL_2639242, EPI_ISL_2639247, EPI_ISL_2639254, EPI_ISL_2651409, EPI_ISL_2651416, EPI_ISL_2651419, EPI_ISL_2651421, EPI_ISL_2651423,
EPI_ISL_2651426, EPI_ISL_2651428, EPI_ISL_2651429, EPI_ISL_2651436, EPI_ISL_2651440, EPI_ISL_2651441, EPI_ISL_2651442, EPI_ISL_2651446, EPI_ISL_2651447, EPI_ISL_2651450, EPI_ISL_2651452, EPI_ISL_2651454, EPI_ISL_2651462, EPI_ISL_2651463, EPI_ISL_2651467, EPI_ISL_2651471, EPI_ISL_2651474, EPI_ISL_2651483, EPI_ISL_2651488, EPI_ISL_2651490, EPI_ISL_2651493, EPI_ISL_2651494,
EPI_ISL_2651498, EPI_ISL_2651501, EPI_ISL_2651504, EPI_ISL_2651506, EPI_ISL_2651507, EPI_ISL_2651511, EPI_ISL_2651520, EPI_ISL_2651523, EPI_ISL_2651525, EPI_ISL_2651530, EPI_ISL_2651536, EPI_ISL_2651537, EPI_ISL_2651541, EPI_ISL_2651544, EPI_ISL_2651545, EPI_ISL_2651550, EPI_ISL_2651553, EPI_ISL_2651554, EPI_ISL_2651559, EPI_ISL_2651567, EPI_ISL_2651569, EPI_ISL_2651570,
EPI_ISL_2651573, EPI_ISL_2651575, EPI_ISL_2651580, EPI_ISL_2651589, EPI_ISL_2651590, EPI_ISL_2651591, EPI_ISL_2651593, EPI_ISL_2651596, EPI_ISL_2651600, EPI_ISL_2651608, EPI_ISL_2651614, EPI_ISL_2651616, EPI_ISL_2651619, EPI_ISL_2651620, EPI_ISL_2651622, EPI_ISL_2651626, EPI_ISL_2651628, EPI_ISL_2651631, EPI_ISL_2651636, EPI_ISL_2651660, EPI_ISL_2651705, EPI_ISL_2664016,
EPI_ISL_2664019, EPI_ISL_2664020, EPI_ISL_2664023, EPI_ISL_2664026, EPI_ISL_2664028, EPI_ISL_2664029, EPI_ISL_2664030, EPI_ISL_2664033, EPI_ISL_2664034, EPI_ISL_2664044, EPI_ISL_2664045, EPI_ISL_2664048, EPI_ISL_2664050, EPI_ISL_2664051, EPI_ISL_2664052, EPI_ISL_2664053, EPI_ISL_2664055, EPI_ISL_2664057, EPI_ISL_2664058, EPI_ISL_2664059, EPI_ISL_2664061, EPI_ISL_2664064,
EPI_ISL_2664065, EPI_ISL_2664066, EPI_ISL_2664072, EPI_ISL_2664073, EPI_ISL_2664076, EPI_ISL_2664077, EPI_ISL_2664079, EPI_ISL_2664080, EPI_ISL_2664081, EPI_ISL_2664083, EPI_ISL_2664084, EPI_ISL_2664086, EPI_ISL_2664091, EPI_ISL_2664101, EPI_ISL_2664102, EPI_ISL_2664103, EPI_ISL_2664112, EPI_ISL_2664113, EPI_ISL_2664115, EPI_ISL_2664117, EPI_ISL_2664118, EPI_ISL_2664120,
EPI_ISL_2664130, EPI_ISL_2664133, EPI_ISL_2664136, EPI_ISL_2664138, EPI_ISL_2664139, EPI_ISL_2664141, EPI_ISL_2664142, EPI_ISL_2664144, EPI_ISL_2664145, EPI_ISL_2664147, EPI_ISL_2664148, EPI_ISL_2664149, EPI_ISL_2664151, EPI_ISL_2664152, EPI_ISL_2664155, EPI_ISL_2664161, EPI_ISL_2664165, EPI_ISL_2664169, EPI_ISL_2664173, EPI_ISL_2664174, EPI_ISL_2664177, EPI_ISL_2664179,
EPI_ISL_2664181, EPI_ISL_2664183, EPI_ISL_2664185, EPI_ISL_2664192, EPI_ISL_2664196, EPI_ISL_2664197, EPI_ISL_2664204, EPI_ISL_2664215, EPI_ISL_2664218, EPI_ISL_2664220, EPI_ISL_2664221, EPI_ISL_2664222, EPI_ISL_2664227, EPI_ISL_2664228, EPI_ISL_2664230, EPI_ISL_2664231, EPI_ISL_2664233, EPI_ISL_2664235, EPI_ISL_2664238, EPI_ISL_2664239, EPI_ISL_2664241, EPI_ISL_2664243,
EPI_ISL_2664244, EPI_ISL_2664245, EPI_ISL_2664247, EPI_ISL_2664249, EPI_ISL_2664252, EPI_ISL_2664255, EPI_ISL_2664256, EPI_ISL_2664257, EPI_ISL_2664261, EPI_ISL_2664262, EPI_ISL_2664268, EPI_ISL_2664271, EPI_ISL_2664278, EPI_ISL_2664279, EPI_ISL_2664284, EPI_ISL_2664287, EPI_ISL_2664288, EPI_ISL_2664290, EPI_ISL_2664298, EPI_ISL_2664303, EPI_ISL_2664305, EPI_ISL_2664306,
EPI_ISL_2664307, EPI_ISL_2664309, EPI_ISL_2664312, EPI_ISL_2664315, EPI_ISL_2664321, EPI_ISL_2664322, EPI_ISL_2664325, EPI_ISL_2664326, EPI_ISL_2664327, EPI_ISL_2664328, EPI_ISL_2664332, EPI_ISL_2664333, EPI_ISL_2664334, EPI_ISL_2664340, EPI_ISL_2664342, EPI_ISL_2664344, EPI_ISL_2664345, EPI_ISL_2664347, EPI_ISL_2664348, EPI_ISL_2664351, EPI_ISL_2664352, EPI_ISL_2664354,
EPI_ISL_2664357, EPI_ISL_2664359, EPI_ISL_2664367, EPI_ISL_2664371, EPI_ISL_2664378, EPI_ISL_2664379, EPI_ISL_2664385, EPI_ISL_2664388, EPI_ISL_2664391, EPI_ISL_2664393, EPI_ISL_2664394, EPI_ISL_2664395, EPI_ISL_2664398, EPI_ISL_2664405, EPI_ISL_2664409, EPI_ISL_2664412, EPI_ISL_2664414, EPI_ISL_2664415, EPI_ISL_2664417, EPI_ISL_2664421, EPI_ISL_2664425, EPI_ISL_2664426,
EPI_ISL_2664430, EPI_ISL_2664431, EPI_ISL_2664432, EPI_ISL_2664433, EPI_ISL_2664438, EPI_ISL_2664443, EPI_ISL_2664444, EPI_ISL_2664452, EPI_ISL_2664459, EPI_ISL_2664460, EPI_ISL_2664461, EPI_ISL_2664463, EPI_ISL_2664469, EPI_ISL_2664475, EPI_ISL_2664476, EPI_ISL_2664480, EPI_ISL_2664481, EPI_ISL_2664490, EPI_ISL_2664494, EPI_ISL_2664497, EPI_ISL_2664498, EPI_ISL_2664509,
EPI_ISL_2664510, EPI_ISL_2664512, EPI_ISL_2664513, EPI_ISL_2664515, EPI_ISL_2664518, EPI_ISL_2664521, EPI_ISL_2664522, EPI_ISL_2664524, EPI_ISL_2664530, EPI_ISL_2664531, EPI_ISL_2664532, EPI_ISL_2664535, EPI_ISL_2664538, EPI_ISL_2664541, EPI_ISL_2664543, EPI_ISL_2664559, EPI_ISL_2664560, EPI_ISL_2664576, EPI_ISL_2664577, EPI_ISL_2664583, EPI_ISL_2664586, EPI_ISL_2664591,
EPI_ISL_2664593, EPI_ISL_2664594, EPI_ISL_2664600, EPI_ISL_2664608, EPI_ISL_2664609, EPI_ISL_2664611, EPI_ISL_2664616, EPI_ISL_2664618, EPI_ISL_2664619, EPI_ISL_2664621, EPI_ISL_2664623, EPI_ISL_2664624, EPI_ISL_2664626, EPI_ISL_2664627, EPI_ISL_2664628, EPI_ISL_2664630, EPI_ISL_2664633, EPI_ISL_2664637, EPI_ISL_2664638, EPI_ISL_2664641, EPI_ISL_2664645, EPI_ISL_2664646,
EPI_ISL_2664655, EPI_ISL_2664661, EPI_ISL_2664663, EPI_ISL_2664666, EPI_ISL_2664667, EPI_ISL_2664671, EPI_ISL_2664673, EPI_ISL_2664674, EPI_ISL_2664675, EPI_ISL_2664679, EPI_ISL_2664687, EPI_ISL_2664690, EPI_ISL_2664692, EPI_ISL_2664696, EPI_ISL_2664697, EPI_ISL_2664698, EPI_ISL_2664699, EPI_ISL_2664703, EPI_ISL_2664713, EPI_ISL_2664714, EPI_ISL_2664720, EPI_ISL_2664725,
EPI_ISL_2664726, EPI_ISL_2664728, EPI_ISL_2664734, EPI_ISL_2664735, EPI_ISL_2664736, EPI_ISL_2664745, EPI_ISL_2664746, EPI_ISL_2664747, EPI_ISL_2664748, EPI_ISL_2664750, EPI_ISL_2664755, EPI_ISL_2664757, EPI_ISL_2664758, EPI_ISL_2664762, EPI_ISL_2664764, EPI_ISL_2664780, EPI_ISL_2664781, EPI_ISL_2664784, EPI_ISL_2664788, EPI_ISL_2664794, EPI_ISL_2664795, EPI_ISL_2664799,
EPI_ISL_2664801, EPI_ISL_2664803, EPI_ISL_2664805, EPI_ISL_2664809, EPI_ISL_2664810, EPI_ISL_2664811, EPI_ISL_2664816, EPI_ISL_2664820, EPI_ISL_2664822, EPI_ISL_2664826, EPI_ISL_2664827, EPI_ISL_2664837, EPI_ISL_2664839, EPI_ISL_2664843, EPI_ISL_2664844, EPI_ISL_2664851, EPI_ISL_2664853, EPI_ISL_2664856, EPI_ISL_2664858, EPI_ISL_2664865, EPI_ISL_2664867, EPI_ISL_2664873,
EPI_ISL_2664878, EPI_ISL_2664884, EPI_ISL_2664885, EPI_ISL_2664887, EPI_ISL_2664888, EPI_ISL_2664890, EPI_ISL_2664892, EPI_ISL_2664894, EPI_ISL_2664895, EPI_ISL_2664899, EPI_ISL_2664900, EPI_ISL_2664901, EPI_ISL_2664915, EPI_ISL_2664917, EPI_ISL_2664930, EPI_ISL_2664931, EPI_ISL_2664933, EPI_ISL_2664940, EPI_ISL_2664944, EPI_ISL_2664946, EPI_ISL_2664950, EPI_ISL_2664957,
EPI_ISL_2664975, EPI_ISL_2664980, EPI_ISL_2664987, EPI_ISL_2664988, EPI_ISL_2664989, EPI_ISL_2664995, EPI_ISL_2664997, EPI_ISL_2664999, EPI_ISL_2665000, EPI_ISL_2665003, EPI_ISL_2665004, EPI_ISL_2665011, EPI_ISL_2665013, EPI_ISL_2665015, EPI_ISL_2665017, EPI_ISL_2665019, EPI_ISL_2665020, EPI_ISL_2665021, EPI_ISL_2665023, EPI_ISL_2665025, EPI_ISL_2665028, EPI_ISL_2665029,
EPI_ISL_2665031, EPI_ISL_2665032, EPI_ISL_2665034, EPI_ISL_2665035, EPI_ISL_2665036, EPI_ISL_2665037, EPI_ISL_2665038, EPI_ISL_2665039, EPI_ISL_2665040, EPI_ISL_2665045, EPI_ISL_2665046, EPI_ISL_2665047, EPI_ISL_2665048, EPI_ISL_2665049, EPI_ISL_2665050, EPI_ISL_2665051, EPI_ISL_2665052, EPI_ISL_2665055, EPI_ISL_2665057, EPI_ISL_2665059, EPI_ISL_2665060, EPI_ISL_2665061,
EPI_ISL_2665062, EPI_ISL_2665063, EPI_ISL_2665064, EPI_ISL_2665065, EPI_ISL_2665066, EPI_ISL_2665067, EPI_ISL_2665068, EPI_ISL_2665069, EPI_ISL_2665071, EPI_ISL_2665072, EPI_ISL_2665073, EPI_ISL_2665074, EPI_ISL_2665076, EPI_ISL_2665078, EPI_ISL_2665079, EPI_ISL_2665080, EPI_ISL_2665081, EPI_ISL_2665082, EPI_ISL_2665083, EPI_ISL_2665084, EPI_ISL_2665086, EPI_ISL_2665087,
EPI_ISL_2665088, EPI_ISL_2665089, EPI_ISL_2665090, EPI_ISL_2665091, EPI_ISL_2665092, EPI_ISL_2665093, EPI_ISL_2665094, EPI_ISL_2665098, EPI_ISL_2665099, EPI_ISL_2665100, EPI_ISL_2665101, EPI_ISL_2665102, EPI_ISL_2665103, EPI_ISL_2665104, EPI_ISL_2665105, EPI_ISL_2665107, EPI_ISL_2665108, EPI_ISL_2665109, EPI_ISL_2665110, EPI_ISL_2665111, EPI_ISL_2665112, EPI_ISL_2665114,
EPI_ISL_2665115, EPI_ISL_2665117, EPI_ISL_2665118, EPI_ISL_2665122, EPI_ISL_2665124, EPI_ISL_2665125, EPI_ISL_2665126, EPI_ISL_2665129, EPI_ISL_2665130, EPI_ISL_2665132, EPI_ISL_2665134, EPI_ISL_2665135, EPI_ISL_2665136, EPI_ISL_2665137, EPI_ISL_2665138, EPI_ISL_2665142, EPI_ISL_2665143, EPI_ISL_2665144, EPI_ISL_2665145, EPI_ISL_2665146, EPI_ISL_2665148, EPI_ISL_2665149,
EPI_ISL_2665151, EPI_ISL_2665152, EPI_ISL_2665153, EPI_ISL_2665154, EPI_ISL_2665155, EPI_ISL_2665156, EPI_ISL_2665159, EPI_ISL_2665161, EPI_ISL_2665162, EPI_ISL_2665163, EPI_ISL_2665165, EPI_ISL_2665166, EPI_ISL_2665169, EPI_ISL_2665170, EPI_ISL_2665171, EPI_ISL_2665173, EPI_ISL_2665174, EPI_ISL_2665175, EPI_ISL_2665176, EPI_ISL_2665177, EPI_ISL_2665178, EPI_ISL_2665181,
EPI_ISL_2665182, EPI_ISL_2665183, EPI_ISL_2665185, EPI_ISL_2665188, EPI_ISL_2665193, EPI_ISL_2665194, EPI_ISL_2665195, EPI_ISL_2665196, EPI_ISL_2665197, EPI_ISL_2665199, EPI_ISL_2665201, EPI_ISL_2665202, EPI_ISL_2665203, EPI_ISL_2665204, EPI_ISL_2665205, EPI_ISL_2665206, EPI_ISL_2665207, EPI_ISL_2665209, EPI_ISL_2665210, EPI_ISL_2665211, EPI_ISL_2665212, EPI_ISL_2665213,
EPI_ISL_2665214, EPI_ISL_2665216, EPI_ISL_2665217, EPI_ISL_2665219, EPI_ISL_2665220, EPI_ISL_2665221, EPI_ISL_2665222, EPI_ISL_2665223, EPI_ISL_2665224, EPI_ISL_2665226, EPI_ISL_2665227, EPI_ISL_2665228, EPI_ISL_2665229, EPI_ISL_2665231, EPI_ISL_2665232, EPI_ISL_2665233, EPI_ISL_2665234, EPI_ISL_2665237, EPI_ISL_2665238, EPI_ISL_2665239, EPI_ISL_2665240, EPI_ISL_2665241,
EPI_ISL_2665242, EPI_ISL_2665243, EPI_ISL_2665244, EPI_ISL_2665245, EPI_ISL_2665249, EPI_ISL_2665250, EPI_ISL_2665252, EPI_ISL_2665253, EPI_ISL_2665254, EPI_ISL_2665256, EPI_ISL_2665257, EPI_ISL_2665258, EPI_ISL_2665259, EPI_ISL_2665261, EPI_ISL_2665263, EPI_ISL_2665264, EPI_ISL_2665265, EPI_ISL_2665267, EPI_ISL_2665268, EPI_ISL_2665269, EPI_ISL_2665270, EPI_ISL_2665271,
EPI_ISL_2665272, EPI_ISL_2665274, EPI_ISL_2665276, EPI_ISL_2665279, EPI_ISL_2665280, EPI_ISL_2665283, EPI_ISL_2665284, EPI_ISL_2665286, EPI_ISL_2665287, EPI_ISL_2665288, EPI_ISL_2665289, EPI_ISL_2665290, EPI_ISL_2665291, EPI_ISL_2665292, EPI_ISL_2665294, EPI_ISL_2665295, EPI_ISL_2665298, EPI_ISL_2665299, EPI_ISL_2665300, EPI_ISL_2665301, EPI_ISL_2665303, EPI_ISL_2665304,
EPI_ISL_2665305, EPI_ISL_2665306, EPI_ISL_2665307, EPI_ISL_2665308, EPI_ISL_2665309, EPI_ISL_2665310, EPI_ISL_2665312, EPI_ISL_2665313, EPI_ISL_2665314, EPI_ISL_2665315, EPI_ISL_2665316, EPI_ISL_2665317, EPI_ISL_2665318, EPI_ISL_2665319, EPI_ISL_2665320, EPI_ISL_2665322, EPI_ISL_2665323, EPI_ISL_2665324, EPI_ISL_2665325, EPI_ISL_2665326, EPI_ISL_2665327, EPI_ISL_2665328,
EPI_ISL_2665329, EPI_ISL_2665331, EPI_ISL_2665333, EPI_ISL_2665334, EPI_ISL_2665335, EPI_ISL_2665336, EPI_ISL_2665337, EPI_ISL_2665338, EPI_ISL_2665339, EPI_ISL_2665340, EPI_ISL_2665341, EPI_ISL_2665342, EPI_ISL_2665343, EPI_ISL_2665344, EPI_ISL_2665345, EPI_ISL_2665346, EPI_ISL_2665348, EPI_ISL_2665349, EPI_ISL_2665350, EPI_ISL_2665351, EPI_ISL_2665352, EPI_ISL_2665354,
EPI_ISL_2665355, EPI_ISL_2665356, EPI_ISL_2665357, EPI_ISL_2665358, EPI_ISL_2665361, EPI_ISL_2665362, EPI_ISL_2665364, EPI_ISL_2665365, EPI_ISL_2665366, EPI_ISL_2665367, EPI_ISL_2665368, EPI_ISL_2665369, EPI_ISL_2665371, EPI_ISL_2665372, EPI_ISL_2665373, EPI_ISL_2665375, EPI_ISL_2665376, EPI_ISL_2665378, EPI_ISL_2665381, EPI_ISL_2665382, EPI_ISL_2665383, EPI_ISL_2665384,
EPI_ISL_2681641, EPI_ISL_2681646, EPI_ISL_2681648, EPI_ISL_2681649, EPI_ISL_2681650, EPI_ISL_2681661, EPI_ISL_2681664, EPI_ISL_2681667, EPI_ISL_2681670, EPI_ISL_2681672, EPI_ISL_2681675, EPI_ISL_2681679, EPI_ISL_2681681, EPI_ISL_2681682, EPI_ISL_2681685, EPI_ISL_2681688, EPI_ISL_2681689, EPI_ISL_2681690, EPI_ISL_2681691, EPI_ISL_2681696, EPI_ISL_2681701, EPI_ISL_2681705,
EPI_ISL_2681706, EPI_ISL_2681713, EPI_ISL_2681715, EPI_ISL_2681716, EPI_ISL_2681719, EPI_ISL_2681730, EPI_ISL_2681731, EPI_ISL_2681733, EPI_ISL_2681735, EPI_ISL_2681736, EPI_ISL_2681738, EPI_ISL_2681741, EPI_ISL_2681743, EPI_ISL_2681746, EPI_ISL_2681748, EPI_ISL_2681753, EPI_ISL_2681756, EPI_ISL_2681761, EPI_ISL_2681762, EPI_ISL_2681764, EPI_ISL_2681769, EPI_ISL_2681770,
EPI_ISL_2681771, EPI_ISL_2681773, EPI_ISL_2681778, EPI_ISL_2681788, EPI_ISL_2681792, EPI_ISL_2681795, EPI_ISL_2681797, EPI_ISL_2681798, EPI_ISL_2681806, EPI_ISL_2681810, EPI_ISL_2681819, EPI_ISL_2681821, EPI_ISL_2681828, EPI_ISL_2681830, EPI_ISL_2681836, EPI_ISL_2681838, EPI_ISL_2681840, EPI_ISL_2681842, EPI_ISL_2681843, EPI_ISL_2681848, EPI_ISL_2681849, EPI_ISL_2681854,
EPI_ISL_2681863, EPI_ISL_2681868, EPI_ISL_2681872, EPI_ISL_2681874, EPI_ISL_2681875, EPI_ISL_2681879, EPI_ISL_2681883, EPI_ISL_2681884, EPI_ISL_2681886, EPI_ISL_2681888, EPI_ISL_2681891, EPI_ISL_2681893, EPI_ISL_2681896, EPI_ISL_2681897, EPI_ISL_2681900, EPI_ISL_2681912, EPI_ISL_2681913, EPI_ISL_2681916, EPI_ISL_2681918, EPI_ISL_2681920, EPI_ISL_2681921, EPI_ISL_2682060,
EPI_ISL_2682061, EPI_ISL_2682066, EPI_ISL_2682067, EPI_ISL_2682070, EPI_ISL_2682072, EPI_ISL_2682081, EPI_ISL_2682084, EPI_ISL_2682086, EPI_ISL_2682091, EPI_ISL_2682094, EPI_ISL_2682099, EPI_ISL_2682108, EPI_ISL_2682109, EPI_ISL_2682110, EPI_ISL_2682112, EPI_ISL_2682116, EPI_ISL_2682118, EPI_ISL_2682119, EPI_ISL_2682122, EPI_ISL_2682125, EPI_ISL_2682126, EPI_ISL_2682127,
EPI_ISL_2682128, EPI_ISL_2682129, EPI_ISL_2682135, EPI_ISL_2682141, EPI_ISL_2682143, EPI_ISL_2682147, EPI_ISL_2682148, EPI_ISL_2682150, EPI_ISL_2682152, EPI_ISL_2682154, EPI_ISL_2682155, EPI_ISL_2682156, EPI_ISL_2682158, EPI_ISL_2682162, EPI_ISL_2682163, EPI_ISL_2682170, EPI_ISL_2682172, EPI_ISL_2682177, EPI_ISL_2682178, EPI_ISL_2682179, EPI_ISL_2682181, EPI_ISL_2682182,
EPI_ISL_2682184, EPI_ISL_2682187, EPI_ISL_2682193, EPI_ISL_2682196, EPI_ISL_2682197, EPI_ISL_2682201, EPI_ISL_2682203, EPI_ISL_2682205, EPI_ISL_2682206, EPI_ISL_2682209, EPI_ISL_2682214, EPI_ISL_2682217, EPI_ISL_2682219, EPI_ISL_2682220, EPI_ISL_2682223, EPI_ISL_2682224, EPI_ISL_2682234, EPI_ISL_2682235, EPI_ISL_2682236, EPI_ISL_2682239, EPI_ISL_2682243, EPI_ISL_2682246,
EPI_ISL_2682247, EPI_ISL_2682250, EPI_ISL_2682253, EPI_ISL_2682255, EPI_ISL_2682257, EPI_ISL_2682259, EPI_ISL_2682263, EPI_ISL_2682264, EPI_ISL_2682267, EPI_ISL_2682272, EPI_ISL_2682275, EPI_ISL_2682277, EPI_ISL_2682281, EPI_ISL_2683113, EPI_ISL_2699610, EPI_ISL_2699623, EPI_ISL_2699625, EPI_ISL_2699639, EPI_ISL_2699657, EPI_ISL_2699691, EPI_ISL_2699694, EPI_ISL_2699696,
EPI_ISL_2699711, EPI_ISL_2699755, EPI_ISL_2699775, EPI_ISL_2699796, EPI_ISL_2699827, EPI_ISL_2699855, EPI_ISL_2699879, EPI_ISL_2699882, EPI_ISL_2699888, EPI_ISL_2699894, EPI_ISL_2699898, EPI_ISL_2699917, EPI_ISL_2699918, EPI_ISL_2699926, EPI_ISL_2699935, EPI_ISL_2699937, EPI_ISL_2699949, EPI_ISL_2699950, EPI_ISL_2699960, EPI_ISL_2699963, EPI_ISL_2699971, EPI_ISL_2699986,
EPI_ISL_2699987, EPI_ISL_2699990, EPI_ISL_2699998, EPI_ISL_2700001, EPI_ISL_2700004, EPI_ISL_2700006, EPI_ISL_2700014, EPI_ISL_2700028, EPI_ISL_2700031, EPI_ISL_2700032, EPI_ISL_2700040, EPI_ISL_2700049, EPI_ISL_2700056, EPI_ISL_2700061, EPI_ISL_2700068, EPI_ISL_2700070, EPI_ISL_2700084, EPI_ISL_2700085, EPI_ISL_2700086, EPI_ISL_2700087, EPI_ISL_2700088, EPI_ISL_2700091,
EPI_ISL_2700095, EPI_ISL_2700096, EPI_ISL_2700108, EPI_ISL_2700115, EPI_ISL_2700125, EPI_ISL_2700130, EPI_ISL_2700131, EPI_ISL_2700174, EPI_ISL_2700175, EPI_ISL_2700176, EPI_ISL_2700192, EPI_ISL_2700197, EPI_ISL_2700206, EPI_ISL_2700232, EPI_ISL_2700233, EPI_ISL_2700246, EPI_ISL_2700279, EPI_ISL_2700318, EPI_ISL_2700321, EPI_ISL_2700328, EPI_ISL_2700329, EPI_ISL_2700338,
EPI_ISL_2700351, EPI_ISL_2700365, EPI_ISL_2700376, EPI_ISL_2700378, EPI_ISL_2700385, EPI_ISL_2700415, EPI_ISL_2700421, EPI_ISL_2700443, EPI_ISL_2700535, EPI_ISL_2700538, EPI_ISL_2700565, EPI_ISL_2702101, EPI_ISL_2702114, EPI_ISL_2702172, EPI_ISL_2702189, EPI_ISL_2702217, EPI_ISL_2702299, EPI_ISL_2702311, EPI_ISL_2702314, EPI_ISL_2702329, EPI_ISL_2702338, EPI_ISL_2702386,
EPI_ISL_2702813, EPI_ISL_2702814, EPI_ISL_2702821, EPI_ISL_2702827, EPI_ISL_2702831, EPI_ISL_2702836, EPI_ISL_2702838, EPI_ISL_2702846, EPI_ISL_2702849, EPI_ISL_2702851, EPI_ISL_2702856, EPI_ISL_2702861, EPI_ISL_2702862, EPI_ISL_2702870, EPI_ISL_2702875, EPI_ISL_2702886, EPI_ISL_2702887, EPI_ISL_2702889, EPI_ISL_2702894, EPI_ISL_2702895, EPI_ISL_2702896, EPI_ISL_2702904,

EPI_ISL_2702912, EPI_ISL_2702922, EPI_ISL_2702932, EPI_ISL_2702933, EPI_ISL_2702935, EPI_ISL_2702938, EPI_ISL_2702941, EPI_ISL_2702944, EPI_ISL_2702949, EPI_ISL_2702951, EPI_ISL_2702955, EPI_ISL_2702956, EPI_ISL_2702957, EPI_ISL_2702966, EPI_ISL_2702974, EPI_ISL_2702978, EPI_ISL_2702979, EPI_ISL_2702980, EPI_ISL_2702985, EPI_ISL_2702995, EPI_ISL_2702996, EPI_ISL_2702999, EPI_ISL_2703002, EPI_ISL_2703012, EPI_ISL_2703015, EPI_ISL_2703026, EPI_ISL_2703027, EPI_ISL_2703034, EPI_ISL_2703037, EPI_ISL_2703038, EPI_ISL_2703040, EPI_ISL_2703042, EPI_ISL_2703043, EPI_ISL_2703049, EPI_ISL_2703052, EPI_ISL_2703053, EPI_ISL_2703055, EPI_ISL_2703057, EPI_ISL_2703058, EPI_ISL_2703060, EPI_ISL_2703062, EPI_ISL_2703065, EPI_ISL_2703069, EPI_ISL_2703072, EPI_ISL_2703073, EPI_ISL_2703074, EPI_ISL_2703075, EPI_ISL_2703077, EPI_ISL_2703082, EPI_ISL_2703091, EPI_ISL_2703098, EPI_ISL_2703099, EPI_ISL_2703105, EPI_ISL_2703111, EPI_ISL_2703112, EPI_ISL_2703117, EPI_ISL_2703118, EPI_ISL_2703124, EPI_ISL_2703127, EPI_ISL_2703131, EPI_ISL_2703147, EPI_ISL_2703148, EPI_ISL_2703149, EPI_ISL_2703150, EPI_ISL_2703151, EPI_ISL_2703159, EPI_ISL_2703167, EPI_ISL_2703169, EPI_ISL_2703180, EPI_ISL_2703184, EPI_ISL_2703185, EPI_ISL_2703195, EPI_ISL_2703221, EPI_ISL_2703248, EPI_ISL_2703261, EPI_ISL_2703267, EPI_ISL_2703285, EPI_ISL_2703286, EPI_ISL_2703306, EPI_ISL_2703308, EPI_ISL_2703317, EPI_ISL_2703322, EPI_ISL_2703330, EPI_ISL_2703341, EPI_ISL_2703348, EPI_ISL_2703366, EPI_ISL_2703373, EPI_ISL_2703382, EPI_ISL_2703398, EPI_ISL_2703420, EPI_ISL_2703445, EPI_ISL_2703454, EPI_ISL_2703469, EPI_ISL_2703473, EPI_ISL_2703475, EPI_ISL_2703484, EPI_ISL_2703489, EPI_ISL_2703502, EPI_ISL_2703541, EPI_ISL_2703571, EPI_ISL_2703634, EPI_ISL_2703641, EPI_ISL_2703649, EPI_ISL_2703658, EPI_ISL_2703716, EPI_ISL_2703717, EPI_ISL_2703799, EPI_ISL_2703818, EPI_ISL_2703831, EPI_ISL_2703834, EPI_ISL_2704535, EPI_ISL_2704666, EPI_ISL_2704675, EPI_ISL_2717552, EPI_ISL_2717835, EPI_ISL_2717858, EPI_ISL_2717871, EPI_ISL_2718074, EPI_ISL_2718135, EPI_ISL_2718143, EPI_ISL_2718149, EPI_ISL_2729412, EPI_ISL_2729581, EPI_ISL_2780934

| | | | |
|---|---|---|---|
| see above | Lighthouse Lab in Milton Keynes | Wellcome Sanger Institute for the COVID-19 Genomics UK (COG-UK) Consortium | Cordelia Langford; David K. Jackson; Dominic Kwiatkowski; Ewan Harrison; Ian Johnston; Jeffrey Barrett; John Sillitoe on behalf of the Wellcome Sanger Institute COVID-19 Surveillance Team; Roberto Amato; Sonia Goncalves; The Lighthouse Lab in Milton Keynes and Alex Alderton |

EPI_ISL_2638395, EPI_ISL_2638396, EPI_ISL_2638398, EPI_ISL_2638401, EPI_ISL_2638402, EPI_ISL_2638407, EPI_ISL_2638412, EPI_ISL_2638413, EPI_ISL_2638415, EPI_ISL_2638417, EPI_ISL_2638421, EPI_ISL_2638425, EPI_ISL_2638426, EPI_ISL_2638431, EPI_ISL_2638433, EPI_ISL_2638438, EPI_ISL_2638439, EPI_ISL_2638442, EPI_ISL_2638445, EPI_ISL_2638448, EPI_ISL_2638451, EPI_ISL_2638455, EPI_ISL_2638459, EPI_ISL_2638461, EPI_ISL_2638463, EPI_ISL_2638464, EPI_ISL_2638466, EPI_ISL_2638467, EPI_ISL_2638468, EPI_ISL_2638469, EPI_ISL_2638470, EPI_ISL_2638471, EPI_ISL_2638472, EPI_ISL_2638474, EPI_ISL_2638475, EPI_ISL_2638481, EPI_ISL_2638483, EPI_ISL_2638484, EPI_ISL_2638488, EPI_ISL_2638489, EPI_ISL_2638491, EPI_ISL_2638492, EPI_ISL_2638493, EPI_ISL_2638495, EPI_ISL_2638496, EPI_ISL_2638497, EPI_ISL_2638499, EPI_ISL_2638500, EPI_ISL_2638502, EPI_ISL_2638507, EPI_ISL_2638511, EPI_ISL_2638518, EPI_ISL_2638519, EPI_ISL_2638522, EPI_ISL_2638524, EPI_ISL_2638525, EPI_ISL_2638526, EPI_ISL_2638529, EPI_ISL_2638531, EPI_ISL_2638532, EPI_ISL_2638533, EPI_ISL_2638541, EPI_ISL_2638543, EPI_ISL_2638546, EPI_ISL_2638547, EPI_ISL_2638549, EPI_ISL_2638550, EPI_ISL_2638551, EPI_ISL_2638552, EPI_ISL_2638553, EPI_ISL_2638555, EPI_ISL_2638556, EPI_ISL_2638558, EPI_ISL_2638559, EPI_ISL_2638569, EPI_ISL_2638575, EPI_ISL_2638583, EPI_ISL_2638584, EPI_ISL_2638587, EPI_ISL_2638594, EPI_ISL_2638596, EPI_ISL_2638598, EPI_ISL_2638599, EPI_ISL_2638602, EPI_ISL_2638916, EPI_ISL_2638917, EPI_ISL_2638918, EPI_ISL_2638919, EPI_ISL_2638920, EPI_ISL_2638922, EPI_ISL_2638926, EPI_ISL_2638927, EPI_ISL_2638928, EPI_ISL_2638929, EPI_ISL_2638931, EPI_ISL_2638932, EPI_ISL_2638933, EPI_ISL_2638935, EPI_ISL_2638936, EPI_ISL_2638937, EPI_ISL_2638938, EPI_ISL_2638940, EPI_ISL_2638941, EPI_ISL_2638942, EPI_ISL_2638944, EPI_ISL_2638946, EPI_ISL_2638948, EPI_ISL_2638949, EPI_ISL_2638951, EPI_ISL_2638952, EPI_ISL_2638955, EPI_ISL_2638957, EPI_ISL_2638958, EPI_ISL_2638963, EPI_ISL_2638964, EPI_ISL_2638965, EPI_ISL_2638966, EPI_ISL_2638967, EPI_ISL_2638969, EPI_ISL_2638970, EPI_ISL_2638971, EPI_ISL_2638973, EPI_ISL_2638974, EPI_ISL_2638977, EPI_ISL_2638983, EPI_ISL_2638985, EPI_ISL_2638987, EPI_ISL_2638989, EPI_ISL_2638993, EPI_ISL_2638996, EPI_ISL_2638999, EPI_ISL_2639000, EPI_ISL_2639003, EPI_ISL_2639006, EPI_ISL_2639007, EPI_ISL_2639008, EPI_ISL_2639009, EPI_ISL_2639013, EPI_ISL_2639015, EPI_ISL_2639017, EPI_ISL_2639021, EPI_ISL_2639024, EPI_ISL_2639025, EPI_ISL_2639026, EPI_ISL_2639027, EPI_ISL_2639032, EPI_ISL_2639035, EPI_ISL_2639039, EPI_ISL_2639040, EPI_ISL_2639041, EPI_ISL_2639044, EPI_ISL_2639046, EPI_ISL_2639047, EPI_ISL_2639049, EPI_ISL_2639050, EPI_ISL_2639052, EPI_ISL_2639055, EPI_ISL_2639056, EPI_ISL_2639058, EPI_ISL_2639065, EPI_ISL_2639067, EPI_ISL_2639068, EPI_ISL_2639070, EPI_ISL_2639071, EPI_ISL_2639075, EPI_ISL_2639076, EPI_ISL_2639077, EPI_ISL_2639078, EPI_ISL_2639079, EPI_ISL_2639080, EPI_ISL_2639081, EPI_ISL_2639082, EPI_ISL_2639083, EPI_ISL_2639085, EPI_ISL_2639087, EPI_ISL_2639089, EPI_ISL_2639090, EPI_ISL_2639091, EPI_ISL_2639093, EPI_ISL_2639094, EPI_ISL_2639095, EPI_ISL_2639096, EPI_ISL_2639097, EPI_ISL_2639100, EPI_ISL_2639102, EPI_ISL_2639103, EPI_ISL_2639105, EPI_ISL_2639106, EPI_ISL_2639107, EPI_ISL_2639109, EPI_ISL_2639111, EPI_ISL_2639112, EPI_ISL_2639114, EPI_ISL_2639115, EPI_ISL_2639117, EPI_ISL_2639122, EPI_ISL_2639123, EPI_ISL_2639124, EPI_ISL_2639125, EPI_ISL_2639126, EPI_ISL_2639127, EPI_ISL_2639128, EPI_ISL_2639131, EPI_ISL_2639132, EPI_ISL_2639134, EPI_ISL_2639135, EPI_ISL_2639136, EPI_ISL_2639138, EPI_ISL_2639139, EPI_ISL_2639142, EPI_ISL_2639147, EPI_ISL_2639148, EPI_ISL_2639149, EPI_ISL_2639151, EPI_ISL_2639152, EPI_ISL_2639153, EPI_ISL_2639154, EPI_ISL_2639155, EPI_ISL_2639158, EPI_ISL_2639159, EPI_ISL_2639163, EPI_ISL_2639165, EPI_ISL_2639167, EPI_ISL_2639171, EPI_ISL_2639172, EPI_ISL_2639173, EPI_ISL_2639174, EPI_ISL_2639177, EPI_ISL_2639178, EPI_ISL_2639179, EPI_ISL_2639180, EPI_ISL_2639181, EPI_ISL_2639184, EPI_ISL_2639185, EPI_ISL_2639186, EPI_ISL_2639188, EPI_ISL_2639189, EPI_ISL_2639190, EPI_ISL_2639191, EPI_ISL_2639192, EPI_ISL_2639194, EPI_ISL_2639196, EPI_ISL_2639199, EPI_ISL_2639201, EPI_ISL_2639205, EPI_ISL_2639206, EPI_ISL_2639209, EPI_ISL_2639210, EPI_ISL_2639211, EPI_ISL_2639212, EPI_ISL_2639214, EPI_ISL_2639216, EPI_ISL_2639217, EPI_ISL_2639218, EPI_ISL_2639220, EPI_ISL_2639221, EPI_ISL_2639222, EPI_ISL_2639224, EPI_ISL_2639226, EPI_ISL_2639229, EPI_ISL_2639231, EPI_ISL_2639232, EPI_ISL_2639234, EPI_ISL_2639235, EPI_ISL_2639236, EPI_ISL_2639237, EPI_ISL_2639239, EPI_ISL_2639240, EPI_ISL_2639243, EPI_ISL_2639244, EPI_ISL_2639246, EPI_ISL_2639248, EPI_ISL_2639250, EPI_ISL_2639251, EPI_ISL_2639253, EPI_ISL_2639255, EPI_ISL_2639620, EPI_ISL_2639626, EPI_ISL_2639628, EPI_ISL_2639629, EPI_ISL_2639631, EPI_ISL_2639633, EPI_ISL_2639636, EPI_ISL_2639648, EPI_ISL_2639655, EPI_ISL_2639656, EPI_ISL_2639658, EPI_ISL_2639661, EPI_ISL_2639668, EPI_ISL_2639671, EPI_ISL_2639682, EPI_ISL_2639684, EPI_ISL_2639686, EPI_ISL_2639690, EPI_ISL_2639693, EPI_ISL_2639694, EPI_ISL_2639695, EPI_ISL_2639696, EPI_ISL_2639698, EPI_ISL_2639699, EPI_ISL_2639703, EPI_ISL_2639704, EPI_ISL_2639717, EPI_ISL_2639720, EPI_ISL_2639721, EPI_ISL_2639732, EPI_ISL_2639738, EPI_ISL_2639740, EPI_ISL_2639748, EPI_ISL_2639755, EPI_ISL_2639757, EPI_ISL_2639759, EPI_ISL_2639762, EPI_ISL_2639764, EPI_ISL_2639773, EPI_ISL_2639774, EPI_ISL_2639781, EPI_ISL_2639782, EPI_ISL_2639788, EPI_ISL_2639790, EPI_ISL_2639791, EPI_ISL_2639798, EPI_ISL_2639799, EPI_ISL_2639805, EPI_ISL_2639811, EPI_ISL_2639813, EPI_ISL_2639820, EPI_ISL_2639822, EPI_ISL_2639828, EPI_ISL_2639832, EPI_ISL_2639836, EPI_ISL_2639838, EPI_ISL_2639839, EPI_ISL_2639840, EPI_ISL_2639841, EPI_ISL_2639844, EPI_ISL_2639847, EPI_ISL_2639858, EPI_ISL_2639860, EPI_ISL_2639867, EPI_ISL_2639872, EPI_ISL_2639880, EPI_ISL_2639885, EPI_ISL_2639889, EPI_ISL_2639894, EPI_ISL_2639898, EPI_ISL_2639901, EPI_ISL_2639917, EPI_ISL_2639920, EPI_ISL_2639922, EPI_ISL_2639924, EPI_ISL_2639927, EPI_ISL_2639928, EPI_ISL_2651411, EPI_ISL_2651415, EPI_ISL_2651417, EPI_ISL_2651420, EPI_ISL_2651422, EPI_ISL_2651424, EPI_ISL_2651435, EPI_ISL_2651439, EPI_ISL_2651443, EPI_ISL_2651444, EPI_ISL_2651448, EPI_ISL_2651455, EPI_ISL_2651459, EPI_ISL_2651460, EPI_ISL_2651464, EPI_ISL_2651470, EPI_ISL_2651473, EPI_ISL_2651475, EPI_ISL_2651479, EPI_ISL_2651484, EPI_ISL_2651485, EPI_ISL_2651487, EPI_ISL_2651489, EPI_ISL_2651491, EPI_ISL_2651495, EPI_ISL_2651496, EPI_ISL_2651497, EPI_ISL_2651505, EPI_ISL_2651508, EPI_ISL_2651509, EPI_ISL_2651512, EPI_ISL_2651513, EPI_ISL_2651514, EPI_ISL_2651517, EPI_ISL_2651518, EPI_ISL_2651519, EPI_ISL_2651524, EPI_ISL_2651526, EPI_ISL_2651527, EPI_ISL_2651529, EPI_ISL_2651534, EPI_ISL_2651535, EPI_ISL_2651540, EPI_ISL_2651542, EPI_ISL_2651546, EPI_ISL_2651548, EPI_ISL_2651549, EPI_ISL_2651555, EPI_ISL_2651558, EPI_ISL_2651560, EPI_ISL_2651562, EPI_ISL_2651563, EPI_ISL_2651564, EPI_ISL_2651568, EPI_ISL_2651578, EPI_ISL_2651579, EPI_ISL_2651581, EPI_ISL_2651583, EPI_ISL_2651584, EPI_ISL_2651585, EPI_ISL_2651587, EPI_ISL_2651597, EPI_ISL_2651599, EPI_ISL_2651601, EPI_ISL_2651602, EPI_ISL_2651603, EPI_ISL_2651604, EPI_ISL_2651607, EPI_ISL_2651610, EPI_ISL_2651612, EPI_ISL_2651617, EPI_ISL_2651618, EPI_ISL_2651621, EPI_ISL_2651624, EPI_ISL_2651625, EPI_ISL_2651629, EPI_ISL_2651630, EPI_ISL_2651633, EPI_ISL_2651635, EPI_ISL_2651637, EPI_ISL_2651639, EPI_ISL_2651945, EPI_ISL_2664468, EPI_ISL_2664478, EPI_ISL_2664479, EPI_ISL_2664484, EPI_ISL_2664485, EPI_ISL_2664487, EPI_ISL_2664489, EPI_ISL_2664493, EPI_ISL_2664496, EPI_ISL_2664499, EPI_ISL_2664501, EPI_ISL_2664502, EPI_ISL_2664504, EPI_ISL_2664505, EPI_ISL_2664506, EPI_ISL_2664507, EPI_ISL_2664511, EPI_ISL_2664520, EPI_ISL_2664523, EPI_ISL_2664525, EPI_ISL_2664527, EPI_ISL_2664529, EPI_ISL_2664533, EPI_ISL_2664534, EPI_ISL_2664537, EPI_ISL_2664544, EPI_ISL_2664550, EPI_ISL_2664551, EPI_ISL_2664553, EPI_ISL_2664554, EPI_ISL_2664555, EPI_ISL_2664561, EPI_ISL_2664563, EPI_ISL_2664565, EPI_ISL_2664578, EPI_ISL_2664580, EPI_ISL_2664588, EPI_ISL_2664589, EPI_ISL_2664590, EPI_ISL_2664595, EPI_ISL_2664602, EPI_ISL_2664603, EPI_ISL_2664604, EPI_ISL_2664606, EPI_ISL_2664607, EPI_ISL_2664613, EPI_ISL_2664614, EPI_ISL_2664622, EPI_ISL_2664625, EPI_ISL_2664629, EPI_ISL_2664631, EPI_ISL_2664634, EPI_ISL_2664635, EPI_ISL_2664639, EPI_ISL_2664642, EPI_ISL_2664649, EPI_ISL_2664654, EPI_ISL_2664656, EPI_ISL_2664657, EPI_ISL_2664658, EPI_ISL_2664660, EPI_ISL_2664670, EPI_ISL_2664672, EPI_ISL_2664676, EPI_ISL_2664677, EPI_ISL_2664678, EPI_ISL_2664682, EPI_ISL_2664688, EPI_ISL_2664695, EPI_ISL_2664701, EPI_ISL_2664702, EPI_ISL_2664704, EPI_ISL_2664707, EPI_ISL_2664716, EPI_ISL_2664727, EPI_ISL_2664729, EPI_ISL_2664732, EPI_ISL_2664737, EPI_ISL_2664740, EPI_ISL_2664741, EPI_ISL_2664743, EPI_ISL_2664749, EPI_ISL_2664751, EPI_ISL_2664754, EPI_ISL_2664765, EPI_ISL_2664769, EPI_ISL_2664770, EPI_ISL_2664771, EPI_ISL_2664772, EPI_ISL_2664773, EPI_ISL_2664774, EPI_ISL_2664779, EPI_ISL_2664783, EPI_ISL_2664787, EPI_ISL_2664790, EPI_ISL_2664792, EPI_ISL_2664793, EPI_ISL_2664796, EPI_ISL_2664800, EPI_ISL_2664802, EPI_ISL_2664812, EPI_ISL_2664818, EPI_ISL_2664819, EPI_ISL_2664823, EPI_ISL_2664824, EPI_ISL_2664825, EPI_ISL_2664830, EPI_ISL_2664833, EPI_ISL_2664834, EPI_ISL_2664836, EPI_ISL_2664840, EPI_ISL_2664845, EPI_ISL_2664850, EPI_ISL_2664852, EPI_ISL_2664855, EPI_ISL_2664857, EPI_ISL_2664866, EPI_ISL_2664868, EPI_ISL_2664869, EPI_ISL_2664874, EPI_ISL_2664875, EPI_ISL_2664876, EPI_ISL_2664877, EPI_ISL_2664880, EPI_ISL_2664882, EPI_ISL_2664886, EPI_ISL_2664891, EPI_ISL_2664896, EPI_ISL_2664897, EPI_ISL_2664898, EPI_ISL_2664902, EPI_ISL_2664903, EPI_ISL_2664905, EPI_ISL_2664908, EPI_ISL_2664912, EPI_ISL_2664914, EPI_ISL_2664918, EPI_ISL_2664919, EPI_ISL_2664922, EPI_ISL_2664923, EPI_ISL_2664926, EPI_ISL_2664928, EPI_ISL_2664942, EPI_ISL_2664951, EPI_ISL_2664955, EPI_ISL_2664958, EPI_ISL_2664961, EPI_ISL_2664966, EPI_ISL_2664967, EPI_ISL_2664970, EPI_ISL_2664972, EPI_ISL_2664974, EPI_ISL_2664979, EPI_ISL_2664982, EPI_ISL_2664986, EPI_ISL_2664993, EPI_ISL_2664994, EPI_ISL_2665002, EPI_ISL_2665012, EPI_ISL_2665014, EPI_ISL_2665024, EPI_ISL_2665027, EPI_ISL_2665385, EPI_ISL_2665386, EPI_ISL_2665387, EPI_ISL_2683428, EPI_ISL_2683460, EPI_ISL_2703507, EPI_ISL_2703573, EPI_ISL_2703586, EPI_ISL_2703694, EPI_ISL_2703805, EPI_ISL_2703808, EPI_ISL_2703826, EPI_ISL_2703835, EPI_ISL_2703849, EPI_ISL_2703851, EPI_ISL_2703858, EPI_ISL_2703878, EPI_ISL_2703891, EPI_ISL_2703909, EPI_ISL_2703933, EPI_ISL_2703938, EPI_ISL_2703940, EPI_ISL_2703967, EPI_ISL_2703981, EPI_ISL_2704025, EPI_ISL_2704029, EPI_ISL_2704037, EPI_ISL_2704095, EPI_ISL_2704671, EPI_ISL_2718298, EPI_ISL_2718299, EPI_ISL_2718300, EPI_ISL_2718301, EPI_ISL_2718302, EPI_ISL_2718307, EPI_ISL_2718310, EPI_ISL_2718313, EPI_ISL_2718317, EPI_ISL_2718319, EPI_ISL_2718324, EPI_ISL_2718327, EPI_ISL_2718329, EPI_ISL_2718331, EPI_ISL_2718332, EPI_ISL_2718333, EPI_ISL_2718334, EPI_ISL_2718338, EPI_ISL_2718339, EPI_ISL_2718345, EPI_ISL_2718349, EPI_ISL_2718350, EPI_ISL_2718355, EPI_ISL_2718359, EPI_ISL_2718360, EPI_ISL_2718361, EPI_ISL_2718364, EPI_ISL_2718367, EPI_ISL_2718369, EPI_ISL_2718374, EPI_ISL_2718375, EPI_ISL_2718384, EPI_ISL_2718386, EPI_ISL_2718387, EPI_ISL_2718392, EPI_ISL_2718396, EPI_ISL_2718399, EPI_ISL_2718402, EPI_ISL_2718405, EPI_ISL_2718407, EPI_ISL_2718409, EPI_ISL_2718411, EPI_ISL_2718413, EPI_ISL_2718419, EPI_ISL_2718420, EPI_ISL_2718422, EPI_ISL_2718427, EPI_ISL_2718428, EPI_ISL_2718430, EPI_ISL_2718431, EPI_ISL_2718432, EPI_ISL_2718433, EPI_ISL_2718437, EPI_ISL_2718441, EPI_ISL_2718444, EPI_ISL_2718450, EPI_ISL_2718452, EPI_ISL_2718457, EPI_ISL_2718459, EPI_ISL_2718460, EPI_ISL_2718464, EPI_ISL_2718468, EPI_ISL_2718473, EPI_ISL_2718481, EPI_ISL_2718482, EPI_ISL_2718483, EPI_ISL_2718485, EPI_ISL_2718491, EPI_ISL_2718492, EPI_ISL_2718496, EPI_ISL_2718497, EPI_ISL_2718498, EPI_ISL_2718500, EPI_ISL_2718501, EPI_ISL_2718503, EPI_ISL_2718505, EPI_ISL_2718508, EPI_ISL_2718510, EPI_ISL_2718514, EPI_ISL_2718516, EPI_ISL_2718517, EPI_ISL_2718523, EPI_ISL_2718524, EPI_ISL_2718526, EPI_ISL_2718529, EPI_ISL_2718531, EPI_ISL_2731187, EPI_ISL_2768009, EPI_ISL_2768010, EPI_ISL_2768011, EPI_ISL_2768012, EPI_ISL_2768013, EPI_ISL_2853263, EPI_ISL_2853264, EPI_ISL_2853265, EPI_ISL_2853268, EPI_ISL_2853275, EPI_ISL_2853281, EPI_ISL_2853285, EPI_ISL_2853288, EPI_ISL_2853290, EPI_ISL_2853295, EPI_ISL_2853296, EPI_ISL_2853299, EPI_ISL_2853300, EPI_ISL_2853301, EPI_ISL_2853307, EPI_ISL_2853311, EPI_ISL_2853312, EPI_ISL_2853314, EPI_ISL_2853315, EPI_ISL_2853322, EPI_ISL_2853327, EPI_ISL_2853331, EPI_ISL_2853335, EPI_ISL_2853336, EPI_ISL_2853346, EPI_ISL_2853347, EPI_ISL_2853348, EPI_ISL_2853356, EPI_ISL_2853361, EPI_ISL_2853363, EPI_ISL_2853368, EPI_ISL_2853371, EPI_ISL_2853372, EPI_ISL_2853374, EPI_ISL_2853377, EPI_ISL_2853383, EPI_ISL_2853394, EPI_ISL_2853399, EPI_ISL_2853401, EPI_ISL_2853403, EPI_ISL_2853411, EPI_ISL_2853417, EPI_ISL_2853420, EPI_ISL_2853424, EPI_ISL_2853428, EPI_ISL_2853429, EPI_ISL_2853433, EPI_ISL_2853438, EPI_ISL_2853447, EPI_ISL_2853454, EPI_ISL_2853456, EPI_ISL_2853457, EPI_ISL_2853458, EPI_ISL_2853459, EPI_ISL_2853460, EPI_ISL_2853461, EPI_ISL_2853467, EPI_ISL_2853473, EPI_ISL_2853476, EPI_ISL_2853479, EPI_ISL_2853487, EPI_ISL_2853490, EPI_ISL_2853491, EPI_ISL_2853493, EPI_ISL_2853495, EPI_ISL_2853501, EPI_ISL_2853502, EPI_ISL_2853508, EPI_ISL_2853514, EPI_ISL_2853516, EPI_ISL_2853533, EPI_ISL_2853536, EPI_ISL_2853540, EPI_ISL_2853543, EPI_ISL_2853544, EPI_ISL_2853545, EPI_ISL_2853547, EPI_ISL_2853557, EPI_ISL_2853582, EPI_ISL_2868228, EPI_ISL_2868234

| | | | |
|---|---|---|---|
| see above | Lighthouse Laboratory Plymouth | Wellcome Sanger Institute for the COVID-19 Genomics UK (COG-UK) Consortium | Cordelia Langford; David K. Jackson; Dominic Kwiatkowski; Ewan Harrison; Ian Johnston; Jeffrey Barrett; John Sillitoe on behalf of the Wellcome Sanger Institute COVID-19 Surveillance Team; Lighthouse Laboratory Plymouth and Alex Alderton; Roberto Amato; Sonia Goncalves |

EPI_ISL_2706158, EPI_ISL_2706160, EPI_ISL_2706161, EPI_ISL_2706164, EPI_ISL_2706165, EPI_ISL_2706166, EPI_ISL_2706167, EPI_ISL_2706169, EPI_ISL_2706171, EPI_ISL_2706172, EPI_ISL_2706173, EPI_ISL_2706174, EPI_ISL_2706175, EPI_ISL_2706178, EPI_ISL_2706179, EPI_ISL_2706180, EPI_ISL_2706184, EPI_ISL_2706185, EPI_ISL_2706186, EPI_ISL_2706188, EPI_ISL_2706189, EPI_ISL_2706190, EPI_ISL_2706191, EPI_ISL_2706192, EPI_ISL_2706193, EPI_ISL_2706194, EPI_ISL_2706195, EPI_ISL_2706196, EPI_ISL_2706197, EPI_ISL_2706198, EPI_ISL_2706199, EPI_ISL_2706200, EPI_ISL_2706201, EPI_ISL_2706202, EPI_ISL_2706204, EPI_ISL_2706205, EPI_ISL_2706206, EPI_ISL_2706207, EPI_ISL_2706208, EPI_ISL_2706209, EPI_ISL_2706210, EPI_ISL_2706211, EPI_ISL_2706212, EPI_ISL_2706215, EPI_ISL_2706216, EPI_ISL_2706217, EPI_ISL_2706219, EPI_ISL_2706220, EPI_ISL_2706221, EPI_ISL_2706223, EPI_ISL_2706224, EPI_ISL_2706225, EPI_ISL_2706226, EPI_ISL_2706227, EPI_ISL_2706228, EPI_ISL_2706229, EPI_ISL_2706230, EPI_ISL_2706231, EPI_ISL_2706232, EPI_ISL_2706233, EPI_ISL_2706234, EPI_ISL_2706235, EPI_ISL_2706236, EPI_ISL_2706237, EPI_ISL_2706238, EPI_ISL_2706239, EPI_ISL_2706240, EPI_ISL_2706241, EPI_ISL_2706242, EPI_ISL_2706243, EPI_ISL_2706244, EPI_ISL_2706245, EPI_ISL_2706246, EPI_ISL_2706247, EPI_ISL_2706248, EPI_ISL_2706249, EPI_ISL_2706251, EPI_ISL_2706252, EPI_ISL_2706253, EPI_ISL_2706254, EPI_ISL_2706255, EPI_ISL_2706256, EPI_ISL_2706258, EPI_ISL_2706259, EPI_ISL_2706260, EPI_ISL_2706261, EPI_ISL_2706262, EPI_ISL_2706264, EPI_ISL_2706265, EPI_ISL_2706266, EPI_ISL_2706267, EPI_ISL_2706268, EPI_ISL_2706269, EPI_ISL_2706270, EPI_ISL_2706271, EPI_ISL_2706272, EPI_ISL_2706273, EPI_ISL_2706274, EPI_ISL_2706275, EPI_ISL_2706276, EPI_ISL_2706277, EPI_ISL_2706278, EPI_ISL_2706279, EPI_ISL_2706280, EPI_ISL_2706281, EPI_ISL_2706282, EPI_ISL_2706283, EPI_ISL_2706284, EPI_ISL_2706286, EPI_ISL_2706287, EPI_ISL_2706288, EPI_ISL_2706289, EPI_ISL_2706290, EPI_ISL_2706291, EPI_ISL_2706292, EPI_ISL_2706293, EPI_ISL_2706294, EPI_ISL_2706295, EPI_ISL_2706296, EPI_ISL_2706297, EPI_ISL_2706298, EPI_ISL_2706299, EPI_ISL_2706300, EPI_ISL_2706301, EPI_ISL_2706303, EPI_ISL_2706304, EPI_ISL_2706305, EPI_ISL_2706306, EPI_ISL_2706307, EPI_ISL_2706308, EPI_ISL_2706309, EPI_ISL_2706310, EPI_ISL_2706311, EPI_ISL_2706312, EPI_ISL_2706313, EPI_ISL_2706314, EPI_ISL_2706315, EPI_ISL_2706317, EPI_ISL_2706318, EPI_ISL_2706319, EPI_ISL_2706320, EPI_ISL_2706321, EPI_ISL_2706322, EPI_ISL_2706324, EPI_ISL_2706325, EPI_ISL_2706327, EPI_ISL_2706328, EPI_ISL_2706329, EPI_ISL_2706330, EPI_ISL_2706332, EPI_ISL_2706333, EPI_ISL_2706334, EPI_ISL_2706335, EPI_ISL_2706336, EPI_ISL_2706337, EPI_ISL_2706339, EPI_ISL_2706340, EPI_ISL_2706341, EPI_ISL_2706342, EPI_ISL_2706344, EPI_ISL_2706346, EPI_ISL_2706347, EPI_ISL_2706348, EPI_ISL_2706349, EPI_ISL_2706351, EPI_ISL_2706352, EPI_ISL_2706353, EPI_ISL_2706354, EPI_ISL_2706355, EPI_ISL_2706356, EPI_ISL_2706357, EPI_ISL_2706358, EPI_ISL_2706359, EPI_ISL_2706360, EPI_ISL_2706361, EPI_ISL_2706362, EPI_ISL_2706363, EPI_ISL_2706364, EPI_ISL_2706365, EPI_ISL_2706366, EPI_ISL_2706367, EPI_ISL_2706368, EPI_ISL_2706369, EPI_ISL_2706370, EPI_ISL_2706371, EPI_ISL_2706373, EPI_ISL_2706374, EPI_ISL_2706375, EPI_ISL_2706376, EPI_ISL_2706377, EPI_ISL_2706378, EPI_ISL_2706379, EPI_ISL_2706382, EPI_ISL_2706383, EPI_ISL_2706384, EPI_ISL_2706385, EPI_ISL_2706386, EPI_ISL_2706388, EPI_ISL_2706389, EPI_ISL_2706390, EPI_ISL_2706391, EPI_ISL_2706392, EPI_ISL_2706393, EPI_ISL_2706395, EPI_ISL_2706396, EPI_ISL_2706397, EPI_ISL_2706398, EPI_ISL_2706400, EPI_ISL_2706401, EPI_ISL_2706402, EPI_ISL_2706403, EPI_ISL_2706404, EPI_ISL_2706405, EPI_ISL_2706406, EPI_ISL_2706407, EPI_ISL_2706408, EPI_ISL_2706409, EPI_ISL_2706410, EPI_ISL_2706411, EPI_ISL_2706412, EPI_ISL_2706413, EPI_ISL_2706414, EPI_ISL_2706416, EPI_ISL_2706417, EPI_ISL_2706419, EPI_ISL_2706420, EPI_ISL_2706421, EPI_ISL_2706422, EPI_ISL_2706423, EPI_ISL_2706424, EPI_ISL_2706425, EPI_ISL_2706426, EPI_ISL_2706427, EPI_ISL_2706428, EPI_ISL_2706429, EPI_ISL_2706431, EPI_ISL_2706432, EPI_ISL_2706434, EPI_ISL_2706435, EPI_ISL_2706436, EPI_ISL_2706437, EPI_ISL_2706438, EPI_ISL_2706439, EPI_ISL_2706440, EPI_ISL_2706441, EPI_ISL_2706442, EPI_ISL_2706443, EPI_ISL_2706445, EPI_ISL_2706446, EPI_ISL_2706447, EPI_ISL_2706448, EPI_ISL_2706449, EPI_ISL_2706451, EPI_ISL_2706452, EPI_ISL_2706453, EPI_ISL_2706454, EPI_ISL_2706455, EPI_ISL_2706456, EPI_ISL_2706457, EPI_ISL_2706458, EPI_ISL_2706459, EPI_ISL_2706461, EPI_ISL_2706462, EPI_ISL_2706464, EPI_ISL_2706465, EPI_ISL_2706466, EPI_ISL_2706467, EPI_ISL_2706469, EPI_ISL_2706470, EPI_ISL_2706471, EPI_ISL_2706472, EPI_ISL_2706473, EPI_ISL_2706474, EPI_ISL_2706476, EPI_ISL_2706478, EPI_ISL_2706479, EPI_ISL_2706480, EPI_ISL_2706481, EPI_ISL_2706482, EPI_ISL_2706483, EPI_ISL_2706484, EPI_ISL_2706485, EPI_ISL_2706486, EPI_ISL_2706488, EPI_ISL_2706490, EPI_ISL_2706492, EPI_ISL_2706493, EPI_ISL_2706495, EPI_ISL_2706496, EPI_ISL_2706498, EPI_ISL_2706499, EPI_ISL_2706500, EPI_ISL_2706501, EPI_ISL_2706502, EPI_ISL_2706503, EPI_ISL_2706504, EPI_ISL_2706505, EPI_ISL_2706506, EPI_ISL_2706507, EPI_ISL_2706508, EPI_ISL_2706509, EPI_ISL_2706510, EPI_ISL_2706511, EPI_ISL_2706512, EPI_ISL_2706513, EPI_ISL_2706514, EPI_ISL_2706515, EPI_ISL_2706516, EPI_ISL_2706517, EPI_ISL_2706518, EPI_ISL_2706519, EPI_ISL_2706520, EPI_ISL_2706521, EPI_ISL_2706522, EPI_ISL_2706523, EPI_ISL_2706524, EPI_ISL_2706525, EPI_ISL_2706526, EPI_ISL_2706527, EPI_ISL_2706528, EPI_ISL_2706529, EPI_ISL_2706530, EPI_ISL_2706531, EPI_ISL_2706532, EPI_ISL_2706533, EPI_ISL_2706534, EPI_ISL_2706535, EPI_ISL_2706536, EPI_ISL_2706537, EPI_ISL_2706538, EPI_ISL_2706539, EPI_ISL_2706540, EPI_ISL_2706541, EPI_ISL_2706542, EPI_ISL_2706543, EPI_ISL_2706544, EPI_ISL_2706545, EPI_ISL_2706546, EPI_ISL_2706547, EPI_ISL_2706548, EPI_ISL_2706549, EPI_ISL_2706550, EPI_ISL_2706551, EPI_ISL_2706552, EPI_ISL_2706553, EPI_ISL_2706554, EPI_ISL_2706555, EPI_ISL_2706556, EPI_ISL_2706557, EPI_ISL_2706558, EPI_ISL_2706559, EPI_ISL_2706560, EPI_ISL_2706561, EPI_ISL_2706562, EPI_ISL_2706563, EPI_ISL_2706564, EPI_ISL_2706565, EPI_ISL_2706566, EPI_ISL_2706567, EPI_ISL_2706568, EPI_ISL_2706569, EPI_ISL_2706570, EPI_ISL_2706572, EPI_ISL_2706581, EPI_ISL_2706601, EPI_ISL_2706614, EPI_ISL_2816673

| | | | |
|---|---|---|---|
| see above | Northumbria University / South Tees Hospitals NHS Foundation Trust / North Cumbria Integrated Care NHS Foundation Trust / North Tees and Hartlepool NHS Foundation Trust / Newcastle Hospitals NHS Foundation Trust | COVID-19 Genomics UK (COG-UK) Consortium | Andrew Nelson; Brendan Payne; Clive Graham; Darren L Smith; Debra Padgett; Edward Barton; Emma Swindells; Garren Scott; Gary Black; Gary Eltringham; Giles S Holt; Greg R Young; Jane Greenaway; Jennifer Collins; John Allan; Joshua Loh; Lynn Dover; Matthew Bashton; Mohammad A Tariq; Paul Baker; Sarah Essex; Steve Liggett; Wen C Yew; Yusri Taha |

EPI_ISL_2707359, EPI_ISL_2707364, EPI_ISL_2707371, EPI_ISL_2707372, EPI_ISL_2707373, EPI_ISL_2707375, EPI_ISL_2707378, EPI_ISL_2707379, EPI_ISL_2707384, EPI_ISL_2707389, EPI_ISL_2707394, EPI_ISL_2707400, EPI_ISL_2707404, EPI_ISL_2707415, EPI_ISL_2707421, EPI_ISL_2707428, EPI_ISL_2707436, EPI_ISL_2707437, EPI_ISL_2707441, EPI_ISL_2707446, EPI_ISL_2707447, EPI_ISL_2707448, EPI_ISL_2707454, EPI_ISL_2707459, EPI_ISL_2707465, EPI_ISL_2707467, EPI_ISL_2707470, EPI_ISL_2707473, EPI_ISL_2707479, EPI_ISL_2707480, EPI_ISL_2707481, EPI_ISL_2707484, EPI_ISL_2707486, EPI_ISL_2707487, EPI_ISL_2707489, EPI_ISL_2707490, EPI_ISL_2707497, EPI_ISL_2707502, EPI_ISL_2707510, EPI_ISL_2707516, EPI_ISL_2707519, EPI_ISL_2707531, EPI_ISL_2707532, EPI_ISL_2707541, EPI_ISL_2707545, EPI_ISL_2707549, EPI_ISL_2707550, EPI_ISL_2707558, EPI_ISL_2707563, EPI_ISL_2707564, EPI_ISL_2707565, EPI_ISL_2707566, EPI_ISL_2707567, EPI_ISL_2707568, EPI_ISL_2707569, EPI_ISL_2707570, EPI_ISL_2707571, EPI_ISL_2707573, EPI_ISL_2707574, EPI_ISL_2707575, EPI_ISL_2707577, EPI_ISL_2707578, EPI_ISL_2707579, EPI_ISL_2707583, EPI_ISL_2707588, EPI_ISL_2707589, EPI_ISL_2707590, EPI_ISL_2707594, EPI_ISL_2707595, EPI_ISL_2707599, EPI_ISL_2707601, EPI_ISL_2707602, EPI_ISL_2707605, EPI_ISL_2707606, EPI_ISL_2707607, EPI_ISL_2707608, EPI_ISL_2707609, EPI_ISL_2707610, EPI_ISL_2707611, EPI_ISL_2707612, EPI_ISL_2707613, EPI_ISL_2707614, EPI_ISL_2707615, EPI_ISL_2707616, EPI_ISL_2707617, EPI_ISL_2707618, EPI_ISL_2707619, EPI_ISL_2707620, EPI_ISL_2707621, EPI_ISL_2707622, EPI_ISL_2707623, EPI_ISL_2707624, EPI_ISL_2707625, EPI_ISL_2707626, EPI_ISL_2707628, EPI_ISL_2707630, EPI_ISL_2707631, EPI_ISL_2707636, EPI_ISL_2707637, EPI_ISL_2707638, EPI_ISL_2707639, EPI_ISL_2707640, EPI_ISL_2707643, EPI_ISL_2707644, EPI_ISL_2707645, EPI_ISL_2707648, EPI_ISL_2707649, EPI_ISL_2707650, EPI_ISL_2707653, EPI_ISL_2707654, EPI_ISL_2707685, EPI_ISL_2707694, EPI_ISL_2707708, EPI_ISL_2707709, EPI_ISL_2707710, EPI_ISL_2707711, EPI_ISL_2707712, EPI_ISL_2707713, EPI_ISL_2707714, EPI_ISL_2707715, EPI_ISL_2707716, EPI_ISL_2707717, EPI_ISL_2707718, EPI_ISL_2707719, EPI_ISL_2707720, EPI_ISL_2707721, EPI_ISL_2707722, EPI_ISL_2707723, EPI_ISL_2707724, EPI_ISL_2707725, EPI_ISL_2707726, EPI_ISL_2707727, EPI_ISL_2707728, EPI_ISL_2707729, EPI_ISL_2707730, EPI_ISL_2707731, EPI_ISL_2707732, EPI_ISL_2707733, EPI_ISL_2707734, EPI_ISL_2707735, EPI_ISL_2707736, EPI_ISL_2707737, EPI_ISL_2707738, EPI_ISL_2707739, EPI_ISL_2707740, EPI_ISL_2707741, EPI_ISL_2707742, EPI_ISL_2707743, EPI_ISL_2707744, EPI_ISL_2707745, EPI_ISL_2707746, EPI_ISL_2707747, EPI_ISL_2707748, EPI_ISL_2707749, EPI_ISL_2707750, EPI_ISL_2707751, EPI_ISL_2707752, EPI_ISL_2707753, EPI_ISL_2707754, EPI_ISL_2707755, EPI_ISL_2707756, EPI_ISL_2707757, EPI_ISL_2707758, EPI_ISL_2707759, EPI_ISL_2707760, EPI_ISL_2707761, EPI_ISL_2707762, EPI_ISL_2707763, EPI_ISL_2707764, EPI_ISL_2707765, EPI_ISL_2707766, EPI_ISL_2707767, EPI_ISL_2707768, EPI_ISL_2707769, EPI_ISL_2707770, EPI_ISL_2707771, EPI_ISL_2707772, EPI_ISL_2707773, EPI_ISL_2707774, EPI_ISL_2707775, EPI_ISL_2707776, EPI_ISL_2707777, EPI_ISL_2707778, EPI_ISL_2707779, EPI_ISL_2707780, EPI_ISL_2707781, EPI_ISL_2707782, EPI_ISL_2707783, EPI_ISL_2707784, EPI_ISL_2707785, EPI_ISL_2707786, EPI_ISL_2707787, EPI_ISL_2707788, EPI_ISL_2707789, EPI_ISL_2707790, EPI_ISL_2707818, EPI_ISL_2707819, EPI_ISL_2707820, EPI_ISL_2707821, EPI_ISL_2707822, EPI_ISL_2707823, EPI_ISL_2707824, EPI_ISL_2707825, EPI_ISL_2707826, EPI_ISL_2707827, EPI_ISL_2707828, EPI_ISL_2707829, EPI_ISL_2707830, EPI_ISL_2707831, EPI_ISL_2707832, EPI_ISL_2707833, EPI_ISL_2707834, EPI_ISL_2707835, EPI_ISL_2707836, EPI_ISL_2707837, EPI_ISL_2707838, EPI_ISL_2707839, EPI_ISL_2707840, EPI_ISL_2707841, EPI_ISL_2707842, EPI_ISL_2707843, EPI_ISL_2707844, EPI_ISL_2707882, EPI_ISL_2707885, EPI_ISL_2707889, EPI_ISL_2707890, EPI_ISL_2707891, EPI_ISL_2707892, EPI_ISL_2707893, EPI_ISL_2707894, EPI_ISL_2707895, EPI_ISL_2707896, EPI_ISL_2707897, EPI_ISL_2707900, EPI_ISL_2707902, EPI_ISL_2707903, EPI_ISL_2707905, EPI_ISL_2707906, EPI_ISL_2707909, EPI_ISL_2707914, EPI_ISL_2707916, EPI_ISL_2707924, EPI_ISL_2707926, EPI_ISL_2707930, EPI_ISL_2707931, EPI_ISL_2707934, EPI_ISL_2707935, EPI_ISL_2707936, EPI_ISL_2707938, EPI_ISL_2707939, EPI_ISL_2707941, EPI_ISL_2707942, EPI_ISL_2707944, EPI_ISL_2707945, EPI_ISL_2707947, EPI_ISL_2707948, EPI_ISL_2707949, EPI_ISL_2707950, EPI_ISL_2707951, EPI_ISL_2707955, EPI_ISL_2707956, EPI_ISL_2707961, EPI_ISL_2707962, EPI_ISL_2707963, EPI_ISL_2707965, EPI_ISL_2707966, EPI_ISL_2707967, EPI_ISL_2707968, EPI_ISL_2707971, EPI_ISL_2707972, EPI_ISL_2707973, EPI_ISL_2707974, EPI_ISL_2707975, EPI_ISL_2707976, EPI_ISL_2707977, EPI_ISL_2707978, EPI_ISL_2707979, EPI_ISL_2707980, EPI_ISL_2707982, EPI_ISL_2708009, EPI_ISL_2708011, EPI_ISL_2708012, EPI_ISL_2708013, EPI_ISL_2708014, EPI_ISL_2708015, EPI_ISL_2708016, EPI_ISL_2708017, EPI_ISL_2708018, EPI_ISL_2708019, EPI_ISL_2708020, EPI_ISL_2708021, EPI_ISL_2708027, EPI_ISL_2708029, EPI_ISL_2708030, EPI_ISL_2708031, EPI_ISL_2708033, EPI_ISL_2708041, EPI_ISL_2708042, EPI_ISL_2708043, EPI_ISL_2708049, EPI_ISL_2708051, EPI_ISL_2708054, EPI_ISL_2708055, EPI_ISL_2708058, EPI_ISL_2708088, EPI_ISL_2708089, EPI_ISL_2708090, EPI_ISL_2708093, EPI_ISL_2708094, EPI_ISL_2708095, EPI_ISL_2708097, EPI_ISL_2708100, EPI_ISL_2708101, EPI_ISL_2708102, EPI_ISL_2708103, EPI_ISL_2708107, EPI_ISL_2708109, EPI_ISL_2708110, EPI_ISL_2708111, EPI_ISL_2708112, EPI_ISL_2708115, EPI_ISL_2708117, EPI_ISL_2749956, EPI_ISL_2749960, EPI_ISL_2749964, EPI_ISL_2749965, EPI_ISL_2749966, EPI_ISL_2749968, EPI_ISL_2749969, EPI_ISL_2749970, EPI_ISL_2749972, EPI_ISL_2749976, EPI_ISL_2749977, EPI_ISL_2749979, EPI_ISL_2749983, EPI_ISL_2749984, EPI_ISL_2749985, EPI_ISL_2749993, EPI_ISL_2749994, EPI_ISL_2749997, EPI_ISL_2750000, EPI_ISL_2750003, EPI_ISL_2750009, EPI_ISL_2750023, EPI_ISL_2750025, EPI_ISL_2750026, EPI_ISL_2750028, EPI_ISL_2750029, EPI_ISL_2750035, EPI_ISL_2750037, EPI_ISL_2750040, EPI_ISL_2750041, EPI_ISL_2750049, EPI_ISL_2750060, EPI_ISL_2750062, EPI_ISL_2750064, EPI_ISL_2750065, EPI_ISL_2750130, EPI_ISL_2750134, EPI_ISL_2750139, EPI_ISL_2750140, EPI_ISL_2750141, EPI_ISL_2750143, EPI_ISL_2750152, EPI_ISL_2750158, EPI_ISL_2750159, EPI_ISL_2750182, EPI_ISL_2750189, EPI_ISL_2750197, EPI_ISL_2750202, EPI_ISL_2750204, EPI_ISL_2750209, EPI_ISL_2750231, EPI_ISL_2750282, EPI_ISL_2750283, EPI_ISL_2750284, EPI_ISL_2750292, EPI_ISL_2750490, EPI_ISL_2750503, EPI_ISL_2750507, EPI_ISL_2750513, EPI_ISL_2750523, EPI_ISL_2750586, EPI_ISL_2750587, EPI_ISL_2750592, EPI_ISL_2750596, EPI_ISL_2750599, EPI_ISL_2750600, EPI_ISL_2750601, EPI_ISL_2750602, EPI_ISL_2750606, EPI_ISL_2750612, EPI_ISL_2750641, EPI_ISL_2750646, EPI_ISL_2750647, EPI_ISL_2750650, EPI_ISL_2750651, EPI_ISL_2750652, EPI_ISL_2750654, EPI_ISL_2750655, EPI_ISL_2750656, EPI_ISL_2750659, EPI_ISL_2750660, EPI_ISL_2750661, EPI_ISL_2750662, EPI_ISL_2750676, EPI_ISL_2750687, EPI_ISL_2750688, EPI_ISL_2750689, EPI_ISL_2818325, EPI_ISL_2818326, EPI_ISL_2818327, EPI_ISL_2818328, EPI_ISL_2818329, EPI_ISL_2818331, EPI_ISL_2818332, EPI_ISL_2818333, EPI_ISL_2818334, EPI_ISL_2818419, EPI_ISL_2818420, EPI_ISL_2818421, EPI_ISL_2818422, EPI_ISL_2818423, EPI_ISL_2818424, EPI_ISL_2818425, EPI_ISL_2818426, EPI_ISL_2818427, EPI_ISL_2818428, EPI_ISL_2818429, EPI_ISL_2818430, EPI_ISL_2818431, EPI_ISL_2818432, EPI_ISL_2818433, EPI_ISL_2818434, EPI_ISL_2818435, EPI_ISL_2818436, EPI_ISL_2818437, EPI_ISL_2818438, EPI_ISL_2818439, EPI_ISL_2818440, EPI_ISL_2818441, EPI_ISL_2818442, EPI_ISL_2818443, EPI_ISL_2818444, EPI_ISL_2818445, EPI_ISL_2818446, EPI_ISL_2818447, EPI_ISL_2818448, EPI_ISL_2818449, EPI_ISL_2818451, EPI_ISL_2818452, EPI_ISL_2818453, EPI_ISL_2818454, EPI_ISL_2818455, EPI_ISL_2818456, EPI_ISL_2818484, EPI_ISL_2818485, EPI_ISL_2818486, EPI_ISL_2818487, EPI_ISL_2818488, EPI_ISL_2818489, EPI_ISL_2818490, EPI_ISL_2818491, EPI_ISL_2818492, EPI_ISL_2818493, EPI_ISL_2818494, EPI_ISL_2818496, EPI_ISL_2818497, EPI_ISL_2818498, EPI_ISL_2818499, EPI_ISL_2818500, EPI_ISL_2818501, EPI_ISL_2818502, EPI_ISL_2818503, EPI_ISL_2818504, EPI_ISL_2818505, EPI_ISL_2818506, EPI_ISL_2818507, EPI_ISL_2818508, EPI_ISL_2818509, EPI_ISL_2818510, EPI_ISL_2818511, EPI_ISL_2818512, EPI_ISL_2818513, EPI_ISL_2818514, EPI_ISL_2818515, EPI_ISL_2818516, EPI_ISL_2818517, EPI_ISL_2818520

| | | | |
|---|---|---|---|
| see above | Originating lab: Wales Specialist Virology Centre Sequencing lab: Pathogen Genomics Unit | Public Health Wales Microbiology Cardiff Wales Specialist Virology Centre | Alec Birchley; Alexander Adams; Amy Gaskin; Angela Marchbank; Bree Gatica-Wilcox; Catherine Moore; Jason Coombes; Joanne Watkins; Joel Southgate; Johnathan Evans; Laura Gifford; Lauren Gilbert; Lee Graham; Malorie Perry; Matthew Bull; Nicole Pacchiarini; Sally Corden; Sara Kumziene-Summerhayes; Sara Rey; Sarah Taylor; Simon Cottrell; Sophie Jones; Tom Connor |

EPI_ISL_2735680, EPI_ISL_2735681, EPI_ISL_2735683, EPI_ISL_2735684, EPI_ISL_2735687, EPI_ISL_2735688, EPI_ISL_2735689, EPI_ISL_2735690, EPI_ISL_2735695, EPI_ISL_2735696, EPI_ISL_2735697, EPI_ISL_2903040, EPI_ISL_2903085, EPI_ISL_2903095

| | | | |
|---|---|---|---|
| see above | Oxford Viromics, NDM, University of Oxford; Oxford University Hospitals; Basingstoke and North Hampshire Hospital | COVID-19 Genomics UK (COG-UK) Consortium | Alex Mobbs; Amy Trebes; Anita Justice; Catrin Moore; Christophe Fraser; David Bonsall; David Buck; Emma Wise; George Macintyre; Jessica Lynch; John Todd; Mariateresa de Cesare; Matilde Mori; Monique Andersson; Nathan Moore; Nick Cortes; Robert Shaw; Stephen Kidd; Tanya Golubchik; Timothy Peto |

EPI_ISL_2706992, EPI_ISL_2706997, EPI_ISL_2706999, EPI_ISL_2707001, EPI_ISL_2707004, EPI_ISL_2707010, EPI_ISL_2707034, EPI_ISL_2707035, EPI_ISL_2707043, EPI_ISL_2707053, EPI_ISL_2707064, EPI_ISL_2707075, EPI_ISL_2707078, EPI_ISL_2707081, EPI_ISL_2707083, EPI_ISL_2707091, EPI_ISL_2707096

| | | | |
|---|---|---|---|
| see above | Quadram Institute Bioscience | COVID-19 Genomics UK (COG-UK) Consortium | Alexander J Trotter; Alison E. Mather; Alp Aydin; Ana P. Tedim; Anastasia Kolyva; Andrew Bell; Andrew J. Page; Claire Stuart; Dave J. Baker; Gemma L. Kay; John Wain; Justin O'Grady; Leonardo de Oliveira Martins; Lizzie Meadows; Maria Diaz; Mark Webber; Muhammed Yasir; Nabil-Fareed Alikhan; Ngozi Elumogo; Nicholas M. Thomson; Rachael Stanley; Rachel Gilroy; Reenesh Prakash; Samir Dervisevic; Samuel Bloomfield; Steven Rudder; Thanh Le-Viet |
| EPI_ISL_2745860, EPI_ISL_2746064, EPI_ISL_2746157, EPI_ISL_2746241, EPI_ISL_2746249, EPI_ISL_2746250, EPI_ISL_2746261, EPI_ISL_2746263, EPI_ISL_2746265, EPI_ISL_2746270, EPI_ISL_2746273, EPI_ISL_2746278, EPI_ISL_2746287, EPI_ISL_2746296, EPI_ISL_2746299, EPI_ISL_2746300, EPI_ISL_2746302, EPI_ISL_2746305, EPI_ISL_2746307, EPI_ISL_2746319, EPI_ISL_2746321, EPI_ISL_2746322, EPI_ISL_2746323, EPI_ISL_2746325, EPI_ISL_2746328, EPI_ISL_2746330, EPI_ISL_2746365, EPI_ISL_2746378, EPI_ISL_2746381, EPI_ISL_2746388, EPI_ISL_2746389, EPI_ISL_2746391, EPI_ISL_2746392, EPI_ISL_2746394, EPI_ISL_2746395, EPI_ISL_2746396, EPI_ISL_2746397, EPI_ISL_2746398, EPI_ISL_2746399, EPI_ISL_2746401, EPI_ISL_2746403, EPI_ISL_2746404, EPI_ISL_2746405, EPI_ISL_2746407, EPI_ISL_2746408, EPI_ISL_2746409, EPI_ISL_2746410, EPI_ISL_2746411, EPI_ISL_2746412, EPI_ISL_2746413, EPI_ISL_2746414, EPI_ISL_2746415, EPI_ISL_2746416, EPI_ISL_2746417, EPI_ISL_2746418, EPI_ISL_2746419, EPI_ISL_2746420, EPI_ISL_2746421, EPI_ISL_2746422, EPI_ISL_2746423, EPI_ISL_2746424, EPI_ISL_2746425, EPI_ISL_2746426, EPI_ISL_2746427, EPI_ISL_2746429, EPI_ISL_2746430, EPI_ISL_2746431, EPI_ISL_2746432, EPI_ISL_2746433, EPI_ISL_2746434, EPI_ISL_2746435, EPI_ISL_2746436, EPI_ISL_2746437, EPI_ISL_2746438, EPI_ISL_2746439, EPI_ISL_2746440, EPI_ISL_2746441, EPI_ISL_2746442, EPI_ISL_2746444, EPI_ISL_2746445, EPI_ISL_2746446, EPI_ISL_2746447, EPI_ISL_2746448, EPI_ISL_2746449, EPI_ISL_2746450, EPI_ISL_2746451, EPI_ISL_2746452, EPI_ISL_2746453, EPI_ISL_2746454, EPI_ISL_2746455, EPI_ISL_2746456, EPI_ISL_2746457, EPI_ISL_2746458, EPI_ISL_2746459, EPI_ISL_2746460, EPI_ISL_2746461, EPI_ISL_2746462, EPI_ISL_2746463, EPI_ISL_2746464, EPI_ISL_2746465, EPI_ISL_2746466, EPI_ISL_2746467, EPI_ISL_2746468, EPI_ISL_2746469, EPI_ISL_2746470, EPI_ISL_2746471, EPI_ISL_2746472, EPI_ISL_2746473, EPI_ISL_2746474, EPI_ISL_2746475, EPI_ISL_2746476, EPI_ISL_2746477, EPI_ISL_2746478, EPI_ISL_2746479, EPI_ISL_2746480, EPI_ISL_2746481, EPI_ISL_2746482, EPI_ISL_2746483, EPI_ISL_2746484, EPI_ISL_2746485, EPI_ISL_2746486, EPI_ISL_2746487, EPI_ISL_2746488, EPI_ISL_2746489, EPI_ISL_2746490, EPI_ISL_2746491, EPI_ISL_2746492, EPI_ISL_2746493, EPI_ISL_2746494, EPI_ISL_2746495, EPI_ISL_2746496, EPI_ISL_2746497, EPI_ISL_2746498, EPI_ISL_2746499, EPI_ISL_2746500, EPI_ISL_2746501, EPI_ISL_2746502, EPI_ISL_2746504, EPI_ISL_2746505, EPI_ISL_2746506, EPI_ISL_2746507, EPI_ISL_2746508, EPI_ISL_2746509, EPI_ISL_2746510, EPI_ISL_2746512, EPI_ISL_2746513, EPI_ISL_2746514, EPI_ISL_2746515, EPI_ISL_2746516, EPI_ISL_2746517, EPI_ISL_2746518, EPI_ISL_2746519, EPI_ISL_2746520, EPI_ISL_2746522, EPI_ISL_2746523, EPI_ISL_2746524, EPI_ISL_2746525, EPI_ISL_2746526, EPI_ISL_2746527, EPI_ISL_2746528, EPI_ISL_2746529, EPI_ISL_2746530, EPI_ISL_2746531, EPI_ISL_2746533, EPI_ISL_2746534, EPI_ISL_2746535, EPI_ISL_2746536, EPI_ISL_2746537, EPI_ISL_2746539, EPI_ISL_2746540, EPI_ISL_2746541, EPI_ISL_2746542, EPI_ISL_2746544, EPI_ISL_2746547, EPI_ISL_2746548, EPI_ISL_2746549, EPI_ISL_2746550, EPI_ISL_2746551, EPI_ISL_2746552, EPI_ISL_2746555, EPI_ISL_2746556, EPI_ISL_2746557, EPI_ISL_2746558, EPI_ISL_2746559, EPI_ISL_2746560, EPI_ISL_2746561, EPI_ISL_2746562, EPI_ISL_2746563, EPI_ISL_2746564, EPI_ISL_2746565, EPI_ISL_2746566, EPI_ISL_2746567, EPI_ISL_2746568, EPI_ISL_2746569, EPI_ISL_2746570, EPI_ISL_2746571, EPI_ISL_2746572, EPI_ISL_2746573, EPI_ISL_2746577, EPI_ISL_2746578, EPI_ISL_2746579, EPI_ISL_2746580, EPI_ISL_2746581, EPI_ISL_2746582, EPI_ISL_2746583, EPI_ISL_2746584, EPI_ISL_2746585, EPI_ISL_2746586, EPI_ISL_2746587, EPI_ISL_2746588, EPI_ISL_2746589, EPI_ISL_2746590, EPI_ISL_2746591, EPI_ISL_2746592, EPI_ISL_2746593, EPI_ISL_2746594, EPI_ISL_2746595, EPI_ISL_2746596, EPI_ISL_2746598, EPI_ISL_2746599, EPI_ISL_2746600, EPI_ISL_2746601, EPI_ISL_2746602, EPI_ISL_2746603, EPI_ISL_2746604, EPI_ISL_2746605, EPI_ISL_2746606, EPI_ISL_2746607, EPI_ISL_2746608, EPI_ISL_2746609, EPI_ISL_2746610, EPI_ISL_2746611, EPI_ISL_2746612, EPI_ISL_2746613, EPI_ISL_2746614, EPI_ISL_2746615, EPI_ISL_2746617, EPI_ISL_2746618, EPI_ISL_2746619, EPI_ISL_2746623, EPI_ISL_2746624, EPI_ISL_2746625, EPI_ISL_2746626, EPI_ISL_2746627, EPI_ISL_2746628, EPI_ISL_2746629, EPI_ISL_2746630, EPI_ISL_2746631, EPI_ISL_2746632, EPI_ISL_2746633, EPI_ISL_2746634, EPI_ISL_2746636, EPI_ISL_2746637, EPI_ISL_2746639, EPI_ISL_2746640, EPI_ISL_2746641, EPI_ISL_2746642, EPI_ISL_2746643, EPI_ISL_2746644, EPI_ISL_2746645, EPI_ISL_2746646, EPI_ISL_2746647, EPI_ISL_2746648, EPI_ISL_2746649, EPI_ISL_2746650, EPI_ISL_2746651, EPI_ISL_2746652, EPI_ISL_2746653, EPI_ISL_2746654, EPI_ISL_2746655, EPI_ISL_2746656, EPI_ISL_2746662, EPI_ISL_2746663, EPI_ISL_2746664, EPI_ISL_2746665, EPI_ISL_2746666, EPI_ISL_2746667, EPI_ISL_2746668, EPI_ISL_2746669, EPI_ISL_2746670, EPI_ISL_2746671, EPI_ISL_2746672, EPI_ISL_2746674, EPI_ISL_2746675, EPI_ISL_2746676, EPI_ISL_2746677, EPI_ISL_2746678, EPI_ISL_2746679, EPI_ISL_2746680, EPI_ISL_2746681, EPI_ISL_2746682, EPI_ISL_2746683, EPI_ISL_2746684, EPI_ISL_2746685, EPI_ISL_2746686, EPI_ISL_2746687, EPI_ISL_2746688, EPI_ISL_2746689, EPI_ISL_2746690, EPI_ISL_2746691, EPI_ISL_2746692, EPI_ISL_2746693, EPI_ISL_2746694, EPI_ISL_2746695, EPI_ISL_2746696, EPI_ISL_2746697, EPI_ISL_2746698, EPI_ISL_2746699, EPI_ISL_2746700, EPI_ISL_2746701, EPI_ISL_2746702, EPI_ISL_2746703, EPI_ISL_2746704, EPI_ISL_2746705, EPI_ISL_2746706, EPI_ISL_2746713, EPI_ISL_2746714, EPI_ISL_2746715, EPI_ISL_2746716, EPI_ISL_2746717, EPI_ISL_2746719, EPI_ISL_2746731, EPI_ISL_2746734, EPI_ISL_2746740, EPI_ISL_2746742, EPI_ISL_2746743, EPI_ISL_2746744, EPI_ISL_2746746, EPI_ISL_2746749, EPI_ISL_2746750, EPI_ISL_2746751, EPI_ISL_2746753, EPI_ISL_2746758, EPI_ISL_2746760, EPI_ISL_2746761, EPI_ISL_2746762, EPI_ISL_2746776, EPI_ISL_2746777, EPI_ISL_2746784, EPI_ISL_2746794, EPI_ISL_2746809, EPI_ISL_2746811, EPI_ISL_2746813, EPI_ISL_2746853, EPI_ISL_2746854, EPI_ISL_2746855, EPI_ISL_2746857, EPI_ISL_2746858, EPI_ISL_2746877, EPI_ISL_2746902, EPI_ISL_2746928, EPI_ISL_2746993, EPI_ISL_2746994, EPI_ISL_2746996, EPI_ISL_2747150, EPI_ISL_2747151, EPI_ISL_2747174, EPI_ISL_2747192, EPI_ISL_2747193, EPI_ISL_2747195, EPI_ISL_2747206, EPI_ISL_2747258, EPI_ISL_2747270, EPI_ISL_2747271, EPI_ISL_2747277, EPI_ISL_2747390, EPI_ISL_2747403, EPI_ISL_2747418, EPI_ISL_2749360, EPI_ISL_2749383, EPI_ISL_2749390, EPI_ISL_2817390, EPI_ISL_2818122, EPI_ISL_2818232, EPI_ISL_2818233, EPI_ISL_2818234, EPI_ISL_2818242, EPI_ISL_2904004 | | | |
| see above | Respiratory Virus Unit, Microbiology Services Colindale, Public Health England | COVID-19 Genomics UK (COG-UK) Consortium | PHE Covid Sequencing Team |
| EPI_ISL_2705562, EPI_ISL_2705563, EPI_ISL_2705564, EPI_ISL_2705565, EPI_ISL_2705566, EPI_ISL_2705567, EPI_ISL_2705568, EPI_ISL_2705569, EPI_ISL_2705570, EPI_ISL_2705572, EPI_ISL_2705573, EPI_ISL_2705576, EPI_ISL_2705578, EPI_ISL_2705579, EPI_ISL_2705580, EPI_ISL_2705581, EPI_ISL_2705582, EPI_ISL_2705583, EPI_ISL_2705584, EPI_ISL_2705585, EPI_ISL_2705586, EPI_ISL_2705588, EPI_ISL_2705589, EPI_ISL_2705593, EPI_ISL_2705594, EPI_ISL_2705595, EPI_ISL_2705597, EPI_ISL_2705600, EPI_ISL_2705601, EPI_ISL_2705602, EPI_ISL_2705604, EPI_ISL_2705605, EPI_ISL_2705606, EPI_ISL_2705607, EPI_ISL_2705608, EPI_ISL_2705609, EPI_ISL_2705610, EPI_ISL_2705611, EPI_ISL_2705612, EPI_ISL_2705614, EPI_ISL_2705616, EPI_ISL_2705617, EPI_ISL_2705618, EPI_ISL_2705619, EPI_ISL_2705620, EPI_ISL_2705621, EPI_ISL_2705622, EPI_ISL_2705623, EPI_ISL_2705624, EPI_ISL_2705625, EPI_ISL_2705626, EPI_ISL_2705627, EPI_ISL_2705628, EPI_ISL_2705629, EPI_ISL_2705630, EPI_ISL_2705633, EPI_ISL_2705637, EPI_ISL_2705640, EPI_ISL_2705649, EPI_ISL_2705653, EPI_ISL_2705655, EPI_ISL_2705666, EPI_ISL_2705667, EPI_ISL_2705671, EPI_ISL_2705675, EPI_ISL_2705677, EPI_ISL_2705681, EPI_ISL_2705682, EPI_ISL_2705683, EPI_ISL_2705687, EPI_ISL_2705688, EPI_ISL_2705690, EPI_ISL_2705691, EPI_ISL_2705693, EPI_ISL_2705698, EPI_ISL_2705699, EPI_ISL_2705701, EPI_ISL_2705703, EPI_ISL_2705704, EPI_ISL_2705708, EPI_ISL_2705710, EPI_ISL_2705711, EPI_ISL_2705712, EPI_ISL_2705713, EPI_ISL_2705714, EPI_ISL_2705715, EPI_ISL_2705716, EPI_ISL_2705717, EPI_ISL_2705718, EPI_ISL_2705719, EPI_ISL_2705720, EPI_ISL_2705721, EPI_ISL_2705722, EPI_ISL_2705723, EPI_ISL_2705724, EPI_ISL_2705725, EPI_ISL_2705726, EPI_ISL_2705728, EPI_ISL_2705729, EPI_ISL_2705731, EPI_ISL_2705732, EPI_ISL_2705733, EPI_ISL_2705734, EPI_ISL_2705735, EPI_ISL_2705737, EPI_ISL_2705738, EPI_ISL_2705739, EPI_ISL_2705740, EPI_ISL_2705741, EPI_ISL_2705742, EPI_ISL_2705743, EPI_ISL_2705744, EPI_ISL_2705745, EPI_ISL_2705746, EPI_ISL_2705747, EPI_ISL_2705748, EPI_ISL_2705749, EPI_ISL_2705750, EPI_ISL_2705751, EPI_ISL_2705752, EPI_ISL_2705753, EPI_ISL_2705754, EPI_ISL_2705755, EPI_ISL_2705756, EPI_ISL_2705757, EPI_ISL_2705758, EPI_ISL_2705759, EPI_ISL_2705760, EPI_ISL_2705761, EPI_ISL_2705762, EPI_ISL_2705763, EPI_ISL_2705764, EPI_ISL_2705765, EPI_ISL_2705766, EPI_ISL_2705767, EPI_ISL_2705768, EPI_ISL_2705769, EPI_ISL_2705771, EPI_ISL_2705772, EPI_ISL_2705773, EPI_ISL_2705775, EPI_ISL_2705776, EPI_ISL_2705777, EPI_ISL_2705778, EPI_ISL_2705779, EPI_ISL_2705780, EPI_ISL_2705782, EPI_ISL_2705783, EPI_ISL_2705784, EPI_ISL_2705786, EPI_ISL_2705787, EPI_ISL_2705788, EPI_ISL_2705789, EPI_ISL_2705791, EPI_ISL_2705792, EPI_ISL_2705793, EPI_ISL_2705794, EPI_ISL_2705795, EPI_ISL_2705796, EPI_ISL_2705797, EPI_ISL_2705798, EPI_ISL_2705800, EPI_ISL_2705802, EPI_ISL_2705803, EPI_ISL_2705805, EPI_ISL_2705806, EPI_ISL_2705807, EPI_ISL_2705808, EPI_ISL_2705809, EPI_ISL_2705810, EPI_ISL_2705811, EPI_ISL_2705813, EPI_ISL_2705814, EPI_ISL_2705815, EPI_ISL_2705816, EPI_ISL_2705817, EPI_ISL_2705819, EPI_ISL_2705820, EPI_ISL_2705821, EPI_ISL_2705822, EPI_ISL_2705823, EPI_ISL_2705825, EPI_ISL_2705826, EPI_ISL_2705827, EPI_ISL_2705829, EPI_ISL_2705830, EPI_ISL_2705831, EPI_ISL_2705832, EPI_ISL_2705834, EPI_ISL_2705835, EPI_ISL_2705836, EPI_ISL_2705837, EPI_ISL_2705838, EPI_ISL_2705839, EPI_ISL_2705840, EPI_ISL_2705841, EPI_ISL_2705843, EPI_ISL_2705844, EPI_ISL_2705845, EPI_ISL_2705847, EPI_ISL_2705850, EPI_ISL_2901205, EPI_ISL_2901213, EPI_ISL_2901234, EPI_ISL_2901249, EPI_ISL_2901251, EPI_ISL_2901255, EPI_ISL_2901256, EPI_ISL_2901262, EPI_ISL_2901278, EPI_ISL_2901288, EPI_ISL_2901292, EPI_ISL_2901294, EPI_ISL_2901301, EPI_ISL_2901319 | | | |
| see above | University College London, Great Ormond Street Hospital for Children NHS Foundation Trust, Imperial College Healthcare NHS Trust | COVID-19 Genomics UK (COG-UK) Consortium | Alison Holmes; Charlotte Williams; Helena Tutill; Jacqueline Findlay; James Price; Judith Breuer; Julianne Brown; Kathryn Harris; Leysa Forrest; Marius Cotic; Mark Kristiansen; Nadua Bayzid; Paola Niola; Paola Resende Silva; Patricia Dyal; Paul Randell; Rachel Williams; Samuel Weeks; Sergi Castellano; Sunando Roy; Tony Brooks; Yasmin Panchbhaya |
| EPI_ISL_2722101, EPI_ISL_2722103, EPI_ISL_2722104, EPI_ISL_2815383, EPI_ISL_2815384, EPI_ISL_2815387, EPI_ISL_2815388, EPI_ISL_2815392, EPI_ISL_2815394, EPI_ISL_2815395, EPI_ISL_2815396, EPI_ISL_2815397, EPI_ISL_2815541, EPI_ISL_2815542, EPI_ISL_2815543, EPI_ISL_2815544, EPI_ISL_2815545, EPI_ISL_2815546, EPI_ISL_2815547, EPI_ISL_2815548 | | | |
| see above | University of Birmingham | COVID-19 Genomics UK (COG-UK) Consortium | Alex Richter; Andrew Bosworth; Andrew Bosworth. Queen Elizabeth Hospital: Anna Casey; Andrew D Beggs PHE Heartlands Lab: Husam Osman; Charlotte Poxon; Institute of Microbiology; Joanne Stockton; Josh Quick; Kasun Wanigasooriya; Mike Kidd; Nicholas Loman. University of Birmingham Testing Laboratory: Celina M Whalley; Oliver Pickles; Radoslaw Poplawski; Samuel Nicholls; University of Birmingham: Claire McMurray; Will Rowe |
| EPI_ISL_2704907, EPI_ISL_2704908, EPI_ISL_2704909, EPI_ISL_2704910, EPI_ISL_2704911, EPI_ISL_2704912, EPI_ISL_2704913, EPI_ISL_2704915, EPI_ISL_2704916, EPI_ISL_2704924, EPI_ISL_2704925, EPI_ISL_2704926, EPI_ISL_2704927, EPI_ISL_2704933, EPI_ISL_2704934, EPI_ISL_2704935, EPI_ISL_2704936, EPI_ISL_2734322, EPI_ISL_2734323, EPI_ISL_2734324, EPI_ISL_2734325, EPI_ISL_2734327, EPI_ISL_2734328, EPI_ISL_2734329, EPI_ISL_2734331, EPI_ISL_2734335, EPI_ISL_2734338, EPI_ISL_2734339, EPI_ISL_2734340, EPI_ISL_2734341, EPI_ISL_2734350, EPI_ISL_2734351, EPI_ISL_2734352, EPI_ISL_2734353, EPI_ISL_2734355, EPI_ISL_2734359, EPI_ISL_2734391, EPI_ISL_2734392, EPI_ISL_2734393, EPI_ISL_2734415, EPI_ISL_2734429, EPI_ISL_2815986, EPI_ISL_2816002 | | | |
| see above | Virology Department, Royal Infirmary of Edinburgh, NHS Lothian / School of Biological Sciences, University of Edinburgh | COVID-19 Genomics UK (COG-UK) Consortium | Colquhoun R; Cotton S; Dewar R; Gallagher A; Hill V; Jackson B; Maloney D; McCrone JT; McHugh M; O'Toole Á; Rambaut A; Rooke S; Scher E; Templeton K; Yu X |
| EPI_ISL_2708501, EPI_ISL_2708554, EPI_ISL_2708558, EPI_ISL_2708562, EPI_ISL_2708572, EPI_ISL_2708588, EPI_ISL_2708615, EPI_ISL_2708667, EPI_ISL_2708679, EPI_ISL_2708707, EPI_ISL_2708756 | | | |
| see above | Virology Department, Sheffield Teaching Hospitals NHS Foundation Trust/Department of Infection, Immunity and Cardiovascular Disease, The Medical School, University of Sheffield | COVID-19 Genomics UK (COG-UK) Consortium | Adri Angyal; Alex Keeley; Benjamin Lindsey; Cariad Evans; Danielle Groves; Dave Partridge; Katie Johnson; Laura Carrilero; Luke Green; Matthew Parker; Matthew Wyles; Mehmet Yavuz; Mohammad Raza; Nikki Smith; Paul Parsons; Rachel Tucker; Rebecca Brown; Thushan de Silva |
| EPI_ISL_2704748, EPI_ISL_2704750, EPI_ISL_2704751, EPI_ISL_2704753, EPI_ISL_2704759, EPI_ISL_2704760, EPI_ISL_2704763, EPI_ISL_2704764, EPI_ISL_2704765, EPI_ISL_2704767, EPI_ISL_2704768, EPI_ISL_2704769, EPI_ISL_2704771, EPI_ISL_2704772, EPI_ISL_2704773, EPI_ISL_2704774, EPI_ISL_2704775, EPI_ISL_2704779, EPI_ISL_2704782, EPI_ISL_2704783, EPI_ISL_2704785, EPI_ISL_2704786, EPI_ISL_2704787, EPI_ISL_2704791, EPI_ISL_2704792, EPI_ISL_2704795, EPI_ISL_2704797, EPI_ISL_2704798, EPI_ISL_2704801, EPI_ISL_2704804, EPI_ISL_2704805, EPI_ISL_2704811, EPI_ISL_2704812, EPI_ISL_2704814, EPI_ISL_2704818, EPI_ISL_2704819, EPI_ISL_2704821, EPI_ISL_2815897, EPI_ISL_2815904, EPI_ISL_2815905, EPI_ISL_2815906, EPI_ISL_2815907, EPI_ISL_2815908, EPI_ISL_2815909, EPI_ISL_2815910, EPI_ISL_2815911, EPI_ISL_2815912, EPI_ISL_2815917, EPI_ISL_2815918, EPI_ISL_2815919, EPI_ISL_2815920, EPI_ISL_2815921, EPI_ISL_2815922, EPI_ISL_2815928, EPI_ISL_2815929, EPI_ISL_2815932 | | | |
| see above | West of Scotland Specialist Virology Centre, NHSGGC / MRC-University of Glasgow Centre for Virus Research | COVID-19 Genomics UK (COG-UK) Consortium | Alasdair MacLean; Alice Broos; Ana da Silva Filipe; Antonia Ho; Daniel Mair; David L Robertson; Emma Thomson; Guy Mollett; Ioulia Tsatsani; James Shepherd; Jenna Nichols; Joseph Hughes; Kathy Li; Kathy Smollett; Kyriaki Nomikou; Lily Tong; Matthew Holden; Natasha Johnson; Rachel Blacow; Richard Orton; Rory Gunson; Sarah McDonald; Sharif Shaaban; Sreenu Vattipally; Stephen Carmichael |

We gratefully acknowledge the following Authors from the Originating laboratories responsible for obtaining the specimens, as well as the Submitting laboratories where the genome data were generated and shared via GISAID, on which this research is based.

All Submitters of data may be contacted directly via www.gisaid.org

Authors are sorted alphabetically.

| Accession ID | Originating Laboratory | Submitting Laboratory | Authors |
|---|---|---|---|
| EPI_ISL_2686195, EPI_ISL_2686196, EPI_ISL_2686198, EPI_ISL_2686201, EPI_ISL_2686202, EPI_ISL_2795922, EPI_ISL_2795943, EPI_ISL_2795944, EPI_ISL_2795946, EPI_ISL_2795947, EPI_ISL_2795948, EPI_ISL_2795949, EPI_ISL_2795951, EPI_ISL_2795952 | | | |
| see above | Amazon Dx, AUV2 Laboratory | Amazon Dx, AUV2 Laboratory | Luke Meredith |
| EPI_ISL_2708168, EPI_ISL_2708170, EPI_ISL_2708171, EPI_ISL_2708172, EPI_ISL_2708173, EPI_ISL_2708178, EPI_ISL_2708179, EPI_ISL_2708180, EPI_ISL_2708181, EPI_ISL_2708183, EPI_ISL_2708185, EPI_ISL_2708186, EPI_ISL_2708187, EPI_ISL_2708188, EPI_ISL_2708189, EPI_ISL_2708190, EPI_ISL_2708191, EPI_ISL_2708192, EPI_ISL_2708193, EPI_ISL_2708195, EPI_ISL_2708196, EPI_ISL_2708197, EPI_ISL_2708199, EPI_ISL_2708201, EPI_ISL_2708202, EPI_ISL_2708203, EPI_ISL_2708204, EPI_ISL_2708205, EPI_ISL_2708206, EPI_ISL_2708207, EPI_ISL_2708208, EPI_ISL_2708209, EPI_ISL_2708210, EPI_ISL_2708211, EPI_ISL_2708212, EPI_ISL_2708213, EPI_ISL_2708214, EPI_ISL_2708215, EPI_ISL_2708216, EPI_ISL_2708217, EPI_ISL_2708218, EPI_ISL_2708219, EPI_ISL_2708220, EPI_ISL_2708221, EPI_ISL_2708222, EPI_ISL_2708223, EPI_ISL_2708224, EPI_ISL_2708225, EPI_ISL_2708226, EPI_ISL_2708227, EPI_ISL_2708228, EPI_ISL_2708229, EPI_ISL_2708230, EPI_ISL_2708231, EPI_ISL_2708232, EPI_ISL_2708233, EPI_ISL_2708234, EPI_ISL_2708235, EPI_ISL_2708236, EPI_ISL_2708237, EPI_ISL_2708238, EPI_ISL_2708239, EPI_ISL_2708240, EPI_ISL_2708242, EPI_ISL_2708243, EPI_ISL_2708244, EPI_ISL_2708245, EPI_ISL_2708246, EPI_ISL_2708247, EPI_ISL_2708248, EPI_ISL_2708249, EPI_ISL_2708250, EPI_ISL_2708251, EPI_ISL_2708252, EPI_ISL_2708253, EPI_ISL_2708254, EPI_ISL_2708255, EPI_ISL_2708256, EPI_ISL_2708257, EPI_ISL_2708258, EPI_ISL_2708259, EPI_ISL_2708260, EPI_ISL_2708261, EPI_ISL_2708262, EPI_ISL_2708263, EPI_ISL_2708264, EPI_ISL_2708265, EPI_ISL_2708266, EPI_ISL_2708267, EPI_ISL_2708268, EPI_ISL_2708269, EPI_ISL_2708270, EPI_ISL_2708271, EPI_ISL_2708272, EPI_ISL_2708273, EPI_ISL_2708274, EPI_ISL_2708275, EPI_ISL_2708276, EPI_ISL_2708277, EPI_ISL_2708278, EPI_ISL_2708279, EPI_ISL_2708280, EPI_ISL_2708281, EPI_ISL_2708282, EPI_ISL_2708283, EPI_ISL_2708284, EPI_ISL_2708285, EPI_ISL_2708286, EPI_ISL_2708287, EPI_ISL_2708288, EPI_ISL_2708289, EPI_ISL_2708300, EPI_ISL_2708301, EPI_ISL_2708303, EPI_ISL_2708304, EPI_ISL_2708305, EPI_ISL_2708306, EPI_ISL_2708307, EPI_ISL_2708308, EPI_ISL_2708309, EPI_ISL_2708310, EPI_ISL_2708311, EPI_ISL_2708312, EPI_ISL_2708313, EPI_ISL_2708314, EPI_ISL_2708315, EPI_ISL_2708316, EPI_ISL_2708317, EPI_ISL_2708318, EPI_ISL_2708319, EPI_ISL_2708320, EPI_ISL_2708322, EPI_ISL_2708323, EPI_ISL_2708324, EPI_ISL_2708325, EPI_ISL_2708326, EPI_ISL_2708327, EPI_ISL_2708328, EPI_ISL_2708329, EPI_ISL_2708330, EPI_ISL_2708331, EPI_ISL_2708332, EPI_ISL_2708333, EPI_ISL_2708334, EPI_ISL_2708335, EPI_ISL_2708336, EPI_ISL_2708337, EPI_ISL_2708338, EPI_ISL_2708339, EPI_ISL_2708340, EPI_ISL_2708341, EPI_ISL_2708342, EPI_ISL_2708343, EPI_ISL_2708344, EPI_ISL_2708345, EPI_ISL_2708350, EPI_ISL_2708352, EPI_ISL_2708353, EPI_ISL_2708354, EPI_ISL_2708355, EPI_ISL_2708357, EPI_ISL_2708358, EPI_ISL_2708359, EPI_ISL_2708365, EPI_ISL_2708366, EPI_ISL_2708367, EPI_ISL_2708368, EPI_ISL_2708369, EPI_ISL_2708370, EPI_ISL_2708373, EPI_ISL_2708374, EPI_ISL_2708375, EPI_ISL_2708379, EPI_ISL_2708380, EPI_ISL_2708381, EPI_ISL_2708405, EPI_ISL_2708407, EPI_ISL_2708413, EPI_ISL_2708414, EPI_ISL_2708415, EPI_ISL_2708416 | | | |
| see above | Centre for Enzyme Innovation, University of Portsmouth / Translational Research Laboratory, Portsmouth Hospitals NHS Trust / Hampshire Hospitals NHS Foundation Trust / Bournemouth University / University Hospitals Dorset NHS Foundation Trust / Brighton and Sussex University Hospitals NHS Trust / Dartford and Gravesham NHS Trust / Isle of Wight NHS Trust / Maidstone and Tunbridge Wells NHS Trust / University Hospital Southampton NHS Foundation Trust | COVID-19 Genomics UK (COG-UK) Consortium | Adhyana Mahanama; Allyson Lloyd; Andrew Butt; Angela Beckett; Anibolina Castigador; Anna Mantzaouratou; Anoop Chauhan; Ben Macklin; Benjamin Cogger; Buddhini Samaraweera; Cassandra Malone; Christopher Fearn; Claire Thomas; Dorian Crudgington; Eleri Wilson-Davies; Emanuela Pelosi; Emily Macnaughton; Helen Umpleby; Helen Wheeler; Jacqui Prieto; Jonnie Seymour; Karen Withell; Kate Cook; Katie Loveson; Kelly Bicknell; Kevin Tucker; Kordo Saeed; Liz Sheridan; Magdalena Barrow; Matthew Harvey; Mohammed Hassan-Ibrahim; Nathan Moore; Nick Cortes; Rachael Jeremiah; Robert Impey; Salman Goudarzi; Samirakhon Raupova; Samuel Robson; Sarah Buchan; Sarah Jeremiah; Sarah Wyllie; Scott Elliott; Sharon Glaysher; Siona Silvieri; Stephen Aplin; Stephen Kidd; Thea Sass |
| EPI_ISL_2722400, EPI_ISL_2722401, EPI_ISL_2722402, EPI_ISL_2722403, EPI_ISL_2722404, EPI_ISL_2722405, EPI_ISL_2722406, EPI_ISL_2722407, EPI_ISL_2722408, EPI_ISL_2722409, EPI_ISL_2722410, EPI_ISL_2722411, EPI_ISL_2722412, EPI_ISL_2722413, EPI_ISL_2722414, EPI_ISL_2722415, EPI_ISL_2722416, EPI_ISL_2722417, EPI_ISL_2722418, EPI_ISL_2722419, EPI_ISL_2722421, EPI_ISL_2722422, EPI_ISL_2722423, EPI_ISL_2722424, EPI_ISL_2722425, EPI_ISL_2722426, EPI_ISL_2722427, EPI_ISL_2722428, EPI_ISL_2722429, EPI_ISL_2722430, EPI_ISL_2722431, EPI_ISL_2722432, EPI_ISL_2722433, EPI_ISL_2722434, EPI_ISL_2722435, EPI_ISL_2722437, EPI_ISL_2722438, EPI_ISL_2722439, EPI_ISL_2722440, EPI_ISL_2722441, EPI_ISL_2722442, EPI_ISL_2722443, EPI_ISL_2722444, EPI_ISL_2722446, EPI_ISL_2722447, EPI_ISL_2722449, EPI_ISL_2722450, EPI_ISL_2722451, EPI_ISL_2722452, EPI_ISL_2722453, EPI_ISL_2722455, EPI_ISL_2722456, EPI_ISL_2722457, EPI_ISL_2722458, EPI_ISL_2722459, EPI_ISL_2722460, EPI_ISL_2722461, EPI_ISL_2722462, EPI_ISL_2722463, EPI_ISL_2722464, EPI_ISL_2722465, EPI_ISL_2722466, EPI_ISL_2722467, EPI_ISL_2722468, EPI_ISL_2722469, EPI_ISL_2722471, EPI_ISL_2722472, EPI_ISL_2722473, EPI_ISL_2722474, EPI_ISL_2722475, EPI_ISL_2722476, EPI_ISL_2722477, EPI_ISL_2722478, EPI_ISL_2722479, EPI_ISL_2722480, EPI_ISL_2722481, EPI_ISL_2722482, EPI_ISL_2722483, EPI_ISL_2722484, EPI_ISL_2722485, EPI_ISL_2722486, EPI_ISL_2722487, EPI_ISL_2722488, EPI_ISL_2722489, EPI_ISL_2722490, EPI_ISL_2722492, EPI_ISL_2722493, EPI_ISL_2722494, EPI_ISL_2722495, EPI_ISL_2722496, EPI_ISL_2722497, EPI_ISL_2722498, EPI_ISL_2722499, EPI_ISL_2722500, EPI_ISL_2722501, EPI_ISL_2722502, EPI_ISL_2722503, EPI_ISL_2722505, EPI_ISL_2722506, EPI_ISL_2722507, EPI_ISL_2722508, EPI_ISL_2722509, EPI_ISL_2722510, EPI_ISL_2722511, EPI_ISL_2722512, EPI_ISL_2722513, EPI_ISL_2722514, EPI_ISL_2722515, EPI_ISL_2722516, EPI_ISL_2722517, EPI_ISL_2722518, EPI_ISL_2722520, EPI_ISL_2722521, EPI_ISL_2722522, EPI_ISL_2722524, EPI_ISL_2722525, EPI_ISL_2722526, EPI_ISL_2722527, EPI_ISL_2722528, EPI_ISL_2722529, EPI_ISL_2722531, EPI_ISL_2722532, EPI_ISL_2722533, EPI_ISL_2722534, EPI_ISL_2722535, EPI_ISL_2722537, EPI_ISL_2722538, EPI_ISL_2722539, EPI_ISL_2722540, EPI_ISL_2722541, EPI_ISL_2722543, EPI_ISL_2722544, EPI_ISL_2722545, EPI_ISL_2722546, EPI_ISL_2722547, EPI_ISL_2722548, EPI_ISL_2722549, EPI_ISL_2722557, EPI_ISL_2722571, EPI_ISL_2722578, EPI_ISL_2722593, EPI_ISL_2722600, EPI_ISL_2722601, EPI_ISL_2722607, EPI_ISL_2722613, EPI_ISL_2733921, EPI_ISL_2733922, EPI_ISL_2733923, EPI_ISL_2733924, EPI_ISL_2733926, EPI_ISL_2733928, EPI_ISL_2733929, EPI_ISL_2733930, EPI_ISL_2733931, EPI_ISL_2733932, EPI_ISL_2733933, EPI_ISL_2733934, EPI_ISL_2733935, EPI_ISL_2733936, EPI_ISL_2733938, EPI_ISL_2733939, EPI_ISL_2733940, EPI_ISL_2733941, EPI_ISL_2733942, EPI_ISL_2733943, EPI_ISL_2733944, EPI_ISL_2733945, EPI_ISL_2733946, EPI_ISL_2733947, EPI_ISL_2733948, EPI_ISL_2733949, EPI_ISL_2733950, EPI_ISL_2733951, EPI_ISL_2733952, EPI_ISL_2733953, EPI_ISL_2733954, EPI_ISL_2733955, EPI_ISL_2733956, EPI_ISL_2733957, EPI_ISL_2733958, EPI_ISL_2733959, EPI_ISL_2733960, EPI_ISL_2733961, EPI_ISL_2733962, EPI_ISL_2733963, EPI_ISL_2733964, EPI_ISL_2733965, EPI_ISL_2733966, EPI_ISL_2733967, EPI_ISL_2733968, EPI_ISL_2733969, EPI_ISL_2733970, EPI_ISL_2733971, EPI_ISL_2733972, EPI_ISL_2733973, EPI_ISL_2733974, EPI_ISL_2733975, EPI_ISL_2733976, EPI_ISL_2733977, EPI_ISL_2733978, EPI_ISL_2733979, EPI_ISL_2733980, EPI_ISL_2733981, EPI_ISL_2733983, EPI_ISL_2733984, EPI_ISL_2733985, EPI_ISL_2733986, EPI_ISL_2733987, EPI_ISL_2733989, EPI_ISL_2733990, EPI_ISL_2733991, EPI_ISL_2733992, EPI_ISL_2733993 | | | |
| see above | Department of Pathology, University of Cambridge | COVID-19 Genomics UK (COG-UK) Consortium | Aminu S. Jahun; Ian Goodfellow; Iliana Georgana; Martin D. Curran; Myra Hosmillo; Rhys Izuagbe; Surendra Parmar; William L. Hamilton; Yasmin Chaudhry |
| EPI_ISL_2682288, EPI_ISL_2682291, EPI_ISL_2682292, EPI_ISL_2682296, EPI_ISL_2682313, EPI_ISL_2682320, EPI_ISL_2682325, EPI_ISL_2682334, EPI_ISL_2682338, EPI_ISL_2682350, EPI_ISL_2682351, EPI_ISL_2682354, EPI_ISL_2682357, EPI_ISL_2682371, EPI_ISL_2682395, EPI_ISL_2682400, EPI_ISL_2682416, EPI_ISL_2682442, EPI_ISL_2682445, EPI_ISL_2682455, EPI_ISL_2682457, EPI_ISL_2682467, EPI_ISL_2682471, EPI_ISL_2682473, EPI_ISL_2682490, EPI_ISL_2682503, EPI_ISL_2682523, EPI_ISL_2682524, EPI_ISL_2682526, EPI_ISL_2682537, EPI_ISL_2682538, EPI_ISL_2682545, EPI_ISL_2682546, EPI_ISL_2682549, EPI_ISL_2682558, EPI_ISL_2682569, EPI_ISL_2682594, EPI_ISL_2682595, EPI_ISL_2682623, EPI_ISL_2682630, EPI_ISL_2682637, EPI_ISL_2682646, EPI_ISL_2682648, EPI_ISL_2682651, EPI_ISL_2704188, EPI_ISL_2704190, EPI_ISL_2704191, EPI_ISL_2704194, EPI_ISL_2704196, EPI_ISL_2704198, EPI_ISL_2704200, EPI_ISL_2704204, EPI_ISL_2704214, EPI_ISL_2704215, EPI_ISL_2704218, EPI_ISL_2704222, EPI_ISL_2704224, EPI_ISL_2704225, EPI_ISL_2704227, EPI_ISL_2704229, EPI_ISL_2704232, EPI_ISL_2704234, EPI_ISL_2704237, EPI_ISL_2704242, EPI_ISL_2704244, EPI_ISL_2704249, EPI_ISL_2704255, EPI_ISL_2704257, EPI_ISL_2704259, EPI_ISL_2704266, EPI_ISL_2704270, EPI_ISL_2704271, EPI_ISL_2704274, EPI_ISL_2704275, EPI_ISL_2704277, EPI_ISL_2704288, EPI_ISL_2704289, EPI_ISL_2704291, EPI_ISL_2704292, EPI_ISL_2704301, EPI_ISL_2704307, EPI_ISL_2704310, EPI_ISL_2704312, EPI_ISL_2704314, EPI_ISL_2704316, EPI_ISL_2704317, EPI_ISL_2704318, EPI_ISL_2704319, EPI_ISL_2704333, EPI_ISL_2704336, EPI_ISL_2704337, EPI_ISL_2704338, EPI_ISL_2704340, EPI_ISL_2704346, EPI_ISL_2704349, EPI_ISL_2704356, EPI_ISL_2704359, EPI_ISL_2704361, EPI_ISL_2704371, EPI_ISL_2704378, EPI_ISL_2704379, EPI_ISL_2704385, EPI_ISL_2704389, EPI_ISL_2704390, EPI_ISL_2704392, EPI_ISL_2704396, EPI_ISL_2704400, EPI_ISL_2704402, EPI_ISL_2704404, EPI_ISL_2704412, EPI_ISL_2704413, EPI_ISL_2704414, EPI_ISL_2704415, EPI_ISL_2704419, EPI_ISL_2704421, EPI_ISL_2704445, EPI_ISL_2704449, EPI_ISL_2704451, EPI_ISL_2704457, EPI_ISL_2704466, EPI_ISL_2704467, EPI_ISL_2704469, EPI_ISL_2704470, EPI_ISL_2704478, EPI_ISL_2704480, EPI_ISL_2704481, EPI_ISL_2704482, EPI_ISL_2704484, EPI_ISL_2704492, EPI_ISL_2704494, EPI_ISL_2704496, EPI_ISL_2704498, EPI_ISL_2704511, EPI_ISL_2704513, EPI_ISL_2704527, EPI_ISL_2704532, EPI_ISL_2781744 | | | |
| see above | Health Services Laboratories | Wellcome Sanger Institute for the COVID-19 Genomics UK (COG-UK) Consortium | Cordelia Langford; David K. Jackson; Dominic Kwiatkowski; Ewan Harrison; Health Services Laboratories and Alex Alderton; Ian Johnston; Jeffrey Barrett; John Sillitoe on behalf of the Wellcome Sanger Institute COVID-19 Surveillance Team; Roberto Amato; Sonia Goncalves |
| EPI_ISL_2700577, EPI_ISL_2700578, EPI_ISL_2700584, EPI_ISL_2700589, EPI_ISL_2700592, EPI_ISL_2700593, EPI_ISL_2700594, EPI_ISL_2700597, EPI_ISL_2700603, EPI_ISL_2700604, EPI_ISL_2700605, EPI_ISL_2700607, EPI_ISL_2700609, EPI_ISL_2700610, EPI_ISL_2700614, EPI_ISL_2700618, EPI_ISL_2700619, EPI_ISL_2700623, EPI_ISL_2700625, EPI_ISL_2700631, EPI_ISL_2700633, EPI_ISL_2700635, EPI_ISL_2700636, EPI_ISL_2700642, EPI_ISL_2700643, EPI_ISL_2700644, EPI_ISL_2700646, EPI_ISL_2700650, EPI_ISL_2700651, EPI_ISL_2700654, EPI_ISL_2700657, EPI_ISL_2700660, EPI_ISL_2700661, EPI_ISL_2700662, EPI_ISL_2700664, EPI_ISL_2700665, EPI_ISL_2700666, EPI_ISL_2700668, EPI_ISL_2700670, EPI_ISL_2700671, EPI_ISL_2700672, EPI_ISL_2700680, EPI_ISL_2700681, EPI_ISL_2700687, EPI_ISL_2700688, EPI_ISL_2700690, EPI_ISL_2700691, EPI_ISL_2700694, EPI_ISL_2700698, EPI_ISL_2700699, EPI_ISL_2700702, EPI_ISL_2700706, EPI_ISL_2700707, EPI_ISL_2700708, EPI_ISL_2700709, EPI_ISL_2700713, EPI_ISL_2700716, EPI_ISL_2700718, EPI_ISL_2700726, EPI_ISL_2700728, EPI_ISL_2700733, EPI_ISL_2700735, EPI_ISL_2700736, EPI_ISL_2700738, EPI_ISL_2700741, EPI_ISL_2700745, EPI_ISL_2700749, EPI_ISL_2700750, EPI_ISL_2700753, EPI_ISL_2700754, EPI_ISL_2700756, EPI_ISL_2700757, EPI_ISL_2700761, EPI_ISL_2700762, EPI_ISL_2700763, EPI_ISL_2700764, EPI_ISL_2700767, EPI_ISL_2700768, EPI_ISL_2700770, EPI_ISL_2700771, EPI_ISL_2700773, EPI_ISL_2700774, EPI_ISL_2700775, EPI_ISL_2700778, EPI_ISL_2700779, EPI_ISL_2700780, EPI_ISL_2700782, EPI_ISL_2700783, EPI_ISL_2700787, EPI_ISL_2700790, EPI_ISL_2700794, EPI_ISL_2700796, EPI_ISL_2700799, EPI_ISL_2700801, EPI_ISL_2700804, EPI_ISL_2700811, EPI_ISL_2700814, EPI_ISL_2700815, EPI_ISL_2700821, EPI_ISL_2700822, EPI_ISL_2700824, EPI_ISL_2700825, EPI_ISL_2700827, EPI_ISL_2700828, EPI_ISL_2700829, EPI_ISL_2700831, EPI_ISL_2700832, EPI_ISL_2700834, EPI_ISL_2700844, EPI_ISL_2700845, EPI_ISL_2700846, EPI_ISL_2700847, EPI_ISL_2700848, EPI_ISL_2700849, EPI_ISL_2700850, EPI_ISL_2700853, EPI_ISL_2700854, EPI_ISL_2700859, EPI_ISL_2700862, EPI_ISL_2700863, EPI_ISL_2700866, EPI_ISL_2700869, EPI_ISL_2700870, EPI_ISL_2700871, EPI_ISL_2700874, EPI_ISL_2700881, EPI_ISL_2700883, EPI_ISL_2700889, EPI_ISL_2700890, EPI_ISL_2700893, EPI_ISL_2700897, EPI_ISL_2700899, EPI_ISL_2700902, EPI_ISL_2700903, EPI_ISL_2700904, EPI_ISL_2700906, EPI_ISL_2700914, EPI_ISL_2700917, EPI_ISL_2700920, EPI_ISL_2700922, EPI_ISL_2700923, EPI_ISL_2700924, EPI_ISL_2700926, EPI_ISL_2700930, EPI_ISL_2700932, EPI_ISL_2700934, EPI_ISL_2700935, EPI_ISL_2700938, EPI_ISL_2702054, EPI_ISL_2702087, EPI_ISL_2702091, EPI_ISL_2702112, EPI_ISL_2702134, EPI_ISL_2702139, EPI_ISL_2702183, EPI_ISL_2702211, EPI_ISL_2702228, EPI_ISL_2702236, EPI_ISL_2702298, EPI_ISL_2702305, EPI_ISL_2702347, EPI_ISL_2702371, EPI_ISL_2702373, EPI_ISL_2702866, EPI_ISL_2702910, EPI_ISL_2702923, EPI_ISL_2702927, EPI_ISL_2702968, EPI_ISL_2702989, EPI_ISL_2703023, EPI_ISL_2703031, EPI_ISL_2703079, EPI_ISL_2703134, EPI_ISL_2716977, EPI_ISL_2716979, EPI_ISL_2716985, EPI_ISL_2716993, EPI_ISL_2716997, EPI_ISL_2717011, EPI_ISL_2717026, EPI_ISL_2717049, EPI_ISL_2717052, EPI_ISL_2717055, EPI_ISL_2717056, EPI_ISL_2717070, EPI_ISL_2717073, EPI_ISL_2717085, EPI_ISL_2717087, EPI_ISL_2717095, EPI_ISL_2717105, EPI_ISL_2717108, EPI_ISL_2717113, EPI_ISL_2717114, EPI_ISL_2717125, EPI_ISL_2717126, EPI_ISL_2717130, EPI_ISL_2717134, EPI_ISL_2717143, EPI_ISL_2717153, EPI_ISL_2717154, EPI_ISL_2717182, EPI_ISL_2717184, EPI_ISL_2717191, EPI_ISL_2717199, EPI_ISL_2717203, EPI_ISL_2717204, EPI_ISL_2717208, EPI_ISL_2717209, EPI_ISL_2717216, EPI_ISL_2717219, EPI_ISL_2717221, EPI_ISL_2717222, EPI_ISL_2717227, EPI_ISL_2717231, EPI_ISL_2717239, EPI_ISL_2717244, EPI_ISL_2717251, EPI_ISL_2717265, EPI_ISL_2717269, EPI_ISL_2717281, EPI_ISL_2717283, EPI_ISL_2717287, EPI_ISL_2717302, EPI_ISL_2717310, EPI_ISL_2717311, EPI_ISL_2717313, EPI_ISL_2717314, EPI_ISL_2717316, EPI_ISL_2717317, EPI_ISL_2717325, EPI_ISL_2717328, EPI_ISL_2717329, EPI_ISL_2717330, EPI_ISL_2717332, EPI_ISL_2717333, EPI_ISL_2717350, EPI_ISL_2717363, EPI_ISL_2717373, EPI_ISL_2717374, EPI_ISL_2717384, EPI_ISL_2717392, EPI_ISL_2717395, EPI_ISL_2717401, EPI_ISL_2717402, EPI_ISL_2717404, EPI_ISL_2717405, EPI_ISL_2717411, EPI_ISL_2717415, EPI_ISL_2717416, EPI_ISL_2717420, EPI_ISL_2717421, EPI_ISL_2717422, EPI_ISL_2717426, EPI_ISL_2717434, EPI_ISL_2717435, EPI_ISL_2717467, EPI_ISL_2717472, EPI_ISL_2717475, EPI_ISL_2717478, EPI_ISL_2717480, EPI_ISL_2717486, EPI_ISL_2717488, EPI_ISL_2717495, EPI_ISL_2717498, EPI_ISL_2717500, EPI_ISL_2717504, EPI_ISL_2717505, EPI_ISL_2717508, EPI_ISL_2717524, EPI_ISL_2717532, EPI_ISL_2717535, EPI_ISL_2717539, EPI_ISL_2717559, EPI_ISL_2717565, EPI_ISL_2717577, EPI_ISL_2717578, EPI_ISL_2717579, EPI_ISL_2717581, EPI_ISL_2717583, EPI_ISL_2717585, EPI_ISL_2717587, EPI_ISL_2717588, EPI_ISL_2717589, EPI_ISL_2717590, EPI_ISL_2717592, EPI_ISL_2717594, EPI_ISL_2717597, EPI_ISL_2717599, EPI_ISL_2717600, EPI_ISL_2717602, EPI_ISL_2717608, EPI_ISL_2717609, EPI_ISL_2717610, EPI_ISL_2717611, EPI_ISL_2717612, EPI_ISL_2717613, EPI_ISL_2717615, EPI_ISL_2717616, EPI_ISL_2717617, EPI_ISL_2717620, EPI_ISL_2717623, EPI_ISL_2717624, EPI_ISL_2717626, EPI_ISL_2717628, EPI_ISL_2717630, EPI_ISL_2717633, EPI_ISL_2717634, EPI_ISL_2717635, EPI_ISL_2717636, EPI_ISL_2717637, EPI_ISL_2717638, EPI_ISL_2717639, EPI_ISL_2717640, EPI_ISL_2717641, EPI_ISL_2717646, EPI_ISL_2717647, EPI_ISL_2717648, EPI_ISL_2717649, EPI_ISL_2717651, EPI_ISL_2717653, EPI_ISL_2717656, EPI_ISL_2717661, EPI_ISL_2717662, EPI_ISL_2717663, EPI_ISL_2717664, EPI_ISL_2717665, EPI_ISL_2717666, EPI_ISL_2717668, EPI_ISL_2717669, EPI_ISL_2717670, EPI_ISL_2717671, EPI_ISL_2717673, EPI_ISL_2717674, EPI_ISL_2717675, EPI_ISL_2717677, EPI_ISL_2717679, EPI_ISL_2717680, EPI_ISL_2717681, EPI_ISL_2717682, EPI_ISL_2717684, EPI_ISL_2717685, EPI_ISL_2717687, EPI_ISL_2717688, EPI_ISL_2717689, EPI_ISL_2717690, EPI_ISL_2717691, EPI_ISL_2717695, EPI_ISL_2717697, EPI_ISL_2717698, EPI_ISL_2717699, EPI_ISL_2717700, EPI_ISL_2717701, EPI_ISL_2717704, EPI_ISL_2717705, EPI_ISL_2717706, EPI_ISL_2717707, EPI_ISL_2717708, EPI_ISL_2717710, EPI_ISL_2717712, EPI_ISL_2717713, EPI_ISL_2717715, EPI_ISL_2717716, EPI_ISL_2717717, EPI_ISL_2717719, EPI_ISL_2717723, EPI_ISL_2717725, EPI_ISL_2717726, EPI_ISL_2717728, EPI_ISL_2717729, EPI_ISL_2717730, EPI_ISL_2717732, EPI_ISL_2717734, EPI_ISL_2717735, EPI_ISL_2717736, EPI_ISL_2717739, EPI_ISL_2717740, EPI_ISL_2717741, EPI_ISL_2717742, EPI_ISL_2717744, EPI_ISL_2717745, EPI_ISL_2717747, EPI_ISL_2717748, EPI_ISL_2717750, EPI_ISL_2717751, EPI_ISL_2717753, EPI_ISL_2717756, EPI_ISL_2717758, EPI_ISL_2717759, EPI_ISL_2717760, EPI_ISL_2717761, EPI_ISL_2717762, EPI_ISL_2717763, EPI_ISL_2717765, EPI_ISL_2717766, EPI_ISL_2717771, EPI_ISL_2717773, EPI_ISL_2717774, EPI_ISL_2717775, EPI_ISL_2717776, EPI_ISL_2717777, EPI_ISL_2717778, EPI_ISL_2717780, EPI_ISL_2717781, EPI_ISL_2717782, EPI_ISL_2717783, EPI_ISL_2717786, EPI_ISL_2717787, EPI_ISL_2717788, EPI_ISL_2717790, EPI_ISL_2717792, EPI_ISL_2717793, EPI_ISL_2717795, EPI_ISL_2717797, EPI_ISL_2717798, EPI_ISL_2717799, EPI_ISL_2717802, EPI_ISL_2717803, EPI_ISL_2717804, EPI_ISL_2717805, EPI_ISL_2717806, EPI_ISL_2717808, EPI_ISL_2717810, EPI_ISL_2717811, EPI_ISL_2717816, EPI_ISL_2717818, EPI_ISL_2717819, EPI_ISL_2717820, EPI_ISL_2717822, EPI_ISL_2717823, EPI_ISL_2717824, EPI_ISL_2717826, EPI_ISL_2717829, EPI_ISL_2717831, EPI_ISL_2717838, EPI_ISL_2717840, EPI_ISL_2717842, EPI_ISL_2717845, EPI_ISL_2717847, EPI_ISL_2717848, EPI_ISL_2717849, EPI_ISL_2717850, EPI_ISL_2717851, EPI_ISL_2717854, EPI_ISL_2717857, EPI_ISL_2717859, EPI_ISL_2717861, EPI_ISL_2717862, EPI_ISL_2717864, EPI_ISL_2717867, EPI_ISL_2717868, EPI_ISL_2717869, EPI_ISL_2717870, EPI_ISL_2717877, EPI_ISL_2717878, EPI_ISL_2717880, EPI_ISL_2717881, EPI_ISL_2717882, EPI_ISL_2717883, EPI_ISL_2717887, EPI_ISL_2717888, EPI_ISL_2717892, EPI_ISL_2717899, EPI_ISL_2717900, EPI_ISL_2717901, EPI_ISL_2717904, EPI_ISL_2717905, EPI_ISL_2717906, EPI_ISL_2717909, EPI_ISL_2717911, EPI_ISL_2717912, EPI_ISL_2717913, EPI_ISL_2717914, EPI_ISL_2717926, EPI_ISL_2717929, EPI_ISL_2717931, EPI_ISL_2717932, EPI_ISL_2717933, EPI_ISL_2717934, EPI_ISL_2717935, EPI_ISL_2717936, EPI_ISL_2717938, EPI_ISL_2717940, EPI_ISL_2717941, EPI_ISL_2717945, EPI_ISL_2717949, EPI_ISL_2717950, EPI_ISL_2717951, EPI_ISL_2717952, EPI_ISL_2717958, EPI_ISL_2717959, EPI_ISL_2717960, EPI_ISL_2717961, EPI_ISL_2717964, EPI_ISL_2717965, EPI_ISL_2717967, EPI_ISL_2717970, EPI_ISL_2717972, EPI_ISL_2717974, EPI_ISL_2717978, EPI_ISL_2717979, EPI_ISL_2717984, EPI_ISL_2717986, EPI_ISL_2717987, EPI_ISL_2717989, EPI_ISL_2717990, EPI_ISL_2717994, EPI_ISL_2717995, EPI_ISL_2717996, EPI_ISL_2717997, EPI_ISL_2718001, EPI_ISL_2718002, EPI_ISL_2718004, EPI_ISL_2718007, EPI_ISL_2718008, EPI_ISL_2718009, EPI_ISL_2718010, EPI_ISL_2718011, EPI_ISL_2718013, EPI_ISL_2718014, EPI_ISL_2718017, EPI_ISL_2718018, EPI_ISL_2718022, EPI_ISL_2718023, EPI_ISL_2718026, EPI_ISL_2718027, EPI_ISL_2718028, EPI_ISL_2718030, EPI_ISL_2718031, EPI_ISL_2718034, EPI_ISL_2718035, EPI_ISL_2718037, EPI_ISL_2718039, EPI_ISL_2718040, EPI_ISL_2718045, EPI_ISL_2718046, EPI_ISL_2718051, EPI_ISL_2718053, EPI_ISL_2718054, EPI_ISL_2718056, EPI_ISL_2718057, EPI_ISL_2718058, EPI_ISL_2718061, EPI_ISL_2718064, EPI_ISL_2718068, EPI_ISL_2718071, EPI_ISL_2718072, EPI_ISL_2718073, EPI_ISL_2718076, EPI_ISL_2718080, EPI_ISL_2718081, EPI_ISL_2718083, EPI_ISL_2718084, EPI_ISL_2718085, EPI_ISL_2718086, EPI_ISL_2718087, EPI_ISL_2718090, EPI_ISL_2718095, EPI_ISL_2718096, EPI_ISL_2718098, EPI_ISL_2718101, EPI_ISL_2718104, EPI_ISL_2718106, EPI_ISL_2718112, EPI_ISL_2718113, EPI_ISL_2718117, EPI_ISL_2718120, EPI_ISL_2718121, EPI_ISL_2718122, EPI_ISL_2718124, EPI_ISL_2718125, EPI_ISL_2718127, EPI_ISL_2718130, EPI_ISL_2718132, EPI_ISL_2718133, EPI_ISL_2718137, EPI_ISL_2718140, EPI_ISL_2718141, EPI_ISL_2718144, EPI_ISL_2718145, EPI_ISL_2718150, EPI_ISL_2718151, EPI_ISL_2718153, EPI_ISL_2718154, EPI_ISL_2718155, EPI_ISL_2718157, EPI_ISL_2718159, EPI_ISL_2718161, EPI_ISL_2718167, EPI_ISL_2729054, EPI_ISL_2729058, EPI_ISL_2729065, EPI_ISL_2729068, EPI_ISL_2729071, EPI_ISL_2729074, EPI_ISL_2729075, EPI_ISL_2729077, EPI_ISL_2729078, EPI_ISL_2729079, EPI_ISL_2729083, EPI_ISL_2729089, EPI_ISL_2729107, EPI_ISL_2729120, EPI_ISL_2729134, EPI_ISL_2729135, EPI_ISL_2729142, EPI_ISL_2729145, EPI_ISL_2729149, EPI_ISL_2729151, EPI_ISL_2729156, EPI_ISL_2729158, EPI_ISL_2729159, EPI_ISL_2729167, EPI_ISL_2729169, EPI_ISL_2729171, EPI_ISL_2729173, EPI_ISL_2729178, EPI_ISL_2729179, EPI_ISL_2729180, EPI_ISL_2729182, EPI_ISL_2729184, EPI_ISL_2729188, EPI_ISL_2729200, EPI_ISL_2729201, EPI_ISL_2729205, EPI_ISL_2729207, EPI_ISL_2729215, EPI_ISL_2729220, EPI_ISL_2729226, EPI_ISL_2729239, EPI_ISL_2729245, EPI_ISL_2729247, EPI_ISL_2729249, EPI_ISL_2729250, EPI_ISL_2729268, EPI_ISL_2729273, EPI_ISL_2729274, EPI_ISL_2729278, EPI_ISL_2729280, EPI_ISL_2729289, EPI_ISL_2729293, EPI_ISL_2729295, EPI_ISL_2729300, EPI_ISL_2729301, EPI_ISL_2729309, EPI_ISL_2729311, EPI_ISL_2729315, EPI_ISL_2729316, EPI_ISL_2729325, EPI_ISL_2729326, EPI_ISL_2729327, EPI_ISL_2729328, EPI_ISL_2729346, EPI_ISL_2729347, EPI_ISL_2729349, EPI_ISL_2729351, EPI_ISL_2729375, EPI_ISL_2729378, EPI_ISL_2729381, EPI_ISL_2729382, EPI_ISL_2729387, EPI_ISL_2729390, EPI_ISL_2729391, EPI_ISL_2729392, EPI_ISL_2729393, EPI_ISL_2729399, EPI_ISL_2729407, EPI_ISL_2729417, EPI_ISL_2729419, EPI_ISL_2729421, EPI_ISL_2729430, EPI_ISL_2729431, EPI_ISL_2729436, EPI_ISL_2729438, EPI_ISL_2729440, EPI_ISL_2729441, EPI_ISL_2729445, EPI_ISL_2729446, EPI_ISL_2729449, EPI_ISL_2729456, EPI_ISL_2729457, EPI_ISL_2729458, EPI_ISL_2729461, EPI_ISL_2729463, EPI_ISL_2729464, EPI_ISL_2729465, EPI_ISL_2729475, EPI_ISL_2729480, EPI_ISL_2729481, EPI_ISL_2729489, EPI_ISL_2729497, EPI_ISL_2729498, EPI_ISL_2729502, EPI_ISL_2729504, EPI_ISL_2729507, EPI_ISL_2729511, EPI_ISL_2729512, EPI_ISL_2729516, EPI_ISL_2729517, EPI_ISL_2729519, EPI_ISL_2729523, EPI_ISL_2729527, EPI_ISL_2729528, EPI_ISL_2729529, EPI_ISL_2729531, EPI_ISL_2729532, EPI_ISL_2729536, EPI_ISL_2729540, EPI_ISL_2729542, EPI_ISL_2729545, EPI_ISL_2729547, EPI_ISL_2729548, EPI_ISL_2729551, EPI_ISL_2729552, EPI_ISL_2729557, EPI_ISL_2729561, EPI_ISL_2729563, EPI_ISL_2729569, EPI_ISL_2729570, EPI_ISL_2729572, EPI_ISL_2729575, EPI_ISL_2729584, EPI_ISL_2729587, EPI_ISL_2729592, EPI_ISL_2729594, EPI_ISL_2729597, EPI_ISL_2729599, EPI_ISL_2729601, EPI_ISL_2729607, EPI_ISL_2729608, EPI_ISL_2729612, EPI_ISL_2729613, EPI_ISL_2729616, EPI_ISL_2729617, EPI_ISL_2729618, EPI_ISL_2729619, EPI_ISL_2729623, EPI_ISL_2729629, EPI_ISL_2729630, EPI_ISL_2729634, EPI_ISL_2729635, EPI_ISL_2729643, EPI_ISL_2729647, EPI_ISL_2729648, EPI_ISL_2729651, EPI_ISL_2729652, EPI_ISL_2729657, EPI_ISL_2729658, EPI_ISL_2729659, EPI_ISL_2729663, EPI_ISL_2729664, EPI_ISL_2729667, EPI_ISL_2729672, EPI_ISL_2729673, EPI_ISL_2729681, EPI_ISL_2729682, EPI_ISL_2729687, EPI_ISL_2729690, EPI_ISL_2729691, EPI_ISL_2729693, EPI_ISL_2729694, EPI_ISL_2729697, EPI_ISL_2729699, EPI_ISL_2729701, EPI_ISL_2729702, EPI_ISL_2729704, EPI_ISL_2729706, EPI_ISL_2729709, EPI_ISL_2729712, EPI_ISL_2729721, EPI_ISL_2729725, EPI_ISL_2729726, EPI_ISL_2729727, EPI_ISL_2729728, EPI_ISL_2729734, EPI_ISL_2729737, EPI_ISL_2729744, EPI_ISL_2729753, EPI_ISL_2729755, EPI_ISL_2729767, EPI_ISL_2729771, EPI_ISL_2729772, EPI_ISL_2729786, EPI_ISL_2729789, EPI_ISL_2729794, EPI_ISL_2729800, EPI_ISL_2729801, EPI_ISL_2729802, EPI_ISL_2729805, EPI_ISL_2729806, EPI_ISL_2729810, EPI_ISL_2729811, EPI_ISL_2729812, EPI_ISL_2729813, EPI_ISL_2729814, EPI_ISL_2729819, EPI_ISL_2729820, EPI_ISL_2729822, EPI_ISL_2729824, EPI_ISL_2729831, EPI_ISL_2729835, EPI_ISL_2729838, EPI_ISL_2729843, EPI_ISL_2729846, EPI_ISL_2729848, EPI_ISL_2729852, EPI_ISL_2729853, EPI_ISL_2729856, EPI_ISL_2729857, EPI_ISL_2729870, EPI_ISL_2729875, EPI_ISL_2729880, EPI_ISL_2729881, EPI_ISL_2729884, EPI_ISL_2729886, EPI_ISL_2729891, EPI_ISL_2729892, EPI_ISL_2729895, EPI_ISL_2729906, EPI_ISL_2729908, EPI_ISL_2729910, EPI_ISL_2729914, EPI_ISL_2729919, EPI_ISL_2729922, EPI_ISL_2729926, EPI_ISL_2729927, EPI_ISL_2729935, EPI_ISL_2729937, EPI_ISL_2729938, EPI_ISL_2729940, EPI_ISL_2729943, EPI_ISL_2729946, EPI_ISL_2729952, EPI_ISL_2729959, EPI_ISL_2729960, EPI_ISL_2729982, EPI_ISL_2729984, EPI_ISL_2729986, EPI_ISL_2729990, EPI_ISL_2730000, EPI_ISL_2730002, EPI_ISL_2730014, EPI_ISL_2730021, EPI_ISL_2730022, EPI_ISL_2730025, EPI_ISL_2730029, EPI_ISL_2730031, EPI_ISL_2730034, EPI_ISL_2730047, EPI_ISL_2730053, EPI_ISL_2730055, EPI_ISL_2730061, EPI_ISL_2730070, EPI_ISL_2730074, EPI_ISL_2730082, EPI_ISL_2730088, EPI_ISL_2730095, EPI_ISL_2730101, EPI_ISL_2730106, EPI_ISL_2730116, EPI_ISL_2730122, EPI_ISL_2730123, EPI_ISL_2730142, EPI_ISL_2730148, EPI_ISL_2730150, EPI_ISL_2730157, EPI_ISL_2730158, EPI_ISL_2730159, EPI_ISL_2730166, EPI_ISL_2730167, EPI_ISL_2730168, EPI_ISL_2730174, EPI_ISL_2730177, EPI_ISL_2730183, EPI_ISL_2730193, EPI_ISL_2730198, EPI_ISL_2730204, EPI_ISL_2730213, EPI_ISL_2730217, EPI_ISL_2730219, EPI_ISL_2730222, EPI_ISL_2730223, EPI_ISL_2730234, EPI_ISL_2730245, EPI_ISL_2730247, EPI_ISL_2730249, EPI_ISL_2730254, EPI_ISL_2730256, EPI_ISL_2730257, EPI_ISL_2730263, EPI_ISL_2730270, EPI_ISL_2730272, EPI_ISL_2730275, EPI_ISL_2730278, EPI_ISL_2730286, EPI_ISL_2730287, EPI_ISL_2730288, EPI_ISL_2730292, EPI_ISL_2730307, EPI_ISL_2730308, EPI_ISL_2730317, EPI_ISL_2730326, EPI_ISL_2730328, EPI_ISL_2730329, EPI_ISL_2730331, EPI_ISL_2730335, EPI_ISL_2730336, EPI_ISL_2730338, EPI_ISL_2730339, EPI_ISL_2730340, EPI_ISL_2730341, EPI_ISL_2730345, EPI_ISL_2730351, EPI_ISL_2730352, EPI_ISL_2730367, EPI_ISL_2730370, EPI_ISL_2730373, EPI_ISL_2730382, EPI_ISL_2730384, EPI_ISL_2730385, EPI_ISL_2730389, EPI_ISL_2730391, EPI_ISL_2730393, EPI_ISL_2730394, EPI_ISL_2730396, EPI_ISL_2730401, EPI_ISL_2730402, EPI_ISL_2730410, EPI_ISL_2730414, EPI_ISL_2730416, EPI_ISL_2730420, EPI_ISL_2730423, EPI_ISL_2730424, EPI_ISL_2730428, EPI_ISL_2730429, EPI_ISL_2730431, EPI_ISL_2730450, EPI_ISL_2730452, EPI_ISL_2730453, EPI_ISL_2730457, EPI_ISL_2730462, EPI_ISL_2730469, EPI_ISL_2730472, EPI_ISL_2730474, EPI_ISL_2730484, EPI_ISL_2730485, EPI_ISL_2730490, EPI_ISL_2730497, EPI_ISL_2730499, EPI_ISL_2730509, EPI_ISL_2730517, EPI_ISL_2730519, EPI_ISL_2730520, EPI_ISL_2730522, EPI_ISL_2730529, EPI_ISL_2730532, EPI_ISL_2730542, EPI_ISL_2730543, EPI_ISL_2730547, EPI_ISL_2730548, EPI_ISL_2730550, EPI_ISL_2730553, EPI_ISL_2730556, EPI_ISL_2730560, EPI_ISL_2730562, EPI_ISL_2730567, EPI_ISL_2730569, EPI_ISL_2730572, EPI_ISL_2730577, EPI_ISL_2730580, EPI_ISL_2730602, EPI_ISL_2730603, EPI_ISL_2730605, EPI_ISL_2730606, EPI_ISL_2730613, EPI_ISL_2730619, EPI_ISL_2730624, EPI_ISL_2730633, EPI_ISL_2730636, EPI_ISL_2730645, EPI_ISL_2765570, EPI_ISL_2765572, EPI_ISL_2765573, EPI_ISL_2765574, EPI_ISL_2765576, EPI_ISL_2765577, EPI_ISL_2765578, EPI_ISL_2765581, EPI_ISL_2765585, EPI_ISL_2765593, EPI_ISL_2765594, EPI_ISL_2765596, EPI_ISL_2765597, EPI_ISL_2765600, EPI_ISL_2765603, EPI_ISL_2765605, EPI_ISL_2765608, EPI_ISL_2765610, EPI_ISL_2765614, EPI_ISL_2765615, EPI_ISL_2765616, EPI_ISL_2765619, EPI_ISL_2765620, EPI_ISL_2765624, EPI_ISL_2765630, EPI_ISL_2765635, EPI_ISL_2765636, EPI_ISL_2765638, EPI_ISL_2765639, EPI_ISL_2765642, EPI_ISL_2765646, EPI_ISL_2765650, EPI_ISL_2765651, EPI_ISL_2765654, EPI_ISL_2765655, EPI_ISL_2765659, EPI_ISL_2765660, EPI_ISL_2765681, EPI_ISL_2765684, EPI_ISL_2765689, EPI_ISL_2765690, EPI_ISL_2765692, EPI_ISL_2765698, EPI_ISL_2765702, EPI_ISL_2765704, EPI_ISL_2765705, EPI_ISL_2765706, EPI_ISL_2765716, EPI_ISL_2765724, EPI_ISL_2765727, EPI_ISL_2765731, EPI_ISL_2765732, EPI_ISL_2765735, EPI_ISL_2765737, EPI_ISL_2765739, EPI_ISL_2765740, EPI_ISL_2765746, EPI_ISL_2765748, EPI_ISL_2765749, EPI_ISL_2765755, EPI_ISL_2765756, EPI_ISL_2765758, EPI_ISL_2765761, EPI_ISL_2765764, EPI_ISL_2765772, EPI_ISL_2765774, EPI_ISL_2765782, EPI_ISL_2765783, EPI_ISL_2765785, EPI_ISL_2765786, EPI_ISL_2765787, EPI_ISL_2765793, EPI_ISL_2765794, EPI_ISL_2765801, EPI_ISL_2765802, EPI_ISL_2765804, EPI_ISL_2765805, EPI_ISL_2765806, EPI_ISL_2765807, EPI_ISL_2765810, EPI_ISL_2765811, EPI_ISL_2765814, EPI_ISL_2765818, EPI_ISL_2765822, EPI_ISL_2765823, EPI_ISL_2765828, EPI_ISL_2765829, EPI_ISL_2765835, EPI_ISL_2765836, EPI_ISL_2765838, EPI_ISL_2765840, EPI_ISL_2765848, EPI_ISL_2765853, EPI_ISL_2765854, EPI_ISL_2765855, EPI_ISL_2765856, EPI_ISL_2765857, EPI_ISL_2765858, EPI_ISL_2765863, EPI_ISL_2765866, EPI_ISL_2765867, EPI_ISL_2765868, EPI_ISL_2765872, EPI_ISL_2765874, EPI_ISL_2765876, EPI_ISL_2765877, EPI_ISL_2765881, EPI_ISL_2765883, EPI_ISL_2765884, EPI_ISL_2765886, EPI_ISL_2765887, EPI_ISL_2765888, EPI_ISL_2765889, EPI_ISL_2765891, EPI_ISL_2765892, EPI_ISL_2765895, EPI_ISL_2765896, EPI_ISL_2765897, EPI_ISL_2765898, EPI_ISL_2765900, EPI_ISL_2765903, EPI_ISL_2765906, EPI_ISL_2765911, EPI_ISL_2765912, EPI_ISL_2765913, EPI_ISL_2765917, EPI_ISL_2765919, EPI_ISL_2765920, EPI_ISL_2765921, EPI_ISL_2765922, EPI_ISL_2765923, EPI_ISL_2765928, EPI_ISL_2765930, EPI_ISL_2765933, EPI_ISL_2765936, EPI_ISL_2765938, EPI_ISL_2765939, EPI_ISL_2765941, EPI_ISL_2765942, EPI_ISL_2765943, EPI_ISL_2765944, EPI_ISL_2765946, EPI_ISL_2765948, EPI_ISL_2765951, EPI_ISL_2765952, EPI_ISL_2765953, EPI_ISL_2765956, EPI_ISL_2765957, EPI_ISL_2765963, EPI_ISL_2765973, EPI_ISL_2765974, EPI_ISL_2765975, EPI_ISL_2765976, EPI_ISL_2765978, EPI_ISL_2765981, EPI_ISL_2765985, EPI_ISL_2765990, EPI_ISL_2765993, EPI_ISL_2765999, EPI_ISL_2766002, EPI_ISL_2766004, EPI_ISL_2766005, EPI_ISL_2766006, EPI_ISL_2766007, | | | |

| | | | | |
|---|---|---|---|---|
| EPI_ISL_2766008, EPI_ISL_2766010, EPI_ISL_2766011, EPI_ISL_2766012, EPI_ISL_2766013, EPI_ISL_2766014, EPI_ISL_2766015, EPI_ISL_2766017, EPI_ISL_2766023, EPI_ISL_2766025, EPI_ISL_2766027, EPI_ISL_2766029, EPI_ISL_2766035, EPI_ISL_2766036, EPI_ISL_2766041, EPI_ISL_2766044, EPI_ISL_2766046, EPI_ISL_2766048, EPI_ISL_2766050, EPI_ISL_2766051, EPI_ISL_2766053, EPI_ISL_2766056, EPI_ISL_2766057, EPI_ISL_2766058, EPI_ISL_2766059, EPI_ISL_2766060, EPI_ISL_2766063, EPI_ISL_2766065, EPI_ISL_2766067, EPI_ISL_2766068, EPI_ISL_2766072, EPI_ISL_2766073, EPI_ISL_2766074, EPI_ISL_2766077, EPI_ISL_2766082, EPI_ISL_2766083, EPI_ISL_2766088, EPI_ISL_2766089, EPI_ISL_2766095, EPI_ISL_2766099, EPI_ISL_2766101, EPI_ISL_2766102, EPI_ISL_2766103, EPI_ISL_2766104, EPI_ISL_2766105, EPI_ISL_2766109, EPI_ISL_2766117, EPI_ISL_2766122, EPI_ISL_2766123, EPI_ISL_2766125, EPI_ISL_2766126, EPI_ISL_2766128, EPI_ISL_2766130, EPI_ISL_2766132, EPI_ISL_2766133, EPI_ISL_2766140, EPI_ISL_2766141, EPI_ISL_2766146, EPI_ISL_2766147, EPI_ISL_2766151, EPI_ISL_2766153, EPI_ISL_2766155, EPI_ISL_2766157, EPI_ISL_2766163, EPI_ISL_2766164, EPI_ISL_2766166, EPI_ISL_2766167, EPI_ISL_2766171, EPI_ISL_2766172, EPI_ISL_2766173, EPI_ISL_2766175, EPI_ISL_2766182, EPI_ISL_2766185, EPI_ISL_2766186, EPI_ISL_2766191, EPI_ISL_2766192, EPI_ISL_2766193, EPI_ISL_2766547, EPI_ISL_2766552, EPI_ISL_2766553, EPI_ISL_2766563, EPI_ISL_2766567, EPI_ISL_2766572, EPI_ISL_2766576, EPI_ISL_2766579, EPI_ISL_2766580, EPI_ISL_2766582, EPI_ISL_2766583, EPI_ISL_2766587, EPI_ISL_2766588, EPI_ISL_2766591, EPI_ISL_2766594, EPI_ISL_2766596, EPI_ISL_2766598, EPI_ISL_2766600, EPI_ISL_2766607, EPI_ISL_2766608, EPI_ISL_2766610, EPI_ISL_2766622, EPI_ISL_2766632, EPI_ISL_2766637, EPI_ISL_2766641, EPI_ISL_2766643, EPI_ISL_2766651, EPI_ISL_2766655, EPI_ISL_2766661, EPI_ISL_2766664, EPI_ISL_2766674, EPI_ISL_2766676, EPI_ISL_2766677, EPI_ISL_2766683, EPI_ISL_2766684, EPI_ISL_2766695, EPI_ISL_2766696, EPI_ISL_2766697, EPI_ISL_2766702, EPI_ISL_2766705, EPI_ISL_2766706, EPI_ISL_2766715, EPI_ISL_2766717, EPI_ISL_2766719, EPI_ISL_2766743, EPI_ISL_2766750, EPI_ISL_2766751, EPI_ISL_2766757, EPI_ISL_2766758, EPI_ISL_2766761, EPI_ISL_2766762, EPI_ISL_2766763, EPI_ISL_2766777, EPI_ISL_2766780, EPI_ISL_2766786, EPI_ISL_2766788, EPI_ISL_2766793, EPI_ISL_2766798, EPI_ISL_2766812, EPI_ISL_2766816, EPI_ISL_2766818, EPI_ISL_2766820, EPI_ISL_2766821, EPI_ISL_2766825, EPI_ISL_2766827, EPI_ISL_2766828, EPI_ISL_2766830, EPI_ISL_2766834, EPI_ISL_2766845, EPI_ISL_2766848, EPI_ISL_2766851, EPI_ISL_2766852, EPI_ISL_2766857, EPI_ISL_2766861, EPI_ISL_2766863, EPI_ISL_2766872, EPI_ISL_2766875, EPI_ISL_2766877, EPI_ISL_2766880, EPI_ISL_2766885, EPI_ISL_2766887, EPI_ISL_2766892, EPI_ISL_2766894, EPI_ISL_2766896, EPI_ISL_2766898, EPI_ISL_2766899, EPI_ISL_2766900, EPI_ISL_2766901, EPI_ISL_2766902, EPI_ISL_2766904, EPI_ISL_2766906, EPI_ISL_2766907, EPI_ISL_2766912, EPI_ISL_2766913, EPI_ISL_2766914, EPI_ISL_2766916, EPI_ISL_2766917, EPI_ISL_2766919, EPI_ISL_2766920, EPI_ISL_2766921, EPI_ISL_2766922, EPI_ISL_2766929, EPI_ISL_2766933, EPI_ISL_2766934, EPI_ISL_2766935, EPI_ISL_2766938, EPI_ISL_2766940, EPI_ISL_2766944, EPI_ISL_2766946, EPI_ISL_2766947, EPI_ISL_2766950, EPI_ISL_2766952, EPI_ISL_2766954, EPI_ISL_2766955, EPI_ISL_2766956, EPI_ISL_2766957, EPI_ISL_2766958, EPI_ISL_2766960, EPI_ISL_2766961, EPI_ISL_2766963, EPI_ISL_2766967, EPI_ISL_2766969, EPI_ISL_2766970, EPI_ISL_2766973, EPI_ISL_2766974, EPI_ISL_2766978, EPI_ISL_2766979, EPI_ISL_2766981, EPI_ISL_2766982, EPI_ISL_2766983, EPI_ISL_2766984, EPI_ISL_2766986, EPI_ISL_2766987, EPI_ISL_2766988, EPI_ISL_2766989, EPI_ISL_2766992, EPI_ISL_2766993, EPI_ISL_2766994, EPI_ISL_2766995, EPI_ISL_2766998, EPI_ISL_2766999, EPI_ISL_2767001, EPI_ISL_2767002, EPI_ISL_2767005, EPI_ISL_2767008, EPI_ISL_2767011, EPI_ISL_2767012, EPI_ISL_2767014, EPI_ISL_2767015, EPI_ISL_2767017, EPI_ISL_2767020, EPI_ISL_2767024, EPI_ISL_2767025, EPI_ISL_2767027, EPI_ISL_2767029, EPI_ISL_2767030, EPI_ISL_2767033, EPI_ISL_2767034, EPI_ISL_2767035, EPI_ISL_2767036, EPI_ISL_2767043, EPI_ISL_2767044, EPI_ISL_2767049, EPI_ISL_2767050, EPI_ISL_2767052, EPI_ISL_2767053, EPI_ISL_2767054, EPI_ISL_2767056, EPI_ISL_2767060, EPI_ISL_2767061, EPI_ISL_2767062, EPI_ISL_2767064, EPI_ISL_2767067, EPI_ISL_2767068, EPI_ISL_2767069, EPI_ISL_2767070, EPI_ISL_2767073, EPI_ISL_2767074, EPI_ISL_2767076, EPI_ISL_2767079, EPI_ISL_2767080, EPI_ISL_2767082, EPI_ISL_2767085, EPI_ISL_2767087, EPI_ISL_2767093, EPI_ISL_2767094, EPI_ISL_2767095, EPI_ISL_2767096, EPI_ISL_2767098, EPI_ISL_2767100, EPI_ISL_2767102, EPI_ISL_2767104, EPI_ISL_2767105, EPI_ISL_2767107, EPI_ISL_2767108, EPI_ISL_2767110, EPI_ISL_2767111, EPI_ISL_2767114, EPI_ISL_2767115, EPI_ISL_2767117, EPI_ISL_2767119, EPI_ISL_2767123, EPI_ISL_2767125, EPI_ISL_2767127, EPI_ISL_2767130, EPI_ISL_2767133, EPI_ISL_2767134, EPI_ISL_2767137, EPI_ISL_2767140, EPI_ISL_2767141, EPI_ISL_2767142, EPI_ISL_2767145, EPI_ISL_2767147, EPI_ISL_2767148, EPI_ISL_2767149, EPI_ISL_2767150, EPI_ISL_2767151, EPI_ISL_2767154, EPI_ISL_2767156, EPI_ISL_2767157, EPI_ISL_2767160, EPI_ISL_2767165, EPI_ISL_2767166, EPI_ISL_2767168, EPI_ISL_2767169, EPI_ISL_2767172, EPI_ISL_2767173, EPI_ISL_2767174, EPI_ISL_2767176, EPI_ISL_2767178, EPI_ISL_2767183, EPI_ISL_2767184, EPI_ISL_2767187, EPI_ISL_2767192, EPI_ISL_2767194, EPI_ISL_2767195, EPI_ISL_2767197, EPI_ISL_2767198, EPI_ISL_2767201, EPI_ISL_2767205, EPI_ISL_2767207, EPI_ISL_2767213, EPI_ISL_2767215, EPI_ISL_2767217, EPI_ISL_2767218, EPI_ISL_2767220, EPI_ISL_2767221, EPI_ISL_2767223, EPI_ISL_2767224, EPI_ISL_2767228, EPI_ISL_2767229, EPI_ISL_2767230, EPI_ISL_2767231, EPI_ISL_2767232, EPI_ISL_2767234, EPI_ISL_2767236, EPI_ISL_2767237, EPI_ISL_2767238, EPI_ISL_2767240, EPI_ISL_2767241, EPI_ISL_2767242, EPI_ISL_2767243, EPI_ISL_2767244, EPI_ISL_2767248, EPI_ISL_2767249, EPI_ISL_2767252, EPI_ISL_2767253, EPI_ISL_2767256, EPI_ISL_2767257, EPI_ISL_2767259, EPI_ISL_2767260, EPI_ISL_2767262, EPI_ISL_2767263, EPI_ISL_2767264, EPI_ISL_2767265, EPI_ISL_2767268, EPI_ISL_2767269, EPI_ISL_2767270, EPI_ISL_2767274, EPI_ISL_2767275, EPI_ISL_2767276, EPI_ISL_2767279, EPI_ISL_2767280, EPI_ISL_2767282, EPI_ISL_2767283, EPI_ISL_2767285, EPI_ISL_2767287, EPI_ISL_2767288, EPI_ISL_2767289, EPI_ISL_2767290, EPI_ISL_2767292, EPI_ISL_2767295, EPI_ISL_2767296, EPI_ISL_2767297, EPI_ISL_2767298, EPI_ISL_2767299, EPI_ISL_2767301, EPI_ISL_2767302, EPI_ISL_2767306, EPI_ISL_2767307, EPI_ISL_2767308, EPI_ISL_2767312, EPI_ISL_2767315, EPI_ISL_2767317, EPI_ISL_2767320, EPI_ISL_2767322, EPI_ISL_2767323, EPI_ISL_2767326, EPI_ISL_2767327, EPI_ISL_2767330, EPI_ISL_2767331, EPI_ISL_2767332, EPI_ISL_2767334, EPI_ISL_2767335, EPI_ISL_2767337, EPI_ISL_2767339, EPI_ISL_2767340, EPI_ISL_2767343, EPI_ISL_2767346, EPI_ISL_2767348, EPI_ISL_2767349, EPI_ISL_2767354, EPI_ISL_2767366, EPI_ISL_2767367, EPI_ISL_2767371, EPI_ISL_2767374, EPI_ISL_2767380, EPI_ISL_2767381, EPI_ISL_2767392, EPI_ISL_2767411, EPI_ISL_2767425, EPI_ISL_2767438, EPI_ISL_2767440, EPI_ISL_2767442, EPI_ISL_2767448, EPI_ISL_2767452, EPI_ISL_2767453, EPI_ISL_2767455, EPI_ISL_2767462, EPI_ISL_2767465, EPI_ISL_2767468, EPI_ISL_2767477, EPI_ISL_2767480, EPI_ISL_2767482, EPI_ISL_2767486, EPI_ISL_2767490, EPI_ISL_2767493, EPI_ISL_2767495, EPI_ISL_2767500, EPI_ISL_2767503, EPI_ISL_2767504, EPI_ISL_2767513, EPI_ISL_2767514, EPI_ISL_2767516, EPI_ISL_2767521, EPI_ISL_2767524, EPI_ISL_2767529, EPI_ISL_2767543, EPI_ISL_2767544, EPI_ISL_2767551, EPI_ISL_2767559, EPI_ISL_2767560, EPI_ISL_2767567, EPI_ISL_2767577, EPI_ISL_2767579, EPI_ISL_2767585, EPI_ISL_2767595, EPI_ISL_2767599, EPI_ISL_2767606, EPI_ISL_2767613, EPI_ISL_2767615, EPI_ISL_2767623, EPI_ISL_2767628, EPI_ISL_2767629, EPI_ISL_2767649, EPI_ISL_2767650, EPI_ISL_2767652, EPI_ISL_2767655, EPI_ISL_2767656, EPI_ISL_2767657, EPI_ISL_2767658, EPI_ISL_2767660, EPI_ISL_2767661, EPI_ISL_2767663, EPI_ISL_2767664, EPI_ISL_2767666, EPI_ISL_2767674, EPI_ISL_2767675, EPI_ISL_2767676, EPI_ISL_2767677, EPI_ISL_2767679, EPI_ISL_2767682, EPI_ISL_2767684, EPI_ISL_2767687, EPI_ISL_2767689, EPI_ISL_2767690, EPI_ISL_2767691, EPI_ISL_2767692, EPI_ISL_2767693, EPI_ISL_2767695, EPI_ISL_2767698, EPI_ISL_2767699, EPI_ISL_2767700, EPI_ISL_2767701, EPI_ISL_2767702, EPI_ISL_2767703, EPI_ISL_2767709, EPI_ISL_2767711, EPI_ISL_2767712, EPI_ISL_2767713, EPI_ISL_2767716, EPI_ISL_2767719, EPI_ISL_2767720, EPI_ISL_2767721, EPI_ISL_2767723, EPI_ISL_2767724, EPI_ISL_2767730, EPI_ISL_2767731, EPI_ISL_2767735, EPI_ISL_2767736, EPI_ISL_2767738, EPI_ISL_2767739, EPI_ISL_2767744, EPI_ISL_2767747, EPI_ISL_2767748, EPI_ISL_2767749, EPI_ISL_2767756, EPI_ISL_2767758, EPI_ISL_2767759, EPI_ISL_2767760, EPI_ISL_2767765, EPI_ISL_2767768, EPI_ISL_2767769, EPI_ISL_2767771, EPI_ISL_2767772, EPI_ISL_2767773, EPI_ISL_2767775, EPI_ISL_2767776, EPI_ISL_2767779, EPI_ISL_2767781, EPI_ISL_2767782, EPI_ISL_2767785, EPI_ISL_2767787, EPI_ISL_2767788, EPI_ISL_2767793, EPI_ISL_2767797, EPI_ISL_2767804, EPI_ISL_2767808, EPI_ISL_2767809, EPI_ISL_2767811, EPI_ISL_2767815, EPI_ISL_2767816, EPI_ISL_2767817, EPI_ISL_2767818, EPI_ISL_2767822, EPI_ISL_2767824, EPI_ISL_2767825, EPI_ISL_2767826, EPI_ISL_2767829, EPI_ISL_2767830, EPI_ISL_2767831, EPI_ISL_2767833, EPI_ISL_2767836, EPI_ISL_2767838, EPI_ISL_2767844, EPI_ISL_2767848, EPI_ISL_2767850, EPI_ISL_2767851, EPI_ISL_2767852, EPI_ISL_2767853, EPI_ISL_2767855, EPI_ISL_2767856, EPI_ISL_2767857, EPI_ISL_2767858, EPI_ISL_2767860, EPI_ISL_2767862, EPI_ISL_2767866, EPI_ISL_2767867, EPI_ISL_2767871, EPI_ISL_2767872, EPI_ISL_2767875, EPI_ISL_2767877, EPI_ISL_2767878, EPI_ISL_2767882, EPI_ISL_2767885, EPI_ISL_2767890, EPI_ISL_2767891, EPI_ISL_2767892, EPI_ISL_2767895, EPI_ISL_2767897, EPI_ISL_2767898, EPI_ISL_2767901, EPI_ISL_2767904, EPI_ISL_2767905, EPI_ISL_2767908, EPI_ISL_2767909, EPI_ISL_2767911, EPI_ISL_2767912, EPI_ISL_2767916, EPI_ISL_2767918, EPI_ISL_2767919, EPI_ISL_2767920, EPI_ISL_2767922, EPI_ISL_2767924, EPI_ISL_2767928, EPI_ISL_2767929, EPI_ISL_2767930, EPI_ISL_2767931, EPI_ISL_2767933, EPI_ISL_2767934, EPI_ISL_2767935, EPI_ISL_2767936, EPI_ISL_2767940, EPI_ISL_2767942, EPI_ISL_2767944, EPI_ISL_2767946, EPI_ISL_2767948, EPI_ISL_2767949, EPI_ISL_2767950, EPI_ISL_2767951, EPI_ISL_2767952, EPI_ISL_2767953, EPI_ISL_2767955, EPI_ISL_2767956, EPI_ISL_2767960, EPI_ISL_2767963, EPI_ISL_2767968, EPI_ISL_2767971, EPI_ISL_2767980, EPI_ISL_2767984, EPI_ISL_2767986, EPI_ISL_2767987, EPI_ISL_2767988, EPI_ISL_2767989, EPI_ISL_2767990, EPI_ISL_2767994, EPI_ISL_2767996, EPI_ISL_2767999, EPI_ISL_2768000, EPI_ISL_2768002, EPI_ISL_2768004, EPI_ISL_2768005, EPI_ISL_2768006, EPI_ISL_2768007, EPI_ISL_2768008, EPI_ISL_2780129, EPI_ISL_2780142, EPI_ISL_2780184, EPI_ISL_2780188, EPI_ISL_2780208, EPI_ISL_2780212, EPI_ISL_2780240, EPI_ISL_2780248, EPI_ISL_2780257, EPI_ISL_2780274, EPI_ISL_2780275, EPI_ISL_2780294, EPI_ISL_2780305, EPI_ISL_2780331, EPI_ISL_2780367, EPI_ISL_2780382, EPI_ISL_2780394, EPI_ISL_2780405, EPI_ISL_2780415, EPI_ISL_2780445, EPI_ISL_2780453, EPI_ISL_2780774, EPI_ISL_2780775, EPI_ISL_2780781, EPI_ISL_2780790, EPI_ISL_2780792, EPI_ISL_2780793, EPI_ISL_2780797, EPI_ISL_2780798, EPI_ISL_2780800, EPI_ISL_2780804, EPI_ISL_2780815, EPI_ISL_2780817, EPI_ISL_2780823, EPI_ISL_2780839, EPI_ISL_2780841, EPI_ISL_2780844, EPI_ISL_2780846, EPI_ISL_2780848, EPI_ISL_2780852, EPI_ISL_2780853, EPI_ISL_2780854, EPI_ISL_2780856, EPI_ISL_2780857, EPI_ISL_2780863, EPI_ISL_2780867, EPI_ISL_2780868, EPI_ISL_2780871, EPI_ISL_2780881, EPI_ISL_2780888, EPI_ISL_2780897, EPI_ISL_2780905, EPI_ISL_2780910, EPI_ISL_2780913, EPI_ISL_2780915, EPI_ISL_2780919, EPI_ISL_2780923, EPI_ISL_2780926, EPI_ISL_2780927, EPI_ISL_2780929, EPI_ISL_2780940, EPI_ISL_2780941, EPI_ISL_2780942, EPI_ISL_2780944, EPI_ISL_2780945, EPI_ISL_2780948, EPI_ISL_2780949, EPI_ISL_2780952, EPI_ISL_2780953, EPI_ISL_2780955, EPI_ISL_2780956, EPI_ISL_2780959, EPI_ISL_2780965, EPI_ISL_2780970, EPI_ISL_2780971, EPI_ISL_2780976, EPI_ISL_2780977, EPI_ISL_2780984, EPI_ISL_2780988, EPI_ISL_2780989, EPI_ISL_2780990, EPI_ISL_2780997, EPI_ISL_2781002, EPI_ISL_2781009, EPI_ISL_2781011, EPI_ISL_2781029, EPI_ISL_2781033, EPI_ISL_2781035, EPI_ISL_2781042, EPI_ISL_2781043, EPI_ISL_2781055, EPI_ISL_2781057, EPI_ISL_2781059, EPI_ISL_2781067, EPI_ISL_2781080, EPI_ISL_2781082, EPI_ISL_2781083, EPI_ISL_2781091, EPI_ISL_2781092, EPI_ISL_2781096, EPI_ISL_2781102, EPI_ISL_2781103, EPI_ISL_2781116, EPI_ISL_2781117, EPI_ISL_2781119, EPI_ISL_2781126, EPI_ISL_2781130, EPI_ISL_2781162, EPI_ISL_2781263, EPI_ISL_2781336, EPI_ISL_2781383, EPI_ISL_2781386, EPI_ISL_2781399, EPI_ISL_2781408, EPI_ISL_2781410, EPI_ISL_2781414, EPI_ISL_2781417, EPI_ISL_2781455, EPI_ISL_2781478, EPI_ISL_2781485, EPI_ISL_2781491, EPI_ISL_2781493, EPI_ISL_2781494, EPI_ISL_2781497, EPI_ISL_2781514, EPI_ISL_2781519, EPI_ISL_2781520, EPI_ISL_2781521, EPI_ISL_2781524, EPI_ISL_2781529, EPI_ISL_2781537, EPI_ISL_2781549, EPI_ISL_2781550, EPI_ISL_2781551, EPI_ISL_2781552, EPI_ISL_2781553, EPI_ISL_2781555, EPI_ISL_2781556, EPI_ISL_2781559, EPI_ISL_2781563, EPI_ISL_2781564, EPI_ISL_2781566, EPI_ISL_2781569, EPI_ISL_2781571, EPI_ISL_2781577, EPI_ISL_2781578, EPI_ISL_2781584, EPI_ISL_2781586, EPI_ISL_2781589, EPI_ISL_2781594, EPI_ISL_2781598, EPI_ISL_2781601, EPI_ISL_2781604, EPI_ISL_2781606, EPI_ISL_2781608, EPI_ISL_2781612, EPI_ISL_2781614, EPI_ISL_2781615, EPI_ISL_2781616, EPI_ISL_2781624, EPI_ISL_2781627, EPI_ISL_2781633, EPI_ISL_2781634, EPI_ISL_2781639, EPI_ISL_2781641, EPI_ISL_2781643, EPI_ISL_2781649, EPI_ISL_2781653, EPI_ISL_2781656, EPI_ISL_2781659, EPI_ISL_2781663, EPI_ISL_2781668, EPI_ISL_2781673, EPI_ISL_2781677, EPI_ISL_2781683, EPI_ISL_2781687, EPI_ISL_2781697, EPI_ISL_2781699, EPI_ISL_2781700, EPI_ISL_2781706, EPI_ISL_2781707, EPI_ISL_2781708, EPI_ISL_2781711, EPI_ISL_2781716, EPI_ISL_2781718, EPI_ISL_2781721, EPI_ISL_2781729, EPI_ISL_2781735, EPI_ISL_2781736, EPI_ISL_2781741, EPI_ISL_2781743, EPI_ISL_2781745, EPI_ISL_2781746, EPI_ISL_2781750, EPI_ISL_2781751, EPI_ISL_2781753, EPI_ISL_2781755, EPI_ISL_2781759, EPI_ISL_2781762, EPI_ISL_2781765, EPI_ISL_2781766, EPI_ISL_2781767, EPI_ISL_2781770, EPI_ISL_2781771, EPI_ISL_2781776, EPI_ISL_2781777, EPI_ISL_2781780, EPI_ISL_2781781, EPI_ISL_2781783, EPI_ISL_2781785, EPI_ISL_2781787, EPI_ISL_2781788, EPI_ISL_2781791, EPI_ISL_2781792, EPI_ISL_2781794, EPI_ISL_2781796, EPI_ISL_2781797, EPI_ISL_2781803, EPI_ISL_2781808, EPI_ISL_2781810, EPI_ISL_2781812, EPI_ISL_2781814, EPI_ISL_2781817, EPI_ISL_2781818, EPI_ISL_2781819, EPI_ISL_2781821, EPI_ISL_2781822, EPI_ISL_2781826, EPI_ISL_2781830, EPI_ISL_2781836, EPI_ISL_2781837, EPI_ISL_2781840, EPI_ISL_2781841, EPI_ISL_2781844, EPI_ISL_2781845, EPI_ISL_2781851, EPI_ISL_2781852, EPI_ISL_2781853, EPI_ISL_2781855, EPI_ISL_2781856, EPI_ISL_2781861, EPI_ISL_2781862, EPI_ISL_2781864, EPI_ISL_2781866, EPI_ISL_2781867, EPI_ISL_2781868, EPI_ISL_2781870, EPI_ISL_2781873, EPI_ISL_2781874, EPI_ISL_2781875, EPI_ISL_2781877, EPI_ISL_2781880, EPI_ISL_2781882, EPI_ISL_2781888, EPI_ISL_2781889, EPI_ISL_2781892, EPI_ISL_2781894, EPI_ISL_2781895, EPI_ISL_2781896, EPI_ISL_2781901, EPI_ISL_2781908, EPI_ISL_2781909, EPI_ISL_2781910, EPI_ISL_2781911, EPI_ISL_2781913, EPI_ISL_2781914, EPI_ISL_2781915, EPI_ISL_2781920, EPI_ISL_2781926, EPI_ISL_2781927, EPI_ISL_2781931, EPI_ISL_2781932, EPI_ISL_2781933, EPI_ISL_2781934, EPI_ISL_2781938, EPI_ISL_2781940, EPI_ISL_2781943, EPI_ISL_2781948, EPI_ISL_2781950, EPI_ISL_2781951, EPI_ISL_2781952, EPI_ISL_2781956, EPI_ISL_2781957, EPI_ISL_2781961, EPI_ISL_2781964, EPI_ISL_2781965, EPI_ISL_2781968, EPI_ISL_2781969, EPI_ISL_2781970, EPI_ISL_2781974, EPI_ISL_2781975, EPI_ISL_2781977, EPI_ISL_2781978, EPI_ISL_2781982, EPI_ISL_2781983, EPI_ISL_2781989, EPI_ISL_2781997, EPI_ISL_2782000, EPI_ISL_2782001, EPI_ISL_2782003, EPI_ISL_2782006, EPI_ISL_2782007, EPI_ISL_2782008, EPI_ISL_2782011, EPI_ISL_2782013, EPI_ISL_2782020, EPI_ISL_2782022, EPI_ISL_2782025, EPI_ISL_2782029, EPI_ISL_2782030, EPI_ISL_2782033, EPI_ISL_2782034, EPI_ISL_2782036, EPI_ISL_2782044, EPI_ISL_2782054, EPI_ISL_2782055, EPI_ISL_2782058, EPI_ISL_2782063, EPI_ISL_2782068, EPI_ISL_2782070, EPI_ISL_2782071, EPI_ISL_2782076, EPI_ISL_2782077, EPI_ISL_2782079, EPI_ISL_2782080, EPI_ISL_2782086, EPI_ISL_2782090, EPI_ISL_2782092, EPI_ISL_2782095, EPI_ISL_2782100, EPI_ISL_2782104, EPI_ISL_2782105, EPI_ISL_2782113, EPI_ISL_2782117, EPI_ISL_2782124, EPI_ISL_2782126, EPI_ISL_2782127, EPI_ISL_2782136, EPI_ISL_2782140, EPI_ISL_2782145, EPI_ISL_2782149, EPI_ISL_2782150, EPI_ISL_2782151, EPI_ISL_2782152, EPI_ISL_2782153, EPI_ISL_2782157, EPI_ISL_2782160, EPI_ISL_2782170, EPI_ISL_2782171, EPI_ISL_2782176, EPI_ISL_2782177, EPI_ISL_2782180, EPI_ISL_2782181, EPI_ISL_2782186, EPI_ISL_2782193, EPI_ISL_2782202, EPI_ISL_2782203, EPI_ISL_2782205, EPI_ISL_2782209, EPI_ISL_2782215, EPI_ISL_2782216, EPI_ISL_2782218, EPI_ISL_2782221, EPI_ISL_2782223, EPI_ISL_2782224, EPI_ISL_2782225, EPI_ISL_2782226, EPI_ISL_2782231, EPI_ISL_2782232, EPI_ISL_2782237, EPI_ISL_2782239, EPI_ISL_2782241, EPI_ISL_2782242, EPI_ISL_2782245, EPI_ISL_2782247, EPI_ISL_2782252, EPI_ISL_2782256, EPI_ISL_2782258, EPI_ISL_2782264, EPI_ISL_2782267, EPI_ISL_2782271, EPI_ISL_2782289, EPI_ISL_2782295, EPI_ISL_2782303, EPI_ISL_2782309, EPI_ISL_2782310, EPI_ISL_2782311, EPI_ISL_2803967, EPI_ISL_2804060, EPI_ISL_2804106, EPI_ISL_2804411, EPI_ISL_2804459, EPI_ISL_2804610, EPI_ISL_2804624, EPI_ISL_2804630, EPI_ISL_2804637, EPI_ISL_2804656, EPI_ISL_2804735, EPI_ISL_2804749, EPI_ISL_2804813, EPI_ISL_2804822, EPI_ISL_2804824, EPI_ISL_2804832, EPI_ISL_2804838, EPI_ISL_2804840, EPI_ISL_2804841, EPI_ISL_2804845, EPI_ISL_2804848, EPI_ISL_2804851, EPI_ISL_2804855, EPI_ISL_2804886, EPI_ISL_2804917, EPI_ISL_2804995, EPI_ISL_2805024, EPI_ISL_2805033, EPI_ISL_2805053, EPI_ISL_2805067, EPI_ISL_2805073, EPI_ISL_2805124, EPI_ISL_2805165, EPI_ISL_2805178, EPI_ISL_2805179, EPI_ISL_2805194, EPI_ISL_2805225, EPI_ISL_2805228, EPI_ISL_2805248, EPI_ISL_2805256, EPI_ISL_2805319, EPI_ISL_2806004, EPI_ISL_2806021, EPI_ISL_2806022, EPI_ISL_2806028, EPI_ISL_2806073, EPI_ISL_2806079, EPI_ISL_2806105, EPI_ISL_2806121, EPI_ISL_2806128, EPI_ISL_2806133, EPI_ISL_2806135, EPI_ISL_2806146, EPI_ISL_2806151, EPI_ISL_2806161, EPI_ISL_2806165, EPI_ISL_2806166, EPI_ISL_2806177, EPI_ISL_2806193, EPI_ISL_2806249, EPI_ISL_2806255, EPI_ISL_2806257, EPI_ISL_2806281, EPI_ISL_2806345, EPI_ISL_2806357, EPI_ISL_2806361, EPI_ISL_2806372, EPI_ISL_2806412, EPI_ISL_2807356, EPI_ISL_2807357, EPI_ISL_2807371, EPI_ISL_2807373, EPI_ISL_2807374, EPI_ISL_2807386, EPI_ISL_2807390, EPI_ISL_2807394, EPI_ISL_2807396, EPI_ISL_2807397, EPI_ISL_2807398, EPI_ISL_2807414, EPI_ISL_2807415, EPI_ISL_2807417, EPI_ISL_2807418, EPI_ISL_2807419, EPI_ISL_2807422, EPI_ISL_2807425, EPI_ISL_2807427, EPI_ISL_2807435, EPI_ISL_2807436, EPI_ISL_2807438, EPI_ISL_2807439, EPI_ISL_2807440, EPI_ISL_2807441, EPI_ISL_2807447, EPI_ISL_2807449, EPI_ISL_2807450, EPI_ISL_2807451, EPI_ISL_2807452, EPI_ISL_2807453, EPI_ISL_2807462, EPI_ISL_2807466, EPI_ISL_2807467, EPI_ISL_2807471, EPI_ISL_2807472, EPI_ISL_2807479, EPI_ISL_2807485, EPI_ISL_2807486, EPI_ISL_2807488, EPI_ISL_2807490, EPI_ISL_2807492, EPI_ISL_2807497, EPI_ISL_2807498, EPI_ISL_2807504, EPI_ISL_2807508, EPI_ISL_2807513, EPI_ISL_2807524, EPI_ISL_2807535, EPI_ISL_2807539, EPI_ISL_2807543, EPI_ISL_2807546, EPI_ISL_2807553, EPI_ISL_2807560, EPI_ISL_2807570, EPI_ISL_2807576, EPI_ISL_2807579, EPI_ISL_2807583, EPI_ISL_2807586, EPI_ISL_2807587, EPI_ISL_2807588, EPI_ISL_2807591, EPI_ISL_2807592, EPI_ISL_2807594, EPI_ISL_2807596, EPI_ISL_2807597, EPI_ISL_2807604, EPI_ISL_2807609, EPI_ISL_2807614, EPI_ISL_2807623, EPI_ISL_2807624, EPI_ISL_2807633, EPI_ISL_2807635, EPI_ISL_2807647, EPI_ISL_2807656, EPI_ISL_2807659, EPI_ISL_2807665, EPI_ISL_2807667, EPI_ISL_2807672, EPI_ISL_2807675, EPI_ISL_2807677, EPI_ISL_2807681, EPI_ISL_2807683, EPI_ISL_2807691, EPI_ISL_2807696, EPI_ISL_2807700, EPI_ISL_2807701, EPI_ISL_2807703, EPI_ISL_2808821, EPI_ISL_2808988, EPI_ISL_2809047, EPI_ISL_2809099, EPI_ISL_2809100, EPI_ISL_2809105, EPI_ISL_2809106, EPI_ISL_2809108, EPI_ISL_2809110, EPI_ISL_2809113, EPI_ISL_2809114, EPI_ISL_2809117, EPI_ISL_2809120, EPI_ISL_2809121, EPI_ISL_2809123, EPI_ISL_2809126, EPI_ISL_2809129, EPI_ISL_2809131, EPI_ISL_2809133, EPI_ISL_2809135, EPI_ISL_2809136, EPI_ISL_2809140, EPI_ISL_2809142, EPI_ISL_2809143, EPI_ISL_2809153, EPI_ISL_2809158, EPI_ISL_2809160, EPI_ISL_2809161, EPI_ISL_2809163, EPI_ISL_2809165, EPI_ISL_2809166, EPI_ISL_2809175, EPI_ISL_2809177, EPI_ISL_2809179, EPI_ISL_2809184, EPI_ISL_2809185, EPI_ISL_2809188, EPI_ISL_2809190, EPI_ISL_2809193, EPI_ISL_2809197, EPI_ISL_2809200, EPI_ISL_2809202, EPI_ISL_2809208, EPI_ISL_2809209, EPI_ISL_2809210, EPI_ISL_2809213, EPI_ISL_2809214, EPI_ISL_2809215, EPI_ISL_2809219, EPI_ISL_2809221, EPI_ISL_2809222, EPI_ISL_2809224, EPI_ISL_2809229, EPI_ISL_2809231, EPI_ISL_2809232, EPI_ISL_2809235, EPI_ISL_2809237, EPI_ISL_2809238, EPI_ISL_2809239, EPI_ISL_2809241, EPI_ISL_2809247, EPI_ISL_2809248, EPI_ISL_2809251, EPI_ISL_2809253, EPI_ISL_2809256, EPI_ISL_2809257, EPI_ISL_2809262, EPI_ISL_2809267, EPI_ISL_2809268, EPI_ISL_2809277, EPI_ISL_2809283, EPI_ISL_2809289, EPI_ISL_2809291, EPI_ISL_2809298, EPI_ISL_2809299, EPI_ISL_2809306, EPI_ISL_2809315, EPI_ISL_2809316, EPI_ISL_2809317, EPI_ISL_2809321, EPI_ISL_2809322, EPI_ISL_2809325, EPI_ISL_2809329, EPI_ISL_2809337, EPI_ISL_2809343, EPI_ISL_2809353, EPI_ISL_2809357, EPI_ISL_2809363, EPI_ISL_2809364, EPI_ISL_2809369, EPI_ISL_2809370, EPI_ISL_2809375, EPI_ISL_2809378, EPI_ISL_2809383, EPI_ISL_2809384, EPI_ISL_2809386, EPI_ISL_2809387, EPI_ISL_2809389, EPI_ISL_2809391, EPI_ISL_2809394, EPI_ISL_2809396, EPI_ISL_2809397, EPI_ISL_2809398, EPI_ISL_2809404, EPI_ISL_2809406, EPI_ISL_2809409, EPI_ISL_2809411, EPI_ISL_2809418, EPI_ISL_2809421, EPI_ISL_2809461, EPI_ISL_2809545, EPI_ISL_2809709, EPI_ISL_2815039, EPI_ISL_2830631, EPI_ISL_2830634, EPI_ISL_2831060, EPI_ISL_2831282, EPI_ISL_2831324, EPI_ISL_2831326, EPI_ISL_2831330, EPI_ISL_2831331, EPI_ISL_2831332, EPI_ISL_2831334, EPI_ISL_2831335, EPI_ISL_2831344, EPI_ISL_2831345, EPI_ISL_2831346, EPI_ISL_2831347, EPI_ISL_2831348, EPI_ISL_2831349, EPI_ISL_2831350, EPI_ISL_2831352, EPI_ISL_2831354, EPI_ISL_2831355, EPI_ISL_2831357, EPI_ISL_2831358, EPI_ISL_2831360, EPI_ISL_2831362, EPI_ISL_2831368, EPI_ISL_2831369, EPI_ISL_2831372, EPI_ISL_2831374, EPI_ISL_2831375, EPI_ISL_2831377, EPI_ISL_2831380, EPI_ISL_2831382, EPI_ISL_2831383, EPI_ISL_2831384, EPI_ISL_2831385, EPI_ISL_2831388, EPI_ISL_2831390, EPI_ISL_2831391, EPI_ISL_2831393, EPI_ISL_2831394, EPI_ISL_2831395, EPI_ISL_2831396, EPI_ISL_2831397, EPI_ISL_2831398, EPI_ISL_2831399, EPI_ISL_2831400, EPI_ISL_2831401, EPI_ISL_2831403, EPI_ISL_2831404, EPI_ISL_2831405, EPI_ISL_2831406, EPI_ISL_2831407, EPI_ISL_2831409, EPI_ISL_2831410, EPI_ISL_2831412, EPI_ISL_2831413, EPI_ISL_2831414, EPI_ISL_2831416, EPI_ISL_2831417, EPI_ISL_2831418, EPI_ISL_2831419, EPI_ISL_2831420, EPI_ISL_2831423, EPI_ISL_2831425, EPI_ISL_2831426, EPI_ISL_2831427, EPI_ISL_2831429, EPI_ISL_2831432, EPI_ISL_2831433, EPI_ISL_2831435, EPI_ISL_2831436, EPI_ISL_2831437, EPI_ISL_2831441, EPI_ISL_2831442, EPI_ISL_2831443, EPI_ISL_2831444, EPI_ISL_2831446, EPI_ISL_2831447, EPI_ISL_2831452, EPI_ISL_2831453, EPI_ISL_2831455, EPI_ISL_2831456, EPI_ISL_2831457, EPI_ISL_2831458, EPI_ISL_2831462, EPI_ISL_2831463, EPI_ISL_2831464, EPI_ISL_2831466, EPI_ISL_2831467, EPI_ISL_2831468, EPI_ISL_2831469, EPI_ISL_2831471, EPI_ISL_2831472, EPI_ISL_2831474, EPI_ISL_2831475, EPI_ISL_2831476, EPI_ISL_2831477, EPI_ISL_2831480, EPI_ISL_2831481, EPI_ISL_2831483, EPI_ISL_2831485, EPI_ISL_2831493, EPI_ISL_2831496, EPI_ISL_2831499, EPI_ISL_2831501, EPI_ISL_2831503, EPI_ISL_2831505, EPI_ISL_2831506, EPI_ISL_2831508, EPI_ISL_2831509, EPI_ISL_2831511, EPI_ISL_2831513, EPI_ISL_2831515, EPI_ISL_2831516, EPI_ISL_2831518, EPI_ISL_2831519, EPI_ISL_2831521, EPI_ISL_2831522, EPI_ISL_2831524, EPI_ISL_2831525, EPI_ISL_2831532, EPI_ISL_2831533, EPI_ISL_2831534, EPI_ISL_2831536, EPI_ISL_2831537, EPI_ISL_2831542, EPI_ISL_2831543, EPI_ISL_2831545, EPI_ISL_2831546, EPI_ISL_2831547, EPI_ISL_2831548, EPI_ISL_2831549, EPI_ISL_2831550, EPI_ISL_2831553, EPI_ISL_2831555, EPI_ISL_2831557, EPI_ISL_2831560, EPI_ISL_2831562, EPI_ISL_2831565, EPI_ISL_2831566, EPI_ISL_2831567, EPI_ISL_2831568, EPI_ISL_2831569, EPI_ISL_2831573, EPI_ISL_2831576, EPI_ISL_2831577, EPI_ISL_2831578, EPI_ISL_2831579, EPI_ISL_2831582, EPI_ISL_2831583, EPI_ISL_2832075, EPI_ISL_2832077, EPI_ISL_2832078, EPI_ISL_2832092, EPI_ISL_2832098, EPI_ISL_2832101, EPI_ISL_2868226, EPI_ISL_2910774, EPI_ISL_2910806 | see above | Lighthouse Lab in Alderley Park | Wellcome Sanger Institute for the COVID-19 Genomics UK (COG-UK) Consortium | Cordelia Langford; David K. Jackson; Dominic Kwiatkowski; Ewan Harrison; Ian Johnston; Jacquelyn Wynn; Jeffrey Barrett; John Sillitoe on behalf of the Wellcome Sanger Institute COVID-19 Surveillance Team; Mairead Hyland; Roberto Amato; Sonia Goncalves; The Lighthouse Lab in Alderley Park and Alex Alderton |
| EPI_ISL_2699521, EPI_ISL_2699525, EPI_ISL_2699526, EPI_ISL_2699528, EPI_ISL_2699529, EPI_ISL_2699530, EPI_ISL_2699532, EPI_ISL_2699533, EPI_ISL_2699539, EPI_ISL_2699541, EPI_ISL_2699548, EPI_ISL_2699549, EPI_ISL_2699553, EPI_ISL_2699554, EPI_ISL_2699556, EPI_ISL_2699557, EPI_ISL_2699558, EPI_ISL_2699559, EPI_ISL_2699560, EPI_ISL_2699564, EPI_ISL_2699565, EPI_ISL_2699572, EPI_ISL_2699573, EPI_ISL_2699576, EPI_ISL_2699579, EPI_ISL_2699580, EPI_ISL_2699582, EPI_ISL_2699584, EPI_ISL_2699585, EPI_ISL_2699588, EPI_ISL_2699590, EPI_ISL_2699591, EPI_ISL_2699592, EPI_ISL_2699598, EPI_ISL_2699600, EPI_ISL_2699601, EPI_ISL_2699602, EPI_ISL_2699603, EPI_ISL_2699613, EPI_ISL_2699618, EPI_ISL_2699619, EPI_ISL_2699621, EPI_ISL_2699626, EPI_ISL_2699627, EPI_ISL_2699631, EPI_ISL_2699640, EPI_ISL_2699641, EPI_ISL_2699645, EPI_ISL_2699652, EPI_ISL_2699655, EPI_ISL_2699656, EPI_ISL_2699658, EPI_ISL_2699659, EPI_ISL_2699661, EPI_ISL_2699671, EPI_ISL_2699672, EPI_ISL_2699673, EPI_ISL_2699677, EPI_ISL_2699679, EPI_ISL_2699684, EPI_ISL_2699687, EPI_ISL_2699695, EPI_ISL_2699700, EPI_ISL_2699705, EPI_ISL_2699706, EPI_ISL_2699707, EPI_ISL_2699709, EPI_ISL_2699713, EPI_ISL_2699718, EPI_ISL_2699719, EPI_ISL_2699725, EPI_ISL_2699727, EPI_ISL_2699731, EPI_ISL_2699733, EPI_ISL_2699747, EPI_ISL_2699754, EPI_ISL_2699768, EPI_ISL_2699769, EPI_ISL_2699770, EPI_ISL_2699780, EPI_ISL_2699781, EPI_ISL_2699784, EPI_ISL_2699786, EPI_ISL_2699787, EPI_ISL_2699788, EPI_ISL_2699790, EPI_ISL_2699791, EPI_ISL_2699793, EPI_ISL_2699795, EPI_ISL_2699797, EPI_ISL_2699798, EPI_ISL_2699807, EPI_ISL_2699808, EPI_ISL_2699813, EPI_ISL_2699817, EPI_ISL_2699820, EPI_ISL_2699824, EPI_ISL_2699831, EPI_ISL_2699836, EPI_ISL_2699841, EPI_ISL_2699847, EPI_ISL_2699851, EPI_ISL_2699852, EPI_ISL_2699856, EPI_ISL_2699858, EPI_ISL_2699859, EPI_ISL_2699862, EPI_ISL_2699865, EPI_ISL_2700214, EPI_ISL_2700216, EPI_ISL_2700217, EPI_ISL_2700218, EPI_ISL_2700219, EPI_ISL_2700220, EPI_ISL_2700221, EPI_ISL_2700222, EPI_ISL_2700223, EPI_ISL_2700224, EPI_ISL_2700226, EPI_ISL_2700228, EPI_ISL_2700229, EPI_ISL_2700234, EPI_ISL_2700242, EPI_ISL_2700245, EPI_ISL_2700247, EPI_ISL_2700254, EPI_ISL_2700256, EPI_ISL_2700257, EPI_ISL_2700258, EPI_ISL_2700259, EPI_ISL_2700260, EPI_ISL_2700264, EPI_ISL_2700265, EPI_ISL_2700266, EPI_ISL_2700267, EPI_ISL_2700268, EPI_ISL_2700269, EPI_ISL_2700270, EPI_ISL_2700273, EPI_ISL_2700274, EPI_ISL_2700285, EPI_ISL_2700286, EPI_ISL_2700287, EPI_ISL_2700288, EPI_ISL_2700289, EPI_ISL_2700290, EPI_ISL_2700295, EPI_ISL_2700297, EPI_ISL_2700299, EPI_ISL_2700301, EPI_ISL_2700304, EPI_ISL_2700308, EPI_ISL_2700309, EPI_ISL_2700311, EPI_ISL_2700312, EPI_ISL_2700313, EPI_ISL_2700314, EPI_ISL_2700315, EPI_ISL_2700317, EPI_ISL_2700323, EPI_ISL_2700327, EPI_ISL_2700330, EPI_ISL_2700331, EPI_ISL_2700332, EPI_ISL_2700334, EPI_ISL_2700335, EPI_ISL_2700336, EPI_ISL_2700339, EPI_ISL_2700340, EPI_ISL_2700341, EPI_ISL_2700342, EPI_ISL_2700345, EPI_ISL_2700350, EPI_ISL_2700353, EPI_ISL_2700354, EPI_ISL_2700357, EPI_ISL_2700358, EPI_ISL_2700366, EPI_ISL_2700367, EPI_ISL_2700368, EPI_ISL_2700369, EPI_ISL_2700370, EPI_ISL_2700372, EPI_ISL_2700373, EPI_ISL_2700374, EPI_ISL_2700375, EPI_ISL_2700377, EPI_ISL_2700382, EPI_ISL_2700383, EPI_ISL_2700386, EPI_ISL_2700387, EPI_ISL_2700390, EPI_ISL_2700391, EPI_ISL_2700392, EPI_ISL_2700393, EPI_ISL_2700394, EPI_ISL_2700397, EPI_ISL_2700398, EPI_ISL_2700399, EPI_ISL_2700400, EPI_ISL_2700401, EPI_ISL_2700403, EPI_ISL_2700405, EPI_ISL_2700409, EPI_ISL_2700411, EPI_ISL_2700414, EPI_ISL_2700416, EPI_ISL_2700417, EPI_ISL_2700426, EPI_ISL_2700428, EPI_ISL_2700432, EPI_ISL_2700437, EPI_ISL_2700438, EPI_ISL_2700439, EPI_ISL_2700440, EPI_ISL_2700447, EPI_ISL_2700449, EPI_ISL_2700450, EPI_ISL_2700451, EPI_ISL_2700452, EPI_ISL_2700453, EPI_ISL_2700457, EPI_ISL_2700458, EPI_ISL_2700459, EPI_ISL_2700462, EPI_ISL_2700464, EPI_ISL_2700466, EPI_ISL_2700468, EPI_ISL_2700469, EPI_ISL_2700470, EPI_ISL_2700473, EPI_ISL_2700478, EPI_ISL_2700479, EPI_ISL_2700480, EPI_ISL_2700481, EPI_ISL_2700482, EPI_ISL_2700483, EPI_ISL_2700484, EPI_ISL_2700487, EPI_ISL_2700490, EPI_ISL_2700492, EPI_ISL_2700493, EPI_ISL_2700495, EPI_ISL_2700499, EPI_ISL_2700503, EPI_ISL_2700506, EPI_ISL_2700508, EPI_ISL_2700510, EPI_ISL_2700512, EPI_ISL_2700514, EPI_ISL_2700516, EPI_ISL_2700518, EPI_ISL_2700520, EPI_ISL_2700521, EPI_ISL_2700523, EPI_ISL_2700525, EPI_ISL_2700527, EPI_ISL_2700531, EPI_ISL_2700532, EPI_ISL_2700533, EPI_ISL_2700536, EPI_ISL_2700539, EPI_ISL_2700540, EPI_ISL_2700541, EPI_ISL_2700542, EPI_ISL_2700543, EPI_ISL_2700545, EPI_ISL_2700546, EPI_ISL_2700549, EPI_ISL_2700554, EPI_ISL_2700557, EPI_ISL_2700560, EPI_ISL_2700561, EPI_ISL_2700562, EPI_ISL_2700563, EPI_ISL_2700564, EPI_ISL_2700566, EPI_ISL_2700567, EPI_ISL_2700569, EPI_ISL_2700571, EPI_ISL_2700573, EPI_ISL_2700575, EPI_ISL_2700950, EPI_ISL_2700956, EPI_ISL_2700966, EPI_ISL_2700973, EPI_ISL_2700974, EPI_ISL_2700983, EPI_ISL_2700987, EPI_ISL_2700989, EPI_ISL_2700996, EPI_ISL_2701001, EPI_ISL_2701004, EPI_ISL_2701008, EPI_ISL_2701017, EPI_ISL_2701024, EPI_ISL_2701034, EPI_ISL_2701038, EPI_ISL_2701042, EPI_ISL_2701047, EPI_ISL_2701055, EPI_ISL_2701059, EPI_ISL_2701061, EPI_ISL_2701068, EPI_ISL_2701070, EPI_ISL_2701072, EPI_ISL_2701084, EPI_ISL_2701086, EPI_ISL_2701089, EPI_ISL_2701091, EPI_ISL_2701092, EPI_ISL_2701093, EPI_ISL_2701099, EPI_ISL_2701115, EPI_ISL_2701118, EPI_ISL_2701120, EPI_ISL_2701123, EPI_ISL_2701130, EPI_ISL_2701132, EPI_ISL_2701134, EPI_ISL_2701144, EPI_ISL_2701147, EPI_ISL_2701153, EPI_ISL_2701158, EPI_ISL_2701165, EPI_ISL_2701172, EPI_ISL_2701174, EPI_ISL_2701186, EPI_ISL_2701200, EPI_ISL_2701203, EPI_ISL_2701204, EPI_ISL_2701208, EPI_ISL_2701211, EPI_ISL_2701214, EPI_ISL_2701217, EPI_ISL_2701219, EPI_ISL_2701220, EPI_ISL_2701221, EPI_ISL_2701224, EPI_ISL_2701232, EPI_ISL_2701233, EPI_ISL_2701234, EPI_ISL_2701236, EPI_ISL_2701239, EPI_ISL_2701241, EPI_ISL_2701245, EPI_ISL_2701252, EPI_ISL_2701254, EPI_ISL_2701259, EPI_ISL_2701260, EPI_ISL_2701261, EPI_ISL_2701273, EPI_ISL_2701274, EPI_ISL_2701275, EPI_ISL_2701282, EPI_ISL_2701288, EPI_ISL_2701295, EPI_ISL_2701297, EPI_ISL_2701298, EPI_ISL_2701300, EPI_ISL_2701303, EPI_ISL_2701305, EPI_ISL_2701307, EPI_ISL_2701308, EPI_ISL_2701314, EPI_ISL_2701316, EPI_ISL_2701317, EPI_ISL_2701321, EPI_ISL_2701324, EPI_ISL_2701327, EPI_ISL_2701329, EPI_ISL_2701330, EPI_ISL_2701333, EPI_ISL_2701337, EPI_ISL_2701338, EPI_ISL_2701339, EPI_ISL_2701340, EPI_ISL_2701341, EPI_ISL_2701345, EPI_ISL_2701346, EPI_ISL_2701347, EPI_ISL_2701348, EPI_ISL_2701349, EPI_ISL_2701350, EPI_ISL_2701351, EPI_ISL_2701354, EPI_ISL_2701356, EPI_ISL_2701360, EPI_ISL_2701364, EPI_ISL_2701366, EPI_ISL_2701369, EPI_ISL_2701370, EPI_ISL_2701372, EPI_ISL_2701374, EPI_ISL_2701375, EPI_ISL_2701379, EPI_ISL_2701380, EPI_ISL_2701383, EPI_ISL_2701385, EPI_ISL_2701389, EPI_ISL_2701390, EPI_ISL_2701391, EPI_ISL_2701397, EPI_ISL_2701399, EPI_ISL_2701400, EPI_ISL_2701401, EPI_ISL_2701404, EPI_ISL_2701405, EPI_ISL_2701407, EPI_ISL_2701409, EPI_ISL_2701411, EPI_ISL_2701415, EPI_ISL_2701423, EPI_ISL_2701425, EPI_ISL_2701426, EPI_ISL_2701430, EPI_ISL_2701431, EPI_ISL_2701434, EPI_ISL_2701438, EPI_ISL_2701439, EPI_ISL_2701441, EPI_ISL_2701443, EPI_ISL_2701444, EPI_ISL_2701448, EPI_ISL_2701452, EPI_ISL_2701454, EPI_ISL_2701455, EPI_ISL_2701456, EPI_ISL_2701457, EPI_ISL_2701458, EPI_ISL_2701459, EPI_ISL_2701460, EPI_ISL_2701462, EPI_ISL_2701464, EPI_ISL_2701468, EPI_ISL_2701470, EPI_ISL_2701471, EPI_ISL_2701474, EPI_ISL_2701475, EPI_ISL_2701476, EPI_ISL_2701478, EPI_ISL_2701479, EPI_ISL_2701480, EPI_ISL_2701486, EPI_ISL_2701488, EPI_ISL_2701489, EPI_ISL_2701490, EPI_ISL_2701491, EPI_ISL_2701492, EPI_ISL_2701498, EPI_ISL_2701505, EPI_ISL_2701506, EPI_ISL_2701507, EPI_ISL_2701508, EPI_ISL_2701512, EPI_ISL_2701513, EPI_ISL_2701514, EPI_ISL_2701516, EPI_ISL_2701517, EPI_ISL_2701518, EPI_ISL_2701519, EPI_ISL_2701522, EPI_ISL_2701523, EPI_ISL_2701525, EPI_ISL_2701527, EPI_ISL_2701528, EPI_ISL_2701529, EPI_ISL_2701530, EPI_ISL_2701534, EPI_ISL_2701536, EPI_ISL_2701537, EPI_ISL_2701539, EPI_ISL_2701540, EPI_ISL_2701541, EPI_ISL_2701542, EPI_ISL_2701543, EPI_ISL_2701545, EPI_ISL_2701546, EPI_ISL_2701547, EPI_ISL_2701548, EPI_ISL_2701556, EPI_ISL_2701558, EPI_ISL_2701562, EPI_ISL_2701564, EPI_ISL_2701565, EPI_ISL_2701567, EPI_ISL_2701568, EPI_ISL_2701569, EPI_ISL_2701574, EPI_ISL_2701576, EPI_ISL_2701579, EPI_ISL_2701580, EPI_ISL_2701582, EPI_ISL_2701585, EPI_ISL_2701589, EPI_ISL_2701590, EPI_ISL_2701591, EPI_ISL_2701597, EPI_ISL_2701599, EPI_ISL_2701600, EPI_ISL_2701602, EPI_ISL_2701604, EPI_ISL_2701605, EPI_ISL_2701614, EPI_ISL_2701615, EPI_ISL_2701621, EPI_ISL_2701624, EPI_ISL_2701627, EPI_ISL_2701629, EPI_ISL_2701630, EPI_ISL_2701633, EPI_ISL_2701635, EPI_ISL_2701636, EPI_ISL_2701638, EPI_ISL_2701640, EPI_ISL_2701643, EPI_ISL_2701646, EPI_ISL_2701647, EPI_ISL_2701651, EPI_ISL_2701659, EPI_ISL_2701661, EPI_ISL_2701662, EPI_ISL_2701663, EPI_ISL_2701667, EPI_ISL_2701670, EPI_ISL_2701671, EPI_ISL_2701672, EPI_ISL_2701674, EPI_ISL_2701678, EPI_ISL_2701679, EPI_ISL_2701680, EPI_ISL_2701682, EPI_ISL_2701683, EPI_ISL_2701687, EPI_ISL_2701688, EPI_ISL_2701691, EPI_ISL_2701694, EPI_ISL_2701697, EPI_ISL_2701699, EPI_ISL_2701701, EPI_ISL_2701708, EPI_ISL_2701710, EPI_ISL_2701711, EPI_ISL_2701713, EPI_ISL_2701714, EPI_ISL_2701715, EPI_ISL_2701718, EPI_ISL_2701719, EPI_ISL_2701724, EPI_ISL_2701725, EPI_ISL_2701727, EPI_ISL_2701728, EPI_ISL_2701729, EPI_ISL_2701730, EPI_ISL_2701732, EPI_ISL_2701734, EPI_ISL_2701737, EPI_ISL_2701738, EPI_ISL_2701740, EPI_ISL_2701742, EPI_ISL_2701747, EPI_ISL_2701750, EPI_ISL_2701756, EPI_ISL_2701757, EPI_ISL_2701758, EPI_ISL_2701759, EPI_ISL_2701760, EPI_ISL_2701761, EPI_ISL_2701762, EPI_ISL_2701763, EPI_ISL_2701764, EPI_ISL_2701766, EPI_ISL_2701767, EPI_ISL_2701774, EPI_ISL_2701775, EPI_ISL_2701776, EPI_ISL_2701783, EPI_ISL_2701784, EPI_ISL_2701787, EPI_ISL_2701788, EPI_ISL_2701789, EPI_ISL_2701791, EPI_ISL_2701792, EPI_ISL_2701793, EPI_ISL_2701794, EPI_ISL_2701797, EPI_ISL_2701798, EPI_ISL_2701799, EPI_ISL_2701800, EPI_ISL_2701804, EPI_ISL_2701806, EPI_ISL_2701807, EPI_ISL_2701814, EPI_ISL_2701823, EPI_ISL_2701825, EPI_ISL_2701828, EPI_ISL_2701832, EPI_ISL_2701833, EPI_ISL_2701835, EPI_ISL_2701838, EPI_ISL_2701840, EPI_ISL_2701841, EPI_ISL_2701842, EPI_ISL_2701843, EPI_ISL_2701844, EPI_ISL_2701848, EPI_ISL_2701849, EPI_ISL_2701852, EPI_ISL_2701855, EPI_ISL_2701857, EPI_ISL_2701859, EPI_ISL_2701865, EPI_ISL_2701869, EPI_ISL_2701871, EPI_ISL_2701872, EPI_ISL_2701873, EPI_ISL_2701874, EPI_ISL_2701875, EPI_ISL_2701876, EPI_ISL_2701882, EPI_ISL_2701886, EPI_ISL_2701887, EPI_ISL_2701889, EPI_ISL_2701891, EPI_ISL_2701893, EPI_ISL_2701896, EPI_ISL_2701898, EPI_ISL_2701899, EPI_ISL_2701900, EPI_ISL_2701903, EPI_ISL_2701905, EPI_ISL_2701907, EPI_ISL_2701908, EPI_ISL_2701909, EPI_ISL_2701914, EPI_ISL_2701917, EPI_ISL_2701918, EPI_ISL_2701920, EPI_ISL_2701921, EPI_ISL_2701922, EPI_ISL_2701924, EPI_ISL_2701925, EPI_ISL_2701928, EPI_ISL_2701930, EPI_ISL_2701934, EPI_ISL_2701935, EPI_ISL_2701942, EPI_ISL_2701947, EPI_ISL_2701949, EPI_ISL_2701954, EPI_ISL_2701958, EPI_ISL_2701959, EPI_ISL_2701961, EPI_ISL_2701962, EPI_ISL_2701971, EPI_ISL_2701972, EPI_ISL_2701974, EPI_ISL_2701975, EPI_ISL_2701976, EPI_ISL_2701977, EPI_ISL_2701980, EPI_ISL_2701983, EPI_ISL_2701989, EPI_ISL_2701991, EPI_ISL_2701992, EPI_ISL_2701994, EPI_ISL_2701998, EPI_ISL_2701999, EPI_ISL_2702000, EPI_ISL_2702001, EPI_ISL_2702002, EPI_ISL_2702005, EPI_ISL_2702006, EPI_ISL_2702007, EPI_ISL_2702008, EPI_ISL_2702009, EPI_ISL_2702010, EPI_ISL_2702012, EPI_ISL_2702013, EPI_ISL_2702016, EPI_ISL_2702018, EPI_ISL_2702019, EPI_ISL_2702021, EPI_ISL_2702025, EPI_ISL_2702026, EPI_ISL_2702029, EPI_ISL_2702031, EPI_ISL_2702038, EPI_ISL_2702042, EPI_ISL_2702043, EPI_ISL_2702044, EPI_ISL_2702046, EPI_ISL_2702047, EPI_ISL_2702049, EPI_ISL_2702050, EPI_ISL_2702052, EPI_ISL_2702053, EPI_ISL_2702057, EPI_ISL_2702059, EPI_ISL_2702060, EPI_ISL_2702063, EPI_ISL_2702065, EPI_ISL_2702066, EPI_ISL_2702068, EPI_ISL_2702070, EPI_ISL_2702075, EPI_ISL_2702076, EPI_ISL_2702079, EPI_ISL_2702080, EPI_ISL_2702083, EPI_ISL_2702092, EPI_ISL_2702093, EPI_ISL_2702094, EPI_ISL_2702096, EPI_ISL_2702097, EPI_ISL_2702102, EPI_ISL_2702103, EPI_ISL_2702104, EPI_ISL_2702105, EPI_ISL_2702106, EPI_ISL_2702108, EPI_ISL_2702110, EPI_ISL_2702115, EPI_ISL_2702118, EPI_ISL_2702121, EPI_ISL_2702122, EPI_ISL_2702124, EPI_ISL_2702126, EPI_ISL_2702128, EPI_ISL_2702132, EPI_ISL_2702133, EPI_ISL_2702135, EPI_ISL_2702137, EPI_ISL_2702138, EPI_ISL_2702141, EPI_ISL_2702142, EPI_ISL_2702147, EPI_ISL_2702152, EPI_ISL_2702153, EPI_ISL_2702155, EPI_ISL_2702157, EPI_ISL_2702158, EPI_ISL_2702161, EPI_ISL_2702162, EPI_ISL_2702164, EPI_ISL_2702165, EPI_ISL_2702167, EPI_ISL_2702169, EPI_ISL_2702171, EPI_ISL_2702173, EPI_ISL_2702174, EPI_ISL_2702175, EPI_ISL_2702177, EPI_ISL_2702178, EPI_ISL_2702179, EPI_ISL_2702181, EPI_ISL_2702185, EPI_ISL_2702187, EPI_ISL_2702188, EPI_ISL_2702190, EPI_ISL_2702191, EPI_ISL_2702195, EPI_ISL_2702199, EPI_ISL_2702202, EPI_ISL_2702203, EPI_ISL_2702204, EPI_ISL_2702206, EPI_ISL_2702208, EPI_ISL_2702209, EPI_ISL_2702210, EPI_ISL_2702212, EPI_ISL_2702218, EPI_ISL_2702220, EPI_ISL_2702221, EPI_ISL_2702227, EPI_ISL_2702230, EPI_ISL_2702232, EPI_ISL_2702234, EPI_ISL_2702235, EPI_ISL_2702238, EPI_ISL_2702239, EPI_ISL_2702240, EPI_ISL_2702243, EPI_ISL_2702244, EPI_ISL_2702245, EPI_ISL_2702246, EPI_ISL_2702247, EPI_ISL_2702248, EPI_ISL_2702249, EPI_ISL_2702250, EPI_ISL_2702251, EPI_ISL_2702252, EPI_ISL_2702253, EPI_ISL_2702254, EPI_ISL_2702259, EPI_ISL_2702262, EPI_ISL_2702263, EPI_ISL_2702264, EPI_ISL_2702265, EPI_ISL_2702266, EPI_ISL_2702267, EPI_ISL_2702268, EPI_ISL_2702269, EPI_ISL_2702270, EPI_ISL_2702271, EPI_ISL_2702272, EPI_ISL_2702273, EPI_ISL_2702276, EPI_ISL_2702278, EPI_ISL_2702279, EPI_ISL_2702280, EPI_ISL_2702284, EPI_ISL_2702286, EPI_ISL_2702289, EPI_ISL_2702290, EPI_ISL_2702291, EPI_ISL_2702292, EPI_ISL_2702293, EPI_ISL_2702294, EPI_ISL_2702296, EPI_ISL_2702297, EPI_ISL_2702302, EPI_ISL_2702307, EPI_ISL_2702310, EPI_ISL_2702319, | | | | |

EPI_ISL_2702321, EPI_ISL_2702322, EPI_ISL_2702323, EPI_ISL_2702324, EPI_ISL_2702325, EPI_ISL_2702327, EPI_ISL_2702328, EPI_ISL_2702332, EPI_ISL_2702333, EPI_ISL_2702335, EPI_ISL_2702336, EPI_ISL_2702337, EPI_ISL_2702342, EPI_ISL_2702343, EPI_ISL_2702344, EPI_ISL_2702346, EPI_ISL_2702348, EPI_ISL_2702350, EPI_ISL_2702351, EPI_ISL_2702353, EPI_ISL_2702355, EPI_ISL_2702356, EPI_ISL_2702358, EPI_ISL_2702360, EPI_ISL_2702363, EPI_ISL_2702364, EPI_ISL_2702367, EPI_ISL_2702369, EPI_ISL_2702370, EPI_ISL_2702374, EPI_ISL_2702377, EPI_ISL_2702378, EPI_ISL_2702380, EPI_ISL_2702382, EPI_ISL_2702383, EPI_ISL_2702387, EPI_ISL_2702388, EPI_ISL_2702389, EPI_ISL_2702390, EPI_ISL_2702392, EPI_ISL_2702393, EPI_ISL_2702394, EPI_ISL_2702395, EPI_ISL_2717693, EPI_ISL_2717785, EPI_ISL_2729122, EPI_ISL_2730319, EPI_ISL_2730614, EPI_ISL_2730785, EPI_ISL_2731100, EPI_ISL_2767294, EPI_ISL_2808866, EPI_ISL_2808894, EPI_ISL_2808906, EPI_ISL_2808918, EPI_ISL_2808920, EPI_ISL_2809446, EPI_ISL_2809454, EPI_ISL_2809985, EPI_ISL_2852526, EPI_ISL_2852547, EPI_ISL_2852566, EPI_ISL_2852568, EPI_ISL_2852573, EPI_ISL_2852578, EPI_ISL_2852582, EPI_ISL_2852588, EPI_ISL_2852596, EPI_ISL_2852598, EPI_ISL_2852601, EPI_ISL_2852607, EPI_ISL_2852612

| | | | |
|---|---|---|---|
| see above | Lighthouse Lab in Glasgow | Wellcome Sanger Institute for the COVID-19 Genomics UK (COG-UK) Consortium | Anna Dominiczak and Alex Alderton; Carol Clugston; Cordelia Langford; David Gray; David K. Jackson; Dominic Kwiatkowski; Ewan Harrison; Harper VanSteenhouse; Ian Johnston; Jeffrey Barrett; John Sillitoe on behalf of the Wellcome Sanger Institute COVID-19 Surveillance Team; Roberto Amato; Sonia Goncalves; Yumi Kasai |

EPI_ISL_2699520, EPI_ISL_2699522, EPI_ISL_2699523, EPI_ISL_2699524, EPI_ISL_2699535, EPI_ISL_2699536, EPI_ISL_2699537, EPI_ISL_2699538, EPI_ISL_2699542, EPI_ISL_2699547, EPI_ISL_2699550, EPI_ISL_2699551, EPI_ISL_2699552, EPI_ISL_2699555, EPI_ISL_2699561, EPI_ISL_2699562, EPI_ISL_2699566, EPI_ISL_2699567, EPI_ISL_2699568, EPI_ISL_2699569, EPI_ISL_2699570, EPI_ISL_2699571, EPI_ISL_2699574, EPI_ISL_2699575, EPI_ISL_2699578, EPI_ISL_2699581, EPI_ISL_2699586, EPI_ISL_2699587, EPI_ISL_2699589, EPI_ISL_2699594, EPI_ISL_2699596, EPI_ISL_2699604, EPI_ISL_2699605, EPI_ISL_2699606, EPI_ISL_2699607, EPI_ISL_2699611, EPI_ISL_2699612, EPI_ISL_2699614, EPI_ISL_2699615, EPI_ISL_2699616, EPI_ISL_2699620, EPI_ISL_2699622, EPI_ISL_2699624, EPI_ISL_2699628, EPI_ISL_2699629, EPI_ISL_2699630, EPI_ISL_2699633, EPI_ISL_2699634, EPI_ISL_2699635, EPI_ISL_2699636, EPI_ISL_2699637, EPI_ISL_2699638, EPI_ISL_2699643, EPI_ISL_2699646, EPI_ISL_2699648, EPI_ISL_2699650, EPI_ISL_2699651, EPI_ISL_2699653, EPI_ISL_2699654, EPI_ISL_2699660, EPI_ISL_2699662, EPI_ISL_2699663, EPI_ISL_2699665, EPI_ISL_2699668, EPI_ISL_2699669, EPI_ISL_2699670, EPI_ISL_2699675, EPI_ISL_2699676, EPI_ISL_2699678, EPI_ISL_2699680, EPI_ISL_2699681, EPI_ISL_2699682, EPI_ISL_2699686, EPI_ISL_2699688, EPI_ISL_2699689, EPI_ISL_2699693, EPI_ISL_2699697, EPI_ISL_2699699, EPI_ISL_2699701, EPI_ISL_2699703, EPI_ISL_2699708, EPI_ISL_2699710, EPI_ISL_2699712, EPI_ISL_2699714, EPI_ISL_2699716, EPI_ISL_2699717, EPI_ISL_2699720, EPI_ISL_2699721, EPI_ISL_2699722, EPI_ISL_2699723, EPI_ISL_2699724, EPI_ISL_2699726, EPI_ISL_2699729, EPI_ISL_2699730, EPI_ISL_2699732, EPI_ISL_2699734, EPI_ISL_2699735, EPI_ISL_2699736, EPI_ISL_2699737, EPI_ISL_2699738, EPI_ISL_2699739, EPI_ISL_2699740, EPI_ISL_2699742, EPI_ISL_2699744, EPI_ISL_2699748, EPI_ISL_2699749, EPI_ISL_2699752, EPI_ISL_2699756, EPI_ISL_2699757, EPI_ISL_2699758, EPI_ISL_2699759, EPI_ISL_2699760, EPI_ISL_2699763, EPI_ISL_2699765, EPI_ISL_2699766, EPI_ISL_2699767, EPI_ISL_2699771, EPI_ISL_2699773, EPI_ISL_2699774, EPI_ISL_2699778, EPI_ISL_2699779, EPI_ISL_2699782, EPI_ISL_2699783, EPI_ISL_2699785, EPI_ISL_2699799, EPI_ISL_2699800, EPI_ISL_2699801, EPI_ISL_2699802, EPI_ISL_2699803, EPI_ISL_2699805, EPI_ISL_2699809, EPI_ISL_2699810, EPI_ISL_2699814, EPI_ISL_2699815, EPI_ISL_2699816, EPI_ISL_2699818, EPI_ISL_2699819, EPI_ISL_2699822, EPI_ISL_2699823, EPI_ISL_2699828, EPI_ISL_2699830, EPI_ISL_2699833, EPI_ISL_2699834, EPI_ISL_2699835, EPI_ISL_2699837, EPI_ISL_2699839, EPI_ISL_2699840, EPI_ISL_2699842, EPI_ISL_2699843, EPI_ISL_2699844, EPI_ISL_2699845, EPI_ISL_2699846, EPI_ISL_2699848, EPI_ISL_2699849, EPI_ISL_2699853, EPI_ISL_2699861, EPI_ISL_2699863, EPI_ISL_2699864, EPI_ISL_2699866, EPI_ISL_2699867, EPI_ISL_2699869, EPI_ISL_2699872, EPI_ISL_2699874, EPI_ISL_2699875, EPI_ISL_2699876, EPI_ISL_2699877, EPI_ISL_2699878, EPI_ISL_2699880, EPI_ISL_2699881, EPI_ISL_2699883, EPI_ISL_2699884, EPI_ISL_2699885, EPI_ISL_2699890, EPI_ISL_2699891, EPI_ISL_2699893, EPI_ISL_2699895, EPI_ISL_2699897, EPI_ISL_2699900, EPI_ISL_2699901, EPI_ISL_2699902, EPI_ISL_2699903, EPI_ISL_2699904, EPI_ISL_2699905, EPI_ISL_2699906, EPI_ISL_2699911, EPI_ISL_2699912, EPI_ISL_2699915, EPI_ISL_2699916, EPI_ISL_2699920, EPI_ISL_2699921, EPI_ISL_2699922, EPI_ISL_2699923, EPI_ISL_2699924, EPI_ISL_2699925, EPI_ISL_2699927, EPI_ISL_2699929, EPI_ISL_2699930, EPI_ISL_2699938, EPI_ISL_2699939, EPI_ISL_2699940, EPI_ISL_2699941, EPI_ISL_2699942, EPI_ISL_2699943, EPI_ISL_2699944, EPI_ISL_2699945, EPI_ISL_2699948, EPI_ISL_2699951, EPI_ISL_2699954, EPI_ISL_2699956, EPI_ISL_2699957, EPI_ISL_2699958, EPI_ISL_2699959, EPI_ISL_2699961, EPI_ISL_2699962, EPI_ISL_2699964, EPI_ISL_2699966, EPI_ISL_2699967, EPI_ISL_2699969, EPI_ISL_2699975, EPI_ISL_2699976, EPI_ISL_2699977, EPI_ISL_2699979, EPI_ISL_2699980, EPI_ISL_2699981, EPI_ISL_2699983, EPI_ISL_2699985, EPI_ISL_2699989, EPI_ISL_2699991, EPI_ISL_2699993, EPI_ISL_2699995, EPI_ISL_2699996, EPI_ISL_2699997, EPI_ISL_2699999, EPI_ISL_2700002, EPI_ISL_2700003, EPI_ISL_2700005, EPI_ISL_2700007, EPI_ISL_2700008, EPI_ISL_2700009, EPI_ISL_2700010, EPI_ISL_2700011, EPI_ISL_2700013, EPI_ISL_2700015, EPI_ISL_2700016, EPI_ISL_2700019, EPI_ISL_2700020, EPI_ISL_2700021, EPI_ISL_2700022, EPI_ISL_2700025, EPI_ISL_2700027, EPI_ISL_2700029, EPI_ISL_2700030, EPI_ISL_2700033, EPI_ISL_2700034, EPI_ISL_2700035, EPI_ISL_2700036, EPI_ISL_2700037, EPI_ISL_2700038, EPI_ISL_2700041, EPI_ISL_2700042, EPI_ISL_2700043, EPI_ISL_2700044, EPI_ISL_2700046, EPI_ISL_2700048, EPI_ISL_2700051, EPI_ISL_2700052, EPI_ISL_2700053, EPI_ISL_2700054, EPI_ISL_2700055, EPI_ISL_2700058, EPI_ISL_2700059, EPI_ISL_2700060, EPI_ISL_2700062, EPI_ISL_2700063, EPI_ISL_2700064, EPI_ISL_2700065, EPI_ISL_2700066, EPI_ISL_2700067, EPI_ISL_2700071, EPI_ISL_2700072, EPI_ISL_2700073, EPI_ISL_2700074, EPI_ISL_2700075, EPI_ISL_2700076, EPI_ISL_2700080, EPI_ISL_2700090, EPI_ISL_2700092, EPI_ISL_2700093, EPI_ISL_2700094, EPI_ISL_2700100, EPI_ISL_2700102, EPI_ISL_2700103, EPI_ISL_2700105, EPI_ISL_2700106, EPI_ISL_2700107, EPI_ISL_2700109, EPI_ISL_2700110, EPI_ISL_2700111, EPI_ISL_2700112, EPI_ISL_2700113, EPI_ISL_2700116, EPI_ISL_2700117, EPI_ISL_2700118, EPI_ISL_2700119, EPI_ISL_2700120, EPI_ISL_2700123, EPI_ISL_2700126, EPI_ISL_2700127, EPI_ISL_2700129, EPI_ISL_2700132, EPI_ISL_2700133, EPI_ISL_2700134, EPI_ISL_2700135, EPI_ISL_2700136, EPI_ISL_2700137, EPI_ISL_2700138, EPI_ISL_2700139, EPI_ISL_2700140, EPI_ISL_2700144, EPI_ISL_2700145, EPI_ISL_2700146, EPI_ISL_2700147, EPI_ISL_2700148, EPI_ISL_2700149, EPI_ISL_2700150, EPI_ISL_2700151, EPI_ISL_2700153, EPI_ISL_2700154, EPI_ISL_2700155, EPI_ISL_2700156, EPI_ISL_2700157, EPI_ISL_2700158, EPI_ISL_2700160, EPI_ISL_2700161, EPI_ISL_2700162, EPI_ISL_2700163, EPI_ISL_2700167, EPI_ISL_2700168, EPI_ISL_2700169, EPI_ISL_2700170, EPI_ISL_2700172, EPI_ISL_2700173, EPI_ISL_2700177, EPI_ISL_2700179, EPI_ISL_2700180, EPI_ISL_2700181, EPI_ISL_2700182, EPI_ISL_2700186, EPI_ISL_2700187, EPI_ISL_2700189, EPI_ISL_2700190, EPI_ISL_2700194, EPI_ISL_2700199, EPI_ISL_2700202, EPI_ISL_2700203, EPI_ISL_2700204, EPI_ISL_2700205, EPI_ISL_2700207, EPI_ISL_2700210, EPI_ISL_2700211, EPI_ISL_2700213, EPI_ISL_2700215, EPI_ISL_2700225, EPI_ISL_2700227, EPI_ISL_2700230, EPI_ISL_2700231, EPI_ISL_2700235, EPI_ISL_2700236, EPI_ISL_2700237, EPI_ISL_2700238, EPI_ISL_2700239, EPI_ISL_2700240, EPI_ISL_2700241, EPI_ISL_2700243, EPI_ISL_2700244, EPI_ISL_2700248, EPI_ISL_2700249, EPI_ISL_2700250, EPI_ISL_2700251, EPI_ISL_2700252, EPI_ISL_2700253, EPI_ISL_2700255, EPI_ISL_2700261, EPI_ISL_2700262, EPI_ISL_2700263, EPI_ISL_2700271, EPI_ISL_2700272, EPI_ISL_2700275, EPI_ISL_2700276, EPI_ISL_2700277, EPI_ISL_2700278, EPI_ISL_2700280, EPI_ISL_2700281, EPI_ISL_2700282, EPI_ISL_2700283, EPI_ISL_2700284, EPI_ISL_2700291, EPI_ISL_2700292, EPI_ISL_2700293, EPI_ISL_2700296, EPI_ISL_2700298, EPI_ISL_2700300, EPI_ISL_2700302, EPI_ISL_2700303, EPI_ISL_2700305, EPI_ISL_2700306, EPI_ISL_2700307, EPI_ISL_2700310, EPI_ISL_2700316, EPI_ISL_2700320, EPI_ISL_2700322, EPI_ISL_2700324, EPI_ISL_2700325, EPI_ISL_2700326, EPI_ISL_2700333, EPI_ISL_2700337, EPI_ISL_2700344, EPI_ISL_2700346, EPI_ISL_2700347, EPI_ISL_2700348, EPI_ISL_2700349, EPI_ISL_2700352, EPI_ISL_2700355, EPI_ISL_2700356, EPI_ISL_2700359, EPI_ISL_2700360, EPI_ISL_2700361, EPI_ISL_2700362, EPI_ISL_2700363, EPI_ISL_2700364, EPI_ISL_2700371, EPI_ISL_2700379, EPI_ISL_2700380, EPI_ISL_2700381, EPI_ISL_2700384, EPI_ISL_2700388, EPI_ISL_2700389, EPI_ISL_2700395, EPI_ISL_2700396, EPI_ISL_2700402, EPI_ISL_2700404, EPI_ISL_2700406, EPI_ISL_2700407, EPI_ISL_2700408, EPI_ISL_2700410, EPI_ISL_2700412, EPI_ISL_2700413, EPI_ISL_2700418, EPI_ISL_2700419, EPI_ISL_2700420, EPI_ISL_2700422, EPI_ISL_2700423, EPI_ISL_2700424, EPI_ISL_2700425, EPI_ISL_2700427, EPI_ISL_2700429, EPI_ISL_2700430, EPI_ISL_2700431, EPI_ISL_2700433, EPI_ISL_2700434, EPI_ISL_2700435, EPI_ISL_2700436, EPI_ISL_2700441, EPI_ISL_2700442, EPI_ISL_2700444, EPI_ISL_2700445, EPI_ISL_2700446, EPI_ISL_2700448, EPI_ISL_2700454, EPI_ISL_2700455, EPI_ISL_2700456, EPI_ISL_2700460, EPI_ISL_2700461, EPI_ISL_2700463, EPI_ISL_2700465, EPI_ISL_2700467, EPI_ISL_2700471, EPI_ISL_2700472, EPI_ISL_2700474, EPI_ISL_2700475, EPI_ISL_2700476, EPI_ISL_2700477, EPI_ISL_2700486, EPI_ISL_2700488, EPI_ISL_2700489, EPI_ISL_2700491, EPI_ISL_2700494, EPI_ISL_2700496, EPI_ISL_2700497, EPI_ISL_2700498, EPI_ISL_2700500, EPI_ISL_2700502, EPI_ISL_2700504, EPI_ISL_2700505, EPI_ISL_2700507, EPI_ISL_2700509, EPI_ISL_2700513, EPI_ISL_2700515, EPI_ISL_2700517, EPI_ISL_2700519, EPI_ISL_2700522, EPI_ISL_2700524, EPI_ISL_2700526, EPI_ISL_2700528, EPI_ISL_2700529, EPI_ISL_2700530, EPI_ISL_2700534, EPI_ISL_2700537, EPI_ISL_2700544, EPI_ISL_2700547, EPI_ISL_2700548, EPI_ISL_2700550, EPI_ISL_2700551, EPI_ISL_2700552, EPI_ISL_2700553, EPI_ISL_2700555, EPI_ISL_2700556, EPI_ISL_2700558, EPI_ISL_2700559, EPI_ISL_2700568, EPI_ISL_2700570, EPI_ISL_2700572, EPI_ISL_2700574, EPI_ISL_2700576, EPI_ISL_2701309, EPI_ISL_2701311, EPI_ISL_2701315, EPI_ISL_2701318, EPI_ISL_2701319, EPI_ISL_2701323, EPI_ISL_2701325, EPI_ISL_2701326, EPI_ISL_2701331, EPI_ISL_2701335, EPI_ISL_2701336, EPI_ISL_2701343, EPI_ISL_2701358, EPI_ISL_2701361, EPI_ISL_2701362, EPI_ISL_2701363, EPI_ISL_2701365, EPI_ISL_2701371, EPI_ISL_2701373, EPI_ISL_2701382, EPI_ISL_2701388, EPI_ISL_2701394, EPI_ISL_2701402, EPI_ISL_2701403, EPI_ISL_2701408, EPI_ISL_2701414, EPI_ISL_2701417, EPI_ISL_2701420, EPI_ISL_2701422, EPI_ISL_2701424, EPI_ISL_2701427, EPI_ISL_2701432, EPI_ISL_2701436, EPI_ISL_2701437, EPI_ISL_2701445, EPI_ISL_2701449, EPI_ISL_2701453, EPI_ISL_2701466, EPI_ISL_2701473, EPI_ISL_2701481, EPI_ISL_2701485, EPI_ISL_2701487, EPI_ISL_2701493, EPI_ISL_2701495, EPI_ISL_2701496, EPI_ISL_2701497, EPI_ISL_2701499, EPI_ISL_2701500, EPI_ISL_2701504, EPI_ISL_2701509, EPI_ISL_2701515, EPI_ISL_2701520, EPI_ISL_2701521, EPI_ISL_2701526, EPI_ISL_2701531, EPI_ISL_2701533, EPI_ISL_2701538, EPI_ISL_2701549, EPI_ISL_2701551, EPI_ISL_2701552, EPI_ISL_2701557, EPI_ISL_2701559, EPI_ISL_2701561, EPI_ISL_2701570, EPI_ISL_2701572, EPI_ISL_2701577, EPI_ISL_2701578, EPI_ISL_2701583, EPI_ISL_2701586, EPI_ISL_2701587, EPI_ISL_2701595, EPI_ISL_2701596, EPI_ISL_2701609, EPI_ISL_2701610, EPI_ISL_2701612, EPI_ISL_2701620, EPI_ISL_2701623, EPI_ISL_2701626, EPI_ISL_2701637, EPI_ISL_2701641, EPI_ISL_2701642, EPI_ISL_2701648, EPI_ISL_2701650, EPI_ISL_2701652, EPI_ISL_2701653, EPI_ISL_2701654, EPI_ISL_2701655, EPI_ISL_2701656, EPI_ISL_2701657, EPI_ISL_2701658, EPI_ISL_2701668, EPI_ISL_2701676, EPI_ISL_2701677, EPI_ISL_2701681, EPI_ISL_2701689, EPI_ISL_2701692, EPI_ISL_2701698, EPI_ISL_2701700, EPI_ISL_2701702, EPI_ISL_2701703, EPI_ISL_2701704, EPI_ISL_2701706, EPI_ISL_2701712, EPI_ISL_2701721, EPI_ISL_2701722, EPI_ISL_2701723, EPI_ISL_2701731, EPI_ISL_2701733, EPI_ISL_2701736, EPI_ISL_2701741, EPI_ISL_2701743, EPI_ISL_2701745, EPI_ISL_2701748, EPI_ISL_2701751, EPI_ISL_2701753, EPI_ISL_2701755, EPI_ISL_2701768, EPI_ISL_2701769, EPI_ISL_2701770, EPI_ISL_2701771, EPI_ISL_2701772, EPI_ISL_2701781, EPI_ISL_2701785, EPI_ISL_2701790, EPI_ISL_2701803, EPI_ISL_2701805, EPI_ISL_2701810, EPI_ISL_2701811, EPI_ISL_2701816, EPI_ISL_2701818, EPI_ISL_2701822, EPI_ISL_2701824, EPI_ISL_2701830, EPI_ISL_2701847, EPI_ISL_2701850, EPI_ISL_2701851, EPI_ISL_2701861, EPI_ISL_2701863, EPI_ISL_2701864, EPI_ISL_2701866, EPI_ISL_2701867, EPI_ISL_2701877, EPI_ISL_2701879, EPI_ISL_2701881, EPI_ISL_2701885, EPI_ISL_2701892, EPI_ISL_2701894, EPI_ISL_2701895, EPI_ISL_2701902, EPI_ISL_2701911, EPI_ISL_2701912, EPI_ISL_2701915, EPI_ISL_2701916, EPI_ISL_2701923, EPI_ISL_2701926, EPI_ISL_2701929, EPI_ISL_2701931, EPI_ISL_2701932, EPI_ISL_2701936, EPI_ISL_2701940, EPI_ISL_2701943, EPI_ISL_2701944, EPI_ISL_2701946, EPI_ISL_2701951, EPI_ISL_2701952, EPI_ISL_2701953, EPI_ISL_2701955, EPI_ISL_2701957, EPI_ISL_2701960, EPI_ISL_2701964, EPI_ISL_2701967, EPI_ISL_2701969, EPI_ISL_2701970, EPI_ISL_2701981, EPI_ISL_2701985, EPI_ISL_2701986, EPI_ISL_2701988, EPI_ISL_2702003, EPI_ISL_2702015, EPI_ISL_2702017, EPI_ISL_2702027, EPI_ISL_2702028, EPI_ISL_2702032, EPI_ISL_2702041, EPI_ISL_2702051, EPI_ISL_2702056, EPI_ISL_2702061, EPI_ISL_2702062, EPI_ISL_2702064, EPI_ISL_2702069, EPI_ISL_2702073, EPI_ISL_2702074, EPI_ISL_2702077, EPI_ISL_2702081, EPI_ISL_2702086, EPI_ISL_2702088, EPI_ISL_2702090, EPI_ISL_2702099, EPI_ISL_2702119, EPI_ISL_2702123, EPI_ISL_2702125, EPI_ISL_2702127, EPI_ISL_2702131, EPI_ISL_2702143, EPI_ISL_2702144, EPI_ISL_2702145, EPI_ISL_2702146, EPI_ISL_2702149, EPI_ISL_2702159, EPI_ISL_2702163, EPI_ISL_2702166, EPI_ISL_2702168, EPI_ISL_2702182, EPI_ISL_2702186, EPI_ISL_2702192, EPI_ISL_2702193, EPI_ISL_2702196, EPI_ISL_2702200, EPI_ISL_2702205, EPI_ISL_2702207, EPI_ISL_2702214, EPI_ISL_2702215, EPI_ISL_2702219, EPI_ISL_2702223, EPI_ISL_2702225, EPI_ISL_2702233, EPI_ISL_2702255, EPI_ISL_2702260, EPI_ISL_2702261, EPI_ISL_2702275, EPI_ISL_2702277, EPI_ISL_2702282, EPI_ISL_2702283, EPI_ISL_2702285, EPI_ISL_2702287, EPI_ISL_2702295, EPI_ISL_2702303, EPI_ISL_2702304, EPI_ISL_2702306, EPI_ISL_2702308, EPI_ISL_2702309, EPI_ISL_2702312, EPI_ISL_2702313, EPI_ISL_2702315, EPI_ISL_2702316, EPI_ISL_2702320, EPI_ISL_2702326, EPI_ISL_2702330, EPI_ISL_2702339, EPI_ISL_2702340, EPI_ISL_2702349, EPI_ISL_2702354, EPI_ISL_2702361, EPI_ISL_2702365, EPI_ISL_2702372, EPI_ISL_2702376, EPI_ISL_2702381, EPI_ISL_2702385, EPI_ISL_2702396, EPI_ISL_2702816, EPI_ISL_2702818, EPI_ISL_2702819, EPI_ISL_2702820, EPI_ISL_2702824, EPI_ISL_2702825, EPI_ISL_2702826, EPI_ISL_2702828, EPI_ISL_2702830, EPI_ISL_2702832, EPI_ISL_2702833, EPI_ISL_2702834, EPI_ISL_2702835, EPI_ISL_2702837, EPI_ISL_2702839, EPI_ISL_2702841, EPI_ISL_2702842, EPI_ISL_2702844, EPI_ISL_2702845, EPI_ISL_2702848, EPI_ISL_2702852, EPI_ISL_2702853, EPI_ISL_2702855, EPI_ISL_2702863, EPI_ISL_2702867, EPI_ISL_2702869, EPI_ISL_2702871, EPI_ISL_2702872, EPI_ISL_2702873, EPI_ISL_2702874, EPI_ISL_2702877, EPI_ISL_2702878, EPI_ISL_2702879, EPI_ISL_2702880, EPI_ISL_2702881, EPI_ISL_2702882, EPI_ISL_2702883, EPI_ISL_2702885, EPI_ISL_2702888, EPI_ISL_2702891, EPI_ISL_2702892, EPI_ISL_2702893, EPI_ISL_2702898, EPI_ISL_2702899, EPI_ISL_2702900, EPI_ISL_2702901, EPI_ISL_2702902, EPI_ISL_2702903, EPI_ISL_2702906, EPI_ISL_2702907, EPI_ISL_2702908, EPI_ISL_2702909, EPI_ISL_2702911, EPI_ISL_2702913, EPI_ISL_2702914, EPI_ISL_2702915, EPI_ISL_2702917, EPI_ISL_2702918, EPI_ISL_2702919, EPI_ISL_2702920, EPI_ISL_2702925, EPI_ISL_2702928, EPI_ISL_2702929, EPI_ISL_2702930, EPI_ISL_2702931, EPI_ISL_2702936, EPI_ISL_2702937, EPI_ISL_2702939, EPI_ISL_2702940, EPI_ISL_2702942, EPI_ISL_2702946, EPI_ISL_2702947, EPI_ISL_2702948, EPI_ISL_2702952, EPI_ISL_2702953, EPI_ISL_2702954, EPI_ISL_2702959, EPI_ISL_2702960, EPI_ISL_2702961, EPI_ISL_2702963, EPI_ISL_2702965, EPI_ISL_2702967, EPI_ISL_2702969, EPI_ISL_2702970, EPI_ISL_2702971, EPI_ISL_2702972, EPI_ISL_2702973, EPI_ISL_2702975, EPI_ISL_2702976, EPI_ISL_2702981, EPI_ISL_2702983, EPI_ISL_2702986, EPI_ISL_2702987, EPI_ISL_2702988, EPI_ISL_2702990, EPI_ISL_2702994, EPI_ISL_2702998, EPI_ISL_2703000, EPI_ISL_2703001, EPI_ISL_2703009, EPI_ISL_2703010, EPI_ISL_2703011, EPI_ISL_2703013, EPI_ISL_2703014, EPI_ISL_2703016, EPI_ISL_2703018, EPI_ISL_2703019, EPI_ISL_2703020, EPI_ISL_2703021, EPI_ISL_2703025, EPI_ISL_2703032, EPI_ISL_2703036, EPI_ISL_2703039, EPI_ISL_2703041, EPI_ISL_2703044, EPI_ISL_2703046, EPI_ISL_2703047, EPI_ISL_2703048, EPI_ISL_2703051, EPI_ISL_2703054, EPI_ISL_2703056, EPI_ISL_2703059, EPI_ISL_2703061, EPI_ISL_2703064, EPI_ISL_2703066, EPI_ISL_2703067, EPI_ISL_2703068, EPI_ISL_2703070, EPI_ISL_2703076, EPI_ISL_2703078, EPI_ISL_2703080, EPI_ISL_2703081, EPI_ISL_2703083, EPI_ISL_2703085, EPI_ISL_2703087, EPI_ISL_2703088, EPI_ISL_2703089, EPI_ISL_2703092, EPI_ISL_2703094, EPI_ISL_2703097, EPI_ISL_2703100, EPI_ISL_2703102, EPI_ISL_2703104, EPI_ISL_2703107, EPI_ISL_2703108, EPI_ISL_2703109, EPI_ISL_2703110, EPI_ISL_2703113, EPI_ISL_2703114, EPI_ISL_2703116, EPI_ISL_2703119, EPI_ISL_2703121, EPI_ISL_2703123, EPI_ISL_2703125, EPI_ISL_2703126, EPI_ISL_2703129, EPI_ISL_2703132, EPI_ISL_2703135, EPI_ISL_2703138, EPI_ISL_2703140, EPI_ISL_2703141, EPI_ISL_2703142, EPI_ISL_2703143, EPI_ISL_2703144, EPI_ISL_2703145, EPI_ISL_2703146, EPI_ISL_2703152, EPI_ISL_2703153, EPI_ISL_2703154, EPI_ISL_2703163, EPI_ISL_2703181, EPI_ISL_2703182, EPI_ISL_2703187, EPI_ISL_2703189, EPI_ISL_2703200, EPI_ISL_2703210, EPI_ISL_2703212, EPI_ISL_2703215, EPI_ISL_2703237, EPI_ISL_2703243, EPI_ISL_2703255, EPI_ISL_2703269, EPI_ISL_2703270, EPI_ISL_2703271, EPI_ISL_2703273, EPI_ISL_2703275, EPI_ISL_2703276, EPI_ISL_2703279, EPI_ISL_2703291, EPI_ISL_2703292, EPI_ISL_2703312, EPI_ISL_2703318, EPI_ISL_2703320, EPI_ISL_2703323, EPI_ISL_2703327, EPI_ISL_2703329, EPI_ISL_2703337, EPI_ISL_2703349, EPI_ISL_2703362, EPI_ISL_2703368, EPI_ISL_2703374, EPI_ISL_2703379, EPI_ISL_2703381, EPI_ISL_2703383, EPI_ISL_2703384, EPI_ISL_2703404, EPI_ISL_2703405, EPI_ISL_2703412, EPI_ISL_2703423, EPI_ISL_2703424, EPI_ISL_2703438, EPI_ISL_2703441, EPI_ISL_2703443, EPI_ISL_2703446, EPI_ISL_2703449, EPI_ISL_2703457, EPI_ISL_2703467, EPI_ISL_2703493, EPI_ISL_2703505, EPI_ISL_2703508, EPI_ISL_2703515, EPI_ISL_2703516, EPI_ISL_2703519, EPI_ISL_2703522, EPI_ISL_2703527, EPI_ISL_2703534, EPI_ISL_2703538, EPI_ISL_2703545, EPI_ISL_2703548, EPI_ISL_2703549, EPI_ISL_2703552, EPI_ISL_2703555, EPI_ISL_2703556, EPI_ISL_2703558, EPI_ISL_2703559, EPI_ISL_2703560, EPI_ISL_2703564, EPI_ISL_2703570, EPI_ISL_2703572, EPI_ISL_2703576, EPI_ISL_2703577, EPI_ISL_2703579, EPI_ISL_2703580, EPI_ISL_2703583, EPI_ISL_2703587, EPI_ISL_2703592, EPI_ISL_2703599, EPI_ISL_2703600, EPI_ISL_2703601, EPI_ISL_2703605, EPI_ISL_2703607, EPI_ISL_2703618, EPI_ISL_2703619, EPI_ISL_2703621, EPI_ISL_2703623, EPI_ISL_2703628, EPI_ISL_2703637, EPI_ISL_2703638, EPI_ISL_2703640, EPI_ISL_2703643, EPI_ISL_2703648, EPI_ISL_2703652, EPI_ISL_2703654, EPI_ISL_2703662, EPI_ISL_2703665, EPI_ISL_2703679, EPI_ISL_2703682, EPI_ISL_2703695, EPI_ISL_2703700, EPI_ISL_2703701, EPI_ISL_2703703, EPI_ISL_2703707, EPI_ISL_2703709, EPI_ISL_2703710, EPI_ISL_2703711, EPI_ISL_2703712, EPI_ISL_2703719, EPI_ISL_2703721, EPI_ISL_2703728, EPI_ISL_2703741, EPI_ISL_2703745, EPI_ISL_2703746, EPI_ISL_2703753, EPI_ISL_2703759, EPI_ISL_2703762, EPI_ISL_2703764, EPI_ISL_2703766, EPI_ISL_2703768, EPI_ISL_2703782, EPI_ISL_2703798, EPI_ISL_2703800, EPI_ISL_2703801, EPI_ISL_2703802, EPI_ISL_2703827, EPI_ISL_2703829, EPI_ISL_2703830, EPI_ISL_2717431, EPI_ISL_2717809, EPI_ISL_2717812, EPI_ISL_2717814, EPI_ISL_2717825, EPI_ISL_2717827, EPI_ISL_2717828, EPI_ISL_2717830, EPI_ISL_2717832, EPI_ISL_2717833, EPI_ISL_2717844, EPI_ISL_2717853, EPI_ISL_2717872, EPI_ISL_2717873, EPI_ISL_2717874, EPI_ISL_2717884, EPI_ISL_2717885, EPI_ISL_2717886, EPI_ISL_2717893, EPI_ISL_2717898, EPI_ISL_2717902, EPI_ISL_2717910, EPI_ISL_2717915, EPI_ISL_2717916, EPI_ISL_2717918, EPI_ISL_2717919, EPI_ISL_2717921, EPI_ISL_2717922, EPI_ISL_2717924, EPI_ISL_2717925, EPI_ISL_2717928, EPI_ISL_2717937, EPI_ISL_2717944, EPI_ISL_2717948, EPI_ISL_2717957, EPI_ISL_2717962, EPI_ISL_2717966, EPI_ISL_2717973, EPI_ISL_2717977, EPI_ISL_2717982, EPI_ISL_2717988, EPI_ISL_2717991, EPI_ISL_2717993, EPI_ISL_2717998, EPI_ISL_2717999, EPI_ISL_2718000, EPI_ISL_2718003, EPI_ISL_2718005, EPI_ISL_2718012, EPI_ISL_2718019, EPI_ISL_2718020, EPI_ISL_2718024, EPI_ISL_2718033, EPI_ISL_2718041, EPI_ISL_2718043, EPI_ISL_2718048, EPI_ISL_2718049, EPI_ISL_2718050, EPI_ISL_2718052, EPI_ISL_2718055, EPI_ISL_2718059, EPI_ISL_2718060, EPI_ISL_2718067, EPI_ISL_2718069, EPI_ISL_2718070, EPI_ISL_2718075, EPI_ISL_2718077, EPI_ISL_2718078, EPI_ISL_2718079, EPI_ISL_2718082, EPI_ISL_2718092, EPI_ISL_2718093, EPI_ISL_2718094, EPI_ISL_2718097, EPI_ISL_2718100, EPI_ISL_2718102, EPI_ISL_2718107, EPI_ISL_2718108, EPI_ISL_2718111, EPI_ISL_2718116, EPI_ISL_2718126, EPI_ISL_2718129, EPI_ISL_2718138, EPI_ISL_2718147, EPI_ISL_2718148, EPI_ISL_2718152, EPI_ISL_2718156, EPI_ISL_2718158, EPI_ISL_2718162, EPI_ISL_2718165, EPI_ISL_2718169, EPI_ISL_2718294, EPI_ISL_2729410, EPI_ISL_2729485, EPI_ISL_2729768, EPI_ISL_2729827, EPI_ISL_2729845, EPI_ISL_2730647, EPI_ISL_2767755, EPI_ISL_2767803, EPI_ISL_2767947, EPI_ISL_2780297, EPI_ISL_2780834, EPI_ISL_2780851, EPI_ISL_2780911, EPI_ISL_2780994, EPI_ISL_2781114, EPI_ISL_2781285, EPI_ISL_2807796

| | | | |
|---|---|---|---|
| see above | Lighthouse Lab in Milton Keynes | Wellcome Sanger Institute for the COVID-19 Genomics UK (COG-UK) Consortium | Cordelia Langford; David K. Jackson; Dominic Kwiatkowski; Ewan Harrison; Ian Johnston; Jeffrey Barrett; John Sillitoe on behalf of the Wellcome Sanger Institute COVID-19 Surveillance Team; Roberto Amato; Sonia Goncalves; The Lighthouse Lab in Milton Keynes and Alex Alderton |

EPI_ISL_2699527, EPI_ISL_2699531, EPI_ISL_2699543, EPI_ISL_2699545, EPI_ISL_2699546, EPI_ISL_2699595, EPI_ISL_2699597, EPI_ISL_2699599, EPI_ISL_2699608, EPI_ISL_2699609, EPI_ISL_2699632, EPI_ISL_2699647, EPI_ISL_2699649, EPI_ISL_2699664, EPI_ISL_2699666, EPI_ISL_2699674, EPI_ISL_2699685, EPI_ISL_2699690, EPI_ISL_2699702, EPI_ISL_2699746, EPI_ISL_2699750, EPI_ISL_2699762, EPI_ISL_2699792, EPI_ISL_2699794, EPI_ISL_2699806, EPI_ISL_2699821, EPI_ISL_2699825, EPI_ISL_2699826, EPI_ISL_2699850, EPI_ISL_2699857, EPI_ISL_2703836, EPI_ISL_2703837, EPI_ISL_2703838, EPI_ISL_2703839, EPI_ISL_2703840, EPI_ISL_2703841, EPI_ISL_2703842, EPI_ISL_2703843, EPI_ISL_2703844, EPI_ISL_2703845, EPI_ISL_2703846, EPI_ISL_2703847, EPI_ISL_2703848, EPI_ISL_2703850, EPI_ISL_2703852, EPI_ISL_2703853, EPI_ISL_2703854, EPI_ISL_2703855, EPI_ISL_2703856, EPI_ISL_2703857, EPI_ISL_2703859, EPI_ISL_2703860, EPI_ISL_2703861, EPI_ISL_2703862, EPI_ISL_2703863, EPI_ISL_2703864, EPI_ISL_2703865, EPI_ISL_2703866, EPI_ISL_2703867, EPI_ISL_2703868, EPI_ISL_2703869, EPI_ISL_2703870, EPI_ISL_2703871, EPI_ISL_2703872, EPI_ISL_2703873, EPI_ISL_2703874, EPI_ISL_2703875, EPI_ISL_2703876, EPI_ISL_2703877, EPI_ISL_2703879, EPI_ISL_2703880, EPI_ISL_2703881, EPI_ISL_2703882, EPI_ISL_2703883, EPI_ISL_2703884, EPI_ISL_2703885, EPI_ISL_2703886, EPI_ISL_2703887, EPI_ISL_2703888, EPI_ISL_2703889, EPI_ISL_2703890, EPI_ISL_2703892, EPI_ISL_2703893, EPI_ISL_2703894, EPI_ISL_2703895, EPI_ISL_2703896, EPI_ISL_2703897, EPI_ISL_2703898, EPI_ISL_2703899, EPI_ISL_2703900, EPI_ISL_2703901, EPI_ISL_2703902, EPI_ISL_2703903, EPI_ISL_2703904, EPI_ISL_2703905, EPI_ISL_2703906, EPI_ISL_2703907, EPI_ISL_2703908, EPI_ISL_2703910, EPI_ISL_2703911, EPI_ISL_2703912, EPI_ISL_2703913, EPI_ISL_2703914, EPI_ISL_2703915, EPI_ISL_2703916, EPI_ISL_2703917, EPI_ISL_2703918, EPI_ISL_2703919, EPI_ISL_2703920, EPI_ISL_2703921, EPI_ISL_2703922, EPI_ISL_2703923, EPI_ISL_2703924, EPI_ISL_2703925, EPI_ISL_2703926, EPI_ISL_2703927, EPI_ISL_2703928, EPI_ISL_2703929, EPI_ISL_2703930, EPI_ISL_2703931, EPI_ISL_2703932, EPI_ISL_2703934, EPI_ISL_2703935, EPI_ISL_2703936, EPI_ISL_2703937, EPI_ISL_2703939, EPI_ISL_2703941, EPI_ISL_2703942, EPI_ISL_2703943, EPI_ISL_2703944, EPI_ISL_2703945, EPI_ISL_2703946, EPI_ISL_2703947, EPI_ISL_2703948, EPI_ISL_2703949, EPI_ISL_2703950, EPI_ISL_2703951, EPI_ISL_2703952, EPI_ISL_2703953, EPI_ISL_2703954, EPI_ISL_2703955, EPI_ISL_2703956, EPI_ISL_2703957, EPI_ISL_2703958, EPI_ISL_2703959, EPI_ISL_2703960, EPI_ISL_2703961, EPI_ISL_2703962, EPI_ISL_2703963, EPI_ISL_2703964, EPI_ISL_2703965, EPI_ISL_2703966, EPI_ISL_2703968, EPI_ISL_2703969, EPI_ISL_2703970, EPI_ISL_2703971, EPI_ISL_2703972, EPI_ISL_2703973, EPI_ISL_2703974, EPI_ISL_2703975, EPI_ISL_2703976, EPI_ISL_2703977, EPI_ISL_2703978, EPI_ISL_2703979, EPI_ISL_2703980, EPI_ISL_2703982, EPI_ISL_2703983, EPI_ISL_2703984, EPI_ISL_2703985, EPI_ISL_2703986, EPI_ISL_2703987, EPI_ISL_2703988, EPI_ISL_2703989, EPI_ISL_2703990, EPI_ISL_2703991, EPI_ISL_2703992, EPI_ISL_2703993, EPI_ISL_2703994, EPI_ISL_2703995, EPI_ISL_2703996, EPI_ISL_2703997, EPI_ISL_2703998, EPI_ISL_2703999, EPI_ISL_2704000, EPI_ISL_2704001, EPI_ISL_2704002, EPI_ISL_2704003, EPI_ISL_2704004, EPI_ISL_2704005, EPI_ISL_2704006, EPI_ISL_2704007, EPI_ISL_2704008, EPI_ISL_2704009, EPI_ISL_2704010, EPI_ISL_2704011, EPI_ISL_2704012, EPI_ISL_2704013, EPI_ISL_2704014, EPI_ISL_2704015, EPI_ISL_2704016, EPI_ISL_2704017, EPI_ISL_2704018, EPI_ISL_2704019, EPI_ISL_2704020, EPI_ISL_2704021, EPI_ISL_2704022, EPI_ISL_2704023, EPI_ISL_2704024, EPI_ISL_2704026, EPI_ISL_2704027, EPI_ISL_2704028, EPI_ISL_2704030, EPI_ISL_2704031, EPI_ISL_2704032, EPI_ISL_2704033, EPI_ISL_2704034, EPI_ISL_2704035, EPI_ISL_2704036, EPI_ISL_2704038, EPI_ISL_2704039, EPI_ISL_2704040, EPI_ISL_2704041, EPI_ISL_2704042, EPI_ISL_2704043, EPI_ISL_2704044, EPI_ISL_2704045, EPI_ISL_2704046, EPI_ISL_2704047, EPI_ISL_2704048, EPI_ISL_2704049, EPI_ISL_2704050, EPI_ISL_2704051, EPI_ISL_2704052, EPI_ISL_2704053, EPI_ISL_2704054, EPI_ISL_2704055, EPI_ISL_2704056, EPI_ISL_2704057, EPI_ISL_2704058, EPI_ISL_2704059, EPI_ISL_2704060, EPI_ISL_2704061, EPI_ISL_2704062, EPI_ISL_2704063, EPI_ISL_2704064, EPI_ISL_2704065, EPI_ISL_2704066, EPI_ISL_2704067, EPI_ISL_2704068, EPI_ISL_2704069, EPI_ISL_2704070, EPI_ISL_2704071, EPI_ISL_2704072, EPI_ISL_2704073, EPI_ISL_2704074, EPI_ISL_2704075, EPI_ISL_2704076, EPI_ISL_2704077, EPI_ISL_2704078, EPI_ISL_2704079, EPI_ISL_2704080, EPI_ISL_2704081, EPI_ISL_2704082, EPI_ISL_2704083, EPI_ISL_2704084, EPI_ISL_2704085, EPI_ISL_2704086, EPI_ISL_2704087, EPI_ISL_2704088, EPI_ISL_2704089, EPI_ISL_2704090, EPI_ISL_2704091, EPI_ISL_2704092, EPI_ISL_2704093, EPI_ISL_2704094, EPI_ISL_2704096, EPI_ISL_2704097, EPI_ISL_2704098, EPI_ISL_2704099, EPI_ISL_2704100, EPI_ISL_2704101, EPI_ISL_2704102, EPI_ISL_2704103, EPI_ISL_2704104, EPI_ISL_2704105, EPI_ISL_2704106, EPI_ISL_2704107, EPI_ISL_2704108, EPI_ISL_2704109, EPI_ISL_2704110, EPI_ISL_2704111, EPI_ISL_2704112, EPI_ISL_2704113, EPI_ISL_2704114, EPI_ISL_2704115, EPI_ISL_2704116, EPI_ISL_2704117, EPI_ISL_2704118, EPI_ISL_2704119, EPI_ISL_2704120, EPI_ISL_2704121, EPI_ISL_2704122, EPI_ISL_2704123, EPI_ISL_2704124, EPI_ISL_2704125, EPI_ISL_2704126, EPI_ISL_2704127, EPI_ISL_2704128, EPI_ISL_2704129, EPI_ISL_2704130, EPI_ISL_2704131, EPI_ISL_2704132, EPI_ISL_2704133, EPI_ISL_2704134, EPI_ISL_2704135, EPI_ISL_2704136, EPI_ISL_2704137, EPI_ISL_2704138, EPI_ISL_2704139, EPI_ISL_2704140, EPI_ISL_2704141, EPI_ISL_2704142, EPI_ISL_2704143, EPI_ISL_2704144, EPI_ISL_2704145, EPI_ISL_2704146, EPI_ISL_2704147, EPI_ISL_2704148, EPI_ISL_2704149, EPI_ISL_2704150, EPI_ISL_2704151, EPI_ISL_2704152, EPI_ISL_2704153, EPI_ISL_2704154, EPI_ISL_2704155, EPI_ISL_2704156, EPI_ISL_2704157, EPI_ISL_2704158, EPI_ISL_2704159, EPI_ISL_2704160, EPI_ISL_2704161, EPI_ISL_2704162, EPI_ISL_2704163, EPI_ISL_2704164, EPI_ISL_2704165, EPI_ISL_2704166, EPI_ISL_2704167, EPI_ISL_2704168, EPI_ISL_2704169, EPI_ISL_2704170, EPI_ISL_2704171, EPI_ISL_2704172, EPI_ISL_2704173, EPI_ISL_2704174, EPI_ISL_2704175, EPI_ISL_2704176, EPI_ISL_2704177, EPI_ISL_2704178, EPI_ISL_2704179, EPI_ISL_2704180, EPI_ISL_2704181, EPI_ISL_2704182, EPI_ISL_2704183, EPI_ISL_2704184, EPI_ISL_2718285, EPI_ISL_2718286, EPI_ISL_2718287

| | | | |
|---|---|---|---|
| see above | Lighthouse Laboratory Plymouth | Wellcome Sanger Institute for the COVID-19 Genomics UK (COG-UK) Consortium | Cordelia Langford; David K. Jackson; Dominic Kwiatkowski; Ewan Harrison; Ian Johnston; Jeffrey Barrett; John Sillitoe on behalf of the Wellcome Sanger Institute COVID-19 Surveillance Team; Lighthouse Laboratory Plymouth and Alex Alderton; Roberto Amato; Sonia Goncalves |

EPI_ISL_2816092, EPI_ISL_2816094, EPI_ISL_2816095, EPI_ISL_2816096, EPI_ISL_2816097, EPI_ISL_2816098, EPI_ISL_2816099, EPI_ISL_2816100, EPI_ISL_2816101, EPI_ISL_2816102, EPI_ISL_2816103, EPI_ISL_2816104, EPI_ISL_2816105, EPI_ISL_2816106, EPI_ISL_2816107, EPI_ISL_2816108, EPI_ISL_2816109, EPI_ISL_2816110, EPI_ISL_2816111, EPI_ISL_2816112, EPI_ISL_2816113, EPI_ISL_2816114, EPI_ISL_2816115, EPI_ISL_2816116, EPI_ISL_2816117, EPI_ISL_2816118, EPI_ISL_2816119, EPI_ISL_2816120, EPI_ISL_2816121, EPI_ISL_2816122, EPI_ISL_2816123, EPI_ISL_2816124, EPI_ISL_2816125, EPI_ISL_2816126, EPI_ISL_2816127, EPI_ISL_2816128, EPI_ISL_2816129, EPI_ISL_2816130, EPI_ISL_2816131, EPI_ISL_2816132, EPI_ISL_2816133, EPI_ISL_2816134, EPI_ISL_2816135, EPI_ISL_2816136, EPI_ISL_2816137, EPI_ISL_2816138, EPI_ISL_2816139, EPI_ISL_2816140, EPI_ISL_2816141, EPI_ISL_2816142, EPI_ISL_2816143, EPI_ISL_2816145, EPI_ISL_2816147, EPI_ISL_2816148

| | | | |
|---|---|---|---|
| see above | Liverpool Clinical Laboratories | COVID-19 Genomics UK (COG-UK) Consortium | A Alrezaihi; Alessandro Gerada; Alistair Darby; Angela Cowell; Anita Lucaci; Anu Chawla; Cassie Olateju; Catherine Hartley; Charlotte Nelson; Ecaterina Vamos; Elaine O'Toole; Eleanor G Bentley; Ghada T Shawli; Hermione Webster; Isabel García-Dorival; James Johnson; James P Stewart; Jenifer Manson; Joanne Watts; Jones Benjamin; Jordan J Clark; Julian Hiscox; L Luu; Lucille Rainbow; M Almsaud; Margaret Hughes; Mark Whitehead; Matthew Gemmell; Miren Iturriza-Gomara; Muhannad Alruwaili; N.P Randle; Neil Swainston; PKF Gilmore; Parul Sharma; Rebekah Penrice-Randal; Richard Eccles; Richard Gregory; Sam Haldenby; Steve Paterson; Stuart D Armstrong; Trevor Ian Robinson; Ximeng Han |

EPI_ISL_2706571, EPI_ISL_2706573, EPI_ISL_2706574, EPI_ISL_2706576, EPI_ISL_2706577, EPI_ISL_2706578, EPI_ISL_2706579, EPI_ISL_2706580, EPI_ISL_2706582, EPI_ISL_2706583, EPI_ISL_2706584, EPI_ISL_2706585, EPI_ISL_2706586, EPI_ISL_2706587, EPI_ISL_2706588, EPI_ISL_2706589, EPI_ISL_2706590, EPI_ISL_2706592, EPI_ISL_2706593, EPI_ISL_2706594, EPI_ISL_2706595, EPI_ISL_2706596, EPI_ISL_2706597, EPI_ISL_2706598, EPI_ISL_2706599, EPI_ISL_2706600, EPI_ISL_2706602, EPI_ISL_2706603, EPI_ISL_2706604, EPI_ISL_2706606, EPI_ISL_2706607, EPI_ISL_2706608, EPI_ISL_2706609, EPI_ISL_2706610, EPI_ISL_2706611, EPI_ISL_2706612, EPI_ISL_2706613, EPI_ISL_2706615, EPI_ISL_2706616, EPI_ISL_2706617, EPI_ISL_2706618, EPI_ISL_2706619, EPI_ISL_2706620, EPI_ISL_2706621, EPI_ISL_2706622, EPI_ISL_2706624, EPI_ISL_2706625, EPI_ISL_2706626, EPI_ISL_2706627, EPI_ISL_2706628, EPI_ISL_2706629, EPI_ISL_2706630, EPI_ISL_2706631, EPI_ISL_2706632, EPI_ISL_2706633, EPI_ISL_2706634, EPI_ISL_2706635, EPI_ISL_2706636, EPI_ISL_2706637, EPI_ISL_2706638, EPI_ISL_2706639, EPI_ISL_2706640, EPI_ISL_2706642, EPI_ISL_2706643, EPI_ISL_2723166, EPI_ISL_2723172, EPI_ISL_2723176, EPI_ISL_2723177, EPI_ISL_2723182, EPI_ISL_2723185, EPI_ISL_2723194, EPI_ISL_2723199, EPI_ISL_2723200, EPI_ISL_2723209, EPI_ISL_2723211, EPI_ISL_2723212, EPI_ISL_2723213, EPI_ISL_2723234, EPI_ISL_2723235, EPI_ISL_2723239, EPI_ISL_2723241, EPI_ISL_2723243, EPI_ISL_2723249, EPI_ISL_2723254, EPI_ISL_2723257, EPI_ISL_2723264, EPI_ISL_2723268, EPI_ISL_2723271, EPI_ISL_2723273, EPI_ISL_2723279, EPI_ISL_2723297, EPI_ISL_2723302, EPI_ISL_2723303, EPI_ISL_2816645, EPI_ISL_2816646, EPI_ISL_2816647, EPI_ISL_2816648, EPI_ISL_2816649, EPI_ISL_2816650, EPI_ISL_2816651, EPI_ISL_2816652, EPI_ISL_2816653, EPI_ISL_2816654, EPI_ISL_2816655, EPI_ISL_2816656, EPI_ISL_2816657, EPI_ISL_2816658, EPI_ISL_2816659, EPI_ISL_2816660, EPI_ISL_2816661, EPI_ISL_2816662, EPI_ISL_2816663, EPI_ISL_2816664, EPI_ISL_2816665, EPI_ISL_2816667, EPI_ISL_2816668, EPI_ISL_2816669, EPI_ISL_2816670, EPI_ISL_2816671, EPI_ISL_2816672, EPI_ISL_2816674, EPI_ISL_2816675, EPI_ISL_2816676, EPI_ISL_2816677, EPI_ISL_2816678, EPI_ISL_2816679, EPI_ISL_2816680, EPI_ISL_2816681, EPI_ISL_2816682, EPI_ISL_2816683, EPI_ISL_2816684, EPI_ISL_2816685, EPI_ISL_2816687, EPI_ISL_2816688, EPI_ISL_2816689, EPI_ISL_2816690, EPI_ISL_2816691, EPI_ISL_2816692, EPI_ISL_2816693, EPI_ISL_2816694, EPI_ISL_2816695, EPI_ISL_2816696, EPI_ISL_2816697, EPI_ISL_2816698, EPI_ISL_2816699, EPI_ISL_2816700, EPI_ISL_2816701, EPI_ISL_2816702, EPI_ISL_2816703, EPI_ISL_2816704, EPI_ISL_2816705, EPI_ISL_2816706, EPI_ISL_2816707, EPI_ISL_2816708, EPI_ISL_2816709, EPI_ISL_2816710, EPI_ISL_2816711, EPI_ISL_2816712, EPI_ISL_2816713, EPI_ISL_2816714, EPI_ISL_2816715, EPI_ISL_2816716, EPI_ISL_2816717, EPI_ISL_2816718, EPI_ISL_2816719, EPI_ISL_2816720, EPI_ISL_2816721, EPI_ISL_2816722 see above | Northumbria University / South Tees Hospitals NHS Foundation Trust / North Cumbria Integrated Care NHS Foundation Trust / North Tees and Hartlepool NHS Foundation Trust / Newcastle Hospitals NHS Foundation Trust | COVID-19 Genomics UK (COG-UK) Consortium | Andrew Nelson; Brendan Payne; Clive Graham; Darren L Smith; Debra Padgett; Edward Barton; Emma Swindells; Garren Scott; Gary Black; Gary Eltringham; Giles S Holt; Greg R Young; Jane Greenaway; Jennifer Collins; John Allan; Joshua Loh; Lynn Dover; Matthew Bashton; Mohammad A Tariq; Paul Baker; Sarah Essex; Steve Liggett; Wen C Yew; Yusri Taha

EPI_ISL_2707360, EPI_ISL_2707361, EPI_ISL_2707362, EPI_ISL_2707363, EPI_ISL_2707367, EPI_ISL_2707369, EPI_ISL_2707370, EPI_ISL_2707374, EPI_ISL_2707377, EPI_ISL_2707381, EPI_ISL_2707383, EPI_ISL_2707385, EPI_ISL_2707387, EPI_ISL_2707388, EPI_ISL_2707390, EPI_ISL_2707391, EPI_ISL_2707393, EPI_ISL_2707395, EPI_ISL_2707396, EPI_ISL_2707398, EPI_ISL_2707399, EPI_ISL_2707410, EPI_ISL_2707414, EPI_ISL_2707416, EPI_ISL_2707420, EPI_ISL_2707423, EPI_ISL_2707424, EPI_ISL_2707426, EPI_ISL_2707429, EPI_ISL_2707431, EPI_ISL_2707433, EPI_ISL_2707435, EPI_ISL_2707438, EPI_ISL_2707439, EPI_ISL_2707442, EPI_ISL_2707443, EPI_ISL_2707444, EPI_ISL_2707445, EPI_ISL_2707450, EPI_ISL_2707451, EPI_ISL_2707452, EPI_ISL_2707453, EPI_ISL_2707455, EPI_ISL_2707456, EPI_ISL_2707457, EPI_ISL_2707458, EPI_ISL_2707461, EPI_ISL_2707462, EPI_ISL_2707463, EPI_ISL_2707464, EPI_ISL_2707466, EPI_ISL_2707469, EPI_ISL_2707474, EPI_ISL_2707482, EPI_ISL_2707483, EPI_ISL_2707485, EPI_ISL_2707493, EPI_ISL_2707496, EPI_ISL_2707504, EPI_ISL_2707505, EPI_ISL_2707513, EPI_ISL_2707514, EPI_ISL_2707518, EPI_ISL_2707523, EPI_ISL_2707524, EPI_ISL_2707525, EPI_ISL_2707533, EPI_ISL_2707536, EPI_ISL_2707542, EPI_ISL_2707546, EPI_ISL_2707547, EPI_ISL_2707548, EPI_ISL_2707554, EPI_ISL_2707555, EPI_ISL_2707559, EPI_ISL_2707560, EPI_ISL_2707561, EPI_ISL_2707572, EPI_ISL_2707576, EPI_ISL_2707580, EPI_ISL_2707581, EPI_ISL_2707582, EPI_ISL_2707584, EPI_ISL_2707585, EPI_ISL_2707586, EPI_ISL_2707591, EPI_ISL_2707592, EPI_ISL_2707597, EPI_ISL_2707598, EPI_ISL_2707600, EPI_ISL_2707603, EPI_ISL_2749952, EPI_ISL_2749953, EPI_ISL_2749954, EPI_ISL_2749955, EPI_ISL_2749957, EPI_ISL_2749958, EPI_ISL_2749959, EPI_ISL_2749961, EPI_ISL_2749962, EPI_ISL_2749963, EPI_ISL_2749967, EPI_ISL_2749971, EPI_ISL_2749973, EPI_ISL_2749974, EPI_ISL_2749975, EPI_ISL_2749978, EPI_ISL_2749980, EPI_ISL_2749981, EPI_ISL_2749982, EPI_ISL_2749986, EPI_ISL_2749987, EPI_ISL_2749989, EPI_ISL_2749990, EPI_ISL_2749991, EPI_ISL_2749992, EPI_ISL_2749995, EPI_ISL_2749996, EPI_ISL_2749998, EPI_ISL_2749999, EPI_ISL_2750001, EPI_ISL_2750002, EPI_ISL_2750004, EPI_ISL_2750005, EPI_ISL_2750006, EPI_ISL_2750007, EPI_ISL_2750008, EPI_ISL_2750010, EPI_ISL_2750011, EPI_ISL_2750012, EPI_ISL_2750013, EPI_ISL_2750014, EPI_ISL_2750015, EPI_ISL_2750016, EPI_ISL_2750017, EPI_ISL_2750018, EPI_ISL_2750019, EPI_ISL_2750020, EPI_ISL_2750021, EPI_ISL_2750022, EPI_ISL_2750024, EPI_ISL_2750027, EPI_ISL_2750030, EPI_ISL_2750031, EPI_ISL_2750038, EPI_ISL_2750044, EPI_ISL_2750045, EPI_ISL_2750046, EPI_ISL_2750047, EPI_ISL_2750048, EPI_ISL_2750050, EPI_ISL_2750051, EPI_ISL_2750052, EPI_ISL_2750053, EPI_ISL_2750054, EPI_ISL_2750055, EPI_ISL_2750056, EPI_ISL_2750057, EPI_ISL_2750058, EPI_ISL_2750059, EPI_ISL_2750063, EPI_ISL_2750074, EPI_ISL_2750088, EPI_ISL_2750090, EPI_ISL_2750091, EPI_ISL_2750094, EPI_ISL_2750095, EPI_ISL_2750097, EPI_ISL_2750099, EPI_ISL_2750103, EPI_ISL_2750109, EPI_ISL_2750115, EPI_ISL_2750117, EPI_ISL_2750118, EPI_ISL_2750121, EPI_ISL_2750122, EPI_ISL_2750123, EPI_ISL_2750124, EPI_ISL_2750126, EPI_ISL_2750129, EPI_ISL_2750131, EPI_ISL_2750132, EPI_ISL_2750133, EPI_ISL_2750135, EPI_ISL_2750136, EPI_ISL_2750137, EPI_ISL_2750138, EPI_ISL_2750144, EPI_ISL_2750145, EPI_ISL_2750146, EPI_ISL_2750147, EPI_ISL_2750148, EPI_ISL_2750149, EPI_ISL_2750150, EPI_ISL_2750151, EPI_ISL_2750153, EPI_ISL_2750155, EPI_ISL_2750156, EPI_ISL_2750157, EPI_ISL_2750186, EPI_ISL_2750191, EPI_ISL_2750193, EPI_ISL_2750196, EPI_ISL_2750207, EPI_ISL_2750210, EPI_ISL_2750211, EPI_ISL_2750212, EPI_ISL_2750213, EPI_ISL_2750214, EPI_ISL_2750215, EPI_ISL_2750216, EPI_ISL_2750217, EPI_ISL_2750218, EPI_ISL_2750219, EPI_ISL_2750220, EPI_ISL_2750221, EPI_ISL_2750222, EPI_ISL_2750223, EPI_ISL_2750224, EPI_ISL_2750225, EPI_ISL_2750226, EPI_ISL_2750227, EPI_ISL_2750228, EPI_ISL_2750229, EPI_ISL_2750230, EPI_ISL_2750232, EPI_ISL_2750233, EPI_ISL_2750234, EPI_ISL_2750288, EPI_ISL_2750293, EPI_ISL_2750298, EPI_ISL_2750306, EPI_ISL_2750359, EPI_ISL_2750393, EPI_ISL_2750394, EPI_ISL_2750395, EPI_ISL_2750396, EPI_ISL_2750397, EPI_ISL_2750398, EPI_ISL_2750399, EPI_ISL_2750400, EPI_ISL_2750401, EPI_ISL_2750402, EPI_ISL_2750403, EPI_ISL_2750404, EPI_ISL_2750405, EPI_ISL_2750406, EPI_ISL_2750489, EPI_ISL_2750491, EPI_ISL_2750492, EPI_ISL_2750494, EPI_ISL_2750495, EPI_ISL_2750496, EPI_ISL_2750504, EPI_ISL_2750505, EPI_ISL_2750506, EPI_ISL_2750509, EPI_ISL_2750517, EPI_ISL_2750518, EPI_ISL_2750521, EPI_ISL_2750555, EPI_ISL_2750563, EPI_ISL_2750565, EPI_ISL_2750567, EPI_ISL_2750569, EPI_ISL_2750570, EPI_ISL_2750571, EPI_ISL_2750572, EPI_ISL_2750574, EPI_ISL_2750575, EPI_ISL_2750576, EPI_ISL_2750577, EPI_ISL_2750578, EPI_ISL_2750579, EPI_ISL_2750580, EPI_ISL_2750581, EPI_ISL_2750583, EPI_ISL_2750584, EPI_ISL_2750585, EPI_ISL_2750590, EPI_ISL_2750591, EPI_ISL_2750593, EPI_ISL_2750603, EPI_ISL_2750604, EPI_ISL_2750607, EPI_ISL_2750608, EPI_ISL_2750609, EPI_ISL_2750626, EPI_ISL_2750642, EPI_ISL_2750643, EPI_ISL_2750644, EPI_ISL_2750645, EPI_ISL_2750648, EPI_ISL_2750649, EPI_ISL_2750653, EPI_ISL_2750657, EPI_ISL_2750658, EPI_ISL_2750664, EPI_ISL_2750665, EPI_ISL_2750666, EPI_ISL_2750668, EPI_ISL_2750669, EPI_ISL_2750675, EPI_ISL_2750684, EPI_ISL_2750686, EPI_ISL_2818335, EPI_ISL_2818340, EPI_ISL_2818450, EPI_ISL_2818682, EPI_ISL_2904738, EPI_ISL_2904917, EPI_ISL_2904925, EPI_ISL_2904928, EPI_ISL_2905010, EPI_ISL_2905017, EPI_ISL_2905027, EPI_ISL_2905030, EPI_ISL_2906002, EPI_ISL_2906004, EPI_ISL_2906007, EPI_ISL_2906009, EPI_ISL_2906010, EPI_ISL_2906013, EPI_ISL_2906014, EPI_ISL_2906015, EPI_ISL_2906018, EPI_ISL_2906023, EPI_ISL_2906030, EPI_ISL_2906031, EPI_ISL_2906032, EPI_ISL_2906033, EPI_ISL_2906036, EPI_ISL_2906037, EPI_ISL_2906038, EPI_ISL_2906039, EPI_ISL_2906040, EPI_ISL_2906043, EPI_ISL_2906047, EPI_ISL_2906050, EPI_ISL_2906051, EPI_ISL_2906052, EPI_ISL_2906053, EPI_ISL_2906056, EPI_ISL_2906062, EPI_ISL_2906066, EPI_ISL_2906068, EPI_ISL_2906070, EPI_ISL_2906071, EPI_ISL_2906074, EPI_ISL_2906100, EPI_ISL_2906106, EPI_ISL_2906113, EPI_ISL_2906117, EPI_ISL_2906119, EPI_ISL_2906121, EPI_ISL_2906122, EPI_ISL_2906123, EPI_ISL_2906124, EPI_ISL_2906125, EPI_ISL_2906127, EPI_ISL_2906297, EPI_ISL_2906298, EPI_ISL_2906300, EPI_ISL_2906301, EPI_ISL_2906302, EPI_ISL_2906303, EPI_ISL_2906304, EPI_ISL_2906305, EPI_ISL_2906306, EPI_ISL_2906307, EPI_ISL_2906308, EPI_ISL_2906309, EPI_ISL_2906310, EPI_ISL_2906311, EPI_ISL_2906312, EPI_ISL_2906313, EPI_ISL_2906314, EPI_ISL_2906315, EPI_ISL_2906316, EPI_ISL_2906317, EPI_ISL_2906322, EPI_ISL_2906323

see above | Originating lab: Wales Specialist Virology Centre Sequencing lab: Pathogen Genomics Unit | Public Health Wales Microbiology Cardiff Wales Specialist Virology Centre | Alec Birchley; Alexander Adams; Amy Gaskin; Angela Marchbank; Bree Gatica-Wilcox; Catherine Moore; Jason Coombes; Joanne Watkins; Joel Southgate; Johnathan Evans; Laura Gifford; Lauren Gilbert; Lee Graham; Malorie Perry; Matthew Bull; Nicole Pacchiarini; Sally Corden; Sara Kumziene-Summerhayes; Sara Rey; Sarah Taylor; Simon Cottrell; Sophie Jones; Tom Connor

EPI_ISL_2735679, EPI_ISL_2735692, EPI_ISL_2735704, EPI_ISL_2903043, EPI_ISL_2903047, EPI_ISL_2903049, EPI_ISL_2903053, EPI_ISL_2903064, EPI_ISL_2903066, EPI_ISL_2903068, EPI_ISL_2903081, EPI_ISL_2903082, EPI_ISL_2903090, EPI_ISL_2903093, EPI_ISL_2903094, EPI_ISL_2903105, EPI_ISL_2903117, EPI_ISL_2903118, EPI_ISL_2903120, EPI_ISL_2903121, EPI_ISL_2903158, EPI_ISL_2903159, EPI_ISL_2903162, EPI_ISL_2903163, EPI_ISL_2903165

see above | Oxford Viromics, NDM, University of Oxford; Oxford University Hospitals; Basingstoke and North Hampshire Hospital | COVID-19 Genomics UK (COG-UK) Consortium | Alex Mobbs; Amy Trebes; Anita Justice; Catrin Moore; Christophe Fraser; David Bonsall; David Buck; Emma Wise; George Macintyre; Jessica Lynch; John Todd; Mariateresa de Cesare; Matilde Mori; Monique Andersson; Nathan Moore; Nick Cortes; Robert Shaw; Stephen Kidd; Tanya Golubchik; Timothy Peto

EPI_ISL_2706993, EPI_ISL_2706994, EPI_ISL_2706995, EPI_ISL_2706996, EPI_ISL_2706998, EPI_ISL_2707000, EPI_ISL_2707002, EPI_ISL_2707003, EPI_ISL_2707005, EPI_ISL_2707006, EPI_ISL_2707007, EPI_ISL_2707008, EPI_ISL_2707009, EPI_ISL_2707011, EPI_ISL_2707012, EPI_ISL_2707013, EPI_ISL_2707014, EPI_ISL_2707015, EPI_ISL_2707016, EPI_ISL_2707017, EPI_ISL_2707018, EPI_ISL_2707019, EPI_ISL_2707020, EPI_ISL_2707021, EPI_ISL_2707022, EPI_ISL_2707023, EPI_ISL_2707024, EPI_ISL_2707025, EPI_ISL_2707026, EPI_ISL_2707027, EPI_ISL_2707028, EPI_ISL_2707029, EPI_ISL_2707030, EPI_ISL_2707031, EPI_ISL_2707032, EPI_ISL_2707036, EPI_ISL_2707037, EPI_ISL_2707038, EPI_ISL_2707039, EPI_ISL_2707040, EPI_ISL_2707041, EPI_ISL_2707042, EPI_ISL_2707044, EPI_ISL_2707045, EPI_ISL_2707046, EPI_ISL_2707047, EPI_ISL_2707048, EPI_ISL_2707049, EPI_ISL_2707050, EPI_ISL_2707051, EPI_ISL_2707052, EPI_ISL_2707054, EPI_ISL_2707055, EPI_ISL_2707056, EPI_ISL_2707057, EPI_ISL_2707058, EPI_ISL_2707059, EPI_ISL_2707060, EPI_ISL_2707061, EPI_ISL_2707062, EPI_ISL_2707063, EPI_ISL_2707065, EPI_ISL_2707066, EPI_ISL_2707067, EPI_ISL_2707068, EPI_ISL_2707069, EPI_ISL_2707070, EPI_ISL_2707071, EPI_ISL_2707072, EPI_ISL_2707073, EPI_ISL_2707074, EPI_ISL_2707076, EPI_ISL_2707077, EPI_ISL_2707079, EPI_ISL_2707082, EPI_ISL_2707084, EPI_ISL_2707085, EPI_ISL_2707087, EPI_ISL_2707088, EPI_ISL_2707089, EPI_ISL_2707090, EPI_ISL_2707092, EPI_ISL_2707093, EPI_ISL_2707094, EPI_ISL_2707095, EPI_ISL_2707097, EPI_ISL_2707098, EPI_ISL_2707099, EPI_ISL_2707100, EPI_ISL_2707101, EPI_ISL_2707102, EPI_ISL_2707103, EPI_ISL_2707104, EPI_ISL_2707105, EPI_ISL_2707106, EPI_ISL_2707107, EPI_ISL_2707108, EPI_ISL_2707109, EPI_ISL_2707110, EPI_ISL_2707111, EPI_ISL_2707112, EPI_ISL_2707113, EPI_ISL_2707114, EPI_ISL_2707115, EPI_ISL_2707116, EPI_ISL_2707117, EPI_ISL_2707118, EPI_ISL_2707119, EPI_ISL_2707120, EPI_ISL_2707121, EPI_ISL_2707122, EPI_ISL_2707123, EPI_ISL_2707124, EPI_ISL_2707125, EPI_ISL_2707126, EPI_ISL_2707127, EPI_ISL_2707128, EPI_ISL_2707129, EPI_ISL_2707130, EPI_ISL_2707131, EPI_ISL_2707132, EPI_ISL_2707133, EPI_ISL_2707134, EPI_ISL_2707135, EPI_ISL_2707136, EPI_ISL_2707137, EPI_ISL_2707138, EPI_ISL_2707139, EPI_ISL_2707140, EPI_ISL_2707141, EPI_ISL_2707142, EPI_ISL_2707143, EPI_ISL_2707144, EPI_ISL_2707145, EPI_ISL_2707146, EPI_ISL_2707147, EPI_ISL_2707148, EPI_ISL_2707149, EPI_ISL_2707150, EPI_ISL_2707151, EPI_ISL_2707152, EPI_ISL_2707153, EPI_ISL_2707154, EPI_ISL_2707155, EPI_ISL_2707156, EPI_ISL_2707157, EPI_ISL_2707158, EPI_ISL_2707159, EPI_ISL_2707160, EPI_ISL_2707161, EPI_ISL_2707162, EPI_ISL_2707163, EPI_ISL_2707164, EPI_ISL_2707165, EPI_ISL_2707166, EPI_ISL_2707167, EPI_ISL_2707168, EPI_ISL_2707169, EPI_ISL_2707170, EPI_ISL_2707171, EPI_ISL_2707172, EPI_ISL_2707173, EPI_ISL_2707174, EPI_ISL_2707175, EPI_ISL_2707176, EPI_ISL_2707177, EPI_ISL_2707178, EPI_ISL_2707179, EPI_ISL_2707180, EPI_ISL_2707181, EPI_ISL_2707182, EPI_ISL_2707183, EPI_ISL_2707184, EPI_ISL_2707185, EPI_ISL_2707186, EPI_ISL_2707187, EPI_ISL_2707188, EPI_ISL_2707189, EPI_ISL_2707190, EPI_ISL_2707191, EPI_ISL_2707192, EPI_ISL_2707193, EPI_ISL_2707194, EPI_ISL_2707195, EPI_ISL_2707196, EPI_ISL_2707197, EPI_ISL_2707198, EPI_ISL_2707199, EPI_ISL_2707200, EPI_ISL_2707201, EPI_ISL_2707202, EPI_ISL_2707203, EPI_ISL_2707204, EPI_ISL_2707205, EPI_ISL_2707206, EPI_ISL_2707207, EPI_ISL_2707208, EPI_ISL_2707209, EPI_ISL_2707210, EPI_ISL_2707211, EPI_ISL_2707212, EPI_ISL_2707213, EPI_ISL_2707214, EPI_ISL_2707215, EPI_ISL_2707216, EPI_ISL_2707217, EPI_ISL_2707218, EPI_ISL_2707219, EPI_ISL_2707220, EPI_ISL_2707221, EPI_ISL_2707222, EPI_ISL_2707223, EPI_ISL_2707224, EPI_ISL_2707225, EPI_ISL_2707226, EPI_ISL_2707227, EPI_ISL_2707228, EPI_ISL_2707229, EPI_ISL_2707230, EPI_ISL_2707231, EPI_ISL_2707232, EPI_ISL_2707233, EPI_ISL_2707234, EPI_ISL_2707235, EPI_ISL_2707236, EPI_ISL_2707237, EPI_ISL_2707238, EPI_ISL_2707239, EPI_ISL_2707240, EPI_ISL_2707241, EPI_ISL_2707242, EPI_ISL_2707243, EPI_ISL_2707244, EPI_ISL_2707245, EPI_ISL_2707246, EPI_ISL_2707247, EPI_ISL_2707248, EPI_ISL_2707249, EPI_ISL_2707250, EPI_ISL_2707251, EPI_ISL_2707252, EPI_ISL_2707253, EPI_ISL_2707254, EPI_ISL_2707255, EPI_ISL_2707256, EPI_ISL_2707257, EPI_ISL_2707258, EPI_ISL_2707259, EPI_ISL_2707260, EPI_ISL_2707261, EPI_ISL_2707262, EPI_ISL_2707263, EPI_ISL_2707264, EPI_ISL_2707265, EPI_ISL_2707266, EPI_ISL_2707267, EPI_ISL_2707268, EPI_ISL_2707269, EPI_ISL_2707270, EPI_ISL_2707271, EPI_ISL_2707272, EPI_ISL_2707273, EPI_ISL_2707274, EPI_ISL_2707275, EPI_ISL_2707276, EPI_ISL_2707277, EPI_ISL_2707278, EPI_ISL_2707279, EPI_ISL_2707280, EPI_ISL_2707281, EPI_ISL_2707282, EPI_ISL_2707283, EPI_ISL_2707284, EPI_ISL_2707285, EPI_ISL_2707286, EPI_ISL_2707287, EPI_ISL_2707288, EPI_ISL_2707289, EPI_ISL_2707290, EPI_ISL_2707291, EPI_ISL_2707292, EPI_ISL_2707293, EPI_ISL_2707294, EPI_ISL_2707295, EPI_ISL_2707296, EPI_ISL_2707297, EPI_ISL_2707298, EPI_ISL_2707299, EPI_ISL_2707300, EPI_ISL_2707301, EPI_ISL_2707302, EPI_ISL_2707303, EPI_ISL_2707304, EPI_ISL_2707305, EPI_ISL_2707306, EPI_ISL_2707307, EPI_ISL_2707308, EPI_ISL_2707309, EPI_ISL_2707310

see above | Quadram Institute Bioscience | COVID-19 Genomics UK (COG-UK) Consortium | Alexander J Trotter; Alison E. Mather; Alp Aydin; Ana P. Tedim; Anastasia Kolyva; Andrew Bell; Andrew J. Page; Claire Stuart; Dave J. Baker; Gemma L. Kay; John Wain; Justin O'Grady; Leonardo de Oliveira Martins; Lizzie Meadows; Maria Diaz; Mark Webber; Muhammed Yasir; Nabil-Fareed Alikhan; Ngozi Elumogo; Nicholas M. Thomson; Rachael Stanley; Rachel Gilroy; Reenesh Prakash; Samir Dervisevic; Samuel Bloomfield; Steven Rudder; Thanh Le-Viet

EPI_ISL_2746503, EPI_ISL_2746538, EPI_ISL_2746546, EPI_ISL_2746553, EPI_ISL_2746554, EPI_ISL_2746620, EPI_ISL_2746621, EPI_ISL_2746622, EPI_ISL_2746635, EPI_ISL_2746657, EPI_ISL_2746658, EPI_ISL_2746659, EPI_ISL_2746660, EPI_ISL_2746661, EPI_ISL_2746673, EPI_ISL_2746709, EPI_ISL_2746710, EPI_ISL_2746711, EPI_ISL_2746712, EPI_ISL_2746718, EPI_ISL_2746720, EPI_ISL_2746721, EPI_ISL_2746722, EPI_ISL_2746723, EPI_ISL_2746724, EPI_ISL_2746725, EPI_ISL_2746726, EPI_ISL_2746727, EPI_ISL_2746728, EPI_ISL_2746729, EPI_ISL_2746730, EPI_ISL_2746732, EPI_ISL_2746733, EPI_ISL_2746735, EPI_ISL_2746736, EPI_ISL_2746737, EPI_ISL_2746738, EPI_ISL_2746739, EPI_ISL_2746741, EPI_ISL_2746745, EPI_ISL_2746748, EPI_ISL_2746752, EPI_ISL_2746754, EPI_ISL_2746755, EPI_ISL_2746756, EPI_ISL_2746757, EPI_ISL_2746759, EPI_ISL_2746763, EPI_ISL_2746764, EPI_ISL_2746765, EPI_ISL_2746766, EPI_ISL_2746767, EPI_ISL_2746768, EPI_ISL_2746769, EPI_ISL_2746770, EPI_ISL_2746771, EPI_ISL_2746772, EPI_ISL_2746773, EPI_ISL_2746774, EPI_ISL_2746775, EPI_ISL_2746778, EPI_ISL_2746779, EPI_ISL_2746780, EPI_ISL_2746781, EPI_ISL_2746782, EPI_ISL_2746783, EPI_ISL_2746785, EPI_ISL_2746786, EPI_ISL_2746787, EPI_ISL_2746788, EPI_ISL_2746789, EPI_ISL_2746790, EPI_ISL_2746791, EPI_ISL_2746792, EPI_ISL_2746795, EPI_ISL_2746796, EPI_ISL_2746797, EPI_ISL_2746798, EPI_ISL_2746799, EPI_ISL_2746800, EPI_ISL_2746801, EPI_ISL_2746802, EPI_ISL_2746803, EPI_ISL_2746804, EPI_ISL_2746805, EPI_ISL_2746806, EPI_ISL_2746807, EPI_ISL_2746808, EPI_ISL_2746810, EPI_ISL_2746812, EPI_ISL_2746814, EPI_ISL_2746815, EPI_ISL_2746816, EPI_ISL_2746817, EPI_ISL_2746818, EPI_ISL_2746819, EPI_ISL_2746820, EPI_ISL_2746821, EPI_ISL_2746822, EPI_ISL_2746823, EPI_ISL_2746825, EPI_ISL_2746826, EPI_ISL_2746827, EPI_ISL_2746828, EPI_ISL_2746829, EPI_ISL_2746830, EPI_ISL_2746831, EPI_ISL_2746832, EPI_ISL_2746833, EPI_ISL_2746834, EPI_ISL_2746835, EPI_ISL_2746836, EPI_ISL_2746837, EPI_ISL_2746838, EPI_ISL_2746839, EPI_ISL_2746840, EPI_ISL_2746841, EPI_ISL_2746842, EPI_ISL_2746843, EPI_ISL_2746844, EPI_ISL_2746845, EPI_ISL_2746846, EPI_ISL_2746847, EPI_ISL_2746848, EPI_ISL_2746849, EPI_ISL_2746850, EPI_ISL_2746851, EPI_ISL_2746852, EPI_ISL_2746859, EPI_ISL_2746860, EPI_ISL_2746861, EPI_ISL_2746862, EPI_ISL_2746863, EPI_ISL_2746864, EPI_ISL_2746865, EPI_ISL_2746867, EPI_ISL_2746869, EPI_ISL_2746870, EPI_ISL_2746871, EPI_ISL_2746872, EPI_ISL_2746873, EPI_ISL_2746874, EPI_ISL_2746875, EPI_ISL_2746876, EPI_ISL_2746878, EPI_ISL_2746879, EPI_ISL_2746880, EPI_ISL_2746881, EPI_ISL_2746882, EPI_ISL_2746883, EPI_ISL_2746884, EPI_ISL_2746885, EPI_ISL_2746886, EPI_ISL_2746887, EPI_ISL_2746888, EPI_ISL_2746889, EPI_ISL_2746890, EPI_ISL_2746891, EPI_ISL_2746892, EPI_ISL_2746893, EPI_ISL_2746894, EPI_ISL_2746895, EPI_ISL_2746896, EPI_ISL_2746897, EPI_ISL_2746898, EPI_ISL_2746900, EPI_ISL_2746901, EPI_ISL_2746903, EPI_ISL_2746904, EPI_ISL_2746905, EPI_ISL_2746906, EPI_ISL_2746907, EPI_ISL_2746908, EPI_ISL_2746909, EPI_ISL_2746910, EPI_ISL_2746911, EPI_ISL_2746912, EPI_ISL_2746913, EPI_ISL_2746914, EPI_ISL_2746915, EPI_ISL_2746916, EPI_ISL_2746917, EPI_ISL_2746918, EPI_ISL_2746919, EPI_ISL_2746920, EPI_ISL_2746921, EPI_ISL_2746922, EPI_ISL_2746923, EPI_ISL_2746924, EPI_ISL_2746925, EPI_ISL_2746926, EPI_ISL_2746927, EPI_ISL_2746929, EPI_ISL_2746930, EPI_ISL_2746931, EPI_ISL_2746932, EPI_ISL_2746933, EPI_ISL_2746934, EPI_ISL_2746935, EPI_ISL_2746936, EPI_ISL_2746937, EPI_ISL_2746938, EPI_ISL_2746939, EPI_ISL_2746940, EPI_ISL_2746942, EPI_ISL_2746943, EPI_ISL_2746944, EPI_ISL_2746945, EPI_ISL_2746946, EPI_ISL_2746947, EPI_ISL_2746948, EPI_ISL_2746949, EPI_ISL_2746950, EPI_ISL_2746951, EPI_ISL_2746952, EPI_ISL_2746953, EPI_ISL_2746954, EPI_ISL_2746955, EPI_ISL_2746957, EPI_ISL_2746958, EPI_ISL_2746959, EPI_ISL_2746960, EPI_ISL_2746961, EPI_ISL_2746962, EPI_ISL_2746963, EPI_ISL_2746964, EPI_ISL_2746965, EPI_ISL_2746966, EPI_ISL_2746967, EPI_ISL_2746968, EPI_ISL_2746969, EPI_ISL_2746972, EPI_ISL_2746973, EPI_ISL_2746974, EPI_ISL_2746975, EPI_ISL_2746976, EPI_ISL_2746977, EPI_ISL_2746978, EPI_ISL_2746979, EPI_ISL_2746980, EPI_ISL_2746981, EPI_ISL_2746982, EPI_ISL_2746983, EPI_ISL_2746984, EPI_ISL_2746985, EPI_ISL_2746986, EPI_ISL_2746987, EPI_ISL_2746988, EPI_ISL_2746989, EPI_ISL_2746990, EPI_ISL_2746991, EPI_ISL_2746995, EPI_ISL_2746997, EPI_ISL_2746998, EPI_ISL_2746999, EPI_ISL_2747000, EPI_ISL_2747001, EPI_ISL_2747002, EPI_ISL_2747014, EPI_ISL_2747015, EPI_ISL_2747022, EPI_ISL_2747023, EPI_ISL_2747025, EPI_ISL_2747075, EPI_ISL_2747086, EPI_ISL_2747097, EPI_ISL_2747110, EPI_ISL_2747111, EPI_ISL_2747133, EPI_ISL_2747152, EPI_ISL_2747153, EPI_ISL_2747160, EPI_ISL_2747161, EPI_ISL_2747162, EPI_ISL_2747163, EPI_ISL_2747164, EPI_ISL_2747165, EPI_ISL_2747166, EPI_ISL_2747167, EPI_ISL_2747168, EPI_ISL_2747169, EPI_ISL_2747170, EPI_ISL_2747171, EPI_ISL_2747172, EPI_ISL_2747175, EPI_ISL_2747176, EPI_ISL_2747177, EPI_ISL_2747178, EPI_ISL_2747179, EPI_ISL_2747180, EPI_ISL_2747181, EPI_ISL_2747182, EPI_ISL_2747183, EPI_ISL_2747184, EPI_ISL_2747185, EPI_ISL_2747186, EPI_ISL_2747187, EPI_ISL_2747189, EPI_ISL_2747190, EPI_ISL_2747191, EPI_ISL_2747194, EPI_ISL_2747196, EPI_ISL_2747197, EPI_ISL_2747198, EPI_ISL_2747199, EPI_ISL_2747200, EPI_ISL_2747201, EPI_ISL_2747202, EPI_ISL_2747203, EPI_ISL_2747204, EPI_ISL_2747207, EPI_ISL_2747208, EPI_ISL_2747209, EPI_ISL_2747210, EPI_ISL_2747211, EPI_ISL_2747212, EPI_ISL_2747213, EPI_ISL_2747214, EPI_ISL_2747215, EPI_ISL_2747216, EPI_ISL_2747217, EPI_ISL_2747218, EPI_ISL_2747219, EPI_ISL_2747220, EPI_ISL_2747221, EPI_ISL_2747222, EPI_ISL_2747223, EPI_ISL_2747224, EPI_ISL_2747225, EPI_ISL_2747226, EPI_ISL_2747227, EPI_ISL_2747228, EPI_ISL_2747229, EPI_ISL_2747230, EPI_ISL_2747231, EPI_ISL_2747232, EPI_ISL_2747233, EPI_ISL_2747234, EPI_ISL_2747235, EPI_ISL_2747236, EPI_ISL_2747237, EPI_ISL_2747238, EPI_ISL_2747239, EPI_ISL_2747240, EPI_ISL_2747241, EPI_ISL_2747242, EPI_ISL_2747243, EPI_ISL_2747244, EPI_ISL_2747246, EPI_ISL_2747247, EPI_ISL_2747248, EPI_ISL_2747249, EPI_ISL_2747251, EPI_ISL_2747252, EPI_ISL_2747253, EPI_ISL_2747254, EPI_ISL_2747255, EPI_ISL_2747256, EPI_ISL_2747257, EPI_ISL_2747259, EPI_ISL_2747260, EPI_ISL_2747261, EPI_ISL_2747262, EPI_ISL_2747263, EPI_ISL_2747264, EPI_ISL_2747265, EPI_ISL_2747266, EPI_ISL_2747267, EPI_ISL_2747268, EPI_ISL_2747269, EPI_ISL_2747272, EPI_ISL_2747273, EPI_ISL_2747274, EPI_ISL_2747275, EPI_ISL_2747276, EPI_ISL_2747278, EPI_ISL_2747279, EPI_ISL_2747280, EPI_ISL_2747281, EPI_ISL_2747282, EPI_ISL_2747283, EPI_ISL_2747287, EPI_ISL_2747289, EPI_ISL_2747292, EPI_ISL_2747295, EPI_ISL_2747296, EPI_ISL_2747300, EPI_ISL_2747318, EPI_ISL_2747320, EPI_ISL_2747321, EPI_ISL_2747323, EPI_ISL_2747333, EPI_ISL_2747335, EPI_ISL_2747357, EPI_ISL_2747358, EPI_ISL_2747359, EPI_ISL_2747360, EPI_ISL_2747361, EPI_ISL_2747362, EPI_ISL_2747364, EPI_ISL_2747365, EPI_ISL_2747366, EPI_ISL_2747367, EPI_ISL_2747368, EPI_ISL_2747369, EPI_ISL_2747370, EPI_ISL_2747371, EPI_ISL_2747372, EPI_ISL_2747373, EPI_ISL_2747374, EPI_ISL_2747375, EPI_ISL_2747376, EPI_ISL_2747377, EPI_ISL_2747378, EPI_ISL_2747380, EPI_ISL_2747381, EPI_ISL_2747382, EPI_ISL_2747383, EPI_ISL_2747384, EPI_ISL_2747385, EPI_ISL_2747386, EPI_ISL_2747387, EPI_ISL_2747388, EPI_ISL_2747389, EPI_ISL_2747391, EPI_ISL_2747392, EPI_ISL_2747393, EPI_ISL_2747394, EPI_ISL_2747395, EPI_ISL_2747396, EPI_ISL_2747397, EPI_ISL_2747398, EPI_ISL_2747399, EPI_ISL_2747400, EPI_ISL_2747401, EPI_ISL_2747402, EPI_ISL_2747404, EPI_ISL_2747405, EPI_ISL_2747406, EPI_ISL_2747407, EPI_ISL_2747408, EPI_ISL_2747409, EPI_ISL_2747410, EPI_ISL_2747411, EPI_ISL_2747414, EPI_ISL_2747415, EPI_ISL_2747416, EPI_ISL_2747417, EPI_ISL_2747419, EPI_ISL_2747420, EPI_ISL_2747421, EPI_ISL_2747422, EPI_ISL_2747423, EPI_ISL_2747426, EPI_ISL_2747427, EPI_ISL_2747428, EPI_ISL_2747429, EPI_ISL_2747431, EPI_ISL_2747444, EPI_ISL_2747449, EPI_ISL_2747453, EPI_ISL_2747456, EPI_ISL_2747459, EPI_ISL_2747461, EPI_ISL_2747465, EPI_ISL_2747466, EPI_ISL_2747467, EPI_ISL_2747469, EPI_ISL_2747487, EPI_ISL_2747489, EPI_ISL_2747490, EPI_ISL_2747495, EPI_ISL_2747501, EPI_ISL_2747503, EPI_ISL_2747507, EPI_ISL_2747515, EPI_ISL_2747523, EPI_ISL_2747525, EPI_ISL_2747535, EPI_ISL_2747539, EPI_ISL_2747541, EPI_ISL_2747543, EPI_ISL_2747545, EPI_ISL_2747804, EPI_ISL_2747812, EPI_ISL_2747816, EPI_ISL_2747818, EPI_ISL_2747825, EPI_ISL_2747831, EPI_ISL_2747865, EPI_ISL_2747873, EPI_ISL_2747888, EPI_ISL_2747973, EPI_ISL_2747977, EPI_ISL_2748021, EPI_ISL_2817397, EPI_ISL_2817503, EPI_ISL_2817508, EPI_ISL_2817517, EPI_ISL_2817520, EPI_ISL_2817524, EPI_ISL_2817527, EPI_ISL_2817529, EPI_ISL_2817530, EPI_ISL_2817534, EPI_ISL_2817535, EPI_ISL_2817536, EPI_ISL_2817537, EPI_ISL_2817538, EPI_ISL_2817539, EPI_ISL_2817540, EPI_ISL_2817548, EPI_ISL_2818076, EPI_ISL_2818077, EPI_ISL_2818078, EPI_ISL_2818079, EPI_ISL_2818080, EPI_ISL_2818081, EPI_ISL_2818082, EPI_ISL_2818083, EPI_ISL_2818084, EPI_ISL_2818085, EPI_ISL_2818086, EPI_ISL_2818087, EPI_ISL_2818244, EPI_ISL_2818250, EPI_ISL_2818251, EPI_ISL_2903901, EPI_ISL_2903994

see above | Respiratory Virus Unit, Microbiology Services Colindale, Public Health England | COVID-19 Genomics UK (COG-UK) Consortium | PHE Covid Sequencing Team

EPI_ISL_2705881 | University College London, Great Ormond Street Hospital for Children NHS Foundation Trust, Imperial College Healthcare NHS Trust | COVID-19 Genomics UK (COG-UK) Consortium | Alison Holmes; Charlotte Williams; Helena Tutill; Jacqueline Findlay; James Price; Judith Breuer; Julianne Brown; Kathryn Harris; Leysa Forrest; Mark Kristiansen; Paola Niola; Paola Resende Silva; Patricia Dyal; Paul Randell; Rachel Williams; Samuel Weeks; Sergi Castellano; Sunando Roy; Tony Brooks; Yasmin Panchbhaya

EPI_ISL_2722036, EPI_ISL_2722037, EPI_ISL_2722039, EPI_ISL_2722041, EPI_ISL_2722044, EPI_ISL_2722046, EPI_ISL_2722048, EPI_ISL_2722049, EPI_ISL_2722050, EPI_ISL_2722051, EPI_ISL_2722052, EPI_ISL_2722053, EPI_ISL_2722054, EPI_ISL_2722055, EPI_ISL_2722056, EPI_ISL_2722057, EPI_ISL_2722058, EPI_ISL_2722065, EPI_ISL_2722066, EPI_ISL_2722067, EPI_ISL_2722068, EPI_ISL_2722070, EPI_ISL_2722071, EPI_ISL_2722072, EPI_ISL_2722073, EPI_ISL_2722074, EPI_ISL_2722075, EPI_ISL_2722076, EPI_ISL_2722077, EPI_ISL_2722078, EPI_ISL_2722079, EPI_ISL_2722080, EPI_ISL_2722081, EPI_ISL_2722082, EPI_ISL_2815378, EPI_ISL_2815379, EPI_ISL_2815380, EPI_ISL_2815381, EPI_ISL_2815382, EPI_ISL_2815385, EPI_ISL_2815386, EPI_ISL_2815389, EPI_ISL_2815390, EPI_ISL_2815391, EPI_ISL_2815393, EPI_ISL_2815398

see above | University of Birmingham | COVID-19 Genomics UK (COG-UK) Consortium | Alex Richter; Andrew Bosworth; Andrew Bosworth. Queen Elizabeth Hospital: Anna Casey; Andrew D Beggs PHE Heartlands Lab: Husam Osman; Charlotte Poxon; Institute of Microbiology; Joanne Stockton; Josh Quick; Kasun Wanigasooriya; Mike Kidd; Nicholas Loman. University of Birmingham Testing Laboratory: Celina M Whalley; Oliver Pickles; Radoslaw Poplawski; Samuel Nicholls; University of Birmingham: Claire McMurray; Will Rowe

EPI_ISL_2704937, EPI_ISL_2704938, EPI_ISL_2704939, EPI_ISL_2704940, EPI_ISL_2734319, EPI_ISL_2734320, EPI_ISL_2734321, EPI_ISL_2734326, EPI_ISL_2734334, EPI_ISL_2734336, EPI_ISL_2734337, EPI_ISL_2734342, EPI_ISL_2734343, EPI_ISL_2734344, EPI_ISL_2734345, EPI_ISL_2734346, EPI_ISL_2734347, EPI_ISL_2734348, EPI_ISL_2734354, EPI_ISL_2734356, EPI_ISL_2734357, EPI_ISL_2734386, EPI_ISL_2734388, EPI_ISL_2734394, EPI_ISL_2734396, EPI_ISL_2734397, EPI_ISL_2734398, EPI_ISL_2734399, EPI_ISL_2734400, EPI_ISL_2734401, EPI_ISL_2734404, EPI_ISL_2734407, EPI_ISL_2734414, EPI_ISL_2734431, EPI_ISL_2734432, EPI_ISL_2734434, EPI_ISL_2734435, EPI_ISL_2815970, EPI_ISL_2815972, EPI_ISL_2815973, EPI_ISL_2815974, EPI_ISL_2815975, EPI_ISL_2815976, EPI_ISL_2815977, EPI_ISL_2815978, EPI_ISL_2815979, EPI_ISL_2815980, EPI_ISL_2815981, EPI_ISL_2815982, EPI_ISL_2815983, EPI_ISL_2815984, EPI_ISL_2815985, EPI_ISL_2815994, EPI_ISL_2816004

see above | Virology Department, Royal Infirmary of Edinburgh, NHS Lothian / School of Biological Sciences, University of Edinburgh | COVID-19 Genomics UK (COG-UK) Consortium | Colquhoun R; Cotton S; Dewar R; Gallagher A; Hill V; Jackson B; Maloney D; McCrone JT; McHugh M; O'Toole Á; Rambaut A; Rooke S; Scher E; Templeton K; Yu X

EPI_ISL_2708437, EPI_ISL_2708445, EPI_ISL_2708446, EPI_ISL_2708447, EPI_ISL_2708448, EPI_ISL_2708449, EPI_ISL_2708450, EPI_ISL_2708451, EPI_ISL_2708452, EPI_ISL_2708453, EPI_ISL_2708455, EPI_ISL_2708457, EPI_ISL_2708458, EPI_ISL_2708460, EPI_ISL_2708464, EPI_ISL_2708466, EPI_ISL_2708467, EPI_ISL_2708468, EPI_ISL_2708469, EPI_ISL_2708470, EPI_ISL_2708471, EPI_ISL_2708472, EPI_ISL_2708473, EPI_ISL_2708474, EPI_ISL_2708475, EPI_ISL_2708476, EPI_ISL_2708479, EPI_ISL_2708480, EPI_ISL_2708484, EPI_ISL_2708485, EPI_ISL_2708486, EPI_ISL_2708487, EPI_ISL_2708488, EPI_ISL_2708489, EPI_ISL_2708490, EPI_ISL_2708491, EPI_ISL_2708493, EPI_ISL_2708494, EPI_ISL_2708495, EPI_ISL_2708497, EPI_ISL_2708498, EPI_ISL_2708499, EPI_ISL_2708500, EPI_ISL_2708502, EPI_ISL_2708503, EPI_ISL_2708504, EPI_ISL_2708505, EPI_ISL_2708507, EPI_ISL_2708508, EPI_ISL_2708509, EPI_ISL_2708513, EPI_ISL_2708514, EPI_ISL_2708515, EPI_ISL_2708516, EPI_ISL_2708518, EPI_ISL_2708519, EPI_ISL_2708520, EPI_ISL_2708521, EPI_ISL_2708522, EPI_ISL_2708523, EPI_ISL_2708525, EPI_ISL_2708526, EPI_ISL_2708527, EPI_ISL_2708529, EPI_ISL_2708532, EPI_ISL_2708533, EPI_ISL_2708534, EPI_ISL_2708535, EPI_ISL_2708536, EPI_ISL_2708537, EPI_ISL_2708538, EPI_ISL_2708539, EPI_ISL_2708540, EPI_ISL_2708543, EPI_ISL_2708544, EPI_ISL_2708546, EPI_ISL_2708549, EPI_ISL_2708552, EPI_ISL_2708555, EPI_ISL_2708556, EPI_ISL_2708557, EPI_ISL_2708559, EPI_ISL_2708560, EPI_ISL_2708561, EPI_ISL_2708564, EPI_ISL_2708569, EPI_ISL_2708570, EPI_ISL_2708571, EPI_ISL_2708573, EPI_ISL_2708575, EPI_ISL_2708578, EPI_ISL_2708579, EPI_ISL_2708581, EPI_ISL_2708582, EPI_ISL_2708583, EPI_ISL_2708584, EPI_ISL_2708586, EPI_ISL_2708587, EPI_ISL_2708589, EPI_ISL_2708590, EPI_ISL_2708591, EPI_ISL_2708592, EPI_ISL_2708593, EPI_ISL_2708594, EPI_ISL_2708595, EPI_ISL_2708596, EPI_ISL_2708597, EPI_ISL_2708598, EPI_ISL_2708602, EPI_ISL_2708603, EPI_ISL_2708604, EPI_ISL_2708605, EPI_ISL_2708606, EPI_ISL_2708607, EPI_ISL_2708610, EPI_ISL_2708612, EPI_ISL_2708613, EPI_ISL_2708614, EPI_ISL_2708616, EPI_ISL_2708617, EPI_ISL_2708619, EPI_ISL_2708620, EPI_ISL_2708621, EPI_ISL_2708622, EPI_ISL_2708624, EPI_ISL_2708625, EPI_ISL_2708627, EPI_ISL_2708628, EPI_ISL_2708630, EPI_ISL_2708633, EPI_ISL_2708635, EPI_ISL_2708636, EPI_ISL_2708637, EPI_ISL_2708638, EPI_ISL_2708639, EPI_ISL_2708641, EPI_ISL_2708642, EPI_ISL_2708646, EPI_ISL_2708647, EPI_ISL_2708650, EPI_ISL_2708651, EPI_ISL_2708652, EPI_ISL_2708653, EPI_ISL_2708655, EPI_ISL_2708656, EPI_ISL_2708658, EPI_ISL_2708659, EPI_ISL_2708660, EPI_ISL_2708662, EPI_ISL_2708663, EPI_ISL_2708665, EPI_ISL_2708666, EPI_ISL_2708668, EPI_ISL_2708669, EPI_ISL_2708670, EPI_ISL_2708671, EPI_ISL_2708672, EPI_ISL_2708673, EPI_ISL_2708674, EPI_ISL_2708675, EPI_ISL_2708676, EPI_ISL_2708678, EPI_ISL_2708680, EPI_ISL_2708681, EPI_ISL_2708682, EPI_ISL_2708683, EPI_ISL_2708685, EPI_ISL_2708687, EPI_ISL_2708688, EPI_ISL_2708689, EPI_ISL_2708690, EPI_ISL_2708694, EPI_ISL_2708695, EPI_ISL_2708696, EPI_ISL_2708697, EPI_ISL_2708698, EPI_ISL_2708699, EPI_ISL_2708700, EPI_ISL_2708701, EPI_ISL_2708702, EPI_ISL_2708703, EPI_ISL_2708704, EPI_ISL_2708706, EPI_ISL_2708708, EPI_ISL_2708709, EPI_ISL_2708710, EPI_ISL_2708711, EPI_ISL_2708712, EPI_ISL_2708715, EPI_ISL_2708716, EPI_ISL_2708717, EPI_ISL_2708718, EPI_ISL_2708720, EPI_ISL_2708721, EPI_ISL_2708723, EPI_ISL_2708724, EPI_ISL_2708725, EPI_ISL_2708726, EPI_ISL_2708728, EPI_ISL_2708729, EPI_ISL_2708732, EPI_ISL_2708734, EPI_ISL_2708735, EPI_ISL_2708736, EPI_ISL_2708739, EPI_ISL_2708740, EPI_ISL_2708741, EPI_ISL_2708742, EPI_ISL_2708743, EPI_ISL_2708745, EPI_ISL_2708746, EPI_ISL_2708747, EPI_ISL_2708749, EPI_ISL_2708750, EPI_ISL_2708752, EPI_ISL_2708753, EPI_ISL_2708754, EPI_ISL_2708755, EPI_ISL_2708757, EPI_ISL_2708758, EPI_ISL_2708759, EPI_ISL_2708760, EPI_ISL_2708761, EPI_ISL_2708762, EPI_ISL_2708764, EPI_ISL_2708765, EPI_ISL_2708766, EPI_ISL_2708767, EPI_ISL_2708768, EPI_ISL_2708769, EPI_ISL_2708773, EPI_ISL_2708775, EPI_ISL_2708776, EPI_ISL_2708777, EPI_ISL_2708778, EPI_ISL_2708780, EPI_ISL_2708781, EPI_ISL_2708782, EPI_ISL_2708783, EPI_ISL_2708785, EPI_ISL_2708786, EPI_ISL_2708787, EPI_ISL_2708788, EPI_ISL_2708789, EPI_ISL_2708790, EPI_ISL_2708791, EPI_ISL_2708792, EPI_ISL_2708793, EPI_ISL_2708794, EPI_ISL_2708795, EPI_ISL_2708796, EPI_ISL_2708797, EPI_ISL_2708798, EPI_ISL_2708800, EPI_ISL_2708803, EPI_ISL_2708804, EPI_ISL_2708806, EPI_ISL_2708807, EPI_ISL_2708809, EPI_ISL_2708810, EPI_ISL_2708812, EPI_ISL_2708813, EPI_ISL_2708814, EPI_ISL_2708815, EPI_ISL_2708818, EPI_ISL_2708819, EPI_ISL_2708820, EPI_ISL_2708821, EPI_ISL_2708822, EPI_ISL_2708823, EPI_ISL_2708824, EPI_ISL_2708825, EPI_ISL_2708827, EPI_ISL_2708828, EPI_ISL_2708831, EPI_ISL_2708832, EPI_ISL_2708833, EPI_ISL_2708834, EPI_ISL_2708835, EPI_ISL_2708837, EPI_ISL_2708838, EPI_ISL_2708841

see above | Virology Department, Sheffield Teaching Hospitals NHS Foundation Trust/Department of Infection, Immunity and | COVID-19 Genomics | Adri Angyal; Alex Keeley; Benjamin Lindsey; Cariad Evans; Danielle Groves; Dave Partridge; Katie Johnson; Laura Carrilero; Luke Green; Matthew Parker; Matthew Wyles; Mehmet Yavuz; Mohammad Raza; Nikki

| | | | |
|---|---|---|---|
| | Cardiovascular Disease, The Medical School, University of Sheffield | UK (COG-UK) Consortium | Smith; Paul Parsons; Rachel Tucker; Rebecca Brown; Thushan de Silva |
| EPI_ISL_2704758, EPI_ISL_2704761, EPI_ISL_2704776, EPI_ISL_2704781, EPI_ISL_2704789, EPI_ISL_2815895, EPI_ISL_2815899, EPI_ISL_2815902, EPI_ISL_2815930, EPI_ISL_2815931, EPI_ISL_2815933, EPI_ISL_2815934, EPI_ISL_2815936, EPI_ISL_2815937, EPI_ISL_2815938, EPI_ISL_2815939, EPI_ISL_2815940, EPI_ISL_2815941, EPI_ISL_2815942, EPI_ISL_2815960, EPI_ISL_2900740, EPI_ISL_2900741, EPI_ISL_2900742, EPI_ISL_2900743, EPI_ISL_2900744 | | | |
| see above | West of Scotland Specialist Virology Centre, NHSGGC / MRC-University of Glasgow Centre for Virus Research | COVID-19 Genomics UK (COG-UK) Consortium | Alasdair MacLean; Alice Broos; Ana da Silva Filipe; Antonia Ho; Daniel Mair; David L Robertson; Emma Thomson; Guy Mollett; Ioulia Tsatsani; James Shepherd; Jenna Nichols; Joseph Hughes; Kathy Li; Kathy Smollett; Kyriaki Nomikou; Lily Tong; Matthew Holden; Natasha Johnson; Rachel Blacow; Richard Orton; Rory Gunson; Sarah McDonald; Sharif Shaaban; Sreenu Vattipally; Stephen Carmichael |

We gratefully acknowledge the following Authors from the Originating laboratories responsible for obtaining the specimens, as well as the Submitting laboratories where the genome data were generated and shared via GISAID, on which this research is based.

All Submitters of data may be contacted directly via www.gisaid.org

Authors are sorted alphabetically.

| Accession ID | Originating Laboratory | Submitting Laboratory | Authors |
|---|---|---|---|
| EPI_ISL_2795921, EPI_ISL_2795923, EPI_ISL_2795924, EPI_ISL_2795925, EPI_ISL_2795926, EPI_ISL_2795927, EPI_ISL_2795928, EPI_ISL_2795929, EPI_ISL_2795930, EPI_ISL_2795931, EPI_ISL_2795932, EPI_ISL_2795933, EPI_ISL_2795934, EPI_ISL_2795945, EPI_ISL_2795950, EPI_ISL_2795953, EPI_ISL_2795956 | | | |
| see above | Amazon Dx, AUV2 Laboratory | Amazon Dx, AUV2 Laboratory | Luke Meredith |
| EPI_ISL_2708290, EPI_ISL_2708291, EPI_ISL_2708292, EPI_ISL_2708293, EPI_ISL_2708294, EPI_ISL_2708295, EPI_ISL_2708296, EPI_ISL_2708297, EPI_ISL_2708298, EPI_ISL_2708299, EPI_ISL_2708321, EPI_ISL_2708346, EPI_ISL_2708347, EPI_ISL_2708348, EPI_ISL_2708349, EPI_ISL_2708351, EPI_ISL_2708356, EPI_ISL_2708360, EPI_ISL_2708361, EPI_ISL_2708362, EPI_ISL_2708363, EPI_ISL_2708364, EPI_ISL_2708371, EPI_ISL_2708372, EPI_ISL_2708376, EPI_ISL_2708377, EPI_ISL_2708378, EPI_ISL_2708382, EPI_ISL_2708383, EPI_ISL_2708384, EPI_ISL_2708385, EPI_ISL_2708386, EPI_ISL_2708387, EPI_ISL_2708388, EPI_ISL_2708389, EPI_ISL_2708390, EPI_ISL_2708391, EPI_ISL_2708392, EPI_ISL_2708393, EPI_ISL_2708394, EPI_ISL_2708395, EPI_ISL_2708396, EPI_ISL_2708397, EPI_ISL_2708398, EPI_ISL_2708399, EPI_ISL_2708400, EPI_ISL_2708401, EPI_ISL_2708402, EPI_ISL_2708403, EPI_ISL_2708404, EPI_ISL_2708406, EPI_ISL_2708408, EPI_ISL_2708409, EPI_ISL_2708410, EPI_ISL_2708411, EPI_ISL_2708412, EPI_ISL_2819861, EPI_ISL_2819864, EPI_ISL_2819870, EPI_ISL_2819873, EPI_ISL_2819877, EPI_ISL_2819879, EPI_ISL_2819880, EPI_ISL_2819884, EPI_ISL_2819889, EPI_ISL_2819915, EPI_ISL_2906737, EPI_ISL_2906740, EPI_ISL_2906742, EPI_ISL_2906743, EPI_ISL_2906744, EPI_ISL_2906746, EPI_ISL_2906748, EPI_ISL_2906750, EPI_ISL_2906753, EPI_ISL_2906757, EPI_ISL_2906759, EPI_ISL_2906762 | | | |
| see above | Centre for Enzyme Innovation, University of Portsmouth / Translational Research Laboratory, Portsmouth Hospitals NHS Trust / Hampshire Hospitals NHS Foundation Trust / Bournemouth University / University Hospitals Dorset NHS Foundation Trust / Brighton and Sussex University Hospitals NHS Trust / Dartford and Gravesham NHS Trust / Isle of Wight NHS Trust / Maidstone and Tunbridge Wells NHS Trust / University Hospital Southampton NHS Foundation Trust | COVID-19 Genomics UK (COG-UK) Consortium | Adhyana Mahanama; Allyson Lloyd; Andrew Butt; Angela Beckett; Anibolina Castigador; Anna Mantzaouratou; Anoop Chauhan; Ben Macklin; Benjamin Cogger; Buddhini Samaraweera; Cassandra Malone; Christopher Fearn; Claire Thomas; Dorian Crudgington; Eleri Wilson-Davies; Emanuela Pelosi; Emily Macnaughton; Helen Umpleby; Helen Wheeler; Jacqui Prieto; Jonnie Seymour; Karen Withell; Kate Cook; Katie Loveson; Kelly Bicknell; Kevin Tucker; Kordo Saeed; Liz Sheridan; Magdalena Barrow; Matthew Harvey; Mohammed Hassan-Ibrahim; Nathan Moore; Nick Cortes; Rachael Jeremiah; Robert Impey; Salman Goudarzi; Samirakhon Raupova; Samuel Robson; Sarah Buchan; Sarah Jeremiah; Sarah Wyllie; Scott Elliott; Sharon Glaysher; Siona Silvieri; Stephen Aplin; Stephen Kidd; Thea Sass |
| EPI_ISL_2722294, EPI_ISL_2722296, EPI_ISL_2722298, EPI_ISL_2722300, EPI_ISL_2722301, EPI_ISL_2722302, EPI_ISL_2722303, EPI_ISL_2722304, EPI_ISL_2722305, EPI_ISL_2722306, EPI_ISL_2722307, EPI_ISL_2722308, EPI_ISL_2722309, EPI_ISL_2722310, EPI_ISL_2722311, EPI_ISL_2722312, EPI_ISL_2722313, EPI_ISL_2722314, EPI_ISL_2722315, EPI_ISL_2722316, EPI_ISL_2722317, EPI_ISL_2722319, EPI_ISL_2722321, EPI_ISL_2722323, EPI_ISL_2722325, EPI_ISL_2722327, EPI_ISL_2722330, EPI_ISL_2722332, EPI_ISL_2722334, EPI_ISL_2722336, EPI_ISL_2722338, EPI_ISL_2722340, EPI_ISL_2722343, EPI_ISL_2722345, EPI_ISL_2722347, EPI_ISL_2722348, EPI_ISL_2722349, EPI_ISL_2722350, EPI_ISL_2722351, EPI_ISL_2722352, EPI_ISL_2722353, EPI_ISL_2722354, EPI_ISL_2722355, EPI_ISL_2722361, EPI_ISL_2722550, EPI_ISL_2722551, EPI_ISL_2722552, EPI_ISL_2722553, EPI_ISL_2722554, EPI_ISL_2722555, EPI_ISL_2722556, EPI_ISL_2722558, EPI_ISL_2722559, EPI_ISL_2722560, EPI_ISL_2722561, EPI_ISL_2722562, EPI_ISL_2722563, EPI_ISL_2722564, EPI_ISL_2722565, EPI_ISL_2722566, EPI_ISL_2722567, EPI_ISL_2722568, EPI_ISL_2722569, EPI_ISL_2722570, EPI_ISL_2722572, EPI_ISL_2722573, EPI_ISL_2722575, EPI_ISL_2722576, EPI_ISL_2722577, EPI_ISL_2722579, EPI_ISL_2722580, EPI_ISL_2722581, EPI_ISL_2722582, EPI_ISL_2722583, EPI_ISL_2722584, EPI_ISL_2722585, EPI_ISL_2722587, EPI_ISL_2722588, EPI_ISL_2722589, EPI_ISL_2722590, EPI_ISL_2722591, EPI_ISL_2722592, EPI_ISL_2722594, EPI_ISL_2722595, EPI_ISL_2722596, EPI_ISL_2722597, EPI_ISL_2722598, EPI_ISL_2722599, EPI_ISL_2722603, EPI_ISL_2722604, EPI_ISL_2722605, EPI_ISL_2722606, EPI_ISL_2722608, EPI_ISL_2722609, EPI_ISL_2722610, EPI_ISL_2722611, EPI_ISL_2722612, EPI_ISL_2722614, EPI_ISL_2722615, EPI_ISL_2722616, EPI_ISL_2722617, EPI_ISL_2722618, EPI_ISL_2722619, EPI_ISL_2722620, EPI_ISL_2722621, EPI_ISL_2722622, EPI_ISL_2722623, EPI_ISL_2722624, EPI_ISL_2722625, EPI_ISL_2722626, EPI_ISL_2722627, EPI_ISL_2722628, EPI_ISL_2722629, EPI_ISL_2722630, EPI_ISL_2722631, EPI_ISL_2815686 | | | |
| see above | Department of Pathology, University of Cambridge | COVID-19 Genomics UK (COG-UK) Consortium | Aminu S. Jahun; Ian Goodfellow; Iliana Georgana; Martin D. Curran; Myra Hosmillo; Rhys Izuagbe; Surendra Parmar; William L. Hamilton; Yasmin Chaudhry |
| EPI_ISL_2716978, EPI_ISL_2716987, EPI_ISL_2717000, EPI_ISL_2717006, EPI_ISL_2717008, EPI_ISL_2717016, EPI_ISL_2717017, EPI_ISL_2717022, EPI_ISL_2717023, EPI_ISL_2717030, EPI_ISL_2717032, EPI_ISL_2717042, EPI_ISL_2717047, EPI_ISL_2717054, EPI_ISL_2717058, EPI_ISL_2717059, EPI_ISL_2717061, EPI_ISL_2717071, EPI_ISL_2717080, EPI_ISL_2717081, EPI_ISL_2717089, EPI_ISL_2717090, EPI_ISL_2717104, EPI_ISL_2717106, EPI_ISL_2717107, EPI_ISL_2717109, EPI_ISL_2717110, EPI_ISL_2717120, EPI_ISL_2717129, EPI_ISL_2717147, EPI_ISL_2717155, EPI_ISL_2717158, EPI_ISL_2717160, EPI_ISL_2717161, EPI_ISL_2717164, EPI_ISL_2717166, EPI_ISL_2717176, EPI_ISL_2717181, EPI_ISL_2717194, EPI_ISL_2717195, EPI_ISL_2717196, EPI_ISL_2717202, EPI_ISL_2717210, EPI_ISL_2717214, EPI_ISL_2717217, EPI_ISL_2717238, EPI_ISL_2717259, EPI_ISL_2717261, EPI_ISL_2717263, EPI_ISL_2717264, EPI_ISL_2717266, EPI_ISL_2717268, EPI_ISL_2717270, EPI_ISL_2717271, EPI_ISL_2717280, EPI_ISL_2717289, EPI_ISL_2717300, EPI_ISL_2717303, EPI_ISL_2717306, EPI_ISL_2717335, EPI_ISL_2717354, EPI_ISL_2717360, EPI_ISL_2717369, EPI_ISL_2717380, EPI_ISL_2717386, EPI_ISL_2717391, EPI_ISL_2717400, EPI_ISL_2717408, EPI_ISL_2717418, EPI_ISL_2717439, EPI_ISL_2717454, EPI_ISL_2717456, EPI_ISL_2717460, EPI_ISL_2717464, EPI_ISL_2717468, EPI_ISL_2717469, EPI_ISL_2717479, EPI_ISL_2717483, EPI_ISL_2717485, EPI_ISL_2717490, EPI_ISL_2717492, EPI_ISL_2717493, EPI_ISL_2717511, EPI_ISL_2717520, EPI_ISL_2717522, EPI_ISL_2717523, EPI_ISL_2717534, EPI_ISL_2717536, EPI_ISL_2717564, EPI_ISL_2717576, EPI_ISL_2781487, EPI_ISL_2781488, EPI_ISL_2781489, EPI_ISL_2781492, EPI_ISL_2781496, EPI_ISL_2781498, EPI_ISL_2781499, EPI_ISL_2781506, EPI_ISL_2781515, EPI_ISL_2781517, EPI_ISL_2781530, EPI_ISL_2781531, EPI_ISL_2781533, EPI_ISL_2781535, EPI_ISL_2781538, EPI_ISL_2781541, EPI_ISL_2781543, EPI_ISL_2781547, EPI_ISL_2781558, EPI_ISL_2781561, EPI_ISL_2781562, EPI_ISL_2781565, EPI_ISL_2781567, EPI_ISL_2781574, EPI_ISL_2781582, EPI_ISL_2781595, EPI_ISL_2781602, EPI_ISL_2781610, EPI_ISL_2781613, EPI_ISL_2781617, EPI_ISL_2781620, EPI_ISL_2781621, EPI_ISL_2781623, EPI_ISL_2781625, EPI_ISL_2781629, EPI_ISL_2781630, EPI_ISL_2781631, EPI_ISL_2781638, EPI_ISL_2781640, EPI_ISL_2781645, EPI_ISL_2781648, EPI_ISL_2781657, EPI_ISL_2781658, EPI_ISL_2781660, EPI_ISL_2781662, EPI_ISL_2781666, EPI_ISL_2781669, EPI_ISL_2781670, EPI_ISL_2781671, EPI_ISL_2781672, EPI_ISL_2781681, EPI_ISL_2781682, EPI_ISL_2781695, EPI_ISL_2781698, EPI_ISL_2781701, EPI_ISL_2781705, EPI_ISL_2781720, EPI_ISL_2781722, EPI_ISL_2781723, EPI_ISL_2781724, EPI_ISL_2781727, EPI_ISL_2781730, EPI_ISL_2781752, EPI_ISL_2781757, EPI_ISL_2781760, EPI_ISL_2781768, EPI_ISL_2809097, EPI_ISL_2809103, EPI_ISL_2809109, EPI_ISL_2809115, EPI_ISL_2809122, EPI_ISL_2809125, EPI_ISL_2809130, EPI_ISL_2809132, EPI_ISL_2809141, EPI_ISL_2809146, EPI_ISL_2809150, EPI_ISL_2809152, EPI_ISL_2809159, EPI_ISL_2809162, EPI_ISL_2809164, EPI_ISL_2809168, EPI_ISL_2809172, EPI_ISL_2809176, EPI_ISL_2809178, EPI_ISL_2809180, EPI_ISL_2809182, EPI_ISL_2809183, EPI_ISL_2809186, EPI_ISL_2809192, EPI_ISL_2809195, EPI_ISL_2809196, EPI_ISL_2809198, EPI_ISL_2809203, EPI_ISL_2809205, EPI_ISL_2809206, EPI_ISL_2809212, EPI_ISL_2809220, EPI_ISL_2809225, EPI_ISL_2809226, EPI_ISL_2809227, EPI_ISL_2809240, EPI_ISL_2809242, EPI_ISL_2809244, EPI_ISL_2809250, EPI_ISL_2809261, EPI_ISL_2809265, EPI_ISL_2809269, EPI_ISL_2809273, EPI_ISL_2809282, EPI_ISL_2809286, EPI_ISL_2809290, EPI_ISL_2809294, EPI_ISL_2809295, EPI_ISL_2809296, EPI_ISL_2809297, EPI_ISL_2809301, EPI_ISL_2809302, EPI_ISL_2809303, EPI_ISL_2809305, EPI_ISL_2809311, EPI_ISL_2809314, EPI_ISL_2809318, EPI_ISL_2809323, EPI_ISL_2809327, EPI_ISL_2809328, EPI_ISL_2809331, EPI_ISL_2809334, EPI_ISL_2809336, EPI_ISL_2809339, EPI_ISL_2809341, EPI_ISL_2809344, EPI_ISL_2809345, EPI_ISL_2809346, EPI_ISL_2809347, EPI_ISL_2809350, EPI_ISL_2809354, EPI_ISL_2809356, EPI_ISL_2809362, EPI_ISL_2809365, EPI_ISL_2809372, EPI_ISL_2809374, EPI_ISL_2809388, EPI_ISL_2809390, EPI_ISL_2809392, EPI_ISL_2809393, EPI_ISL_2809395, EPI_ISL_2809401, EPI_ISL_2809402, EPI_ISL_2809407, EPI_ISL_2809408, EPI_ISL_2809413, EPI_ISL_2809417, EPI_ISL_2809420, EPI_ISL_2815082, EPI_ISL_2815091, EPI_ISL_2815094, EPI_ISL_2815095 | | | |
| see above | Health Services Laboratories | Wellcome Sanger Institute for the COVID-19 Genomics UK (COG-UK) Consortium | Cordelia Langford; David K. Jackson; Dominic Kwiatkowski; Ewan Harrison; Health Services Laboratories and Alex Alderton; Ian Johnston; Jeffrey Barrett; John Sillitoe on behalf of the Wellcome Sanger Institute COVID-19 Surveillance Team; Roberto Amato; Sonia Goncalves |
| EPI_ISL_2729055, EPI_ISL_2729056, EPI_ISL_2729061, EPI_ISL_2729063, EPI_ISL_2729066, EPI_ISL_2729067, EPI_ISL_2729070, EPI_ISL_2729080, EPI_ISL_2729084, EPI_ISL_2729086, EPI_ISL_2729093, EPI_ISL_2729096, EPI_ISL_2729097, EPI_ISL_2729098, EPI_ISL_2729099, EPI_ISL_2729100, EPI_ISL_2729102, EPI_ISL_2729104, EPI_ISL_2729108, EPI_ISL_2729110, EPI_ISL_2729112, EPI_ISL_2729114, EPI_ISL_2729115, EPI_ISL_2729118, EPI_ISL_2729121, EPI_ISL_2729123, EPI_ISL_2729125, EPI_ISL_2729130, EPI_ISL_2729131, EPI_ISL_2729132, EPI_ISL_2729141, EPI_ISL_2729143, EPI_ISL_2729147, EPI_ISL_2729152, EPI_ISL_2729153, EPI_ISL_2729157, EPI_ISL_2729162, EPI_ISL_2729168, EPI_ISL_2729181, EPI_ISL_2729183, EPI_ISL_2729187, EPI_ISL_2729189, EPI_ISL_2729192, EPI_ISL_2729193, EPI_ISL_2729196, EPI_ISL_2729202, EPI_ISL_2729203, EPI_ISL_2729208, EPI_ISL_2729212, EPI_ISL_2729217, EPI_ISL_2729218, EPI_ISL_2729228, EPI_ISL_2729229, EPI_ISL_2729232, EPI_ISL_2729233, EPI_ISL_2729234, EPI_ISL_2729237, EPI_ISL_2729240, EPI_ISL_2729244, EPI_ISL_2729246, EPI_ISL_2729251, EPI_ISL_2729252, EPI_ISL_2729256, EPI_ISL_2729265, EPI_ISL_2729267, EPI_ISL_2729269, EPI_ISL_2729270, EPI_ISL_2729272, EPI_ISL_2729276, EPI_ISL_2729284, EPI_ISL_2729291, EPI_ISL_2729294, EPI_ISL_2729297, EPI_ISL_2729302, EPI_ISL_2729308, EPI_ISL_2729312, EPI_ISL_2729314, EPI_ISL_2729324, EPI_ISL_2729329, EPI_ISL_2729341, EPI_ISL_2729342, EPI_ISL_2729343, EPI_ISL_2729355, EPI_ISL_2729362, EPI_ISL_2729366, EPI_ISL_2729368, EPI_ISL_2729371, EPI_ISL_2729372, EPI_ISL_2729374, EPI_ISL_2729941, EPI_ISL_2729987, EPI_ISL_2729992, EPI_ISL_2729994, EPI_ISL_2730039, EPI_ISL_2730069, EPI_ISL_2730131, EPI_ISL_2730154, EPI_ISL_2730165, EPI_ISL_2730194, EPI_ISL_2730221, EPI_ISL_2730300, EPI_ISL_2765571, EPI_ISL_2765579, EPI_ISL_2765586, EPI_ISL_2765589, EPI_ISL_2765592, EPI_ISL_2765599, EPI_ISL_2765601, EPI_ISL_2765607, EPI_ISL_2765609, EPI_ISL_2765617, EPI_ISL_2765623, EPI_ISL_2765626, EPI_ISL_2765628, EPI_ISL_2765634, EPI_ISL_2765643, EPI_ISL_2765647, EPI_ISL_2765661, EPI_ISL_2765662, EPI_ISL_2765667, EPI_ISL_2765668, EPI_ISL_2765669, EPI_ISL_2765671, EPI_ISL_2765672, EPI_ISL_2765677, EPI_ISL_2765679, EPI_ISL_2765680, EPI_ISL_2765683, EPI_ISL_2765686, EPI_ISL_2765688, EPI_ISL_2765699, EPI_ISL_2765710, EPI_ISL_2765715, EPI_ISL_2765717, EPI_ISL_2765721, EPI_ISL_2765723, EPI_ISL_2765729, EPI_ISL_2765733, EPI_ISL_2765734, EPI_ISL_2765747, EPI_ISL_2765754, EPI_ISL_2765760, EPI_ISL_2765762, EPI_ISL_2765765, EPI_ISL_2765766, EPI_ISL_2765769, EPI_ISL_2765773, EPI_ISL_2765775, EPI_ISL_2765776, EPI_ISL_2765779, EPI_ISL_2765780, EPI_ISL_2765791, EPI_ISL_2765796, EPI_ISL_2765800, EPI_ISL_2765808, EPI_ISL_2765809, EPI_ISL_2765812, EPI_ISL_2765816, EPI_ISL_2765820, EPI_ISL_2765825, EPI_ISL_2765831, EPI_ISL_2765841, EPI_ISL_2765843, EPI_ISL_2765845, EPI_ISL_2765846, EPI_ISL_2765847, EPI_ISL_2765861, EPI_ISL_2765865, EPI_ISL_2765869, EPI_ISL_2765870, EPI_ISL_2765873, EPI_ISL_2765875, EPI_ISL_2765878, EPI_ISL_2765880, EPI_ISL_2765885, EPI_ISL_2765890, EPI_ISL_2765893, EPI_ISL_2765894, EPI_ISL_2765901, EPI_ISL_2765902, EPI_ISL_2765905, EPI_ISL_2765907, EPI_ISL_2765909, EPI_ISL_2765915, EPI_ISL_2765918, EPI_ISL_2765931, EPI_ISL_2765940, EPI_ISL_2765947, EPI_ISL_2765958, EPI_ISL_2765959, EPI_ISL_2765962, EPI_ISL_2765968, EPI_ISL_2765970, EPI_ISL_2765972, EPI_ISL_2765979, EPI_ISL_2765980, EPI_ISL_2765982, EPI_ISL_2765986, EPI_ISL_2765988, EPI_ISL_2765996, EPI_ISL_2766000, EPI_ISL_2766016, EPI_ISL_2766018, EPI_ISL_2766019, EPI_ISL_2766021, EPI_ISL_2766022, EPI_ISL_2766024, EPI_ISL_2766026, EPI_ISL_2766030, EPI_ISL_2766034, EPI_ISL_2766037, EPI_ISL_2766043, EPI_ISL_2766045, EPI_ISL_2766047, EPI_ISL_2766049, EPI_ISL_2766052, EPI_ISL_2766054, EPI_ISL_2766055, EPI_ISL_2766062, EPI_ISL_2766064, EPI_ISL_2766066, EPI_ISL_2766070, EPI_ISL_2766071, EPI_ISL_2766076, EPI_ISL_2766078, EPI_ISL_2766080, EPI_ISL_2766084, EPI_ISL_2766087, EPI_ISL_2766090, EPI_ISL_2766092, EPI_ISL_2766093, EPI_ISL_2766094, EPI_ISL_2766096, EPI_ISL_2766100, EPI_ISL_2766108, EPI_ISL_2766114, EPI_ISL_2766115, EPI_ISL_2766116, EPI_ISL_2766120, EPI_ISL_2766121, EPI_ISL_2766124, EPI_ISL_2766127, EPI_ISL_2766131, EPI_ISL_2766134, EPI_ISL_2766136, EPI_ISL_2766138, EPI_ISL_2766139, EPI_ISL_2766150, EPI_ISL_2766154, EPI_ISL_2766158, EPI_ISL_2766161, EPI_ISL_2766165, EPI_ISL_2766168, EPI_ISL_2766176, EPI_ISL_2766177, EPI_ISL_2766179, EPI_ISL_2766180, EPI_ISL_2766181, EPI_ISL_2766183, EPI_ISL_2766188, EPI_ISL_2766189, EPI_ISL_2766190, EPI_ISL_2766194, EPI_ISL_2766550, EPI_ISL_2766551, EPI_ISL_2766556, EPI_ISL_2766557, EPI_ISL_2766562, EPI_ISL_2766564, EPI_ISL_2766565, EPI_ISL_2766570, EPI_ISL_2766573, EPI_ISL_2766575, EPI_ISL_2766603, EPI_ISL_2766604, EPI_ISL_2766612, EPI_ISL_2766613, EPI_ISL_2766614, EPI_ISL_2766619, EPI_ISL_2766621, EPI_ISL_2766624, EPI_ISL_2766626, EPI_ISL_2766628, EPI_ISL_2766630, EPI_ISL_2766633, EPI_ISL_2766636, EPI_ISL_2766640, EPI_ISL_2766645, EPI_ISL_2766647, EPI_ISL_2766652, EPI_ISL_2766653, EPI_ISL_2766657, EPI_ISL_2766658, EPI_ISL_2766659, EPI_ISL_2766662, EPI_ISL_2766668, EPI_ISL_2766672, EPI_ISL_2766673, EPI_ISL_2766682, EPI_ISL_2766685, EPI_ISL_2766686, EPI_ISL_2766688, EPI_ISL_2766690, EPI_ISL_2766693, EPI_ISL_2766699, EPI_ISL_2766707, EPI_ISL_2766711, EPI_ISL_2766714, EPI_ISL_2766716, EPI_ISL_2766718, EPI_ISL_2766721, EPI_ISL_2766728, EPI_ISL_2766730, EPI_ISL_2766735, EPI_ISL_2766737, EPI_ISL_2766741, EPI_ISL_2766745, EPI_ISL_2766755, EPI_ISL_2766756, EPI_ISL_2766759, EPI_ISL_2766760, EPI_ISL_2766768, EPI_ISL_2766772, EPI_ISL_2766773, EPI_ISL_2766779, EPI_ISL_2766783, EPI_ISL_2766784, EPI_ISL_2766785, EPI_ISL_2766787, EPI_ISL_2766801, EPI_ISL_2766802, EPI_ISL_2766804, EPI_ISL_2766806, EPI_ISL_2766807, EPI_ISL_2766811, EPI_ISL_2766822, EPI_ISL_2766826, EPI_ISL_2766832, EPI_ISL_2766835, EPI_ISL_2766838, EPI_ISL_2766847, EPI_ISL_2766856, EPI_ISL_2766865, EPI_ISL_2766868, EPI_ISL_2766870, EPI_ISL_2766879, EPI_ISL_2766881, EPI_ISL_2766882, EPI_ISL_2766884, EPI_ISL_2766891, EPI_ISL_2766903, EPI_ISL_2766909, EPI_ISL_2766911, EPI_ISL_2766915, EPI_ISL_2766925, EPI_ISL_2766932, EPI_ISL_2766936, EPI_ISL_2766937, EPI_ISL_2766939, EPI_ISL_2766943, EPI_ISL_2766945, EPI_ISL_2766951, EPI_ISL_2766953, EPI_ISL_2766964, EPI_ISL_2766968, EPI_ISL_2766972, EPI_ISL_2766975, EPI_ISL_2766976, EPI_ISL_2766980, EPI_ISL_2766990, EPI_ISL_2766991, EPI_ISL_2766997, EPI_ISL_2767000, EPI_ISL_2767003, EPI_ISL_2767006, EPI_ISL_2767007, EPI_ISL_2767016, EPI_ISL_2767019, EPI_ISL_2767022, EPI_ISL_2767026, EPI_ISL_2767037, EPI_ISL_2767041, EPI_ISL_2767048, EPI_ISL_2767055, EPI_ISL_2767057, EPI_ISL_2767059, EPI_ISL_2767063, EPI_ISL_2767065, EPI_ISL_2767075, EPI_ISL_2767077, EPI_ISL_2767086, EPI_ISL_2767090, EPI_ISL_2767097, EPI_ISL_2767103, EPI_ISL_2767106, EPI_ISL_2767112, EPI_ISL_2767120, EPI_ISL_2767121, EPI_ISL_2767122, EPI_ISL_2767126, EPI_ISL_2767128, EPI_ISL_2767129, EPI_ISL_2767132, EPI_ISL_2767144, EPI_ISL_2767146, EPI_ISL_2767155, EPI_ISL_2767158, EPI_ISL_2767161, EPI_ISL_2767162, EPI_ISL_2767163, EPI_ISL_2767181, EPI_ISL_2767185, EPI_ISL_2767186, EPI_ISL_2767188, EPI_ISL_2767189, EPI_ISL_2767190, EPI_ISL_2767199, EPI_ISL_2767200, EPI_ISL_2767203, EPI_ISL_2767206, EPI_ISL_2767214, EPI_ISL_2767226, EPI_ISL_2767254, EPI_ISL_2767281, EPI_ISL_2767300, EPI_ISL_2767309, EPI_ISL_2767310, EPI_ISL_2767311, EPI_ISL_2767313, EPI_ISL_2767321, EPI_ISL_2767324, EPI_ISL_2767325, EPI_ISL_2767328, EPI_ISL_2767341, EPI_ISL_2767345, EPI_ISL_2767641, EPI_ISL_2768001, EPI_ISL_2780124, EPI_ISL_2780126, EPI_ISL_2780127, EPI_ISL_2780131, EPI_ISL_2780133, EPI_ISL_2780134, EPI_ISL_2780135, EPI_ISL_2780136, EPI_ISL_2780139, EPI_ISL_2780145, EPI_ISL_2780147, EPI_ISL_2780148, EPI_ISL_2780149, EPI_ISL_2780150, EPI_ISL_2780152, EPI_ISL_2780153, EPI_ISL_2780154, EPI_ISL_2780155, EPI_ISL_2780157, EPI_ISL_2780158, EPI_ISL_2780160, EPI_ISL_2780163, EPI_ISL_2780164, EPI_ISL_2780166, EPI_ISL_2780169, EPI_ISL_2780170, EPI_ISL_2780172, EPI_ISL_2780173, EPI_ISL_2780174, EPI_ISL_2780175, EPI_ISL_2780176, EPI_ISL_2780180, EPI_ISL_2780181, EPI_ISL_2780182, EPI_ISL_2780190, EPI_ISL_2780191, EPI_ISL_2780192, EPI_ISL_2780194, EPI_ISL_2780195, EPI_ISL_2780196, EPI_ISL_2780200, EPI_ISL_2780201, EPI_ISL_2780204, EPI_ISL_2780205, EPI_ISL_2780209, EPI_ISL_2780214, EPI_ISL_2780215, EPI_ISL_2780216, EPI_ISL_2780218, EPI_ISL_2780222, EPI_ISL_2780225, EPI_ISL_2780229, EPI_ISL_2780230, EPI_ISL_2780233, EPI_ISL_2780234, EPI_ISL_2780237, EPI_ISL_2780238, EPI_ISL_2780239, EPI_ISL_2780241, EPI_ISL_2780243, EPI_ISL_2780244, EPI_ISL_2780245, EPI_ISL_2780253, EPI_ISL_2780254, EPI_ISL_2780255, EPI_ISL_2780263, EPI_ISL_2780264, EPI_ISL_2780265, EPI_ISL_2780266, EPI_ISL_2780268, EPI_ISL_2780270, EPI_ISL_2780278, EPI_ISL_2780280, EPI_ISL_2780282, EPI_ISL_2780285, EPI_ISL_2780287, EPI_ISL_2780290, EPI_ISL_2780295, EPI_ISL_2780296, EPI_ISL_2780300, EPI_ISL_2780301, EPI_ISL_2780302, EPI_ISL_2780303, EPI_ISL_2780304, EPI_ISL_2780306, EPI_ISL_2780307, EPI_ISL_2780308, EPI_ISL_2780309, EPI_ISL_2780318, EPI_ISL_2780321, EPI_ISL_2780323, EPI_ISL_2780325, EPI_ISL_2780333, EPI_ISL_2780336, EPI_ISL_2780337, EPI_ISL_2780338, EPI_ISL_2780340, EPI_ISL_2780341, EPI_ISL_2780342, EPI_ISL_2780344, EPI_ISL_2780345, EPI_ISL_2780346, EPI_ISL_2780348, EPI_ISL_2780351, EPI_ISL_2780352, EPI_ISL_2780356, EPI_ISL_2780360, EPI_ISL_2780361, EPI_ISL_2780364, EPI_ISL_2780366, EPI_ISL_2780372, EPI_ISL_2780373, EPI_ISL_2780378, EPI_ISL_2780380, EPI_ISL_2780384, EPI_ISL_2780387, EPI_ISL_2780390, EPI_ISL_2780393, EPI_ISL_2780396, EPI_ISL_2780399, EPI_ISL_2780400, EPI_ISL_2780408, EPI_ISL_2780409, EPI_ISL_2780410, EPI_ISL_2780412, EPI_ISL_2780413, EPI_ISL_2780416, EPI_ISL_2780417, EPI_ISL_2780422, EPI_ISL_2780424, EPI_ISL_2780425, EPI_ISL_2780427, EPI_ISL_2780429, EPI_ISL_2780430, EPI_ISL_2780431, EPI_ISL_2780432, EPI_ISL_2780435, EPI_ISL_2780436, EPI_ISL_2780438, EPI_ISL_2780442, EPI_ISL_2780443, EPI_ISL_2780448, EPI_ISL_2780452, EPI_ISL_2780455, EPI_ISL_2780456, EPI_ISL_2780457, EPI_ISL_2780460, EPI_ISL_2780461, EPI_ISL_2780464, EPI_ISL_2780466, EPI_ISL_2781135, EPI_ISL_2781154, EPI_ISL_2781155, EPI_ISL_2781161, EPI_ISL_2781164, EPI_ISL_2781172, EPI_ISL_2781179, EPI_ISL_2781181, EPI_ISL_2781184, EPI_ISL_2781186, EPI_ISL_2781191, EPI_ISL_2781193, EPI_ISL_2781201, EPI_ISL_2781210, EPI_ISL_2781211, EPI_ISL_2781218, EPI_ISL_2781222, EPI_ISL_2781227, EPI_ISL_2781229, EPI_ISL_2781231, EPI_ISL_2781232, EPI_ISL_2781233, EPI_ISL_2781236, EPI_ISL_2781238, EPI_ISL_2781249, EPI_ISL_2781250, EPI_ISL_2781253, EPI_ISL_2781259, EPI_ISL_2781268, EPI_ISL_2781269, EPI_ISL_2781271, EPI_ISL_2781278, EPI_ISL_2781279, EPI_ISL_2781281, EPI_ISL_2781284, EPI_ISL_2781289, EPI_ISL_2781293, EPI_ISL_2781295, EPI_ISL_2781299, EPI_ISL_2781306, EPI_ISL_2781311, EPI_ISL_2781315, EPI_ISL_2781322, EPI_ISL_2781323, EPI_ISL_2781325, EPI_ISL_2781326, EPI_ISL_2781327, EPI_ISL_2781330, EPI_ISL_2781338, EPI_ISL_2781354, EPI_ISL_2781374, EPI_ISL_2781378, EPI_ISL_2781380, EPI_ISL_2781384, EPI_ISL_2781389, EPI_ISL_2781390, EPI_ISL_2781398, EPI_ISL_2781402, EPI_ISL_2781403, EPI_ISL_2781407, EPI_ISL_2781409, EPI_ISL_2781428, EPI_ISL_2781434, EPI_ISL_2781448, EPI_ISL_2781450, EPI_ISL_2781452, EPI_ISL_2781456, EPI_ISL_2781457, EPI_ISL_2781461, EPI_ISL_2781466, EPI_ISL_2781474, EPI_ISL_2781476, EPI_ISL_2781772, EPI_ISL_2781774, EPI_ISL_2781775, EPI_ISL_2803925, EPI_ISL_2803927, EPI_ISL_2803932, EPI_ISL_2803938, EPI_ISL_2803949, EPI_ISL_2803955, EPI_ISL_2803961, EPI_ISL_2803968, EPI_ISL_2803973, EPI_ISL_2803981, EPI_ISL_2803982, EPI_ISL_2803984, EPI_ISL_2803985, EPI_ISL_2803992, EPI_ISL_2804004, EPI_ISL_2804005, EPI_ISL_2804011, EPI_ISL_2804015, EPI_ISL_2804017, EPI_ISL_2804020, EPI_ISL_2804021, EPI_ISL_2804031, EPI_ISL_2804047, EPI_ISL_2804059, EPI_ISL_2804071, EPI_ISL_2804076, EPI_ISL_2804077, EPI_ISL_2804085, EPI_ISL_2804087, EPI_ISL_2804104, EPI_ISL_2804108, EPI_ISL_2804111, EPI_ISL_2804114, EPI_ISL_2804119, EPI_ISL_2804126, EPI_ISL_2804132, EPI_ISL_2804137, EPI_ISL_2804142, EPI_ISL_2804143, EPI_ISL_2804147, EPI_ISL_2804148, EPI_ISL_2804149, EPI_ISL_2804166, EPI_ISL_2804171, EPI_ISL_2804172, EPI_ISL_2804176, EPI_ISL_2804177, EPI_ISL_2804187, EPI_ISL_2804194, EPI_ISL_2804196, EPI_ISL_2804215, EPI_ISL_2804216, EPI_ISL_2804231, EPI_ISL_2804246, EPI_ISL_2804248, EPI_ISL_2804254, EPI_ISL_2804256, EPI_ISL_2804261, EPI_ISL_2804266, EPI_ISL_2804267, EPI_ISL_2804268, EPI_ISL_2804270, EPI_ISL_2804272, EPI_ISL_2804277, EPI_ISL_2804280, EPI_ISL_2804287, EPI_ISL_2804288, EPI_ISL_2804292, EPI_ISL_2804295, EPI_ISL_2804296, EPI_ISL_2804300, EPI_ISL_2804302, EPI_ISL_2804304, EPI_ISL_2804306, EPI_ISL_2804307, EPI_ISL_2804308, EPI_ISL_2804310, EPI_ISL_2804311, EPI_ISL_2804314, EPI_ISL_2804317, EPI_ISL_2804318, EPI_ISL_2804319, EPI_ISL_2804320, EPI_ISL_2804327, EPI_ISL_2804328, EPI_ISL_2804330, EPI_ISL_2804331, EPI_ISL_2804336, EPI_ISL_2804340, EPI_ISL_2804341, EPI_ISL_2804347, EPI_ISL_2804349, EPI_ISL_2804350, EPI_ISL_2804352, EPI_ISL_2804359, EPI_ISL_2804360, EPI_ISL_2804362, EPI_ISL_2804368, EPI_ISL_2804369, EPI_ISL_2804371, EPI_ISL_2804374, EPI_ISL_2804377, EPI_ISL_2804381, EPI_ISL_2804383, EPI_ISL_2804384, EPI_ISL_2804387, EPI_ISL_2804390, EPI_ISL_2804391, EPI_ISL_2804394, EPI_ISL_2804397, EPI_ISL_2804399, EPI_ISL_2804401, EPI_ISL_2804402, EPI_ISL_2804406, EPI_ISL_2804407, EPI_ISL_2804418, EPI_ISL_2804419, EPI_ISL_2804420, EPI_ISL_2804431, EPI_ISL_2804432, EPI_ISL_2804433, EPI_ISL_2804435, EPI_ISL_2804437, EPI_ISL_2804438, EPI_ISL_2804448, EPI_ISL_2804451, EPI_ISL_2804460, EPI_ISL_2804461, EPI_ISL_2804462, EPI_ISL_2804465, EPI_ISL_2804467, EPI_ISL_2804468, EPI_ISL_2804470, EPI_ISL_2804472, EPI_ISL_2804476, EPI_ISL_2804478, EPI_ISL_2804480, EPI_ISL_2804481, EPI_ISL_2804484, EPI_ISL_2804491, EPI_ISL_2804497, EPI_ISL_2804498, EPI_ISL_2804503, EPI_ISL_2804506, EPI_ISL_2804508, EPI_ISL_2804512, EPI_ISL_2804519, EPI_ISL_2804523, EPI_ISL_2804524, EPI_ISL_2804530, EPI_ISL_2804537, EPI_ISL_2804546, EPI_ISL_2804547, EPI_ISL_2804553, EPI_ISL_2804554, EPI_ISL_2804557, EPI_ISL_2804561, EPI_ISL_2804562, EPI_ISL_2804563, EPI_ISL_2804564, EPI_ISL_2804567, EPI_ISL_2804568, EPI_ISL_2804571, EPI_ISL_2804572, EPI_ISL_2804576, EPI_ISL_2804580, EPI_ISL_2804586, EPI_ISL_2804587, EPI_ISL_2804591, EPI_ISL_2804594, EPI_ISL_2804596, EPI_ISL_2804597, EPI_ISL_2804598, EPI_ISL_2804599, EPI_ISL_2804609, EPI_ISL_2804611, EPI_ISL_2804612, EPI_ISL_2804613, EPI_ISL_2804615, EPI_ISL_2804616, EPI_ISL_2804618, EPI_ISL_2804627, EPI_ISL_2804628, EPI_ISL_2804632, EPI_ISL_2804633, EPI_ISL_2804634, EPI_ISL_2804636, EPI_ISL_2804640, EPI_ISL_2804642, EPI_ISL_2804643, EPI_ISL_2804644, EPI_ISL_2804646, EPI_ISL_2804649, EPI_ISL_2804652, EPI_ISL_2804653, EPI_ISL_2804654, EPI_ISL_2804655, EPI_ISL_2804658, EPI_ISL_2804659, EPI_ISL_2804661, EPI_ISL_2804662, EPI_ISL_2804669, EPI_ISL_2804670, EPI_ISL_2804671, EPI_ISL_2804672, EPI_ISL_2804675, EPI_ISL_2804676, EPI_ISL_2804677, EPI_ISL_2804678, EPI_ISL_2804679, EPI_ISL_2804681, EPI_ISL_2804683, EPI_ISL_2804684, EPI_ISL_2804687, EPI_ISL_2804688, EPI_ISL_2804695, EPI_ISL_2804696, EPI_ISL_2804704, EPI_ISL_2804706, EPI_ISL_2804707, EPI_ISL_2804708, EPI_ISL_2804710, EPI_ISL_2804711, EPI_ISL_2804712, EPI_ISL_2804714, EPI_ISL_2804715, EPI_ISL_2804719, EPI_ISL_2804720, EPI_ISL_2804721, EPI_ISL_2804728, EPI_ISL_2804729, EPI_ISL_2804730, EPI_ISL_2804731, EPI_ISL_2804733, EPI_ISL_2804734, EPI_ISL_2804738, EPI_ISL_2804740, EPI_ISL_2804741, EPI_ISL_2804743, EPI_ISL_2804745, EPI_ISL_2804747, EPI_ISL_2804752, EPI_ISL_2804753, EPI_ISL_2804754, EPI_ISL_2804755, EPI_ISL_2804758, EPI_ISL_2804760, EPI_ISL_2804761, EPI_ISL_2804762, EPI_ISL_2804764, EPI_ISL_2804765, EPI_ISL_2804766, EPI_ISL_2804767, EPI_ISL_2804768, EPI_ISL_2804769, EPI_ISL_2804771, EPI_ISL_2804772, EPI_ISL_2804776, EPI_ISL_2804777, EPI_ISL_2804779, EPI_ISL_2804780, EPI_ISL_2804781, EPI_ISL_2804782, EPI_ISL_2804783, EPI_ISL_2804784, EPI_ISL_2804786, EPI_ISL_2804787, EPI_ISL_2804789, EPI_ISL_2804792, EPI_ISL_2804793, EPI_ISL_2804795, EPI_ISL_2804796, EPI_ISL_2804800, EPI_ISL_2804801, EPI_ISL_2804803, EPI_ISL_2804809, EPI_ISL_2804811, EPI_ISL_2804812, EPI_ISL_2804814, EPI_ISL_2804815, EPI_ISL_2804817, EPI_ISL_2804819, EPI_ISL_2804821, EPI_ISL_2804825, EPI_ISL_2804826, EPI_ISL_2804828, EPI_ISL_2804830, EPI_ISL_2804833, EPI_ISL_2804835, EPI_ISL_2804836, EPI_ISL_2804837, EPI_ISL_2804843, EPI_ISL_2804846, EPI_ISL_2804847, EPI_ISL_2804849, EPI_ISL_2804852, EPI_ISL_2804854, EPI_ISL_2804857, EPI_ISL_2804858, EPI_ISL_2804859, EPI_ISL_2804860, EPI_ISL_2804861, EPI_ISL_2804862, EPI_ISL_2804863, EPI_ISL_2804864, EPI_ISL_2804866, EPI_ISL_2804867, EPI_ISL_2804868, EPI_ISL_2804870, EPI_ISL_2804871, EPI_ISL_2804874, EPI_ISL_2804878, EPI_ISL_2804879, EPI_ISL_2804880, EPI_ISL_2804881, EPI_ISL_2804882, EPI_ISL_2804883, EPI_ISL_2804884, EPI_ISL_2804887, EPI_ISL_2804888, EPI_ISL_2804893, EPI_ISL_2804894, EPI_ISL_2804897, EPI_ISL_2804898, EPI_ISL_2804900, EPI_ISL_2804903, EPI_ISL_2804904, EPI_ISL_2804906, EPI_ISL_2804912, EPI_ISL_2804913, EPI_ISL_2804918, EPI_ISL_2804924, EPI_ISL_2804927, EPI_ISL_2804928, EPI_ISL_2804930, EPI_ISL_2804932, EPI_ISL_2804933, EPI_ISL_2804934, EPI_ISL_2804938, EPI_ISL_2804942, EPI_ISL_2804947, EPI_ISL_2804951, EPI_ISL_2804952, EPI_ISL_2804956, EPI_ISL_2804961, EPI_ISL_2804962, EPI_ISL_2804964, EPI_ISL_2804967, EPI_ISL_2804968, EPI_ISL_2804972, EPI_ISL_2804973, EPI_ISL_2804980, EPI_ISL_2804981, EPI_ISL_2804983, EPI_ISL_2804984, EPI_ISL_2804986, EPI_ISL_2804988, EPI_ISL_2804989, EPI_ISL_2804994, EPI_ISL_2804996, EPI_ISL_2804998, EPI_ISL_2805004, EPI_ISL_2805006, EPI_ISL_2805008, EPI_ISL_2805011, EPI_ISL_2805014, EPI_ISL_2805019, EPI_ISL_2805020, EPI_ISL_2805021, EPI_ISL_2805023, EPI_ISL_2805026, EPI_ISL_2805028, EPI_ISL_2805030, EPI_ISL_2805036, EPI_ISL_2805038, EPI_ISL_2805045, EPI_ISL_2805051, EPI_ISL_2805052, EPI_ISL_2805054, EPI_ISL_2805065, EPI_ISL_2805066, EPI_ISL_2805071, EPI_ISL_2805072, EPI_ISL_2805074, EPI_ISL_2805075, EPI_ISL_2805083, EPI_ISL_2805096, EPI_ISL_2805098, EPI_ISL_2805101, EPI_ISL_2805105, EPI_ISL_2805106, EPI_ISL_2805107, EPI_ISL_2805114, EPI_ISL_2805121, EPI_ISL_2805131, EPI_ISL_2805132, EPI_ISL_2805136, EPI_ISL_2805143, EPI_ISL_2805145, EPI_ISL_2805148, EPI_ISL_2805149, EPI_ISL_2805151, EPI_ISL_2805153, EPI_ISL_2805154, EPI_ISL_2805156, EPI_ISL_2805164, EPI_ISL_2805167, EPI_ISL_2805173, EPI_ISL_2805174, EPI_ISL_2805192, EPI_ISL_2805193, EPI_ISL_2805196, EPI_ISL_2805197, EPI_ISL_2805203, EPI_ISL_2805205, EPI_ISL_2805206, EPI_ISL_2805210, EPI_ISL_2805212, EPI_ISL_2805213, EPI_ISL_2805214, EPI_ISL_2805215, EPI_ISL_2805216, EPI_ISL_2805218, EPI_ISL_2805220, EPI_ISL_2805222, EPI_ISL_2805224, EPI_ISL_2805230, EPI_ISL_2805232, EPI_ISL_2805233, EPI_ISL_2805238, EPI_ISL_2805239, EPI_ISL_2805243, EPI_ISL_2805245, EPI_ISL_2805246, EPI_ISL_2805247, EPI_ISL_2805250, EPI_ISL_2805252, EPI_ISL_2805254, EPI_ISL_2805258, EPI_ISL_2805259, EPI_ISL_2805261, EPI_ISL_2805262, EPI_ISL_2805263, EPI_ISL_2805267, EPI_ISL_2805269, EPI_ISL_2805271, EPI_ISL_2805273, EPI_ISL_2805277, EPI_ISL_2805278, EPI_ISL_2805279, EPI_ISL_2805281, EPI_ISL_2805287, EPI_ISL_2805292, EPI_ISL_2805294, EPI_ISL_2805296, EPI_ISL_2805299, EPI_ISL_2805304, EPI_ISL_2805305, EPI_ISL_2805306, EPI_ISL_2805308, EPI_ISL_2805309, EPI_ISL_2805310, EPI_ISL_2805311, EPI_ISL_2805313, EPI_ISL_2805314, EPI_ISL_2805318, EPI_ISL_2805325, EPI_ISL_2805331, EPI_ISL_2805334, EPI_ISL_2805337, EPI_ISL_2805340, EPI_ISL_2805342, EPI_ISL_2805343, EPI_ISL_2805344, EPI_ISL_2805345, EPI_ISL_2805346, EPI_ISL_2805349, EPI_ISL_2805356, EPI_ISL_2805359, EPI_ISL_2805360, EPI_ISL_2805363, EPI_ISL_2805364, EPI_ISL_2805366, EPI_ISL_2805368, EPI_ISL_2805371, EPI_ISL_2805373, EPI_ISL_2805378, EPI_ISL_2805384, EPI_ISL_2805385, EPI_ISL_2805386, EPI_ISL_2805391, EPI_ISL_2805393, EPI_ISL_2805395, EPI_ISL_2805396, EPI_ISL_2805398, EPI_ISL_2805399, EPI_ISL_2805400, EPI_ISL_2805401, EPI_ISL_2805403, EPI_ISL_2805405, EPI_ISL_2805407, EPI_ISL_2805411, EPI_ISL_2805412, EPI_ISL_2805413, EPI_ISL_2805418, EPI_ISL_2805419, EPI_ISL_2805421, EPI_ISL_2805427, EPI_ISL_2805435, EPI_ISL_2805436, EPI_ISL_2805437, EPI_ISL_2805438, EPI_ISL_2805445, EPI_ISL_2805448, EPI_ISL_2805449, EPI_ISL_2805452, EPI_ISL_2805456, EPI_ISL_2805458, EPI_ISL_2805460, EPI_ISL_2805461, EPI_ISL_2805462, EPI_ISL_2805465, EPI_ISL_2805467, EPI_ISL_2805470, EPI_ISL_2805472, EPI_ISL_2805476, EPI_ISL_2805478, EPI_ISL_2805481, EPI_ISL_2805482, EPI_ISL_2805489, EPI_ISL_2805492, EPI_ISL_2805495, EPI_ISL_2805498, EPI_ISL_2805499, EPI_ISL_2805500, EPI_ISL_2805501, EPI_ISL_2805503, EPI_ISL_2805504, EPI_ISL_2805505, EPI_ISL_2805508, EPI_ISL_2805516, EPI_ISL_2805519, EPI_ISL_2805520, EPI_ISL_2805524, EPI_ISL_2805526, EPI_ISL_2805530, EPI_ISL_2805531, EPI_ISL_2805535, EPI_ISL_2805537, EPI_ISL_2805540, EPI_ISL_2805543, EPI_ISL_2805548, EPI_ISL_2805558, EPI_ISL_2805561, EPI_ISL_2805563, EPI_ISL_2805564, EPI_ISL_2805567, EPI_ISL_2805569, EPI_ISL_2805572, EPI_ISL_2805573, EPI_ISL_2805574, EPI_ISL_2805580, EPI_ISL_2805583, EPI_ISL_2805586, EPI_ISL_2805589, EPI_ISL_2805590, EPI_ISL_2805592, EPI_ISL_2805597, EPI_ISL_2805598, EPI_ISL_2805606, EPI_ISL_2805607, EPI_ISL_2805608, EPI_ISL_2805610, EPI_ISL_2805611, EPI_ISL_2805612, EPI_ISL_2805614, EPI_ISL_2805617, EPI_ISL_2805622, EPI_ISL_2805624, EPI_ISL_2805629, EPI_ISL_2805632, EPI_ISL_2805634, EPI_ISL_2805636, EPI_ISL_2805638, EPI_ISL_2805640, EPI_ISL_2805642, EPI_ISL_2805645, EPI_ISL_2805646, EPI_ISL_2805653, EPI_ISL_2805654, EPI_ISL_2805655, EPI_ISL_2805657, | | | |

EPI_ISL_2805669, EPI_ISL_2805671, EPI_ISL_2805672, EPI_ISL_2805680, EPI_ISL_2805682, EPI_ISL_2805685, EPI_ISL_2805686, EPI_ISL_2805687, EPI_ISL_2805688, EPI_ISL_2805690, EPI_ISL_2805693, EPI_ISL_2805992, EPI_ISL_2805993, EPI_ISL_2805994, EPI_ISL_2805995, EPI_ISL_2805996, EPI_ISL_2805998, EPI_ISL_2805999, EPI_ISL_2806000, EPI_ISL_2806001, EPI_ISL_2806002, EPI_ISL_2806003, EPI_ISL_2806005, EPI_ISL_2806006, EPI_ISL_2806007, EPI_ISL_2806009, EPI_ISL_2806010, EPI_ISL_2806011, EPI_ISL_2806012, EPI_ISL_2806014, EPI_ISL_2806015, EPI_ISL_2806016, EPI_ISL_2806017, EPI_ISL_2806020, EPI_ISL_2806023, EPI_ISL_2806026, EPI_ISL_2806027, EPI_ISL_2806029, EPI_ISL_2806030, EPI_ISL_2806032, EPI_ISL_2806033, EPI_ISL_2806035, EPI_ISL_2806036, EPI_ISL_2806038, EPI_ISL_2806040, EPI_ISL_2806041, EPI_ISL_2806042, EPI_ISL_2806044, EPI_ISL_2806046, EPI_ISL_2806047, EPI_ISL_2806048, EPI_ISL_2806049, EPI_ISL_2806050, EPI_ISL_2806051, EPI_ISL_2806052, EPI_ISL_2806053, EPI_ISL_2806054, EPI_ISL_2806055, EPI_ISL_2806056, EPI_ISL_2806059, EPI_ISL_2806060, EPI_ISL_2806061, EPI_ISL_2806062, EPI_ISL_2806063, EPI_ISL_2806065, EPI_ISL_2806066, EPI_ISL_2806067, EPI_ISL_2806069, EPI_ISL_2806070, EPI_ISL_2806071, EPI_ISL_2806072, EPI_ISL_2806075, EPI_ISL_2806077, EPI_ISL_2806078, EPI_ISL_2806080, EPI_ISL_2806081, EPI_ISL_2806082, EPI_ISL_2806084, EPI_ISL_2806086, EPI_ISL_2806087, EPI_ISL_2806088, EPI_ISL_2806089, EPI_ISL_2806090, EPI_ISL_2806091, EPI_ISL_2806092, EPI_ISL_2806093, EPI_ISL_2806097, EPI_ISL_2806099, EPI_ISL_2806101, EPI_ISL_2806106, EPI_ISL_2806107, EPI_ISL_2806109, EPI_ISL_2806112, EPI_ISL_2806113, EPI_ISL_2806114, EPI_ISL_2806115, EPI_ISL_2806118, EPI_ISL_2806119, EPI_ISL_2806123, EPI_ISL_2806126, EPI_ISL_2806127, EPI_ISL_2806129, EPI_ISL_2806130, EPI_ISL_2806131, EPI_ISL_2806132, EPI_ISL_2806134, EPI_ISL_2806136, EPI_ISL_2806137, EPI_ISL_2806138, EPI_ISL_2806139, EPI_ISL_2806140, EPI_ISL_2806141, EPI_ISL_2806142, EPI_ISL_2806143, EPI_ISL_2806145, EPI_ISL_2806147, EPI_ISL_2806148, EPI_ISL_2806149, EPI_ISL_2806150, EPI_ISL_2806153, EPI_ISL_2806156, EPI_ISL_2806157, EPI_ISL_2806158, EPI_ISL_2806159, EPI_ISL_2806160, EPI_ISL_2806167, EPI_ISL_2806168, EPI_ISL_2806169, EPI_ISL_2806170, EPI_ISL_2806172, EPI_ISL_2806173, EPI_ISL_2806174, EPI_ISL_2806175, EPI_ISL_2806178, EPI_ISL_2806180, EPI_ISL_2806182, EPI_ISL_2806183, EPI_ISL_2806184, EPI_ISL_2806185, EPI_ISL_2806186, EPI_ISL_2806188, EPI_ISL_2806191, EPI_ISL_2806195, EPI_ISL_2806196, EPI_ISL_2806197, EPI_ISL_2806199, EPI_ISL_2806200, EPI_ISL_2806202, EPI_ISL_2806204, EPI_ISL_2806205, EPI_ISL_2806206, EPI_ISL_2806208, EPI_ISL_2806214, EPI_ISL_2806215, EPI_ISL_2806216, EPI_ISL_2806217, EPI_ISL_2806220, EPI_ISL_2806225, EPI_ISL_2806227, EPI_ISL_2806230, EPI_ISL_2806232, EPI_ISL_2806233, EPI_ISL_2806234, EPI_ISL_2806237, EPI_ISL_2806240, EPI_ISL_2806247, EPI_ISL_2806252, EPI_ISL_2806253, EPI_ISL_2806254, EPI_ISL_2806262, EPI_ISL_2806264, EPI_ISL_2806266, EPI_ISL_2806268, EPI_ISL_2806273, EPI_ISL_2806275, EPI_ISL_2806277, EPI_ISL_2806282, EPI_ISL_2806283, EPI_ISL_2806284, EPI_ISL_2806285, EPI_ISL_2806287, EPI_ISL_2806291, EPI_ISL_2806299, EPI_ISL_2806303, EPI_ISL_2806304, EPI_ISL_2806307, EPI_ISL_2806311, EPI_ISL_2806312, EPI_ISL_2806319, EPI_ISL_2806324, EPI_ISL_2806327, EPI_ISL_2806333, EPI_ISL_2806334, EPI_ISL_2806341, EPI_ISL_2806346, EPI_ISL_2806349, EPI_ISL_2806351, EPI_ISL_2806352, EPI_ISL_2806353, EPI_ISL_2806354, EPI_ISL_2806355, EPI_ISL_2806358, EPI_ISL_2806360, EPI_ISL_2806366, EPI_ISL_2806371, EPI_ISL_2806381, EPI_ISL_2806387, EPI_ISL_2806391, EPI_ISL_2806392, EPI_ISL_2806394, EPI_ISL_2806402, EPI_ISL_2806406, EPI_ISL_2806407, EPI_ISL_2806408, EPI_ISL_2806413, EPI_ISL_2806414, EPI_ISL_2806417, EPI_ISL_2806422, EPI_ISL_2806429, EPI_ISL_2806430, EPI_ISL_2806432, EPI_ISL_2806433, EPI_ISL_2806436, EPI_ISL_2806437, EPI_ISL_2806438, EPI_ISL_2806441, EPI_ISL_2807406, EPI_ISL_2807491, EPI_ISL_2807611, EPI_ISL_2807664, EPI_ISL_2808794, EPI_ISL_2808797, EPI_ISL_2808804, EPI_ISL_2808806, EPI_ISL_2809096, EPI_ISL_2809111, EPI_ISL_2809116, EPI_ISL_2809118, EPI_ISL_2809128, EPI_ISL_2809138, EPI_ISL_2809144, EPI_ISL_2809145, EPI_ISL_2809147, EPI_ISL_2809155, EPI_ISL_2809170, EPI_ISL_2809174, EPI_ISL_2809204, EPI_ISL_2809217, EPI_ISL_2809223, EPI_ISL_2809228, EPI_ISL_2809233, EPI_ISL_2809236, EPI_ISL_2809249, EPI_ISL_2809260, EPI_ISL_2809281, EPI_ISL_2809293, EPI_ISL_2809304, EPI_ISL_2809310, EPI_ISL_2809313, EPI_ISL_2809319, EPI_ISL_2809349, EPI_ISL_2809352, EPI_ISL_2809359, EPI_ISL_2809366, EPI_ISL_2809373, EPI_ISL_2809399, EPI_ISL_2809415, EPI_ISL_2809419, EPI_ISL_2813660, EPI_ISL_2813670, EPI_ISL_2813672, EPI_ISL_2813684, EPI_ISL_2813685, EPI_ISL_2813689, EPI_ISL_2813692, EPI_ISL_2813693, EPI_ISL_2813698, EPI_ISL_2813701, EPI_ISL_2813703, EPI_ISL_2813704, EPI_ISL_2813706, EPI_ISL_2813708, EPI_ISL_2813714, EPI_ISL_2813715, EPI_ISL_2813716, EPI_ISL_2813720, EPI_ISL_2813721, EPI_ISL_2813724, EPI_ISL_2813728, EPI_ISL_2813730, EPI_ISL_2813732, EPI_ISL_2813733, EPI_ISL_2813737, EPI_ISL_2813740, EPI_ISL_2813751, EPI_ISL_2813755, EPI_ISL_2813757, EPI_ISL_2813759, EPI_ISL_2813761, EPI_ISL_2813769, EPI_ISL_2813770, EPI_ISL_2813772, EPI_ISL_2813773, EPI_ISL_2813789, EPI_ISL_2813790, EPI_ISL_2813791, EPI_ISL_2813793, EPI_ISL_2813795, EPI_ISL_2813797, EPI_ISL_2813801, EPI_ISL_2813802, EPI_ISL_2813803, EPI_ISL_2813809, EPI_ISL_2813811, EPI_ISL_2813825, EPI_ISL_2813831, EPI_ISL_2813833, EPI_ISL_2813838, EPI_ISL_2813839, EPI_ISL_2813845, EPI_ISL_2813848, EPI_ISL_2813850, EPI_ISL_2813857, EPI_ISL_2813861, EPI_ISL_2813871, EPI_ISL_2813872, EPI_ISL_2813884, EPI_ISL_2813888, EPI_ISL_2813894, EPI_ISL_2813895, EPI_ISL_2813899, EPI_ISL_2813906, EPI_ISL_2813910, EPI_ISL_2813928, EPI_ISL_2813937, EPI_ISL_2813941, EPI_ISL_2813946, EPI_ISL_2813948, EPI_ISL_2813949, EPI_ISL_2813953, EPI_ISL_2813955, EPI_ISL_2813977, EPI_ISL_2813978, EPI_ISL_2813979, EPI_ISL_2813980, EPI_ISL_2814000, EPI_ISL_2814001, EPI_ISL_2814003, EPI_ISL_2814004, EPI_ISL_2814007, EPI_ISL_2814008, EPI_ISL_2814009, EPI_ISL_2814013, EPI_ISL_2814015, EPI_ISL_2814016, EPI_ISL_2814028, EPI_ISL_2814031, EPI_ISL_2814034, EPI_ISL_2814046, EPI_ISL_2814049, EPI_ISL_2814053, EPI_ISL_2814063, EPI_ISL_2814065, EPI_ISL_2814076, EPI_ISL_2814088, EPI_ISL_2814097, EPI_ISL_2814099, EPI_ISL_2814104, EPI_ISL_2814112, EPI_ISL_2814117, EPI_ISL_2814120, EPI_ISL_2814126, EPI_ISL_2814134, EPI_ISL_2814137, EPI_ISL_2814143, EPI_ISL_2814147, EPI_ISL_2814150, EPI_ISL_2814154, EPI_ISL_2814166, EPI_ISL_2814169, EPI_ISL_2814173, EPI_ISL_2814174, EPI_ISL_2814185, EPI_ISL_2814194, EPI_ISL_2814200, EPI_ISL_2814202, EPI_ISL_2814203, EPI_ISL_2814207, EPI_ISL_2814730, EPI_ISL_2814791, EPI_ISL_2814802, EPI_ISL_2814811, EPI_ISL_2814814, EPI_ISL_2814816, EPI_ISL_2814834, EPI_ISL_2814843, EPI_ISL_2814845, EPI_ISL_2814877, EPI_ISL_2814879, EPI_ISL_2814886, EPI_ISL_2815040, EPI_ISL_2830336, EPI_ISL_2830338, EPI_ISL_2830340, EPI_ISL_2830344, EPI_ISL_2830349, EPI_ISL_2830353, EPI_ISL_2830354, EPI_ISL_2830356, EPI_ISL_2830359, EPI_ISL_2830360, EPI_ISL_2830364, EPI_ISL_2830365, EPI_ISL_2830367, EPI_ISL_2830372, EPI_ISL_2830374, EPI_ISL_2830378, EPI_ISL_2830380, EPI_ISL_2830382, EPI_ISL_2830383, EPI_ISL_2830386, EPI_ISL_2830390, EPI_ISL_2830392, EPI_ISL_2830393, EPI_ISL_2830400, EPI_ISL_2830404, EPI_ISL_2830406, EPI_ISL_2830409, EPI_ISL_2830410, EPI_ISL_2830418, EPI_ISL_2830421, EPI_ISL_2830422, EPI_ISL_2830423, EPI_ISL_2830424, EPI_ISL_2830428, EPI_ISL_2830431, EPI_ISL_2830434, EPI_ISL_2830437, EPI_ISL_2830446, EPI_ISL_2830447, EPI_ISL_2830450, EPI_ISL_2830452, EPI_ISL_2830453, EPI_ISL_2830454, EPI_ISL_2830456, EPI_ISL_2830457, EPI_ISL_2830459, EPI_ISL_2830461, EPI_ISL_2830466, EPI_ISL_2830467, EPI_ISL_2830470, EPI_ISL_2830471, EPI_ISL_2830477, EPI_ISL_2830478, EPI_ISL_2830480, EPI_ISL_2830481, EPI_ISL_2830488, EPI_ISL_2830492, EPI_ISL_2830493, EPI_ISL_2830502, EPI_ISL_2830503, EPI_ISL_2830504, EPI_ISL_2830505, EPI_ISL_2830506, EPI_ISL_2830507, EPI_ISL_2830509, EPI_ISL_2830510, EPI_ISL_2830513, EPI_ISL_2830515, EPI_ISL_2830516, EPI_ISL_2830521, EPI_ISL_2830522, EPI_ISL_2830523, EPI_ISL_2830524, EPI_ISL_2830529, EPI_ISL_2830530, EPI_ISL_2830536, EPI_ISL_2830538, EPI_ISL_2830539, EPI_ISL_2830545, EPI_ISL_2830549, EPI_ISL_2830552, EPI_ISL_2830555, EPI_ISL_2830561, EPI_ISL_2830564, EPI_ISL_2830568, EPI_ISL_2830572, EPI_ISL_2830575, EPI_ISL_2830589, EPI_ISL_2830595, EPI_ISL_2830600, EPI_ISL_2830603, EPI_ISL_2830644, EPI_ISL_2830692, EPI_ISL_2830710, EPI_ISL_2830716, EPI_ISL_2830723, EPI_ISL_2830734, EPI_ISL_2830751, EPI_ISL_2830813, EPI_ISL_2830822, EPI_ISL_2830823, EPI_ISL_2830829, EPI_ISL_2830850, EPI_ISL_2830855, EPI_ISL_2830872, EPI_ISL_2830884, EPI_ISL_2830886, EPI_ISL_2830890, EPI_ISL_2830897, EPI_ISL_2830900, EPI_ISL_2830904, EPI_ISL_2830928, EPI_ISL_2830929, EPI_ISL_2830931, EPI_ISL_2830960, EPI_ISL_2830973, EPI_ISL_2830982, EPI_ISL_2830997, EPI_ISL_2830998, EPI_ISL_2831002, EPI_ISL_2831014, EPI_ISL_2831037, EPI_ISL_2831040, EPI_ISL_2831045, EPI_ISL_2831047, EPI_ISL_2831056, EPI_ISL_2831058, EPI_ISL_2831062, EPI_ISL_2831070, EPI_ISL_2831074, EPI_ISL_2831075, EPI_ISL_2831078, EPI_ISL_2831079, EPI_ISL_2831082, EPI_ISL_2831085, EPI_ISL_2831087, EPI_ISL_2831092, EPI_ISL_2831094, EPI_ISL_2831095, EPI_ISL_2831096, EPI_ISL_2831102, EPI_ISL_2831119, EPI_ISL_2831130, EPI_ISL_2831134, EPI_ISL_2831142, EPI_ISL_2831148, EPI_ISL_2831150, EPI_ISL_2831151, EPI_ISL_2831154, EPI_ISL_2831157, EPI_ISL_2831161, EPI_ISL_2831163, EPI_ISL_2831166, EPI_ISL_2831168, EPI_ISL_2831176, EPI_ISL_2831188, EPI_ISL_2831196, EPI_ISL_2831203, EPI_ISL_2831205, EPI_ISL_2831211, EPI_ISL_2831222, EPI_ISL_2831223, EPI_ISL_2831231, EPI_ISL_2831239, EPI_ISL_2831240, EPI_ISL_2831242, EPI_ISL_2831246, EPI_ISL_2831248, EPI_ISL_2831256, EPI_ISL_2831258, EPI_ISL_2831260, EPI_ISL_2831275, EPI_ISL_2831288, EPI_ISL_2831295, EPI_ISL_2831297, EPI_ISL_2831298, EPI_ISL_2831316, EPI_ISL_2831321, EPI_ISL_2832065, EPI_ISL_2832069, EPI_ISL_2832070, EPI_ISL_2832071, EPI_ISL_2832073, EPI_ISL_2832079, EPI_ISL_2832080, EPI_ISL_2832081, EPI_ISL_2832082, EPI_ISL_2832086, EPI_ISL_2852490, EPI_ISL_2852852, EPI_ISL_2852853, EPI_ISL_2852854, EPI_ISL_2868224, EPI_ISL_2868225, EPI_ISL_2910608, EPI_ISL_2910618, EPI_ISL_2910624, EPI_ISL_2910634, EPI_ISL_2910638, EPI_ISL_2910650, EPI_ISL_2910662, EPI_ISL_2910665, EPI_ISL_2910668, EPI_ISL_2910679, EPI_ISL_2910687, EPI_ISL_2910688, EPI_ISL_2910689, EPI_ISL_2910691, EPI_ISL_2910696, EPI_ISL_2910697, EPI_ISL_2910700, EPI_ISL_2910712, EPI_ISL_2910721, EPI_ISL_2910722, EPI_ISL_2910725, EPI_ISL_2910733, EPI_ISL_2910735, EPI_ISL_2910738, EPI_ISL_2910741, EPI_ISL_2910750, EPI_ISL_2910752, EPI_ISL_2910757, EPI_ISL_2910763, EPI_ISL_2910765, EPI_ISL_2910772, EPI_ISL_2910775, EPI_ISL_2910779, EPI_ISL_2910780, EPI_ISL_2910783, EPI_ISL_2910796, EPI_ISL_2910802, EPI_ISL_2910803, EPI_ISL_2910809, EPI_ISL_2910813, EPI_ISL_2910817, EPI_ISL_2910823, EPI_ISL_2910836, EPI_ISL_2910838, EPI_ISL_2910844

| | | | |
|---|---|---|---|
| see above | Lighthouse Lab in Alderley Park | Wellcome Sanger Institute for the COVID-19 Genomics UK (COG-UK) Consortium | Cordelia Langford; David K. Jackson; Dominic Kwiatkowski; Ewan Harrison; Ian Johnston; Jacquelyn Wynn; Jeffrey Barrett; John Sillitoe on behalf of the Wellcome Sanger Institute COVID-19 Surveillance Team; Mairead Hyland; Roberto Amato; Sonia Goncalves; The Lighthouse Lab in Alderley Park and Alex Alderton |

EPI_ISL_2716974, EPI_ISL_2716980, EPI_ISL_2716983, EPI_ISL_2717001, EPI_ISL_2717015, EPI_ISL_2717025, EPI_ISL_2717031, EPI_ISL_2717038, EPI_ISL_2717063, EPI_ISL_2717111, EPI_ISL_2717171, EPI_ISL_2717189, EPI_ISL_2717193, EPI_ISL_2717197, EPI_ISL_2717229, EPI_ISL_2717240, EPI_ISL_2717255, EPI_ISL_2717257, EPI_ISL_2717274, EPI_ISL_2717284, EPI_ISL_2717318, EPI_ISL_2717319, EPI_ISL_2717321, EPI_ISL_2717323, EPI_ISL_2717346, EPI_ISL_2717352, EPI_ISL_2717353, EPI_ISL_2717355, EPI_ISL_2717362, EPI_ISL_2717364, EPI_ISL_2717370, EPI_ISL_2717371, EPI_ISL_2717372, EPI_ISL_2717377, EPI_ISL_2717389, EPI_ISL_2717393, EPI_ISL_2717394, EPI_ISL_2717409, EPI_ISL_2717417, EPI_ISL_2717423, EPI_ISL_2717427, EPI_ISL_2717430, EPI_ISL_2717432, EPI_ISL_2717442, EPI_ISL_2717443, EPI_ISL_2717447, EPI_ISL_2717449, EPI_ISL_2717450, EPI_ISL_2717453, EPI_ISL_2717459, EPI_ISL_2717474, EPI_ISL_2717476, EPI_ISL_2717482, EPI_ISL_2717484, EPI_ISL_2717497, EPI_ISL_2717499, EPI_ISL_2717515, EPI_ISL_2717518, EPI_ISL_2717525, EPI_ISL_2717542, EPI_ISL_2717545, EPI_ISL_2717548, EPI_ISL_2717549, EPI_ISL_2717554, EPI_ISL_2717555, EPI_ISL_2717557, EPI_ISL_2717558, EPI_ISL_2717561, EPI_ISL_2717584, EPI_ISL_2717586, EPI_ISL_2717591, EPI_ISL_2717593, EPI_ISL_2717595, EPI_ISL_2717598, EPI_ISL_2717604, EPI_ISL_2717605, EPI_ISL_2717607, EPI_ISL_2717614, EPI_ISL_2717618, EPI_ISL_2717619, EPI_ISL_2717621, EPI_ISL_2717625, EPI_ISL_2717631, EPI_ISL_2717632, EPI_ISL_2717642, EPI_ISL_2717652, EPI_ISL_2717654, EPI_ISL_2717655, EPI_ISL_2717667, EPI_ISL_2717683, EPI_ISL_2717686, EPI_ISL_2717692, EPI_ISL_2717694, EPI_ISL_2717703, EPI_ISL_2717711, EPI_ISL_2717718, EPI_ISL_2717722, EPI_ISL_2717727, EPI_ISL_2717737, EPI_ISL_2717738, EPI_ISL_2717754, EPI_ISL_2717767, EPI_ISL_2717768, EPI_ISL_2717779, EPI_ISL_2717784, EPI_ISL_2717789, EPI_ISL_2717794, EPI_ISL_2717796, EPI_ISL_2717800, EPI_ISL_2717801, EPI_ISL_2718170, EPI_ISL_2718171, EPI_ISL_2718172, EPI_ISL_2718173, EPI_ISL_2718174, EPI_ISL_2718175, EPI_ISL_2718176, EPI_ISL_2718177, EPI_ISL_2718178, EPI_ISL_2718179, EPI_ISL_2718180, EPI_ISL_2718181, EPI_ISL_2718182, EPI_ISL_2718183, EPI_ISL_2718184, EPI_ISL_2718185, EPI_ISL_2718186, EPI_ISL_2718187, EPI_ISL_2718188, EPI_ISL_2718189, EPI_ISL_2718190, EPI_ISL_2718191, EPI_ISL_2718192, EPI_ISL_2718193, EPI_ISL_2718194, EPI_ISL_2718195, EPI_ISL_2718196, EPI_ISL_2718197, EPI_ISL_2718198, EPI_ISL_2718199, EPI_ISL_2718200, EPI_ISL_2718201, EPI_ISL_2718202, EPI_ISL_2718203, EPI_ISL_2718204, EPI_ISL_2718205, EPI_ISL_2718206, EPI_ISL_2718207, EPI_ISL_2718208, EPI_ISL_2718209, EPI_ISL_2718210, EPI_ISL_2718211, EPI_ISL_2718212, EPI_ISL_2718213, EPI_ISL_2718214, EPI_ISL_2718215, EPI_ISL_2718216, EPI_ISL_2718217, EPI_ISL_2718218, EPI_ISL_2718219, EPI_ISL_2718220, EPI_ISL_2718221, EPI_ISL_2718222, EPI_ISL_2718223, EPI_ISL_2718224, EPI_ISL_2718225, EPI_ISL_2718226, EPI_ISL_2718227, EPI_ISL_2718228, EPI_ISL_2718229, EPI_ISL_2718230, EPI_ISL_2718231, EPI_ISL_2718232, EPI_ISL_2718233, EPI_ISL_2718234, EPI_ISL_2718235, EPI_ISL_2718236, EPI_ISL_2718237, EPI_ISL_2718238, EPI_ISL_2718239, EPI_ISL_2718240, EPI_ISL_2718242, EPI_ISL_2718243, EPI_ISL_2718244, EPI_ISL_2718245, EPI_ISL_2718246, EPI_ISL_2718247, EPI_ISL_2718248, EPI_ISL_2718249, EPI_ISL_2718250, EPI_ISL_2718251, EPI_ISL_2718252, EPI_ISL_2718253, EPI_ISL_2718254, EPI_ISL_2718255, EPI_ISL_2718256, EPI_ISL_2718257, EPI_ISL_2718258, EPI_ISL_2718259, EPI_ISL_2718260, EPI_ISL_2718261, EPI_ISL_2718262, EPI_ISL_2718263, EPI_ISL_2718264, EPI_ISL_2718265, EPI_ISL_2718266, EPI_ISL_2718267, EPI_ISL_2718268, EPI_ISL_2718269, EPI_ISL_2718270, EPI_ISL_2718271, EPI_ISL_2718272, EPI_ISL_2718273, EPI_ISL_2718274, EPI_ISL_2718275, EPI_ISL_2718276, EPI_ISL_2718277, EPI_ISL_2718278, EPI_ISL_2718279, EPI_ISL_2718280, EPI_ISL_2728694, EPI_ISL_2728709, EPI_ISL_2728719, EPI_ISL_2728728, EPI_ISL_2728735, EPI_ISL_2728753, EPI_ISL_2728771, EPI_ISL_2728772, EPI_ISL_2728786, EPI_ISL_2728788, EPI_ISL_2728799, EPI_ISL_2728821, EPI_ISL_2728825, EPI_ISL_2728828, EPI_ISL_2728854, EPI_ISL_2728859, EPI_ISL_2728874, EPI_ISL_2728878, EPI_ISL_2728889, EPI_ISL_2728890, EPI_ISL_2728898, EPI_ISL_2728915, EPI_ISL_2728921, EPI_ISL_2728928, EPI_ISL_2728936, EPI_ISL_2728939, EPI_ISL_2728945, EPI_ISL_2728953, EPI_ISL_2728969, EPI_ISL_2728994, EPI_ISL_2729009, EPI_ISL_2729015, EPI_ISL_2729024, EPI_ISL_2729028, EPI_ISL_2729052, EPI_ISL_2729064, EPI_ISL_2729073, EPI_ISL_2729095, EPI_ISL_2729103, EPI_ISL_2729127, EPI_ISL_2729129, EPI_ISL_2729146, EPI_ISL_2729161, EPI_ISL_2729164, EPI_ISL_2729166, EPI_ISL_2729176, EPI_ISL_2729185, EPI_ISL_2729211, EPI_ISL_2729225, EPI_ISL_2729253, EPI_ISL_2729258, EPI_ISL_2729288, EPI_ISL_2729317, EPI_ISL_2729322, EPI_ISL_2729353, EPI_ISL_2729356, EPI_ISL_2729359, EPI_ISL_2729360, EPI_ISL_2729369, EPI_ISL_2729377, EPI_ISL_2729394, EPI_ISL_2729395, EPI_ISL_2729396, EPI_ISL_2729397, EPI_ISL_2729398, EPI_ISL_2729400, EPI_ISL_2729404, EPI_ISL_2729405, EPI_ISL_2729409, EPI_ISL_2729411, EPI_ISL_2729413, EPI_ISL_2729415, EPI_ISL_2729416, EPI_ISL_2729418, EPI_ISL_2729422, EPI_ISL_2729424, EPI_ISL_2729426, EPI_ISL_2729428, EPI_ISL_2729429, EPI_ISL_2729434, EPI_ISL_2729439, EPI_ISL_2729443, EPI_ISL_2729447, EPI_ISL_2729448, EPI_ISL_2729450, EPI_ISL_2729452, EPI_ISL_2729455, EPI_ISL_2729462, EPI_ISL_2729466, EPI_ISL_2729467, EPI_ISL_2729469, EPI_ISL_2729470, EPI_ISL_2729471, EPI_ISL_2729472, EPI_ISL_2729473, EPI_ISL_2729476, EPI_ISL_2729478, EPI_ISL_2729479, EPI_ISL_2729483, EPI_ISL_2729488, EPI_ISL_2729492, EPI_ISL_2729495, EPI_ISL_2729499, EPI_ISL_2729500, EPI_ISL_2729501, EPI_ISL_2729503, EPI_ISL_2729506, EPI_ISL_2729508, EPI_ISL_2729510, EPI_ISL_2729513, EPI_ISL_2729514, EPI_ISL_2729515, EPI_ISL_2729520, EPI_ISL_2729530, EPI_ISL_2729535, EPI_ISL_2729538, EPI_ISL_2729549, EPI_ISL_2729554, EPI_ISL_2729556, EPI_ISL_2729558, EPI_ISL_2729562, EPI_ISL_2729565, EPI_ISL_2729568, EPI_ISL_2729574, EPI_ISL_2729576, EPI_ISL_2729578, EPI_ISL_2729579, EPI_ISL_2729580, EPI_ISL_2729582, EPI_ISL_2729583, EPI_ISL_2729588, EPI_ISL_2729591, EPI_ISL_2729593, EPI_ISL_2729605, EPI_ISL_2729611, EPI_ISL_2729624, EPI_ISL_2729626, EPI_ISL_2729628, EPI_ISL_2729633, EPI_ISL_2729636, EPI_ISL_2729637, EPI_ISL_2729638, EPI_ISL_2729641, EPI_ISL_2729650, EPI_ISL_2729654, EPI_ISL_2729674, EPI_ISL_2729683, EPI_ISL_2729685, EPI_ISL_2729720, EPI_ISL_2729729, EPI_ISL_2729730, EPI_ISL_2729733, EPI_ISL_2729739, EPI_ISL_2729740, EPI_ISL_2729742, EPI_ISL_2729754, EPI_ISL_2729760, EPI_ISL_2729780, EPI_ISL_2729782, EPI_ISL_2729823, EPI_ISL_2729832, EPI_ISL_2729854, EPI_ISL_2729865, EPI_ISL_2729867, EPI_ISL_2729874, EPI_ISL_2729885, EPI_ISL_2729917, EPI_ISL_2729918, EPI_ISL_2729920, EPI_ISL_2729931, EPI_ISL_2729939, EPI_ISL_2729958, EPI_ISL_2729961, EPI_ISL_2729965, EPI_ISL_2729967, EPI_ISL_2729973, EPI_ISL_2729975, EPI_ISL_2729983, EPI_ISL_2729988, EPI_ISL_2729993, EPI_ISL_2730011, EPI_ISL_2730012, EPI_ISL_2730013, EPI_ISL_2730016, EPI_ISL_2730017, EPI_ISL_2730026, EPI_ISL_2730037, EPI_ISL_2730040, EPI_ISL_2730041, EPI_ISL_2730044, EPI_ISL_2730052, EPI_ISL_2730058, EPI_ISL_2730063, EPI_ISL_2730086, EPI_ISL_2730096, EPI_ISL_2730097, EPI_ISL_2730100, EPI_ISL_2730110, EPI_ISL_2730111, EPI_ISL_2730117, EPI_ISL_2730120, EPI_ISL_2730126, EPI_ISL_2730134, EPI_ISL_2730145, EPI_ISL_2730155, EPI_ISL_2730164, EPI_ISL_2730173, EPI_ISL_2730179, EPI_ISL_2730181, EPI_ISL_2730188, EPI_ISL_2730189, EPI_ISL_2730196, EPI_ISL_2730208, EPI_ISL_2730214, EPI_ISL_2730218, EPI_ISL_2730226, EPI_ISL_2730227, EPI_ISL_2730236, EPI_ISL_2730241, EPI_ISL_2730251, EPI_ISL_2730267, EPI_ISL_2730309, EPI_ISL_2730314, EPI_ISL_2730324, EPI_ISL_2730354, EPI_ISL_2730360, EPI_ISL_2730388, EPI_ISL_2730434, EPI_ISL_2730449, EPI_ISL_2730465, EPI_ISL_2730466, EPI_ISL_2730473, EPI_ISL_2730489, EPI_ISL_2730508, EPI_ISL_2730515, EPI_ISL_2730518, EPI_ISL_2730533, EPI_ISL_2730552, EPI_ISL_2730554, EPI_ISL_2730561, EPI_ISL_2730566, EPI_ISL_2730595, EPI_ISL_2730626, EPI_ISL_2730648, EPI_ISL_2730649, EPI_ISL_2730650, EPI_ISL_2730651, EPI_ISL_2730652, EPI_ISL_2730653, EPI_ISL_2730654, EPI_ISL_2730655, EPI_ISL_2730656, EPI_ISL_2730657, EPI_ISL_2730658, EPI_ISL_2730659, EPI_ISL_2730660, EPI_ISL_2730661, EPI_ISL_2730662, EPI_ISL_2730663, EPI_ISL_2730664, EPI_ISL_2730665, EPI_ISL_2730666, EPI_ISL_2730667, EPI_ISL_2730668, EPI_ISL_2730669, EPI_ISL_2730670, EPI_ISL_2730671, EPI_ISL_2730672, EPI_ISL_2730673, EPI_ISL_2730674, EPI_ISL_2730675, EPI_ISL_2730676, EPI_ISL_2730677, EPI_ISL_2730678, EPI_ISL_2730679, EPI_ISL_2730680, EPI_ISL_2730681, EPI_ISL_2730682, EPI_ISL_2730683, EPI_ISL_2730684, EPI_ISL_2730685, EPI_ISL_2730686, EPI_ISL_2730687, EPI_ISL_2730688, EPI_ISL_2730689, EPI_ISL_2730690, EPI_ISL_2730691, EPI_ISL_2730692, EPI_ISL_2730693, EPI_ISL_2730694, EPI_ISL_2730695, EPI_ISL_2730696, EPI_ISL_2730697, EPI_ISL_2730698, EPI_ISL_2730699, EPI_ISL_2730700, EPI_ISL_2730701, EPI_ISL_2730702, EPI_ISL_2730703, EPI_ISL_2730704, EPI_ISL_2730705, EPI_ISL_2730706, EPI_ISL_2730707, EPI_ISL_2730708, EPI_ISL_2730709, EPI_ISL_2730710, EPI_ISL_2730711, EPI_ISL_2730712, EPI_ISL_2730713, EPI_ISL_2730714, EPI_ISL_2730715, EPI_ISL_2730716, EPI_ISL_2730717, EPI_ISL_2730718, EPI_ISL_2730719, EPI_ISL_2730720, EPI_ISL_2730721, EPI_ISL_2730722, EPI_ISL_2730723, EPI_ISL_2730724, EPI_ISL_2730725, EPI_ISL_2730726, EPI_ISL_2730727, EPI_ISL_2730728, EPI_ISL_2730729, EPI_ISL_2730730, EPI_ISL_2730731, EPI_ISL_2730732, EPI_ISL_2730733, EPI_ISL_2730734, EPI_ISL_2730735, EPI_ISL_2730736, EPI_ISL_2730737, EPI_ISL_2730738, EPI_ISL_2730739, EPI_ISL_2730740, EPI_ISL_2730741, EPI_ISL_2730742, EPI_ISL_2730743, EPI_ISL_2730744, EPI_ISL_2730745, EPI_ISL_2730746, EPI_ISL_2730747, EPI_ISL_2730748, EPI_ISL_2730749, EPI_ISL_2730750, EPI_ISL_2730751, EPI_ISL_2730752, EPI_ISL_2730753, EPI_ISL_2730754, EPI_ISL_2730755, EPI_ISL_2730756, EPI_ISL_2730757, EPI_ISL_2730758, EPI_ISL_2730759, EPI_ISL_2730760, EPI_ISL_2730761, EPI_ISL_2730762, EPI_ISL_2730763, EPI_ISL_2730764, EPI_ISL_2730765, EPI_ISL_2730766, EPI_ISL_2730767, EPI_ISL_2730768, EPI_ISL_2730769, EPI_ISL_2730770, EPI_ISL_2730771, EPI_ISL_2730772, EPI_ISL_2730773, EPI_ISL_2730774, EPI_ISL_2730775, EPI_ISL_2730776, EPI_ISL_2730777, EPI_ISL_2730778, EPI_ISL_2730779, EPI_ISL_2730780, EPI_ISL_2730781, EPI_ISL_2730782, EPI_ISL_2730783, EPI_ISL_2730784, EPI_ISL_2730786, EPI_ISL_2730787, EPI_ISL_2730788, EPI_ISL_2730789, EPI_ISL_2730790, EPI_ISL_2730791, EPI_ISL_2730792, EPI_ISL_2730793, EPI_ISL_2730794, EPI_ISL_2730795, EPI_ISL_2730796, EPI_ISL_2730797, EPI_ISL_2730798, EPI_ISL_2730799, EPI_ISL_2730800, EPI_ISL_2730801, EPI_ISL_2730802, EPI_ISL_2730803, EPI_ISL_2730804, EPI_ISL_2730805, EPI_ISL_2730806, EPI_ISL_2730807, EPI_ISL_2730808, EPI_ISL_2730809, EPI_ISL_2730810, EPI_ISL_2730811, EPI_ISL_2730812, EPI_ISL_2730813, EPI_ISL_2730814, EPI_ISL_2730815, EPI_ISL_2730816, EPI_ISL_2730817, EPI_ISL_2730818, EPI_ISL_2730819, EPI_ISL_2730820, EPI_ISL_2730821, EPI_ISL_2730822, EPI_ISL_2730823, EPI_ISL_2730824, EPI_ISL_2730825, EPI_ISL_2730826, EPI_ISL_2730827, EPI_ISL_2730828, EPI_ISL_2730829, EPI_ISL_2730830, EPI_ISL_2730831, EPI_ISL_2730832, EPI_ISL_2730833, EPI_ISL_2730834, EPI_ISL_2730835, EPI_ISL_2730836, EPI_ISL_2730837, EPI_ISL_2730838, EPI_ISL_2730839, EPI_ISL_2730840, EPI_ISL_2730841, EPI_ISL_2730842, EPI_ISL_2730843, EPI_ISL_2730844, EPI_ISL_2730845, EPI_ISL_2730846, EPI_ISL_2730847, EPI_ISL_2730848, EPI_ISL_2730849, EPI_ISL_2730850, EPI_ISL_2730851, EPI_ISL_2730852, EPI_ISL_2730853, EPI_ISL_2730854, EPI_ISL_2730855, EPI_ISL_2730856, EPI_ISL_2730857, EPI_ISL_2730858, EPI_ISL_2730859, EPI_ISL_2730860, EPI_ISL_2730861, EPI_ISL_2730862, EPI_ISL_2730863, EPI_ISL_2730864, EPI_ISL_2730865, EPI_ISL_2730866, EPI_ISL_2730867, EPI_ISL_2730868, EPI_ISL_2730869, EPI_ISL_2730870, EPI_ISL_2730871, EPI_ISL_2730872, EPI_ISL_2730873, EPI_ISL_2730874, EPI_ISL_2730875, EPI_ISL_2730876, EPI_ISL_2730877, EPI_ISL_2730878, EPI_ISL_2730879, EPI_ISL_2730880, EPI_ISL_2730881, EPI_ISL_2730882, EPI_ISL_2730883, EPI_ISL_2730884, EPI_ISL_2730885, EPI_ISL_2730886, EPI_ISL_2730887, EPI_ISL_2730888, EPI_ISL_2730889, EPI_ISL_2730890, EPI_ISL_2730891, EPI_ISL_2730892, EPI_ISL_2730893, EPI_ISL_2730894, EPI_ISL_2730895, EPI_ISL_2730896, EPI_ISL_2730897, EPI_ISL_2730898, EPI_ISL_2730899, EPI_ISL_2730900, EPI_ISL_2730901, EPI_ISL_2730902, EPI_ISL_2730903, EPI_ISL_2730904, EPI_ISL_2730905, EPI_ISL_2730906, EPI_ISL_2730907, EPI_ISL_2730908, EPI_ISL_2730909, EPI_ISL_2730910, EPI_ISL_2730911, EPI_ISL_2730913, EPI_ISL_2730914, EPI_ISL_2730915, EPI_ISL_2730916, EPI_ISL_2730917, EPI_ISL_2730918, EPI_ISL_2730919, EPI_ISL_2730920, EPI_ISL_2730921, EPI_ISL_2730922, EPI_ISL_2730924, EPI_ISL_2730925, EPI_ISL_2730927, EPI_ISL_2730928, EPI_ISL_2730929, EPI_ISL_2730930, EPI_ISL_2730931, EPI_ISL_2730932, EPI_ISL_2730933, EPI_ISL_2730934, EPI_ISL_2730935, EPI_ISL_2730936, EPI_ISL_2730937, EPI_ISL_2730938, EPI_ISL_2730939, EPI_ISL_2730940, EPI_ISL_2730941, EPI_ISL_2730942, EPI_ISL_2730943, EPI_ISL_2730944, EPI_ISL_2730945, EPI_ISL_2730946, EPI_ISL_2730947, EPI_ISL_2730948, EPI_ISL_2730949, EPI_ISL_2730950, EPI_ISL_2730951, EPI_ISL_2730952, EPI_ISL_2730953, EPI_ISL_2730954, EPI_ISL_2730955, EPI_ISL_2730956, EPI_ISL_2730957, EPI_ISL_2730958, EPI_ISL_2730959, EPI_ISL_2730960, EPI_ISL_2730961, EPI_ISL_2730962, EPI_ISL_2730963, EPI_ISL_2730964, EPI_ISL_2730965, EPI_ISL_2730966, EPI_ISL_2730967, EPI_ISL_2730968, EPI_ISL_2730969, EPI_ISL_2730970, EPI_ISL_2730971, EPI_ISL_2730972, EPI_ISL_2730973, EPI_ISL_2730974, EPI_ISL_2730976, EPI_ISL_2730977, EPI_ISL_2730978, EPI_ISL_2730979, EPI_ISL_2730980, EPI_ISL_2730981, EPI_ISL_2730983, EPI_ISL_2730984, EPI_ISL_2730985, EPI_ISL_2730986, EPI_ISL_2730987, EPI_ISL_2730988, EPI_ISL_2730989, EPI_ISL_2730990, EPI_ISL_2730991, EPI_ISL_2730992, EPI_ISL_2730993, EPI_ISL_2730994, EPI_ISL_2730995, EPI_ISL_2730996, EPI_ISL_2730997, EPI_ISL_2730998, EPI_ISL_2730999, EPI_ISL_2731000, EPI_ISL_2731001, EPI_ISL_2731003, EPI_ISL_2731004, EPI_ISL_2731005, EPI_ISL_2731006, EPI_ISL_2731007, EPI_ISL_2731008, EPI_ISL_2731009, EPI_ISL_2731010, EPI_ISL_2731011, EPI_ISL_2731012, EPI_ISL_2731013, EPI_ISL_2731014, EPI_ISL_2731015, EPI_ISL_2731016, EPI_ISL_2731017, EPI_ISL_2731018, EPI_ISL_2731019, EPI_ISL_2731020, EPI_ISL_2731021, EPI_ISL_2731022, EPI_ISL_2731023, EPI_ISL_2731024, EPI_ISL_2731025, EPI_ISL_2731026, EPI_ISL_2731027, EPI_ISL_2731028, EPI_ISL_2731029, EPI_ISL_2731030, EPI_ISL_2731031, EPI_ISL_2731032, EPI_ISL_2731033, EPI_ISL_2731034, EPI_ISL_2731035, EPI_ISL_2731036, EPI_ISL_2731037, EPI_ISL_2731038, EPI_ISL_2731039, EPI_ISL_2731040, EPI_ISL_2731041, EPI_ISL_2731042, EPI_ISL_2731043, EPI_ISL_2731044, EPI_ISL_2731045, EPI_ISL_2731046, EPI_ISL_2731047, EPI_ISL_2731048, EPI_ISL_2731049, EPI_ISL_2731050, EPI_ISL_2731051, EPI_ISL_2731052, EPI_ISL_2731053, EPI_ISL_2731054, EPI_ISL_2731055, EPI_ISL_2731056, EPI_ISL_2731058, EPI_ISL_2731060, EPI_ISL_2731061, EPI_ISL_2731062, EPI_ISL_2731063, EPI_ISL_2731064, EPI_ISL_2731065, EPI_ISL_2731066, EPI_ISL_2731067, EPI_ISL_2731068, EPI_ISL_2731069, EPI_ISL_2731070, EPI_ISL_2731071, EPI_ISL_2731072, EPI_ISL_2731073, EPI_ISL_2731074, EPI_ISL_2731075, EPI_ISL_2731076, EPI_ISL_2731077, EPI_ISL_2731078, EPI_ISL_2731079, EPI_ISL_2731081, EPI_ISL_2731083, EPI_ISL_2731084, EPI_ISL_2731085, EPI_ISL_2731086, EPI_ISL_2731087, EPI_ISL_2731088, EPI_ISL_2731089, EPI_ISL_2731090, EPI_ISL_2731091, EPI_ISL_2731092, EPI_ISL_2731093, EPI_ISL_2731094, EPI_ISL_2731095, EPI_ISL_2731096, EPI_ISL_2731097, EPI_ISL_2731098, EPI_ISL_2731099, EPI_ISL_2731101, EPI_ISL_2731102, EPI_ISL_2731103, EPI_ISL_2731104, EPI_ISL_2731105, EPI_ISL_2731106, EPI_ISL_2731107, EPI_ISL_2731108, EPI_ISL_2731109, EPI_ISL_2731110, EPI_ISL_2731111, EPI_ISL_2731112, EPI_ISL_2731113, EPI_ISL_2731114, EPI_ISL_2731115, EPI_ISL_2731116, EPI_ISL_2731117, EPI_ISL_2731118, EPI_ISL_2731119, EPI_ISL_2731120, EPI_ISL_2731121, EPI_ISL_2731122, EPI_ISL_2731123, EPI_ISL_2731124, EPI_ISL_2731125, EPI_ISL_2731126, EPI_ISL_2731127, EPI_ISL_2731128, EPI_ISL_2731129, EPI_ISL_2731130, EPI_ISL_2731131, EPI_ISL_2731132, EPI_ISL_2731133, EPI_ISL_2731134, EPI_ISL_2731135, EPI_ISL_2731136, EPI_ISL_2731137, EPI_ISL_2731138, EPI_ISL_2731139, EPI_ISL_2731140, EPI_ISL_2731141, EPI_ISL_2731142, EPI_ISL_2731143, EPI_ISL_2731144, EPI_ISL_2731145, EPI_ISL_2731146, EPI_ISL_2731147, EPI_ISL_2767239, EPI_ISL_2767246, EPI_ISL_2767250, EPI_ISL_2767272, EPI_ISL_2767273, EPI_ISL_2767277, EPI_ISL_2767291, EPI_ISL_2767293, EPI_ISL_2767303, EPI_ISL_2767316, EPI_ISL_2767319, EPI_ISL_2767333, EPI_ISL_2767338, EPI_ISL_2767344, EPI_ISL_2767347, EPI_ISL_2767384, EPI_ISL_2767388, EPI_ISL_2767391, EPI_ISL_2767398, EPI_ISL_2767401, EPI_ISL_2767416, EPI_ISL_2767427, EPI_ISL_2767429, EPI_ISL_2767459, EPI_ISL_2767464, EPI_ISL_2767469, EPI_ISL_2767479, EPI_ISL_2767483, EPI_ISL_2767488, EPI_ISL_2767508, EPI_ISL_2767518, EPI_ISL_2767527, EPI_ISL_2767528, EPI_ISL_2767536, EPI_ISL_2767540, EPI_ISL_2767552, EPI_ISL_2767565, EPI_ISL_2767592, EPI_ISL_2767598, EPI_ISL_2767602, EPI_ISL_2767603, EPI_ISL_2767604, EPI_ISL_2767620, EPI_ISL_2767625, EPI_ISL_2767627, EPI_ISL_2767635, EPI_ISL_2767639, EPI_ISL_2781509, EPI_ISL_2781510, EPI_ISL_2781511, EPI_ISL_2781528, EPI_ISL_2781532, EPI_ISL_2781534, EPI_ISL_2781573, EPI_ISL_2781579, EPI_ISL_2781585, EPI_ISL_2781588, EPI_ISL_2781592, EPI_ISL_2781644, EPI_ISL_2781651, EPI_ISL_2781686, EPI_ISL_2781693, EPI_ISL_2781710, EPI_ISL_2781733, EPI_ISL_2781809, EPI_ISL_2781815, EPI_ISL_2781816, EPI_ISL_2781834, EPI_ISL_2781865, EPI_ISL_2781899, EPI_ISL_2781921, EPI_ISL_2781937, EPI_ISL_2781966, EPI_ISL_2781971, EPI_ISL_2781985, EPI_ISL_2781995, EPI_ISL_2782009, EPI_ISL_2782012, EPI_ISL_2782032, EPI_ISL_2782094, EPI_ISL_2782158, EPI_ISL_2808859, EPI_ISL_2808860, EPI_ISL_2808861, EPI_ISL_2808862, EPI_ISL_2808863, EPI_ISL_2808864, EPI_ISL_2808865, EPI_ISL_2808867, EPI_ISL_2808868, EPI_ISL_2808869, EPI_ISL_2808870, EPI_ISL_2808871, EPI_ISL_2808872, EPI_ISL_2808873, EPI_ISL_2808874, EPI_ISL_2808875, EPI_ISL_2808876, EPI_ISL_2808877, EPI_ISL_2808878, EPI_ISL_2808879, EPI_ISL_2808880, EPI_ISL_2808881, EPI_ISL_2808882, EPI_ISL_2808883, EPI_ISL_2808884, EPI_ISL_2808885, EPI_ISL_2808886, EPI_ISL_2808887, EPI_ISL_2808888, EPI_ISL_2808889, EPI_ISL_2808890, EPI_ISL_2808891, EPI_ISL_2808892, EPI_ISL_2808893, EPI_ISL_2808895, EPI_ISL_2808896, EPI_ISL_2808897, EPI_ISL_2808898, EPI_ISL_2808899, EPI_ISL_2808900, EPI_ISL_2808902, EPI_ISL_2808903, EPI_ISL_2808904, EPI_ISL_2808905, EPI_ISL_2808907, EPI_ISL_2808908, EPI_ISL_2808909, EPI_ISL_2808910, EPI_ISL_2808911, EPI_ISL_2808912, EPI_ISL_2808913, EPI_ISL_2808914, EPI_ISL_2808915, EPI_ISL_2808916, EPI_ISL_2808917, EPI_ISL_2808919, EPI_ISL_2808921, EPI_ISL_2808922, EPI_ISL_2808923, EPI_ISL_2808924, EPI_ISL_2808925, EPI_ISL_2808926, EPI_ISL_2808927, EPI_ISL_2808928, EPI_ISL_2808929, EPI_ISL_2808930, EPI_ISL_2808931, EPI_ISL_2808932, EPI_ISL_2808933, EPI_ISL_2808934, EPI_ISL_2808935, EPI_ISL_2808936, EPI_ISL_2808937, EPI_ISL_2808938, EPI_ISL_2808939, EPI_ISL_2808940, EPI_ISL_2808941, EPI_ISL_2808942, EPI_ISL_2808943, EPI_ISL_2808944, EPI_ISL_2808945, EPI_ISL_2808946, EPI_ISL_2808947, EPI_ISL_2808948, EPI_ISL_2808949, EPI_ISL_2808950, EPI_ISL_2808951, EPI_ISL_2808952, EPI_ISL_2808953, EPI_ISL_2808954, EPI_ISL_2808955, EPI_ISL_2808956, EPI_ISL_2808957, EPI_ISL_2808958, EPI_ISL_2808959, EPI_ISL_2808960, EPI_ISL_2808961, EPI_ISL_2808962, EPI_ISL_2808963, EPI_ISL_2808964, EPI_ISL_2808965, EPI_ISL_2808966, EPI_ISL_2808967, EPI_ISL_2808968, EPI_ISL_2808969, EPI_ISL_2808970, EPI_ISL_2808971, EPI_ISL_2808972, EPI_ISL_2808973, EPI_ISL_2808974, EPI_ISL_2808975, EPI_ISL_2808976, EPI_ISL_2808977, EPI_ISL_2808978, EPI_ISL_2808979, EPI_ISL_2808980, EPI_ISL_2808981, EPI_ISL_2808982, EPI_ISL_2808983, EPI_ISL_2808984, EPI_ISL_2808985, EPI_ISL_2808986, EPI_ISL_2808987, EPI_ISL_2808989, EPI_ISL_2808990, EPI_ISL_2808991, EPI_ISL_2808992, EPI_ISL_2808993, EPI_ISL_2808994, EPI_ISL_2808995, EPI_ISL_2808996, EPI_ISL_2808997, EPI_ISL_2808998, EPI_ISL_2808999, EPI_ISL_2809000, EPI_ISL_2809001, EPI_ISL_2809002, EPI_ISL_2809003, EPI_ISL_2809004, EPI_ISL_2809005, EPI_ISL_2809006, EPI_ISL_2809007, EPI_ISL_2809008, EPI_ISL_2809009, EPI_ISL_2809010, EPI_ISL_2809011, EPI_ISL_2809012, EPI_ISL_2809013, EPI_ISL_2809014, EPI_ISL_2809015, EPI_ISL_2809016, EPI_ISL_2809017, EPI_ISL_2809018, EPI_ISL_2809019, EPI_ISL_2809020, EPI_ISL_2809021, EPI_ISL_2809022, EPI_ISL_2809023, EPI_ISL_2809024, EPI_ISL_2809025, EPI_ISL_2809026, EPI_ISL_2809027, EPI_ISL_2809028, EPI_ISL_2809029, EPI_ISL_2809030, EPI_ISL_2809031, EPI_ISL_2809032, EPI_ISL_2809033, EPI_ISL_2809034, EPI_ISL_2809035, EPI_ISL_2809036, EPI_ISL_2809037, EPI_ISL_2809038, EPI_ISL_2809039, EPI_ISL_2809040, EPI_ISL_2809041, EPI_ISL_2809042, EPI_ISL_2809043, EPI_ISL_2809044, EPI_ISL_2809045, EPI_ISL_2809046, EPI_ISL_2809048, EPI_ISL_2809049, EPI_ISL_2809050, EPI_ISL_2809052, EPI_ISL_2809053, EPI_ISL_2809054, EPI_ISL_2809055, EPI_ISL_2809056, EPI_ISL_2809057, EPI_ISL_2809058, EPI_ISL_2809059, EPI_ISL_2809060, EPI_ISL_2809061, EPI_ISL_2809062, EPI_ISL_2809063, EPI_ISL_2809064, EPI_ISL_2809065, EPI_ISL_2809066, EPI_ISL_2809067, EPI_ISL_2809068, EPI_ISL_2809069, EPI_ISL_2809070, EPI_ISL_2809071, EPI_ISL_2809072, EPI_ISL_2809073, EPI_ISL_2809074, EPI_ISL_2809076, EPI_ISL_2809077, EPI_ISL_2809078, EPI_ISL_2809079, EPI_ISL_2809080, EPI_ISL_2809081, EPI_ISL_2809082, EPI_ISL_2809083, EPI_ISL_2809084, EPI_ISL_2809085, EPI_ISL_2809086, EPI_ISL_2809087, EPI_ISL_2809088, EPI_ISL_2809089, EPI_ISL_2809090, EPI_ISL_2809091, EPI_ISL_2809092, EPI_ISL_2809093, EPI_ISL_2809094, EPI_ISL_2809300, EPI_ISL_2809422, EPI_ISL_2809423, EPI_ISL_2809424, EPI_ISL_2809425, EPI_ISL_2809426, EPI_ISL_2809427, EPI_ISL_2809428, EPI_ISL_2809429, EPI_ISL_2809430, EPI_ISL_2809431, EPI_ISL_2809432, EPI_ISL_2809433, EPI_ISL_2809434, EPI_ISL_2809436, EPI_ISL_2809437, EPI_ISL_2809438, EPI_ISL_2809439, EPI_ISL_2809440, EPI_ISL_2809441, EPI_ISL_2809442, EPI_ISL_2809443, EPI_ISL_2809444, EPI_ISL_2809445, EPI_ISL_2809447, EPI_ISL_2809448, EPI_ISL_2809449, EPI_ISL_2809450, EPI_ISL_2809451, EPI_ISL_2809452, EPI_ISL_2809453, EPI_ISL_2809455, EPI_ISL_2809456, EPI_ISL_2809457, EPI_ISL_2809458, EPI_ISL_2809459, EPI_ISL_2809460, EPI_ISL_2809462, EPI_ISL_2809463, EPI_ISL_2809464, EPI_ISL_2809465, EPI_ISL_2809466, EPI_ISL_2809467, EPI_ISL_2809468, EPI_ISL_2809469, EPI_ISL_2809470, EPI_ISL_2809471, EPI_ISL_2809472, EPI_ISL_2809473, EPI_ISL_2809474, EPI_ISL_2809475, EPI_ISL_2809476, EPI_ISL_2809477, EPI_ISL_2809478, EPI_ISL_2809479, EPI_ISL_2809480, EPI_ISL_2809481, EPI_ISL_2809482, EPI_ISL_2809483, EPI_ISL_2809484, EPI_ISL_2809485, EPI_ISL_2809486, EPI_ISL_2809487, EPI_ISL_2809489, EPI_ISL_2809490, EPI_ISL_2809491, EPI_ISL_2809492, EPI_ISL_2809493, EPI_ISL_2809494, EPI_ISL_2809495, EPI_ISL_2809496, EPI_ISL_2809497, EPI_ISL_2809498, EPI_ISL_2809499, EPI_ISL_2809526, EPI_ISL_2809574, EPI_ISL_2809576, EPI_ISL_2809582, EPI_ISL_2809583, EPI_ISL_2809587, EPI_ISL_2809588, EPI_ISL_2809589, EPI_ISL_2809591, EPI_ISL_2809594, EPI_ISL_2809598, EPI_ISL_2809599, EPI_ISL_2809600, EPI_ISL_2809601, EPI_ISL_2809602, EPI_ISL_2809605, EPI_ISL_2809606, EPI_ISL_2809612, EPI_ISL_2809615, EPI_ISL_2809616, EPI_ISL_2809620, EPI_ISL_2809621, EPI_ISL_2809622, EPI_ISL_2809624, EPI_ISL_2809625, EPI_ISL_2809626, EPI_ISL_2809628, EPI_ISL_2809629, EPI_ISL_2809634, EPI_ISL_2809635, EPI_ISL_2809637, EPI_ISL_2809638, EPI_ISL_2809641, EPI_ISL_2809643, EPI_ISL_2809644, EPI_ISL_2809645, EPI_ISL_2809646, EPI_ISL_2809649, EPI_ISL_2809651, EPI_ISL_2809652, EPI_ISL_2809660, EPI_ISL_2809662, EPI_ISL_2809665, EPI_ISL_2809666, EPI_ISL_2809671, EPI_ISL_2809672, EPI_ISL_2809675, EPI_ISL_2809679, EPI_ISL_2809680, EPI_ISL_2809683, EPI_ISL_2809685, EPI_ISL_2809686, EPI_ISL_2809689, EPI_ISL_2809690, EPI_ISL_2809696, EPI_ISL_2809701, EPI_ISL_2809702, EPI_ISL_2809707, EPI_ISL_2809710, EPI_ISL_2809711, EPI_ISL_2809713, EPI_ISL_2809719, EPI_ISL_2809721, EPI_ISL_2809723, EPI_ISL_2809727, EPI_ISL_2809733, EPI_ISL_2809735, EPI_ISL_2809738, EPI_ISL_2809746, EPI_ISL_2809747, EPI_ISL_2809756, EPI_ISL_2809761, EPI_ISL_2809764, EPI_ISL_2809765, EPI_ISL_2809767, EPI_ISL_2809770, EPI_ISL_2809771, EPI_ISL_2809772, EPI_ISL_2809773, EPI_ISL_2809778, EPI_ISL_2809780, EPI_ISL_2809786, EPI_ISL_2809788, EPI_ISL_2809789, EPI_ISL_2809794, EPI_ISL_2809795, EPI_ISL_2809798, EPI_ISL_2809800, EPI_ISL_2809801, EPI_ISL_2809804, EPI_ISL_2809805, EPI_ISL_2809806, EPI_ISL_2809807, EPI_ISL_2809809, EPI_ISL_2809810, EPI_ISL_2809820, EPI_ISL_2809821, EPI_ISL_2809822, EPI_ISL_2809823, EPI_ISL_2809824, EPI_ISL_2809825, EPI_ISL_2809826, EPI_ISL_2809827, EPI_ISL_2809828, EPI_ISL_2809829, EPI_ISL_2809830, EPI_ISL_2809831, EPI_ISL_2809832, EPI_ISL_2809833, EPI_ISL_2809834, EPI_ISL_2809836, EPI_ISL_2809837, EPI_ISL_2809838, EPI_ISL_2809839, EPI_ISL_2809840, EPI_ISL_2809841, EPI_ISL_2809842, EPI_ISL_2809843, EPI_ISL_2809844, EPI_ISL_2809845, EPI_ISL_2809846, EPI_ISL_2809847, EPI_ISL_2809848, EPI_ISL_2809849, EPI_ISL_2809850, EPI_ISL_2809851, EPI_ISL_2809852, EPI_ISL_2809853, EPI_ISL_2809854, EPI_ISL_2809856, EPI_ISL_2809857, EPI_ISL_2809858, EPI_ISL_2809859, EPI_ISL_2809860, EPI_ISL_2809861, EPI_ISL_2809862, EPI_ISL_2809863, EPI_ISL_2809864, EPI_ISL_2809865, EPI_ISL_2809866, EPI_ISL_2809867, EPI_ISL_2809868, EPI_ISL_2809869, EPI_ISL_2809870, EPI_ISL_2809871, EPI_ISL_2809872, EPI_ISL_2809873, EPI_ISL_2809874, EPI_ISL_2809875, EPI_ISL_2809876, EPI_ISL_2809877, EPI_ISL_2809878, EPI_ISL_2809879, EPI_ISL_2809880, EPI_ISL_2809881, EPI_ISL_2809885, EPI_ISL_2809893, EPI_ISL_2809895, EPI_ISL_2809906, EPI_ISL_2809909, EPI_ISL_2809913, EPI_ISL_2809916, EPI_ISL_2809922, EPI_ISL_2809923, EPI_ISL_2809924, EPI_ISL_2809925, EPI_ISL_2809928, EPI_ISL_2809933, EPI_ISL_2809934, EPI_ISL_2809936, EPI_ISL_2809937, EPI_ISL_2809939, EPI_ISL_2809949, EPI_ISL_2809955, EPI_ISL_2809956, EPI_ISL_2809957, EPI_ISL_2809958, EPI_ISL_2809963, EPI_ISL_2809966, EPI_ISL_2809969, EPI_ISL_2809972, EPI_ISL_2809973, EPI_ISL_2809975, EPI_ISL_2809978, EPI_ISL_2809979, EPI_ISL_2809983, EPI_ISL_2809984, EPI_ISL_2809986, EPI_ISL_2809990, EPI_ISL_2809992, EPI_ISL_2809996, EPI_ISL_2809998, EPI_ISL_2809999, EPI_ISL_2810003, EPI_ISL_2810004, EPI_ISL_2810007, EPI_ISL_2810008, EPI_ISL_2810012, EPI_ISL_2810016, EPI_ISL_2810017, EPI_ISL_2810019, EPI_ISL_2814665, EPI_ISL_2815083, EPI_ISL_2815084, EPI_ISL_2815085, EPI_ISL_2815086, EPI_ISL_2815087, EPI_ISL_2815088, EPI_ISL_2815089, EPI_ISL_2815090, EPI_ISL_2815092, EPI_ISL_2815093, EPI_ISL_2815096, EPI_ISL_2815097, EPI_ISL_2815098, EPI_ISL_2815099, EPI_ISL_2815101, EPI_ISL_2815102, EPI_ISL_2815103, EPI_ISL_2815104, EPI_ISL_2815105, EPI_ISL_2815106, EPI_ISL_2815107, EPI_ISL_2815108, EPI_ISL_2815109, EPI_ISL_2815110, EPI_ISL_2815111, EPI_ISL_2815112, EPI_ISL_2815113, EPI_ISL_2815114, EPI_ISL_2815115, EPI_ISL_2815116, EPI_ISL_2815117, EPI_ISL_2815118, EPI_ISL_2815119, EPI_ISL_2815120, EPI_ISL_2815121, EPI_ISL_2815122, EPI_ISL_2815123, EPI_ISL_2815124, EPI_ISL_2815125, EPI_ISL_2815126, EPI_ISL_2815127, EPI_ISL_2815128, EPI_ISL_2815129, EPI_ISL_2815130, EPI_ISL_2815131, EPI_ISL_2815132, EPI_ISL_2815133, EPI_ISL_2815134, EPI_ISL_2815135, EPI_ISL_2815136, EPI_ISL_2815137, EPI_ISL_2815138, EPI_ISL_2815139, EPI_ISL_2815140, EPI_ISL_2815141, EPI_ISL_2815142, EPI_ISL_2815143, EPI_ISL_2815144, EPI_ISL_2815145, EPI_ISL_2815146, EPI_ISL_2815147, EPI_ISL_2815148, EPI_ISL_2815149, EPI_ISL_2815150, EPI_ISL_2815151, EPI_ISL_2815152, EPI_ISL_2815153, EPI_ISL_2815154, EPI_ISL_2815155, EPI_ISL_2815156, EPI_ISL_2815157, EPI_ISL_2815158, EPI_ISL_2815159, EPI_ISL_2815160, EPI_ISL_2815161, EPI_ISL_2815162, EPI_ISL_2815163,

EPI_ISL_2815164, EPI_ISL_2815165, EPI_ISL_2815166, EPI_ISL_2815167, EPI_ISL_2815168, EPI_ISL_2815169, EPI_ISL_2815170, EPI_ISL_2815171, EPI_ISL_2815172, EPI_ISL_2815173, EPI_ISL_2815174, EPI_ISL_2815175, EPI_ISL_2815176, EPI_ISL_2815177, EPI_ISL_2815178, EPI_ISL_2815179, EPI_ISL_2815180, EPI_ISL_2815181, EPI_ISL_2815182, EPI_ISL_2815183, EPI_ISL_2815184, EPI_ISL_2815185, EPI_ISL_2815186, EPI_ISL_2815187, EPI_ISL_2815188, EPI_ISL_2815189, EPI_ISL_2815190, EPI_ISL_2815191, EPI_ISL_2815192, EPI_ISL_2815193, EPI_ISL_2815194, EPI_ISL_2815195, EPI_ISL_2815196, EPI_ISL_2815197, EPI_ISL_2815198, EPI_ISL_2815199, EPI_ISL_2815200, EPI_ISL_2815201, EPI_ISL_2815202, EPI_ISL_2815203, EPI_ISL_2815204, EPI_ISL_2815205, EPI_ISL_2815206, EPI_ISL_2815207, EPI_ISL_2815208, EPI_ISL_2815209, EPI_ISL_2815210, EPI_ISL_2815211, EPI_ISL_2815212, EPI_ISL_2815213, EPI_ISL_2815214, EPI_ISL_2815215, EPI_ISL_2815216, EPI_ISL_2815217, EPI_ISL_2815218, EPI_ISL_2815219, EPI_ISL_2815220, EPI_ISL_2815221, EPI_ISL_2815222, EPI_ISL_2815223, EPI_ISL_2815224, EPI_ISL_2815225, EPI_ISL_2815226, EPI_ISL_2815227, EPI_ISL_2815228, EPI_ISL_2815229, EPI_ISL_2815230, EPI_ISL_2815231, EPI_ISL_2815232, EPI_ISL_2815233, EPI_ISL_2815234, EPI_ISL_2815235, EPI_ISL_2815236, EPI_ISL_2815237, EPI_ISL_2815238, EPI_ISL_2815239, EPI_ISL_2815240, EPI_ISL_2815241, EPI_ISL_2815242, EPI_ISL_2815243, EPI_ISL_2815244, EPI_ISL_2815245, EPI_ISL_2815246, EPI_ISL_2815247, EPI_ISL_2815248, EPI_ISL_2815249, EPI_ISL_2815250, EPI_ISL_2815251, EPI_ISL_2815252, EPI_ISL_2815253, EPI_ISL_2815254, EPI_ISL_2815255, EPI_ISL_2815256, EPI_ISL_2815257, EPI_ISL_2815258, EPI_ISL_2815259, EPI_ISL_2815260, EPI_ISL_2815261, EPI_ISL_2815262, EPI_ISL_2815263, EPI_ISL_2815264, EPI_ISL_2815265, EPI_ISL_2815266, EPI_ISL_2815267, EPI_ISL_2815268, EPI_ISL_2815269, EPI_ISL_2815270, EPI_ISL_2815271, EPI_ISL_2815272, EPI_ISL_2815273, EPI_ISL_2815274, EPI_ISL_2815275, EPI_ISL_2815276, EPI_ISL_2815277, EPI_ISL_2815278, EPI_ISL_2815279, EPI_ISL_2815280, EPI_ISL_2815281, EPI_ISL_2815282, EPI_ISL_2815283, EPI_ISL_2815284, EPI_ISL_2815285, EPI_ISL_2815286, EPI_ISL_2815287, EPI_ISL_2815288, EPI_ISL_2815289, EPI_ISL_2815290, EPI_ISL_2815291, EPI_ISL_2815292, EPI_ISL_2815293, EPI_ISL_2815294, EPI_ISL_2815295, EPI_ISL_2815296, EPI_ISL_2815297, EPI_ISL_2815298, EPI_ISL_2815299, EPI_ISL_2815300, EPI_ISL_2815301, EPI_ISL_2815302, EPI_ISL_2815303, EPI_ISL_2815304, EPI_ISL_2815305, EPI_ISL_2815306, EPI_ISL_2815307, EPI_ISL_2815308, EPI_ISL_2815309, EPI_ISL_2815310, EPI_ISL_2815311, EPI_ISL_2815312, EPI_ISL_2815313, EPI_ISL_2815314, EPI_ISL_2815315, EPI_ISL_2815316, EPI_ISL_2815317, EPI_ISL_2815318, EPI_ISL_2815319, EPI_ISL_2815320, EPI_ISL_2815321, EPI_ISL_2815322, EPI_ISL_2815323, EPI_ISL_2815324, EPI_ISL_2815325, EPI_ISL_2815326, EPI_ISL_2832099, EPI_ISL_2852524, EPI_ISL_2852525, EPI_ISL_2852527, EPI_ISL_2852528, EPI_ISL_2852529, EPI_ISL_2852530, EPI_ISL_2852531, EPI_ISL_2852532, EPI_ISL_2852533, EPI_ISL_2852534, EPI_ISL_2852535, EPI_ISL_2852536, EPI_ISL_2852537, EPI_ISL_2852538, EPI_ISL_2852539, EPI_ISL_2852540, EPI_ISL_2852541, EPI_ISL_2852542, EPI_ISL_2852543, EPI_ISL_2852544, EPI_ISL_2852545, EPI_ISL_2852546, EPI_ISL_2852548, EPI_ISL_2852549, EPI_ISL_2852550, EPI_ISL_2852551, EPI_ISL_2852552, EPI_ISL_2852553, EPI_ISL_2852554, EPI_ISL_2852555, EPI_ISL_2852556, EPI_ISL_2852557, EPI_ISL_2852558, EPI_ISL_2852559, EPI_ISL_2852560, EPI_ISL_2852561, EPI_ISL_2852562, EPI_ISL_2852563, EPI_ISL_2852564, EPI_ISL_2852565, EPI_ISL_2852567, EPI_ISL_2852569, EPI_ISL_2852570, EPI_ISL_2852571, EPI_ISL_2852572, EPI_ISL_2852574, EPI_ISL_2852575, EPI_ISL_2852577, EPI_ISL_2852579, EPI_ISL_2852580, EPI_ISL_2852581, EPI_ISL_2852583, EPI_ISL_2852584, EPI_ISL_2852585, EPI_ISL_2852586, EPI_ISL_2852587, EPI_ISL_2852589, EPI_ISL_2852590, EPI_ISL_2852591, EPI_ISL_2852592, EPI_ISL_2852593, EPI_ISL_2852594, EPI_ISL_2852595, EPI_ISL_2852597, EPI_ISL_2852599, EPI_ISL_2852600, EPI_ISL_2852602, EPI_ISL_2852603, EPI_ISL_2852604, EPI_ISL_2852605, EPI_ISL_2852606, EPI_ISL_2852608, EPI_ISL_2852609, EPI_ISL_2852610, EPI_ISL_2852611

see above | Lighthouse Lab in Glasgow | Wellcome Sanger Institute for the COVID-19 Genomics UK (COG-UK) Consortium | Anna Dominiczak and Alex Alderton; Carol Clugston; Cordelia Langford; David Gray; David K. Jackson; Dominic Kwiatkowski; Ewan Harrison; Harper VanSteenhouse; Ian Johnston; Jeffrey Barrett; John Sillitoe on behalf of the Wellcome Sanger Institute COVID-19 Surveillance Team; Roberto Amato; Sonia Goncalves; Yumi Kasai

EPI_ISL_2700039, EPI_ISL_2716975, EPI_ISL_2716976, EPI_ISL_2716982, EPI_ISL_2716986, EPI_ISL_2716988, EPI_ISL_2716992, EPI_ISL_2716995, EPI_ISL_2716996, EPI_ISL_2716998, EPI_ISL_2716999, EPI_ISL_2717002, EPI_ISL_2717003, EPI_ISL_2717005, EPI_ISL_2717007, EPI_ISL_2717009, EPI_ISL_2717010, EPI_ISL_2717012, EPI_ISL_2717014, EPI_ISL_2717018, EPI_ISL_2717024, EPI_ISL_2717027, EPI_ISL_2717028, EPI_ISL_2717029, EPI_ISL_2717033, EPI_ISL_2717035, EPI_ISL_2717036, EPI_ISL_2717039, EPI_ISL_2717041, EPI_ISL_2717043, EPI_ISL_2717044, EPI_ISL_2717045, EPI_ISL_2717046, EPI_ISL_2717048, EPI_ISL_2717050, EPI_ISL_2717051, EPI_ISL_2717057, EPI_ISL_2717060, EPI_ISL_2717062, EPI_ISL_2717064, EPI_ISL_2717066, EPI_ISL_2717067, EPI_ISL_2717069, EPI_ISL_2717072, EPI_ISL_2717075, EPI_ISL_2717077, EPI_ISL_2717078, EPI_ISL_2717079, EPI_ISL_2717082, EPI_ISL_2717083, EPI_ISL_2717084, EPI_ISL_2717086, EPI_ISL_2717088, EPI_ISL_2717092, EPI_ISL_2717093, EPI_ISL_2717094, EPI_ISL_2717096, EPI_ISL_2717097, EPI_ISL_2717099, EPI_ISL_2717100, EPI_ISL_2717102, EPI_ISL_2717103, EPI_ISL_2717115, EPI_ISL_2717116, EPI_ISL_2717119, EPI_ISL_2717121, EPI_ISL_2717122, EPI_ISL_2717124, EPI_ISL_2717127, EPI_ISL_2717128, EPI_ISL_2717131, EPI_ISL_2717132, EPI_ISL_2717133, EPI_ISL_2717137, EPI_ISL_2717138, EPI_ISL_2717139, EPI_ISL_2717140, EPI_ISL_2717142, EPI_ISL_2717144, EPI_ISL_2717145, EPI_ISL_2717146, EPI_ISL_2717148, EPI_ISL_2717149, EPI_ISL_2717150, EPI_ISL_2717151, EPI_ISL_2717152, EPI_ISL_2717156, EPI_ISL_2717157, EPI_ISL_2717159, EPI_ISL_2717170, EPI_ISL_2717173, EPI_ISL_2717177, EPI_ISL_2717178, EPI_ISL_2717179, EPI_ISL_2717180, EPI_ISL_2717183, EPI_ISL_2717185, EPI_ISL_2717186, EPI_ISL_2717187, EPI_ISL_2717188, EPI_ISL_2717190, EPI_ISL_2717192, EPI_ISL_2717198, EPI_ISL_2717200, EPI_ISL_2717205, EPI_ISL_2717206, EPI_ISL_2717207, EPI_ISL_2717211, EPI_ISL_2717213, EPI_ISL_2717215, EPI_ISL_2717224, EPI_ISL_2717225, EPI_ISL_2717226, EPI_ISL_2717230, EPI_ISL_2717232, EPI_ISL_2717233, EPI_ISL_2717234, EPI_ISL_2717235, EPI_ISL_2717236, EPI_ISL_2717237, EPI_ISL_2717241, EPI_ISL_2717242, EPI_ISL_2717243, EPI_ISL_2717246, EPI_ISL_2717247, EPI_ISL_2717248, EPI_ISL_2717249, EPI_ISL_2717250, EPI_ISL_2717252, EPI_ISL_2717253, EPI_ISL_2717254, EPI_ISL_2717258, EPI_ISL_2717260, EPI_ISL_2717262, EPI_ISL_2717267, EPI_ISL_2717272, EPI_ISL_2717273, EPI_ISL_2717275, EPI_ISL_2717276, EPI_ISL_2717277, EPI_ISL_2717282, EPI_ISL_2717285, EPI_ISL_2717286, EPI_ISL_2717288, EPI_ISL_2717290, EPI_ISL_2717292, EPI_ISL_2717293, EPI_ISL_2717294, EPI_ISL_2717295, EPI_ISL_2717296, EPI_ISL_2717297, EPI_ISL_2717298, EPI_ISL_2717299, EPI_ISL_2717304, EPI_ISL_2717305, EPI_ISL_2717308, EPI_ISL_2717309, EPI_ISL_2717312, EPI_ISL_2717315, EPI_ISL_2717320, EPI_ISL_2717324, EPI_ISL_2717331, EPI_ISL_2717336, EPI_ISL_2717337, EPI_ISL_2717338, EPI_ISL_2717339, EPI_ISL_2717340, EPI_ISL_2717341, EPI_ISL_2717342, EPI_ISL_2717344, EPI_ISL_2717345, EPI_ISL_2717347, EPI_ISL_2717348, EPI_ISL_2717349, EPI_ISL_2717351, EPI_ISL_2717356, EPI_ISL_2717357, EPI_ISL_2717365, EPI_ISL_2717366, EPI_ISL_2717367, EPI_ISL_2717375, EPI_ISL_2717376, EPI_ISL_2717378, EPI_ISL_2717379, EPI_ISL_2717381, EPI_ISL_2717382, EPI_ISL_2717385, EPI_ISL_2717387, EPI_ISL_2717388, EPI_ISL_2717390, EPI_ISL_2717396, EPI_ISL_2717397, EPI_ISL_2717398, EPI_ISL_2717399, EPI_ISL_2717406, EPI_ISL_2717407, EPI_ISL_2717410, EPI_ISL_2717412, EPI_ISL_2717414, EPI_ISL_2717419, EPI_ISL_2717424, EPI_ISL_2717428, EPI_ISL_2717429, EPI_ISL_2717433, EPI_ISL_2717436, EPI_ISL_2717437, EPI_ISL_2717438, EPI_ISL_2717440, EPI_ISL_2717441, EPI_ISL_2717448, EPI_ISL_2717452, EPI_ISL_2717455, EPI_ISL_2717461, EPI_ISL_2717463, EPI_ISL_2717465, EPI_ISL_2717466, EPI_ISL_2717470, EPI_ISL_2717471, EPI_ISL_2717473, EPI_ISL_2717477, EPI_ISL_2717481, EPI_ISL_2717487, EPI_ISL_2717489, EPI_ISL_2717491, EPI_ISL_2717496, EPI_ISL_2717501, EPI_ISL_2717503, EPI_ISL_2717506, EPI_ISL_2717507, EPI_ISL_2717509, EPI_ISL_2717512, EPI_ISL_2717513, EPI_ISL_2717514, EPI_ISL_2717516, EPI_ISL_2717519, EPI_ISL_2717521, EPI_ISL_2717526, EPI_ISL_2717528, EPI_ISL_2717529, EPI_ISL_2717530, EPI_ISL_2717531, EPI_ISL_2717537, EPI_ISL_2717538, EPI_ISL_2717540, EPI_ISL_2717543, EPI_ISL_2717544, EPI_ISL_2717546, EPI_ISL_2717547, EPI_ISL_2717551, EPI_ISL_2717553, EPI_ISL_2717556, EPI_ISL_2717560, EPI_ISL_2717562, EPI_ISL_2717563, EPI_ISL_2717566, EPI_ISL_2717567, EPI_ISL_2717568, EPI_ISL_2717570, EPI_ISL_2717571, EPI_ISL_2717572, EPI_ISL_2717573, EPI_ISL_2717574, EPI_ISL_2717575, EPI_ISL_2717813, EPI_ISL_2717817, EPI_ISL_2717821, EPI_ISL_2717834, EPI_ISL_2717836, EPI_ISL_2717841, EPI_ISL_2717843, EPI_ISL_2717846, EPI_ISL_2717852, EPI_ISL_2717855, EPI_ISL_2717856, EPI_ISL_2717860, EPI_ISL_2717865, EPI_ISL_2717875, EPI_ISL_2717876, EPI_ISL_2717879, EPI_ISL_2717889, EPI_ISL_2717890, EPI_ISL_2717891, EPI_ISL_2717894, EPI_ISL_2717896, EPI_ISL_2717903, EPI_ISL_2717907, EPI_ISL_2717908, EPI_ISL_2717917, EPI_ISL_2717920, EPI_ISL_2717923, EPI_ISL_2717927, EPI_ISL_2717930, EPI_ISL_2717939, EPI_ISL_2717942, EPI_ISL_2717943, EPI_ISL_2717946, EPI_ISL_2717947, EPI_ISL_2717953, EPI_ISL_2717954, EPI_ISL_2717955, EPI_ISL_2717963, EPI_ISL_2717969, EPI_ISL_2717971, EPI_ISL_2717975, EPI_ISL_2717980, EPI_ISL_2717983, EPI_ISL_2717985, EPI_ISL_2717992, EPI_ISL_2718006, EPI_ISL_2718015, EPI_ISL_2718016, EPI_ISL_2718021, EPI_ISL_2718025, EPI_ISL_2718029, EPI_ISL_2718032, EPI_ISL_2718038, EPI_ISL_2718042, EPI_ISL_2718044, EPI_ISL_2718062, EPI_ISL_2718063, EPI_ISL_2718065, EPI_ISL_2718066, EPI_ISL_2718089, EPI_ISL_2718091, EPI_ISL_2718099, EPI_ISL_2718103, EPI_ISL_2718105, EPI_ISL_2718109, EPI_ISL_2718110, EPI_ISL_2718114, EPI_ISL_2718115, EPI_ISL_2718118, EPI_ISL_2718119, EPI_ISL_2718123, EPI_ISL_2718131, EPI_ISL_2718134, EPI_ISL_2718139, EPI_ISL_2718142, EPI_ISL_2718146, EPI_ISL_2718160, EPI_ISL_2718163, EPI_ISL_2718164, EPI_ISL_2718166, EPI_ISL_2718168, EPI_ISL_2728696, EPI_ISL_2728698, EPI_ISL_2728704, EPI_ISL_2728706, EPI_ISL_2728713, EPI_ISL_2728717, EPI_ISL_2728720, EPI_ISL_2728724, EPI_ISL_2728725, EPI_ISL_2728739, EPI_ISL_2728742, EPI_ISL_2728750, EPI_ISL_2728762, EPI_ISL_2728764, EPI_ISL_2728767, EPI_ISL_2728787, EPI_ISL_2728794, EPI_ISL_2728796, EPI_ISL_2728812, EPI_ISL_2728843, EPI_ISL_2728849, EPI_ISL_2728857, EPI_ISL_2728870, EPI_ISL_2728873, EPI_ISL_2728882, EPI_ISL_2728892, EPI_ISL_2728894, EPI_ISL_2728900, EPI_ISL_2728902, EPI_ISL_2728911, EPI_ISL_2728916, EPI_ISL_2728919, EPI_ISL_2728946, EPI_ISL_2728957, EPI_ISL_2728959, EPI_ISL_2728970, EPI_ISL_2728975, EPI_ISL_2728987, EPI_ISL_2729004, EPI_ISL_2729005, EPI_ISL_2729021, EPI_ISL_2729022, EPI_ISL_2729030, EPI_ISL_2729032, EPI_ISL_2729037, EPI_ISL_2729039, EPI_ISL_2729044, EPI_ISL_2729049, EPI_ISL_2729062, EPI_ISL_2729214, EPI_ISL_2729221, EPI_ISL_2729224, EPI_ISL_2729231, EPI_ISL_2729279, EPI_ISL_2729384, EPI_ISL_2729385, EPI_ISL_2729386, EPI_ISL_2729388, EPI_ISL_2729389, EPI_ISL_2729401, EPI_ISL_2729402, EPI_ISL_2729403, EPI_ISL_2729406, EPI_ISL_2729414, EPI_ISL_2729420, EPI_ISL_2729423, EPI_ISL_2729425, EPI_ISL_2729427, EPI_ISL_2729432, EPI_ISL_2729433, EPI_ISL_2729435, EPI_ISL_2729437, EPI_ISL_2729442, EPI_ISL_2729444, EPI_ISL_2729451, EPI_ISL_2729453, EPI_ISL_2729454, EPI_ISL_2729459, EPI_ISL_2729460, EPI_ISL_2729468, EPI_ISL_2729474, EPI_ISL_2729477, EPI_ISL_2729482, EPI_ISL_2729484, EPI_ISL_2729486, EPI_ISL_2729487, EPI_ISL_2729493, EPI_ISL_2729494, EPI_ISL_2729496, EPI_ISL_2729505, EPI_ISL_2729509, EPI_ISL_2729518, EPI_ISL_2729521, EPI_ISL_2729522, EPI_ISL_2729524, EPI_ISL_2729526, EPI_ISL_2729533, EPI_ISL_2729534, EPI_ISL_2729537, EPI_ISL_2729539, EPI_ISL_2729541, EPI_ISL_2729543, EPI_ISL_2729544, EPI_ISL_2729546, EPI_ISL_2729550, EPI_ISL_2729553, EPI_ISL_2729555, EPI_ISL_2729559, EPI_ISL_2729560, EPI_ISL_2729564, EPI_ISL_2729566, EPI_ISL_2729567, EPI_ISL_2729571, EPI_ISL_2729573, EPI_ISL_2729577, EPI_ISL_2729585, EPI_ISL_2729586, EPI_ISL_2729589, EPI_ISL_2729590, EPI_ISL_2729595, EPI_ISL_2729596, EPI_ISL_2729598, EPI_ISL_2729600, EPI_ISL_2729602, EPI_ISL_2729604, EPI_ISL_2729606, EPI_ISL_2729609, EPI_ISL_2729610, EPI_ISL_2729615, EPI_ISL_2729620, EPI_ISL_2729621, EPI_ISL_2729625, EPI_ISL_2729631, EPI_ISL_2729632, EPI_ISL_2729640, EPI_ISL_2729642, EPI_ISL_2729644, EPI_ISL_2729645, EPI_ISL_2729646, EPI_ISL_2729649, EPI_ISL_2729653, EPI_ISL_2729655, EPI_ISL_2729661, EPI_ISL_2729669, EPI_ISL_2729670, EPI_ISL_2729671, EPI_ISL_2729678, EPI_ISL_2729679, EPI_ISL_2729686, EPI_ISL_2729688, EPI_ISL_2729692, EPI_ISL_2729695, EPI_ISL_2729696, EPI_ISL_2729711, EPI_ISL_2729713, EPI_ISL_2729714, EPI_ISL_2729717, EPI_ISL_2729723, EPI_ISL_2729724, EPI_ISL_2729731, EPI_ISL_2729736, EPI_ISL_2729743, EPI_ISL_2729750, EPI_ISL_2729751, EPI_ISL_2729756, EPI_ISL_2729759, EPI_ISL_2729764, EPI_ISL_2729769, EPI_ISL_2729770, EPI_ISL_2729773, EPI_ISL_2729774, EPI_ISL_2729776, EPI_ISL_2729777, EPI_ISL_2729778, EPI_ISL_2729783, EPI_ISL_2729787, EPI_ISL_2729791, EPI_ISL_2729795, EPI_ISL_2729799, EPI_ISL_2729803, EPI_ISL_2729804, EPI_ISL_2729807, EPI_ISL_2729815, EPI_ISL_2729817, EPI_ISL_2729818, EPI_ISL_2729821, EPI_ISL_2729825, EPI_ISL_2729830, EPI_ISL_2729837, EPI_ISL_2729840, EPI_ISL_2729842, EPI_ISL_2729847, EPI_ISL_2729849, EPI_ISL_2729850, EPI_ISL_2729851, EPI_ISL_2729855, EPI_ISL_2729860, EPI_ISL_2729863, EPI_ISL_2729864, EPI_ISL_2729866, EPI_ISL_2729868, EPI_ISL_2729869, EPI_ISL_2729873, EPI_ISL_2729876, EPI_ISL_2729877, EPI_ISL_2729879, EPI_ISL_2729883, EPI_ISL_2729889, EPI_ISL_2729890, EPI_ISL_2729899, EPI_ISL_2729900, EPI_ISL_2729902, EPI_ISL_2729903, EPI_ISL_2729905, EPI_ISL_2729909, EPI_ISL_2729913, EPI_ISL_2729916, EPI_ISL_2729921, EPI_ISL_2729925, EPI_ISL_2729932, EPI_ISL_2729933, EPI_ISL_2729936, EPI_ISL_2730299, EPI_ISL_2730320, EPI_ISL_2730332, EPI_ISL_2730342, EPI_ISL_2730368, EPI_ISL_2730372, EPI_ISL_2730395, EPI_ISL_2730406, EPI_ISL_2730407, EPI_ISL_2730417, EPI_ISL_2730464, EPI_ISL_2730475, EPI_ISL_2730482, EPI_ISL_2730487, EPI_ISL_2730498, EPI_ISL_2730503, EPI_ISL_2730505, EPI_ISL_2730510, EPI_ISL_2730511, EPI_ISL_2730524, EPI_ISL_2730530, EPI_ISL_2730535, EPI_ISL_2730545, EPI_ISL_2730546, EPI_ISL_2730551, EPI_ISL_2730555, EPI_ISL_2730575, EPI_ISL_2730578, EPI_ISL_2730581, EPI_ISL_2730583, EPI_ISL_2730607, EPI_ISL_2730608, EPI_ISL_2730625, EPI_ISL_2730628, EPI_ISL_2730630, EPI_ISL_2730643, EPI_ISL_2730646, EPI_ISL_2730926, EPI_ISL_2730975, EPI_ISL_2765850, EPI_ISL_2767351, EPI_ISL_2767353, EPI_ISL_2767355, EPI_ISL_2767357, EPI_ISL_2767358, EPI_ISL_2767363, EPI_ISL_2767365, EPI_ISL_2767368, EPI_ISL_2767370, EPI_ISL_2767377, EPI_ISL_2767379, EPI_ISL_2767382, EPI_ISL_2767385, EPI_ISL_2767386, EPI_ISL_2767387, EPI_ISL_2767389, EPI_ISL_2767393, EPI_ISL_2767395, EPI_ISL_2767396, EPI_ISL_2767399, EPI_ISL_2767403, EPI_ISL_2767404, EPI_ISL_2767405, EPI_ISL_2767408, EPI_ISL_2767410, EPI_ISL_2767418, EPI_ISL_2767419, EPI_ISL_2767422, EPI_ISL_2767428, EPI_ISL_2767430, EPI_ISL_2767433, EPI_ISL_2767435, EPI_ISL_2767447, EPI_ISL_2767451, EPI_ISL_2767456, EPI_ISL_2767458, EPI_ISL_2767472, EPI_ISL_2767474, EPI_ISL_2767487, EPI_ISL_2767491, EPI_ISL_2767492, EPI_ISL_2767496, EPI_ISL_2767498, EPI_ISL_2767505, EPI_ISL_2767510, EPI_ISL_2767511, EPI_ISL_2767512, EPI_ISL_2767515, EPI_ISL_2767519, EPI_ISL_2767522, EPI_ISL_2767523, EPI_ISL_2767525, EPI_ISL_2767532, EPI_ISL_2767534, EPI_ISL_2767538, EPI_ISL_2767539, EPI_ISL_2767541, EPI_ISL_2767546, EPI_ISL_2767547, EPI_ISL_2767549, EPI_ISL_2767554, EPI_ISL_2767555, EPI_ISL_2767556, EPI_ISL_2767562, EPI_ISL_2767563, EPI_ISL_2767564, EPI_ISL_2767566, EPI_ISL_2767569, EPI_ISL_2767570, EPI_ISL_2767575, EPI_ISL_2767583, EPI_ISL_2767584, EPI_ISL_2767586, EPI_ISL_2767588, EPI_ISL_2767589, EPI_ISL_2767590, EPI_ISL_2767594, EPI_ISL_2767601, EPI_ISL_2767607, EPI_ISL_2767611, EPI_ISL_2767614, EPI_ISL_2767617, EPI_ISL_2767618, EPI_ISL_2767619, EPI_ISL_2767622, EPI_ISL_2767624, EPI_ISL_2767630, EPI_ISL_2767637, EPI_ISL_2767638, EPI_ISL_2767643, EPI_ISL_2767645, EPI_ISL_2767646, EPI_ISL_2767647, EPI_ISL_2767648, EPI_ISL_2767651, EPI_ISL_2767654, EPI_ISL_2767659, EPI_ISL_2767662, EPI_ISL_2767665, EPI_ISL_2767668, EPI_ISL_2767669, EPI_ISL_2767670, EPI_ISL_2767671, EPI_ISL_2767672, EPI_ISL_2767673, EPI_ISL_2767678, EPI_ISL_2767680, EPI_ISL_2767681, EPI_ISL_2767683, EPI_ISL_2767685, EPI_ISL_2767686, EPI_ISL_2767688, EPI_ISL_2767694, EPI_ISL_2767696, EPI_ISL_2767697, EPI_ISL_2767704, EPI_ISL_2767705, EPI_ISL_2767706, EPI_ISL_2767707, EPI_ISL_2767708, EPI_ISL_2767710, EPI_ISL_2767714, EPI_ISL_2767715, EPI_ISL_2767717, EPI_ISL_2767718, EPI_ISL_2767722, EPI_ISL_2767725, EPI_ISL_2767726, EPI_ISL_2767727, EPI_ISL_2767728, EPI_ISL_2767732, EPI_ISL_2767733, EPI_ISL_2767734, EPI_ISL_2767737, EPI_ISL_2767741, EPI_ISL_2767742, EPI_ISL_2767745, EPI_ISL_2767746, EPI_ISL_2767750, EPI_ISL_2767751, EPI_ISL_2767753, EPI_ISL_2767754, EPI_ISL_2767757, EPI_ISL_2767761, EPI_ISL_2767762, EPI_ISL_2767763, EPI_ISL_2767764, EPI_ISL_2767766, EPI_ISL_2767767, EPI_ISL_2767770, EPI_ISL_2767774, EPI_ISL_2767777, EPI_ISL_2767778, EPI_ISL_2767780, EPI_ISL_2767783, EPI_ISL_2767784, EPI_ISL_2767786, EPI_ISL_2767789, EPI_ISL_2767790, EPI_ISL_2767791, EPI_ISL_2767792, EPI_ISL_2767794, EPI_ISL_2767795, EPI_ISL_2767796, EPI_ISL_2767798, EPI_ISL_2767799, EPI_ISL_2767801, EPI_ISL_2767802, EPI_ISL_2767805, EPI_ISL_2767807, EPI_ISL_2767810, EPI_ISL_2767814, EPI_ISL_2767819, EPI_ISL_2767820, EPI_ISL_2767821, EPI_ISL_2767823, EPI_ISL_2767827, EPI_ISL_2767828, EPI_ISL_2767832, EPI_ISL_2767834, EPI_ISL_2767835, EPI_ISL_2767837, EPI_ISL_2767839, EPI_ISL_2767840, EPI_ISL_2767841, EPI_ISL_2767842, EPI_ISL_2767843, EPI_ISL_2767845, EPI_ISL_2767846, EPI_ISL_2767847, EPI_ISL_2767849, EPI_ISL_2767854, EPI_ISL_2767859, EPI_ISL_2767861, EPI_ISL_2767863, EPI_ISL_2767864, EPI_ISL_2767865, EPI_ISL_2767868, EPI_ISL_2767869, EPI_ISL_2767870, EPI_ISL_2767873, EPI_ISL_2767874, EPI_ISL_2767876, EPI_ISL_2767879, EPI_ISL_2767881, EPI_ISL_2767882, EPI_ISL_2767884, EPI_ISL_2767886, EPI_ISL_2767887, EPI_ISL_2767888, EPI_ISL_2767893, EPI_ISL_2767894, EPI_ISL_2767896, EPI_ISL_2767899, EPI_ISL_2767900, EPI_ISL_2767902, EPI_ISL_2767903, EPI_ISL_2767906, EPI_ISL_2767907, EPI_ISL_2767910, EPI_ISL_2767913, EPI_ISL_2767914, EPI_ISL_2767915, EPI_ISL_2767917, EPI_ISL_2767921, EPI_ISL_2767923, EPI_ISL_2767925, EPI_ISL_2767926, EPI_ISL_2767927, EPI_ISL_2767937, EPI_ISL_2767938, EPI_ISL_2767939, EPI_ISL_2767943, EPI_ISL_2767945, EPI_ISL_2767954, EPI_ISL_2767957, EPI_ISL_2767958, EPI_ISL_2767959, EPI_ISL_2767961, EPI_ISL_2767962, EPI_ISL_2767964, EPI_ISL_2767965, EPI_ISL_2767966, EPI_ISL_2767967, EPI_ISL_2767969, EPI_ISL_2767970, EPI_ISL_2767973, EPI_ISL_2767974, EPI_ISL_2767975, EPI_ISL_2767976, EPI_ISL_2767978, EPI_ISL_2767979, EPI_ISL_2767981, EPI_ISL_2767982, EPI_ISL_2767983, EPI_ISL_2767985, EPI_ISL_2767991, EPI_ISL_2767992, EPI_ISL_2767993, EPI_ISL_2767995, EPI_ISL_2767997, EPI_ISL_2767998, EPI_ISL_2780434, EPI_ISL_2780514, EPI_ISL_2780563, EPI_ISL_2780568, EPI_ISL_2780583, EPI_ISL_2780588, EPI_ISL_2780671, EPI_ISL_2780688, EPI_ISL_2780725, EPI_ISL_2780749, EPI_ISL_2780754, EPI_ISL_2780763, EPI_ISL_2781125, EPI_ISL_2781165, EPI_ISL_2781169, EPI_ISL_2781198, EPI_ISL_2781202, EPI_ISL_2781205, EPI_ISL_2781209, EPI_ISL_2781235, EPI_ISL_2781251, EPI_ISL_2781287, EPI_ISL_2781288, EPI_ISL_2781296, EPI_ISL_2781310, EPI_ISL_2781314, EPI_ISL_2781316, EPI_ISL_2781339, EPI_ISL_2781344, EPI_ISL_2781361, EPI_ISL_2781363, EPI_ISL_2781373, EPI_ISL_2781381, EPI_ISL_2781388, EPI_ISL_2781406, EPI_ISL_2781451, EPI_ISL_2781458, EPI_ISL_2781490, EPI_ISL_2781500, EPI_ISL_2781502, EPI_ISL_2781503, EPI_ISL_2781504, EPI_ISL_2781505, EPI_ISL_2781507, EPI_ISL_2781508, EPI_ISL_2781512, EPI_ISL_2781513, EPI_ISL_2781516, EPI_ISL_2781518, EPI_ISL_2781523, EPI_ISL_2781525, EPI_ISL_2781526, EPI_ISL_2781527, EPI_ISL_2781536, EPI_ISL_2781539, EPI_ISL_2781540, EPI_ISL_2781542, EPI_ISL_2781544, EPI_ISL_2781545, EPI_ISL_2781546, EPI_ISL_2781548, EPI_ISL_2781557, EPI_ISL_2781568, EPI_ISL_2781570, EPI_ISL_2781575, EPI_ISL_2781576, EPI_ISL_2781580, EPI_ISL_2781581, EPI_ISL_2781587, EPI_ISL_2781590, EPI_ISL_2781593, EPI_ISL_2781596, EPI_ISL_2781597, EPI_ISL_2781603, EPI_ISL_2781605, EPI_ISL_2781607, EPI_ISL_2781609, EPI_ISL_2781618, EPI_ISL_2781619, EPI_ISL_2781622, EPI_ISL_2781626, EPI_ISL_2781628, EPI_ISL_2781632, EPI_ISL_2781636, EPI_ISL_2781637, EPI_ISL_2781646, EPI_ISL_2781650, EPI_ISL_2781654, EPI_ISL_2781655, EPI_ISL_2781661, EPI_ISL_2781664, EPI_ISL_2781667, EPI_ISL_2781674, EPI_ISL_2781675, EPI_ISL_2781676, EPI_ISL_2781678, EPI_ISL_2781685, EPI_ISL_2781688, EPI_ISL_2781692, EPI_ISL_2781696, EPI_ISL_2781702, EPI_ISL_2781703, EPI_ISL_2781704, EPI_ISL_2781709, EPI_ISL_2781712, EPI_ISL_2781713, EPI_ISL_2781714, EPI_ISL_2781715, EPI_ISL_2781717, EPI_ISL_2781725, EPI_ISL_2781726, EPI_ISL_2781728, EPI_ISL_2781732, EPI_ISL_2781734, EPI_ISL_2781737, EPI_ISL_2781738, EPI_ISL_2781739, EPI_ISL_2781740, EPI_ISL_2781742, EPI_ISL_2781748, EPI_ISL_2781749, EPI_ISL_2781754, EPI_ISL_2781758, EPI_ISL_2781763, EPI_ISL_2781782, EPI_ISL_2781784, EPI_ISL_2781786, EPI_ISL_2781789, EPI_ISL_2781790, EPI_ISL_2781793, EPI_ISL_2781795, EPI_ISL_2781798, EPI_ISL_2781799, EPI_ISL_2781802, EPI_ISL_2781807, EPI_ISL_2781813, EPI_ISL_2781820, EPI_ISL_2781824, EPI_ISL_2781825, EPI_ISL_2781827, EPI_ISL_2781831, EPI_ISL_2781832, EPI_ISL_2781833, EPI_ISL_2781839, EPI_ISL_2781846, EPI_ISL_2781847, EPI_ISL_2781849, EPI_ISL_2781850, EPI_ISL_2781854, EPI_ISL_2781858, EPI_ISL_2781859, EPI_ISL_2781860, EPI_ISL_2781863, EPI_ISL_2781869, EPI_ISL_2781876, EPI_ISL_2781878, EPI_ISL_2781879, EPI_ISL_2781883, EPI_ISL_2781884, EPI_ISL_2781886, EPI_ISL_2781887, EPI_ISL_2781890, EPI_ISL_2781893, EPI_ISL_2781898, EPI_ISL_2781903, EPI_ISL_2781906, EPI_ISL_2781916, EPI_ISL_2781917, EPI_ISL_2781919, EPI_ISL_2781922, EPI_ISL_2781923, EPI_ISL_2781925, EPI_ISL_2781928, EPI_ISL_2781929, EPI_ISL_2781930, EPI_ISL_2781935, EPI_ISL_2781941, EPI_ISL_2781945, EPI_ISL_2781946, EPI_ISL_2781947, EPI_ISL_2781949, EPI_ISL_2781953, EPI_ISL_2781959, EPI_ISL_2781960, EPI_ISL_2781962, EPI_ISL_2781963, EPI_ISL_2781973, EPI_ISL_2781979, EPI_ISL_2781980, EPI_ISL_2781981, EPI_ISL_2781987, EPI_ISL_2781988, EPI_ISL_2781990, EPI_ISL_2781992, EPI_ISL_2781993, EPI_ISL_2781994, EPI_ISL_2781996, EPI_ISL_2781999, EPI_ISL_2782004, EPI_ISL_2782005, EPI_ISL_2782016, EPI_ISL_2782017, EPI_ISL_2782021, EPI_ISL_2782024, EPI_ISL_2782027, EPI_ISL_2782028, EPI_ISL_2782035, EPI_ISL_2782043, EPI_ISL_2782045, EPI_ISL_2782051, EPI_ISL_2782053, EPI_ISL_2782059, EPI_ISL_2782061, EPI_ISL_2782062, EPI_ISL_2782065, EPI_ISL_2782081, EPI_ISL_2782088, EPI_ISL_2782091, EPI_ISL_2782093, EPI_ISL_2782103, EPI_ISL_2782106, EPI_ISL_2782111, EPI_ISL_2782116, EPI_ISL_2782119, EPI_ISL_2782120, EPI_ISL_2782128, EPI_ISL_2782139, EPI_ISL_2782142, EPI_ISL_2782164, EPI_ISL_2782179, EPI_ISL_2782183, EPI_ISL_2782192, EPI_ISL_2782197, EPI_ISL_2782199, EPI_ISL_2782201, EPI_ISL_2782204, EPI_ISL_2782207, EPI_ISL_2782210, EPI_ISL_2782217, EPI_ISL_2782219, EPI_ISL_2782234, EPI_ISL_2782238, EPI_ISL_2782240, EPI_ISL_2782248, EPI_ISL_2782249, EPI_ISL_2782250, EPI_ISL_2782254, EPI_ISL_2782261, EPI_ISL_2782262, EPI_ISL_2782263, EPI_ISL_2782268, EPI_ISL_2782274, EPI_ISL_2782279, EPI_ISL_2782282, EPI_ISL_2782290, EPI_ISL_2782294, EPI_ISL_2782298, EPI_ISL_2782301, EPI_ISL_2782308, EPI_ISL_2782313, EPI_ISL_2807366, EPI_ISL_2807383, EPI_ISL_2807384, EPI_ISL_2807391, EPI_ISL_2807400, EPI_ISL_2807446, EPI_ISL_2807478, EPI_ISL_2807483, EPI_ISL_2807506, EPI_ISL_2807510, EPI_ISL_2807523, EPI_ISL_2807544, EPI_ISL_2807568, EPI_ISL_2807578, EPI_ISL_2807605, EPI_ISL_2807628, EPI_ISL_2807630, EPI_ISL_2807651, EPI_ISL_2807715, EPI_ISL_2808688, EPI_ISL_2808901, EPI_ISL_2809102, EPI_ISL_2809104, EPI_ISL_2809137, EPI_ISL_2809187, EPI_ISL_2809230, EPI_ISL_2809234, EPI_ISL_2809285, EPI_ISL_2809348, EPI_ISL_2809403, EPI_ISL_2809435, EPI_ISL_2809573, EPI_ISL_2809883, EPI_ISL_2809884, EPI_ISL_2809886, EPI_ISL_2809887, EPI_ISL_2809888, EPI_ISL_2809889, EPI_ISL_2809890, EPI_ISL_2809891, EPI_ISL_2809892, EPI_ISL_2809894, EPI_ISL_2809896, EPI_ISL_2809897, EPI_ISL_2809898, EPI_ISL_2809899, EPI_ISL_2809900, EPI_ISL_2809901, EPI_ISL_2809902, EPI_ISL_2809903, EPI_ISL_2809904, EPI_ISL_2809905, EPI_ISL_2809907, EPI_ISL_2809908, EPI_ISL_2809910, EPI_ISL_2809911, EPI_ISL_2809912, EPI_ISL_2809914, EPI_ISL_2809915, EPI_ISL_2809917, EPI_ISL_2809918, EPI_ISL_2809919, EPI_ISL_2809920, EPI_ISL_2809921, EPI_ISL_2809926, EPI_ISL_2809927, EPI_ISL_2809929, EPI_ISL_2809930, EPI_ISL_2809931, EPI_ISL_2809932, EPI_ISL_2809935, EPI_ISL_2809938, EPI_ISL_2809940, EPI_ISL_2809941, EPI_ISL_2809942, EPI_ISL_2809943, EPI_ISL_2809944, EPI_ISL_2809945, EPI_ISL_2809946, EPI_ISL_2809947, EPI_ISL_2809948, EPI_ISL_2809950, EPI_ISL_2809951, EPI_ISL_2809952, EPI_ISL_2809954, EPI_ISL_2809959, EPI_ISL_2809960, EPI_ISL_2809961, EPI_ISL_2809962, EPI_ISL_2809964, EPI_ISL_2809965, EPI_ISL_2809967, EPI_ISL_2809968, EPI_ISL_2809970, EPI_ISL_2809971, EPI_ISL_2809974, EPI_ISL_2809976, EPI_ISL_2809977, EPI_ISL_2809980, EPI_ISL_2809981, EPI_ISL_2809982, EPI_ISL_2809987, EPI_ISL_2809988, EPI_ISL_2809989, EPI_ISL_2809993, EPI_ISL_2809994, EPI_ISL_2809995, EPI_ISL_2809997, EPI_ISL_2810000, EPI_ISL_2810001, EPI_ISL_2810002, EPI_ISL_2810005, EPI_ISL_2810006, EPI_ISL_2810009, EPI_ISL_2810010, EPI_ISL_2810011, EPI_ISL_2810013, EPI_ISL_2810014, EPI_ISL_2810015, EPI_ISL_2810018, EPI_ISL_2810020, EPI_ISL_2810021, EPI_ISL_2813359, EPI_ISL_2813362, EPI_ISL_2813644, EPI_ISL_2832094, EPI_ISL_2832100

see above | Lighthouse Lab in Milton Keynes | Wellcome Sanger Institute for the COVID-19 Genomics UK (COG-UK) Consortium | Cordelia Langford; David K. Jackson; Dominic Kwiatkowski; Ewan Harrison; Ian Johnston; Jeffrey Barrett; John Sillitoe on behalf of the Wellcome Sanger Institute COVID-19 Surveillance Team; Roberto Amato; Sonia Goncalves; The Lighthouse Lab in Milton Keynes and Alex Alderton

EPI_ISL_2728912, EPI_ISL_2729010, EPI_ISL_2729656, EPI_ISL_2729660, EPI_ISL_2729662, EPI_ISL_2729665, EPI_ISL_2729666, EPI_ISL_2729668, EPI_ISL_2729675, EPI_ISL_2729676, EPI_ISL_2729680, EPI_ISL_2729684, EPI_ISL_2729689, EPI_ISL_2729700, EPI_ISL_2729703, EPI_ISL_2729705, EPI_ISL_2729707, EPI_ISL_2729708, EPI_ISL_2729710, EPI_ISL_2729715, EPI_ISL_2729716, EPI_ISL_2729718, EPI_ISL_2729719, EPI_ISL_2729722, EPI_ISL_2729732, EPI_ISL_2729735, EPI_ISL_2729738, EPI_ISL_2729741, EPI_ISL_2729745, EPI_ISL_2729746, EPI_ISL_2729747, EPI_ISL_2729748, EPI_ISL_2729749, EPI_ISL_2729752, EPI_ISL_2729757, EPI_ISL_2729758, EPI_ISL_2729761, EPI_ISL_2729762, EPI_ISL_2729763, EPI_ISL_2729765, EPI_ISL_2729766, EPI_ISL_2729775, EPI_ISL_2729779, EPI_ISL_2729781, EPI_ISL_2729784, EPI_ISL_2729785, EPI_ISL_2729790, EPI_ISL_2729792, EPI_ISL_2729793, EPI_ISL_2729796, EPI_ISL_2729797, EPI_ISL_2729798, EPI_ISL_2729808, EPI_ISL_2729809, EPI_ISL_2729816, EPI_ISL_2729826, EPI_ISL_2729828, EPI_ISL_2729829, EPI_ISL_2729833, EPI_ISL_2729834, EPI_ISL_2729836, EPI_ISL_2729839, EPI_ISL_2729841, EPI_ISL_2729844, EPI_ISL_2729858, EPI_ISL_2729859, EPI_ISL_2729861, EPI_ISL_2729862, EPI_ISL_2729871, EPI_ISL_2729872, EPI_ISL_2729878, EPI_ISL_2729882, EPI_ISL_2729887, EPI_ISL_2729888, EPI_ISL_2729893, EPI_ISL_2729894, EPI_ISL_2729896, EPI_ISL_2729897, EPI_ISL_2729898, EPI_ISL_2729901, EPI_ISL_2729904, EPI_ISL_2729907, EPI_ISL_2729911, EPI_ISL_2729912, EPI_ISL_2729915, EPI_ISL_2729923, EPI_ISL_2729924, EPI_ISL_2729928, EPI_ISL_2729929, EPI_ISL_2729930, EPI_ISL_2729934, EPI_ISL_2729954, EPI_ISL_2729971, EPI_ISL_2730054, EPI_ISL_2730076, EPI_ISL_2730107, EPI_ISL_2730129, EPI_ISL_2730138, EPI_ISL_2730146, EPI_ISL_2730203, EPI_ISL_2730239, EPI_ISL_2730240, EPI_ISL_2767352, EPI_ISL_2767356, EPI_ISL_2767359, EPI_ISL_2767361, EPI_ISL_2767362, EPI_ISL_2767364, EPI_ISL_2767369, EPI_ISL_2767373, EPI_ISL_2767378, EPI_ISL_2767383, EPI_ISL_2767390, EPI_ISL_2767394, EPI_ISL_2767400, EPI_ISL_2767402, EPI_ISL_2767412, EPI_ISL_2767413, EPI_ISL_2767414, EPI_ISL_2767415, EPI_ISL_2767417, EPI_ISL_2767420, EPI_ISL_2767423, EPI_ISL_2767426, EPI_ISL_2767431, EPI_ISL_2767434, EPI_ISL_2767439, EPI_ISL_2767441, EPI_ISL_2767443, EPI_ISL_2767444, EPI_ISL_2767445, EPI_ISL_2767446, EPI_ISL_2767450, EPI_ISL_2767454, EPI_ISL_2767457, EPI_ISL_2767460, EPI_ISL_2767463, EPI_ISL_2767467, EPI_ISL_2767470, EPI_ISL_2767471, EPI_ISL_2767473, EPI_ISL_2767476, EPI_ISL_2767478, EPI_ISL_2767481, EPI_ISL_2767485, EPI_ISL_2767497, EPI_ISL_2767501, EPI_ISL_2767502, EPI_ISL_2767506, EPI_ISL_2767507, EPI_ISL_2767509, EPI_ISL_2767520, EPI_ISL_2767526, EPI_ISL_2767531, EPI_ISL_2767533, EPI_ISL_2767535, EPI_ISL_2767545, EPI_ISL_2767548, EPI_ISL_2767550, EPI_ISL_2767553, EPI_ISL_2767557, EPI_ISL_2767561, EPI_ISL_2767568, EPI_ISL_2767571, EPI_ISL_2767576, EPI_ISL_2767578, EPI_ISL_2767580, EPI_ISL_2767587, EPI_ISL_2767591, EPI_ISL_2767593, EPI_ISL_2767608, EPI_ISL_2767612, EPI_ISL_2767616, EPI_ISL_2767621, EPI_ISL_2767631, EPI_ISL_2767632, EPI_ISL_2767636, EPI_ISL_2780315, EPI_ISL_2781495, EPI_ISL_2781501, EPI_ISL_2781522, EPI_ISL_2781554, EPI_ISL_2781572, EPI_ISL_2781583, EPI_ISL_2781591, EPI_ISL_2781599, EPI_ISL_2781600, EPI_ISL_2781611, EPI_ISL_2781635, EPI_ISL_2781642, EPI_ISL_2781647, EPI_ISL_2781665, EPI_ISL_2781680, EPI_ISL_2781684, EPI_ISL_2781689, EPI_ISL_2781690, EPI_ISL_2781691, EPI_ISL_2781694, EPI_ISL_2781719, EPI_ISL_2781756, EPI_ISL_2781764, EPI_ISL_2781779, EPI_ISL_2806919, EPI_ISL_2809075, EPI_ISL_2809855, EPI_ISL_2813370, EPI_ISL_2813419, EPI_ISL_2813474, EPI_ISL_2813492, EPI_ISL_2813499, EPI_ISL_2831325, EPI_ISL_2831327, EPI_ISL_2831328, EPI_ISL_2831329, EPI_ISL_2831333, EPI_ISL_2831337, EPI_ISL_2831338, EPI_ISL_2831339, EPI_ISL_2831340, EPI_ISL_2831343, EPI_ISL_2831351, EPI_ISL_2831353, EPI_ISL_2831356, EPI_ISL_2831359, EPI_ISL_2831363, EPI_ISL_2831364, EPI_ISL_2831365, EPI_ISL_2831366, EPI_ISL_2831367, EPI_ISL_2831370, EPI_ISL_2831371, EPI_ISL_2831376, EPI_ISL_2831378, EPI_ISL_2831379, EPI_ISL_2831381, EPI_ISL_2831386, EPI_ISL_2831387, EPI_ISL_2831389, EPI_ISL_2831408, EPI_ISL_2831421, EPI_ISL_2831424, EPI_ISL_2831428, EPI_ISL_2831431, EPI_ISL_2831434, EPI_ISL_2831438, EPI_ISL_2831439, EPI_ISL_2831445, EPI_ISL_2831448, EPI_ISL_2831451, EPI_ISL_2831454, EPI_ISL_2831459, EPI_ISL_2831460, EPI_ISL_2831461, EPI_ISL_2831465, EPI_ISL_2831470, EPI_ISL_2831478, EPI_ISL_2831479, EPI_ISL_2831482, EPI_ISL_2831484, EPI_ISL_2831486, EPI_ISL_2831488, EPI_ISL_2831489, EPI_ISL_2831490, EPI_ISL_2831491, EPI_ISL_2831492, EPI_ISL_2831494, EPI_ISL_2831498, EPI_ISL_2831502, EPI_ISL_2831507, EPI_ISL_2831510, EPI_ISL_2831512, EPI_ISL_2831517, EPI_ISL_2831520, EPI_ISL_2831526, EPI_ISL_2831527, EPI_ISL_2831528, EPI_ISL_2831529, EPI_ISL_2831530, EPI_ISL_2831531, EPI_ISL_2831535, EPI_ISL_2831539, EPI_ISL_2831540, EPI_ISL_2831541, EPI_ISL_2831544, EPI_ISL_2831551, EPI_ISL_2831552, EPI_ISL_2831558, EPI_ISL_2831559, EPI_ISL_2831564, EPI_ISL_2831570, EPI_ISL_2831572, EPI_ISL_2831574, EPI_ISL_2831575, EPI_ISL_2831580, EPI_ISL_2831581

see above | Lighthouse Laboratory Plymouth | Wellcome Sanger Institute for the COVID-19 Genomics UK (COG-UK) Consortium | Cordelia Langford; David K. Jackson; Dominic Kwiatkowski; Ewan Harrison; Ian Johnston; Jeffrey Barrett; John Sillitoe on behalf of the Wellcome Sanger Institute COVID-19 Surveillance Team; Lighthouse Laboratory Plymouth and Alex Alderton; Roberto Amato; Sonia Goncalves

EPI_ISL_2723164, EPI_ISL_2723165, EPI_ISL_2723167, EPI_ISL_2723168, EPI_ISL_2723169, EPI_ISL_2723170, EPI_ISL_2723171, EPI_ISL_2723173, EPI_ISL_2723174, EPI_ISL_2723175, EPI_ISL_2723178, EPI_ISL_2723180, EPI_ISL_2723181, EPI_ISL_2723183, EPI_ISL_2723184, EPI_ISL_2723186, EPI_ISL_2723187, EPI_ISL_2723188, EPI_ISL_2723189, EPI_ISL_2723190, EPI_ISL_2723191, EPI_ISL_2723193, EPI_ISL_2723195, EPI_ISL_2723196, EPI_ISL_2723197, EPI_ISL_2723198, EPI_ISL_2723201, EPI_ISL_2723202, EPI_ISL_2723205, EPI_ISL_2723207, EPI_ISL_2723208, EPI_ISL_2723210, EPI_ISL_2723215, EPI_ISL_2723216, EPI_ISL_2723217, EPI_ISL_2723218, EPI_ISL_2723219, EPI_ISL_2723220, EPI_ISL_2723221, EPI_ISL_2723222, EPI_ISL_2723223, EPI_ISL_2723224, EPI_ISL_2723225, EPI_ISL_2723226, EPI_ISL_2723227, EPI_ISL_2723228, EPI_ISL_2723229, EPI_ISL_2723230, EPI_ISL_2723231, EPI_ISL_2723232, EPI_ISL_2723233, EPI_ISL_2723236, EPI_ISL_2723237, EPI_ISL_2723238, EPI_ISL_2723242, EPI_ISL_2723244, EPI_ISL_2723245, EPI_ISL_2723246, EPI_ISL_2723251, EPI_ISL_2723252, EPI_ISL_2723253, EPI_ISL_2723255, EPI_ISL_2723256, EPI_ISL_2723258, EPI_ISL_2723259, EPI_ISL_2723260, EPI_ISL_2723261, EPI_ISL_2723262, EPI_ISL_2723263, EPI_ISL_2723265, EPI_ISL_2723266, EPI_ISL_2723267, EPI_ISL_2723269, EPI_ISL_2723270, EPI_ISL_2723272, EPI_ISL_2723274, EPI_ISL_2723277, EPI_ISL_2723278, EPI_ISL_2723280, EPI_ISL_2723281, EPI_ISL_2723282, EPI_ISL_2723283, EPI_ISL_2723284, EPI_ISL_2723287, EPI_ISL_2723288, EPI_ISL_2723289, EPI_ISL_2723290, EPI_ISL_2723291, EPI_ISL_2723292, EPI_ISL_2723294, EPI_ISL_2723295, EPI_ISL_2723296, EPI_ISL_2723298, EPI_ISL_2723299, EPI_ISL_2723300, EPI_ISL_2723301, EPI_ISL_2723304, EPI_ISL_2723305, EPI_ISL_2723306, EPI_ISL_2723307, EPI_ISL_2723308, EPI_ISL_2723310, EPI_ISL_2723311, EPI_ISL_2723313, EPI_ISL_2723314, EPI_ISL_2723315, EPI_ISL_2723316, EPI_ISL_2723317, EPI_ISL_2723318, EPI_ISL_2723319, EPI_ISL_2723320, EPI_ISL_2723321, EPI_ISL_2723322, EPI_ISL_2723323, EPI_ISL_2723325, EPI_ISL_2723326, EPI_ISL_2723327, EPI_ISL_2723328, EPI_ISL_2723329, EPI_ISL_2723330, EPI_ISL_2723331, EPI_ISL_2723333, EPI_ISL_2723334, EPI_ISL_2723335, EPI_ISL_2723336, EPI_ISL_2723337, EPI_ISL_2723338, EPI_ISL_2723339, EPI_ISL_2723340, EPI_ISL_2723341, EPI_ISL_2723342, EPI_ISL_2723344, EPI_ISL_2723345, EPI_ISL_2723346, EPI_ISL_2723347, EPI_ISL_2723348, EPI_ISL_2723349, EPI_ISL_2723350, EPI_ISL_2723351, EPI_ISL_2723352, EPI_ISL_2723353, EPI_ISL_2723354, EPI_ISL_2723355, EPI_ISL_2723356, EPI_ISL_2723358, EPI_ISL_2723359, EPI_ISL_2723360, EPI_ISL_2723361, EPI_ISL_2723362, EPI_ISL_2723363, EPI_ISL_2723364, EPI_ISL_2723365, EPI_ISL_2723366, EPI_ISL_2723367, EPI_ISL_2723368, EPI_ISL_2723369, EPI_ISL_2723370, EPI_ISL_2723371, EPI_ISL_2723372, EPI_ISL_2723373, EPI_ISL_2723374, EPI_ISL_2723375, EPI_ISL_2723376, EPI_ISL_2723377, EPI_ISL_2723378, EPI_ISL_2723379, EPI_ISL_2723380, EPI_ISL_2723381, EPI_ISL_2723382, EPI_ISL_2723383, EPI_ISL_2723384, EPI_ISL_2723385, EPI_ISL_2723386, EPI_ISL_2723387, EPI_ISL_2723388, EPI_ISL_2723389, EPI_ISL_2723390, EPI_ISL_2723391, EPI_ISL_2723392, EPI_ISL_2723393, EPI_ISL_2723394, EPI_ISL_2723396, EPI_ISL_2723397, EPI_ISL_2723398, EPI_ISL_2723399, EPI_ISL_2723400, EPI_ISL_2723401, EPI_ISL_2723402, EPI_ISL_2723403, EPI_ISL_2723404, EPI_ISL_2723405, EPI_ISL_2723406, EPI_ISL_2723407, EPI_ISL_2723408, EPI_ISL_2723409, EPI_ISL_2723412, EPI_ISL_2723413, EPI_ISL_2723414, EPI_ISL_2723415, EPI_ISL_2723416, EPI_ISL_2723417, EPI_ISL_2723418, EPI_ISL_2723419, EPI_ISL_2723420, EPI_ISL_2723421, EPI_ISL_2723422, EPI_ISL_2723423, EPI_ISL_2723424, EPI_ISL_2723425, EPI_ISL_2723426, EPI_ISL_2723427, EPI_ISL_2723428, EPI_ISL_2723429, EPI_ISL_2723430, EPI_ISL_2723431, EPI_ISL_2723432, EPI_ISL_2723433, EPI_ISL_2723434, EPI_ISL_2723435,

EPI_ISL_2723436, EPI_ISL_2723437, EPI_ISL_2723438, EPI_ISL_2723439, EPI_ISL_2723440, EPI_ISL_2723441, EPI_ISL_2723442, EPI_ISL_2723443, EPI_ISL_2723444, EPI_ISL_2723445, EPI_ISL_2723446, EPI_ISL_2723447, EPI_ISL_2723448, EPI_ISL_2723449, EPI_ISL_2723450, EPI_ISL_2723451, EPI_ISL_2723452, EPI_ISL_2723453, EPI_ISL_2723454, EPI_ISL_2723455, EPI_ISL_2723456, EPI_ISL_2723457, EPI_ISL_2723458, EPI_ISL_2723459, EPI_ISL_2723460, EPI_ISL_2723461, EPI_ISL_2723462, EPI_ISL_2723463, EPI_ISL_2723464, EPI_ISL_2723465, EPI_ISL_2723466, EPI_ISL_2723467, EPI_ISL_2723468, EPI_ISL_2723469, EPI_ISL_2723470, EPI_ISL_2723471, EPI_ISL_2723472, EPI_ISL_2723473, EPI_ISL_2723474, EPI_ISL_2723475, EPI_ISL_2723476, EPI_ISL_2723477, EPI_ISL_2723478, EPI_ISL_2723479, EPI_ISL_2723480, EPI_ISL_2723481, EPI_ISL_2723482, EPI_ISL_2723483, EPI_ISL_2723484, EPI_ISL_2723485, EPI_ISL_2723486, EPI_ISL_2723487, EPI_ISL_2723488, EPI_ISL_2723489, EPI_ISL_2723490, EPI_ISL_2723491, EPI_ISL_2723492, EPI_ISL_2723493, EPI_ISL_2723494, EPI_ISL_2723495, EPI_ISL_2723496, EPI_ISL_2723497, EPI_ISL_2723498, EPI_ISL_2723499, EPI_ISL_2723500, EPI_ISL_2723501, EPI_ISL_2723502, EPI_ISL_2723503, EPI_ISL_2723504, EPI_ISL_2723505, EPI_ISL_2723506, EPI_ISL_2723507, EPI_ISL_2723508, EPI_ISL_2723509, EPI_ISL_2723510, EPI_ISL_2723511, EPI_ISL_2723512, EPI_ISL_2723513, EPI_ISL_2723514, EPI_ISL_2723515, EPI_ISL_2723516, EPI_ISL_2723517, EPI_ISL_2723518, EPI_ISL_2723519, EPI_ISL_2723520, EPI_ISL_2723521, EPI_ISL_2723522, EPI_ISL_2723523, EPI_ISL_2723524, EPI_ISL_2723525, EPI_ISL_2723526, EPI_ISL_2723527, EPI_ISL_2723528, EPI_ISL_2723529, EPI_ISL_2723530, EPI_ISL_2723531, EPI_ISL_2723532, EPI_ISL_2723533, EPI_ISL_2723534, EPI_ISL_2723535, EPI_ISL_2723536, EPI_ISL_2723537, EPI_ISL_2723538, EPI_ISL_2723539, EPI_ISL_2723540, EPI_ISL_2723541, EPI_ISL_2723542, EPI_ISL_2723543, EPI_ISL_2723544, EPI_ISL_2723545, EPI_ISL_2723546, EPI_ISL_2723547

| | | | |
|---|---|---|---|
| see above | Northumbria University / South Tees Hospitals NHS Foundation Trust / North Cumbria Integrated Care NHS Foundation Trust / North Tees and Hartlepool NHS Foundation Trust / Newcastle Hospitals NHS Foundation Trust | COVID-19 Genomics UK (COG-UK) Consortium | Andrew Nelson; Brendan Payne; Clive Graham; Darren L Smith; Debra Padgett; Edward Barton; Emma Swindells; Garren Scott; Gary Black; Gary Eltringham; Giles S Holt; Greg R Young; Jane Greenaway; Jennifer Collins; John Allan; Joshua Loh; Lynn Dover; Matthew Bashton; Mohammad A Tariq; Paul Baker; Sarah Essex; Steve Liggett; Wen C Yew; Yusri Taha |

EPI_ISL_2750066, EPI_ISL_2750067, EPI_ISL_2750068, EPI_ISL_2750069, EPI_ISL_2750070, EPI_ISL_2750071, EPI_ISL_2750072, EPI_ISL_2750073, EPI_ISL_2750075, EPI_ISL_2750076, EPI_ISL_2750077, EPI_ISL_2750078, EPI_ISL_2750079, EPI_ISL_2750080, EPI_ISL_2750081, EPI_ISL_2750082, EPI_ISL_2750083, EPI_ISL_2750084, EPI_ISL_2750085, EPI_ISL_2750086, EPI_ISL_2750087, EPI_ISL_2750089, EPI_ISL_2750093, EPI_ISL_2750096, EPI_ISL_2750098, EPI_ISL_2750100, EPI_ISL_2750101, EPI_ISL_2750102, EPI_ISL_2750104, EPI_ISL_2750105, EPI_ISL_2750106, EPI_ISL_2750107, EPI_ISL_2750108, EPI_ISL_2750110, EPI_ISL_2750111, EPI_ISL_2750112, EPI_ISL_2750113, EPI_ISL_2750114, EPI_ISL_2750116, EPI_ISL_2750119, EPI_ISL_2750120, EPI_ISL_2750125, EPI_ISL_2750127, EPI_ISL_2750128, EPI_ISL_2750142, EPI_ISL_2750154, EPI_ISL_2750160, EPI_ISL_2750161, EPI_ISL_2750164, EPI_ISL_2750165, EPI_ISL_2750167, EPI_ISL_2750168, EPI_ISL_2750170, EPI_ISL_2750171, EPI_ISL_2750172, EPI_ISL_2750173, EPI_ISL_2750174, EPI_ISL_2750175, EPI_ISL_2750176, EPI_ISL_2750177, EPI_ISL_2750179, EPI_ISL_2750180, EPI_ISL_2750181, EPI_ISL_2750183, EPI_ISL_2750184, EPI_ISL_2750185, EPI_ISL_2750187, EPI_ISL_2750188, EPI_ISL_2750190, EPI_ISL_2750192, EPI_ISL_2750194, EPI_ISL_2750195, EPI_ISL_2750198, EPI_ISL_2750199, EPI_ISL_2750200, EPI_ISL_2750201, EPI_ISL_2750203, EPI_ISL_2750205, EPI_ISL_2750206, EPI_ISL_2750208, EPI_ISL_2750235, EPI_ISL_2750236, EPI_ISL_2750237, EPI_ISL_2750238, EPI_ISL_2750239, EPI_ISL_2750240, EPI_ISL_2750241, EPI_ISL_2750242, EPI_ISL_2750244, EPI_ISL_2750245, EPI_ISL_2750246, EPI_ISL_2750247, EPI_ISL_2750248, EPI_ISL_2750249, EPI_ISL_2750250, EPI_ISL_2750251, EPI_ISL_2750252, EPI_ISL_2750253, EPI_ISL_2750255, EPI_ISL_2750256, EPI_ISL_2750257, EPI_ISL_2750258, EPI_ISL_2750259, EPI_ISL_2750261, EPI_ISL_2750262, EPI_ISL_2750263, EPI_ISL_2750264, EPI_ISL_2750265, EPI_ISL_2750266, EPI_ISL_2750267, EPI_ISL_2750268, EPI_ISL_2750269, EPI_ISL_2750270, EPI_ISL_2750271, EPI_ISL_2750272, EPI_ISL_2750273, EPI_ISL_2750274, EPI_ISL_2750275, EPI_ISL_2750276, EPI_ISL_2750277, EPI_ISL_2750278, EPI_ISL_2750279, EPI_ISL_2750280, EPI_ISL_2750281, EPI_ISL_2750285, EPI_ISL_2750287, EPI_ISL_2750289, EPI_ISL_2750290, EPI_ISL_2750291, EPI_ISL_2750294, EPI_ISL_2750295, EPI_ISL_2750296, EPI_ISL_2750297, EPI_ISL_2750299, EPI_ISL_2750300, EPI_ISL_2750301, EPI_ISL_2750302, EPI_ISL_2750303, EPI_ISL_2750304, EPI_ISL_2750305, EPI_ISL_2750307, EPI_ISL_2750308, EPI_ISL_2750309, EPI_ISL_2750310, EPI_ISL_2750311, EPI_ISL_2750312, EPI_ISL_2750313, EPI_ISL_2750314, EPI_ISL_2750315, EPI_ISL_2750316, EPI_ISL_2750317, EPI_ISL_2750318, EPI_ISL_2750319, EPI_ISL_2750320, EPI_ISL_2750321, EPI_ISL_2750322, EPI_ISL_2750323, EPI_ISL_2750324, EPI_ISL_2750325, EPI_ISL_2750326, EPI_ISL_2750327, EPI_ISL_2750328, EPI_ISL_2750329, EPI_ISL_2750330, EPI_ISL_2750331, EPI_ISL_2750332, EPI_ISL_2750333, EPI_ISL_2750334, EPI_ISL_2750335, EPI_ISL_2750336, EPI_ISL_2750337, EPI_ISL_2750338, EPI_ISL_2750339, EPI_ISL_2750340, EPI_ISL_2750341, EPI_ISL_2750342, EPI_ISL_2750343, EPI_ISL_2750344, EPI_ISL_2750345, EPI_ISL_2750346, EPI_ISL_2750347, EPI_ISL_2750348, EPI_ISL_2750349, EPI_ISL_2750350, EPI_ISL_2750351, EPI_ISL_2750352, EPI_ISL_2750353, EPI_ISL_2750354, EPI_ISL_2750355, EPI_ISL_2750356, EPI_ISL_2750357, EPI_ISL_2750358, EPI_ISL_2750360, EPI_ISL_2750361, EPI_ISL_2750362, EPI_ISL_2750363, EPI_ISL_2750364, EPI_ISL_2750367, EPI_ISL_2750368, EPI_ISL_2750369, EPI_ISL_2750370, EPI_ISL_2750371, EPI_ISL_2750373, EPI_ISL_2750374, EPI_ISL_2750375, EPI_ISL_2750376, EPI_ISL_2750377, EPI_ISL_2750378, EPI_ISL_2750379, EPI_ISL_2750380, EPI_ISL_2750381, EPI_ISL_2750382, EPI_ISL_2750383, EPI_ISL_2750384, EPI_ISL_2750385, EPI_ISL_2750386, EPI_ISL_2750387, EPI_ISL_2750388, EPI_ISL_2750389, EPI_ISL_2750390, EPI_ISL_2750391, EPI_ISL_2750392, EPI_ISL_2750407, EPI_ISL_2750408, EPI_ISL_2750410, EPI_ISL_2750411, EPI_ISL_2750412, EPI_ISL_2750413, EPI_ISL_2750414, EPI_ISL_2750415, EPI_ISL_2750416, EPI_ISL_2750417, EPI_ISL_2750418, EPI_ISL_2750419, EPI_ISL_2750420, EPI_ISL_2750421, EPI_ISL_2750422, EPI_ISL_2750423, EPI_ISL_2750424, EPI_ISL_2750425, EPI_ISL_2750426, EPI_ISL_2750427, EPI_ISL_2750428, EPI_ISL_2750429, EPI_ISL_2750430, EPI_ISL_2750431, EPI_ISL_2750432, EPI_ISL_2750433, EPI_ISL_2750434, EPI_ISL_2750435, EPI_ISL_2750436, EPI_ISL_2750437, EPI_ISL_2750438, EPI_ISL_2750439, EPI_ISL_2750440, EPI_ISL_2750441, EPI_ISL_2750442, EPI_ISL_2750443, EPI_ISL_2750444, EPI_ISL_2750445, EPI_ISL_2750446, EPI_ISL_2750449, EPI_ISL_2750450, EPI_ISL_2750451, EPI_ISL_2750452, EPI_ISL_2750453, EPI_ISL_2750454, EPI_ISL_2750455, EPI_ISL_2750456, EPI_ISL_2750457, EPI_ISL_2750458, EPI_ISL_2750459, EPI_ISL_2750460, EPI_ISL_2750461, EPI_ISL_2750462, EPI_ISL_2750463, EPI_ISL_2750464, EPI_ISL_2750465, EPI_ISL_2750466, EPI_ISL_2750467, EPI_ISL_2750468, EPI_ISL_2750469, EPI_ISL_2750470, EPI_ISL_2750471, EPI_ISL_2750472, EPI_ISL_2750473, EPI_ISL_2750474, EPI_ISL_2750475, EPI_ISL_2750476, EPI_ISL_2750477, EPI_ISL_2750478, EPI_ISL_2750479, EPI_ISL_2750480, EPI_ISL_2750481, EPI_ISL_2750482, EPI_ISL_2750483, EPI_ISL_2750484, EPI_ISL_2750485, EPI_ISL_2750486, EPI_ISL_2750487, EPI_ISL_2750497, EPI_ISL_2750498, EPI_ISL_2750499, EPI_ISL_2750500, EPI_ISL_2750501, EPI_ISL_2750502, EPI_ISL_2750508, EPI_ISL_2750510, EPI_ISL_2750511, EPI_ISL_2750512, EPI_ISL_2750514, EPI_ISL_2750515, EPI_ISL_2750516, EPI_ISL_2750519, EPI_ISL_2750520, EPI_ISL_2750522, EPI_ISL_2750524, EPI_ISL_2750525, EPI_ISL_2750526, EPI_ISL_2750527, EPI_ISL_2750528, EPI_ISL_2750529, EPI_ISL_2750530, EPI_ISL_2750531, EPI_ISL_2750532, EPI_ISL_2750533, EPI_ISL_2750534, EPI_ISL_2750535, EPI_ISL_2750536, EPI_ISL_2750537, EPI_ISL_2750538, EPI_ISL_2750539, EPI_ISL_2750540, EPI_ISL_2750541, EPI_ISL_2750542, EPI_ISL_2750543, EPI_ISL_2750544, EPI_ISL_2750545, EPI_ISL_2750546, EPI_ISL_2750547, EPI_ISL_2750548, EPI_ISL_2750549, EPI_ISL_2750550, EPI_ISL_2750551, EPI_ISL_2750552, EPI_ISL_2750553, EPI_ISL_2750554, EPI_ISL_2750556, EPI_ISL_2750557, EPI_ISL_2750558, EPI_ISL_2750559, EPI_ISL_2750560, EPI_ISL_2750561, EPI_ISL_2750562, EPI_ISL_2750564, EPI_ISL_2750566, EPI_ISL_2750568, EPI_ISL_2750573, EPI_ISL_2750582, EPI_ISL_2750588, EPI_ISL_2750589, EPI_ISL_2750594, EPI_ISL_2750595, EPI_ISL_2750597, EPI_ISL_2750598, EPI_ISL_2750605, EPI_ISL_2750610, EPI_ISL_2750611, EPI_ISL_2750613, EPI_ISL_2750614, EPI_ISL_2750615, EPI_ISL_2750616, EPI_ISL_2750617, EPI_ISL_2750618, EPI_ISL_2750619, EPI_ISL_2750620, EPI_ISL_2750621, EPI_ISL_2750622, EPI_ISL_2750623, EPI_ISL_2750624, EPI_ISL_2750625, EPI_ISL_2750627, EPI_ISL_2750628, EPI_ISL_2750629, EPI_ISL_2750630, EPI_ISL_2750631, EPI_ISL_2750632, EPI_ISL_2750633, EPI_ISL_2750634, EPI_ISL_2750635, EPI_ISL_2750636, EPI_ISL_2750637, EPI_ISL_2750638, EPI_ISL_2750639, EPI_ISL_2750640, EPI_ISL_2750663, EPI_ISL_2750667, EPI_ISL_2750670, EPI_ISL_2750671, EPI_ISL_2750672, EPI_ISL_2750673, EPI_ISL_2750674, EPI_ISL_2750677, EPI_ISL_2750678, EPI_ISL_2750679, EPI_ISL_2750680, EPI_ISL_2750681, EPI_ISL_2750682, EPI_ISL_2750683, EPI_ISL_2750685, EPI_ISL_2818336, EPI_ISL_2818338, EPI_ISL_2818339, EPI_ISL_2818495, EPI_ISL_2818522, EPI_ISL_2818525, EPI_ISL_2818527, EPI_ISL_2818532, EPI_ISL_2818538, EPI_ISL_2818540, EPI_ISL_2818543, EPI_ISL_2818544, EPI_ISL_2818546, EPI_ISL_2818587, EPI_ISL_2818831, EPI_ISL_2818838, EPI_ISL_2818839, EPI_ISL_2818840, EPI_ISL_2818841, EPI_ISL_2818844, EPI_ISL_2818845, EPI_ISL_2818846, EPI_ISL_2818847, EPI_ISL_2818848, EPI_ISL_2818849, EPI_ISL_2818851, EPI_ISL_2818854, EPI_ISL_2818855, EPI_ISL_2818959, EPI_ISL_2818967, EPI_ISL_2818977, EPI_ISL_2818986, EPI_ISL_2818995, EPI_ISL_2819001, EPI_ISL_2819043, EPI_ISL_2819131, EPI_ISL_2819201, EPI_ISL_2819203, EPI_ISL_2819207, EPI_ISL_2819222, EPI_ISL_2819244, EPI_ISL_2819255, EPI_ISL_2819359, EPI_ISL_2819365, EPI_ISL_2819373, EPI_ISL_2819386, EPI_ISL_2819396, EPI_ISL_2819403, EPI_ISL_2819427, EPI_ISL_2819447, EPI_ISL_2819460, EPI_ISL_2819520, EPI_ISL_2819542, EPI_ISL_2819567, EPI_ISL_2819572, EPI_ISL_2905487, EPI_ISL_2905495, EPI_ISL_2906025, EPI_ISL_2906028, EPI_ISL_2906034, EPI_ISL_2906042, EPI_ISL_2906045, EPI_ISL_2906076, EPI_ISL_2906078, EPI_ISL_2906086, EPI_ISL_2906087, EPI_ISL_2906104, EPI_ISL_2906107, EPI_ISL_2906108, EPI_ISL_2906109, EPI_ISL_2906110, EPI_ISL_2906111, EPI_ISL_2906114, EPI_ISL_2906120, EPI_ISL_2906126, EPI_ISL_2906216, EPI_ISL_2906253, EPI_ISL_2906274, EPI_ISL_2906278, EPI_ISL_2906463, EPI_ISL_2906498, EPI_ISL_2906512

| | | | |
|---|---|---|---|
| see above | Originating lab: Wales Specialist Virology Centre Sequencing lab: Pathogen Genomics Unit | Public Health Wales Microbiology Cardiff Wales Specialist Virology Centre | Alec Birchley; Alexander Adams; Amy Gaskin; Angela Marchbank; Bree Gatica-Wilcox; Catherine Moore; Jason Coombes; Joanne Watkins; Joel Southgate; Johnathan Evans; Laura Gifford; Lauren Gilbert; Lee Graham; Malorie Perry; Matthew Bull; Nicole Pacchiarini; Sally Corden; Sara Kumziene-Summerhayes; Sara Rey; Sarah Taylor; Simon Cottrell; Sophie Jones; Tom Connor |

EPI_ISL_2903042, EPI_ISL_2903048, EPI_ISL_2903050, EPI_ISL_2903056, EPI_ISL_2903061, EPI_ISL_2903062, EPI_ISL_2903069, EPI_ISL_2903072, EPI_ISL_2903073, EPI_ISL_2903075, EPI_ISL_2903078, EPI_ISL_2903079, EPI_ISL_2903080, EPI_ISL_2903084, EPI_ISL_2903088, EPI_ISL_2903089, EPI_ISL_2903092, EPI_ISL_2903096, EPI_ISL_2903098, EPI_ISL_2903099, EPI_ISL_2903107, EPI_ISL_2903110, EPI_ISL_2903113, EPI_ISL_2903122, EPI_ISL_2903128, EPI_ISL_2903130, EPI_ISL_2903133, EPI_ISL_2903134, EPI_ISL_2903135, EPI_ISL_2903136, EPI_ISL_2903139, EPI_ISL_2903142, EPI_ISL_2903152, EPI_ISL_2903157

| | | | |
|---|---|---|---|
| see above | Oxford Viromics, NDM, University of Oxford; Oxford University Hospitals; Basingstoke and North Hampshire Hospital | COVID-19 Genomics UK (COG-UK) Consortium | Alex Mobbs; Amy Trebes; Anita Justice; Catrin Moore; Christophe Fraser; David Bonsall; David Buck; Emma Wise; George Macintyre; Jessica Lynch; John Todd; Mariateresa de Cesare; Matilde Mori; Monique Andersson; Nathan Moore; Nick Cortes; Robert Shaw; Stephen Kidd; Tanya Golubchik; Timothy Peto |

EPI_ISL_2817223, EPI_ISL_2817231, EPI_ISL_2817236, EPI_ISL_2817238, EPI_ISL_2817259, EPI_ISL_2817273, EPI_ISL_2817280

| | | | |
|---|---|---|---|
| see above | Queens Medical Centre, Clinical Microbiology Department / DeepSeq Nottingham | COVID-19 Genomics UK (COG-UK) Consortium | Christopher Moore; Emily Park; Fei Sang; Gemma Clark; Hannah Howson-Wells; Jack Hill; Jayasree Dey; Johnny Debebe; Jonathan Ball; Joseph Chappell; Manjinder Khakh; Matthew Carlisle; Matthew Loose; Michelle M Lister; Nadine Holmes; Patrick McClure; Theocharis Tsoleridis; Timothy Byaruhanga; Vicki M Fleming; Victoria Wright; Wendy Smith |

EPI_ISL_2746824, EPI_ISL_2746856, EPI_ISL_2746868, EPI_ISL_2746899, EPI_ISL_2746941, EPI_ISL_2746956, EPI_ISL_2746970, EPI_ISL_2746971, EPI_ISL_2746992, EPI_ISL_2747007, EPI_ISL_2747008, EPI_ISL_2747043, EPI_ISL_2747051, EPI_ISL_2747052, EPI_ISL_2747061, EPI_ISL_2747074, EPI_ISL_2747082, EPI_ISL_2747083, EPI_ISL_2747098, EPI_ISL_2747105, EPI_ISL_2747107, EPI_ISL_2747108, EPI_ISL_2747123, EPI_ISL_2747126, EPI_ISL_2747137, EPI_ISL_2747173, EPI_ISL_2747188, EPI_ISL_2747205, EPI_ISL_2747245, EPI_ISL_2747250, EPI_ISL_2747285, EPI_ISL_2747286, EPI_ISL_2747288, EPI_ISL_2747290, EPI_ISL_2747291, EPI_ISL_2747293, EPI_ISL_2747294, EPI_ISL_2747299, EPI_ISL_2747303, EPI_ISL_2747308, EPI_ISL_2747311, EPI_ISL_2747312, EPI_ISL_2747314, EPI_ISL_2747326, EPI_ISL_2747328, EPI_ISL_2747337, EPI_ISL_2747338, EPI_ISL_2747351, EPI_ISL_2747363, EPI_ISL_2747379, EPI_ISL_2747412, EPI_ISL_2747413, EPI_ISL_2747424, EPI_ISL_2747425, EPI_ISL_2747430, EPI_ISL_2747432, EPI_ISL_2747433, EPI_ISL_2747434, EPI_ISL_2747435, EPI_ISL_2747436, EPI_ISL_2747437, EPI_ISL_2747438, EPI_ISL_2747439, EPI_ISL_2747440, EPI_ISL_2747441, EPI_ISL_2747442, EPI_ISL_2747448, EPI_ISL_2747450, EPI_ISL_2747451, EPI_ISL_2747452, EPI_ISL_2747454, EPI_ISL_2747455, EPI_ISL_2747457, EPI_ISL_2747458, EPI_ISL_2747464, EPI_ISL_2747471, EPI_ISL_2747473, EPI_ISL_2747475, EPI_ISL_2747477, EPI_ISL_2747479, EPI_ISL_2747481, EPI_ISL_2747483, EPI_ISL_2747485, EPI_ISL_2747492, EPI_ISL_2747497, EPI_ISL_2747499, EPI_ISL_2747503, EPI_ISL_2747505, EPI_ISL_2747509, EPI_ISL_2747511, EPI_ISL_2747513, EPI_ISL_2747516, EPI_ISL_2747518, EPI_ISL_2747520, EPI_ISL_2747527, EPI_ISL_2747529, EPI_ISL_2747530, EPI_ISL_2747533, EPI_ISL_2747537, EPI_ISL_2747547, EPI_ISL_2747549, EPI_ISL_2747551, EPI_ISL_2747557, EPI_ISL_2747558, EPI_ISL_2747560, EPI_ISL_2747562, EPI_ISL_2747564, EPI_ISL_2747567, EPI_ISL_2747568, EPI_ISL_2747570, EPI_ISL_2747573, EPI_ISL_2747575, EPI_ISL_2747579, EPI_ISL_2747581, EPI_ISL_2747583, EPI_ISL_2747585, EPI_ISL_2747587, EPI_ISL_2747589, EPI_ISL_2747592, EPI_ISL_2747596, EPI_ISL_2747598, EPI_ISL_2747600, EPI_ISL_2747602, EPI_ISL_2747604, EPI_ISL_2747612, EPI_ISL_2747616, EPI_ISL_2747618, EPI_ISL_2747619, EPI_ISL_2747624, EPI_ISL_2747625, EPI_ISL_2747630, EPI_ISL_2747632, EPI_ISL_2747634, EPI_ISL_2747636, EPI_ISL_2747638, EPI_ISL_2747642, EPI_ISL_2747644, EPI_ISL_2747647, EPI_ISL_2747649, EPI_ISL_2747651, EPI_ISL_2747653, EPI_ISL_2747655, EPI_ISL_2747657, EPI_ISL_2747659, EPI_ISL_2747660, EPI_ISL_2747662, EPI_ISL_2747665, EPI_ISL_2747667, EPI_ISL_2747669, EPI_ISL_2747673, EPI_ISL_2747675, EPI_ISL_2747677, EPI_ISL_2747679, EPI_ISL_2747681, EPI_ISL_2747687, EPI_ISL_2747689, EPI_ISL_2747691, EPI_ISL_2747693, EPI_ISL_2747695, EPI_ISL_2747697, EPI_ISL_2747699, EPI_ISL_2747701, EPI_ISL_2747703, EPI_ISL_2747705, EPI_ISL_2747707, EPI_ISL_2747709, EPI_ISL_2747711, EPI_ISL_2747713, EPI_ISL_2747715, EPI_ISL_2747717, EPI_ISL_2747719, EPI_ISL_2747721, EPI_ISL_2747723, EPI_ISL_2747725, EPI_ISL_2747729, EPI_ISL_2747731, EPI_ISL_2747733, EPI_ISL_2747735, EPI_ISL_2747741, EPI_ISL_2747743, EPI_ISL_2747745, EPI_ISL_2747747, EPI_ISL_2747749, EPI_ISL_2747750, EPI_ISL_2747752, EPI_ISL_2747755, EPI_ISL_2747756, EPI_ISL_2747758, EPI_ISL_2747760, EPI_ISL_2747762, EPI_ISL_2747764, EPI_ISL_2747766, EPI_ISL_2747768, EPI_ISL_2747770, EPI_ISL_2747772, EPI_ISL_2747774, EPI_ISL_2747776, EPI_ISL_2747778, EPI_ISL_2747780, EPI_ISL_2747782, EPI_ISL_2747784, EPI_ISL_2747786, EPI_ISL_2747788, EPI_ISL_2747790, EPI_ISL_2747792, EPI_ISL_2747794, EPI_ISL_2747796, EPI_ISL_2747800, EPI_ISL_2747802, EPI_ISL_2747806, EPI_ISL_2747808, EPI_ISL_2747810, EPI_ISL_2747814, EPI_ISL_2747820, EPI_ISL_2747822, EPI_ISL_2747827, EPI_ISL_2747829, EPI_ISL_2747832, EPI_ISL_2747834, EPI_ISL_2747837, EPI_ISL_2747839, EPI_ISL_2747841, EPI_ISL_2747843, EPI_ISL_2747845, EPI_ISL_2747847, EPI_ISL_2747849, EPI_ISL_2747851, EPI_ISL_2747853, EPI_ISL_2747855, EPI_ISL_2747857, EPI_ISL_2747859, EPI_ISL_2747861, EPI_ISL_2747863, EPI_ISL_2747867, EPI_ISL_2747869, EPI_ISL_2747871, EPI_ISL_2747875, EPI_ISL_2747877, EPI_ISL_2747878, EPI_ISL_2747880, EPI_ISL_2747882, EPI_ISL_2747884, EPI_ISL_2747886, EPI_ISL_2747890, EPI_ISL_2747892, EPI_ISL_2747894, EPI_ISL_2747896, EPI_ISL_2747898, EPI_ISL_2747900, EPI_ISL_2747902, EPI_ISL_2747904, EPI_ISL_2747906, EPI_ISL_2747908, EPI_ISL_2747910, EPI_ISL_2747912, EPI_ISL_2747914, EPI_ISL_2747916, EPI_ISL_2747918, EPI_ISL_2747920, EPI_ISL_2747922, EPI_ISL_2747924, EPI_ISL_2747926, EPI_ISL_2747928, EPI_ISL_2747930, EPI_ISL_2747932, EPI_ISL_2747936, EPI_ISL_2747942, EPI_ISL_2747944, EPI_ISL_2747946, EPI_ISL_2747948, EPI_ISL_2747951, EPI_ISL_2747957, EPI_ISL_2747959, EPI_ISL_2747961, EPI_ISL_2747963, EPI_ISL_2747965, EPI_ISL_2747967, EPI_ISL_2747971, EPI_ISL_2747975, EPI_ISL_2747979, EPI_ISL_2747987, EPI_ISL_2747990, EPI_ISL_2747992, EPI_ISL_2747996, EPI_ISL_2747998, EPI_ISL_2748000, EPI_ISL_2748002, EPI_ISL_2748004, EPI_ISL_2748006, EPI_ISL_2748008, EPI_ISL_2748009, EPI_ISL_2748011, EPI_ISL_2748013, EPI_ISL_2748015, EPI_ISL_2748017, EPI_ISL_2748019, EPI_ISL_2748023, EPI_ISL_2748026, EPI_ISL_2748027, EPI_ISL_2748029, EPI_ISL_2748069, EPI_ISL_2748071, EPI_ISL_2748073, EPI_ISL_2748075, EPI_ISL_2748085, EPI_ISL_2748092, EPI_ISL_2748093, EPI_ISL_2748096, EPI_ISL_2748097, EPI_ISL_2748139, EPI_ISL_2748142, EPI_ISL_2748144, EPI_ISL_2748146, EPI_ISL_2748148, EPI_ISL_2748150, EPI_ISL_2748152, EPI_ISL_2817299, EPI_ISL_2817300, EPI_ISL_2817301, EPI_ISL_2817302, EPI_ISL_2817303, EPI_ISL_2817304, EPI_ISL_2817309, EPI_ISL_2817310, EPI_ISL_2817311, EPI_ISL_2817392, EPI_ISL_2817393, EPI_ISL_2817395, EPI_ISL_2817496, EPI_ISL_2817502, EPI_ISL_2817505, EPI_ISL_2817507, EPI_ISL_2817509, EPI_ISL_2817510, EPI_ISL_2817511, EPI_ISL_2817512, EPI_ISL_2817514, EPI_ISL_2817515, EPI_ISL_2817516, EPI_ISL_2817518, EPI_ISL_2817519, EPI_ISL_2817521, EPI_ISL_2817523, EPI_ISL_2817525, EPI_ISL_2817526, EPI_ISL_2817544, EPI_ISL_2817545, EPI_ISL_2817546, EPI_ISL_2817547, EPI_ISL_2817549, EPI_ISL_2817550, EPI_ISL_2817551, EPI_ISL_2817552, EPI_ISL_2817553, EPI_ISL_2817554, EPI_ISL_2817555, EPI_ISL_2817556, EPI_ISL_2817557, EPI_ISL_2817558, EPI_ISL_2817559, EPI_ISL_2817560, EPI_ISL_2817591, EPI_ISL_2817593, EPI_ISL_2817595, EPI_ISL_2817601, EPI_ISL_2817732, EPI_ISL_2817734, EPI_ISL_2817735, EPI_ISL_2817736, EPI_ISL_2817737, EPI_ISL_2817738, EPI_ISL_2817739, EPI_ISL_2817740, EPI_ISL_2817741, EPI_ISL_2817742, EPI_ISL_2817744, EPI_ISL_2817745, EPI_ISL_2817746, EPI_ISL_2817747, EPI_ISL_2817755, EPI_ISL_2817756, EPI_ISL_2817762, EPI_ISL_2817887, EPI_ISL_2817910, EPI_ISL_2817911, EPI_ISL_2818168, EPI_ISL_2818174, EPI_ISL_2818175, EPI_ISL_2818179, EPI_ISL_2818235, EPI_ISL_2818236, EPI_ISL_2818237, EPI_ISL_2818238, EPI_ISL_2818239, EPI_ISL_2818240, EPI_ISL_2818241, EPI_ISL_2818243, EPI_ISL_2818245, EPI_ISL_2818246, EPI_ISL_2818247, EPI_ISL_2818248, EPI_ISL_2818249, EPI_ISL_2818252, EPI_ISL_2818253, EPI_ISL_2818254, EPI_ISL_2818255, EPI_ISL_2818256, EPI_ISL_2818257, EPI_ISL_2818258, EPI_ISL_2818259, EPI_ISL_2818260, EPI_ISL_2818261, EPI_ISL_2818262, EPI_ISL_2818263, EPI_ISL_2818264, EPI_ISL_2818269, EPI_ISL_2818273, EPI_ISL_2818274, EPI_ISL_2818276, EPI_ISL_2818280, EPI_ISL_2904275, EPI_ISL_2904279

| | | | |
|---|---|---|---|
| see above | Respiratory Virus Unit, Microbiology Services Colindale, Public Health England | COVID-19 Genomics UK (COG-UK) Consortium | PHE Covid Sequencing Team |

EPI_ISL_2705851, EPI_ISL_2705852, EPI_ISL_2705853, EPI_ISL_2705854, EPI_ISL_2705855, EPI_ISL_2705856, EPI_ISL_2705857, EPI_ISL_2705858, EPI_ISL_2705859, EPI_ISL_2705860, EPI_ISL_2705861, EPI_ISL_2705862, EPI_ISL_2705863, EPI_ISL_2705864, EPI_ISL_2705865, EPI_ISL_2705866, EPI_ISL_2705867, EPI_ISL_2705868, EPI_ISL_2705869, EPI_ISL_2705870, EPI_ISL_2705871, EPI_ISL_2705872, EPI_ISL_2705873, EPI_ISL_2705874, EPI_ISL_2705875, EPI_ISL_2705876, EPI_ISL_2705877, EPI_ISL_2705878, EPI_ISL_2705879, EPI_ISL_2705880, EPI_ISL_2705882, EPI_ISL_2705883, EPI_ISL_2705884, EPI_ISL_2705885, EPI_ISL_2705886, EPI_ISL_2705887, EPI_ISL_2705888, EPI_ISL_2705889, EPI_ISL_2705890, EPI_ISL_2705891, EPI_ISL_2705892, EPI_ISL_2705893, EPI_ISL_2705894, EPI_ISL_2705895, EPI_ISL_2705896, EPI_ISL_2705897, EPI_ISL_2705898, EPI_ISL_2705899, EPI_ISL_2705900, EPI_ISL_2705901, EPI_ISL_2705902, EPI_ISL_2705903, EPI_ISL_2705904, EPI_ISL_2705905, EPI_ISL_2705906, EPI_ISL_2705907, EPI_ISL_2705908, EPI_ISL_2705909, EPI_ISL_2705910, EPI_ISL_2705911, EPI_ISL_2705912, EPI_ISL_2705913, EPI_ISL_2705914, EPI_ISL_2705915, EPI_ISL_2705916, EPI_ISL_2705917, EPI_ISL_2705918, EPI_ISL_2705919, EPI_ISL_2705920, EPI_ISL_2705921, EPI_ISL_2705922, EPI_ISL_2705923, EPI_ISL_2705924, EPI_ISL_2705925, EPI_ISL_2705926, EPI_ISL_2705927, EPI_ISL_2705928, EPI_ISL_2705929, EPI_ISL_2722999, EPI_ISL_2723000, EPI_ISL_2723001, EPI_ISL_2723002, EPI_ISL_2723003, EPI_ISL_2723004, EPI_ISL_2723005, EPI_ISL_2723006, EPI_ISL_2723007, EPI_ISL_2723008, EPI_ISL_2723009, EPI_ISL_2723010, EPI_ISL_2723011, EPI_ISL_2723012, EPI_ISL_2723013, EPI_ISL_2723014, EPI_ISL_2723015, EPI_ISL_2723016, EPI_ISL_2723017, EPI_ISL_2723018, EPI_ISL_2723019, EPI_ISL_2723020, EPI_ISL_2723021, EPI_ISL_2723022, EPI_ISL_2723023, EPI_ISL_2723024, EPI_ISL_2723025, EPI_ISL_2723026, EPI_ISL_2723027, EPI_ISL_2723028, EPI_ISL_2723029, EPI_ISL_2723030, EPI_ISL_2723031, EPI_ISL_2723032, EPI_ISL_2723033, EPI_ISL_2723034, EPI_ISL_2723035, EPI_ISL_2723036, EPI_ISL_2723037, EPI_ISL_2723038, EPI_ISL_2723039, EPI_ISL_2723040, EPI_ISL_2723041, EPI_ISL_2723042, EPI_ISL_2723043, EPI_ISL_2723044, EPI_ISL_2723045, EPI_ISL_2723046, EPI_ISL_2723047, EPI_ISL_2723048, EPI_ISL_2723049, EPI_ISL_2723050, EPI_ISL_2723051, EPI_ISL_2723052, EPI_ISL_2723053, EPI_ISL_2723054, EPI_ISL_2723055, EPI_ISL_2723056, EPI_ISL_2723057, EPI_ISL_2723058, EPI_ISL_2723059, EPI_ISL_2723060, EPI_ISL_2723061, EPI_ISL_2723062, EPI_ISL_2723063, EPI_ISL_2723064, EPI_ISL_2723065, EPI_ISL_2723066, EPI_ISL_2723067, EPI_ISL_2723068, EPI_ISL_2723069, EPI_ISL_2723070, EPI_ISL_2723071, EPI_ISL_2723072, EPI_ISL_2723073, EPI_ISL_2723074, EPI_ISL_2723075, EPI_ISL_2723076, EPI_ISL_2723077, EPI_ISL_2723078, EPI_ISL_2723079, EPI_ISL_2723080, EPI_ISL_2723081, EPI_ISL_2723082, EPI_ISL_2723083, EPI_ISL_2723084, EPI_ISL_2723085, EPI_ISL_2723086, EPI_ISL_2723087, EPI_ISL_2723088, EPI_ISL_2723089, EPI_ISL_2723090, EPI_ISL_2723091, EPI_ISL_2723092, EPI_ISL_2723093, EPI_ISL_2723094, EPI_ISL_2723095, EPI_ISL_2723096, EPI_ISL_2723097, EPI_ISL_2723098, EPI_ISL_2723099, EPI_ISL_2723100, EPI_ISL_2723101, EPI_ISL_2723102, EPI_ISL_2723103, EPI_ISL_2723104, EPI_ISL_2723105, EPI_ISL_2723106, EPI_ISL_2723107, EPI_ISL_2723108, EPI_ISL_2723109, EPI_ISL_2723110, EPI_ISL_2723111, EPI_ISL_2723112, EPI_ISL_2723113, EPI_ISL_2723114, EPI_ISL_2723115, EPI_ISL_2723116, EPI_ISL_2723117, EPI_ISL_2723118, EPI_ISL_2723119, EPI_ISL_2723120, EPI_ISL_2723121, EPI_ISL_2723122, EPI_ISL_2723123, EPI_ISL_2723124, EPI_ISL_2723125, EPI_ISL_2723126, EPI_ISL_2723127, EPI_ISL_2723128, EPI_ISL_2723129, EPI_ISL_2723130, EPI_ISL_2723131, EPI_ISL_2723132, EPI_ISL_2723133, EPI_ISL_2723134, EPI_ISL_2723135, EPI_ISL_2723136, EPI_ISL_2723137, EPI_ISL_2723138, EPI_ISL_2723139, EPI_ISL_2723140, EPI_ISL_2723141, EPI_ISL_2723142, EPI_ISL_2723143, EPI_ISL_2723144, EPI_ISL_2723145, EPI_ISL_2723146, EPI_ISL_2723147, EPI_ISL_2723148, EPI_ISL_2723149, EPI_ISL_2723150, EPI_ISL_2723151, EPI_ISL_2723152, EPI_ISL_2723153, EPI_ISL_2723154, EPI_ISL_2723155, EPI_ISL_2723156, EPI_ISL_2723157, EPI_ISL_2723158, EPI_ISL_2723159, EPI_ISL_2723160, EPI_ISL_2723161, EPI_ISL_2723162, EPI_ISL_2723163, EPI_ISL_2734990, EPI_ISL_2734991, EPI_ISL_2734992, EPI_ISL_2734993, EPI_ISL_2734994, EPI_ISL_2734995, EPI_ISL_2734996, EPI_ISL_2734997, EPI_ISL_2734998, EPI_ISL_2734999, EPI_ISL_2735000, EPI_ISL_2735001, EPI_ISL_2735002, EPI_ISL_2735003, EPI_ISL_2735004, EPI_ISL_2735005, EPI_ISL_2735006, EPI_ISL_2735007, EPI_ISL_2735008, EPI_ISL_2735009, EPI_ISL_2735010, EPI_ISL_2735011, EPI_ISL_2735012, EPI_ISL_2735013, EPI_ISL_2735014, EPI_ISL_2735015, EPI_ISL_2735016, EPI_ISL_2735017, EPI_ISL_2735018, EPI_ISL_2735019, EPI_ISL_2735020, EPI_ISL_2735021, EPI_ISL_2735022, EPI_ISL_2735023, EPI_ISL_2735024, EPI_ISL_2735025, EPI_ISL_2735026, EPI_ISL_2735027, EPI_ISL_2735028, EPI_ISL_2735029, EPI_ISL_2735030, EPI_ISL_2735031, EPI_ISL_2735032, EPI_ISL_2735033, EPI_ISL_2735034, EPI_ISL_2735035, EPI_ISL_2735036, EPI_ISL_2735037, EPI_ISL_2735038, EPI_ISL_2735039, EPI_ISL_2735040, EPI_ISL_2735041, EPI_ISL_2735042, EPI_ISL_2735043, EPI_ISL_2735044, EPI_ISL_2735045, EPI_ISL_2735046, EPI_ISL_2735047, EPI_ISL_2735048, EPI_ISL_2735049, EPI_ISL_2735050, EPI_ISL_2735051, EPI_ISL_2735052, EPI_ISL_2735053, EPI_ISL_2735054, EPI_ISL_2735055, EPI_ISL_2735056, EPI_ISL_2735057, EPI_ISL_2735058, EPI_ISL_2735059, EPI_ISL_2735060, EPI_ISL_2735061, EPI_ISL_2816284, EPI_ISL_2816286, EPI_ISL_2816287, EPI_ISL_2816289, EPI_ISL_2816291, EPI_ISL_2816293, EPI_ISL_2816295, EPI_ISL_2816296, EPI_ISL_2816297, EPI_ISL_2816299, EPI_ISL_2816300, EPI_ISL_2816302, EPI_ISL_2816303, EPI_ISL_2816305, EPI_ISL_2816307, EPI_ISL_2816308, EPI_ISL_2816310, EPI_ISL_2816312, EPI_ISL_2816313, EPI_ISL_2816315, EPI_ISL_2816317, EPI_ISL_2816319, EPI_ISL_2816320, EPI_ISL_2816322, EPI_ISL_2816324, EPI_ISL_2816325, EPI_ISL_2816327, EPI_ISL_2816329, EPI_ISL_2816330, EPI_ISL_2816332, EPI_ISL_2816333, EPI_ISL_2816335, EPI_ISL_2816337, EPI_ISL_2816338, EPI_ISL_2816340, EPI_ISL_2816342, EPI_ISL_2816343, EPI_ISL_2816345, EPI_ISL_2816347, EPI_ISL_2816348, EPI_ISL_2816350, EPI_ISL_2816352, EPI_ISL_2816353, EPI_ISL_2816355, EPI_ISL_2816357, EPI_ISL_2816358, EPI_ISL_2816360, EPI_ISL_2816362, EPI_ISL_2816364, EPI_ISL_2816365, EPI_ISL_2816367, EPI_ISL_2816369, EPI_ISL_2816370, EPI_ISL_2816372, EPI_ISL_2816374, EPI_ISL_2816375, EPI_ISL_2816377, EPI_ISL_2816379, EPI_ISL_2816381, EPI_ISL_2816382, EPI_ISL_2816384, EPI_ISL_2816386, EPI_ISL_2816387, EPI_ISL_2816389, EPI_ISL_2816391, EPI_ISL_2816392, EPI_ISL_2816394, EPI_ISL_2816396, EPI_ISL_2816397, EPI_ISL_2816399, EPI_ISL_2816400, EPI_ISL_2816402, EPI_ISL_2816404, EPI_ISL_2816405, EPI_ISL_2816407, EPI_ISL_2816409, EPI_ISL_2816411, EPI_ISL_2816412, EPI_ISL_2816426, EPI_ISL_2816438, EPI_ISL_2816472, EPI_ISL_2816474, EPI_ISL_2816498, EPI_ISL_2816503, EPI_ISL_2816508, EPI_ISL_2816538, EPI_ISL_2816544, EPI_ISL_2816552

| | | | |
|---|---|---|---|
| see above | University College London, Great Ormond Street Hospital for Children NHS Foundation Trust, Imperial College Healthcare NHS Trust | COVID-19 Genomics UK (COG-UK) Consortium | Alison Holmes; Charlotte Williams; Helena Tutill; Jacqueline Findlay; James Price; Judith Breuer; Julianne Brown; Kathryn Harris; Leysa Forrest; Marius Cotic; Mark Kristiansen; Nadua Bayzid; Paola Niola; Paola Resende Silva; Patricia Dyal; Paul Randell; Rachel Williams; Samuel Weeks; Sergi Castellano; Sunando Roy; Tony Brooks; Yasmin Panchbhaya |

EPI_ISL_2815420, EPI_ISL_2815424, EPI_ISL_2815511, EPI_ISL_2815522, EPI_ISL_2815524, EPI_ISL_2815531, EPI_ISL_2815533, EPI_ISL_2815534, EPI_ISL_2815536, EPI_ISL_2815538, EPI_ISL_2815539, EPI_ISL_2815540

| | | | |
|---|---|---|---|
| see above | University of Birmingham | COVID-19 Genomics UK (COG-UK) Consortium | Alex Richter; Andrew Bosworth; Andrew Bosworth. Queen Elizabeth Hospital: Anna Casey; Andrew D Beggs PHE Heartlands Lab: Husam Osman; Charlotte Poxon; Institute of Microbiology; Joanne Stockton; Josh Quick; Kasun Wanigasooriya; Mike Kidd; Nicholas Loman. University of Birmingham Testing Laboratory: Celina M Whalley; Oliver Pickles; Radoslaw Poplawski; Samuel Nicholls; University of Birmingham: Claire McMurray; Will Rowe |

EPI_ISL_2734349, EPI_ISL_2734377, EPI_ISL_2734378, EPI_ISL_2734379, EPI_ISL_2734380, EPI_ISL_2734381, EPI_ISL_2734382, EPI_ISL_2734383, EPI_ISL_2734384, EPI_ISL_2734385, EPI_ISL_2734387, EPI_ISL_2734389, EPI_ISL_2734390, EPI_ISL_2734402, EPI_ISL_2734405, EPI_ISL_2734406, EPI_ISL_2734408, EPI_ISL_2734409, EPI_ISL_2734410, EPI_ISL_2734411, EPI_ISL_2734412, EPI_ISL_2734413, EPI_ISL_2815988, EPI_ISL_2815989, EPI_ISL_2815990, EPI_ISL_2815991, EPI_ISL_2815993, EPI_ISL_2815995, EPI_ISL_2816003, EPI_ISL_2816027, EPI_ISL_2816028, EPI_ISL_2816030, EPI_ISL_2816038, EPI_ISL_2816039, EPI_ISL_2816040, EPI_ISL_2816041, EPI_ISL_2816042, EPI_ISL_2816043, EPI_ISL_2816044, EPI_ISL_2816049, EPI_ISL_2816072

| | | | |
|---|---|---|---|
| see above | Virology Department, Royal Infirmary of Edinburgh, NHS Lothian / School of Biological Sciences, University of Edinburgh | COVID-19 Genomics UK (COG-UK) Consortium | Colquhoun R; Cotton S; Dewar R; Gallagher A; Hill V; Jackson B; Maloney D; McCrone JT; McHugh M; O'Toole Á; Rambaut A; Rooke S; Scher E; Templeton K; Yu X |

EPI_ISL_2708438, EPI_ISL_2708439, EPI_ISL_2708443, EPI_ISL_2708454, EPI_ISL_2708456, EPI_ISL_2708459, EPI_ISL_2708461, EPI_ISL_2708462, EPI_ISL_2708463, EPI_ISL_2708477, EPI_ISL_2708481, EPI_ISL_2708482, EPI_ISL_2708483, EPI_ISL_2708492, EPI_ISL_2708496, EPI_ISL_2708506, EPI_ISL_2708511, EPI_ISL_2708512, EPI_ISL_2708517, EPI_ISL_2708524, EPI_ISL_2708528, EPI_ISL_2708530, EPI_ISL_2708531, EPI_ISL_2708541, EPI_ISL_2708545, EPI_ISL_2708548, EPI_ISL_2708550, EPI_ISL_2708551, EPI_ISL_2708565, EPI_ISL_2708566, EPI_ISL_2708567, EPI_ISL_2708568, EPI_ISL_2708574, EPI_ISL_2708576, EPI_ISL_2708577, EPI_ISL_2708580, EPI_ISL_2708585, EPI_ISL_2708600, EPI_ISL_2708608, EPI_ISL_2708618, EPI_ISL_2708626, EPI_ISL_2708629, EPI_ISL_2708631, EPI_ISL_2708632, EPI_ISL_2708634, EPI_ISL_2708640, EPI_ISL_2708643, EPI_ISL_2708644, EPI_ISL_2708645, EPI_ISL_2708648, EPI_ISL_2708654, EPI_ISL_2708657, EPI_ISL_2708661, EPI_ISL_2708664, EPI_ISL_2708686, EPI_ISL_2708705, EPI_ISL_2708714, EPI_ISL_2708719, EPI_ISL_2708727, EPI_ISL_2708733, EPI_ISL_2708737, EPI_ISL_2708738, EPI_ISL_2708744, EPI_ISL_2708748, EPI_ISL_2708763, EPI_ISL_2708772,

| | | | |
|---|---|---|---|
| EPI_ISL_2708784, EPI_ISL_2708802, EPI_ISL_2708805, EPI_ISL_2708811, EPI_ISL_2708816, EPI_ISL_2708817, EPI_ISL_2708826, EPI_ISL_2708829, EPI_ISL_2708830, EPI_ISL_2708839, EPI_ISL_2708840, EPI_ISL_2820142 | | | |
| see above | Virology Department, Sheffield Teaching Hospitals NHS Foundation Trust/Department of Infection, Immunity and Cardiovascular Disease, The Medical School, University of Sheffield | COVID-19 Genomics UK (COG-UK) Consortium | Adri Angyal; Alex Keeley; Benjamin Lindsey; Cariad Evans; Danielle Groves; Dave Partridge; Katie Johnson; Laura Carrilero; Luke Green; Matthew Parker; Matthew Wyles; Mehmet Yavuz; Mohammad Raza; Nikki Smith; Paul Parsons; Rachel Tucker; Rebecca Brown; Thushan de Silva |
| EPI_ISL_2815900, EPI_ISL_2815901, EPI_ISL_2815923, EPI_ISL_2815927, EPI_ISL_2815943, EPI_ISL_2815944, EPI_ISL_2815945, EPI_ISL_2815946, EPI_ISL_2815947, EPI_ISL_2815948, EPI_ISL_2815949, EPI_ISL_2815950, EPI_ISL_2815951, EPI_ISL_2815952, EPI_ISL_2815953, EPI_ISL_2815954, EPI_ISL_2815957, EPI_ISL_2815958, EPI_ISL_2815959, EPI_ISL_2815961, EPI_ISL_2815962, EPI_ISL_2815963, EPI_ISL_2815964, EPI_ISL_2815965, EPI_ISL_2900692, EPI_ISL_2900696, EPI_ISL_2900713, EPI_ISL_2900717, EPI_ISL_2900718, EPI_ISL_2900720, EPI_ISL_2900722, EPI_ISL_2900727, EPI_ISL_2900729, EPI_ISL_2900730, EPI_ISL_2900731, EPI_ISL_2900732, EPI_ISL_2900733, EPI_ISL_2900734, EPI_ISL_2900735, EPI_ISL_2900736, EPI_ISL_2900737, EPI_ISL_2900738, EPI_ISL_2900739 | | | |
| see above | West of Scotland Specialist Virology Centre, NHSGGC / MRC-University of Glasgow Centre for Virus Research | COVID-19 Genomics UK (COG-UK) Consortium | Alasdair MacLean; Alice Broos; Ana da Silva Filipe; Antonia Ho; Daniel Mair; David L Robertson; Emma Thomson; Guy Mollett; Ioulia Tsatsani; James Shepherd; Jenna Nichols; Joseph Hughes; Kathy Li; Kathy Smollett; Kyriaki Nomikou; Lily Tong; Matthew Holden; Natasha Johnson; Rachel Blacow; Richard Orton; Rory Gunson; Sarah McDonald; Sharif Shaaban; Sreenu Vattipally; Stephen Carmichael |

We gratefully acknowledge the following Authors from the Originating laboratories responsible for obtaining the specimens, as well as the Submitting laboratories where the genome data were generated and shared via GISAID, on which this research is based.

All Submitters of data may be contacted directly via www.gisaid.org

Authors are sorted alphabetically.

| Accession ID | Originating Laboratory | Submitting Laboratory | Authors |
|---|---|---|---|
| EPI_ISL_2795935, EPI_ISL_2795936, EPI_ISL_2795937, EPI_ISL_2795938, EPI_ISL_2795939, EPI_ISL_2795940, EPI_ISL_2795941, EPI_ISL_2795942, EPI_ISL_2795954, EPI_ISL_2795955, EPI_ISL_2795957, EPI_ISL_2795958, EPI_ISL_2795959, EPI_ISL_2795960, EPI_ISL_2795961, EPI_ISL_2795962, EPI_ISL_2795963, EPI_ISL_2795964, EPI_ISL_2795965 | | | |
| see above | Amazon Dx, AUV2 Laboratory | Amazon Dx, AUV2 Laboratory | Luke Meredith |
| EPI_ISL_2819851, EPI_ISL_2819852, EPI_ISL_2819853, EPI_ISL_2819854, EPI_ISL_2819855, EPI_ISL_2819856, EPI_ISL_2819857, EPI_ISL_2819858, EPI_ISL_2819859, EPI_ISL_2819860, EPI_ISL_2819862, EPI_ISL_2819863, EPI_ISL_2819865, EPI_ISL_2819871, EPI_ISL_2819872, EPI_ISL_2819874, EPI_ISL_2819875, EPI_ISL_2819876, EPI_ISL_2819881, EPI_ISL_2819882, EPI_ISL_2819883, EPI_ISL_2819885, EPI_ISL_2819886, EPI_ISL_2819887, EPI_ISL_2819888, EPI_ISL_2819890, EPI_ISL_2819891, EPI_ISL_2819892, EPI_ISL_2819893, EPI_ISL_2819894, EPI_ISL_2819895, EPI_ISL_2819896, EPI_ISL_2819897, EPI_ISL_2819898, EPI_ISL_2819899, EPI_ISL_2819900, EPI_ISL_2819901, EPI_ISL_2819902, EPI_ISL_2819903, EPI_ISL_2819904, EPI_ISL_2819905, EPI_ISL_2819906, EPI_ISL_2819907, EPI_ISL_2819908, EPI_ISL_2819909, EPI_ISL_2819910, EPI_ISL_2819912, EPI_ISL_2819913, EPI_ISL_2819914, EPI_ISL_2819916, EPI_ISL_2819917, EPI_ISL_2819918, EPI_ISL_2906736, EPI_ISL_2906738, EPI_ISL_2906739, EPI_ISL_2906741, EPI_ISL_2906745, EPI_ISL_2906747, EPI_ISL_2906749, EPI_ISL_2906751, EPI_ISL_2906752, EPI_ISL_2906754, EPI_ISL_2906755, EPI_ISL_2906756, EPI_ISL_2906758, EPI_ISL_2906760, EPI_ISL_2906763, EPI_ISL_2906764, EPI_ISL_2906765 | | | |
| see above | Centre for Enzyme Innovation, University of Portsmouth / Translational Research Laboratory, Portsmouth Hospitals NHS Trust / Hampshire Hospitals NHS Foundation Trust / Bournemouth University / University Hospitals Dorset NHS Foundation Trust / Brighton and Sussex University Hospitals NHS Trust / Dartford and Gravesham NHS Trust / Isle of Wight NHS Trust / Maidstone and Tunbridge Wells NHS Trust / University Hospital Southampton NHS Foundation Trust | COVID-19 Genomics UK (COG-UK) Consortium | Adhyana Mahanama; Allyson Lloyd; Andrew Butt; Angela Beckett; Anibolina Castigador; Anna Mantzaouratou; Anoop Chauhan; Ben Macklin; Benjamin Cogger; Buddhini Samaraweera; Cassandra Malone; Christopher Fearn; Claire Thomas; Dorian Crudgington; Eleri Wilson-Davies; Emanuela Pelosi; Emily Macnaughton; Helen Umpleby; Helen Wheeler; Jacqui Prieto; Jonnie Seymour; Karen Withell; Kate Cook; Katie Loveson; Kelly Bicknell; Kevin Tucker; Kordo Saeed; Liz Sheridan; Magdalena Barrow; Matthew Harvey; Mohammed Hassan-Ibrahim; Nathan Moore; Nick Cortes; Rachael Jeremiah; Robert Impey; Salman Goudarzi; Samirakhon Raupova; Samuel Robson; Sarah Buchan; Sarah Jeremiah; Sarah Wyllie; Scott Elliott; Sharon Glaysher; Siona Silvieri; Stephen Aplin; Stephen Kidd; Thea Sass |
| EPI_ISL_2722297, EPI_ISL_2722299, EPI_ISL_2722356, EPI_ISL_2722357, EPI_ISL_2722358, EPI_ISL_2722359, EPI_ISL_2722360, EPI_ISL_2722362, EPI_ISL_2722363, EPI_ISL_2815638, EPI_ISL_2815639, EPI_ISL_2815645, EPI_ISL_2815646, EPI_ISL_2815649, EPI_ISL_2815660, EPI_ISL_2815687, EPI_ISL_2815688, EPI_ISL_2815689, EPI_ISL_2815690, EPI_ISL_2815691, EPI_ISL_2815692, EPI_ISL_2815693, EPI_ISL_2815694, EPI_ISL_2815695, EPI_ISL_2815696, EPI_ISL_2815697, EPI_ISL_2815698, EPI_ISL_2815699, EPI_ISL_2815700, EPI_ISL_2815701, EPI_ISL_2815702, EPI_ISL_2815703, EPI_ISL_2815704, EPI_ISL_2815841, EPI_ISL_2815849 | | | |
| see above | Department of Pathology, University of Cambridge | COVID-19 Genomics UK (COG-UK) Consortium | Aminu S. Jahun; Ian Goodfellow; Iliana Georgana; Martin D. Curran; Myra Hosmillo; Rhys Izuagbe; Surendra Parmar; William L. Hamilton; Yasmin Chaudhry |
| EPI_ISL_2765871, EPI_ISL_2765879, EPI_ISL_2765882, EPI_ISL_2765899, EPI_ISL_2765904, EPI_ISL_2765908, EPI_ISL_2765916, EPI_ISL_2765924, EPI_ISL_2765925, EPI_ISL_2765926, EPI_ISL_2765927, EPI_ISL_2765929, EPI_ISL_2765932, EPI_ISL_2765934, EPI_ISL_2765935, EPI_ISL_2765937, EPI_ISL_2765945, EPI_ISL_2765949, EPI_ISL_2765950, EPI_ISL_2765954, EPI_ISL_2765955, EPI_ISL_2765960, EPI_ISL_2765961, EPI_ISL_2765964, EPI_ISL_2765965, EPI_ISL_2765966, EPI_ISL_2765967, EPI_ISL_2765969, EPI_ISL_2765971, EPI_ISL_2765977, EPI_ISL_2765983, EPI_ISL_2765984, EPI_ISL_2765987, EPI_ISL_2765989, EPI_ISL_2765991, EPI_ISL_2765992, EPI_ISL_2765994, EPI_ISL_2765995, EPI_ISL_2765997, EPI_ISL_2765998, EPI_ISL_2766001, EPI_ISL_2766003, EPI_ISL_2766009, EPI_ISL_2766020, EPI_ISL_2766028, EPI_ISL_2766031, EPI_ISL_2766032, EPI_ISL_2766033, EPI_ISL_2766038, EPI_ISL_2766039, EPI_ISL_2766040, EPI_ISL_2766042, EPI_ISL_2766061, EPI_ISL_2766069, EPI_ISL_2766075, EPI_ISL_2766081, EPI_ISL_2766085, EPI_ISL_2766086, EPI_ISL_2766091, EPI_ISL_2766097, EPI_ISL_2766098, EPI_ISL_2766106, EPI_ISL_2766110, EPI_ISL_2766111, EPI_ISL_2766112, EPI_ISL_2766113, EPI_ISL_2766118, EPI_ISL_2766119, EPI_ISL_2766129, EPI_ISL_2766135, EPI_ISL_2766137, EPI_ISL_2766142, EPI_ISL_2766143, EPI_ISL_2766144, EPI_ISL_2766145, EPI_ISL_2766148, EPI_ISL_2766149, EPI_ISL_2766156, EPI_ISL_2766159, EPI_ISL_2766160, EPI_ISL_2766162, EPI_ISL_2766169, EPI_ISL_2766170, EPI_ISL_2766174, EPI_ISL_2766178, EPI_ISL_2766184, EPI_ISL_2766187, EPI_ISL_2766197, EPI_ISL_2766199, EPI_ISL_2766201, EPI_ISL_2766202, EPI_ISL_2766212, EPI_ISL_2766214, EPI_ISL_2766220, EPI_ISL_2766221, EPI_ISL_2766222, EPI_ISL_2766225, EPI_ISL_2766228, EPI_ISL_2766243, EPI_ISL_2766244, EPI_ISL_2766250, EPI_ISL_2766255, EPI_ISL_2766256, EPI_ISL_2766261, EPI_ISL_2766265, EPI_ISL_2766273, EPI_ISL_2766274, EPI_ISL_2766280, EPI_ISL_2766282, EPI_ISL_2766283, EPI_ISL_2766284, EPI_ISL_2766285, EPI_ISL_2766288, EPI_ISL_2766297, EPI_ISL_2766300, EPI_ISL_2766311, EPI_ISL_2766313, EPI_ISL_2766315, EPI_ISL_2766317, EPI_ISL_2766318, EPI_ISL_2766319, EPI_ISL_2766320, EPI_ISL_2766321, EPI_ISL_2766322, EPI_ISL_2766324, EPI_ISL_2766327, EPI_ISL_2766341, EPI_ISL_2766344, EPI_ISL_2766349, EPI_ISL_2766355, EPI_ISL_2766357, EPI_ISL_2766358, EPI_ISL_2766360, EPI_ISL_2766361, EPI_ISL_2766367, EPI_ISL_2766368, EPI_ISL_2766371, EPI_ISL_2766381, EPI_ISL_2766388, EPI_ISL_2766396, EPI_ISL_2766402, EPI_ISL_2766403, EPI_ISL_2766410, EPI_ISL_2766412, EPI_ISL_2766415, EPI_ISL_2766417, EPI_ISL_2766420, EPI_ISL_2766422, EPI_ISL_2766424, EPI_ISL_2766428, EPI_ISL_2766435, EPI_ISL_2766438, EPI_ISL_2766449, EPI_ISL_2766450, EPI_ISL_2766452, EPI_ISL_2766453, EPI_ISL_2766466, EPI_ISL_2766471, EPI_ISL_2766475, EPI_ISL_2766476, EPI_ISL_2766478, EPI_ISL_2766481, EPI_ISL_2766484, EPI_ISL_2766496, EPI_ISL_2766499, EPI_ISL_2766508, EPI_ISL_2766511, EPI_ISL_2766512, EPI_ISL_2766519, EPI_ISL_2766521, EPI_ISL_2766522, EPI_ISL_2766526, EPI_ISL_2766531, EPI_ISL_2766532, EPI_ISL_2766536, EPI_ISL_2766540, EPI_ISL_2766544, EPI_ISL_2779685, EPI_ISL_2781769, EPI_ISL_2804046, EPI_ISL_2804346, EPI_ISL_2806219, EPI_ISL_2806245, EPI_ISL_2806256, EPI_ISL_2806313, EPI_ISL_2806340, EPI_ISL_2806362, EPI_ISL_2806377, EPI_ISL_2806378, EPI_ISL_2806389, EPI_ISL_2806423, EPI_ISL_2806443, EPI_ISL_2806448, EPI_ISL_2806449, EPI_ISL_2806450, EPI_ISL_2806452, EPI_ISL_2806454, EPI_ISL_2806457, EPI_ISL_2806468, EPI_ISL_2806473, EPI_ISL_2806474, EPI_ISL_2806475, EPI_ISL_2806480, EPI_ISL_2806483, EPI_ISL_2806491, EPI_ISL_2806494, EPI_ISL_2806497, EPI_ISL_2806500, EPI_ISL_2806505, EPI_ISL_2806508, EPI_ISL_2806509, EPI_ISL_2806513, EPI_ISL_2806514, EPI_ISL_2806518, EPI_ISL_2806528, EPI_ISL_2806530, EPI_ISL_2806535, EPI_ISL_2806537, EPI_ISL_2806538, EPI_ISL_2806539, EPI_ISL_2806540, EPI_ISL_2806549, EPI_ISL_2806551, EPI_ISL_2806553, EPI_ISL_2806557, EPI_ISL_2806558, EPI_ISL_2806565, EPI_ISL_2806567, EPI_ISL_2806569, EPI_ISL_2806571, EPI_ISL_2806574, EPI_ISL_2806576, EPI_ISL_2806578, EPI_ISL_2806580, EPI_ISL_2806585, EPI_ISL_2806588, EPI_ISL_2806602, EPI_ISL_2806608, EPI_ISL_2806617, EPI_ISL_2806623, EPI_ISL_2806625, EPI_ISL_2806627, EPI_ISL_2806632, EPI_ISL_2806633, EPI_ISL_2806634, EPI_ISL_2806638, EPI_ISL_2806640, EPI_ISL_2806645, EPI_ISL_2806647, EPI_ISL_2806655, EPI_ISL_2806661, EPI_ISL_2806664, EPI_ISL_2806670, EPI_ISL_2806677, EPI_ISL_2806685, EPI_ISL_2806693, EPI_ISL_2806694, EPI_ISL_2806695, EPI_ISL_2806696, EPI_ISL_2806700, EPI_ISL_2806703, EPI_ISL_2806705, EPI_ISL_2806706, EPI_ISL_2806708, EPI_ISL_2806722, EPI_ISL_2806723, EPI_ISL_2806725, EPI_ISL_2806727, EPI_ISL_2806729, EPI_ISL_2806731, EPI_ISL_2806736, EPI_ISL_2806739, EPI_ISL_2806743, EPI_ISL_2806754, EPI_ISL_2806755, EPI_ISL_2806759, EPI_ISL_2806760, EPI_ISL_2806763, EPI_ISL_2806765, EPI_ISL_2806768, EPI_ISL_2806774, EPI_ISL_2806786, EPI_ISL_2806789, EPI_ISL_2806790, EPI_ISL_2806791, EPI_ISL_2806792, EPI_ISL_2806793, EPI_ISL_2808072, EPI_ISL_2808073, EPI_ISL_2808077, EPI_ISL_2808079, EPI_ISL_2808080, EPI_ISL_2808084, EPI_ISL_2808089, EPI_ISL_2808093, EPI_ISL_2808099, EPI_ISL_2808100, EPI_ISL_2808104, EPI_ISL_2808117, EPI_ISL_2808118, EPI_ISL_2808119, EPI_ISL_2808121, EPI_ISL_2808123, EPI_ISL_2808124, EPI_ISL_2808125, EPI_ISL_2808126, EPI_ISL_2808129, EPI_ISL_2808144, EPI_ISL_2808148, EPI_ISL_2808150, EPI_ISL_2808156, EPI_ISL_2808158, EPI_ISL_2808168, EPI_ISL_2808172, EPI_ISL_2808174, EPI_ISL_2808178, EPI_ISL_2808187, EPI_ISL_2808189, EPI_ISL_2808198, EPI_ISL_2808200, EPI_ISL_2808210, EPI_ISL_2808216, EPI_ISL_2808220, EPI_ISL_2808224, EPI_ISL_2808225, EPI_ISL_2808233, EPI_ISL_2808237, EPI_ISL_2808238, EPI_ISL_2808240, EPI_ISL_2808241, EPI_ISL_2808256, EPI_ISL_2808261, EPI_ISL_2808264, EPI_ISL_2808266, EPI_ISL_2808272, EPI_ISL_2808277, EPI_ISL_2808290, EPI_ISL_2808293, EPI_ISL_2808301, EPI_ISL_2808310, EPI_ISL_2808315, EPI_ISL_2808326, EPI_ISL_2808329, EPI_ISL_2808332, EPI_ISL_2808341, EPI_ISL_2808344, EPI_ISL_2808350, EPI_ISL_2808352, EPI_ISL_2808353, EPI_ISL_2808359, EPI_ISL_2808360, EPI_ISL_2808361, EPI_ISL_2808382, EPI_ISL_2808391, EPI_ISL_2808397, EPI_ISL_2808399, EPI_ISL_2808400, EPI_ISL_2808423, EPI_ISL_2808463, EPI_ISL_2808477, EPI_ISL_2808517, EPI_ISL_2808522, EPI_ISL_2808528, EPI_ISL_2808562, EPI_ISL_2808567, EPI_ISL_2808570, EPI_ISL_2808586, EPI_ISL_2808600, EPI_ISL_2808613, EPI_ISL_2808617, EPI_ISL_2808623, EPI_ISL_2808630, EPI_ISL_2808648, EPI_ISL_2808654, EPI_ISL_2808661, EPI_ISL_2808665, EPI_ISL_2808684, EPI_ISL_2808704, EPI_ISL_2808717, EPI_ISL_2808795, EPI_ISL_2808796, EPI_ISL_2808801, EPI_ISL_2809171, EPI_ISL_2829602, EPI_ISL_2829667, EPI_ISL_2829725, EPI_ISL_2829730, EPI_ISL_2829760, EPI_ISL_2829823, EPI_ISL_2829893, EPI_ISL_2829936, EPI_ISL_2829947, EPI_ISL_2832089, EPI_ISL_2846990, EPI_ISL_2847040, EPI_ISL_2847041, EPI_ISL_2847090, EPI_ISL_2847135, EPI_ISL_2847137, EPI_ISL_2847138, EPI_ISL_2851426, EPI_ISL_2851438, EPI_ISL_2851481, EPI_ISL_2851559, EPI_ISL_2851674, EPI_ISL_2851688, EPI_ISL_2851710, EPI_ISL_2851716, EPI_ISL_2851741 | | | |
| see above | Health Services Laboratories | Wellcome Sanger Institute for the COVID-19 Genomics UK (COG-UK) Consortium | Cordelia Langford; David K. Jackson; Dominic Kwiatkowski; Ewan Harrison; Health Services Laboratories and Alex Alderton; Ian Johnston; Jeffrey Barrett; John Sillitoe on behalf of the Wellcome Sanger Institute COVID-19 Surveillance Team; Roberto Amato; Sonia Goncalves |
| EPI_ISL_2803934, EPI_ISL_2804010, EPI_ISL_2804043, EPI_ISL_2804045, EPI_ISL_2804072, EPI_ISL_2804086, EPI_ISL_2804099, EPI_ISL_2804102, EPI_ISL_2804182, EPI_ISL_2804195, EPI_ISL_2804198, EPI_ISL_2804211, EPI_ISL_2804257, EPI_ISL_2804258, EPI_ISL_2804259, EPI_ISL_2804260, EPI_ISL_2804264, EPI_ISL_2804265, EPI_ISL_2804271, EPI_ISL_2804274, EPI_ISL_2804278, EPI_ISL_2804279, EPI_ISL_2804281, EPI_ISL_2804282, EPI_ISL_2804283, EPI_ISL_2804284, EPI_ISL_2804285, EPI_ISL_2804289, EPI_ISL_2804294, EPI_ISL_2804297, EPI_ISL_2804298, EPI_ISL_2804299, EPI_ISL_2804303, EPI_ISL_2804305, EPI_ISL_2804312, EPI_ISL_2804313, EPI_ISL_2804316, EPI_ISL_2804323, EPI_ISL_2804324, EPI_ISL_2804325, EPI_ISL_2804326, EPI_ISL_2804329, EPI_ISL_2804333, EPI_ISL_2804334, EPI_ISL_2804337, EPI_ISL_2804338, EPI_ISL_2804343, EPI_ISL_2804344, EPI_ISL_2804345, EPI_ISL_2804348, EPI_ISL_2804354, EPI_ISL_2804355, EPI_ISL_2804356, EPI_ISL_2804357, EPI_ISL_2804361, EPI_ISL_2804363, EPI_ISL_2804364, EPI_ISL_2804365, EPI_ISL_2804367, EPI_ISL_2804372, EPI_ISL_2804375, EPI_ISL_2804376, EPI_ISL_2804382, EPI_ISL_2804385, EPI_ISL_2804386, EPI_ISL_2804388, EPI_ISL_2804393, EPI_ISL_2804395, EPI_ISL_2804396, EPI_ISL_2804398, EPI_ISL_2804403, EPI_ISL_2804404, EPI_ISL_2804405, EPI_ISL_2804408, EPI_ISL_2804409, EPI_ISL_2804412, EPI_ISL_2804413, EPI_ISL_2804415, EPI_ISL_2804416, EPI_ISL_2804422, EPI_ISL_2804426, EPI_ISL_2804428, EPI_ISL_2804430, EPI_ISL_2804436, EPI_ISL_2804441, EPI_ISL_2804444, EPI_ISL_2804445, EPI_ISL_2804446, EPI_ISL_2804447, EPI_ISL_2804449, EPI_ISL_2804452, EPI_ISL_2804454, EPI_ISL_2804458, EPI_ISL_2804473, EPI_ISL_2804474, EPI_ISL_2804477, EPI_ISL_2804482, EPI_ISL_2804483, EPI_ISL_2804485, EPI_ISL_2804486, EPI_ISL_2804489, EPI_ISL_2804490, EPI_ISL_2804492, EPI_ISL_2804494, EPI_ISL_2804495, EPI_ISL_2804496, EPI_ISL_2804500, EPI_ISL_2804501, EPI_ISL_2804502, EPI_ISL_2804504, EPI_ISL_2804505, EPI_ISL_2804509, EPI_ISL_2804510, EPI_ISL_2804511, EPI_ISL_2804513, EPI_ISL_2804514, EPI_ISL_2804516, EPI_ISL_2804518, EPI_ISL_2804520, EPI_ISL_2804521, EPI_ISL_2804522, EPI_ISL_2804525, EPI_ISL_2804526, EPI_ISL_2804527, EPI_ISL_2804528, EPI_ISL_2804529, EPI_ISL_2804536, EPI_ISL_2804538, EPI_ISL_2804539, EPI_ISL_2804540, EPI_ISL_2804542, EPI_ISL_2804544, EPI_ISL_2804548, EPI_ISL_2804549, EPI_ISL_2804552, EPI_ISL_2804555, EPI_ISL_2804556, EPI_ISL_2804558, EPI_ISL_2804559, EPI_ISL_2804565, EPI_ISL_2804569, EPI_ISL_2804578, EPI_ISL_2804579, EPI_ISL_2804581, EPI_ISL_2804582, EPI_ISL_2804584, EPI_ISL_2804585, EPI_ISL_2804588, EPI_ISL_2804589, EPI_ISL_2804592, EPI_ISL_2804593, EPI_ISL_2804595, EPI_ISL_2804603, EPI_ISL_2804605, EPI_ISL_2804606, EPI_ISL_2804607, EPI_ISL_2804608, EPI_ISL_2805348, EPI_ISL_2805365, EPI_ISL_2805367, EPI_ISL_2805370, EPI_ISL_2805372, EPI_ISL_2805375, EPI_ISL_2805388, EPI_ISL_2805397, EPI_ISL_2805406, EPI_ISL_2805414, EPI_ISL_2805415, EPI_ISL_2805420, EPI_ISL_2805423, EPI_ISL_2805426, EPI_ISL_2805429, EPI_ISL_2805440, EPI_ISL_2805442, EPI_ISL_2805443, EPI_ISL_2805446, EPI_ISL_2805471, EPI_ISL_2805473, EPI_ISL_2805485, EPI_ISL_2805522, EPI_ISL_2805525, EPI_ISL_2805527, EPI_ISL_2805539, EPI_ISL_2805552, EPI_ISL_2805560, EPI_ISL_2805565, EPI_ISL_2805581, EPI_ISL_2805585, EPI_ISL_2805588, EPI_ISL_2805601, EPI_ISL_2805602, EPI_ISL_2805615, EPI_ISL_2805616, EPI_ISL_2805631, EPI_ISL_2805647, EPI_ISL_2805649, EPI_ISL_2805650, EPI_ISL_2805660, EPI_ISL_2805664, EPI_ISL_2805665, EPI_ISL_2805666, EPI_ISL_2805668, EPI_ISL_2805675, EPI_ISL_2805679, EPI_ISL_2805683, EPI_ISL_2805689, EPI_ISL_2805694, EPI_ISL_2814010, EPI_ISL_2814012, EPI_ISL_2814014, EPI_ISL_2814020, EPI_ISL_2814036, EPI_ISL_2814037, EPI_ISL_2814038, EPI_ISL_2814040, EPI_ISL_2814043, EPI_ISL_2814048, EPI_ISL_2814052, EPI_ISL_2814055, EPI_ISL_2814064, EPI_ISL_2814068, EPI_ISL_2814070, EPI_ISL_2814074, EPI_ISL_2814075, EPI_ISL_2814080, EPI_ISL_2814089, EPI_ISL_2814092, EPI_ISL_2814096, EPI_ISL_2814098, EPI_ISL_2814100, EPI_ISL_2814101, EPI_ISL_2814111, EPI_ISL_2814115, EPI_ISL_2814116, EPI_ISL_2814118, EPI_ISL_2814119, EPI_ISL_2814125, EPI_ISL_2814132, EPI_ISL_2814141, EPI_ISL_2814144, EPI_ISL_2814149, EPI_ISL_2814152, EPI_ISL_2814157, EPI_ISL_2814160, EPI_ISL_2814161, EPI_ISL_2814162, EPI_ISL_2814167, EPI_ISL_2814180, EPI_ISL_2814181, EPI_ISL_2814186, EPI_ISL_2814187, EPI_ISL_2814188, EPI_ISL_2814189, EPI_ISL_2814192, EPI_ISL_2814193, EPI_ISL_2814210, EPI_ISL_2814211, EPI_ISL_2814215, EPI_ISL_2814216, EPI_ISL_2814224, EPI_ISL_2814225, EPI_ISL_2829036, EPI_ISL_2829047, EPI_ISL_2829053, EPI_ISL_2829061, EPI_ISL_2829063, EPI_ISL_2829068, EPI_ISL_2829101, EPI_ISL_2829121, EPI_ISL_2829124, EPI_ISL_2829136, EPI_ISL_2829179, EPI_ISL_2829190, EPI_ISL_2829193, EPI_ISL_2829285, EPI_ISL_2829300, EPI_ISL_2829305, EPI_ISL_2829308, EPI_ISL_2829310, EPI_ISL_2829312, EPI_ISL_2829316, EPI_ISL_2829322, EPI_ISL_2829323, EPI_ISL_2829326, EPI_ISL_2829327, EPI_ISL_2829333, EPI_ISL_2829356, EPI_ISL_2829369, EPI_ISL_2829377, EPI_ISL_2829385, EPI_ISL_2829386, EPI_ISL_2829391, EPI_ISL_2829397, EPI_ISL_2829405, EPI_ISL_2829408, EPI_ISL_2829409, EPI_ISL_2829419, EPI_ISL_2829431, EPI_ISL_2829444, EPI_ISL_2829456, EPI_ISL_2829458, EPI_ISL_2829474, EPI_ISL_2829482, EPI_ISL_2829487, EPI_ISL_2829490, EPI_ISL_2829494, EPI_ISL_2829503, EPI_ISL_2829504, EPI_ISL_2829506, EPI_ISL_2829507, EPI_ISL_2829524, EPI_ISL_2829526, EPI_ISL_2829532, EPI_ISL_2829538, EPI_ISL_2829540, EPI_ISL_2829542, EPI_ISL_2829559, EPI_ISL_2829562, EPI_ISL_2829569, EPI_ISL_2829571, EPI_ISL_2829575, EPI_ISL_2829578, EPI_ISL_2829581, EPI_ISL_2829585, EPI_ISL_2829613, EPI_ISL_2829614, EPI_ISL_2829672, EPI_ISL_2829677, EPI_ISL_2829688, EPI_ISL_2829746, EPI_ISL_2829765, EPI_ISL_2829799, EPI_ISL_2829878, EPI_ISL_2829914, EPI_ISL_2829915, EPI_ISL_2829940, EPI_ISL_2829962, EPI_ISL_2829971, EPI_ISL_2829990, EPI_ISL_2830006, EPI_ISL_2830012, EPI_ISL_2830028, EPI_ISL_2830038, EPI_ISL_2830054, EPI_ISL_2830059, EPI_ISL_2830062, EPI_ISL_2830064, EPI_ISL_2830065, EPI_ISL_2830097, EPI_ISL_2830104, EPI_ISL_2830123, EPI_ISL_2830125, EPI_ISL_2830147, EPI_ISL_2830170, EPI_ISL_2830171, EPI_ISL_2830179, EPI_ISL_2830181, EPI_ISL_2830182, EPI_ISL_2830187, EPI_ISL_2830192, EPI_ISL_2830193, EPI_ISL_2830196, EPI_ISL_2830197, EPI_ISL_2830198, EPI_ISL_2830199, EPI_ISL_2830202, EPI_ISL_2830203, EPI_ISL_2830204, EPI_ISL_2830205, EPI_ISL_2830207, EPI_ISL_2830209, EPI_ISL_2830211, EPI_ISL_2830212, EPI_ISL_2830216, EPI_ISL_2830217, EPI_ISL_2830218, EPI_ISL_2830220, EPI_ISL_2830226, EPI_ISL_2830230, EPI_ISL_2830231, EPI_ISL_2830233, EPI_ISL_2830239, EPI_ISL_2830242, EPI_ISL_2830245, EPI_ISL_2830246, EPI_ISL_2830248, EPI_ISL_2830252, EPI_ISL_2830260, EPI_ISL_2830262, EPI_ISL_2830263, EPI_ISL_2830264, EPI_ISL_2830270, EPI_ISL_2830272, EPI_ISL_2830273, EPI_ISL_2830276, EPI_ISL_2830277, EPI_ISL_2830279, EPI_ISL_2830281, EPI_ISL_2830282, EPI_ISL_2830287, EPI_ISL_2830289, EPI_ISL_2830291, EPI_ISL_2830292, EPI_ISL_2830293, EPI_ISL_2830296, EPI_ISL_2830297, EPI_ISL_2830299, EPI_ISL_2830301, EPI_ISL_2830306, EPI_ISL_2830308, EPI_ISL_2830311, EPI_ISL_2830312, EPI_ISL_2830315, EPI_ISL_2830317, EPI_ISL_2830319, EPI_ISL_2830321, EPI_ISL_2830615, EPI_ISL_2830635, EPI_ISL_2830639, EPI_ISL_2830640, EPI_ISL_2830661, EPI_ISL_2830674, EPI_ISL_2830697, EPI_ISL_2830727, EPI_ISL_2830761, EPI_ISL_2830799, EPI_ISL_2830801, EPI_ISL_2830811, EPI_ISL_2830814, EPI_ISL_2830818, EPI_ISL_2830826, EPI_ISL_2830837, EPI_ISL_2830842, EPI_ISL_2830854, EPI_ISL_2830856, EPI_ISL_2830860, EPI_ISL_2830864, EPI_ISL_2830866, EPI_ISL_2830867, EPI_ISL_2830869, EPI_ISL_2830870, EPI_ISL_2830871, EPI_ISL_2830874, EPI_ISL_2830876, EPI_ISL_2830877, EPI_ISL_2830880, EPI_ISL_2830883, EPI_ISL_2830885, EPI_ISL_2830887, EPI_ISL_2830891, EPI_ISL_2830898, EPI_ISL_2830902, EPI_ISL_2830909, EPI_ISL_2830914, EPI_ISL_2830915, EPI_ISL_2830918, EPI_ISL_2830919, EPI_ISL_2830925, EPI_ISL_2830932, EPI_ISL_2830933, EPI_ISL_2830935, EPI_ISL_2830940, EPI_ISL_2830941, EPI_ISL_2830942, EPI_ISL_2830944, EPI_ISL_2830946, EPI_ISL_2830949, EPI_ISL_2830951, EPI_ISL_2830956, EPI_ISL_2830957, EPI_ISL_2830959, EPI_ISL_2830961, EPI_ISL_2830962, EPI_ISL_2830968, EPI_ISL_2830977, EPI_ISL_2830979, EPI_ISL_2830985, EPI_ISL_2830988, EPI_ISL_2830992, EPI_ISL_2830993, EPI_ISL_2831001, EPI_ISL_2831003, EPI_ISL_2831005, EPI_ISL_2831006, EPI_ISL_2831008, EPI_ISL_2831009, EPI_ISL_2831010, EPI_ISL_2831011, EPI_ISL_2831012, EPI_ISL_2831015, EPI_ISL_2831021, EPI_ISL_2831024, EPI_ISL_2831025, EPI_ISL_2831026, EPI_ISL_2831028, EPI_ISL_2831031, EPI_ISL_2831033, EPI_ISL_2831034, EPI_ISL_2831035, EPI_ISL_2831038, EPI_ISL_2831039, EPI_ISL_2831042, EPI_ISL_2831584, EPI_ISL_2831585, EPI_ISL_2831586, EPI_ISL_2831587, EPI_ISL_2831588, EPI_ISL_2831592, EPI_ISL_2831593, EPI_ISL_2831596, EPI_ISL_2831597, EPI_ISL_2831602, EPI_ISL_2831603, EPI_ISL_2831605, EPI_ISL_2831606, EPI_ISL_2831607, EPI_ISL_2831608, EPI_ISL_2831609, EPI_ISL_2831611, EPI_ISL_2831614, EPI_ISL_2831615, EPI_ISL_2831617, EPI_ISL_2831618, EPI_ISL_2831621, EPI_ISL_2831624, EPI_ISL_2831625, EPI_ISL_2831630, EPI_ISL_2831632, EPI_ISL_2831637, EPI_ISL_2831638, EPI_ISL_2831640, EPI_ISL_2831643, EPI_ISL_2831644, EPI_ISL_2831645, EPI_ISL_2831648, EPI_ISL_2831651, EPI_ISL_2831654, EPI_ISL_2831655, EPI_ISL_2831657, EPI_ISL_2831658, EPI_ISL_2831660, EPI_ISL_2831661, EPI_ISL_2831662, EPI_ISL_2831664, EPI_ISL_2831667, EPI_ISL_2831669, EPI_ISL_2831674, EPI_ISL_2831676, EPI_ISL_2831681, EPI_ISL_2831682, EPI_ISL_2831683, EPI_ISL_2831685, EPI_ISL_2831686, EPI_ISL_2831691, EPI_ISL_2831692, EPI_ISL_2831694, EPI_ISL_2831697, EPI_ISL_2831698, EPI_ISL_2831699, EPI_ISL_2831700, EPI_ISL_2831701, EPI_ISL_2831703, EPI_ISL_2831705, EPI_ISL_2831711, EPI_ISL_2831715, EPI_ISL_2831718, EPI_ISL_2831720, EPI_ISL_2831721, EPI_ISL_2831725, EPI_ISL_2831726, EPI_ISL_2831727, EPI_ISL_2831728, EPI_ISL_2831729, EPI_ISL_2831733, EPI_ISL_2831734, EPI_ISL_2831736, EPI_ISL_2831740, EPI_ISL_2831742, EPI_ISL_2831743, EPI_ISL_2831746, EPI_ISL_2831748, EPI_ISL_2831750, EPI_ISL_2831751, EPI_ISL_2831752, EPI_ISL_2831753, EPI_ISL_2831754, EPI_ISL_2831758, EPI_ISL_2831759, EPI_ISL_2831760, EPI_ISL_2831761, EPI_ISL_2831762, EPI_ISL_2832074, EPI_ISL_2847150, EPI_ISL_2847278, EPI_ISL_2847291, EPI_ISL_2847523, EPI_ISL_2848064, EPI_ISL_2848074, EPI_ISL_2848088, EPI_ISL_2848113, EPI_ISL_2848115, EPI_ISL_2848228, EPI_ISL_2848232, EPI_ISL_2849594, EPI_ISL_2849752, EPI_ISL_2850391, EPI_ISL_2850392, EPI_ISL_2850393, EPI_ISL_2850395, EPI_ISL_2850407, EPI_ISL_2850421, EPI_ISL_2850423, EPI_ISL_2850424, EPI_ISL_2850438, EPI_ISL_2850447, EPI_ISL_2850448, EPI_ISL_2850453, EPI_ISL_2850486, EPI_ISL_2850507, EPI_ISL_2850517, EPI_ISL_2850526, EPI_ISL_2850628, EPI_ISL_2850631, EPI_ISL_2850645, EPI_ISL_2850653, EPI_ISL_2850669, EPI_ISL_2850670, EPI_ISL_2850673, EPI_ISL_2850686, EPI_ISL_2850693, EPI_ISL_2850696, EPI_ISL_2850717, EPI_ISL_2850722, EPI_ISL_2851455, EPI_ISL_2851468, EPI_ISL_2851483, EPI_ISL_2851518, EPI_ISL_2851534, EPI_ISL_2851541, EPI_ISL_2851562, EPI_ISL_2851575, EPI_ISL_2851581, EPI_ISL_2851591, EPI_ISL_2851693, EPI_ISL_2851704, EPI_ISL_2851706, EPI_ISL_2851719, EPI_ISL_2851731, EPI_ISL_2851737, EPI_ISL_2851770, EPI_ISL_2851790, EPI_ISL_2851873, EPI_ISL_2851874, EPI_ISL_2851939, EPI_ISL_2851947, EPI_ISL_2851977, EPI_ISL_2851989, EPI_ISL_2851991, EPI_ISL_2851994, EPI_ISL_2851996, EPI_ISL_2851999, EPI_ISL_2852023, EPI_ISL_2852044, EPI_ISL_2852045, EPI_ISL_2852090, EPI_ISL_2852092, EPI_ISL_2852129, EPI_ISL_2852678, EPI_ISL_2852698, EPI_ISL_2853100, EPI_ISL_2867711 | | | |
| see above | Lighthouse Lab in Alderley Park | Wellcome Sanger Institute for the COVID-19 Genomics UK (COG-UK) Consortium | Cordelia Langford; David K. Jackson; Dominic Kwiatkowski; Ewan Harrison; Ian Johnston; Jacquelyn Wynn; Jeffrey Barrett; John Sillitoe on behalf of the Wellcome Sanger Institute COVID-19 Surveillance Team; Mairead Hyland; Roberto Amato; Sonia Goncalves; The Lighthouse Lab in Alderley Park and Alex Alderton |
| EPI_ISL_2728684, EPI_ISL_2728688, EPI_ISL_2728690, EPI_ISL_2728691, EPI_ISL_2728692, EPI_ISL_2728693, EPI_ISL_2728695, EPI_ISL_2728697, EPI_ISL_2728699, EPI_ISL_2728701, EPI_ISL_2728703, EPI_ISL_2728707, EPI_ISL_2728708, EPI_ISL_2728710, EPI_ISL_2728722, EPI_ISL_2728723, EPI_ISL_2728729, EPI_ISL_2728730, EPI_ISL_2728731, EPI_ISL_2728732, EPI_ISL_2728733, EPI_ISL_2728734, EPI_ISL_2728737, EPI_ISL_2728738, EPI_ISL_2728740, EPI_ISL_2728741, EPI_ISL_2728744, EPI_ISL_2728747, EPI_ISL_2728748, EPI_ISL_2728752, EPI_ISL_2728754, EPI_ISL_2728763, EPI_ISL_2728765, EPI_ISL_2728766, EPI_ISL_2728768, EPI_ISL_2728770, EPI_ISL_2728773, EPI_ISL_2728775, EPI_ISL_2728777, EPI_ISL_2728780, EPI_ISL_2728782, EPI_ISL_2728783, EPI_ISL_2728785, EPI_ISL_2728791, EPI_ISL_2728800, EPI_ISL_2728801, EPI_ISL_2728802, EPI_ISL_2728803, EPI_ISL_2728804, EPI_ISL_2728806, EPI_ISL_2728808, EPI_ISL_2728810, EPI_ISL_2728813, EPI_ISL_2728814, EPI_ISL_2728815, EPI_ISL_2728820, EPI_ISL_2728824, EPI_ISL_2728826, EPI_ISL_2728827, EPI_ISL_2728830, EPI_ISL_2728831, EPI_ISL_2728837, EPI_ISL_2728839, EPI_ISL_2728840, EPI_ISL_2728841, EPI_ISL_2728845, EPI_ISL_2728852, EPI_ISL_2728855, EPI_ISL_2728862, EPI_ISL_2728864, EPI_ISL_2728865, EPI_ISL_2728868, EPI_ISL_2728869, EPI_ISL_2728871, EPI_ISL_2728875, EPI_ISL_2728876, EPI_ISL_2728879, EPI_ISL_2728880, EPI_ISL_2728881, EPI_ISL_2728883, EPI_ISL_2728885, EPI_ISL_2728886, EPI_ISL_2728887, EPI_ISL_2728888, EPI_ISL_2728899, EPI_ISL_2728903, EPI_ISL_2728906, EPI_ISL_2728910, EPI_ISL_2728913, EPI_ISL_2728918, EPI_ISL_2728920, EPI_ISL_2728925, EPI_ISL_2728926, EPI_ISL_2728931, EPI_ISL_2728934, EPI_ISL_2728935, EPI_ISL_2728940, EPI_ISL_2728941, EPI_ISL_2728943, EPI_ISL_2728944, EPI_ISL_2728948, EPI_ISL_2728950, EPI_ISL_2728951, EPI_ISL_2728952, EPI_ISL_2728954, EPI_ISL_2728958, EPI_ISL_2728960, EPI_ISL_2728962, EPI_ISL_2728964, EPI_ISL_2728965, EPI_ISL_2728966, EPI_ISL_2728967, EPI_ISL_2728971, EPI_ISL_2728972, EPI_ISL_2728973, EPI_ISL_2728974, EPI_ISL_2728976, EPI_ISL_2728977, EPI_ISL_2728978, EPI_ISL_2728981, EPI_ISL_2728982, EPI_ISL_2728983, EPI_ISL_2728984, EPI_ISL_2728986, EPI_ISL_2728992, EPI_ISL_2728993, EPI_ISL_2728997, EPI_ISL_2728998, EPI_ISL_2729000, EPI_ISL_2729002, EPI_ISL_2729003, EPI_ISL_2729006, EPI_ISL_2729007, EPI_ISL_2729008, EPI_ISL_2729011, EPI_ISL_2729012, EPI_ISL_2729013, EPI_ISL_2729018, EPI_ISL_2729019, EPI_ISL_2729023, EPI_ISL_2729025, EPI_ISL_2729026, EPI_ISL_2729027, EPI_ISL_2729033, EPI_ISL_2729034, EPI_ISL_2729035, EPI_ISL_2729038, EPI_ISL_2729040, EPI_ISL_2729042, EPI_ISL_2729043, EPI_ISL_2729045, EPI_ISL_2729047, EPI_ISL_2729050, EPI_ISL_2729053, EPI_ISL_2729059, EPI_ISL_2729072, EPI_ISL_2729076, EPI_ISL_2729087, EPI_ISL_2729090, EPI_ISL_2729101, EPI_ISL_2729105, EPI_ISL_2729106, EPI_ISL_2729111, EPI_ISL_2729113, EPI_ISL_2729116, EPI_ISL_2729119, EPI_ISL_2729124, EPI_ISL_2729126, EPI_ISL_2729128, EPI_ISL_2729137, EPI_ISL_2729139, EPI_ISL_2729144, EPI_ISL_2729148, EPI_ISL_2729150, EPI_ISL_2729154, EPI_ISL_2729160, EPI_ISL_2729165, EPI_ISL_2729170, EPI_ISL_2729172, EPI_ISL_2729191, EPI_ISL_2729197, EPI_ISL_2729198, EPI_ISL_2729199, EPI_ISL_2729204, EPI_ISL_2729209, EPI_ISL_2729213, EPI_ISL_2729222, EPI_ISL_2729223, EPI_ISL_2729242, EPI_ISL_2729257, EPI_ISL_2729261, EPI_ISL_2729264, EPI_ISL_2729275, EPI_ISL_2729282, EPI_ISL_2729287, EPI_ISL_2729292, EPI_ISL_2729296, EPI_ISL_2729299, EPI_ISL_2729306, EPI_ISL_2729313, EPI_ISL_2729320, EPI_ISL_2729321, EPI_ISL_2729332, EPI_ISL_2729340, EPI_ISL_2729350, EPI_ISL_2729352, EPI_ISL_2729354, EPI_ISL_2729358, EPI_ISL_2729361, EPI_ISL_2729363, EPI_ISL_2729364, EPI_ISL_2729367, EPI_ISL_2729373, EPI_ISL_2729944, EPI_ISL_2729945, EPI_ISL_2729953, EPI_ISL_2729955, EPI_ISL_2729966, EPI_ISL_2729977, EPI_ISL_2729979, EPI_ISL_2729981, EPI_ISL_2729989, EPI_ISL_2730003, EPI_ISL_2730005, EPI_ISL_2730007, EPI_ISL_2730015, EPI_ISL_2730020, EPI_ISL_2730032, EPI_ISL_2730056, EPI_ISL_2730072, EPI_ISL_2730073, EPI_ISL_2730077, EPI_ISL_2730084, EPI_ISL_2730090, EPI_ISL_2730109, EPI_ISL_2730130, EPI_ISL_2730132, EPI_ISL_2730144, EPI_ISL_2730147, EPI_ISL_2730149, EPI_ISL_2730161, EPI_ISL_2730162, EPI_ISL_2730187, EPI_ISL_2730199, EPI_ISL_2730201, EPI_ISL_2730211, EPI_ISL_2730212, EPI_ISL_2730220, EPI_ISL_2730228, EPI_ISL_2730258, EPI_ISL_2730282, EPI_ISL_2730289, EPI_ISL_2730290, EPI_ISL_2730291, EPI_ISL_2730294, EPI_ISL_2730295, EPI_ISL_2730296, EPI_ISL_2730297, EPI_ISL_2730298, EPI_ISL_2730301, EPI_ISL_2730302, EPI_ISL_2730303, EPI_ISL_2730304, EPI_ISL_2730305, EPI_ISL_2730306, EPI_ISL_2730310, EPI_ISL_2730312, EPI_ISL_2730313, EPI_ISL_2730315, EPI_ISL_2730316, EPI_ISL_2730321, EPI_ISL_2730322, EPI_ISL_2730323, EPI_ISL_2730325, EPI_ISL_2730327, EPI_ISL_2730333, EPI_ISL_2730337, EPI_ISL_2730343, EPI_ISL_2730344, EPI_ISL_2730346, EPI_ISL_2730348, EPI_ISL_2730350, EPI_ISL_2730353, EPI_ISL_2730355, EPI_ISL_2730356, EPI_ISL_2730358, EPI_ISL_2730361, EPI_ISL_2730362, EPI_ISL_2730363, EPI_ISL_2730364, EPI_ISL_2730366, EPI_ISL_2730369, EPI_ISL_2730371, EPI_ISL_2730374, EPI_ISL_2730375, EPI_ISL_2730377, EPI_ISL_2730378, EPI_ISL_2730379, EPI_ISL_2730380, EPI_ISL_2730381, EPI_ISL_2730383, EPI_ISL_2730386, EPI_ISL_2730387, EPI_ISL_2730390, EPI_ISL_2730392, EPI_ISL_2730403, EPI_ISL_2730404, EPI_ISL_2730405, EPI_ISL_2730409, EPI_ISL_2730411, EPI_ISL_2730415, EPI_ISL_2730418, EPI_ISL_2730419, EPI_ISL_2730421, EPI_ISL_2730422, EPI_ISL_2730426, EPI_ISL_2730430, EPI_ISL_2730432, EPI_ISL_2730433, EPI_ISL_2730435, EPI_ISL_2730436, EPI_ISL_2730438, EPI_ISL_2730439, EPI_ISL_2730440, EPI_ISL_2730442, EPI_ISL_2730443, EPI_ISL_2730446, EPI_ISL_2730448, EPI_ISL_2730455, EPI_ISL_2730456, EPI_ISL_2730458, EPI_ISL_2730459, EPI_ISL_2730461, EPI_ISL_2730467, EPI_ISL_2730468, EPI_ISL_2730471, EPI_ISL_2730477, EPI_ISL_2730478, EPI_ISL_2730479, EPI_ISL_2730480, EPI_ISL_2730481, EPI_ISL_2730483, EPI_ISL_2730486, EPI_ISL_2730488, EPI_ISL_2730491, EPI_ISL_2730492, EPI_ISL_2730494, EPI_ISL_2730495, EPI_ISL_2730500, EPI_ISL_2730501, EPI_ISL_2730502, EPI_ISL_2730504, EPI_ISL_2730506, EPI_ISL_2730507, EPI_ISL_2730512, EPI_ISL_2730513, EPI_ISL_2730514, EPI_ISL_2730516, EPI_ISL_2730523, EPI_ISL_2730525, EPI_ISL_2730526, EPI_ISL_2730528, EPI_ISL_2730534, EPI_ISL_2730537, EPI_ISL_2730538, EPI_ISL_2730539, EPI_ISL_2730540, EPI_ISL_2730541, EPI_ISL_2730549, EPI_ISL_2730557, EPI_ISL_2730559, | | | |

EPI_ISL_2730563, EPI_ISL_2730570, EPI_ISL_2730571, EPI_ISL_2730574, EPI_ISL_2730576, EPI_ISL_2730579, EPI_ISL_2730582, EPI_ISL_2730585, EPI_ISL_2730587, EPI_ISL_2730588, EPI_ISL_2730589, EPI_ISL_2730591, EPI_ISL_2730592, EPI_ISL_2730594, EPI_ISL_2730596, EPI_ISL_2730597, EPI_ISL_2730598, EPI_ISL_2730599, EPI_ISL_2730600, EPI_ISL_2730609, EPI_ISL_2730610, EPI_ISL_2730612,
EPI_ISL_2730615, EPI_ISL_2730616, EPI_ISL_2730618, EPI_ISL_2730620, EPI_ISL_2730621, EPI_ISL_2730623, EPI_ISL_2730627, EPI_ISL_2730629, EPI_ISL_2730631, EPI_ISL_2730634, EPI_ISL_2730637, EPI_ISL_2730639, EPI_ISL_2730640, EPI_ISL_2730641, EPI_ISL_2730642, EPI_ISL_2730644, EPI_ISL_2765587, EPI_ISL_2765611, EPI_ISL_2765632, EPI_ISL_2765771, EPI_ISL_2765815, EPI_ISL_2767245,
EPI_ISL_2767247, EPI_ISL_2767251, EPI_ISL_2767255, EPI_ISL_2767258, EPI_ISL_2767261, EPI_ISL_2767266, EPI_ISL_2767271, EPI_ISL_2767278, EPI_ISL_2767284, EPI_ISL_2767286, EPI_ISL_2767304, EPI_ISL_2767305, EPI_ISL_2767318, EPI_ISL_2767329, EPI_ISL_2767342, EPI_ISL_2779640, EPI_ISL_2779650, EPI_ISL_2779652, EPI_ISL_2779659, EPI_ISL_2779677, EPI_ISL_2779686, EPI_ISL_2779688,
EPI_ISL_2779691, EPI_ISL_2779694, EPI_ISL_2779705, EPI_ISL_2779711, EPI_ISL_2779715, EPI_ISL_2779716, EPI_ISL_2779718, EPI_ISL_2779724, EPI_ISL_2779728, EPI_ISL_2779735, EPI_ISL_2779740, EPI_ISL_2779754, EPI_ISL_2779768, EPI_ISL_2779781, EPI_ISL_2779782, EPI_ISL_2779784, EPI_ISL_2779785, EPI_ISL_2779818, EPI_ISL_2779819, EPI_ISL_2779821, EPI_ISL_2779822, EPI_ISL_2779835,
EPI_ISL_2779836, EPI_ISL_2779841, EPI_ISL_2779854, EPI_ISL_2779867, EPI_ISL_2779868, EPI_ISL_2779874, EPI_ISL_2779878, EPI_ISL_2779881, EPI_ISL_2779882, EPI_ISL_2779885, EPI_ISL_2779886, EPI_ISL_2779887, EPI_ISL_2779888, EPI_ISL_2779892, EPI_ISL_2779894, EPI_ISL_2779895, EPI_ISL_2779907, EPI_ISL_2779913, EPI_ISL_2779916, EPI_ISL_2779918, EPI_ISL_2779919, EPI_ISL_2779921,
EPI_ISL_2779922, EPI_ISL_2779930, EPI_ISL_2779933, EPI_ISL_2779937, EPI_ISL_2779938, EPI_ISL_2779945, EPI_ISL_2779947, EPI_ISL_2779953, EPI_ISL_2779956, EPI_ISL_2779959, EPI_ISL_2779962, EPI_ISL_2779965, EPI_ISL_2779968, EPI_ISL_2779969, EPI_ISL_2779974, EPI_ISL_2779975, EPI_ISL_2779980, EPI_ISL_2779981, EPI_ISL_2779992, EPI_ISL_2780000, EPI_ISL_2780001, EPI_ISL_2780009,
EPI_ISL_2780012, EPI_ISL_2780014, EPI_ISL_2780016, EPI_ISL_2780024, EPI_ISL_2780026, EPI_ISL_2780032, EPI_ISL_2780036, EPI_ISL_2780041, EPI_ISL_2780042, EPI_ISL_2780044, EPI_ISL_2780045, EPI_ISL_2780047, EPI_ISL_2780051, EPI_ISL_2780055, EPI_ISL_2780056, EPI_ISL_2780057, EPI_ISL_2780059, EPI_ISL_2780065, EPI_ISL_2780070, EPI_ISL_2780072, EPI_ISL_2780074, EPI_ISL_2780077,
EPI_ISL_2780080, EPI_ISL_2780090, EPI_ISL_2780092, EPI_ISL_2780093, EPI_ISL_2780101, EPI_ISL_2780105, EPI_ISL_2780106, EPI_ISL_2780109, EPI_ISL_2780112, EPI_ISL_2780113, EPI_ISL_2780120, EPI_ISL_2780121, EPI_ISL_2780477, EPI_ISL_2780480, EPI_ISL_2780481, EPI_ISL_2780484, EPI_ISL_2780489, EPI_ISL_2780491, EPI_ISL_2780506, EPI_ISL_2780508, EPI_ISL_2780512, EPI_ISL_2780515,
EPI_ISL_2780536, EPI_ISL_2780538, EPI_ISL_2780539, EPI_ISL_2780543, EPI_ISL_2780545, EPI_ISL_2780555, EPI_ISL_2780556, EPI_ISL_2780560, EPI_ISL_2780585, EPI_ISL_2780587, EPI_ISL_2780594, EPI_ISL_2780597, EPI_ISL_2780609, EPI_ISL_2780618, EPI_ISL_2780620, EPI_ISL_2780623, EPI_ISL_2780624, EPI_ISL_2780634, EPI_ISL_2780637, EPI_ISL_2780656, EPI_ISL_2780674, EPI_ISL_2780680,
EPI_ISL_2780699, EPI_ISL_2780701, EPI_ISL_2780729, EPI_ISL_2780739, EPI_ISL_2780751, EPI_ISL_2782049, EPI_ISL_2782087, EPI_ISL_2782097, EPI_ISL_2782101, EPI_ISL_2782118, EPI_ISL_2782131, EPI_ISL_2782155, EPI_ISL_2782161, EPI_ISL_2782163, EPI_ISL_2782182, EPI_ISL_2782191, EPI_ISL_2782198, EPI_ISL_2782208, EPI_ISL_2782213, EPI_ISL_2782214, EPI_ISL_2782230, EPI_ISL_2782251,
EPI_ISL_2782253, EPI_ISL_2782275, EPI_ISL_2782284, EPI_ISL_2782293, EPI_ISL_2803767, EPI_ISL_2803785, EPI_ISL_2803795, EPI_ISL_2803803, EPI_ISL_2803811, EPI_ISL_2803822, EPI_ISL_2803825, EPI_ISL_2803834, EPI_ISL_2803841, EPI_ISL_2803851, EPI_ISL_2803861, EPI_ISL_2803873, EPI_ISL_2803874, EPI_ISL_2803877, EPI_ISL_2803886, EPI_ISL_2803898, EPI_ISL_2803915, EPI_ISL_2804619,
EPI_ISL_2804620, EPI_ISL_2804626, EPI_ISL_2804635, EPI_ISL_2804638, EPI_ISL_2804641, EPI_ISL_2804651, EPI_ISL_2804660, EPI_ISL_2804664, EPI_ISL_2804666, EPI_ISL_2804667, EPI_ISL_2804691, EPI_ISL_2804697, EPI_ISL_2804702, EPI_ISL_2804709, EPI_ISL_2804713, EPI_ISL_2804722, EPI_ISL_2804723, EPI_ISL_2804724, EPI_ISL_2804725, EPI_ISL_2804742, EPI_ISL_2804757, EPI_ISL_2804778,
EPI_ISL_2804788, EPI_ISL_2804791, EPI_ISL_2804797, EPI_ISL_2804798, EPI_ISL_2804799, EPI_ISL_2804802, EPI_ISL_2804806, EPI_ISL_2804807, EPI_ISL_2804816, EPI_ISL_2804818, EPI_ISL_2804827, EPI_ISL_2804834, EPI_ISL_2804842, EPI_ISL_2804850, EPI_ISL_2804873, EPI_ISL_2804875, EPI_ISL_2804877, EPI_ISL_2804891, EPI_ISL_2804895, EPI_ISL_2804899, EPI_ISL_2804909, EPI_ISL_2804911,
EPI_ISL_2804915, EPI_ISL_2804919, EPI_ISL_2804920, EPI_ISL_2804921, EPI_ISL_2804926, EPI_ISL_2804929, EPI_ISL_2804936, EPI_ISL_2804939, EPI_ISL_2804940, EPI_ISL_2804941, EPI_ISL_2804944, EPI_ISL_2804946, EPI_ISL_2804950, EPI_ISL_2804953, EPI_ISL_2804954, EPI_ISL_2804957, EPI_ISL_2804960, EPI_ISL_2804970, EPI_ISL_2804971, EPI_ISL_2804974, EPI_ISL_2804982, EPI_ISL_2804987,
EPI_ISL_2804990, EPI_ISL_2804991, EPI_ISL_2804992, EPI_ISL_2804993, EPI_ISL_2804997, EPI_ISL_2804999, EPI_ISL_2805007, EPI_ISL_2805012, EPI_ISL_2805015, EPI_ISL_2805017, EPI_ISL_2805018, EPI_ISL_2805029, EPI_ISL_2805031, EPI_ISL_2805032, EPI_ISL_2805039, EPI_ISL_2805040, EPI_ISL_2805041, EPI_ISL_2805042, EPI_ISL_2805044, EPI_ISL_2805046, EPI_ISL_2805050, EPI_ISL_2805056,
EPI_ISL_2805057, EPI_ISL_2805059, EPI_ISL_2805062, EPI_ISL_2805064, EPI_ISL_2805070, EPI_ISL_2805076, EPI_ISL_2805078, EPI_ISL_2805081, EPI_ISL_2805084, EPI_ISL_2805088, EPI_ISL_2805089, EPI_ISL_2805091, EPI_ISL_2805093, EPI_ISL_2805095, EPI_ISL_2805097, EPI_ISL_2805102, EPI_ISL_2805103, EPI_ISL_2805113, EPI_ISL_2805115, EPI_ISL_2805116, EPI_ISL_2805117, EPI_ISL_2805120,
EPI_ISL_2805122, EPI_ISL_2805123, EPI_ISL_2805129, EPI_ISL_2805133, EPI_ISL_2805135, EPI_ISL_2805141, EPI_ISL_2805142, EPI_ISL_2805150, EPI_ISL_2805159, EPI_ISL_2805160, EPI_ISL_2805171, EPI_ISL_2805185, EPI_ISL_2805187, EPI_ISL_2805189, EPI_ISL_2805190, EPI_ISL_2805198, EPI_ISL_2805199, EPI_ISL_2805204, EPI_ISL_2805208, EPI_ISL_2805219, EPI_ISL_2805223, EPI_ISL_2805229,
EPI_ISL_2805242, EPI_ISL_2805264, EPI_ISL_2805270, EPI_ISL_2805275, EPI_ISL_2805285, EPI_ISL_2805290, EPI_ISL_2805293, EPI_ISL_2805295, EPI_ISL_2805315, EPI_ISL_2805322, EPI_ISL_2805324, EPI_ISL_2805328, EPI_ISL_2805329, EPI_ISL_2805338, EPI_ISL_2805341, EPI_ISL_2805754, EPI_ISL_2805997, EPI_ISL_2806019, EPI_ISL_2806037, EPI_ISL_2806039, EPI_ISL_2806043, EPI_ISL_2806068,
EPI_ISL_2806074, EPI_ISL_2806076, EPI_ISL_2806102, EPI_ISL_2806108, EPI_ISL_2806201, EPI_ISL_2806210, EPI_ISL_2806212, EPI_ISL_2806221, EPI_ISL_2806226, EPI_ISL_2806228, EPI_ISL_2806239, EPI_ISL_2806241, EPI_ISL_2806242, EPI_ISL_2806243, EPI_ISL_2806244, EPI_ISL_2806250, EPI_ISL_2806251, EPI_ISL_2806259, EPI_ISL_2806265, EPI_ISL_2806272, EPI_ISL_2806280, EPI_ISL_2806289,
EPI_ISL_2806290, EPI_ISL_2806292, EPI_ISL_2806295, EPI_ISL_2806298, EPI_ISL_2806300, EPI_ISL_2806309, EPI_ISL_2806310, EPI_ISL_2806314, EPI_ISL_2806315, EPI_ISL_2806318, EPI_ISL_2806320, EPI_ISL_2806323, EPI_ISL_2806329, EPI_ISL_2806330, EPI_ISL_2806335, EPI_ISL_2806337, EPI_ISL_2806338, EPI_ISL_2806342, EPI_ISL_2806343, EPI_ISL_2806350, EPI_ISL_2806363, EPI_ISL_2806367,
EPI_ISL_2806369, EPI_ISL_2806374, EPI_ISL_2806380, EPI_ISL_2806386, EPI_ISL_2806393, EPI_ISL_2806395, EPI_ISL_2806396, EPI_ISL_2806398, EPI_ISL_2806399, EPI_ISL_2806400, EPI_ISL_2806401, EPI_ISL_2806403, EPI_ISL_2806404, EPI_ISL_2806405, EPI_ISL_2806415, EPI_ISL_2806416, EPI_ISL_2806421, EPI_ISL_2806425, EPI_ISL_2806442, EPI_ISL_2807149, EPI_ISL_2807152, EPI_ISL_2807155,
EPI_ISL_2807158, EPI_ISL_2807161, EPI_ISL_2807164, EPI_ISL_2807165, EPI_ISL_2807166, EPI_ISL_2807167, EPI_ISL_2807169, EPI_ISL_2807173, EPI_ISL_2807175, EPI_ISL_2807186, EPI_ISL_2807192, EPI_ISL_2807193, EPI_ISL_2807196, EPI_ISL_2807200, EPI_ISL_2807202, EPI_ISL_2807203, EPI_ISL_2807204, EPI_ISL_2807220, EPI_ISL_2807221, EPI_ISL_2807223, EPI_ISL_2807224, EPI_ISL_2807226,
EPI_ISL_2807229, EPI_ISL_2807231, EPI_ISL_2807232, EPI_ISL_2807235, EPI_ISL_2807236, EPI_ISL_2807239, EPI_ISL_2807240, EPI_ISL_2807241, EPI_ISL_2807250, EPI_ISL_2807257, EPI_ISL_2807265, EPI_ISL_2807266, EPI_ISL_2807270, EPI_ISL_2807271, EPI_ISL_2807273, EPI_ISL_2807274, EPI_ISL_2807275, EPI_ISL_2807276, EPI_ISL_2807277, EPI_ISL_2807278, EPI_ISL_2807279, EPI_ISL_2807280,
EPI_ISL_2807281, EPI_ISL_2807282, EPI_ISL_2807283, EPI_ISL_2807284, EPI_ISL_2807285, EPI_ISL_2807286, EPI_ISL_2807287, EPI_ISL_2807288, EPI_ISL_2807289, EPI_ISL_2807290, EPI_ISL_2807291, EPI_ISL_2807293, EPI_ISL_2807294, EPI_ISL_2807295, EPI_ISL_2807297, EPI_ISL_2807298, EPI_ISL_2807299, EPI_ISL_2807300, EPI_ISL_2807301, EPI_ISL_2807302, EPI_ISL_2807304, EPI_ISL_2807305,
EPI_ISL_2807306, EPI_ISL_2807307, EPI_ISL_2807308, EPI_ISL_2807309, EPI_ISL_2807310, EPI_ISL_2807311, EPI_ISL_2807312, EPI_ISL_2807313, EPI_ISL_2807314, EPI_ISL_2807315, EPI_ISL_2807316, EPI_ISL_2807317, EPI_ISL_2807318, EPI_ISL_2807319, EPI_ISL_2807320, EPI_ISL_2807321, EPI_ISL_2807322, EPI_ISL_2807323, EPI_ISL_2807324, EPI_ISL_2807325, EPI_ISL_2807326, EPI_ISL_2807328,
EPI_ISL_2807329, EPI_ISL_2807330, EPI_ISL_2807332, EPI_ISL_2807333, EPI_ISL_2807334, EPI_ISL_2807335, EPI_ISL_2807336, EPI_ISL_2807337, EPI_ISL_2807338, EPI_ISL_2807339, EPI_ISL_2807340, EPI_ISL_2807341, EPI_ISL_2807342, EPI_ISL_2807343, EPI_ISL_2807345, EPI_ISL_2807346, EPI_ISL_2807347, EPI_ISL_2807348, EPI_ISL_2807349, EPI_ISL_2807350, EPI_ISL_2807351, EPI_ISL_2807352,
EPI_ISL_2807353, EPI_ISL_2807359, EPI_ISL_2807360, EPI_ISL_2807363, EPI_ISL_2807365, EPI_ISL_2807368, EPI_ISL_2807369, EPI_ISL_2807372, EPI_ISL_2807375, EPI_ISL_2807379, EPI_ISL_2807380, EPI_ISL_2807381, EPI_ISL_2807385, EPI_ISL_2807389, EPI_ISL_2807392, EPI_ISL_2807393, EPI_ISL_2807399, EPI_ISL_2807403, EPI_ISL_2807411, EPI_ISL_2807412, EPI_ISL_2807413, EPI_ISL_2807429,
EPI_ISL_2807433, EPI_ISL_2807434, EPI_ISL_2807437, EPI_ISL_2807443, EPI_ISL_2807444, EPI_ISL_2807454, EPI_ISL_2807455, EPI_ISL_2807457, EPI_ISL_2807464, EPI_ISL_2807465, EPI_ISL_2807468, EPI_ISL_2807469, EPI_ISL_2807470, EPI_ISL_2807474, EPI_ISL_2807475, EPI_ISL_2807481, EPI_ISL_2807489, EPI_ISL_2807493, EPI_ISL_2807500, EPI_ISL_2807501, EPI_ISL_2807505, EPI_ISL_2807520,
EPI_ISL_2807527, EPI_ISL_2807530, EPI_ISL_2807532, EPI_ISL_2807533, EPI_ISL_2807538, EPI_ISL_2807540, EPI_ISL_2807542, EPI_ISL_2807545, EPI_ISL_2807547, EPI_ISL_2807551, EPI_ISL_2807559, EPI_ISL_2807562, EPI_ISL_2807564, EPI_ISL_2807565, EPI_ISL_2807567, EPI_ISL_2807584, EPI_ISL_2807585, EPI_ISL_2807600, EPI_ISL_2807603, EPI_ISL_2807613, EPI_ISL_2807615, EPI_ISL_2807616,
EPI_ISL_2807617, EPI_ISL_2807618, EPI_ISL_2807620, EPI_ISL_2807622, EPI_ISL_2807625, EPI_ISL_2807627, EPI_ISL_2807629, EPI_ISL_2807631, EPI_ISL_2807632, EPI_ISL_2807638, EPI_ISL_2807641, EPI_ISL_2807644, EPI_ISL_2807646, EPI_ISL_2807648, EPI_ISL_2807650, EPI_ISL_2807655, EPI_ISL_2807657, EPI_ISL_2807674, EPI_ISL_2807676, EPI_ISL_2807682, EPI_ISL_2807685, EPI_ISL_2807686,
EPI_ISL_2807688, EPI_ISL_2807712, EPI_ISL_2807714, EPI_ISL_2807716, EPI_ISL_2807717, EPI_ISL_2807718, EPI_ISL_2807719, EPI_ISL_2807720, EPI_ISL_2807721, EPI_ISL_2807722, EPI_ISL_2807723, EPI_ISL_2807724, EPI_ISL_2807725, EPI_ISL_2807726, EPI_ISL_2807729, EPI_ISL_2807730, EPI_ISL_2807731, EPI_ISL_2807732, EPI_ISL_2807737, EPI_ISL_2807739, EPI_ISL_2807741, EPI_ISL_2807742,
EPI_ISL_2807746, EPI_ISL_2807747, EPI_ISL_2807750, EPI_ISL_2807751, EPI_ISL_2807753, EPI_ISL_2807755, EPI_ISL_2807757, EPI_ISL_2807759, EPI_ISL_2807760, EPI_ISL_2807761, EPI_ISL_2807765, EPI_ISL_2807767, EPI_ISL_2807768, EPI_ISL_2807769, EPI_ISL_2807771, EPI_ISL_2807772, EPI_ISL_2807773, EPI_ISL_2807776, EPI_ISL_2807777, EPI_ISL_2807778, EPI_ISL_2807781, EPI_ISL_2807782,
EPI_ISL_2807783, EPI_ISL_2807784, EPI_ISL_2807785, EPI_ISL_2807787, EPI_ISL_2807788, EPI_ISL_2807790, EPI_ISL_2807791, EPI_ISL_2807792, EPI_ISL_2807793, EPI_ISL_2807797, EPI_ISL_2807800, EPI_ISL_2807802, EPI_ISL_2807803, EPI_ISL_2807806, EPI_ISL_2807808, EPI_ISL_2807810, EPI_ISL_2807813, EPI_ISL_2807817, EPI_ISL_2807819, EPI_ISL_2807822, EPI_ISL_2807823, EPI_ISL_2807825,
EPI_ISL_2807827, EPI_ISL_2807829, EPI_ISL_2807830, EPI_ISL_2807832, EPI_ISL_2807833, EPI_ISL_2807837, EPI_ISL_2807838, EPI_ISL_2807839, EPI_ISL_2807842, EPI_ISL_2807850, EPI_ISL_2807852, EPI_ISL_2807854, EPI_ISL_2807855, EPI_ISL_2807858, EPI_ISL_2807861, EPI_ISL_2807862, EPI_ISL_2807863, EPI_ISL_2807866, EPI_ISL_2807868, EPI_ISL_2807869, EPI_ISL_2807871, EPI_ISL_2807877,
EPI_ISL_2807878, EPI_ISL_2807880, EPI_ISL_2807882, EPI_ISL_2807887, EPI_ISL_2807888, EPI_ISL_2807890, EPI_ISL_2807891, EPI_ISL_2807893, EPI_ISL_2807894, EPI_ISL_2807898, EPI_ISL_2807902, EPI_ISL_2807904, EPI_ISL_2807905, EPI_ISL_2807906, EPI_ISL_2807907, EPI_ISL_2807908, EPI_ISL_2807909, EPI_ISL_2807912, EPI_ISL_2807914, EPI_ISL_2807915, EPI_ISL_2807916, EPI_ISL_2807917,
EPI_ISL_2807920, EPI_ISL_2807921, EPI_ISL_2807924, EPI_ISL_2807925, EPI_ISL_2807926, EPI_ISL_2807931, EPI_ISL_2807932, EPI_ISL_2807933, EPI_ISL_2807935, EPI_ISL_2807937, EPI_ISL_2807938, EPI_ISL_2807942, EPI_ISL_2807943, EPI_ISL_2807949, EPI_ISL_2807950, EPI_ISL_2807951, EPI_ISL_2807952, EPI_ISL_2807953, EPI_ISL_2807954, EPI_ISL_2807956, EPI_ISL_2807959, EPI_ISL_2807960,
EPI_ISL_2807961, EPI_ISL_2807964, EPI_ISL_2807968, EPI_ISL_2807972, EPI_ISL_2807973, EPI_ISL_2807978, EPI_ISL_2807979, EPI_ISL_2807980, EPI_ISL_2807981, EPI_ISL_2807987, EPI_ISL_2807989, EPI_ISL_2807994, EPI_ISL_2807995, EPI_ISL_2807997, EPI_ISL_2807998, EPI_ISL_2807999, EPI_ISL_2808000, EPI_ISL_2808003, EPI_ISL_2808004, EPI_ISL_2808007, EPI_ISL_2808008, EPI_ISL_2808009,
EPI_ISL_2808010, EPI_ISL_2808011, EPI_ISL_2808012, EPI_ISL_2808013, EPI_ISL_2808014, EPI_ISL_2808019, EPI_ISL_2808022, EPI_ISL_2808023, EPI_ISL_2808026, EPI_ISL_2808029, EPI_ISL_2808030, EPI_ISL_2808032, EPI_ISL_2808035, EPI_ISL_2808038, EPI_ISL_2808040, EPI_ISL_2808041, EPI_ISL_2808043, EPI_ISL_2808044, EPI_ISL_2808046, EPI_ISL_2808047, EPI_ISL_2808048, EPI_ISL_2808049,
EPI_ISL_2808050, EPI_ISL_2808054, EPI_ISL_2808056, EPI_ISL_2808060, EPI_ISL_2808062, EPI_ISL_2808063, EPI_ISL_2808064, EPI_ISL_2808066, EPI_ISL_2808067, EPI_ISL_2808430, EPI_ISL_2808432, EPI_ISL_2808434, EPI_ISL_2808435, EPI_ISL_2808439, EPI_ISL_2808442, EPI_ISL_2808452, EPI_ISL_2808460, EPI_ISL_2808467, EPI_ISL_2808472, EPI_ISL_2808473, EPI_ISL_2808481, EPI_ISL_2808482,
EPI_ISL_2808483, EPI_ISL_2808500, EPI_ISL_2808505, EPI_ISL_2808508, EPI_ISL_2808510, EPI_ISL_2808524, EPI_ISL_2808538, EPI_ISL_2808544, EPI_ISL_2808551, EPI_ISL_2808553, EPI_ISL_2808558, EPI_ISL_2808559, EPI_ISL_2808568, EPI_ISL_2808584, EPI_ISL_2808591, EPI_ISL_2808595, EPI_ISL_2808607, EPI_ISL_2808614, EPI_ISL_2808618, EPI_ISL_2808626, EPI_ISL_2808643, EPI_ISL_2808646,
EPI_ISL_2808656, EPI_ISL_2808657, EPI_ISL_2808660, EPI_ISL_2808672, EPI_ISL_2808676, EPI_ISL_2808682, EPI_ISL_2808702, EPI_ISL_2808707, EPI_ISL_2808716, EPI_ISL_2808724, EPI_ISL_2808726, EPI_ISL_2808727, EPI_ISL_2808733, EPI_ISL_2808734, EPI_ISL_2808735, EPI_ISL_2808736, EPI_ISL_2808737, EPI_ISL_2808738, EPI_ISL_2808739, EPI_ISL_2808740, EPI_ISL_2808741, EPI_ISL_2808742,
EPI_ISL_2808743, EPI_ISL_2808744, EPI_ISL_2808745, EPI_ISL_2808746, EPI_ISL_2808747, EPI_ISL_2808748, EPI_ISL_2808749, EPI_ISL_2808750, EPI_ISL_2808751, EPI_ISL_2808752, EPI_ISL_2808753, EPI_ISL_2808754, EPI_ISL_2808755, EPI_ISL_2808756, EPI_ISL_2808757, EPI_ISL_2808758, EPI_ISL_2808759, EPI_ISL_2808760, EPI_ISL_2808761, EPI_ISL_2808762, EPI_ISL_2808764, EPI_ISL_2808765,
EPI_ISL_2808766, EPI_ISL_2808767, EPI_ISL_2808768, EPI_ISL_2808769, EPI_ISL_2808770, EPI_ISL_2808771, EPI_ISL_2808772, EPI_ISL_2808773, EPI_ISL_2808774, EPI_ISL_2808775, EPI_ISL_2808776, EPI_ISL_2808777, EPI_ISL_2808778, EPI_ISL_2808779, EPI_ISL_2808780, EPI_ISL_2808781, EPI_ISL_2808782, EPI_ISL_2808783, EPI_ISL_2808784, EPI_ISL_2808785, EPI_ISL_2808786, EPI_ISL_2808787,
EPI_ISL_2808788, EPI_ISL_2808789, EPI_ISL_2808790, EPI_ISL_2808791, EPI_ISL_2808792, EPI_ISL_2808807, EPI_ISL_2808808, EPI_ISL_2808809, EPI_ISL_2808811, EPI_ISL_2808812, EPI_ISL_2808813, EPI_ISL_2808814, EPI_ISL_2808815, EPI_ISL_2808816, EPI_ISL_2808817, EPI_ISL_2808818, EPI_ISL_2808819, EPI_ISL_2808820, EPI_ISL_2808822, EPI_ISL_2808823, EPI_ISL_2808824, EPI_ISL_2808825,
EPI_ISL_2808826, EPI_ISL_2808827, EPI_ISL_2808828, EPI_ISL_2808829, EPI_ISL_2808830, EPI_ISL_2808831, EPI_ISL_2808832, EPI_ISL_2808833, EPI_ISL_2808834, EPI_ISL_2808835, EPI_ISL_2808836, EPI_ISL_2808837, EPI_ISL_2808838, EPI_ISL_2808839, EPI_ISL_2808840, EPI_ISL_2808841, EPI_ISL_2808842, EPI_ISL_2808843, EPI_ISL_2808844, EPI_ISL_2808845, EPI_ISL_2808846, EPI_ISL_2808847,
EPI_ISL_2808848, EPI_ISL_2808849, EPI_ISL_2808850, EPI_ISL_2808851, EPI_ISL_2808852, EPI_ISL_2808853, EPI_ISL_2808854, EPI_ISL_2808855, EPI_ISL_2808856, EPI_ISL_2808857, EPI_ISL_2808858, EPI_ISL_2809098, EPI_ISL_2809258, EPI_ISL_2809287, EPI_ISL_2809326, EPI_ISL_2809342, EPI_ISL_2809500, EPI_ISL_2809501, EPI_ISL_2809502, EPI_ISL_2809503, EPI_ISL_2809504, EPI_ISL_2809505,
EPI_ISL_2809506, EPI_ISL_2809507, EPI_ISL_2809508, EPI_ISL_2809509, EPI_ISL_2809510, EPI_ISL_2809511, EPI_ISL_2809512, EPI_ISL_2809513, EPI_ISL_2809514, EPI_ISL_2809515, EPI_ISL_2809516, EPI_ISL_2809517, EPI_ISL_2809518, EPI_ISL_2809519, EPI_ISL_2809520, EPI_ISL_2809521, EPI_ISL_2809522, EPI_ISL_2809524, EPI_ISL_2809525, EPI_ISL_2809527, EPI_ISL_2809528, EPI_ISL_2809529,
EPI_ISL_2809530, EPI_ISL_2809531, EPI_ISL_2809532, EPI_ISL_2809533, EPI_ISL_2809534, EPI_ISL_2809535, EPI_ISL_2809536, EPI_ISL_2809537, EPI_ISL_2809538, EPI_ISL_2809539, EPI_ISL_2809540, EPI_ISL_2809541, EPI_ISL_2809542, EPI_ISL_2809543, EPI_ISL_2809544, EPI_ISL_2809546, EPI_ISL_2809547, EPI_ISL_2809548, EPI_ISL_2809549, EPI_ISL_2809550, EPI_ISL_2809551, EPI_ISL_2809552,
EPI_ISL_2809553, EPI_ISL_2809554, EPI_ISL_2809555, EPI_ISL_2809556, EPI_ISL_2809557, EPI_ISL_2809558, EPI_ISL_2809560, EPI_ISL_2809561, EPI_ISL_2809562, EPI_ISL_2809563, EPI_ISL_2809564, EPI_ISL_2809565, EPI_ISL_2809566, EPI_ISL_2809567, EPI_ISL_2809568, EPI_ISL_2809569, EPI_ISL_2809570, EPI_ISL_2809571, EPI_ISL_2809572, EPI_ISL_2809575, EPI_ISL_2809577, EPI_ISL_2809578,
EPI_ISL_2809579, EPI_ISL_2809580, EPI_ISL_2809581, EPI_ISL_2809584, EPI_ISL_2809585, EPI_ISL_2809586, EPI_ISL_2809590, EPI_ISL_2809592, EPI_ISL_2809593, EPI_ISL_2809595, EPI_ISL_2809596, EPI_ISL_2809597, EPI_ISL_2809603, EPI_ISL_2809604, EPI_ISL_2809607, EPI_ISL_2809608, EPI_ISL_2809609, EPI_ISL_2809610, EPI_ISL_2809611, EPI_ISL_2809613, EPI_ISL_2809614, EPI_ISL_2809617,
EPI_ISL_2809618, EPI_ISL_2809619, EPI_ISL_2809623, EPI_ISL_2809627, EPI_ISL_2809630, EPI_ISL_2809631, EPI_ISL_2809632, EPI_ISL_2809633, EPI_ISL_2809636, EPI_ISL_2809639, EPI_ISL_2809640, EPI_ISL_2809642, EPI_ISL_2809647, EPI_ISL_2809648, EPI_ISL_2809650, EPI_ISL_2809653, EPI_ISL_2809654, EPI_ISL_2809655, EPI_ISL_2809656, EPI_ISL_2809657, EPI_ISL_2809658, EPI_ISL_2809659,
EPI_ISL_2809661, EPI_ISL_2809663, EPI_ISL_2809664, EPI_ISL_2809667, EPI_ISL_2809668, EPI_ISL_2809669, EPI_ISL_2809670, EPI_ISL_2809673, EPI_ISL_2809674, EPI_ISL_2809676, EPI_ISL_2809677, EPI_ISL_2809678, EPI_ISL_2809681, EPI_ISL_2809682, EPI_ISL_2809684, EPI_ISL_2809687, EPI_ISL_2809688, EPI_ISL_2809691, EPI_ISL_2809692, EPI_ISL_2809693, EPI_ISL_2809694, EPI_ISL_2809695,
EPI_ISL_2809697, EPI_ISL_2809698, EPI_ISL_2809699, EPI_ISL_2809700, EPI_ISL_2809703, EPI_ISL_2809704, EPI_ISL_2809705, EPI_ISL_2809706, EPI_ISL_2809712, EPI_ISL_2809714, EPI_ISL_2809715, EPI_ISL_2809716, EPI_ISL_2809717, EPI_ISL_2809718, EPI_ISL_2809720, EPI_ISL_2809722, EPI_ISL_2809724, EPI_ISL_2809725, EPI_ISL_2809726, EPI_ISL_2809728, EPI_ISL_2809729, EPI_ISL_2809730,
EPI_ISL_2809731, EPI_ISL_2809732, EPI_ISL_2809734, EPI_ISL_2809736, EPI_ISL_2809737, EPI_ISL_2809739, EPI_ISL_2809740, EPI_ISL_2809741, EPI_ISL_2809742, EPI_ISL_2809743, EPI_ISL_2809744, EPI_ISL_2809745, EPI_ISL_2809748, EPI_ISL_2809749, EPI_ISL_2809750, EPI_ISL_2809751, EPI_ISL_2809752, EPI_ISL_2809753, EPI_ISL_2809754, EPI_ISL_2809755, EPI_ISL_2809757, EPI_ISL_2809758,
EPI_ISL_2809759, EPI_ISL_2809760, EPI_ISL_2809762, EPI_ISL_2809763, EPI_ISL_2809766, EPI_ISL_2809768, EPI_ISL_2809769, EPI_ISL_2809774, EPI_ISL_2809775, EPI_ISL_2809776, EPI_ISL_2809777, EPI_ISL_2809779, EPI_ISL_2809781, EPI_ISL_2809782, EPI_ISL_2809783, EPI_ISL_2809784, EPI_ISL_2809785, EPI_ISL_2809787, EPI_ISL_2809790, EPI_ISL_2809791, EPI_ISL_2809792, EPI_ISL_2809793,
EPI_ISL_2809796, EPI_ISL_2809797, EPI_ISL_2809799, EPI_ISL_2809802, EPI_ISL_2809803, EPI_ISL_2809808, EPI_ISL_2809811, EPI_ISL_2809812, EPI_ISL_2809813, EPI_ISL_2809814, EPI_ISL_2809815, EPI_ISL_2809816, EPI_ISL_2809817, EPI_ISL_2809818, EPI_ISL_2809819, EPI_ISL_2814062, EPI_ISL_2814085, EPI_ISL_2814129, EPI_ISL_2814131, EPI_ISL_2814142, EPI_ISL_2814146, EPI_ISL_2814148,
EPI_ISL_2814153, EPI_ISL_2814178, EPI_ISL_2814182, EPI_ISL_2814183, EPI_ISL_2814204, EPI_ISL_2814236, EPI_ISL_2814252, EPI_ISL_2814254, EPI_ISL_2814267, EPI_ISL_2814323, EPI_ISL_2814369, EPI_ISL_2814374, EPI_ISL_2814380, EPI_ISL_2814394, EPI_ISL_2814407, EPI_ISL_2814425, EPI_ISL_2814426, EPI_ISL_2814427, EPI_ISL_2814428, EPI_ISL_2814429, EPI_ISL_2814430, EPI_ISL_2814431,
EPI_ISL_2814432, EPI_ISL_2814433, EPI_ISL_2814434, EPI_ISL_2814435, EPI_ISL_2814436, EPI_ISL_2814437, EPI_ISL_2814438, EPI_ISL_2814439, EPI_ISL_2814440, EPI_ISL_2814441, EPI_ISL_2814442, EPI_ISL_2814443, EPI_ISL_2814444, EPI_ISL_2814445, EPI_ISL_2814446, EPI_ISL_2814447, EPI_ISL_2814448, EPI_ISL_2814449, EPI_ISL_2814450, EPI_ISL_2814451, EPI_ISL_2814452, EPI_ISL_2814453,
EPI_ISL_2814454, EPI_ISL_2814455, EPI_ISL_2814456, EPI_ISL_2814457, EPI_ISL_2814458, EPI_ISL_2814459, EPI_ISL_2814460, EPI_ISL_2814461, EPI_ISL_2814462, EPI_ISL_2814463, EPI_ISL_2814464, EPI_ISL_2814465, EPI_ISL_2814466, EPI_ISL_2814467, EPI_ISL_2814468, EPI_ISL_2814469, EPI_ISL_2814470, EPI_ISL_2814471, EPI_ISL_2814472, EPI_ISL_2814473, EPI_ISL_2814474, EPI_ISL_2814476,
EPI_ISL_2814477, EPI_ISL_2814478, EPI_ISL_2814480, EPI_ISL_2814481, EPI_ISL_2814482, EPI_ISL_2814483, EPI_ISL_2814484, EPI_ISL_2814485, EPI_ISL_2814488, EPI_ISL_2814490, EPI_ISL_2814491, EPI_ISL_2814492, EPI_ISL_2814494, EPI_ISL_2814495, EPI_ISL_2814496, EPI_ISL_2814497, EPI_ISL_2814498, EPI_ISL_2814499, EPI_ISL_2814500, EPI_ISL_2814501, EPI_ISL_2814502, EPI_ISL_2814503,
EPI_ISL_2814504, EPI_ISL_2814507, EPI_ISL_2814509, EPI_ISL_2814511, EPI_ISL_2814512, EPI_ISL_2814513, EPI_ISL_2814515, EPI_ISL_2814516, EPI_ISL_2814518, EPI_ISL_2814519, EPI_ISL_2814520, EPI_ISL_2814521, EPI_ISL_2814524, EPI_ISL_2814525, EPI_ISL_2814526, EPI_ISL_2814527, EPI_ISL_2814528, EPI_ISL_2814531, EPI_ISL_2814532, EPI_ISL_2814533, EPI_ISL_2814534, EPI_ISL_2814537,
EPI_ISL_2814538, EPI_ISL_2814539, EPI_ISL_2814543, EPI_ISL_2814545, EPI_ISL_2814549, EPI_ISL_2814550, EPI_ISL_2814551, EPI_ISL_2814553, EPI_ISL_2814554, EPI_ISL_2814555, EPI_ISL_2814556, EPI_ISL_2814557, EPI_ISL_2814558, EPI_ISL_2814560, EPI_ISL_2814561, EPI_ISL_2814564, EPI_ISL_2814565, EPI_ISL_2814567, EPI_ISL_2814569, EPI_ISL_2814570, EPI_ISL_2814571, EPI_ISL_2814573,
EPI_ISL_2814574, EPI_ISL_2814575, EPI_ISL_2814576, EPI_ISL_2814577, EPI_ISL_2814578, EPI_ISL_2814579, EPI_ISL_2814580, EPI_ISL_2814581, EPI_ISL_2814582, EPI_ISL_2814583, EPI_ISL_2814584, EPI_ISL_2814585, EPI_ISL_2814586, EPI_ISL_2814587, EPI_ISL_2814588, EPI_ISL_2814589, EPI_ISL_2814590, EPI_ISL_2814591, EPI_ISL_2814592, EPI_ISL_2814594, EPI_ISL_2814595, EPI_ISL_2814596,
EPI_ISL_2814600, EPI_ISL_2814602, EPI_ISL_2814603, EPI_ISL_2814604, EPI_ISL_2814605, EPI_ISL_2814606, EPI_ISL_2814610, EPI_ISL_2814611, EPI_ISL_2814612, EPI_ISL_2814614, EPI_ISL_2814615, EPI_ISL_2814616, EPI_ISL_2814618, EPI_ISL_2814620, EPI_ISL_2814621, EPI_ISL_2814622, EPI_ISL_2814624, EPI_ISL_2814625, EPI_ISL_2814626, EPI_ISL_2814627, EPI_ISL_2814628, EPI_ISL_2814629,
EPI_ISL_2814631, EPI_ISL_2814634, EPI_ISL_2814635, EPI_ISL_2814637, EPI_ISL_2814638, EPI_ISL_2814639, EPI_ISL_2814641, EPI_ISL_2814642, EPI_ISL_2814643, EPI_ISL_2814644, EPI_ISL_2814645, EPI_ISL_2814646, EPI_ISL_2814647, EPI_ISL_2814648, EPI_ISL_2814649, EPI_ISL_2814650, EPI_ISL_2814651, EPI_ISL_2814654, EPI_ISL_2814657, EPI_ISL_2814658, EPI_ISL_2814661, EPI_ISL_2814664,
EPI_ISL_2814666, EPI_ISL_2814667, EPI_ISL_2814668, EPI_ISL_2814669, EPI_ISL_2814670, EPI_ISL_2814671, EPI_ISL_2814672, EPI_ISL_2814673, EPI_ISL_2814674, EPI_ISL_2814675, EPI_ISL_2814677, EPI_ISL_2814678, EPI_ISL_2814679, EPI_ISL_2814680, EPI_ISL_2814681, EPI_ISL_2814683, EPI_ISL_2814684, EPI_ISL_2814685, EPI_ISL_2814687, EPI_ISL_2814689, EPI_ISL_2814691, EPI_ISL_2814692,
EPI_ISL_2814693, EPI_ISL_2814695, EPI_ISL_2814696, EPI_ISL_2814699, EPI_ISL_2814700, EPI_ISL_2814702, EPI_ISL_2814703, EPI_ISL_2814705, EPI_ISL_2814707, EPI_ISL_2814708, EPI_ISL_2814709, EPI_ISL_2814711, EPI_ISL_2814712, EPI_ISL_2814713, EPI_ISL_2814714, EPI_ISL_2814715, EPI_ISL_2814717, EPI_ISL_2814722, EPI_ISL_2814724, EPI_ISL_2814725, EPI_ISL_2814726, EPI_ISL_2814727,
EPI_ISL_2814728, EPI_ISL_2814729, EPI_ISL_2814740, EPI_ISL_2814795, EPI_ISL_2814812, EPI_ISL_2814825, EPI_ISL_2814827, EPI_ISL_2814830, EPI_ISL_2814856, EPI_ISL_2814896, EPI_ISL_2814914, EPI_ISL_2814921, EPI_ISL_2814923, EPI_ISL_2814924, EPI_ISL_2814926, EPI_ISL_2814927, EPI_ISL_2814930, EPI_ISL_2814935, EPI_ISL_2814936, EPI_ISL_2814939, EPI_ISL_2814940, EPI_ISL_2814941,
EPI_ISL_2814949, EPI_ISL_2814950, EPI_ISL_2814951, EPI_ISL_2814952, EPI_ISL_2814956, EPI_ISL_2814957, EPI_ISL_2814961, EPI_ISL_2814962, EPI_ISL_2814965, EPI_ISL_2814966, EPI_ISL_2814967, EPI_ISL_2814973, EPI_ISL_2814982, EPI_ISL_2814983, EPI_ISL_2814988, EPI_ISL_2814989, EPI_ISL_2814990, EPI_ISL_2814992, EPI_ISL_2814993, EPI_ISL_2814995, EPI_ISL_2814996, EPI_ISL_2814998,
EPI_ISL_2814999, EPI_ISL_2815001, EPI_ISL_2815004, EPI_ISL_2815005, EPI_ISL_2815008, EPI_ISL_2815010, EPI_ISL_2815014, EPI_ISL_2815016, EPI_ISL_2815018, EPI_ISL_2815021, EPI_ISL_2815023, EPI_ISL_2815025, EPI_ISL_2815030, EPI_ISL_2815031, EPI_ISL_2815032, EPI_ISL_2815033, EPI_ISL_2815035, EPI_ISL_2815037, EPI_ISL_2815041, EPI_ISL_2815043, EPI_ISL_2815044, EPI_ISL_2815045,
EPI_ISL_2815046, EPI_ISL_2815047, EPI_ISL_2815048, EPI_ISL_2815049, EPI_ISL_2815050, EPI_ISL_2815051, EPI_ISL_2815052, EPI_ISL_2815053, EPI_ISL_2815054, EPI_ISL_2815055, EPI_ISL_2815056, EPI_ISL_2815057, EPI_ISL_2815058, EPI_ISL_2815059, EPI_ISL_2815060, EPI_ISL_2815061, EPI_ISL_2815062, EPI_ISL_2815063, EPI_ISL_2815064, EPI_ISL_2815065, EPI_ISL_2815066, EPI_ISL_2815067,
EPI_ISL_2815068, EPI_ISL_2815069, EPI_ISL_2815070, EPI_ISL_2815071, EPI_ISL_2815072, EPI_ISL_2815073, EPI_ISL_2815074, EPI_ISL_2815075, EPI_ISL_2815076, EPI_ISL_2815077, EPI_ISL_2815078, EPI_ISL_2815079, EPI_ISL_2815080, EPI_ISL_2815081, EPI_ISL_2815100, EPI_ISL_2828759, EPI_ISL_2830613, EPI_ISL_2830789, EPI_ISL_2832076, EPI_ISL_2832091, EPI_ISL_2850762, EPI_ISL_2851795,
EPI_ISL_2852620, EPI_ISL_2852856, EPI_ISL_2852868, EPI_ISL_2852873, EPI_ISL_2852910, EPI_ISL_2852917, EPI_ISL_2852918, EPI_ISL_2852926, EPI_ISL_2852935, EPI_ISL_2852941, EPI_ISL_2852947, EPI_ISL_2852971, EPI_ISL_2853022

| | | | |
|---|---|---|---|
| see above | Lighthouse Lab in Glasgow | Wellcome Sanger Institute for the COVID-19 Genomics UK (COG-UK) Consortium | Anna Dominiczak and Alex Alderton; Carol Clugston; Cordelia Langford; David Gray; David K. Jackson; Dominic Kwiatkowski; Ewan Harrison; Harper VanSteenhouse; Ian Johnston; Jeffrey Barrett; John Sillitoe on behalf of the Wellcome Sanger Institute COVID-19 Surveillance Team; Roberto Amato; Sonia Goncalves; Yumi Kasai |

EPI_ISL_2717201, EPI_ISL_2728686, EPI_ISL_2728687, EPI_ISL_2728700, EPI_ISL_2728702, EPI_ISL_2728721, EPI_ISL_2728745, EPI_ISL_2728758, EPI_ISL_2728761, EPI_ISL_2728778, EPI_ISL_2728781, EPI_ISL_2728784, EPI_ISL_2728792, EPI_ISL_2728793, EPI_ISL_2728809, EPI_ISL_2728816, EPI_ISL_2728818, EPI_ISL_2728822, EPI_ISL_2728832, EPI_ISL_2728833, EPI_ISL_2728838, EPI_ISL_2728845,
EPI_ISL_2728848, EPI_ISL_2728850, EPI_ISL_2728867, EPI_ISL_2728872, EPI_ISL_2728884, EPI_ISL_2728897, EPI_ISL_2728901, EPI_ISL_2728907, EPI_ISL_2728908, EPI_ISL_2728923, EPI_ISL_2728924, EPI_ISL_2728929, EPI_ISL_2728930, EPI_ISL_2728932, EPI_ISL_2728956, EPI_ISL_2728979, EPI_ISL_2728980, EPI_ISL_2728985, EPI_ISL_2728990, EPI_ISL_2728996, EPI_ISL_2729020, EPI_ISL_2729048,
EPI_ISL_2729051, EPI_ISL_2729057, EPI_ISL_2729060, EPI_ISL_2729069, EPI_ISL_2729082, EPI_ISL_2729085, EPI_ISL_2729088, EPI_ISL_2729091, EPI_ISL_2729092, EPI_ISL_2729109, EPI_ISL_2729117, EPI_ISL_2729133, EPI_ISL_2729136, EPI_ISL_2729138, EPI_ISL_2729140, EPI_ISL_2729155, EPI_ISL_2729163, EPI_ISL_2729174, EPI_ISL_2729177, EPI_ISL_2729186, EPI_ISL_2729190, EPI_ISL_2729194,
EPI_ISL_2729195, EPI_ISL_2729206, EPI_ISL_2729210, EPI_ISL_2729216, EPI_ISL_2729219, EPI_ISL_2729227, EPI_ISL_2729230, EPI_ISL_2729235, EPI_ISL_2729236, EPI_ISL_2729238, EPI_ISL_2729241, EPI_ISL_2729243, EPI_ISL_2729248, EPI_ISL_2729254, EPI_ISL_2729255, EPI_ISL_2729259, EPI_ISL_2729260, EPI_ISL_2729262, EPI_ISL_2729263, EPI_ISL_2729266, EPI_ISL_2729271, EPI_ISL_2729277,
EPI_ISL_2729281, EPI_ISL_2729283, EPI_ISL_2729285, EPI_ISL_2729286, EPI_ISL_2729290, EPI_ISL_2729298, EPI_ISL_2729303, EPI_ISL_2729304, EPI_ISL_2729307, EPI_ISL_2729310, EPI_ISL_2729318, EPI_ISL_2729319, EPI_ISL_2729323, EPI_ISL_2729330, EPI_ISL_2729331, EPI_ISL_2729333, EPI_ISL_2729334, EPI_ISL_2729335, EPI_ISL_2729336, EPI_ISL_2729337, EPI_ISL_2729338, EPI_ISL_2729339,
EPI_ISL_2729344, EPI_ISL_2729345, EPI_ISL_2729348, EPI_ISL_2729357, EPI_ISL_2729365, EPI_ISL_2729370, EPI_ISL_2729376, EPI_ISL_2729379, EPI_ISL_2729380, EPI_ISL_2729942, EPI_ISL_2729948, EPI_ISL_2729951, EPI_ISL_2729956, EPI_ISL_2729963, EPI_ISL_2729964, EPI_ISL_2729968, EPI_ISL_2729970, EPI_ISL_2729972, EPI_ISL_2729976, EPI_ISL_2729985, EPI_ISL_2729991, EPI_ISL_2729996,
EPI_ISL_2729997, EPI_ISL_2729998, EPI_ISL_2729999, EPI_ISL_2730004, EPI_ISL_2730006, EPI_ISL_2730009, EPI_ISL_2730018, EPI_ISL_2730019, EPI_ISL_2730024, EPI_ISL_2730028, EPI_ISL_2730030, EPI_ISL_2730033, EPI_ISL_2730035, EPI_ISL_2730038, EPI_ISL_2730042, EPI_ISL_2730045, EPI_ISL_2730048, EPI_ISL_2730059, EPI_ISL_2730062, EPI_ISL_2730064, EPI_ISL_2730065, EPI_ISL_2730066,
EPI_ISL_2730067, EPI_ISL_2730068, EPI_ISL_2730075, EPI_ISL_2730079, EPI_ISL_2730080, EPI_ISL_2730091, EPI_ISL_2730092, EPI_ISL_2730113, EPI_ISL_2730114, EPI_ISL_2730115, EPI_ISL_2730118, EPI_ISL_2730119, EPI_ISL_2730121, EPI_ISL_2730124, EPI_ISL_2730125, EPI_ISL_2730133, EPI_ISL_2730135, EPI_ISL_2730136, EPI_ISL_2730137, EPI_ISL_2730139, EPI_ISL_2730140, EPI_ISL_2730152,
EPI_ISL_2730153, EPI_ISL_2730156, EPI_ISL_2730163, EPI_ISL_2730169, EPI_ISL_2730170, EPI_ISL_2730171, EPI_ISL_2730172, EPI_ISL_2730176, EPI_ISL_2730178, EPI_ISL_2730180, EPI_ISL_2730182, EPI_ISL_2730184, EPI_ISL_2730186, EPI_ISL_2730191, EPI_ISL_2730192, EPI_ISL_2730195, EPI_ISL_2730197, EPI_ISL_2730200, EPI_ISL_2730205, EPI_ISL_2730207, EPI_ISL_2730210, EPI_ISL_2730215,
EPI_ISL_2730216, EPI_ISL_2730224, EPI_ISL_2730225, EPI_ISL_2730229, EPI_ISL_2730242, EPI_ISL_2730243, EPI_ISL_2730244, EPI_ISL_2730248, EPI_ISL_2730250, EPI_ISL_2730253, EPI_ISL_2730266, EPI_ISL_2730271, EPI_ISL_2730277, EPI_ISL_2730280, EPI_ISL_2730285, EPI_ISL_2730293, EPI_ISL_2730311, EPI_ISL_2730318, EPI_ISL_2730330, EPI_ISL_2730334, EPI_ISL_2730349, EPI_ISL_2730357,
EPI_ISL_2730359, EPI_ISL_2730365, EPI_ISL_2730376, EPI_ISL_2730397, EPI_ISL_2730398, EPI_ISL_2730399, EPI_ISL_2730400, EPI_ISL_2730408, EPI_ISL_2730413, EPI_ISL_2730425, EPI_ISL_2730427, EPI_ISL_2730437, EPI_ISL_2730441, EPI_ISL_2730445, EPI_ISL_2730451, EPI_ISL_2730454, EPI_ISL_2730460, EPI_ISL_2730463, EPI_ISL_2730470, EPI_ISL_2730476, EPI_ISL_2730493, EPI_ISL_2730496,
EPI_ISL_2730521, EPI_ISL_2730531, EPI_ISL_2730536, EPI_ISL_2730544, EPI_ISL_2730558, EPI_ISL_2730564, EPI_ISL_2730565, EPI_ISL_2730568, EPI_ISL_2730573, EPI_ISL_2730584, EPI_ISL_2730586, EPI_ISL_2730590, EPI_ISL_2730593, EPI_ISL_2730601, EPI_ISL_2730604, EPI_ISL_2730611, EPI_ISL_2730617, EPI_ISL_2730622, EPI_ISL_2730632, EPI_ISL_2730635, EPI_ISL_2730638, EPI_ISL_2765575,
EPI_ISL_2765580, EPI_ISL_2765584, EPI_ISL_2765588, EPI_ISL_2765590, EPI_ISL_2765604, EPI_ISL_2765606, EPI_ISL_2765612, EPI_ISL_2765613, EPI_ISL_2765629, EPI_ISL_2765631, EPI_ISL_2765633, EPI_ISL_2765637, EPI_ISL_2765641, EPI_ISL_2765644, EPI_ISL_2765645, EPI_ISL_2765648, EPI_ISL_2765649, EPI_ISL_2765653, EPI_ISL_2765656, EPI_ISL_2765657, EPI_ISL_2765658, EPI_ISL_2765663,
EPI_ISL_2765670, EPI_ISL_2765673, EPI_ISL_2765674, EPI_ISL_2765676, EPI_ISL_2765682, EPI_ISL_2765685, EPI_ISL_2765687, EPI_ISL_2765691, EPI_ISL_2765694, EPI_ISL_2765695, EPI_ISL_2765696, EPI_ISL_2765697, EPI_ISL_2765701, EPI_ISL_2765703, EPI_ISL_2765707, EPI_ISL_2765708, EPI_ISL_2765709, EPI_ISL_2765711, EPI_ISL_2765712, EPI_ISL_2765718, EPI_ISL_2765719, EPI_ISL_2765722,
EPI_ISL_2765725, EPI_ISL_2765726, EPI_ISL_2765728, EPI_ISL_2765736, EPI_ISL_2765738, EPI_ISL_2765741, EPI_ISL_2765744, EPI_ISL_2765750, EPI_ISL_2765751, EPI_ISL_2765752, EPI_ISL_2765757, EPI_ISL_2765763, EPI_ISL_2765768, EPI_ISL_2765770, EPI_ISL_2765777, EPI_ISL_2765784, EPI_ISL_2765789, EPI_ISL_2765790, EPI_ISL_2765792, EPI_ISL_2765797, EPI_ISL_2765803, EPI_ISL_2765817,
EPI_ISL_2765819, EPI_ISL_2765830, EPI_ISL_2765839, EPI_ISL_2765842, EPI_ISL_2765844, EPI_ISL_2765851, EPI_ISL_2765852, EPI_ISL_2765860, EPI_ISL_2765862, EPI_ISL_2765864, EPI_ISL_2766555, EPI_ISL_2766559, EPI_ISL_2766560, EPI_ISL_2766561, EPI_ISL_2766566, EPI_ISL_2766569, EPI_ISL_2766571, EPI_ISL_2766574, EPI_ISL_2766577, EPI_ISL_2766578, EPI_ISL_2766586, EPI_ISL_2766589,
EPI_ISL_2766595, EPI_ISL_2766597, EPI_ISL_2766599, EPI_ISL_2766601, EPI_ISL_2766611, EPI_ISL_2766615, EPI_ISL_2766618, EPI_ISL_2766620, EPI_ISL_2766625, EPI_ISL_2766627, EPI_ISL_2766629, EPI_ISL_2766631, EPI_ISL_2766634, EPI_ISL_2766635, EPI_ISL_2766642, EPI_ISL_2766644, EPI_ISL_2766646, EPI_ISL_2766648, EPI_ISL_2766649, EPI_ISL_2766654, EPI_ISL_2766656, EPI_ISL_2766665,
EPI_ISL_2766666, EPI_ISL_2766667, EPI_ISL_2766671, EPI_ISL_2766675, EPI_ISL_2766678, EPI_ISL_2766681, EPI_ISL_2766687, EPI_ISL_2766691, EPI_ISL_2766692, EPI_ISL_2766698, EPI_ISL_2766700, EPI_ISL_2766701, EPI_ISL_2766709, EPI_ISL_2766710, EPI_ISL_2766712, EPI_ISL_2766713, EPI_ISL_2766720, EPI_ISL_2766722, EPI_ISL_2766723, EPI_ISL_2766725, EPI_ISL_2766726, EPI_ISL_2766727,
EPI_ISL_2766729, EPI_ISL_2766731, EPI_ISL_2766732, EPI_ISL_2766733, EPI_ISL_2766739, EPI_ISL_2766740, EPI_ISL_2766744, EPI_ISL_2766746, EPI_ISL_2766747, EPI_ISL_2766749, EPI_ISL_2766752, EPI_ISL_2766753, EPI_ISL_2766764, EPI_ISL_2766765, EPI_ISL_2766766, EPI_ISL_2766769, EPI_ISL_2766771, EPI_ISL_2766774, EPI_ISL_2766776, EPI_ISL_2766778, EPI_ISL_2766789, EPI_ISL_2766791,
EPI_ISL_2766792, EPI_ISL_2766795, EPI_ISL_2766797, EPI_ISL_2766799, EPI_ISL_2766800, EPI_ISL_2766803, EPI_ISL_2766805, EPI_ISL_2766808, EPI_ISL_2766813, EPI_ISL_2766817, EPI_ISL_2766819, EPI_ISL_2766823, EPI_ISL_2766824, EPI_ISL_2766831, EPI_ISL_2766833, EPI_ISL_2766836, EPI_ISL_2766839, EPI_ISL_2766841, EPI_ISL_2766843, EPI_ISL_2766850, EPI_ISL_2766853, EPI_ISL_2766854,
EPI_ISL_2766855, EPI_ISL_2766858, EPI_ISL_2766859, EPI_ISL_2766860, EPI_ISL_2766866, EPI_ISL_2766867, EPI_ISL_2766869, EPI_ISL_2766874, EPI_ISL_2766876, EPI_ISL_2766878, EPI_ISL_2766883, EPI_ISL_2766886, EPI_ISL_2766888, EPI_ISL_2766889, EPI_ISL_2766890, EPI_ISL_2766893, EPI_ISL_2766895, EPI_ISL_2766897, EPI_ISL_2766905, EPI_ISL_2766908, EPI_ISL_2766918, EPI_ISL_2766923,
EPI_ISL_2766924, EPI_ISL_2766926, EPI_ISL_2766927, EPI_ISL_2766928, EPI_ISL_2766930, EPI_ISL_2766931, EPI_ISL_2766941, EPI_ISL_2766942, EPI_ISL_2766948, EPI_ISL_2766949, EPI_ISL_2766959, EPI_ISL_2766962, EPI_ISL_2766965, EPI_ISL_2766966, EPI_ISL_2766971, EPI_ISL_2766977, EPI_ISL_2766985, EPI_ISL_2766996, EPI_ISL_2767004, EPI_ISL_2767009, EPI_ISL_2767010, EPI_ISL_2767013,
EPI_ISL_2767018, EPI_ISL_2767021, EPI_ISL_2767023, EPI_ISL_2767028, EPI_ISL_2767031, EPI_ISL_2767032, EPI_ISL_2767039, EPI_ISL_2767040, EPI_ISL_2767045, EPI_ISL_2767046, EPI_ISL_2767047, EPI_ISL_2767051, EPI_ISL_2767058, EPI_ISL_2767066, EPI_ISL_2767072, EPI_ISL_2767081, EPI_ISL_2767083, EPI_ISL_2767084, EPI_ISL_2767088, EPI_ISL_2767089, EPI_ISL_2767091, EPI_ISL_2767092,
EPI_ISL_2767099, EPI_ISL_2767101, EPI_ISL_2767109, EPI_ISL_2767113, EPI_ISL_2767116, EPI_ISL_2767118, EPI_ISL_2767124, EPI_ISL_2767131, EPI_ISL_2767135, EPI_ISL_2767136, EPI_ISL_2767138, EPI_ISL_2767139, EPI_ISL_2767143, EPI_ISL_2767152, EPI_ISL_2767153, EPI_ISL_2767159, EPI_ISL_2767164, EPI_ISL_2767167, EPI_ISL_2767170, EPI_ISL_2767171, EPI_ISL_2767177, EPI_ISL_2767179,
EPI_ISL_2767180, EPI_ISL_2767191, EPI_ISL_2767193, EPI_ISL_2767196, EPI_ISL_2767202, EPI_ISL_2767204, EPI_ISL_2767208, EPI_ISL_2767209, EPI_ISL_2767210, EPI_ISL_2767211, EPI_ISL_2767212, EPI_ISL_2767216, EPI_ISL_2767219, EPI_ISL_2767225, EPI_ISL_2767227, EPI_ISL_2767233, EPI_ISL_2779654, EPI_ISL_2779658, EPI_ISL_2779665, EPI_ISL_2779668, EPI_ISL_2779676, EPI_ISL_2779690,
EPI_ISL_2779703, EPI_ISL_2779706, EPI_ISL_2779712, EPI_ISL_2779739, EPI_ISL_2779744, EPI_ISL_2779747, EPI_ISL_2779760, EPI_ISL_2779769, EPI_ISL_2779776, EPI_ISL_2779783, EPI_ISL_2779786, EPI_ISL_2779788, EPI_ISL_2779791, EPI_ISL_2779792, EPI_ISL_2779797, EPI_ISL_2779800, EPI_ISL_2779801, EPI_ISL_2779811, EPI_ISL_2779830, EPI_ISL_2779834, EPI_ISL_2779839, EPI_ISL_2779844,
EPI_ISL_2779852, EPI_ISL_2779857, EPI_ISL_2779861, EPI_ISL_2779871, EPI_ISL_2779883, EPI_ISL_2779890, EPI_ISL_2779900, EPI_ISL_2779902, EPI_ISL_2779905, EPI_ISL_2779912, EPI_ISL_2779929, EPI_ISL_2779935, EPI_ISL_2779936, EPI_ISL_2779941, EPI_ISL_2779950, EPI_ISL_2779958, EPI_ISL_2779966, EPI_ISL_2779967, EPI_ISL_2779973, EPI_ISL_2779984, EPI_ISL_2779985, EPI_ISL_2779988,
EPI_ISL_2779993, EPI_ISL_2779995, EPI_ISL_2780003, EPI_ISL_2780007, EPI_ISL_2780010, EPI_ISL_2780017, EPI_ISL_2780018, EPI_ISL_2780020, EPI_ISL_2780021, EPI_ISL_2780022, EPI_ISL_2780030, EPI_ISL_2780033, EPI_ISL_2780049, EPI_ISL_2780053, EPI_ISL_2780073, EPI_ISL_2780075, EPI_ISL_2780079, EPI_ISL_2780088, EPI_ISL_2780100, EPI_ISL_2780103, EPI_ISL_2780110, EPI_ISL_2780111,
EPI_ISL_2780116, EPI_ISL_2780118, EPI_ISL_2780125, EPI_ISL_2780128, EPI_ISL_2780130, EPI_ISL_2780132, EPI_ISL_2780137, EPI_ISL_2780144, EPI_ISL_2780151, EPI_ISL_2780159, EPI_ISL_2780161, EPI_ISL_2780162, EPI_ISL_2780165, EPI_ISL_2780167, EPI_ISL_2780171, EPI_ISL_2780177, EPI_ISL_2780178, EPI_ISL_2780185, EPI_ISL_2780186, EPI_ISL_2780187, EPI_ISL_2780206, EPI_ISL_2780210,
EPI_ISL_2780211, EPI_ISL_2780213, EPI_ISL_2780219, EPI_ISL_2780223, EPI_ISL_2780226, EPI_ISL_2780235, EPI_ISL_2780242, EPI_ISL_2780246, EPI_ISL_2780247, EPI_ISL_2780249, EPI_ISL_2780250, EPI_ISL_2780251, EPI_ISL_2780252, EPI_ISL_2780256, EPI_ISL_2780272, EPI_ISL_2780279, EPI_ISL_2780286, EPI_ISL_2780288, EPI_ISL_2780289, EPI_ISL_2780291, EPI_ISL_2780299, EPI_ISL_2780310,
EPI_ISL_2780311, EPI_ISL_2780312, EPI_ISL_2780314, EPI_ISL_2780327, EPI_ISL_2780328, EPI_ISL_2780332, EPI_ISL_2780334, EPI_ISL_2780343, EPI_ISL_2780347, EPI_ISL_2780350, EPI_ISL_2780353, EPI_ISL_2780355, EPI_ISL_2780357, EPI_ISL_2780358, EPI_ISL_2780368, EPI_ISL_2780370, EPI_ISL_2780376, EPI_ISL_2780377, EPI_ISL_2780379, EPI_ISL_2780386, EPI_ISL_2780388, EPI_ISL_2780398,

EPI_ISL_2780401, EPI_ISL_2780402, EPI_ISL_2780406, EPI_ISL_2780423, EPI_ISL_2780426, EPI_ISL_2780428, EPI_ISL_2780433, EPI_ISL_2780437, EPI_ISL_2780439, EPI_ISL_2780440, EPI_ISL_2780441, EPI_ISL_2780444, EPI_ISL_2780446, EPI_ISL_2780449, EPI_ISL_2780450, EPI_ISL_2780451, EPI_ISL_2780458, EPI_ISL_2780459, EPI_ISL_2780467, EPI_ISL_2780468, EPI_ISL_2780471, EPI_ISL_2780472,
EPI_ISL_2780473, EPI_ISL_2780474, EPI_ISL_2780475, EPI_ISL_2780476, EPI_ISL_2780478, EPI_ISL_2780479, EPI_ISL_2780482, EPI_ISL_2780483, EPI_ISL_2780485, EPI_ISL_2780486, EPI_ISL_2780487, EPI_ISL_2780488, EPI_ISL_2780490, EPI_ISL_2780492, EPI_ISL_2780493, EPI_ISL_2780494, EPI_ISL_2780495, EPI_ISL_2780496, EPI_ISL_2780497, EPI_ISL_2780498, EPI_ISL_2780499, EPI_ISL_2780500,
EPI_ISL_2780501, EPI_ISL_2780502, EPI_ISL_2780503, EPI_ISL_2780504, EPI_ISL_2780505, EPI_ISL_2780507, EPI_ISL_2780509, EPI_ISL_2780510, EPI_ISL_2780511, EPI_ISL_2780513, EPI_ISL_2780516, EPI_ISL_2780517, EPI_ISL_2780518, EPI_ISL_2780519, EPI_ISL_2780520, EPI_ISL_2780521, EPI_ISL_2780522, EPI_ISL_2780523, EPI_ISL_2780524, EPI_ISL_2780525, EPI_ISL_2780526, EPI_ISL_2780527,
EPI_ISL_2780528, EPI_ISL_2780529, EPI_ISL_2780530, EPI_ISL_2780531, EPI_ISL_2780532, EPI_ISL_2780533, EPI_ISL_2780534, EPI_ISL_2780535, EPI_ISL_2780537, EPI_ISL_2780540, EPI_ISL_2780541, EPI_ISL_2780542, EPI_ISL_2780544, EPI_ISL_2780546, EPI_ISL_2780547, EPI_ISL_2780548, EPI_ISL_2780549, EPI_ISL_2780550, EPI_ISL_2780551, EPI_ISL_2780552, EPI_ISL_2780553, EPI_ISL_2780554,
EPI_ISL_2780557, EPI_ISL_2780558, EPI_ISL_2780559, EPI_ISL_2780561, EPI_ISL_2780562, EPI_ISL_2780564, EPI_ISL_2780565, EPI_ISL_2780566, EPI_ISL_2780567, EPI_ISL_2780569, EPI_ISL_2780570, EPI_ISL_2780571, EPI_ISL_2780572, EPI_ISL_2780573, EPI_ISL_2780574, EPI_ISL_2780575, EPI_ISL_2780576, EPI_ISL_2780577, EPI_ISL_2780578, EPI_ISL_2780579, EPI_ISL_2780580, EPI_ISL_2780581,
EPI_ISL_2780582, EPI_ISL_2780584, EPI_ISL_2780586, EPI_ISL_2780589, EPI_ISL_2780590, EPI_ISL_2780591, EPI_ISL_2780592, EPI_ISL_2780593, EPI_ISL_2780595, EPI_ISL_2780596, EPI_ISL_2780598, EPI_ISL_2780599, EPI_ISL_2780600, EPI_ISL_2780601, EPI_ISL_2780602, EPI_ISL_2780603, EPI_ISL_2780604, EPI_ISL_2780605, EPI_ISL_2780606, EPI_ISL_2780607, EPI_ISL_2780608, EPI_ISL_2780610,
EPI_ISL_2780611, EPI_ISL_2780612, EPI_ISL_2780613, EPI_ISL_2780614, EPI_ISL_2780615, EPI_ISL_2780616, EPI_ISL_2780617, EPI_ISL_2780619, EPI_ISL_2780621, EPI_ISL_2780622, EPI_ISL_2780625, EPI_ISL_2780626, EPI_ISL_2780627, EPI_ISL_2780628, EPI_ISL_2780629, EPI_ISL_2780630, EPI_ISL_2780631, EPI_ISL_2780632, EPI_ISL_2780633, EPI_ISL_2780635, EPI_ISL_2780636, EPI_ISL_2780638,
EPI_ISL_2780639, EPI_ISL_2780640, EPI_ISL_2780641, EPI_ISL_2780642, EPI_ISL_2780643, EPI_ISL_2780644, EPI_ISL_2780645, EPI_ISL_2780646, EPI_ISL_2780647, EPI_ISL_2780648, EPI_ISL_2780649, EPI_ISL_2780650, EPI_ISL_2780651, EPI_ISL_2780652, EPI_ISL_2780653, EPI_ISL_2780654, EPI_ISL_2780655, EPI_ISL_2780657, EPI_ISL_2780658, EPI_ISL_2780659, EPI_ISL_2780660, EPI_ISL_2780661,
EPI_ISL_2780662, EPI_ISL_2780663, EPI_ISL_2780664, EPI_ISL_2780665, EPI_ISL_2780666, EPI_ISL_2780667, EPI_ISL_2780668, EPI_ISL_2780669, EPI_ISL_2780670, EPI_ISL_2780672, EPI_ISL_2780673, EPI_ISL_2780675, EPI_ISL_2780676, EPI_ISL_2780677, EPI_ISL_2780678, EPI_ISL_2780679, EPI_ISL_2780681, EPI_ISL_2780682, EPI_ISL_2780683, EPI_ISL_2780684, EPI_ISL_2780685, EPI_ISL_2780686,
EPI_ISL_2780687, EPI_ISL_2780689, EPI_ISL_2780690, EPI_ISL_2780691, EPI_ISL_2780692, EPI_ISL_2780693, EPI_ISL_2780694, EPI_ISL_2780695, EPI_ISL_2780696, EPI_ISL_2780697, EPI_ISL_2780698, EPI_ISL_2780700, EPI_ISL_2780702, EPI_ISL_2780703, EPI_ISL_2780704, EPI_ISL_2780705, EPI_ISL_2780706, EPI_ISL_2780707, EPI_ISL_2780708, EPI_ISL_2780709, EPI_ISL_2780710, EPI_ISL_2780711,
EPI_ISL_2780712, EPI_ISL_2780713, EPI_ISL_2780714, EPI_ISL_2780715, EPI_ISL_2780716, EPI_ISL_2780717, EPI_ISL_2780718, EPI_ISL_2780719, EPI_ISL_2780720, EPI_ISL_2780721, EPI_ISL_2780722, EPI_ISL_2780723, EPI_ISL_2780724, EPI_ISL_2780726, EPI_ISL_2780727, EPI_ISL_2780728, EPI_ISL_2780730, EPI_ISL_2780731, EPI_ISL_2780732, EPI_ISL_2780733, EPI_ISL_2780734, EPI_ISL_2780735,
EPI_ISL_2780736, EPI_ISL_2780737, EPI_ISL_2780738, EPI_ISL_2780740, EPI_ISL_2780741, EPI_ISL_2780742, EPI_ISL_2780743, EPI_ISL_2780744, EPI_ISL_2780745, EPI_ISL_2780746, EPI_ISL_2780747, EPI_ISL_2780748, EPI_ISL_2780750, EPI_ISL_2780752, EPI_ISL_2780753, EPI_ISL_2780755, EPI_ISL_2780756, EPI_ISL_2780757, EPI_ISL_2780758, EPI_ISL_2780759, EPI_ISL_2780760, EPI_ISL_2780761,
EPI_ISL_2780762, EPI_ISL_2780764, EPI_ISL_2780765, EPI_ISL_2780766, EPI_ISL_2780767, EPI_ISL_2780768, EPI_ISL_2780769, EPI_ISL_2780770, EPI_ISL_2780771, EPI_ISL_2780772, EPI_ISL_2780773, EPI_ISL_2780776, EPI_ISL_2780777, EPI_ISL_2780778, EPI_ISL_2780779, EPI_ISL_2780780, EPI_ISL_2780782, EPI_ISL_2780783, EPI_ISL_2780784, EPI_ISL_2780785, EPI_ISL_2780786, EPI_ISL_2780787,
EPI_ISL_2780788, EPI_ISL_2780789, EPI_ISL_2780791, EPI_ISL_2780794, EPI_ISL_2780795, EPI_ISL_2780796, EPI_ISL_2780799, EPI_ISL_2780801, EPI_ISL_2780802, EPI_ISL_2780803, EPI_ISL_2780805, EPI_ISL_2780806, EPI_ISL_2780807, EPI_ISL_2780808, EPI_ISL_2780809, EPI_ISL_2780810, EPI_ISL_2780811, EPI_ISL_2780812, EPI_ISL_2780813, EPI_ISL_2780814, EPI_ISL_2780816, EPI_ISL_2780818,
EPI_ISL_2780819, EPI_ISL_2780820, EPI_ISL_2780821, EPI_ISL_2780822, EPI_ISL_2780824, EPI_ISL_2780825, EPI_ISL_2780826, EPI_ISL_2780827, EPI_ISL_2780828, EPI_ISL_2780829, EPI_ISL_2780830, EPI_ISL_2780831, EPI_ISL_2780832, EPI_ISL_2780833, EPI_ISL_2780835, EPI_ISL_2780836, EPI_ISL_2780837, EPI_ISL_2780838, EPI_ISL_2780840, EPI_ISL_2780842, EPI_ISL_2780843, EPI_ISL_2780845,
EPI_ISL_2780847, EPI_ISL_2780849, EPI_ISL_2780850, EPI_ISL_2780855, EPI_ISL_2780858, EPI_ISL_2780859, EPI_ISL_2780860, EPI_ISL_2780861, EPI_ISL_2780862, EPI_ISL_2780864, EPI_ISL_2780865, EPI_ISL_2780866, EPI_ISL_2780869, EPI_ISL_2780870, EPI_ISL_2780872, EPI_ISL_2780873, EPI_ISL_2780874, EPI_ISL_2780875, EPI_ISL_2780876, EPI_ISL_2780877, EPI_ISL_2780878, EPI_ISL_2780879,
EPI_ISL_2780880, EPI_ISL_2780882, EPI_ISL_2780883, EPI_ISL_2780884, EPI_ISL_2780885, EPI_ISL_2780886, EPI_ISL_2780887, EPI_ISL_2780889, EPI_ISL_2780890, EPI_ISL_2780891, EPI_ISL_2780892, EPI_ISL_2780893, EPI_ISL_2780894, EPI_ISL_2780895, EPI_ISL_2780896, EPI_ISL_2780898, EPI_ISL_2780899, EPI_ISL_2780900, EPI_ISL_2780901, EPI_ISL_2780902, EPI_ISL_2780903, EPI_ISL_2780904,
EPI_ISL_2780906, EPI_ISL_2780907, EPI_ISL_2780908, EPI_ISL_2780909, EPI_ISL_2780912, EPI_ISL_2780914, EPI_ISL_2780916, EPI_ISL_2780917, EPI_ISL_2780920, EPI_ISL_2780921, EPI_ISL_2780922, EPI_ISL_2780924, EPI_ISL_2780925, EPI_ISL_2780928, EPI_ISL_2780930, EPI_ISL_2780931, EPI_ISL_2780932, EPI_ISL_2780933, EPI_ISL_2780935, EPI_ISL_2780936, EPI_ISL_2780937, EPI_ISL_2780938,
EPI_ISL_2780939, EPI_ISL_2780943, EPI_ISL_2780946, EPI_ISL_2780947, EPI_ISL_2780954, EPI_ISL_2780957, EPI_ISL_2780958, EPI_ISL_2780960, EPI_ISL_2780961, EPI_ISL_2780962, EPI_ISL_2780963, EPI_ISL_2780964, EPI_ISL_2780966, EPI_ISL_2780967, EPI_ISL_2780968, EPI_ISL_2780969, EPI_ISL_2780972, EPI_ISL_2780973, EPI_ISL_2780974, EPI_ISL_2780975, EPI_ISL_2780978, EPI_ISL_2780979,
EPI_ISL_2780980, EPI_ISL_2780981, EPI_ISL_2780983, EPI_ISL_2780985, EPI_ISL_2780986, EPI_ISL_2780987, EPI_ISL_2780991, EPI_ISL_2780992, EPI_ISL_2780993, EPI_ISL_2780995, EPI_ISL_2780996, EPI_ISL_2780998, EPI_ISL_2780999, EPI_ISL_2781000, EPI_ISL_2781001, EPI_ISL_2781003, EPI_ISL_2781004, EPI_ISL_2781005, EPI_ISL_2781006, EPI_ISL_2781007, EPI_ISL_2781008, EPI_ISL_2781010,
EPI_ISL_2781012, EPI_ISL_2781013, EPI_ISL_2781014, EPI_ISL_2781015, EPI_ISL_2781016, EPI_ISL_2781017, EPI_ISL_2781018, EPI_ISL_2781019, EPI_ISL_2781020, EPI_ISL_2781021, EPI_ISL_2781022, EPI_ISL_2781023, EPI_ISL_2781024, EPI_ISL_2781025, EPI_ISL_2781026, EPI_ISL_2781027, EPI_ISL_2781028, EPI_ISL_2781030, EPI_ISL_2781031, EPI_ISL_2781032, EPI_ISL_2781034, EPI_ISL_2781036,
EPI_ISL_2781037, EPI_ISL_2781038, EPI_ISL_2781039, EPI_ISL_2781040, EPI_ISL_2781044, EPI_ISL_2781045, EPI_ISL_2781046, EPI_ISL_2781047, EPI_ISL_2781048, EPI_ISL_2781049, EPI_ISL_2781050, EPI_ISL_2781051, EPI_ISL_2781052, EPI_ISL_2781053, EPI_ISL_2781054, EPI_ISL_2781056, EPI_ISL_2781058, EPI_ISL_2781060, EPI_ISL_2781061, EPI_ISL_2781062, EPI_ISL_2781063, EPI_ISL_2781064,
EPI_ISL_2781066, EPI_ISL_2781068, EPI_ISL_2781069, EPI_ISL_2781070, EPI_ISL_2781071, EPI_ISL_2781072, EPI_ISL_2781073, EPI_ISL_2781074, EPI_ISL_2781075, EPI_ISL_2781076, EPI_ISL_2781077, EPI_ISL_2781078, EPI_ISL_2781079, EPI_ISL_2781081, EPI_ISL_2781084, EPI_ISL_2781085, EPI_ISL_2781086, EPI_ISL_2781087, EPI_ISL_2781088, EPI_ISL_2781089, EPI_ISL_2781090, EPI_ISL_2781093,
EPI_ISL_2781094, EPI_ISL_2781095, EPI_ISL_2781097, EPI_ISL_2781098, EPI_ISL_2781099, EPI_ISL_2781100, EPI_ISL_2781101, EPI_ISL_2781104, EPI_ISL_2781105, EPI_ISL_2781106, EPI_ISL_2781107, EPI_ISL_2781108, EPI_ISL_2781109, EPI_ISL_2781110, EPI_ISL_2781111, EPI_ISL_2781112, EPI_ISL_2781113, EPI_ISL_2781115, EPI_ISL_2781118, EPI_ISL_2781120, EPI_ISL_2781121, EPI_ISL_2781122,
EPI_ISL_2781123, EPI_ISL_2781124, EPI_ISL_2781127, EPI_ISL_2781128, EPI_ISL_2781129, EPI_ISL_2781131, EPI_ISL_2781132, EPI_ISL_2781133, EPI_ISL_2781134, EPI_ISL_2781136, EPI_ISL_2781137, EPI_ISL_2781138, EPI_ISL_2781139, EPI_ISL_2781140, EPI_ISL_2781141, EPI_ISL_2781142, EPI_ISL_2781143, EPI_ISL_2781144, EPI_ISL_2781145, EPI_ISL_2781146, EPI_ISL_2781147, EPI_ISL_2781148,
EPI_ISL_2781149, EPI_ISL_2781150, EPI_ISL_2781151, EPI_ISL_2781152, EPI_ISL_2781153, EPI_ISL_2781156, EPI_ISL_2781157, EPI_ISL_2781158, EPI_ISL_2781159, EPI_ISL_2781160, EPI_ISL_2781163, EPI_ISL_2781166, EPI_ISL_2781167, EPI_ISL_2781168, EPI_ISL_2781170, EPI_ISL_2781171, EPI_ISL_2781173, EPI_ISL_2781174, EPI_ISL_2781175, EPI_ISL_2781176, EPI_ISL_2781177, EPI_ISL_2781178,
EPI_ISL_2781180, EPI_ISL_2781182, EPI_ISL_2781183, EPI_ISL_2781185, EPI_ISL_2781187, EPI_ISL_2781188, EPI_ISL_2781189, EPI_ISL_2781190, EPI_ISL_2781192, EPI_ISL_2781194, EPI_ISL_2781195, EPI_ISL_2781196, EPI_ISL_2781197, EPI_ISL_2781199, EPI_ISL_2781200, EPI_ISL_2781203, EPI_ISL_2781204, EPI_ISL_2781206, EPI_ISL_2781207, EPI_ISL_2781208, EPI_ISL_2781212, EPI_ISL_2781213,
EPI_ISL_2781214, EPI_ISL_2781215, EPI_ISL_2781216, EPI_ISL_2781217, EPI_ISL_2781219, EPI_ISL_2781220, EPI_ISL_2781221, EPI_ISL_2781223, EPI_ISL_2781224, EPI_ISL_2781225, EPI_ISL_2781226, EPI_ISL_2781228, EPI_ISL_2781230, EPI_ISL_2781234, EPI_ISL_2781237, EPI_ISL_2781239, EPI_ISL_2781240, EPI_ISL_2781241, EPI_ISL_2781242, EPI_ISL_2781243, EPI_ISL_2781244, EPI_ISL_2781245,
EPI_ISL_2781246, EPI_ISL_2781247, EPI_ISL_2781248, EPI_ISL_2781252, EPI_ISL_2781254, EPI_ISL_2781255, EPI_ISL_2781256, EPI_ISL_2781257, EPI_ISL_2781258, EPI_ISL_2781260, EPI_ISL_2781261, EPI_ISL_2781262, EPI_ISL_2781264, EPI_ISL_2781265, EPI_ISL_2781266, EPI_ISL_2781267, EPI_ISL_2781270, EPI_ISL_2781272, EPI_ISL_2781273, EPI_ISL_2781274, EPI_ISL_2781275, EPI_ISL_2781276,
EPI_ISL_2781277, EPI_ISL_2781280, EPI_ISL_2781282, EPI_ISL_2781283, EPI_ISL_2781286, EPI_ISL_2781290, EPI_ISL_2781291, EPI_ISL_2781292, EPI_ISL_2781294, EPI_ISL_2781297, EPI_ISL_2781298, EPI_ISL_2781300, EPI_ISL_2781301, EPI_ISL_2781302, EPI_ISL_2781303, EPI_ISL_2781304, EPI_ISL_2781305, EPI_ISL_2781307, EPI_ISL_2781308, EPI_ISL_2781309, EPI_ISL_2781312, EPI_ISL_2781313,
EPI_ISL_2781317, EPI_ISL_2781318, EPI_ISL_2781319, EPI_ISL_2781320, EPI_ISL_2781321, EPI_ISL_2781324, EPI_ISL_2781328, EPI_ISL_2781329, EPI_ISL_2781331, EPI_ISL_2781332, EPI_ISL_2781333, EPI_ISL_2781334, EPI_ISL_2781335, EPI_ISL_2781337, EPI_ISL_2781340, EPI_ISL_2781341, EPI_ISL_2781342, EPI_ISL_2781343, EPI_ISL_2781345, EPI_ISL_2781346, EPI_ISL_2781347, EPI_ISL_2781348,
EPI_ISL_2781349, EPI_ISL_2781350, EPI_ISL_2781351, EPI_ISL_2781352, EPI_ISL_2781353, EPI_ISL_2781355, EPI_ISL_2781356, EPI_ISL_2781357, EPI_ISL_2781358, EPI_ISL_2781359, EPI_ISL_2781360, EPI_ISL_2781362, EPI_ISL_2781364, EPI_ISL_2781365, EPI_ISL_2781366, EPI_ISL_2781367, EPI_ISL_2781368, EPI_ISL_2781369, EPI_ISL_2781370, EPI_ISL_2781371, EPI_ISL_2781372, EPI_ISL_2781375,
EPI_ISL_2781376, EPI_ISL_2781377, EPI_ISL_2781379, EPI_ISL_2781382, EPI_ISL_2781385, EPI_ISL_2781387, EPI_ISL_2781391, EPI_ISL_2781392, EPI_ISL_2781393, EPI_ISL_2781394, EPI_ISL_2781395, EPI_ISL_2781396, EPI_ISL_2781397, EPI_ISL_2781400, EPI_ISL_2781401, EPI_ISL_2781404, EPI_ISL_2781405, EPI_ISL_2781411, EPI_ISL_2781412, EPI_ISL_2781413, EPI_ISL_2781415, EPI_ISL_2781416,
EPI_ISL_2781418, EPI_ISL_2781419, EPI_ISL_2781420, EPI_ISL_2781421, EPI_ISL_2781422, EPI_ISL_2781423, EPI_ISL_2781424, EPI_ISL_2781425, EPI_ISL_2781426, EPI_ISL_2781427, EPI_ISL_2781429, EPI_ISL_2781430, EPI_ISL_2781431, EPI_ISL_2781432, EPI_ISL_2781433, EPI_ISL_2781435, EPI_ISL_2781436, EPI_ISL_2781437, EPI_ISL_2781438, EPI_ISL_2781439, EPI_ISL_2781440, EPI_ISL_2781441,
EPI_ISL_2781442, EPI_ISL_2781443, EPI_ISL_2781444, EPI_ISL_2781445, EPI_ISL_2781446, EPI_ISL_2781447, EPI_ISL_2781449, EPI_ISL_2781453, EPI_ISL_2781454, EPI_ISL_2781459, EPI_ISL_2781460, EPI_ISL_2781462, EPI_ISL_2781463, EPI_ISL_2781464, EPI_ISL_2781465, EPI_ISL_2781467, EPI_ISL_2781468, EPI_ISL_2781469, EPI_ISL_2781470, EPI_ISL_2781471, EPI_ISL_2781472, EPI_ISL_2781473,
EPI_ISL_2781475, EPI_ISL_2781477, EPI_ISL_2781479, EPI_ISL_2781480, EPI_ISL_2781481, EPI_ISL_2781482, EPI_ISL_2781483, EPI_ISL_2781484, EPI_ISL_2782039, EPI_ISL_2782040, EPI_ISL_2782041, EPI_ISL_2782067, EPI_ISL_2782069, EPI_ISL_2782072, EPI_ISL_2782082, EPI_ISL_2782089, EPI_ISL_2782107, EPI_ISL_2782114, EPI_ISL_2782115, EPI_ISL_2782122, EPI_ISL_2782130, EPI_ISL_2782135,
EPI_ISL_2782137, EPI_ISL_2782144, EPI_ISL_2782172, EPI_ISL_2782178, EPI_ISL_2782184, EPI_ISL_2782189, EPI_ISL_2782190, EPI_ISL_2782200, EPI_ISL_2782206, EPI_ISL_2782244, EPI_ISL_2782246, EPI_ISL_2782260, EPI_ISL_2782266, EPI_ISL_2782280, EPI_ISL_2782297, EPI_ISL_2782306, EPI_ISL_2782307, EPI_ISL_2803769, EPI_ISL_2803843, EPI_ISL_2803887, EPI_ISL_2803931, EPI_ISL_2804067,
EPI_ISL_2804202, EPI_ISL_2804625, EPI_ISL_2804631, EPI_ISL_2804639, EPI_ISL_2804647, EPI_ISL_2804648, EPI_ISL_2804650, EPI_ISL_2804657, EPI_ISL_2804663, EPI_ISL_2804665, EPI_ISL_2804668, EPI_ISL_2804673, EPI_ISL_2804680, EPI_ISL_2804685, EPI_ISL_2804686, EPI_ISL_2804692, EPI_ISL_2804693, EPI_ISL_2804701, EPI_ISL_2804703, EPI_ISL_2804716, EPI_ISL_2804717, EPI_ISL_2804718,
EPI_ISL_2804726, EPI_ISL_2804732, EPI_ISL_2804739, EPI_ISL_2804748, EPI_ISL_2804751, EPI_ISL_2804763, EPI_ISL_2804774, EPI_ISL_2804775, EPI_ISL_2804794, EPI_ISL_2804804, EPI_ISL_2804808, EPI_ISL_2804820, EPI_ISL_2804829, EPI_ISL_2804831, EPI_ISL_2804853, EPI_ISL_2804856, EPI_ISL_2804865, EPI_ISL_2804869, EPI_ISL_2804872, EPI_ISL_2804885, EPI_ISL_2804892, EPI_ISL_2804902,
EPI_ISL_2805049, EPI_ISL_2805061, EPI_ISL_2805063, EPI_ISL_2805085, EPI_ISL_2805086, EPI_ISL_2805140, EPI_ISL_2805236, EPI_ISL_2805266, EPI_ISL_2805297, EPI_ISL_2805317, EPI_ISL_2805339, EPI_ISL_2805713, EPI_ISL_2805927, EPI_ISL_2805957, EPI_ISL_2805961, EPI_ISL_2806493, EPI_ISL_2806560, EPI_ISL_2806681, EPI_ISL_2806800, EPI_ISL_2806801, EPI_ISL_2806803, EPI_ISL_2806804,
EPI_ISL_2806805, EPI_ISL_2806809, EPI_ISL_2806810, EPI_ISL_2806815, EPI_ISL_2806816, EPI_ISL_2806817, EPI_ISL_2806820, EPI_ISL_2806821, EPI_ISL_2806823, EPI_ISL_2806824, EPI_ISL_2806826, EPI_ISL_2806828, EPI_ISL_2806830, EPI_ISL_2806833, EPI_ISL_2806834, EPI_ISL_2806838, EPI_ISL_2806839, EPI_ISL_2806840, EPI_ISL_2806844, EPI_ISL_2806845, EPI_ISL_2806847, EPI_ISL_2806849,
EPI_ISL_2806850, EPI_ISL_2806851, EPI_ISL_2806852, EPI_ISL_2806854, EPI_ISL_2806855, EPI_ISL_2806856, EPI_ISL_2806857, EPI_ISL_2806858, EPI_ISL_2806860, EPI_ISL_2806864, EPI_ISL_2806868, EPI_ISL_2806870, EPI_ISL_2806873, EPI_ISL_2806874, EPI_ISL_2806875, EPI_ISL_2806876, EPI_ISL_2806878, EPI_ISL_2806880, EPI_ISL_2806885, EPI_ISL_2806887, EPI_ISL_2806888, EPI_ISL_2806889,
EPI_ISL_2806892, EPI_ISL_2806893, EPI_ISL_2806894, EPI_ISL_2806896, EPI_ISL_2806897, EPI_ISL_2806899, EPI_ISL_2806901, EPI_ISL_2806902, EPI_ISL_2806904, EPI_ISL_2806907, EPI_ISL_2806908, EPI_ISL_2806910, EPI_ISL_2806912, EPI_ISL_2806915, EPI_ISL_2806916, EPI_ISL_2806918, EPI_ISL_2806920, EPI_ISL_2806921, EPI_ISL_2806922, EPI_ISL_2806925, EPI_ISL_2806926, EPI_ISL_2806927,
EPI_ISL_2806928, EPI_ISL_2806931, EPI_ISL_2806933, EPI_ISL_2806937, EPI_ISL_2806938, EPI_ISL_2806939, EPI_ISL_2806940, EPI_ISL_2806944, EPI_ISL_2806945, EPI_ISL_2806946, EPI_ISL_2806947, EPI_ISL_2806948, EPI_ISL_2806949, EPI_ISL_2806950, EPI_ISL_2806951, EPI_ISL_2806955, EPI_ISL_2806958, EPI_ISL_2806959, EPI_ISL_2806960, EPI_ISL_2806961, EPI_ISL_2806962, EPI_ISL_2806966,
EPI_ISL_2806967, EPI_ISL_2806968, EPI_ISL_2806970, EPI_ISL_2806976, EPI_ISL_2806978, EPI_ISL_2806980, EPI_ISL_2806981, EPI_ISL_2806983, EPI_ISL_2806987, EPI_ISL_2806988, EPI_ISL_2806989, EPI_ISL_2806992, EPI_ISL_2806993, EPI_ISL_2806994, EPI_ISL_2806995, EPI_ISL_2806996, EPI_ISL_2806999, EPI_ISL_2807001, EPI_ISL_2807002, EPI_ISL_2807003, EPI_ISL_2807010, EPI_ISL_2807011,
EPI_ISL_2807012, EPI_ISL_2807013, EPI_ISL_2807014, EPI_ISL_2807015, EPI_ISL_2807018, EPI_ISL_2807020, EPI_ISL_2807022, EPI_ISL_2807024, EPI_ISL_2807031, EPI_ISL_2807033, EPI_ISL_2807037, EPI_ISL_2807038, EPI_ISL_2807044, EPI_ISL_2807047, EPI_ISL_2807048, EPI_ISL_2807049, EPI_ISL_2807054, EPI_ISL_2807059, EPI_ISL_2807061, EPI_ISL_2807062, EPI_ISL_2807063, EPI_ISL_2807068,
EPI_ISL_2807070, EPI_ISL_2807071, EPI_ISL_2807072, EPI_ISL_2807074, EPI_ISL_2807076, EPI_ISL_2807077, EPI_ISL_2807078, EPI_ISL_2807079, EPI_ISL_2807084, EPI_ISL_2807085, EPI_ISL_2807086, EPI_ISL_2807088, EPI_ISL_2807089, EPI_ISL_2807093, EPI_ISL_2807096, EPI_ISL_2807097, EPI_ISL_2807098, EPI_ISL_2807100, EPI_ISL_2807102, EPI_ISL_2807104, EPI_ISL_2807105, EPI_ISL_2807107,
EPI_ISL_2807108, EPI_ISL_2807109, EPI_ISL_2807110, EPI_ISL_2807112, EPI_ISL_2807113, EPI_ISL_2807116, EPI_ISL_2807118, EPI_ISL_2807119, EPI_ISL_2807122, EPI_ISL_2807123, EPI_ISL_2807127, EPI_ISL_2807129, EPI_ISL_2807131, EPI_ISL_2807132, EPI_ISL_2807134, EPI_ISL_2807137, EPI_ISL_2807138, EPI_ISL_2807139, EPI_ISL_2807354, EPI_ISL_2807355, EPI_ISL_2807358, EPI_ISL_2807361,
EPI_ISL_2807362, EPI_ISL_2807367, EPI_ISL_2807370, EPI_ISL_2807376, EPI_ISL_2807377, EPI_ISL_2807378, EPI_ISL_2807382, EPI_ISL_2807387, EPI_ISL_2807388, EPI_ISL_2807395, EPI_ISL_2807401, EPI_ISL_2807402, EPI_ISL_2807404, EPI_ISL_2807405, EPI_ISL_2807407, EPI_ISL_2807408, EPI_ISL_2807409, EPI_ISL_2807410, EPI_ISL_2807416, EPI_ISL_2807420, EPI_ISL_2807421, EPI_ISL_2807423,
EPI_ISL_2807424, EPI_ISL_2807426, EPI_ISL_2807428, EPI_ISL_2807430, EPI_ISL_2807431, EPI_ISL_2807432, EPI_ISL_2807442, EPI_ISL_2807445, EPI_ISL_2807448, EPI_ISL_2807458, EPI_ISL_2807459, EPI_ISL_2807460, EPI_ISL_2807461, EPI_ISL_2807463, EPI_ISL_2807473, EPI_ISL_2807476, EPI_ISL_2807477, EPI_ISL_2807480, EPI_ISL_2807482, EPI_ISL_2807484, EPI_ISL_2807487, EPI_ISL_2807494,
EPI_ISL_2807495, EPI_ISL_2807496, EPI_ISL_2807499, EPI_ISL_2807502, EPI_ISL_2807503, EPI_ISL_2807507, EPI_ISL_2807509, EPI_ISL_2807511, EPI_ISL_2807512, EPI_ISL_2807514, EPI_ISL_2807515, EPI_ISL_2807516, EPI_ISL_2807517, EPI_ISL_2807518, EPI_ISL_2807519, EPI_ISL_2807521, EPI_ISL_2807522, EPI_ISL_2807525, EPI_ISL_2807526, EPI_ISL_2807528, EPI_ISL_2807529, EPI_ISL_2807531,
EPI_ISL_2807534, EPI_ISL_2807536, EPI_ISL_2807537, EPI_ISL_2807541, EPI_ISL_2807548, EPI_ISL_2807549, EPI_ISL_2807550, EPI_ISL_2807552, EPI_ISL_2807554, EPI_ISL_2807555, EPI_ISL_2807556, EPI_ISL_2807557, EPI_ISL_2807558, EPI_ISL_2807561, EPI_ISL_2807563, EPI_ISL_2807566, EPI_ISL_2807569, EPI_ISL_2807571, EPI_ISL_2807572, EPI_ISL_2807573, EPI_ISL_2807574, EPI_ISL_2807575,
EPI_ISL_2807577, EPI_ISL_2807580, EPI_ISL_2807581, EPI_ISL_2807582, EPI_ISL_2807589, EPI_ISL_2807590, EPI_ISL_2807593, EPI_ISL_2807595, EPI_ISL_2807598, EPI_ISL_2807599, EPI_ISL_2807601, EPI_ISL_2807602, EPI_ISL_2807606, EPI_ISL_2807607, EPI_ISL_2807608, EPI_ISL_2807610, EPI_ISL_2807612, EPI_ISL_2807619, EPI_ISL_2807621, EPI_ISL_2807626, EPI_ISL_2807634, EPI_ISL_2807636,
EPI_ISL_2807637, EPI_ISL_2807639, EPI_ISL_2807640, EPI_ISL_2807642, EPI_ISL_2807643, EPI_ISL_2807645, EPI_ISL_2807649, EPI_ISL_2807652, EPI_ISL_2807653, EPI_ISL_2807654, EPI_ISL_2807658, EPI_ISL_2807660, EPI_ISL_2807661, EPI_ISL_2807662, EPI_ISL_2807663, EPI_ISL_2807666, EPI_ISL_2807668, EPI_ISL_2807669, EPI_ISL_2807670, EPI_ISL_2807671, EPI_ISL_2807673, EPI_ISL_2807678,
EPI_ISL_2807679, EPI_ISL_2807680, EPI_ISL_2807684, EPI_ISL_2807687, EPI_ISL_2807689, EPI_ISL_2807690, EPI_ISL_2807692, EPI_ISL_2807693, EPI_ISL_2807694, EPI_ISL_2807695, EPI_ISL_2807697, EPI_ISL_2807698, EPI_ISL_2807699, EPI_ISL_2807702, EPI_ISL_2807704, EPI_ISL_2807705, EPI_ISL_2807706, EPI_ISL_2807707, EPI_ISL_2807708, EPI_ISL_2807709, EPI_ISL_2807710, EPI_ISL_2807711,
EPI_ISL_2807713, EPI_ISL_2807727, EPI_ISL_2807728, EPI_ISL_2807733, EPI_ISL_2807734, EPI_ISL_2807735, EPI_ISL_2807736, EPI_ISL_2807738, EPI_ISL_2807740, EPI_ISL_2807743, EPI_ISL_2807744, EPI_ISL_2807745, EPI_ISL_2807748, EPI_ISL_2807749, EPI_ISL_2807752, EPI_ISL_2807754, EPI_ISL_2807756, EPI_ISL_2807758, EPI_ISL_2807762, EPI_ISL_2807763, EPI_ISL_2807764, EPI_ISL_2807766,
EPI_ISL_2807770, EPI_ISL_2807774, EPI_ISL_2807775, EPI_ISL_2807779, EPI_ISL_2807780, EPI_ISL_2807786, EPI_ISL_2807789, EPI_ISL_2807794, EPI_ISL_2807795, EPI_ISL_2807798, EPI_ISL_2807799, EPI_ISL_2807801, EPI_ISL_2807804, EPI_ISL_2807805, EPI_ISL_2807807, EPI_ISL_2807809, EPI_ISL_2807811, EPI_ISL_2807812, EPI_ISL_2807814, EPI_ISL_2807815, EPI_ISL_2807816, EPI_ISL_2807818,
EPI_ISL_2807820, EPI_ISL_2807821, EPI_ISL_2807824, EPI_ISL_2807826, EPI_ISL_2807828, EPI_ISL_2807831, EPI_ISL_2807834, EPI_ISL_2807835, EPI_ISL_2807836, EPI_ISL_2807840, EPI_ISL_2807841, EPI_ISL_2807843, EPI_ISL_2807844, EPI_ISL_2807845, EPI_ISL_2807846, EPI_ISL_2807847, EPI_ISL_2807848, EPI_ISL_2807849, EPI_ISL_2807851, EPI_ISL_2807853, EPI_ISL_2807856, EPI_ISL_2807857,
EPI_ISL_2807859, EPI_ISL_2807860, EPI_ISL_2807864, EPI_ISL_2807865, EPI_ISL_2807867, EPI_ISL_2807870, EPI_ISL_2807872, EPI_ISL_2807873, EPI_ISL_2807874, EPI_ISL_2807875, EPI_ISL_2807876, EPI_ISL_2807879, EPI_ISL_2807881, EPI_ISL_2807883, EPI_ISL_2807884, EPI_ISL_2807885, EPI_ISL_2807886, EPI_ISL_2807889, EPI_ISL_2807892, EPI_ISL_2807895, EPI_ISL_2807896, EPI_ISL_2807897,
EPI_ISL_2807899, EPI_ISL_2807900, EPI_ISL_2807901, EPI_ISL_2807903, EPI_ISL_2807910, EPI_ISL_2807911, EPI_ISL_2807913, EPI_ISL_2807918, EPI_ISL_2807919, EPI_ISL_2807922, EPI_ISL_2807923, EPI_ISL_2807927, EPI_ISL_2807928, EPI_ISL_2807929, EPI_ISL_2807930, EPI_ISL_2807934, EPI_ISL_2807936, EPI_ISL_2807939, EPI_ISL_2807940, EPI_ISL_2807941, EPI_ISL_2807944, EPI_ISL_2807945,
EPI_ISL_2807946, EPI_ISL_2807947, EPI_ISL_2807948, EPI_ISL_2807955, EPI_ISL_2807957, EPI_ISL_2807958, EPI_ISL_2807962, EPI_ISL_2807963, EPI_ISL_2807965, EPI_ISL_2807966, EPI_ISL_2807967, EPI_ISL_2807969, EPI_ISL_2807970, EPI_ISL_2807971, EPI_ISL_2807974, EPI_ISL_2807975, EPI_ISL_2807976, EPI_ISL_2807977, EPI_ISL_2807982, EPI_ISL_2807983, EPI_ISL_2807984, EPI_ISL_2807985,
EPI_ISL_2807986, EPI_ISL_2807988, EPI_ISL_2807990, EPI_ISL_2807991, EPI_ISL_2807992, EPI_ISL_2807993, EPI_ISL_2807996, EPI_ISL_2808001, EPI_ISL_2808002, EPI_ISL_2808005, EPI_ISL_2808006, EPI_ISL_2808015, EPI_ISL_2808017, EPI_ISL_2808018, EPI_ISL_2808020, EPI_ISL_2808021, EPI_ISL_2808024, EPI_ISL_2808025, EPI_ISL_2808027, EPI_ISL_2808028, EPI_ISL_2808031, EPI_ISL_2808033,
EPI_ISL_2808034, EPI_ISL_2808036, EPI_ISL_2808037, EPI_ISL_2808039, EPI_ISL_2808042, EPI_ISL_2808045, EPI_ISL_2808051, EPI_ISL_2808052, EPI_ISL_2808053, EPI_ISL_2808055, EPI_ISL_2808057, EPI_ISL_2808058, EPI_ISL_2808059, EPI_ISL_2808061, EPI_ISL_2808065, EPI_ISL_2808068, EPI_ISL_2808069, EPI_ISL_2808070, EPI_ISL_2808071, EPI_ISL_2808155, EPI_ISL_2808206, EPI_ISL_2808302,
EPI_ISL_2808303, EPI_ISL_2808375, EPI_ISL_2808388, EPI_ISL_2808407, EPI_ISL_2808414, EPI_ISL_2808494, EPI_ISL_2808537, EPI_ISL_2808605, EPI_ISL_2808763, EPI_ISL_2808793, EPI_ISL_2808802, EPI_ISL_2808803, EPI_ISL_2808810, EPI_ISL_2809095, EPI_ISL_2809101, EPI_ISL_2809107, EPI_ISL_2809112, EPI_ISL_2809119, EPI_ISL_2809124, EPI_ISL_2809127, EPI_ISL_2809134, EPI_ISL_2809139,
EPI_ISL_2809148, EPI_ISL_2809151, EPI_ISL_2809154, EPI_ISL_2809156, EPI_ISL_2809157, EPI_ISL_2809169, EPI_ISL_2809173, EPI_ISL_2809181, EPI_ISL_2809189, EPI_ISL_2809191, EPI_ISL_2809194, EPI_ISL_2809199, EPI_ISL_2809207, EPI_ISL_2809218, EPI_ISL_2809243, EPI_ISL_2809245, EPI_ISL_2809246, EPI_ISL_2809252, EPI_ISL_2809254, EPI_ISL_2809255, EPI_ISL_2809259, EPI_ISL_2809263,
EPI_ISL_2809264, EPI_ISL_2809266, EPI_ISL_2809270, EPI_ISL_2809271, EPI_ISL_2809274, EPI_ISL_2809276, EPI_ISL_2809278, EPI_ISL_2809279, EPI_ISL_2809280, EPI_ISL_2809284, EPI_ISL_2809288, EPI_ISL_2809292, EPI_ISL_2809307, EPI_ISL_2809308, EPI_ISL_2809309, EPI_ISL_2809312, EPI_ISL_2809320, EPI_ISL_2809324, EPI_ISL_2809330, EPI_ISL_2809332, EPI_ISL_2809335, EPI_ISL_2809338,
EPI_ISL_2809340, EPI_ISL_2809351, EPI_ISL_2809355, EPI_ISL_2809358, EPI_ISL_2809360, EPI_ISL_2809361, EPI_ISL_2809367, EPI_ISL_2809368, EPI_ISL_2809371, EPI_ISL_2809376, EPI_ISL_2809377, EPI_ISL_2809379, EPI_ISL_2809381, EPI_ISL_2809382, EPI_ISL_2809400, EPI_ISL_2809405, EPI_ISL_2809410, EPI_ISL_2809412, EPI_ISL_2809414, EPI_ISL_2809416, EPI_ISL_2809523, EPI_ISL_2809882,
EPI_ISL_2812988, EPI_ISL_2813002, EPI_ISL_2813008, EPI_ISL_2813009, EPI_ISL_2813013, EPI_ISL_2813019, EPI_ISL_2813020, EPI_ISL_2813026, EPI_ISL_2813034, EPI_ISL_2813040, EPI_ISL_2813044, EPI_ISL_2813045, EPI_ISL_2813046, EPI_ISL_2813058, EPI_ISL_2813062, EPI_ISL_2813068, EPI_ISL_2813087, EPI_ISL_2813099, EPI_ISL_2813101, EPI_ISL_2813103, EPI_ISL_2813112, EPI_ISL_2813113,
EPI_ISL_2813120, EPI_ISL_2813130, EPI_ISL_2813139, EPI_ISL_2813141, EPI_ISL_2813145, EPI_ISL_2813147, EPI_ISL_2813152, EPI_ISL_2813153, EPI_ISL_2813155, EPI_ISL_2813160, EPI_ISL_2813161, EPI_ISL_2813162, EPI_ISL_2813166, EPI_ISL_2813167, EPI_ISL_2813175, EPI_ISL_2813178, EPI_ISL_2813181, EPI_ISL_2813189, EPI_ISL_2813203, EPI_ISL_2813217, EPI_ISL_2813218, EPI_ISL_2813221,
EPI_ISL_2813235, EPI_ISL_2813240, EPI_ISL_2813249, EPI_ISL_2813250, EPI_ISL_2813255, EPI_ISL_2813256, EPI_ISL_2813258, EPI_ISL_2813265, EPI_ISL_2813280, EPI_ISL_2813286, EPI_ISL_2813288, EPI_ISL_2813317, EPI_ISL_2813318, EPI_ISL_2813319, EPI_ISL_2813323, EPI_ISL_2813332, EPI_ISL_2813338, EPI_ISL_2813342, EPI_ISL_2813352, EPI_ISL_2813357, EPI_ISL_2813360, EPI_ISL_2813363,
EPI_ISL_2813367, EPI_ISL_2813382, EPI_ISL_2813384, EPI_ISL_2813391, EPI_ISL_2813397, EPI_ISL_2813400, EPI_ISL_2813401, EPI_ISL_2813403, EPI_ISL_2813405, EPI_ISL_2813407, EPI_ISL_2813408, EPI_ISL_2813411, EPI_ISL_2813417, EPI_ISL_2813418, EPI_ISL_2813423, EPI_ISL_2813426, EPI_ISL_2813428, EPI_ISL_2813431, EPI_ISL_2813433, EPI_ISL_2813435, EPI_ISL_2813439, EPI_ISL_2813447,
EPI_ISL_2813453, EPI_ISL_2813454, EPI_ISL_2813456, EPI_ISL_2813465, EPI_ISL_2813473, EPI_ISL_2813476, EPI_ISL_2813477, EPI_ISL_2813493, EPI_ISL_2813495, EPI_ISL_2813496, EPI_ISL_2813497, EPI_ISL_2813500, EPI_ISL_2813507, EPI_ISL_2813510, EPI_ISL_2813511, EPI_ISL_2813512, EPI_ISL_2813515, EPI_ISL_2813516, EPI_ISL_2813527, EPI_ISL_2813530, EPI_ISL_2813531, EPI_ISL_2813535,
EPI_ISL_2813546, EPI_ISL_2813547, EPI_ISL_2813552, EPI_ISL_2813553, EPI_ISL_2813556, EPI_ISL_2813557, EPI_ISL_2813566, EPI_ISL_2813573, EPI_ISL_2813575, EPI_ISL_2813581, EPI_ISL_2813584, EPI_ISL_2813587, EPI_ISL_2813588, EPI_ISL_2813590, EPI_ISL_2813605, EPI_ISL_2813606, EPI_ISL_2813607, EPI_ISL_2813618, EPI_ISL_2813620, EPI_ISL_2813624, EPI_ISL_2813626, EPI_ISL_2813637,
EPI_ISL_2813642, EPI_ISL_2813643, EPI_ISL_2813646, EPI_ISL_2813656, EPI_ISL_2813681, EPI_ISL_2813695, EPI_ISL_2813717, EPI_ISL_2813753, EPI_ISL_2813756, EPI_ISL_2813758, EPI_ISL_2813763, EPI_ISL_2813764, EPI_ISL_2813766, EPI_ISL_2813784, EPI_ISL_2813794, EPI_ISL_2813830, EPI_ISL_2813837, EPI_ISL_2813867, EPI_ISL_2813882, EPI_ISL_2813956, EPI_ISL_2813966, EPI_ISL_2813995,
EPI_ISL_2814069, EPI_ISL_2814094, EPI_ISL_2814191, EPI_ISL_2830092, EPI_ISL_2830593, EPI_ISL_2830605, EPI_ISL_2830617, EPI_ISL_2830620, EPI_ISL_2830625, EPI_ISL_2830653, EPI_ISL_2830666, EPI_ISL_2830682, EPI_ISL_2830685, EPI_ISL_2830702, EPI_ISL_2830705, EPI_ISL_2830713, EPI_ISL_2830730, EPI_ISL_2830738, EPI_ISL_2830790, EPI_ISL_2831061, EPI_ISL_2831068, EPI_ISL_2831089,
EPI_ISL_2831140, EPI_ISL_2831158, EPI_ISL_2831162, EPI_ISL_2831192, EPI_ISL_2831198, EPI_ISL_2831254, EPI_ISL_2831263, EPI_ISL_2831272, EPI_ISL_2831314, EPI_ISL_2832090, EPI_ISL_2847649, EPI_ISL_2848359, EPI_ISL_2848367, EPI_ISL_2852480, EPI_ISL_2865209, EPI_ISL_2910756

| | | | |
|---|---|---|---|
| see above | Lighthouse Lab in Milton Keynes | Wellcome Sanger Institute for the COVID-19 Genomics UK (COG-UK) Consortium | Cordelia Langford; David K. Jackson; Dominic Kwiatkowski; Ewan Harrison; Ian Johnston; Jeffrey Barrett; John Sillitoe on behalf of the Wellcome Sanger Institute COVID-19 Surveillance Team; Roberto Amato; Sonia Goncalves; The Lighthouse Lab in Milton Keynes and Alex Alderton |

EPI_ISL_2728683, EPI_ISL_2728685, EPI_ISL_2728689, EPI_ISL_2728705, EPI_ISL_2728711, EPI_ISL_2728712, EPI_ISL_2728714, EPI_ISL_2728715, EPI_ISL_2728716, EPI_ISL_2728718, EPI_ISL_2728726, EPI_ISL_2728727, EPI_ISL_2728736, EPI_ISL_2728743, EPI_ISL_2728746, EPI_ISL_2728749, EPI_ISL_2728751, EPI_ISL_2728755, EPI_ISL_2728756, EPI_ISL_2728757, EPI_ISL_2728759, EPI_ISL_2728760,
EPI_ISL_2728769, EPI_ISL_2728774, EPI_ISL_2728776, EPI_ISL_2728779, EPI_ISL_2728789, EPI_ISL_2728790, EPI_ISL_2728795, EPI_ISL_2728797, EPI_ISL_2728798, EPI_ISL_2728805, EPI_ISL_2728807, EPI_ISL_2728811, EPI_ISL_2728817, EPI_ISL_2728819, EPI_ISL_2728823, EPI_ISL_2728829, EPI_ISL_2728834, EPI_ISL_2728835, EPI_ISL_2728836, EPI_ISL_2728842, EPI_ISL_2728844, EPI_ISL_2728846,
EPI_ISL_2728847, EPI_ISL_2728853, EPI_ISL_2728856, EPI_ISL_2728858, EPI_ISL_2728860, EPI_ISL_2728861, EPI_ISL_2728863, EPI_ISL_2728866, EPI_ISL_2728877, EPI_ISL_2728891, EPI_ISL_2728893, EPI_ISL_2728895, EPI_ISL_2728896, EPI_ISL_2728904, EPI_ISL_2728905, EPI_ISL_2728909, EPI_ISL_2728914, EPI_ISL_2728917, EPI_ISL_2728922, EPI_ISL_2728927, EPI_ISL_2728933, EPI_ISL_2728937,
EPI_ISL_2728938, EPI_ISL_2728942, EPI_ISL_2728947, EPI_ISL_2728949, EPI_ISL_2728955, EPI_ISL_2728963, EPI_ISL_2728968, EPI_ISL_2728988, EPI_ISL_2728989, EPI_ISL_2728991, EPI_ISL_2728995, EPI_ISL_2728999, EPI_ISL_2729001, EPI_ISL_2729014, EPI_ISL_2729016, EPI_ISL_2729017, EPI_ISL_2729029, EPI_ISL_2729031, EPI_ISL_2729036, EPI_ISL_2729041, EPI_ISL_2729947, EPI_ISL_2729949,
EPI_ISL_2729950, EPI_ISL_2729957, EPI_ISL_2729962, EPI_ISL_2729969, EPI_ISL_2729974, EPI_ISL_2729978, EPI_ISL_2729980, EPI_ISL_2729995, EPI_ISL_2730001, EPI_ISL_2730008, EPI_ISL_2730023, EPI_ISL_2730027, EPI_ISL_2730036, EPI_ISL_2730043, EPI_ISL_2730046, EPI_ISL_2730049, EPI_ISL_2730050, EPI_ISL_2730051, EPI_ISL_2730057, EPI_ISL_2730060, EPI_ISL_2730071, EPI_ISL_2730078,
EPI_ISL_2730081, EPI_ISL_2730083, EPI_ISL_2730085, EPI_ISL_2730087, EPI_ISL_2730089, EPI_ISL_2730093, EPI_ISL_2730094, EPI_ISL_2730098, EPI_ISL_2730099, EPI_ISL_2730102, EPI_ISL_2730103, EPI_ISL_2730104, EPI_ISL_2730105, EPI_ISL_2730108, EPI_ISL_2730112, EPI_ISL_2730128, EPI_ISL_2730141, EPI_ISL_2730151, EPI_ISL_2730160, EPI_ISL_2730175, EPI_ISL_2730185, EPI_ISL_2730190,
EPI_ISL_2730202, EPI_ISL_2730206, EPI_ISL_2730209, EPI_ISL_2730230, EPI_ISL_2730231, EPI_ISL_2730232, EPI_ISL_2730233, EPI_ISL_2730235, EPI_ISL_2730237, EPI_ISL_2730238, EPI_ISL_2730246, EPI_ISL_2730252, EPI_ISL_2730255, EPI_ISL_2730259, EPI_ISL_2730260, EPI_ISL_2730261, EPI_ISL_2730262, EPI_ISL_2730264, EPI_ISL_2730265, EPI_ISL_2730268, EPI_ISL_2730269, EPI_ISL_2730273,
EPI_ISL_2730274, EPI_ISL_2730276, EPI_ISL_2730279, EPI_ISL_2730281, EPI_ISL_2730283, EPI_ISL_2730284, EPI_ISL_2765582, EPI_ISL_2765583, EPI_ISL_2765591, EPI_ISL_2765595, EPI_ISL_2765602, EPI_ISL_2765618, EPI_ISL_2765621, EPI_ISL_2765622, EPI_ISL_2765625, EPI_ISL_2765627, EPI_ISL_2765640, EPI_ISL_2765652, EPI_ISL_2765664, EPI_ISL_2765665, EPI_ISL_2765675, EPI_ISL_2765700,
EPI_ISL_2765713, EPI_ISL_2765714, EPI_ISL_2765720, EPI_ISL_2765730, EPI_ISL_2765742, EPI_ISL_2765743, EPI_ISL_2765745, EPI_ISL_2765753, EPI_ISL_2765759, EPI_ISL_2765767, EPI_ISL_2765781, EPI_ISL_2765788, EPI_ISL_2765795, EPI_ISL_2765798, EPI_ISL_2765799, EPI_ISL_2765813, EPI_ISL_2765821, EPI_ISL_2765827, EPI_ISL_2765832, EPI_ISL_2765833, EPI_ISL_2765834, EPI_ISL_2765837,
EPI_ISL_2765849, EPI_ISL_2766195, EPI_ISL_2766196, EPI_ISL_2766198, EPI_ISL_2766200, EPI_ISL_2766203, EPI_ISL_2766204, EPI_ISL_2766205, EPI_ISL_2766206, EPI_ISL_2766207, EPI_ISL_2766208, EPI_ISL_2766209, EPI_ISL_2766210, EPI_ISL_2766211, EPI_ISL_2766213, EPI_ISL_2766215, EPI_ISL_2766216, EPI_ISL_2766217, EPI_ISL_2766218, EPI_ISL_2766219, EPI_ISL_2766223, EPI_ISL_2766224,
EPI_ISL_2766226, EPI_ISL_2766227, EPI_ISL_2766229, EPI_ISL_2766230, EPI_ISL_2766231, EPI_ISL_2766232, EPI_ISL_2766233, EPI_ISL_2766234, EPI_ISL_2766235, EPI_ISL_2766236, EPI_ISL_2766237, EPI_ISL_2766238, EPI_ISL_2766239, EPI_ISL_2766240, EPI_ISL_2766241, EPI_ISL_2766242, EPI_ISL_2766245, EPI_ISL_2766246, EPI_ISL_2766247, EPI_ISL_2766248, EPI_ISL_2766249, EPI_ISL_2766251,
EPI_ISL_2766252, EPI_ISL_2766253, EPI_ISL_2766254, EPI_ISL_2766257, EPI_ISL_2766258, EPI_ISL_2766259, EPI_ISL_2766260, EPI_ISL_2766262, EPI_ISL_2766263, EPI_ISL_2766264, EPI_ISL_2766266, EPI_ISL_2766267, EPI_ISL_2766268, EPI_ISL_2766269, EPI_ISL_2766270, EPI_ISL_2766271, EPI_ISL_2766272, EPI_ISL_2766275, EPI_ISL_2766276, EPI_ISL_2766277, EPI_ISL_2766278, EPI_ISL_2766279,
EPI_ISL_2766281, EPI_ISL_2766286, EPI_ISL_2766287, EPI_ISL_2766289, EPI_ISL_2766290, EPI_ISL_2766291, EPI_ISL_2766292, EPI_ISL_2766293, EPI_ISL_2766294, EPI_ISL_2766295, EPI_ISL_2766296, EPI_ISL_2766298, EPI_ISL_2766299, EPI_ISL_2766301, EPI_ISL_2766302, EPI_ISL_2766303, EPI_ISL_2766304, EPI_ISL_2766305, EPI_ISL_2766306, EPI_ISL_2766307, EPI_ISL_2766308, EPI_ISL_2766309,
EPI_ISL_2766310, EPI_ISL_2766312, EPI_ISL_2766314, EPI_ISL_2766316, EPI_ISL_2766323, EPI_ISL_2766325, EPI_ISL_2766326, EPI_ISL_2766328, EPI_ISL_2766329, EPI_ISL_2766330, EPI_ISL_2766331, EPI_ISL_2766332, EPI_ISL_2766333, EPI_ISL_2766334, EPI_ISL_2766335, EPI_ISL_2766336, EPI_ISL_2766337, EPI_ISL_2766338, EPI_ISL_2766339, EPI_ISL_2766340, EPI_ISL_2766342, EPI_ISL_2766343,
EPI_ISL_2766345, EPI_ISL_2766346, EPI_ISL_2766347, EPI_ISL_2766348, EPI_ISL_2766350, EPI_ISL_2766351, EPI_ISL_2766352, EPI_ISL_2766353, EPI_ISL_2766354, EPI_ISL_2766356, EPI_ISL_2766359, EPI_ISL_2766362, EPI_ISL_2766363, EPI_ISL_2766364, EPI_ISL_2766365, EPI_ISL_2766366, EPI_ISL_2766369, EPI_ISL_2766370, EPI_ISL_2766372, EPI_ISL_2766373, EPI_ISL_2766374, EPI_ISL_2766375,
EPI_ISL_2766376, EPI_ISL_2766377, EPI_ISL_2766378, EPI_ISL_2766379, EPI_ISL_2766380, EPI_ISL_2766382, EPI_ISL_2766383, EPI_ISL_2766384, EPI_ISL_2766385, EPI_ISL_2766386, EPI_ISL_2766387, EPI_ISL_2766389, EPI_ISL_2766390, EPI_ISL_2766391, EPI_ISL_2766392, EPI_ISL_2766393, EPI_ISL_2766394, EPI_ISL_2766395, EPI_ISL_2766397, EPI_ISL_2766398, EPI_ISL_2766399, EPI_ISL_2766400,
EPI_ISL_2766401, EPI_ISL_2766404, EPI_ISL_2766405, EPI_ISL_2766406, EPI_ISL_2766407, EPI_ISL_2766408, EPI_ISL_2766409, EPI_ISL_2766411, EPI_ISL_2766413, EPI_ISL_2766414, EPI_ISL_2766416, EPI_ISL_2766418, EPI_ISL_2766419, EPI_ISL_2766421, EPI_ISL_2766423, EPI_ISL_2766425, EPI_ISL_2766426, EPI_ISL_2766427, EPI_ISL_2766429, EPI_ISL_2766430, EPI_ISL_2766431, EPI_ISL_2766432,
EPI_ISL_2766433, EPI_ISL_2766434, EPI_ISL_2766436, EPI_ISL_2766437, EPI_ISL_2766439, EPI_ISL_2766440, EPI_ISL_2766441, EPI_ISL_2766442, EPI_ISL_2766443, EPI_ISL_2766444, EPI_ISL_2766445, EPI_ISL_2766446, EPI_ISL_2766447, EPI_ISL_2766448, EPI_ISL_2766451, EPI_ISL_2766454, EPI_ISL_2766455, EPI_ISL_2766456, EPI_ISL_2766457, EPI_ISL_2766458, EPI_ISL_2766459, EPI_ISL_2766460,
EPI_ISL_2766461, EPI_ISL_2766462, EPI_ISL_2766463, EPI_ISL_2766464, EPI_ISL_2766465, EPI_ISL_2766467, EPI_ISL_2766468, EPI_ISL_2766469, EPI_ISL_2766470, EPI_ISL_2766472, EPI_ISL_2766473, EPI_ISL_2766474, EPI_ISL_2766477, EPI_ISL_2766479, EPI_ISL_2766480, EPI_ISL_2766482, EPI_ISL_2766483, EPI_ISL_2766485, EPI_ISL_2766486, EPI_ISL_2766487, EPI_ISL_2766488, EPI_ISL_2766489,
EPI_ISL_2766490, EPI_ISL_2766491, EPI_ISL_2766492, EPI_ISL_2766493, EPI_ISL_2766494, EPI_ISL_2766495, EPI_ISL_2766497, EPI_ISL_2766498, EPI_ISL_2766500, EPI_ISL_2766501, EPI_ISL_2766502, EPI_ISL_2766503, EPI_ISL_2766504, EPI_ISL_2766505, EPI_ISL_2766506, EPI_ISL_2766507, EPI_ISL_2766509, EPI_ISL_2766510, EPI_ISL_2766514, EPI_ISL_2766515, EPI_ISL_2766516, EPI_ISL_2766517,
EPI_ISL_2766518, EPI_ISL_2766520, EPI_ISL_2766523, EPI_ISL_2766524, EPI_ISL_2766525, EPI_ISL_2766527, EPI_ISL_2766528, EPI_ISL_2766529, EPI_ISL_2766530, EPI_ISL_2766533, EPI_ISL_2766534, EPI_ISL_2766535, EPI_ISL_2766537, EPI_ISL_2766538, EPI_ISL_2766539, EPI_ISL_2766541, EPI_ISL_2766542, EPI_ISL_2766543, EPI_ISL_2766545, EPI_ISL_2766546, EPI_ISL_2766548, EPI_ISL_2766549,
EPI_ISL_2766554, EPI_ISL_2766558, EPI_ISL_2766568, EPI_ISL_2766581, EPI_ISL_2766584, EPI_ISL_2766585, EPI_ISL_2766590, EPI_ISL_2766592, EPI_ISL_2766593, EPI_ISL_2766602, EPI_ISL_2766606, EPI_ISL_2766609, EPI_ISL_2766616, EPI_ISL_2766623, EPI_ISL_2766638, EPI_ISL_2766639, EPI_ISL_2766660, EPI_ISL_2766663, EPI_ISL_2766669, EPI_ISL_2766670, EPI_ISL_2766679, EPI_ISL_2766680,
EPI_ISL_2766689, EPI_ISL_2766694, EPI_ISL_2766703, EPI_ISL_2766704, EPI_ISL_2766708, EPI_ISL_2766724, EPI_ISL_2766734, EPI_ISL_2766736, EPI_ISL_2766738, EPI_ISL_2766742, EPI_ISL_2766748, EPI_ISL_2766754, EPI_ISL_2766770, EPI_ISL_2766775, EPI_ISL_2766781, EPI_ISL_2766782, EPI_ISL_2766790, EPI_ISL_2766794, EPI_ISL_2766796, EPI_ISL_2766809, EPI_ISL_2766810, EPI_ISL_2766814,
EPI_ISL_2766815, EPI_ISL_2766829, EPI_ISL_2766840, EPI_ISL_2766842, EPI_ISL_2766846, EPI_ISL_2766849, EPI_ISL_2766862, EPI_ISL_2766871, EPI_ISL_2766873, EPI_ISL_2767640, EPI_ISL_2767642, EPI_ISL_2779873, EPI_ISL_2779875, EPI_ISL_2779877, EPI_ISL_2779879, EPI_ISL_2779884, EPI_ISL_2779910, EPI_ISL_2779915, EPI_ISL_2779917, EPI_ISL_2779920, EPI_ISL_2779924, EPI_ISL_2779925,
EPI_ISL_2779926, EPI_ISL_2779928, EPI_ISL_2779942, EPI_ISL_2779943, EPI_ISL_2779944, EPI_ISL_2779948, EPI_ISL_2779954, EPI_ISL_2779963, EPI_ISL_2779986, EPI_ISL_2779989, EPI_ISL_2779998, EPI_ISL_2780019, EPI_ISL_2780027, EPI_ISL_2780031, EPI_ISL_2780034, EPI_ISL_2780039, EPI_ISL_2780052, EPI_ISL_2780062, EPI_ISL_2780063, EPI_ISL_2780078, EPI_ISL_2780084, EPI_ISL_2780085,
EPI_ISL_2780087, EPI_ISL_2780091, EPI_ISL_2780096, EPI_ISL_2780115, EPI_ISL_2780138, EPI_ISL_2780140, EPI_ISL_2780141, EPI_ISL_2780143, EPI_ISL_2780146, EPI_ISL_2780156, EPI_ISL_2780168, EPI_ISL_2780179, EPI_ISL_2780183, EPI_ISL_2780189, EPI_ISL_2780193, EPI_ISL_2780197, EPI_ISL_2780198, EPI_ISL_2780199, EPI_ISL_2780202, EPI_ISL_2780203, EPI_ISL_2780207, EPI_ISL_2780217,
EPI_ISL_2780220, EPI_ISL_2780221, EPI_ISL_2780224, EPI_ISL_2780227, EPI_ISL_2780228, EPI_ISL_2780231, EPI_ISL_2780232, EPI_ISL_2780236, EPI_ISL_2780258, EPI_ISL_2780259, EPI_ISL_2780260, EPI_ISL_2780261, EPI_ISL_2780262, EPI_ISL_2780267, EPI_ISL_2780269, EPI_ISL_2780271, EPI_ISL_2780273, EPI_ISL_2780276, EPI_ISL_2780277, EPI_ISL_2780281, EPI_ISL_2780283, EPI_ISL_2780284,
EPI_ISL_2780292, EPI_ISL_2780293, EPI_ISL_2780298, EPI_ISL_2780313, EPI_ISL_2780316, EPI_ISL_2780317, EPI_ISL_2780319, EPI_ISL_2780320, EPI_ISL_2780322, EPI_ISL_2780324, EPI_ISL_2780326, EPI_ISL_2780329, EPI_ISL_2780330, EPI_ISL_2780335, EPI_ISL_2780339, EPI_ISL_2780349, EPI_ISL_2780354, EPI_ISL_2780359, EPI_ISL_2780362, EPI_ISL_2780363, EPI_ISL_2780365, EPI_ISL_2780369,

EPI_ISL_2780371, EPI_ISL_2780374, EPI_ISL_2780375, EPI_ISL_2780381, EPI_ISL_2780383, EPI_ISL_2780385, EPI_ISL_2780389, EPI_ISL_2780391, EPI_ISL_2780392, EPI_ISL_2780395, EPI_ISL_2780397, EPI_ISL_2780403, EPI_ISL_2780404, EPI_ISL_2780407, EPI_ISL_2780411, EPI_ISL_2780414, EPI_ISL_2780418, EPI_ISL_2780419, EPI_ISL_2780420, EPI_ISL_2780421, EPI_ISL_2780447, EPI_ISL_2780454, EPI_ISL_2780462, EPI_ISL_2780463, EPI_ISL_2780465, EPI_ISL_2780469, EPI_ISL_2780470, EPI_ISL_2780951, EPI_ISL_2780982, EPI_ISL_2781041, EPI_ISL_2781065, EPI_ISL_2781773, EPI_ISL_2782037, EPI_ISL_2782046, EPI_ISL_2782047, EPI_ISL_2782048, EPI_ISL_2782050, EPI_ISL_2782052, EPI_ISL_2782056, EPI_ISL_2782057, EPI_ISL_2782060, EPI_ISL_2782064, EPI_ISL_2782073, EPI_ISL_2782074, EPI_ISL_2782075, EPI_ISL_2782078, EPI_ISL_2782083, EPI_ISL_2782084, EPI_ISL_2782085, EPI_ISL_2782098, EPI_ISL_2782099, EPI_ISL_2782102, EPI_ISL_2782108, EPI_ISL_2782109, EPI_ISL_2782110, EPI_ISL_2782112, EPI_ISL_2782121, EPI_ISL_2782123, EPI_ISL_2782125, EPI_ISL_2782129, EPI_ISL_2782132, EPI_ISL_2782133, EPI_ISL_2782134, EPI_ISL_2782138, EPI_ISL_2782143, EPI_ISL_2782146, EPI_ISL_2782148, EPI_ISL_2782154, EPI_ISL_2782156, EPI_ISL_2782159, EPI_ISL_2782162, EPI_ISL_2782165, EPI_ISL_2782166, EPI_ISL_2782167, EPI_ISL_2782168, EPI_ISL_2782169, EPI_ISL_2782173, EPI_ISL_2782174, EPI_ISL_2782175, EPI_ISL_2782185, EPI_ISL_2782188, EPI_ISL_2782194, EPI_ISL_2782195, EPI_ISL_2782211, EPI_ISL_2782212, EPI_ISL_2782220, EPI_ISL_2782222, EPI_ISL_2782227, EPI_ISL_2782228, EPI_ISL_2782229, EPI_ISL_2782233, EPI_ISL_2782235, EPI_ISL_2782236, EPI_ISL_2782243, EPI_ISL_2782255, EPI_ISL_2782265, EPI_ISL_2782269, EPI_ISL_2782270, EPI_ISL_2782272, EPI_ISL_2782273, EPI_ISL_2782276, EPI_ISL_2782277, EPI_ISL_2782278, EPI_ISL_2782281, EPI_ISL_2782283, EPI_ISL_2782285, EPI_ISL_2782286, EPI_ISL_2782288, EPI_ISL_2782292, EPI_ISL_2782296, EPI_ISL_2782299, EPI_ISL_2782304, EPI_ISL_2782305, EPI_ISL_2782312, EPI_ISL_2804189, EPI_ISL_2804286, EPI_ISL_2804358, EPI_ISL_2804389, EPI_ISL_2804493, EPI_ISL_2804517, EPI_ISL_2804532, EPI_ISL_2804551, EPI_ISL_2804566, EPI_ISL_2804573, EPI_ISL_2806456, EPI_ISL_2806461, EPI_ISL_2806465, EPI_ISL_2806502, EPI_ISL_2806531, EPI_ISL_2806547, EPI_ISL_2806575, EPI_ISL_2806589, EPI_ISL_2806600, EPI_ISL_2806613, EPI_ISL_2806657, EPI_ISL_2806675, EPI_ISL_2806686, EPI_ISL_2806707, EPI_ISL_2806724, EPI_ISL_2806728, EPI_ISL_2806737, EPI_ISL_2806745, EPI_ISL_2806747, EPI_ISL_2806751, EPI_ISL_2806753, EPI_ISL_2806756, EPI_ISL_2806766, EPI_ISL_2806773, EPI_ISL_2806782, EPI_ISL_2806785, EPI_ISL_2806798, EPI_ISL_2806799, EPI_ISL_2806802, EPI_ISL_2806806, EPI_ISL_2806807, EPI_ISL_2806808, EPI_ISL_2806811, EPI_ISL_2806812, EPI_ISL_2806813, EPI_ISL_2806814, EPI_ISL_2806818, EPI_ISL_2806819, EPI_ISL_2806822, EPI_ISL_2806825, EPI_ISL_2806827, EPI_ISL_2806829, EPI_ISL_2806831, EPI_ISL_2806832, EPI_ISL_2806835, EPI_ISL_2806836, EPI_ISL_2806837, EPI_ISL_2806841, EPI_ISL_2806842, EPI_ISL_2806843, EPI_ISL_2806846, EPI_ISL_2806848, EPI_ISL_2806853, EPI_ISL_2806859, EPI_ISL_2806861, EPI_ISL_2806862, EPI_ISL_2806863, EPI_ISL_2806865, EPI_ISL_2806866, EPI_ISL_2806867, EPI_ISL_2806869, EPI_ISL_2806871, EPI_ISL_2806872, EPI_ISL_2806877, EPI_ISL_2806879, EPI_ISL_2806881, EPI_ISL_2806882, EPI_ISL_2806883, EPI_ISL_2806884, EPI_ISL_2806886, EPI_ISL_2806890, EPI_ISL_2806891, EPI_ISL_2806895, EPI_ISL_2806898, EPI_ISL_2806900, EPI_ISL_2806903, EPI_ISL_2806905, EPI_ISL_2806906, EPI_ISL_2806909, EPI_ISL_2806911, EPI_ISL_2806913, EPI_ISL_2806914, EPI_ISL_2806917, EPI_ISL_2806923, EPI_ISL_2806924, EPI_ISL_2806929, EPI_ISL_2806930, EPI_ISL_2806932, EPI_ISL_2806934, EPI_ISL_2806935, EPI_ISL_2806936, EPI_ISL_2806941, EPI_ISL_2806942, EPI_ISL_2806943, EPI_ISL_2806952, EPI_ISL_2806953, EPI_ISL_2806954, EPI_ISL_2806956, EPI_ISL_2806957, EPI_ISL_2806963, EPI_ISL_2806964, EPI_ISL_2806965, EPI_ISL_2806969, EPI_ISL_2806971, EPI_ISL_2806972, EPI_ISL_2806973, EPI_ISL_2806974, EPI_ISL_2806975, EPI_ISL_2806977, EPI_ISL_2806979, EPI_ISL_2806982, EPI_ISL_2806984, EPI_ISL_2806985, EPI_ISL_2806986, EPI_ISL_2806990, EPI_ISL_2806991, EPI_ISL_2806997, EPI_ISL_2806998, EPI_ISL_2807000, EPI_ISL_2807004, EPI_ISL_2807005, EPI_ISL_2807006, EPI_ISL_2807007, EPI_ISL_2807008, EPI_ISL_2807009, EPI_ISL_2807016, EPI_ISL_2807017, EPI_ISL_2807019, EPI_ISL_2807021, EPI_ISL_2807023, EPI_ISL_2807025, EPI_ISL_2807026, EPI_ISL_2807027, EPI_ISL_2807028, EPI_ISL_2807029, EPI_ISL_2807030, EPI_ISL_2807032, EPI_ISL_2807034, EPI_ISL_2807035, EPI_ISL_2807036, EPI_ISL_2807039, EPI_ISL_2807040, EPI_ISL_2807041, EPI_ISL_2807042, EPI_ISL_2807043, EPI_ISL_2807045, EPI_ISL_2807046, EPI_ISL_2807051, EPI_ISL_2807052, EPI_ISL_2807053, EPI_ISL_2807055, EPI_ISL_2807056, EPI_ISL_2807057, EPI_ISL_2807058, EPI_ISL_2807060, EPI_ISL_2807064, EPI_ISL_2807065, EPI_ISL_2807066, EPI_ISL_2807067, EPI_ISL_2807069, EPI_ISL_2807073, EPI_ISL_2807075, EPI_ISL_2807080, EPI_ISL_2807081, EPI_ISL_2807082, EPI_ISL_2807083, EPI_ISL_2807087, EPI_ISL_2807090, EPI_ISL_2807091, EPI_ISL_2807092, EPI_ISL_2807094, EPI_ISL_2807095, EPI_ISL_2807099, EPI_ISL_2807101, EPI_ISL_2807103, EPI_ISL_2807106, EPI_ISL_2807111, EPI_ISL_2807114, EPI_ISL_2807115, EPI_ISL_2807117, EPI_ISL_2807120, EPI_ISL_2807121, EPI_ISL_2807124, EPI_ISL_2807125, EPI_ISL_2807126, EPI_ISL_2807128, EPI_ISL_2807130, EPI_ISL_2807133, EPI_ISL_2807135, EPI_ISL_2807136, EPI_ISL_2807140, EPI_ISL_2808120, EPI_ISL_2808130, EPI_ISL_2808132, EPI_ISL_2808175, EPI_ISL_2808180, EPI_ISL_2808221, EPI_ISL_2808260, EPI_ISL_2808269, EPI_ISL_2808282, EPI_ISL_2808300, EPI_ISL_2808309, EPI_ISL_2808327, EPI_ISL_2808356, EPI_ISL_2808358, EPI_ISL_2808363, EPI_ISL_2808371, EPI_ISL_2808392, EPI_ISL_2808420, EPI_ISL_2808421, EPI_ISL_2808798, EPI_ISL_2808799, EPI_ISL_2808800, EPI_ISL_2808805, EPI_ISL_2809559, EPI_ISL_2813320, EPI_ISL_2813321, EPI_ISL_2813322, EPI_ISL_2813324, EPI_ISL_2813325, EPI_ISL_2813326, EPI_ISL_2813327, EPI_ISL_2813328, EPI_ISL_2813329, EPI_ISL_2813330, EPI_ISL_2813331, EPI_ISL_2813333, EPI_ISL_2813334, EPI_ISL_2813335, EPI_ISL_2813336, EPI_ISL_2813337, EPI_ISL_2813339, EPI_ISL_2813340, EPI_ISL_2813341, EPI_ISL_2813343, EPI_ISL_2813344, EPI_ISL_2813345, EPI_ISL_2813346, EPI_ISL_2813347, EPI_ISL_2813348, EPI_ISL_2813349, EPI_ISL_2813350, EPI_ISL_2813351, EPI_ISL_2813353, EPI_ISL_2813354, EPI_ISL_2813355, EPI_ISL_2813356, EPI_ISL_2813358, EPI_ISL_2813361, EPI_ISL_2813364, EPI_ISL_2813365, EPI_ISL_2813366, EPI_ISL_2813368, EPI_ISL_2813369, EPI_ISL_2813371, EPI_ISL_2813372, EPI_ISL_2813373, EPI_ISL_2813374, EPI_ISL_2813375, EPI_ISL_2813376, EPI_ISL_2813377, EPI_ISL_2813378, EPI_ISL_2813379, EPI_ISL_2813380, EPI_ISL_2813381, EPI_ISL_2813383, EPI_ISL_2813385, EPI_ISL_2813386, EPI_ISL_2813387, EPI_ISL_2813388, EPI_ISL_2813389, EPI_ISL_2813390, EPI_ISL_2813392, EPI_ISL_2813393, EPI_ISL_2813394, EPI_ISL_2813395, EPI_ISL_2813396, EPI_ISL_2813398, EPI_ISL_2813399, EPI_ISL_2813402, EPI_ISL_2813404, EPI_ISL_2813406, EPI_ISL_2813409, EPI_ISL_2813410, EPI_ISL_2813412, EPI_ISL_2813413, EPI_ISL_2813414, EPI_ISL_2813415, EPI_ISL_2813416, EPI_ISL_2813420, EPI_ISL_2813421, EPI_ISL_2813422, EPI_ISL_2813424, EPI_ISL_2813425, EPI_ISL_2813427, EPI_ISL_2813429, EPI_ISL_2813430, EPI_ISL_2813432, EPI_ISL_2813434, EPI_ISL_2813436, EPI_ISL_2813437, EPI_ISL_2813438, EPI_ISL_2813440, EPI_ISL_2813441, EPI_ISL_2813442, EPI_ISL_2813443, EPI_ISL_2813444, EPI_ISL_2813445, EPI_ISL_2813446, EPI_ISL_2813448, EPI_ISL_2813449, EPI_ISL_2813450, EPI_ISL_2813451, EPI_ISL_2813452, EPI_ISL_2813455, EPI_ISL_2813457, EPI_ISL_2813458, EPI_ISL_2813459, EPI_ISL_2813460, EPI_ISL_2813461, EPI_ISL_2813462, EPI_ISL_2813463, EPI_ISL_2813464, EPI_ISL_2813466, EPI_ISL_2813467, EPI_ISL_2813468, EPI_ISL_2813469, EPI_ISL_2813470, EPI_ISL_2813471, EPI_ISL_2813472, EPI_ISL_2813475, EPI_ISL_2813478, EPI_ISL_2813479, EPI_ISL_2813480, EPI_ISL_2813481, EPI_ISL_2813482, EPI_ISL_2813483, EPI_ISL_2813484, EPI_ISL_2813485, EPI_ISL_2813486, EPI_ISL_2813487, EPI_ISL_2813488, EPI_ISL_2813489, EPI_ISL_2813490, EPI_ISL_2813491, EPI_ISL_2813494, EPI_ISL_2813498, EPI_ISL_2813501, EPI_ISL_2813502, EPI_ISL_2813503, EPI_ISL_2813504, EPI_ISL_2813505, EPI_ISL_2813506, EPI_ISL_2813508, EPI_ISL_2813509, EPI_ISL_2813513, EPI_ISL_2813514, EPI_ISL_2813517, EPI_ISL_2813518, EPI_ISL_2813519, EPI_ISL_2813520, EPI_ISL_2813521, EPI_ISL_2813522, EPI_ISL_2813523, EPI_ISL_2813524, EPI_ISL_2813525, EPI_ISL_2813526, EPI_ISL_2813528, EPI_ISL_2813529, EPI_ISL_2813532, EPI_ISL_2813533, EPI_ISL_2813534, EPI_ISL_2813536, EPI_ISL_2813537, EPI_ISL_2813538, EPI_ISL_2813539, EPI_ISL_2813540, EPI_ISL_2813541, EPI_ISL_2813542, EPI_ISL_2813543, EPI_ISL_2813544, EPI_ISL_2813545, EPI_ISL_2813548, EPI_ISL_2813549, EPI_ISL_2813550, EPI_ISL_2813551, EPI_ISL_2813554, EPI_ISL_2813555, EPI_ISL_2813558, EPI_ISL_2813559, EPI_ISL_2813560, EPI_ISL_2813561, EPI_ISL_2813562, EPI_ISL_2813563, EPI_ISL_2813564, EPI_ISL_2813565, EPI_ISL_2813567, EPI_ISL_2813568, EPI_ISL_2813569, EPI_ISL_2813570, EPI_ISL_2813571, EPI_ISL_2813572, EPI_ISL_2813574, EPI_ISL_2813576, EPI_ISL_2813577, EPI_ISL_2813578, EPI_ISL_2813579, EPI_ISL_2813580, EPI_ISL_2813582, EPI_ISL_2813583, EPI_ISL_2813585, EPI_ISL_2813586, EPI_ISL_2813589, EPI_ISL_2813591, EPI_ISL_2813592, EPI_ISL_2813593, EPI_ISL_2813594, EPI_ISL_2813595, EPI_ISL_2813596, EPI_ISL_2813597, EPI_ISL_2813598, EPI_ISL_2813599, EPI_ISL_2813600, EPI_ISL_2813601, EPI_ISL_2813602, EPI_ISL_2813603, EPI_ISL_2813604, EPI_ISL_2813608, EPI_ISL_2813609, EPI_ISL_2813610, EPI_ISL_2813611, EPI_ISL_2813612, EPI_ISL_2813613, EPI_ISL_2813614, EPI_ISL_2813615, EPI_ISL_2813616, EPI_ISL_2813617, EPI_ISL_2813619, EPI_ISL_2813621, EPI_ISL_2813622, EPI_ISL_2813623, EPI_ISL_2813625, EPI_ISL_2813627, EPI_ISL_2813628, EPI_ISL_2813629, EPI_ISL_2813630, EPI_ISL_2813631, EPI_ISL_2813632, EPI_ISL_2813633, EPI_ISL_2813634, EPI_ISL_2813635, EPI_ISL_2813636, EPI_ISL_2813638, EPI_ISL_2813639, EPI_ISL_2813640, EPI_ISL_2813641, EPI_ISL_2813645, EPI_ISL_2813647, EPI_ISL_2813648, EPI_ISL_2813649, EPI_ISL_2813650, EPI_ISL_2813651, EPI_ISL_2813652, EPI_ISL_2813653, EPI_ISL_2813654, EPI_ISL_2813655, EPI_ISL_2813657, EPI_ISL_2813658, EPI_ISL_2813659, EPI_ISL_2829412, EPI_ISL_2832095, EPI_ISL_2846783, EPI_ISL_2847925, EPI_ISL_2852481, EPI_ISL_2852482

| | | | |
|---|---|---|---|
| see above | Lighthouse Laboratory Plymouth | Wellcome Sanger Institute for the COVID-19 Genomics UK (COG-UK) Consortium | Cordelia Langford; David K. Jackson; Dominic Kwiatkowski; Ewan Harrison; Ian Johnston; Jeffrey Barrett; John Sillitoe on behalf of the Wellcome Sanger Institute COVID-19 Surveillance Team; Lighthouse Laboratory Plymouth and Alex Alderton; Roberto Amato; Sonia Goncalves |
| EPI_ISL_2901163 | Liverpool Clinical Laboratories | COVID-19 Genomics UK (COG-UK) Consortium | A Alrezaihi; Alessandro Gerada; Alistair Darby; Angela Cowell; Anita Lucaci; Anu Chawla; Cassie Olateju; Catherine Hartley; Charlotte Nelson; Ecaterina Vamos; Elaine O'Toole; Eleanor G Bentley; Ghada T Shawli; Hermione Webster; Isabel García-Dorival; James Johnson; James P Stewart; Jenifer Manson; Joanne Watts; Jones Benjamin; Jordan J Clark; Julian Hiscox; L Luu; Lucille Rainbow; M Almsaud; Margaret Hughes; Mark Whitehead; Matthew Gemmell; Miren Iturriza-Gomara; Muhannad Alruwaili; N.P Randle; Neil Swainston; PKF Gilmore; Parul Sharma; Rebekah Penrice-Randal; Richard Eccles; Richard Gregory; Sam Haldenby; Steve Paterson; Stuart D Armstrong; Trevor Ian Robinson; Ximeng Han |

EPI_ISL_2816744, EPI_ISL_2816748, EPI_ISL_2816756, EPI_ISL_2816771, EPI_ISL_2816782, EPI_ISL_2816790, EPI_ISL_2816796, EPI_ISL_2816797, EPI_ISL_2816799, EPI_ISL_2816803, EPI_ISL_2816804

| | | | |
|---|---|---|---|
| see above | Northumbria University / South Tees Hospitals NHS Foundation Trust / North Cumbria Integrated Care NHS Foundation Trust / North Tees and Hartlepool NHS Foundation Trust / Newcastle Hospitals NHS Foundation Trust | COVID-19 Genomics UK (COG-UK) Consortium | Andrew Nelson; Brendan Payne; Clive Graham; Darren L Smith; Debra Padgett; Edward Barton; Emma Swindells; Garren Scott; Gary Black; Gary Eltringham; Giles S Holt; Greg R Young; Jane Greenaway; Jennifer Collins; John Allan; Joshua Loh; Lynn Dover; Matthew Bashton; Mohammad A Tariq; Paul Baker; Sarah Essex; Steve Liggett; Wen C Yew; Yusri Taha |

EPI_ISL_2750162, EPI_ISL_2750163, EPI_ISL_2750169, EPI_ISL_2750178, EPI_ISL_2750243, EPI_ISL_2750254, EPI_ISL_2750260, EPI_ISL_2750365, EPI_ISL_2750366, EPI_ISL_2750372, EPI_ISL_2750409, EPI_ISL_2750447, EPI_ISL_2750448, EPI_ISL_2818337, EPI_ISL_2818342, EPI_ISL_2818347, EPI_ISL_2818350, EPI_ISL_2818351, EPI_ISL_2818354, EPI_ISL_2818360, EPI_ISL_2818364, EPI_ISL_2818366, EPI_ISL_2818367, EPI_ISL_2818368, EPI_ISL_2818369, EPI_ISL_2818370, EPI_ISL_2818371, EPI_ISL_2818372, EPI_ISL_2818373, EPI_ISL_2818374, EPI_ISL_2818376, EPI_ISL_2818377, EPI_ISL_2818378, EPI_ISL_2818379, EPI_ISL_2818380, EPI_ISL_2818381, EPI_ISL_2818382, EPI_ISL_2818383, EPI_ISL_2818384, EPI_ISL_2818385, EPI_ISL_2818386, EPI_ISL_2818388, EPI_ISL_2818389, EPI_ISL_2818391, EPI_ISL_2818392, EPI_ISL_2818393, EPI_ISL_2818394, EPI_ISL_2818395, EPI_ISL_2818396, EPI_ISL_2818397, EPI_ISL_2818398, EPI_ISL_2818399, EPI_ISL_2818402, EPI_ISL_2818403, EPI_ISL_2818404, EPI_ISL_2818410, EPI_ISL_2818413, EPI_ISL_2818414, EPI_ISL_2818415, EPI_ISL_2818416, EPI_ISL_2818418, EPI_ISL_2818457, EPI_ISL_2818458, EPI_ISL_2818459, EPI_ISL_2818460, EPI_ISL_2818461, EPI_ISL_2818462, EPI_ISL_2818463, EPI_ISL_2818464, EPI_ISL_2818465, EPI_ISL_2818466, EPI_ISL_2818468, EPI_ISL_2818469, EPI_ISL_2818470, EPI_ISL_2818471, EPI_ISL_2818472, EPI_ISL_2818473, EPI_ISL_2818474, EPI_ISL_2818475, EPI_ISL_2818476, EPI_ISL_2818477, EPI_ISL_2818478, EPI_ISL_2818479, EPI_ISL_2818480, EPI_ISL_2818481, EPI_ISL_2818482, EPI_ISL_2818483, EPI_ISL_2818523, EPI_ISL_2818524, EPI_ISL_2818526, EPI_ISL_2818528, EPI_ISL_2818529, EPI_ISL_2818530, EPI_ISL_2818531, EPI_ISL_2818533, EPI_ISL_2818534, EPI_ISL_2818535, EPI_ISL_2818536, EPI_ISL_2818539, EPI_ISL_2818541, EPI_ISL_2818542, EPI_ISL_2818545, EPI_ISL_2818547, EPI_ISL_2818553, EPI_ISL_2818554, EPI_ISL_2818555, EPI_ISL_2818557, EPI_ISL_2818558, EPI_ISL_2818560, EPI_ISL_2818562, EPI_ISL_2818563, EPI_ISL_2818567, EPI_ISL_2818568, EPI_ISL_2818571, EPI_ISL_2818573, EPI_ISL_2818574, EPI_ISL_2818576, EPI_ISL_2818577, EPI_ISL_2818578, EPI_ISL_2818579, EPI_ISL_2818580, EPI_ISL_2818581, EPI_ISL_2818583, EPI_ISL_2818584, EPI_ISL_2818585, EPI_ISL_2818588, EPI_ISL_2818589, EPI_ISL_2818590, EPI_ISL_2818591, EPI_ISL_2818592, EPI_ISL_2818593, EPI_ISL_2818594, EPI_ISL_2818595, EPI_ISL_2818596, EPI_ISL_2818598, EPI_ISL_2818599, EPI_ISL_2818601, EPI_ISL_2818602, EPI_ISL_2818603, EPI_ISL_2818604, EPI_ISL_2818605, EPI_ISL_2818607, EPI_ISL_2818608, EPI_ISL_2818609, EPI_ISL_2818610, EPI_ISL_2818611, EPI_ISL_2818612, EPI_ISL_2818613, EPI_ISL_2818614, EPI_ISL_2818615, EPI_ISL_2818616, EPI_ISL_2818617, EPI_ISL_2818618, EPI_ISL_2818619, EPI_ISL_2818620, EPI_ISL_2818621, EPI_ISL_2818623, EPI_ISL_2818625, EPI_ISL_2818626, EPI_ISL_2818627, EPI_ISL_2818628, EPI_ISL_2818629, EPI_ISL_2818630, EPI_ISL_2818631, EPI_ISL_2818633, EPI_ISL_2818636, EPI_ISL_2818637, EPI_ISL_2818638, EPI_ISL_2818640, EPI_ISL_2818641, EPI_ISL_2818644, EPI_ISL_2818647, EPI_ISL_2818648, EPI_ISL_2818652, EPI_ISL_2818654, EPI_ISL_2818655, EPI_ISL_2818656, EPI_ISL_2818659, EPI_ISL_2818661, EPI_ISL_2818662, EPI_ISL_2818663, EPI_ISL_2818665, EPI_ISL_2818666, EPI_ISL_2818667, EPI_ISL_2818668, EPI_ISL_2818669, EPI_ISL_2818670, EPI_ISL_2818671, EPI_ISL_2818672, EPI_ISL_2818674, EPI_ISL_2818675, EPI_ISL_2818676, EPI_ISL_2818677, EPI_ISL_2818678, EPI_ISL_2818680, EPI_ISL_2818683, EPI_ISL_2818689, EPI_ISL_2818693, EPI_ISL_2818694, EPI_ISL_2818695, EPI_ISL_2818696, EPI_ISL_2818697, EPI_ISL_2818698, EPI_ISL_2818699, EPI_ISL_2818701, EPI_ISL_2818702, EPI_ISL_2818703, EPI_ISL_2818705, EPI_ISL_2818706, EPI_ISL_2818707, EPI_ISL_2818708, EPI_ISL_2818710, EPI_ISL_2818711, EPI_ISL_2818712, EPI_ISL_2818713, EPI_ISL_2818714, EPI_ISL_2818715, EPI_ISL_2818716, EPI_ISL_2818717, EPI_ISL_2818719, EPI_ISL_2818722, EPI_ISL_2818725, EPI_ISL_2818726, EPI_ISL_2818727, EPI_ISL_2818728, EPI_ISL_2818729, EPI_ISL_2818730, EPI_ISL_2818731, EPI_ISL_2818733, EPI_ISL_2818734, EPI_ISL_2818735, EPI_ISL_2818736, EPI_ISL_2818737, EPI_ISL_2818740, EPI_ISL_2818742, EPI_ISL_2818743, EPI_ISL_2818744, EPI_ISL_2818746, EPI_ISL_2818747, EPI_ISL_2818748, EPI_ISL_2818751, EPI_ISL_2818753, EPI_ISL_2818754, EPI_ISL_2818756, EPI_ISL_2818759, EPI_ISL_2818762, EPI_ISL_2818765, EPI_ISL_2818766, EPI_ISL_2818768, EPI_ISL_2818775, EPI_ISL_2818777, EPI_ISL_2818778, EPI_ISL_2818779, EPI_ISL_2818780, EPI_ISL_2818781, EPI_ISL_2818782, EPI_ISL_2818783, EPI_ISL_2818784, EPI_ISL_2818785, EPI_ISL_2818786, EPI_ISL_2818787, EPI_ISL_2818788, EPI_ISL_2818789, EPI_ISL_2818790, EPI_ISL_2818791, EPI_ISL_2818792, EPI_ISL_2818793, EPI_ISL_2818794, EPI_ISL_2818795, EPI_ISL_2818796, EPI_ISL_2818797, EPI_ISL_2818798, EPI_ISL_2818799, EPI_ISL_2818800, EPI_ISL_2818801, EPI_ISL_2818802, EPI_ISL_2818803, EPI_ISL_2818805, EPI_ISL_2818806, EPI_ISL_2818807, EPI_ISL_2818808, EPI_ISL_2818809, EPI_ISL_2818810, EPI_ISL_2818813, EPI_ISL_2818814, EPI_ISL_2818815, EPI_ISL_2818816, EPI_ISL_2818817, EPI_ISL_2818818, EPI_ISL_2818819, EPI_ISL_2818820, EPI_ISL_2818821, EPI_ISL_2818822, EPI_ISL_2818823, EPI_ISL_2818827, EPI_ISL_2818828, EPI_ISL_2818830, EPI_ISL_2818832, EPI_ISL_2818833, EPI_ISL_2818834, EPI_ISL_2818835, EPI_ISL_2818836, EPI_ISL_2818837, EPI_ISL_2818842, EPI_ISL_2818843, EPI_ISL_2818850, EPI_ISL_2818852, EPI_ISL_2818853, EPI_ISL_2818856, EPI_ISL_2818857, EPI_ISL_2818858, EPI_ISL_2818859, EPI_ISL_2818860, EPI_ISL_2818861, EPI_ISL_2818862, EPI_ISL_2818863, EPI_ISL_2818864, EPI_ISL_2818865, EPI_ISL_2818866, EPI_ISL_2818867, EPI_ISL_2818868, EPI_ISL_2818869, EPI_ISL_2818870, EPI_ISL_2818871, EPI_ISL_2818874, EPI_ISL_2818875, EPI_ISL_2818878, EPI_ISL_2818879, EPI_ISL_2818880, EPI_ISL_2818881, EPI_ISL_2818882, EPI_ISL_2818883, EPI_ISL_2818884, EPI_ISL_2818885, EPI_ISL_2818886, EPI_ISL_2818887, EPI_ISL_2818889, EPI_ISL_2818890, EPI_ISL_2818891, EPI_ISL_2818893, EPI_ISL_2818894, EPI_ISL_2818899, EPI_ISL_2818900, EPI_ISL_2818901, EPI_ISL_2818902, EPI_ISL_2818903, EPI_ISL_2818904, EPI_ISL_2818905, EPI_ISL_2818906, EPI_ISL_2818908, EPI_ISL_2818909, EPI_ISL_2818911, EPI_ISL_2818913, EPI_ISL_2818914, EPI_ISL_2818915, EPI_ISL_2818918, EPI_ISL_2818919, EPI_ISL_2818921, EPI_ISL_2818922, EPI_ISL_2818923, EPI_ISL_2818924, EPI_ISL_2818926, EPI_ISL_2818927, EPI_ISL_2818928, EPI_ISL_2818929, EPI_ISL_2818934, EPI_ISL_2818936, EPI_ISL_2818938, EPI_ISL_2818939, EPI_ISL_2818940, EPI_ISL_2818941, EPI_ISL_2818942, EPI_ISL_2818943, EPI_ISL_2818944, EPI_ISL_2818945, EPI_ISL_2818946, EPI_ISL_2818949, EPI_ISL_2818951, EPI_ISL_2818952, EPI_ISL_2818953, EPI_ISL_2818954, EPI_ISL_2818955, EPI_ISL_2818958, EPI_ISL_2818960, EPI_ISL_2818961, EPI_ISL_2818962, EPI_ISL_2818963, EPI_ISL_2818964, EPI_ISL_2818965, EPI_ISL_2818966, EPI_ISL_2818968, EPI_ISL_2818969, EPI_ISL_2818970, EPI_ISL_2818971, EPI_ISL_2818972, EPI_ISL_2818973, EPI_ISL_2818974, EPI_ISL_2818976, EPI_ISL_2818978, EPI_ISL_2818979, EPI_ISL_2818980, EPI_ISL_2818981, EPI_ISL_2818982, EPI_ISL_2818983, EPI_ISL_2818984, EPI_ISL_2818985, EPI_ISL_2818987, EPI_ISL_2818988, EPI_ISL_2818989, EPI_ISL_2818990, EPI_ISL_2818992, EPI_ISL_2818993, EPI_ISL_2818994, EPI_ISL_2818996, EPI_ISL_2818997, EPI_ISL_2818998, EPI_ISL_2818999, EPI_ISL_2819000, EPI_ISL_2819002, EPI_ISL_2819003, EPI_ISL_2819004, EPI_ISL_2819005, EPI_ISL_2819006, EPI_ISL_2819007, EPI_ISL_2819008, EPI_ISL_2819009, EPI_ISL_2819010, EPI_ISL_2819011, EPI_ISL_2819012, EPI_ISL_2819013, EPI_ISL_2819014, EPI_ISL_2819015, EPI_ISL_2819016, EPI_ISL_2819017, EPI_ISL_2819018, EPI_ISL_2819019, EPI_ISL_2819020, EPI_ISL_2819021, EPI_ISL_2819022, EPI_ISL_2819023, EPI_ISL_2819025, EPI_ISL_2819026, EPI_ISL_2819027, EPI_ISL_2819028, EPI_ISL_2819029, EPI_ISL_2819030, EPI_ISL_2819031, EPI_ISL_2819032, EPI_ISL_2819033, EPI_ISL_2819034, EPI_ISL_2819035, EPI_ISL_2819036, EPI_ISL_2819037, EPI_ISL_2819038, EPI_ISL_2819039, EPI_ISL_2819040, EPI_ISL_2819041, EPI_ISL_2819042, EPI_ISL_2819044, EPI_ISL_2819046, EPI_ISL_2819048, EPI_ISL_2819049, EPI_ISL_2819051, EPI_ISL_2819053, EPI_ISL_2819059, EPI_ISL_2819065, EPI_ISL_2819066, EPI_ISL_2819068, EPI_ISL_2819070, EPI_ISL_2819072, EPI_ISL_2819076, EPI_ISL_2819078, EPI_ISL_2819079, EPI_ISL_2819081, EPI_ISL_2819085, EPI_ISL_2819087, EPI_ISL_2819089, EPI_ISL_2819091, EPI_ISL_2819093, EPI_ISL_2819097, EPI_ISL_2819101, EPI_ISL_2819103, EPI_ISL_2819105, EPI_ISL_2819107, EPI_ISL_2819109, EPI_ISL_2819111, EPI_ISL_2819113, EPI_ISL_2819115, EPI_ISL_2819117, EPI_ISL_2819120, EPI_ISL_2819121, EPI_ISL_2819123, EPI_ISL_2819125, EPI_ISL_2819127, EPI_ISL_2819129, EPI_ISL_2819133, EPI_ISL_2819135, EPI_ISL_2819137, EPI_ISL_2819139, EPI_ISL_2819141, EPI_ISL_2819143, EPI_ISL_2819145, EPI_ISL_2819147, EPI_ISL_2819149, EPI_ISL_2819151, EPI_ISL_2819153, EPI_ISL_2819155, EPI_ISL_2819156, EPI_ISL_2819158, EPI_ISL_2819160, EPI_ISL_2819162, EPI_ISL_2819164, EPI_ISL_2819166, EPI_ISL_2819168, EPI_ISL_2819170, EPI_ISL_2819172, EPI_ISL_2819174, EPI_ISL_2819176, EPI_ISL_2819178, EPI_ISL_2819180, EPI_ISL_2819182, EPI_ISL_2819184, EPI_ISL_2819185, EPI_ISL_2819187, EPI_ISL_2819189, EPI_ISL_2819191, EPI_ISL_2819193, EPI_ISL_2819195, EPI_ISL_2819197, EPI_ISL_2819199, EPI_ISL_2819205, EPI_ISL_2819209, EPI_ISL_2819211, EPI_ISL_2819212, EPI_ISL_2819214, EPI_ISL_2819216, EPI_ISL_2819218, EPI_ISL_2819220, EPI_ISL_2819224, EPI_ISL_2819226, EPI_ISL_2819228, EPI_ISL_2819230, EPI_ISL_2819232, EPI_ISL_2819233, EPI_ISL_2819235, EPI_ISL_2819237, EPI_ISL_2819239, EPI_ISL_2819242, EPI_ISL_2819245, EPI_ISL_2819247, EPI_ISL_2819249, EPI_ISL_2819251, EPI_ISL_2819253, EPI_ISL_2819257, EPI_ISL_2819259, EPI_ISL_2819260, EPI_ISL_2819262, EPI_ISL_2819264, EPI_ISL_2819266, EPI_ISL_2819268, EPI_ISL_2819270, EPI_ISL_2819271, EPI_ISL_2819273, EPI_ISL_2819275, EPI_ISL_2819277, EPI_ISL_2819279, EPI_ISL_2819280, EPI_ISL_2819282, EPI_ISL_2819284, EPI_ISL_2819286, EPI_ISL_2819288, EPI_ISL_2819290, EPI_ISL_2819294, EPI_ISL_2819296, EPI_ISL_2819298, EPI_ISL_2819300, EPI_ISL_2819301, EPI_ISL_2819303, EPI_ISL_2819305, EPI_ISL_2819307, EPI_ISL_2819309, EPI_ISL_2819312, EPI_ISL_2819314, EPI_ISL_2819316, EPI_ISL_2819320, EPI_ISL_2819322, EPI_ISL_2819327, EPI_ISL_2819333, EPI_ISL_2819337, EPI_ISL_2819339, EPI_ISL_2819343, EPI_ISL_2819344, EPI_ISL_2819346, EPI_ISL_2819348, EPI_ISL_2819356, EPI_ISL_2819358, EPI_ISL_2819361, EPI_ISL_2819363, EPI_ISL_2819367, EPI_ISL_2819369, EPI_ISL_2819371, EPI_ISL_2819375, EPI_ISL_2819377, EPI_ISL_2819379, EPI_ISL_2819381, EPI_ISL_2819382, EPI_ISL_2819384, EPI_ISL_2819388, EPI_ISL_2819390, EPI_ISL_2819392, EPI_ISL_2819394, EPI_ISL_2819398, EPI_ISL_2819400, EPI_ISL_2819401, EPI_ISL_2819405, EPI_ISL_2819407, EPI_ISL_2819409, EPI_ISL_2819411, EPI_ISL_2819413, EPI_ISL_2819414, EPI_ISL_2819418, EPI_ISL_2819420, EPI_ISL_2819423, EPI_ISL_2819425, EPI_ISL_2819429, EPI_ISL_2819439, EPI_ISL_2819441, EPI_ISL_2819449, EPI_ISL_2819451, EPI_ISL_2819455, EPI_ISL_2819457, EPI_ISL_2819458, EPI_ISL_2819462, EPI_ISL_2819464, EPI_ISL_2819469, EPI_ISL_2819471, EPI_ISL_2819473, EPI_ISL_2819475, EPI_ISL_2819477, EPI_ISL_2819479, EPI_ISL_2819481, EPI_ISL_2819482, EPI_ISL_2819484, EPI_ISL_2819486, EPI_ISL_2819488, EPI_ISL_2819491, EPI_ISL_2819493, EPI_ISL_2819495, EPI_ISL_2819497, EPI_ISL_2819499, EPI_ISL_2819500, EPI_ISL_2819503, EPI_ISL_2819505, EPI_ISL_2819506, EPI_ISL_2819508, EPI_ISL_2819510, EPI_ISL_2819512, EPI_ISL_2819514, EPI_ISL_2819516, EPI_ISL_2819518, EPI_ISL_2819522, EPI_ISL_2819524, EPI_ISL_2819526, EPI_ISL_2819528, EPI_ISL_2819529, EPI_ISL_2819531, EPI_ISL_2819533, EPI_ISL_2819534, EPI_ISL_2819536, EPI_ISL_2819538, EPI_ISL_2819540, EPI_ISL_2819544, EPI_ISL_2819546, EPI_ISL_2819548, EPI_ISL_2819550, EPI_ISL_2819552, EPI_ISL_2819554, EPI_ISL_2819556, EPI_ISL_2819558, EPI_ISL_2819559, EPI_ISL_2819561, EPI_ISL_2819563, EPI_ISL_2819565, EPI_ISL_2819569, EPI_ISL_2819574, EPI_ISL_2819576, EPI_ISL_2819578, EPI_ISL_2819581, EPI_ISL_2819584, EPI_ISL_2819586, EPI_ISL_2819588, EPI_ISL_2819592, EPI_ISL_2819594, EPI_ISL_2819595, EPI_ISL_2819597, EPI_ISL_2819599, EPI_ISL_2819601, EPI_ISL_2819605, EPI_ISL_2819607, EPI_ISL_2819609, EPI_ISL_2819611, EPI_ISL_2819616, EPI_ISL_2819618, EPI_ISL_2819620, EPI_ISL_2819622, EPI_ISL_2819624, EPI_ISL_2819625, EPI_ISL_2819627, EPI_ISL_2819629, EPI_ISL_2819631, EPI_ISL_2819633, EPI_ISL_2819636, EPI_ISL_2819640, EPI_ISL_2819643, EPI_ISL_2819647, EPI_ISL_2819649, EPI_ISL_2819651, EPI_ISL_2819653, EPI_ISL_2819655, EPI_ISL_2819657, EPI_ISL_2819659, EPI_ISL_2819663, EPI_ISL_2819665, EPI_ISL_2819667, EPI_ISL_2819671, EPI_ISL_2819674, EPI_ISL_2819676, EPI_ISL_2819678, EPI_ISL_2819680, EPI_ISL_2819682, EPI_ISL_2819684, EPI_ISL_2819686, EPI_ISL_2819688, EPI_ISL_2819689, EPI_ISL_2819691, EPI_ISL_2819692, EPI_ISL_2819693, EPI_ISL_2819694, EPI_ISL_2819695, EPI_ISL_2819696, EPI_ISL_2819697, EPI_ISL_2819698, EPI_ISL_2819699, EPI_ISL_2819700, EPI_ISL_2819701, EPI_ISL_2819702, EPI_ISL_2819703, EPI_ISL_2819704, EPI_ISL_2819705, EPI_ISL_2819707, EPI_ISL_2819708, EPI_ISL_2819709, EPI_ISL_2819711, EPI_ISL_2819712, EPI_ISL_2819713, EPI_ISL_2819715, EPI_ISL_2819716, EPI_ISL_2819717, EPI_ISL_2819718, EPI_ISL_2819719, EPI_ISL_2819720, EPI_ISL_2819721, EPI_ISL_2819722, EPI_ISL_2819723, EPI_ISL_2819724, EPI_ISL_2819725, EPI_ISL_2819726, EPI_ISL_2819727, EPI_ISL_2819729, EPI_ISL_2819731, EPI_ISL_2819732, EPI_ISL_2819733, EPI_ISL_2819735, EPI_ISL_2819736, EPI_ISL_2819737, EPI_ISL_2819740, EPI_ISL_2819741, EPI_ISL_2819744, EPI_ISL_2819745, EPI_ISL_2819746, EPI_ISL_2819748, EPI_ISL_2819750, EPI_ISL_2819751, EPI_ISL_2819752, EPI_ISL_2819754, EPI_ISL_2819755, EPI_ISL_2819757, EPI_ISL_2819758, EPI_ISL_2819759, EPI_ISL_2819760, EPI_ISL_2819761, EPI_ISL_2819762, EPI_ISL_2819763, EPI_ISL_2819764, EPI_ISL_2819765, EPI_ISL_2819767, EPI_ISL_2819770, EPI_ISL_2819771, EPI_ISL_2819772, EPI_ISL_2819773, EPI_ISL_2819774, EPI_ISL_2819775, EPI_ISL_2819776, EPI_ISL_2819777, EPI_ISL_2819778, EPI_ISL_2819779, EPI_ISL_2819780, EPI_ISL_2819781, EPI_ISL_2819782, EPI_ISL_2819783, EPI_ISL_2819784, EPI_ISL_2819785, EPI_ISL_2819786, EPI_ISL_2819787, EPI_ISL_2819788, EPI_ISL_2819789, EPI_ISL_2819790, EPI_ISL_2819791, EPI_ISL_2819792, EPI_ISL_2819794, EPI_ISL_2819795, EPI_ISL_2819796, EPI_ISL_2819797, EPI_ISL_2819798, EPI_ISL_2819800, EPI_ISL_2819801, EPI_ISL_2819802, EPI_ISL_2819803, EPI_ISL_2819804, EPI_ISL_2819806, EPI_ISL_2819807, EPI_ISL_2819808, EPI_ISL_2819809, EPI_ISL_2819810, EPI_ISL_2819811, EPI_ISL_2819812, EPI_ISL_2819813, EPI_ISL_2819814, EPI_ISL_2819816, EPI_ISL_2819817, EPI_ISL_2819818, EPI_ISL_2819819, EPI_ISL_2819820, EPI_ISL_2819821, EPI_ISL_2819822, EPI_ISL_2819824, EPI_ISL_2819825, EPI_ISL_2819826, EPI_ISL_2819827, EPI_ISL_2819828, EPI_ISL_2819830, EPI_ISL_2819831, EPI_ISL_2819832, EPI_ISL_2819833, EPI_ISL_2819834, EPI_ISL_2819835, EPI_ISL_2819837, EPI_ISL_2819838, EPI_ISL_2819839, EPI_ISL_2819840, EPI_ISL_2819841, EPI_ISL_2819842, EPI_ISL_2819843, EPI_ISL_2819844, EPI_ISL_2819846, EPI_ISL_2819849, EPI_ISL_2905020, EPI_ISL_2905386, EPI_ISL_2905387, EPI_ISL_2905388, EPI_ISL_2905389, EPI_ISL_2905390, EPI_ISL_2905391, EPI_ISL_2905392, EPI_ISL_2905393, EPI_ISL_2905394, EPI_ISL_2905395, EPI_ISL_2905396, EPI_ISL_2905397, EPI_ISL_2905398, EPI_ISL_2905399, EPI_ISL_2905400, EPI_ISL_2905401, EPI_ISL_2905402, EPI_ISL_2905403, EPI_ISL_2905404, EPI_ISL_2905405, EPI_ISL_2905406, EPI_ISL_2905464, EPI_ISL_2905473, EPI_ISL_2905484, EPI_ISL_2905485, EPI_ISL_2905486, EPI_ISL_2905488, EPI_ISL_2905489, EPI_ISL_2905490, EPI_ISL_2905491, EPI_ISL_2905492, EPI_ISL_2905493, EPI_ISL_2905494, EPI_ISL_2905496, EPI_ISL_2905497, EPI_ISL_2905498, EPI_ISL_2905499, EPI_ISL_2905500, EPI_ISL_2905501, EPI_ISL_2905502, EPI_ISL_2905503, EPI_ISL_2905504, EPI_ISL_2905505, EPI_ISL_2905506, EPI_ISL_2905507, EPI_ISL_2905508, EPI_ISL_2905509, EPI_ISL_2905510, EPI_ISL_2905511, EPI_ISL_2905512, EPI_ISL_2905513, EPI_ISL_2905514, EPI_ISL_2905515, EPI_ISL_2905516, EPI_ISL_2905517, EPI_ISL_2905518, EPI_ISL_2905519, EPI_ISL_2905520, EPI_ISL_2905521, EPI_ISL_2905522, EPI_ISL_2905523, EPI_ISL_2905524, EPI_ISL_2905525, EPI_ISL_2905526, EPI_ISL_2905527, EPI_ISL_2905528, EPI_ISL_2905529, EPI_ISL_2905530, EPI_ISL_2905531, EPI_ISL_2905532, EPI_ISL_2905533, EPI_ISL_2905534, EPI_ISL_2905535, EPI_ISL_2905595, EPI_ISL_2905596, EPI_ISL_2905597, EPI_ISL_2905598, EPI_ISL_2905599, EPI_ISL_2905600, EPI_ISL_2905601, EPI_ISL_2905602, EPI_ISL_2905603, EPI_ISL_2905604, EPI_ISL_2905605, EPI_ISL_2905606, EPI_ISL_2905607, EPI_ISL_2905608, EPI_ISL_2905609, EPI_ISL_2905610, EPI_ISL_2905611, EPI_ISL_2905612, EPI_ISL_2905613, EPI_ISL_2905614, EPI_ISL_2905615, EPI_ISL_2905616, EPI_ISL_2905617, EPI_ISL_2905618, EPI_ISL_2905619, EPI_ISL_2905620, EPI_ISL_2905621, EPI_ISL_2905622, EPI_ISL_2905623, EPI_ISL_2905624, EPI_ISL_2905625, EPI_ISL_2905626, EPI_ISL_2905627, EPI_ISL_2905628, EPI_ISL_2905629, EPI_ISL_2905630, EPI_ISL_2905631, EPI_ISL_2905632, EPI_ISL_2905633, EPI_ISL_2905634, EPI_ISL_2905635, EPI_ISL_2905636, EPI_ISL_2905637, EPI_ISL_2905638, EPI_ISL_2905639, EPI_ISL_2905640, EPI_ISL_2905641, EPI_ISL_2905642, EPI_ISL_2905643, EPI_ISL_2905644, EPI_ISL_2905645, EPI_ISL_2905646, EPI_ISL_2905647, EPI_ISL_2905648, EPI_ISL_2905649, EPI_ISL_2905650, EPI_ISL_2905651, EPI_ISL_2905652, EPI_ISL_2905653, EPI_ISL_2905654, EPI_ISL_2905664, EPI_ISL_2905730, EPI_ISL_2905736, EPI_ISL_2905739, EPI_ISL_2905740, EPI_ISL_2905741, EPI_ISL_2905742, EPI_ISL_2905743, EPI_ISL_2905744, EPI_ISL_2905745, EPI_ISL_2905746, EPI_ISL_2905747, EPI_ISL_2905748, EPI_ISL_2905749, EPI_ISL_2905750, EPI_ISL_2905972, EPI_ISL_2905978, EPI_ISL_2905981, EPI_ISL_2905982, EPI_ISL_2905986, EPI_ISL_2905987, EPI_ISL_2905988, EPI_ISL_2905989, EPI_ISL_2905991, EPI_ISL_2905993, EPI_ISL_2905995, EPI_ISL_2905996, EPI_ISL_2905997, EPI_ISL_2905998, EPI_ISL_2905999, EPI_ISL_2906077, EPI_ISL_2906079, EPI_ISL_2906080, EPI_ISL_2906081, EPI_ISL_2906082, EPI_ISL_2906083, EPI_ISL_2906084, EPI_ISL_2906085, EPI_ISL_2906088, EPI_ISL_2906089, EPI_ISL_2906090, EPI_ISL_2906091, EPI_ISL_2906092, EPI_ISL_2906093, EPI_ISL_2906094, EPI_ISL_2906095, EPI_ISL_2906096, EPI_ISL_2906097, EPI_ISL_2906098, EPI_ISL_2906099, EPI_ISL_2906155, EPI_ISL_2906156, EPI_ISL_2906157, EPI_ISL_2906158, EPI_ISL_2906159, EPI_ISL_2906160, EPI_ISL_2906165, EPI_ISL_2906178, EPI_ISL_2906182, EPI_ISL_2906183, EPI_ISL_2906184, EPI_ISL_2906185, EPI_ISL_2906186, EPI_ISL_2906187, EPI_ISL_2906188, EPI_ISL_2906191, EPI_ISL_2906192, EPI_ISL_2906193, EPI_ISL_2906194, EPI_ISL_2906195, EPI_ISL_2906197, EPI_ISL_2906199, EPI_ISL_2906200, EPI_ISL_2906201, EPI_ISL_2906209, EPI_ISL_2906217, EPI_ISL_2906218, EPI_ISL_2906220, EPI_ISL_2906226, EPI_ISL_2906233, EPI_ISL_2906236, EPI_ISL_2906237, EPI_ISL_2906238, EPI_ISL_2906241, EPI_ISL_2906242, EPI_ISL_2906244, EPI_ISL_2906245, EPI_ISL_2906250, EPI_ISL_2906251, EPI_ISL_2906254, EPI_ISL_2906255, EPI_ISL_2906256, EPI_ISL_2906257, EPI_ISL_2906258, EPI_ISL_2906259, EPI_ISL_2906260, EPI_ISL_2906261, EPI_ISL_2906262, EPI_ISL_2906263, EPI_ISL_2906264, EPI_ISL_2906265, EPI_ISL_2906267, EPI_ISL_2906268, EPI_ISL_2906269, EPI_ISL_2906270, EPI_ISL_2906271, EPI_ISL_2906272, EPI_ISL_2906273, EPI_ISL_2906275, EPI_ISL_2906276, EPI_ISL_2906277, EPI_ISL_2906279, EPI_ISL_2906280, EPI_ISL_2906281, EPI_ISL_2906282, EPI_ISL_2906283, EPI_ISL_2906287, EPI_ISL_2906288, EPI_ISL_2906289, EPI_ISL_2906290, EPI_ISL_2906291, EPI_ISL_2906292, EPI_ISL_2906293, EPI_ISL_2906294, EPI_ISL_2906295, EPI_ISL_2906353, EPI_ISL_2906354, EPI_ISL_2906355, EPI_ISL_2906356, EPI_ISL_2906357, EPI_ISL_2906358, EPI_ISL_2906359, EPI_ISL_2906365, EPI_ISL_2906367, EPI_ISL_2906368, EPI_ISL_2906374, EPI_ISL_2906379, EPI_ISL_2906380, EPI_ISL_2906381, EPI_ISL_2906382, EPI_ISL_2906383, EPI_ISL_2906384, EPI_ISL_2906385, EPI_ISL_2906386, EPI_ISL_2906388, EPI_ISL_2906389, EPI_ISL_2906390, EPI_ISL_2906391, EPI_ISL_2906392, EPI_ISL_2906394, EPI_ISL_2906395, EPI_ISL_2906397, EPI_ISL_2906398, EPI_ISL_2906399, EPI_ISL_2906400, EPI_ISL_2906401, EPI_ISL_2906402, EPI_ISL_2906403, EPI_ISL_2906404, EPI_ISL_2906405, EPI_ISL_2906406, EPI_ISL_2906407, EPI_ISL_2906408, EPI_ISL_2906410, EPI_ISL_2906411, EPI_ISL_2906412, EPI_ISL_2906413, EPI_ISL_2906414, EPI_ISL_2906415, EPI_ISL_2906416, EPI_ISL_2906417, EPI_ISL_2906418, EPI_ISL_2906419, EPI_ISL_2906420, EPI_ISL_2906421, EPI_ISL_2906422, EPI_ISL_2906423, EPI_ISL_2906424, EPI_ISL_2906425, EPI_ISL_2906426, EPI_ISL_2906427, EPI_ISL_2906428, EPI_ISL_2906429, EPI_ISL_2906430, EPI_ISL_2906457, EPI_ISL_2906458, EPI_ISL_2906459, EPI_ISL_2906460, EPI_ISL_2906461, EPI_ISL_2906462, EPI_ISL_2906464, EPI_ISL_2906465, EPI_ISL_2906466, EPI_ISL_2906467, EPI_ISL_2906468, EPI_ISL_2906469, EPI_ISL_2906470, EPI_ISL_2906471, EPI_ISL_2906472, EPI_ISL_2906473, EPI_ISL_2906474, EPI_ISL_2906475, EPI_ISL_2906476, EPI_ISL_2906477, EPI_ISL_2906478, EPI_ISL_2906479, EPI_ISL_2906480, EPI_ISL_2906508, EPI_ISL_2906509, EPI_ISL_2906510, EPI_ISL_2906511, EPI_ISL_2906513, EPI_ISL_2906514, EPI_ISL_2906516, EPI_ISL_2906520, EPI_ISL_2906521, EPI_ISL_2906523, EPI_ISL_2906524, EPI_ISL_2906526, EPI_ISL_2906528, EPI_ISL_2906529, EPI_ISL_2906530, EPI_ISL_2906531

| | | | |
|---|---|---|---|
| see above | Originating lab: Wales Specialist Virology Centre Sequencing lab: Pathogen Genomics Unit | Public Health Wales Microbiology Cardiff Wales Specialist Virology Centre | Alec Birchley; Alexander Adams; Amy Gaskin; Angela Marchbank; Bree Gatica-Wilcox; Catherine Moore; Jason Coombes; Joanne Watkins; Joel Southgate; Johnathan Evans; Laura Gifford; Lauren Gilbert; Lee Graham; Malorie Perry; Matthew Bull; Nicole Pacchiarini; Sally Corden; Sara Kumziene-Summerhayes; Sara Rey; Sarah Taylor; Simon Cottrell; Sophie Jones; Tom Connor |

EPI_ISL_2903045, EPI_ISL_2903046, EPI_ISL_2903059, EPI_ISL_2903060, EPI_ISL_2903063, EPI_ISL_2903070, EPI_ISL_2903071, EPI_ISL_2903074, EPI_ISL_2903077, EPI_ISL_2903091, EPI_ISL_2903097, EPI_ISL_2903100, EPI_ISL_2903101, EPI_ISL_2903102, EPI_ISL_2903103, EPI_ISL_2903106, EPI_ISL_2903108, EPI_ISL_2903111, EPI_ISL_2903114, EPI_ISL_2903116, EPI_ISL_2903119, EPI_ISL_2903124, EPI_ISL_2903126, EPI_ISL_2903127, EPI_ISL_2903131, EPI_ISL_2903137, EPI_ISL_2903138, EPI_ISL_2903141, EPI_ISL_2903143, EPI_ISL_2903144, EPI_ISL_2903145, EPI_ISL_2903147, EPI_ISL_2903148, EPI_ISL_2903149, EPI_ISL_2903150, EPI_ISL_2903151, EPI_ISL_2903153, EPI_ISL_2903154, EPI_ISL_2903155, EPI_ISL_2903156, EPI_ISL_2903160, EPI_ISL_2903164, EPI_ISL_2903167, EPI_ISL_2903168, EPI_ISL_2903169

| | | | |
|---|---|---|---|
| see above | Oxford Viromics, NDM, University of Oxford; Oxford University Hospitals; Basingstoke and North Hampshire Hospital | COVID-19 Genomics UK (COG-UK) | Alex Mobbs; Amy Trebes; Anita Justice; Catrin Moore; Christophe Fraser; David Bonsall; David Buck; Emma Wise; George Macintyre; Jessica Lynch; John Todd; Mariateresa de Cesare; Matilde Mori; Monique Andersson; Nathan Moore; Nick Cortes; Robert Shaw; Stephen Kidd; Tanya Golubchik; Timothy Peto |

| | | | |
|---|---|---|---|
| | | Consortium | |
| EPI_ISL_2816883, EPI_ISL_2816884, EPI_ISL_2816885, EPI_ISL_2816886, EPI_ISL_2816888, EPI_ISL_2816889, EPI_ISL_2816890, EPI_ISL_2816891, EPI_ISL_2816892, EPI_ISL_2816893, EPI_ISL_2816894, EPI_ISL_2816896, EPI_ISL_2816897, EPI_ISL_2816898, EPI_ISL_2816899, EPI_ISL_2816900, EPI_ISL_2816901, EPI_ISL_2816902, EPI_ISL_2816903, EPI_ISL_2816904, EPI_ISL_2816905, EPI_ISL_2816906, EPI_ISL_2816907, EPI_ISL_2816908, EPI_ISL_2816909, EPI_ISL_2816910, EPI_ISL_2816911, EPI_ISL_2816912, EPI_ISL_2816913, EPI_ISL_2816914, EPI_ISL_2816915, EPI_ISL_2816916, EPI_ISL_2816917, EPI_ISL_2816918, EPI_ISL_2816919, EPI_ISL_2816920, EPI_ISL_2816921, EPI_ISL_2816922, EPI_ISL_2816924, EPI_ISL_2816925, EPI_ISL_2816926, EPI_ISL_2816927, EPI_ISL_2816928, EPI_ISL_2816929, EPI_ISL_2816930, EPI_ISL_2816931, EPI_ISL_2816932, EPI_ISL_2816933, EPI_ISL_2816934, EPI_ISL_2816935, EPI_ISL_2816936, EPI_ISL_2816937, EPI_ISL_2816938, EPI_ISL_2816939, EPI_ISL_2816943, EPI_ISL_2816947, EPI_ISL_2816954, EPI_ISL_2816957, EPI_ISL_2816959, EPI_ISL_2816961, EPI_ISL_2816962, EPI_ISL_2816964, EPI_ISL_2816966, EPI_ISL_2816968, EPI_ISL_2816971, EPI_ISL_2816972, EPI_ISL_2816975, EPI_ISL_2816976, EPI_ISL_2816994, EPI_ISL_2816995, EPI_ISL_2816996, EPI_ISL_2816997, EPI_ISL_2816998, EPI_ISL_2816999, EPI_ISL_2817000, EPI_ISL_2817001, EPI_ISL_2817002, EPI_ISL_2817003, EPI_ISL_2817004, EPI_ISL_2817005, EPI_ISL_2817006, EPI_ISL_2817007, EPI_ISL_2817008, EPI_ISL_2817009, EPI_ISL_2817010, EPI_ISL_2817011, EPI_ISL_2817012, EPI_ISL_2817013, EPI_ISL_2817014, EPI_ISL_2817015, EPI_ISL_2817016, EPI_ISL_2817017, EPI_ISL_2817018, EPI_ISL_2817019, EPI_ISL_2817021, EPI_ISL_2817022, EPI_ISL_2817023, EPI_ISL_2817024, EPI_ISL_2817025, EPI_ISL_2817026, EPI_ISL_2817027, EPI_ISL_2817028, EPI_ISL_2817029, EPI_ISL_2817030, EPI_ISL_2817031, EPI_ISL_2817032, EPI_ISL_2817033, EPI_ISL_2817034, EPI_ISL_2817035, EPI_ISL_2817036, EPI_ISL_2817037, EPI_ISL_2817038, EPI_ISL_2817039, EPI_ISL_2817040, EPI_ISL_2817041, EPI_ISL_2817042, EPI_ISL_2817043, EPI_ISL_2817044, EPI_ISL_2817045, EPI_ISL_2817046, EPI_ISL_2817047, EPI_ISL_2817048, EPI_ISL_2817049, EPI_ISL_2817050, EPI_ISL_2817051, EPI_ISL_2817052, EPI_ISL_2817053, EPI_ISL_2817054, EPI_ISL_2817055, EPI_ISL_2817056, EPI_ISL_2817057, EPI_ISL_2817058, EPI_ISL_2817059, EPI_ISL_2817060, EPI_ISL_2817061, EPI_ISL_2817062, EPI_ISL_2817063, EPI_ISL_2817064, EPI_ISL_2817065, EPI_ISL_2817066, EPI_ISL_2817067, EPI_ISL_2817068, EPI_ISL_2817069, EPI_ISL_2817070, EPI_ISL_2817071, EPI_ISL_2817072, EPI_ISL_2817073, EPI_ISL_2817074, EPI_ISL_2817075, EPI_ISL_2817076, EPI_ISL_2817077, EPI_ISL_2817078, EPI_ISL_2817079, EPI_ISL_2817080, EPI_ISL_2817081, EPI_ISL_2817082, EPI_ISL_2817083, EPI_ISL_2817084, EPI_ISL_2817085, EPI_ISL_2817086, EPI_ISL_2817087, EPI_ISL_2817088, EPI_ISL_2817089, EPI_ISL_2817090, EPI_ISL_2817091, EPI_ISL_2817092, EPI_ISL_2817093, EPI_ISL_2817094, EPI_ISL_2817095, EPI_ISL_2817096, EPI_ISL_2817097, EPI_ISL_2817098, EPI_ISL_2817099, EPI_ISL_2817100, EPI_ISL_2817101, EPI_ISL_2817102, EPI_ISL_2817103, EPI_ISL_2817104, EPI_ISL_2817105, EPI_ISL_2817106, EPI_ISL_2817107, EPI_ISL_2817108, EPI_ISL_2817109, EPI_ISL_2817110, EPI_ISL_2817111, EPI_ISL_2817112, EPI_ISL_2817113, EPI_ISL_2817114, EPI_ISL_2817115, EPI_ISL_2817116, EPI_ISL_2817117, EPI_ISL_2817118, EPI_ISL_2817133, EPI_ISL_2817170, EPI_ISL_2817171, EPI_ISL_2817172, EPI_ISL_2902456, EPI_ISL_2902501, EPI_ISL_2902507, EPI_ISL_2902573 | | | |
| see above | Quadram Institute Bioscience | COVID-19 Genomics UK (COG-UK) Consortium | Alexander J Trotter; Alison E. Mather; Alp Aydin; Ana P. Tedim; Anastasia Kolyva; Andrew Bell; Andrew J. Page; Christopher Jeanes; Claire Stuart; Dave J. Baker; Ebenezer Foster-Nyarko; Gemma L. Kay; John Wain; Justin O'Grady; Leonardo de Oliveira Martins; Lewis G. Spurgin; Lindsay Coupland; Lizzie Meadows; Luke Bedford; Maria Diaz; Mark Webber; Martin Lott; Muhammed Yasir; Nabil-Fareed Alikhan; Ngozi Elumogo; Nicholas M. Thomson; Rachael Stanley; Rachel Gilroy; Reenesh Prakash; Rose K Davidson; Samir Dervisevic; Samuel Bloomfield; Sophie J. Prosolek; Steven Rudder; Thanh Le-Viet |
| EPI_ISL_2817173, EPI_ISL_2817174, EPI_ISL_2817175, EPI_ISL_2817176, EPI_ISL_2817177, EPI_ISL_2817178, EPI_ISL_2817179, EPI_ISL_2817180, EPI_ISL_2817181, EPI_ISL_2817182, EPI_ISL_2817183, EPI_ISL_2817184, EPI_ISL_2817185, EPI_ISL_2817186, EPI_ISL_2817187, EPI_ISL_2817188, EPI_ISL_2817189, EPI_ISL_2817190, EPI_ISL_2817191, EPI_ISL_2817192, EPI_ISL_2817193, EPI_ISL_2817194, EPI_ISL_2817195, EPI_ISL_2817196, EPI_ISL_2817197, EPI_ISL_2817198, EPI_ISL_2817199, EPI_ISL_2817200, EPI_ISL_2817201, EPI_ISL_2817202, EPI_ISL_2817203, EPI_ISL_2817204, EPI_ISL_2817205, EPI_ISL_2817206, EPI_ISL_2817207, EPI_ISL_2817208, EPI_ISL_2817209, EPI_ISL_2817210, EPI_ISL_2817211, EPI_ISL_2817212, EPI_ISL_2817213, EPI_ISL_2817214, EPI_ISL_2817215, EPI_ISL_2817216, EPI_ISL_2817217, EPI_ISL_2817218, EPI_ISL_2817219, EPI_ISL_2817220, EPI_ISL_2817221, EPI_ISL_2817222, EPI_ISL_2817224, EPI_ISL_2817225, EPI_ISL_2817226, EPI_ISL_2817227, EPI_ISL_2817228, EPI_ISL_2817229, EPI_ISL_2817230, EPI_ISL_2817232, EPI_ISL_2817233, EPI_ISL_2817234, EPI_ISL_2817235, EPI_ISL_2817237, EPI_ISL_2817239, EPI_ISL_2817240, EPI_ISL_2817241, EPI_ISL_2817242, EPI_ISL_2817243, EPI_ISL_2817244, EPI_ISL_2817245, EPI_ISL_2817246, EPI_ISL_2817247, EPI_ISL_2817248, EPI_ISL_2817249, EPI_ISL_2817250, EPI_ISL_2817251, EPI_ISL_2817252, EPI_ISL_2817253, EPI_ISL_2817254, EPI_ISL_2817255, EPI_ISL_2817256, EPI_ISL_2817257, EPI_ISL_2817258, EPI_ISL_2817260, EPI_ISL_2817261, EPI_ISL_2817262, EPI_ISL_2817263, EPI_ISL_2817264, EPI_ISL_2817265, EPI_ISL_2817266, EPI_ISL_2817267, EPI_ISL_2817268, EPI_ISL_2817269, EPI_ISL_2817270, EPI_ISL_2817271, EPI_ISL_2817272, EPI_ISL_2817274, EPI_ISL_2817275, EPI_ISL_2817276, EPI_ISL_2817277, EPI_ISL_2817278, EPI_ISL_2817279, EPI_ISL_2817281 | | | |
| see above | Queens Medical Centre, Clinical Microbiology Department / DeepSeq Nottingham | COVID-19 Genomics UK (COG-UK) Consortium | Christopher Moore; Emily Park; Fei Sang; Gemma Clark; Hannah Howson-Wells; Jack Hill; Jayasree Dey; Johnny Debebe; Jonathan Ball; Joseph Chappell; Manjinder Khakh; Matthew Carlisle; Matthew Loose; Michelle M Lister; Nadine Holmes; Patrick McClure; Theocharis Tsoleridis; Timothy Byaruhanga; Vicki M Fleming; Victoria Wright; Wendy Smith |
| EPI_ISL_2747284, EPI_ISL_2747443, EPI_ISL_2747445, EPI_ISL_2747446, EPI_ISL_2747447, EPI_ISL_2747577, EPI_ISL_2747594, EPI_ISL_2747606, EPI_ISL_2747608, EPI_ISL_2747610, EPI_ISL_2747614, EPI_ISL_2747622, EPI_ISL_2747627, EPI_ISL_2747640, EPI_ISL_2747671, EPI_ISL_2747683, EPI_ISL_2747685, EPI_ISL_2747727, EPI_ISL_2747737, EPI_ISL_2747739, EPI_ISL_2747798, EPI_ISL_2747934, EPI_ISL_2747938, EPI_ISL_2747940, EPI_ISL_2747953, EPI_ISL_2747955, EPI_ISL_2747969, EPI_ISL_2747994, EPI_ISL_2748036, EPI_ISL_2748038, EPI_ISL_2748040, EPI_ISL_2748043, EPI_ISL_2748045, EPI_ISL_2748048, EPI_ISL_2748050, EPI_ISL_2748052, EPI_ISL_2748054, EPI_ISL_2748056, EPI_ISL_2748058, EPI_ISL_2748060, EPI_ISL_2748062, EPI_ISL_2748064, EPI_ISL_2748066, EPI_ISL_2748067, EPI_ISL_2748078, EPI_ISL_2748080, EPI_ISL_2748082, EPI_ISL_2748084, EPI_ISL_2748088, EPI_ISL_2748089, EPI_ISL_2748099, EPI_ISL_2748101, EPI_ISL_2748104, EPI_ISL_2748106, EPI_ISL_2748108, EPI_ISL_2748110, EPI_ISL_2748112, EPI_ISL_2748114, EPI_ISL_2748116, EPI_ISL_2748118, EPI_ISL_2748120, EPI_ISL_2748122, EPI_ISL_2748124, EPI_ISL_2748126, EPI_ISL_2748128, EPI_ISL_2748130, EPI_ISL_2748132, EPI_ISL_2748134, EPI_ISL_2748136, EPI_ISL_2748138, EPI_ISL_2748154, EPI_ISL_2748156, EPI_ISL_2748158, EPI_ISL_2748160, EPI_ISL_2748162, EPI_ISL_2748163, EPI_ISL_2748165, EPI_ISL_2748167, EPI_ISL_2748169, EPI_ISL_2748171, EPI_ISL_2748173, EPI_ISL_2748175, EPI_ISL_2748177, EPI_ISL_2748179, EPI_ISL_2817305, EPI_ISL_2817306, EPI_ISL_2817307, EPI_ISL_2817308, EPI_ISL_2817312, EPI_ISL_2817313, EPI_ISL_2817314, EPI_ISL_2817315, EPI_ISL_2817316, EPI_ISL_2817317, EPI_ISL_2817318, EPI_ISL_2817319, EPI_ISL_2817320, EPI_ISL_2817321, EPI_ISL_2817323, EPI_ISL_2817325, EPI_ISL_2817326, EPI_ISL_2817327, EPI_ISL_2817328, EPI_ISL_2817329, EPI_ISL_2817330, EPI_ISL_2817331, EPI_ISL_2817332, EPI_ISL_2817333, EPI_ISL_2817334, EPI_ISL_2817335, EPI_ISL_2817336, EPI_ISL_2817337, EPI_ISL_2817338, EPI_ISL_2817339, EPI_ISL_2817340, EPI_ISL_2817341, EPI_ISL_2817342, EPI_ISL_2817344, EPI_ISL_2817345, EPI_ISL_2817346, EPI_ISL_2817347, EPI_ISL_2817348, EPI_ISL_2817349, EPI_ISL_2817350, EPI_ISL_2817351, EPI_ISL_2817352, EPI_ISL_2817353, EPI_ISL_2817354, EPI_ISL_2817396, EPI_ISL_2817398, EPI_ISL_2817399, EPI_ISL_2817400, EPI_ISL_2817401, EPI_ISL_2817402, EPI_ISL_2817403, EPI_ISL_2817404, EPI_ISL_2817405, EPI_ISL_2817406, EPI_ISL_2817407, EPI_ISL_2817408, EPI_ISL_2817418, EPI_ISL_2817421, EPI_ISL_2817423, EPI_ISL_2817425, EPI_ISL_2817426, EPI_ISL_2817428, EPI_ISL_2817429, EPI_ISL_2817430, EPI_ISL_2817431, EPI_ISL_2817432, EPI_ISL_2817434, EPI_ISL_2817435, EPI_ISL_2817436, EPI_ISL_2817437, EPI_ISL_2817438, EPI_ISL_2817439, EPI_ISL_2817440, EPI_ISL_2817441, EPI_ISL_2817442, EPI_ISL_2817445, EPI_ISL_2817446, EPI_ISL_2817447, EPI_ISL_2817448, EPI_ISL_2817449, EPI_ISL_2817450, EPI_ISL_2817451, EPI_ISL_2817452, EPI_ISL_2817453, EPI_ISL_2817454, EPI_ISL_2817455, EPI_ISL_2817456, EPI_ISL_2817457, EPI_ISL_2817458, EPI_ISL_2817459, EPI_ISL_2817460, EPI_ISL_2817461, EPI_ISL_2817462, EPI_ISL_2817463, EPI_ISL_2817464, EPI_ISL_2817465, EPI_ISL_2817466, EPI_ISL_2817467, EPI_ISL_2817468, EPI_ISL_2817469, EPI_ISL_2817470, EPI_ISL_2817471, EPI_ISL_2817472, EPI_ISL_2817473, EPI_ISL_2817474, EPI_ISL_2817475, EPI_ISL_2817476, EPI_ISL_2817477, EPI_ISL_2817478, EPI_ISL_2817481, EPI_ISL_2817482, EPI_ISL_2817483, EPI_ISL_2817484, EPI_ISL_2817485, EPI_ISL_2817486, EPI_ISL_2817487, EPI_ISL_2817488, EPI_ISL_2817489, EPI_ISL_2817490, EPI_ISL_2817491, EPI_ISL_2817494, EPI_ISL_2817495, EPI_ISL_2817497, EPI_ISL_2817498, EPI_ISL_2817499, EPI_ISL_2817500, EPI_ISL_2817501, EPI_ISL_2817504, EPI_ISL_2817506, EPI_ISL_2817513, EPI_ISL_2817522, EPI_ISL_2817528, EPI_ISL_2817531, EPI_ISL_2817532, EPI_ISL_2817533, EPI_ISL_2817541, EPI_ISL_2817542, EPI_ISL_2817543, EPI_ISL_2817561, EPI_ISL_2817562, EPI_ISL_2817563, EPI_ISL_2817564, EPI_ISL_2817565, EPI_ISL_2817566, EPI_ISL_2817567, EPI_ISL_2817568, EPI_ISL_2817569, EPI_ISL_2817570, EPI_ISL_2817571, EPI_ISL_2817572, EPI_ISL_2817573, EPI_ISL_2817574, EPI_ISL_2817575, EPI_ISL_2817576, EPI_ISL_2817577, EPI_ISL_2817578, EPI_ISL_2817579, EPI_ISL_2817580, EPI_ISL_2817581, EPI_ISL_2817582, EPI_ISL_2817583, EPI_ISL_2817584, EPI_ISL_2817585, EPI_ISL_2817586, EPI_ISL_2817587, EPI_ISL_2817588, EPI_ISL_2817589, EPI_ISL_2817590, EPI_ISL_2817592, EPI_ISL_2817594, EPI_ISL_2817596, EPI_ISL_2817597, EPI_ISL_2817598, EPI_ISL_2817599, EPI_ISL_2817600, EPI_ISL_2817602, EPI_ISL_2817603, EPI_ISL_2817604, EPI_ISL_2817605, EPI_ISL_2817606, EPI_ISL_2817607, EPI_ISL_2817608, EPI_ISL_2817609, EPI_ISL_2817610, EPI_ISL_2817611, EPI_ISL_2817612, EPI_ISL_2817613, EPI_ISL_2817614, EPI_ISL_2817615, EPI_ISL_2817616, EPI_ISL_2817617, EPI_ISL_2817618, EPI_ISL_2817619, EPI_ISL_2817620, EPI_ISL_2817621, EPI_ISL_2817622, EPI_ISL_2817623, EPI_ISL_2817624, EPI_ISL_2817625, EPI_ISL_2817626, EPI_ISL_2817627, EPI_ISL_2817628, EPI_ISL_2817629, EPI_ISL_2817630, EPI_ISL_2817631, EPI_ISL_2817651, EPI_ISL_2817652, EPI_ISL_2817653, EPI_ISL_2817654, EPI_ISL_2817655, EPI_ISL_2817656, EPI_ISL_2817657, EPI_ISL_2817658, EPI_ISL_2817659, EPI_ISL_2817660, EPI_ISL_2817661, EPI_ISL_2817662, EPI_ISL_2817663, EPI_ISL_2817665, EPI_ISL_2817666, EPI_ISL_2817667, EPI_ISL_2817669, EPI_ISL_2817670, EPI_ISL_2817671, EPI_ISL_2817673, EPI_ISL_2817683, EPI_ISL_2817684, EPI_ISL_2817685, EPI_ISL_2817686, EPI_ISL_2817687, EPI_ISL_2817690, EPI_ISL_2817691, EPI_ISL_2817692, EPI_ISL_2817693, EPI_ISL_2817695, EPI_ISL_2817696, EPI_ISL_2817697, EPI_ISL_2817698, EPI_ISL_2817699, EPI_ISL_2817700, EPI_ISL_2817701, EPI_ISL_2817702, EPI_ISL_2817703, EPI_ISL_2817704, EPI_ISL_2817705, EPI_ISL_2817706, EPI_ISL_2817707, EPI_ISL_2817708, EPI_ISL_2817709, EPI_ISL_2817710, EPI_ISL_2817711, EPI_ISL_2817713, EPI_ISL_2817714, EPI_ISL_2817715, EPI_ISL_2817716, EPI_ISL_2817717, EPI_ISL_2817718, EPI_ISL_2817720, EPI_ISL_2817721, EPI_ISL_2817722, EPI_ISL_2817723, EPI_ISL_2817724, EPI_ISL_2817725, EPI_ISL_2817726, EPI_ISL_2817727, EPI_ISL_2817729, EPI_ISL_2817730, EPI_ISL_2817731, EPI_ISL_2817733, EPI_ISL_2817748, EPI_ISL_2817749, EPI_ISL_2817750, EPI_ISL_2817751, EPI_ISL_2817752, EPI_ISL_2817753, EPI_ISL_2817754, EPI_ISL_2817757, EPI_ISL_2817758, EPI_ISL_2817759, EPI_ISL_2817760, EPI_ISL_2817761, EPI_ISL_2817763, EPI_ISL_2817764, EPI_ISL_2817765, EPI_ISL_2817766, EPI_ISL_2817767, EPI_ISL_2817768, EPI_ISL_2817770, EPI_ISL_2817771, EPI_ISL_2817772, EPI_ISL_2817773, EPI_ISL_2817774, EPI_ISL_2817776, EPI_ISL_2817777, EPI_ISL_2817778, EPI_ISL_2817780, EPI_ISL_2817781, EPI_ISL_2817782, EPI_ISL_2817784, EPI_ISL_2817785, EPI_ISL_2817786, EPI_ISL_2817787, EPI_ISL_2817788, EPI_ISL_2817789, EPI_ISL_2817790, EPI_ISL_2817791, EPI_ISL_2817792, EPI_ISL_2817793, EPI_ISL_2817794, EPI_ISL_2817795, EPI_ISL_2817796, EPI_ISL_2817797, EPI_ISL_2817798, EPI_ISL_2817799, EPI_ISL_2817800, EPI_ISL_2817801, EPI_ISL_2817802, EPI_ISL_2817803, EPI_ISL_2817804, EPI_ISL_2817805, EPI_ISL_2817806, EPI_ISL_2817807, EPI_ISL_2817808, EPI_ISL_2817809, EPI_ISL_2817810, EPI_ISL_2817811, EPI_ISL_2817815, EPI_ISL_2817825, EPI_ISL_2817843, EPI_ISL_2817855, EPI_ISL_2817856, EPI_ISL_2817860, EPI_ISL_2817875, EPI_ISL_2817876, EPI_ISL_2817878, EPI_ISL_2817879, EPI_ISL_2817880, EPI_ISL_2817881, EPI_ISL_2817882, EPI_ISL_2817883, EPI_ISL_2817884, EPI_ISL_2817885, EPI_ISL_2817886, EPI_ISL_2817888, EPI_ISL_2817890, EPI_ISL_2817891, EPI_ISL_2817892, EPI_ISL_2817893, EPI_ISL_2817894, EPI_ISL_2817895, EPI_ISL_2817896, EPI_ISL_2817897, EPI_ISL_2817898, EPI_ISL_2817899, EPI_ISL_2817900, EPI_ISL_2817901, EPI_ISL_2817902, EPI_ISL_2817905, EPI_ISL_2817906, EPI_ISL_2817907, EPI_ISL_2817908, EPI_ISL_2817909, EPI_ISL_2817912, EPI_ISL_2817916, EPI_ISL_2817917, EPI_ISL_2817918, EPI_ISL_2817919, EPI_ISL_2817952, EPI_ISL_2817956, EPI_ISL_2817968, EPI_ISL_2817969, EPI_ISL_2817970, EPI_ISL_2817971, EPI_ISL_2817972, EPI_ISL_2817973, EPI_ISL_2817974, EPI_ISL_2817975, EPI_ISL_2817976, EPI_ISL_2817977, EPI_ISL_2817978, EPI_ISL_2817979, EPI_ISL_2817980, EPI_ISL_2817981, EPI_ISL_2817982, EPI_ISL_2817983, EPI_ISL_2817984, EPI_ISL_2817985, EPI_ISL_2817986, EPI_ISL_2817987, EPI_ISL_2817988, EPI_ISL_2817993, EPI_ISL_2817994, EPI_ISL_2817995, EPI_ISL_2817996, EPI_ISL_2818042, EPI_ISL_2818043, EPI_ISL_2818044, EPI_ISL_2818045, EPI_ISL_2818046, EPI_ISL_2818047, EPI_ISL_2818048, EPI_ISL_2818049, EPI_ISL_2818051, EPI_ISL_2818060, EPI_ISL_2818063, EPI_ISL_2818064, EPI_ISL_2818065, EPI_ISL_2818066, EPI_ISL_2818071, EPI_ISL_2818072, EPI_ISL_2818074, EPI_ISL_2818124, EPI_ISL_2818125, EPI_ISL_2818126, EPI_ISL_2818127, EPI_ISL_2818128, EPI_ISL_2818129, EPI_ISL_2818130, EPI_ISL_2818131, EPI_ISL_2818132, EPI_ISL_2818133, EPI_ISL_2818134, EPI_ISL_2818135, EPI_ISL_2818136, EPI_ISL_2818137, EPI_ISL_2818138, EPI_ISL_2818139, EPI_ISL_2818140, EPI_ISL_2818141, EPI_ISL_2818142, EPI_ISL_2818144, EPI_ISL_2818159, EPI_ISL_2818160, EPI_ISL_2818161, EPI_ISL_2818164, EPI_ISL_2818166, EPI_ISL_2818167, EPI_ISL_2818169, EPI_ISL_2818171, EPI_ISL_2818172, EPI_ISL_2818173, EPI_ISL_2818176, EPI_ISL_2818177, EPI_ISL_2818178, EPI_ISL_2818180, EPI_ISL_2818181, EPI_ISL_2818182, EPI_ISL_2818183, EPI_ISL_2818185, EPI_ISL_2818265, EPI_ISL_2818266, EPI_ISL_2818267, EPI_ISL_2818268, EPI_ISL_2818270, EPI_ISL_2818271, EPI_ISL_2818272, EPI_ISL_2818275, EPI_ISL_2818277, EPI_ISL_2818278, EPI_ISL_2818279, EPI_ISL_2818281, EPI_ISL_2818282, EPI_ISL_2818283, EPI_ISL_2818284, EPI_ISL_2818285, EPI_ISL_2818286, EPI_ISL_2818287, EPI_ISL_2818288, EPI_ISL_2818289, EPI_ISL_2818290, EPI_ISL_2818291, EPI_ISL_2818292, EPI_ISL_2818293, EPI_ISL_2818294, EPI_ISL_2903415, EPI_ISL_2903418, EPI_ISL_2903419, EPI_ISL_2903450, EPI_ISL_2903453, EPI_ISL_2903454, EPI_ISL_2903455, EPI_ISL_2903456, EPI_ISL_2903457, EPI_ISL_2903465, EPI_ISL_2903466, EPI_ISL_2903467, EPI_ISL_2903472, EPI_ISL_2903474, EPI_ISL_2903475, EPI_ISL_2903488, EPI_ISL_2903493, EPI_ISL_2903494, EPI_ISL_2903496, EPI_ISL_2903634, EPI_ISL_2903635, EPI_ISL_2903636, EPI_ISL_2903637, EPI_ISL_2903638, EPI_ISL_2903639, EPI_ISL_2903640, EPI_ISL_2903641, EPI_ISL_2903642, EPI_ISL_2903643, EPI_ISL_2903644, EPI_ISL_2903645, EPI_ISL_2903646, EPI_ISL_2903647, EPI_ISL_2903648, EPI_ISL_2903732, EPI_ISL_2903790, EPI_ISL_2903930, EPI_ISL_2903932, EPI_ISL_2903949, EPI_ISL_2903953, EPI_ISL_2903955, EPI_ISL_2903987, EPI_ISL_2903989, EPI_ISL_2903991, EPI_ISL_2903992, EPI_ISL_2904042, EPI_ISL_2904044, EPI_ISL_2904055, EPI_ISL_2904339 | | | |
| see above | Respiratory Virus Unit, Microbiology Services Colindale, Public Health England | COVID-19 Genomics UK (COG-UK) Consortium | PHE Covid Sequencing Team |
| EPI_ISL_2816413, EPI_ISL_2816414, EPI_ISL_2816415, EPI_ISL_2816416, EPI_ISL_2816417, EPI_ISL_2816418, EPI_ISL_2816419, EPI_ISL_2816420, EPI_ISL_2816421, EPI_ISL_2816422, EPI_ISL_2816423, EPI_ISL_2816424, EPI_ISL_2816425, EPI_ISL_2816427, EPI_ISL_2816428, EPI_ISL_2816429, EPI_ISL_2816430, EPI_ISL_2816431, EPI_ISL_2816432, EPI_ISL_2816433, EPI_ISL_2816434, EPI_ISL_2816435, EPI_ISL_2816436, EPI_ISL_2816437, EPI_ISL_2816439, EPI_ISL_2816440, EPI_ISL_2816441, EPI_ISL_2816442, EPI_ISL_2816443, EPI_ISL_2816444, EPI_ISL_2816445, EPI_ISL_2816446, EPI_ISL_2816447, EPI_ISL_2816448, EPI_ISL_2816449, EPI_ISL_2816450, EPI_ISL_2816451, EPI_ISL_2816452, EPI_ISL_2816453, EPI_ISL_2816454, EPI_ISL_2816455, EPI_ISL_2816456, EPI_ISL_2816457, EPI_ISL_2816458, EPI_ISL_2816459, EPI_ISL_2816460, EPI_ISL_2816461, EPI_ISL_2816462, EPI_ISL_2816463, EPI_ISL_2816464, EPI_ISL_2816465, EPI_ISL_2816466, EPI_ISL_2816467, EPI_ISL_2816468, EPI_ISL_2816469, EPI_ISL_2816470, EPI_ISL_2816471, EPI_ISL_2816473, EPI_ISL_2816475, EPI_ISL_2816476, EPI_ISL_2816477, EPI_ISL_2816478, EPI_ISL_2816479, EPI_ISL_2816480, EPI_ISL_2816481, EPI_ISL_2816482, EPI_ISL_2816483, EPI_ISL_2816484, EPI_ISL_2816485, EPI_ISL_2816486, EPI_ISL_2816487, EPI_ISL_2816488, EPI_ISL_2816489, EPI_ISL_2816490, EPI_ISL_2816491, EPI_ISL_2816492, EPI_ISL_2816493, EPI_ISL_2816494, EPI_ISL_2816495, EPI_ISL_2816496, EPI_ISL_2816497, EPI_ISL_2816499, EPI_ISL_2816500, EPI_ISL_2816501, EPI_ISL_2816502, EPI_ISL_2816504, EPI_ISL_2816505, EPI_ISL_2816506, EPI_ISL_2816507, EPI_ISL_2816509, EPI_ISL_2816510, EPI_ISL_2816511, EPI_ISL_2816512, EPI_ISL_2816513, EPI_ISL_2816514, EPI_ISL_2816515, EPI_ISL_2816516, EPI_ISL_2816517, EPI_ISL_2816518, EPI_ISL_2816519, EPI_ISL_2816520, EPI_ISL_2816521, EPI_ISL_2816522, EPI_ISL_2816523, EPI_ISL_2816524, EPI_ISL_2816525, EPI_ISL_2816526, EPI_ISL_2816527, EPI_ISL_2816528, EPI_ISL_2816529, EPI_ISL_2816530, EPI_ISL_2816531, EPI_ISL_2816532, EPI_ISL_2816533, EPI_ISL_2816534, EPI_ISL_2816535, EPI_ISL_2816536, EPI_ISL_2816537, EPI_ISL_2816539, EPI_ISL_2816540, EPI_ISL_2816541, EPI_ISL_2816542, EPI_ISL_2816543, EPI_ISL_2816545, EPI_ISL_2816546, EPI_ISL_2816547, EPI_ISL_2816548, EPI_ISL_2816549, EPI_ISL_2816550, EPI_ISL_2816551, EPI_ISL_2816553, EPI_ISL_2816554, EPI_ISL_2816555, EPI_ISL_2816556, EPI_ISL_2816557, EPI_ISL_2816558, EPI_ISL_2816559, EPI_ISL_2816560, EPI_ISL_2816561, EPI_ISL_2816562, EPI_ISL_2816563 | | | |
| see above | University College London, Great Ormond Street Hospital for Children NHS Foundation Trust, Imperial College Healthcare NHS Trust | COVID-19 Genomics UK (COG-UK) Consortium | Charlotte Williams; Helena Tutill; Judith Breuer; Marius Cotic; Mark Kristiansen; Nadua Bayzid; Patricia Dyal; Rachel Williams; Sergi Castellano; Sunando Roy |
| EPI_ISL_2815400, EPI_ISL_2815401, EPI_ISL_2815402, EPI_ISL_2815403, EPI_ISL_2815404, EPI_ISL_2815405, EPI_ISL_2815406, EPI_ISL_2815407, EPI_ISL_2815408, EPI_ISL_2815409, EPI_ISL_2815410, EPI_ISL_2815411, EPI_ISL_2815412, EPI_ISL_2815413, EPI_ISL_2815414, EPI_ISL_2815415, EPI_ISL_2815416, EPI_ISL_2815417, EPI_ISL_2815418, EPI_ISL_2815419, EPI_ISL_2815421, EPI_ISL_2815422, EPI_ISL_2815423, EPI_ISL_2815425, EPI_ISL_2815426, EPI_ISL_2815427, EPI_ISL_2815428, EPI_ISL_2815429, EPI_ISL_2815430, EPI_ISL_2815431, EPI_ISL_2815432, EPI_ISL_2815433, EPI_ISL_2815434, EPI_ISL_2815435, EPI_ISL_2815436, EPI_ISL_2815437, EPI_ISL_2815438, EPI_ISL_2815439, EPI_ISL_2815440, EPI_ISL_2815441, EPI_ISL_2815442, EPI_ISL_2815443, EPI_ISL_2815444, EPI_ISL_2815445, EPI_ISL_2815446, EPI_ISL_2815447, EPI_ISL_2815450, EPI_ISL_2815451, EPI_ISL_2815452, EPI_ISL_2815453, EPI_ISL_2815454, EPI_ISL_2815455, EPI_ISL_2815456, EPI_ISL_2815457, EPI_ISL_2815458, EPI_ISL_2815459, EPI_ISL_2815460, EPI_ISL_2815461, EPI_ISL_2815462, EPI_ISL_2815463, EPI_ISL_2815464, EPI_ISL_2815465, EPI_ISL_2815467, EPI_ISL_2815468, EPI_ISL_2815469, EPI_ISL_2815471, EPI_ISL_2815472, EPI_ISL_2815474, EPI_ISL_2815475, EPI_ISL_2815476, EPI_ISL_2815477, EPI_ISL_2815478, EPI_ISL_2815479, EPI_ISL_2815480, EPI_ISL_2815481, EPI_ISL_2815482, EPI_ISL_2815483, EPI_ISL_2815484, EPI_ISL_2815486, EPI_ISL_2815487, EPI_ISL_2815488, EPI_ISL_2815489, EPI_ISL_2815491, EPI_ISL_2815493, EPI_ISL_2815494, EPI_ISL_2815495, EPI_ISL_2815496, EPI_ISL_2815497, EPI_ISL_2815498, EPI_ISL_2815499, EPI_ISL_2815500, EPI_ISL_2815501, EPI_ISL_2815503, EPI_ISL_2815504, EPI_ISL_2815505, EPI_ISL_2815506, EPI_ISL_2815507, EPI_ISL_2815508, EPI_ISL_2815509, EPI_ISL_2815510, EPI_ISL_2815512, EPI_ISL_2815514, EPI_ISL_2815515, EPI_ISL_2815517, EPI_ISL_2815518, EPI_ISL_2815519, EPI_ISL_2815520, EPI_ISL_2815521, EPI_ISL_2815523, EPI_ISL_2815525, EPI_ISL_2815526, EPI_ISL_2815527, EPI_ISL_2815528, EPI_ISL_2815529, EPI_ISL_2815530, EPI_ISL_2815535, EPI_ISL_2815537 | | | |
| see above | University of Birmingham | COVID-19 Genomics UK (COG-UK) Consortium | Alex Richter; Andrew Bosworth; Andrew Bosworth. Queen Elizabeth Hospital: Anna Casey; Andrew D Beggs PHE Heartlands Lab: Husam Osman; Charlotte Poxon; Institute of Microbiology; Joanne Stockton; Josh Quick; Kasun Wanigasooriya; Mike Kidd; Nicholas Loman. University of Birmingham Testing Laboratory: Celina M Whalley; Oliver Pickles; Radoslaw Poplawski; Samuel Nicholls; University of Birmingham: Claire McMurray; Will Rowe |
| EPI_ISL_2734403, EPI_ISL_2815992, EPI_ISL_2815998, EPI_ISL_2815999, EPI_ISL_2816000, EPI_ISL_2816001, EPI_ISL_2816005, EPI_ISL_2816006, EPI_ISL_2816007, EPI_ISL_2816008, EPI_ISL_2816009, EPI_ISL_2816010, EPI_ISL_2816011, EPI_ISL_2816012, EPI_ISL_2816013, EPI_ISL_2816014, EPI_ISL_2816015, EPI_ISL_2816016, EPI_ISL_2816017, EPI_ISL_2816018, EPI_ISL_2816019, EPI_ISL_2816020, EPI_ISL_2816021, EPI_ISL_2816022, EPI_ISL_2816023, EPI_ISL_2816024, EPI_ISL_2816025, EPI_ISL_2816026, EPI_ISL_2816031, EPI_ISL_2816032, EPI_ISL_2816033, EPI_ISL_2816034, EPI_ISL_2816035, EPI_ISL_2816036, EPI_ISL_2816037, EPI_ISL_2816045, EPI_ISL_2816046, EPI_ISL_2816047, EPI_ISL_2816048, EPI_ISL_2816050, EPI_ISL_2816051, EPI_ISL_2816052, EPI_ISL_2816053, EPI_ISL_2816054, EPI_ISL_2816055, EPI_ISL_2816056, EPI_ISL_2816057, EPI_ISL_2816058, EPI_ISL_2816059, EPI_ISL_2816060, EPI_ISL_2816062, EPI_ISL_2816063, EPI_ISL_2816065, EPI_ISL_2816066, EPI_ISL_2816067, EPI_ISL_2816068, EPI_ISL_2816074, EPI_ISL_2816075, EPI_ISL_2816076, EPI_ISL_2816077, EPI_ISL_2816078, EPI_ISL_2816079, EPI_ISL_2816080, EPI_ISL_2816081, EPI_ISL_2816082, EPI_ISL_2816083, EPI_ISL_2816084, EPI_ISL_2816089, EPI_ISL_2900756, EPI_ISL_2900757, EPI_ISL_2900758, EPI_ISL_2900759, EPI_ISL_2900760, EPI_ISL_2900761, EPI_ISL_2900762, EPI_ISL_2900763, EPI_ISL_2900764, EPI_ISL_2900765, EPI_ISL_2900780, EPI_ISL_2900785, EPI_ISL_2900786, EPI_ISL_2900789, EPI_ISL_2900790, EPI_ISL_2900818 | | | |
| see above | Virology Department, Royal Infirmary of Edinburgh, NHS Lothian / School of Biological Sciences, University of Edinburgh | COVID-19 Genomics UK (COG-UK) Consortium | Colquhoun R; Cotton S; Dewar R; Gallagher A; Hill V; Jackson B; Maloney D; McCrone JT; McHugh M; O'Toole Á; Rambaut A; Rooke S; Scher E; Templeton K; Yu X |
| EPI_ISL_2708440, EPI_ISL_2708770, EPI_ISL_2708799, EPI_ISL_2819921, EPI_ISL_2819922, EPI_ISL_2819923, EPI_ISL_2819924, EPI_ISL_2819926, EPI_ISL_2819927, EPI_ISL_2819928, EPI_ISL_2819929, EPI_ISL_2819931, EPI_ISL_2819934, EPI_ISL_2819935, EPI_ISL_2819937, EPI_ISL_2819938, EPI_ISL_2819939, EPI_ISL_2819940, EPI_ISL_2819941, EPI_ISL_2819942, EPI_ISL_2819944, EPI_ISL_2819946, EPI_ISL_2819947, EPI_ISL_2819948, EPI_ISL_2819949, EPI_ISL_2819950, EPI_ISL_2819952, EPI_ISL_2819953, EPI_ISL_2819954, EPI_ISL_2819960, EPI_ISL_2819962, EPI_ISL_2819965, EPI_ISL_2819966, EPI_ISL_2819968, EPI_ISL_2819970, EPI_ISL_2819972, EPI_ISL_2819973, EPI_ISL_2819977, EPI_ISL_2819978, EPI_ISL_2819979, EPI_ISL_2819980, EPI_ISL_2819981, EPI_ISL_2819982, EPI_ISL_2819983, EPI_ISL_2819984, EPI_ISL_2819985, EPI_ISL_2819986, EPI_ISL_2819990, EPI_ISL_2819993, EPI_ISL_2819996, EPI_ISL_2819998, EPI_ISL_2819999, EPI_ISL_2820002, EPI_ISL_2820003, EPI_ISL_2820004, EPI_ISL_2820006, EPI_ISL_2820009, EPI_ISL_2820011, EPI_ISL_2820013, EPI_ISL_2820017, EPI_ISL_2820018, EPI_ISL_2820019, EPI_ISL_2820020, EPI_ISL_2820022, EPI_ISL_2820023, EPI_ISL_2820024, EPI_ISL_2820026, EPI_ISL_2820029, EPI_ISL_2820030, EPI_ISL_2820032, EPI_ISL_2820033, EPI_ISL_2820034, EPI_ISL_2820039, EPI_ISL_2820041, EPI_ISL_2820042, EPI_ISL_2820043, EPI_ISL_2820045, EPI_ISL_2820046, EPI_ISL_2820049, EPI_ISL_2820053, EPI_ISL_2820054, EPI_ISL_2820059, EPI_ISL_2820061, EPI_ISL_2820062, EPI_ISL_2820063, EPI_ISL_2820064, EPI_ISL_2820066, EPI_ISL_2820067, EPI_ISL_2820068, EPI_ISL_2820070, EPI_ISL_2820071, EPI_ISL_2820073, EPI_ISL_2820075, EPI_ISL_2820077, EPI_ISL_2820078, EPI_ISL_2820079, EPI_ISL_2820080, EPI_ISL_2820081, EPI_ISL_2820082, EPI_ISL_2820083, EPI_ISL_2820085, EPI_ISL_2820086, EPI_ISL_2820087, EPI_ISL_2820090, EPI_ISL_2820091, EPI_ISL_2820092, EPI_ISL_2820093, EPI_ISL_2820094, EPI_ISL_2820096, EPI_ISL_2820098, EPI_ISL_2820099, EPI_ISL_2820106, EPI_ISL_2820109, EPI_ISL_2820110, EPI_ISL_2820111, EPI_ISL_2820114, EPI_ISL_2820115, EPI_ISL_2820116, EPI_ISL_2820117, EPI_ISL_2820118, EPI_ISL_2820120, EPI_ISL_2820126, EPI_ISL_2820127, EPI_ISL_2820128, EPI_ISL_2820129, EPI_ISL_2820135, EPI_ISL_2820136, EPI_ISL_2820137, EPI_ISL_2820139, EPI_ISL_2820140, EPI_ISL_2820143, EPI_ISL_2820147, EPI_ISL_2820148, EPI_ISL_2820149, EPI_ISL_2820150, EPI_ISL_2820151, EPI_ISL_2820152, EPI_ISL_2820154, EPI_ISL_2820156, EPI_ISL_2820157, EPI_ISL_2820159, EPI_ISL_2820160, EPI_ISL_2820161, EPI_ISL_2820162, EPI_ISL_2820163, EPI_ISL_2820164, EPI_ISL_2820165, EPI_ISL_2820166, EPI_ISL_2820167, EPI_ISL_2820168, EPI_ISL_2820169, EPI_ISL_2820170, EPI_ISL_2820173, EPI_ISL_2820174, EPI_ISL_2820176, EPI_ISL_2820177, EPI_ISL_2820179, EPI_ISL_2820180, EPI_ISL_2820182, EPI_ISL_2820184, EPI_ISL_2820185, EPI_ISL_2820188, EPI_ISL_2820189, EPI_ISL_2820190, EPI_ISL_2820191, EPI_ISL_2820194, EPI_ISL_2820195, EPI_ISL_2820196, EPI_ISL_2820199, EPI_ISL_2820200, EPI_ISL_2820201, EPI_ISL_2820204, EPI_ISL_2820205, EPI_ISL_2820206, EPI_ISL_2820207, EPI_ISL_2820208, EPI_ISL_2820209, EPI_ISL_2820210, EPI_ISL_2820211, EPI_ISL_2820214, EPI_ISL_2820217, EPI_ISL_2820219, EPI_ISL_2820221 | | | |
| see above | Virology Department, Sheffield Teaching Hospitals NHS Foundation Trust/Department of Infection, Immunity and Cardiovascular Disease, The Medical School, University of Sheffield | COVID-19 Genomics UK (COG-UK) Consortium | Adri Angyal; Alex Keeley; Benjamin Lindsey; Cariad Evans; Danielle Groves; Dave Partridge; Katie Johnson; Laura Carrilero; Luke Green; Matthew Parker; Matthew Wyles; Mehmet Yavuz; Mohammad Raza; Nikki Smith; Paul Parsons; Rachel Tucker; Rebecca Brown; Thushan de Silva |
| EPI_ISL_2815903, EPI_ISL_2815924, EPI_ISL_2815925, EPI_ISL_2815926, EPI_ISL_2815955, EPI_ISL_2815956, EPI_ISL_2815966, EPI_ISL_2815967, EPI_ISL_2900693, EPI_ISL_2900694, EPI_ISL_2900695, EPI_ISL_2900697, EPI_ISL_2900698, EPI_ISL_2900699, EPI_ISL_2900700, EPI_ISL_2900701, EPI_ISL_2900702, EPI_ISL_2900703, EPI_ISL_2900704, EPI_ISL_2900705, EPI_ISL_2900706, EPI_ISL_2900707, EPI_ISL_2900709, EPI_ISL_2900711, EPI_ISL_2900712, EPI_ISL_2900714, EPI_ISL_2900715, EPI_ISL_2900716, EPI_ISL_2900719, EPI_ISL_2900721, EPI_ISL_2900723, EPI_ISL_2900745, EPI_ISL_2900746 | | | |
| see above | West of Scotland Specialist Virology Centre, NHSGGC / MRC-University of Glasgow Centre for Virus Research | COVID-19 Genomics UK (COG-UK) Consortium | Alasdair MacLean; Alice Broos; Ana da Silva Filipe; Antonia Ho; Daniel Mair; David L Robertson; Emma Thomson; Guy Mollett; Ioulia Tsatsani; James Shepherd; Jenna Nichols; Joseph Hughes; Kathy Li; Kathy Smollett; Kyriaki Nomikou; Lily Tong; Matthew Holden; Natasha Johnson; Rachel Blacow; Richard Orton; Rory Gunson; Sarah McDonald; Sharif Shaaban; Sreenu Vattipally; Stephen Carmichael |

We gratefully acknowledge the following Authors from the Originating laboratories responsible for obtaining the specimens, as well as the Submitting laboratories where the genome data were generated and shared via GISAID, on which this research is based.

All Submitters of data may be contacted directly via www.gisaid.org

Authors are sorted alphabetically.

| Accession ID | Originating Laboratory | Submitting Laboratory | Authors |
|---|---|---|---|
| EPI_ISL_2846370, EPI_ISL_2918432 | Berkshire and Surrey Pathology Services Lighthouse Laboratory | Wellcome Sanger Institute for the COVID-19 Genomics UK (COG-UK) Consortium | Berkshire and Surrey Pathology Services Lighthouse Laboratory and Alex Alderton; Cordelia Langford; David K. Jackson; Dominic Kwiatkowski; Ewan Harrison; Ian Johnston; Jeffrey Barrett; John Sillitoe on behalf of the Wellcome Sanger Institute COVID-19 Surveillance Team; Roberto Amato; Sonia Goncalves |
| EPI_ISL_2815549, EPI_ISL_2815550, EPI_ISL_2815551, EPI_ISL_2815552, EPI_ISL_2815553, EPI_ISL_2815554, EPI_ISL_2815555, EPI_ISL_2815556, EPI_ISL_2815557, EPI_ISL_2815558, EPI_ISL_2815559, EPI_ISL_2815560, EPI_ISL_2815561, EPI_ISL_2815562, EPI_ISL_2815563, EPI_ISL_2815564, EPI_ISL_2815565, EPI_ISL_2815566, EPI_ISL_2815567, EPI_ISL_2815568, EPI_ISL_2815569, EPI_ISL_2815570, EPI_ISL_2815571, EPI_ISL_2815572, EPI_ISL_2815573, EPI_ISL_2815574, EPI_ISL_2815575, EPI_ISL_2815576, EPI_ISL_2815577, EPI_ISL_2815578, EPI_ISL_2815579, EPI_ISL_2815580, EPI_ISL_2815581, EPI_ISL_2815582, EPI_ISL_2815583, EPI_ISL_2815584, EPI_ISL_2815585, EPI_ISL_2815586, EPI_ISL_2815587, EPI_ISL_2815588, EPI_ISL_2815589, EPI_ISL_2815590, EPI_ISL_2815591, EPI_ISL_2815592, EPI_ISL_2815593, EPI_ISL_2815594, EPI_ISL_2815595, EPI_ISL_2815596, EPI_ISL_2815597, EPI_ISL_2815598, EPI_ISL_2815600, EPI_ISL_2815601, EPI_ISL_2815602, EPI_ISL_2815604, EPI_ISL_2815605, EPI_ISL_2815606, EPI_ISL_2815607, EPI_ISL_2815608, EPI_ISL_2815609, EPI_ISL_2815610, EPI_ISL_2815611, EPI_ISL_2815612, EPI_ISL_2815613, EPI_ISL_2815614, EPI_ISL_2815615, EPI_ISL_2815616, EPI_ISL_2815617, EPI_ISL_2815618, EPI_ISL_2815619, EPI_ISL_2815620, EPI_ISL_2815621, EPI_ISL_2815622, EPI_ISL_2815623, EPI_ISL_2815624, EPI_ISL_2815626, EPI_ISL_2815627, EPI_ISL_2815628, EPI_ISL_2815629, EPI_ISL_2815630, EPI_ISL_2815631, EPI_ISL_2815632, EPI_ISL_2815633, EPI_ISL_2815634, EPI_ISL_2815635, EPI_ISL_2815636, EPI_ISL_2815637, EPI_ISL_2815640, EPI_ISL_2815641, EPI_ISL_2815642, EPI_ISL_2815643, EPI_ISL_2815644, EPI_ISL_2815647, EPI_ISL_2815648, EPI_ISL_2815650, EPI_ISL_2815651, EPI_ISL_2815652, EPI_ISL_2815653, EPI_ISL_2815654, EPI_ISL_2815655, EPI_ISL_2815657, EPI_ISL_2815658, EPI_ISL_2815659, EPI_ISL_2815661, EPI_ISL_2815662, EPI_ISL_2815663, EPI_ISL_2815664, EPI_ISL_2815665, EPI_ISL_2815666, EPI_ISL_2815667, EPI_ISL_2815668, EPI_ISL_2815669, EPI_ISL_2815670, EPI_ISL_2815671, EPI_ISL_2815672, EPI_ISL_2815673, EPI_ISL_2815674, EPI_ISL_2815675, EPI_ISL_2815676, EPI_ISL_2815677, EPI_ISL_2815678, EPI_ISL_2815680, EPI_ISL_2815681, EPI_ISL_2815682, EPI_ISL_2815683, EPI_ISL_2815684, EPI_ISL_2815685, EPI_ISL_2815710, EPI_ISL_2815835, EPI_ISL_2815848, EPI_ISL_2815855, EPI_ISL_2815859, EPI_ISL_2815865 | | | |
| see above | Department of Pathology, University of Cambridge | COVID-19 Genomics UK (COG-UK) Consortium | Aminu S. Jahun; Ian Goodfellow; Iliana Georgana; Martin D. Curran; Myra Hosmillo; Rhys Izuagbe; Surendra Parmar; William L. Hamilton; Yasmin Chaudhry |
| EPI_ISL_2779641, EPI_ISL_2779642, EPI_ISL_2779643, EPI_ISL_2779645, EPI_ISL_2779646, EPI_ISL_2779647, EPI_ISL_2779648, EPI_ISL_2779649, EPI_ISL_2779653, EPI_ISL_2779655, EPI_ISL_2779660, EPI_ISL_2779662, EPI_ISL_2779663, EPI_ISL_2779664, EPI_ISL_2779666, EPI_ISL_2779670, EPI_ISL_2779671, EPI_ISL_2779673, EPI_ISL_2779674, EPI_ISL_2779675, EPI_ISL_2779682, EPI_ISL_2779683, EPI_ISL_2779684, EPI_ISL_2779692, EPI_ISL_2779699, EPI_ISL_2779704, EPI_ISL_2779709, EPI_ISL_2779710, EPI_ISL_2779719, EPI_ISL_2779720, EPI_ISL_2779721, EPI_ISL_2779723, EPI_ISL_2779725, EPI_ISL_2779726, EPI_ISL_2779732, EPI_ISL_2779733, EPI_ISL_2779734, EPI_ISL_2779737, EPI_ISL_2779738, EPI_ISL_2779741, EPI_ISL_2779742, EPI_ISL_2779745, EPI_ISL_2779746, EPI_ISL_2779748, EPI_ISL_2779749, EPI_ISL_2779750, EPI_ISL_2779752, EPI_ISL_2779756, EPI_ISL_2779758, EPI_ISL_2779761, EPI_ISL_2779762, EPI_ISL_2779763, EPI_ISL_2779764, EPI_ISL_2779766, EPI_ISL_2779770, EPI_ISL_2779772, EPI_ISL_2779774, EPI_ISL_2779777, EPI_ISL_2779780, EPI_ISL_2779787, EPI_ISL_2779793, EPI_ISL_2779794, EPI_ISL_2779796, EPI_ISL_2779798, EPI_ISL_2779802, EPI_ISL_2779803, EPI_ISL_2779804, EPI_ISL_2779805, EPI_ISL_2779806, EPI_ISL_2779809, EPI_ISL_2779810, EPI_ISL_2779812, EPI_ISL_2779813, EPI_ISL_2779814, EPI_ISL_2779816, EPI_ISL_2779820, EPI_ISL_2779828, EPI_ISL_2779829, EPI_ISL_2779832, EPI_ISL_2779833, EPI_ISL_2779837, EPI_ISL_2779840, EPI_ISL_2779842, EPI_ISL_2779845, EPI_ISL_2779848, EPI_ISL_2779850, EPI_ISL_2779851, EPI_ISL_2779853, EPI_ISL_2779855, EPI_ISL_2779859, EPI_ISL_2779860, EPI_ISL_2779862, EPI_ISL_2779864, EPI_ISL_2803933, EPI_ISL_2803962, EPI_ISL_2803974, EPI_ISL_2803978, EPI_ISL_2803993, EPI_ISL_2803997, EPI_ISL_2804001, EPI_ISL_2804009, EPI_ISL_2804039, EPI_ISL_2804040, EPI_ISL_2804062, EPI_ISL_2804068, EPI_ISL_2804081, EPI_ISL_2804082, EPI_ISL_2804083, EPI_ISL_2804090, EPI_ISL_2804093, EPI_ISL_2804101, EPI_ISL_2804115, EPI_ISL_2804118, EPI_ISL_2804125, EPI_ISL_2804135, EPI_ISL_2804151, EPI_ISL_2804152, EPI_ISL_2804155, EPI_ISL_2804160, EPI_ISL_2804164, EPI_ISL_2804167, EPI_ISL_2804181, EPI_ISL_2804183, EPI_ISL_2804204, EPI_ISL_2804208, EPI_ISL_2804212, EPI_ISL_2804214, EPI_ISL_2804218, EPI_ISL_2804226, EPI_ISL_2804230, EPI_ISL_2804234, EPI_ISL_2804244, EPI_ISL_2804249, EPI_ISL_2804309, EPI_ISL_2804351, EPI_ISL_2804353, EPI_ISL_2804378, EPI_ISL_2804379, EPI_ISL_2804417, EPI_ISL_2804424, EPI_ISL_2804450, EPI_ISL_2804456, EPI_ISL_2804487, EPI_ISL_2804488, EPI_ISL_2804499, EPI_ISL_2804531, EPI_ISL_2804541, EPI_ISL_2804550, EPI_ISL_2804574, EPI_ISL_2804604, EPI_ISL_2806209, EPI_ISL_2806211, EPI_ISL_2806213, EPI_ISL_2806218, EPI_ISL_2806222, EPI_ISL_2806223, EPI_ISL_2806229, EPI_ISL_2806231, EPI_ISL_2806235, EPI_ISL_2806236, EPI_ISL_2806238, EPI_ISL_2806246, EPI_ISL_2806248, EPI_ISL_2806258, EPI_ISL_2806260, EPI_ISL_2806261, EPI_ISL_2806263, EPI_ISL_2806267, EPI_ISL_2806269, EPI_ISL_2806270, EPI_ISL_2806271, EPI_ISL_2806274, EPI_ISL_2806276, EPI_ISL_2806278, EPI_ISL_2806279, EPI_ISL_2806286, EPI_ISL_2806288, EPI_ISL_2806293, EPI_ISL_2806294, EPI_ISL_2806296, EPI_ISL_2806297, EPI_ISL_2806301, EPI_ISL_2806302, EPI_ISL_2806305, EPI_ISL_2806306, EPI_ISL_2806308, EPI_ISL_2806317, EPI_ISL_2806321, EPI_ISL_2806322, EPI_ISL_2806325, EPI_ISL_2806326, EPI_ISL_2806328, EPI_ISL_2806331, EPI_ISL_2806332, EPI_ISL_2806336, EPI_ISL_2806339, EPI_ISL_2806344, EPI_ISL_2806347, EPI_ISL_2806348, EPI_ISL_2806356, EPI_ISL_2806359, EPI_ISL_2806364, EPI_ISL_2806365, EPI_ISL_2806368, EPI_ISL_2806370, EPI_ISL_2806373, EPI_ISL_2806375, EPI_ISL_2806376, EPI_ISL_2806379, EPI_ISL_2806382, EPI_ISL_2806383, EPI_ISL_2806384, EPI_ISL_2806385, EPI_ISL_2806388, EPI_ISL_2806390, EPI_ISL_2806409, EPI_ISL_2806410, EPI_ISL_2806411, EPI_ISL_2806418, EPI_ISL_2806419, EPI_ISL_2806420, EPI_ISL_2806424, EPI_ISL_2806426, EPI_ISL_2806427, EPI_ISL_2806428, EPI_ISL_2806431, EPI_ISL_2806434, EPI_ISL_2806435, EPI_ISL_2806439, EPI_ISL_2806440, EPI_ISL_2806447, EPI_ISL_2806453, EPI_ISL_2806458, EPI_ISL_2806460, EPI_ISL_2806469, EPI_ISL_2806471, EPI_ISL_2806477, EPI_ISL_2806484, EPI_ISL_2806485, EPI_ISL_2806489, EPI_ISL_2806496, EPI_ISL_2806498, EPI_ISL_2806501, EPI_ISL_2806503, EPI_ISL_2806511, EPI_ISL_2806512, EPI_ISL_2806520, EPI_ISL_2806525, EPI_ISL_2806527, EPI_ISL_2806536, EPI_ISL_2806545, EPI_ISL_2806552, EPI_ISL_2806556, EPI_ISL_2806568, EPI_ISL_2806572, EPI_ISL_2806579, EPI_ISL_2806584, EPI_ISL_2806590, EPI_ISL_2806599, EPI_ISL_2806601, EPI_ISL_2806605, EPI_ISL_2806609, EPI_ISL_2806611, EPI_ISL_2806614, EPI_ISL_2806619, EPI_ISL_2806621, EPI_ISL_2806622, EPI_ISL_2806626, EPI_ISL_2806629, EPI_ISL_2806630, EPI_ISL_2806631, EPI_ISL_2806643, EPI_ISL_2806648, EPI_ISL_2806649, EPI_ISL_2806653, EPI_ISL_2806660, EPI_ISL_2806666, EPI_ISL_2806667, EPI_ISL_2806668, EPI_ISL_2806669, EPI_ISL_2806672, EPI_ISL_2806673, EPI_ISL_2806678, EPI_ISL_2806680, EPI_ISL_2806683, EPI_ISL_2806688, EPI_ISL_2806689, EPI_ISL_2806711, EPI_ISL_2806712, EPI_ISL_2806715, EPI_ISL_2806730, EPI_ISL_2806733, EPI_ISL_2806735, EPI_ISL_2806740, EPI_ISL_2806741, EPI_ISL_2806748, EPI_ISL_2806750, EPI_ISL_2806758, EPI_ISL_2806767, EPI_ISL_2806770, EPI_ISL_2806772, EPI_ISL_2806777, EPI_ISL_2806794, EPI_ISL_2806795, EPI_ISL_2829105, EPI_ISL_2829267, EPI_ISL_2851415, EPI_ISL_2851417, EPI_ISL_2851418, EPI_ISL_2851441, EPI_ISL_2851443, EPI_ISL_2851466, EPI_ISL_2851474, EPI_ISL_2851475, EPI_ISL_2851478, EPI_ISL_2851500, EPI_ISL_2851516, EPI_ISL_2851537, EPI_ISL_2851567, EPI_ISL_2851580, EPI_ISL_2851621, EPI_ISL_2851627, EPI_ISL_2851632, EPI_ISL_2851633, EPI_ISL_2851644, EPI_ISL_2851651, EPI_ISL_2851654, EPI_ISL_2851680, EPI_ISL_2851717, EPI_ISL_2851725, EPI_ISL_2851755, EPI_ISL_2851760, EPI_ISL_2851765 | | | |
| see above | Health Services Laboratories | Wellcome Sanger Institute for the COVID-19 Genomics UK (COG-UK) Consortium | Cordelia Langford; David K. Jackson; Dominic Kwiatkowski; Ewan Harrison; Health Services Laboratories and Alex Alderton; Ian Johnston; Jeffrey Barrett; John Sillitoe on behalf of the Wellcome Sanger Institute COVID-19 Surveillance Team; Roberto Amato; Sonia Goncalves |
| EPI_ISL_2829026, EPI_ISL_2829028, EPI_ISL_2829038, EPI_ISL_2829040, EPI_ISL_2829042, EPI_ISL_2829043, EPI_ISL_2829044, EPI_ISL_2829051, EPI_ISL_2829058, EPI_ISL_2829060, EPI_ISL_2829066, EPI_ISL_2829072, EPI_ISL_2829076, EPI_ISL_2829080, EPI_ISL_2829081, EPI_ISL_2829083, EPI_ISL_2829084, EPI_ISL_2829088, EPI_ISL_2829092, EPI_ISL_2829093, EPI_ISL_2829095, EPI_ISL_2829098, EPI_ISL_2829106, EPI_ISL_2829109, EPI_ISL_2829115, EPI_ISL_2829119, EPI_ISL_2829120, EPI_ISL_2829123, EPI_ISL_2829126, EPI_ISL_2829127, EPI_ISL_2829128, EPI_ISL_2829129, EPI_ISL_2829133, EPI_ISL_2829135, EPI_ISL_2829137, EPI_ISL_2829146, EPI_ISL_2829153, EPI_ISL_2829161, EPI_ISL_2829164, EPI_ISL_2829170, EPI_ISL_2829171, EPI_ISL_2829172, EPI_ISL_2829173, EPI_ISL_2829184, EPI_ISL_2829185, EPI_ISL_2829187, EPI_ISL_2829188, EPI_ISL_2829199, EPI_ISL_2829200, EPI_ISL_2829210, EPI_ISL_2829212, EPI_ISL_2829215, EPI_ISL_2829221, EPI_ISL_2829223, EPI_ISL_2829226, EPI_ISL_2829227, EPI_ISL_2829229, EPI_ISL_2829234, EPI_ISL_2829236, EPI_ISL_2829240, EPI_ISL_2829252, EPI_ISL_2829253, EPI_ISL_2829254, EPI_ISL_2829255, EPI_ISL_2829261, EPI_ISL_2829264, EPI_ISL_2829268, EPI_ISL_2829274, EPI_ISL_2829275, EPI_ISL_2829276, EPI_ISL_2829288, EPI_ISL_2829289, EPI_ISL_2829294, EPI_ISL_2829297, EPI_ISL_2829299, EPI_ISL_2829301, EPI_ISL_2829303, EPI_ISL_2829304, EPI_ISL_2829307, EPI_ISL_2829313, EPI_ISL_2829314, EPI_ISL_2829318, EPI_ISL_2829320, EPI_ISL_2829324, EPI_ISL_2829328, EPI_ISL_2829331, EPI_ISL_2829335, EPI_ISL_2829337, EPI_ISL_2829339, EPI_ISL_2829341, EPI_ISL_2829343, EPI_ISL_2829344, EPI_ISL_2829348, EPI_ISL_2829349, EPI_ISL_2829350, EPI_ISL_2829352, EPI_ISL_2829355, EPI_ISL_2829362, EPI_ISL_2829364, EPI_ISL_2829366, EPI_ISL_2829367, EPI_ISL_2829368, EPI_ISL_2829370, EPI_ISL_2829372, EPI_ISL_2829375, EPI_ISL_2829378, EPI_ISL_2829384, EPI_ISL_2829387, EPI_ISL_2829389, EPI_ISL_2829393, EPI_ISL_2829395, EPI_ISL_2829396, EPI_ISL_2829400, EPI_ISL_2829403, EPI_ISL_2829410, EPI_ISL_2829411, EPI_ISL_2829413, EPI_ISL_2829414, EPI_ISL_2829416, EPI_ISL_2829418, EPI_ISL_2829420, EPI_ISL_2829422, EPI_ISL_2829423, EPI_ISL_2829424, EPI_ISL_2829427, EPI_ISL_2829429, EPI_ISL_2829430, EPI_ISL_2829434, EPI_ISL_2829435, EPI_ISL_2829436, EPI_ISL_2829437, EPI_ISL_2829439, EPI_ISL_2829440, EPI_ISL_2829441, EPI_ISL_2829442, EPI_ISL_2829443, EPI_ISL_2829448, EPI_ISL_2829449, EPI_ISL_2829452, EPI_ISL_2829455, EPI_ISL_2829460, EPI_ISL_2829462, EPI_ISL_2829464, EPI_ISL_2829468, EPI_ISL_2829469, EPI_ISL_2829470, EPI_ISL_2829471, EPI_ISL_2829472, EPI_ISL_2829477, EPI_ISL_2829479, EPI_ISL_2829480, EPI_ISL_2829483, EPI_ISL_2829484, EPI_ISL_2829485, EPI_ISL_2829488, EPI_ISL_2829492, EPI_ISL_2829495, EPI_ISL_2829496, EPI_ISL_2829498, EPI_ISL_2829500, EPI_ISL_2829501, EPI_ISL_2829505, EPI_ISL_2829508, EPI_ISL_2829509, EPI_ISL_2829510, EPI_ISL_2829511, EPI_ISL_2829512, EPI_ISL_2829513, EPI_ISL_2829514, EPI_ISL_2829516, EPI_ISL_2829517, EPI_ISL_2829520, EPI_ISL_2829523, EPI_ISL_2829525, EPI_ISL_2829528, EPI_ISL_2829529, EPI_ISL_2829530, EPI_ISL_2829535, EPI_ISL_2829536, EPI_ISL_2829539, EPI_ISL_2829541, EPI_ISL_2829543, EPI_ISL_2829544, EPI_ISL_2829547, EPI_ISL_2829552, EPI_ISL_2829553, EPI_ISL_2829554, EPI_ISL_2829555, EPI_ISL_2829557, EPI_ISL_2829560, EPI_ISL_2829561, EPI_ISL_2829564, EPI_ISL_2829568, EPI_ISL_2829570, EPI_ISL_2829573, EPI_ISL_2829574, EPI_ISL_2829576, EPI_ISL_2829577, EPI_ISL_2829579, EPI_ISL_2829580, EPI_ISL_2829582, EPI_ISL_2829587, EPI_ISL_2829590, EPI_ISL_2829598, EPI_ISL_2829599, EPI_ISL_2829600, EPI_ISL_2829605, EPI_ISL_2829619, EPI_ISL_2829623, EPI_ISL_2829628, EPI_ISL_2829632, EPI_ISL_2829633, EPI_ISL_2829641, EPI_ISL_2829642, EPI_ISL_2829646, EPI_ISL_2829655, EPI_ISL_2829657, EPI_ISL_2829662, EPI_ISL_2829663, EPI_ISL_2829664, EPI_ISL_2829665, EPI_ISL_2829676, EPI_ISL_2829680, EPI_ISL_2829691, EPI_ISL_2829701, EPI_ISL_2829703, EPI_ISL_2829707, EPI_ISL_2829717, EPI_ISL_2829718, EPI_ISL_2829720, EPI_ISL_2829726, EPI_ISL_2829731, EPI_ISL_2829733, EPI_ISL_2829738, EPI_ISL_2829740, EPI_ISL_2829742, EPI_ISL_2829744, EPI_ISL_2829750, EPI_ISL_2829751, EPI_ISL_2829762, EPI_ISL_2829769, EPI_ISL_2829773, EPI_ISL_2829776, EPI_ISL_2829782, EPI_ISL_2829784, EPI_ISL_2829791, EPI_ISL_2829794, EPI_ISL_2829798, EPI_ISL_2829807, EPI_ISL_2829810, EPI_ISL_2829812, EPI_ISL_2829819, EPI_ISL_2829831, EPI_ISL_2829833, EPI_ISL_2829840, EPI_ISL_2829841, EPI_ISL_2829846, EPI_ISL_2829852, EPI_ISL_2829855, EPI_ISL_2829858, EPI_ISL_2829862, EPI_ISL_2829865, EPI_ISL_2829868, EPI_ISL_2829869, EPI_ISL_2829875, EPI_ISL_2829883, EPI_ISL_2829892, EPI_ISL_2829900, EPI_ISL_2829908, EPI_ISL_2829919, EPI_ISL_2829920, EPI_ISL_2829921, EPI_ISL_2829925, EPI_ISL_2829927, EPI_ISL_2829928, EPI_ISL_2829938, EPI_ISL_2829946, EPI_ISL_2829948, EPI_ISL_2829957, EPI_ISL_2829959, EPI_ISL_2829963, EPI_ISL_2829964, EPI_ISL_2829965, EPI_ISL_2829968, EPI_ISL_2829969, EPI_ISL_2829970, EPI_ISL_2829985, EPI_ISL_2829986, EPI_ISL_2829991, EPI_ISL_2829995, EPI_ISL_2829996, EPI_ISL_2829998, EPI_ISL_2829999, EPI_ISL_2830008, EPI_ISL_2830010, EPI_ISL_2830014, EPI_ISL_2830020, EPI_ISL_2830021, EPI_ISL_2830029, EPI_ISL_2830033, EPI_ISL_2830035, EPI_ISL_2830045, EPI_ISL_2830047, EPI_ISL_2830048, EPI_ISL_2830049, EPI_ISL_2830056, EPI_ISL_2830060, EPI_ISL_2830067, EPI_ISL_2830068, EPI_ISL_2830077, EPI_ISL_2830078, EPI_ISL_2830079, EPI_ISL_2830081, EPI_ISL_2830082, EPI_ISL_2830084, EPI_ISL_2830087, EPI_ISL_2830088, EPI_ISL_2830089, EPI_ISL_2830090, EPI_ISL_2830099, EPI_ISL_2830100, EPI_ISL_2830105, EPI_ISL_2830107, EPI_ISL_2830108, EPI_ISL_2830109, EPI_ISL_2830114, EPI_ISL_2830115, EPI_ISL_2830116, EPI_ISL_2830118, EPI_ISL_2830119, EPI_ISL_2830128, EPI_ISL_2830134, EPI_ISL_2830135, EPI_ISL_2830143, EPI_ISL_2830144, EPI_ISL_2830148, EPI_ISL_2830149, EPI_ISL_2830154, EPI_ISL_2830162, EPI_ISL_2830163, EPI_ISL_2830164, EPI_ISL_2830165, EPI_ISL_2830167, EPI_ISL_2830169, EPI_ISL_2830173, EPI_ISL_2830174, EPI_ISL_2830176, EPI_ISL_2830178, EPI_ISL_2830184, EPI_ISL_2830185, EPI_ISL_2830186, EPI_ISL_2830188, EPI_ISL_2830191, EPI_ISL_2830214, EPI_ISL_2830219, EPI_ISL_2830225, EPI_ISL_2830243, EPI_ISL_2830256, EPI_ISL_2830266, EPI_ISL_2830267, EPI_ISL_2830274, EPI_ISL_2830284, EPI_ISL_2830286, EPI_ISL_2830300, EPI_ISL_2830320, EPI_ISL_2830328, EPI_ISL_2830329, EPI_ISL_2830330, EPI_ISL_2846027, EPI_ISL_2846087, EPI_ISL_2846094, EPI_ISL_2846125, EPI_ISL_2846188, EPI_ISL_2846192, EPI_ISL_2846195, EPI_ISL_2846263, EPI_ISL_2846340, EPI_ISL_2846356, EPI_ISL_2846589, EPI_ISL_2846596, EPI_ISL_2846717, EPI_ISL_2846847, EPI_ISL_2846878, EPI_ISL_2846930, EPI_ISL_2846933, EPI_ISL_2846939, EPI_ISL_2846940, EPI_ISL_2846941, EPI_ISL_2846948, EPI_ISL_2846957, EPI_ISL_2846962, EPI_ISL_2846963, EPI_ISL_2846968, EPI_ISL_2846976, EPI_ISL_2846977, EPI_ISL_2846979, EPI_ISL_2846982, EPI_ISL_2846985, EPI_ISL_2846986, EPI_ISL_2846996, EPI_ISL_2847000, EPI_ISL_2847002, EPI_ISL_2847006, EPI_ISL_2847013, EPI_ISL_2847015, EPI_ISL_2847017, EPI_ISL_2847021, EPI_ISL_2847026, EPI_ISL_2847034, EPI_ISL_2847042, EPI_ISL_2847045, EPI_ISL_2847051, EPI_ISL_2847057, EPI_ISL_2847058, EPI_ISL_2847061, EPI_ISL_2847063, EPI_ISL_2847064, EPI_ISL_2847065, EPI_ISL_2847069, EPI_ISL_2847073, EPI_ISL_2847076, EPI_ISL_2847092, EPI_ISL_2847093, EPI_ISL_2847094, EPI_ISL_2847096, EPI_ISL_2847099, EPI_ISL_2847101, EPI_ISL_2847105, EPI_ISL_2847106, EPI_ISL_2847108, EPI_ISL_2847109, EPI_ISL_2847120, EPI_ISL_2847127, EPI_ISL_2847130, EPI_ISL_2847131, EPI_ISL_2847134, EPI_ISL_2847140, EPI_ISL_2847142, EPI_ISL_2847145, EPI_ISL_2847148, EPI_ISL_2847158, EPI_ISL_2847161, EPI_ISL_2847162, EPI_ISL_2847163, EPI_ISL_2847170, EPI_ISL_2847183, EPI_ISL_2847190, EPI_ISL_2847192, EPI_ISL_2847195, EPI_ISL_2847196, EPI_ISL_2847197, EPI_ISL_2847199, EPI_ISL_2847208, EPI_ISL_2847212, EPI_ISL_2847217, EPI_ISL_2847218, EPI_ISL_2847221, EPI_ISL_2847222, EPI_ISL_2847224, EPI_ISL_2847227, EPI_ISL_2847233, EPI_ISL_2847235, EPI_ISL_2847240, EPI_ISL_2847247, EPI_ISL_2847249, EPI_ISL_2847251, EPI_ISL_2847253, EPI_ISL_2847256, EPI_ISL_2847257, EPI_ISL_2847258, EPI_ISL_2847260, EPI_ISL_2847272, EPI_ISL_2847280, EPI_ISL_2847308, EPI_ISL_2847324, EPI_ISL_2847329, EPI_ISL_2847330, EPI_ISL_2847335, EPI_ISL_2847337, EPI_ISL_2847340, EPI_ISL_2847341, EPI_ISL_2847343, EPI_ISL_2847351, EPI_ISL_2847359, EPI_ISL_2847362, EPI_ISL_2847370, EPI_ISL_2847374, EPI_ISL_2847377, EPI_ISL_2847379, EPI_ISL_2847384, EPI_ISL_2847389, EPI_ISL_2847394, EPI_ISL_2847398, EPI_ISL_2847404, EPI_ISL_2847408, EPI_ISL_2847411, EPI_ISL_2847414, EPI_ISL_2847416, EPI_ISL_2847417, EPI_ISL_2847423, EPI_ISL_2847426, EPI_ISL_2847428, EPI_ISL_2847431, EPI_ISL_2847436, EPI_ISL_2847449, EPI_ISL_2847459, EPI_ISL_2847462, EPI_ISL_2847464, EPI_ISL_2847465, EPI_ISL_2847479, EPI_ISL_2847486, EPI_ISL_2847487, EPI_ISL_2847490, EPI_ISL_2847508, EPI_ISL_2847522, EPI_ISL_2847524, EPI_ISL_2847528, EPI_ISL_2847530, EPI_ISL_2847537, EPI_ISL_2847542, EPI_ISL_2847549, EPI_ISL_2847553, EPI_ISL_2847555, EPI_ISL_2847558, EPI_ISL_2847561, EPI_ISL_2847573, EPI_ISL_2847575, EPI_ISL_2847580, EPI_ISL_2847581, EPI_ISL_2847592, EPI_ISL_2847600, EPI_ISL_2847612, EPI_ISL_2847613, EPI_ISL_2847614, EPI_ISL_2847615, EPI_ISL_2847957, EPI_ISL_2847958, EPI_ISL_2847961, EPI_ISL_2847966, EPI_ISL_2847974, EPI_ISL_2847975, EPI_ISL_2847981, EPI_ISL_2847983, EPI_ISL_2847986, EPI_ISL_2847996, EPI_ISL_2847998, EPI_ISL_2848002, EPI_ISL_2848005, EPI_ISL_2848006, EPI_ISL_2848010, EPI_ISL_2848014, EPI_ISL_2848022, EPI_ISL_2848029, EPI_ISL_2848036, EPI_ISL_2848038, EPI_ISL_2848039, EPI_ISL_2848042, EPI_ISL_2848051, EPI_ISL_2848055, EPI_ISL_2848062, EPI_ISL_2848063, EPI_ISL_2848070, EPI_ISL_2848073, EPI_ISL_2848077, EPI_ISL_2848081, EPI_ISL_2848084, EPI_ISL_2848089, EPI_ISL_2848100, EPI_ISL_2848104, EPI_ISL_2848106, EPI_ISL_2848107, EPI_ISL_2848111, EPI_ISL_2848119, EPI_ISL_2848137, EPI_ISL_2848147, EPI_ISL_2848153, EPI_ISL_2848158, EPI_ISL_2848159, EPI_ISL_2848162, EPI_ISL_2848164, EPI_ISL_2848166, EPI_ISL_2848171, EPI_ISL_2848173, EPI_ISL_2848174, EPI_ISL_2848178, EPI_ISL_2848179, EPI_ISL_2848181, EPI_ISL_2848183, EPI_ISL_2848187, EPI_ISL_2848188, EPI_ISL_2848190, EPI_ISL_2848193, EPI_ISL_2848197, EPI_ISL_2848205, EPI_ISL_2848211, EPI_ISL_2848215, EPI_ISL_2848222, EPI_ISL_2848223, EPI_ISL_2848226, EPI_ISL_2848227, EPI_ISL_2848231, EPI_ISL_2848236, EPI_ISL_2848239, EPI_ISL_2848248, EPI_ISL_2848249, EPI_ISL_2848253, EPI_ISL_2848276, EPI_ISL_2848280, EPI_ISL_2848281, EPI_ISL_2848283, EPI_ISL_2848287, EPI_ISL_2848293, EPI_ISL_2848297, EPI_ISL_2848298, EPI_ISL_2848299, EPI_ISL_2849040, EPI_ISL_2849092, EPI_ISL_2849417, EPI_ISL_2849517, EPI_ISL_2849572, EPI_ISL_2849631, EPI_ISL_2849647, EPI_ISL_2849720, EPI_ISL_2849738, EPI_ISL_2849747, EPI_ISL_2849749, EPI_ISL_2849768, EPI_ISL_2849806, EPI_ISL_2849808, EPI_ISL_2849813, EPI_ISL_2849922, EPI_ISL_2849951, EPI_ISL_2849990, EPI_ISL_2850010, EPI_ISL_2850011, EPI_ISL_2850374, EPI_ISL_2850377, EPI_ISL_2850378, EPI_ISL_2850382, EPI_ISL_2850383, EPI_ISL_2850384, EPI_ISL_2850385, EPI_ISL_2850388, EPI_ISL_2850397, EPI_ISL_2850400, EPI_ISL_2850403, EPI_ISL_2850413, EPI_ISL_2850417, EPI_ISL_2850418, EPI_ISL_2850419, EPI_ISL_2850420, EPI_ISL_2850425, EPI_ISL_2850429, EPI_ISL_2850430, EPI_ISL_2850431, EPI_ISL_2850432, EPI_ISL_2850433, EPI_ISL_2850434, EPI_ISL_2850437, EPI_ISL_2850440, EPI_ISL_2850441, EPI_ISL_2850446, EPI_ISL_2850451, EPI_ISL_2850454, EPI_ISL_2850455, EPI_ISL_2850456, EPI_ISL_2850457, EPI_ISL_2850459, EPI_ISL_2850465, EPI_ISL_2850466, EPI_ISL_2850468, EPI_ISL_2850471, EPI_ISL_2850472, EPI_ISL_2850473, EPI_ISL_2850475, EPI_ISL_2850476, EPI_ISL_2850480, EPI_ISL_2850482, EPI_ISL_2850483, EPI_ISL_2850484, EPI_ISL_2850487, EPI_ISL_2850488, EPI_ISL_2850490, EPI_ISL_2850491, EPI_ISL_2850495, EPI_ISL_2850500, EPI_ISL_2850501, EPI_ISL_2850506, EPI_ISL_2850508, EPI_ISL_2850512, EPI_ISL_2850514, EPI_ISL_2850518, EPI_ISL_2850519, EPI_ISL_2850520, EPI_ISL_2850521, EPI_ISL_2850523, EPI_ISL_2850525, EPI_ISL_2850529, EPI_ISL_2850533, EPI_ISL_2850534, EPI_ISL_2850539, EPI_ISL_2850542, EPI_ISL_2850544, EPI_ISL_2850545, EPI_ISL_2850548, EPI_ISL_2850550, EPI_ISL_2850553, EPI_ISL_2850557, EPI_ISL_2850558, EPI_ISL_2850559, EPI_ISL_2850560, EPI_ISL_2850566, EPI_ISL_2850567, EPI_ISL_2850568, EPI_ISL_2850569, EPI_ISL_2850570, EPI_ISL_2850571, EPI_ISL_2850576, EPI_ISL_2850582, EPI_ISL_2850583, EPI_ISL_2850584, EPI_ISL_2850585, EPI_ISL_2850587, EPI_ISL_2850589, EPI_ISL_2850590, EPI_ISL_2850592, EPI_ISL_2850594, EPI_ISL_2850596, EPI_ISL_2850599, EPI_ISL_2850600, EPI_ISL_2850601, EPI_ISL_2850603, EPI_ISL_2850606, EPI_ISL_2850611, EPI_ISL_2850616, EPI_ISL_2850617, EPI_ISL_2850618, EPI_ISL_2850619, EPI_ISL_2850621, EPI_ISL_2850622, EPI_ISL_2850623, EPI_ISL_2850626, EPI_ISL_2850627, EPI_ISL_2850630, EPI_ISL_2850635, EPI_ISL_2850637, EPI_ISL_2850639, EPI_ISL_2850642, EPI_ISL_2850643, EPI_ISL_2850644, EPI_ISL_2850646, EPI_ISL_2850647, EPI_ISL_2850648, EPI_ISL_2850649, EPI_ISL_2850650, EPI_ISL_2850652, EPI_ISL_2850666, EPI_ISL_2850667, EPI_ISL_2850672, EPI_ISL_2850674, EPI_ISL_2850678, EPI_ISL_2850679, EPI_ISL_2850681, EPI_ISL_2850682, EPI_ISL_2850687, EPI_ISL_2850688, EPI_ISL_2850690, EPI_ISL_2850692, EPI_ISL_2850695, EPI_ISL_2850700, EPI_ISL_2850704, EPI_ISL_2850706, EPI_ISL_2850710, EPI_ISL_2850711, EPI_ISL_2850712, EPI_ISL_2850713, EPI_ISL_2850714, EPI_ISL_2850716, EPI_ISL_2850720, EPI_ISL_2850724, EPI_ISL_2850725, EPI_ISL_2850726, EPI_ISL_2850727, EPI_ISL_2850728, EPI_ISL_2850730, EPI_ISL_2850731, EPI_ISL_2850732, EPI_ISL_2850735, EPI_ISL_2850736, EPI_ISL_2850737, EPI_ISL_2850922, EPI_ISL_2851016, EPI_ISL_2851079, EPI_ISL_2851377, EPI_ISL_2851411, EPI_ISL_2851414, EPI_ISL_2851425, EPI_ISL_2851427, EPI_ISL_2851430, EPI_ISL_2851431, EPI_ISL_2851434, EPI_ISL_2851437, EPI_ISL_2851444, EPI_ISL_2851451, EPI_ISL_2851456, EPI_ISL_2851458, EPI_ISL_2851459, EPI_ISL_2851460, EPI_ISL_2851464, EPI_ISL_2851465, EPI_ISL_2851467, EPI_ISL_2851470, EPI_ISL_2851487, EPI_ISL_2851493, EPI_ISL_2851496, EPI_ISL_2851497, EPI_ISL_2851498, EPI_ISL_2851504, EPI_ISL_2851506, EPI_ISL_2851509, EPI_ISL_2851510, EPI_ISL_2851515, EPI_ISL_2851521, EPI_ISL_2851538, EPI_ISL_2851544, EPI_ISL_2851545, EPI_ISL_2851546, EPI_ISL_2851549, EPI_ISL_2851556, EPI_ISL_2851560, EPI_ISL_2851568, EPI_ISL_2851570, EPI_ISL_2851578, EPI_ISL_2851583, EPI_ISL_2851584, EPI_ISL_2851589, EPI_ISL_2851593, EPI_ISL_2851598, EPI_ISL_2851600, EPI_ISL_2851602, EPI_ISL_2851604, EPI_ISL_2851606, EPI_ISL_2851610, EPI_ISL_2851620, EPI_ISL_2851622, EPI_ISL_2851628, EPI_ISL_2851629, EPI_ISL_2851635, EPI_ISL_2851642, EPI_ISL_2851660, EPI_ISL_2851668, EPI_ISL_2851672, EPI_ISL_2851681, EPI_ISL_2851684, EPI_ISL_2851685, EPI_ISL_2851713, EPI_ISL_2851721, EPI_ISL_2851722, EPI_ISL_2851726, EPI_ISL_2851728, EPI_ISL_2851730, EPI_ISL_2851738, EPI_ISL_2851751, EPI_ISL_2851758, EPI_ISL_2851759, EPI_ISL_2851766, EPI_ISL_2851773, EPI_ISL_2851784, EPI_ISL_2851791, EPI_ISL_2851793, EPI_ISL_2851796, EPI_ISL_2851802, EPI_ISL_2851804, EPI_ISL_2851813, EPI_ISL_2851824, EPI_ISL_2851826, EPI_ISL_2851835, EPI_ISL_2851840, EPI_ISL_2851846, EPI_ISL_2851863, EPI_ISL_2851869, EPI_ISL_2851876, EPI_ISL_2851881, EPI_ISL_2851882, EPI_ISL_2851892, EPI_ISL_2851893, EPI_ISL_2851896, EPI_ISL_2851897, EPI_ISL_2851901, EPI_ISL_2851906, EPI_ISL_2851909, EPI_ISL_2851935, EPI_ISL_2851943, EPI_ISL_2851958, EPI_ISL_2851965, EPI_ISL_2851966, EPI_ISL_2851973, EPI_ISL_2851978, EPI_ISL_2852000, EPI_ISL_2852001, EPI_ISL_2852002, EPI_ISL_2852003, EPI_ISL_2852004, EPI_ISL_2852005, EPI_ISL_2852008, EPI_ISL_2852011, EPI_ISL_2852014, EPI_ISL_2852020, EPI_ISL_2852022, EPI_ISL_2852025, EPI_ISL_2852026, EPI_ISL_2852028, EPI_ISL_2852037, EPI_ISL_2852039, EPI_ISL_2852040, EPI_ISL_2852048, EPI_ISL_2852049, EPI_ISL_2852052, EPI_ISL_2852055, EPI_ISL_2852056, EPI_ISL_2852061, EPI_ISL_2852063, EPI_ISL_2852064, EPI_ISL_2852065, EPI_ISL_2852067, EPI_ISL_2852068, EPI_ISL_2852069, EPI_ISL_2852070, EPI_ISL_2852075, EPI_ISL_2852076, EPI_ISL_2852080, EPI_ISL_2852101, EPI_ISL_2852102, EPI_ISL_2852103, EPI_ISL_2852111, EPI_ISL_2852115, EPI_ISL_2852121, EPI_ISL_2852123, EPI_ISL_2852127, EPI_ISL_2852136, EPI_ISL_2852139, EPI_ISL_2865079, EPI_ISL_2865099, EPI_ISL_2865224, EPI_ISL_2865228, EPI_ISL_2865326, EPI_ISL_2865676, EPI_ISL_2865711, EPI_ISL_2865713, EPI_ISL_2865753, EPI_ISL_2865760, EPI_ISL_2865858, EPI_ISL_2865878, EPI_ISL_2865896, EPI_ISL_2865938, EPI_ISL_2865960, EPI_ISL_2865985, EPI_ISL_2865999, EPI_ISL_2866016, EPI_ISL_2867132, EPI_ISL_2867221, EPI_ISL_2867283, EPI_ISL_2867323, EPI_ISL_2867324, EPI_ISL_2867694, EPI_ISL_2867733, EPI_ISL_2867752, EPI_ISL_2867781, EPI_ISL_2867823, EPI_ISL_2867841, EPI_ISL_2867843, EPI_ISL_2867856, EPI_ISL_2867881, EPI_ISL_2867897, EPI_ISL_2867901, EPI_ISL_2867958, EPI_ISL_2867965, EPI_ISL_2867991, EPI_ISL_2867995, EPI_ISL_2868014, EPI_ISL_2868016, EPI_ISL_2868057, EPI_ISL_2868059, EPI_ISL_2868061, EPI_ISL_2868083, EPI_ISL_2868214, EPI_ISL_2868215, EPI_ISL_2868222, EPI_ISL_2907829, EPI_ISL_2909625, EPI_ISL_2909713, EPI_ISL_2909728, EPI_ISL_2909747, EPI_ISL_2909859, EPI_ISL_2913349, EPI_ISL_2913582, EPI_ISL_2915741, EPI_ISL_2915748, EPI_ISL_2915878, EPI_ISL_2916020, EPI_ISL_2918172, EPI_ISL_2918193 | | | |
| see above | Lighthouse Lab in Alderley Park | Wellcome Sanger Institute for the COVID-19 Genomics UK (COG-UK) Consortium | Cordelia Langford; David K. Jackson; Dominic Kwiatkowski; Ewan Harrison; Ian Johnston; Jacquelyn Wynn; Jeffrey Barrett; John Sillitoe on behalf of the Wellcome Sanger Institute COVID-19 Surveillance Team; Mairead Hyland; Roberto Amato; Sonia Goncalves; The Lighthouse Lab in Alderley Park and Alex Alderton |
| EPI_ISL_2779651, EPI_ISL_2779672, EPI_ISL_2779680, EPI_ISL_2779681, EPI_ISL_2779693, EPI_ISL_2779696, EPI_ISL_2779701, EPI_ISL_2779713, EPI_ISL_2779727, EPI_ISL_2779730, EPI_ISL_2779743, EPI_ISL_2779755, EPI_ISL_2779757, EPI_ISL_2779759, EPI_ISL_2779767, EPI_ISL_2779773, EPI_ISL_2779807, EPI_ISL_2779824, EPI_ISL_2779831, EPI_ISL_2779838, EPI_ISL_2779856, EPI_ISL_2779858, EPI_ISL_2780082, EPI_ISL_2803766, EPI_ISL_2803768, EPI_ISL_2803773, EPI_ISL_2803778, EPI_ISL_2803789, EPI_ISL_2803790, EPI_ISL_2803794, EPI_ISL_2803798, EPI_ISL_2803799, EPI_ISL_2803801, EPI_ISL_2803808, EPI_ISL_2803809, EPI_ISL_2803810, EPI_ISL_2803812, EPI_ISL_2803813, EPI_ISL_2803814, EPI_ISL_2803817, EPI_ISL_2803818, EPI_ISL_2803819, EPI_ISL_2803821, EPI_ISL_2803826, EPI_ISL_2803827, EPI_ISL_2803830, EPI_ISL_2803831, EPI_ISL_2803832, EPI_ISL_2803833, EPI_ISL_2803839, EPI_ISL_2803842, EPI_ISL_2803844, EPI_ISL_2803845, EPI_ISL_2803847, EPI_ISL_2803849, EPI_ISL_2803853, EPI_ISL_2803860, EPI_ISL_2803865, EPI_ISL_2803866, EPI_ISL_2803867, EPI_ISL_2803868, EPI_ISL_2803870, EPI_ISL_2803875, EPI_ISL_2803878, EPI_ISL_2803880, EPI_ISL_2803881, EPI_ISL_2803884, EPI_ISL_2803888, EPI_ISL_2803891, EPI_ISL_2803892, EPI_ISL_2803897, EPI_ISL_2803899, EPI_ISL_2803901, EPI_ISL_2803907, EPI_ISL_2803911, EPI_ISL_2803912, EPI_ISL_2803916, EPI_ISL_2803917, EPI_ISL_2803918, EPI_ISL_2803919, EPI_ISL_2803921, EPI_ISL_2805698, EPI_ISL_2805707, EPI_ISL_2805717, EPI_ISL_2805720, EPI_ISL_2805769, EPI_ISL_2805775, EPI_ISL_2805778, EPI_ISL_2805784, EPI_ISL_2805791, EPI_ISL_2805808, EPI_ISL_2805811, EPI_ISL_2805813, EPI_ISL_2805814, EPI_ISL_2805816, EPI_ISL_2805836, EPI_ISL_2805838, EPI_ISL_2805840, EPI_ISL_2805845, EPI_ISL_2805861, EPI_ISL_2805875, EPI_ISL_2805885, EPI_ISL_2805887, EPI_ISL_2805898, EPI_ISL_2805899, EPI_ISL_2805906, EPI_ISL_2805913, EPI_ISL_2805922, EPI_ISL_2805930, EPI_ISL_2805945, EPI_ISL_2805948, EPI_ISL_2805949, EPI_ISL_2805956, EPI_ISL_2805959, EPI_ISL_2805963, EPI_ISL_2805968, EPI_ISL_2805971, EPI_ISL_2805972, EPI_ISL_2805973, EPI_ISL_2805978, EPI_ISL_2805983, EPI_ISL_2805985, EPI_ISL_2805988, EPI_ISL_2806008, EPI_ISL_2806013, EPI_ISL_2806018, EPI_ISL_2806024, EPI_ISL_2806025, EPI_ISL_2806031, EPI_ISL_2806034, EPI_ISL_2806045, EPI_ISL_2806057, EPI_ISL_2806058, EPI_ISL_2806064, EPI_ISL_2806083, EPI_ISL_2806085, EPI_ISL_2806094, EPI_ISL_2806096, EPI_ISL_2806098, EPI_ISL_2806100, EPI_ISL_2806103, EPI_ISL_2806104, EPI_ISL_2806110, EPI_ISL_2806111, EPI_ISL_2806116, EPI_ISL_2806117, EPI_ISL_2806120, EPI_ISL_2806122, EPI_ISL_2806124, EPI_ISL_2806125, EPI_ISL_2806144, EPI_ISL_2806152, EPI_ISL_2806154, EPI_ISL_2806155, EPI_ISL_2806162, EPI_ISL_2806163, EPI_ISL_2806171, EPI_ISL_2806176, EPI_ISL_2806179, EPI_ISL_2806181, EPI_ISL_2806187, EPI_ISL_2806189, EPI_ISL_2806190, EPI_ISL_2806192, EPI_ISL_2806194, EPI_ISL_2806198, EPI_ISL_2806203, EPI_ISL_2806207, EPI_ISL_2806224, EPI_ISL_2806316, EPI_ISL_2806397, EPI_ISL_2807141, EPI_ISL_2807142, EPI_ISL_2807143, EPI_ISL_2807144, EPI_ISL_2807145, EPI_ISL_2807146, EPI_ISL_2807147, EPI_ISL_2807148, EPI_ISL_2807150, EPI_ISL_2807151, EPI_ISL_2807153, EPI_ISL_2807154, EPI_ISL_2807156, EPI_ISL_2807157, EPI_ISL_2807159, EPI_ISL_2807160, EPI_ISL_2807162, EPI_ISL_2807163, EPI_ISL_2807168, EPI_ISL_2807170, EPI_ISL_2807171, EPI_ISL_2807172, EPI_ISL_2807174, EPI_ISL_2807176, EPI_ISL_2807177, EPI_ISL_2807178, EPI_ISL_2807179, EPI_ISL_2807180, EPI_ISL_2807181, EPI_ISL_2807182, EPI_ISL_2807183, EPI_ISL_2807184, EPI_ISL_2807185, EPI_ISL_2807187, EPI_ISL_2807188, EPI_ISL_2807189, EPI_ISL_2807190, EPI_ISL_2807191, EPI_ISL_2807194, EPI_ISL_2807195, EPI_ISL_2807197, EPI_ISL_2807198, EPI_ISL_2807199, EPI_ISL_2807201, EPI_ISL_2807205, EPI_ISL_2807206, EPI_ISL_2807207, EPI_ISL_2807208, EPI_ISL_2807209, EPI_ISL_2807210, EPI_ISL_2807211, EPI_ISL_2807212, EPI_ISL_2807213, EPI_ISL_2807214, EPI_ISL_2807215, EPI_ISL_2807216, EPI_ISL_2807217, EPI_ISL_2807218, EPI_ISL_2807219, EPI_ISL_2807222, EPI_ISL_2807225, EPI_ISL_2807227, EPI_ISL_2807228, EPI_ISL_2807230, EPI_ISL_2807233, EPI_ISL_2807234, EPI_ISL_2807237, EPI_ISL_2807238, EPI_ISL_2807242, EPI_ISL_2807243, EPI_ISL_2807244, EPI_ISL_2807245, EPI_ISL_2807246, EPI_ISL_2807247, EPI_ISL_2807248, EPI_ISL_2807249, EPI_ISL_2807251, EPI_ISL_2807252, EPI_ISL_2807253, EPI_ISL_2807254, EPI_ISL_2807255, EPI_ISL_2807256, EPI_ISL_2807258, EPI_ISL_2807259, EPI_ISL_2807260, EPI_ISL_2807261, EPI_ISL_2807262, EPI_ISL_2807263, EPI_ISL_2807264, EPI_ISL_2807267, EPI_ISL_2807268, EPI_ISL_2807269, EPI_ISL_2807272, EPI_ISL_2807292, EPI_ISL_2807296, EPI_ISL_2807303, EPI_ISL_2807327, EPI_ISL_2807331, EPI_ISL_2807344, EPI_ISL_2808571, EPI_ISL_2814019, EPI_ISL_2814023, EPI_ISL_2814029, EPI_ISL_2814032, EPI_ISL_2814050, EPI_ISL_2814060, EPI_ISL_2814102, EPI_ISL_2814103, EPI_ISL_2814124, EPI_ISL_2814130, EPI_ISL_2814133, EPI_ISL_2814139, EPI_ISL_2814156, EPI_ISL_2814175, EPI_ISL_2814177, EPI_ISL_2814205, EPI_ISL_2814206, EPI_ISL_2814213, EPI_ISL_2814218, EPI_ISL_2814220, EPI_ISL_2814227, EPI_ISL_2814228, EPI_ISL_2814229, EPI_ISL_2814230, EPI_ISL_2814231, EPI_ISL_2814232, EPI_ISL_2814233, EPI_ISL_2814234, EPI_ISL_2814235, EPI_ISL_2814237, EPI_ISL_2814238, EPI_ISL_2814239, EPI_ISL_2814240, EPI_ISL_2814241, EPI_ISL_2814242, EPI_ISL_2814243, EPI_ISL_2814244, EPI_ISL_2814245, EPI_ISL_2814246, EPI_ISL_2814247, EPI_ISL_2814248, EPI_ISL_2814249, EPI_ISL_2814250, EPI_ISL_2814251, EPI_ISL_2814253, EPI_ISL_2814255, EPI_ISL_2814256, EPI_ISL_2814257, EPI_ISL_2814258, EPI_ISL_2814259, EPI_ISL_2814260, EPI_ISL_2814261, EPI_ISL_2814262, EPI_ISL_2814263, EPI_ISL_2814264, EPI_ISL_2814265, EPI_ISL_2814266, EPI_ISL_2814268, EPI_ISL_2814269, EPI_ISL_2814270, EPI_ISL_2814271, EPI_ISL_2814272, EPI_ISL_2814273, EPI_ISL_2814274, EPI_ISL_2814275, EPI_ISL_2814276, EPI_ISL_2814277, EPI_ISL_2814278, EPI_ISL_2814279, EPI_ISL_2814280, EPI_ISL_2814281, EPI_ISL_2814282, EPI_ISL_2814283, EPI_ISL_2814284, EPI_ISL_2814285, EPI_ISL_2814286, EPI_ISL_2814287, EPI_ISL_2814288, EPI_ISL_2814289, EPI_ISL_2814290, EPI_ISL_2814291, EPI_ISL_2814292, EPI_ISL_2814293, EPI_ISL_2814294, EPI_ISL_2814295, EPI_ISL_2814296, EPI_ISL_2814297, EPI_ISL_2814298, EPI_ISL_2814299, EPI_ISL_2814300, EPI_ISL_2814301, EPI_ISL_2814302, EPI_ISL_2814303, EPI_ISL_2814304, EPI_ISL_2814305, EPI_ISL_2814306, EPI_ISL_2814307, EPI_ISL_2814308, EPI_ISL_2814309, EPI_ISL_2814310, EPI_ISL_2814311, EPI_ISL_2814312, EPI_ISL_2814313, | | | |

| | | | |
|---|---|---|---|
| EPI_ISL_2814314, EPI_ISL_2814315, EPI_ISL_2814316, EPI_ISL_2814317, EPI_ISL_2814318, EPI_ISL_2814319, EPI_ISL_2814320, EPI_ISL_2814321, EPI_ISL_2814322, EPI_ISL_2814324, EPI_ISL_2814325, EPI_ISL_2814326, EPI_ISL_2814327, EPI_ISL_2814328, EPI_ISL_2814329, EPI_ISL_2814330, EPI_ISL_2814331, EPI_ISL_2814332, EPI_ISL_2814333, EPI_ISL_2814334, EPI_ISL_2814335, EPI_ISL_2814336, EPI_ISL_2814337, EPI_ISL_2814338, EPI_ISL_2814339, EPI_ISL_2814340, EPI_ISL_2814341, EPI_ISL_2814342, EPI_ISL_2814343, EPI_ISL_2814344, EPI_ISL_2814345, EPI_ISL_2814346, EPI_ISL_2814347, EPI_ISL_2814348, EPI_ISL_2814349, EPI_ISL_2814350, EPI_ISL_2814351, EPI_ISL_2814352, EPI_ISL_2814353, EPI_ISL_2814354, EPI_ISL_2814355, EPI_ISL_2814356, EPI_ISL_2814357, EPI_ISL_2814358, EPI_ISL_2814359, EPI_ISL_2814360, EPI_ISL_2814361, EPI_ISL_2814362, EPI_ISL_2814363, EPI_ISL_2814364, EPI_ISL_2814365, EPI_ISL_2814366, EPI_ISL_2814367, EPI_ISL_2814368, EPI_ISL_2814370, EPI_ISL_2814371, EPI_ISL_2814372, EPI_ISL_2814373, EPI_ISL_2814375, EPI_ISL_2814376, EPI_ISL_2814377, EPI_ISL_2814378, EPI_ISL_2814379, EPI_ISL_2814381, EPI_ISL_2814382, EPI_ISL_2814383, EPI_ISL_2814384, EPI_ISL_2814385, EPI_ISL_2814386, EPI_ISL_2814387, EPI_ISL_2814388, EPI_ISL_2814390, EPI_ISL_2814391, EPI_ISL_2814392, EPI_ISL_2814393, EPI_ISL_2814395, EPI_ISL_2814396, EPI_ISL_2814397, EPI_ISL_2814398, EPI_ISL_2814399, EPI_ISL_2814400, EPI_ISL_2814401, EPI_ISL_2814402, EPI_ISL_2814403, EPI_ISL_2814404, EPI_ISL_2814405, EPI_ISL_2814406, EPI_ISL_2814408, EPI_ISL_2814409, EPI_ISL_2814410, EPI_ISL_2814411, EPI_ISL_2814412, EPI_ISL_2814413, EPI_ISL_2814414, EPI_ISL_2814415, EPI_ISL_2814416, EPI_ISL_2814417, EPI_ISL_2814418, EPI_ISL_2814419, EPI_ISL_2814420, EPI_ISL_2814421, EPI_ISL_2814422, EPI_ISL_2814423, EPI_ISL_2814475, EPI_ISL_2814479, EPI_ISL_2814486, EPI_ISL_2814487, EPI_ISL_2814489, EPI_ISL_2814493, EPI_ISL_2814505, EPI_ISL_2814506, EPI_ISL_2814508, EPI_ISL_2814510, EPI_ISL_2814514, EPI_ISL_2814517, EPI_ISL_2814522, EPI_ISL_2814523, EPI_ISL_2814529, EPI_ISL_2814530, EPI_ISL_2814535, EPI_ISL_2814536, EPI_ISL_2814540, EPI_ISL_2814541, EPI_ISL_2814542, EPI_ISL_2814544, EPI_ISL_2814546, EPI_ISL_2814547, EPI_ISL_2814548, EPI_ISL_2814552, EPI_ISL_2814559, EPI_ISL_2814562, EPI_ISL_2814563, EPI_ISL_2814566, EPI_ISL_2814568, EPI_ISL_2814572, EPI_ISL_2814593, EPI_ISL_2814597, EPI_ISL_2814598, EPI_ISL_2814599, EPI_ISL_2814601, EPI_ISL_2814607, EPI_ISL_2814608, EPI_ISL_2814609, EPI_ISL_2814613, EPI_ISL_2814617, EPI_ISL_2814619, EPI_ISL_2814623, EPI_ISL_2814630, EPI_ISL_2814632, EPI_ISL_2814633, EPI_ISL_2814636, EPI_ISL_2814640, EPI_ISL_2814652, EPI_ISL_2814653, EPI_ISL_2814655, EPI_ISL_2814656, EPI_ISL_2814660, EPI_ISL_2814662, EPI_ISL_2814663, EPI_ISL_2814676, EPI_ISL_2814682, EPI_ISL_2814686, EPI_ISL_2814688, EPI_ISL_2814690, EPI_ISL_2814694, EPI_ISL_2814697, EPI_ISL_2814698, EPI_ISL_2814701, EPI_ISL_2814704, EPI_ISL_2814706, EPI_ISL_2814710, EPI_ISL_2814716, EPI_ISL_2814718, EPI_ISL_2814719, EPI_ISL_2814720, EPI_ISL_2814721, EPI_ISL_2814723, EPI_ISL_2814731, EPI_ISL_2814732, EPI_ISL_2814733, EPI_ISL_2814734, EPI_ISL_2814735, EPI_ISL_2814736, EPI_ISL_2814737, EPI_ISL_2814738, EPI_ISL_2814739, EPI_ISL_2814741, EPI_ISL_2814742, EPI_ISL_2814743, EPI_ISL_2814744, EPI_ISL_2814745, EPI_ISL_2814746, EPI_ISL_2814747, EPI_ISL_2814748, EPI_ISL_2814749, EPI_ISL_2814750, EPI_ISL_2814751, EPI_ISL_2814752, EPI_ISL_2814753, EPI_ISL_2814754, EPI_ISL_2814755, EPI_ISL_2814756, EPI_ISL_2814757, EPI_ISL_2814758, EPI_ISL_2814759, EPI_ISL_2814760, EPI_ISL_2814761, EPI_ISL_2814762, EPI_ISL_2814763, EPI_ISL_2814764, EPI_ISL_2814765, EPI_ISL_2814766, EPI_ISL_2814767, EPI_ISL_2814769, EPI_ISL_2814770, EPI_ISL_2814771, EPI_ISL_2814772, EPI_ISL_2814773, EPI_ISL_2814774, EPI_ISL_2814775, EPI_ISL_2814776, EPI_ISL_2814777, EPI_ISL_2814778, EPI_ISL_2814779, EPI_ISL_2814780, EPI_ISL_2814781, EPI_ISL_2814782, EPI_ISL_2814783, EPI_ISL_2814784, EPI_ISL_2814785, EPI_ISL_2814786, EPI_ISL_2814787, EPI_ISL_2814788, EPI_ISL_2814789, EPI_ISL_2814790, EPI_ISL_2814792, EPI_ISL_2814793, EPI_ISL_2814794, EPI_ISL_2814796, EPI_ISL_2814797, EPI_ISL_2814798, EPI_ISL_2814799, EPI_ISL_2814800, EPI_ISL_2814801, EPI_ISL_2814803, EPI_ISL_2814804, EPI_ISL_2814805, EPI_ISL_2814806, EPI_ISL_2814807, EPI_ISL_2814808, EPI_ISL_2814809, EPI_ISL_2814810, EPI_ISL_2814813, EPI_ISL_2814815, EPI_ISL_2814817, EPI_ISL_2814818, EPI_ISL_2814819, EPI_ISL_2814820, EPI_ISL_2814821, EPI_ISL_2814822, EPI_ISL_2814823, EPI_ISL_2814824, EPI_ISL_2814826, EPI_ISL_2814828, EPI_ISL_2814829, EPI_ISL_2814831, EPI_ISL_2814832, EPI_ISL_2814833, EPI_ISL_2814835, EPI_ISL_2814836, EPI_ISL_2814837, EPI_ISL_2814838, EPI_ISL_2814839, EPI_ISL_2814840, EPI_ISL_2814841, EPI_ISL_2814842, EPI_ISL_2814844, EPI_ISL_2814846, EPI_ISL_2814847, EPI_ISL_2814848, EPI_ISL_2814849, EPI_ISL_2814850, EPI_ISL_2814851, EPI_ISL_2814852, EPI_ISL_2814853, EPI_ISL_2814854, EPI_ISL_2814855, EPI_ISL_2814857, EPI_ISL_2814858, EPI_ISL_2814859, EPI_ISL_2814860, EPI_ISL_2814861, EPI_ISL_2814862, EPI_ISL_2814863, EPI_ISL_2814864, EPI_ISL_2814865, EPI_ISL_2814866, EPI_ISL_2814867, EPI_ISL_2814868, EPI_ISL_2814869, EPI_ISL_2814870, EPI_ISL_2814871, EPI_ISL_2814872, EPI_ISL_2814873, EPI_ISL_2814874, EPI_ISL_2814875, EPI_ISL_2814876, EPI_ISL_2814878, EPI_ISL_2814880, EPI_ISL_2814881, EPI_ISL_2814882, EPI_ISL_2814883, EPI_ISL_2814884, EPI_ISL_2814885, EPI_ISL_2814887, EPI_ISL_2814888, EPI_ISL_2814889, EPI_ISL_2814890, EPI_ISL_2814891, EPI_ISL_2814892, EPI_ISL_2814893, EPI_ISL_2814894, EPI_ISL_2814895, EPI_ISL_2814897, EPI_ISL_2814898, EPI_ISL_2814899, EPI_ISL_2814900, EPI_ISL_2814901, EPI_ISL_2814902, EPI_ISL_2814903, EPI_ISL_2814904, EPI_ISL_2814905, EPI_ISL_2814906, EPI_ISL_2814907, EPI_ISL_2814908, EPI_ISL_2814909, EPI_ISL_2814910, EPI_ISL_2814911, EPI_ISL_2814912, EPI_ISL_2814913, EPI_ISL_2814915, EPI_ISL_2814917, EPI_ISL_2814918, EPI_ISL_2814919, EPI_ISL_2814920, EPI_ISL_2814922, EPI_ISL_2814925, EPI_ISL_2814928, EPI_ISL_2814929, EPI_ISL_2814931, EPI_ISL_2814932, EPI_ISL_2814933, EPI_ISL_2814934, EPI_ISL_2814937, EPI_ISL_2814938, EPI_ISL_2814942, EPI_ISL_2814943, EPI_ISL_2814944, EPI_ISL_2814945, EPI_ISL_2814946, EPI_ISL_2814947, EPI_ISL_2814948, EPI_ISL_2814953, EPI_ISL_2814954, EPI_ISL_2814955, EPI_ISL_2814958, EPI_ISL_2814959, EPI_ISL_2814960, EPI_ISL_2814963, EPI_ISL_2814964, EPI_ISL_2814968, EPI_ISL_2814969, EPI_ISL_2814970, EPI_ISL_2814971, EPI_ISL_2814972, EPI_ISL_2814974, EPI_ISL_2814975, EPI_ISL_2814976, EPI_ISL_2814977, EPI_ISL_2814978, EPI_ISL_2814979, EPI_ISL_2814980, EPI_ISL_2814981, EPI_ISL_2814984, EPI_ISL_2814985, EPI_ISL_2814986, EPI_ISL_2814987, EPI_ISL_2814991, EPI_ISL_2814994, EPI_ISL_2814997, EPI_ISL_2815000, EPI_ISL_2815002, EPI_ISL_2815003, EPI_ISL_2815006, EPI_ISL_2815007, EPI_ISL_2815009, EPI_ISL_2815011, EPI_ISL_2815012, EPI_ISL_2815013, EPI_ISL_2815015, EPI_ISL_2815017, EPI_ISL_2815019, EPI_ISL_2815020, EPI_ISL_2815022, EPI_ISL_2815024, EPI_ISL_2815026, EPI_ISL_2815027, EPI_ISL_2815028, EPI_ISL_2815029, EPI_ISL_2815034, EPI_ISL_2815036, EPI_ISL_2815038, EPI_ISL_2829302, EPI_ISL_2829306, EPI_ISL_2829309, EPI_ISL_2829311, EPI_ISL_2829329, EPI_ISL_2829330, EPI_ISL_2829334, EPI_ISL_2829358, EPI_ISL_2829365, EPI_ISL_2829382, EPI_ISL_2829399, EPI_ISL_2829406, EPI_ISL_2829407, EPI_ISL_2829417, EPI_ISL_2829446, EPI_ISL_2829457, EPI_ISL_2829466, EPI_ISL_2829467, EPI_ISL_2829491, EPI_ISL_2829499, EPI_ISL_2829515, EPI_ISL_2829518, EPI_ISL_2829527, EPI_ISL_2829534, EPI_ISL_2829550, EPI_ISL_2829558, EPI_ISL_2829563, EPI_ISL_2829566, EPI_ISL_2830258, EPI_ISL_2830271, EPI_ISL_2830611, EPI_ISL_2830618, EPI_ISL_2830621, EPI_ISL_2830622, EPI_ISL_2830623, EPI_ISL_2830624, EPI_ISL_2830629, EPI_ISL_2830637, EPI_ISL_2830643, EPI_ISL_2830645, EPI_ISL_2830646, EPI_ISL_2830647, EPI_ISL_2830649, EPI_ISL_2830651, EPI_ISL_2830652, EPI_ISL_2830657, EPI_ISL_2830658, EPI_ISL_2830660, EPI_ISL_2830664, EPI_ISL_2830667, EPI_ISL_2830669, EPI_ISL_2830671, EPI_ISL_2830675, EPI_ISL_2830676, EPI_ISL_2830677, EPI_ISL_2830679, EPI_ISL_2830691, EPI_ISL_2830693, EPI_ISL_2830694, EPI_ISL_2830699, EPI_ISL_2830707, EPI_ISL_2830708, EPI_ISL_2830712, EPI_ISL_2830715, EPI_ISL_2830717, EPI_ISL_2830719, EPI_ISL_2830721, EPI_ISL_2830729, EPI_ISL_2830733, EPI_ISL_2830739, EPI_ISL_2830740, EPI_ISL_2830742, EPI_ISL_2830746, EPI_ISL_2830748, EPI_ISL_2830752, EPI_ISL_2830756, EPI_ISL_2830764, EPI_ISL_2830765, EPI_ISL_2830767, EPI_ISL_2830768, EPI_ISL_2830771, EPI_ISL_2830772, EPI_ISL_2830782, EPI_ISL_2830785, EPI_ISL_2830786, EPI_ISL_2830788, EPI_ISL_2830791, EPI_ISL_2830792, EPI_ISL_2830793, EPI_ISL_2830794, EPI_ISL_2830797, EPI_ISL_2830804, EPI_ISL_2830807, EPI_ISL_2830809, EPI_ISL_2830815, EPI_ISL_2830819, EPI_ISL_2830820, EPI_ISL_2830821, EPI_ISL_2830824, EPI_ISL_2830825, EPI_ISL_2830827, EPI_ISL_2830828, EPI_ISL_2830836, EPI_ISL_2830845, EPI_ISL_2830846, EPI_ISL_2830847, EPI_ISL_2830851, EPI_ISL_2830857, EPI_ISL_2830858, EPI_ISL_2830859, EPI_ISL_2830861, EPI_ISL_2830862, EPI_ISL_2830863, EPI_ISL_2830865, EPI_ISL_2830868, EPI_ISL_2830873, EPI_ISL_2830875, EPI_ISL_2830878, EPI_ISL_2830879, EPI_ISL_2830881, EPI_ISL_2830882, EPI_ISL_2830888, EPI_ISL_2830889, EPI_ISL_2830892, EPI_ISL_2830893, EPI_ISL_2830894, EPI_ISL_2830895, EPI_ISL_2830896, EPI_ISL_2830899, EPI_ISL_2830901, EPI_ISL_2830903, EPI_ISL_2830905, EPI_ISL_2830906, EPI_ISL_2830907, EPI_ISL_2830908, EPI_ISL_2830910, EPI_ISL_2830911, EPI_ISL_2830912, EPI_ISL_2830913, EPI_ISL_2830916, EPI_ISL_2830917, EPI_ISL_2830920, EPI_ISL_2830921, EPI_ISL_2830922, EPI_ISL_2830923, EPI_ISL_2830924, EPI_ISL_2830926, EPI_ISL_2830927, EPI_ISL_2830930, EPI_ISL_2830936, EPI_ISL_2830937, EPI_ISL_2830938, EPI_ISL_2830939, EPI_ISL_2830943, EPI_ISL_2830945, EPI_ISL_2830947, EPI_ISL_2830948, EPI_ISL_2830950, EPI_ISL_2830952, EPI_ISL_2830953, EPI_ISL_2830954, EPI_ISL_2830955, EPI_ISL_2830958, EPI_ISL_2830963, EPI_ISL_2830964, EPI_ISL_2830966, EPI_ISL_2830967, EPI_ISL_2830969, EPI_ISL_2830970, EPI_ISL_2830971, EPI_ISL_2830972, EPI_ISL_2830974, EPI_ISL_2830975, EPI_ISL_2830976, EPI_ISL_2830978, EPI_ISL_2830980, EPI_ISL_2830981, EPI_ISL_2830983, EPI_ISL_2830984, EPI_ISL_2830986, EPI_ISL_2830987, EPI_ISL_2830989, EPI_ISL_2830990, EPI_ISL_2830991, EPI_ISL_2830994, EPI_ISL_2830995, EPI_ISL_2830996, EPI_ISL_2830999, EPI_ISL_2831000, EPI_ISL_2831004, EPI_ISL_2831007, EPI_ISL_2831013, EPI_ISL_2831016, EPI_ISL_2831017, EPI_ISL_2831018, EPI_ISL_2831019, EPI_ISL_2831020, EPI_ISL_2831022, EPI_ISL_2831023, EPI_ISL_2831027, EPI_ISL_2831029, EPI_ISL_2831030, EPI_ISL_2831032, EPI_ISL_2831036, EPI_ISL_2831041, EPI_ISL_2831043, EPI_ISL_2831044, EPI_ISL_2831051, EPI_ISL_2831052, EPI_ISL_2831054, EPI_ISL_2831067, EPI_ISL_2831069, EPI_ISL_2831076, EPI_ISL_2831080, EPI_ISL_2831088, EPI_ISL_2831091, EPI_ISL_2831104, EPI_ISL_2831108, EPI_ISL_2831113, EPI_ISL_2831114, EPI_ISL_2831121, EPI_ISL_2831122, EPI_ISL_2831128, EPI_ISL_2831137, EPI_ISL_2831159, EPI_ISL_2831160, EPI_ISL_2831165, EPI_ISL_2831170, EPI_ISL_2831182, EPI_ISL_2831193, EPI_ISL_2831199, EPI_ISL_2831201, EPI_ISL_2831204, EPI_ISL_2831206, EPI_ISL_2831208, EPI_ISL_2831212, EPI_ISL_2831220, EPI_ISL_2831225, EPI_ISL_2831227, EPI_ISL_2831229, EPI_ISL_2831233, EPI_ISL_2831234, EPI_ISL_2831237, EPI_ISL_2831241, EPI_ISL_2831245, EPI_ISL_2831251, EPI_ISL_2831252, EPI_ISL_2831255, EPI_ISL_2831265, EPI_ISL_2831267, EPI_ISL_2831270, EPI_ISL_2831278, EPI_ISL_2831286, EPI_ISL_2831290, EPI_ISL_2831294, EPI_ISL_2831296, EPI_ISL_2831299, EPI_ISL_2831303, EPI_ISL_2831308, EPI_ISL_2831322, EPI_ISL_2831687, EPI_ISL_2831755, EPI_ISL_2831765, EPI_ISL_2831773, EPI_ISL_2831793, EPI_ISL_2831811, EPI_ISL_2831816, EPI_ISL_2831844, EPI_ISL_2831846, EPI_ISL_2831854, EPI_ISL_2831855, EPI_ISL_2831882, EPI_ISL_2831898, EPI_ISL_2831907, EPI_ISL_2831919, EPI_ISL_2831936, EPI_ISL_2831950, EPI_ISL_2831952, EPI_ISL_2831986, EPI_ISL_2832016, EPI_ISL_2832017, EPI_ISL_2832022, EPI_ISL_2832024, EPI_ISL_2832029, EPI_ISL_2832030, EPI_ISL_2832039, EPI_ISL_2832046, EPI_ISL_2832053, EPI_ISL_2850750, EPI_ISL_2850758, EPI_ISL_2850765, EPI_ISL_2850776, EPI_ISL_2850780, EPI_ISL_2850782, EPI_ISL_2850793, EPI_ISL_2850796, EPI_ISL_2850810, EPI_ISL_2850821, EPI_ISL_2851142, EPI_ISL_2851169, EPI_ISL_2851346, EPI_ISL_2851347, EPI_ISL_2851348, EPI_ISL_2851349, EPI_ISL_2851352, EPI_ISL_2851353, EPI_ISL_2851354, EPI_ISL_2851355, EPI_ISL_2851358, EPI_ISL_2851359, EPI_ISL_2851360, EPI_ISL_2851361, EPI_ISL_2851362, EPI_ISL_2851364, EPI_ISL_2851365, EPI_ISL_2851366, EPI_ISL_2851368, EPI_ISL_2851369, EPI_ISL_2851370, EPI_ISL_2851371, EPI_ISL_2851372, EPI_ISL_2851373, EPI_ISL_2851374, EPI_ISL_2851376, EPI_ISL_2851379, EPI_ISL_2851381, EPI_ISL_2851382, EPI_ISL_2851383, EPI_ISL_2851386, EPI_ISL_2851387, EPI_ISL_2851388, EPI_ISL_2851389, EPI_ISL_2851390, EPI_ISL_2851391, EPI_ISL_2851392, EPI_ISL_2851393, EPI_ISL_2851394, EPI_ISL_2851396, EPI_ISL_2851397, EPI_ISL_2851398, EPI_ISL_2851399, EPI_ISL_2851401, EPI_ISL_2851402, EPI_ISL_2851403, EPI_ISL_2851404, EPI_ISL_2851406, EPI_ISL_2851407, EPI_ISL_2851408, EPI_ISL_2851409, EPI_ISL_2852140, EPI_ISL_2852148, EPI_ISL_2852178, EPI_ISL_2852188, EPI_ISL_2852194, EPI_ISL_2852196, EPI_ISL_2852230, EPI_ISL_2852232, EPI_ISL_2852289, EPI_ISL_2852290, EPI_ISL_2852301, EPI_ISL_2852343, EPI_ISL_2852381, EPI_ISL_2852384, EPI_ISL_2852427, EPI_ISL_2852454, EPI_ISL_2852457, EPI_ISL_2852478, EPI_ISL_2852483, EPI_ISL_2852484, EPI_ISL_2852485, EPI_ISL_2852486, EPI_ISL_2852487, EPI_ISL_2852488, EPI_ISL_2852489, EPI_ISL_2852491, EPI_ISL_2852492, EPI_ISL_2852493, EPI_ISL_2852494, EPI_ISL_2852495, EPI_ISL_2852496, EPI_ISL_2852497, EPI_ISL_2852498, EPI_ISL_2852499, EPI_ISL_2852500, EPI_ISL_2852501, EPI_ISL_2852502, EPI_ISL_2852503, EPI_ISL_2852504, EPI_ISL_2852505, EPI_ISL_2852506, EPI_ISL_2852507, EPI_ISL_2852508, EPI_ISL_2852509, EPI_ISL_2852510, EPI_ISL_2852511, EPI_ISL_2852512, EPI_ISL_2852513, EPI_ISL_2852514, EPI_ISL_2852515, EPI_ISL_2852516, EPI_ISL_2852517, EPI_ISL_2852518, EPI_ISL_2852519, EPI_ISL_2852520, EPI_ISL_2852521, EPI_ISL_2852522, EPI_ISL_2852523, EPI_ISL_2852640, EPI_ISL_2852645, EPI_ISL_2852661, EPI_ISL_2852685, EPI_ISL_2852692, EPI_ISL_2852703, EPI_ISL_2852720, EPI_ISL_2852747, EPI_ISL_2852788, EPI_ISL_2852797, EPI_ISL_2852798, EPI_ISL_2852799, EPI_ISL_2852801, EPI_ISL_2852803, EPI_ISL_2852807, EPI_ISL_2852808, EPI_ISL_2852811, EPI_ISL_2852814, EPI_ISL_2852818, EPI_ISL_2852821, EPI_ISL_2852822, EPI_ISL_2852824, EPI_ISL_2852825, EPI_ISL_2852829, EPI_ISL_2852833, EPI_ISL_2852835, EPI_ISL_2852837, EPI_ISL_2852838, EPI_ISL_2852839, EPI_ISL_2852840, EPI_ISL_2852841, EPI_ISL_2852844, EPI_ISL_2852851, EPI_ISL_2852857, EPI_ISL_2852858, EPI_ISL_2852859, EPI_ISL_2852860, EPI_ISL_2852861, EPI_ISL_2852862, EPI_ISL_2852863, EPI_ISL_2852864, EPI_ISL_2852865, EPI_ISL_2852866, EPI_ISL_2852867, EPI_ISL_2852869, EPI_ISL_2852870, EPI_ISL_2852871, EPI_ISL_2852872, EPI_ISL_2852874, EPI_ISL_2852875, EPI_ISL_2852876, EPI_ISL_2852877, EPI_ISL_2852878, EPI_ISL_2852879, EPI_ISL_2852880, EPI_ISL_2852881, EPI_ISL_2852882, EPI_ISL_2852883, EPI_ISL_2852884, EPI_ISL_2852885, EPI_ISL_2852886, EPI_ISL_2852887, EPI_ISL_2852888, EPI_ISL_2852889, EPI_ISL_2852890, EPI_ISL_2852891, EPI_ISL_2852892, EPI_ISL_2852893, EPI_ISL_2852894, EPI_ISL_2852895, EPI_ISL_2852896, EPI_ISL_2852897, EPI_ISL_2852898, EPI_ISL_2852899, EPI_ISL_2852900, EPI_ISL_2852901, EPI_ISL_2852902, EPI_ISL_2852903, EPI_ISL_2852904, EPI_ISL_2852905, EPI_ISL_2852906, EPI_ISL_2852907, EPI_ISL_2852908, EPI_ISL_2852909, EPI_ISL_2852911, EPI_ISL_2852912, EPI_ISL_2852913, EPI_ISL_2852914, EPI_ISL_2852915, EPI_ISL_2852916, EPI_ISL_2852919, EPI_ISL_2852920, EPI_ISL_2852921, EPI_ISL_2852922, EPI_ISL_2852923, EPI_ISL_2852924, EPI_ISL_2852925, EPI_ISL_2852927, EPI_ISL_2852928, EPI_ISL_2852929, EPI_ISL_2852930, EPI_ISL_2852931, EPI_ISL_2852932, EPI_ISL_2852933, EPI_ISL_2852934, EPI_ISL_2852936, EPI_ISL_2852937, EPI_ISL_2852938, EPI_ISL_2852939, EPI_ISL_2852940, EPI_ISL_2852942, EPI_ISL_2852943, EPI_ISL_2852944, EPI_ISL_2852945, EPI_ISL_2852946, EPI_ISL_2852948, EPI_ISL_2852949, EPI_ISL_2852950, EPI_ISL_2852951, EPI_ISL_2852952, EPI_ISL_2852953, EPI_ISL_2852954, EPI_ISL_2852955, EPI_ISL_2852956, EPI_ISL_2852957, EPI_ISL_2852958, EPI_ISL_2852959, EPI_ISL_2852960, EPI_ISL_2852961, EPI_ISL_2852962, EPI_ISL_2852963, EPI_ISL_2852964, EPI_ISL_2852965, EPI_ISL_2852966, EPI_ISL_2852967, EPI_ISL_2852968, EPI_ISL_2852969, EPI_ISL_2852970, EPI_ISL_2852972, EPI_ISL_2852973, EPI_ISL_2852974, EPI_ISL_2852975, EPI_ISL_2852976, EPI_ISL_2852977, EPI_ISL_2852978, EPI_ISL_2852979, EPI_ISL_2852980, EPI_ISL_2852981, EPI_ISL_2852982, EPI_ISL_2852983, EPI_ISL_2852984, EPI_ISL_2852985, EPI_ISL_2852986, EPI_ISL_2852987, EPI_ISL_2852988, EPI_ISL_2852989, EPI_ISL_2852990, EPI_ISL_2852991, EPI_ISL_2852992, EPI_ISL_2852993, EPI_ISL_2852994, EPI_ISL_2852995, EPI_ISL_2852996, EPI_ISL_2852997, EPI_ISL_2852998, EPI_ISL_2852999, EPI_ISL_2853000, EPI_ISL_2853001, EPI_ISL_2853002, EPI_ISL_2853003, EPI_ISL_2853004, EPI_ISL_2853005, EPI_ISL_2853006, EPI_ISL_2853007, EPI_ISL_2853008, EPI_ISL_2853009, EPI_ISL_2853010, EPI_ISL_2853011, EPI_ISL_2853012, EPI_ISL_2853013, EPI_ISL_2853015, EPI_ISL_2853016, EPI_ISL_2853017, EPI_ISL_2853018, EPI_ISL_2853020, EPI_ISL_2853021, EPI_ISL_2853023, EPI_ISL_2853024, EPI_ISL_2853025, EPI_ISL_2853026, EPI_ISL_2853027, EPI_ISL_2853028, EPI_ISL_2853029, EPI_ISL_2853030, EPI_ISL_2853031, EPI_ISL_2853032, EPI_ISL_2853033, EPI_ISL_2853034, EPI_ISL_2853035, EPI_ISL_2853036, EPI_ISL_2853037, EPI_ISL_2853038, EPI_ISL_2853039, EPI_ISL_2853040, EPI_ISL_2853041, EPI_ISL_2853042, EPI_ISL_2853043, EPI_ISL_2853044, EPI_ISL_2853045, EPI_ISL_2853046, EPI_ISL_2853047, EPI_ISL_2853048, EPI_ISL_2853049, EPI_ISL_2853050, EPI_ISL_2853051, EPI_ISL_2853052, EPI_ISL_2853053, EPI_ISL_2853054, EPI_ISL_2853055, EPI_ISL_2853056, EPI_ISL_2853057, EPI_ISL_2853058, EPI_ISL_2853059, EPI_ISL_2853060, EPI_ISL_2853061, EPI_ISL_2853062, EPI_ISL_2853064, EPI_ISL_2853065, EPI_ISL_2853066, EPI_ISL_2853067, EPI_ISL_2853068, EPI_ISL_2853069, EPI_ISL_2853070, EPI_ISL_2853071, EPI_ISL_2853072, EPI_ISL_2853073, EPI_ISL_2853074, EPI_ISL_2853075, EPI_ISL_2853076, EPI_ISL_2853077, EPI_ISL_2853078, EPI_ISL_2853079, EPI_ISL_2853080, EPI_ISL_2853081, EPI_ISL_2853082, EPI_ISL_2853083, EPI_ISL_2853084, EPI_ISL_2853085, EPI_ISL_2853086, EPI_ISL_2853087, EPI_ISL_2853088, EPI_ISL_2853089, EPI_ISL_2853090, EPI_ISL_2853091, EPI_ISL_2853092, EPI_ISL_2853093, EPI_ISL_2853094, EPI_ISL_2853095, EPI_ISL_2853096, EPI_ISL_2853097, EPI_ISL_2853098, EPI_ISL_2853099, EPI_ISL_2853101, EPI_ISL_2853102, EPI_ISL_2853103, EPI_ISL_2853104, EPI_ISL_2853105, EPI_ISL_2853106, EPI_ISL_2853107, EPI_ISL_2853108, EPI_ISL_2853109, EPI_ISL_2853110, EPI_ISL_2853111, EPI_ISL_2853112, EPI_ISL_2853113, EPI_ISL_2853114, EPI_ISL_2853115, EPI_ISL_2853116, EPI_ISL_2853117, EPI_ISL_2853118, EPI_ISL_2853119, EPI_ISL_2853120, EPI_ISL_2853121, EPI_ISL_2853122, EPI_ISL_2853123, EPI_ISL_2853124, EPI_ISL_2853125, EPI_ISL_2853126, EPI_ISL_2853127, EPI_ISL_2853128, EPI_ISL_2853129, EPI_ISL_2853130, EPI_ISL_2853131, EPI_ISL_2853132, EPI_ISL_2853133, EPI_ISL_2853134, EPI_ISL_2853135, EPI_ISL_2853136, EPI_ISL_2853137, EPI_ISL_2853138, EPI_ISL_2853139, EPI_ISL_2853140, EPI_ISL_2853141, EPI_ISL_2853142, EPI_ISL_2853143, EPI_ISL_2853144, EPI_ISL_2853145, EPI_ISL_2853146, EPI_ISL_2853147, EPI_ISL_2853148, EPI_ISL_2853149, EPI_ISL_2853150, EPI_ISL_2853151, EPI_ISL_2853152, EPI_ISL_2853153, EPI_ISL_2853154, EPI_ISL_2853155, EPI_ISL_2853156, EPI_ISL_2853157, EPI_ISL_2853158, EPI_ISL_2853159, EPI_ISL_2853160, EPI_ISL_2853161, EPI_ISL_2853162, EPI_ISL_2853163, EPI_ISL_2853164, EPI_ISL_2853165, EPI_ISL_2853166, EPI_ISL_2853167, EPI_ISL_2853168, EPI_ISL_2853169, EPI_ISL_2853170, EPI_ISL_2853171, EPI_ISL_2853172, EPI_ISL_2853173, EPI_ISL_2853174, EPI_ISL_2853175, EPI_ISL_2853176, EPI_ISL_2853177, EPI_ISL_2853178, EPI_ISL_2853179, EPI_ISL_2853180, EPI_ISL_2853181, EPI_ISL_2853182, EPI_ISL_2853183, EPI_ISL_2853184, EPI_ISL_2853185, EPI_ISL_2853186, EPI_ISL_2853187, EPI_ISL_2853188, EPI_ISL_2853189, EPI_ISL_2853190, EPI_ISL_2853191, EPI_ISL_2853192, EPI_ISL_2853193, EPI_ISL_2853194, EPI_ISL_2853195, EPI_ISL_2853196, EPI_ISL_2853197, EPI_ISL_2853198, EPI_ISL_2853199, EPI_ISL_2853200, EPI_ISL_2853201, EPI_ISL_2853202, EPI_ISL_2853203, EPI_ISL_2853204, EPI_ISL_2853205, EPI_ISL_2853206, EPI_ISL_2853207, EPI_ISL_2853208, EPI_ISL_2853209, EPI_ISL_2853210, EPI_ISL_2853211, EPI_ISL_2853212, EPI_ISL_2853213, EPI_ISL_2853214, EPI_ISL_2853215, EPI_ISL_2853216, EPI_ISL_2853217, EPI_ISL_2853218, EPI_ISL_2853219, EPI_ISL_2853220, EPI_ISL_2853221, EPI_ISL_2853222, EPI_ISL_2853223, EPI_ISL_2853224, EPI_ISL_2853225, EPI_ISL_2853226, EPI_ISL_2853227, EPI_ISL_2853228, EPI_ISL_2853229, EPI_ISL_2853230, EPI_ISL_2853231, EPI_ISL_2853232, EPI_ISL_2853233, EPI_ISL_2853234, EPI_ISL_2853235, EPI_ISL_2853236, EPI_ISL_2853237, EPI_ISL_2853238, EPI_ISL_2853239, EPI_ISL_2853240, EPI_ISL_2853242, EPI_ISL_2853244, EPI_ISL_2853245, EPI_ISL_2853246, EPI_ISL_2853247, EPI_ISL_2853248, EPI_ISL_2853249, EPI_ISL_2853250, EPI_ISL_2853251, EPI_ISL_2853252, EPI_ISL_2853253, EPI_ISL_2853254, EPI_ISL_2853255, EPI_ISL_2853256, EPI_ISL_2853257, EPI_ISL_2853258, EPI_ISL_2937661, EPI_ISL_2937706 | | | |
| see above | Lighthouse Lab in Glasgow | Wellcome Sanger Institute for the COVID-19 Genomics UK (COG-UK) Consortium | Anna Dominiczak and Alex Alderton; Carol Clugston; Cordelia Langford; David Gray; David K. Jackson; Dominic Kwiatkowski; Ewan Harrison; Harper VanSteenhouse; Ian Johnston; Jeffrey Barrett; John Sillitoe on behalf of the Wellcome Sanger Institute COVID-19 Surveillance Team; Roberto Amato; Sonia Goncalves; Yumi Kasai |
| EPI_ISL_2779644, EPI_ISL_2779656, EPI_ISL_2779657, EPI_ISL_2779661, EPI_ISL_2779667, EPI_ISL_2779669, EPI_ISL_2779678, EPI_ISL_2779679, EPI_ISL_2779687, EPI_ISL_2779689, EPI_ISL_2779695, EPI_ISL_2779697, EPI_ISL_2779698, EPI_ISL_2779700, EPI_ISL_2779702, EPI_ISL_2779707, EPI_ISL_2779708, EPI_ISL_2779714, EPI_ISL_2779717, EPI_ISL_2779722, EPI_ISL_2779729, EPI_ISL_2779731, EPI_ISL_2779736, EPI_ISL_2779751, EPI_ISL_2779765, EPI_ISL_2779771, EPI_ISL_2779775, EPI_ISL_2779778, EPI_ISL_2779779, EPI_ISL_2779789, EPI_ISL_2779790, EPI_ISL_2779795, EPI_ISL_2779799, EPI_ISL_2779808, EPI_ISL_2779815, EPI_ISL_2779817, EPI_ISL_2779823, EPI_ISL_2779825, EPI_ISL_2779826, EPI_ISL_2779827, EPI_ISL_2779843, EPI_ISL_2779846, EPI_ISL_2779847, EPI_ISL_2779849, EPI_ISL_2779863, EPI_ISL_2779865, EPI_ISL_2779866, EPI_ISL_2779880, EPI_ISL_2779899, EPI_ISL_2779904, EPI_ISL_2779908, EPI_ISL_2779932, EPI_ISL_2779934, EPI_ISL_2779940, EPI_ISL_2779977, EPI_ISL_2779979, EPI_ISL_2779987, EPI_ISL_2779990, EPI_ISL_2779997, EPI_ISL_2780002, EPI_ISL_2780004, EPI_ISL_2780005, EPI_ISL_2780025, EPI_ISL_2780029, EPI_ISL_2780037, EPI_ISL_2780038, EPI_ISL_2780043, EPI_ISL_2780046, EPI_ISL_2780060, EPI_ISL_2780066, EPI_ISL_2780067, EPI_ISL_2780068, EPI_ISL_2780069, EPI_ISL_2780071, EPI_ISL_2780076, EPI_ISL_2780081, EPI_ISL_2780089, EPI_ISL_2780095, EPI_ISL_2780099, EPI_ISL_2780104, EPI_ISL_2780114, EPI_ISL_2780117, EPI_ISL_2780119, EPI_ISL_2803770, EPI_ISL_2803771, EPI_ISL_2803772, EPI_ISL_2803774, EPI_ISL_2803775, EPI_ISL_2803776, EPI_ISL_2803777, EPI_ISL_2803779, EPI_ISL_2803780, EPI_ISL_2803781, EPI_ISL_2803782, EPI_ISL_2803783, EPI_ISL_2803784, EPI_ISL_2803786, EPI_ISL_2803787, EPI_ISL_2803788, EPI_ISL_2803791, EPI_ISL_2803792, EPI_ISL_2803793, EPI_ISL_2803796, EPI_ISL_2803797, EPI_ISL_2803800, EPI_ISL_2803802, EPI_ISL_2803804, EPI_ISL_2803805, EPI_ISL_2803806, EPI_ISL_2803807, EPI_ISL_2803815, EPI_ISL_2803816, EPI_ISL_2803820, EPI_ISL_2803823, EPI_ISL_2803824, EPI_ISL_2803828, EPI_ISL_2803829, EPI_ISL_2803835, EPI_ISL_2803836, EPI_ISL_2803837, EPI_ISL_2803838, EPI_ISL_2803840, EPI_ISL_2803846, EPI_ISL_2803848, EPI_ISL_2803850, EPI_ISL_2803852, EPI_ISL_2803854, EPI_ISL_2803855, EPI_ISL_2803856, EPI_ISL_2803857, EPI_ISL_2803858, EPI_ISL_2803859, EPI_ISL_2803862, EPI_ISL_2803863, EPI_ISL_2803864, EPI_ISL_2803869, EPI_ISL_2803871, EPI_ISL_2803872, EPI_ISL_2803876, EPI_ISL_2803879, EPI_ISL_2803882, EPI_ISL_2803883, EPI_ISL_2803885, EPI_ISL_2803889, EPI_ISL_2803890, EPI_ISL_2803893, EPI_ISL_2803894, EPI_ISL_2803895, EPI_ISL_2803896, EPI_ISL_2803900, EPI_ISL_2803902, EPI_ISL_2803903, EPI_ISL_2803904, EPI_ISL_2803905, EPI_ISL_2803906, EPI_ISL_2803908, EPI_ISL_2803909, EPI_ISL_2803910, EPI_ISL_2803913, EPI_ISL_2803914, EPI_ISL_2803920, EPI_ISL_2803922, EPI_ISL_2803923, EPI_ISL_2803935, EPI_ISL_2803939, EPI_ISL_2803948, EPI_ISL_2803953, EPI_ISL_2803959, EPI_ISL_2803969, EPI_ISL_2803972, EPI_ISL_2803975, EPI_ISL_2803996, EPI_ISL_2803998, EPI_ISL_2803999, EPI_ISL_2804002, EPI_ISL_2804007, EPI_ISL_2804012, EPI_ISL_2804013, EPI_ISL_2804022, EPI_ISL_2804036, EPI_ISL_2804044, EPI_ISL_2804049, EPI_ISL_2804054, EPI_ISL_2804058, EPI_ISL_2804065, EPI_ISL_2804070, EPI_ISL_2804084, EPI_ISL_2804107, EPI_ISL_2804123, EPI_ISL_2804141, EPI_ISL_2804146, EPI_ISL_2804173, EPI_ISL_2804191, EPI_ISL_2804199, EPI_ISL_2804207, EPI_ISL_2804210, EPI_ISL_2804219, EPI_ISL_2804225, EPI_ISL_2804229, EPI_ISL_2804245, EPI_ISL_2804614, EPI_ISL_2804617, EPI_ISL_2804621, EPI_ISL_2804622, EPI_ISL_2804623, EPI_ISL_2804629, EPI_ISL_2804645, EPI_ISL_2804674, EPI_ISL_2804682, EPI_ISL_2804689, EPI_ISL_2804690, EPI_ISL_2804694, EPI_ISL_2804698, EPI_ISL_2804699, EPI_ISL_2804700, EPI_ISL_2804705, EPI_ISL_2804727, EPI_ISL_2804736, EPI_ISL_2804737, EPI_ISL_2804744, EPI_ISL_2804746, EPI_ISL_2804750, EPI_ISL_2804756, EPI_ISL_2804759, EPI_ISL_2804770, EPI_ISL_2804773, EPI_ISL_2804790, EPI_ISL_2804805, EPI_ISL_2804810, EPI_ISL_2804823, EPI_ISL_2804844, EPI_ISL_2804876, EPI_ISL_2804889, EPI_ISL_2804890, EPI_ISL_2804896, EPI_ISL_2804901, EPI_ISL_2804905, EPI_ISL_2804907, EPI_ISL_2804908, EPI_ISL_2804910, EPI_ISL_2804914, EPI_ISL_2804916, EPI_ISL_2804922, EPI_ISL_2804923, EPI_ISL_2804925, EPI_ISL_2804931, EPI_ISL_2804935, EPI_ISL_2804937, EPI_ISL_2804943, EPI_ISL_2804945, EPI_ISL_2804948, EPI_ISL_2804949, EPI_ISL_2804955, EPI_ISL_2804958, EPI_ISL_2804959, EPI_ISL_2804963, EPI_ISL_2804965, EPI_ISL_2804966, EPI_ISL_2804969, EPI_ISL_2804975, EPI_ISL_2804976, EPI_ISL_2804977, EPI_ISL_2804978, EPI_ISL_2804979, EPI_ISL_2804985, EPI_ISL_2805000, EPI_ISL_2805001, EPI_ISL_2805002, EPI_ISL_2805003, EPI_ISL_2805005, EPI_ISL_2805009, EPI_ISL_2805010, EPI_ISL_2805013, EPI_ISL_2805016, EPI_ISL_2805022, EPI_ISL_2805025, EPI_ISL_2805027, EPI_ISL_2805034, EPI_ISL_2805035, EPI_ISL_2805037, EPI_ISL_2805043, EPI_ISL_2805048, EPI_ISL_2805055, EPI_ISL_2805058, EPI_ISL_2805060, EPI_ISL_2805068, EPI_ISL_2805069, EPI_ISL_2805077, EPI_ISL_2805079, EPI_ISL_2805080, EPI_ISL_2805082, EPI_ISL_2805087, EPI_ISL_2805090, EPI_ISL_2805092, EPI_ISL_2805094, EPI_ISL_2805099, EPI_ISL_2805100, EPI_ISL_2805104, EPI_ISL_2805108, EPI_ISL_2805109, EPI_ISL_2805110, EPI_ISL_2805111, EPI_ISL_2805112, EPI_ISL_2805118, EPI_ISL_2805119, EPI_ISL_2805125, EPI_ISL_2805126, EPI_ISL_2805127, EPI_ISL_2805128, EPI_ISL_2805130, EPI_ISL_2805134, EPI_ISL_2805137, EPI_ISL_2805139, EPI_ISL_2805144, EPI_ISL_2805146, EPI_ISL_2805147, EPI_ISL_2805152, EPI_ISL_2805155, EPI_ISL_2805157, EPI_ISL_2805158, EPI_ISL_2805161, EPI_ISL_2805162, EPI_ISL_2805163, EPI_ISL_2805166, EPI_ISL_2805168, EPI_ISL_2805169, EPI_ISL_2805170, EPI_ISL_2805172, EPI_ISL_2805175, EPI_ISL_2805176, EPI_ISL_2805177, EPI_ISL_2805180, EPI_ISL_2805181, EPI_ISL_2805182, EPI_ISL_2805183, EPI_ISL_2805184, EPI_ISL_2805186, EPI_ISL_2805188, EPI_ISL_2805191, EPI_ISL_2805195, EPI_ISL_2805200, EPI_ISL_2805201, EPI_ISL_2805202, EPI_ISL_2805207, EPI_ISL_2805209, EPI_ISL_2805211, EPI_ISL_2805217, EPI_ISL_2805221, EPI_ISL_2805226, EPI_ISL_2805227, EPI_ISL_2805231, EPI_ISL_2805234, EPI_ISL_2805235, EPI_ISL_2805237, EPI_ISL_2805240, EPI_ISL_2805241, EPI_ISL_2805244, EPI_ISL_2805249, EPI_ISL_2805251, EPI_ISL_2805253, EPI_ISL_2805255, EPI_ISL_2805257, EPI_ISL_2805260, EPI_ISL_2805265, EPI_ISL_2805268, EPI_ISL_2805272, EPI_ISL_2805274, EPI_ISL_2805276, EPI_ISL_2805280, EPI_ISL_2805282, EPI_ISL_2805283, EPI_ISL_2805284, EPI_ISL_2805286, EPI_ISL_2805288, EPI_ISL_2805289, EPI_ISL_2805291, EPI_ISL_2805298, EPI_ISL_2805300, EPI_ISL_2805301, EPI_ISL_2805302, EPI_ISL_2805303, EPI_ISL_2805307, EPI_ISL_2805312, EPI_ISL_2805316, EPI_ISL_2805320, EPI_ISL_2805321, EPI_ISL_2805323, EPI_ISL_2805326, EPI_ISL_2805327, EPI_ISL_2805330, EPI_ISL_2805332, EPI_ISL_2805333, EPI_ISL_2805335, EPI_ISL_2805336, EPI_ISL_2805695, EPI_ISL_2805697, EPI_ISL_2805699, EPI_ISL_2805700, EPI_ISL_2805702, EPI_ISL_2805703, EPI_ISL_2805704, EPI_ISL_2805709, EPI_ISL_2805711, EPI_ISL_2805712, EPI_ISL_2805714, EPI_ISL_2805715, EPI_ISL_2805718, EPI_ISL_2805721, EPI_ISL_2805722, EPI_ISL_2805723, EPI_ISL_2805724, EPI_ISL_2805726, EPI_ISL_2805727, EPI_ISL_2805728, EPI_ISL_2805729, EPI_ISL_2805730, EPI_ISL_2805731, EPI_ISL_2805732, EPI_ISL_2805733, EPI_ISL_2805736, EPI_ISL_2805737, EPI_ISL_2805738, EPI_ISL_2805740, EPI_ISL_2805742, EPI_ISL_2805743, EPI_ISL_2805745, EPI_ISL_2805748, EPI_ISL_2805749, EPI_ISL_2805753, EPI_ISL_2805755, EPI_ISL_2805758, EPI_ISL_2805759, EPI_ISL_2805760, EPI_ISL_2805761, EPI_ISL_2805763, EPI_ISL_2805764, EPI_ISL_2805765, EPI_ISL_2805766, EPI_ISL_2805768, EPI_ISL_2805771, EPI_ISL_2805773, EPI_ISL_2805776, EPI_ISL_2805777, EPI_ISL_2805779, EPI_ISL_2805780, EPI_ISL_2805782, EPI_ISL_2805783, EPI_ISL_2805787, EPI_ISL_2805788, EPI_ISL_2805792, EPI_ISL_2805793, EPI_ISL_2805794, EPI_ISL_2805795, EPI_ISL_2805797, EPI_ISL_2805798, EPI_ISL_2805799, EPI_ISL_2805801, EPI_ISL_2805802, EPI_ISL_2805803, EPI_ISL_2805804, EPI_ISL_2805806, EPI_ISL_2805809, EPI_ISL_2805810, EPI_ISL_2805812, EPI_ISL_2805815, EPI_ISL_2805818, EPI_ISL_2805819, EPI_ISL_2805820, EPI_ISL_2805821, EPI_ISL_2805822, EPI_ISL_2805825, EPI_ISL_2805826, EPI_ISL_2805827, EPI_ISL_2805829, EPI_ISL_2805830, EPI_ISL_2805831, EPI_ISL_2805832, EPI_ISL_2805833, EPI_ISL_2805834, EPI_ISL_2805835, EPI_ISL_2805837, EPI_ISL_2805839, EPI_ISL_2805841, EPI_ISL_2805843, EPI_ISL_2805846, EPI_ISL_2805849, EPI_ISL_2805850, EPI_ISL_2805854, EPI_ISL_2805857, EPI_ISL_2805858, EPI_ISL_2805859, EPI_ISL_2805860, EPI_ISL_2805862, EPI_ISL_2805863, EPI_ISL_2805864, EPI_ISL_2805866, EPI_ISL_2805867, EPI_ISL_2805871, EPI_ISL_2805872, EPI_ISL_2805876, EPI_ISL_2805877, EPI_ISL_2805878, EPI_ISL_2805879, EPI_ISL_2805880, EPI_ISL_2805883, EPI_ISL_2805884, EPI_ISL_2805888, EPI_ISL_2805894, EPI_ISL_2805895, EPI_ISL_2805896, EPI_ISL_2805897, EPI_ISL_2805900, EPI_ISL_2805901, EPI_ISL_2805904, EPI_ISL_2805905, EPI_ISL_2805907, EPI_ISL_2805909, EPI_ISL_2805910, EPI_ISL_2805911, EPI_ISL_2805914, EPI_ISL_2805915, EPI_ISL_2805916, EPI_ISL_2805919, EPI_ISL_2805923, EPI_ISL_2805924, EPI_ISL_2805928, EPI_ISL_2805929, EPI_ISL_2805931, EPI_ISL_2805932, EPI_ISL_2805933, EPI_ISL_2805935, EPI_ISL_2805936, EPI_ISL_2805937, EPI_ISL_2805939, EPI_ISL_2805940, EPI_ISL_2805942, EPI_ISL_2805943, EPI_ISL_2805947, EPI_ISL_2805950, EPI_ISL_2805951, EPI_ISL_2805952, EPI_ISL_2805953, EPI_ISL_2805958, EPI_ISL_2805960, EPI_ISL_2805962, EPI_ISL_2805964, EPI_ISL_2805966, EPI_ISL_2805967, EPI_ISL_2805969, EPI_ISL_2805970, EPI_ISL_2805974, EPI_ISL_2805976, EPI_ISL_2805977, EPI_ISL_2805979, EPI_ISL_2805980, EPI_ISL_2805987, EPI_ISL_2805989, EPI_ISL_2805991, EPI_ISL_2806445, EPI_ISL_2806446, EPI_ISL_2806451, EPI_ISL_2806455, EPI_ISL_2806463, EPI_ISL_2806464, EPI_ISL_2806466, EPI_ISL_2806476, EPI_ISL_2806487, EPI_ISL_2806488, EPI_ISL_2806490, EPI_ISL_2806495, EPI_ISL_2806499, EPI_ISL_2806504, EPI_ISL_2806507, EPI_ISL_2806515, EPI_ISL_2806516, EPI_ISL_2806519, EPI_ISL_2806522, EPI_ISL_2806526, EPI_ISL_2806529, EPI_ISL_2806532, EPI_ISL_2806542, EPI_ISL_2806546, EPI_ISL_2806550, EPI_ISL_2806555, EPI_ISL_2806563, EPI_ISL_2806566, EPI_ISL_2806573, EPI_ISL_2806581, EPI_ISL_2806582, EPI_ISL_2806587, EPI_ISL_2806591, EPI_ISL_2806592, EPI_ISL_2806596, EPI_ISL_2806597, EPI_ISL_2806598, EPI_ISL_2806603, EPI_ISL_2806604, EPI_ISL_2806607, EPI_ISL_2806610, EPI_ISL_2806615, EPI_ISL_2806616, EPI_ISL_2806620, EPI_ISL_2806635, EPI_ISL_2806637, EPI_ISL_2806641, EPI_ISL_2806642, EPI_ISL_2806646, EPI_ISL_2806650, EPI_ISL_2806654, EPI_ISL_2806656, EPI_ISL_2806674, EPI_ISL_2806679, EPI_ISL_2806682, EPI_ISL_2806684, EPI_ISL_2806691, EPI_ISL_2806692, EPI_ISL_2806697, EPI_ISL_2806698, EPI_ISL_2806699, EPI_ISL_2806701, EPI_ISL_2806702, EPI_ISL_2806704, EPI_ISL_2806714, EPI_ISL_2806716, EPI_ISL_2806717, EPI_ISL_2806718, EPI_ISL_2806719, EPI_ISL_2806721, EPI_ISL_2806726, EPI_ISL_2806738, EPI_ISL_2806744, EPI_ISL_2806752, EPI_ISL_2806761, EPI_ISL_2806762, EPI_ISL_2806775, EPI_ISL_2806776, EPI_ISL_2806778, EPI_ISL_2806783, EPI_ISL_2806787, EPI_ISL_2806796, EPI_ISL_2806797, EPI_ISL_2808074, EPI_ISL_2808090, EPI_ISL_2808092, EPI_ISL_2808112, EPI_ISL_2808116, EPI_ISL_2808133, EPI_ISL_2808136, EPI_ISL_2808142, EPI_ISL_2808171, EPI_ISL_2808182, EPI_ISL_2808183, EPI_ISL_2808259, EPI_ISL_2808267, EPI_ISL_2808281, EPI_ISL_2808304, EPI_ISL_2808319, EPI_ISL_2808323, EPI_ISL_2808324, EPI_ISL_2808328, EPI_ISL_2808338, EPI_ISL_2808347, EPI_ISL_2808351, EPI_ISL_2808370, EPI_ISL_2808379, EPI_ISL_2808385, EPI_ISL_2808390, EPI_ISL_2808403, EPI_ISL_2808404, EPI_ISL_2808426, EPI_ISL_2808429, EPI_ISL_2808433, EPI_ISL_2808440, EPI_ISL_2808443, EPI_ISL_2808450, EPI_ISL_2808464, EPI_ISL_2808471, EPI_ISL_2808474, EPI_ISL_2808475, EPI_ISL_2808476, EPI_ISL_2808480, EPI_ISL_2808484, EPI_ISL_2808485, EPI_ISL_2808486, EPI_ISL_2808488, EPI_ISL_2808490, EPI_ISL_2808492, EPI_ISL_2808495, EPI_ISL_2808498, EPI_ISL_2808502, EPI_ISL_2808503, EPI_ISL_2808506, EPI_ISL_2808507, EPI_ISL_2808511, EPI_ISL_2808513, EPI_ISL_2808516, EPI_ISL_2808518, EPI_ISL_2808519, EPI_ISL_2808530, EPI_ISL_2808531, EPI_ISL_2808535, EPI_ISL_2808540, EPI_ISL_2808543, EPI_ISL_2808550, EPI_ISL_2808554, EPI_ISL_2808556, EPI_ISL_2808574, EPI_ISL_2808581, EPI_ISL_2808583, EPI_ISL_2808588, EPI_ISL_2808597, EPI_ISL_2808598, EPI_ISL_2808601, EPI_ISL_2808616, EPI_ISL_2808620, EPI_ISL_2808621, EPI_ISL_2808625, EPI_ISL_2808627, EPI_ISL_2808632, EPI_ISL_2808633, EPI_ISL_2808636, EPI_ISL_2808637, EPI_ISL_2808641, EPI_ISL_2808647, EPI_ISL_2808649, EPI_ISL_2808652, EPI_ISL_2808653, EPI_ISL_2808662, EPI_ISL_2808668, EPI_ISL_2808669, EPI_ISL_2808673, EPI_ISL_2808675, EPI_ISL_2808677, EPI_ISL_2808679, EPI_ISL_2808680, EPI_ISL_2808685, EPI_ISL_2808686, EPI_ISL_2808687, EPI_ISL_2808690, EPI_ISL_2808692, EPI_ISL_2808695, EPI_ISL_2808698, EPI_ISL_2808709, EPI_ISL_2808711, EPI_ISL_2808712, EPI_ISL_2808713, EPI_ISL_2808715, EPI_ISL_2808720, EPI_ISL_2808721, EPI_ISL_2808722, EPI_ISL_2808723, EPI_ISL_2808725, EPI_ISL_2808729, EPI_ISL_2808730, EPI_ISL_2812984, EPI_ISL_2812985, EPI_ISL_2812986, EPI_ISL_2812987, EPI_ISL_2812989, EPI_ISL_2812990, EPI_ISL_2812991, EPI_ISL_2812992, EPI_ISL_2812993, EPI_ISL_2812994, EPI_ISL_2812995, EPI_ISL_2812996, EPI_ISL_2812997, EPI_ISL_2812998, EPI_ISL_2812999, EPI_ISL_2813000, EPI_ISL_2813001, EPI_ISL_2813003, EPI_ISL_2813004, EPI_ISL_2813005, EPI_ISL_2813006, EPI_ISL_2813007, EPI_ISL_2813010, EPI_ISL_2813011, EPI_ISL_2813012, EPI_ISL_2813014, EPI_ISL_2813015, EPI_ISL_2813016, EPI_ISL_2813017, EPI_ISL_2813018, EPI_ISL_2813021, EPI_ISL_2813022, EPI_ISL_2813024, EPI_ISL_2813025, EPI_ISL_2813027, EPI_ISL_2813028, EPI_ISL_2813029, EPI_ISL_2813030, EPI_ISL_2813031, EPI_ISL_2813032, EPI_ISL_2813035, EPI_ISL_2813036, EPI_ISL_2813037, EPI_ISL_2813038, EPI_ISL_2813039, EPI_ISL_2813041, EPI_ISL_2813042, EPI_ISL_2813043, EPI_ISL_2813047, EPI_ISL_2813048, EPI_ISL_2813049, EPI_ISL_2813050, EPI_ISL_2813051, EPI_ISL_2813052, EPI_ISL_2813053, EPI_ISL_2813054, EPI_ISL_2813055, EPI_ISL_2813056, EPI_ISL_2813057, EPI_ISL_2813059, EPI_ISL_2813060, EPI_ISL_2813061, EPI_ISL_2813063, EPI_ISL_2813064, EPI_ISL_2813065, EPI_ISL_2813066, EPI_ISL_2813067, EPI_ISL_2813069, EPI_ISL_2813070, EPI_ISL_2813071, EPI_ISL_2813072, EPI_ISL_2813073, EPI_ISL_2813074, EPI_ISL_2813075, EPI_ISL_2813076, EPI_ISL_2813077, EPI_ISL_2813078, EPI_ISL_2813079, EPI_ISL_2813080, EPI_ISL_2813081, EPI_ISL_2813082, EPI_ISL_2813083, EPI_ISL_2813084, EPI_ISL_2813085, EPI_ISL_2813086, EPI_ISL_2813088, EPI_ISL_2813089, EPI_ISL_2813090, EPI_ISL_2813091, EPI_ISL_2813092, EPI_ISL_2813093, EPI_ISL_2813094, EPI_ISL_2813095, EPI_ISL_2813096, EPI_ISL_2813097, EPI_ISL_2813098, EPI_ISL_2813100, EPI_ISL_2813102, EPI_ISL_2813104, EPI_ISL_2813105, EPI_ISL_2813106, EPI_ISL_2813107, EPI_ISL_2813108, EPI_ISL_2813109, EPI_ISL_2813110, EPI_ISL_2813111, EPI_ISL_2813114, EPI_ISL_2813115, EPI_ISL_2813116, EPI_ISL_2813117, EPI_ISL_2813118, EPI_ISL_2813119, EPI_ISL_2813121, EPI_ISL_2813122, EPI_ISL_2813123, EPI_ISL_2813124, EPI_ISL_2813125, EPI_ISL_2813126, EPI_ISL_2813127, EPI_ISL_2813128, EPI_ISL_2813129, EPI_ISL_2813131, EPI_ISL_2813132, EPI_ISL_2813133, EPI_ISL_2813134, EPI_ISL_2813135, EPI_ISL_2813136, EPI_ISL_2813137, EPI_ISL_2813138, EPI_ISL_2813140, EPI_ISL_2813142, EPI_ISL_2813143, EPI_ISL_2813144, EPI_ISL_2813146, EPI_ISL_2813148, EPI_ISL_2813149, EPI_ISL_2813150, EPI_ISL_2813151, EPI_ISL_2813154, EPI_ISL_2813156, EPI_ISL_2813157, EPI_ISL_2813158, EPI_ISL_2813159, EPI_ISL_2813163, EPI_ISL_2813164, EPI_ISL_2813165, EPI_ISL_2813168, EPI_ISL_2813169, EPI_ISL_2813170, EPI_ISL_2813171, EPI_ISL_2813172, EPI_ISL_2813173, EPI_ISL_2813174, EPI_ISL_2813176, EPI_ISL_2813177, EPI_ISL_2813179, EPI_ISL_2813180, EPI_ISL_2813182, EPI_ISL_2813183, EPI_ISL_2813184, EPI_ISL_2813186, EPI_ISL_2813187, EPI_ISL_2813188, EPI_ISL_2813190, EPI_ISL_2813192, EPI_ISL_2813193, EPI_ISL_2813194, EPI_ISL_2813195, EPI_ISL_2813196, EPI_ISL_2813197, EPI_ISL_2813199, EPI_ISL_2813200, EPI_ISL_2813201, EPI_ISL_2813202, EPI_ISL_2813204, EPI_ISL_2813205, EPI_ISL_2813206, EPI_ISL_2813207, EPI_ISL_2813208, EPI_ISL_2813209, EPI_ISL_2813210, EPI_ISL_2813211, EPI_ISL_2813212, EPI_ISL_2813213, EPI_ISL_2813214, EPI_ISL_2813215, EPI_ISL_2813216, EPI_ISL_2813219, EPI_ISL_2813220, EPI_ISL_2813222, EPI_ISL_2813223, EPI_ISL_2813224, EPI_ISL_2813225, EPI_ISL_2813226, EPI_ISL_2813227, EPI_ISL_2813228, EPI_ISL_2813229, EPI_ISL_2813230, EPI_ISL_2813231, EPI_ISL_2813232, EPI_ISL_2813233, EPI_ISL_2813236, EPI_ISL_2813237, EPI_ISL_2813238, EPI_ISL_2813239, EPI_ISL_2813241, EPI_ISL_2813242, EPI_ISL_2813244, EPI_ISL_2813245, EPI_ISL_2813246, EPI_ISL_2813247, EPI_ISL_2813248, EPI_ISL_2813251, EPI_ISL_2813252, EPI_ISL_2813253, EPI_ISL_2813254, EPI_ISL_2813257, EPI_ISL_2813259, EPI_ISL_2813260, EPI_ISL_2813261, EPI_ISL_2813262, EPI_ISL_2813263, EPI_ISL_2813264, EPI_ISL_2813266, EPI_ISL_2813267, EPI_ISL_2813268, EPI_ISL_2813269, EPI_ISL_2813270, EPI_ISL_2813271, EPI_ISL_2813272, EPI_ISL_2813273, EPI_ISL_2813274, EPI_ISL_2813275, EPI_ISL_2813276, EPI_ISL_2813277, EPI_ISL_2813278, EPI_ISL_2813279, EPI_ISL_2813281, EPI_ISL_2813282, EPI_ISL_2813283, EPI_ISL_2813284, EPI_ISL_2813285, EPI_ISL_2813288, EPI_ISL_2813289, EPI_ISL_2813290, EPI_ISL_2813291, EPI_ISL_2813292, EPI_ISL_2813293, EPI_ISL_2813294, EPI_ISL_2813295, EPI_ISL_2813297, EPI_ISL_2813298, EPI_ISL_2813300, EPI_ISL_2813301, EPI_ISL_2813302, EPI_ISL_2813303, EPI_ISL_2813304, EPI_ISL_2813305, EPI_ISL_2813306, EPI_ISL_2813307, EPI_ISL_2813308, EPI_ISL_2813309, EPI_ISL_2813310, EPI_ISL_2813311, EPI_ISL_2813312, EPI_ISL_2813313, EPI_ISL_2813314, EPI_ISL_2813315, EPI_ISL_2813316, EPI_ISL_2813661, EPI_ISL_2813662, EPI_ISL_2813663, EPI_ISL_2813664, EPI_ISL_2813665, EPI_ISL_2813666, EPI_ISL_2813667, EPI_ISL_2813668, EPI_ISL_2813669, EPI_ISL_2813671, EPI_ISL_2813673, EPI_ISL_2813674, EPI_ISL_2813675, EPI_ISL_2813676, EPI_ISL_2813677, EPI_ISL_2813678, EPI_ISL_2813679, EPI_ISL_2813680, EPI_ISL_2813682, EPI_ISL_2813683, EPI_ISL_2813686, EPI_ISL_2813687, EPI_ISL_2813688, EPI_ISL_2813690, EPI_ISL_2813691, EPI_ISL_2813694, EPI_ISL_2813696, EPI_ISL_2813697, EPI_ISL_2813699, EPI_ISL_2813700, EPI_ISL_2813702, | | | |

EPI_ISL_2813705, EPI_ISL_2813707, EPI_ISL_2813709, EPI_ISL_2813710, EPI_ISL_2813711, EPI_ISL_2813712, EPI_ISL_2813713, EPI_ISL_2813718, EPI_ISL_2813719, EPI_ISL_2813722, EPI_ISL_2813723, EPI_ISL_2813725, EPI_ISL_2813726, EPI_ISL_2813727, EPI_ISL_2813729, EPI_ISL_2813731, EPI_ISL_2813734, EPI_ISL_2813735, EPI_ISL_2813736, EPI_ISL_2813738, EPI_ISL_2813739, EPI_ISL_2813741, EPI_ISL_2813742, EPI_ISL_2813743, EPI_ISL_2813744, EPI_ISL_2813745, EPI_ISL_2813746, EPI_ISL_2813747, EPI_ISL_2813748, EPI_ISL_2813749, EPI_ISL_2813750, EPI_ISL_2813752, EPI_ISL_2813754, EPI_ISL_2813760, EPI_ISL_2813762, EPI_ISL_2813765, EPI_ISL_2813767, EPI_ISL_2813768, EPI_ISL_2813771, EPI_ISL_2813774, EPI_ISL_2813775, EPI_ISL_2813776, EPI_ISL_2813777, EPI_ISL_2813778, EPI_ISL_2813779, EPI_ISL_2813780, EPI_ISL_2813781, EPI_ISL_2813782, EPI_ISL_2813783, EPI_ISL_2813785, EPI_ISL_2813786, EPI_ISL_2813787, EPI_ISL_2813788, EPI_ISL_2813792, EPI_ISL_2813796, EPI_ISL_2813798, EPI_ISL_2813799, EPI_ISL_2813800, EPI_ISL_2813804, EPI_ISL_2813805, EPI_ISL_2813806, EPI_ISL_2813807, EPI_ISL_2813808, EPI_ISL_2813810, EPI_ISL_2813812, EPI_ISL_2813813, EPI_ISL_2813814, EPI_ISL_2813815, EPI_ISL_2813816, EPI_ISL_2813817, EPI_ISL_2813818, EPI_ISL_2813819, EPI_ISL_2813820, EPI_ISL_2813821, EPI_ISL_2813822, EPI_ISL_2813823, EPI_ISL_2813824, EPI_ISL_2813826, EPI_ISL_2813827, EPI_ISL_2813828, EPI_ISL_2813829, EPI_ISL_2813832, EPI_ISL_2813834, EPI_ISL_2813835, EPI_ISL_2813836, EPI_ISL_2813840, EPI_ISL_2813841, EPI_ISL_2813842, EPI_ISL_2813843, EPI_ISL_2813844, EPI_ISL_2813846, EPI_ISL_2813847, EPI_ISL_2813849, EPI_ISL_2813852, EPI_ISL_2813853, EPI_ISL_2813854, EPI_ISL_2813855, EPI_ISL_2813856, EPI_ISL_2813858, EPI_ISL_2813859, EPI_ISL_2813860, EPI_ISL_2813862, EPI_ISL_2813863, EPI_ISL_2813864, EPI_ISL_2813865, EPI_ISL_2813866, EPI_ISL_2813868, EPI_ISL_2813869, EPI_ISL_2813870, EPI_ISL_2813873, EPI_ISL_2813874, EPI_ISL_2813875, EPI_ISL_2813876, EPI_ISL_2813877, EPI_ISL_2813878, EPI_ISL_2813879, EPI_ISL_2813880, EPI_ISL_2813881, EPI_ISL_2813883, EPI_ISL_2813885, EPI_ISL_2813886, EPI_ISL_2813887, EPI_ISL_2813889, EPI_ISL_2813890, EPI_ISL_2813891, EPI_ISL_2813892, EPI_ISL_2813893, EPI_ISL_2813896, EPI_ISL_2813897, EPI_ISL_2813898, EPI_ISL_2813900, EPI_ISL_2813901, EPI_ISL_2813902, EPI_ISL_2813903, EPI_ISL_2813904, EPI_ISL_2813905, EPI_ISL_2813907, EPI_ISL_2813908, EPI_ISL_2813909, EPI_ISL_2813911, EPI_ISL_2813912, EPI_ISL_2813913, EPI_ISL_2813914, EPI_ISL_2813915, EPI_ISL_2813916, EPI_ISL_2813917, EPI_ISL_2813918, EPI_ISL_2813919, EPI_ISL_2813920, EPI_ISL_2813921, EPI_ISL_2813922, EPI_ISL_2813923, EPI_ISL_2813924, EPI_ISL_2813925, EPI_ISL_2813926, EPI_ISL_2813927, EPI_ISL_2813929, EPI_ISL_2813930, EPI_ISL_2813931, EPI_ISL_2813932, EPI_ISL_2813933, EPI_ISL_2813934, EPI_ISL_2813935, EPI_ISL_2813936, EPI_ISL_2813938, EPI_ISL_2813939, EPI_ISL_2813940, EPI_ISL_2813942, EPI_ISL_2813943, EPI_ISL_2813944, EPI_ISL_2813945, EPI_ISL_2813947, EPI_ISL_2813950, EPI_ISL_2813951, EPI_ISL_2813952, EPI_ISL_2813954, EPI_ISL_2813957, EPI_ISL_2813958, EPI_ISL_2813959, EPI_ISL_2813960, EPI_ISL_2813961, EPI_ISL_2813962, EPI_ISL_2813963, EPI_ISL_2813964, EPI_ISL_2813965, EPI_ISL_2813967, EPI_ISL_2813968, EPI_ISL_2813969, EPI_ISL_2813970, EPI_ISL_2813971, EPI_ISL_2813972, EPI_ISL_2813973, EPI_ISL_2813974, EPI_ISL_2813975, EPI_ISL_2813976, EPI_ISL_2813981, EPI_ISL_2813982, EPI_ISL_2813983, EPI_ISL_2813984, EPI_ISL_2813985, EPI_ISL_2813986, EPI_ISL_2813987, EPI_ISL_2813988, EPI_ISL_2813989, EPI_ISL_2813990, EPI_ISL_2813991, EPI_ISL_2813992, EPI_ISL_2813993, EPI_ISL_2813994, EPI_ISL_2813996, EPI_ISL_2813997, EPI_ISL_2813998, EPI_ISL_2813999, EPI_ISL_2814002, EPI_ISL_2814005, EPI_ISL_2814006, EPI_ISL_2814011, EPI_ISL_2814017, EPI_ISL_2814018, EPI_ISL_2814021, EPI_ISL_2814022, EPI_ISL_2814024, EPI_ISL_2814025, EPI_ISL_2814026, EPI_ISL_2814027, EPI_ISL_2814030, EPI_ISL_2814033, EPI_ISL_2814035, EPI_ISL_2814039, EPI_ISL_2814041, EPI_ISL_2814042, EPI_ISL_2814044, EPI_ISL_2814045, EPI_ISL_2814047, EPI_ISL_2814051, EPI_ISL_2814054, EPI_ISL_2814056, EPI_ISL_2814057, EPI_ISL_2814058, EPI_ISL_2814059, EPI_ISL_2814061, EPI_ISL_2814066, EPI_ISL_2814067, EPI_ISL_2814071, EPI_ISL_2814072, EPI_ISL_2814073, EPI_ISL_2814077, EPI_ISL_2814078, EPI_ISL_2814079, EPI_ISL_2814081, EPI_ISL_2814082, EPI_ISL_2814083, EPI_ISL_2814084, EPI_ISL_2814086, EPI_ISL_2814087, EPI_ISL_2814090, EPI_ISL_2814091, EPI_ISL_2814093, EPI_ISL_2814095, EPI_ISL_2814105, EPI_ISL_2814106, EPI_ISL_2814107, EPI_ISL_2814108, EPI_ISL_2814109, EPI_ISL_2814110, EPI_ISL_2814113, EPI_ISL_2814114, EPI_ISL_2814121, EPI_ISL_2814122, EPI_ISL_2814123, EPI_ISL_2814127, EPI_ISL_2814128, EPI_ISL_2814135, EPI_ISL_2814136, EPI_ISL_2814138, EPI_ISL_2814140, EPI_ISL_2814145, EPI_ISL_2814151, EPI_ISL_2814155, EPI_ISL_2814158, EPI_ISL_2814159, EPI_ISL_2814163, EPI_ISL_2814164, EPI_ISL_2814165, EPI_ISL_2814170, EPI_ISL_2814171, EPI_ISL_2814172, EPI_ISL_2814176, EPI_ISL_2814179, EPI_ISL_2814184, EPI_ISL_2814190, EPI_ISL_2814195, EPI_ISL_2814196, EPI_ISL_2814197, EPI_ISL_2814198, EPI_ISL_2814199, EPI_ISL_2814201, EPI_ISL_2814208, EPI_ISL_2814209, EPI_ISL_2814212, EPI_ISL_2814214, EPI_ISL_2814217, EPI_ISL_2814221, EPI_ISL_2814222, EPI_ISL_2814223, EPI_ISL_2814226, EPI_ISL_2814768, EPI_ISL_2829675, EPI_ISL_2829689, EPI_ISL_2829715, EPI_ISL_2829931, EPI_ISL_2829975, EPI_ISL_2830069, EPI_ISL_2830098, EPI_ISL_2830124, EPI_ISL_2830137, EPI_ISL_2830346, EPI_ISL_2830363, EPI_ISL_2830399, EPI_ISL_2830412, EPI_ISL_2830430, EPI_ISL_2830442, EPI_ISL_2830473, EPI_ISL_2830486, EPI_ISL_2830489, EPI_ISL_2830491, EPI_ISL_2830517, EPI_ISL_2830525, EPI_ISL_2830531, EPI_ISL_2830533, EPI_ISL_2830535, EPI_ISL_2830553, EPI_ISL_2830565, EPI_ISL_2830567, EPI_ISL_2830576, EPI_ISL_2830580, EPI_ISL_2830594, EPI_ISL_2830606, EPI_ISL_2830609, EPI_ISL_2830612, EPI_ISL_2830616, EPI_ISL_2830619, EPI_ISL_2830626, EPI_ISL_2830633, EPI_ISL_2830636, EPI_ISL_2830650, EPI_ISL_2830654, EPI_ISL_2830655, EPI_ISL_2830656, EPI_ISL_2830662, EPI_ISL_2830665, EPI_ISL_2830670, EPI_ISL_2830683, EPI_ISL_2830684, EPI_ISL_2830686, EPI_ISL_2830687, EPI_ISL_2830688, EPI_ISL_2830689, EPI_ISL_2830690, EPI_ISL_2830695, EPI_ISL_2830698, EPI_ISL_2830700, EPI_ISL_2830701, EPI_ISL_2830703, EPI_ISL_2830704, EPI_ISL_2830706, EPI_ISL_2830709, EPI_ISL_2830711, EPI_ISL_2830714, EPI_ISL_2830724, EPI_ISL_2830725, EPI_ISL_2830726, EPI_ISL_2830732, EPI_ISL_2830735, EPI_ISL_2830737, EPI_ISL_2830741, EPI_ISL_2830745, EPI_ISL_2830747, EPI_ISL_2830750, EPI_ISL_2830755, EPI_ISL_2830757, EPI_ISL_2830758, EPI_ISL_2830759, EPI_ISL_2830760, EPI_ISL_2830762, EPI_ISL_2830763, EPI_ISL_2830770, EPI_ISL_2830773, EPI_ISL_2830776, EPI_ISL_2830778, EPI_ISL_2830779, EPI_ISL_2830780, EPI_ISL_2830784, EPI_ISL_2830787, EPI_ISL_2830795, EPI_ISL_2830796, EPI_ISL_2830798, EPI_ISL_2830802, EPI_ISL_2830803, EPI_ISL_2830805, EPI_ISL_2830806, EPI_ISL_2830808, EPI_ISL_2830810, EPI_ISL_2830817, EPI_ISL_2830831, EPI_ISL_2830832, EPI_ISL_2830838, EPI_ISL_2830840, EPI_ISL_2830843, EPI_ISL_2830849, EPI_ISL_2831046, EPI_ISL_2831053, EPI_ISL_2831055, EPI_ISL_2831059, EPI_ISL_2831064, EPI_ISL_2831066, EPI_ISL_2831073, EPI_ISL_2831077, EPI_ISL_2831081, EPI_ISL_2831086, EPI_ISL_2831093, EPI_ISL_2831097, EPI_ISL_2831103, EPI_ISL_2831105, EPI_ISL_2831109, EPI_ISL_2831110, EPI_ISL_2831112, EPI_ISL_2831115, EPI_ISL_2831120, EPI_ISL_2831123, EPI_ISL_2831126, EPI_ISL_2831129, EPI_ISL_2831132, EPI_ISL_2831143, EPI_ISL_2831146, EPI_ISL_2831147, EPI_ISL_2831155, EPI_ISL_2831156, EPI_ISL_2831164, EPI_ISL_2831175, EPI_ISL_2831178, EPI_ISL_2831183, EPI_ISL_2831184, EPI_ISL_2831185, EPI_ISL_2831186, EPI_ISL_2831191, EPI_ISL_2831194, EPI_ISL_2831195, EPI_ISL_2831197, EPI_ISL_2831202, EPI_ISL_2831207, EPI_ISL_2831216, EPI_ISL_2831217, EPI_ISL_2831219, EPI_ISL_2831224, EPI_ISL_2831226, EPI_ISL_2831232, EPI_ISL_2831235, EPI_ISL_2831236, EPI_ISL_2831238, EPI_ISL_2831244, EPI_ISL_2831247, EPI_ISL_2831249, EPI_ISL_2831257, EPI_ISL_2831266, EPI_ISL_2831274, EPI_ISL_2831279, EPI_ISL_2831280, EPI_ISL_2831287, EPI_ISL_2831300, EPI_ISL_2831301, EPI_ISL_2831302, EPI_ISL_2831306, EPI_ISL_2831309, EPI_ISL_2831310, EPI_ISL_2831313, EPI_ISL_2831315, EPI_ISL_2831320, EPI_ISL_2831784, EPI_ISL_2831791, EPI_ISL_2831792, EPI_ISL_2831794, EPI_ISL_2831801, EPI_ISL_2831815, EPI_ISL_2831821, EPI_ISL_2831874, EPI_ISL_2831880, EPI_ISL_2831900, EPI_ISL_2831903, EPI_ISL_2831941, EPI_ISL_2831944, EPI_ISL_2831953, EPI_ISL_2831963, EPI_ISL_2831972, EPI_ISL_2831974, EPI_ISL_2831976, EPI_ISL_2832000, EPI_ISL_2832003, EPI_ISL_2832008, EPI_ISL_2832027, EPI_ISL_2832036, EPI_ISL_2832051, EPI_ISL_2832057, EPI_ISL_2832083, EPI_ISL_2832088, EPI_ISL_2832092, EPI_ISL_2846172, EPI_ISL_2846223, EPI_ISL_2846839, EPI_ISL_2847738, EPI_ISL_2847879, EPI_ISL_2847999, EPI_ISL_2848087, EPI_ISL_2848112, EPI_ISL_2848140, EPI_ISL_2848145, EPI_ISL_2848169, EPI_ISL_2848189, EPI_ISL_2848213, EPI_ISL_2848294, EPI_ISL_2848373, EPI_ISL_2848436, EPI_ISL_2848457, EPI_ISL_2848458, EPI_ISL_2848494, EPI_ISL_2848498, EPI_ISL_2848507, EPI_ISL_2848530, EPI_ISL_2848538, EPI_ISL_2848558, EPI_ISL_2848564, EPI_ISL_2848569, EPI_ISL_2848572, EPI_ISL_2848602, EPI_ISL_2848628, EPI_ISL_2848762, EPI_ISL_2848959, EPI_ISL_2848964, EPI_ISL_2850406, EPI_ISL_2850439, EPI_ISL_2850504, EPI_ISL_2850586, EPI_ISL_2851539, EPI_ISL_2851563, EPI_ISL_2851634, EPI_ISL_2851807, EPI_ISL_2852855, EPI_ISL_2853019, EPI_ISL_2865065, EPI_ISL_2865092, EPI_ISL_2865126, EPI_ISL_2865194, EPI_ISL_2865246, EPI_ISL_2865251, EPI_ISL_2865266, EPI_ISL_2865278, EPI_ISL_2865295, EPI_ISL_2865315, EPI_ISL_2865323, EPI_ISL_2865335, EPI_ISL_2865337, EPI_ISL_2867384, EPI_ISL_2867659, EPI_ISL_2910648, EPI_ISL_2910652, EPI_ISL_2910670, EPI_ISL_2910705, EPI_ISL_2910719, EPI_ISL_2910726, EPI_ISL_2910753, EPI_ISL_2910754, EPI_ISL_2910764, EPI_ISL_2910766, EPI_ISL_2910821

| | | | |
|---|---|---|---|
| see above | Lighthouse Lab in Milton Keynes | Wellcome Sanger Institute for the COVID-19 Genomics UK (COG-UK) Consortium | Cordelia Langford; David K. Jackson; Dominic Kwiatkowski; Ewan Harrison; Ian Johnston; Jeffrey Barrett; John Sillitoe on behalf of the Wellcome Sanger Institute COVID-19 Surveillance Team; Roberto Amato; Sonia Goncalves; The Lighthouse Lab in Milton Keynes and Alex Alderton |

EPI_ISL_2779869, EPI_ISL_2779870, EPI_ISL_2779872, EPI_ISL_2779876, EPI_ISL_2779889, EPI_ISL_2779891, EPI_ISL_2779893, EPI_ISL_2779896, EPI_ISL_2779897, EPI_ISL_2779898, EPI_ISL_2779901, EPI_ISL_2779903, EPI_ISL_2779906, EPI_ISL_2779909, EPI_ISL_2779911, EPI_ISL_2779914, EPI_ISL_2779923, EPI_ISL_2779927, EPI_ISL_2779931, EPI_ISL_2779939, EPI_ISL_2779946, EPI_ISL_2779949, EPI_ISL_2779951, EPI_ISL_2779952, EPI_ISL_2779955, EPI_ISL_2779957, EPI_ISL_2779960, EPI_ISL_2779961, EPI_ISL_2779964, EPI_ISL_2779970, EPI_ISL_2779971, EPI_ISL_2779972, EPI_ISL_2779976, EPI_ISL_2779978, EPI_ISL_2779982, EPI_ISL_2779983, EPI_ISL_2779991, EPI_ISL_2779994, EPI_ISL_2779996, EPI_ISL_2779999, EPI_ISL_2780006, EPI_ISL_2780008, EPI_ISL_2780011, EPI_ISL_2780013, EPI_ISL_2780015, EPI_ISL_2780023, EPI_ISL_2780028, EPI_ISL_2780035, EPI_ISL_2780040, EPI_ISL_2780048, EPI_ISL_2780050, EPI_ISL_2780054, EPI_ISL_2780058, EPI_ISL_2780061, EPI_ISL_2780064, EPI_ISL_2780083, EPI_ISL_2780086, EPI_ISL_2780094, EPI_ISL_2780097, EPI_ISL_2780098, EPI_ISL_2780102, EPI_ISL_2780107, EPI_ISL_2780108, EPI_ISL_2780122, EPI_ISL_2780123, EPI_ISL_2803924, EPI_ISL_2803926, EPI_ISL_2803928, EPI_ISL_2803929, EPI_ISL_2803930, EPI_ISL_2803936, EPI_ISL_2803937, EPI_ISL_2803940, EPI_ISL_2803941, EPI_ISL_2803942, EPI_ISL_2803943, EPI_ISL_2803944, EPI_ISL_2803945, EPI_ISL_2803946, EPI_ISL_2803947, EPI_ISL_2803950, EPI_ISL_2803951, EPI_ISL_2803952, EPI_ISL_2803954, EPI_ISL_2803956, EPI_ISL_2803957, EPI_ISL_2803958, EPI_ISL_2803960, EPI_ISL_2803963, EPI_ISL_2803964, EPI_ISL_2803965, EPI_ISL_2803966, EPI_ISL_2803970, EPI_ISL_2803971, EPI_ISL_2803976, EPI_ISL_2803977, EPI_ISL_2803979, EPI_ISL_2803980, EPI_ISL_2803983, EPI_ISL_2803986, EPI_ISL_2803987, EPI_ISL_2803988, EPI_ISL_2803989, EPI_ISL_2803990, EPI_ISL_2803991, EPI_ISL_2803994, EPI_ISL_2803995, EPI_ISL_2804000, EPI_ISL_2804003, EPI_ISL_2804006, EPI_ISL_2804008, EPI_ISL_2804014, EPI_ISL_2804016, EPI_ISL_2804018, EPI_ISL_2804019, EPI_ISL_2804023, EPI_ISL_2804024, EPI_ISL_2804025, EPI_ISL_2804026, EPI_ISL_2804027, EPI_ISL_2804028, EPI_ISL_2804029, EPI_ISL_2804030, EPI_ISL_2804032, EPI_ISL_2804033, EPI_ISL_2804034, EPI_ISL_2804035, EPI_ISL_2804037, EPI_ISL_2804038, EPI_ISL_2804041, EPI_ISL_2804042, EPI_ISL_2804048, EPI_ISL_2804050, EPI_ISL_2804051, EPI_ISL_2804052, EPI_ISL_2804053, EPI_ISL_2804055, EPI_ISL_2804056, EPI_ISL_2804057, EPI_ISL_2804061, EPI_ISL_2804063, EPI_ISL_2804064, EPI_ISL_2804066, EPI_ISL_2804069, EPI_ISL_2804073, EPI_ISL_2804074, EPI_ISL_2804075, EPI_ISL_2804078, EPI_ISL_2804079, EPI_ISL_2804080, EPI_ISL_2804088, EPI_ISL_2804089, EPI_ISL_2804091, EPI_ISL_2804092, EPI_ISL_2804094, EPI_ISL_2804095, EPI_ISL_2804096, EPI_ISL_2804097, EPI_ISL_2804098, EPI_ISL_2804100, EPI_ISL_2804103, EPI_ISL_2804105, EPI_ISL_2804109, EPI_ISL_2804110, EPI_ISL_2804112, EPI_ISL_2804113, EPI_ISL_2804116, EPI_ISL_2804117, EPI_ISL_2804120, EPI_ISL_2804121, EPI_ISL_2804122, EPI_ISL_2804124, EPI_ISL_2804127, EPI_ISL_2804128, EPI_ISL_2804129, EPI_ISL_2804130, EPI_ISL_2804131, EPI_ISL_2804133, EPI_ISL_2804134, EPI_ISL_2804136, EPI_ISL_2804138, EPI_ISL_2804139, EPI_ISL_2804140, EPI_ISL_2804144, EPI_ISL_2804145, EPI_ISL_2804150, EPI_ISL_2804153, EPI_ISL_2804154, EPI_ISL_2804156, EPI_ISL_2804157, EPI_ISL_2804158, EPI_ISL_2804159, EPI_ISL_2804161, EPI_ISL_2804162, EPI_ISL_2804163, EPI_ISL_2804165, EPI_ISL_2804168, EPI_ISL_2804169, EPI_ISL_2804170, EPI_ISL_2804174, EPI_ISL_2804175, EPI_ISL_2804178, EPI_ISL_2804179, EPI_ISL_2804180, EPI_ISL_2804184, EPI_ISL_2804185, EPI_ISL_2804186, EPI_ISL_2804188, EPI_ISL_2804190, EPI_ISL_2804192, EPI_ISL_2804193, EPI_ISL_2804197, EPI_ISL_2804200, EPI_ISL_2804201, EPI_ISL_2804203, EPI_ISL_2804205, EPI_ISL_2804206, EPI_ISL_2804209, EPI_ISL_2804213, EPI_ISL_2804217, EPI_ISL_2804220, EPI_ISL_2804221, EPI_ISL_2804222, EPI_ISL_2804223, EPI_ISL_2804224, EPI_ISL_2804227, EPI_ISL_2804228, EPI_ISL_2804232, EPI_ISL_2804233, EPI_ISL_2804235, EPI_ISL_2804236, EPI_ISL_2804237, EPI_ISL_2804238, EPI_ISL_2804239, EPI_ISL_2804240, EPI_ISL_2804241, EPI_ISL_2804242, EPI_ISL_2804243, EPI_ISL_2804247, EPI_ISL_2804250, EPI_ISL_2804251, EPI_ISL_2804252, EPI_ISL_2804253, EPI_ISL_2804255, EPI_ISL_2804262, EPI_ISL_2804263, EPI_ISL_2804269, EPI_ISL_2804273, EPI_ISL_2804275, EPI_ISL_2804276, EPI_ISL_2804290, EPI_ISL_2804291, EPI_ISL_2804293, EPI_ISL_2804301, EPI_ISL_2804315, EPI_ISL_2804321, EPI_ISL_2804322, EPI_ISL_2804332, EPI_ISL_2804335, EPI_ISL_2804339, EPI_ISL_2804342, EPI_ISL_2804366, EPI_ISL_2804370, EPI_ISL_2804373, EPI_ISL_2804380, EPI_ISL_2804392, EPI_ISL_2804400, EPI_ISL_2804410, EPI_ISL_2804414, EPI_ISL_2804421, EPI_ISL_2804423, EPI_ISL_2804425, EPI_ISL_2804427, EPI_ISL_2804429, EPI_ISL_2804434, EPI_ISL_2804439, EPI_ISL_2804440, EPI_ISL_2804442, EPI_ISL_2804453, EPI_ISL_2804455, EPI_ISL_2804457, EPI_ISL_2804463, EPI_ISL_2804464, EPI_ISL_2804466, EPI_ISL_2804469, EPI_ISL_2804471, EPI_ISL_2804475, EPI_ISL_2804479, EPI_ISL_2804507, EPI_ISL_2804515, EPI_ISL_2804533, EPI_ISL_2804534, EPI_ISL_2804535, EPI_ISL_2804543, EPI_ISL_2804545, EPI_ISL_2804560, EPI_ISL_2804570, EPI_ISL_2804575, EPI_ISL_2804577, EPI_ISL_2804583, EPI_ISL_2804590, EPI_ISL_2804600, EPI_ISL_2804601, EPI_ISL_2804602, EPI_ISL_2805347, EPI_ISL_2805350, EPI_ISL_2805351, EPI_ISL_2805352, EPI_ISL_2805353, EPI_ISL_2805354, EPI_ISL_2805355, EPI_ISL_2805357, EPI_ISL_2805358, EPI_ISL_2805361, EPI_ISL_2805362, EPI_ISL_2805369, EPI_ISL_2805374, EPI_ISL_2805376, EPI_ISL_2805377, EPI_ISL_2805379, EPI_ISL_2805380, EPI_ISL_2805381, EPI_ISL_2805382, EPI_ISL_2805383, EPI_ISL_2805387, EPI_ISL_2805389, EPI_ISL_2805390, EPI_ISL_2805392, EPI_ISL_2805394, EPI_ISL_2805402, EPI_ISL_2805404, EPI_ISL_2805408, EPI_ISL_2805409, EPI_ISL_2805410, EPI_ISL_2805416, EPI_ISL_2805417, EPI_ISL_2805422, EPI_ISL_2805424, EPI_ISL_2805425, EPI_ISL_2805428, EPI_ISL_2805430, EPI_ISL_2805431, EPI_ISL_2805432, EPI_ISL_2805433, EPI_ISL_2805434, EPI_ISL_2805439, EPI_ISL_2805441, EPI_ISL_2805444, EPI_ISL_2805447, EPI_ISL_2805450, EPI_ISL_2805451, EPI_ISL_2805453, EPI_ISL_2805454, EPI_ISL_2805455, EPI_ISL_2805457, EPI_ISL_2805459, EPI_ISL_2805463, EPI_ISL_2805464, EPI_ISL_2805466, EPI_ISL_2805468, EPI_ISL_2805469, EPI_ISL_2805474, EPI_ISL_2805475, EPI_ISL_2805477, EPI_ISL_2805479, EPI_ISL_2805480, EPI_ISL_2805483, EPI_ISL_2805484, EPI_ISL_2805486, EPI_ISL_2805487, EPI_ISL_2805488, EPI_ISL_2805490, EPI_ISL_2805491, EPI_ISL_2805493, EPI_ISL_2805494, EPI_ISL_2805496, EPI_ISL_2805497, EPI_ISL_2805502, EPI_ISL_2805506, EPI_ISL_2805507, EPI_ISL_2805509, EPI_ISL_2805510, EPI_ISL_2805511, EPI_ISL_2805512, EPI_ISL_2805513, EPI_ISL_2805514, EPI_ISL_2805515, EPI_ISL_2805517, EPI_ISL_2805518, EPI_ISL_2805521, EPI_ISL_2805523, EPI_ISL_2805528, EPI_ISL_2805529, EPI_ISL_2805532, EPI_ISL_2805533, EPI_ISL_2805534, EPI_ISL_2805536, EPI_ISL_2805538, EPI_ISL_2805541, EPI_ISL_2805542, EPI_ISL_2805544, EPI_ISL_2805545, EPI_ISL_2805546, EPI_ISL_2805547, EPI_ISL_2805549, EPI_ISL_2805550, EPI_ISL_2805551, EPI_ISL_2805553, EPI_ISL_2805554, EPI_ISL_2805555, EPI_ISL_2805556, EPI_ISL_2805557, EPI_ISL_2805559, EPI_ISL_2805562, EPI_ISL_2805566, EPI_ISL_2805568, EPI_ISL_2805570, EPI_ISL_2805571, EPI_ISL_2805575, EPI_ISL_2805576, EPI_ISL_2805577, EPI_ISL_2805578, EPI_ISL_2805579, EPI_ISL_2805582, EPI_ISL_2805584, EPI_ISL_2805587, EPI_ISL_2805591, EPI_ISL_2805593, EPI_ISL_2805594, EPI_ISL_2805595, EPI_ISL_2805596, EPI_ISL_2805599, EPI_ISL_2805600, EPI_ISL_2805603, EPI_ISL_2805604, EPI_ISL_2805605, EPI_ISL_2805609, EPI_ISL_2805613, EPI_ISL_2805618, EPI_ISL_2805619, EPI_ISL_2805620, EPI_ISL_2805621, EPI_ISL_2805623, EPI_ISL_2805625, EPI_ISL_2805626, EPI_ISL_2805627, EPI_ISL_2805628, EPI_ISL_2805630, EPI_ISL_2805633, EPI_ISL_2805635, EPI_ISL_2805637, EPI_ISL_2805639, EPI_ISL_2805641, EPI_ISL_2805643, EPI_ISL_2805644, EPI_ISL_2805648, EPI_ISL_2805651, EPI_ISL_2805652, EPI_ISL_2805656, EPI_ISL_2805658, EPI_ISL_2805659, EPI_ISL_2805661, EPI_ISL_2805662, EPI_ISL_2805663, EPI_ISL_2805667, EPI_ISL_2805670, EPI_ISL_2805673, EPI_ISL_2805674, EPI_ISL_2805676, EPI_ISL_2805677, EPI_ISL_2805678, EPI_ISL_2805681, EPI_ISL_2805684, EPI_ISL_2805691, EPI_ISL_2805692, EPI_ISL_2805696, EPI_ISL_2805701, EPI_ISL_2805705, EPI_ISL_2805706, EPI_ISL_2805708, EPI_ISL_2805716, EPI_ISL_2805719, EPI_ISL_2805725, EPI_ISL_2805734, EPI_ISL_2805735, EPI_ISL_2805739, EPI_ISL_2805741, EPI_ISL_2805744, EPI_ISL_2805746, EPI_ISL_2805747, EPI_ISL_2805750, EPI_ISL_2805751, EPI_ISL_2805752, EPI_ISL_2805756, EPI_ISL_2805757, EPI_ISL_2805762, EPI_ISL_2805770, EPI_ISL_2805772, EPI_ISL_2805774, EPI_ISL_2805781, EPI_ISL_2805785, EPI_ISL_2805786, EPI_ISL_2805789, EPI_ISL_2805790, EPI_ISL_2805800, EPI_ISL_2805805, EPI_ISL_2805807, EPI_ISL_2805817, EPI_ISL_2805823, EPI_ISL_2805824, EPI_ISL_2805828, EPI_ISL_2805842, EPI_ISL_2805844, EPI_ISL_2805848, EPI_ISL_2805851, EPI_ISL_2805852, EPI_ISL_2805853, EPI_ISL_2805855, EPI_ISL_2805856, EPI_ISL_2805865, EPI_ISL_2805868, EPI_ISL_2805869, EPI_ISL_2805870, EPI_ISL_2805873, EPI_ISL_2805874, EPI_ISL_2805881, EPI_ISL_2805882, EPI_ISL_2805886, EPI_ISL_2805889, EPI_ISL_2805890, EPI_ISL_2805891, EPI_ISL_2805892, EPI_ISL_2805893, EPI_ISL_2805902, EPI_ISL_2805903, EPI_ISL_2805908, EPI_ISL_2805912, EPI_ISL_2805917, EPI_ISL_2805918, EPI_ISL_2805920, EPI_ISL_2805925, EPI_ISL_2805926, EPI_ISL_2805934, EPI_ISL_2805938, EPI_ISL_2805941, EPI_ISL_2805944, EPI_ISL_2805946, EPI_ISL_2805954, EPI_ISL_2805955, EPI_ISL_2805965, EPI_ISL_2805975, EPI_ISL_2805981, EPI_ISL_2805982, EPI_ISL_2805984, EPI_ISL_2805986, EPI_ISL_2805990, EPI_ISL_2806444, EPI_ISL_2806459, EPI_ISL_2806462, EPI_ISL_2806467, EPI_ISL_2806470, EPI_ISL_2806472, EPI_ISL_2806479, EPI_ISL_2806481, EPI_ISL_2806482, EPI_ISL_2806486, EPI_ISL_2806492, EPI_ISL_2806510, EPI_ISL_2806517, EPI_ISL_2806521, EPI_ISL_2806523, EPI_ISL_2806524, EPI_ISL_2806534, EPI_ISL_2806541, EPI_ISL_2806543, EPI_ISL_2806544, EPI_ISL_2806548, EPI_ISL_2806554, EPI_ISL_2806559, EPI_ISL_2806561, EPI_ISL_2806562, EPI_ISL_2806564, EPI_ISL_2806570, EPI_ISL_2806577, EPI_ISL_2806583, EPI_ISL_2806586, EPI_ISL_2806593, EPI_ISL_2806594, EPI_ISL_2806595, EPI_ISL_2806606, EPI_ISL_2806612, EPI_ISL_2806618, EPI_ISL_2806624, EPI_ISL_2806628, EPI_ISL_2806636, EPI_ISL_2806639, EPI_ISL_2806644, EPI_ISL_2806651, EPI_ISL_2806652, EPI_ISL_2806658, EPI_ISL_2806659, EPI_ISL_2806662, EPI_ISL_2806663, EPI_ISL_2806665, EPI_ISL_2806671, EPI_ISL_2806676, EPI_ISL_2806687, EPI_ISL_2806690, EPI_ISL_2806709, EPI_ISL_2806710, EPI_ISL_2806713, EPI_ISL_2806720, EPI_ISL_2806732, EPI_ISL_2806734, EPI_ISL_2806742, EPI_ISL_2806746, EPI_ISL_2806749, EPI_ISL_2806757, EPI_ISL_2806764, EPI_ISL_2806769, EPI_ISL_2806771, EPI_ISL_2806779, EPI_ISL_2806780, EPI_ISL_2806781, EPI_ISL_2806784, EPI_ISL_2806788, EPI_ISL_2808075, EPI_ISL_2808076, EPI_ISL_2808078, EPI_ISL_2808081, EPI_ISL_2808082, EPI_ISL_2808083, EPI_ISL_2808085, EPI_ISL_2808086, EPI_ISL_2808087, EPI_ISL_2808088, EPI_ISL_2808091, EPI_ISL_2808094, EPI_ISL_2808095, EPI_ISL_2808096, EPI_ISL_2808097, EPI_ISL_2808098, EPI_ISL_2808101, EPI_ISL_2808102, EPI_ISL_2808103, EPI_ISL_2808105, EPI_ISL_2808106, EPI_ISL_2808107, EPI_ISL_2808108, EPI_ISL_2808109, EPI_ISL_2808110, EPI_ISL_2808111, EPI_ISL_2808113, EPI_ISL_2808114, EPI_ISL_2808115, EPI_ISL_2808122, EPI_ISL_2808127, EPI_ISL_2808128, EPI_ISL_2808131, EPI_ISL_2808134, EPI_ISL_2808135, EPI_ISL_2808137, EPI_ISL_2808138, EPI_ISL_2808139, EPI_ISL_2808140, EPI_ISL_2808141, EPI_ISL_2808143, EPI_ISL_2808145, EPI_ISL_2808146, EPI_ISL_2808147, EPI_ISL_2808149, EPI_ISL_2808151, EPI_ISL_2808152, EPI_ISL_2808153, EPI_ISL_2808154, EPI_ISL_2808157, EPI_ISL_2808159, EPI_ISL_2808160, EPI_ISL_2808161, EPI_ISL_2808162, EPI_ISL_2808163, EPI_ISL_2808164, EPI_ISL_2808165, EPI_ISL_2808166, EPI_ISL_2808167, EPI_ISL_2808169, EPI_ISL_2808170, EPI_ISL_2808173, EPI_ISL_2808176, EPI_ISL_2808179, EPI_ISL_2808181, EPI_ISL_2808184, EPI_ISL_2808185, EPI_ISL_2808186, EPI_ISL_2808188, EPI_ISL_2808190, EPI_ISL_2808191, EPI_ISL_2808192, EPI_ISL_2808193, EPI_ISL_2808194, EPI_ISL_2808195, EPI_ISL_2808196, EPI_ISL_2808197, EPI_ISL_2808199, EPI_ISL_2808201, EPI_ISL_2808202, EPI_ISL_2808203, EPI_ISL_2808204, EPI_ISL_2808205, EPI_ISL_2808207, EPI_ISL_2808208, EPI_ISL_2808209, EPI_ISL_2808211, EPI_ISL_2808212, EPI_ISL_2808213, EPI_ISL_2808214, EPI_ISL_2808215, EPI_ISL_2808217, EPI_ISL_2808218, EPI_ISL_2808219, EPI_ISL_2808222, EPI_ISL_2808223, EPI_ISL_2808226, EPI_ISL_2808227, EPI_ISL_2808228, EPI_ISL_2808229, EPI_ISL_2808230, EPI_ISL_2808231, EPI_ISL_2808232, EPI_ISL_2808234, EPI_ISL_2808235, EPI_ISL_2808236, EPI_ISL_2808239, EPI_ISL_2808242, EPI_ISL_2808243, EPI_ISL_2808244, EPI_ISL_2808245, EPI_ISL_2808246, EPI_ISL_2808247, EPI_ISL_2808248, EPI_ISL_2808249, EPI_ISL_2808250, EPI_ISL_2808251, EPI_ISL_2808252, EPI_ISL_2808253, EPI_ISL_2808254, EPI_ISL_2808255, EPI_ISL_2808257, EPI_ISL_2808258, EPI_ISL_2808262, EPI_ISL_2808263, EPI_ISL_2808265, EPI_ISL_2808268, EPI_ISL_2808270, EPI_ISL_2808271, EPI_ISL_2808273, EPI_ISL_2808274, EPI_ISL_2808275, EPI_ISL_2808276, EPI_ISL_2808278, EPI_ISL_2808279, EPI_ISL_2808280, EPI_ISL_2808283, EPI_ISL_2808284, EPI_ISL_2808285, EPI_ISL_2808286, EPI_ISL_2808287, EPI_ISL_2808288, EPI_ISL_2808289, EPI_ISL_2808291, EPI_ISL_2808292, EPI_ISL_2808294, EPI_ISL_2808295, EPI_ISL_2808296, EPI_ISL_2808297, EPI_ISL_2808298, EPI_ISL_2808299, EPI_ISL_2808305, EPI_ISL_2808306, EPI_ISL_2808307, EPI_ISL_2808308, EPI_ISL_2808311, EPI_ISL_2808312, EPI_ISL_2808313, EPI_ISL_2808314, EPI_ISL_2808316, EPI_ISL_2808317, EPI_ISL_2808318, EPI_ISL_2808320, EPI_ISL_2808321, EPI_ISL_2808322, EPI_ISL_2808325, EPI_ISL_2808330, EPI_ISL_2808331, EPI_ISL_2808333, EPI_ISL_2808334, EPI_ISL_2808335, EPI_ISL_2808336, EPI_ISL_2808337, EPI_ISL_2808339, EPI_ISL_2808340, EPI_ISL_2808342, EPI_ISL_2808343, EPI_ISL_2808345, EPI_ISL_2808346, EPI_ISL_2808348, EPI_ISL_2808349, EPI_ISL_2808354, EPI_ISL_2808355, EPI_ISL_2808357, EPI_ISL_2808362, EPI_ISL_2808364, EPI_ISL_2808365, EPI_ISL_2808366, EPI_ISL_2808367, EPI_ISL_2808368, EPI_ISL_2808369, EPI_ISL_2808372, EPI_ISL_2808373, EPI_ISL_2808374, EPI_ISL_2808376, EPI_ISL_2808377, EPI_ISL_2808378, EPI_ISL_2808380, EPI_ISL_2808381, EPI_ISL_2808383, EPI_ISL_2808384, EPI_ISL_2808386, EPI_ISL_2808387, EPI_ISL_2808389, EPI_ISL_2808393, EPI_ISL_2808394, EPI_ISL_2808395, EPI_ISL_2808396, EPI_ISL_2808398, EPI_ISL_2808401, EPI_ISL_2808402, EPI_ISL_2808405, EPI_ISL_2808406, EPI_ISL_2808408, EPI_ISL_2808409, EPI_ISL_2808410, EPI_ISL_2808411, EPI_ISL_2808412, EPI_ISL_2808413, EPI_ISL_2808415, EPI_ISL_2808416, EPI_ISL_2808417, EPI_ISL_2808418, EPI_ISL_2808419, EPI_ISL_2808422, EPI_ISL_2808424, EPI_ISL_2808425, EPI_ISL_2808427, EPI_ISL_2808428, EPI_ISL_2808431, EPI_ISL_2808436, EPI_ISL_2808437, EPI_ISL_2808438, EPI_ISL_2808441, EPI_ISL_2808444, EPI_ISL_2808445, EPI_ISL_2808446, EPI_ISL_2808447, EPI_ISL_2808448, EPI_ISL_2808449, EPI_ISL_2808451, EPI_ISL_2808453, EPI_ISL_2808454, EPI_ISL_2808455, EPI_ISL_2808456, EPI_ISL_2808457, EPI_ISL_2808458, EPI_ISL_2808459, EPI_ISL_2808461, EPI_ISL_2808462, EPI_ISL_2808465, EPI_ISL_2808466, EPI_ISL_2808468, EPI_ISL_2808469, EPI_ISL_2808470, EPI_ISL_2808478, EPI_ISL_2808479, EPI_ISL_2808487, EPI_ISL_2808489, EPI_ISL_2808491, EPI_ISL_2808493, EPI_ISL_2808496, EPI_ISL_2808497, EPI_ISL_2808499, EPI_ISL_2808501, EPI_ISL_2808509, EPI_ISL_2808514, EPI_ISL_2808515, EPI_ISL_2808520, EPI_ISL_2808521, EPI_ISL_2808523, EPI_ISL_2808525, EPI_ISL_2808526, EPI_ISL_2808527, EPI_ISL_2808529, EPI_ISL_2808532, EPI_ISL_2808533, EPI_ISL_2808534, EPI_ISL_2808536, EPI_ISL_2808539, EPI_ISL_2808541, EPI_ISL_2808542, EPI_ISL_2808545, EPI_ISL_2808546, EPI_ISL_2808547, EPI_ISL_2808548, EPI_ISL_2808549, EPI_ISL_2808552, EPI_ISL_2808555, EPI_ISL_2808557, EPI_ISL_2808560, EPI_ISL_2808561, EPI_ISL_2808563, EPI_ISL_2808564, EPI_ISL_2808565, EPI_ISL_2808566, EPI_ISL_2808569, EPI_ISL_2808572, EPI_ISL_2808573, EPI_ISL_2808576, EPI_ISL_2808577, EPI_ISL_2808578, EPI_ISL_2808579, EPI_ISL_2808580, EPI_ISL_2808585, EPI_ISL_2808587, EPI_ISL_2808589, EPI_ISL_2808590, EPI_ISL_2808592, EPI_ISL_2808593, EPI_ISL_2808594, EPI_ISL_2808596, EPI_ISL_2808599, EPI_ISL_2808602, EPI_ISL_2808603, EPI_ISL_2808604, EPI_ISL_2808606, EPI_ISL_2808608, EPI_ISL_2808609, EPI_ISL_2808610, EPI_ISL_2808611, EPI_ISL_2808612, EPI_ISL_2808615, EPI_ISL_2808619, EPI_ISL_2808622, EPI_ISL_2808624, EPI_ISL_2808628, EPI_ISL_2808629, EPI_ISL_2808631, EPI_ISL_2808634, EPI_ISL_2808635, EPI_ISL_2808638, EPI_ISL_2808639, EPI_ISL_2808640, EPI_ISL_2808642, EPI_ISL_2808644, EPI_ISL_2808645, EPI_ISL_2808650, EPI_ISL_2808651, EPI_ISL_2808655, EPI_ISL_2808658, EPI_ISL_2808659, EPI_ISL_2808663, EPI_ISL_2808664, EPI_ISL_2808666, EPI_ISL_2808667, EPI_ISL_2808670, EPI_ISL_2808671, EPI_ISL_2808674, EPI_ISL_2808678, EPI_ISL_2808681, EPI_ISL_2808683, EPI_ISL_2808689, EPI_ISL_2808691, EPI_ISL_2808693, EPI_ISL_2808694, EPI_ISL_2808696, EPI_ISL_2808697, EPI_ISL_2808699, EPI_ISL_2808700, EPI_ISL_2808701, EPI_ISL_2808703, EPI_ISL_2808705, EPI_ISL_2808706, EPI_ISL_2808708, EPI_ISL_2808710, EPI_ISL_2808714, EPI_ISL_2808718, EPI_ISL_2808719, EPI_ISL_2808728, EPI_ISL_2808731, EPI_ISL_2808732, EPI_ISL_2814424, EPI_ISL_2815042, EPI_ISL_2829315, EPI_ISL_2829319, EPI_ISL_2829325, EPI_ISL_2829332, EPI_ISL_2829336, EPI_ISL_2829342, EPI_ISL_2829347, EPI_ISL_2829354, EPI_ISL_2829363, EPI_ISL_2829376, EPI_ISL_2829381, EPI_ISL_2829390, EPI_ISL_2829394, EPI_ISL_2829404, EPI_ISL_2829415, EPI_ISL_2829425, EPI_ISL_2829432, EPI_ISL_2829433, EPI_ISL_2829445, EPI_ISL_2829463, EPI_ISL_2829465, EPI_ISL_2829475, EPI_ISL_2829489, EPI_ISL_2829497, EPI_ISL_2829502, EPI_ISL_2829522, EPI_ISL_2829546, EPI_ISL_2829548, EPI_ISL_2829572, EPI_ISL_2830607, EPI_ISL_2830608, EPI_ISL_2830610, EPI_ISL_2830614, EPI_ISL_2830627, EPI_ISL_2830628, EPI_ISL_2830630, EPI_ISL_2830632, EPI_ISL_2830638, EPI_ISL_2830641, EPI_ISL_2830642, EPI_ISL_2830648, EPI_ISL_2830659, EPI_ISL_2830663, EPI_ISL_2830668, EPI_ISL_2830672, EPI_ISL_2830673, EPI_ISL_2830678, EPI_ISL_2830680, EPI_ISL_2830681, EPI_ISL_2830696, EPI_ISL_2830718, EPI_ISL_2830720, EPI_ISL_2830722, EPI_ISL_2830728, EPI_ISL_2830731, EPI_ISL_2830736, EPI_ISL_2830743, EPI_ISL_2830744, EPI_ISL_2830749, EPI_ISL_2830753, EPI_ISL_2830754, EPI_ISL_2830766, EPI_ISL_2830769, EPI_ISL_2830774, EPI_ISL_2830775, EPI_ISL_2830777, EPI_ISL_2830781, EPI_ISL_2830783, EPI_ISL_2830800, EPI_ISL_2830812, EPI_ISL_2830816, EPI_ISL_2830833, EPI_ISL_2830834, EPI_ISL_2830835, EPI_ISL_2830839, EPI_ISL_2830841, EPI_ISL_2830844, EPI_ISL_2830848, EPI_ISL_2830852, EPI_ISL_2831048, EPI_ISL_2831049, EPI_ISL_2831050, EPI_ISL_2831057, EPI_ISL_2831063, EPI_ISL_2831065, EPI_ISL_2831071, EPI_ISL_2831072, EPI_ISL_2831083, EPI_ISL_2831084, EPI_ISL_2831090, EPI_ISL_2831098, EPI_ISL_2831099, EPI_ISL_2831100, EPI_ISL_2831101, EPI_ISL_2831106, EPI_ISL_2831107, EPI_ISL_2831111, EPI_ISL_2831116, EPI_ISL_2831117, EPI_ISL_2831118, EPI_ISL_2831124, EPI_ISL_2831125, EPI_ISL_2831127, EPI_ISL_2831131, EPI_ISL_2831133, EPI_ISL_2831135, EPI_ISL_2831136, EPI_ISL_2831139, EPI_ISL_2831141, EPI_ISL_2831144, EPI_ISL_2831145, EPI_ISL_2831149, EPI_ISL_2831152, EPI_ISL_2831153, EPI_ISL_2831167, EPI_ISL_2831169, EPI_ISL_2831171, EPI_ISL_2831172, EPI_ISL_2831173, EPI_ISL_2831174, EPI_ISL_2831177, EPI_ISL_2831179, EPI_ISL_2831180, EPI_ISL_2831181, EPI_ISL_2831187, EPI_ISL_2831189, EPI_ISL_2831190, EPI_ISL_2831200, EPI_ISL_2831209, EPI_ISL_2831210, EPI_ISL_2831213, EPI_ISL_2831214, EPI_ISL_2831215, EPI_ISL_2831218, EPI_ISL_2831221, EPI_ISL_2831228, EPI_ISL_2831230, EPI_ISL_2831243, EPI_ISL_2831250, EPI_ISL_2831253, EPI_ISL_2831259, EPI_ISL_2831261, EPI_ISL_2831262, EPI_ISL_2831264, EPI_ISL_2831268, EPI_ISL_2831269, EPI_ISL_2831271, EPI_ISL_2831273, EPI_ISL_2831276, EPI_ISL_2831277, EPI_ISL_2831281, EPI_ISL_2831283, EPI_ISL_2831284, EPI_ISL_2831285, EPI_ISL_2831289, EPI_ISL_2831291, EPI_ISL_2831292, EPI_ISL_2831293, EPI_ISL_2831304, EPI_ISL_2831305, EPI_ISL_2831307, EPI_ISL_2831311, EPI_ISL_2831317, EPI_ISL_2831318, EPI_ISL_2831319, EPI_ISL_2831964, EPI_ISL_2832066, EPI_ISL_2832067, EPI_ISL_2832068, EPI_ISL_2832072, EPI_ISL_2832084, EPI_ISL_2832085, EPI_ISL_2832093, EPI_ISL_2846194, EPI_ISL_2846269, EPI_ISL_2847623, EPI_ISL_2847696, EPI_ISL_2847707, EPI_ISL_2847764, EPI_ISL_2847769, EPI_ISL_2847777, EPI_ISL_2847828, EPI_ISL_2847889, EPI_ISL_2848099, EPI_ISL_2852170, EPI_ISL_2852183, EPI_ISL_2852206, EPI_ISL_2852211, EPI_ISL_2852231, EPI_ISL_2852237, EPI_ISL_2852286, EPI_ISL_2852319, EPI_ISL_2852361, EPI_ISL_2852371, EPI_ISL_2852428, EPI_ISL_2852446, EPI_ISL_2853014, EPI_ISL_2853063

| | | | |
|---|---|---|---|
| see above | Lighthouse Laboratory Plymouth | Wellcome Sanger Institute for the COVID-19 Genomics UK (COG-UK) Consortium | Cordelia Langford; David K. Jackson; Dominic Kwiatkowski; Ewan Harrison; Ian Johnston; Jeffrey Barrett; John Sillitoe on behalf of the Wellcome Sanger Institute COVID-19 Surveillance Team; Lighthouse Laboratory Plymouth and Alex Alderton; Roberto Amato; Sonia Goncalves |

EPI_ISL_2901129, EPI_ISL_2901130, EPI_ISL_2901131, EPI_ISL_2901132, EPI_ISL_2901133, EPI_ISL_2901134, EPI_ISL_2901135, EPI_ISL_2901136, EPI_ISL_2901137, EPI_ISL_2901138, EPI_ISL_2901139, EPI_ISL_2901140, EPI_ISL_2901141, EPI_ISL_2901142, EPI_ISL_2901143, EPI_ISL_2901144, EPI_ISL_2901145, EPI_ISL_2901146, EPI_ISL_2901147, EPI_ISL_2901148, EPI_ISL_2901149, EPI_ISL_2901150, EPI_ISL_2901151, EPI_ISL_2901152, EPI_ISL_2901153, EPI_ISL_2901154, EPI_ISL_2901155, EPI_ISL_2901156, EPI_ISL_2901157, EPI_ISL_2901158, EPI_ISL_2901159, EPI_ISL_2901160, EPI_ISL_2901162, EPI_ISL_2901164, EPI_ISL_2901166, EPI_ISL_2901167, EPI_ISL_2901168, EPI_ISL_2901169, EPI_ISL_2901170, EPI_ISL_2901171, EPI_ISL_2901172, EPI_ISL_2901173, EPI_ISL_2901174, EPI_ISL_2901175, EPI_ISL_2901176, EPI_ISL_2901178, EPI_ISL_2901179, EPI_ISL_2901180, EPI_ISL_2901181, EPI_ISL_2901182, EPI_ISL_2901183, EPI_ISL_2901184, EPI_ISL_2901185

| | | | |
|---|---|---|---|
| see above | Liverpool Clinical Laboratories | COVID-19 Genomics UK (COG-UK) Consortium | A Alrezaihi; Alessandro Gerada; Alistair Darby; Angela Cowell; Anita Lucaci; Anu Chawla; Cassie Olateju; Catherine Hartley; Charlotte Nelson; Ecaterina Vamos; Elaine O'Toole; Eleanor G Bentley; Ghada T Shawli; Hermione Webster; Isabel García-Dorival; James Johnson; James P Stewart; Jenifer Manson; Joanne Watts; Jones Benjamin; Jordan J Clark; Julian Hiscox; L Luu; Lucille Rainbow; M Almsaud; Margaret Hughes; Mark Whitehead; Matthew Gemmell; Miren Iturriza-Gomara; Muhannad Alruwaili; N.P Randle; Neil Swainston; PKF Gilmore; Parul Sharma; Rebekah Penrice-Randal; Richard Eccles; Richard Gregory; Sam Haldenby; Steve Paterson; Stuart D Armstrong; Trevor Ian Robinson; Ximeng Han |

EPI_ISL_2816564, EPI_ISL_2816565, EPI_ISL_2816566, EPI_ISL_2816567, EPI_ISL_2816568, EPI_ISL_2816569, EPI_ISL_2816570, EPI_ISL_2816571, EPI_ISL_2816572, EPI_ISL_2816573, EPI_ISL_2816574, EPI_ISL_2816575, EPI_ISL_2816576, EPI_ISL_2816577, EPI_ISL_2816578, EPI_ISL_2816579, EPI_ISL_2816580, EPI_ISL_2816581, EPI_ISL_2816582, EPI_ISL_2816583, EPI_ISL_2816585, EPI_ISL_2816586, EPI_ISL_2816587, EPI_ISL_2816588, EPI_ISL_2816589, EPI_ISL_2816590, EPI_ISL_2816591, EPI_ISL_2816592, EPI_ISL_2816593, EPI_ISL_2816594, EPI_ISL_2816595, EPI_ISL_2816596, EPI_ISL_2816597, EPI_ISL_2816598, EPI_ISL_2816599, EPI_ISL_2816600, EPI_ISL_2816601, EPI_ISL_2816602, EPI_ISL_2816603, EPI_ISL_2816604, EPI_ISL_2816605, EPI_ISL_2816606, EPI_ISL_2816607, EPI_ISL_2816608, EPI_ISL_2816609, EPI_ISL_2816610, EPI_ISL_2816611, EPI_ISL_2816612, EPI_ISL_2816613, EPI_ISL_2816614, EPI_ISL_2816615, EPI_ISL_2816616, EPI_ISL_2816617, EPI_ISL_2816618, EPI_ISL_2816619, EPI_ISL_2816620, EPI_ISL_2816621, EPI_ISL_2816622, EPI_ISL_2816623, EPI_ISL_2816625, EPI_ISL_2816626, EPI_ISL_2816627, EPI_ISL_2816628, EPI_ISL_2816629, EPI_ISL_2816630, EPI_ISL_2816631, EPI_ISL_2816632, EPI_ISL_2816633, EPI_ISL_2816634, EPI_ISL_2816635, EPI_ISL_2816636, EPI_ISL_2816637, EPI_ISL_2816638, EPI_ISL_2816639, EPI_ISL_2816640, EPI_ISL_2816641, EPI_ISL_2816642, EPI_ISL_2816643, EPI_ISL_2816644, EPI_ISL_2816723, EPI_ISL_2816724, EPI_ISL_2816725, EPI_ISL_2816726, EPI_ISL_2816727, EPI_ISL_2816728, EPI_ISL_2816729, EPI_ISL_2816730, EPI_ISL_2816731, EPI_ISL_2816732, EPI_ISL_2816733, EPI_ISL_2816734, EPI_ISL_2816735, EPI_ISL_2816736, EPI_ISL_2816737, EPI_ISL_2816738, EPI_ISL_2816739, EPI_ISL_2816740, EPI_ISL_2816741, EPI_ISL_2816742, EPI_ISL_2816743, EPI_ISL_2816745, EPI_ISL_2816747, EPI_ISL_2816749, EPI_ISL_2816750, EPI_ISL_2816751, EPI_ISL_2816752, EPI_ISL_2816753, EPI_ISL_2816754, EPI_ISL_2816755, EPI_ISL_2816757, EPI_ISL_2816758, EPI_ISL_2816759, EPI_ISL_2816760, EPI_ISL_2816761, EPI_ISL_2816762, EPI_ISL_2816763, EPI_ISL_2816764, EPI_ISL_2816765, EPI_ISL_2816766, EPI_ISL_2816767, EPI_ISL_2816768, EPI_ISL_2816769, EPI_ISL_2816770, EPI_ISL_2816772, EPI_ISL_2816773, EPI_ISL_2816774, EPI_ISL_2816775, EPI_ISL_2816776, EPI_ISL_2816777, EPI_ISL_2816778, EPI_ISL_2816779, EPI_ISL_2816780, EPI_ISL_2816781, EPI_ISL_2816783, EPI_ISL_2816784, EPI_ISL_2816785, EPI_ISL_2816786, EPI_ISL_2816787, EPI_ISL_2816788, EPI_ISL_2816789, EPI_ISL_2816791, EPI_ISL_2816792, EPI_ISL_2816793, EPI_ISL_2816794, EPI_ISL_2816795, EPI_ISL_2816798, EPI_ISL_2816800, EPI_ISL_2816801, EPI_ISL_2816802, EPI_ISL_2816805, EPI_ISL_2816806, EPI_ISL_2816807, EPI_ISL_2816808, EPI_ISL_2816809, EPI_ISL_2816810, EPI_ISL_2816811, EPI_ISL_2816812, EPI_ISL_2816814, EPI_ISL_2816815, EPI_ISL_2816816, EPI_ISL_2816817, EPI_ISL_2816818, EPI_ISL_2816819, EPI_ISL_2816820, EPI_ISL_2816821, EPI_ISL_2816822, EPI_ISL_2816823, EPI_ISL_2816824, EPI_ISL_2816825, EPI_ISL_2816826, EPI_ISL_2816827, EPI_ISL_2816828, EPI_ISL_2816829, EPI_ISL_2816830, EPI_ISL_2816831, EPI_ISL_2816832, EPI_ISL_2816833, EPI_ISL_2816834, EPI_ISL_2816835, EPI_ISL_2816836, EPI_ISL_2816837, EPI_ISL_2816838, EPI_ISL_2816839, EPI_ISL_2816840, EPI_ISL_2816841, EPI_ISL_2816842, EPI_ISL_2816843, EPI_ISL_2816844, EPI_ISL_2816845, EPI_ISL_2816846, EPI_ISL_2816847, EPI_ISL_2816848, EPI_ISL_2816849, EPI_ISL_2816850, EPI_ISL_2816852, EPI_ISL_2816853, EPI_ISL_2816854, EPI_ISL_2816855, EPI_ISL_2816856, EPI_ISL_2816857, EPI_ISL_2816858, EPI_ISL_2816859, EPI_ISL_2816860, EPI_ISL_2816861, EPI_ISL_2816862, EPI_ISL_2816863, EPI_ISL_2816864, EPI_ISL_2816865, EPI_ISL_2816866, EPI_ISL_2816867, EPI_ISL_2816868, EPI_ISL_2816869, EPI_ISL_2816870, EPI_ISL_2816871, EPI_ISL_2816872, EPI_ISL_2816873, EPI_ISL_2816874, EPI_ISL_2816875, EPI_ISL_2816876, EPI_ISL_2816877, EPI_ISL_2816879, EPI_ISL_2816880, EPI_ISL_2816881, EPI_ISL_2816882, EPI_ISL_2901725, EPI_ISL_2901774

| | | | |
|---|---|---|---|
| see above | Northumbria University / South Tees Hospitals NHS Foundation Trust / North Cumbria Integrated Care NHS Foundation Trust / North Tees and Hartlepool NHS | COVID-19 Genomics UK (COG-UK) Consortium | Andrew Nelson; Brendan Payne; Clive Graham; Darren L Smith; Debra Padgett; Edward Barton; Emma Swindells; Garren Scott; Gary Black; Gary Eltringham; Giles S Holt; Greg R Young; Jane Greenaway; Jennifer Collins; John Allan; Joshua Loh; Lynn Dover; Matthew Bashton; Mohammad A Tariq; Paul Baker; Sarah Essex; Steve Liggett; Wen C Yew; Yusri Taha |

Foundation Trust / Newcastle Hospitals NHS Foundation Trust

EPI_ISL_2818341, EPI_ISL_2818343, EPI_ISL_2818344, EPI_ISL_2818345, EPI_ISL_2818346, EPI_ISL_2818348, EPI_ISL_2818349, EPI_ISL_2818352, EPI_ISL_2818353, EPI_ISL_2818355, EPI_ISL_2818356, EPI_ISL_2818357, EPI_ISL_2818358, EPI_ISL_2818359, EPI_ISL_2818361, EPI_ISL_2818362, EPI_ISL_2818363, EPI_ISL_2818365, EPI_ISL_2818375, EPI_ISL_2818387, EPI_ISL_2818390, EPI_ISL_2818400, EPI_ISL_2818401, EPI_ISL_2818405, EPI_ISL_2818406, EPI_ISL_2818407, EPI_ISL_2818408, EPI_ISL_2818409, EPI_ISL_2818411, EPI_ISL_2818412, EPI_ISL_2818417, EPI_ISL_2818548, EPI_ISL_2818549, EPI_ISL_2818550, EPI_ISL_2818551, EPI_ISL_2818552, EPI_ISL_2818556, EPI_ISL_2818559, EPI_ISL_2818561, EPI_ISL_2818564, EPI_ISL_2818565, EPI_ISL_2818566, EPI_ISL_2818569, EPI_ISL_2818570, EPI_ISL_2818572, EPI_ISL_2818575, EPI_ISL_2818582, EPI_ISL_2818586, EPI_ISL_2818597, EPI_ISL_2818600, EPI_ISL_2818606, EPI_ISL_2818622, EPI_ISL_2818624, EPI_ISL_2818632, EPI_ISL_2818634, EPI_ISL_2818635, EPI_ISL_2818639, EPI_ISL_2818642, EPI_ISL_2818643, EPI_ISL_2818645, EPI_ISL_2818646, EPI_ISL_2818649, EPI_ISL_2818650, EPI_ISL_2818651, EPI_ISL_2818653, EPI_ISL_2818657, EPI_ISL_2818658, EPI_ISL_2818660, EPI_ISL_2818664, EPI_ISL_2818673, EPI_ISL_2818679, EPI_ISL_2818681, EPI_ISL_2818684, EPI_ISL_2818685, EPI_ISL_2818686, EPI_ISL_2818687, EPI_ISL_2818688, EPI_ISL_2818690, EPI_ISL_2818691, EPI_ISL_2818692, EPI_ISL_2818700, EPI_ISL_2818704, EPI_ISL_2818709, EPI_ISL_2818718, EPI_ISL_2818720, EPI_ISL_2818721, EPI_ISL_2818723, EPI_ISL_2818724, EPI_ISL_2818732, EPI_ISL_2818738, EPI_ISL_2818739, EPI_ISL_2818741, EPI_ISL_2818745, EPI_ISL_2818749, EPI_ISL_2818750, EPI_ISL_2818752, EPI_ISL_2818755, EPI_ISL_2818757, EPI_ISL_2818758, EPI_ISL_2818760, EPI_ISL_2818761, EPI_ISL_2818763, EPI_ISL_2818764, EPI_ISL_2818767, EPI_ISL_2818769, EPI_ISL_2818770, EPI_ISL_2818771, EPI_ISL_2818772, EPI_ISL_2818773, EPI_ISL_2818774, EPI_ISL_2818776, EPI_ISL_2818804, EPI_ISL_2818811, EPI_ISL_2818812, EPI_ISL_2818824, EPI_ISL_2818825, EPI_ISL_2818826, EPI_ISL_2818829, EPI_ISL_2818872, EPI_ISL_2818876, EPI_ISL_2818877, EPI_ISL_2818888, EPI_ISL_2818892, EPI_ISL_2818895, EPI_ISL_2818896, EPI_ISL_2818897, EPI_ISL_2818898, EPI_ISL_2818907, EPI_ISL_2818910, EPI_ISL_2818912, EPI_ISL_2818916, EPI_ISL_2818917, EPI_ISL_2818920, EPI_ISL_2818925, EPI_ISL_2818930, EPI_ISL_2818931, EPI_ISL_2818932, EPI_ISL_2818933, EPI_ISL_2818935, EPI_ISL_2818937, EPI_ISL_2818947, EPI_ISL_2818948, EPI_ISL_2818950, EPI_ISL_2818956, EPI_ISL_2818957, EPI_ISL_2819055, EPI_ISL_2819057, EPI_ISL_2819061, EPI_ISL_2819063, EPI_ISL_2819074, EPI_ISL_2819084, EPI_ISL_2819095, EPI_ISL_2819099, EPI_ISL_2819310, EPI_ISL_2819323, EPI_ISL_2819325, EPI_ISL_2819329, EPI_ISL_2819331, EPI_ISL_2819335, EPI_ISL_2819341, EPI_ISL_2819350, EPI_ISL_2819352, EPI_ISL_2819354, EPI_ISL_2819422, EPI_ISL_2819431, EPI_ISL_2819433, EPI_ISL_2819435, EPI_ISL_2819437, EPI_ISL_2819442, EPI_ISL_2819444, EPI_ISL_2819446, EPI_ISL_2819453, EPI_ISL_2819466, EPI_ISL_2819468, EPI_ISL_2819580, EPI_ISL_2819590, EPI_ISL_2819603, EPI_ISL_2819638, EPI_ISL_2819641, EPI_ISL_2819661, EPI_ISL_2819669, EPI_ISL_2819673, EPI_ISL_2819706, EPI_ISL_2819710, EPI_ISL_2819728, EPI_ISL_2819730, EPI_ISL_2819734, EPI_ISL_2819738, EPI_ISL_2819739, EPI_ISL_2819742, EPI_ISL_2819743, EPI_ISL_2819747, EPI_ISL_2819749, EPI_ISL_2819753, EPI_ISL_2819756, EPI_ISL_2819766, EPI_ISL_2819768, EPI_ISL_2819769, EPI_ISL_2819793, EPI_ISL_2819799, EPI_ISL_2819805, EPI_ISL_2819815, EPI_ISL_2819823, EPI_ISL_2819829, EPI_ISL_2819836, EPI_ISL_2819845, EPI_ISL_2819847, EPI_ISL_2819848, EPI_ISL_2904674, EPI_ISL_2904675, EPI_ISL_2904676, EPI_ISL_2904677, EPI_ISL_2904678, EPI_ISL_2904679, EPI_ISL_2904680, EPI_ISL_2904681, EPI_ISL_2904682, EPI_ISL_2904684, EPI_ISL_2904685, EPI_ISL_2904686, EPI_ISL_2904687, EPI_ISL_2904689, EPI_ISL_2904690, EPI_ISL_2904691, EPI_ISL_2904692, EPI_ISL_2904693, EPI_ISL_2904694, EPI_ISL_2904695, EPI_ISL_2904696, EPI_ISL_2904697, EPI_ISL_2904698, EPI_ISL_2904722, EPI_ISL_2904723, EPI_ISL_2904724, EPI_ISL_2904725, EPI_ISL_2904726, EPI_ISL_2904727, EPI_ISL_2904729, EPI_ISL_2904730, EPI_ISL_2904731, EPI_ISL_2904732, EPI_ISL_2904736, EPI_ISL_2904737, EPI_ISL_2904740, EPI_ISL_2904745, EPI_ISL_2904746, EPI_ISL_2904748, EPI_ISL_2904908, EPI_ISL_2904909, EPI_ISL_2904910, EPI_ISL_2904913, EPI_ISL_2904914, EPI_ISL_2904915, EPI_ISL_2904916, EPI_ISL_2904918, EPI_ISL_2904921, EPI_ISL_2904922, EPI_ISL_2904923, EPI_ISL_2904924, EPI_ISL_2904926, EPI_ISL_2904927, EPI_ISL_2904929, EPI_ISL_2904930, EPI_ISL_2904931, EPI_ISL_2904933, EPI_ISL_2905015, EPI_ISL_2905016, EPI_ISL_2905019, EPI_ISL_2905021, EPI_ISL_2905022, EPI_ISL_2905023, EPI_ISL_2905024, EPI_ISL_2905025, EPI_ISL_2905026, EPI_ISL_2905028, EPI_ISL_2905029, EPI_ISL_2905031, EPI_ISL_2905032, EPI_ISL_2905033, EPI_ISL_2905034, EPI_ISL_2905170, EPI_ISL_2905171, EPI_ISL_2905172, EPI_ISL_2905173, EPI_ISL_2905174, EPI_ISL_2905176, EPI_ISL_2905177, EPI_ISL_2905178, EPI_ISL_2905180, EPI_ISL_2905181, EPI_ISL_2905182, EPI_ISL_2905183, EPI_ISL_2905184, EPI_ISL_2905185, EPI_ISL_2905186, EPI_ISL_2905188, EPI_ISL_2905189, EPI_ISL_2905192, EPI_ISL_2905193, EPI_ISL_2905194, EPI_ISL_2905195, EPI_ISL_2905196, EPI_ISL_2905197, EPI_ISL_2905465, EPI_ISL_2905470, EPI_ISL_2905474, EPI_ISL_2905475, EPI_ISL_2905476, EPI_ISL_2905477, EPI_ISL_2905478, EPI_ISL_2905480, EPI_ISL_2905481, EPI_ISL_2905482, EPI_ISL_2905483, EPI_ISL_2905571, EPI_ISL_2905578, EPI_ISL_2905588, EPI_ISL_2905670, EPI_ISL_2905672, EPI_ISL_2905673, EPI_ISL_2905682, EPI_ISL_2905687, EPI_ISL_2905689, EPI_ISL_2905691, EPI_ISL_2905693, EPI_ISL_2905694, EPI_ISL_2905696, EPI_ISL_2905698, EPI_ISL_2905710, EPI_ISL_2905722, EPI_ISL_2905735, EPI_ISL_2905778, EPI_ISL_2905780, EPI_ISL_2905811, EPI_ISL_2905818, EPI_ISL_2905851, EPI_ISL_2905854, EPI_ISL_2905857, EPI_ISL_2905863, EPI_ISL_2905883, EPI_ISL_2905884, EPI_ISL_2905887, EPI_ISL_2905888, EPI_ISL_2905920, EPI_ISL_2905973, EPI_ISL_2905974, EPI_ISL_2905975, EPI_ISL_2905976, EPI_ISL_2905977, EPI_ISL_2905979, EPI_ISL_2905983, EPI_ISL_2905984, EPI_ISL_2905985, EPI_ISL_2905992, EPI_ISL_2905994, EPI_ISL_2906000, EPI_ISL_2906001, EPI_ISL_2906003, EPI_ISL_2906005, EPI_ISL_2906006, EPI_ISL_2906008, EPI_ISL_2906011, EPI_ISL_2906012, EPI_ISL_2906016, EPI_ISL_2906017, EPI_ISL_2906019, EPI_ISL_2906020, EPI_ISL_2906021, EPI_ISL_2906022, EPI_ISL_2906024, EPI_ISL_2906048, EPI_ISL_2906054, EPI_ISL_2906055, EPI_ISL_2906057, EPI_ISL_2906058, EPI_ISL_2906059, EPI_ISL_2906060, EPI_ISL_2906061, EPI_ISL_2906063, EPI_ISL_2906064, EPI_ISL_2906065, EPI_ISL_2906067, EPI_ISL_2906069, EPI_ISL_2906072, EPI_ISL_2906073, EPI_ISL_2906075, EPI_ISL_2906101, EPI_ISL_2906102, EPI_ISL_2906103, EPI_ISL_2906105, EPI_ISL_2906112, EPI_ISL_2906154, EPI_ISL_2906161, EPI_ISL_2906162, EPI_ISL_2906163, EPI_ISL_2906164, EPI_ISL_2906166, EPI_ISL_2906167, EPI_ISL_2906168, EPI_ISL_2906169, EPI_ISL_2906170, EPI_ISL_2906171, EPI_ISL_2906172, EPI_ISL_2906173, EPI_ISL_2906174, EPI_ISL_2906175, EPI_ISL_2906176, EPI_ISL_2906177, EPI_ISL_2906179, EPI_ISL_2906180, EPI_ISL_2906189, EPI_ISL_2906190, EPI_ISL_2906196, EPI_ISL_2906198, EPI_ISL_2906202, EPI_ISL_2906203, EPI_ISL_2906204, EPI_ISL_2906205, EPI_ISL_2906206, EPI_ISL_2906207, EPI_ISL_2906208, EPI_ISL_2906210, EPI_ISL_2906211, EPI_ISL_2906212, EPI_ISL_2906213, EPI_ISL_2906214, EPI_ISL_2906215, EPI_ISL_2906219, EPI_ISL_2906221, EPI_ISL_2906222, EPI_ISL_2906223, EPI_ISL_2906224, EPI_ISL_2906225, EPI_ISL_2906227, EPI_ISL_2906228, EPI_ISL_2906229, EPI_ISL_2906230, EPI_ISL_2906231, EPI_ISL_2906232, EPI_ISL_2906234, EPI_ISL_2906235, EPI_ISL_2906239, EPI_ISL_2906240, EPI_ISL_2906243, EPI_ISL_2906247, EPI_ISL_2906248, EPI_ISL_2906249, EPI_ISL_2906252, EPI_ISL_2906296, EPI_ISL_2906299, EPI_ISL_2906301, EPI_ISL_2906318, EPI_ISL_2906319, EPI_ISL_2906320, EPI_ISL_2906321, EPI_ISL_2906352, EPI_ISL_2906360, EPI_ISL_2906361, EPI_ISL_2906362, EPI_ISL_2906363, EPI_ISL_2906364, EPI_ISL_2906366, EPI_ISL_2906369, EPI_ISL_2906370, EPI_ISL_2906371, EPI_ISL_2906372, EPI_ISL_2906373, EPI_ISL_2906375, EPI_ISL_2906376, EPI_ISL_2906377, EPI_ISL_2906378, EPI_ISL_2906409, EPI_ISL_2906515, EPI_ISL_2906517, EPI_ISL_2906518, EPI_ISL_2906519, EPI_ISL_2906522, EPI_ISL_2906525, EPI_ISL_2906527, EPI_ISL_2906532, EPI_ISL_2906682, EPI_ISL_2906685, EPI_ISL_2906691, EPI_ISL_2906692, EPI_ISL_2906693, EPI_ISL_2906695, EPI_ISL_2906699

| | | | |
|---|---|---|---|
| see above | Originating lab: Wales Specialist Virology Centre Sequencing lab: Pathogen Genomics Unit | Public Health Wales Microbiology Cardiff Wales Specialist Virology Centre | Alec Birchley; Alexander Adams; Amy Gaskin; Angela Marchbank; Bree Gatica-Wilcox; Catherine Moore; Jason Coombes; Joanne Watkins; Joel Southgate; Johnathan Evans; Laura Gifford; Lauren Gilbert; Lee Graham; Malorie Perry; Matthew Bull; Nicole Pacchiarini; Sally Corden; Sara Kumziene-Summerhayes; Sara Rey; Sarah Taylor; Simon Cottrell; Sophie Jones; Tom Connor |
| EPI_ISL_2903109, EPI_ISL_2903125 | Oxford Viromics, NDM, University of Oxford; Oxford University Hospitals; Basingstoke and North Hampshire Hospital | COVID-19 Genomics UK (COG-UK) Consortium | Alex Mobbs; Amy Trebes; Anita Justice; Catrin Moore; Christophe Fraser; David Bonsall; David Buck; Emma Wise; George Macintyre; Jessica Lynch; John Todd; Mariateresa de Cesare; Matilde Mori; Monique Andersson; Nathan Moore; Nick Cortes; Robert Shaw; Stephen Kidd; Tanya Golubchik; Timothy Peto |

EPI_ISL_2816887, EPI_ISL_2816895, EPI_ISL_2816940, EPI_ISL_2816941, EPI_ISL_2816942, EPI_ISL_2816944, EPI_ISL_2816945, EPI_ISL_2816946, EPI_ISL_2816948, EPI_ISL_2816949, EPI_ISL_2816950, EPI_ISL_2816951, EPI_ISL_2816952, EPI_ISL_2816953, EPI_ISL_2816955, EPI_ISL_2816956, EPI_ISL_2816958, EPI_ISL_2816960, EPI_ISL_2816963, EPI_ISL_2816965, EPI_ISL_2816967, EPI_ISL_2816969, EPI_ISL_2816970, EPI_ISL_2816973, EPI_ISL_2816974, EPI_ISL_2816977, EPI_ISL_2816978, EPI_ISL_2816979, EPI_ISL_2816980, EPI_ISL_2816981, EPI_ISL_2816982, EPI_ISL_2816983, EPI_ISL_2816984, EPI_ISL_2816985, EPI_ISL_2816986, EPI_ISL_2816987, EPI_ISL_2816988, EPI_ISL_2816989, EPI_ISL_2816990, EPI_ISL_2816991, EPI_ISL_2816992, EPI_ISL_2816993, EPI_ISL_2817020, EPI_ISL_2817119, EPI_ISL_2817120, EPI_ISL_2817121, EPI_ISL_2817122, EPI_ISL_2817123, EPI_ISL_2817124, EPI_ISL_2817125, EPI_ISL_2817126, EPI_ISL_2817127, EPI_ISL_2817128, EPI_ISL_2817129, EPI_ISL_2817130, EPI_ISL_2817131, EPI_ISL_2817132, EPI_ISL_2817134, EPI_ISL_2817135, EPI_ISL_2817136, EPI_ISL_2817137, EPI_ISL_2817138, EPI_ISL_2817139, EPI_ISL_2817140, EPI_ISL_2817141, EPI_ISL_2817142, EPI_ISL_2817143, EPI_ISL_2817144, EPI_ISL_2817145, EPI_ISL_2817146, EPI_ISL_2817147, EPI_ISL_2817148, EPI_ISL_2817149, EPI_ISL_2817150, EPI_ISL_2817151, EPI_ISL_2817152, EPI_ISL_2817153, EPI_ISL_2817154, EPI_ISL_2817155, EPI_ISL_2817156, EPI_ISL_2817157, EPI_ISL_2817158, EPI_ISL_2817159, EPI_ISL_2817160, EPI_ISL_2817161, EPI_ISL_2817162, EPI_ISL_2817163, EPI_ISL_2817164, EPI_ISL_2817165, EPI_ISL_2817166, EPI_ISL_2817167, EPI_ISL_2817168, EPI_ISL_2817169, EPI_ISL_2902410, EPI_ISL_2902411, EPI_ISL_2902412, EPI_ISL_2902413, EPI_ISL_2902414, EPI_ISL_2902415, EPI_ISL_2902416, EPI_ISL_2902417, EPI_ISL_2902418, EPI_ISL_2902419, EPI_ISL_2902420, EPI_ISL_2902421, EPI_ISL_2902422, EPI_ISL_2902423, EPI_ISL_2902424, EPI_ISL_2902425, EPI_ISL_2902426, EPI_ISL_2902427, EPI_ISL_2902428, EPI_ISL_2902429, EPI_ISL_2902430, EPI_ISL_2902431, EPI_ISL_2902432, EPI_ISL_2902434, EPI_ISL_2902435, EPI_ISL_2902436, EPI_ISL_2902437, EPI_ISL_2902438, EPI_ISL_2902439, EPI_ISL_2902440, EPI_ISL_2902441, EPI_ISL_2902442, EPI_ISL_2902443, EPI_ISL_2902444, EPI_ISL_2902445, EPI_ISL_2902446, EPI_ISL_2902447, EPI_ISL_2902448, EPI_ISL_2902449, EPI_ISL_2902450, EPI_ISL_2902451, EPI_ISL_2902452, EPI_ISL_2902453, EPI_ISL_2902454, EPI_ISL_2902455, EPI_ISL_2902457, EPI_ISL_2902458, EPI_ISL_2902459, EPI_ISL_2902460, EPI_ISL_2902461, EPI_ISL_2902462, EPI_ISL_2902463, EPI_ISL_2902464, EPI_ISL_2902466, EPI_ISL_2902467, EPI_ISL_2902468, EPI_ISL_2902469, EPI_ISL_2902470, EPI_ISL_2902471, EPI_ISL_2902472, EPI_ISL_2902473, EPI_ISL_2902474, EPI_ISL_2902475, EPI_ISL_2902476, EPI_ISL_2902477, EPI_ISL_2902478, EPI_ISL_2902479, EPI_ISL_2902480, EPI_ISL_2902481, EPI_ISL_2902482, EPI_ISL_2902483, EPI_ISL_2902484, EPI_ISL_2902485, EPI_ISL_2902486, EPI_ISL_2902487, EPI_ISL_2902488, EPI_ISL_2902489, EPI_ISL_2902490, EPI_ISL_2902491, EPI_ISL_2902492, EPI_ISL_2902493, EPI_ISL_2902494, EPI_ISL_2902495, EPI_ISL_2902496, EPI_ISL_2902497, EPI_ISL_2902498, EPI_ISL_2902499, EPI_ISL_2902502, EPI_ISL_2902503, EPI_ISL_2902504, EPI_ISL_2902505, EPI_ISL_2902508, EPI_ISL_2902509, EPI_ISL_2902510, EPI_ISL_2902511, EPI_ISL_2902512, EPI_ISL_2902513, EPI_ISL_2902514, EPI_ISL_2902515, EPI_ISL_2902516, EPI_ISL_2902517, EPI_ISL_2902518, EPI_ISL_2902519, EPI_ISL_2902520, EPI_ISL_2902521, EPI_ISL_2902522, EPI_ISL_2902523, EPI_ISL_2902524, EPI_ISL_2902525, EPI_ISL_2902526, EPI_ISL_2902527, EPI_ISL_2902528, EPI_ISL_2902529, EPI_ISL_2902530, EPI_ISL_2902531, EPI_ISL_2902532, EPI_ISL_2902533, EPI_ISL_2902534, EPI_ISL_2902535, EPI_ISL_2902536, EPI_ISL_2902537, EPI_ISL_2902538, EPI_ISL_2902539, EPI_ISL_2902540, EPI_ISL_2902541, EPI_ISL_2902542, EPI_ISL_2902543, EPI_ISL_2902544, EPI_ISL_2902545, EPI_ISL_2902546, EPI_ISL_2902547, EPI_ISL_2902548, EPI_ISL_2902549, EPI_ISL_2902550, EPI_ISL_2902551, EPI_ISL_2902552, EPI_ISL_2902553, EPI_ISL_2902554, EPI_ISL_2902555, EPI_ISL_2902556, EPI_ISL_2902557, EPI_ISL_2902558, EPI_ISL_2902559, EPI_ISL_2902560, EPI_ISL_2902561, EPI_ISL_2902562, EPI_ISL_2902563, EPI_ISL_2902564, EPI_ISL_2902565, EPI_ISL_2902566, EPI_ISL_2902567, EPI_ISL_2902568, EPI_ISL_2902569, EPI_ISL_2902570, EPI_ISL_2902571, EPI_ISL_2902572, EPI_ISL_2902574, EPI_ISL_2902575, EPI_ISL_2902576, EPI_ISL_2902577, EPI_ISL_2902578, EPI_ISL_2902579, EPI_ISL_2902580, EPI_ISL_2902581, EPI_ISL_2902582, EPI_ISL_2902583, EPI_ISL_2902584, EPI_ISL_2902585, EPI_ISL_2902586, EPI_ISL_2902587, EPI_ISL_2902588, EPI_ISL_2902589, EPI_ISL_2902590, EPI_ISL_2902592, EPI_ISL_2902593, EPI_ISL_2902594, EPI_ISL_2902595, EPI_ISL_2902596, EPI_ISL_2902597, EPI_ISL_2902598, EPI_ISL_2902599, EPI_ISL_2902600, EPI_ISL_2902601, EPI_ISL_2902602, EPI_ISL_2902603, EPI_ISL_2902604, EPI_ISL_2902605, EPI_ISL_2902606, EPI_ISL_2902607, EPI_ISL_2902608, EPI_ISL_2902609, EPI_ISL_2902610, EPI_ISL_2902611, EPI_ISL_2902612, EPI_ISL_2902613, EPI_ISL_2902614, EPI_ISL_2902615, EPI_ISL_2902616, EPI_ISL_2902617, EPI_ISL_2902618, EPI_ISL_2902619, EPI_ISL_2902620, EPI_ISL_2902622, EPI_ISL_2902623, EPI_ISL_2902624, EPI_ISL_2902625, EPI_ISL_2902626, EPI_ISL_2902627, EPI_ISL_2902628, EPI_ISL_2902629, EPI_ISL_2902630, EPI_ISL_2902631, EPI_ISL_2902632, EPI_ISL_2902633, EPI_ISL_2902634, EPI_ISL_2902635, EPI_ISL_2902636, EPI_ISL_2902637, EPI_ISL_2902638, EPI_ISL_2902639

| | | | |
|---|---|---|---|
| see above | Quadram Institute Bioscience | COVID-19 Genomics UK (COG-UK) Consortium | Alexander J Trotter; Alison E. Mather; Alp Aydin; Ana P. Tedim; Anastasia Kolyva; Andrew Bell; Andrew J. Page; Christopher Jeanes; Claire Stuart; Dave J. Baker; Ebenezer Foster-Nyarko; Gemma L. Kay; John Wain; Justin O'Grady; Leonardo de Oliveira Martins; Lewis G. Spurgin; Lindsay Coupland; Lizzie Meadows; Luke Bedford; Maria Diaz; Mark Webber; Martin Lott; Muhammed Yasir; Nabil-Fareed Alikhan; Ngozi Elumogo; Nicholas M. Thomson; Rachael Stanley; Rachel Gilroy; Reenesh Prakash; Rose K Davidson; Samir Dervisevic; Samuel Bloomfield; Sophie J. Prosolek; Steven Rudder; Thanh Le-Viet |

EPI_ISL_2817282, EPI_ISL_2817283, EPI_ISL_2817284, EPI_ISL_2817285, EPI_ISL_2817286, EPI_ISL_2817287, EPI_ISL_2817288, EPI_ISL_2817289, EPI_ISL_2817290, EPI_ISL_2817291, EPI_ISL_2817292, EPI_ISL_2817293, EPI_ISL_2817294, EPI_ISL_2817295, EPI_ISL_2817296, EPI_ISL_2817297, EPI_ISL_2817298, EPI_ISL_2817409, EPI_ISL_2817410, EPI_ISL_2817411, EPI_ISL_2817412, EPI_ISL_2817413, EPI_ISL_2817414, EPI_ISL_2817415, EPI_ISL_2817416, EPI_ISL_2817417, EPI_ISL_2817419, EPI_ISL_2817420, EPI_ISL_2817422, EPI_ISL_2817424, EPI_ISL_2817427, EPI_ISL_2817433, EPI_ISL_2817443, EPI_ISL_2817444, EPI_ISL_2817479, EPI_ISL_2817480, EPI_ISL_2817492, EPI_ISL_2817493, EPI_ISL_2817632, EPI_ISL_2817633, EPI_ISL_2817634, EPI_ISL_2817635, EPI_ISL_2817636, EPI_ISL_2817637, EPI_ISL_2817638, EPI_ISL_2817639, EPI_ISL_2817640, EPI_ISL_2817641, EPI_ISL_2817642, EPI_ISL_2817643, EPI_ISL_2817644, EPI_ISL_2817645, EPI_ISL_2817646, EPI_ISL_2817647, EPI_ISL_2817648, EPI_ISL_2817649, EPI_ISL_2817650, EPI_ISL_2817664, EPI_ISL_2817668, EPI_ISL_2817672, EPI_ISL_2817674, EPI_ISL_2817675, EPI_ISL_2817676, EPI_ISL_2817677, EPI_ISL_2817678, EPI_ISL_2817679, EPI_ISL_2817680, EPI_ISL_2817681, EPI_ISL_2817682, EPI_ISL_2817688, EPI_ISL_2817689, EPI_ISL_2817694, EPI_ISL_2817712, EPI_ISL_2817719, EPI_ISL_2817728, EPI_ISL_2817813, EPI_ISL_2817814, EPI_ISL_2817816, EPI_ISL_2817817, EPI_ISL_2817818, EPI_ISL_2817819, EPI_ISL_2817820, EPI_ISL_2817822, EPI_ISL_2817824, EPI_ISL_2817827, EPI_ISL_2817828, EPI_ISL_2817829, EPI_ISL_2817830, EPI_ISL_2817831, EPI_ISL_2817832, EPI_ISL_2817833, EPI_ISL_2817834, EPI_ISL_2817835, EPI_ISL_2817836, EPI_ISL_2817837, EPI_ISL_2817838, EPI_ISL_2817840, EPI_ISL_2817841, EPI_ISL_2817842, EPI_ISL_2817844, EPI_ISL_2817845, EPI_ISL_2817846, EPI_ISL_2817847, EPI_ISL_2817848, EPI_ISL_2817849, EPI_ISL_2817850, EPI_ISL_2817851, EPI_ISL_2817852, EPI_ISL_2817857, EPI_ISL_2817858, EPI_ISL_2817859, EPI_ISL_2817862, EPI_ISL_2817863, EPI_ISL_2817864, EPI_ISL_2817865, EPI_ISL_2817866, EPI_ISL_2817867, EPI_ISL_2817868, EPI_ISL_2817869, EPI_ISL_2817870, EPI_ISL_2817871, EPI_ISL_2817872, EPI_ISL_2817873, EPI_ISL_2817874, EPI_ISL_2817877, EPI_ISL_2817903, EPI_ISL_2817913, EPI_ISL_2817914, EPI_ISL_2817915, EPI_ISL_2817920, EPI_ISL_2817921, EPI_ISL_2817922, EPI_ISL_2817923, EPI_ISL_2817924, EPI_ISL_2817925, EPI_ISL_2817926, EPI_ISL_2817927, EPI_ISL_2817928, EPI_ISL_2817929, EPI_ISL_2817930, EPI_ISL_2817931, EPI_ISL_2817932, EPI_ISL_2817933, EPI_ISL_2817935, EPI_ISL_2817936, EPI_ISL_2817937, EPI_ISL_2817938, EPI_ISL_2817939, EPI_ISL_2817940, EPI_ISL_2817941, EPI_ISL_2817942, EPI_ISL_2817943, EPI_ISL_2817944, EPI_ISL_2817945, EPI_ISL_2817946, EPI_ISL_2817947, EPI_ISL_2817948, EPI_ISL_2817949, EPI_ISL_2817950, EPI_ISL_2817951, EPI_ISL_2817953, EPI_ISL_2817954, EPI_ISL_2817955, EPI_ISL_2817957, EPI_ISL_2817958, EPI_ISL_2817959, EPI_ISL_2817960, EPI_ISL_2817961, EPI_ISL_2817962, EPI_ISL_2817963, EPI_ISL_2817964, EPI_ISL_2817965, EPI_ISL_2817966, EPI_ISL_2817967, EPI_ISL_2817989, EPI_ISL_2817990, EPI_ISL_2817991, EPI_ISL_2817992, EPI_ISL_2817997, EPI_ISL_2817998, EPI_ISL_2817999, EPI_ISL_2818000, EPI_ISL_2818001, EPI_ISL_2818002, EPI_ISL_2818003, EPI_ISL_2818004, EPI_ISL_2818005, EPI_ISL_2818006, EPI_ISL_2818007, EPI_ISL_2818008, EPI_ISL_2818009, EPI_ISL_2818010, EPI_ISL_2818013, EPI_ISL_2818014, EPI_ISL_2818015, EPI_ISL_2818016, EPI_ISL_2818017, EPI_ISL_2818018, EPI_ISL_2818019, EPI_ISL_2818020, EPI_ISL_2818021, EPI_ISL_2818022, EPI_ISL_2818024, EPI_ISL_2818025, EPI_ISL_2818026, EPI_ISL_2818027, EPI_ISL_2818028, EPI_ISL_2818029, EPI_ISL_2818030, EPI_ISL_2818031, EPI_ISL_2818034, EPI_ISL_2818035, EPI_ISL_2818036, EPI_ISL_2818037, EPI_ISL_2818038, EPI_ISL_2818039, EPI_ISL_2818040, EPI_ISL_2818050, EPI_ISL_2818052, EPI_ISL_2818053, EPI_ISL_2818054, EPI_ISL_2818055, EPI_ISL_2818056, EPI_ISL_2818057, EPI_ISL_2818058, EPI_ISL_2818059, EPI_ISL_2818061, EPI_ISL_2818067, EPI_ISL_2818068, EPI_ISL_2818069, EPI_ISL_2818070, EPI_ISL_2818073, EPI_ISL_2818158, EPI_ISL_2818162, EPI_ISL_2818163, EPI_ISL_2818165, EPI_ISL_2818295, EPI_ISL_2818296, EPI_ISL_2818297, EPI_ISL_2818298, EPI_ISL_2818299, EPI_ISL_2818300, EPI_ISL_2818301, EPI_ISL_2818302, EPI_ISL_2818303, EPI_ISL_2818304, EPI_ISL_2818305, EPI_ISL_2818306, EPI_ISL_2818307, EPI_ISL_2818308, EPI_ISL_2818309, EPI_ISL_2818310, EPI_ISL_2818311, EPI_ISL_2818312, EPI_ISL_2818313, EPI_ISL_2818314, EPI_ISL_2903412, EPI_ISL_2903413, EPI_ISL_2903414, EPI_ISL_2903416, EPI_ISL_2903417, EPI_ISL_2903425, EPI_ISL_2903426, EPI_ISL_2903427, EPI_ISL_2903428, EPI_ISL_2903429, EPI_ISL_2903430, EPI_ISL_2903431, EPI_ISL_2903432, EPI_ISL_2903433, EPI_ISL_2903434, EPI_ISL_2903435, EPI_ISL_2903436, EPI_ISL_2903437, EPI_ISL_2903439, EPI_ISL_2903440, EPI_ISL_2903441, EPI_ISL_2903442, EPI_ISL_2903443, EPI_ISL_2903444, EPI_ISL_2903445, EPI_ISL_2903446, EPI_ISL_2903447, EPI_ISL_2903448, EPI_ISL_2903449, EPI_ISL_2903451, EPI_ISL_2903458, EPI_ISL_2903459, EPI_ISL_2903461, EPI_ISL_2903462, EPI_ISL_2903463, EPI_ISL_2903464, EPI_ISL_2903468, EPI_ISL_2903469, EPI_ISL_2903470, EPI_ISL_2903471, EPI_ISL_2903473, EPI_ISL_2903476, EPI_ISL_2903477, EPI_ISL_2903479, EPI_ISL_2903480, EPI_ISL_2903481, EPI_ISL_2903482, EPI_ISL_2903483, EPI_ISL_2903484, EPI_ISL_2903485, EPI_ISL_2903491, EPI_ISL_2903492, EPI_ISL_2903497, EPI_ISL_2903527, EPI_ISL_2903556, EPI_ISL_2903654, EPI_ISL_2903656, EPI_ISL_2903657, EPI_ISL_2903661, EPI_ISL_2903664, EPI_ISL_2903671, EPI_ISL_2903687, EPI_ISL_2903697, EPI_ISL_2903698, EPI_ISL_2903699, EPI_ISL_2903700, EPI_ISL_2903701, EPI_ISL_2903702, EPI_ISL_2903703, EPI_ISL_2903704, EPI_ISL_2903705, EPI_ISL_2903706, EPI_ISL_2903707, EPI_ISL_2903750, EPI_ISL_2903756, EPI_ISL_2903933, EPI_ISL_2903934, EPI_ISL_2903935, EPI_ISL_2903936, EPI_ISL_2903937, EPI_ISL_2903938, EPI_ISL_2903939, EPI_ISL_2903945, EPI_ISL_2903946, EPI_ISL_2903947, EPI_ISL_2903951, EPI_ISL_2903952, EPI_ISL_2903956, EPI_ISL_2903957, EPI_ISL_2903958, EPI_ISL_2903959, EPI_ISL_2903960, EPI_ISL_2903962, EPI_ISL_2903963, EPI_ISL_2903964, EPI_ISL_2903965, EPI_ISL_2903966, EPI_ISL_2903968, EPI_ISL_2903969, EPI_ISL_2903970, EPI_ISL_2903971, EPI_ISL_2903972, EPI_ISL_2903973, EPI_ISL_2903974, EPI_ISL_2903975, EPI_ISL_2903976, EPI_ISL_2903977, EPI_ISL_2903978, EPI_ISL_2903979, EPI_ISL_2903980, EPI_ISL_2903981, EPI_ISL_2903982, EPI_ISL_2903983, EPI_ISL_2903984, EPI_ISL_2903985, EPI_ISL_2903986, EPI_ISL_2903990, EPI_ISL_2904043, EPI_ISL_2904046, EPI_ISL_2904047, EPI_ISL_2904058, EPI_ISL_2904065, EPI_ISL_2904066, EPI_ISL_2904067, EPI_ISL_2904068, EPI_ISL_2904069, EPI_ISL_2904070, EPI_ISL_2904071, EPI_ISL_2904072, EPI_ISL_2904073, EPI_ISL_2904075, EPI_ISL_2904078, EPI_ISL_2904082, EPI_ISL_2904084, EPI_ISL_2904087, EPI_ISL_2904089, EPI_ISL_2904091, EPI_ISL_2904092, EPI_ISL_2904109, EPI_ISL_2904340, EPI_ISL_2904341, EPI_ISL_2904342, EPI_ISL_2904344, EPI_ISL_2904351, EPI_ISL_2904352, EPI_ISL_2904353, EPI_ISL_2904374, EPI_ISL_2904401

| | | | |
|---|---|---|---|
| see above | Respiratory Virus Unit, Microbiology Services Colindale, Public Health England | COVID-19 Genomics UK (COG-UK) Consortium | PHE Covid Sequencing Team |

EPI_ISL_2816061, EPI_ISL_2816064, EPI_ISL_2816069, EPI_ISL_2816070, EPI_ISL_2816071, EPI_ISL_2816085, EPI_ISL_2816086, EPI_ISL_2816087, EPI_ISL_2816088, EPI_ISL_2816090, EPI_ISL_2816091, EPI_ISL_2900747, EPI_ISL_2900748, EPI_ISL_2900749, EPI_ISL_2900750, EPI_ISL_2900751, EPI_ISL_2900752, EPI_ISL_2900753, EPI_ISL_2900754, EPI_ISL_2900766, EPI_ISL_2900767, EPI_ISL_2900769, EPI_ISL_2900770, EPI_ISL_2900771, EPI_ISL_2900772, EPI_ISL_2900774, EPI_ISL_2900775, EPI_ISL_2900776, EPI_ISL_2900778, EPI_ISL_2900779, EPI_ISL_2900784, EPI_ISL_2900787, EPI_ISL_2900788, EPI_ISL_2900791, EPI_ISL_2900798, EPI_ISL_2900801, EPI_ISL_2900809, EPI_ISL_2900811, EPI_ISL_2900814, EPI_ISL_2900816, EPI_ISL_2900820, EPI_ISL_2900821

| | | | |
|---|---|---|---|
| see above | Virology Department, Royal Infirmary of Edinburgh, NHS Lothian / School of Biological Sciences, University of Edinburgh | COVID-19 Genomics UK (COG-UK) Consortium | Colquhoun R; Cotton S; Dewar R; Gallagher A; Hill V; Jackson B; Maloney D; McCrone JT; McHugh M; O'Toole Á; Rambaut A; Rooke S; Scher E; Templeton K; Yu X |

EPI_ISL_2819943, EPI_ISL_2819987, EPI_ISL_2819989, EPI_ISL_2820069, EPI_ISL_2820095, EPI_ISL_2820103, EPI_ISL_2820138, EPI_ISL_2820171

| | | | |
|---|---|---|---|
| see above | Virology Department, Sheffield Teaching Hospitals NHS Foundation Trust/Department of Infection, Immunity and Cardiovascular Disease, The Medical School, University of Sheffield | COVID-19 Genomics UK (COG-UK) Consortium | Adri Angyal; Alex Keeley; Benjamin Lindsey; Cariad Evans; Danielle Groves; Dave Partridge; Katie Johnson; Laura Carrilero; Luke Green; Matthew Parker; Matthew Wyles; Mehmet Yavuz; Mohammad Raza; Nikki Smith; Paul Parsons; Rachel Tucker; Rebecca Brown; Thushan de Silva |
| EPI_ISL_2900708, EPI_ISL_2900710, EPI_ISL_2900724, EPI_ISL_2900725, EPI_ISL_2900726 | West of Scotland Specialist Virology Centre, NHSGGC / MRC-University of Glasgow Centre for Virus Research | COVID-19 Genomics UK (COG-UK) Consortium | Alasdair MacLean; Alice Broos; Ana da Silva Filipe; Antonia Ho; Daniel Mair; David L Robertson; Emma Thomson; Guy Mollett; Ioulia Tsatsani; James Shepherd; Jenna Nichols; Joseph Hughes; Kathy Li; Kathy Smollett; Kyriaki Nomikou; Lily Tong; Matthew Holden; Natasha Johnson; Rachel Blacow; Richard Orton; Rory Gunson; Sarah McDonald; Sharif Shaaban; Sreenu Vattipally; Stephen Carmichael |

We gratefully acknowledge the following Authors from the Originating laboratories responsible for obtaining the specimens, as well as the Submitting laboratories where the genome data were generated and shared via GISAID, on which this research is based.

All Submitters of data may be contacted directly via www.gisaid.org

Authors are sorted alphabetically.

| Accession ID | Originating Laboratory | Submitting Laboratory | Authors |
|---|---|---|---|
| EPI_ISL_2846363, EPI_ISL_2846364, EPI_ISL_2846369, EPI_ISL_2846372, EPI_ISL_2846377, EPI_ISL_2846379, EPI_ISL_2846386, EPI_ISL_2846390, EPI_ISL_2846392, EPI_ISL_2846393, EPI_ISL_2846394, EPI_ISL_2846398, EPI_ISL_2846399, EPI_ISL_2846401, EPI_ISL_2846402, EPI_ISL_2846404, EPI_ISL_2846408, EPI_ISL_2846411, EPI_ISL_2846413, EPI_ISL_2846416, EPI_ISL_2846418, EPI_ISL_2846423, EPI_ISL_2846431, EPI_ISL_2846432, EPI_ISL_2846436, EPI_ISL_2846437, EPI_ISL_2846438, EPI_ISL_2846439, EPI_ISL_2846442, EPI_ISL_2846444, EPI_ISL_2846445, EPI_ISL_2846448, EPI_ISL_2846453, EPI_ISL_2846455, EPI_ISL_2846459, EPI_ISL_2846461, EPI_ISL_2846462, EPI_ISL_2846464, EPI_ISL_2846466, EPI_ISL_2846467, EPI_ISL_2846470, EPI_ISL_2846475, EPI_ISL_2846476, EPI_ISL_2846477, EPI_ISL_2846478, EPI_ISL_2846482, EPI_ISL_2846487, EPI_ISL_2846493, EPI_ISL_2846504, EPI_ISL_2846506, EPI_ISL_2846508, EPI_ISL_2846510, EPI_ISL_2846517, EPI_ISL_2846519, EPI_ISL_2846520, EPI_ISL_2846523, EPI_ISL_2846526, EPI_ISL_2846527, EPI_ISL_2846529, EPI_ISL_2846530, EPI_ISL_2846540, EPI_ISL_2846547, EPI_ISL_2846550, EPI_ISL_2846552, EPI_ISL_2846553, EPI_ISL_2846558, EPI_ISL_2846561, EPI_ISL_2846562, EPI_ISL_2846563, EPI_ISL_2846565, EPI_ISL_2846567, EPI_ISL_2846571, EPI_ISL_2846577, EPI_ISL_2864810, EPI_ISL_2864853, EPI_ISL_2864885, EPI_ISL_2864894, EPI_ISL_2864895, EPI_ISL_2864955, EPI_ISL_2864961, EPI_ISL_2864990, EPI_ISL_2865005, EPI_ISL_2865023, EPI_ISL_2865056, EPI_ISL_2865112, EPI_ISL_2865145, EPI_ISL_2865231, EPI_ISL_2865237, EPI_ISL_2865289, EPI_ISL_2867032, EPI_ISL_2867046, EPI_ISL_2867047, EPI_ISL_2867055, EPI_ISL_2867065, EPI_ISL_2867066, EPI_ISL_2867067, EPI_ISL_2867069, EPI_ISL_2867070, EPI_ISL_2867074, EPI_ISL_2867078, EPI_ISL_2867080, EPI_ISL_2867083, EPI_ISL_2867084, EPI_ISL_2867092, EPI_ISL_2867093, EPI_ISL_2867096, EPI_ISL_2867101, EPI_ISL_2867102, EPI_ISL_2867105, EPI_ISL_2867109, EPI_ISL_2867114, EPI_ISL_2867115, EPI_ISL_2867123, EPI_ISL_2867126, EPI_ISL_2867127, EPI_ISL_2867133, EPI_ISL_2867134, EPI_ISL_2867135, EPI_ISL_2867145, EPI_ISL_2867153, EPI_ISL_2867161, EPI_ISL_2867167, EPI_ISL_2867169, EPI_ISL_2867174, EPI_ISL_2867185, EPI_ISL_2867193, EPI_ISL_2867198, EPI_ISL_2867201, EPI_ISL_2867206, EPI_ISL_2867207, EPI_ISL_2867208, EPI_ISL_2867211, EPI_ISL_2867215, EPI_ISL_2867231, EPI_ISL_2867234, EPI_ISL_2867235, EPI_ISL_2867240, EPI_ISL_2867243, EPI_ISL_2867248, EPI_ISL_2867251, EPI_ISL_2867252, EPI_ISL_2867256, EPI_ISL_2867258, EPI_ISL_2867263, EPI_ISL_2867274, EPI_ISL_2867279, EPI_ISL_2867288, EPI_ISL_2867292, EPI_ISL_2867299, EPI_ISL_2867300, EPI_ISL_2867304, EPI_ISL_2867311, EPI_ISL_2867312, EPI_ISL_2867315, EPI_ISL_2867317, EPI_ISL_2867319, EPI_ISL_2867325, EPI_ISL_2867328, EPI_ISL_2867359, EPI_ISL_2867365, EPI_ISL_2867366, EPI_ISL_2867367, EPI_ISL_2867371, EPI_ISL_2867373, EPI_ISL_2867377, EPI_ISL_2867388, EPI_ISL_2867397, EPI_ISL_2867398, EPI_ISL_2867400, EPI_ISL_2867403, EPI_ISL_2867405, EPI_ISL_2867407, EPI_ISL_2867409, EPI_ISL_2867411, EPI_ISL_2867417, EPI_ISL_2867418, EPI_ISL_2867427, EPI_ISL_2867428, EPI_ISL_2867429, EPI_ISL_2867432, EPI_ISL_2867434, EPI_ISL_2867436, EPI_ISL_2867440, EPI_ISL_2867442, EPI_ISL_2867444, EPI_ISL_2867445, EPI_ISL_2867446, EPI_ISL_2867449, EPI_ISL_2867450, EPI_ISL_2867455, EPI_ISL_2867456, EPI_ISL_2867458, EPI_ISL_2867459, EPI_ISL_2867461, EPI_ISL_2867465, EPI_ISL_2867466, EPI_ISL_2867470, EPI_ISL_2867476, EPI_ISL_2867482, EPI_ISL_2867485, EPI_ISL_2867492, EPI_ISL_2867503, EPI_ISL_2867504, EPI_ISL_2867508, EPI_ISL_2867509, EPI_ISL_2867514, EPI_ISL_2867515, EPI_ISL_2867521, EPI_ISL_2867524, EPI_ISL_2867527, EPI_ISL_2867528, EPI_ISL_2867533, EPI_ISL_2867534, EPI_ISL_2867535, EPI_ISL_2867536, EPI_ISL_2867540, EPI_ISL_2867541, EPI_ISL_2867548, EPI_ISL_2867561, EPI_ISL_2867564, EPI_ISL_2867565, EPI_ISL_2867566, EPI_ISL_2867568, EPI_ISL_2867569, EPI_ISL_2867570, EPI_ISL_2867571, EPI_ISL_2867575, EPI_ISL_2867576, EPI_ISL_2867581, EPI_ISL_2867585, EPI_ISL_2867589, EPI_ISL_2867590, EPI_ISL_2867591, EPI_ISL_2867593, EPI_ISL_2867594, EPI_ISL_2867601, EPI_ISL_2867603, EPI_ISL_2867604, EPI_ISL_2867607, EPI_ISL_2867610, EPI_ISL_2867612, EPI_ISL_2867617, EPI_ISL_2867619, EPI_ISL_2867620, EPI_ISL_2867623, EPI_ISL_2867624, EPI_ISL_2867625, EPI_ISL_2867626, EPI_ISL_2867629, EPI_ISL_2867630, EPI_ISL_2867633, EPI_ISL_2867637, EPI_ISL_2867639, EPI_ISL_2867644, EPI_ISL_2867655, EPI_ISL_2867666, EPI_ISL_2867672, EPI_ISL_2867673, EPI_ISL_2867683, EPI_ISL_2867685, EPI_ISL_2867688, EPI_ISL_2868210, EPI_ISL_2913623, EPI_ISL_2914640, EPI_ISL_2915556, EPI_ISL_2915782, EPI_ISL_2915932, EPI_ISL_2915941, EPI_ISL_2916691, EPI_ISL_2916694, EPI_ISL_2916698, EPI_ISL_2916725, EPI_ISL_2916732, EPI_ISL_2916748, EPI_ISL_2916763, EPI_ISL_2916825, EPI_ISL_2916849, EPI_ISL_2916903, EPI_ISL_2916934, EPI_ISL_2916968, EPI_ISL_2917011, EPI_ISL_2917016, EPI_ISL_2917018, EPI_ISL_2917023, EPI_ISL_2917047, EPI_ISL_2917052, EPI_ISL_2917054, EPI_ISL_2917067, EPI_ISL_2917068, EPI_ISL_2917069, EPI_ISL_2917071, EPI_ISL_2917073, EPI_ISL_2917075, EPI_ISL_2917079, EPI_ISL_2917083, EPI_ISL_2917086, EPI_ISL_2917092, EPI_ISL_2917094, EPI_ISL_2917121, EPI_ISL_2917133, EPI_ISL_2917143, EPI_ISL_2917146, EPI_ISL_2917151, EPI_ISL_2917153, EPI_ISL_2917154, EPI_ISL_2917155, EPI_ISL_2917156, EPI_ISL_2917158, EPI_ISL_2917159, EPI_ISL_2917161, EPI_ISL_2917164, EPI_ISL_2917168, EPI_ISL_2917172, EPI_ISL_2917180, EPI_ISL_2917181, EPI_ISL_2917184, EPI_ISL_2917188, EPI_ISL_2917189, EPI_ISL_2917194, EPI_ISL_2917196, EPI_ISL_2917210, EPI_ISL_2917211, EPI_ISL_2917213, EPI_ISL_2917218, EPI_ISL_2917221, EPI_ISL_2917222, EPI_ISL_2917228, EPI_ISL_2917229, EPI_ISL_2917232, EPI_ISL_2917239, EPI_ISL_2917242, EPI_ISL_2917258, EPI_ISL_2917264, EPI_ISL_2917265, EPI_ISL_2917274, EPI_ISL_2917281, EPI_ISL_2917282, EPI_ISL_2917284, EPI_ISL_2917295, EPI_ISL_2917296, EPI_ISL_2917297, EPI_ISL_2917311, EPI_ISL_2917315, EPI_ISL_2917329, EPI_ISL_2917332, EPI_ISL_2917335, EPI_ISL_2917337, EPI_ISL_2917344, EPI_ISL_2917604, EPI_ISL_2918109, EPI_ISL_2918213, EPI_ISL_2918285, EPI_ISL_2918322, EPI_ISL_2918371, EPI_ISL_2918418, EPI_ISL_2918441, EPI_ISL_2918445, EPI_ISL_2918446, EPI_ISL_2918448, EPI_ISL_2918461, EPI_ISL_2918464, EPI_ISL_2918475, EPI_ISL_2918485, EPI_ISL_2918492, EPI_ISL_2918514, EPI_ISL_2918566, EPI_ISL_2918570, EPI_ISL_2918591, EPI_ISL_2918594, EPI_ISL_2918598 | | | |
| see above | Berkshire and Surrey Pathology Services Lighthouse Laboratory | Wellcome Sanger Institute for the COVID-19 Genomics UK (COG-UK) Consortium | Berkshire and Surrey Pathology Services Lighthouse Laboratory and Alex Alderton; Cordelia Langford; David K. Jackson; Dominic Kwiatkowski; Ewan Harrison; Ian Johnston; Jeffrey Barrett; John Sillitoe on behalf of the Wellcome Sanger Institute COVID-19 Surveillance Team; Roberto Amato; Sonia Goncalves |
| EPI_ISL_2815705, EPI_ISL_2815706, EPI_ISL_2815707, EPI_ISL_2815708, EPI_ISL_2815709, EPI_ISL_2815711, EPI_ISL_2815712, EPI_ISL_2815713, EPI_ISL_2815714, EPI_ISL_2815715, EPI_ISL_2815716, EPI_ISL_2815717, EPI_ISL_2815718, EPI_ISL_2815719, EPI_ISL_2815720, EPI_ISL_2815721, EPI_ISL_2815722, EPI_ISL_2815723, EPI_ISL_2815724, EPI_ISL_2815725, EPI_ISL_2815726, EPI_ISL_2815727, EPI_ISL_2815728, EPI_ISL_2815729, EPI_ISL_2815730, EPI_ISL_2815731, EPI_ISL_2815732, EPI_ISL_2815733, EPI_ISL_2815734, EPI_ISL_2815744, EPI_ISL_2815745, EPI_ISL_2815746, EPI_ISL_2815747, EPI_ISL_2815748, EPI_ISL_2815749, EPI_ISL_2815750, EPI_ISL_2815751, EPI_ISL_2815752, EPI_ISL_2815753, EPI_ISL_2815754, EPI_ISL_2815755, EPI_ISL_2815756, EPI_ISL_2815757, EPI_ISL_2815758, EPI_ISL_2815759, EPI_ISL_2815760, EPI_ISL_2815761, EPI_ISL_2815762, EPI_ISL_2815763, EPI_ISL_2815764, EPI_ISL_2815765, EPI_ISL_2815766, EPI_ISL_2815767, EPI_ISL_2815768, EPI_ISL_2815769, EPI_ISL_2815770, EPI_ISL_2815772, EPI_ISL_2815773, EPI_ISL_2815775, EPI_ISL_2815777, EPI_ISL_2815779, EPI_ISL_2815780, EPI_ISL_2815781, EPI_ISL_2815782, EPI_ISL_2815783, EPI_ISL_2815784, EPI_ISL_2815785, EPI_ISL_2815786, EPI_ISL_2815787, EPI_ISL_2815788, EPI_ISL_2815789, EPI_ISL_2815790, EPI_ISL_2815791, EPI_ISL_2815792, EPI_ISL_2815793, EPI_ISL_2815794, EPI_ISL_2815795, EPI_ISL_2815796, EPI_ISL_2815797, EPI_ISL_2815798, EPI_ISL_2815799, EPI_ISL_2815800, EPI_ISL_2815801, EPI_ISL_2815802, EPI_ISL_2815803, EPI_ISL_2815804, EPI_ISL_2815805, EPI_ISL_2815806, EPI_ISL_2815807, EPI_ISL_2815808, EPI_ISL_2815809, EPI_ISL_2815810, EPI_ISL_2815811, EPI_ISL_2815812, EPI_ISL_2815813, EPI_ISL_2815814, EPI_ISL_2815815, EPI_ISL_2815816, EPI_ISL_2815817, EPI_ISL_2815818, EPI_ISL_2815819, EPI_ISL_2815820, EPI_ISL_2815822, EPI_ISL_2815823, EPI_ISL_2815824, EPI_ISL_2815825, EPI_ISL_2815826, EPI_ISL_2815827, EPI_ISL_2815828, EPI_ISL_2815829, EPI_ISL_2815830, EPI_ISL_2815831, EPI_ISL_2815832, EPI_ISL_2815833, EPI_ISL_2815834, EPI_ISL_2815836, EPI_ISL_2815837, EPI_ISL_2815838, EPI_ISL_2815840, EPI_ISL_2815842, EPI_ISL_2815843, EPI_ISL_2815844, EPI_ISL_2815845, EPI_ISL_2815846, EPI_ISL_2815847, EPI_ISL_2815850, EPI_ISL_2815851, EPI_ISL_2815852, EPI_ISL_2815853, EPI_ISL_2815854, EPI_ISL_2815856, EPI_ISL_2815858, EPI_ISL_2815860, EPI_ISL_2815861, EPI_ISL_2815862, EPI_ISL_2815863, EPI_ISL_2815864, EPI_ISL_2815866, EPI_ISL_2815867, EPI_ISL_2815868, EPI_ISL_2815869, EPI_ISL_2815870, EPI_ISL_2815871, EPI_ISL_2815872, EPI_ISL_2815873, EPI_ISL_2815874, EPI_ISL_2815875, EPI_ISL_2815876, EPI_ISL_2815878, EPI_ISL_2815879, EPI_ISL_2815880, EPI_ISL_2815881, EPI_ISL_2815882, EPI_ISL_2815883, EPI_ISL_2815884, EPI_ISL_2815885, EPI_ISL_2815886, EPI_ISL_2815887, EPI_ISL_2815888, EPI_ISL_2815889, EPI_ISL_2815890, EPI_ISL_2815891, EPI_ISL_2815892, EPI_ISL_2815893, EPI_ISL_2815894 | | | |
| see above | Department of Pathology, University of Cambridge | COVID-19 Genomics UK (COG-UK) Consortium | Aminu S. Jahun; Ian Goodfellow; Iliana Georgana; Martin D. Curran; Myra Hosmillo; Rhys Izuagbe; Surendra Parmar; William L. Hamilton; Yasmin Chaudhry |
| EPI_ISL_2829027, EPI_ISL_2829030, EPI_ISL_2829032, EPI_ISL_2829035, EPI_ISL_2829037, EPI_ISL_2829039, EPI_ISL_2829041, EPI_ISL_2829057, EPI_ISL_2829062, EPI_ISL_2829067, EPI_ISL_2829071, EPI_ISL_2829074, EPI_ISL_2829075, EPI_ISL_2829086, EPI_ISL_2829091, EPI_ISL_2829104, EPI_ISL_2829107, EPI_ISL_2829116, EPI_ISL_2829142, EPI_ISL_2829143, EPI_ISL_2829152, EPI_ISL_2829168, EPI_ISL_2829183, EPI_ISL_2829186, EPI_ISL_2829192, EPI_ISL_2829194, EPI_ISL_2829195, EPI_ISL_2829196, EPI_ISL_2829197, EPI_ISL_2829201, EPI_ISL_2829202, EPI_ISL_2829211, EPI_ISL_2829217, EPI_ISL_2829228, EPI_ISL_2829235, EPI_ISL_2829238, EPI_ISL_2829241, EPI_ISL_2829247, EPI_ISL_2829249, EPI_ISL_2829256, EPI_ISL_2829259, EPI_ISL_2829265, EPI_ISL_2829270, EPI_ISL_2829272, EPI_ISL_2829279, EPI_ISL_2829283, EPI_ISL_2829291, EPI_ISL_2829296, EPI_ISL_2829591, EPI_ISL_2829592, EPI_ISL_2829594, EPI_ISL_2829597, EPI_ISL_2829607, EPI_ISL_2829608, EPI_ISL_2829609, EPI_ISL_2829611, EPI_ISL_2829612, EPI_ISL_2829615, EPI_ISL_2829616, EPI_ISL_2829617, EPI_ISL_2829618, EPI_ISL_2829620, EPI_ISL_2829622, EPI_ISL_2829624, EPI_ISL_2829625, EPI_ISL_2829630, EPI_ISL_2829636, EPI_ISL_2829643, EPI_ISL_2829644, EPI_ISL_2829645, EPI_ISL_2829649, EPI_ISL_2829650, EPI_ISL_2829652, EPI_ISL_2829669, EPI_ISL_2829673, EPI_ISL_2829683, EPI_ISL_2829686, EPI_ISL_2829693, EPI_ISL_2829694, EPI_ISL_2829695, EPI_ISL_2829700, EPI_ISL_2829705, EPI_ISL_2829713, EPI_ISL_2829714, EPI_ISL_2829716, EPI_ISL_2829719, EPI_ISL_2829722, EPI_ISL_2829735, EPI_ISL_2829739, EPI_ISL_2829745, EPI_ISL_2829747, EPI_ISL_2829752, EPI_ISL_2829753, EPI_ISL_2829754, EPI_ISL_2829756, EPI_ISL_2829757, EPI_ISL_2829759, EPI_ISL_2829763, EPI_ISL_2829778, EPI_ISL_2829779, EPI_ISL_2829780, EPI_ISL_2829787, EPI_ISL_2829789, EPI_ISL_2829796, EPI_ISL_2829805, EPI_ISL_2829820, EPI_ISL_2829824, EPI_ISL_2829832, EPI_ISL_2829837, EPI_ISL_2829842, EPI_ISL_2829843, EPI_ISL_2829853, EPI_ISL_2829856, EPI_ISL_2829860, EPI_ISL_2829861, EPI_ISL_2829864, EPI_ISL_2829867, EPI_ISL_2829873, EPI_ISL_2829877, EPI_ISL_2829880, EPI_ISL_2829884, EPI_ISL_2829905, EPI_ISL_2829906, EPI_ISL_2829907, EPI_ISL_2829909, EPI_ISL_2829913, EPI_ISL_2829916, EPI_ISL_2829917, EPI_ISL_2829923, EPI_ISL_2829929, EPI_ISL_2829934, EPI_ISL_2829941, EPI_ISL_2829952, EPI_ISL_2846583, EPI_ISL_2846593, EPI_ISL_2846600, EPI_ISL_2846603, EPI_ISL_2846604, EPI_ISL_2846605, EPI_ISL_2846606, EPI_ISL_2846607, EPI_ISL_2846615, EPI_ISL_2846617, EPI_ISL_2846620, EPI_ISL_2846622, EPI_ISL_2846623, EPI_ISL_2846627, EPI_ISL_2846635, EPI_ISL_2846641, EPI_ISL_2846655, EPI_ISL_2846660, EPI_ISL_2846666, EPI_ISL_2846667, EPI_ISL_2846675, EPI_ISL_2846679, EPI_ISL_2846684, EPI_ISL_2846685, EPI_ISL_2846687, EPI_ISL_2846690, EPI_ISL_2846697, EPI_ISL_2846707, EPI_ISL_2846709, EPI_ISL_2846710, EPI_ISL_2846716, EPI_ISL_2846723, EPI_ISL_2846724, EPI_ISL_2846726, EPI_ISL_2846729, EPI_ISL_2846730, EPI_ISL_2846731, EPI_ISL_2846736, EPI_ISL_2846738, EPI_ISL_2846739, EPI_ISL_2846740, EPI_ISL_2846741, EPI_ISL_2846742, EPI_ISL_2846744, EPI_ISL_2846745, EPI_ISL_2846746, EPI_ISL_2846749, EPI_ISL_2846754, EPI_ISL_2846755, EPI_ISL_2846764, EPI_ISL_2846765, EPI_ISL_2846770, EPI_ISL_2846776, EPI_ISL_2846779, EPI_ISL_2846788, EPI_ISL_2846792, EPI_ISL_2846795, EPI_ISL_2846798, EPI_ISL_2846809, EPI_ISL_2846811, EPI_ISL_2846812, EPI_ISL_2846814, EPI_ISL_2846815, EPI_ISL_2846816, EPI_ISL_2846818, EPI_ISL_2846820, EPI_ISL_2846825, EPI_ISL_2846827, EPI_ISL_2846832, EPI_ISL_2846837, EPI_ISL_2846843, EPI_ISL_2846844, EPI_ISL_2846849, EPI_ISL_2846854, EPI_ISL_2846859, EPI_ISL_2846865, EPI_ISL_2846866, EPI_ISL_2846872, EPI_ISL_2846879, EPI_ISL_2846881, EPI_ISL_2846885, EPI_ISL_2846892, EPI_ISL_2846895, EPI_ISL_2846897, EPI_ISL_2846899, EPI_ISL_2846904, EPI_ISL_2846905, EPI_ISL_2846906, EPI_ISL_2846907, EPI_ISL_2846914, EPI_ISL_2846916, EPI_ISL_2846921, EPI_ISL_2846934, EPI_ISL_2846935, EPI_ISL_2846936, EPI_ISL_2846938, EPI_ISL_2846942, EPI_ISL_2846946, EPI_ISL_2846949, EPI_ISL_2846950, EPI_ISL_2846951, EPI_ISL_2846952, EPI_ISL_2846966, EPI_ISL_2846984, EPI_ISL_2846987, EPI_ISL_2846988, EPI_ISL_2846993, EPI_ISL_2846994, EPI_ISL_2846995, EPI_ISL_2846997, EPI_ISL_2847003, EPI_ISL_2847010, EPI_ISL_2847011, EPI_ISL_2847014, EPI_ISL_2847016, EPI_ISL_2847019, EPI_ISL_2847028, EPI_ISL_2847035, EPI_ISL_2847038, EPI_ISL_2847050, EPI_ISL_2847053, EPI_ISL_2847055, EPI_ISL_2847071, EPI_ISL_2847074, EPI_ISL_2847077, EPI_ISL_2847078, EPI_ISL_2847080, EPI_ISL_2847081, EPI_ISL_2847084, EPI_ISL_2847097, EPI_ISL_2847098, EPI_ISL_2847102, EPI_ISL_2847107, EPI_ISL_2847117, EPI_ISL_2847124, EPI_ISL_2847125, EPI_ISL_2847129, EPI_ISL_2847132, EPI_ISL_2847153, EPI_ISL_2847154, EPI_ISL_2847156, EPI_ISL_2847166, EPI_ISL_2847172, EPI_ISL_2847173, EPI_ISL_2847174, EPI_ISL_2847178, EPI_ISL_2847179, EPI_ISL_2847181, EPI_ISL_2847182, EPI_ISL_2847187, EPI_ISL_2847188, EPI_ISL_2847194, EPI_ISL_2847201, EPI_ISL_2847204, EPI_ISL_2847213, EPI_ISL_2847216, EPI_ISL_2847228, EPI_ISL_2847229, EPI_ISL_2847237, EPI_ISL_2847238, EPI_ISL_2847239, EPI_ISL_2847246, EPI_ISL_2847248, EPI_ISL_2847250, EPI_ISL_2847261, EPI_ISL_2847265, EPI_ISL_2847270, EPI_ISL_2847273, EPI_ISL_2847275, EPI_ISL_2847281, EPI_ISL_2847283, EPI_ISL_2851412, EPI_ISL_2851413, EPI_ISL_2851416, EPI_ISL_2851419, EPI_ISL_2851420, EPI_ISL_2851421, EPI_ISL_2851428, EPI_ISL_2851429, EPI_ISL_2851432, EPI_ISL_2851433, EPI_ISL_2851435, EPI_ISL_2851439, EPI_ISL_2851440, EPI_ISL_2851442, EPI_ISL_2851445, EPI_ISL_2851448, EPI_ISL_2851449, EPI_ISL_2851450, EPI_ISL_2851457, EPI_ISL_2851461, EPI_ISL_2851462, EPI_ISL_2851463, EPI_ISL_2851469, EPI_ISL_2851471, EPI_ISL_2851472, EPI_ISL_2851476, EPI_ISL_2851477, EPI_ISL_2851480, EPI_ISL_2851482, EPI_ISL_2851484, EPI_ISL_2851485, EPI_ISL_2851486, EPI_ISL_2851488, EPI_ISL_2851489, EPI_ISL_2851490, EPI_ISL_2851491, EPI_ISL_2851492, EPI_ISL_2851494, EPI_ISL_2851495, EPI_ISL_2851499, EPI_ISL_2851505, EPI_ISL_2851507, EPI_ISL_2851508, EPI_ISL_2851513, EPI_ISL_2851517, EPI_ISL_2851520, EPI_ISL_2851522, EPI_ISL_2851525, EPI_ISL_2851527, EPI_ISL_2851528, EPI_ISL_2851532, EPI_ISL_2851535, EPI_ISL_2851540, EPI_ISL_2851542, EPI_ISL_2851543, EPI_ISL_2851547, EPI_ISL_2851548, EPI_ISL_2851550, EPI_ISL_2851551, EPI_ISL_2851552, EPI_ISL_2851553, EPI_ISL_2851554, EPI_ISL_2851557, EPI_ISL_2851564, EPI_ISL_2851565, EPI_ISL_2851566, EPI_ISL_2851572, EPI_ISL_2851576, EPI_ISL_2851577, EPI_ISL_2851582, EPI_ISL_2851585, EPI_ISL_2851586, EPI_ISL_2851587, EPI_ISL_2851590, EPI_ISL_2851594, EPI_ISL_2851595, EPI_ISL_2851596, EPI_ISL_2851597, EPI_ISL_2851607, EPI_ISL_2851609, EPI_ISL_2851612, EPI_ISL_2851617, EPI_ISL_2851619, EPI_ISL_2851624, EPI_ISL_2851625, EPI_ISL_2851630, EPI_ISL_2851631, EPI_ISL_2851637, EPI_ISL_2851638, EPI_ISL_2851643, EPI_ISL_2851645, EPI_ISL_2851646, EPI_ISL_2851647, EPI_ISL_2851648, EPI_ISL_2851650, EPI_ISL_2851652, EPI_ISL_2851658, EPI_ISL_2851659, EPI_ISL_2851662, EPI_ISL_2851663, EPI_ISL_2851665, EPI_ISL_2851667, EPI_ISL_2851671, EPI_ISL_2851673, EPI_ISL_2851676, EPI_ISL_2851677, EPI_ISL_2851679, EPI_ISL_2851682, EPI_ISL_2851683, EPI_ISL_2851687, EPI_ISL_2851689, EPI_ISL_2851690, EPI_ISL_2851692, EPI_ISL_2851694, EPI_ISL_2851696, EPI_ISL_2851697, EPI_ISL_2851698, EPI_ISL_2851700, EPI_ISL_2851702, EPI_ISL_2851707, EPI_ISL_2851708, EPI_ISL_2851709, EPI_ISL_2851714, EPI_ISL_2851715, EPI_ISL_2851718, EPI_ISL_2851720, EPI_ISL_2851723, EPI_ISL_2851724, EPI_ISL_2851727, EPI_ISL_2851732, EPI_ISL_2851733, EPI_ISL_2851734, EPI_ISL_2851735, EPI_ISL_2851736, EPI_ISL_2851739, EPI_ISL_2851740, EPI_ISL_2851744, EPI_ISL_2851746, EPI_ISL_2851747, EPI_ISL_2851748, EPI_ISL_2851749, EPI_ISL_2851750, EPI_ISL_2851752, EPI_ISL_2851761, EPI_ISL_2851762, EPI_ISL_2851764, EPI_ISL_2851769, EPI_ISL_2851771, EPI_ISL_2868053, EPI_ISL_2868058, EPI_ISL_2868220, EPI_ISL_2868221 | | | |
| see above | Health Services Laboratories | Wellcome Sanger Institute for the COVID-19 Genomics UK (COG-UK) Consortium | Cordelia Langford; David K. Jackson; Dominic Kwiatkowski; Ewan Harrison; Health Services Laboratories and Alex Alderton; Ian Johnston; Jeffrey Barrett; John Sillitoe on behalf of the Wellcome Sanger Institute COVID-19 Surveillance Team; Roberto Amato; Sonia Goncalves |
| EPI_ISL_2828760, EPI_ISL_2828767, EPI_ISL_2828768, EPI_ISL_2828775, EPI_ISL_2828778, EPI_ISL_2828779, EPI_ISL_2828788, EPI_ISL_2828789, EPI_ISL_2828790, EPI_ISL_2828796, EPI_ISL_2828798, EPI_ISL_2828801, EPI_ISL_2828806, EPI_ISL_2828807, EPI_ISL_2828812, EPI_ISL_2828814, EPI_ISL_2828816, EPI_ISL_2828818, EPI_ISL_2828822, EPI_ISL_2828828, EPI_ISL_2828829, EPI_ISL_2828834, EPI_ISL_2828838, EPI_ISL_2828839, EPI_ISL_2828840, EPI_ISL_2828842, EPI_ISL_2828843, EPI_ISL_2828848, EPI_ISL_2828849, EPI_ISL_2828851, EPI_ISL_2828852, EPI_ISL_2828855, EPI_ISL_2828859, EPI_ISL_2828860, EPI_ISL_2828861, EPI_ISL_2828864, EPI_ISL_2828867, EPI_ISL_2828871, EPI_ISL_2828872, EPI_ISL_2828876, EPI_ISL_2828882, EPI_ISL_2828886, EPI_ISL_2828887, EPI_ISL_2828888, EPI_ISL_2828890, EPI_ISL_2828891, EPI_ISL_2828896, EPI_ISL_2828906, EPI_ISL_2828912, EPI_ISL_2828913, EPI_ISL_2828916, EPI_ISL_2828917, EPI_ISL_2828918, EPI_ISL_2828931, EPI_ISL_2828932, EPI_ISL_2828934, EPI_ISL_2828938, EPI_ISL_2828945, EPI_ISL_2828948, EPI_ISL_2828951, EPI_ISL_2828952, EPI_ISL_2828954, EPI_ISL_2828955, EPI_ISL_2828959, EPI_ISL_2828961, EPI_ISL_2828963, EPI_ISL_2828965, EPI_ISL_2828966, EPI_ISL_2828967, EPI_ISL_2828969, EPI_ISL_2828971, EPI_ISL_2828973, EPI_ISL_2828975, EPI_ISL_2828977, EPI_ISL_2828981, EPI_ISL_2828983, EPI_ISL_2828984, EPI_ISL_2828989, EPI_ISL_2828990, EPI_ISL_2828992, EPI_ISL_2828995, EPI_ISL_2828998, EPI_ISL_2829001, EPI_ISL_2829006, EPI_ISL_2829007, EPI_ISL_2829009, EPI_ISL_2829019, EPI_ISL_2829023, EPI_ISL_2830041, EPI_ISL_2846025, EPI_ISL_2846029, EPI_ISL_2846032, EPI_ISL_2846034, EPI_ISL_2846035, EPI_ISL_2846037, EPI_ISL_2846038, EPI_ISL_2846041, EPI_ISL_2846042, EPI_ISL_2846043, EPI_ISL_2846044, EPI_ISL_2846046, EPI_ISL_2846049, EPI_ISL_2846052, EPI_ISL_2846054, EPI_ISL_2846057, EPI_ISL_2846058, EPI_ISL_2846059, EPI_ISL_2846060, EPI_ISL_2846061, EPI_ISL_2846062, EPI_ISL_2846063, EPI_ISL_2846065, EPI_ISL_2846072, EPI_ISL_2846073, EPI_ISL_2846077, EPI_ISL_2846079, EPI_ISL_2846082, EPI_ISL_2846083, EPI_ISL_2846085, EPI_ISL_2846088, EPI_ISL_2846090, EPI_ISL_2846091, EPI_ISL_2846092, EPI_ISL_2846093, EPI_ISL_2846095, EPI_ISL_2846098, EPI_ISL_2846099, EPI_ISL_2846101, EPI_ISL_2846102, EPI_ISL_2846107, EPI_ISL_2846109, EPI_ISL_2846111, EPI_ISL_2846116, EPI_ISL_2846117, EPI_ISL_2846119, EPI_ISL_2846120, EPI_ISL_2846121, EPI_ISL_2846122, EPI_ISL_2846123, EPI_ISL_2846126, EPI_ISL_2846127, EPI_ISL_2846128, EPI_ISL_2846129, EPI_ISL_2846131, EPI_ISL_2846132, EPI_ISL_2846133, EPI_ISL_2846137, EPI_ISL_2846138, EPI_ISL_2846140, EPI_ISL_2846144, EPI_ISL_2846146, EPI_ISL_2846149, EPI_ISL_2846151, EPI_ISL_2846153, EPI_ISL_2846155, EPI_ISL_2846156, EPI_ISL_2846157, EPI_ISL_2846160, EPI_ISL_2846161, EPI_ISL_2846165, EPI_ISL_2846168, EPI_ISL_2846169, EPI_ISL_2846173, EPI_ISL_2846174, EPI_ISL_2846175, EPI_ISL_2846177, EPI_ISL_2846181, EPI_ISL_2846182, EPI_ISL_2846184, EPI_ISL_2846187, EPI_ISL_2846198, EPI_ISL_2846200, EPI_ISL_2846201, EPI_ISL_2846202, EPI_ISL_2846204, EPI_ISL_2846205, EPI_ISL_2846206, EPI_ISL_2846207, EPI_ISL_2846208, EPI_ISL_2846212, EPI_ISL_2846215, EPI_ISL_2846218, EPI_ISL_2846222, EPI_ISL_2846224, EPI_ISL_2846225, EPI_ISL_2846230, EPI_ISL_2846231, EPI_ISL_2846233, EPI_ISL_2846234, EPI_ISL_2846235, EPI_ISL_2846237, EPI_ISL_2846238, EPI_ISL_2846239, EPI_ISL_2846242, EPI_ISL_2846244, EPI_ISL_2846245, EPI_ISL_2846248, EPI_ISL_2846249, EPI_ISL_2846254, EPI_ISL_2846256, EPI_ISL_2846257, EPI_ISL_2846258, EPI_ISL_2846260, EPI_ISL_2846261, EPI_ISL_2846265, EPI_ISL_2846272, EPI_ISL_2846273, EPI_ISL_2846275, EPI_ISL_2846279, EPI_ISL_2846282, EPI_ISL_2846283, EPI_ISL_2846284, EPI_ISL_2846287, EPI_ISL_2846288, EPI_ISL_2846289, EPI_ISL_2846290, EPI_ISL_2846295, EPI_ISL_2846296, EPI_ISL_2846297, EPI_ISL_2846298, EPI_ISL_2846305, EPI_ISL_2846306, EPI_ISL_2846307, EPI_ISL_2846310, EPI_ISL_2846311, EPI_ISL_2846314, EPI_ISL_2846316, EPI_ISL_2846319, EPI_ISL_2846326, EPI_ISL_2846327, EPI_ISL_2846328, EPI_ISL_2846330, EPI_ISL_2846331, EPI_ISL_2846333, EPI_ISL_2846334, EPI_ISL_2846336, EPI_ISL_2846337, EPI_ISL_2846338, EPI_ISL_2846341, EPI_ISL_2846343, EPI_ISL_2846344, EPI_ISL_2846345, EPI_ISL_2846346, EPI_ISL_2846352, EPI_ISL_2846353, EPI_ISL_2846357, EPI_ISL_2846359, EPI_ISL_2846581, EPI_ISL_2846588, EPI_ISL_2846597, EPI_ISL_2846599, EPI_ISL_2846601, EPI_ISL_2846611, EPI_ISL_2846613, EPI_ISL_2846618, EPI_ISL_2846619, EPI_ISL_2846625, EPI_ISL_2846626, EPI_ISL_2846633, EPI_ISL_2846634, EPI_ISL_2846637, EPI_ISL_2846639, EPI_ISL_2846643, EPI_ISL_2846645, EPI_ISL_2846649, EPI_ISL_2846650, EPI_ISL_2846654, EPI_ISL_2846658, EPI_ISL_2846663, EPI_ISL_2846668, EPI_ISL_2846669, EPI_ISL_2846673, EPI_ISL_2846676, EPI_ISL_2846677, EPI_ISL_2846680, EPI_ISL_2846682, EPI_ISL_2846683, EPI_ISL_2846688, EPI_ISL_2846698, EPI_ISL_2846700, EPI_ISL_2846701, EPI_ISL_2846703, EPI_ISL_2846704, EPI_ISL_2846708, EPI_ISL_2846713, EPI_ISL_2846715, EPI_ISL_2846718, EPI_ISL_2846720, EPI_ISL_2846721, EPI_ISL_2846722, EPI_ISL_2846728, EPI_ISL_2846748, EPI_ISL_2846753, EPI_ISL_2846761, EPI_ISL_2846763, EPI_ISL_2846766, EPI_ISL_2846771, EPI_ISL_2846772, EPI_ISL_2846780, EPI_ISL_2846782, EPI_ISL_2846787, EPI_ISL_2846790, EPI_ISL_2846800, EPI_ISL_2846802, EPI_ISL_2846803, EPI_ISL_2846808, EPI_ISL_2846828, EPI_ISL_2846831, EPI_ISL_2846833, EPI_ISL_2846841, EPI_ISL_2846845, EPI_ISL_2846846, EPI_ISL_2846853, EPI_ISL_2846856, EPI_ISL_2846857, EPI_ISL_2846858, EPI_ISL_2846860, EPI_ISL_2846864, EPI_ISL_2846868, EPI_ISL_2846870, EPI_ISL_2846883, EPI_ISL_2846887, EPI_ISL_2846889, EPI_ISL_2846896, EPI_ISL_2846901, EPI_ISL_2846903, EPI_ISL_2846909, EPI_ISL_2846911, EPI_ISL_2846917, EPI_ISL_2846920, EPI_ISL_2847306, EPI_ISL_2847311, EPI_ISL_2847334, EPI_ISL_2847357, EPI_ISL_2847360, EPI_ISL_2847371, EPI_ISL_2847386, EPI_ISL_2847395, EPI_ISL_2847399, EPI_ISL_2847406, EPI_ISL_2847409, EPI_ISL_2847421, EPI_ISL_2847433, EPI_ISL_2847435, EPI_ISL_2847444, EPI_ISL_2847445, EPI_ISL_2847451, EPI_ISL_2847453, EPI_ISL_2847467, EPI_ISL_2847475, EPI_ISL_2847501, EPI_ISL_2847502, EPI_ISL_2847518, EPI_ISL_2847521, EPI_ISL_2847536, EPI_ISL_2847547, EPI_ISL_2847604, EPI_ISL_2849009, EPI_ISL_2849011, EPI_ISL_2849012, EPI_ISL_2849013, EPI_ISL_2849016, EPI_ISL_2849017, EPI_ISL_2849020, EPI_ISL_2849021, EPI_ISL_2849022, EPI_ISL_2849023, EPI_ISL_2849025, EPI_ISL_2849027, EPI_ISL_2849028, EPI_ISL_2849029, EPI_ISL_2849032, EPI_ISL_2849035, EPI_ISL_2849039, EPI_ISL_2849042, EPI_ISL_2849043, EPI_ISL_2849044, EPI_ISL_2849045, EPI_ISL_2849048, EPI_ISL_2849049, EPI_ISL_2849050, EPI_ISL_2849052, EPI_ISL_2849054, EPI_ISL_2849056, EPI_ISL_2849057, EPI_ISL_2849060, EPI_ISL_2849062, EPI_ISL_2849065, EPI_ISL_2849067, EPI_ISL_2849068, EPI_ISL_2849069, EPI_ISL_2849070, EPI_ISL_2849071, EPI_ISL_2849078, EPI_ISL_2849079, EPI_ISL_2849083, EPI_ISL_2849084, EPI_ISL_2849086, EPI_ISL_2849090, EPI_ISL_2849091, EPI_ISL_2849094, EPI_ISL_2849097, EPI_ISL_2849101, EPI_ISL_2849103, EPI_ISL_2849104, EPI_ISL_2849105, EPI_ISL_2849106, EPI_ISL_2849107, EPI_ISL_2849108, EPI_ISL_2849110, EPI_ISL_2849114, EPI_ISL_2849119, EPI_ISL_2849120, EPI_ISL_2849121, EPI_ISL_2849124, EPI_ISL_2849126, EPI_ISL_2849127, EPI_ISL_2849128, EPI_ISL_2849130, EPI_ISL_2849132, EPI_ISL_2849138, EPI_ISL_2849139, EPI_ISL_2849140, EPI_ISL_2849141, EPI_ISL_2849143, EPI_ISL_2849144, EPI_ISL_2849146, EPI_ISL_2849147, EPI_ISL_2849148, EPI_ISL_2849149, EPI_ISL_2849152, EPI_ISL_2849154, EPI_ISL_2849156, EPI_ISL_2849157, EPI_ISL_2849159, EPI_ISL_2849162, EPI_ISL_2849163, EPI_ISL_2849166, EPI_ISL_2849168, EPI_ISL_2849169, EPI_ISL_2849170, EPI_ISL_2849175, EPI_ISL_2849180, EPI_ISL_2849183, EPI_ISL_2849184, EPI_ISL_2849187, EPI_ISL_2849188, EPI_ISL_2849190, EPI_ISL_2849191, EPI_ISL_2849193, EPI_ISL_2849194, EPI_ISL_2849195, EPI_ISL_2849196, EPI_ISL_2849199, EPI_ISL_2849201, EPI_ISL_2849202, EPI_ISL_2849203, EPI_ISL_2849204, EPI_ISL_2849207, EPI_ISL_2849208, EPI_ISL_2849209, EPI_ISL_2849210, EPI_ISL_2849211, EPI_ISL_2849214, EPI_ISL_2849217, EPI_ISL_2849218, EPI_ISL_2849219, EPI_ISL_2849220, EPI_ISL_2849221, EPI_ISL_2849223, EPI_ISL_2849224, EPI_ISL_2849225, EPI_ISL_2849227, EPI_ISL_2849232, EPI_ISL_2849234, EPI_ISL_2849238, EPI_ISL_2849239, EPI_ISL_2849240, EPI_ISL_2849241, EPI_ISL_2849242, EPI_ISL_2849249, EPI_ISL_2849251, EPI_ISL_2849253, EPI_ISL_2849259, EPI_ISL_2849261, EPI_ISL_2849263, EPI_ISL_2849267, EPI_ISL_2849272, EPI_ISL_2849279, EPI_ISL_2849280, EPI_ISL_2849281, EPI_ISL_2849283, EPI_ISL_2849284, EPI_ISL_2849285, EPI_ISL_2849287, EPI_ISL_2849294, EPI_ISL_2849295, EPI_ISL_2849296, EPI_ISL_2849300, EPI_ISL_2849303, EPI_ISL_2849305, EPI_ISL_2849308, EPI_ISL_2849309, EPI_ISL_2849312, EPI_ISL_2849313, EPI_ISL_2849315, EPI_ISL_2849317, EPI_ISL_2849319, EPI_ISL_2849320, EPI_ISL_2849321, EPI_ISL_2849323, EPI_ISL_2849324, EPI_ISL_2849325, EPI_ISL_2849326, EPI_ISL_2849327, EPI_ISL_2849330, EPI_ISL_2849331, EPI_ISL_2849332, EPI_ISL_2849333, EPI_ISL_2849337, EPI_ISL_2849339, EPI_ISL_2849340, EPI_ISL_2849341, EPI_ISL_2849343, EPI_ISL_2849347, EPI_ISL_2849349, EPI_ISL_2849350, EPI_ISL_2849351, EPI_ISL_2849354, EPI_ISL_2849356, EPI_ISL_2849357, EPI_ISL_2849359, EPI_ISL_2849360, EPI_ISL_2849368, EPI_ISL_2849375, EPI_ISL_2849381, EPI_ISL_2849387, EPI_ISL_2849394, EPI_ISL_2849399, EPI_ISL_2849401, EPI_ISL_2849404, EPI_ISL_2849406, EPI_ISL_2849408, EPI_ISL_2849409, EPI_ISL_2849411, EPI_ISL_2849414, EPI_ISL_2849415, EPI_ISL_2849416, EPI_ISL_2849435, EPI_ISL_2849436, EPI_ISL_2849446, EPI_ISL_2849447, EPI_ISL_2849451, EPI_ISL_2849452, EPI_ISL_2849453, EPI_ISL_2849454, EPI_ISL_2849455, EPI_ISL_2849459, EPI_ISL_2849460, EPI_ISL_2849461, EPI_ISL_2849462, EPI_ISL_2849468, EPI_ISL_2849470, EPI_ISL_2849472, EPI_ISL_2849478, EPI_ISL_2849481, EPI_ISL_2849483, EPI_ISL_2849488, EPI_ISL_2849502, EPI_ISL_2849505, EPI_ISL_2849508, EPI_ISL_2849509, EPI_ISL_2849511, EPI_ISL_2849527, EPI_ISL_2849532, EPI_ISL_2849533, EPI_ISL_2849535, EPI_ISL_2849537, EPI_ISL_2849538, EPI_ISL_2849546, EPI_ISL_2849554, EPI_ISL_2849556, EPI_ISL_2849558, EPI_ISL_2849560, EPI_ISL_2849563, EPI_ISL_2849566, EPI_ISL_2849568, EPI_ISL_2849569, EPI_ISL_2849579, EPI_ISL_2849583, EPI_ISL_2849584, EPI_ISL_2849587, EPI_ISL_2849589, EPI_ISL_2849607, EPI_ISL_2849616, EPI_ISL_2849623, EPI_ISL_2849626, EPI_ISL_2849630, EPI_ISL_2849633, EPI_ISL_2849638, EPI_ISL_2849639, EPI_ISL_2849641, EPI_ISL_2849642, EPI_ISL_2849644, EPI_ISL_2849655, EPI_ISL_2849664, EPI_ISL_2849666, EPI_ISL_2849669, EPI_ISL_2849673, EPI_ISL_2849675, EPI_ISL_2849676, EPI_ISL_2849681, EPI_ISL_2849689, EPI_ISL_2849690, EPI_ISL_2849693, EPI_ISL_2849695, EPI_ISL_2849696, EPI_ISL_2849698, EPI_ISL_2849699, EPI_ISL_2849700, EPI_ISL_2849701, EPI_ISL_2849702, EPI_ISL_2849704, EPI_ISL_2849706, EPI_ISL_2849707, EPI_ISL_2849708, EPI_ISL_2849710, EPI_ISL_2849711, EPI_ISL_2849715, EPI_ISL_2849717, EPI_ISL_2849718, EPI_ISL_2849719, EPI_ISL_2849721, EPI_ISL_2849725, EPI_ISL_2849726, EPI_ISL_2849728, EPI_ISL_2849729, EPI_ISL_2849730, EPI_ISL_2849731, EPI_ISL_2849732, EPI_ISL_2849735, EPI_ISL_2849736, EPI_ISL_2849740, EPI_ISL_2849743, EPI_ISL_2849745, EPI_ISL_2849746, EPI_ISL_2849748, EPI_ISL_2849750, EPI_ISL_2849751, EPI_ISL_2849753, EPI_ISL_2849754, EPI_ISL_2849757, EPI_ISL_2849758, EPI_ISL_2849760, EPI_ISL_2849761, EPI_ISL_2849765, EPI_ISL_2849767, EPI_ISL_2849770, EPI_ISL_2849771, EPI_ISL_2849772, EPI_ISL_2849773, EPI_ISL_2849774, EPI_ISL_2849776, EPI_ISL_2849777, EPI_ISL_2849779, EPI_ISL_2849780, EPI_ISL_2849783, EPI_ISL_2849784, EPI_ISL_2849787, EPI_ISL_2849790, EPI_ISL_2849791, EPI_ISL_2849793, EPI_ISL_2849795, EPI_ISL_2849796, EPI_ISL_2849797, EPI_ISL_2849801, EPI_ISL_2849805, EPI_ISL_2849810, EPI_ISL_2849812, EPI_ISL_2849814, EPI_ISL_2849815, EPI_ISL_2849818, EPI_ISL_2849819, EPI_ISL_2849821, EPI_ISL_2849822, EPI_ISL_2849824, EPI_ISL_2849826, EPI_ISL_2849827, EPI_ISL_2849834, EPI_ISL_2849835, EPI_ISL_2849838, EPI_ISL_2849839, EPI_ISL_2849842, EPI_ISL_2849845, EPI_ISL_2849847, EPI_ISL_2849851, EPI_ISL_2849853, EPI_ISL_2849854, EPI_ISL_2849855, EPI_ISL_2849858, EPI_ISL_2849860, EPI_ISL_2849862, EPI_ISL_2849863, EPI_ISL_2849867, EPI_ISL_2849868, EPI_ISL_2849869, EPI_ISL_2849871, EPI_ISL_2849878, EPI_ISL_2849879, EPI_ISL_2849881, EPI_ISL_2849885, EPI_ISL_2849886, EPI_ISL_2849892, EPI_ISL_2849893, EPI_ISL_2849894, EPI_ISL_2849897, EPI_ISL_2849901, EPI_ISL_2849902, EPI_ISL_2849904, EPI_ISL_2849905, EPI_ISL_2849908, EPI_ISL_2849909, EPI_ISL_2849910, EPI_ISL_2849915, EPI_ISL_2849917, EPI_ISL_2849919, EPI_ISL_2849920, EPI_ISL_2849923, EPI_ISL_2849926, EPI_ISL_2849927, EPI_ISL_2849928, EPI_ISL_2849932, EPI_ISL_2849933, EPI_ISL_2849934, EPI_ISL_2849939, EPI_ISL_2849940, EPI_ISL_2849941, EPI_ISL_2849942, EPI_ISL_2849945, EPI_ISL_2849947, EPI_ISL_2849948, EPI_ISL_2849952, EPI_ISL_2849956, EPI_ISL_2849957, EPI_ISL_2849958, EPI_ISL_2849959, EPI_ISL_2849960, EPI_ISL_2849961, EPI_ISL_2849963, EPI_ISL_2849964, EPI_ISL_2849965, EPI_ISL_2849968, EPI_ISL_2849969, EPI_ISL_2849972, EPI_ISL_2849973, EPI_ISL_2849977, EPI_ISL_2849986, EPI_ISL_2849987, EPI_ISL_2849988, EPI_ISL_2849992, EPI_ISL_2849994, EPI_ISL_2849995, EPI_ISL_2849997, EPI_ISL_2849998, EPI_ISL_2850004, EPI_ISL_2850005, EPI_ISL_2850006, EPI_ISL_2850008, EPI_ISL_2850013, EPI_ISL_2850020, EPI_ISL_2850021, EPI_ISL_2850027, EPI_ISL_2850028, EPI_ISL_2850029, EPI_ISL_2850031, EPI_ISL_2850033, EPI_ISL_2850034, EPI_ISL_2850035, EPI_ISL_2850036, EPI_ISL_2851189, EPI_ISL_2851245, EPI_ISL_2865063, EPI_ISL_2865067, EPI_ISL_2865070, EPI_ISL_2865072, EPI_ISL_2865074, EPI_ISL_2865075, EPI_ISL_2865078, EPI_ISL_2865080, EPI_ISL_2865081, EPI_ISL_2865083, EPI_ISL_2865087, EPI_ISL_2865089, EPI_ISL_2865091, EPI_ISL_2865093, EPI_ISL_2865095, EPI_ISL_2865098, EPI_ISL_2865101, EPI_ISL_2865103, EPI_ISL_2865104, EPI_ISL_2865107, EPI_ISL_2865113, EPI_ISL_2865115, EPI_ISL_2865118, EPI_ISL_2865120, EPI_ISL_2865121, EPI_ISL_2865124, EPI_ISL_2865127, EPI_ISL_2865133, EPI_ISL_2865135, EPI_ISL_2865136, EPI_ISL_2865140, EPI_ISL_2865141, EPI_ISL_2865152, EPI_ISL_2865156, EPI_ISL_2865167, EPI_ISL_2865168, EPI_ISL_2865169, EPI_ISL_2865172, EPI_ISL_2865175, EPI_ISL_2865196, EPI_ISL_2865201, EPI_ISL_2865202, EPI_ISL_2865205, EPI_ISL_2865212, EPI_ISL_2865213, EPI_ISL_2865214, EPI_ISL_2865225, EPI_ISL_2865226, EPI_ISL_2865227, EPI_ISL_2865234, EPI_ISL_2865239, EPI_ISL_2865243, EPI_ISL_2865249, EPI_ISL_2865253, EPI_ISL_2865258, EPI_ISL_2865260, EPI_ISL_2865263, EPI_ISL_2865265, EPI_ISL_2865274, EPI_ISL_2865280, EPI_ISL_2865282, EPI_ISL_2865284, EPI_ISL_2865285, EPI_ISL_2865288, EPI_ISL_2865293, EPI_ISL_2865297, EPI_ISL_2865298, EPI_ISL_2865299, EPI_ISL_2865306, EPI_ISL_2865307, EPI_ISL_2865309, EPI_ISL_2865310, EPI_ISL_2865313, EPI_ISL_2865318, EPI_ISL_2865320, EPI_ISL_2865321, EPI_ISL_2865330, EPI_ISL_2865332, EPI_ISL_2865336, EPI_ISL_2865341, EPI_ISL_2865347, EPI_ISL_2865352, EPI_ISL_2865356, EPI_ISL_2865360, EPI_ISL_2865371, EPI_ISL_2865384, EPI_ISL_2865433, EPI_ISL_2865453, EPI_ISL_2865461, EPI_ISL_2865470, EPI_ISL_2865476, EPI_ISL_2865496, EPI_ISL_2865499, EPI_ISL_2865527, EPI_ISL_2865547, EPI_ISL_2865608, EPI_ISL_2865647, EPI_ISL_2865662, EPI_ISL_2865715, EPI_ISL_2865717, EPI_ISL_2865723, EPI_ISL_2865724, EPI_ISL_2865725, EPI_ISL_2865730, EPI_ISL_2865731, EPI_ISL_2865733, EPI_ISL_2865749, EPI_ISL_2865761, EPI_ISL_2865774, EPI_ISL_2865778, EPI_ISL_2865779, EPI_ISL_2865791, EPI_ISL_2865793, EPI_ISL_2865795, EPI_ISL_2865798, EPI_ISL_2865800, EPI_ISL_2865805, EPI_ISL_2865811, EPI_ISL_2865815, EPI_ISL_2865818, EPI_ISL_2865819, EPI_ISL_2865828, EPI_ISL_2865838, EPI_ISL_2865854, EPI_ISL_2865863, EPI_ISL_2865864, EPI_ISL_2865865, EPI_ISL_2865868, EPI_ISL_2865879, EPI_ISL_2865882, EPI_ISL_2865883, EPI_ISL_2865884, EPI_ISL_2865887, EPI_ISL_2865890, EPI_ISL_2865893, EPI_ISL_2865894, EPI_ISL_2865900, EPI_ISL_2865901, EPI_ISL_2865903, EPI_ISL_2865906, EPI_ISL_2865920, EPI_ISL_2865926, EPI_ISL_2865935, | | | |

EPI_ISL_2865948, EPI_ISL_2865953, EPI_ISL_2865955, EPI_ISL_2865956, EPI_ISL_2865957, EPI_ISL_2865963, EPI_ISL_2865968, EPI_ISL_2865996, EPI_ISL_2865998, EPI_ISL_2866009, EPI_ISL_2866012, EPI_ISL_2866015, EPI_ISL_2866017, EPI_ISL_2866018, EPI_ISL_2866048, EPI_ISL_2866058, EPI_ISL_2866060, EPI_ISL_2866061, EPI_ISL_2866063, EPI_ISL_2867031, EPI_ISL_2867033, EPI_ISL_2867038,
EPI_ISL_2867040, EPI_ISL_2867041, EPI_ISL_2867042, EPI_ISL_2867044, EPI_ISL_2867049, EPI_ISL_2867052, EPI_ISL_2867054, EPI_ISL_2867062, EPI_ISL_2867063, EPI_ISL_2867068, EPI_ISL_2867073, EPI_ISL_2867076, EPI_ISL_2867077, EPI_ISL_2867081, EPI_ISL_2867082, EPI_ISL_2867086, EPI_ISL_2867097, EPI_ISL_2867100, EPI_ISL_2867106, EPI_ISL_2867108, EPI_ISL_2867116, EPI_ISL_2867120,
EPI_ISL_2867124, EPI_ISL_2867139, EPI_ISL_2867140, EPI_ISL_2867141, EPI_ISL_2867143, EPI_ISL_2867147, EPI_ISL_2867149, EPI_ISL_2867150, EPI_ISL_2867155, EPI_ISL_2867156, EPI_ISL_2867160, EPI_ISL_2867163, EPI_ISL_2867164, EPI_ISL_2867165, EPI_ISL_2867176, EPI_ISL_2867177, EPI_ISL_2867187, EPI_ISL_2867190, EPI_ISL_2867196, EPI_ISL_2867199, EPI_ISL_2867202, EPI_ISL_2867205,
EPI_ISL_2867214, EPI_ISL_2867219, EPI_ISL_2867223, EPI_ISL_2867233, EPI_ISL_2867241, EPI_ISL_2867244, EPI_ISL_2867246, EPI_ISL_2867253, EPI_ISL_2867255, EPI_ISL_2867261, EPI_ISL_2867262, EPI_ISL_2867266, EPI_ISL_2867269, EPI_ISL_2867270, EPI_ISL_2867271, EPI_ISL_2867272, EPI_ISL_2867275, EPI_ISL_2867278, EPI_ISL_2867284, EPI_ISL_2867285, EPI_ISL_2867287, EPI_ISL_2867294,
EPI_ISL_2867296, EPI_ISL_2867301, EPI_ISL_2867306, EPI_ISL_2867313, EPI_ISL_2867316, EPI_ISL_2867318, EPI_ISL_2867322, EPI_ISL_2867330, EPI_ISL_2867333, EPI_ISL_2867338, EPI_ISL_2867339, EPI_ISL_2867340, EPI_ISL_2867344, EPI_ISL_2867352, EPI_ISL_2867355, EPI_ISL_2867356, EPI_ISL_2867358, EPI_ISL_2867695, EPI_ISL_2867697, EPI_ISL_2867699, EPI_ISL_2867700, EPI_ISL_2867702,
EPI_ISL_2867704, EPI_ISL_2867706, EPI_ISL_2867709, EPI_ISL_2867710, EPI_ISL_2867717, EPI_ISL_2867720, EPI_ISL_2867724, EPI_ISL_2867725, EPI_ISL_2867726, EPI_ISL_2867729, EPI_ISL_2867730, EPI_ISL_2867731, EPI_ISL_2867732, EPI_ISL_2867734, EPI_ISL_2867737, EPI_ISL_2867738, EPI_ISL_2867740, EPI_ISL_2867743, EPI_ISL_2867744, EPI_ISL_2867746, EPI_ISL_2867748, EPI_ISL_2867751,
EPI_ISL_2867753, EPI_ISL_2867754, EPI_ISL_2867756, EPI_ISL_2867757, EPI_ISL_2867759, EPI_ISL_2867762, EPI_ISL_2867765, EPI_ISL_2867766, EPI_ISL_2867768, EPI_ISL_2867769, EPI_ISL_2867770, EPI_ISL_2867771, EPI_ISL_2867773, EPI_ISL_2867779, EPI_ISL_2867780, EPI_ISL_2867783, EPI_ISL_2867789, EPI_ISL_2867790, EPI_ISL_2867791, EPI_ISL_2867792, EPI_ISL_2867794, EPI_ISL_2867801,
EPI_ISL_2867805, EPI_ISL_2867807, EPI_ISL_2867809, EPI_ISL_2867810, EPI_ISL_2867812, EPI_ISL_2867813, EPI_ISL_2867815, EPI_ISL_2867818, EPI_ISL_2867820, EPI_ISL_2867822, EPI_ISL_2867826, EPI_ISL_2867827, EPI_ISL_2867830, EPI_ISL_2867832, EPI_ISL_2867836, EPI_ISL_2867842, EPI_ISL_2867844, EPI_ISL_2867845, EPI_ISL_2867848, EPI_ISL_2867852, EPI_ISL_2867853, EPI_ISL_2867857,
EPI_ISL_2867859, EPI_ISL_2867861, EPI_ISL_2867862, EPI_ISL_2867865, EPI_ISL_2867867, EPI_ISL_2867869, EPI_ISL_2867873, EPI_ISL_2867874, EPI_ISL_2867875, EPI_ISL_2867876, EPI_ISL_2867877, EPI_ISL_2867879, EPI_ISL_2867880, EPI_ISL_2867883, EPI_ISL_2867888, EPI_ISL_2867889, EPI_ISL_2867890, EPI_ISL_2867892, EPI_ISL_2867895, EPI_ISL_2867898, EPI_ISL_2867903, EPI_ISL_2867905,
EPI_ISL_2867906, EPI_ISL_2867908, EPI_ISL_2867910, EPI_ISL_2867911, EPI_ISL_2867912, EPI_ISL_2867913, EPI_ISL_2867915, EPI_ISL_2867919, EPI_ISL_2867920, EPI_ISL_2867921, EPI_ISL_2867922, EPI_ISL_2867924, EPI_ISL_2867925, EPI_ISL_2867926, EPI_ISL_2867927, EPI_ISL_2867929, EPI_ISL_2867935, EPI_ISL_2867936, EPI_ISL_2867939, EPI_ISL_2867940, EPI_ISL_2867941, EPI_ISL_2867945,
EPI_ISL_2867947, EPI_ISL_2867954, EPI_ISL_2867955, EPI_ISL_2867956, EPI_ISL_2867957, EPI_ISL_2867960, EPI_ISL_2867962, EPI_ISL_2867963, EPI_ISL_2867964, EPI_ISL_2867966, EPI_ISL_2867967, EPI_ISL_2867969, EPI_ISL_2867970, EPI_ISL_2867971, EPI_ISL_2867974, EPI_ISL_2867979, EPI_ISL_2867980, EPI_ISL_2867981, EPI_ISL_2867982, EPI_ISL_2867984, EPI_ISL_2867987, EPI_ISL_2867989,
EPI_ISL_2867990, EPI_ISL_2867992, EPI_ISL_2867993, EPI_ISL_2867994, EPI_ISL_2867997, EPI_ISL_2868000, EPI_ISL_2868001, EPI_ISL_2868007, EPI_ISL_2868008, EPI_ISL_2868012, EPI_ISL_2868017, EPI_ISL_2868019, EPI_ISL_2868022, EPI_ISL_2868023, EPI_ISL_2868024, EPI_ISL_2868025, EPI_ISL_2868028, EPI_ISL_2868029, EPI_ISL_2868033, EPI_ISL_2868036, EPI_ISL_2868037, EPI_ISL_2868038,
EPI_ISL_2868043, EPI_ISL_2868045, EPI_ISL_2868047, EPI_ISL_2868048, EPI_ISL_2868049, EPI_ISL_2868050, EPI_ISL_2868052, EPI_ISL_2868056, EPI_ISL_2868074, EPI_ISL_2868075, EPI_ISL_2868080, EPI_ISL_2868216, EPI_ISL_2868217, EPI_ISL_2868218, EPI_ISL_2907584, EPI_ISL_2907598, EPI_ISL_2907603, EPI_ISL_2907605, EPI_ISL_2907615, EPI_ISL_2907624, EPI_ISL_2907625, EPI_ISL_2907628,
EPI_ISL_2907629, EPI_ISL_2907639, EPI_ISL_2907650, EPI_ISL_2907666, EPI_ISL_2907673, EPI_ISL_2907678, EPI_ISL_2907686, EPI_ISL_2907706, EPI_ISL_2907715, EPI_ISL_2907748, EPI_ISL_2907749, EPI_ISL_2907756, EPI_ISL_2907757, EPI_ISL_2907767, EPI_ISL_2907779, EPI_ISL_2907784, EPI_ISL_2907794, EPI_ISL_2907796, EPI_ISL_2907800, EPI_ISL_2907806, EPI_ISL_2907813, EPI_ISL_2907820,
EPI_ISL_2907825, EPI_ISL_2907834, EPI_ISL_2907850, EPI_ISL_2907854, EPI_ISL_2907862, EPI_ISL_2907872, EPI_ISL_2907877, EPI_ISL_2907888, EPI_ISL_2907905, EPI_ISL_2907922, EPI_ISL_2907931, EPI_ISL_2909546, EPI_ISL_2909549, EPI_ISL_2909553, EPI_ISL_2909557, EPI_ISL_2909558, EPI_ISL_2909560, EPI_ISL_2909563, EPI_ISL_2909567, EPI_ISL_2909570, EPI_ISL_2909575, EPI_ISL_2909578,
EPI_ISL_2909579, EPI_ISL_2909585, EPI_ISL_2909587, EPI_ISL_2909588, EPI_ISL_2909591, EPI_ISL_2909594, EPI_ISL_2909595, EPI_ISL_2909598, EPI_ISL_2909600, EPI_ISL_2909602, EPI_ISL_2909608, EPI_ISL_2909616, EPI_ISL_2909618, EPI_ISL_2909619, EPI_ISL_2909621, EPI_ISL_2909627, EPI_ISL_2909633, EPI_ISL_2909635, EPI_ISL_2909637, EPI_ISL_2909640, EPI_ISL_2909648, EPI_ISL_2909651,
EPI_ISL_2909653, EPI_ISL_2909655, EPI_ISL_2909657, EPI_ISL_2909661, EPI_ISL_2909663, EPI_ISL_2909667, EPI_ISL_2909668, EPI_ISL_2909670, EPI_ISL_2909676, EPI_ISL_2909677, EPI_ISL_2909679, EPI_ISL_2909683, EPI_ISL_2909684, EPI_ISL_2909685, EPI_ISL_2909689, EPI_ISL_2909702, EPI_ISL_2909704, EPI_ISL_2909707, EPI_ISL_2909708, EPI_ISL_2909710, EPI_ISL_2909719, EPI_ISL_2909720,
EPI_ISL_2909722, EPI_ISL_2909727, EPI_ISL_2909729, EPI_ISL_2909730, EPI_ISL_2909732, EPI_ISL_2909734, EPI_ISL_2909735, EPI_ISL_2909739, EPI_ISL_2909741, EPI_ISL_2909742, EPI_ISL_2909753, EPI_ISL_2909755, EPI_ISL_2909761, EPI_ISL_2909762, EPI_ISL_2909770, EPI_ISL_2909774, EPI_ISL_2909787, EPI_ISL_2909788, EPI_ISL_2909793, EPI_ISL_2909794, EPI_ISL_2909796, EPI_ISL_2909798,
EPI_ISL_2909799, EPI_ISL_2909804, EPI_ISL_2909805, EPI_ISL_2909806, EPI_ISL_2909809, EPI_ISL_2909810, EPI_ISL_2909822, EPI_ISL_2909823, EPI_ISL_2909826, EPI_ISL_2909831, EPI_ISL_2909836, EPI_ISL_2909837, EPI_ISL_2909838, EPI_ISL_2909845, EPI_ISL_2909849, EPI_ISL_2909852, EPI_ISL_2909853, EPI_ISL_2909855, EPI_ISL_2909860, EPI_ISL_2909864, EPI_ISL_2909867, EPI_ISL_2909871,
EPI_ISL_2909872, EPI_ISL_2909876, EPI_ISL_2909883, EPI_ISL_2909884, EPI_ISL_2909885, EPI_ISL_2909888, EPI_ISL_2909889, EPI_ISL_2909890, EPI_ISL_2909894, EPI_ISL_2909895, EPI_ISL_2909899, EPI_ISL_2909971, EPI_ISL_2909973, EPI_ISL_2909988, EPI_ISL_2910332, EPI_ISL_2910517, EPI_ISL_2911893, EPI_ISL_2913329, EPI_ISL_2913333, EPI_ISL_2913345, EPI_ISL_2913356, EPI_ISL_2913360,
EPI_ISL_2913362, EPI_ISL_2913363, EPI_ISL_2913366, EPI_ISL_2913370, EPI_ISL_2913371, EPI_ISL_2913373, EPI_ISL_2913377, EPI_ISL_2913379, EPI_ISL_2913382, EPI_ISL_2913383, EPI_ISL_2913384, EPI_ISL_2913388, EPI_ISL_2913390, EPI_ISL_2913408, EPI_ISL_2913413, EPI_ISL_2913414, EPI_ISL_2913416, EPI_ISL_2913418, EPI_ISL_2913423, EPI_ISL_2913427, EPI_ISL_2913434, EPI_ISL_2913436,
EPI_ISL_2913442, EPI_ISL_2913453, EPI_ISL_2913456, EPI_ISL_2913460, EPI_ISL_2913461, EPI_ISL_2913464, EPI_ISL_2913466, EPI_ISL_2913470, EPI_ISL_2913471, EPI_ISL_2913472, EPI_ISL_2913480, EPI_ISL_2913483, EPI_ISL_2913486, EPI_ISL_2913487, EPI_ISL_2913488, EPI_ISL_2913489, EPI_ISL_2913490, EPI_ISL_2913491, EPI_ISL_2913492, EPI_ISL_2913500, EPI_ISL_2913505, EPI_ISL_2913506,
EPI_ISL_2913509, EPI_ISL_2913511, EPI_ISL_2913514, EPI_ISL_2913515, EPI_ISL_2913517, EPI_ISL_2913518, EPI_ISL_2913523, EPI_ISL_2913525, EPI_ISL_2913527, EPI_ISL_2913532, EPI_ISL_2913534, EPI_ISL_2913541, EPI_ISL_2913543, EPI_ISL_2913546, EPI_ISL_2913548, EPI_ISL_2913549, EPI_ISL_2913551, EPI_ISL_2913552, EPI_ISL_2913556, EPI_ISL_2913557, EPI_ISL_2913558, EPI_ISL_2913561,
EPI_ISL_2913562, EPI_ISL_2913569, EPI_ISL_2913571, EPI_ISL_2913573, EPI_ISL_2913575, EPI_ISL_2913576, EPI_ISL_2913577, EPI_ISL_2913584, EPI_ISL_2913587, EPI_ISL_2913588, EPI_ISL_2913590, EPI_ISL_2913591, EPI_ISL_2913595, EPI_ISL_2914280, EPI_ISL_2914281, EPI_ISL_2914282, EPI_ISL_2914283, EPI_ISL_2914296, EPI_ISL_2914308, EPI_ISL_2914311, EPI_ISL_2914316, EPI_ISL_2914317,
EPI_ISL_2914327, EPI_ISL_2914337, EPI_ISL_2914339, EPI_ISL_2914341, EPI_ISL_2914342, EPI_ISL_2914345, EPI_ISL_2914346, EPI_ISL_2914350, EPI_ISL_2914351, EPI_ISL_2914352, EPI_ISL_2914360, EPI_ISL_2914362, EPI_ISL_2914372, EPI_ISL_2914376, EPI_ISL_2914383, EPI_ISL_2914384, EPI_ISL_2914389, EPI_ISL_2914394, EPI_ISL_2914395, EPI_ISL_2914397, EPI_ISL_2914398, EPI_ISL_2914401,
EPI_ISL_2914405, EPI_ISL_2914406, EPI_ISL_2914411, EPI_ISL_2914412, EPI_ISL_2914413, EPI_ISL_2914414, EPI_ISL_2914420, EPI_ISL_2914423, EPI_ISL_2914424, EPI_ISL_2914429, EPI_ISL_2914438, EPI_ISL_2914439, EPI_ISL_2914445, EPI_ISL_2914449, EPI_ISL_2914452, EPI_ISL_2914454, EPI_ISL_2914461, EPI_ISL_2914462, EPI_ISL_2914466, EPI_ISL_2914471, EPI_ISL_2914475, EPI_ISL_2914483,
EPI_ISL_2914485, EPI_ISL_2914490, EPI_ISL_2914497, EPI_ISL_2914498, EPI_ISL_2914507, EPI_ISL_2914508, EPI_ISL_2914513, EPI_ISL_2914516, EPI_ISL_2914520, EPI_ISL_2914526, EPI_ISL_2914531, EPI_ISL_2914538, EPI_ISL_2914540, EPI_ISL_2914543, EPI_ISL_2914545, EPI_ISL_2914548, EPI_ISL_2914549, EPI_ISL_2914552, EPI_ISL_2914555, EPI_ISL_2914556, EPI_ISL_2914557, EPI_ISL_2914559,
EPI_ISL_2914561, EPI_ISL_2914575, EPI_ISL_2914578, EPI_ISL_2914579, EPI_ISL_2914581, EPI_ISL_2914586, EPI_ISL_2914597, EPI_ISL_2914599, EPI_ISL_2914604, EPI_ISL_2914605, EPI_ISL_2914606, EPI_ISL_2914612, EPI_ISL_2914615, EPI_ISL_2915692, EPI_ISL_2915699, EPI_ISL_2915707, EPI_ISL_2915708, EPI_ISL_2915713, EPI_ISL_2915714, EPI_ISL_2915721, EPI_ISL_2915722, EPI_ISL_2915724,
EPI_ISL_2915726, EPI_ISL_2915728, EPI_ISL_2915730, EPI_ISL_2915731, EPI_ISL_2915736, EPI_ISL_2915740, EPI_ISL_2915745, EPI_ISL_2915747, EPI_ISL_2915749, EPI_ISL_2915750, EPI_ISL_2915753, EPI_ISL_2915754, EPI_ISL_2915755, EPI_ISL_2915757, EPI_ISL_2915758, EPI_ISL_2915759, EPI_ISL_2915764, EPI_ISL_2915767, EPI_ISL_2915768, EPI_ISL_2915769, EPI_ISL_2915776, EPI_ISL_2915778,
EPI_ISL_2915781, EPI_ISL_2915783, EPI_ISL_2915785, EPI_ISL_2915786, EPI_ISL_2915787, EPI_ISL_2915793, EPI_ISL_2915795, EPI_ISL_2915796, EPI_ISL_2915797, EPI_ISL_2915798, EPI_ISL_2915799, EPI_ISL_2915803, EPI_ISL_2915806, EPI_ISL_2915807, EPI_ISL_2915808, EPI_ISL_2915810, EPI_ISL_2915811, EPI_ISL_2915812, EPI_ISL_2915813, EPI_ISL_2915818, EPI_ISL_2915819, EPI_ISL_2915820,
EPI_ISL_2915822, EPI_ISL_2915824, EPI_ISL_2915827, EPI_ISL_2915830, EPI_ISL_2915835, EPI_ISL_2915838, EPI_ISL_2915841, EPI_ISL_2915842, EPI_ISL_2915844, EPI_ISL_2915848, EPI_ISL_2915849, EPI_ISL_2915850, EPI_ISL_2915851, EPI_ISL_2915856, EPI_ISL_2915857, EPI_ISL_2915861, EPI_ISL_2915863, EPI_ISL_2915864, EPI_ISL_2915865, EPI_ISL_2915866, EPI_ISL_2915867, EPI_ISL_2915868,
EPI_ISL_2915870, EPI_ISL_2915872, EPI_ISL_2915873, EPI_ISL_2915875, EPI_ISL_2915879, EPI_ISL_2915880, EPI_ISL_2915882, EPI_ISL_2915883, EPI_ISL_2915884, EPI_ISL_2915885, EPI_ISL_2915887, EPI_ISL_2915888, EPI_ISL_2915891, EPI_ISL_2915892, EPI_ISL_2915893, EPI_ISL_2915894, EPI_ISL_2915895, EPI_ISL_2915897, EPI_ISL_2915898, EPI_ISL_2915899, EPI_ISL_2915904, EPI_ISL_2915905,
EPI_ISL_2915909, EPI_ISL_2915910, EPI_ISL_2915911, EPI_ISL_2915913, EPI_ISL_2915915, EPI_ISL_2915916, EPI_ISL_2915917, EPI_ISL_2915919, EPI_ISL_2915922, EPI_ISL_2915923, EPI_ISL_2915925, EPI_ISL_2915926, EPI_ISL_2915928, EPI_ISL_2915929, EPI_ISL_2915931, EPI_ISL_2915933, EPI_ISL_2915934, EPI_ISL_2915937, EPI_ISL_2915938, EPI_ISL_2915939, EPI_ISL_2915940, EPI_ISL_2915942,
EPI_ISL_2915943, EPI_ISL_2915944, EPI_ISL_2915946, EPI_ISL_2915948, EPI_ISL_2915950, EPI_ISL_2915952, EPI_ISL_2915955, EPI_ISL_2915957, EPI_ISL_2915958, EPI_ISL_2915959, EPI_ISL_2915962, EPI_ISL_2915963, EPI_ISL_2915965, EPI_ISL_2915967, EPI_ISL_2915970, EPI_ISL_2915973, EPI_ISL_2915977, EPI_ISL_2915980, EPI_ISL_2915982, EPI_ISL_2915983, EPI_ISL_2915984, EPI_ISL_2915985,
EPI_ISL_2915986, EPI_ISL_2915987, EPI_ISL_2915990, EPI_ISL_2915991, EPI_ISL_2915992, EPI_ISL_2915993, EPI_ISL_2915995, EPI_ISL_2915996, EPI_ISL_2915997, EPI_ISL_2916000, EPI_ISL_2916001, EPI_ISL_2916002, EPI_ISL_2916004, EPI_ISL_2916009, EPI_ISL_2916014, EPI_ISL_2916017, EPI_ISL_2916022, EPI_ISL_2916024, EPI_ISL_2916025, EPI_ISL_2916028, EPI_ISL_2916031, EPI_ISL_2916033,
EPI_ISL_2916043, EPI_ISL_2916045, EPI_ISL_2916046, EPI_ISL_2916053, EPI_ISL_2916068, EPI_ISL_2916071, EPI_ISL_2916074, EPI_ISL_2916076, EPI_ISL_2916091, EPI_ISL_2916095, EPI_ISL_2916106, EPI_ISL_2916112, EPI_ISL_2916116, EPI_ISL_2916120, EPI_ISL_2916126, EPI_ISL_2916127, EPI_ISL_2916128, EPI_ISL_2916137, EPI_ISL_2916142, EPI_ISL_2916143, EPI_ISL_2916150, EPI_ISL_2916160,
EPI_ISL_2916162, EPI_ISL_2916169, EPI_ISL_2916180, EPI_ISL_2916181, EPI_ISL_2916183, EPI_ISL_2916184, EPI_ISL_2916185, EPI_ISL_2916190, EPI_ISL_2916193, EPI_ISL_2916194, EPI_ISL_2916201, EPI_ISL_2916206, EPI_ISL_2916207, EPI_ISL_2916210, EPI_ISL_2916221, EPI_ISL_2916222, EPI_ISL_2916230, EPI_ISL_2916237, EPI_ISL_2916241, EPI_ISL_2916245, EPI_ISL_2916253, EPI_ISL_2916261,
EPI_ISL_2916262, EPI_ISL_2916264, EPI_ISL_2916265, EPI_ISL_2916267, EPI_ISL_2916273, EPI_ISL_2916274, EPI_ISL_2916280, EPI_ISL_2916283, EPI_ISL_2916286, EPI_ISL_2916298, EPI_ISL_2916356, EPI_ISL_2916360, EPI_ISL_2916418, EPI_ISL_2916643, EPI_ISL_2918068, EPI_ISL_2918073, EPI_ISL_2918074, EPI_ISL_2918078, EPI_ISL_2918081, EPI_ISL_2918082, EPI_ISL_2918086, EPI_ISL_2918087,
EPI_ISL_2918089, EPI_ISL_2918092, EPI_ISL_2918094, EPI_ISL_2918098, EPI_ISL_2918101, EPI_ISL_2918103, EPI_ISL_2918107, EPI_ISL_2918108, EPI_ISL_2918112, EPI_ISL_2918113, EPI_ISL_2918116, EPI_ISL_2918118, EPI_ISL_2918119, EPI_ISL_2918127, EPI_ISL_2918128, EPI_ISL_2918130, EPI_ISL_2918132, EPI_ISL_2918133, EPI_ISL_2918134, EPI_ISL_2918136, EPI_ISL_2918140, EPI_ISL_2918142,
EPI_ISL_2918165, EPI_ISL_2918168, EPI_ISL_2918173, EPI_ISL_2918174, EPI_ISL_2918175, EPI_ISL_2918177, EPI_ISL_2918188, EPI_ISL_2918197, EPI_ISL_2918207, EPI_ISL_2918210, EPI_ISL_2918221, EPI_ISL_2918224, EPI_ISL_2918229, EPI_ISL_2918233, EPI_ISL_2918235, EPI_ISL_2918241, EPI_ISL_2918245, EPI_ISL_2918246, EPI_ISL_2918254, EPI_ISL_2918258, EPI_ISL_2918262, EPI_ISL_2918266,
EPI_ISL_2918276, EPI_ISL_2918279, EPI_ISL_2918280, EPI_ISL_2918286, EPI_ISL_2918288, EPI_ISL_2918293, EPI_ISL_2918305, EPI_ISL_2918332, EPI_ISL_2918333, EPI_ISL_2918334, EPI_ISL_2918338, EPI_ISL_2918339, EPI_ISL_2918341, EPI_ISL_2918351, EPI_ISL_2918356, EPI_ISL_2918359, EPI_ISL_2918361, EPI_ISL_2918363, EPI_ISL_2918374, EPI_ISL_2918375, EPI_ISL_2918379, EPI_ISL_2918380,
EPI_ISL_2918382, EPI_ISL_2918384, EPI_ISL_2918388, EPI_ISL_2918389, EPI_ISL_2918391, EPI_ISL_2918392, EPI_ISL_2925225, EPI_ISL_2925286, EPI_ISL_2937159

| | | | |
|---|---|---|---|
| see above | Lighthouse Lab in Alderley Park | Wellcome Sanger Institute for the COVID-19 Genomics UK (COG-UK) Consortium | Cordelia Langford; David K. Jackson; Dominic Kwiatkowski; Ewan Harrison; Ian Johnston; Jacquelyn Wynn; Jeffrey Barrett; John Sillitoe on behalf of the Wellcome Sanger Institute COVID-19 Surveillance Team; Mairead Hyland; Roberto Amato; Sonia Goncalves; The Lighthouse Lab in Alderley Park and Alex Alderton |

EPI_ISL_2829090, EPI_ISL_2829169, EPI_ISL_2829260, EPI_ISL_2829266, EPI_ISL_2829280, EPI_ISL_2829956, EPI_ISL_2829966, EPI_ISL_2829973, EPI_ISL_2829993, EPI_ISL_2830001, EPI_ISL_2830004, EPI_ISL_2830005, EPI_ISL_2830007, EPI_ISL_2830009, EPI_ISL_2830013, EPI_ISL_2830017, EPI_ISL_2830022, EPI_ISL_2830023, EPI_ISL_2830024, EPI_ISL_2830025, EPI_ISL_2830026, EPI_ISL_2830030,
EPI_ISL_2830032, EPI_ISL_2830034, EPI_ISL_2830044, EPI_ISL_2830051, EPI_ISL_2830055, EPI_ISL_2830071, EPI_ISL_2830076, EPI_ISL_2830083, EPI_ISL_2830086, EPI_ISL_2830094, EPI_ISL_2830101, EPI_ISL_2830102, EPI_ISL_2830106, EPI_ISL_2830113, EPI_ISL_2830121, EPI_ISL_2830129, EPI_ISL_2830131, EPI_ISL_2830139, EPI_ISL_2830141, EPI_ISL_2830146, EPI_ISL_2830152, EPI_ISL_2830158,
EPI_ISL_2830160, EPI_ISL_2830161, EPI_ISL_2830177, EPI_ISL_2830180, EPI_ISL_2830183, EPI_ISL_2830189, EPI_ISL_2830190, EPI_ISL_2830194, EPI_ISL_2830195, EPI_ISL_2830200, EPI_ISL_2830201, EPI_ISL_2830206, EPI_ISL_2830208, EPI_ISL_2830210, EPI_ISL_2830213, EPI_ISL_2830215, EPI_ISL_2830221, EPI_ISL_2830222, EPI_ISL_2830223, EPI_ISL_2830224, EPI_ISL_2830227, EPI_ISL_2830228,
EPI_ISL_2830229, EPI_ISL_2830232, EPI_ISL_2830234, EPI_ISL_2830235, EPI_ISL_2830236, EPI_ISL_2830237, EPI_ISL_2830238, EPI_ISL_2830240, EPI_ISL_2830241, EPI_ISL_2830244, EPI_ISL_2830247, EPI_ISL_2830249, EPI_ISL_2830250, EPI_ISL_2830251, EPI_ISL_2830253, EPI_ISL_2830254, EPI_ISL_2830255, EPI_ISL_2830257, EPI_ISL_2830259, EPI_ISL_2830261, EPI_ISL_2830265, EPI_ISL_2830268,
EPI_ISL_2830269, EPI_ISL_2830275, EPI_ISL_2830278, EPI_ISL_2830280, EPI_ISL_2830283, EPI_ISL_2830285, EPI_ISL_2830288, EPI_ISL_2830290, EPI_ISL_2830294, EPI_ISL_2830295, EPI_ISL_2830298, EPI_ISL_2830302, EPI_ISL_2830303, EPI_ISL_2830304, EPI_ISL_2830305, EPI_ISL_2830307, EPI_ISL_2830309, EPI_ISL_2830310, EPI_ISL_2830313, EPI_ISL_2830314, EPI_ISL_2830316, EPI_ISL_2830318,
EPI_ISL_2830322, EPI_ISL_2830323, EPI_ISL_2830324, EPI_ISL_2830325, EPI_ISL_2830326, EPI_ISL_2830327, EPI_ISL_2830331, EPI_ISL_2830379, EPI_ISL_2830487, EPI_ISL_2830520, EPI_ISL_2830544, EPI_ISL_2830546, EPI_ISL_2830577, EPI_ISL_2830583, EPI_ISL_2831589, EPI_ISL_2831590, EPI_ISL_2831591, EPI_ISL_2831594, EPI_ISL_2831595, EPI_ISL_2831598, EPI_ISL_2831599, EPI_ISL_2831600,
EPI_ISL_2831601, EPI_ISL_2831610, EPI_ISL_2831612, EPI_ISL_2831613, EPI_ISL_2831616, EPI_ISL_2831619, EPI_ISL_2831620, EPI_ISL_2831622, EPI_ISL_2831623, EPI_ISL_2831626, EPI_ISL_2831627, EPI_ISL_2831628, EPI_ISL_2831629, EPI_ISL_2831631, EPI_ISL_2831633, EPI_ISL_2831634, EPI_ISL_2831635, EPI_ISL_2831636, EPI_ISL_2831639, EPI_ISL_2831641, EPI_ISL_2831642, EPI_ISL_2831646,
EPI_ISL_2831647, EPI_ISL_2831649, EPI_ISL_2831650, EPI_ISL_2831652, EPI_ISL_2831653, EPI_ISL_2831656, EPI_ISL_2831659, EPI_ISL_2831663, EPI_ISL_2831665, EPI_ISL_2831666, EPI_ISL_2831668, EPI_ISL_2831670, EPI_ISL_2831671, EPI_ISL_2831672, EPI_ISL_2831673, EPI_ISL_2831675, EPI_ISL_2831677, EPI_ISL_2831678, EPI_ISL_2831679, EPI_ISL_2831680, EPI_ISL_2831684, EPI_ISL_2831688,
EPI_ISL_2831689, EPI_ISL_2831690, EPI_ISL_2831693, EPI_ISL_2831695, EPI_ISL_2831696, EPI_ISL_2831702, EPI_ISL_2831704, EPI_ISL_2831706, EPI_ISL_2831707, EPI_ISL_2831708, EPI_ISL_2831709, EPI_ISL_2831710, EPI_ISL_2831712, EPI_ISL_2831713, EPI_ISL_2831714, EPI_ISL_2831716, EPI_ISL_2831717, EPI_ISL_2831719, EPI_ISL_2831722, EPI_ISL_2831723, EPI_ISL_2831724, EPI_ISL_2831730,
EPI_ISL_2831731, EPI_ISL_2831732, EPI_ISL_2831735, EPI_ISL_2831737, EPI_ISL_2831738, EPI_ISL_2831739, EPI_ISL_2831741, EPI_ISL_2831744, EPI_ISL_2831745, EPI_ISL_2831747, EPI_ISL_2831749, EPI_ISL_2831756, EPI_ISL_2831757, EPI_ISL_2831790, EPI_ISL_2831823, EPI_ISL_2831831, EPI_ISL_2831840, EPI_ISL_2831876, EPI_ISL_2831886, EPI_ISL_2831916, EPI_ISL_2831935, EPI_ISL_2831945,
EPI_ISL_2831968, EPI_ISL_2831983, EPI_ISL_2846937, EPI_ISL_2846945, EPI_ISL_2846956, EPI_ISL_2846958, EPI_ISL_2846964, EPI_ISL_2846965, EPI_ISL_2846969, EPI_ISL_2846972, EPI_ISL_2846973, EPI_ISL_2846975, EPI_ISL_2846991, EPI_ISL_2846992, EPI_ISL_2846998, EPI_ISL_2846999, EPI_ISL_2847004, EPI_ISL_2847007, EPI_ISL_2847012, EPI_ISL_2847022, EPI_ISL_2847024, EPI_ISL_2847025,
EPI_ISL_2847030, EPI_ISL_2847032, EPI_ISL_2847036, EPI_ISL_2847043, EPI_ISL_2847044, EPI_ISL_2847047, EPI_ISL_2847049, EPI_ISL_2847052, EPI_ISL_2847054, EPI_ISL_2847056, EPI_ISL_2847059, EPI_ISL_2847062, EPI_ISL_2847067, EPI_ISL_2847068, EPI_ISL_2847075, EPI_ISL_2847079, EPI_ISL_2847082, EPI_ISL_2847083, EPI_ISL_2847085, EPI_ISL_2847086, EPI_ISL_2847087, EPI_ISL_2847089,
EPI_ISL_2847091, EPI_ISL_2847095, EPI_ISL_2847100, EPI_ISL_2847103, EPI_ISL_2847110, EPI_ISL_2847111, EPI_ISL_2847113, EPI_ISL_2847118, EPI_ISL_2847119, EPI_ISL_2847123, EPI_ISL_2847133, EPI_ISL_2847146, EPI_ISL_2847149, EPI_ISL_2847152, EPI_ISL_2847157, EPI_ISL_2847159, EPI_ISL_2847168, EPI_ISL_2847171, EPI_ISL_2847176, EPI_ISL_2847177, EPI_ISL_2847180, EPI_ISL_2847185,
EPI_ISL_2847191, EPI_ISL_2847193, EPI_ISL_2847200, EPI_ISL_2847202, EPI_ISL_2847205, EPI_ISL_2847206, EPI_ISL_2847207, EPI_ISL_2847211, EPI_ISL_2847214, EPI_ISL_2847215, EPI_ISL_2847223, EPI_ISL_2847225, EPI_ISL_2847226, EPI_ISL_2847231, EPI_ISL_2847232, EPI_ISL_2847244, EPI_ISL_2847245, EPI_ISL_2847252, EPI_ISL_2847255, EPI_ISL_2847262, EPI_ISL_2847263, EPI_ISL_2847266,
EPI_ISL_2847267, EPI_ISL_2847271, EPI_ISL_2847277, EPI_ISL_2847282, EPI_ISL_2847284, EPI_ISL_2847307, EPI_ISL_2847312, EPI_ISL_2847317, EPI_ISL_2847333, EPI_ISL_2847338, EPI_ISL_2847356, EPI_ISL_2847366, EPI_ISL_2847372, EPI_ISL_2847422, EPI_ISL_2847439, EPI_ISL_2847452, EPI_ISL_2847470, EPI_ISL_2847473, EPI_ISL_2847491, EPI_ISL_2847510, EPI_ISL_2847534, EPI_ISL_2847535,
EPI_ISL_2847568, EPI_ISL_2847576, EPI_ISL_2847594, EPI_ISL_2848679, EPI_ISL_2848694, EPI_ISL_2848703, EPI_ISL_2848726, EPI_ISL_2848730, EPI_ISL_2848853, EPI_ISL_2850037, EPI_ISL_2850038, EPI_ISL_2850040, EPI_ISL_2850041, EPI_ISL_2850042, EPI_ISL_2850044, EPI_ISL_2850045, EPI_ISL_2850046, EPI_ISL_2850047, EPI_ISL_2850048, EPI_ISL_2850049, EPI_ISL_2850051, EPI_ISL_2850052,
EPI_ISL_2850053, EPI_ISL_2850054, EPI_ISL_2850055, EPI_ISL_2850056, EPI_ISL_2850057, EPI_ISL_2850058, EPI_ISL_2850059, EPI_ISL_2850061, EPI_ISL_2850062, EPI_ISL_2850063, EPI_ISL_2850066, EPI_ISL_2850067, EPI_ISL_2850068, EPI_ISL_2850071, EPI_ISL_2850072, EPI_ISL_2850073, EPI_ISL_2850074, EPI_ISL_2850076, EPI_ISL_2850078, EPI_ISL_2850080, EPI_ISL_2850082, EPI_ISL_2850083,
EPI_ISL_2850084, EPI_ISL_2850085, EPI_ISL_2850086, EPI_ISL_2850087, EPI_ISL_2850088, EPI_ISL_2850091, EPI_ISL_2850093, EPI_ISL_2850094, EPI_ISL_2850095, EPI_ISL_2850096, EPI_ISL_2850097, EPI_ISL_2850098, EPI_ISL_2850100, EPI_ISL_2850103, EPI_ISL_2850106, EPI_ISL_2850107, EPI_ISL_2850108, EPI_ISL_2850109, EPI_ISL_2850110, EPI_ISL_2850111, EPI_ISL_2850112, EPI_ISL_2850113,
EPI_ISL_2850115, EPI_ISL_2850116, EPI_ISL_2850117, EPI_ISL_2850119, EPI_ISL_2850121, EPI_ISL_2850124, EPI_ISL_2850125, EPI_ISL_2850126, EPI_ISL_2850127, EPI_ISL_2850128, EPI_ISL_2850129, EPI_ISL_2850130, EPI_ISL_2850132, EPI_ISL_2850133, EPI_ISL_2850134, EPI_ISL_2850137, EPI_ISL_2850138, EPI_ISL_2850141, EPI_ISL_2850142, EPI_ISL_2850143, EPI_ISL_2850144, EPI_ISL_2850145,
EPI_ISL_2850147, EPI_ISL_2850148, EPI_ISL_2850149, EPI_ISL_2850151, EPI_ISL_2850152, EPI_ISL_2850153, EPI_ISL_2850154, EPI_ISL_2850156, EPI_ISL_2850157, EPI_ISL_2850158, EPI_ISL_2850159, EPI_ISL_2850160, EPI_ISL_2850161, EPI_ISL_2850162, EPI_ISL_2850163, EPI_ISL_2850164, EPI_ISL_2850165, EPI_ISL_2850166, EPI_ISL_2850167, EPI_ISL_2850169, EPI_ISL_2850171, EPI_ISL_2850172,
EPI_ISL_2850173, EPI_ISL_2850174, EPI_ISL_2850175, EPI_ISL_2850177, EPI_ISL_2850179, EPI_ISL_2850180, EPI_ISL_2850183, EPI_ISL_2850184, EPI_ISL_2850185, EPI_ISL_2850186, EPI_ISL_2850188, EPI_ISL_2850189, EPI_ISL_2850190, EPI_ISL_2850191, EPI_ISL_2850192, EPI_ISL_2850194, EPI_ISL_2850195, EPI_ISL_2850196, EPI_ISL_2850197, EPI_ISL_2850199, EPI_ISL_2850200, EPI_ISL_2850201,
EPI_ISL_2850202, EPI_ISL_2850203, EPI_ISL_2850204, EPI_ISL_2850205, EPI_ISL_2850206, EPI_ISL_2850207, EPI_ISL_2850208, EPI_ISL_2850209, EPI_ISL_2850210, EPI_ISL_2850211, EPI_ISL_2850212, EPI_ISL_2850214, EPI_ISL_2850215, EPI_ISL_2850216, EPI_ISL_2850217, EPI_ISL_2850219, EPI_ISL_2850220, EPI_ISL_2850222, EPI_ISL_2850223, EPI_ISL_2850224, EPI_ISL_2850225, EPI_ISL_2850226,
EPI_ISL_2850227, EPI_ISL_2850228, EPI_ISL_2850229, EPI_ISL_2850233, EPI_ISL_2850234, EPI_ISL_2850235, EPI_ISL_2850236, EPI_ISL_2850237, EPI_ISL_2850238, EPI_ISL_2850239, EPI_ISL_2850240, EPI_ISL_2850241, EPI_ISL_2850243, EPI_ISL_2850245, EPI_ISL_2850247, EPI_ISL_2850248, EPI_ISL_2850249, EPI_ISL_2850250, EPI_ISL_2850251, EPI_ISL_2850252, EPI_ISL_2850253, EPI_ISL_2850254,
EPI_ISL_2850255, EPI_ISL_2850256, EPI_ISL_2850258, EPI_ISL_2850259, EPI_ISL_2850260, EPI_ISL_2850261, EPI_ISL_2850264, EPI_ISL_2850265, EPI_ISL_2850266, EPI_ISL_2850267, EPI_ISL_2850268, EPI_ISL_2850269, EPI_ISL_2850270, EPI_ISL_2850271, EPI_ISL_2850272, EPI_ISL_2850273, EPI_ISL_2850274, EPI_ISL_2850275, EPI_ISL_2850278, EPI_ISL_2850279, EPI_ISL_2850280, EPI_ISL_2850281,
EPI_ISL_2850282, EPI_ISL_2850283, EPI_ISL_2850284, EPI_ISL_2850285, EPI_ISL_2850286, EPI_ISL_2850287, EPI_ISL_2850288, EPI_ISL_2850291, EPI_ISL_2850292, EPI_ISL_2850294, EPI_ISL_2850295, EPI_ISL_2850296, EPI_ISL_2850297, EPI_ISL_2850298, EPI_ISL_2850299, EPI_ISL_2850303, EPI_ISL_2850304, EPI_ISL_2850305, EPI_ISL_2850307, EPI_ISL_2850308, EPI_ISL_2850309, EPI_ISL_2850310,
EPI_ISL_2850311, EPI_ISL_2850313, EPI_ISL_2850314, EPI_ISL_2850315, EPI_ISL_2850316, EPI_ISL_2850317, EPI_ISL_2850318, EPI_ISL_2850319, EPI_ISL_2850320, EPI_ISL_2850321, EPI_ISL_2850322, EPI_ISL_2850323, EPI_ISL_2850324, EPI_ISL_2850325, EPI_ISL_2850327, EPI_ISL_2850328, EPI_ISL_2850330, EPI_ISL_2850332, EPI_ISL_2850334, EPI_ISL_2850335, EPI_ISL_2850336, EPI_ISL_2850338,
EPI_ISL_2850339, EPI_ISL_2850340, EPI_ISL_2850342, EPI_ISL_2850343, EPI_ISL_2850345, EPI_ISL_2850347, EPI_ISL_2850348, EPI_ISL_2850349, EPI_ISL_2850350, EPI_ISL_2850352, EPI_ISL_2850353, EPI_ISL_2850354, EPI_ISL_2850355, EPI_ISL_2850356, EPI_ISL_2850357, EPI_ISL_2850359, EPI_ISL_2850360, EPI_ISL_2850361, EPI_ISL_2850362, EPI_ISL_2850364, EPI_ISL_2850366, EPI_ISL_2850367,
EPI_ISL_2850369, EPI_ISL_2850370, EPI_ISL_2850371, EPI_ISL_2850373, EPI_ISL_2850739, EPI_ISL_2850740, EPI_ISL_2850741, EPI_ISL_2850742, EPI_ISL_2850743, EPI_ISL_2850744, EPI_ISL_2850745, EPI_ISL_2850746, EPI_ISL_2850747, EPI_ISL_2850748, EPI_ISL_2850749, EPI_ISL_2850751, EPI_ISL_2850752, EPI_ISL_2850753, EPI_ISL_2850754, EPI_ISL_2850755, EPI_ISL_2850756, EPI_ISL_2850757,
EPI_ISL_2850759, EPI_ISL_2850760, EPI_ISL_2850761, EPI_ISL_2850763, EPI_ISL_2850764, EPI_ISL_2850766, EPI_ISL_2850767, EPI_ISL_2850768, EPI_ISL_2850769, EPI_ISL_2850770, EPI_ISL_2850771, EPI_ISL_2850772, EPI_ISL_2850773, EPI_ISL_2850774, EPI_ISL_2850775, EPI_ISL_2850777, EPI_ISL_2850778, EPI_ISL_2850779, EPI_ISL_2850781, EPI_ISL_2850783, EPI_ISL_2850784, EPI_ISL_2850785,
EPI_ISL_2850786, EPI_ISL_2850787, EPI_ISL_2850788, EPI_ISL_2850789, EPI_ISL_2850790, EPI_ISL_2850791, EPI_ISL_2850792, EPI_ISL_2850794, EPI_ISL_2850795, EPI_ISL_2850797, EPI_ISL_2850798, EPI_ISL_2850799, EPI_ISL_2850800, EPI_ISL_2850801, EPI_ISL_2850802, EPI_ISL_2850803, EPI_ISL_2850804, EPI_ISL_2850805, EPI_ISL_2850806, EPI_ISL_2850807, EPI_ISL_2850808, EPI_ISL_2850809,
EPI_ISL_2850811, EPI_ISL_2850812, EPI_ISL_2850813, EPI_ISL_2850814, EPI_ISL_2850815, EPI_ISL_2850816, EPI_ISL_2850817, EPI_ISL_2850818, EPI_ISL_2850819, EPI_ISL_2850820, EPI_ISL_2850822, EPI_ISL_2850823, EPI_ISL_2850824, EPI_ISL_2850825, EPI_ISL_2850826, EPI_ISL_2850827, EPI_ISL_2850828, EPI_ISL_2850829, EPI_ISL_2850830, EPI_ISL_2850831, EPI_ISL_2850832, EPI_ISL_2850833,
EPI_ISL_2850834, EPI_ISL_2850835, EPI_ISL_2850836, EPI_ISL_2850837, EPI_ISL_2850838, EPI_ISL_2850839, EPI_ISL_2850840, EPI_ISL_2850841, EPI_ISL_2850842, EPI_ISL_2850843, EPI_ISL_2850844, EPI_ISL_2850845, EPI_ISL_2850846, EPI_ISL_2850847, EPI_ISL_2850848, EPI_ISL_2850849, EPI_ISL_2850850, EPI_ISL_2850851, EPI_ISL_2850852, EPI_ISL_2850853, EPI_ISL_2850854, EPI_ISL_2850855,
EPI_ISL_2850857, EPI_ISL_2850858, EPI_ISL_2850859, EPI_ISL_2850860, EPI_ISL_2850861, EPI_ISL_2850862, EPI_ISL_2850863, EPI_ISL_2850864, EPI_ISL_2850865, EPI_ISL_2850866, EPI_ISL_2850867, EPI_ISL_2850868, EPI_ISL_2850869, EPI_ISL_2850870, EPI_ISL_2850871, EPI_ISL_2850872, EPI_ISL_2850873, EPI_ISL_2850874, EPI_ISL_2850875, EPI_ISL_2850876, EPI_ISL_2850877, EPI_ISL_2850878,
EPI_ISL_2850879, EPI_ISL_2850880, EPI_ISL_2850881, EPI_ISL_2850882, EPI_ISL_2850883, EPI_ISL_2850884, EPI_ISL_2850885, EPI_ISL_2850886, EPI_ISL_2850887, EPI_ISL_2850888, EPI_ISL_2850889, EPI_ISL_2850890, EPI_ISL_2850891, EPI_ISL_2850892, EPI_ISL_2850893, EPI_ISL_2850894, EPI_ISL_2850895, EPI_ISL_2850896, EPI_ISL_2850897, EPI_ISL_2850898, EPI_ISL_2850899, EPI_ISL_2850900,
EPI_ISL_2850901, EPI_ISL_2850902, EPI_ISL_2850903, EPI_ISL_2850904, EPI_ISL_2850905, EPI_ISL_2850906, EPI_ISL_2850907, EPI_ISL_2850908, EPI_ISL_2850909, EPI_ISL_2850910, EPI_ISL_2850911, EPI_ISL_2850912, EPI_ISL_2850913, EPI_ISL_2850914, EPI_ISL_2850915, EPI_ISL_2850916, EPI_ISL_2850917, EPI_ISL_2850918, EPI_ISL_2850919, EPI_ISL_2850920, EPI_ISL_2850921, EPI_ISL_2850923,
EPI_ISL_2850924, EPI_ISL_2850925, EPI_ISL_2850926, EPI_ISL_2850927, EPI_ISL_2850928, EPI_ISL_2850929, EPI_ISL_2850930, EPI_ISL_2850931, EPI_ISL_2850932, EPI_ISL_2850933, EPI_ISL_2850934, EPI_ISL_2850935, EPI_ISL_2850936, EPI_ISL_2850937, EPI_ISL_2850938, EPI_ISL_2850940, EPI_ISL_2850941, EPI_ISL_2850942, EPI_ISL_2850943, EPI_ISL_2850944, EPI_ISL_2850945, EPI_ISL_2850946,
EPI_ISL_2850947, EPI_ISL_2850948, EPI_ISL_2850949, EPI_ISL_2850950, EPI_ISL_2850951, EPI_ISL_2850952, EPI_ISL_2850953, EPI_ISL_2850954, EPI_ISL_2850955, EPI_ISL_2850956, EPI_ISL_2850957, EPI_ISL_2850958, EPI_ISL_2850959, EPI_ISL_2850960, EPI_ISL_2850961, EPI_ISL_2850962, EPI_ISL_2850963, EPI_ISL_2850964, EPI_ISL_2850965, EPI_ISL_2850966, EPI_ISL_2850967, EPI_ISL_2850968,
EPI_ISL_2850969, EPI_ISL_2850970, EPI_ISL_2850971, EPI_ISL_2850972, EPI_ISL_2850973, EPI_ISL_2850974, EPI_ISL_2850975, EPI_ISL_2850976, EPI_ISL_2850977, EPI_ISL_2850978, EPI_ISL_2850979, EPI_ISL_2850980, EPI_ISL_2850981, EPI_ISL_2850982, EPI_ISL_2850983, EPI_ISL_2850984, EPI_ISL_2850985, EPI_ISL_2850986, EPI_ISL_2850987, EPI_ISL_2850988, EPI_ISL_2850989, EPI_ISL_2850990,
EPI_ISL_2850991, EPI_ISL_2850992, EPI_ISL_2850993, EPI_ISL_2850994, EPI_ISL_2850995, EPI_ISL_2850996, EPI_ISL_2850997, EPI_ISL_2850998, EPI_ISL_2850999, EPI_ISL_2851000, EPI_ISL_2851001, EPI_ISL_2851002, EPI_ISL_2851003, EPI_ISL_2851004, EPI_ISL_2851005, EPI_ISL_2851006, EPI_ISL_2851007, EPI_ISL_2851008, EPI_ISL_2851009, EPI_ISL_2851010, EPI_ISL_2851011, EPI_ISL_2851012,
EPI_ISL_2851013, EPI_ISL_2851014, EPI_ISL_2851015, EPI_ISL_2851017, EPI_ISL_2851018, EPI_ISL_2851019, EPI_ISL_2851020, EPI_ISL_2851021, EPI_ISL_2851022, EPI_ISL_2851023, EPI_ISL_2851024, EPI_ISL_2851025, EPI_ISL_2851026, EPI_ISL_2851027, EPI_ISL_2851028, EPI_ISL_2851029, EPI_ISL_2851030, EPI_ISL_2851031, EPI_ISL_2851032, EPI_ISL_2851033, EPI_ISL_2851034, EPI_ISL_2851035,
EPI_ISL_2851037, EPI_ISL_2851038, EPI_ISL_2851039, EPI_ISL_2851040, EPI_ISL_2851041, EPI_ISL_2851042, EPI_ISL_2851043, EPI_ISL_2851044, EPI_ISL_2851045, EPI_ISL_2851046, EPI_ISL_2851047, EPI_ISL_2851048, EPI_ISL_2851049, EPI_ISL_2851050, EPI_ISL_2851051, EPI_ISL_2851052, EPI_ISL_2851053, EPI_ISL_2851054, EPI_ISL_2851055, EPI_ISL_2851056, EPI_ISL_2851057, EPI_ISL_2851058,
EPI_ISL_2851059, EPI_ISL_2851060, EPI_ISL_2851061, EPI_ISL_2851062, EPI_ISL_2851063, EPI_ISL_2851064, EPI_ISL_2851065, EPI_ISL_2851066, EPI_ISL_2851067, EPI_ISL_2851068, EPI_ISL_2851069, EPI_ISL_2851070, EPI_ISL_2851071, EPI_ISL_2851072, EPI_ISL_2851073, EPI_ISL_2851074, EPI_ISL_2851075, EPI_ISL_2851076, EPI_ISL_2851077, EPI_ISL_2851078, EPI_ISL_2851080, EPI_ISL_2851081,
EPI_ISL_2851082, EPI_ISL_2851083, EPI_ISL_2851084, EPI_ISL_2851085, EPI_ISL_2851086, EPI_ISL_2851087, EPI_ISL_2851088, EPI_ISL_2851089, EPI_ISL_2851090, EPI_ISL_2851091, EPI_ISL_2851092, EPI_ISL_2851093, EPI_ISL_2851094, EPI_ISL_2851095, EPI_ISL_2851096, EPI_ISL_2851097, EPI_ISL_2851098, EPI_ISL_2851099, EPI_ISL_2851100, EPI_ISL_2851101, EPI_ISL_2851102, EPI_ISL_2851103,
EPI_ISL_2851104, EPI_ISL_2851105, EPI_ISL_2851106, EPI_ISL_2851107, EPI_ISL_2851108, EPI_ISL_2851109, EPI_ISL_2851110, EPI_ISL_2851111, EPI_ISL_2851112, EPI_ISL_2851113, EPI_ISL_2851114, EPI_ISL_2851115, EPI_ISL_2851116, EPI_ISL_2851117, EPI_ISL_2851118, EPI_ISL_2851119, EPI_ISL_2851120, EPI_ISL_2851121, EPI_ISL_2851122, EPI_ISL_2851123, EPI_ISL_2851124, EPI_ISL_2851125,
EPI_ISL_2851126, EPI_ISL_2851127, EPI_ISL_2851128, EPI_ISL_2851129, EPI_ISL_2851130, EPI_ISL_2851131, EPI_ISL_2851132, EPI_ISL_2851134, EPI_ISL_2851135, EPI_ISL_2851136, EPI_ISL_2851137, EPI_ISL_2851138, EPI_ISL_2851139, EPI_ISL_2851140, EPI_ISL_2851141, EPI_ISL_2851143, EPI_ISL_2851144, EPI_ISL_2851145, EPI_ISL_2851146, EPI_ISL_2851147, EPI_ISL_2851148, EPI_ISL_2851149,
EPI_ISL_2851150, EPI_ISL_2851151, EPI_ISL_2851152, EPI_ISL_2851153, EPI_ISL_2851154, EPI_ISL_2851155, EPI_ISL_2851156, EPI_ISL_2851157, EPI_ISL_2851158, EPI_ISL_2851159, EPI_ISL_2851160, EPI_ISL_2851161, EPI_ISL_2851162, EPI_ISL_2851163, EPI_ISL_2851164, EPI_ISL_2851165, EPI_ISL_2851166, EPI_ISL_2851167, EPI_ISL_2851168, EPI_ISL_2851170, EPI_ISL_2851171, EPI_ISL_2851172,
EPI_ISL_2851173, EPI_ISL_2851174, EPI_ISL_2851184, EPI_ISL_2851264, EPI_ISL_2851265, EPI_ISL_2851266, EPI_ISL_2851267, EPI_ISL_2851268, EPI_ISL_2851269, EPI_ISL_2851270, EPI_ISL_2851271, EPI_ISL_2851272, EPI_ISL_2851273, EPI_ISL_2851274, EPI_ISL_2851275, EPI_ISL_2851276, EPI_ISL_2851277, EPI_ISL_2851278, EPI_ISL_2851279, EPI_ISL_2851280, EPI_ISL_2851281, EPI_ISL_2851282,
EPI_ISL_2851283, EPI_ISL_2851284, EPI_ISL_2851285, EPI_ISL_2851286, EPI_ISL_2851287, EPI_ISL_2851288, EPI_ISL_2851289, EPI_ISL_2851290, EPI_ISL_2851291, EPI_ISL_2851292, EPI_ISL_2851293, EPI_ISL_2851294, EPI_ISL_2851295, EPI_ISL_2851296, EPI_ISL_2851297, EPI_ISL_2851298, EPI_ISL_2851299, EPI_ISL_2851300, EPI_ISL_2851301, EPI_ISL_2851302, EPI_ISL_2851303, EPI_ISL_2851304,
EPI_ISL_2851305, EPI_ISL_2851306, EPI_ISL_2851307, EPI_ISL_2851308, EPI_ISL_2851309, EPI_ISL_2851310, EPI_ISL_2851311, EPI_ISL_2851312, EPI_ISL_2851313, EPI_ISL_2851314, EPI_ISL_2851315, EPI_ISL_2851316, EPI_ISL_2851317, EPI_ISL_2851318, EPI_ISL_2851319, EPI_ISL_2851320, EPI_ISL_2851321, EPI_ISL_2851322, EPI_ISL_2851323, EPI_ISL_2851324, EPI_ISL_2851325, EPI_ISL_2851326,
EPI_ISL_2851327, EPI_ISL_2851328, EPI_ISL_2851329, EPI_ISL_2851330, EPI_ISL_2851331, EPI_ISL_2851332, EPI_ISL_2851333, EPI_ISL_2851334, EPI_ISL_2851335, EPI_ISL_2851336, EPI_ISL_2851337, EPI_ISL_2851338, EPI_ISL_2851339, EPI_ISL_2851340, EPI_ISL_2851341, EPI_ISL_2851342, EPI_ISL_2851343, EPI_ISL_2851344, EPI_ISL_2851345, EPI_ISL_2851350, EPI_ISL_2851351, EPI_ISL_2851356,
EPI_ISL_2851363, EPI_ISL_2851367, EPI_ISL_2851375, EPI_ISL_2851378, EPI_ISL_2851380, EPI_ISL_2851384, EPI_ISL_2851385, EPI_ISL_2851395, EPI_ISL_2851400, EPI_ISL_2851405, EPI_ISL_2851774, EPI_ISL_2851775, EPI_ISL_2851776, EPI_ISL_2851778, EPI_ISL_2851779, EPI_ISL_2851780, EPI_ISL_2851781, EPI_ISL_2851782, EPI_ISL_2851783, EPI_ISL_2851785, EPI_ISL_2851787, EPI_ISL_2851788,
EPI_ISL_2851789, EPI_ISL_2851794, EPI_ISL_2851797, EPI_ISL_2851799, EPI_ISL_2851805, EPI_ISL_2851806, EPI_ISL_2851809, EPI_ISL_2851810, EPI_ISL_2851811, EPI_ISL_2851815, EPI_ISL_2851817, EPI_ISL_2851818, EPI_ISL_2851821, EPI_ISL_2851823, EPI_ISL_2851827, EPI_ISL_2851829, EPI_ISL_2851830, EPI_ISL_2851831, EPI_ISL_2851832, EPI_ISL_2851833, EPI_ISL_2851834, EPI_ISL_2851836,
EPI_ISL_2851838, EPI_ISL_2851839, EPI_ISL_2851841, EPI_ISL_2851842, EPI_ISL_2851843, EPI_ISL_2851844, EPI_ISL_2851845, EPI_ISL_2851847, EPI_ISL_2851848, EPI_ISL_2851849, EPI_ISL_2851850, EPI_ISL_2851851, EPI_ISL_2851852, EPI_ISL_2851853, EPI_ISL_2851854, EPI_ISL_2851855, EPI_ISL_2851856, EPI_ISL_2851857, EPI_ISL_2851858, EPI_ISL_2851859, EPI_ISL_2851860, EPI_ISL_2851861,
EPI_ISL_2851867, EPI_ISL_2851868, EPI_ISL_2851870, EPI_ISL_2851871, EPI_ISL_2851875, EPI_ISL_2851883, EPI_ISL_2851886, EPI_ISL_2851888, EPI_ISL_2851889, EPI_ISL_2851890, EPI_ISL_2851891, EPI_ISL_2851894, EPI_ISL_2851895, EPI_ISL_2851898, EPI_ISL_2851899, EPI_ISL_2851900, EPI_ISL_2851903, EPI_ISL_2851908, EPI_ISL_2851910, EPI_ISL_2851911, EPI_ISL_2851912, EPI_ISL_2851915,
EPI_ISL_2851916, EPI_ISL_2851917, EPI_ISL_2851919, EPI_ISL_2851921, EPI_ISL_2851923, EPI_ISL_2851924, EPI_ISL_2851925, EPI_ISL_2851926, EPI_ISL_2851927, EPI_ISL_2851928, EPI_ISL_2851930, EPI_ISL_2851932, EPI_ISL_2851933, EPI_ISL_2851934, EPI_ISL_2851937, EPI_ISL_2851938, EPI_ISL_2851940, EPI_ISL_2851941, EPI_ISL_2851942, EPI_ISL_2851944, EPI_ISL_2851946, EPI_ISL_2851948,
EPI_ISL_2851949, EPI_ISL_2851951, EPI_ISL_2851952, EPI_ISL_2851954, EPI_ISL_2851956, EPI_ISL_2851959, EPI_ISL_2851963, EPI_ISL_2851964, EPI_ISL_2851968, EPI_ISL_2851970, EPI_ISL_2851971, EPI_ISL_2851972, EPI_ISL_2851974, EPI_ISL_2851975, EPI_ISL_2851979, EPI_ISL_2851980, EPI_ISL_2851981, EPI_ISL_2851982, EPI_ISL_2851985, EPI_ISL_2851986, EPI_ISL_2851987, EPI_ISL_2851992,
EPI_ISL_2851993, EPI_ISL_2851995, EPI_ISL_2851997, EPI_ISL_2852006, EPI_ISL_2852009, EPI_ISL_2852010, EPI_ISL_2852012, EPI_ISL_2852015, EPI_ISL_2852016, EPI_ISL_2852017, EPI_ISL_2852018, EPI_ISL_2852019, EPI_ISL_2852021, EPI_ISL_2852024, EPI_ISL_2852027, EPI_ISL_2852030, EPI_ISL_2852031, EPI_ISL_2852033, EPI_ISL_2852034, EPI_ISL_2852036, EPI_ISL_2852041, EPI_ISL_2852042,
EPI_ISL_2852043, EPI_ISL_2852046, EPI_ISL_2852047, EPI_ISL_2852050, EPI_ISL_2852051, EPI_ISL_2852057, EPI_ISL_2852059, EPI_ISL_2852060, EPI_ISL_2852062, EPI_ISL_2852066, EPI_ISL_2852071, EPI_ISL_2852072, EPI_ISL_2852073, EPI_ISL_2852078, EPI_ISL_2852083, EPI_ISL_2852084, EPI_ISL_2852086, EPI_ISL_2852087, EPI_ISL_2852088, EPI_ISL_2852089, EPI_ISL_2852091, EPI_ISL_2852093,
EPI_ISL_2852094, EPI_ISL_2852099, EPI_ISL_2852104, EPI_ISL_2852106, EPI_ISL_2852112, EPI_ISL_2852114, EPI_ISL_2852116, EPI_ISL_2852117, EPI_ISL_2852120, EPI_ISL_2852122, EPI_ISL_2852124, EPI_ISL_2852125, EPI_ISL_2852128, EPI_ISL_2852130, EPI_ISL_2852131, EPI_ISL_2852132, EPI_ISL_2852134, EPI_ISL_2852135, EPI_ISL_2852141, EPI_ISL_2852143, EPI_ISL_2852144, EPI_ISL_2852145,
EPI_ISL_2852146, EPI_ISL_2852147, EPI_ISL_2852149, EPI_ISL_2852150, EPI_ISL_2852151, EPI_ISL_2852153, EPI_ISL_2852154, EPI_ISL_2852156, EPI_ISL_2852160, EPI_ISL_2852161, EPI_ISL_2852162, EPI_ISL_2852163, EPI_ISL_2852164, EPI_ISL_2852165, EPI_ISL_2852166, EPI_ISL_2852168, EPI_ISL_2852171, EPI_ISL_2852172, EPI_ISL_2852173, EPI_ISL_2852174, EPI_ISL_2852175, EPI_ISL_2852176,
EPI_ISL_2852177, EPI_ISL_2852179, EPI_ISL_2852181, EPI_ISL_2852182, EPI_ISL_2852184, EPI_ISL_2852185, EPI_ISL_2852189, EPI_ISL_2852190, EPI_ISL_2852191, EPI_ISL_2852198, EPI_ISL_2852199, EPI_ISL_2852200, EPI_ISL_2852201, EPI_ISL_2852202, EPI_ISL_2852203, EPI_ISL_2852204, EPI_ISL_2852205, EPI_ISL_2852207, EPI_ISL_2852208, EPI_ISL_2852209, EPI_ISL_2852210, EPI_ISL_2852212,
EPI_ISL_2852213, EPI_ISL_2852214, EPI_ISL_2852215, EPI_ISL_2852216, EPI_ISL_2852217, EPI_ISL_2852218, EPI_ISL_2852219, EPI_ISL_2852220, EPI_ISL_2852221, EPI_ISL_2852222, EPI_ISL_2852223, EPI_ISL_2852224, EPI_ISL_2852226, EPI_ISL_2852228, EPI_ISL_2852229, EPI_ISL_2852233, EPI_ISL_2852234, EPI_ISL_2852235, EPI_ISL_2852236, EPI_ISL_2852239, EPI_ISL_2852240, EPI_ISL_2852241,
EPI_ISL_2852242, EPI_ISL_2852243, EPI_ISL_2852244, EPI_ISL_2852245, EPI_ISL_2852246, EPI_ISL_2852247, EPI_ISL_2852252, EPI_ISL_2852253, EPI_ISL_2852254, EPI_ISL_2852256, EPI_ISL_2852257, EPI_ISL_2852258, EPI_ISL_2852260, EPI_ISL_2852261, EPI_ISL_2852262, EPI_ISL_2852264, EPI_ISL_2852265, EPI_ISL_2852267, EPI_ISL_2852268, EPI_ISL_2852269, EPI_ISL_2852270, EPI_ISL_2852271,
EPI_ISL_2852272, EPI_ISL_2852273, EPI_ISL_2852274, EPI_ISL_2852275, EPI_ISL_2852276, EPI_ISL_2852277, EPI_ISL_2852278, EPI_ISL_2852279, EPI_ISL_2852280, EPI_ISL_2852281, EPI_ISL_2852283, EPI_ISL_2852284, EPI_ISL_2852285, EPI_ISL_2852287, EPI_ISL_2852288, EPI_ISL_2852291, EPI_ISL_2852292, EPI_ISL_2852293, EPI_ISL_2852294, EPI_ISL_2852295, EPI_ISL_2852296, EPI_ISL_2852297,

EPI_ISL_2852298, EPI_ISL_2852299, EPI_ISL_2852300, EPI_ISL_2852304, EPI_ISL_2852305, EPI_ISL_2852306, EPI_ISL_2852307, EPI_ISL_2852308, EPI_ISL_2852309, EPI_ISL_2852311, EPI_ISL_2852312, EPI_ISL_2852314, EPI_ISL_2852315, EPI_ISL_2852317, EPI_ISL_2852318, EPI_ISL_2852322, EPI_ISL_2852325, EPI_ISL_2852326, EPI_ISL_2852327, EPI_ISL_2852328, EPI_ISL_2852329, EPI_ISL_2852330, EPI_ISL_2852331, EPI_ISL_2852333, EPI_ISL_2852334, EPI_ISL_2852335, EPI_ISL_2852336, EPI_ISL_2852337, EPI_ISL_2852338, EPI_ISL_2852339, EPI_ISL_2852340, EPI_ISL_2852342, EPI_ISL_2852346, EPI_ISL_2852347, EPI_ISL_2852348, EPI_ISL_2852351, EPI_ISL_2852352, EPI_ISL_2852353, EPI_ISL_2852354, EPI_ISL_2852356, EPI_ISL_2852359, EPI_ISL_2852360, EPI_ISL_2852362, EPI_ISL_2852363, EPI_ISL_2852364, EPI_ISL_2852365, EPI_ISL_2852367, EPI_ISL_2852368, EPI_ISL_2852369, EPI_ISL_2852370, EPI_ISL_2852372, EPI_ISL_2852373, EPI_ISL_2852375, EPI_ISL_2852376, EPI_ISL_2852377, EPI_ISL_2852379, EPI_ISL_2852382, EPI_ISL_2852383, EPI_ISL_2852385, EPI_ISL_2852386, EPI_ISL_2852389, EPI_ISL_2852391, EPI_ISL_2852392, EPI_ISL_2852393, EPI_ISL_2852394, EPI_ISL_2852395, EPI_ISL_2852396, EPI_ISL_2852397, EPI_ISL_2852398, EPI_ISL_2852399, EPI_ISL_2852400, EPI_ISL_2852401, EPI_ISL_2852402, EPI_ISL_2852403, EPI_ISL_2852405, EPI_ISL_2852407, EPI_ISL_2852408, EPI_ISL_2852409, EPI_ISL_2852411, EPI_ISL_2852412, EPI_ISL_2852413, EPI_ISL_2852414, EPI_ISL_2852415, EPI_ISL_2852416, EPI_ISL_2852417, EPI_ISL_2852418, EPI_ISL_2852419, EPI_ISL_2852420, EPI_ISL_2852421, EPI_ISL_2852422, EPI_ISL_2852423, EPI_ISL_2852425, EPI_ISL_2852426, EPI_ISL_2852429, EPI_ISL_2852430, EPI_ISL_2852431, EPI_ISL_2852432, EPI_ISL_2852433, EPI_ISL_2852434, EPI_ISL_2852435, EPI_ISL_2852436, EPI_ISL_2852438, EPI_ISL_2852439, EPI_ISL_2852441, EPI_ISL_2852442, EPI_ISL_2852443, EPI_ISL_2852444, EPI_ISL_2852445, EPI_ISL_2852447, EPI_ISL_2852449, EPI_ISL_2852450, EPI_ISL_2852453, EPI_ISL_2852455, EPI_ISL_2852456, EPI_ISL_2852458, EPI_ISL_2852459, EPI_ISL_2852461, EPI_ISL_2852462, EPI_ISL_2852464, EPI_ISL_2852465, EPI_ISL_2852467, EPI_ISL_2852468, EPI_ISL_2852469, EPI_ISL_2852470, EPI_ISL_2852471, EPI_ISL_2852472, EPI_ISL_2852473, EPI_ISL_2852474, EPI_ISL_2852475, EPI_ISL_2852476, EPI_ISL_2852477, EPI_ISL_2852479, EPI_ISL_2852613, EPI_ISL_2852614, EPI_ISL_2852615, EPI_ISL_2852616, EPI_ISL_2852617, EPI_ISL_2852618, EPI_ISL_2852619, EPI_ISL_2852621, EPI_ISL_2852622, EPI_ISL_2852623, EPI_ISL_2852624, EPI_ISL_2852625, EPI_ISL_2852626, EPI_ISL_2852627, EPI_ISL_2852628, EPI_ISL_2852629, EPI_ISL_2852630, EPI_ISL_2852631, EPI_ISL_2852632, EPI_ISL_2852633, EPI_ISL_2852634, EPI_ISL_2852635, EPI_ISL_2852636, EPI_ISL_2852637, EPI_ISL_2852638, EPI_ISL_2852639, EPI_ISL_2852641, EPI_ISL_2852642, EPI_ISL_2852643, EPI_ISL_2852644, EPI_ISL_2852646, EPI_ISL_2852647, EPI_ISL_2852648, EPI_ISL_2852649, EPI_ISL_2852650, EPI_ISL_2852651, EPI_ISL_2852652, EPI_ISL_2852653, EPI_ISL_2852654, EPI_ISL_2852655, EPI_ISL_2852656, EPI_ISL_2852657, EPI_ISL_2852658, EPI_ISL_2852659, EPI_ISL_2852660, EPI_ISL_2852662, EPI_ISL_2852663, EPI_ISL_2852664, EPI_ISL_2852665, EPI_ISL_2852666, EPI_ISL_2852667, EPI_ISL_2852668, EPI_ISL_2852669, EPI_ISL_2852670, EPI_ISL_2852671, EPI_ISL_2852672, EPI_ISL_2852673, EPI_ISL_2852674, EPI_ISL_2852675, EPI_ISL_2852676, EPI_ISL_2852677, EPI_ISL_2852678, EPI_ISL_2852679, EPI_ISL_2852680, EPI_ISL_2852681, EPI_ISL_2852682, EPI_ISL_2852683, EPI_ISL_2852684, EPI_ISL_2852686, EPI_ISL_2852687, EPI_ISL_2852688, EPI_ISL_2852689, EPI_ISL_2852690, EPI_ISL_2852691, EPI_ISL_2852693, EPI_ISL_2852694, EPI_ISL_2852695, EPI_ISL_2852696, EPI_ISL_2852697, EPI_ISL_2852699, EPI_ISL_2852700, EPI_ISL_2852701, EPI_ISL_2852702, EPI_ISL_2852704, EPI_ISL_2852705, EPI_ISL_2852706, EPI_ISL_2852707, EPI_ISL_2852708, EPI_ISL_2852709, EPI_ISL_2852710, EPI_ISL_2852711, EPI_ISL_2852712, EPI_ISL_2852713, EPI_ISL_2852714, EPI_ISL_2852715, EPI_ISL_2852716, EPI_ISL_2852717, EPI_ISL_2852718, EPI_ISL_2852719, EPI_ISL_2852721, EPI_ISL_2852722, EPI_ISL_2852723, EPI_ISL_2852724, EPI_ISL_2852725, EPI_ISL_2852726, EPI_ISL_2852727, EPI_ISL_2852728, EPI_ISL_2852729, EPI_ISL_2852730, EPI_ISL_2852731, EPI_ISL_2852732, EPI_ISL_2852733, EPI_ISL_2852734, EPI_ISL_2852735, EPI_ISL_2852736, EPI_ISL_2852737, EPI_ISL_2852738, EPI_ISL_2852739, EPI_ISL_2852740, EPI_ISL_2852741, EPI_ISL_2852742, EPI_ISL_2852743, EPI_ISL_2852744, EPI_ISL_2852745, EPI_ISL_2852746, EPI_ISL_2852748, EPI_ISL_2852749, EPI_ISL_2852750, EPI_ISL_2852751, EPI_ISL_2852752, EPI_ISL_2852753, EPI_ISL_2852754, EPI_ISL_2852755, EPI_ISL_2852756, EPI_ISL_2852757, EPI_ISL_2852758, EPI_ISL_2852759, EPI_ISL_2852760, EPI_ISL_2852761, EPI_ISL_2852762, EPI_ISL_2852763, EPI_ISL_2852764, EPI_ISL_2852765, EPI_ISL_2852766, EPI_ISL_2852767, EPI_ISL_2852768, EPI_ISL_2852769, EPI_ISL_2852770, EPI_ISL_2852771, EPI_ISL_2852772, EPI_ISL_2852773, EPI_ISL_2852774, EPI_ISL_2852775, EPI_ISL_2852776, EPI_ISL_2852777, EPI_ISL_2852778, EPI_ISL_2852779, EPI_ISL_2852780, EPI_ISL_2852781, EPI_ISL_2852782, EPI_ISL_2852783, EPI_ISL_2852784, EPI_ISL_2852785, EPI_ISL_2852786, EPI_ISL_2852787, EPI_ISL_2852789, EPI_ISL_2852790, EPI_ISL_2852791, EPI_ISL_2852792, EPI_ISL_2852793, EPI_ISL_2852794, EPI_ISL_2852795, EPI_ISL_2852796, EPI_ISL_2852800, EPI_ISL_2852802, EPI_ISL_2852804, EPI_ISL_2852805, EPI_ISL_2852806, EPI_ISL_2852809, EPI_ISL_2852810, EPI_ISL_2852812, EPI_ISL_2852813, EPI_ISL_2852815, EPI_ISL_2852816, EPI_ISL_2852817, EPI_ISL_2852820, EPI_ISL_2852823, EPI_ISL_2852826, EPI_ISL_2852827, EPI_ISL_2852828, EPI_ISL_2852830, EPI_ISL_2852831, EPI_ISL_2852832, EPI_ISL_2852834, EPI_ISL_2852836, EPI_ISL_2852843, EPI_ISL_2852845, EPI_ISL_2852846, EPI_ISL_2852847, EPI_ISL_2852848, EPI_ISL_2852849, EPI_ISL_2852850, EPI_ISL_2866763, EPI_ISL_2866927, EPI_ISL_2866970, EPI_ISL_2868062, EPI_ISL_2868219, EPI_ISL_2910607, EPI_ISL_2910609, EPI_ISL_2910611, EPI_ISL_2910612, EPI_ISL_2910622, EPI_ISL_2910625, EPI_ISL_2910639, EPI_ISL_2910641, EPI_ISL_2910642, EPI_ISL_2910644, EPI_ISL_2910649, EPI_ISL_2910651, EPI_ISL_2910664, EPI_ISL_2910666, EPI_ISL_2910669, EPI_ISL_2910672, EPI_ISL_2910678, EPI_ISL_2910684, EPI_ISL_2910685, EPI_ISL_2910686, EPI_ISL_2910690, EPI_ISL_2910702, EPI_ISL_2910703, EPI_ISL_2910710, EPI_ISL_2910714, EPI_ISL_2910720, EPI_ISL_2910731, EPI_ISL_2910747, EPI_ISL_2910755, EPI_ISL_2910758, EPI_ISL_2910778, EPI_ISL_2910792, EPI_ISL_2910800, EPI_ISL_2910816, EPI_ISL_2910819, EPI_ISL_2910820, EPI_ISL_2910825, EPI_ISL_2910827, EPI_ISL_2910828, EPI_ISL_2910834, EPI_ISL_2910835, EPI_ISL_2910841, EPI_ISL_2910842, EPI_ISL_2910843, EPI_ISL_2913763, EPI_ISL_2914464, EPI_ISL_2916742, EPI_ISL_2937642, EPI_ISL_2937643, EPI_ISL_2937644, EPI_ISL_2937645, EPI_ISL_2937646, EPI_ISL_2937647, EPI_ISL_2937648, EPI_ISL_2937649, EPI_ISL_2937650, EPI_ISL_2937651, EPI_ISL_2937652, EPI_ISL_2937653, EPI_ISL_2937654, EPI_ISL_2937655, EPI_ISL_2937656, EPI_ISL_2937657, EPI_ISL_2937658, EPI_ISL_2937659, EPI_ISL_2937660, EPI_ISL_2937662, EPI_ISL_2937663, EPI_ISL_2937664, EPI_ISL_2937665, EPI_ISL_2937666, EPI_ISL_2937667, EPI_ISL_2937668, EPI_ISL_2937669, EPI_ISL_2937670, EPI_ISL_2937671, EPI_ISL_2937672, EPI_ISL_2937673, EPI_ISL_2937674, EPI_ISL_2937675, EPI_ISL_2937676, EPI_ISL_2937677, EPI_ISL_2937678, EPI_ISL_2937679, EPI_ISL_2937680, EPI_ISL_2937681, EPI_ISL_2937683, EPI_ISL_2937684, EPI_ISL_2937685, EPI_ISL_2937686, EPI_ISL_2937687, EPI_ISL_2937688, EPI_ISL_2937689, EPI_ISL_2937690, EPI_ISL_2937691, EPI_ISL_2937692, EPI_ISL_2937693, EPI_ISL_2937694, EPI_ISL_2937695, EPI_ISL_2937696, EPI_ISL_2937697, EPI_ISL_2937698, EPI_ISL_2937699, EPI_ISL_2937700, EPI_ISL_2937701, EPI_ISL_2937702, EPI_ISL_2937703, EPI_ISL_2937704, EPI_ISL_2937705, EPI_ISL_2937707, EPI_ISL_2937708, EPI_ISL_2937709, EPI_ISL_2937710, EPI_ISL_2937712, EPI_ISL_2937713

| | | | |
|---|---|---|---|
| see above | Lighthouse Lab in Glasgow | Wellcome Sanger Institute for the COVID-19 Genomics UK (COG-UK) Consortium | Anna Dominiczak and Alex Alderton; Carol Clugston; Cordelia Langford; David Gray; David K. Jackson; Dominic Kwiatkowski; Ewan Harrison; Harper VanSteenhouse; Ian Johnston; Jeffrey Barrett; John Sillitoe on behalf of the Wellcome Sanger Institute COVID-19 Surveillance Team; Roberto Amato; Sonia Goncalves; Yumi Kasai |

EPI_ISL_2807456, EPI_ISL_2828765, EPI_ISL_2828878, EPI_ISL_2828936, EPI_ISL_2828968, EPI_ISL_2829025, EPI_ISL_2829031, EPI_ISL_2829033, EPI_ISL_2829034, EPI_ISL_2829045, EPI_ISL_2829048, EPI_ISL_2829049, EPI_ISL_2829050, EPI_ISL_2829052, EPI_ISL_2829054, EPI_ISL_2829055, EPI_ISL_2829059, EPI_ISL_2829064, EPI_ISL_2829065, EPI_ISL_2829069, EPI_ISL_2829070, EPI_ISL_2829073, EPI_ISL_2829077, EPI_ISL_2829079, EPI_ISL_2829085, EPI_ISL_2829087, EPI_ISL_2829094, EPI_ISL_2829097, EPI_ISL_2829100, EPI_ISL_2829102, EPI_ISL_2829103, EPI_ISL_2829110, EPI_ISL_2829113, EPI_ISL_2829114, EPI_ISL_2829122, EPI_ISL_2829125, EPI_ISL_2829130, EPI_ISL_2829134, EPI_ISL_2829139, EPI_ISL_2829140, EPI_ISL_2829141, EPI_ISL_2829147, EPI_ISL_2829148, EPI_ISL_2829149, EPI_ISL_2829150, EPI_ISL_2829151, EPI_ISL_2829154, EPI_ISL_2829156, EPI_ISL_2829160, EPI_ISL_2829163, EPI_ISL_2829165, EPI_ISL_2829166, EPI_ISL_2829167, EPI_ISL_2829175, EPI_ISL_2829177, EPI_ISL_2829178, EPI_ISL_2829180, EPI_ISL_2829181, EPI_ISL_2829198, EPI_ISL_2829203, EPI_ISL_2829206, EPI_ISL_2829207, EPI_ISL_2829208, EPI_ISL_2829209, EPI_ISL_2829214, EPI_ISL_2829218, EPI_ISL_2829219, EPI_ISL_2829222, EPI_ISL_2829224, EPI_ISL_2829225, EPI_ISL_2829230, EPI_ISL_2829231, EPI_ISL_2829232, EPI_ISL_2829233, EPI_ISL_2829237, EPI_ISL_2829239, EPI_ISL_2829242, EPI_ISL_2829244, EPI_ISL_2829245, EPI_ISL_2829246, EPI_ISL_2829251, EPI_ISL_2829258, EPI_ISL_2829262, EPI_ISL_2829269, EPI_ISL_2829273, EPI_ISL_2829281, EPI_ISL_2829284, EPI_ISL_2829286, EPI_ISL_2829287, EPI_ISL_2829290, EPI_ISL_2829293, EPI_ISL_2829295, EPI_ISL_2829593, EPI_ISL_2829595, EPI_ISL_2829596, EPI_ISL_2829601, EPI_ISL_2829603, EPI_ISL_2829604, EPI_ISL_2829606, EPI_ISL_2829610, EPI_ISL_2829621, EPI_ISL_2829626, EPI_ISL_2829627, EPI_ISL_2829629, EPI_ISL_2829631, EPI_ISL_2829634, EPI_ISL_2829635, EPI_ISL_2829637, EPI_ISL_2829638, EPI_ISL_2829639, EPI_ISL_2829640, EPI_ISL_2829647, EPI_ISL_2829648, EPI_ISL_2829651, EPI_ISL_2829653, EPI_ISL_2829654, EPI_ISL_2829656, EPI_ISL_2829658, EPI_ISL_2829659, EPI_ISL_2829660, EPI_ISL_2829661, EPI_ISL_2829666, EPI_ISL_2829668, EPI_ISL_2829670, EPI_ISL_2829671, EPI_ISL_2829674, EPI_ISL_2829678, EPI_ISL_2829679, EPI_ISL_2829681, EPI_ISL_2829682, EPI_ISL_2829684, EPI_ISL_2829685, EPI_ISL_2829687, EPI_ISL_2829690, EPI_ISL_2829692, EPI_ISL_2829696, EPI_ISL_2829697, EPI_ISL_2829698, EPI_ISL_2829699, EPI_ISL_2829702, EPI_ISL_2829704, EPI_ISL_2829706, EPI_ISL_2829708, EPI_ISL_2829709, EPI_ISL_2829710, EPI_ISL_2829711, EPI_ISL_2829712, EPI_ISL_2829721, EPI_ISL_2829723, EPI_ISL_2829724, EPI_ISL_2829727, EPI_ISL_2829728, EPI_ISL_2829729, EPI_ISL_2829732, EPI_ISL_2829734, EPI_ISL_2829736, EPI_ISL_2829737, EPI_ISL_2829741, EPI_ISL_2829743, EPI_ISL_2829748, EPI_ISL_2829749, EPI_ISL_2829755, EPI_ISL_2829758, EPI_ISL_2829761, EPI_ISL_2829764, EPI_ISL_2829766, EPI_ISL_2829767, EPI_ISL_2829770, EPI_ISL_2829771, EPI_ISL_2829772, EPI_ISL_2829774, EPI_ISL_2829775, EPI_ISL_2829777, EPI_ISL_2829781, EPI_ISL_2829783, EPI_ISL_2829785, EPI_ISL_2829786, EPI_ISL_2829788, EPI_ISL_2829790, EPI_ISL_2829792, EPI_ISL_2829793, EPI_ISL_2829795, EPI_ISL_2829797, EPI_ISL_2829800, EPI_ISL_2829801, EPI_ISL_2829802, EPI_ISL_2829803, EPI_ISL_2829804, EPI_ISL_2829806, EPI_ISL_2829808, EPI_ISL_2829809, EPI_ISL_2829811, EPI_ISL_2829813, EPI_ISL_2829814, EPI_ISL_2829815, EPI_ISL_2829816, EPI_ISL_2829817, EPI_ISL_2829818, EPI_ISL_2829821, EPI_ISL_2829822, EPI_ISL_2829825, EPI_ISL_2829826, EPI_ISL_2829827, EPI_ISL_2829828, EPI_ISL_2829829, EPI_ISL_2829830, EPI_ISL_2829834, EPI_ISL_2829835, EPI_ISL_2829836, EPI_ISL_2829838, EPI_ISL_2829839, EPI_ISL_2829844, EPI_ISL_2829845, EPI_ISL_2829847, EPI_ISL_2829848, EPI_ISL_2829849, EPI_ISL_2829850, EPI_ISL_2829851, EPI_ISL_2829854, EPI_ISL_2829857, EPI_ISL_2829859, EPI_ISL_2829863, EPI_ISL_2829866, EPI_ISL_2829870, EPI_ISL_2829871, EPI_ISL_2829872, EPI_ISL_2829874, EPI_ISL_2829876, EPI_ISL_2829879, EPI_ISL_2829881, EPI_ISL_2829882, EPI_ISL_2829885, EPI_ISL_2829886, EPI_ISL_2829887, EPI_ISL_2829888, EPI_ISL_2829889, EPI_ISL_2829890, EPI_ISL_2829891, EPI_ISL_2829894, EPI_ISL_2829895, EPI_ISL_2829896, EPI_ISL_2829897, EPI_ISL_2829898, EPI_ISL_2829899, EPI_ISL_2829901, EPI_ISL_2829902, EPI_ISL_2829903, EPI_ISL_2829904, EPI_ISL_2829910, EPI_ISL_2829911, EPI_ISL_2829912, EPI_ISL_2829918, EPI_ISL_2829922, EPI_ISL_2829924, EPI_ISL_2829926, EPI_ISL_2829930, EPI_ISL_2829932, EPI_ISL_2829933, EPI_ISL_2829935, EPI_ISL_2829937, EPI_ISL_2829939, EPI_ISL_2829942, EPI_ISL_2829943, EPI_ISL_2829944, EPI_ISL_2829945, EPI_ISL_2829949, EPI_ISL_2829950, EPI_ISL_2829951, EPI_ISL_2829953, EPI_ISL_2829954, EPI_ISL_2829955, EPI_ISL_2829958, EPI_ISL_2829960, EPI_ISL_2829961, EPI_ISL_2829967, EPI_ISL_2829972, EPI_ISL_2829974, EPI_ISL_2829976, EPI_ISL_2829977, EPI_ISL_2829978, EPI_ISL_2829979, EPI_ISL_2829980, EPI_ISL_2829982, EPI_ISL_2829983, EPI_ISL_2829984, EPI_ISL_2829987, EPI_ISL_2829988, EPI_ISL_2829989, EPI_ISL_2829992, EPI_ISL_2829994, EPI_ISL_2829997, EPI_ISL_2830000, EPI_ISL_2830002, EPI_ISL_2830003, EPI_ISL_2830011, EPI_ISL_2830015, EPI_ISL_2830016, EPI_ISL_2830019, EPI_ISL_2830027, EPI_ISL_2830031, EPI_ISL_2830036, EPI_ISL_2830037, EPI_ISL_2830039, EPI_ISL_2830040, EPI_ISL_2830042, EPI_ISL_2830043, EPI_ISL_2830046, EPI_ISL_2830050, EPI_ISL_2830052, EPI_ISL_2830053, EPI_ISL_2830057, EPI_ISL_2830058, EPI_ISL_2830061, EPI_ISL_2830063, EPI_ISL_2830066, EPI_ISL_2830070, EPI_ISL_2830072, EPI_ISL_2830073, EPI_ISL_2830074, EPI_ISL_2830075, EPI_ISL_2830080, EPI_ISL_2830085, EPI_ISL_2830091, EPI_ISL_2830093, EPI_ISL_2830095, EPI_ISL_2830096, EPI_ISL_2830103, EPI_ISL_2830110, EPI_ISL_2830111, EPI_ISL_2830112, EPI_ISL_2830117, EPI_ISL_2830120, EPI_ISL_2830122, EPI_ISL_2830126, EPI_ISL_2830127, EPI_ISL_2830130, EPI_ISL_2830132, EPI_ISL_2830133, EPI_ISL_2830136, EPI_ISL_2830138, EPI_ISL_2830140, EPI_ISL_2830142, EPI_ISL_2830145, EPI_ISL_2830150, EPI_ISL_2830151, EPI_ISL_2830153, EPI_ISL_2830155, EPI_ISL_2830156, EPI_ISL_2830157, EPI_ISL_2830159, EPI_ISL_2830166, EPI_ISL_2830168, EPI_ISL_2830172, EPI_ISL_2830175, EPI_ISL_2830332, EPI_ISL_2830333, EPI_ISL_2830334, EPI_ISL_2830335, EPI_ISL_2830337, EPI_ISL_2830339, EPI_ISL_2830341, EPI_ISL_2830342, EPI_ISL_2830343, EPI_ISL_2830345, EPI_ISL_2830347, EPI_ISL_2830348, EPI_ISL_2830350, EPI_ISL_2830351, EPI_ISL_2830352, EPI_ISL_2830355, EPI_ISL_2830357, EPI_ISL_2830358, EPI_ISL_2830361, EPI_ISL_2830362, EPI_ISL_2830366, EPI_ISL_2830368, EPI_ISL_2830369, EPI_ISL_2830370, EPI_ISL_2830371, EPI_ISL_2830373, EPI_ISL_2830375, EPI_ISL_2830376, EPI_ISL_2830377, EPI_ISL_2830381, EPI_ISL_2830384, EPI_ISL_2830385, EPI_ISL_2830387, EPI_ISL_2830388, EPI_ISL_2830389, EPI_ISL_2830391, EPI_ISL_2830394, EPI_ISL_2830395, EPI_ISL_2830396, EPI_ISL_2830397, EPI_ISL_2830398, EPI_ISL_2830401, EPI_ISL_2830402, EPI_ISL_2830403, EPI_ISL_2830405, EPI_ISL_2830407, EPI_ISL_2830408, EPI_ISL_2830411, EPI_ISL_2830413, EPI_ISL_2830414, EPI_ISL_2830415, EPI_ISL_2830416, EPI_ISL_2830417, EPI_ISL_2830419, EPI_ISL_2830420, EPI_ISL_2830425, EPI_ISL_2830426, EPI_ISL_2830427, EPI_ISL_2830429, EPI_ISL_2830432, EPI_ISL_2830433, EPI_ISL_2830435, EPI_ISL_2830436, EPI_ISL_2830438, EPI_ISL_2830439, EPI_ISL_2830440, EPI_ISL_2830441, EPI_ISL_2830443, EPI_ISL_2830444, EPI_ISL_2830445, EPI_ISL_2830448, EPI_ISL_2830449, EPI_ISL_2830451, EPI_ISL_2830455, EPI_ISL_2830458, EPI_ISL_2830460, EPI_ISL_2830462, EPI_ISL_2830463, EPI_ISL_2830464, EPI_ISL_2830465, EPI_ISL_2830468, EPI_ISL_2830469, EPI_ISL_2830472, EPI_ISL_2830474, EPI_ISL_2830475, EPI_ISL_2830476, EPI_ISL_2830479, EPI_ISL_2830482, EPI_ISL_2830483, EPI_ISL_2830484, EPI_ISL_2830485, EPI_ISL_2830490, EPI_ISL_2830494, EPI_ISL_2830495, EPI_ISL_2830496, EPI_ISL_2830497, EPI_ISL_2830498, EPI_ISL_2830499, EPI_ISL_2830500, EPI_ISL_2830501, EPI_ISL_2830508, EPI_ISL_2830511, EPI_ISL_2830512, EPI_ISL_2830514, EPI_ISL_2830518, EPI_ISL_2830519, EPI_ISL_2830526, EPI_ISL_2830527, EPI_ISL_2830528, EPI_ISL_2830532, EPI_ISL_2830534, EPI_ISL_2830537, EPI_ISL_2830540, EPI_ISL_2830541, EPI_ISL_2830542, EPI_ISL_2830543, EPI_ISL_2830547, EPI_ISL_2830548, EPI_ISL_2830550, EPI_ISL_2830554, EPI_ISL_2830556, EPI_ISL_2830557, EPI_ISL_2830558, EPI_ISL_2830559, EPI_ISL_2830560, EPI_ISL_2830562, EPI_ISL_2830563, EPI_ISL_2830566, EPI_ISL_2830569, EPI_ISL_2830570, EPI_ISL_2830571, EPI_ISL_2830573, EPI_ISL_2830574, EPI_ISL_2830578, EPI_ISL_2830579, EPI_ISL_2830581, EPI_ISL_2830582, EPI_ISL_2830584, EPI_ISL_2830585, EPI_ISL_2830586, EPI_ISL_2830587, EPI_ISL_2830588, EPI_ISL_2830590, EPI_ISL_2830591, EPI_ISL_2830592, EPI_ISL_2830596, EPI_ISL_2830597, EPI_ISL_2830598, EPI_ISL_2830599, EPI_ISL_2830601, EPI_ISL_2830602, EPI_ISL_2830604, EPI_ISL_2831763, EPI_ISL_2831764, EPI_ISL_2831766, EPI_ISL_2831768, EPI_ISL_2831769, EPI_ISL_2831770, EPI_ISL_2831772, EPI_ISL_2831774, EPI_ISL_2831776, EPI_ISL_2831777, EPI_ISL_2831779, EPI_ISL_2831781, EPI_ISL_2831782, EPI_ISL_2831783, EPI_ISL_2831785, EPI_ISL_2831787, EPI_ISL_2831796, EPI_ISL_2831798, EPI_ISL_2831799, EPI_ISL_2831802, EPI_ISL_2831804, EPI_ISL_2831806, EPI_ISL_2831807, EPI_ISL_2831809, EPI_ISL_2831810, EPI_ISL_2831813, EPI_ISL_2831814, EPI_ISL_2831817, EPI_ISL_2831818, EPI_ISL_2831824, EPI_ISL_2831825, EPI_ISL_2831826, EPI_ISL_2831827, EPI_ISL_2831828, EPI_ISL_2831832, EPI_ISL_2831833, EPI_ISL_2831834, EPI_ISL_2831836, EPI_ISL_2831837, EPI_ISL_2831838, EPI_ISL_2831842, EPI_ISL_2831843, EPI_ISL_2831845, EPI_ISL_2831847, EPI_ISL_2831848, EPI_ISL_2831849, EPI_ISL_2831851, EPI_ISL_2831852, EPI_ISL_2831853, EPI_ISL_2831858, EPI_ISL_2831861, EPI_ISL_2831863, EPI_ISL_2831864, EPI_ISL_2831867, EPI_ISL_2831868, EPI_ISL_2831869, EPI_ISL_2831870, EPI_ISL_2831871, EPI_ISL_2831872, EPI_ISL_2831873, EPI_ISL_2831875, EPI_ISL_2831877, EPI_ISL_2831884, EPI_ISL_2831887, EPI_ISL_2831888, EPI_ISL_2831889, EPI_ISL_2831891, EPI_ISL_2831895, EPI_ISL_2831896, EPI_ISL_2831897, EPI_ISL_2831899, EPI_ISL_2831901, EPI_ISL_2831902, EPI_ISL_2831910, EPI_ISL_2831913, EPI_ISL_2831915, EPI_ISL_2831917, EPI_ISL_2831918, EPI_ISL_2831921, EPI_ISL_2831922, EPI_ISL_2831923, EPI_ISL_2831924, EPI_ISL_2831927, EPI_ISL_2831928, EPI_ISL_2831929, EPI_ISL_2831930, EPI_ISL_2831937, EPI_ISL_2831938, EPI_ISL_2831939, EPI_ISL_2831940, EPI_ISL_2831942, EPI_ISL_2831947, EPI_ISL_2831949, EPI_ISL_2831954, EPI_ISL_2831956, EPI_ISL_2831957, EPI_ISL_2831959, EPI_ISL_2831960, EPI_ISL_2831966, EPI_ISL_2831967, EPI_ISL_2831970, EPI_ISL_2831971, EPI_ISL_2831973, EPI_ISL_2831975, EPI_ISL_2831977, EPI_ISL_2831978, EPI_ISL_2831981, EPI_ISL_2831982, EPI_ISL_2831984, EPI_ISL_2831985, EPI_ISL_2831987, EPI_ISL_2831988, EPI_ISL_2831989, EPI_ISL_2831990, EPI_ISL_2831991, EPI_ISL_2831992, EPI_ISL_2831995, EPI_ISL_2831996, EPI_ISL_2831999, EPI_ISL_2832001, EPI_ISL_2832004, EPI_ISL_2832005, EPI_ISL_2832006, EPI_ISL_2832007, EPI_ISL_2832009, EPI_ISL_2832011, EPI_ISL_2832012, EPI_ISL_2832013, EPI_ISL_2832019, EPI_ISL_2832023, EPI_ISL_2832025, EPI_ISL_2832028, EPI_ISL_2832031, EPI_ISL_2832032, EPI_ISL_2832033, EPI_ISL_2832034, EPI_ISL_2832035, EPI_ISL_2832037, EPI_ISL_2832040, EPI_ISL_2832043, EPI_ISL_2832044, EPI_ISL_2832045, EPI_ISL_2832047, EPI_ISL_2832048, EPI_ISL_2832049, EPI_ISL_2832050, EPI_ISL_2832052, EPI_ISL_2832054, EPI_ISL_2832056, EPI_ISL_2832058, EPI_ISL_2832059, EPI_ISL_2832060, EPI_ISL_2832062, EPI_ISL_2832064, EPI_ISL_2846081, EPI_ISL_2846096, EPI_ISL_2846124, EPI_ISL_2846203, EPI_ISL_2846241, EPI_ISL_2846313, EPI_ISL_2846321, EPI_ISL_2846322, EPI_ISL_2846360, EPI_ISL_2846366, EPI_ISL_2846368, EPI_ISL_2846373, EPI_ISL_2846374, EPI_ISL_2846376, EPI_ISL_2846380, EPI_ISL_2846381, EPI_ISL_2846382, EPI_ISL_2846383, EPI_ISL_2846388, EPI_ISL_2846389, EPI_ISL_2846391, EPI_ISL_2846395, EPI_ISL_2846396, EPI_ISL_2846400, EPI_ISL_2846406, EPI_ISL_2846407, EPI_ISL_2846409, EPI_ISL_2846412, EPI_ISL_2846420, EPI_ISL_2846421, EPI_ISL_2846422, EPI_ISL_2846426, EPI_ISL_2846429, EPI_ISL_2846433, EPI_ISL_2846435, EPI_ISL_2846440, EPI_ISL_2846441, EPI_ISL_2846447, EPI_ISL_2846449, EPI_ISL_2846452, EPI_ISL_2846457, EPI_ISL_2846458, EPI_ISL_2846468, EPI_ISL_2846469, EPI_ISL_2846471, EPI_ISL_2846472, EPI_ISL_2846474, EPI_ISL_2846480, EPI_ISL_2846481, EPI_ISL_2846483, EPI_ISL_2846486, EPI_ISL_2846488, EPI_ISL_2846489, EPI_ISL_2846490, EPI_ISL_2846492, EPI_ISL_2846495, EPI_ISL_2846496, EPI_ISL_2846497, EPI_ISL_2846502, EPI_ISL_2846505, EPI_ISL_2846507, EPI_ISL_2846511, EPI_ISL_2846512, EPI_ISL_2846513, EPI_ISL_2846514, EPI_ISL_2846516, EPI_ISL_2846522, EPI_ISL_2846525, EPI_ISL_2846528, EPI_ISL_2846531, EPI_ISL_2846532, EPI_ISL_2846535, EPI_ISL_2846536, EPI_ISL_2846537, EPI_ISL_2846541, EPI_ISL_2846542, EPI_ISL_2846544, EPI_ISL_2846545, EPI_ISL_2846548, EPI_ISL_2846549, EPI_ISL_2846551, EPI_ISL_2846554, EPI_ISL_2846555, EPI_ISL_2846557, EPI_ISL_2846559, EPI_ISL_2846560, EPI_ISL_2846564, EPI_ISL_2846566, EPI_ISL_2846568, EPI_ISL_2846570, EPI_ISL_2846572, EPI_ISL_2846573, EPI_ISL_2846579, EPI_ISL_2846582, EPI_ISL_2846585, EPI_ISL_2846586, EPI_ISL_2846590, EPI_ISL_2846595, EPI_ISL_2846598, EPI_ISL_2846612, EPI_ISL_2846614, EPI_ISL_2846616, EPI_ISL_2846621, EPI_ISL_2846626, EPI_ISL_2846632, EPI_ISL_2846638, EPI_ISL_2846640, EPI_ISL_2846646, EPI_ISL_2846647, EPI_ISL_2846648, EPI_ISL_2846659, EPI_ISL_2846661, EPI_ISL_2846664, EPI_ISL_2846665, EPI_ISL_2846670, EPI_ISL_2846672, EPI_ISL_2846674, EPI_ISL_2846678, EPI_ISL_2846686, EPI_ISL_2846691, EPI_ISL_2846695, EPI_ISL_2846702, EPI_ISL_2846706, EPI_ISL_2846711, EPI_ISL_2846727, EPI_ISL_2846733, EPI_ISL_2846737, EPI_ISL_2846750, EPI_ISL_2846751, EPI_ISL_2846752, EPI_ISL_2846756, EPI_ISL_2846758, EPI_ISL_2846762, EPI_ISL_2846773, EPI_ISL_2846774, EPI_ISL_2846775, EPI_ISL_2846786, EPI_ISL_2846794, EPI_ISL_2846801, EPI_ISL_2846804, EPI_ISL_2846806, EPI_ISL_2846817, EPI_ISL_2846821, EPI_ISL_2846826, EPI_ISL_2846829, EPI_ISL_2846834, EPI_ISL_2846852, EPI_ISL_2846863, EPI_ISL_2846867, EPI_ISL_2846873, EPI_ISL_2846874, EPI_ISL_2846882, EPI_ISL_2846886, EPI_ISL_2846893, EPI_ISL_2846898, EPI_ISL_2846902, EPI_ISL_2846908, EPI_ISL_2846913, EPI_ISL_2846919, EPI_ISL_2846922, EPI_ISL_2846925, EPI_ISL_2846926, EPI_ISL_2846928, EPI_ISL_2846929, EPI_ISL_2846931, EPI_ISL_2846932, EPI_ISL_2846943, EPI_ISL_2846944, EPI_ISL_2846947, EPI_ISL_2846953, EPI_ISL_2846954, EPI_ISL_2846955, EPI_ISL_2846959, EPI_ISL_2846960, EPI_ISL_2846961, EPI_ISL_2846967, EPI_ISL_2846970, EPI_ISL_2846971, EPI_ISL_2846974, EPI_ISL_2846978, EPI_ISL_2846980, EPI_ISL_2846981, EPI_ISL_2846983, EPI_ISL_2846989, EPI_ISL_2847001, EPI_ISL_2847005, EPI_ISL_2847008, EPI_ISL_2847009, EPI_ISL_2847018, EPI_ISL_2847020, EPI_ISL_2847023, EPI_ISL_2847027, EPI_ISL_2847029, EPI_ISL_2847031, EPI_ISL_2847033, EPI_ISL_2847037, EPI_ISL_2847039, EPI_ISL_2847046, EPI_ISL_2847048, EPI_ISL_2847060, EPI_ISL_2847066, EPI_ISL_2847070, EPI_ISL_2847072, EPI_ISL_2847088, EPI_ISL_2847104, EPI_ISL_2847112, EPI_ISL_2847114, EPI_ISL_2847116, EPI_ISL_2847121, EPI_ISL_2847122, EPI_ISL_2847126, EPI_ISL_2847128, EPI_ISL_2847136, EPI_ISL_2847139, EPI_ISL_2847141, EPI_ISL_2847143, EPI_ISL_2847144, EPI_ISL_2847147, EPI_ISL_2847151, EPI_ISL_2847155, EPI_ISL_2847160, EPI_ISL_2847164, EPI_ISL_2847165, EPI_ISL_2847167, EPI_ISL_2847169, EPI_ISL_2847175, EPI_ISL_2847184, EPI_ISL_2847186, EPI_ISL_2847189, EPI_ISL_2847198, EPI_ISL_2847203, EPI_ISL_2847209, EPI_ISL_2847210, EPI_ISL_2847219, EPI_ISL_2847220, EPI_ISL_2847230, EPI_ISL_2847234, EPI_ISL_2847236, EPI_ISL_2847241, EPI_ISL_2847242, EPI_ISL_2847243, EPI_ISL_2847254, EPI_ISL_2847259, EPI_ISL_2847264, EPI_ISL_2847268, EPI_ISL_2847269, EPI_ISL_2847274, EPI_ISL_2847276, EPI_ISL_2847279, EPI_ISL_2847286, EPI_ISL_2847287, EPI_ISL_2847290, EPI_ISL_2847292, EPI_ISL_2847293, EPI_ISL_2847294, EPI_ISL_2847298, EPI_ISL_2847300, EPI_ISL_2847301, EPI_ISL_2847304, EPI_ISL_2847313, EPI_ISL_2847315, EPI_ISL_2847321, EPI_ISL_2847332, EPI_ISL_2847336, EPI_ISL_2847344, EPI_ISL_2847347, EPI_ISL_2847348, EPI_ISL_2847350, EPI_ISL_2847352, EPI_ISL_2847354, EPI_ISL_2847358, EPI_ISL_2847361, EPI_ISL_2847367, EPI_ISL_2847376, EPI_ISL_2847378, EPI_ISL_2847388, EPI_ISL_2847390, EPI_ISL_2847392, EPI_ISL_2847402, EPI_ISL_2847403, EPI_ISL_2847405, EPI_ISL_2847412, EPI_ISL_2847413, EPI_ISL_2847418, EPI_ISL_2847425, EPI_ISL_2847427, EPI_ISL_2847430, EPI_ISL_2847437, EPI_ISL_2847446, EPI_ISL_2847448, EPI_ISL_2847450, EPI_ISL_2847454, EPI_ISL_2847457, EPI_ISL_2847460, EPI_ISL_2847463, EPI_ISL_2847472, EPI_ISL_2847476, EPI_ISL_2847480, EPI_ISL_2847481, EPI_ISL_2847492, EPI_ISL_2847493, EPI_ISL_2847495, EPI_ISL_2847497, EPI_ISL_2847500, EPI_ISL_2847503, EPI_ISL_2847509, EPI_ISL_2847511, EPI_ISL_2847512, EPI_ISL_2847513, EPI_ISL_2847514, EPI_ISL_2847516, EPI_ISL_2847519, EPI_ISL_2847525, EPI_ISL_2847526, EPI_ISL_2847527, EPI_ISL_2847531, EPI_ISL_2847538, EPI_ISL_2847541, EPI_ISL_2847550, EPI_ISL_2847556, EPI_ISL_2847559, EPI_ISL_2847562, EPI_ISL_2847564, EPI_ISL_2847571, EPI_ISL_2847574, EPI_ISL_2847579, EPI_ISL_2847585, EPI_ISL_2847593, EPI_ISL_2847595, EPI_ISL_2847596, EPI_ISL_2847597, EPI_ISL_2847602, EPI_ISL_2847603, EPI_ISL_2847605, EPI_ISL_2847606, EPI_ISL_2847611, EPI_ISL_2847618, EPI_ISL_2847626, EPI_ISL_2847628, EPI_ISL_2847629, EPI_ISL_2847635, EPI_ISL_2847636, EPI_ISL_2847647, EPI_ISL_2847648, EPI_ISL_2847653, EPI_ISL_2847661, EPI_ISL_2847675, EPI_ISL_2847677, EPI_ISL_2847684, EPI_ISL_2847686, EPI_ISL_2847687, EPI_ISL_2847688, EPI_ISL_2847692, EPI_ISL_2847693, EPI_ISL_2847702, EPI_ISL_2847703, EPI_ISL_2847704, EPI_ISL_2847706, EPI_ISL_2847711, EPI_ISL_2847715, EPI_ISL_2847716, EPI_ISL_2847717, EPI_ISL_2847718, EPI_ISL_2847721, EPI_ISL_2847722, EPI_ISL_2847732, EPI_ISL_2847733, EPI_ISL_2847735, EPI_ISL_2847736, EPI_ISL_2847742, EPI_ISL_2847743, EPI_ISL_2847751, EPI_ISL_2847758, EPI_ISL_2847765, EPI_ISL_2847771, EPI_ISL_2847774, EPI_ISL_2847782, EPI_ISL_2847787, EPI_ISL_2847788, EPI_ISL_2847789, EPI_ISL_2847796, EPI_ISL_2847799, EPI_ISL_2847802, EPI_ISL_2847806, EPI_ISL_2847808, EPI_ISL_2847810, EPI_ISL_2847811, EPI_ISL_2847818, EPI_ISL_2847824, EPI_ISL_2847832, EPI_ISL_2847835, EPI_ISL_2847837, EPI_ISL_2847838, EPI_ISL_2847840, EPI_ISL_2847844, EPI_ISL_2847845, EPI_ISL_2847847, EPI_ISL_2847849, EPI_ISL_2847850, EPI_ISL_2847854, EPI_ISL_2847858, EPI_ISL_2847859, EPI_ISL_2847864, EPI_ISL_2847869, EPI_ISL_2847871, EPI_ISL_2847874, EPI_ISL_2847878, EPI_ISL_2847885, EPI_ISL_2847891, EPI_ISL_2847898, EPI_ISL_2847900, EPI_ISL_2847904, EPI_ISL_2847914, EPI_ISL_2847922, EPI_ISL_2847926, EPI_ISL_2847928, EPI_ISL_2847940, EPI_ISL_2847941, EPI_ISL_2847943, EPI_ISL_2847950, EPI_ISL_2847951, EPI_ISL_2847962, EPI_ISL_2847967, EPI_ISL_2847971, EPI_ISL_2847979, EPI_ISL_2847988, EPI_ISL_2847989, EPI_ISL_2847990, EPI_ISL_2847991, EPI_ISL_2847992, EPI_ISL_2847995, EPI_ISL_2847997, EPI_ISL_2848004, EPI_ISL_2848011, EPI_ISL_2848012, EPI_ISL_2848016, EPI_ISL_2848018, EPI_ISL_2848019, EPI_ISL_2848024, EPI_ISL_2848025, EPI_ISL_2848026, EPI_ISL_2848028, EPI_ISL_2848033, EPI_ISL_2848060, EPI_ISL_2848061, EPI_ISL_2848066, EPI_ISL_2848069, EPI_ISL_2848085, EPI_ISL_2848086, EPI_ISL_2848090, EPI_ISL_2848114, EPI_ISL_2848116, EPI_ISL_2848124, EPI_ISL_2848148, EPI_ISL_2848152, EPI_ISL_2848156, EPI_ISL_2848160, EPI_ISL_2848168, EPI_ISL_2848170, EPI_ISL_2848172, EPI_ISL_2848176, EPI_ISL_2848180, EPI_ISL_2848184, EPI_ISL_2848186, EPI_ISL_2848192, EPI_ISL_2848195, EPI_ISL_2848198, EPI_ISL_2848199, EPI_ISL_2848201, EPI_ISL_2848202, EPI_ISL_2848208, EPI_ISL_2848209, EPI_ISL_2848210, EPI_ISL_2848217, EPI_ISL_2848218, EPI_ISL_2848219, EPI_ISL_2848220, EPI_ISL_2848225, EPI_ISL_2848233, EPI_ISL_2848235, EPI_ISL_2848238, EPI_ISL_2848240, EPI_ISL_2848242, EPI_ISL_2848243, EPI_ISL_2848244, EPI_ISL_2848257, EPI_ISL_2848258, EPI_ISL_2848260, EPI_ISL_2848262, EPI_ISL_2848263, EPI_ISL_2848267, EPI_ISL_2848270, EPI_ISL_2848277, EPI_ISL_2848279, EPI_ISL_2848288, EPI_ISL_2848289, EPI_ISL_2848290, EPI_ISL_2848291, EPI_ISL_2848296, EPI_ISL_2848303, EPI_ISL_2848304, EPI_ISL_2848310, EPI_ISL_2848315, EPI_ISL_2848320, EPI_ISL_2848324, EPI_ISL_2848325, EPI_ISL_2848326, EPI_ISL_2848327, EPI_ISL_2848328, EPI_ISL_2848334, EPI_ISL_2848340, EPI_ISL_2848345, EPI_ISL_2848347, EPI_ISL_2848348, EPI_ISL_2848349, EPI_ISL_2848351, EPI_ISL_2848352, EPI_ISL_2848360, EPI_ISL_2848365, EPI_ISL_2848368, EPI_ISL_2848376, EPI_ISL_2848377, EPI_ISL_2848379, EPI_ISL_2848381, EPI_ISL_2848387, EPI_ISL_2848389, EPI_ISL_2848390, EPI_ISL_2848395, EPI_ISL_2848398, EPI_ISL_2848400, EPI_ISL_2848404, EPI_ISL_2848405, EPI_ISL_2848409, EPI_ISL_2848418, EPI_ISL_2848423, EPI_ISL_2848424, EPI_ISL_2848430, EPI_ISL_2848434, EPI_ISL_2848435, EPI_ISL_2848438, EPI_ISL_2848439, EPI_ISL_2848440, EPI_ISL_2848443, EPI_ISL_2848447, EPI_ISL_2848459, EPI_ISL_2848460, EPI_ISL_2848463, EPI_ISL_2848464, EPI_ISL_2848465, EPI_ISL_2848468, EPI_ISL_2848475, EPI_ISL_2848481, EPI_ISL_2848495, EPI_ISL_2848496, EPI_ISL_2848499, EPI_ISL_2848500, EPI_ISL_2848502, EPI_ISL_2848504, EPI_ISL_2848508, EPI_ISL_2848509, EPI_ISL_2848510, EPI_ISL_2848518, EPI_ISL_2848524, EPI_ISL_2848536, EPI_ISL_2848542, EPI_ISL_2848550, EPI_ISL_2848571, EPI_ISL_2848573, EPI_ISL_2848574, EPI_ISL_2848580, EPI_ISL_2848582, EPI_ISL_2848586, EPI_ISL_2848590, EPI_ISL_2848597, EPI_ISL_2848608, EPI_ISL_2848613, EPI_ISL_2848614, EPI_ISL_2848619, EPI_ISL_2848627, EPI_ISL_2848634, EPI_ISL_2848636, EPI_ISL_2848637, EPI_ISL_2848642, EPI_ISL_2848650, EPI_ISL_2848651, EPI_ISL_2848658, EPI_ISL_2848668, EPI_ISL_2848669, EPI_ISL_2848671, EPI_ISL_2848693, EPI_ISL_2848695, EPI_ISL_2848697, EPI_ISL_2848716, EPI_ISL_2848720, EPI_ISL_2848722, EPI_ISL_2848737, EPI_ISL_2848742, EPI_ISL_2848745, EPI_ISL_2848747, EPI_ISL_2848748, EPI_ISL_2848749, EPI_ISL_2848755, EPI_ISL_2848768, EPI_ISL_2848781, EPI_ISL_2848784, EPI_ISL_2848791, EPI_ISL_2848817, EPI_ISL_2848828, EPI_ISL_2848830, EPI_ISL_2848834, EPI_ISL_2848837, EPI_ISL_2848843, EPI_ISL_2848856, EPI_ISL_2848861, EPI_ISL_2848863, EPI_ISL_2848871, EPI_ISL_2848875, EPI_ISL_2848878, EPI_ISL_2848879, EPI_ISL_2848894, EPI_ISL_2848897, EPI_ISL_2848904, EPI_ISL_2848908, EPI_ISL_2848909, EPI_ISL_2848921, EPI_ISL_2848932, EPI_ISL_2848938, EPI_ISL_2848940, EPI_ISL_2848943, EPI_ISL_2848954, EPI_ISL_2848956, EPI_ISL_2848972, EPI_ISL_2848976, EPI_ISL_2848977, EPI_ISL_2848978, EPI_ISL_2848980, EPI_ISL_2848983, EPI_ISL_2848996, EPI_ISL_2849002, EPI_ISL_2850376, EPI_ISL_2850379, EPI_ISL_2850389, EPI_ISL_2850396, EPI_ISL_2850402, EPI_ISL_2850409, EPI_ISL_2850411, EPI_ISL_2850415, EPI_ISL_2850422, EPI_ISL_2850427, EPI_ISL_2850436, EPI_ISL_2850442, EPI_ISL_2850445, EPI_ISL_2850449, EPI_ISL_2850458, EPI_ISL_2850460, EPI_ISL_2850461, EPI_ISL_2850463, EPI_ISL_2850464, EPI_ISL_2850469, EPI_ISL_2850474, EPI_ISL_2850477, EPI_ISL_2850478, EPI_ISL_2850479, EPI_ISL_2850481, EPI_ISL_2850492, EPI_ISL_2850494, EPI_ISL_2850496, EPI_ISL_2850497, EPI_ISL_2850498, EPI_ISL_2850522, EPI_ISL_2850524, EPI_ISL_2850528, EPI_ISL_2850531, EPI_ISL_2850535, EPI_ISL_2850537, EPI_ISL_2850540, EPI_ISL_2850546, EPI_ISL_2850547, EPI_ISL_2850549, EPI_ISL_2850551, EPI_ISL_2850554, EPI_ISL_2850556, EPI_ISL_2850561, EPI_ISL_2850562, EPI_ISL_2850563, EPI_ISL_2850572, EPI_ISL_2850573, EPI_ISL_2850578, EPI_ISL_2850579, EPI_ISL_2850580, EPI_ISL_2850581, EPI_ISL_2850595, EPI_ISL_2850597, EPI_ISL_2850605, EPI_ISL_2850609, EPI_ISL_2850612, EPI_ISL_2850613, EPI_ISL_2850615, EPI_ISL_2850620, EPI_ISL_2850625, EPI_ISL_2850629, EPI_ISL_2850632, EPI_ISL_2850636, EPI_ISL_2850638, EPI_ISL_2850641, EPI_ISL_2850656, EPI_ISL_2850657, EPI_ISL_2850658, EPI_ISL_2850660, EPI_ISL_2850661, EPI_ISL_2850677, EPI_ISL_2850691, EPI_ISL_2850694, EPI_ISL_2850698, EPI_ISL_2850699, EPI_ISL_2850701, EPI_ISL_2850705, EPI_ISL_2850707, EPI_ISL_2850719, EPI_ISL_2850723, EPI_ISL_2850729, EPI_ISL_2850733, EPI_ISL_2850738, EPI_ISL_2850856, EPI_ISL_2851410, EPI_ISL_2851422, EPI_ISL_2851423, EPI_ISL_2851424, EPI_ISL_2851436, EPI_ISL_2851446, EPI_ISL_2851447, EPI_ISL_2851452, EPI_ISL_2851454, EPI_ISL_2851473, EPI_ISL_2851479, EPI_ISL_2851501, EPI_ISL_2851502, EPI_ISL_2851503, EPI_ISL_2851511, EPI_ISL_2851512, EPI_ISL_2851514, EPI_ISL_2851519, EPI_ISL_2851523, EPI_ISL_2851524, EPI_ISL_2851526, EPI_ISL_2851529, EPI_ISL_2851530, EPI_ISL_2851531, EPI_ISL_2851533, EPI_ISL_2851536, EPI_ISL_2851555, EPI_ISL_2851558, EPI_ISL_2851561, EPI_ISL_2851569, EPI_ISL_2851571, EPI_ISL_2851573, EPI_ISL_2851574, EPI_ISL_2851579, EPI_ISL_2851588, EPI_ISL_2851592, EPI_ISL_2851599, EPI_ISL_2851601, EPI_ISL_2851603, EPI_ISL_2851605, EPI_ISL_2851608, EPI_ISL_2851611, EPI_ISL_2851613, EPI_ISL_2851614, EPI_ISL_2851615, EPI_ISL_2851618, EPI_ISL_2851623, EPI_ISL_2851626, EPI_ISL_2851635, EPI_ISL_2851639, EPI_ISL_2851640, EPI_ISL_2851641, EPI_ISL_2851649, EPI_ISL_2851653, EPI_ISL_2851656, EPI_ISL_2851657, EPI_ISL_2851661, EPI_ISL_2851664, EPI_ISL_2851666, EPI_ISL_2851669, EPI_ISL_2851670, EPI_ISL_2851675, EPI_ISL_2851678, EPI_ISL_2851686, EPI_ISL_2851691, EPI_ISL_2851695, EPI_ISL_2851699, EPI_ISL_2851701, EPI_ISL_2851703, EPI_ISL_2851705, EPI_ISL_2851711, EPI_ISL_2851712, EPI_ISL_2851729, EPI_ISL_2851742, EPI_ISL_2851743, EPI_ISL_2851745, EPI_ISL_2851753, EPI_ISL_2851754, EPI_ISL_2851756, EPI_ISL_2851763, EPI_ISL_2851767, EPI_ISL_2851768, EPI_ISL_2851772, EPI_ISL_2851777, EPI_ISL_2851786, EPI_ISL_2851792, EPI_ISL_2851798, EPI_ISL_2851800, EPI_ISL_2851801, EPI_ISL_2851803, EPI_ISL_2851808, EPI_ISL_2851812, EPI_ISL_2851814, EPI_ISL_2851816, EPI_ISL_2851819, EPI_ISL_2851820, EPI_ISL_2851822, EPI_ISL_2851825, EPI_ISL_2851828, EPI_ISL_2851837, EPI_ISL_2851862, EPI_ISL_2851864, EPI_ISL_2851865, EPI_ISL_2851866, EPI_ISL_2851872, EPI_ISL_2851877, EPI_ISL_2851878, EPI_ISL_2851880, EPI_ISL_2851885, EPI_ISL_2851887, EPI_ISL_2851902, EPI_ISL_2851904, EPI_ISL_2851905, EPI_ISL_2851907, EPI_ISL_2851913, EPI_ISL_2851914, EPI_ISL_2851918, EPI_ISL_2851920, EPI_ISL_2851922, EPI_ISL_2851929, EPI_ISL_2851931, EPI_ISL_2851936, EPI_ISL_2851945, EPI_ISL_2851950, EPI_ISL_2851953, EPI_ISL_2851955, EPI_ISL_2851957, EPI_ISL_2851960, EPI_ISL_2851961, EPI_ISL_2851962, EPI_ISL_2851967, EPI_ISL_2851969, EPI_ISL_2851976, EPI_ISL_2851983, EPI_ISL_2851984, EPI_ISL_2851988, EPI_ISL_2851990, EPI_ISL_2851998, EPI_ISL_2852007, EPI_ISL_2852013, EPI_ISL_2852029, EPI_ISL_2852032, EPI_ISL_2852035, EPI_ISL_2852038, EPI_ISL_2852053, EPI_ISL_2852054, EPI_ISL_2852058, EPI_ISL_2852074, EPI_ISL_2852077, EPI_ISL_2852079, EPI_ISL_2852081, EPI_ISL_2852082, EPI_ISL_2852085, EPI_ISL_2852095, EPI_ISL_2852096, EPI_ISL_2852097, EPI_ISL_2852098, EPI_ISL_2852100, EPI_ISL_2852105, EPI_ISL_2852107, EPI_ISL_2852108, EPI_ISL_2852109, EPI_ISL_2852113, EPI_ISL_2852118, EPI_ISL_2852119, EPI_ISL_2852126, EPI_ISL_2852133, EPI_ISL_2852137, EPI_ISL_2852138, EPI_ISL_2865062, EPI_ISL_2865066, EPI_ISL_2865069, EPI_ISL_2865071, EPI_ISL_2865084, EPI_ISL_2865086, EPI_ISL_2865090, EPI_ISL_2865094, EPI_ISL_2865096, EPI_ISL_2865105, EPI_ISL_2865117, EPI_ISL_2865119, EPI_ISL_2865122, EPI_ISL_2865131, EPI_ISL_2865137, EPI_ISL_2865148, EPI_ISL_2865150, EPI_ISL_2865153, EPI_ISL_2865157, EPI_ISL_2865158, EPI_ISL_2865163, EPI_ISL_2865166, EPI_ISL_2865171, EPI_ISL_2865173, EPI_ISL_2865176, EPI_ISL_2865180, EPI_ISL_2865181, EPI_ISL_2865183, EPI_ISL_2865185, EPI_ISL_2865187, EPI_ISL_2865192, EPI_ISL_2865195, EPI_ISL_2865198, EPI_ISL_2865204, EPI_ISL_2865206, EPI_ISL_2865207, EPI_ISL_2865218, EPI_ISL_2865221, EPI_ISL_2865222, EPI_ISL_2865230, EPI_ISL_2865232, EPI_ISL_2865236, EPI_ISL_2865240, EPI_ISL_2865242, EPI_ISL_2865245, EPI_ISL_2865248, EPI_ISL_2865250, EPI_ISL_2865254, EPI_ISL_2865255, EPI_ISL_2865257, EPI_ISL_2865262, EPI_ISL_2865267, EPI_ISL_2865271, EPI_ISL_2865275, EPI_ISL_2865277, EPI_ISL_2865279, EPI_ISL_2865283, EPI_ISL_2865290, EPI_ISL_2865291, EPI_ISL_2865292, EPI_ISL_2865294, EPI_ISL_2865303, EPI_ISL_2865305, EPI_ISL_2865316, EPI_ISL_2865329, EPI_ISL_2865331, EPI_ISL_2865338, EPI_ISL_2865342, EPI_ISL_2865343, EPI_ISL_2865344, EPI_ISL_2865349, EPI_ISL_2865350, EPI_ISL_2865358, EPI_ISL_2865362, EPI_ISL_2865380, EPI_ISL_2865382, EPI_ISL_2865388, EPI_ISL_2865400, EPI_ISL_2865404, EPI_ISL_2865422, EPI_ISL_2865436, EPI_ISL_2865440, EPI_ISL_2865443, EPI_ISL_2865448, EPI_ISL_2865449, EPI_ISL_2865464, EPI_ISL_2865465, EPI_ISL_2865475, EPI_ISL_2865492, EPI_ISL_2865511, EPI_ISL_2865534, EPI_ISL_2865548, EPI_ISL_2865553, EPI_ISL_2865558, EPI_ISL_2865566, EPI_ISL_2865576, EPI_ISL_2865583, EPI_ISL_2865590, EPI_ISL_2865595, EPI_ISL_2865624, EPI_ISL_2865649, EPI_ISL_2865650, EPI_ISL_2865654, EPI_ISL_2865658, EPI_ISL_2865697, EPI_ISL_2865703, EPI_ISL_2865720, EPI_ISL_2865952, EPI_ISL_2866034, EPI_ISL_2866065, EPI_ISL_2866088, EPI_ISL_2866096, EPI_ISL_2866107, EPI_ISL_2866109, EPI_ISL_2866128, EPI_ISL_2866130, EPI_ISL_2866134, EPI_ISL_2866138, EPI_ISL_2866142, EPI_ISL_2866146, EPI_ISL_2866149, EPI_ISL_2866154, EPI_ISL_2866169, EPI_ISL_2866173, EPI_ISL_2866181, EPI_ISL_2866192, EPI_ISL_2866203, EPI_ISL_2866205, EPI_ISL_2866207, EPI_ISL_2866211, EPI_ISL_2866214, EPI_ISL_2866218, EPI_ISL_2866224, EPI_ISL_2866226, EPI_ISL_2866230, EPI_ISL_2866231, EPI_ISL_2866232, EPI_ISL_2866238, EPI_ISL_2866241, EPI_ISL_2866244, EPI_ISL_2866250, EPI_ISL_2866252, EPI_ISL_2866262, EPI_ISL_2866264, EPI_ISL_2866278, EPI_ISL_2866281, EPI_ISL_2866283, EPI_ISL_2866291, EPI_ISL_2866310, EPI_ISL_2866316, EPI_ISL_2866335, EPI_ISL_2866337, EPI_ISL_2866370, EPI_ISL_2866374, EPI_ISL_2866383, EPI_ISL_2866397, EPI_ISL_2866406, EPI_ISL_2866436, EPI_ISL_2866449, EPI_ISL_2866460, EPI_ISL_2866477, EPI_ISL_2866496, EPI_ISL_2866509, EPI_ISL_2866583, EPI_ISL_2866587, EPI_ISL_2866614, EPI_ISL_2866617, EPI_ISL_2866631, EPI_ISL_2866639, EPI_ISL_2866642, EPI_ISL_2866649, EPI_ISL_2866662, EPI_ISL_2866701, EPI_ISL_2866723, EPI_ISL_2866731, EPI_ISL_2866758, EPI_ISL_2866801, EPI_ISL_2866805, EPI_ISL_2866814, EPI_ISL_2866835, EPI_ISL_2866849, EPI_ISL_2866871, EPI_ISL_2866888, EPI_ISL_2866940, EPI_ISL_2866978, EPI_ISL_2867000, EPI_ISL_2867023, EPI_ISL_2867364, EPI_ISL_2867369, EPI_ISL_2867370, EPI_ISL_2867372, EPI_ISL_2867376, EPI_ISL_2867382, EPI_ISL_2867392, EPI_ISL_2867406, EPI_ISL_2867421, EPI_ISL_2867422, EPI_ISL_2867430, EPI_ISL_2867441, EPI_ISL_2867452, EPI_ISL_2867454, EPI_ISL_2867462, EPI_ISL_2867464, EPI_ISL_2867475, EPI_ISL_2867479, EPI_ISL_2867480, EPI_ISL_2867493, EPI_ISL_2867499, EPI_ISL_2867539, EPI_ISL_2867547, EPI_ISL_2867551, EPI_ISL_2867552, EPI_ISL_2867553, EPI_ISL_2867558, EPI_ISL_2867562, EPI_ISL_2867574, EPI_ISL_2867586, EPI_ISL_2867587, EPI_ISL_2867595, EPI_ISL_2867597, EPI_ISL_2867642, EPI_ISL_2867647, EPI_ISL_2867653, EPI_ISL_2867654, EPI_ISL_2867657, EPI_ISL_2867665, EPI_ISL_2867668, EPI_ISL_2867677, EPI_ISL_2867678, EPI_ISL_2867686, EPI_ISL_2867690, EPI_ISL_2868065, EPI_ISL_2868066, EPI_ISL_2868067, EPI_ISL_2868068, EPI_ISL_2868069, EPI_ISL_2868223, EPI_ISL_2910394, EPI_ISL_2910443, EPI_ISL_2910514, EPI_ISL_2910604, EPI_ISL_2910606, EPI_ISL_2910610, EPI_ISL_2910613, EPI_ISL_2910614, EPI_ISL_2910615, EPI_ISL_2910616, EPI_ISL_2910617, EPI_ISL_2910619, EPI_ISL_2910620, EPI_ISL_2910621, EPI_ISL_2910623, EPI_ISL_2910626, EPI_ISL_2910627, EPI_ISL_2910628, EPI_ISL_2910629, EPI_ISL_2910630, EPI_ISL_2910631, EPI_ISL_2910632, EPI_ISL_2910633, EPI_ISL_2910635, EPI_ISL_2910636, EPI_ISL_2910637, EPI_ISL_2910640, EPI_ISL_2910643, EPI_ISL_2910645, EPI_ISL_2910646, EPI_ISL_2910647, EPI_ISL_2910653, EPI_ISL_2910654, EPI_ISL_2910655, EPI_ISL_2910656, EPI_ISL_2910657, EPI_ISL_2910658, EPI_ISL_2910659, EPI_ISL_2910660, EPI_ISL_2910661, EPI_ISL_2910663, EPI_ISL_2910667, EPI_ISL_2910671, EPI_ISL_2910673, EPI_ISL_2910674, EPI_ISL_2910675, EPI_ISL_2910676, EPI_ISL_2910677, EPI_ISL_2910680, EPI_ISL_2910681, EPI_ISL_2910682, EPI_ISL_2910683, EPI_ISL_2910692, EPI_ISL_2910693, EPI_ISL_2910694, EPI_ISL_2910695, EPI_ISL_2910698,

EPI_ISL_2910699, EPI_ISL_2910701, EPI_ISL_2910704, EPI_ISL_2910706, EPI_ISL_2910707, EPI_ISL_2910708, EPI_ISL_2910709, EPI_ISL_2910711, EPI_ISL_2910713, EPI_ISL_2910715, EPI_ISL_2910716, EPI_ISL_2910717, EPI_ISL_2910718, EPI_ISL_2910723, EPI_ISL_2910724, EPI_ISL_2910727, EPI_ISL_2910728, EPI_ISL_2910729, EPI_ISL_2910730, EPI_ISL_2910732, EPI_ISL_2910734, EPI_ISL_2910736, EPI_ISL_2910737, EPI_ISL_2910739, EPI_ISL_2910740, EPI_ISL_2910742, EPI_ISL_2910743, EPI_ISL_2910744, EPI_ISL_2910745, EPI_ISL_2910746, EPI_ISL_2910748, EPI_ISL_2910749, EPI_ISL_2910751, EPI_ISL_2910759, EPI_ISL_2910760, EPI_ISL_2910761, EPI_ISL_2910762, EPI_ISL_2910767, EPI_ISL_2910768, EPI_ISL_2910769, EPI_ISL_2910770, EPI_ISL_2910771, EPI_ISL_2910773, EPI_ISL_2910776, EPI_ISL_2910777, EPI_ISL_2910781, EPI_ISL_2910782, EPI_ISL_2910784, EPI_ISL_2910785, EPI_ISL_2910786, EPI_ISL_2910787, EPI_ISL_2910788, EPI_ISL_2910789, EPI_ISL_2910790, EPI_ISL_2910791, EPI_ISL_2910793, EPI_ISL_2910794, EPI_ISL_2910795, EPI_ISL_2910797, EPI_ISL_2910798, EPI_ISL_2910799, EPI_ISL_2910801, EPI_ISL_2910804, EPI_ISL_2910805, EPI_ISL_2910807, EPI_ISL_2910808, EPI_ISL_2910810, EPI_ISL_2910811, EPI_ISL_2910812, EPI_ISL_2910814, EPI_ISL_2910815, EPI_ISL_2910818, EPI_ISL_2910822, EPI_ISL_2910824, EPI_ISL_2910826, EPI_ISL_2910829, EPI_ISL_2910830, EPI_ISL_2910831, EPI_ISL_2910832, EPI_ISL_2910833, EPI_ISL_2910837, EPI_ISL_2910839, EPI_ISL_2910840, EPI_ISL_2910845, EPI_ISL_2910846, EPI_ISL_2910847, EPI_ISL_2910848, EPI_ISL_2911165, EPI_ISL_2911944, EPI_ISL_2912071, EPI_ISL_2912206, EPI_ISL_2916328, EPI_ISL_2916408, EPI_ISL_2916543, EPI_ISL_2916567, EPI_ISL_2916614, EPI_ISL_2935924, EPI_ISL_2935938, EPI_ISL_2935939, EPI_ISL_2935958, EPI_ISL_2935967, EPI_ISL_2935968, EPI_ISL_2935998, EPI_ISL_2936015, EPI_ISL_2936039, EPI_ISL_2936094, EPI_ISL_2936110, EPI_ISL_2936118, EPI_ISL_2936142, EPI_ISL_2936147, EPI_ISL_2937682

| | | | |
|---|---|---|---|
| see above | Lighthouse Lab in Milton Keynes | Wellcome Sanger Institute for the COVID-19 Genomics UK (COG-UK) Consortium | Cordelia Langford; David K. Jackson; Dominic Kwiatkowski; Ewan Harrison; Ian Johnston; Jeffrey Barrett; John Sillitoe on behalf of the Wellcome Sanger Institute COVID-19 Surveillance Team; Roberto Amato; Sonia Goncalves; The Lighthouse Lab in Milton Keynes and Alex Alderton |

EPI_ISL_2829029, EPI_ISL_2829046, EPI_ISL_2829056, EPI_ISL_2829078, EPI_ISL_2829082, EPI_ISL_2829096, EPI_ISL_2829099, EPI_ISL_2829108, EPI_ISL_2829111, EPI_ISL_2829112, EPI_ISL_2829117, EPI_ISL_2829118, EPI_ISL_2829131, EPI_ISL_2829144, EPI_ISL_2829145, EPI_ISL_2829157, EPI_ISL_2829158, EPI_ISL_2829159, EPI_ISL_2829162, EPI_ISL_2829174, EPI_ISL_2829176, EPI_ISL_2829182, EPI_ISL_2829189, EPI_ISL_2829191, EPI_ISL_2829204, EPI_ISL_2829205, EPI_ISL_2829213, EPI_ISL_2829216, EPI_ISL_2829220, EPI_ISL_2829243, EPI_ISL_2829250, EPI_ISL_2829257, EPI_ISL_2829263, EPI_ISL_2829271, EPI_ISL_2829277, EPI_ISL_2829278, EPI_ISL_2829282, EPI_ISL_2829292, EPI_ISL_2829298, EPI_ISL_2829317, EPI_ISL_2829321, EPI_ISL_2829338, EPI_ISL_2829340, EPI_ISL_2829345, EPI_ISL_2829346, EPI_ISL_2829351, EPI_ISL_2829357, EPI_ISL_2829359, EPI_ISL_2829360, EPI_ISL_2829361, EPI_ISL_2829371, EPI_ISL_2829373, EPI_ISL_2829374, EPI_ISL_2829379, EPI_ISL_2829380, EPI_ISL_2829383, EPI_ISL_2829388, EPI_ISL_2829392, EPI_ISL_2829398, EPI_ISL_2829401, EPI_ISL_2829402, EPI_ISL_2829421, EPI_ISL_2829426, EPI_ISL_2829428, EPI_ISL_2829438, EPI_ISL_2829447, EPI_ISL_2829450, EPI_ISL_2829451, EPI_ISL_2829453, EPI_ISL_2829454, EPI_ISL_2829459, EPI_ISL_2829461, EPI_ISL_2829473, EPI_ISL_2829476, EPI_ISL_2829478, EPI_ISL_2829481, EPI_ISL_2829486, EPI_ISL_2829493, EPI_ISL_2829519, EPI_ISL_2829521, EPI_ISL_2829531, EPI_ISL_2829533, EPI_ISL_2829537, EPI_ISL_2829545, EPI_ISL_2829549, EPI_ISL_2829551, EPI_ISL_2829556, EPI_ISL_2829565, EPI_ISL_2829567, EPI_ISL_2829583, EPI_ISL_2829584, EPI_ISL_2829586, EPI_ISL_2829588, EPI_ISL_2829589, EPI_ISL_2831767, EPI_ISL_2831771, EPI_ISL_2831775, EPI_ISL_2831778, EPI_ISL_2831780, EPI_ISL_2831786, EPI_ISL_2831788, EPI_ISL_2831789, EPI_ISL_2831795, EPI_ISL_2831797, EPI_ISL_2831800, EPI_ISL_2831803, EPI_ISL_2831805, EPI_ISL_2831808, EPI_ISL_2831812, EPI_ISL_2831819, EPI_ISL_2831820, EPI_ISL_2831822, EPI_ISL_2831829, EPI_ISL_2831830, EPI_ISL_2831835, EPI_ISL_2831839, EPI_ISL_2831841, EPI_ISL_2831850, EPI_ISL_2831856, EPI_ISL_2831857, EPI_ISL_2831859, EPI_ISL_2831860, EPI_ISL_2831862, EPI_ISL_2831865, EPI_ISL_2831866, EPI_ISL_2831878, EPI_ISL_2831879, EPI_ISL_2831881, EPI_ISL_2831883, EPI_ISL_2831885, EPI_ISL_2831890, EPI_ISL_2831892, EPI_ISL_2831893, EPI_ISL_2831894, EPI_ISL_2831904, EPI_ISL_2831905, EPI_ISL_2831906, EPI_ISL_2831908, EPI_ISL_2831909, EPI_ISL_2831911, EPI_ISL_2831912, EPI_ISL_2831914, EPI_ISL_2831920, EPI_ISL_2831925, EPI_ISL_2831926, EPI_ISL_2831931, EPI_ISL_2831932, EPI_ISL_2831933, EPI_ISL_2831934, EPI_ISL_2831943, EPI_ISL_2831946, EPI_ISL_2831948, EPI_ISL_2831951, EPI_ISL_2831955, EPI_ISL_2831958, EPI_ISL_2831961, EPI_ISL_2831962, EPI_ISL_2831965, EPI_ISL_2831969, EPI_ISL_2831979, EPI_ISL_2831980, EPI_ISL_2831993, EPI_ISL_2831994, EPI_ISL_2831997, EPI_ISL_2831998, EPI_ISL_2832002, EPI_ISL_2832010, EPI_ISL_2832014, EPI_ISL_2832015, EPI_ISL_2832018, EPI_ISL_2832020, EPI_ISL_2832021, EPI_ISL_2832026, EPI_ISL_2832038, EPI_ISL_2832041, EPI_ISL_2832042, EPI_ISL_2832055, EPI_ISL_2832061, EPI_ISL_2832063, EPI_ISL_2846031, EPI_ISL_2846045, EPI_ISL_2846048, EPI_ISL_2846066, EPI_ISL_2846074, EPI_ISL_2846078, EPI_ISL_2846080, EPI_ISL_2846089, EPI_ISL_2846105, EPI_ISL_2846112, EPI_ISL_2846154, EPI_ISL_2846158, EPI_ISL_2846162, EPI_ISL_2846163, EPI_ISL_2846167, EPI_ISL_2846178, EPI_ISL_2846183, EPI_ISL_2846185, EPI_ISL_2846190, EPI_ISL_2846193, EPI_ISL_2846197, EPI_ISL_2846211, EPI_ISL_2846243, EPI_ISL_2846246, EPI_ISL_2846247, EPI_ISL_2846251, EPI_ISL_2846252, EPI_ISL_2846266, EPI_ISL_2846270, EPI_ISL_2846274, EPI_ISL_2846286, EPI_ISL_2846303, EPI_ISL_2846308, EPI_ISL_2846323, EPI_ISL_2846335, EPI_ISL_2846348, EPI_ISL_2846350, EPI_ISL_2846354, EPI_ISL_2846656, EPI_ISL_2846768, EPI_ISL_2846876, EPI_ISL_2846880, EPI_ISL_2846910, EPI_ISL_2847285, EPI_ISL_2847288, EPI_ISL_2847297, EPI_ISL_2847305, EPI_ISL_2847316, EPI_ISL_2847319, EPI_ISL_2847320, EPI_ISL_2847323, EPI_ISL_2847325, EPI_ISL_2847326, EPI_ISL_2847328, EPI_ISL_2847339, EPI_ISL_2847345, EPI_ISL_2847346, EPI_ISL_2847353, EPI_ISL_2847355, EPI_ISL_2847363, EPI_ISL_2847368, EPI_ISL_2847369, EPI_ISL_2847373, EPI_ISL_2847375, EPI_ISL_2847382, EPI_ISL_2847383, EPI_ISL_2847391, EPI_ISL_2847393, EPI_ISL_2847396, EPI_ISL_2847397, EPI_ISL_2847401, EPI_ISL_2847407, EPI_ISL_2847419, EPI_ISL_2847424, EPI_ISL_2847429, EPI_ISL_2847432, EPI_ISL_2847434, EPI_ISL_2847438, EPI_ISL_2847441, EPI_ISL_2847447, EPI_ISL_2847455, EPI_ISL_2847456, EPI_ISL_2847458, EPI_ISL_2847468, EPI_ISL_2847474, EPI_ISL_2847477, EPI_ISL_2847478, EPI_ISL_2847483, EPI_ISL_2847485, EPI_ISL_2847489, EPI_ISL_2847494, EPI_ISL_2847496, EPI_ISL_2847498, EPI_ISL_2847499, EPI_ISL_2847504, EPI_ISL_2847506, EPI_ISL_2847515, EPI_ISL_2847517, EPI_ISL_2847520, EPI_ISL_2847529, EPI_ISL_2847533, EPI_ISL_2847539, EPI_ISL_2847540, EPI_ISL_2847544, EPI_ISL_2847545, EPI_ISL_2847546, EPI_ISL_2847551, EPI_ISL_2847552, EPI_ISL_2847557, EPI_ISL_2847560, EPI_ISL_2847563, EPI_ISL_2847565, EPI_ISL_2847566, EPI_ISL_2847567, EPI_ISL_2847569, EPI_ISL_2847570, EPI_ISL_2847572, EPI_ISL_2847577, EPI_ISL_2847578, EPI_ISL_2847583, EPI_ISL_2847586, EPI_ISL_2847588, EPI_ISL_2847589, EPI_ISL_2847590, EPI_ISL_2847591, EPI_ISL_2847598, EPI_ISL_2847608, EPI_ISL_2847609, EPI_ISL_2847610, EPI_ISL_2847616, EPI_ISL_2847619, EPI_ISL_2847620, EPI_ISL_2847621, EPI_ISL_2847622, EPI_ISL_2847624, EPI_ISL_2847625, EPI_ISL_2847627, EPI_ISL_2847630, EPI_ISL_2847631, EPI_ISL_2847632, EPI_ISL_2847633, EPI_ISL_2847634, EPI_ISL_2847637, EPI_ISL_2847638, EPI_ISL_2847639, EPI_ISL_2847640, EPI_ISL_2847641, EPI_ISL_2847642, EPI_ISL_2847643, EPI_ISL_2847644, EPI_ISL_2847645, EPI_ISL_2847646, EPI_ISL_2847650, EPI_ISL_2847651, EPI_ISL_2847652, EPI_ISL_2847654, EPI_ISL_2847655, EPI_ISL_2847656, EPI_ISL_2847657, EPI_ISL_2847658, EPI_ISL_2847659, EPI_ISL_2847660, EPI_ISL_2847662, EPI_ISL_2847663, EPI_ISL_2847664, EPI_ISL_2847665, EPI_ISL_2847666, EPI_ISL_2847667, EPI_ISL_2847668, EPI_ISL_2847669, EPI_ISL_2847670, EPI_ISL_2847672, EPI_ISL_2847673, EPI_ISL_2847674, EPI_ISL_2847676, EPI_ISL_2847678, EPI_ISL_2847679, EPI_ISL_2847680, EPI_ISL_2847681, EPI_ISL_2847682, EPI_ISL_2847683, EPI_ISL_2847685, EPI_ISL_2847689, EPI_ISL_2847690, EPI_ISL_2847691, EPI_ISL_2847694, EPI_ISL_2847695, EPI_ISL_2847697, EPI_ISL_2847698, EPI_ISL_2847699, EPI_ISL_2847700, EPI_ISL_2847701, EPI_ISL_2847705, EPI_ISL_2847708, EPI_ISL_2847709, EPI_ISL_2847712, EPI_ISL_2847713, EPI_ISL_2847714, EPI_ISL_2847719, EPI_ISL_2847720, EPI_ISL_2847723, EPI_ISL_2847724, EPI_ISL_2847725, EPI_ISL_2847726, EPI_ISL_2847727, EPI_ISL_2847728, EPI_ISL_2847729, EPI_ISL_2847730, EPI_ISL_2847731, EPI_ISL_2847734, EPI_ISL_2847737, EPI_ISL_2847739, EPI_ISL_2847740, EPI_ISL_2847741, EPI_ISL_2847744, EPI_ISL_2847745, EPI_ISL_2847746, EPI_ISL_2847747, EPI_ISL_2847748, EPI_ISL_2847749, EPI_ISL_2847750, EPI_ISL_2847752, EPI_ISL_2847753, EPI_ISL_2847754, EPI_ISL_2847755, EPI_ISL_2847756, EPI_ISL_2847757, EPI_ISL_2847759, EPI_ISL_2847760, EPI_ISL_2847761, EPI_ISL_2847762, EPI_ISL_2847763, EPI_ISL_2847766, EPI_ISL_2847767, EPI_ISL_2847768, EPI_ISL_2847770, EPI_ISL_2847772, EPI_ISL_2847773, EPI_ISL_2847775, EPI_ISL_2847776, EPI_ISL_2847778, EPI_ISL_2847779, EPI_ISL_2847780, EPI_ISL_2847781, EPI_ISL_2847783, EPI_ISL_2847784, EPI_ISL_2847785, EPI_ISL_2847786, EPI_ISL_2847790, EPI_ISL_2847791, EPI_ISL_2847792, EPI_ISL_2847793, EPI_ISL_2847794, EPI_ISL_2847795, EPI_ISL_2847797, EPI_ISL_2847798, EPI_ISL_2847800, EPI_ISL_2847801, EPI_ISL_2847803, EPI_ISL_2847804, EPI_ISL_2847805, EPI_ISL_2847807, EPI_ISL_2847809, EPI_ISL_2847812, EPI_ISL_2847813, EPI_ISL_2847814, EPI_ISL_2847815, EPI_ISL_2847816, EPI_ISL_2847817, EPI_ISL_2847819, EPI_ISL_2847820, EPI_ISL_2847821, EPI_ISL_2847822, EPI_ISL_2847823, EPI_ISL_2847825, EPI_ISL_2847826, EPI_ISL_2847827, EPI_ISL_2847829, EPI_ISL_2847830, EPI_ISL_2847831, EPI_ISL_2847833, EPI_ISL_2847834, EPI_ISL_2847836, EPI_ISL_2847839, EPI_ISL_2847841, EPI_ISL_2847842, EPI_ISL_2847843, EPI_ISL_2847846, EPI_ISL_2847848, EPI_ISL_2847851, EPI_ISL_2847852, EPI_ISL_2847853, EPI_ISL_2847855, EPI_ISL_2847856, EPI_ISL_2847857, EPI_ISL_2847860, EPI_ISL_2847861, EPI_ISL_2847862, EPI_ISL_2847863, EPI_ISL_2847865, EPI_ISL_2847866, EPI_ISL_2847870, EPI_ISL_2847872, EPI_ISL_2847873, EPI_ISL_2847875, EPI_ISL_2847876, EPI_ISL_2847877, EPI_ISL_2847880, EPI_ISL_2847881, EPI_ISL_2847882, EPI_ISL_2847883, EPI_ISL_2847884, EPI_ISL_2847886, EPI_ISL_2847887, EPI_ISL_2847888, EPI_ISL_2847890, EPI_ISL_2847892, EPI_ISL_2847893, EPI_ISL_2847894, EPI_ISL_2847895, EPI_ISL_2847896, EPI_ISL_2847897, EPI_ISL_2847899, EPI_ISL_2847901, EPI_ISL_2847902, EPI_ISL_2847903, EPI_ISL_2847905, EPI_ISL_2847906, EPI_ISL_2847907, EPI_ISL_2847908, EPI_ISL_2847909, EPI_ISL_2847910, EPI_ISL_2847911, EPI_ISL_2847912, EPI_ISL_2847913, EPI_ISL_2847915, EPI_ISL_2847916, EPI_ISL_2847917, EPI_ISL_2847918, EPI_ISL_2847919, EPI_ISL_2847920, EPI_ISL_2847921, EPI_ISL_2847923, EPI_ISL_2847924, EPI_ISL_2847927, EPI_ISL_2847929, EPI_ISL_2847930, EPI_ISL_2847931, EPI_ISL_2847932, EPI_ISL_2847933, EPI_ISL_2847934, EPI_ISL_2847935, EPI_ISL_2847936, EPI_ISL_2847937, EPI_ISL_2847938, EPI_ISL_2847939, EPI_ISL_2847942, EPI_ISL_2847944, EPI_ISL_2847945, EPI_ISL_2847946, EPI_ISL_2847947, EPI_ISL_2847948, EPI_ISL_2847949, EPI_ISL_2847952, EPI_ISL_2847953, EPI_ISL_2847954, EPI_ISL_2847955, EPI_ISL_2847956, EPI_ISL_2847959, EPI_ISL_2847960, EPI_ISL_2847963, EPI_ISL_2847964, EPI_ISL_2847965, EPI_ISL_2847968, EPI_ISL_2847969, EPI_ISL_2847970, EPI_ISL_2847972, EPI_ISL_2847973, EPI_ISL_2847976, EPI_ISL_2847977, EPI_ISL_2847978, EPI_ISL_2847980, EPI_ISL_2847982, EPI_ISL_2847984, EPI_ISL_2847985, EPI_ISL_2847987, EPI_ISL_2847993, EPI_ISL_2847994, EPI_ISL_2848000, EPI_ISL_2848001, EPI_ISL_2848003, EPI_ISL_2848007, EPI_ISL_2848008, EPI_ISL_2848009, EPI_ISL_2848013, EPI_ISL_2848015, EPI_ISL_2848017, EPI_ISL_2848020, EPI_ISL_2848021, EPI_ISL_2848023, EPI_ISL_2848027, EPI_ISL_2848030, EPI_ISL_2848031, EPI_ISL_2848032, EPI_ISL_2848034, EPI_ISL_2848035, EPI_ISL_2848037, EPI_ISL_2848040, EPI_ISL_2848041, EPI_ISL_2848043, EPI_ISL_2848044, EPI_ISL_2848045, EPI_ISL_2848046, EPI_ISL_2848047, EPI_ISL_2848048, EPI_ISL_2848049, EPI_ISL_2848050, EPI_ISL_2848052, EPI_ISL_2848053, EPI_ISL_2848054, EPI_ISL_2848056, EPI_ISL_2848057, EPI_ISL_2848058, EPI_ISL_2848059, EPI_ISL_2848065, EPI_ISL_2848067, EPI_ISL_2848068, EPI_ISL_2848071, EPI_ISL_2848072, EPI_ISL_2848075, EPI_ISL_2848076, EPI_ISL_2848078, EPI_ISL_2848079, EPI_ISL_2848080, EPI_ISL_2848082, EPI_ISL_2848083, EPI_ISL_2848091, EPI_ISL_2848092, EPI_ISL_2848093, EPI_ISL_2848094, EPI_ISL_2848095, EPI_ISL_2848096, EPI_ISL_2848097, EPI_ISL_2848098, EPI_ISL_2848101, EPI_ISL_2848102, EPI_ISL_2848103, EPI_ISL_2848105, EPI_ISL_2848108, EPI_ISL_2848109, EPI_ISL_2848110, EPI_ISL_2848117, EPI_ISL_2848118, EPI_ISL_2848120, EPI_ISL_2848121, EPI_ISL_2848122, EPI_ISL_2848123, EPI_ISL_2848125, EPI_ISL_2848126, EPI_ISL_2848127, EPI_ISL_2848128, EPI_ISL_2848129, EPI_ISL_2848130, EPI_ISL_2848131, EPI_ISL_2848132, EPI_ISL_2848133, EPI_ISL_2848134, EPI_ISL_2848135, EPI_ISL_2848136, EPI_ISL_2848138, EPI_ISL_2848139, EPI_ISL_2848141, EPI_ISL_2848142, EPI_ISL_2848143, EPI_ISL_2848144, EPI_ISL_2848146, EPI_ISL_2848150, EPI_ISL_2848151, EPI_ISL_2848154, EPI_ISL_2848155, EPI_ISL_2848157, EPI_ISL_2848161, EPI_ISL_2848163, EPI_ISL_2848165, EPI_ISL_2848167, EPI_ISL_2848175, EPI_ISL_2848177, EPI_ISL_2848182, EPI_ISL_2848185, EPI_ISL_2848191, EPI_ISL_2848194, EPI_ISL_2848196, EPI_ISL_2848200, EPI_ISL_2848204, EPI_ISL_2848206, EPI_ISL_2848207, EPI_ISL_2848212, EPI_ISL_2848214, EPI_ISL_2848216, EPI_ISL_2848221, EPI_ISL_2848224, EPI_ISL_2848229, EPI_ISL_2848230, EPI_ISL_2848234, EPI_ISL_2848237, EPI_ISL_2848241, EPI_ISL_2848245, EPI_ISL_2848246, EPI_ISL_2848247, EPI_ISL_2848250, EPI_ISL_2848251, EPI_ISL_2848252, EPI_ISL_2848254, EPI_ISL_2848255, EPI_ISL_2848256, EPI_ISL_2848259, EPI_ISL_2848261, EPI_ISL_2848264, EPI_ISL_2848265, EPI_ISL_2848266, EPI_ISL_2848268, EPI_ISL_2848269, EPI_ISL_2848271, EPI_ISL_2848272, EPI_ISL_2848273, EPI_ISL_2848274, EPI_ISL_2848275, EPI_ISL_2848278, EPI_ISL_2848282, EPI_ISL_2848284, EPI_ISL_2848285, EPI_ISL_2848286, EPI_ISL_2848292, EPI_ISL_2848295, EPI_ISL_2848300, EPI_ISL_2849031, EPI_ISL_2849037, EPI_ISL_2849059, EPI_ISL_2849061, EPI_ISL_2849072, EPI_ISL_2849077, EPI_ISL_2849080, EPI_ISL_2849082, EPI_ISL_2849085, EPI_ISL_2849087, EPI_ISL_2849095, EPI_ISL_2849100, EPI_ISL_2849111, EPI_ISL_2849115, EPI_ISL_2849131, EPI_ISL_2849134, EPI_ISL_2849155, EPI_ISL_2849172, EPI_ISL_2849189, EPI_ISL_2849216, EPI_ISL_2849222, EPI_ISL_2849231, EPI_ISL_2849233, EPI_ISL_2849246, EPI_ISL_2849250, EPI_ISL_2849254, EPI_ISL_2849266, EPI_ISL_2849282, EPI_ISL_2849286, EPI_ISL_2849291, EPI_ISL_2849292, EPI_ISL_2849299, EPI_ISL_2849306, EPI_ISL_2849314, EPI_ISL_2849318, EPI_ISL_2849322, EPI_ISL_2849329, EPI_ISL_2849342, EPI_ISL_2849344, EPI_ISL_2849352, EPI_ISL_2849386, EPI_ISL_2849395, EPI_ISL_2849407, EPI_ISL_2849437, EPI_ISL_2849440, EPI_ISL_2849503, EPI_ISL_2849523, EPI_ISL_2849542, EPI_ISL_2849552, EPI_ISL_2849561, EPI_ISL_2849610, EPI_ISL_2849613, EPI_ISL_2849657, EPI_ISL_2850039, EPI_ISL_2850043, EPI_ISL_2850050, EPI_ISL_2850060, EPI_ISL_2850065, EPI_ISL_2850069, EPI_ISL_2850070, EPI_ISL_2850075, EPI_ISL_2850077, EPI_ISL_2850079, EPI_ISL_2850081, EPI_ISL_2850089, EPI_ISL_2850090, EPI_ISL_2850092, EPI_ISL_2850099, EPI_ISL_2850101, EPI_ISL_2850102, EPI_ISL_2850104, EPI_ISL_2850105, EPI_ISL_2850114, EPI_ISL_2850118, EPI_ISL_2850120, EPI_ISL_2850122, EPI_ISL_2850123, EPI_ISL_2850131, EPI_ISL_2850135, EPI_ISL_2850136, EPI_ISL_2850139, EPI_ISL_2850140, EPI_ISL_2850146, EPI_ISL_2850150, EPI_ISL_2850168, EPI_ISL_2850176, EPI_ISL_2850178, EPI_ISL_2850181, EPI_ISL_2850182, EPI_ISL_2850187, EPI_ISL_2850198, EPI_ISL_2850213, EPI_ISL_2850218, EPI_ISL_2850221, EPI_ISL_2850230, EPI_ISL_2850231, EPI_ISL_2850232, EPI_ISL_2850242, EPI_ISL_2850244, EPI_ISL_2850246, EPI_ISL_2850257, EPI_ISL_2850262, EPI_ISL_2850263, EPI_ISL_2850276, EPI_ISL_2850277, EPI_ISL_2850289, EPI_ISL_2850290, EPI_ISL_2850293, EPI_ISL_2850300, EPI_ISL_2850301, EPI_ISL_2850302, EPI_ISL_2850306, EPI_ISL_2850312, EPI_ISL_2850326, EPI_ISL_2850329, EPI_ISL_2850331, EPI_ISL_2850333, EPI_ISL_2850337, EPI_ISL_2850341, EPI_ISL_2850344, EPI_ISL_2850346, EPI_ISL_2850351, EPI_ISL_2850358, EPI_ISL_2850363, EPI_ISL_2850365, EPI_ISL_2850368, EPI_ISL_2850372, EPI_ISL_2850375, EPI_ISL_2850380, EPI_ISL_2850381, EPI_ISL_2850386, EPI_ISL_2850387, EPI_ISL_2850390, EPI_ISL_2850394, EPI_ISL_2850398, EPI_ISL_2850399, EPI_ISL_2850401, EPI_ISL_2850404, EPI_ISL_2850405, EPI_ISL_2850408, EPI_ISL_2850410, EPI_ISL_2850414, EPI_ISL_2850416, EPI_ISL_2850426, EPI_ISL_2850428, EPI_ISL_2850435, EPI_ISL_2850443, EPI_ISL_2850444, EPI_ISL_2850450, EPI_ISL_2850452, EPI_ISL_2850462, EPI_ISL_2850467, EPI_ISL_2850470, EPI_ISL_2850485, EPI_ISL_2850489, EPI_ISL_2850493, EPI_ISL_2850499, EPI_ISL_2850502, EPI_ISL_2850503, EPI_ISL_2850505, EPI_ISL_2850509, EPI_ISL_2850510, EPI_ISL_2850511, EPI_ISL_2850513, EPI_ISL_2850515, EPI_ISL_2850516, EPI_ISL_2850527, EPI_ISL_2850530, EPI_ISL_2850532, EPI_ISL_2850536, EPI_ISL_2850538, EPI_ISL_2850541, EPI_ISL_2850543, EPI_ISL_2850552, EPI_ISL_2850555, EPI_ISL_2850564, EPI_ISL_2850565, EPI_ISL_2850574, EPI_ISL_2850575, EPI_ISL_2850577, EPI_ISL_2850588, EPI_ISL_2850591, EPI_ISL_2850593, EPI_ISL_2850602, EPI_ISL_2850604, EPI_ISL_2850607, EPI_ISL_2850608, EPI_ISL_2850610, EPI_ISL_2850614, EPI_ISL_2850624, EPI_ISL_2850634, EPI_ISL_2850640, EPI_ISL_2850651, EPI_ISL_2850654, EPI_ISL_2850655, EPI_ISL_2850659, EPI_ISL_2850662, EPI_ISL_2850663, EPI_ISL_2850664, EPI_ISL_2850665, EPI_ISL_2850668, EPI_ISL_2850675, EPI_ISL_2850676, EPI_ISL_2850680, EPI_ISL_2850683, EPI_ISL_2850684, EPI_ISL_2850685, EPI_ISL_2850689, EPI_ISL_2850697, EPI_ISL_2850702, EPI_ISL_2850703, EPI_ISL_2850708, EPI_ISL_2850709, EPI_ISL_2850715, EPI_ISL_2850718, EPI_ISL_2850721, EPI_ISL_2850734, EPI_ISL_2852142, EPI_ISL_2852152, EPI_ISL_2852155, EPI_ISL_2852157, EPI_ISL_2852158, EPI_ISL_2852159, EPI_ISL_2852167, EPI_ISL_2852169, EPI_ISL_2852180, EPI_ISL_2852186, EPI_ISL_2852187, EPI_ISL_2852192, EPI_ISL_2852193, EPI_ISL_2852195, EPI_ISL_2852197, EPI_ISL_2852225, EPI_ISL_2852227, EPI_ISL_2852238, EPI_ISL_2852248, EPI_ISL_2852249, EPI_ISL_2852250, EPI_ISL_2852251, EPI_ISL_2852255, EPI_ISL_2852259, EPI_ISL_2852263, EPI_ISL_2852266, EPI_ISL_2852282, EPI_ISL_2852302, EPI_ISL_2852303, EPI_ISL_2852310, EPI_ISL_2852313, EPI_ISL_2852316, EPI_ISL_2852320, EPI_ISL_2852321, EPI_ISL_2852323, EPI_ISL_2852324, EPI_ISL_2852332, EPI_ISL_2852344, EPI_ISL_2852345, EPI_ISL_2852349, EPI_ISL_2852350, EPI_ISL_2852355, EPI_ISL_2852357, EPI_ISL_2852358, EPI_ISL_2852366, EPI_ISL_2852374, EPI_ISL_2852380, EPI_ISL_2852387, EPI_ISL_2852388, EPI_ISL_2852390, EPI_ISL_2852404, EPI_ISL_2852406, EPI_ISL_2852410, EPI_ISL_2852424, EPI_ISL_2852437, EPI_ISL_2852440, EPI_ISL_2852448, EPI_ISL_2852451, EPI_ISL_2852452, EPI_ISL_2852460, EPI_ISL_2852463, EPI_ISL_2852466, EPI_ISL_2852819, EPI_ISL_2866068, EPI_ISL_2866075, EPI_ISL_2866083, EPI_ISL_2866093, EPI_ISL_2866113, EPI_ISL_2866115, EPI_ISL_2866117, EPI_ISL_2866122, EPI_ISL_2866140, EPI_ISL_2866141, EPI_ISL_2866151, EPI_ISL_2866152, EPI_ISL_2866160, EPI_ISL_2866162, EPI_ISL_2866165, EPI_ISL_2866166, EPI_ISL_2866172, EPI_ISL_2866174, EPI_ISL_2866180, EPI_ISL_2866183, EPI_ISL_2866185, EPI_ISL_2866193, EPI_ISL_2866201, EPI_ISL_2866204, EPI_ISL_2866208, EPI_ISL_2866219, EPI_ISL_2866220, EPI_ISL_2866240, EPI_ISL_2866247, EPI_ISL_2866253, EPI_ISL_2866255, EPI_ISL_2866268, EPI_ISL_2866275, EPI_ISL_2866280, EPI_ISL_2866284, EPI_ISL_2866293, EPI_ISL_2866297, EPI_ISL_2866302, EPI_ISL_2866307, EPI_ISL_2866834, EPI_ISL_2866921, EPI_ISL_2867019, EPI_ISL_2868060, EPI_ISL_2868063, EPI_ISL_2868064, EPI_ISL_2868079, EPI_ISL_2868227, EPI_ISL_2914035, EPI_ISL_2914054, EPI_ISL_2914075

| | | | |
|---|---|---|---|
| see above | Lighthouse Laboratory Plymouth | Wellcome Sanger Institute for the COVID-19 Genomics UK (COG-UK) Consortium | Cordelia Langford; David K. Jackson; Dominic Kwiatkowski; Ewan Harrison; Ian Johnston; Jeffrey Barrett; John Sillitoe on behalf of the Wellcome Sanger Institute COVID-19 Surveillance Team; Lighthouse Laboratory Plymouth and Alex Alderton; Roberto Amato; Sonia Goncalves |

EPI_ISL_2900845, EPI_ISL_2900846, EPI_ISL_2900847, EPI_ISL_2900848, EPI_ISL_2900849, EPI_ISL_2900850, EPI_ISL_2900851, EPI_ISL_2900852, EPI_ISL_2900853, EPI_ISL_2900854, EPI_ISL_2900855, EPI_ISL_2900856, EPI_ISL_2900857, EPI_ISL_2900858, EPI_ISL_2900859, EPI_ISL_2900860, EPI_ISL_2900861, EPI_ISL_2900862, EPI_ISL_2900863, EPI_ISL_2900864, EPI_ISL_2900865, EPI_ISL_2900866, EPI_ISL_2900867, EPI_ISL_2900868, EPI_ISL_2900869, EPI_ISL_2900870, EPI_ISL_2900871, EPI_ISL_2900872, EPI_ISL_2900873, EPI_ISL_2900874, EPI_ISL_2900875, EPI_ISL_2900876, EPI_ISL_2900877, EPI_ISL_2900878, EPI_ISL_2900879, EPI_ISL_2900880, EPI_ISL_2900881, EPI_ISL_2900882, EPI_ISL_2900883, EPI_ISL_2900884, EPI_ISL_2900885, EPI_ISL_2900886, EPI_ISL_2900887, EPI_ISL_2900888, EPI_ISL_2900889, EPI_ISL_2900890, EPI_ISL_2900891, EPI_ISL_2900892, EPI_ISL_2900894, EPI_ISL_2900895, EPI_ISL_2900896, EPI_ISL_2900897, EPI_ISL_2900898, EPI_ISL_2900899, EPI_ISL_2900900, EPI_ISL_2900901, EPI_ISL_2900902, EPI_ISL_2900903, EPI_ISL_2900904, EPI_ISL_2900905, EPI_ISL_2900906, EPI_ISL_2900907, EPI_ISL_2900908, EPI_ISL_2900909, EPI_ISL_2900910, EPI_ISL_2900911, EPI_ISL_2900912, EPI_ISL_2900913, EPI_ISL_2900914, EPI_ISL_2900915, EPI_ISL_2900916, EPI_ISL_2900917, EPI_ISL_2900918, EPI_ISL_2900919, EPI_ISL_2900920, EPI_ISL_2900921, EPI_ISL_2900922, EPI_ISL_2900923, EPI_ISL_2900924, EPI_ISL_2900925, EPI_ISL_2900926, EPI_ISL_2900927, EPI_ISL_2900929, EPI_ISL_2900930, EPI_ISL_2900931, EPI_ISL_2900932, EPI_ISL_2900933, EPI_ISL_2900934, EPI_ISL_2900935, EPI_ISL_2900936, EPI_ISL_2900937, EPI_ISL_2900938, EPI_ISL_2900939, EPI_ISL_2900940, EPI_ISL_2900941, EPI_ISL_2900942, EPI_ISL_2900943, EPI_ISL_2900944, EPI_ISL_2900945, EPI_ISL_2900946, EPI_ISL_2900947, EPI_ISL_2900948, EPI_ISL_2900949, EPI_ISL_2900950, EPI_ISL_2900951, EPI_ISL_2900952, EPI_ISL_2900953, EPI_ISL_2900954, EPI_ISL_2900955, EPI_ISL_2900956, EPI_ISL_2900957, EPI_ISL_2900958, EPI_ISL_2900959, EPI_ISL_2900960, EPI_ISL_2900961, EPI_ISL_2900962, EPI_ISL_2900963, EPI_ISL_2900964, EPI_ISL_2900965, EPI_ISL_2900966, EPI_ISL_2900967, EPI_ISL_2900968, EPI_ISL_2900969, EPI_ISL_2900970, EPI_ISL_2900971, EPI_ISL_2900972, EPI_ISL_2900973, EPI_ISL_2900974, EPI_ISL_2900975, EPI_ISL_2900976, EPI_ISL_2900977, EPI_ISL_2900978, EPI_ISL_2900979, EPI_ISL_2900980, EPI_ISL_2900981, EPI_ISL_2900982, EPI_ISL_2900983, EPI_ISL_2900984, EPI_ISL_2900985, EPI_ISL_2900986, EPI_ISL_2900987, EPI_ISL_2900988, EPI_ISL_2900989, EPI_ISL_2900990, EPI_ISL_2900991, EPI_ISL_2900992, EPI_ISL_2900993, EPI_ISL_2900994, EPI_ISL_2900995, EPI_ISL_2900996, EPI_ISL_2900997, EPI_ISL_2900998, EPI_ISL_2900999, EPI_ISL_2901000, EPI_ISL_2901001, EPI_ISL_2901002, EPI_ISL_2901003, EPI_ISL_2901004, EPI_ISL_2901005, EPI_ISL_2901006, EPI_ISL_2901007, EPI_ISL_2901008, EPI_ISL_2901009, EPI_ISL_2901010, EPI_ISL_2901011, EPI_ISL_2901012, EPI_ISL_2901013, EPI_ISL_2901014, EPI_ISL_2901015, EPI_ISL_2901016, EPI_ISL_2901017, EPI_ISL_2901018, EPI_ISL_2901019, EPI_ISL_2901020, EPI_ISL_2901021, EPI_ISL_2901022, EPI_ISL_2901023, EPI_ISL_2901024, EPI_ISL_2901025, EPI_ISL_2901026, EPI_ISL_2901027, EPI_ISL_2901028, EPI_ISL_2901029, EPI_ISL_2901030, EPI_ISL_2901031, EPI_ISL_2901032, EPI_ISL_2901033, EPI_ISL_2901034, EPI_ISL_2901035, EPI_ISL_2901036, EPI_ISL_2901037, EPI_ISL_2901038, EPI_ISL_2901039, EPI_ISL_2901040, EPI_ISL_2901041, EPI_ISL_2901043, EPI_ISL_2901044, EPI_ISL_2901045, EPI_ISL_2901046, EPI_ISL_2901047, EPI_ISL_2901048, EPI_ISL_2901049, EPI_ISL_2901050, EPI_ISL_2901051, EPI_ISL_2901052, EPI_ISL_2901053, EPI_ISL_2901055, EPI_ISL_2901056, EPI_ISL_2901057, EPI_ISL_2901058, EPI_ISL_2901059, EPI_ISL_2901060, EPI_ISL_2901061, EPI_ISL_2901062, EPI_ISL_2901063, EPI_ISL_2901064, EPI_ISL_2901065, EPI_ISL_2901066, EPI_ISL_2901067, EPI_ISL_2901068, EPI_ISL_2901069, EPI_ISL_2901070, EPI_ISL_2901071, EPI_ISL_2901072, EPI_ISL_2901073, EPI_ISL_2901075, EPI_ISL_2901076, EPI_ISL_2901077, EPI_ISL_2901078, EPI_ISL_2901079, EPI_ISL_2901080, EPI_ISL_2901081, EPI_ISL_2901082, EPI_ISL_2901083, EPI_ISL_2901084, EPI_ISL_2901085, EPI_ISL_2901086, EPI_ISL_2901087, EPI_ISL_2901088, EPI_ISL_2901089, EPI_ISL_2901090, EPI_ISL_2901091, EPI_ISL_2901092, EPI_ISL_2901093, EPI_ISL_2901094, EPI_ISL_2901095, EPI_ISL_2901096, EPI_ISL_2901097, EPI_ISL_2901098, EPI_ISL_2901099, EPI_ISL_2901100, EPI_ISL_2901101, EPI_ISL_2901102, EPI_ISL_2901103, EPI_ISL_2901104, EPI_ISL_2901105, EPI_ISL_2901106, EPI_ISL_2901107, EPI_ISL_2901108, EPI_ISL_2901109, EPI_ISL_2901110, EPI_ISL_2901111, EPI_ISL_2901112, EPI_ISL_2901113, EPI_ISL_2901114, EPI_ISL_2901115, EPI_ISL_2901116, EPI_ISL_2901117, EPI_ISL_2901118, EPI_ISL_2901119, EPI_ISL_2901120, EPI_ISL_2901121, EPI_ISL_2901122, EPI_ISL_2901123, EPI_ISL_2901124, EPI_ISL_2901125, EPI_ISL_2901126, EPI_ISL_2901127, EPI_ISL_2901128, EPI_ISL_2901161, EPI_ISL_2901177

| | | | |
|---|---|---|---|
| see above | Liverpool Clinical Laboratories | COVID-19 Genomics UK (COG-UK) Consortium | A Alrezaihi; Alessandro Gerada; Alistair Darby; Angela Cowell; Anita Lucaci; Anu Chawla; Cassie Olateju; Catherine Hartley; Charlotte Nelson; Ecaterina Vamos; Elaine O'Toole; Eleanor G Bentley; Ghada T Shawli; Hermione Webster; Isabel García-Dorival; James Johnson; James P Stewart; Jenifer Manson; Joanne Watts; Jones Benjamin; Jordan J Clark; Julian Hiscox; L Luu; Lucille Rainbow; M Almsaud; Margaret Hughes; Mark Whitehead; Matthew Gemmell; Miren Iturriza-Gomara; Muhannad Alruwaili; N.P Randle; Neil Swainston; PKF Gilmore; Parul Sharma; Rebekah Penrice-Randal; Richard Eccles; Richard Gregory; Sam Haldenby; Steve Paterson; Stuart D Armstrong; Trevor Ian Robinson; Ximeng Han |

EPI_ISL_2901563, EPI_ISL_2901564, EPI_ISL_2901565, EPI_ISL_2901566, EPI_ISL_2901567, EPI_ISL_2901568, EPI_ISL_2901569, EPI_ISL_2901570, EPI_ISL_2901571, EPI_ISL_2901572, EPI_ISL_2901573, EPI_ISL_2901574, EPI_ISL_2901575, EPI_ISL_2901576, EPI_ISL_2901577, EPI_ISL_2901578, EPI_ISL_2901579, EPI_ISL_2901580, EPI_ISL_2901581, EPI_ISL_2901582, EPI_ISL_2901583, EPI_ISL_2901584, EPI_ISL_2901585, EPI_ISL_2901586, EPI_ISL_2901587, EPI_ISL_2901588, EPI_ISL_2901589, EPI_ISL_2901590, EPI_ISL_2901591, EPI_ISL_2901592, EPI_ISL_2901593, EPI_ISL_2901595, EPI_ISL_2901596, EPI_ISL_2901597, EPI_ISL_2901598, EPI_ISL_2901599, EPI_ISL_2901600, EPI_ISL_2901601, EPI_ISL_2901602, EPI_ISL_2901603, EPI_ISL_2901604, EPI_ISL_2901605, EPI_ISL_2901606, EPI_ISL_2901607, EPI_ISL_2901608, EPI_ISL_2901609, EPI_ISL_2901610, EPI_ISL_2901611, EPI_ISL_2901612, EPI_ISL_2901613, EPI_ISL_2901614, EPI_ISL_2901615, EPI_ISL_2901616, EPI_ISL_2901617, EPI_ISL_2901618, EPI_ISL_2901619, EPI_ISL_2901620, EPI_ISL_2901622, EPI_ISL_2901623, EPI_ISL_2901624, EPI_ISL_2901625, EPI_ISL_2901626, EPI_ISL_2901627, EPI_ISL_2901628, EPI_ISL_2901629, EPI_ISL_2901630, EPI_ISL_2901631, EPI_ISL_2901632, EPI_ISL_2901633, EPI_ISL_2901634, EPI_ISL_2901635, EPI_ISL_2901636, EPI_ISL_2901637, EPI_ISL_2901639, EPI_ISL_2901640, EPI_ISL_2901647, EPI_ISL_2901652, EPI_ISL_2901657, EPI_ISL_2901658, EPI_ISL_2901663, EPI_ISL_2901664, EPI_ISL_2901665, EPI_ISL_2901670, EPI_ISL_2901671, EPI_ISL_2901672, EPI_ISL_2901677, EPI_ISL_2901678, EPI_ISL_2901682, EPI_ISL_2901687, EPI_ISL_2901688, EPI_ISL_2901692, EPI_ISL_2901698, EPI_ISL_2901699, EPI_ISL_2901706, EPI_ISL_2901712, EPI_ISL_2901713, EPI_ISL_2901714, EPI_ISL_2901715, EPI_ISL_2901716, EPI_ISL_2901717, EPI_ISL_2901718, EPI_ISL_2901719, EPI_ISL_2901720, EPI_ISL_2901721, EPI_ISL_2901722, EPI_ISL_2901723, EPI_ISL_2901724, EPI_ISL_2901726, EPI_ISL_2901727, EPI_ISL_2901729, EPI_ISL_2901730, EPI_ISL_2901731, EPI_ISL_2901732, EPI_ISL_2901733, EPI_ISL_2901734, EPI_ISL_2901735, EPI_ISL_2901736, EPI_ISL_2901737, EPI_ISL_2901738, EPI_ISL_2901739, EPI_ISL_2901740, EPI_ISL_2901741, EPI_ISL_2901742, EPI_ISL_2901743, EPI_ISL_2901744, EPI_ISL_2901745, EPI_ISL_2901746, EPI_ISL_2901747, EPI_ISL_2901748, EPI_ISL_2901749, EPI_ISL_2901750, EPI_ISL_2901751, EPI_ISL_2901752, EPI_ISL_2901753, EPI_ISL_2901754, EPI_ISL_2901755, EPI_ISL_2901756, EPI_ISL_2901757, EPI_ISL_2901758, EPI_ISL_2901759, EPI_ISL_2901760, EPI_ISL_2901761, EPI_ISL_2901762, EPI_ISL_2901763, EPI_ISL_2901764, EPI_ISL_2901765, EPI_ISL_2901766, EPI_ISL_2901767, EPI_ISL_2901768, EPI_ISL_2901769, EPI_ISL_2901770, EPI_ISL_2901771, EPI_ISL_2901772, EPI_ISL_2901773, EPI_ISL_2901775, EPI_ISL_2901776, EPI_ISL_2901777, EPI_ISL_2901778, EPI_ISL_2901779, EPI_ISL_2901780, EPI_ISL_2901781, EPI_ISL_2901782, EPI_ISL_2901783, EPI_ISL_2901784, EPI_ISL_2901785, EPI_ISL_2901786, EPI_ISL_2901787, EPI_ISL_2901789, EPI_ISL_2901790, EPI_ISL_2901791, EPI_ISL_2901792, EPI_ISL_2901793, EPI_ISL_2901794, EPI_ISL_2901795, EPI_ISL_2901796, EPI_ISL_2901797, EPI_ISL_2901798, EPI_ISL_2901799, EPI_ISL_2901800, EPI_ISL_2901801, EPI_ISL_2901802, EPI_ISL_2901803, EPI_ISL_2901804, EPI_ISL_2901805, EPI_ISL_2901806, EPI_ISL_2901807, EPI_ISL_2901808, EPI_ISL_2901809, EPI_ISL_2901810, EPI_ISL_2901811, EPI_ISL_2901812, EPI_ISL_2901813, EPI_ISL_2901815, EPI_ISL_2901816, EPI_ISL_2901817, EPI_ISL_2901818, EPI_ISL_2901819, EPI_ISL_2901820, EPI_ISL_2901821, EPI_ISL_2901822, EPI_ISL_2901823, EPI_ISL_2901824, EPI_ISL_2901825, EPI_ISL_2901826, EPI_ISL_2901827, EPI_ISL_2901828, EPI_ISL_2901829, EPI_ISL_2901830, EPI_ISL_2901831, EPI_ISL_2901832, EPI_ISL_2901833, EPI_ISL_2901834, EPI_ISL_2901835, EPI_ISL_2901836, EPI_ISL_2901837, EPI_ISL_2901838, EPI_ISL_2901839, EPI_ISL_2901840, EPI_ISL_2901841, EPI_ISL_2901842, EPI_ISL_2901843, EPI_ISL_2901844, EPI_ISL_2901845, EPI_ISL_2901846, EPI_ISL_2901848, EPI_ISL_2901849, EPI_ISL_2901850, EPI_ISL_2901851, EPI_ISL_2901852, EPI_ISL_2901853, EPI_ISL_2901854, EPI_ISL_2901855, EPI_ISL_2901856, EPI_ISL_2901857, EPI_ISL_2901858, EPI_ISL_2901859, EPI_ISL_2901860, EPI_ISL_2901861, EPI_ISL_2901862, EPI_ISL_2901863, EPI_ISL_2901864, EPI_ISL_2901865, EPI_ISL_2901866, EPI_ISL_2901867, EPI_ISL_2901868, EPI_ISL_2901869, EPI_ISL_2901870, EPI_ISL_2901871, EPI_ISL_2901872, EPI_ISL_2901873, EPI_ISL_2901874, EPI_ISL_2901875, EPI_ISL_2901876, EPI_ISL_2901928, EPI_ISL_2901942, EPI_ISL_2901962, EPI_ISL_2901963, EPI_ISL_2901964, EPI_ISL_2901965, EPI_ISL_2901966, EPI_ISL_2901967, EPI_ISL_2901968, EPI_ISL_2901969, EPI_ISL_2901970, EPI_ISL_2901971, EPI_ISL_2901972, EPI_ISL_2901973, EPI_ISL_2901974, EPI_ISL_2901975, EPI_ISL_2901976, EPI_ISL_2901977, EPI_ISL_2901978, EPI_ISL_2901979, EPI_ISL_2901980, EPI_ISL_2901981, EPI_ISL_2901982, EPI_ISL_2901983, EPI_ISL_2901984, EPI_ISL_2901985, EPI_ISL_2901986, EPI_ISL_2901987, EPI_ISL_2901988, EPI_ISL_2901989, EPI_ISL_2901990, EPI_ISL_2901991, EPI_ISL_2901992, EPI_ISL_2901993, EPI_ISL_2901994, EPI_ISL_2901995, EPI_ISL_2901996, EPI_ISL_2901997, EPI_ISL_2901998, EPI_ISL_2901999, EPI_ISL_2902000, EPI_ISL_2902001, EPI_ISL_2902002, EPI_ISL_2902003, EPI_ISL_2902004, EPI_ISL_2902005, EPI_ISL_2902006, EPI_ISL_2902007, EPI_ISL_2902008, EPI_ISL_2902009, EPI_ISL_2902010, EPI_ISL_2902011, EPI_ISL_2902012, EPI_ISL_2902013, EPI_ISL_2902014, EPI_ISL_2902015, EPI_ISL_2902016, EPI_ISL_2902017, EPI_ISL_2902018, EPI_ISL_2902019, EPI_ISL_2902020, EPI_ISL_2902021, EPI_ISL_2902022, EPI_ISL_2902023, EPI_ISL_2902024, EPI_ISL_2902025, EPI_ISL_2902026, EPI_ISL_2902027, EPI_ISL_2902028, EPI_ISL_2902029, EPI_ISL_2902030, EPI_ISL_2902031, EPI_ISL_2902032, EPI_ISL_2902033, EPI_ISL_2902034, EPI_ISL_2902035, EPI_ISL_2902036, EPI_ISL_2902037, EPI_ISL_2902038, EPI_ISL_2902039, EPI_ISL_2902040, EPI_ISL_2902041, EPI_ISL_2902152, EPI_ISL_2902289, EPI_ISL_2902394

| | | | |
|---|---|---|---|
| see above | Northumbria University / South Tees Hospitals NHS Foundation Trust / North Cumbria Integrated Care NHS Foundation Trust / North Tees and Hartlepool NHS Foundation Trust / Newcastle Hospitals NHS Foundation Trust | COVID-19 Genomics UK (COG-UK) Consortium | Andrew Nelson; Brendan Payne; Clive Graham; Darren L Smith; Debra Padgett; Edward Barton; Emma Swindells; Garren Scott; Gary Black; Gary Eltringham; Giles S Holt; Greg R Young; Jane Greenaway; Jennifer Collins; John Allan; Joshua Loh; Lynn Dover; Matthew Bashton; Mohammad A Tariq; Paul Baker; Sarah Essex; Steve Liggett; Wen C Yew; Yusri Taha |

EPI_ISL_2904580, EPI_ISL_2904582, EPI_ISL_2904584, EPI_ISL_2904589, EPI_ISL_2904590, EPI_ISL_2904595, EPI_ISL_2904597, EPI_ISL_2904619, EPI_ISL_2904620, EPI_ISL_2904621, EPI_ISL_2904622, EPI_ISL_2904623, EPI_ISL_2904624, EPI_ISL_2904625, EPI_ISL_2904626, EPI_ISL_2904627, EPI_ISL_2904628, EPI_ISL_2904629, EPI_ISL_2904630, EPI_ISL_2904631, EPI_ISL_2904632, EPI_ISL_2904633, EPI_ISL_2904634, EPI_ISL_2904635, EPI_ISL_2904636, EPI_ISL_2904637, EPI_ISL_2904638, EPI_ISL_2904639, EPI_ISL_2904640, EPI_ISL_2904641, EPI_ISL_2904642, EPI_ISL_2904683, EPI_ISL_2904688, EPI_ISL_2904699, EPI_ISL_2904700, EPI_ISL_2904701, EPI_ISL_2904702, EPI_ISL_2904703, EPI_ISL_2904704, EPI_ISL_2904705, EPI_ISL_2904706, EPI_ISL_2904707, EPI_ISL_2904708, EPI_ISL_2904709, EPI_ISL_2904710, EPI_ISL_2904711, EPI_ISL_2904712, EPI_ISL_2904713, EPI_ISL_2904714, EPI_ISL_2904715, EPI_ISL_2904717, EPI_ISL_2904718, EPI_ISL_2904719, EPI_ISL_2904720, EPI_ISL_2904721, EPI_ISL_2904728, EPI_ISL_2904733, EPI_ISL_2904734, EPI_ISL_2904735, EPI_ISL_2904739, EPI_ISL_2904741, EPI_ISL_2904742, EPI_ISL_2904743, EPI_ISL_2904744, EPI_ISL_2904747, EPI_ISL_2904749, EPI_ISL_2904758, EPI_ISL_2904760, EPI_ISL_2904763, EPI_ISL_2904766, EPI_ISL_2904768, EPI_ISL_2904769, EPI_ISL_2904771, EPI_ISL_2904772, EPI_ISL_2904775, EPI_ISL_2904833, EPI_ISL_2904835, EPI_ISL_2904836, EPI_ISL_2904837, EPI_ISL_2904838, EPI_ISL_2904839, EPI_ISL_2904840, EPI_ISL_2904841, EPI_ISL_2904842, EPI_ISL_2904843, EPI_ISL_2904844, EPI_ISL_2904845, EPI_ISL_2904846, EPI_ISL_2904847, EPI_ISL_2904848, EPI_ISL_2904849, EPI_ISL_2904850, EPI_ISL_2904851, EPI_ISL_2904852, EPI_ISL_2904853, EPI_ISL_2904854, EPI_ISL_2904855, EPI_ISL_2904856, EPI_ISL_2904857, EPI_ISL_2904858, EPI_ISL_2904859, EPI_ISL_2904861, EPI_ISL_2904862, EPI_ISL_2904863, EPI_ISL_2904864, EPI_ISL_2904866, EPI_ISL_2904867, EPI_ISL_2904868, EPI_ISL_2904869, EPI_ISL_2904870, EPI_ISL_2904871, EPI_ISL_2904872, EPI_ISL_2904873, EPI_ISL_2904874, EPI_ISL_2904875, EPI_ISL_2904876, EPI_ISL_2904877, EPI_ISL_2904878, EPI_ISL_2904879, EPI_ISL_2904880, EPI_ISL_2904881, EPI_ISL_2904882, EPI_ISL_2904883, EPI_ISL_2904884, EPI_ISL_2904885, EPI_ISL_2904886, EPI_ISL_2904887, EPI_ISL_2904888, EPI_ISL_2904889, EPI_ISL_2904890, EPI_ISL_2904891, EPI_ISL_2904892, EPI_ISL_2904893, EPI_ISL_2904894, EPI_ISL_2904895, EPI_ISL_2904896, EPI_ISL_2904898, EPI_ISL_2904899, EPI_ISL_2904900, EPI_ISL_2904901, EPI_ISL_2904902, EPI_ISL_2904903, EPI_ISL_2904904, EPI_ISL_2904905, EPI_ISL_2904906, EPI_ISL_2904907, EPI_ISL_2904911, EPI_ISL_2904912, EPI_ISL_2904919, EPI_ISL_2904920, EPI_ISL_2904932, EPI_ISL_2904934, EPI_ISL_2904935, EPI_ISL_2904936, EPI_ISL_2904937, EPI_ISL_2904938, EPI_ISL_2904939, EPI_ISL_2904940, EPI_ISL_2904941, EPI_ISL_2904942, EPI_ISL_2904943, EPI_ISL_2904944, EPI_ISL_2904945, EPI_ISL_2904946, EPI_ISL_2904947, EPI_ISL_2904948, EPI_ISL_2904949, EPI_ISL_2904950, EPI_ISL_2904951, EPI_ISL_2904952, EPI_ISL_2904953, EPI_ISL_2904954, EPI_ISL_2904955, EPI_ISL_2904956, EPI_ISL_2904957, EPI_ISL_2904958, EPI_ISL_2904959, EPI_ISL_2904960, EPI_ISL_2904961, EPI_ISL_2904962, EPI_ISL_2904963, EPI_ISL_2904964, EPI_ISL_2904965, EPI_ISL_2904966, EPI_ISL_2904967, EPI_ISL_2904968, EPI_ISL_2904969, EPI_ISL_2904970, EPI_ISL_2904971, EPI_ISL_2904972, EPI_ISL_2904973, EPI_ISL_2904974, EPI_ISL_2904975, EPI_ISL_2904976, EPI_ISL_2904977, EPI_ISL_2904978, EPI_ISL_2904979, EPI_ISL_2904980, EPI_ISL_2904981, EPI_ISL_2904982, EPI_ISL_2904983, EPI_ISL_2904984, EPI_ISL_2904986, EPI_ISL_2904987, EPI_ISL_2904988, EPI_ISL_2904989, EPI_ISL_2904990, EPI_ISL_2904991, EPI_ISL_2904992, EPI_ISL_2904993, EPI_ISL_2904994, EPI_ISL_2904995, EPI_ISL_2904996, EPI_ISL_2904997, EPI_ISL_2904998, EPI_ISL_2904999, EPI_ISL_2905000, EPI_ISL_2905001, EPI_ISL_2905002, EPI_ISL_2905003, EPI_ISL_2905004, EPI_ISL_2905005, EPI_ISL_2905006, EPI_ISL_2905007, EPI_ISL_2905008, EPI_ISL_2905009, EPI_ISL_2905012, EPI_ISL_2905013, EPI_ISL_2905014, EPI_ISL_2905060, EPI_ISL_2905066, EPI_ISL_2905069, EPI_ISL_2905070, EPI_ISL_2905071, EPI_ISL_2905072, EPI_ISL_2905073, EPI_ISL_2905075, EPI_ISL_2905076, EPI_ISL_2905077, EPI_ISL_2905078, EPI_ISL_2905079, EPI_ISL_2905080, EPI_ISL_2905081, EPI_ISL_2905141, EPI_ISL_2905142, EPI_ISL_2905143, EPI_ISL_2905144, EPI_ISL_2905145, EPI_ISL_2905146, EPI_ISL_2905147, EPI_ISL_2905148, EPI_ISL_2905149, EPI_ISL_2905150, EPI_ISL_2905151, EPI_ISL_2905152, EPI_ISL_2905153, EPI_ISL_2905154, EPI_ISL_2905155, EPI_ISL_2905156, EPI_ISL_2905158, EPI_ISL_2905159,

| | | | |
|---|---|---|---|
| EPI_ISL_2905160, EPI_ISL_2905161, EPI_ISL_2905162, EPI_ISL_2905163, EPI_ISL_2905164, EPI_ISL_2905165, EPI_ISL_2905166, EPI_ISL_2905167, EPI_ISL_2905168, EPI_ISL_2905169, EPI_ISL_2905179, EPI_ISL_2905187, EPI_ISL_2905190, EPI_ISL_2905191, EPI_ISL_2905224, EPI_ISL_2905225, EPI_ISL_2905226, EPI_ISL_2905227, EPI_ISL_2905228, EPI_ISL_2905229, EPI_ISL_2905230, EPI_ISL_2905231, EPI_ISL_2905232, EPI_ISL_2905233, EPI_ISL_2905234, EPI_ISL_2905235, EPI_ISL_2905236, EPI_ISL_2905237, EPI_ISL_2905238, EPI_ISL_2905239, EPI_ISL_2905240, EPI_ISL_2905241, EPI_ISL_2905242, EPI_ISL_2905243, EPI_ISL_2905244, EPI_ISL_2905245, EPI_ISL_2905246, EPI_ISL_2905247, EPI_ISL_2905248, EPI_ISL_2905249, EPI_ISL_2905250, EPI_ISL_2905251, EPI_ISL_2905252, EPI_ISL_2905253, EPI_ISL_2905254, EPI_ISL_2905255, EPI_ISL_2905256, EPI_ISL_2905257, EPI_ISL_2905258, EPI_ISL_2905259, EPI_ISL_2905260, EPI_ISL_2905261, EPI_ISL_2905262, EPI_ISL_2905263, EPI_ISL_2905264, EPI_ISL_2905265, EPI_ISL_2905266, EPI_ISL_2905267, EPI_ISL_2905268, EPI_ISL_2905269, EPI_ISL_2905270, EPI_ISL_2905271, EPI_ISL_2905272, EPI_ISL_2905273, EPI_ISL_2905274, EPI_ISL_2905275, EPI_ISL_2905276, EPI_ISL_2905277, EPI_ISL_2905278, EPI_ISL_2905279, EPI_ISL_2905280, EPI_ISL_2905281, EPI_ISL_2905282, EPI_ISL_2905283, EPI_ISL_2905284, EPI_ISL_2905285, EPI_ISL_2905286, EPI_ISL_2905287, EPI_ISL_2905288, EPI_ISL_2905289, EPI_ISL_2905290, EPI_ISL_2905291, EPI_ISL_2905292, EPI_ISL_2905293, EPI_ISL_2905294, EPI_ISL_2905295, EPI_ISL_2905296, EPI_ISL_2905297, EPI_ISL_2905298, EPI_ISL_2905299, EPI_ISL_2905300, EPI_ISL_2905301, EPI_ISL_2905302, EPI_ISL_2905303, EPI_ISL_2905304, EPI_ISL_2905305, EPI_ISL_2905306, EPI_ISL_2905307, EPI_ISL_2905308, EPI_ISL_2905309, EPI_ISL_2905310, EPI_ISL_2905311, EPI_ISL_2905312, EPI_ISL_2905313, EPI_ISL_2905314, EPI_ISL_2905315, EPI_ISL_2905316, EPI_ISL_2905317, EPI_ISL_2905318, EPI_ISL_2905319, EPI_ISL_2905320, EPI_ISL_2905321, EPI_ISL_2905322, EPI_ISL_2905323, EPI_ISL_2905324, EPI_ISL_2905325, EPI_ISL_2905326, EPI_ISL_2905327, EPI_ISL_2905328, EPI_ISL_2905329, EPI_ISL_2905330, EPI_ISL_2905331, EPI_ISL_2905332, EPI_ISL_2905333, EPI_ISL_2905334, EPI_ISL_2905335, EPI_ISL_2905336, EPI_ISL_2905337, EPI_ISL_2905338, EPI_ISL_2905339, EPI_ISL_2905340, EPI_ISL_2905341, EPI_ISL_2905342, EPI_ISL_2905343, EPI_ISL_2905344, EPI_ISL_2905345, EPI_ISL_2905346, EPI_ISL_2905347, EPI_ISL_2905348, EPI_ISL_2905349, EPI_ISL_2905350, EPI_ISL_2905351, EPI_ISL_2905352, EPI_ISL_2905353, EPI_ISL_2905354, EPI_ISL_2905355, EPI_ISL_2905356, EPI_ISL_2905357, EPI_ISL_2905358, EPI_ISL_2905359, EPI_ISL_2905360, EPI_ISL_2905361, EPI_ISL_2905362, EPI_ISL_2905363, EPI_ISL_2905364, EPI_ISL_2905365, EPI_ISL_2905366, EPI_ISL_2905367, EPI_ISL_2905368, EPI_ISL_2905369, EPI_ISL_2905370, EPI_ISL_2905371, EPI_ISL_2905372, EPI_ISL_2905373, EPI_ISL_2905374, EPI_ISL_2905375, EPI_ISL_2905376, EPI_ISL_2905377, EPI_ISL_2905378, EPI_ISL_2905379, EPI_ISL_2905380, EPI_ISL_2905381, EPI_ISL_2905382, EPI_ISL_2905383, EPI_ISL_2905384, EPI_ISL_2905385, EPI_ISL_2905407, EPI_ISL_2905408, EPI_ISL_2905409, EPI_ISL_2905410, EPI_ISL_2905411, EPI_ISL_2905412, EPI_ISL_2905413, EPI_ISL_2905414, EPI_ISL_2905415, EPI_ISL_2905416, EPI_ISL_2905417, EPI_ISL_2905418, EPI_ISL_2905419, EPI_ISL_2905420, EPI_ISL_2905421, EPI_ISL_2905422, EPI_ISL_2905423, EPI_ISL_2905424, EPI_ISL_2905425, EPI_ISL_2905426, EPI_ISL_2905427, EPI_ISL_2905428, EPI_ISL_2905429, EPI_ISL_2905430, EPI_ISL_2905431, EPI_ISL_2905432, EPI_ISL_2905433, EPI_ISL_2905434, EPI_ISL_2905435, EPI_ISL_2905436, EPI_ISL_2905437, EPI_ISL_2905438, EPI_ISL_2905439, EPI_ISL_2905440, EPI_ISL_2905441, EPI_ISL_2905442, EPI_ISL_2905443, EPI_ISL_2905444, EPI_ISL_2905445, EPI_ISL_2905446, EPI_ISL_2905447, EPI_ISL_2905448, EPI_ISL_2905449, EPI_ISL_2905450, EPI_ISL_2905451, EPI_ISL_2905452, EPI_ISL_2905453, EPI_ISL_2905454, EPI_ISL_2905455, EPI_ISL_2905456, EPI_ISL_2905457, EPI_ISL_2905458, EPI_ISL_2905459, EPI_ISL_2905460, EPI_ISL_2905461, EPI_ISL_2905462, EPI_ISL_2905463, EPI_ISL_2905466, EPI_ISL_2905467, EPI_ISL_2905468, EPI_ISL_2905469, EPI_ISL_2905471, EPI_ISL_2905472, EPI_ISL_2905479, EPI_ISL_2905536, EPI_ISL_2905537, EPI_ISL_2905538, EPI_ISL_2905539, EPI_ISL_2905540, EPI_ISL_2905541, EPI_ISL_2905542, EPI_ISL_2905543, EPI_ISL_2905544, EPI_ISL_2905545, EPI_ISL_2905546, EPI_ISL_2905547, EPI_ISL_2905548, EPI_ISL_2905549, EPI_ISL_2905550, EPI_ISL_2905551, EPI_ISL_2905552, EPI_ISL_2905553, EPI_ISL_2905554, EPI_ISL_2905555, EPI_ISL_2905556, EPI_ISL_2905557, EPI_ISL_2905558, EPI_ISL_2905559, EPI_ISL_2905560, EPI_ISL_2905561, EPI_ISL_2905562, EPI_ISL_2905563, EPI_ISL_2905564, EPI_ISL_2905565, EPI_ISL_2905566, EPI_ISL_2905567, EPI_ISL_2905568, EPI_ISL_2905569, EPI_ISL_2905570, EPI_ISL_2905572, EPI_ISL_2905573, EPI_ISL_2905574, EPI_ISL_2905575, EPI_ISL_2905576, EPI_ISL_2905577, EPI_ISL_2905579, EPI_ISL_2905580, EPI_ISL_2905581, EPI_ISL_2905582, EPI_ISL_2905583, EPI_ISL_2905584, EPI_ISL_2905585, EPI_ISL_2905586, EPI_ISL_2905587, EPI_ISL_2905589, EPI_ISL_2905590, EPI_ISL_2905591, EPI_ISL_2905592, EPI_ISL_2905593, EPI_ISL_2905594, EPI_ISL_2905655, EPI_ISL_2905656, EPI_ISL_2905657, EPI_ISL_2905658, EPI_ISL_2905659, EPI_ISL_2905660, EPI_ISL_2905661, EPI_ISL_2905662, EPI_ISL_2905663, EPI_ISL_2905665, EPI_ISL_2905666, EPI_ISL_2905667, EPI_ISL_2905668, EPI_ISL_2905669, EPI_ISL_2905671, EPI_ISL_2905674, EPI_ISL_2905675, EPI_ISL_2905676, EPI_ISL_2905677, EPI_ISL_2905678, EPI_ISL_2905679, EPI_ISL_2905680, EPI_ISL_2905681, EPI_ISL_2905683, EPI_ISL_2905684, EPI_ISL_2905685, EPI_ISL_2905686, EPI_ISL_2905688, EPI_ISL_2905690, EPI_ISL_2905692, EPI_ISL_2905697, EPI_ISL_2905699, EPI_ISL_2905700, EPI_ISL_2905701, EPI_ISL_2905702, EPI_ISL_2905703, EPI_ISL_2905704, EPI_ISL_2905705, EPI_ISL_2905706, EPI_ISL_2905707, EPI_ISL_2905708, EPI_ISL_2905709, EPI_ISL_2905711, EPI_ISL_2905712, EPI_ISL_2905713, EPI_ISL_2905714, EPI_ISL_2905715, EPI_ISL_2905716, EPI_ISL_2905717, EPI_ISL_2905718, EPI_ISL_2905719, EPI_ISL_2905720, EPI_ISL_2905721, EPI_ISL_2905723, EPI_ISL_2905724, EPI_ISL_2905725, EPI_ISL_2905726, EPI_ISL_2905727, EPI_ISL_2905728, EPI_ISL_2905729, EPI_ISL_2905731, EPI_ISL_2905732, EPI_ISL_2905733, EPI_ISL_2905734, EPI_ISL_2905737, EPI_ISL_2905738, EPI_ISL_2905751, EPI_ISL_2905752, EPI_ISL_2905753, EPI_ISL_2905754, EPI_ISL_2905755, EPI_ISL_2905756, EPI_ISL_2905757, EPI_ISL_2905758, EPI_ISL_2905759, EPI_ISL_2905760, EPI_ISL_2905761, EPI_ISL_2905762, EPI_ISL_2905763, EPI_ISL_2905764, EPI_ISL_2905765, EPI_ISL_2905766, EPI_ISL_2905767, EPI_ISL_2905768, EPI_ISL_2905769, EPI_ISL_2905770, EPI_ISL_2905771, EPI_ISL_2905772, EPI_ISL_2905773, EPI_ISL_2905774, EPI_ISL_2905775, EPI_ISL_2905776, EPI_ISL_2905777, EPI_ISL_2905779, EPI_ISL_2905781, EPI_ISL_2905782, EPI_ISL_2905783, EPI_ISL_2905784, EPI_ISL_2905785, EPI_ISL_2905786, EPI_ISL_2905787, EPI_ISL_2905788, EPI_ISL_2905789, EPI_ISL_2905790, EPI_ISL_2905791, EPI_ISL_2905792, EPI_ISL_2905793, EPI_ISL_2905794, EPI_ISL_2905795, EPI_ISL_2905796, EPI_ISL_2905797, EPI_ISL_2905798, EPI_ISL_2905799, EPI_ISL_2905800, EPI_ISL_2905801, EPI_ISL_2905802, EPI_ISL_2905803, EPI_ISL_2905804, EPI_ISL_2905805, EPI_ISL_2905806, EPI_ISL_2905807, EPI_ISL_2905808, EPI_ISL_2905809, EPI_ISL_2905810, EPI_ISL_2905812, EPI_ISL_2905813, EPI_ISL_2905814, EPI_ISL_2905815, EPI_ISL_2905816, EPI_ISL_2905817, EPI_ISL_2905819, EPI_ISL_2905820, EPI_ISL_2905821, EPI_ISL_2905822, EPI_ISL_2905823, EPI_ISL_2905824, EPI_ISL_2905825, EPI_ISL_2905826, EPI_ISL_2905827, EPI_ISL_2905828, EPI_ISL_2905829, EPI_ISL_2905830, EPI_ISL_2905831, EPI_ISL_2905832, EPI_ISL_2905833, EPI_ISL_2905834, EPI_ISL_2905835, EPI_ISL_2905836, EPI_ISL_2905837, EPI_ISL_2905838, EPI_ISL_2905839, EPI_ISL_2905840, EPI_ISL_2905841, EPI_ISL_2905842, EPI_ISL_2905843, EPI_ISL_2905844, EPI_ISL_2905845, EPI_ISL_2905846, EPI_ISL_2905847, EPI_ISL_2905848, EPI_ISL_2905849, EPI_ISL_2905850, EPI_ISL_2905852, EPI_ISL_2905853, EPI_ISL_2905855, EPI_ISL_2905856, EPI_ISL_2905858, EPI_ISL_2905859, EPI_ISL_2905860, EPI_ISL_2905861, EPI_ISL_2905862, EPI_ISL_2905864, EPI_ISL_2905865, EPI_ISL_2905866, EPI_ISL_2905867, EPI_ISL_2905868, EPI_ISL_2905869, EPI_ISL_2905870, EPI_ISL_2905871, EPI_ISL_2905872, EPI_ISL_2905873, EPI_ISL_2905874, EPI_ISL_2905875, EPI_ISL_2905876, EPI_ISL_2905877, EPI_ISL_2905878, EPI_ISL_2905879, EPI_ISL_2905880, EPI_ISL_2905881, EPI_ISL_2905882, EPI_ISL_2905885, EPI_ISL_2905886, EPI_ISL_2905889, EPI_ISL_2905890, EPI_ISL_2905891, EPI_ISL_2905892, EPI_ISL_2905893, EPI_ISL_2905894, EPI_ISL_2905895, EPI_ISL_2905896, EPI_ISL_2905897, EPI_ISL_2905898, EPI_ISL_2905899, EPI_ISL_2905900, EPI_ISL_2905901, EPI_ISL_2905902, EPI_ISL_2905903, EPI_ISL_2905904, EPI_ISL_2905905, EPI_ISL_2905906, EPI_ISL_2905907, EPI_ISL_2905908, EPI_ISL_2905909, EPI_ISL_2905910, EPI_ISL_2905911, EPI_ISL_2905912, EPI_ISL_2905913, EPI_ISL_2905914, EPI_ISL_2905915, EPI_ISL_2905916, EPI_ISL_2905917, EPI_ISL_2905918, EPI_ISL_2905919, EPI_ISL_2905990, EPI_ISL_2906026, EPI_ISL_2906027, EPI_ISL_2906029, EPI_ISL_2906035, EPI_ISL_2906041, EPI_ISL_2906044, EPI_ISL_2906046, EPI_ISL_2906049, EPI_ISL_2906115, EPI_ISL_2906116, EPI_ISL_2906118, EPI_ISL_2906181, EPI_ISL_2906246, EPI_ISL_2906266, EPI_ISL_2906284, EPI_ISL_2906285, EPI_ISL_2906286, EPI_ISL_2906387, EPI_ISL_2906393, EPI_ISL_2906396, EPI_ISL_2906534, EPI_ISL_2906538, EPI_ISL_2906643, EPI_ISL_2906677, EPI_ISL_2906678, EPI_ISL_2906679, EPI_ISL_2906680, EPI_ISL_2906681, EPI_ISL_2906683, EPI_ISL_2906684, EPI_ISL_2906686, EPI_ISL_2906687, EPI_ISL_2906688, EPI_ISL_2906689, EPI_ISL_2906690, EPI_ISL_2906694, EPI_ISL_2906696, EPI_ISL_2906697, EPI_ISL_2906698, EPI_ISL_2906700, EPI_ISL_2906701, EPI_ISL_2906702 | | | |
| see above | Originating lab: Wales Specialist Virology Centre Sequencing lab: Pathogen Genomics Unit | Public Health Wales Microbiology Cardiff Wales Specialist Virology Centre | Alec Birchley; Alexander Adams; Amy Gaskin; Angela Marchbank; Bree Gatica-Wilcox; Catherine Moore; Jason Coombes; Joanne Watkins; Joel Southgate; Johnathan Evans; Laura Gifford; Lauren Gilbert; Lee Graham; Malorie Perry; Matthew Bull; Nicole Pacchiarini; Sally Corden; Sara Kumziene-Summerhayes; Sara Rey; Sarah Taylor; Simon Cottrell; Sophie Jones; Tom Connor |
| EPI_ISL_2902433, EPI_ISL_2902465, EPI_ISL_2902500, EPI_ISL_2902506, EPI_ISL_2902591, EPI_ISL_2902621 | Quadram Institute Bioscience | COVID-19 Genomics UK (COG-UK) Consortium | Alexander J Trotter; Alison E. Mather; Alp Aydin; Ana P. Tedim; Anastasia Kolyva; Andrew Bell; Andrew J. Page; Christopher Jeanes; Claire Stuart; Dave J. Baker; Ebenezer Foster-Nyarko; Gemma L. Kay; John Wain; Justin O'Grady; Leonardo de Oliveira Martins; Lewis G. Spurgin; Lindsay Coupland; Lizzie Meadows; Luke Bedford; Maria Diaz; Mark Webber; Martin Lott; Muhammed Yasir; Nabil-Fareed Alikhan; Ngozi Elumogo; Nicholas M. Thomson; Rachael Stanley; Rachel Gilroy; Reenesh Prakash; Rose K Davidson; Samir Dervisevic; Samuel Bloomfield; Sophie J. Prosolek; Steven Rudder; Thanh Le-Viet |
| EPI_ISL_2918091, EPI_ISL_2918111, EPI_ISL_2918294, EPI_ISL_2918303, EPI_ISL_2918410, EPI_ISL_2918420, EPI_ISL_2918426, EPI_ISL_2918428, EPI_ISL_2918457, EPI_ISL_2918468, EPI_ISL_2918473, EPI_ISL_2918478, EPI_ISL_2918487, EPI_ISL_2918497, EPI_ISL_2918512, EPI_ISL_2918518, EPI_ISL_2918521, EPI_ISL_2918527, EPI_ISL_2918530, EPI_ISL_2918534, EPI_ISL_2918536, EPI_ISL_2918539, EPI_ISL_2918544, EPI_ISL_2918550, EPI_ISL_2918560, EPI_ISL_2918567, EPI_ISL_2918575, EPI_ISL_2918585, EPI_ISL_2918586, EPI_ISL_2937745 | | | |
| see above | Randox Laboratories | Wellcome Sanger Institute for the COVID-19 Genomics UK (COG-UK) Consortium | Cordelia Langford; David K. Jackson; Dominic Kwiatkowski; Ewan Harrison; Ian Johnston; Jeffrey Barrett; John Sillitoe on behalf of the Wellcome Sanger Institute COVID-19 Surveillance Team; Randox Laboratories and Alex Alderton; Roberto Amato; Sonia Goncalves |
| EPI_ISL_2817812, EPI_ISL_2817821, EPI_ISL_2817823, EPI_ISL_2817826, EPI_ISL_2817853, EPI_ISL_2817854, EPI_ISL_2817861, EPI_ISL_2818011, EPI_ISL_2818012, EPI_ISL_2818023, EPI_ISL_2818032, EPI_ISL_2818033, EPI_ISL_2818041, EPI_ISL_2818062, EPI_ISL_2818075, EPI_ISL_2818315, EPI_ISL_2818316, EPI_ISL_2818317, EPI_ISL_2818318, EPI_ISL_2818319, EPI_ISL_2818320, EPI_ISL_2818321, EPI_ISL_2818322, EPI_ISL_2818323, EPI_ISL_2818324, EPI_ISL_2903420, EPI_ISL_2903421, EPI_ISL_2903422, EPI_ISL_2903423, EPI_ISL_2903424, EPI_ISL_2903460, EPI_ISL_2903486, EPI_ISL_2903487, EPI_ISL_2903489, EPI_ISL_2903490, EPI_ISL_2903498, EPI_ISL_2903499, EPI_ISL_2903500, EPI_ISL_2903501, EPI_ISL_2903502, EPI_ISL_2903503, EPI_ISL_2903504, EPI_ISL_2903505, EPI_ISL_2903506, EPI_ISL_2903507, EPI_ISL_2903508, EPI_ISL_2903509, EPI_ISL_2903510, EPI_ISL_2903511, EPI_ISL_2903512, EPI_ISL_2903513, EPI_ISL_2903514, EPI_ISL_2903525, EPI_ISL_2903526, EPI_ISL_2903528, EPI_ISL_2903529, EPI_ISL_2903530, EPI_ISL_2903531, EPI_ISL_2903532, EPI_ISL_2903533, EPI_ISL_2903534, EPI_ISL_2903535, EPI_ISL_2903536, EPI_ISL_2903537, EPI_ISL_2903538, EPI_ISL_2903539, EPI_ISL_2903540, EPI_ISL_2903541, EPI_ISL_2903542, EPI_ISL_2903543, EPI_ISL_2903544, EPI_ISL_2903545, EPI_ISL_2903546, EPI_ISL_2903547, EPI_ISL_2903548, EPI_ISL_2903549, EPI_ISL_2903550, EPI_ISL_2903551, EPI_ISL_2903552, EPI_ISL_2903553, EPI_ISL_2903555, EPI_ISL_2903557, EPI_ISL_2903558, EPI_ISL_2903559, EPI_ISL_2903560, EPI_ISL_2903561, EPI_ISL_2903562, EPI_ISL_2903563, EPI_ISL_2903564, EPI_ISL_2903565, EPI_ISL_2903566, EPI_ISL_2903572, EPI_ISL_2903573, EPI_ISL_2903574, EPI_ISL_2903575, EPI_ISL_2903576, EPI_ISL_2903577, EPI_ISL_2903578, EPI_ISL_2903579, EPI_ISL_2903580, EPI_ISL_2903581, EPI_ISL_2903582, EPI_ISL_2903583, EPI_ISL_2903584, EPI_ISL_2903585, EPI_ISL_2903587, EPI_ISL_2903589, EPI_ISL_2903591, EPI_ISL_2903592, EPI_ISL_2903593, EPI_ISL_2903594, EPI_ISL_2903595, EPI_ISL_2903596, EPI_ISL_2903597, EPI_ISL_2903598, EPI_ISL_2903599, EPI_ISL_2903600, EPI_ISL_2903601, EPI_ISL_2903603, EPI_ISL_2903604, EPI_ISL_2903605, EPI_ISL_2903606, EPI_ISL_2903607, EPI_ISL_2903608, EPI_ISL_2903609, EPI_ISL_2903610, EPI_ISL_2903612, EPI_ISL_2903613, EPI_ISL_2903617, EPI_ISL_2903618, EPI_ISL_2903619, EPI_ISL_2903620, EPI_ISL_2903621, EPI_ISL_2903622, EPI_ISL_2903623, EPI_ISL_2903624, EPI_ISL_2903625, EPI_ISL_2903626, EPI_ISL_2903627, EPI_ISL_2903628, EPI_ISL_2903631, EPI_ISL_2903633, EPI_ISL_2903653, EPI_ISL_2903655, EPI_ISL_2903658, EPI_ISL_2903659, EPI_ISL_2903660, EPI_ISL_2903662, EPI_ISL_2903663, EPI_ISL_2903665, EPI_ISL_2903666, EPI_ISL_2903667, EPI_ISL_2903668, EPI_ISL_2903669, EPI_ISL_2903670, EPI_ISL_2903672, EPI_ISL_2903673, EPI_ISL_2903674, EPI_ISL_2903675, EPI_ISL_2903676, EPI_ISL_2903677, EPI_ISL_2903678, EPI_ISL_2903679, EPI_ISL_2903680, EPI_ISL_2903681, EPI_ISL_2903682, EPI_ISL_2903683, EPI_ISL_2903684, EPI_ISL_2903685, EPI_ISL_2903686, EPI_ISL_2903688, EPI_ISL_2903689, EPI_ISL_2903690, EPI_ISL_2903691, EPI_ISL_2903692, EPI_ISL_2903693, EPI_ISL_2903694, EPI_ISL_2903695, EPI_ISL_2903696, EPI_ISL_2903708, EPI_ISL_2903709, EPI_ISL_2903710, EPI_ISL_2903711, EPI_ISL_2903712, EPI_ISL_2903713, EPI_ISL_2903714, EPI_ISL_2903715, EPI_ISL_2903716, EPI_ISL_2903718, EPI_ISL_2903719, EPI_ISL_2903720, EPI_ISL_2903721, EPI_ISL_2903722, EPI_ISL_2903723, EPI_ISL_2903724, EPI_ISL_2903725, EPI_ISL_2903726, EPI_ISL_2903727, EPI_ISL_2903728, EPI_ISL_2903729, EPI_ISL_2903730, EPI_ISL_2903731, EPI_ISL_2903733, EPI_ISL_2903734, EPI_ISL_2903735, EPI_ISL_2903736, EPI_ISL_2903737, EPI_ISL_2903738, EPI_ISL_2903739, EPI_ISL_2903740, EPI_ISL_2903741, EPI_ISL_2903742, EPI_ISL_2903743, EPI_ISL_2903745, EPI_ISL_2903747, EPI_ISL_2903748, EPI_ISL_2903749, EPI_ISL_2903751, EPI_ISL_2903752, EPI_ISL_2903753, EPI_ISL_2903754, EPI_ISL_2903755, EPI_ISL_2903757, EPI_ISL_2903758, EPI_ISL_2903759, EPI_ISL_2903760, EPI_ISL_2903761, EPI_ISL_2903762, EPI_ISL_2903763, EPI_ISL_2903764, EPI_ISL_2903765, EPI_ISL_2903768, EPI_ISL_2903769, EPI_ISL_2903770, EPI_ISL_2903771, EPI_ISL_2903772, EPI_ISL_2903773, EPI_ISL_2903774, EPI_ISL_2903775, EPI_ISL_2903776, EPI_ISL_2903777, EPI_ISL_2903779, EPI_ISL_2903785, EPI_ISL_2903786, EPI_ISL_2903787, EPI_ISL_2903788, EPI_ISL_2903789, EPI_ISL_2903791, EPI_ISL_2903793, EPI_ISL_2903794, EPI_ISL_2903795, EPI_ISL_2903796, EPI_ISL_2903797, EPI_ISL_2903798, EPI_ISL_2903799, EPI_ISL_2903800, EPI_ISL_2903801, EPI_ISL_2903824, EPI_ISL_2903825, EPI_ISL_2903826, EPI_ISL_2903844, EPI_ISL_2903847, EPI_ISL_2903853, EPI_ISL_2903854, EPI_ISL_2903855, EPI_ISL_2903856, EPI_ISL_2903857, EPI_ISL_2903858, EPI_ISL_2903859, EPI_ISL_2903860, EPI_ISL_2903861, EPI_ISL_2903862, EPI_ISL_2903863, EPI_ISL_2903864, EPI_ISL_2903865, EPI_ISL_2903866, EPI_ISL_2903867, EPI_ISL_2903868, EPI_ISL_2903869, EPI_ISL_2903870, EPI_ISL_2903871, EPI_ISL_2903872, EPI_ISL_2903873, EPI_ISL_2903874, EPI_ISL_2903875, EPI_ISL_2903876, EPI_ISL_2903877, EPI_ISL_2903878, EPI_ISL_2903879, EPI_ISL_2903880, EPI_ISL_2903881, EPI_ISL_2903882, EPI_ISL_2903883, EPI_ISL_2903884, EPI_ISL_2903885, EPI_ISL_2903886, EPI_ISL_2903887, EPI_ISL_2903888, EPI_ISL_2903931, EPI_ISL_2903942, EPI_ISL_2903943, EPI_ISL_2903950, EPI_ISL_2903954, EPI_ISL_2904027, EPI_ISL_2904029, EPI_ISL_2904030, EPI_ISL_2904031, EPI_ISL_2904032, EPI_ISL_2904033, EPI_ISL_2904034, EPI_ISL_2904035, EPI_ISL_2904036, EPI_ISL_2904037, EPI_ISL_2904038, EPI_ISL_2904039, EPI_ISL_2904040, EPI_ISL_2904041, EPI_ISL_2904045, EPI_ISL_2904048, EPI_ISL_2904049, EPI_ISL_2904050, EPI_ISL_2904051, EPI_ISL_2904053, EPI_ISL_2904054, EPI_ISL_2904056, EPI_ISL_2904057, EPI_ISL_2904059, EPI_ISL_2904060, EPI_ISL_2904061, EPI_ISL_2904062, EPI_ISL_2904063, EPI_ISL_2904064, EPI_ISL_2904074, EPI_ISL_2904076, EPI_ISL_2904077, EPI_ISL_2904079, EPI_ISL_2904080, EPI_ISL_2904081, EPI_ISL_2904083, EPI_ISL_2904085, EPI_ISL_2904086, EPI_ISL_2904088, EPI_ISL_2904090, EPI_ISL_2904093, EPI_ISL_2904094, EPI_ISL_2904095, EPI_ISL_2904096, EPI_ISL_2904097, EPI_ISL_2904098, EPI_ISL_2904099, EPI_ISL_2904100, EPI_ISL_2904101, EPI_ISL_2904102, EPI_ISL_2904103, EPI_ISL_2904104, EPI_ISL_2904105, EPI_ISL_2904106, EPI_ISL_2904107, EPI_ISL_2904108, EPI_ISL_2904110, EPI_ISL_2904111, EPI_ISL_2904112, EPI_ISL_2904189, EPI_ISL_2904192, EPI_ISL_2904194, EPI_ISL_2904195, EPI_ISL_2904196, EPI_ISL_2904197, EPI_ISL_2904219, EPI_ISL_2904220, EPI_ISL_2904221, EPI_ISL_2904222, EPI_ISL_2904223, EPI_ISL_2904224, EPI_ISL_2904226, EPI_ISL_2904227, EPI_ISL_2904228, EPI_ISL_2904229, EPI_ISL_2904231, EPI_ISL_2904232, EPI_ISL_2904233, EPI_ISL_2904234, EPI_ISL_2904237, EPI_ISL_2904240, EPI_ISL_2904241, EPI_ISL_2904245, EPI_ISL_2904246, EPI_ISL_2904248, EPI_ISL_2904249, EPI_ISL_2904250, EPI_ISL_2904251, EPI_ISL_2904252, EPI_ISL_2904253, EPI_ISL_2904254, EPI_ISL_2904255, EPI_ISL_2904257, EPI_ISL_2904262, EPI_ISL_2904264, EPI_ISL_2904266, EPI_ISL_2904270, EPI_ISL_2904272, EPI_ISL_2904278, EPI_ISL_2904281, EPI_ISL_2904283, EPI_ISL_2904285, EPI_ISL_2904286, EPI_ISL_2904292, EPI_ISL_2904297, EPI_ISL_2904309, EPI_ISL_2904310, EPI_ISL_2904320, EPI_ISL_2904343, EPI_ISL_2904345, EPI_ISL_2904346, EPI_ISL_2904347, EPI_ISL_2904348, EPI_ISL_2904349, EPI_ISL_2904350, EPI_ISL_2904355, EPI_ISL_2904357, EPI_ISL_2904358, EPI_ISL_2904360, EPI_ISL_2904362, EPI_ISL_2904363, EPI_ISL_2904364, EPI_ISL_2904365, EPI_ISL_2904367, EPI_ISL_2904369, EPI_ISL_2904371, EPI_ISL_2904400, EPI_ISL_2904410 | | | |
| see above | Respiratory Virus Unit, Microbiology Services Colindale, Public Health England | COVID-19 Genomics UK (COG-UK) Consortium | PHE Covid Sequencing Team |
| EPI_ISL_2900755, EPI_ISL_2900777, EPI_ISL_2900781, EPI_ISL_2900782, EPI_ISL_2900783, EPI_ISL_2900792, EPI_ISL_2900793, EPI_ISL_2900794, EPI_ISL_2900795, EPI_ISL_2900796, EPI_ISL_2900797, EPI_ISL_2900799, EPI_ISL_2900800, EPI_ISL_2900802, EPI_ISL_2900803, EPI_ISL_2900804, EPI_ISL_2900805, EPI_ISL_2900806, EPI_ISL_2900807, EPI_ISL_2900808, EPI_ISL_2900810, EPI_ISL_2900812, EPI_ISL_2900813, EPI_ISL_2900815, EPI_ISL_2900817, EPI_ISL_2900819, EPI_ISL_2900822, EPI_ISL_2900823, EPI_ISL_2900824, EPI_ISL_2900825, EPI_ISL_2900826, EPI_ISL_2900827, EPI_ISL_2900828, EPI_ISL_2900829, EPI_ISL_2900831, EPI_ISL_2900832, EPI_ISL_2900833, EPI_ISL_2900834, EPI_ISL_2900835, EPI_ISL_2900840, EPI_ISL_2900841, EPI_ISL_2900844 | | | |
| see above | Virology Department, Royal Infirmary of Edinburgh, NHS Lothian / School of Biological Sciences, University of Edinburgh | COVID-19 Genomics UK (COG-UK) Consortium | Colquhoun R; Cotton S; Dewar R; Gallagher A; Hill V; Jackson B; Maloney D; McCrone JT; McHugh M; O'Toole Á; Rambaut A; Rooke S; Scher E; Templeton K; Yu X |
| EPI_ISL_2819920, EPI_ISL_2819925, EPI_ISL_2819930, EPI_ISL_2819932, EPI_ISL_2819933, EPI_ISL_2819936, EPI_ISL_2819956, EPI_ISL_2819961, EPI_ISL_2819964, EPI_ISL_2819967, EPI_ISL_2819969, EPI_ISL_2819971, EPI_ISL_2819974, EPI_ISL_2819975, EPI_ISL_2819976, EPI_ISL_2819988, EPI_ISL_2819991, EPI_ISL_2819992, EPI_ISL_2819994, EPI_ISL_2819995, EPI_ISL_2819997, EPI_ISL_2820001, EPI_ISL_2820005, EPI_ISL_2820007, EPI_ISL_2820008, EPI_ISL_2820010, EPI_ISL_2820012, EPI_ISL_2820014, EPI_ISL_2820015, EPI_ISL_2820016, EPI_ISL_2820021, EPI_ISL_2820025, EPI_ISL_2820027, EPI_ISL_2820028, EPI_ISL_2820031, EPI_ISL_2820035, EPI_ISL_2820036, EPI_ISL_2820037, EPI_ISL_2820040, EPI_ISL_2820044, EPI_ISL_2820047, EPI_ISL_2820048, EPI_ISL_2820050, EPI_ISL_2820051, EPI_ISL_2820052, EPI_ISL_2820055, EPI_ISL_2820056, EPI_ISL_2820057, EPI_ISL_2820060, EPI_ISL_2820065, EPI_ISL_2820072, EPI_ISL_2820074, EPI_ISL_2820076, EPI_ISL_2820084, EPI_ISL_2820088, EPI_ISL_2820089, EPI_ISL_2820097, EPI_ISL_2820100, EPI_ISL_2820101, EPI_ISL_2820102, EPI_ISL_2820104, EPI_ISL_2820105, EPI_ISL_2820107, EPI_ISL_2820108, EPI_ISL_2820112, EPI_ISL_2820113, EPI_ISL_2820119, EPI_ISL_2820121, EPI_ISL_2820122, EPI_ISL_2820123, EPI_ISL_2820125, EPI_ISL_2820130, EPI_ISL_2820131, EPI_ISL_2820133, EPI_ISL_2820134, EPI_ISL_2820144, EPI_ISL_2820145, EPI_ISL_2820146, EPI_ISL_2820153, EPI_ISL_2820155, EPI_ISL_2820158, EPI_ISL_2820172, EPI_ISL_2820175, EPI_ISL_2820178, EPI_ISL_2820181, EPI_ISL_2820183, EPI_ISL_2820186, EPI_ISL_2820187, EPI_ISL_2820192, EPI_ISL_2820193, EPI_ISL_2820197, EPI_ISL_2820198, EPI_ISL_2820202, EPI_ISL_2820203, EPI_ISL_2820212, EPI_ISL_2820213, EPI_ISL_2820216, EPI_ISL_2820218, EPI_ISL_2820220, EPI_ISL_2820222, EPI_ISL_2907050, EPI_ISL_2907074, EPI_ISL_2907076, EPI_ISL_2907084, EPI_ISL_2907102, EPI_ISL_2907110, EPI_ISL_2907122, EPI_ISL_2907126, EPI_ISL_2907129, EPI_ISL_2907146, EPI_ISL_2907147, EPI_ISL_2907148, EPI_ISL_2907159, EPI_ISL_2907201, EPI_ISL_2907211, EPI_ISL_2907212, EPI_ISL_2907226, EPI_ISL_2907230, EPI_ISL_2907235, EPI_ISL_2907255, EPI_ISL_2907267, EPI_ISL_2907270, EPI_ISL_2907311, EPI_ISL_2907319, EPI_ISL_2907357, EPI_ISL_2907362, EPI_ISL_2907365, EPI_ISL_2907366, EPI_ISL_2907367, EPI_ISL_2907373, EPI_ISL_2907374, EPI_ISL_2907376, EPI_ISL_2907379, EPI_ISL_2907392, EPI_ISL_2907395, EPI_ISL_2907398, EPI_ISL_2907420, EPI_ISL_2907431, EPI_ISL_2907432, EPI_ISL_2907442, EPI_ISL_2907448, EPI_ISL_2907450, EPI_ISL_2907452, EPI_ISL_2907469, EPI_ISL_2907478 | | | |
| see above | Virology Department, Sheffield Teaching Hospitals NHS Foundation Trust/Department of Infection, Immunity and Cardiovascular Disease, The Medical School, University of Sheffield | COVID-19 Genomics UK (COG-UK) Consortium | Adri Angyal; Alex Keeley; Benjamin Lindsey; Cariad Evans; Danielle Groves; Dave Partridge; Katie Johnson; Laura Carrilero; Luke Green; Matthew Parker; Matthew Wyles; Mehmet Yavuz; Mohammad Raza; Nikki Smith; Paul Parsons; Rachel Tucker; Rebecca Brown; Thushan de Silva |

We gratefully acknowledge the following Authors from the Originating laboratories responsible for obtaining the specimens, as well as the Submitting laboratories where the genome data were generated and shared via GISAID, on which this research is based.

All Submitters of data may be contacted directly via www.gisaid.org

Authors are sorted alphabetically.

| Accession ID | Originating Laboratory | Submitting Laboratory | Authors |
|---|---|---|---|
| EPI_ISL_2846415, EPI_ISL_2846546, EPI_ISL_2864790, EPI_ISL_2864802, EPI_ISL_2864841, EPI_ISL_2864864, EPI_ISL_2864919, EPI_ISL_2864992, EPI_ISL_2865029, EPI_ISL_2865060, EPI_ISL_2865061, EPI_ISL_2865068, EPI_ISL_2865076, EPI_ISL_2865077, EPI_ISL_2865082, EPI_ISL_2865085, EPI_ISL_2865088, EPI_ISL_2865097, EPI_ISL_2865100, EPI_ISL_2865102, EPI_ISL_2865106, EPI_ISL_2865109, EPI_ISL_2865130, EPI_ISL_2865134, EPI_ISL_2865138, EPI_ISL_2865139, EPI_ISL_2865143, EPI_ISL_2865147, EPI_ISL_2865149, EPI_ISL_2865154, EPI_ISL_2865159, EPI_ISL_2865160, EPI_ISL_2865161, EPI_ISL_2865162, EPI_ISL_2865165, EPI_ISL_2865170, EPI_ISL_2865174, EPI_ISL_2865177, EPI_ISL_2865182, EPI_ISL_2865184, EPI_ISL_2865186, EPI_ISL_2865189, EPI_ISL_2865190, EPI_ISL_2865193, EPI_ISL_2865199, EPI_ISL_2865203, EPI_ISL_2865211, EPI_ISL_2865215, EPI_ISL_2865216, EPI_ISL_2865219, EPI_ISL_2865223, EPI_ISL_2865229, EPI_ISL_2865233, EPI_ISL_2865238, EPI_ISL_2865241, EPI_ISL_2865256, EPI_ISL_2865264, EPI_ISL_2865269, EPI_ISL_2865276, EPI_ISL_2865287, EPI_ISL_2865296, EPI_ISL_2865300, EPI_ISL_2865304, EPI_ISL_2865308, EPI_ISL_2865314, EPI_ISL_2865319, EPI_ISL_2865322, EPI_ISL_2865340, EPI_ISL_2865345, EPI_ISL_2865351, EPI_ISL_2865353, EPI_ISL_2865354, EPI_ISL_2865355, EPI_ISL_2865359, EPI_ISL_2867030, EPI_ISL_2867035, EPI_ISL_2867043, EPI_ISL_2867045, EPI_ISL_2867048, EPI_ISL_2867056, EPI_ISL_2867064, EPI_ISL_2867072, EPI_ISL_2867089, EPI_ISL_2867090, EPI_ISL_2867095, EPI_ISL_2867099, EPI_ISL_2867117, EPI_ISL_2867118, EPI_ISL_2867119, EPI_ISL_2867121, EPI_ISL_2867138, EPI_ISL_2867142, EPI_ISL_2867144, EPI_ISL_2867148, EPI_ISL_2867154, EPI_ISL_2867157, EPI_ISL_2867158, EPI_ISL_2867182, EPI_ISL_2867183, EPI_ISL_2867184, EPI_ISL_2867189, EPI_ISL_2867192, EPI_ISL_2867200, EPI_ISL_2867213, EPI_ISL_2867217, EPI_ISL_2867218, EPI_ISL_2867220, EPI_ISL_2867222, EPI_ISL_2867224, EPI_ISL_2867226, EPI_ISL_2867227, EPI_ISL_2867228, EPI_ISL_2867229, EPI_ISL_2867230, EPI_ISL_2867236, EPI_ISL_2867238, EPI_ISL_2867242, EPI_ISL_2867245, EPI_ISL_2867249, EPI_ISL_2867250, EPI_ISL_2867260, EPI_ISL_2867273, EPI_ISL_2867286, EPI_ISL_2867289, EPI_ISL_2867291, EPI_ISL_2867293, EPI_ISL_2867298, EPI_ISL_2867303, EPI_ISL_2867307, EPI_ISL_2867308, EPI_ISL_2867310, EPI_ISL_2867314, EPI_ISL_2867320, EPI_ISL_2867326, EPI_ISL_2867329, EPI_ISL_2867342, EPI_ISL_2867345, EPI_ISL_2867347, EPI_ISL_2867349, EPI_ISL_2867375, EPI_ISL_2867390, EPI_ISL_2867416, EPI_ISL_2867419, EPI_ISL_2867435, EPI_ISL_2867453, EPI_ISL_2867463, EPI_ISL_2867478, EPI_ISL_2867487, EPI_ISL_2867488, EPI_ISL_2867491, EPI_ISL_2867511, EPI_ISL_2867517, EPI_ISL_2867537, EPI_ISL_2867554, EPI_ISL_2867572, EPI_ISL_2867583, EPI_ISL_2867584, EPI_ISL_2867596, EPI_ISL_2867627, EPI_ISL_2867640, EPI_ISL_2867643, EPI_ISL_2867645, EPI_ISL_2867651, EPI_ISL_2867656, EPI_ISL_2867661, EPI_ISL_2867663, EPI_ISL_2867670, EPI_ISL_2867691, EPI_ISL_2913634, EPI_ISL_2913733, EPI_ISL_2913771, EPI_ISL_2913792, EPI_ISL_2913837, EPI_ISL_2913911, EPI_ISL_2914431, EPI_ISL_2914624, EPI_ISL_2914629, EPI_ISL_2914650, EPI_ISL_2914651, EPI_ISL_2914654, EPI_ISL_2914659, EPI_ISL_2914660, EPI_ISL_2914661, EPI_ISL_2914662, EPI_ISL_2914670, EPI_ISL_2914671, EPI_ISL_2914672, EPI_ISL_2914674, EPI_ISL_2914677, EPI_ISL_2914680, EPI_ISL_2914683, EPI_ISL_2914684, EPI_ISL_2914688, EPI_ISL_2914691, EPI_ISL_2914694, EPI_ISL_2914698, EPI_ISL_2914701, EPI_ISL_2914705, EPI_ISL_2914706, EPI_ISL_2914715, EPI_ISL_2914721, EPI_ISL_2914730, EPI_ISL_2914740, EPI_ISL_2914741, EPI_ISL_2914750, EPI_ISL_2914754, EPI_ISL_2914755, EPI_ISL_2914771, EPI_ISL_2914796, EPI_ISL_2914799, EPI_ISL_2914801, EPI_ISL_2914813, EPI_ISL_2914814, EPI_ISL_2914818, EPI_ISL_2914819, EPI_ISL_2914822, EPI_ISL_2914827, EPI_ISL_2914842, EPI_ISL_2914844, EPI_ISL_2914851, EPI_ISL_2914864, EPI_ISL_2914872, EPI_ISL_2914878, EPI_ISL_2914886, EPI_ISL_2914889, EPI_ISL_2914890, EPI_ISL_2914891, EPI_ISL_2914908, EPI_ISL_2914920, EPI_ISL_2914924, EPI_ISL_2914944, EPI_ISL_2914946, EPI_ISL_2914948, EPI_ISL_2914961, EPI_ISL_2915688, EPI_ISL_2915689, EPI_ISL_2915690, EPI_ISL_2915735, EPI_ISL_2915739, EPI_ISL_2915766, EPI_ISL_2915801, EPI_ISL_2915809, EPI_ISL_2915823, EPI_ISL_2915829, EPI_ISL_2915852, EPI_ISL_2915854, EPI_ISL_2915862, EPI_ISL_2915886, EPI_ISL_2915896, EPI_ISL_2915906, EPI_ISL_2915908, EPI_ISL_2915921, EPI_ISL_2915968, EPI_ISL_2915981, EPI_ISL_2916010, EPI_ISL_2916672, EPI_ISL_2916674, EPI_ISL_2916676, EPI_ISL_2916677, EPI_ISL_2916678, EPI_ISL_2916682, EPI_ISL_2916683, EPI_ISL_2916687, EPI_ISL_2916689, EPI_ISL_2916690, EPI_ISL_2916692, EPI_ISL_2916695, EPI_ISL_2916697, EPI_ISL_2916699, EPI_ISL_2916702, EPI_ISL_2916704, EPI_ISL_2916707, EPI_ISL_2916708, EPI_ISL_2916711, EPI_ISL_2916717, EPI_ISL_2916718, EPI_ISL_2916720, EPI_ISL_2916722, EPI_ISL_2916724, EPI_ISL_2916727, EPI_ISL_2916729, EPI_ISL_2916735, EPI_ISL_2916736, EPI_ISL_2916737, EPI_ISL_2916738, EPI_ISL_2916741, EPI_ISL_2916749, EPI_ISL_2916754, EPI_ISL_2916755, EPI_ISL_2916761, EPI_ISL_2916767, EPI_ISL_2916771, EPI_ISL_2916774, EPI_ISL_2916778, EPI_ISL_2916780, EPI_ISL_2916781, EPI_ISL_2916782, EPI_ISL_2916784, EPI_ISL_2916785, EPI_ISL_2916786, EPI_ISL_2916788, EPI_ISL_2916791, EPI_ISL_2916793, EPI_ISL_2916795, EPI_ISL_2916797, EPI_ISL_2916799, EPI_ISL_2916801, EPI_ISL_2916802, EPI_ISL_2916807, EPI_ISL_2916809, EPI_ISL_2916810, EPI_ISL_2916814, EPI_ISL_2916815, EPI_ISL_2916817, EPI_ISL_2916820, EPI_ISL_2916824, EPI_ISL_2916828, EPI_ISL_2916830, EPI_ISL_2916831, EPI_ISL_2916832, EPI_ISL_2916837, EPI_ISL_2916843, EPI_ISL_2916844, EPI_ISL_2916846, EPI_ISL_2916848, EPI_ISL_2916850, EPI_ISL_2916856, EPI_ISL_2916857, EPI_ISL_2916859, EPI_ISL_2916863, EPI_ISL_2916866, EPI_ISL_2916868, EPI_ISL_2916869, EPI_ISL_2916871, EPI_ISL_2916873, EPI_ISL_2916876, EPI_ISL_2916877, EPI_ISL_2916878, EPI_ISL_2916879, EPI_ISL_2916880, EPI_ISL_2916881, EPI_ISL_2916882, EPI_ISL_2916886, EPI_ISL_2916887, EPI_ISL_2916888, EPI_ISL_2916890, EPI_ISL_2916891, EPI_ISL_2916892, EPI_ISL_2916894, EPI_ISL_2916895, EPI_ISL_2916901, EPI_ISL_2916902, EPI_ISL_2916904, EPI_ISL_2916908, EPI_ISL_2916910, EPI_ISL_2916913, EPI_ISL_2916914, EPI_ISL_2916920, EPI_ISL_2916921, EPI_ISL_2916924, EPI_ISL_2916925, EPI_ISL_2916926, EPI_ISL_2916927, EPI_ISL_2916930, EPI_ISL_2916931, EPI_ISL_2916933, EPI_ISL_2916939, EPI_ISL_2916940, EPI_ISL_2916941, EPI_ISL_2916944, EPI_ISL_2916945, EPI_ISL_2916948, EPI_ISL_2916949, EPI_ISL_2916952, EPI_ISL_2916953, EPI_ISL_2916954, EPI_ISL_2916957, EPI_ISL_2916958, EPI_ISL_2916959, EPI_ISL_2916961, EPI_ISL_2916967, EPI_ISL_2916969, EPI_ISL_2916970, EPI_ISL_2916973, EPI_ISL_2916976, EPI_ISL_2916979, EPI_ISL_2916981, EPI_ISL_2916983, EPI_ISL_2916984, EPI_ISL_2916985, EPI_ISL_2916986, EPI_ISL_2916988, EPI_ISL_2916989, EPI_ISL_2916990, EPI_ISL_2916991, EPI_ISL_2916992, EPI_ISL_2916994, EPI_ISL_2916998, EPI_ISL_2916999, EPI_ISL_2917000, EPI_ISL_2917001, EPI_ISL_2917002, EPI_ISL_2917003, EPI_ISL_2917007, EPI_ISL_2917009, EPI_ISL_2917012, EPI_ISL_2917013, EPI_ISL_2917022, EPI_ISL_2917045, EPI_ISL_2917066, EPI_ISL_2917078, EPI_ISL_2917080, EPI_ISL_2917082, EPI_ISL_2917091, EPI_ISL_2917093, EPI_ISL_2917103, EPI_ISL_2917111, EPI_ISL_2917129, EPI_ISL_2917138, EPI_ISL_2917141, EPI_ISL_2917147, EPI_ISL_2917170, EPI_ISL_2917182, EPI_ISL_2917183, EPI_ISL_2917198, EPI_ISL_2917202, EPI_ISL_2917244, EPI_ISL_2917256, EPI_ISL_2917261, EPI_ISL_2917294, EPI_ISL_2917301, EPI_ISL_2917321, EPI_ISL_2917324, EPI_ISL_2917483, EPI_ISL_2917506, EPI_ISL_2917616, EPI_ISL_2917695, EPI_ISL_2917701, EPI_ISL_2917719, EPI_ISL_2917791, EPI_ISL_2917862, EPI_ISL_2917865, EPI_ISL_2917945, EPI_ISL_2918070, EPI_ISL_2918075, EPI_ISL_2918084, EPI_ISL_2918114, EPI_ISL_2918141, EPI_ISL_2918145, EPI_ISL_2918148, EPI_ISL_2918149, EPI_ISL_2918169, EPI_ISL_2918186, EPI_ISL_2918192, EPI_ISL_2918200, EPI_ISL_2918205, EPI_ISL_2918206, EPI_ISL_2918214, EPI_ISL_2918216, EPI_ISL_2918234, EPI_ISL_2918236, EPI_ISL_2918248, EPI_ISL_2918295, EPI_ISL_2918299, EPI_ISL_2918300, EPI_ISL_2918302, EPI_ISL_2918313, EPI_ISL_2918326, EPI_ISL_2918350, EPI_ISL_2918354, EPI_ISL_2918370, EPI_ISL_2918404, EPI_ISL_2918409, EPI_ISL_2918417, EPI_ISL_2918419, EPI_ISL_2918439, EPI_ISL_2918456, EPI_ISL_2918459, EPI_ISL_2918466, EPI_ISL_2918471, EPI_ISL_2918476, EPI_ISL_2918482, EPI_ISL_2918498, EPI_ISL_2918508, EPI_ISL_2918516, EPI_ISL_2918520, EPI_ISL_2918522, EPI_ISL_2918524, EPI_ISL_2918535, EPI_ISL_2918540, EPI_ISL_2918542, EPI_ISL_2918568, EPI_ISL_2918572, EPI_ISL_2918592, EPI_ISL_2918593, EPI_ISL_2918595 | | | |
| see above | Berkshire and Surrey Pathology Services Lighthouse Laboratory | Wellcome Sanger Institute for the COVID-19 Genomics UK (COG-UK) Consortium | Berkshire and Surrey Pathology Services Lighthouse Laboratory and Alex Alderton; Cordelia Langford; David K. Jackson; Dominic Kwiatkowski; Ewan Harrison; Ian Johnston; Jeffrey Barrett; John Sillitoe on behalf of the Wellcome Sanger Institute COVID-19 Surveillance Team; Roberto Amato; Sonia Goncalves |
| EPI_ISL_2815735, EPI_ISL_2815736, EPI_ISL_2815737, EPI_ISL_2815738, EPI_ISL_2815739, EPI_ISL_2815740, EPI_ISL_2815741, EPI_ISL_2815742, EPI_ISL_2815743, EPI_ISL_2900469, EPI_ISL_2900472, EPI_ISL_2900500 | | | |
| see above | Department of Pathology, University of Cambridge | COVID-19 Genomics UK (COG-UK) Consortium | Aminu S. Jahun; Ian Goodfellow; Iliana Georgana; Martin D. Curran; Myra Hosmillo; Rhys Izuagbe; Surendra Parmar; William L. Hamilton; Yasmin Chaudhry |
| EPI_ISL_2918077, EPI_ISL_2918124, EPI_ISL_2918164, EPI_ISL_2918183, EPI_ISL_2918337 | Health Services Laboratories | Wellcome Sanger Institute for the COVID-19 Genomics UK (COG-UK) Consortium | Cordelia Langford; David K. Jackson; Dominic Kwiatkowski; Ewan Harrison; Health Services Laboratories and Alex Alderton; Ian Johnston; Jeffrey Barrett; John Sillitoe on behalf of the Wellcome Sanger Institute COVID-19 Surveillance Team; Roberto Amato; Sonia Goncalves |
| EPI_ISL_2907579, EPI_ISL_2907580, EPI_ISL_2907586, EPI_ISL_2907587, EPI_ISL_2907595, EPI_ISL_2907596, EPI_ISL_2907597, EPI_ISL_2907599, EPI_ISL_2907609, EPI_ISL_2907612, EPI_ISL_2907619, EPI_ISL_2907621, EPI_ISL_2907627, EPI_ISL_2907630, EPI_ISL_2907631, EPI_ISL_2907634, EPI_ISL_2907636, EPI_ISL_2907645, EPI_ISL_2907651, EPI_ISL_2907652, EPI_ISL_2907653, EPI_ISL_2907658, EPI_ISL_2907661, EPI_ISL_2907671, EPI_ISL_2907676, EPI_ISL_2907690, EPI_ISL_2907691, EPI_ISL_2907694, EPI_ISL_2907707, EPI_ISL_2907712, EPI_ISL_2907713, EPI_ISL_2907720, EPI_ISL_2907724, EPI_ISL_2907732, EPI_ISL_2907733, EPI_ISL_2907736, EPI_ISL_2907743, EPI_ISL_2907745, EPI_ISL_2907746, EPI_ISL_2907751, EPI_ISL_2907777, EPI_ISL_2907780, EPI_ISL_2907781, EPI_ISL_2907782, EPI_ISL_2907783, EPI_ISL_2907787, EPI_ISL_2907789, EPI_ISL_2907798, EPI_ISL_2907799, EPI_ISL_2907807, EPI_ISL_2907811, EPI_ISL_2907821, EPI_ISL_2907823, EPI_ISL_2907833, EPI_ISL_2907836, EPI_ISL_2907838, EPI_ISL_2907839, EPI_ISL_2907843, EPI_ISL_2907844, EPI_ISL_2907849, EPI_ISL_2907851, EPI_ISL_2907853, EPI_ISL_2907855, EPI_ISL_2907867, EPI_ISL_2907882, EPI_ISL_2907884, EPI_ISL_2907893, EPI_ISL_2907908, EPI_ISL_2907909, EPI_ISL_2907911, EPI_ISL_2907914, EPI_ISL_2907915, EPI_ISL_2907917, EPI_ISL_2907918, EPI_ISL_2907925, EPI_ISL_2907927, EPI_ISL_2907928, EPI_ISL_2907940, EPI_ISL_2909550, EPI_ISL_2909552, EPI_ISL_2909561, EPI_ISL_2909562, EPI_ISL_2909564, EPI_ISL_2909565, EPI_ISL_2909569, EPI_ISL_2909573, EPI_ISL_2909574, EPI_ISL_2909580, EPI_ISL_2909582, EPI_ISL_2909583, EPI_ISL_2909586, EPI_ISL_2909597, EPI_ISL_2909601, EPI_ISL_2909612, EPI_ISL_2909613, EPI_ISL_2909615, EPI_ISL_2909620, EPI_ISL_2909622, EPI_ISL_2909623, EPI_ISL_2909626, EPI_ISL_2909628, EPI_ISL_2909630, EPI_ISL_2909632, EPI_ISL_2909636, EPI_ISL_2909638, EPI_ISL_2909639, EPI_ISL_2909641, EPI_ISL_2909643, EPI_ISL_2909644, EPI_ISL_2909645, EPI_ISL_2909649, EPI_ISL_2909654, EPI_ISL_2909656, EPI_ISL_2909660, EPI_ISL_2909664, EPI_ISL_2909669, EPI_ISL_2909671, EPI_ISL_2909672, EPI_ISL_2909674, EPI_ISL_2909675, EPI_ISL_2909680, EPI_ISL_2909681, EPI_ISL_2909682, EPI_ISL_2909686, EPI_ISL_2909687, EPI_ISL_2909688, EPI_ISL_2909690, EPI_ISL_2909691, EPI_ISL_2909692, EPI_ISL_2909693, EPI_ISL_2909694, EPI_ISL_2909698, EPI_ISL_2909701, EPI_ISL_2909705, EPI_ISL_2909706, EPI_ISL_2909709, EPI_ISL_2909711, EPI_ISL_2909714, EPI_ISL_2909716, EPI_ISL_2909717, EPI_ISL_2909721, EPI_ISL_2909723, EPI_ISL_2909725, EPI_ISL_2909731, EPI_ISL_2909736, EPI_ISL_2909740, EPI_ISL_2909743, EPI_ISL_2909745, EPI_ISL_2909746, EPI_ISL_2909749, EPI_ISL_2909751, EPI_ISL_2909754, EPI_ISL_2909756, EPI_ISL_2909760, EPI_ISL_2909764, EPI_ISL_2909765, EPI_ISL_2909766, EPI_ISL_2909767, EPI_ISL_2909768, EPI_ISL_2909773, EPI_ISL_2909775, EPI_ISL_2909777, EPI_ISL_2909778, EPI_ISL_2909780, EPI_ISL_2909781, EPI_ISL_2909782, EPI_ISL_2909783, EPI_ISL_2909785, EPI_ISL_2909786, EPI_ISL_2909789, EPI_ISL_2909791, EPI_ISL_2909795, EPI_ISL_2909797, EPI_ISL_2909801, EPI_ISL_2909803, EPI_ISL_2909807, EPI_ISL_2909808, EPI_ISL_2909812, EPI_ISL_2909819, EPI_ISL_2909820, EPI_ISL_2909825, EPI_ISL_2909827, EPI_ISL_2909830, EPI_ISL_2909834, EPI_ISL_2909835, EPI_ISL_2909839, EPI_ISL_2909842, EPI_ISL_2909843, EPI_ISL_2909857, EPI_ISL_2909858, EPI_ISL_2909861, EPI_ISL_2909863, EPI_ISL_2909870, EPI_ISL_2909877, EPI_ISL_2909879, EPI_ISL_2909881, EPI_ISL_2909886, EPI_ISL_2909891, EPI_ISL_2909893, EPI_ISL_2909896, EPI_ISL_2909897, EPI_ISL_2909904, EPI_ISL_2909906, EPI_ISL_2909907, EPI_ISL_2909911, EPI_ISL_2909919, EPI_ISL_2909920, EPI_ISL_2909921, EPI_ISL_2909926, EPI_ISL_2909930, EPI_ISL_2909935, EPI_ISL_2909936, EPI_ISL_2909955, EPI_ISL_2909963, EPI_ISL_2909966, EPI_ISL_2909970, EPI_ISL_2909972, EPI_ISL_2909982, EPI_ISL_2909989, EPI_ISL_2909990, EPI_ISL_2909991, EPI_ISL_2909993, EPI_ISL_2909995, EPI_ISL_2909996, EPI_ISL_2910002, EPI_ISL_2910004, EPI_ISL_2910012, EPI_ISL_2910013, EPI_ISL_2910018, EPI_ISL_2910025, EPI_ISL_2910032, EPI_ISL_2910033, EPI_ISL_2910038, EPI_ISL_2910039, EPI_ISL_2910041, EPI_ISL_2910059, EPI_ISL_2910060, EPI_ISL_2910063, EPI_ISL_2910064, EPI_ISL_2910066, EPI_ISL_2910067, EPI_ISL_2910069, EPI_ISL_2910071, EPI_ISL_2910072, EPI_ISL_2910074, EPI_ISL_2910075, EPI_ISL_2910077, EPI_ISL_2910079, EPI_ISL_2910084, EPI_ISL_2910085, EPI_ISL_2910090, EPI_ISL_2910091, EPI_ISL_2910094, EPI_ISL_2910097, EPI_ISL_2910098, EPI_ISL_2910113, EPI_ISL_2910115, EPI_ISL_2910119, EPI_ISL_2910121, EPI_ISL_2910122, EPI_ISL_2910126, EPI_ISL_2910131, EPI_ISL_2910134, EPI_ISL_2910135, EPI_ISL_2910137, EPI_ISL_2910145, EPI_ISL_2910148, EPI_ISL_2910151, EPI_ISL_2910157, EPI_ISL_2910165, EPI_ISL_2910166, EPI_ISL_2910176, EPI_ISL_2910177, EPI_ISL_2910178, EPI_ISL_2910182, EPI_ISL_2910184, EPI_ISL_2910186, EPI_ISL_2910190, EPI_ISL_2910191, EPI_ISL_2910195, EPI_ISL_2910201, EPI_ISL_2910211, EPI_ISL_2910218, EPI_ISL_2910219, EPI_ISL_2910223, EPI_ISL_2910225, EPI_ISL_2910230, EPI_ISL_2910234, EPI_ISL_2910237, EPI_ISL_2910238, EPI_ISL_2910241, EPI_ISL_2910252, EPI_ISL_2910290, EPI_ISL_2910291, EPI_ISL_2910300, EPI_ISL_2910322, EPI_ISL_2910342, EPI_ISL_2910349, EPI_ISL_2910408, EPI_ISL_2910487, EPI_ISL_2910518, EPI_ISL_2910529, EPI_ISL_2911764, EPI_ISL_2913326, EPI_ISL_2913398, EPI_ISL_2913446, EPI_ISL_2913458, EPI_ISL_2913519, EPI_ISL_2913553, EPI_ISL_2913578, EPI_ISL_2915333, EPI_ISL_2915334, EPI_ISL_2915341, EPI_ISL_2915345, EPI_ISL_2915346, EPI_ISL_2915351, EPI_ISL_2915353, EPI_ISL_2915354, EPI_ISL_2915356, EPI_ISL_2915360, EPI_ISL_2915365, EPI_ISL_2915369, EPI_ISL_2915372, EPI_ISL_2915374, EPI_ISL_2915379, EPI_ISL_2915383, EPI_ISL_2915384, EPI_ISL_2915386, EPI_ISL_2915389, EPI_ISL_2915394, EPI_ISL_2915396, EPI_ISL_2915398, EPI_ISL_2915404, EPI_ISL_2915406, EPI_ISL_2915410, EPI_ISL_2915411, EPI_ISL_2915413, EPI_ISL_2915417, EPI_ISL_2915418, EPI_ISL_2915425, EPI_ISL_2915426, EPI_ISL_2915433, EPI_ISL_2915439, EPI_ISL_2915440, EPI_ISL_2915444, EPI_ISL_2915447, EPI_ISL_2915450, EPI_ISL_2915451, EPI_ISL_2915454, EPI_ISL_2915462, EPI_ISL_2915467, EPI_ISL_2915471, EPI_ISL_2915474, EPI_ISL_2915478, EPI_ISL_2915480, EPI_ISL_2915487, EPI_ISL_2915494, EPI_ISL_2915498, EPI_ISL_2915503, EPI_ISL_2915509, EPI_ISL_2915510, EPI_ISL_2915516, EPI_ISL_2915527, EPI_ISL_2915535, EPI_ISL_2915536, EPI_ISL_2915538, EPI_ISL_2915539, EPI_ISL_2915540, EPI_ISL_2915545, EPI_ISL_2915547, EPI_ISL_2915548, EPI_ISL_2915553, EPI_ISL_2915555, EPI_ISL_2915559, EPI_ISL_2915562, EPI_ISL_2915565, EPI_ISL_2915568, EPI_ISL_2915569, EPI_ISL_2915571, EPI_ISL_2915576, EPI_ISL_2915591, EPI_ISL_2915595, EPI_ISL_2915598, EPI_ISL_2915601, EPI_ISL_2915610, EPI_ISL_2915622, EPI_ISL_2915628, EPI_ISL_2915632, EPI_ISL_2915633, EPI_ISL_2915634, EPI_ISL_2915638, EPI_ISL_2915649, EPI_ISL_2915650, EPI_ISL_2915658, EPI_ISL_2915661, EPI_ISL_2915663, EPI_ISL_2915666, EPI_ISL_2915669, EPI_ISL_2915674, EPI_ISL_2915677, EPI_ISL_2915678, EPI_ISL_2915684, EPI_ISL_2915686, EPI_ISL_2915698, EPI_ISL_2915702, EPI_ISL_2915709, EPI_ISL_2915733, EPI_ISL_2915800, EPI_ISL_2915815, EPI_ISL_2915816, EPI_ISL_2915817, EPI_ISL_2915832, EPI_ISL_2915876, EPI_ISL_2915901, EPI_ISL_2915971, EPI_ISL_2915976, EPI_ISL_2916008, EPI_ISL_2916038, EPI_ISL_2916049, EPI_ISL_2916059, EPI_ISL_2916063, EPI_ISL_2916069, EPI_ISL_2916070, EPI_ISL_2916073, EPI_ISL_2916077, EPI_ISL_2916080, EPI_ISL_2916099, EPI_ISL_2916125, EPI_ISL_2916130, EPI_ISL_2916131, EPI_ISL_2916139, EPI_ISL_2916144, EPI_ISL_2916147, EPI_ISL_2916153, EPI_ISL_2916164, EPI_ISL_2916165, EPI_ISL_2916172, EPI_ISL_2916173, EPI_ISL_2916177, EPI_ISL_2916189, EPI_ISL_2916197, EPI_ISL_2916199, EPI_ISL_2916213, EPI_ISL_2916214, EPI_ISL_2916223, EPI_ISL_2916232, EPI_ISL_2916248, EPI_ISL_2916250, EPI_ISL_2916257, EPI_ISL_2916269, EPI_ISL_2916270, EPI_ISL_2916277, EPI_ISL_2916282, EPI_ISL_2916292, EPI_ISL_2916303, EPI_ISL_2916311, EPI_ISL_2916312, EPI_ISL_2916314, EPI_ISL_2916315, EPI_ISL_2916319, EPI_ISL_2916327, EPI_ISL_2916336, EPI_ISL_2916337, EPI_ISL_2916338, EPI_ISL_2916339, EPI_ISL_2916341, EPI_ISL_2916342, EPI_ISL_2916355, EPI_ISL_2916365, EPI_ISL_2916371, EPI_ISL_2916372, EPI_ISL_2916375, EPI_ISL_2916376, EPI_ISL_2916378, EPI_ISL_2916379, EPI_ISL_2916380, EPI_ISL_2916391, EPI_ISL_2916394, EPI_ISL_2916399, EPI_ISL_2916400, EPI_ISL_2916401, EPI_ISL_2916409, EPI_ISL_2916417, EPI_ISL_2916427, EPI_ISL_2916432, EPI_ISL_2916438, EPI_ISL_2916446, EPI_ISL_2916453, EPI_ISL_2916456, EPI_ISL_2916461, EPI_ISL_2916462, EPI_ISL_2916464, EPI_ISL_2916465, EPI_ISL_2916473, EPI_ISL_2916477, EPI_ISL_2916482, EPI_ISL_2916484, EPI_ISL_2916485, EPI_ISL_2916486, EPI_ISL_2916491, EPI_ISL_2916493, EPI_ISL_2916496, EPI_ISL_2916501, EPI_ISL_2916503, EPI_ISL_2916509, EPI_ISL_2916510, EPI_ISL_2916512, EPI_ISL_2916513, EPI_ISL_2916527, EPI_ISL_2916532, EPI_ISL_2916536, EPI_ISL_2916548, EPI_ISL_2916550, EPI_ISL_2916551, EPI_ISL_2916556, EPI_ISL_2916563, EPI_ISL_2916568, EPI_ISL_2916570, EPI_ISL_2916574, EPI_ISL_2916576, EPI_ISL_2916577, EPI_ISL_2916580, EPI_ISL_2916596, EPI_ISL_2916603, EPI_ISL_2916606, EPI_ISL_2916608, EPI_ISL_2916610, EPI_ISL_2916615, EPI_ISL_2916620, EPI_ISL_2916622, EPI_ISL_2916626, EPI_ISL_2916627, EPI_ISL_2916631, EPI_ISL_2916634, EPI_ISL_2916640, EPI_ISL_2916650, EPI_ISL_2916652, EPI_ISL_2916657, EPI_ISL_2916661, EPI_ISL_2918069, EPI_ISL_2918102, EPI_ISL_2918135, EPI_ISL_2918170, EPI_ISL_2918215, EPI_ISL_2918259, EPI_ISL_2918274, EPI_ISL_2918312, EPI_ISL_2918355, EPI_ISL_2924317, EPI_ISL_2924506, EPI_ISL_2924642, EPI_ISL_2924689, EPI_ISL_2924707, EPI_ISL_2924740, EPI_ISL_2924757, EPI_ISL_2924793, EPI_ISL_2924806, EPI_ISL_2924830, EPI_ISL_2925136, EPI_ISL_2925146, EPI_ISL_2925248, EPI_ISL_2925325, EPI_ISL_2925348, EPI_ISL_2925393, EPI_ISL_2925401, EPI_ISL_2925449, EPI_ISL_2925472, EPI_ISL_2936749, EPI_ISL_2936796, EPI_ISL_2936917, EPI_ISL_2936994, EPI_ISL_2936997, EPI_ISL_2937157, EPI_ISL_2937158, EPI_ISL_2937160 | | | |
| see above | Lighthouse Lab in Alderley Park | Wellcome Sanger Institute for the COVID-19 Genomics UK (COG-UK) Consortium | Cordelia Langford; David K. Jackson; Dominic Kwiatkowski; Ewan Harrison; Ian Johnston; Jacquelyn Wynn; Jeffrey Barrett; John Sillitoe on behalf of the Wellcome Sanger Institute COVID-19 Surveillance Team; Mairead Hyland; Roberto Amato; Sonia Goncalves; The Lighthouse Lab in Alderley Park and Alex Alderton |
| EPI_ISL_2846361, EPI_ISL_2846362, EPI_ISL_2846365, EPI_ISL_2846371, EPI_ISL_2846375, EPI_ISL_2846384, EPI_ISL_2846385, EPI_ISL_2846387, EPI_ISL_2846397, EPI_ISL_2846403, EPI_ISL_2846405, EPI_ISL_2846410, EPI_ISL_2846414, EPI_ISL_2846417, EPI_ISL_2846419, EPI_ISL_2846424, EPI_ISL_2846425, EPI_ISL_2846428, EPI_ISL_2846430, EPI_ISL_2846434, EPI_ISL_2846443, EPI_ISL_2846446, EPI_ISL_2846450, EPI_ISL_2846451, EPI_ISL_2846454, EPI_ISL_2846456, EPI_ISL_2846460, EPI_ISL_2846463, EPI_ISL_2846465, EPI_ISL_2846473, EPI_ISL_2846485, EPI_ISL_2846491, EPI_ISL_2846494, EPI_ISL_2846498, EPI_ISL_2846500, EPI_ISL_2846503, EPI_ISL_2846509, EPI_ISL_2846515, EPI_ISL_2846521, EPI_ISL_2846524, EPI_ISL_2846533, EPI_ISL_2846534, EPI_ISL_2846538, EPI_ISL_2846539, EPI_ISL_2846543, EPI_ISL_2846556, EPI_ISL_2846569, EPI_ISL_2846574, EPI_ISL_2846575, EPI_ISL_2846576, EPI_ISL_2846578, EPI_ISL_2846580, EPI_ISL_2847289, EPI_ISL_2847295, EPI_ISL_2847296, EPI_ISL_2847299, EPI_ISL_2847302, EPI_ISL_2847303, EPI_ISL_2847309, EPI_ISL_2847310, EPI_ISL_2847314, EPI_ISL_2847318, EPI_ISL_2847327, EPI_ISL_2847331, EPI_ISL_2847342, EPI_ISL_2847349, EPI_ISL_2847364, EPI_ISL_2847365, EPI_ISL_2847380, EPI_ISL_2847381, EPI_ISL_2847385, EPI_ISL_2847387, EPI_ISL_2847400, EPI_ISL_2847410, EPI_ISL_2847415, EPI_ISL_2847420, EPI_ISL_2847440, EPI_ISL_2847442, EPI_ISL_2847443, EPI_ISL_2847461, EPI_ISL_2847466, EPI_ISL_2847469, EPI_ISL_2847471, EPI_ISL_2847482, EPI_ISL_2847484, EPI_ISL_2847488, EPI_ISL_2847505, EPI_ISL_2847507, EPI_ISL_2847532, EPI_ISL_2847543, EPI_ISL_2847548, EPI_ISL_2847554, EPI_ISL_2847582, EPI_ISL_2847584, EPI_ISL_2847587, EPI_ISL_2847599, EPI_ISL_2847601, EPI_ISL_2847607, EPI_ISL_2847617, EPI_ISL_2848657, EPI_ISL_2848666, EPI_ISL_2848667, EPI_ISL_2848670, EPI_ISL_2848682, EPI_ISL_2848691, EPI_ISL_2848692, EPI_ISL_2848699, EPI_ISL_2848706, EPI_ISL_2848714, EPI_ISL_2848715, EPI_ISL_2848717, EPI_ISL_2848724, EPI_ISL_2848727, EPI_ISL_2848728, EPI_ISL_2848729, EPI_ISL_2848732, EPI_ISL_2848735, EPI_ISL_2848736, EPI_ISL_2848754, EPI_ISL_2848760, EPI_ISL_2848761, EPI_ISL_2848763, EPI_ISL_2848766, EPI_ISL_2848767, EPI_ISL_2848772, EPI_ISL_2848783, EPI_ISL_2848796, EPI_ISL_2848799, EPI_ISL_2848802, EPI_ISL_2848805, EPI_ISL_2848808, EPI_ISL_2848809, EPI_ISL_2848811, EPI_ISL_2848816, EPI_ISL_2848821, EPI_ISL_2848828, EPI_ISL_2848833, EPI_ISL_2848835, EPI_ISL_2848838, EPI_ISL_2848839, EPI_ISL_2848842, EPI_ISL_2848845, EPI_ISL_2848846, EPI_ISL_2848855, EPI_ISL_2848860, EPI_ISL_2848869, EPI_ISL_2848870, EPI_ISL_2848877, EPI_ISL_2848884, EPI_ISL_2848886, EPI_ISL_2848888, EPI_ISL_2848891, EPI_ISL_2848892, EPI_ISL_2848896, EPI_ISL_2848917, EPI_ISL_2848918, EPI_ISL_2848922, EPI_ISL_2848923, EPI_ISL_2848924, EPI_ISL_2848928, EPI_ISL_2848930, EPI_ISL_2848933, EPI_ISL_2848949, EPI_ISL_2848955, EPI_ISL_2848957, EPI_ISL_2848965, EPI_ISL_2848966, EPI_ISL_2848969, EPI_ISL_2848993, EPI_ISL_2849003, EPI_ISL_2850939, EPI_ISL_2851175, EPI_ISL_2851176, EPI_ISL_2851177, EPI_ISL_2851178, EPI_ISL_2851179, EPI_ISL_2851180, EPI_ISL_2851181, EPI_ISL_2851182, EPI_ISL_2851183, EPI_ISL_2851185, EPI_ISL_2851186, EPI_ISL_2851187, EPI_ISL_2851188, EPI_ISL_2851190, EPI_ISL_2851191, EPI_ISL_2851192, EPI_ISL_2851193, EPI_ISL_2851194, EPI_ISL_2851195, EPI_ISL_2851196, EPI_ISL_2851197, EPI_ISL_2851198, EPI_ISL_2851199, EPI_ISL_2851200, EPI_ISL_2851201, EPI_ISL_2851202, EPI_ISL_2851203, EPI_ISL_2851204, EPI_ISL_2851205, EPI_ISL_2851206, EPI_ISL_2851207, EPI_ISL_2851208, EPI_ISL_2851209, EPI_ISL_2851210, EPI_ISL_2851211, EPI_ISL_2851212, EPI_ISL_2851213, EPI_ISL_2851214, EPI_ISL_2851215, EPI_ISL_2851216, EPI_ISL_2851217, EPI_ISL_2851218, EPI_ISL_2851219, EPI_ISL_2851220, EPI_ISL_2851221, EPI_ISL_2851222, EPI_ISL_2851223, EPI_ISL_2851224, EPI_ISL_2851225, EPI_ISL_2851226, EPI_ISL_2851227, EPI_ISL_2851228, EPI_ISL_2851229, EPI_ISL_2851230, EPI_ISL_2851231, EPI_ISL_2851232, EPI_ISL_2851233, EPI_ISL_2851234, EPI_ISL_2851235, EPI_ISL_2851236, EPI_ISL_2851237, EPI_ISL_2851238, EPI_ISL_2851239, EPI_ISL_2851240, EPI_ISL_2851241, EPI_ISL_2851242, EPI_ISL_2851243, EPI_ISL_2851246, EPI_ISL_2851247, EPI_ISL_2851248, EPI_ISL_2851249, EPI_ISL_2851250, EPI_ISL_2851251, EPI_ISL_2851252, EPI_ISL_2851253, EPI_ISL_2851254, EPI_ISL_2851255, EPI_ISL_2851256, EPI_ISL_2851257, EPI_ISL_2851258, EPI_ISL_2851259, EPI_ISL_2851260, EPI_ISL_2851261, EPI_ISL_2851262, EPI_ISL_2851263, EPI_ISL_2864898, EPI_ISL_2864988, EPI_ISL_2865010, EPI_ISL_2865403, EPI_ISL_2865431, EPI_ISL_2865452, EPI_ISL_2865455, EPI_ISL_2865639, EPI_ISL_2865832, EPI_ISL_2866674, EPI_ISL_2866677, EPI_ISL_2866678, EPI_ISL_2866680, EPI_ISL_2866682, EPI_ISL_2866685, EPI_ISL_2866686, EPI_ISL_2866689, EPI_ISL_2866690, EPI_ISL_2866691, EPI_ISL_2866692, EPI_ISL_2866693, EPI_ISL_2866697, EPI_ISL_2866698, EPI_ISL_2866699, EPI_ISL_2866700, EPI_ISL_2866702, EPI_ISL_2866703, EPI_ISL_2866705, EPI_ISL_2866706, EPI_ISL_2866712, EPI_ISL_2866713, EPI_ISL_2866714, EPI_ISL_2866716, EPI_ISL_2866717, EPI_ISL_2866718, EPI_ISL_2866721, EPI_ISL_2866722, EPI_ISL_2866726, EPI_ISL_2866729, EPI_ISL_2866730, EPI_ISL_2866732, EPI_ISL_2866734, EPI_ISL_2866735, EPI_ISL_2866737, EPI_ISL_2866738, EPI_ISL_2866739, EPI_ISL_2866741, EPI_ISL_2866742, EPI_ISL_2866744, EPI_ISL_2866745, EPI_ISL_2866746, EPI_ISL_2866749, EPI_ISL_2866751, EPI_ISL_2866752, EPI_ISL_2866753, EPI_ISL_2866756, EPI_ISL_2866757, EPI_ISL_2866759, EPI_ISL_2866761, EPI_ISL_2866762, EPI_ISL_2866764, EPI_ISL_2866766, EPI_ISL_2866767, EPI_ISL_2866769, EPI_ISL_2866770, EPI_ISL_2866771, EPI_ISL_2866772, EPI_ISL_2866773, EPI_ISL_2866776, EPI_ISL_2866778, EPI_ISL_2866780, EPI_ISL_2866781, EPI_ISL_2866782, EPI_ISL_2866784, EPI_ISL_2866785, EPI_ISL_2866786, EPI_ISL_2866787, EPI_ISL_2866788, EPI_ISL_2866791, EPI_ISL_2866792, EPI_ISL_2866794, EPI_ISL_2866795, EPI_ISL_2866799, EPI_ISL_2866800, EPI_ISL_2866802, EPI_ISL_2866803, EPI_ISL_2866806, EPI_ISL_2866807, EPI_ISL_2866811, EPI_ISL_2866812, EPI_ISL_2866818, EPI_ISL_2866819, EPI_ISL_2866820, EPI_ISL_2866821, EPI_ISL_2866822, EPI_ISL_2866823, EPI_ISL_2866827, EPI_ISL_2866829, EPI_ISL_2866831, EPI_ISL_2866838, EPI_ISL_2866841, EPI_ISL_2866842, EPI_ISL_2866844, EPI_ISL_2866846, EPI_ISL_2866848, EPI_ISL_2866850, EPI_ISL_2866851, EPI_ISL_2866854, EPI_ISL_2866856, EPI_ISL_2866858, EPI_ISL_2866859, EPI_ISL_2866860, EPI_ISL_2866862, EPI_ISL_2866864, EPI_ISL_2866866, EPI_ISL_2866869, EPI_ISL_2866870, EPI_ISL_2866872, EPI_ISL_2866874, EPI_ISL_2866878, EPI_ISL_2866880, EPI_ISL_2866884, EPI_ISL_2866886, EPI_ISL_2866887, EPI_ISL_2866892, EPI_ISL_2866894, EPI_ISL_2866895, EPI_ISL_2866898, EPI_ISL_2866899, EPI_ISL_2866900, EPI_ISL_2866901, EPI_ISL_2866903, EPI_ISL_2866904, EPI_ISL_2866906, EPI_ISL_2866907, EPI_ISL_2866911, EPI_ISL_2866913, EPI_ISL_2866915, EPI_ISL_2866916, EPI_ISL_2866923, EPI_ISL_2866925, EPI_ISL_2866928, EPI_ISL_2866929, EPI_ISL_2866930, EPI_ISL_2866931, EPI_ISL_2866933, EPI_ISL_2866934, EPI_ISL_2866935, EPI_ISL_2866936, EPI_ISL_2866941, EPI_ISL_2866947, EPI_ISL_2866949, EPI_ISL_2866951, EPI_ISL_2866955, EPI_ISL_2866957, EPI_ISL_2866958, EPI_ISL_2866961, EPI_ISL_2866962, EPI_ISL_2866964, EPI_ISL_2866965, EPI_ISL_2866966, EPI_ISL_2866968, EPI_ISL_2866972, EPI_ISL_2866977, EPI_ISL_2866982, EPI_ISL_2866983, EPI_ISL_2866985, EPI_ISL_2866987, EPI_ISL_2866988, EPI_ISL_2866991, EPI_ISL_2866992, EPI_ISL_2866993, EPI_ISL_2866998, EPI_ISL_2867004, EPI_ISL_2867005, EPI_ISL_2867006, EPI_ISL_2867007, EPI_ISL_2867008, EPI_ISL_2867009, EPI_ISL_2867011, EPI_ISL_2867012, EPI_ISL_2867013, EPI_ISL_2867014, EPI_ISL_2867015, EPI_ISL_2867016, EPI_ISL_2867018, EPI_ISL_2867021, EPI_ISL_2867022, EPI_ISL_2867024, EPI_ISL_2867025, EPI_ISL_2867028, EPI_ISL_2868085, EPI_ISL_2868086, EPI_ISL_2868087, EPI_ISL_2868088, EPI_ISL_2868089, EPI_ISL_2868090, EPI_ISL_2868091, EPI_ISL_2868092, EPI_ISL_2868093, EPI_ISL_2868094, EPI_ISL_2868095, EPI_ISL_2868096, EPI_ISL_2868097, EPI_ISL_2868098, EPI_ISL_2868099, EPI_ISL_2868100, EPI_ISL_2868101, EPI_ISL_2868102, EPI_ISL_2868103, EPI_ISL_2868104, EPI_ISL_2868105, EPI_ISL_2868106, EPI_ISL_2868107, EPI_ISL_2868108, EPI_ISL_2868109, EPI_ISL_2868110, EPI_ISL_2868111, EPI_ISL_2868112, EPI_ISL_2868113, EPI_ISL_2868114, EPI_ISL_2868115, EPI_ISL_2868116, EPI_ISL_2868117, EPI_ISL_2868118, EPI_ISL_2868119, EPI_ISL_2868120, EPI_ISL_2868121, EPI_ISL_2868123, EPI_ISL_2868124, EPI_ISL_2868125, EPI_ISL_2868126, EPI_ISL_2868127, EPI_ISL_2868128, EPI_ISL_2868129, EPI_ISL_2868130, EPI_ISL_2868131, EPI_ISL_2868132, EPI_ISL_2868133, EPI_ISL_2868134, EPI_ISL_2868135, EPI_ISL_2868136, EPI_ISL_2868137, EPI_ISL_2868138, EPI_ISL_2868139, EPI_ISL_2868140, EPI_ISL_2868141, EPI_ISL_2868142, EPI_ISL_2868143, EPI_ISL_2868144, EPI_ISL_2868145, EPI_ISL_2868146, EPI_ISL_2868147, EPI_ISL_2868148, EPI_ISL_2868149, EPI_ISL_2868150, EPI_ISL_2868151, EPI_ISL_2868152, EPI_ISL_2868153, EPI_ISL_2868154, EPI_ISL_2868155, EPI_ISL_2868156, EPI_ISL_2868157, EPI_ISL_2868158, EPI_ISL_2868159, EPI_ISL_2868160, EPI_ISL_2868161, EPI_ISL_2868162, EPI_ISL_2868163, EPI_ISL_2868164, EPI_ISL_2868165, EPI_ISL_2868166, EPI_ISL_2868167, EPI_ISL_2868169, EPI_ISL_2868170, EPI_ISL_2868171, EPI_ISL_2868172, EPI_ISL_2868173, EPI_ISL_2868174, EPI_ISL_2868175, EPI_ISL_2868176, EPI_ISL_2868177, EPI_ISL_2868178, EPI_ISL_2868179, EPI_ISL_2868180, EPI_ISL_2868181, EPI_ISL_2868182, EPI_ISL_2868183, EPI_ISL_2868184, EPI_ISL_2868185, EPI_ISL_2868186, EPI_ISL_2868187, EPI_ISL_2868188, EPI_ISL_2868189, EPI_ISL_2868190, EPI_ISL_2868191, EPI_ISL_2868192, EPI_ISL_2868193, EPI_ISL_2868194, EPI_ISL_2868195, EPI_ISL_2868196, EPI_ISL_2868197, EPI_ISL_2868198, EPI_ISL_2868199, EPI_ISL_2868200, EPI_ISL_2868201, EPI_ISL_2868202, EPI_ISL_2868203, EPI_ISL_2868204, EPI_ISL_2868205, EPI_ISL_2868206, EPI_ISL_2868207, EPI_ISL_2868208, EPI_ISL_2868209, EPI_ISL_2868211, EPI_ISL_2868212, EPI_ISL_2868213, EPI_ISL_2908293, EPI_ISL_2913677, EPI_ISL_2913790, EPI_ISL_2913854, EPI_ISL_2916709, EPI_ISL_2916839, EPI_ISL_2917107, EPI_ISL_2917186, EPI_ISL_2935979, EPI_ISL_2935994, EPI_ISL_2936069 | | | |
| see above | Lighthouse Lab in Glasgow | Wellcome Sanger Institute for the COVID-19 Genomics UK (COG-UK) Consortium | Anna Dominiczak and Alex Alderton; Carol Clugston; Cordelia Langford; David Gray; David K. Jackson; Dominic Kwiatkowski; Ewan Harrison; Harper VanSteenhouse; Ian Johnston; Jeffrey Barrett; John Sillitoe on behalf of the Wellcome Sanger Institute COVID-19 Surveillance Team; Roberto Amato; Sonia Goncalves; Yumi Kasai |
| EPI_ISL_2828762, EPI_ISL_2828763, EPI_ISL_2828764, EPI_ISL_2828773, EPI_ISL_2828777, EPI_ISL_2828781, EPI_ISL_2828782, EPI_ISL_2828792, EPI_ISL_2828793, EPI_ISL_2828794, EPI_ISL_2828795, EPI_ISL_2828797, EPI_ISL_2828800, EPI_ISL_2828803, EPI_ISL_2828809, EPI_ISL_2828811, EPI_ISL_2828813, EPI_ISL_2828817, EPI_ISL_2828819, EPI_ISL_2828820, EPI_ISL_2828821, EPI_ISL_2828823, EPI_ISL_2828825, EPI_ISL_2828826, EPI_ISL_2828827, EPI_ISL_2828831, EPI_ISL_2828833, EPI_ISL_2828835, EPI_ISL_2828845, EPI_ISL_2828847, EPI_ISL_2828850, EPI_ISL_2828854, EPI_ISL_2828856, EPI_ISL_2828857, EPI_ISL_2828863, EPI_ISL_2828868, EPI_ISL_2828873, EPI_ISL_2828874, EPI_ISL_2828875, EPI_ISL_2828883, EPI_ISL_2828884, EPI_ISL_2828889, EPI_ISL_2828893, EPI_ISL_2828894, EPI_ISL_2828900, EPI_ISL_2828901, EPI_ISL_2828902, EPI_ISL_2828904, EPI_ISL_2828905, EPI_ISL_2828907, EPI_ISL_2828908, EPI_ISL_2828914, EPI_ISL_2828919, EPI_ISL_2828920, EPI_ISL_2828921, EPI_ISL_2828922, EPI_ISL_2828924, EPI_ISL_2828925, EPI_ISL_2828926, EPI_ISL_2828927, EPI_ISL_2828935, EPI_ISL_2828940, EPI_ISL_2828943, EPI_ISL_2828946, EPI_ISL_2828947, EPI_ISL_2828950, | | | |

EPI_ISL_2828953, EPI_ISL_2828956, EPI_ISL_2828957, EPI_ISL_2828964, EPI_ISL_2828970, EPI_ISL_2828976, EPI_ISL_2828978, EPI_ISL_2828980, EPI_ISL_2828982, EPI_ISL_2828986, EPI_ISL_2828993, EPI_ISL_2828996, EPI_ISL_2829002, EPI_ISL_2829003, EPI_ISL_2829004, EPI_ISL_2829012, EPI_ISL_2829013, EPI_ISL_2829018, EPI_ISL_2829022, EPI_ISL_2829024, EPI_ISL_2846026, EPI_ISL_2846028, EPI_ISL_2846030, EPI_ISL_2846033, EPI_ISL_2846039, EPI_ISL_2846040, EPI_ISL_2846047, EPI_ISL_2846050, EPI_ISL_2846051, EPI_ISL_2846053, EPI_ISL_2846055, EPI_ISL_2846056, EPI_ISL_2846064, EPI_ISL_2846071, EPI_ISL_2846075, EPI_ISL_2846076, EPI_ISL_2846084, EPI_ISL_2846086, EPI_ISL_2846104, EPI_ISL_2846108, EPI_ISL_2846113, EPI_ISL_2846114, EPI_ISL_2846118, EPI_ISL_2846130, EPI_ISL_2846135, EPI_ISL_2846136, EPI_ISL_2846139, EPI_ISL_2846142, EPI_ISL_2846143, EPI_ISL_2846148, EPI_ISL_2846159, EPI_ISL_2846164, EPI_ISL_2846166, EPI_ISL_2846170, EPI_ISL_2846171, EPI_ISL_2846176, EPI_ISL_2846180, EPI_ISL_2846186, EPI_ISL_2846189, EPI_ISL_2846196, EPI_ISL_2846209, EPI_ISL_2846213, EPI_ISL_2846214, EPI_ISL_2846216, EPI_ISL_2846217, EPI_ISL_2846219, EPI_ISL_2846220, EPI_ISL_2846221, EPI_ISL_2846227, EPI_ISL_2846228, EPI_ISL_2846232, EPI_ISL_2846236, EPI_ISL_2846240, EPI_ISL_2846250, EPI_ISL_2846253, EPI_ISL_2846255, EPI_ISL_2846259, EPI_ISL_2846262, EPI_ISL_2846264, EPI_ISL_2846267, EPI_ISL_2846268, EPI_ISL_2846271, EPI_ISL_2846276, EPI_ISL_2846277, EPI_ISL_2846280, EPI_ISL_2846293, EPI_ISL_2846294, EPI_ISL_2846301, EPI_ISL_2846304, EPI_ISL_2846309, EPI_ISL_2846312, EPI_ISL_2846315, EPI_ISL_2846317, EPI_ISL_2846320, EPI_ISL_2846329, EPI_ISL_2846347, EPI_ISL_2846349, EPI_ISL_2846351, EPI_ISL_2846358, EPI_ISL_2846587, EPI_ISL_2846610, EPI_ISL_2846693, EPI_ISL_2846714, EPI_ISL_2846732, EPI_ISL_2846767, EPI_ISL_2846796, EPI_ISL_2846805, EPI_ISL_2846824, EPI_ISL_2846835, EPI_ISL_2846840, EPI_ISL_2846861, EPI_ISL_2846871, EPI_ISL_2848301, EPI_ISL_2848302, EPI_ISL_2848305, EPI_ISL_2848306, EPI_ISL_2848307, EPI_ISL_2848308, EPI_ISL_2848309, EPI_ISL_2848311, EPI_ISL_2848312, EPI_ISL_2848313, EPI_ISL_2848314, EPI_ISL_2848316, EPI_ISL_2848317, EPI_ISL_2848318, EPI_ISL_2848319, EPI_ISL_2848321, EPI_ISL_2848322, EPI_ISL_2848323, EPI_ISL_2848329, EPI_ISL_2848330, EPI_ISL_2848331, EPI_ISL_2848332, EPI_ISL_2848333, EPI_ISL_2848335, EPI_ISL_2848336, EPI_ISL_2848337, EPI_ISL_2848338, EPI_ISL_2848339, EPI_ISL_2848341, EPI_ISL_2848342, EPI_ISL_2848343, EPI_ISL_2848344, EPI_ISL_2848346, EPI_ISL_2848350, EPI_ISL_2848353, EPI_ISL_2848354, EPI_ISL_2848355, EPI_ISL_2848356, EPI_ISL_2848357, EPI_ISL_2848358, EPI_ISL_2848362, EPI_ISL_2848363, EPI_ISL_2848364, EPI_ISL_2848366, EPI_ISL_2848369, EPI_ISL_2848370, EPI_ISL_2848371, EPI_ISL_2848372, EPI_ISL_2848374, EPI_ISL_2848375, EPI_ISL_2848378, EPI_ISL_2848380, EPI_ISL_2848382, EPI_ISL_2848383, EPI_ISL_2848384, EPI_ISL_2848385, EPI_ISL_2848386, EPI_ISL_2848388, EPI_ISL_2848391, EPI_ISL_2848392, EPI_ISL_2848393, EPI_ISL_2848394, EPI_ISL_2848396, EPI_ISL_2848397, EPI_ISL_2848399, EPI_ISL_2848401, EPI_ISL_2848402, EPI_ISL_2848403, EPI_ISL_2848406, EPI_ISL_2848407, EPI_ISL_2848408, EPI_ISL_2848410, EPI_ISL_2848411, EPI_ISL_2848412, EPI_ISL_2848413, EPI_ISL_2848414, EPI_ISL_2848415, EPI_ISL_2848416, EPI_ISL_2848417, EPI_ISL_2848419, EPI_ISL_2848420, EPI_ISL_2848421, EPI_ISL_2848422, EPI_ISL_2848425, EPI_ISL_2848426, EPI_ISL_2848427, EPI_ISL_2848428, EPI_ISL_2848429, EPI_ISL_2848431, EPI_ISL_2848432, EPI_ISL_2848433, EPI_ISL_2848437, EPI_ISL_2848441, EPI_ISL_2848442, EPI_ISL_2848444, EPI_ISL_2848445, EPI_ISL_2848446, EPI_ISL_2848448, EPI_ISL_2848449, EPI_ISL_2848450, EPI_ISL_2848451, EPI_ISL_2848452, EPI_ISL_2848453, EPI_ISL_2848454, EPI_ISL_2848455, EPI_ISL_2848456, EPI_ISL_2848461, EPI_ISL_2848462, EPI_ISL_2848466, EPI_ISL_2848467, EPI_ISL_2848469, EPI_ISL_2848470, EPI_ISL_2848471, EPI_ISL_2848472, EPI_ISL_2848473, EPI_ISL_2848474, EPI_ISL_2848476, EPI_ISL_2848477, EPI_ISL_2848478, EPI_ISL_2848479, EPI_ISL_2848480, EPI_ISL_2848482, EPI_ISL_2848483, EPI_ISL_2848484, EPI_ISL_2848485, EPI_ISL_2848486, EPI_ISL_2848487, EPI_ISL_2848489, EPI_ISL_2848490, EPI_ISL_2848491, EPI_ISL_2848492, EPI_ISL_2848493, EPI_ISL_2848497, EPI_ISL_2848501, EPI_ISL_2848503, EPI_ISL_2848505, EPI_ISL_2848506, EPI_ISL_2848512, EPI_ISL_2848513, EPI_ISL_2848514, EPI_ISL_2848515, EPI_ISL_2848516, EPI_ISL_2848517, EPI_ISL_2848519, EPI_ISL_2848520, EPI_ISL_2848521, EPI_ISL_2848522, EPI_ISL_2848523, EPI_ISL_2848525, EPI_ISL_2848526, EPI_ISL_2848527, EPI_ISL_2848528, EPI_ISL_2848529, EPI_ISL_2848531, EPI_ISL_2848532, EPI_ISL_2848533, EPI_ISL_2848534, EPI_ISL_2848535, EPI_ISL_2848537, EPI_ISL_2848539, EPI_ISL_2848540, EPI_ISL_2848541, EPI_ISL_2848543, EPI_ISL_2848544, EPI_ISL_2848545, EPI_ISL_2848546, EPI_ISL_2848547, EPI_ISL_2848548, EPI_ISL_2848549, EPI_ISL_2848551, EPI_ISL_2848552, EPI_ISL_2848553, EPI_ISL_2848554, EPI_ISL_2848555, EPI_ISL_2848556, EPI_ISL_2848557, EPI_ISL_2848559, EPI_ISL_2848560, EPI_ISL_2848561, EPI_ISL_2848562, EPI_ISL_2848563, EPI_ISL_2848565, EPI_ISL_2848566, EPI_ISL_2848567, EPI_ISL_2848568, EPI_ISL_2848570, EPI_ISL_2848575, EPI_ISL_2848576, EPI_ISL_2848577, EPI_ISL_2848578, EPI_ISL_2848579, EPI_ISL_2848581, EPI_ISL_2848583, EPI_ISL_2848584, EPI_ISL_2848585, EPI_ISL_2848587, EPI_ISL_2848588, EPI_ISL_2848589, EPI_ISL_2848591, EPI_ISL_2848592, EPI_ISL_2848593, EPI_ISL_2848594, EPI_ISL_2848595, EPI_ISL_2848596, EPI_ISL_2848598, EPI_ISL_2848599, EPI_ISL_2848600, EPI_ISL_2848601, EPI_ISL_2848603, EPI_ISL_2848604, EPI_ISL_2848605, EPI_ISL_2848606, EPI_ISL_2848607, EPI_ISL_2848609, EPI_ISL_2848610, EPI_ISL_2848611, EPI_ISL_2848612, EPI_ISL_2848615, EPI_ISL_2848616, EPI_ISL_2848617, EPI_ISL_2848618, EPI_ISL_2848620, EPI_ISL_2848621, EPI_ISL_2848622, EPI_ISL_2848623, EPI_ISL_2848624, EPI_ISL_2848625, EPI_ISL_2848626, EPI_ISL_2848629, EPI_ISL_2848630, EPI_ISL_2848631, EPI_ISL_2848632, EPI_ISL_2848633, EPI_ISL_2848635, EPI_ISL_2848638, EPI_ISL_2848639, EPI_ISL_2848640, EPI_ISL_2848641, EPI_ISL_2848643, EPI_ISL_2848644, EPI_ISL_2848645, EPI_ISL_2848646, EPI_ISL_2848647, EPI_ISL_2848648, EPI_ISL_2848649, EPI_ISL_2848652, EPI_ISL_2848653, EPI_ISL_2848654, EPI_ISL_2848655, EPI_ISL_2848656, EPI_ISL_2848659, EPI_ISL_2848660, EPI_ISL_2848661, EPI_ISL_2848662, EPI_ISL_2848664, EPI_ISL_2848665, EPI_ISL_2848672, EPI_ISL_2848673, EPI_ISL_2848674, EPI_ISL_2848675, EPI_ISL_2848676, EPI_ISL_2848677, EPI_ISL_2848678, EPI_ISL_2848680, EPI_ISL_2848681, EPI_ISL_2848683, EPI_ISL_2848684, EPI_ISL_2848685, EPI_ISL_2848687, EPI_ISL_2848688, EPI_ISL_2848689, EPI_ISL_2848696, EPI_ISL_2848698, EPI_ISL_2848700, EPI_ISL_2848701, EPI_ISL_2848702, EPI_ISL_2848704, EPI_ISL_2848705, EPI_ISL_2848707, EPI_ISL_2848708, EPI_ISL_2848710, EPI_ISL_2848711, EPI_ISL_2848712, EPI_ISL_2848713, EPI_ISL_2848719, EPI_ISL_2848721, EPI_ISL_2848723, EPI_ISL_2848725, EPI_ISL_2848731, EPI_ISL_2848733, EPI_ISL_2848734, EPI_ISL_2848738, EPI_ISL_2848740, EPI_ISL_2848741, EPI_ISL_2848743, EPI_ISL_2848744, EPI_ISL_2848746, EPI_ISL_2848750, EPI_ISL_2848751, EPI_ISL_2848752, EPI_ISL_2848753, EPI_ISL_2848756, EPI_ISL_2848757, EPI_ISL_2848758, EPI_ISL_2848759, EPI_ISL_2848764, EPI_ISL_2848765, EPI_ISL_2848769, EPI_ISL_2848770, EPI_ISL_2848771, EPI_ISL_2848773, EPI_ISL_2848774, EPI_ISL_2848775, EPI_ISL_2848776, EPI_ISL_2848777, EPI_ISL_2848778, EPI_ISL_2848779, EPI_ISL_2848780, EPI_ISL_2848782, EPI_ISL_2848785, EPI_ISL_2848786, EPI_ISL_2848787, EPI_ISL_2848788, EPI_ISL_2848789, EPI_ISL_2848790, EPI_ISL_2848792, EPI_ISL_2848793, EPI_ISL_2848794, EPI_ISL_2848795, EPI_ISL_2848797, EPI_ISL_2848798, EPI_ISL_2848800, EPI_ISL_2848801, EPI_ISL_2848803, EPI_ISL_2848804, EPI_ISL_2848806, EPI_ISL_2848807, EPI_ISL_2848810, EPI_ISL_2848813, EPI_ISL_2848814, EPI_ISL_2848815, EPI_ISL_2848818, EPI_ISL_2848819, EPI_ISL_2848820, EPI_ISL_2848823, EPI_ISL_2848824, EPI_ISL_2848825, EPI_ISL_2848826, EPI_ISL_2848827, EPI_ISL_2848829, EPI_ISL_2848831, EPI_ISL_2848832, EPI_ISL_2848836, EPI_ISL_2848840, EPI_ISL_2848841, EPI_ISL_2848844, EPI_ISL_2848847, EPI_ISL_2848848, EPI_ISL_2848849, EPI_ISL_2848850, EPI_ISL_2848851, EPI_ISL_2848852, EPI_ISL_2848854, EPI_ISL_2848857, EPI_ISL_2848858, EPI_ISL_2848859, EPI_ISL_2848862, EPI_ISL_2848864, EPI_ISL_2848866, EPI_ISL_2848867, EPI_ISL_2848868, EPI_ISL_2848872, EPI_ISL_2848873, EPI_ISL_2848874, EPI_ISL_2848876, EPI_ISL_2848880, EPI_ISL_2848881, EPI_ISL_2848882, EPI_ISL_2848883, EPI_ISL_2848885, EPI_ISL_2848887, EPI_ISL_2848889, EPI_ISL_2848890, EPI_ISL_2848893, EPI_ISL_2848895, EPI_ISL_2848898, EPI_ISL_2848899, EPI_ISL_2848900, EPI_ISL_2848902, EPI_ISL_2848903, EPI_ISL_2848905, EPI_ISL_2848906, EPI_ISL_2848907, EPI_ISL_2848910, EPI_ISL_2848911, EPI_ISL_2848912, EPI_ISL_2848913, EPI_ISL_2848914, EPI_ISL_2848916, EPI_ISL_2848919, EPI_ISL_2848920, EPI_ISL_2848925, EPI_ISL_2848926, EPI_ISL_2848927, EPI_ISL_2848929, EPI_ISL_2848931, EPI_ISL_2848934, EPI_ISL_2848935, EPI_ISL_2848936, EPI_ISL_2848937, EPI_ISL_2848939, EPI_ISL_2848941, EPI_ISL_2848942, EPI_ISL_2848944, EPI_ISL_2848945, EPI_ISL_2848946, EPI_ISL_2848947, EPI_ISL_2848948, EPI_ISL_2848950, EPI_ISL_2848951, EPI_ISL_2848952, EPI_ISL_2848953, EPI_ISL_2848958, EPI_ISL_2848960, EPI_ISL_2848961, EPI_ISL_2848962, EPI_ISL_2848963, EPI_ISL_2848967, EPI_ISL_2848968, EPI_ISL_2848970, EPI_ISL_2848971, EPI_ISL_2848973, EPI_ISL_2848974, EPI_ISL_2848975, EPI_ISL_2848979, EPI_ISL_2848981, EPI_ISL_2848982, EPI_ISL_2848985, EPI_ISL_2848986, EPI_ISL_2848987, EPI_ISL_2848988, EPI_ISL_2848989, EPI_ISL_2848990, EPI_ISL_2848991, EPI_ISL_2848992, EPI_ISL_2848994, EPI_ISL_2848995, EPI_ISL_2848997, EPI_ISL_2848998, EPI_ISL_2848999, EPI_ISL_2849000, EPI_ISL_2849001, EPI_ISL_2849004, EPI_ISL_2849005, EPI_ISL_2849006, EPI_ISL_2849007, EPI_ISL_2849008, EPI_ISL_2849441, EPI_ISL_2849495, EPI_ISL_2851655, EPI_ISL_2851757, EPI_ISL_2865365, EPI_ISL_2865370, EPI_ISL_2865376, EPI_ISL_2865385, EPI_ISL_2865390, EPI_ISL_2865394, EPI_ISL_2865402, EPI_ISL_2865406, EPI_ISL_2865407, EPI_ISL_2865411, EPI_ISL_2865418, EPI_ISL_2865419, EPI_ISL_2865430, EPI_ISL_2865441, EPI_ISL_2865442, EPI_ISL_2865456, EPI_ISL_2865458, EPI_ISL_2865459, EPI_ISL_2865462, EPI_ISL_2865463, EPI_ISL_2865468, EPI_ISL_2865479, EPI_ISL_2865487, EPI_ISL_2865504, EPI_ISL_2865509, EPI_ISL_2865516, EPI_ISL_2865523, EPI_ISL_2865526, EPI_ISL_2865536, EPI_ISL_2865550, EPI_ISL_2865560, EPI_ISL_2865565, EPI_ISL_2865592, EPI_ISL_2865600, EPI_ISL_2865607, EPI_ISL_2865609, EPI_ISL_2865613, EPI_ISL_2865616, EPI_ISL_2865622, EPI_ISL_2865651, EPI_ISL_2865653, EPI_ISL_2865660, EPI_ISL_2865661, EPI_ISL_2865664, EPI_ISL_2865665, EPI_ISL_2865677, EPI_ISL_2865695, EPI_ISL_2865705, EPI_ISL_2865757, EPI_ISL_2865804, EPI_ISL_2865855, EPI_ISL_2865866, EPI_ISL_2865919, EPI_ISL_2866005, EPI_ISL_2866066, EPI_ISL_2866067, EPI_ISL_2866071, EPI_ISL_2866074, EPI_ISL_2866076, EPI_ISL_2866078, EPI_ISL_2866080, EPI_ISL_2866081, EPI_ISL_2866090, EPI_ISL_2866097, EPI_ISL_2866106, EPI_ISL_2866108, EPI_ISL_2866110, EPI_ISL_2866111, EPI_ISL_2866114, EPI_ISL_2866116, EPI_ISL_2866118, EPI_ISL_2866119, EPI_ISL_2866137, EPI_ISL_2866139, EPI_ISL_2866145, EPI_ISL_2866147, EPI_ISL_2866155, EPI_ISL_2866157, EPI_ISL_2866159, EPI_ISL_2866167, EPI_ISL_2866171, EPI_ISL_2866189, EPI_ISL_2866190, EPI_ISL_2866197, EPI_ISL_2866199, EPI_ISL_2866200, EPI_ISL_2866206, EPI_ISL_2866210, EPI_ISL_2866212, EPI_ISL_2866223, EPI_ISL_2866228, EPI_ISL_2866234, EPI_ISL_2866261, EPI_ISL_2866266, EPI_ISL_2866276, EPI_ISL_2866277, EPI_ISL_2866287, EPI_ISL_2866288, EPI_ISL_2866296, EPI_ISL_2866298, EPI_ISL_2866299, EPI_ISL_2866300, EPI_ISL_2866304, EPI_ISL_2866308, EPI_ISL_2866321, EPI_ISL_2866322, EPI_ISL_2866327, EPI_ISL_2866331, EPI_ISL_2866338, EPI_ISL_2866342, EPI_ISL_2866344, EPI_ISL_2866346, EPI_ISL_2866348, EPI_ISL_2866351, EPI_ISL_2866353, EPI_ISL_2866367, EPI_ISL_2866369, EPI_ISL_2866384, EPI_ISL_2866388, EPI_ISL_2866391, EPI_ISL_2866393, EPI_ISL_2866398, EPI_ISL_2866412, EPI_ISL_2866416, EPI_ISL_2866417, EPI_ISL_2866425, EPI_ISL_2866427, EPI_ISL_2866429, EPI_ISL_2866431, EPI_ISL_2866438, EPI_ISL_2866440, EPI_ISL_2866443, EPI_ISL_2866453, EPI_ISL_2866456, EPI_ISL_2866461, EPI_ISL_2866464, EPI_ISL_2866468, EPI_ISL_2866472, EPI_ISL_2866473, EPI_ISL_2866479, EPI_ISL_2866480, EPI_ISL_2866487, EPI_ISL_2866488, EPI_ISL_2866491, EPI_ISL_2866510, EPI_ISL_2866513, EPI_ISL_2866518, EPI_ISL_2866519, EPI_ISL_2866529, EPI_ISL_2866532, EPI_ISL_2866539, EPI_ISL_2866542, EPI_ISL_2866546, EPI_ISL_2866548, EPI_ISL_2866549, EPI_ISL_2866550, EPI_ISL_2866551, EPI_ISL_2866554, EPI_ISL_2866556, EPI_ISL_2866564, EPI_ISL_2866568, EPI_ISL_2866572, EPI_ISL_2866578, EPI_ISL_2866584, EPI_ISL_2866591, EPI_ISL_2866592, EPI_ISL_2866598, EPI_ISL_2866600, EPI_ISL_2866602, EPI_ISL_2866604, EPI_ISL_2866606, EPI_ISL_2866610, EPI_ISL_2866615, EPI_ISL_2866616, EPI_ISL_2866622, EPI_ISL_2866625, EPI_ISL_2866629, EPI_ISL_2866630, EPI_ISL_2866643, EPI_ISL_2866648, EPI_ISL_2866654, EPI_ISL_2866655, EPI_ISL_2866660, EPI_ISL_2866664, EPI_ISL_2866670, EPI_ISL_2866675, EPI_ISL_2866683, EPI_ISL_2866684, EPI_ISL_2866706, EPI_ISL_2866707, EPI_ISL_2866724, EPI_ISL_2866728, EPI_ISL_2866733, EPI_ISL_2866736, EPI_ISL_2866743, EPI_ISL_2866747, EPI_ISL_2866748, EPI_ISL_2866750, EPI_ISL_2866754, EPI_ISL_2866755, EPI_ISL_2866768, EPI_ISL_2866783, EPI_ISL_2866789, EPI_ISL_2866797, EPI_ISL_2866798, EPI_ISL_2866804, EPI_ISL_2866809, EPI_ISL_2866810, EPI_ISL_2866815, EPI_ISL_2866817, EPI_ISL_2866825, EPI_ISL_2866833, EPI_ISL_2866836, EPI_ISL_2866837, EPI_ISL_2866839, EPI_ISL_2866843, EPI_ISL_2866845, EPI_ISL_2866853, EPI_ISL_2866855, EPI_ISL_2866867, EPI_ISL_2866873, EPI_ISL_2866879, EPI_ISL_2866883, EPI_ISL_2866885, EPI_ISL_2866890, EPI_ISL_2866891, EPI_ISL_2866897, EPI_ISL_2866905, EPI_ISL_2866908, EPI_ISL_2866909, EPI_ISL_2866910, EPI_ISL_2866912, EPI_ISL_2866918, EPI_ISL_2866924, EPI_ISL_2866926, EPI_ISL_2866937, EPI_ISL_2866944, EPI_ISL_2866946, EPI_ISL_2866953, EPI_ISL_2866954, EPI_ISL_2866960, EPI_ISL_2866969, EPI_ISL_2866971, EPI_ISL_2866973, EPI_ISL_2866975, EPI_ISL_2866980, EPI_ISL_2866981, EPI_ISL_2866984, EPI_ISL_2866986, EPI_ISL_2866990, EPI_ISL_2866995, EPI_ISL_2866996, EPI_ISL_2866997, EPI_ISL_2867001, EPI_ISL_2867003, EPI_ISL_2867010, EPI_ISL_2867026, EPI_ISL_2867027, EPI_ISL_2867363, EPI_ISL_2867368, EPI_ISL_2867380, EPI_ISL_2867386, EPI_ISL_2867391, EPI_ISL_2867393, EPI_ISL_2867395, EPI_ISL_2867401, EPI_ISL_2867408, EPI_ISL_2867410, EPI_ISL_2867415, EPI_ISL_2867426, EPI_ISL_2867431, EPI_ISL_2867451, EPI_ISL_2867457, EPI_ISL_2867467, EPI_ISL_2867468, EPI_ISL_2867471, EPI_ISL_2867473, EPI_ISL_2867484, EPI_ISL_2867486, EPI_ISL_2867489, EPI_ISL_2867490, EPI_ISL_2867494, EPI_ISL_2867496, EPI_ISL_2867497, EPI_ISL_2867498, EPI_ISL_2867500, EPI_ISL_2867502, EPI_ISL_2867506, EPI_ISL_2867513, EPI_ISL_2867518, EPI_ISL_2867519, EPI_ISL_2867520, EPI_ISL_2867522, EPI_ISL_2867526, EPI_ISL_2867529, EPI_ISL_2867531, EPI_ISL_2867532, EPI_ISL_2867543, EPI_ISL_2867544, EPI_ISL_2867546, EPI_ISL_2867549, EPI_ISL_2867557, EPI_ISL_2867559, EPI_ISL_2867563, EPI_ISL_2867567, EPI_ISL_2867578, EPI_ISL_2867580, EPI_ISL_2867599, EPI_ISL_2867600, EPI_ISL_2867605, EPI_ISL_2867606, EPI_ISL_2867609, EPI_ISL_2867611, EPI_ISL_2867614, EPI_ISL_2867615, EPI_ISL_2867622, EPI_ISL_2867631, EPI_ISL_2867632, EPI_ISL_2867635, EPI_ISL_2867636, EPI_ISL_2867638, EPI_ISL_2867646, EPI_ISL_2867649, EPI_ISL_2867652, EPI_ISL_2867658, EPI_ISL_2867660, EPI_ISL_2867667, EPI_ISL_2867671, EPI_ISL_2867679, EPI_ISL_2867680, EPI_ISL_2867684, EPI_ISL_2867687, EPI_ISL_2867689, EPI_ISL_2867693, EPI_ISL_2868084, EPI_ISL_2908540, EPI_ISL_2910262, EPI_ISL_2910266, EPI_ISL_2910328, EPI_ISL_2910347, EPI_ISL_2910432, EPI_ISL_2910465, EPI_ISL_2910541, EPI_ISL_2911097, EPI_ISL_2912134, EPI_ISL_2914931, EPI_ISL_2916041, EPI_ISL_2916096, EPI_ISL_2916397, EPI_ISL_2916425, EPI_ISL_2916443, EPI_ISL_2916463, EPI_ISL_2916578, EPI_ISL_2916638, EPI_ISL_2917354, EPI_ISL_2917456, EPI_ISL_2918151, EPI_ISL_2918247, EPI_ISL_2918309, EPI_ISL_2924842, EPI_ISL_2924846, EPI_ISL_2924872, EPI_ISL_2924885, EPI_ISL_2925124, EPI_ISL_2935914, EPI_ISL_2935926, EPI_ISL_2935935, EPI_ISL_2935943, EPI_ISL_2935951, EPI_ISL_2935955, EPI_ISL_2935965, EPI_ISL_2935966, EPI_ISL_2935991, EPI_ISL_2935995, EPI_ISL_2935996, EPI_ISL_2935997, EPI_ISL_2936003, EPI_ISL_2936006, EPI_ISL_2936009, EPI_ISL_2936012, EPI_ISL_2936021, EPI_ISL_2936040, EPI_ISL_2936041, EPI_ISL_2936045, EPI_ISL_2936050, EPI_ISL_2936051, EPI_ISL_2936061, EPI_ISL_2936064, EPI_ISL_2936065, EPI_ISL_2936072, EPI_ISL_2936076, EPI_ISL_2936085, EPI_ISL_2936100, EPI_ISL_2936108, EPI_ISL_2936112, EPI_ISL_2936113, EPI_ISL_2936121, EPI_ISL_2936123, EPI_ISL_2936127, EPI_ISL_2936145, EPI_ISL_2936148, EPI_ISL_2936159, EPI_ISL_2936162, EPI_ISL_2936176, EPI_ISL_2936184, EPI_ISL_2936666

| | | | |
|---|---|---|---|
| see above | Lighthouse Lab in Milton Keynes | Wellcome Sanger Institute for the COVID-19 Genomics UK (COG-UK) Consortium | Cordelia Langford; David K. Jackson; Dominic Kwiatkowski; Ewan Harrison; Ian Johnston; Jeffrey Barrett; John Sillitoe on behalf of the Wellcome Sanger Institute COVID-19 Surveillance Team; Roberto Amato; Sonia Goncalves; The Lighthouse Lab in Milton Keynes and Alex Alderton |

EPI_ISL_2828761, EPI_ISL_2828766, EPI_ISL_2828769, EPI_ISL_2828770, EPI_ISL_2828771, EPI_ISL_2828772, EPI_ISL_2828774, EPI_ISL_2828776, EPI_ISL_2828780, EPI_ISL_2828783, EPI_ISL_2828784, EPI_ISL_2828785, EPI_ISL_2828786, EPI_ISL_2828787, EPI_ISL_2828791, EPI_ISL_2828799, EPI_ISL_2828802, EPI_ISL_2828804, EPI_ISL_2828805, EPI_ISL_2828808, EPI_ISL_2828810, EPI_ISL_2828815, EPI_ISL_2828824, EPI_ISL_2828830, EPI_ISL_2828832, EPI_ISL_2828836, EPI_ISL_2828841, EPI_ISL_2828844, EPI_ISL_2828846, EPI_ISL_2828853, EPI_ISL_2828858, EPI_ISL_2828862, EPI_ISL_2828865, EPI_ISL_2828869, EPI_ISL_2828870, EPI_ISL_2828877, EPI_ISL_2828880, EPI_ISL_2828881, EPI_ISL_2828892, EPI_ISL_2828895, EPI_ISL_2828897, EPI_ISL_2828898, EPI_ISL_2828899, EPI_ISL_2828903, EPI_ISL_2828909, EPI_ISL_2828910, EPI_ISL_2828911, EPI_ISL_2828915, EPI_ISL_2828923, EPI_ISL_2828928, EPI_ISL_2828929, EPI_ISL_2828930, EPI_ISL_2828933, EPI_ISL_2828937, EPI_ISL_2828939, EPI_ISL_2828941, EPI_ISL_2828942, EPI_ISL_2828944, EPI_ISL_2828949, EPI_ISL_2828958, EPI_ISL_2828960, EPI_ISL_2828962, EPI_ISL_2828972, EPI_ISL_2828974, EPI_ISL_2828979, EPI_ISL_2828985, EPI_ISL_2828987, EPI_ISL_2828988, EPI_ISL_2828991, EPI_ISL_2828994, EPI_ISL_2828997, EPI_ISL_2828999, EPI_ISL_2829000, EPI_ISL_2829005, EPI_ISL_2829008, EPI_ISL_2829010, EPI_ISL_2829011, EPI_ISL_2829014, EPI_ISL_2829015, EPI_ISL_2829016, EPI_ISL_2829017, EPI_ISL_2829020, EPI_ISL_2829021, EPI_ISL_2846036, EPI_ISL_2846067, EPI_ISL_2846068, EPI_ISL_2846069, EPI_ISL_2846070, EPI_ISL_2846097, EPI_ISL_2846100, EPI_ISL_2846103, EPI_ISL_2846106, EPI_ISL_2846110, EPI_ISL_2846115, EPI_ISL_2846134, EPI_ISL_2846141, EPI_ISL_2846145, EPI_ISL_2846147, EPI_ISL_2846150, EPI_ISL_2846152, EPI_ISL_2846179, EPI_ISL_2846191, EPI_ISL_2846199, EPI_ISL_2846210, EPI_ISL_2846226, EPI_ISL_2846229, EPI_ISL_2846278, EPI_ISL_2846281, EPI_ISL_2846285, EPI_ISL_2846291, EPI_ISL_2846292, EPI_ISL_2846299, EPI_ISL_2846300, EPI_ISL_2846302, EPI_ISL_2846318, EPI_ISL_2846324, EPI_ISL_2846325, EPI_ISL_2846332, EPI_ISL_2846339, EPI_ISL_2846355, EPI_ISL_2846367, EPI_ISL_2846378, EPI_ISL_2846479, EPI_ISL_2846484, EPI_ISL_2846499, EPI_ISL_2846501, EPI_ISL_2846518, EPI_ISL_2846584, EPI_ISL_2846591, EPI_ISL_2846592, EPI_ISL_2846594, EPI_ISL_2846602, EPI_ISL_2846608, EPI_ISL_2846609, EPI_ISL_2846624, EPI_ISL_2846629, EPI_ISL_2846630, EPI_ISL_2846631, EPI_ISL_2846636, EPI_ISL_2846642, EPI_ISL_2846644, EPI_ISL_2846651, EPI_ISL_2846652, EPI_ISL_2846653, EPI_ISL_2846657, EPI_ISL_2846662, EPI_ISL_2846671, EPI_ISL_2846689, EPI_ISL_2846692, EPI_ISL_2846694, EPI_ISL_2846696, EPI_ISL_2846699, EPI_ISL_2846705, EPI_ISL_2846712, EPI_ISL_2846719, EPI_ISL_2846725, EPI_ISL_2846734, EPI_ISL_2846735, EPI_ISL_2846743, EPI_ISL_2846747, EPI_ISL_2846757, EPI_ISL_2846759, EPI_ISL_2846760, EPI_ISL_2846769, EPI_ISL_2846777, EPI_ISL_2846778, EPI_ISL_2846781, EPI_ISL_2846784, EPI_ISL_2846785, EPI_ISL_2846789, EPI_ISL_2846791, EPI_ISL_2846793, EPI_ISL_2846797, EPI_ISL_2846799, EPI_ISL_2846807, EPI_ISL_2846810, EPI_ISL_2846813, EPI_ISL_2846819, EPI_ISL_2846822, EPI_ISL_2846823, EPI_ISL_2846830, EPI_ISL_2846836, EPI_ISL_2846838, EPI_ISL_2846842, EPI_ISL_2846848, EPI_ISL_2846850, EPI_ISL_2846851, EPI_ISL_2846855, EPI_ISL_2846862, EPI_ISL_2846869, EPI_ISL_2846875, EPI_ISL_2846877, EPI_ISL_2846884, EPI_ISL_2846888, EPI_ISL_2846890, EPI_ISL_2846891, EPI_ISL_2846894, EPI_ISL_2846900, EPI_ISL_2846912, EPI_ISL_2846915, EPI_ISL_2846918, EPI_ISL_2846923, EPI_ISL_2846924, EPI_ISL_2846927, EPI_ISL_2848149, EPI_ISL_2849010, EPI_ISL_2849014, EPI_ISL_2849015, EPI_ISL_2849018, EPI_ISL_2849019, EPI_ISL_2849024, EPI_ISL_2849026, EPI_ISL_2849030, EPI_ISL_2849033, EPI_ISL_2849034, EPI_ISL_2849036, EPI_ISL_2849038, EPI_ISL_2849041, EPI_ISL_2849046, EPI_ISL_2849047, EPI_ISL_2849051, EPI_ISL_2849053, EPI_ISL_2849055, EPI_ISL_2849058, EPI_ISL_2849063, EPI_ISL_2849064, EPI_ISL_2849066, EPI_ISL_2849073, EPI_ISL_2849074, EPI_ISL_2849075, EPI_ISL_2849076, EPI_ISL_2849081, EPI_ISL_2849088, EPI_ISL_2849089, EPI_ISL_2849093, EPI_ISL_2849096, EPI_ISL_2849098, EPI_ISL_2849099, EPI_ISL_2849102, EPI_ISL_2849109, EPI_ISL_2849112, EPI_ISL_2849113, EPI_ISL_2849116, EPI_ISL_2849117, EPI_ISL_2849118, EPI_ISL_2849122, EPI_ISL_2849123, EPI_ISL_2849125, EPI_ISL_2849129, EPI_ISL_2849135, EPI_ISL_2849136, EPI_ISL_2849137, EPI_ISL_2849142, EPI_ISL_2849145, EPI_ISL_2849150, EPI_ISL_2849151, EPI_ISL_2849153, EPI_ISL_2849158, EPI_ISL_2849160, EPI_ISL_2849161, EPI_ISL_2849164, EPI_ISL_2849165, EPI_ISL_2849167, EPI_ISL_2849171, EPI_ISL_2849173, EPI_ISL_2849174, EPI_ISL_2849176, EPI_ISL_2849177, EPI_ISL_2849178, EPI_ISL_2849179, EPI_ISL_2849181, EPI_ISL_2849182, EPI_ISL_2849185, EPI_ISL_2849186, EPI_ISL_2849192, EPI_ISL_2849197, EPI_ISL_2849198, EPI_ISL_2849200, EPI_ISL_2849205, EPI_ISL_2849206, EPI_ISL_2849212, EPI_ISL_2849213, EPI_ISL_2849215, EPI_ISL_2849226, EPI_ISL_2849228, EPI_ISL_2849229, EPI_ISL_2849230, EPI_ISL_2849235, EPI_ISL_2849236, EPI_ISL_2849237, EPI_ISL_2849243, EPI_ISL_2849244, EPI_ISL_2849245, EPI_ISL_2849247, EPI_ISL_2849248, EPI_ISL_2849252, EPI_ISL_2849255, EPI_ISL_2849256, EPI_ISL_2849257, EPI_ISL_2849258, EPI_ISL_2849260, EPI_ISL_2849264, EPI_ISL_2849265, EPI_ISL_2849268, EPI_ISL_2849269, EPI_ISL_2849270, EPI_ISL_2849271, EPI_ISL_2849273, EPI_ISL_2849274, EPI_ISL_2849275, EPI_ISL_2849276, EPI_ISL_2849277, EPI_ISL_2849278, EPI_ISL_2849288, EPI_ISL_2849289, EPI_ISL_2849290, EPI_ISL_2849293, EPI_ISL_2849297, EPI_ISL_2849298, EPI_ISL_2849301, EPI_ISL_2849302, EPI_ISL_2849304, EPI_ISL_2849307, EPI_ISL_2849310, EPI_ISL_2849311, EPI_ISL_2849316, EPI_ISL_2849328, EPI_ISL_2849334, EPI_ISL_2849335, EPI_ISL_2849336, EPI_ISL_2849338, EPI_ISL_2849345, EPI_ISL_2849346, EPI_ISL_2849348, EPI_ISL_2849353, EPI_ISL_2849355, EPI_ISL_2849358, EPI_ISL_2849363, EPI_ISL_2849364, EPI_ISL_2849371, EPI_ISL_2849383, EPI_ISL_2849388, EPI_ISL_2849393, EPI_ISL_2849398, EPI_ISL_2849426, EPI_ISL_2849427, EPI_ISL_2849430, EPI_ISL_2849443, EPI_ISL_2849444, EPI_ISL_2849449, EPI_ISL_2849456, EPI_ISL_2849457, EPI_ISL_2849465, EPI_ISL_2849474, EPI_ISL_2849477, EPI_ISL_2849493, EPI_ISL_2849496, EPI_ISL_2849506, EPI_ISL_2849519, EPI_ISL_2849521, EPI_ISL_2849522, EPI_ISL_2849529, EPI_ISL_2849530, EPI_ISL_2849539, EPI_ISL_2849541, EPI_ISL_2849543, EPI_ISL_2849545, EPI_ISL_2849547, EPI_ISL_2849548, EPI_ISL_2849549, EPI_ISL_2849550, EPI_ISL_2849555, EPI_ISL_2849557, EPI_ISL_2849575, EPI_ISL_2849577, EPI_ISL_2849578, EPI_ISL_2849580, EPI_ISL_2849582, EPI_ISL_2849593, EPI_ISL_2849596, EPI_ISL_2849603, EPI_ISL_2849604, EPI_ISL_2849609, EPI_ISL_2849612, EPI_ISL_2849621, EPI_ISL_2849629, EPI_ISL_2849646, EPI_ISL_2849649, EPI_ISL_2849654, EPI_ISL_2849659, EPI_ISL_2849662, EPI_ISL_2849668, EPI_ISL_2849671, EPI_ISL_2849680, EPI_ISL_2849682, EPI_ISL_2849685, EPI_ISL_2849799, EPI_ISL_2849954, EPI_ISL_2850018, EPI_ISL_2850022, EPI_ISL_2864852, EPI_ISL_2864856, EPI_ISL_2864877, EPI_ISL_2864892, EPI_ISL_2864909, EPI_ISL_2864963, EPI_ISL_2865012, EPI_ISL_2865048, EPI_ISL_2865059, EPI_ISL_2866014, EPI_ISL_2866064, EPI_ISL_2866069, EPI_ISL_2866070, EPI_ISL_2866072, EPI_ISL_2866073, EPI_ISL_2866077, EPI_ISL_2866079, EPI_ISL_2866082, EPI_ISL_2866084, EPI_ISL_2866085, EPI_ISL_2866086, EPI_ISL_2866087, EPI_ISL_2866089, EPI_ISL_2866091, EPI_ISL_2866092, EPI_ISL_2866094, EPI_ISL_2866095, EPI_ISL_2866098, EPI_ISL_2866099, EPI_ISL_2866100, EPI_ISL_2866101, EPI_ISL_2866102, EPI_ISL_2866103, EPI_ISL_2866104, EPI_ISL_2866105, EPI_ISL_2866112, EPI_ISL_2866120, EPI_ISL_2866121, EPI_ISL_2866123, EPI_ISL_2866124, EPI_ISL_2866125, EPI_ISL_2866126, EPI_ISL_2866127, EPI_ISL_2866129, EPI_ISL_2866131, EPI_ISL_2866132, EPI_ISL_2866133, EPI_ISL_2866135, EPI_ISL_2866136, EPI_ISL_2866143, EPI_ISL_2866144, EPI_ISL_2866148, EPI_ISL_2866150, EPI_ISL_2866153, EPI_ISL_2866158, EPI_ISL_2866161, EPI_ISL_2866163, EPI_ISL_2866164, EPI_ISL_2866168, EPI_ISL_2866170, EPI_ISL_2866175, EPI_ISL_2866176, EPI_ISL_2866177, EPI_ISL_2866178, EPI_ISL_2866179, EPI_ISL_2866182, EPI_ISL_2866184, EPI_ISL_2866186, EPI_ISL_2866187, EPI_ISL_2866188, EPI_ISL_2866191, EPI_ISL_2866194, EPI_ISL_2866195, EPI_ISL_2866196, EPI_ISL_2866198, EPI_ISL_2866202, EPI_ISL_2866209, EPI_ISL_2866213, EPI_ISL_2866215, EPI_ISL_2866216, EPI_ISL_2866217, EPI_ISL_2866222, EPI_ISL_2866225, EPI_ISL_2866227, EPI_ISL_2866229, EPI_ISL_2866233, EPI_ISL_2866235, EPI_ISL_2866236, EPI_ISL_2866237, EPI_ISL_2866239, EPI_ISL_2866242, EPI_ISL_2866245, EPI_ISL_2866246, EPI_ISL_2866248, EPI_ISL_2866249, EPI_ISL_2866251, EPI_ISL_2866254, EPI_ISL_2866256, EPI_ISL_2866257, EPI_ISL_2866258, EPI_ISL_2866259, EPI_ISL_2866260, EPI_ISL_2866263, EPI_ISL_2866265, EPI_ISL_2866267, EPI_ISL_2866269, EPI_ISL_2866270, EPI_ISL_2866271, EPI_ISL_2866272, EPI_ISL_2866273, EPI_ISL_2866274, EPI_ISL_2866279, EPI_ISL_2866282, EPI_ISL_2866285, EPI_ISL_2866286, EPI_ISL_2866289, EPI_ISL_2866290, EPI_ISL_2866292, EPI_ISL_2866294, EPI_ISL_2866295, EPI_ISL_2866301, EPI_ISL_2866303, EPI_ISL_2866305, EPI_ISL_2866306, EPI_ISL_2866309, EPI_ISL_2866311, EPI_ISL_2866312, EPI_ISL_2866313, EPI_ISL_2866314, EPI_ISL_2866315, EPI_ISL_2866526, EPI_ISL_2866671, EPI_ISL_2866672, EPI_ISL_2866673, EPI_ISL_2866676, EPI_ISL_2866679, EPI_ISL_2866681, EPI_ISL_2866687, EPI_ISL_2866688, EPI_ISL_2866694, EPI_ISL_2866695, EPI_ISL_2866696, EPI_ISL_2866704, EPI_ISL_2866709, EPI_ISL_2866710, EPI_ISL_2866711, EPI_ISL_2866715, EPI_ISL_2866719, EPI_ISL_2866720, EPI_ISL_2866725, EPI_ISL_2866727, EPI_ISL_2866740, EPI_ISL_2866760, EPI_ISL_2866765, EPI_ISL_2866774, EPI_ISL_2866775, EPI_ISL_2866777, EPI_ISL_2866779, EPI_ISL_2866790, EPI_ISL_2866793, EPI_ISL_2866796, EPI_ISL_2866808, EPI_ISL_2866813, EPI_ISL_2866816, EPI_ISL_2866824, EPI_ISL_2866826, EPI_ISL_2866828, EPI_ISL_2866830, EPI_ISL_2866840, EPI_ISL_2866852, EPI_ISL_2866857, EPI_ISL_2866863, EPI_ISL_2866865, EPI_ISL_2866868, EPI_ISL_2866875, EPI_ISL_2866876, EPI_ISL_2866877, EPI_ISL_2866881, EPI_ISL_2866882, EPI_ISL_2866889, EPI_ISL_2866893, EPI_ISL_2866896, EPI_ISL_2866902, EPI_ISL_2866914, EPI_ISL_2866917, EPI_ISL_2866919, EPI_ISL_2866920, EPI_ISL_2866922, EPI_ISL_2866932, EPI_ISL_2866938, EPI_ISL_2866939, EPI_ISL_2866942, EPI_ISL_2866943, EPI_ISL_2866945, EPI_ISL_2866948, EPI_ISL_2866950, EPI_ISL_2866952, EPI_ISL_2866956, EPI_ISL_2866959, EPI_ISL_2866963, EPI_ISL_2866967, EPI_ISL_2866974, EPI_ISL_2866976, EPI_ISL_2866979, EPI_ISL_2866989, EPI_ISL_2866994, EPI_ISL_2866999, EPI_ISL_2867002, EPI_ISL_2867017, EPI_ISL_2867020, EPI_ISL_2867796, EPI_ISL_2867950, EPI_ISL_2868054, EPI_ISL_2868055, EPI_ISL_2868070, EPI_ISL_2868071, EPI_ISL_2868072, EPI_ISL_2868073, EPI_ISL_2868076, EPI_ISL_2868077, EPI_ISL_2868168, EPI_ISL_2913719, EPI_ISL_2913742, EPI_ISL_2913806, EPI_ISL_2913921, EPI_ISL_2914126, EPI_ISL_2914131, EPI_ISL_2914181, EPI_ISL_2914192, EPI_ISL_2914240, EPI_ISL_2917411, EPI_ISL_2935554

| | | | |
|---|---|---|---|
| see above | Lighthouse Laboratory Plymouth | Wellcome Sanger Institute for the COVID-19 Genomics UK (COG-UK) Consortium | Cordelia Langford; David K. Jackson; Dominic Kwiatkowski; Ewan Harrison; Ian Johnston; Jeffrey Barrett; John Sillitoe on behalf of the Wellcome Sanger Institute COVID-19 Surveillance Team; Lighthouse Laboratory Plymouth and Alex Alderton; Roberto Amato; Sonia Goncalves |

EPI_ISL_2901638, EPI_ISL_2901641, EPI_ISL_2901642, EPI_ISL_2901643, EPI_ISL_2901644, EPI_ISL_2901645, EPI_ISL_2901646, EPI_ISL_2901648, EPI_ISL_2901649, EPI_ISL_2901650, EPI_ISL_2901651, EPI_ISL_2901653, EPI_ISL_2901654, EPI_ISL_2901655, EPI_ISL_2901656, EPI_ISL_2901659, EPI_ISL_2901660, EPI_ISL_2901661, EPI_ISL_2901662, EPI_ISL_2901666, EPI_ISL_2901667, EPI_ISL_2901668, EPI_ISL_2901669, EPI_ISL_2901673, EPI_ISL_2901674, EPI_ISL_2901675, EPI_ISL_2901676, EPI_ISL_2901679, EPI_ISL_2901680, EPI_ISL_2901681, EPI_ISL_2901683, EPI_ISL_2901684, EPI_ISL_2901685, EPI_ISL_2901686, EPI_ISL_2901689, EPI_ISL_2901690, EPI_ISL_2901691, EPI_ISL_2901693, EPI_ISL_2901694, EPI_ISL_2901695, EPI_ISL_2901696, EPI_ISL_2901697, EPI_ISL_2901700, EPI_ISL_2901701, EPI_ISL_2901702, EPI_ISL_2901703, EPI_ISL_2901707, EPI_ISL_2901708, EPI_ISL_2901709, EPI_ISL_2901710, EPI_ISL_2901711, EPI_ISL_2901877, EPI_ISL_2901878, EPI_ISL_2901879, EPI_ISL_2901880, EPI_ISL_2901881, EPI_ISL_2901882, EPI_ISL_2901883, EPI_ISL_2901884, EPI_ISL_2901885, EPI_ISL_2901886, EPI_ISL_2901887, EPI_ISL_2901888, EPI_ISL_2901889, EPI_ISL_2901890, EPI_ISL_2901891, EPI_ISL_2901892, EPI_ISL_2901893, EPI_ISL_2901894, EPI_ISL_2901895, EPI_ISL_2901896, EPI_ISL_2901897, EPI_ISL_2901898, EPI_ISL_2901899, EPI_ISL_2901900, EPI_ISL_2901901, EPI_ISL_2901902, EPI_ISL_2901903, EPI_ISL_2901904, EPI_ISL_2901905, EPI_ISL_2901906, EPI_ISL_2901907, EPI_ISL_2901908, EPI_ISL_2901909, EPI_ISL_2901910, EPI_ISL_2901911, EPI_ISL_2901912, EPI_ISL_2901913, EPI_ISL_2901914, EPI_ISL_2901915, EPI_ISL_2901916, EPI_ISL_2901917, EPI_ISL_2901918, EPI_ISL_2901919, EPI_ISL_2901920, EPI_ISL_2901921, EPI_ISL_2901922, EPI_ISL_2901923, EPI_ISL_2901924, EPI_ISL_2901925, EPI_ISL_2901926, EPI_ISL_2901927, EPI_ISL_2901929, EPI_ISL_2901930, EPI_ISL_2901931, EPI_ISL_2901932, EPI_ISL_2901933, EPI_ISL_2901934, EPI_ISL_2901935, EPI_ISL_2901936, EPI_ISL_2901937, EPI_ISL_2901938, EPI_ISL_2901939, EPI_ISL_2901940, EPI_ISL_2901941, EPI_ISL_2901943, EPI_ISL_2901944, EPI_ISL_2901945, EPI_ISL_2901946, EPI_ISL_2901947, EPI_ISL_2901948, EPI_ISL_2901949, EPI_ISL_2901950, EPI_ISL_2901951, EPI_ISL_2901952, EPI_ISL_2901953, EPI_ISL_2901954, EPI_ISL_2901955, EPI_ISL_2901956, EPI_ISL_2901957, EPI_ISL_2901958, EPI_ISL_2901959, EPI_ISL_2901960, EPI_ISL_2901961, EPI_ISL_2902043, EPI_ISL_2902044, EPI_ISL_2902045, EPI_ISL_2902046, EPI_ISL_2902047, EPI_ISL_2902048, EPI_ISL_2902049, EPI_ISL_2902050, EPI_ISL_2902051, EPI_ISL_2902052, EPI_ISL_2902053, EPI_ISL_2902054, EPI_ISL_2902055, EPI_ISL_2902056, EPI_ISL_2902057, EPI_ISL_2902058, EPI_ISL_2902059, EPI_ISL_2902060, EPI_ISL_2902061, EPI_ISL_2902062, EPI_ISL_2902063, EPI_ISL_2902064, EPI_ISL_2902065, EPI_ISL_2902066, EPI_ISL_2902067, EPI_ISL_2902068, EPI_ISL_2902069, EPI_ISL_2902070, EPI_ISL_2902071, EPI_ISL_2902072, EPI_ISL_2902073, EPI_ISL_2902074, EPI_ISL_2902075, EPI_ISL_2902076, EPI_ISL_2902077, EPI_ISL_2902078, EPI_ISL_2902079, EPI_ISL_2902080, EPI_ISL_2902081, EPI_ISL_2902082, EPI_ISL_2902083, EPI_ISL_2902084, EPI_ISL_2902086, EPI_ISL_2902087, EPI_ISL_2902088, EPI_ISL_2902089, EPI_ISL_2902090, EPI_ISL_2902091, EPI_ISL_2902092, EPI_ISL_2902093, EPI_ISL_2902094, EPI_ISL_2902095, EPI_ISL_2902096, EPI_ISL_2902097, EPI_ISL_2902098, EPI_ISL_2902099, EPI_ISL_2902100, EPI_ISL_2902101, EPI_ISL_2902102, EPI_ISL_2902103, EPI_ISL_2902104, EPI_ISL_2902105, EPI_ISL_2902106, EPI_ISL_2902107, EPI_ISL_2902108, EPI_ISL_2902109, EPI_ISL_2902110, EPI_ISL_2902111, EPI_ISL_2902112, EPI_ISL_2902113, EPI_ISL_2902114, EPI_ISL_2902115, EPI_ISL_2902116, EPI_ISL_2902117, EPI_ISL_2902118, EPI_ISL_2902119, EPI_ISL_2902120, EPI_ISL_2902121, EPI_ISL_2902168, EPI_ISL_2902175, EPI_ISL_2902222, EPI_ISL_2902317, EPI_ISL_2902323, EPI_ISL_2902325, EPI_ISL_2902381, EPI_ISL_2902385, EPI_ISL_2902399

| | | | |
|---|---|---|---|
| see above | Northumbria University / South Tees Hospitals NHS Foundation Trust / North Cumbria Integrated Care NHS Foundation Trust / North Tees and Hartlepool NHS Foundation Trust / Newcastle Hospitals NHS Foundation Trust | COVID-19 Genomics UK (COG-UK) Consortium | Andrew Nelson; Brendan Payne; Clive Graham; Darren L Smith; Debra Padgett; Edward Barton; Emma Swindells; Garren Scott; Gary Black; Gary Eltringham; Giles S Holt; Greg R Young; Jane Greenaway; Jennifer Collins; John Allan; Joshua Loh; Lynn Dover; Matthew Bashton; Mohammad A Tariq; Paul Baker; Sarah Essex; Steve Liggett; Wen C Yew; Yusri Taha |

EPI_ISL_2904578, EPI_ISL_2904579, EPI_ISL_2904581, EPI_ISL_2904583, EPI_ISL_2904585, EPI_ISL_2904586, EPI_ISL_2904587, EPI_ISL_2904588, EPI_ISL_2904591, EPI_ISL_2904592, EPI_ISL_2904593, EPI_ISL_2904594, EPI_ISL_2904596, EPI_ISL_2904599, EPI_ISL_2904605, EPI_ISL_2904750, EPI_ISL_2904751, EPI_ISL_2904752, EPI_ISL_2904753, EPI_ISL_2904754, EPI_ISL_2904755, EPI_ISL_2904756, EPI_ISL_2904757, EPI_ISL_2904759, EPI_ISL_2904761, EPI_ISL_2904762, EPI_ISL_2904764, EPI_ISL_2904765, EPI_ISL_2904767, EPI_ISL_2904770, EPI_ISL_2904773, EPI_ISL_2904774, EPI_ISL_2904776, EPI_ISL_2904834, EPI_ISL_2904860, EPI_ISL_2904897, EPI_ISL_2905011, EPI_ISL_2905018, EPI_ISL_2905157, EPI_ISL_2905205, EPI_ISL_2905209, EPI_ISL_2905921, EPI_ISL_2905922, EPI_ISL_2905923, EPI_ISL_2905924, EPI_ISL_2905925, EPI_ISL_2905926, EPI_ISL_2905927, EPI_ISL_2905928, EPI_ISL_2905929, EPI_ISL_2905930, EPI_ISL_2905931, EPI_ISL_2905932, EPI_ISL_2905933, EPI_ISL_2905934, EPI_ISL_2905935, EPI_ISL_2905936, EPI_ISL_2905937, EPI_ISL_2905938, EPI_ISL_2905939, EPI_ISL_2905940, EPI_ISL_2905941, EPI_ISL_2905942, EPI_ISL_2905943, EPI_ISL_2905944, EPI_ISL_2905945, EPI_ISL_2905946, EPI_ISL_2905947, EPI_ISL_2905948, EPI_ISL_2905949, EPI_ISL_2905950, EPI_ISL_2905951, EPI_ISL_2905952, EPI_ISL_2905953, EPI_ISL_2905954, EPI_ISL_2905955, EPI_ISL_2905956, EPI_ISL_2905957, EPI_ISL_2905958, EPI_ISL_2905959, EPI_ISL_2905960, EPI_ISL_2905962, EPI_ISL_2905963, EPI_ISL_2905964, EPI_ISL_2905965, EPI_ISL_2905966, EPI_ISL_2905967, EPI_ISL_2905968, EPI_ISL_2905969, EPI_ISL_2905970, EPI_ISL_2905971, EPI_ISL_2905980, EPI_ISL_2906128, EPI_ISL_2906129, EPI_ISL_2906130, EPI_ISL_2906131, EPI_ISL_2906132, EPI_ISL_2906133, EPI_ISL_2906134, EPI_ISL_2906135, EPI_ISL_2906136, EPI_ISL_2906137, EPI_ISL_2906138, EPI_ISL_2906139, EPI_ISL_2906140, EPI_ISL_2906141, EPI_ISL_2906142, EPI_ISL_2906143, EPI_ISL_2906144, EPI_ISL_2906145, EPI_ISL_2906146, EPI_ISL_2906147, EPI_ISL_2906148, EPI_ISL_2906149, EPI_ISL_2906150, EPI_ISL_2906151, EPI_ISL_2906152, EPI_ISL_2906153, EPI_ISL_2906324, EPI_ISL_2906325, EPI_ISL_2906326, EPI_ISL_2906327, EPI_ISL_2906328, EPI_ISL_2906329, EPI_ISL_2906330, EPI_ISL_2906331, EPI_ISL_2906332, EPI_ISL_2906333, EPI_ISL_2906334, EPI_ISL_2906335, EPI_ISL_2906336, EPI_ISL_2906337, EPI_ISL_2906338, EPI_ISL_2906339, EPI_ISL_2906340, EPI_ISL_2906341, EPI_ISL_2906342, EPI_ISL_2906343, EPI_ISL_2906344, EPI_ISL_2906346, EPI_ISL_2906347, EPI_ISL_2906348, EPI_ISL_2906349, EPI_ISL_2906350, EPI_ISL_2906351, EPI_ISL_2906430, EPI_ISL_2906431, EPI_ISL_2906432, EPI_ISL_2906433, EPI_ISL_2906434, EPI_ISL_2906435, EPI_ISL_2906436, EPI_ISL_2906437, EPI_ISL_2906438, EPI_ISL_2906439, EPI_ISL_2906440, EPI_ISL_2906441, EPI_ISL_2906442, EPI_ISL_2906443, EPI_ISL_2906444, EPI_ISL_2906445, EPI_ISL_2906446, EPI_ISL_2906447, EPI_ISL_2906448, EPI_ISL_2906449, EPI_ISL_2906450, EPI_ISL_2906451, EPI_ISL_2906452, EPI_ISL_2906453, EPI_ISL_2906454, EPI_ISL_2906455, EPI_ISL_2906456, EPI_ISL_2906481, EPI_ISL_2906482, EPI_ISL_2906483, EPI_ISL_2906484, EPI_ISL_2906485, EPI_ISL_2906487, EPI_ISL_2906488, EPI_ISL_2906490, EPI_ISL_2906493, EPI_ISL_2906494, EPI_ISL_2906501, EPI_ISL_2906502, EPI_ISL_2906505, EPI_ISL_2906507, EPI_ISL_2906533, EPI_ISL_2906535, EPI_ISL_2906536, EPI_ISL_2906537, EPI_ISL_2906539, EPI_ISL_2906540, EPI_ISL_2906541, EPI_ISL_2906542, EPI_ISL_2906543, EPI_ISL_2906544, EPI_ISL_2906545, EPI_ISL_2906546, EPI_ISL_2906547, EPI_ISL_2906548, EPI_ISL_2906549, EPI_ISL_2906550, EPI_ISL_2906551, EPI_ISL_2906552, EPI_ISL_2906553, EPI_ISL_2906554, EPI_ISL_2906555, EPI_ISL_2906556, EPI_ISL_2906557, EPI_ISL_2906558, EPI_ISL_2906559, EPI_ISL_2906560, EPI_ISL_2906561, EPI_ISL_2906562, EPI_ISL_2906563, EPI_ISL_2906564, EPI_ISL_2906565, EPI_ISL_2906566, EPI_ISL_2906567, EPI_ISL_2906568, EPI_ISL_2906569, EPI_ISL_2906570, EPI_ISL_2906571, EPI_ISL_2906572, EPI_ISL_2906574, EPI_ISL_2906575, EPI_ISL_2906576, EPI_ISL_2906577, EPI_ISL_2906578, EPI_ISL_2906579, EPI_ISL_2906580, EPI_ISL_2906581, EPI_ISL_2906582, EPI_ISL_2906583, EPI_ISL_2906584, EPI_ISL_2906585, EPI_ISL_2906586, EPI_ISL_2906587, EPI_ISL_2906588, EPI_ISL_2906589, EPI_ISL_2906590, EPI_ISL_2906591, EPI_ISL_2906592, EPI_ISL_2906593, EPI_ISL_2906594, EPI_ISL_2906595, EPI_ISL_2906596, EPI_ISL_2906597, EPI_ISL_2906598, EPI_ISL_2906599, EPI_ISL_2906600, EPI_ISL_2906601, EPI_ISL_2906602, EPI_ISL_2906603, EPI_ISL_2906604, EPI_ISL_2906605, EPI_ISL_2906606, EPI_ISL_2906607, EPI_ISL_2906608, EPI_ISL_2906609, EPI_ISL_2906610, EPI_ISL_2906611, EPI_ISL_2906612, EPI_ISL_2906613, EPI_ISL_2906615, EPI_ISL_2906616, EPI_ISL_2906617, EPI_ISL_2906618, EPI_ISL_2906619, EPI_ISL_2906620, EPI_ISL_2906622, EPI_ISL_2906623, EPI_ISL_2906624, EPI_ISL_2906626, EPI_ISL_2906627, EPI_ISL_2906628, EPI_ISL_2906629, EPI_ISL_2906630, EPI_ISL_2906632, EPI_ISL_2906633, EPI_ISL_2906634, EPI_ISL_2906635, EPI_ISL_2906636, EPI_ISL_2906637, EPI_ISL_2906638, EPI_ISL_2906639, EPI_ISL_2906640, EPI_ISL_2906641, EPI_ISL_2906642, EPI_ISL_2906644, EPI_ISL_2906645, EPI_ISL_2906646, EPI_ISL_2906647, EPI_ISL_2906648, EPI_ISL_2906649, EPI_ISL_2906650, EPI_ISL_2906651, EPI_ISL_2906652, EPI_ISL_2906653, EPI_ISL_2906654, EPI_ISL_2906655, EPI_ISL_2906656, EPI_ISL_2906657,

| | | | |
|---|---|---|---|
| EPI_ISL_2906658, EPI_ISL_2906659, EPI_ISL_2906660, EPI_ISL_2906661, EPI_ISL_2906662, EPI_ISL_2906663, EPI_ISL_2906664, EPI_ISL_2906665, EPI_ISL_2906666, EPI_ISL_2906667, EPI_ISL_2906668, EPI_ISL_2906669, EPI_ISL_2906670, EPI_ISL_2906671, EPI_ISL_2906672, EPI_ISL_2906673, EPI_ISL_2906674, EPI_ISL_2906675, EPI_ISL_2906676, EPI_ISL_2906703, EPI_ISL_2906704, EPI_ISL_2906705, EPI_ISL_2906706, EPI_ISL_2906707, EPI_ISL_2906708, EPI_ISL_2906709, EPI_ISL_2906710, EPI_ISL_2906711, EPI_ISL_2906712, EPI_ISL_2906713, EPI_ISL_2906714, EPI_ISL_2906715, EPI_ISL_2906716, EPI_ISL_2906717, EPI_ISL_2906718, EPI_ISL_2906719, EPI_ISL_2906720, EPI_ISL_2906721, EPI_ISL_2906722, EPI_ISL_2906723, EPI_ISL_2906724, EPI_ISL_2906725, EPI_ISL_2906726, EPI_ISL_2906727, EPI_ISL_2906728, EPI_ISL_2906729, EPI_ISL_2906730 | | | |
| see above | Originating lab: Wales Specialist Virology Centre Sequencing lab: Pathogen Genomics Unit | Public Health Wales Microbiology Cardiff Wales Specialist Virology Centre | Alec Birchley; Alexander Adams; Amy Gaskin; Angela Marchbank; Bree Gatica-Wilcox; Catherine Moore; Jason Coombes; Joanne Watkins; Joel Southgate; Johnathan Evans; Laura Gifford; Lauren Gilbert; Lee Graham; Malorie Perry; Matthew Bull; Nicole Pacchiarini; Sally Corden; Sara Kumziene-Summerhayes; Sara Rey; Sarah Taylor; Simon Cottrell; Sophie Jones; Tom Connor |
| EPI_ISL_2918272, EPI_ISL_2918298, EPI_ISL_2918422, EPI_ISL_2918435, EPI_ISL_2918458, EPI_ISL_2918495 | Randox Laboratories | Wellcome Sanger Institute for the COVID-19 Genomics UK (COG-UK) Consortium | Cordelia Langford; David K. Jackson; Dominic Kwiatkowski; Ewan Harrison; Ian Johnston; Jeffrey Barrett; John Sillitoe on behalf of the Wellcome Sanger Institute COVID-19 Surveillance Team; Randox Laboratories and Alex Alderton; Roberto Amato; Sonia Goncalves |
| EPI_ISL_2903515, EPI_ISL_2903516, EPI_ISL_2903517, EPI_ISL_2903518, EPI_ISL_2903519, EPI_ISL_2903520, EPI_ISL_2903521, EPI_ISL_2903522, EPI_ISL_2903523, EPI_ISL_2903524, EPI_ISL_2903554, EPI_ISL_2903567, EPI_ISL_2903568, EPI_ISL_2903569, EPI_ISL_2903570, EPI_ISL_2903571, EPI_ISL_2903616, EPI_ISL_2903629, EPI_ISL_2903630, EPI_ISL_2903632, EPI_ISL_2903717, EPI_ISL_2903744, EPI_ISL_2903746, EPI_ISL_2903766, EPI_ISL_2903767, EPI_ISL_2903780, EPI_ISL_2903781, EPI_ISL_2903782, EPI_ISL_2903783, EPI_ISL_2903784, EPI_ISL_2903792, EPI_ISL_2903805, EPI_ISL_2903806, EPI_ISL_2903807, EPI_ISL_2903809, EPI_ISL_2903810, EPI_ISL_2903811, EPI_ISL_2903812, EPI_ISL_2903813, EPI_ISL_2903814, EPI_ISL_2903815, EPI_ISL_2903816, EPI_ISL_2903817, EPI_ISL_2903818, EPI_ISL_2903819, EPI_ISL_2903820, EPI_ISL_2903821, EPI_ISL_2903822, EPI_ISL_2903823, EPI_ISL_2903828, EPI_ISL_2903833, EPI_ISL_2903838, EPI_ISL_2903842, EPI_ISL_2903843, EPI_ISL_2903845, EPI_ISL_2903846, EPI_ISL_2903895, EPI_ISL_2903898, EPI_ISL_2903903, EPI_ISL_2903904, EPI_ISL_2903906, EPI_ISL_2903993, EPI_ISL_2903995, EPI_ISL_2903996, EPI_ISL_2903998, EPI_ISL_2903999, EPI_ISL_2904000, EPI_ISL_2904002, EPI_ISL_2904003, EPI_ISL_2904118, EPI_ISL_2904120, EPI_ISL_2904123, EPI_ISL_2904124, EPI_ISL_2904125, EPI_ISL_2904133, EPI_ISL_2904187, EPI_ISL_2904191, EPI_ISL_2904193, EPI_ISL_2904198, EPI_ISL_2904199, EPI_ISL_2904200, EPI_ISL_2904201, EPI_ISL_2904202, EPI_ISL_2904203, EPI_ISL_2904204, EPI_ISL_2904205, EPI_ISL_2904206, EPI_ISL_2904207, EPI_ISL_2904208, EPI_ISL_2904209, EPI_ISL_2904210, EPI_ISL_2904211, EPI_ISL_2904212, EPI_ISL_2904213, EPI_ISL_2904214, EPI_ISL_2904215, EPI_ISL_2904216, EPI_ISL_2904217, EPI_ISL_2904218, EPI_ISL_2904225, EPI_ISL_2904230, EPI_ISL_2904235, EPI_ISL_2904236, EPI_ISL_2904238, EPI_ISL_2904239, EPI_ISL_2904242, EPI_ISL_2904243, EPI_ISL_2904244, EPI_ISL_2904247, EPI_ISL_2904256, EPI_ISL_2904258, EPI_ISL_2904259, EPI_ISL_2904260, EPI_ISL_2904261, EPI_ISL_2904263, EPI_ISL_2904274, EPI_ISL_2904300, EPI_ISL_2904308, EPI_ISL_2904312, EPI_ISL_2904315, EPI_ISL_2904318, EPI_ISL_2904319, EPI_ISL_2904356, EPI_ISL_2904359, EPI_ISL_2904361, EPI_ISL_2904368, EPI_ISL_2904373, EPI_ISL_2904375, EPI_ISL_2904396, EPI_ISL_2904397, EPI_ISL_2904398, EPI_ISL_2904399, EPI_ISL_2904402, EPI_ISL_2904403, EPI_ISL_2904406, EPI_ISL_2904409, EPI_ISL_2904411, EPI_ISL_2904412, EPI_ISL_2904414 | | | |
| see above | Respiratory Virus Unit, Microbiology Services Colindale, Public Health England | COVID-19 Genomics UK (COG-UK) Consortium | PHE Covid Sequencing Team |
| EPI_ISL_2900830, EPI_ISL_2900836, EPI_ISL_2900837, EPI_ISL_2900838, EPI_ISL_2900839, EPI_ISL_2900842, EPI_ISL_2900843 | | | |
| see above | Virology Department, Royal Infirmary of Edinburgh, NHS Lothian / School of Biological Sciences, University of Edinburgh | COVID-19 Genomics UK (COG-UK) Consortium | Colquhoun R; Cotton S; Dewar R; Gallagher A; Hill V; Jackson B; Maloney D; McCrone JT; McHugh M; O’Toole Á; Rambaut A; Rooke S; Scher E; Templeton K; Yu X |
| EPI_ISL_2907123, EPI_ISL_2907260, EPI_ISL_2907320 | Virology Department, Sheffield Teaching Hospitals NHS Foundation Trust/Department of Infection, Immunity and Cardiovascular Disease, The Medical School, University of Sheffield | COVID-19 Genomics UK (COG-UK) Consortium | Adri Angyal; Alex Keeley; Benjamin Lindsey; Cariad Evans; Danielle Groves; Dave Partridge; Katie Johnson; Laura Carrilero; Luke Green; Matthew Parker; Matthew Wyles; Mehmet Yavuz; Mohammad Raza; Nikki Smith; Paul Parsons; Rachel Tucker; Rebecca Brown; Thushan de Silva |
